# A YOUNG CHILD FORMULA SUPPLEMENTED WITH L. REUTERI AND GALACTO-OLIGOSACCHARIDES MODULATES THE COMPOSITION AND FUNCTION OF THE GUT MICROBIOME SUPPORTING BONE AND MUSCLE DEVELOPMENT IN TODDLERS

**DOI:** 10.1101/2024.12.11.24318836

**Authors:** Nicolas Bonnet, Maria Rosario Capeding, Léa Siegwald, Marc Garcia-Garcera, Thibaut Desgeorges, Hanne L.P. Tytgat, Laura-Florina Krattinger, Jowena Lebumfacil, Loudhie Cyd Phee, Janne Marie Moll, Alexander Gudjonsson, Paula Rodriguez-Garcia, Jerome N Feige, Ivana Jankovic, Yipu Chen, Delphine Egli, Marie-Noëlle Horcajada

## Abstract

**Key Points:** This section will be completed further

**Importance:** Toddlerhood is a key window of opportunity for development of musculoskeletal system and microbiome. In this study we tested the efficacy of a synbiotic-based young child formula on bone and muscle strength and microbiome maturation in young children during motor-skill development.

**Intervention:** In this randomized, double-blind controlled trial, children aged 2-3 years received either an experimental young child formula (EYCF) containing a combination of *Limosilactobacillus reuteri* DSM 17938 and galacto-oligosaccharides (GOS) or a minimally fortified milk (CM) for 6 months. A third arm remained on their habitual diet.

**Main outcomes and measures:** Bone quality (tibia speed of sound), muscle strength (handgrip), microbiota composition (shotgun metagenomics) and functionality (fecal metabolome) were evaluated at baseline, and after 3 months and 6 months of intervention. Microbiota and metabolomic features were associated to each other and to clinical bone and muscle readouts at the same timepoints.

**Results:** Tibial speed of sound was significantly increased after 6 months (primary end point, p<0.01) and 3 months (p<0.05) of EYCF vs CM feeding. These effects on bone strength were paralleled by significantly higher muscle strength after 6 months in EYCF vs CM. The intervention significantly remodeled microbiome composition, with enrichment of *L. reuteri*, and higher bifidobacteria presence in the stools of EYCF vs CM children at both 3 and 6 months. Increased *L. reuteri* abundance after 6 months of EYCF consumption was associated with higher bone quality and muscle strength. Stool metabolomics were significantly modulated by EYCF consumption with 45 metabolites significantly modified and associated to microbiome compositional changes such as *Bifidobacterium* spp. and *L. reuteri* expansion. Pairing of metagenomic and metabolomic signatures induced by EYCF revealed an enrichment of tryptophane and indole metabolism which significantly associated to bone and muscle strength clinical outcomes.

**Conclusions and relevance:** Consumption of an experimental young child formula containing a *L. reuteri* + GOS synbiotic improves musculoskeletal development in toddlers that was associated with a modulation of microbiota composition and functionality. These results provide novel mechanistic insights on gut-musculoskeletal crosstalk during early life and demonstrate that nutritional interventions targeting the microbiome can support healthy bone and muscle development and may contribute to functional motorskills acquisition during childhood.

**Trial registration:** The trial was registered at clinicaltrial.gov as NCT04799028

## Introduction

Bone is a “dynamic” and highly specialized connective tissue, providing mechanical support for muscular activity and physical protection to tissues and internal organs, as well as a repository for the systemic mineral homeostasis (1).

Bone tissue is constantly renewed in adults as well as in children and adolescents. “Strong bones” are defined by two different aspects of bone geometry (length & width) and by the amount of mineral deposited in the bone tissue. It is interesting to note that osteoblasts first synthesize and excrete the bone matrix proteins (osteoids) which later on, at a short distance from the bone-forming cells, mineralize until about 70% of the bone, with the remaining 30% at longer distance (6-12 months later). Therefore, bone could rapidly be under-mineralized if the mineralization rate is reduced compared to velocity growth (2).

Most data seem to confirm that an adequate intake of calcium is important in the mineralization process to reach skeletal maturity. Prospective randomized clinical trials have demonstrated that calcium supplementation may increase bone mass acquisition in children, adolescence and adulthood up to the third decade of life (3, 4). When calcium supplementation ceases, the beneficial effect on bone accretion seems to disappear (5–7). Inadequate intake of calcium may contribute to failure to develop strong bones during early life (5). Therefore, daily recommended calcium intakes have been established by various authorities to meet the needs for growth and bone development in children (5). In addition, nutrients (such as vitamin D) which help calcium incorporation into bone may also contribute to bone development (5, 8).

Early life and toddlerhood are crucial windows for bone growth, but they are also critical for microbiome development (9). The influence of the microbiome on bone (re)modeling has been demonstrated through different mechanisms of action such as modulation of mineral absorption, immune cell activity, and production of microbial metabolites (10). Given the link between the microbiome functionality and bone development, modulation of the gut microbiome by nutritional supplements has been explored to provide novel strategies to maximize bone gain during childhood through the gut-bone axis (11). Pre-clinical growth models documented an effect of the prebiotic GOS on mineral absorption (12), on gut hormones known to impact bone metabolism (13, 14) and on bone strength and mineral density (15–17). In a pre-clinical model of bone loss, a positive effect of *Limosilactobacillus reuteri* (*L. reuteri*) ATCC PTA 6475 on trabecular bone microarchitecture was reported (18) (19). We have shown previously *in vitro* that the administration of a synbiotic consisting of pre-cultivated *L. reuteri* DSM 17938 and GOS stimulate microbiome metabolite production, microbial engraftment and microbiome profiles compared to *L. reuteri* DSM 17938 alone (20). In this study, we also confirmed the ability of specific metabolites (e.g. short chain fatty acids (SCFAs), known to regulate bone homeostasis *in vivo* (21), to improve osteoblastogenesis (20). The impact of the intervention on muscle progenitor cells remained to be evaluated.

In clinical settings, GOS was shown to positively impact calcium absorption in adolescents (22) and post-menopausal women (23). Additionally, formula containing a fat blend with low levels of sn-1,3 long-chain saturated fatty acids (LCSFAs) positively impacted calcium absorption in infants by decreasing calcium soaps formation (24–27). Finally, one year of supplementation with *L. reuteri ATCC PTA* 6475 in post-menopausal women reduced loss of tibia bone mineral density compared to placebo, suggesting that probiotics are a novel candidate approach to prevent age-associated bone loss and osteoporosis (20, 21, 28).

Skeletal muscle mass and strength drastically increase from birth to adulthood (29) and is highly dependent on nutrition (30). Muscle mass and strength increase through muscle fiber hypertrophy due to an increase of protein synthesis. The mechanism is driven by both an increase in the myonuclear protein synthesis rate and muscle progenitor cell activity, which insure myonuclear accretion. The role of muscle progenitor cells in muscle development is important especially during the first years of life (31–34).

Another potential driver of muscle development is the gut microbiome, as more and more studies underline the existence of a gut-muscle axis (35), similarly to growing evidence for the gut-bone axis. It remains however unknown whether modulating the microbiome can impact bone and muscle development in children.

In this current study we assessed the effect of an experimental young child formula (EYCF), containing synbiotic blend of pre-cultivated *L. reuteri* DSM 17938 and GOS (4g/L), in toddlers through a double-blind randomized controlled trial, compared to minimally fortified milk (control milk, CM). The primary endpoint of this clinical study was tibia quality (ultrasound) after 6 months of intervention. Evaluation of muscle force (handgrip) was also performed. To evaluate the potential contribution of the microbiome on bone and muscle development, the fecal microbiota composition and fecal metabolome were analyzed. Additionally, we investigated the effects of product digestion of this experimental formula, obtained via *in vitro* fermentation, on muscle progenitor cells activities.

## Material & methods

### *In vitro* experiments with human myoblasts

*In vitro* assays were performed on primary human skeletal muscle myoblasts (Lonza). Myoblasts were seeded at 90-110 cells per mm^2^ on well plates pre-coated with 20 µg/ml human fibronectin. To evaluate the differentiation potential of myoblasts, cells were cultured for 3 days in growth medium (SkM-M, AMSBIO) with the product generated after the *in vitro* digestion of experimental blends (milk matrix+/- GOS+/- *L. reuteri*) as previously described (20). To evaluate the myogenesis process, cells were cultured for 3 days in growth medium (SkM-M, AMSBIO) followed by 3 days of culture in differentiation medium (DMEM/F12, Horse serum 2%, Pen/Strep 1%). Cells were exposed to different treatments at each step (6 days of treatment, range of fusion index in milk matrix control group is 46.46% ±3.832(SD)). At the end of the treatment period, cells were fixed with PFA 4%, permeabilized with Triton 0.5%, blocked with 4% Bovin Serum Albumin and stained for Troponin T assay overnight at 4°C. The next day, the cells were washed and stained with Hoechst 33342 (Sigma) and secondary antibodies coupled with fluorescent molecules (ThermoFisher Scientific).

Image acquisition was performed with ImageXpress Micro confocal microscope (Molecular devices) and image analysis was performed with MetaXpress Software to quantify the myotube area and/or cell fusion index (myogenesis).

### Study population

Healthy Filipino toddlers between 24 months (±1 week) to 36 months (±1 week) of age, not consuming pre- or probiotics in the past month and without history of bone malabsorption, metabolic, congenital, or chromosomal abnormalities were eligible for this trial.

Children who fulfilled all of the following inclusion criteria were included:

1. Written informed consent has been obtained from the parent(s)/legally acceptable representative (LAR).
2. Singleton, full-term gestational birth (≥ 37 completed weeks of gestation), with a birth weight of ≥ 2.5 kg and ≤ 4.5 kg.
3. Child is between 24 months ±1 week to 36 months ±1 week at inclusion.
4. Child is not currently consuming nor has consumed any formulas or taking any supplements with pre- or probiotics at enrolment or in the past month.
5. Child’s parent(s)/guardian is of legal age of consent, must understand the informed consent and other study documents, and is willing and able to fulfill the requirements of the study protocol.

Exclusion criteria, that rendered children ineligible for inclusion:

1. Chronic infectious, metabolic, genetic illness or other disease including any condition that impacts feeding or growth.
2. History of bone malabsorption, metabolic, congenital, or chromosomal abnormality known to affect feeding or growth.
3. Use of systemic antibiotics or anti-mycotic medication in the 4 weeks preceding enrollment.
4. Known or suspected cows’ milk protein intolerance / allergy, or lactose intolerance, or severe food allergies that impact diet.
5. Current breast milk feeding in place of all other milk, and/or milk alternatives.
6. Clinical signs of severe micronutrient deficiencies.
7. Parent(s) not willing / not able to comply with the requirements of study protocol.
8. Child’s participation in another interventional clinical trial.

### Study design

The study is a randomized, controlled, double-blind intervention conducted in healthy male and female children, which were exposed to either minimally fortified cow milk (CM) or to experimental young child formula (EYCF). A non-randomized habitual diet reference group was also included from enrollment to 6 months (REF). The study was conducted at the Asian Hospital and Medical Center in Muntinlupa City, Philippines from June 2021 to August 2022. The Asian Hospital and Medical Center Research Ethics Committee approved the study (REF:QF-REC-007). Written informed consent from all parents or legally acceptable representatives was obtained prior to screening and enrolment. The trial was registered at clinicaltrial.gov as NCT04799028.

Children completed three visits (enrollment, after 3 months, and after 6 months). Stool samples were collected at all three visits. The primary endpoint was tibia bone mass index measured by speed of sound (SOS) at 6 months of intervention. Secondary endpoints included additional bone mass index parameters, muscle force (Handgrip strength) gastrointestinal (GI) tolerance and stooling patterns, excretion of fecal calcium fatty acid soaps, serum vitamin D, urine and blood bone turnover markers, gut microbiota, fecal metabolism and safety endpoints including growth, medication use, and standard adverse events (AEs). A schematic of the trial design is presented in Fig. 1A and 1B.

**Figure 1.**
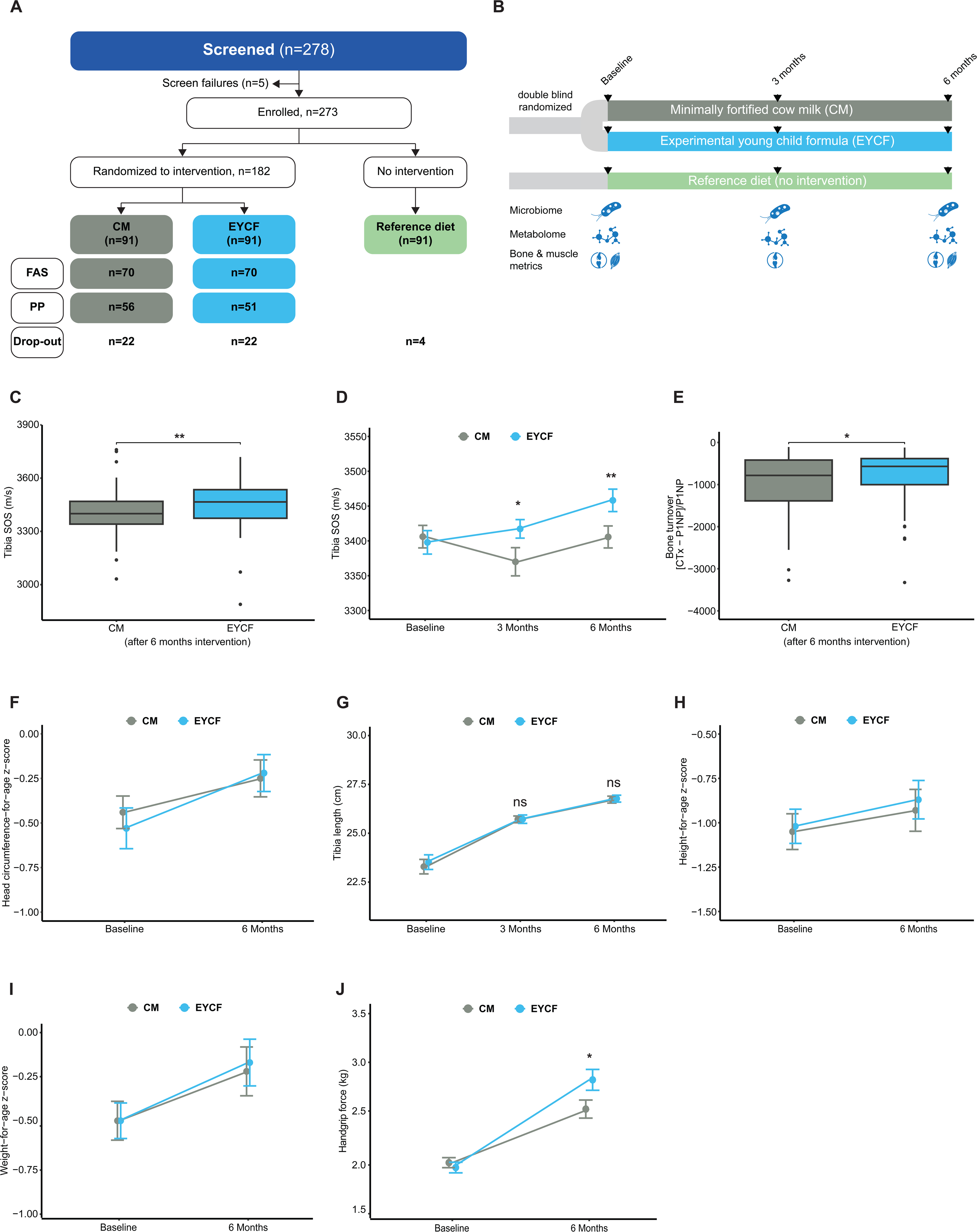
Subjects and design of the clinical trial and effect of the experimental blend on bone and muscle clinical outcomes. (A) A total of 273 toddlers were enrolled into the study; 182 toddlers were randomized to one of the feeding groups and 91 toddlers were enrolled in parallel as a reference population. Randomized toddlers were assigned to either a minimally fortified cow milk (CM, n=91) or experimental young child formula group (EYCF, n=91). Overall, 225 (82%) subjects completed the study. In the FAS, the retention rate was 77% in both CM & EYCF groups. Across all groups, 48 toddlers (22 each in CM and EYCF, 4 in REF) withdrew from the study with the common reasons of issues with taste (n=16; 33%) and transfer to province/other country (n=15; 31%). Other reasons for withdrawal included relatives’ disapproval (n=9; 19%), unable to comply (n=4; 8%), adverse events (n=2; 4%), and preference to solid food (n=2; 4%). (B) Schematic of experimental design. (C) Tibia speed of sound (SOS) measured by quantitative ultrasound after 6 months of intervention. (D) Longitudinal tibia SOS. (E) Bone turnover marker (CTx-P1NP)/P1NP. CTX: carboxy-terminal collagen crosslinks, P1NP: Procollagen type I N-terminal propeptide. C(D-H) Anthropometric outcomes: (F) Head circumference, (G) Tibia length, (H) Height for age, (I) Weight for age. (J) Muscle force (right arm) measured by handgrip. For all outcome, EYCF and CM groups n=69. Data are presented as mean ± SD, * P<0.05, ** P<0.01 compared to CM group.

### Product allocation, product description, groups and timing

Toddlers were randomly assigned to the control (CM) or experimental (EYCF) formula. Children were fed the assigned formulas for 6 months from enrollment. The nutritional composition of EYCF and CM are within regulatory limits for children’s formula and fortified milks, respectively.

- CM: Powdered cow’s milk, fortified with the key nutrients (vitamins A, D, E, and C), with nutritional composition all within regulatory limits and an average of nutrient levels found in commonly consumed children’s milk in Philippines (Department of Science and Technology - Food and Nutrition Research Institute (DOST-FNRI). 2016. Philippine Nutrition Facts and Figures 2015: Dietary Survey).
- EYCF: Powdered cow’s milk-based toddler formula containing calcium (120 mg / 100 mL), Vitamin D (61IU/100 mL), GOS (4 g/L), pre-cultivated *L. reuteri* DSM 17938 (at concentration that guarantees 10^8^ cfu/day)) and fat blend low in sn-1, −3 Long-Chain Saturated Fatty Acids (LCSFAs).

Since EYCF is a toddler formula tailored to the needs of young children, nutritional profile of EYCF is slightly different from that found in CM, especially pertaining to protein levels. The nutritional composition of the CM and EYCF is summarized in Supplementary table S1.

Children randomized to CM or EYCF received the feeding orally, ad libitum, every day for 6 months from enrollment. Recommended consumption in line with dietary guidelines was at least 2 servings per day (∼ 235mL of reconstituted product per serving). Children in the reference group continued their habitual diet.

### Clinical readouts

#### Anthropometry

Anthropometric parameters were collected at baseline and 6 months after intervention, including weight (kg), height (cm) and head circumference (cm); BMI was calculated (kg/m^2^). Children were weighted without clothing or diaper on a calibrated electronic weighing scale and measurements were recorded to the nearest gram. Height was measured using a stadiometer to the nearest 0.1 cm. Head circumference was measured using a pediatric non-elastic tape measure to the nearest 0.1 cm. All anthropometric measures were repeated until reproduced within a pre-defined acceptable range (i.e. 10 grams for weight, 0.5 cm for height and 0.2 cm for head circumference). For the baseline and 6 months timepoint, corresponding WHO child growth standard z-scores including weight-for-age, height-for-age, head circumference-for-age and BMI-for-age were computed. Radius and tibia length were determined using a rubber, as previously described (36).

#### Speed of sound of the tibia and radius

Speed of sound (SOS) readings were measured using the Sunlight Omnisense® 9000S Bone Sonometer (BeamMed Ltd) as previously described (37, 38) at baseline, 3 and 6 months after intervention. This device uses the axial transmission technique to measure the speed at which ultrasound waves travel across the long axis of the bone surface (39). A handheld ultrasound probe contains transmitting and receiving transducers. The probe emits an array of ultrasound waves with a center frequency of 1.25 MHz. The signal travels through the soft tissues until it encounters bone. The waves that encounter the bone at a “critical angle” (angle of incidence that results in 90 degrees angle of refraction) are propagated along the surface of the long axis of the bone, and then exit the bone at the same “critical angle” back into the soft tissue. The first signal to be detected by the receiving transducers is used to calculate SOS in meters per second (m/s). The device uses proprietary algorithms to account for differences in soft tissue thickness, normalizing the SOS measurements for comparison to children of similar age. The device is designed to measure SOS of bones that are close to the surface of the skin, such as the tibia or radius. Ultrasound transmission gel (Aquasonic 100 Ultrasound Transmission Gel; Parker Laboratories Inc.) was used to obtain good acoustic contact between the probe surface and the soft tissue overlying the tibia.

#### Muscle force as measured by Handgrip digital dynamometers

Force was measured with the Jamar hand dynamometer at baseline and 6 months after the intervention as previously described (40). The child was seated upright on a chair in front of the instrument, which was placed on a table. The most used posture was as follows: shoulders adducted and neutrally rotated, elbow flexed at 90°, forearm in neutral and wrist between 0 and 30° of dorsiflexion. The child grasped the handle and was allowed to become familiar with the instrument by obtaining a good grip, squeezing lightly and watching the corresponding increase in grip strength on the digital display. When ready, the child was asked to squeeze as hard as possible for 10 s on a verbal “go” signal. Three trials for each hand were conducted, alternating hands, and always starting with the dominant hand. There was always a break of at least 1 min between the tests on the same hand. The results of each of the three tests per hand were noted in a test protocol. Some systematic verbal instruction was given like: ‘I want you to hold the handle like this and squeeze as hard as you can’. The examiner demonstrated and then gave the dynamometer to the subject. After the child was positioned appropriately, the examiner said, ‘Are you ready? Squeeze as hard as you can’. As the child began to squeeze, the examiner said, ‘Harder!… Harder!… Relax’(41).

#### Gastrointestinal tolerance and stooling patterns

Gastrointestinal (GI) tolerance was assessed using the Toddler Gut Comfort Questionnaire (TGCQ) at baseline, after 3 and 6 months of intervention. It consists of 10 questions pertaining to GI-related symptoms and behaviors including constipation, diarrhea, gassiness, abdominal pain, difficult to pass / hard stools, bloating, fussiness, and sleep problems that are recalled retrospectively over the past week from a parent’s perspective. Possible scores for each individual question on the TGCQ range from 1 (Never/None/Not a problem at all) to 6 (Always/Very strong). Scores for each of the 10 questions were summed to provide a total score with a possible total score of 10 to 60. Stooling patterns, which includes stool frequency and consistency were recorded via the GI symptom and Behavior Diary at baseline, after 3 and 6 months of intervention. Stool consistency was rated by parents on a 5-point stool scale: 1=watery, 2=runny, 3=mushy soft, 4=formed, and 5=hard.

### Blood markers

Subject’s blood bone turnover marker levels including C-terminal cross-linked telopeptides of type I collagen (CTX) and procollagen type I N-propeptide (P1NP) were measured at baseline and 6 months after the intervention as described in (42). Bone turnover index define as (CTX-P1NP)/P1NP was calculated as described in (42)

### Blood vitamins

Sample preparation including calibration curves, quality controls (QC) and study samples, was automated and performed on a Microlab Star M liquid handler (Hamilton, Reno, NV, USA). Briefly, samples were thawed at room temperature, vortexed, and transferred to polypropylene plates containing internal standards (IS) and Ascorbic Acid. Samples were precipitated using 7.5% trichloroacetic acid (TCA), vortexed during 10min and finally centrifuged at 2500 rpm for 10 minutes. Supernatant was transferred and filtered onto an AcroPrep Advance 96 filter plate with a 0.2 µm membrane (Pall, Port Washington, NY, USA) prior to LC-MSMS analysis.

Water-soluble B vitamins (B3, B6 and B9) analyses in CM and EYCF groups were performed on an Acquity I-class UPLC system (Waters, Milford, MA, USA) composed of a binary solvent pump, a sample manager with a fixed-loop injection system (SM-FL) set at 6°C, and a column oven equipped with an active preheater set at 40°C. Separations were performed on an ACE Excel, 1.7μm, C18-AR 100 x 3.0 mm column (ACE, UK) in gradient mode using solutions containing 1% Acetic acid, 0.4% Formic acid and 0.2% Heptafluorobutyric acid (HFBA) in Milli-Q water (Merck®, DE), and Methanol as mobile phases. A constant flow rate of 450 µL/min was used and a volume of 2 µL was systematically injected. The UPLC system was hyphenated to a Xevo TQ-XS triple quadrupole mass spectrometer (Waters, Milford, MA, USA) equipped with an Electrospray Ionization (ESI) source. Argon was used as collision gas and Multiple Reaction Monitoring (MRM) transitions were experimentally determined for each compound. Data were acquired using MassLynx software (Waters, Wilmslow, UK), and chromatographic peaks were integrated with TargetLynx (Waters, Wilmslow, UK). A calibration curve is built using response (ratio of analyte area and internal standard area) and theoretical concentration of each calibration point, then weighted. QCs and samples are quantified using the calibration curve and reported in ng/ml.

For vitamin D (25-hydroxycholecalciferol) levels, blood samples were centrifuged and sent on ice to Asian Hospital and Medical Center in Muntinlupa City. Concentrations of serum 25(OH)D were measured using the DiaSorin radioimmunoassay (RIA) method. The intra-assay coefficient of variation (CV) was less than 2%. All laboratory technicians were blinded to the case status. Deficiency was defined as levels below 20ng/mL and insufficiency as levels between 20 to 30 ng/mL.

### Measurement of Fatty acid and mineral excretion in feces

A 27g stool sample was collected by the parents at home and stored in the freezer until picked up by study staff. The samples were stored locally in a −80 °C freezer at the study site in the Philippines, and shipped on dry ice to Eurofins (Madison, WI, USA) for analysis of total fatty acid soaps, and mineral content (total calcium, magnesium and phosphorus), in CM and EYCF groups. The dried stool sample was extracted and neutral lipids including non-soap free fatty acids were obtained by solvent reflux. The remaining sample was treated by acetic acid to release the fatty acid soaps, which were obtained by a second solvent reflux step, as described previously (43). Total fatty acid soaps were calculated based on the sum of all measured individual fatty acid soaps and was normalized to gram of dry stool weight in the acid form and expressed as mg/g. The limit of quantification for major fatty acids is a standard 0.05 mg/g based on a 0.5 gram sample weight. Mineral analyses were performed by ICP.

### Statistical analysis on primary and secondary outcomes

The primary outcome was tibia SOS measurements after 6 months of product intake. The estimated effect for the primary endpoint was obtained using a mixed model for repeated measurements (MMRM) for the tibia SOS measurements at 3 months and 6 months. The fixed covariates of the MMRM were: randomized treatment (EYCF or CM), visit (3 months or 6 months), visit in interaction with treatment, baseline tibia SOS measurement (at enrollment), and sex. Participant identification number was used as a random effect to account for the intra-participant correlation. No imputation methods were used, assuming that the data was missing at random, and that the resulting estimator of the mixed linear model is unbiased under this assumption. Significance of the estimated effect for the primary endpoint was tested at 5% level and no multiplicity adjustment was needed.

All conclusions for the primary outcome were based on the intention-to-treat analysis population (ITT), which quantified the average treatment effect in all randomized participants regardless of adherence to treatment. A supportive analysis for the primary outcome was carried out on the per-protocol set (PP), consisting of all children with at least 70% compliance for the product intake, no consumption of other fortified milk and no intake of vitamin supplements. No difference on the primary outcome between ITT and PP analysis was observed.

Continuous secondary endpoints were analyzed using MMRM, when repeated measurements were performed at both 3 months and 6 months visits, with same covariates as for the primary endpoint. For continuous secondary outcomes measured only at baseline and at end of the study (6-months visit), an analysis of covariance model was used to estimate the treatment effect. The factors of the analysis of covariance model were randomized treatment, sex and baseline endpoint. When necessary, a natural logarithm transformation was performed to meet the assumptions of homoskedasticity of residuals. Estimated effects of secondary outcomes were tested at 5% without adjustment for multiplicity. No imputation for missing data was performed on secondary endpoints.

Association between different clinical outcomes were performed using both Pearson and Spearman correlation coefficient, significance of the correlation was tested at 5% level without adjustment for multiplicity.

Subjects in the formula group are randomized to receive the experimental young child formula or the minimally fortified milk. Subjects belonging to the reference group (REF) are not randomized, they are recruited in the trial and they continue their habitual diet, which may or may not consist of cow’s milk-based products.

In this situation, confounding can occur if some covariates are related to both the treatment assignment (exposed to either EYCF or CM) and the outcome. In the presence of confounding, propensity score methods are used to remove the effects of confounding when estimating the effect of treatment. For this analysis gender, gestational age (GA), delivery mode, breastfeeding history, age at enrollment (months), BMI at baseline were used as variables to build the propensity scores.

First a comparative model without adjustment was carried out. After which, a model including the computed propensity scores as a covariate was carried out. Then a model including the computed propensity score as a weight (inverse probability of treatment weighing method) was carried out. The three models included as covariates by default the gender, gestational age (GA), weight at birth, delivery mode, breastfeeding history, age at enrollment (months), BMI at baseline. The comparison between the first and the two next models highlight a part of the bias introduced by the lack of randomization. The results were reported when the three models lead to similar conclusions, for a robust interpretation of the results.

### Microbiome analysis

Out of 278 screened subjects, a total of 220 participants (81%) provided stool samples at all timepoints, while 41 participants (6%) provided samples only at baseline. Stool samples were subjected to DNA extraction, library preparation and shotgun metagenomics sequencing similarly to Capeding *et al*. (44) with the exception that DNA extraction was performed using the NucleoSpin 96 Stool (Macherey-Nagel) kit. After removing low-quality reads and reads mapping to the human genome (GRCh38), the remaining reads were aligned to the Clinical Microbiomics in-house extended infant fecal microbiome gene catalog (20,992,486 microbial genes) allowing for taxonomical profiling of corresponding set of 1472 metagenomics species (MGS) (45). All MGS relative abundances are shared in Supplementary Table S2.

### Metabolome analysis

Stool metabolomics were performed untargeted for semi-polar metabolites following an analytical protocol adapted to a UHPLC system (Vanquish, Thermo Fisher Scientific) coupled with a high-resolution quadrupole-orbitrap mass spectrometer (Orbitrap Exploris 240 MS, Thermo Fisher Scientific). Data was processed with Compound Discoverer (3.2, Thermo Fisher Scientific) and MS-Omics software, with compound annotations based on the MS-Omics internal spectral library, mzCloud, the Human metabolome database (HMDB, version 5.0) and the FooDB.

Short chain fatty acid analysis of Acetic acid, Formic acid, Propanoic acid, 2-Methylpropanoic acid, Butanoic acid, 3-Methylbutanoic acid, Pentanoic acid, 4-Methylpentanoic acid, Hexanoic acid, Heptanoic acid, and p-Cresol was performed in a targeted manner on a high-polarity column (ZebronTM ZB-FFAP, GC Cap. Column 30 m x 0.25 mm x 0.25 μm) installed in a GC (7890B, Agilent) coupled with a time of flight MS (Pegasus® BT, LECO). Raw GC-MS data were processed and quantified in Skyline 22.2 (Adams et al. 2020, PMID: 31984744) and the PARADISe software (v2.6) developed by MS-Omics and collaborators. Finally, clusters of co-abundant compounds were identified using the weighted correlation network analysis (WGCNA) framework as implemented in the WGCNA R package (Langfelder & Horvath, 2008), applied on the merged data set including the SCFA processed peak areas and the semi-polar processed PQN peak areas. A signed, weighted compound co-abundance correlation network using biweight midcorrelation, with <5% of the individuals regarded as outliers was calculated across all included samples using all pairwise observations. A scale-free topology criterion (R2-cutoff for scale-free topology = 0.8) was used to choose the soft threshold, resulting in β=4I. Clusters of positively correlated compounds were identified with the dynamic hybrid tree-cutting algorithm (46), using a deepSplit of 2, a minimum cluster size of 10 and the partitioning around medoids option turned on. The profile of each metabolite module was summarized by the module eigenvector, i.e. the first principal component of the metabolite abundances across samples. Pairs of modules were subsequently merged if the correlation between the modules’ eigenvectors exceeded 0.85. The resulting metabolite modules were named ‘MXX’, where ‘XX’ is an integer. Compounds that did not fit the clustering criteria were assigned to a leftover group named ‘M0’. Complete metabolite data is available upon request.

### Integration of microbiome, metabolome & clinical outcomes

Microbial abundances for taxa present in at least 10% of samples were centered log-ratio (CLR) transformed after replacing all zeroes with half the lowest observed non-zero abundance per taxon (47). Semi-polar metabolite intensities were power transformed (Box-Cox). SCFA quantifications were log2 transformed. To be robust against outliers in the data, winsorization was performed, capping the transformed abundances at their 3rd and 97th percentile.

The intervention effect on the microbial abundances and the metabolite intensities was estimated with cross-sectional linear models with the covariates: intervention group (CM or EYCF) and sex.

Correlations between microbial abundances, metabolite intensities and clinical outcomes were estimated on samples at all time points and the two intervention groups collectively using linear mixed models adjusting for sex as a fixed effect and participant as a random effect, with effect sizes reported as partial Pearson correlation coefficients (obtaining by scaling the T-values of the linear model).

All models including metabolite intensities have the additional fixed effect covariate: storage time of metabolome sample. All models including microbial abundances used a compositional bias correction based on LinDA (48).

P-values were adjusted for multiple testing by the FDR method (49). These adjusted P-values are referred to as Q-values. Adjustments were performed separately for each visit, each microbial taxonomy level, and for the set of WGCNA modules, as well as for each annotation level of the metabolites (SCFA, 1, 2a, 2b, 3), where relevant.

The alpha diversity of the microbial composition was estimated with richness and Shannon index, both estimated at gene-level and species-level, and with species-level Faith’s PD. Weighted UniFrac distances were used for beta diversity analysis.

Procrustes analysis was performed on the first two principal co-ordinates of the weighted UniFrac distance matrix (microbial composition) and the first two dimensions of a redundancy analysis (for metabolite intensities) using protest from the vegan package with 999 permutations.

### Association of *L. reuteri* increase vs. non-increase groups with clinical outcomes

Statistical comparisons between samples displaying an increase in *L. reuteri,* or a lack thereof, between baseline and after 6 months of intervention, were run using the same approach as in the section “Statistical analysis on primary and secondary outcomes”.

## Results

We have previously shown *in vitro* that administration of a synbiotic consisting of *L. reuteri* and GOS improves osteoblastogenesis (20). Prior to assessing the clinical efficacy of this synbiotic on muscle strength in toddlers, we evaluated its impact on muscle progenitor cells, key drivers of muscle development.

### Experimental formula positively impacts myogenesis *in vitro*

Human myoblasts exposed to the product of *in vitro* fermentation of the experimental formula, containing GOS + *L. reuteri*, showed a statistically significant increase in the fusion index of myotubes (+10% as compared to milk matrix alone versus non-statistical significant increase by GOS and *L. reuteri* separately, by 4.5% and 2.4%, respectively, Fig.S1A, C) and the myotube total area (+18% as compared to milk matrix alone versus non-signficant increase by GOS and *L. reuteri* alone by 5.8% and 3.8%, respectively, Fig.S1B, C).

### Clinical trial population

A total of 273 of 278 screened toddlers were eligible for the study (Fig. 1A). Of the 273 toddlers, 182 were randomized to experimental young child formula (EYCF) or a minimally fortified control milk (CM), whereas 91 remained under their habitual diet (REF). Study participants were included between June 2021 and March 2022 and completed their 6-month visit between November 2021 and August 2022. The FAS analysis included all study subjects (n = 138) who were randomized to the EYCF or CM. The study groups were well balanced with regard to baseline characteristics (Supplementary table S3). All study participants were Asian. A total of 107 (59%) subjects completed the 6 months intervention period in the study, of whom 51 had been randomized to EYCF and 56 to CM. Four participants dropped out in the REF group. Four participants had a major protocol deviation defined as a compliance rate of <70%.

### Experimental young child formula increases bone and muscle strength

The mean relative change in tibia SOS at 6 months of intervention in EYCF was 58.91m/s (95% confidence interval [CI]: 21.34, 96.48; p=0.002 compared to CM) (Fig. 1C).

After 3 months of intervention, the difference was already significant reaching 53.00 m/s (95% CI: 7.05, 98.95; p=0.024 in EYCF compared to CM) (Fig. 1D). Tibia SOS was significantly greater in EYCF compared to REF after 6 months of intervention (p=0.007; Supplementary table S4, S5) while no difference was observed between the CM and REF. Bone turnover index was also significantly higher in EYCF compared to CM (+25%, p=0.02) (Fig. 1E) after 6 months of intervention, while no difference was observed between CM and REF (Supplementary table S4, S5).

Secondary outcomes, including bone growth (head circumference, tibia & radius length, Fig. 1F-G, Supplementary table S4, S5) and body growth (height & weight for age Z score, figure 1H-I) were not significantly different between EYCF and CM. However, both CM and EYCF groups showed higher bone and body growth than REF after 6 months of intervention (supplementary table S4, S5). Finally, muscle force of the right arm was higher in EYCF than CM group by 11% (95% CI: [2%, 20%]; p=0.015) (Fig. 1J), after 6 months of intervention, while no difference was observed between CM and REF (Supplementary table S4, S5).

### Vitamins and minerals levels in experimental young child formula do not drive effect on bone and muscle strength

To determine whether the observed improvement of bone and muscle strength in EYCF was associated to higher vitamin levels, and/or lower intakes of sn-1,3 palmitate, we measured vitamins D, B3, B6 concentrations and excretion of fecal calcium fatty acid soaps. After 6 months of intervention, Vitamin D levels were 31.3, 36.5, and 32.7 ng/mL in the CM, EYCF, and REF groups, respectively. Vitamin D levels were 17% (95% CI: [11%, 23%]) higher in the EYCF compared to CM (p<0.0001) (Fig. 2A) and 10% (95% CI: [10%, 15%]) higher compared to REF (p<0.001) (Supplementary table S6).

**Figure 2.**
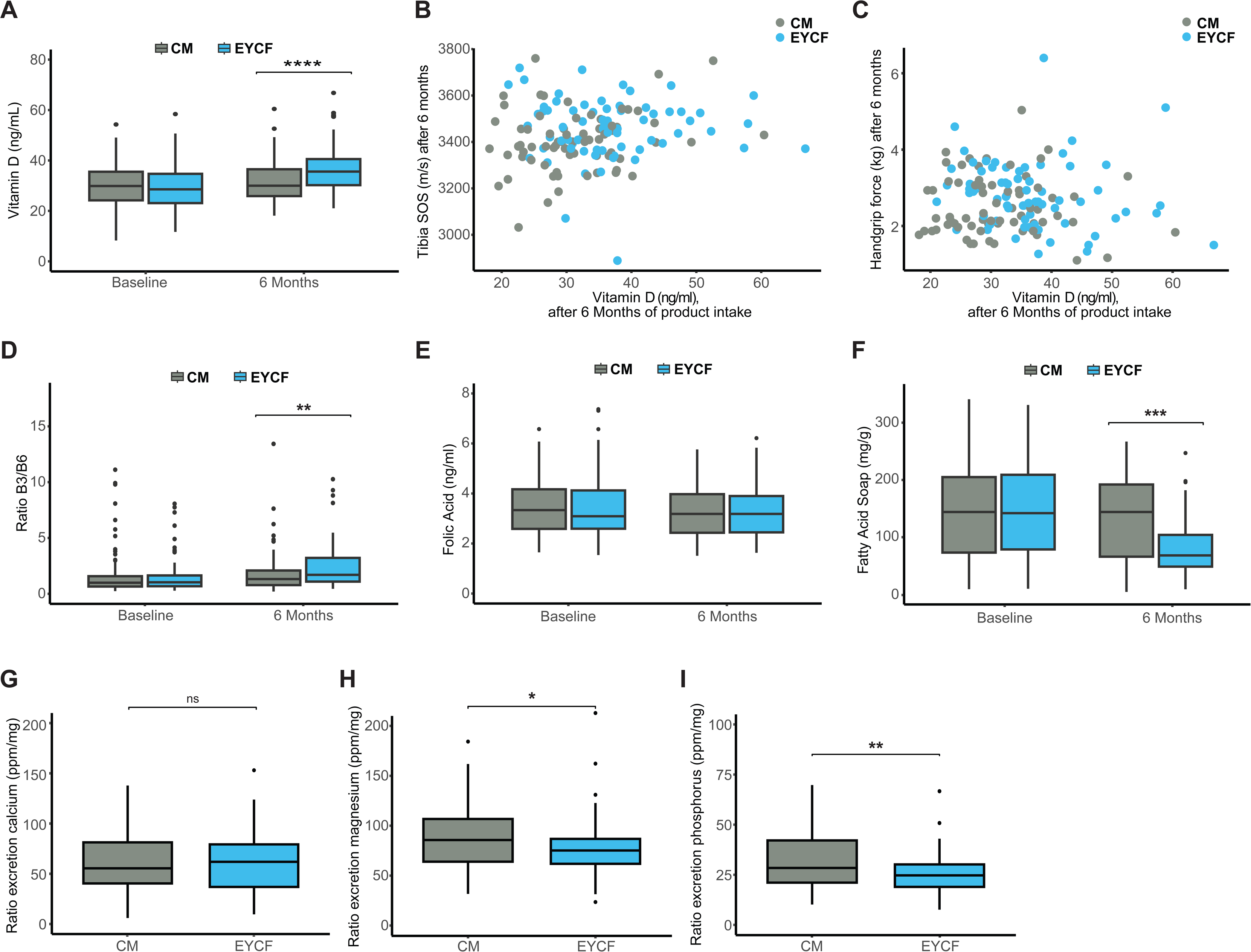
Influence of «non synbiotic elements» in the blend on bone & muscle readouts. (A) 25 hydroxy vitamin D3 levels in blood at baseline and after 6 months of intervention. (B) Lack of association between 25 hydroxy vitamin D3 and tibia SOS after 6 months of intervention. (C) Lack of association between 25 hydroxy vitamin D3 and handgrip force after 6 months of intervention. Blood vitamins measure showing the specific ratio of vitamin B3 and B6 (D), Folic acid (E) at baseline and after 6 months of intervention. (F) Soap Fatty Acids levels in Infant Stool at baseline and after 6 months of intervention. (G-I) Respectively calcium, magnesium and phosphorus excretion levels in feces adjust by product intake. For all outcome, EYCF n=68 and CM n=69. Data are presented as mean ± SD, * P<0.05, ** P<0.01, *** P<0.001 and **** P<0.0001 compared to CM group.

This effect totally abolished deficiency and reduced insufficiency of vitamin D by 52% reported at baseline (Supplementary Fig. S1D) in the EYCF. However, blood vitamin D levels were not associated to tibia SOS and muscle force (Fig. 2B-2C).

A specific ratio of vitamins B3/B6, previously described to stimulate muscle regeneration (50), was increased in EYCF after 6 months of intervention (Fig. 2D). However, no association was found with tibia SOS, handgrip, radius SOS and radius length, and only a mild correlation was found with the tibia length (Supplementary table S7). Finally, folic acid level was not different between CM and EYCF (Fig. 2E).

Total fecal fatty acids soaps levels did not differ at baseline between groups, whereas they significantly lowered after 6 months of intervention in EYCF compared to CM (p<0.001, Fig. 2F). However, calcium excretion was not significantly different between groups after adjustment by intake of the product (Fig. 2G). Magnesium and phosphorus excretion appeared lower in EYCF compared to CM (Fig. 2H, 2I, p<0.01). However, none of the calcium and magnesium minerals nor total fatty acids soaps were associated with tibia/radius SOS, muscle force or tibia/radius length and only phosphorus was mildly associated with handgrip (Supplementary table S7).

### The experimental young child formula induces a bifidogenic effect

Given the important link between microbiome and musculoskeletal development in early life, we investigated whether the gut microbiome and metabolome of toddlers were impacted by the nutritional intervention.

The gut microbiome composition of all children was overall not significantly different between groups at baseline, as determined by alpha and beta diversity (Supplementary Fig. S2, Supplementary table S8). After 6 months of intervention, toddlers consuming EYCF showed a distinct microbial community compared to baseline (P=0.0199, Supplementary Fig. S2, Supplementary table S8). Looking at the 16 species significantly higher in EYCF compared to CM after 6 months of intervention (Fig. 3A), a notably significant increase was observed in *L. reuteri* (Q=6.10e-14, effect size=4.23), and in several bifidobacteria such as *B. breve* (Q=0.047, effect size=3.57), *B. bifidum* (Q=0.038, effect size=3.40), *Bifidobacterium longum* spp. (Q=0.021, effect size=2.56), *B. longum* subsp*. longum* (Q=0.038, effect size=1.85), overall suggesting a bifidogenic effect of the intervention (Fig. 3A). Other taxa significantly enriched in EYCF include 3 species belonging to the *Bacteroides* genus (Q=0.202, effect size=1.04). In CM, 21 species were significantly higher than in EYCF, including 8 species belonging to the *Streptococcus* genus (Q=3.84e-05, effect size=-2.17) (Statistics for all taxonomic levels are reported in Supplementary table S9).

**Figure 3.**
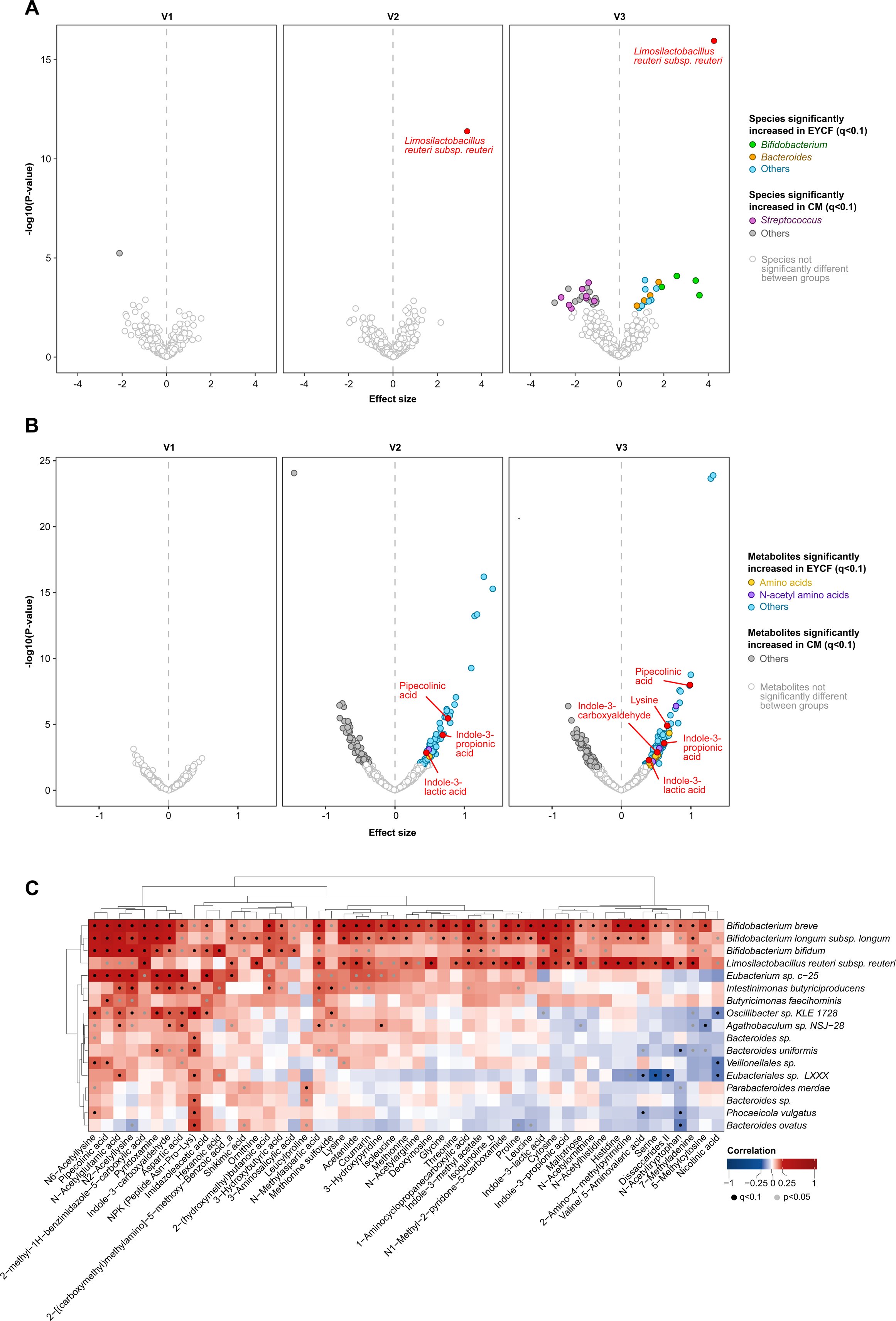
EYCF consumption impacts both the fecal microbiome and metabolome. Volcano plots of (A) the fecal microbiome and (B) the fecal metabolome depicting the difference between control-fed and EYCF-fed infants at each visit (421 samples in total, 208 EYCF and 213 CM). Group comparison was performed using a linear model at each visit with intervention group and sex as confounders, adding storage time as confounder for the fecal metabolome, filtering species prevalence at 10% minimum (C) Associations between the fecal metagenome and metabolome features significantly increased in the EYCF group at V3 using a linear model with sex and storage time as confounders.

### The experimental young child formula altered the microbial metabolism of amino acids

Given the changes in microbiome composition, we further investigated the impact of the intervention on its functionality by exploring the fecal metabolome (Fig. 3B).

Within it, 266 metabolites were found to significantly differ between EYCF and CM at 6-month visit, of which 69 had an unambiguous annotation (Fig. 3B). Of note, metabolites like pipecolinic acid (Q=2.83e-06, effect size=0.99), indole-3-carboxyaldehyde (Q=0.013, effect size=0.52), IPA (indole-3-propionic acid, (Q=0.007, effect size=0.61), ILA (indole-3-lactic acid, Q=0.04, effect size=0.39) and lysine (Q=0.001, effect size=0.64) were all significantly increased in EYCF compared to CM. Other metabolites strongly associated with the EYCF group included several amino acids (Q<0.06, effect size 0.41-0.71), and N-acetylated amino acids (Q<0.09, effect size 0.35-0.79) (Fig. 3B, Statistics for all metabolites are reported in Supplementary table S10).

### The experimental young child formula promotes microbiome & metabolites changes that are associated to clinical outcomes

We then assessed whether the metabolomic changes observed were associated with changes in microbiota composition. In this analysis, we found a significant correlation between microbiome and metabolome at all timepoints, suggesting that children with similar microbiota compositions have similar metabolomic profiles (all p<0.02, correlation between 0.27 and 0.47 depending on sample subset, see Supplementary table S11). Looking at correlations between species and metabolites significantly increased in EYCF at 6 months (Fig. 3C), we observed a majority of these metabolites correlating with the most enriched species, namely: 42 out of 45 significantly correlated with *Bifidobacterium breve*, 27 with *L. reuteri* and 26 with *B. longum* subsp*. longum* – suggesting that the increase of *L. reuteri* and bifidobacteria caused by the EYCF intervention is associated with a shift in metabolomic profiles. Of note, abovementioned metabolites known to have an impact on bone and muscle development (i.e. pipecolinic acid, lysine, indole-3-carboxyaldehyde, IPA, and ILA) all significantly correlated positively with *B. breve, B. longum* subsp. *longum* and some to *L. reuteri* and *B. bifidum* too, although with a low effect size (Q<0.07, effect size 0.18-0.41).

We further determined if the observed taxonomic and metabolic differences were associated to clinical outcomes related to bone and muscle strength. 37 species were significantly associated to at least one clinical outcome and one metabolite (Fig. 4A), including several which were significantly increased after 6 months of intervention in EYCF compared to CM. Notably, *L. reuteri* showed significant associations with 4 of the 5 measured clinical outcomes (tibia length, radius length, radius SOS and handgrip strength, Q<0.07). *Bifidobacterium longum* subsp. *longum* was significantly associated with tibia length and radius length (Q<0.004), and nominally also with handgrip (P=0.02, Q=0.43). Worth noting, tibia length was the outcome associated with the highest number of species (32 out of 37, 12 of which were positively correlated to tibia length and significantly higher in EYCF compared to CM after 6 months of intervention). All statistical associations between microbiome species and clinical outcomes can be found in the Supplementary table S12.

**Figure 4.**
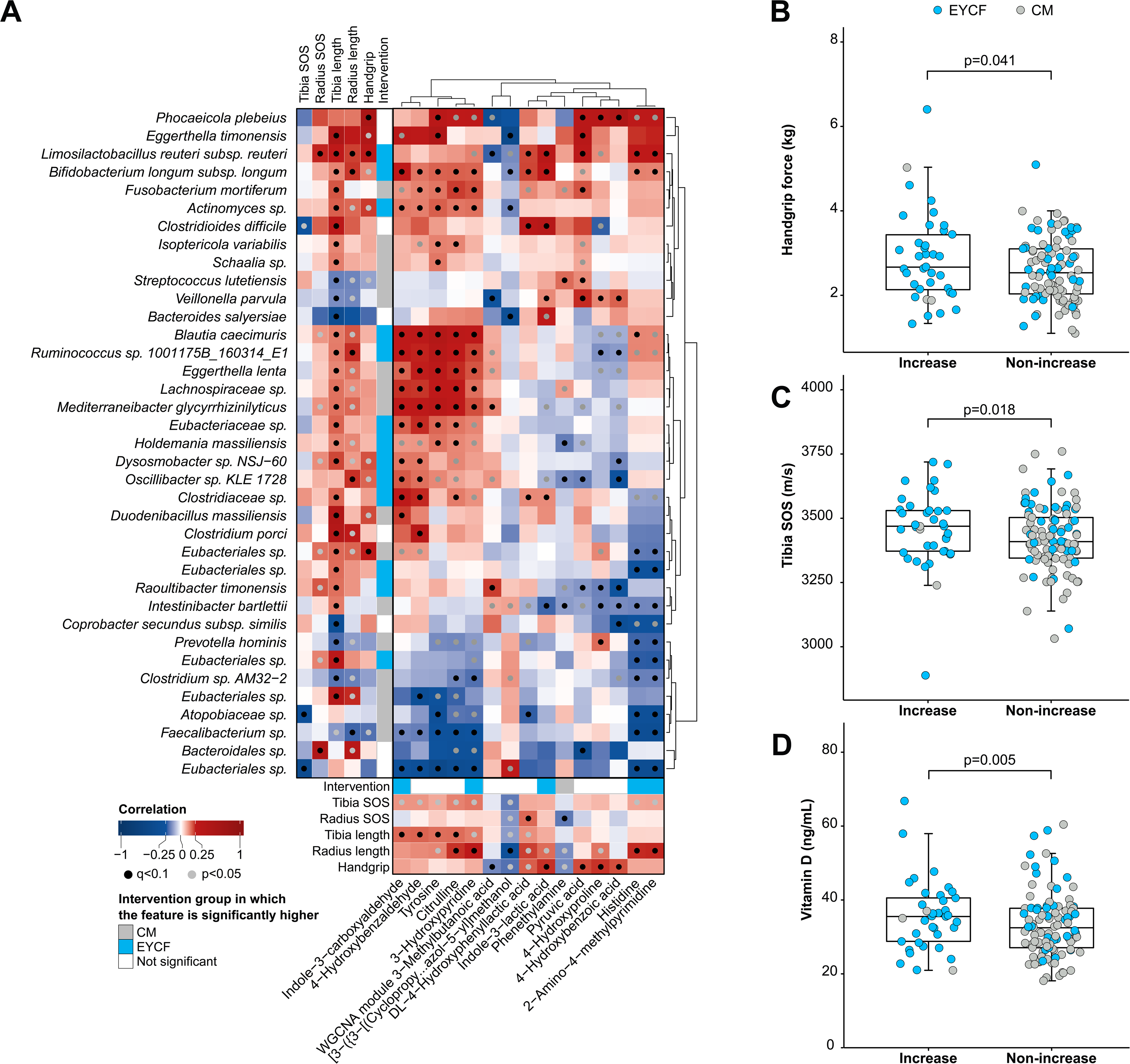
Changes in fecal microbiome and metabolome are associated to the nutritional intervention and clinical outcomes. (A) Correlations between metagenome, metabolome and clinical outcomes (see methods for more details on the statistical approach) (B-D) Comparison of in handgrip force (B), tibia SOS (C), and Vitamin D (D) between samples showing an increase in *L. reuteri* after 6 months compared to baseline (“Increase”, n=37) and samples not showing an increase (“Non-increase”, n=101). Non-significant associations are shown in Supplementary table 14.

We also assessed the relationship between clinical outcomes and fecal metabolites, with 15 metabolites significantly associated with at least one clinical outcome (Fig. 4A). Of interest, several metabolites that were significantly increased in the EYCF group at 6 months compared to CM were associated with at least one clinical outcome. For example, indole-3-carboxyldehyde was significantly associated with tibia length (Q=0.058) and ILA with handgrip strength (Q=0.012) and nominally to radius length (P=0.046, Q=0.012). Of note, 8 metabolites such as indole-3-carboxyaldehyde, tyrosine, citrulline, 3-hydroxypyridine or 4-hydroxybenzaldehyde positively correlated with tibia SOS, although only nominally significant (P<0.05, Q<=1). All statistical associations between fecal metabolites and clinical outcomes can be found in the Supplemental Table S13.

### *L. reuteri* abundance increases are associated with clinical outcomes

While there was an association between EYCF and tibia SOS as well as handgrip strength, this relationship was not observed in all the children in the EYCF group. To better understand this effect, we investigated whether it was driven by an increased abundance of *L. reuteri* throughout the intervention period. For this, we classified the overall study population into two categories: “increase” when *L. reuteri* abundance (derived from the associated rarefied species abundance) was increased between baseline and 6 months, and “non-increase” when *L. reuteri* did not increase. Notably, *L. reuteri* abundance increased between baseline and 6 months in half of the children in the EYCF group. Participants in which *L. reuteri* increased in abundance over time showed a significant increase of handgrip force (mean increase=0.3kg, Fig. 4B), tibia SOS (mean increase = 55 m/s, Fig. 4C) and vitamin D (mean increase = 3ng/mL, Fig. 4D), with most subjects indeed belonging to the EYCF group. However, in this stratified analysis we did not see associations with bone length or bone radius, despite their association with overall *L. reuteri* abundance. All of these trends persist when including samples from the reference group, except for handgrip force which was no longer significant (P<0.1, Supplementary table S14).

### Consumption of EYCF is safe and well tolerated

Mean total TGCQ scores were low at baseline and remained low after 3 and 6 months of intervention, indicating maintenance of good GI tolerance in these healthy children. No significant difference in total TGCQ scores was observed between the EYCF and CM groups. After 3 months of intervention, mean (± SD) scores were 10.64 ± 2.28 in EYCF *vs*. 10.85 ± 1.96 in CM; P=0.236, and after 6 months scores were 10.62 ± 1.63 in EYCF *vs*. 10.57 ± 1.44 in CM; P=1, Supplementary table S15, S16). The mean number of stools per day was approximately 1.3 after 3 and 6 months of intervention and not significantly different between treatment groups. Mean stool consistency scores ranged from 3.5 – 3.87 or between “mushy-soft” and “formed” at Visits 2 and 3 in both intervention groups. After 3 months of intervention, stool consistency scores were significantly lower, indicating softer stools, in EYCF *vs* CM (3.5 ± 0.62 *vs* 3.78 ± 0.39, P<0.001, Supplementary table S15, S16). There was no significant difference in mean stool consistency score after 6 months of intervention (3.72 ± 0.40 *vs* 3.87 ± 0.37, P=0.079, Supplementary table S15, S16).

The overall incidence of AEs was comparable between the groups. In the EYCF group, a total of 42 events occurred in 35 participants, while a total of 35 events occurred in 31 participants in the CM group. No AE was considered to be related to the study products and no serious adverse event was reported in this trial.

## Discussion

Toddlerhood is a critical window for bone and muscle development, coinciding with the establishment of a more adult-type microbiome (51). Previous reports highlight the existence of a gut-musculoskeletal axis in elderly people (11), while our study is-to the best of our knowledge-the first one exploring this axis in early life in a controlled randomized trial.

In this study, an experimental young child formula (EYCF) containing a synbiotic composed of GOS and *L. reuteri* was shown to improve bone quality and muscle strength after 6 months of intake. We found that this formula improved tibia SOS compared to a minimally fortified control milk (CM) after 6 months of intervention and, more importantly, exhibited a gain in tibia SOS by 4 times compared to the habitual diet group. Handgrip strength after 6 months was increased 1.7 times compared to control and 1.4 times compared to habitual diet. The results of this clinical study have shown that this experimental young child formula containing a synbiotic of GOS and *L. reuteri* is efficient in improving bone and muscle strength acquisition, within normal physiological growth rates represented by the habitual diet reference group. Furthermore, the latter are consistent with previous studies in toddlers reporting increases of tibia SOS at a rate of 2.1% per year (39) and handgrip force at a rate of 30% per year (52).

Interestingly, EYCF containing 520IU/l Vitamin D significantly increased circulating levels of vitamin D by 17%, effectively reduced insufficiency by 52% and abolished deficiency. Meanwhile, in a previous study supplementation of 400IU and 1000IU, respectively increased vitamin D levels by 8.2% and 18% in 12-30 months old toddlers (53). The similar increase in our study to the latter higher dose suggests a potential improvement of Vitamin D absorption when co-administered with a synbiotic, which is further supported by another previous RCT combining a probiotic and vitamin D intervention (54). However, in our study, vitamin D levels or even B vitamin blood levels did not associate to muscle strength or bone length and quality.

Fecal soap fatty acids have been shown to decrease mineral absorption in infants via formation of fatty acid mineral soaps (55). In our study, excretion of fatty acid mineral soaps was significantly decreased in EYCF compared to CM, but did not impact fecal calcium levels and was not associated with bone quality or muscle strength.

We also demonstrated that the supplementation with a young child formula with *L. reuteri* and GOS had a significant impact on microbiome composition both compositionally and functionally. Indeed, supplementation of EYCF with *L. reuteri* and GOS resulted in a bifidogenic effect previously reported with GOS alone (56) and L. reuteri alone (57), with not only *L. reuteri* being significantly increased in EYCF subjects but also several other members of the *Bifidobacterium* genus, some being reported in clinical studies to positively impact bone mass (58) and muscle function (59) in adults.

The analysis of the fecal metabolome showed that the supplementation with a young child formula with *L. reuteri* and GOS also promoted significant increases in metabolites such as pipecolinic acid, lysine, indole-3-carboxyaldehyde, known to be positively associated to bone (10) and muscle outcomes (60). These metabolites are part of the tryptophane metabolism pathway, i.e. either direct (indoles) and indirect (pipecolinic acid, lysine) derivatives. Interestingly, an increasing number of studies indicate that amino acid metabolism is crucial for bone and muscle development and homeostasis, particularly through autophagy mediated effects (61). Tryptophan metabolism through the kynurenine pathway is one major known factor in promoting bone-aging phenotypes but also to stimulate bone anabolism during growth through activation of the NAD signaling pathway (62, 63). The multiple faces of tryptophan in muscle biology also indicate the ability of tryptophan metabolites to improve muscle growth, protein synthesis as well as antioxidant capacity, which might be partly related to myogenic regulatory factors, IGF/PIK3Ca/AKT/TOR and Keap1/Nrf2 signaling pathways (64).

The correlation between some specific features of the microbiome and the metabolome affected by our intervention further supports the importance of using a holistic approach, including microbiome ecology and functionality to more comprehensively assess the impact of a nutritional intervention. This analysis allowed us to show that several of the taxa and metabolites that were significantly impacted by the nutritional intervention are strongly linked to muscle and bone metabolism. Some examples stand out, like ILA and indole-3-carboxyaldehyde, as well as precursors and metabolites related to amino acid metabolism, such as citrulline, which has been clinically shown to improve muscle performance, recovery and protein synthesis (65, 66), as well as fracture healing and mineralization in preclinical models (67, 68). In our study, we too could observe some associations to health outcomes; for example citrulline was associated with tibia SOS, and 4-hydroxyproline, a metabolite found in collagen, was positively associated with handgrip, consistent with a previous intervention with collagen peptide in sarcopenic men (69). Also, we report for the first time, to our knowledge, two metabolite associations, 4−Hydroxybenzaldehyde with bone quality and 4−Hydroxybenzoic acid with muscle strength, both being present as aromatic compounds in plants.

It is interesting to note that, in our study, different metabolites associate to muscle and tibia quality. However, radius quality (a non-weight bearing bone) and muscle strength shared associations with the same metabolites, namely positively and negatively with DL−4−hydroxyphenyllactic acid and phenethylamine, respectively. This highlights the importance of diverse microbes and metabolites to impact the gut-musculoskeletal axis.

In addition, we could also show a link between *L. reuteri* increase from baseline to 6-months and key clinical outcomes like handgrip strength, tibia SOS and vitamin D, especially in EYCF. Further analysis using qPCR is needed to confirm the origin of the *L. reuteri* strain, i.e. from the synbiotic intervention or from other sources, as some samples in the control group (3 out of 69 participants) also showed an increase in *L. reuteri* abundance.

Here we provide further *in vitro* evidence for a synergistic effect of *L. reuteri* and GOS on muscle stem cell fusion and osteocalcin secretion from osteoblast, two key cell processes for muscle and bone development. Even if we showed an effect of EYCF supplemented with the *L. reuteri* and GOS synbiotic on clinical outcomes and the microbiome, the synergistic effects was not assessed.

In conclusion, we have shown clinically that a young child formula supplemented with a synbiotic consisting of *L. reuteri* and GOS is safe and well tolerated, can mitigate vitamin D insufficiency and supports healthy and strong muscle and bone development. We further substantiated the presence of a gut-musculoskeletal axis in toddlers, as the nutritional intervention significantly affected the microbiome and the metabolome of the toddlers in the ECYF group in a correlated manner. Furthermore, several metabolites and taxa further correlated to muscle and bone outcomes. Our study additionally provides novel mechanistic insights as to how microbiome-modulating interventions can benefit bone and muscle development in early life.

## Supplemental figures & tables

**Supplemental Figure S1. EYCF impacts muscle development in vitro.** (A) Fusion index of myotube after 6 days of treatment with final product of ex vivo colonic incubation (1% final concentration). (B) Troponin T positive area of myotube after 6 days of treatment with final product of ex vivo colonic incubation (1% final concentration). (C) Representative images, scale bar: 200µm. MM=Milk matrix, GOS=Galacto-oligosaccharide. Experiments performed with 2 different donors (10-12 replicates per donor). Statistical analysis: One-way ANOVA Dunnett’s test. **p<0.01, ***p<0.001.D) Vitamin D insufficiency (%) and deficiency (%) among study participants (at baseline n=91 per group, at 6 months n= 87 (REF), 69 (CM), 69 (EYCF)

**Supplemental Figure S2. Alpha and beta diversity of subjects grouped by intervention at baseline.** (A) Alpha-diversity is represented by boxplots showing differences in species (MGS) richness, species (MGS) diversity (Shannon and Faith’s) as well as gene richness and diversity (Shannon). Intervention groups were compared by Kruskal-Wallis and Dunn’s test. No significant differences were observed for any alpha-diversity metric at baseline between intervention groups. (B) Beta diversity is represented in a Principal Coordinates Analysis (PCoA) based on weighted UniFrac distance between samples (color-coded by intervention group).

**Supplemental table S1. Complete formulation of the control milk (CM) and the experimental blend (EYCF).**

**Supplemental table S2. Relative species abundance for each sample. Supplemental table S3. Demographics and baseline characteristics of subjects.** Characteristics of the subjects in the ITT population.

**Supplemental table S4. Descriptive statistics of subjects at all visits.** Characteristics of the subjects in the Reference group (REF), Control milk group (CM) and Experimental young child formula group (EYCF) at baseline (V1), 3 months, if recorded, (V2) and 6 months (V3).

**Supplemental table S5. Clinical outcomes comparisons between the experimental and control blends versus the habitual diet arm**. Comparison between Experimental young child formula group and Reference group (EYCF – REF) or Control milk group and Reference group (CM – REF) at 3 months, if recorded, (V2) and 6 months (V3) with ANCOVA models

**Supplemental table S6. Comparisons of the blood vitamin D level between Experimental group and Habitual diet group.** Comparison between Experimental group (EYCF) and Habitual diet group (REF) at 6 months (V3) with ANCOVA models. Three models are proposed to evaluate the differences between the Experimental group and the Reference group, by taking into account that the Reference group was not randomized in this trial. The first model, Model 1 is a model computed without propensity scores but includes as covariates: Arm (Experimental or Reference), Visit (Visit 2 or Visit 3), Arm in interaction with Visit, baseline Vitamin D level, measurement, sex, gestational age, delivery method, duration of breastfeeding, age at baseline (measured in months), BMI (kg/m2) at enrolment and vitamin D (ng/mL) at enrolment. The other two models have additional corrections: the propensity scores included either as covariate in Model 2 or as weights in Model 3.

**Supplemental table S7. Correlation of the vitamin B3/B6 ratio and mineral excretion (adjusted to intake) and the clinical outcome.** Spearman correlation of ratio of vitamin B3/B6, the calcium, magnesium and phosphorus excretion adjusted to intake and the clinical outcomes tibia SOS, Radius SOS, Tibia length, Radius length and Handgrip at 6 months (V3) after the beginning of the trial. n=222 for the ratio B3/B6 (participants of the 3 groups were measured), n=137 for the calcium, magnesium and phosphorus (only participants from the CM and EYCF were measured)

**Supplemental table S8. PERMANOVA analysis on microbiome beta diversity.** P-values for tests comparing beta-diversity between intervention groups, overall and pairwise

**Supplemental table S9. Comparison of microbiome taxonomic composition between EYCF and CM.** Results of the linear model at each visit and taxonomic level.

**Supplemental table S10. Comparison of metabolome intensities between EYCF and CM.** Results of the linear model at each visit comparing metabolites intensity between EYCF and CM.

**Supplemental table S11. Procrustes analysis of microbiome-metabolome associations.** Results from a Procrustes analysis stratified by intervention group and visit. P-values are derived from 10 000 permutations (thus the smallest possible P-value being 0.0001)

**Supplemental table S12. Associations between microbiome taxonomic composition and clinical outcomes.** Results of the linear model at species (MGS) level considering EYCF and CM samples.

**Supplemental table S13. Associations between metabolome intensities and clinical outcomes.** Results of the linear model for metabolites considering EYCF and CM samples.

**Supplemental table S14. Associations between *L. reuteri* “Increased” and “Non-increased” groups and clinical outcomes.** The comparisons were run considering EYCF+CM samples, and EYCF+CM+REF samples.

**Supplemental table S15. Stool consistency and Descriptive statistics for the TCGQ questionnaire.** Average consistency of stools averaged over 3 days and Descriptive statistics of the questionnaire “Toddler Gut Comfort Questionnaire” in the ITT population.

**Supplemental table S16. Comparison of stool consistency and Gastrointestinal total score.** Stool consistency at baseline is extracted from “24h Recall GI Symptom and Behavior Recall”, from question 3: “If yes, thinking about the past 24 hours, mark the one picture that looks the most like your child’s stool”. The stool consistency was transformed using a 5-point stool scale: 1=watery, 2=runny, 3=mushy soft, 4=formed, and 5=hard. The effect of treatment group on stool consistency at V2 and V3 is evaluated based on a linear mixed model with a log transformation of the response and the following explanatory variables: stool consistency at baseline, treatment group, mode of delivery, gender, visit and interaction between treatment and visit as covariates. A subject specific random effect is be included in order to consider the repeated measurements.

Effect of treatment group on the log-transformed scores of GI TOTAL at V2 and V3 is evaluated based on a linear mixed model, including GI TOTAL at baseline with log transformation, treatment group, mode of delivery, gender, visit and interaction between treatment and visit as covariates. A subject specific random effect is be included in order to consider the repeated measurements.

## Author contributions

NB, YC and MNH designed the study. MRC, JL and LP were responsible for recruiting participants. MRC, JL and LP were investigators in the study and were responsible for collecting data. NB, TD and MNH, were responsible for analyses of bone, muscles outcomes, circulating levels of vitamin and fecal fatty soap and minerals. LFK had access to raw data and were responsible for statistical analyses. NB, LS, TD, HLPT, LFK and MNH drafted the manuscript and designed figures. NB, LFK, TD, LS and MNH have directly accessed and verified the underlying data reported in the manuscript. JMM, AG and PRG generated, analyzed and integrated microbiome and metabolome data. MGG oversaw the microbiome analysis. MGG, LS and HLPT interpreted microbiome, metabolome and clinical integration results. All authors had full access to all the data in the study and had final responsibility for the decision to submit for publication.

## Supporting information

Supplemental Figure S1. EYCF impacts muscle development in vitro.

Supplemental Figure S2. Alpha and beta diversity of subjects grouped by intervention at baseline.

Supplemental table S1. Complete formulation of the control milk (CM) and the experimental blend (EYCF).

Supplemental table S2. Relative species abundance for each sample.

Supplemental table S3. Demographics and baseline characteristics of subjects.

Supplemental table S4. Descriptive statistics of subjects at all visits.

Supplemental table S5. Clinical outcomes comparisons between the experimental and control blends versus the habitual diet arm.

Supplemental table S6. Comparisons of the blood vitamin D level between Experimental group and Habitual diet group.

Supplemental table S7. Correlation of the vitamin B3/B6 ratio and mineral excretion (adjusted to intake) and the clinical outcome.

Supplemental table S8. PERMANOVA analysis on microbiome beta diversity.

Supplemental table S9. Comparison of microbiome taxonomic composition between EYCF and CM.

Supplemental table S10. Comparison of metabolome intensities between EYCF and CM.

Supplemental table S11. Procrustes analysis of microbiome-metabolome associations.

Supplemental table S12. Associations between microbiome taxonomic composition and clinical outcomes.

Supplemental table S13. Associations between metabolome intensities and clinical outcomes.

Supplemental table S15. Stool consistency and Descriptive statistics for the TCGQ questionnaire.

Supplemental table S16. Comparison of stool consistency and Gastrointestinal total score.

## Data Availability

All clinical outcomes, microbiome and metabolome data necessary to interpret the results are included in supplementary data files or are available upon request in a deidentified and anonymized format. No expiration date of data request is currently set once data are made available.

## Acknowledgements

Authors would like to thank Nestlé Research colleagues: Olivier Ciclet and Adrien Frezal for technical expertise on B vitamins analyses, Dustin Larue and Francesca Giuffrida for fatty acid soap analysis and interpretation, Héloise Denurra for metabolomics results interpretation, Ute Haeberlein Schwan for experimental product development, Flavien Bermont and Aurelie Hermant for technical support on bone biomarkers analyses, Michael Baruchet for sample preparation, and Eugenia Migliavacca for her support on statistical analysis. We would like to thank Eline van der Beek, Irma Silva-Zolezzi and Laurence Biehl for critical review of the manuscript. We thank Helle Pedersen and Thorsten Gravert (Clinical Microbiomics) for their expert insights on metabolomics and statistical models associated, and Claire Boulangé for supporting metabolomics interpretation.

## References

1. Ma NS, Gordon CM. Pediatric osteoporosis: where are we now?. J Pediatr Endocrinol Metab. 2012;161:983–90.

2. Shea JE, Miller SC. Skeletal function and structure: implications for tissue-targeted therapeutics. Adv Drug Deliv Rev 2005;57(7):945–57.

3. Dibba B, Prentice A, Ceesay M, Stirling DM, Cole TJ, Poskitt EM. EFFECT OF CALCIUM SUPPLEMENTATION ON BONE MINERAL ACCRETION IN GAMBIAN CHILDREN ACCUSTOMED TO A LOW-CALCIUM DIE. Am J Clin Nutr 2000;71(2):544–9.

4. Uusi-Rasi K, Kärkkäinen MU, Lamberg-Allardt CJ. Calcium intake in health maintenance - a systematic review. Food Nutr Res 2013;57:doi: 10.3402.

5. Lloyd T, Andon MB, Rollings N, Martel JK, Landis JR, Demers LM, et al. Calcium supplementation and bone mineral density in adolescent girls. JAMA. 1993;270:841–4.

6. Mitchell DM, Jüppner H. Regulation of calcium homeostasis and bone metabolism in the fetus and neonate. Curr Opin Endocrinol Diabetes Obes. 2010;17:25–30.

7. Lee WT, Leung SS, Leung DM, Wang SH, Xu YC, Zeng WP, Cheng JC. Bone mineral acquisition in low calcium intake children following the withdrawal of calcium supplement. Acta Paediatr 1997;86:570–6.

8. Rizzoli R, Bianchi ML, Garabédian M, McKay HA, Moreno LA. Maximizing bone mineral mass gain during growth for the prevention of fractures in the adolescents and the elderly. Bone. 2010;46(2):294–305.

9. He J, Xu S, Zhang B, Xiao C, Chen Z, Si F, et al. Gut microbiota and metabolite alterations associated with reduced bone mineral density or bone metabolic indexes in postmenopausal osteoporosis. Aging (Albany NY). 2020;12(9):8583–604.

10. Lu L, Chen X, Liu Y, Yu X. Gut microbiota and bone metabolism. FASEB J. 2021;35(7):e21740.

11. Wang Y, Li Y, Bo L, Zhou E, Chen Y, Naranmandakh S, et al. Progress of linking gut microbiota and musculoskeletal health: casualty, mechanisms, and translational values. Gut Microbes 2023;15(2):2263207.

12. Chonan O, Matsumoto K, Watanuki M. Effect of galactooligosaccharides on calcium absorption and preventing bone loss in ovariectomized rats. Biosci Biotechnol Biochem 1995;59:236–9.

13. Adam CL, Williams PA, Garden KE, Thomson LM, Ross AW. Dose-dependent effects of a soluble dietary fibre (pectin) on food intake, adiposity, gut hypertrophy and gut satiety hormone secretion in rats. PLoS One 2015;10(1):e0115438.

14. Singh A, Zapata RC, Pezeshki A, Reidelberger RD, Chelikani PK. Inulin fiber dose-dependently modulates energy balance, glucose tolerance, gut microbiota, hormones and diet preference in high-fat-fed male rats. J Nutr Biochem 2018;59:142–52.

15. Weaver CM, Martin BR, Nakatsu CH, Armstrong AP, Clavijo A, McCabe LD, et al. Galactooligosaccharides improve mineral absorption and bone properties in growing rats through gut fermentation. Journal of agricultural and food chemistry. 2011;59(12):6501–10.

16. Lobo AR, Filho JM, Alvares EP, Cocato ML, Colli C. Effects of dietary lipid composition and inulin-type fructans on mineral bioavailability in growing rats. Nutrition. 2009;25:216–25.

17. Takahara S, Morohashi T, Sano T, Ohta A, Yamada S, Sasa R. Fructooligosaccharide consumption enhances femoral bone volume and mineral concentrations in rats. J Nutr 2000;130:1792–5.

18. Ribeiro JL, Santos TA, Garcia MT, Carvalho BFDC, Esteves JECS, Moraes RM, Anbinder AL. Heat-killed Limosilactobacillus reuteri ATCC PTA 6475 prevents bone loss in ovariectomized mice: A preliminary study. PLoS One 2024 19(5):e0304358.

19. Britton RA, Irwin R, Quach D, Schaefer L, Zhang J, Lee T, et al. Probiotic L. reuteri Treatment Prevents Bone Loss in a Menopausal Ovariectomized Mouse Model. Journal of Cellular Physiology. 2014;229(11):1822–30.

20. De Bruyn F, Bonnet N, Baruchet M, Sabatier M, Breton I, Bourqui B, et al. Galacto-oligosaccharide preconditioning improves metabolic activity and engraftment of Limosilactobacillus reuteri and stimulates osteoblastogenesis ex vivo. Sci Rep. 2024;14(4329):doi: 10.1038/s41598-024-54887-z.

21. Lucas S, Omata Y, Hofmann J, Bottcher M, Iljazovic A, Sarter K, et al. Short-chain fatty acids regulate systemic bone mass and protect from pathological bone loss. Nature communications. 2018;9(1):55.

22. Scholz-Ahrens KE, Adolphi B, Rochat F, Barclay DV, de Vrese M, Açil Y, Schrezenmeir J. Effects of probiotics, prebiotics, and synbiotics on mineral metabolism in ovariectomized rats - impact of bacterial mass, intestinal absorptive area and reduction of bone turn-over. NFS Journal. 2016;3:41–50.

23. van den Heuvel E, Schoterman M, Muijs T. Trans-galactooligosaccharides stimulate calcium absorption in postmenopausal women. J Nutr 2000;130:2938–42.

24. Leite ME, Lasekan J, Baggs G, Ribeiro T, Menezes-Filho J, Pontes M, et al. Calcium and fat metabolic balance, and gastrointestinal tolerance in term infants fed milk-based formulas with and without palm olein and palm kernel oils: a randomized blinded crossover study. BMC Pediatr. 2013;13(215):doi: 10.1186/471-2431-13-215.

25. Nelson SE, Frantz JA, Ziegler EE. Absorption of fat and calcium by infants fed a milk-based formula containing palm olein. J Am Coll Nutr. 1998;17(4):327–32.

26. Nelson SE, Rogers RR, Frantz JA, Ziegler EE. Palm olein in infant formula: absorption of fat and minerals by normal infants. Am J Clin Nutr 1996;64(3):291–6.

27. Ostrom KM, Borschel MW, Westcott JE, Richardson KS, Krebs NF. Lower calcium absorption in infants fed casein hydrolysate- and soy protein-based infant formulas containing palm olein versus formulas without palm olein. J Am Coll Nutr. 2002;21(6):564–9.

28. Nilsson A, Sundh D, Bäckhed F, Lorentzon M. Lactobacillus reuteri reduces bone loss in older women with low bone mineral density: a randomized, placebo-controlled, double-blind, clinical trial. Journal of internal medicine. 2018;284(3):307–17.

29. Fiorotto ML, Davis TA. Critical Windows for the Programming Effects of Early-Life Nutrition on Skeletal Muscle Mass. Nestle Nutr Inst Workshop Ser. 2018;89:25–35.

30. Orsso CE, Tibaes JRB, Oliveira CLP, Rubin DA, Field CJ, Heymsfield SB, et al. Low muscle mass and strength in pediatrics patients: Why should we care?. Clin Nutr 2019;38(5):2002–15.

31. Fiorotto M. L, Davis T. A. Critical Windows for the Programming Effects of Early-Life Nutrition on Skeletal Muscle Mass. Nestle Nutr Inst Workshop. 2018;89:25–35.

32. Bachman JF, J.V C. Insights into muscle stem cell dynamics during postnatal development. FEBS J. 2022;289(10):2710–22.

33. Verdijk LB, Snijders T, Drost M, Delhaas T, Kadi F, van Loon LJ. Satellite cells in human skeletal muscle; from birth to old age. Age. 2014;36(2):545–7.

34. Davis TA, Fiorotto ML,. Regulation of muscle growth in neonates. Current opinion in clinical nutrition and metabolic care. 2009;12(1):78–85.

35. Giron M, Thomas M, Dardevet D, Chassard C, Savary-Auzeloux I. Gut microbes and muscle function: can probiotics make our muscles stronger? J Cachexia Sarcopenia Muscle. 2022;13:1460–76.

36. Chen HL, Lee WT, Lee PL, Liu PL, Yang RC. Postnatal Changes in Tibial Bone Speed of Sound of Preterm and Term Infants during Infancy. PLoS One. 2016;11(11):e0166434.

37. Soto Martinez ME, Love JC, Crowder CM, Wiersema JM, Pinto DC, Derrick SM, et al. The first step in an investigation of quantitative ultrasound as a technique for evaluating infant bone strength. J Forensic Sci. 2021;66(2):456–69.

38. Wu Z, Yuan Y, Tian J, Long F, Luo W. The associations between serum trace elements and bone mineral density in children under 3 years of age. Sci Rep. 2021;11(1):1890.

39. Zadik Z, Price D, Diamond G. Pediatric reference curves for multi-site quantitative ultrasound and its modulators. Osteoporos Int 2003;14(10):857–62.

40. Häger-Ross C, Rösblad B. Norms for grip strength in children aged 4-16 years. Acta Paediatr 2002;91(6):617–25.

41. Roberts HC, Denison HJ, Martin HJ, Patel HP, Syddall H, Cooper C, Sayer AA. A review of the measurement of grip strength in clinical and epidemiological studies: towards a standardised approach. Age Ageing 2011;40(4):423–9.

42. Greenblatt MB, Tsai JN, Wein MN. Bone Turnover Markers in the Diagnosis and Monitoring of Metabolic Bone Disease. Clin Chem. 2017;63(2):464–74.

43. Quinlan PT, Lockton S, Irwin J, Lucas AL. The relationship between stool hardness and stool composition in breast- and formula-fed infants. J Pediatr Gastroenterol Nutr. 1995;20(1):81–90.

44. Capeding MRZ, Phee LCM, Ming C, Noti M, Vidal K, Le Carrou G, et al. Safety, efficacy, and impact on gut microbial ecology of a Bifidobacterium longum subspecies infantis LMG11588 supplementation in healthy term infants: a randomized, double-blind, controlled trial in the Philippines. Front Nutr 2023;10:1319873.

45. Nielsen HB, Almeida M, Juncker AS, Rasmussen S, Li J, Sunagawa S, et al. Identification and assembly of genomes and genetic elements in complex metagenomic samples without using reference genomes. Nat Biotechnol 2014;32(8):822–8.

46. Langfelder P, Zhang B, Horvath S. Defining clusters from a hierarchical cluster tree: the Dynamic Tree Cut package for R. Bioinformatics. Bioinformatics. 2008;24(5):719–20.

47. Aitchison J. The Statistical Analysis of Compositional Data. Journal of the Royal Statistical Society. 1982;44(2):139–77.

48. Zhou H, He K, Chen J, Zhang X. LinDA: linear models for differential abundance analysis of microbiome compositional data. Genome Biol. 2022;23(1):95. doi: 10.1186.

49. Benjamini Y, Hochberg Y. Controlling the False Discovery Rate: A Practical and Powerful Approach to Multiple Testing Get access Arrow. Journal of the Royal Statistical Society:. 1995;57(1):289–300.

50. Ancel S, Michaud J, Migliavacca E, Jomard C, Fessard A, Garcia P, et al. The combination of nicotinamide and pyridoxine stimulates muscle stem cell expansion and enhances regenerative capacity. Journal clinical investigation. 2024;Under review.

51. Lui JC. Gut microbiota in regulation of childhood bone growth. Exp Physiol. 2024;109(5):662–71.

52. Bohannon RW, Wang YC, Bubela D, Gershon RC. Handgrip Strength: A Population-Based Study of Norms and Age Trajectories for 3-to 17-Year-Olds. Pediatr Phys Ther 2017;29(2):118–23.

53. Flores-Aldana M, Rivera-Pasquel M, García-Guerra A, Pérez-Cortés JG, Bárcena-Echegollén JE. Effect of Vitamin D Supplementation on (25(OH)D) Status in Children 12-30 Months of Age: A Randomized Clinical Trial. Nutrients. 2023;15(12):2756. doi: 10.3390.

54. Jones ML, Martoni CJ, Prakash S. Oral supplementation with probiotic L. reuteri NCIMB 30242 increases mean circulating 25-hydroxyvitamin D: a post hoc analysis of a randomized controlled trial. J Clin Endocrinol Metab. 2013;98(7):2944–51.

55. Havlicekova Z, Jesenak M, Banovcin P, Kuchta M. Beta-palmitate - a natural component of human milk in supplemental milk formulas. Nutr J 2016;15(28):doi: 10.1186.

56. Depeint F, Tzortzis G, Vulevic J, I’anson K, Gibson GR. Prebiotic evaluation of a novel galactooligosaccharide mixture produced by the enzymatic activity of Bifidobacterium bifidum NCIMB 41171, in healthy humans: a randomized, double-blind, crossover, placebo-controlled intervention study. Am J Clin Nutr 2008;87:785–91.

57. Knysh O. V. Bifidogenic properties of cell-free extracts derived from probiotic strains of Bifidobacterium bifidum and Lactobacillus reuteri. Regulatory Mechanisms in Biosystems. 2019;10:124–8.

58. Yu J, Cao G, Yuan S, Luo C, Yu J, Cai M. Probiotic supplements and bone health in postmenopausal women: a meta-analysis of randomised controlled trials. BMJ Open. 2021;11(3):e041393.

59. Prokopidis K, Giannos P, Reginster JY, Bruyere O, Petrovic M, Cherubini A, et al. Sarcopenia is associated with a greater risk of polypharmacy and number of medications: a systematic review and meta-analysis. J Cachexia Sarcopenia Muscle. 2023;14(2):671–83.

60. Du L, Qi R, Wang J, Liu Z, Wu Z. Indole-3-Propionic Acid, a Functional Metabolite of Clostridium sporogenes, Promotes Muscle Tissue Development and Reduces Muscle Cell Inflammation. Int J Mol Sci 2021;22(22):12435.

61. Suzuki A, Iwata J. Amino acid metabolism and autophagy in skeletal development and homeostasis. Bone. 2021;146: doi: 10.1016/j.bone.2021.

62. Michalowska M, Znorko B, Kaminski T, Oksztulska-Kolanek E, Pawlak D. New insights into tryptophan and its metabolites in the regulation of bone metabolism. J Physiol Pharmacol. 2015;66(6):779–91.

63. Al Saedi A, Sharma S, Summers MA, Nurgali K, Duque G. The multiple faces of tryptophan in bone biology. Exp Gerontol. 2020;129:110778. doi: 10.1016.

64. Jiang Q, Zhao Y, Zhou XQ, Wu XY, Xu SX, Feng L, et al. Effects of dietary tryptophan on muscle growth, protein synthesis and antioxidant capacity in hybrid catfish Pelteobagrus vachellilZl × Leiocassis longirostrislZl. Br J Nutr 2022;127(12):1761–73.

65. Jourdan M, Nair KS, Carter RE, Schimke J, Ford GC, Marc J, et al. Citrulline stimulates muscle protein synthesis in the post-absorptive state in healthy people fed a low-protein diet - A pilot study. Clin Nutr. 2015;34(3):449–56.

66. Suzuki T, Morita M, Kobayashi Y, Kamimura A. Oral L-citrulline supplementation enhances cycling time trial performance in healthy trained men: Double-blind randomized placebo-controlled 2-way crossover study. J Int Soc Sports Nutr 2016;13(6):doi: 10.1186.

67. Nauta S, Greven J, Hofman M, Mohren R, Meesters DM, Möckel D, et al. Mass Spectrometry Reveals Molecular Effects of Citrulline Supplementation during Bone Fracture Healing in a Rat Model. J Am Soc Mass Spectrom. 2024;35(6):1184–96.

68. Dao HT, Moss AF, Bradbury EJ, Swick RA. Effects of L-arginine, guanidinoacetic acid and L-citrulline supplementation in reduced-protein diets on bone morphology and mineralization of laying hens. Anim Nutr. 2023;14(16):225–34.

69. Zdzieblik D, Oesser S, Baumstark MW, Gollhofer A, König D. Collagen peptide supplementation in combination with resistance training improves body composition and increases muscle strength in elderly sarcopenic men: a randomised controlled trial. Br J Nutr 2015;114(8):1237–45.

