## Supplementary figures and images for "A YOUNG CHILD FORMULA SUPPLEMENTED WITH L. REUTERI AND GALACTO-OLIGOSACCHARIDES MODULATES THE COMPOSITION AND FUNCTION OF THE GUT MICROBIOME SUPPORTING BONE AND MUSCLE DEVELOPMENT IN TODDLERS"

### Supplemental Figure S1. EYCF impacts muscle development in vitro.

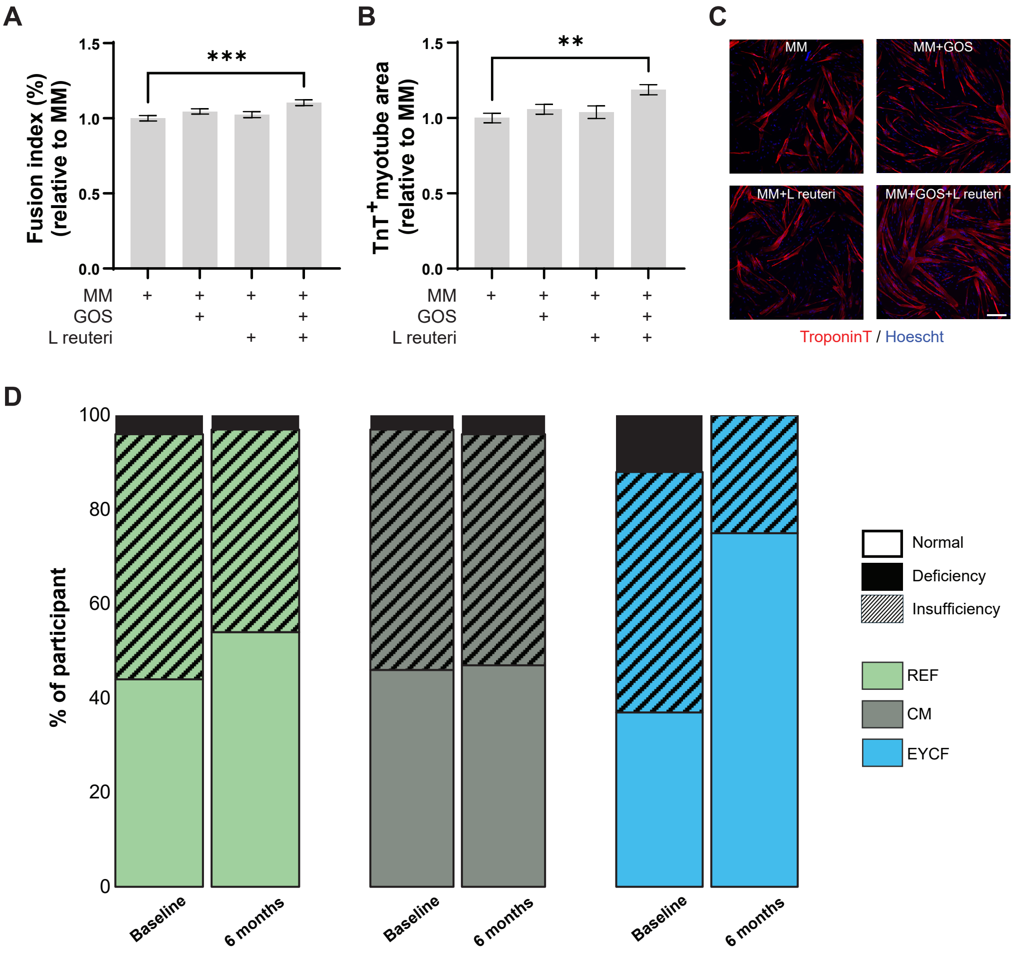

### Supplemental Figure S2. Alpha and beta diversity of subjects grouped by intervention at baseline.

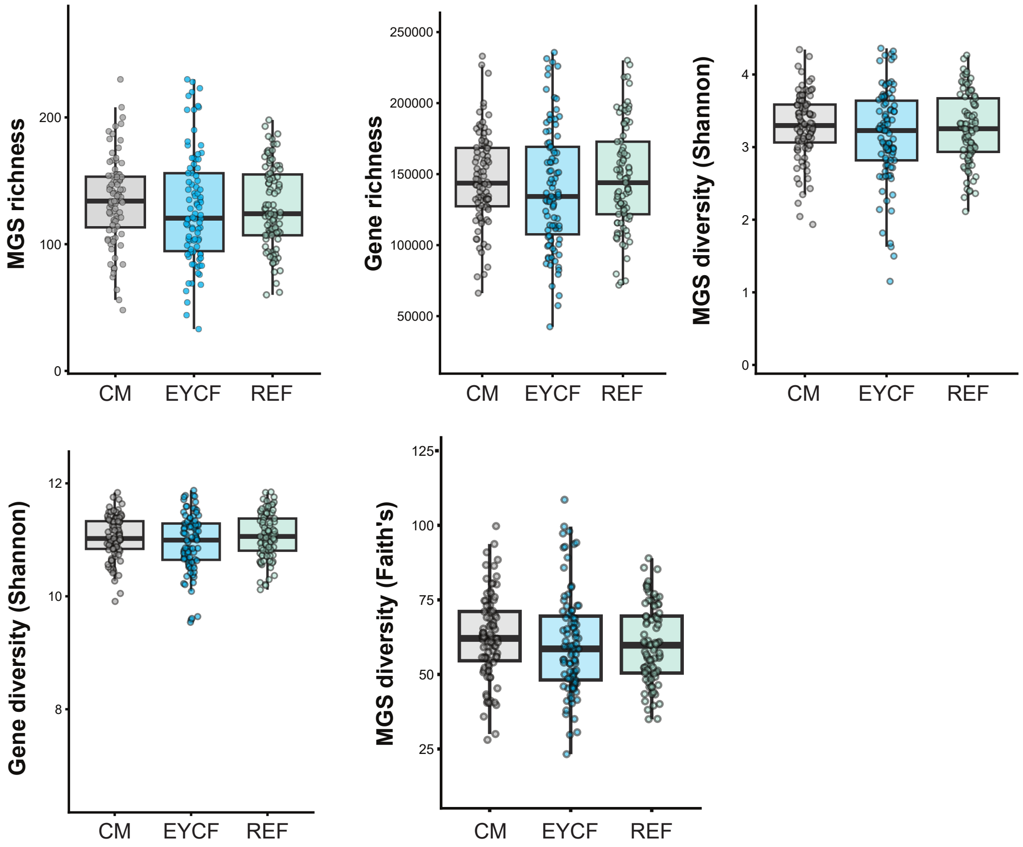

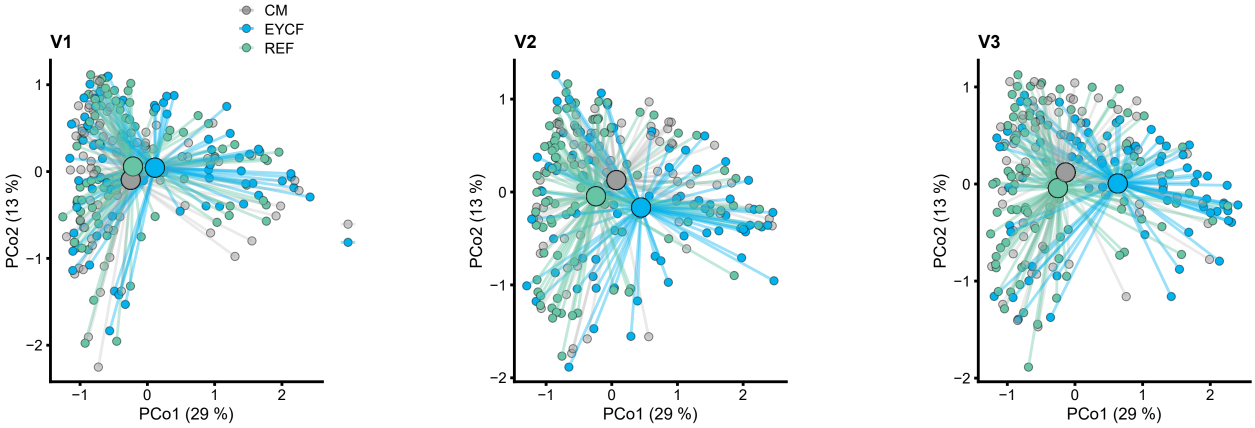
