## Supplemental table S1. Complete formulation of the control milk (CM) and the experimental blend (EYCF). for "A YOUNG CHILD FORMULA SUPPLEMENTED WITH L. REUTERI AND GALACTO-OLIGOSACCHARIDES MODULATES THE COMPOSITION AND FUNCTION OF THE GUT MICROBIOME SUPPORTING BONE AND MUSCLE DEVELOPMENT IN TODDLERS"

| Parameter | UoM | Oper. | /100g |
|---|---|---|---|
| ENERGY (KCAL) 449 | kcal | = | 480,2 |
| Protein | g | = | 10,6 |
| Fat | g | = | 22,2 |
| Available Carbohydrates | g | = | 59,5 |
| Sum of Fibers (incl. GOS) | g | = | 2,92 |
| Na (Sodium) | mg | = | 240 |
| K (Potassium) | mg | = | 715 |
| Cl (Chloride) | mg | = | 360 |
| Ca (Calcium) | mg | = | 865 |
| P (Phosphorus) | mg | = | 473 |
| Mg (Magnesium) | mg | = | 68 |
| Fe (Iron) | mg | = | 7 |
| Cu (Copper) | mg | = | 0,4 |
| Zn (Zinc) | mg | = | 4,4 |
| I (Iodine) | µg | = | 112 |
| Mn (Manganese) | µg | = | 80 |
| Se (Selenium) | µg | = | 15 |
| Vit A | µgRE | = | 620 |
| Total Vitamin D [Sum of Vita | µgD | = | 10 |
| Vit E | mgTE | = | 10 |
| Vit K1 (Phytomenadione) | µg | = | 40 |
| Vit C | mg | = | 110 |
| Vit B1 | mg | = | 0,6 |
| Vit B2 | mg | = | 1,2 |
| Niacin | mg | = | 5,8 |
| Vit B6 | mg | = | 0,48 |
| Total Folic Acid | µg | = | 126 |
| Pantothenic Acid | mg | = | 3,5 |
| Vit B12 | µg | = | 1,2 |
| Biotin | µg | = | 18 |
| Choline | mg | = | 64 |
| Taurine | mg | = | 1,461 |
| L.reuteri | CFU | = | |

# CM

| Parameter | UoM | Oper. | /100g |
|---|---|---|---|
| ENERGY (KCAL) 449 | kcal | = | 478 |
| Protein | g | = | 15,75 |
| Fat | g | = | 21 |
| Available Carbohydrates | g | = | 56,5 |
| Sum of Fibers | g | = | 0 |
| Na (Sodium) | mg | = | 310 |
| K (Potassium) | mg | = | 860 |
| Cl (Chloride) | mg | = | 480 |
| Ca (Calcium) | mg | = | 720 |
| P (Phosphorus) | mg | = | 542 |
| Mg (Magnesium) | mg | = | 58 |
| Fe (Iron) | mg | = | 7,5 |
| Cu (Copper) | mg | = | 0,5 |
| Zn (Zinc) | mg | = | 5,1 |
| I (Iodine) | µg | = | 112 |
| Mn (Manganese) | µg | = | 66 |
| Se (Selenium) | µg | = | 6,77 |
| Vit A | µgRE | = | 612 |
| Total Vitamin D [Sum of Vitamin D2+D3] | IUD | = | 122 |
| Vit E | mgTE | = | 7,37 |
| Vit K1 | µg | = | 22 |
| Vit C | mg | = | 80 |
| Vit B1 | mg | = | 1 |
| Vit B2 | mg | = | 1,1 |
| Niacin | mg | = | 1,3 |
| Vit B6 | mg | = | 1,5 |
| Total Folic Acid | µg | = | 30 |
| Pantothenic Acid | mg | = | 2 |
| Vit B12 | µg | = | 3,2 |
| Biotin | µg | = | 9,4 |
