## Supplemental table S2. Relative species abundance for each sample. for "A YOUNG CHILD FORMULA SUPPLEMENTED WITH L. REUTERI AND GALACTO-OLIGOSACCHARIDES MODULATES THE COMPOSITION AND FUNCTION OF THE GUT MICROBIOME SUPPORTING BONE AND MUSCLE DEVELOPMENT IN TODDLERS"

| MGS Group | Anotation | Level |
|---|---|---|
| Hgn3C.0001 | Phocaeicola vulgatus | species |
| Hgn3C.0010 | Anaerostipes hadrus | species |
| Hgn3C.0100 | Bifidobacterium bifidum | species |
| Hgn3C.1000 | Bacteroidales sp. | order |
| Hgn3C.1001 | Archaea sp. | superkingdom |
| Hgn3C.1002 | Eggerthellales sp. | order |
| Hgn3C.1003 | Veillonellaceae sp. | family |
| Hgn3C.1004 | Bacteroidales sp. | order |
| Hgn3C.1005 | Barnesiella sp. An22 | species |
| Hgn3C.1006 | Eubacteriales sp. | order |
| Hgn3C.1007 | Firmicutes sp. | phylum |
| Hgn3C.1008 | Oscillospiraceae sp. | family |
| Hgn3C.1009 | Prevotellaceae sp. | family |
| Hgn3C.0101 | Bifidobacterium pseudocatenulatum | species |
| Hgn3C.1010 | Parabacteroides massiliensis | species |
| Hgn3C.1011 | Eubacteriales sp. | order |
| Hgn3C.1012 | Erysipelotrichaceae sp. | family |
| Hgn3C.1013 | Oscillospiraceae sp. | family |
| Hgn3C.1014 | Burkholderiales sp. | order |
| Hgn3C.1015 | Bacteria sp. | superkingdom |
| Hgn3C.1016 | Eubacteriales sp. | order |
| Hgn3C.1017 | Emergencia timonensis | species |
| Hgn3C.1018 | Bacteroidia sp. | class |
| Hgn3C.1019 | Prevotellaceae sp. | family |
| Hgn3C.0102 | Eubacteriales sp. | order |
| Hgn3C.1020 | Phocaeicola barnesiae | species |
| Hgn3C.1021 | Bacteroides stercorirosoris | species |
| Hgn3C.1022 | Eubacteriales sp. | order |
| Hgn3C.1023 | Firmicutes sp. | phylum |
| Hgn3C.1024 | Prevotellaceae sp. | family |
| Hgn3C.1025 | Eubacteriales sp. | order |
| Hgn3C.1026 | Bacteroidales sp. | order |
| Hgn3C.1027 | Lachnospiraceae sp. | family |
| Hgn3C.1028 | Bacteroidales sp. | order |
| Hgn3C.1029 | Eubacteriales sp. | order |
| Hgn3C.0103 | Holdemanella porci | species |
| Hgn3C.1030 | Lachnospiraceae sp. | family |
| Hgn3C.1031 | Eubacteriales sp. | order |
| Hgn3C.1032 | Bacteroidales sp. | order |
| Hgn3C.1033 | Hafnia paralvei | species |
| Hgn3C.1034 | Odoribacter sp. AF15-53 | species |
| Hgn3C.1035 | Klebsiella sp. | genus |
| Hgn3C.1036 | Escherichia coli | species |
| Hgn3C.1037 | Rothia sp. | genus |
| Hgn3C.1038 | Veillonella sp. | genus |
| Hgn3C.1039 | Prevotellaceae sp. | family |

| | | |
|---|---|---|
| Hgn3C.0104 | Clostridium sp. AM32-2 | species |
| Hgn3C.1040 | Lachnospiraceae sp. | family |
| Hgn3C.1041 | Bacteroides stercoris | species |
| Hgn3C.1042 | Dorea formicigenerans | species |
| Hgn3C.1043 | Clostridium phoceensis | species |
| Hgn3C.1044 | Eubacteriales sp. | order |
| Hgn3C.1045 | Lactococcus lactis subsp. lactis | subspecies |
| Hgn3C.1046 | Bacteroidales sp. | order |
| Hgn3C.1047 | Eubacteriales sp. | order |
| Hgn3C.1048 | Akkermansia sp. BIOML-A59 | species |
| Hgn3C.1049 | Enterococcus faecium | species |
| Hgn3C.0105 | Vescimonas coprocola | species |
| Hgn3C.1050 | Staphylococcus epidermidis | species |
| Hgn3C.1051 | Holdemania massiliensis | species |
| Hgn3C.1052 | Enterococcus gallinarum | species |
| Hgn3C.1053 | Eubacteriales sp. | order |
| Hgn3C.1054 | Streptococcus lutetiensis | species |
| Hgn3C.1055 | Veillonella seminalis | species |
| Hgn3C.1056 | Muribaculum sp. NM65_B17 | species |
| Hgn3C.1057 | Eubacteriales sp. | order |
| Hgn3C.1058 | Mediterraneibacter glycyrrhizinilyticus | species |
| Hgn3C.1059 | Faecalicatena sp. | genus |
| Hgn3C.0106 | Faecalibacterium sp. CLA-AA-H233 | species |
| Hgn3C.1060 | Eubacteriales sp. | order |
| Hgn3C.1061 | Acidaminococcales sp. | order |
| Hgn3C.1062 | Eubacteriales sp. | order |
| Hgn3C.1063 | Enterococcus avium | species |
| Hgn3C.1064 | Eubacteriales sp. | order |
| Hgn3C.1065 | Klebsiella variicola subsp. variicola | subspecies |
| Hgn3C.1066 | Lacrimispora saccharolytica | species |
| Hgn3C.1067 | Lachnospiraceae sp. | family |
| Hgn3C.1068 | Eubacteriales sp. | order |
| Hgn3C.1069 | Bacteroidales sp. | order |
| Hgn3C.0107 | Parasutterella excrementihominis | species |
| Hgn3C.1070 | Raoultella planticola | species |
| Hgn3C.1071 | Lactobacillus gasseri | species |
| Hgn3C.1072 | Eubacteriales sp. | order |
| Hgn3C.1073 | Streptococcus gallolyticus subsp. gallolyticus | subspecies |
| Hgn3C.1074 | Eubacteriales sp. | order |
| Hgn3C.1075 | Brachyspira aalborgi | species |
| Hgn3C.1076 | Clostridia sp. | class |
| Hgn3C.1077 | Eubacteriales sp. | order |
| Hgn3C.1078 | Eubacteriales sp. | order |
| Hgn3C.1079 | Eubacteriales sp. | order |
| Hgn3C.0108 | Eubacteriales sp. | order |
| Hgn3C.1080 | Bacteroidaceae sp. | family |
| Hgn3C.1081 | Lacticaseibacillus rhamnosus | species |
| Hgn3C.1082 | Lacticaseibacillus paracasei subsp. paracasei | subspecies |

| | | |
|---|---|---|
| Hgn3C.1083 | Firmicutes sp. | phylum |
| Hgn3C.1084 | Massilimicrobiota timonensis | species |
| Hgn3C.1085 | Eubacteriales sp. | order |
| Hgn3C.1086 | Bacteroidales sp. | order |
| Hgn3C.1087 | Ligilactobacillus salivarius | species |
| Hgn3C.1088 | Enterococcus casseliflavus | species |
| Hgn3C.1089 | Eubacteriales sp. | order |
| Hgn3C.0109 | Bacteroides eggerthii | species |
| Hgn3C.1090 | Turicibacter sanguinis | species |
| Hgn3C.1091 | Eubacteriales sp. | order |
| Hgn3C.1092 | Eubacteriales sp. | order |
| Hgn3C.1093 | Enterobacter kobei | species |
| Hgn3C.1094 | Eubacteriales sp. | order |
| Hgn3C.1095 | Clostridia sp. | class |
| Hgn3C.1096 | Clostridia sp. | class |
| Hgn3C.1097 | Alistipes provencensis | species |
| Hgn3C.1098 | Bacteroidales sp. | order |
| Hgn3C.1099 | Eubacteriales sp. | order |
| Hgn3C.0011 | Roseburia faecis | species |
| Hgn3C.0110 | Eubacteriales sp. | order |
| Hgn3C.1100 | Bacteria sp. | superkingdom |
| Hgn3C.1101 | Firmicutes sp. | phylum |
| Hgn3C.1102 | Eubacteriales sp. | order |
| Hgn3C.1103 | Eubacteriales sp. | order |
| Hgn3C.1104 | Bacteroidales sp. | order |
| Hgn3C.1105 | Barnesiellaceae sp. | family |
| Hgn3C.1106 | Clostridium butyricum | species |
| Hgn3C.1107 | Eubacteriales sp. | order |
| Hgn3C.1108 | Eubacteriales sp. | order |
| Hgn3C.1109 | Eubacteriales sp. | order |
| Hgn3C.0111 | Eubacteriales sp. | order |
| Hgn3C.1110 | Porphyromonas sp. | genus |
| Hgn3C.1111 | Eubacteriales sp. | order |
| Hgn3C.1112 | Eubacteriales sp. | order |
| Hgn3C.1113 | Firmicutes sp. | phylum |
| Hgn3C.1114 | Bacteria sp. | superkingdom |
| Hgn3C.1115 | Bacteria sp. | superkingdom |
| Hgn3C.1116 | Bacteroidales sp. | order |
| Hgn3C.1117 | Atopobiaceae sp. | family |
| Hgn3C.1118 | Rhodospirillales sp. | order |
| Hgn3C.1119 | Bacteroidales sp. | order |
| Hgn3C.0112 | Clostridium sp. | genus |
| Hgn3C.1120 | Actinomycetaceae sp. | family |
| Hgn3C.1121 | Roseburia sp. | genus |
| Hgn3C.1122 | Ruminococcus sp. | genus |
| Hgn3C.1123 | Eubacteriales sp. | order |
| Hgn3C.1124 | Clostridium porci | species |
| Hgn3C.1125 | Bacteroidales sp. | order |

| | | |
|---|---|---|
| Hgn3C.1126 | Phocaeicola sartorii | species |
| Hgn3C.1127 | Eubacteriales sp. | order |
| Hgn3C.1128 | Actinobacteria sp. | phylum |
| Hgn3C.1129 | Eubacteriales sp. | order |
| Hgn3C.0113 | Clostridium sp. MCC345 | species |
| Hgn3C.1130 | Bacteroidales sp. | order |
| Hgn3C.1131 | Lachnospiraceae sp. | family |
| Hgn3C.1132 | Bacteroidales sp. | order |
| Hgn3C.1133 | Lachnospiraceae sp. | family |
| Hgn3C.1134 | Eubacteriales sp. | order |
| Hgn3C.1135 | Eubacteriales sp. | order |
| Hgn3C.1136 | Bacteria sp. | superkingdom |
| Hgn3C.1137 | Eubacteriales sp. | order |
| Hgn3C.1138 | Eubacteriales sp. | order |
| Hgn3C.1139 | Firmicutes sp. | phylum |
| Hgn3C.0114 | Eubacteriales sp. | order |
| Hgn3C.1140 | Eubacteriales sp. | order |
| Hgn3C.1141 | Archaea sp. | superkingdom |
| Hgn3C.1142 | Prevotella sp. | genus |
| Hgn3C.1143 | Gemmiger sp. | genus |
| Hgn3C.1144 | Culturomica massiliensis | species |
| Hgn3C.1145 | Lachnospiraceae sp. | family |
| Hgn3C.1146 | Bacteria sp. | superkingdom |
| Hgn3C.1147 | Methanomassiliicoccales sp. | order |
| Hgn3C.1148 | Eggerthellales sp. | order |
| Hgn3C.1149 | Bacteria sp. | superkingdom |
| Hgn3C.0115 | Eubacteriales sp. | order |
| Hgn3C.1150 | Oscillospiraceae sp. | family |
| Hgn3C.1151 | Lachnospiraceae sp. | family |
| Hgn3C.1152 | Acidaminococcus fermentans | species |
| Hgn3C.1153 | Eubacterium sp. OM08-24 | species |
| Hgn3C.1154 | Lactococcus garvieae | species |
| Hgn3C.1155 | Firmicutes sp. | phylum |
| Hgn3C.1156 | Bacteria sp. | superkingdom |
| Hgn3C.1157 | Eubacteriales sp. | order |
| Hgn3C.1158 | Bacteroidales sp. | order |
| Hgn3C.1159 | Firmicutes sp. | phylum |
| Hgn3C.0116 | Anaerostipes hadrus | species |
| Hgn3C.1160 | Deltaproteobacteria sp. | class |
| Hgn3C.1161 | Eubacteriales sp. | order |
| Hgn3C.1162 | Bacteria sp. | superkingdom |
| Hgn3C.1163 | Bacteria sp. | superkingdom |
| Hgn3C.1164 | Eubacteriales sp. | order |
| Hgn3C.1165 | Desulfovibrio legallii | species |
| Hgn3C.1166 | Eubacteriales sp. | order |
| Hgn3C.1167 | Firmicutes sp. | phylum |
| Hgn3C.1168 | Bacteria sp. | superkingdom |
| Hgn3C.1169 | Oscillospiraceae sp. | family |

| | | |
|---|---|---|
| Hgn3C.0117 | Methanobrevibacter smithii | species |
| Hgn3C.1170 | Bacteria sp. | superkingdom |
| Hgn3C.1171 | Clostridia sp. | class |
| Hgn3C.1172 | Treponemataceae sp. | family |
| Hgn3C.1173 | Negativicutes sp. | class |
| Hgn3C.1174 | Sutterella parvirubra | species |
| Hgn3C.1175 | Firmicutes sp. | phylum |
| Hgn3C.1176 | Eubacteriales sp. | order |
| Hgn3C.1177 | Bacteroidaceae sp. | family |
| Hgn3C.1178 | Clostridium sp. 12(A) | species |
| Hgn3C.1179 | Lacrimispora celerecrescens | species |
| Hgn3C.0118 | Eubacteriales sp. | order |
| Hgn3C.1180 | Enterocloster citroniae | species |
| Hgn3C.1181 | Acidovorax delafieldii | species |
| Hgn3C.1182 | Comamonas kerstersii | species |
| Hgn3C.1183 | Acinetobacter bereziniae | species |
| Hgn3C.1184 | Acinetobacter johnsonii | species |
| Hgn3C.1185 | Acinetobacter ursingii | species |
| Hgn3C.1186 | Acinetobacter guillouiae | species |
| Hgn3C.1187 | Actinomyces sp. HMSC065F12 | species |
| Hgn3C.1188 | Actinomyces oris | species |
| Hgn3C.1189 | Aeromonas caviae | species |
| Hgn3C.0119 | Bacteroides faecis | species |
| Hgn3C.1190 | Fenollaria massiliensis | species |
| Hgn3C.1191 | Aliarcobacter cryaerophilus | species |
| Hgn3C.1192 | Methylobacterium sp. UNCCL125 | species |
| Hgn3C.1193 | Atopobium sp. | genus |
| Hgn3C.1194 | Bacteroides gallinaceum | species |
| Hgn3C.1195 | Bifidobacterium pullorum subsp. saeculare | subspecies |
| Hgn3C.1196 | Blautia marasmi | species |
| Hgn3C.1197 | Blautia hansenii | species |
| Hgn3C.1198 | Turicimonas muris | species |
| Hgn3C.1199 | Butyricimonas synergistica | species |
| Hgn3C.0012 | Lachnospira sp. | genus |
| Hgn3C.0120 | Flavonifractor plautii | species |
| Hgn3C.1200 | Megasphaera sp. DJF_B143 | species |
| Hgn3C.1201 | Caulobacter sp. CCH5-E12 | species |
| Hgn3C.1202 | Chryseobacterium gambrini | species |
| Hgn3C.1203 | Citrobacter sp. CRE-46 | species |
| Hgn3C.1204 | Citrobacter amalonaticus | species |
| Hgn3C.1205 | Citrobacter braakii | species |
| Hgn3C.1206 | Citrobacter pasteurii | species |
| Hgn3C.1207 | Citrobacter sp. | genus |
| Hgn3C.1208 | Citrobacter sp. S-77 | species |
| Hgn3C.1209 | Cloacibacillus evryensis | species |
| Hgn3C.0121 | Oscillospiraceae sp. | family |
| Hgn3C.1210 | Dielma sp. | genus |
| Hgn3C.1211 | Lachnoclostridium pacaense | species |

| | | |
|---|---|---|
| Hgn3C.1212 | Clostridium jeddahense | species |
| Hgn3C.1213 | Rubneribacter badeniensis | species |
| Hgn3C.1214 | Corynebacterium tuberculostearicum | species |
| Hgn3C.1215 | Cutibacterium acnes subsp. acnes | subspecies |
| Hgn3C.1216 | Cutibacterium avidum | species |
| Hgn3C.1217 | Cutibacterium granulosum | species |
| Hgn3C.1218 | Enterobacter bugandensis | species |
| Hgn3C.1219 | Enterobacter sp. NFIX58 | species |
| Hgn3C.0122 | Bacteroides finegoldii | species |
| Hgn3C.1220 | Enterobacter ludwigii | species |
| Hgn3C.1221 | Phytobacter diazotrophicus | species |
| Hgn3C.1222 | Enterococcus durans | species |
| Hgn3C.1223 | Enterococcus gilvus | species |
| Hgn3C.1224 | Enterococcus raffinosus | species |
| Hgn3C.1225 | Enterococcus faecium | species |
| Hgn3C.1226 | Enterococcus thailandicus | species |
| Hgn3C.1227 | Erysipelatoclostridium sp. An15 | species |
| Hgn3C.1228 | Escherichia marmotae | species |
| Hgn3C.1229 | Escherichia sp. 93.0750 | species |
| Hgn3C.0123 | Eubacteriales sp. | order |
| Hgn3C.1230 | Eubacterium limosum | species |
| Hgn3C.1231 | Flavobacterium sp. Leaf359 | species |
| Hgn3C.1232 | Sanguibacteroides justesenii | species |
| Hgn3C.1233 | Gardnerella vaginalis | species |
| Hgn3C.1234 | Granulicatella adiacens | species |
| Hgn3C.1235 | Haemophilus sp. CCUG 66565 | species |
| Hgn3C.1236 | Hafnia sp. | genus |
| Hgn3C.1237 | Kosakonia cowanii | species |
| Hgn3C.1238 | Klebsiella quasivariicola | species |
| Hgn3C.1239 | Lactobacillus amylovorus | species |
| Hgn3C.0124 | Ruthenibacterium lactatiformans | species |
| Hgn3C.1240 | Lactobacillus helveticus | species |
| Hgn3C.1241 | Limosilactobacillus reuteri subsp. reuteri | subspecies |
| Hgn3C.1242 | Limosilactobacillus vaginalis | species |
| Hgn3C.1243 | Lysinibacillus fusiformis | species |
| Hgn3C.1244 | Megasphaera genomosp. type_1 | species |
| Hgn3C.1245 | Megasphaera massiliensis | species |
| Hgn3C.1246 | Morganella morganii subsp. morganii | subspecies |
| Hgn3C.1247 | Mycobacteroides chelonae | species |
| Hgn3C.1248 | Neisseria sp. HMSC061E12 | species |
| Hgn3C.1249 | Pantoea sp. 3.5.1 | species |
| Hgn3C.0125 | Sutterella sp. KLE1602 | species |
| Hgn3C.1250 | Parabacteroides gordonii | species |
| Hgn3C.1251 | Prevotella jejuni | species |
| Hgn3C.1252 | Prevotella sp. | genus |
| Hgn3C.1253 | Pseudoflavonifractor sp. An176 | species |
| Hgn3C.1254 | Pseudomonas extremaustralis | species |
| Hgn3C.1255 | Pseudomonas fulva | species |

| | | |
|---|---|---|
| Hgn3C.1256 | Pseudomonas nitroreducens | species |
| Hgn3C.1257 | Robinsoniella peoriensis | species |
| Hgn3C.1258 | Rothia sp. HMSC072E10 | species |
| Hgn3C.1259 | Selenomonas artemidis | species |
| Hgn3C.0126 | Ruminococcus sp. BSD2780120874_150323_B10 | species |
| Hgn3C.1260 | Serratia marcescens | species |
| Hgn3C.1261 | Shewanella xiamenensis | species |
| Hgn3C.1262 | Sphingomonas sp. CCH9-F2 | species |
| Hgn3C.1263 | Staphylococcus borealis | species |
| Hgn3C.1264 | Staphylococcus hominis subsp. hominis | subspecies |
| Hgn3C.1265 | Staphylococcus lugdunensis | species |
| Hgn3C.1266 | Stenotrophomonas bentonitica | species |
| Hgn3C.1267 | Stenotrophomonas maltophilia | species |
| Hgn3C.1268 | Streptococcus sp. | genus |
| Hgn3C.1269 | Streptococcus constellatus subsp. constellatus | subspecies |
| Hgn3C.0127 | Faecalibacterium sp. | genus |
| Hgn3C.1270 | Streptococcus sp. | genus |
| Hgn3C.1271 | Streptococcus agalactiae | species |
| Hgn3C.1272 | Veillonella sp. CNR 79/14 | species |
| Hgn3C.1273 | Citrobacter koseri | species |
| Hgn3C.1274 | Kluyvera cryocrescens | species |
| Hgn3C.1275 | Lachnospiraceae sp. | family |
| Hgn3C.1276 | Citrobacter sp. MGH106 | species |
| Hgn3C.1277 | Lachnospiraceae sp. | family |
| Hgn3C.1278 | Clostridium sp. NSJ-6 | species |
| Hgn3C.1279 | Enterococcus malodoratus | species |
| Hgn3C.0128 | Eubacteriales sp. | order |
| Hgn3C.1280 | Clostridium sp. | genus |
| Hgn3C.1281 | Atlantibacter sp. RC6 | species |
| Hgn3C.1282 | Terrisporobacter othiniensis | species |
| Hgn3C.1283 | Clostridium baratii | species |
| Hgn3C.1284 | Enterobacteriaceae sp. | family |
| Hgn3C.1285 | Enterobacteriaceae sp. | family |
| Hgn3C.1286 | Corynebacterium variabile | species |
| Hgn3C.1287 | Macrococcus caseolyticus subsp. caseolyticus | subspecies |
| Hgn3C.1288 | Enterobacter hormaechei | species |
| Hgn3C.1289 | Anaerosporobacter mobilis | species |
| Hgn3C.0129 | Haemophilus parainfluenzae | species |
| Hgn3C.1290 | Veillonella ratti | species |
| Hgn3C.1291 | Lachnoclostridium edouardi | species |
| Hgn3C.1292 | Limosilactobacillus portuensis | species |
| Hgn3C.1293 | Peptostreptococcaceae sp. | family |
| Hgn3C.1294 | Eubacteriales sp. | order |
| Hgn3C.1295 | Butyricicoccus pullicaecorum | species |
| Hgn3C.1296 | Raoultella planticola | species |
| Hgn3C.1297 | Clostridium sp. | genus |
| Hgn3C.1298 | Firmicutes sp. | phylum |
| Hgn3C.1299 | Erysipelotrichaceae sp. | family |

| | | |
|---|---|---|
| Hgn3C.0013 | Ruminococcus bromii | species |
| Hgn3C.0130 | Phocaeicola plebeius | species |
| Hgn3C.1300 | Rothia sp. | genus |
| Hgn3C.1301 | Enterobacteriaceae sp. | family |
| Hgn3C.1302 | Schaalia sp. | genus |
| Hgn3C.1303 | Clostridium paraputrificum | species |
| Hgn3C.1304 | Citrobacter sp. RHBSTW-00671 | species |
| Hgn3C.1305 | Isoptericola variabilis | species |
| Hgn3C.1306 | Peptostreptococcaceae sp. | family |
| Hgn3C.1307 | Bifidobacterium longum spp. | subspecies |
| Hgn3C.1308 | Eisenbergiella sp. | genus |
| Hgn3C.1309 | Enterobacter cloacae subsp. cloacae | subspecies |
| Hgn3C.0131 | Streptococcus thermophilus | species |
| Hgn3C.1310 | Erysipelotrichaceae sp. | family |
| Hgn3C.1311 | Lachnospiraceae sp. | family |
| Hgn3C.1312 | Candida tropicalis | species |
| Hgn3C.1313 | Klebsiella aerogenes | species |
| Hgn3C.1314 | Eubacteriales sp. | order |
| Hgn3C.1315 | Eubacteriales sp. | order |
| Hgn3C.1316 | Eubacteriales sp. | order |
| Hgn3C.1317 | Lachnospiraceae sp. | family |
| Hgn3C.1318 | Eubacterium maltosivorans | species |
| Hgn3C.1319 | Atlantibacter hermannii | species |
| Hgn3C.0132 | Eubacteriales sp. | order |
| Hgn3C.1320 | Proteus mirabilis | species |
| Hgn3C.1321 | Veillonellaceae sp. | family |
| Hgn3C.1322 | Enterococcus hirae | species |
| Hgn3C.1323 | Lachnospiraceae sp. | family |
| Hgn3C.1324 | Eubacterium sp. c-25 | species |
| Hgn3C.1325 | Luxibacter massiliensis | species |
| Hgn3C.1326 | Eggerthella guodeyinii | species |
| Hgn3C.1327 | Erysipelotrichaceae sp. | family |
| Hgn3C.1328 | Lacrimispora saccharolytica | species |
| Hgn3C.1329 | Eubacteriales sp. | order |
| Hgn3C.0133 | Dialister hominis | species |
| Hgn3C.1330 | Lachnospiraceae sp. | family |
| Hgn3C.1331 | Lachnospiraceae sp. | family |
| Hgn3C.1332 | Erysipelotrichales sp. | order |
| Hgn3C.1333 | Tannerellaceae sp. | family |
| Hgn3C.1334 | Eubacteriales sp. | order |
| Hgn3C.1335 | Eubacteriales sp. | order |
| Hgn3C.1336 | Eubacteriales sp. | order |
| Hgn3C.1337 | Lachnospiraceae sp. | family |
| Hgn3C.1338 | Erysipelatoclostridium sp. | genus |
| Hgn3C.1339 | Eubacteriales sp. | order |
| Hgn3C.0134 | Eubacteriales sp. | order |
| Hgn3C.1340 | Clostridiaceae sp. | family |
| Hgn3C.1341 | Lachnospiraceae sp. | family |

| | | |
|---|---|---|
| Hgn3C.1342 | Eubacteriales sp. | order |
| Hgn3C.1343 | Enterococcus italicus | species |
| Hgn3C.1344 | Eubacteriaceae sp. | family |
| Hgn3C.1345 | Limosilactobacillus mucosae | species |
| Hgn3C.1346 | Erysipelotrichales sp. | order |
| Hgn3C.1347 | [Clostridium] hylemonae | species |
| Hgn3C.1348 | Eubacteriales sp. | order |
| Hgn3C.1349 | Corynebacterium falsenii | species |
| Hgn3C.0135 | Firmicutes sp. | phylum |
| Hgn3C.1350 | Enterococcus asini | species |
| Hgn3C.1351 | Actinomyces sp. ICM47 | species |
| Hgn3C.1352 | Akkermansia sp. | genus |
| Hgn3C.1353 | Beduini sp. | genus |
| Hgn3C.1354 | Actinomyces sp. | genus |
| Hgn3C.1355 | Collinsella stercoris | species |
| Hgn3C.1356 | Eubacteriales sp. | order |
| Hgn3C.1357 | Bacteroidales sp. | order |
| Hgn3C.1358 | Lachnospiraceae sp. | family |
| Hgn3C.1359 | Extibacter muris | species |
| Hgn3C.0136 | Alistipes ihumii | species |
| Hgn3C.1360 | Bifidobacterium crudilactis | species |
| Hgn3C.1361 | Senegalimassilia anaerobia | species |
| Hgn3C.1362 | Christensenella intestinihominis | species |
| Hgn3C.1363 | Oscillospiraceae sp. | family |
| Hgn3C.1364 | Clostridia sp. | class |
| Hgn3C.1365 | Erysipelotrichales sp. | order |
| Hgn3C.1366 | Lachnospiraceae sp. | family |
| Hgn3C.1367 | Oscillospiraceae sp. | family |
| Hgn3C.1368 | Lachnospiraceae sp. | family |
| Hgn3C.1369 | Lachnospiraceae sp. | family |
| Hgn3C.0137 | Clostridium sp. OF03-18AA | species |
| Hgn3C.1370 | Kluyvera genomosp. 3 | species |
| Hgn3C.1371 | Lachnospiraceae sp. | family |
| Hgn3C.1372 | Eubacteriales sp. | order |
| Hgn3C.1373 | Eubacteriales sp. | order |
| Hgn3C.1374 | Eubacterium sp. An3 | species |
| Hgn3C.1375 | Atopobiaceae sp. | family |
| Hgn3C.1376 | Kluyvera sp. CRP | species |
| Hgn3C.1377 | Collinsella tanakaei | species |
| Hgn3C.1378 | Collinsella sp. AF08-23 | species |
| Hgn3C.1379 | Gordonibacter urolithinfaciens | species |
| Hgn3C.0138 | Anaerobutyricum soehngenii | species |
| Hgn3C.1380 | Kluyvera georgiana | species |
| Hgn3C.1381 | Eubacteriales sp. | order |
| Hgn3C.1382 | Oscillospiraceae sp. | family |
| Hgn3C.1383 | Sellimonas monacensis | species |
| Hgn3C.1384 | Anaerococcus sp. | genus |
| Hgn3C.1385 | Eubacteriales sp. | order |

| | | |
|---|---|---|
| Hgn3C.1386 | Veillonella sp. | genus |
| Hgn3C.1387 | Alistipes sp. | genus |
| Hgn3C.1388 | Streptococcus infantarius subsp. infantarius | subspecies |
| Hgn3C.1389 | Eubacteriales sp. | order |
| Hgn3C.0139 | Ruminococcus callidus | species |
| Hgn3C.1390 | Erysipelatoclostridium sp. | genus |
| Hgn3C.1391 | Clostridium sp. | genus |
| Hgn3C.1392 | Enterobacter sp. JBIWA005 | species |
| Hgn3C.1393 | Anaerofustis stercorihominis | species |
| Hgn3C.1394 | Eubacteriales sp. | order |
| Hgn3C.1395 | Bilophila sp. | genus |
| Hgn3C.1396 | Anaerostipes sp. | genus |
| Hgn3C.1397 | Erysipelotrichales sp. | order |
| Hgn3C.1398 | Actinomyces sp. | genus |
| Hgn3C.1399 | Clostridium sp. | genus |
| Hgn3C.0014 | Lacrimispora saccharolytica | species |
| Hgn3C.0140 | Veillonella parvula | species |
| Hgn3C.1400 | Eubacterium sp. | genus |
| Hgn3C.1401 | Eubacteriales sp. | order |
| Hgn3C.1402 | Collinsella phocaeensis | species |
| Hgn3C.1403 | Eubacteriaceae sp. | family |
| Hgn3C.1404 | Clostridium sp. | genus |
| Hgn3C.1405 | Eubacteriales sp. | order |
| Hgn3C.1406 | Slackia piriformis | species |
| Hgn3C.1407 | Christensenella minuta | species |
| Hgn3C.1408 | Eubacterium callanderi | species |
| Hgn3C.1409 | Eggerthella timonensis | species |
| Hgn3C.0141 | Adlercreutzia equolifaciens subsp. celatus | subspecies |
| Hgn3C.1410 | Erysipelatoclostridium sp. | genus |
| Hgn3C.1411 | Eubacteriales sp. | order |
| Hgn3C.1412 | Christensenella hongkongensis | species |
| Hgn3C.1413 | Streptococcus sp. | genus |
| Hgn3C.1414 | Collinsella sp. | genus |
| Hgn3C.1415 | Klebsiella quasipneumoniae subsp. quasipneumoniae | subspecies |
| Hgn3C.1416 | Limosilactobacillus fermentum | species |
| Hgn3C.1417 | Oscillospiraceae sp. | family |
| Hgn3C.1418 | Carnobacteriaceae sp. | family |
| Hgn3C.1419 | Agathobaculum sp. NSJ-28 | species |
| Hgn3C.0142 | Eubacterium ramulus | species |
| Hgn3C.1420 | Eubacteriales sp. | order |
| Hgn3C.1421 | Bilophila sp. | genus |
| Hgn3C.1422 | Eubacteriales sp. | order |
| Hgn3C.1423 | Lachnospiraceae sp. | family |
| Hgn3C.1424 | Mogibacterium sp. NSJ-24 | species |
| Hgn3C.1425 | Eubacteriales sp. | order |
| Hgn3C.1426 | Limosilactobacillus oris | species |
| Hgn3C.1427 | Erysipelotrichales sp. | order |
| Hgn3C.1428 | Mediterraneibacter glycyrrhizinilyticus | species |

| | | |
|---|---|---|
| Hgn3C.1429 | Lachnospiraceae sp. | family |
| Hgn3C.0143 | Streptococcus salivarius | species |
| Hgn3C.1430 | Corynebacterium provencense | species |
| Hgn3C.1431 | Bacteroidales sp. | order |
| Hgn3C.1432 | Eggerthella timonensis | species |
| Hgn3C.1433 | Robinsoniella sp. | genus |
| Hgn3C.1434 | Blautia sp. | genus |
| Hgn3C.1435 | Acinetobacter baumannii | species |
| Hgn3C.1436 | Bacteria sp. | superkingdom |
| Hgn3C.1437 | Candidatus Stoquefichus sp. SB1 | species |
| Hgn3C.1438 | Dielma fastidiosa | species |
| Hgn3C.1439 | Streptococcus sp. IMAU 99161 | species |
| Hgn3C.0144 | Eubacteriales sp. | order |
| Hgn3C.1440 | Eubacteriales sp. | order |
| Hgn3C.1441 | Lancefieldella sp. | genus |
| Hgn3C.1442 | Oscillospiraceae sp. | family |
| Hgn3C.1443 | Streptococcus sp. | genus |
| Hgn3C.1444 | Ruminococcus sp. 1001175B_160314_E1 | species |
| Hgn3C.1445 | Oscillospiraceae sp. | family |
| Hgn3C.1446 | Eubacteriales sp. | order |
| Hgn3C.1447 | Bacteria sp. | superkingdom |
| Hgn3C.1448 | Leuconostoc falkenbergense | species |
| Hgn3C.1449 | Bifidobacterium scardovii | species |
| Hgn3C.0145 | Faecalibacterium sp. | genus |
| Hgn3C.1450 | Lactococcus sp. NH2-7C | species |
| Hgn3C.1451 | Faecalicatena orotica | species |
| Hgn3C.1452 | Raoultibacter timonensis | species |
| Hgn3C.1453 | Collinsella sp. | genus |
| Hgn3C.1454 | Escherichia fergusonii | species |
| Hgn3C.1455 | Enterocloster sp. | genus |
| Hgn3C.1456 | Eubacteriales sp. | order |
| Hgn3C.1457 | Eisenbergiella sp. OF01-20 | species |
| Hgn3C.1458 | Intestinimonas timonensis | species |
| Hgn3C.1459 | Eggerthellaceae sp. | family |
| Hgn3C.0146 | Phascolarctobacterium succinatutens | species |
| Hgn3C.1460 | Blautia sp. | genus |
| Hgn3C.1461 | Eubacteriales sp. | order |
| Hgn3C.1462 | Eubacteriales sp. | order |
| Hgn3C.1463 | Enterobacter sp. | genus |
| Hgn3C.1464 | Oscillospiraceae sp. | family |
| Hgn3C.1465 | Clostridia sp. | class |
| Hgn3C.1466 | Lachnospiraceae sp. | family |
| Hgn3C.1467 | Eubacteriales sp. | order |
| Hgn3C.1468 | Staphylococcus aureus | species |
| Hgn3C.1469 | Lachnospiraceae sp. | family |
| Hgn3C.0147 | Oscillospiraceae sp. | family |
| Hgn3C.1470 | Erysipelotrichales sp. | order |
| Hgn3C.1471 | Eggerthella sp. | genus |

| | | |
|---|---|---|
| Hgn3C.1472 | Amedibacillus dolichus | species |
| Hgn3C.0148 | Agathobaculum sp. M2 | species |
| Hgn3C.0149 | Eggerthellales sp. | order |
| Hgn3C.0015 | Faecalibacterium sp. Marseille-Q4896 | species |
| Hgn3C.0150 | Eubacteriales sp. | order |
| Hgn3C.0151 | Clostridium sp. AF34-13 | species |
| Hgn3C.0152 | Bacteroides salyersiae | species |
| Hgn3C.0153 | Lachnospiraceae sp. | family |
| Hgn3C.0154 | Prevotella copri | species |
| Hgn3C.0155 | Blautia sp. | genus |
| Hgn3C.0156 | Eubacteriales sp. | order |
| Hgn3C.0157 | Eubacteriales sp. | order |
| Hgn3C.0158 | Eubacteriales sp. | order |
| Hgn3C.0159 | Firmicutes sp. | phylum |
| Hgn3C.0016 | Parabacteroides merdae | species |
| Hgn3C.0160 | Eubacteriales sp. | order |
| Hgn3C.0161 | Blautia sp. | genus |
| Hgn3C.0162 | Prevotella sp. | genus |
| Hgn3C.0163 | Bacteria sp. | superkingdom |
| Hgn3C.0164 | Kineothrix sp. | genus |
| Hgn3C.0165 | Butyrivibrio crossotus | species |
| Hgn3C.0166 | Eubacteriales sp. | order |
| Hgn3C.0167 | Lachnospiraceae sp. | family |
| Hgn3C.0168 | Evtepia gabavorous | species |
| Hgn3C.0169 | Bacteroides intestinalis | species |
| Hgn3C.0017 | Subdoligranulum sp. APC924/74 | species |
| Hgn3C.0170 | Oscillospiraceae sp. | family |
| Hgn3C.0171 | Catenibacterium mitsuokai | species |
| Hgn3C.0172 | Eubacteriales sp. | order |
| Hgn3C.0173 | Proteobacteria sp. | phylum |
| Hgn3C.0174 | Lachnospiraceae sp. | family |
| Hgn3C.0175 | Oscillospiraceae sp. | family |
| Hgn3C.0176 | Alistipes indistinctus | species |
| Hgn3C.0177 | Clostridia sp. | class |
| Hgn3C.0178 | Clostridia sp. | class |
| Hgn3C.0179 | Bacteroidales sp. | order |
| Hgn3C.0018 | Faecalibacterium prausnitzii | species |
| Hgn3C.0180 | Bacteria sp. | superkingdom |
| Hgn3C.0181 | Alistipes onderdonkii | species |
| Hgn3C.0182 | Butyricimonas faecihominis | species |
| Hgn3C.0183 | Intestinibacter bartlettii | species |
| Hgn3C.0184 | Eubacteriales sp. | order |
| Hgn3C.0185 | Bifidobacterium catenulatum subsp. kashiwanohense | subspecies |
| Hgn3C.0186 | Butyricimonas virosa | species |
| Hgn3C.0187 | Bacteria sp. | superkingdom |
| Hgn3C.0188 | Eubacteriales sp. | order |
| Hgn3C.0189 | Eubacteriales sp. | order |
| Hgn3C.0019 | Phocaeicola dorei | species |

| | | |
|---|---|---|
| Hgn3C.0190 | Eubacteriales sp. | order |
| Hgn3C.0191 | Prevotella sp. | genus |
| Hgn3C.0192 | Erysipelatoclostridium ramosum | species |
| Hgn3C.0193 | Eubacteriales sp. | order |
| Hgn3C.0194 | Faecalibacillus faecis | species |
| Hgn3C.0195 | Eubacteriales sp. | order |
| Hgn3C.0196 | Phocaeicola coprocola | species |
| Hgn3C.0197 | Bacteroides clarus | species |
| Hgn3C.0198 | Oscillospiraceae sp. | family |
| Hgn3C.0199 | Bacteroidales sp. | order |
| Hgn3C.0002 | Bacteroides uniformis | species |
| Hgn3C.0020 | Collinsella aerofaciens | species |
| Hgn3C.0200 | Bacteroidales sp. | order |
| Hgn3C.0201 | Veillonella dispar | species |
| Hgn3C.0202 | Eggerthella lenta | species |
| Hgn3C.0203 | Faecalibacterium sp. Marseille-Q3530 | species |
| Hgn3C.0204 | Coprobacter fastidiosus | species |
| Hgn3C.0205 | Proteobacteria sp. | phylum |
| Hgn3C.0206 | Gemmiger formicilis | species |
| Hgn3C.0207 | Eubacteriales sp. | order |
| Hgn3C.0208 | Romboutsia timonensis | species |
| Hgn3C.0209 | Bifidobacterium breve | species |
| Hgn3C.0021 | Bifidobacterium longum subsp. longum | subspecies |
| Hgn3C.0210 | Firmicutes sp. | phylum |
| Hgn3C.0211 | Eubacteriales sp. | order |
| Hgn3C.0212 | Holdemanella biformis | species |
| Hgn3C.0213 | Akkermansia sp. GGCC_0220 | species |
| Hgn3C.0214 | Duodenibacillus massiliensis | species |
| Hgn3C.0215 | Alistipes sp. cv1 | species |
| Hgn3C.0216 | Eggerthellales sp. | order |
| Hgn3C.0217 | Eubacteriales sp. | order |
| Hgn3C.0218 | Eubacteriales sp. | order |
| Hgn3C.0219 | Tyzzerella nexilis | species |
| Hgn3C.0022 | Alistipes shahii | species |
| Hgn3C.0220 | Clostridium sp. MCC334 | species |
| Hgn3C.0221 | Clostridium sp. | genus |
| Hgn3C.0222 | Parabacteroides merdae | species |
| Hgn3C.0223 | Firmicutes sp. | phylum |
| Hgn3C.0224 | Veillonellales sp. | order |
| Hgn3C.0225 | Eubacteriales sp. | order |
| Hgn3C.0226 | Eubacteriales sp. | order |
| Hgn3C.0227 | Lachnospiraceae sp. | family |
| Hgn3C.0228 | Coprococcus eutactus | species |
| Hgn3C.0229 | Eubacteriales sp. | order |
| Hgn3C.0023 | Bacteroides xylanisolvens | species |
| Hgn3C.0230 | Gemmiger sp. | genus |
| Hgn3C.0231 | Firmicutes sp. | phylum |
| Hgn3C.0232 | Megamonas funiformis | species |

| | | |
|---|---|---|
| Hgn3C.0233 | Eubacteriales sp. | order |
| Hgn3C.0234 | Eubacteriales sp. | order |
| Hgn3C.0235 | Parasutterella sp. | genus |
| Hgn3C.0236 | Oscillospiraceae sp. | family |
| Hgn3C.0237 | Pseudoruminococcus massiliensis | species |
| Hgn3C.0238 | Oscillospiraceae sp. | family |
| Hgn3C.0239 | Blautia sp. BIOML-A1 | species |
| Hgn3C.0024 | Bacteroides caccae | species |
| Hgn3C.0240 | Lachnospiraceae sp. | family |
| Hgn3C.0241 | Eubacteriales sp. | order |
| Hgn3C.0242 | Enterocloster bolteae | species |
| Hgn3C.0243 | Acidaminococcales sp. | order |
| Hgn3C.0244 | Desulfovibrio piger | species |
| Hgn3C.0245 | Blautia stercoris | species |
| Hgn3C.0246 | Roseburia sp. AM59-24XD | species |
| Hgn3C.0247 | Eubacteriales sp. | order |
| Hgn3C.0248 | Coprococcus catus | species |
| Hgn3C.0249 | Clostridium sp. AM33-3 | species |
| Hgn3C.0025 | Ruminococcus bicirculans | species |
| Hgn3C.0250 | Eubacteriales sp. | order |
| Hgn3C.0251 | Alistipes senegalensis | species |
| Hgn3C.0252 | Firmicutes sp. | phylum |
| Hgn3C.0253 | Waltera sp. | genus |
| Hgn3C.0254 | Acidaminococcus intestini | species |
| Hgn3C.0255 | Firmicutes sp. | phylum |
| Hgn3C.0256 | Bacteria sp. | superkingdom |
| Hgn3C.0257 | Eubacteriales sp. | order |
| Hgn3C.0258 | Eubacteriales sp. | order |
| Hgn3C.0259 | Clostridiaceae sp. | family |
| Hgn3C.0026 | Dorea longicatena | species |
| Hgn3C.0260 | Hydrogenoanaerobacterium saccharovorans | species |
| Hgn3C.0261 | Firmicutes sp. | phylum |
| Hgn3C.0262 | Oscillospiraceae sp. | family |
| Hgn3C.0263 | Eubacterium sp. AF22-9 | species |
| Hgn3C.0264 | Clostridium sp. AF27-2AA | species |
| Hgn3C.0265 | Eubacteriales sp. | order |
| Hgn3C.0266 | Dorea sp. AF36-15AT | species |
| Hgn3C.0267 | Oscillospiraceae sp. | family |
| Hgn3C.0268 | Eubacteriales sp. | order |
| Hgn3C.0269 | Flintibacter sp. NSJ-23 | species |
| Hgn3C.0027 | Barnesiella intestinihominis | species |
| Hgn3C.0270 | Eubacteriales sp. | order |
| Hgn3C.0271 | Oscillibacter sp. | genus |
| Hgn3C.0272 | Blautia faecicola | species |
| Hgn3C.0273 | Slackia isoflavoniconvertens | species |
| Hgn3C.0274 | Eubacteriales sp. | order |
| Hgn3C.0275 | Lachnospiraceae sp. | family |
| Hgn3C.0276 | Eubacteriales sp. | order |

| | | |
|---|---|---|
| Hgn3C.0277 | Faecalibacterium sp. | genus |
| Hgn3C.0278 | Firmicutes sp. | phylum |
| Hgn3C.0279 | Oscillospiraceae sp. | family |
| Hgn3C.0028 | Oscillibacter sp. ER4 | species |
| Hgn3C.0280 | Clostridia sp. | class |
| Hgn3C.0281 | Eubacteriales sp. | order |
| Hgn3C.0282 | Eubacteriales sp. | order |
| Hgn3C.0283 | Eubacteriales sp. | order |
| Hgn3C.0284 | Erysipelotrichaceae sp. | family |
| Hgn3C.0285 | Colidextribacter sp. | genus |
| Hgn3C.0286 | Clostridia sp. | class |
| Hgn3C.0287 | Ruminococcus sp. 1001095IJ_161003_G2 | species |
| Hgn3C.0288 | Butyricimonas paravirosa | species |
| Hgn3C.0289 | Bacteroides cellulosilyticus | species |
| Hgn3C.0029 | Anaerobutyricum hallii | species |
| Hgn3C.0290 | Sellimonas intestinalis | species |
| Hgn3C.0291 | Parabacteroides johnsonii | species |
| Hgn3C.0292 | Clostridium sp. OM07-9AC | species |
| Hgn3C.0293 | Eubacteriales sp. | order |
| Hgn3C.0294 | Firmicutes sp. | phylum |
| Hgn3C.0295 | Eubacteriales sp. | order |
| Hgn3C.0296 | Firmicutes sp. | phylum |
| Hgn3C.0297 | Eubacteriales sp. | order |
| Hgn3C.0298 | Monoglobus pectinilyticus | species |
| Hgn3C.0299 | Escherichia coli | species |
| Hgn3C.0003 | Lachnospiraceae sp. | family |
| Hgn3C.0030 | Faecalibacterium prausnitzii | species |
| Hgn3C.0300 | Clostridia sp. | class |
| Hgn3C.0301 | Prevotella hominis | species |
| Hgn3C.0302 | Lachnospiraceae sp. | family |
| Hgn3C.0303 | Firmicutes sp. | phylum |
| Hgn3C.0304 | Eubacteriales sp. | order |
| Hgn3C.0305 | Candidatus Borkfalkia ceftriaxoniphila | species |
| Hgn3C.0306 | Bacteroides nordii | species |
| Hgn3C.0307 | Veillonella atypica | species |
| Hgn3C.0308 | [Clostridium] symbiosum | species |
| Hgn3C.0309 | Eubacteriales sp. | order |
| Hgn3C.0031 | Blautia massiliensis | species |
| Hgn3C.0310 | Ligilactobacillus ruminis | species |
| Hgn3C.0311 | Mesosutterella multiformis | species |
| Hgn3C.0312 | Bifidobacterium angulatum | species |
| Hgn3C.0313 | Ruminococcus bromii | species |
| Hgn3C.0314 | Ruminococcus sp. | genus |
| Hgn3C.0315 | Bacteroidia sp. | class |
| Hgn3C.0316 | Firmicutes sp. | phylum |
| Hgn3C.0317 | Intestinimonas butyriciproducens | species |
| Hgn3C.0318 | Eubacterium sp. AF34-35BH | species |
| Hgn3C.0319 | Clostridiaceae sp. | family |

| | | |
|---|---|---|
| Hgn3C.0032 | Alistipes onderdonkii subsp. vulgaris | subspecies |
| Hgn3C.0320 | Eubacteriales sp. | order |
| Hgn3C.0321 | Blautia luti | species |
| Hgn3C.0322 | Eubacteriales sp. | order |
| Hgn3C.0323 | Bacteroides sp. | genus |
| Hgn3C.0324 | Prevotella stercorea | species |
| Hgn3C.0325 | Roseburia sp. BX0805 | species |
| Hgn3C.0326 | Eubacteriales sp. | order |
| Hgn3C.0327 | Ruminococcus sp. 1001270H_150608_F2 | species |
| Hgn3C.0328 | Eubacteriales sp. | order |
| Hgn3C.0329 | Eubacteriales sp. | order |
| Hgn3C.0033 | Gemmiger formicilis | species |
| Hgn3C.0330 | Faecalibacillus intestinalis | species |
| Hgn3C.0331 | Clostridia sp. | class |
| Hgn3C.0332 | Klebsiella pneumoniae subsp. pneumoniae | subspecies |
| Hgn3C.0333 | Eubacteriales sp. | order |
| Hgn3C.0334 | Bacteroides cellulosilyticus | species |
| Hgn3C.0335 | Alistipes sp. Marseille-P2263 | species |
| Hgn3C.0336 | Parabacteroides goldsteinii | species |
| Hgn3C.0337 | Eubacteriales sp. | order |
| Hgn3C.0338 | Eubacteriales sp. | order |
| Hgn3C.0339 | Eubacteriales sp. | order |
| Hgn3C.0034 | Roseburia inulinivorans | species |
| Hgn3C.0340 | Eubacterium sp. AF19-12LB | species |
| Hgn3C.0341 | Eubacteriales sp. | order |
| Hgn3C.0342 | Eubacteriales sp. | order |
| Hgn3C.0343 | Bacteroidales sp. | order |
| Hgn3C.0344 | Akkermansia sp. BIOML-A8 | species |
| Hgn3C.0345 | Oscillospiraceae sp. | family |
| Hgn3C.0346 | Eubacteriales sp. | order |
| Hgn3C.0347 | unclassified sp. | unclassified |
| Hgn3C.0348 | Streptococcus sp. | genus |
| Hgn3C.0349 | Eubacteriales sp. | order |
| Hgn3C.0035 | Odoribacter splanchnicus | species |
| Hgn3C.0350 | [Clostridium] symbiosum | species |
| Hgn3C.0351 | Prevotella sp. | genus |
| Hgn3C.0352 | Eubacteriales sp. | order |
| Hgn3C.0353 | Prevotella sp. P4-67 | species |
| Hgn3C.0354 | Sutterella seckii | species |
| Hgn3C.0355 | Oscillospiraceae sp. | family |
| Hgn3C.0356 | Blautia sp. M29 | species |
| Hgn3C.0357 | Eubacteriales sp. | order |
| Hgn3C.0358 | Firmicutes sp. | phylum |
| Hgn3C.0359 | Eubacterium sp. BX4 | species |
| Hgn3C.0036 | Bacteroides thetaiotaomicron | species |
| Hgn3C.0360 | Eubacteriales sp. | order |
| Hgn3C.0361 | Dorea sp. AF24-7LB | species |
| Hgn3C.0362 | Dysosmobacter sp. NSJ-60 | species |

| | | |
|---|---|---|
| Hgn3C.0363 | Anaerostipes sp. | genus |
| Hgn3C.0364 | Eubacteriales sp. | order |
| Hgn3C.0365 | Eubacteriales sp. | order |
| Hgn3C.0366 | Eubacteriales sp. | order |
| Hgn3C.0367 | Lachnospiraceae sp. | family |
| Hgn3C.0368 | Eubacteriales sp. | order |
| Hgn3C.0369 | Prevotella sp. | genus |
| Hgn3C.0037 | Roseburia intestinalis | species |
| Hgn3C.0370 | Ruminococcus sp. AF41-9 | species |
| Hgn3C.0371 | Lachnospiraceae sp. | family |
| Hgn3C.0372 | Eubacteriales sp. | order |
| Hgn3C.0373 | Enterocloster clostridioformis | species |
| Hgn3C.0374 | Bacteroidia sp. | class |
| Hgn3C.0375 | Ellagibacter isourolithinifaciens | species |
| Hgn3C.0376 | Eubacteriales sp. | order |
| Hgn3C.0377 | Butyricicoccus sp. MSJd-7 | species |
| Hgn3C.0378 | Firmicutes sp. | phylum |
| Hgn3C.0379 | Eubacteriales sp. | order |
| Hgn3C.0038 | Bifidobacterium adolescentis | species |
| Hgn3C.0380 | Victivallis lenta | species |
| Hgn3C.0381 | Clostridium sp. AM30-24 | species |
| Hgn3C.0382 | Eubacteriales sp. | order |
| Hgn3C.0383 | Eubacteriales sp. | order |
| Hgn3C.0384 | Eggerthellales sp. | order |
| Hgn3C.0385 | Veillonella rogosae | species |
| Hgn3C.0386 | Clostridium sp. AT4 | species |
| Hgn3C.0387 | Eubacteriales sp. | order |
| Hgn3C.0388 | Clostridium sp. | genus |
| Hgn3C.0389 | Vescimonas fastidiosa | species |
| Hgn3C.0039 | Coprococcus comes | species |
| Hgn3C.0390 | Bacteria sp. | superkingdom |
| Hgn3C.0391 | Firmicutes sp. | phylum |
| Hgn3C.0392 | Clostridia sp. | class |
| Hgn3C.0393 | Rikenellaceae sp. | family |
| Hgn3C.0394 | Hungatella hathewayi | species |
| Hgn3C.0395 | Bacteria sp. | superkingdom |
| Hgn3C.0396 | Bacteroidales sp. | order |
| Hgn3C.0397 | Parabacteroides sp. | genus |
| Hgn3C.0398 | Eubacteriales sp. | order |
| Hgn3C.0399 | Butyricimonas faecalis | species |
| Hgn3C.0004 | Blautia wexlerae | species |
| Hgn3C.0040 | Eubacteriales sp. | order |
| Hgn3C.0400 | Prevotellamassilia timonensis | species |
| Hgn3C.0401 | Eubacteriales sp. | order |
| Hgn3C.0402 | Firmicutes sp. | phylum |
| Hgn3C.0403 | Desulfovibrionales sp. | order |
| Hgn3C.0404 | Eubacteriales sp. | order |
| Hgn3C.0405 | Bacteroides ovatus | species |

| | | |
|---|---|---|
| Hgn3C.0406 | Phocaeicola coprophilus | species |
| Hgn3C.0407 | Eubacteriales sp. | order |
| Hgn3C.0408 | Eubacteriales sp. | order |
| Hgn3C.0409 | Eubacteriales sp. | order |
| Hgn3C.0041 | Ruminococcus sp. | genus |
| Hgn3C.0410 | Eubacteriales sp. | order |
| Hgn3C.0411 | Eubacteriales sp. | order |
| Hgn3C.0412 | Eubacteriales sp. | order |
| Hgn3C.0413 | Blautia caecimuris | species |
| Hgn3C.0414 | Eubacteriales sp. | order |
| Hgn3C.0415 | Clostridium sp. C5-48 | species |
| Hgn3C.0416 | Firmicutes sp. | phylum |
| Hgn3C.0417 | Eubacteriales sp. | order |
| Hgn3C.0418 | Blautia sp. | genus |
| Hgn3C.0419 | Prevotella sp. 885 | species |
| Hgn3C.0042 | Blautia faecis | species |
| Hgn3C.0420 | Clostridium sp. 1001270J_160509_D11 | species |
| Hgn3C.0421 | Bacteroidales sp. | order |
| Hgn3C.0422 | Bacteroidia sp. | class |
| Hgn3C.0423 | Eubacteriales sp. | order |
| Hgn3C.0424 | Eubacteriales sp. | order |
| Hgn3C.0425 | Eubacteriales sp. | order |
| Hgn3C.0426 | Bifidobacterium animalis subsp. lactis | subspecies |
| Hgn3C.0427 | Bacteria sp. | superkingdom |
| Hgn3C.0428 | Eubacteriales sp. | order |
| Hgn3C.0429 | Firmicutes sp. | phylum |
| Hgn3C.0043 | Lachnoclostridium sp. 210928-DFI.6.3 | species |
| Hgn3C.0430 | Enterocloster aldenensis | species |
| Hgn3C.0431 | Bacteria sp. | superkingdom |
| Hgn3C.0432 | Eubacteriales sp. | order |
| Hgn3C.0433 | Firmicutes sp. | phylum |
| Hgn3C.0434 | Firmicutes sp. | phylum |
| Hgn3C.0435 | Eubacteriales sp. | order |
| Hgn3C.0436 | Eubacteriales sp. | order |
| Hgn3C.0437 | Prevotella sp. | genus |
| Hgn3C.0438 | Coprobacillus cateniformis | species |
| Hgn3C.0439 | Enterococcus faecalis | species |
| Hgn3C.0044 | Lachnospira pectinoschiza | species |
| Hgn3C.0440 | Bacteria sp. | superkingdom |
| Hgn3C.0441 | Firmicutes sp. | phylum |
| Hgn3C.0442 | Holdemanella sp. | genus |
| Hgn3C.0443 | Prevotella stercorea | species |
| Hgn3C.0444 | Eubacteriales sp. | order |
| Hgn3C.0445 | Aeromonadales sp. | order |
| Hgn3C.0446 | Akkermansia sp. KLE1798 | species |
| Hgn3C.0447 | Bacteria sp. | superkingdom |
| Hgn3C.0448 | Firmicutes sp. | phylum |
| Hgn3C.0449 | Coprococcus sp. OM04-5BH | species |

| | | |
|---|---|---|
| Hgn3C.0045 | Faecalibacterium sp. | genus |
| Hgn3C.0450 | Eubacteriales sp. | order |
| Hgn3C.0451 | Roseburia sp. AM16-25 | species |
| Hgn3C.0452 | Oscillospiraceae sp. | family |
| Hgn3C.0453 | Blautia sp. Marseille-P3087 | species |
| Hgn3C.0454 | Eubacteriales sp. | order |
| Hgn3C.0455 | Peptostreptococcaceae sp. | family |
| Hgn3C.0456 | Alistipes sp. | genus |
| Hgn3C.0457 | Coprobacter secundus subsp. similis | subspecies |
| Hgn3C.0458 | Eubacteriales sp. | order |
| Hgn3C.0459 | Eubacteriales sp. | order |
| Hgn3C.0046 | Clostridium sp. AF37-5 | species |
| Hgn3C.0460 | Sutterella sp. | genus |
| Hgn3C.0461 | Dialister succinatiphilus | species |
| Hgn3C.0462 | Eubacteriales sp. | order |
| Hgn3C.0463 | Eubacteriales sp. | order |
| Hgn3C.0464 | Bifidobacterium longum subsp. infantis | subspecies |
| Hgn3C.0465 | Eisenbergiella massiliensis | species |
| Hgn3C.0466 | Prevotella pectinovora | species |
| Hgn3C.0467 | Eubacteriales sp. | order |
| Hgn3C.0468 | Clostridium sp. NSJ-42 | species |
| Hgn3C.0469 | Firmicutes sp. | phylum |
| Hgn3C.0047 | Dysosmobacter sp. BX15 | species |
| Hgn3C.0470 | Eubacteriales sp. | order |
| Hgn3C.0471 | Mitsuokella jalaludinii | species |
| Hgn3C.0472 | Parabacteroides goldsteinii | species |
| Hgn3C.0473 | Bacteroidales sp. | order |
| Hgn3C.0474 | Oscillospiraceae sp. | family |
| Hgn3C.0475 | Coriobacteriia sp. | class |
| Hgn3C.0476 | Streptococcus sp. | genus |
| Hgn3C.0477 | Eubacteriales sp. | order |
| Hgn3C.0478 | Burkholderiales sp. | order |
| Hgn3C.0479 | Eubacteriales sp. | order |
| Hgn3C.0048 | Akkermansia muciniphila | species |
| Hgn3C.0480 | Eubacteriales sp. | order |
| Hgn3C.0481 | Firmicutes sp. | phylum |
| Hgn3C.0482 | Lachnospiraceae sp. | family |
| Hgn3C.0483 | Bacteroidia sp. | class |
| Hgn3C.0484 | Bacteroidales sp. | order |
| Hgn3C.0485 | Prevotella sp. | genus |
| Hgn3C.0486 | Rikenellaceae sp. | family |
| Hgn3C.0487 | Bacteria sp. | superkingdom |
| Hgn3C.0488 | Clostridia sp. | class |
| Hgn3C.0489 | Eubacteriales sp. | order |
| Hgn3C.0049 | Eubacteriales sp. | order |
| Hgn3C.0490 | Mogibacterium kristiansenii | species |
| Hgn3C.0491 | Eubacteriales sp. | order |
| Hgn3C.0492 | Prevotellaceae sp. | family |

| | | |
|---|---|---|
| Hgn3C.0493 | Eisenbergiella tayi | species |
| Hgn3C.0494 | Prevotellaceae sp. | family |
| Hgn3C.0495 | Ruminococcus sp. | genus |
| Hgn3C.0496 | Sutterella sp. AM11-39 | species |
| Hgn3C.0497 | Odoribacter laneus | species |
| Hgn3C.0498 | Prevotella sp. | genus |
| Hgn3C.0499 | Bacteroides sp. | genus |
| Hgn3C.0005 | Fusicatenibacter saccharivorans | species |
| Hgn3C.0050 | Escherichia coli | species |
| Hgn3C.0500 | Anaerotignum sp. | genus |
| Hgn3C.0501 | Oscillospiraceae sp. | family |
| Hgn3C.0502 | Lachnospiraceae sp. | family |
| Hgn3C.0503 | Eubacteriales sp. | order |
| Hgn3C.0504 | Megasphaera sp. BL7 | species |
| Hgn3C.0505 | Bacteroides sp. | genus |
| Hgn3C.0506 | Lachnospiraceae sp. | family |
| Hgn3C.0507 | [Clostridium] spiroforme | species |
| Hgn3C.0508 | Erysipelotrichales sp. | order |
| Hgn3C.0509 | Bacteroidales sp. | order |
| Hgn3C.0051 | Alistipes communis | species |
| Hgn3C.0510 | Methanosphaera stadtmanae | species |
| Hgn3C.0511 | Clostridia sp. | class |
| Hgn3C.0512 | Eubacteriales sp. | order |
| Hgn3C.0513 | Eubacteriales sp. | order |
| Hgn3C.0514 | Blautia glucerasea | species |
| Hgn3C.0515 | Eubacteriales sp. | order |
| Hgn3C.0516 | Eubacteriales sp. | order |
| Hgn3C.0517 | Ruminococcus callidus | species |
| Hgn3C.0518 | Eubacteriales sp. | order |
| Hgn3C.0519 | Bacteroidales sp. | order |
| Hgn3C.0052 | Phocaeicola massiliensis | species |
| Hgn3C.0520 | Bacteroidales sp. | order |
| Hgn3C.0521 | Firmicutes sp. | phylum |
| Hgn3C.0522 | Roseburia sp. BX1005 | species |
| Hgn3C.0523 | Phascolarctobacterium succinatutens | species |
| Hgn3C.0524 | Butyricicoccus sp. | genus |
| Hgn3C.0525 | Firmicutes sp. | phylum |
| Hgn3C.0526 | Anaerostipes sp. | genus |
| Hgn3C.0527 | Sutterella sp. | genus |
| Hgn3C.0528 | Coprococcus sp. AM27-12LB | species |
| Hgn3C.0529 | Bacteria sp. | superkingdom |
| Hgn3C.0053 | Eubacteriales sp. | order |
| Hgn3C.0530 | Eubacteriales sp. | order |
| Hgn3C.0531 | Clostridia sp. | class |
| Hgn3C.0532 | Paraprevotella xylaniphila | species |
| Hgn3C.0533 | Flintibacter sp. KGMB00164 | species |
| Hgn3C.0534 | Eubacteriales sp. | order |
| Hgn3C.0535 | Victivallis vadensis | species |

| | | |
|---|---|---|
| Hgn3C.0536 | Bacteroidaceae sp. | family |
| Hgn3C.0537 | Bifidobacterium dentium | species |
| Hgn3C.0538 | Eubacteriales sp. | order |
| Hgn3C.0539 | Eubacteriales sp. | order |
| Hgn3C.0054 | Prevotella copri | species |
| Hgn3C.0540 | Firmicutes sp. | phylum |
| Hgn3C.0541 | Phocaeicola plebeius | species |
| Hgn3C.0542 | [Clostridium] innocuum | species |
| Hgn3C.0543 | Oscillospiraceae sp. | family |
| Hgn3C.0544 | Eubacteriales sp. | order |
| Hgn3C.0545 | Eubacteriales sp. | order |
| Hgn3C.0546 | Bacteroidia sp. | class |
| Hgn3C.0547 | Eubacteriales sp. | order |
| Hgn3C.0548 | Bacteria sp. | superkingdom |
| Hgn3C.0549 | Longicatena caecimuris | species |
| Hgn3C.0055 | Alistipes finegoldii | species |
| Hgn3C.0550 | Eubacteriales sp. | order |
| Hgn3C.0551 | Eubacteriales sp. | order |
| Hgn3C.0552 | Firmicutes sp. | phylum |
| Hgn3C.0553 | Firmicutes sp. | phylum |
| Hgn3C.0554 | Eubacteriales sp. | order |
| Hgn3C.0555 | Bacteria sp. | superkingdom |
| Hgn3C.0556 | Firmicutes sp. | phylum |
| Hgn3C.0557 | Enterocloster bolteae | species |
| Hgn3C.0558 | Olsenella sp. | genus |
| Hgn3C.0559 | Oscillospiraceae sp. | family |
| Hgn3C.0056 | Bacteroides fragilis | species |
| Hgn3C.0560 | Oscillospiraceae sp. | family |
| Hgn3C.0561 | Enterocloster clostridioformis | species |
| Hgn3C.0562 | Eubacteriales sp. | order |
| Hgn3C.0563 | Parasutterella sp. | genus |
| Hgn3C.0564 | Neglecta timonensis | species |
| Hgn3C.0565 | Ruminococcus champanellensis | species |
| Hgn3C.0566 | Eubacteriales sp. | order |
| Hgn3C.0567 | Prevotella rara | species |
| Hgn3C.0568 | Eubacteriales sp. | order |
| Hgn3C.0569 | Bacteria sp. | superkingdom |
| Hgn3C.0057 | Oscillibacter sp. MSJ-31 | species |
| Hgn3C.0570 | Eubacteriales sp. | order |
| Hgn3C.0571 | Eubacterium sp. | genus |
| Hgn3C.0572 | Eubacteriales sp. | order |
| Hgn3C.0573 | Prevotella sp. Marseille-P4119 | species |
| Hgn3C.0574 | Eubacteriales sp. | order |
| Hgn3C.0575 | Bacteria sp. | superkingdom |
| Hgn3C.0576 | Eubacteriales sp. | order |
| Hgn3C.0577 | Eubacteriales sp. | order |
| Hgn3C.0578 | Firmicutes sp. | phylum |
| Hgn3C.0579 | Eubacteriales sp. | order |

| | | |
|---|---|---|
| Hgn3C.0058 | Lachnospiraceae sp. | family |
| Hgn3C.0580 | Methanobrevibacter smithii | species |
| Hgn3C.0581 | Negativibacillus massiliensis | species |
| Hgn3C.0582 | Bacteroides sp. | genus |
| Hgn3C.0583 | Clostridia sp. | class |
| Hgn3C.0584 | Bacteroides fragilis | species |
| Hgn3C.0585 | Bacteria sp. | superkingdom |
| Hgn3C.0586 | Lawsonibacter sp. NSJ-51 | species |
| Hgn3C.0587 | Eubacteriales sp. | order |
| Hgn3C.0588 | Eubacteriales sp. | order |
| Hgn3C.0589 | Erysipelotrichales sp. | order |
| Hgn3C.0059 | Bacteroides cellulosilyticus | species |
| Hgn3C.0590 | Clostridia sp. | class |
| Hgn3C.0591 | Eubacteriales sp. | order |
| Hgn3C.0592 | Clostridia sp. | class |
| Hgn3C.0593 | Eubacteriales sp. | order |
| Hgn3C.0594 | Eubacteriales sp. | order |
| Hgn3C.0595 | Eubacteriales sp. | order |
| Hgn3C.0596 | Butyricimonas sp. | genus |
| Hgn3C.0597 | Butyricimonas sp. | genus |
| Hgn3C.0598 | Eubacteriales sp. | order |
| Hgn3C.0599 | Rhodospirillales sp. | order |
| Hgn3C.0006 | Bacteroides ovatus | species |
| Hgn3C.0060 | Eubacteriales sp. | order |
| Hgn3C.0600 | Eubacteriales sp. | order |
| Hgn3C.0601 | Bacteroidales sp. | order |
| Hgn3C.0602 | Eubacteriales sp. | order |
| Hgn3C.0603 | Clostridia sp. | class |
| Hgn3C.0604 | Megasphaera elsdenii | species |
| Hgn3C.0605 | Oscillospiraceae sp. | family |
| Hgn3C.0606 | Oscillospiraceae sp. | family |
| Hgn3C.0607 | Eubacteriales sp. | order |
| Hgn3C.0608 | Lachnospiraceae sp. | family |
| Hgn3C.0609 | Eubacteriaceae sp. | family |
| Hgn3C.0061 | Faecalibacillus sp. TM498 | species |
| Hgn3C.0610 | Eubacteriales sp. | order |
| Hgn3C.0611 | Clostridia sp. | class |
| Hgn3C.0612 | Bacteroidales sp. | order |
| Hgn3C.0613 | Clostridiaceae sp. | family |
| Hgn3C.0614 | Eubacteriales sp. | order |
| Hgn3C.0615 | Firmicutes sp. | phylum |
| Hgn3C.0616 | Lachnospiraceae sp. | family |
| Hgn3C.0617 | Prevotella sp. | genus |
| Hgn3C.0618 | Eubacteriales sp. | order |
| Hgn3C.0619 | Erysipelotrichales sp. | order |
| Hgn3C.0062 | Waltera intestinalis | species |
| Hgn3C.0620 | Alistipes sp. An31A | species |
| Hgn3C.0621 | Eubacteriales sp. | order |

| | | |
|---|---|---|
| Hgn3C.0622 | Lachnospiraceae sp. | family |
| Hgn3C.0623 | Bacteroidales sp. | order |
| Hgn3C.0624 | Eubacteriales sp. | order |
| Hgn3C.0625 | Eubacteriales sp. | order |
| Hgn3C.0626 | Oscillospiraceae sp. | family |
| Hgn3C.0627 | Clostridia sp. | class |
| Hgn3C.0628 | Eubacteriales sp. | order |
| Hgn3C.0629 | Collinsella intestinalis | species |
| Hgn3C.0063 | Eubacteriales sp. | order |
| Hgn3C.0630 | Allisonella histaminiformans | species |
| Hgn3C.0631 | Firmicutes sp. | phylum |
| Hgn3C.0632 | Eubacteriales sp. | order |
| Hgn3C.0633 | Oscillospiraceae sp. | family |
| Hgn3C.0634 | Blautia schinkii | species |
| Hgn3C.0635 | Eubacteriales sp. | order |
| Hgn3C.0636 | Anaerostipes caccae | species |
| Hgn3C.0637 | Veillonellales sp. | order |
| Hgn3C.0638 | Blautia sp. | genus |
| Hgn3C.0639 | Veillonellaceae sp. | family |
| Hgn3C.0064 | Blautia sp. DFI.9.9 | species |
| Hgn3C.0640 | Eubacterium sp. AF22-8LB | species |
| Hgn3C.0641 | Eubacteriales sp. | order |
| Hgn3C.0642 | Eubacteriales sp. | order |
| Hgn3C.0643 | Eubacteriales sp. | order |
| Hgn3C.0644 | Bacteroides cellulosilyticus | species |
| Hgn3C.0645 | Eubacteriales sp. | order |
| Hgn3C.0646 | Lachnospiraceae sp. | family |
| Hgn3C.0647 | Mitsuokella multacida | species |
| Hgn3C.0648 | Eubacteriales sp. | order |
| Hgn3C.0649 | Lachnospiraceae sp. | family |
| Hgn3C.0065 | Blautia obeum | species |
| Hgn3C.0650 | Eubacteriales sp. | order |
| Hgn3C.0651 | Eubacteriales sp. | order |
| Hgn3C.0652 | Firmicutes sp. | phylum |
| Hgn3C.0653 | Clostridia sp. | class |
| Hgn3C.0654 | Firmicutes sp. | phylum |
| Hgn3C.0655 | Eubacteriales sp. | order |
| Hgn3C.0656 | Eubacteriales sp. | order |
| Hgn3C.0657 | Parolsenella catena | species |
| Hgn3C.0658 | Eubacteriales sp. | order |
| Hgn3C.0659 | Oscillospiraceae sp. | family |
| Hgn3C.0066 | Oscillospiraceae sp. | family |
| Hgn3C.0660 | Eubacteriales sp. | order |
| Hgn3C.0661 | Alistipes sp. | genus |
| Hgn3C.0662 | Eubacteriales sp. | order |
| Hgn3C.0663 | Prevotella lascolaii | species |
| Hgn3C.0664 | Blautia hansenii | species |
| Hgn3C.0665 | Desulfovibrionales sp. | order |

| | | |
|---|---|---|
| Hgn3C.0666 | Burkholderiales sp. | order |
| Hgn3C.0667 | Oscillospiraceae sp. | family |
| Hgn3C.0668 | Oscillospiraceae sp. | family |
| Hgn3C.0669 | Burkholderiales sp. | order |
| Hgn3C.0067 | Roseburia hominis | species |
| Hgn3C.0670 | Eubacteriales sp. | order |
| Hgn3C.0671 | Coriobacteriia sp. | class |
| Hgn3C.0672 | Clostridium sp. | genus |
| Hgn3C.0673 | Eubacteriales sp. | order |
| Hgn3C.0674 | Eubacteriales sp. | order |
| Hgn3C.0675 | Eubacteriales sp. | order |
| Hgn3C.0676 | Bacteroides sp. | genus |
| Hgn3C.0677 | Bacteroides sp. | genus |
| Hgn3C.0678 | Bacteroidales sp. | order |
| Hgn3C.0679 | Holdemania filiformis | species |
| Hgn3C.0068 | [Ruminococcus] lactaris | species |
| Hgn3C.0680 | Eubacteriales sp. | order |
| Hgn3C.0681 | Pseudoflavonifractor sp. | genus |
| Hgn3C.0682 | [Clostridium] scindens | species |
| Hgn3C.0683 | Clostridium disporicum | species |
| Hgn3C.0684 | Prevotella buccae | species |
| Hgn3C.0685 | Lachnospiraceae sp. | family |
| Hgn3C.0686 | Eubacteriales sp. | order |
| Hgn3C.0687 | Bacteroidales sp. | order |
| Hgn3C.0688 | Firmicutes sp. | phylum |
| Hgn3C.0689 | Eubacteriales sp. | order |
| Hgn3C.0069 | Oscillibacter sp. KLE 1728 | species |
| Hgn3C.0690 | Eubacteriales sp. | order |
| Hgn3C.0691 | Firmicutes sp. | phylum |
| Hgn3C.0692 | Clostridia sp. | class |
| Hgn3C.0693 | Hungatella hathewayi | species |
| Hgn3C.0694 | Anaeromassilibacillus sp. An250 | species |
| Hgn3C.0695 | Eubacteriales sp. | order |
| Hgn3C.0696 | Eubacteriales sp. | order |
| Hgn3C.0697 | Eubacteriales sp. | order |
| Hgn3C.0698 | Clostridia sp. | class |
| Hgn3C.0699 | Firmicutes sp. | phylum |
| Hgn3C.0007 | Parabacteroides distasonis | species |
| Hgn3C.0070 | Faecalibacterium sp. OF04-11AC | species |
| Hgn3C.0700 | Eubacteriales sp. | order |
| Hgn3C.0701 | Bacteroides sp. | genus |
| Hgn3C.0702 | Eubacteriales sp. | order |
| Hgn3C.0703 | Firmicutes sp. | phylum |
| Hgn3C.0704 | [Clostridium] scindens | species |
| Hgn3C.0705 | Bacteria sp. | superkingdom |
| Hgn3C.0706 | Eubacteriales sp. | order |
| Hgn3C.0707 | Bacteroidales sp. | order |
| Hgn3C.0708 | Eubacteriales sp. | order |

| | | |
|---|---|---|
| Hgn3C.0709 | Firmicutes sp. | phylum |
| Hgn3C.0071 | Faecalicatena fissicatena | species |
| Hgn3C.0710 | Oscillospiraceae sp. | family |
| Hgn3C.0711 | Enterocloster asparagiformis | species |
| Hgn3C.0712 | Megasphaera micronuciformis | species |
| Hgn3C.0713 | Veillonellales sp. | order |
| Hgn3C.0714 | Intestinimonas sp. MSJ-38 | species |
| Hgn3C.0715 | Desulfovibrio fairfieldensis | species |
| Hgn3C.0716 | Eubacteriales sp. | order |
| Hgn3C.0717 | Eubacteriales sp. | order |
| Hgn3C.0718 | Eubacteriales sp. | order |
| Hgn3C.0719 | Eubacteriales sp. | order |
| Hgn3C.0072 | Clostridium fessum | species |
| Hgn3C.0720 | Eubacteriales sp. | order |
| Hgn3C.0721 | Desulfovibrionales sp. | order |
| Hgn3C.0722 | Eubacteriales sp. | order |
| Hgn3C.0723 | Bacteroidales sp. | order |
| Hgn3C.0724 | Eubacteriales sp. | order |
| Hgn3C.0725 | Sutterellaceae sp. | family |
| Hgn3C.0726 | Oscillospiraceae sp. | family |
| Hgn3C.0727 | Firmicutes sp. | phylum |
| Hgn3C.0728 | Firmicutes sp. | phylum |
| Hgn3C.0729 | Eubacteriales sp. | order |
| Hgn3C.0073 | Blautia sp. | genus |
| Hgn3C.0730 | Eubacteriales sp. | order |
| Hgn3C.0731 | Eubacteriales sp. | order |
| Hgn3C.0732 | Eubacterium ramulus | species |
| Hgn3C.0733 | Desulfovibrio piger | species |
| Hgn3C.0734 | Bacteria sp. | superkingdom |
| Hgn3C.0735 | Bacteroidales sp. | order |
| Hgn3C.0736 | Coprococcus sp. | genus |
| Hgn3C.0737 | Alistipes timonensis | species |
| Hgn3C.0738 | Sutterella seckii | species |
| Hgn3C.0739 | Clostridia sp. | class |
| Hgn3C.0074 | Clostridium sp. D43t1_170807_D5 | species |
| Hgn3C.0740 | Anaerostipes sp. NSJ-7 | species |
| Hgn3C.0741 | Blautia sp. OF01-4LB | species |
| Hgn3C.0742 | Eubacteriales sp. | order |
| Hgn3C.0743 | Enterobacter sp. | genus |
| Hgn3C.0744 | Oscillospiraceae sp. | family |
| Hgn3C.0745 | Eubacteriales sp. | order |
| Hgn3C.0746 | Bacteroidia sp. | class |
| Hgn3C.0747 | Eubacteriales sp. | order |
| Hgn3C.0748 | Eubacteriales sp. | order |
| Hgn3C.0749 | Eubacteriales sp. | order |
| Hgn3C.0075 | Agathobaculum butyriciproducens | species |
| Hgn3C.0750 | Merdimonas faecis | species |
| Hgn3C.0751 | Eubacteriales sp. | order |

| | | |
|---|---|---|
| Hgn3C.0752 | Oscillospiraceae sp. | family |
| Hgn3C.0753 | Firmicutes sp. | phylum |
| Hgn3C.0754 | Collinsella bouchesdurhonensis | species |
| Hgn3C.0755 | Oscillibacter sp. | genus |
| Hgn3C.0756 | Oscillibacter valericigenes | species |
| Hgn3C.0757 | Oxalobacter formigenes | species |
| Hgn3C.0758 | Bacteroides sp. | genus |
| Hgn3C.0759 | Thermoanaerobacterales sp. | order |
| Hgn3C.0076 | Lachnospira pectinoschiza | species |
| Hgn3C.0760 | Klebsiella oxytoca | species |
| Hgn3C.0761 | Bacteroides sp. | genus |
| Hgn3C.0762 | Enterocloster lavalensis | species |
| Hgn3C.0763 | Prevotellaceae sp. | family |
| Hgn3C.0764 | Bacteroides sp. | genus |
| Hgn3C.0765 | Marseilla massiliensis | species |
| Hgn3C.0766 | Eubacteriales sp. | order |
| Hgn3C.0767 | Citrobacter freundii | species |
| Hgn3C.0768 | Eubacteriales sp. | order |
| Hgn3C.0769 | Rikenellaceae sp. | family |
| Hgn3C.0077 | Sutterella wadsworthensis | species |
| Hgn3C.0770 | Eubacteriales sp. | order |
| Hgn3C.0771 | Eubacteriales sp. | order |
| Hgn3C.0772 | Eubacteriales sp. | order |
| Hgn3C.0773 | Eubacteriales sp. | order |
| Hgn3C.0774 | Eubacteriales sp. | order |
| Hgn3C.0775 | Clostridiaceae sp. | family |
| Hgn3C.0776 | Oscillospiraceae sp. | family |
| Hgn3C.0777 | Holdemanella sp. | genus |
| Hgn3C.0778 | Oscillibacter sp. | genus |
| Hgn3C.0779 | Clostridium neonatale | species |
| Hgn3C.0078 | Clostridium sp. MCC328 | species |
| Hgn3C.0780 | Eubacteriales sp. | order |
| Hgn3C.0781 | Collinsella sp. | genus |
| Hgn3C.0782 | Streptococcus sp. | genus |
| Hgn3C.0783 | Clostridia sp. | class |
| Hgn3C.0784 | Eubacteriales sp. | order |
| Hgn3C.0785 | Eubacteriales sp. | order |
| Hgn3C.0786 | Hallella sp. CLA-AA-H145 | species |
| Hgn3C.0787 | Bacteroides mediterraneensis | species |
| Hgn3C.0788 | Eubacteriales sp. | order |
| Hgn3C.0789 | Eubacteriales sp. | order |
| Hgn3C.0079 | [Ruminococcus] torques | species |
| Hgn3C.0790 | Oscillospiraceae sp. | family |
| Hgn3C.0791 | Eubacteriales sp. | order |
| Hgn3C.0792 | Clostridia sp. | class |
| Hgn3C.0793 | Prevotellaceae sp. | family |
| Hgn3C.0794 | Bacteroides sp. | genus |
| Hgn3C.0795 | Oscillospiraceae sp. | family |

| | | |
|---|---|---|
| Hgn3C.0796 | Eubacteriales sp. | order |
| Hgn3C.0797 | Senegalimassilia sp. | genus |
| Hgn3C.0798 | Streptococcus vestibularis | species |
| Hgn3C.0799 | Bacteroidales sp. | order |
| Hgn3C.0008 | Alistipes putredinis | species |
| Hgn3C.0080 | Phascolarctobacterium faecium | species |
| Hgn3C.0800 | Archaea sp. | superkingdom |
| Hgn3C.0801 | Marseilla massiliensis | species |
| Hgn3C.0802 | Prevotellaceae sp. | family |
| Hgn3C.0803 | Bacteroidales sp. | order |
| Hgn3C.0804 | Lachnospiraceae sp. | family |
| Hgn3C.0805 | Eubacteriales sp. | order |
| Hgn3C.0806 | Bacteroidia sp. | class |
| Hgn3C.0807 | Enorma massiliensis | species |
| Hgn3C.0808 | Bacteria sp. | superkingdom |
| Hgn3C.0809 | Frisingicoccus sp. | genus |
| Hgn3C.0081 | Coprococcus eutactus | species |
| Hgn3C.0810 | Eubacteriales sp. | order |
| Hgn3C.0811 | Eubacteriales sp. | order |
| Hgn3C.0812 | Clostridia sp. | class |
| Hgn3C.0813 | Clostridia sp. | class |
| Hgn3C.0814 | Massilimicrobiota timonensis | species |
| Hgn3C.0815 | Eubacteriales sp. | order |
| Hgn3C.0816 | Lachnospiraceae sp. | family |
| Hgn3C.0817 | Prevotella sp. | genus |
| Hgn3C.0818 | Rothia sp. | genus |
| Hgn3C.0819 | Archaea sp. | superkingdom |
| Hgn3C.0082 | Roseburia sp. CLA-AA-H204 | species |
| Hgn3C.0820 | Eubacteriales sp. | order |
| Hgn3C.0821 | Bacteroidales sp. | order |
| Hgn3C.0822 | Bacteroidales sp. | order |
| Hgn3C.0823 | Anaerotignum lactatifermentans | species |
| Hgn3C.0824 | Eubacteriales sp. | order |
| Hgn3C.0825 | Bacteroidales sp. | order |
| Hgn3C.0826 | Bacteria sp. | superkingdom |
| Hgn3C.0827 | Eubacteriales sp. | order |
| Hgn3C.0828 | Bacteroides congonensis | species |
| Hgn3C.0829 | Dorea phocaeensis | species |
| Hgn3C.0083 | Eubacteriales sp. | order |
| Hgn3C.0830 | Dakarella massiliensis | species |
| Hgn3C.0831 | Eubacteriales sp. | order |
| Hgn3C.0832 | Veillonellales sp. | order |
| Hgn3C.0833 | Eisenbergiella tayi | species |
| Hgn3C.0834 | Olsenella sp. AF21-51 | species |
| Hgn3C.0835 | Eubacteriales sp. | order |
| Hgn3C.0836 | Lachnospiraceae sp. | family |
| Hgn3C.0837 | Anaerotruncus colihominis | species |
| Hgn3C.0838 | Eubacteriales sp. | order |

| | | |
|---|---|---|
| Hgn3C.0839 | Eubacteriales sp. | order |
| Hgn3C.0084 | Flavonifractor plautii | species |
| Hgn3C.0840 | Eubacteriales sp. | order |
| Hgn3C.0841 | Barnesiella sp. GGCC_0306 | species |
| Hgn3C.0842 | Ruminococcus sp. 210702-SL.1.03 | species |
| Hgn3C.0843 | Eubacteriales sp. | order |
| Hgn3C.0844 | Bacteroidales sp. | order |
| Hgn3C.0845 | Eubacteriales sp. | order |
| Hgn3C.0846 | Eubacteriales sp. | order |
| Hgn3C.0847 | Clostridia sp. | class |
| Hgn3C.0848 | Eubacteriales sp. | order |
| Hgn3C.0849 | Clostridia sp. | class |
| Hgn3C.0085 | Faecalibacterium sp. | genus |
| Hgn3C.0850 | Clostridia sp. | class |
| Hgn3C.0851 | Oscillospiraceae sp. | family |
| Hgn3C.0852 | Bacteria sp. | superkingdom |
| Hgn3C.0853 | Eubacteriales sp. | order |
| Hgn3C.0854 | Eubacteriales sp. | order |
| Hgn3C.0855 | Eubacteriales sp. | order |
| Hgn3C.0856 | Bilophila sp. | genus |
| Hgn3C.0857 | Blautia glucerasea | species |
| Hgn3C.0858 | Turicibacter sanguinis | species |
| Hgn3C.0859 | Hydrogeniiclostidium mannosilyticum | species |
| Hgn3C.0086 | Dialister invisus | species |
| Hgn3C.0860 | Bacteria sp. | superkingdom |
| Hgn3C.0861 | Firmicutes sp. | phylum |
| Hgn3C.0862 | Eubacteriales sp. | order |
| Hgn3C.0863 | Eubacteriales sp. | order |
| Hgn3C.0864 | Eubacteriales sp. | order |
| Hgn3C.0865 | Eubacteriales sp. | order |
| Hgn3C.0866 | Victivallales sp. | order |
| Hgn3C.0867 | Bacteria sp. | superkingdom |
| Hgn3C.0868 | Blastocystis sp. subtype 4 | species |
| Hgn3C.0869 | Bacteroidales sp. | order |
| Hgn3C.0087 | Eubacteriales sp. | order |
| Hgn3C.0870 | Eubacteriales sp. | order |
| Hgn3C.0871 | Anaerotruncus massiliensis | species |
| Hgn3C.0872 | Eubacteriales sp. | order |
| Hgn3C.0873 | Clostridium perfringens | species |
| Hgn3C.0874 | Methanomassiliicoccales sp. | order |
| Hgn3C.0875 | Lachnospiraceae sp. | family |
| Hgn3C.0876 | Clostridiaceae sp. | family |
| Hgn3C.0877 | Bacteroidales sp. | order |
| Hgn3C.0878 | Eubacterium sp. | genus |
| Hgn3C.0879 | Massilistercora sp. | genus |
| Hgn3C.0088 | [Ruminococcus] gnavus | species |
| Hgn3C.0880 | Firmicutes sp. | phylum |
| Hgn3C.0881 | Oscillospiraceae sp. | family |

| | | |
|---|---|---|
| Hgn3C.0882 | Eubacteriales sp. | order |
| Hgn3C.0883 | Oscillospiraceae sp. | family |
| Hgn3C.0884 | Eggerthellales sp. | order |
| Hgn3C.0885 | Eubacteriales sp. | order |
| Hgn3C.0886 | Bacteroides sp. | genus |
| Hgn3C.0887 | Eubacteriales sp. | order |
| Hgn3C.0888 | Eubacteriales sp. | order |
| Hgn3C.0889 | Clostridium paraputrificum | species |
| Hgn3C.0089 | Paraprevotella clara | species |
| Hgn3C.0890 | Enterobacter roggenkampii | species |
| Hgn3C.0891 | Eubacteriales sp. | order |
| Hgn3C.0892 | Eggerthellales sp. | order |
| Hgn3C.0893 | Phocea massiliensis | species |
| Hgn3C.0894 | Eubacteriales sp. | order |
| Hgn3C.0895 | Eubacteriales sp. | order |
| Hgn3C.0896 | Lachnospiraceae sp. | family |
| Hgn3C.0897 | Prevotella sp. | genus |
| Hgn3C.0898 | Bacteria sp. | superkingdom |
| Hgn3C.0899 | Eubacteriales sp. | order |
| Hgn3C.0009 | Faecalibacterium longum CLA-AA-H243 | subspecies |
| Hgn3C.0090 | Clostridium sp. AM49-4BH | species |
| Hgn3C.0900 | Bacteria sp. | superkingdom |
| Hgn3C.0901 | Firmicutes sp. | phylum |
| Hgn3C.0902 | Candidatus Methanomassiliicoccus intestinalis | species |
| Hgn3C.0903 | Archaea sp. | superkingdom |
| Hgn3C.0904 | Bacteroidia sp. | class |
| Hgn3C.0905 | Eubacteriales sp. | order |
| Hgn3C.0906 | Bacteria sp. | superkingdom |
| Hgn3C.0907 | Klebsiella sp. | genus |
| Hgn3C.0908 | Eubacteriales sp. | order |
| Hgn3C.0909 | Eubacteriales sp. | order |
| Hgn3C.0091 | Lachnospira sp. NSJ-43 | species |
| Hgn3C.0910 | Firmicutes sp. | phylum |
| Hgn3C.0911 | Bacteroides sp. | genus |
| Hgn3C.0912 | Clostridia sp. | class |
| Hgn3C.0913 | Erysipelotrichaceae sp. | family |
| Hgn3C.0914 | Eubacteriales sp. | order |
| Hgn3C.0915 | Firmicutes sp. | phylum |
| Hgn3C.0916 | Firmicutes sp. | phylum |
| Hgn3C.0917 | Firmicutes sp. | phylum |
| Hgn3C.0918 | Eubacteriales sp. | order |
| Hgn3C.0919 | Eubacteriales sp. | order |
| Hgn3C.0092 | Lachnospiraceae sp. | family |
| Hgn3C.0920 | Eubacteriales sp. | order |
| Hgn3C.0921 | Bacteroides sp. | genus |
| Hgn3C.0922 | Eubacteriales sp. | order |
| Hgn3C.0923 | Eubacteriales sp. | order |
| Hgn3C.0924 | Clostridioides difficile | species |

| | | |
|---|---|---|
| Hgn3C.0925 | Bacteroides sp. Marseille-P3684 | species |
| Hgn3C.0926 | Victivallales sp. | order |
| Hgn3C.0927 | Eubacteriales sp. | order |
| Hgn3C.0928 | Eubacteriales sp. | order |
| Hgn3C.0929 | Lachnospiraceae sp. | family |
| Hgn3C.0093 | Bilophila wadsworthia | species |
| Hgn3C.0930 | Eubacteriales sp. | order |
| Hgn3C.0931 | Bacteroides sp. | genus |
| Hgn3C.0932 | Eubacteriales sp. | order |
| Hgn3C.0933 | Oscillospiraceae sp. | family |
| Hgn3C.0934 | Phocaeicola coprophilus | species |
| Hgn3C.0935 | Bacteria sp. | superkingdom |
| Hgn3C.0936 | Lactobacillus acidophilus | species |
| Hgn3C.0937 | Elusimicrobiaceae sp. | family |
| Hgn3C.0938 | Firmicutes sp. | phylum |
| Hgn3C.0939 | Eubacteriales sp. | order |
| Hgn3C.0094 | Eubacteriales sp. | order |
| Hgn3C.0940 | Eubacteriales sp. | order |
| Hgn3C.0941 | Bacteria sp. | superkingdom |
| Hgn3C.0942 | Eubacteriales sp. | order |
| Hgn3C.0943 | Lactiplantibacillus plantarum subsp. plantarum | subspecies |
| Hgn3C.0944 | Bacteria sp. | superkingdom |
| Hgn3C.0945 | Eubacteriales sp. | order |
| Hgn3C.0946 | Eubacteriales sp. | order |
| Hgn3C.0947 | Eubacteriales sp. | order |
| Hgn3C.0948 | Enterobacter sp. | genus |
| Hgn3C.0949 | Eubacteriales sp. | order |
| Hgn3C.0095 | Eubacteriales sp. | order |
| Hgn3C.0950 | Clostridia sp. | class |
| Hgn3C.0951 | Eubacteriales sp. | order |
| Hgn3C.0952 | Bacteroidales sp. | order |
| Hgn3C.0953 | Lachnospiraceae sp. | family |
| Hgn3C.0954 | Bacteria sp. | superkingdom |
| Hgn3C.0955 | Eubacteriales sp. | order |
| Hgn3C.0956 | Erysipelotrichales sp. | order |
| Hgn3C.0957 | Alistipes sp. | genus |
| Hgn3C.0958 | Firmicutes sp. | phylum |
| Hgn3C.0959 | Eubacteriales sp. | order |
| Hgn3C.0096 | Eubacteriales sp. | order |
| Hgn3C.0960 | Clostridia sp. | class |
| Hgn3C.0961 | Eubacteriales sp. | order |
| Hgn3C.0962 | Clostridia sp. | class |
| Hgn3C.0963 | Eubacteriales sp. | order |
| Hgn3C.0964 | Bacteria sp. | superkingdom |
| Hgn3C.0965 | Lactobacillus delbrueckii subsp. bulgaricus | subspecies |
| Hgn3C.0966 | Lachnospiraceae sp. | family |
| Hgn3C.0967 | Firmicutes sp. | phylum |
| Hgn3C.0968 | Bacteria sp. | superkingdom |

| | | |
|---|---|---|
| Hgn3C.0969 | Bacteroides ndongoniae | species |
| Hgn3C.0097 | Eubacterium ventriosum | species |
| Hgn3C.0970 | Eubacteriales sp. | order |
| Hgn3C.0971 | Bacteroides cutis | species |
| Hgn3C.0972 | Eubacteriales sp. | order |
| Hgn3C.0973 | Desulfovibrio piger | species |
| Hgn3C.0974 | Bacteroides finegoldii | species |
| Hgn3C.0975 | Eubacteriales sp. | order |
| Hgn3C.0976 | Eubacteriales sp. | order |
| Hgn3C.0977 | Eubacteriales sp. | order |
| Hgn3C.0978 | Veillonella sp. | genus |
| Hgn3C.0979 | Eubacteriales sp. | order |
| Hgn3C.0098 | Coprococcus catus | species |
| Hgn3C.0980 | Firmicutes sp. | phylum |
| Hgn3C.0981 | Rikenellaceae sp. | family |
| Hgn3C.0982 | Acidaminococcales sp. | order |
| Hgn3C.0983 | Bacteroidales sp. | order |
| Hgn3C.0984 | Firmicutes sp. | phylum |
| Hgn3C.0985 | Firmicutes sp. | phylum |
| Hgn3C.0986 | Desulfovibrio sp. | genus |
| Hgn3C.0987 | Fusobacterium mortiferum | species |
| Hgn3C.0988 | Eubacteriales sp. | order |
| Hgn3C.0989 | Bacteria sp. | superkingdom |
| Hgn3C.0099 | Dorea longicatena | species |
| Hgn3C.0990 | Raoultella ornithinolytica | species |
| Hgn3C.0991 | Eubacteriales sp. | order |
| Hgn3C.0992 | Eubacteriales sp. | order |
| Hgn3C.0993 | Faecalitalea cylindroides | species |
| Hgn3C.0994 | Bacteroides sp. | genus |
| Hgn3C.0995 | Bacteria sp. | superkingdom |
| Hgn3C.0996 | Oscillospiraceae sp. | family |
| Hgn3C.0997 | Mediterranea massiliensis | species |
| Hgn3C.0998 | Eubacteriales sp. | order |
| Hgn3C.0999 | Burkholderiales sp. | order |

| subspecies | species |
|---|---|
| unclassified | Phocaeicola vulgatus |
| unclassified | Anaerostipes hadrus |
| unclassified | Bifidobacterium bifidum |
| unclassified | unclassified |
| unclassified | unclassified |
| unclassified | unclassified |
| unclassified | unclassified |
| unclassified | unclassified |
| unclassified | Barnesiella sp. An22 |
| unclassified | unclassified |
| unclassified | unclassified |
| unclassified | unclassified |
| unclassified | unclassified |
| unclassified | Bifidobacterium pseudocatenulatum |
| unclassified | Parabacteroides massiliensis |
| unclassified | unclassified |
| unclassified | unclassified |
| unclassified | unclassified |
| unclassified | unclassified |
| unclassified | unclassified |
| unclassified | unclassified |
| unclassified | Emergencia timonensis |
| unclassified | unclassified |
| unclassified | unclassified |
| unclassified | unclassified |
| unclassified | Phocaeicola barnesiae |
| unclassified | Bacteroides stercorirosoris |
| unclassified | unclassified |
| unclassified | unclassified |
| unclassified | unclassified |
| unclassified | unclassified |
| unclassified | unclassified |
| unclassified | unclassified |
| unclassified | unclassified |
| unclassified | unclassified |
| unclassified | Holdemanella porci |
| unclassified | unclassified |
| unclassified | unclassified |
| unclassified | unclassified |
| unclassified | Hafnia paralvei |
| unclassified | Odoribacter sp. AF15-53 |
| unclassified | unclassified |
| unclassified | Escherichia coli |
| unclassified | unclassified |
| unclassified | unclassified |
| unclassified | unclassified |

| | |
|---|---|
| unclassified | Clostridium sp. AM32-2 |
| unclassified | unclassified |
| unclassified | Bacteroides stercoris |
| unclassified | Dorea formicigenerans |
| unclassified | Clostridium phoceensis |
| unclassified | unclassified |
| Lactococcus lactis subsp. lactis | Lactococcus lactis |
| unclassified | unclassified |
| unclassified | unclassified |
| unclassified | Akkermansia sp. BIOML-A59 |
| unclassified | Enterococcus faecium |
| unclassified | Vescimonas coprocola |
| unclassified | Staphylococcus epidermidis |
| unclassified | Holdemania massiliensis |
| unclassified | Enterococcus gallinarum |
| unclassified | unclassified |
| unclassified | Streptococcus lutetiensis |
| unclassified | Veillonella seminalis |
| unclassified | Muribaculum sp. NM65_B17 |
| unclassified | unclassified |
| unclassified | Mediterraneibacter glycyrrhizinilyticus |
| unclassified | unclassified |
| unclassified | Faecalibacterium sp. CLA-AA-H233 |
| unclassified | unclassified |
| unclassified | unclassified |
| unclassified | unclassified |
| unclassified | Enterococcus avium |
| unclassified | unclassified |
| Klebsiella variicola subsp. variicola | Klebsiella variicola |
| unclassified | Lacrimispora saccharolytica |
| unclassified | unclassified |
| unclassified | unclassified |
| unclassified | unclassified |
| unclassified | Parasutterella excrementihominis |
| unclassified | Raoultella planticola |
| unclassified | Lactobacillus gasseri |
| unclassified | unclassified |
| Streptococcus gallolyticus subsp. gallolyticus | Streptococcus gallolyticus |
| unclassified | unclassified |
| unclassified | Brachyspira aalborgi |
| unclassified | unclassified |
| unclassified | unclassified |
| unclassified | unclassified |
| unclassified | unclassified |
| unclassified | unclassified |
| unclassified | unclassified |
| unclassified | Lacticaseibacillus rhamnosus |
| Lacticaseibacillus paracasei subsp. paracasei | Lacticaseibacillus paracasei |

| | |
|---|---|
| unclassified | unclassified |
| unclassified | Massilimicrobiota timonensis |
| unclassified | unclassified |
| unclassified | unclassified |
| unclassified | Ligilactobacillus salivarius |
| unclassified | Enterococcus casseliflavus |
| unclassified | unclassified |
| unclassified | Bacteroides eggerthii |
| unclassified | Turicibacter sanguinis |
| unclassified | unclassified |
| unclassified | unclassified |
| unclassified | Enterobacter kobei |
| unclassified | unclassified |
| unclassified | unclassified |
| unclassified | unclassified |
| unclassified | Alistipes provencensis |
| unclassified | unclassified |
| unclassified | unclassified |
| unclassified | Roseburia faecis |
| unclassified | unclassified |
| unclassified | unclassified |
| unclassified | unclassified |
| unclassified | unclassified |
| unclassified | unclassified |
| unclassified | unclassified |
| unclassified | unclassified |
| unclassified | Clostridium butyricum |
| unclassified | unclassified |
| unclassified | unclassified |
| unclassified | unclassified |
| unclassified | unclassified |
| unclassified | unclassified |
| unclassified | unclassified |
| unclassified | unclassified |
| unclassified | unclassified |
| unclassified | unclassified |
| unclassified | unclassified |
| unclassified | unclassified |
| unclassified | unclassified |
| unclassified | unclassified |
| unclassified | unclassified |
| unclassified | unclassified |
| unclassified | unclassified |
| unclassified | unclassified |
| unclassified | unclassified |
| unclassified | unclassified |
| unclassified | Clostridium porci |
| unclassified | unclassified |

| | |
|---|---|
| unclassified | Phocaeicola sartorii |
| unclassified | unclassified |
| unclassified | unclassified |
| unclassified | unclassified |
| unclassified | Clostridium sp. MCC345 |
| unclassified | unclassified |
| unclassified | unclassified |
| unclassified | unclassified |
| unclassified | unclassified |
| unclassified | unclassified |
| unclassified | unclassified |
| unclassified | unclassified |
| unclassified | unclassified |
| unclassified | unclassified |
| unclassified | unclassified |
| unclassified | unclassified |
| unclassified | unclassified |
| unclassified | unclassified |
| unclassified | unclassified |
| unclassified | unclassified |
| unclassified | Culturomica massiliensis |
| unclassified | unclassified |
| unclassified | unclassified |
| unclassified | unclassified |
| unclassified | unclassified |
| unclassified | unclassified |
| unclassified | unclassified |
| unclassified | unclassified |
| unclassified | unclassified |
| unclassified | Acidaminococcus fermentans |
| unclassified | Eubacterium sp. OM08-24 |
| unclassified | Lactococcus garvieae |
| unclassified | unclassified |
| unclassified | unclassified |
| unclassified | unclassified |
| unclassified | unclassified |
| unclassified | unclassified |
| unclassified | Anaerostipes hadrus |
| unclassified | unclassified |
| unclassified | unclassified |
| unclassified | unclassified |
| unclassified | unclassified |
| unclassified | unclassified |
| unclassified | Desulfovibrio legallii |
| unclassified | unclassified |
| unclassified | unclassified |
| unclassified | unclassified |
| unclassified | unclassified |

| | |
|---|---|
| unclassified | Methanobrevibacter smithii |
| unclassified | unclassified |
| unclassified | unclassified |
| unclassified | unclassified |
| unclassified | unclassified |
| unclassified | Sutterella parvirubra |
| unclassified | unclassified |
| unclassified | unclassified |
| unclassified | unclassified |
| unclassified | Clostridium sp. 12(A) |
| unclassified | Lacrimispora celerecrescens |
| unclassified | unclassified |
| unclassified | Enterocloster citroniae |
| unclassified | Acidovorax delafieldii |
| unclassified | Comamonas kerstersii |
| unclassified | Acinetobacter bereziniae |
| unclassified | Acinetobacter johnsonii |
| unclassified | Acinetobacter ursingii |
| unclassified | Acinetobacter guillouiae |
| unclassified | Actinomyces sp. HMSC065F12 |
| unclassified | Actinomyces oris |
| unclassified | Aeromonas caviae |
| unclassified | Bacteroides faecis |
| unclassified | Fenollaria massiliensis |
| unclassified | Aliarcobacter cryaerophilus |
| unclassified | Methylobacterium sp. UNCCL125 |
| unclassified | unclassified |
| unclassified | Bacteroides gallinaceum |
| Bifidobacterium pullorum subsp. saeculare | Bifidobacterium pullorum |
| unclassified | Blautia marasmi |
| unclassified | Blautia hansenii |
| unclassified | Turicimonas muris |
| unclassified | Butyricimonas synergistica |
| unclassified | unclassified |
| unclassified | Flavonifractor plautii |
| unclassified | Megasphaera sp. DJF_B143 |
| unclassified | Caulobacter sp. CCH5-E12 |
| unclassified | Chryseobacterium gambrini |
| unclassified | Citrobacter sp. CRE-46 |
| unclassified | Citrobacter amalonaticus |
| unclassified | Citrobacter braakii |
| unclassified | Citrobacter pasteurii |
| unclassified | unclassified |
| unclassified | Citrobacter sp. S-77 |
| unclassified | Cloacibacillus evryensis |
| unclassified | unclassified |
| unclassified | unclassified |
| unclassified | Lachnoclostridium pacaense |

| | |
|---|---|
| unclassified | Clostridium jeddahense |
| unclassified | Rubneribacter badeniensis |
| unclassified | Corynebacterium tuberculostearicum |
| Cutibacterium acnes subsp. acnes | Cutibacterium acnes |
| unclassified | Cutibacterium avidum |
| unclassified | Cutibacterium granulosum |
| unclassified | Enterobacter bugandensis |
| unclassified | Enterobacter sp. NFIX58 |
| unclassified | Bacteroides finegoldii |
| unclassified | Enterobacter ludwigii |
| unclassified | Phytobacter diazotrophicus |
| unclassified | Enterococcus durans |
| unclassified | Enterococcus gilvus |
| unclassified | Enterococcus raffinosus |
| unclassified | Enterococcus faecium |
| unclassified | Enterococcus thailandicus |
| unclassified | Erysipelatoclostridium sp. An15 |
| unclassified | Escherichia marmotae |
| unclassified | Escherichia sp. 93.0750 |
| unclassified | unclassified |
| unclassified | Eubacterium limosum |
| unclassified | Flavobacterium sp. Leaf359 |
| unclassified | Sanguibacteroides justesenii |
| unclassified | Gardnerella vaginalis |
| unclassified | Granulicatella adiacens |
| unclassified | Haemophilus sp. CCUG 66565 |
| unclassified | unclassified |
| unclassified | Kosakonia cowanii |
| unclassified | Klebsiella quasivariicola |
| unclassified | Lactobacillus amylovorus |
| unclassified | Ruthenibacterium lactatiformans |
| unclassified | Lactobacillus helveticus |
| Limosilactobacillus reuteri subsp. reuteri | Limosilactobacillus reuteri |
| unclassified | Limosilactobacillus vaginalis |
| unclassified | Lysinibacillus fusiformis |
| unclassified | Megasphaera genomosp. type_1 |
| unclassified | Megasphaera massiliensis |
| Morganella morganii subsp. morganii | Morganella morganii |
| unclassified | Mycobacteroides chelonae |
| unclassified | Neisseria sp. HMSC061E12 |
| unclassified | Pantoea sp. 3.5.1 |
| unclassified | Sutterella sp. KLE1602 |
| unclassified | Parabacteroides gordonii |
| unclassified | Prevotella jejuni |
| unclassified | unclassified |
| unclassified | Pseudoflavonifractor sp. An176 |
| unclassified | Pseudomonas extremaustralis |
| unclassified | Pseudomonas fulva |

| | |
|---|---|
| unclassified | Pseudomonas nitroreducens |
| unclassified | Robinsoniella peoriensis |
| unclassified | Rothia sp. HMSC072E10 |
| unclassified | Selenomonas artemidis |
| unclassified | Ruminococcus sp. BSD2780120874_150323_B10 |
| unclassified | Serratia marcescens |
| unclassified | Shewanella xiamenensis |
| unclassified | Sphingomonas sp. CCH9-F2 |
| unclassified | Staphylococcus borealis |
| Staphylococcus hominis subsp. hominis | Staphylococcus hominis |
| unclassified | Staphylococcus lugdunensis |
| unclassified | Stenotrophomonas bentonitica |
| unclassified | Stenotrophomonas maltophilia |
| unclassified | unclassified |
| Streptococcus constellatus subsp. constellatus | Streptococcus constellatus |
| unclassified | unclassified |
| unclassified | unclassified |
| unclassified | Streptococcus agalactiae |
| unclassified | Veillonella sp. CNR 79/14 |
| unclassified | Citrobacter koseri |
| unclassified | Kluyvera cryocrescens |
| unclassified | unclassified |
| unclassified | Citrobacter sp. MGH106 |
| unclassified | unclassified |
| unclassified | Clostridium sp. NSJ-6 |
| unclassified | Enterococcus malodoratus |
| unclassified | unclassified |
| unclassified | unclassified |
| unclassified | Atlantibacter sp. RC6 |
| unclassified | Terrisporobacter othiniensis |
| unclassified | Clostridium baratii |
| unclassified | unclassified |
| unclassified | unclassified |
| unclassified | Corynebacterium variabile |
| Macrococcus caseolyticus subsp. caseolyticus | Macrococcus caseolyticus |
| unclassified | Enterobacter hormaechei |
| unclassified | Anaerosporobacter mobilis |
| unclassified | Haemophilus parainfluenzae |
| unclassified | Veillonella ratti |
| unclassified | Lachnoclostridium edouardi |
| unclassified | Limosilactobacillus portuensis |
| unclassified | unclassified |
| unclassified | unclassified |
| unclassified | Butyricicoccus pullicaecorum |
| unclassified | Raoultella planticola |
| unclassified | unclassified |
| unclassified | unclassified |
| unclassified | unclassified |

| | |
|---|---|
| unclassified | Ruminococcus bromii |
| unclassified | Phocaeicola plebeius |
| unclassified | unclassified |
| unclassified | unclassified |
| unclassified | unclassified |
| unclassified | Clostridium paraputrificum |
| unclassified | Citrobacter sp. RHBSTW-00671 |
| unclassified | Isoptericola variabilis |
| unclassified | unclassified |
| Bifidobacterium longum spp. | Bifidobacterium longum |
| unclassified | unclassified |
| Enterobacter cloacae subsp. cloacae | Enterobacter cloacae |
| unclassified | Streptococcus thermophilus |
| unclassified | unclassified |
| unclassified | unclassified |
| unclassified | Candida tropicalis |
| unclassified | Klebsiella aerogenes |
| unclassified | unclassified |
| unclassified | unclassified |
| unclassified | unclassified |
| unclassified | unclassified |
| unclassified | Eubacterium maltosivorans |
| unclassified | Atlantibacter hermannii |
| unclassified | unclassified |
| unclassified | Proteus mirabilis |
| unclassified | unclassified |
| unclassified | Enterococcus hirae |
| unclassified | unclassified |
| unclassified | Eubacterium sp. c-25 |
| unclassified | Luxibacter massiliensis |
| unclassified | Eggerthella guodeyinii |
| unclassified | unclassified |
| unclassified | Lacrimispora saccharolytica |
| unclassified | unclassified |
| unclassified | Dialister hominis |
| unclassified | unclassified |
| unclassified | unclassified |
| unclassified | unclassified |
| unclassified | unclassified |
| unclassified | unclassified |
| unclassified | unclassified |
| unclassified | unclassified |
| unclassified | unclassified |
| unclassified | unclassified |
| unclassified | unclassified |
| unclassified | unclassified |
| unclassified | unclassified |
| unclassified | unclassified |

| | |
|---|---|
| unclassified | unclassified |
| unclassified | Enterococcus italicus |
| unclassified | unclassified |
| unclassified | Limosilactobacillus mucosae |
| unclassified | unclassified |
| unclassified | [Clostridium] hylemonae |
| unclassified | unclassified |
| unclassified | Corynebacterium falsenii |
| unclassified | unclassified |
| unclassified | Enterococcus asini |
| unclassified | Actinomyces sp. ICM47 |
| unclassified | unclassified |
| unclassified | unclassified |
| unclassified | unclassified |
| unclassified | Collinsella stercoris |
| unclassified | unclassified |
| unclassified | unclassified |
| unclassified | unclassified |
| unclassified | Extibacter muris |
| unclassified | Alistipes ihumii |
| unclassified | Bifidobacterium crudilactis |
| unclassified | Senegalimassilia anaerobia |
| unclassified | Christensenella intestinihominis |
| unclassified | unclassified |
| unclassified | unclassified |
| unclassified | unclassified |
| unclassified | unclassified |
| unclassified | unclassified |
| unclassified | unclassified |
| unclassified | unclassified |
| unclassified | Clostridium sp. OF03-18AA |
| unclassified | Kluyvera genomosp. 3 |
| unclassified | unclassified |
| unclassified | unclassified |
| unclassified | unclassified |
| unclassified | Eubacterium sp. An3 |
| unclassified | unclassified |
| unclassified | Kluyvera sp. CRP |
| unclassified | Collinsella tanakaei |
| unclassified | Collinsella sp. AF08-23 |
| unclassified | Gordonibacter urolithinfaciens |
| unclassified | Anaerobutyricum soehngenii |
| unclassified | Kluyvera georgiana |
| unclassified | unclassified |
| unclassified | unclassified |
| unclassified | Sellimonas monacensis |
| unclassified | unclassified |
| unclassified | unclassified |

| | |
|---|---|
| unclassified | unclassified |
| unclassified | unclassified |
| Streptococcus infantarius subsp. infantarius | Streptococcus infantarius |
| unclassified | unclassified |
| unclassified | Ruminococcus callidus |
| unclassified | unclassified |
| unclassified | unclassified |
| unclassified | Enterobacter sp. JBIWA005 |
| unclassified | Anaerofustis stercorihominis |
| unclassified | unclassified |
| unclassified | unclassified |
| unclassified | unclassified |
| unclassified | unclassified |
| unclassified | unclassified |
| unclassified | unclassified |
| unclassified | Lacrimispora saccharolytica |
| unclassified | Veillonella parvula |
| unclassified | unclassified |
| unclassified | unclassified |
| unclassified | Collinsella phocaeensis |
| unclassified | unclassified |
| unclassified | unclassified |
| unclassified | unclassified |
| unclassified | Slackia piriformis |
| unclassified | Christensenella minuta |
| unclassified | Eubacterium callanderi |
| unclassified | Eggerthella timonensis |
| Adlercreutzia equolifaciens subsp. celatus | Adlercreutzia equolifaciens |
| unclassified | unclassified |
| unclassified | unclassified |
| unclassified | Christensenella hongkongensis |
| unclassified | unclassified |
| unclassified | unclassified |
| Klebsiella quasipneumoniae subsp. quasipneumoniae | Klebsiella quasipneumoniae |
| unclassified | Limosilactobacillus fermentum |
| unclassified | unclassified |
| unclassified | unclassified |
| unclassified | Agathobaculum sp. NSJ-28 |
| unclassified | Eubacterium ramulus |
| unclassified | unclassified |
| unclassified | unclassified |
| unclassified | unclassified |
| unclassified | unclassified |
| unclassified | Mogibacterium sp. NSJ-24 |
| unclassified | unclassified |
| unclassified | Limosilactobacillus oris |
| unclassified | unclassified |
| unclassified | Mediterraneibacter glycyrrhizinilyticus |

| | |
|---|---|
| unclassified | unclassified |
| unclassified | Streptococcus salivarius |
| unclassified | Corynebacterium provencense |
| unclassified | unclassified |
| unclassified | Eggerthella timonensis |
| unclassified | unclassified |
| unclassified | unclassified |
| unclassified | Acinetobacter baumannii |
| unclassified | unclassified |
| unclassified | Candidatus Stoquefichus sp. SB1 |
| unclassified | Dielma fastidiosa |
| unclassified | Streptococcus sp. IMAU 99161 |
| unclassified | unclassified |
| unclassified | unclassified |
| unclassified | unclassified |
| unclassified | unclassified |
| unclassified | unclassified |
| unclassified | Ruminococcus sp. 1001175B_160314_E1 |
| unclassified | unclassified |
| unclassified | unclassified |
| unclassified | unclassified |
| unclassified | Leuconostoc falkenbergense |
| unclassified | Bifidobacterium scardovii |
| unclassified | unclassified |
| unclassified | Lactococcus sp. NH2-7C |
| unclassified | Faecalicatena orotica |
| unclassified | Raoultibacter timonensis |
| unclassified | unclassified |
| unclassified | Escherichia fergusonii |
| unclassified | unclassified |
| unclassified | unclassified |
| unclassified | Eisenbergiella sp. OF01-20 |
| unclassified | Intestinimonas timonensis |
| unclassified | unclassified |
| unclassified | Phascolarctobacterium succinatutens |
| unclassified | unclassified |
| unclassified | unclassified |
| unclassified | unclassified |
| unclassified | unclassified |
| unclassified | unclassified |
| unclassified | unclassified |
| unclassified | unclassified |
| unclassified | unclassified |
| unclassified | Staphylococcus aureus |
| unclassified | unclassified |
| unclassified | unclassified |
| unclassified | unclassified |
| unclassified | unclassified |

| | |
|---|---|
| unclassified | Amedibacillus dolichus |
| unclassified | Agathobaculum sp. M2 |
| unclassified | unclassified |
| unclassified | Faecalibacterium sp. Marseille-Q4896 |
| unclassified | unclassified |
| unclassified | Clostridium sp. AF34-13 |
| unclassified | Bacteroides salyersiae |
| unclassified | unclassified |
| unclassified | Prevotella copri |
| unclassified | unclassified |
| unclassified | unclassified |
| unclassified | unclassified |
| unclassified | unclassified |
| unclassified | unclassified |
| unclassified | Parabacteroides merdae |
| unclassified | unclassified |
| unclassified | unclassified |
| unclassified | unclassified |
| unclassified | unclassified |
| unclassified | unclassified |
| unclassified | Butyrivibrio crossotus |
| unclassified | unclassified |
| unclassified | unclassified |
| unclassified | Evtepia gabavorous |
| unclassified | Bacteroides intestinalis |
| unclassified | Subdoligranulum sp. APC924/74 |
| unclassified | unclassified |
| unclassified | Catenibacterium mitsuokai |
| unclassified | unclassified |
| unclassified | unclassified |
| unclassified | unclassified |
| unclassified | unclassified |
| unclassified | Alistipes indistinctus |
| unclassified | unclassified |
| unclassified | unclassified |
| unclassified | unclassified |
| unclassified | Faecalibacterium prausnitzii |
| unclassified | unclassified |
| unclassified | Alistipes onderdonkii |
| unclassified | Butyricimonas faecihominis |
| unclassified | Intestinibacter bartlettii |
| unclassified | unclassified |
| Bifidobacterium catenulatum subsp. kashiwanohense | Bifidobacterium catenulatum |
| unclassified | Butyricimonas virosa |
| unclassified | unclassified |
| unclassified | unclassified |
| unclassified | unclassified |
| unclassified | Phocaeicola dorei |

| | |
|---|---|
| unclassified | unclassified |
| unclassified | unclassified |
| unclassified | Erysipelatoclostridium ramosum |
| unclassified | unclassified |
| unclassified | Faecalibacillus faecis |
| unclassified | unclassified |
| unclassified | Phocaeicola coprocola |
| unclassified | Bacteroides clarus |
| unclassified | unclassified |
| unclassified | unclassified |
| unclassified | Bacteroides uniformis |
| unclassified | Collinsella aerofaciens |
| unclassified | unclassified |
| unclassified | Veillonella dispar |
| unclassified | Eggerthella lenta |
| unclassified | Faecalibacterium sp. Marseille-Q3530 |
| unclassified | Coprobacter fastidiosus |
| unclassified | unclassified |
| unclassified | Gemmiger formicilis |
| unclassified | unclassified |
| unclassified | Romboutsia timonensis |
| unclassified | Bifidobacterium breve |
| Bifidobacterium longum subsp. longum | Bifidobacterium longum |
| unclassified | unclassified |
| unclassified | unclassified |
| unclassified | Holdemanella biformis |
| unclassified | Akkermansia sp. GGCC_0220 |
| unclassified | Duodenibacillus massiliensis |
| unclassified | Alistipes sp. cv1 |
| unclassified | unclassified |
| unclassified | unclassified |
| unclassified | unclassified |
| unclassified | Tyzzerella nexilis |
| unclassified | Alistipes shahii |
| unclassified | Clostridium sp. MCC334 |
| unclassified | unclassified |
| unclassified | Parabacteroides merdae |
| unclassified | unclassified |
| unclassified | unclassified |
| unclassified | unclassified |
| unclassified | unclassified |
| unclassified | unclassified |
| unclassified | Coprococcus eutactus |
| unclassified | unclassified |
| unclassified | Bacteroides xylanisolvens |
| unclassified | unclassified |
| unclassified | unclassified |
| unclassified | Megamonas funiformis |

| | |
|---|---|
| unclassified | unclassified |
| unclassified | unclassified |
| unclassified | unclassified |
| unclassified | unclassified |
| unclassified | Pseudoruminococcus massiliensis |
| unclassified | unclassified |
| unclassified | Blautia sp. BIOML-A1 |
| unclassified | Bacteroides caccae |
| unclassified | unclassified |
| unclassified | unclassified |
| unclassified | Enterocloster bolteae |
| unclassified | unclassified |
| unclassified | Desulfovibrio piger |
| unclassified | Blautia stercoris |
| unclassified | Roseburia sp. AM59-24XD |
| unclassified | unclassified |
| unclassified | Coprococcus catus |
| unclassified | Clostridium sp. AM33-3 |
| unclassified | Ruminococcus bicirculans |
| unclassified | unclassified |
| unclassified | Alistipes senegalensis |
| unclassified | unclassified |
| unclassified | unclassified |
| unclassified | Acidaminococcus intestini |
| unclassified | unclassified |
| unclassified | unclassified |
| unclassified | unclassified |
| unclassified | unclassified |
| unclassified | unclassified |
| unclassified | Dorea longicatena |
| unclassified | Hydrogenoanaerobacterium saccharovorans |
| unclassified | unclassified |
| unclassified | unclassified |
| unclassified | Eubacterium sp. AF22-9 |
| unclassified | Clostridium sp. AF27-2AA |
| unclassified | unclassified |
| unclassified | Dorea sp. AF36-15AT |
| unclassified | unclassified |
| unclassified | unclassified |
| unclassified | Flintibacter sp. NSJ-23 |
| unclassified | Barnesiella intestinihominis |
| unclassified | unclassified |
| unclassified | unclassified |
| unclassified | Blautia faecicola |
| unclassified | Slackia isoflavoniconvertens |
| unclassified | unclassified |
| unclassified | unclassified |
| unclassified | unclassified |

| | |
|---|---|
| unclassified | unclassified |
| unclassified | unclassified |
| unclassified | unclassified |
| unclassified | Oscillibacter sp. ER4 |
| unclassified | unclassified |
| unclassified | unclassified |
| unclassified | unclassified |
| unclassified | unclassified |
| unclassified | unclassified |
| unclassified | unclassified |
| unclassified | unclassified |
| unclassified | Ruminococcus sp. 1001095IJ_161003_G2 |
| unclassified | Butyricimonas paravirosa |
| unclassified | Bacteroides cellulosilyticus |
| unclassified | Anaerobutyricum hallii |
| unclassified | Sellimonas intestinalis |
| unclassified | Parabacteroides johnsonii |
| unclassified | Clostridium sp. OM07-9AC |
| unclassified | unclassified |
| unclassified | unclassified |
| unclassified | unclassified |
| unclassified | unclassified |
| unclassified | unclassified |
| unclassified | Monoglobus pectinilyticus |
| unclassified | Escherichia coli |
| unclassified | unclassified |
| unclassified | Faecalibacterium prausnitzii |
| unclassified | unclassified |
| unclassified | Prevotella hominis |
| unclassified | unclassified |
| unclassified | unclassified |
| unclassified | unclassified |
| unclassified | Candidatus Borkfalkia ceftriaxoniphila |
| unclassified | Bacteroides nordii |
| unclassified | Veillonella atypica |
| unclassified | [Clostridium] symbiosum |
| unclassified | unclassified |
| unclassified | Blautia massiliensis |
| unclassified | Ligilactobacillus ruminis |
| unclassified | Mesosutterella multiformis |
| unclassified | Bifidobacterium angulatum |
| unclassified | Ruminococcus bromii |
| unclassified | unclassified |
| unclassified | unclassified |
| unclassified | unclassified |
| unclassified | Intestinimonas butyriciproducens |
| unclassified | Eubacterium sp. AF34-35BH |
| unclassified | unclassified |

| | |
|---|---|
| Alistipes onderdonkii subsp. vulgaris | Alistipes onderdonkii |
| unclassified | unclassified |
| unclassified | Blautia luti |
| unclassified | unclassified |
| unclassified | unclassified |
| unclassified | Prevotella stercorea |
| unclassified | Roseburia sp. BX0805 |
| unclassified | unclassified |
| unclassified | Ruminococcus sp. 1001270H_150608_F2 |
| unclassified | unclassified |
| unclassified | unclassified |
| unclassified | Gemmiger formicilis |
| unclassified | Faecalibacillus intestinalis |
| unclassified | unclassified |
| Klebsiella pneumoniae subsp. pneumoniae | Klebsiella pneumoniae |
| unclassified | unclassified |
| unclassified | Bacteroides cellulosilyticus |
| unclassified | Alistipes sp. Marseille-P2263 |
| unclassified | Parabacteroides goldsteinii |
| unclassified | unclassified |
| unclassified | unclassified |
| unclassified | unclassified |
| unclassified | Roseburia inulinivorans |
| unclassified | Eubacterium sp. AF19-12LB |
| unclassified | unclassified |
| unclassified | unclassified |
| unclassified | unclassified |
| unclassified | Akkermansia sp. BIOML-A8 |
| unclassified | unclassified |
| unclassified | unclassified |
| unclassified | unclassified |
| unclassified | unclassified |
| unclassified | unclassified |
| unclassified | Odoribacter splanchnicus |
| unclassified | [Clostridium] symbiosum |
| unclassified | unclassified |
| unclassified | unclassified |
| unclassified | Prevotella sp. P4-67 |
| unclassified | Sutterella seckii |
| unclassified | unclassified |
| unclassified | Blautia sp. M29 |
| unclassified | unclassified |
| unclassified | unclassified |
| unclassified | Eubacterium sp. BX4 |
| unclassified | Bacteroides thetaiotaomicron |
| unclassified | unclassified |
| unclassified | Dorea sp. AF24-7LB |
| unclassified | Dysosmobacter sp. NSJ-60 |

| | |
|---|---|
| unclassified | unclassified |
| unclassified | unclassified |
| unclassified | unclassified |
| unclassified | unclassified |
| unclassified | unclassified |
| unclassified | unclassified |
| unclassified | unclassified |
| unclassified | Roseburia intestinalis |
| unclassified | Ruminococcus sp. AF41-9 |
| unclassified | unclassified |
| unclassified | unclassified |
| unclassified | Enterocloster clostridioformis |
| unclassified | unclassified |
| unclassified | Ellagibacter isourolithinifaciens |
| unclassified | unclassified |
| unclassified | Butyricicoccus sp. MSJd-7 |
| unclassified | unclassified |
| unclassified | unclassified |
| unclassified | Bifidobacterium adolescentis |
| unclassified | Victivallis lenta |
| unclassified | Clostridium sp. AM30-24 |
| unclassified | unclassified |
| unclassified | unclassified |
| unclassified | unclassified |
| unclassified | Veillonella rogosae |
| unclassified | Clostridium sp. AT4 |
| unclassified | unclassified |
| unclassified | unclassified |
| unclassified | Vescimonas fastidiosa |
| unclassified | Coprococcus comes |
| unclassified | unclassified |
| unclassified | unclassified |
| unclassified | unclassified |
| unclassified | unclassified |
| unclassified | Hungatella hathewayi |
| unclassified | unclassified |
| unclassified | unclassified |
| unclassified | unclassified |
| unclassified | unclassified |
| unclassified | Butyricimonas faecalis |
| unclassified | Blautia wexlerae |
| unclassified | unclassified |
| unclassified | Prevotellamassilia timonensis |
| unclassified | unclassified |
| unclassified | unclassified |
| unclassified | unclassified |
| unclassified | unclassified |
| unclassified | Bacteroides ovatus |

| | |
|---|---|
| unclassified | Phocaeicola coprophilus |
| unclassified | unclassified |
| unclassified | unclassified |
| unclassified | unclassified |
| unclassified | unclassified |
| unclassified | unclassified |
| unclassified | unclassified |
| unclassified | unclassified |
| unclassified | Blautia caecimuris |
| unclassified | unclassified |
| unclassified | Clostridium sp. C5-48 |
| unclassified | unclassified |
| unclassified | unclassified |
| unclassified | unclassified |
| unclassified | Prevotella sp. 885 |
| unclassified | Blautia faecis |
| unclassified | Clostridium sp. 1001270J_160509_D11 |
| unclassified | unclassified |
| unclassified | unclassified |
| unclassified | unclassified |
| unclassified | unclassified |
| unclassified | unclassified |
| Bifidobacterium animalis subsp. lactis | Bifidobacterium animalis |
| unclassified | unclassified |
| unclassified | unclassified |
| unclassified | unclassified |
| unclassified | Lachnoclostridium sp. 210928-DFI.6.3 |
| unclassified | Enterocloster aldenensis |
| unclassified | unclassified |
| unclassified | unclassified |
| unclassified | unclassified |
| unclassified | unclassified |
| unclassified | unclassified |
| unclassified | unclassified |
| unclassified | unclassified |
| unclassified | Coprobacillus cateniformis |
| unclassified | Enterococcus faecalis |
| unclassified | Lachnospira pectinoschiza |
| unclassified | unclassified |
| unclassified | unclassified |
| unclassified | unclassified |
| unclassified | Prevotella stercorea |
| unclassified | unclassified |
| unclassified | unclassified |
| unclassified | Akkermansia sp. KLE1798 |
| unclassified | unclassified |
| unclassified | unclassified |
| unclassified | Coprococcus sp. OM04-5BH |

| | |
|---|---|
| unclassified | unclassified |
| unclassified | unclassified |
| unclassified | Roseburia sp. AM16-25 |
| unclassified | unclassified |
| unclassified | Blautia sp. Marseille-P3087 |
| unclassified | unclassified |
| unclassified | unclassified |
| unclassified | unclassified |
| Coprobacter secundus subsp. similis | Coprobacter secundus |
| unclassified | unclassified |
| unclassified | unclassified |
| unclassified | Clostridium sp. AF37-5 |
| unclassified | unclassified |
| unclassified | Dialister succinatiphilus |
| unclassified | unclassified |
| unclassified | unclassified |
| Bifidobacterium longum subsp. infantis | Bifidobacterium longum |
| unclassified | Eisenbergiella massiliensis |
| unclassified | Prevotella pectinovora |
| unclassified | unclassified |
| unclassified | Clostridium sp. NSJ-42 |
| unclassified | unclassified |
| unclassified | Dysosmobacter sp. BX15 |
| unclassified | unclassified |
| unclassified | Mitsuokella jalaludinii |
| unclassified | Parabacteroides goldsteinii |
| unclassified | unclassified |
| unclassified | unclassified |
| unclassified | unclassified |
| unclassified | unclassified |
| unclassified | unclassified |
| unclassified | unclassified |
| unclassified | unclassified |
| unclassified | Akkermansia muciniphila |
| unclassified | unclassified |
| unclassified | unclassified |
| unclassified | unclassified |
| unclassified | unclassified |
| unclassified | unclassified |
| unclassified | unclassified |
| unclassified | unclassified |
| unclassified | unclassified |
| unclassified | unclassified |
| unclassified | unclassified |
| unclassified | unclassified |
| unclassified | Mogibacterium kristiansenii |
| unclassified | unclassified |
| unclassified | unclassified |

| | |
|---|---|
| unclassified | Eisenbergiella tayi |
| unclassified | unclassified |
| unclassified | unclassified |
| unclassified | Sutterella sp. AM11-39 |
| unclassified | Odoribacter laneus |
| unclassified | unclassified |
| unclassified | unclassified |
| unclassified | Fusicatenibacter saccharivorans |
| unclassified | Escherichia coli |
| unclassified | unclassified |
| unclassified | unclassified |
| unclassified | unclassified |
| unclassified | unclassified |
| unclassified | Megasphaera sp. BL7 |
| unclassified | unclassified |
| unclassified | unclassified |
| unclassified | [Clostridium] spiroforme |
| unclassified | unclassified |
| unclassified | unclassified |
| unclassified | Alistipes communis |
| unclassified | Methanosphaera stadtmanae |
| unclassified | unclassified |
| unclassified | unclassified |
| unclassified | unclassified |
| unclassified | Blautia glucerasea |
| unclassified | unclassified |
| unclassified | unclassified |
| unclassified | Ruminococcus callidus |
| unclassified | unclassified |
| unclassified | unclassified |
| unclassified | Phocaeicola massiliensis |
| unclassified | unclassified |
| unclassified | unclassified |
| unclassified | Roseburia sp. BX1005 |
| unclassified | Phascolarctobacterium succinatutens |
| unclassified | unclassified |
| unclassified | unclassified |
| unclassified | unclassified |
| unclassified | unclassified |
| unclassified | Coprococcus sp. AM27-12LB |
| unclassified | unclassified |
| unclassified | unclassified |
| unclassified | unclassified |
| unclassified | unclassified |
| unclassified | Paraprevotella xylaniphila |
| unclassified | Flintibacter sp. KGMB00164 |
| unclassified | unclassified |
| unclassified | Victivallis vadensis |

| | |
|---|---|
| unclassified | unclassified |
| unclassified | Bifidobacterium dentium |
| unclassified | unclassified |
| unclassified | unclassified |
| unclassified | Prevotella copri |
| unclassified | unclassified |
| unclassified | Phocaeicola plebeius |
| unclassified | [Clostridium] innocuum |
| unclassified | unclassified |
| unclassified | unclassified |
| unclassified | unclassified |
| unclassified | unclassified |
| unclassified | unclassified |
| unclassified | unclassified |
| unclassified | Longicatena caecimuris |
| unclassified | Alistipes finegoldii |
| unclassified | unclassified |
| unclassified | unclassified |
| unclassified | unclassified |
| unclassified | unclassified |
| unclassified | unclassified |
| unclassified | unclassified |
| unclassified | unclassified |
| unclassified | Enterocloster bolteae |
| unclassified | unclassified |
| unclassified | unclassified |
| unclassified | Bacteroides fragilis |
| unclassified | unclassified |
| unclassified | Enterocloster clostridioformis |
| unclassified | unclassified |
| unclassified | unclassified |
| unclassified | Neglecta timonensis |
| unclassified | Ruminococcus champanellensis |
| unclassified | unclassified |
| unclassified | Prevotella rara |
| unclassified | unclassified |
| unclassified | unclassified |
| unclassified | Oscillibacter sp. MSJ-31 |
| unclassified | unclassified |
| unclassified | unclassified |
| unclassified | unclassified |
| unclassified | Prevotella sp. Marseille-P4119 |
| unclassified | unclassified |
| unclassified | unclassified |
| unclassified | unclassified |
| unclassified | unclassified |
| unclassified | unclassified |
| unclassified | unclassified |

| | |
|---|---|
| unclassified | unclassified |
| unclassified | Methanobrevibacter smithii |
| unclassified | Negativibacillus massiliensis |
| unclassified | unclassified |
| unclassified | unclassified |
| unclassified | Bacteroides fragilis |
| unclassified | unclassified |
| unclassified | Lawsonibacter sp. NSJ-51 |
| unclassified | unclassified |
| unclassified | unclassified |
| unclassified | unclassified |
| unclassified | Bacteroides cellulosilyticus |
| unclassified | unclassified |
| unclassified | unclassified |
| unclassified | unclassified |
| unclassified | unclassified |
| unclassified | unclassified |
| unclassified | unclassified |
| unclassified | unclassified |
| unclassified | unclassified |
| unclassified | unclassified |
| unclassified | unclassified |
| unclassified | Bacteroides ovatus |
| unclassified | unclassified |
| unclassified | unclassified |
| unclassified | unclassified |
| unclassified | unclassified |
| unclassified | unclassified |
| unclassified | Megasphaera elsdenii |
| unclassified | unclassified |
| unclassified | unclassified |
| unclassified | unclassified |
| unclassified | unclassified |
| unclassified | unclassified |
| unclassified | Faecalibacillus sp. TM498 |
| unclassified | unclassified |
| unclassified | unclassified |
| unclassified | unclassified |
| unclassified | unclassified |
| unclassified | unclassified |
| unclassified | unclassified |
| unclassified | unclassified |
| unclassified | unclassified |
| unclassified | unclassified |
| unclassified | unclassified |
| unclassified | Waltera intestinalis |
| unclassified | Alistipes sp. An31A |
| unclassified | unclassified |

| | |
|---|---|
| unclassified | unclassified |
| unclassified | unclassified |
| unclassified | unclassified |
| unclassified | unclassified |
| unclassified | unclassified |
| unclassified | unclassified |
| unclassified | unclassified |
| unclassified | Collinsella intestinalis |
| unclassified | unclassified |
| unclassified | Allisonella histaminiformans |
| unclassified | unclassified |
| unclassified | unclassified |
| unclassified | unclassified |
| unclassified | Blautia schinkii |
| unclassified | unclassified |
| unclassified | Anaerostipes caccae |
| unclassified | unclassified |
| unclassified | unclassified |
| unclassified | unclassified |
| unclassified | Blautia sp. DFI.9.9 |
| unclassified | Eubacterium sp. AF22-8LB |
| unclassified | unclassified |
| unclassified | unclassified |
| unclassified | unclassified |
| unclassified | Bacteroides cellulosilyticus |
| unclassified | unclassified |
| unclassified | unclassified |
| unclassified | Mitsuokella multacida |
| unclassified | unclassified |
| unclassified | unclassified |
| unclassified | Blautia obeum |
| unclassified | unclassified |
| unclassified | unclassified |
| unclassified | unclassified |
| unclassified | unclassified |
| unclassified | unclassified |
| unclassified | unclassified |
| unclassified | unclassified |
| unclassified | Parolsenella catena |
| unclassified | unclassified |
| unclassified | unclassified |
| unclassified | unclassified |
| unclassified | unclassified |
| unclassified | unclassified |
| unclassified | unclassified |
| unclassified | Prevotella lascolaii |
| unclassified | Blautia hansenii |
| unclassified | unclassified |

| | |
|---|---|
| unclassified | unclassified |
| unclassified | unclassified |
| unclassified | unclassified |
| unclassified | unclassified |
| unclassified | Roseburia hominis |
| unclassified | unclassified |
| unclassified | unclassified |
| unclassified | unclassified |
| unclassified | unclassified |
| unclassified | unclassified |
| unclassified | unclassified |
| unclassified | unclassified |
| unclassified | unclassified |
| unclassified | unclassified |
| unclassified | Holdemania filiformis |
| unclassified | [Ruminococcus] lactaris |
| unclassified | unclassified |
| unclassified | unclassified |
| unclassified | [Clostridium] scindens |
| unclassified | Clostridium disporicum |
| unclassified | Prevotella buccae |
| unclassified | unclassified |
| unclassified | unclassified |
| unclassified | unclassified |
| unclassified | unclassified |
| unclassified | unclassified |
| unclassified | Oscillibacter sp. KLE 1728 |
| unclassified | unclassified |
| unclassified | unclassified |
| unclassified | unclassified |
| unclassified | Hungatella hathewayi |
| unclassified | Anaeromassilibacillus sp. An250 |
| unclassified | unclassified |
| unclassified | unclassified |
| unclassified | unclassified |
| unclassified | unclassified |
| unclassified | unclassified |
| unclassified | Parabacteroides distasonis |
| unclassified | Faecalibacterium sp. OF04-11AC |
| unclassified | unclassified |
| unclassified | unclassified |
| unclassified | unclassified |
| unclassified | unclassified |
| unclassified | [Clostridium] scindens |
| unclassified | unclassified |
| unclassified | unclassified |
| unclassified | unclassified |
| unclassified | unclassified |

| | |
|---|---|
| unclassified | unclassified |
| unclassified | Faecalicatena fissicatena |
| unclassified | unclassified |
| unclassified | Enterocloster asparagiformis |
| unclassified | Megasphaera micronuciformis |
| unclassified | unclassified |
| unclassified | Intestinimonas sp. MSJ-38 |
| unclassified | Desulfovibrio fairfieldensis |
| unclassified | unclassified |
| unclassified | unclassified |
| unclassified | unclassified |
| unclassified | unclassified |
| unclassified | Clostridium fessum |
| unclassified | unclassified |
| unclassified | unclassified |
| unclassified | unclassified |
| unclassified | unclassified |
| unclassified | unclassified |
| unclassified | unclassified |
| unclassified | unclassified |
| unclassified | unclassified |
| unclassified | unclassified |
| unclassified | unclassified |
| unclassified | unclassified |
| unclassified | unclassified |
| unclassified | unclassified |
| unclassified | Eubacterium ramulus |
| unclassified | Desulfovibrio piger |
| unclassified | unclassified |
| unclassified | unclassified |
| unclassified | unclassified |
| unclassified | Alistipes timonensis |
| unclassified | Sutterella seckii |
| unclassified | unclassified |
| unclassified | Clostridium sp. D43t1_170807_D5 |
| unclassified | Anaerostipes sp. NSJ-7 |
| unclassified | Blautia sp. OF01-4LB |
| unclassified | unclassified |
| unclassified | unclassified |
| unclassified | unclassified |
| unclassified | unclassified |
| unclassified | unclassified |
| unclassified | unclassified |
| unclassified | unclassified |
| unclassified | unclassified |
| unclassified | Agathobaculum butyriciprodicens |
| unclassified | Merdimonas faecis |
| unclassified | unclassified |

| | |
|---|---|
| unclassified | unclassified |
| unclassified | unclassified |
| unclassified | Collinsella bouchesdurhonensis |
| unclassified | unclassified |
| unclassified | Oscillibacter valericigenes |
| unclassified | Oxalobacter formigenes |
| unclassified | unclassified |
| unclassified | unclassified |
| unclassified | Lachnospira pectinoschiza |
| unclassified | Klebsiella oxytoca |
| unclassified | unclassified |
| unclassified | Enterocloster lavalensis |
| unclassified | unclassified |
| unclassified | unclassified |
| unclassified | Marseilla massiliensis |
| unclassified | unclassified |
| unclassified | Citrobacter freundii |
| unclassified | unclassified |
| unclassified | unclassified |
| unclassified | Sutterella wadsworthensis |
| unclassified | unclassified |
| unclassified | unclassified |
| unclassified | unclassified |
| unclassified | unclassified |
| unclassified | unclassified |
| unclassified | unclassified |
| unclassified | unclassified |
| unclassified | unclassified |
| unclassified | unclassified |
| unclassified | Clostridium neonatale |
| unclassified | Clostridium sp. MCC328 |
| unclassified | unclassified |
| unclassified | unclassified |
| unclassified | unclassified |
| unclassified | unclassified |
| unclassified | unclassified |
| unclassified | unclassified |
| unclassified | Hallella sp. CLA-AA-H145 |
| unclassified | Bacteroides mediterraneensis |
| unclassified | unclassified |
| unclassified | unclassified |
| unclassified | [Ruminococcus] torques |
| unclassified | unclassified |
| unclassified | unclassified |
| unclassified | unclassified |
| unclassified | unclassified |
| unclassified | unclassified |
| unclassified | unclassified |

| | |
|---|---|
| unclassified | unclassified |
| unclassified | unclassified |
| unclassified | Streptococcus vestibularis |
| unclassified | unclassified |
| unclassified | Alistipes putredinis |
| unclassified | Phascolarctobacterium faecium |
| unclassified | unclassified |
| unclassified | Marseilla massiliensis |
| unclassified | unclassified |
| unclassified | unclassified |
| unclassified | unclassified |
| unclassified | unclassified |
| unclassified | unclassified |
| unclassified | Enorma massiliensis |
| unclassified | unclassified |
| unclassified | unclassified |
| unclassified | Coprococcus eutactus |
| unclassified | unclassified |
| unclassified | unclassified |
| unclassified | unclassified |
| unclassified | unclassified |
| unclassified | Massilimicrobiota timonensis |
| unclassified | unclassified |
| unclassified | unclassified |
| unclassified | unclassified |
| unclassified | unclassified |
| unclassified | unclassified |
| unclassified | Roseburia sp. CLA-AA-H204 |
| unclassified | unclassified |
| unclassified | unclassified |
| unclassified | unclassified |
| unclassified | Anaerotignum lactatifermentans |
| unclassified | unclassified |
| unclassified | unclassified |
| unclassified | unclassified |
| unclassified | unclassified |
| unclassified | Bacteroides congonensis |
| unclassified | Dorea phocaeensis |
| unclassified | unclassified |
| unclassified | Dakarella massiliensis |
| unclassified | unclassified |
| unclassified | unclassified |
| unclassified | Eisenbergiella tayi |
| unclassified | Olsenella sp. AF21-51 |
| unclassified | unclassified |
| unclassified | unclassified |
| unclassified | Anaerotruncus colihominis |
| unclassified | unclassified |

| | |
|---|---|
| unclassified | unclassified |
| unclassified | Flavonifractor plautii |
| unclassified | unclassified |
| unclassified | Barnesiella sp. GGCC_0306 |
| unclassified | Ruminococcus sp. 210702-SL.1.03 |
| unclassified | unclassified |
| unclassified | unclassified |
| unclassified | unclassified |
| unclassified | unclassified |
| unclassified | unclassified |
| unclassified | unclassified |
| unclassified | unclassified |
| unclassified | unclassified |
| unclassified | unclassified |
| unclassified | unclassified |
| unclassified | unclassified |
| unclassified | unclassified |
| unclassified | unclassified |
| unclassified | unclassified |
| unclassified | unclassified |
| unclassified | Blautia glucerasea |
| unclassified | Turicibacter sanguinis |
| unclassified | Hydrogeniiclostidium mannosilyticum |
| unclassified | Dialister invisus |
| unclassified | unclassified |
| unclassified | unclassified |
| unclassified | unclassified |
| unclassified | unclassified |
| unclassified | unclassified |
| unclassified | unclassified |
| unclassified | unclassified |
| unclassified | unclassified |
| unclassified | Blastocystis sp. subtype 4 |
| unclassified | unclassified |
| unclassified | unclassified |
| unclassified | unclassified |
| unclassified | Anaerotruncus massiliensis |
| unclassified | unclassified |
| unclassified | Clostridium perfringens |
| unclassified | unclassified |
| unclassified | unclassified |
| unclassified | unclassified |
| unclassified | unclassified |
| unclassified | unclassified |
| unclassified | unclassified |
| unclassified | [Ruminococcus] gnavus |
| unclassified | unclassified |
| unclassified | unclassified |

| | |
|---|---|
| unclassified | unclassified |
| unclassified | unclassified |
| unclassified | unclassified |
| unclassified | unclassified |
| unclassified | unclassified |
| unclassified | unclassified |
| unclassified | unclassified |
| unclassified | Clostridium paraputrificum |
| unclassified | Paraprevotella clara |
| unclassified | Enterobacter roggenkampii |
| unclassified | unclassified |
| unclassified | unclassified |
| unclassified | Phocea massiliensis |
| unclassified | unclassified |
| unclassified | unclassified |
| unclassified | unclassified |
| unclassified | unclassified |
| unclassified | unclassified |
| unclassified | unclassified |
| Faecalibacterium longum CLA-AA-H243 | Faecalibacterium longum |
| unclassified | Clostridium sp. AM49-4BH |
| unclassified | unclassified |
| unclassified | unclassified |
| unclassified | Candidatus Methanomassiliicoccus intestinalis |
| unclassified | unclassified |
| unclassified | unclassified |
| unclassified | unclassified |
| unclassified | unclassified |
| unclassified | unclassified |
| unclassified | unclassified |
| unclassified | unclassified |
| unclassified | Lachnospira sp. NSJ-43 |
| unclassified | unclassified |
| unclassified | unclassified |
| unclassified | unclassified |
| unclassified | unclassified |
| unclassified | unclassified |
| unclassified | unclassified |
| unclassified | unclassified |
| unclassified | unclassified |
| unclassified | unclassified |
| unclassified | unclassified |
| unclassified | unclassified |
| unclassified | unclassified |
| unclassified | unclassified |
| unclassified | unclassified |
| unclassified | unclassified |
| unclassified | Clostridioides difficile |

| | |
|---|---|
| unclassified | Bacteroides sp. Marseille-P3684 |
| unclassified | unclassified |
| unclassified | unclassified |
| unclassified | unclassified |
| unclassified | unclassified |
| unclassified | Bilophila wadsworthia |
| unclassified | unclassified |
| unclassified | unclassified |
| unclassified | unclassified |
| unclassified | unclassified |
| unclassified | Phocaeicola coprophilus |
| unclassified | unclassified |
| unclassified | Lactobacillus acidophilus |
| unclassified | unclassified |
| unclassified | unclassified |
| unclassified | unclassified |
| unclassified | unclassified |
| unclassified | unclassified |
| unclassified | unclassified |
| unclassified | unclassified |
| Lactiplantibacillus plantarum subsp. plantarum | Lactiplantibacillus plantarum |
| unclassified | unclassified |
| unclassified | unclassified |
| unclassified | unclassified |
| unclassified | unclassified |
| unclassified | unclassified |
| unclassified | unclassified |
| unclassified | unclassified |
| unclassified | unclassified |
| unclassified | unclassified |
| unclassified | unclassified |
| unclassified | unclassified |
| unclassified | unclassified |
| unclassified | unclassified |
| unclassified | unclassified |
| unclassified | unclassified |
| unclassified | unclassified |
| unclassified | unclassified |
| unclassified | unclassified |
| unclassified | unclassified |
| unclassified | unclassified |
| unclassified | unclassified |
| unclassified | unclassified |
| Lactobacillus delbrueckii subsp. bulgaricus | Lactobacillus delbrueckii |
| unclassified | unclassified |
| unclassified | unclassified |
| unclassified | unclassified |

| | |
|---|---|
| unclassified | Bacteroides ndongoniae |
| unclassified | Eubacterium ventriosum |
| unclassified | unclassified |
| unclassified | Bacteroides cutis |
| unclassified | unclassified |
| unclassified | Desulfovibrio piger |
| unclassified | Bacteroides finegoldii |
| unclassified | unclassified |
| unclassified | unclassified |
| unclassified | unclassified |
| unclassified | unclassified |
| unclassified | unclassified |
| unclassified | Coprococcus catus |
| unclassified | unclassified |
| unclassified | unclassified |
| unclassified | unclassified |
| unclassified | unclassified |
| unclassified | unclassified |
| unclassified | unclassified |
| unclassified | unclassified |
| unclassified | Fusobacterium mortiferum |
| unclassified | unclassified |
| unclassified | unclassified |
| unclassified | Dorea longicatena |
| unclassified | Raoultella ornithinolytica |
| unclassified | unclassified |
| unclassified | unclassified |
| unclassified | Faecalitalea cylindroides |
| unclassified | unclassified |
| unclassified | unclassified |
| unclassified | unclassified |
| unclassified | Mediterranea massiliensis |
| unclassified | unclassified |
| unclassified | unclassified |

| genus | family | order |
|---|---|---|
| Phocaeicola | Bacteroidaceae | Bacteroidales |
| Anaerostipes | Lachnospiraceae | Eubacteriales |
| Bifidobacterium | Bifidobacteriaceae | Bifidobacteriales |
| unclassified | unclassified | Bacteroidales |
| unclassified | unclassified | unclassified |
| unclassified | unclassified | Eggerthellales |
| unclassified | Veillonellaceae | Veillonellales |
| unclassified | unclassified | Bacteroidales |
| Barnesiella | Barnesiellaceae | Bacteroidales |
| unclassified | unclassified | Eubacteriales |
| unclassified | unclassified | unclassified |
| unclassified | Oscillospiraceae | Eubacteriales |
| unclassified | Prevotellaceae | Bacteroidales |
| Bifidobacterium | Bifidobacteriaceae | Bifidobacteriales |
| Parabacteroides | Tannerellaceae | Bacteroidales |
| unclassified | unclassified | Eubacteriales |
| unclassified | Erysipelotrichaceae | Erysipelotrichales |
| unclassified | Oscillospiraceae | Eubacteriales |
| unclassified | unclassified | Burkholderiales |
| unclassified | unclassified | unclassified |
| unclassified | unclassified | Eubacteriales |
| Emergencia | Eubacteriales Family XIII. Incertae Sedis | Eubacteriales |
| unclassified | unclassified | unclassified |
| unclassified | Prevotellaceae | Bacteroidales |
| unclassified | unclassified | Eubacteriales |
| Phocaeicola | Bacteroidaceae | Bacteroidales |
| Bacteroides | Bacteroidaceae | Bacteroidales |
| unclassified | unclassified | Eubacteriales |
| unclassified | unclassified | unclassified |
| unclassified | Prevotellaceae | Bacteroidales |
| unclassified | unclassified | Eubacteriales |
| unclassified | unclassified | Bacteroidales |
| unclassified | unclassified | unclassified |
| unclassified | unclassified | Bacteroidales |
| unclassified | unclassified | unclassified |
| Holdemanella | Erysipelotrichaceae | Erysipelotrichales |
| unclassified | Lachnospiraceae | Eubacteriales |
| unclassified | unclassified | Eubacteriales |
| unclassified | unclassified | Bacteroidales |
| Hafnia | Hafniaceae | Enterobacterales |
| Odoribacter | Odoribacteraceae | Bacteroidales |
| Klebsiella | Enterobacteriaceae | Enterobacterales |
| Escherichia | Enterobacteriaceae | Enterobacterales |
| Rothia | Micrococcaceae | Micrococcales |
| Veillonella | Veillonellaceae | Veillonellales |
| unclassified | Prevotellaceae | Bacteroidales |

| | | |
|---|---|---|
| Clostridium | Clostridiaceae | Eubacteriales |
| unclassified | Lachnospiraceae | Eubacteriales |
| Bacteroides | Bacteroidaceae | Bacteroidales |
| Dorea | Lachnospiraceae | Eubacteriales |
| Clostridium | Clostridiaceae | Eubacteriales |
| unclassified | unclassified | Eubacteriales |
| Lactococcus | Streptococcaceae | Lactobacillales |
| unclassified | unclassified | Bacteroidales |
| unclassified | unclassified | Eubacteriales |
| Akkermansia | Akkermansiaceae | Verrucomicrobiales |
| Enterococcus | Enterococcaceae | Lactobacillales |
| Vescimonas | Oscillospiraceae | Eubacteriales |
| Staphylococcus | Staphylococcaceae | Bacillales |
| Holdemania | Erysipelotrichaceae | Erysipelotrichales |
| Enterococcus | Enterococcaceae | Lactobacillales |
| unclassified | unclassified | Eubacteriales |
| Streptococcus | Streptococcaceae | Lactobacillales |
| Veillonella | Veillonellaceae | Veillonellales |
| Muribaculum | Muribaculaceae | Bacteroidales |
| unclassified | unclassified | Eubacteriales |
| Mediterraneibacter | Lachnospiraceae | Eubacteriales |
| Faecalicatena | Lachnospiraceae | Eubacteriales |
| Faecalibacterium | Oscillospiraceae | Eubacteriales |
| unclassified | unclassified | unclassified |
| unclassified | unclassified | Acidaminococcales |
| unclassified | unclassified | Eubacteriales |
| Enterococcus | Enterococcaceae | Lactobacillales |
| unclassified | unclassified | Eubacteriales |
| Klebsiella | Enterobacteriaceae | Enterobacterales |
| Lacrimispora | Lachnospiraceae | Eubacteriales |
| unclassified | Lachnospiraceae | Eubacteriales |
| unclassified | unclassified | Eubacteriales |
| unclassified | unclassified | Bacteroidales |
| Parasutterella | Sutterellaceae | Burkholderiales |
| Raoultella | Enterobacteriaceae | Enterobacterales |
| Lactobacillus | Lactobacillaceae | Lactobacillales |
| unclassified | unclassified | unclassified |
| Streptococcus | Streptococcaceae | Lactobacillales |
| unclassified | unclassified | Eubacteriales |
| Brachyspira | Brachyspiraceae | Brachyspirales |
| unclassified | unclassified | unclassified |
| unclassified | unclassified | Eubacteriales |
| unclassified | unclassified | unclassified |
| unclassified | unclassified | Eubacteriales |
| unclassified | unclassified | unclassified |
| unclassified | Bacteroidaceae | Bacteroidales |
| Lacticaseibacillus | Lactobacillaceae | Lactobacillales |
| Lacticaseibacillus | Lactobacillaceae | Lactobacillales |

| | | |
|---|---|---|
| unclassified | unclassified | unclassified |
| Massilimicrobiota | Erysipelotrichaceae | Erysipelotrichales |
| unclassified | unclassified | unclassified |
| unclassified | unclassified | Bacteroidales |
| Ligilactobacillus | Lactobacillaceae | Lactobacillales |
| Enterococcus | Enterococcaceae | Lactobacillales |
| unclassified | unclassified | Eubacteriales |
| Bacteroides | Bacteroidaceae | Bacteroidales |
| Turicibacter | Turicibacteraceae | Erysipelotrichales |
| unclassified | unclassified | unclassified |
| unclassified | unclassified | Eubacteriales |
| Enterobacter | Enterobacteriaceae | Enterobacterales |
| unclassified | unclassified | unclassified |
| unclassified | unclassified | unclassified |
| unclassified | unclassified | unclassified |
| Alistipes | Rikenellaceae | Bacteroidales |
| unclassified | unclassified | Bacteroidales |
| unclassified | unclassified | unclassified |
| Roseburia | Lachnospiraceae | Eubacteriales |
| unclassified | unclassified | Eubacteriales |
| unclassified | unclassified | unclassified |
| unclassified | unclassified | unclassified |
| unclassified | unclassified | Eubacteriales |
| unclassified | unclassified | Eubacteriales |
| unclassified | unclassified | Bacteroidales |
| unclassified | Barnesiellaceae | Bacteroidales |
| Clostridium | Clostridiaceae | Eubacteriales |
| unclassified | unclassified | Eubacteriales |
| unclassified | unclassified | Eubacteriales |
| unclassified | unclassified | Eubacteriales |
| unclassified | unclassified | Eubacteriales |
| Porphyromonas | Porphyromonadaceae | Bacteroidales |
| unclassified | unclassified | unclassified |
| unclassified | unclassified | unclassified |
| unclassified | unclassified | unclassified |
| unclassified | unclassified | unclassified |
| unclassified | unclassified | unclassified |
| unclassified | unclassified | Bacteroidales |
| unclassified | Atopobiaceae | Coriobacteriales |
| unclassified | unclassified | unclassified |
| unclassified | unclassified | Bacteroidales |
| Clostridium | Clostridiaceae | Eubacteriales |
| unclassified | Actinomycetaceae | Actinomycetales |
| Roseburia | Lachnospiraceae | Eubacteriales |
| Ruminococcus | Oscillospiraceae | Eubacteriales |
| unclassified | unclassified | Eubacteriales |
| Clostridium | Clostridiaceae | Eubacteriales |
| unclassified | unclassified | Bacteroidales |

| | | |
|---|---|---|
| Phocaeicola | Bacteroidaceae | Bacteroidales |
| unclassified | unclassified | Eubacteriales |
| unclassified | unclassified | unclassified |
| unclassified | unclassified | Eubacteriales |
| Clostridium | Clostridiaceae | Eubacteriales |
| unclassified | unclassified | Bacteroidales |
| unclassified | Lachnospiraceae | Eubacteriales |
| unclassified | unclassified | Bacteroidales |
| unclassified | Lachnospiraceae | Eubacteriales |
| unclassified | unclassified | unclassified |
| unclassified | unclassified | Eubacteriales |
| unclassified | unclassified | unclassified |
| unclassified | unclassified | Eubacteriales |
| unclassified | unclassified | Eubacteriales |
| unclassified | unclassified | unclassified |
| unclassified | unclassified | Eubacteriales |
| unclassified | unclassified | unclassified |
| unclassified | unclassified | unclassified |
| unclassified | unclassified | unclassified |
| Gemmiger | unclassified | Eubacteriales |
| Culturomica | Odoribacteraceae | Bacteroidales |
| unclassified | Lachnospiraceae | Eubacteriales |
| unclassified | unclassified | unclassified |
| unclassified | unclassified | Methanomassiliicoccales |
| unclassified | unclassified | Eggerthellales |
| unclassified | unclassified | unclassified |
| unclassified | unclassified | Eubacteriales |
| unclassified | Oscillospiraceae | Eubacteriales |
| unclassified | Lachnospiraceae | Eubacteriales |
| Acidaminococcus | Acidaminococcaceae | Acidaminococcales |
| Eubacterium | Eubacteriaceae | Eubacteriales |
| Lactococcus | Streptococcaceae | Lactobacillales |
| unclassified | unclassified | unclassified |
| unclassified | unclassified | unclassified |
| unclassified | unclassified | Eubacteriales |
| unclassified | unclassified | Bacteroidales |
| unclassified | unclassified | unclassified |
| Anaerostipes | Lachnospiraceae | Eubacteriales |
| unclassified | unclassified | unclassified |
| unclassified | unclassified | Eubacteriales |
| unclassified | unclassified | unclassified |
| unclassified | unclassified | unclassified |
| unclassified | unclassified | Eubacteriales |
| Desulfovibrio | Desulfovibrionaceae | Desulfovibrionales |
| unclassified | unclassified | Eubacteriales |
| unclassified | unclassified | unclassified |
| unclassified | unclassified | unclassified |
| unclassified | Oscillospiraceae | Eubacteriales |

| | | |
|---|---|---|
| Methanobrevibacter | Methanobacteriaceae | Methanobacteriales |
| unclassified | unclassified | unclassified |
| unclassified | unclassified | unclassified |
| unclassified | Treponemataceae | Spirochaetales |
| unclassified | unclassified | unclassified |
| Sutterella | Sutterellaceae | Burkholderiales |
| unclassified | unclassified | unclassified |
| unclassified | unclassified | unclassified |
| unclassified | Bacteroidaceae | Bacteroidales |
| Clostridium | Clostridiaceae | Eubacteriales |
| Lacrimispora | Lachnospiraceae | Eubacteriales |
| unclassified | unclassified | Eubacteriales |
| Enterocloster | Lachnospiraceae | Eubacteriales |
| Acidovorax | Comamonadaceae | Burkholderiales |
| Comamonas | Comamonadaceae | Burkholderiales |
| Acinetobacter | Moraxellaceae | Moraxellales |
| Acinetobacter | Moraxellaceae | Moraxellales |
| Acinetobacter | Moraxellaceae | Moraxellales |
| Acinetobacter | Moraxellaceae | Moraxellales |
| Actinomyces | Actinomycetaceae | Actinomycetales |
| Actinomyces | Actinomycetaceae | Actinomycetales |
| Aeromonas | Aeromonadaceae | Aeromonadales |
| Bacteroides | Bacteroidaceae | Bacteroidales |
| Fenollaria | unclassified | Eubacteriales |
| Aliarcobacter | Arcobacteraceae | Campylobacterales |
| Methylobacterium | Methylobacteriaceae | Hyphomicrobiales |
| Atopobium | Atopobiaceae | Coriobacteriales |
| Bacteroides | Bacteroidaceae | Bacteroidales |
| Bifidobacterium | Bifidobacteriaceae | Bifidobacteriales |
| Blautia | Lachnospiraceae | Eubacteriales |
| Blautia | Lachnospiraceae | Eubacteriales |
| Turicimonas | Sutterellaceae | Burkholderiales |
| Butyricimonas | Odoribacteraceae | Bacteroidales |
| Lachnospira | Lachnospiraceae | Eubacteriales |
| Flavonifractor | Oscillospiraceae | Eubacteriales |
| Megasphaera | Veillonellaceae | Veillonellales |
| Caulobacter | Caulobacteraceae | Caulobacterales |
| Chryseobacterium | Weeksellaceae | Flavobacteriales |
| Citrobacter | Enterobacteriaceae | Enterobacterales |
| Citrobacter | Enterobacteriaceae | Enterobacterales |
| Citrobacter | Enterobacteriaceae | Enterobacterales |
| Citrobacter | Enterobacteriaceae | Enterobacterales |
| Citrobacter | Enterobacteriaceae | Enterobacterales |
| Citrobacter | Enterobacteriaceae | Enterobacterales |
| Cloacibacillus | Synergistaceae | Synergistales |
| unclassified | Oscillospiraceae | Eubacteriales |
| Dielma | Erysipelotrichaceae | Erysipelotrichales |
| Lachnoclostridium | Lachnospiraceae | Eubacteriales |

| | | |
|---|---|---|
| Clostridium | Clostridiaceae | Eubacteriales |
| Rubneribacter | Eggerthellaceae | Eggerthellales |
| Corynebacterium | Corynebacteriaceae | Corynebacteriales |
| Cutibacterium | Propionibacteriaceae | Propionibacteriales |
| Cutibacterium | Propionibacteriaceae | Propionibacteriales |
| Cutibacterium | Propionibacteriaceae | Propionibacteriales |
| Enterobacter | Enterobacteriaceae | Enterobacterales |
| Enterobacter | Enterobacteriaceae | Enterobacterales |
| Bacteroides | Bacteroidaceae | Bacteroidales |
| Enterobacter | Enterobacteriaceae | Enterobacterales |
| Phytobacter | Enterobacteriaceae | Enterobacterales |
| Enterococcus | Enterococcaceae | Lactobacillales |
| Enterococcus | Enterococcaceae | Lactobacillales |
| Enterococcus | Enterococcaceae | Lactobacillales |
| Enterococcus | Enterococcaceae | Lactobacillales |
| Enterococcus | Enterococcaceae | Lactobacillales |
| Erysipelatoclostridium | Erysipelotrichaceae | Erysipelotrichales |
| Escherichia | Enterobacteriaceae | Enterobacterales |
| Escherichia | Enterobacteriaceae | Enterobacterales |
| unclassified | unclassified | Eubacteriales |
| Eubacterium | Eubacteriaceae | Eubacteriales |
| Flavobacterium | Flavobacteriaceae | Flavobacteriales |
| Sanguibacteroides | Porphyromonadaceae | Bacteroidales |
| Gardnerella | Bifidobacteriaceae | Bifidobacteriales |
| Granulicatella | Carnobacteriaceae | Lactobacillales |
| Haemophilus | Pasteurellaceae | Pasteurellales |
| Hafnia | Hafniaceae | Enterobacterales |
| Kosakonia | Enterobacteriaceae | Enterobacterales |
| Klebsiella | Enterobacteriaceae | Enterobacterales |
| Lactobacillus | Lactobacillaceae | Lactobacillales |
| Ruthenibacterium | Oscillospiraceae | Eubacteriales |
| Lactobacillus | Lactobacillaceae | Lactobacillales |
| Limosilactobacillus | Lactobacillaceae | Lactobacillales |
| Limosilactobacillus | Lactobacillaceae | Lactobacillales |
| Lysinibacillus | Bacillaceae | Bacillales |
| Megasphaera | Veillonellaceae | Veillonellales |
| Megasphaera | Veillonellaceae | Veillonellales |
| Morganella | Morganellaceae | Enterobacterales |
| Mycobacteroides | Mycobacteriaceae | Corynebacteriales |
| Neisseria | Neisseriaceae | Neisseriales |
| Pantoea | Erwiniaceae | Enterobacterales |
| Sutterella | Sutterellaceae | Burkholderiales |
| Parabacteroides | Tannerellaceae | Bacteroidales |
| Prevotella | Prevotellaceae | Bacteroidales |
| Prevotella | Prevotellaceae | Bacteroidales |
| Pseudoflavonifractor | Oscillospiraceae | Eubacteriales |
| Pseudomonas | Pseudomonadaceae | Pseudomonadales |
| Pseudomonas | Pseudomonadaceae | Pseudomonadales |

| | | |
|---|---|---|
| Pseudomonas | Pseudomonadaceae | Pseudomonadales |
| Robinsoniella | Lachnospiraceae | Eubacteriales |
| Rothia | Micrococcaceae | Micrococcales |
| Selenomonas | Selenomonadaceae | Selenomonadales |
| Ruminococcus | Oscillospiraceae | Eubacteriales |
| Serratia | Yersiniaceae | Enterobacterales |
| Shewanella | Shewanellaceae | Alteromonadales |
| Sphingomonas | Sphingomonadaceae | Sphingomonadales |
| Staphylococcus | Staphylococcaceae | Bacillales |
| Staphylococcus | Staphylococcaceae | Bacillales |
| Staphylococcus | Staphylococcaceae | Bacillales |
| Stenotrophomonas | Xanthomonadaceae | Xanthomonadales |
| Stenotrophomonas | Xanthomonadaceae | Xanthomonadales |
| Streptococcus | Streptococcaceae | Lactobacillales |
| Streptococcus | Streptococcaceae | Lactobacillales |
| Faecalibacterium | Oscillospiraceae | Eubacteriales |
| Streptococcus | Streptococcaceae | Lactobacillales |
| Streptococcus | Streptococcaceae | Lactobacillales |
| Veillonella | Veillonellaceae | Veillonellales |
| Citrobacter | Enterobacteriaceae | Enterobacterales |
| Kluyvera | Enterobacteriaceae | Enterobacterales |
| unclassified | Lachnospiraceae | Eubacteriales |
| Citrobacter | Enterobacteriaceae | Enterobacterales |
| unclassified | Lachnospiraceae | Eubacteriales |
| Clostridium | Clostridiaceae | Eubacteriales |
| Enterococcus | Enterococcaceae | Lactobacillales |
| unclassified | unclassified | Eubacteriales |
| Clostridium | Clostridiaceae | Eubacteriales |
| Atlantibacter | Enterobacteriaceae | Enterobacterales |
| Terrisporobacter | Peptostreptococcaceae | Eubacteriales |
| Clostridium | Clostridiaceae | Eubacteriales |
| unclassified | Enterobacteriaceae | Enterobacterales |
| unclassified | Enterobacteriaceae | Enterobacterales |
| Corynebacterium | Corynebacteriaceae | Corynebacteriales |
| Macrococcus | Staphylococcaceae | Bacillales |
| Enterobacter | Enterobacteriaceae | Enterobacterales |
| Anaerosporobacter | Lachnospiraceae | Eubacteriales |
| Haemophilus | Pasteurellaceae | Pasteurellales |
| Veillonella | Veillonellaceae | Veillonellales |
| Lachnoclostridium | Lachnospiraceae | Eubacteriales |
| Limosilactobacillus | Lactobacillaceae | Lactobacillales |
| unclassified | Peptostreptococcaceae | Eubacteriales |
| unclassified | unclassified | Eubacteriales |
| Butyricicoccus | Clostridiaceae | Eubacteriales |
| Raoultella | Enterobacteriaceae | Enterobacterales |
| Clostridium | Clostridiaceae | Eubacteriales |
| unclassified | unclassified | unclassified |
| unclassified | Erysipelotrichaceae | Erysipelotrichales |

| | | |
|---|---|---|
| Ruminococcus | Oscillospiraceae | Eubacteriales |
| Phocaeicola | Bacteroidaceae | Bacteroidales |
| Rothia | Micrococcaceae | Micrococcales |
| unclassified | Enterobacteriaceae | Enterobacterales |
| Schaalia | Actinomycetaceae | Actinomycetales |
| Clostridium | Clostridiaceae | Eubacteriales |
| Citrobacter | Enterobacteriaceae | Enterobacterales |
| Isoptericola | Promicromonosporaceae | Micrococcales |
| unclassified | Peptostreptococcaceae | Eubacteriales |
| Bifidobacterium | Bifidobacteriaceae | Bifidobacteriales |
| Eisenbergiella | Lachnospiraceae | Eubacteriales |
| Enterobacter | Enterobacteriaceae | Enterobacterales |
| Streptococcus | Streptococcaceae | Lactobacillales |
| unclassified | Erysipelotrichaceae | Erysipelotrichales |
| unclassified | Lachnospiraceae | Eubacteriales |
| Candida | Debaryomycetaceae | Saccharomycetales |
| Klebsiella | Enterobacteriaceae | Enterobacterales |
| unclassified | unclassified | Eubacteriales |
| unclassified | unclassified | Eubacteriales |
| unclassified | unclassified | Eubacteriales |
| unclassified | Lachnospiraceae | Eubacteriales |
| Eubacterium | Eubacteriaceae | Eubacteriales |
| Atlantibacter | Enterobacteriaceae | Enterobacterales |
| unclassified | unclassified | Eubacteriales |
| Proteus | Morganellaceae | Enterobacterales |
| unclassified | Veillonellaceae | Veillonellales |
| Enterococcus | Enterococcaceae | Lactobacillales |
| unclassified | unclassified | unclassified |
| Eubacterium | Eubacteriaceae | Eubacteriales |
| Luxibacter | Lachnospiraceae | Eubacteriales |
| Eggerthella | Eggerthellaceae | Eggerthellales |
| unclassified | Erysipelotrichaceae | Erysipelotrichales |
| Lacrimispora | Lachnospiraceae | Eubacteriales |
| unclassified | unclassified | Eubacteriales |
| Dialister | Veillonellaceae | Veillonellales |
| unclassified | unclassified | unclassified |
| unclassified | Lachnospiraceae | Eubacteriales |
| unclassified | unclassified | Erysipelotrichales |
| unclassified | Tannerellaceae | Bacteroidales |
| unclassified | unclassified | Eubacteriales |
| unclassified | unclassified | Eubacteriales |
| unclassified | unclassified | Eubacteriales |
| unclassified | unclassified | unclassified |
| Erysipelatoclostridium | Erysipelotrichaceae | Erysipelotrichales |
| unclassified | unclassified | Eubacteriales |
| unclassified | unclassified | Eubacteriales |
| unclassified | Clostridiaceae | Eubacteriales |
| unclassified | Lachnospiraceae | Eubacteriales |

| | | |
|---|---|---|
| unclassified | unclassified | Eubacteriales |
| Enterococcus | Enterococcaceae | Lactobacillales |
| unclassified | Eubacteriaceae | Eubacteriales |
| Limosilactobacillus | Lactobacillaceae | Lactobacillales |
| unclassified | unclassified | Erysipelotrichales |
| Lachnoclostridium | Lachnospiraceae | Eubacteriales |
| unclassified | unclassified | Eubacteriales |
| Corynebacterium | Corynebacteriaceae | Corynebacteriales |
| unclassified | unclassified | unclassified |
| Enterococcus | Enterococcaceae | Lactobacillales |
| Actinomyces | Actinomycetaceae | Actinomycetales |
| Akkermansia | Akkermansiaceae | Verrucomicrobiales |
| Beduini | Clostridiaceae | Eubacteriales |
| Actinomyces | Actinomycetaceae | Actinomycetales |
| Collinsella | Coriobacteriaceae | Coriobacteriales |
| unclassified | unclassified | Eubacteriales |
| unclassified | unclassified | Bacteroidales |
| unclassified | Lachnospiraceae | Eubacteriales |
| Extibacter | Lachnospiraceae | Eubacteriales |
| Alistipes | Rikenellaceae | Bacteroidales |
| Bifidobacterium | Bifidobacteriaceae | Bifidobacteriales |
| Senegalimassilia | Coriobacteriaceae | Coriobacteriales |
| Christensenella | Christensenellaceae | Eubacteriales |
| unclassified | Oscillospiraceae | Eubacteriales |
| unclassified | unclassified | unclassified |
| unclassified | unclassified | Erysipelotrichales |
| unclassified | Lachnospiraceae | Eubacteriales |
| unclassified | Oscillospiraceae | Eubacteriales |
| unclassified | Lachnospiraceae | Eubacteriales |
| unclassified | Lachnospiraceae | Eubacteriales |
| Clostridium | Clostridiaceae | Eubacteriales |
| Kluyvera | Enterobacteriaceae | Enterobacterales |
| unclassified | Lachnospiraceae | Eubacteriales |
| unclassified | unclassified | Eubacteriales |
| unclassified | unclassified | Eubacteriales |
| Eubacterium | Eubacteriaceae | Eubacteriales |
| unclassified | Atopobiaceae | Coriobacteriales |
| Kluyvera | Enterobacteriaceae | Enterobacterales |
| Collinsella | Coriobacteriaceae | Coriobacteriales |
| Collinsella | Coriobacteriaceae | Coriobacteriales |
| Gordonibacter | Eggerthellaceae | Eggerthellales |
| Anaerobutyricum | Lachnospiraceae | Eubacteriales |
| Kluyvera | Enterobacteriaceae | Enterobacterales |
| unclassified | unclassified | Eubacteriales |
| unclassified | Oscillospiraceae | Eubacteriales |
| Sellimonas | Lachnospiraceae | Eubacteriales |
| Anaerococcus | Peptoniphilaceae | Tissierellales |
| unclassified | unclassified | Eubacteriales |

| | | |
|---|---|---|
| Veillonella | Veillonellaceae | Veillonellales |
| Alistipes | Rikenellaceae | Bacteroidales |
| Streptococcus | Streptococcaceae | Lactobacillales |
| unclassified | unclassified | Eubacteriales |
| Ruminococcus | Oscillospiraceae | Eubacteriales |
| Erysipelatoclostridium | Erysipelotrichaceae | Erysipelotrichales |
| Clostridium | Clostridiaceae | Eubacteriales |
| Enterobacter | Enterobacteriaceae | Enterobacterales |
| Anaerofustis | Eubacteriaceae | Eubacteriales |
| unclassified | unclassified | Eubacteriales |
| Bilophila | Desulfovibrionaceae | Desulfovibrionales |
| Anaerostipes | Lachnospiraceae | Eubacteriales |
| unclassified | unclassified | Erysipelotrichales |
| Actinomyces | Actinomycetaceae | Actinomycetales |
| Clostridium | Clostridiaceae | Eubacteriales |
| Lacrimispora | Lachnospiraceae | Eubacteriales |
| Veillonella | Veillonellaceae | Veillonellales |
| Eubacterium | Eubacteriaceae | Eubacteriales |
| unclassified | unclassified | Eubacteriales |
| Collinsella | Coriobacteriaceae | Coriobacteriales |
| unclassified | Eubacteriaceae | Eubacteriales |
| Clostridium | Clostridiaceae | Eubacteriales |
| unclassified | unclassified | Eubacteriales |
| Slackia | Eggerthellaceae | Eggerthellales |
| Christensenella | Christensenellaceae | Eubacteriales |
| Eubacterium | Eubacteriaceae | Eubacteriales |
| Eggerthella | Eggerthellaceae | Eggerthellales |
| Adlercreutzia | Eggerthellaceae | Eggerthellales |
| Erysipelatoclostridium | Erysipelotrichaceae | Erysipelotrichales |
| unclassified | unclassified | Eubacteriales |
| Christensenella | Christensenellaceae | Eubacteriales |
| Streptococcus | Streptococcaceae | Lactobacillales |
| Collinsella | Coriobacteriaceae | Coriobacteriales |
| Klebsiella | Enterobacteriaceae | Enterobacterales |
| Limosilactobacillus | Lactobacillaceae | Lactobacillales |
| unclassified | Oscillospiraceae | Eubacteriales |
| unclassified | Carnobacteriaceae | Lactobacillales |
| Agathobaculum | Oscillospiraceae | Eubacteriales |
| Eubacterium | Eubacteriaceae | Eubacteriales |
| unclassified | unclassified | Eubacteriales |
| Bilophila | Desulfovibrionaceae | Desulfovibrionales |
| unclassified | unclassified | Eubacteriales |
| unclassified | Lachnospiraceae | Eubacteriales |
| Mogibacterium | Eubacteriales Family XIII. Incertae Sedis | Eubacteriales |
| unclassified | unclassified | Eubacteriales |
| Limosilactobacillus | Lactobacillaceae | Lactobacillales |
| unclassified | unclassified | Erysipelotrichales |
| Mediterraneibacter | Lachnospiraceae | Eubacteriales |

| | | |
|---|---|---|
| unclassified | Lachnospiraceae | Eubacteriales |
| Streptococcus | Streptococcaceae | Lactobacillales |
| Corynebacterium | Corynebacteriaceae | Corynebacteriales |
| unclassified | unclassified | Bacteroidales |
| Eggerthella | Eggerthellaceae | Eggerthellales |
| Robinsoniella | Lachnospiraceae | Eubacteriales |
| Blautia | Lachnospiraceae | Eubacteriales |
| Acinetobacter | Moraxellaceae | Moraxellales |
| unclassified | unclassified | unclassified |
| Candidatus Stoquefichus | Erysipelotrichaceae | Erysipelotrichales |
| Dielma | Erysipelotrichaceae | Erysipelotrichales |
| Streptococcus | Streptococcaceae | Lactobacillales |
| unclassified | unclassified | Eubacteriales |
| unclassified | unclassified | Eubacteriales |
| Lancefieldella | Atopobiaceae | Coriobacteriales |
| unclassified | Oscillospiraceae | Eubacteriales |
| Streptococcus | Streptococcaceae | Lactobacillales |
| Ruminococcus | Oscillospiraceae | Eubacteriales |
| unclassified | Oscillospiraceae | Eubacteriales |
| unclassified | unclassified | Eubacteriales |
| unclassified | unclassified | unclassified |
| Leuconostoc | Lactobacillaceae | Lactobacillales |
| Bifidobacterium | Bifidobacteriaceae | Bifidobacteriales |
| Faecalibacterium | Oscillospiraceae | Eubacteriales |
| Lactococcus | Streptococcaceae | Lactobacillales |
| Faecalicatena | Lachnospiraceae | Eubacteriales |
| Raoultibacter | Eggerthellaceae | Eggerthellales |
| Collinsella | Coriobacteriaceae | Coriobacteriales |
| Escherichia | Enterobacteriaceae | Enterobacterales |
| Enterocloster | Lachnospiraceae | Eubacteriales |
| unclassified | unclassified | Eubacteriales |
| Eisenbergiella | Lachnospiraceae | Eubacteriales |
| Intestinimonas | unclassified | Eubacteriales |
| unclassified | Eggerthellaceae | Eggerthellales |
| Phascolarctobacterium | Acidaminococcaceae | Acidaminococcales |
| Blautia | Lachnospiraceae | Eubacteriales |
| unclassified | unclassified | Eubacteriales |
| unclassified | unclassified | Eubacteriales |
| Enterobacter | Enterobacteriaceae | Enterobacterales |
| unclassified | Oscillospiraceae | Eubacteriales |
| unclassified | unclassified | unclassified |
| unclassified | Lachnospiraceae | Eubacteriales |
| unclassified | unclassified | Eubacteriales |
| Staphylococcus | Staphylococcaceae | Bacillales |
| unclassified | Lachnospiraceae | Eubacteriales |
| unclassified | Oscillospiraceae | Eubacteriales |
| unclassified | unclassified | Erysipelotrichales |
| Eggerthella | Eggerthellaceae | Eggerthellales |

| | | |
|---|---|---|
| Amedibacillus | Erysipelotrichaceae | Erysipelotrichales |
| Agathobaculum | Oscillospiraceae | Eubacteriales |
| unclassified | unclassified | Eggerthellales |
| Faecalibacterium | Oscillospiraceae | Eubacteriales |
| unclassified | unclassified | Eubacteriales |
| Clostridium | Clostridiaceae | Eubacteriales |
| Bacteroides | Bacteroidaceae | Bacteroidales |
| unclassified | unclassified | unclassified |
| Prevotella | Prevotellaceae | Bacteroidales |
| Blautia | Lachnospiraceae | Eubacteriales |
| unclassified | unclassified | Eubacteriales |
| unclassified | unclassified | Eubacteriales |
| unclassified | unclassified | Eubacteriales |
| unclassified | unclassified | unclassified |
| Parabacteroides | Tannerellaceae | Bacteroidales |
| unclassified | unclassified | Eubacteriales |
| Blautia | Lachnospiraceae | Eubacteriales |
| unclassified | unclassified | unclassified |
| unclassified | unclassified | unclassified |
| Kineothrix | Lachnospiraceae | Eubacteriales |
| Butyrivibrio | Lachnospiraceae | Eubacteriales |
| unclassified | unclassified | Eubacteriales |
| unclassified | unclassified | unclassified |
| Evtepia | unclassified | Eubacteriales |
| Bacteroides | Bacteroidaceae | Bacteroidales |
| Subdoligranulum | Oscillospiraceae | Eubacteriales |
| unclassified | Oscillospiraceae | Eubacteriales |
| Catenibacterium | Coprobacillaceae | Erysipelotrichales |
| unclassified | unclassified | Eubacteriales |
| unclassified | unclassified | unclassified |
| unclassified | Lachnospiraceae | Eubacteriales |
| unclassified | Oscillospiraceae | Eubacteriales |
| Alistipes | Rikenellaceae | Bacteroidales |
| unclassified | unclassified | unclassified |
| unclassified | unclassified | unclassified |
| unclassified | unclassified | Bacteroidales |
| Faecalibacterium | Oscillospiraceae | Eubacteriales |
| unclassified | unclassified | unclassified |
| Alistipes | Rikenellaceae | Bacteroidales |
| Butyricimonas | Odoribacteraceae | Bacteroidales |
| Intestinibacter | Peptostreptococcaceae | Eubacteriales |
| unclassified | unclassified | Eubacteriales |
| Bifidobacterium | Bifidobacteriaceae | Bifidobacteriales |
| Butyricimonas | Odoribacteraceae | Bacteroidales |
| unclassified | unclassified | unclassified |
| unclassified | unclassified | Eubacteriales |
| unclassified | unclassified | Eubacteriales |
| Phocaeicola | Bacteroidaceae | Bacteroidales |

| | | |
|---|---|---|
| unclassified | unclassified | Eubacteriales |
| unclassified | unclassified | unclassified |
| Erysipelatoclostridium | Erysipelotrichaceae | Erysipelotrichales |
| unclassified | unclassified | Eubacteriales |
| Faecalibacillus | Erysipelotrichaceae | Erysipelotrichales |
| unclassified | unclassified | Eubacteriales |
| Phocaeicola | Bacteroidaceae | Bacteroidales |
| Bacteroides | Bacteroidaceae | Bacteroidales |
| unclassified | Oscillospiraceae | Eubacteriales |
| unclassified | unclassified | Bacteroidales |
| Bacteroides | Bacteroidaceae | Bacteroidales |
| Collinsella | Coriobacteriaceae | Coriobacteriales |
| unclassified | unclassified | Bacteroidales |
| Veillonella | Veillonellaceae | Veillonellales |
| Eggerthella | Eggerthellaceae | Eggerthellales |
| Faecalibacterium | Oscillospiraceae | Eubacteriales |
| Coprobacter | Barnesiellaceae | Bacteroidales |
| unclassified | unclassified | unclassified |
| Gemmiger | unclassified | Eubacteriales |
| unclassified | unclassified | Eubacteriales |
| Romboutsia | Peptostreptococcaceae | Eubacteriales |
| Bifidobacterium | Bifidobacteriaceae | Bifidobacteriales |
| Bifidobacterium | Bifidobacteriaceae | Bifidobacteriales |
| unclassified | unclassified | unclassified |
| unclassified | unclassified | Eubacteriales |
| Holdemanella | Erysipelotrichaceae | Erysipelotrichales |
| Akkermansia | Akkermansiaceae | Verrucomicrobiales |
| Duodenibacillus | Sutterellaceae | Burkholderiales |
| Alistipes | Rikenellaceae | Bacteroidales |
| unclassified | unclassified | Eggerthellales |
| unclassified | unclassified | Eubacteriales |
| unclassified | unclassified | Eubacteriales |
| Tyzzerella | Lachnospiraceae | Eubacteriales |
| Alistipes | Rikenellaceae | Bacteroidales |
| Clostridium | Clostridiaceae | Eubacteriales |
| Clostridium | Clostridiaceae | Eubacteriales |
| Parabacteroides | Tannerellaceae | Bacteroidales |
| unclassified | unclassified | unclassified |
| unclassified | unclassified | Veillonellales |
| unclassified | unclassified | Eubacteriales |
| unclassified | unclassified | Eubacteriales |
| unclassified | Lachnospiraceae | Eubacteriales |
| Coprococcus | Lachnospiraceae | Eubacteriales |
| unclassified | unclassified | Eubacteriales |
| Bacteroides | Bacteroidaceae | Bacteroidales |
| Gemmiger | unclassified | Eubacteriales |
| unclassified | unclassified | unclassified |
| Megamonas | Selenomonadaceae | Selenomonadales |

| | | |
|---|---|---|
| unclassified | unclassified | Eubacteriales |
| unclassified | unclassified | unclassified |
| Parasutterella | Sutterellaceae | Burkholderiales |
| unclassified | Oscillospiraceae | Eubacteriales |
| Pseudoruminococcus | Oscillospiraceae | Eubacteriales |
| unclassified | Oscillospiraceae | Eubacteriales |
| Blautia | Lachnospiraceae | Eubacteriales |
| Bacteroides | Bacteroidaceae | Bacteroidales |
| unclassified | Lachnospiraceae | Eubacteriales |
| unclassified | unclassified | Eubacteriales |
| Enterocloster | Lachnospiraceae | Eubacteriales |
| unclassified | unclassified | Acidaminococcales |
| Desulfovibrio | Desulfovibrionaceae | Desulfovibrionales |
| Blautia | Lachnospiraceae | Eubacteriales |
| Roseburia | Lachnospiraceae | Eubacteriales |
| unclassified | unclassified | Eubacteriales |
| Coprococcus | Lachnospiraceae | Eubacteriales |
| Clostridium | Clostridiaceae | Eubacteriales |
| Ruminococcus | Oscillospiraceae | Eubacteriales |
| unclassified | unclassified | Eubacteriales |
| Alistipes | Rikenellaceae | Bacteroidales |
| unclassified | unclassified | unclassified |
| Waltera | Lachnospiraceae | Eubacteriales |
| Acidaminococcus | Acidaminococcaceae | Acidaminococcales |
| unclassified | unclassified | unclassified |
| unclassified | unclassified | unclassified |
| unclassified | unclassified | Eubacteriales |
| unclassified | unclassified | Eubacteriales |
| unclassified | Clostridiaceae | Eubacteriales |
| Dorea | Lachnospiraceae | Eubacteriales |
| Hydrogenoanaerobacterium | Oscillospiraceae | Eubacteriales |
| unclassified | unclassified | unclassified |
| unclassified | Oscillospiraceae | Eubacteriales |
| Eubacterium | Eubacteriaceae | Eubacteriales |
| Clostridium | Clostridiaceae | Eubacteriales |
| unclassified | unclassified | Eubacteriales |
| Dorea | Lachnospiraceae | Eubacteriales |
| unclassified | Oscillospiraceae | Eubacteriales |
| unclassified | unclassified | unclassified |
| Flintibacter | unclassified | Eubacteriales |
| Barnesiella | Barnesiellaceae | Bacteroidales |
| unclassified | unclassified | Eubacteriales |
| Oscillibacter | Oscillospiraceae | Eubacteriales |
| Blautia | Lachnospiraceae | Eubacteriales |
| Slackia | Eggerthellaceae | Eggerthellales |
| unclassified | unclassified | Eubacteriales |
| unclassified | unclassified | unclassified |
| unclassified | unclassified | Eubacteriales |

| | | |
|---|---|---|
| Faecalibacterium | Oscillospiraceae | Eubacteriales |
| unclassified | unclassified | unclassified |
| unclassified | Oscillospiraceae | Eubacteriales |
| Oscillibacter | Oscillospiraceae | Eubacteriales |
| unclassified | unclassified | unclassified |
| unclassified | unclassified | Eubacteriales |
| unclassified | unclassified | unclassified |
| unclassified | unclassified | Eubacteriales |
| unclassified | Erysipelotrichaceae | Erysipelotrichales |
| Colidextribacter | unclassified | Eubacteriales |
| unclassified | unclassified | unclassified |
| Ruminococcus | Oscillospiraceae | Eubacteriales |
| Butyricimonas | Odoribacteraceae | Bacteroidales |
| Bacteroides | Bacteroidaceae | Bacteroidales |
| Anaerobutyricum | Lachnospiraceae | Eubacteriales |
| Sellimonas | Lachnospiraceae | Eubacteriales |
| Parabacteroides | Tannerellaceae | Bacteroidales |
| Clostridium | Clostridiaceae | Eubacteriales |
| unclassified | unclassified | Eubacteriales |
| unclassified | unclassified | unclassified |
| unclassified | unclassified | unclassified |
| unclassified | unclassified | unclassified |
| unclassified | unclassified | Eubacteriales |
| Monoglobus | Oscillospiraceae | Eubacteriales |
| Escherichia | Enterobacteriaceae | Enterobacterales |
| unclassified | unclassified | unclassified |
| Faecalibacterium | Oscillospiraceae | Eubacteriales |
| unclassified | unclassified | unclassified |
| Prevotella | Prevotellaceae | Bacteroidales |
| unclassified | unclassified | unclassified |
| unclassified | unclassified | unclassified |
| unclassified | unclassified | Eubacteriales |
| Candidatus Borkfalkia | Candidatus Borkfalkiaceae | Candidatus Borkfalkiales |
| Bacteroides | Bacteroidaceae | Bacteroidales |
| Veillonella | Veillonellaceae | Veillonellales |
| Lachnoclostridium | Lachnospiraceae | Eubacteriales |
| unclassified | unclassified | Eubacteriales |
| Blautia | Lachnospiraceae | Eubacteriales |
| Ligilactobacillus | Lactobacillaceae | Lactobacillales |
| Mesosutterella | Sutterellaceae | Burkholderiales |
| Bifidobacterium | Bifidobacteriaceae | Bifidobacteriales |
| Ruminococcus | Oscillospiraceae | Eubacteriales |
| Ruminococcus | Oscillospiraceae | Eubacteriales |
| unclassified | unclassified | unclassified |
| unclassified | unclassified | unclassified |
| Intestinimonas | unclassified | Eubacteriales |
| Eubacterium | Eubacteriaceae | Eubacteriales |
| unclassified | Clostridiaceae | Eubacteriales |

| | | |
|---|---|---|
| Alistipes | Rikenellaceae | Bacteroidales |
| unclassified | unclassified | Eubacteriales |
| Blautia | Lachnospiraceae | Eubacteriales |
| unclassified | unclassified | Eubacteriales |
| Bacteroides | Bacteroidaceae | Bacteroidales |
| Prevotella | Prevotellaceae | Bacteroidales |
| Roseburia | Lachnospiraceae | Eubacteriales |
| unclassified | unclassified | Eubacteriales |
| Ruminococcus | Oscillospiraceae | Eubacteriales |
| unclassified | unclassified | Eubacteriales |
| unclassified | unclassified | Eubacteriales |
| Gemmiger | unclassified | Eubacteriales |
| Faecalibacillus | Erysipelotrichaceae | Erysipelotrichales |
| unclassified | unclassified | unclassified |
| Klebsiella | Enterobacteriaceae | Enterobacterales |
| unclassified | unclassified | unclassified |
| Bacteroides | Bacteroidaceae | Bacteroidales |
| Alistipes | Rikenellaceae | Bacteroidales |
| Parabacteroides | Tannerellaceae | Bacteroidales |
| unclassified | unclassified | Eubacteriales |
| unclassified | unclassified | Eubacteriales |
| unclassified | unclassified | Eubacteriales |
| Roseburia | Lachnospiraceae | Eubacteriales |
| Eubacterium | Eubacteriaceae | Eubacteriales |
| unclassified | unclassified | Eubacteriales |
| unclassified | unclassified | Eubacteriales |
| unclassified | unclassified | Bacteroidales |
| Akkermansia | Akkermansiaceae | Verrucomicrobiales |
| unclassified | Oscillospiraceae | Eubacteriales |
| unclassified | unclassified | Eubacteriales |
| unclassified | unclassified | unclassified |
| Streptococcus | Streptococcaceae | Lactobacillales |
| unclassified | unclassified | Eubacteriales |
| Odoribacter | Odoribacteraceae | Bacteroidales |
| Lachnoclostridium | Lachnospiraceae | Eubacteriales |
| Prevotella | Prevotellaceae | Bacteroidales |
| unclassified | unclassified | Eubacteriales |
| Prevotella | Prevotellaceae | Bacteroidales |
| Sutterella | Sutterellaceae | Burkholderiales |
| unclassified | Oscillospiraceae | Eubacteriales |
| Blautia | Lachnospiraceae | Eubacteriales |
| unclassified | unclassified | Eubacteriales |
| unclassified | unclassified | unclassified |
| Eubacterium | Eubacteriaceae | Eubacteriales |
| Bacteroides | Bacteroidaceae | Bacteroidales |
| unclassified | unclassified | unclassified |
| Dorea | Lachnospiraceae | Eubacteriales |
| Dysosmobacter | Oscillospiraceae | Eubacteriales |

| | | |
|---|---|---|
| Anaerostipes | Lachnospiraceae | Eubacteriales |
| unclassified | unclassified | Eubacteriales |
| unclassified | unclassified | unclassified |
| unclassified | unclassified | Eubacteriales |
| unclassified | Lachnospiraceae | Eubacteriales |
| unclassified | unclassified | Eubacteriales |
| unclassified | unclassified | unclassified |
| Roseburia | Lachnospiraceae | Eubacteriales |
| Ruminococcus | Oscillospiraceae | Eubacteriales |
| unclassified | Lachnospiraceae | Eubacteriales |
| unclassified | unclassified | Eubacteriales |
| Enterocloster | Lachnospiraceae | Eubacteriales |
| unclassified | unclassified | unclassified |
| Ellagibacter | Eggerthellaceae | Eggerthellales |
| unclassified | unclassified | Eubacteriales |
| Butyricicoccus | Clostridiaceae | Eubacteriales |
| unclassified | unclassified | unclassified |
| unclassified | unclassified | unclassified |
| Bifidobacterium | Bifidobacteriaceae | Bifidobacteriales |
| Victivallis | Victivallaceae | Victivallales |
| Clostridium | Clostridiaceae | Eubacteriales |
| unclassified | unclassified | Eubacteriales |
| unclassified | unclassified | unclassified |
| unclassified | unclassified | Eggerthellales |
| Veillonella | Veillonellaceae | Veillonellales |
| Clostridium | Clostridiaceae | Eubacteriales |
| unclassified | unclassified | Eubacteriales |
| Clostridium | Clostridiaceae | Eubacteriales |
| Vescimonas | Oscillospiraceae | Eubacteriales |
| Coprococcus | Lachnospiraceae | Eubacteriales |
| unclassified | unclassified | unclassified |
| unclassified | unclassified | unclassified |
| unclassified | unclassified | unclassified |
| unclassified | Rikenellaceae | Bacteroidales |
| Hungatella | Clostridiaceae | Eubacteriales |
| unclassified | unclassified | unclassified |
| unclassified | unclassified | Bacteroidales |
| Parabacteroides | Tannerellaceae | Bacteroidales |
| unclassified | unclassified | unclassified |
| Butyricimonas | Odoribacteraceae | Bacteroidales |
| Blautia | Lachnospiraceae | Eubacteriales |
| unclassified | unclassified | Eubacteriales |
| Prevotellamassilia | Prevotellaceae | Bacteroidales |
| unclassified | unclassified | Eubacteriales |
| unclassified | unclassified | unclassified |
| unclassified | unclassified | Desulfovibrionales |
| unclassified | unclassified | Eubacteriales |
| Bacteroides | Bacteroidaceae | Bacteroidales |

| | | |
|---|---|---|
| Phocaeicola | Bacteroidaceae | Bacteroidales |
| unclassified | unclassified | Eubacteriales |
| unclassified | unclassified | unclassified |
| unclassified | unclassified | Eubacteriales |
| Ruminococcus | Oscillospiraceae | Eubacteriales |
| unclassified | unclassified | Eubacteriales |
| unclassified | unclassified | Eubacteriales |
| unclassified | unclassified | Eubacteriales |
| Blautia | Lachnospiraceae | Eubacteriales |
| unclassified | unclassified | Eubacteriales |
| Clostridium | Clostridiaceae | Eubacteriales |
| unclassified | unclassified | unclassified |
| unclassified | unclassified | Eubacteriales |
| Blautia | Lachnospiraceae | Eubacteriales |
| Prevotella | Prevotellaceae | Bacteroidales |
| Blautia | Lachnospiraceae | Eubacteriales |
| Clostridium | Clostridiaceae | Eubacteriales |
| unclassified | unclassified | Bacteroidales |
| unclassified | unclassified | unclassified |
| unclassified | unclassified | Eubacteriales |
| unclassified | unclassified | unclassified |
| unclassified | unclassified | Eubacteriales |
| Bifidobacterium | Bifidobacteriaceae | Bifidobacteriales |
| unclassified | unclassified | unclassified |
| unclassified | unclassified | unclassified |
| unclassified | unclassified | unclassified |
| Lachnoclostridium | Lachnospiraceae | Eubacteriales |
| Enterocloster | Lachnospiraceae | Eubacteriales |
| unclassified | unclassified | unclassified |
| unclassified | unclassified | Eubacteriales |
| unclassified | unclassified | unclassified |
| unclassified | unclassified | unclassified |
| unclassified | unclassified | Eubacteriales |
| unclassified | unclassified | Eubacteriales |
| Prevotella | Prevotellaceae | Bacteroidales |
| Coprobacillus | Coprobacillaceae | Erysipelotrichales |
| Enterococcus | Enterococcaceae | Lactobacillales |
| Lachnospira | Lachnospiraceae | Eubacteriales |
| unclassified | unclassified | unclassified |
| unclassified | unclassified | unclassified |
| Holdemanella | Erysipelotrichaceae | Erysipelotrichales |
| Prevotella | Prevotellaceae | Bacteroidales |
| unclassified | unclassified | Eubacteriales |
| unclassified | unclassified | Aeromonadales |
| Akkermansia | Akkermansiaceae | Verrucomicrobiales |
| unclassified | unclassified | unclassified |
| unclassified | unclassified | unclassified |
| Coprococcus | Lachnospiraceae | Eubacteriales |

| | | |
|---|---|---|
| Faecalibacterium | Oscillospiraceae | Eubacteriales |
| unclassified | unclassified | Eubacteriales |
| Roseburia | Lachnospiraceae | Eubacteriales |
| unclassified | Oscillospiraceae | Eubacteriales |
| Blautia | Lachnospiraceae | Eubacteriales |
| unclassified | unclassified | Eubacteriales |
| unclassified | Peptostreptococcaceae | Eubacteriales |
| Alistipes | Rikenellaceae | Bacteroidales |
| Coprobacter | Barnesiellaceae | Bacteroidales |
| unclassified | unclassified | Eubacteriales |
| unclassified | unclassified | unclassified |
| Clostridium | Clostridiaceae | Eubacteriales |
| Sutterella | Sutterellaceae | Burkholderiales |
| Dialister | Veillonellaceae | Veillonellales |
| unclassified | unclassified | Eubacteriales |
| unclassified | unclassified | Eubacteriales |
| Bifidobacterium | Bifidobacteriaceae | Bifidobacteriales |
| Eisenbergiella | Lachnospiraceae | Eubacteriales |
| Prevotella | Prevotellaceae | Bacteroidales |
| unclassified | unclassified | unclassified |
| Clostridium | Clostridiaceae | Eubacteriales |
| unclassified | unclassified | unclassified |
| Dysosmobacter | Oscillospiraceae | Eubacteriales |
| unclassified | unclassified | Eubacteriales |
| Mitsuokella | Selenomonadaceae | Selenomonadales |
| Parabacteroides | Tannerellaceae | Bacteroidales |
| unclassified | unclassified | Bacteroidales |
| unclassified | Oscillospiraceae | Eubacteriales |
| unclassified | unclassified | unclassified |
| Streptococcus | Streptococcaceae | Lactobacillales |
| unclassified | unclassified | unclassified |
| unclassified | unclassified | Burkholderiales |
| unclassified | unclassified | Eubacteriales |
| Akkermansia | Akkermansiaceae | Verrucomicrobiales |
| unclassified | unclassified | Eubacteriales |
| unclassified | unclassified | unclassified |
| unclassified | Lachnospiraceae | Eubacteriales |
| unclassified | unclassified | unclassified |
| unclassified | unclassified | Bacteroidales |
| unclassified | unclassified | unclassified |
| unclassified | Rikenellaceae | Bacteroidales |
| unclassified | unclassified | unclassified |
| unclassified | unclassified | unclassified |
| unclassified | unclassified | Eubacteriales |
| unclassified | unclassified | unclassified |
| Mogibacterium | Eubacteriales Family XIII. Incertae Sedis | Eubacteriales |
| unclassified | unclassified | unclassified |
| unclassified | Prevotellaceae | Bacteroidales |

| | | |
|---|---|---|
| Eisenbergiella | Lachnospiraceae | Eubacteriales |
| unclassified | Prevotellaceae | Bacteroidales |
| Ruminococcus | Oscillospiraceae | Eubacteriales |
| Sutterella | Sutterellaceae | Burkholderiales |
| Odoribacter | Odoribacteraceae | Bacteroidales |
| unclassified | unclassified | unclassified |
| Bacteroides | Bacteroidaceae | Bacteroidales |
| Fusicatenibacter | Lachnospiraceae | Eubacteriales |
| Escherichia | Enterobacteriaceae | Enterobacterales |
| Anaerotignum | Lachnospiraceae | Eubacteriales |
| unclassified | Oscillospiraceae | Eubacteriales |
| unclassified | unclassified | unclassified |
| unclassified | unclassified | Eubacteriales |
| Megasphaera | Veillonellaceae | Veillonellales |
| Bacteroides | Bacteroidaceae | Bacteroidales |
| unclassified | unclassified | unclassified |
| Erysipelatoclostridium | Erysipelotrichaceae | Erysipelotrichales |
| unclassified | unclassified | Erysipelotrichales |
| unclassified | unclassified | Bacteroidales |
| Alistipes | Rikenellaceae | Bacteroidales |
| Methanosphaera | Methanobacteriaceae | Methanobacteriales |
| unclassified | unclassified | unclassified |
| unclassified | unclassified | Eubacteriales |
| unclassified | unclassified | Eubacteriales |
| Blautia | Lachnospiraceae | Eubacteriales |
| unclassified | unclassified | unclassified |
| unclassified | unclassified | Eubacteriales |
| Ruminococcus | Oscillospiraceae | Eubacteriales |
| unclassified | unclassified | Eubacteriales |
| unclassified | unclassified | Bacteroidales |
| Phocaeicola | Bacteroidaceae | Bacteroidales |
| unclassified | unclassified | Bacteroidales |
| unclassified | unclassified | unclassified |
| Roseburia | Lachnospiraceae | Eubacteriales |
| Phascolarctobacterium | Acidaminococcaceae | Acidaminococcales |
| Butyricicoccus | Clostridiaceae | Eubacteriales |
| unclassified | unclassified | unclassified |
| Anaerostipes | Lachnospiraceae | Eubacteriales |
| Sutterella | Sutterellaceae | Burkholderiales |
| Coprococcus | Lachnospiraceae | Eubacteriales |
| unclassified | unclassified | unclassified |
| unclassified | unclassified | Eubacteriales |
| unclassified | unclassified | Eubacteriales |
| unclassified | unclassified | unclassified |
| Paraprevotella | Prevotellaceae | Bacteroidales |
| Flintibacter | unclassified | Eubacteriales |
| unclassified | unclassified | Eubacteriales |
| Victivallis | Victivallaceae | Victivallales |

| | | |
|---|---|---|
| unclassified | Bacteroidaceae | Bacteroidales |
| Bifidobacterium | Bifidobacteriaceae | Bifidobacteriales |
| unclassified | unclassified | Eubacteriales |
| unclassified | unclassified | Eubacteriales |
| Prevotella | Prevotellaceae | Bacteroidales |
| unclassified | unclassified | unclassified |
| Phocaeicola | Bacteroidaceae | Bacteroidales |
| Erysipelatoclostridium | Erysipelotrichaceae | Erysipelotrichales |
| unclassified | Oscillospiraceae | Eubacteriales |
| unclassified | unclassified | Eubacteriales |
| unclassified | unclassified | Eubacteriales |
| unclassified | unclassified | unclassified |
| unclassified | unclassified | unclassified |
| unclassified | unclassified | unclassified |
| Longicatena | Erysipelotrichaceae | Erysipelotrichales |
| Alistipes | Rikenellaceae | Bacteroidales |
| unclassified | unclassified | Eubacteriales |
| unclassified | unclassified | unclassified |
| unclassified | unclassified | unclassified |
| unclassified | unclassified | unclassified |
| unclassified | unclassified | Eubacteriales |
| unclassified | unclassified | unclassified |
| unclassified | unclassified | unclassified |
| Enterocloster | Lachnospiraceae | Eubacteriales |
| Olsenella | Atopobiaceae | Coriobacteriales |
| unclassified | Oscillospiraceae | Eubacteriales |
| Bacteroides | Bacteroidaceae | Bacteroidales |
| unclassified | Oscillospiraceae | Eubacteriales |
| Enterocloster | Lachnospiraceae | Eubacteriales |
| unclassified | unclassified | unclassified |
| Parasutterella | Sutterellaceae | Burkholderiales |
| Neglecta | unclassified | Eubacteriales |
| Ruminococcus | Oscillospiraceae | Eubacteriales |
| unclassified | unclassified | unclassified |
| Prevotella | Prevotellaceae | Bacteroidales |
| unclassified | unclassified | Eubacteriales |
| unclassified | unclassified | unclassified |
| Oscillibacter | Oscillospiraceae | Eubacteriales |
| unclassified | unclassified | Eubacteriales |
| Eubacterium | Eubacteriaceae | Eubacteriales |
| unclassified | unclassified | unclassified |
| Prevotella | Prevotellaceae | Bacteroidales |
| unclassified | unclassified | Eubacteriales |
| unclassified | unclassified | unclassified |
| unclassified | unclassified | Eubacteriales |
| unclassified | unclassified | Eubacteriales |
| unclassified | unclassified | unclassified |
| unclassified | unclassified | Eubacteriales |

| | | |
|---|---|---|
| unclassified | unclassified | unclassified |
| Methanobrevibacter | Methanobacteriaceae | Methanobacteriales |
| Negativibacillus | unclassified | unclassified |
| Bacteroides | Bacteroidaceae | Bacteroidales |
| unclassified | unclassified | unclassified |
| Bacteroides | Bacteroidaceae | Bacteroidales |
| unclassified | unclassified | unclassified |
| Lawsonibacter | unclassified | Eubacteriales |
| unclassified | unclassified | Eubacteriales |
| unclassified | unclassified | unclassified |
| unclassified | unclassified | Erysipelotrichales |
| Bacteroides | Bacteroidaceae | Bacteroidales |
| unclassified | unclassified | unclassified |
| unclassified | unclassified | unclassified |
| unclassified | unclassified | unclassified |
| unclassified | unclassified | unclassified |
| unclassified | unclassified | Eubacteriales |
| unclassified | unclassified | Eubacteriales |
| Butyricimonas | Odoribacteraceae | Bacteroidales |
| Butyricimonas | Odoribacteraceae | Bacteroidales |
| unclassified | unclassified | Eubacteriales |
| unclassified | unclassified | unclassified |
| Bacteroides | Bacteroidaceae | Bacteroidales |
| unclassified | unclassified | Eubacteriales |
| unclassified | unclassified | Eubacteriales |
| unclassified | unclassified | Bacteroidales |
| unclassified | unclassified | unclassified |
| unclassified | unclassified | unclassified |
| Megasphaera | Veillonellaceae | Veillonellales |
| unclassified | Oscillospiraceae | Eubacteriales |
| unclassified | Oscillospiraceae | Eubacteriales |
| unclassified | unclassified | Eubacteriales |
| unclassified | Lachnospiraceae | Eubacteriales |
| unclassified | Eubacteriaceae | Eubacteriales |
| Faecalibacillus | Erysipelotrichaceae | Erysipelotrichales |
| unclassified | unclassified | Eubacteriales |
| unclassified | unclassified | unclassified |
| unclassified | unclassified | Bacteroidales |
| unclassified | Clostridiaceae | Eubacteriales |
| unclassified | unclassified | Eubacteriales |
| unclassified | unclassified | unclassified |
| unclassified | Lachnospiraceae | Eubacteriales |
| Prevotella | Prevotellaceae | Bacteroidales |
| unclassified | unclassified | Eubacteriales |
| unclassified | unclassified | Erysipelotrichales |
| Waltera | Lachnospiraceae | Eubacteriales |
| Alistipes | Rikenellaceae | Bacteroidales |
| unclassified | unclassified | Eubacteriales |

| | | |
|---|---|---|
| unclassified | unclassified | unclassified |
| unclassified | unclassified | Bacteroidales |
| unclassified | unclassified | Eubacteriales |
| unclassified | unclassified | unclassified |
| unclassified | Oscillospiraceae | Eubacteriales |
| unclassified | unclassified | unclassified |
| unclassified | unclassified | Eubacteriales |
| Collinsella | Coriobacteriaceae | Coriobacteriales |
| unclassified | unclassified | unclassified |
| Allisonella | Veillonellaceae | Veillonellales |
| unclassified | unclassified | unclassified |
| unclassified | unclassified | Eubacteriales |
| unclassified | Oscillospiraceae | Eubacteriales |
| Blautia | Lachnospiraceae | Eubacteriales |
| unclassified | unclassified | Eubacteriales |
| Anaerostipes | Lachnospiraceae | Eubacteriales |
| unclassified | unclassified | Veillonellales |
| Blautia | Lachnospiraceae | Eubacteriales |
| unclassified | Veillonellaceae | Veillonellales |
| Blautia | Lachnospiraceae | Eubacteriales |
| Eubacterium | Eubacteriaceae | Eubacteriales |
| unclassified | unclassified | Eubacteriales |
| unclassified | unclassified | Eubacteriales |
| unclassified | unclassified | Eubacteriales |
| Bacteroides | Bacteroidaceae | Bacteroidales |
| unclassified | unclassified | Eubacteriales |
| unclassified | unclassified | unclassified |
| Mitsuokella | Selenomonadaceae | Selenomonadales |
| unclassified | unclassified | Eubacteriales |
| unclassified | Lachnospiraceae | Eubacteriales |
| Blautia | Lachnospiraceae | Eubacteriales |
| unclassified | unclassified | Eubacteriales |
| unclassified | unclassified | Eubacteriales |
| unclassified | unclassified | unclassified |
| unclassified | unclassified | unclassified |
| unclassified | unclassified | unclassified |
| unclassified | unclassified | unclassified |
| unclassified | unclassified | Eubacteriales |
| Parolsenella | Atopobiaceae | Coriobacteriales |
| unclassified | unclassified | Eubacteriales |
| unclassified | Oscillospiraceae | Eubacteriales |
| unclassified | Oscillospiraceae | Eubacteriales |
| unclassified | unclassified | Eubacteriales |
| Alistipes | Rikenellaceae | Bacteroidales |
| unclassified | unclassified | Eubacteriales |
| Prevotella | Prevotellaceae | Bacteroidales |
| Blautia | Lachnospiraceae | Eubacteriales |
| unclassified | unclassified | Desulfovibrionales |

| | | |
|---|---|---|
| unclassified | unclassified | Burkholderiales |
| unclassified | Oscillospiraceae | Eubacteriales |
| unclassified | Oscillospiraceae | Eubacteriales |
| unclassified | unclassified | Burkholderiales |
| Roseburia | Lachnospiraceae | Eubacteriales |
| unclassified | unclassified | unclassified |
| unclassified | unclassified | unclassified |
| Clostridium | Clostridiaceae | Eubacteriales |
| unclassified | unclassified | Eubacteriales |
| unclassified | unclassified | unclassified |
| unclassified | unclassified | Eubacteriales |
| Bacteroides | Bacteroidaceae | Bacteroidales |
| Bacteroides | Bacteroidaceae | Bacteroidales |
| unclassified | unclassified | Bacteroidales |
| Holdemania | Erysipelotrichaceae | Erysipelotrichales |
| Mediterraneibacter | Lachnospiraceae | Eubacteriales |
| unclassified | unclassified | Eubacteriales |
| Pseudoflavonifractor | Oscillospiraceae | Eubacteriales |
| Lachnoclostridium | Lachnospiraceae | Eubacteriales |
| Clostridium | Clostridiaceae | Eubacteriales |
| Prevotella | Prevotellaceae | Bacteroidales |
| unclassified | Lachnospiraceae | Eubacteriales |
| unclassified | unclassified | Eubacteriales |
| unclassified | unclassified | Bacteroidales |
| unclassified | unclassified | unclassified |
| unclassified | unclassified | unclassified |
| Oscillibacter | Oscillospiraceae | Eubacteriales |
| unclassified | unclassified | Eubacteriales |
| unclassified | unclassified | unclassified |
| unclassified | unclassified | unclassified |
| Hungatella | Clostridiaceae | Eubacteriales |
| Anaeromassilibacillus | Oscillospiraceae | Eubacteriales |
| unclassified | unclassified | Eubacteriales |
| unclassified | unclassified | Eubacteriales |
| unclassified | unclassified | Eubacteriales |
| unclassified | unclassified | unclassified |
| unclassified | unclassified | unclassified |
| Parabacteroides | Tannerellaceae | Bacteroidales |
| Faecalibacterium | Oscillospiraceae | Eubacteriales |
| unclassified | unclassified | Eubacteriales |
| Bacteroides | Bacteroidaceae | Bacteroidales |
| unclassified | unclassified | unclassified |
| unclassified | unclassified | unclassified |
| Lachnoclostridium | Lachnospiraceae | Eubacteriales |
| unclassified | unclassified | unclassified |
| unclassified | unclassified | Eubacteriales |
| unclassified | unclassified | Bacteroidales |
| unclassified | unclassified | unclassified |

| | | |
|---|---|---|
| unclassified | unclassified | unclassified |
| Faecalicatena | Lachnospiraceae | Eubacteriales |
| unclassified | Oscillospiraceae | Eubacteriales |
| Enterocloster | Lachnospiraceae | Eubacteriales |
| Megasphaera | Veillonellaceae | Veillonellales |
| unclassified | unclassified | Veillonellales |
| Intestinimonas | unclassified | Eubacteriales |
| Desulfovibrio | Desulfovibrionaceae | Desulfovibrionales |
| unclassified | unclassified | Eubacteriales |
| unclassified | unclassified | Eubacteriales |
| unclassified | unclassified | Eubacteriales |
| unclassified | unclassified | Eubacteriales |
| Clostridium | Clostridiaceae | Eubacteriales |
| unclassified | unclassified | Eubacteriales |
| unclassified | unclassified | Desulfovibrionales |
| unclassified | unclassified | Eubacteriales |
| unclassified | unclassified | Bacteroidales |
| unclassified | unclassified | Eubacteriales |
| unclassified | Sutterellaceae | Burkholderiales |
| unclassified | Oscillospiraceae | Eubacteriales |
| unclassified | unclassified | unclassified |
| unclassified | unclassified | unclassified |
| unclassified | unclassified | Eubacteriales |
| Blautia | Lachnospiraceae | Eubacteriales |
| unclassified | unclassified | unclassified |
| unclassified | unclassified | Eubacteriales |
| Eubacterium | Eubacteriaceae | Eubacteriales |
| Desulfovibrio | Desulfovibrionaceae | Desulfovibrionales |
| unclassified | unclassified | unclassified |
| unclassified | unclassified | Bacteroidales |
| Coprococcus | Lachnospiraceae | Eubacteriales |
| Alistipes | Rikenellaceae | Bacteroidales |
| Sutterella | Sutterellaceae | Burkholderiales |
| unclassified | unclassified | unclassified |
| Clostridium | Clostridiaceae | Eubacteriales |
| Anaerostipes | Lachnospiraceae | Eubacteriales |
| Blautia | Lachnospiraceae | Eubacteriales |
| unclassified | unclassified | Eubacteriales |
| Enterobacter | Enterobacteriaceae | Enterobacterales |
| unclassified | Oscillospiraceae | Eubacteriales |
| unclassified | unclassified | Eubacteriales |
| unclassified | unclassified | unclassified |
| unclassified | unclassified | Eubacteriales |
| unclassified | unclassified | unclassified |
| unclassified | unclassified | Eubacteriales |
| Agathobaculum | Oscillospiraceae | Eubacteriales |
| Merdimonas | Lachnospiraceae | Eubacteriales |
| unclassified | unclassified | Eubacteriales |

| | | |
|---|---|---|
| unclassified | Oscillospiraceae | Eubacteriales |
| unclassified | unclassified | unclassified |
| Collinsella | Coriobacteriaceae | Coriobacteriales |
| Oscillibacter | Oscillospiraceae | Eubacteriales |
| Oscillibacter | Oscillospiraceae | Eubacteriales |
| Oxalobacter | Oxalobacteraceae | Burkholderiales |
| Bacteroides | Bacteroidaceae | Bacteroidales |
| unclassified | unclassified | unclassified |
| Lachnospira | Lachnospiraceae | Eubacteriales |
| Klebsiella | Enterobacteriaceae | Enterobacterales |
| Bacteroides | Bacteroidaceae | Bacteroidales |
| Enterocloster | Lachnospiraceae | Eubacteriales |
| unclassified | Prevotellaceae | Bacteroidales |
| Bacteroides | Bacteroidaceae | Bacteroidales |
| Marseilla | Prevotellaceae | Bacteroidales |
| unclassified | unclassified | Eubacteriales |
| Citrobacter | Enterobacteriaceae | Enterobacterales |
| unclassified | unclassified | Eubacteriales |
| unclassified | Rikenellaceae | Bacteroidales |
| Sutterella | Sutterellaceae | Burkholderiales |
| unclassified | unclassified | Eubacteriales |
| unclassified | unclassified | unclassified |
| unclassified | unclassified | unclassified |
| unclassified | unclassified | Eubacteriales |
| unclassified | unclassified | Eubacteriales |
| unclassified | Clostridiaceae | Eubacteriales |
| unclassified | Oscillospiraceae | Eubacteriales |
| Holdemanella | Erysipelotrichaceae | Erysipelotrichales |
| Oscillibacter | Oscillospiraceae | Eubacteriales |
| Clostridium | Clostridiaceae | Eubacteriales |
| Clostridium | Clostridiaceae | Eubacteriales |
| unclassified | unclassified | Eubacteriales |
| Collinsella | Coriobacteriaceae | Coriobacteriales |
| Streptococcus | Streptococcaceae | Lactobacillales |
| unclassified | unclassified | unclassified |
| unclassified | unclassified | Eubacteriales |
| unclassified | unclassified | Eubacteriales |
| Hallella | Prevotellaceae | Bacteroidales |
| Bacteroides | Bacteroidaceae | Bacteroidales |
| unclassified | unclassified | Eubacteriales |
| unclassified | unclassified | Eubacteriales |
| Mediterraneibacter | Lachnospiraceae | Eubacteriales |
| unclassified | Oscillospiraceae | Eubacteriales |
| unclassified | unclassified | unclassified |
| unclassified | unclassified | unclassified |
| unclassified | Prevotellaceae | Bacteroidales |
| Bacteroides | Bacteroidaceae | Bacteroidales |
| unclassified | Oscillospiraceae | Eubacteriales |

| | | |
|---|---|---|
| unclassified | unclassified | Eubacteriales |
| Senegalimassilia | Coriobacteriaceae | Coriobacteriales |
| Streptococcus | Streptococcaceae | Lactobacillales |
| unclassified | unclassified | Bacteroidales |
| Alistipes | Rikenellaceae | Bacteroidales |
| Phascolarctobacterium | Acidaminococcaceae | Acidaminococcales |
| unclassified | unclassified | unclassified |
| Marseilla | Prevotellaceae | Bacteroidales |
| unclassified | Prevotellaceae | Bacteroidales |
| unclassified | unclassified | Bacteroidales |
| unclassified | Lachnospiraceae | Eubacteriales |
| unclassified | unclassified | unclassified |
| unclassified | unclassified | unclassified |
| Enorma | Coriobacteriaceae | Coriobacteriales |
| unclassified | unclassified | unclassified |
| Frisingicoccus | Lachnospiraceae | Eubacteriales |
| Coprococcus | Lachnospiraceae | Eubacteriales |
| unclassified | unclassified | Eubacteriales |
| unclassified | unclassified | Eubacteriales |
| unclassified | unclassified | unclassified |
| unclassified | unclassified | unclassified |
| Massilimicrobiota | Erysipelotrichaceae | Erysipelotrichales |
| unclassified | unclassified | Eubacteriales |
| unclassified | Lachnospiraceae | Eubacteriales |
| unclassified | unclassified | unclassified |
| Rothia | Micrococcaceae | Micrococcales |
| unclassified | unclassified | unclassified |
| Roseburia | Lachnospiraceae | Eubacteriales |
| unclassified | unclassified | Eubacteriales |
| unclassified | unclassified | Bacteroidales |
| unclassified | unclassified | Bacteroidales |
| Anaerotignum | Lachnospiraceae | Eubacteriales |
| unclassified | unclassified | unclassified |
| unclassified | unclassified | Bacteroidales |
| unclassified | unclassified | unclassified |
| unclassified | unclassified | unclassified |
| Bacteroides | Bacteroidaceae | Bacteroidales |
| Dorea | Lachnospiraceae | Eubacteriales |
| unclassified | unclassified | Eubacteriales |
| Dakarella | Sutterellaceae | Burkholderiales |
| unclassified | unclassified | Eubacteriales |
| unclassified | unclassified | Veillonellales |
| Eisenbergiella | Lachnospiraceae | Eubacteriales |
| Olsenella | Atopobiaceae | Coriobacteriales |
| unclassified | unclassified | Eubacteriales |
| unclassified | Lachnospiraceae | Eubacteriales |
| Anaerotruncus | Oscillospiraceae | Eubacteriales |
| unclassified | unclassified | unclassified |

| | | |
|---|---|---|
| unclassified | unclassified | Eubacteriales |
| Flavonifractor | Oscillospiraceae | Eubacteriales |
| unclassified | unclassified | Eubacteriales |
| Barnesiella | Barnesiellaceae | Bacteroidales |
| Ruminococcus | Oscillospiraceae | Eubacteriales |
| unclassified | unclassified | Eubacteriales |
| unclassified | unclassified | Bacteroidales |
| unclassified | unclassified | Eubacteriales |
| unclassified | unclassified | Eubacteriales |
| unclassified | unclassified | unclassified |
| unclassified | unclassified | Eubacteriales |
| unclassified | unclassified | unclassified |
| Faecalibacterium | Oscillospiraceae | Eubacteriales |
| unclassified | unclassified | unclassified |
| unclassified | Oscillospiraceae | Eubacteriales |
| unclassified | unclassified | unclassified |
| unclassified | unclassified | Eubacteriales |
| unclassified | unclassified | unclassified |
| unclassified | unclassified | unclassified |
| Bilophila | Desulfovibrionaceae | Desulfovibrionales |
| Blautia | Lachnospiraceae | Eubacteriales |
| Turicibacter | Turicibacteraceae | Erysipelotrichales |
| Hydrogeniiclostidium | Oscillospiraceae | Eubacteriales |
| Dialister | Veillonellaceae | Veillonellales |
| unclassified | unclassified | unclassified |
| unclassified | unclassified | unclassified |
| unclassified | unclassified | Eubacteriales |
| unclassified | unclassified | Eubacteriales |
| unclassified | unclassified | Eubacteriales |
| unclassified | unclassified | unclassified |
| unclassified | unclassified | Victivallales |
| unclassified | unclassified | unclassified |
| Blastocystis | Blastocystidae | Opalinata |
| unclassified | unclassified | Bacteroidales |
| unclassified | unclassified | Eubacteriales |
| unclassified | unclassified | unclassified |
| Anaerotruncus | Oscillospiraceae | Eubacteriales |
| unclassified | unclassified | Eubacteriales |
| Clostridium | Clostridiaceae | Eubacteriales |
| unclassified | unclassified | Methanomassiliicoccales |
| unclassified | Lachnospiraceae | Eubacteriales |
| unclassified | Clostridiaceae | Eubacteriales |
| unclassified | unclassified | Bacteroidales |
| Eubacterium | Eubacteriaceae | Eubacteriales |
| Massilistercora | unclassified | Eubacteriales |
| Mediterraneibacter | Lachnospiraceae | Eubacteriales |
| unclassified | unclassified | unclassified |
| unclassified | Oscillospiraceae | Eubacteriales |

| | | |
|---|---|---|
| unclassified | unclassified | Eubacteriales |
| unclassified | Oscillospiraceae | Eubacteriales |
| unclassified | unclassified | Eggerthellales |
| unclassified | unclassified | Eubacteriales |
| Bacteroides | Bacteroidaceae | Bacteroidales |
| unclassified | unclassified | Eubacteriales |
| unclassified | unclassified | Eubacteriales |
| Clostridium | Clostridiaceae | Eubacteriales |
| Paraprevotella | Prevotellaceae | Bacteroidales |
| Enterobacter | Enterobacteriaceae | Enterobacterales |
| unclassified | unclassified | unclassified |
| unclassified | unclassified | Eggerthellales |
| Phocea | Oscillospiraceae | Eubacteriales |
| unclassified | unclassified | unclassified |
| unclassified | unclassified | Eubacteriales |
| unclassified | unclassified | unclassified |
| unclassified | unclassified | unclassified |
| unclassified | unclassified | unclassified |
| unclassified | unclassified | Eubacteriales |
| Faecalibacterium | Oscillospiraceae | Eubacteriales |
| Clostridium | Clostridiaceae | Eubacteriales |
| unclassified | unclassified | unclassified |
| unclassified | unclassified | unclassified |
| Methanomassiliicoccus | Methanomassiliicoccaceae | Methanomassiliicoccales |
| unclassified | unclassified | unclassified |
| unclassified | unclassified | unclassified |
| unclassified | unclassified | Eubacteriales |
| unclassified | unclassified | unclassified |
| Klebsiella | Enterobacteriaceae | Enterobacterales |
| unclassified | unclassified | Eubacteriales |
| unclassified | unclassified | Eubacteriales |
| Lachnospira | Lachnospiraceae | Eubacteriales |
| unclassified | unclassified | unclassified |
| Bacteroides | Bacteroidaceae | Bacteroidales |
| unclassified | unclassified | unclassified |
| unclassified | Erysipelotrichaceae | Erysipelotrichales |
| unclassified | unclassified | Eubacteriales |
| unclassified | unclassified | unclassified |
| unclassified | unclassified | unclassified |
| unclassified | unclassified | unclassified |
| unclassified | unclassified | unclassified |
| unclassified | unclassified | Eubacteriales |
| unclassified | unclassified | unclassified |
| unclassified | unclassified | Eubacteriales |
| Bacteroides | Bacteroidaceae | Bacteroidales |
| unclassified | unclassified | Eubacteriales |
| unclassified | unclassified | Eubacteriales |
| Clostridioides | Peptostreptococcaceae | Eubacteriales |

| | | |
|---|---|---|
| Bacteroides | Bacteroidaceae | Bacteroidales |
| unclassified | unclassified | Victivallales |
| unclassified | unclassified | unclassified |
| unclassified | unclassified | unclassified |
| unclassified | unclassified | unclassified |
| Bilophila | Desulfovibrionaceae | Desulfovibrionales |
| unclassified | unclassified | unclassified |
| Bacteroides | Bacteroidaceae | Bacteroidales |
| unclassified | unclassified | Eubacteriales |
| unclassified | Oscillospiraceae | Eubacteriales |
| Phocaeicola | Bacteroidaceae | Bacteroidales |
| unclassified | unclassified | unclassified |
| Lactobacillus | Lactobacillaceae | Lactobacillales |
| unclassified | Elusimicrobiaceae | Elusimicrobiales |
| unclassified | unclassified | unclassified |
| unclassified | unclassified | unclassified |
| unclassified | unclassified | Eubacteriales |
| unclassified | unclassified | Eubacteriales |
| unclassified | unclassified | unclassified |
| unclassified | unclassified | Eubacteriales |
| Lactiplantibacillus | Lactobacillaceae | Lactobacillales |
| unclassified | unclassified | unclassified |
| unclassified | unclassified | Eubacteriales |
| unclassified | unclassified | Eubacteriales |
| unclassified | unclassified | unclassified |
| Enterobacter | Enterobacteriaceae | Enterobacterales |
| unclassified | unclassified | Eubacteriales |
| unclassified | unclassified | Eubacteriales |
| unclassified | unclassified | unclassified |
| unclassified | unclassified | Eubacteriales |
| unclassified | unclassified | Bacteroidales |
| unclassified | Lachnospiraceae | Eubacteriales |
| unclassified | unclassified | unclassified |
| unclassified | unclassified | Eubacteriales |
| unclassified | unclassified | Erysipelotrichales |
| Alistipes | Rikenellaceae | Bacteroidales |
| unclassified | unclassified | unclassified |
| unclassified | unclassified | Eubacteriales |
| unclassified | unclassified | Eubacteriales |
| unclassified | unclassified | unclassified |
| unclassified | unclassified | unclassified |
| unclassified | unclassified | unclassified |
| unclassified | unclassified | Eubacteriales |
| unclassified | unclassified | unclassified |
| Lactobacillus | Lactobacillaceae | Lactobacillales |
| unclassified | Lachnospiraceae | Eubacteriales |
| unclassified | unclassified | unclassified |
| unclassified | unclassified | unclassified |

| | | |
|---|---|---|
| Bacteroides | Bacteroidaceae | Bacteroidales |
| Eubacterium | Eubacteriaceae | Eubacteriales |
| unclassified | unclassified | Eubacteriales |
| Bacteroides | Bacteroidaceae | Bacteroidales |
| unclassified | unclassified | Eubacteriales |
| Desulfovibrio | Desulfovibrionaceae | Desulfovibrionales |
| Bacteroides | Bacteroidaceae | Bacteroidales |
| unclassified | unclassified | Eubacteriales |
| unclassified | unclassified | Eubacteriales |
| unclassified | unclassified | Eubacteriales |
| Veillonella | Veillonellaceae | Veillonellales |
| unclassified | unclassified | Eubacteriales |
| Coprococcus | Lachnospiraceae | Eubacteriales |
| unclassified | unclassified | unclassified |
| unclassified | Rikenellaceae | Bacteroidales |
| unclassified | unclassified | Acidaminococcales |
| unclassified | unclassified | Bacteroidales |
| unclassified | unclassified | unclassified |
| unclassified | unclassified | unclassified |
| Desulfovibrio | Desulfovibrionaceae | Desulfovibrionales |
| Fusobacterium | Fusobacteriaceae | Fusobacteriales |
| unclassified | unclassified | unclassified |
| unclassified | unclassified | unclassified |
| Dorea | Lachnospiraceae | Eubacteriales |
| Raoultella | Enterobacteriaceae | Enterobacterales |
| unclassified | unclassified | Eubacteriales |
| unclassified | unclassified | Eubacteriales |
| Faecalitalea | Erysipelotrichaceae | Erysipelotrichales |
| Bacteroides | Bacteroidaceae | Bacteroidales |
| unclassified | unclassified | unclassified |
| unclassified | Oscillospiraceae | Eubacteriales |
| Mediterranea | Bacteroidaceae | Bacteroidales |
| unclassified | unclassified | Eubacteriales |
| unclassified | unclassified | Burkholderiales |

| class | phylum | superkingdom | PHL018-0047_V3 EYCF |
|---|---|---|---|
| Bacteroidia | Bacteroidetes | Bacteria | 0.121883321 |
| Clostridia | Firmicutes | Bacteria | 2.030872148 |
| Actinomycetia | Actinobacteria | Bacteria | 7.616228544 |
| Bacteroidia | Bacteroidetes | Bacteria | 0 |
| unclassified | unclassified | unclassified | 0 |
| Coriobacteriia | Actinobacteria | Bacteria | 0 |
| Negativicutes | Firmicutes | Bacteria | 0 |
| Bacteroidia | Bacteroidetes | Bacteria | 0 |
| Bacteroidia | Bacteroidetes | Bacteria | 0 |
| Clostridia | Firmicutes | Bacteria | 0 |
| unclassified | Firmicutes | Bacteria | 0 |
| Clostridia | Firmicutes | Bacteria | 0 |
| Bacteroidia | Bacteroidetes | Bacteria | 0 |
| Actinomycetia | Actinobacteria | Bacteria | 0.632164511 |
| Bacteroidia | Bacteroidetes | Bacteria | 0 |
| Clostridia | Firmicutes | Bacteria | 0 |
| Erysipelotrichia | Firmicutes | Bacteria | 0 |
| Clostridia | Firmicutes | Bacteria | 0 |
| Betaproteobacteria | Proteobacteria | Bacteria | 0 |
| unclassified | unclassified | unclassified | 0 |
| Clostridia | Firmicutes | Bacteria | 0 |
| Clostridia | Firmicutes | Bacteria | 0 |
| Bacteroidia | Bacteroidetes | Bacteria | 0 |
| Bacteroidia | Bacteroidetes | Bacteria | 0 |
| Clostridia | Firmicutes | Bacteria | 0 |
| Bacteroidia | Bacteroidetes | Bacteria | 0 |
| Bacteroidia | Bacteroidetes | Bacteria | 0 |
| Clostridia | Firmicutes | Bacteria | 0 |
| unclassified | Firmicutes | Bacteria | 0 |
| Bacteroidia | Bacteroidetes | Bacteria | 0 |
| Clostridia | Firmicutes | Bacteria | 0 |
| Bacteroidia | Bacteroidetes | Bacteria | 0 |
| unclassified | unclassified | unclassified | 0 |
| Bacteroidia | Bacteroidetes | Bacteria | 0 |
| unclassified | unclassified | unclassified | 0 |
| Erysipelotrichia | Firmicutes | Bacteria | 5.910912985 |
| Clostridia | Firmicutes | Bacteria | 0 |
| Clostridia | Firmicutes | Bacteria | 0 |
| Bacteroidia | Bacteroidetes | Bacteria | 0 |
| Gammaproteobacteria | Proteobacteria | Bacteria | 0 |
| Bacteroidia | Bacteroidetes | Bacteria | 0 |
| Gammaproteobacteria | Proteobacteria | Bacteria | 0.001688946 |
| Gammaproteobacteria | Proteobacteria | Bacteria | 0.240978237 |
| Actinomycetia | Actinobacteria | Bacteria | 0.011869077 |
| Negativicutes | Firmicutes | Bacteria | 0.001140958 |
| Bacteroidia | Bacteroidetes | Bacteria | 0 |

| | | | |
|---|---|---|---|
| Clostridia | Firmicutes | Bacteria | 0 |
| Clostridia | Firmicutes | Bacteria | 0.008138359 |
| Bacteroidia | Bacteroidetes | Bacteria | 0.005257099 |
| Clostridia | Firmicutes | Bacteria | 0.56691284 |
| Clostridia | Firmicutes | Bacteria | 0 |
| Clostridia | Firmicutes | Bacteria | 0.00454935 |
| Bacilli | Firmicutes | Bacteria | 0 |
| Bacteroidia | Bacteroidetes | Bacteria | 0 |
| Clostridia | Firmicutes | Bacteria | 0 |
| Verrucomicrobiae | Verrucomicrobia | Bacteria | 0 |
| Bacilli | Firmicutes | Bacteria | 0 |
| Clostridia | Firmicutes | Bacteria | 1.743315985 |
| Bacilli | Firmicutes | Bacteria | 0 |
| Erysipelotrichia | Firmicutes | Bacteria | 0.014641743 |
| Bacilli | Firmicutes | Bacteria | 0 |
| Clostridia | Firmicutes | Bacteria | 0 |
| Bacilli | Firmicutes | Bacteria | 0 |
| Negativicutes | Firmicutes | Bacteria | 0 |
| Bacteroidia | Bacteroidetes | Bacteria | 0 |
| Clostridia | Firmicutes | Bacteria | 0 |
| Clostridia | Firmicutes | Bacteria | 0.005982435 |
| Clostridia | Firmicutes | Bacteria | 0.010071724 |
| Clostridia | Firmicutes | Bacteria | 0 |
| unclassified | unclassified | unclassified | 0 |
| Negativicutes | Firmicutes | Bacteria | 0 |
| Clostridia | Firmicutes | Bacteria | 0 |
| Bacilli | Firmicutes | Bacteria | 0 |
| Clostridia | Firmicutes | Bacteria | 0 |
| Gammaproteobacteria | Proteobacteria | Bacteria | 0 |
| Clostridia | Firmicutes | Bacteria | 0 |
| Clostridia | Firmicutes | Bacteria | 0 |
| Clostridia | Firmicutes | Bacteria | 0 |
| Bacteroidia | Bacteroidetes | Bacteria | 0 |
| Betaproteobacteria | Proteobacteria | Bacteria | 0 |
| Gammaproteobacteria | Proteobacteria | Bacteria | 0 |
| Bacilli | Firmicutes | Bacteria | 0 |
| unclassified | unclassified | unclassified | 0 |
| Bacilli | Firmicutes | Bacteria | 0 |
| Clostridia | Firmicutes | Bacteria | 0 |
| Spirochaetia | Spirochaetes | Bacteria | 0 |
| Clostridia | Firmicutes | Bacteria | 0 |
| Clostridia | Firmicutes | Bacteria | 0 |
| unclassified | unclassified | unclassified | 0 |
| Clostridia | Firmicutes | Bacteria | 0 |
| unclassified | unclassified | unclassified | 0.028731044 |
| Bacteroidia | Bacteroidetes | Bacteria | 0 |
| Bacilli | Firmicutes | Bacteria | 0 |
| Bacilli | Firmicutes | Bacteria | 1.462561322 |

| | | | |
|---|---|---|---|
| unclassified | Firmicutes | Bacteria | 0 |
| Erysipelotrichia | Firmicutes | Bacteria | 0 |
| unclassified | unclassified | unclassified | 0 |
| Bacteroidia | Bacteroidetes | Bacteria | 0 |
| Bacilli | Firmicutes | Bacteria | 0 |
| Bacilli | Firmicutes | Bacteria | 0 |
| Clostridia | Firmicutes | Bacteria | 0 |
| Bacteroidia | Bacteroidetes | Bacteria | 0 |
| Erysipelotrichia | Firmicutes | Bacteria | 0 |
| unclassified | unclassified | unclassified | 0 |
| Clostridia | Firmicutes | Bacteria | 0 |
| Gammaproteobacteria | Proteobacteria | Bacteria | 0 |
| unclassified | unclassified | unclassified | 0 |
| Clostridia | Firmicutes | Bacteria | 0 |
| unclassified | unclassified | unclassified | 0 |
| Bacteroidia | Bacteroidetes | Bacteria | 0 |
| Bacteroidia | Bacteroidetes | Bacteria | 0 |
| unclassified | unclassified | unclassified | 0 |
| Clostridia | Firmicutes | Bacteria | 0.028241153 |
| Clostridia | Firmicutes | Bacteria | 0 |
| unclassified | unclassified | Bacteria | 0 |
| unclassified | Firmicutes | Bacteria | 0 |
| Clostridia | Firmicutes | Bacteria | 0 |
| Clostridia | Firmicutes | Bacteria | 0 |
| Bacteroidia | Bacteroidetes | Bacteria | 0 |
| Bacteroidia | Bacteroidetes | Bacteria | 0 |
| Clostridia | Firmicutes | Bacteria | 0.004389511 |
| Clostridia | Firmicutes | Bacteria | 0 |
| Clostridia | Firmicutes | Bacteria | 0 |
| Clostridia | Firmicutes | Bacteria | 0 |
| Clostridia | Firmicutes | Bacteria | 0.015591786 |
| Bacteroidia | Bacteroidetes | Bacteria | 0.003089682 |
| unclassified | unclassified | unclassified | 0 |
| unclassified | unclassified | unclassified | 0 |
| unclassified | Firmicutes | Bacteria | 0 |
| unclassified | unclassified | Bacteria | 0 |
| unclassified | unclassified | Bacteria | 0 |
| Bacteroidia | Bacteroidetes | Bacteria | 0 |
| Coriobacteriia | Actinobacteria | Bacteria | 0 |
| unclassified | unclassified | unclassified | 0 |
| Bacteroidia | Bacteroidetes | Bacteria | 0 |
| Clostridia | Firmicutes | Bacteria | 0 |
| Actinomycetia | Actinobacteria | Bacteria | 0 |
| Clostridia | Firmicutes | Bacteria | 0 |
| Clostridia | Firmicutes | Bacteria | 0 |
| Clostridia | Firmicutes | Bacteria | 0 |
| Clostridia | Firmicutes | Bacteria | 0 |
| Bacteroidia | Bacteroidetes | Bacteria | 0 |

| | | | |
|---|---|---|---|
| Bacteroidia | Bacteroidetes | Bacteria | 0 |
| Clostridia | Firmicutes | Bacteria | 0 |
| unclassified | Actinobacteria | Bacteria | 0 |
| Clostridia | Firmicutes | Bacteria | 0 |
| Clostridia | Firmicutes | Bacteria | 0 |
| Bacteroidia | Bacteroidetes | Bacteria | 0 |
| Clostridia | Firmicutes | Bacteria | 0 |
| Bacteroidia | Bacteroidetes | Bacteria | 0 |
| Clostridia | Firmicutes | Bacteria | 0 |
| unclassified | unclassified | unclassified | 0 |
| Clostridia | Firmicutes | Bacteria | 0 |
| unclassified | unclassified | Bacteria | 0 |
| Clostridia | Firmicutes | Bacteria | 0 |
| Clostridia | Firmicutes | Bacteria | 0 |
| unclassified | Firmicutes | Bacteria | 0 |
| Clostridia | Firmicutes | Bacteria | 0 |
| unclassified | unclassified | unclassified | 0 |
| unclassified | unclassified | unclassified | 0 |
| unclassified | unclassified | unclassified | 0 |
| Clostridia | Firmicutes | Bacteria | 0 |
| Bacteroidia | Bacteroidetes | Bacteria | 0 |
| Clostridia | Firmicutes | Bacteria | 0 |
| unclassified | unclassified | Bacteria | 0 |
| Thermoplasmata | Candidatus Thermoplasmatota | Archaea | 0 |
| Coriobacteriia | Actinobacteria | Bacteria | 0 |
| unclassified | unclassified | Bacteria | 0 |
| Clostridia | Firmicutes | Bacteria | 0 |
| Clostridia | Firmicutes | Bacteria | 0 |
| Clostridia | Firmicutes | Bacteria | 0 |
| Negativicutes | Firmicutes | Bacteria | 0 |
| Clostridia | Firmicutes | Bacteria | 0 |
| Bacilli | Firmicutes | Bacteria | 0 |
| unclassified | Firmicutes | Bacteria | 0 |
| unclassified | unclassified | Bacteria | 0 |
| Clostridia | Firmicutes | Bacteria | 0 |
| Bacteroidia | Bacteroidetes | Bacteria | 0 |
| unclassified | Firmicutes | Bacteria | 0 |
| Clostridia | Firmicutes | Bacteria | 0 |
| Deltaproteobacteria | Proteobacteria | Bacteria | 0 |
| Clostridia | Firmicutes | Bacteria | 0 |
| unclassified | unclassified | Bacteria | 0 |
| unclassified | unclassified | Bacteria | 0 |
| Clostridia | Firmicutes | Bacteria | 0 |
| Deltaproteobacteria | Proteobacteria | Bacteria | 0 |
| Clostridia | Firmicutes | Bacteria | 0 |
| unclassified | Firmicutes | Bacteria | 0 |
| unclassified | unclassified | Bacteria | 0 |
| Clostridia | Firmicutes | Bacteria | 0 |

| | | | |
|---|---|---|---|
| Methanobacteria | Euryarchaeota | Archaea | 0 |
| unclassified | unclassified | Bacteria | 0 |
| Clostridia | Firmicutes | Bacteria | 0 |
| Spirochaetia | Spirochaetes | Bacteria | 0 |
| Negativicutes | Firmicutes | Bacteria | 0 |
| Betaproteobacteria | Proteobacteria | Bacteria | 0 |
| unclassified | Firmicutes | Bacteria | 0 |
| unclassified | unclassified | unclassified | 0 |
| Bacteroidia | Bacteroidetes | Bacteria | 0.005535507 |
| Clostridia | Firmicutes | Bacteria | 0 |
| Clostridia | Firmicutes | Bacteria | 0 |
| Clostridia | Firmicutes | Bacteria | 0 |
| Clostridia | Firmicutes | Bacteria | 0 |
| Betaproteobacteria | Proteobacteria | Bacteria | 0 |
| Betaproteobacteria | Proteobacteria | Bacteria | 0 |
| Gammaproteobacteria | Proteobacteria | Bacteria | 0 |
| Gammaproteobacteria | Proteobacteria | Bacteria | 0 |
| Gammaproteobacteria | Proteobacteria | Bacteria | 0 |
| Gammaproteobacteria | Proteobacteria | Bacteria | 0 |
| Actinomycetia | Actinobacteria | Bacteria | 0 |
| Actinomycetia | Actinobacteria | Bacteria | 0 |
| Gammaproteobacteria | Proteobacteria | Bacteria | 0 |
| Bacteroidia | Bacteroidetes | Bacteria | 0 |
| Clostridia | Firmicutes | Bacteria | 0 |
| Epsilonproteobacteria | Proteobacteria | Bacteria | 0 |
| Alphaproteobacteria | Proteobacteria | Bacteria | 0 |
| Coriobacteriia | Actinobacteria | Bacteria | 0.001310985 |
| Bacteroidia | Bacteroidetes | Bacteria | 0 |
| Actinomycetia | Actinobacteria | Bacteria | 0 |
| Clostridia | Firmicutes | Bacteria | 0 |
| Clostridia | Firmicutes | Bacteria | 0 |
| Betaproteobacteria | Proteobacteria | Bacteria | 0 |
| Bacteroidia | Bacteroidetes | Bacteria | 0 |
| Clostridia | Firmicutes | Bacteria | 0 |
| Clostridia | Firmicutes | Bacteria | 0.001220569 |
| Negativicutes | Firmicutes | Bacteria | 0 |
| Alphaproteobacteria | Proteobacteria | Bacteria | 0 |
| Flavobacteriia | Bacteroidetes | Bacteria | 0 |
| Gammaproteobacteria | Proteobacteria | Bacteria | 0 |
| Gammaproteobacteria | Proteobacteria | Bacteria | 0 |
| Gammaproteobacteria | Proteobacteria | Bacteria | 0 |
| Gammaproteobacteria | Proteobacteria | Bacteria | 0 |
| Gammaproteobacteria | Proteobacteria | Bacteria | 0 |
| Gammaproteobacteria | Proteobacteria | Bacteria | 0 |
| Synergistia | Synergistetes | Bacteria | 0 |
| Clostridia | Firmicutes | Bacteria | 0.141777998 |
| Erysipelotrichia | Firmicutes | Bacteria | 0 |
| Clostridia | Firmicutes | Bacteria | 0.009567916 |

| | | | |
|---|---|---|---|
| Clostridia | Firmicutes | Bacteria | 0 |
| Coriobacteriia | Actinobacteria | Bacteria | 0 |
| Actinomycetia | Actinobacteria | Bacteria | 0 |
| Actinomycetia | Actinobacteria | Bacteria | 0 |
| Actinomycetia | Actinobacteria | Bacteria | 0 |
| Actinomycetia | Actinobacteria | Bacteria | 0 |
| Gammaproteobacteria | Proteobacteria | Bacteria | 0 |
| Gammaproteobacteria | Proteobacteria | Bacteria | 0 |
| Bacteroidia | Bacteroidetes | Bacteria | 0 |
| Gammaproteobacteria | Proteobacteria | Bacteria | 0 |
| Gammaproteobacteria | Proteobacteria | Bacteria | 0 |
| Bacilli | Firmicutes | Bacteria | 0 |
| Bacilli | Firmicutes | Bacteria | 0 |
| Bacilli | Firmicutes | Bacteria | 0 |
| Bacilli | Firmicutes | Bacteria | 0 |
| Bacilli | Firmicutes | Bacteria | 0 |
| Erysipelotrichia | Firmicutes | Bacteria | 0 |
| Gammaproteobacteria | Proteobacteria | Bacteria | 0 |
| Gammaproteobacteria | Proteobacteria | Bacteria | 0.001946101 |
| Clostridia | Firmicutes | Bacteria | 0.015611 |
| Clostridia | Firmicutes | Bacteria | 0.005741396 |
| Flavobacteriia | Bacteroidetes | Bacteria | 0 |
| Bacteroidia | Bacteroidetes | Bacteria | 0 |
| Actinomycetia | Actinobacteria | Bacteria | 0 |
| Bacilli | Firmicutes | Bacteria | 0.002073908 |
| Gammaproteobacteria | Proteobacteria | Bacteria | 0 |
| Gammaproteobacteria | Proteobacteria | Bacteria | 0 |
| Gammaproteobacteria | Proteobacteria | Bacteria | 0 |
| Gammaproteobacteria | Proteobacteria | Bacteria | 0 |
| Bacilli | Firmicutes | Bacteria | 0 |
| Clostridia | Firmicutes | Bacteria | 0.010170329 |
| Bacilli | Firmicutes | Bacteria | 0 |
| Bacilli | Firmicutes | Bacteria | 0.00429487 |
| Bacilli | Firmicutes | Bacteria | 0 |
| Bacilli | Firmicutes | Bacteria | 0 |
| Negativicutes | Firmicutes | Bacteria | 0 |
| Negativicutes | Firmicutes | Bacteria | 0 |
| Gammaproteobacteria | Proteobacteria | Bacteria | 0 |
| Actinomycetia | Actinobacteria | Bacteria | 0 |
| Betaproteobacteria | Proteobacteria | Bacteria | 0 |
| Gammaproteobacteria | Proteobacteria | Bacteria | 0 |
| Betaproteobacteria | Proteobacteria | Bacteria | 0.007237311 |
| Bacteroidia | Bacteroidetes | Bacteria | 0 |
| Bacteroidia | Bacteroidetes | Bacteria | 0 |
| Bacteroidia | Bacteroidetes | Bacteria | 0 |
| Clostridia | Firmicutes | Bacteria | 0 |
| Gammaproteobacteria | Proteobacteria | Bacteria | 0 |
| Gammaproteobacteria | Proteobacteria | Bacteria | 0 |

| | | | |
|---|---|---|---|
| Gammaproteobacteria | Proteobacteria | Bacteria | 0 |
| Clostridia | Firmicutes | Bacteria | 0 |
| Actinomycetia | Actinobacteria | Bacteria | 0 |
| Negativicutes | Firmicutes | Bacteria | 0 |
| Clostridia | Firmicutes | Bacteria | 0 |
| Gammaproteobacteria | Proteobacteria | Bacteria | 0 |
| Gammaproteobacteria | Proteobacteria | Bacteria | 0 |
| Alphaproteobacteria | Proteobacteria | Bacteria | 0 |
| Bacilli | Firmicutes | Bacteria | 0 |
| Bacilli | Firmicutes | Bacteria | 0 |
| Bacilli | Firmicutes | Bacteria | 0 |
| Gammaproteobacteria | Proteobacteria | Bacteria | 0 |
| Gammaproteobacteria | Proteobacteria | Bacteria | 0 |
| Bacilli | Firmicutes | Bacteria | 0.000814336 |
| Bacilli | Firmicutes | Bacteria | 0.001568298 |
| Clostridia | Firmicutes | Bacteria | 0 |
| Bacilli | Firmicutes | Bacteria | 0.002042211 |
| Bacilli | Firmicutes | Bacteria | 0 |
| Negativicutes | Firmicutes | Bacteria | 0 |
| Gammaproteobacteria | Proteobacteria | Bacteria | 0 |
| Gammaproteobacteria | Proteobacteria | Bacteria | 0 |
| Clostridia | Firmicutes | Bacteria | 0.005300161 |
| Gammaproteobacteria | Proteobacteria | Bacteria | 0 |
| Clostridia | Firmicutes | Bacteria | 0 |
| Clostridia | Firmicutes | Bacteria | 0 |
| Bacilli | Firmicutes | Bacteria | 0 |
| Clostridia | Firmicutes | Bacteria | 0 |
| Clostridia | Firmicutes | Bacteria | 0 |
| Gammaproteobacteria | Proteobacteria | Bacteria | 0 |
| Clostridia | Firmicutes | Bacteria | 0 |
| Clostridia | Firmicutes | Bacteria | 0 |
| Gammaproteobacteria | Proteobacteria | Bacteria | 0 |
| Gammaproteobacteria | Proteobacteria | Bacteria | 0 |
| Actinomycetia | Actinobacteria | Bacteria | 0 |
| Bacilli | Firmicutes | Bacteria | 0 |
| Gammaproteobacteria | Proteobacteria | Bacteria | 0 |
| Clostridia | Firmicutes | Bacteria | 0 |
| Gammaproteobacteria | Proteobacteria | Bacteria | 0.042849882 |
| Negativicutes | Firmicutes | Bacteria | 0 |
| Clostridia | Firmicutes | Bacteria | 0 |
| Bacilli | Firmicutes | Bacteria | 0 |
| Clostridia | Firmicutes | Bacteria | 0 |
| Clostridia | Firmicutes | Bacteria | 0 |
| Clostridia | Firmicutes | Bacteria | 0 |
| Gammaproteobacteria | Proteobacteria | Bacteria | 0 |
| Clostridia | Firmicutes | Bacteria | 0 |
| unclassified | Firmicutes | Bacteria | 0 |
| Erysipelotrichia | Firmicutes | Bacteria | 0 |

| | | | |
|---|---|---|---|
| Clostridia | Firmicutes | Bacteria | 0.212464784 |
| Bacteroidia | Bacteroidetes | Bacteria | 0 |
| Actinomycetia | Actinobacteria | Bacteria | 0 |
| Gammaproteobacteria | Proteobacteria | Bacteria | 0 |
| Actinomycetia | Actinobacteria | Bacteria | 0.002671254 |
| Clostridia | Firmicutes | Bacteria | 0 |
| Gammaproteobacteria | Proteobacteria | Bacteria | 0 |
| Actinomycetia | Actinobacteria | Bacteria | 0.007488701 |
| Clostridia | Firmicutes | Bacteria | 0 |
| Actinomycetia | Actinobacteria | Bacteria | 0.792226137 |
| Clostridia | Firmicutes | Bacteria | 0 |
| Gammaproteobacteria | Proteobacteria | Bacteria | 0 |
| Bacilli | Firmicutes | Bacteria | 0.427182811 |
| Erysipelotrichia | Firmicutes | Bacteria | 0 |
| Clostridia | Firmicutes | Bacteria | 0 |
| Saccharomycetes | Ascomycota | Eukaryota | 0 |
| Gammaproteobacteria | Proteobacteria | Bacteria | 0 |
| Clostridia | Firmicutes | Bacteria | 0 |
| Clostridia | Firmicutes | Bacteria | 0 |
| Clostridia | Firmicutes | Bacteria | 0 |
| Clostridia | Firmicutes | Bacteria | 0 |
| Clostridia | Firmicutes | Bacteria | 0 |
| Gammaproteobacteria | Proteobacteria | Bacteria | 0 |
| Clostridia | Firmicutes | Bacteria | 0 |
| Gammaproteobacteria | Proteobacteria | Bacteria | 0 |
| Negativicutes | Firmicutes | Bacteria | 0 |
| Bacilli | Firmicutes | Bacteria | 0 |
| unclassified | unclassified | unclassified | 0 |
| Clostridia | Firmicutes | Bacteria | 0 |
| Clostridia | Firmicutes | Bacteria | 0 |
| Coriobacteriia | Actinobacteria | Bacteria | 0 |
| Erysipelotrichia | Firmicutes | Bacteria | 0 |
| Clostridia | Firmicutes | Bacteria | 0 |
| Clostridia | Firmicutes | Bacteria | 0.002179431 |
| Negativicutes | Firmicutes | Bacteria | 0 |
| unclassified | unclassified | unclassified | 0 |
| Clostridia | Firmicutes | Bacteria | 0 |
| Erysipelotrichia | Firmicutes | Bacteria | 0 |
| Bacteroidia | Bacteroidetes | Bacteria | 0 |
| Clostridia | Firmicutes | Bacteria | 0 |
| Clostridia | Firmicutes | Bacteria | 0 |
| Clostridia | Firmicutes | Bacteria | 0 |
| unclassified | unclassified | unclassified | 0 |
| Erysipelotrichia | Firmicutes | Bacteria | 0.004603075 |
| Clostridia | Firmicutes | Bacteria | 0 |
| Clostridia | Firmicutes | Bacteria | 0.000522537 |
| Clostridia | Firmicutes | Bacteria | 0 |
| Clostridia | Firmicutes | Bacteria | 0 |

| | | | |
|---|---|---|---|
| Clostridia | Firmicutes | Bacteria | 0 |
| Bacilli | Firmicutes | Bacteria | 0 |
| Clostridia | Firmicutes | Bacteria | 0 |
| Bacilli | Firmicutes | Bacteria | 0 |
| Erysipelotrichia | Firmicutes | Bacteria | 0 |
| Clostridia | Firmicutes | Bacteria | 0.005596292 |
| Clostridia | Firmicutes | Bacteria | 0 |
| Actinomycetia | Actinobacteria | Bacteria | 0 |
| unclassified | Firmicutes | Bacteria | 0 |
| Bacilli | Firmicutes | Bacteria | 0 |
| Actinomycetia | Actinobacteria | Bacteria | 0.00594986 |
| Verrucomicrobiae | Verrucomicrobia | Bacteria | 0 |
| Clostridia | Firmicutes | Bacteria | 0 |
| Actinomycetia | Actinobacteria | Bacteria | 0 |
| Coriobacteriia | Actinobacteria | Bacteria | 0.002068208 |
| Clostridia | Firmicutes | Bacteria | 0 |
| Bacteroidia | Bacteroidetes | Bacteria | 0 |
| Clostridia | Firmicutes | Bacteria | 0 |
| Clostridia | Firmicutes | Bacteria | 0 |
| Bacteroidia | Bacteroidetes | Bacteria | 0 |
| Actinomycetia | Actinobacteria | Bacteria | 0 |
| Coriobacteriia | Actinobacteria | Bacteria | 0 |
| Clostridia | Firmicutes | Bacteria | 0 |
| Clostridia | Firmicutes | Bacteria | 0 |
| Clostridia | Firmicutes | Bacteria | 0.001898916 |
| Erysipelotrichia | Firmicutes | Bacteria | 0 |
| Clostridia | Firmicutes | Bacteria | 0 |
| Clostridia | Firmicutes | Bacteria | 0 |
| Clostridia | Firmicutes | Bacteria | 0 |
| Clostridia | Firmicutes | Bacteria | 0.064417103 |
| Clostridia | Firmicutes | Bacteria | 0 |
| Gammaproteobacteria | Proteobacteria | Bacteria | 0 |
| Clostridia | Firmicutes | Bacteria | 0 |
| Clostridia | Firmicutes | Bacteria | 0 |
| Clostridia | Firmicutes | Bacteria | 0 |
| Clostridia | Firmicutes | Bacteria | 0 |
| Coriobacteriia | Actinobacteria | Bacteria | 0 |
| Gammaproteobacteria | Proteobacteria | Bacteria | 0 |
| Coriobacteriia | Actinobacteria | Bacteria | 0.002999228 |
| Coriobacteriia | Actinobacteria | Bacteria | 0 |
| Coriobacteriia | Actinobacteria | Bacteria | 0.012183109 |
| Clostridia | Firmicutes | Bacteria | 0.199262422 |
| Gammaproteobacteria | Proteobacteria | Bacteria | 0 |
| Clostridia | Firmicutes | Bacteria | 0 |
| Clostridia | Firmicutes | Bacteria | 0 |
| Clostridia | Firmicutes | Bacteria | 0 |
| Tissierellia | Firmicutes | Bacteria | 0 |
| Clostridia | Firmicutes | Bacteria | 0 |

| | | | |
|---|---|---|---|
| Negativicutes | Firmicutes | Bacteria | 0 |
| Bacteroidia | Bacteroidetes | Bacteria | 0 |
| Bacilli | Firmicutes | Bacteria | 0.009266149 |
| Clostridia | Firmicutes | Bacteria | 0 |
| Clostridia | Firmicutes | Bacteria | 0 |
| Erysipelotrichia | Firmicutes | Bacteria | 0 |
| Clostridia | Firmicutes | Bacteria | 0 |
| Gammaproteobacteria | Proteobacteria | Bacteria | 0 |
| Clostridia | Firmicutes | Bacteria | 0.043482114 |
| Clostridia | Firmicutes | Bacteria | 0 |
| Deltaproteobacteria | Proteobacteria | Bacteria | 0 |
| Clostridia | Firmicutes | Bacteria | 0 |
| Erysipelotrichia | Firmicutes | Bacteria | 0 |
| Actinomycetia | Actinobacteria | Bacteria | 0 |
| Clostridia | Firmicutes | Bacteria | 0 |
| Clostridia | Firmicutes | Bacteria | 0.020577566 |
| Negativicutes | Firmicutes | Bacteria | 0.003197395 |
| Clostridia | Firmicutes | Bacteria | 0 |
| Clostridia | Firmicutes | Bacteria | 0 |
| Coriobacteriia | Actinobacteria | Bacteria | 0 |
| Clostridia | Firmicutes | Bacteria | 0 |
| Clostridia | Firmicutes | Bacteria | 0 |
| Clostridia | Firmicutes | Bacteria | 0 |
| Coriobacteriia | Actinobacteria | Bacteria | 0 |
| Clostridia | Firmicutes | Bacteria | 0.011156227 |
| Clostridia | Firmicutes | Bacteria | 0 |
| Coriobacteriia | Actinobacteria | Bacteria | 0.001623648 |
| Coriobacteriia | Actinobacteria | Bacteria | 0 |
| Erysipelotrichia | Firmicutes | Bacteria | 0 |
| Clostridia | Firmicutes | Bacteria | 0 |
| Clostridia | Firmicutes | Bacteria | 0.01502315 |
| Bacilli | Firmicutes | Bacteria | 0.003681809 |
| Coriobacteriia | Actinobacteria | Bacteria | 0.016651506 |
| Gammaproteobacteria | Proteobacteria | Bacteria | 0 |
| Bacilli | Firmicutes | Bacteria | 0 |
| Clostridia | Firmicutes | Bacteria | 0 |
| Bacilli | Firmicutes | Bacteria | 0 |
| Clostridia | Firmicutes | Bacteria | 0.047505132 |
| Clostridia | Firmicutes | Bacteria | 0 |
| Clostridia | Firmicutes | Bacteria | 0.000506613 |
| Deltaproteobacteria | Proteobacteria | Bacteria | 0 |
| Clostridia | Firmicutes | Bacteria | 0 |
| Clostridia | Firmicutes | Bacteria | 0.001853318 |
| Clostridia | Firmicutes | Bacteria | 0 |
| Clostridia | Firmicutes | Bacteria | 0 |
| Bacilli | Firmicutes | Bacteria | 0 |
| Erysipelotrichia | Firmicutes | Bacteria | 0 |
| Clostridia | Firmicutes | Bacteria | 0 |

| | | | |
|---|---|---|---|
| Clostridia | Firmicutes | Bacteria | 0 |
| Bacilli | Firmicutes | Bacteria | 0.13762537 |
| Actinomycetia | Actinobacteria | Bacteria | 0 |
| Bacteroidia | Bacteroidetes | Bacteria | 0 |
| Coriobacteriia | Actinobacteria | Bacteria | 0 |
| Clostridia | Firmicutes | Bacteria | 0 |
| Clostridia | Firmicutes | Bacteria | 0 |
| Gammaproteobacteria | Proteobacteria | Bacteria | 0 |
| unclassified | unclassified | Bacteria | 0 |
| Erysipelotrichia | Firmicutes | Bacteria | 0 |
| Erysipelotrichia | Firmicutes | Bacteria | 0 |
| Bacilli | Firmicutes | Bacteria | 0.002803714 |
| Clostridia | Firmicutes | Bacteria | 0 |
| Clostridia | Firmicutes | Bacteria | 0 |
| Coriobacteriia | Actinobacteria | Bacteria | 0 |
| Clostridia | Firmicutes | Bacteria | 0 |
| Bacilli | Firmicutes | Bacteria | 0.028032309 |
| Clostridia | Firmicutes | Bacteria | 0.00369584 |
| Clostridia | Firmicutes | Bacteria | 0 |
| Clostridia | Firmicutes | Bacteria | 0.001846066 |
| unclassified | unclassified | Bacteria | 0.02145544 |
| Bacilli | Firmicutes | Bacteria | 0 |
| Actinomycetia | Actinobacteria | Bacteria | 0 |
| Clostridia | Firmicutes | Bacteria | 0 |
| Bacilli | Firmicutes | Bacteria | 0 |
| Clostridia | Firmicutes | Bacteria | 0 |
| Coriobacteriia | Actinobacteria | Bacteria | 0.033661225 |
| Coriobacteriia | Actinobacteria | Bacteria | 0.028324926 |
| Gammaproteobacteria | Proteobacteria | Bacteria | 0 |
| Clostridia | Firmicutes | Bacteria | 0 |
| Clostridia | Firmicutes | Bacteria | 0 |
| Clostridia | Firmicutes | Bacteria | 0 |
| Clostridia | Firmicutes | Bacteria | 0 |
| Coriobacteriia | Actinobacteria | Bacteria | 0 |
| Negativicutes | Firmicutes | Bacteria | 0 |
| Clostridia | Firmicutes | Bacteria | 0.643916289 |
| Clostridia | Firmicutes | Bacteria | 0 |
| Clostridia | Firmicutes | Bacteria | 0 |
| Gammaproteobacteria | Proteobacteria | Bacteria | 0 |
| Clostridia | Firmicutes | Bacteria | 0 |
| Clostridia | Firmicutes | Bacteria | 0 |
| Clostridia | Firmicutes | Bacteria | 0 |
| Clostridia | Firmicutes | Bacteria | 0.001079201 |
| Bacilli | Firmicutes | Bacteria | 0.001389264 |
| Clostridia | Firmicutes | Bacteria | 0 |
| Clostridia | Firmicutes | Bacteria | 0 |
| Erysipelotrichia | Firmicutes | Bacteria | 0 |
| Coriobacteriia | Actinobacteria | Bacteria | 0 |

| | | | |
|---|---|---|---|
| Erysipelotrichia | Firmicutes | Bacteria | 0 |
| Clostridia | Firmicutes | Bacteria | 0.043391514 |
| Coriobacteriia | Actinobacteria | Bacteria | 0 |
| Clostridia | Firmicutes | Bacteria | 0.103068052 |
| Clostridia | Firmicutes | Bacteria | 0 |
| Clostridia | Firmicutes | Bacteria | 0 |
| Bacteroidia | Bacteroidetes | Bacteria | 0 |
| unclassified | unclassified | unclassified | 0 |
| Bacteroidia | Bacteroidetes | Bacteria | 0 |
| Clostridia | Firmicutes | Bacteria | 0 |
| Clostridia | Firmicutes | Bacteria | 0 |
| Clostridia | Firmicutes | Bacteria | 0.002207831 |
| Clostridia | Firmicutes | Bacteria | 0 |
| unclassified | Firmicutes | Bacteria | 0 |
| Bacteroidia | Bacteroidetes | Bacteria | 0.024347316 |
| Clostridia | Firmicutes | Bacteria | 0.442468306 |
| Clostridia | Firmicutes | Bacteria | 0 |
| unclassified | unclassified | unclassified | 0 |
| unclassified | unclassified | unclassified | 0 |
| Clostridia | Firmicutes | Bacteria | 0.001792412 |
| Clostridia | Firmicutes | Bacteria | 0.000634721 |
| Clostridia | Firmicutes | Bacteria | 0 |
| unclassified | unclassified | unclassified | 0 |
| Clostridia | Firmicutes | Bacteria | 0.247609519 |
| Bacteroidia | Bacteroidetes | Bacteria | 0 |
| Clostridia | Firmicutes | Bacteria | 0 |
| Clostridia | Firmicutes | Bacteria | 0.002683116 |
| Erysipelotrichia | Firmicutes | Bacteria | 0 |
| Clostridia | Firmicutes | Bacteria | 0 |
| unclassified | Proteobacteria | Bacteria | 0 |
| Clostridia | Firmicutes | Bacteria | 0 |
| Clostridia | Firmicutes | Bacteria | 0 |
| Bacteroidia | Bacteroidetes | Bacteria | 0 |
| unclassified | unclassified | unclassified | 0 |
| Clostridia | Firmicutes | Bacteria | 0 |
| Bacteroidia | Bacteroidetes | Bacteria | 0 |
| Clostridia | Firmicutes | Bacteria | 0.002991428 |
| unclassified | unclassified | Bacteria | 0 |
| Bacteroidia | Bacteroidetes | Bacteria | 0.003551999 |
| Bacteroidia | Bacteroidetes | Bacteria | 0 |
| Clostridia | Firmicutes | Bacteria | 0.16231731 |
| Clostridia | Firmicutes | Bacteria | 0 |
| Actinomycetia | Actinobacteria | Bacteria | 8.093218493 |
| Bacteroidia | Bacteroidetes | Bacteria | 0 |
| unclassified | unclassified | Bacteria | 0 |
| Clostridia | Firmicutes | Bacteria | 0 |
| Clostridia | Firmicutes | Bacteria | 0 |
| Bacteroidia | Bacteroidetes | Bacteria | 0.000925855 |

| | | | |
|---|---|---|---|
| Clostridia | Firmicutes | Bacteria | 0 |
| unclassified | unclassified | unclassified | 0 |
| Erysipelotrichia | Firmicutes | Bacteria | 0.013756822 |
| Clostridia | Firmicutes | Bacteria | 0 |
| Erysipelotrichia | Firmicutes | Bacteria | 0 |
| Clostridia | Firmicutes | Bacteria | 0.003670629 |
| Bacteroidia | Bacteroidetes | Bacteria | 0 |
| Bacteroidia | Bacteroidetes | Bacteria | 0 |
| Clostridia | Firmicutes | Bacteria | 0 |
| Bacteroidia | Bacteroidetes | Bacteria | 0 |
| Bacteroidia | Bacteroidetes | Bacteria | 0.001307106 |
| Coriobacteriia | Actinobacteria | Bacteria | 1.468938081 |
| Bacteroidia | Bacteroidetes | Bacteria | 0 |
| Negativicutes | Firmicutes | Bacteria | 0.02774341 |
| Coriobacteriia | Actinobacteria | Bacteria | 0.134141685 |
| Clostridia | Firmicutes | Bacteria | 0 |
| Bacteroidia | Bacteroidetes | Bacteria | 0 |
| unclassified | Proteobacteria | Bacteria | 0 |
| Clostridia | Firmicutes | Bacteria | 0 |
| Clostridia | Firmicutes | Bacteria | 0 |
| Clostridia | Firmicutes | Bacteria | 0.00310405 |
| Actinomycetia | Actinobacteria | Bacteria | 0.030398326 |
| Actinomycetia | Actinobacteria | Bacteria | 13.58102237 |
| unclassified | Firmicutes | Bacteria | 0 |
| Clostridia | Firmicutes | Bacteria | 0 |
| Erysipelotrichia | Firmicutes | Bacteria | 0.011377549 |
| Verrucomicrobiae | Verrucomicrobia | Bacteria | 0 |
| Betaproteobacteria | Proteobacteria | Bacteria | 0 |
| Bacteroidia | Bacteroidetes | Bacteria | 0.001249608 |
| Coriobacteriia | Actinobacteria | Bacteria | 0 |
| Clostridia | Firmicutes | Bacteria | 0.003423687 |
| Clostridia | Firmicutes | Bacteria | 0.162554418 |
| Clostridia | Firmicutes | Bacteria | 0 |
| Bacteroidia | Bacteroidetes | Bacteria | 0.001876798 |
| Clostridia | Firmicutes | Bacteria | 0.001772043 |
| Clostridia | Firmicutes | Bacteria | 0.612118888 |
| Bacteroidia | Bacteroidetes | Bacteria | 0 |
| unclassified | Firmicutes | Bacteria | 0 |
| Negativicutes | Firmicutes | Bacteria | 0.017094397 |
| Clostridia | Firmicutes | Bacteria | 0 |
| Clostridia | Firmicutes | Bacteria | 0.007324846 |
| Clostridia | Firmicutes | Bacteria | 0 |
| Clostridia | Firmicutes | Bacteria | 0 |
| Clostridia | Firmicutes | Bacteria | 0 |
| Bacteroidia | Bacteroidetes | Bacteria | 0.004164713 |
| Clostridia | Firmicutes | Bacteria | 0 |
| unclassified | Firmicutes | Bacteria | 0 |
| Negativicutes | Firmicutes | Bacteria | 0.374829919 |

| | | | |
|---|---|---|---|
| Clostridia | Firmicutes | Bacteria | 0 |
| unclassified | unclassified | unclassified | 0 |
| Betaproteobacteria | Proteobacteria | Bacteria | 0 |
| Clostridia | Firmicutes | Bacteria | 0.005324351 |
| Clostridia | Firmicutes | Bacteria | 0 |
| Clostridia | Firmicutes | Bacteria | 0 |
| Clostridia | Firmicutes | Bacteria | 0 |
| Bacteroidia | Bacteroidetes | Bacteria | 0.010524566 |
| Clostridia | Firmicutes | Bacteria | 0 |
| Clostridia | Firmicutes | Bacteria | 0 |
| Clostridia | Firmicutes | Bacteria | 0.003294399 |
| Negativicutes | Firmicutes | Bacteria | 0 |
| Deltaproteobacteria | Proteobacteria | Bacteria | 0 |
| Clostridia | Firmicutes | Bacteria | 0.373328016 |
| Clostridia | Firmicutes | Bacteria | 0 |
| Clostridia | Firmicutes | Bacteria | 0 |
| Clostridia | Firmicutes | Bacteria | 0 |
| Clostridia | Firmicutes | Bacteria | 0 |
| Clostridia | Firmicutes | Bacteria | 0 |
| Clostridia | Firmicutes | Bacteria | 0 |
| Bacteroidia | Bacteroidetes | Bacteria | 0 |
| unclassified | Firmicutes | Bacteria | 0 |
| Clostridia | Firmicutes | Bacteria | 0 |
| Negativicutes | Firmicutes | Bacteria | 0 |
| unclassified | Firmicutes | Bacteria | 0 |
| unclassified | unclassified | Bacteria | 0 |
| Clostridia | Firmicutes | Bacteria | 0 |
| Clostridia | Firmicutes | Bacteria | 0 |
| Clostridia | Firmicutes | Bacteria | 0 |
| Clostridia | Firmicutes | Bacteria | 0 |
| Clostridia | Firmicutes | Bacteria | 0 |
| unclassified | Firmicutes | Bacteria | 0 |
| Clostridia | Firmicutes | Bacteria | 0 |
| Clostridia | Firmicutes | Bacteria | 0 |
| Clostridia | Firmicutes | Bacteria | 0.001921 |
| Clostridia | Firmicutes | Bacteria | 0 |
| Clostridia | Firmicutes | Bacteria | 0 |
| Clostridia | Firmicutes | Bacteria | 0.003532437 |
| unclassified | unclassified | unclassified | 0 |
| Clostridia | Firmicutes | Bacteria | 0 |
| Bacteroidia | Bacteroidetes | Bacteria | 0 |
| Clostridia | Firmicutes | Bacteria | 0 |
| Clostridia | Firmicutes | Bacteria | 0 |
| Clostridia | Firmicutes | Bacteria | 0.001805482 |
| Coriobacteriia | Actinobacteria | Bacteria | 0 |
| Clostridia | Firmicutes | Bacteria | 0 |
| unclassified | unclassified | unclassified | 0 |
| Clostridia | Firmicutes | Bacteria | 0 |

| | | | |
|---|---|---|---|
| Clostridia | Firmicutes | Bacteria | 0 |
| unclassified | Firmicutes | Bacteria | 0 |
| Clostridia | Firmicutes | Bacteria | 0 |
| Clostridia | Firmicutes | Bacteria | 0 |
| Clostridia | Firmicutes | Bacteria | 0 |
| Clostridia | Firmicutes | Bacteria | 0 |
| unclassified | unclassified | unclassified | 0 |
| Clostridia | Firmicutes | Bacteria | 0 |
| Erysipelotrichia | Firmicutes | Bacteria | 0.011282111 |
| Clostridia | Firmicutes | Bacteria | 0 |
| unclassified | unclassified | unclassified | 0 |
| Clostridia | Firmicutes | Bacteria | 0 |
| Bacteroidia | Bacteroidetes | Bacteria | 0 |
| Bacteroidia | Bacteroidetes | Bacteria | 0 |
| Clostridia | Firmicutes | Bacteria | 2.899949319 |
| Clostridia | Firmicutes | Bacteria | 0 |
| Bacteroidia | Bacteroidetes | Bacteria | 0 |
| Clostridia | Firmicutes | Bacteria | 0 |
| Clostridia | Firmicutes | Bacteria | 0 |
| unclassified | Firmicutes | Bacteria | 0 |
| unclassified | unclassified | unclassified | 0.027644947 |
| unclassified | Firmicutes | Bacteria | 0 |
| Clostridia | Firmicutes | Bacteria | 0 |
| Clostridia | Firmicutes | Bacteria | 0 |
| Gammaproteobacteria | Proteobacteria | Bacteria | 0.012965909 |
| unclassified | unclassified | unclassified | 3.727093195 |
| Clostridia | Firmicutes | Bacteria | 0 |
| Clostridia | Firmicutes | Bacteria | 0 |
| Bacteroidia | Bacteroidetes | Bacteria | 0 |
| unclassified | unclassified | unclassified | 0 |
| unclassified | Firmicutes | Bacteria | 0 |
| Clostridia | Firmicutes | Bacteria | 0.001359321 |
| Clostridia | Firmicutes | Bacteria | 0 |
| Bacteroidia | Bacteroidetes | Bacteria | 0.001127265 |
| Negativicutes | Firmicutes | Bacteria | 0.006368148 |
| Clostridia | Firmicutes | Bacteria | 0 |
| Clostridia | Firmicutes | Bacteria | 0 |
| Clostridia | Firmicutes | Bacteria | 2.862777612 |
| Bacilli | Firmicutes | Bacteria | 0 |
| Betaproteobacteria | Proteobacteria | Bacteria | 0 |
| Actinomycetia | Actinobacteria | Bacteria | 0 |
| Clostridia | Firmicutes | Bacteria | 0.001276187 |
| Clostridia | Firmicutes | Bacteria | 5.460129436 |
| Bacteroidia | Bacteroidetes | Bacteria | 0 |
| unclassified | Firmicutes | Bacteria | 0 |
| Clostridia | Firmicutes | Bacteria | 0 |
| Clostridia | Firmicutes | Bacteria | 0 |
| Clostridia | Firmicutes | Bacteria | 0.124840528 |

| | | | |
|---|---|---|---|
| Bacteroidia | Bacteroidetes | Bacteria | 0.001899872 |
| Clostridia | Firmicutes | Bacteria | 0 |
| Clostridia | Firmicutes | Bacteria | 0 |
| Clostridia | Firmicutes | Bacteria | 0 |
| Bacteroidia | Bacteroidetes | Bacteria | 0 |
| Bacteroidia | Bacteroidetes | Bacteria | 0 |
| Clostridia | Firmicutes | Bacteria | 0 |
| Clostridia | Firmicutes | Bacteria | 0 |
| Clostridia | Firmicutes | Bacteria | 0 |
| Clostridia | Firmicutes | Bacteria | 0 |
| Clostridia | Firmicutes | Bacteria | 0 |
| Clostridia | Firmicutes | Bacteria | 2.327145982 |
| Erysipelotrichia | Firmicutes | Bacteria | 0.240993222 |
| unclassified | unclassified | unclassified | 0 |
| Gammaproteobacteria | Proteobacteria | Bacteria | 0.001458379 |
| unclassified | unclassified | unclassified | 0 |
| Bacteroidia | Bacteroidetes | Bacteria | 0 |
| Bacteroidia | Bacteroidetes | Bacteria | 0 |
| Bacteroidia | Bacteroidetes | Bacteria | 0 |
| Clostridia | Firmicutes | Bacteria | 0.003071014 |
| Clostridia | Firmicutes | Bacteria | 0.131756546 |
| Clostridia | Firmicutes | Bacteria | 0 |
| Clostridia | Firmicutes | Bacteria | 0.390489754 |
| Clostridia | Firmicutes | Bacteria | 0 |
| Clostridia | Firmicutes | Bacteria | 0 |
| Clostridia | Firmicutes | Bacteria | 0 |
| Bacteroidia | Bacteroidetes | Bacteria | 0 |
| Verrucomicrobiae | Verrucomicrobia | Bacteria | 0 |
| Clostridia | Firmicutes | Bacteria | 0 |
| Clostridia | Firmicutes | Bacteria | 0 |
| unclassified | unclassified | unclassified | 0 |
| Bacilli | Firmicutes | Bacteria | 0.015770425 |
| Clostridia | Firmicutes | Bacteria | 0 |
| Bacteroidia | Bacteroidetes | Bacteria | 0.187511449 |
| Clostridia | Firmicutes | Bacteria | 0 |
| Bacteroidia | Bacteroidetes | Bacteria | 0.002368166 |
| Clostridia | Firmicutes | Bacteria | 0 |
| Bacteroidia | Bacteroidetes | Bacteria | 0 |
| Betaproteobacteria | Proteobacteria | Bacteria | 0 |
| Clostridia | Firmicutes | Bacteria | 0 |
| Clostridia | Firmicutes | Bacteria | 0 |
| Clostridia | Firmicutes | Bacteria | 0 |
| unclassified | Firmicutes | Bacteria | 0 |
| Clostridia | Firmicutes | Bacteria | 0.029910371 |
| Bacteroidia | Bacteroidetes | Bacteria | 0.004423631 |
| unclassified | unclassified | unclassified | 0 |
| Clostridia | Firmicutes | Bacteria | 0 |
| Clostridia | Firmicutes | Bacteria | 0 |

| | | | |
|---|---|---|---|
| Clostridia | Firmicutes | Bacteria | 0 |
| Clostridia | Firmicutes | Bacteria | 0.003052252 |
| unclassified | unclassified | unclassified | 0 |
| Clostridia | Firmicutes | Bacteria | 0 |
| Clostridia | Firmicutes | Bacteria | 0 |
| Clostridia | Firmicutes | Bacteria | 0 |
| unclassified | unclassified | unclassified | 0 |
| Clostridia | Firmicutes | Bacteria | 0 |
| Clostridia | Firmicutes | Bacteria | 0 |
| Clostridia | Firmicutes | Bacteria | 0 |
| Clostridia | Firmicutes | Bacteria | 0 |
| Clostridia | Firmicutes | Bacteria | 0 |
| Bacteroidia | Bacteroidetes | Bacteria | 0 |
| Coriobacteriia | Actinobacteria | Bacteria | 0 |
| Clostridia | Firmicutes | Bacteria | 0 |
| Clostridia | Firmicutes | Bacteria | 0 |
| unclassified | Firmicutes | Bacteria | 0 |
| unclassified | unclassified | unclassified | 0 |
| Actinomycetia | Actinobacteria | Bacteria | 0.012888361 |
| Lentisphaeria | Lentisphaerae | Bacteria | 0 |
| Clostridia | Firmicutes | Bacteria | 0 |
| Clostridia | Firmicutes | Bacteria | 0 |
| unclassified | unclassified | unclassified | 0 |
| Coriobacteriia | Actinobacteria | Bacteria | 0 |
| Negativicutes | Firmicutes | Bacteria | 0 |
| Clostridia | Firmicutes | Bacteria | 0.043054256 |
| Clostridia | Firmicutes | Bacteria | 0 |
| Clostridia | Firmicutes | Bacteria | 0.017955966 |
| Clostridia | Firmicutes | Bacteria | 0.178211852 |
| Clostridia | Firmicutes | Bacteria | 0 |
| unclassified | unclassified | unclassified | 0 |
| unclassified | Firmicutes | Bacteria | 0 |
| Clostridia | Firmicutes | Bacteria | 0 |
| Bacteroidia | Bacteroidetes | Bacteria | 0 |
| Clostridia | Firmicutes | Bacteria | 0.00098083 |
| unclassified | unclassified | Bacteria | 0 |
| Bacteroidia | Bacteroidetes | Bacteria | 0 |
| Bacteroidia | Bacteroidetes | Bacteria | 0 |
| unclassified | unclassified | unclassified | 0 |
| Bacteroidia | Bacteroidetes | Bacteria | 0 |
| Clostridia | Firmicutes | Bacteria | 1.786907598 |
| Clostridia | Firmicutes | Bacteria | 1.392953949 |
| Bacteroidia | Bacteroidetes | Bacteria | 0 |
| Clostridia | Firmicutes | Bacteria | 0 |
| unclassified | Firmicutes | Bacteria | 0 |
| Deltaproteobacteria | Proteobacteria | Bacteria | 0 |
| Clostridia | Firmicutes | Bacteria | 0 |
| Bacteroidia | Bacteroidetes | Bacteria | 0 |

| | | | |
|---|---|---|---|
| Bacteroidia | Bacteroidetes | Bacteria | 0 |
| Clostridia | Firmicutes | Bacteria | 0 |
| unclassified | unclassified | unclassified | 0 |
| Clostridia | Firmicutes | Bacteria | 0 |
| Clostridia | Firmicutes | Bacteria | 3.59705331 |
| Clostridia | Firmicutes | Bacteria | 0 |
| Clostridia | Firmicutes | Bacteria | 0 |
| Clostridia | Firmicutes | Bacteria | 0 |
| Clostridia | Firmicutes | Bacteria | 0.038647161 |
| Clostridia | Firmicutes | Bacteria | 0 |
| Clostridia | Firmicutes | Bacteria | 0 |
| unclassified | Firmicutes | Bacteria | 0 |
| Clostridia | Firmicutes | Bacteria | 0.021893714 |
| Clostridia | Firmicutes | Bacteria | 0.002387718 |
| Bacteroidia | Bacteroidetes | Bacteria | 0 |
| Clostridia | Firmicutes | Bacteria | 0.365328625 |
| Clostridia | Firmicutes | Bacteria | 0.000751345 |
| Bacteroidia | Bacteroidetes | Bacteria | 0 |
| Bacteroidia | Bacteroidetes | Bacteria | 0 |
| Clostridia | Firmicutes | Bacteria | 0 |
| unclassified | unclassified | unclassified | 0 |
| Clostridia | Firmicutes | Bacteria | 0 |
| Actinomycetia | Actinobacteria | Bacteria | 0 |
| unclassified | unclassified | Bacteria | 0 |
| unclassified | unclassified | unclassified | 0.014796694 |
| unclassified | Firmicutes | Bacteria | 0 |
| Clostridia | Firmicutes | Bacteria | 0.040669521 |
| Clostridia | Firmicutes | Bacteria | 0 |
| unclassified | unclassified | unclassified | 0 |
| Clostridia | Firmicutes | Bacteria | 0 |
| unclassified | Firmicutes | Bacteria | 0 |
| unclassified | Firmicutes | Bacteria | 0 |
| Clostridia | Firmicutes | Bacteria | 0 |
| Clostridia | Firmicutes | Bacteria | 0.012171403 |
| Bacteroidia | Bacteroidetes | Bacteria | 0 |
| Erysipelotrichia | Firmicutes | Bacteria | 0 |
| Bacilli | Firmicutes | Bacteria | 0 |
| Clostridia | Firmicutes | Bacteria | 0 |
| unclassified | unclassified | Bacteria | 0 |
| unclassified | Firmicutes | Bacteria | 0 |
| Erysipelotrichia | Firmicutes | Bacteria | 6.084977804 |
| Bacteroidia | Bacteroidetes | Bacteria | 0 |
| Clostridia | Firmicutes | Bacteria | 0 |
| Gammaproteobacteria | Proteobacteria | Bacteria | 0 |
| Verrucomicrobiae | Verrucomicrobia | Bacteria | 0 |
| unclassified | unclassified | Bacteria | 0 |
| unclassified | Firmicutes | Bacteria | 0 |
| Clostridia | Firmicutes | Bacteria | 0 |

| | | | |
|---|---|---|---|
| Clostridia | Firmicutes | Bacteria | 0.07970807 |
| Clostridia | Firmicutes | Bacteria | 0 |
| Clostridia | Firmicutes | Bacteria | 0 |
| Clostridia | Firmicutes | Bacteria | 0 |
| Clostridia | Firmicutes | Bacteria | 0 |
| Clostridia | Firmicutes | Bacteria | 0 |
| Clostridia | Firmicutes | Bacteria | 0.000749518 |
| Bacteroidia | Bacteroidetes | Bacteria | 0 |
| Bacteroidia | Bacteroidetes | Bacteria | 0 |
| Clostridia | Firmicutes | Bacteria | 0 |
| unclassified | unclassified | unclassified | 0 |
| Clostridia | Firmicutes | Bacteria | 0.000819335 |
| Betaproteobacteria | Proteobacteria | Bacteria | 0 |
| Negativicutes | Firmicutes | Bacteria | 0 |
| Clostridia | Firmicutes | Bacteria | 0.004050553 |
| Clostridia | Firmicutes | Bacteria | 0 |
| Actinomycetia | Actinobacteria | Bacteria | 0 |
| Clostridia | Firmicutes | Bacteria | 0 |
| Bacteroidia | Bacteroidetes | Bacteria | 0 |
| unclassified | unclassified | unclassified | 0 |
| Clostridia | Firmicutes | Bacteria | 0.58121356 |
| unclassified | Firmicutes | Bacteria | 0 |
| Clostridia | Firmicutes | Bacteria | 0 |
| Clostridia | Firmicutes | Bacteria | 0.000874849 |
| Negativicutes | Firmicutes | Bacteria | 0 |
| Bacteroidia | Bacteroidetes | Bacteria | 0 |
| Bacteroidia | Bacteroidetes | Bacteria | 0 |
| Clostridia | Firmicutes | Bacteria | 0 |
| Coriobacteriia | Actinobacteria | Bacteria | 0 |
| Bacilli | Firmicutes | Bacteria | 0 |
| unclassified | unclassified | unclassified | 0 |
| Betaproteobacteria | Proteobacteria | Bacteria | 0 |
| Clostridia | Firmicutes | Bacteria | 0 |
| Verrucomicrobiae | Verrucomicrobia | Bacteria | 1.96237527 |
| Clostridia | Firmicutes | Bacteria | 0 |
| unclassified | Firmicutes | Bacteria | 0 |
| Clostridia | Firmicutes | Bacteria | 0.000656798 |
| Bacteroidia | Bacteroidetes | Bacteria | 0 |
| Bacteroidia | Bacteroidetes | Bacteria | 0 |
| unclassified | unclassified | unclassified | 0 |
| Bacteroidia | Bacteroidetes | Bacteria | 0 |
| unclassified | unclassified | Bacteria | 0 |
| Clostridia | Firmicutes | Bacteria | 0 |
| Clostridia | Firmicutes | Bacteria | 0 |
| unclassified | unclassified | unclassified | 0.016783461 |
| Clostridia | Firmicutes | Bacteria | 0 |
| unclassified | unclassified | unclassified | 0 |
| Bacteroidia | Bacteroidetes | Bacteria | 0 |

| | | | |
|---|---|---|---|
| Clostridia | Firmicutes | Bacteria | 0.000941369 |
| Bacteroidia | Bacteroidetes | Bacteria | 0 |
| Clostridia | Firmicutes | Bacteria | 0 |
| Betaproteobacteria | Proteobacteria | Bacteria | 0 |
| Bacteroidia | Bacteroidetes | Bacteria | 0 |
| unclassified | unclassified | unclassified | 0 |
| Bacteroidia | Bacteroidetes | Bacteria | 0 |
| Clostridia | Firmicutes | Bacteria | 1.960597123 |
| Gammaproteobacteria | Proteobacteria | Bacteria | 0.338670518 |
| Clostridia | Firmicutes | Bacteria | 0.004937639 |
| Clostridia | Firmicutes | Bacteria | 0 |
| unclassified | unclassified | unclassified | 0 |
| Clostridia | Firmicutes | Bacteria | 0.008908612 |
| Negativicutes | Firmicutes | Bacteria | 0 |
| Bacteroidia | Bacteroidetes | Bacteria | 0 |
| unclassified | unclassified | unclassified | 0 |
| Erysipelotrichia | Firmicutes | Bacteria | 0 |
| Erysipelotrichia | Firmicutes | Bacteria | 0 |
| Bacteroidia | Bacteroidetes | Bacteria | 0 |
| Bacteroidia | Bacteroidetes | Bacteria | 0 |
| Methanobacteria | Euryarchaeota | Archaea | 0 |
| unclassified | unclassified | unclassified | 0 |
| Clostridia | Firmicutes | Bacteria | 0 |
| Clostridia | Firmicutes | Bacteria | 0 |
| Clostridia | Firmicutes | Bacteria | 0 |
| unclassified | unclassified | unclassified | 0 |
| Clostridia | Firmicutes | Bacteria | 0 |
| Clostridia | Firmicutes | Bacteria | 0 |
| Clostridia | Firmicutes | Bacteria | 0 |
| Bacteroidia | Bacteroidetes | Bacteria | 0 |
| Bacteroidia | Bacteroidetes | Bacteria | 0 |
| Bacteroidia | Bacteroidetes | Bacteria | 0 |
| unclassified | Firmicutes | Bacteria | 0 |
| Clostridia | Firmicutes | Bacteria | 0 |
| Negativicutes | Firmicutes | Bacteria | 0 |
| Clostridia | Firmicutes | Bacteria | 0 |
| unclassified | Firmicutes | Bacteria | 0 |
| Clostridia | Firmicutes | Bacteria | 0 |
| Betaproteobacteria | Proteobacteria | Bacteria | 0 |
| Clostridia | Firmicutes | Bacteria | 0.033644163 |
| unclassified | unclassified | unclassified | 0 |
| Clostridia | Firmicutes | Bacteria | 0.17820347 |
| Clostridia | Firmicutes | Bacteria | 0 |
| Clostridia | Firmicutes | Bacteria | 0 |
| Bacteroidia | Bacteroidetes | Bacteria | 0 |
| Clostridia | Firmicutes | Bacteria | 0 |
| Clostridia | Firmicutes | Bacteria | 0 |
| Lentisphaeria | Lentisphaerae | Bacteria | 0 |

| | | | |
|---|---|---|---|
| Bacteroidia | Bacteroidetes | Bacteria | 0 |
| Actinomycetia | Actinobacteria | Bacteria | 0 |
| Clostridia | Firmicutes | Bacteria | 0 |
| Clostridia | Firmicutes | Bacteria | 0 |
| Bacteroidia | Bacteroidetes | Bacteria | 0.139696757 |
| unclassified | Firmicutes | Bacteria | 0 |
| Bacteroidia | Bacteroidetes | Bacteria | 0 |
| Erysipelotrichia | Firmicutes | Bacteria | 0.008142929 |
| Clostridia | Firmicutes | Bacteria | 0 |
| Clostridia | Firmicutes | Bacteria | 0 |
| Clostridia | Firmicutes | Bacteria | 0 |
| Bacteroidia | Bacteroidetes | Bacteria | 0 |
| unclassified | unclassified | unclassified | 0 |
| unclassified | unclassified | Bacteria | 0 |
| Erysipelotrichia | Firmicutes | Bacteria | 0 |
| Bacteroidia | Bacteroidetes | Bacteria | 0 |
| Clostridia | Firmicutes | Bacteria | 0 |
| unclassified | unclassified | unclassified | 0 |
| unclassified | Firmicutes | Bacteria | 0 |
| unclassified | Firmicutes | Bacteria | 0 |
| Clostridia | Firmicutes | Bacteria | 0 |
| unclassified | unclassified | Bacteria | 0 |
| unclassified | Firmicutes | Bacteria | 0 |
| Clostridia | Firmicutes | Bacteria | 0.003523517 |
| Coriobacteriia | Actinobacteria | Bacteria | 0 |
| Clostridia | Firmicutes | Bacteria | 0 |
| Bacteroidia | Bacteroidetes | Bacteria | 0.000758272 |
| Clostridia | Firmicutes | Bacteria | 0 |
| Clostridia | Firmicutes | Bacteria | 0 |
| unclassified | unclassified | unclassified | 0 |
| Betaproteobacteria | Proteobacteria | Bacteria | 0 |
| Clostridia | Firmicutes | Bacteria | 0.001457631 |
| Clostridia | Firmicutes | Bacteria | 0 |
| unclassified | unclassified | unclassified | 0 |
| Bacteroidia | Bacteroidetes | Bacteria | 0 |
| Clostridia | Firmicutes | Bacteria | 0 |
| unclassified | unclassified | unclassified | 0 |
| Clostridia | Firmicutes | Bacteria | 0.000583239 |
| Clostridia | Firmicutes | Bacteria | 0 |
| Clostridia | Firmicutes | Bacteria | 0 |
| unclassified | unclassified | unclassified | 0 |
| Bacteroidia | Bacteroidetes | Bacteria | 0 |
| Clostridia | Firmicutes | Bacteria | 0 |
| unclassified | unclassified | Bacteria | 0 |
| Clostridia | Firmicutes | Bacteria | 0 |
| Clostridia | Firmicutes | Bacteria | 0 |
| unclassified | Firmicutes | Bacteria | 0 |
| Clostridia | Firmicutes | Bacteria | 0 |

| | | | |
|---|---|---|---|
| unclassified | unclassified | unclassified | 0.025136407 |
| Methanobacteria | Euryarchaeota | Archaea | 0 |
| unclassified | Firmicutes | Bacteria | 0 |
| Bacteroidia | Bacteroidetes | Bacteria | 0 |
| Clostridia | Firmicutes | Bacteria | 0 |
| Bacteroidia | Bacteroidetes | Bacteria | 0 |
| unclassified | unclassified | Bacteria | 0 |
| Clostridia | Firmicutes | Bacteria | 0 |
| Clostridia | Firmicutes | Bacteria | 0 |
| unclassified | unclassified | unclassified | 0.012088521 |
| Erysipelotrichia | Firmicutes | Bacteria | 0 |
| Bacteroidia | Bacteroidetes | Bacteria | 0 |
| unclassified | unclassified | unclassified | 0 |
| unclassified | unclassified | unclassified | 0 |
| unclassified | unclassified | unclassified | 0 |
| unclassified | unclassified | unclassified | 0 |
| Clostridia | Firmicutes | Bacteria | 0 |
| Clostridia | Firmicutes | Bacteria | 0 |
| Bacteroidia | Bacteroidetes | Bacteria | 0 |
| Bacteroidia | Bacteroidetes | Bacteria | 0 |
| Clostridia | Firmicutes | Bacteria | 0.002476791 |
| unclassified | unclassified | unclassified | 0 |
| Bacteroidia | Bacteroidetes | Bacteria | 0 |
| Clostridia | Firmicutes | Bacteria | 0.100679186 |
| Clostridia | Firmicutes | Bacteria | 0 |
| Bacteroidia | Bacteroidetes | Bacteria | 0 |
| unclassified | unclassified | unclassified | 0 |
| Clostridia | Firmicutes | Bacteria | 0 |
| Negativicutes | Firmicutes | Bacteria | 0 |
| Clostridia | Firmicutes | Bacteria | 0 |
| Clostridia | Firmicutes | Bacteria | 0 |
| Clostridia | Firmicutes | Bacteria | 0 |
| Clostridia | Firmicutes | Bacteria | 0 |
| Clostridia | Firmicutes | Bacteria | 0 |
| Erysipelotrichia | Firmicutes | Bacteria | 1.57654949 |
| Clostridia | Firmicutes | Bacteria | 0 |
| Clostridia | Firmicutes | Bacteria | 0 |
| Bacteroidia | Bacteroidetes | Bacteria | 0 |
| Clostridia | Firmicutes | Bacteria | 0.005664476 |
| Clostridia | Firmicutes | Bacteria | 0 |
| unclassified | Firmicutes | Bacteria | 0 |
| Clostridia | Firmicutes | Bacteria | 0 |
| Bacteroidia | Bacteroidetes | Bacteria | 0 |
| Clostridia | Firmicutes | Bacteria | 0 |
| Erysipelotrichia | Firmicutes | Bacteria | 0 |
| Clostridia | Firmicutes | Bacteria | 0.01514933 |
| Bacteroidia | Bacteroidetes | Bacteria | 0 |
| Clostridia | Firmicutes | Bacteria | 0 |

| | | | |
|---|---|---|---|
| unclassified | unclassified | unclassified | 0 |
| Bacteroidia | Bacteroidetes | Bacteria | 0 |
| Clostridia | Firmicutes | Bacteria | 0 |
| unclassified | unclassified | unclassified | 0 |
| Clostridia | Firmicutes | Bacteria | 0 |
| unclassified | unclassified | unclassified | 0 |
| Clostridia | Firmicutes | Bacteria | 0 |
| Coriobacteriia | Actinobacteria | Bacteria | 0.098406962 |
| unclassified | unclassified | unclassified | 0.012025064 |
| Negativicutes | Firmicutes | Bacteria | 0 |
| unclassified | Firmicutes | Bacteria | 0 |
| Clostridia | Firmicutes | Bacteria | 0 |
| Clostridia | Firmicutes | Bacteria | 0 |
| Clostridia | Firmicutes | Bacteria | 0.000559191 |
| Clostridia | Firmicutes | Bacteria | 0 |
| Clostridia | Firmicutes | Bacteria | 0 |
| Negativicutes | Firmicutes | Bacteria | 0 |
| Clostridia | Firmicutes | Bacteria | 0 |
| Negativicutes | Firmicutes | Bacteria | 0 |
| Clostridia | Firmicutes | Bacteria | 1.186618511 |
| Clostridia | Firmicutes | Bacteria | 0.00601288 |
| Clostridia | Firmicutes | Bacteria | 0 |
| Clostridia | Firmicutes | Bacteria | 0 |
| Clostridia | Firmicutes | Bacteria | 0.008520636 |
| Bacteroidia | Bacteroidetes | Bacteria | 0 |
| Clostridia | Firmicutes | Bacteria | 0 |
| unclassified | unclassified | unclassified | 0 |
| Negativicutes | Firmicutes | Bacteria | 0 |
| Clostridia | Firmicutes | Bacteria | 0 |
| Clostridia | Firmicutes | Bacteria | 0 |
| Clostridia | Firmicutes | Bacteria | 0 |
| Clostridia | Firmicutes | Bacteria | 0 |
| Clostridia | Firmicutes | Bacteria | 0 |
| unclassified | Firmicutes | Bacteria | 0 |
| unclassified | unclassified | unclassified | 0 |
| unclassified | Firmicutes | Bacteria | 0 |
| unclassified | unclassified | unclassified | 0 |
| Clostridia | Firmicutes | Bacteria | 0 |
| Coriobacteriia | Actinobacteria | Bacteria | 0 |
| Clostridia | Firmicutes | Bacteria | 0 |
| Clostridia | Firmicutes | Bacteria | 0 |
| Clostridia | Firmicutes | Bacteria | 0 |
| Clostridia | Firmicutes | Bacteria | 0 |
| Bacteroidia | Bacteroidetes | Bacteria | 0 |
| Clostridia | Firmicutes | Bacteria | 0 |
| Bacteroidia | Bacteroidetes | Bacteria | 0 |
| Clostridia | Firmicutes | Bacteria | 0 |
| Deltaproteobacteria | Proteobacteria | Bacteria | 0 |

| | | | |
|---|---|---|---|
| Betaproteobacteria | Proteobacteria | Bacteria | 0 |
| Clostridia | Firmicutes | Bacteria | 0 |
| Clostridia | Firmicutes | Bacteria | 0 |
| Betaproteobacteria | Proteobacteria | Bacteria | 0 |
| Clostridia | Firmicutes | Bacteria | 0.078273487 |
| unclassified | unclassified | unclassified | 0 |
| Coriobacteriia | Actinobacteria | Bacteria | 0 |
| Clostridia | Firmicutes | Bacteria | 0 |
| Clostridia | Firmicutes | Bacteria | 0 |
| unclassified | unclassified | unclassified | 0 |
| Clostridia | Firmicutes | Bacteria | 0 |
| Bacteroidia | Bacteroidetes | Bacteria | 0.001358392 |
| Bacteroidia | Bacteroidetes | Bacteria | 0 |
| Bacteroidia | Bacteroidetes | Bacteria | 0 |
| Erysipelotrichia | Firmicutes | Bacteria | 0 |
| Clostridia | Firmicutes | Bacteria | 0 |
| Clostridia | Firmicutes | Bacteria | 0.001915402 |
| Clostridia | Firmicutes | Bacteria | 0 |
| Clostridia | Firmicutes | Bacteria | 0.049029079 |
| Clostridia | Firmicutes | Bacteria | 0.000895343 |
| Bacteroidia | Bacteroidetes | Bacteria | 0 |
| Clostridia | Firmicutes | Bacteria | 0 |
| Clostridia | Firmicutes | Bacteria | 0 |
| Bacteroidia | Bacteroidetes | Bacteria | 0 |
| unclassified | Firmicutes | Bacteria | 0 |
| unclassified | unclassified | unclassified | 0 |
| Clostridia | Firmicutes | Bacteria | 0.002511178 |
| Clostridia | Firmicutes | Bacteria | 0 |
| unclassified | Firmicutes | Bacteria | 0 |
| Clostridia | Firmicutes | Bacteria | 0 |
| Clostridia | Firmicutes | Bacteria | 0 |
| Clostridia | Firmicutes | Bacteria | 0 |
| Clostridia | Firmicutes | Bacteria | 0 |
| Clostridia | Firmicutes | Bacteria | 0 |
| Clostridia | Firmicutes | Bacteria | 0 |
| Clostridia | Firmicutes | Bacteria | 0 |
| unclassified | Firmicutes | Bacteria | 0 |
| Bacteroidia | Bacteroidetes | Bacteria | 0.004714062 |
| Clostridia | Firmicutes | Bacteria | 0 |
| Clostridia | Firmicutes | Bacteria | 0 |
| Bacteroidia | Bacteroidetes | Bacteria | 0 |
| unclassified | unclassified | unclassified | 0 |
| unclassified | Firmicutes | Bacteria | 0 |
| Clostridia | Firmicutes | Bacteria | 0 |
| unclassified | unclassified | unclassified | 0 |
| Clostridia | Firmicutes | Bacteria | 0 |
| Bacteroidia | Bacteroidetes | Bacteria | 0 |
| unclassified | unclassified | unclassified | 0 |

| | | | |
|---|---|---|---|
| unclassified | Firmicutes | Bacteria | 0 |
| Clostridia | Firmicutes | Bacteria | 0.037477689 |
| Clostridia | Firmicutes | Bacteria | 0 |
| Clostridia | Firmicutes | Bacteria | 0 |
| Negativicutes | Firmicutes | Bacteria | 0 |
| Negativicutes | Firmicutes | Bacteria | 0 |
| Clostridia | Firmicutes | Bacteria | 0 |
| Deltaproteobacteria | Proteobacteria | Bacteria | 0 |
| Clostridia | Firmicutes | Bacteria | 0 |
| Clostridia | Firmicutes | Bacteria | 0 |
| Clostridia | Firmicutes | Bacteria | 0.026415386 |
| Clostridia | Firmicutes | Bacteria | 0 |
| Clostridia | Firmicutes | Bacteria | 0.04862583 |
| Clostridia | Firmicutes | Bacteria | 0 |
| Deltaproteobacteria | Proteobacteria | Bacteria | 0 |
| Clostridia | Firmicutes | Bacteria | 0 |
| Bacteroidia | Bacteroidetes | Bacteria | 0 |
| Clostridia | Firmicutes | Bacteria | 0 |
| Betaproteobacteria | Proteobacteria | Bacteria | 0 |
| Clostridia | Firmicutes | Bacteria | 0 |
| unclassified | Firmicutes | Bacteria | 0 |
| unclassified | Firmicutes | Bacteria | 0 |
| Clostridia | Firmicutes | Bacteria | 0.013054459 |
| Clostridia | Firmicutes | Bacteria | 0.066998265 |
| unclassified | unclassified | unclassified | 0 |
| Clostridia | Firmicutes | Bacteria | 0 |
| Clostridia | Firmicutes | Bacteria | 0 |
| Deltaproteobacteria | Proteobacteria | Bacteria | 0 |
| unclassified | unclassified | Bacteria | 0 |
| Bacteroidia | Bacteroidetes | Bacteria | 0 |
| Clostridia | Firmicutes | Bacteria | 0 |
| Bacteroidia | Bacteroidetes | Bacteria | 0 |
| Betaproteobacteria | Proteobacteria | Bacteria | 0 |
| Clostridia | Firmicutes | Bacteria | 0 |
| Clostridia | Firmicutes | Bacteria | 0 |
| Clostridia | Firmicutes | Bacteria | 0 |
| Clostridia | Firmicutes | Bacteria | 0 |
| Clostridia | Firmicutes | Bacteria | 0 |
| Gammaproteobacteria | Proteobacteria | Bacteria | 0 |
| Clostridia | Firmicutes | Bacteria | 0 |
| Clostridia | Firmicutes | Bacteria | 0 |
| Bacteroidia | Bacteroidetes | Bacteria | 0 |
| Clostridia | Firmicutes | Bacteria | 0 |
| unclassified | unclassified | unclassified | 0 |
| Clostridia | Firmicutes | Bacteria | 1.916164768 |
| Clostridia | Firmicutes | Bacteria | 0.101988982 |
| Clostridia | Firmicutes | Bacteria | 0 |
| Clostridia | Firmicutes | Bacteria | 0 |

| | | | |
|---|---|---|---|
| Clostridia | Firmicutes | Bacteria | 0 |
| unclassified | Firmicutes | Bacteria | 0 |
| Coriobacteriia | Actinobacteria | Bacteria | 0 |
| Clostridia | Firmicutes | Bacteria | 0 |
| Clostridia | Firmicutes | Bacteria | 0 |
| Betaproteobacteria | Proteobacteria | Bacteria | 0 |
| Bacteroidia | Bacteroidetes | Bacteria | 0 |
| unclassified | unclassified | unclassified | 0 |
| Clostridia | Firmicutes | Bacteria | 0 |
| Gammaproteobacteria | Proteobacteria | Bacteria | 0 |
| Bacteroidia | Bacteroidetes | Bacteria | 0 |
| Clostridia | Firmicutes | Bacteria | 0 |
| Bacteroidia | Bacteroidetes | Bacteria | 0 |
| Bacteroidia | Bacteroidetes | Bacteria | 0 |
| Bacteroidia | Bacteroidetes | Bacteria | 0 |
| Clostridia | Firmicutes | Bacteria | 0 |
| Gammaproteobacteria | Proteobacteria | Bacteria | 0 |
| Clostridia | Firmicutes | Bacteria | 0 |
| Bacteroidia | Bacteroidetes | Bacteria | 0 |
| Betaproteobacteria | Proteobacteria | Bacteria | 0.000630932 |
| Clostridia | Firmicutes | Bacteria | 0 |
| unclassified | unclassified | unclassified | 0 |
| unclassified | unclassified | unclassified | 0 |
| Clostridia | Firmicutes | Bacteria | 0 |
| Clostridia | Firmicutes | Bacteria | 0 |
| Clostridia | Firmicutes | Bacteria | 0 |
| Clostridia | Firmicutes | Bacteria | 0 |
| Erysipelotrichia | Firmicutes | Bacteria | 0 |
| Clostridia | Firmicutes | Bacteria | 0 |
| Clostridia | Firmicutes | Bacteria | 0 |
| Clostridia | Firmicutes | Bacteria | 0.000722715 |
| Clostridia | Firmicutes | Bacteria | 0 |
| Coriobacteriia | Actinobacteria | Bacteria | 0.322651459 |
| Bacilli | Firmicutes | Bacteria | 0.020580378 |
| unclassified | unclassified | unclassified | 0 |
| Clostridia | Firmicutes | Bacteria | 0 |
| Clostridia | Firmicutes | Bacteria | 0 |
| Bacteroidia | Bacteroidetes | Bacteria | 0 |
| Bacteroidia | Bacteroidetes | Bacteria | 0 |
| Clostridia | Firmicutes | Bacteria | 0 |
| Clostridia | Firmicutes | Bacteria | 0 |
| Clostridia | Firmicutes | Bacteria | 0.00276143 |
| Clostridia | Firmicutes | Bacteria | 0.107444137 |
| unclassified | unclassified | unclassified | 0 |
| Clostridia | Firmicutes | Bacteria | 0 |
| Bacteroidia | Bacteroidetes | Bacteria | 0 |
| Bacteroidia | Bacteroidetes | Bacteria | 0 |
| Clostridia | Firmicutes | Bacteria | 0 |

| | | | |
|---|---|---|---|
| Clostridia | Firmicutes | Bacteria | 0 |
| Coriobacteriia | Actinobacteria | Bacteria | 0 |
| Bacilli | Firmicutes | Bacteria | 0.006690003 |
| Bacteroidia | Bacteroidetes | Bacteria | 0 |
| Bacteroidia | Bacteroidetes | Bacteria | 0.068825042 |
| Negativicutes | Firmicutes | Bacteria | 0.002999726 |
| unclassified | unclassified | unclassified | 0 |
| Bacteroidia | Bacteroidetes | Bacteria | 0 |
| Bacteroidia | Bacteroidetes | Bacteria | 0 |
| Bacteroidia | Bacteroidetes | Bacteria | 0 |
| Clostridia | Firmicutes | Bacteria | 0 |
| unclassified | unclassified | unclassified | 0 |
| Bacteroidia | Bacteroidetes | Bacteria | 0 |
| Coriobacteriia | Actinobacteria | Bacteria | 0 |
| unclassified | unclassified | unclassified | 0 |
| Clostridia | Firmicutes | Bacteria | 0 |
| Clostridia | Firmicutes | Bacteria | 0 |
| Clostridia | Firmicutes | Bacteria | 0 |
| Clostridia | Firmicutes | Bacteria | 0 |
| Clostridia | Firmicutes | Bacteria | 0 |
| Clostridia | Firmicutes | Bacteria | 0 |
| Erysipelotrichia | Firmicutes | Bacteria | 0 |
| Clostridia | Firmicutes | Bacteria | 0 |
| Clostridia | Firmicutes | Bacteria | 0 |
| unclassified | unclassified | unclassified | 0 |
| Actinomycetia | Actinobacteria | Bacteria | 0 |
| unclassified | unclassified | unclassified | 0 |
| Clostridia | Firmicutes | Bacteria | 0 |
| Clostridia | Firmicutes | Bacteria | 0 |
| Bacteroidia | Bacteroidetes | Bacteria | 0 |
| Bacteroidia | Bacteroidetes | Bacteria | 0 |
| Clostridia | Firmicutes | Bacteria | 0 |
| unclassified | unclassified | unclassified | 0 |
| Bacteroidia | Bacteroidetes | Bacteria | 0 |
| unclassified | unclassified | Bacteria | 0 |
| unclassified | unclassified | unclassified | 0 |
| Bacteroidia | Bacteroidetes | Bacteria | 0 |
| Clostridia | Firmicutes | Bacteria | 0.002548167 |
| Clostridia | Firmicutes | Bacteria | 0 |
| Betaproteobacteria | Proteobacteria | Bacteria | 0 |
| Clostridia | Firmicutes | Bacteria | 0 |
| Negativicutes | Firmicutes | Bacteria | 1.780351553 |
| Clostridia | Firmicutes | Bacteria | 0.000642285 |
| Coriobacteriia | Actinobacteria | Bacteria | 0 |
| Clostridia | Firmicutes | Bacteria | 0 |
| Clostridia | Firmicutes | Bacteria | 0 |
| Clostridia | Firmicutes | Bacteria | 0 |
| unclassified | unclassified | unclassified | 0 |

| | | | |
|---|---|---|---|
| Clostridia | Firmicutes | Bacteria | 0 |
| Clostridia | Firmicutes | Bacteria | 0.001239823 |
| Clostridia | Firmicutes | Bacteria | 0 |
| Bacteroidia | Bacteroidetes | Bacteria | 0 |
| Clostridia | Firmicutes | Bacteria | 0 |
| Clostridia | Firmicutes | Bacteria | 0 |
| Bacteroidia | Bacteroidetes | Bacteria | 0 |
| Clostridia | Firmicutes | Bacteria | 0 |
| Clostridia | Firmicutes | Bacteria | 0 |
| Clostridia | Firmicutes | Bacteria | 0 |
| Clostridia | Firmicutes | Bacteria | 0 |
| unclassified | unclassified | unclassified | 0 |
| Clostridia | Firmicutes | Bacteria | 0.031454475 |
| Clostridia | Firmicutes | Bacteria | 0 |
| Clostridia | Firmicutes | Bacteria | 0 |
| unclassified | unclassified | unclassified | 0 |
| Clostridia | Firmicutes | Bacteria | 0 |
| unclassified | unclassified | unclassified | 0 |
| unclassified | unclassified | unclassified | 0.009120723 |
| Deltaproteobacteria | Proteobacteria | Bacteria | 0 |
| Clostridia | Firmicutes | Bacteria | 0 |
| Erysipelotrichia | Firmicutes | Bacteria | 0 |
| Clostridia | Firmicutes | Bacteria | 0 |
| Negativicutes | Firmicutes | Bacteria | 0 |
| unclassified | unclassified | unclassified | 0 |
| unclassified | Firmicutes | Bacteria | 0 |
| Clostridia | Firmicutes | Bacteria | 0 |
| Clostridia | Firmicutes | Bacteria | 0 |
| Clostridia | Firmicutes | Bacteria | 0 |
| unclassified | unclassified | unclassified | 0 |
| Lentisphaeria | Lentisphaerae | Bacteria | 0 |
| unclassified | unclassified | Bacteria | 0 |
| Bigyra | unclassified | Eukaryota | 0 |
| Bacteroidia | Bacteroidetes | Bacteria | 0 |
| Clostridia | Firmicutes | Bacteria | 0 |
| unclassified | unclassified | unclassified | 0 |
| Clostridia | Firmicutes | Bacteria | 0.001112835 |
| Clostridia | Firmicutes | Bacteria | 0 |
| Clostridia | Firmicutes | Bacteria | 0 |
| Thermoplasmata | Candidatus Thermoplasmatota | Archaea | 0 |
| Clostridia | Firmicutes | Bacteria | 0.026054444 |
| Clostridia | Firmicutes | Bacteria | 0 |
| Bacteroidia | Bacteroidetes | Bacteria | 0 |
| Clostridia | Firmicutes | Bacteria | 0 |
| Clostridia | Firmicutes | Bacteria | 0 |
| Clostridia | Firmicutes | Bacteria | 0.167310559 |
| unclassified | Firmicutes | Bacteria | 0 |
| Clostridia | Firmicutes | Bacteria | 0 |

| | | | |
|---|---|---|---|
| Clostridia | Firmicutes | Bacteria | 0 |
| Clostridia | Firmicutes | Bacteria | 0 |
| Coriobacteriia | Actinobacteria | Bacteria | 0 |
| Clostridia | Firmicutes | Bacteria | 0 |
| Bacteroidia | Bacteroidetes | Bacteria | 0 |
| Clostridia | Firmicutes | Bacteria | 0 |
| Clostridia | Firmicutes | Bacteria | 0 |
| Clostridia | Firmicutes | Bacteria | 0 |
| Bacteroidia | Bacteroidetes | Bacteria | 0 |
| Gammaproteobacteria | Proteobacteria | Bacteria | 0.00076107 |
| unclassified | unclassified | unclassified | 0 |
| Coriobacteriia | Actinobacteria | Bacteria | 0 |
| Clostridia | Firmicutes | Bacteria | 0 |
| unclassified | unclassified | unclassified | 0 |
| Clostridia | Firmicutes | Bacteria | 0 |
| unclassified | unclassified | unclassified | 0 |
| unclassified | unclassified | unclassified | 0 |
| unclassified | unclassified | unclassified | 0 |
| Clostridia | Firmicutes | Bacteria | 0 |
| Clostridia | Firmicutes | Bacteria | 0.889209833 |
| Clostridia | Firmicutes | Bacteria | 0 |
| unclassified | unclassified | Bacteria | 0 |
| unclassified | Firmicutes | Bacteria | 0 |
| Thermoplasmata | Candidatus Thermoplasmatota | Archaea | 0 |
| unclassified | unclassified | unclassified | 0 |
| Bacteroidia | Bacteroidetes | Bacteria | 0 |
| Clostridia | Firmicutes | Bacteria | 0 |
| unclassified | unclassified | Bacteria | 0 |
| Gammaproteobacteria | Proteobacteria | Bacteria | 0 |
| Clostridia | Firmicutes | Bacteria | 0 |
| Clostridia | Firmicutes | Bacteria | 0 |
| Clostridia | Firmicutes | Bacteria | 0 |
| unclassified | Firmicutes | Bacteria | 0 |
| Bacteroidia | Bacteroidetes | Bacteria | 0 |
| unclassified | unclassified | unclassified | 0 |
| Erysipelotrichia | Firmicutes | Bacteria | 0 |
| Clostridia | Firmicutes | Bacteria | 0 |
| unclassified | Firmicutes | Bacteria | 0 |
| unclassified | Firmicutes | Bacteria | 0 |
| unclassified | Firmicutes | Bacteria | 0 |
| unclassified | unclassified | unclassified | 0 |
| Clostridia | Firmicutes | Bacteria | 0 |
| unclassified | unclassified | unclassified | 0.007521717 |
| Clostridia | Firmicutes | Bacteria | 0 |
| Bacteroidia | Bacteroidetes | Bacteria | 0.002072918 |
| Clostridia | Firmicutes | Bacteria | 0 |
| Clostridia | Firmicutes | Bacteria | 0 |
| Clostridia | Firmicutes | Bacteria | 0 |

| | | | |
|---|---|---|---|
| Bacteroidia | Bacteroidetes | Bacteria | 0 |
| Lentisphaeria | Lentisphaerae | Bacteria | 0 |
| unclassified | unclassified | unclassified | 0 |
| unclassified | unclassified | unclassified | 0 |
| unclassified | unclassified | unclassified | 0 |
| Deltaproteobacteria | Proteobacteria | Bacteria | 0.027236241 |
| unclassified | unclassified | unclassified | 0 |
| Bacteroidia | Bacteroidetes | Bacteria | 0 |
| Clostridia | Firmicutes | Bacteria | 0 |
| Clostridia | Firmicutes | Bacteria | 0.009000854 |
| Bacteroidia | Bacteroidetes | Bacteria | 0 |
| unclassified | unclassified | Bacteria | 0 |
| Bacilli | Firmicutes | Bacteria | 0 |
| Elusimicrobia | Elusimicrobia | Bacteria | 0 |
| unclassified | Firmicutes | Bacteria | 0 |
| unclassified | unclassified | unclassified | 0 |
| Clostridia | Firmicutes | Bacteria | 0.006382219 |
| Clostridia | Firmicutes | Bacteria | 0 |
| unclassified | unclassified | Bacteria | 0 |
| Clostridia | Firmicutes | Bacteria | 0 |
| Bacilli | Firmicutes | Bacteria | 0 |
| unclassified | unclassified | unclassified | 0 |
| Clostridia | Firmicutes | Bacteria | 0 |
| Clostridia | Firmicutes | Bacteria | 0 |
| unclassified | unclassified | unclassified | 0 |
| Gammaproteobacteria | Proteobacteria | Bacteria | 0 |
| Clostridia | Firmicutes | Bacteria | 0 |
| Clostridia | Firmicutes | Bacteria | 0 |
| Clostridia | Firmicutes | Bacteria | 0 |
| Clostridia | Firmicutes | Bacteria | 0 |
| Bacteroidia | Bacteroidetes | Bacteria | 0 |
| Clostridia | Firmicutes | Bacteria | 0 |
| unclassified | unclassified | unclassified | 0 |
| Clostridia | Firmicutes | Bacteria | 0 |
| Erysipelotrichia | Firmicutes | Bacteria | 0 |
| Bacteroidia | Bacteroidetes | Bacteria | 0 |
| unclassified | Firmicutes | Bacteria | 0 |
| Clostridia | Firmicutes | Bacteria | 0 |
| Clostridia | Firmicutes | Bacteria | 1.992222309 |
| unclassified | unclassified | unclassified | 0 |
| unclassified | unclassified | unclassified | 0 |
| unclassified | unclassified | unclassified | 0 |
| Clostridia | Firmicutes | Bacteria | 0 |
| unclassified | unclassified | unclassified | 0 |
| Bacilli | Firmicutes | Bacteria | 0 |
| Clostridia | Firmicutes | Bacteria | 0 |
| unclassified | Firmicutes | Bacteria | 0 |
| unclassified | unclassified | Bacteria | 0 |

| | | | |
|---|---|---|---|
| Bacteroidia | Bacteroidetes | Bacteria | 0 |
| Clostridia | Firmicutes | Bacteria | 0.010741911 |
| Clostridia | Firmicutes | Bacteria | 0 |
| Bacteroidia | Bacteroidetes | Bacteria | 0 |
| Clostridia | Firmicutes | Bacteria | 0 |
| Deltaproteobacteria | Proteobacteria | Bacteria | 0 |
| Bacteroidia | Bacteroidetes | Bacteria | 0 |
| Clostridia | Firmicutes | Bacteria | 0 |
| Clostridia | Firmicutes | Bacteria | 0 |
| Clostridia | Firmicutes | Bacteria | 0 |
| Negativicutes | Firmicutes | Bacteria | 0.000680597 |
| Clostridia | Firmicutes | Bacteria | 0 |
| Clostridia | Firmicutes | Bacteria | 0.684935321 |
| unclassified | Firmicutes | Bacteria | 0 |
| Bacteroidia | Bacteroidetes | Bacteria | 0 |
| Negativicutes | Firmicutes | Bacteria | 0 |
| Bacteroidia | Bacteroidetes | Bacteria | 0 |
| unclassified | Firmicutes | Bacteria | 0 |
| unclassified | Firmicutes | Bacteria | 0 |
| Deltaproteobacteria | Proteobacteria | Bacteria | 0 |
| Fusobacteriia | Fusobacteria | Bacteria | 0 |
| unclassified | unclassified | unclassified | 0 |
| unclassified | unclassified | Bacteria | 0 |
| Clostridia | Firmicutes | Bacteria | 1.45804161 |
| Gammaproteobacteria | Proteobacteria | Bacteria | 0 |
| Clostridia | Firmicutes | Bacteria | 0 |
| Clostridia | Firmicutes | Bacteria | 0 |
| Erysipelotrichia | Firmicutes | Bacteria | 0 |
| Bacteroidia | Bacteroidetes | Bacteria | 0 |
| unclassified | unclassified | Bacteria | 0 |
| Clostridia | Firmicutes | Bacteria | 0 |
| Bacteroidia | Bacteroidetes | Bacteria | 0 |
| Clostridia | Firmicutes | Bacteria | 0 |
| Betaproteobacteria | Proteobacteria | Bacteria | 0 |

| PHL018-0056_V2 | PHL018-0247_V1 | PHL018-0046_V3 | PHL018-0097_V2 | PHL018-0234_V1 |
|---|---|---|---|---|
| REF | REF | CTRL | CTRL | CTRL |
| 0.546842458 | 0.977717031 | 0.000538193 | 0.387206409 | 0.01245673 |
| 3.025926573 | 1.945555306 | 0.039779184 | 3.555190428 | 0.043354332 |
| 0.008250213 | 2.097812698 | 0.004147746 | 0 | 0 |
| 0 | 0 | 0 | 0 | 0 |
| 0 | 0 | 0 | 0 | 0 |
| 0 | 0 | 0 | 0 | 0 |
| 0.010423819 | 0 | 0 | 0 | 0 |
| 0 | 0 | 0 | 0 | 0 |
| 0 | 0 | 0 | 0 | 0 |
| 0 | 0 | 0 | 0 | 0 |
| 0 | 0 | 0 | 0 | 0 |
| 0 | 0 | 0 | 0 | 0 |
| 0 | 0 | 0 | 0 | 0 |
| 0.219898814 | 11.15617075 | 0.649029382 | 2.411935164 | 0.34758004 |
| 0 | 0 | 0 | 0 | 0 |
| 0 | 0 | 0 | 0 | 0 |
| 0 | 0.005728779 | 0 | 0 | 0 |
| 0 | 0 | 0 | 0 | 0 |
| 0 | 0 | 0 | 0 | 0 |
| 0 | 0 | 0 | 0 | 0 |
| 0 | 0 | 0 | 0 | 0 |
| 0 | 0 | 0 | 0 | 0 |
| 0 | 0 | 0 | 0 | 0 |
| 0 | 0 | 0 | 0 | 0 |
| 0 | 0 | 0 | 0 | 0 |
| 0 | 0 | 0 | 0 | 0 |
| 0 | 0 | 0 | 0 | 0 |
| 0 | 0 | 0 | 0 | 0 |
| 0 | 0 | 0 | 0 | 0 |
| 0 | 0 | 0 | 0 | 0 |
| 0 | 0 | 0 | 0 | 0 |
| 0 | 0 | 0 | 0 | 0 |
| 0 | 0 | 0 | 0 | 0 |
| 0 | 0 | 0 | 0 | 0 |
| 0 | 0 | 0 | 0 | 0 |
| 0 | 0 | 2.345745919 | 0 | 0.336720511 |
| 0 | 0 | 0 | 0 | 0 |
| 0 | 0 | 0 | 0 | 0 |
| 0 | 0 | 0 | 0 | 0 |
| 0 | 0 | 0 | 0 | 0 |
| 0 | 0 | 0 | 0 | 0 |
| 0.013961058 | 0.854668847 | 0.189313681 | 0 | 0.553782791 |
| 0.10385799 | 0.101038512 | 6.361958862 | 0.141734765 | 0.122987807 |
| 0.049151898 | 0.012660339 | 0.003037137 | 0.009407475 | 0.054976502 |
| 0.006472472 | 0.003439407 | 0.001843931 | 0 | 0.004251192 |
| 0 | 0 | 0 | 0 | 0 |

| | | | | |
|---|---|---|---|---|
| 0 | 0 | 0 | 0.007768095 | 0 |
| 0.030586036 | 0.423902007 | 0 | 0.043034544 | 0.003567459 |
| 0 | 0 | 0 | 0.006976961 | 0 |
| 0.001137067 | 0 | 0.22392157 | 0.842565043 | 0.957564254 |
| 0.004654329 | 0 | 0 | 0.003128942 | 0.013393822 |
| 0.007831312 | 0 | 0.009127019 | 0 | 0 |
| 0.002164935 | 0.001517044 | 0.035463962 | 1.288214204 | 0 |
| 0 | 0 | 0 | 0 | 0 |
| 0 | 0 | 0 | 0 | 0 |
| 0 | 0 | 0 | 0 | 0 |
| 0.013355761 | 0 | 0 | 0 | 0.001537059 |
| 0.003492227 | 0 | 0.207684269 | 0.364884225 | 0.021925401 |
| 0 | 0 | 0 | 0 | 0 |
| 0.099672306 | 0.137980282 | 0 | 0.026988606 | 0.009071091 |
| 0.003342806 | 0 | 0 | 0 | 0 |
| 0 | 0 | 0 | 0 | 0 |
| 0.003036448 | 0.08688284 | 1.275081648 | 0.045582177 | 0.000938529 |
| 0 | 0 | 0 | 0 | 0 |
| 0 | 0 | 0 | 0 | 0 |
| 0 | 0 | 0 | 0 | 0 |
| 0.094556589 | 0.55584875 | 0.000500088 | 0.113390132 | 0.002351124 |
| 0.012965312 | 0.026768084 | 0.000782556 | 0.006212085 | 0 |
| 0 | 0 | 0 | 0 | 0.002666399 |
| 0 | 0 | 0 | 0 | 0 |
| 0 | 0 | 0 | 0 | 0 |
| 0 | 0 | 0 | 0 | 0 |
| 0 | 0.017804354 | 0 | 0 | 0 |
| 0 | 0 | 0 | 0 | 0 |
| 0 | 0.018845758 | 0.006807587 | 0 | 0.004127385 |
| 0 | 0 | 0 | 0 | 0 |
| 0.129232132 | 0.602959682 | 0 | 0.349764376 | 0 |
| 0 | 0 | 0 | 0 | 0 |
| 0 | 0 | 0 | 0 | 0 |
| 0.001828622 | 0 | 0 | 0 | 0 |
| 0 | 0 | 0 | 0 | 0 |
| 0 | 0 | 0 | 0 | 0 |
| 0 | 0 | 0 | 0 | 0 |
| 0 | 5.879021295 | 54.52732704 | 0.004675535 | 0 |
| 0 | 0 | 0 | 0 | 0 |
| 0 | 0 | 0 | 0 | 0 |
| 0 | 0 | 0 | 0 | 0 |
| 0 | 0 | 0 | 0 | 0 |
| 0 | 0 | 0 | 0 | 0 |
| 0 | 0 | 0 | 0 | 0 |
| 0.030182702 | 0 | 0 | 0 | 0 |
| 0 | 0 | 0 | 0 | 0 |
| 0 | 0 | 0 | 0 | 0 |
| 0 | 0 | 0 | 0 | 0 |

| | | | | |
|---|---|---|---|---|
| 0 | 0 | 0 | 0 | 0 |
| 0.003230882 | 0 | 0 | 0 | 0 |
| 0 | 0 | 0 | 0 | 0 |
| 0 | 0 | 0 | 0 | 0 |
| 0 | 0 | 0 | 0 | 0 |
| 0.013505132 | 0 | 0 | 0 | 0 |
| 0 | 0 | 0 | 0 | 0 |
| 0 | 0 | 0 | 0 | 0 |
| 0.023364727 | 0.007987777 | 0.002117925 | 0.004141012 | 0.010504361 |
| 0 | 0 | 0 | 0 | 0 |
| 0 | 0 | 0 | 0 | 0 |
| 0.009124195 | 0.011692864 | 0.004917739 | 0 | 0.008003202 |
| 0 | 0 | 0 | 0 | 0 |
| 0 | 0 | 0 | 0 | 0 |
| 0 | 0 | 0 | 0 | 0 |
| 0 | 0 | 0 | 0 | 0 |
| 0 | 0 | 0 | 0 | 0 |
| 0 | 0 | 0 | 0 | 0 |
| 5.57983038 | 0 | 0.00719006 | 0 | 0.165682357 |
| 0 | 0 | 0 | 0 | 0 |
| 0 | 0 | 0 | 0 | 0 |
| 0 | 0 | 0 | 0 | 0 |
| 0 | 0 | 0 | 0 | 0 |
| 0 | 0 | 0 | 0 | 0 |
| 0 | 0 | 0 | 0 | 0 |
| 0 | 0 | 0 | 0 | 0 |
| 0.00124505 | 0.002290178 | 0 | 0 | 0.007937107 |
| 0.001143748 | 0 | 0 | 0 | 0.000662847 |
| 0 | 0 | 0 | 0 | 0 |
| 0 | 0 | 0 | 0 | 0 |
| 0.001572439 | 0 | 0.000559962 | 0.121665093 | 0.082471771 |
| 0.00061861 | 0 | 0 | 0 | 0.028999361 |
| 0 | 0 | 0 | 0 | 0 |
| 0 | 0 | 0 | 0 | 0 |
| 0 | 0 | 0 | 0 | 0 |
| 0 | 0 | 0 | 0 | 0 |
| 0 | 0 | 0 | 0 | 0 |
| 0 | 0 | 0 | 0 | 0 |
| 0 | 0 | 0 | 0 | 0 |
| 0 | 0 | 0 | 0 | 0 |
| 0 | 0 | 0 | 0 | 0 |
| 0 | 0 | 0 | 0 | 0 |
| 0 | 0 | 0 | 0.011616418 | 0.003130865 |
| 0 | 0 | 0 | 0 | 0 |
| 0 | 0 | 0 | 0 | 0.004319684 |
| 0 | 0 | 0 | 0 | 0 |
| 0 | 0 | 0 | 0 | 0 |
| 0 | 0 | 0 | 0 | 0 |

| | | | | |
|---|---|---|---|---|
| 0.003703399 | 0 | 0 | 0 | 0 |
| 0 | 0 | 0 | 0 | 0 |
| 0 | 0 | 0 | 0 | 0 |
| 0 | 0 | 0 | 0 | 0 |
| 0.001435812 | 0 | 0 | 0 | 0.002080274 |
| 0 | 0 | 0 | 0 | 0 |
| 0 | 0 | 0 | 0 | 0 |
| 0 | 0 | 0 | 0 | 0 |
| 0 | 0 | 0 | 0 | 0 |
| 0 | 0 | 0 | 0 | 0 |
| 0 | 0 | 0 | 0 | 0 |
| 0 | 0 | 0 | 0 | 0 |
| 0 | 0 | 0 | 0 | 0 |
| 0 | 0 | 0 | 0 | 0 |
| 0 | 0 | 0 | 0 | 0 |
| 0 | 0 | 0.224444004 | 0 | 3.581799711 |
| 0 | 0 | 0 | 0 | 0 |
| 0 | 0 | 0 | 0 | 0 |
| 0 | 0 | 0 | 0 | 0 |
| 0 | 0 | 0 | 0 | 0 |
| 0 | 0 | 0 | 0 | 0 |
| 0 | 0 | 0 | 0 | 0 |
| 0 | 0 | 0 | 0 | 0 |
| 0 | 0 | 0 | 0 | 0 |
| 0 | 0 | 0 | 0 | 0 |
| 0 | 0 | 0 | 0 | 0 |
| 0 | 0 | 0 | 0 | 0 |
| 0 | 0 | 0 | 0 | 0 |
| 0 | 0 | 0 | 0 | 0 |
| 0 | 0 | 0 | 0 | 0 |
| 0 | 0 | 0 | 0 | 0.007518461 |
| 0 | 0 | 0.010844626 | 0 | 0 |
| 0 | 0 | 0 | 0 | 0 |
| 0 | 0 | 0 | 0 | 0 |
| 0 | 0 | 0 | 0 | 0 |
| 0 | 0 | 0 | 0 | 0 |
| 0 | 0 | 0 | 0 | 0 |
| 0 | 0 | 0 | 0 | 0 |
| 0 | 0 | 0 | 0 | 0 |
| 0 | 0 | 0 | 0 | 0 |
| 0 | 0 | 0 | 0 | 0 |
| 0 | 0 | 0 | 0 | 0 |
| 0 | 0 | 0 | 0 | 0 |
| 0 | 0 | 0 | 0 | 0 |
| 0 | 0 | 0 | 0 | 0 |
| 0 | 0 | 0 | 0 | 0 |
| 0 | 0 | 0 | 0 | 0 |
| 0 | 0.087753719 | 0 | 0 | 0 |

| | | | | |
|---|---|---|---|---|
| 0 | 0 | 0 | 0 | 0 |
| 0 | 0 | 0 | 0 | 0 |
| 0 | 0 | 0 | 0 | 0 |
| 0 | 0 | 0 | 0 | 0 |
| 0 | 0 | 0 | 0 | 0 |
| 0 | 0 | 0 | 0 | 0 |
| 0 | 0 | 0 | 0 | 0 |
| 0 | 0 | 0 | 0 | 0 |
| 0 | 0 | 0 | 0 | 0 |
| 0 | 0 | 0 | 0 | 0 |
| 0 | 0 | 0 | 0 | 0 |
| 0 | 0 | 0 | 0 | 0 |
| 0.028685204 | 0 | 0 | 0 | 0.001055505 |
| 0 | 0 | 0 | 0 | 0 |
| 0 | 0 | 0 | 0.001204066 | 0 |
| 0 | 0 | 0 | 0 | 0 |
| 0 | 0 | 0 | 0 | 0 |
| 0 | 0 | 0 | 0 | 0 |
| 0 | 0 | 0 | 0 | 0 |
| 0 | 0 | 0 | 0 | 0 |
| 0.011067424 | 0.045239378 | 0.052145432 | 0.106223746 | 0.012828016 |
| 0 | 0 | 0 | 0 | 0 |
| 0 | 0 | 0 | 0 | 0 |
| 0 | 0 | 0 | 0.002596172 | 0.009604029 |
| 0 | 0 | 0 | 0 | 0 |
| 0 | 0 | 0 | 0 | 0 |
| 0.002726903 | 0.009322554 | 0.000706239 | 0.00314144 | 0.000957786 |
| 0 | 0 | 0 | 0 | 0 |
| 0 | 0 | 0 | 0 | 0 |
| 0 | 0 | 0 | 0 | 0 |
| 0.002965374 | 0.061818672 | 0 | 0.064556714 | 0.001336651 |
| 0 | 0 | 0 | 0 | 0 |
| 0 | 0 | 0 | 0 | 0 |
| 0 | 0 | 0.000741117 | 0 | 0.609686374 |
| 0.012186402 | 0.040982786 | 0 | 0.242171886 | 0.001926136 |
| 0 | 0 | 0 | 0 | 0 |
| 0 | 0 | 0 | 0 | 0 |
| 0 | 0 | 0 | 0 | 0 |
| 0 | 0 | 0.004206331 | 0 | 0.001037189 |
| 0.003894809 | 0 | 0 | 0 | 0 |
| 0 | 0 | 0 | 0 | 0 |
| 0 | 0 | 0 | 0 | 0 |
| 0 | 0 | 0 | 0 | 0 |
| 0 | 0 | 0 | 0 | 0.002619384 |
| 0 | 0 | 0 | 0 | 0 |
| 0.005010292 | 0 | 0.001502499 | 0 | 0 |
| 0.001799268 | 0 | 0 | 0.008184857 | 0 |
| 0.210398352 | 0.001290848 | 0 | 0 | 0.001372617 |

| | | | | |
|---|---|---|---|---|
| 0 | 0 | 0 | 0 | 0 |
| 0 | 0 | 0 | 0 | 0 |
| 0 | 0.00117955 | 0 | 0 | 0.002229811 |
| 0 | 0 | 0 | 0 | 0 |
| 0 | 0 | 0 | 0 | 0 |
| 0 | 0 | 0 | 0 | 0 |
| 0.00224765 | 0.002067194 | 0 | 0 | 0.043637159 |
| 0 | 0 | 0.002020753 | 0 | 0.10041201 |
| 0 | 0 | 0 | 0 | 0 |
| 0 | 0 | 0 | 0 | 0 |
| 0 | 0 | 0 | 0 | 0 |
| 0 | 0 | 0 | 0 | 0 |
| 0 | 0 | 0 | 0 | 0 |
| 0 | 0 | 0 | 0 | 0 |
| 0.012136407 | 0 | 0 | 0 | 0 |
| 0 | 0 | 0 | 0 | 0 |
| 0 | 0 | 0 | 0 | 0 |
| 0 | 0 | 0.043628843 | 0 | 0 |
| 0 | 0 | 0.032738092 | 0.001760979 | 0.001085733 |
| 0 | 0 | 0.019823049 | 0 | 0.046924112 |
| 0.321258774 | 0.259973815 | 0 | 0.05368422 | 0 |
| 0 | 0 | 0 | 0 | 0 |
| 0 | 0 | 0 | 0 | 0 |
| 0 | 0 | 0 | 0 | 0 |
| 0.005882474 | 0.006251777 | 0.002513772 | 0.006880971 | 0 |
| 0 | 0 | 0 | 0 | 0 |
| 0 | 0 | 0 | 0 | 0 |
| 0 | 0 | 0 | 0 | 0 |
| 0 | 0.103719419 | 0.037783136 | 0 | 0.011061484 |
| 0 | 0 | 0 | 0 | 0 |
| 0.301753847 | 1.096759349 | 0.006512586 | 1.461174115 | 0.053161635 |
| 0 | 0 | 0 | 0 | 0 |
| 0 | 0 | 0 | 0 | 0 |
| 0.000845026 | 0 | 0 | 0 | 0 |
| 0 | 0 | 0 | 0 | 0 |
| 0 | 0 | 0 | 0 | 0 |
| 0 | 0 | 0 | 0 | 0 |
| 0 | 0 | 0 | 0 | 0 |
| 0 | 0 | 0 | 0 | 0 |
| 0 | 0 | 0 | 0 | 0 |
| 0 | 0 | 0 | 0 | 0 |
| 0.053927744 | 0.029572882 | 0 | 0.034179956 | 0.002400798 |
| 0 | 0.00563234 | 0 | 0.000726032 | 0 |
| 0 | 0 | 0 | 0 | 0 |
| 0 | 0 | 0 | 0 | 0 |
| 0 | 0 | 0 | 0 | 0 |
| 0 | 0 | 0 | 0 | 0 |
| 0 | 0 | 0 | 0 | 0 |

| | | | | |
|---|---|---|---|---|
| 0 | 0 | 0 | 0 | 0 |
| 0 | 0 | 0 | 0 | 0 |
| 0.061305903 | 0 | 0.001984697 | 0.004346183 | 0.075149605 |
| 0 | 0 | 0 | 0 | 0 |
| 0.369276883 | 0 | 0.096969711 | 0 | 0.830814765 |
| 0 | 0 | 0 | 0 | 0 |
| 0 | 0 | 0 | 0 | 0 |
| 0 | 0 | 0 | 0 | 0 |
| 0 | 0 | 0 | 0 | 0 |
| 0 | 0 | 0 | 0 | 0 |
| 0 | 0 | 0 | 0 | 0 |
| 0 | 0 | 0 | 0 | 0 |
| 0 | 0 | 0 | 0 | 0 |
| 0 | 0.001510651 | 0 | 0.006604561 | 0 |
| 0.005338022 | 0.003324919 | 0.002896648 | 0.005698982 | 0.001571349 |
| 0.009525095 | 0 | 0 | 0 | 0.010304313 |
| 0.004923683 | 0.006866561 | 0 | 0.00762278 | 0 |
| 0 | 0 | 0 | 0 | 0 |
| 0 | 0 | 0 | 0 | 0 |
| 0 | 0 | 0 | 0 | 0 |
| 0 | 0 | 0 | 0 | 0 |
| 0.193474352 | 0.002351172 | 0 | 0 | 0.000808115 |
| 0 | 0 | 0 | 0 | 0.001007632 |
| 0 | 0.005632955 | 0 | 0 | 0 |
| 0 | 0.002741794 | 0 | 0.002043258 | 0.001619707 |
| 0 | 0 | 0 | 0 | 0 |
| 0 | 0 | 0 | 0 | 0.00638916 |
| 0 | 0 | 0 | 0 | 0 |
| 0 | 0 | 0 | 0 | 0 |
| 0.012858544 | 0.008966252 | 0 | 0.081472978 | 0 |
| 0 | 0 | 0.001141574 | 0 | 0.001548179 |
| 0 | 0 | 0 | 0 | 0 |
| 0 | 0 | 0 | 0 | 0.01212522 |
| 0 | 0 | 0.003005816 | 0.000881555 | 0 |
| 0 | 0 | 0 | 0 | 0 |
| 0.013577534 | 0.011953791 | 0 | 0 | 0.016544478 |
| 0 | 0 | 0 | 0 | 0 |
| 0.090662453 | 0.051560804 | 0.006316795 | 0 | 0.138844082 |
| 0 | 0 | 0 | 0 | 0 |
| 0 | 0 | 0 | 0 | 0 |
| 0 | 0 | 0 | 0 | 0 |
| 0 | 0 | 0 | 0 | 0 |
| 0 | 0 | 0 | 0 | 0 |
| 0.030194398 | 0.026686075 | 0 | 0.001193231 | 0 |
| 0 | 0 | 0 | 0 | 0 |
| 0 | 0.025185986 | 0.001798513 | 0 | 0.011707711 |
| 0 | 0.004948596 | 0.006086378 | 0 | 0 |
| 0 | 0.013011055 | 0 | 0.009308338 | 0.001537249 |

| | | | | |
|---|---|---|---|---|
| 0 | 0.001501067 | 1.346764187 | 0.02173348 | 0 |
| 0 | 0 | 0 | 0 | 0 |
| 0.000821551 | 0.015671533 | 0.000780168 | 0 | 0 |
| 0 | 0 | 0 | 0 | 0 |
| 0.002755199 | 0.009685488 | 0.009615319 | 0 | 0.002128995 |
| 0.004043342 | 0.015058506 | 0 | 0 | 0.023606333 |
| 0 | 0 | 0 | 0 | 0 |
| 0.004634422 | 0.010419054 | 0.002475548 | 0.220984003 | 0 |
| 0 | 0.005474245 | 0 | 0 | 0 |
| 0.027466593 | 0.005206858 | 0.003685646 | 0.584759641 | 0.006459467 |
| 0.001799518 | 0 | 0 | 0 | 0 |
| 0.059615152 | 0.005748439 | 0.060531503 | 0.001490049 | 0.089769189 |
| 0.004869869 | 0.006066637 | 0 | 0.003616816 | 0.002508694 |
| 0.033810626 | 0 | 0 | 0 | 0 |
| 0.000954009 | 0.006239398 | 0 | 0 | 0 |
| 0 | 0 | 0.013636533 | 0 | 0 |
| 0 | 0 | 0.202658004 | 0 | 0 |
| 0 | 0 | 0 | 0 | 0 |
| 0.318012798 | 0 | 0 | 0.068278969 | 0 |
| 0 | 0.280065551 | 0 | 0 | 0 |
| 0 | 0 | 0 | 0 | 0.023292153 |
| 0 | 0 | 0 | 0 | 0 |
| 0.129216222 | 0 | 0 | 0 | 0 |
| 0 | 0 | 0 | 0 | 0 |
| 0 | 0 | 0 | 0 | 0 |
| 0.005402053 | 0 | 0 | 0 | 0 |
| 0 | 0 | 3.368060375 | 0 | 0 |
| 0.005879325 | 1.566913926 | 0 | 0.00222875 | 0 |
| 0.041205375 | 0 | 0 | 0 | 0 |
| 0.001389508 | 0.003635056 | 0 | 0.028714773 | 0 |
| 0.006666201 | 0.00284336 | 0 | 0 | 0 |
| 0 | 0 | 0 | 0 | 0 |
| 0 | 0 | 0 | 0 | 0 |
| 0 | 0 | 0 | 0.095381415 | 0 |
| 0 | 0 | 0 | 0 | 0 |
| 0 | 0 | 0 | 0 | 0 |
| 0 | 0 | 0 | 0.005372427 | 0 |
| 0 | 0 | 0 | 0 | 0 |
| 0 | 0 | 0 | 0.000703129 | 0 |
| 0.344587633 | 0 | 0 | 0 | 0 |
| 0 | 0 | 0 | 0.002962382 | 0 |
| 0 | 0 | 0 | 0.036796727 | 0 |
| 0 | 0 | 0 | 0 | 0 |
| 0 | 0.027929747 | 0 | 0.054301983 | 0 |
| 0 | 0 | 0 | 0 | 0 |
| 0 | 0 | 0 | 0 | 0.047796056 |
| 0.004030728 | 0.003569814 | 0 | 0 | 0 |
| 0 | 0.036390199 | 0 | 0 | 0 |

| | | | | |
|---|---|---|---|---|
| 0 | 0 | 0 | 0 | 0 |
| 0.00600616 | 0 | 0.002138856 | 0.004756951 | 0 |
| 0 | 0 | 0 | 0 | 0 |
| 0 | 0 | 0 | 0 | 0 |
| 0 | 0 | 0 | 0 | 0 |
| 0.050265866 | 0.085214962 | 0 | 0.002578861 | 0 |
| 0 | 0 | 0 | 0 | 0 |
| 0 | 0 | 0 | 0 | 0 |
| 0 | 0 | 0 | 0 | 0 |
| 0 | 0 | 0 | 0 | 0 |
| 0.005829994 | 0.031857985 | 0.006556173 | 0 | 0 |
| 0 | 0 | 0 | 0 | 0 |
| 0 | 0.061383328 | 0 | 0 | 0 |
| 0 | 0.02275111 | 0.003303436 | 0.001978052 | 0.013440153 |
| 0 | 0.023259827 | 0 | 0 | 0 |
| 0 | 0 | 0 | 0 | 0 |
| 0 | 0 | 0 | 0 | 0 |
| 0 | 0 | 0 | 0.006874873 | 0 |
| 0 | 0 | 0 | 0 | 0 |
| 0 | 0 | 0 | 0.006947207 | 0.091203057 |
| 0 | 0 | 0 | 0 | 0 |
| 0 | 0 | 0.150095995 | 0 | 0.463211018 |
| 0 | 0 | 0 | 0 | 0 |
| 0 | 0 | 0 | 0 | 0 |
| 0 | 0 | 0 | 0 | 0 |
| 0 | 0 | 0 | 0 | 0 |
| 0 | 0.032120087 | 0 | 0.031864596 | 0 |
| 0 | 0 | 0 | 0 | 0 |
| 0 | 0 | 0 | 0 | 0 |
| 0.266847324 | 0.088581587 | 0.191264737 | 0.103136169 | 0.151365116 |
| 0.004719995 | 0 | 0 | 0.008243415 | 0 |
| 0 | 0.004288855 | 0.018485457 | 0 | 0 |
| 0.014233885 | 0 | 0 | 0 | 0 |
| 0 | 0 | 0 | 0.001086093 | 0 |
| 0 | 0 | 0 | 0.00178636 | 0 |
| 0 | 0 | 0 | 0 | 0 |
| 0 | 0 | 0 | 0 | 0.307469809 |
| 0 | 0.00362335 | 0.029251519 | 0 | 0.008904413 |
| 0 | 0 | 0 | 0 | 0.010039642 |
| 0 | 0 | 0 | 0 | 0 |
| 0 | 0.120406279 | 0.005261831 | 0.016029432 | 0 |
| 0 | 0 | 0.682751012 | 0 | 0.493672378 |
| 0 | 0.001106408 | 0.032579654 | 0 | 0.007320401 |
| 0.004025139 | 0 | 0 | 0 | 0.00599844 |
| 0.004032562 | 0 | 0 | 0.149897443 | 0 |
| 0 | 0 | 0 | 0 | 0 |
| 0.001750938 | 0.006543691 | 0 | 0.018043172 | 0.003092533 |
| 0 | 0 | 0 | 0 | 0 |

| | | | | |
|---|---|---|---|---|
| 0.001543117 | 0.005298442 | 0 | 0 | 0.005485021 |
| 0 | 0 | 0 | 0 | 0 |
| 0.001682092 | 0.005844384 | 0.003893568 | 0.114367441 | 0 |
| 0.019821102 | 0 | 0 | 0 | 0 |
| 0.065357569 | 0 | 0 | 0 | 0.078151863 |
| 0.012786126 | 0 | 0.000929239 | 0 | 0 |
| 0 | 0 | 0 | 0 | 0.002202922 |
| 0 | 0 | 0.00073908 | 0 | 0.064148834 |
| 0.118038572 | 0 | 0 | 0 | 0 |
| 0 | 0 | 0 | 0.008560317 | 0 |
| 0 | 0 | 0 | 0 | 0 |
| 0 | 0 | 0 | 0 | 0 |
| 0 | 0 | 0 | 0 | 0 |
| 0.006421687 | 0.002282556 | 0.000795419 | 0 | 0 |
| 0 | 0 | 0 | 0 | 0 |
| 0.6705069 | 0 | 0 | 0.03998591 | 0.13707331 |
| 0.073520613 | 0.053975696 | 0.010334784 | 0.003771933 | 0.048961926 |
| 0.325858776 | 0.019457317 | 0 | 0.101966219 | 0 |
| 0.004996134 | 0 | 0 | 0 | 0 |
| 0 | 0 | 0 | 0 | 0 |
| 0 | 0 | 0.001085397 | 0.026739612 | 0.016957363 |
| 0.005891059 | 0.005733425 | 0 | 0.00191417 | 0 |
| 0 | 0 | 0 | 0 | 0.001711359 |
| 0 | 0 | 0.128533549 | 0 | 0 |
| 0 | 0 | 0.000482943 | 0 | 0.020783965 |
| 0.009239023 | 0.749842042 | 0 | 0 | 0 |
| 0 | 0.01305192 | 0 | 0.002693531 | 0 |
| 0.021372712 | 0 | 0 | 0 | 0 |
| 0 | 0 | 0 | 0 | 0 |
| 0 | 0 | 0 | 0 | 0.006901965 |
| 0.009806587 | 0.030159612 | 0 | 0.019573816 | 0.039873333 |
| 0 | 0.0196363 | 0 | 0 | 0 |
| 0.028172673 | 0.583924183 | 0 | 0.026816102 | 0.00922005 |
| 0 | 0.00543642 | 0.004397875 | 0.002315066 | 0.031931975 |
| 0 | 0 | 0 | 0.002974262 | 0 |
| 0 | 0 | 0 | 0 | 0 |
| 0 | 0.000734906 | 0 | 0.035343087 | 0.000926173 |
| 0.034682059 | 0.141958512 | 0 | 0.05093971 | 0.000543233 |
| 0 | 0 | 0.002355723 | 0 | 0.023944049 |
| 0 | 0.131728958 | 0 | 0.002054406 | 0 |
| 0.038371335 | 0 | 0 | 0 | 0 |
| 0 | 0 | 0 | 0 | 0 |
| 0.018924433 | 0.001719018 | 0 | 0 | 0 |
| 0.036585976 | 0 | 0 | 0.018366665 | 0.043819559 |
| 0.270629533 | 0 | 0 | 0 | 0 |
| 0 | 0 | 0 | 0 | 0 |
| 0 | 0 | 0 | 0 | 0 |
| 0 | 0 | 0 | 0 | 0 |

| | | | | |
|---|---|---|---|---|
| 0.003127013 | 0 | 0 | 0.004064216 | 0 |
| 0.441294131 | 2.83419686 | 0.083464307 | 0 | 0.267714263 |
| 0 | 0 | 0 | 0 | 0 |
| 0 | 0 | 0 | 0 | 0 |
| 0 | 0 | 0 | 0.001221457 | 0 |
| 0 | 0 | 0 | 0 | 0 |
| 0 | 0.001207516 | 0 | 0.066518648 | 0 |
| 0 | 0 | 0.00174685 | 0 | 0 |
| 0 | 0 | 0.000832749 | 0.0031343 | 0.005872654 |
| 0 | 0 | 0 | 0.004976164 | 0 |
| 0.019457484 | 0.01144524 | 0 | 0.024055512 | 0 |
| 0.012370568 | 0.014158517 | 0.00679673 | 0.200862378 | 0.023214653 |
| 0 | 0 | 0.001118972 | 0 | 0.744888775 |
| 0 | 0 | 0 | 0 | 0 |
| 0 | 0 | 0 | 0.001592348 | 0.006942467 |
| 0 | 0 | 0 | 0 | 0 |
| 0.022611052 | 0.023969637 | 0.012564706 | 0.010657136 | 0.004991998 |
| 0.039351394 | 0.084118406 | 0 | 0.032385153 | 0 |
| 0 | 0 | 0 | 0 | 0 |
| 0.019897624 | 0 | 0 | 0.039812612 | 0 |
| 0.019217884 | 0.034983305 | 0.009307415 | 0 | 0.085932153 |
| 0.004463866 | 0 | 0 | 0.00124323 | 0 |
| 0 | 0 | 0 | 0 | 0 |
| 0 | 0 | 0 | 0 | 0 |
| 0 | 0 | 0 | 0.017206562 | 0 |
| 0.031017077 | 0 | 0 | 0 | 0 |
| 0.025292015 | 0 | 0 | 0.005423936 | 0.007622013 |
| 0.029957801 | 0.116667153 | 0.006788908 | 0.489007265 | 0.03724879 |
| 0 | 0 | 0 | 0 | 0 |
| 0.01445445 | 0 | 0 | 0 | 0 |
| 0 | 0.001911205 | 0 | 0 | 0 |
| 0 | 0 | 0 | 0 | 0 |
| 0 | 0 | 0 | 0 | 0 |
| 0 | 0 | 0 | 0 | 0 |
| 0 | 0 | 0.004901879 | 0 | 0.012103627 |
| 0.469222028 | 0.088552034 | 0.163812025 | 0.090309848 | 0.047726224 |
| 0 | 0 | 0 | 0 | 0 |
| 0 | 0 | 0 | 0 | 0 |
| 0.030908904 | 0.032959233 | 0.015314061 | 0.001309979 | 0.047508195 |
| 0 | 0 | 0 | 0 | 0 |
| 0.113421496 | 0 | 0 | 0 | 0 |
| 0 | 0 | 0 | 0 | 0 |
| 0.007754701 | 0.318317066 | 0.008284589 | 0.014919394 | 0 |
| 0.077431531 | 0 | 0 | 0.052047878 | 0.000608985 |
| 0 | 0 | 0 | 0 | 0 |
| 0.001424985 | 0 | 0.186307367 | 0 | 0.387749162 |
| 0 | 0 | 0 | 0 | 0 |
| 0.001910871 | 0.00364509 | 0 | 0 | 0 |

| | | | | |
|---|---|---|---|---|
| 0.001425459 | 0 | 0 | 0 | 0 |
| 0 | 0 | 0 | 0 | 0 |
| 0 | 0 | 0 | 0 | 0 |
| 5.579734491 | 0.55319337 | 0.041304168 | 0 | 0.343257787 |
| 0 | 0 | 0 | 0 | 0 |
| 0 | 0 | 0 | 0 | 0 |
| 0 | 0 | 0 | 0 | 0 |
| 0 | 0 | 0 | 0.006839813 | 0.004271858 |
| 0 | 0 | 0.057765991 | 0.014261126 | 6.156415091 |
| 0.070525398 | 0 | 0 | 0 | 0.064345058 |
| 0 | 0 | 0 | 0 | 0 |
| 0 | 0 | 0.00373032 | 0 | 0.020059966 |
| 0 | 0 | 0 | 0 | 0 |
| 0 | 0 | 0.039622179 | 0 | 0 |
| 0.145447141 | 0 | 0.00307827 | 0 | 0.062003152 |
| 0 | 0 | 0 | 0 | 0 |
| 0 | 0 | 0.001396638 | 0 | 0.243580082 |
| 0 | 0 | 0.093677813 | 0 | 1.793025482 |
| 0 | 0 | 0 | 0 | 0 |
| 0 | 0 | 0 | 0 | 0.068984977 |
| 0 | 0 | 0.056760276 | 0 | 0 |
| 0 | 0 | 0 | 0 | 0.000816851 |
| 0 | 0 | 0 | 0 | 0.000810168 |
| 0.156031884 | 0.357078314 | 0.020498299 | 0.42488462 | 0.012028908 |
| 2.263814298 | 0 | 0 | 0.001039146 | 0 |
| 0 | 0 | 0 | 0.287014199 | 1.742213984 |
| 0 | 0 | 0 | 0 | 0.266397296 |
| 0 | 0 | 0.647922215 | 0 | 1.229876579 |
| 0 | 0 | 0 | 0 | 0 |
| 0 | 0 | 0 | 0 | 0 |
| 0.005755982 | 0 | 0 | 0 | 0.732169353 |
| 0.387133271 | 0 | 0 | 0 | 0 |
| 0.018533838 | 0 | 0 | 0.023650678 | 0 |
| 0 | 0 | 0 | 0 | 0 |
| 0 | 0 | 0.000582315 | 0 | 0 |
| 0 | 0 | 0.001203211 | 0 | 0.703329022 |
| 0.006303101 | 0.000594569 | 0.457380351 | 0 | 1.101863023 |
| 0 | 0 | 0 | 0 | 0 |
| 0 | 0 | 0 | 0.091045628 | 0 |
| 0.001303975 | 0 | 0 | 0 | 0 |
| 0.122554875 | 0.084832332 | 0.084641091 | 0.129899059 | 0.154795642 |
| 0 | 0 | 0.2193202 | 0 | 0.326356538 |
| 0.070496037 | 0.23074282 | 0.170411078 | 20.06577776 | 0.018427885 |
| 0 | 0 | 0 | 0.007420266 | 0.002941052 |
| 0 | 0 | 0 | 0 | 0 |
| 0 | 0 | 0 | 0 | 0 |
| 0 | 0 | 0 | 0 | 0 |
| 0.452742128 | 0.010734547 | 0 | 0.013311452 | 0.0008117 |

| | | | | |
|---|---|---|---|---|
| 0 | 0 | 0 | 0 | 0 |
| 0 | 0 | 0 | 0 | 0.020335735 |
| 0.175716415 | 2.065052398 | 0 | 0.497906046 | 0.000583579 |
| 0.001904853 | 0 | 0 | 0 | 0.002943832 |
| 0 | 0 | 0 | 0 | 0 |
| 0.028953728 | 0 | 0 | 0 | 0.027736514 |
| 0 | 0 | 0 | 0 | 0 |
| 0 | 0 | 0 | 0 | 0 |
| 0 | 0 | 0 | 0 | 0 |
| 0 | 0 | 0 | 0 | 0 |
| 0.101832682 | 0.117803607 | 0.001056224 | 0.035658258 | 0.005729713 |
| 0.267527159 | 7.628904493 | 0.356904005 | 6.490554891 | 0.728845522 |
| 0 | 0 | 0 | 0 | 0 |
| 0.077091439 | 0.082214747 | 0.011969155 | 0.001820182 | 0.129240167 |
| 0.078408743 | 0.618496964 | 0.016319122 | 0.100406119 | 0.017542888 |
| 0 | 0 | 0 | 0 | 6.204996625 |
| 0 | 0 | 0 | 0 | 0 |
| 0 | 0 | 0 | 0 | 0 |
| 0 | 0 | 0 | 0 | 0.006494114 |
| 0 | 0 | 0 | 0.002381596 | 0.004877102 |
| 0.51894105 | 1.050200192 | 0.0258204 | 0.030896563 | 0.190492956 |
| 0 | 0.586574023 | 0.002953869 | 1.681177457 | 0 |
| 0.492696103 | 0.691686266 | 0.442328522 | 12.07623605 | 0.188089887 |
| 0 | 0 | 0 | 0 | 0 |
| 0 | 0 | 0 | 0 | 0.036085208 |
| 0 | 0 | 0.00424328 | 0 | 1.869301467 |
| 0 | 0.011951171 | 0 | 0 | 0.003637757 |
| 0 | 0 | 0 | 0 | 0 |
| 0 | 0 | 0 | 0 | 0.024588652 |
| 0 | 0 | 0 | 0 | 0 |
| 0 | 0 | 0.001301908 | 0 | 0 |
| 0.000782308 | 0 | 0.082462003 | 0 | 0.213087675 |
| 3.825468917 | 3.344688659 | 0.001265564 | 0.888082524 | 0.024126715 |
| 0.002710088 | 0 | 0.002068051 | 0 | 0.008226966 |
| 0.008544639 | 0.02794176 | 0.000715961 | 0.029723734 | 0.000647314 |
| 1.191475748 | 0 | 0 | 0.002243879 | 0.159019404 |
| 0.016899565 | 0 | 0 | 0 | 0.003264654 |
| 0 | 0 | 0 | 0 | 0 |
| 5.379888952 | 0 | 0.00383704 | 0.017067643 | 0.477076507 |
| 0 | 0 | 0 | 0 | 0 |
| 0 | 0 | 0 | 0 | 0.002506033 |
| 0 | 0 | 0 | 0 | 0 |
| 0 | 0 | 0 | 0 | 0 |
| 0 | 0 | 0 | 0 | 0 |
| 6.448515823 | 0.110951611 | 0 | 0.009212 | 0.055376731 |
| 0.312057433 | 0 | 0.018641572 | 0 | 0.274527595 |
| 0 | 0 | 0 | 0 | 0 |
| 0 | 0.356987524 | 0 | 0.002160704 | 0.137269241 |

| | | | | |
|---|---|---|---|---|
| 0 | 0 | 0 | 0 | 0 |
| 0 | 0 | 0 | 0 | 0 |
| 0 | 0 | 0 | 0 | 0 |
| 0 | 0 | 0 | 0.003053303 | 0.004322103 |
| 0 | 0 | 0 | 0 | 0 |
| 0.000988006 | 0 | 0 | 0 | 0 |
| 0.02155533 | 0 | 0.312276757 | 0 | 0.000757101 |
| 0.54083746 | 0.69090752 | 0.001173036 | 0.095526108 | 0 |
| 0 | 0 | 0 | 0 | 0 |
| 0 | 0 | 0 | 0 | 0 |
| 1.714616225 | 0.015278359 | 0 | 0.003400571 | 0.000770188 |
| 0 | 0 | 0 | 0 | 0 |
| 0.302612432 | 0 | 0 | 0.129307086 | 0.011295534 |
| 0 | 0 | 0.00075721 | 0 | 0.086424948 |
| 0 | 0 | 0 | 0 | 0 |
| 0 | 0 | 0 | 0 | 0 |
| 0 | 0 | 0.023494663 | 0 | 0.071888591 |
| 0.00539338 | 0 | 0 | 0 | 0.007052808 |
| 0 | 0 | 0 | 0 | 0.913509865 |
| 0.010289095 | 0 | 0 | 0 | 0 |
| 0 | 0 | 0 | 0 | 0 |
| 0 | 0 | 0 | 0 | 0 |
| 0.004783217 | 0 | 0.002214359 | 0 | 0.013490703 |
| 0 | 0 | 0 | 0 | 0 |
| 0 | 0 | 0 | 0 | 0 |
| 0 | 0 | 0 | 0 | 0 |
| 0 | 0 | 0 | 0 | 0 |
| 0 | 0 | 0 | 0 | 0 |
| 0 | 0 | 0 | 0 | 0 |
| 0 | 0 | 0.010172809 | 0 | 0.017620348 |
| 0 | 0.035297999 | 0 | 0 | 0 |
| 0 | 0 | 0 | 0 | 0 |
| 0 | 0 | 0 | 0 | 0.512311807 |
| 0 | 0 | 0 | 0 | 0 |
| 0.205071586 | 0 | 0 | 0 | 0 |
| 0 | 0 | 0 | 0 | 0 |
| 0 | 0 | 0.014742506 | 0 | 0 |
| 0.004640598 | 0.01431296 | 0 | 0.042974026 | 0 |
| 0 | 0 | 0 | 0 | 0 |
| 0 | 0 | 0 | 0 | 0 |
| 0 | 0 | 0 | 0 | 0.002599796 |
| 0 | 0 | 0 | 0 | 0 |
| 0 | 0 | 0 | 0 | 0 |
| 0 | 0 | 0.008899549 | 0 | 0.365326349 |
| 0 | 0 | 0.015182354 | 0.323131297 | 0.175454493 |
| 0 | 0 | 0 | 0 | 0 |
| 0 | 0 | 0 | 0 | 0.004181546 |
| 0 | 0 | 0 | 0 | 0 |

| | | | | |
|---|---|---|---|---|
| 0 | 0 | 0 | 0 | 0 |
| 0 | 0 | 0 | 0 | 0 |
| 0 | 0 | 0 | 0 | 0 |
| 0 | 0 | 0.001195504 | 0 | 1.778585443 |
| 0 | 0 | 0 | 0 | 0 |
| 0 | 0 | 0 | 0 | 0 |
| 0 | 0 | 0 | 0 | 0 |
| 0 | 0 | 0 | 0 | 0 |
| 7.051813024 | 0.535119476 | 0 | 0.597504457 | 0 |
| 0 | 0 | 0 | 0 | 0 |
| 0 | 0 | 0 | 0 | 0.002732522 |
| 0 | 0 | 0 | 0 | 0 |
| 0 | 0 | 0 | 0 | 0.000655711 |
| 0.01919581 | 0 | 0 | 0 | 0 |
| 0.001254361 | 0.001196381 | 0.0248658 | 1.968189803 | 0.621624415 |
| 0 | 1.32179701 | 0 | 0.309267357 | 0 |
| 0 | 0 | 0 | 0 | 0 |
| 0 | 0 | 0 | 0 | 0 |
| 0 | 0 | 0 | 0 | 0 |
| 0 | 0 | 0 | 0 | 0 |
| 0 | 0 | 0.000633726 | 0 | 0.033862188 |
| 0 | 0 | 0 | 0 | 0 |
| 0 | 0 | 0 | 0 | 0 |
| 0 | 0 | 0 | 0 | 0 |
| 0.003421094 | 0.013984117 | 0.190052769 | 0.010671449 | 0.010309824 |
| 0.008061309 | 0.002636123 | 4.667553975 | 0.351725673 | 2.402200698 |
| 0.330326528 | 0 | 0 | 0 | 0.123691845 |
| 0 | 0 | 0 | 0 | 0 |
| 0.491423641 | 0 | 0.005751708 | 0 | 0.062171655 |
| 0 | 0 | 0 | 0 | 0.006044154 |
| 0 | 0 | 0 | 0 | 0 |
| 0.000881281 | 0 | 0 | 0 | 0.032687181 |
| 0 | 0 | 0 | 0.000807447 | 0 |
| 0.005115834 | 0 | 0 | 0.035442142 | 0.001129458 |
| 0.008128233 | 0.013290038 | 0.003644987 | 0.003228 | 0.013492178 |
| 0.087887558 | 0.06123787 | 0 | 0.014935449 | 0 |
| 0 | 0 | 0 | 0.061437811 | 0.002715108 |
| 2.228940664 | 0.000589331 | 0.283408401 | 0.084557748 | 0.427986787 |
| 0 | 0 | 0 | 0 | 0.107991601 |
| 0 | 0 | 0 | 0 | 0 |
| 0 | 0 | 0 | 0 | 0 |
| 0 | 0 | 0.00378121 | 0 | 0 |
| 1.602592296 | 0 | 0.02381799 | 0.153922075 | 0.293016378 |
| 0 | 0 | 0 | 0 | 0 |
| 0 | 0 | 0 | 0 | 0 |
| 0 | 0 | 0 | 0.015918146 | 0 |
| 0 | 0 | 0 | 0 | 0 |
| 0.9889109 | 1.28843822 | 0.102022733 | 0 | 0.592657058 |

| | | | | |
|---|---|---|---|---|
| 0 | 0 | 0 | 0.067004287 | 0 |
| 0 | 0 | 0 | 0 | 0.55455488 |
| 0 | 0 | 0 | 0 | 0 |
| 0 | 0 | 0 | 0 | 0 |
| 0.003099898 | 0.003119063 | 0 | 0 | 0.001842577 |
| 0.440654893 | 0 | 0 | 0 | 0.053408672 |
| 0 | 0 | 0 | 0 | 0 |
| 0 | 0 | 0 | 0 | 0 |
| 0 | 0 | 0 | 0 | 0 |
| 0 | 0 | 0 | 0 | 0 |
| 0 | 0 | 0 | 0 | 0.000563958 |
| 0.004839753 | 0.365087176 | 0.082570651 | 0.630243682 | 1.981057822 |
| 0.009816671 | 0 | 0.003290182 | 0.73654194 | 0.024764523 |
| 0 | 0 | 0 | 0 | 0 |
| 0.222547754 | 3.980315237 | 1.772210309 | 0.013440893 | 0.139363515 |
| 0 | 0 | 0 | 0 | 0 |
| 0.027241144 | 0 | 0 | 0.000748007 | 0 |
| 0 | 0 | 0 | 0 | 0 |
| 0.019337032 | 0 | 0 | 0.005890447 | 0.005416509 |
| 0.000804063 | 0 | 0 | 0 | 0.053640187 |
| 0.710011985 | 0 | 0.002565483 | 0 | 0.113501027 |
| 0 | 0 | 0 | 0 | 0 |
| 0.019977801 | 0 | 0.003013116 | 0.020848681 | 0.244634676 |
| 0 | 0 | 0 | 0 | 0 |
| 0 | 0 | 0 | 0 | 0 |
| 0 | 0 | 0 | 0 | 0 |
| 0 | 0 | 0 | 0 | 0 |
| 0 | 0 | 0 | 0 | 0 |
| 0 | 0 | 0 | 0 | 0 |
| 0 | 0 | 0 | 0 | 0 |
| 0 | 0 | 0 | 0 | 0 |
| 0.04294228 | 0.047393516 | 0.029310033 | 0.018487925 | 0.010784254 |
| 0 | 0 | 0.118384318 | 0 | 0 |
| 0.007687405 | 0 | 0 | 0 | 0.001519587 |
| 0.082906633 | 0.053200123 | 0 | 0.017156484 | 0 |
| 0 | 0.016840275 | 0.002976752 | 0 | 0.010842253 |
| 0 | 0 | 0 | 0 | 0 |
| 0 | 0 | 0 | 0 | 0.134497332 |
| 0 | 0 | 0 | 0 | 0 |
| 0 | 0 | 0 | 0 | 0 |
| 0.005327353 | 0 | 0.002670029 | 0 | 0.014484149 |
| 0.008923141 | 0 | 0 | 0 | 0.678993145 |
| 0 | 0 | 0 | 0 | 0 |
| 0 | 0 | 0 | 0 | 0 |
| 0.023041357 | 0.148643205 | 0 | 0.006671391 | 0.006875004 |
| 0 | 0 | 0 | 0 | 0 |
| 0 | 0 | 0.154342699 | 0 | 0 |
| 0.001941951 | 0 | 0 | 0 | 0.00116712 |

| | | | | |
|---|---|---|---|---|
| 0 | 0 | 0 | 0 | 0 |
| 0 | 0 | 0 | 0 | 0 |
| 0.003539187 | 0 | 0 | 0.001379977 | 0 |
| 0 | 0 | 0 | 0 | 0 |
| 0 | 0 | 0 | 0 | 0 |
| 0 | 0 | 0 | 0 | 0 |
| 0 | 0 | 0.057529095 | 0.019241307 | 0 |
| 0.084229279 | 0 | 0.000634813 | 0.0458313 | 0.026344161 |
| 0 | 0 | 0 | 0 | 0 |
| 0 | 0 | 0.004060422 | 0 | 0 |
| 0 | 0 | 0 | 0 | 0 |
| 0.143081218 | 0 | 0 | 0 | 0 |
| 0 | 0 | 0 | 0 | 0 |
| 0 | 0 | 0 | 0 | 0.504230047 |
| 0 | 0 | 0 | 0 | 0 |
| 0 | 0 | 0 | 0 | 0 |
| 0.003117011 | 0 | 0.001009091 | 0 | 0.44317764 |
| 0 | 0 | 0 | 0 | 0 |
| 0.001764811 | 0.042664884 | 0 | 0 | 0.278585084 |
| 0.000669568 | 0 | 0 | 0 | 0 |
| 0 | 0 | 0 | 0 | 0 |
| 0 | 0 | 0 | 0 | 0 |
| 0 | 0 | 0 | 0 | 0 |
| 0 | 0 | 0 | 0 | 0 |
| 0 | 0.020784658 | 0 | 0 | 0 |
| 0.536478184 | 0.070666437 | 0 | 0.006831894 | 0 |
| 0 | 0 | 0 | 0 | 0 |
| 0.598279274 | 10.2330124 | 0.002665019 | 0 | 2.341386942 |
| 0 | 0 | 0 | 0 | 0.789354134 |
| 0 | 0 | 0.001653551 | 0 | 0.715361212 |
| 0 | 0 | 0 | 0 | 0 |
| 0 | 0 | 0 | 0 | 0 |
| 0 | 0 | 0 | 0 | 0.000678458 |
| 0 | 0 | 0 | 0 | 0 |
| 0.018027661 | 0.005640481 | 0 | 0 | 0 |
| 0 | 0 | 0 | 0.005669064 | 0.508503433 |
| 0 | 0 | 0 | 0 | 0 |
| 0.00707125 | 0.003096916 | 0 | 0.004677353 | 0 |
| 0 | 0 | 0 | 0 | 0 |
| 0 | 0 | 0 | 0 | 0 |
| 3.18232544 | 2.418633568 | 1.958620622 | 2.5890164 | 1.605750603 |
| 0.090213542 | 0.138737754 | 0.019222921 | 0.032977023 | 0.026569692 |
| 0 | 0 | 0.002472584 | 0 | 1.291846044 |
| 0 | 0 | 0 | 0 | 0 |
| 0 | 0 | 0 | 0 | 0 |
| 0 | 0 | 0 | 0 | 0 |
| 0 | 0 | 0 | 0 | 0 |
| 0.206646828 | 0.003441038 | 0 | 0.025203928 | 0.002942696 |

| | | | | |
|---|---|---|---|---|
| 0 | 0 | 0 | 0 | 0 |
| 0.003092434 | 0.005898986 | 0 | 0.019493481 | 0.006531527 |
| 0.000963144 | 0 | 0 | 0 | 0 |
| 0 | 0 | 0 | 0 | 0 |
| 1.828831545 | 0.001687139 | 0.056655054 | 2.33556269 | 1.753274077 |
| 0 | 0 | 0 | 0 | 0 |
| 0 | 0 | 0 | 0 | 0 |
| 0 | 0 | 0 | 0 | 0 |
| 2.061877121 | 1.044095729 | 0.001026316 | 0.240800925 | 0.001590707 |
| 0 | 0 | 0 | 0 | 0 |
| 0 | 0 | 0 | 0 | 0 |
| 0 | 0 | 0 | 0 | 0 |
| 0.291868867 | 0 | 0.012697928 | 0 | 0.127239426 |
| 0.00837336 | 0.008657437 | 0.00964713 | 0.010322805 | 0.00589935 |
| 0 | 0 | 0 | 0 | 2.193022946 |
| 0.336482706 | 0 | 0.028786783 | 0 | 0.423350125 |
| 0.064147031 | 0.025262605 | 0.011231982 | 0.006232177 | 0.078056679 |
| 0 | 0 | 0 | 0 | 0 |
| 0 | 0 | 0 | 0 | 0 |
| 0 | 0 | 0 | 0 | 0 |
| 0 | 0 | 0 | 0 | 0 |
| 0 | 0 | 0 | 0 | 0 |
| 0 | 0 | 0 | 0 | 0 |
| 0 | 0 | 0 | 0 | 0 |
| 0 | 0 | 0 | 0 | 0 |
| 0 | 0 | 0 | 0 | 0 |
| 0 | 0 | 0.00778694 | 0 | 0.083624722 |
| 0.054575686 | 0.0230777 | 0 | 0.007337864 | 0 |
| 0 | 0 | 0 | 0 | 0 |
| 0 | 0 | 0 | 0 | 0 |
| 0 | 0 | 0 | 0 | 0 |
| 0 | 0 | 0 | 0 | 0 |
| 0.018707786 | 0 | 0 | 0 | 0.108157729 |
| 0 | 0 | 0.005151802 | 0 | 0.07113798 |
| 0 | 0 | 0 | 0 | 2.24185768 |
| 0.005805309 | 0.034408319 | 0 | 0 | 0.00934557 |
| 0 | 0.001414863 | 0 | 0 | 0 |
| 0.001142054 | 0 | 0 | 0 | 0.003000455 |
| 0 | 0 | 0 | 0 | 0 |
| 0 | 0 | 0 | 0 | 0 |
| 0 | 0 | 2.52350864 | 0 | 0.372221282 |
| 0 | 0 | 0.002511859 | 0 | 0 |
| 0 | 0 | 0.009339135 | 0 | 0.143443444 |
| 0 | 0 | 0 | 0 | 0.003559619 |
| 0 | 0 | 0 | 0 | 0 |
| 0 | 0 | 0 | 0 | 0 |
| 0 | 0 | 0 | 0 | 0 |
| 0 | 0 | 0 | 0 | 0 |

| | | | | |
|---|---|---|---|---|
| 0 | 0 | 0.004728771 | 0 | 0.427773254 |
| 0 | 0 | 0 | 0 | 0 |
| 0 | 0 | 0 | 0 | 0 |
| 0 | 0 | 0 | 0 | 0 |
| 0.548695448 | 0 | 0.003712607 | 0 | 0.069876497 |
| 0.007975983 | 0 | 0 | 0 | 0 |
| 0.021684675 | 0.086379206 | 0.034825328 | 0 | 0.659568594 |
| 0 | 0 | 0 | 0 | 0 |
| 0 | 0 | 0 | 0 | 0 |
| 0 | 0 | 0 | 0 | 0 |
| 0 | 0 | 0 | 0 | 0.003785022 |
| 0 | 0 | 0.001721393 | 0 | 0.02241136 |
| 0 | 0 | 0 | 0 | 0 |
| 0 | 0 | 0 | 0 | 0 |
| 0 | 0 | 0 | 0 | 0 |
| 0 | 0 | 0 | 0 | 0 |
| 0 | 0 | 0 | 0 | 0 |
| 0 | 0 | 0 | 0 | 0 |
| 0.002128076 | 0 | 0 | 0 | 0 |
| 0 | 0 | 0 | 0 | 0 |
| 1.404063734 | 0.289073379 | 0.651868966 | 0.000986492 | 0.870678885 |
| 0 | 0 | 0 | 0 | 0 |
| 0.004464977 | 0 | 0 | 0 | 0.029671491 |
| 0.003374758 | 0 | 0 | 0.003096151 | 0.00306793 |
| 0 | 0 | 0 | 0 | 0 |
| 0.006791896 | 0 | 0 | 0.006164037 | 0.001990707 |
| 0 | 0 | 0 | 0 | 0 |
| 0 | 0 | 0 | 0 | 0 |
| 0 | 0 | 0.021111361 | 0 | 0.04444182 |
| 0.020856824 | 0.00677366 | 0 | 0.365956206 | 0.339991215 |
| 0 | 0 | 0 | 0 | 0.117657867 |
| 0 | 0 | 0 | 0 | 0 |
| 0.011548293 | 0 | 0 | 0 | 0 |
| 0 | 0 | 0 | 6.29041558 | 0.013269275 |
| 0 | 0 | 0 | 0 | 0 |
| 0 | 0 | 0 | 0 | 0 |
| 0.101096329 | 0.002639881 | 0.00406896 | 1.082081003 | 0.018809997 |
| 0 | 0 | 0 | 0 | 0 |
| 0 | 0 | 0 | 0 | 0 |
| 0 | 0 | 0 | 0 | 0 |
| 0 | 0 | 0 | 0 | 0 |
| 0 | 0 | 0 | 0 | 0 |
| 0 | 0 | 0 | 0 | 0 |
| 0 | 0 | 0 | 0 | 0 |
| 0 | 0 | 0.050899694 | 0 | 0 |
| 0 | 0 | 0 | 0 | 0 |
| 0 | 0 | 0 | 0 | 0 |
| 0 | 0 | 0 | 0 | 0 |

| | | | | |
|---|---|---|---|---|
| 0.851660686 | 0.048023424 | 0 | 0.016241398 | 0.001815661 |
| 0 | 0 | 0 | 0 | 0.181745568 |
| 0.002030828 | 0 | 0 | 0 | 0 |
| 0 | 0 | 0 | 0 | 0 |
| 0 | 0 | 0 | 0 | 0 |
| 0 | 0 | 0 | 0 | 0 |
| 0.002463192 | 0 | 0 | 0 | 0 |
| 0.376367841 | 0.339785236 | 0.005154942 | 0.266606122 | 0.211679756 |
| 0.162281347 | 0.11348113 | 8.206667713 | 0.215712652 | 0.169442619 |
| 0.061622949 | 0.751283808 | 0 | 0.333337837 | 0.000541105 |
| 0 | 0 | 0 | 0 | 0.676462207 |
| 0 | 0 | 0 | 0 | 0 |
| 0 | 0 | 0 | 0 | 0 |
| 0 | 0 | 0 | 0 | 0 |
| 0 | 0 | 0 | 0 | 0 |
| 0 | 0 | 0 | 0 | 0 |
| 0 | 0 | 0 | 0 | 0.000963773 |
| 0 | 0 | 0 | 0 | 0 |
| 0 | 0 | 0 | 0 | 0 |
| 0 | 0 | 0 | 0 | 0.016452231 |
| 0 | 0 | 0 | 0 | 0 |
| 0 | 0 | 0 | 0 | 0 |
| 0 | 0 | 0 | 0 | 0 |
| 0 | 0 | 0 | 0 | 0 |
| 0 | 0 | 0.003714699 | 0 | 0.040302383 |
| 0 | 0 | 0 | 0 | 0 |
| 0 | 0 | 0 | 0 | 0 |
| 0 | 0 | 0 | 0 | 0 |
| 0 | 0 | 0 | 0 | 0 |
| 0.0005994 | 0 | 0 | 0 | 0 |
| 0 | 0 | 0 | 0 | 0 |
| 0 | 0 | 0 | 0 | 0 |
| 0 | 0 | 0 | 0 | 0 |
| 0 | 0 | 0 | 0 | 0 |
| 0 | 0 | 0 | 0 | 0 |
| 0 | 0 | 0 | 0 | 0 |
| 0 | 0 | 0 | 0 | 0 |
| 0 | 0 | 0 | 0 | 0 |
| 0 | 0 | 0 | 0 | 0 |
| 0.007890216 | 1.330532827 | 0 | 0 | 0.007415174 |
| 0 | 0 | 0 | 0 | 0 |
| 0.899597466 | 0 | 0.070041148 | 0.230540918 | 0.206721835 |
| 0 | 0 | 0 | 0 | 0 |
| 0 | 0 | 0 | 0 | 0 |
| 0.001662451 | 0 | 0 | 0 | 0 |
| 0 | 0 | 0 | 0 | 0.005870878 |
| 0 | 0 | 0 | 0 | 0 |
| 0 | 0 | 0 | 0 | 0 |

| | | | | |
|---|---|---|---|---|
| 0 | 0 | 0 | 0 | 0 |
| 0.004434212 | 0 | 0 | 0.005637912 | 0 |
| 0 | 0 | 0 | 0 | 0 |
| 0 | 0 | 0 | 0 | 0 |
| 0.005930102 | 0.919576614 | 0.00230375 | 0.009459113 | 0.224116278 |
| 0 | 0 | 0 | 0 | 0.383749093 |
| 0 | 0 | 0 | 0 | 0 |
| 0.637470672 | 0.054202846 | 0 | 0.375327592 | 0 |
| 0 | 0 | 0 | 0.001057871 | 0.006918306 |
| 0 | 0 | 0.005473283 | 0 | 0 |
| 0 | 0 | 0 | 0 | 0 |
| 0 | 0 | 0 | 0 | 0 |
| 0 | 0 | 0 | 0 | 0 |
| 0 | 0 | 0 | 0 | 0 |
| 0 | 0 | 0 | 0.018086864 | 0 |
| 0 | 0 | 0 | 0.019756258 | 0.026040871 |
| 0 | 0 | 0 | 0 | 0 |
| 0 | 0 | 0 | 0 | 0 |
| 0 | 0 | 0 | 0 | 0 |
| 0 | 0 | 0 | 0 | 0 |
| 0 | 0 | 0 | 0 | 0 |
| 0 | 0 | 0 | 0 | 0 |
| 0 | 0 | 0 | 0 | 0 |
| 0.708763259 | 0.012111518 | 0 | 0 | 0.000726841 |
| 0.000627671 | 0 | 0 | 0 | 0.10670303 |
| 0 | 0 | 0 | 0 | 0 |
| 0 | 0.001406649 | 0 | 10.04055913 | 0.122651702 |
| 0 | 0 | 0 | 0.000662278 | 0 |
| 0.117644662 | 0 | 0 | 0 | 0 |
| 0 | 0 | 0 | 0 | 0 |
| 0 | 0 | 0 | 0 | 0 |
| 0 | 0 | 0 | 0 | 0 |
| 0 | 0 | 0 | 0 | 0 |
| 0 | 0 | 0 | 0 | 0 |
| 0 | 0 | 0 | 0 | 0 |
| 0 | 0 | 0 | 0.031160058 | 0 |
| 0 | 0 | 0 | 0 | 0 |
| 0 | 0 | 0.000628392 | 0 | 0 |
| 0 | 0 | 0.002040399 | 0 | 0 |
| 0 | 0 | 0 | 0 | 0 |
| 0 | 0 | 0 | 0 | 0 |
| 0 | 0 | 0 | 0 | 0.080324295 |
| 0 | 0 | 0 | 0 | 0 |
| 0 | 0 | 0 | 0 | 0 |
| 0 | 0 | 0 | 0 | 0 |
| 0 | 0 | 0 | 0 | 0 |
| 0 | 0 | 0 | 0 | 0 |
| 0 | 0 | 0 | 0 | 0.002354933 |

| | | | | |
|---|---|---|---|---|
| 0.011528941 | 0 | 0.005474101 | 0 | 0.143163297 |
| 0 | 0 | 0 | 0 | 0 |
| 0 | 0 | 0 | 0 | 0 |
| 0.062384293 | 0 | 0 | 0 | 0 |
| 0 | 0 | 0 | 0 | 0 |
| 1.237977908 | 0.619735873 | 0 | 0.019872842 | 0 |
| 0 | 0 | 0 | 0 | 0 |
| 0 | 0 | 0 | 0 | 0 |
| 0 | 0 | 0 | 0 | 0 |
| 0 | 0 | 0 | 0 | 0 |
| 0 | 0 | 0 | 0 | 0 |
| 0.019509954 | 0 | 0 | 0.001056555 | 0 |
| 0 | 0 | 0 | 0 | 0 |
| 0 | 0 | 0 | 0 | 0 |
| 0 | 0 | 0 | 0 | 0 |
| 0 | 0 | 0 | 0 | 0 |
| 0 | 0 | 0 | 0 | 0 |
| 0 | 0 | 0 | 0 | 0 |
| 0 | 0 | 0 | 0 | 0.000397523 |
| 0 | 0 | 0 | 0 | 0 |
| 0.003980958 | 0.262467893 | 0 | 0.004565382 | 0 |
| 0 | 0 | 0 | 0 | 0 |
| 0.064472345 | 0.00265956 | 0 | 0.061405181 | 0.003427914 |
| 0.049805297 | 0 | 0 | 0 | 0.156661108 |
| 0 | 0 | 0 | 0 | 0.006568457 |
| 0 | 0 | 0 | 0 | 0 |
| 0 | 0 | 0 | 0 | 0 |
| 0 | 0 | 0 | 0 | 0 |
| 0 | 0 | 0 | 0 | 0 |
| 0 | 0 | 0 | 0 | 0 |
| 0 | 0 | 0 | 0 | 0 |
| 0 | 0 | 0.046382849 | 0 | 0 |
| 0.056777557 | 0.149892843 | 0 | 0.004859637 | 0.002996212 |
| 0 | 0 | 0 | 0 | 0 |
| 0.011643907 | 0.004455779 | 0.002470265 | 0.713136668 | 0.033118347 |
| 0.168244579 | 0.009266494 | 0 | 0.012348906 | 0.000686953 |
| 0 | 0 | 0 | 0 | 0 |
| 0 | 0 | 0 | 0 | 0 |
| 0.170694022 | 0.211841397 | 0 | 0.002756458 | 0.057010361 |
| 0 | 0 | 0 | 0 | 0 |
| 0 | 0 | 0 | 0 | 0 |
| 0 | 0 | 0.007225969 | 0 | 0.038373595 |
| 0 | 0 | 0.005138092 | 0.001465057 | 0.812489056 |
| 0 | 0 | 0 | 0 | 0 |
| 0 | 0 | 0 | 0 | 0 |
| 0.017187943 | 0 | 0.004250559 | 0 | 0.016048422 |
| 0 | 0 | 0 | 0 | 0 |
| 0 | 0 | 0 | 0 | 0 |

| | | | | |
|---|---|---|---|---|
| 0 | 0 | 0 | 0 | 0 |
| 0 | 0 | 0 | 0 | 0 |
| 0 | 0 | 0 | 0 | 0 |
| 0 | 0 | 0 | 0 | 0 |
| 0 | 0 | 0 | 0 | 0.007929069 |
| 0 | 0 | 0 | 0 | 0 |
| 0 | 0 | 0 | 0 | 0 |
| 0.002537486 | 1.301813809 | 0 | 0 | 0 |
| 0.034667276 | 0 | 0 | 0 | 0.001901089 |
| 0 | 0.000804052 | 0 | 0 | 0.000759986 |
| 0 | 0 | 0 | 0 | 0 |
| 0 | 0 | 0 | 0 | 0 |
| 0 | 0 | 0 | 0 | 0 |
| 0.006132917 | 0 | 0.003313648 | 0 | 0 |
| 0 | 0 | 0 | 0 | 0 |
| 0.240467656 | 0.009295403 | 0 | 0.001179095 | 0 |
| 0 | 0 | 0 | 0 | 0 |
| 0 | 0 | 0 | 0 | 0 |
| 0 | 0 | 0 | 0 | 0 |
| 0.001215254 | 0 | 0.529126919 | 0 | 0.52383352 |
| 0 | 0 | 0.004858784 | 0 | 0.003514337 |
| 0 | 0 | 0 | 0 | 0 |
| 0 | 0 | 0 | 0 | 0 |
| 0.008700515 | 0 | 0 | 0 | 0 |
| 0.004250483 | 0 | 0 | 0 | 0 |
| 0 | 0 | 0 | 0 | 0.102612539 |
| 0 | 0 | 0 | 0 | 0 |
| 0 | 0 | 0 | 0 | 0 |
| 0 | 0 | 0 | 0 | 0 |
| 0 | 0 | 0.072144531 | 0 | 0.538415609 |
| 0.058201032 | 0 | 0.017224979 | 0.086058497 | 0.666391221 |
| 0 | 0 | 0 | 0 | 0 |
| 0 | 0 | 0 | 0 | 0 |
| 0 | 0 | 0 | 0 | 0 |
| 0 | 0 | 0 | 0 | 0 |
| 0 | 0 | 0 | 0 | 0 |
| 0 | 0 | 0 | 0 | 0 |
| 0 | 0 | 0 | 0 | 0 |
| 0 | 0 | 0 | 0 | 0 |
| 0 | 0 | 0 | 0 | 0 |
| 0 | 0 | 0 | 0 | 0 |
| 0 | 0 | 0 | 0 | 0 |
| 0 | 0 | 0 | 0 | 0 |
| 0 | 0 | 0 | 0.002015107 | 0 |
| 0 | 0 | 0 | 0 | 0 |
| 0 | 0 | 0 | 0 | 0 |
| 0.335319307 | 0.153667398 | 0 | 0.071214815 | 0.002870469 |
| 0 | 0 | 0 | 0 | 0 |

| | | | | |
|---|---|---|---|---|
| 0 | 0 | 0 | 0 | 0 |
| 0 | 0 | 0 | 0 | 0 |
| 0 | 0 | 0 | 0 | 0 |
| 0 | 0 | 0 | 0 | 0 |
| 0 | 0 | 0.000790624 | 0.04238188 | 0.153257016 |
| 0 | 0 | 0 | 0 | 0 |
| 0 | 0 | 0 | 0 | 0 |
| 0 | 0 | 0 | 0 | 0 |
| 0 | 0 | 0 | 0 | 0 |
| 0 | 0 | 0 | 0 | 0 |
| 0 | 0 | 0 | 0 | 0.089667982 |
| 0.019264846 | 0.023729183 | 0 | 0.29495653 | 0.006351495 |
| 0.002216094 | 0 | 0 | 0 | 0 |
| 0 | 0 | 0 | 0 | 0 |
| 0 | 0 | 0 | 0.00133569 | 0 |
| 0 | 0 | 0.017214081 | 3.577050698 | 0.004321789 |
| 0 | 0 | 0 | 0.005199599 | 0.076786928 |
| 0 | 0 | 0 | 0 | 0 |
| 0.00914147 | 0.005247255 | 0 | 0.347906611 | 0.011873177 |
| 0.058918001 | 1.87767592 | 0.268054339 | 0.00396085 | 0.257659646 |
| 0 | 0 | 0 | 0 | 0 |
| 0 | 0 | 0.000475759 | 0 | 0.259806095 |
| 0 | 0 | 0 | 0 | 0 |
| 0 | 0 | 0 | 0 | 0 |
| 0 | 0 | 0 | 0 | 0 |
| 0 | 0 | 0 | 0 | 0 |
| 0.005087683 | 0 | 0 | 0.08133403 | 0.001100778 |
| 0 | 0 | 0 | 0 | 0 |
| 0 | 0 | 0 | 0 | 0 |
| 0 | 0 | 0 | 0 | 0 |
| 0.109806638 | 0.001320139 | 0 | 0.000899477 | 0 |
| 0 | 0 | 0 | 0 | 0 |
| 0 | 0 | 0 | 0 | 0 |
| 0 | 0 | 0 | 0 | 0 |
| 0 | 0 | 0 | 0 | 0.115949042 |
| 0 | 0 | 0 | 0 | 0 |
| 0 | 0 | 0 | 0 | 0 |
| 0.046839224 | 0.034714683 | 0 | 0.094633981 | 0.0063871 |
| 0.016959878 | 0 | 0.053592672 | 0 | 0.100473373 |
| 0 | 0 | 0 | 0 | 0 |
| 0.08235698 | 0.011111226 | 0 | 0.161506881 | 0 |
| 0 | 0 | 0 | 0 | 0 |
| 0 | 0 | 0 | 0 | 0 |
| 0.78709502 | 0.251885955 | 0 | 0.006793949 | 0.002042821 |
| 0 | 0 | 0 | 0 | 0 |
| 0 | 0 | 0 | 0 | 0.047815709 |
| 0 | 0 | 0 | 0 | 0 |
| 0 | 0 | 0 | 0 | 0.002518057 |

| | | | | |
|---|---|---|---|---|
| 0 | 0 | 0 | 0 | 0 |
| 0 | 0 | 0.000731505 | 0 | 0.024761627 |
| 0 | 0 | 0 | 0 | 0.014980173 |
| 0.095711168 | 0.004324126 | 0 | 0.006874569 | 0 |
| 0 | 0.003437621 | 0 | 0 | 0.000541537 |
| 0.001205968 | 0.002711243 | 0 | 0 | 0.000931874 |
| 0 | 0 | 0 | 0 | 0.095001446 |
| 0 | 0 | 0 | 0 | 0 |
| 0 | 0 | 0 | 0 | 0 |
| 0 | 0 | 0 | 0.001134852 | 0.017992125 |
| 0 | 0 | 0 | 0 | 0.036185046 |
| 0 | 0 | 0 | 0 | 0 |
| 0.686848892 | 0 | 0.013606218 | 0.005569259 | 0.623865885 |
| 0.001260172 | 0 | 0 | 0 | 0 |
| 0 | 0 | 0 | 0 | 0 |
| 0 | 0 | 0 | 0 | 0 |
| 0 | 0 | 0 | 0 | 0 |
| 0 | 0 | 0 | 0 | 0 |
| 0 | 0 | 0 | 0 | 0 |
| 0 | 0 | 0 | 0 | 0 |
| 0 | 0 | 0 | 0 | 0 |
| 0 | 0 | 0 | 0 | 0 |
| 0.062771146 | 0.027099918 | 0 | 0.051104848 | 0 |
| 0.018864807 | 0 | 0.001383108 | 0 | 0.046304039 |
| 0 | 0 | 0 | 0 | 0 |
| 0 | 0 | 0 | 0 | 0.002533035 |
| 0 | 0 | 0 | 0 | 0 |
| 0.001540969 | 0 | 0 | 0.001502112 | 0 |
| 0 | 0 | 0 | 0 | 0 |
| 0 | 0 | 0 | 0 | 0 |
| 0 | 0 | 0 | 0 | 0.014989092 |
| 0 | 0 | 0 | 0 | 0 |
| 0 | 0 | 0 | 0 | 0 |
| 0 | 0 | 0 | 0 | 0 |
| 0 | 0 | 0 | 0 | 0.056209693 |
| 0 | 0 | 0 | 0 | 0 |
| 0 | 0.10245509 | 0 | 0.064339509 | 0 |
| 0 | 0 | 0 | 0 | 0 |
| 0.022400065 | 0.021757511 | 0 | 0 | 0.029476642 |
| 0 | 0 | 0 | 0 | 0 |
| 0 | 0 | 0 | 0 | 0 |
| 0 | 0 | 0 | 0 | 0 |
| 0 | 0 | 0.144272858 | 0 | 0 |
| 0 | 0 | 0 | 0 | 0 |
| 0 | 0 | 0 | 0 | 0 |
| 0.687646019 | 0 | 0.030019082 | 0 | 0.087906148 |
| 0.054577322 | 0 | 0 | 0 | 0 |
| 0 | 0 | 0 | 0 | 0 |

| | | | | |
|---|---|---|---|---|
| 0 | 0 | 0 | 0 | 0 |
| 0 | 0 | 0 | 0 | 0 |
| 0 | 0 | 0 | 0 | 0.076247389 |
| 0.000883825 | 0 | 0.009232355 | 0 | 0 |
| 0 | 0 | 0 | 0 | 0 |
| 0.006108061 | 0 | 0 | 0 | 0 |
| 0.056810492 | 0 | 0 | 0 | 0 |
| 0 | 0 | 0 | 0 | 0 |
| 0 | 0 | 0 | 0 | 0 |
| 0 | 0 | 0 | 0 | 0 |
| 0.056496538 | 0.000735465 | 0 | 0.025782081 | 0.000556127 |
| 0.014505967 | 0 | 0 | 0 | 0 |
| 0 | 0 | 0 | 0 | 0 |
| 0 | 0 | 0 | 0 | 0 |
| 0 | 0 | 0 | 0 | 0 |
| 0 | 0 | 0 | 0 | 0 |
| 0 | 0 | 0 | 0 | 0.096462926 |
| 0 | 0 | 0 | 0 | 0 |
| 0 | 0 | 0 | 0 | 0 |
| 0.000715835 | 0 | 0 | 0.00279114 | 0 |
| 0 | 0 | 0 | 0 | 0 |
| 0 | 0 | 0 | 0 | 0 |
| 0 | 0 | 0 | 0 | 0 |
| 0 | 0 | 0 | 0 | 0 |
| 0 | 0 | 0 | 0 | 0 |
| 0 | 0 | 0 | 0 | 0 |
| 0 | 0 | 0 | 0 | 0 |
| 0 | 0 | 0 | 0 | 0.002652618 |
| 0 | 0 | 0 | 0 | 0 |
| 0 | 0 | 0 | 0 | 0 |
| 0.001229954 | 0.002513789 | 0 | 0.000999116 | 0 |
| 0 | 0 | 0 | 0 | 0 |
| 0.004760338 | 0.097097574 | 0.017630139 | 0.138048971 | 0.011219132 |
| 0.00097291 | 0.035593088 | 0.000866158 | 0 | 0 |
| 0 | 0 | 0 | 0 | 0 |
| 0 | 0 | 0 | 0 | 0 |
| 0 | 0 | 0 | 0 | 0 |
| 0 | 0 | 0 | 0 | 0 |
| 0 | 0 | 0 | 0 | 0 |
| 0 | 0 | 0 | 0 | 0 |
| 0 | 0 | 0 | 0 | 0 |
| 0.324855976 | 1.228955286 | 0 | 0.115098649 | 0 |
| 0.011689296 | 0.033934512 | 0.010704034 | 0.025812122 | 0.100095322 |
| 0 | 0 | 0 | 0 | 0 |
| 0 | 0 | 0 | 0 | 0 |
| 0.001378926 | 0 | 0 | 0 | 0.130526525 |
| 0 | 0 | 0 | 0.003745382 | 0 |
| 0 | 0 | 0 | 0 | 0 |

| | | | | |
|---|---|---|---|---|
| 0 | 0 | 0 | 0 | 0 |
| 0 | 0 | 0.001601985 | 0 | 0.021049872 |
| 0.005254797 | 0.029832764 | 0 | 0 | 0.008797711 |
| 0 | 0 | 0 | 0 | 0 |
| 0 | 0 | 0 | 0 | 0.026837378 |
| 0 | 0 | 0 | 0.030300502 | 0 |
| 0 | 0 | 0 | 0 | 0 |
| 0 | 0 | 0 | 0 | 0 |
| 0 | 0 | 0 | 0 | 0 |
| 0 | 0 | 0 | 0 | 0 |
| 0.006997798 | 0 | 0 | 0 | 0 |
| 0 | 0 | 0 | 0 | 0 |
| 0 | 0 | 0 | 0 | 0 |
| 0.001472264 | 0.00240722 | 0 | 0 | 0 |
| 0 | 0 | 0 | 0 | 0 |
| 0 | 0 | 0 | 0 | 0 |
| 2.345514433 | 0 | 0.020837774 | 0 | 0 |
| 0 | 0 | 0 | 0 | 0 |
| 0 | 0 | 0 | 0 | 0 |
| 0 | 0 | 0 | 0 | 0 |
| 0 | 0 | 0 | 0 | 0 |
| 0.004696529 | 0.066807674 | 0 | 0 | 0 |
| 0 | 0 | 0 | 0 | 0 |
| 0 | 0 | 0 | 0 | 0 |
| 0 | 0 | 0.003084408 | 0 | 0.013757455 |
| 0.004356573 | 0 | 0 | 0 | 0.002805339 |
| 0 | 0 | 0 | 0 | 0 |
| 0.492567009 | 0 | 0 | 0 | 0.057509655 |
| 0 | 0 | 0 | 0 | 0 |
| 0 | 0 | 0 | 0 | 0 |
| 0 | 0 | 0 | 0 | 0 |
| 0 | 0 | 0 | 0.001034478 | 0 |
| 0 | 0 | 0 | 0 | 0 |
| 0 | 0 | 0 | 0 | 0 |
| 0 | 0 | 0 | 0 | 0 |
| 0 | 0 | 0 | 0 | 0 |
| 0 | 0 | 0 | 0.004313944 | 0 |
| 0.005541214 | 0.196959598 | 0 | 0.040628578 | 0 |
| 0.016920433 | 0 | 0.023632644 | 3.225067384 | 0.138388505 |
| 0 | 0 | 0 | 0 | 0 |
| 0 | 0 | 0 | 0 | 0 |
| 0.081395838 | 0 | 0 | 0 | 0.004260705 |
| 0.734788704 | 0.035148794 | 0 | 0.013496476 | 0.001126186 |
| 0 | 0 | 0 | 0 | 0 |
| 0 | 0 | 0 | 0 | 0 |
| 0 | 0 | 0 | 0 | 0.010365093 |
| 0.016721895 | 0.007898532 | 0 | 0.065925404 | 0.000861422 |
| 0 | 0 | 0 | 0 | 0 |

| | | | | |
|---|---|---|---|---|
| 0 | 0 | 0 | 0 | 0 |
| 0.047826565 | 0.040632632 | 0 | 0.094612345 | 0.001449275 |
| 0 | 0 | 0.021184514 | 0 | 0.343685935 |
| 0 | 0 | 0 | 0 | 0 |
| 0 | 0 | 0 | 0 | 0 |
| 0 | 0 | 0.00061125 | 0 | 0 |
| 0 | 0 | 0 | 0 | 0 |
| 0 | 0 | 0 | 0 | 0 |
| 0 | 0 | 0 | 0 | 0 |
| 0 | 0 | 0 | 0 | 0 |
| 0 | 0 | 0 | 0 | 0 |
| 0 | 0 | 0 | 0 | 0 |
| 0.039404656 | 0 | 0.04571564 | 0 | 0.492733249 |
| 0 | 0 | 0 | 0 | 0 |
| 0.002923163 | 0 | 0 | 0.006268796 | 0.003388176 |
| 0 | 0 | 0 | 0 | 0 |
| 0 | 0 | 0 | 0 | 0 |
| 0 | 0 | 0 | 0 | 0 |
| 0.000970132 | 0 | 0 | 0 | 0 |
| 0 | 0 | 0 | 0 | 0 |
| 0 | 0 | 0 | 0 | 0 |
| 0.022104979 | 0.304161643 | 0 | 0.002798387 | 0.036158353 |
| 0 | 0.016771813 | 0 | 0 | 0 |
| 0 | 0 | 0 | 0 | 0 |
| 0 | 0 | 0 | 0 | 0 |
| 0 | 0 | 0 | 0 | 0 |
| 0 | 0 | 0 | 0 | 0 |
| 0 | 0 | 0 | 0 | 0 |
| 0 | 0 | 0 | 0 | 0 |
| 0 | 0 | 0 | 0 | 0 |
| 0 | 0 | 0 | 0 | 0 |
| 0 | 0 | 0 | 0 | 0 |
| 0 | 0 | 0 | 0 | 0 |
| 0 | 0 | 0 | 0 | 0 |
| 6.147182042 | 0.001917791 | 0.00519794 | 0 | 15.80683065 |
| 0 | 0 | 0 | 0 | 0 |
| 0 | 0 | 0 | 0 | 0 |
| 0 | 0 | 0 | 0 | 0 |
| 0.008027089 | 0.216088204 | 0.079834654 | 0.006427415 | 0.174118245 |
| 0 | 0 | 0 | 0 | 0 |
| 0.076343292 | 0.014499853 | 0.064076251 | 0.01328002 | 0.039526574 |
| 0 | 0 | 0 | 0 | 0 |
| 0 | 0 | 0 | 0 | 0 |
| 0 | 0 | 0 | 0 | 0 |
| 0 | 0 | 0 | 0 | 0 |
| 4.370982272 | 9.591968128 | 0.081118459 | 0.166965034 | 0.234690476 |
| 0 | 0 | 0 | 0 | 0 |
| 0 | 0 | 0 | 0 | 0 |

| | | | | |
|---|---|---|---|---|
| 0 | 0 | 0 | 0 | 0 |
| 0 | 0 | 0 | 0 | 0 |
| 0 | 0 | 0 | 0 | 0 |
| 0 | 0 | 0 | 0 | 0.011804643 |
| 0.209590674 | 0 | 0 | 0 | 0 |
| 0 | 0 | 0 | 0 | 0 |
| 0 | 0 | 0 | 0 | 0 |
| 0.004453393 | 0.012482579 | 0.0015859 | 0 | 0.026546572 |
| 0.048446858 | 0 | 0 | 0 | 0.00246491 |
| 0.023026273 | 0.020001043 | 0.021524723 | 0 | 2.067305779 |
| 0 | 0 | 0.009728955 | 0 | 0.04620904 |
| 0 | 0 | 0 | 0 | 0 |
| 0.005345233 | 0.008279719 | 0 | 0 | 0.000652163 |
| 0 | 0 | 0 | 0 | 0 |
| 0 | 0 | 0 | 0 | 0 |
| 0 | 0 | 0 | 0 | 0 |
| 0 | 0 | 0 | 0 | 0 |
| 0 | 0 | 0 | 0 | 0 |
| 0 | 0 | 0 | 0 | 0 |
| 1.531958254 | 2.011268647 | 0.05243236 | 0.295102032 | 0.372855743 |
| 0 | 0 | 0 | 0 | 0 |
| 0 | 0 | 0 | 0 | 0 |
| 0 | 0 | 0 | 0 | 0 |
| 0 | 0 | 0 | 0 | 0 |
| 0 | 0 | 0 | 0 | 0 |
| 0 | 0 | 0 | 0 | 0 |
| 0 | 0 | 0 | 0 | 0 |
| 0 | 0 | 0 | 0 | 0 |
| 0 | 0 | 0 | 0 | 0 |
| 0 | 0 | 0.004766245 | 0.036791652 | 0 |
| 0 | 0 | 0 | 0 | 0 |
| 0 | 0 | 0 | 0 | 0.160957151 |
| 0 | 0 | 0 | 0 | 0 |
| 0 | 0 | 0 | 0 | 0 |
| 0 | 0 | 0 | 0 | 0 |
| 0 | 0 | 0 | 0.003791591 | 0 |
| 0 | 0 | 0 | 0 | 0 |
| 0 | 0 | 0 | 0 | 0 |
| 0 | 0 | 0 | 0 | 0 |
| 0 | 0 | 0 | 0 | 0.030954475 |
| 0 | 0 | 0 | 0 | 0 |
| 0 | 0 | 0 | 0 | 0 |
| 0.000624432 | 0 | 0 | 0 | 0 |
| 0 | 0 | 0 | 0 | 0 |
| 0.235656953 | 0.011151688 | 0 | 0.009170246 | 0 |
| 0 | 0 | 0 | 0 | 0 |
| 0 | 0 | 0 | 0 | 0.006142554 |
| 0 | 0 | 0 | 0 | 0 |

| | | | | |
|---|---|---|---|---|
| 0 | 0 | 0 | 0 | 0 |
| 0 | 0 | 0 | 0 | 0 |
| 0 | 0 | 0 | 0 | 0 |
| 0 | 0 | 0 | 0 | 0 |
| 0 | 0 | 0 | 0 | 0 |
| 0.012114739 | 0.008612243 | 0 | 0.013349592 | 0.003256101 |
| 0 | 0 | 0 | 0 | 0 |
| 0 | 0 | 0 | 0 | 0 |
| 0 | 0 | 0 | 0 | 0 |
| 0 | 0 | 0 | 0 | 0.579319951 |
| 0 | 0 | 0 | 0 | 0 |
| 0 | 0 | 0 | 0 | 0 |
| 0 | 0 | 0 | 0 | 0 |
| 0 | 0 | 0 | 0 | 0.012489174 |
| 0 | 0 | 0 | 0 | 0 |
| 0 | 0 | 0 | 0 | 0 |
| 0 | 0 | 0 | 0 | 0.068131077 |
| 0.005405778 | 0.001237418 | 0 | 0 | 0 |
| 0 | 0 | 0 | 0 | 0 |
| 0 | 0 | 0 | 0 | 0 |
| 0 | 0 | 0 | 0 | 0 |
| 0 | 0 | 0 | 0 | 0 |
| 0 | 0 | 0 | 0 | 0 |
| 0 | 0 | 0.021145954 | 0 | 0 |
| 0 | 0 | 0 | 0 | 0 |
| 0 | 0 | 0.001564779 | 0 | 0.016764749 |
| 0 | 0 | 0 | 0 | 0 |
| 0 | 0 | 0 | 0 | 0 |
| 0 | 0 | 0 | 0 | 0 |
| 0 | 0 | 0 | 0 | 0 |
| 0 | 0 | 0 | 0 | 0 |
| 0.006450193 | 0 | 0 | 0 | 0 |
| 0 | 0 | 0 | 0 | 0 |
| 0 | 0 | 0 | 0 | 0 |
| 0 | 0 | 0 | 0 | 0 |
| 0 | 0 | 0 | 0 | 0 |
| 0 | 0 | 0 | 0 | 0 |
| 0.119863938 | 0 | 0.00059491 | 0 | 0 |
| 0.003965459 | 0 | 0.073784363 | 0 | 0.002106628 |
| 0 | 0 | 0 | 0 | 0 |
| 0 | 0 | 0 | 0 | 0 |
| 0 | 0 | 0 | 0 | 0 |
| 0 | 0 | 0 | 0 | 0 |
| 0 | 0 | 0 | 0 | 0 |
| 0 | 0 | 0 | 0 | 0 |
| 0 | 0 | 0 | 0 | 0 |
| 0 | 0 | 0 | 0 | 0 |
| 0 | 0 | 0 | 0 | 0 |

| | | | | |
|---|---|---|---|---|
| 0 | 0 | 0 | 0 | 0 |
| 0.001179428 | 0 | 0 | 0 | 0.001518945 |
| 0.000721986 | 0 | 0 | 0 | 0 |
| 0 | 0 | 0 | 0 | 0 |
| 0 | 0 | 0 | 0 | 0 |
| 0.005391157 | 0 | 0 | 0 | 0 |
| 0 | 0 | 0 | 0 | 0 |
| 0 | 0 | 0 | 0 | 0 |
| 0 | 0 | 0 | 0 | 0 |
| 0 | 0 | 0 | 0 | 0 |
| 0.001286973 | 0.002104261 | 0.001283253 | 0 | 0.010740264 |
| 0 | 0 | 0 | 0 | 0 |
| 0 | 0 | 0 | 0 | 0.014796028 |
| 0 | 0 | 0 | 0 | 0 |
| 0 | 0 | 0 | 0 | 0 |
| 0 | 0 | 0 | 0 | 0 |
| 0 | 0 | 0 | 0 | 0 |
| 0 | 0 | 0 | 0 | 0 |
| 0 | 0 | 0.004632331 | 0 | 0 |
| 0.002570179 | 0 | 0 | 0.001461466 | 0 |
| 0.170067303 | 0.190504289 | 0 | 0.633321254 | 0 |
| 0 | 0 | 0.001151062 | 0 | 0 |
| 0 | 0 | 0 | 0 | 0 |
| 0.005193869 | 1.476230149 | 0.428026058 | 1.125651541 | 1.162715875 |
| 0 | 0 | 0 | 0 | 0 |
| 0 | 0 | 0 | 0 | 0 |
| 0 | 0 | 0 | 0 | 0 |
| 0 | 0 | 0 | 0 | 0.002782174 |
| 0.00879039 | 0 | 0 | 0 | 0 |
| 0 | 0 | 0 | 0 | 0 |
| 0 | 0 | 0 | 0 | 0 |
| 0 | 0 | 0 | 0 | 0 |
| 0 | 0 | 0 | 0 | 0.028878637 |
| 0 | 0 | 0 | 0 | 0 |

| PHL018-0099_V2 | PHL018-0038_V3 | PHL018-0229_V1 | PHL018-0219_V1 | PHL018-0237_V1 |
|---|---|---|---|---|
| EYCF | CTRL | REF | REF | CTRL |
| 1.832785982 | 0.072620604 | 0.015211216 | 0.098557984 | 0.017623724 |
| 2.3462854 | 1.944871547 | 0.061926585 | 0.195969569 | 0.169582546 |
| 1.79906268 | 14.35775995 | 0 | 18.1583808 | 2.163872044 |
| 0 | 0 | 0 | 0 | 0 |
| 0 | 0 | 0 | 0 | 0 |
| 0 | 0 | 0 | 0 | 0 |
| 0 | 0 | 0 | 0.105821604 | 0 |
| 0 | 0 | 0 | 0 | 0 |
| 0 | 0 | 0 | 0.01901673 | 0 |
| 0 | 0 | 0 | 0 | 0 |
| 0 | 0 | 0 | 0 | 0 |
| 0 | 0 | 0 | 0 | 0 |
| 0 | 0 | 0 | 0 | 0 |
| 0.519463505 | 6.747604331 | 0.042753723 | 1.492602487 | 0.517928358 |
| 0 | 0 | 0 | 0 | 0 |
| 0 | 0 | 0 | 0 | 0 |
| 0.001667377 | 0 | 0 | 0.001483711 | 0 |
| 0 | 0 | 0 | 0 | 0 |
| 0 | 0 | 0 | 0 | 0 |
| 0 | 0 | 0 | 0 | 0 |
| 0 | 0 | 0 | 0 | 0 |
| 0 | 0 | 0 | 0 | 0 |
| 0 | 0 | 0 | 0 | 0 |
| 0 | 0 | 0 | 0 | 0 |
| 0 | 0 | 0 | 0 | 0 |
| 0 | 0 | 0 | 0 | 0 |
| 0 | 0 | 0 | 0 | 0 |
| 0 | 0 | 0 | 0 | 0 |
| 0 | 0 | 0 | 0 | 0 |
| 0 | 0 | 0.012661191 | 0 | 0 |
| 0 | 0 | 0 | 0 | 0 |
| 0 | 0 | 0 | 0 | 0 |
| 0 | 0 | 0 | 0 | 0 |
| 0 | 0 | 0 | 0 | 0 |
| 0 | 0 | 0 | 0 | 0 |
| 0 | 0 | 1.935908988 | 0.447082565 | 0.967629013 |
| 0 | 0 | 0 | 0 | 0 |
| 0 | 0 | 0 | 0 | 0 |
| 0 | 0 | 0 | 0 | 0 |
| 0 | 0 | 0 | 0 | 0 |
| 0 | 0 | 0 | 0 | 0 |
| 0.023883141 | 0 | 0.344404196 | 0.001092978 | 0.0049111 |
| 0.001585701 | 0 | 1.529552373 | 0.597816781 | 2.035132237 |
| 0.285885636 | 0 | 0.000531517 | 0.00433118 | 0.008321417 |
| 0 | 0 | 0.012454173 | 0 | 0.01006359 |
| 0 | 0 | 0 | 0 | 0 |

| | | | | |
|---|---|---|---|---|
| 0.003659281 | 0 | 0 | 0 | 0 |
| 0.0196361 | 0.013401541 | 0 | 0 | 0 |
| 0 | 0.004599608 | 0.001553784 | 0.005874223 | 0 |
| 0.538182061 | 1.212990065 | 0.490878864 | 0.411116215 | 0.341576187 |
| 0.023664052 | 0 | 0 | 0.0013884 | 0 |
| 0.100157815 | 0 | 0 | 0 | 0 |
| 0.011332855 | 0 | 0.004943925 | 0.000823226 | 0 |
| 0 | 0 | 0 | 0 | 0.038527746 |
| 0 | 0 | 0 | 0 | 0.013232083 |
| 0 | 0 | 0 | 0 | 0 |
| 0 | 0 | 0 | 0 | 0 |
| 0.715239363 | 0 | 0.360147864 | 0 | 0.348891907 |
| 0 | 0 | 0 | 0.001782919 | 0 |
| 0.084833974 | 0.027173876 | 0 | 0.04878677 | 0 |
| 0 | 0 | 0 | 0 | 0 |
| 0 | 0 | 0 | 0 | 0 |
| 0 | 0 | 43.39945796 | 0.002222648 | 0 |
| 0 | 0 | 0 | 0 | 0 |
| 0 | 0 | 0 | 0 | 0 |
| 0 | 0 | 0 | 0 | 0 |
| 0.035122238 | 0.349700593 | 0.01856849 | 0.033485823 | 0 |
| 0.007477198 | 0.020797745 | 0 | 0.007604075 | 0 |
| 0 | 0 | 0 | 0 | 0 |
| 0 | 0 | 0 | 0 | 0 |
| 0 | 0 | 0 | 0 | 0 |
| 0 | 0 | 0 | 0 | 0 |
| 0 | 0 | 0 | 0 | 0 |
| 0 | 0 | 0 | 0 | 0 |
| 0 | 0 | 0 | 0 | 0 |
| 0 | 0 | 0 | 0 | 0 |
| 0.848986937 | 0 | 0 | 0.020380759 | 0 |
| 0 | 0 | 0 | 0 | 0 |
| 0 | 0 | 0 | 0 | 0 |
| 0 | 0 | 0 | 0 | 0 |
| 0 | 0 | 0 | 0 | 0 |
| 0 | 0 | 0 | 0 | 0 |
| 0 | 0 | 0 | 0 | 0 |
| 0 | 0 | 3.176132635 | 0 | 0 |
| 0 | 0 | 0 | 0 | 0 |
| 0 | 0 | 0 | 0 | 0 |
| 0 | 0 | 0 | 0 | 0 |
| 0 | 0 | 0 | 0 | 0 |
| 0 | 0 | 0 | 0 | 0 |
| 0 | 0 | 0 | 0 | 0 |
| 0 | 0 | 0 | 0 | 0 |
| 0 | 0 | 0 | 0 | 0 |
| 0 | 0 | 0 | 0 | 0 |
| 0 | 0 | 0 | 0 | 0 |

| | | | | |
|---|---|---|---|---|
| 0 | 0 | 0 | 0 | 0 |
| 0 | 0.048536206 | 0 | 0.009060603 | 0 |
| 0 | 0 | 0 | 0 | 0 |
| 0 | 0 | 0 | 0 | 0 |
| 0.049756621 | 0 | 0 | 0 | 0 |
| 0 | 0 | 0 | 0.000539215 | 0 |
| 0 | 0 | 0 | 0 | 0 |
| 0.07525352 | 0 | 0 | 0.001359423 | 0 |
| 0.068751983 | 0.002047492 | 0 | 0 | 0 |
| 0 | 0 | 0 | 0 | 0 |
| 0 | 0 | 0 | 0 | 0 |
| 0 | 0 | 0 | 0 | 0 |
| 0 | 0 | 0 | 0 | 0 |
| 0 | 0 | 0 | 0 | 0 |
| 0 | 0 | 0 | 0 | 0 |
| 0 | 0 | 0 | 0 | 0 |
| 0 | 0 | 0 | 0 | 0 |
| 0 | 0 | 0 | 0 | 0 |
| 0.012254153 | 0 | 0 | 0.105440951 | 1.455462416 |
| 0 | 0 | 0 | 0 | 0 |
| 0 | 0 | 0 | 0 | 0 |
| 0 | 0 | 0 | 0 | 0 |
| 0 | 0 | 0 | 0 | 0 |
| 0 | 0.005085254 | 0 | 0 | 0 |
| 0 | 0 | 0 | 0 | 0 |
| 0 | 0 | 0 | 0 | 0 |
| 0 | 0 | 0 | 0 | 0 |
| 0.011475495 | 0 | 0 | 0.002283303 | 0 |
| 0 | 0 | 0 | 0 | 0 |
| 0 | 0 | 0 | 0 | 0 |
| 0.51942925 | 0 | 0 | 0.222615534 | 0.005994644 |
| 0 | 0 | 0 | 0.003018771 | 0 |
| 0 | 0 | 0 | 0 | 0 |
| 0 | 0 | 0 | 0 | 0 |
| 0 | 0 | 0 | 0 | 0 |
| 0 | 0 | 0 | 0 | 0 |
| 0 | 0 | 0 | 0 | 0 |
| 0 | 0 | 0 | 0 | 0 |
| 0 | 0 | 0 | 0.235990693 | 0 |
| 0 | 0 | 0 | 0 | 0 |
| 0 | 0 | 0 | 0 | 0 |
| 0 | 0 | 0 | 0 | 0 |
| 0 | 0.009816422 | 0 | 0 | 0 |
| 0 | 0 | 0 | 0 | 0 |
| 0 | 0 | 0 | 0.000507272 | 0 |
| 0 | 0 | 0 | 0 | 0 |
| 0 | 0 | 0 | 0 | 0 |
| 0 | 0 | 0 | 0 | 0 |

| | | | | |
|---|---|---|---|---|
| 0 | 0 | 0 | 0 | 0 |
| 0 | 0 | 0 | 0 | 0 |
| 0 | 0 | 0 | 0 | 0 |
| 0 | 0 | 0 | 0 | 0 |
| 0.014719018 | 0 | 0 | 0 | 0.03123274 |
| 0 | 0 | 0 | 0 | 0 |
| 0 | 0 | 0 | 0 | 0 |
| 0 | 0 | 0 | 0 | 0 |
| 0 | 0 | 0 | 0 | 0 |
| 0 | 0 | 0 | 0 | 0 |
| 0 | 0 | 0 | 0 | 0 |
| 0 | 0 | 0 | 0 | 0 |
| 0 | 0 | 0 | 0 | 0 |
| 0 | 0 | 0 | 0 | 0 |
| 0 | 0 | 0 | 0 | 0 |
| 0 | 0 | 9.155734585 | 0 | 0.571902171 |
| 0 | 0 | 0 | 0 | 0 |
| 0 | 0 | 0 | 0 | 0 |
| 0 | 0 | 0 | 0 | 0 |
| 0 | 0 | 0 | 0 | 0 |
| 0 | 0 | 0 | 0 | 0 |
| 0 | 0 | 0 | 0 | 0 |
| 0 | 0 | 0 | 0 | 0 |
| 0 | 0 | 0 | 0 | 0 |
| 0.128889938 | 0 | 0 | 0 | 0.013302453 |
| 0 | 0 | 0 | 0 | 0 |
| 0 | 0 | 0 | 0 | 0 |
| 0 | 0 | 0 | 0 | 0 |
| 0 | 0 | 0 | 0 | 0 |
| 0 | 0 | 0 | 0 | 0 |
| 0 | 0 | 0 | 0.022841287 | 0 |
| 0 | 0 | 0.174804697 | 0 | 0 |
| 0 | 0 | 0 | 0 | 0 |
| 0 | 0 | 0 | 0 | 0 |
| 0 | 0 | 0 | 0 | 0 |
| 0 | 0 | 0 | 0 | 0 |
| 0 | 0 | 0 | 0 | 0 |
| 0 | 0 | 0 | 0 | 0 |
| 0 | 0 | 0 | 0 | 0 |
| 0 | 0 | 0 | 0 | 0 |
| 0 | 0 | 0 | 0 | 0 |
| 0 | 0 | 0 | 0 | 0 |
| 0 | 0 | 0 | 0 | 0 |
| 0 | 0 | 0 | 0 | 0 |
| 0 | 0 | 0 | 0 | 0 |
| 0 | 0 | 0 | 0 | 0 |
| 0 | 0 | 0 | 0 | 0 |
| 0.019340805 | 0 | 0 | 0 | 0 |

| | | | | |
|---|---|---|---|---|
| 0 | 0 | 0 | 0 | 0 |
| 0 | 0 | 0 | 0 | 0 |
| 0 | 0 | 0 | 0 | 0 |
| 0 | 0 | 0 | 0 | 0 |
| 0 | 0 | 0 | 0 | 0 |
| 0 | 0 | 0 | 0 | 0 |
| 0 | 0 | 0 | 0 | 0 |
| 0 | 0 | 0 | 0 | 0 |
| 0 | 0 | 0 | 0 | 0 |
| 0 | 0 | 0 | 0 | 0 |
| 0 | 0 | 0 | 0 | 0 |
| 0 | 0 | 0 | 0.04810168 | 0 |
| 0.000595871 | 0.000798011 | 0 | 0.000757477 | 0 |
| 0 | 0 | 0 | 0 | 0 |
| 0 | 0 | 0 | 0.006904518 | 0 |
| 0 | 0 | 0 | 0 | 0 |
| 0 | 0 | 0 | 0 | 0 |
| 0 | 0 | 0 | 0 | 0 |
| 0 | 0 | 0 | 0 | 0 |
| 0 | 0 | 0 | 0 | 0 |
| 0.116612781 | 0.032452937 | 0.002520197 | 0.047021164 | 0.011964112 |
| 0 | 0 | 0 | 0 | 0 |
| 0.001955168 | 0 | 0 | 0 | 0 |
| 0 | 0.003111898 | 0.001051224 | 0.002363066 | 0 |
| 0 | 0 | 0 | 0 | 0 |
| 0 | 0 | 0 | 0 | 0 |
| 0.004974485 | 0.001303436 | 0.001320933 | 0.002309495 | 0.001185977 |
| 0 | 0 | 0 | 0 | 0 |
| 0 | 0 | 0 | 0 | 0 |
| 0 | 0.029285059 | 0 | 0 | 0 |
| 0 | 0.08683686 | 0 | 0 | 0 |
| 0 | 0 | 0 | 0 | 0 |
| 0 | 0 | 0 | 0 | 0 |
| 0 | 0 | 0.017866182 | 0.021638867 | 0.01291219 |
| 0.153198813 | 0.056793712 | 0 | 0.000737216 | 0.006293842 |
| 0 | 0 | 0 | 0 | 0 |
| 0 | 0 | 0 | 0 | 0 |
| 0 | 0 | 0 | 0 | 0 |
| 0 | 0 | 0 | 0 | 0 |
| 0 | 0 | 0 | 0 | 0 |
| 0 | 0 | 0 | 0 | 0 |
| 0 | 0 | 0 | 0 | 0 |
| 0 | 0 | 0 | 0.001915634 | 0 |
| 0 | 0 | 0 | 0 | 0 |
| 0 | 0 | 0 | 0 | 0 |
| 0 | 0 | 0 | 0 | 0 |
| 0.001046521 | 0 | 0 | 0.001795971 | 0 |
| 0 | 0 | 0 | 0 | 0.002242581 |

| | | | | |
|---|---|---|---|---|
| 0 | 0 | 0 | 0 | 0 |
| 0 | 0 | 0 | 0 | 0 |
| 0 | 0 | 0 | 0 | 0 |
| 0 | 0 | 0 | 0.000834752 | 0 |
| 0 | 0 | 0 | 0 | 0 |
| 0 | 0 | 0 | 0 | 0 |
| 0 | 0 | 0 | 0 | 0 |
| 0 | 0 | 0 | 0 | 0 |
| 0.015778735 | 0 | 0 | 0 | 0.00034888 |
| 0 | 0 | 0 | 0 | 0 |
| 0 | 0 | 0 | 0 | 0 |
| 0 | 0 | 0 | 0 | 0 |
| 0 | 0 | 0 | 0 | 0 |
| 0 | 0 | 0 | 0 | 0 |
| 0 | 0 | 0 | 0 | 0 |
| 0 | 0 | 0 | 0 | 0 |
| 0 | 0 | 0 | 0 | 0 |
| 0 | 0 | 0 | 0 | 0 |
| 0 | 0 | 0.008199997 | 0.002537868 | 0.003361017 |
| 0 | 0 | 0 | 0 | 0 |
| 0 | 0.001556819 | 0 | 0.007487226 | 0 |
| 0 | 0 | 0 | 0 | 0 |
| 0 | 0 | 0 | 0 | 0 |
| 0 | 0 | 0 | 0 | 0 |
| 0.005132197 | 0.00309295 | 0 | 0.007828925 | 0.002814229 |
| 0 | 0 | 0 | 0 | 0 |
| 0 | 0 | 0 | 0 | 0 |
| 0 | 0 | 0 | 0 | 0 |
| 0 | 0 | 0.00415454 | 0 | 0 |
| 0 | 0 | 0 | 0 | 0 |
| 0.285490635 | 1.196622569 | 0 | 0.379579683 | 0.050646402 |
| 0 | 0 | 0 | 0 | 0 |
| 0.021965153 | 0 | 0 | 0 | 0 |
| 0 | 0 | 0 | 0 | 0 |
| 0 | 0 | 0 | 0 | 0 |
| 0 | 0 | 0 | 0 | 0 |
| 0 | 0 | 0 | 0 | 0 |
| 0 | 0 | 0 | 0 | 0 |
| 0 | 0 | 0 | 0 | 0 |
| 0 | 0 | 0 | 0 | 0 |
| 0 | 0 | 0 | 0 | 0 |
| 0.037949438 | 0 | 0.000788349 | 0.00118143 | 0.031585834 |
| 0 | 0 | 0 | 0 | 0 |
| 0 | 0 | 0 | 0 | 0 |
| 0 | 0 | 0 | 0 | 0 |
| 0 | 0 | 0 | 0 | 0 |
| 0 | 0 | 0 | 0 | 0 |
| 0 | 0 | 0 | 0 | 0 |

| | | | | |
|---|---|---|---|---|
| 0 | 0 | 0 | 0 | 0 |
| 0 | 0 | 0 | 0 | 0 |
| 0.295149191 | 0 | 0 | 0.00482131 | 0.010331912 |
| 0 | 0 | 0 | 0 | 0 |
| 0 | 0 | 0 | 2.922891868 | 0 |
| 0 | 0 | 0 | 0 | 0 |
| 0 | 0 | 0 | 0 | 0 |
| 0 | 0 | 0 | 0 | 0 |
| 0 | 0 | 0 | 0 | 0 |
| 0 | 0 | 0 | 0 | 0 |
| 0 | 0 | 0 | 0 | 0 |
| 0 | 0 | 0 | 0 | 0 |
| 0 | 0 | 0 | 0 | 0 |
| 0.025794516 | 0.000809647 | 0.00109402 | 0.001639513 | 0.001657543 |
| 0.008649061 | 0.001559268 | 0.002257427 | 0.002706407 | 0.000456028 |
| 0 | 0 | 0.016434739 | 0.001992077 | 0.020506014 |
| 0.003284943 | 0.002791872 | 0 | 0.002312781 | 0.00155881 |
| 0 | 0 | 0 | 0 | 0 |
| 0 | 0 | 0 | 0 | 0 |
| 0.001544483 | 0 | 0 | 0 | 0 |
| 0 | 0 | 0 | 0 | 0 |
| 0 | 0 | 0 | 0 | 0.002032565 |
| 0 | 0 | 0 | 0 | 0 |
| 0 | 0 | 0 | 0 | 0 |
| 0 | 0 | 0 | 0 | 0 |
| 0 | 0 | 0 | 0 | 0 |
| 0 | 0 | 0 | 0 | 0 |
| 0 | 0 | 0 | 0 | 0 |
| 0 | 0 | 0 | 0 | 0 |
| 0 | 0.003314167 | 0 | 0 | 0 |
| 0 | 0 | 0 | 0 | 0 |
| 0 | 0 | 0 | 0 | 0 |
| 0 | 0 | 0 | 0.001440264 | 0 |
| 0 | 0 | 0 | 0 | 0 |
| 0 | 0 | 0 | 0 | 0 |
| 0 | 0 | 0 | 0 | 0 |
| 0 | 0 | 0 | 0 | 0 |
| 0 | 0 | 0 | 0.011148094 | 0.012768537 |
| 0 | 0 | 0 | 0 | 0 |
| 0 | 0 | 0 | 0 | 0 |
| 0 | 0 | 0 | 0 | 0 |
| 0 | 0 | 0 | 0 | 0 |
| 0 | 0 | 0 | 0 | 0 |
| 0 | 0 | 0 | 0 | 0 |
| 0 | 0 | 0 | 0 | 0 |
| 0 | 0 | 0 | 0 | 0 |
| 0 | 0 | 0 | 0 | 0 |
| 0.030200562 | 0.003486696 | 0 | 0 | 0 |

| | | | | |
|---|---|---|---|---|
| 0.006407716 | 0 | 0 | 0 | 0 |
| 0 | 0 | 0 | 0 | 0 |
| 0.001911378 | 0 | 0 | 0 | 0 |
| 0 | 0 | 0 | 0 | 0 |
| 0 | 0.007001846 | 0 | 0.005683643 | 0 |
| 0 | 0 | 0 | 0.008071863 | 0 |
| 0 | 0 | 0 | 0 | 0 |
| 0.048182977 | 0.025721094 | 0.00297248 | 0.002312964 | 0.001385718 |
| 0 | 0 | 0 | 0 | 0 |
| 0.03229847 | 0.191858013 | 0 | 0.034170984 | 0.0635589 |
| 0 | 0 | 0 | 0 | 0 |
| 0 | 0 | 0.003077034 | 0 | 0 |
| 0.012139268 | 0.002845035 | 0 | 0.006481258 | 0.014145149 |
| 0.017514596 | 0 | 0 | 0 | 0 |
| 0 | 0 | 0 | 0 | 0 |
| 0 | 0 | 0.000824419 | 0 | 0 |
| 0 | 0 | 0 | 0 | 0 |
| 0 | 0 | 0 | 0 | 0 |
| 0.143791391 | 0.123091972 | 0 | 0 | 0 |
| 0 | 0 | 0 | 0 | 0 |
| 0 | 0 | 0 | 0 | 0 |
| 0 | 0 | 0 | 0 | 0 |
| 0 | 0 | 0 | 0 | 0 |
| 0 | 0 | 0 | 0 | 0 |
| 0 | 0 | 0 | 0 | 0 |
| 0 | 0 | 0.023187727 | 0 | 0.013406528 |
| 0 | 0 | 0.818790503 | 0 | 0 |
| 0 | 0.001144924 | 0 | 0.006955319 | 0 |
| 0 | 0.22295488 | 0 | 0.046926684 | 0 |
| 0 | 0.002435301 | 0 | 0 | 0 |
| 0 | 0.021588926 | 0 | 0.000771476 | 0 |
| 0 | 0 | 0 | 0 | 0 |
| 0 | 0 | 0 | 0 | 0 |
| 0 | 0.00278599 | 0 | 0 | 0 |
| 0 | 0 | 0 | 0 | 0 |
| 0.064998282 | 0 | 0 | 0 | 0 |
| 0.045413171 | 0.245780052 | 0 | 0 | 0 |
| 0 | 0 | 0 | 0 | 0 |
| 0 | 0 | 0 | 0 | 0 |
| 0 | 0 | 0 | 0 | 0 |
| 0 | 0 | 0 | 0 | 0 |
| 0 | 0 | 0 | 0 | 0 |
| 0 | 0 | 0 | 0 | 0 |
| 0.056385448 | 0.019069041 | 0 | 0 | 0 |
| 0 | 0 | 0 | 0 | 0 |
| 0 | 0 | 0 | 0 | 0 |
| 0 | 0 | 0 | 0 | 0 |
| 0.003723941 | 0.011755625 | 0 | 0 | 0 |

| | | | | |
|---|---|---|---|---|
| 0.014956998 | 0 | 0 | 0 | 0 |
| 0 | 0 | 0.001333489 | 0 | 0 |
| 0 | 0.052967637 | 0 | 0 | 0 |
| 0.002207767 | 0 | 0 | 0 | 0 |
| 0 | 0 | 0 | 0.0008837 | 0 |
| 0.02054248 | 0.011823643 | 0 | 0.003697013 | 0 |
| 0 | 0 | 0 | 0 | 0 |
| 0.002388238 | 0 | 0 | 0 | 0 |
| 0 | 0 | 0 | 0 | 0 |
| 0 | 0 | 0 | 0 | 0 |
| 0 | 0.007260054 | 0 | 0.014293041 | 0 |
| 0 | 0 | 0 | 0 | 0 |
| 0 | 0 | 0 | 0 | 0 |
| 0.003793725 | 0.001016138 | 0.003947485 | 0.003279384 | 0.041432258 |
| 0.0056299 | 0.52281396 | 0 | 0.060897764 | 0 |
| 0 | 0 | 0 | 0 | 0 |
| 0.1976777 | 0.001242064 | 0 | 0 | 0 |
| 0.046764158 | 0.005438767 | 0 | 0 | 0 |
| 0 | 0 | 0 | 0 | 0 |
| 0.224906708 | 0 | 0 | 0 | 0 |
| 0 | 0 | 0 | 0 | 0 |
| 0.314542618 | 0 | 0.434275115 | 0 | 0.098391287 |
| 0 | 0 | 0 | 0 | 0 |
| 0 | 0 | 0 | 0 | 0 |
| 0 | 0 | 0 | 0 | 0 |
| 0 | 0 | 0 | 0 | 0 |
| 0.021389692 | 0.002754407 | 0 | 0 | 0 |
| 0 | 0 | 0 | 0 | 0 |
| 0 | 0 | 0 | 0 | 0 |
| 0.319931723 | 1.302520505 | 0.346919591 | 0.115648126 | 2.632270129 |
| 0.001201079 | 0 | 0.002328743 | 0 | 0.036458716 |
| 0 | 0 | 0 | 0 | 0 |
| 0 | 0 | 0 | 0 | 0 |
| 0 | 0 | 0 | 0 | 0 |
| 0.009593015 | 0 | 0 | 0 | 0 |
| 0 | 0 | 0 | 0 | 0 |
| 0 | 0 | 0.384361006 | 8.733967976 | 0.386370921 |
| 0 | 0 | 0 | 0 | 0 |
| 0 | 0 | 0 | 0.195973025 | 0 |
| 0 | 2.341279374 | 0 | 0 | 0 |
| 0.014138808 | 0.036756556 | 0 | 0 | 0 |
| 2.063261134 | 11.03822241 | 0.698372357 | 1.74637743 | 2.952994448 |
| 0 | 0 | 0.000641012 | 0 | 0 |
| 0 | 0 | 0 | 0 | 0 |
| 0.016887507 | 0.00706761 | 0 | 0 | 0 |
| 0 | 0 | 0 | 0 | 0 |
| 0 | 0.005041527 | 0.001184743 | 0 | 0.005684153 |
| 0 | 0 | 0 | 0 | 0 |

| | | | | |
|---|---|---|---|---|
| 0 | 0 | 0.072357803 | 0 | 0.004798567 |
| 0 | 0.002419353 | 0 | 0 | 0 |
| 0.001100663 | 0 | 2.362000644 | 0 | 0.000754433 |
| 0 | 0 | 0 | 0 | 0 |
| 0 | 0 | 0 | 0.064136729 | 0 |
| 0 | 0.003430012 | 0 | 0 | 0 |
| 0 | 0 | 0 | 0 | 0 |
| 0 | 0 | 0 | 0 | 0 |
| 0.073462998 | 0.009959143 | 0 | 0 | 0 |
| 0 | 0 | 0 | 0.088261158 | 0 |
| 0 | 0 | 0 | 0 | 0 |
| 0 | 0 | 0 | 0 | 0 |
| 0 | 0 | 0 | 0 | 0 |
| 0 | 0 | 0 | 0.058339543 | 0 |
| 0 | 0 | 0 | 0 | 0 |
| 1.240409453 | 0.030784218 | 0.004004642 | 0.012631997 | 0.005088792 |
| 0.011182907 | 0 | 2.716931399 | 0 | 0.159666581 |
| 0 | 0 | 0 | 0 | 0 |
| 0 | 0 | 0 | 0 | 0 |
| 0 | 0 | 0 | 0 | 0 |
| 0 | 0.036859105 | 0 | 0 | 0 |
| 0.00244747 | 0.001229152 | 0 | 0 | 0 |
| 0 | 0 | 0 | 0 | 0 |
| 0 | 0 | 0 | 0.273049342 | 0 |
| 0 | 0 | 0 | 0.004361841 | 0 |
| 0 | 0 | 0 | 0.021412211 | 0 |
| 0 | 0.003497646 | 0 | 0 | 0 |
| 0 | 0 | 0 | 0 | 0 |
| 0 | 0 | 0 | 0 | 0 |
| 0.004584009 | 0 | 0 | 0 | 0 |
| 0.072316975 | 0.014732032 | 0 | 0.020354104 | 0 |
| 0.102956436 | 0 | 0 | 0 | 0.000832683 |
| 0.02139854 | 0.062156199 | 0.966444028 | 0.020732375 | 1.09826384 |
| 0 | 0 | 0.00253098 | 0 | 0 |
| 0.002662037 | 0 | 0 | 0 | 0 |
| 0 | 0 | 0 | 0 | 0 |
| 0.002091433 | 0.001312932 | 0 | 0.001196393 | 0.008780438 |
| 0.033938649 | 0.002053547 | 0 | 0.01528204 | 0 |
| 0 | 0 | 0.061530665 | 0 | 0.153032148 |
| 0.018888912 | 0 | 0 | 0.001529955 | 0 |
| 0 | 0 | 0 | 0 | 0 |
| 0 | 0 | 0 | 0 | 0 |
| 0.025377598 | 0 | 0 | 0.00069962 | 0 |
| 0.049635035 | 0.08287752 | 0 | 0.118929965 | 0 |
| 4.144144471 | 0.048309848 | 0 | 0 | 0 |
| 0 | 0 | 0 | 0 | 0 |
| 0.003180671 | 0 | 0 | 0 | 0 |
| 0 | 0 | 0 | 0 | 0 |

| | | | | |
|---|---|---|---|---|
| 0.000909393 | 0.003349202 | 0 | 0 | 0 |
| 0.407853774 | 0.102414806 | 0.017959948 | 0.394163227 | 0.030361739 |
| 0 | 0 | 0 | 0 | 0 |
| 0 | 0 | 0 | 0 | 0.001850387 |
| 0 | 0.001098074 | 0 | 0 | 0 |
| 0 | 0 | 0 | 0 | 0 |
| 0 | 0.090605089 | 0 | 0 | 0 |
| 0 | 0 | 0 | 0 | 0 |
| 0.0040804 | 0 | 0.001730616 | 0 | 0 |
| 0 | 0 | 0 | 0 | 0 |
| 0.003312349 | 0.001663506 | 0 | 0.008210853 | 0.001844699 |
| 0.032069947 | 0.002168109 | 0.005649978 | 0.007055941 | 0.052841 |
| 0 | 0 | 0.27480111 | 0 | 0.158781813 |
| 0 | 0 | 0 | 0 | 0 |
| 0 | 0 | 0.000805953 | 0 | 0.012373746 |
| 0 | 0 | 0 | 0 | 0 |
| 0.018209665 | 0.02395155 | 0 | 0.018022621 | 0.003417564 |
| 0.316307798 | 0.057379648 | 0 | 0.011375965 | 0 |
| 0 | 0 | 0 | 0 | 0 |
| 0.101052265 | 0.057632681 | 0 | 0.024251519 | 0 |
| 0.011401389 | 0.004490925 | 0.216789317 | 0.011253829 | 0.105424646 |
| 0 | 0 | 0 | 0 | 0 |
| 0 | 0 | 0 | 0 | 0 |
| 0 | 0 | 0 | 0 | 0 |
| 0.010892889 | 0 | 0 | 0 | 0 |
| 0 | 0 | 0 | 0 | 0 |
| 0.082968757 | 0 | 0 | 0 | 0.028132336 |
| 0.016394933 | 0.034605633 | 0.032087189 | 0.340893031 | 0.013192031 |
| 0 | 0 | 0 | 0 | 0 |
| 0 | 0 | 0 | 0 | 0.002946792 |
| 0 | 0 | 0 | 0 | 0 |
| 0 | 0 | 0 | 0 | 0 |
| 0.000865362 | 0.001738386 | 0 | 0 | 0 |
| 0 | 0.001084532 | 0 | 0 | 0 |
| 0 | 0 | 0.005620453 | 0 | 0.004541605 |
| 0.545699759 | 0.830049505 | 0.236597996 | 0.097843815 | 0.082967217 |
| 0 | 0 | 0 | 0 | 0 |
| 0 | 0 | 0 | 0 | 0 |
| 0.002638041 | 0.005299438 | 0.001392372 | 0.011401921 | 0 |
| 0 | 0 | 0 | 0 | 0 |
| 2.450704912 | 0.015888142 | 0 | 0 | 0 |
| 0 | 0 | 0 | 0 | 0 |
| 0.001246301 | 0.001430649 | 0 | 0.001493779 | 0 |
| 0.008766753 | 0.001553923 | 0 | 0.009439954 | 0 |
| 0 | 0 | 0 | 0.008956936 | 0 |
| 0 | 0 | 0.203444556 | 0 | 0.407452893 |
| 0 | 0 | 0 | 0 | 0 |
| 0.001389292 | 0.015070772 | 0 | 0 | 0 |

| | | | | |
|---|---|---|---|---|
| 0 | 0.000999324 | 0 | 0 | 0 |
| 0 | 0 | 0 | 0 | 0 |
| 0.002610643 | 0 | 0 | 0 | 0 |
| 0.394613121 | 0 | 0.004502988 | 3.548833784 | 0.421143884 |
| 0 | 0 | 0 | 0 | 0 |
| 0 | 0 | 0 | 0 | 0 |
| 0.003858347 | 0 | 0 | 0 | 0 |
| 0 | 0 | 0 | 0 | 0 |
| 0.371949055 | 0.018668532 | 0.185544267 | 0.118362967 | 0.098694037 |
| 0 | 0 | 0.014085973 | 0.144184879 | 0.005716489 |
| 0 | 0 | 0 | 0 | 0 |
| 0 | 0.002394674 | 0.001483057 | 0.000606144 | 0.041398697 |
| 0 | 0 | 0 | 0 | 0 |
| 0 | 0 | 0 | 0 | 0 |
| 0.569721144 | 0 | 0.012349477 | 0.020632899 | 0.002921776 |
| 0 | 0 | 0 | 0 | 0 |
| 0.027587488 | 0 | 0.001741493 | 0.015332715 | 0.107153394 |
| 0 | 0 | 0.01452843 | 0 | 0.405019404 |
| 0 | 0 | 0 | 0 | 0 |
| 0 | 0 | 0 | 0 | 0 |
| 0 | 0 | 0 | 0 | 0 |
| 0 | 0 | 0 | 0 | 0 |
| 0 | 0 | 0 | 0 | 0 |
| 1.121421559 | 0.318674696 | 0 | 0.168393507 | 0.115282957 |
| 0.200893152 | 0 | 0 | 0 | 0 |
| 0 | 0 | 0 | 2.834006326 | 0 |
| 0 | 0 | 0 | 0 | 0.015322142 |
| 0 | 0 | 0.398548507 | 0 | 1.497884271 |
| 0 | 0 | 0 | 0 | 0 |
| 0 | 0 | 0 | 0 | 0 |
| 0.004828691 | 0 | 0.007733204 | 0.004124927 | 0.014430521 |
| 0 | 0 | 0 | 0 | 0.044406029 |
| 0.666201907 | 0 | 0 | 0 | 0 |
| 0 | 0 | 0 | 0 | 0 |
| 0 | 0 | 0 | 0 | 0 |
| 0 | 0 | 0.005667822 | 0 | 0.004826836 |
| 0.188664018 | 0 | 0.219170103 | 0.676099583 | 0.086549521 |
| 0 | 0 | 0 | 0 | 0 |
| 0.176626395 | 0 | 0 | 0.001735239 | 0 |
| 0.063993373 | 0 | 0 | 0 | 0 |
| 0.154953567 | 0.02680139 | 0.000905372 | 0.002471319 | 0.000653201 |
| 0 | 0 | 0 | 0 | 0 |
| 0.01851999 | 30.43627196 | 0 | 0.050747832 | 0.861014558 |
| 0 | 0 | 0 | 0 | 0 |
| 0 | 0 | 0 | 0 | 0 |
| 0 | 0 | 0 | 0 | 0 |
| 0 | 0 | 0 | 0 | 0 |
| 0.459379196 | 0 | 0 | 0.001048521 | 0 |

| | | | | |
|---|---|---|---|---|
| 0 | 0 | 0 | 0 | 0 |
| 0.163854542 | 0 | 0.007648929 | 0 | 0.031046645 |
| 0.029211365 | 0.471436268 | 0 | 0.005025627 | 0 |
| 0 | 0 | 0.000644443 | 0 | 0.001301856 |
| 0 | 0 | 0 | 0 | 0 |
| 0.002422272 | 0 | 0.000587061 | 0 | 0.000711561 |
| 0 | 0 | 0 | 0 | 0 |
| 0.448516656 | 0 | 0 | 0 | 0 |
| 0 | 0 | 0 | 0 | 0.131282089 |
| 0 | 0 | 0 | 0 | 0 |
| 1.799747782 | 0.015594951 | 0 | 0.012993589 | 0.001625893 |
| 1.380692531 | 1.658955603 | 1.046480756 | 0.861485428 | 0.421451107 |
| 0 | 0 | 0 | 0 | 0 |
| 0.000698189 | 0 | 0.015003524 | 0 | 0.097148257 |
| 0.103070712 | 1.052033028 | 0 | 0.028396432 | 0.001005669 |
| 0 | 0 | 2.059691716 | 0 | 8.812288763 |
| 0.002532655 | 0 | 0 | 0 | 0 |
| 0 | 0 | 0 | 0 | 0 |
| 0.002122514 | 0 | 0 | 0 | 0 |
| 0.039079088 | 0 | 0 | 0.319202149 | 0 |
| 0.292075381 | 0.206955371 | 0 | 0.270609275 | 0 |
| 0 | 3.428047655 | 0 | 0 | 0.105734688 |
| 0.257776694 | 1.868666327 | 0.014365027 | 3.604139309 | 2.311935938 |
| 0 | 0 | 0 | 0 | 0 |
| 0 | 0 | 0 | 0 | 0 |
| 0 | 0 | 5.67300601 | 0 | 3.180578243 |
| 0.031345102 | 0 | 0 | 0 | 0 |
| 0 | 0 | 0.007291607 | 0 | 0.001292101 |
| 0 | 0 | 0 | 0 | 0 |
| 0 | 0 | 0 | 0.085531736 | 0 |
| 0 | 0 | 0 | 0 | 0 |
| 0 | 0 | 0 | 0 | 0.151927405 |
| 0.019710822 | 0.178182684 | 0 | 0.014020439 | 0 |
| 0 | 0 | 0.001146085 | 0.00437972 | 0.001041855 |
| 0.015786672 | 0.004551419 | 0 | 0.019176267 | 0 |
| 0.014459948 | 0 | 0.003543445 | 0.002144525 | 0.280683949 |
| 0.0372483 | 0 | 0 | 0 | 0 |
| 0 | 0 | 0 | 0 | 0 |
| 0 | 0.017940184 | 0.00255172 | 0 | 8.968371558 |
| 0 | 0 | 0 | 0 | 0 |
| 0 | 0 | 0 | 0 | 0.018545817 |
| 0 | 0 | 0 | 0 | 0 |
| 0 | 0 | 0 | 0 | 0 |
| 0 | 0 | 0 | 0 | 0 |
| 0.372626601 | 0 | 0.000466257 | 0 | 0.008634061 |
| 0 | 0 | 0 | 0 | 0 |
| 0 | 0 | 0 | 0 | 0 |
| 0.39865603 | 0.01193215 | 0.000562434 | 0.001896458 | 1.888151491 |

| | | | | |
|---|---|---|---|---|
| 0 | 0 | 0 | 0 | 0 |
| 0 | 0 | 0 | 0 | 0 |
| 0 | 0 | 0 | 0 | 0 |
| 0.000585596 | 0 | 0 | 0.003870961 | 0.002006939 |
| 0 | 0 | 0 | 0 | 0 |
| 0 | 0 | 0 | 0 | 0.513793159 |
| 0 | 0 | 0.423753766 | 0 | 0 |
| 0.429107537 | 0 | 0.00036567 | 0.003150987 | 0.037427403 |
| 0 | 0 | 0 | 0 | 0 |
| 0 | 0 | 0 | 0 | 0 |
| 0.006304587 | 0.001528534 | 0 | 0 | 0.000745066 |
| 0 | 0 | 0 | 0 | 0 |
| 0 | 0 | 0 | 0 | 0.025572653 |
| 0 | 0 | 0 | 0 | 0 |
| 0 | 0 | 0 | 0 | 0 |
| 0 | 0 | 0 | 0 | 0 |
| 0 | 0 | 0.006473631 | 0 | 0 |
| 0.003619604 | 0.003272064 | 0 | 0 | 0.105070435 |
| 0 | 0 | 0 | 0.121431478 | 0 |
| 0.044394249 | 0 | 0 | 0.06834377 | 0 |
| 0 | 0 | 0 | 0 | 0 |
| 0 | 0 | 0 | 0 | 0 |
| 0.006885687 | 0 | 0.004672673 | 0.00190978 | 0.007580107 |
| 0 | 0 | 0 | 0 | 0 |
| 0 | 0 | 0 | 0 | 0 |
| 0 | 0 | 0 | 0 | 0 |
| 0 | 0 | 0 | 0 | 0 |
| 0 | 0 | 0 | 0 | 0 |
| 0 | 0 | 0 | 0 | 0 |
| 0 | 0 | 0.002670455 | 1.397024013 | 0.203797924 |
| 0.111125913 | 0.001719843 | 0 | 0 | 0.006220319 |
| 0 | 0 | 0 | 0 | 0 |
| 0 | 0 | 0 | 0 | 0 |
| 0 | 0 | 0 | 0 | 0 |
| 0 | 0 | 0 | 0 | 0.001895809 |
| 0 | 0 | 0 | 0 | 0 |
| 0 | 0 | 0 | 0 | 0.115994574 |
| 0.018771319 | 0.004066638 | 0 | 0 | 0 |
| 0 | 0 | 0 | 0 | 0 |
| 0.007079261 | 0 | 0 | 0 | 0 |
| 0 | 0 | 0 | 0 | 0.001893643 |
| 0 | 0 | 0 | 0 | 0 |
| 0 | 0 | 0 | 0 | 0 |
| 0 | 0 | 0.04438804 | 0 | 0.006124956 |
| 0 | 0 | 0.173003813 | 0 | 0.030508686 |
| 0 | 0 | 0 | 0 | 0 |
| 0.009617395 | 0 | 0 | 0 | 0 |
| 0 | 0 | 0 | 0 | 0 |

| | | | | |
|---|---|---|---|---|
| 0 | 0 | 0 | 0 | 0 |
| 0 | 0 | 0 | 0 | 0 |
| 0 | 0 | 0 | 0 | 0 |
| 0 | 0 | 0.024472257 | 0.002094353 | 0.797767573 |
| 0 | 0 | 0 | 0 | 0 |
| 0 | 0 | 0 | 0 | 0 |
| 0 | 0 | 0 | 0 | 0 |
| 0 | 0 | 0 | 0 | 0 |
| 0.184013413 | 0.321983101 | 0.006717291 | 0.078403469 | 0.006262966 |
| 0 | 0 | 0 | 0 | 0 |
| 0 | 0 | 0 | 0 | 0 |
| 0 | 0 | 0 | 0 | 0 |
| 0 | 0 | 0 | 0 | 0 |
| 0 | 0 | 0 | 0 | 0 |
| 1.66691446 | 0 | 0.000866425 | 0 | 0.059759733 |
| 0.195424042 | 0.429124581 | 0 | 0.080719516 | 0 |
| 0 | 0 | 0 | 0 | 0 |
| 0 | 0 | 0 | 0 | 0 |
| 0 | 0 | 0 | 0 | 0 |
| 0 | 0 | 0 | 0 | 0 |
| 0 | 0 | 0 | 0 | 0.006917352 |
| 0 | 0 | 0 | 0 | 0 |
| 0 | 0 | 0 | 0 | 0 |
| 0 | 0 | 0 | 0.022364061 | 0 |
| 0 | 0 | 0.086082577 | 0.015480535 | 2.413901773 |
| 3.672983555 | 0 | 2.912892808 | 0.363917946 | 0.728705165 |
| 0 | 0 | 0.04188658 | 0.001524135 | 1.271980754 |
| 0.002038052 | 0 | 0 | 0 | 0 |
| 0.249731727 | 0.013367501 | 0.034531421 | 0 | 0.065286105 |
| 0 | 0 | 0 | 0 | 0 |
| 0 | 0 | 0 | 0 | 0 |
| 0.049016021 | 0 | 0 | 0 | 0 |
| 0 | 0 | 0 | 0 | 0 |
| 0 | 0 | 0 | 0 | 0 |
| 0.001838543 | 0 | 0.037964122 | 0 | 0.003600575 |
| 0.003898881 | 0.000932414 | 0 | 0 | 0.002799687 |
| 2.007936531 | 0 | 0 | 0 | 0 |
| 2.529137241 | 1.87366405 | 0.841214979 | 0.892724549 | 0.392579734 |
| 0 | 0 | 0 | 16.18852221 | 7.200008522 |
| 0 | 0.021550653 | 0 | 0.001893449 | 0 |
| 0 | 0 | 0 | 0.252191256 | 0 |
| 0 | 0 | 0 | 0 | 0 |
| 5.188096137 | 0.375877401 | 0.72190301 | 0.042378818 | 0.00347802 |
| 0 | 0 | 0 | 0 | 0 |
| 0 | 0 | 0 | 0 | 0 |
| 0 | 0 | 0 | 0 | 0 |
| 0 | 0 | 0 | 0 | 0 |
| 0.036708114 | 0.272167024 | 0 | 0.05492786 | 0.008835166 |

| | | | | |
|---|---|---|---|---|
| 0.147314221 | 0 | 0 | 0 | 0 |
| 0 | 0 | 0 | 0 | 0 |
| 0 | 0 | 0 | 0 | 0 |
| 0 | 0 | 0 | 0 | 0 |
| 3.386063075 | 0 | 0 | 0.015709101 | 0 |
| 0 | 0 | 0.001223566 | 0 | 0 |
| 0 | 0 | 0 | 0 | 0 |
| 0 | 0 | 0 | 0 | 0 |
| 0 | 0 | 0 | 0 | 0 |
| 0.01983714 | 0 | 0 | 0.002866796 | 0 |
| 0 | 0 | 0 | 0 | 0.000982013 |
| 0.854282222 | 0 | 0.005079564 | 0.103058832 | 0.79512305 |
| 0.999792139 | 0 | 0.034188234 | 0.095296967 | 0.001899273 |
| 0 | 0 | 0 | 0 | 0 |
| 0.028631226 | 0.010874862 | 0.056328752 | 0.005688837 | 0.936385074 |
| 0 | 0 | 0 | 0 | 0 |
| 0.002343195 | 0 | 0 | 0 | 0 |
| 0 | 0 | 0 | 0.001715285 | 0 |
| 0.000632651 | 0 | 0 | 0 | 0.002601847 |
| 0 | 0 | 0.001904195 | 0 | 0 |
| 0 | 0 | 0 | 0 | 0 |
| 0 | 0 | 0 | 0 | 0 |
| 0.553855715 | 0 | 0.026160615 | 0.039100368 | 0.141957893 |
| 0 | 0 | 0 | 0 | 0 |
| 0 | 0 | 0 | 0 | 0 |
| 0 | 0 | 0 | 0 | 0 |
| 0 | 0 | 0 | 0 | 0 |
| 0 | 0 | 0 | 0 | 0 |
| 0 | 0 | 0 | 0 | 0 |
| 0 | 0 | 0 | 0 | 0 |
| 0 | 0 | 0 | 0 | 0 |
| 0.111771268 | 0.009721362 | 0.003178019 | 0.020717397 | 0.010913987 |
| 0 | 0 | 0 | 0 | 0 |
| 0.092961015 | 0 | 0 | 0.05829341 | 0.00363496 |
| 0.005455233 | 0 | 0 | 0 | 0.002631293 |
| 0.10444501 | 0 | 0.003358265 | 0 | 0.010176169 |
| 0 | 0 | 0 | 0 | 0.00049173 |
| 0 | 0 | 0 | 0 | 0.006488958 |
| 0 | 0 | 0 | 0 | 0.000636414 |
| 0 | 0 | 0 | 0 | 0 |
| 0 | 0 | 0.059109812 | 0 | 0 |
| 0 | 0 | 0 | 0 | 0.012806734 |
| 0 | 0 | 0 | 0 | 0 |
| 0 | 0 | 0 | 0.055200693 | 0 |
| 0.617805379 | 0.010795481 | 0 | 0.004706092 | 0.012960438 |
| 0 | 0 | 0 | 0 | 0 |
| 3.18934723 | 0 | 0.00738673 | 0 | 0.538114313 |
| 0 | 0.004348956 | 0 | 0 | 0 |

| | | | | |
|---|---|---|---|---|
| 0 | 0 | 0 | 0 | 0 |
| 0 | 0 | 0 | 0 | 0 |
| 0.0366417 | 0.00413527 | 0 | 0.003140179 | 0 |
| 0 | 0 | 0 | 0 | 0 |
| 0 | 0 | 0 | 0 | 0 |
| 0 | 0 | 0 | 0 | 0 |
| 0 | 0 | 0 | 0 | 0.017861576 |
| 0.003499348 | 0.005662795 | 0 | 0.021500639 | 0.010660331 |
| 0 | 0 | 0 | 0 | 0 |
| 0 | 0 | 0 | 0 | 0 |
| 0 | 0 | 0 | 0 | 0 |
| 0 | 0 | 0 | 0 | 0.000861384 |
| 0 | 0 | 0 | 0 | 0 |
| 0 | 0 | 0.16116171 | 0.188586802 | 0.061596759 |
| 0 | 0 | 0 | 0 | 0 |
| 0 | 0 | 0 | 0 | 0 |
| 0 | 0 | 0 | 0 | 0 |
| 0 | 0 | 0 | 0 | 0 |
| 1.281046298 | 0 | 0 | 17.11380086 | 0.002261522 |
| 0 | 0 | 0 | 0 | 0 |
| 0.000935176 | 0 | 0.000507693 | 0 | 0 |
| 0 | 0 | 0 | 0 | 0 |
| 0 | 0 | 0 | 0 | 0 |
| 0 | 0 | 0 | 0 | 0 |
| 0 | 0 | 0 | 0 | 0 |
| 0.010191186 | 0.005583436 | 0 | 0.001130631 | 0 |
| 0 | 0 | 0 | 0 | 0 |
| 0.620635301 | 0.011112315 | 0 | 0.002213325 | 0.000372944 |
| 0 | 0 | 0 | 0 | 0 |
| 0 | 0 | 0.621416636 | 0 | 1.351369269 |
| 0 | 0 | 0 | 0 | 0 |
| 0 | 0 | 0 | 0 | 0 |
| 0 | 0 | 0 | 0 | 0 |
| 0 | 0 | 0 | 0 | 0 |
| 0.004077716 | 0.001365256 | 0 | 0.010219168 | 0.001996434 |
| 0 | 0 | 0 | 0 | 0 |
| 0 | 0 | 0 | 0 | 0 |
| 0.022474064 | 0.000885237 | 0 | 0 | 0.000906147 |
| 0 | 0 | 0 | 0 | 0 |
| 0 | 0 | 0 | 0 | 0.003358528 |
| 6.714392543 | 6.612878616 | 1.425437627 | 2.097059348 | 1.897992196 |
| 0.911907978 | 0.15713944 | 0.212112877 | 0.08032989 | 0.003758722 |
| 0 | 0 | 0.129876016 | 0 | 0.004281932 |
| 0 | 0 | 0 | 0 | 0 |
| 0 | 0 | 0 | 0 | 0 |
| 0 | 0 | 0 | 0 | 0 |
| 0 | 0 | 0 | 0 | 0 |
| 0.015388492 | 0 | 0 | 0.000800264 | 0 |

| | | | | |
|---|---|---|---|---|
| 0 | 0 | 0 | 0 | 0 |
| 0.015828334 | 0.007226546 | 0 | 0.000548759 | 0 |
| 0.060711678 | 0 | 0 | 0 | 0 |
| 0 | 0 | 0 | 0 | 0 |
| 2.86296556 | 0.252857636 | 3.893985078 | 0.090762129 | 0.003365804 |
| 0 | 0 | 0 | 0 | 0 |
| 0 | 0 | 0 | 0 | 0 |
| 0 | 0 | 0 | 0.001454566 | 0 |
| 0.457446922 | 0.738907317 | 0 | 0.012876813 | 0 |
| 0.001526432 | 0 | 0 | 0 | 0 |
| 0 | 0.000816449 | 0 | 0 | 0 |
| 0 | 0 | 0 | 0 | 0 |
| 0.561542002 | 0 | 0.09962512 | 0.089579694 | 0.007307477 |
| 0.024064794 | 0.018991753 | 0.019684117 | 0.002622122 | 0.148011787 |
| 0 | 0 | 0.027810125 | 0 | 0 |
| 0.934651302 | 0.095632129 | 0.056833378 | 0.327192019 | 0.063810456 |
| 0.004648292 | 0.002427812 | 0 | 0 | 0 |
| 0 | 0 | 0 | 0 | 0 |
| 0 | 0 | 0 | 0 | 0 |
| 0 | 0 | 0 | 0 | 0 |
| 0 | 0 | 0 | 0 | 0 |
| 0 | 0 | 0 | 0 | 0 |
| 0 | 0 | 0 | 0 | 0 |
| 0 | 0 | 0 | 0 | 0 |
| 0 | 0 | 0 | 0 | 0 |
| 0 | 0 | 0 | 0 | 0 |
| 0 | 0 | 0.11146929 | 0.028855468 | 0.122896959 |
| 0 | 0 | 0 | 0 | 0 |
| 0 | 0 | 0 | 0 | 0 |
| 0 | 0 | 0 | 0 | 0 |
| 0 | 0 | 0 | 0 | 0 |
| 0 | 0 | 0 | 0 | 0 |
| 0 | 0 | 0 | 0 | 0.241676263 |
| 0 | 0 | 0 | 0 | 0 |
| 0.63693253 | 0 | 0.003683749 | 0 | 0.050541077 |
| 0 | 0 | 0 | 0 | 0 |
| 0.001941342 | 0 | 0.102757769 | 0 | 0.000887109 |
| 0 | 0 | 0 | 0 | 0 |
| 0 | 0 | 0 | 0 | 0 |
| 0 | 0 | 0 | 0 | 0 |
| 0 | 0 | 0.294841125 | 0.489691617 | 0.277257148 |
| 0 | 0 | 0.072733932 | 0 | 0 |
| 0 | 0 | 0 | 0 | 0 |
| 0 | 0 | 0 | 0 | 0 |
| 0 | 0 | 0 | 0 | 0 |
| 0 | 0 | 0 | 0 | 0 |
| 0 | 0 | 0 | 0 | 0 |
| 0 | 0 | 0 | 0 | 0 |

| | | | | |
|---|---|---|---|---|
| 0.079299322 | 0 | 0.422642463 | 0 | 0.213036821 |
| 0 | 0 | 0 | 0 | 0 |
| 0 | 0 | 0 | 0 | 0 |
| 0.001653281 | 0 | 0 | 0 | 0 |
| 0 | 0 | 0.000536733 | 0.019304503 | 0 |
| 0 | 0 | 0 | 0 | 0 |
| 0.030047689 | 0.005589019 | 0 | 0 | 0 |
| 0 | 0 | 0 | 0 | 0 |
| 0.000654593 | 0 | 0 | 0 | 0 |
| 0 | 0 | 0 | 0.085641351 | 0 |
| 0 | 0 | 0 | 0 | 0 |
| 0 | 0 | 0 | 0.024125074 | 0 |
| 0 | 0 | 0 | 0 | 0 |
| 0 | 0 | 0 | 0 | 0 |
| 0 | 0 | 0 | 0 | 0 |
| 0 | 0 | 0 | 0 | 0 |
| 0 | 0 | 0 | 0 | 0 |
| 0.027996103 | 0 | 0 | 0 | 0 |
| 0 | 0 | 0 | 0 | 0 |
| 0 | 0 | 0 | 0 | 0 |
| 0.046089175 | 0.014366855 | 0.001677662 | 0.518681464 | 0.123822673 |
| 0 | 0 | 0 | 0 | 0 |
| 0 | 0 | 0.002643508 | 0.012033345 | 0.002803612 |
| 0.004849479 | 0.003827172 | 0.00070519 | 0.002377817 | 0.011515291 |
| 0 | 0 | 0 | 0 | 0 |
| 0 | 0 | 0 | 0 | 0.000840337 |
| 0 | 0 | 0 | 0 | 0 |
| 0 | 0 | 0 | 0 | 0 |
| 0 | 0 | 0 | 0 | 0 |
| 0.091980333 | 0 | 0.009811031 | 0 | 0.710939167 |
| 0 | 0 | 0 | 0 | 0 |
| 0 | 0 | 0 | 0 | 0 |
| 0 | 0 | 0 | 0 | 0 |
| 0 | 1.600506175 | 0 | 2.471225232 | 0 |
| 0 | 0 | 0 | 0 | 0 |
| 0 | 0 | 0 | 0 | 0 |
| 0.025788806 | 0 | 0 | 0.014380445 | 0 |
| 0 | 0 | 0 | 0 | 0 |
| 0 | 0 | 0 | 0 | 0 |
| 0 | 0 | 0 | 0 | 0.028991156 |
| 0 | 0 | 0 | 0 | 0 |
| 0 | 0 | 0 | 0 | 0 |
| 0 | 0 | 0 | 0 | 0 |
| 0.003722333 | 0 | 0 | 0 | 0 |
| 0 | 0 | 0 | 0 | 0 |
| 0 | 0 | 0 | 0 | 0 |
| 0 | 0 | 0 | 0 | 0 |
| 0 | 0 | 0 | 0 | 0 |

| | | | | |
|---|---|---|---|---|
| 0.077154988 | 0.001310328 | 0 | 0.011371622 | 0 |
| 0 | 0 | 0.00659681 | 0 | 0 |
| 0.039422713 | 0 | 0 | 0 | 0 |
| 0 | 0 | 0 | 0 | 0 |
| 0 | 0 | 0 | 0 | 0 |
| 0 | 0 | 0 | 0 | 0 |
| 0 | 0 | 0 | 0 | 0 |
| 3.908796973 | 0 | 0.132451459 | 0.529802412 | 0.61055434 |
| 0.004972966 | 0 | 2.031958674 | 0.820629359 | 2.625658013 |
| 0.125854956 | 0.000613651 | 0.079601707 | 0 | 0 |
| 0 | 0 | 0.007459922 | 0 | 0 |
| 0 | 0 | 0 | 0 | 0 |
| 0 | 0 | 0 | 0 | 0 |
| 0.016675363 | 0 | 0 | 0 | 1.182027002 |
| 0.007833719 | 0 | 0 | 0 | 0 |
| 0 | 0 | 0 | 0 | 0 |
| 0.025027893 | 0 | 0 | 0.00269742 | 0 |
| 0 | 0 | 0 | 0 | 0 |
| 0 | 0 | 0 | 0 | 0 |
| 0.074640752 | 0 | 0 | 0.002060832 | 0.002662243 |
| 0 | 0 | 0 | 0 | 0 |
| 0 | 0 | 0 | 0 | 0 |
| 0 | 0 | 0 | 0.263005407 | 0 |
| 0 | 0 | 0 | 0 | 0 |
| 0 | 0 | 0 | 0 | 0.004024545 |
| 0 | 0 | 0 | 0 | 0 |
| 0 | 0 | 0 | 0 | 0 |
| 0 | 0 | 0 | 0 | 0 |
| 0 | 0 | 0 | 0 | 0 |
| 0 | 0 | 0 | 0 | 0 |
| 0 | 0 | 0 | 0.015041072 | 0 |
| 0 | 0 | 0 | 0 | 0 |
| 0 | 0 | 0 | 0 | 0 |
| 0 | 0 | 0 | 0 | 0 |
| 0 | 0 | 0 | 0 | 0 |
| 0 | 0 | 0.008337331 | 0 | 0 |
| 0 | 0 | 0 | 0 | 0 |
| 0.028349763 | 0 | 0 | 0 | 0 |
| 0 | 0 | 0.000751904 | 0 | 0.00197462 |
| 0 | 0 | 0 | 0 | 0 |
| 0 | 0 | 0 | 0 | 0 |
| 0.768735494 | 0 | 0.211793684 | 0 | 0.107357079 |
| 0 | 0 | 0 | 0 | 0 |
| 0 | 0 | 0 | 0 | 0 |
| 0 | 0 | 0 | 0 | 0 |
| 0.002320025 | 0 | 0 | 0.010490871 | 0 |
| 0 | 0 | 0 | 0 | 0 |
| 0 | 0 | 0 | 0 | 0 |

| | | | | |
|---|---|---|---|---|
| 0 | 0 | 0 | 0 | 0 |
| 0.072326988 | 0 | 0.000570716 | 0 | 0 |
| 0 | 0 | 0 | 0 | 0 |
| 0 | 0 | 0 | 0 | 0 |
| 0.277030801 | 0.005196653 | 0.117297338 | 0 | 0.135241769 |
| 0 | 0 | 0 | 0 | 0 |
| 0 | 0 | 0 | 0 | 0 |
| 0.281164309 | 0.021430696 | 0 | 0.057500471 | 0.0147871 |
| 0 | 0 | 0 | 0 | 0.002433688 |
| 0 | 0 | 0 | 0 | 0 |
| 0 | 0 | 0 | 0 | 0 |
| 0 | 0 | 0 | 0 | 0 |
| 0 | 0 | 0 | 0 | 0 |
| 0 | 0 | 0 | 0 | 0 |
| 0.027870379 | 0 | 0 | 0 | 0 |
| 0 | 0.019619276 | 0 | 0 | 0.082162903 |
| 0 | 0 | 0 | 0 | 0 |
| 0 | 0 | 0 | 0 | 0 |
| 0 | 0 | 0 | 0 | 0 |
| 0 | 0 | 0 | 0 | 0 |
| 0 | 0 | 0 | 0 | 0 |
| 0 | 0 | 0 | 0 | 0 |
| 0 | 0 | 0 | 0 | 0 |
| 0.006360085 | 0 | 0 | 0 | 0 |
| 0 | 0 | 0.051778419 | 0 | 0.200189156 |
| 0 | 0 | 0 | 0 | 0 |
| 0.582077278 | 0.008104488 | 0.000382013 | 0.008873589 | 0.038328448 |
| 0 | 0 | 0 | 0 | 0 |
| 0 | 0 | 0 | 0 | 0 |
| 0 | 0 | 0 | 0 | 0 |
| 0 | 0 | 0 | 0 | 0 |
| 0 | 0 | 0 | 0 | 0 |
| 0 | 0 | 0 | 0 | 0 |
| 0 | 0 | 0 | 0 | 0 |
| 0 | 0 | 0 | 0 | 0 |
| 0 | 0 | 0 | 0.008476687 | 0 |
| 0 | 0 | 0 | 0 | 0 |
| 0 | 0 | 0 | 0 | 0 |
| 0 | 0 | 0.028834392 | 0 | 0.014795882 |
| 0 | 0 | 0 | 0 | 0 |
| 0 | 0 | 0 | 0 | 0 |
| 0 | 0 | 0.014824498 | 0 | 0.083943261 |
| 0 | 0 | 0 | 0 | 0 |
| 0 | 0 | 0 | 0 | 0.055550586 |
| 0 | 0 | 0 | 0 | 0 |
| 0 | 0 | 0 | 0 | 0 |
| 0 | 0 | 0 | 0 | 0 |
| 0 | 0 | 0 | 0 | 0 |

| | | | | |
|---|---|---|---|---|
| 0 | 0 | 0.14717288 | 0 | 0.145992733 |
| 0 | 0 | 0 | 0 | 0 |
| 0.004020356 | 0 | 0 | 0 | 0 |
| 0.004389316 | 0 | 0 | 0 | 0 |
| 0 | 0 | 0 | 0 | 0 |
| 0 | 0 | 0.049529261 | 0 | 0 |
| 0 | 0 | 0 | 0 | 0 |
| 0 | 0 | 0 | 0 | 0 |
| 0 | 0 | 0 | 0 | 0 |
| 0 | 0 | 0 | 0 | 0 |
| 0 | 0 | 0 | 0 | 0 |
| 0.000756514 | 0 | 0 | 0 | 0 |
| 0 | 0 | 0 | 0 | 0 |
| 0 | 0 | 0 | 0 | 0 |
| 0 | 0 | 0 | 0 | 0 |
| 0 | 0 | 0 | 0 | 0 |
| 0 | 0 | 0 | 0 | 0 |
| 0 | 0 | 0 | 0 | 0 |
| 0 | 0 | 0 | 0 | 0 |
| 0 | 0 | 0 | 0 | 0 |
| 0.019000495 | 0.001846897 | 0 | 0.005454046 | 0 |
| 0 | 0 | 0 | 0 | 0 |
| 0.010578988 | 0.004146661 | 0 | 0.015153738 | 0.000884293 |
| 0 | 0 | 0.002403866 | 0 | 0.001294962 |
| 0 | 0 | 0 | 0 | 0 |
| 0 | 0 | 0 | 0 | 0 |
| 0 | 0 | 0 | 0 | 0 |
| 0 | 0 | 0 | 0 | 0 |
| 0 | 0 | 0 | 0.045147494 | 0.002143449 |
| 0 | 0 | 0 | 0 | 0 |
| 0 | 0 | 0 | 0 | 0 |
| 0 | 0 | 0 | 0 | 0 |
| 0.010873733 | 0.065531206 | 0.026892306 | 0 | 0.56578329 |
| 0 | 0 | 0 | 0 | 0 |
| 0.857012926 | 0 | 0.042389674 | 0.082099839 | 0.0025186 |
| 0 | 0.025319207 | 0 | 0.033572458 | 0 |
| 0 | 0 | 0 | 0 | 0 |
| 0 | 0 | 0 | 0 | 0.076216123 |
| 0.031175134 | 0.000675823 | 0 | 0.000855327 | 0 |
| 0 | 0 | 0 | 0 | 0 |
| 0 | 0 | 0 | 0 | 0 |
| 0 | 0 | 0 | 0 | 0 |
| 0.05297497 | 0.002107306 | 0.054813574 | 0.011601553 | 0.020492282 |
| 0 | 0 | 0 | 0 | 0 |
| 0 | 0 | 0 | 0 | 0 |
| 0.869751522 | 0 | 0.001696032 | 0.021286697 | 0.001284823 |
| 0 | 0 | 0 | 0 | 0 |
| 0 | 0 | 0 | 0 | 0 |

| | | | | |
|---|---|---|---|---|
| 0 | 0 | 0 | 0 | 0 |
| 0 | 0 | 0 | 0 | 0 |
| 0 | 0 | 0 | 0 | 0 |
| 0 | 0 | 0 | 0 | 0 |
| 0 | 0 | 0 | 0 | 0 |
| 0 | 0 | 0 | 0 | 0 |
| 0 | 0 | 0 | 0 | 0.152860521 |
| 0 | 0 | 0 | 0 | 0 |
| 1.338028836 | 0 | 0 | 0 | 0.010566339 |
| 0.003432316 | 0 | 0.00160132 | 0.003599637 | 0.331867673 |
| 0 | 0 | 0 | 0 | 0 |
| 0 | 0 | 0 | 0 | 0 |
| 0 | 0 | 0 | 0 | 0 |
| 0.221039148 | 0 | 0.001126868 | 0 | 0 |
| 0.177001813 | 0 | 0 | 0 | 0 |
| 0.010131058 | 0.005511949 | 0 | 0.010624936 | 0 |
| 0 | 0 | 0 | 0 | 0 |
| 0 | 0 | 0 | 0 | 0 |
| 0 | 0 | 0 | 0 | 0 |
| 0 | 0 | 1.542360041 | 0 | 0.274100531 |
| 0 | 0 | 0.00703086 | 0 | 0 |
| 0 | 0 | 0 | 0 | 0 |
| 0 | 0 | 0 | 0 | 0 |
| 0 | 0 | 0 | 0 | 0 |
| 0 | 0 | 0 | 0 | 0 |
| 0 | 0 | 0 | 0 | 0 |
| 0 | 0 | 0 | 0 | 0 |
| 0 | 0 | 0 | 0 | 0.000570261 |
| 0 | 0 | 0 | 0 | 0 |
| 0 | 0 | 0 | 0.005005423 | 0.02653934 |
| 0.007891147 | 0 | 0.064841869 | 0.718852377 | 0.055499668 |
| 0 | 0 | 0 | 0 | 0 |
| 0 | 0 | 0 | 0 | 0 |
| 0 | 0 | 0 | 0 | 0 |
| 0 | 0 | 0 | 0 | 0 |
| 0 | 0 | 0 | 0 | 0 |
| 0 | 0 | 0 | 0 | 0 |
| 0 | 0 | 0 | 0 | 0 |
| 0 | 0 | 0 | 0.102930832 | 0.004903307 |
| 0 | 0 | 0 | 0 | 0 |
| 0 | 0 | 0 | 0 | 0 |
| 0 | 0 | 0 | 0 | 0 |
| 0 | 0 | 0 | 0 | 0 |
| 0.007665171 | 0 | 0 | 0 | 0 |
| 0 | 0 | 0 | 0 | 0 |
| 0 | 0 | 0 | 0 | 0 |
| 0.20180414 | 0.08485515 | 0 | 0.006591921 | 0 |
| 0 | 0 | 0 | 0 | 0 |

| | | | | |
|---|---|---|---|---|
| 0 | 0 | 0 | 0 | 0 |
| 0 | 0 | 0 | 0 | 0 |
| 0 | 0 | 0 | 0 | 0 |
| 0 | 0 | 0 | 0 | 0.000796376 |
| 0.081838032 | 0 | 0 | 0.083472936 | 0.186207618 |
| 0 | 0 | 0 | 0 | 0 |
| 0 | 0 | 0 | 0 | 0 |
| 0 | 0 | 0 | 0 | 0 |
| 0 | 0 | 0 | 0 | 0 |
| 0 | 0 | 0 | 0 | 0 |
| 0 | 0.050639038 | 0 | 0 | 0 |
| 0.117654052 | 0.003826615 | 0 | 0.001880222 | 0.003379375 |
| 0.023781359 | 0 | 0 | 0.001032282 | 0 |
| 0 | 0 | 0 | 0 | 0 |
| 0 | 0 | 0 | 0 | 0 |
| 0.312120824 | 0 | 0.012535772 | 0 | 0 |
| 0.005343215 | 0 | 0 | 0 | 0 |
| 0 | 0 | 0 | 0 | 0 |
| 0.090106474 | 1.735793743 | 0 | 0.143018604 | 0 |
| 0.055169959 | 0.018693941 | 0 | 0.007604744 | 0 |
| 0 | 0 | 0 | 0 | 0 |
| 0 | 0 | 0 | 0 | 0.038215719 |
| 0 | 0 | 0 | 0 | 0 |
| 0 | 0 | 0 | 0 | 0 |
| 0 | 0 | 0 | 0 | 0 |
| 0 | 0 | 0 | 0 | 0 |
| 0.070310224 | 0.005171774 | 0 | 0.010969254 | 0 |
| 0.013019778 | 0 | 0 | 0 | 0 |
| 0 | 0 | 0 | 0 | 0 |
| 0 | 0 | 0 | 0 | 0 |
| 0.006641694 | 0.001212926 | 0 | 0.000614036 | 0 |
| 0.008912907 | 0 | 0 | 0.221353088 | 0 |
| 0 | 0 | 0 | 0 | 0 |
| 0 | 0 | 0 | 0 | 0 |
| 0 | 0 | 0 | 0 | 0 |
| 0 | 0 | 0 | 0 | 0 |
| 0 | 0 | 0 | 0 | 0 |
| 0.131645139 | 0.007385446 | 0 | 0.002157019 | 0.002326122 |
| 0 | 0 | 0 | 0 | 0.259034107 |
| 0.0024673 | 0 | 0 | 0.008382898 | 0 |
| 0 | 0 | 0.003161243 | 0 | 0 |
| 0 | 0 | 0 | 0 | 0 |
| 0 | 0 | 0 | 0 | 0 |
| 0.001467767 | 0.026115545 | 0 | 0.002718798 | 0 |
| 0 | 0 | 0 | 0 | 0 |
| 0 | 0 | 0 | 0 | 0.108701673 |
| 0 | 0 | 0 | 0 | 0 |
| 0 | 0 | 0 | 0 | 0 |

| | | | | |
|---|---|---|---|---|
| 0 | 0 | 0 | 0 | 0 |
| 0 | 0 | 0.005715999 | 0 | 0.011301346 |
| 0 | 0 | 0 | 0 | 0 |
| 0.002197467 | 0 | 0 | 0 | 0.004970526 |
| 0.000815246 | 0 | 0.002212925 | 0 | 0.007124675 |
| 0 | 0 | 0 | 0 | 0 |
| 0 | 0 | 0.004630756 | 0 | 0.015493725 |
| 0.003609079 | 0 | 0 | 0 | 0 |
| 0 | 0 | 0 | 0 | 0 |
| 0 | 0 | 0 | 0 | 0 |
| 0 | 0 | 0 | 0 | 0 |
| 0 | 0 | 0 | 0 | 0 |
| 0.056855853 | 0 | 0.066172067 | 0.001663333 | 0.471281897 |
| 0 | 0 | 0 | 0 | 0 |
| 0 | 0 | 0 | 0 | 0 |
| 0 | 0 | 0 | 0 | 0 |
| 0 | 0 | 0 | 0 | 0 |
| 0 | 0 | 0 | 0 | 0 |
| 0 | 0 | 0 | 0 | 0 |
| 0 | 0 | 0 | 0 | 0 |
| 0 | 0 | 0 | 0 | 0 |
| 0 | 0 | 0 | 0 | 0 |
| 0.128195303 | 0.004120409 | 0 | 0.000938668 | 0 |
| 0.029770894 | 0.044003132 | 0.011168985 | 0.026214639 | 0.00331805 |
| 0 | 0 | 0 | 0 | 0 |
| 0 | 0 | 0 | 0 | 0 |
| 0 | 0 | 0 | 0 | 0 |
| 0 | 0 | 0 | 0 | 0 |
| 0 | 0 | 0 | 0 | 0 |
| 0 | 0 | 0 | 0 | 0 |
| 0 | 0 | 0.020175608 | 0 | 0.036838269 |
| 0 | 0 | 0 | 0 | 0 |
| 0 | 0 | 0 | 0 | 0 |
| 0 | 0 | 0 | 0 | 0 |
| 0 | 0 | 0 | 0 | 0 |
| 0 | 0 | 0 | 0 | 0 |
| 0 | 0 | 0 | 0 | 0 |
| 0.041015296 | 0 | 0 | 0 | 0 |
| 0 | 0.001166113 | 0.001444382 | 0 | 0 |
| 0 | 0 | 0 | 0 | 0 |
| 0 | 0 | 0 | 0 | 0 |
| 0 | 0 | 0 | 0 | 0 |
| 0 | 0 | 0 | 0 | 0 |
| 0 | 0 | 0 | 0 | 0 |
| 0 | 0 | 0 | 0 | 0 |
| 0.107764581 | 0.101401711 | 0.297486743 | 0.116085676 | 0.293128525 |
| 0 | 0 | 0 | 0 | 0 |
| 0 | 0 | 0 | 0 | 0 |

| | | | | |
|---|---|---|---|---|
| 0 | 0 | 0 | 0 | 0 |
| 0 | 0 | 0 | 0 | 0 |
| 0 | 0 | 0 | 0.342961769 | 0.057691635 |
| 0 | 0 | 0.000872121 | 0 | 0 |
| 0 | 0 | 0 | 0 | 0 |
| 0 | 0 | 0 | 0.009085528 | 0.009561453 |
| 0 | 0 | 0 | 0 | 0 |
| 0 | 0 | 0 | 0 | 0 |
| 0 | 0 | 0 | 0.001502101 | 0.001227168 |
| 0 | 0 | 0 | 0 | 0 |
| 0.003610467 | 0 | 0 | 0 | 0 |
| 0 | 0 | 0 | 0 | 0.00048193 |
| 0 | 0 | 0 | 0 | 0 |
| 0 | 0 | 0 | 0 | 0 |
| 0 | 0 | 0 | 0 | 0 |
| 0 | 0 | 0 | 0 | 0 |
| 0 | 0 | 0 | 0 | 0 |
| 0 | 0 | 0 | 0 | 0 |
| 0 | 0 | 0 | 0 | 0 |
| 0.003955386 | 0 | 0 | 0.001429047 | 0.000856155 |
| 0 | 0 | 0 | 0 | 0 |
| 0 | 0 | 0 | 0 | 0 |
| 0 | 0 | 0 | 0 | 0 |
| 0 | 0 | 0 | 0 | 0 |
| 0 | 0 | 0 | 0 | 0 |
| 0 | 0 | 0 | 0 | 0 |
| 0 | 0 | 0 | 0 | 0 |
| 0 | 0 | 0.034707558 | 0 | 0.007106106 |
| 0 | 0 | 0 | 0 | 0 |
| 0 | 0 | 0 | 0 | 0 |
| 0.005007706 | 0.006646618 | 0 | 0 | 0.069874985 |
| 0 | 0 | 0 | 0 | 0 |
| 0.024919098 | 0.038587003 | 0.07158678 | 0.027717745 | 0.058073412 |
| 0 | 0.008952069 | 0 | 0.007930868 | 0 |
| 0 | 0 | 0 | 0 | 0 |
| 0 | 0 | 0 | 0 | 0 |
| 0 | 0 | 0 | 0 | 0 |
| 0 | 0 | 0 | 0 | 0 |
| 0 | 0 | 0 | 0 | 0 |
| 0 | 0 | 0 | 0 | 0 |
| 0 | 0 | 0 | 0 | 0 |
| 0.223799479 | 0 | 0 | 0.038309271 | 0.003630075 |
| 0.041417956 | 0 | 0.005225831 | 0.008096075 | 0.036235597 |
| 0 | 0 | 0 | 0 | 0 |
| 0 | 0 | 0 | 0 | 0 |
| 0.063761745 | 0 | 0.030410856 | 0 | 0.01525535 |
| 0 | 0 | 0 | 0 | 0 |
| 0 | 0 | 0 | 0 | 0 |

| | | | | |
|---|---|---|---|---|
| 0 | 0 | 0 | 0 | 0 |
| 0.007413504 | 0 | 0.014648658 | 0 | 0.003885832 |
| 0.026488664 | 0.001023305 | 0.058074307 | 0.006022228 | 0.002967849 |
| 0 | 0 | 0 | 0 | 0 |
| 1.722701887 | 0 | 0 | 0.061269075 | 0.017702935 |
| 3.742121038 | 0.001789472 | 0 | 0 | 0 |
| 0 | 0 | 0 | 0 | 0 |
| 0 | 0 | 0 | 0 | 0 |
| 0 | 0 | 0 | 0 | 0 |
| 0 | 0 | 0 | 0 | 0 |
| 0 | 0 | 0 | 0 | 0 |
| 0 | 0 | 0 | 0 | 0 |
| 0 | 0 | 0 | 0 | 0 |
| 0 | 0 | 0 | 0 | 0 |
| 0 | 0 | 0 | 0 | 0 |
| 0 | 0 | 0 | 0 | 0 |
| 0 | 0 | 0 | 0 | 0.005440025 |
| 0 | 0 | 0 | 0 | 0 |
| 0 | 0 | 0 | 0 | 0 |
| 0 | 0 | 0 | 0 | 0 |
| 0 | 0 | 0 | 0 | 0 |
| 0.017209554 | 0 | 0 | 0.011407277 | 0 |
| 0.005183929 | 0 | 0 | 0 | 0 |
| 0 | 0 | 0 | 0 | 0 |
| 0.00167926 | 0 | 0 | 0 | 0.163157552 |
| 0.013725531 | 0 | 0 | 0 | 0.0130264 |
| 0 | 0 | 0 | 0 | 0 |
| 0 | 0 | 0 | 0.004218283 | 0.057002681 |
| 0 | 0 | 0 | 0 | 0 |
| 0 | 0 | 0 | 0 | 0 |
| 0 | 0 | 0 | 0 | 0 |
| 0 | 0.014135719 | 0 | 0 | 0 |
| 0 | 0 | 0 | 0 | 0 |
| 0 | 0 | 0 | 0 | 0 |
| 0 | 0 | 0 | 0 | 0 |
| 0 | 0 | 0 | 0 | 0 |
| 0 | 0 | 0 | 0 | 0 |
| 0 | 0 | 0 | 0.003206411 | 0 |
| 0 | 0 | 0.019385114 | 0 | 2.528338559 |
| 0 | 0 | 0 | 0 | 0 |
| 0 | 0 | 0 | 0 | 0 |
| 0 | 0 | 0 | 0 | 0.112110975 |
| 0.061881844 | 0 | 0 | 0.005980685 | 0 |
| 0 | 0 | 0 | 0 | 0 |
| 0 | 0 | 0 | 0 | 0 |
| 0 | 0 | 0 | 0 | 0 |
| 0.039552697 | 0.003256376 | 0 | 0.004451 | 0.002222196 |
| 0 | 0 | 0 | 0 | 0 |

| | | | | |
|---|---|---|---|---|
| 0.002361298 | 0 | 0 | 0 | 0 |
| 0.103702753 | 0.041294921 | 0 | 0.005460333 | 0.003505003 |
| 0 | 0 | 0 | 0 | 0 |
| 0 | 0 | 0 | 0 | 0 |
| 0 | 0 | 0 | 0 | 0 |
| 0 | 0 | 0 | 0 | 0 |
| 0 | 0 | 0 | 0 | 0 |
| 0 | 0 | 0 | 0 | 0 |
| 0 | 0 | 0 | 0 | 0 |
| 0 | 0 | 0 | 0 | 0 |
| 0 | 0 | 0 | 0 | 0 |
| 0 | 0 | 0 | 0 | 0 |
| 0.149191064 | 0 | 0.003081279 | 0.016986337 | 0.167618688 |
| 0 | 0 | 0 | 0 | 0 |
| 0 | 0 | 0 | 0.007975335 | 0 |
| 0 | 0 | 0 | 0 | 0 |
| 0 | 0 | 0 | 0 | 0 |
| 0 | 0 | 0 | 0 | 0 |
| 0 | 0 | 0 | 0 | 0 |
| 0 | 0 | 0 | 0 | 0 |
| 0 | 0 | 0 | 0 | 0 |
| 0.044582322 | 0.001257856 | 0 | 0.000955172 | 0 |
| 0.004234118 | 0 | 0 | 0.004991005 | 0 |
| 0 | 0 | 0 | 0 | 0 |
| 0 | 0 | 0 | 0 | 0 |
| 0 | 0 | 0 | 0 | 0 |
| 0 | 0 | 0 | 0 | 0 |
| 0 | 0 | 0 | 0 | 0 |
| 0 | 0 | 0 | 0 | 0 |
| 0 | 0 | 0 | 0 | 0 |
| 0 | 0 | 0 | 0 | 0 |
| 0 | 0 | 0 | 0 | 0 |
| 0 | 0 | 0 | 0 | 0 |
| 0 | 0 | 0 | 0 | 0 |
| 0 | 0 | 0 | 0.126964344 | 17.05613435 |
| 0 | 0 | 0 | 0 | 0 |
| 0 | 0 | 0 | 0 | 0 |
| 0 | 0 | 0 | 0 | 0 |
| 0.014506791 | 0.002763467 | 0.091485062 | 0.001907709 | 0.015943822 |
| 0 | 0 | 0 | 0 | 0 |
| 0.04260998 | 0.052259346 | 0.39461928 | 0.039512863 | 0.148798723 |
| 0 | 0 | 0 | 0 | 0 |
| 0 | 0 | 0 | 0 | 0 |
| 0 | 0 | 0 | 0 | 0 |
| 0 | 0 | 0 | 0 | 0 |
| 0.621630328 | 0.832251574 | 0.014984902 | 0.00724616 | 0.047518999 |
| 0 | 0 | 0 | 0 | 0 |
| 0 | 0 | 0 | 0 | 0 |

| | | | | |
|---|---|---|---|---|
| 0 | 0 | 0 | 0.003314175 | 0 |
| 0 | 0 | 0 | 0.003030326 | 0 |
| 0 | 0 | 0 | 0.009255861 | 0 |
| 0 | 0 | 0 | 0 | 0 |
| 0.017721115 | 0 | 0 | 0 | 0 |
| 0 | 0 | 0 | 0 | 0 |
| 0 | 0 | 0 | 0 | 0 |
| 0 | 0 | 0 | 0.015240794 | 0 |
| 0 | 0 | 0 | 0 | 0 |
| 0 | 0 | 0 | 0 | 0 |
| 0 | 0 | 0 | 0 | 0 |
| 0 | 0 | 0 | 0 | 0 |
| 0.009817848 | 0 | 0 | 0.0022465 | 0 |
| 0 | 0 | 0 | 0 | 0 |
| 0 | 0 | 0 | 0 | 0 |
| 0 | 0 | 0 | 0 | 0 |
| 0 | 0 | 0 | 0 | 0 |
| 0 | 0 | 0 | 0 | 0 |
| 0 | 0 | 0 | 0 | 0 |
| 0.997945568 | 0.127928577 | 0.033653068 | 0.07731567 | 0.248573576 |
| 0 | 0 | 0 | 0 | 0 |
| 0 | 0 | 0 | 0 | 0 |
| 0 | 0 | 0 | 0 | 0 |
| 0 | 0 | 0 | 0 | 0 |
| 0 | 0 | 0 | 0 | 0 |
| 0 | 0 | 0 | 0 | 0 |
| 0 | 0 | 0 | 0 | 0 |
| 0 | 0 | 0 | 0 | 0 |
| 0 | 0 | 0 | 0 | 0 |
| 0 | 0 | 0 | 0 | 0 |
| 0 | 0 | 0 | 0 | 0 |
| 0 | 0 | 0 | 0 | 0 |
| 0 | 0 | 0 | 0 | 0 |
| 0 | 0 | 0 | 0 | 0 |
| 0 | 0 | 0 | 0 | 0 |
| 0.00706993 | 0 | 0 | 0 | 0 |
| 0 | 0 | 0 | 0 | 0 |
| 0 | 0 | 0 | 0 | 0 |
| 0 | 0 | 0 | 0 | 0 |
| 0 | 0 | 0 | 0 | 0 |
| 0 | 0 | 0 | 0 | 0 |
| 0 | 0 | 0 | 0 | 0 |
| 0.007990235 | 0 | 0 | 0 | 0.000746834 |
| 0 | 0 | 0 | 0 | 0 |
| 0.038165309 | 0 | 0 | 0 | 0.000843865 |
| 0.153332067 | 0 | 0 | 0 | 0 |
| 0 | 0 | 0 | 0 | 0 |
| 0 | 0.002888351 | 0 | 0 | 0 |

| | | | | |
|---|---|---|---|---|
| 0 | 0 | 0 | 0 | 0 |
| 0 | 0 | 0 | 0 | 0 |
| 0 | 0 | 0 | 0 | 0 |
| 0 | 0 | 0 | 0 | 0 |
| 0 | 0 | 0 | 0 | 0 |
| 0.012407757 | 0 | 0 | 0.047669128 | 0.034858802 |
| 0 | 0 | 0 | 0 | 0 |
| 0 | 0 | 0 | 0 | 0 |
| 0 | 0 | 0 | 0 | 0 |
| 0 | 0 | 0 | 0 | 0 |
| 0 | 0 | 0 | 0 | 0 |
| 0 | 0 | 0 | 0 | 0 |
| 0 | 0 | 0 | 0 | 0 |
| 0 | 0 | 0 | 0 | 0 |
| 0 | 0 | 0 | 0 | 0 |
| 0 | 0 | 0 | 0 | 0 |
| 0 | 0 | 0 | 0 | 0 |
| 0 | 0 | 0 | 0 | 0 |
| 0 | 0 | 0 | 0 | 0 |
| 0 | 0 | 0 | 0 | 0 |
| 0 | 0 | 0 | 0 | 0 |
| 0 | 0 | 0 | 0 | 0 |
| 0 | 0 | 0 | 0 | 0 |
| 0 | 0 | 0 | 0 | 0 |
| 0 | 0 | 0 | 0 | 0 |
| 0 | 0 | 0 | 0 | 0 |
| 0 | 0 | 0 | 0 | 0 |
| 1.449312912 | 0 | 0 | 0.506484092 | 0 |
| 0 | 0 | 0 | 0 | 0 |
| 0 | 0 | 0 | 0.00971603 | 0 |
| 0 | 0 | 0 | 0 | 0 |
| 0 | 0 | 0 | 0 | 0 |
| 0 | 0 | 0 | 0 | 0 |
| 0 | 0 | 0 | 0 | 0 |
| 0 | 0 | 0 | 0 | 0 |
| 0 | 0 | 0 | 0 | 0 |
| 0 | 0 | 0 | 0 | 0 |
| 0.237391063 | 0 | 0 | 0 | 0 |
| 0 | 0 | 0 | 0 | 0.479168818 |
| 0 | 0 | 0 | 0 | 0 |
| 0 | 0 | 0 | 0 | 0 |
| 0 | 0 | 0 | 0 | 0 |
| 0 | 0 | 0 | 0 | 0 |
| 0 | 0 | 0 | 0 | 0 |
| 0 | 0 | 0 | 0 | 0 |
| 0 | 0 | 0 | 0 | 0 |
| 0 | 0 | 0 | 0 | 0 |
| 0 | 0 | 0 | 0 | 0 |

| | | | | |
|---|---|---|---|---|
| 0 | 0 | 0 | 0 | 0 |
| 0 | 0 | 0 | 0.024722604 | 0.008463739 |
| 0 | 0 | 0 | 0 | 0 |
| 0 | 0 | 0 | 0 | 0 |
| 0 | 0 | 0 | 0 | 0 |
| 0 | 0 | 0 | 0 | 0 |
| 0 | 0 | 0 | 0 | 0 |
| 0 | 0 | 0 | 0 | 0 |
| 0 | 0 | 0 | 0 | 0 |
| 0 | 0 | 0 | 0 | 0 |
| 0 | 0 | 0.011429366 | 0 | 0.018624916 |
| 0 | 0 | 0 | 0 | 0 |
| 0 | 0 | 0.00549996 | 0 | 0.192281444 |
| 0 | 0 | 0 | 0 | 0 |
| 0 | 0 | 0 | 0 | 0 |
| 0 | 0 | 0 | 0 | 0 |
| 0 | 0 | 0 | 0 | 0 |
| 0 | 0 | 0 | 0 | 0 |
| 0 | 0 | 0 | 0 | 0 |
| 0 | 0 | 0 | 0 | 0 |
| 0 | 0 | 0.000773341 | 0 | 0.000781123 |
| 0 | 0 | 0 | 0 | 0 |
| 0 | 0 | 0 | 0 | 0 |
| 2.112396636 | 0 | 0.519171085 | 0.051267459 | 0.661188095 |
| 0 | 0 | 0 | 0 | 0 |
| 0 | 0 | 0 | 0 | 0 |
| 0 | 0 | 0 | 0 | 0 |
| 0 | 0 | 0 | 0.189478383 | 0 |
| 0 | 0 | 0 | 0 | 0 |
| 0 | 0 | 0 | 0 | 0 |
| 0.002460741 | 0 | 0 | 0 | 0 |
| 0 | 0 | 0 | 0 | 0 |
| 0.001918004 | 0 | 0 | 0 | 0 |
| 0 | 0 | 0 | 0 | 0 |

| PHL018-0029_V3 | PHL018-0058_V2 | PHL018-0102_V2 | PHL018-0243_V1 | PHL018-0217_V1 |
|---|---|---|---|---|
| EYCF | EYCF | CTRL | REF | REF |
| 1.061200361 | 0.013717676 | 0.03865884 | 0.909487982 | 0.036246467 |
| 1.99655061 | 0.20075359 | 0.652832967 | 22.83567174 | 1.676590255 |
| 5.082994643 | 2.749759714 | 0 | 0.033717616 | 0.103008143 |
| 0 | 0 | 0 | 0 | 0 |
| 0 | 0 | 0 | 0 | 0 |
| 0 | 0 | 0 | 0 | 0 |
| 0 | 0 | 0 | 0 | 0 |
| 0 | 0 | 0 | 0 | 0 |
| 0 | 0 | 0 | 0 | 0 |
| 0 | 0 | 0 | 0 | 0 |
| 0 | 0 | 0 | 0 | 0 |
| 0 | 0 | 0 | 0 | 0 |
| 0 | 0 | 0 | 0 | 0 |
| 5.052572284 | 0.682259871 | 37.62770525 | 9.741686287 | 9.264426392 |
| 0 | 0 | 0 | 0 | 0 |
| 0 | 0 | 0 | 0 | 0 |
| 0 | 0 | 0 | 0 | 0 |
| 0 | 0 | 0 | 0 | 0 |
| 0 | 0 | 0 | 0 | 0 |
| 0 | 0 | 0 | 0 | 0 |
| 0 | 0 | 0 | 0 | 0 |
| 0 | 0 | 0 | 0 | 0 |
| 0 | 0 | 0 | 0 | 0 |
| 0 | 0 | 0 | 0 | 0 |
| 0.014851107 | 0 | 0 | 0 | 0.051546281 |
| 0 | 0 | 0 | 0 | 0 |
| 0 | 0 | 0 | 0 | 0 |
| 0 | 0 | 0 | 0 | 0 |
| 0 | 0 | 0 | 0 | 0 |
| 0 | 0 | 0 | 0 | 0 |
| 0 | 0 | 0 | 0 | 0 |
| 0 | 0 | 0 | 0 | 0 |
| 0 | 0 | 0 | 0 | 0 |
| 0 | 0 | 0 | 0 | 0 |
| 0 | 0 | 0 | 0 | 0 |
| 0.67367999 | 0.662976547 | 1.440814567 | 0.046621529 | 2.061771808 |
| 0 | 0 | 0 | 0 | 0 |
| 0 | 0 | 0 | 0 | 0 |
| 0 | 0 | 0 | 0 | 0 |
| 0 | 0 | 0 | 0 | 0 |
| 0 | 0 | 0 | 0 | 0 |
| 0.001441628 | 0 | 0 | 0.002270931 | 0.007878462 |
| 0.595488088 | 24.40301521 | 0.149123531 | 0.132467951 | 0.666931786 |
| 0 | 0 | 0.000866574 | 0.037935829 | 0.04106307 |
| 0 | 0 | 0 | 0 | 0.002993766 |
| 0 | 0 | 0 | 0 | 0 |

| | | | | |
|---|---|---|---|---|
| 0.001104403 | 0 | 0 | 0 | 0 |
| 0.002026101 | 0.009742876 | 0.004456425 | 0.024236402 | 0 |
| 0 | 0.001039453 | 0.016350997 | 0.009896061 | 0.000735687 |
| 0.383905627 | 0.167196845 | 2.352841766 | 1.492352137 | 0.819488147 |
| 0 | 0 | 0.001549203 | 0 | 0 |
| 0 | 0 | 0 | 0 | 0 |
| 0 | 0 | 0.063886606 | 0.006093491 | 0 |
| 0.025048147 | 0 | 0 | 0 | 0 |
| 0 | 0 | 0 | 0 | 0 |
| 0 | 0 | 0 | 0 | 0 |
| 0 | 0 | 0 | 0 | 0 |
| 0.625871832 | 0 | 0.159066515 | 0 | 0.58374753 |
| 0 | 0 | 0 | 0 | 0 |
| 0.017691193 | 0.010132565 | 0.033780679 | 0.050759818 | 0.023904878 |
| 0.000800191 | 0 | 0 | 0 | 0 |
| 0 | 0 | 0 | 0 | 0 |
| 0 | 0 | 0 | 0.289680674 | 0.273716346 |
| 0 | 0 | 0 | 0 | 0 |
| 0 | 0 | 0 | 0 | 0 |
| 0 | 0 | 0 | 0 | 0 |
| 0.026089629 | 0.113953085 | 0.284888428 | 0.32595094 | 0.001684006 |
| 0.012344249 | 0.007180593 | 0.008060534 | 0.07200851 | 0 |
| 0 | 0 | 0 | 0 | 0 |
| 0 | 0 | 0 | 0 | 0 |
| 0 | 0 | 0 | 0 | 0 |
| 0 | 0 | 0 | 0 | 0 |
| 0 | 0.125178662 | 0 | 0 | 0 |
| 0 | 0 | 0 | 0 | 0 |
| 0 | 0 | 0 | 0.010203132 | 0 |
| 0 | 0 | 0 | 0 | 0 |
| 0 | 0 | 0 | 0 | 0 |
| 0 | 0 | 0 | 0 | 0 |
| 0 | 0 | 0 | 0 | 0 |
| 0 | 0 | 0 | 0 | 0 |
| 0 | 0 | 0 | 0 | 0 |
| 0 | 0 | 0 | 0.024105783 | 0 |
| 0 | 0 | 0 | 0 | 0 |
| 0 | 0 | 0 | 0.001381612 | 0 |
| 0 | 0 | 0 | 0 | 0 |
| 0 | 0 | 0 | 0 | 0 |
| 0 | 0 | 0 | 0 | 0 |
| 0 | 0 | 0 | 0 | 0 |
| 0 | 0 | 0 | 0 | 0 |
| 0 | 0 | 0 | 0 | 0 |
| 0 | 0 | 0 | 0 | 0 |
| 0 | 0 | 0 | 0 | 0 |
| 0 | 0 | 0 | 0.001972118 | 0 |
| 0 | 0 | 0 | 0 | 0 |

| | | | | |
|---|---|---|---|---|
| 0 | 0 | 0 | 0 | 0 |
| 0 | 0 | 0 | 0 | 0 |
| 0 | 0 | 0 | 0 | 0 |
| 0 | 0 | 0 | 0 | 0 |
| 0 | 0 | 0.031629572 | 0.145253481 | 0.110040293 |
| 0.00071122 | 0 | 0 | 0 | 0 |
| 0 | 0 | 0 | 0 | 0 |
| 0 | 0 | 0 | 0 | 0 |
| 0 | 0 | 0 | 0.00704829 | 0.01026127 |
| 0 | 0 | 0 | 0 | 0 |
| 0 | 0 | 0 | 0 | 0 |
| 0 | 0 | 0 | 0.000767149 | 0 |
| 0 | 0 | 0 | 0 | 0 |
| 0 | 0 | 0 | 0 | 0 |
| 0 | 0 | 0 | 0 | 0 |
| 0 | 0 | 0 | 0 | 0 |
| 0 | 0 | 0 | 0 | 0 |
| 0 | 0 | 0 | 0 | 0 |
| 0.158145537 | 0.01984021 | 0.001482885 | 0.02730908 | 0.008346358 |
| 0 | 0 | 0 | 0 | 0 |
| 0 | 0 | 0 | 0 | 0 |
| 0 | 0 | 0 | 0 | 0 |
| 0 | 0 | 0 | 0 | 0 |
| 0 | 0 | 0 | 0 | 0 |
| 0 | 0 | 0 | 0 | 0 |
| 0 | 0 | 0 | 0 | 0 |
| 0 | 0 | 0 | 0 | 0 |
| 0 | 0 | 0 | 0 | 0 |
| 0 | 0 | 0 | 0 | 0 |
| 0 | 0 | 0 | 0 | 0 |
| 0.233418553 | 0 | 0.059764553 | 0.002445856 | 0.037577857 |
| 0.000904939 | 0 | 0.00099521 | 0.028652727 | 0.402914347 |
| 0 | 0 | 0 | 0 | 0 |
| 0 | 0 | 0 | 0 | 0 |
| 0 | 0 | 0 | 0 | 0 |
| 0 | 0 | 0 | 0 | 0 |
| 0 | 0 | 0 | 0 | 0 |
| 0 | 0 | 0 | 0 | 0 |
| 0.163156122 | 0 | 0.0473394 | 0 | 0 |
| 0 | 0 | 0 | 0 | 0 |
| 0 | 0 | 0 | 0 | 0 |
| 0 | 0 | 0 | 0 | 0 |
| 0.010878046 | 0 | 0.024846592 | 0.028175033 | 0 |
| 0 | 0 | 0 | 0 | 0 |
| 0 | 0 | 0 | 0 | 0.001305909 |
| 0 | 0 | 0 | 0 | 0 |
| 1.439382593 | 0 | 0 | 0 | 0.007448234 |
| 0 | 0 | 0 | 0 | 0 |

| | | | | |
|---|---|---|---|---|
| 0 | 0 | 0 | 0 | 0 |
| 0 | 0 | 0 | 0 | 0 |
| 0 | 0 | 0 | 0 | 0 |
| 0 | 0 | 0 | 0 | 0 |
| 0.000630117 | 0 | 0.007449471 | 0.001675005 | 0.000737909 |
| 0 | 0 | 0 | 0 | 0 |
| 0 | 0 | 0 | 0 | 0 |
| 0 | 0 | 0 | 0 | 0 |
| 0 | 0 | 0 | 0 | 0 |
| 0 | 0 | 0 | 0 | 0 |
| 0 | 0 | 0 | 0 | 0 |
| 0 | 0 | 0 | 0 | 0 |
| 0 | 0 | 0 | 0 | 0 |
| 0 | 0 | 0 | 0 | 0 |
| 0 | 0 | 0 | 0 | 0 |
| 2.283204056 | 0 | 0 | 0 | 0.689815117 |
| 0 | 0 | 0 | 0 | 0 |
| 0 | 0 | 0 | 0 | 0 |
| 0 | 0 | 0 | 0 | 0 |
| 0 | 0 | 0 | 0 | 0 |
| 0 | 0 | 0 | 0 | 0 |
| 0 | 0 | 0 | 0 | 0 |
| 0.001613442 | 0 | 0 | 0 | 0 |
| 0 | 0 | 0 | 0 | 0 |
| 0 | 0 | 0 | 0 | 0 |
| 0 | 0 | 0 | 0 | 0 |
| 0 | 0 | 0 | 0 | 0.017958171 |
| 0 | 0 | 0 | 0 | 0 |
| 0 | 0 | 0 | 0 | 0 |
| 0 | 0 | 0 | 0 | 0 |
| 0 | 0 | 0 | 0 | 0 |
| 0 | 0 | 0 | 0 | 0 |
| 0 | 0 | 0 | 0 | 0 |
| 0 | 0 | 0 | 0 | 0 |
| 0 | 0 | 0 | 0 | 0 |
| 0 | 0 | 0 | 0 | 0 |
| 0 | 0 | 0 | 0 | 0 |
| 0 | 0 | 0 | 0 | 0 |
| 0 | 0 | 0 | 0 | 0 |
| 0 | 0 | 0 | 0 | 0 |
| 0 | 0 | 0 | 0 | 0 |
| 0 | 0 | 0 | 0 | 0 |
| 0 | 0 | 0 | 0 | 0 |
| 0 | 0 | 0 | 0 | 0 |
| 0 | 0 | 0 | 0 | 0 |
| 0 | 0 | 0 | 0 | 0 |
| 0 | 0 | 0 | 0 | 0 |
| 0 | 0 | 0 | 0 | 0 |

| | | | | |
|---|---|---|---|---|
| 0 | 0 | 0 | 0 | 0 |
| 0 | 0 | 0 | 0 | 0 |
| 0 | 0 | 0 | 0 | 0 |
| 0 | 0 | 0 | 0 | 0 |
| 0 | 0 | 0 | 0 | 0 |
| 0 | 0 | 0 | 0 | 0 |
| 0 | 0 | 0 | 0 | 0 |
| 0 | 0 | 0 | 0 | 0 |
| 0 | 0 | 0 | 0 | 0 |
| 0 | 0 | 0 | 0 | 0 |
| 0 | 0 | 0 | 0 | 0 |
| 0 | 0 | 0 | 0 | 0.001186886 |
| 0.000999105 | 0.001487808 | 0 | 0.00141646 | 0 |
| 0 | 0 | 0 | 0 | 0 |
| 0 | 0 | 0 | 0 | 0.01218845 |
| 0 | 0 | 0 | 0 | 0 |
| 0 | 0 | 0 | 0 | 0 |
| 0 | 0 | 0 | 0 | 0 |
| 0 | 0 | 0 | 0 | 0 |
| 0 | 0 | 0 | 0 | 0 |
| 0.013450222 | 0.021327462 | 0.024653225 | 0.112117322 | 0.053816265 |
| 0 | 0 | 0 | 0 | 0 |
| 0 | 0 | 0 | 0 | 0 |
| 0.001558431 | 0 | 0 | 0 | 0 |
| 0 | 0 | 0 | 0 | 0 |
| 0 | 0 | 0 | 0 | 0 |
| 0 | 0 | 0 | 0 | 0.022423061 |
| 0 | 0 | 0 | 0 | 0 |
| 0 | 0 | 0 | 0 | 0 |
| 0 | 0.001444416 | 0 | 0.039879326 | 0 |
| 0 | 0 | 0 | 0 | 0 |
| 0 | 0 | 0 | 0 | 0 |
| 0 | 0 | 0 | 0 | 0 |
| 0 | 0 | 0 | 0 | 0.008556529 |
| 0.027955961 | 0 | 0.006148936 | 0.011200908 | 0.001138723 |
| 0 | 0 | 0 | 0 | 0 |
| 0 | 0 | 0 | 0 | 0 |
| 0 | 0 | 0 | 0 | 0 |
| 0 | 0 | 0 | 0 | 0 |
| 0 | 0 | 0 | 0 | 0 |
| 0 | 0 | 0 | 0 | 0 |
| 0 | 0 | 0 | 0 | 0 |
| 0 | 0 | 0 | 0 | 0 |
| 0 | 0 | 0 | 0 | 0 |
| 0 | 0 | 0 | 0 | 0 |
| 0 | 0 | 0 | 0.015586555 | 0 |
| 0.005264152 | 0 | 0 | 0 | 0 |
| 0 | 0.002794709 | 0.001333621 | 0 | 0 |

| | | | | |
|---|---|---|---|---|
| 0 | 0 | 0 | 0 | 0 |
| 0 | 0 | 0 | 0 | 0 |
| 0 | 0 | 0 | 0 | 0 |
| 0 | 0 | 0 | 0 | 0 |
| 0 | 0 | 0 | 0 | 0 |
| 0 | 0 | 0 | 0 | 0 |
| 0 | 0 | 0 | 0.001165372 | 0 |
| 0 | 0 | 0 | 0 | 0 |
| 0 | 0.005083529 | 0 | 0 | 0 |
| 0 | 0 | 0 | 0 | 0 |
| 0 | 0 | 0 | 0 | 0 |
| 0 | 0 | 0 | 0 | 0 |
| 0 | 0 | 0 | 0 | 0 |
| 0 | 0.01704348 | 0 | 0 | 0 |
| 0 | 0 | 0 | 0 | 0 |
| 0.011921315 | 0 | 0 | 0 | 0 |
| 0 | 0 | 0 | 0 | 0 |
| 0 | 0.181518031 | 0 | 0 | 0 |
| 0.002995063 | 0.124619571 | 0 | 0 | 0.00309478 |
| 0 | 0 | 0 | 0 | 0.00141859 |
| 0 | 0.057405486 | 0.026294346 | 0 | 0.001521705 |
| 0 | 0 | 0 | 0 | 0 |
| 0 | 0 | 0 | 0 | 0 |
| 0 | 0 | 0 | 0 | 0 |
| 0 | 0.00192216 | 0 | 0 | 0.006046378 |
| 0 | 0 | 0 | 0 | 0.001760196 |
| 0 | 0 | 0 | 0 | 0 |
| 0 | 0 | 0 | 0 | 0 |
| 0 | 0 | 0 | 0 | 0 |
| 0 | 0 | 0 | 0 | 0 |
| 0.850743205 | 0.184966178 | 0.008616338 | 0.098207496 | 0.070491277 |
| 0 | 0 | 0 | 0 | 0 |
| 0.038017338 | 0.026537415 | 0 | 0 | 0 |
| 0 | 0 | 0 | 0.011975633 | 0 |
| 0 | 0 | 0 | 0 | 0 |
| 0 | 0 | 0 | 0 | 0 |
| 0 | 0 | 0 | 0 | 0 |
| 0 | 0 | 0 | 0 | 0 |
| 0 | 0 | 0 | 0 | 0 |
| 0 | 0 | 0 | 0 | 0.003129377 |
| 0 | 0 | 0 | 0 | 0 |
| 0 | 0.00290065 | 0.008782924 | 0.004372455 | 0.005930818 |
| 0 | 0 | 0 | 0 | 0 |
| 0 | 0 | 0 | 0 | 0 |
| 0 | 0 | 0 | 0 | 0 |
| 0 | 0 | 0 | 0 | 0 |
| 0 | 0 | 0 | 0 | 0 |
| 0 | 0 | 0 | 0 | 0 |

| | | | | |
|---|---|---|---|---|
| 0 | 0 | 0 | 0 | 0 |
| 0 | 0 | 0 | 0 | 0 |
| 0 | 0 | 0 | 0.035253743 | 0.038954152 |
| 0 | 0 | 0 | 0 | 0 |
| 0 | 0 | 0 | 0 | 0 |
| 0 | 0 | 0 | 0 | 0 |
| 0 | 0 | 0 | 0 | 0 |
| 0 | 0 | 0 | 0 | 0 |
| 0 | 0 | 0 | 0 | 0 |
| 0 | 0 | 0 | 0 | 0 |
| 0 | 0 | 0 | 0 | 0 |
| 0 | 0 | 0 | 0 | 0 |
| 0 | 0 | 0 | 0 | 0 |
| 0.002432815 | 0 | 0 | 0.004151662 | 0.002532434 |
| 0.001115539 | 0 | 0 | 0.012915848 | 0.017505362 |
| 0 | 0 | 0.002101554 | 0 | 0.002517555 |
| 0 | 0.001892782 | 0 | 0 | 0.009228649 |
| 0 | 0 | 0 | 0 | 0 |
| 0 | 0 | 0 | 0 | 0 |
| 0 | 0 | 0 | 0 | 0 |
| 0 | 0 | 0 | 0 | 0 |
| 0 | 0 | 0.001009488 | 0 | 0 |
| 0 | 0 | 0 | 0 | 0 |
| 0 | 0.008387819 | 0 | 0 | 0 |
| 0 | 0 | 0 | 0.001738884 | 0.004739939 |
| 0 | 0 | 0 | 0 | 0 |
| 0 | 0 | 0.006207645 | 0 | 0 |
| 0 | 0 | 0 | 0 | 0 |
| 0 | 0 | 0 | 0 | 0 |
| 0 | 0 | 0.001825291 | 0.003921739 | 0 |
| 0 | 0 | 0 | 0 | 0.002883121 |
| 0 | 0 | 0 | 0 | 0 |
| 0 | 0 | 0 | 0 | 0 |
| 0 | 0 | 0 | 0 | 0 |
| 0 | 0 | 0 | 0 | 0 |
| 0 | 0 | 0 | 0.075812598 | 0 |
| 0 | 0 | 0 | 0 | 0 |
| 0.002306546 | 0 | 0 | 0.001021893 | 0.432178932 |
| 0 | 0 | 0 | 0 | 0 |
| 0 | 0 | 0 | 0 | 0 |
| 0 | 0 | 0 | 0.002458134 | 0 |
| 0 | 0 | 0 | 0 | 0 |
| 0 | 0 | 0 | 0 | 0 |
| 0 | 0 | 0 | 0 | 0 |
| 0 | 0 | 0 | 0 | 0 |
| 0 | 0 | 0.001462513 | 0.004189718 | 0.090845276 |
| 0 | 0 | 0 | 0 | 0.032573038 |
| 0 | 0 | 0 | 0.009489561 | 0 |

| | | | | |
|---|---|---|---|---|
| 0.001611587 | 0 | 0.012912828 | 0 | 5.969990132 |
| 0 | 0 | 0.007076897 | 0.013370006 | 0 |
| 0 | 0 | 0 | 0 | 0.006755537 |
| 0 | 0 | 0 | 0 | 0 |
| 0.003627421 | 0 | 0.001329757 | 0 | 0.006796721 |
| 0 | 0 | 0 | 0 | 0.007388447 |
| 0 | 0 | 0 | 0 | 0 |
| 0.022372369 | 0 | 0 | 0.001201438 | 0.012305839 |
| 0 | 0 | 0 | 0 | 0 |
| 0.009433508 | 0.356831896 | 0.02151756 | 0.071824191 | 0.04091579 |
| 0 | 0 | 0 | 0 | 0 |
| 0 | 0 | 0 | 0.009510626 | 0.010684057 |
| 0 | 0 | 0 | 0.019718669 | 0.008819316 |
| 0 | 0 | 0 | 0 | 0 |
| 0 | 0 | 0 | 0 | 0 |
| 0 | 0 | 0 | 0 | 0 |
| 0 | 0 | 0 | 0 | 0 |
| 0 | 0 | 0 | 0 | 0 |
| 0 | 0 | 0 | 0 | 0 |
| 0 | 0.008829699 | 0 | 0 | 0 |
| 0 | 0 | 0 | 0 | 0 |
| 0 | 0 | 0 | 0 | 0 |
| 0 | 0 | 0 | 0 | 0 |
| 0.067823616 | 0 | 0 | 0 | 0 |
| 0 | 0 | 0 | 0 | 0 |
| 0 | 0 | 0 | 0.001120352 | 0 |
| 0 | 2.345450033 | 0 | 0 | 0 |
| 0 | 0 | 0 | 0 | 0 |
| 0 | 0 | 0.286161811 | 0 | 0 |
| 0.015366869 | 0 | 0 | 0 | 0 |
| 0 | 0 | 0 | 0 | 0 |
| 0 | 0 | 0 | 0 | 0 |
| 0 | 0 | 0 | 0 | 0 |
| 0 | 0 | 0 | 0 | 0 |
| 0 | 0 | 0 | 0 | 0 |
| 0 | 0 | 0 | 0 | 0 |
| 0 | 0 | 0.214573218 | 0 | 0 |
| 0 | 0 | 0.008143163 | 0 | 0 |
| 0 | 0 | 0 | 0 | 0 |
| 0 | 0 | 0.006855327 | 0.006775362 | 0 |
| 0 | 0 | 0 | 0 | 0 |
| 0 | 0 | 0 | 0 | 0 |
| 0 | 0 | 0 | 0 | 0 |
| 0 | 0 | 0 | 0 | 0 |
| 0 | 0 | 0 | 0 | 0 |
| 0 | 0 | 0 | 0 | 0 |
| 0.025649282 | 0 | 0.003566516 | 0 | 0.00345253 |
| 0 | 0 | 0 | 0 | 0 |

| | | | | |
|---|---|---|---|---|
| 0 | 0 | 0 | 0 | 0 |
| 0.001317925 | 0 | 0 | 0 | 0 |
| 0 | 0 | 0 | 0 | 0 |
| 0 | 0.030318713 | 0 | 0 | 0 |
| 0 | 0 | 0.001281865 | 0 | 0 |
| 0 | 0 | 0.20480626 | 0.171735349 | 0 |
| 0 | 0 | 0 | 0.001220193 | 0 |
| 0 | 0 | 0 | 0 | 0 |
| 0 | 0 | 0.004658212 | 0 | 0 |
| 0.000908733 | 0 | 0 | 0 | 0 |
| 0.001346599 | 0 | 0.005035154 | 0 | 0.029331394 |
| 0.01893105 | 0 | 0 | 0 | 0 |
| 0 | 0 | 0 | 0 | 0 |
| 0 | 0 | 0.001119285 | 0 | 0.005661352 |
| 0.025744735 | 0.031947936 | 0.007078219 | 0 | 0 |
| 0 | 0 | 0.001426783 | 0 | 0 |
| 0 | 0 | 0 | 0 | 0 |
| 0 | 0 | 0 | 0 | 0 |
| 0 | 0 | 0 | 0 | 0 |
| 0 | 0 | 0 | 0.002163042 | 0 |
| 0 | 0 | 0 | 0 | 0 |
| 0.008917825 | 0.094958377 | 0.261003752 | 0.002039203 | 0 |
| 0 | 0 | 0 | 0 | 0.016682567 |
| 0 | 0 | 0 | 0 | 0 |
| 0 | 0 | 0 | 0 | 0 |
| 0 | 0 | 0 | 0 | 0 |
| 0.00082764 | 0 | 0.016383613 | 0 | 0 |
| 0 | 0 | 0 | 0 | 0 |
| 0 | 0 | 0 | 0 | 0 |
| 0.516170342 | 1.401879277 | 0.799810518 | 0.613349815 | 0.527600724 |
| 0.006904687 | 0 | 0 | 0 | 0 |
| 0 | 0 | 0 | 0 | 0 |
| 0 | 0 | 0 | 0 | 0 |
| 0 | 0 | 0 | 0.000866534 | 0 |
| 0 | 0 | 0 | 0 | 0 |
| 0 | 0 | 0 | 0 | 0 |
| 2.595508059 | 0 | 0.211982686 | 0.032532606 | 0 |
| 0 | 0 | 0 | 0 | 0 |
| 0.146077487 | 0.028303144 | 0.237390691 | 0.192149776 | 0 |
| 0 | 0.160278551 | 0 | 0 | 0 |
| 0 | 0.008046934 | 0.000920174 | 0.015816354 | 0 |
| 0.146439594 | 0 | 6.729570711 | 7.014546141 | 0.113554505 |
| 0 | 0 | 0 | 0 | 0 |
| 0 | 0.001252625 | 0 | 0 | 0 |
| 0 | 0 | 0 | 0 | 0 |
| 0 | 0 | 0 | 0 | 0.007988351 |
| 0.012294603 | 0 | 0.021971687 | 0.032941414 | 0.001028414 |
| 0 | 0 | 0.003389942 | 0 | 0 |

| | | | | |
|---|---|---|---|---|
| 0 | 0 | 0 | 0 | 0 |
| 0 | 0 | 0.001903526 | 0 | 0 |
| 0 | 0 | 0 | 0.024855953 | 0.010806001 |
| 0 | 0 | 0 | 0 | 0 |
| 0 | 0 | 0 | 0.005790797 | 0.044006253 |
| 0 | 0 | 0 | 0 | 0 |
| 0 | 0 | 0 | 0.001591835 | 0 |
| 0 | 0 | 0 | 0 | 0 |
| 0 | 0 | 0 | 0 | 0 |
| 0 | 0 | 0 | 0 | 0 |
| 0 | 0 | 0 | 0 | 0 |
| 0 | 0 | 0 | 0 | 0 |
| 0 | 0 | 0 | 0 | 0 |
| 0 | 0 | 0.006468176 | 0 | 0 |
| 0 | 0 | 0 | 0 | 0 |
| 0.368850794 | 0 | 0.017481073 | 0.003167631 | 0.003364087 |
| 0 | 0.002107335 | 0 | 0.002257064 | 0.015909288 |
| 0 | 0.006400826 | 0 | 0 | 0 |
| 0 | 0 | 0 | 0 | 0 |
| 1.429319961 | 0.237830277 | 0 | 0 | 0.002652135 |
| 0.004012815 | 0 | 0.014710365 | 0.016751196 | 0 |
| 0 | 0 | 0 | 0 | 0 |
| 0 | 0 | 0 | 0 | 0 |
| 0.002108878 | 0 | 0 | 0.453732546 | 0.187966734 |
| 0.025591941 | 0 | 0.039708223 | 0 | 0.000929298 |
| 0 | 0.270664185 | 0 | 0 | 0 |
| 0 | 0 | 0.001778163 | 0.001273494 | 0 |
| 0 | 0 | 0 | 0 | 0 |
| 0 | 0 | 0 | 0 | 0 |
| 0 | 0 | 0 | 0 | 0 |
| 0.026232055 | 0 | 0.043949355 | 0 | 0.017519701 |
| 0 | 0 | 0 | 0.007580461 | 0.010376329 |
| 0.034618626 | 0.43718112 | 0.020475682 | 0.021309173 | 0.084147519 |
| 0 | 0 | 0 | 0 | 0.001464675 |
| 0 | 0 | 0 | 0.011249781 | 0.001393874 |
| 0 | 0 | 0 | 0 | 0 |
| 0 | 0 | 0 | 0 | 0 |
| 0.010284115 | 0 | 0.016964994 | 0.048843302 | 0.010357307 |
| 0.035448483 | 0 | 0 | 0.682540456 | 0.004473332 |
| 0 | 0 | 0 | 0 | 0 |
| 0 | 0 | 0 | 0 | 0 |
| 0 | 0 | 0.003982351 | 0 | 0 |
| 0.00246078 | 0 | 0 | 0.001453633 | 0.001080651 |
| 0.051700443 | 0.081292961 | 0.214894112 | 0 | 0.001128161 |
| 0 | 0 | 0.006219782 | 0 | 0 |
| 0 | 0 | 0 | 0.010595659 | 0 |
| 0 | 0 | 0 | 0 | 0 |
| 0 | 0 | 0 | 0 | 0 |

| | | | | |
|---|---|---|---|---|
| 0 | 0.002270631 | 0 | 0 | 0 |
| 0.020179312 | 0.002782391 | 0.01972647 | 0.139401677 | 1.244910675 |
| 0 | 0 | 0 | 0 | 0 |
| 0 | 0 | 0.019414036 | 0 | 0 |
| 0 | 0 | 0 | 0 | 0 |
| 0 | 0 | 0 | 0 | 0 |
| 0 | 0 | 0 | 0 | 0 |
| 0 | 0 | 0 | 0 | 0 |
| 0 | 0 | 0 | 0 | 0 |
| 0 | 0 | 0 | 0 | 0 |
| 0.038321667 | 0 | 0 | 0.005905407 | 0 |
| 0.005894255 | 0.001924863 | 0 | 0.030053929 | 0.027246963 |
| 0 | 0 | 0 | 0 | 0.45862552 |
| 0 | 0 | 0 | 0 | 0 |
| 0 | 0 | 0 | 0 | 0.001679054 |
| 0 | 0 | 0 | 0 | 0 |
| 0 | 0.001082549 | 0.008154692 | 0 | 0.025284235 |
| 0 | 0 | 0.322870476 | 0.06823328 | 0 |
| 0.0023116 | 0 | 0 | 0 | 0 |
| 0 | 0 | 0.034976232 | 0.010208014 | 0 |
| 0.004947898 | 0.009489238 | 0.006925508 | 0.003719959 | 0.097186491 |
| 0 | 0 | 0 | 0 | 0 |
| 0 | 0 | 0 | 0 | 0 |
| 0 | 0 | 0 | 0 | 0 |
| 0 | 0 | 0 | 0 | 0 |
| 0 | 0 | 0 | 0 | 0 |
| 0 | 0.008539909 | 0.052733486 | 0 | 0.008318953 |
| 0.101353056 | 0.027883882 | 0.042587442 | 0.112399948 | 0.036951019 |
| 0 | 0 | 0 | 0 | 0 |
| 0 | 0 | 0 | 0 | 0 |
| 0.001025961 | 0 | 0 | 0 | 0.015318729 |
| 0 | 0 | 0.001088581 | 0 | 0 |
| 0 | 0 | 0 | 0 | 0 |
| 0 | 0 | 0 | 0 | 0 |
| 0.06415853 | 0 | 0.002978123 | 0 | 0.007074295 |
| 0.008900349 | 0.043677523 | 0.195763888 | 0.447375562 | 0.506458144 |
| 0 | 0 | 0.000824237 | 0.001549554 | 0 |
| 0 | 0 | 0 | 0 | 0 |
| 0 | 0 | 0.001297194 | 0.019509656 | 0.021755625 |
| 0 | 0 | 0.00240055 | 0 | 0 |
| 0 | 0 | 0.002034991 | 0 | 0 |
| 0 | 0 | 0 | 0 | 0 |
| 0 | 0 | 0 | 0 | 0 |
| 0 | 0 | 0.003613501 | 0 | 0.001620132 |
| 0.018629947 | 0 | 0.015741056 | 0 | 0 |
| 0.121499626 | 0 | 0.041657906 | 0.012032419 | 0 |
| 0 | 0 | 0 | 0 | 0 |
| 0 | 0 | 0 | 0 | 0 |

| | | | | |
|---|---|---|---|---|
| 0 | 0 | 0 | 0 | 0 |
| 0 | 0 | 0 | 0 | 0.446710732 |
| 0 | 0 | 0 | 0 | 0 |
| 0 | 0 | 0.006653294 | 0.00640809 | 1.796831489 |
| 0 | 0 | 0 | 0 | 0 |
| 0 | 0 | 0 | 0 | 0.006143531 |
| 0 | 0 | 0 | 0 | 0 |
| 0.004105805 | 0 | 0 | 0 | 0 |
| 0.549234711 | 0 | 0.054505772 | 0.005056965 | 2.814682944 |
| 0.013051465 | 0 | 0 | 0 | 0.158081581 |
| 0 | 0 | 0 | 0 | 0 |
| 0.01858833 | 0.002728378 | 0 | 0 | 0 |
| 0 | 0 | 0 | 0 | 0 |
| 0 | 0 | 0 | 0 | 0 |
| 0.698178137 | 0.037456177 | 0 | 0 | 0.001158913 |
| 0 | 0 | 0 | 0 | 0 |
| 0.0718587 | 0 | 0 | 0.034568763 | 0 |
| 0 | 0 | 0.007106044 | 0.002290159 | 0 |
| 0 | 0 | 0 | 0 | 0 |
| 0 | 0 | 0 | 0 | 0 |
| 0 | 0 | 0 | 0 | 0 |
| 0 | 0 | 0 | 0 | 0.003259701 |
| 0 | 0 | 0 | 0 | 0 |
| 0.244352154 | 0.01419537 | 0.906823439 | 0.200091706 | 0.046141638 |
| 0 | 0 | 0.001234806 | 0 | 0 |
| 0 | 0 | 0.113897376 | 0 | 1.971694511 |
| 0 | 0 | 0 | 0 | 0 |
| 1.053769504 | 0.834505727 | 0.095795719 | 2.346569335 | 0.679087582 |
| 0 | 0 | 0 | 0 | 0 |
| 0 | 0 | 0 | 0 | 0 |
| 0 | 0 | 0 | 0 | 0.007054123 |
| 0.005492341 | 0 | 0 | 0.001297775 | 0 |
| 0.000887315 | 0 | 0.036756183 | 0 | 0.000692736 |
| 0 | 0 | 0 | 0 | 0 |
| 0 | 0 | 0 | 0 | 0 |
| 0.292440014 | 0 | 0 | 0 | 0.014470422 |
| 0 | 0 | 0.006786224 | 0.007290289 | 5.943233005 |
| 0 | 0 | 0 | 0 | 0 |
| 0.005409804 | 0 | 0 | 0 | 0.001949304 |
| 0.006008724 | 0.012605082 | 0 | 0 | 0 |
| 0.087243459 | 0.001189723 | 0.013495749 | 0.199349981 | 0.041989875 |
| 0 | 0 | 0 | 0 | 0 |
| 30.84549284 | 10.04019897 | 0.876273344 | 0.200011992 | 0.257516795 |
| 0.002004408 | 0 | 0 | 0 | 0.000782431 |
| 0 | 0 | 0 | 0 | 0 |
| 0 | 0 | 0 | 0 | 0 |
| 0 | 0 | 0 | 0 | 0 |
| 0.031808741 | 0.022882937 | 0.00202793 | 0.007080318 | 0 |

| | | | | |
|---|---|---|---|---|
| 0 | 0 | 0 | 0 | 0 |
| 0.083996128 | 0.165473094 | 0.000692813 | 0 | 0.041559228 |
| 0.01303655 | 0.013161527 | 0.007289999 | 0.112251244 | 0.016042936 |
| 0 | 0 | 0 | 0 | 0.004195559 |
| 0 | 0 | 0 | 0 | 0.498188439 |
| 0.003307189 | 0 | 0 | 0.006991918 | 0.098250306 |
| 0 | 0 | 0 | 0 | 0 |
| 0 | 0 | 0 | 0 | 0 |
| 0 | 0 | 0 | 0 | 0.006824473 |
| 0 | 0 | 0 | 0 | 0.036820906 |
| 0.033626016 | 0.0169605 | 0.012644892 | 0.204530769 | 0.006097284 |
| 1.447622389 | 1.718384917 | 1.538282803 | 3.13532251 | 0.791654212 |
| 0 | 0 | 0 | 0 | 0 |
| 0.001873062 | 0 | 0 | 0 | 0.027418491 |
| 0.001476044 | 0.023583107 | 0.016029945 | 0.094386384 | 0.040836894 |
| 0 | 0 | 0.020280876 | 0 | 0 |
| 0 | 0 | 0.000659315 | 0 | 0 |
| 0 | 0 | 0 | 0 | 0 |
| 0 | 0 | 0 | 0 | 0.006137813 |
| 0.002323141 | 0 | 0.052276845 | 0.001689018 | 0.005231831 |
| 0.006909516 | 0.004964107 | 0.020596661 | 0.196990565 | 0.638163967 |
| 11.09316232 | 8.221359123 | 0.038137386 | 0.0857883 | 0.041956563 |
| 0.139798192 | 0.387884945 | 0.095570357 | 9.455701445 | 3.114166322 |
| 0 | 0 | 0 | 0 | 0 |
| 0 | 0 | 0 | 0 | 0 |
| 0.072120309 | 0.007570804 | 0.002635833 | 0 | 0.054093731 |
| 0.6791788 | 0 | 0 | 0 | 0.001018717 |
| 0.015740513 | 0 | 0 | 0 | 0 |
| 0.059454385 | 0.006176925 | 0 | 0 | 0.01101047 |
| 0 | 0 | 0 | 0 | 0 |
| 0.004612721 | 0 | 0 | 0 | 0 |
| 0.171832528 | 0 | 0.512739306 | 0.159203645 | 0.129661938 |
| 0 | 0.005806292 | 0.91249478 | 6.602889237 | 0.000782759 |
| 0.057428281 | 0.013283241 | 0 | 0 | 0.019698166 |
| 0 | 0 | 0.001132064 | 0.002258565 | 0.004305254 |
| 0.033135119 | 0.011533388 | 0.003555169 | 0.02410894 | 0.076423813 |
| 0.036106667 | 0.000726593 | 0 | 0 | 0 |
| 0 | 0 | 0 | 0 | 0 |
| 0.049414202 | 0 | 0.008580547 | 0.00251397 | 0.000830631 |
| 0 | 0 | 0 | 0 | 0 |
| 0 | 0 | 0 | 0.001050947 | 0.066044952 |
| 0 | 0 | 0 | 0 | 0 |
| 0 | 0 | 0 | 0 | 0 |
| 0 | 0 | 0 | 0 | 0 |
| 0.217965564 | 0.031165754 | 0.057266512 | 0.051992675 | 0.005396451 |
| 0 | 0 | 1.324741329 | 0 | 0.057143126 |
| 0 | 0 | 0.00456579 | 0 | 0 |
| 0.558370404 | 0.053115046 | 0.011615064 | 0.029881053 | 0 |

| | | | | |
|---|---|---|---|---|
| 0 | 0 | 0 | 0 | 0 |
| 0 | 0 | 0 | 0 | 0 |
| 0 | 0 | 0 | 0 | 0 |
| 0.006087638 | 0 | 0.005183152 | 0 | 0 |
| 0 | 0 | 0 | 0 | 0 |
| 0 | 0 | 0.002098125 | 0.00163925 | 0.117598876 |
| 0.012039648 | 0 | 0.015841493 | 0 | 0 |
| 0.016624488 | 0.018163513 | 0 | 0 | 0.001058064 |
| 0 | 0 | 0 | 0 | 0 |
| 0 | 0 | 0 | 0 | 0 |
| 0.003280654 | 0.000814226 | 0 | 0.001808753 | 0.001792869 |
| 0 | 0 | 0 | 0 | 0 |
| 0.036193106 | 0 | 0 | 0 | 0 |
| 0 | 0 | 1.452957531 | 0 | 0.003715488 |
| 0 | 0 | 0 | 0 | 0 |
| 0 | 0 | 0 | 0 | 0 |
| 0 | 0 | 0 | 0.00057653 | 0 |
| 0.004551782 | 0 | 0 | 0 | 0 |
| 0 | 0 | 0 | 0 | 0.135738654 |
| 0 | 0 | 0 | 0 | 0 |
| 0 | 0 | 0 | 0 | 0.002651978 |
| 0 | 0 | 0 | 0 | 0 |
| 0.043032608 | 0 | 0 | 0 | 0 |
| 0 | 0 | 0 | 0 | 0 |
| 0 | 0 | 0 | 0 | 0 |
| 0 | 0 | 0 | 0 | 0 |
| 0 | 0 | 0 | 0 | 0 |
| 0 | 0 | 0 | 0 | 0 |
| 0 | 0 | 0 | 0 | 0 |
| 0.028592263 | 0 | 0.602282259 | 0.005196934 | 2.747187185 |
| 0 | 0 | 0 | 0 | 0.002622442 |
| 0 | 0 | 0 | 0 | 0 |
| 0 | 0 | 0 | 0 | 0 |
| 0 | 0 | 0 | 0 | 0 |
| 0 | 0 | 0 | 0.001849156 | 0 |
| 0 | 0 | 0 | 0 | 0 |
| 0.125454135 | 0 | 0 | 0 | 0.04842146 |
| 0.006479972 | 0.00551405 | 0.001425275 | 0.010280517 | 0 |
| 0 | 0 | 0 | 0 | 0 |
| 0 | 0 | 0 | 0 | 0 |
| 0.011812216 | 0.0030179 | 0 | 0 | 0.001423974 |
| 0 | 0 | 0 | 0 | 0 |
| 0 | 0 | 0 | 0 | 0 |
| 0.038116511 | 0 | 1.6099186 | 0 | 0.040004825 |
| 0.260050648 | 0 | 0 | 0.000745112 | 0.410398092 |
| 0 | 0 | 0 | 0 | 0 |
| 0 | 0 | 0 | 0 | 0.107124807 |
| 0 | 0 | 0 | 0 | 0 |

| | | | | |
|---|---|---|---|---|
| 0 | 0 | 0 | 0 | 0 |
| 0 | 0 | 0 | 0 | 0 |
| 0.164966364 | 0 | 0 | 0 | 0 |
| 0.159852678 | 0 | 0.069873875 | 0 | 0.066209462 |
| 0 | 0 | 0 | 0 | 0 |
| 0 | 0 | 0 | 0 | 0 |
| 0 | 0 | 0 | 0 | 0 |
| 0 | 0 | 0 | 0 | 0 |
| 0.02834294 | 0.026616785 | 0.03903303 | 0.292922814 | 0 |
| 0 | 0 | 0 | 0 | 0 |
| 0 | 0 | 0 | 0 | 0.004276203 |
| 0 | 0 | 0 | 0 | 0 |
| 0 | 0.001293978 | 0 | 0.000879947 | 0 |
| 0 | 0 | 0 | 0 | 0 |
| 1.209234416 | 0 | 0.208862743 | 0 | 1.349049507 |
| 0 | 0.039319068 | 0 | 0.003022584 | 0.020280895 |
| 0 | 0 | 0 | 0.026870212 | 0 |
| 0 | 0 | 0 | 0 | 0 |
| 0 | 0 | 0 | 0 | 0 |
| 0 | 0 | 0 | 0 | 0 |
| 0 | 0 | 0 | 0.000922684 | 0.005030198 |
| 0 | 0 | 0 | 0 | 0 |
| 0 | 0 | 1.138104953 | 0 | 0 |
| 0 | 0 | 0 | 0 | 0 |
| 0.011410438 | 0.715441851 | 0.010237077 | 0.006030866 | 0.097053075 |
| 0 | 0 | 0.518860289 | 5.298141831 | 0.872010331 |
| 0 | 0 | 0.068290902 | 0 | 0.009416868 |
| 0 | 0 | 0 | 0 | 0 |
| 0.043054255 | 0 | 0 | 0 | 0 |
| 0 | 0 | 0 | 0 | 0.016714468 |
| 0 | 0 | 0 | 0 | 0 |
| 0 | 0 | 0 | 0 | 0 |
| 0 | 0 | 0 | 0 | 0 |
| 0.00080183 | 0.001393043 | 0.002116357 | 0.02652481 | 0 |
| 0.001321162 | 0 | 0 | 0 | 0.002166036 |
| 0.065746669 | 0.006489982 | 0.002875773 | 0.007502777 | 0 |
| 0 | 0 | 0.309066699 | 0 | 0 |
| 0.57894041 | 0 | 9.618108578 | 0.006976885 | 0.072120023 |
| 0 | 0 | 0 | 0.041525707 | 0 |
| 0 | 0 | 0 | 0 | 0 |
| 0 | 0 | 0 | 0 | 0 |
| 0 | 0 | 0 | 0 | 0.015874816 |
| 0.004083818 | 0 | 1.520269514 | 0 | 0.066953903 |
| 0 | 0 | 0.009491415 | 0 | 0 |
| 0 | 0 | 0.007588799 | 0 | 0 |
| 0.006901085 | 0 | 0.027049741 | 0 | 0.000828885 |
| 0 | 0 | 0 | 0 | 0.002018661 |
| 0 | 0.004547775 | 0.003410275 | 0.033305294 | 1.586926589 |

| | | | | |
|---|---|---|---|---|
| 0.002522593 | 0.032869336 | 0 | 0 | 0.001723239 |
| 0 | 0 | 0 | 0 | 0 |
| 0 | 0 | 0 | 0 | 0 |
| 0 | 0 | 0 | 0 | 0 |
| 0 | 0 | 0.021636211 | 0.153554114 | 0 |
| 0 | 0 | 0 | 0 | 0 |
| 0 | 0 | 0 | 0 | 0 |
| 0 | 0 | 0 | 0 | 0 |
| 0 | 0 | 0 | 0 | 0 |
| 0 | 0 | 0 | 0 | 0 |
| 0 | 0 | 0 | 0 | 0 |
| 0.150994447 | 0 | 0.025694202 | 0.437993331 | 0.077784696 |
| 0.003801284 | 0 | 1.149073859 | 0 | 0.742816271 |
| 0 | 0 | 0 | 0 | 0 |
| 0.001936393 | 0 | 0.002661944 | 0.06691621 | 0.030896652 |
| 0 | 0 | 0 | 0 | 0 |
| 0 | 0 | 0 | 0 | 0 |
| 0 | 0 | 0 | 0 | 0 |
| 0 | 0 | 0.000933274 | 0 | 0 |
| 0 | 0 | 0 | 0 | 0.15427378 |
| 0 | 0 | 0 | 0 | 0.107165509 |
| 0 | 0 | 0 | 0 | 0 |
| 0 | 0 | 0.056490757 | 0.01023632 | 0.053389551 |
| 0 | 0 | 0 | 0 | 0 |
| 0 | 0 | 0 | 0 | 0 |
| 0 | 0 | 0 | 0 | 0 |
| 0 | 0 | 0.000921687 | 0 | 0 |
| 0 | 0 | 0 | 0 | 0 |
| 0 | 0 | 0 | 0 | 0 |
| 0 | 0 | 0 | 0 | 0 |
| 0 | 0 | 0 | 0 | 0 |
| 0 | 0.002338639 | 0.005526792 | 0.096481266 | 0.129841582 |
| 0 | 0 | 0.004643131 | 0 | 0 |
| 0.043779344 | 0.01518847 | 0 | 0.010752392 | 0.160788355 |
| 0.063621564 | 0.006053766 | 0.002235402 | 0.011046632 | 0 |
| 0.057384934 | 0.004552356 | 0.001440852 | 0.022289397 | 0.49158404 |
| 0.001082586 | 0 | 0 | 0 | 0 |
| 0.014577548 | 0 | 0 | 0 | 0.048710038 |
| 0.007706163 | 0 | 0 | 0 | 0 |
| 0 | 0 | 0 | 0 | 0 |
| 0.003117267 | 0 | 0 | 0.000818415 | 0.023119999 |
| 0 | 0 | 0 | 0 | 0.006420236 |
| 0 | 0 | 0 | 0 | 0 |
| 0 | 0 | 0 | 0 | 0.001356479 |
| 0.241884119 | 0.003727229 | 0.017176338 | 0.175058897 | 0.006565683 |
| 0 | 0 | 0 | 0 | 0 |
| 0 | 0 | 0 | 0 | 0 |
| 0.008901173 | 0 | 0.001457951 | 0 | 0 |

| | | | | |
|---|---|---|---|---|
| 0 | 0 | 0 | 0 | 0 |
| 0 | 0 | 0 | 0 | 0 |
| 0 | 0 | 0.006377047 | 0.001957348 | 0 |
| 0 | 0 | 0 | 0 | 0 |
| 0 | 0 | 0 | 0 | 0 |
| 0 | 0 | 0 | 0.000631458 | 0.003755476 |
| 0 | 0 | 0 | 0.00269326 | 0 |
| 0 | 0 | 0 | 0 | 0 |
| 0 | 0 | 0 | 0 | 0 |
| 0 | 0 | 0 | 0 | 0 |
| 0 | 0 | 0 | 0 | 0 |
| 0 | 0 | 0.002780782 | 0 | 0 |
| 0 | 0 | 0 | 0 | 0 |
| 0.291533529 | 0 | 0 | 0.001967046 | 0.524878653 |
| 0 | 0 | 0 | 0 | 0 |
| 0 | 0 | 0 | 0 | 0 |
| 0 | 0 | 0 | 0 | 2.136392412 |
| 0 | 0 | 0 | 0 | 0 |
| 0 | 0.00109842 | 0.018738731 | 0.045751359 | 0.784159345 |
| 0 | 0 | 0 | 0 | 0 |
| 0 | 0 | 0.001655462 | 0 | 0 |
| 0 | 0 | 0 | 0 | 0 |
| 0 | 0 | 0 | 0 | 0 |
| 0 | 0 | 0 | 0 | 0 |
| 0 | 0 | 0 | 0 | 0.002305793 |
| 0 | 0 | 0.003485114 | 0 | 0.0008732 |
| 0 | 0 | 0 | 0 | 0 |
| 0 | 0.003849198 | 0 | 0.59258916 | 0.14657926 |
| 0 | 0 | 0 | 0 | 0.295797517 |
| 0.131266838 | 0 | 2.566006819 | 0.197616775 | 0.070795723 |
| 0 | 0 | 0 | 0 | 0 |
| 0 | 0 | 0 | 0 | 0 |
| 0 | 0 | 0 | 0 | 0 |
| 0 | 0 | 0 | 0 | 0 |
| 0.070715837 | 0.0014545 | 0.01890542 | 0.002538709 | 0.000686295 |
| 0 | 0 | 0.004102348 | 0 | 0 |
| 0 | 0 | 0.00066556 | 0 | 0 |
| 0.001108312 | 0 | 0 | 0 | 0.001297906 |
| 0 | 0 | 0 | 0 | 0 |
| 0 | 0 | 0 | 0 | 0 |
| 0.550744465 | 1.602452238 | 3.810315418 | 7.782380811 | 6.106830986 |
| 0 | 0.011477172 | 0 | 0.512217202 | 0.849935656 |
| 0 | 0 | 0.015289335 | 0.001124999 | 0 |
| 0 | 0 | 0 | 0 | 0 |
| 0 | 0 | 0.002446375 | 0 | 0 |
| 0 | 0 | 0 | 0 | 0 |
| 0 | 0 | 0 | 0 | 0 |
| 0.014601649 | 0.002183123 | 0.004062924 | 0.001870589 | 0 |

| | | | | |
|---|---|---|---|---|
| 0 | 0 | 0 | 0 | 0 |
| 0.001809518 | 0 | 0.006169077 | 0 | 0.00932389 |
| 0 | 0 | 0 | 0 | 0.00148497 |
| 0 | 0 | 0 | 0 | 0 |
| 0.044789977 | 0 | 4.677513688 | 0 | 1.854333645 |
| 0 | 0 | 0 | 0 | 0 |
| 0 | 0 | 0 | 0 | 0 |
| 0 | 0 | 0 | 0 | 0.016850696 |
| 0.023669741 | 0 | 0.372778271 | 0.418398703 | 0.002750724 |
| 0 | 0 | 0 | 0 | 0 |
| 0 | 0 | 0 | 0 | 0 |
| 0 | 0 | 0 | 0 | 0 |
| 0 | 0 | 0.213474785 | 0.00166182 | 0.043239708 |
| 0.001080798 | 0.081545978 | 0.011648398 | 0.015067411 | 0.02025098 |
| 0.064689694 | 0 | 0.003256534 | 0.001345547 | 5.376269773 |
| 0.05268157 | 0 | 1.756244803 | 0.004191257 | 0.131488406 |
| 0.018705263 | 0 | 0 | 0.061877661 | 0.001533357 |
| 0 | 0 | 0 | 0 | 0 |
| 0 | 0 | 0 | 0 | 0 |
| 0 | 0 | 0 | 0 | 0.00604713 |
| 0 | 0 | 0 | 0 | 0 |
| 0 | 0 | 0 | 0 | 0 |
| 0 | 0 | 0 | 0 | 0 |
| 0 | 0 | 0 | 0 | 0 |
| 0 | 0 | 0 | 0 | 0 |
| 0 | 0 | 0 | 0 | 0 |
| 0.009271047 | 0 | 0.009480371 | 0 | 0.003047582 |
| 0 | 0 | 0 | 0.001517828 | 0 |
| 0 | 0 | 0 | 0 | 0 |
| 0 | 0 | 0 | 0 | 0 |
| 0 | 0 | 0 | 0 | 0 |
| 0 | 0 | 0 | 0 | 0 |
| 0 | 0 | 0.008702691 | 0 | 0 |
| 0.002501081 | 0 | 0 | 0 | 0.012616934 |
| 0.005461129 | 0 | 0.017767451 | 0.001075334 | 0.059156939 |
| 0 | 0 | 0 | 0 | 0 |
| 0.4548427 | 0 | 0.007636871 | 0.005896711 | 0 |
| 0 | 0 | 0 | 0 | 0 |
| 0 | 0 | 0 | 0 | 0 |
| 0 | 0 | 0 | 0 | 0 |
| 0.17109552 | 0.665035948 | 1.542192928 | 0.035942696 | 2.207355767 |
| 0.011178288 | 0 | 0 | 0 | 0 |
| 0 | 0 | 0 | 0 | 0 |
| 0.265509801 | 0 | 0 | 0 | 0 |
| 0 | 0 | 0 | 0 | 0 |
| 0 | 0 | 0 | 0 | 0 |
| 0 | 0 | 0 | 0 | 0 |
| 0 | 0 | 0 | 0 | 0 |

| | | | | |
|---|---|---|---|---|
| 0 | 0 | 0 | 0.003323759 | 5.369680956 |
| 0 | 0 | 0 | 0 | 0 |
| 0 | 0 | 0 | 0 | 0 |
| 0 | 0 | 0 | 0 | 0 |
| 0.002387105 | 0 | 0 | 0 | 0.008153421 |
| 0 | 0 | 0 | 0 | 0.166508639 |
| 0.002798967 | 0 | 0.00123127 | 1.415757434 | 0.266373919 |
| 0 | 0 | 0 | 0 | 0 |
| 0 | 0 | 0 | 0 | 0 |
| 0 | 0 | 0.004090708 | 0.005319726 | 0 |
| 0 | 0 | 0 | 0 | 0.028088869 |
| 0 | 0 | 0 | 0 | 0 |
| 0 | 0 | 0 | 0 | 0 |
| 0 | 0 | 0 | 0 | 0 |
| 0 | 0 | 0 | 0 | 0 |
| 0 | 0 | 0 | 0 | 0 |
| 0 | 0 | 0 | 0 | 0 |
| 0 | 0 | 0 | 0 | 0 |
| 0 | 0 | 0 | 0 | 0 |
| 0 | 0 | 0 | 0 | 0 |
| 0.00124356 | 0.014991054 | 0 | 1.015632393 | 0.045145046 |
| 0 | 0 | 0 | 0 | 0 |
| 0.026061215 | 0 | 0 | 0 | 0.001835754 |
| 0.015681587 | 0.004756906 | 0.001341347 | 0 | 0 |
| 0 | 0 | 0 | 0 | 0 |
| 0 | 0 | 0.000904279 | 0 | 0.001203645 |
| 0 | 0 | 0 | 0 | 0 |
| 0 | 0 | 0 | 0 | 0 |
| 0 | 0 | 0 | 0 | 0.128918464 |
| 0 | 0.014314998 | 0 | 0.272866824 | 0.005286072 |
| 0 | 0 | 0 | 0 | 0 |
| 0 | 0 | 0 | 0 | 0 |
| 0 | 0 | 0 | 0 | 0 |
| 0 | 1.40649195 | 0.050955276 | 0 | 0 |
| 0 | 0 | 0 | 0 | 0 |
| 0 | 0 | 0 | 0 | 0 |
| 0 | 0 | 0.528687282 | 0.013393994 | 0.002042519 |
| 0 | 0 | 0 | 0 | 0 |
| 0 | 0 | 0 | 0 | 0 |
| 0 | 0 | 0 | 0 | 0 |
| 0 | 0 | 0 | 0 | 0 |
| 0 | 0 | 0 | 0 | 0 |
| 0 | 0 | 0 | 0 | 0 |
| 0 | 0 | 0 | 0 | 0 |
| 0 | 0 | 0.004765358 | 0 | 0 |
| 0 | 0 | 0 | 0 | 0 |
| 0 | 0 | 0 | 0 | 0 |
| 0.02659918 | 0 | 0 | 0 | 0 |

| | | | | |
|---|---|---|---|---|
| 0.006937066 | 0 | 0.000824764 | 0 | 0.001097806 |
| 0 | 0 | 0 | 0 | 0 |
| 0 | 0 | 0 | 0 | 0 |
| 0 | 0 | 0 | 0 | 0 |
| 0 | 0 | 0 | 0 | 0 |
| 0.007333814 | 0 | 0 | 0 | 0 |
| 0 | 0 | 0 | 0 | 0 |
| 0.012705534 | 0.635694176 | 0.254053839 | 1.254997078 | 2.435876329 |
| 0.910830667 | 32.90967888 | 0.2116656 | 0.172105297 | 0.814715456 |
| 0.301988065 | 0.035339588 | 0.07187511 | 0.012344529 | 0.000959696 |
| 0 | 0 | 0 | 0 | 0.0042232 |
| 0 | 0 | 0 | 0 | 0 |
| 0 | 0 | 0 | 0 | 0.017245819 |
| 0 | 0.644007847 | 0 | 0 | 0 |
| 0 | 0 | 0 | 0 | 0 |
| 0 | 0 | 0.002434773 | 0 | 0 |
| 0 | 0 | 0.023175717 | 0 | 0 |
| 0 | 0 | 0 | 0 | 0 |
| 0 | 0 | 0 | 0 | 0 |
| 0 | 0 | 0 | 0 | 0 |
| 0 | 0 | 0 | 0 | 0 |
| 0 | 0 | 0 | 0 | 0 |
| 0 | 0 | 0 | 0 | 0 |
| 0 | 0 | 0 | 0 | 0 |
| 0.022446297 | 0 | 0 | 0 | 0.042369047 |
| 0 | 0 | 0 | 0 | 0 |
| 0 | 0 | 0 | 0 | 0 |
| 0 | 0 | 0 | 0 | 0 |
| 0 | 0 | 0 | 0 | 0 |
| 0 | 0 | 0 | 0 | 0 |
| 0 | 0.005736524 | 0 | 0 | 0 |
| 0 | 0 | 0 | 0 | 0 |
| 0 | 0 | 0.008287341 | 0 | 0 |
| 0 | 0 | 0 | 0 | 0 |
| 0.003631695 | 0 | 0 | 0 | 0 |
| 0 | 0 | 0 | 0 | 0.003735332 |
| 0 | 0 | 0 | 0 | 0 |
| 0.019088645 | 0 | 0 | 0 | 0 |
| 0.021402068 | 0 | 0 | 0 | 0 |
| 0 | 0 | 0 | 0 | 0 |
| 0 | 0 | 0 | 0 | 0 |
| 0.420574306 | 0 | 0.000637091 | 0 | 0.155806501 |
| 0 | 0 | 0 | 0 | 0 |
| 0 | 0 | 0 | 0 | 0 |
| 0 | 0 | 0 | 0 | 0 |
| 0 | 0 | 0.00128342 | 0 | 0.001366642 |
| 0 | 0 | 0 | 0 | 0 |
| 0 | 0 | 0 | 0 | 0 |

| | | | | |
|---|---|---|---|---|
| 0 | 0 | 0 | 0 | 0 |
| 0 | 0 | 0 | 0 | 0 |
| 0 | 0 | 0 | 0 | 0 |
| 0 | 0 | 0 | 0 | 0 |
| 0.27278628 | 0.164706395 | 0.045012679 | 1.336920684 | 0.377356482 |
| 0 | 0 | 0 | 0 | 0 |
| 0 | 0 | 0 | 0 | 0 |
| 0.037205792 | 0.040843168 | 0.007868696 | 0.022259996 | 0.000837893 |
| 0.003095717 | 0 | 0 | 0 | 0 |
| 0 | 0 | 0 | 0 | 0 |
| 0 | 0 | 0 | 0 | 0 |
| 0.263872587 | 0 | 0 | 0.000841998 | 0 |
| 0 | 0 | 0 | 0 | 0 |
| 0 | 0 | 0 | 0 | 0 |
| 0 | 0 | 0 | 0.002380291 | 0 |
| 0.046333976 | 0.001540129 | 0.032984895 | 0 | 0.021558724 |
| 0 | 0 | 0 | 0 | 0 |
| 0 | 0 | 0 | 0 | 0 |
| 0 | 0 | 0 | 0 | 0.02196894 |
| 0 | 0 | 0 | 0 | 0 |
| 0 | 0 | 0 | 0 | 0 |
| 0 | 0 | 0 | 0 | 0 |
| 0 | 0 | 0 | 0 | 0 |
| 0.001238406 | 0.0007684 | 0 | 0 | 0.001691964 |
| 0.234139716 | 0 | 0 | 0.003688297 | 1.203223995 |
| 0 | 0 | 0 | 0 | 0 |
| 11.37079959 | 0.002108367 | 0 | 0.113298735 | 0 |
| 0.001391433 | 0 | 0 | 0 | 0 |
| 0 | 0 | 0.001851505 | 0 | 0 |
| 0 | 0 | 0 | 0 | 0 |
| 0 | 0 | 0 | 0 | 0 |
| 0 | 0 | 0 | 0 | 0 |
| 0 | 0 | 0 | 0 | 0 |
| 0 | 0 | 0.13323294 | 0 | 0 |
| 0 | 0 | 0 | 0 | 0 |
| 0 | 0 | 0 | 0 | 0 |
| 0 | 0 | 0 | 0 | 0 |
| 0.070858323 | 0 | 0 | 0 | 0.008388668 |
| 0 | 0 | 0 | 0 | 0 |
| 0 | 0 | 0 | 0 | 0 |
| 0 | 0 | 0 | 0 | 0 |
| 0 | 0 | 0 | 0 | 0 |
| 0 | 0 | 0 | 0 | 0 |
| 0 | 0 | 0 | 0 | 0 |
| 0 | 0 | 0 | 0 | 0 |
| 0 | 0 | 0 | 0 | 0 |
| 0 | 0 | 0 | 0 | 0 |
| 0 | 0 | 0 | 0 | 0 |

| | | | | |
|---|---|---|---|---|
| 0.003373043 | 0 | 0.002538091 | 0.325515794 | 0.092306602 |
| 0 | 0 | 0 | 0 | 0 |
| 0 | 0 | 0.000580181 | 0 | 0.000617803 |
| 0 | 0 | 0 | 0 | 0 |
| 0 | 0 | 0 | 0 | 0 |
| 0 | 0.027310157 | 0.071477385 | 0 | 0.150538485 |
| 0 | 0 | 0 | 0 | 0 |
| 0 | 0 | 0 | 0 | 0 |
| 0 | 0 | 0 | 0 | 0 |
| 0 | 0 | 0 | 0 | 0 |
| 0 | 0 | 0 | 0 | 0 |
| 0 | 0.000944456 | 0 | 0 | 0 |
| 0 | 0 | 0 | 0 | 0 |
| 0 | 0 | 0 | 0 | 0 |
| 0 | 0 | 0 | 0 | 0 |
| 0 | 0 | 0 | 0 | 0 |
| 0 | 0 | 0 | 0 | 0 |
| 0 | 0 | 0 | 0 | 0 |
| 0 | 0.00130745 | 0 | 0.000533465 | 0 |
| 0.098795774 | 0 | 0 | 0 | 0 |
| 0 | 0 | 0.001356248 | 0.000971325 | 0.002647687 |
| 0 | 0 | 0 | 0 | 0 |
| 0.045426447 | 0 | 0.017413902 | 0.000920034 | 0 |
| 0 | 0 | 0 | 0.007017195 | 0.011407141 |
| 0 | 0 | 0 | 0 | 0 |
| 0.019099155 | 0 | 0 | 0 | 0 |
| 0 | 0 | 0 | 0 | 0 |
| 0 | 0 | 0 | 0 | 0 |
| 0 | 0.006692603 | 0 | 0 | 0 |
| 0 | 0 | 0.005438334 | 0 | 0 |
| 0 | 0 | 0 | 0 | 0 |
| 0 | 0 | 0 | 0 | 0 |
| 0.10015527 | 0.335456153 | 0.006950971 | 0.084150414 | 0 |
| 0 | 0 | 0 | 0 | 0 |
| 0.006088533 | 0 | 0.934076423 | 0 | 0.743819701 |
| 0 | 0 | 0 | 0.004839832 | 0 |
| 0 | 0 | 0 | 0 | 0 |
| 0 | 0 | 0 | 0 | 0 |
| 0 | 0 | 0.009181238 | 0 | 0 |
| 0 | 0 | 0 | 0 | 0 |
| 0 | 0 | 0 | 0 | 0 |
| 0 | 0 | 0 | 0 | 0 |
| 0.28045492 | 0 | 0.005512885 | 0 | 0.219057197 |
| 0 | 0 | 0 | 0 | 0 |
| 0 | 0 | 0 | 0 | 0 |
| 0.049658489 | 0 | 0.019356195 | 0 | 0 |
| 0 | 0 | 0 | 0 | 0 |
| 0 | 0 | 0 | 0 | 0 |

| | | | | |
|---|---|---|---|---|
| 0 | 0 | 0 | 0 | 0 |
| 0 | 0 | 0 | 0 | 0 |
| 0 | 0 | 0 | 0 | 0 |
| 0 | 0 | 0 | 0 | 0 |
| 0 | 0 | 0 | 0 | 0 |
| 0 | 0 | 0 | 0 | 0 |
| 0 | 0 | 0 | 0 | 0 |
| 0 | 0 | 0 | 0 | 0 |
| 0 | 0 | 0 | 0 | 0.030575594 |
| 0.001294878 | 0 | 0 | 0 | 0 |
| 0 | 0 | 0 | 0 | 0 |
| 0 | 0 | 0 | 0 | 0 |
| 0 | 0 | 0 | 0 | 0 |
| 0 | 0 | 0 | 0 | 0 |
| 0 | 0 | 0 | 0 | 0 |
| 0 | 0 | 0 | 0.020069015 | 0 |
| 0 | 0 | 0 | 0 | 0 |
| 0 | 0 | 0 | 0 | 0 |
| 0 | 0 | 0 | 0 | 0 |
| 0 | 0 | 0.032063361 | 0 | 3.181703915 |
| 0 | 0.002476851 | 0.004633958 | 0 | 0.003635905 |
| 0 | 0 | 0 | 0 | 0 |
| 0 | 0 | 0 | 0 | 0 |
| 0 | 0 | 0 | 0.00233908 | 0 |
| 0 | 0 | 0 | 0 | 0 |
| 0 | 0 | 0 | 0 | 0 |
| 0 | 0 | 0 | 0 | 0 |
| 0 | 0 | 0 | 0 | 0 |
| 0 | 0 | 0 | 0 | 0 |
| 0 | 0 | 0.038481664 | 0 | 0.048450752 |
| 0.121669533 | 0 | 0.024483752 | 1.183809449 | 0.345395185 |
| 0 | 0 | 0 | 0 | 0 |
| 0 | 0 | 0 | 0 | 0 |
| 0 | 0 | 0 | 0 | 0 |
| 0 | 0 | 0 | 0 | 0 |
| 0 | 0 | 0 | 0 | 0 |
| 0 | 0 | 0 | 0 | 0 |
| 0 | 0 | 0 | 0 | 0 |
| 0.031704385 | 0 | 0.003636438 | 0 | 0.005238911 |
| 0 | 0 | 0 | 0 | 0 |
| 0 | 0 | 0 | 0 | 0 |
| 0.026435344 | 0 | 0.000726809 | 0 | 0 |
| 0 | 0 | 0 | 0 | 0.001092965 |
| 0 | 0 | 0 | 0 | 0 |
| 0 | 0 | 0 | 0 | 0 |
| 0 | 0 | 0 | 0 | 0 |
| 0 | 0.005664575 | 0 | 0 | 0.00203641 |
| 0 | 0 | 0 | 0 | 0 |

| | | | | |
|---|---|---|---|---|
| 0 | 0 | 0 | 0 | 0 |
| 0 | 0 | 0 | 0 | 0 |
| 0 | 0 | 0 | 0 | 0 |
| 0 | 0 | 0 | 0 | 0 |
| 0.172457537 | 0.009068273 | 0.00446471 | 0.114344682 | 0.00456405 |
| 0 | 0 | 0 | 0 | 0 |
| 0 | 0 | 0 | 0 | 0 |
| 0 | 0 | 0 | 0 | 0 |
| 0 | 0 | 0 | 0 | 0 |
| 0 | 0 | 0 | 0 | 0 |
| 0.019793097 | 0 | 0 | 0 | 0 |
| 0.399730023 | 0.001678664 | 0.003967104 | 0.201635018 | 0.001320108 |
| 0 | 0 | 0 | 0 | 0 |
| 0 | 0 | 0 | 0 | 0 |
| 0.002920799 | 0 | 0 | 0 | 0.002414435 |
| 0 | 0 | 0 | 0 | 0 |
| 0.002427613 | 0 | 0 | 0 | 0.00101532 |
| 0 | 0 | 0 | 0 | 0 |
| 0 | 0.294681883 | 0.088052219 | 0.001411829 | 0.021591219 |
| 0 | 0 | 0.001470823 | 0.260975336 | 1.023771053 |
| 0 | 0 | 0 | 0 | 0 |
| 0 | 0 | 0 | 0 | 0 |
| 0 | 0 | 0 | 0 | 0 |
| 0 | 0 | 0 | 0 | 0 |
| 0 | 0 | 0 | 0 | 0 |
| 0 | 0 | 0 | 0 | 0 |
| 0.037867759 | 0.001773283 | 0.008054028 | 0 | 0.000627533 |
| 0 | 0 | 0 | 0 | 0 |
| 0 | 0 | 0 | 0 | 0 |
| 0 | 0 | 0 | 0 | 0 |
| 0.011136227 | 0 | 0 | 0 | 0.015886627 |
| 0 | 0 | 0 | 0 | 0 |
| 0 | 0 | 0 | 0 | 0 |
| 0 | 0 | 0 | 0 | 0 |
| 0 | 0 | 0 | 0 | 0 |
| 0 | 0 | 0 | 0 | 0 |
| 0 | 0 | 0 | 0 | 0 |
| 0.043387581 | 0.004766326 | 0.007196461 | 0.021176208 | 0.019490947 |
| 0 | 0 | 0 | 0 | 0.00360671 |
| 0 | 0 | 0.009926481 | 0 | 0 |
| 0 | 0.002467282 | 0.007809117 | 0 | 0.005987153 |
| 0 | 0 | 0 | 0 | 0 |
| 0 | 0 | 0 | 0 | 0 |
| 0 | 0.012303307 | 0 | 0.114391654 | 0 |
| 0 | 0 | 0 | 0 | 0 |
| 0 | 0 | 0 | 0 | 0 |
| 0 | 0 | 0 | 0 | 0 |
| 0 | 0 | 0 | 0 | 0 |

| | | | | |
|---|---|---|---|---|
| 0 | 0 | 0 | 0 | 0 |
| 0 | 0 | 0 | 0 | 0 |
| 0 | 0 | 0 | 0 | 0 |
| 0 | 0 | 0 | 0.001305915 | 0 |
| 0 | 0 | 0 | 0 | 0 |
| 0.00317549 | 0 | 0 | 0.010317049 | 0 |
| 0.016948115 | 0 | 0 | 0 | 0 |
| 0 | 0.000858227 | 0 | 0 | 0.018222652 |
| 0 | 0 | 0 | 0 | 0 |
| 0 | 0 | 0 | 0 | 0 |
| 0.033064143 | 0 | 0 | 0 | 0 |
| 0 | 0 | 0 | 0 | 0 |
| 0.004231132 | 0 | 0.009651089 | 0.006479968 | 0.147179941 |
| 0 | 0 | 0 | 0 | 0 |
| 0 | 0 | 0 | 0 | 0 |
| 0 | 0 | 0 | 0 | 0 |
| 0 | 0 | 0 | 0 | 0 |
| 0 | 0 | 0 | 0 | 0 |
| 0 | 0 | 0 | 0 | 0 |
| 0 | 0 | 0 | 0 | 0 |
| 0 | 0 | 0 | 0 | 0 |
| 0 | 0 | 0 | 0 | 0 |
| 0.093476135 | 0 | 0 | 0.000731368 | 0 |
| 0.007061471 | 0 | 0.084353539 | 0 | 0.00085546 |
| 0 | 0 | 0 | 0 | 1.098617488 |
| 0 | 0 | 0 | 0 | 0 |
| 0 | 0 | 0 | 0.001397517 | 0.000692623 |
| 0 | 0 | 0 | 0 | 0 |
| 0 | 0 | 0 | 0 | 0 |
| 0 | 0 | 0 | 0 | 0 |
| 0.002300783 | 0 | 0.05516044 | 0.01440667 | 0.001616622 |
| 0 | 0 | 0.001565354 | 0 | 0 |
| 0 | 0 | 0 | 0 | 0 |
| 0 | 0 | 0 | 0 | 0 |
| 0 | 0 | 0 | 0 | 0.02100776 |
| 0 | 0 | 0 | 0 | 0 |
| 0 | 0 | 0 | 0 | 0 |
| 0 | 0 | 0 | 0 | 0 |
| 0 | 0 | 0 | 0.165127017 | 0.001823699 |
| 0 | 0 | 0 | 0 | 0 |
| 0 | 0 | 0 | 0 | 0 |
| 0 | 0 | 0 | 0 | 0 |
| 0 | 0 | 0 | 0 | 0 |
| 0 | 0 | 0 | 0 | 0 |
| 0 | 0 | 0.042882535 | 0 | 0 |
| 0.016270706 | 0.08544039 | 0.023167727 | 0.061715559 | 0.300452683 |
| 0 | 0 | 0 | 0 | 0 |
| 0 | 0 | 0 | 0 | 0 |

| | | | | |
|---|---|---|---|---|
| 0 | 0 | 0 | 0 | 0 |
| 0 | 0 | 0 | 0 | 0 |
| 0.102769918 | 0.085457581 | 0.578245689 | 0.001245298 | 0 |
| 0 | 0 | 0 | 0 | 0 |
| 0 | 0 | 0 | 0 | 0 |
| 0 | 0 | 0 | 0.000931452 | 0 |
| 0 | 0 | 0 | 0 | 0 |
| 0 | 0 | 0 | 0 | 0 |
| 0 | 0 | 0 | 0 | 0 |
| 0 | 0 | 0 | 0 | 0 |
| 0.001895078 | 0 | 0.001215737 | 0 | 0 |
| 0 | 0 | 0 | 0.00194992 | 0 |
| 0 | 0 | 0 | 0 | 0 |
| 0 | 0 | 0 | 0 | 0 |
| 0 | 0 | 0 | 0 | 0 |
| 0 | 0 | 0 | 0 | 0 |
| 0 | 0 | 0 | 0 | 0 |
| 0 | 0 | 0 | 0 | 0 |
| 0 | 0 | 0 | 0 | 0 |
| 0.000837733 | 0.004678134 | 0 | 0 | 0 |
| 0 | 0 | 0 | 0 | 0 |
| 0 | 0 | 0 | 0 | 0 |
| 0 | 0 | 0 | 0 | 0 |
| 0 | 0 | 0 | 0 | 0 |
| 0 | 0 | 0 | 0 | 0 |
| 0 | 0 | 0 | 0 | 0 |
| 0 | 0 | 0 | 0 | 0 |
| 0.005099014 | 0.289632532 | 0 | 0 | 0 |
| 0 | 0 | 0 | 0 | 0 |
| 0 | 0 | 0 | 0 | 0 |
| 0.001439399 | 0.008261279 | 0 | 0.003613701 | 0.00210704 |
| 0 | 0 | 0 | 0 | 0 |
| 0.064762441 | 0.166955899 | 0.046179913 | 0.229785829 | 0.018593305 |
| 0 | 0 | 0.000939122 | 0 | 0.013000234 |
| 0 | 0 | 0 | 0 | 0 |
| 0 | 0 | 0 | 0 | 0 |
| 0 | 0 | 0 | 0 | 0 |
| 0 | 0 | 0 | 0 | 0 |
| 0 | 0 | 0 | 0 | 0 |
| 0 | 0 | 0 | 0 | 0 |
| 0 | 0 | 0 | 0 | 0 |
| 0 | 0.002985937 | 0.229273873 | 0.041422894 | 0.002817784 |
| 0.009841106 | 0 | 0.002302722 | 0.022263867 | 0.043891514 |
| 0 | 0 | 0 | 0 | 0 |
| 0 | 0 | 0 | 0 | 0 |
| 0.091680592 | 0.012015425 | 0 | 0.034675167 | 0.086103845 |
| 0 | 0.00221559 | 0 | 0 | 0 |
| 0 | 0 | 0 | 0 | 0 |

| | | | | |
|---|---|---|---|---|
| 0 | 0 | 0 | 0 | 0 |
| 0 | 0.004627627 | 0.395393518 | 0 | 0 |
| 0 | 0 | 0 | 0.024823502 | 0.007501682 |
| 0 | 0 | 0 | 0 | 0 |
| 0.037440099 | 0.040114129 | 0 | 0 | 0.003593839 |
| 0 | 0.08847325 | 0.007446446 | 0 | 0 |
| 0 | 0 | 0 | 0 | 0 |
| 0 | 0 | 0 | 0 | 0 |
| 0 | 0 | 0 | 0 | 0.077973518 |
| 0 | 0 | 0 | 0 | 0 |
| 0 | 0 | 0 | 0 | 0 |
| 0 | 0 | 0 | 0 | 0 |
| 0 | 0 | 0 | 0 | 0 |
| 0 | 0 | 0 | 0 | 0.003531001 |
| 0 | 0 | 0 | 0 | 0 |
| 0 | 0 | 0 | 0 | 0 |
| 0 | 0 | 0 | 0 | 0 |
| 0 | 0 | 0 | 0 | 0 |
| 0 | 0 | 0 | 0 | 0 |
| 0 | 0 | 0.104093534 | 0 | 0 |
| 0 | 0 | 0 | 0 | 0 |
| 0.000824443 | 0 | 0 | 0 | 0.000965478 |
| 0 | 0 | 0 | 0 | 0 |
| 0 | 0 | 0 | 0 | 0 |
| 0 | 0 | 0 | 0 | 0.003956762 |
| 0 | 0 | 0 | 0.007278406 | 0.00447796 |
| 0 | 0 | 0 | 0 | 0 |
| 0 | 0 | 0.000906503 | 0.006086472 | 0.174233746 |
| 0 | 0 | 0 | 0 | 0 |
| 0 | 0 | 0 | 0 | 0 |
| 0 | 0 | 0 | 0 | 0 |
| 0.001304051 | 0 | 0 | 0 | 0 |
| 0 | 0 | 0 | 0 | 0 |
| 0 | 0 | 0 | 0 | 0 |
| 0 | 0 | 0 | 0 | 0 |
| 0 | 0 | 0 | 0 | 0 |
| 0 | 0 | 0.014061485 | 0 | 0 |
| 0 | 0 | 0.013023101 | 0 | 0 |
| 0 | 0.749875692 | 0.156328271 | 0.197107273 | 0.59426046 |
| 0 | 0 | 0 | 0 | 0 |
| 0 | 0 | 0 | 0 | 0 |
| 0.002074134 | 0 | 0.010644839 | 0.051187527 | 0 |
| 0.005330045 | 0 | 0 | 0 | 0.001997387 |
| 0 | 0 | 0 | 0 | 0 |
| 0 | 0 | 0 | 0 | 0 |
| 0 | 0 | 0 | 0 | 0 |
| 0.020874049 | 0.003237957 | 0 | 0 | 0 |
| 0 | 0 | 0 | 0 | 0 |

| | | | | |
|---|---|---|---|---|
| 0 | 0 | 0 | 0 | 0 |
| 0.162356536 | 0.011235692 | 0.006562757 | 0.007779556 | 0.002409763 |
| 0 | 0 | 0.00107332 | 0 | 0 |
| 0 | 0 | 0 | 0 | 0 |
| 0 | 0 | 0 | 0 | 0 |
| 0 | 0 | 0 | 0 | 0 |
| 0 | 0 | 0 | 0 | 0 |
| 0 | 0 | 0 | 0 | 0 |
| 0 | 0 | 0 | 0 | 0 |
| 0 | 0 | 0 | 0 | 0 |
| 0 | 0 | 0 | 0 | 0 |
| 0 | 0 | 0 | 0 | 0 |
| 0 | 0 | 0.001435327 | 0 | 0.204805544 |
| 0 | 0 | 0 | 0 | 0 |
| 0.019242788 | 0 | 0.001692987 | 0.003940598 | 0 |
| 0 | 0 | 0 | 0 | 0 |
| 0 | 0 | 0 | 0 | 0 |
| 0 | 0 | 0 | 0 | 0 |
| 0 | 0 | 0 | 0 | 0 |
| 0 | 0 | 0 | 0 | 0 |
| 0 | 0 | 0 | 0 | 0 |
| 0 | 0 | 0 | 0.022624488 | 0.233995661 |
| 0.09003341 | 0 | 0.004471627 | 0 | 0.003627874 |
| 0 | 0 | 0 | 0 | 0 |
| 0 | 0 | 0 | 0 | 0 |
| 0 | 0 | 0 | 0 | 0 |
| 0 | 0 | 0 | 0 | 0 |
| 0 | 0 | 0 | 0 | 0 |
| 0 | 0 | 0 | 0 | 0 |
| 0 | 0 | 0 | 0 | 0 |
| 0 | 0 | 0 | 0 | 0 |
| 0 | 0 | 0 | 0 | 0 |
| 0 | 0 | 0 | 0 | 0 |
| 0 | 0 | 0 | 0 | 0 |
| 0 | 0 | 0.018115086 | 0.584629765 | 0.007501565 |
| 0 | 0 | 0 | 0 | 0 |
| 0 | 0 | 0 | 0 | 0 |
| 0 | 0 | 0 | 0 | 0 |
| 0.000754875 | 0 | 0.001106902 | 0.010404804 | 0.480900763 |
| 0 | 0 | 0 | 0 | 0 |
| 0.004512302 | 0.0097992 | 0.02009781 | 0.070102899 | 0.184418759 |
| 0 | 0 | 0 | 0 | 0 |
| 0 | 0 | 0 | 0 | 0 |
| 0 | 0 | 0 | 0 | 0 |
| 0 | 0 | 0 | 0 | 0 |
| 0.001808198 | 1.351138339 | 0.333796103 | 2.721314062 | 0.008470077 |
| 0 | 0 | 0 | 0 | 0 |
| 0 | 0 | 0 | 0 | 0 |

| | | | | |
|---|---|---|---|---|
| 0 | 0 | 0 | 0 | 0 |
| 0 | 0 | 0 | 0 | 0 |
| 0 | 0 | 0 | 0 | 0 |
| 0 | 0 | 0 | 0 | 0 |
| 0 | 0 | 0 | 0 | 0 |
| 0 | 0 | 0 | 0 | 0 |
| 0 | 0 | 0 | 0 | 0 |
| 0 | 0 | 0 | 0 | 0.009481919 |
| 0 | 0 | 0 | 0 | 0 |
| 0 | 0 | 0 | 0.003880099 | 0.004437723 |
| 0.047692033 | 0 | 0 | 0 | 0 |
| 0 | 0 | 0 | 0 | 0 |
| 0 | 0 | 0 | 0.002333828 | 0 |
| 0 | 0 | 0 | 0 | 0 |
| 0 | 0 | 0 | 0 | 0 |
| 0 | 0 | 0 | 0 | 0 |
| 0 | 0 | 0 | 0 | 0 |
| 0 | 0 | 0 | 0 | 0 |
| 0 | 0 | 0 | 0 | 0 |
| 0.529673505 | 0.180552531 | 0.038672899 | 0.646291061 | 0.114790925 |
| 0 | 0 | 0 | 0 | 0 |
| 0 | 0 | 0 | 0 | 0 |
| 0 | 0 | 0 | 0 | 0 |
| 0 | 0 | 0 | 0 | 0 |
| 0 | 0 | 0 | 0 | 0 |
| 0 | 0 | 0 | 0 | 0 |
| 0 | 0 | 0 | 0 | 0 |
| 0 | 0 | 0 | 0 | 0 |
| 0 | 0 | 0 | 0 | 0 |
| 0 | 0 | 0.012919361 | 0.000832739 | 0 |
| 0 | 0 | 0 | 0 | 0 |
| 0 | 0 | 0 | 0 | 0 |
| 0 | 0 | 0 | 0 | 0 |
| 0 | 0 | 0 | 0 | 0 |
| 0 | 0 | 0 | 0 | 0 |
| 0 | 0 | 0.005006118 | 0 | 0.002998538 |
| 0 | 0 | 0 | 0 | 0 |
| 0 | 0 | 0 | 0 | 0 |
| 0 | 0 | 0 | 0 | 0 |
| 0 | 0 | 0 | 0 | 0 |
| 0 | 0 | 0 | 0 | 0 |
| 0 | 0 | 0 | 0 | 0 |
| 0 | 0 | 0 | 0 | 0.002353377 |
| 0 | 0 | 0 | 0 | 0 |
| 0.017546286 | 0.003073986 | 0 | 0.016096159 | 0 |
| 0.012929012 | 0 | 0 | 0 | 0 |
| 0 | 0 | 0 | 0 | 0 |
| 0 | 0 | 0 | 0 | 0 |

| | | | | |
|---|---|---|---|---|
| 0 | 0 | 0 | 0 | 0 |
| 0 | 0 | 0 | 0 | 0 |
| 0 | 0 | 0 | 0 | 0 |
| 0 | 0 | 0 | 0 | 0 |
| 0 | 0 | 0 | 0 | 0 |
| 0.006318334 | 0.001147425 | 0.000508435 | 0.007282672 | 0.00812106 |
| 0 | 0 | 0 | 0 | 0 |
| 0 | 0 | 0 | 0 | 0 |
| 0 | 0 | 0 | 0 | 0 |
| 0 | 0 | 0 | 0 | 0 |
| 0 | 0 | 0 | 0 | 0 |
| 0 | 0 | 0 | 0 | 0 |
| 0 | 0 | 0 | 0 | 0 |
| 0.015368373 | 0 | 0 | 0 | 0.004153242 |
| 0 | 0 | 0 | 0 | 0 |
| 0 | 0 | 0 | 0 | 0 |
| 0 | 0 | 0 | 0 | 0 |
| 0 | 0 | 0 | 0 | 0 |
| 0 | 0 | 0 | 0 | 0 |
| 0 | 0 | 0 | 0 | 0 |
| 0 | 0 | 0 | 0.001266625 | 0 |
| 0 | 0 | 0 | 0 | 0 |
| 0 | 0 | 0 | 0 | 0 |
| 0 | 0 | 0 | 0.021674601 | 0.002685533 |
| 0 | 0 | 0 | 0 | 0 |
| 0 | 0 | 0 | 0 | 0 |
| 0 | 0 | 0 | 0 | 0 |
| 0 | 0 | 0 | 0 | 0.083873692 |
| 0 | 0 | 0 | 0 | 0 |
| 0 | 0 | 0 | 0 | 0 |
| 0 | 0 | 0 | 0 | 0 |
| 0 | 0 | 0 | 0 | 0 |
| 0 | 0 | 0 | 0 | 0 |
| 0 | 0 | 0 | 0 | 0 |
| 0 | 0 | 0 | 0 | 0 |
| 0 | 0 | 0 | 0 | 0 |
| 0 | 0 | 0 | 0 | 0 |
| 0 | 0 | 0 | 0 | 0 |
| 0 | 0 | 0.413156946 | 0 | 0.006878163 |
| 0 | 0 | 0 | 0 | 0 |
| 0 | 0 | 0 | 0 | 0 |
| 0 | 0 | 0 | 0 | 0 |
| 0 | 0 | 0 | 0 | 0 |
| 0 | 0 | 0 | 0 | 0 |
| 0 | 0 | 0 | 0 | 0 |
| 0 | 0 | 0 | 0 | 0 |
| 0 | 0 | 0 | 0 | 0 |
| 0 | 0 | 0 | 0 | 0 |

| | | | | |
|---|---|---|---|---|
| 0 | 0 | 0 | 0 | 0 |
| 0 | 0 | 0 | 0.113945697 | 0 |
| 0 | 0 | 0 | 0 | 0 |
| 0 | 0 | 0 | 0 | 0 |
| 0 | 0 | 0 | 0 | 0 |
| 0 | 0 | 0 | 0 | 0 |
| 0 | 0 | 0 | 0 | 0 |
| 0 | 0 | 0 | 0 | 0 |
| 0 | 0 | 0 | 0 | 0 |
| 0 | 0 | 0 | 0 | 0.001097334 |
| 0 | 0 | 0 | 0 | 0.001851965 |
| 0 | 0 | 0 | 0 | 0 |
| 0.289204891 | 0 | 0 | 0.098689654 | 0.086715626 |
| 0 | 0 | 0 | 0 | 0 |
| 0 | 0 | 0 | 0 | 0 |
| 0 | 0 | 0 | 0 | 0 |
| 0 | 0 | 0 | 0 | 0 |
| 0 | 0 | 0 | 0 | 0 |
| 0 | 0 | 0 | 0 | 0 |
| 0 | 0 | 0 | 0 | 0 |
| 0.134137266 | 0 | 0 | 0 | 0 |
| 0 | 0 | 0.003788647 | 0 | 0 |
| 0 | 0 | 0 | 0 | 0 |
| 1.368571345 | 0 | 0.9569501 | 0.091111152 | 0.945596931 |
| 0 | 0 | 0 | 0 | 0 |
| 0 | 0 | 0 | 0 | 0 |
| 0 | 0 | 0 | 0 | 0 |
| 0 | 0 | 0 | 0 | 0.04255942 |
| 0 | 0 | 0 | 0 | 0 |
| 0 | 0 | 0 | 0 | 0 |
| 0.001575367 | 0 | 0 | 0 | 0 |
| 0 | 0 | 0 | 0 | 0 |
| 0.000723588 | 0 | 0 | 0 | 0 |
| 0 | 0 | 0 | 0 | 0 |

| PHL018-0262_V1 | PHL018-0244_V1 | PHL018-0258_V1 | PHL018-0264_V1 | PHL018-0030_V3 |
|---|---|---|---|---|
| REF | REF | REF | REF | CTRL |
| 0.028126862 | 0.10197194 | 0.11110906 | 0.012985722 | 0.027372903 |
| 0.509422584 | 3.599791855 | 0 | 4.671229371 | 0.875846781 |
| 0.008210908 | 8.067993652 | 0 | 0 | 9.886192779 |
| 0 | 0 | 0 | 0 | 0 |
| 0 | 0 | 0 | 0 | 0 |
| 0 | 0 | 0 | 0 | 0 |
| 0 | 0 | 0 | 0 | 0 |
| 0 | 0 | 0 | 0 | 0 |
| 0 | 0 | 0 | 0 | 0 |
| 0 | 0 | 0 | 0 | 0 |
| 0 | 0 | 0 | 0 | 0 |
| 0 | 0 | 0 | 0 | 0 |
| 0 | 0 | 0 | 0 | 0 |
| 1.353662591 | 17.32448435 | 0 | 4.450002489 | 3.185537692 |
| 0 | 0 | 0 | 0 | 0 |
| 0 | 0 | 0 | 0 | 0 |
| 0 | 0 | 0 | 0 | 0 |
| 0 | 0 | 0 | 0 | 0 |
| 0 | 0 | 0 | 0 | 0 |
| 0 | 0 | 0 | 0 | 0 |
| 0 | 0 | 0 | 0 | 0 |
| 0 | 0 | 0 | 0 | 0 |
| 0 | 0 | 0 | 0 | 0 |
| 0 | 0 | 0 | 0 | 0 |
| 0.005978387 | 0 | 0 | 0 | 0 |
| 0 | 0 | 0 | 0 | 0 |
| 0 | 0 | 0 | 0 | 0 |
| 0 | 0 | 0 | 0 | 0 |
| 0 | 0 | 0 | 0 | 0 |
| 0 | 0 | 0 | 0 | 0 |
| 0 | 0 | 0 | 0 | 0 |
| 0 | 0 | 0 | 0 | 0 |
| 0 | 0 | 0 | 0 | 0 |
| 0 | 0 | 0 | 0 | 0 |
| 0 | 0 | 0 | 0 | 0 |
| 0.005201155 | 2.161816994 | 0 | 0.413692816 | 3.941821847 |
| 0 | 0 | 0 | 0 | 0 |
| 0 | 0 | 0 | 0 | 0 |
| 0 | 0 | 0 | 0 | 0 |
| 0 | 0 | 0 | 0 | 0 |
| 0 | 0 | 0 | 0 | 0 |
| 0.004631515 | 0.001288629 | 0.021195873 | 0.019093729 | 0 |
| 0.513094906 | 0.213282478 | 0.130371575 | 0.001837264 | 0.008030762 |
| 0.003037813 | 0.000704344 | 0.108129664 | 0.026544562 | 0.0011257 |
| 0.002190156 | 0 | 0.000759328 | 0 | 0.00081159 |
| 0 | 0 | 0 | 0 | 0 |

| | | | | |
|---|---|---|---|---|
| 0 | 0 | 0 | 0.001059951 | 0.000876532 |
| 0 | 0.050030884 | 0.052582498 | 0.004375242 | 0.000723625 |
| 0 | 0.00074873 | 0 | 0 | 0.004188234 |
| 1.863526058 | 0.192842821 | 0 | 0.296167863 | 0.568886245 |
| 0 | 0 | 0.001506289 | 0 | 0 |
| 0 | 0 | 0 | 0.050197206 | 0 |
| 0 | 0 | 0.000967553 | 0.127555865 | 0.061014612 |
| 0 | 0 | 0 | 0.001966908 | 0 |
| 0 | 0 | 0 | 0 | 0 |
| 0 | 0 | 0 | 0 | 0 |
| 0 | 0 | 0 | 0 | 0 |
| 0.022156886 | 1.962950698 | 0 | 0.152238723 | 0.629472443 |
| 0 | 0 | 0 | 0.001354201 | 0 |
| 0 | 0.021026933 | 0.001732327 | 0.019927165 | 0.068322455 |
| 0 | 0.016927989 | 0 | 0 | 0 |
| 0 | 0 | 0 | 0 | 0 |
| 0 | 0 | 0 | 0 | 0 |
| 0 | 0 | 0 | 0 | 0 |
| 0 | 0 | 0 | 0 | 0 |
| 0 | 0 | 0 | 0 | 0 |
| 0.005807869 | 0.055272041 | 0.092625147 | 0.159937625 | 0.103500204 |
| 0 | 0.005689492 | 0.002062434 | 0.093298305 | 0.107096334 |
| 0 | 0 | 0 | 0.043602055 | 0 |
| 0 | 0 | 0 | 0 | 0 |
| 0 | 0 | 0 | 0 | 0 |
| 0 | 0 | 0 | 0 | 0 |
| 0 | 0 | 0.001436528 | 0 | 0 |
| 0 | 0 | 0 | 0 | 0 |
| 0 | 0 | 0 | 0 | 0 |
| 0 | 0 | 0 | 0 | 0 |
| 0 | 0 | 0 | 0.00453074 | 0 |
| 0 | 0 | 0 | 0 | 0 |
| 0 | 0 | 0 | 0 | 0 |
| 0 | 0 | 0 | 0 | 0 |
| 0 | 0 | 0 | 0 | 0 |
| 0 | 0.007816408 | 0 | 0 | 0 |
| 0 | 0 | 0 | 0 | 0 |
| 0 | 0 | 0 | 0 | 0 |
| 0 | 0 | 0 | 0 | 0 |
| 0 | 0 | 0 | 0 | 0 |
| 0 | 0 | 0 | 0 | 0 |
| 0 | 0 | 0 | 0 | 0 |
| 0 | 0 | 0 | 0 | 0 |
| 0 | 0 | 0 | 0 | 0 |
| 0 | 0 | 0 | 0 | 0 |
| 0 | 0 | 0 | 0 | 0 |
| 0 | 0 | 0 | 0.005492786 | 0 |
| 0 | 0.001916076 | 0 | 0 | 0 |

| | | | | |
|---|---|---|---|---|
| 0 | 0 | 0 | 0 | 0 |
| 0 | 0 | 0 | 0 | 0 |
| 0 | 0 | 0 | 0 | 0 |
| 0 | 0 | 0 | 0 | 0 |
| 0 | 0.288089453 | 0 | 0.059925904 | 0 |
| 0 | 0 | 0 | 0 | 0 |
| 0 | 0 | 0 | 0 | 0 |
| 0 | 0 | 0 | 0 | 0 |
| 0 | 0 | 0.001495128 | 0.000858858 | 0 |
| 0 | 0 | 0 | 0 | 0 |
| 0 | 0 | 0 | 0 | 0 |
| 0 | 0 | 0.000771473 | 0 | 0 |
| 0 | 0 | 0 | 0 | 0 |
| 0 | 0 | 0 | 0 | 0 |
| 0 | 0 | 0 | 0 | 0 |
| 0 | 0.00107669 | 0 | 0 | 0 |
| 0 | 0 | 0 | 0 | 0 |
| 0 | 0 | 0 | 0 | 0 |
| 0.016832655 | 0 | 0 | 0.02440317 | 0 |
| 0 | 0 | 0 | 0 | 0 |
| 0 | 0 | 0 | 0 | 0 |
| 0 | 0 | 0 | 0 | 0 |
| 0 | 0 | 0 | 0 | 0 |
| 0 | 0 | 0 | 0 | 0 |
| 0 | 0 | 0 | 0 | 0 |
| 0 | 0 | 0 | 0 | 0 |
| 0 | 0 | 0.00316474 | 0 | 0 |
| 0 | 0 | 0 | 0 | 0 |
| 0 | 0 | 0 | 0 | 0 |
| 0 | 0 | 0 | 0 | 0 |
| 0 | 0.009098387 | 0 | 0 | 0.341103583 |
| 0 | 0.004368051 | 0 | 0 | 0.005688325 |
| 0 | 0 | 0 | 0 | 0 |
| 0 | 0 | 0 | 0 | 0 |
| 0 | 0 | 0 | 0 | 0 |
| 0 | 0 | 0 | 0 | 0 |
| 0 | 0 | 0 | 0 | 0 |
| 0 | 0 | 0 | 0 | 0 |
| 0.002974022 | 0 | 0 | 0 | 0 |
| 0 | 0 | 0 | 0 | 0 |
| 0 | 0 | 0 | 0 | 0 |
| 0 | 0 | 0 | 0 | 0 |
| 0 | 0 | 0 | 0 | 0 |
| 0 | 0 | 0 | 0 | 0 |
| 0 | 0 | 0 | 0 | 0 |
| 0 | 0 | 0 | 0 | 0 |
| 0 | 0 | 0 | 0 | 0 |
| 0 | 0 | 0 | 0 | 0 |

| | | | | |
|---|---|---|---|---|
| 0 | 0 | 0 | 0 | 0 |
| 0 | 0 | 0 | 0 | 0 |
| 0 | 0 | 0 | 0 | 0 |
| 0 | 0 | 0 | 0 | 0 |
| 0.263371432 | 0 | 0 | 0.021952631 | 0.087768496 |
| 0 | 0 | 0 | 0 | 0 |
| 0 | 0 | 0 | 0 | 0 |
| 0 | 0 | 0 | 0 | 0 |
| 0 | 0 | 0 | 0 | 0 |
| 0 | 0 | 0 | 0 | 0 |
| 0 | 0 | 0 | 0 | 0 |
| 0 | 0 | 0 | 0 | 0 |
| 0 | 0 | 0 | 0 | 0 |
| 0 | 0 | 0 | 0 | 0 |
| 0 | 0 | 0 | 0 | 0 |
| 0.014536101 | 0 | 0 | 0 | 0.326037724 |
| 0 | 0 | 0 | 0 | 0 |
| 0 | 0 | 0 | 0 | 0 |
| 0 | 0 | 0 | 0 | 0 |
| 0 | 0 | 0 | 0 | 0 |
| 0 | 0 | 0 | 0 | 0 |
| 0 | 0 | 0 | 0 | 0 |
| 0 | 0 | 0 | 0 | 0 |
| 0 | 0 | 0 | 0 | 0 |
| 0 | 0 | 0 | 0 | 0 |
| 0 | 0 | 0 | 0 | 0 |
| 0.003284424 | 0 | 0 | 0 | 0 |
| 0 | 0 | 0 | 0 | 0 |
| 0 | 0 | 0 | 0 | 0 |
| 0 | 0 | 0 | 0 | 0 |
| 0 | 0 | 0 | 0 | 0 |
| 0 | 0 | 0.014699922 | 0.001202496 | 0 |
| 0 | 0 | 0 | 0 | 0 |
| 0 | 0 | 0 | 0 | 0 |
| 0 | 0 | 0 | 0 | 0 |
| 0 | 0 | 0 | 0 | 0 |
| 0 | 0 | 0 | 0 | 0 |
| 0 | 0 | 0 | 0 | 0 |
| 0 | 0 | 0 | 0 | 0 |
| 0 | 0 | 0 | 0 | 0 |
| 0 | 0 | 0 | 0 | 0 |
| 0 | 0 | 0 | 0 | 0 |
| 0 | 0 | 0 | 0 | 0 |
| 0 | 0 | 0 | 0 | 0 |
| 0 | 0 | 0 | 0 | 0 |
| 0 | 0 | 0 | 0 | 0 |
| 0 | 0 | 0 | 0 | 0 |
| 0 | 0 | 0 | 0 | 0 |

| | | | | |
|---|---|---|---|---|
| 0.027789603 | 0 | 0 | 0 | 0 |
| 0 | 0 | 0 | 0 | 0 |
| 0 | 0 | 0 | 0 | 0 |
| 0 | 0 | 0 | 0 | 0 |
| 0 | 0 | 0 | 0 | 0 |
| 0 | 0 | 0 | 0 | 0 |
| 0 | 0 | 0 | 0 | 0 |
| 0 | 0 | 0 | 0 | 0 |
| 0 | 0 | 0 | 0 | 0 |
| 0 | 0 | 0 | 0 | 0 |
| 0 | 0 | 0 | 0 | 0 |
| 0 | 0 | 0 | 0 | 0 |
| 0 | 0 | 0 | 0.000690402 | 0 |
| 0 | 0 | 0 | 0 | 0 |
| 0 | 0 | 0 | 0 | 0 |
| 0 | 0 | 0 | 0 | 0 |
| 0 | 0 | 0 | 0 | 0 |
| 0 | 0 | 0 | 0 | 0 |
| 0 | 0 | 0 | 0 | 0 |
| 0 | 0 | 0 | 0 | 0 |
| 0.001920511 | 0.035400357 | 0.042613886 | 0.146730738 | 0.017791736 |
| 0 | 0 | 0 | 0 | 0 |
| 0 | 0 | 0 | 0 | 0 |
| 0 | 0 | 0 | 0 | 0 |
| 0 | 0 | 0 | 0 | 0 |
| 0 | 0 | 0 | 0 | 0 |
| 0 | 0.002139431 | 0.005622675 | 0.001754157 | 0.006424122 |
| 0 | 0 | 0 | 0 | 0 |
| 0 | 0 | 0 | 0 | 0 |
| 0 | 0 | 0.000622305 | 0 | 0 |
| 0 | 0 | 0.016621285 | 0 | 0 |
| 0 | 0 | 0 | 0 | 0 |
| 0 | 0 | 0 | 0 | 0 |
| 0 | 0 | 0 | 0.006311276 | 0.004349281 |
| 0.004998348 | 0.069969261 | 0.008231413 | 0.003639651 | 0.006251176 |
| 0 | 0 | 0 | 0 | 0 |
| 0 | 0 | 0 | 0 | 0 |
| 0 | 0 | 0 | 0 | 0 |
| 0 | 0 | 0 | 0 | 0 |
| 0 | 0 | 0 | 0 | 0 |
| 0 | 0 | 0 | 0 | 0 |
| 0 | 0 | 0 | 0 | 0 |
| 0 | 0 | 0 | 0 | 0 |
| 0 | 0 | 0 | 0 | 0 |
| 0 | 0 | 0 | 0 | 0 |
| 0 | 0 | 0 | 0 | 0 |
| 0.001014728 | 0 | 0 | 0 | 0.0025068 |
| 0 | 0.002322763 | 0.003859163 | 0 | 0 |

| | | | | |
|---|---|---|---|---|
| 0 | 0 | 0 | 0 | 0 |
| 0 | 0 | 0 | 0 | 0 |
| 0 | 0 | 0 | 0 | 0 |
| 0 | 0 | 0 | 0 | 0 |
| 0 | 0 | 0 | 0 | 0 |
| 0 | 0 | 0 | 0 | 0 |
| 0 | 0 | 0 | 0 | 0 |
| 0 | 0 | 0 | 0 | 0 |
| 0 | 0 | 0 | 0 | 0 |
| 0 | 0 | 0 | 0 | 0 |
| 0 | 0 | 0 | 0 | 0 |
| 0 | 0 | 0 | 0 | 0 |
| 0 | 0 | 0 | 0 | 0 |
| 0 | 0 | 0 | 0 | 0 |
| 0 | 0 | 0 | 0 | 0 |
| 0 | 0 | 0 | 0 | 0 |
| 0 | 0 | 0 | 0 | 0 |
| 0 | 0 | 0 | 0 | 0 |
| 0.001584838 | 0 | 0.000470969 | 0 | 0 |
| 0.001556702 | 0 | 0 | 0 | 0 |
| 0 | 0.007898284 | 0.063220201 | 0 | 0 |
| 0 | 0 | 0 | 0 | 0 |
| 0 | 0 | 0 | 0 | 0 |
| 0 | 0 | 0 | 0 | 0 |
| 0 | 0 | 0.007361189 | 0.010703506 | 0.002458698 |
| 0.00676048 | 0 | 0 | 0 | 0 |
| 0 | 0 | 0 | 0 | 0 |
| 0 | 0 | 0 | 0 | 0 |
| 0 | 0 | 0 | 0 | 0 |
| 0 | 0 | 0 | 0 | 0 |
| 0 | 0.082331343 | 0.107785728 | 0.04291585 | 0.189459077 |
| 0 | 0 | 0 | 0 | 0 |
| 0.029007832 | 0 | 0 | 0 | 0 |
| 0 | 0.320881002 | 0 | 0.004840511 | 0 |
| 0 | 0 | 0 | 0 | 0 |
| 0 | 0 | 0 | 0 | 0 |
| 0 | 0 | 0 | 0 | 0 |
| 0 | 0 | 0 | 0 | 0 |
| 0 | 0 | 0 | 0 | 0 |
| 0.000981157 | 0 | 0 | 0 | 0 |
| 0 | 0 | 0 | 0 | 0 |
| 0.044556355 | 0.018804349 | 0.008678486 | 0.004935398 | 0.012800595 |
| 0 | 0 | 0 | 0 | 0 |
| 0 | 0 | 0 | 0 | 0 |
| 0 | 0 | 0 | 0 | 0 |
| 0 | 0 | 0 | 0 | 0 |
| 0 | 0 | 0 | 0 | 0 |
| 0 | 0 | 0 | 0 | 0 |

| | | | | |
|---|---|---|---|---|
| 0 | 0 | 0 | 0 | 0 |
| 0 | 0 | 0 | 0 | 0 |
| 0.003143137 | 0 | 0.125318567 | 0.027887419 | 0 |
| 0 | 0 | 0 | 0 | 0 |
| 0 | 0 | 0 | 0 | 0 |
| 0 | 0 | 0 | 0 | 0 |
| 0 | 0 | 0 | 0 | 0 |
| 0 | 0 | 0 | 0 | 0 |
| 0 | 0 | 0 | 0 | 0 |
| 0 | 0 | 0 | 0 | 0 |
| 0 | 0 | 0 | 0 | 0 |
| 0 | 0 | 0 | 0 | 0 |
| 0 | 0 | 0 | 0 | 0 |
| 0 | 0 | 0.001445214 | 0 | 0 |
| 0 | 0 | 0.00516895 | 0.020556281 | 0.002337377 |
| 0 | 0 | 0 | 0 | 0 |
| 0.000980045 | 0 | 0.009287376 | 0.004684408 | 0.003389568 |
| 0 | 0 | 0 | 0 | 0 |
| 0 | 0 | 0 | 0 | 0 |
| 0 | 0 | 0 | 0 | 0 |
| 0 | 0 | 0 | 0 | 0 |
| 0 | 0.00389739 | 0.003067262 | 0 | 0 |
| 0 | 0 | 0 | 0 | 0 |
| 0 | 0.002225942 | 0.176567378 | 0 | 0 |
| 0 | 0 | 0 | 0 | 0 |
| 0 | 0 | 0 | 0 | 0 |
| 0 | 0 | 0 | 0 | 0 |
| 0 | 0 | 0 | 0 | 0 |
| 0 | 0 | 0 | 0 | 0 |
| 0 | 0 | 0.001183153 | 0 | 0 |
| 0 | 0 | 0 | 0 | 0 |
| 0 | 0 | 0 | 0 | 0 |
| 0 | 0 | 0 | 0 | 0 |
| 0 | 0 | 0 | 0 | 0 |
| 0 | 0 | 0 | 0 | 0 |
| 0 | 0 | 0 | 0.016364501 | 0 |
| 0 | 0 | 0 | 0 | 0 |
| 0.017414055 | 0 | 0.007707395 | 0.003320564 | 0 |
| 0 | 0 | 0 | 0 | 0 |
| 0 | 0 | 0 | 0 | 0 |
| 0 | 0.00409158 | 0 | 0 | 0 |
| 0 | 0 | 0 | 0 | 0 |
| 0 | 0 | 0 | 0 | 0 |
| 0 | 0 | 0 | 0 | 0 |
| 0 | 0 | 0 | 0 | 0 |
| 0 | 0 | 0 | 0 | 0 |
| 0 | 0 | 0 | 0 | 0 |
| 0 | 0.001872976 | 0.004356608 | 0 | 0 |

| | | | | |
|---|---|---|---|---|
| 0 | 0 | 1.264950112 | 0 | 0 |
| 0 | 0 | 0 | 0 | 0 |
| 0 | 0 | 0.001070906 | 0 | 0 |
| 0 | 0 | 0 | 0 | 0 |
| 0 | 0 | 0 | 0.001114055 | 0.013358451 |
| 0 | 0 | 0 | 0 | 0 |
| 0 | 0 | 0 | 0 | 0 |
| 0.006534152 | 0 | 0 | 0.044114997 | 0.022276066 |
| 0 | 0 | 0 | 0 | 0 |
| 0.087104604 | 0.105560387 | 0.430260726 | 0.023741121 | 0.035272535 |
| 0 | 0 | 0 | 0 | 0 |
| 0 | 0 | 0.002391057 | 0.021323802 | 0 |
| 0 | 0.095700074 | 0.017955649 | 0.019691135 | 0 |
| 0 | 0.001934232 | 0.005509091 | 0.006052355 | 0 |
| 0 | 0 | 0 | 0 | 0 |
| 0 | 0 | 0 | 0 | 0 |
| 0 | 0 | 0 | 0 | 0 |
| 0 | 0 | 0 | 0 | 0 |
| 0 | 0 | 0.368540748 | 0 | 0 |
| 0 | 0 | 0.03099673 | 0 | 0 |
| 0 | 0 | 0 | 0 | 0 |
| 0 | 0 | 0 | 0 | 0 |
| 0 | 0.001689641 | 0.001497205 | 0 | 0 |
| 0.000869414 | 0 | 0 | 0 | 0 |
| 0 | 0 | 0 | 0 | 0 |
| 0.001827948 | 0.00148339 | 0 | 0 | 0 |
| 0 | 0 | 0 | 0 | 0 |
| 0 | 0.008712895 | 1.065523722 | 0 | 0 |
| 0 | 0 | 0 | 0 | 0 |
| 0 | 0 | 0.003042896 | 0 | 0 |
| 0.004903718 | 0 | 0 | 0 | 0 |
| 0 | 0 | 0 | 0 | 0 |
| 0 | 0 | 0 | 0 | 0 |
| 0 | 0 | 0.005525525 | 0 | 0 |
| 0 | 0 | 0 | 0 | 0 |
| 0 | 0 | 0 | 0 | 0 |
| 0 | 0.107620794 | 0 | 0 | 0.002047645 |
| 0 | 0 | 0 | 0 | 0 |
| 0 | 0 | 0 | 0 | 0 |
| 0 | 0 | 0.266172797 | 0 | 0 |
| 0 | 0 | 0 | 0 | 0 |
| 0 | 0 | 0 | 0 | 0 |
| 0 | 0 | 0 | 0 | 0 |
| 0 | 0.164066249 | 0 | 0 | 0 |
| 0 | 0 | 0 | 0 | 0 |
| 0.004903796 | 0 | 0 | 0 | 0 |
| 0 | 0.016338887 | 0.001050824 | 0 | 0.001123149 |
| 0 | 0 | 0 | 0 | 0 |

| | | | | |
|---|---|---|---|---|
| 0 | 0 | 0 | 0 | 0 |
| 0 | 0 | 0 | 0 | 0.004079391 |
| 0 | 0 | 0 | 0 | 0 |
| 0 | 0 | 0 | 0 | 0 |
| 0 | 0.068504204 | 0.004414177 | 0 | 0 |
| 0 | 0 | 0.026691762 | 0 | 0 |
| 0 | 0 | 0 | 0 | 0 |
| 0 | 0 | 0 | 0 | 0 |
| 0 | 0 | 0 | 0 | 0 |
| 0 | 0 | 0 | 0 | 0 |
| 0 | 0 | 0.029038077 | 0 | 0.005899531 |
| 0 | 0 | 0 | 0 | 0 |
| 0 | 0 | 0 | 0 | 0 |
| 0 | 0 | 0.003174149 | 0.001758232 | 0.011631819 |
| 0.003639243 | 0 | 0.475097489 | 0 | 0 |
| 0 | 0 | 0 | 0 | 0 |
| 0 | 0 | 0 | 0 | 0 |
| 0 | 0 | 0 | 0 | 0 |
| 0 | 0 | 0 | 0 | 0 |
| 0.00137246 | 0 | 0 | 0 | 0 |
| 0 | 0 | 0 | 0.002841634 | 0 |
| 0.353045405 | 0 | 0 | 0.479739624 | 0.369598811 |
| 0 | 0.026546625 | 0 | 0.025583324 | 0 |
| 0 | 0 | 0 | 0 | 0 |
| 0 | 0 | 0 | 0.003266788 | 0 |
| 0 | 0 | 0 | 0 | 0 |
| 0 | 0.00197281 | 0 | 0 | 0 |
| 0 | 0 | 0 | 0 | 0 |
| 0 | 0 | 0 | 0 | 0 |
| 0.062785331 | 2.683994436 | 0.177132553 | 0.405908863 | 0.11188938 |
| 0 | 0 | 0 | 0.00119282 | 0 |
| 0 | 0 | 0 | 0 | 0 |
| 0 | 0 | 0.019659464 | 0 | 0 |
| 0 | 0 | 0 | 0 | 0 |
| 0 | 0 | 0 | 0 | 0 |
| 0 | 0 | 0 | 0 | 0 |
| 1.25429125 | 0 | 0 | 0 | 0 |
| 0 | 0 | 0 | 0 | 0 |
| 0.056176777 | 0 | 0 | 0 | 0 |
| 0 | 0 | 2.676297479 | 0 | 0 |
| 0 | 0.006918165 | 0.008946843 | 0.025054599 | 0.004980532 |
| 0.001155937 | 0.310199552 | 0 | 4.655406664 | 0.136728282 |
| 0 | 0 | 0 | 0 | 0 |
| 0 | 0 | 0 | 0 | 0 |
| 0 | 0.001476442 | 0 | 0 | 0 |
| 0 | 0 | 0 | 0 | 0 |
| 0 | 0.000981231 | 0.001956324 | 0 | 0 |
| 0 | 0 | 0 | 0 | 0 |

| | | | | |
|---|---|---|---|---|
| 0.001392427 | 0.001937079 | 0 | 0 | 0 |
| 0.0022243 | 0 | 0.001233865 | 0 | 0 |
| 0 | 0.002969346 | 0.002631163 | 0.002975646 | 0.002460724 |
| 0 | 0 | 0 | 0 | 0 |
| 0 | 0 | 0 | 0.004390577 | 0.046163122 |
| 0 | 0 | 0 | 0.018464368 | 0 |
| 0 | 0 | 0 | 0 | 0 |
| 0 | 0 | 0 | 0 | 0 |
| 0 | 0.046000461 | 0.001212068 | 0 | 0 |
| 0 | 0 | 0 | 0 | 0 |
| 0 | 0 | 0 | 0 | 0 |
| 0 | 0 | 0.019335749 | 0 | 0 |
| 0 | 0 | 0 | 0 | 0 |
| 0 | 0 | 0 | 0 | 0.003500964 |
| 0 | 0 | 0 | 0 | 0 |
| 0.025102937 | 0 | 0.021331366 | 0.120207264 | 0.013862111 |
| 0.014184779 | 0.000758969 | 0 | 0.006845215 | 0.000808669 |
| 0 | 0.008068531 | 0.159946625 | 0 | 0 |
| 0 | 0 | 0.003907528 | 0 | 0.001461764 |
| 0 | 0 | 0 | 0.378526315 | 0 |
| 0 | 0 | 0 | 0.004929668 | 0 |
| 0 | 0 | 0 | 0 | 0.000938014 |
| 0.361505144 | 0 | 0 | 0 | 0 |
| 0.001806717 | 0 | 0 | 0 | 0.089490031 |
| 0 | 0 | 0 | 0 | 0 |
| 0.006468212 | 0 | 0.002740872 | 0 | 0 |
| 0 | 0.001445276 | 0.005522895 | 0.004034669 | 0 |
| 0 | 0 | 0 | 0 | 0 |
| 0 | 0 | 0 | 0 | 0 |
| 0 | 0 | 0.00678038 | 0 | 0 |
| 0 | 0.064482199 | 0 | 0.014633689 | 0 |
| 0 | 0.004915997 | 0.012796062 | 0.005278315 | 0 |
| 0.015701534 | 0.016902504 | 0.008295204 | 0.003015402 | 1.675144855 |
| 0 | 0 | 0.004829433 | 0 | 0 |
| 0 | 0.22129938 | 0 | 0.004797885 | 0 |
| 0 | 0 | 0 | 0 | 0 |
| 0 | 0.000940371 | 0.001171787 | 0.001514517 | 0 |
| 0.024317262 | 0.064348701 | 0 | 0.009001612 | 0.009011064 |
| 0.021206225 | 0 | 0 | 0.447326874 | 0.003201502 |
| 0 | 0 | 0 | 0 | 0 |
| 0 | 0 | 0 | 0 | 0 |
| 0 | 0 | 0 | 0 | 0 |
| 0 | 0 | 0 | 0 | 0 |
| 0 | 0 | 0.0204592 | 0.093013535 | 0.389789006 |
| 0 | 0 | 15.23771318 | 0 | 0 |
| 0 | 0.00420872 | 0 | 0.004389799 | 0 |
| 0 | 0 | 0 | 0 | 0 |
| 0 | 0 | 0 | 0 | 0 |

| | | | | |
|---|---|---|---|---|
| 0 | 0 | 0 | 0 | 0 |
| 0 | 0.003256807 | 0.776191858 | 0.743374336 | 0.464455875 |
| 0 | 0 | 0 | 0 | 0 |
| 0 | 0.001820719 | 0 | 0 | 0 |
| 0 | 0.000983102 | 0 | 0 | 0 |
| 0 | 0 | 0 | 0 | 0 |
| 0 | 0 | 0.012014185 | 0 | 0 |
| 0 | 0 | 0 | 0 | 0.002050294 |
| 0 | 0 | 0 | 0 | 0 |
| 0 | 0 | 0 | 0.006098604 | 0 |
| 0 | 0 | 0.002969346 | 0 | 0.017852146 |
| 0.004385286 | 0.009150907 | 0.652381403 | 0.096467025 | 0.009750135 |
| 0 | 0 | 0 | 0.009075282 | 0 |
| 0 | 0 | 0 | 0 | 0 |
| 0 | 0.01495219 | 0 | 0.006880369 | 0.00091036 |
| 0 | 0 | 0 | 0 | 0 |
| 0 | 0 | 0.036729003 | 0.074347198 | 0.017032105 |
| 0.002182907 | 0.021257302 | 0.033804386 | 0.008477487 | 0.012133551 |
| 0 | 0 | 0 | 0 | 0 |
| 0 | 0.006408713 | 0.039642492 | 0 | 0.005602769 |
| 0.032659266 | 0.008041422 | 0.007815862 | 0.426869399 | 0.019063797 |
| 0 | 0 | 0.000997496 | 0 | 0 |
| 0 | 0 | 0 | 0 | 0 |
| 0 | 0 | 0 | 0 | 0 |
| 0 | 0 | 0 | 0.164072829 | 0 |
| 0 | 0 | 0 | 0 | 0 |
| 0 | 0.111518862 | 0.138270196 | 0.064939759 | 0 |
| 0.034097607 | 0.189953623 | 0.003834049 | 0.011562721 | 0.021172654 |
| 0.011679666 | 0 | 0 | 0 | 0 |
| 0 | 0 | 0.001284656 | 0 | 0 |
| 0 | 0 | 0 | 0 | 0.020764015 |
| 0 | 0 | 0 | 0 | 0 |
| 0 | 0 | 0 | 0 | 0 |
| 0 | 0 | 0 | 0 | 0 |
| 0 | 0 | 0 | 0.005997682 | 0 |
| 0.026514563 | 0.116805075 | 0.152128656 | 0.197089959 | 0.062419564 |
| 0 | 0 | 0 | 0 | 0 |
| 0 | 0 | 0 | 0 | 0 |
| 0 | 0 | 0.006831831 | 0.010867741 | 0 |
| 0 | 0 | 0 | 0 | 0 |
| 0 | 0 | 14.6938887 | 0 | 0 |
| 0 | 0 | 0 | 0 | 0 |
| 0.003913038 | 0 | 0 | 0 | 0.118390283 |
| 0 | 0.000772901 | 0.01433099 | 0.009958383 | 0 |
| 0 | 0.07250047 | 0 | 0 | 0 |
| 0.215835966 | 0 | 0 | 0 | 0 |
| 0 | 0 | 0 | 0 | 0 |
| 0.003233005 | 0 | 0 | 0 | 0 |

| | | | | |
|---|---|---|---|---|
| 0 | 0 | 0 | 0 | 0 |
| 0 | 0 | 0 | 0 | 0 |
| 0 | 0 | 0 | 0 | 0 |
| 0.041553232 | 0 | 0 | 0.047331381 | 0.007748705 |
| 0 | 0 | 0 | 0 | 0 |
| 0 | 0 | 0 | 0 | 0 |
| 0 | 0 | 0 | 0 | 0 |
| 0.018227165 | 0 | 0 | 0 | 0 |
| 4.772179967 | 0.035240102 | 0 | 0.07705849 | 0.367753189 |
| 0 | 0.000999971 | 0 | 0.051750914 | 0.047945332 |
| 0 | 0 | 0 | 0 | 0 |
| 0.162588963 | 0 | 0.004096373 | 0.004373718 | 0.016561437 |
| 0 | 0 | 0 | 0 | 0 |
| 0 | 0 | 0 | 0 | 0 |
| 0.151125392 | 0 | 0 | 0.001519665 | 0.18441975 |
| 0 | 0 | 0 | 0 | 0 |
| 0 | 0 | 0 | 0 | 0 |
| 0 | 0 | 0 | 0 | 0 |
| 0 | 0 | 0 | 0 | 0 |
| 0 | 0 | 0 | 0 | 0 |
| 0 | 0 | 0 | 0 | 0 |
| 0 | 0 | 0 | 0 | 0 |
| 0 | 0 | 0 | 0 | 0 |
| 0 | 0.494780768 | 0.032419741 | 0.574185928 | 0.069302144 |
| 0 | 0 | 0 | 0 | 0 |
| 0.018092421 | 0 | 0 | 6.021839806 | 0 |
| 0 | 0 | 0 | 0 | 0 |
| 0.656144179 | 0 | 0 | 2.134495296 | 1.886440695 |
| 0 | 0 | 0 | 0 | 0 |
| 0 | 0 | 0 | 0 | 0 |
| 0 | 0 | 0 | 0 | 0 |
| 0.000705809 | 0 | 0 | 0 | 0 |
| 0.003990944 | 0.004362297 | 0 | 0 | 0 |
| 0 | 0 | 0 | 0 | 0 |
| 0.000922204 | 0 | 0 | 0 | 0 |
| 0.481670193 | 0 | 0 | 0 | 0 |
| 0.118126776 | 0 | 0 | 1.342689964 | 4.155428388 |
| 0 | 0 | 0 | 0 | 0 |
| 0 | 0 | 0 | 0 | 0 |
| 0 | 0 | 0 | 0 | 0 |
| 0 | 0.082269251 | 0.004954886 | 0.285535278 | 0.008948267 |
| 0 | 0 | 0 | 0 | 0 |
| 10.53143498 | 9.328661903 | 0 | 2.316678441 | 23.37501775 |
| 0 | 0 | 0 | 0.00057712 | 0 |
| 0 | 0 | 0 | 0 | 0 |
| 0 | 0 | 0 | 0 | 0 |
| 0 | 0 | 0 | 0 | 0 |
| 0.0035545 | 0 | 0.19327302 | 0 | 0 |

| | | | | |
|---|---|---|---|---|
| 0 | 0 | 0 | 0 | 0 |
| 0.069892242 | 0 | 0 | 0 | 0 |
| 0.011074017 | 0.14832878 | 1.684398674 | 0.004580606 | 0.086070698 |
| 0 | 0 | 0 | 0 | 0 |
| 0 | 0 | 0 | 0 | 0 |
| 0.0085 | 0 | 0 | 0.003007021 | 0.004641782 |
| 0 | 0 | 0 | 0 | 0 |
| 0 | 0 | 0 | 0 | 0 |
| 0 | 0 | 0 | 0 | 0 |
| 0 | 0 | 0 | 0 | 0 |
| 0.011709084 | 0.019779655 | 0.58592748 | 0.001998819 | 0.075828268 |
| 0.849962564 | 1.477599843 | 1.779544653 | 0.289718749 | 0.617318751 |
| 0 | 0 | 0 | 0 | 0 |
| 0.02286681 | 0 | 0.011057378 | 0 | 0.001783912 |
| 0 | 0.290266491 | 0.108180633 | 0.066935996 | 0.288890045 |
| 0 | 0 | 0 | 0 | 0 |
| 0 | 0 | 0 | 0 | 0 |
| 0 | 0 | 0 | 0 | 0 |
| 0 | 0 | 0 | 0 | 0 |
| 0 | 0.00063895 | 0 | 0.005762753 | 0.004935732 |
| 0.002803982 | 0.0081266 | 0.263450363 | 0.043557922 | 0.020088309 |
| 0.20312374 | 2.781577327 | 0 | 0 | 0.977506842 |
| 0.755243658 | 10.54128915 | 9.831594068 | 1.11438116 | 5.025465101 |
| 0 | 0 | 0 | 0 | 0 |
| 0 | 0 | 0 | 0 | 0 |
| 0.899643085 | 0.005842863 | 0 | 11.60796818 | 0.004150314 |
| 0 | 0 | 0 | 0 | 0 |
| 0 | 0 | 0 | 0 | 0.001083711 |
| 0 | 0 | 0 | 0 | 0 |
| 0 | 0 | 0 | 0 | 0 |
| 0 | 0 | 0 | 0 | 0 |
| 0.063090979 | 0 | 0 | 0.341431361 | 0.250631015 |
| 0 | 0 | 0.16458639 | 0.077238074 | 0 |
| 0.017904117 | 0 | 0 | 0.000782724 | 0.038674826 |
| 0.002078739 | 0.004732108 | 0.005896632 | 0.002032349 | 0.005462145 |
| 0.262238223 | 0.011377846 | 0.284096512 | 0.003955791 | 0.037330841 |
| 0.007775124 | 0 | 0 | 0 | 0.006691747 |
| 0 | 0 | 0 | 0 | 0 |
| 0.136725005 | 0 | 0 | 0.012253433 | 0.096826741 |
| 0 | 0 | 0 | 0 | 0 |
| 0 | 0 | 0 | 0 | 0 |
| 0 | 0 | 0 | 0 | 0 |
| 0.001779743 | 0 | 0 | 0 | 0 |
| 0 | 0 | 0 | 0 | 0 |
| 0.093861166 | 0.022243101 | 0.051738223 | 0 | 0 |
| 0.001373135 | 0 | 0 | 0 | 0.006105986 |
| 0 | 0 | 0 | 0 | 0 |
| 4.276013771 | 0.068071896 | 0.686762448 | 0.01632497 | 0.095558871 |

| | | | | |
|---|---|---|---|---|
| 0 | 0 | 0 | 0 | 0 |
| 0.009477179 | 0 | 0 | 0 | 0 |
| 0 | 0 | 0 | 0 | 0 |
| 0.194568024 | 0 | 0 | 0 | 0.001309202 |
| 0 | 0 | 0 | 0 | 0 |
| 0 | 0 | 0 | 0.005193453 | 0 |
| 0 | 0 | 0 | 0 | 0 |
| 0 | 0 | 0.083729404 | 0 | 0.036313177 |
| 0 | 0 | 0 | 0 | 0 |
| 0 | 0 | 0 | 0 | 0 |
| 0 | 0.000586496 | 0.007795488 | 0 | 0.003957711 |
| 0 | 0 | 0 | 0 | 0 |
| 0 | 0 | 0 | 0.000882193 | 0.002735752 |
| 0 | 0 | 0 | 0 | 0.021685361 |
| 0 | 0 | 0 | 0 | 0 |
| 0 | 0 | 0 | 0 | 0 |
| 0 | 0 | 0 | 0 | 0 |
| 0.014272539 | 0 | 0 | 0 | 0.001213838 |
| 0 | 0 | 0 | 0 | 0 |
| 0.003376122 | 0 | 0 | 0 | 0 |
| 0 | 0 | 0 | 0 | 0.007394741 |
| 0 | 0 | 0 | 0 | 0 |
| 0.002427819 | 0 | 0 | 0 | 0.056378598 |
| 0 | 0 | 0 | 0 | 0 |
| 0 | 0 | 0 | 0 | 0 |
| 0 | 0 | 0 | 0 | 0 |
| 0 | 0 | 0 | 0 | 0 |
| 0 | 0 | 0 | 0 | 0 |
| 0 | 0 | 0 | 0 | 0 |
| 0.295077172 | 0.012778897 | 0 | 0.015451647 | 0.12924439 |
| 0 | 0 | 0.005065348 | 0 | 0.012632612 |
| 0 | 0 | 0 | 0 | 0 |
| 0 | 0 | 0 | 0 | 0 |
| 0 | 0 | 0 | 0 | 0 |
| 0 | 0 | 0 | 0.002002897 | 0 |
| 0 | 0 | 0 | 0 | 0 |
| 0.283792889 | 0 | 0 | 0.170640993 | 0.085472901 |
| 0.003330911 | 0.01870072 | 0.003299509 | 0.007036534 | 0.004760912 |
| 0 | 0 | 0 | 0 | 0 |
| 0 | 0 | 0 | 0.000841266 | 0 |
| 0.009822116 | 0.015837895 | 0 | 0 | 0 |
| 0 | 0 | 0 | 0 | 0 |
| 0 | 0 | 0 | 0 | 0 |
| 0 | 0 | 0 | 0.002898979 | 0 |
| 0 | 0 | 0 | 0.091278682 | 0 |
| 0 | 0 | 0 | 0 | 0 |
| 0.000678198 | 0 | 0 | 0 | 0 |
| 0.002542914 | 0 | 0 | 0 | 0 |

| | | | | |
|---|---|---|---|---|
| 0 | 0 | 0 | 0 | 0 |
| 0 | 0 | 0 | 0 | 0 |
| 0 | 0 | 0 | 0 | 0 |
| 0.027926218 | 0 | 0 | 0 | 0.246432436 |
| 0 | 0 | 0 | 0 | 0 |
| 1.162448109 | 0 | 0 | 0 | 0 |
| 0 | 0 | 0 | 0 | 0 |
| 0 | 0 | 0 | 0 | 0 |
| 0 | 0.103165665 | 0.335832324 | 0.002646697 | 0 |
| 0.002378207 | 0 | 0 | 0.001184096 | 0 |
| 0 | 0 | 0 | 0 | 0 |
| 0 | 0 | 0 | 0 | 0 |
| 0 | 0 | 0 | 0 | 0 |
| 0 | 0 | 0 | 0 | 0 |
| 0.002593867 | 2.759984269 | 0 | 8.126975727 | 1.589108794 |
| 0 | 0.001583216 | 0.089610328 | 0.012692604 | 0.001499458 |
| 0 | 0 | 0.003609057 | 0 | 0 |
| 0 | 0 | 0 | 0 | 0 |
| 0 | 0 | 0 | 0 | 0 |
| 0 | 0 | 0 | 0 | 0 |
| 0 | 0 | 0 | 0 | 0 |
| 0 | 0 | 0 | 0 | 0 |
| 0 | 0 | 0 | 0 | 0 |
| 0 | 0 | 0 | 0 | 0 |
| 0.026625499 | 0.003220882 | 0.004013509 | 0 | 0 |
| 0.035643086 | 0 | 0 | 4.081621782 | 2.322719355 |
| 0 | 0 | 0 | 0 | 0 |
| 0 | 0 | 0 | 0 | 0 |
| 0.239530329 | 0 | 0 | 0.018141085 | 0.006250771 |
| 0 | 0 | 0 | 0 | 0 |
| 0 | 0 | 0 | 0 | 0 |
| 0 | 0 | 0 | 0 | 0 |
| 0 | 0 | 0 | 0 | 0 |
| 0 | 0 | 0.006144618 | 0 | 0.014509657 |
| 0.040068039 | 0 | 0.002354509 | 0.001217267 | 0.005033123 |
| 0 | 0.020368814 | 0.001664352 | 0 | 0.001956794 |
| 0 | 0.003962797 | 0 | 0 | 0 |
| 0.042010045 | 0.629292674 | 0 | 1.238072048 | 0.008034759 |
| 0.00242515 | 19.38032423 | 0 | 0 | 0 |
| 0 | 0 | 0 | 0 | 0 |
| 0.036893565 | 0 | 0 | 0 | 0 |
| 0 | 0 | 0.006039645 | 0 | 0 |
| 0.125624717 | 0.107230683 | 0 | 0.45269623 | 1.241705578 |
| 0.156997332 | 0 | 0 | 0 | 0 |
| 0 | 0 | 0 | 0 | 0 |
| 0 | 0.011704674 | 0 | 0.000611384 | 0 |
| 0 | 0 | 0 | 0 | 0 |
| 0.00724538 | 0.000755957 | 0.025370955 | 0.244475852 | 0.045642718 |

| | | | | |
|---|---|---|---|---|
| 0 | 0 | 0 | 0 | 0 |
| 0 | 0 | 0 | 0 | 0 |
| 0 | 0 | 0 | 0 | 0 |
| 0 | 0 | 0 | 0 | 0 |
| 0 | 0 | 0.009511351 | 0 | 0 |
| 0.139603424 | 0 | 0 | 0 | 0 |
| 0 | 0 | 0 | 0 | 0 |
| 0 | 0 | 0 | 0 | 0 |
| 0 | 0 | 0 | 0 | 0 |
| 0 | 0 | 0 | 0 | 0 |
| 0 | 0 | 0 | 0 | 0 |
| 0.57939006 | 0.238354156 | 0 | 2.482640405 | 3.597584744 |
| 0 | 0 | 0 | 0.168988597 | 0.035962236 |
| 0 | 0 | 0 | 0 | 0 |
| 0.001866314 | 0.01016895 | 0.186566538 | 0.025089136 | 0.003688457 |
| 0.001891689 | 0 | 0 | 0 | 0 |
| 0 | 0 | 0 | 0 | 0.001122562 |
| 0 | 0 | 0 | 0 | 0 |
| 0.000817909 | 0.000568918 | 0.001323323 | 0.000977357 | 0.001414402 |
| 0 | 0 | 0 | 0 | 0 |
| 0 | 0 | 0 | 0 | 0.756797639 |
| 0 | 0 | 0 | 0 | 0 |
| 0.380420874 | 0.02065263 | 0 | 0.249703707 | 0.148533925 |
| 0 | 0 | 0 | 0 | 0 |
| 0 | 0 | 0 | 0 | 0 |
| 0 | 0 | 0 | 0 | 0 |
| 0 | 0 | 0 | 0.017567069 | 0 |
| 0 | 0 | 0 | 0 | 0 |
| 0 | 0 | 0 | 0 | 0 |
| 0 | 0 | 0 | 0 | 0 |
| 0 | 0 | 0 | 0 | 0 |
| 0 | 0 | 0.114470907 | 0.070539436 | 0.02722202 |
| 0 | 0 | 0 | 0 | 0 |
| 0.122803932 | 0.015148291 | 0.007969923 | 0 | 0.01733582 |
| 0.00130605 | 0.021439608 | 0.003984712 | 0 | 0.002129482 |
| 0.045122017 | 0.003513337 | 0 | 0 | 0.001497361 |
| 0 | 0 | 0 | 0 | 0 |
| 0 | 0 | 0 | 0 | 0 |
| 0.62749205 | 0 | 0 | 0 | 0 |
| 0 | 0 | 0 | 0 | 0 |
| 0 | 0 | 0 | 0.12445886 | 0 |
| 0 | 0 | 0 | 0 | 0 |
| 0 | 0 | 0 | 0 | 0 |
| 0 | 0 | 0 | 0 | 0 |
| 0.011064679 | 0.008770237 | 0.035863337 | 0.002536722 | 0.008200309 |
| 0 | 0 | 0 | 0 | 0 |
| 0 | 0 | 0 | 0.108851371 | 0 |
| 0 | 0 | 0 | 0 | 0.008477516 |

| | | | | |
|---|---|---|---|---|
| 0 | 0 | 0 | 0 | 0 |
| 0 | 0 | 0 | 0 | 0 |
| 0 | 0 | 0.000738143 | 0.007393797 | 0 |
| 0 | 0 | 0 | 0 | 0 |
| 0 | 0.004881059 | 0 | 0 | 0 |
| 0 | 0 | 0 | 0 | 0 |
| 0.02079958 | 0 | 0 | 0 | 0 |
| 0 | 0 | 0 | 0.016218 | 0.002980344 |
| 0 | 0 | 0 | 0 | 0 |
| 0 | 0 | 0 | 0 | 0 |
| 0 | 0 | 0 | 0 | 0 |
| 0 | 0 | 0 | 0 | 0.001404784 |
| 0 | 0 | 0 | 0 | 0 |
| 0 | 0 | 0 | 0 | 0 |
| 0 | 0 | 0 | 0 | 0 |
| 0 | 0 | 0 | 0 | 0 |
| 0 | 0 | 0 | 0 | 0 |
| 0 | 0 | 0 | 0 | 0 |
| 0.012227912 | 1.37689356 | 0 | 0 | 0 |
| 0.004342565 | 0 | 0 | 0 | 0 |
| 0 | 0 | 0 | 0 | 0 |
| 0 | 0 | 0 | 0 | 0 |
| 0 | 0 | 0 | 0 | 0 |
| 0 | 0 | 0 | 0 | 0 |
| 0 | 0 | 0 | 0 | 0.000833447 |
| 0 | 0.073316204 | 0.026577043 | 0 | 0.001597851 |
| 0 | 0 | 0 | 0 | 0 |
| 0.000703425 | 0.001467859 | 0.047149721 | 0.038875717 | 0.022590801 |
| 0 | 0 | 0 | 0 | 0 |
| 0.029242215 | 0 | 1.334319525 | 0.965558134 | 0.306644528 |
| 0 | 0 | 0 | 0 | 0 |
| 0 | 0 | 0 | 0 | 0 |
| 0 | 0 | 0 | 0 | 0 |
| 0 | 0 | 0 | 0 | 0 |
| 0 | 0.000873078 | 0.000696277 | 0 | 0.002976799 |
| 0 | 0 | 0 | 0 | 0 |
| 0 | 0 | 0 | 0 | 0.044958044 |
| 0 | 0.001386963 | 0.003357802 | 0 | 0.000844449 |
| 0 | 0 | 0 | 0 | 0 |
| 0 | 0 | 0 | 0 | 0 |
| 0.173833412 | 2.857503171 | 19.41476039 | 9.154359352 | 0.907002223 |
| 0 | 0.031038038 | 0 | 0.147629286 | 0.460359933 |
| 0 | 0 | 0 | 0 | 0 |
| 0 | 0 | 0 | 0 | 0 |
| 0 | 0 | 0 | 0 | 0 |
| 0 | 0 | 0 | 0 | 0 |
| 0 | 0 | 0 | 0 | 0 |
| 0.002938983 | 0 | 0.000783805 | 0 | 0.001172852 |

| | | | | |
|---|---|---|---|---|
| 0 | 0 | 0 | 0 | 0 |
| 0 | 0.010675341 | 0 | 0 | 0.011719073 |
| 0 | 0 | 0 | 0 | 0 |
| 0 | 0 | 0 | 0 | 0 |
| 2.347420417 | 1.756010883 | 0 | 3.092614389 | 2.884915384 |
| 0 | 0 | 0 | 0 | 0 |
| 0 | 0 | 0 | 0 | 0 |
| 0 | 0 | 0 | 0 | 0 |
| 0.002321946 | 0.026810513 | 2.459334754 | 0.136510426 | 0.015659755 |
| 0 | 0 | 0 | 0 | 0 |
| 0 | 0 | 0 | 0 | 0 |
| 0 | 0 | 0 | 0 | 0 |
| 0.011975338 | 0 | 0 | 0.02105984 | 0.04420865 |
| 0 | 0.140856512 | 0.070111648 | 0.023152468 | 0.002264585 |
| 0.004098026 | 0 | 0 | 0 | 0.086558757 |
| 0.006154545 | 0 | 0 | 0.94115302 | 0.204244286 |
| 0 | 0.000668804 | 0 | 0.002585145 | 0 |
| 0 | 0 | 0 | 0 | 0 |
| 0 | 0 | 0 | 0 | 0 |
| 0 | 0 | 0 | 0 | 0 |
| 0 | 0 | 0 | 0 | 0 |
| 0 | 0 | 0 | 0 | 0 |
| 0 | 0 | 0 | 0 | 0 |
| 0 | 0 | 0 | 0 | 0 |
| 0 | 0 | 0 | 0 | 0 |
| 0 | 0 | 0 | 0 | 0 |
| 0 | 0 | 0 | 0.003933806 | 0.005576706 |
| 0 | 0.001148379 | 0 | 0 | 0 |
| 0 | 0 | 0 | 0 | 0 |
| 0 | 0 | 0 | 0 | 0 |
| 0 | 0 | 0 | 0 | 0 |
| 0 | 0 | 0 | 0 | 0 |
| 0 | 0 | 0 | 0 | 0 |
| 0 | 0 | 0 | 0 | 0 |
| 1.110011208 | 0 | 0 | 0 | 0.033807821 |
| 0 | 0 | 0 | 0 | 0.01556969 |
| 0 | 0 | 0 | 0 | 0 |
| 0 | 0 | 0 | 0 | 0 |
| 0 | 0 | 0 | 0 | 0 |
| 0 | 0 | 0 | 0 | 0 |
| 0.00367566 | 2.242460834 | 0 | 0.542339159 | 1.338782852 |
| 7.948699354 | 0 | 0 | 0 | 0 |
| 0 | 0 | 0 | 0 | 0 |
| 0.109981082 | 0 | 0 | 0 | 0 |
| 0 | 0 | 0 | 0 | 0 |
| 0.001042118 | 0 | 0 | 0 | 0 |
| 0 | 0 | 0 | 0 | 0 |
| 0 | 0 | 0 | 0 | 0 |

| | | | | |
|---|---|---|---|---|
| 0 | 0 | 0 | 0.002545786 | 0.134161773 |
| 0 | 0 | 0 | 0 | 0 |
| 0 | 0 | 0 | 0 | 0 |
| 0 | 0.000743365 | 0 | 0 | 0 |
| 0 | 0.017603553 | 0 | 0.037114627 | 0.005115351 |
| 0.002481761 | 0 | 0 | 3.061026389 | 0 |
| 0 | 0.001000766 | 0.050546874 | 0.018696724 | 0 |
| 0 | 0 | 0 | 0 | 0 |
| 0 | 0 | 0 | 0 | 0 |
| 0 | 0 | 0 | 0 | 0 |
| 0.003780247 | 0 | 0 | 0 | 0 |
| 0 | 0 | 0 | 0.007893412 | 0 |
| 0 | 0 | 0 | 0 | 0 |
| 0 | 0 | 0 | 0 | 0 |
| 0 | 0 | 0 | 0 | 0 |
| 0 | 0 | 0 | 0 | 0 |
| 0 | 0 | 0 | 0 | 0 |
| 0 | 0 | 0 | 0 | 0 |
| 0 | 0 | 0 | 0 | 0 |
| 0 | 0 | 0 | 0 | 0 |
| 0.007077184 | 0.003493545 | 0.034034671 | 0.052582565 | 0.134849255 |
| 0 | 0 | 0 | 0 | 0 |
| 0.085363585 | 0 | 0 | 0.009026986 | 0.005225434 |
| 0.027989005 | 0.005606926 | 0.000931565 | 0.001204033 | 0 |
| 0.000803073 | 0 | 0 | 0 | 0 |
| 0 | 0.002021223 | 0 | 0 | 0 |
| 0.001297368 | 0 | 0 | 0 | 0 |
| 0 | 0 | 0 | 0 | 0 |
| 0 | 0 | 0 | 0 | 0 |
| 0.023847382 | 0.277507377 | 0.010279006 | 0.364772575 | 0 |
| 0 | 0 | 0 | 0 | 0 |
| 0 | 0 | 0 | 0 | 0 |
| 0 | 0 | 0 | 0 | 0 |
| 0.001434747 | 0 | 0 | 0 | 1.085125278 |
| 0 | 0 | 0 | 0 | 0 |
| 0 | 0 | 0 | 0 | 0 |
| 0 | 0 | 0 | 0.065786349 | 0 |
| 0 | 0 | 0 | 0 | 0 |
| 0 | 0 | 0 | 0 | 0 |
| 0 | 0 | 0 | 0 | 0 |
| 0 | 0 | 0.000726634 | 0 | 0 |
| 0 | 0 | 0 | 0 | 0 |
| 0 | 0 | 0 | 0 | 0 |
| 0 | 0 | 0 | 0 | 0 |
| 0 | 0 | 0 | 0 | 0 |
| 0.002245511 | 0 | 0 | 0 | 0 |
| 0 | 0 | 0 | 0 | 0 |
| 0 | 0 | 0 | 0 | 0 |

| | | | | |
|---|---|---|---|---|
| 0 | 0.002513855 | 0.006682644 | 0.002375235 | 0 |
| 0.139387382 | 0 | 0 | 0 | 0.072989825 |
| 0 | 0 | 0 | 0 | 0 |
| 0 | 0 | 0 | 0 | 0 |
| 0 | 0 | 0 | 0 | 0 |
| 0 | 0 | 0 | 0 | 0 |
| 0 | 0 | 0 | 0 | 0 |
| 0.059867934 | 0.175518934 | 0 | 3.959328718 | 0.199846628 |
| 0.614554339 | 0.283576723 | 0.18094238 | 0.001694676 | 0.009249373 |
| 0.02790796 | 0.196318808 | 0.01478739 | 0 | 0.162734526 |
| 0 | 0 | 0 | 0 | 0 |
| 0 | 0 | 0 | 0 | 0 |
| 0 | 0 | 0 | 0 | 0 |
| 0.382730845 | 0.012463781 | 0 | 0 | 0 |
| 0 | 0 | 0 | 0 | 0 |
| 0 | 0 | 0 | 0 | 0 |
| 0 | 0.002446368 | 0 | 0 | 0 |
| 0 | 0 | 0 | 0 | 0 |
| 0 | 0 | 0 | 0 | 0 |
| 0 | 0 | 0 | 0 | 0 |
| 0 | 0 | 0 | 0 | 0 |
| 0 | 0 | 0 | 0 | 0 |
| 0 | 0 | 0 | 0.004076806 | 0 |
| 0 | 0 | 0 | 0 | 0 |
| 0 | 0 | 0 | 0 | 0 |
| 0 | 0 | 0 | 0 | 0 |
| 0 | 0 | 0 | 0 | 0 |
| 0 | 0 | 0 | 0 | 0 |
| 0 | 0 | 0 | 0 | 0 |
| 0 | 0 | 0 | 0 | 0 |
| 0 | 0.004304256 | 0 | 0 | 0 |
| 0 | 0 | 0 | 0 | 0 |
| 0 | 0 | 0 | 0 | 0 |
| 0 | 0 | 0 | 0 | 0 |
| 0 | 0 | 0 | 0.002091314 | 0 |
| 0.256955901 | 0 | 0 | 0 | 0 |
| 0 | 0 | 0 | 0 | 0 |
| 0 | 0 | 0 | 0 | 0 |
| 0.022059983 | 0 | 0 | 0 | 0 |
| 0 | 0.192738387 | 0 | 0 | 0 |
| 0 | 0 | 0 | 0 | 0 |
| 0.014640867 | 0.010183843 | 0 | 0.36828579 | 0.165151512 |
| 0 | 0 | 0 | 0 | 0 |
| 0 | 0 | 0 | 0 | 0 |
| 0 | 0 | 0 | 0 | 0 |
| 0 | 0 | 0 | 0 | 0 |
| 0 | 0 | 0 | 0 | 0 |
| 0 | 0 | 0 | 0 | 0 |

| | | | | |
|---|---|---|---|---|
| 0 | 0 | 0 | 0 | 0 |
| 0 | 0 | 0 | 0 | 0 |
| 0 | 0 | 0 | 0 | 0 |
| 0 | 0 | 0 | 0 | 0 |
| 0.463044843 | 0.048851738 | 0 | 0.0051771 | 0.039432346 |
| 0 | 0 | 0 | 0 | 0 |
| 0 | 0 | 0 | 0 | 0 |
| 0 | 0.022171451 | 0.346407886 | 0 | 0.00318006 |
| 0 | 0 | 0 | 0 | 0 |
| 0 | 0 | 0 | 0 | 0 |
| 0 | 0 | 0 | 0 | 0 |
| 1.562457333 | 0 | 0 | 0 | 0 |
| 0 | 0 | 0 | 0 | 0 |
| 0 | 0 | 0 | 0 | 0 |
| 0 | 0 | 0.017503985 | 0 | 0 |
| 0.00212653 | 0.00166406 | 0 | 0.001429363 | 0.005319082 |
| 0 | 0 | 0 | 0 | 0 |
| 0 | 0 | 0 | 0 | 0 |
| 0 | 0 | 0 | 0 | 0 |
| 0 | 0 | 0 | 0 | 0 |
| 0 | 0 | 0 | 0 | 0 |
| 0 | 0 | 0 | 0 | 0 |
| 0 | 0 | 0 | 0 | 0 |
| 0 | 0 | 0.000735675 | 0 | 0 |
| 0.207199809 | 0 | 0 | 0.149105543 | 0 |
| 0.003384405 | 0 | 0 | 0 | 0 |
| 0.007035213 | 0.006580952 | 0 | 0 | 0 |
| 0 | 0 | 0 | 0 | 0 |
| 0 | 0 | 0 | 0 | 0.001870672 |
| 0 | 0 | 0 | 0 | 0 |
| 0 | 0 | 0 | 0 | 0 |
| 0 | 0 | 0 | 0 | 0 |
| 0 | 0 | 0 | 0 | 0 |
| 0 | 0 | 0 | 0 | 0 |
| 0 | 0 | 0 | 0 | 0 |
| 0 | 0 | 0 | 0 | 0 |
| 0 | 0 | 0 | 0 | 0 |
| 0.011693313 | 0 | 0 | 0 | 0.071911069 |
| 0.000881278 | 0 | 0 | 0 | 0 |
| 0 | 0 | 0 | 0 | 0 |
| 0 | 0 | 0 | 0 | 0 |
| 9.592447078 | 0 | 0 | 0 | 0.05222224 |
| 0 | 0 | 0 | 0 | 0 |
| 0 | 0 | 0 | 0 | 0 |
| 0 | 0 | 0 | 0 | 0 |
| 0 | 0 | 0 | 0 | 0 |
| 0 | 0 | 0 | 0 | 0 |
| 0 | 0 | 0 | 0 | 0 |

| | | | | |
|---|---|---|---|---|
| 0.015285267 | 0 | 0 | 1.092224602 | 0 |
| 0 | 0 | 0 | 0 | 0 |
| 0 | 0 | 0 | 0 | 0 |
| 0 | 0 | 0 | 0 | 0 |
| 0 | 0 | 0 | 0 | 0 |
| 0 | 0 | 0 | 0.018002773 | 0 |
| 0 | 0 | 0 | 0 | 0 |
| 0 | 0 | 0 | 0 | 0 |
| 0 | 0 | 0 | 0 | 0 |
| 0 | 0 | 0 | 0 | 0 |
| 0 | 0 | 0 | 0.187973053 | 0 |
| 0 | 0 | 0.000508631 | 0 | 0 |
| 0 | 0 | 0 | 0 | 0 |
| 0 | 0 | 0 | 0 | 0 |
| 0 | 0 | 0 | 0 | 0 |
| 0 | 0 | 0 | 0 | 0 |
| 0 | 0 | 0 | 0 | 0 |
| 0 | 0 | 0 | 0 | 0 |
| 0 | 0 | 0 | 0 | 0 |
| 0 | 0 | 0 | 0 | 0 |
| 0 | 0.012493271 | 0.245420834 | 0.001893748 | 0 |
| 0 | 0 | 0 | 0 | 0 |
| 0.004002966 | 0.001160154 | 0.002775659 | 0 | 0.01384459 |
| 0 | 0 | 0 | 0 | 0 |
| 0 | 0 | 0 | 0 | 0 |
| 5.905371669 | 0 | 0 | 0 | 0 |
| 0 | 0 | 0 | 0 | 0 |
| 0 | 0 | 0 | 0 | 0 |
| 12.2821722 | 0 | 0 | 0 | 0 |
| 0 | 0 | 0 | 0 | 0 |
| 0 | 0 | 0 | 0 | 0 |
| 0 | 0 | 0 | 0 | 0 |
| 0.001561984 | 0.536720731 | 0.744076428 | 0.108349603 | 0.00138915 |
| 0 | 0 | 0 | 0 | 0 |
| 0 | 0 | 0 | 0.122212104 | 0.053638392 |
| 0.138628975 | 0 | 0.002085904 | 0 | 0.004087361 |
| 0 | 0 | 0 | 0 | 0 |
| 0 | 0 | 0 | 0 | 0 |
| 0 | 0.000605062 | 0.050867304 | 0 | 0.001719156 |
| 0 | 0 | 0 | 0 | 0 |
| 0 | 0 | 0 | 0 | 0 |
| 0 | 0 | 0 | 0 | 0 |
| 0.510943624 | 0.030422461 | 0 | 0.012154289 | 0.0346761 |
| 0 | 0 | 0 | 0 | 0 |
| 0 | 0 | 0 | 0 | 0 |
| 0 | 0 | 0 | 0.036921247 | 0.002394681 |
| 0 | 0 | 0 | 0 | 0 |
| 0 | 0 | 0 | 0 | 0 |

| | | | | |
|---|---|---|---|---|
| 0 | 0 | 0 | 0 | 0 |
| 0 | 0 | 0 | 0 | 0 |
| 0 | 0 | 0 | 0 | 0 |
| 0 | 0 | 0 | 0 | 0 |
| 0 | 0 | 0 | 0 | 0 |
| 0 | 0 | 0 | 0 | 0 |
| 0 | 0 | 0 | 0 | 0 |
| 0 | 0 | 0 | 0.184217779 | 0.003663866 |
| 0 | 0 | 0 | 0 | 0.000934831 |
| 0 | 0 | 0 | 0 | 0 |
| 0 | 0 | 0 | 0 | 0 |
| 0 | 0 | 0 | 0 | 0 |
| 0 | 0 | 0 | 0 | 0 |
| 0 | 0 | 0 | 0 | 0 |
| 0 | 0 | 0 | 0 | 0 |
| 0 | 0.081234896 | 6.080181639 | 0 | 0 |
| 0 | 0 | 0 | 0 | 0 |
| 0 | 0 | 0 | 0 | 0 |
| 0 | 0 | 0 | 0 | 0 |
| 0.003655271 | 0 | 0 | 0.409486343 | 0.355388567 |
| 0 | 0.002775274 | 0 | 0 | 0.002323363 |
| 0 | 0 | 0 | 0 | 0 |
| 0 | 0 | 0 | 0 | 0 |
| 0 | 0 | 0 | 0 | 0 |
| 0 | 0 | 0 | 0 | 0 |
| 0.001190328 | 0 | 0 | 0 | 0 |
| 0 | 0 | 0 | 0 | 0 |
| 0 | 0 | 0 | 0 | 0 |
| 0 | 0 | 0 | 0 | 0 |
| 0 | 0 | 0 | 0.988728151 | 0 |
| 0.005988064 | 0.070653394 | 0 | 0.905557869 | 0.466472957 |
| 0 | 0 | 0 | 0 | 0 |
| 0 | 0 | 0 | 0 | 0 |
| 0 | 0 | 0 | 0 | 0 |
| 0 | 0 | 0 | 0 | 0 |
| 0 | 0 | 0 | 0 | 0 |
| 0 | 0 | 0 | 0 | 0 |
| 0 | 0 | 0 | 0 | 0 |
| 0.018746618 | 0 | 0 | 0 | 0 |
| 0 | 0 | 0 | 0 | 0 |
| 0 | 0 | 0 | 0 | 0 |
| 0.023213867 | 0 | 0 | 0 | 0 |
| 0 | 0 | 0 | 0 | 0 |
| 0 | 0 | 0 | 0 | 0 |
| 0 | 0 | 0 | 0 | 0 |
| 0 | 0 | 0 | 0 | 0 |
| 0 | 0.011657884 | 1.628935377 | 0 | 0 |
| 0 | 0 | 0 | 0 | 0 |

| | | | | |
|---|---|---|---|---|
| 0 | 0 | 0 | 0 | 0 |
| 0 | 0 | 0 | 0 | 0 |
| 0 | 0 | 0 | 0 | 0 |
| 0 | 0 | 0 | 0 | 0 |
| 0.003964982 | 0 | 0 | 0.003553457 | 0.02582828 |
| 0 | 0 | 0 | 0 | 0 |
| 0 | 0 | 0 | 0 | 0 |
| 0 | 0 | 0 | 0 | 0 |
| 0 | 0 | 0 | 0 | 0 |
| 0 | 0 | 0 | 0 | 0 |
| 0 | 0 | 0 | 0 | 0 |
| 0.002317811 | 0.001612215 | 0.006629583 | 0 | 0.001717787 |
| 0 | 0 | 0 | 0 | 0.002408279 |
| 0 | 0 | 0 | 0 | 0 |
| 0 | 0 | 0.014850421 | 0 | 0 |
| 0 | 0.044977398 | 0 | 2.21545623 | 0 |
| 0 | 0.002634967 | 0 | 0 | 0 |
| 0 | 0 | 0 | 0 | 0 |
| 0 | 0.305652321 | 0 | 0.003735642 | 0.013982722 |
| 0 | 0 | 0.017876006 | 0 | 0 |
| 0 | 0 | 0 | 0 | 0 |
| 0 | 0 | 0 | 0 | 0 |
| 0 | 0 | 0 | 0 | 0 |
| 0 | 0 | 0 | 0 | 0 |
| 0 | 0 | 0 | 0 | 0 |
| 0 | 0 | 0 | 0 | 0 |
| 0 | 0.003193292 | 0 | 0.002674342 | 0.007145035 |
| 0 | 0 | 0 | 0 | 0 |
| 0 | 0 | 0 | 0 | 0 |
| 0 | 0 | 0 | 0 | 0 |
| 0 | 0 | 0.058095899 | 0 | 0.001157039 |
| 0 | 0 | 0 | 0 | 0 |
| 0 | 0 | 0 | 0 | 0 |
| 0 | 0 | 0 | 0 | 0 |
| 0 | 0 | 0 | 0 | 0 |
| 0 | 0 | 0 | 0 | 0 |
| 0 | 0 | 0 | 0 | 0 |
| 0.006215477 | 0.010426878 | 0.063886684 | 0 | 0.00406451 |
| 0.000659642 | 0 | 0 | 0 | 0 |
| 0 | 0 | 0 | 0 | 0 |
| 0 | 0 | 0 | 0.001635596 | 0 |
| 0 | 0 | 0 | 0 | 0 |
| 0 | 0 | 0 | 0 | 0 |
| 0 | 0.008107984 | 0 | 0 | 0 |
| 0 | 0 | 0 | 0 | 0 |
| 0 | 0 | 0 | 0 | 0 |
| 0 | 0 | 0 | 0 | 0 |
| 0 | 0 | 0 | 0 | 0 |

| | | | | |
|---|---|---|---|---|
| 0 | 0 | 0 | 0 | 0 |
| 0 | 0 | 0 | 0 | 0.018373496 |
| 0 | 0 | 0 | 0 | 0 |
| 0 | 0.001185656 | 0.001181948 | 0 | 0 |
| 0 | 0 | 0 | 0 | 0.001757531 |
| 0 | 0 | 0.0014148 | 0 | 0 |
| 0.008546454 | 0 | 0 | 0 | 0 |
| 0.005776847 | 0.000772738 | 0 | 0 | 0 |
| 0 | 0 | 0 | 0 | 0 |
| 0 | 0 | 0 | 0 | 0 |
| 0 | 0 | 0 | 0 | 0 |
| 0 | 0 | 0 | 0 | 0 |
| 0.084379456 | 0.015128342 | 0 | 0.014979959 | 0.011044494 |
| 0 | 0 | 0 | 0 | 0 |
| 0 | 0 | 0 | 0 | 0 |
| 0 | 0 | 0 | 0 | 0 |
| 0 | 0 | 0 | 0 | 0 |
| 0 | 0 | 0 | 0 | 0 |
| 0 | 0 | 0 | 0 | 0 |
| 0 | 0 | 0 | 0 | 0 |
| 0 | 0 | 0 | 0 | 0 |
| 0 | 0 | 0 | 0 | 0 |
| 0.044814564 | 0 | 0.00441294 | 0 | 0.039502079 |
| 0.003129155 | 0.013494715 | 0 | 0.020191542 | 0 |
| 0 | 0 | 0 | 0 | 0 |
| 0 | 0 | 0 | 0 | 0 |
| 0 | 0 | 0 | 0 | 0 |
| 0 | 0 | 0 | 0 | 0 |
| 0 | 0 | 0 | 0 | 0 |
| 0 | 0 | 0 | 0 | 0 |
| 0.003252358 | 0 | 0.02603716 | 0.031002828 | 0.014024165 |
| 0 | 0 | 0 | 0 | 0 |
| 0.006520084 | 0 | 0 | 0 | 0 |
| 0 | 0 | 0 | 0 | 0 |
| 0 | 0 | 0 | 0 | 0 |
| 0 | 0.00128995 | 0.064510087 | 0 | 0 |
| 0 | 0 | 0.008059176 | 0 | 0 |
| 0 | 0 | 0 | 0 | 0 |
| 0 | 0 | 0 | 0.029817625 | 0.00129778 |
| 0 | 0 | 0 | 0 | 0 |
| 0 | 0 | 0 | 0 | 0 |
| 0 | 0 | 0 | 0 | 0 |
| 0 | 0 | 0 | 0 | 0 |
| 0 | 0 | 0 | 0 | 0 |
| 0 | 0 | 0 | 0 | 0 |
| 0.54165696 | 0.077771652 | 0 | 0.293511743 | 0.040616592 |
| 0 | 0 | 0 | 0 | 0 |
| 0 | 0 | 0 | 0 | 0 |

| | | | | |
|---|---|---|---|---|
| 0 | 0 | 0 | 0 | 0 |
| 0 | 0 | 0 | 0 | 0 |
| 0.327121028 | 0 | 0 | 0 | 0 |
| 0 | 0 | 0 | 0 | 0 |
| 0 | 0 | 0 | 0 | 0 |
| 0 | 0 | 0 | 0 | 0 |
| 0 | 0 | 0 | 0 | 0 |
| 0 | 0 | 0 | 0 | 0 |
| 0 | 0 | 0 | 0 | 0 |
| 0 | 0 | 0 | 0 | 0 |
| 0 | 0 | 0 | 0 | 0.004060982 |
| 0 | 0 | 0 | 0 | 0 |
| 0 | 0 | 0 | 0 | 0 |
| 0 | 0 | 0 | 0 | 0 |
| 0 | 0 | 0 | 0 | 0 |
| 0 | 0 | 0 | 0 | 0 |
| 0 | 0 | 0 | 0 | 0 |
| 0 | 0 | 0 | 0 | 0 |
| 0 | 0 | 0 | 0 | 0 |
| 0.399401374 | 0 | 0 | 0 | 0.004986629 |
| 0 | 0 | 0 | 0 | 0 |
| 0 | 0 | 0 | 0 | 0 |
| 0 | 0 | 0 | 0 | 0 |
| 0 | 0 | 0 | 0 | 0 |
| 0 | 0 | 0 | 0 | 0 |
| 0 | 0 | 0 | 0 | 0 |
| 0 | 0 | 0 | 0 | 0 |
| 0.005361248 | 0 | 0 | 0.025358693 | 0.009491903 |
| 0 | 0 | 0 | 0 | 0 |
| 0 | 0 | 0 | 0 | 0 |
| 0.282085634 | 0 | 0 | 0.001864977 | 0.005312198 |
| 0 | 0 | 0 | 0 | 0 |
| 0.022014321 | 0.037161173 | 0.193601657 | 0.008180507 | 0.065062481 |
| 0 | 0 | 0.004375305 | 0.002704573 | 0.010979472 |
| 0.001331933 | 0 | 0 | 0 | 0 |
| 0 | 0 | 0 | 0 | 0 |
| 0 | 0 | 0 | 0 | 0 |
| 0 | 0 | 0 | 0 | 0 |
| 0 | 0 | 0 | 0 | 0 |
| 0 | 0 | 0 | 0 | 0 |
| 0 | 0 | 0 | 0 | 0 |
| 0 | 0.009064108 | 0.037827653 | 0.042359592 | 0.068258326 |
| 0.196156859 | 0.010106822 | 0 | 0.117198506 | 0.18765365 |
| 0 | 0 | 0 | 0 | 0 |
| 0 | 0 | 0 | 0 | 0 |
| 0.110428978 | 0.002434177 | 0 | 0 | 0.025071215 |
| 0 | 0 | 0 | 0 | 0 |
| 0 | 0 | 0 | 0 | 0 |

| | | | | |
|---|---|---|---|---|
| 0 | 0 | 0 | 0 | 0 |
| 0.012121418 | 0 | 0 | 0.011873881 | 0.012535096 |
| 0 | 0.095738863 | 0.011872843 | 0.014460157 | 0.004148656 |
| 0 | 0 | 0 | 0 | 0 |
| 0.014986171 | 0.001828777 | 0 | 0.025683379 | 0.002727943 |
| 0.025080176 | 0.00178012 | 0.020584806 | 0 | 0 |
| 0 | 0 | 0 | 0 | 0 |
| 0 | 0 | 0 | 0 | 0 |
| 0 | 0 | 0 | 0 | 0 |
| 0 | 0 | 0 | 0 | 0 |
| 0 | 0 | 0 | 0 | 0 |
| 0 | 0 | 0 | 0 | 0 |
| 0.161770999 | 0 | 0 | 0 | 0 |
| 0 | 0 | 0 | 0.004960872 | 0 |
| 0 | 0 | 0 | 0 | 0 |
| 0 | 0 | 0 | 0 | 0 |
| 0 | 0 | 0 | 0.00298239 | 0 |
| 0 | 0 | 0 | 0 | 0 |
| 0 | 0 | 0 | 0 | 0 |
| 0 | 0 | 0 | 0 | 0 |
| 0 | 0 | 0 | 0 | 0 |
| 0 | 0 | 0 | 0 | 0 |
| 0 | 0 | 0 | 0 | 0 |
| 0 | 0 | 0 | 0 | 0 |
| 0.014925575 | 0 | 0 | 0 | 0 |
| 0 | 0.014051827 | 0.004353798 | 0.00146797 | 0 |
| 0 | 0 | 0 | 0 | 0 |
| 0 | 0 | 0 | 0.143584886 | 0.017271005 |
| 0 | 0 | 0 | 0 | 0 |
| 0 | 0 | 0 | 0 | 0 |
| 0 | 0 | 0 | 0 | 0 |
| 0 | 0.001332175 | 0 | 0 | 0 |
| 0 | 0 | 0 | 0 | 0 |
| 0 | 0 | 0 | 0 | 0 |
| 0 | 0 | 0 | 0 | 0 |
| 0 | 0 | 0 | 0 | 0 |
| 0 | 0 | 0 | 0 | 0 |
| 0 | 0 | 0 | 0 | 0 |
| 0.010410072 | 0 | 0 | 1.546272022 | 1.429325908 |
| 0 | 0 | 0 | 0 | 0 |
| 0 | 0 | 0 | 0 | 0 |
| 0.001480792 | 0 | 0 | 0 | 0.001975414 |
| 0 | 0.001905749 | 0.00455949 | 0.00466535 | 0 |
| 0 | 0 | 0 | 0 | 0 |
| 0 | 0 | 0 | 0 | 0 |
| 0 | 0 | 0 | 0 | 0 |
| 0.003353101 | 0.001166169 | 0 | 0 | 0.013978501 |
| 0 | 0 | 0 | 0 | 0 |

| | | | | |
|---|---|---|---|---|
| 0 | 0 | 0 | 0 | 0 |
| 0.003173248 | 0.080564128 | 0.007701162 | 0.002606905 | 0.031552953 |
| 0 | 0 | 0 | 0 | 0 |
| 0 | 0 | 0 | 0 | 0 |
| 0 | 0 | 0 | 0 | 0 |
| 0 | 0 | 0 | 0 | 0 |
| 0 | 0 | 0 | 0 | 0 |
| 0 | 0 | 0 | 0 | 0 |
| 0 | 0 | 0 | 0 | 0 |
| 0 | 0 | 0 | 0 | 0 |
| 0 | 0 | 0 | 0 | 0 |
| 0 | 0 | 0 | 0 | 0 |
| 0.003633937 | 0 | 0 | 0.027056256 | 0.003521879 |
| 0 | 0 | 0 | 0 | 0 |
| 0.002637708 | 0.002752091 | 0 | 0.004136895 | 0 |
| 0 | 0 | 0 | 0 | 0 |
| 0 | 0 | 0 | 0 | 0 |
| 0 | 0 | 0 | 0 | 0 |
| 0 | 0 | 0 | 0 | 0 |
| 0 | 0 | 0 | 0 | 0 |
| 0 | 0 | 0 | 0 | 0 |
| 0 | 0 | 0.000898105 | 0 | 0 |
| 0 | 0 | 0 | 0 | 0 |
| 0 | 0 | 0 | 0 | 0 |
| 0 | 0 | 0 | 0 | 0 |
| 0 | 0 | 0 | 0 | 0 |
| 0 | 0 | 0 | 0 | 0 |
| 0 | 0 | 0 | 0 | 0 |
| 0 | 0 | 0 | 0 | 0 |
| 0 | 0 | 0 | 0 | 0 |
| 0 | 0 | 0 | 0 | 0 |
| 0 | 0 | 0 | 0 | 0 |
| 0 | 0 | 0 | 0 | 0 |
| 0 | 0 | 0 | 0 | 0 |
| 1.808373009 | 0 | 0 | 0 | 14.36558695 |
| 0 | 0 | 0 | 0 | 0 |
| 0 | 0 | 0 | 0 | 0 |
| 0 | 0 | 0 | 0 | 0 |
| 0.004527013 | 0.000674761 | 0.001121084 | 0.001738781 | 0 |
| 0 | 0 | 0 | 0 | 0 |
| 0.0034792 | 0.023797154 | 0.158620531 | 0.061582355 | 0.009454579 |
| 0 | 0 | 0 | 0 | 0 |
| 0 | 0 | 0 | 0 | 0 |
| 0 | 0 | 0 | 0 | 0 |
| 0 | 0 | 0 | 0 | 0 |
| 0.068382505 | 0.024475329 | 9.477541105 | 0.251982878 | 0.022387758 |
| 0 | 0 | 0 | 0 | 0 |
| 0 | 0 | 0 | 0 | 0 |

| | | | | |
|---|---|---|---|---|
| 0 | 0 | 0 | 0 | 0 |
| 0 | 0 | 0 | 0 | 0 |
| 0 | 0 | 0 | 0 | 0 |
| 0 | 0 | 0 | 0 | 0 |
| 0 | 0 | 0 | 0 | 0 |
| 0 | 0 | 0 | 0 | 0 |
| 0 | 0 | 0 | 0 | 0 |
| 0 | 0 | 0 | 0 | 0 |
| 0 | 0 | 0 | 0 | 0 |
| 0 | 0 | 0.007203633 | 0 | 0 |
| 0.008559783 | 0 | 0 | 0 | 0 |
| 0 | 0 | 0 | 0 | 0 |
| 0 | 0.003752239 | 0.002200295 | 0 | 0 |
| 0 | 0 | 0 | 0 | 0 |
| 0 | 0 | 0 | 0 | 0 |
| 0 | 0 | 0 | 0 | 0 |
| 0 | 0 | 0 | 0 | 0 |
| 0 | 0 | 0 | 0 | 0 |
| 0 | 0 | 0 | 0 | 0 |
| 12.64892792 | 0.087423068 | 0.038384995 | 0.010884273 | 0.080797862 |
| 0 | 0 | 0 | 0 | 0 |
| 0 | 0 | 0 | 0 | 0 |
| 0 | 0 | 0 | 0 | 0 |
| 0 | 0 | 0 | 0 | 0 |
| 0 | 0 | 0 | 0 | 0 |
| 0 | 0 | 0 | 0 | 0 |
| 0 | 0 | 0 | 0 | 0 |
| 0 | 0 | 0 | 0 | 0 |
| 0 | 0 | 0 | 0 | 0 |
| 0.004528941 | 0.006090435 | 0.237412046 | 0.018670861 | 0 |
| 0 | 0 | 0 | 0 | 0 |
| 0 | 0 | 0 | 0 | 0 |
| 0 | 0 | 0 | 0 | 0 |
| 0 | 0 | 0 | 0 | 0 |
| 0 | 0 | 0 | 0 | 0 |
| 0 | 0 | 0 | 0 | 0 |
| 0 | 0 | 0 | 0 | 0 |
| 0 | 0 | 0 | 0 | 0 |
| 0 | 0 | 0 | 0 | 0 |
| 0 | 0 | 0 | 0 | 0 |
| 0 | 0 | 0 | 0 | 0 |
| 0 | 0 | 0 | 0 | 0 |
| 0 | 0 | 0 | 0.001052028 | 0 |
| 0 | 0 | 0 | 0 | 0 |
| 0 | 0.002952304 | 0.003310953 | 0 | 0 |
| 0 | 0 | 0 | 0 | 0 |
| 0 | 0 | 0 | 0 | 0 |
| 0 | 0 | 0 | 0 | 0 |

| | | | | |
|---|---|---|---|---|
| 0 | 0 | 0 | 0 | 0 |
| 0 | 0 | 0 | 0 | 0 |
| 0 | 0 | 0 | 0 | 0 |
| 0 | 0 | 0 | 0 | 0 |
| 0.001889541 | 0 | 0 | 0 | 0 |
| 0.108128777 | 0.003168264 | 0 | 0.001952321 | 0.00308219 |
| 0 | 0 | 0 | 0 | 0 |
| 0 | 0 | 0 | 0 | 0 |
| 0 | 0 | 0 | 0 | 0 |
| 0 | 0 | 0 | 0 | 0 |
| 0 | 0 | 0 | 0 | 0 |
| 0 | 0 | 0 | 0 | 0 |
| 0 | 0 | 0 | 0 | 0 |
| 0 | 0 | 0 | 0 | 0.083693071 |
| 0 | 0 | 0 | 0 | 0 |
| 0 | 0 | 0 | 0 | 0 |
| 0 | 0 | 0 | 0 | 0 |
| 0 | 0 | 0 | 0 | 0 |
| 0 | 0 | 0 | 0 | 0 |
| 0 | 0 | 0 | 0 | 0 |
| 0 | 0.004472165 | 0 | 0 | 0 |
| 0 | 0 | 0 | 0 | 0 |
| 0 | 0 | 0 | 0 | 0 |
| 0 | 0 | 0 | 0 | 0 |
| 0 | 0 | 0 | 0 | 0 |
| 0 | 0 | 0 | 0 | 0 |
| 0 | 0 | 0 | 0 | 0 |
| 0 | 0 | 0 | 0 | 0 |
| 0 | 0 | 0 | 0 | 0 |
| 0 | 0 | 0 | 0 | 0 |
| 0 | 0 | 0 | 0 | 0 |
| 0 | 0 | 0 | 0 | 0 |
| 0 | 0 | 0 | 0 | 0 |
| 0 | 0 | 0 | 0 | 0 |
| 0 | 0 | 0 | 0 | 0 |
| 0 | 0 | 0 | 0 | 0 |
| 0 | 0 | 0 | 0 | 0 |
| 0 | 0 | 0 | 0 | 0 |
| 0 | 0 | 0 | 0.13553898 | 0 |
| 0 | 0 | 0 | 0 | 0 |
| 0 | 0 | 0 | 0 | 0 |
| 0 | 0 | 0 | 0 | 0 |
| 0 | 0 | 0 | 0 | 0 |
| 0 | 0 | 0 | 0 | 0 |
| 0 | 0 | 0 | 0 | 0 |
| 0 | 0 | 0 | 0 | 0 |
| 0 | 0 | 0 | 0 | 0 |
| 0 | 0 | 0 | 0 | 0 |

| | | | | |
|---|---|---|---|---|
| 0 | 0 | 0 | 0 | 0 |
| 0 | 0 | 0 | 0 | 0 |
| 0 | 0 | 0 | 0 | 0 |
| 0 | 0 | 0 | 0 | 0 |
| 0 | 0 | 0 | 0 | 0 |
| 0 | 0 | 0 | 0 | 0 |
| 0 | 0 | 0 | 0 | 0 |
| 0 | 0 | 0 | 0 | 0 |
| 0 | 0 | 0 | 0 | 0 |
| 0 | 0 | 0 | 0 | 0 |
| 0.002032269 | 0 | 0.001409177 | 0 | 0 |
| 0 | 0 | 0 | 0 | 0 |
| 0 | 0 | 0 | 0 | 0.645493135 |
| 0 | 0 | 0 | 0 | 0 |
| 0 | 0 | 0 | 0 | 0 |
| 0 | 0 | 0 | 0 | 0 |
| 0 | 0 | 0 | 0 | 0 |
| 0 | 0 | 0 | 0 | 0 |
| 0 | 0 | 0 | 0 | 0 |
| 0 | 0 | 0 | 0 | 0 |
| 0.007955863 | 0.003484314 | 0 | 0 | 0.000655143 |
| 0 | 0 | 0 | 0 | 0 |
| 0 | 0 | 0 | 0 | 0 |
| 2.087278004 | 0.597825327 | 0.00389292 | 3.82244023 | 0.796351589 |
| 0 | 0 | 0 | 0 | 0 |
| 0 | 0 | 0 | 0 | 0 |
| 0 | 0 | 0 | 0 | 0 |
| 0 | 0 | 0 | 0 | 0 |
| 0 | 0 | 0 | 0 | 0 |
| 0 | 0 | 0 | 0 | 0 |
| 0 | 0 | 0 | 0 | 0 |
| 0 | 0 | 0 | 0 | 0 |
| 0 | 0 | 0 | 0 | 0 |
| 0 | 0 | 0 | 0 | 0 |

| PHL018-0260_V1 | PHL018-0041_V3 | PHL018-0122_V2 | PHL018-0105_V2 | PHL018-0218_V1 |
|---|---|---|---|---|
| REF | EYCF | CTRL | CTRL | REF |
| 8.097828946 | 0.212515807 | 0.025619503 | 0.008729296 | 1.104637958 |
| 0.347423552 | 2.346958249 | 0.630984331 | 2.645176338 | 0.519886983 |
| 0 | 9.099618512 | 26.02033471 | 10.89512463 | 9.39126418 |
| 0 | 0 | 0 | 0 | 0 |
| 0 | 0 | 0 | 0 | 0 |
| 0 | 0 | 0 | 0 | 0 |
| 0 | 0 | 0 | 0 | 1.401167234 |
| 0 | 0 | 0 | 0 | 0 |
| 0 | 0 | 0 | 0 | 0 |
| 0 | 0 | 0 | 0 | 0 |
| 0 | 0 | 0 | 0 | 0 |
| 0 | 0 | 0 | 0 | 0 |
| 0 | 0 | 0 | 0 | 0 |
| 0.477090289 | 7.934515795 | 15.97029689 | 4.086615263 | 2.176589041 |
| 0 | 0 | 0 | 0 | 0 |
| 0 | 0 | 0 | 0 | 0 |
| 0 | 0 | 0.001705116 | 0 | 0 |
| 0 | 0 | 0 | 0 | 0 |
| 0 | 0 | 0 | 0 | 0 |
| 0 | 0 | 0 | 0 | 0 |
| 0 | 0 | 0 | 0 | 0 |
| 0 | 0 | 0 | 0 | 0 |
| 0 | 0 | 0 | 0 | 0 |
| 0 | 0 | 0 | 0 | 0 |
| 0 | 0 | 0 | 0 | 0.145719652 |
| 0 | 0 | 0 | 0 | 0 |
| 0 | 0 | 0 | 0 | 0 |
| 0 | 0 | 0 | 0 | 0 |
| 0 | 0 | 0 | 0 | 0 |
| 0 | 0 | 0 | 0 | 0 |
| 0 | 0 | 0 | 0 | 0 |
| 0 | 0 | 0 | 0 | 0 |
| 0 | 0 | 0 | 0 | 0 |
| 0 | 0 | 0 | 0 | 0 |
| 0 | 0 | 0 | 0 | 0 |
| 0.86976966 | 3.010199748 | 0 | 4.785323972 | 0 |
| 0 | 0 | 0 | 0 | 0 |
| 0 | 0 | 0 | 0 | 0 |
| 0 | 0 | 0 | 0 | 0 |
| 0 | 0 | 0 | 0 | 0 |
| 0 | 0 | 0 | 0 | 0 |
| 0.00415473 | 0.208046371 | 0 | 0.053588243 | 0.009128462 |
| 0.377870408 | 0.014818232 | 0.003772682 | 0.041343185 | 7.57817601 |
| 0 | 0.00512357 | 0 | 0.062855379 | 0.121858201 |
| 0.00463886 | 0 | 0 | 0 | 0 |
| 0 | 0 | 0 | 0 | 0 |

| | | | | |
|---|---|---|---|---|
| 0.023517778 | 0 | 0 | 0 | 0 |
| 0 | 0 | 0 | 0.235402227 | 0.009868751 |
| 0 | 0.003857898 | 0 | 0 | 0 |
| 0.405873607 | 0.311670095 | 0.094637826 | 0.221703447 | 0.308198341 |
| 0.000811956 | 0.031753212 | 0 | 0.001052282 | 0.007273105 |
| 0 | 0.001692961 | 0 | 0 | 0.07533068 |
| 0 | 0.032065533 | 0.011116326 | 0.024333306 | 0 |
| 0 | 0 | 0 | 0 | 0 |
| 0 | 0.037933006 | 0 | 0 | 0 |
| 0 | 0 | 0 | 0 | 0 |
| 0.001198017 | 0.002702937 | 0 | 0 | 0.00145783 |
| 0 | 0.017646829 | 0 | 0 | 0.392470173 |
| 0 | 0 | 0 | 0 | 0 |
| 0.023345016 | 0.088697076 | 0.019479441 | 0.009128297 | 0.121206755 |
| 0 | 0 | 0 | 0 | 0 |
| 0 | 0 | 0 | 0 | 0 |
| 0 | 0 | 0 | 0.10636839 | 0.854434237 |
| 0 | 0 | 0 | 0 | 0 |
| 0 | 0 | 0 | 0 | 0 |
| 0 | 0 | 0 | 0 | 0 |
| 0.006512482 | 0 | 0.274032624 | 0.688140042 | 0.058435685 |
| 0.001482324 | 0.005852672 | 0.005377712 | 0.056602906 | 0.00751581 |
| 0.020782489 | 0.204904756 | 0 | 0 | 0 |
| 0 | 0 | 0 | 0 | 0 |
| 0 | 0 | 0 | 0 | 0 |
| 0 | 0 | 0 | 0 | 0 |
| 0 | 0 | 0 | 0 | 0 |
| 0 | 0 | 0 | 0 | 0 |
| 0.001422241 | 0.209108484 | 0.000644967 | 0 | 0 |
| 0 | 0 | 0 | 0 | 0 |
| 0.001049773 | 0 | 0.107017824 | 0.079063846 | 0 |
| 0 | 0 | 0 | 0 | 0 |
| 0 | 0 | 0 | 0 | 0 |
| 0.038375287 | 0 | 0 | 0 | 0 |
| 0 | 0 | 0 | 0 | 0 |
| 0 | 0 | 0 | 0 | 0 |
| 0 | 0 | 0 | 0 | 0 |
| 0 | 0 | 0 | 0 | 0.002050585 |
| 0 | 0 | 0 | 0 | 0 |
| 0 | 0 | 0 | 0 | 0 |
| 0 | 0 | 0 | 0 | 0 |
| 0 | 0 | 0 | 0 | 0 |
| 0 | 0 | 0 | 0 | 0 |
| 0 | 0 | 0 | 0 | 0 |
| 0 | 0 | 0 | 0 | 0 |
| 0 | 0 | 0 | 0 | 0 |
| 0 | 0 | 0 | 0.001017898 | 0 |
| 0 | 0.002956546 | 0 | 0 | 0 |

| | | | | |
|---|---|---|---|---|
| 0 | 0 | 0 | 0 | 0 |
| 0 | 0 | 0.06953539 | 0 | 0.001775908 |
| 0 | 0 | 0 | 0 | 0 |
| 0 | 0 | 0 | 0 | 0 |
| 0 | 0.005710172 | 0 | 0 | 0 |
| 0 | 0 | 0 | 0 | 0 |
| 0 | 0 | 0 | 0 | 0 |
| 0 | 0 | 0 | 0 | 0 |
| 0.682720719 | 0.125869725 | 0.000649748 | 0 | 0.018706395 |
| 0 | 0 | 0 | 0 | 0 |
| 0 | 0 | 0 | 0 | 0 |
| 0 | 0.191637334 | 0 | 0 | 0 |
| 0 | 0 | 0 | 0 | 0 |
| 0 | 0 | 0 | 0 | 0 |
| 0 | 0 | 0 | 0 | 0 |
| 0 | 0 | 0.002273897 | 0 | 0 |
| 0 | 0 | 0 | 0 | 0 |
| 0 | 0 | 0 | 0 | 0 |
| 0 | 0.101034713 | 0 | 0 | 0.119156742 |
| 0 | 0 | 0 | 0 | 0 |
| 0 | 0 | 0 | 0 | 0 |
| 0 | 0 | 0 | 0 | 0 |
| 0 | 0 | 0 | 0 | 0 |
| 0 | 0 | 0 | 0 | 0 |
| 0 | 0 | 0 | 0 | 0 |
| 0 | 0 | 0 | 0 | 0 |
| 0 | 0 | 0 | 0 | 0 |
| 0 | 0 | 0 | 0 | 0 |
| 0 | 0 | 0 | 0 | 0 |
| 0 | 0 | 0 | 0 | 0 |
| 0.001657319 | 0.16396405 | 0.032768531 | 0 | 0.149238768 |
| 0.006780819 | 0.001176825 | 0 | 0 | 0 |
| 0 | 0 | 0 | 0 | 0 |
| 0 | 0 | 0 | 0 | 0 |
| 0 | 0 | 0 | 0 | 0 |
| 0 | 0 | 0 | 0 | 0 |
| 0 | 0 | 0 | 0 | 0 |
| 0 | 0 | 0 | 0 | 0 |
| 0 | 0.248792565 | 0 | 0 | 0.155784203 |
| 0 | 0 | 0 | 0 | 0 |
| 0 | 0 | 0 | 0 | 0 |
| 0 | 0 | 0 | 0 | 0 |
| 0 | 0 | 0.000729072 | 0.001562669 | 0 |
| 0 | 0 | 0 | 0 | 0 |
| 0 | 0 | 0 | 0 | 0 |
| 0 | 0 | 0 | 0 | 0 |
| 0 | 0 | 0 | 0 | 0 |
| 0 | 0 | 0 | 0 | 0 |

| | | | | |
|---|---|---|---|---|
| 0.015613228 | 0 | 0 | 0 | 0.003166543 |
| 0 | 0 | 0 | 0 | 0 |
| 0 | 0 | 0 | 0 | 0 |
| 0 | 0 | 0 | 0 | 0 |
| 0.002118644 | 0.004950749 | 0 | 0 | 0.129519451 |
| 0 | 0 | 0 | 0 | 0 |
| 0 | 0 | 0 | 0 | 0 |
| 0 | 0 | 0 | 0 | 0 |
| 0 | 0 | 0 | 0 | 0 |
| 0 | 0 | 0 | 0 | 0 |
| 0 | 0 | 0 | 0 | 0 |
| 0 | 0 | 0 | 0 | 0 |
| 0 | 0 | 0 | 0 | 0 |
| 0 | 0 | 0 | 0 | 0 |
| 0 | 0 | 0 | 0 | 0 |
| 0 | 0 | 0 | 0 | 0 |
| 0 | 0 | 0 | 0 | 0 |
| 0 | 0 | 0 | 0 | 0 |
| 0 | 0 | 0 | 0 | 0 |
| 0 | 0 | 0 | 0 | 0 |
| 0 | 0 | 0 | 0 | 0 |
| 0 | 0 | 0 | 0 | 0 |
| 0 | 0 | 0 | 0 | 0 |
| 0 | 0 | 0 | 0 | 0 |
| 0 | 0 | 0 | 0 | 0.281369805 |
| 0 | 0 | 0 | 0 | 0 |
| 0.018697761 | 0 | 0 | 0 | 0 |
| 0 | 0 | 0 | 0 | 0 |
| 0 | 0 | 0 | 0 | 0 |
| 0 | 0 | 0 | 0 | 0 |
| 0 | 0 | 0 | 0 | 0 |
| 0.001404236 | 0.001357802 | 0 | 0 | 0.000813701 |
| 0 | 0 | 0 | 0 | 0 |
| 0 | 0 | 0 | 0 | 0 |
| 0 | 0 | 0 | 0 | 0 |
| 0 | 0 | 0 | 0 | 0 |
| 0 | 0 | 0 | 0 | 0 |
| 0 | 0 | 0 | 0 | 0 |
| 0 | 0 | 0 | 0 | 0 |
| 0 | 0 | 0 | 0 | 0 |
| 0 | 0 | 0 | 0 | 0 |
| 0 | 0 | 0 | 0 | 0 |
| 0 | 0 | 0 | 0 | 0 |
| 0 | 0 | 0 | 0 | 0 |
| 0 | 0 | 0 | 0 | 0 |
| 0 | 0 | 0 | 0 | 0 |
| 0 | 0 | 0 | 0 | 0 |
| 0 | 0 | 0 | 0 | 0 |

| | | | | |
|---|---|---|---|---|
| 0 | 0 | 0 | 0 | 0 |
| 0 | 0 | 0 | 0 | 0 |
| 0 | 0 | 0 | 0 | 0 |
| 0 | 0 | 0 | 0 | 0 |
| 0 | 0 | 0 | 0 | 0 |
| 0 | 0 | 0 | 0 | 0 |
| 0 | 0 | 0 | 0 | 0 |
| 0 | 0 | 0 | 0 | 0 |
| 0 | 0 | 0 | 0 | 0 |
| 0 | 0 | 0 | 0 | 0 |
| 0 | 0 | 0 | 0 | 0 |
| 0 | 0 | 0 | 0 | 0 |
| 0 | 0 | 0 | 0 | 0 |
| 0 | 0 | 0 | 0 | 0 |
| 0 | 0 | 0 | 0 | 0.008660455 |
| 0 | 0.005244197 | 0 | 0 | 0 |
| 0 | 0 | 0 | 0 | 0 |
| 0 | 0 | 0 | 0 | 0 |
| 0 | 0 | 0 | 0 | 0 |
| 0 | 0 | 0 | 0 | 0 |
| 0.011126462 | 0.021459238 | 0.017253044 | 0.092498837 | 0.102637762 |
| 0 | 0 | 0 | 0 | 0 |
| 0 | 0 | 0 | 0 | 0.000958067 |
| 0 | 0 | 0 | 0.005404956 | 0.005693114 |
| 0 | 0 | 0 | 0 | 0 |
| 0 | 0 | 0 | 0 | 0 |
| 0.001254153 | 0.028295896 | 0.000758322 | 0.010680951 | 0.001695711 |
| 0 | 0 | 0 | 0 | 0 |
| 0 | 0 | 0 | 0 | 0 |
| 0 | 0 | 0.010547123 | 0 | 0.005442646 |
| 0 | 0 | 0.091655086 | 0.623780845 | 0 |
| 0 | 0 | 0 | 0 | 0 |
| 0 | 0 | 0 | 0 | 0 |
| 2.735494617 | 0 | 0 | 0 | 0.084079298 |
| 0.015413063 | 0.010010866 | 0.004871552 | 0.007004251 | 0.006393983 |
| 0 | 0 | 0 | 0 | 0 |
| 0 | 0 | 0 | 0 | 0 |
| 0 | 0 | 0 | 0 | 0 |
| 0 | 0.0048516 | 0 | 0 | 0 |
| 0 | 0 | 0 | 0 | 0 |
| 0 | 0 | 0 | 0 | 0 |
| 0 | 0 | 0 | 0 | 0 |
| 0 | 0 | 0 | 0 | 0 |
| 0 | 0 | 0 | 0 | 0 |
| 0 | 0 | 0 | 0 | 0 |
| 0 | 0.0027591 | 0 | 0 | 0.870550833 |
| 0.001264261 | 0 | 0 | 0 | 0 |
| 0 | 0 | 0 | 0 | 0.029533032 |

| | | | | |
|---|---|---|---|---|
| 0 | 0 | 0 | 0 | 0 |
| 0 | 0 | 0 | 0 | 0 |
| 0 | 0 | 0 | 0 | 0.001850514 |
| 0 | 0 | 0 | 0 | 0 |
| 0 | 0 | 0 | 0 | 0 |
| 0 | 0 | 0 | 0 | 0 |
| 0 | 0.001336208 | 0 | 0 | 0 |
| 0 | 0 | 0 | 0.003454753 | 0 |
| 0 | 0 | 0 | 0 | 0 |
| 0 | 0 | 0 | 0 | 0 |
| 0 | 0 | 0 | 0 | 0 |
| 0 | 0 | 0 | 0 | 0 |
| 0 | 0 | 0 | 0 | 0 |
| 0 | 0 | 0 | 0 | 0 |
| 0 | 0.002959993 | 0 | 0 | 0 |
| 0 | 0 | 0 | 0 | 0 |
| 0 | 0 | 0 | 0 | 0 |
| 0 | 0 | 0 | 0 | 0.056951845 |
| 0.002030985 | 0 | 0 | 0 | 0.032266063 |
| 0 | 0.000656382 | 0 | 0 | 0 |
| 0 | 0 | 0.001358605 | 0.001941326 | 0.004557038 |
| 0 | 0 | 0 | 0 | 0 |
| 0 | 0 | 0 | 0 | 0 |
| 0 | 0 | 0 | 0 | 0 |
| 0 | 0.003916727 | 0 | 0.002754895 | 0 |
| 0 | 0 | 0 | 0 | 0 |
| 0 | 0 | 0 | 0 | 0 |
| 0 | 0 | 0 | 0 | 0 |
| 0 | 0.064610359 | 0 | 0 | 0 |
| 0 | 0 | 0 | 0 | 0 |
| 0.061313798 | 0.193420133 | 0.366126752 | 0.356863982 | 0.747039567 |
| 0 | 0 | 0 | 0 | 0 |
| 0 | 0.022659789 | 0 | 0 | 0 |
| 0 | 0 | 0 | 0.005012767 | 0 |
| 0 | 0 | 0 | 0 | 0 |
| 0 | 0 | 0 | 0 | 0 |
| 0 | 0 | 0 | 0 | 0 |
| 0 | 0 | 0 | 0 | 0 |
| 0 | 0 | 0 | 0 | 0 |
| 0 | 0 | 0 | 0 | 0 |
| 0 | 0 | 0 | 0 | 0 |
| 0 | 0.004221834 | 0.00084858 | 0 | 0.005882373 |
| 0 | 0 | 0 | 0 | 0 |
| 0 | 0 | 0 | 0 | 0 |
| 0 | 0 | 0 | 0 | 0 |
| 0 | 0 | 0 | 0 | 0 |
| 0 | 0 | 0 | 0 | 0 |
| 0 | 0 | 0 | 0 | 0 |

| | | | | |
|---|---|---|---|---|
| 0 | 0 | 0 | 0 | 0 |
| 0 | 0 | 0 | 0 | 0 |
| 0 | 0 | 0 | 0.077258524 | 0.156779893 |
| 0 | 0 | 0 | 0 | 0 |
| 0 | 0 | 0 | 0 | 0 |
| 0 | 0 | 0 | 0 | 0 |
| 0 | 0 | 0 | 0 | 0 |
| 0 | 0 | 0 | 0 | 0 |
| 0 | 0 | 0 | 0 | 0 |
| 0 | 0 | 0 | 0 | 0 |
| 0 | 0 | 0 | 0 | 0 |
| 0 | 0 | 0 | 0 | 0 |
| 0 | 0 | 0 | 0 | 0 |
| 0 | 0.002636456 | 0 | 0.004038462 | 0.010006479 |
| 0.036221791 | 0.033849659 | 0 | 0.001190436 | 0.007607009 |
| 0 | 0.012684185 | 0 | 0 | 0.022338673 |
| 0 | 0.000826471 | 0 | 0.002712791 | 0.001238215 |
| 0.001136627 | 0 | 0 | 0 | 0 |
| 0 | 0 | 0 | 0 | 0 |
| 0 | 0 | 0 | 0 | 0 |
| 0 | 0 | 0 | 0 | 0 |
| 0 | 0 | 0 | 0 | 0.03263844 |
| 0 | 0 | 0 | 0 | 0 |
| 0 | 0 | 0 | 0 | 0 |
| 0.00212089 | 0 | 0 | 0 | 0 |
| 0 | 0 | 0 | 0 | 0 |
| 0 | 0 | 0 | 0 | 0 |
| 0 | 0 | 0 | 0 | 0 |
| 0 | 0 | 0 | 0 | 0 |
| 0 | 0.00287785 | 0.008098182 | 0.001062696 | 0 |
| 0 | 0.001524596 | 0 | 0 | 0 |
| 0 | 0 | 0 | 0 | 0 |
| 0 | 0 | 0 | 0 | 0 |
| 0 | 0 | 0 | 0 | 0 |
| 0 | 0 | 0 | 0 | 0 |
| 0.001761285 | 0 | 0 | 0.003423897 | 0 |
| 0 | 0 | 0 | 0 | 0 |
| 0.054841048 | 0.001249809 | 0 | 0 | 0.074149319 |
| 0 | 0 | 0 | 0 | 0 |
| 0 | 0 | 0 | 0 | 0 |
| 0 | 0 | 0 | 0 | 0 |
| 0 | 0 | 0 | 0 | 0 |
| 0 | 0 | 0 | 0 | 0 |
| 0 | 0 | 0 | 0 | 0 |
| 0 | 0 | 0 | 0 | 0 |
| 0 | 0.003362732 | 0 | 0 | 0 |
| 0 | 0 | 0 | 0 | 0 |
| 0 | 0.006055328 | 0.436333211 | 0 | 0 |

| | | | | |
|---|---|---|---|---|
| 18.50542413 | 0 | 0 | 0.95816787 | 0 |
| 0 | 0 | 0 | 0 | 0 |
| 0 | 0 | 0 | 0 | 0 |
| 0 | 0 | 0 | 0 | 0 |
| 0.007666358 | 0.011531104 | 0.00337124 | 0 | 0 |
| 0.001515235 | 0 | 0 | 0 | 0 |
| 0 | 0 | 0 | 0 | 0 |
| 0 | 0.002571447 | 0.003248805 | 0.031470861 | 0 |
| 0 | 0 | 0 | 0 | 0 |
| 0.054038557 | 0.878812155 | 0.309898964 | 0.021625132 | 0.064668704 |
| 0 | 0 | 0 | 0 | 0 |
| 0.001031108 | 0.100615145 | 0 | 0.016035598 | 0 |
| 0.000586599 | 0.001764628 | 0.00053203 | 0.003258099 | 0.024388659 |
| 0 | 0 | 0 | 0 | 0 |
| 0 | 0 | 0 | 0 | 0 |
| 0 | 0 | 0 | 0 | 0 |
| 0 | 0 | 0 | 0 | 0 |
| 0 | 0 | 0 | 0 | 0 |
| 0 | 0 | 1.324845677 | 0.012279482 | 0 |
| 0 | 0 | 0 | 0.032340513 | 0 |
| 0 | 0 | 0 | 0 | 0 |
| 0 | 0 | 0 | 0 | 0 |
| 0 | 0.005917821 | 0 | 0 | 0 |
| 0 | 0 | 0 | 0 | 0 |
| 0 | 0 | 0 | 0 | 0 |
| 0 | 0 | 0 | 0.001517942 | 0.002309477 |
| 0 | 0 | 0 | 0 | 0 |
| 0 | 0 | 0 | 0 | 0 |
| 0 | 0 | 0.041662994 | 0 | 0.007330172 |
| 0 | 0 | 0 | 0.002602952 | 0 |
| 0 | 0 | 0 | 0.002443251 | 0 |
| 0 | 0 | 0 | 0 | 0 |
| 0 | 0 | 0 | 0.002249957 | 0 |
| 0 | 0 | 0.002701421 | 0 | 0 |
| 2.60959172 | 0 | 0 | 0 | 0 |
| 0 | 0 | 0 | 0.056696602 | 0 |
| 0 | 0 | 0.008195478 | 0.739058991 | 0 |
| 0 | 0 | 0 | 0 | 0 |
| 0 | 0 | 0 | 0 | 0 |
| 0 | 0 | 0 | 0.018093512 | 0 |
| 0 | 0 | 0 | 0 | 0 |
| 0 | 0 | 0.555285373 | 0 | 0 |
| 0 | 0 | 0 | 0 | 0 |
| 0 | 0 | 0 | 0.031591778 | 0 |
| 0 | 0 | 0 | 0 | 0 |
| 0 | 0 | 0 | 0 | 0 |
| 0 | 0.00127799 | 0 | 0.001573067 | 0.004595228 |
| 0 | 0 | 0.021683586 | 0 | 0 |

| | | | | |
|---|---|---|---|---|
| 0 | 0 | 0 | 0 | 0 |
| 0 | 0.035706077 | 0 | 0 | 0 |
| 0.001029594 | 0 | 0 | 0 | 0 |
| 0 | 0 | 0 | 0 | 0 |
| 0 | 0 | 0 | 0 | 0 |
| 0.000892281 | 0 | 0.007485795 | 0.015363365 | 0.044562614 |
| 0 | 0 | 0 | 0 | 0 |
| 0 | 0 | 0 | 0 | 0 |
| 0 | 0 | 0 | 0 | 0 |
| 0 | 0 | 0 | 0 | 0 |
| 0.009314063 | 0.020138579 | 0.001173279 | 0.036500038 | 0 |
| 0 | 0 | 0.002419185 | 0 | 0 |
| 0 | 0 | 0 | 0 | 0 |
| 0.001222147 | 0.003584593 | 0 | 0.001357614 | 0.01611126 |
| 0 | 0 | 0.008523833 | 0.062913702 | 0 |
| 0 | 0 | 0.021929408 | 0 | 0 |
| 0 | 0 | 0 | 0 | 0 |
| 0 | 0 | 0.003020375 | 0 | 0 |
| 0 | 0 | 0 | 0 | 0 |
| 0 | 0 | 0 | 0.002605032 | 0 |
| 0 | 0.002970963 | 0 | 0 | 0 |
| 0 | 0 | 0 | 0 | 0 |
| 0 | 0 | 0 | 0 | 0 |
| 0 | 0 | 0 | 0 | 0 |
| 0 | 0 | 0 | 0 | 0 |
| 0 | 0 | 0 | 0 | 0 |
| 0 | 0 | 0.023796795 | 1.873872872 | 0 |
| 0 | 0 | 0 | 0 | 0 |
| 0 | 0 | 0 | 0 | 0 |
| 0.094944815 | 0.574735497 | 0.090661742 | 0.205763735 | 0.828367241 |
| 0.0044773 | 0.001309464 | 0 | 0 | 0.001345257 |
| 0 | 0.041758996 | 0 | 0 | 0 |
| 0 | 0 | 0 | 0 | 0 |
| 0 | 0 | 0.001065087 | 0 | 0 |
| 0 | 0.001743114 | 0.007407657 | 0 | 0 |
| 0 | 0 | 0 | 0 | 0 |
| 0 | 0 | 0 | 0 | 0 |
| 0 | 0.05508588 | 0 | 0.031273215 | 0 |
| 0.001304186 | 0 | 0 | 0.005215487 | 0 |
| 0 | 0.005872834 | 0 | 0 | 0 |
| 0.02150139 | 0.057351705 | 0.022781759 | 0.05067124 | 0.030565867 |
| 0.74573295 | 1.769067294 | 0 | 0.067384896 | 0.003855564 |
| 0 | 0.012615557 | 0 | 0.030676485 | 0 |
| 0 | 0 | 0 | 0 | 0 |
| 0 | 0 | 0 | 0 | 0 |
| 0 | 0 | 0 | 0 | 0 |
| 0 | 0.019747678 | 0.002104176 | 0.004920023 | 0.009624321 |
| 0 | 0 | 0 | 0 | 0 |

| | | | | |
|---|---|---|---|---|
| 0.002342036 | 0 | 0 | 0 | 0.012138682 |
| 0.000831384 | 0 | 0 | 0 | 0 |
| 0 | 0 | 0 | 0.247357161 | 0.058249156 |
| 0 | 0 | 0 | 0 | 0 |
| 0 | 0 | 0 | 0 | 0.078342495 |
| 0 | 0.003723054 | 0.045099111 | 0 | 0 |
| 0 | 0 | 0 | 0 | 0 |
| 0 | 0 | 0 | 0 | 0 |
| 0.000653358 | 0 | 0 | 0.0573365 | 0 |
| 0.02195857 | 0 | 0 | 0.072043073 | 0 |
| 0 | 0 | 0 | 0 | 0 |
| 0 | 0 | 0 | 0 | 0 |
| 0 | 0 | 0 | 0 | 0.003244967 |
| 0.00094168 | 0.011685282 | 0 | 0 | 0 |
| 0 | 0 | 0 | 0 | 0 |
| 0.041026808 | 0.013697769 | 0 | 0 | 0.039364796 |
| 0.027529066 | 0.002300386 | 0 | 0 | 0.016749638 |
| 0 | 0 | 0.001572941 | 0 | 0 |
| 0 | 0 | 0 | 0 | 0 |
| 0 | 0.019327519 | 3.594725675 | 0.003323059 | 0 |
| 0.028783549 | 0.008987338 | 0.029369147 | 0 | 0 |
| 0.000709606 | 0.001334164 | 0 | 0.001051016 | 0.000959442 |
| 0 | 0 | 0 | 0 | 0 |
| 0 | 0.525642831 | 0 | 0.566617747 | 0 |
| 0 | 0 | 0 | 0 | 0.042903982 |
| 0 | 0 | 0 | 0.004774447 | 0.024788732 |
| 0.001294384 | 0.006132753 | 0.002347944 | 0.003738432 | 0 |
| 0.040137951 | 0 | 0 | 0 | 0.491749032 |
| 0 | 0 | 0 | 0 | 0 |
| 0 | 0 | 0 | 0 | 0 |
| 0.001771876 | 0.00266511 | 0 | 0.062110071 | 0.064284953 |
| 0.006750882 | 0 | 0 | 0.002282357 | 0.325025922 |
| 0 | 0.017970046 | 0.002027756 | 0.037563805 | 0.043926407 |
| 0 | 0.038855145 | 0 | 0.004004077 | 0 |
| 0 | 0 | 0 | 0.012701744 | 0.002608889 |
| 0 | 0 | 0 | 0 | 0 |
| 0 | 0 | 0 | 0.018523783 | 0 |
| 0.003161443 | 0.031428778 | 0 | 0.001975426 | 0.039672833 |
| 0.197864549 | 0.035269617 | 0 | 0.230558118 | 0 |
| 0 | 0 | 0 | 0 | 0 |
| 0 | 0 | 0 | 0 | 0 |
| 0 | 0 | 0 | 0 | 0 |
| 0 | 0 | 0 | 0 | 0.005693346 |
| 0.006323983 | 0.006438009 | 0.223271958 | 0 | 0.009384708 |
| 0 | 0 | 0.344937639 | 0.947546722 | 0 |
| 0 | 0 | 0 | 0 | 0 |
| 0 | 0 | 0.001393998 | 0 | 0 |
| 0 | 0 | 0 | 0 | 0 |

| | | | | |
|---|---|---|---|---|
| 0 | 0 | 0.005314149 | 0 | 0 |
| 0.507520551 | 0.591662817 | 0.005128095 | 0.080154883 | 0.067437571 |
| 0 | 0 | 0 | 0 | 0 |
| 0.000978378 | 0 | 0 | 0 | 0 |
| 0 | 0 | 0 | 0 | 0 |
| 0 | 0 | 0 | 0 | 0 |
| 0 | 0 | 0.16807866 | 0.527930128 | 0 |
| 0 | 0 | 0 | 0 | 0 |
| 0 | 0 | 0 | 0 | 0 |
| 0 | 0 | 0 | 0 | 0 |
| 0.003201216 | 0 | 0.001088781 | 0 | 0 |
| 0.006258396 | 0.004706682 | 0 | 0.051644168 | 0.052886462 |
| 0 | 0.030742155 | 0 | 0 | 0 |
| 0 | 0 | 0.029727219 | 0 | 0 |
| 0 | 0 | 0 | 0.001275038 | 0.026072404 |
| 0 | 0 | 0 | 0 | 0 |
| 0.002304593 | 0.005435912 | 0.000570056 | 0.017687565 | 0 |
| 0.004351535 | 0.001840843 | 0.298224443 | 0.14380776 | 0 |
| 0 | 0 | 0 | 0 | 0 |
| 0.007770563 | 0 | 0.060065561 | 0.041197608 | 0.003581727 |
| 0.020525326 | 0.03265986 | 0 | 0.018240342 | 0.00197184 |
| 0 | 0 | 0 | 0 | 0 |
| 0 | 0 | 0 | 0 | 0 |
| 0 | 0 | 0 | 0 | 0 |
| 0 | 0 | 0 | 0 | 0 |
| 0 | 0 | 0 | 0 | 0.006429272 |
| 0 | 0 | 0 | 0 | 0.033952202 |
| 0.037889891 | 0.105174025 | 0.067688868 | 0.04719153 | 0.060311742 |
| 0 | 0 | 0 | 0 | 0 |
| 0 | 0 | 0 | 0 | 0.011797311 |
| 0 | 0 | 0 | 0 | 0 |
| 0 | 0 | 0 | 0 | 0 |
| 0 | 0 | 0 | 0 | 0.00169617 |
| 0 | 0 | 0 | 0 | 0 |
| 0 | 0 | 0 | 0 | 0 |
| 0.070344696 | 0.429657014 | 3.295785003 | 0.246945847 | 0.212030213 |
| 0 | 0 | 0 | 0 | 0 |
| 0 | 0 | 0 | 0 | 0 |
| 0.046458138 | 0.041224055 | 0.001284643 | 0.008811075 | 0.010628754 |
| 0 | 0 | 0 | 0 | 0 |
| 0 | 0 | 0.188632103 | 0.908334923 | 0 |
| 0 | 0 | 0 | 0 | 0 |
| 0.004990013 | 0 | 0 | 0.017967366 | 0 |
| 0.012127473 | 0.002998544 | 0 | 0.011995372 | 0.00488549 |
| 0 | 0 | 0.172812408 | 0 | 0 |
| 0 | 0.078421026 | 0 | 0 | 0.363261682 |
| 0 | 0 | 0 | 0 | 0 |
| 0 | 0 | 0 | 0 | 0 |

| | | | | |
|---|---|---|---|---|
| 0 | 0 | 0 | 0 | 0 |
| 0 | 0 | 0 | 0 | 0 |
| 0 | 0 | 0 | 0 | 0.001918891 |
| 0.4847831 | 0.289829558 | 0 | 0 | 1.74691068 |
| 0 | 0 | 0 | 0 | 0 |
| 0.012859933 | 0 | 0 | 0 | 0 |
| 0 | 0.000876359 | 0 | 0 | 0 |
| 0 | 0 | 0 | 0 | 0 |
| 0 | 0.258542356 | 0 | 0 | 0 |
| 0.023105711 | 0 | 0 | 0 | 0.071199737 |
| 0 | 0 | 0 | 0 | 0 |
| 0.009408533 | 0.000649816 | 0 | 0 | 0.00077884 |
| 0 | 0 | 0 | 0 | 0 |
| 0.06807658 | 0 | 0 | 0 | 0 |
| 0.004278092 | 0.088120381 | 0.00143708 | 0.005632346 | 1.192049966 |
| 0 | 0 | 0 | 0 | 0 |
| 0.014467695 | 0.009093037 | 0 | 0 | 0.069582829 |
| 0 | 0 | 0 | 0 | 0 |
| 0 | 0 | 0 | 0 | 0 |
| 0 | 0 | 0 | 0 | 0 |
| 0 | 0 | 0 | 0 | 0 |
| 0 | 0 | 0 | 0 | 0 |
| 0 | 0 | 0 | 0 | 0 |
| 0 | 0 | 0.440615099 | 0.693602502 | 0.330901434 |
| 0.261567175 | 0 | 0.000489143 | 0 | 0 |
| 0 | 0 | 0 | 0 | 0 |
| 0 | 0.167702182 | 0 | 0 | 0.008025559 |
| 0 | 2.025397542 | 0 | 0 | 0.819739312 |
| 0 | 0 | 0 | 0 | 0 |
| 0 | 0 | 0 | 0 | 0 |
| 0.159227221 | 0 | 0 | 0 | 0.027068676 |
| 0 | 0 | 0 | 0 | 0 |
| 0.006393039 | 0 | 0 | 0 | 0.014838662 |
| 0 | 0 | 0 | 0 | 0 |
| 0.009996162 | 0 | 0 | 0 | 0 |
| 0 | 0 | 0 | 0 | 0 |
| 0.390832781 | 8.116182223 | 0 | 0 | 2.109529289 |
| 0 | 0 | 0 | 0 | 0.022487555 |
| 0 | 0 | 0 | 0 | 0.006080798 |
| 0.016629788 | 0.060310649 | 0 | 0 | 0.002369261 |
| 0.185305318 | 0.225456461 | 0.153198219 | 0.060157551 | 0.03791828 |
| 0 | 0.014618262 | 0 | 0 | 0 |
| 0.017235711 | 4.781581129 | 0.241299022 | 13.13550603 | 1.046215524 |
| 0 | 0 | 0 | 0 | 0.110485602 |
| 0 | 0 | 0 | 0 | 0 |
| 0 | 0 | 0 | 0 | 0 |
| 0 | 0 | 0 | 0 | 0 |
| 0.063771745 | 0.001998338 | 0 | 0.014020503 | 1.104748979 |

| | | | | |
|---|---|---|---|---|
| 0 | 0 | 0 | 0 | 0 |
| 0 | 0 | 0 | 0 | 0 |
| 0.004882103 | 0 | 0.592188395 | 0.578165885 | 0.463862268 |
| 0 | 0 | 0 | 0 | 0.001628719 |
| 0 | 0 | 0 | 0 | 0 |
| 0.071734999 | 0.006602143 | 0 | 0 | 0.025773897 |
| 0 | 0 | 0 | 0 | 0 |
| 0.246558454 | 0 | 0 | 0 | 0 |
| 0.185827397 | 0 | 0 | 0 | 0 |
| 0.017000729 | 0 | 0 | 0 | 0 |
| 1.049537252 | 0.060891258 | 0.006993722 | 0.01389045 | 0.068895749 |
| 1.251882491 | 2.24906625 | 2.213847648 | 3.001326203 | 1.199871168 |
| 0 | 0 | 0 | 0 | 0 |
| 0.053306335 | 0 | 0 | 0.001249262 | 0.001140416 |
| 0.218013276 | 0.10557347 | 0.162847747 | 0.094705635 | 0.630521796 |
| 0 | 0.051045846 | 0 | 0 | 0 |
| 0.006479158 | 0.007633909 | 0 | 0 | 0 |
| 0 | 0 | 0 | 0 | 0 |
| 0 | 0 | 0 | 0 | 0 |
| 0.003605121 | 0.007165481 | 0.038614176 | 0.042717078 | 0.019149436 |
| 2.21576112 | 0.182073811 | 0.040557101 | 0.032598278 | 0.018421649 |
| 0 | 0.006228874 | 16.21811555 | 1.089337252 | 0.291533821 |
| 2.160649084 | 7.000646326 | 8.599659321 | 0.86065 | 0.93590275 |
| 0 | 0 | 0 | 0 | 0 |
| 0.003595184 | 0 | 0 | 0 | 0 |
| 0 | 0.006139237 | 0 | 0.010230659 | 0 |
| 0 | 0 | 0.098342761 | 0.025760628 | 0 |
| 0 | 0 | 0 | 0 | 0.000738979 |
| 0 | 0 | 0 | 0 | 0 |
| 0 | 0 | 0 | 0 | 0 |
| 0 | 0 | 0 | 0 | 0 |
| 0 | 0.293927254 | 0 | 0 | 0.299500809 |
| 0.004066747 | 0 | 0.029313321 | 0.069427154 | 0.147158712 |
| 0.157182368 | 0 | 0 | 0 | 0.068854695 |
| 0 | 0 | 0.001793776 | 0.002040055 | 0.0047274 |
| 0.030861017 | 0.022333476 | 0.002112462 | 0.008193126 | 0.296612228 |
| 0 | 0.003172622 | 0 | 0 | 1.199329371 |
| 0 | 0 | 0 | 0 | 0 |
| 0 | 0 | 0 | 0.005298417 | 0 |
| 0 | 0 | 0 | 0 | 0 |
| 0 | 0.005591243 | 0 | 0 | 0 |
| 0 | 0 | 0 | 0 | 0 |
| 0 | 0 | 0 | 0 | 0 |
| 0 | 0 | 0 | 0 | 0 |
| 0.195666933 | 0.069664644 | 0.004617294 | 0.008113944 | 0.089557252 |
| 0 | 0 | 0 | 0 | 0 |
| 0 | 0 | 0 | 0 | 0 |
| 0.286223746 | 0 | 0.008717819 | 0.023431003 | 0.031678122 |

| | | | | |
|---|---|---|---|---|
| 0 | 0 | 0 | 0 | 0 |
| 0 | 0 | 0 | 0 | 0 |
| 0 | 0 | 0 | 0 | 0 |
| 0.003207041 | 0.002766572 | 0 | 0 | 0.031182137 |
| 0.064716588 | 0 | 0 | 0 | 0 |
| 0 | 0 | 0 | 0 | 0.173349099 |
| 0 | 0.051024803 | 0 | 0 | 0.057176757 |
| 0 | 0.188385287 | 0 | 0.003278164 | 0.048408792 |
| 0 | 0 | 0 | 0 | 0 |
| 0 | 0 | 0 | 0 | 0 |
| 0 | 0.000711053 | 0 | 0 | 0.047725254 |
| 0 | 0 | 0 | 0 | 0 |
| 0 | 0 | 0 | 0 | 0.004290649 |
| 0 | 0 | 0 | 0 | 0 |
| 0 | 0 | 0 | 0 | 0 |
| 0 | 0 | 0 | 0 | 0 |
| 0.170326906 | 0.058348245 | 0 | 0 | 0 |
| 0.008395588 | 0.00118387 | 0 | 0 | 0.000709468 |
| 0.005337441 | 0 | 0 | 0 | 0 |
| 0.002050596 | 0 | 0 | 0 | 0.029111914 |
| 0 | 0 | 0 | 0 | 0 |
| 0 | 0 | 0 | 0 | 0 |
| 0 | 0.000909945 | 0 | 0 | 0.003271861 |
| 0 | 0 | 0 | 0 | 0 |
| 0 | 0 | 0 | 0 | 0 |
| 0 | 0 | 0 | 0 | 0 |
| 0 | 0 | 0 | 0 | 0 |
| 0 | 0 | 0 | 0 | 0 |
| 0 | 0 | 0 | 0 | 0 |
| 0.139449685 | 0 | 0 | 0.002816487 | 0.054421438 |
| 0 | 0 | 0 | 0 | 0 |
| 0 | 0 | 0 | 0 | 0 |
| 0.057931484 | 0 | 0 | 0 | 0 |
| 0 | 0 | 0 | 0 | 0 |
| 0 | 0 | 0 | 0 | 0 |
| 0 | 0 | 0 | 0 | 0 |
| 0 | 0 | 0 | 0.316725488 | 0.068201056 |
| 0 | 0 | 0.001129187 | 0.001975725 | 0.007935766 |
| 0 | 0 | 0 | 0 | 0 |
| 0 | 0.009697088 | 0 | 0.002728244 | 0 |
| 0.314725971 | 0.247359905 | 0.004843233 | 0 | 0 |
| 0 | 0 | 0 | 0 | 0 |
| 0 | 0 | 0 | 0 | 0 |
| 0 | 0.00818348 | 0 | 0.002110529 | 0 |
| 0 | 0 | 0 | 0 | 0 |
| 0 | 0 | 0 | 0 | 0 |
| 0.00067598 | 0 | 0 | 0 | 0 |
| 0 | 0 | 0 | 0 | 0 |

| | | | | |
|---|---|---|---|---|
| 0 | 0 | 0 | 0 | 0 |
| 0.183323967 | 0 | 0 | 0 | 0 |
| 0 | 0 | 0 | 0 | 0 |
| 0 | 0.137908078 | 0 | 0 | 0 |
| 0 | 0 | 0 | 0 | 0 |
| 0 | 0.429492138 | 0 | 0 | 0 |
| 0 | 0 | 0 | 0 | 0 |
| 0.001642811 | 0 | 0 | 0 | 0 |
| 0.003434143 | 0.002213724 | 1.283246456 | 0.174935635 | 0.049002615 |
| 0 | 0 | 0 | 0 | 0 |
| 0 | 0 | 0 | 0 | 0 |
| 0 | 0 | 0 | 0 | 0 |
| 0 | 0 | 0 | 0 | 0 |
| 0.013289224 | 0 | 0 | 0 | 0 |
| 0.027499076 | 0.675311637 | 2.507202627 | 2.741217105 | 0.505159301 |
| 0.001512451 | 0 | 0.141633471 | 0 | 0.321696266 |
| 0.223875311 | 0 | 0 | 0 | 0 |
| 0 | 0 | 0 | 0 | 0 |
| 0 | 0 | 0 | 0 | 0 |
| 0 | 0 | 0 | 0 | 0 |
| 0 | 0.005712893 | 0 | 0 | 0.00874923 |
| 0 | 0 | 0 | 0 | 0 |
| 0 | 0 | 0 | 0 | 0 |
| 0 | 0 | 0 | 0 | 0 |
| 0.01673074 | 0.002277866 | 0.001831385 | 0.007690442 | 0.242495883 |
| 0.142740435 | 3.458791147 | 0 | 0 | 0.958087582 |
| 0 | 1.134079686 | 0 | 0 | 0.274172672 |
| 0 | 0 | 0 | 0 | 0 |
| 0 | 0.016785555 | 0 | 0 | 0 |
| 0.368296667 | 0 | 0 | 0 | 0 |
| 0 | 0 | 0 | 0 | 0 |
| 0 | 0.006286965 | 0 | 0 | 0.077236625 |
| 0 | 0 | 0 | 0 | 0 |
| 0.006470382 | 0.001390316 | 0 | 0.001369063 | 0.021558691 |
| 0.008630451 | 0 | 0 | 0.004793541 | 0 |
| 0.006638978 | 0 | 0 | 0 | 0.018559334 |
| 0 | 0.012115424 | 0 | 0 | 0 |
| 0.64754016 | 1.130007812 | 0 | 1.735270681 | 1.177478636 |
| 0 | 0 | 0 | 0 | 0 |
| 0 | 0 | 0 | 0 | 0 |
| 0 | 0 | 0 | 0 | 0 |
| 0.120254995 | 0 | 0 | 0.007233001 | 0 |
| 0.329054115 | 0.175919901 | 0.145218675 | 0.074630832 | 0.655424854 |
| 0 | 0 | 0 | 0 | 0 |
| 0 | 0 | 0 | 0 | 0 |
| 0 | 0 | 0 | 0 | 0 |
| 0 | 0 | 0 | 0 | 0 |
| 2.148631019 | 0.025967588 | 0 | 5.319594217 | 0.091723116 |

| | | | | |
|---|---|---|---|---|
| 0 | 0 | 0 | 0.016151759 | 0.003686121 |
| 0 | 0 | 0 | 0 | 0 |
| 0 | 0 | 0 | 0 | 0 |
| 0 | 0 | 0 | 0 | 0 |
| 0.02312193 | 0.025856762 | 0 | 0 | 0 |
| 0 | 0 | 0 | 0 | 0 |
| 0 | 0 | 0 | 0 | 0 |
| 0.004507894 | 0 | 0 | 0 | 0 |
| 0 | 0 | 0 | 0 | 0 |
| 0 | 0 | 0 | 0 | 0 |
| 0 | 0 | 0 | 0 | 0 |
| 4.605182956 | 0.809379417 | 0 | 0 | 1.218125892 |
| 0.181856191 | 0.207896134 | 0 | 0 | 0.057767828 |
| 0 | 0 | 0 | 0 | 0 |
| 0.502489068 | 2.441319449 | 0.010755646 | 0.486895902 | 0.005659079 |
| 0 | 0 | 0 | 0 | 0 |
| 0.095921037 | 0 | 0 | 0 | 0 |
| 0 | 0.018563815 | 0 | 0 | 0 |
| 0.033220763 | 0 | 0 | 0 | 0 |
| 0 | 0 | 0 | 0 | 0.045604405 |
| 0 | 0.006471817 | 0 | 0 | 0 |
| 0 | 0 | 0 | 0 | 0 |
| 0.53012064 | 0.028168552 | 0.001617662 | 0 | 0.170616609 |
| 0 | 0 | 0 | 0 | 0 |
| 0 | 0 | 0 | 0 | 0 |
| 0 | 0 | 0 | 0 | 0 |
| 0 | 0 | 0 | 0 | 0 |
| 0 | 0 | 0 | 0 | 0 |
| 0 | 0 | 0 | 0 | 0 |
| 0 | 0 | 0 | 0 | 0 |
| 0 | 0 | 0 | 0 | 0 |
| 0.042544668 | 0.01463651 | 0 | 0.013742407 | 0.012545055 |
| 0 | 0 | 0 | 0 | 0 |
| 0.225660356 | 0.117164417 | 0.003691405 | 0.001004703 | 0.265519228 |
| 0.006639071 | 0 | 0 | 0 | 0.019801182 |
| 0 | 0.002271722 | 0 | 0 | 0.104912372 |
| 0.003466646 | 0 | 0 | 0 | 0 |
| 0 | 0 | 0 | 0 | 0 |
| 0 | 0 | 0 | 0 | 0 |
| 0 | 0 | 0 | 0 | 0 |
| 0.116623991 | 0 | 0 | 0 | 0 |
| 0 | 0 | 0 | 0 | 0 |
| 0 | 0 | 0 | 0 | 0 |
| 0.025752355 | 0.003227881 | 0 | 0 | 0.032723569 |
| 0.065593865 | 0.041229255 | 0 | 0.001495753 | 0.319315655 |
| 0 | 0 | 0 | 0 | 0 |
| 0.053247055 | 0.338860319 | 0 | 0.231922199 | 0 |
| 0.000909678 | 0.001847155 | 0.000990065 | 0 | 0.00055348 |

| | | | | |
|---|---|---|---|---|
| 0 | 0 | 0 | 0 | 0 |
| 0 | 0 | 0 | 0 | 0 |
| 0 | 0.002244283 | 0.001263069 | 0.001988978 | 0.006455757 |
| 0 | 0 | 0 | 0 | 0 |
| 0 | 0 | 0.006587662 | 0 | 0.030484797 |
| 0.00171151 | 0 | 0 | 0 | 0 |
| 0 | 0 | 0 | 0 | 0 |
| 0.455058879 | 0.002119515 | 0.003748958 | 0 | 0.022291639 |
| 0 | 0 | 0 | 0 | 0 |
| 0 | 0.004588504 | 0 | 0 | 0 |
| 0.040153202 | 0 | 0 | 0 | 0 |
| 0 | 0 | 0 | 0 | 0 |
| 0 | 0 | 0 | 0 | 0 |
| 0 | 0 | 0 | 0 | 0 |
| 0 | 0 | 0 | 0 | 0 |
| 0 | 0 | 0 | 0 | 0 |
| 0.275962343 | 0 | 0 | 0 | 0.926748663 |
| 0 | 0 | 0 | 0 | 0 |
| 7.527456528 | 0 | 0 | 0.341462909 | 2.219276273 |
| 0 | 0 | 0 | 0 | 0 |
| 0.007230398 | 0 | 0 | 0 | 0 |
| 0 | 0 | 0 | 0 | 0 |
| 0 | 0 | 0 | 0 | 0 |
| 0 | 0 | 0 | 0 | 0 |
| 0.004203343 | 0 | 0 | 0 | 0 |
| 0.001969852 | 0 | 0.003573206 | 0.004774249 | 0 |
| 0 | 0 | 0 | 0 | 0 |
| 1.42363391 | 0.580345325 | 0 | 0.305889407 | 0.10238127 |
| 0 | 0 | 0 | 0 | 0 |
| 0.186625067 | 0.536997527 | 0 | 0.831600848 | 0 |
| 0 | 0 | 0 | 0 | 0 |
| 0 | 0 | 0 | 0 | 0 |
| 0 | 0 | 0 | 0 | 0 |
| 0 | 0 | 0 | 0 | 0 |
| 0.001501297 | 0 | 0 | 0 | 0.150146852 |
| 0 | 0 | 0 | 0 | 0 |
| 0 | 0 | 0 | 0 | 0 |
| 0.006814128 | 0 | 0 | 0 | 0 |
| 0 | 0 | 0 | 0 | 0 |
| 0 | 0 | 0 | 0 | 0 |
| 1.873039493 | 11.42927763 | 2.475409544 | 9.31804723 | 1.674646053 |
| 1.352208198 | 0.127835519 | 0.15565242 | 1.01466951 | 0.150952442 |
| 0 | 0 | 0 | 0 | 0 |
| 0 | 0 | 0 | 0 | 0 |
| 0 | 0 | 0 | 0 | 0 |
| 0 | 0 | 0 | 0 | 0 |
| 0 | 0 | 0 | 0 | 0 |
| 0.033124423 | 0.001715844 | 0.001686085 | 0 | 0.003941693 |

| | | | | |
|---|---|---|---|---|
| 0 | 0 | 0 | 0 | 0 |
| 0.005910311 | 0.003529774 | 0.001261293 | 0 | 0.004759462 |
| 0.021926889 | 0 | 0 | 0 | 0 |
| 0 | 0 | 0 | 0 | 0 |
| 3.138341005 | 1.895525786 | 0.45725335 | 2.529383991 | 1.134931076 |
| 0 | 0 | 0 | 0 | 0 |
| 0 | 0 | 0 | 0 | 0 |
| 0 | 0 | 0 | 0 | 0 |
| 0.001735764 | 0.001370666 | 1.171745061 | 1.332360027 | 0.014433351 |
| 0 | 0 | 0 | 0 | 0 |
| 0 | 0 | 0 | 0 | 0 |
| 0 | 0 | 0 | 0 | 0 |
| 0.021451324 | 0.097177028 | 0 | 0 | 0 |
| 0.00353014 | 0.086673786 | 0.004331769 | 0.030448776 | 0.022531467 |
| 0 | 0 | 0 | 0 | 0 |
| 0.512734888 | 0.135712055 | 0 | 0.010032276 | 0.209428601 |
| 1.204844542 | 0.095476433 | 0 | 0.021558022 | 0.00400883 |
| 0 | 0 | 0 | 0 | 0 |
| 0 | 0 | 0 | 0 | 0 |
| 0 | 0 | 0 | 0 | 0 |
| 0 | 0 | 0 | 0 | 0 |
| 0 | 0 | 0 | 0 | 0 |
| 0 | 0 | 0 | 0 | 0 |
| 0 | 0 | 0 | 0 | 0 |
| 0 | 0 | 0 | 0 | 0 |
| 0 | 0 | 0 | 0 | 0 |
| 1.377351114 | 0.002115177 | 0 | 0 | 0.001742921 |
| 0 | 0 | 0 | 0 | 0.009833457 |
| 0 | 0 | 0 | 0 | 0 |
| 0 | 0 | 0 | 0 | 0 |
| 0 | 0 | 0 | 0 | 0 |
| 0 | 0 | 0 | 0 | 0 |
| 0.183017556 | 0 | 0 | 0 | 2.0727149 |
| 0 | 0.012926694 | 0 | 0 | 0 |
| 0 | 0 | 0 | 0 | 0 |
| 0 | 0 | 0 | 0.009742214 | 0.019234546 |
| 0.003543948 | 0 | 0 | 0 | 0.001902583 |
| 0 | 0 | 0 | 0 | 0 |
| 0 | 0 | 0 | 0 | 0 |
| 0 | 0 | 0 | 0 | 0 |
| 0.928232717 | 3.159701339 | 0 | 4.985788119 | 0 |
| 0 | 0 | 0 | 0 | 0 |
| 0 | 0 | 0 | 0 | 0 |
| 0 | 0 | 0 | 0 | 0 |
| 0 | 0 | 0 | 0 | 0 |
| 0 | 0 | 0 | 0 | 0 |
| 0 | 0 | 0 | 0 | 0 |
| 0 | 0 | 0 | 0 | 0 |

| | | | | |
|---|---|---|---|---|
| 0 | 0.339941187 | 0 | 0 | 0 |
| 0.000736938 | 0 | 0 | 0 | 0 |
| 0 | 0 | 0 | 0 | 0 |
| 0 | 0 | 0 | 0.008874594 | 0 |
| 0.01337694 | 0 | 0 | 0 | 0.012402276 |
| 0.270168264 | 0 | 0 | 0 | 0.820958013 |
| 0.879432415 | 1.357281466 | 0 | 0.019514342 | 0.052896947 |
| 0 | 0 | 0 | 0 | 0 |
| 0 | 0 | 0 | 0 | 0 |
| 0.005265939 | 0 | 0.107461478 | 0 | 0 |
| 0 | 0.019862933 | 0 | 0 | 0 |
| 0.124156455 | 0 | 0 | 0 | 0.017486363 |
| 0 | 0 | 0 | 0 | 0 |
| 0 | 0 | 0 | 0 | 0 |
| 0 | 0 | 0 | 0 | 0 |
| 0 | 0 | 0 | 0 | 0 |
| 0 | 0 | 0 | 0 | 0.011841373 |
| 0 | 0 | 0 | 0 | 0 |
| 0 | 0 | 0 | 0 | 0.228539737 |
| 0 | 0 | 0 | 0 | 0 |
| 0.008703771 | 0.030418475 | 0 | 0.346739977 | 0.098471886 |
| 0.004139383 | 0 | 0 | 0 | 0 |
| 0.002823597 | 0.006582884 | 0 | 0 | 0 |
| 0 | 0.008497115 | 0.001669949 | 0 | 0.00152764 |
| 0 | 0 | 0 | 0 | 0 |
| 0.006714197 | 0 | 0 | 0 | 0 |
| 0 | 0 | 0 | 0 | 0 |
| 0 | 0 | 0 | 0 | 0 |
| 0 | 0 | 0 | 0 | 0 |
| 0 | 0 | 0 | 0.464301273 | 8.408548575 |
| 0 | 0 | 0 | 0 | 0 |
| 0 | 0 | 0 | 0 | 0 |
| 0 | 0 | 0 | 0 | 0 |
| 0 | 0 | 0 | 0 | 0 |
| 0 | 0 | 0 | 0 | 0 |
| 0 | 0 | 0 | 0 | 0 |
| 0.006911576 | 0 | 0.024314626 | 0.078870712 | 0.143785306 |
| 0 | 0 | 0 | 0 | 0 |
| 0 | 0 | 0 | 0 | 0 |
| 0 | 0 | 0 | 0 | 0 |
| 0 | 0 | 0 | 0 | 0 |
| 0 | 0 | 0 | 0 | 0 |
| 0 | 0 | 0 | 0 | 0 |
| 0 | 0 | 0 | 0 | 0 |
| 0.530835196 | 0 | 0 | 0 | 0 |
| 0 | 0 | 0 | 0 | 0 |
| 0 | 0 | 0 | 0 | 0 |
| 0 | 0 | 0 | 0 | 0 |

| | | | | |
|---|---|---|---|---|
| 0.001621008 | 0 | 0.002450351 | 0 | 0.005296674 |
| 0 | 0 | 0 | 0 | 0 |
| 0 | 0.013521861 | 0 | 0 | 0.00998449 |
| 0 | 0 | 0 | 0 | 0 |
| 0.000458552 | 0 | 0 | 0 | 0 |
| 0 | 0 | 0 | 0 | 0 |
| 0 | 0 | 0 | 0 | 0 |
| 1.377945236 | 0.609802577 | 0 | 0.080118888 | 1.384563736 |
| 0.472130312 | 0.012438072 | 0.005641084 | 0.054330591 | 11.17506191 |
| 0 | 0 | 0.022491879 | 0 | 0.001197498 |
| 0 | 0 | 0 | 0 | 0.00398152 |
| 0 | 0 | 0 | 0 | 0 |
| 0.022158278 | 0 | 0 | 0 | 0 |
| 0 | 0 | 0 | 0 | 0 |
| 0 | 0 | 0 | 0 | 0 |
| 0 | 0 | 0 | 0 | 0 |
| 0.001840405 | 0.035294363 | 0.030760935 | 0.002920574 | 0 |
| 0 | 0 | 0 | 0 | 0 |
| 0 | 0 | 0 | 0 | 0 |
| 0.00522255 | 0.042161023 | 0 | 0 | 0 |
| 0 | 0 | 0 | 0 | 0 |
| 0 | 0 | 0 | 0 | 0 |
| 0 | 0 | 0 | 0 | 0 |
| 0 | 0 | 0 | 0 | 0 |
| 0.055326621 | 0 | 0 | 0 | 0 |
| 0 | 0 | 0 | 0 | 0 |
| 0 | 0 | 0 | 0 | 0 |
| 0 | 0 | 0 | 0 | 0 |
| 0 | 0 | 0.004786157 | 0 | 0 |
| 0 | 0 | 0 | 0 | 0.116339481 |
| 0 | 0.003131021 | 0 | 0 | 0 |
| 0 | 0 | 0 | 0 | 0 |
| 0 | 0 | 0 | 0 | 0 |
| 0 | 0 | 0 | 0 | 0 |
| 0 | 0 | 0 | 0 | 0 |
| 0 | 0 | 0 | 0 | 0 |
| 0.004553834 | 0 | 0 | 0 | 0 |
| 0 | 0 | 0 | 0.023725354 | 0 |
| 0 | 0 | 0 | 0.002616969 | 0 |
| 0 | 0 | 0 | 0.69429177 | 0 |
| 0 | 0 | 0 | 0 | 0 |
| 0.37453258 | 0.164691453 | 0 | 0 | 0 |
| 0 | 0 | 0 | 0 | 0 |
| 0 | 0 | 0 | 0 | 0 |
| 0 | 0 | 0 | 0 | 0 |
| 0.016656237 | 0.003794075 | 0 | 0 | 0 |
| 0 | 0 | 0 | 0 | 0 |
| 0 | 0 | 0 | 0 | 0 |

| | | | | |
|---|---|---|---|---|
| 0 | 0 | 0 | 0 | 0 |
| 0 | 0.004951281 | 0 | 0.022030459 | 0 |
| 0 | 0 | 0 | 0 | 0 |
| 0 | 0 | 0 | 0 | 0 |
| 0 | 0.179218096 | 0 | 0 | 1.084616607 |
| 0.39901085 | 0 | 0 | 0 | 0 |
| 0 | 0 | 0 | 0 | 0 |
| 0.004353194 | 0.003618474 | 0.169773893 | 0.058283117 | 0.012081487 |
| 0 | 0.000774195 | 0 | 0 | 0 |
| 0 | 0 | 0 | 0 | 0 |
| 0 | 0 | 0 | 0 | 0 |
| 0 | 0 | 0 | 0 | 0 |
| 0 | 0 | 0 | 0 | 0 |
| 0 | 0 | 0 | 0 | 0 |
| 0.019354703 | 0 | 0.000438855 | 0.001791671 | 0 |
| 0.175063612 | 0.019950465 | 0 | 0.002207359 | 0 |
| 0 | 0 | 0 | 0 | 0 |
| 0 | 0 | 0 | 0 | 0 |
| 0 | 0 | 0 | 0 | 0 |
| 0 | 0 | 0 | 0 | 0 |
| 0 | 0 | 0 | 0 | 0 |
| 0 | 0 | 0 | 0 | 0 |
| 0 | 0 | 0 | 0 | 0 |
| 0 | 0 | 0 | 0 | 0.053886205 |
| 0 | 0.437027439 | 0 | 0 | 0.418433296 |
| 0 | 0 | 0 | 0 | 0 |
| 0.316455869 | 0 | 0.003454073 | 0 | 0.006068682 |
| 0.000572867 | 0 | 0 | 0 | 0 |
| 0 | 0 | 0 | 0 | 0 |
| 0 | 0 | 0 | 0 | 0 |
| 0 | 0 | 0 | 0 | 0 |
| 0 | 0 | 0 | 0 | 0 |
| 0 | 0 | 0 | 0 | 0 |
| 0 | 0 | 0 | 0 | 0 |
| 0 | 0 | 0 | 0 | 0 |
| 0 | 0 | 0 | 0 | 0 |
| 0 | 0 | 0 | 0 | 0 |
| 0 | 0.002307884 | 0 | 0 | 0 |
| 0 | 0 | 0 | 0 | 0 |
| 0 | 0 | 0 | 0 | 0 |
| 0 | 0 | 0 | 0 | 0 |
| 0 | 0 | 0 | 0 | 0 |
| 0 | 0 | 0 | 0 | 0 |
| 0 | 0 | 0 | 0 | 0 |
| 0 | 0 | 0 | 0 | 0 |
| 0 | 0 | 0 | 0 | 0 |
| 0 | 0 | 0 | 0 | 0 |
| 0 | 0 | 0 | 0 | 0 |

| | | | | |
|---|---|---|---|---|
| 0.222389519 | 0.108892057 | 0 | 0 | 1.147121478 |
| 0 | 0 | 0 | 0 | 0 |
| 0 | 0 | 0 | 0 | 0 |
| 0.009332509 | 0 | 0 | 0 | 0 |
| 0 | 0 | 0 | 0 | 0 |
| 0 | 0 | 0 | 0.003731267 | 0.637955232 |
| 0 | 0 | 0 | 0 | 0 |
| 0 | 0 | 0 | 0 | 0 |
| 0 | 0 | 0 | 0 | 0 |
| 0 | 0 | 0 | 0 | 0 |
| 0 | 0 | 0 | 0 | 0 |
| 0.075854961 | 0 | 0 | 0 | 0 |
| 0 | 0 | 0 | 0 | 0 |
| 0 | 0 | 0 | 0 | 0 |
| 0 | 0 | 0 | 0 | 0 |
| 0 | 0 | 0 | 0 | 0 |
| 0 | 0 | 0 | 0 | 0 |
| 0 | 0 | 0 | 0 | 0 |
| 0.00144591 | 0.001631113 | 0 | 0 | 0 |
| 0 | 0 | 0 | 0 | 0 |
| 0 | 0 | 0.001969917 | 0.011844248 | 0 |
| 0 | 0 | 0 | 0 | 0 |
| 0.328915196 | 0.001687848 | 0.024652452 | 0 | 0 |
| 0 | 0 | 0 | 0 | 0.005091729 |
| 0 | 0 | 0 | 0 | 0 |
| 0 | 0 | 0 | 0 | 0 |
| 0 | 0 | 0 | 0 | 0 |
| 0 | 0 | 0 | 0 | 0 |
| 0 | 0 | 0 | 0 | 0.001532352 |
| 0 | 0 | 0 | 0 | 0 |
| 0 | 0 | 0 | 0 | 0 |
| 0 | 0 | 0 | 0 | 0.00076279 |
| 0 | 0.002897882 | 0.001694453 | 0.046176172 | 0.354510825 |
| 0 | 0 | 0 | 0 | 0 |
| 0.201790249 | 0.179564743 | 0 | 0 | 0.333630924 |
| 0 | 0 | 0 | 0 | 0 |
| 0 | 0 | 0 | 0 | 0 |
| 0 | 0 | 0 | 0 | 0 |
| 0.004551889 | 0 | 0 | 0.001926262 | 0.00527529 |
| 0 | 0 | 0 | 0 | 0 |
| 0 | 0 | 0 | 0 | 0 |
| 0 | 0 | 0 | 0 | 0 |
| 0 | 0.022873428 | 0 | 0 | 0 |
| 0 | 0 | 0 | 0 | 0 |
| 0 | 0 | 0 | 0 | 0 |
| 0 | 0.002043615 | 0 | 0 | 0.037965508 |
| 0 | 0 | 0 | 0 | 0 |
| 0 | 0 | 0 | 0 | 0 |

| | | | | |
|---|---|---|---|---|
| 0 | 0 | 0 | 0 | 0 |
| 0 | 0 | 0 | 0 | 0 |
| 0 | 0 | 0.087426221 | 0 | 0 |
| 0 | 0 | 0 | 0 | 0 |
| 0 | 0 | 0 | 0 | 0 |
| 0 | 0 | 0 | 0 | 0 |
| 0 | 0 | 0 | 0 | 0 |
| 0 | 0 | 0.392692623 | 1.944592145 | 0 |
| 0.478820675 | 0 | 0 | 0 | 0 |
| 0 | 0 | 0 | 0 | 0.0039945 |
| 0 | 0 | 0 | 0 | 0 |
| 0 | 0 | 0 | 0 | 0 |
| 0 | 0 | 0 | 0 | 0 |
| 0 | 0.087905436 | 0 | 0 | 0.0790197 |
| 0 | 0 | 0 | 0 | 0 |
| 0 | 0 | 0.001110039 | 0.014275322 | 0 |
| 0 | 0 | 0 | 0 | 0 |
| 0 | 0 | 0 | 0 | 0 |
| 0 | 0 | 0 | 0 | 0 |
| 0 | 2.035211359 | 0 | 1.887617841 | 1.01778597 |
| 0.00127827 | 0.008411675 | 0 | 0.00757311 | 0 |
| 0 | 0 | 0 | 0 | 0 |
| 0 | 0 | 0 | 0 | 0 |
| 0 | 0 | 0 | 0 | 1.109276593 |
| 0.039974689 | 0 | 0 | 0 | 0 |
| 0 | 0 | 0 | 0 | 0 |
| 0 | 0 | 0 | 0 | 0 |
| 0 | 0 | 0 | 0 | 0 |
| 0 | 0 | 0 | 0 | 0 |
| 0 | 0.022179696 | 0 | 0 | 0 |
| 1.61696035 | 0.415012803 | 0 | 0 | 0.240918431 |
| 0 | 0 | 0 | 0 | 0 |
| 0 | 0 | 0 | 0 | 0 |
| 0 | 0 | 0 | 0 | 0 |
| 0 | 0 | 0 | 0 | 0 |
| 0 | 0 | 0 | 0 | 0 |
| 0 | 0 | 0 | 0 | 0 |
| 0 | 0 | 0 | 0 | 0 |
| 0 | 0 | 0 | 0 | 0 |
| 0 | 0 | 0 | 0 | 0 |
| 0 | 0 | 0 | 0 | 0 |
| 0 | 0 | 0 | 0 | 0 |
| 0.00376567 | 0 | 0 | 0 | 0.026002896 |
| 0 | 0 | 0 | 0 | 0 |
| 0 | 0 | 0 | 0 | 0 |
| 0 | 0 | 0 | 0 | 0 |
| 0 | 0 | 0.265145695 | 0.941297752 | 0 |
| 0 | 0 | 0 | 0 | 0 |

| | | | | |
|---|---|---|---|---|
| 0 | 0 | 0 | 0 | 0 |
| 0 | 0 | 0 | 0 | 0 |
| 0 | 0 | 0 | 0 | 0 |
| 0 | 0 | 0 | 0 | 0 |
| 0.002652009 | 0.013198677 | 0.003395719 | 0 | 0.060746324 |
| 0 | 0 | 0 | 0 | 0 |
| 0 | 0 | 0 | 0 | 0 |
| 0 | 0 | 0 | 0 | 0 |
| 0 | 0 | 0 | 0 | 0 |
| 0 | 0 | 0 | 0 | 0 |
| 0 | 0 | 0 | 0 | 0 |
| 0.035736367 | 0.00537517 | 0 | 0 | 0.059519385 |
| 0.205356346 | 0 | 0 | 0 | 0 |
| 0 | 0 | 0 | 0 | 0 |
| 0 | 0.00055858 | 0 | 0 | 0.076321827 |
| 0 | 0 | 0 | 0 | 0.09683933 |
| 0 | 0.001315409 | 0.001510825 | 0.004996162 | 0 |
| 0 | 0 | 0 | 0 | 0 |
| 0.010823913 | 0 | 0.050423678 | 0.034977961 | 0 |
| 2.116055414 | 1.451534748 | 0.003690033 | 0.006898157 | 0.041908507 |
| 0 | 0 | 0 | 0 | 0 |
| 0 | 0 | 0 | 0 | 0 |
| 0 | 0 | 0 | 0 | 0 |
| 0 | 0 | 0 | 0 | 0 |
| 0 | 0 | 0 | 0 | 0 |
| 0 | 0 | 0 | 0 | 0 |
| 0.002230722 | 0.001742159 | 0.004824567 | 0.001143685 | 0 |
| 0 | 0 | 0 | 0 | 0 |
| 0 | 0 | 0 | 0 | 0 |
| 0 | 0 | 0 | 0 | 0 |
| 0.000583533 | 0 | 0.001234913 | 0 | 0.00453664 |
| 0 | 0 | 0 | 0 | 0 |
| 0 | 0 | 0 | 0 | 0 |
| 0 | 0 | 0 | 0 | 0 |
| 0 | 0 | 0 | 0 | 0 |
| 0 | 0 | 0 | 0 | 0 |
| 0 | 0 | 0 | 0 | 0 |
| 0.170685432 | 0.005241505 | 0.000495779 | 0.002277079 | 0.006513202 |
| 0.020875139 | 0.113109357 | 0 | 0 | 0 |
| 0 | 0 | 0 | 0 | 0 |
| 0.021964973 | 0 | 0 | 0 | 0.039290344 |
| 0 | 0 | 0 | 0 | 0 |
| 0 | 0 | 0 | 0 | 0 |
| 0 | 0 | 0.364648286 | 0 | 0 |
| 0 | 0 | 0 | 0 | 0 |
| 0 | 0 | 0 | 0 | 0 |
| 0 | 0 | 0 | 0 | 0 |
| 0 | 0 | 0 | 0 | 0 |

| | | | | |
|---|---|---|---|---|
| 0 | 0 | 0 | 0 | 0 |
| 0.509216449 | 0.002540045 | 0 | 0 | 0.001405102 |
| 0 | 0 | 0 | 0 | 0.001657601 |
| 0 | 0 | 0 | 0 | 0.043287244 |
| 0 | 0.001777627 | 0 | 0.00196926 | 0 |
| 0 | 0 | 0 | 0 | 0.002320081 |
| 0.007625432 | 0 | 0 | 0 | 0 |
| 0.004650654 | 0 | 0 | 0 | 0 |
| 0 | 0.014597057 | 0 | 0 | 0 |
| 0 | 0 | 0 | 0 | 0 |
| 0.018777203 | 0.0075227 | 0 | 0 | 0 |
| 0 | 0 | 0 | 0 | 0 |
| 0.321409764 | 0.005094781 | 0 | 0.009532108 | 0.024883501 |
| 0 | 0 | 0 | 0 | 0 |
| 0 | 0 | 0 | 0 | 0 |
| 0 | 0 | 0 | 0 | 0 |
| 0 | 0 | 0 | 0 | 0 |
| 0 | 0 | 0 | 0 | 0 |
| 0 | 0 | 0 | 0 | 0 |
| 0 | 0 | 0 | 0 | 0 |
| 0 | 0 | 0 | 0 | 0 |
| 0 | 0 | 0 | 0 | 0 |
| 0 | 0 | 0.005933069 | 0 | 0.004422383 |
| 0.064561664 | 0.007124796 | 0 | 0.04677255 | 0.010437128 |
| 0 | 0 | 0 | 0 | 0 |
| 0 | 0 | 0 | 0 | 0 |
| 0 | 0 | 0 | 0 | 0 |
| 0 | 0 | 0 | 0 | 0 |
| 0 | 0 | 0 | 0 | 0 |
| 0 | 0 | 0 | 0 | 0 |
| 0.006409765 | 0.014710897 | 0 | 0.018659956 | 0 |
| 0 | 0.001349698 | 0 | 0 | 0 |
| 0 | 0 | 0 | 0 | 0 |
| 0 | 0 | 0 | 0 | 0 |
| 0 | 0 | 0 | 0 | 0 |
| 0 | 0 | 0 | 0 | 0 |
| 0 | 0 | 0 | 0.004434503 | 0 |
| 0 | 0 | 0 | 0 | 0 |
| 0.004301084 | 0 | 0 | 0.005608759 | 0 |
| 0 | 0 | 0 | 0 | 0 |
| 0 | 0 | 0 | 0 | 0 |
| 0 | 0 | 0 | 0 | 0 |
| 0 | 0 | 0 | 0 | 0 |
| 0 | 0 | 0 | 0 | 0 |
| 0.119954623 | 0 | 0 | 0 | 0 |
| 0.184276191 | 0.184864529 | 0 | 0 | 0.013347621 |
| 0 | 0 | 0 | 0 | 0 |
| 0 | 0 | 0 | 0 | 0 |

| | | | | |
|---|---|---|---|---|
| 0 | 0 | 0 | 0 | 0 |
| 0 | 0 | 0 | 0 | 0 |
| 0 | 0 | 0 | 0 | 0 |
| 0 | 0.001120909 | 0 | 0 | 0.088417286 |
| 0 | 0 | 0 | 0 | 0 |
| 0 | 0 | 0 | 0 | 0.005376224 |
| 0.301856603 | 0 | 0 | 0 | 0 |
| 0 | 0 | 0 | 0 | 0 |
| 0 | 0 | 0 | 0 | 0.019125194 |
| 0 | 0 | 0 | 0 | 0 |
| 0.002427361 | 0 | 0 | 0 | 0 |
| 0 | 0 | 0 | 0 | 0.004364067 |
| 0 | 0 | 0 | 0 | 0 |
| 0 | 0 | 0 | 0 | 0 |
| 0 | 0 | 0 | 0 | 0 |
| 0 | 0 | 0 | 0 | 0 |
| 0 | 0 | 0 | 0 | 0 |
| 0 | 0 | 0 | 0 | 0 |
| 0 | 0 | 0 | 0 | 0 |
| 0.026758748 | 0.001134821 | 0 | 0 | 0 |
| 0 | 0 | 0 | 0 | 0 |
| 0 | 0 | 0 | 0 | 0 |
| 0 | 0 | 0 | 0 | 0 |
| 0 | 0 | 0 | 0 | 0 |
| 0 | 0 | 0 | 0 | 0 |
| 0 | 0 | 0 | 0 | 0 |
| 0 | 0 | 0 | 0 | 0 |
| 0 | 0 | 0 | 0 | 0 |
| 0 | 0 | 0 | 0 | 0 |
| 0 | 0 | 0 | 0 | 0 |
| 0.065681524 | 0.002534814 | 0.000627067 | 0.002880075 | 0 |
| 0 | 0 | 0 | 0 | 0 |
| 0.012242208 | 0.041883668 | 1.483117076 | 0.034332353 | 0.016077544 |
| 0.002255939 | 0.001850835 | 0 | 0.002278178 | 0 |
| 0 | 0 | 0 | 0 | 0 |
| 0 | 0 | 0 | 0 | 0 |
| 0 | 0 | 0 | 0 | 0 |
| 0 | 0 | 0 | 0 | 0 |
| 0 | 0 | 0 | 0 | 0 |
| 0 | 0 | 0 | 0 | 0 |
| 0 | 0 | 0 | 0 | 0 |
| 0.052504166 | 0.017694296 | 0.011530598 | 0.004218396 | 0.070319962 |
| 0.20557266 | 0.112321451 | 0 | 0 | 0.077918487 |
| 0 | 0 | 0 | 0 | 0 |
| 0 | 0 | 0 | 0 | 0 |
| 0 | 0.005246458 | 0 | 0 | 0 |
| 0.180091312 | 0 | 0 | 0 | 0 |
| 0 | 0 | 0 | 0 | 0 |

| | | | | |
|---|---|---|---|---|
| 0 | 0 | 0 | 0 | 0 |
| 0 | 0 | 0 | 0 | 0 |
| 0.009353824 | 0.002221461 | 0 | 0 | 0.051170791 |
| 0 | 0 | 0 | 0 | 0 |
| 0.891905432 | 0 | 0 | 0.015282882 | 0.05859551 |
| 0 | 0 | 0.013187154 | 0 | 0 |
| 0 | 0 | 0 | 0 | 0 |
| 0 | 0 | 0 | 0 | 0 |
| 0 | 0 | 0 | 0 | 0 |
| 0 | 0 | 0 | 0 | 0 |
| 0 | 0 | 0 | 0 | 0 |
| 0 | 0 | 0 | 0 | 0 |
| 0 | 0 | 0 | 0 | 0 |
| 0 | 0 | 0 | 0 | 0 |
| 0 | 0 | 0 | 0 | 0 |
| 0 | 0 | 0 | 0 | 0 |
| 0 | 0 | 0 | 0 | 0.30818292 |
| 0 | 0 | 0 | 0 | 0 |
| 0 | 0 | 0 | 0 | 0 |
| 0 | 0 | 0 | 0 | 0 |
| 0 | 0 | 0 | 0 | 0 |
| 0 | 0 | 0 | 0 | 0 |
| 0 | 0 | 0 | 0 | 0 |
| 0 | 0 | 0 | 0 | 0 |
| 0 | 0 | 0 | 0 | 0 |
| 0 | 0 | 0 | 0.002493677 | 0.006001438 |
| 0 | 0 | 0 | 0 | 0 |
| 2.064744489 | 0.139127538 | 0 | 0 | 0.029710257 |
| 0 | 0 | 0 | 0 | 0 |
| 0 | 0 | 0 | 0 | 0 |
| 0 | 0 | 0 | 0 | 0 |
| 0 | 0 | 0 | 0 | 0 |
| 0 | 0 | 0 | 0 | 0 |
| 0 | 0 | 0 | 0 | 0 |
| 0 | 0 | 0 | 0 | 0 |
| 0 | 0 | 0 | 0 | 0 |
| 0 | 0 | 0 | 0 | 0 |
| 0 | 0 | 0.00442186 | 0 | 0 |
| 1.627368104 | 1.225896992 | 0 | 0.123288524 | 3.82282702 |
| 0 | 0 | 0 | 0 | 0 |
| 0 | 0 | 0 | 0 | 0 |
| 0 | 0.023976012 | 0 | 0 | 0 |
| 0.000614441 | 0.001617336 | 0.002414889 | 0 | 0.003115396 |
| 0.02823106 | 0 | 0 | 0 | 0 |
| 0 | 0 | 0 | 0 | 0 |
| 0.021442424 | 0 | 0 | 0 | 0 |
| 0.002819926 | 0.001767292 | 0 | 0 | 0.018428323 |
| 0 | 0 | 0 | 0 | 0 |

| | | | | |
|---|---|---|---|---|
| 0 | 0 | 0 | 0 | 0 |
| 0.004744303 | 0.010480985 | 0.005378695 | 0.005270179 | 0.005412372 |
| 0 | 0 | 0 | 0 | 0 |
| 0 | 0 | 0 | 0 | 0 |
| 0 | 0 | 0 | 0 | 0 |
| 0 | 0 | 0 | 0 | 0 |
| 0 | 0 | 0 | 0 | 0 |
| 0 | 0 | 0 | 0 | 0 |
| 0 | 0 | 0 | 0 | 0 |
| 0 | 0 | 0 | 0 | 0 |
| 0 | 0 | 0 | 0 | 0 |
| 0 | 0 | 0 | 0 | 0 |
| 0.023195064 | 0.263075152 | 0 | 0 | 0.479958277 |
| 0 | 0 | 0 | 0 | 0 |
| 0.002218287 | 0.001112188 | 0.002682569 | 0 | 0.000999765 |
| 0 | 0 | 0 | 0 | 0 |
| 0 | 0 | 0 | 0 | 0 |
| 0 | 0 | 0 | 0 | 0 |
| 0.001908666 | 0 | 0 | 0 | 0.145162333 |
| 0 | 0 | 0 | 0 | 0 |
| 0 | 0 | 0 | 0 | 0 |
| 0.57779525 | 0.179130044 | 0.013392013 | 0 | 0.027982666 |
| 0 | 0.005875167 | 0.003205289 | 0.005578736 | 0 |
| 0.156572836 | 0 | 0 | 0 | 0.06649289 |
| 0 | 0 | 0 | 0 | 0 |
| 0 | 0 | 0 | 0 | 0 |
| 0 | 0 | 0 | 0 | 0 |
| 0 | 0 | 0 | 0 | 0 |
| 0 | 0 | 0 | 0 | 0 |
| 0 | 0 | 0 | 0 | 0 |
| 0 | 0 | 0 | 0 | 0 |
| 0 | 0 | 0 | 0 | 0 |
| 0 | 0 | 0 | 0 | 0 |
| 0 | 0 | 0 | 0 | 0 |
| 0.042197042 | 0 | 4.324888803 | 0 | 0 |
| 0 | 0 | 0 | 0 | 0 |
| 0 | 0 | 0 | 0 | 0 |
| 0 | 0 | 0 | 0 | 0 |
| 0.139233816 | 0.139616019 | 0.001753908 | 0.00590742 | 0.000980494 |
| 0 | 0 | 0 | 0 | 0 |
| 0.021023661 | 0.267972484 | 0.019854203 | 0 | 0.538255006 |
| 0 | 0 | 0 | 0 | 0 |
| 0 | 0 | 0 | 0 | 0 |
| 0 | 0 | 0 | 0 | 0 |
| 0 | 0 | 0 | 0 | 0 |
| 0.039952334 | 0.001399682 | 0.055366346 | 3.040222192 | 0.113237814 |
| 0 | 0 | 0 | 0 | 0 |
| 0 | 0 | 0 | 0 | 0 |

| | | | | |
|---|---|---|---|---|
| 0 | 0 | 0 | 0 | 0 |
| 0 | 0 | 0 | 0 | 0 |
| 0 | 0 | 0 | 0 | 0 |
| 0 | 0 | 0 | 0 | 0 |
| 0.058343197 | 0 | 0 | 0 | 0 |
| 0 | 0 | 0 | 0 | 0 |
| 0 | 0 | 0 | 0 | 0 |
| 0.004559677 | 0 | 0 | 0 | 0 |
| 0 | 0 | 0 | 0 | 0 |
| 0.006309997 | 0.031758272 | 0 | 0.069824608 | 0 |
| 0 | 0 | 0 | 0 | 0 |
| 0 | 0 | 0 | 0 | 0 |
| 0 | 0.000802785 | 0 | 0 | 0.002646004 |
| 0 | 0 | 0 | 0 | 0 |
| 0 | 0 | 0 | 0 | 0 |
| 0 | 0 | 0 | 0 | 0 |
| 0 | 0 | 0 | 0 | 0 |
| 0 | 0 | 0 | 0 | 0 |
| 0 | 0 | 0 | 0 | 0 |
| 0.071363531 | 0.056210141 | 0.017617405 | 0.024860989 | 0.044961585 |
| 0 | 0 | 0 | 0 | 0 |
| 0 | 0 | 0 | 0 | 0 |
| 0 | 0 | 0 | 0 | 0 |
| 0 | 0 | 0 | 0 | 0 |
| 0 | 0 | 0 | 0 | 0 |
| 0 | 0 | 0 | 0 | 0 |
| 0 | 0 | 0 | 0 | 0 |
| 0 | 0 | 0 | 0 | 0 |
| 0 | 0 | 0 | 0 | 0 |
| 0 | 0.001018467 | 0 | 0.025072452 | 0 |
| 0 | 0 | 0 | 0 | 0 |
| 0 | 0 | 0 | 0 | 0 |
| 0 | 0 | 0 | 0 | 0 |
| 0 | 0 | 0 | 0 | 0 |
| 0 | 0 | 0 | 0 | 0 |
| 0.000819927 | 0.0018499 | 0 | 0 | 0 |
| 0 | 0 | 0 | 0 | 0 |
| 0 | 0 | 0 | 0 | 0 |
| 0 | 0 | 0 | 0 | 0 |
| 0 | 0 | 0 | 0 | 0 |
| 0 | 0 | 0 | 0 | 0 |
| 0 | 0 | 0 | 0 | 0 |
| 0 | 0 | 0 | 0 | 0.09966358 |
| 0 | 0 | 0 | 0 | 0 |
| 0.091220519 | 0.004474121 | 0 | 0 | 0.017294016 |
| 0 | 0 | 0 | 0 | 0 |
| 0 | 0 | 0 | 0 | 0 |
| 0 | 0 | 0 | 0 | 0 |

| | | | | |
|---|---|---|---|---|
| 0 | 0 | 0 | 0 | 0 |
| 0 | 0 | 0 | 0 | 0 |
| 0 | 0 | 0 | 0 | 0 |
| 0 | 0 | 0 | 0 | 0 |
| 0 | 0 | 0 | 0 | 0 |
| 0.011103206 | 0.001503047 | 0.000537083 | 0.001480071 | 0.196962551 |
| 0 | 0 | 0 | 0 | 0 |
| 0.004055892 | 0 | 0 | 0 | 0 |
| 0 | 0 | 0 | 0 | 0 |
| 0 | 0 | 0 | 0 | 0 |
| 0 | 0 | 0 | 0 | 0 |
| 0 | 0 | 0 | 0 | 0 |
| 0 | 0 | 0 | 0 | 0 |
| 0 | 0 | 0 | 0 | 0 |
| 0 | 0 | 0 | 0 | 0 |
| 0 | 0 | 0 | 0 | 0 |
| 0 | 0 | 0 | 0 | 0 |
| 0 | 0 | 0 | 0 | 0 |
| 0 | 0 | 0 | 0 | 0 |
| 0 | 0 | 0 | 0 | 0 |
| 0 | 0 | 0 | 0 | 0 |
| 0 | 0 | 0 | 0 | 0 |
| 0 | 0 | 0 | 0 | 0 |
| 0 | 0 | 0 | 0 | 0 |
| 0 | 0 | 0 | 0 | 0 |
| 0 | 0.001044897 | 0 | 0.000771693 | 0 |
| 0 | 0 | 0 | 0 | 0 |
| 0 | 0 | 0 | 0 | 0 |
| 0 | 0 | 0 | 0 | 0 |
| 0 | 0 | 0.002184631 | 0 | 0 |
| 0 | 0 | 0 | 0 | 0 |
| 0 | 0 | 0 | 0 | 0 |
| 0 | 0 | 0 | 0 | 0 |
| 0 | 0 | 0 | 0 | 0 |
| 0 | 0 | 0 | 0 | 0 |
| 0 | 0 | 0 | 0 | 0 |
| 0 | 0 | 0 | 0 | 0 |
| 0 | 0 | 0 | 0 | 0 |
| 0.796823998 | 1.242365153 | 0 | 0 | 0.441203567 |
| 0 | 0 | 0 | 0 | 0 |
| 0 | 0 | 0 | 0 | 0 |
| 0 | 0 | 0 | 0 | 0 |
| 0 | 0 | 0 | 0 | 0 |
| 0 | 0 | 0 | 0 | 0 |
| 0 | 0 | 0 | 0 | 0 |
| 0 | 0 | 0 | 0 | 0 |
| 0 | 0 | 0 | 0 | 0 |
| 0 | 0 | 0 | 0 | 0 |

| | | | | |
|---|---|---|---|---|
| 0 | 0 | 0 | 0 | 0 |
| 0.14320436 | 0.045809086 | 0 | 0.097214117 | 0.086222886 |
| 0 | 0 | 0 | 0 | 0 |
| 0 | 0 | 0 | 0 | 0 |
| 0 | 0 | 0 | 0 | 0 |
| 0 | 0 | 0 | 0 | 0 |
| 0 | 0 | 0 | 0 | 0 |
| 0 | 0 | 0 | 0 | 0 |
| 0 | 0 | 0 | 0 | 0 |
| 0 | 0 | 0 | 0 | 0 |
| 0.001953279 | 0 | 0 | 0 | 0 |
| 0 | 0 | 0 | 0 | 0 |
| 0.056588417 | 0.128935756 | 0 | 0 | 0 |
| 0 | 0 | 0 | 0 | 0 |
| 0 | 0 | 0 | 0 | 0 |
| 0 | 0 | 0 | 0 | 0 |
| 0 | 0 | 0 | 0 | 0 |
| 0 | 0 | 0 | 0 | 0 |
| 0 | 0 | 0 | 0 | 0 |
| 0 | 0 | 0 | 0 | 0 |
| 0 | 0 | 0 | 0.006117232 | 0.038196257 |
| 0 | 0 | 0 | 0 | 0 |
| 0 | 0 | 0 | 0 | 0 |
| 0.794675893 | 1.176004949 | 0 | 4.096171445 | 2.50506546 |
| 0 | 0 | 0 | 0 | 0 |
| 0 | 0 | 0 | 0 | 0 |
| 0 | 0 | 0 | 0 | 0 |
| 0 | 0 | 0 | 0 | 0 |
| 0 | 0 | 0 | 0 | 0 |
| 0 | 0 | 0 | 0 | 0 |
| 0 | 0 | 0.002549118 | 0 | 0 |
| 0 | 0 | 0 | 0 | 0 |
| 0 | 0 | 0 | 0 | 0 |
| 0 | 0 | 0 | 0 | 0 |

| PHL018-0022_V3 | PHL018-0001_V3 | PHL018-0235_V1 | PHL018-0240_V1 | PHL018-0067_V2 |
|---|---|---|---|---|
| CTRL | EYCF | EYCF | REF | EYCF |
| 0.082035874 | 0.213691486 | 0 | 0.547707929 | 0.023498432 |
| 0.966535588 | 1.148171572 | 0 | 5.326191263 | 2.164278086 |
| 0.006841335 | 4.057316733 | 0 | 0 | 6.833401224 |
| 0 | 0 | 0 | 0 | 0 |
| 0 | 0 | 0 | 0 | 0 |
| 0 | 0 | 0 | 0 | 0 |
| 0 | 0 | 0 | 0 | 0 |
| 0 | 0 | 0 | 0 | 0 |
| 0 | 0 | 0 | 0 | 0 |
| 0 | 0 | 0 | 0 | 0 |
| 0 | 0 | 0 | 0 | 0 |
| 0 | 0 | 0 | 0 | 0 |
| 0 | 0 | 0 | 0 | 0 |
| 1.420409414 | 0.652376645 | 0 | 0.179728143 | 5.340543495 |
| 0 | 0 | 0 | 0 | 0 |
| 0 | 0 | 0 | 0 | 0 |
| 0.001548339 | 0 | 0 | 0 | 0 |
| 0 | 0 | 0 | 0 | 0 |
| 0 | 0 | 0 | 0 | 0 |
| 0 | 0 | 0 | 0 | 0 |
| 0 | 0 | 0 | 0 | 0 |
| 0.001973144 | 0 | 0 | 0 | 0.002195935 |
| 0 | 0 | 0 | 0 | 0 |
| 0 | 0 | 0 | 0 | 0 |
| 0 | 0.061677738 | 0 | 0 | 0 |
| 0 | 0 | 0 | 0 | 0 |
| 0 | 0 | 0 | 0 | 0 |
| 0 | 0 | 0 | 0 | 0 |
| 0 | 0 | 0 | 0 | 0 |
| 0 | 0 | 0 | 0 | 0 |
| 0 | 0 | 0 | 0 | 0 |
| 0 | 0 | 0 | 0 | 0 |
| 0 | 0 | 0 | 0 | 0 |
| 0 | 0 | 0 | 0 | 0 |
| 0 | 0 | 0 | 0 | 0 |
| 0 | 0 | 0.001440632 | 6.988329182 | 0.238374238 |
| 0 | 0 | 0 | 0 | 0 |
| 0 | 0 | 0 | 0 | 0 |
| 0 | 0 | 0 | 0 | 0 |
| 0 | 0 | 0 | 0 | 0 |
| 0 | 0 | 0 | 0 | 0 |
| 0 | 0.819396169 | 0.240890946 | 0.00543003 | 0.000789831 |
| 0.177708901 | 0.142171171 | 0.598920626 | 0.398528028 | 0.001748005 |
| 0.016146727 | 0.000782759 | 0.118967841 | 0.208320554 | 0.007123196 |
| 0 | 0 | 0 | 0.000885434 | 0.003734962 |
| 0 | 0 | 0 | 0 | 0 |

| | | | | |
|---|---|---|---|---|
| 0 | 0.002194197 | 0 | 0.00731558 | 0 |
| 0.002244212 | 0 | 0.038462979 | 0.004736796 | 0 |
| 0.027138076 | 0.103802811 | 0 | 0 | 0 |
| 0.236936786 | 1.219282698 | 0 | 0.967526017 | 0.16677109 |
| 0 | 0.011614732 | 0 | 0.015478694 | 0.001080493 |
| 0 | 0 | 0 | 0 | 0 |
| 0.001503396 | 0.002157291 | 0 | 0.017135148 | 0 |
| 0 | 0 | 0 | 0 | 0 |
| 0 | 0 | 0 | 0 | 0 |
| 0 | 0 | 0 | 0 | 0 |
| 0 | 0 | 6.139919013 | 0.001166214 | 0 |
| 0 | 0.00913473 | 0 | 0.070018284 | 1.503844217 |
| 0 | 0 | 0 | 0.00054979 | 0 |
| 0.134585605 | 0.072227997 | 0 | 0.271339909 | 0.025136728 |
| 0 | 0 | 0 | 0 | 0 |
| 0 | 0 | 0 | 0 | 0 |
| 0 | 0 | 0 | 0.004361554 | 0.420818312 |
| 0 | 0 | 0 | 0 | 0 |
| 0 | 0 | 0 | 0 | 0 |
| 0 | 0 | 0 | 0 | 0 |
| 0.112179616 | 0.016937936 | 0 | 0.481713386 | 0.015531889 |
| 0.024996167 | 0.08602987 | 0 | 0.149858784 | 0.003804243 |
| 0 | 0 | 0 | 0 | 0.00071115 |
| 0 | 0 | 0 | 0 | 0 |
| 0 | 0 | 0 | 0 | 0.000700247 |
| 0 | 0 | 0 | 0 | 0 |
| 0 | 0 | 0 | 0 | 0 |
| 0 | 0 | 0 | 0 | 0 |
| 0 | 0.007108364 | 0.070262977 | 0.001538317 | 0 |
| 0 | 0 | 0 | 0 | 0 |
| 0.056404749 | 0 | 0 | 0.010219049 | 0 |
| 0 | 0 | 0 | 0 | 0 |
| 0 | 0 | 0 | 0 | 0 |
| 0 | 0 | 0 | 0 | 0 |
| 0 | 0 | 0 | 0 | 0 |
| 0 | 0 | 0 | 0 | 0 |
| 0 | 0 | 0 | 0 | 0 |
| 0 | 0 | 0 | 0 | 0 |
| 0 | 0 | 0 | 0 | 0 |
| 0 | 0 | 0 | 0 | 0 |
| 0 | 0 | 0 | 0 | 0 |
| 0 | 0 | 0 | 0 | 0 |
| 0 | 0 | 0 | 0 | 0 |
| 0 | 0 | 0 | 0 | 0 |
| 0 | 0.017895301 | 0 | 0 | 0 |
| 0 | 0 | 0 | 0 | 0 |
| 0 | 0 | 0 | 0 | 0 |
| 0 | 0 | 0.001156051 | 0.00106303 | 0 |

| | | | | |
|---|---|---|---|---|
| 0 | 0 | 0 | 0 | 0 |
| 0.01346173 | 0 | 0 | 0 | 0 |
| 0 | 0 | 0 | 0 | 0 |
| 0 | 0 | 0 | 0 | 0 |
| 0.001459085 | 0 | 0.003646837 | 0 | 0 |
| 0 | 0 | 0 | 0 | 0 |
| 0 | 0 | 0 | 0 | 0 |
| 0 | 0 | 0 | 0 | 0 |
| 0.001652017 | 0.001666796 | 0 | 0 | 0 |
| 0 | 0 | 0 | 0 | 0 |
| 0 | 0 | 0 | 0 | 0 |
| 0 | 0 | 0 | 0.00118072 | 0 |
| 0 | 0 | 0 | 0 | 0 |
| 0 | 0 | 0 | 0 | 0 |
| 0 | 0 | 0 | 0 | 0 |
| 0 | 0.01116788 | 0 | 0 | 0 |
| 0 | 0 | 0 | 0 | 0 |
| 0 | 0 | 0 | 0 | 0 |
| 0.034777971 | 0.013394609 | 0 | 0 | 0 |
| 0 | 0 | 0 | 0 | 0 |
| 0 | 0 | 0 | 0 | 0 |
| 0 | 0 | 0 | 0 | 0 |
| 0 | 0.003443628 | 0 | 0 | 0 |
| 0 | 0 | 0 | 0 | 0 |
| 0 | 0 | 0 | 0 | 0 |
| 0 | 0 | 0 | 0 | 0 |
| 0 | 0 | 0.008751974 | 0 | 0 |
| 0 | 0.003988984 | 0 | 0 | 0 |
| 0 | 0 | 0 | 0 | 0 |
| 0 | 0.001715672 | 0 | 0 | 0 |
| 0 | 0.039245628 | 0 | 0.010083265 | 0.01776955 |
| 0.001403298 | 0.09061454 | 0 | 0 | 0.000594951 |
| 0 | 0 | 0 | 0 | 0 |
| 0 | 0 | 0 | 0 | 0 |
| 0 | 0 | 0 | 0 | 0 |
| 0 | 0 | 0 | 0 | 0 |
| 0 | 0 | 0 | 0 | 0 |
| 0 | 0 | 0 | 0 | 0 |
| 0 | 0 | 0 | 0 | 0 |
| 0 | 0 | 0 | 0 | 0 |
| 0 | 0 | 0 | 0 | 0 |
| 0 | 0 | 0 | 0 | 0 |
| 0 | 0.013299819 | 0 | 0 | 0 |
| 0 | 0 | 0 | 0 | 0 |
| 0 | 0 | 0 | 0 | 0 |
| 0 | 0 | 0 | 0 | 0 |
| 0 | 0 | 0 | 0 | 0 |
| 0 | 0 | 0 | 0 | 0 |

| | | | | |
|---|---|---|---|---|
| 0 | 0 | 0 | 0 | 0 |
| 0 | 0 | 0 | 0 | 0 |
| 0 | 0 | 0 | 0 | 0 |
| 0 | 0 | 0 | 0 | 0 |
| 0.047111621 | 0.026915851 | 0 | 0.016081816 | 0 |
| 0 | 0 | 0 | 0 | 0 |
| 0 | 0 | 0 | 0 | 0 |
| 0 | 0 | 0 | 0 | 0 |
| 0 | 0 | 0 | 0 | 0 |
| 0 | 0 | 0 | 0 | 0 |
| 0 | 0 | 0 | 0 | 0 |
| 0 | 0 | 0 | 0 | 0 |
| 0 | 0 | 0 | 0 | 0 |
| 0 | 0 | 0 | 0 | 0 |
| 0 | 0 | 0 | 0 | 0 |
| 0 | 4.154374364 | 0 | 11.05196415 | 0 |
| 0 | 0 | 0 | 0 | 0 |
| 0 | 0 | 0 | 0 | 0 |
| 0 | 0 | 0 | 0 | 0 |
| 0 | 0 | 0 | 0 | 0 |
| 0 | 0 | 0 | 0.002089151 | 0 |
| 0 | 0 | 0 | 0 | 0 |
| 0 | 0 | 0 | 0 | 0 |
| 0 | 0 | 0 | 0 | 0 |
| 0 | 0 | 0 | 0 | 0 |
| 0 | 0 | 0 | 0 | 0 |
| 0 | 0 | 0 | 0 | 0 |
| 0 | 0 | 0 | 0 | 0 |
| 0 | 0 | 0 | 0 | 0 |
| 0 | 0 | 0 | 0 | 0 |
| 0 | 0.020030679 | 0 | 0 | 0 |
| 0.000925202 | 0.015972859 | 0 | 0.039055959 | 0 |
| 0 | 0 | 0 | 0 | 0 |
| 0 | 0 | 0 | 0 | 0 |
| 0 | 0 | 0 | 0 | 0 |
| 0 | 0 | 0 | 0 | 0 |
| 0 | 0 | 0 | 0 | 0 |
| 0 | 0 | 0 | 0 | 0 |
| 0 | 0 | 0 | 0 | 0 |
| 0 | 0 | 0 | 0 | 0 |
| 0 | 0 | 0 | 0 | 0 |
| 0 | 0 | 0 | 0 | 0 |
| 0 | 0 | 0 | 0 | 0 |
| 0 | 0 | 0 | 0 | 0 |
| 0 | 0 | 0 | 0 | 0 |
| 0 | 0 | 0 | 0 | 0 |
| 0 | 0 | 0 | 0 | 0 |
| 0 | 0 | 0 | 0 | 0 |

| | | | | |
|---|---|---|---|---|
| 0 | 0 | 0 | 0 | 0 |
| 0 | 0 | 0 | 0 | 0 |
| 0 | 0 | 0 | 0 | 0 |
| 0 | 0 | 0 | 0 | 0 |
| 0 | 0 | 0 | 0 | 0 |
| 0 | 0 | 0 | 0 | 0 |
| 0 | 0 | 0 | 0 | 0 |
| 0 | 0 | 0 | 0 | 0 |
| 0 | 0 | 0 | 0 | 0 |
| 0 | 0 | 0 | 0 | 0 |
| 0 | 0 | 0 | 0 | 0 |
| 0 | 0 | 0 | 0 | 0 |
| 0 | 0 | 0.000711253 | 0.001245757 | 0.001313722 |
| 0 | 0 | 0 | 0 | 0 |
| 0 | 0.001507794 | 0 | 0 | 0 |
| 0 | 0 | 0 | 0 | 0 |
| 0 | 0 | 0 | 0 | 0 |
| 0 | 0 | 0 | 0 | 0 |
| 0 | 0 | 0 | 0 | 0 |
| 0 | 0 | 0 | 0 | 0 |
| 0.050902008 | 0.048249024 | 0.024381 | 0.021836879 | 0.003275125 |
| 0 | 0 | 0 | 0 | 0 |
| 0 | 0 | 0 | 0 | 0 |
| 0 | 0.01199796 | 0 | 0 | 0 |
| 0 | 0 | 0 | 0 | 0 |
| 0 | 0 | 0 | 0 | 0 |
| 0.013014496 | 0 | 0 | 0.017125942 | 0 |
| 0 | 0 | 0 | 0 | 0 |
| 0 | 0 | 0 | 0 | 0 |
| 0 | 0 | 0 | 0 | 0 |
| 0 | 0 | 0 | 0 | 0.001188319 |
| 0 | 0 | 0 | 0 | 0 |
| 0 | 0 | 0 | 0 | 0 |
| 0 | 0.002721859 | 0 | 0 | 0 |
| 0.005385294 | 0.018111569 | 0 | 0.003978306 | 0.016515027 |
| 0 | 0 | 0 | 0 | 0 |
| 0 | 0 | 0 | 0 | 0 |
| 0 | 0 | 0 | 0 | 0 |
| 0 | 0 | 0 | 0 | 0 |
| 0 | 0 | 0 | 0 | 0 |
| 0 | 0 | 0 | 0 | 0 |
| 0 | 0 | 0 | 0 | 0 |
| 0 | 0 | 0 | 0 | 0 |
| 0 | 0 | 0 | 0 | 0 |
| 0 | 0 | 0 | 0 | 0 |
| 0 | 0.72172669 | 0 | 0 | 0 |
| 0.002623879 | 0.001568801 | 0 | 0.001640933 | 0.004037731 |
| 0.000767541 | 0 | 0.00102771 | 0.000540009 | 0.009680996 |

| | | | | |
|---|---|---|---|---|
| 0 | 0 | 0 | 0 | 0 |
| 0 | 0 | 0 | 0 | 0 |
| 0 | 0.001887033 | 0 | 0 | 0 |
| 0 | 0 | 0 | 0 | 0 |
| 0 | 0 | 0 | 0 | 0 |
| 0 | 0 | 0 | 0 | 0 |
| 0 | 0 | 0 | 0 | 0 |
| 0 | 0.02647002 | 0 | 0 | 0 |
| 0 | 0 | 0 | 0 | 0.001496238 |
| 0 | 0 | 0 | 0 | 0 |
| 0 | 0 | 0 | 0 | 0 |
| 0 | 0 | 0 | 0 | 0 |
| 0 | 0 | 0 | 0 | 0 |
| 0 | 0 | 0.008147714 | 0 | 0 |
| 0 | 0 | 0.027546096 | 0.001064269 | 0 |
| 0 | 0 | 0 | 0 | 0 |
| 0 | 0 | 0 | 0 | 0 |
| 0 | 0 | 0 | 0 | 0 |
| 0 | 0 | 0 | 0.002745929 | 0 |
| 0 | 0 | 0 | 0 | 0 |
| 0.025043851 | 0.009552102 | 0.015725739 | 0 | 0.006549646 |
| 0 | 0 | 0 | 0 | 0 |
| 0 | 0 | 0 | 0 | 0 |
| 0 | 0 | 0 | 0 | 0 |
| 0.009722223 | 0.004103194 | 0.018990542 | 0.010059031 | 0 |
| 0 | 0 | 0 | 0 | 0 |
| 0 | 0 | 0 | 0 | 0 |
| 0 | 0 | 0 | 0 | 0 |
| 0 | 0.082475214 | 1.448374438 | 0 | 0 |
| 0 | 0 | 0 | 0 | 0 |
| 1.090490885 | 0.265190503 | 0 | 0.036484783 | 0.101972554 |
| 0 | 0 | 0 | 0 | 0 |
| 0 | 0.029504616 | 0 | 0.02851575 | 0.013278313 |
| 0 | 0 | 0 | 0 | 0 |
| 0 | 0 | 0 | 0 | 0 |
| 0 | 0 | 0 | 0 | 0 |
| 0 | 0 | 0 | 0 | 0 |
| 0 | 0 | 0 | 0 | 0 |
| 0 | 0 | 0 | 0 | 0 |
| 0 | 0 | 0 | 0 | 0.008366029 |
| 0 | 0 | 0 | 0 | 0 |
| 0.009277501 | 0.219427475 | 0 | 0.125991186 | 0.029240937 |
| 0 | 0 | 0 | 0 | 0 |
| 0 | 0 | 0 | 0 | 0 |
| 0 | 0 | 0 | 0 | 0 |
| 0 | 0 | 0 | 0 | 0 |
| 0 | 0 | 0 | 0 | 0 |
| 0 | 0 | 0 | 0 | 0 |

| | | | | |
|---|---|---|---|---|
| 0 | 0 | 0 | 0 | 0 |
| 0 | 0 | 0 | 0 | 0 |
| 0.010836679 | 0.000728908 | 0.12565597 | 0.188699566 | 0.007504165 |
| 0 | 0 | 0 | 0 | 0 |
| 0 | 1.759800515 | 0 | 0 | 0 |
| 0 | 0 | 0 | 0 | 0 |
| 0 | 0 | 0 | 0 | 0 |
| 0 | 0 | 0 | 0 | 0 |
| 0 | 0 | 0 | 0 | 0 |
| 0 | 0 | 0 | 0 | 0 |
| 0 | 0 | 0 | 0 | 0 |
| 0 | 0 | 0 | 0 | 0 |
| 0 | 0 | 0 | 0 | 0 |
| 0.005688831 | 0 | 0 | 0.003581112 | 0 |
| 0.02273555 | 0.005097543 | 0 | 0.006085345 | 0 |
| 0 | 0.004983019 | 0 | 0 | 0 |
| 0.002815774 | 0.00075759 | 0.012969561 | 0.003764011 | 0 |
| 0 | 0 | 0 | 0 | 0 |
| 0 | 0 | 0 | 0 | 0 |
| 0 | 0.001029012 | 0 | 0 | 0 |
| 0 | 0 | 0 | 0 | 0 |
| 0.000762552 | 0 | 0.001633648 | 0 | 0.007543583 |
| 0 | 0 | 0 | 0 | 0 |
| 0.002758303 | 0 | 0 | 0 | 0 |
| 0 | 0 | 0.003056036 | 0 | 0 |
| 0 | 0 | 0 | 0 | 0 |
| 0 | 0 | 0 | 0 | 0 |
| 0 | 0 | 0 | 0 | 0 |
| 0 | 0 | 0 | 0 | 0 |
| 0 | 0.015168502 | 0 | 0 | 0 |
| 0 | 0 | 0 | 0.015074687 | 0 |
| 0 | 0 | 0 | 0 | 0 |
| 0 | 0 | 0 | 0 | 0 |
| 0 | 0.001051361 | 0 | 0 | 0 |
| 0 | 0 | 0 | 0 | 0 |
| 0.002030786 | 0.03414922 | 0.00598213 | 0 | 0 |
| 0 | 0 | 0 | 0 | 0 |
| 0 | 0 | 0 | 0.002022167 | 0.00505479 |
| 0 | 0 | 0 | 0 | 0 |
| 0 | 0 | 0 | 0 | 0 |
| 0 | 0 | 0 | 0 | 0 |
| 0 | 0.005382625 | 0 | 0 | 0 |
| 0 | 0 | 0 | 0 | 0 |
| 0 | 0 | 0 | 0 | 0 |
| 0 | 0 | 0 | 0 | 0 |
| 0 | 0 | 0 | 0 | 0 |
| 0 | 0 | 0 | 0 | 0 |
| 0 | 0 | 0 | 0 | 0 |

| | | | | |
|---|---|---|---|---|
| 2.048246464 | 0.037507467 | 0.001638962 | 0.256563013 | 0.282543087 |
| 0 | 0 | 0 | 0 | 0 |
| 0.022896465 | 0 | 0 | 0.002247764 | 0 |
| 0 | 0 | 0 | 0 | 0 |
| 0.019018136 | 0.006966634 | 0 | 0.035618112 | 0 |
| 0 | 0.002350289 | 0 | 0 | 0 |
| 0 | 0 | 0 | 0 | 0 |
| 0.014267717 | 0.002020401 | 0 | 0.028793661 | 0 |
| 0 | 0 | 0 | 0 | 0 |
| 0.026703082 | 0.087116696 | 0 | 0.000816833 | 0.563927397 |
| 0 | 0 | 0 | 0 | 0 |
| 0 | 0.060242427 | 0 | 0.003764009 | 0.00176416 |
| 0.002479975 | 0 | 0.019078343 | 0.003489609 | 0.003791505 |
| 0.143124789 | 0 | 0 | 0.157067261 | 0.00812939 |
| 0 | 0 | 0 | 0 | 0.001376286 |
| 0 | 0 | 0 | 0 | 0 |
| 0 | 0 | 0 | 0 | 0 |
| 0 | 0.001316662 | 0 | 0 | 0.00203327 |
| 0.016751409 | 0 | 0 | 0 | 0 |
| 0 | 0 | 0 | 0 | 0 |
| 0 | 0 | 0 | 0.000702882 | 0 |
| 0 | 0 | 0 | 0 | 0 |
| 0 | 0.046526472 | 0 | 0 | 0 |
| 0 | 0 | 0 | 0 | 0 |
| 0 | 0 | 0 | 0 | 0 |
| 0 | 0 | 0 | 0 | 0.020781819 |
| 0 | 0 | 0.083501576 | 0.005682115 | 0 |
| 0.001270198 | 0 | 0 | 0.010500474 | 0 |
| 0 | 0 | 0 | 0 | 0 |
| 0 | 0.001453832 | 0 | 0.003991779 | 0.00080182 |
| 0.003381337 | 0 | 0 | 0 | 0 |
| 0 | 0 | 0 | 0 | 0 |
| 0 | 0 | 0 | 0.034504405 | 0 |
| 0 | 0 | 0 | 0 | 0 |
| 0 | 0 | 0 | 0 | 0 |
| 0 | 0 | 0 | 0 | 0 |
| 0.001488388 | 0 | 0 | 0 | 0 |
| 0 | 0 | 0 | 0 | 0 |
| 0 | 0 | 0 | 0 | 0 |
| 0 | 0 | 0 | 0.004080508 | 0 |
| 0 | 0 | 0 | 0.577458653 | 0 |
| 0.123975665 | 0 | 0 | 0 | 0 |
| 0 | 0 | 0 | 0 | 0 |
| 0 | 0 | 0 | 0.00119073 | 0.000837128 |
| 0.004456385 | 0 | 0 | 0 | 0 |
| 0 | 0 | 0 | 0 | 0 |
| 0 | 0.003807304 | 0 | 0.001148757 | 0.001292192 |
| 0 | 0 | 0 | 0 | 0 |

| | | | | |
|---|---|---|---|---|
| 0 | 0 | 0 | 0 | 0 |
| 0 | 0 | 0 | 0 | 0 |
| 0 | 0 | 0 | 0 | 0 |
| 0 | 0 | 0.013552831 | 0 | 0 |
| 0 | 0 | 0 | 0 | 0 |
| 0.031121573 | 0 | 0 | 0.129384338 | 0.002798833 |
| 0 | 0.002308433 | 0 | 0 | 0 |
| 0 | 0.00179006 | 0 | 0 | 0 |
| 0 | 0 | 0 | 0 | 0 |
| 0 | 0 | 0 | 0 | 0 |
| 0.01312574 | 0.002140309 | 0 | 0.020778099 | 0 |
| 0 | 0.002005957 | 0 | 0 | 0 |
| 0 | 0 | 0 | 0 | 0 |
| 0.007327591 | 0 | 0 | 0.005551948 | 0 |
| 0.065302041 | 0 | 0 | 0 | 0.007898733 |
| 0 | 0 | 0 | 0 | 0 |
| 0 | 0 | 0 | 0 | 0 |
| 0 | 0 | 0 | 0 | 0 |
| 0 | 0.002421189 | 0 | 0 | 0 |
| 0 | 0.13428381 | 0 | 0 | 0 |
| 0 | 0 | 0 | 0 | 0 |
| 0 | 0.254048859 | 0 | 0.471453518 | 0 |
| 0 | 0 | 0 | 0 | 0 |
| 0 | 0 | 0 | 0 | 0 |
| 0 | 0 | 0 | 0 | 0 |
| 0 | 0 | 0 | 0 | 0 |
| 0.049809346 | 0 | 0 | 0.008384696 | 0.002116067 |
| 0 | 0 | 0 | 0 | 0 |
| 0 | 0 | 0.155758528 | 0 | 0 |
| 1.599127091 | 0.624644263 | 0 | 2.264687325 | 0.139763595 |
| 0 | 0.000857378 | 0 | 0.004842714 | 0 |
| 0 | 0 | 0 | 0 | 0 |
| 0 | 0 | 0 | 0 | 0 |
| 0 | 0 | 0 | 0 | 0 |
| 0 | 0 | 0 | 0 | 0 |
| 0 | 0 | 0 | 0 | 0 |
| 0 | 0 | 0 | 0 | 0 |
| 0 | 0 | 0 | 0 | 0 |
| 0.242403512 | 0 | 0 | 0.74671525 | 0 |
| 0.02434386 | 0 | 0 | 0 | 0.049154241 |
| 0.096153543 | 0.045299047 | 0 | 0.048088221 | 0.002750468 |
| 0 | 0.191399945 | 0 | 1.960785032 | 0 |
| 0 | 0 | 0 | 0 | 0 |
| 0 | 0 | 0 | 0.028711894 | 0 |
| 0 | 0 | 0 | 0 | 0 |
| 0 | 0 | 0 | 0 | 0 |
| 0 | 0.006106646 | 0 | 0.007185849 | 0.000962271 |
| 0 | 0 | 0 | 0 | 0 |

| | | | | |
|---|---|---|---|---|
| 0 | 0 | 0.011891761 | 0 | 0.005936406 |
| 0 | 0 | 0 | 0 | 0 |
| 0.002044167 | 0.001833292 | 0.002627584 | 0.002013464 | 0.017795377 |
| 0 | 0 | 0 | 0 | 0 |
| 0 | 0 | 0 | 0 | 0 |
| 0.004312692 | 0.389282495 | 0 | 0.05068942 | 0 |
| 0 | 0 | 0 | 0 | 0 |
| 0 | 0 | 0 | 0 | 0 |
| 0.009039967 | 0 | 0 | 0 | 0 |
| 0 | 0 | 0 | 0 | 0 |
| 0.104128729 | 0 | 0 | 0 | 0 |
| 0 | 0 | 0 | 0 | 0 |
| 0 | 0 | 0 | 0 | 0 |
| 0.064060437 | 0.001947526 | 0 | 0.003246582 | 0.001074103 |
| 0 | 0 | 0 | 0 | 0 |
| 0.094608934 | 0.079712548 | 0 | 0.017610788 | 0.004826511 |
| 0 | 0.004849929 | 0.006044528 | 0.000992528 | 0.003954114 |
| 0.00085699 | 0 | 0.007037871 | 0 | 0.001695563 |
| 0 | 0 | 0 | 0 | 0 |
| 0 | 0 | 0 | 0.163693088 | 0 |
| 0 | 0.002923269 | 0 | 0.194718456 | 0.007328396 |
| 0.001363643 | 0 | 0 | 0.000959401 | 0 |
| 0 | 0 | 0 | 0.20623879 | 0 |
| 0 | 0 | 0 | 0 | 0 |
| 0 | 0.103464425 | 0 | 0 | 0 |
| 0 | 0.042778027 | 0 | 0 | 0 |
| 0.008059189 | 0.004550843 | 0 | 0.005460098 | 0 |
| 0.008311439 | 0.00067764 | 0 | 0 | 0 |
| 0 | 0 | 0 | 0 | 0 |
| 0 | 0 | 0 | 0 | 0 |
| 0.057884913 | 0.004682399 | 0.002674711 | 0.073784761 | 0.044238417 |
| 0.002030572 | 0.001214066 | 0.012325514 | 0.008333638 | 0 |
| 0.012889193 | 0.006068766 | 0 | 0.022897422 | 0 |
| 0 | 0.009525432 | 0 | 0 | 0 |
| 0 | 0 | 0 | 0 | 0 |
| 0 | 0 | 0 | 0 | 0 |
| 0 | 0 | 0.027772584 | 0.004509096 | 0 |
| 0.044881358 | 0.00408644 | 0 | 0.063313505 | 0.052512638 |
| 0 | 0.112927792 | 0 | 0 | 0 |
| 0 | 0.003341082 | 0 | 0 | 0.001474144 |
| 0 | 0 | 0 | 0 | 0 |
| 0 | 0 | 0 | 0 | 0 |
| 0.135262132 | 0.001833379 | 0.002919676 | 0 | 0.012807886 |
| 0 | 0.010526901 | 0 | 0.397268813 | 0 |
| 0 | 0.053685106 | 0 | 0 | 0.051642755 |
| 0 | 0 | 0 | 0 | 0 |
| 0 | 0 | 0 | 0 | 0 |
| 0 | 0 | 0 | 0 | 0 |

| | | | | |
|---|---|---|---|---|
| 0.00135115 | 0 | 0 | 0 | 0 |
| 0.509741346 | 0.16760562 | 0 | 1.16872183 | 0.365146138 |
| 0 | 0 | 0 | 0 | 0 |
| 0.000940069 | 0.023775206 | 0 | 0 | 0 |
| 0 | 0 | 0 | 0 | 0 |
| 0 | 0 | 0.156365503 | 0 | 0 |
| 0 | 0 | 0 | 0 | 0.001420554 |
| 0 | 0 | 0 | 0 | 0 |
| 0 | 0 | 0 | 0 | 0 |
| 0 | 0 | 0 | 0 | 0 |
| 0.00645933 | 0.006827453 | 0 | 0.010549753 | 0.016659441 |
| 0.013057553 | 0.001232689 | 0 | 0.033120606 | 0.002039565 |
| 0 | 0.275733521 | 0 | 0 | 0 |
| 0 | 0 | 0 | 0 | 0 |
| 0 | 0 | 0.005670532 | 0.002420904 | 0 |
| 0.004185157 | 0 | 0 | 0 | 0 |
| 0.042274066 | 0.002166463 | 0.003363863 | 0.06254366 | 0.006213231 |
| 0.115399707 | 0 | 0 | 0.185546335 | 0.006514549 |
| 0 | 0 | 0 | 0 | 0 |
| 0.027285946 | 0.004930754 | 0 | 0.007736196 | 0.012690622 |
| 0.004484081 | 0 | 0 | 0.058889745 | 0.00394302 |
| 0 | 0 | 0 | 0 | 0 |
| 0 | 0 | 0 | 0 | 0 |
| 0 | 0 | 0 | 0 | 0 |
| 0 | 0.001126147 | 0 | 0 | 0 |
| 0.001827568 | 0 | 0 | 0 | 0 |
| 0.04180122 | 0.064613465 | 0 | 0 | 0 |
| 0.132916092 | 0.176907194 | 0 | 0.292278107 | 0.017025323 |
| 0 | 0 | 0 | 0 | 0 |
| 0 | 0 | 0 | 0 | 0 |
| 0 | 0 | 0 | 0 | 0 |
| 0 | 0 | 0 | 0 | 0 |
| 0 | 0 | 0 | 0 | 0 |
| 0 | 0 | 0 | 0 | 0 |
| 0.069450126 | 0 | 0 | 0 | 0.086507528 |
| 0.197169571 | 0.296591672 | 0 | 0.091429156 | 0.168896379 |
| 0.00539601 | 0.004094845 | 0 | 0 | 0 |
| 0.00708992 | 0 | 0 | 0 | 0 |
| 0 | 0.04891975 | 0.001399495 | 0.006204626 | 0 |
| 0 | 0 | 0 | 0 | 0 |
| 0 | 0.008823204 | 0 | 0 | 0.047850871 |
| 0 | 0 | 0 | 0 | 0 |
| 0.056146737 | 0.059251274 | 0 | 0.00753757 | 0.019822919 |
| 0 | 0 | 0 | 0 | 0 |
| 0.035482952 | 0 | 0 | 0.067987857 | 0.002240533 |
| 0 | 0.090167072 | 0 | 0 | 0 |
| 0.106104479 | 0 | 0 | 0 | 0 |
| 0 | 0 | 0 | 0 | 0 |

| | | | | |
|---|---|---|---|---|
| 0.002217334 | 0 | 0 | 0.001560022 | 0 |
| 0 | 0 | 0 | 0 | 0 |
| 0 | 0 | 0 | 0 | 0 |
| 13.9474645 | 0.014713628 | 0 | 0.005202301 | 0.101950425 |
| 0 | 0 | 0 | 0 | 0 |
| 0 | 0 | 0 | 0 | 0 |
| 0 | 0 | 0 | 0 | 0 |
| 0 | 0.230072973 | 0 | 0.00145438 | 0 |
| 0.81971428 | 0.053709826 | 0.003207505 | 0.239991101 | 1.48481008 |
| 0 | 0.025930296 | 0 | 0 | 0 |
| 0 | 0.163611163 | 0 | 0 | 0 |
| 0.005091997 | 0.026010395 | 0 | 0.002647943 | 0 |
| 0 | 0 | 0 | 0 | 0 |
| 0 | 5.675702765 | 0 | 0 | 0 |
| 0 | 0.296397661 | 0.028180088 | 0.076349601 | 0.104823093 |
| 0 | 0 | 0 | 0 | 0 |
| 0 | 0 | 0 | 0.001173636 | 0 |
| 0 | 0 | 0 | 0 | 0 |
| 0 | 0 | 0 | 0 | 0 |
| 0 | 0 | 0 | 0 | 0 |
| 0 | 0 | 0 | 0 | 0 |
| 0 | 0 | 0 | 0 | 0 |
| 0 | 0 | 0 | 0 | 0 |
| 0.253410835 | 0.610628111 | 0 | 0.620792576 | 0.12360305 |
| 0 | 0.016916536 | 0 | 0.011666596 | 0 |
| 0 | 0 | 0 | 0 | 0 |
| 0 | 0.014156003 | 0 | 0 | 0 |
| 0 | 0 | 0.00227501 | 0 | 0 |
| 0 | 0 | 0 | 0 | 0 |
| 0 | 0 | 0 | 0 | 0 |
| 0 | 0.024321404 | 0 | 0 | 0 |
| 0 | 0 | 0 | 0 | 0 |
| 0.047503681 | 0.078595635 | 0 | 0 | 0 |
| 0 | 0 | 0 | 0 | 0 |
| 0 | 0.350735862 | 0 | 0 | 0 |
| 0 | 0 | 0 | 0 | 0 |
| 3.63973024 | 0.543866974 | 0.001262312 | 0 | 0 |
| 0 | 0 | 0 | 0 | 0 |
| 0 | 0.0031004 | 0 | 0 | 0 |
| 0 | 0.001705424 | 0.003225139 | 0 | 0 |
| 0.064777498 | 0.168571045 | 0.034125153 | 0.538827249 | 0.069334026 |
| 0 | 0.398854669 | 0 | 0 | 0 |
| 6.407007147 | 7.445870106 | 0 | 0.093929024 | 0.09527281 |
| 0 | 0 | 0 | 0 | 0 |
| 0 | 0 | 0 | 0 | 0 |
| 0 | 0 | 0 | 0 | 0 |
| 0 | 0 | 0 | 0 | 0 |
| 0 | 0.00183179 | 0 | 0.003233272 | 0.026014508 |

| | | | | |
|---|---|---|---|---|
| 0 | 0 | 0 | 0 | 0 |
| 0 | 0.029204215 | 0 | 0 | 0 |
| 0.379843965 | 0.002194972 | 0 | 0.176324624 | 0.004115955 |
| 0 | 0.001423596 | 0 | 0 | 0 |
| 0 | 0 | 0 | 0 | 0 |
| 0 | 0.001556203 | 0 | 0.00718925 | 0.045774923 |
| 0 | 0 | 0 | 0.013719835 | 0 |
| 0 | 0 | 0 | 0 | 0 |
| 0 | 0 | 0 | 0 | 0 |
| 0 | 0 | 0 | 0 | 0 |
| 0.055988971 | 0.120468569 | 0.001801788 | 0.075232601 | 0.137398678 |
| 1.067319943 | 3.521332904 | 0 | 2.879111283 | 1.074199125 |
| 0 | 0 | 0 | 0 | 0 |
| 0.001556024 | 0 | 0 | 0.007115886 | 0.076195532 |
| 0.096665923 | 0.098974062 | 0.041155618 | 0.469599836 | 0.11746177 |
| 0 | 0 | 0.002372561 | 0 | 0 |
| 0 | 0.004019941 | 0 | 0 | 0 |
| 0 | 0 | 0 | 0 | 0 |
| 0 | 0 | 0 | 0 | 0 |
| 0 | 0.010651277 | 0 | 0.030637774 | 0.000783255 |
| 0.059212613 | 0.087965269 | 0 | 0.163520498 | 0 |
| 0.011035271 | 1.514032564 | 0 | 0.202459818 | 13.60723956 |
| 2.936777461 | 6.984626959 | 0 | 0.058621052 | 30.58329507 |
| 0 | 0 | 0 | 0 | 0 |
| 0 | 0 | 0 | 0 | 0 |
| 0 | 0 | 0 | 0.014602607 | 0 |
| 0 | 0.714654062 | 0.022964871 | 0 | 0.005957488 |
| 0 | 0 | 0 | 0 | 0 |
| 0 | 0 | 0 | 0 | 0 |
| 0 | 0.032651489 | 0 | 0 | 0 |
| 0 | 0.038450757 | 0 | 0 | 0 |
| 0 | 0.204971745 | 0 | 0.4149688 | 0.033105105 |
| 0.004195435 | 0.502201375 | 0 | 0.013022333 | 0 |
| 0 | 0.04438979 | 0 | 0 | 0 |
| 0.002606154 | 0.01460816 | 0 | 0.01650221 | 0.106832111 |
| 0.068257029 | 0.007024751 | 0 | 0.0198389 | 0.802755867 |
| 0 | 0.015704312 | 0 | 0.003346114 | 0.138580358 |
| 0 | 0 | 0 | 0 | 0 |
| 0 | 0.19681933 | 0 | 0.04956329 | 0 |
| 0 | 0 | 0 | 0 | 0 |
| 0 | 0.016082683 | 0 | 0 | 0 |
| 0 | 0 | 0 | 0 | 0 |
| 0 | 0 | 0 | 0 | 0 |
| 0 | 0 | 0 | 0 | 0 |
| 0.000765633 | 0.029754889 | 0 | 0.026215126 | 0.351689067 |
| 0 | 0 | 0 | 0 | 0 |
| 0 | 0 | 0 | 0 | 0 |
| 0.01200632 | 0.002760964 | 16.07438625 | 0.008880321 | 0.87222086 |

| | | | | |
|---|---|---|---|---|
| 0 | 0 | 0 | 0 | 0 |
| 0 | 0 | 0 | 0 | 0 |
| 0 | 0 | 0 | 0 | 0 |
| 0 | 0.000975383 | 0 | 0 | 0 |
| 0 | 0 | 0 | 0 | 0 |
| 0 | 0 | 0 | 0 | 0 |
| 0 | 1.067645617 | 0.00076526 | 0 | 0 |
| 0.028021497 | 0.02566934 | 0 | 0.041682571 | 0 |
| 0 | 0 | 0 | 0 | 0 |
| 0 | 0 | 0 | 0 | 0 |
| 0.00581411 | 0.015860257 | 0.011417832 | 0 | 0.004313727 |
| 0 | 0 | 0 | 0 | 0 |
| 0 | 0 | 0 | 0 | 0 |
| 0 | 0.741589692 | 0 | 0 | 0 |
| 0 | 0 | 0 | 0 | 0 |
| 0 | 0 | 0 | 0 | 0 |
| 0 | 0.486861797 | 0 | 0 | 0 |
| 0 | 0.00253214 | 0 | 0.003121523 | 0.004189592 |
| 0 | 0 | 0 | 0 | 0 |
| 0.001636868 | 0 | 0 | 0.005758152 | 0 |
| 0 | 0 | 0 | 0 | 0 |
| 0 | 0 | 0 | 0 | 0 |
| 0 | 0.573448911 | 0 | 0 | 0 |
| 0 | 0 | 0 | 0 | 0 |
| 0 | 0 | 0 | 0 | 0 |
| 0 | 0 | 0 | 0 | 0 |
| 0 | 0 | 0 | 0 | 0 |
| 0 | 0 | 0 | 0 | 0 |
| 0 | 0 | 0 | 0 | 0 |
| 0 | 0.394367044 | 0 | 0 | 0 |
| 0 | 0.036961796 | 0 | 0 | 0 |
| 0 | 0 | 0 | 0 | 0 |
| 0 | 0 | 0 | 0 | 0 |
| 0 | 0 | 0 | 0 | 0 |
| 0 | 0 | 0 | 0 | 0.02572549 |
| 0 | 0 | 0 | 0 | 0 |
| 0 | 0.117467478 | 0 | 0 | 0 |
| 0.001435509 | 0.001655258 | 0 | 0.020199259 | 0.001014346 |
| 0 | 0 | 0 | 0 | 0 |
| 0 | 0 | 0 | 0 | 0 |
| 0 | 0.263499464 | 0 | 0.09448878 | 0 |
| 0 | 0 | 0 | 0 | 0 |
| 0 | 0 | 0 | 0 | 0 |
| 0 | 0.008573077 | 0 | 0.022558255 | 0 |
| 0 | 0 | 0 | 0 | 0 |
| 0 | 0 | 0 | 0 | 0 |
| 0 | 0 | 0 | 0 | 0 |
| 0 | 0 | 0 | 0 | 0 |

| | | | | |
|---|---|---|---|---|
| 0 | 0 | 0 | 0 | 0 |
| 0 | 0 | 0 | 0 | 0 |
| 0 | 0 | 0 | 0 | 0 |
| 0 | 1.034183022 | 0 | 0 | 0 |
| 0 | 0 | 0 | 0 | 0 |
| 0 | 0 | 0 | 0 | 0 |
| 0 | 0 | 0 | 0 | 0 |
| 0 | 0 | 0 | 0 | 0 |
| 0.010747523 | 0.062398659 | 0.030295895 | 0.060690974 | 0.002518114 |
| 0 | 0 | 0 | 0 | 0 |
| 0 | 0 | 0 | 0 | 0 |
| 0 | 0 | 0 | 0 | 0 |
| 0 | 0.007398822 | 0.001590665 | 0 | 0 |
| 0 | 0 | 0 | 0 | 0 |
| 3.157876217 | 0.093692846 | 0 | 2.497445519 | 0.990043164 |
| 0.704744197 | 0 | 0 | 0 | 0.081728431 |
| 0.019670432 | 0.000773739 | 0 | 0 | 0 |
| 0 | 0 | 0 | 0 | 0 |
| 0 | 0 | 0 | 0 | 0 |
| 0 | 0 | 0 | 0 | 0 |
| 0 | 0.205979317 | 0 | 0 | 0 |
| 0 | 0 | 0 | 0 | 0 |
| 0 | 0 | 0 | 0 | 0 |
| 0 | 0 | 0 | 0 | 0.012613612 |
| 0.033592549 | 0.203134677 | 0 | 0.2080285 | 0 |
| 6.630289849 | 0.436625378 | 0 | 6.642527298 | 0 |
| 0 | 0.039718585 | 0 | 0 | 0 |
| 0 | 0 | 0 | 0 | 0 |
| 0 | 0 | 0 | 0.025368586 | 0 |
| 0 | 0 | 0 | 0 | 0 |
| 0 | 0 | 0 | 0 | 0 |
| 0 | 0.022091459 | 0 | 0 | 0 |
| 0 | 0 | 0 | 0 | 0 |
| 0 | 0.005257077 | 0 | 0 | 0 |
| 0 | 0 | 0 | 0.001647322 | 0.040824067 |
| 0.001655099 | 0.002783175 | 0.017507336 | 0.003638928 | 0.020875759 |
| 0 | 0.019530688 | 0 | 1.107955717 | 0.016263008 |
| 1.263137817 | 0.605909878 | 0 | 2.517488972 | 0.123639232 |
| 0 | 0 | 0 | 0 | 0 |
| 0 | 0 | 0 | 0 | 0 |
| 0 | 0 | 0 | 0 | 0 |
| 0.007507581 | 0 | 0 | 0.000825315 | 0.001856728 |
| 0.716965882 | 0.107758998 | 0 | 0.007359536 | 0.040553224 |
| 0 | 0 | 0.000910217 | 0 | 0 |
| 0 | 0 | 0 | 0 | 0 |
| 0 | 0.007558567 | 0 | 0 | 0.000969469 |
| 0 | 0 | 0 | 0 | 0 |
| 0.024667842 | 0.052087366 | 0 | 0.553390551 | 0 |

| | | | | |
|---|---|---|---|---|
| 0 | 0.180425524 | 0 | 0 | 0 |
| 0 | 0 | 0 | 0 | 0 |
| 0 | 0 | 0 | 0 | 0 |
| 0 | 0 | 0 | 0 | 0 |
| 0.001159126 | 0.001871193 | 0 | 0.017615051 | 0.002064008 |
| 0 | 0 | 0 | 0 | 0 |
| 0 | 0 | 0 | 0 | 0 |
| 0 | 0 | 0 | 0 | 0 |
| 0 | 0 | 0 | 0 | 0 |
| 0 | 0 | 0 | 0 | 0 |
| 0 | 0 | 0 | 0 | 0 |
| 3.599705692 | 0.973630262 | 0 | 3.490957454 | 1.351120668 |
| 0 | 0.136696415 | 0 | 0.47140462 | 0 |
| 0 | 0 | 0 | 0 | 0 |
| 0.00321727 | 3.347761267 | 66.2664697 | 0.041875379 | 0.0053045 |
| 0 | 0 | 0 | 0 | 0 |
| 0 | 0.001003599 | 0 | 0 | 0 |
| 0 | 0 | 0 | 0 | 0 |
| 0 | 0.00210752 | 0 | 0 | 0 |
| 0 | 0.001168452 | 0 | 0 | 0 |
| 0 | 0.042573943 | 0 | 0.005201418 | 0 |
| 0 | 0 | 0 | 0 | 0 |
| 0.357601803 | 1.249444381 | 0 | 0.291623244 | 0.1598133 |
| 0 | 0 | 0 | 0.050917638 | 0 |
| 0 | 0 | 0 | 0 | 0 |
| 0 | 0 | 0 | 0 | 0 |
| 0 | 0 | 0 | 0 | 0 |
| 0 | 0 | 0 | 0 | 0 |
| 0 | 0 | 0 | 0 | 0 |
| 0 | 0 | 0 | 0 | 0 |
| 0 | 0 | 0 | 0 | 0 |
| 0.075495347 | 0.002808137 | 0 | 0.113573741 | 0.012389994 |
| 0 | 0 | 0 | 0 | 0 |
| 0.104979582 | 0.13109293 | 0 | 0 | 0.021492457 |
| 0.001125726 | 0.003028792 | 0.019534707 | 0.004435273 | 0.022718056 |
| 0.002031677 | 0.00390448 | 0 | 0.001429401 | 0 |
| 0 | 0 | 0 | 0 | 0 |
| 0 | 0 | 0 | 0 | 0 |
| 0 | 0 | 0 | 0 | 0 |
| 0 | 0 | 0 | 0 | 0 |
| 0 | 0.378135434 | 0 | 0 | 0 |
| 0 | 0 | 0 | 0 | 0 |
| 0 | 0 | 0 | 0 | 0 |
| 0 | 0 | 0 | 0 | 0 |
| 0.064762876 | 0.125512797 | 0.001425455 | 0.014199863 | 0.116066555 |
| 0 | 0 | 0 | 0 | 0 |
| 0 | 0.660733988 | 0 | 0 | 0.249216071 |
| 0 | 0 | 0 | 0.000885529 | 0 |

| | | | | |
|---|---|---|---|---|
| 0 | 0 | 0 | 0 | 0 |
| 0 | 0 | 0 | 0 | 0 |
| 0 | 0.011520526 | 0 | 0.009199056 | 0.007488434 |
| 0 | 0 | 0 | 0 | 0 |
| 0.002326316 | 0 | 0 | 0.004255412 | 0 |
| 0 | 0.036104334 | 0 | 0 | 0 |
| 0 | 0.26880369 | 0 | 0 | 0 |
| 0.082537794 | 0 | 0 | 0.049230008 | 0 |
| 0 | 0 | 0 | 0 | 0 |
| 0 | 0.006691497 | 0 | 0 | 0 |
| 0 | 0 | 0 | 0 | 0 |
| 0.001867166 | 0 | 0 | 0 | 0 |
| 0 | 0 | 0 | 0 | 0 |
| 0 | 0 | 0 | 0 | 0 |
| 0 | 0 | 0 | 0 | 0 |
| 0 | 0 | 0 | 0 | 0 |
| 0 | 0 | 0 | 0 | 0 |
| 0 | 0 | 0 | 0 | 0 |
| 0 | 0 | 0 | 0.44474144 | 0 |
| 0 | 0 | 0 | 0 | 0 |
| 0 | 0 | 0 | 0 | 0 |
| 0 | 0 | 0 | 0 | 0 |
| 0 | 0 | 0 | 0 | 0 |
| 0 | 0 | 0 | 0 | 0 |
| 0 | 0 | 0 | 0 | 0 |
| 0.04872898 | 0.000740714 | 0 | 0.005084437 | 0.019608966 |
| 0 | 0 | 0 | 0 | 0 |
| 0.04021822 | 0.048032039 | 0.065378617 | 0.06114173 | 0 |
| 0 | 0 | 0 | 0 | 0 |
| 0.066209613 | 0.61015794 | 0 | 0.095017221 | 0 |
| 0 | 0 | 0 | 0 | 0 |
| 0 | 0 | 0 | 0 | 0 |
| 0 | 0 | 0 | 0 | 0 |
| 0 | 0 | 0 | 0 | 0 |
| 0.008438701 | 0.001552445 | 0.883764989 | 0.003653606 | 0.002140519 |
| 0 | 0 | 0 | 0 | 0 |
| 0 | 0 | 0 | 0 | 0 |
| 0 | 0.000880785 | 0.145175335 | 0 | 0 |
| 0 | 0 | 0 | 0 | 0 |
| 0 | 0 | 0 | 0 | 0 |
| 9.555838485 | 10.92824414 | 0 | 1.715830231 | 5.384096411 |
| 1.437081163 | 0.386682159 | 0 | 0.093444221 | 0.139212561 |
| 0 | 0 | 0.000903841 | 0 | 0 |
| 0 | 0 | 0 | 0 | 0 |
| 0 | 0 | 0 | 0 | 0 |
| 0 | 0 | 0 | 0 | 0 |
| 0 | 0.00074963 | 0 | 0 | 0 |
| 0.004676683 | 0.01188368 | 0 | 0 | 0.116817377 |

| | | | | |
|---|---|---|---|---|
| 0 | 0 | 0 | 0 | 0 |
| 0.031668166 | 0.006291428 | 0 | 0.008601915 | 0.001387944 |
| 0.002809111 | 0 | 0 | 0 | 0 |
| 0 | 0 | 0 | 0 | 0 |
| 2.90483518 | 1.120359307 | 0 | 0.040166003 | 0.381080606 |
| 0 | 0 | 0 | 0 | 0 |
| 0 | 0 | 0 | 0 | 0 |
| 0 | 0 | 0 | 0 | 0 |
| 0.206740416 | 0 | 0 | 0.36736573 | 0.007523446 |
| 0 | 0.002397181 | 0 | 0 | 0 |
| 0 | 0 | 0 | 0 | 0 |
| 0 | 0 | 0 | 0 | 0 |
| 0 | 0.128027767 | 0 | 0.126880623 | 0 |
| 0.007901169 | 0.025552875 | 0 | 0.1066303 | 0 |
| 0 | 0 | 0 | 0 | 0 |
| 0.278993096 | 0.478046652 | 0 | 0.722670266 | 0 |
| 0 | 0.220005603 | 0 | 0.036150794 | 0 |
| 0 | 0 | 0 | 0 | 0 |
| 0 | 0 | 0 | 0 | 0 |
| 0 | 0 | 0 | 0 | 0 |
| 0 | 0 | 0 | 0 | 0 |
| 0 | 0 | 0 | 0 | 0 |
| 0 | 0 | 0 | 0 | 0 |
| 0 | 0 | 0 | 0 | 0 |
| 0 | 0 | 0 | 0 | 0 |
| 0 | 0 | 0 | 0 | 0 |
| 0 | 0.005170376 | 0 | 0 | 0 |
| 0 | 0 | 0 | 0 | 0.002011053 |
| 0 | 0 | 0 | 0 | 0 |
| 0 | 0 | 0 | 0 | 0 |
| 0 | 0 | 0 | 0 | 0 |
| 0 | 0 | 0 | 0 | 0 |
| 0 | 0.020045479 | 0.001173627 | 0.288298245 | 0 |
| 0 | 0.017200658 | 0 | 0 | 0 |
| 0 | 0 | 0 | 0 | 0 |
| 0.008230738 | 0.370559433 | 0 | 0.180341937 | 0 |
| 0 | 0 | 0 | 0 | 0 |
| 0 | 0 | 0 | 0 | 0 |
| 0 | 0 | 0 | 0 | 0 |
| 0 | 0 | 0 | 0 | 0 |
| 0 | 0 | 0 | 7.340868828 | 0.256143557 |
| 0 | 0 | 0 | 0 | 0 |
| 0 | 0 | 0 | 0 | 0 |
| 0 | 0 | 0 | 0 | 0 |
| 0 | 0 | 0 | 0 | 0 |
| 0 | 0 | 0 | 0 | 0 |
| 0 | 0 | 0 | 0 | 0 |
| 0 | 0 | 0 | 0 | 0 |

| | | | | |
|---|---|---|---|---|
| 0 | 0 | 0 | 0 | 0 |
| 0 | 0 | 0 | 0 | 0 |
| 0 | 0 | 0 | 0 | 0 |
| 0.00598747 | 0 | 0 | 0 | 0 |
| 0 | 0.028060596 | 0 | 0 | 0 |
| 0 | 0 | 0 | 0 | 0 |
| 0.053324779 | 0.076555215 | 0.001549767 | 0.323111865 | 0 |
| 0 | 0 | 0 | 0 | 0 |
| 0 | 0 | 0 | 0 | 0 |
| 0.610422612 | 0.008168022 | 0 | 0 | 0 |
| 0 | 0.015060519 | 0 | 0 | 0 |
| 0 | 0.024801923 | 0 | 0 | 0 |
| 0 | 0 | 0 | 0 | 0 |
| 0 | 0 | 0 | 0 | 0 |
| 0 | 0 | 0 | 0 | 0 |
| 0 | 0.008643293 | 0 | 0 | 0 |
| 0.166882624 | 0 | 0 | 0 | 0 |
| 0.110878013 | 0.000712831 | 0 | 0 | 0 |
| 0 | 0 | 0 | 0 | 0 |
| 0 | 0 | 0 | 0 | 0 |
| 0.091697532 | 0.052942977 | 0 | 0.540481749 | 0.012069008 |
| 0 | 0 | 0 | 0 | 0 |
| 0 | 0.02413698 | 0 | 0.005955387 | 0.026838858 |
| 0 | 0.00207705 | 0 | 0.000950491 | 0.031502314 |
| 0 | 0 | 0 | 0 | 0 |
| 0 | 0.002042044 | 0 | 0 | 0 |
| 0 | 0 | 0 | 0 | 0 |
| 0 | 0 | 0 | 0 | 0 |
| 0 | 0 | 0 | 0 | 0 |
| 0 | 0 | 2.515079974 | 0.040843946 | 0 |
| 0 | 0 | 0 | 0 | 0 |
| 0 | 0 | 0 | 0 | 0 |
| 0 | 0 | 0 | 0 | 0 |
| 0 | 0 | 0 | 8.772062897 | 0 |
| 0 | 0 | 0 | 0 | 0 |
| 0 | 0 | 0 | 0 | 0 |
| 0.004829783 | 0.668357577 | 0 | 0.00934458 | 0 |
| 0 | 0 | 0 | 0 | 0 |
| 0 | 0 | 0 | 0 | 0 |
| 0 | 0 | 0 | 0 | 0 |
| 0 | 0 | 0 | 0 | 0 |
| 0 | 0 | 0 | 0 | 0 |
| 0 | 0 | 0 | 0 | 0 |
| 0 | 0.007026682 | 0 | 0 | 0 |
| 0 | 0 | 0.001224088 | 0 | 0 |
| 0 | 0 | 0 | 0 | 0 |
| 0 | 0 | 0 | 0 | 0 |
| 0 | 0 | 0 | 0 | 0 |

| | | | | |
|---|---|---|---|---|
| 0.119826474 | 0.005773698 | 0.009342977 | 0.004675483 | 0.034924809 |
| 0 | 0 | 0 | 0 | 0 |
| 0 | 0.171757974 | 0 | 0 | 0 |
| 0 | 0 | 0 | 0 | 0 |
| 0 | 0 | 0 | 0 | 0 |
| 0 | 0 | 0 | 0 | 0 |
| 0 | 0 | 0 | 0 | 0.001579326 |
| 1.017755033 | 1.311297852 | 0 | 1.195965387 | 0.384333593 |
| 0.227202247 | 0.172775716 | 0.002095029 | 0.46349564 | 0.002257283 |
| 0.105750168 | 0.089753029 | 0 | 0.220809027 | 0 |
| 0 | 0.017434518 | 0 | 0 | 0 |
| 0 | 0.001425195 | 0 | 0 | 0 |
| 0 | 0.018154308 | 0 | 0 | 0 |
| 0 | 0 | 0 | 0 | 0 |
| 0 | 0 | 0 | 0 | 0 |
| 0 | 0 | 0 | 0 | 0 |
| 0.117316942 | 0 | 0 | 0.434450493 | 0 |
| 0 | 0 | 0 | 0 | 0 |
| 0 | 0 | 0 | 0 | 0 |
| 0.079975317 | 0.016538971 | 0 | 0 | 0.007073898 |
| 0 | 0 | 0 | 0 | 0 |
| 0 | 0 | 0 | 0 | 0 |
| 0 | 0 | 0 | 0 | 0 |
| 0 | 0 | 0 | 0 | 0 |
| 0 | 0.071147737 | 0 | 0.005869129 | 0 |
| 0 | 0 | 0 | 0 | 0 |
| 0 | 0 | 0 | 0 | 0 |
| 0 | 0 | 0 | 0 | 0 |
| 0.001170102 | 0 | 0 | 0 | 0 |
| 0 | 0 | 0 | 0 | 0 |
| 0 | 0.120160324 | 0 | 0 | 0 |
| 0 | 0 | 0 | 0 | 0 |
| 0 | 0 | 0 | 0 | 0 |
| 0 | 0 | 0 | 0 | 0 |
| 0.003016987 | 0 | 0 | 0 | 0.004725568 |
| 0 | 0 | 0 | 0 | 0 |
| 0 | 0 | 0 | 0 | 0 |
| 0 | 0 | 0 | 0 | 0 |
| 0 | 0 | 0 | 0 | 0 |
| 0 | 0 | 0 | 0.001021037 | 0 |
| 0 | 0 | 0 | 0 | 0 |
| 0 | 0.267978218 | 0 | 0.626008947 | 0.202280377 |
| 0 | 0 | 0 | 0 | 0 |
| 0 | 0 | 0 | 0 | 0 |
| 0 | 0 | 0 | 0 | 0 |
| 0.049304806 | 0 | 0 | 0.004936977 | 0 |
| 0 | 0 | 0 | 0 | 0 |
| 0 | 0.000520323 | 0 | 0 | 0 |

| | | | | |
|---|---|---|---|---|
| 0 | 0 | 0 | 0 | 0 |
| 0.000749731 | 0.001176681 | 0 | 0.001714305 | 0 |
| 0 | 0 | 0 | 0 | 0 |
| 0 | 0 | 0 | 0 | 0 |
| 0.235851464 | 0.107170754 | 0 | 0.033002633 | 0 |
| 0 | 0 | 0 | 0 | 0 |
| 0 | 0 | 0 | 0 | 0 |
| 0.020605512 | 0.0388551 | 0.007640321 | 0.021188155 | 0.001829342 |
| 0 | 0.002602129 | 0 | 0 | 0 |
| 0 | 0 | 0 | 0 | 0 |
| 0 | 0 | 0 | 0 | 0 |
| 0 | 0 | 0 | 0 | 0 |
| 0 | 0 | 0 | 0 | 0 |
| 0 | 0 | 0 | 0 | 0 |
| 0 | 0.005670665 | 0 | 0 | 0 |
| 0 | 0.058767311 | 0 | 0 | 0 |
| 0 | 0 | 0 | 0 | 0 |
| 0 | 0 | 0 | 0 | 0 |
| 0 | 0 | 0 | 0 | 0 |
| 0 | 0 | 0 | 0 | 0 |
| 0 | 0 | 0 | 0 | 0 |
| 0 | 0 | 0 | 0 | 0 |
| 0 | 0 | 0 | 0 | 0 |
| 0.001600341 | 0.004100718 | 0 | 0 | 0.003844781 |
| 0 | 0 | 0 | 0 | 0.061372772 |
| 0 | 0 | 0 | 0 | 0 |
| 0 | 0.352928839 | 0 | 0 | 0.033303216 |
| 0.001321047 | 0 | 0 | 0 | 0 |
| 0 | 0 | 0 | 0 | 0 |
| 0 | 0 | 0 | 0 | 0 |
| 0 | 0 | 0 | 0 | 0 |
| 0 | 0.000901218 | 0 | 0 | 0 |
| 0 | 0 | 0 | 0 | 0 |
| 0 | 0 | 0.010787796 | 0 | 0 |
| 0 | 0 | 0 | 0 | 0 |
| 0 | 0 | 0 | 0 | 0 |
| 0 | 0 | 0 | 0 | 0 |
| 0 | 0.554847749 | 0 | 0 | 0 |
| 0 | 1.276873584 | 0 | 0 | 0 |
| 0 | 0 | 0 | 0 | 0 |
| 0 | 0 | 0 | 0 | 0 |
| 0 | 0 | 0 | 0 | 0 |
| 0 | 0 | 0 | 0 | 0 |
| 0 | 0 | 0 | 0 | 0 |
| 0 | 0 | 0 | 0 | 0 |
| 0 | 0 | 0 | 0 | 0 |
| 0 | 0 | 0 | 0 | 0 |
| 0 | 0 | 0 | 0 | 0 |

| | | | | |
|---|---|---|---|---|
| 0 | 0.13579296 | 0 | 0.12907802 | 0.005252222 |
| 0 | 0 | 0 | 0 | 0 |
| 0 | 0 | 0 | 0 | 0 |
| 0 | 0 | 0 | 0 | 0 |
| 0 | 0 | 0 | 0 | 0 |
| 0.018681113 | 0 | 0 | 0.014746067 | 0 |
| 0 | 0 | 0 | 0 | 0 |
| 0 | 0.000992256 | 0 | 0 | 0 |
| 0 | 0 | 0 | 0 | 0 |
| 0 | 0 | 0 | 0 | 0 |
| 0 | 0 | 0 | 0 | 0 |
| 0 | 0 | 0 | 0 | 0 |
| 0 | 0 | 0 | 0 | 0 |
| 0 | 0 | 0 | 0 | 0 |
| 0 | 0 | 0 | 0 | 0 |
| 0 | 0 | 0 | 0 | 0 |
| 0 | 0 | 0 | 0 | 0 |
| 0 | 0 | 0 | 0 | 0 |
| 0 | 0.001495171 | 0 | 0 | 0 |
| 0 | 0 | 0 | 0 | 0 |
| 0.055550131 | 0.000612536 | 0 | 0.006246825 | 0.015765271 |
| 0 | 0 | 0 | 0 | 0 |
| 0.037090551 | 0.084579048 | 0 | 0.000809157 | 0.065135418 |
| 0 | 0.03020923 | 0 | 0 | 0 |
| 0 | 0 | 0 | 0 | 0 |
| 0 | 0 | 0 | 0 | 0 |
| 0 | 0 | 0 | 0 | 0 |
| 0 | 0 | 0 | 0 | 0 |
| 0 | 0 | 0 | 0 | 0 |
| 0 | 0 | 0 | 0 | 0 |
| 0 | 0 | 0 | 0 | 0 |
| 0 | 0 | 0 | 0 | 0 |
| 0.016694412 | 0 | 0.095758326 | 0.444812548 | 0.019711483 |
| 0 | 0 | 0 | 0 | 0 |
| 0 | 0.16049506 | 0 | 2.495409206 | 0.009054182 |
| 0.001512516 | 0 | 0.00601775 | 0 | 0.000855008 |
| 0 | 0 | 0 | 0 | 0 |
| 0 | 0 | 0 | 0 | 0 |
| 0.010496781 | 0 | 0 | 0.001934189 | 0.009889515 |
| 0 | 0 | 0 | 0 | 0 |
| 0 | 0 | 0 | 0 | 0 |
| 0 | 0.019089023 | 0 | 0 | 0 |
| 0.101405644 | 0.011269807 | 0 | 0.028990203 | 0.146571239 |
| 0 | 0 | 0 | 0 | 0 |
| 0 | 0 | 0 | 0 | 0 |
| 0 | 0.031429707 | 0 | 0.005551706 | 0.008724484 |
| 0 | 0 | 0 | 0 | 0 |
| 0 | 0 | 0 | 0 | 0 |

| | | | | |
|---|---|---|---|---|
| 0 | 0 | 0 | 0 | 0 |
| 0 | 0 | 0 | 0 | 0 |
| 0 | 0 | 0 | 0 | 0 |
| 0 | 0 | 0 | 0 | 0 |
| 0 | 0 | 0 | 0 | 0 |
| 0 | 0 | 0 | 0 | 0 |
| 0 | 0 | 0 | 0 | 0 |
| 0 | 0 | 0 | 0.04465329 | 0.002440441 |
| 0.079801433 | 0.007410433 | 0 | 0.002447733 | 0 |
| 0.01171323 | 0 | 0 | 0.003700009 | 0 |
| 0 | 0 | 0 | 0 | 0 |
| 0 | 0 | 0 | 0 | 0 |
| 0 | 0 | 0 | 0 | 0 |
| 0 | 1.253862656 | 0 | 0 | 0 |
| 0 | 0.002598794 | 0 | 0 | 0 |
| 0.010113357 | 0 | 0 | 0.001985673 | 0 |
| 0 | 0 | 0 | 0 | 0 |
| 0 | 0 | 0 | 0 | 0 |
| 0 | 0 | 0 | 0 | 0 |
| 0.221723079 | 1.617822119 | 0 | 0.917959324 | 0 |
| 0 | 0 | 0 | 0.009159699 | 0.000972017 |
| 0 | 0 | 0 | 0 | 0 |
| 0 | 0 | 0 | 0 | 0 |
| 0 | 0.043549672 | 0 | 0 | 0 |
| 0 | 0 | 0 | 0 | 0 |
| 0 | 0 | 0 | 0 | 0 |
| 0 | 0 | 0 | 0 | 0 |
| 0 | 0 | 0 | 0 | 0 |
| 0 | 0 | 0 | 0 | 0 |
| 0 | 0.014428572 | 0 | 0 | 0 |
| 0.186571539 | 0.176068391 | 0 | 0.5027295 | 0.07015807 |
| 0 | 0 | 0 | 0 | 0 |
| 0 | 0.460017668 | 0 | 0 | 0 |
| 0 | 0 | 0 | 0 | 0 |
| 0 | 0 | 0 | 0 | 0 |
| 0 | 0 | 0 | 0 | 0 |
| 0 | 0 | 0 | 0 | 0 |
| 0 | 0 | 0 | 0 | 0 |
| 0 | 0 | 0 | 0 | 0 |
| 0 | 0 | 0 | 0 | 0 |
| 0 | 0 | 0 | 0 | 0 |
| 0 | 0 | 0 | 0 | 0 |
| 0 | 0 | 0 | 0 | 0 |
| 0 | 0 | 0 | 0 | 0 |
| 0 | 0 | 0 | 0 | 0 |
| 0 | 0 | 0 | 0 | 0 |
| 0.030818143 | 0 | 0.110640163 | 1.066330243 | 0.04692139 |
| 0 | 0 | 0 | 0 | 0 |

| | | | | |
|---|---|---|---|---|
| 0 | 0 | 0 | 0 | 0 |
| 0 | 0 | 0 | 0 | 0 |
| 0 | 0 | 0 | 0 | 0 |
| 0 | 0 | 0 | 0 | 0 |
| 0.142097883 | 0.845454132 | 0 | 0.126549377 | 0.356053905 |
| 0 | 0 | 0 | 0 | 0 |
| 0 | 0 | 0 | 0 | 0 |
| 0 | 0 | 0 | 0 | 0 |
| 0 | 0 | 0 | 0 | 0 |
| 0 | 0 | 0 | 0 | 0 |
| 0 | 0.041372535 | 0 | 0.074713285 | 0 |
| 0.012236493 | 0.034714264 | 0.002407478 | 0.002986822 | 0.044220339 |
| 0.079500751 | 0.004637367 | 0 | 0 | 0 |
| 0 | 0 | 0 | 0 | 0 |
| 0.00361584 | 0 | 0 | 0 | 0 |
| 0 | 1.257630842 | 0 | 0.761018387 | 0 |
| 0 | 0.003445082 | 0 | 0.018648024 | 0.01235025 |
| 0 | 0 | 0 | 0 | 0 |
| 0.075069804 | 0.032560523 | 0 | 0.290420993 | 0.027685007 |
| 0.044935337 | 0.056464069 | 0 | 0.025882271 | 0 |
| 0 | 0 | 0 | 0 | 0 |
| 0 | 0 | 0 | 0 | 0 |
| 0 | 0 | 0 | 0 | 0 |
| 0 | 0 | 0 | 0 | 0 |
| 0 | 0 | 0 | 0 | 0 |
| 0 | 0 | 0 | 0 | 0 |
| 0.022950629 | 0.004613445 | 0 | 0.009047935 | 0.079268341 |
| 0 | 0 | 0 | 0 | 0 |
| 0 | 0 | 0 | 0 | 0 |
| 0 | 0 | 0 | 0 | 0 |
| 0.016147703 | 0.000804551 | 0.000720707 | 0 | 0 |
| 0.108766506 | 0 | 0 | 0 | 0 |
| 0 | 0 | 0 | 0 | 0 |
| 0 | 0 | 0 | 0 | 0 |
| 0 | 0 | 0 | 0 | 0 |
| 0 | 0 | 0 | 0 | 0 |
| 0 | 0 | 0 | 0 | 0 |
| 0.021901973 | 0.013566101 | 2.547265781 | 0.004212613 | 0.004052751 |
| 0 | 0.026005601 | 0 | 0 | 0 |
| 0.010497646 | 0 | 0 | 0 | 0 |
| 0.001573038 | 0.007335977 | 0 | 0.000774706 | 0.001400522 |
| 0 | 0 | 0 | 0 | 0 |
| 0 | 0 | 0 | 0 | 0 |
| 0.001635575 | 0.002514601 | 0 | 0.005917993 | 0.001155715 |
| 0 | 0 | 0 | 0 | 0 |
| 0 | 0 | 0 | 0 | 0 |
| 0 | 0 | 0 | 0 | 0 |
| 0 | 0 | 0 | 0 | 0 |

| | | | | |
|---|---|---|---|---|
| 0 | 0 | 0 | 0 | 0 |
| 0 | 0 | 0 | 0.728373292 | 0 |
| 0 | 0 | 0 | 0 | 0 |
| 0.00073461 | 0 | 0.002885276 | 0 | 0 |
| 0 | 0 | 0 | 0.003195741 | 0.002696072 |
| 0 | 0.020504177 | 0 | 0 | 0 |
| 0 | 0.006606543 | 0 | 0 | 0 |
| 0.000574529 | 0 | 0 | 0 | 0 |
| 0 | 0.028009828 | 0 | 0 | 0 |
| 0 | 0 | 0 | 0 | 0 |
| 0 | 0 | 0 | 0 | 0 |
| 0 | 0 | 0 | 0 | 0 |
| 0.010414702 | 0.009807347 | 0 | 0.00781583 | 0.466544854 |
| 0 | 0 | 0 | 0 | 0 |
| 0 | 0 | 0 | 0 | 0 |
| 0 | 0 | 0 | 0 | 0 |
| 0 | 0 | 0 | 0 | 0 |
| 0 | 0 | 0 | 0 | 0 |
| 0 | 0 | 0 | 0 | 0 |
| 0 | 0 | 0 | 0 | 0 |
| 0 | 0 | 0 | 0 | 0 |
| 0 | 0 | 0 | 0 | 0 |
| 0.030170279 | 0.032592517 | 0 | 0.004663402 | 0.037533722 |
| 0 | 0.035799516 | 0 | 0.069071827 | 0.005069461 |
| 0 | 0.081362349 | 0 | 0 | 0 |
| 0 | 0 | 0 | 0 | 0 |
| 0 | 0 | 0 | 0 | 0 |
| 0 | 0 | 0 | 0 | 0 |
| 0 | 0 | 0 | 0 | 0 |
| 0 | 0 | 0 | 0 | 0 |
| 0 | 0.027655358 | 0 | 0 | 0 |
| 0 | 0 | 0 | 0 | 0 |
| 0 | 0 | 0 | 0 | 0 |
| 0 | 0 | 0 | 0 | 0 |
| 0 | 0 | 0 | 0 | 0 |
| 0 | 0 | 0 | 0 | 0 |
| 0 | 0 | 0 | 0 | 0 |
| 0 | 0 | 0 | 0 | 0 |
| 0.005174826 | 0.065554057 | 0.004850235 | 0.002578891 | 0.000853204 |
| 0 | 0 | 0 | 0 | 0 |
| 0 | 0 | 0 | 0 | 0 |
| 0 | 0 | 0 | 0 | 0 |
| 0 | 0 | 0 | 0 | 0 |
| 0 | 0 | 0 | 0 | 0 |
| 0 | 0 | 0.005615908 | 0 | 0 |
| 0.187810915 | 0.037940747 | 0 | 0.039774195 | 0.228956835 |
| 0 | 0.006074878 | 0 | 0 | 0 |
| 0 | 0 | 0 | 0 | 0 |

| | | | | |
|---|---|---|---|---|
| 0 | 0 | 0 | 0 | 0 |
| 0 | 0 | 0 | 0 | 0 |
| 0 | 0 | 0 | 0 | 0 |
| 0 | 0.020036035 | 0 | 0 | 0 |
| 0 | 0 | 0 | 0 | 0 |
| 0 | 0 | 0 | 0 | 0 |
| 0 | 0.017297473 | 0 | 0 | 0 |
| 0 | 0 | 0 | 0 | 0 |
| 0 | 0 | 0 | 0.01684348 | 0 |
| 0 | 0 | 0 | 0 | 0 |
| 0 | 0.022590542 | 0 | 0.000615344 | 0.007613943 |
| 0.002350455 | 0 | 0 | 0 | 0.000775066 |
| 0 | 0 | 0 | 0 | 0 |
| 0 | 0 | 0 | 0 | 0 |
| 0 | 0 | 0 | 0 | 0 |
| 0 | 0 | 0 | 0 | 0 |
| 0 | 0 | 0 | 0 | 0 |
| 0 | 0 | 0 | 0 | 0 |
| 0 | 0 | 0 | 0 | 0 |
| 0 | 0.002704627 | 0 | 0 | 0.001606404 |
| 0 | 0 | 0 | 0 | 0 |
| 0 | 0 | 0 | 0 | 0 |
| 0 | 0 | 0 | 0 | 0 |
| 0 | 0 | 0 | 0 | 0 |
| 0 | 0 | 0 | 0 | 0 |
| 0 | 0 | 0 | 0 | 0 |
| 0 | 0 | 0 | 0 | 0 |
| 0 | 0 | 0 | 0 | 0 |
| 0 | 0 | 0 | 0 | 0 |
| 0 | 0 | 0 | 0 | 0 |
| 0.008968227 | 0.018230963 | 0 | 0.002103221 | 0.003943049 |
| 0 | 0 | 0 | 0 | 0 |
| 0.02568546 | 0.026356241 | 0 | 0.128940409 | 0.019915525 |
| 0.00567519 | 0.004453524 | 0 | 0.0123112 | 0.000935701 |
| 0 | 0 | 0 | 0 | 0 |
| 0 | 0 | 0 | 0 | 0 |
| 0 | 0 | 0 | 0 | 0 |
| 0 | 0 | 0 | 0 | 0 |
| 0 | 0 | 0 | 0 | 0 |
| 0 | 0 | 0 | 0 | 0 |
| 0 | 0 | 0 | 0 | 0 |
| 1.366674303 | 0.186439998 | 0 | 0.010581023 | 0.008662975 |
| 0.181597829 | 0.0468001 | 0 | 0.14685573 | 0.084202135 |
| 0 | 0 | 0 | 0 | 0 |
| 0 | 0 | 0 | 0 | 0 |
| 0.005697529 | 0.013225305 | 0 | 0.001886371 | 0 |
| 0 | 0.00177359 | 0 | 0 | 0 |
| 0 | 0 | 0 | 0 | 0 |

| | | | | |
|---|---|---|---|---|
| 0 | 0 | 0 | 0 | 0 |
| 0 | 0.006319333 | 0 | 0.01743643 | 0 |
| 0.009933632 | 0.001272698 | 0 | 0.044529146 | 0.007299977 |
| 0 | 0 | 0 | 0 | 0 |
| 1.403653616 | 0.202831156 | 0 | 0.016421978 | 0.023090542 |
| 0 | 0.007517551 | 0 | 0 | 0 |
| 0 | 0 | 0 | 0 | 0 |
| 0 | 0 | 0 | 0 | 0 |
| 0 | 0 | 0 | 0 | 0 |
| 0 | 0 | 0 | 0 | 0 |
| 0 | 0 | 0 | 0 | 0 |
| 0 | 0 | 0 | 0 | 0 |
| 0 | 0 | 0 | 0 | 0 |
| 0 | 0 | 0 | 0 | 0 |
| 0 | 0 | 0 | 0 | 0 |
| 0 | 0 | 0 | 0 | 0 |
| 0 | 0 | 0 | 0 | 0 |
| 0 | 0 | 0 | 0 | 0 |
| 0 | 0 | 0 | 0 | 0 |
| 0 | 0 | 0 | 0 | 0 |
| 0 | 0 | 0 | 0 | 0 |
| 0.00182639 | 0 | 0 | 0 | 0 |
| 0 | 0 | 0 | 0 | 0 |
| 0 | 0 | 0 | 0 | 0 |
| 0.00233906 | 0.011118151 | 0 | 0 | 0.00092557 |
| 0 | 0 | 0.009829982 | 0.005628692 | 0 |
| 0 | 0 | 0 | 0 | 0 |
| 0 | 0.27512565 | 0.000733494 | 0.020876577 | 0 |
| 0 | 0 | 0 | 0 | 0 |
| 0 | 0 | 0 | 0 | 0 |
| 0 | 0 | 0 | 0 | 0 |
| 0 | 0 | 0 | 0 | 0 |
| 0 | 0 | 0 | 0 | 0 |
| 0 | 0 | 0 | 0 | 0 |
| 0 | 0 | 0 | 0 | 0 |
| 0 | 0 | 0 | 0 | 0 |
| 0 | 0 | 0 | 0 | 0 |
| 0 | 0 | 0 | 0.122769207 | 0 |
| 0 | 0.002873982 | 0 | 0 | 0 |
| 0 | 0 | 0 | 0 | 0 |
| 0 | 0 | 0 | 0 | 0 |
| 0 | 0.002060416 | 0 | 0 | 0 |
| 0.106741211 | 0.005506586 | 0.006576973 | 0.002658356 | 0.031070875 |
| 0 | 0 | 0 | 0 | 0 |
| 0 | 0 | 0 | 0 | 0 |
| 0 | 0 | 0 | 0 | 0 |
| 0.002528876 | 0.003887999 | 0 | 0.004320938 | 0.006432951 |
| 0 | 0 | 0 | 0 | 0 |

| | | | | |
|---|---|---|---|---|
| 0 | 0 | 0 | 0 | 0 |
| 0.008433296 | 0.021872316 | 0 | 0.002084676 | 0.016459879 |
| 0 | 0 | 0 | 0 | 0 |
| 0 | 0 | 0 | 0 | 0 |
| 0 | 0 | 0 | 0 | 0 |
| 0 | 0 | 0 | 0 | 0 |
| 0 | 0 | 0 | 0 | 0 |
| 0 | 0 | 0 | 0 | 0 |
| 0 | 0 | 0 | 0 | 0 |
| 0 | 0 | 0 | 0 | 0 |
| 0 | 0 | 0 | 0 | 0 |
| 0 | 0 | 0 | 0 | 0 |
| 0.09444807 | 0.061367826 | 0 | 0 | 0 |
| 0 | 0 | 0 | 0 | 0 |
| 0.084972965 | 0.017586283 | 0 | 0.073379567 | 0.004498189 |
| 0 | 0 | 0 | 0 | 0 |
| 0 | 0 | 0 | 0 | 0 |
| 0 | 0 | 0 | 0 | 0 |
| 0.00188633 | 0 | 0 | 0.012386651 | 0 |
| 0 | 0 | 0 | 0 | 0 |
| 0 | 0 | 0 | 0 | 0 |
| 0.003070073 | 0.017521424 | 0 | 0.002749056 | 0 |
| 0 | 0.005385512 | 0 | 0 | 0.002758069 |
| 0 | 0 | 0 | 0 | 0 |
| 0 | 0 | 0 | 0 | 0 |
| 0 | 0 | 0 | 0 | 0 |
| 0 | 0 | 0 | 0 | 0 |
| 0 | 0 | 0 | 0 | 0 |
| 0 | 0 | 0 | 0 | 0 |
| 0 | 0 | 0 | 0 | 0 |
| 0 | 0 | 0 | 0 | 0 |
| 0 | 0 | 0 | 0 | 0 |
| 0 | 0 | 0 | 0 | 0 |
| 0 | 0 | 0 | 0 | 0 |
| 17.70920961 | 1.469261552 | 0.001085761 | 0 | 12.99912129 |
| 0 | 0 | 0 | 0 | 0 |
| 0 | 0 | 0 | 0 | 0 |
| 0 | 0 | 0 | 0 | 0 |
| 0 | 0.126480206 | 1.101645988 | 0.145499022 | 0.001378589 |
| 0 | 0 | 0 | 0 | 0 |
| 0.28638855 | 0.288222166 | 0 | 0.040790452 | 0.030902185 |
| 0 | 0 | 0 | 0 | 0 |
| 0 | 0 | 0 | 0 | 0 |
| 0 | 0 | 0 | 0 | 0 |
| 0 | 0 | 0 | 0 | 0 |
| 0.423460027 | 0.006158534 | 0.446549158 | 0.025968084 | 0.014718454 |
| 0 | 0 | 0 | 0 | 0 |
| 0 | 0 | 0 | 0 | 0 |

| | | | | |
|---|---|---|---|---|
| 0 | 0 | 0 | 0 | 0 |
| 0 | 0.010239806 | 0 | 0 | 0 |
| 0 | 0.002385768 | 0 | 0 | 0 |
| 0 | 0 | 0 | 0 | 0 |
| 0 | 0 | 0 | 0.000610692 | 0 |
| 0 | 0 | 0 | 0 | 0 |
| 0 | 0 | 0 | 0 | 0 |
| 0 | 0.001386772 | 0 | 0 | 0 |
| 0 | 0 | 0 | 0 | 0.002109195 |
| 0 | 0.495898252 | 0 | 0.002362496 | 0 |
| 0 | 0.050552705 | 0 | 0 | 0 |
| 0 | 0 | 0 | 0 | 0 |
| 0.006564186 | 0.003434101 | 0 | 0 | 0 |
| 0 | 0 | 0 | 0 | 0 |
| 0 | 0 | 0 | 0 | 0 |
| 0 | 0 | 0 | 0 | 0 |
| 0 | 0 | 0 | 0 | 0 |
| 0 | 0 | 0 | 0 | 0 |
| 0 | 0 | 0 | 0 | 0 |
| 0.253422277 | 0.529882054 | 0 | 0.26821639 | 5.592687197 |
| 0 | 0 | 0 | 0 | 0 |
| 0 | 0 | 0 | 0 | 0 |
| 0 | 0 | 0 | 0 | 0 |
| 0 | 0 | 0 | 0 | 0 |
| 0 | 0 | 0 | 0 | 0 |
| 0 | 0 | 0 | 0 | 0 |
| 0 | 0 | 0 | 0 | 0 |
| 0 | 0 | 0 | 0 | 0 |
| 0 | 0 | 0 | 0 | 0 |
| 0.038255657 | 0 | 0 | 0.030393864 | 0 |
| 0 | 0 | 0 | 0 | 0 |
| 0 | 0 | 0 | 0 | 0 |
| 0 | 0 | 0 | 0 | 0 |
| 0 | 0 | 0 | 0 | 0 |
| 0 | 0 | 0 | 0 | 0 |
| 0 | 0.001695723 | 0 | 0 | 0 |
| 0 | 0 | 0 | 0 | 0 |
| 0 | 0 | 0 | 0 | 0 |
| 0 | 0 | 0 | 0 | 0 |
| 0 | 0 | 0 | 0 | 0 |
| 0 | 0 | 0 | 0 | 0 |
| 0 | 0 | 0 | 0 | 0 |
| 0.102595463 | 0.170956837 | 0 | 0 | 0 |
| 0 | 0 | 0 | 0 | 0 |
| 0 | 0.003486049 | 0 | 0.004826029 | 0 |
| 0 | 0.021479655 | 0 | 0 | 0 |
| 0 | 0 | 0 | 0 | 0 |
| 0 | 0 | 0.017407419 | 0 | 0 |

| | | | | |
|---|---|---|---|---|
| 0 | 0 | 0 | 0 | 0 |
| 0 | 0 | 0 | 0 | 0 |
| 0 | 0 | 0 | 0 | 0 |
| 0 | 0 | 0 | 0 | 0 |
| 0 | 0 | 0 | 0 | 0 |
| 0 | 0.030004957 | 0 | 0.088148946 | 0.005572418 |
| 0 | 0 | 0 | 0 | 0 |
| 0 | 0 | 0 | 0 | 0 |
| 0 | 0 | 0 | 0 | 0 |
| 0 | 0 | 0 | 0 | 0 |
| 0 | 0 | 0 | 0 | 0 |
| 0 | 0 | 0 | 0 | 0 |
| 0 | 0 | 0 | 0 | 0 |
| 0 | 0 | 0 | 0 | 0 |
| 0 | 0 | 0 | 0 | 0 |
| 0 | 0 | 0 | 0 | 0 |
| 0 | 0.194606126 | 0 | 0 | 0 |
| 0 | 0 | 0 | 0 | 0 |
| 0 | 0 | 0 | 0 | 0 |
| 0 | 0 | 0 | 0 | 0 |
| 0 | 0 | 0 | 0 | 0 |
| 0 | 0 | 0 | 0 | 0 |
| 0 | 0 | 0 | 0 | 0 |
| 0 | 0 | 0 | 0 | 0 |
| 0 | 0 | 0 | 0 | 0 |
| 0 | 0.003831248 | 0 | 0 | 0 |
| 0 | 0 | 0 | 0 | 0 |
| 0 | 0.167744326 | 0 | 0 | 0 |
| 0 | 0 | 0 | 0 | 0 |
| 0 | 0 | 0 | 0 | 0 |
| 0 | 0 | 0 | 0 | 0 |
| 0 | 0 | 0 | 0 | 0 |
| 0 | 0 | 0 | 0 | 0 |
| 0 | 0 | 0 | 0 | 0 |
| 0 | 0 | 0 | 0 | 0 |
| 0 | 0 | 0 | 0 | 0 |
| 0 | 0 | 0 | 0 | 0 |
| 0 | 0.094132408 | 0 | 0 | 0 |
| 0 | 0 | 0.000774302 | 1.566226233 | 0.006197429 |
| 0 | 0 | 0 | 0 | 0 |
| 0 | 0 | 0 | 0 | 0 |
| 0 | 0 | 0 | 0 | 0 |
| 0 | 0 | 0 | 0 | 0 |
| 0 | 0 | 0 | 0 | 0 |
| 0 | 0 | 0 | 0 | 0 |
| 0 | 0 | 0 | 0 | 0 |
| 0 | 0 | 0 | 0 | 0 |
| 0 | 0 | 0 | 0 | 0 |

| | | | | |
|---|---|---|---|---|
| 0 | 0 | 0 | 0 | 0 |
| 0 | 0.016967877 | 0 | 0 | 0.00415918 |
| 0 | 0 | 0 | 0 | 0 |
| 0 | 0 | 0 | 0 | 0 |
| 0 | 0 | 0 | 0 | 0 |
| 0 | 0 | 0 | 0 | 0 |
| 0 | 0 | 0 | 0 | 0 |
| 0 | 0 | 0 | 0 | 0 |
| 0 | 0 | 0 | 0 | 0 |
| 0 | 0 | 0 | 0 | 0 |
| 0 | 0 | 0 | 0.001056348 | 0.005198566 |
| 0 | 0 | 0 | 0 | 0 |
| 0 | 0 | 0 | 0.197625589 | 0 |
| 0 | 0 | 0 | 0 | 0 |
| 0 | 0 | 0 | 0 | 0 |
| 0 | 0 | 0 | 0 | 0 |
| 0 | 0 | 0 | 0 | 0 |
| 0 | 0 | 0 | 0 | 0 |
| 0 | 2.88147991 | 0 | 0 | 0 |
| 0 | 0 | 0 | 0 | 0 |
| 0 | 0 | 0 | 0.001250818 | 0.006532478 |
| 0.007407241 | 0 | 0 | 0 | 0 |
| 0 | 0 | 0 | 0 | 0 |
| 0.771520055 | 1.270457964 | 0 | 2.87346617 | 0 |
| 0 | 0 | 0 | 0 | 0 |
| 0 | 0 | 0 | 0 | 0 |
| 0 | 0 | 0 | 0 | 0 |
| 0 | 0 | 0 | 0 | 0 |
| 0 | 0 | 0 | 0 | 0 |
| 0 | 0 | 0 | 0 | 0 |
| 0.001246397 | 0 | 0 | 0 | 0 |
| 0 | 0 | 0 | 0 | 0 |
| 0 | 0 | 0 | 0 | 0.00132145 |
| 0 | 0 | 0 | 0 | 0 |

| | | | | |
|---|---|---|---|---|
| PHL018-0021_V3 | PHL018-0228_V1 | PHL018-0037_V3 | PHL018-0256_V1 | PHL018-0088_V2 |
| EYCF | REF | CTRL | REF | EYCF |
| 2.188907648 | 0.035634512 | 0.040458178 | 0.10884487 | 2.118541257 |
| 0.042070814 | 0.230860609 | 0.393533851 | 1.134292678 | 0.353996039 |
| 0.007506118 | 0.088483055 | 2.447429105 | 3.392689832 | 0.179856078 |
| 0 | 0 | 0 | 0 | 0 |
| 0 | 0 | 0 | 0 | 0 |
| 0 | 0 | 0 | 0 | 0 |
| 0 | 0 | 0 | 0 | 0 |
| 0 | 0 | 0 | 0 | 0 |
| 0 | 0 | 0 | 0 | 0 |
| 0 | 0 | 0 | 0 | 0 |
| 0 | 0 | 0 | 0 | 0 |
| 0 | 0 | 0 | 0 | 0 |
| 0 | 0 | 0 | 0 | 0 |
| 6.440923356 | 0.07295886 | 0.373799759 | 1.326369622 | 0.037486673 |
| 0 | 0 | 0 | 0 | 0 |
| 0 | 0 | 0 | 0 | 0 |
| 0 | 0 | 0 | 0 | 0 |
| 0 | 0 | 0 | 0 | 0 |
| 0 | 0 | 0 | 0 | 0 |
| 0 | 0 | 0 | 0 | 0 |
| 0 | 0 | 0 | 0 | 0 |
| 0 | 0 | 0 | 0 | 0 |
| 0 | 0 | 0 | 0 | 0 |
| 0 | 0 | 0 | 0 | 0 |
| 0 | 0 | 0.111862065 | 0 | 0 |
| 0 | 0 | 0 | 0 | 0 |
| 0 | 0 | 0 | 0 | 0 |
| 0 | 0 | 0 | 0 | 0 |
| 0 | 0 | 0 | 0 | 0 |
| 0 | 0 | 0 | 0 | 0 |
| 0 | 0 | 0 | 0 | 0 |
| 0 | 0 | 0 | 0 | 0 |
| 0 | 0 | 0 | 0 | 0 |
| 0 | 0 | 0 | 0 | 0 |
| 0 | 0 | 0 | 0 | 0 |
| 0 | 0.11599374 | 1.322737139 | 0 | 0 |
| 0 | 0 | 0 | 0 | 0 |
| 0 | 0 | 0 | 0 | 0 |
| 0 | 0 | 0 | 0 | 0 |
| 0 | 0 | 0 | 0 | 0 |
| 0 | 0 | 0 | 0 | 0 |
| 0.008064696 | 0.023816218 | 0 | 0.051167971 | 0 |
| 0.456661162 | 3.635320868 | 0.002394522 | 21.80191622 | 0.000737358 |
| 0.067773493 | 0.09185869 | 0 | 0.004510901 | 0.027619551 |
| 0.002621879 | 0.058514685 | 0 | 0.009431381 | 0.006126991 |
| 0 | 0 | 0 | 0 | 0 |

| | | | | |
|---|---|---|---|---|
| 0 | 0.125970767 | 0 | 0 | 0.153851332 |
| 0 | 0.001091476 | 0 | 0 | 0.00191202 |
| 0.29678782 | 0 | 0.000916778 | 0 | 0.126022439 |
| 0.290599915 | 0.638861785 | 0.423387067 | 0.140168312 | 0.708971783 |
| 0 | 0.008256478 | 0.024668727 | 0 | 0.00470975 |
| 0 | 0 | 0 | 0 | 0 |
| 0.000683357 | 0.109969288 | 0 | 0.008702457 | 0 |
| 0 | 0.023225393 | 0 | 0 | 0 |
| 0 | 0 | 0 | 0 | 0 |
| 0 | 0 | 0 | 0 | 0 |
| 0 | 0 | 0 | 0 | 0 |
| 0.000643017 | 0.390352351 | 0.128825363 | 0.150248519 | 0.228721629 |
| 0 | 0 | 0 | 0 | 0 |
| 0 | 0.098026763 | 0 | 0 | 0.049482438 |
| 0.001566738 | 0 | 0 | 0 | 0 |
| 0 | 0 | 0 | 0 | 0 |
| 0 | 0.889463338 | 1.063414456 | 0.00671314 | 0 |
| 0 | 0 | 0 | 0 | 0 |
| 0 | 0 | 0 | 0 | 0 |
| 0 | 0 | 0 | 0 | 0 |
| 0 | 0.053474185 | 0.022484233 | 0 | 0.017281527 |
| 0 | 0.01895233 | 0.005910941 | 0 | 0.006032621 |
| 0 | 0 | 0 | 0 | 0.0506305 |
| 0 | 0 | 0 | 0 | 0 |
| 0 | 0 | 0 | 0 | 0 |
| 0 | 0 | 0 | 0 | 0 |
| 0.110154629 | 0 | 0 | 0.001845794 | 0 |
| 0 | 0 | 0 | 0 | 0 |
| 0 | 0.163763538 | 0 | 0.003107631 | 0 |
| 0 | 0 | 0 | 0 | 0 |
| 0 | 0 | 0 | 0 | 0 |
| 0 | 0 | 0.002628185 | 0 | 0 |
| 0 | 0 | 0 | 0 | 0 |
| 0 | 0 | 0 | 0 | 0.003077362 |
| 0 | 0 | 0 | 0 | 0 |
| 0 | 0 | 0 | 0 | 0 |
| 0 | 0 | 0 | 0 | 0 |
| 0 | 18.25875325 | 0.00552932 | 0 | 0 |
| 0 | 0 | 0 | 0 | 0 |
| 0 | 0 | 0 | 0 | 0 |
| 0 | 0 | 0 | 0 | 0 |
| 0 | 0 | 0 | 0 | 0 |
| 0 | 0 | 0 | 0 | 0 |
| 0 | 0 | 0 | 0 | 0 |
| 0 | 0 | 0.153558456 | 0 | 0 |
| 0 | 0 | 0 | 0 | 0 |
| 0 | 0 | 0 | 0 | 0 |
| 0 | 0 | 0 | 0.005354757 | 0 |

| | | | | |
|---|---|---|---|---|
| 0 | 0 | 0 | 0 | 0 |
| 0 | 0 | 0 | 0 | 0 |
| 0 | 0 | 0 | 0 | 0 |
| 0 | 0 | 0 | 0 | 0 |
| 0 | 0 | 0.073817883 | 0 | 0 |
| 0 | 0 | 0 | 0.034472252 | 0 |
| 0 | 0 | 0 | 0 | 0 |
| 0 | 0 | 0.002442225 | 0 | 0 |
| 0 | 0 | 0 | 0 | 0.00402138 |
| 0 | 0 | 0 | 0 | 0 |
| 0 | 0 | 0 | 0 | 0 |
| 0 | 0.002052165 | 0 | 0 | 0 |
| 0 | 0 | 0 | 0 | 0 |
| 0 | 0 | 0 | 0 | 0 |
| 0 | 0 | 0 | 0 | 0 |
| 0 | 0 | 0 | 0 | 0 |
| 0 | 0 | 0.000876732 | 0 | 0 |
| 0 | 0 | 0 | 0 | 0 |
| 0.009051653 | 0 | 0.36717753 | 0 | 0.005816948 |
| 0 | 0 | 0 | 0 | 0 |
| 0 | 0 | 0 | 0 | 0.056884285 |
| 0 | 0 | 0 | 0 | 0 |
| 0 | 0 | 0 | 0 | 0 |
| 0 | 0 | 0 | 0 | 0 |
| 0 | 0 | 0 | 0 | 0 |
| 0 | 0 | 0 | 0 | 0 |
| 0 | 0 | 0 | 0 | 0 |
| 0 | 0 | 0.001977746 | 0 | 0 |
| 0 | 0 | 0 | 0 | 0 |
| 0 | 0 | 0 | 0 | 0 |
| 0 | 0.008475893 | 0.098942473 | 0 | 0.01265231 |
| 0.00273368 | 0.013103927 | 0 | 0 | 0.04430965 |
| 0 | 0 | 0 | 0 | 0 |
| 0 | 0 | 0 | 0 | 0 |
| 0 | 0 | 0 | 0 | 0 |
| 0 | 0 | 0 | 0 | 0 |
| 0 | 0 | 0 | 0 | 0 |
| 0 | 0 | 0 | 0 | 0 |
| 0 | 0.607399752 | 0.465253902 | 0 | 0 |
| 0 | 0 | 0 | 0 | 0 |
| 0 | 0 | 0 | 0 | 0 |
| 0 | 0 | 0 | 0 | 0 |
| 0 | 0 | 0 | 0 | 0 |
| 0 | 0 | 0 | 0 | 0 |
| 0 | 0 | 0 | 0 | 0 |
| 0 | 0 | 0 | 0 | 0 |
| 0 | 0.004536889 | 0 | 0 | 0.029093924 |
| 0 | 0 | 0 | 0 | 0 |

| | | | | |
|---|---|---|---|---|
| 0 | 0 | 0 | 0 | 0 |
| 0 | 0 | 0 | 0 | 0 |
| 0 | 0 | 0 | 0 | 0 |
| 0 | 0 | 0 | 0 | 0 |
| 0 | 0 | 0 | 0 | 0.0202177 |
| 0 | 0 | 0 | 0 | 0 |
| 0 | 0 | 0 | 0 | 0 |
| 0 | 0 | 0 | 0 | 0 |
| 0 | 0 | 0 | 0 | 0 |
| 0 | 0 | 0 | 0 | 0 |
| 0 | 0 | 0 | 0 | 0 |
| 0 | 0 | 0 | 0 | 0 |
| 0 | 0 | 0 | 0 | 0 |
| 0 | 0 | 0 | 0 | 0 |
| 0 | 0 | 0 | 0 | 0 |
| 0 | 0 | 0 | 0 | 1.457492679 |
| 0 | 0 | 0 | 0 | 0 |
| 0 | 0 | 0 | 0 | 0 |
| 0 | 0 | 0 | 0 | 0 |
| 0 | 0 | 0 | 0 | 0 |
| 0 | 0 | 0 | 0 | 0 |
| 0 | 0 | 0 | 0 | 0 |
| 0 | 0 | 0.020989033 | 0 | 0 |
| 0 | 0 | 0 | 0 | 0 |
| 0 | 0.002743801 | 0 | 0 | 0 |
| 0 | 0 | 0 | 0 | 0 |
| 0 | 0 | 0 | 0 | 0.001484251 |
| 0 | 0 | 0 | 0 | 0 |
| 0 | 0 | 0 | 0 | 0 |
| 0 | 0 | 0 | 0 | 0 |
| 0 | 0 | 0 | 0 | 0 |
| 0 | 0 | 0 | 0 | 0 |
| 0 | 0 | 0 | 0 | 0 |
| 0 | 0 | 0 | 0 | 0 |
| 0 | 0 | 0 | 0 | 0 |
| 0 | 0 | 0 | 0 | 0 |
| 0 | 0 | 0 | 0 | 0 |
| 0 | 0 | 0 | 0 | 0 |
| 0 | 0 | 0 | 0 | 0 |
| 0 | 0 | 0 | 0 | 0 |
| 0 | 0 | 0 | 0 | 0 |
| 0 | 0 | 0 | 0 | 0 |
| 0 | 0 | 0 | 0 | 0 |
| 0 | 0 | 0 | 0 | 0 |
| 0 | 0 | 0 | 0 | 0 |
| 0 | 0 | 0 | 0 | 0 |
| 0 | 0 | 0 | 0 | 0 |
| 0 | 0 | 0 | 0 | 0 |
| 0 | 0 | 0 | 0 | 0 |

| | | | | |
|---|---|---|---|---|
| 0 | 0 | 0 | 0 | 0 |
| 0 | 0 | 0 | 0 | 0 |
| 0 | 0 | 0 | 0 | 0 |
| 0 | 0 | 0 | 0 | 0 |
| 0 | 0 | 0 | 0 | 0 |
| 0 | 0 | 0 | 0 | 0 |
| 0 | 0 | 0 | 0 | 0 |
| 0 | 0 | 0 | 0 | 0 |
| 0 | 0 | 0 | 0 | 0 |
| 0 | 0 | 0 | 0 | 0 |
| 0 | 0 | 0 | 0 | 0 |
| 0 | 0 | 0 | 0 | 0 |
| 0 | 0.004994732 | 0 | 0 | 0.006249748 |
| 0 | 0 | 0 | 0 | 0 |
| 0 | 0 | 0 | 0 | 0 |
| 0 | 0 | 0 | 0 | 0 |
| 0.004499748 | 0 | 0 | 0 | 0 |
| 0.003024792 | 0 | 0 | 0 | 0 |
| 0 | 0 | 0 | 0 | 0 |
| 0 | 0 | 0 | 0 | 0 |
| 0.031978141 | 0.013203342 | 0.007851309 | 0.0042777 | 0.008864882 |
| 0.032406646 | 0 | 0 | 0 | 0 |
| 0 | 0 | 0 | 0 | 0 |
| 0 | 0 | 0.008869614 | 0 | 0 |
| 0 | 0 | 0 | 0 | 0 |
| 0 | 0 | 0 | 0 | 0 |
| 0.014241359 | 0.006564049 | 0 | 0.000747369 | 0.003989358 |
| 0 | 0 | 0 | 0 | 0 |
| 0 | 0 | 0 | 0 | 0 |
| 0 | 0 | 0 | 0 | 0 |
| 0 | 0 | 0 | 0 | 0 |
| 0 | 0 | 0 | 0 | 0 |
| 0 | 0 | 0 | 0 | 0 |
| 0 | 0 | 0.017393593 | 0 | 0.009357786 |
| 0.022030605 | 0.012362368 | 0.001489973 | 0.019483097 | 0.041162295 |
| 0 | 0 | 0 | 0 | 0 |
| 0 | 0 | 0 | 0 | 0 |
| 0.000647036 | 0 | 0 | 0 | 0 |
| 0.000790872 | 0 | 0 | 0 | 0 |
| 0 | 0 | 0 | 0 | 0 |
| 0 | 0 | 0 | 0 | 0 |
| 0.000611165 | 0 | 0 | 0 | 0 |
| 0 | 0 | 0 | 0 | 0 |
| 0 | 0 | 0 | 0 | 0 |
| 0 | 0 | 0 | 0 | 0 |
| 0 | 0.159597566 | 0.664520712 | 0 | 0.031702265 |
| 0 | 0.001588069 | 0 | 0 | 0 |
| 0.000581467 | 0.000895908 | 0 | 0 | 0.001307857 |

| | | | | |
|---|---|---|---|---|
| 0 | 0 | 0 | 0 | 0 |
| 0 | 0 | 0 | 0 | 0 |
| 0 | 0 | 0 | 0 | 0 |
| 0 | 0 | 0 | 0 | 0 |
| 0 | 0 | 0 | 0 | 0 |
| 0 | 0 | 0 | 0 | 0 |
| 0 | 0 | 0 | 0 | 0 |
| 0 | 0.001931825 | 0 | 0 | 0 |
| 0 | 0 | 0 | 0 | 0 |
| 0 | 0 | 0 | 0 | 0 |
| 0 | 0 | 0 | 0 | 0 |
| 0 | 0 | 0 | 0 | 0 |
| 0 | 0 | 0 | 0 | 0 |
| 0 | 0 | 0 | 0 | 0 |
| 0 | 0 | 0 | 0 | 0 |
| 0 | 0 | 0 | 0 | 0 |
| 0 | 0 | 0 | 0 | 0 |
| 0 | 0 | 0 | 0 | 0 |
| 0.002217555 | 0.016248521 | 0 | 0.097832265 | 0 |
| 0 | 0 | 0 | 0 | 0.41240843 |
| 0.009377208 | 0.007840073 | 0 | 0.004165723 | 0 |
| 0 | 0 | 0 | 0 | 0 |
| 0 | 0 | 0 | 0 | 0 |
| 0 | 0 | 0.008454543 | 0 | 0 |
| 0.004765764 | 0 | 0 | 0.006502642 | 0.005011637 |
| 0 | 0 | 0 | 0.00200775 | 0 |
| 0 | 0 | 0 | 0 | 0 |
| 0 | 0 | 0 | 0 | 0 |
| 0 | 0.005521051 | 0 | 0.006769708 | 0 |
| 0 | 0 | 0 | 0 | 0 |
| 0 | 0.564621228 | 0.103068968 | 0.02209775 | 0.097894949 |
| 0 | 0 | 0 | 0 | 0 |
| 0.163493942 | 0 | 0 | 0 | 0.032801601 |
| 0 | 0 | 0.002597692 | 0 | 0 |
| 0 | 0 | 0 | 0 | 0 |
| 0 | 0 | 0 | 0 | 0 |
| 0 | 0 | 0 | 0 | 0 |
| 0 | 0 | 0 | 0 | 0 |
| 0 | 0 | 0 | 0 | 0 |
| 0 | 0 | 0 | 0.000655621 | 0.001235159 |
| 0 | 0 | 0 | 0 | 0 |
| 0 | 0.015949842 | 0 | 0 | 0.354403619 |
| 0 | 0 | 0 | 0 | 0 |
| 0 | 0 | 0 | 0 | 0 |
| 0 | 0 | 0 | 0 | 0 |
| 0 | 0 | 0 | 0 | 0 |
| 0 | 0 | 0 | 0 | 0 |
| 0.000663314 | 0 | 0 | 0 | 0 |

| | | | | |
|---|---|---|---|---|
| 0 | 0 | 0 | 0 | 0 |
| 0 | 0 | 0 | 0 | 0 |
| 0.05603019 | 0.075050988 | 0 | 0.001400189 | 0.022158277 |
| 0 | 0 | 0 | 0 | 0 |
| 0 | 0 | 0 | 0 | 0 |
| 0 | 0 | 0 | 0 | 0 |
| 0 | 0 | 0 | 0 | 0 |
| 0 | 0 | 0 | 0 | 0 |
| 0 | 0 | 0 | 0 | 0 |
| 0 | 0 | 0 | 0 | 0 |
| 0 | 0 | 0 | 0 | 0 |
| 0 | 0 | 0 | 0 | 0 |
| 0.002501174 | 0 | 0 | 0 | 0 |
| 0.00113413 | 0 | 0.000946739 | 0 | 0 |
| 0.005990883 | 0 | 0.000976762 | 0.002043563 | 0 |
| 0 | 0 | 0.006065365 | 0 | 0.028338121 |
| 0.002773093 | 0.1220459 | 0 | 0.003201629 | 0.001370845 |
| 0 | 0 | 0 | 0 | 0 |
| 0 | 0 | 0 | 0 | 0 |
| 0 | 0 | 0 | 0 | 0 |
| 0 | 0 | 0 | 0 | 0 |
| 0.001347937 | 0.001186779 | 0 | 0 | 0.001484978 |
| 0 | 0 | 0 | 0 | 0 |
| 0 | 0 | 0 | 0 | 0 |
| 0 | 0 | 0 | 0 | 0 |
| 0 | 0 | 0 | 0 | 0 |
| 0 | 0 | 0 | 0 | 0 |
| 0 | 0 | 0 | 0 | 0 |
| 0 | 0 | 0 | 0 | 0 |
| 0 | 0 | 0 | 0 | 0 |
| 0 | 0 | 0 | 0 | 0 |
| 0 | 0 | 0 | 0 | 0 |
| 0.018331906 | 0 | 0 | 0 | 0 |
| 0.000666072 | 0 | 0 | 0 | 0 |
| 0 | 0 | 0 | 0 | 0 |
| 0 | 0 | 0 | 0.002099156 | 0 |
| 0 | 0 | 0 | 0 | 0 |
| 0.001935478 | 0.011182984 | 0 | 0.305679612 | 0.009950513 |
| 0 | 0 | 0 | 0 | 0 |
| 0 | 0 | 0 | 0 | 0 |
| 0.001396721 | 0 | 0 | 0 | 0 |
| 0 | 0 | 0 | 0 | 0 |
| 0 | 0 | 0 | 0 | 0 |
| 0 | 0.004630338 | 0 | 0 | 0 |
| 0 | 0 | 0 | 0 | 0 |
| 0 | 0 | 0.001940692 | 0 | 0 |
| 0 | 0 | 0 | 0 | 0 |
| 0 | 0 | 0 | 0 | 0 |

| | | | | |
|---|---|---|---|---|
| 0 | 0 | 1.670136846 | 0 | 0 |
| 0 | 0 | 0.001942913 | 0.481693797 | 0 |
| 0 | 0.000621529 | 0 | 0.004403221 | 0 |
| 0 | 0 | 0 | 0 | 0 |
| 0 | 0.002084394 | 0.004234875 | 0.001938153 | 0 |
| 0 | 0 | 0 | 0 | 0 |
| 0 | 0 | 0 | 0 | 0 |
| 0.011946575 | 0.006135647 | 0 | 0 | 0.003655875 |
| 0 | 0 | 0 | 0 | 0 |
| 2.86056511 | 0.001204599 | 0.028450918 | 0.055204171 | 0 |
| 0 | 0 | 0 | 0 | 0 |
| 0.031973503 | 0.001387714 | 0 | 0 | 0 |
| 0.003757513 | 0.009298237 | 0 | 0.00279651 | 0.001975684 |
| 0 | 0 | 0 | 0 | 0 |
| 0 | 0 | 0 | 0 | 0 |
| 0 | 0 | 0 | 0 | 0 |
| 0 | 0 | 0 | 0 | 0 |
| 0 | 0 | 0 | 0 | 0 |
| 0 | 0.011618433 | 0 | 0 | 0 |
| 0 | 0 | 0 | 0 | 0 |
| 0 | 0 | 0 | 0 | 0 |
| 0 | 0 | 0 | 0 | 0 |
| 0 | 0 | 0 | 0 | 0 |
| 0 | 0 | 0 | 0 | 0 |
| 0 | 0 | 0 | 0 | 0 |
| 0.016710442 | 0.095018591 | 0 | 0.001357175 | 0.002556854 |
| 0 | 3.866695869 | 0 | 1.857285214 | 0 |
| 0 | 0.013096572 | 0 | 0 | 0 |
| 0 | 0 | 0 | 0 | 0 |
| 0 | 0 | 0 | 0 | 0 |
| 0 | 0 | 0 | 0 | 0 |
| 0 | 0 | 0 | 0 | 0 |
| 0 | 0 | 0 | 0 | 0 |
| 0 | 0 | 0.000814431 | 0 | 0 |
| 0 | 0 | 0 | 0 | 0 |
| 0 | 0 | 0 | 0 | 0 |
| 0 | 0 | 0 | 0 | 0 |
| 0 | 0 | 0 | 0 | 0 |
| 0 | 0 | 0 | 0 | 0 |
| 0 | 0 | 0 | 0 | 0 |
| 0 | 0 | 0 | 0 | 0.0014384 |
| 0 | 0 | 0 | 0 | 0 |
| 0 | 0 | 0 | 0 | 0 |
| 0 | 0 | 0 | 0 | 0 |
| 0 | 0.009587421 | 0 | 0 | 0 |
| 0 | 0 | 0.017161772 | 0 | 0 |
| 0 | 0.003049371 | 0 | 0 | 0.00381558 |
| 0 | 0 | 0.001640164 | 0 | 0 |

| | | | | |
|---|---|---|---|---|
| 0 | 0 | 0 | 0 | 0 |
| 0.007741316 | 0 | 0 | 0 | 0 |
| 0 | 0 | 0 | 0 | 0 |
| 0.010117549 | 0 | 0 | 0 | 0 |
| 0 | 0.028884014 | 0 | 0 | 0 |
| 0 | 0.01601166 | 0.001829868 | 0 | 0 |
| 0 | 0.012685411 | 0 | 0 | 0 |
| 0 | 0.001294319 | 0 | 0 | 0 |
| 0 | 0 | 0 | 0 | 0 |
| 0 | 0 | 0 | 0 | 0 |
| 0.016723457 | 0 | 0.005659569 | 0.002466844 | 0.005518807 |
| 0 | 0 | 0 | 0 | 0 |
| 0 | 0 | 0 | 0 | 0 |
| 0 | 0 | 0.001336719 | 0.006700331 | 0 |
| 0.022899203 | 0 | 0.015779395 | 0 | 0 |
| 0 | 0 | 0 | 0 | 0 |
| 0 | 0 | 0 | 0 | 0 |
| 0 | 0 | 0 | 0 | 0 |
| 0 | 0 | 0 | 0 | 0 |
| 0 | 0.002761621 | 0.302983263 | 0 | 0 |
| 0 | 0 | 0 | 0 | 0 |
| 0 | 0.108851605 | 0.275309568 | 0.030301801 | 0.043435881 |
| 0 | 0 | 0.012473412 | 0 | 0 |
| 0 | 0 | 0 | 0 | 0 |
| 0 | 0 | 0 | 0 | 0 |
| 0 | 0 | 0 | 0 | 0 |
| 0 | 0.003091269 | 0.008213026 | 0 | 0 |
| 0 | 0 | 0 | 0 | 0 |
| 0 | 0 | 0 | 0 | 0 |
| 0.064781856 | 0.118941041 | 0.155441791 | 2.825966935 | 0.098547675 |
| 0.001303625 | 0.032137406 | 0 | 0 | 0.002420197 |
| 0 | 0 | 0 | 0 | 0 |
| 0 | 0 | 0 | 0 | 0.002293468 |
| 0 | 0 | 0 | 0 | 0 |
| 0 | 0 | 0.001408375 | 0 | 0 |
| 0 | 0 | 0 | 0 | 0 |
| 0 | 0 | 0 | 0.146400187 | 0 |
| 0 | 0 | 0 | 0 | 0 |
| 0 | 0 | 0 | 0 | 0 |
| 0 | 0.002075997 | 0.044550678 | 0 | 0 |
| 0 | 0 | 0 | 0 | 0 |
| 0 | 0.424406438 | 0.169177372 | 0.006385268 | 0.030073858 |
| 0 | 0 | 0 | 0 | 0 |
| 0 | 0 | 0.003977251 | 0 | 0 |
| 0 | 0 | 0 | 0 | 0 |
| 0 | 0 | 0 | 0 | 0 |
| 0.003131854 | 0 | 0.010188406 | 0 | 0 |
| 0 | 0 | 0 | 0 | 0 |

| | | | | |
|---|---|---|---|---|
| 0.001591134 | 0.048097539 | 0 | 0.004652187 | 0.0029215 |
| 0 | 0 | 0.000909324 | 0 | 0 |
| 0 | 0.926101888 | 0.063020523 | 0.00169039 | 0 |
| 0 | 0 | 0 | 0 | 0 |
| 0 | 0.019089539 | 0 | 0 | 0 |
| 0 | 0 | 0 | 0 | 0.010868576 |
| 0.002261216 | 0 | 0 | 0 | 0 |
| 0 | 0 | 0 | 0 | 0 |
| 0 | 0 | 0 | 0 | 0 |
| 0 | 0 | 0.067257219 | 0 | 0 |
| 0 | 0 | 0 | 0 | 0 |
| 0 | 0 | 0 | 0 | 0 |
| 0 | 0 | 0 | 0 | 0 |
| 0 | 0 | 0 | 0 | 0 |
| 0 | 0 | 0 | 0 | 0 |
| 0 | 0.307194806 | 0.032531198 | 0.000657808 | 0.124547523 |
| 0.030993072 | 0.25962394 | 0 | 0.157731103 | 0.005723381 |
| 0 | 0 | 0 | 0 | 0 |
| 0 | 0.007181489 | 0 | 0 | 0 |
| 0 | 0.124442097 | 0 | 0 | 0 |
| 0 | 0.000691755 | 0 | 0 | 0 |
| 0 | 0.004668997 | 0 | 0 | 0.003186636 |
| 0 | 0 | 0 | 0 | 0 |
| 0 | 0 | 0 | 0 | 0 |
| 0 | 0 | 0 | 0 | 0 |
| 0 | 0 | 0 | 0 | 0 |
| 0 | 0 | 0 | 0 | 0 |
| 0 | 0 | 0.000895934 | 0 | 0.001103558 |
| 0 | 0 | 0 | 0 | 0 |
| 0 | 0 | 0 | 0 | 0.004476327 |
| 0 | 0.012188338 | 0.009689916 | 0 | 0 |
| 0.006665975 | 0.001316762 | 0 | 0 | 0 |
| 0.018796616 | 0.002758218 | 0.051453879 | 1.329981304 | 0.012550072 |
| 0.002099017 | 0 | 0 | 0.013842673 | 0 |
| 0.000998776 | 0.001795367 | 0 | 0 | 0 |
| 0 | 0 | 0 | 0 | 0 |
| 0.000662082 | 0.000906771 | 0 | 0.004215759 | 0 |
| 0 | 0.020033112 | 0.084824577 | 0 | 0.00776405 |
| 0 | 0 | 0 | 0 | 0 |
| 0 | 0.029134533 | 0 | 0 | 0.002176424 |
| 0 | 0 | 0 | 0 | 0 |
| 0 | 0 | 0 | 0 | 0 |
| 0 | 0.081924614 | 0 | 0 | 0 |
| 0 | 0 | 0.113734127 | 0 | 0.00277065 |
| 0 | 0 | 0 | 0 | 0 |
| 0 | 0.002125796 | 0.002159497 | 0 | 0 |
| 0 | 0 | 0.00180114 | 0 | 0 |
| 0 | 0 | 0.001097726 | 0 | 0 |

| | | | | |
|---|---|---|---|---|
| 0 | 0 | 0 | 0 | 0 |
| 0.284567509 | 0.037443687 | 0.003190223 | 0.037223328 | 0.136928989 |
| 0 | 0.005753624 | 0 | 0 | 0 |
| 0 | 0 | 0 | 0 | 0 |
| 0 | 0 | 0 | 0 | 0 |
| 0 | 0 | 0 | 0 | 0 |
| 0 | 0 | 0 | 0 | 0 |
| 0.001083857 | 0 | 0 | 0 | 0 |
| 0 | 0 | 0 | 0 | 0 |
| 0 | 0 | 0 | 0 | 0 |
| 0 | 0.005744465 | 0 | 0 | 0 |
| 0.006247592 | 0.025669632 | 0 | 0.013141969 | 0.007695317 |
| 0 | 0 | 0 | 0 | 0 |
| 0 | 0 | 0 | 0 | 0 |
| 0.001604161 | 0.001235823 | 0 | 0 | 0 |
| 0 | 0 | 0 | 0 | 0 |
| 0.009333174 | 0.007331128 | 0 | 0.011735844 | 0.020463302 |
| 0.000712691 | 0.003416293 | 0.008924018 | 0 | 0.001221342 |
| 0 | 0 | 0 | 0 | 0 |
| 0.001234092 | 0.000633818 | 0.008209287 | 0 | 0 |
| 0.004340622 | 0.02209917 | 0.005120063 | 0.00206002 | 0.005336347 |
| 0 | 0 | 0 | 0 | 0 |
| 0 | 0 | 0 | 0 | 0 |
| 0 | 0 | 0 | 0 | 0 |
| 0 | 0.001954249 | 0 | 0 | 0 |
| 0 | 0 | 0 | 0 | 0 |
| 0 | 0 | 0.182518404 | 0 | 0 |
| 0.011633893 | 0.005562984 | 0.174349167 | 0.02846346 | 0.006960783 |
| 0 | 0 | 0 | 0 | 0 |
| 0 | 0 | 0 | 0 | 0 |
| 0.015926812 | 0.005305855 | 0 | 0 | 0.002489642 |
| 0 | 0 | 0 | 0 | 0 |
| 0 | 0 | 0 | 0 | 0 |
| 0 | 0 | 0 | 0 | 0 |
| 0 | 0 | 0 | 0 | 0 |
| 1.981867147 | 0.155870863 | 0.12398133 | 0.168141278 | 0.071556906 |
| 0 | 0 | 0.007710779 | 0 | 0 |
| 0 | 0 | 0 | 0 | 0 |
| 0.017815914 | 0 | 0.00154919 | 0.002700989 | 0.010177072 |
| 0 | 0 | 0 | 0 | 0 |
| 0 | 0 | 0 | 0 | 0 |
| 0 | 0 | 0 | 0 | 0 |
| 0.000601203 | 0 | 0.048147916 | 0 | 0 |
| 0.001451126 | 0.001788683 | 0.001968459 | 0 | 0.00074604 |
| 0 | 0 | 0.008355087 | 0 | 0 |
| 0 | 0.675164547 | 0.254227939 | 0 | 0 |
| 0 | 0 | 0 | 0 | 0 |
| 0 | 0 | 0 | 0 | 0 |

| | | | | |
|---|---|---|---|---|
| 0 | 0 | 0.0015337 | 0 | 0 |
| 0 | 0 | 0 | 0 | 0 |
| 0 | 0 | 0 | 0 | 0.012038441 |
| 0 | 0.611896805 | 1.046467841 | 0 | 0.958239708 |
| 0 | 0 | 0 | 0 | 0 |
| 0 | 0 | 0 | 0 | 0 |
| 0 | 0 | 0 | 0 | 0 |
| 0 | 0 | 0 | 0 | 0.071655402 |
| 0.012854621 | 0.67204589 | 0 | 0 | 8.670040925 |
| 0 | 0 | 0.247983548 | 0 | 0 |
| 0 | 0 | 0 | 0 | 0 |
| 0 | 0 | 0.001750094 | 0 | 0.002155662 |
| 0 | 0 | 0 | 0 | 0 |
| 0 | 0 | 0 | 0 | 0 |
| 0 | 0.031276181 | 0.003754873 | 0.179589876 | 0.537215162 |
| 0 | 0 | 0 | 0 | 0 |
| 0 | 0.283168527 | 0 | 0 | 0 |
| 0 | 0.769500086 | 0 | 0 | 0 |
| 0 | 0 | 0 | 0 | 0 |
| 0 | 0 | 0 | 0 | 0 |
| 0 | 0 | 0 | 0 | 0 |
| 0 | 0 | 0 | 0 | 0 |
| 0 | 0 | 0 | 0 | 0 |
| 0 | 0.1251639 | 0.134381921 | 0 | 0.080705677 |
| 0 | 0 | 0.003113216 | 0 | 0 |
| 0 | 0 | 0.064001033 | 0 | 0 |
| 0 | 0.004797948 | 0.021835561 | 0 | 0.000960563 |
| 0 | 1.460752409 | 0 | 0 | 2.8399175 |
| 0 | 0 | 0 | 0 | 0 |
| 0 | 0 | 0 | 0 | 0 |
| 0 | 0.639955764 | 0.023309036 | 0 | 0 |
| 0 | 0 | 0 | 0 | 0 |
| 0.000372283 | 0.00305922 | 0.007769294 | 0 | 0.005741856 |
| 0 | 0 | 0 | 0 | 0 |
| 0 | 0 | 0.188347525 | 0 | 0 |
| 0 | 0.005396288 | 0 | 0.132798893 | 0 |
| 0 | 0 | 0.91385558 | 0 | 0 |
| 0 | 0 | 0 | 0 | 0 |
| 0 | 0 | 0 | 0 | 0 |
| 0 | 0 | 0 | 0 | 0 |
| 0 | 0.002479053 | 0.14623237 | 0.003732104 | 0.077755777 |
| 0 | 0 | 0 | 0 | 0 |
| 0.184522849 | 0.272411829 | 2.64243244 | 0.07690107 | 0.11461237 |
| 0 | 0.002591495 | 0.002632578 | 0 | 0.007025751 |
| 0 | 0 | 0 | 0 | 0 |
| 0 | 0 | 0 | 0 | 0 |
| 0 | 0 | 0 | 0 | 0 |
| 0.018181149 | 0.002384085 | 0.004238289 | 0.016362228 | 0.014418455 |

| | | | | |
|---|---|---|---|---|
| 0 | 0 | 0 | 0 | 0 |
| 0 | 0.016470763 | 0 | 0 | 0 |
| 0.001668704 | 0 | 0 | 0.075136414 | 0 |
| 0 | 0 | 0.014011861 | 0 | 0 |
| 0 | 0 | 0 | 0 | 0 |
| 0 | 0.001800361 | 0 | 0 | 0.070022491 |
| 1.042534454 | 0 | 0 | 0 | 0.532373625 |
| 0 | 0 | 0 | 0 | 0 |
| 0 | 0 | 0.1804016 | 0 | 0 |
| 0 | 0 | 0.187750178 | 0 | 0 |
| 0.036954373 | 0.003552797 | 0.01253694 | 0.063338424 | 0.178755912 |
| 1.562735142 | 0.197861446 | 1.315779076 | 0.374735975 | 0.312254445 |
| 0 | 0 | 0 | 0 | 0 |
| 0.046955454 | 0.506332462 | 0 | 0.162449372 | 0.043684916 |
| 0.004335044 | 0.000636125 | 0.001130866 | 0.036334669 | 0.025271817 |
| 0 | 0 | 0 | 0 | 6.713879562 |
| 0 | 0 | 0 | 0 | 0 |
| 0 | 0 | 0 | 0 | 0 |
| 0 | 0 | 0.066798215 | 0 | 0 |
| 0 | 0.018329567 | 0.014551877 | 0 | 0 |
| 0.000457732 | 0 | 0.137556675 | 0.158428043 | 0.233756471 |
| 1.138889115 | 0 | 0 | 0.384540491 | 0 |
| 3.684970057 | 0.073126898 | 0.224963793 | 0.058597187 | 0.047047246 |
| 0 | 0 | 0.012494304 | 0 | 0 |
| 0 | 0 | 0.029647696 | 0 | 0 |
| 0 | 0.496175661 | 0 | 0 | 0 |
| 0 | 0 | 0 | 0 | 0 |
| 0 | 0.000653844 | 0 | 0 | 0 |
| 0 | 0 | 0 | 0 | 0.029525956 |
| 0 | 0 | 0.210602607 | 0 | 0 |
| 0 | 0.017286315 | 0.007199745 | 0 | 0.00454226 |
| 0 | 0.066138125 | 0 | 0 | 0.021846238 |
| 0 | 0.005377201 | 0.047016046 | 0 | 0.081460714 |
| 0 | 0.002196714 | 0.004909383 | 0 | 0.077329448 |
| 0 | 0.004182773 | 0.00090132 | 0 | 0.079299532 |
| 4.62571439 | 0.075580102 | 0.002830535 | 0.12537973 | 0.151444266 |
| 0 | 0.000672897 | 0 | 0.009608763 | 0.025048745 |
| 0 | 0 | 0 | 0 | 0 |
| 0 | 4.990767554 | 0 | 2.424663266 | 0.816743681 |
| 0 | 0 | 0 | 0 | 0 |
| 0 | 0 | 0.007633063 | 0 | 0 |
| 0 | 0 | 0 | 0 | 0 |
| 0 | 0 | 0 | 0 | 0 |
| 0 | 0 | 0.004050367 | 0 | 0 |
| 0.007346939 | 0.01012838 | 0.000806976 | 0.01556439 | 0.060881617 |
| 0 | 0 | 0.001715254 | 0 | 0 |
| 0 | 0 | 0 | 0 | 0 |
| 0.74560906 | 0.031622008 | 0.002676942 | 0.27207706 | 0.01378871 |

| | | | | |
|---|---|---|---|---|
| 0 | 0 | 0 | 0 | 0 |
| 0 | 0 | 0 | 0 | 0 |
| 0 | 0 | 0 | 0 | 0 |
| 0 | 0 | 0.020805416 | 0 | 0.010166038 |
| 0 | 0 | 0 | 0 | 0 |
| 0 | 0 | 0.073652711 | 0 | 0 |
| 0 | 0 | 0.006965164 | 0 | 0.677531218 |
| 0 | 0.009968125 | 0.0030062 | 0 | 0.83879541 |
| 0 | 0 | 0 | 0 | 0 |
| 0 | 0 | 0 | 0 | 0 |
| 0 | 0.008671615 | 0 | 0 | 0.001179404 |
| 0 | 0 | 0 | 0 | 0 |
| 0 | 0.003961397 | 0.001676749 | 0.010962697 | 0 |
| 0 | 0 | 0 | 0 | 0 |
| 0 | 0 | 0 | 0 | 0 |
| 0 | 0 | 0 | 0 | 0 |
| 0 | 0.687982596 | 0 | 0 | 0 |
| 0 | 0.004551059 | 0 | 0.000833844 | 0.055375021 |
| 0 | 0 | 0.234183327 | 0 | 0 |
| 0 | 0 | 0.000575085 | 0 | 0 |
| 0 | 0.000836536 | 0 | 0 | 0.008722756 |
| 0 | 0 | 0 | 0 | 0 |
| 0 | 0.19703562 | 0 | 0 | 0.024224283 |
| 0 | 0 | 0 | 0 | 0 |
| 0 | 0 | 0 | 0 | 0 |
| 0 | 0 | 0 | 0 | 0 |
| 0 | 0 | 0 | 0 | 0 |
| 0 | 0 | 0 | 0 | 0 |
| 0 | 0 | 0 | 0 | 0 |
| 0 | 0.032796261 | 0 | 0 | 0 |
| 0 | 0 | 0 | 0 | 0 |
| 0 | 0 | 0 | 0 | 0 |
| 0 | 0 | 0 | 0 | 0 |
| 0 | 0 | 0 | 0 | 0 |
| 0 | 0 | 0 | 0 | 0 |
| 0 | 0 | 0 | 0 | 0 |
| 0 | 0.034694341 | 0.036341447 | 0 | 0.041047062 |
| 0 | 0.005425716 | 0 | 0 | 0 |
| 0 | 0 | 0 | 0 | 0 |
| 0 | 0 | 0.000959376 | 0 | 0.001378654 |
| 0 | 0 | 0.033613879 | 0 | 0.049084809 |
| 0 | 0 | 0 | 0 | 0 |
| 0 | 0 | 0 | 0 | 0 |
| 0 | 0 | 0 | 0 | 0.002714743 |
| 0 | 0.056534804 | 0 | 0.00385116 | 0 |
| 0 | 0 | 0 | 0 | 0 |
| 0 | 0 | 0 | 0 | 0.005502076 |
| 0 | 0 | 0 | 0 | 0 |

| | | | | |
|---|---|---|---|---|
| 0 | 0 | 0 | 0 | 0 |
| 0 | 0 | 0 | 0 | 0 |
| 0 | 0 | 0 | 0 | 0 |
| 0 | 0 | 0.111263614 | 0 | 0.348975786 |
| 0 | 0 | 0 | 0 | 0 |
| 0 | 0 | 0 | 0 | 0 |
| 0 | 0 | 0 | 0 | 0 |
| 0 | 0 | 0.03570602 | 0 | 0 |
| 0.001928371 | 0.001760698 | 0.104857294 | 0.007308787 | 0.000826164 |
| 0 | 0 | 0 | 0 | 0 |
| 0 | 0 | 0.015666675 | 0 | 0.002953667 |
| 0 | 0 | 0 | 0 | 0 |
| 0 | 0 | 0 | 0 | 0 |
| 0 | 0 | 0 | 0 | 0 |
| 0 | 0 | 2.874508407 | 0 | 0.208587864 |
| 0 | 0 | 0.317062338 | 0 | 0 |
| 0 | 0.002819672 | 0 | 0 | 0 |
| 0 | 0 | 0 | 0 | 0 |
| 0 | 0 | 0 | 0 | 0 |
| 0 | 0 | 0 | 0 | 0 |
| 0 | 0 | 0.286351567 | 0 | 0.048642534 |
| 0 | 0 | 0 | 0 | 0 |
| 0 | 0 | 0 | 0 | 0 |
| 0 | 0 | 0.022836018 | 0 | 0 |
| 0.034015635 | 0.172630535 | 0 | 0.820987238 | 0.000971546 |
| 0 | 6.689954224 | 1.746086732 | 0.095592686 | 4.323736423 |
| 0 | 0 | 0.295913868 | 0 | 1.165739392 |
| 0 | 0 | 0 | 0 | 0 |
| 0 | 0.375850315 | 0 | 0 | 0 |
| 0 | 0 | 0 | 0 | 0 |
| 0 | 0 | 0 | 0 | 0 |
| 0 | 0.058671074 | 0 | 0 | 0.011470817 |
| 0 | 0 | 0.000636595 | 0 | 0 |
| 0 | 0 | 0 | 0.002386926 | 0.005707547 |
| 0.045675095 | 0.406990021 | 0 | 0.028135359 | 0.003989675 |
| 0 | 0.00579571 | 0 | 0 | 0.000805776 |
| 0 | 0 | 0 | 0 | 0 |
| 0 | 0.592628006 | 0.330189238 | 0 | 0.116007224 |
| 0 | 0 | 11.40004599 | 0.125319905 | 0.03689012 |
| 0 | 0 | 0 | 0 | 0 |
| 0 | 0 | 0 | 0 | 0 |
| 0 | 0 | 0.009273018 | 0 | 0 |
| 0.130433686 | 0.844353706 | 0.030841064 | 0 | 0.156907819 |
| 0 | 0 | 0 | 0 | 0 |
| 0 | 0 | 0 | 0 | 0 |
| 0 | 0.000610079 | 0 | 0 | 0.005725288 |
| 0 | 0 | 0 | 0 | 0 |
| 3.082987075 | 0.001700875 | 0.608692819 | 0.029048726 | 0.588613093 |

| | | | | |
|---|---|---|---|---|
| 0 | 0.002174304 | 0.000552193 | 0 | 0 |
| 0 | 0 | 0 | 0 | 0 |
| 0 | 0 | 0 | 0 | 0 |
| 0 | 0 | 0 | 0 | 0 |
| 0.000702496 | 0 | 0 | 0.00311519 | 0.003160161 |
| 0 | 0.002953244 | 0.009176659 | 0 | 0 |
| 0 | 0 | 0 | 0.03128576 | 0 |
| 0 | 0 | 0.069114954 | 0 | 0 |
| 0 | 0 | 0 | 0 | 0 |
| 0 | 0 | 0.002759059 | 0 | 0 |
| 0 | 0 | 0 | 0 | 0.001036319 |
| 0 | 0 | 1.429120913 | 0.019233434 | 0.532277874 |
| 0.026793886 | 0.108122809 | 0.233652978 | 0 | 0.780859052 |
| 0 | 0 | 0 | 0 | 0 |
| 0.067229133 | 0.010014234 | 0 | 0.011639547 | 0 |
| 0 | 0 | 0 | 0 | 0 |
| 0 | 0 | 0 | 0 | 0 |
| 0 | 0 | 0 | 0 | 0 |
| 0 | 0.000914317 | 0.001857622 | 0 | 0 |
| 0 | 0 | 0.00617942 | 0 | 0 |
| 0 | 0 | 0.023069116 | 0 | 0 |
| 0 | 0 | 0 | 0 | 0 |
| 0 | 2.435637897 | 0.143960952 | 0 | 4.446192568 |
| 0 | 0 | 0 | 0 | 0 |
| 0 | 0 | 0 | 0 | 0 |
| 0 | 0 | 0 | 0 | 0 |
| 0 | 0 | 0 | 0 | 0 |
| 0 | 0 | 0 | 0 | 0 |
| 0 | 0 | 0 | 0 | 0 |
| 0 | 0 | 0 | 0 | 0 |
| 0 | 0 | 0 | 0 | 0 |
| 0.028728317 | 0.014889954 | 0 | 0.012226999 | 0.039972651 |
| 0 | 0 | 0 | 0 | 0 |
| 0.000526685 | 0.023533545 | 0 | 0.016348947 | 0.042477809 |
| 0 | 0.006131985 | 0 | 0 | 0 |
| 0.05474938 | 0.002484389 | 0 | 0.876032016 | 0.365405969 |
| 0 | 0 | 0 | 0 | 0.005254097 |
| 0 | 0 | 0 | 0.042282452 | 0 |
| 0 | 0.003421734 | 0 | 0 | 0 |
| 0 | 0 | 0 | 0 | 0 |
| 0 | 0 | 0.0896175 | 0 | 0 |
| 0 | 0 | 0 | 0 | 0 |
| 0 | 0 | 0 | 0 | 0 |
| 0 | 0 | 0.015213415 | 0 | 0.013920392 |
| 0.007392974 | 0.012944186 | 0.001051951 | 0.04332969 | 0.1872332 |
| 0 | 0 | 0 | 0 | 0 |
| 0 | 0 | 2.277773736 | 0 | 0.092121834 |
| 0 | 0.001305908 | 0 | 0 | 0.000612765 |

| | | | | |
|---|---|---|---|---|
| 0 | 0 | 0 | 0 | 0 |
| 0 | 0 | 0 | 0 | 0.015922412 |
| 0 | 0 | 0.013055768 | 0 | 0 |
| 0 | 0 | 0 | 0 | 0 |
| 0 | 0 | 0 | 0 | 0 |
| 0 | 0 | 0.009203793 | 0 | 0 |
| 0.127207878 | 0.738014539 | 0 | 0 | 0 |
| 0 | 0 | 0.118847044 | 0 | 0.004218697 |
| 0 | 0 | 0 | 0 | 0 |
| 0 | 0 | 0 | 0 | 0.089254225 |
| 0 | 0 | 0 | 0 | 0 |
| 0 | 0.002179437 | 0 | 0.001206267 | 0 |
| 0 | 0 | 0 | 0 | 0 |
| 0 | 0.161015719 | 0 | 0 | 0 |
| 0 | 0 | 0 | 0 | 0 |
| 0 | 0 | 0 | 0 | 0 |
| 0 | 0 | 0 | 0 | 0 |
| 0 | 0 | 0 | 0 | 0 |
| 0 | 0 | 7.499371399 | 0 | 0.000715976 |
| 0 | 0 | 0 | 0 | 0 |
| 0 | 0 | 0 | 0 | 0 |
| 0 | 0 | 0 | 0 | 0 |
| 0 | 0 | 0 | 0 | 0 |
| 0 | 0 | 0 | 0 | 0 |
| 0 | 0.003394235 | 0 | 0 | 0.00070785 |
| 0.0014078 | 0.01092583 | 0 | 0.017928135 | 0.066948376 |
| 0 | 0 | 0 | 0 | 0 |
| 0 | 0 | 0.163755036 | 0.003133585 | 0.172776541 |
| 0 | 0 | 0 | 0 | 0 |
| 0 | 0.584597087 | 0.648879652 | 0.118212245 | 0.75145052 |
| 0 | 0 | 0 | 0 | 0 |
| 0 | 0 | 0 | 0 | 0 |
| 0 | 0 | 0 | 0 | 0.01371395 |
| 0 | 0 | 0 | 0 | 0 |
| 0 | 0.002020517 | 0 | 0.000670985 | 0.000632052 |
| 0 | 0 | 0 | 0 | 0 |
| 0 | 0 | 0 | 0 | 0 |
| 0 | 0 | 0 | 0.002791695 | 0.001195323 |
| 0 | 0 | 0 | 0 | 0 |
| 0 | 0 | 0 | 0 | 0 |
| 1.375618154 | 1.411794284 | 3.675944527 | 1.720931387 | 1.85228682 |
| 0 | 0 | 0.449232748 | 0.076911159 | 0.316743401 |
| 0 | 0 | 0 | 0 | 0 |
| 0 | 0 | 0.135416978 | 0 | 0 |
| 0 | 0 | 0 | 0 | 0 |
| 0 | 0 | 0 | 0 | 0 |
| 0 | 0 | 0.006827678 | 0 | 0 |
| 0.003838162 | 0.001819609 | 0.000616152 | 0.001208533 | 0.06849429 |

| | | | | |
|---|---|---|---|---|
| 0 | 0 | 0 | 0 | 0 |
| 0 | 0.022771356 | 0.009664885 | 0 | 0.00097579 |
| 0 | 0 | 0 | 0 | 0.015955346 |
| 0 | 0 | 0 | 0 | 0 |
| 0.087881029 | 5.134303316 | 0.485610309 | 0.008107067 | 0.63046884 |
| 0 | 0 | 0 | 0 | 0 |
| 0 | 0 | 0 | 0 | 0 |
| 0 | 0 | 0 | 0 | 0 |
| 0.001971023 | 0.023204998 | 0.013131193 | 0 | 0.007015329 |
| 0 | 0 | 0.001536681 | 0 | 0 |
| 0.000857744 | 0 | 0 | 0 | 0 |
| 0 | 0 | 0 | 0 | 0 |
| 0 | 0 | 0.081902561 | 0 | 0.017825149 |
| 0.002358001 | 0.009129431 | 0.002460491 | 0.120280216 | 0.004196338 |
| 0 | 0 | 0 | 17.46417153 | 8.296783179 |
| 0 | 0.096625642 | 0.350229471 | 0 | 0.202974268 |
| 0 | 0 | 0.097286849 | 0 | 0.002017377 |
| 0 | 0 | 0 | 0 | 0 |
| 0 | 0 | 0 | 0 | 0 |
| 0 | 0 | 0 | 0 | 0 |
| 0 | 0 | 0 | 0 | 0 |
| 0 | 0 | 0 | 0 | 0 |
| 0 | 0 | 0 | 0 | 0 |
| 0 | 0 | 0 | 0 | 0 |
| 0 | 0 | 0 | 0 | 0 |
| 0 | 0 | 0 | 0 | 0 |
| 0 | 0 | 0.003133145 | 0 | 0.154368946 |
| 0 | 0.017084775 | 0 | 0.000840535 | 0.005146466 |
| 0 | 0 | 0 | 0 | 0 |
| 0 | 0 | 0 | 0 | 0 |
| 0 | 0 | 0 | 0 | 0 |
| 0 | 0 | 0.002818914 | 0 | 0 |
| 0 | 0 | 0 | 0 | 0 |
| 0 | 0 | 0 | 0 | 0.032576717 |
| 0.004200684 | 0.518568024 | 0 | 0 | 0 |
| 0 | 0 | 0.004833315 | 0 | 0 |
| 0 | 0 | 0 | 0 | 0 |
| 0 | 0 | 0.006319412 | 0 | 0.002594627 |
| 0 | 0 | 0 | 0 | 0 |
| 0 | 0 | 0 | 0 | 0 |
| 0 | 0.147024476 | 1.409995437 | 0 | 0 |
| 0 | 0.093532887 | 0 | 0 | 0 |
| 0 | 0 | 0 | 0 | 0 |
| 0 | 0 | 0.00255688 | 0 | 0 |
| 0 | 0 | 0 | 0 | 0 |
| 0 | 0 | 0 | 0 | 0 |
| 0 | 0 | 0 | 0 | 0 |
| 0 | 0 | 0 | 0 | 0 |

| | | | | |
|---|---|---|---|---|
| 0 | 0 | 0.003167129 | 0 | 0 |
| 0 | 0 | 0 | 0 | 0 |
| 0 | 0 | 0 | 0 | 0 |
| 0 | 0 | 0 | 0 | 0 |
| 0 | 0 | 0.037448348 | 0 | 0 |
| 0 | 0 | 0 | 0 | 0.042177258 |
| 0 | 0 | 0.064046655 | 0.010254892 | 0.050915541 |
| 0 | 0 | 0 | 0 | 0 |
| 0 | 0 | 0 | 0 | 0 |
| 0 | 0.009280793 | 0.077308952 | 0 | 0 |
| 0 | 0 | 0.009708382 | 0 | 0.046856675 |
| 0 | 0 | 0.041293094 | 0 | 0.273143431 |
| 0 | 0 | 0 | 0 | 0 |
| 0 | 0 | 0.837221041 | 0 | 0 |
| 0 | 0 | 0 | 0 | 0.001977421 |
| 0 | 0 | 0 | 0 | 0 |
| 57.98604524 | 0 | 0 | 0.010621081 | 0 |
| 0 | 0 | 0 | 0 | 0 |
| 0 | 0 | 0 | 0 | 0 |
| 0 | 0 | 0 | 0 | 0 |
| 0 | 0 | 0.001399959 | 0.69379945 | 0.196963309 |
| 0 | 0 | 0 | 0 | 0 |
| 0 | 0 | 0 | 0 | 0.058750444 |
| 0 | 0.003003659 | 0.000762819 | 0.010573207 | 0.022926127 |
| 0 | 0 | 0 | 0.001192496 | 0 |
| 0 | 0 | 0.000539973 | 0 | 0 |
| 0 | 0 | 0 | 0 | 0 |
| 0 | 0 | 0 | 0 | 0 |
| 0 | 0 | 0.194312476 | 0 | 0 |
| 0.329321047 | 0.314712136 | 0 | 0.152747478 | 0 |
| 0 | 0 | 0 | 0 | 0 |
| 0 | 0 | 0 | 0 | 0 |
| 0 | 0 | 0 | 0 | 0 |
| 0 | 0 | 0 | 0.006178394 | 0.056794207 |
| 0 | 0 | 0 | 0 | 0 |
| 0 | 0 | 0 | 0 | 0 |
| 0 | 0.007328792 | 0.056505448 | 0 | 0.069600086 |
| 0 | 0 | 0 | 0 | 0 |
| 0 | 0 | 0 | 0 | 0 |
| 0 | 0 | 0 | 0 | 0 |
| 0 | 0 | 0 | 0 | 0 |
| 0 | 0 | 0 | 0 | 0 |
| 0 | 0 | 0 | 0 | 0 |
| 0 | 0 | 0 | 0 | 0 |
| 0 | 0 | 0.146342213 | 0 | 0.30955027 |
| 0 | 0 | 0 | 0 | 0 |
| 0 | 0 | 0 | 0 | 0 |
| 0 | 0 | 0 | 0 | 0 |

| | | | | |
|---|---|---|---|---|
| 0 | 0.059954402 | 0 | 0.006010568 | 0.003437529 |
| 0 | 0 | 0 | 0 | 0 |
| 0 | 0 | 0 | 0 | 0 |
| 0 | 0 | 0 | 0 | 0 |
| 0 | 0 | 0 | 0 | 0 |
| 0 | 0 | 0 | 0 | 0 |
| 0 | 0 | 0 | 0 | 0.005440668 |
| 0.105161324 | 0.191316019 | 0.614567401 | 0 | 0.104225991 |
| 0.681078764 | 4.372064217 | 0.003092161 | 28.74250746 | 0.001269581 |
| 0 | 0.317155089 | 0 | 0 | 0 |
| 0 | 0.010982919 | 0 | 0 | 0 |
| 0 | 0 | 0 | 0 | 0.025724152 |
| 0 | 0 | 0.023303405 | 0 | 0 |
| 1.042616213 | 0 | 0.028042468 | 0.143182607 | 0 |
| 0 | 0 | 0 | 0 | 0 |
| 0 | 0 | 0 | 0 | 0 |
| 0 | 0 | 0 | 0 | 0.122494998 |
| 0 | 0 | 0 | 0 | 0 |
| 0 | 0.00579865 | 0 | 0 | 0 |
| 0 | 0 | 0.014131611 | 0.071580605 | 0.003114845 |
| 0 | 0 | 0 | 0 | 0 |
| 0 | 0 | 0 | 0 | 0 |
| 0 | 0 | 0 | 0 | 0 |
| 0 | 0 | 0 | 0 | 0 |
| 0 | 0.01641965 | 0.095036918 | 0 | 0.002070465 |
| 0 | 0 | 0 | 0 | 0 |
| 0 | 0 | 0 | 0 | 0 |
| 0 | 0 | 0 | 0 | 0 |
| 0 | 0 | 0.004254832 | 0 | 0 |
| 0 | 0 | 0 | 0 | 0 |
| 0 | 0 | 0 | 0 | 0 |
| 0 | 0 | 0 | 0 | 0 |
| 0 | 0 | 0 | 0 | 0 |
| 0 | 0 | 0 | 0 | 0 |
| 0 | 0 | 0 | 0 | 0 |
| 0 | 0 | 0 | 0 | 0 |
| 0 | 0 | 0 | 0 | 0 |
| 0 | 0 | 0 | 0 | 0 |
| 0 | 0.003074522 | 0 | 0 | 0 |
| 0 | 0 | 0 | 0 | 0.003027998 |
| 0 | 0 | 0 | 0 | 0 |
| 0 | 0 | 0.214560793 | 0 | 0.339882151 |
| 0 | 0 | 0 | 0 | 0 |
| 0 | 0 | 0 | 0 | 0 |
| 0 | 0 | 0 | 0 | 0.008458714 |
| 0 | 0 | 0.004963156 | 0 | 0 |
| 0 | 0 | 0 | 0 | 0 |
| 0 | 0 | 0 | 0 | 0 |

| | | | | |
|---|---|---|---|---|
| 0 | 0 | 0 | 0 | 0 |
| 0.001135948 | 0 | 0.084009677 | 0 | 0 |
| 0 | 0 | 0 | 0 | 0 |
| 0 | 0 | 0 | 0 | 0 |
| 1.781977208 | 0.077898597 | 0.109378263 | 1.810560381 | 19.81488581 |
| 0 | 0 | 0 | 0 | 0 |
| 0 | 0 | 0 | 0.042286215 | 0 |
| 0.001801169 | 0.002877975 | 0.012320883 | 0.001365335 | 0 |
| 0 | 0 | 0 | 0 | 0.004879719 |
| 0 | 0 | 0 | 0 | 0 |
| 0 | 0 | 0 | 0 | 0 |
| 0 | 0 | 0 | 0 | 0 |
| 0 | 0 | 0 | 0 | 0 |
| 0 | 0 | 0 | 0 | 0 |
| 0 | 0 | 0 | 0 | 0.000724303 |
| 0 | 0.002317757 | 0.000905577 | 0 | 0.039263369 |
| 0 | 0 | 0 | 0 | 0 |
| 0 | 0 | 0 | 0 | 0 |
| 0 | 0 | 0 | 0 | 0 |
| 0 | 0 | 0 | 0 | 0 |
| 0 | 0 | 0 | 0 | 0 |
| 0 | 0 | 0 | 0 | 0 |
| 0 | 0 | 0 | 0 | 0 |
| 0 | 0.00195694 | 0 | 0 | 0 |
| 0 | 0.137549094 | 0 | 0.171358585 | 0 |
| 0 | 0 | 0 | 0 | 0.00101442 |
| 0.00966285 | 0.00162713 | 0.000826463 | 0.123631371 | 0.275874741 |
| 0 | 0 | 0.000522143 | 0 | 0 |
| 0 | 0.003627793 | 0 | 0 | 0 |
| 0 | 0 | 0 | 0 | 0 |
| 0 | 0 | 0 | 0 | 0 |
| 0 | 0 | 0 | 0 | 0 |
| 0 | 0 | 0 | 0 | 0 |
| 0 | 0 | 0 | 0 | 0 |
| 0 | 0 | 0 | 0 | 0 |
| 0 | 0 | 0 | 0 | 0 |
| 0 | 0 | 0 | 0 | 0 |
| 0 | 0 | 0 | 0 | 0.040298644 |
| 0 | 0 | 0 | 0 | 0 |
| 0 | 0 | 0 | 0 | 0.000716523 |
| 0 | 0 | 0 | 0 | 0 |
| 0 | 0.113274446 | 0 | 0 | 0 |
| 0 | 0 | 0 | 0 | 0 |
| 0 | 0 | 0 | 0 | 0 |
| 0 | 0 | 0 | 0 | 0 |
| 0 | 0 | 0 | 0 | 0 |
| 0 | 0 | 0 | 0 | 0 |
| 0 | 0 | 0 | 0 | 0 |

| | | | | |
|---|---|---|---|---|
| 0 | 0 | 0.030777786 | 0 | 0.098030587 |
| 0 | 0 | 0 | 0 | 0 |
| 0 | 0 | 0 | 0 | 0 |
| 0 | 0 | 0 | 0 | 0 |
| 0 | 0 | 0 | 0 | 0 |
| 0 | 0.048929206 | 0 | 0 | 0 |
| 0 | 0 | 0 | 0 | 0 |
| 0 | 0 | 0 | 0 | 0 |
| 0 | 0 | 0 | 0 | 0 |
| 0 | 0 | 0 | 0 | 0 |
| 0 | 0 | 0 | 0 | 0 |
| 0 | 0 | 0 | 0 | 0 |
| 0 | 0 | 0 | 0 | 0 |
| 0 | 0 | 0 | 0 | 0 |
| 0 | 0 | 0 | 0 | 0.009018618 |
| 0 | 0 | 0 | 0 | 0 |
| 0 | 0 | 0 | 0 | 0 |
| 0 | 0 | 0 | 0 | 0 |
| 0 | 0 | 0 | 0 | 0 |
| 0 | 0 | 0 | 0 | 0 |
| 0 | 0.04163258 | 0.000539905 | 0 | 0 |
| 0 | 0 | 0 | 0 | 0 |
| 0.060118189 | 0.005146022 | 0.002045583 | 0.028531546 | 0.558237769 |
| 0 | 0 | 0.799231705 | 0 | 0 |
| 0 | 0 | 0.025314599 | 0 | 0 |
| 0 | 0 | 0 | 0 | 0 |
| 0 | 0 | 0 | 0 | 0 |
| 0 | 0 | 0 | 0 | 0 |
| 0 | 0 | 0 | 0 | 0 |
| 0 | 0 | 0 | 0 | 0 |
| 0 | 0 | 0 | 0 | 0 |
| 0 | 0 | 0 | 0 | 0 |
| 0 | 0.701302452 | 0.00830128 | 0.392166903 | 0.008651947 |
| 0 | 0 | 0 | 0 | 0 |
| 0.029924339 | 0.131384975 | 0.193061851 | 0 | 0.743659641 |
| 0.013258978 | 0 | 0.001195641 | 0 | 0 |
| 0 | 0 | 0 | 0 | 0 |
| 0 | 0 | 0 | 0 | 0 |
| 0 | 0.011085413 | 0 | 0 | 0.00973391 |
| 0 | 0 | 0 | 0 | 0 |
| 0 | 0 | 0 | 0 | 0 |
| 0 | 0 | 0 | 0 | 0.023757614 |
| 0.065309732 | 0.084140401 | 0 | 0.115392217 | 0.897743986 |
| 0 | 0 | 0 | 0 | 0 |
| 0 | 0 | 0 | 0 | 0 |
| 0 | 0.071156476 | 0.004036191 | 0 | 0.2652996 |
| 0 | 0 | 0 | 0 | 0 |
| 0 | 0 | 0 | 0 | 0 |

| | | | | |
|---|---|---|---|---|
| 0 | 0 | 0 | 0 | 0 |
| 0 | 0 | 0 | 0 | 0 |
| 0 | 0 | 0 | 0 | 0 |
| 0 | 0 | 0.10493215 | 0 | 0 |
| 0 | 0 | 0 | 0 | 0 |
| 0 | 0 | 0 | 0 | 0 |
| 0 | 0 | 0 | 0 | 0 |
| 0 | 0 | 0 | 0 | 0 |
| 0 | 0.008798693 | 0 | 0 | 0.380040225 |
| 0 | 0.221359285 | 0.001133791 | 0 | 0 |
| 0 | 0 | 0 | 0 | 0 |
| 0 | 0 | 0.012907652 | 0 | 0 |
| 0 | 0 | 0 | 0 | 0 |
| 0 | 0 | 0 | 0 | 0 |
| 0 | 0 | 0.003206901 | 0 | 0 |
| 0 | 0 | 0.001115525 | 0 | 0 |
| 0 | 0 | 0 | 0 | 0 |
| 0 | 0 | 0 | 0 | 0 |
| 0 | 0 | 0 | 0 | 0 |
| 0 | 0 | 0 | 0 | 0.134020173 |
| 0 | 0 | 0.002524354 | 0 | 0 |
| 0 | 0 | 0 | 0 | 0 |
| 0 | 0 | 0 | 0 | 0 |
| 0 | 0.009263852 | 0.031946888 | 0 | 0.001525205 |
| 0 | 0 | 0 | 0 | 0 |
| 0 | 0 | 1.724535139 | 0 | 0 |
| 0 | 0 | 0 | 0 | 0 |
| 0 | 0 | 0.015756528 | 0.36535192 | 0 |
| 0 | 0 | 0 | 0 | 0 |
| 0 | 0 | 0 | 0 | 0 |
| 0.044024293 | 0.007288898 | 0.14808896 | 0.008298992 | 0.329449864 |
| 0 | 0 | 0 | 0 | 0 |
| 0 | 0 | 0 | 0 | 0 |
| 0 | 0 | 0 | 0 | 0 |
| 0 | 0 | 0 | 0 | 0 |
| 0 | 0 | 0 | 0 | 0 |
| 0 | 0 | 0.000762026 | 0 | 0 |
| 0 | 0 | 0 | 0 | 0 |
| 0 | 0.002850058 | 0 | 0.001781579 | 0 |
| 0 | 0 | 0 | 0 | 0 |
| 0 | 0 | 0 | 0 | 0 |
| 0 | 0 | 0 | 0 | 0 |
| 0 | 0 | 0.008008562 | 0 | 0.001811842 |
| 0 | 0 | 0 | 0 | 0 |
| 0 | 0 | 0 | 0 | 0 |
| 0 | 0 | 0 | 0 | 0 |
| 0.05362497 | 0 | 0 | 0 | 0.000703296 |
| 0 | 0 | 0 | 0 | 0 |

| | | | | |
|---|---|---|---|---|
| 0 | 0 | 0 | 0 | 0 |
| 0 | 0 | 0 | 0 | 0 |
| 0 | 0 | 0 | 0 | 0 |
| 0 | 0 | 0 | 0 | 0 |
| 0 | 0 | 0.20361256 | 0.00873854 | 0.006129839 |
| 0 | 0 | 0 | 0 | 0 |
| 0 | 0 | 0 | 0 | 0 |
| 0 | 0 | 0 | 0 | 0 |
| 0 | 0 | 0 | 0 | 0 |
| 0 | 0 | 0 | 0 | 0 |
| 0 | 0 | 0 | 0 | 0 |
| 0.003594494 | 0.002914891 | 0.00118444 | 0.014197245 | 0.079268196 |
| 0.021055124 | 0 | 0 | 0 | 0.007342322 |
| 0 | 0 | 0.066586302 | 0 | 0.063145901 |
| 0 | 0 | 0.000451313 | 0 | 0 |
| 0 | 0 | 0 | 0 | 0.00226904 |
| 0 | 0.003736495 | 0.002732925 | 0 | 0.004114314 |
| 0 | 0 | 0 | 0 | 0 |
| 0 | 0.002207172 | 0.001494775 | 0 | 0 |
| 0 | 0 | 0.017760658 | 0.001786468 | 0.008894854 |
| 0 | 0 | 0 | 0.000764198 | 0 |
| 0 | 0 | 0 | 0 | 0 |
| 0 | 0 | 0 | 0 | 0 |
| 0 | 0 | 0 | 0 | 0 |
| 0 | 0 | 0 | 0 | 0 |
| 0 | 0 | 0 | 0 | 0 |
| 0 | 0 | 0.031749264 | 0 | 0.000770579 |
| 0 | 0 | 0 | 0 | 0 |
| 0 | 0 | 0 | 0 | 0 |
| 0 | 0 | 0.000447892 | 0 | 0 |
| 0 | 0.00261782 | 0 | 0 | 0 |
| 0 | 0 | 0.013978891 | 0 | 0 |
| 0 | 0 | 0 | 0 | 0 |
| 0 | 0 | 0 | 0 | 0 |
| 0 | 0 | 0 | 0 | 0 |
| 0 | 0 | 0 | 0 | 0 |
| 0 | 0 | 0 | 0 | 0 |
| 0 | 0.005885453 | 0.000747344 | 0.118245758 | 0.036514545 |
| 0 | 0 | 0 | 0 | 1.046871454 |
| 0 | 0 | 0 | 0 | 0 |
| 0 | 0.002570566 | 0 | 0.002601595 | 0.005207612 |
| 0 | 0 | 0 | 0 | 0 |
| 0 | 0 | 0 | 0 | 0 |
| 0 | 0 | 0 | 0 | 0 |
| 0 | 0 | 0 | 0 | 0 |
| 0 | 0 | 0 | 0 | 0 |
| 0 | 0 | 0 | 0 | 0 |
| 0 | 0 | 0 | 0 | 0 |

| | | | | |
|---|---|---|---|---|
| 0 | 0 | 0 | 0 | 0 |
| 0 | 0 | 0.002683734 | 0 | 1.254597368 |
| 0 | 0 | 0 | 0 | 0 |
| 0 | 0.001143292 | 0 | 0 | 0.005006975 |
| 0.001204379 | 0 | 0 | 0.000704277 | 0.002211376 |
| 0.002442577 | 0 | 0.001853631 | 0 | 0 |
| 0 | 0 | 0 | 0 | 0.047200633 |
| 0 | 0 | 0 | 0.001583656 | 0 |
| 0 | 0 | 0.107874938 | 0 | 0 |
| 0 | 0 | 0.001118403 | 0 | 0 |
| 0 | 0 | 0 | 0 | 0.0157613 |
| 0 | 0 | 0 | 0 | 0 |
| 0.003418946 | 0.428854083 | 0.002607055 | 0.001794222 | 0.179659162 |
| 0 | 0 | 0 | 0 | 0 |
| 0 | 0 | 0 | 0 | 0 |
| 0 | 0 | 0 | 0 | 0 |
| 0 | 0 | 0 | 0 | 0 |
| 0 | 0 | 0 | 0 | 0 |
| 0 | 0 | 0 | 0 | 0 |
| 0 | 0 | 0 | 0 | 0 |
| 0 | 0 | 0 | 0 | 0 |
| 0 | 0 | 0 | 0 | 0 |
| 0 | 0.029702436 | 0 | 0 | 0 |
| 0 | 0 | 0.006822618 | 0 | 0.074582836 |
| 0 | 0 | 0 | 0 | 0 |
| 0 | 0 | 0.001889463 | 0 | 0.015515533 |
| 0 | 0 | 0 | 0 | 0.032744469 |
| 0 | 0 | 0 | 0 | 0 |
| 0 | 0 | 0 | 0 | 0 |
| 0 | 0 | 0 | 0 | 0 |
| 0 | 0.008329099 | 0.022160127 | 0.003687967 | 0.054839224 |
| 0 | 0 | 0 | 0 | 0 |
| 0 | 0 | 0 | 0 | 0 |
| 0 | 0 | 0 | 0 | 0 |
| 0 | 0 | 0 | 0 | 0 |
| 0 | 0 | 0 | 0 | 0 |
| 0 | 0 | 0 | 0 | 0 |
| 0 | 0.004899334 | 0 | 0 | 0 |
| 0 | 0 | 0 | 0.006463429 | 0 |
| 0 | 0 | 0 | 0 | 0 |
| 0 | 0 | 0 | 0 | 0 |
| 0 | 0 | 0.001242875 | 0 | 0 |
| 0 | 0 | 0 | 0 | 0 |
| 0 | 0 | 0 | 0 | 0 |
| 0 | 0 | 0 | 0 | 6.693649983 |
| 0.001293455 | 0.418070079 | 0.212348852 | 0.048827612 | 0.10639681 |
| 0 | 0 | 0 | 0 | 0 |
| 0 | 0 | 0 | 0 | 0 |

| | | | | |
|---|---|---|---|---|
| 0 | 0 | 0 | 0 | 0 |
| 0 | 0 | 0 | 0 | 0 |
| 0.322834801 | 0 | 0.279645545 | 0 | 0 |
| 0 | 0 | 0 | 0 | 0 |
| 0 | 0 | 0 | 0 | 0 |
| 0 | 0 | 0 | 0 | 0 |
| 0 | 0 | 0 | 0 | 0 |
| 0 | 0 | 0 | 0 | 0 |
| 0 | 0 | 0 | 0 | 0.013792085 |
| 0 | 0 | 0 | 0.003052324 | 0 |
| 0.00185524 | 0 | 0.000829662 | 0 | 0.011581857 |
| 0 | 0 | 0 | 0 | 0 |
| 0 | 0 | 0 | 0 | 0 |
| 0 | 0 | 0 | 0 | 0 |
| 0 | 0 | 0 | 0 | 0 |
| 0 | 0 | 0 | 0 | 0 |
| 0.107721337 | 0 | 0 | 0 | 0 |
| 0 | 0 | 0 | 0 | 0 |
| 0 | 0 | 0 | 0 | 0 |
| 0 | 0 | 0 | 0 | 0.001355253 |
| 0 | 0 | 0 | 0 | 0 |
| 0 | 0 | 0 | 0 | 0 |
| 0 | 0 | 0 | 0 | 0 |
| 0 | 0 | 0 | 0 | 0 |
| 0 | 0 | 0 | 0 | 0 |
| 0 | 0 | 0 | 0 | 0 |
| 0 | 0 | 0 | 0 | 0 |
| 0 | 0 | 0 | 0 | 0 |
| 0 | 0 | 0 | 0 | 0 |
| 0 | 0 | 0 | 0 | 0 |
| 0 | 0.002636411 | 0 | 0.001236021 | 0 |
| 0 | 0 | 0 | 0 | 0 |
| 0.009466324 | 0.003781411 | 0.013719133 | 0.085869576 | 0.01487061 |
| 0 | 0.001840091 | 0 | 0.002199846 | 0.00391416 |
| 0 | 0 | 0 | 0 | 0 |
| 0 | 0 | 0 | 0 | 0 |
| 0 | 0 | 0 | 0 | 0 |
| 0 | 0 | 0 | 0 | 0 |
| 0 | 0 | 0 | 0 | 0 |
| 0 | 0 | 0 | 0 | 0 |
| 0 | 0 | 0 | 0 | 0 |
| 0.001442196 | 0 | 0.004966107 | 0 | 0.025765369 |
| 0 | 0.008482365 | 0.071868071 | 0.011507226 | 0.032066955 |
| 0 | 0 | 0 | 0 | 0 |
| 0 | 0 | 0 | 0 | 0 |
| 0.085310039 | 0.044075225 | 0 | 0.066514965 | 0.610891337 |
| 0 | 0 | 0 | 0 | 0 |
| 0 | 0 | 0 | 0 | 0 |

| | | | | |
|---|---|---|---|---|
| 0 | 0 | 0 | 0 | 0 |
| 0 | 0.003025158 | 0.00691451 | 0 | 0 |
| 0.005590319 | 0 | 0 | 0 | 0.003039857 |
| 0 | 0.897845008 | 0 | 0 | 0 |
| 0 | 0.002645149 | 0 | 0.01218071 | 0.034421821 |
| 0.001503984 | 0.009612484 | 0 | 0 | 0 |
| 0 | 0 | 0 | 0 | 0 |
| 0 | 0 | 0 | 0 | 0 |
| 0 | 0 | 0 | 0.107301218 | 0 |
| 0 | 0 | 0 | 0 | 0 |
| 0 | 0 | 0 | 0 | 0 |
| 0 | 0 | 0 | 0 | 0 |
| 0 | 0 | 0 | 0 | 0 |
| 0 | 0 | 0.049407568 | 0 | 0 |
| 0 | 0 | 0 | 0 | 0 |
| 0 | 0 | 0 | 0 | 0 |
| 0 | 0 | 0 | 0 | 0 |
| 0 | 0 | 0 | 0 | 0 |
| 0 | 0 | 0 | 0 | 0 |
| 0 | 0 | 0 | 0 | 0 |
| 0 | 0 | 0 | 0 | 0 |
| 0 | 0 | 0.001082819 | 0 | 0 |
| 0 | 0 | 0 | 0 | 0 |
| 0 | 0 | 0 | 0 | 0 |
| 2.404608589 | 0.027848573 | 0 | 0 | 0.040312067 |
| 0.001426075 | 0.002197255 | 0 | 0.009242579 | 0 |
| 0 | 0 | 0 | 0 | 0 |
| 0 | 0 | 0.023095504 | 0 | 0.031781396 |
| 0 | 0 | 0 | 0 | 0 |
| 0 | 0 | 0 | 0 | 0 |
| 0 | 0 | 0 | 0 | 0 |
| 0 | 0 | 0 | 0 | 0 |
| 0 | 0 | 0 | 0 | 0 |
| 0 | 0 | 0 | 0 | 0 |
| 0 | 0 | 0 | 0 | 0 |
| 0 | 0 | 0 | 0 | 0 |
| 0 | 0 | 0 | 0 | 0 |
| 0 | 0 | 0 | 0 | 0 |
| 0 | 0 | 0 | 0.801391662 | 0.092567354 |
| 0 | 0 | 0 | 0 | 0 |
| 0 | 0 | 0 | 0 | 0 |
| 0 | 0.061578485 | 0 | 0.025594659 | 0.007208014 |
| 0 | 0.055680936 | 0 | 0.00610259 | 0.003908977 |
| 0 | 0 | 0 | 0 | 0 |
| 0 | 0 | 0 | 0 | 0 |
| 0 | 0 | 0 | 0 | 0 |
| 0 | 0 | 0.000856746 | 0.013443507 | 0 |
| 0 | 0 | 0 | 0 | 0 |

| | | | | |
|---|---|---|---|---|
| 0 | 0 | 0.030784124 | 0 | 0 |
| 0.020202482 | 0.011351305 | 0.001261231 | 0.022853244 | 0.029294767 |
| 0 | 0 | 0 | 0 | 0 |
| 0 | 0 | 0 | 0 | 0 |
| 0 | 0 | 0 | 0 | 0 |
| 0 | 0 | 0 | 0 | 0 |
| 0 | 0 | 0 | 0 | 0 |
| 0 | 0 | 0 | 0 | 0 |
| 0 | 0 | 0 | 0 | 0 |
| 0 | 0 | 0 | 0 | 0 |
| 0 | 0 | 0 | 0 | 0 |
| 0 | 0 | 0 | 0 | 0 |
| 0 | 0.001499917 | 0.007999399 | 0 | 0 |
| 0 | 0 | 0 | 0 | 0 |
| 0 | 0.002653756 | 0.001347913 | 0 | 0.110685331 |
| 0 | 0 | 0 | 0 | 0 |
| 0 | 0 | 0 | 0 | 0 |
| 0 | 0 | 0 | 0 | 0 |
| 0 | 0 | 0.014911408 | 0 | 0 |
| 0 | 0 | 0 | 0 | 0 |
| 0 | 0 | 0 | 0 | 0 |
| 0 | 0 | 0.03618269 | 0 | 0.01005491 |
| 0 | 0 | 0.005764127 | 0 | 0.017540961 |
| 0 | 0 | 0 | 0.015451657 | 0 |
| 0 | 0 | 0 | 0 | 0 |
| 0 | 0 | 0 | 0 | 0 |
| 0 | 0 | 0 | 0 | 0 |
| 0 | 0 | 0 | 0 | 0 |
| 0 | 0 | 0 | 0 | 0 |
| 0 | 0 | 0 | 0 | 0 |
| 0 | 0 | 0 | 0 | 0 |
| 0 | 0 | 0 | 0 | 0 |
| 0 | 0 | 0 | 0 | 0 |
| 0 | 0 | 0 | 0 | 0 |
| 0 | 19.72710864 | 31.07266023 | 0.001571618 | 0 |
| 0 | 0 | 0 | 0 | 0 |
| 0 | 0 | 0 | 0 | 0 |
| 0 | 0 | 0 | 0 | 0 |
| 0.041595479 | 0 | 0.054199192 | 0.014404798 | 0.015740016 |
| 0 | 0 | 0 | 0 | 0 |
| 0 | 0.007195201 | 0.055313367 | 0.038747421 | 0.046475588 |
| 0 | 0 | 0 | 0 | 0 |
| 0 | 0 | 0 | 0 | 0 |
| 0 | 0 | 0 | 0 | 0 |
| 0 | 0 | 0 | 0 | 0 |
| 0.000650272 | 0.018257225 | 0.017868119 | 0.094937161 | 0.102243865 |
| 0 | 0 | 0 | 0 | 0 |
| 0 | 0 | 0 | 0 | 0 |

| | | | | |
|---|---|---|---|---|
| 0 | 0 | 0 | 0 | 0 |
| 0 | 0 | 0 | 0 | 0 |
| 0 | 0 | 0.155087149 | 0 | 0 |
| 0 | 0 | 0 | 0 | 0 |
| 0 | 0 | 0 | 0 | 0 |
| 0 | 0 | 0 | 0 | 0 |
| 0 | 0 | 0 | 0 | 0 |
| 0 | 0 | 0.000977869 | 0 | 0 |
| 0 | 0.002262442 | 0 | 0 | 0.23232422 |
| 0.001271934 | 0.114537107 | 0 | 0.010702186 | 0 |
| 0 | 0 | 0.099848249 | 0 | 0 |
| 0 | 0 | 0 | 0 | 0 |
| 0 | 0.002554001 | 0.000648622 | 0 | 0 |
| 0 | 0 | 0 | 0 | 0 |
| 0 | 0 | 0 | 0 | 0 |
| 0 | 0 | 0 | 0 | 0 |
| 0 | 0 | 0 | 0 | 0 |
| 0 | 0 | 0 | 0 | 0 |
| 0 | 0 | 0 | 0 | 0 |
| 0.742121928 | 0.198150853 | 0.070240558 | 3.242343222 | 1.783696881 |
| 0 | 0 | 0.01501501 | 0 | 0 |
| 0 | 0 | 0 | 0 | 0 |
| 0 | 0 | 0 | 0 | 0 |
| 0 | 0 | 0 | 0 | 0 |
| 0 | 0 | 0 | 0 | 0 |
| 0 | 0 | 0 | 0 | 0 |
| 0 | 0 | 0 | 0 | 0 |
| 0 | 0 | 0 | 0 | 0 |
| 0 | 0 | 0 | 0 | 0 |
| 0 | 0 | 0.00761169 | 0 | 0 |
| 0 | 0 | 0 | 0 | 0 |
| 0 | 0 | 0 | 0 | 0 |
| 0 | 0 | 0 | 0 | 0 |
| 0 | 0 | 0 | 0 | 0 |
| 0 | 0 | 0 | 0 | 0 |
| 0 | 0 | 0.001494654 | 0 | 0.002454703 |
| 0 | 0 | 0 | 0 | 0 |
| 0 | 0 | 0 | 0 | 0 |
| 0 | 0 | 0 | 0 | 0 |
| 0 | 0 | 0 | 0 | 0 |
| 0 | 0 | 0 | 0 | 0 |
| 0 | 0 | 0 | 0 | 0 |
| 0 | 0 | 0.949544054 | 0 | 0 |
| 0 | 0 | 0 | 0 | 0 |
| 0.036722161 | 0.001245482 | 0.001265226 | 0.008272136 | 0.106195962 |
| 0 | 0 | 0 | 0 | 0 |
| 0 | 0 | 0 | 0 | 0 |
| 0 | 0 | 0 | 0 | 0 |

| | | | | |
|---|---|---|---|---|
| 0 | 0 | 0 | 0 | 0 |
| 0 | 0 | 0 | 0 | 0 |
| 0 | 0 | 0 | 0 | 0 |
| 0 | 0 | 0 | 0 | 0 |
| 0 | 0 | 0 | 0 | 0 |
| 0 | 0.002258087 | 0.001889081 | 0.014997573 | 0.073628471 |
| 0 | 0 | 0 | 0 | 0 |
| 0 | 0 | 0 | 0 | 0 |
| 0 | 0 | 0 | 0 | 0 |
| 0 | 0 | 0 | 0 | 0 |
| 0 | 0 | 0 | 0 | 0 |
| 0 | 0 | 0 | 0 | 0 |
| 0 | 0 | 0 | 0 | 0 |
| 0 | 0.000679307 | 0 | 0 | 0 |
| 0 | 0 | 0 | 0 | 0 |
| 0 | 0 | 0 | 0 | 0 |
| 0 | 0 | 0.272517952 | 0 | 0.000806418 |
| 0 | 0 | 0 | 0 | 0 |
| 0 | 0 | 0 | 0 | 0 |
| 0 | 0 | 0 | 0 | 0 |
| 0 | 0 | 0 | 0 | 0 |
| 0 | 0 | 0 | 0 | 0 |
| 0 | 0 | 0 | 0 | 0 |
| 0 | 0 | 0 | 0 | 0 |
| 0 | 0 | 0 | 0 | 0 |
| 0 | 0 | 0 | 0 | 0 |
| 0 | 0 | 0 | 0 | 0 |
| 0 | 0 | 0.328722723 | 0 | 0 |
| 0 | 0 | 0 | 0 | 0 |
| 0 | 0 | 0.095135189 | 0 | 0 |
| 0 | 0 | 0 | 0 | 0 |
| 0 | 0 | 0 | 0 | 0 |
| 0 | 0 | 0 | 0 | 0 |
| 0 | 0 | 0 | 0 | 0 |
| 0 | 0 | 0 | 0 | 0 |
| 0 | 0 | 0 | 0 | 0 |
| 0 | 0 | 0 | 0 | 0 |
| 0 | 0 | 0.047343057 | 0 | 0 |
| 0 | 1.862808046 | 0 | 0 | 0.928829546 |
| 0 | 0 | 0 | 0 | 0 |
| 0 | 0 | 0 | 0 | 0 |
| 0 | 0 | 0 | 0 | 0 |
| 0 | 0 | 0 | 0 | 0 |
| 0 | 0 | 0 | 0 | 0 |
| 0 | 0.000553207 | 0 | 0 | 0 |
| 0 | 0 | 0 | 0 | 0 |
| 0 | 0 | 0 | 0 | 0 |
| 0 | 0 | 0 | 0 | 0 |

| | | | | |
|---|---|---|---|---|
| 0 | 0 | 0 | 0 | 0 |
| 0 | 0.006692057 | 0.062996151 | 0 | 0.132302186 |
| 0 | 0.001638615 | 0 | 0 | 0 |
| 0 | 0 | 0 | 0 | 0 |
| 0 | 0 | 0 | 0 | 0 |
| 0 | 0 | 0 | 0 | 0 |
| 0 | 0 | 0 | 0 | 0 |
| 0 | 0 | 0 | 0 | 0 |
| 0 | 0 | 0.08798649 | 0 | 0 |
| 0 | 0 | 0 | 0 | 0.004042413 |
| 0.001516594 | 0.051797403 | 0 | 0.098551134 | 0.006335049 |
| 0 | 0 | 0 | 0 | 0 |
| 0 | 0.013482076 | 0 | 0 | 0 |
| 0 | 0 | 0 | 0 | 0 |
| 0 | 0 | 0 | 0 | 0 |
| 0 | 0 | 0 | 0 | 0 |
| 0 | 0 | 0 | 0 | 0 |
| 0 | 0 | 0 | 0 | 0 |
| 0 | 0 | 0 | 0 | 0 |
| 0 | 0 | 0 | 0 | 0 |
| 0.011159575 | 0.007905451 | 0 | 0 | 0.571995432 |
| 0 | 0 | 0 | 0 | 0 |
| 0 | 0 | 0 | 0 | 0 |
| 0 | 1.357090007 | 1.081727506 | 0.028272237 | 1.4594214 |
| 0 | 0 | 0 | 0 | 0 |
| 0 | 0 | 0 | 0 | 0 |
| 0 | 0 | 0 | 0 | 0 |
| 0 | 0.001134954 | 0.059953198 | 0 | 0 |
| 0 | 0 | 0 | 0 | 0 |
| 0 | 0 | 0 | 0 | 0 |
| 0 | 0.001357859 | 0 | 0 | 0 |
| 0 | 0 | 0 | 0 | 0 |
| 0 | 0 | 0 | 0 | 0.001300658 |
| 0 | 0 | 0 | 0 | 0 |

| PHL018-0251_V1 | PHL018-0042_V3 | PHL018-0248_V1 | PHL018-0261_V1 | PHL018-0055_V3 |
|---|---|---|---|---|
| REF | CTRL | REF | REF | EYCF |
| 0.077231429 | 0.045404955 | 0.083258236 | 0.363388413 | 0.015676306 |
| 26.48355036 | 0.100679742 | 0.311379608 | 2.917898435 | 0.193658013 |
| 0 | 0 | 2.830300359 | 19.16276109 | 0 |
| 0 | 0 | 0 | 0 | 0 |
| 0 | 0 | 0 | 0 | 0 |
| 0 | 0 | 0 | 0 | 0.303410181 |
| 0 | 0 | 0 | 0 | 0 |
| 0 | 0 | 0 | 0 | 0 |
| 0 | 0 | 0 | 0 | 0 |
| 0 | 0 | 0 | 0 | 0 |
| 0 | 0 | 0 | 0 | 0 |
| 0 | 0 | 0 | 0 | 0 |
| 0 | 0 | 0 | 0 | 0 |
| 0 | 0.012162785 | 0.081086937 | 0.173031293 | 0.025860353 |
| 0 | 0 | 0 | 0 | 0 |
| 0 | 0 | 0 | 0 | 0 |
| 0 | 0 | 0 | 0 | 0 |
| 0 | 0 | 0 | 0 | 0 |
| 0 | 0 | 0 | 0 | 0 |
| 0 | 0 | 0 | 0 | 0 |
| 0 | 0 | 0 | 0 | 0 |
| 0 | 0 | 0 | 0.048377836 | 0 |
| 0 | 0 | 0 | 0 | 0 |
| 0 | 0 | 0 | 0 | 0 |
| 0 | 0.037381081 | 0 | 0 | 0.020745408 |
| 0 | 0 | 0 | 0 | 0 |
| 0 | 0 | 0 | 0 | 0 |
| 0 | 0 | 0 | 0 | 0.096316218 |
| 0 | 0 | 0 | 0 | 0 |
| 0 | 0 | 0 | 0 | 0 |
| 0 | 0 | 0 | 0 | 0 |
| 0 | 0 | 0 | 0 | 0 |
| 0 | 0 | 0 | 0 | 0 |
| 0 | 0 | 0 | 0 | 0 |
| 0 | 0 | 0 | 0 | 0 |
| 0 | 3.566371639 | 3.976728655 | 0 | 1.878580689 |
| 0 | 0 | 0 | 0 | 0 |
| 0 | 0 | 0 | 0 | 0 |
| 0 | 0 | 0 | 0 | 0 |
| 0 | 0 | 0 | 0 | 0 |
| 0 | 0 | 0 | 0 | 0 |
| 0.001951962 | 0 | 0.001822943 | 0.002733059 | 0.246573977 |
| 0.319060042 | 2.791925128 | 0.091796248 | 0.495901835 | 0.268811344 |
| 0.14216587 | 0.006556875 | 0.006114217 | 0.032939298 | 0.043980658 |
| 0 | 0.001609285 | 0 | 0 | 0 |
| 0 | 0 | 0 | 0 | 0 |

| | | | | |
|---|---|---|---|---|
| 0 | 0.017467474 | 0.006601733 | 0 | 0.000529119 |
| 0.114191433 | 0 | 0.009956905 | 0.002016583 | 0 |
| 0.001984754 | 0 | 0.001733205 | 0 | 0 |
| 1.925405466 | 0.291903347 | 0.179204172 | 1.069962011 | 1.051774584 |
| 0 | 0 | 0 | 0.003204719 | 0.004184373 |
| 0 | 0.051444343 | 0 | 0 | 0.069605635 |
| 0 | 0 | 0.005804176 | 0 | 0.002861206 |
| 0 | 0 | 0 | 0 | 0.012909692 |
| 0 | 0.009275796 | 0 | 0 | 0.003462182 |
| 0 | 0 | 0 | 0 | 0 |
| 0 | 0 | 0.012730184 | 0 | 0.004301827 |
| 0.014525882 | 0.023878005 | 0.111653256 | 0 | 0.025148536 |
| 0 | 0 | 0.000486601 | 0 | 0.000901341 |
| 0.162412374 | 0.027810975 | 0.024940612 | 0.045996645 | 0 |
| 0 | 0 | 0 | 0 | 0 |
| 0 | 0 | 0 | 0 | 0 |
| 0.001804296 | 0 | 0.041543827 | 0 | 0 |
| 0 | 0 | 0 | 0 | 0 |
| 0 | 0 | 0 | 0 | 0 |
| 0 | 0 | 0 | 0 | 0 |
| 0.637802833 | 0 | 0.200067843 | 0.054212476 | 0 |
| 0.021153777 | 0.007102924 | 0.038047764 | 0.07525336 | 0 |
| 0 | 0.006741102 | 0.005670106 | 0 | 0 |
| 0 | 0 | 0 | 0 | 0 |
| 0 | 0 | 0 | 0 | 0 |
| 0 | 0 | 0 | 0.001016816 | 0 |
| 0 | 0 | 0 | 0 | 0 |
| 0 | 0 | 0 | 0 | 0 |
| 0 | 0 | 0 | 0 | 0.00709301 |
| 0 | 0 | 0 | 0 | 0 |
| 1.368632499 | 0 | 0 | 0 | 0 |
| 0 | 0 | 0 | 0 | 0 |
| 0 | 0 | 0 | 0 | 0 |
| 0 | 0 | 0 | 0 | 0 |
| 0 | 0 | 0 | 0 | 0 |
| 0 | 0 | 0 | 0 | 0 |
| 0 | 0 | 0 | 0 | 0 |
| 0 | 0 | 0.002581086 | 0 | 0 |
| 0 | 0 | 0 | 0 | 0 |
| 0 | 0 | 0 | 0 | 0 |
| 0 | 0 | 0 | 0 | 0 |
| 0 | 0 | 0 | 0 | 0 |
| 0 | 0 | 0 | 0 | 0 |
| 0 | 0 | 0 | 0 | 0 |
| 0 | 0 | 0 | 0 | 0 |
| 0 | 0 | 0 | 0 | 0 |
| 0 | 0 | 0 | 0 | 0 |
| 0 | 0 | 0 | 0 | 0 |

| | | | | |
|---|---|---|---|---|
| 0 | 0 | 0 | 0 | 0 |
| 0 | 0 | 0.000558837 | 0 | 0 |
| 0 | 0 | 0 | 0 | 0 |
| 0 | 0 | 0 | 0 | 0 |
| 0 | 0.005130852 | 0 | 0 | 0 |
| 0 | 0 | 0 | 0 | 0 |
| 0 | 0 | 0 | 0 | 0 |
| 0 | 0.001410921 | 0 | 0 | 0 |
| 0 | 0 | 0.011572913 | 0 | 0.005537826 |
| 0 | 0 | 0 | 0 | 0 |
| 0 | 0 | 0 | 0 | 0 |
| 0 | 0.001226269 | 0 | 0 | 0.00207397 |
| 0 | 0 | 0 | 0 | 0 |
| 0 | 0 | 0 | 0 | 0 |
| 0 | 0 | 0 | 0 | 0 |
| 0 | 0 | 0 | 0 | 0 |
| 0 | 0 | 0 | 0 | 0 |
| 0 | 0 | 0 | 0 | 0 |
| 0 | 0.035740792 | 0.349951587 | 0 | 1.781135535 |
| 0 | 0.053284383 | 0 | 0 | 0 |
| 0 | 0 | 0 | 0 | 0 |
| 0 | 0 | 0 | 0 | 0 |
| 0 | 0.000722052 | 0 | 0 | 0 |
| 0 | 0 | 0 | 0 | 0 |
| 0 | 0 | 0 | 0 | 0 |
| 0 | 0 | 0 | 0 | 0 |
| 0 | 0 | 0 | 0 | 0 |
| 0 | 0 | 0 | 0 | 0 |
| 0 | 0 | 0 | 0 | 0 |
| 0 | 0 | 0 | 0 | 0 |
| 0 | 0.011851093 | 0.257616627 | 0.045331139 | 0.002149002 |
| 0.031612369 | 0.001281735 | 0.007489948 | 0.096553727 | 0.004682403 |
| 0 | 0 | 0 | 0 | 0 |
| 0 | 0 | 0 | 0 | 0 |
| 0 | 0 | 0 | 0 | 0 |
| 0 | 0 | 0 | 0 | 0 |
| 0 | 0 | 0 | 0 | 0 |
| 0 | 0 | 0 | 0 | 0 |
| 0 | 0 | 0.078055144 | 0 | 0 |
| 0 | 0 | 0 | 0 | 0 |
| 0 | 0 | 0 | 0 | 0 |
| 0 | 0 | 0 | 0 | 0.001657821 |
| 0 | 0 | 0 | 0 | 0 |
| 0 | 0 | 0 | 0 | 0 |
| 0 | 0 | 0 | 0 | 0 |
| 0 | 0 | 0 | 0 | 0 |
| 0 | 0 | 0 | 0.775606858 | 0 |
| 0 | 0 | 0 | 0 | 0 |

| | | | | |
|---|---|---|---|---|
| 0 | 0 | 0 | 0 | 0 |
| 0 | 0 | 0 | 0 | 0 |
| 0 | 0 | 0 | 0 | 0 |
| 0 | 0 | 0 | 0 | 0 |
| 0 | 0.089359978 | 0 | 0 | 0.000754723 |
| 0 | 0 | 0 | 0 | 0 |
| 0 | 0 | 0 | 0 | 0 |
| 0 | 0 | 0 | 0 | 0 |
| 0 | 0 | 0 | 0 | 0 |
| 0 | 0 | 0 | 0 | 0 |
| 0 | 0 | 0 | 0 | 0 |
| 0 | 0 | 0 | 0 | 0 |
| 0 | 0 | 0 | 0 | 0 |
| 0 | 0 | 0 | 0 | 0 |
| 0 | 0 | 0 | 0 | 0 |
| 0 | 0.008766358 | 0 | 0 | 6.872092249 |
| 0 | 0 | 0 | 0 | 0 |
| 0 | 0 | 0 | 0 | 0 |
| 0 | 0 | 0 | 0 | 0 |
| 0 | 0 | 0 | 0 | 0 |
| 0 | 0 | 0 | 0 | 0 |
| 0 | 0 | 0 | 0 | 0 |
| 0 | 0 | 0 | 0 | 0 |
| 0 | 0 | 0 | 0 | 0 |
| 0 | 0 | 0 | 0 | 0.301974713 |
| 0 | 0 | 0 | 0 | 0 |
| 0 | 0.043300757 | 0 | 0 | 0.01902669 |
| 0 | 0 | 0 | 0 | 0 |
| 0 | 0 | 0 | 0 | 0 |
| 0 | 0 | 0 | 0 | 0 |
| 0 | 0 | 0 | 0 | 0 |
| 0 | 0.03149941 | 0.000432089 | 0 | 0.001400644 |
| 0 | 0 | 0 | 0 | 0 |
| 0 | 0 | 0 | 0 | 0 |
| 0 | 0 | 0 | 0 | 0 |
| 0 | 0 | 0 | 0 | 0 |
| 0 | 0 | 0 | 0 | 0 |
| 0 | 0 | 0 | 0 | 0.099221413 |
| 0 | 0 | 0 | 0 | 0 |
| 0 | 0 | 0 | 0 | 0 |
| 0 | 0 | 0 | 0 | 0 |
| 0 | 0 | 0 | 0 | 0 |
| 0 | 0 | 0 | 0 | 0 |
| 0 | 0 | 0 | 0 | 0 |
| 0 | 0 | 0 | 0 | 0 |
| 0 | 0 | 0 | 0 | 0 |
| 0 | 0 | 0 | 0 | 0 |
| 0.046450929 | 0 | 0 | 0 | 0 |

| | | | | |
|---|---|---|---|---|
| 0 | 0 | 0 | 0 | 0.071883561 |
| 0 | 0 | 0 | 0 | 0 |
| 0 | 0 | 0 | 0 | 0 |
| 0 | 0 | 0 | 0 | 0 |
| 0 | 0 | 0 | 0 | 0 |
| 0 | 0 | 0 | 0 | 0 |
| 0 | 0 | 0 | 0 | 0 |
| 0 | 0 | 0 | 0 | 0 |
| 0 | 0 | 0 | 0 | 0 |
| 0 | 0 | 0 | 0 | 0 |
| 0 | 0 | 0 | 0 | 0 |
| 0 | 0.000430653 | 0 | 0 | 0.000776915 |
| 0.003517246 | 0.002122665 | 0.001240401 | 0 | 0 |
| 0 | 0 | 0 | 0 | 0 |
| 0 | 0 | 0 | 0 | 0 |
| 0 | 0 | 0 | 0 | 0 |
| 0 | 0 | 0 | 0 | 0 |
| 0 | 0 | 0 | 0 | 0 |
| 0 | 0 | 0 | 0 | 0 |
| 0 | 0 | 0 | 0 | 0 |
| 0.158339585 | 0.011377439 | 0.002576946 | 0.194643322 | 0.013961994 |
| 0 | 0 | 0 | 0 | 0 |
| 0 | 0 | 0 | 0 | 0 |
| 0.002110113 | 0 | 0 | 0 | 0.001119965 |
| 0 | 0 | 0 | 0 | 0 |
| 0 | 0 | 0 | 0 | 0 |
| 0.025041935 | 0 | 0.001350677 | 0.006930042 | 0.005629248 |
| 0 | 0 | 0 | 0 | 0 |
| 0 | 0 | 0 | 0 | 0 |
| 0 | 0 | 0 | 0.015887885 | 0 |
| 0 | 0 | 0 | 0.00814166 | 0 |
| 0 | 0 | 0 | 0 | 0 |
| 0 | 0 | 0 | 0 | 0 |
| 0 | 0.0413957 | 0 | 0 | 0.003719383 |
| 0.020736495 | 0.001549417 | 0.025653489 | 0.029863962 | 0.003028139 |
| 0 | 0 | 0 | 0 | 0 |
| 0 | 0 | 0 | 0 | 0 |
| 0 | 0 | 0 | 0 | 0 |
| 0 | 0 | 0 | 0 | 0.00225775 |
| 0 | 0 | 0 | 0 | 0 |
| 0 | 0 | 0 | 0 | 0 |
| 0 | 0 | 0 | 0 | 0.006513661 |
| 0 | 0 | 0 | 0 | 0.002905302 |
| 0 | 0 | 0 | 0 | 0 |
| 0 | 0 | 0 | 0 | 0 |
| 0 | 0 | 2.110440393 | 0 | 0 |
| 0 | 0 | 0.001815419 | 0 | 0 |
| 0.010789838 | 0.000490736 | 0.006930192 | 0.010246825 | 0 |

| | | | | |
|---|---|---|---|---|
| 0 | 0 | 0 | 0 | 0 |
| 0 | 0 | 0 | 0 | 0 |
| 0 | 0 | 0 | 0 | 0 |
| 0 | 0 | 0 | 0 | 0 |
| 0 | 0 | 0 | 0 | 0 |
| 0 | 0 | 0 | 0 | 0 |
| 0 | 0 | 0 | 0 | 0.021738831 |
| 0 | 0 | 0 | 0 | 0.002386207 |
| 0 | 0 | 0 | 0 | 0 |
| 0 | 0 | 0 | 0 | 0 |
| 0 | 0 | 0 | 0 | 0 |
| 0 | 0 | 0 | 0 | 0 |
| 0 | 0 | 0 | 0 | 0 |
| 0 | 0 | 0 | 0 | 0 |
| 0 | 0 | 0.010361446 | 0 | 0.002878908 |
| 0 | 0 | 0 | 0 | 0 |
| 0 | 0 | 0 | 0 | 0 |
| 0 | 0.018789053 | 0 | 0 | 0 |
| 0 | 0.007236592 | 0 | 0.002404835 | 0.006752629 |
| 0 | 0.025736283 | 0 | 0 | 0.087635141 |
| 0.058764317 | 0 | 0.021330664 | 0.007685983 | 0 |
| 0 | 0 | 0 | 0 | 0 |
| 0 | 0 | 0 | 0.000984844 | 0 |
| 0 | 0 | 0 | 0 | 0 |
| 0.003495441 | 0 | 0.001780583 | 0.001761903 | 0.00296839 |
| 0 | 0 | 0 | 0 | 0 |
| 0 | 0 | 0 | 0 | 0 |
| 0 | 0 | 0 | 0 | 0 |
| 0 | 0 | 0 | 0 | 0.014399951 |
| 0 | 0 | 0 | 0 | 0 |
| 0.668063557 | 0.053184488 | 0.467106015 | 1.031398811 | 0.004577614 |
| 0 | 0 | 0 | 0 | 0 |
| 0 | 0 | 0 | 0 | 0 |
| 0 | 0.00087543 | 0 | 0 | 0.015398265 |
| 0 | 0 | 0 | 0 | 0 |
| 0 | 0 | 0 | 0 | 0 |
| 0 | 0 | 0 | 0 | 0 |
| 0 | 0 | 0 | 0 | 0 |
| 0 | 0 | 0 | 0 | 0 |
| 0 | 0 | 0 | 0 | 0 |
| 0 | 0 | 0 | 0 | 0 |
| 0.016088212 | 0 | 0.015450267 | 0.08042914 | 0 |
| 0 | 0 | 0.007327107 | 0.002788554 | 0 |
| 0 | 0 | 0 | 0 | 0 |
| 0 | 0 | 0 | 0 | 0 |
| 0 | 0 | 0 | 0 | 0 |
| 0 | 0 | 0 | 0 | 0 |
| 0 | 0 | 0 | 0 | 0 |

| | | | | |
|---|---|---|---|---|
| 0 | 0 | 0 | 0 | 0 |
| 0 | 0 | 0 | 0 | 0 |
| 0.136773393 | 0.011258896 | 0.006073154 | 0.033385792 | 0.014296149 |
| 0 | 0 | 0 | 0 | 0 |
| 0 | 0.035625771 | 0 | 0 | 0 |
| 0 | 0 | 0 | 0 | 0 |
| 0 | 0 | 0 | 0 | 0 |
| 0 | 0 | 0 | 0 | 0 |
| 0 | 0 | 0 | 0 | 0 |
| 0 | 0 | 0 | 0 | 0 |
| 0 | 0 | 0 | 0 | 0 |
| 0 | 0 | 0 | 0 | 0 |
| 0 | 0 | 0 | 0 | 0 |
| 0 | 0 | 0 | 0.001537389 | 0.073818859 |
| 0.017521085 | 0.000474007 | 0.001538839 | 0.002030261 | 0.069906638 |
| 0 | 0.035524128 | 0 | 0 | 0.008926383 |
| 0.002409413 | 0 | 0 | 0 | 0.010717747 |
| 0 | 0 | 0 | 0 | 0 |
| 0 | 0 | 0 | 0 | 0 |
| 0 | 0 | 0 | 0 | 0 |
| 0 | 0 | 0 | 0 | 0 |
| 0.010253629 | 0 | 0.005539772 | 0.009397125 | 0 |
| 0 | 0 | 0 | 0 | 0 |
| 0.002408404 | 0 | 0 | 0 | 0 |
| 0 | 0 | 0 | 0 | 0 |
| 0 | 0 | 0 | 0 | 0 |
| 0 | 1.047753957 | 0 | 0 | 0.039019398 |
| 0 | 0 | 0 | 0 | 0 |
| 0 | 0 | 0 | 0 | 0 |
| 0 | 0 | 0 | 0 | 0.002862629 |
| 0 | 0 | 0 | 0 | 0.009773208 |
| 0 | 0 | 0 | 0 | 0 |
| 0 | 0 | 0 | 0 | 0.016837643 |
| 0 | 0 | 0 | 0 | 0 |
| 0 | 0 | 0 | 0 | 0 |
| 0 | 0 | 0 | 0 | 0.006587922 |
| 0 | 0 | 0 | 0 | 0 |
| 0 | 0.026543891 | 0 | 0.04427443 | 0.002762664 |
| 0 | 0 | 0 | 0 | 0 |
| 0 | 0 | 0 | 0 | 0 |
| 0 | 0 | 0 | 0 | 0 |
| 0 | 0 | 0 | 0 | 0 |
| 0 | 0 | 0 | 0 | 0 |
| 0 | 0 | 0 | 0 | 0 |
| 0 | 0 | 0 | 0 | 0 |
| 0 | 0 | 0 | 0 | 0 |
| 0 | 0 | 0 | 0 | 0 |
| 0.010402725 | 0 | 0.003372195 | 0.196633946 | 0 |

| | | | | |
|---|---|---|---|---|
| 0 | 4.70544919 | 0 | 0 | 0 |
| 0 | 0 | 0 | 0 | 0 |
| 0 | 0 | 0 | 0 | 0.026716588 |
| 0 | 0 | 0 | 0 | 0 |
| 0.006221263 | 0.00137008 | 0.002501937 | 0.006601841 | 0.014830049 |
| 0 | 0 | 0 | 0 | 0 |
| 0 | 0 | 0 | 0 | 0 |
| 0.014228134 | 0.004080995 | 0 | 0 | 0.001299224 |
| 0 | 0 | 0 | 0 | 0 |
| 0.016557536 | 0 | 0.00506066 | 0.022255869 | 0.027787112 |
| 0.037211522 | 0 | 0 | 0 | 0 |
| 0 | 0.284666725 | 0 | 0 | 0.008999113 |
| 0.007165447 | 0 | 0.006879014 | 0.001852204 | 0.015992687 |
| 0.084129827 | 0 | 0.003837838 | 0.105488243 | 0 |
| 0 | 0 | 0 | 0.022859393 | 0 |
| 0 | 0 | 0 | 0 | 0 |
| 0 | 0 | 0 | 0 | 0.002341991 |
| 0 | 0 | 0 | 0.008744421 | 0 |
| 0.018251255 | 0 | 0 | 0 | 0 |
| 0 | 0 | 0 | 0 | 0 |
| 0 | 0 | 0 | 0 | 0 |
| 0 | 0 | 0 | 0 | 0 |
| 0 | 0 | 0 | 0 | 0 |
| 0 | 0.04791215 | 0.072444694 | 0 | 0.037599358 |
| 0 | 0 | 0 | 0 | 0 |
| 0 | 0.001511034 | 0 | 0 | 0 |
| 0 | 0 | 0.008527522 | 0 | 0.011543016 |
| 0.188652775 | 0 | 0.00118642 | 0 | 0 |
| 0 | 0 | 0 | 0 | 0 |
| 0 | 0 | 0 | 0 | 0 |
| 0 | 0 | 0 | 0 | 0 |
| 0 | 0 | 0 | 0 | 0 |
| 0 | 0 | 0 | 0 | 0 |
| 0 | 0 | 0.000855397 | 0 | 0 |
| 0 | 0 | 0 | 0 | 0 |
| 0 | 0 | 0 | 0 | 0 |
| 0.536727171 | 0 | 0 | 0 | 0 |
| 0 | 0 | 0 | 0 | 0 |
| 0 | 0 | 0 | 0 | 0 |
| 0.409854334 | 0 | 0 | 0 | 0 |
| 0 | 0 | 0 | 0 | 0 |
| 0 | 0 | 0 | 0 | 0 |
| 0 | 0 | 0 | 0 | 0 |
| 0.108614682 | 0 | 0 | 0 | 0 |
| 0 | 0 | 0 | 0 | 0 |
| 0 | 0 | 0 | 0 | 0.001163412 |
| 0.002794295 | 0 | 0.00264349 | 0.002012122 | 0 |
| 0 | 0 | 0 | 0 | 0 |

| | | | | |
|---|---|---|---|---|
| 0 | 0 | 0 | 0 | 0 |
| 0 | 0 | 0 | 0 | 0 |
| 0 | 0 | 0 | 0 | 0 |
| 0 | 0 | 0 | 0 | 0 |
| 0 | 0 | 0 | 0.08187111 | 0 |
| 0 | 0 | 0.088087527 | 0.088484092 | 0 |
| 0 | 0 | 0.126620678 | 0 | 0 |
| 0 | 0 | 0 | 0 | 0 |
| 0 | 0.441950685 | 0.00233718 | 0 | 0.123111807 |
| 0 | 0 | 0 | 0 | 0 |
| 0.077307626 | 0 | 0 | 0.007658705 | 0.018580467 |
| 0 | 0 | 0 | 0 | 0 |
| 0.089232349 | 0 | 0 | 0 | 0 |
| 0.004478645 | 0 | 0.000877472 | 0.379143323 | 0.004876081 |
| 0 | 0 | 0 | 0 | 0 |
| 0 | 0 | 0 | 0 | 0 |
| 0 | 0 | 0 | 0.009501299 | 0 |
| 0.011063744 | 0 | 0 | 0 | 0 |
| 0.023467464 | 0 | 0 | 0 | 0 |
| 0.017146205 | 0 | 0.291704645 | 0.060915231 | 0 |
| 0 | 0 | 0 | 0 | 0 |
| 0 | 0.136496389 | 0.168057958 | 0 | 0.177516611 |
| 0 | 0.010772562 | 0.006161827 | 0.212577555 | 0 |
| 0 | 0 | 0 | 0 | 0 |
| 0 | 0 | 0 | 0 | 0 |
| 0 | 0 | 0 | 0 | 0 |
| 0.006723743 | 0 | 0 | 0 | 0 |
| 0 | 0 | 0 | 0 | 0 |
| 0 | 0 | 0 | 0 | 0 |
| 1.659873042 | 0.252501492 | 0.313830538 | 0.367385427 | 0.143660999 |
| 0 | 0.001589192 | 0.005476716 | 0.0007854 | 0 |
| 0 | 0 | 0 | 0 | 0.002089969 |
| 0 | 0 | 0 | 0 | 0 |
| 0 | 0 | 0 | 0 | 0 |
| 0 | 0 | 0 | 0 | 0 |
| 0 | 0 | 0 | 0 | 0 |
| 0 | 0 | 2.27366286 | 0 | 1.596182388 |
| 0 | 0 | 0 | 0 | 0.008697545 |
| 0 | 0 | 0 | 0 | 0 |
| 0 | 0 | 0 | 0 | 0 |
| 0.04729868 | 0.002814602 | 0.001587031 | 0.00214143 | 0 |
| 1.975726034 | 0.576248487 | 1.55699434 | 0 | 0.288566183 |
| 0 | 0 | 0 | 0 | 0.005691081 |
| 0 | 0 | 0 | 0 | 0 |
| 0 | 0 | 0 | 0.00375767 | 0 |
| 0 | 0 | 0 | 0 | 0 |
| 0.002378125 | 0 | 0 | 0.00149839 | 0 |
| 0 | 0 | 0.003401693 | 0 | 0 |

| | | | | |
|---|---|---|---|---|
| 0 | 0.00057551 | 0 | 0 | 0 |
| 0 | 0 | 0 | 0 | 0 |
| 0 | 0 | 0.005855308 | 0 | 0 |
| 0 | 0 | 0 | 0 | 0 |
| 0 | 0.03522591 | 0.740244855 | 0 | 0.02243924 |
| 0 | 0 | 0 | 0 | 0 |
| 0 | 0 | 0 | 0 | 0 |
| 0 | 0 | 0 | 0 | 0.000872744 |
| 0 | 0 | 0 | 0.06163203 | 0 |
| 0 | 0 | 0 | 0.041117165 | 0 |
| 0 | 0 | 0 | 0 | 0 |
| 0 | 0 | 0 | 0 | 0 |
| 0.143424876 | 0 | 0 | 0 | 0 |
| 0.003649942 | 0 | 0.005577857 | 0 | 0.002582995 |
| 0 | 0 | 0 | 0 | 0 |
| 0.001815142 | 0.144713137 | 0 | 0 | 0.014313469 |
| 0.011496549 | 0.025856327 | 0 | 0.018350587 | 0.001423781 |
| 0.185772518 | 0 | 0.001067287 | 0 | 0 |
| 0.002078137 | 0 | 0.008015159 | 0.000997619 | 0 |
| 0 | 0 | 0 | 0.375787232 | 0 |
| 0.09924912 | 0 | 0.009594882 | 0 | 0 |
| 0.00466739 | 0 | 0.006113765 | 0.002520678 | 0 |
| 0 | 0 | 0 | 0 | 0 |
| 0 | 0.029205907 | 0.067071102 | 0.02105635 | 0 |
| 0.036531763 | 0 | 0 | 0 | 0 |
| 0.003786169 | 0 | 0 | 0 | 0 |
| 0.002554118 | 0 | 0 | 0.002758763 | 0 |
| 0 | 0 | 0 | 0.000581954 | 0 |
| 0 | 0 | 0 | 0 | 0 |
| 0 | 0 | 0 | 0 | 0 |
| 0.110716782 | 0.000435404 | 0.024877884 | 0.541058926 | 0 |
| 0.004550668 | 0 | 0.012644274 | 0 | 0.03161869 |
| 0.012604658 | 0 | 0.004414322 | 0.027796443 | 0.004738792 |
| 0 | 0 | 0 | 0 | 0.052731292 |
| 0 | 0 | 0 | 0.001692385 | 0.009979435 |
| 0 | 0 | 0 | 0 | 0 |
| 0 | 0 | 0 | 0 | 0.002268103 |
| 0.055698707 | 0.013546558 | 0.149242001 | 0.002105652 | 0 |
| 0 | 0.024779598 | 0.070082524 | 0 | 0.024303769 |
| 0 | 0.00512102 | 0 | 0.005738617 | 0 |
| 0 | 0 | 0 | 0 | 0 |
| 0 | 0 | 0 | 0 | 0 |
| 0 | 0 | 0.138327612 | 0.03848776 | 0 |
| 0.486755311 | 0 | 0 | 0.052964328 | 0 |
| 0.014889955 | 0 | 0 | 0 | 0 |
| 0 | 0 | 0 | 0 | 0 |
| 0 | 0 | 0 | 0 | 0 |
| 0 | 0 | 0 | 0 | 0 |

| | | | | |
|---|---|---|---|---|
| 0 | 0 | 0 | 0 | 0 |
| 0.491809676 | 0.105592869 | 1.750027166 | 0 | 0.444454928 |
| 0 | 0 | 0 | 0 | 0 |
| 0 | 0 | 0 | 0 | 0 |
| 0 | 0 | 0 | 0.002502077 | 0 |
| 0 | 0 | 0 | 0 | 0 |
| 0 | 0 | 0 | 0 | 0 |
| 0 | 0 | 0 | 0 | 0 |
| 0 | 0 | 0 | 0 | 0 |
| 0.043018983 | 0 | 0 | 0 | 0 |
| 0.005357947 | 0.00132745 | 0.000718249 | 0.000947619 | 0 |
| 0.022262269 | 0.001318183 | 0.004065438 | 0.00564602 | 0.023780513 |
| 0 | 0.028041775 | 0 | 0 | 0.011514526 |
| 0 | 0 | 0 | 0 | 0 |
| 0.00097067 | 0 | 0 | 0.043490961 | 0 |
| 0 | 0 | 0 | 0 | 0 |
| 0.011221087 | 0.002007824 | 0.008874902 | 0.001736514 | 0.009194787 |
| 0.336179974 | 0 | 0 | 0.039932159 | 0 |
| 0 | 0 | 0 | 0 | 0 |
| 0.061979623 | 0 | 0.000887581 | 0 | 0 |
| 0.007308487 | 0.051764319 | 0.065306823 | 0.001790784 | 0.065728505 |
| 0 | 0 | 0 | 0 | 0 |
| 0 | 0 | 0 | 0 | 0 |
| 0 | 0 | 0 | 0 | 0.014254485 |
| 0 | 0 | 0 | 0 | 0 |
| 0 | 0 | 0 | 0 | 0 |
| 0.021942103 | 0.015878434 | 0.02740745 | 0.559316966 | 0 |
| 0.072176209 | 0.011511423 | 0.019290135 | 0.020936727 | 0.050161554 |
| 0 | 0 | 0 | 0 | 0 |
| 0.00156164 | 0.000612594 | 0.001392128 | 0 | 0 |
| 0.032992321 | 0 | 0 | 0 | 0.003686533 |
| 0 | 0 | 0 | 0 | 0 |
| 0 | 0 | 0 | 0 | 0 |
| 0 | 0 | 0 | 0 | 0 |
| 0 | 0.190596668 | 0.021263944 | 0 | 0 |
| 0.578998662 | 0.077929585 | 0.131984799 | 0.53427353 | 0.12676168 |
| 0 | 0 | 0 | 0 | 0 |
| 0 | 0 | 0 | 0.008154617 | 0 |
| 0 | 0.110681363 | 0.009152515 | 0.005366815 | 0.066118236 |
| 0 | 0 | 0 | 0 | 0 |
| 0.016535944 | 0 | 0 | 0 | 0 |
| 0 | 0 | 0 | 0 | 0 |
| 0.006790652 | 0.004820232 | 0.027920429 | 0.001833685 | 0 |
| 0.021776105 | 0 | 0.00083494 | 0 | 0.001657047 |
| 0 | 0 | 0 | 0 | 0 |
| 0 | 0.101229039 | 0 | 0 | 0.606073313 |
| 0 | 0 | 0 | 0 | 0 |
| 0 | 0 | 0 | 0 | 0 |

| | | | | |
|---|---|---|---|---|
| 0 | 0 | 0 | 0 | 0 |
| 0 | 0 | 0 | 0 | 0 |
| 0 | 0 | 0 | 0 | 0.002831173 |
| 0 | 0.21362918 | 0 | 0 | 1.014660294 |
| 0 | 0 | 0 | 0 | 0 |
| 0 | 0 | 0 | 0 | 0 |
| 0 | 0 | 0 | 0 | 0 |
| 0 | 0.00581537 | 0 | 0 | 0 |
| 0.175392222 | 4.567350945 | 0.587035874 | 0 | 0.053318399 |
| 0 | 0.023239297 | 0.002314793 | 0 | 0.064673394 |
| 0 | 0 | 0 | 0 | 0.008142243 |
| 0 | 0.004529565 | 0.008547272 | 0 | 0.001340637 |
| 0 | 0 | 0 | 0 | 0 |
| 0 | 0 | 0 | 0 | 0.001199689 |
| 0.054491155 | 0.034925149 | 0.030146872 | 0 | 0 |
| 0 | 0 | 0 | 0 | 0 |
| 0 | 0.049975349 | 0.483016866 | 0 | 0 |
| 0 | 0.637277548 | 0.164005559 | 0 | 0.138068012 |
| 0 | 0 | 0 | 0 | 0 |
| 0 | 0.002907361 | 0 | 0 | 0 |
| 0 | 0.041927692 | 0 | 0 | 0 |
| 0 | 0.03968817 | 0 | 0 | 0.023648987 |
| 0 | 0.022809955 | 0 | 0 | 0 |
| 0.314718573 | 0.079772862 | 0.13555263 | 0.744386871 | 0 |
| 0 | 0 | 0.003614001 | 0 | 0 |
| 0 | 0.078944309 | 0 | 0 | 0.741229039 |
| 0 | 0 | 0.045765385 | 0 | 0.012801167 |
| 0 | 3.947440669 | 0.708300084 | 0 | 3.628382266 |
| 0 | 0 | 0 | 0 | 0 |
| 0 | 0 | 0 | 0 | 0 |
| 0 | 0.007430922 | 0 | 0 | 0.017874215 |
| 0 | 0.06521588 | 0.156453756 | 0 | 0.001403402 |
| 0 | 0 | 0.001326014 | 0.038690232 | 0 |
| 0 | 0 | 0 | 0 | 0 |
| 0 | 0 | 0 | 0 | 0 |
| 0.019407782 | 0.394020301 | 0.060227207 | 0 | 0.006946195 |
| 0 | 1.541556358 | 0.803961246 | 0 | 0.738684316 |
| 0 | 0 | 0 | 0.099590362 | 0 |
| 0 | 0.001178819 | 0.000861069 | 0.071949676 | 0 |
| 0.00968548 | 0 | 0.00049338 | 0.009601344 | 0 |
| 0.023106262 | 0.001357909 | 0.103023749 | 0.117995602 | 0.042441364 |
| 0 | 0 | 0.001033761 | 0 | 0.035265284 |
| 0 | 0 | 0.074407623 | 0 | 0.002865269 |
| 0 | 0.007807245 | 0.007258114 | 0 | 0 |
| 0 | 0 | 0 | 0 | 0 |
| 0 | 0.000926923 | 0 | 0 | 0 |
| 0 | 0 | 0 | 0 | 0 |
| 0 | 0.031341352 | 0.00397453 | 0.00524378 | 0 |

| | | | | |
|---|---|---|---|---|
| 0 | 0 | 0 | 0 | 0 |
| 0 | 0.279877505 | 0.044754444 | 0 | 0.052516485 |
| 0.974720167 | 0.013457309 | 0.011430108 | 1.181036127 | 0.013550225 |
| 0 | 0 | 0 | 0 | 0.002059755 |
| 0 | 0.042301641 | 0 | 0 | 0 |
| 0 | 0.01343634 | 0.015967445 | 0 | 0.005337172 |
| 0 | 0 | 0 | 0 | 0 |
| 0 | 0 | 0 | 0 | 0 |
| 0 | 0 | 0 | 0 | 0 |
| 0 | 0 | 0 | 7.051597763 | 0 |
| 0.027611486 | 0.016439045 | 0.078556374 | 0.325488453 | 0.000623621 |
| 4.780113838 | 0.220343474 | 1.116644639 | 1.219653514 | 0.610627269 |
| 0 | 0 | 0 | 0 | 0 |
| 0.015216736 | 0.008124698 | 0.002422321 | 0.001331616 | 0.001794766 |
| 0.616306528 | 0.007186562 | 0.028760489 | 0.158034338 | 0 |
| 0 | 4.820209132 | 0 | 0 | 0.06791405 |
| 0 | 0 | 0 | 0 | 0 |
| 0 | 0 | 0 | 0 | 0 |
| 0 | 0.023920308 | 0 | 0 | 0.048733274 |
| 0 | 0.002657667 | 0.379877205 | 0.085171302 | 0.009760294 |
| 0.056379048 | 0.043009026 | 0.168680446 | 0.106930516 | 2.631900068 |
| 0.932077945 | 0 | 0.002378635 | 6.315711184 | 0 |
| 1.4715053 | 0.007297557 | 0.190950412 | 15.00598842 | 0.570705567 |
| 0 | 0 | 0 | 0 | 0 |
| 0 | 0 | 0 | 0 | 0.023735814 |
| 0 | 0.123906541 | 0.006011294 | 0 | 0.798785963 |
| 0 | 0 | 0 | 0 | 0 |
| 0 | 0 | 0 | 0 | 0 |
| 0 | 0 | 0 | 0 | 0 |
| 0 | 0 | 0 | 0 | 0.002132469 |
| 0 | 0.016475418 | 0 | 0 | 0.002498208 |
| 0 | 0.055556721 | 0.178783182 | 0 | 0.258570941 |
| 0.376192244 | 0 | 0.002458802 | 0.001013753 | 0.001067459 |
| 0.016793847 | 0.015040706 | 0.028008215 | 0 | 0 |
| 0.033649777 | 0.021244991 | 0.207216369 | 0.237359573 | 0 |
| 0.012584059 | 0.042210044 | 0.077063412 | 0.483086027 | 0.029814071 |
| 0.112575396 | 0.001935053 | 0.001480767 | 0 | 0 |
| 0 | 0 | 0 | 0 | 0 |
| 0.029131651 | 0.030808633 | 0.231564649 | 0.081757229 | 0.209330902 |
| 0 | 0 | 0 | 0 | 0 |
| 0 | 0.019402992 | 0.189953914 | 0 | 0.014149231 |
| 0 | 0 | 0 | 0 | 0 |
| 0 | 0 | 0 | 0 | 0.367589515 |
| 0 | 0 | 0 | 0 | 0 |
| 0.010919003 | 0.025291672 | 0.005562157 | 0.101951455 | 0 |
| 0 | 0.025097711 | 0 | 0 | 0.052728607 |
| 0 | 0.283234451 | 0 | 0 | 0 |
| 0.016558208 | 0 | 0.088948545 | 0 | 0 |

| | | | | |
|---|---|---|---|---|
| 0 | 0 | 0 | 0 | 0 |
| 0 | 0 | 0 | 0 | 0 |
| 0 | 0 | 0 | 0 | 0 |
| 0 | 0.001251638 | 0.001760796 | 0 | 0.003010669 |
| 0 | 0 | 0 | 0 | 0 |
| 0 | 0.079591598 | 0.219033163 | 0 | 0.027587165 |
| 0 | 1.526672461 | 0 | 0 | 0.026986895 |
| 0.009052747 | 0 | 0.053343637 | 0.208217373 | 0 |
| 0 | 0 | 0 | 0 | 0 |
| 0 | 0 | 0 | 0 | 0 |
| 0.015695068 | 0.002633099 | 0.019007197 | 0.009453656 | 0 |
| 0 | 0 | 0 | 0 | 0 |
| 0.009075077 | 0.007458918 | 0.067097475 | 0 | 0.024037606 |
| 0 | 0.48480143 | 0 | 0 | 0 |
| 0 | 0 | 0 | 0 | 0 |
| 0 | 0 | 0 | 0 | 0 |
| 0 | 0.026149494 | 0.055535662 | 0 | 0.006078673 |
| 0 | 0.00606022 | 0.005274353 | 0.004556288 | 0 |
| 0 | 0 | 0 | 0 | 0.001101473 |
| 0 | 0.002441949 | 0.045300882 | 0 | 0 |
| 0.001533122 | 0.008362427 | 0 | 0 | 0 |
| 0 | 0 | 0 | 0 | 0.689797564 |
| 0 | 0.03909747 | 0 | 0 | 0.006235358 |
| 0 | 0 | 0 | 0 | 0 |
| 0 | 0 | 0 | 0 | 0 |
| 0 | 0 | 0 | 0 | 0 |
| 0 | 0 | 0 | 0 | 0 |
| 0 | 0 | 0 | 0 | 0 |
| 0 | 0 | 0 | 0 | 0 |
| 0 | 0 | 0.637440484 | 0 | 1.808207806 |
| 0 | 0 | 0 | 0.003722903 | 0 |
| 0 | 0 | 0 | 0 | 0 |
| 0 | 0.057660691 | 0 | 0 | 0.004010195 |
| 0 | 0 | 0 | 0 | 0 |
| 0 | 0 | 0 | 0 | 0 |
| 0 | 0 | 0 | 0 | 0 |
| 0 | 0.024734687 | 0.005293305 | 0 | 0.080440197 |
| 0.012032739 | 0 | 0.083897466 | 0.034537905 | 0 |
| 0 | 0.009928821 | 0 | 0 | 0.001235308 |
| 0 | 0 | 0 | 0 | 0 |
| 0 | 0.010948675 | 0.006589669 | 0.186329338 | 0.000665792 |
| 0 | 0 | 0 | 0 | 0 |
| 0 | 0.026901306 | 0 | 0 | 0.007799618 |
| 0 | 0.347225422 | 0 | 0 | 0.205873787 |
| 0 | 0 | 0.050314844 | 0 | 0.535224108 |
| 0 | 0 | 0 | 0 | 0.011732952 |
| 0 | 0.0333879 | 0 | 0 | 0.001685625 |
| 0 | 0 | 0 | 0 | 2.504016605 |

| | | | | |
|---|---|---|---|---|
| 0 | 0 | 0 | 0 | 0 |
| 0 | 0 | 0 | 0 | 0 |
| 0 | 0.145806594 | 0 | 0 | 0 |
| 0 | 0.586766463 | 0.118384411 | 0 | 0.227814994 |
| 0 | 0 | 0 | 0 | 0 |
| 0 | 0 | 0 | 0 | 0.65163484 |
| 0 | 0 | 0 | 0 | 0 |
| 0 | 0 | 0 | 0.033367818 | 0 |
| 0.036301906 | 0 | 0.038041189 | 0.065060443 | 0 |
| 0 | 0 | 0 | 0 | 0 |
| 0 | 0 | 0 | 0 | 0 |
| 0 | 0 | 0 | 0 | 0.001342455 |
| 0 | 0 | 0.00493167 | 0.001863864 | 0 |
| 0 | 0 | 0 | 0 | 0 |
| 0 | 0.125271194 | 0 | 3.786877542 | 2.119044507 |
| 0 | 0 | 0.324820973 | 3.28621583 | 0 |
| 0 | 0 | 0 | 0.027288143 | 0 |
| 0 | 0 | 0 | 0 | 0 |
| 0 | 0 | 0 | 0 | 0 |
| 0 | 0 | 0 | 0 | 0.011586059 |
| 0 | 0.078122911 | 0.130626418 | 0 | 0.064731078 |
| 0 | 0.004744754 | 0 | 0 | 0 |
| 0 | 0 | 0 | 0 | 0 |
| 0 | 0 | 0 | 0 | 0 |
| 0.010570859 | 2.078665429 | 0.008077217 | 0.062163748 | 0.012084367 |
| 0 | 0.143309319 | 0 | 0 | 11.73687228 |
| 0 | 0.102030807 | 0 | 0 | 0.306921654 |
| 0 | 0 | 0 | 0.005724583 | 0 |
| 0 | 0 | 0.000724287 | 0 | 0.285260183 |
| 0 | 0 | 0 | 0 | 0 |
| 0 | 0 | 0 | 0 | 0 |
| 0 | 0.000821691 | 0 | 0 | 0 |
| 0 | 0.001235719 | 0 | 0 | 0 |
| 0 | 0.000794986 | 0 | 0.003101046 | 0 |
| 0.014668585 | 0.008607818 | 0 | 0 | 0 |
| 0.002276102 | 0 | 0.026925005 | 0.054814805 | 0 |
| 0 | 0 | 0 | 0 | 0 |
| 0 | 0.040925235 | 0.496339629 | 0 | 0.717827916 |
| 0 | 0 | 3.38049247 | 0 | 4.973209459 |
| 0 | 0 | 0 | 0 | 0 |
| 0 | 0 | 0 | 0 | 0 |
| 0 | 0.013800161 | 0 | 0 | 0 |
| 0 | 0.247998811 | 0.000528136 | 0 | 0.24408034 |
| 0 | 1.48168863 | 0 | 0 | 0.078654314 |
| 0 | 1.04903802 | 0 | 0 | 0.005142673 |
| 0 | 0.000501259 | 0 | 0.002817915 | 0 |
| 0 | 0 | 0 | 0 | 0 |
| 0 | 0.211020822 | 0.042776211 | 0.262622663 | 5.642541266 |

| | | | | |
|---|---|---|---|---|
| 0 | 0 | 0.001449921 | 0.042084847 | 0 |
| 0 | 0 | 0 | 0 | 0.016625467 |
| 0 | 0 | 0 | 0 | 0 |
| 0 | 0 | 0 | 0 | 0 |
| 0 | 0 | 0 | 0.004523332 | 0 |
| 0 | 0 | 0 | 0 | 0 |
| 0 | 0.014631548 | 0 | 0 | 0 |
| 0 | 0 | 0 | 0 | 0 |
| 0 | 0 | 0 | 0 | 0 |
| 0 | 0 | 0 | 0 | 0.01006449 |
| 0 | 0.001360972 | 0.013917711 | 0 | 0.000767264 |
| 0 | 0.026254548 | 2.187582569 | 0 | 0.422762345 |
| 0 | 0.101758624 | 0.003670598 | 0 | 0.015055203 |
| 0 | 0 | 0 | 0 | 0 |
| 0.014748035 | 0.002571247 | 0.007512671 | 0.002753281 | 0.515351275 |
| 0 | 0 | 0 | 0 | 0 |
| 0 | 0.002027163 | 0 | 0.003447548 | 0 |
| 0 | 0 | 0 | 0.018382179 | 0.000619392 |
| 0 | 0 | 0 | 0 | 0 |
| 0 | 0.002998777 | 0.001947073 | 0 | 0.00247954 |
| 0 | 0.124265452 | 0.002665244 | 0 | 0.110406725 |
| 0 | 0 | 0 | 0 | 0 |
| 0.636009711 | 0.019430467 | 0.016078272 | 0 | 0.621471066 |
| 0 | 0 | 0 | 0 | 0 |
| 0 | 0 | 0 | 0 | 0.006787318 |
| 0 | 0 | 0 | 0 | 0 |
| 0 | 0.210937929 | 0 | 0 | 0.004015263 |
| 0 | 0 | 0 | 0 | 0 |
| 0 | 0.000463209 | 0 | 0 | 0.002089114 |
| 0 | 0 | 0 | 0 | 0 |
| 0 | 0 | 0 | 0 | 0 |
| 0.135664586 | 0.006005794 | 0.017764372 | 0.001429107 | 0.112259293 |
| 0 | 0.804131549 | 0 | 0 | 0.003371243 |
| 0 | 0.032337552 | 0.009740606 | 0.102631332 | 0.001202848 |
| 0.003027408 | 0.000719746 | 0.025095245 | 0.047398101 | 0 |
| 0 | 0.023752692 | 0.070304283 | 0.022652411 | 0.142110469 |
| 0 | 0 | 0 | 0 | 0 |
| 0 | 0 | 0.018232261 | 0 | 0.008567161 |
| 0 | 0 | 0 | 0 | 0 |
| 0 | 0 | 0 | 0 | 0 |
| 0 | 0.004415227 | 0.122076135 | 0 | 0.341841316 |
| 0 | 0.032863131 | 0 | 0 | 0.019699566 |
| 0 | 0 | 0 | 0 | 0 |
| 0 | 0 | 0 | 0 | 0 |
| 0.033889772 | 0.057005344 | 0.0034527 | 0.061952153 | 0 |
| 0 | 0 | 0 | 0 | 0 |
| 0 | 0.161904986 | 0 | 0 | 0.125270516 |
| 0 | 0.000536486 | 0 | 0 | 0 |

| | | | | |
|---|---|---|---|---|
| 0 | 0 | 0 | 0 | 0 |
| 0 | 0.022140539 | 0 | 0 | 0 |
| 0 | 0.001173289 | 0 | 0.005418025 | 0 |
| 0 | 0 | 0 | 0 | 0 |
| 0 | 0 | 0 | 0 | 0 |
| 0 | 0 | 0.027648519 | 0 | 0.002902122 |
| 0 | 0.252978944 | 0.277516878 | 0 | 0.138924635 |
| 0 | 0 | 0.012545454 | 0 | 0.060906587 |
| 0 | 0 | 0 | 0 | 0 |
| 0 | 0.004770393 | 0 | 0 | 0.002028549 |
| 0 | 0 | 0 | 0 | 0 |
| 0 | 0 | 0 | 0.004074611 | 0 |
| 0 | 0 | 0 | 0 | 0 |
| 0 | 0 | 0.321083467 | 0 | 0 |
| 0 | 0 | 0 | 0 | 0 |
| 0 | 0 | 0 | 0 | 0 |
| 0 | 0.145488466 | 0 | 0 | 0 |
| 0 | 0.039664264 | 0 | 0 | 0.000662554 |
| 0 | 0.029775332 | 3.180753658 | 0 | 5.034584819 |
| 0 | 0 | 0 | 0 | 0 |
| 0 | 0 | 0 | 0 | 0 |
| 0 | 0 | 0 | 0 | 0 |
| 0 | 0 | 0 | 0 | 0 |
| 0 | 0 | 0 | 0 | 0.006529859 |
| 0 | 0 | 0 | 0 | 0.002934812 |
| 0.012872439 | 0 | 0.010543013 | 0.14906472 | 0 |
| 0 | 0 | 0 | 0 | 0 |
| 0.000988201 | 0.012792376 | 0.049206456 | 0.001037729 | 0.496699842 |
| 0 | 0.058942922 | 0 | 0 | 0.025165182 |
| 0 | 0.495715636 | 2.170230812 | 0 | 0.214785685 |
| 0 | 0 | 0 | 0 | 0.036321189 |
| 0 | 0 | 0 | 0 | 0 |
| 0 | 0.000818645 | 0 | 0 | 0.081227642 |
| 0 | 0 | 0 | 0 | 0 |
| 0.002909504 | 0 | 0.192808495 | 0.004221908 | 0 |
| 0 | 0.853552107 | 0 | 0 | 0 |
| 0 | 0.100075423 | 0 | 0 | 0.080315079 |
| 0 | 0 | 0 | 0.085474912 | 0 |
| 0 | 0 | 0 | 0 | 0 |
| 0 | 0 | 0 | 0 | 0.000372895 |
| 27.37884794 | 0.959111571 | 1.229977488 | 17.336107 | 1.406875536 |
| 0.214204448 | 0.099970994 | 0 | 0 | 0 |
| 0 | 2.634439629 | 0.129253917 | 0 | 0 |
| 0 | 0.020544055 | 0 | 0 | 0.031889443 |
| 0 | 0.241378318 | 0 | 0 | 0 |
| 0 | 0 | 0 | 0 | 0 |
| 0 | 0.000712552 | 0 | 0 | 0 |
| 0.003334802 | 0.004609721 | 0.002305451 | 0.002401332 | 0 |

| | | | | |
|---|---|---|---|---|
| 0 | 0 | 0 | 0 | 0 |
| 0.00171506 | 0.004613331 | 0.05653789 | 0.006792412 | 0 |
| 0 | 0 | 0 | 0 | 0 |
| 0 | 0 | 0 | 0 | 0.00487813 |
| 0 | 0.41165722 | 0 | 0 | 1.000203925 |
| 0 | 0.001576871 | 0 | 0 | 0 |
| 0 | 0 | 0 | 0 | 0 |
| 0 | 0 | 0 | 0 | 0.005331249 |
| 3.49356341 | 0 | 0.006106561 | 0.277872763 | 0 |
| 0 | 0 | 0 | 0 | 0 |
| 0 | 0 | 0 | 0 | 0 |
| 0 | 0 | 0 | 0 | 0 |
| 0 | 0.027767446 | 0.137522398 | 0 | 0.066716465 |
| 0.022243667 | 0.007807174 | 0.00775276 | 0.052126373 | 0.013877309 |
| 0 | 1.716517762 | 0.057972434 | 0 | 0.214330569 |
| 0 | 0.071602175 | 0.012234344 | 0 | 0.199198352 |
| 0 | 0 | 0 | 0.001489392 | 0.006810881 |
| 0 | 0 | 0 | 0 | 0 |
| 0 | 0 | 0 | 0 | 0.007198717 |
| 0 | 0 | 0 | 0 | 0.000927737 |
| 0 | 0.002537318 | 0 | 0 | 0 |
| 0 | 0 | 0 | 0 | 0 |
| 0 | 0 | 0 | 0 | 0 |
| 0 | 0 | 0 | 0 | 0 |
| 0 | 0 | 0 | 0 | 0.005936409 |
| 0 | 0 | 0 | 0 | 0 |
| 0 | 0.204456717 | 0.014696152 | 0 | 0.076522877 |
| 0 | 0 | 0.001772225 | 0.008559401 | 0 |
| 0 | 0 | 0 | 0 | 0 |
| 0 | 0.003709346 | 0 | 0 | 0 |
| 0 | 0 | 0 | 0 | 0 |
| 0 | 0 | 0 | 0 | 0 |
| 0 | 1.361379935 | 0 | 0 | 0 |
| 0 | 0.013216156 | 0.022027785 | 0 | 0.038897586 |
| 0 | 0.270081192 | 0.227100543 | 0 | 0.426259293 |
| 0.003162123 | 0 | 0 | 0.060133199 | 0 |
| 0 | 0.354848508 | 0 | 0 | 0 |
| 0 | 0 | 0 | 0 | 0.058590336 |
| 0 | 0 | 0 | 0 | 0 |
| 0 | 0 | 0 | 0 | 0.003961428 |
| 0 | 0.413407346 | 3.262120358 | 0 | 0.214023656 |
| 0 | 0 | 0.002253689 | 0 | 0 |
| 0 | 0.137762799 | 0.007315632 | 0 | 0 |
| 0 | 0 | 0.049578282 | 0 | 0.259577031 |
| 0 | 0 | 0 | 0 | 0 |
| 0 | 0 | 0 | 0 | 0 |
| 0 | 0 | 0 | 0 | 0 |
| 0 | 0 | 0 | 0 | 0 |

| | | | | |
|---|---|---|---|---|
| 0 | 0.110148725 | 0.296745742 | 0 | 0.134015344 |
| 0 | 0 | 0 | 0 | 0 |
| 0 | 0 | 0 | 0 | 0 |
| 0 | 0 | 0.002150982 | 0.001655436 | 0 |
| 0 | 0.003521919 | 0.002058069 | 0 | 0.01753614 |
| 0 | 0 | 0 | 0 | 1.097338298 |
| 0 | 0.000792879 | 0.005276777 | 0.060279816 | 0.032720013 |
| 0 | 0 | 0 | 0 | 0 |
| 0 | 0 | 0 | 0 | 0 |
| 0 | 0 | 0.432768666 | 0 | 0 |
| 0 | 0.071310914 | 0.006867173 | 0 | 0.003252326 |
| 0 | 0.014445574 | 0 | 0 | 0 |
| 0 | 0 | 0 | 0 | 0 |
| 0 | 0 | 0 | 0 | 0 |
| 0 | 0 | 0 | 0 | 0 |
| 0 | 0 | 0 | 0 | 0 |
| 0 | 0 | 0 | 0.03674987 | 0 |
| 0 | 0 | 0 | 0 | 0 |
| 0 | 0 | 0 | 0 | 0.005548282 |
| 0 | 0 | 0 | 0 | 0 |
| 0.001202699 | 0.025917028 | 0.015806547 | 0.001212459 | 3.455919121 |
| 0 | 0 | 0.003090857 | 0 | 0.002477275 |
| 0 | 0.016374729 | 0.053384911 | 0 | 0.003755164 |
| 0.00471841 | 0.004318816 | 0.004086061 | 0.001387371 | 0 |
| 0 | 0 | 0 | 0 | 0 |
| 0 | 0 | 0 | 0 | 0 |
| 0 | 0 | 0 | 0 | 0.157921928 |
| 0 | 0.024292995 | 0 | 0 | 0 |
| 0 | 0 | 0 | 0 | 0 |
| 0.592845475 | 0 | 0.005361906 | 0.163733682 | 0 |
| 0 | 0.005458112 | 0 | 0 | 0.10920339 |
| 0 | 0 | 0 | 0 | 0.001550276 |
| 0 | 0 | 0 | 0 | 0.002466646 |
| 0 | 0 | 0 | 0 | 0 |
| 0 | 0.254266172 | 0 | 0 | 0 |
| 0 | 0 | 0 | 0 | 0 |
| 0.010922314 | 0 | 0.004962346 | 0.000743983 | 0.002715776 |
| 0 | 0 | 0 | 0 | 0 |
| 0 | 0.001565038 | 0 | 0 | 0 |
| 0 | 0 | 0.041197498 | 0 | 0.06837599 |
| 0 | 0 | 0 | 0 | 0 |
| 0 | 0 | 0 | 0 | 0 |
| 0 | 0 | 0 | 0 | 0 |
| 0 | 0 | 0 | 0 | 0.000797387 |
| 0 | 0.602919096 | 0 | 0 | 0.005140554 |
| 0 | 0 | 0 | 0 | 1.531860138 |
| 0 | 0 | 0 | 0 | 0 |
| 0 | 0 | 0 | 0 | 0 |

| | | | | |
|---|---|---|---|---|
| 0 | 0 | 0.13552325 | 0.185327969 | 0 |
| 0 | 0 | 0.009934068 | 0 | 0.186225748 |
| 0 | 0 | 0 | 0 | 0.029035662 |
| 0 | 0 | 0 | 0 | 0 |
| 0 | 0 | 0 | 0 | 0 |
| 0 | 0 | 0 | 0 | 0 |
| 0 | 0 | 0 | 0 | 0 |
| 0 | 0.076345967 | 0.002257233 | 0 | 0.505976854 |
| 0.407634408 | 3.200191092 | 0.110421668 | 0.687916766 | 0.281707462 |
| 0.110961035 | 0 | 0.026142218 | 0.128640716 | 0 |
| 0 | 0 | 0 | 0 | 0.191667311 |
| 0 | 0 | 0 | 0 | 0 |
| 0 | 0 | 0 | 0 | 0.031962295 |
| 0 | 0 | 0 | 0 | 0.027139025 |
| 0 | 0 | 0 | 0 | 0 |
| 0 | 0.567999035 | 0 | 0 | 0 |
| 0 | 0 | 0 | 0 | 0 |
| 0 | 0 | 0 | 0 | 0 |
| 0 | 0 | 0 | 0 | 0 |
| 0 | 0 | 0 | 0 | 0 |
| 0 | 0 | 0 | 0 | 0.197251242 |
| 0 | 0 | 0 | 0 | 0 |
| 0 | 0 | 0 | 0 | 0 |
| 0 | 0 | 0 | 0 | 0 |
| 0 | 0.012758807 | 0.001731524 | 0 | 0.01542352 |
| 0 | 0 | 0 | 0 | 0 |
| 0 | 0 | 0 | 0 | 0 |
| 0 | 0 | 0 | 0 | 0 |
| 0 | 0 | 0 | 0 | 0 |
| 0 | 0 | 0 | 0 | 0.006385404 |
| 0 | 0 | 0 | 0 | 0 |
| 0 | 0 | 0 | 0 | 0 |
| 0 | 1.010151722 | 0 | 0 | 0 |
| 0 | 0.007179252 | 0 | 0 | 0 |
| 0 | 0.011412922 | 0.001722108 | 0 | 0 |
| 0 | 0 | 0 | 0 | 0.007831906 |
| 0 | 0 | 0.003405775 | 0 | 0.003595895 |
| 0 | 0 | 0 | 0.031675606 | 0.002797405 |
| 0 | 0 | 0 | 0 | 0 |
| 0.497991213 | 0 | 0 | 0 | 0 |
| 0 | 0 | 0 | 0 | 0 |
| 0 | 0.08259826 | 0 | 0.43402913 | 0.067165671 |
| 0 | 0 | 0 | 0 | 0.002991725 |
| 0 | 0 | 0 | 0 | 0 |
| 0 | 0 | 0.000377409 | 0 | 0 |
| 0 | 0 | 0 | 0 | 0 |
| 0 | 0 | 0 | 0 | 0 |
| 0 | 0 | 0 | 0 | 0 |

| | | | | |
|---|---|---|---|---|
| 0 | 0 | 0 | 0 | 0 |
| 0 | 0.001198371 | 0 | 0 | 0.01718716 |
| 0 | 0 | 0 | 0 | 0 |
| 0 | 0 | 0 | 0 | 0.012271242 |
| 0.178109144 | 0.28114753 | 0.42786259 | 0.854983053 | 0.943293414 |
| 0 | 0 | 0 | 0 | 0 |
| 0 | 0 | 0 | 0 | 0 |
| 0.007750244 | 0 | 0.082085209 | 0.101733572 | 0 |
| 0 | 0.001180497 | 0 | 0 | 0.002738136 |
| 0 | 0 | 0 | 0 | 0.014395153 |
| 0 | 0 | 0 | 0 | 0 |
| 0 | 0 | 0.26421416 | 0 | 0.007170416 |
| 0 | 0 | 0 | 0 | 0 |
| 0 | 0 | 0 | 0 | 0 |
| 0 | 0 | 0.002663441 | 0 | 0 |
| 0.037529654 | 0.0011719 | 0.005706765 | 0.291285775 | 0 |
| 0 | 0 | 0 | 0 | 0 |
| 0 | 0 | 0 | 0 | 0 |
| 0 | 0 | 0 | 0 | 0 |
| 0 | 0 | 0 | 0 | 0 |
| 0 | 0 | 0 | 0 | 0.020558085 |
| 0 | 0.036869389 | 0 | 0 | 0 |
| 0 | 0 | 0 | 0 | 0 |
| 0.013414397 | 0 | 0.018649252 | 0.00187823 | 0 |
| 0 | 0.00598235 | 0.307786515 | 0 | 0.455039705 |
| 0 | 0 | 0 | 0 | 0 |
| 0 | 0.015909091 | 0.274081558 | 0.183595694 | 0 |
| 0 | 0 | 0 | 0 | 0 |
| 0 | 0 | 0 | 0.006064274 | 0 |
| 0 | 0 | 0 | 0 | 0 |
| 0 | 0 | 0 | 0 | 0 |
| 0 | 0 | 0 | 0 | 0 |
| 0 | 0 | 0 | 0 | 0 |
| 0 | 19.14269311 | 0 | 0 | 0 |
| 0 | 0 | 0 | 0 | 0 |
| 0 | 0 | 0 | 0 | 0 |
| 0 | 0 | 0 | 0 | 0 |
| 0 | 0.042228884 | 0.018828135 | 0 | 0.002040602 |
| 0 | 0 | 0 | 0 | 0 |
| 0 | 0 | 0.001260138 | 0 | 0 |
| 0 | 0 | 0 | 0 | 0 |
| 0 | 0.354041908 | 0.001990485 | 0 | 0 |
| 0 | 0 | 0 | 0 | 0 |
| 0 | 0 | 0.060058991 | 0 | 0 |
| 0 | 0 | 0 | 0 | 0 |
| 0 | 0 | 0 | 0 | 0 |
| 0 | 0 | 0 | 0 | 0 |
| 0 | 0 | 0 | 0 | 0.002929264 |

| | | | | |
|---|---|---|---|---|
| 0 | 0.907348462 | 0 | 0 | 0.024665628 |
| 0 | 0 | 0 | 0 | 0 |
| 0 | 0 | 0 | 0 | 0 |
| 0 | 0 | 0 | 0 | 0 |
| 0 | 0 | 0 | 0 | 0.003466351 |
| 0.007797624 | 0 | 0 | 0 | 0 |
| 0 | 0 | 0 | 0 | 0 |
| 0 | 0 | 0 | 0 | 0 |
| 0 | 0 | 0 | 0 | 0 |
| 0 | 0 | 0 | 0 | 0 |
| 0 | 0 | 0 | 0 | 0 |
| 0 | 0.000404239 | 0 | 0.004545 | 0 |
| 0 | 0 | 0 | 0 | 0.011251074 |
| 0 | 0 | 0 | 0 | 0 |
| 0 | 0 | 0 | 0 | 0.000890287 |
| 0 | 0 | 0 | 0 | 0 |
| 0 | 0 | 0 | 0 | 0 |
| 0 | 0 | 0 | 0 | 0 |
| 0 | 0 | 0.005502081 | 0 | 0 |
| 0 | 0 | 0 | 0 | 0 |
| 0.37653766 | 0 | 0 | 0.23543388 | 0 |
| 0 | 0 | 0 | 0 | 0 |
| 0.023899997 | 0.024633415 | 0.020231476 | 0.031593779 | 0 |
| 0 | 0 | 0.46013681 | 0 | 0.144557642 |
| 0 | 0 | 0 | 0 | 0.02077707 |
| 0 | 0 | 0 | 0 | 0.283662371 |
| 0 | 0 | 0 | 0 | 0 |
| 0 | 0 | 0 | 0 | 0 |
| 0 | 0 | 0 | 0 | 0 |
| 0 | 0.63620149 | 0 | 0 | 0.000592292 |
| 0 | 0 | 0 | 0 | 0 |
| 0 | 0.021626558 | 0 | 0 | 0 |
| 0.536186511 | 0 | 0.01257524 | 0.006359918 | 0 |
| 0 | 0 | 0 | 0 | 0 |
| 0 | 0.145524306 | 0.012448331 | 0 | 0.017502085 |
| 0 | 0 | 0.000403644 | 0.04082854 | 0 |
| 0 | 0 | 0 | 0 | 0 |
| 0 | 0 | 0 | 0 | 0 |
| 0.103872625 | 0.001438122 | 0.014628852 | 0.056977584 | 0 |
| 0 | 0 | 0 | 0 | 0.010461326 |
| 0 | 0 | 0 | 0 | 0 |
| 0 | 0 | 0 | 0 | 0.000449094 |
| 0.015003663 | 0.442633174 | 0.066784284 | 0 | 0.028314161 |
| 0 | 0 | 0 | 0 | 0 |
| 0 | 0 | 0 | 0 | 0 |
| 0 | 0.023890219 | 0 | 0 | 0.001405397 |
| 0 | 0 | 0 | 0 | 0 |
| 0 | 0 | 0 | 0 | 0 |

| | | | | |
|---|---|---|---|---|
| 0 | 0 | 0 | 0 | 0 |
| 0 | 0 | 0 | 0 | 0 |
| 0 | 0 | 0 | 0 | 0 |
| 0 | 0 | 0 | 0 | 0 |
| 0 | 0 | 0 | 0 | 0.001473412 |
| 0 | 0 | 0 | 0 | 0.000606383 |
| 0 | 0 | 0 | 0 | 0 |
| 0 | 0 | 0.000977555 | 0.001151547 | 0 |
| 0 | 0.005560974 | 0.019768469 | 0.008262102 | 0 |
| 0 | 0 | 0.02024394 | 0 | 0.0384633 |
| 0 | 0 | 0 | 0 | 0 |
| 0 | 0 | 0 | 0 | 0 |
| 0 | 0 | 0 | 0 | 0 |
| 0 | 0 | 0 | 0 | 0.17981663 |
| 0.07533058 | 0 | 0 | 0 | 0 |
| 0 | 0 | 0 | 0.013042621 | 0 |
| 0 | 0 | 0 | 0 | 0 |
| 0 | 0 | 0 | 0 | 0 |
| 0 | 0 | 0 | 0 | 0 |
| 0 | 0.1183441 | 0 | 0 | 0.058087128 |
| 0 | 0.940603055 | 0.004588837 | 0 | 0.0019125 |
| 0 | 0 | 0 | 0 | 0 |
| 0 | 0 | 0 | 0 | 0 |
| 0 | 0 | 0.093248376 | 0 | 0 |
| 0 | 0.000609704 | 0 | 0.00228504 | 0 |
| 0 | 0.020225832 | 0 | 0 | 0.590072635 |
| 0 | 0 | 0 | 0 | 0 |
| 0 | 0 | 0 | 0 | 0.030275478 |
| 0 | 0 | 0 | 0 | 0 |
| 0 | 0.006701604 | 0 | 0 | 0.058200422 |
| 0 | 0.097531458 | 0.913229079 | 0 | 0.227379493 |
| 0 | 0 | 0 | 0 | 0 |
| 0 | 0 | 0 | 0 | 0 |
| 0 | 0 | 0 | 0 | 0 |
| 0 | 0 | 0 | 0 | 0 |
| 0 | 0 | 0 | 0 | 0 |
| 0 | 0 | 0 | 0 | 0 |
| 0 | 0 | 0 | 0 | 0.000635592 |
| 0 | 0 | 0.04306408 | 0 | 0.013046248 |
| 0 | 0 | 0 | 0 | 0 |
| 0 | 0 | 0 | 0 | 0 |
| 0 | 0.1493017 | 0 | 0 | 0.022797296 |
| 0 | 0.001189724 | 0 | 0 | 0.040958677 |
| 0 | 0 | 0 | 0.001031954 | 0 |
| 0 | 0 | 0 | 0.000905443 | 0 |
| 0 | 0 | 0 | 0 | 0 |
| 1.248188356 | 0 | 0 | 0.343186493 | 0 |
| 0 | 0.018515446 | 0 | 0 | 0 |

| | | | | |
|---|---|---|---|---|
| 0 | 0 | 0 | 0 | 0 |
| 0 | 0 | 0 | 0 | 0 |
| 0 | 0 | 0 | 0 | 0 |
| 0 | 0 | 0 | 0 | 0.019786682 |
| 0 | 0.018170373 | 0 | 0 | 0.005912854 |
| 0 | 0 | 0 | 0 | 0 |
| 0 | 0 | 0 | 0 | 0 |
| 0 | 0 | 0 | 0 | 0 |
| 0 | 0 | 0 | 0 | 0 |
| 0 | 0 | 0 | 0 | 0 |
| 0 | 0.004526404 | 0 | 0 | 0.010273105 |
| 0.00824214 | 0.012294132 | 0.012284671 | 0.016925794 | 0 |
| 0 | 0.007713991 | 0 | 0 | 0 |
| 0 | 0 | 0 | 0 | 0.005373747 |
| 0 | 0 | 0 | 0 | 0 |
| 0 | 0 | 0 | 0 | 0 |
| 0 | 0.006754033 | 0.07271657 | 0 | 0 |
| 0 | 0 | 0 | 0 | 0 |
| 0.046229599 | 0.00084629 | 0.082658486 | 0.569743143 | 0 |
| 0 | 0.001104997 | 0.073027391 | 0 | 0.016018834 |
| 0 | 0 | 0 | 0 | 0 |
| 0 | 0.027542709 | 0 | 0 | 0 |
| 0 | 0 | 0 | 0 | 0.027437156 |
| 0 | 0 | 0 | 0 | 0 |
| 0 | 0 | 0 | 0 | 0 |
| 0 | 0 | 0 | 0 | 0 |
| 0.008706718 | 0.001897466 | 0 | 0.037791468 | 0 |
| 0 | 0 | 0 | 0 | 0 |
| 0 | 0 | 0 | 0 | 0 |
| 0 | 0 | 0 | 0 | 0 |
| 0.003015687 | 0 | 0.017317113 | 0.001381891 | 0 |
| 0 | 0 | 0 | 0.00348274 | 0 |
| 0 | 0 | 0 | 0 | 0 |
| 0 | 0 | 0 | 0 | 0 |
| 0 | 0 | 0 | 0 | 0.026134781 |
| 0 | 0.005782925 | 0 | 0 | 0 |
| 0 | 0 | 0 | 0 | 0 |
| 0.043530359 | 0.00544012 | 0.00932111 | 0.168608859 | 0 |
| 0 | 0.042047077 | 0 | 0 | 0.218218295 |
| 0 | 0 | 0 | 0 | 0 |
| 0 | 0 | 0 | 0 | 0 |
| 0 | 0 | 0 | 0 | 0.001397916 |
| 0 | 0 | 0 | 0 | 0 |
| 0 | 0 | 0.001600445 | 0.220991879 | 0 |
| 0 | 0 | 0 | 0 | 0 |
| 0 | 0 | 0 | 0 | 0.009396571 |
| 0 | 0 | 0 | 0 | 0 |
| 0 | 0 | 0 | 0 | 0 |

| | | | | |
|---|---|---|---|---|
| 0 | 0 | 0 | 0 | 0 |
| 0 | 0.071758291 | 0 | 0 | 0.005528312 |
| 0 | 0 | 0 | 0 | 0.007919279 |
| 0.006884602 | 0 | 0.005794217 | 0.002263197 | 0 |
| 0.007495855 | 0.001887694 | 0 | 0.001399386 | 0 |
| 0 | 0 | 0 | 0 | 0.000760688 |
| 0 | 0 | 0 | 0 | 0.002055655 |
| 0 | 0 | 0 | 0 | 0 |
| 0 | 0 | 0 | 0 | 0 |
| 0 | 0 | 0 | 0 | 0.000734348 |
| 0 | 0.009185148 | 0 | 0 | 0.021004667 |
| 0 | 0 | 0 | 0 | 0 |
| 0 | 0.02885461 | 0.317017503 | 0.043137523 | 0.031383034 |
| 0 | 0 | 0.008544368 | 0 | 0 |
| 0 | 0 | 0.003607684 | 0 | 0 |
| 0 | 0 | 0 | 0 | 0 |
| 0 | 0 | 0 | 0 | 0 |
| 0 | 0 | 0 | 0 | 0 |
| 0 | 0 | 0 | 0 | 0 |
| 0 | 0.026211663 | 0 | 0 | 0 |
| 0 | 0 | 0 | 0 | 0 |
| 0 | 0 | 0 | 0 | 0 |
| 0.008661287 | 0 | 0 | 0.005867994 | 0 |
| 0 | 0 | 0.011252536 | 0 | 0.499772722 |
| 0 | 0 | 0 | 0 | 0 |
| 0 | 0.020248814 | 0 | 0 | 0.000861548 |
| 0 | 0.000837711 | 0.020260893 | 0 | 0.007178494 |
| 0 | 0 | 0 | 0 | 0 |
| 0 | 0 | 0 | 0 | 0.035151834 |
| 0 | 0 | 0 | 0 | 0 |
| 0 | 0.035031864 | 0.040625064 | 0 | 0.007936592 |
| 0 | 0 | 0 | 0.002023831 | 0 |
| 0 | 0 | 0 | 0 | 0 |
| 0 | 0 | 0 | 0 | 0 |
| 0 | 0.014616661 | 0 | 0 | 0.000739318 |
| 0.005210568 | 0 | 0 | 0 | 0 |
| 0.01522477 | 0 | 0 | 0 | 0 |
| 0 | 0 | 0 | 0 | 0 |
| 0.000790717 | 0 | 0 | 0 | 0.015668117 |
| 0 | 0 | 0 | 0 | 0 |
| 0 | 0 | 0 | 0 | 0 |
| 0 | 0 | 0 | 0 | 0 |
| 0 | 0 | 0 | 0 | 0 |
| 0 | 0 | 0 | 0 | 0 |
| 0 | 8.830829025 | 0 | 0 | 0.001727765 |
| 0 | 0.153798158 | 0.494138918 | 0.080654796 | 0.129155643 |
| 0 | 0 | 0 | 0 | 0 |
| 0 | 0 | 0 | 0 | 0 |

| | | | | |
|---|---|---|---|---|
| 0 | 0.011387473 | 0 | 0 | 0 |
| 0 | 0 | 0 | 0 | 0 |
| 0 | 0 | 0.149036852 | 0 | 0.077432601 |
| 0 | 0 | 0 | 0 | 0.000991094 |
| 0 | 0 | 0 | 0 | 0 |
| 0 | 0 | 0.059054973 | 0 | 0 |
| 0 | 0 | 0.000545909 | 0 | 0 |
| 0 | 0 | 0 | 0 | 0 |
| 0 | 0 | 0.012795695 | 0 | 0 |
| 0 | 0 | 0 | 0 | 0 |
| 0 | 0.000671037 | 0 | 0 | 0 |
| 0.001276982 | 0 | 0.000975745 | 0 | 0 |
| 0 | 0 | 0 | 0 | 0 |
| 0 | 0 | 0 | 0 | 0 |
| 0 | 0 | 0 | 0 | 0 |
| 0 | 0 | 0 | 0 | 0 |
| 0 | 0 | 0 | 0 | 0.053384801 |
| 0 | 0 | 0 | 0 | 0 |
| 0 | 0 | 0 | 0 | 0 |
| 0.002552151 | 0 | 0 | 0 | 0 |
| 0 | 0 | 0 | 0 | 0 |
| 0 | 0 | 0 | 0 | 0 |
| 0 | 0 | 0 | 0 | 0 |
| 0 | 0 | 0 | 0 | 0 |
| 0 | 0 | 0 | 0 | 0 |
| 0 | 0.010363193 | 0 | 0 | 0 |
| 0 | 0 | 0 | 0 | 0 |
| 0 | 0.014772537 | 0.004316241 | 0 | 0.014657614 |
| 0 | 0 | 0 | 0 | 0 |
| 0 | 0 | 0 | 0 | 0 |
| 0.005846833 | 0.00152905 | 0.013154534 | 0 | 0.00706856 |
| 0 | 0 | 0 | 0 | 0 |
| 0.3394384 | 0.006361786 | 0.127809621 | 0.12095571 | 0.014412851 |
| 0.003179635 | 0 | 0.008540292 | 0 | 0.049503688 |
| 0 | 0 | 0 | 0 | 0.007945081 |
| 0 | 0 | 0 | 0 | 0 |
| 0 | 0 | 0 | 0 | 0 |
| 0 | 0 | 0 | 0 | 0 |
| 0 | 0 | 0 | 0 | 0 |
| 0 | 0 | 0 | 0 | 0 |
| 0 | 0 | 0 | 0 | 0 |
| 1.719499684 | 0 | 0.045757674 | 0.373403743 | 0.003129015 |
| 0 | 0.033808818 | 0.098493785 | 0 | 0.027486692 |
| 0 | 0 | 0 | 0 | 0 |
| 0 | 0 | 0 | 0 | 0 |
| 0.006145316 | 0.479561116 | 0.034643462 | 0 | 0.038270539 |
| 0.000805809 | 0.003090753 | 0 | 0.006543914 | 0 |
| 0 | 0 | 0 | 0 | 0 |

| | | | | |
|---|---|---|---|---|
| 0 | 0 | 0 | 0 | 0 |
| 0 | 0.005281814 | 0.003025962 | 0 | 0.006095494 |
| 0.00451023 | 0.003084858 | 0.056377557 | 0 | 0.033391193 |
| 0 | 0 | 0 | 0 | 0 |
| 0.165655283 | 0.011880887 | 0 | 0 | 0 |
| 0 | 0 | 0 | 0 | 0 |
| 0 | 0 | 0 | 0 | 0 |
| 0 | 0 | 0 | 0 | 0 |
| 0 | 0 | 0 | 0 | 0 |
| 0 | 0 | 0 | 0 | 0 |
| 0 | 0 | 0 | 0 | 0 |
| 0 | 0 | 0 | 0 | 0 |
| 0 | 0 | 0 | 0 | 0.00074458 |
| 0 | 0 | 0 | 0 | 0 |
| 0 | 0 | 0 | 0 | 0 |
| 0 | 0 | 0 | 0 | 0 |
| 0 | 0.185528772 | 0 | 0 | 0 |
| 0 | 0 | 0 | 0 | 0 |
| 0 | 0 | 0 | 0 | 0 |
| 0 | 0 | 0 | 0 | 0 |
| 0 | 0 | 0 | 0 | 0 |
| 0 | 0 | 0 | 0 | 0 |
| 0 | 0.001025926 | 0 | 0 | 0.005552427 |
| 0 | 0 | 0 | 0 | 0 |
| 0 | 0.014356838 | 0.006699999 | 0 | 0.005058647 |
| 0.005177462 | 0 | 0 | 0.002658067 | 0.017098633 |
| 0 | 0 | 0 | 0 | 0 |
| 0 | 1.216523187 | 0.24418285 | 0 | 0.010267691 |
| 0 | 0 | 0 | 0 | 0 |
| 0 | 0 | 0 | 0 | 0 |
| 0 | 0 | 0 | 0 | 0 |
| 0 | 0 | 0 | 0 | 0 |
| 0 | 0 | 0 | 0 | 0 |
| 0 | 0 | 0 | 0 | 0 |
| 0 | 0 | 0 | 0 | 0 |
| 0 | 0 | 0 | 0 | 0 |
| 0 | 0 | 0 | 0 | 0 |
| 0 | 0 | 0 | 0.030307384 | 0 |
| 0 | 0.085369672 | 0 | 0 | 4.648763179 |
| 0 | 0 | 0 | 0 | 0 |
| 0 | 0.001094351 | 0 | 0 | 0 |
| 0 | 0 | 0.002384317 | 0 | 0.003533215 |
| 0 | 0 | 0.103671259 | 0.142113565 | 0 |
| 0 | 0 | 0 | 0 | 0.09954742 |
| 0 | 0 | 0 | 0 | 0 |
| 0 | 0 | 0 | 0 | 0 |
| 0.012806868 | 0 | 0.006973757 | 0.015210996 | 0 |
| 0 | 0 | 0 | 0 | 0 |

| | | | | |
|---|---|---|---|---|
| 0 | 0 | 0 | 0 | 0 |
| 0.010308909 | 0.002914552 | 0.015612192 | 0.020831947 | 0.002563256 |
| 0 | 0.31396258 | 0.014136164 | 0 | 0.001636544 |
| 0 | 0 | 0 | 0 | 0 |
| 0 | 0 | 0 | 0 | 0 |
| 0 | 0 | 0 | 0 | 0 |
| 0 | 0 | 0 | 0 | 0 |
| 0 | 0 | 0 | 0 | 0 |
| 0 | 0 | 0 | 0 | 0 |
| 0 | 0 | 0 | 0 | 0 |
| 0 | 0 | 0 | 0 | 0 |
| 0 | 0 | 0 | 0 | 0 |
| 0 | 0.615725891 | 0 | 0 | 0.039184823 |
| 0 | 0 | 0 | 0 | 0 |
| 0.009032299 | 0.024711252 | 0.121751294 | 0 | 0 |
| 0 | 0 | 0 | 0 | 0 |
| 0 | 0 | 0 | 0 | 0 |
| 0 | 0 | 0 | 0 | 0 |
| 0 | 0 | 0 | 0 | 0 |
| 0 | 0 | 0 | 0 | 0 |
| 0 | 0 | 0 | 0 | 0 |
| 0.021834905 | 0 | 0.063086751 | 0.001433079 | 0.005070237 |
| 0 | 0 | 0.00707791 | 0 | 0 |
| 0 | 0 | 0 | 0.000885639 | 0 |
| 0 | 0 | 0 | 0 | 0 |
| 0 | 0 | 0 | 0 | 0 |
| 0 | 0 | 0 | 0 | 0 |
| 0 | 0 | 0 | 0 | 0.003358993 |
| 0 | 0 | 0 | 0 | 0 |
| 0 | 0 | 0 | 0 | 0 |
| 0 | 0 | 0 | 0 | 0 |
| 0 | 0 | 0 | 0 | 0 |
| 0 | 0 | 0 | 0 | 0 |
| 0 | 0 | 0 | 0 | 0 |
| 0 | 1.294680441 | 42.94035156 | 1.022448546 | 0.060064628 |
| 0 | 0 | 0 | 0 | 0 |
| 0 | 0 | 0 | 0 | 0 |
| 0 | 0 | 0 | 0 | 0 |
| 0.0020442 | 0 | 0.019785047 | 0.003148427 | 4.493268728 |
| 0 | 0 | 0 | 0 | 0 |
| 0.369939354 | 0.183425271 | 0.752701326 | 0.102910486 | 0.294442949 |
| 0 | 0 | 0 | 0 | 0 |
| 0 | 0 | 0 | 0 | 0 |
| 0 | 0 | 0 | 0 | 0 |
| 0 | 0 | 0 | 0 | 0 |
| 9.038063318 | 0 | 0.641577888 | 0.83917615 | 0 |
| 0 | 0 | 0 | 0 | 0 |
| 0 | 0 | 0 | 0 | 0 |

| | | | | |
|---|---|---|---|---|
| 0 | 0 | 0 | 0 | 0.000661364 |
| 0 | 0 | 0 | 0 | 0 |
| 0 | 0 | 0 | 0 | 0 |
| 0 | 0.006281873 | 0 | 0 | 0.002174303 |
| 0 | 0 | 0.000405378 | 0 | 0 |
| 0 | 0 | 0 | 0 | 0 |
| 0 | 0 | 0 | 0 | 0 |
| 0 | 0 | 0 | 0 | 0 |
| 0 | 0 | 0.00772452 | 0 | 0 |
| 0 | 0 | 0 | 0 | 0.027354055 |
| 0 | 0.033020219 | 0 | 0 | 0.016339119 |
| 0 | 0 | 0 | 0 | 0 |
| 0.007355423 | 0.002972794 | 0.032188963 | 0.009549767 | 0 |
| 0 | 0 | 0 | 0 | 0 |
| 0 | 0 | 0 | 0 | 0 |
| 0 | 0 | 0 | 0 | 0 |
| 0 | 0 | 0 | 0 | 0 |
| 0 | 0 | 0 | 0 | 0 |
| 0 | 0 | 0 | 0 | 0 |
| 0.124092482 | 0.042491587 | 1.078105764 | 1.365994572 | 0.013899239 |
| 0 | 0 | 0 | 0 | 0 |
| 0 | 0 | 0 | 0 | 0 |
| 0 | 0 | 0 | 0 | 0 |
| 0 | 0 | 0 | 0 | 0 |
| 0 | 0 | 0 | 0 | 0 |
| 0 | 0.023488841 | 0 | 0 | 0 |
| 0 | 0 | 0 | 0 | 0 |
| 0 | 0 | 0 | 0 | 0 |
| 0 | 0 | 0 | 0 | 0 |
| 0 | 0 | 0 | 0.04302773 | 0 |
| 0 | 0 | 0 | 0 | 0 |
| 0 | 0 | 0 | 0 | 0 |
| 0 | 0 | 0 | 0 | 0 |
| 0 | 0 | 0 | 0 | 0 |
| 0 | 0 | 0 | 0 | 0 |
| 0 | 0.002417775 | 0 | 0 | 0 |
| 0 | 0 | 0 | 0 | 0 |
| 0 | 0 | 0 | 0 | 0 |
| 0 | 0 | 0 | 0 | 0 |
| 0 | 0.019458515 | 0 | 0 | 0 |
| 0 | 0 | 0 | 0 | 0 |
| 0 | 0 | 0 | 0 | 0 |
| 0 | 0.008150927 | 0.03893626 | 0 | 0 |
| 0 | 0 | 0 | 0 | 0 |
| 0.00167701 | 0.002485217 | 0.003417086 | 0.010331563 | 0 |
| 0 | 0.016575848 | 0 | 0 | 0.013456551 |
| 0 | 0 | 0 | 0 | 0 |
| 0 | 0 | 0 | 0 | 0 |

| | | | | |
|---|---|---|---|---|
| 0 | 0 | 0 | 0 | 0 |
| 0 | 0 | 0 | 0 | 0 |
| 0 | 0 | 0 | 0 | 0.009703867 |
| 0 | 0 | 0 | 0 | 0 |
| 0 | 0 | 0.008789351 | 0 | 0.000939271 |
| 0.009180992 | 0.015169896 | 0.136690426 | 0.17424173 | 0 |
| 0 | 0 | 0 | 0 | 0 |
| 0 | 0 | 0 | 0 | 0 |
| 0 | 0 | 0 | 0 | 0 |
| 0 | 0 | 0 | 0 | 0 |
| 0 | 0 | 0 | 0 | 0 |
| 0 | 0 | 0 | 0 | 0 |
| 0 | 0 | 0 | 0 | 0 |
| 0 | 0 | 0 | 0 | 0.001887944 |
| 0 | 0 | 0 | 0 | 0 |
| 0 | 0 | 0 | 0 | 0 |
| 0 | 0.026740989 | 0 | 0 | 0.031345191 |
| 0 | 0 | 0 | 0 | 0 |
| 0 | 0 | 0 | 0 | 0 |
| 0 | 0.102670478 | 0 | 0 | 0 |
| 0 | 0 | 0 | 0 | 0 |
| 0 | 0 | 0 | 0 | 0 |
| 0 | 0 | 0 | 0 | 0 |
| 0 | 0 | 0 | 0 | 0 |
| 0 | 0 | 0 | 0 | 0 |
| 0 | 0 | 0 | 0 | 0.002540684 |
| 0 | 0 | 0 | 0 | 0 |
| 0.002686981 | 0 | 0 | 0.001436478 | 0.036128998 |
| 0 | 0 | 0 | 0 | 0 |
| 0 | 0 | 0 | 0 | 0 |
| 0 | 0 | 0 | 0 | 0 |
| 0 | 0 | 0 | 0 | 0 |
| 0 | 0 | 0 | 0 | 0 |
| 0 | 0 | 0 | 0 | 0 |
| 0 | 0 | 0 | 0 | 0.555586197 |
| 0 | 0 | 0 | 0 | 0 |
| 0 | 0 | 0 | 0 | 0 |
| 0 | 0 | 0 | 0 | 0 |
| 0 | 0.456311414 | 0 | 0 | 0.001667528 |
| 0 | 0 | 0 | 0 | 0 |
| 0 | 0 | 0 | 0 | 0 |
| 0 | 0 | 0 | 0 | 0 |
| 0 | 0 | 0 | 0 | 0 |
| 0 | 0 | 0 | 0 | 0 |
| 0 | 0 | 0 | 0 | 0 |
| 0 | 0 | 0 | 0 | 0 |
| 0 | 0 | 0 | 0 | 0 |
| 0 | 0 | 0 | 0 | 0 |

| | | | | |
|---|---|---|---|---|
| 0 | 0 | 0 | 0 | 0 |
| 0 | 0.004276527 | 0 | 0 | 0 |
| 0.001144036 | 0 | 0 | 0 | 0 |
| 0 | 0 | 0 | 0 | 0 |
| 0 | 0 | 0 | 0 | 0 |
| 0 | 0 | 0 | 0 | 0 |
| 0 | 0 | 0 | 0 | 0 |
| 0 | 0 | 0 | 0 | 0 |
| 0 | 0 | 0 | 0 | 0 |
| 0 | 0 | 0 | 0 | 0.002244674 |
| 0 | 0.001759929 | 0 | 0 | 0 |
| 0 | 0 | 0 | 0 | 0 |
| 0 | 0 | 0 | 0 | 0.040020771 |
| 0 | 0 | 0 | 0 | 0 |
| 0 | 0 | 0 | 0 | 0 |
| 0 | 0 | 0 | 0 | 0 |
| 0 | 0 | 0 | 0 | 0 |
| 0 | 0 | 0 | 0 | 0 |
| 0 | 0 | 0 | 0 | 0 |
| 0 | 0 | 0.000388987 | 0 | 0 |
| 0.012108111 | 0 | 0.001265208 | 0 | 0 |
| 0 | 0.00330078 | 0 | 0 | 0 |
| 0 | 0 | 0 | 0 | 0 |
| 0.817345283 | 0.194648512 | 1.736043186 | 0 | 0.583516104 |
| 0 | 0 | 0 | 0 | 0 |
| 0 | 0 | 0 | 0 | 0 |
| 0 | 0 | 0 | 0 | 0 |
| 0 | 0 | 0 | 0 | 0 |
| 0 | 0 | 0 | 0 | 0 |
| 0 | 0 | 0 | 0 | 0 |
| 0 | 0 | 0.007295581 | 0 | 0.002587741 |
| 0 | 0 | 0 | 0 | 0 |
| 0 | 0.000854061 | 0 | 0.003566648 | 0 |
| 0 | 0 | 0 | 0 | 0 |

| PHL018-0059_V2 | PHL018-0245_V1 | PHL018-0027_V3 | PHL018-0048_V3 | PHL018-0254_V1 |
|---|---|---|---|---|
| CTRL | REF | EYCF | CTRL | REF |
| 0.12365609 | 0.004603559 | 0.163829071 | 0.003883267 | 0.397303528 |
| 5.877302843 | 0.12192661 | 1.526201917 | 5.511339193 | 1.517436271 |
| 0 | 0 | 2.969561322 | 7.986912789 | 0 |
| 0 | 0 | 0 | 0 | 0 |
| 0 | 0 | 0 | 0 | 0 |
| 0 | 0 | 0 | 0 | 0 |
| 0 | 0 | 0 | 0 | 0 |
| 0 | 0 | 0 | 0 | 0 |
| 0 | 0 | 0 | 0 | 0 |
| 0 | 0 | 0 | 0 | 0 |
| 0 | 0 | 0 | 0 | 0 |
| 0 | 0 | 0 | 0 | 0 |
| 0 | 0 | 0 | 0 | 0 |
| 0.088461536 | 2.517529401 | 5.226607122 | 6.330177372 | 2.737212599 |
| 0 | 0 | 0 | 0 | 0 |
| 0 | 0 | 0 | 0 | 0 |
| 0 | 0 | 0 | 0 | 0 |
| 0 | 0 | 0.001574787 | 0 | 0 |
| 0 | 0 | 0 | 0 | 0 |
| 0 | 0 | 0 | 0 | 0 |
| 0 | 0 | 0 | 0 | 0 |
| 0 | 0 | 0 | 0 | 0 |
| 0 | 0 | 0 | 0 | 0 |
| 0 | 0 | 0 | 0 | 0 |
| 0 | 0 | 0 | 0 | 0 |
| 0 | 0 | 0 | 0 | 0 |
| 0 | 0 | 0 | 0 | 0 |
| 0 | 0 | 0 | 0 | 0 |
| 0 | 0 | 0 | 0 | 0 |
| 0 | 0 | 0 | 0 | 0 |
| 0 | 0 | 0 | 0 | 0 |
| 0 | 0 | 0 | 0 | 0 |
| 0 | 0 | 0 | 0 | 0 |
| 0 | 0 | 0 | 0 | 0 |
| 0 | 0 | 0 | 0 | 0 |
| 0 | 1.806564193 | 0 | 0 | 1.133318274 |
| 0 | 0 | 0 | 0 | 0 |
| 0 | 0 | 0 | 0 | 0 |
| 0 | 0 | 0 | 0 | 0 |
| 0 | 0 | 0 | 0 | 0 |
| 0 | 0 | 0 | 0 | 0 |
| 0.39361315 | 0.004447203 | 0.001094556 | 0 | 0.036924075 |
| 0.087224401 | 0.023726912 | 0.002941491 | 0.073653356 | 0.277555595 |
| 0.130943155 | 0.253635557 | 0.0008974 | 0.002306746 | 0.03377413 |
| 0.003939029 | 0 | 0 | 0.004324014 | 0 |
| 0 | 0 | 0 | 0 | 0 |

| | | | | |
|---|---|---|---|---|
| 0 | 0 | 0 | 0 | 0 |
| 0.030672301 | 0.001464894 | 0.002115187 | 0 | 0.005030545 |
| 0 | 0 | 0 | 0 | 0 |
| 0.898214089 | 3.540364174 | 0.345533185 | 0.407279446 | 0.28639977 |
| 0.005079038 | 0 | 0.006738115 | 0 | 0 |
| 0 | 0 | 0 | 0 | 0 |
| 23.83793625 | 0.673585672 | 0 | 0.001907223 | 0.005391933 |
| 0 | 0.006585488 | 0 | 0 | 0 |
| 0 | 0 | 0 | 0 | 0 |
| 0 | 0 | 0 | 0 | 0 |
| 0 | 0 | 0 | 0 | 0 |
| 0 | 0.008421412 | 0.002472693 | 0.181482428 | 0.05767919 |
| 0 | 0 | 0 | 0 | 0.000983385 |
| 0.09687428 | 0 | 0 | 0 | 0.082921337 |
| 0 | 0.004936933 | 0.00364527 | 0 | 0 |
| 0 | 0 | 0 | 0 | 0 |
| 0.002463903 | 0.004008003 | 0.001011753 | 0 | 0.018388825 |
| 0 | 0 | 0 | 0 | 0 |
| 0 | 0 | 0 | 0 | 0 |
| 0 | 0 | 0 | 0 | 0 |
| 0.343373353 | 0.01568459 | 0.084225372 | 0.082510285 | 0.075595725 |
| 0.042795679 | 0.004685641 | 0.000732217 | 0.006775725 | 0.002016393 |
| 0 | 0 | 0 | 0 | 0.049755628 |
| 0 | 0 | 0 | 0 | 0 |
| 0 | 0 | 0 | 0 | 0 |
| 0 | 0 | 0 | 0 | 0 |
| 0 | 0 | 0 | 0 | 0 |
| 0 | 0 | 0 | 0 | 0 |
| 0.005474805 | 0 | 0.001311404 | 0 | 0.005417053 |
| 0 | 0 | 0 | 0 | 0 |
| 0 | 0.035648483 | 0.178381727 | 0 | 0.002513275 |
| 0 | 0 | 0 | 0 | 0 |
| 0 | 0 | 0 | 0 | 0 |
| 0 | 0 | 0 | 0 | 0 |
| 0 | 0 | 0 | 0 | 0 |
| 0 | 0 | 0 | 0 | 0 |
| 0 | 0 | 0 | 0 | 0 |
| 0 | 0 | 0 | 0 | 0.002689597 |
| 0 | 0 | 0 | 0 | 0 |
| 0 | 0 | 0 | 0 | 0 |
| 0 | 0 | 0 | 0 | 0 |
| 0 | 0 | 0 | 0 | 0 |
| 0 | 0 | 0 | 0 | 0 |
| 0 | 0 | 0 | 0 | 0 |
| 0 | 0 | 0 | 0 | 0 |
| 0 | 0 | 0 | 0 | 0 |
| 0 | 0 | 0 | 0 | 0 |
| 0.007746716 | 0.326421544 | 0 | 0 | 0 |

| | | | | |
|---|---|---|---|---|
| 0 | 0 | 0 | 0 | 0 |
| 0 | 0 | 0 | 0 | 0 |
| 0 | 0 | 0 | 0 | 0 |
| 0 | 0 | 0 | 0 | 0 |
| 0.457086975 | 0 | 0.014835476 | 0 | 0 |
| 0 | 0 | 0 | 0 | 0 |
| 0 | 0 | 0 | 0 | 0 |
| 0 | 0 | 0 | 0.000933176 | 0 |
| 0 | 0.611636001 | 0 | 0 | 0 |
| 0 | 0 | 0 | 0 | 0 |
| 0 | 0 | 0 | 0 | 0 |
| 0.123862896 | 0.002754256 | 0 | 0 | 0 |
| 0 | 0 | 0 | 0 | 0 |
| 0 | 0 | 0 | 0 | 0 |
| 0 | 0 | 0 | 0 | 0 |
| 0 | 0 | 0 | 0 | 0 |
| 0 | 0 | 0 | 0 | 0 |
| 0 | 0 | 0 | 0 | 0 |
| 0 | 0.001114163 | 0 | 0 | 0.013089334 |
| 0 | 0 | 0 | 0 | 0 |
| 0 | 0 | 0 | 0 | 0 |
| 0 | 0 | 0 | 0 | 0 |
| 0 | 0 | 0 | 0 | 0 |
| 0 | 0 | 0 | 0 | 0 |
| 0 | 0 | 0 | 0 | 0 |
| 0 | 0 | 0 | 0 | 0 |
| 0 | 0 | 0 | 0 | 0 |
| 0 | 0 | 0 | 0 | 0 |
| 0 | 0 | 0 | 0 | 0 |
| 0 | 0 | 0 | 0 | 0 |
| 0.244821717 | 0 | 0 | 0 | 0.032103174 |
| 0 | 0 | 0 | 0 | 0.010028024 |
| 0 | 0 | 0 | 0 | 0 |
| 0 | 0 | 0 | 0 | 0 |
| 0 | 0 | 0 | 0 | 0 |
| 0 | 0 | 0 | 0 | 0 |
| 0 | 0 | 0 | 0 | 0 |
| 0 | 0 | 0 | 0 | 0 |
| 0 | 0 | 0 | 0 | 0 |
| 0 | 0 | 0 | 0 | 0 |
| 0 | 0 | 0 | 0 | 0 |
| 0 | 0 | 0 | 0 | 0 |
| 0 | 0 | 0.00095298 | 0.001224806 | 0.001312167 |
| 0 | 0 | 0 | 0 | 0 |
| 0 | 0 | 0 | 0 | 0 |
| 0 | 0 | 0 | 0 | 0 |
| 0 | 0 | 0 | 0 | 0 |
| 0 | 0 | 0 | 0 | 0 |

| | | | | |
|---|---|---|---|---|
| 0 | 0 | 0 | 0 | 0.001061927 |
| 0 | 0 | 0 | 0 | 0 |
| 0 | 0 | 0 | 0 | 0 |
| 0 | 0 | 0 | 0 | 0 |
| 0 | 0 | 0 | 0.002951421 | 0.010869155 |
| 0 | 0 | 0 | 0 | 0 |
| 0 | 0 | 0 | 0 | 0 |
| 0 | 0 | 0 | 0 | 0 |
| 0 | 0 | 0 | 0 | 0 |
| 0 | 0 | 0 | 0 | 0 |
| 0 | 0 | 0 | 0 | 0 |
| 0 | 0 | 0 | 0 | 0 |
| 0 | 0 | 0 | 0 | 0 |
| 0 | 0 | 0 | 0 | 0 |
| 0 | 0 | 0 | 0 | 0 |
| 0 | 0 | 0 | 0 | 0 |
| 0 | 0 | 0 | 0 | 0 |
| 0 | 0 | 0 | 0 | 0 |
| 0 | 0 | 0 | 0 | 0 |
| 0 | 0 | 0 | 0 | 0 |
| 0 | 0 | 0 | 0 | 0 |
| 0 | 0 | 0 | 0 | 0 |
| 0 | 0 | 0 | 0 | 0 |
| 0 | 0 | 0 | 0 | 0 |
| 0 | 0 | 0 | 0 | 0 |
| 0 | 0 | 0 | 0 | 0 |
| 0 | 0 | 0 | 0 | 0 |
| 0 | 0 | 0 | 0 | 0 |
| 0 | 0 | 0 | 0 | 0 |
| 0 | 0 | 0 | 0 | 0 |
| 0.011404227 | 0 | 0 | 0 | 0 |
| 0 | 0 | 0 | 0 | 0.003056273 |
| 0 | 0 | 0 | 0 | 0 |
| 0 | 0 | 0 | 0 | 0 |
| 0 | 0 | 0 | 0 | 0 |
| 0 | 0 | 0 | 0 | 0 |
| 0 | 0 | 0 | 0 | 0 |
| 0 | 0 | 0 | 0 | 0 |
| 0 | 0 | 0 | 0 | 0 |
| 0 | 0 | 0 | 0 | 0 |
| 0 | 0 | 0 | 0 | 0 |
| 0 | 0 | 0 | 0 | 0 |
| 0 | 0 | 0 | 0 | 0 |
| 0 | 0 | 0 | 0 | 0 |
| 0 | 0 | 0 | 0 | 0 |
| 0 | 0 | 0 | 0 | 0 |
| 0 | 0 | 0 | 0 | 0 |
| 0 | 0 | 0 | 0 | 0 |

| | | | | |
|---|---|---|---|---|
| 0 | 0 | 0 | 0 | 0 |
| 0 | 0 | 0 | 0 | 0 |
| 0 | 0 | 0 | 0 | 0 |
| 0 | 0 | 0 | 0 | 0 |
| 0 | 0 | 0 | 0 | 0 |
| 0 | 0 | 0 | 0 | 0 |
| 0 | 0 | 0 | 0 | 0 |
| 0 | 0 | 0 | 0 | 0 |
| 0 | 0 | 0 | 0 | 0 |
| 0 | 0 | 0 | 0 | 0 |
| 0 | 0 | 0 | 0 | 0 |
| 0 | 0 | 0 | 0 | 0 |
| 0.002586266 | 0 | 0 | 0 | 0.00376014 |
| 0 | 0 | 0 | 0 | 0 |
| 0 | 0 | 0 | 0 | 0 |
| 0 | 0 | 0 | 0 | 0 |
| 0 | 0 | 0 | 0 | 0 |
| 0 | 0 | 0 | 0 | 0 |
| 0 | 0 | 0 | 0 | 0 |
| 0 | 0 | 0 | 0 | 0 |
| 0.054919688 | 0.092492341 | 0.039997336 | 0.004812485 | 0.033590448 |
| 0 | 0 | 0 | 0 | 0 |
| 0 | 0 | 0 | 0 | 0 |
| 0 | 0.002704231 | 0 | 0 | 0 |
| 0 | 0 | 0 | 0 | 0 |
| 0 | 0 | 0 | 0 | 0 |
| 0.011063624 | 0.010823413 | 0 | 0.019873535 | 0.004776837 |
| 0 | 0 | 0 | 0 | 0 |
| 0 | 0 | 0 | 0 | 0 |
| 0.004304292 | 0 | 0 | 0 | 0 |
| 0.002606749 | 0.011040015 | 0 | 0 | 0 |
| 0 | 0 | 0 | 0 | 0 |
| 0 | 0 | 0 | 0 | 0 |
| 0 | 0 | 0 | 0 | 0.021483205 |
| 0.054836585 | 0.001687302 | 0.008121089 | 0.00227728 | 0.044473881 |
| 0 | 0 | 0 | 0 | 0 |
| 0 | 0 | 0 | 0 | 0 |
| 0 | 0 | 0 | 0 | 0 |
| 0.067528269 | 0.001362872 | 0 | 0 | 0 |
| 0.0474062 | 0 | 0 | 0 | 0 |
| 0.000593237 | 0 | 0 | 0 | 0 |
| 0 | 0 | 0 | 0 | 0 |
| 0 | 0 | 0 | 0 | 0 |
| 0 | 0 | 0 | 0 | 0 |
| 0 | 0 | 0 | 0 | 0 |
| 0 | 0 | 0 | 0 | 0 |
| 0.001460003 | 0 | 0.000799362 | 0 | 0.001650973 |
| 0.000800779 | 0.001202417 | 0 | 0 | 0.001267732 |

| | | | | |
|---|---|---|---|---|
| 0 | 0 | 0 | 0 | 0 |
| 0 | 0 | 0 | 0 | 0 |
| 0 | 0.006592462 | 0 | 0 | 0 |
| 0 | 0.001273691 | 0 | 0 | 0 |
| 0 | 0 | 0 | 0 | 0 |
| 0 | 0 | 0 | 0 | 0 |
| 0 | 0 | 0 | 0 | 0 |
| 0 | 0 | 0 | 0.009914732 | 0 |
| 0 | 0 | 0 | 0 | 0 |
| 0 | 0 | 0 | 0 | 0 |
| 0 | 0 | 0 | 0 | 0 |
| 0 | 0 | 0 | 0 | 0 |
| 0 | 0 | 0 | 0 | 0 |
| 0 | 0 | 0 | 0 | 0 |
| 0 | 0 | 0 | 0 | 0 |
| 0 | 0 | 0 | 0 | 0 |
| 0 | 0 | 0 | 0 | 0 |
| 0 | 0 | 0 | 0 | 0 |
| 0 | 0 | 0 | 0 | 0.002762731 |
| 0 | 0 | 0 | 0 | 0 |
| 0.175630674 | 0 | 0.001775851 | 0.030127562 | 0.003531935 |
| 0 | 0 | 0 | 0 | 0 |
| 0 | 0 | 0 | 0 | 0 |
| 0 | 0 | 0 | 0 | 0 |
| 0 | 0.012542905 | 0 | 0 | 0.011874815 |
| 0 | 0 | 0 | 0 | 0 |
| 0 | 0 | 0 | 0 | 0 |
| 0 | 0 | 0 | 0 | 0 |
| 0.035986233 | 0 | 0 | 0 | 0.002833077 |
| 0.00218282 | 0 | 0 | 0 | 0 |
| 1.258375521 | 0.32805169 | 0.076605906 | 0.174537087 | 0.436101381 |
| 0 | 0 | 0 | 0 | 0 |
| 0.041736816 | 0 | 0.05285844 | 0 | 0 |
| 0.021942079 | 0 | 0.001501681 | 0 | 0 |
| 0 | 0 | 0 | 0 | 0 |
| 0 | 0 | 0 | 0 | 0 |
| 0 | 0 | 0 | 0 | 0 |
| 0 | 0 | 0 | 0 | 0 |
| 0 | 0 | 0 | 0 | 0 |
| 0 | 0 | 0 | 0.001788085 | 0 |
| 0 | 0 | 0 | 0 | 0 |
| 0.006843012 | 0.004957326 | 0 | 0 | 0.007534455 |
| 0 | 0 | 0 | 0 | 0 |
| 0 | 0 | 0 | 0 | 0 |
| 0 | 0 | 0 | 0 | 0 |
| 0 | 0 | 0 | 0 | 0 |
| 0 | 0 | 0 | 0 | 0 |
| 0 | 0 | 0 | 0 | 0 |

| | | | | |
|---|---|---|---|---|
| 0 | 0 | 0 | 0 | 0 |
| 0 | 0 | 0 | 0 | 0 |
| 0.142455064 | 0.28379147 | 0.000557108 | 0.00085922 | 0.021989847 |
| 0 | 0 | 0 | 0 | 0 |
| 1.356751128 | 0 | 0 | 0 | 0 |
| 0 | 0 | 0 | 0 | 0 |
| 0 | 0 | 0 | 0 | 0 |
| 0 | 0 | 0 | 0 | 0 |
| 0 | 0 | 0 | 0 | 0 |
| 0 | 0 | 0 | 0 | 0 |
| 0 | 0 | 0 | 0 | 0 |
| 0 | 0 | 0 | 0 | 0 |
| 0 | 0 | 0 | 0 | 0 |
| 0.029488494 | 0 | 0.001641881 | 0 | 0.003391082 |
| 0.006806639 | 0.002580951 | 0.003218499 | 0.00104502 | 0.007930199 |
| 0 | 0.006631798 | 0 | 0 | 0 |
| 0.002115151 | 0 | 0 | 0.003572119 | 0.010896053 |
| 0.088418655 | 0.00068436 | 0.007186616 | 0 | 0 |
| 0 | 0 | 0 | 0 | 0 |
| 0 | 0 | 0 | 0 | 0 |
| 0 | 0 | 0 | 0 | 0 |
| 0.001272919 | 0 | 0 | 0 | 0.001679322 |
| 0 | 0 | 0 | 0 | 0 |
| 0 | 0 | 0 | 0 | 0 |
| 0 | 0 | 0 | 0 | 0 |
| 0 | 0 | 0 | 0 | 0 |
| 0 | 0 | 0 | 0 | 0 |
| 0 | 0 | 0 | 0 | 0 |
| 0 | 0 | 0 | 0 | 0 |
| 0 | 0 | 0 | 0 | 0 |
| 0 | 0.001627452 | 0 | 0 | 0 |
| 0 | 0 | 0 | 0 | 0 |
| 0 | 0 | 0 | 0 | 0 |
| 0 | 0.00122433 | 0 | 0 | 0 |
| 0 | 0 | 0 | 0 | 0.00304447 |
| 0 | 0 | 0 | 0.004293793 | 0 |
| 0 | 0 | 0 | 0 | 0 |
| 0 | 0.003335317 | 0 | 0.002025691 | 0 |
| 0 | 0 | 0 | 0 | 0 |
| 0 | 0 | 0 | 0 | 0 |
| 0 | 0 | 0 | 0 | 0 |
| 0 | 0 | 0 | 0 | 0 |
| 0 | 0 | 0 | 0 | 0 |
| 0 | 0 | 0 | 0 | 0 |
| 0 | 0 | 0 | 0 | 0 |
| 0 | 0.006153598 | 0 | 0 | 0 |
| 0 | 0.001446145 | 0 | 0 | 0 |
| 0 | 0.006059859 | 0 | 0 | 0 |

| | | | | |
|---|---|---|---|---|
| 0 | 0 | 0 | 0 | 0 |
| 0 | 0 | 0 | 0 | 0 |
| 0 | 0 | 0 | 0 | 0.004774316 |
| 0 | 0 | 0 | 0 | 0 |
| 0 | 0.008112782 | 0 | 0 | 0.000758435 |
| 0 | 0 | 0 | 0 | 0 |
| 0 | 0 | 0 | 0 | 0 |
| 0.054528214 | 0.216065132 | 0.002316304 | 0.001984668 | 0.044650773 |
| 0 | 0 | 0 | 0 | 0 |
| 0.102393572 | 0.01374211 | 1.06109541 | 0.075688686 | 0.016801872 |
| 0 | 0 | 0 | 0 | 0 |
| 0.054079948 | 0.004035378 | 0 | 0 | 0 |
| 0.010913993 | 0.01106661 | 0 | 0.002383425 | 0.005106853 |
| 0 | 0 | 0 | 0 | 0.004201162 |
| 0.00212884 | 0.00096866 | 0 | 0 | 0 |
| 0 | 0.004948499 | 0 | 0 | 0 |
| 0 | 0 | 0 | 0 | 0 |
| 0 | 0 | 0 | 0 | 0 |
| 0.108204329 | 0.006465537 | 0.086118161 | 0 | 0 |
| 0 | 0 | 0 | 0.074060862 | 0 |
| 0 | 0 | 0 | 0 | 0 |
| 0 | 0 | 0 | 0 | 0 |
| 0.000582509 | 0 | 0 | 0 | 0 |
| 0 | 0 | 0 | 0 | 0 |
| 0 | 0.053121214 | 0 | 0 | 0 |
| 0.005917664 | 0 | 0 | 0.000832825 | 0.000991364 |
| 0 | 0 | 0 | 0 | 0 |
| 0.003710571 | 0.001326581 | 0 | 0 | 0 |
| 0 | 0 | 0.204564055 | 0 | 0 |
| 0 | 0.001693016 | 0 | 0 | 0 |
| 0 | 0 | 0 | 0 | 0 |
| 0 | 0 | 0 | 0 | 0 |
| 0 | 0 | 0.009604818 | 0 | 0 |
| 0.019204725 | 0 | 0 | 0 | 0 |
| 0 | 0 | 0 | 0 | 0 |
| 0 | 0 | 0 | 0 | 0 |
| 0 | 0 | 0.027546227 | 0 | 0 |
| 0 | 0 | 0 | 0 | 0 |
| 0 | 0 | 0 | 0 | 0 |
| 0 | 0 | 0 | 0 | 0 |
| 0 | 0 | 0 | 0 | 0 |
| 0 | 0.016426414 | 0.00105697 | 0 | 0 |
| 0 | 0 | 0 | 0 | 0 |
| 0.008475518 | 0 | 0 | 0 | 0 |
| 0 | 0 | 0 | 0 | 0 |
| 0 | 0 | 0 | 0 | 0 |
| 0 | 0 | 0.0026861 | 0.001726139 | 0 |
| 0 | 0 | 0 | 0 | 0.001119014 |

| | | | | |
|---|---|---|---|---|
| 0 | 0 | 0 | 0 | 0 |
| 0.090771795 | 0.092619406 | 0 | 0 | 0 |
| 0 | 0 | 0 | 0 | 0 |
| 0 | 0 | 0.001927261 | 0 | 0 |
| 0 | 0 | 0 | 0 | 0 |
| 0.319003986 | 0 | 0.003173444 | 0 | 0.02136222 |
| 0 | 0 | 0 | 0 | 0 |
| 0 | 0 | 0 | 0 | 0 |
| 0 | 0 | 0 | 0 | 3.202082161 |
| 0 | 0 | 0 | 0 | 0 |
| 0 | 0.086300454 | 0.001226887 | 0.075373026 | 0.123319788 |
| 0 | 0 | 0 | 0 | 0 |
| 0 | 0 | 0 | 0 | 0 |
| 0.00352842 | 0.025313287 | 0 | 0.001191777 | 0.00771389 |
| 0 | 0 | 0 | 0 | 0 |
| 0.001439287 | 0 | 0 | 0 | 0 |
| 0 | 0 | 0 | 0 | 0 |
| 0.072566004 | 0.012030517 | 0.001315991 | 0 | 0 |
| 0 | 0 | 0 | 0 | 0 |
| 0 | 0 | 0 | 0 | 0 |
| 0.125203573 | 2.406457277 | 0 | 0 | 0 |
| 0 | 1.407343106 | 0 | 2.673643067 | 0 |
| 0 | 0 | 0 | 0 | 0 |
| 0 | 0 | 0 | 0 | 0 |
| 0 | 0 | 0 | 0 | 0 |
| 0 | 0 | 0 | 0 | 0 |
| 0 | 0.011489151 | 0.400491467 | 0 | 0 |
| 0 | 0 | 0 | 0 | 0 |
| 0 | 0 | 0 | 0 | 0 |
| 0.322728106 | 0.455170872 | 0.352542359 | 1.027174106 | 1.412978451 |
| 0 | 0 | 0 | 0.001617051 | 0.001624116 |
| 0 | 0 | 0 | 0 | 0 |
| 0 | 0 | 0 | 0 | 0 |
| 0 | 0 | 0 | 0 | 0.001022357 |
| 0 | 0 | 0.003663706 | 0.021256568 | 0.019697926 |
| 0 | 0 | 0 | 0 | 0 |
| 0 | 0.005992445 | 0 | 0.937327314 | 0 |
| 0.0011988 | 0.000900034 | 0 | 0 | 0.000813363 |
| 0 | 0 | 0 | 0 | 0 |
| 0 | 0 | 0 | 0 | 0 |
| 0.063042863 | 0 | 0.009529072 | 0 | 0.005248268 |
| 3.806842539 | 5.482111965 | 0 | 0 | 3.106763946 |
| 0 | 0 | 0 | 0 | 0 |
| 0 | 0 | 0.002490778 | 0 | 0 |
| 0 | 0 | 0.001433239 | 0 | 0 |
| 0 | 0 | 0.000647397 | 0 | 0 |
| 0 | 0.006857334 | 0.00233367 | 0 | 0.001606626 |
| 0 | 0 | 0 | 0 | 0 |

| | | | | |
|---|---|---|---|---|
| 0.005008602 | 0 | 0 | 0.002220396 | 0 |
| 0.016481798 | 0 | 0 | 0 | 0 |
| 0.011943058 | 0.003415848 | 0 | 0 | 0.030869094 |
| 0 | 0 | 0 | 0 | 0 |
| 0 | 0 | 0 | 0 | 0 |
| 0 | 0 | 0.033561313 | 0 | 0.010535125 |
| 0 | 0 | 0 | 0 | 0 |
| 0.014876113 | 0 | 0 | 0 | 0 |
| 0 | 0 | 0 | 0 | 0.023463191 |
| 0 | 0 | 0 | 0 | 0 |
| 0 | 0 | 0 | 0 | 0 |
| 0 | 0 | 0 | 0 | 0 |
| 0 | 0.003853401 | 0 | 0 | 0 |
| 0 | 0.005102848 | 0 | 0.200586642 | 0 |
| 0 | 0 | 0 | 0 | 0 |
| 0.007082041 | 0.005981681 | 0 | 0.003363841 | 0.028629968 |
| 0.111662034 | 0 | 0.004190324 | 0.054435622 | 0.000887646 |
| 0.006199104 | 0 | 0 | 0.000905988 | 0 |
| 0 | 0 | 0 | 0 | 0 |
| 0 | 0 | 0 | 0 | 0 |
| 0.033883046 | 0.00495157 | 0.001421808 | 0 | 0 |
| 0 | 0.002563512 | 0 | 0.002883219 | 0.002574057 |
| 0 | 0 | 0 | 0 | 0 |
| 0 | 0 | 0 | 0 | 0 |
| 0 | 0 | 0 | 0 | 0 |
| 0.00155109 | 0 | 0.010403103 | 0.116568602 | 0 |
| 0.002491315 | 0 | 0 | 0 | 0.001408592 |
| 0 | 0 | 0 | 0 | 0.063290633 |
| 0 | 0 | 0 | 0 | 0 |
| 0 | 0 | 0 | 0 | 0 |
| 0.208220759 | 0.004741518 | 0.012447936 | 0 | 0.004927658 |
| 0.088412276 | 0.040646872 | 0 | 0.005008897 | 0.008304777 |
| 0.08418044 | 0.104325292 | 5.420951415 | 0.120931937 | 0.023417686 |
| 0.009250258 | 0.003375994 | 0 | 0 | 0.003050893 |
| 0.056670004 | 0 | 0 | 0 | 0 |
| 0 | 0 | 0 | 0 | 0 |
| 0.002431467 | 0.001216997 | 0 | 0 | 0.004674162 |
| 0.008746992 | 0 | 0.031389035 | 0 | 0.004730532 |
| 0.008192975 | 0.224303651 | 0 | 0 | 0.597089404 |
| 0.013370307 | 0 | 0 | 0 | 0 |
| 0 | 0 | 0 | 0 | 0.001228213 |
| 0 | 0 | 0 | 0 | 0 |
| 0 | 0 | 0 | 0.015128015 | 0 |
| 0.253827393 | 0 | 0.017920112 | 0 | 0 |
| 0.820596343 | 0 | 0.25883433 | 0 | 0 |
| 0.030055775 | 0 | 0.000851159 | 0 | 0 |
| 0 | 0 | 0 | 0 | 0.001911532 |
| 0 | 0 | 0 | 0 | 0 |

| | | | | |
|---|---|---|---|---|
| 0 | 0 | 0 | 0 | 0 |
| 0.060112643 | 0.68053637 | 0 | 0.369540494 | 0.517433984 |
| 0 | 0 | 0 | 0 | 0 |
| 0.000470774 | 0 | 0 | 0 | 0 |
| 0 | 0 | 0 | 0 | 0 |
| 0 | 0 | 0 | 0 | 0 |
| 0.001917656 | 0 | 0 | 0 | 0 |
| 0 | 0 | 0 | 0 | 0.011252657 |
| 0 | 0.003561553 | 0 | 0 | 0.001341077 |
| 0.010920671 | 0 | 0 | 0 | 0.017459089 |
| 0.011745199 | 0 | 0 | 0 | 0.006314151 |
| 0.974260027 | 0.025121088 | 0.00306199 | 0.002179598 | 0.010702363 |
| 0 | 0 | 0 | 0 | 0 |
| 0 | 0 | 0 | 0 | 0 |
| 0.090135545 | 0.001382188 | 0 | 0 | 0 |
| 0 | 0 | 0 | 0 | 0 |
| 0.000604865 | 0.047682605 | 0 | 0.008682841 | 0.13451649 |
| 0.005234654 | 0.017357811 | 0.24396914 | 0.001326062 | 0 |
| 0 | 0 | 0 | 0 | 0 |
| 0.007817955 | 0.021479185 | 0.02163456 | 0 | 0 |
| 0 | 0.042408284 | 0.000853794 | 0.006847329 | 0.049139952 |
| 0 | 0 | 0 | 0 | 0 |
| 0 | 0 | 0.212360652 | 0 | 0 |
| 0 | 0 | 0 | 0 | 0 |
| 0.179216048 | 0 | 0 | 0 | 0 |
| 0 | 0 | 0 | 0 | 0 |
| 0 | 0 | 0 | 0 | 0.030169696 |
| 0.034253636 | 0.057240891 | 0.007078164 | 0.207134529 | 0.030737355 |
| 0 | 0 | 0 | 0 | 0 |
| 0 | 0 | 0 | 0 | 0 |
| 0.001659866 | 0 | 0 | 0.003003453 | 0.028690969 |
| 0 | 0 | 0 | 0 | 0 |
| 0 | 0 | 0 | 0 | 0 |
| 0 | 0 | 0 | 0 | 0 |
| 0 | 0.033013328 | 0 | 0 | 0 |
| 0.01608262 | 0.141805161 | 3.863807467 | 0.623907096 | 0.233883663 |
| 0 | 0 | 0 | 0 | 0 |
| 0 | 0 | 0 | 0 | 0 |
| 0.022082006 | 0.043322977 | 0 | 0 | 0 |
| 0 | 0 | 0 | 0 | 0 |
| 0.807786911 | 0 | 0.233778822 | 0 | 0 |
| 0 | 0 | 0 | 0 | 0 |
| 0.0534862 | 0 | 0 | 0.002726644 | 0.009362582 |
| 0.000479628 | 0.001000263 | 0 | 0 | 0 |
| 0 | 0.185815628 | 0.299110645 | 0 | 0 |
| 0 | 0 | 0 | 0 | 0 |
| 0.061851036 | 0 | 0 | 0 | 0 |
| 0.002067416 | 0 | 0 | 0 | 0 |

| | | | | |
|---|---|---|---|---|
| 0 | 0.002605229 | 0 | 0 | 0 |
| 0 | 0 | 0 | 0 | 0 |
| 0 | 0 | 0 | 0 | 0 |
| 0 | 0.000965313 | 0 | 0 | 0.286568822 |
| 0 | 0 | 0 | 0 | 0 |
| 0 | 0 | 0 | 0 | 0 |
| 0 | 0 | 0 | 0 | 0 |
| 0 | 0 | 0 | 0 | 0.002341247 |
| 0 | 0.937670988 | 0.019841169 | 0.511771142 | 0.325714316 |
| 0 | 0.012078536 | 0 | 0.000654986 | 0.073678946 |
| 0 | 0 | 0 | 0 | 0 |
| 0 | 0 | 0.000455264 | 0.029724252 | 0 |
| 0 | 0 | 0 | 0 | 0 |
| 0 | 0 | 0 | 0 | 0 |
| 0 | 0 | 0.026423201 | 0.020665768 | 0 |
| 0 | 0 | 0 | 0 | 0 |
| 0 | 0 | 0 | 0 | 0.165764211 |
| 0 | 0 | 0 | 0 | 0 |
| 0 | 0 | 0 | 0 | 0 |
| 0 | 0 | 0 | 0 | 0 |
| 0 | 0 | 0 | 0 | 0 |
| 0 | 0 | 0 | 0 | 0 |
| 0 | 0 | 0 | 0 | 0 |
| 0.019681972 | 0.151995381 | 0.154402669 | 0.024929549 | 0.119918842 |
| 0 | 0 | 0 | 0 | 0 |
| 0 | 0 | 0 | 0 | 0 |
| 0 | 0 | 0 | 0 | 0 |
| 0 | 4.556984706 | 0 | 7.055323613 | 1.939809244 |
| 0 | 0 | 0 | 0 | 0 |
| 0 | 0 | 0 | 0 | 0 |
| 0 | 0.018657051 | 0 | 0 | 0.004265889 |
| 0 | 0 | 0 | 0 | 0 |
| 0.006972694 | 0 | 0 | 0 | 0.039269062 |
| 0 | 0 | 0 | 0 | 0 |
| 0 | 0 | 0 | 0 | 0 |
| 0 | 0.022299203 | 0 | 0 | 0 |
| 0 | 0.003446099 | 0 | 0 | 0 |
| 0.006344627 | 0 | 0 | 0 | 0 |
| 0 | 0 | 0 | 0 | 0 |
| 0 | 0 | 0.005105236 | 0 | 0 |
| 0.086683078 | 0.069427394 | 0.004658611 | 0.002694341 | 0.017238931 |
| 0 | 0 | 0 | 0 | 0 |
| 0.831895939 | 9.07984481 | 24.07802983 | 17.85059073 | 0.065166719 |
| 0.002161906 | 0 | 0 | 0 | 0 |
| 0 | 0 | 0 | 0 | 0 |
| 0 | 0 | 0 | 0 | 0 |
| 0 | 0 | 0 | 0 | 0 |
| 0.008949951 | 0.002133155 | 0 | 0 | 0.218075244 |

| | | | | |
|---|---|---|---|---|
| 0 | 0 | 0 | 0 | 0 |
| 0 | 0 | 0 | 0.026310727 | 0 |
| 1.46750966 | 0.056745141 | 0.136391154 | 0.017335455 | 0.023099419 |
| 0 | 0 | 0 | 0 | 0 |
| 0 | 0 | 0 | 0 | 0 |
| 0.000643679 | 0 | 0 | 0 | 0.144118906 |
| 0 | 0 | 0 | 0 | 0 |
| 0.046313842 | 0 | 0 | 0 | 0 |
| 0 | 0 | 0 | 0 | 0 |
| 0 | 0 | 0 | 0 | 0 |
| 0.05395106 | 0 | 0.016471332 | 0.006096839 | 0.160797889 |
| 3.823251035 | 0.812879639 | 0.340154276 | 1.296375939 | 1.595197858 |
| 0 | 0 | 0 | 0 | 0 |
| 0.009091077 | 0.005958676 | 0 | 0.021933191 | 0.017133674 |
| 1.438964112 | 0.016861707 | 0.101982531 | 1.058440954 | 0.2825774 |
| 0 | 0.447740386 | 0 | 0 | 0 |
| 0 | 0 | 0 | 0 | 0 |
| 0 | 0 | 0 | 0 | 0 |
| 0 | 0 | 0 | 0 | 0 |
| 0 | 0 | 0.000814083 | 0 | 0.001494557 |
| 0.109265079 | 2.406119259 | 0 | 0.002129187 | 0.000760352 |
| 2.132766666 | 0.069815588 | 27.22179905 | 1.311102965 | 0 |
| 2.194265508 | 1.527012439 | 12.51560773 | 1.72469264 | 0.355494374 |
| 0 | 0 | 0 | 0 | 0 |
| 0 | 0 | 0 | 0 | 0 |
| 0 | 0.417150368 | 0 | 0 | 0.002050041 |
| 0 | 0 | 0 | 0.016298285 | 0 |
| 0 | 0.022377214 | 0 | 0 | 0 |
| 0 | 0 | 0 | 0 | 0 |
| 0 | 0 | 0 | 0 | 0 |
| 0 | 0 | 0 | 0 | 0 |
| 0.200754742 | 0 | 0 | 0 | 0 |
| 1.603981416 | 0 | 0 | 0 | 0.018168128 |
| 0 | 0.005699965 | 0 | 0 | 0.084370985 |
| 0.003126865 | 0 | 0.001116509 | 0.001721976 | 0.018140518 |
| 0.002988606 | 0.005375731 | 0.016260562 | 0.171290407 | 0.422651261 |
| 0 | 0 | 0.001926389 | 0 | 0 |
| 0 | 0 | 0 | 0 | 0 |
| 0.011147556 | 0.231388042 | 0.010411624 | 0.088594196 | 0.042760424 |
| 0 | 0 | 0 | 0 | 0 |
| 0 | 0 | 0 | 0 | 0 |
| 0 | 0 | 0 | 0 | 0 |
| 0 | 0 | 0 | 0 | 0 |
| 0 | 0 | 0 | 0 | 0 |
| 0.160183793 | 0.017858155 | 0.001399496 | 0.001079211 | 0.045284022 |
| 0 | 0 | 0 | 0 | 0 |
| 0 | 0 | 0 | 0 | 0 |
| 0 | 0.140182561 | 0.001899197 | 0.200480878 | 0.09908028 |

| | | | | |
|---|---|---|---|---|
| 0 | 0 | 0 | 0 | 0 |
| 0 | 0 | 0 | 0 | 0 |
| 0 | 0 | 0 | 0 | 0 |
| 0 | 0.004089072 | 0 | 0 | 0.004105892 |
| 0 | 0 | 0 | 0 | 0 |
| 0 | 0 | 0 | 0 | 0 |
| 0 | 1.311937863 | 0 | 0 | 0 |
| 0 | 0.001045191 | 0.081769567 | 0.064537192 | 0.323410856 |
| 0 | 0 | 0 | 0 | 0 |
| 0 | 0 | 0 | 0 | 0 |
| 0 | 0 | 0 | 0 | 0.007773891 |
| 0 | 0 | 0 | 0 | 0 |
| 0 | 0.004209037 | 0 | 0 | 0 |
| 0 | 0.188695674 | 0 | 0 | 0.208375719 |
| 0 | 0 | 0 | 0 | 0 |
| 0 | 0 | 0 | 0 | 0 |
| 0 | 0 | 0 | 0 | 0 |
| 0 | 0 | 0 | 0.013005321 | 0.007994313 |
| 0 | 0 | 0 | 0 | 0 |
| 0 | 0 | 0.001745345 | 0 | 0 |
| 0 | 0 | 0 | 0 | 0 |
| 0 | 0 | 0 | 0 | 0 |
| 0 | 0.011898849 | 0 | 0 | 0.038623077 |
| 0 | 0 | 0 | 0 | 0 |
| 0 | 0 | 0 | 0 | 0 |
| 0 | 0 | 0 | 0 | 0 |
| 0 | 0 | 0 | 0 | 0 |
| 0 | 0 | 0 | 0 | 0 |
| 0 | 0 | 0 | 0 | 0 |
| 0 | 0 | 0 | 0 | 0.737155469 |
| 0 | 0 | 0.005100711 | 0 | 0.003421567 |
| 0 | 0 | 0 | 0 | 0 |
| 0 | 0 | 0 | 0 | 0 |
| 0 | 0 | 0 | 0 | 0 |
| 0 | 0 | 0 | 0 | 0 |
| 0 | 0 | 0 | 0 | 0 |
| 0 | 0.225572868 | 0 | 0 | 0 |
| 0.007017669 | 0.044976888 | 0.001967968 | 0.008671912 | 0.00387102 |
| 0 | 0 | 0 | 0 | 0 |
| 0.000506476 | 0 | 0 | 0 | 0 |
| 0 | 0 | 0 | 0 | 0 |
| 0 | 0 | 0 | 0 | 0 |
| 0 | 0 | 0 | 0 | 0 |
| 1.248638841 | 0 | 0 | 0 | 0.022179394 |
| 0 | 0.002918331 | 0 | 0 | 0 |
| 0 | 0 | 0 | 0 | 0 |
| 0 | 0 | 0 | 0 | 0 |
| 0 | 0 | 0 | 0 | 0 |

| | | | | |
|---|---|---|---|---|
| 0 | 0 | 0 | 0 | 0 |
| 0 | 0 | 0 | 0 | 0 |
| 0 | 0 | 0 | 0 | 0 |
| 0 | 0 | 0 | 0 | 0 |
| 0 | 0 | 0 | 0 | 0 |
| 0 | 0 | 0 | 0 | 0 |
| 0 | 0 | 0 | 0 | 0 |
| 0 | 0 | 0 | 0 | 0 |
| 0.069638262 | 0.037809152 | 0.055446349 | 0.013455016 | 0.041108633 |
| 0 | 0 | 0 | 0 | 0 |
| 0 | 0 | 0 | 0 | 0 |
| 0 | 0 | 0 | 0 | 0 |
| 0 | 0 | 0 | 0 | 0 |
| 0 | 0 | 0 | 0 | 0 |
| 0 | 0 | 0.911842297 | 0 | 0.533477941 |
| 0 | 0 | 0.165407642 | 0 | 0 |
| 0 | 0 | 0 | 0 | 0 |
| 0 | 0 | 0 | 0 | 0 |
| 0 | 0 | 0 | 0 | 0 |
| 0 | 0 | 0 | 0 | 0 |
| 0 | 0 | 0 | 0 | 0 |
| 0 | 0 | 0 | 0 | 0 |
| 0 | 0 | 0 | 0 | 0 |
| 0 | 0 | 0 | 0 | 0 |
| 0.003331225 | 0 | 0 | 0.088958825 | 0.524862317 |
| 0.109370338 | 1.16850583 | 0 | 0 | 14.09405145 |
| 0 | 0.137984025 | 0 | 0 | 0 |
| 0 | 0 | 0 | 0 | 0 |
| 0 | 0.154884319 | 0 | 0.004918606 | 0 |
| 0 | 0 | 0 | 0 | 0 |
| 0 | 0 | 0 | 0 | 0 |
| 0.003217991 | 0 | 0 | 0 | 0 |
| 0 | 0 | 0 | 0 | 0 |
| 0 | 0 | 0.003165706 | 0 | 0 |
| 0.007572719 | 0 | 0.000802474 | 0.021349368 | 0.004419738 |
| 0.005698349 | 0.009291046 | 0 | 0.003062026 | 0 |
| 0 | 0 | 0.033220794 | 0.008125525 | 0.195058507 |
| 0.619264228 | 0.803674439 | 0 | 0 | 4.934707278 |
| 0 | 0 | 0 | 0 | 0 |
| 0 | 0 | 0 | 0 | 0 |
| 0 | 0 | 0 | 0 | 0 |
| 0 | 0 | 0 | 0 | 0 |
| 1.138066485 | 1.259303669 | 0 | 0.653948419 | 0.082717705 |
| 0 | 0 | 0 | 0 | 0 |
| 0 | 0 | 0 | 0 | 0 |
| 0.02470209 | 0 | 0 | 0 | 0.00258983 |
| 0 | 0 | 0 | 0 | 0 |
| 0 | 7.263484019 | 0 | 0 | 0.003536493 |

| | | | | |
|---|---|---|---|---|
| 0 | 0 | 0 | 0 | 0 |
| 0 | 0 | 0 | 0 | 0 |
| 0 | 0 | 0 | 0 | 0 |
| 0 | 0 | 0 | 0 | 0 |
| 0.00135444 | 0 | 0.020816806 | 0 | 0.001312803 |
| 0 | 0.017719685 | 0 | 0 | 0 |
| 0 | 0 | 0 | 0 | 0 |
| 0 | 0 | 0 | 0 | 0 |
| 0 | 0 | 0 | 0 | 0 |
| 0 | 0 | 0 | 0 | 0 |
| 0 | 0 | 0 | 0 | 0 |
| 0 | 2.17514913 | 0.002638832 | 1.077827258 | 1.367785554 |
| 0 | 0.16651465 | 0 | 0 | 0.438457454 |
| 0 | 0 | 0 | 0 | 0 |
| 1.487418202 | 0.018200438 | 0.000918879 | 0.035429354 | 0.142969128 |
| 0 | 0 | 0 | 0 | 0 |
| 0 | 0 | 0 | 0 | 0.003344527 |
| 0 | 0 | 0.049201606 | 0 | 0 |
| 0.006472492 | 0 | 0 | 0 | 0 |
| 0 | 0 | 0 | 0 | 0 |
| 0 | 0 | 0.004267473 | 0 | 0 |
| 0 | 0 | 0 | 0 | 0 |
| 0.035663839 | 0.014795827 | 0 | 0.599078832 | 3.496522126 |
| 0 | 0 | 0 | 0 | 0 |
| 0 | 0 | 0 | 0 | 0 |
| 0 | 0 | 0 | 0 | 0 |
| 0 | 0 | 0 | 0 | 0 |
| 0 | 0 | 0 | 0 | 0 |
| 0 | 0 | 0 | 0 | 0 |
| 0 | 0 | 0 | 0 | 0 |
| 0 | 0 | 0 | 0 | 0 |
| 0.045298746 | 0.113727891 | 0 | 0.015079649 | 0.041044787 |
| 0 | 0 | 0 | 0 | 0 |
| 0 | 0.004538059 | 0.007624827 | 0 | 0.140584118 |
| 0.006201231 | 0.009170429 | 0 | 0 | 0 |
| 0 | 0.099424327 | 0 | 0.015341703 | 0.027119355 |
| 0 | 0 | 0 | 0 | 0 |
| 0 | 0 | 0 | 0 | 0 |
| 0 | 0 | 0 | 0.007929979 | 0 |
| 0 | 0 | 0 | 0 | 0 |
| 0 | 1.18620996 | 0 | 0 | 0 |
| 0 | 0 | 0 | 0 | 0 |
| 0 | 0 | 0 | 0 | 0 |
| 0 | 0 | 0 | 0 | 0 |
| 0.015919992 | 0 | 0.014138664 | 0.001641301 | 0.002511956 |
| 0 | 0 | 0 | 0 | 0 |
| 0 | 0 | 0 | 0 | 0 |
| 0 | 0 | 0 | 0 | 0.004949711 |

| | | | | |
|---|---|---|---|---|
| 0 | 0 | 0 | 0 | 0 |
| 0 | 0 | 0 | 0 | 0 |
| 0 | 0 | 0.001257885 | 0 | 0.004329986 |
| 0 | 0 | 0 | 0 | 0 |
| 0 | 0.007774652 | 0.011481098 | 0 | 0 |
| 0 | 0 | 0 | 0 | 0 |
| 0 | 0 | 0 | 0.150821937 | 0 |
| 0.014645832 | 0 | 0 | 0 | 0.178700948 |
| 0 | 0 | 0 | 0 | 0 |
| 0 | 0 | 0 | 0 | 0 |
| 0 | 0 | 0 | 0 | 0 |
| 0 | 0 | 0 | 0 | 0 |
| 0 | 0 | 0 | 0 | 0 |
| 0 | 0 | 0 | 0 | 0 |
| 0 | 0 | 0 | 0 | 0 |
| 0 | 0 | 0 | 0 | 0 |
| 0 | 0 | 0 | 0 | 0 |
| 0 | 0 | 0 | 0 | 0 |
| 0 | 0 | 0 | 0 | 0 |
| 0 | 0 | 0 | 0 | 0 |
| 0 | 0.002829712 | 0 | 0 | 0 |
| 0 | 0 | 0 | 0 | 0 |
| 0 | 0 | 0 | 0 | 0 |
| 0 | 0 | 0 | 0 | 0 |
| 0.000606767 | 0 | 0 | 0.008453959 | 0 |
| 0.002585043 | 0.003665949 | 0.000707665 | 0.00785823 | 0 |
| 0 | 0 | 0 | 0 | 0 |
| 0.018555112 | 1.341577761 | 0 | 0 | 0.004577924 |
| 0 | 0 | 0 | 0 | 0 |
| 0.181483585 | 0.074059696 | 0 | 0.543456704 | 1.213046206 |
| 0 | 0 | 0 | 0 | 0 |
| 0 | 0 | 0 | 0 | 0 |
| 0 | 0 | 0 | 0 | 0 |
| 0 | 0 | 0 | 0 | 0 |
| 0.001986574 | 0 | 0 | 0 | 0.002042201 |
| 0 | 0 | 0 | 0.139777308 | 0 |
| 0 | 0 | 0 | 0 | 0 |
| 0 | 0 | 0 | 0 | 0.00208557 |
| 0 | 0 | 0 | 0 | 0 |
| 0 | 0 | 0 | 0 | 0 |
| 1.84254071 | 4.284898297 | 8.851973413 | 16.90794091 | 23.46637711 |
| 0.255160801 | 0 | 0 | 0.000570002 | 0.989945612 |
| 0 | 0.607616429 | 0 | 0 | 0 |
| 0 | 0 | 0 | 0 | 0 |
| 0 | 0 | 0 | 0 | 0 |
| 0 | 0 | 0 | 0 | 0 |
| 0 | 0 | 0 | 0 | 0 |
| 0.008619929 | 0 | 0 | 0 | 0.008276119 |

| | | | | |
|---|---|---|---|---|
| 0 | 0 | 0 | 0 | 0 |
| 0.017398039 | 0.00251194 | 0.000549551 | 0 | 0.000756682 |
| 0 | 0 | 0 | 0 | 0 |
| 0 | 0 | 0 | 0 | 0 |
| 15.79184432 | 16.60748255 | 0 | 3.601571916 | 1.982285786 |
| 0 | 0 | 0 | 0 | 0 |
| 0 | 0 | 0 | 0 | 0 |
| 0 | 0 | 0 | 0 | 0 |
| 0.163868396 | 0.039922766 | 0.236918813 | 0.014598932 | 0.01076682 |
| 0 | 0 | 0 | 0 | 0 |
| 0 | 0 | 0.038804884 | 0 | 0 |
| 0 | 0 | 0 | 0 | 0 |
| 0 | 0.014644667 | 0.001868938 | 0 | 0.168555022 |
| 0.00219822 | 0.010002294 | 0.008041844 | 0.068088951 | 0.042935702 |
| 0 | 0.382866544 | 0 | 0.153367872 | 0 |
| 0.157590354 | 0.012927301 | 0.002828177 | 0.295359976 | 0.659222856 |
| 0.011586242 | 0.609126165 | 0 | 0.001095175 | 0.001759935 |
| 0 | 0 | 0 | 0 | 0 |
| 0 | 0 | 0 | 0 | 0 |
| 0 | 0 | 0 | 0 | 0 |
| 0 | 0 | 0 | 0 | 0 |
| 0 | 0 | 0 | 0 | 0 |
| 0 | 0 | 0 | 0 | 0 |
| 0 | 0 | 0 | 0 | 0 |
| 0 | 0 | 0 | 0 | 0 |
| 0 | 0 | 0 | 0 | 0 |
| 0 | 0.037443122 | 0 | 0 | 0.120046367 |
| 0.000509023 | 0 | 0 | 0.002578953 | 0.016116923 |
| 0 | 0 | 0 | 0 | 0 |
| 0 | 0 | 0 | 0 | 0 |
| 0 | 0 | 0 | 0 | 0 |
| 0 | 0 | 0 | 0 | 0 |
| 0 | 0 | 0 | 0 | 0 |
| 0 | 0 | 0 | 0 | 0 |
| 0 | 0.467760668 | 0 | 0 | 0 |
| 0 | 0 | 0 | 0 | 0 |
| 0.001003099 | 0 | 0 | 0 | 0.001134306 |
| 0 | 0 | 0 | 0 | 0 |
| 0 | 0 | 0 | 0 | 0 |
| 0 | 0 | 0 | 0 | 0 |
| 0 | 0.178675183 | 0 | 0 | 1.249890893 |
| 0 | 0 | 0 | 0 | 0 |
| 0 | 0 | 0 | 0 | 0 |
| 0 | 0 | 0 | 0 | 0 |
| 0 | 0 | 0 | 0 | 0 |
| 0 | 0 | 0 | 0 | 0 |
| 0 | 0 | 0 | 0 | 0 |
| 0 | 0 | 0 | 0 | 0 |

| | | | | |
|---|---|---|---|---|
| 0 | 0.01743475 | 0 | 0 | 0.107349669 |
| 0 | 0 | 0 | 0 | 0 |
| 0 | 0 | 0 | 0 | 0 |
| 0 | 0 | 0 | 0 | 0 |
| 0 | 0.088826524 | 0 | 0.013044521 | 0.040968205 |
| 0 | 0 | 0 | 0 | 0 |
| 0.003105153 | 4.224589987 | 0 | 0.042389473 | 0.002340875 |
| 0 | 0 | 0 | 0 | 0 |
| 0 | 0 | 0 | 0 | 0.005782979 |
| 0 | 0 | 0.004459186 | 0 | 0 |
| 0.00522987 | 0 | 0 | 0 | 0 |
| 0 | 0 | 0 | 0 | 0 |
| 0 | 0 | 0 | 0 | 0 |
| 0 | 0 | 0 | 0 | 0 |
| 0 | 0 | 0 | 0 | 0 |
| 0 | 0 | 0 | 0 | 0 |
| 0 | 0 | 0 | 0 | 0 |
| 0 | 0 | 0 | 0 | 0 |
| 0 | 0 | 0 | 0 | 0 |
| 0 | 0 | 0 | 0 | 0 |
| 0.001313902 | 0.389236802 | 0 | 0.005200663 | 0.005757328 |
| 0 | 0 | 0 | 0 | 0 |
| 0 | 0 | 0 | 0 | 0.007374556 |
| 0.004510343 | 0.000604691 | 0 | 0.001224189 | 0.004189534 |
| 0 | 0 | 0 | 0 | 0 |
| 0.001900425 | 0 | 0 | 0 | 0 |
| 0 | 0 | 0 | 0 | 0 |
| 0 | 0 | 0 | 0 | 0 |
| 0 | 0 | 0 | 0 | 0 |
| 7.132238316 | 0.056569028 | 0.041697323 | 0 | 0.000786485 |
| 0 | 0 | 0 | 0 | 0 |
| 0 | 0 | 0 | 0 | 0 |
| 0 | 0 | 0 | 0 | 0 |
| 0.30122041 | 0 | 0 | 0 | 0.007392107 |
| 0 | 0 | 0 | 0 | 0 |
| 0 | 0 | 0 | 0 | 0 |
| 0.045148854 | 0 | 0 | 0 | 0 |
| 0 | 0 | 0 | 0 | 0 |
| 0 | 0 | 0 | 0 | 0 |
| 0 | 0 | 0 | 0 | 0 |
| 0 | 0 | 0 | 0 | 0 |
| 0 | 0 | 0 | 0 | 0 |
| 0 | 0 | 0 | 0 | 0 |
| 0 | 0 | 0 | 0 | 0 |
| 0 | 0 | 0 | 0 | 0 |
| 0 | 0 | 0 | 0 | 0 |
| 0 | 0 | 0 | 0 | 0 |
| 0 | 0.043751678 | 0 | 0 | 0 |

| | | | | |
|---|---|---|---|---|
| 0.03189304 | 0 | 0 | 0.017783159 | 0 |
| 0 | 0.074873843 | 0 | 0 | 0 |
| 0 | 0.162383745 | 0 | 0 | 0 |
| 0 | 0 | 0 | 0 | 0 |
| 0 | 0 | 0 | 0 | 0 |
| 0 | 0 | 0 | 0 | 0 |
| 0.000799496 | 0 | 0 | 0 | 0 |
| 0.326757551 | 0.406867092 | 0 | 0.284258753 | 1.13324782 |
| 0.097673202 | 0.039491172 | 0.006926657 | 0.092699856 | 0.418106424 |
| 0.144707033 | 0.052852095 | 0.00062221 | 0 | 0 |
| 0 | 0 | 0 | 0 | 0 |
| 0 | 0 | 0 | 0 | 0 |
| 0 | 0 | 0 | 0 | 0 |
| 0 | 0.01108952 | 0 | 0.01403161 | 0 |
| 0 | 0 | 0 | 0 | 0 |
| 0 | 0 | 0 | 0 | 0 |
| 0 | 0 | 0 | 0 | 0.090983875 |
| 0 | 0 | 0 | 0 | 0 |
| 0 | 0 | 0 | 0 | 0 |
| 0 | 0 | 0 | 0.007161706 | 0 |
| 0 | 0 | 0 | 0 | 0 |
| 0 | 0 | 0 | 0 | 0 |
| 0 | 0 | 0 | 0 | 0 |
| 0 | 0 | 0 | 0 | 0 |
| 0 | 0 | 0 | 0 | 0 |
| 0 | 0 | 0 | 0 | 0 |
| 0 | 0 | 0 | 0 | 0 |
| 0 | 0 | 0 | 0 | 0 |
| 0 | 0 | 0 | 0 | 0 |
| 0 | 0 | 0 | 0 | 0 |
| 0 | 0 | 0 | 0 | 0 |
| 0 | 0 | 0 | 0 | 0 |
| 0 | 0 | 0 | 0 | 0 |
| 0 | 0 | 0 | 0 | 0 |
| 0 | 0.002100605 | 0 | 0 | 0 |
| 0 | 0 | 0 | 0 | 0 |
| 0 | 0 | 0 | 0 | 0 |
| 0.067475096 | 0 | 0 | 0 | 0.015494961 |
| 0 | 0.001031596 | 0 | 0.014880213 | 0 |
| 0 | 0.003464383 | 0 | 0 | 0.032481748 |
| 0 | 0 | 0 | 0 | 0 |
| 0.027492205 | 0 | 0.004105139 | 0.004748473 | 0 |
| 0 | 0 | 0 | 0 | 0 |
| 0 | 0 | 0 | 0 | 0 |
| 0 | 0 | 0 | 0 | 0 |
| 0 | 0 | 0 | 0 | 0 |
| 0 | 0 | 0 | 0 | 0 |
| 0 | 0 | 0 | 0 | 0 |

| | | | | |
|---|---|---|---|---|
| 0 | 0 | 0 | 0 | 0 |
| 0.07164928 | 0 | 0.024410741 | 0 | 0 |
| 0 | 0 | 0 | 0 | 0 |
| 0 | 0 | 0 | 0 | 0 |
| 0 | 0.819700287 | 0.001437038 | 0.186171105 | 0.312630023 |
| 0 | 0 | 0 | 0 | 0 |
| 0 | 0 | 0 | 0 | 0 |
| 0.046743992 | 0.001379498 | 0.008148609 | 0.020387248 | 0.054104835 |
| 0 | 0 | 0 | 0 | 0.003983165 |
| 0 | 0 | 0 | 0 | 0 |
| 0 | 0 | 0 | 0 | 0 |
| 0 | 0 | 0 | 0 | 0 |
| 0 | 0 | 0 | 0 | 0 |
| 0 | 0 | 0 | 0 | 0 |
| 0 | 0 | 0 | 0 | 0 |
| 0 | 0 | 0.003298039 | 0 | 0 |
| 0 | 0 | 0 | 0 | 0 |
| 0 | 0 | 0 | 0 | 0 |
| 0 | 0 | 0 | 0 | 0 |
| 0 | 0 | 0 | 0 | 0 |
| 0 | 0 | 0 | 0 | 0 |
| 0 | 0 | 0 | 0 | 0 |
| 0 | 0 | 0 | 0 | 0 |
| 0 | 0 | 0 | 0 | 0.001510428 |
| 0 | 0.274680906 | 0 | 0.301267876 | 0 |
| 0 | 0 | 0 | 0 | 0 |
| 0.033508463 | 0.004149234 | 0 | 0 | 0.013025177 |
| 0 | 0 | 0 | 0 | 0 |
| 0 | 0 | 0 | 0 | 0 |
| 0 | 0 | 0 | 0 | 0 |
| 0 | 0 | 0 | 0 | 0 |
| 0 | 0 | 0 | 0 | 0 |
| 0 | 0 | 0 | 0 | 0 |
| 0 | 0 | 0 | 0 | 0 |
| 0 | 0 | 0 | 0 | 0 |
| 0 | 0 | 0 | 0 | 0 |
| 0 | 0 | 0 | 0 | 0 |
| 0 | 0 | 0 | 0 | 0.02185868 |
| 0 | 0 | 0 | 0 | 0 |
| 0 | 0 | 0 | 0 | 0 |
| 0 | 0 | 0 | 0 | 0 |
| 0 | 0.174113291 | 0 | 0.145571046 | 0 |
| 0 | 0 | 0 | 0 | 0 |
| 0 | 0 | 0 | 0 | 0 |
| 0 | 0 | 0 | 0 | 0 |
| 0 | 0 | 0 | 0 | 0 |
| 0 | 0 | 0 | 0 | 0 |
| 0 | 0 | 0 | 0 | 0 |

| | | | | |
|---|---|---|---|---|
| 0 | 0 | 0 | 0 | 1.898974908 |
| 0 | 0 | 0 | 0 | 0 |
| 0 | 0 | 0 | 0 | 0 |
| 0 | 0 | 0 | 0 | 0 |
| 0 | 0 | 0 | 0 | 0 |
| 0 | 0 | 0.005309455 | 0 | 0.026662331 |
| 0 | 0 | 0 | 0 | 0 |
| 0 | 0 | 0 | 0 | 0 |
| 0 | 0 | 0 | 0 | 0 |
| 0 | 0 | 0 | 0 | 0 |
| 0 | 0 | 0 | 0 | 0 |
| 0 | 0 | 0 | 0 | 0.001392374 |
| 0 | 0 | 0 | 0 | 0 |
| 0 | 0 | 0 | 0 | 0 |
| 0 | 0 | 0 | 0 | 0 |
| 0 | 0 | 0 | 0 | 0 |
| 0 | 0 | 0 | 0 | 0 |
| 0 | 0 | 0 | 0 | 0 |
| 0 | 0 | 0 | 0 | 0 |
| 0 | 0 | 0 | 0 | 0 |
| 0.007790768 | 0 | 0 | 0 | 0 |
| 0 | 0 | 0 | 0 | 0 |
| 0.002879758 | 0 | 0.01320476 | 0 | 0.078968539 |
| 0 | 0 | 0 | 0 | 0 |
| 0 | 0 | 0 | 0 | 0 |
| 0 | 0 | 0 | 0 | 0 |
| 0 | 0 | 0 | 0 | 0 |
| 0 | 0 | 0 | 0 | 0 |
| 0 | 0 | 0 | 0.427331844 | 0 |
| 0 | 0 | 0 | 0 | 0 |
| 0 | 0 | 0 | 0 | 0 |
| 0 | 0 | 0 | 0 | 0 |
| 0.143384185 | 0.014904503 | 0.021594714 | 0.003700579 | 0.206612945 |
| 0 | 0 | 0 | 0 | 0.088775718 |
| 0.001206174 | 0.15973254 | 0 | 0 | 0.518753048 |
| 0.001442756 | 0 | 0.018809951 | 0 | 0.268988762 |
| 0 | 0 | 0 | 0 | 0 |
| 0 | 0 | 0 | 0 | 0 |
| 0 | 0.002088137 | 0 | 0 | 0 |
| 0 | 0 | 0 | 0 | 0 |
| 0 | 0 | 0 | 0 | 0 |
| 0 | 0 | 0 | 0 | 0 |
| 0 | 0.086068444 | 0.001402209 | 0.056845673 | 0.034477058 |
| 0 | 0 | 0 | 0 | 0 |
| 0 | 0 | 0 | 0 | 0 |
| 0 | 0 | 0 | 0 | 0.271617178 |
| 0 | 0 | 0 | 0 | 0 |
| 0 | 0 | 0 | 0 | 0 |

| | | | | |
|---|---|---|---|---|
| 0 | 0 | 0 | 0 | 0 |
| 0 | 0 | 0 | 0 | 0 |
| 0 | 0 | 0 | 0 | 0 |
| 0 | 0 | 0 | 0 | 0 |
| 0 | 0 | 0 | 0 | 0 |
| 0 | 0 | 0 | 0 | 0 |
| 0 | 0 | 0 | 0 | 0 |
| 0 | 0 | 0 | 0 | 0 |
| 0 | 0 | 0 | 0 | 0 |
| 0 | 0.009736601 | 0 | 0.007581389 | 0 |
| 0 | 0 | 0 | 0 | 0 |
| 0 | 0 | 0 | 0 | 0 |
| 0 | 0 | 0 | 0 | 0 |
| 0 | 0.92998524 | 0 | 0 | 0 |
| 0.00612703 | 0 | 0 | 0 | 0 |
| 0.027286185 | 0.001228172 | 0.002257029 | 0 | 0 |
| 0 | 0 | 0 | 0 | 0 |
| 0 | 0 | 0 | 0 | 0 |
| 0 | 0 | 0 | 0 | 0 |
| 0.030569377 | 0.001028264 | 0 | 0 | 1.237010436 |
| 0 | 0 | 0 | 0 | 0 |
| 0 | 0 | 0 | 0 | 0 |
| 0 | 0 | 0 | 0 | 0 |
| 0 | 0 | 0 | 0 | 0 |
| 0 | 0 | 0 | 0 | 0.002700111 |
| 0 | 0 | 0 | 0 | 0 |
| 0 | 0 | 0 | 0 | 0 |
| 0 | 0 | 0 | 0 | 0 |
| 0 | 0 | 0 | 0 | 0 |
| 0 | 0.012474476 | 0 | 0 | 2.690616486 |
| 0 | 0.000598934 | 0 | 0.152779227 | 2.478420629 |
| 0 | 0 | 0 | 0 | 0 |
| 0 | 0 | 0 | 0 | 0 |
| 0 | 0 | 0 | 0 | 0 |
| 0 | 0 | 0 | 0 | 0 |
| 0 | 0 | 0 | 0 | 0 |
| 0 | 0 | 0 | 0 | 0 |
| 0 | 0 | 0 | 0 | 0 |
| 0 | 0 | 0 | 0 | 0 |
| 0 | 0 | 0 | 0 | 0 |
| 0 | 0 | 0 | 0 | 0 |
| 0 | 0 | 0 | 0 | 0 |
| 0 | 0 | 0 | 0 | 0 |
| 0 | 0 | 0 | 0 | 0 |
| 0 | 0 | 0 | 0 | 0 |
| 0 | 0 | 0 | 0 | 0 |
| 0.048831931 | 0.005280536 | 0.082022883 | 0.005599717 | 0 |
| 0 | 0 | 0 | 0 | 0 |

| | | | | |
|---|---|---|---|---|
| 0 | 0 | 0 | 0 | 0 |
| 0 | 0 | 0 | 0 | 0 |
| 0 | 0 | 0 | 0 | 0 |
| 0 | 0 | 0 | 0 | 0 |
| 0 | 0.003193535 | 0.01787766 | 0 | 0 |
| 0 | 0 | 0 | 0 | 0 |
| 0 | 0 | 0 | 0 | 0 |
| 0 | 0 | 0 | 0 | 0 |
| 0 | 0 | 0 | 0 | 0 |
| 0 | 0 | 0 | 0 | 0 |
| 0 | 0 | 0 | 0 | 0.059489731 |
| 0.00479391 | 0 | 0.004108225 | 0.001584014 | 0.01013484 |
| 0 | 0 | 0 | 0 | 0 |
| 0 | 0 | 0 | 0 | 0 |
| 0.028273319 | 0 | 0 | 0.017704565 | 0 |
| 1.096015653 | 0 | 0 | 0 | 1.608513142 |
| 0 | 0 | 0 | 0 | 0 |
| 0 | 0 | 0 | 0 | 0 |
| 0.373571867 | 0 | 0.027737716 | 0 | 0.055860736 |
| 0.041214283 | 0.763772231 | 0 | 0 | 0.001864207 |
| 0 | 0 | 0 | 0 | 0 |
| 0 | 0 | 0 | 0 | 0 |
| 0 | 0 | 0 | 0 | 0 |
| 0 | 0 | 0 | 0 | 0 |
| 0 | 0 | 0 | 0 | 0 |
| 0 | 0 | 0 | 0 | 0 |
| 0.006935648 | 0 | 0.005695974 | 0 | 0 |
| 0 | 0 | 0 | 0 | 0 |
| 0 | 0 | 0 | 0 | 0 |
| 0 | 0 | 0 | 0 | 0 |
| 0.002059075 | 0 | 0 | 0 | 0 |
| 0 | 0 | 0 | 0 | 0 |
| 0 | 0 | 0 | 0 | 0 |
| 0 | 0 | 0 | 0 | 0 |
| 0 | 0 | 0 | 0 | 0 |
| 0 | 0 | 0 | 0 | 0 |
| 0 | 0 | 0 | 0 | 0 |
| 0.027486274 | 0.00065825 | 0.002700167 | 0.002665234 | 0.044912055 |
| 0 | 0.025929466 | 0 | 0 | 0 |
| 0 | 0 | 0 | 0 | 0 |
| 0 | 0 | 0 | 0 | 0.001484659 |
| 0 | 0 | 0 | 0 | 0 |
| 0 | 0 | 0 | 0 | 0 |
| 0.014626313 | 0 | 0.006566568 | 0 | 0.010364727 |
| 0 | 0 | 0 | 0 | 0 |
| 0 | 0 | 0 | 0 | 0 |
| 0 | 0 | 0 | 0 | 0 |
| 0 | 0 | 0 | 0 | 0 |

| | | | | |
|---|---|---|---|---|
| 0 | 0 | 0 | 0 | 0 |
| 0 | 0 | 0 | 0 | 0 |
| 0 | 0 | 0 | 0 | 0 |
| 0.000817517 | 0.001790176 | 0 | 0 | 0 |
| 0 | 0 | 0 | 0.003841572 | 0.007502355 |
| 0 | 0 | 0 | 0 | 0.003043074 |
| 0 | 0 | 0 | 0 | 0 |
| 0 | 0 | 0 | 0 | 0 |
| 0 | 0 | 0 | 0 | 0 |
| 0 | 0 | 0 | 0 | 0 |
| 0.011821721 | 0 | 0 | 0 | 0 |
| 0 | 0 | 0 | 0 | 0 |
| 0.004346283 | 0.044051843 | 0 | 0.082025725 | 0.084206628 |
| 0 | 0 | 0 | 0 | 0 |
| 0 | 0 | 0 | 0 | 0 |
| 0 | 0 | 0 | 0 | 0 |
| 0 | 0 | 0 | 0 | 0 |
| 0 | 0 | 0 | 0 | 0 |
| 0 | 0 | 0 | 0 | 0 |
| 0 | 0 | 0 | 0 | 0 |
| 0 | 0 | 0 | 0 | 0 |
| 0 | 0 | 0 | 0 | 0 |
| 0.029950645 | 0.00143225 | 0.003760095 | 0.003866093 | 0 |
| 0.018684406 | 0.039999135 | 0 | 0 | 0.028001435 |
| 0 | 0 | 0 | 0 | 0 |
| 0 | 0 | 0 | 0 | 0 |
| 0 | 0 | 0 | 0 | 0 |
| 0 | 0 | 0 | 0 | 0 |
| 0 | 0 | 0 | 0 | 0 |
| 0 | 0 | 0 | 0 | 0 |
| 0.004679531 | 0 | 0 | 0.058194136 | 0.048586705 |
| 0.00361871 | 0 | 0 | 0 | 0.005952062 |
| 0 | 0 | 0 | 0 | 0 |
| 0 | 0 | 0 | 0 | 0 |
| 0 | 0 | 0 | 0 | 0 |
| 0 | 0 | 0 | 0 | 0 |
| 0 | 0.002262193 | 0 | 0 | 0 |
| 0 | 0 | 0 | 0 | 0 |
| 0 | 0.001351133 | 0.000443392 | 0.001823566 | 0 |
| 0 | 0 | 0 | 0 | 0 |
| 0 | 0 | 0 | 0 | 0 |
| 0 | 0 | 0 | 0 | 0 |
| 0 | 0 | 0 | 0 | 0 |
| 0 | 0 | 0 | 0 | 0 |
| 0 | 0 | 0 | 0 | 0 |
| 0.078535939 | 0.011491476 | 0.004421267 | 0.438612852 | 0.292925024 |
| 0 | 0 | 0.165757268 | 0 | 0 |
| 0 | 0 | 0 | 0 | 0 |

| | | | | |
|---|---|---|---|---|
| 0 | 0 | 0 | 0 | 0 |
| 0 | 0 | 0 | 0 | 0 |
| 0 | 0.001219345 | 0 | 0 | 0 |
| 0 | 0 | 0 | 0 | 0 |
| 0 | 0 | 0.001645742 | 0 | 0 |
| 0 | 0 | 0 | 0 | 0 |
| 0 | 0 | 0 | 0 | 0 |
| 0 | 0 | 0 | 0 | 0 |
| 0 | 0 | 0 | 0 | 0.002447834 |
| 0 | 0 | 0 | 0 | 0 |
| 0.00437997 | 0 | 0 | 0 | 0 |
| 0.000871908 | 0 | 0 | 0 | 0 |
| 0 | 0 | 0 | 0 | 0 |
| 0 | 0 | 0 | 0 | 0 |
| 0 | 0 | 0 | 0 | 0 |
| 0 | 0 | 0 | 0 | 0 |
| 0 | 0 | 0 | 0 | 0 |
| 0 | 0 | 0 | 0 | 0 |
| 0 | 0 | 0 | 0 | 0 |
| 0.000968099 | 0.002665038 | 0.002544197 | 0.000980969 | 0.002846292 |
| 0 | 0 | 0 | 0 | 0 |
| 0 | 0 | 0 | 0 | 0 |
| 0 | 0 | 0 | 0 | 0 |
| 0 | 0 | 0 | 0 | 0 |
| 0 | 0 | 0 | 0 | 0 |
| 0 | 0 | 0 | 0 | 0 |
| 0 | 0 | 0 | 0 | 0 |
| 0 | 0.004826152 | 0 | 0 | 0 |
| 0 | 0 | 0 | 0 | 0 |
| 0 | 0 | 0 | 0 | 0 |
| 0.001330715 | 0.00603607 | 0 | 0.034342238 | 0.004326228 |
| 0 | 0 | 0 | 0 | 0 |
| 0.049443074 | 0.011116918 | 0.34641696 | 0.005626511 | 0.054163109 |
| 0 | 0.005680142 | 0 | 0.002249876 | 0.007141784 |
| 0 | 0 | 0 | 0 | 0 |
| 0 | 0 | 0 | 0 | 0 |
| 0 | 0 | 0 | 0 | 0 |
| 0 | 0 | 0 | 0 | 0 |
| 0 | 0 | 0 | 0 | 0 |
| 0 | 0 | 0 | 0 | 0 |
| 0 | 0 | 0 | 0 | 0 |
| 0.092951858 | 0 | 0.037973114 | 0.025760719 | 0.452422652 |
| 0 | 0.058375166 | 0 | 0.042907566 | 0.061071788 |
| 0 | 0 | 0 | 0 | 0 |
| 0 | 0 | 0 | 0 | 0 |
| 0 | 0.17956314 | 0 | 0.031179363 | 0 |
| 0 | 0 | 0 | 0 | 0.006636405 |
| 0 | 0 | 0 | 0 | 0 |

| | | | | |
|---|---|---|---|---|
| 0 | 0 | 0 | 0 | 0 |
| 0 | 0.037556214 | 0 | 0.054455368 | 0 |
| 0.009949252 | 0.019859898 | 0 | 0.005100771 | 0.006964676 |
| 0 | 0 | 0 | 0 | 0 |
| 0 | 0 | 0 | 0 | 0.861093 |
| 0.068489105 | 0 | 0 | 0 | 0 |
| 0 | 0 | 0 | 0 | 0 |
| 0 | 0 | 0 | 0 | 0 |
| 0 | 0 | 0 | 0 | 0 |
| 0 | 0 | 0 | 0 | 0 |
| 0 | 0 | 0 | 0 | 0 |
| 0 | 0 | 0 | 0 | 0 |
| 0 | 0 | 0 | 0 | 0 |
| 0 | 0 | 0 | 0 | 0 |
| 0 | 0 | 0 | 0 | 0 |
| 0 | 0 | 0 | 0 | 0 |
| 0 | 0 | 0 | 0 | 0 |
| 0 | 0 | 0 | 0 | 0 |
| 0 | 0 | 0 | 0 | 0 |
| 0 | 0 | 0 | 0 | 0 |
| 0 | 0 | 0 | 0 | 0 |
| 0 | 0 | 0 | 0 | 0 |
| 0 | 0 | 0 | 0 | 0.003331809 |
| 0 | 0 | 0 | 0 | 0 |
| 0 | 0.001221444 | 0 | 0.00568743 | 0.000883057 |
| 0.084646204 | 0.00393198 | 0 | 0 | 0.001110418 |
| 0 | 0 | 0 | 0 | 0 |
| 0 | 0 | 0 | 0 | 0.055365229 |
| 0 | 0 | 0 | 0 | 0 |
| 0 | 0 | 0 | 0 | 0 |
| 0 | 0 | 0 | 0 | 0 |
| 0 | 0 | 0 | 0 | 0 |
| 0 | 0 | 0 | 0 | 0 |
| 0 | 0 | 0 | 0 | 0 |
| 0 | 0 | 0 | 0 | 0 |
| 0 | 0 | 0 | 0 | 0 |
| 0 | 0 | 0 | 0 | 0 |
| 0 | 0 | 0 | 0 | 0 |
| 1.516357943 | 0 | 0 | 11.37165717 | 1.734460149 |
| 0 | 0 | 0 | 0 | 0 |
| 0 | 0 | 0 | 0 | 0 |
| 0 | 0.002132796 | 0 | 0 | 0 |
| 0.029762662 | 0 | 0 | 0.01248276 | 0 |
| 0 | 0 | 0 | 0 | 0 |
| 0 | 0 | 0 | 0 | 0 |
| 0 | 0 | 0 | 0 | 0 |
| 0.005126012 | 0 | 0 | 0 | 0.00477359 |
| 0 | 0 | 0 | 0 | 0 |

| | | | | |
|---|---|---|---|---|
| 0 | 0 | 0 | 0 | 0 |
| 0.049081346 | 0.006903285 | 0.009999027 | 0.012047934 | 0.024738935 |
| 0 | 0 | 0 | 0 | 0 |
| 0 | 0 | 0 | 0 | 0 |
| 0 | 0 | 0 | 0 | 0 |
| 0 | 0 | 0 | 0 | 0 |
| 0 | 0 | 0 | 0 | 0 |
| 0 | 0 | 0 | 0 | 0 |
| 0 | 0 | 0 | 0 | 0 |
| 0 | 0 | 0 | 0 | 0 |
| 0 | 0 | 0 | 0 | 0 |
| 0 | 0 | 0 | 0 | 0 |
| 0 | 0.011323527 | 0 | 0 | 0 |
| 0.001190826 | 0 | 0 | 0 | 0 |
| 0.004269561 | 0 | 0 | 0 | 0 |
| 0 | 0 | 0 | 0 | 0 |
| 0 | 0 | 0 | 0 | 0 |
| 0 | 0 | 0 | 0 | 0 |
| 0.001312011 | 0 | 0 | 0 | 0 |
| 0 | 0 | 0 | 0 | 0 |
| 0 | 0 | 0 | 0 | 0 |
| 0 | 1.176438771 | 0 | 0 | 0.00079025 |
| 0.009129036 | 0 | 0.002940125 | 0.063256438 | 0 |
| 0 | 0 | 0 | 0 | 0 |
| 0 | 0 | 0 | 0 | 0 |
| 0 | 0 | 0 | 0 | 0 |
| 0 | 0 | 0 | 0 | 0 |
| 0 | 0 | 0 | 0 | 0 |
| 0 | 0 | 0 | 0 | 0 |
| 0 | 0 | 0 | 0 | 0 |
| 0 | 0 | 0 | 0 | 0 |
| 0 | 0 | 0 | 0 | 0 |
| 0 | 0 | 0 | 0 | 0 |
| 0 | 0 | 0 | 0 | 0 |
| 0.596015191 | 0 | 0 | 0 | 0 |
| 0 | 0 | 0 | 0 | 0 |
| 0 | 0 | 0 | 0 | 0 |
| 0 | 0 | 0 | 0 | 0 |
| 0.00162838 | 0.036676629 | 0 | 0 | 0 |
| 0 | 0 | 0 | 0 | 0 |
| 0.006048813 | 0.046457221 | 0.038542129 | 0.127868429 | 0 |
| 0 | 0 | 0 | 0 | 0 |
| 0 | 0 | 0 | 0 | 0 |
| 0 | 0 | 0 | 0 | 0 |
| 0 | 0 | 0 | 0 | 0 |
| 2.122301187 | 0.03884689 | 0.299875052 | 0.040834892 | 0.038886668 |
| 0 | 0 | 0 | 0 | 0 |
| 0 | 0 | 0 | 0 | 0 |

| | | | | |
|---|---|---|---|---|
| 0 | 0 | 0 | 0 | 0 |
| 0 | 0 | 0 | 0 | 0 |
| 0 | 0 | 0 | 0 | 0 |
| 0 | 0 | 0 | 0 | 0 |
| 0 | 0 | 0 | 0 | 0 |
| 0 | 0 | 0 | 0 | 0 |
| 0 | 0 | 0 | 0 | 0 |
| 0 | 0.001291938 | 0 | 0 | 0 |
| 0 | 0 | 0 | 0 | 0 |
| 0.018217383 | 0.003214735 | 0.002877158 | 0.314463658 | 0.001584634 |
| 0 | 0 | 0 | 0 | 0 |
| 0 | 0 | 0 | 0 | 0 |
| 0 | 0 | 0 | 0 | 0 |
| 0 | 0 | 0 | 0 | 0 |
| 0 | 0 | 0 | 0 | 0 |
| 0 | 0 | 0 | 0 | 0 |
| 0 | 0 | 0 | 0 | 0 |
| 0 | 0 | 0 | 0 | 0 |
| 0 | 0 | 0 | 0 | 0 |
| 0.082290597 | 0.024661983 | 0.022694212 | 0.133055104 | 0.136479695 |
| 0 | 0 | 0 | 0 | 0 |
| 0 | 0 | 0 | 0 | 0 |
| 0 | 0 | 0 | 0 | 0 |
| 0 | 0 | 0 | 0 | 0 |
| 0 | 0 | 0 | 0 | 0 |
| 0 | 0 | 0 | 0 | 0 |
| 0 | 0 | 0 | 0 | 0 |
| 0 | 0 | 0 | 0 | 0 |
| 0 | 0 | 0 | 0 | 0 |
| 0 | 0 | 0.000713544 | 0 | 0 |
| 0 | 0 | 0 | 0 | 0 |
| 0 | 0 | 0 | 0 | 0 |
| 0 | 0 | 0 | 0 | 0 |
| 0 | 0 | 0 | 0 | 0 |
| 0 | 0 | 0 | 0 | 0 |
| 0 | 0 | 0 | 0 | 0.00148712 |
| 0 | 0 | 0 | 0 | 0 |
| 0 | 0 | 0 | 0 | 0 |
| 0 | 0 | 0 | 0 | 0 |
| 0 | 0 | 0 | 0 | 0 |
| 0 | 0 | 0 | 0 | 0 |
| 0 | 0 | 0 | 0 | 0 |
| 0 | 0 | 0 | 0 | 0 |
| 0 | 0 | 0 | 0 | 0 |
| 0 | 0.005969959 | 0 | 0 | 0.00496346 |
| 0 | 0 | 0 | 0 | 0.103702147 |
| 0 | 0 | 0 | 0 | 0 |
| 0 | 0 | 0.000941348 | 0 | 0 |

| | | | | |
|---|---|---|---|---|
| 0 | 0 | 0 | 0 | 0 |
| 0 | 0 | 0 | 0 | 0 |
| 0 | 0 | 0 | 0 | 0 |
| 0 | 0 | 0 | 0 | 0 |
| 0 | 0 | 0 | 0 | 0 |
| 0.001852107 | 0.003743717 | 0 | 0.004691825 | 0 |
| 0 | 0 | 0 | 0 | 0 |
| 0 | 0 | 0 | 0 | 0 |
| 0 | 0 | 0 | 0 | 0 |
| 0 | 0 | 0 | 0 | 0 |
| 0 | 0 | 0 | 0 | 0 |
| 0 | 0 | 0 | 0 | 0 |
| 0 | 0 | 0 | 0 | 0 |
| 0 | 0 | 0 | 0 | 0 |
| 0 | 0 | 0 | 0 | 0 |
| 0 | 0 | 0 | 0 | 0 |
| 0 | 0 | 0 | 0 | 0 |
| 0 | 0 | 0 | 0 | 0 |
| 0 | 0 | 0 | 0 | 0 |
| 0 | 0 | 0 | 0 | 0 |
| 0.000495576 | 0 | 0 | 0 | 0 |
| 0 | 0 | 0 | 0 | 0 |
| 0 | 0 | 0 | 0 | 0 |
| 0 | 0 | 0 | 0 | 0 |
| 0 | 0 | 0 | 0 | 0 |
| 0.002897006 | 0.001394239 | 0 | 0.002258093 | 0 |
| 0 | 0 | 0 | 0 | 0 |
| 0 | 0 | 0 | 0 | 0 |
| 0 | 0 | 0 | 0 | 0 |
| 0 | 0 | 0 | 0 | 0.009611187 |
| 0 | 0 | 0 | 0 | 0 |
| 0 | 0 | 0 | 0 | 0 |
| 0 | 0 | 0 | 0 | 0 |
| 0 | 0 | 0 | 0 | 0 |
| 0 | 0 | 0 | 0 | 0 |
| 0 | 0 | 0 | 0 | 0 |
| 0 | 0 | 0 | 0 | 0 |
| 0 | 0 | 0 | 0 | 0 |
| 0 | 0 | 0 | 0.001273641 | 0 |
| 0 | 0 | 0 | 0 | 0 |
| 0 | 0 | 0 | 0 | 0 |
| 0 | 0 | 0 | 0 | 0 |
| 0 | 0 | 0 | 0 | 0 |
| 0 | 0 | 0 | 0 | 0 |
| 0 | 0.003464865 | 0 | 0 | 0 |
| 0 | 0 | 0 | 0 | 0 |
| 0 | 0 | 0 | 0 | 0 |
| 0 | 0 | 0 | 0 | 0 |

| | | | | |
|---|---|---|---|---|
| 0 | 0 | 0 | 0 | 0 |
| 0.132390296 | 0 | 0 | 0 | 0 |
| 0 | 0 | 0 | 0 | 0 |
| 0 | 0 | 0 | 0 | 0 |
| 0 | 0 | 0 | 0 | 0 |
| 0 | 0 | 0 | 0 | 0 |
| 0 | 0 | 0 | 0 | 0 |
| 0 | 0 | 0 | 0 | 0 |
| 0 | 0 | 0 | 0 | 0 |
| 0 | 0 | 0 | 0 | 0 |
| 0.000626583 | 0 | 0 | 0.005026396 | 0 |
| 0 | 0 | 0 | 0 | 0 |
| 0.577656663 | 0 | 0 | 0 | 0 |
| 0 | 0 | 0 | 0 | 0 |
| 0 | 0 | 0 | 0 | 0 |
| 0 | 0 | 0 | 0 | 0 |
| 0 | 0 | 0 | 0 | 0 |
| 0 | 0 | 0 | 0 | 0 |
| 0 | 0 | 0 | 0 | 0 |
| 0 | 0 | 0 | 0 | 0 |
| 0.01950228 | 0.006896551 | 0.002089103 | 0.005369989 | 0.001198544 |
| 0 | 0 | 0 | 0 | 0 |
| 0 | 0 | 0 | 0 | 0 |
| 1.075230811 | 4.372719094 | 0 | 1.819879868 | 0.638417565 |
| 0 | 0 | 0 | 0 | 0 |
| 0 | 0 | 0 | 0 | 0 |
| 0 | 0 | 0 | 0 | 0 |
| 0 | 0 | 0 | 0 | 0 |
| 0 | 0 | 0 | 0 | 0 |
| 0 | 0 | 0 | 0 | 0 |
| 0 | 0 | 0 | 0 | 0 |
| 0 | 0 | 0 | 0 | 0 |
| 0 | 0 | 0 | 0 | 0 |
| 0 | 0 | 0 | 0 | 0 |

| PHL018-0062_V2 | PHL018-0131_V2 | PHL018-0252_V1 | PHL018-0044_V3 | PHL018-0031_V3 |
|---|---|---|---|---|
| EYCF | EYCF | REF | CTRL | CTRL |
| 0.008854071 | 0 | 0.216805811 | 0.016968487 | 0.180267464 |
| 2.24963331 | 0.12446868 | 7.320441836 | 2.573517137 | 2.636154334 |
| 0 | 0.019356662 | 0 | 0 | 14.49174452 |
| 0 | 0 | 0 | 0 | 0 |
| 0 | 0 | 0 | 0 | 0 |
| 0 | 0 | 0 | 0 | 0 |
| 0 | 0 | 0 | 0 | 0 |
| 0 | 0 | 0 | 0 | 0 |
| 0 | 0 | 0 | 0 | 0 |
| 0 | 0 | 0 | 0 | 0 |
| 0 | 0 | 0 | 0 | 0 |
| 0 | 0 | 0 | 0 | 0 |
| 0 | 0 | 0 | 0 | 0 |
| 2.986305611 | 3.36210968 | 3.605195637 | 1.973765349 | 18.69039985 |
| 0 | 0 | 0 | 0 | 0 |
| 0 | 0 | 0 | 0 | 0 |
| 0 | 0 | 0 | 0 | 0 |
| 0 | 0 | 0 | 0 | 0 |
| 0 | 0 | 0 | 0 | 0 |
| 0 | 0 | 0 | 0 | 0 |
| 0 | 0 | 0 | 0 | 0 |
| 0 | 0 | 0.215814156 | 0 | 0 |
| 0 | 0 | 0 | 0 | 0 |
| 0 | 0 | 0 | 0 | 0 |
| 0 | 0.004019795 | 0 | 0 | 0 |
| 0 | 0 | 0 | 0 | 0 |
| 0 | 0 | 0 | 0 | 0 |
| 0 | 0 | 0 | 0 | 0 |
| 0 | 0 | 0 | 0 | 0 |
| 0 | 0.008139659 | 0 | 0 | 0 |
| 0 | 0 | 0 | 0 | 0 |
| 0 | 0 | 0 | 0 | 0 |
| 0 | 0 | 0 | 0 | 0 |
| 0 | 0 | 0 | 0 | 0 |
| 0 | 0 | 0 | 0 | 0 |
| 0 | 2.152490621 | 0 | 0.750219275 | 0 |
| 0 | 0 | 0 | 0 | 0 |
| 0 | 0 | 0 | 0 | 0 |
| 0 | 0 | 0 | 0 | 0 |
| 0 | 0 | 0 | 0 | 0 |
| 0 | 0 | 0 | 0 | 0 |
| 0 | 0.020855532 | 0.003340769 | 0.008781517 | 0.002487783 |
| 0.026306779 | 3.188965131 | 0.695058945 | 0.401187995 | 0.098711278 |
| 0.002784448 | 0.003017461 | 0.001933422 | 0.190793453 | 0.005779074 |
| 0 | 0 | 0 | 0.007613126 | 0.00196071 |
| 0 | 0 | 0 | 0 | 0 |

| | | | | |
|---|---|---|---|---|
| 0 | 0 | 0 | 0 | 0.009529209 |
| 0.07994914 | 0 | 0.008492793 | 0.007096518 | 0.016607875 |
| 0 | 0 | 0.253653571 | 0.001785808 | 0.00090342 |
| 0 | 0.071680589 | 0.040813657 | 0.791935189 | 0.326081671 |
| 0 | 0.00129466 | 0 | 0.003260715 | 0.001458557 |
| 0 | 0 | 0 | 0 | 0.004987314 |
| 0 | 0 | 0 | 0 | 0.269122442 |
| 0 | 0.006200862 | 0 | 0 | 0 |
| 0 | 0 | 0 | 0 | 0 |
| 0 | 0 | 0 | 0 | 0 |
| 0 | 0 | 0 | 0 | 0 |
| 0 | 0.079621641 | 0.836706443 | 0.031040902 | 0.051738043 |
| 0 | 0 | 0 | 0.001145987 | 0 |
| 0.006182832 | 0.003771984 | 0.057718887 | 0.083369043 | 0.012245259 |
| 0 | 0 | 0 | 0 | 0 |
| 0 | 0 | 0 | 0 | 0 |
| 0 | 0.084913098 | 0.010463006 | 0 | 2.325108084 |
| 0 | 0 | 0 | 0 | 0 |
| 0 | 0 | 0 | 0 | 0 |
| 0 | 0 | 0 | 0 | 0 |
| 0.067995226 | 0 | 0.088378272 | 0.246516971 | 0.085081435 |
| 0.000908767 | 0 | 0.190882296 | 0.010809085 | 0.019637947 |
| 0 | 0 | 0 | 0 | 0 |
| 0 | 0 | 0 | 0 | 0 |
| 0 | 0 | 0 | 0 | 0 |
| 0 | 0 | 0 | 0 | 0 |
| 0 | 0 | 0 | 0 | 0 |
| 0 | 0 | 0 | 0 | 0 |
| 0 | 0 | 0 | 0 | 0 |
| 0 | 0 | 0 | 0 | 0 |
| 2.964602578 | 0 | 0.001430021 | 0 | 0.029983575 |
| 0 | 0 | 0 | 0 | 0 |
| 0 | 0 | 0 | 0 | 0 |
| 0 | 0 | 0 | 0 | 0 |
| 0 | 0 | 0 | 0 | 0 |
| 0 | 0 | 0 | 0 | 0 |
| 0 | 0 | 0 | 0 | 0 |
| 0 | 4.244878764 | 0 | 0 | 0.05105685 |
| 0 | 0 | 0 | 0 | 0 |
| 0 | 0 | 0 | 0 | 0 |
| 0 | 0 | 0 | 0 | 0 |
| 0 | 0 | 0 | 0 | 0 |
| 0 | 0 | 0 | 0 | 0 |
| 0 | 0 | 0 | 0 | 0 |
| 0 | 0 | 0 | 0 | 0 |
| 0 | 0 | 0 | 0 | 0.021840755 |
| 0 | 0 | 0 | 0 | 0 |
| 0.005967968 | 0 | 0.071403386 | 0 | 0.045566313 |

| | | | | |
|---|---|---|---|---|
| 0 | 0 | 0 | 0 | 0 |
| 0 | 0 | 0 | 0 | 0 |
| 0 | 0 | 0 | 0 | 0 |
| 0 | 0 | 0 | 0 | 0 |
| 0 | 0 | 0 | 0 | 0.024058004 |
| 0 | 0 | 0 | 0 | 0 |
| 0 | 0 | 0 | 0 | 0 |
| 0 | 0 | 0 | 0 | 0 |
| 0 | 0 | 0 | 0 | 0 |
| 0 | 0 | 0 | 0 | 0 |
| 0 | 0 | 0 | 0 | 0 |
| 0 | 0.005083641 | 0 | 0 | 0 |
| 0 | 0 | 0 | 0 | 0 |
| 0 | 0 | 0 | 0 | 0 |
| 0 | 0 | 0 | 0 | 0 |
| 0 | 0 | 0 | 0 | 0 |
| 0 | 0 | 0 | 0 | 0 |
| 0 | 0 | 0 | 0 | 0 |
| 0.012252239 | 0.012392382 | 0 | 0 | 0.007645414 |
| 0 | 0 | 0 | 0 | 0 |
| 0 | 0 | 0 | 0 | 0 |
| 0 | 0 | 0 | 0 | 0 |
| 0 | 0 | 0 | 0 | 0 |
| 0 | 0 | 0 | 0 | 0 |
| 0 | 0 | 0 | 0 | 0 |
| 0 | 0 | 0 | 0 | 0 |
| 0 | 0 | 0 | 0 | 0 |
| 0 | 0 | 0 | 0 | 0 |
| 0 | 0 | 0 | 0 | 0 |
| 0 | 0 | 0 | 0 | 0 |
| 0.01056693 | 0.044219288 | 0 | 0 | 0.020988726 |
| 0 | 0 | 0 | 0 | 0.016709455 |
| 0 | 0 | 0 | 0 | 0 |
| 0 | 0 | 0 | 0 | 0 |
| 0 | 0 | 0 | 0 | 0 |
| 0 | 0 | 0 | 0 | 0 |
| 0 | 0 | 0 | 0 | 0 |
| 0 | 0 | 0 | 0 | 0 |
| 0 | 1.530694771 | 0 | 0 | 0 |
| 0 | 0 | 0 | 0 | 0 |
| 0 | 0 | 0 | 0 | 0 |
| 0 | 0 | 0 | 0 | 0 |
| 0 | 0 | 0.002395362 | 0 | 0.005414996 |
| 0 | 0 | 0 | 0 | 0 |
| 0 | 0 | 0.000729654 | 0 | 0 |
| 0 | 0 | 0 | 0 | 0 |
| 0 | 0 | 0 | 0 | 0.051472129 |
| 0 | 0 | 0 | 0 | 0 |

| | | | | |
|---|---|---|---|---|
| 0 | 0 | 0 | 0 | 0 |
| 0 | 0 | 0 | 0 | 0 |
| 0 | 0 | 0 | 0 | 0 |
| 0 | 0 | 0 | 0 | 0 |
| 0 | 0.002573842 | 0 | 0.010555299 | 0.045262137 |
| 0 | 0 | 0 | 0 | 0 |
| 0 | 0 | 0 | 0 | 0 |
| 0 | 0 | 0 | 0 | 0 |
| 0 | 0 | 0 | 0 | 0 |
| 0 | 0 | 0 | 0 | 0 |
| 0 | 0 | 0 | 0 | 0 |
| 0 | 0 | 0 | 0 | 0 |
| 0 | 0 | 0 | 0 | 0 |
| 0 | 0 | 0 | 0 | 0 |
| 0 | 0 | 0 | 0 | 0 |
| 0 | 0 | 0 | 0 | 1.568036522 |
| 0 | 0 | 0 | 0 | 0 |
| 0 | 0 | 0 | 0 | 0 |
| 0 | 0 | 0 | 0 | 0 |
| 0 | 0 | 0 | 0 | 0 |
| 0 | 0 | 0 | 0 | 0 |
| 0 | 0 | 0 | 0 | 0 |
| 0 | 0 | 0 | 0 | 0 |
| 0 | 0 | 0 | 0 | 0 |
| 0 | 0 | 0 | 0 | 0 |
| 0 | 0 | 0 | 0 | 0 |
| 0 | 0 | 0 | 0 | 0.003901604 |
| 0 | 0 | 0 | 0 | 0 |
| 0 | 0 | 0 | 0 | 0 |
| 0 | 0 | 0 | 0 | 0 |
| 0 | 0 | 0.039580773 | 0 | 0.002456231 |
| 0 | 0.000852971 | 0 | 0.091584637 | 0.000900893 |
| 0 | 0 | 0 | 0 | 0 |
| 0 | 0 | 0 | 0 | 0 |
| 0 | 0 | 0 | 0 | 0 |
| 0 | 0 | 0 | 0 | 0 |
| 0 | 0 | 0 | 0 | 0 |
| 0 | 0 | 0 | 0 | 0 |
| 0 | 0 | 0 | 0 | 0 |
| 0 | 0 | 0 | 0 | 0 |
| 0 | 0 | 0 | 0 | 0 |
| 0 | 0 | 0 | 0 | 0 |
| 0 | 0 | 0 | 0 | 0 |
| 0 | 0 | 0 | 0 | 0 |
| 0 | 0 | 0 | 0 | 0 |
| 0 | 0 | 0 | 0 | 0 |
| 0 | 0 | 0 | 0 | 0 |
| 0 | 0 | 0 | 0 | 0 |

| | | | | |
|---|---|---|---|---|
| 0 | 0 | 0 | 0 | 0 |
| 0 | 0 | 0 | 0 | 0 |
| 0 | 0 | 0 | 0 | 0 |
| 0 | 0 | 0 | 0 | 0 |
| 0 | 0 | 0 | 0 | 0 |
| 0 | 0 | 0 | 0 | 0 |
| 0 | 0 | 0 | 0 | 0 |
| 0 | 0 | 0 | 0 | 0 |
| 0 | 0 | 0 | 0 | 0 |
| 0 | 0 | 0 | 0 | 0 |
| 0 | 0 | 0 | 0 | 0 |
| 0 | 0 | 0 | 0 | 0 |
| 0 | 0 | 0.002288046 | 0 | 0 |
| 0 | 0 | 0 | 0 | 0 |
| 0 | 0 | 0 | 0 | 0 |
| 0 | 0 | 0 | 0 | 0 |
| 0 | 0 | 0 | 0 | 0 |
| 0 | 0 | 0 | 0 | 0 |
| 0 | 0 | 0 | 0 | 0 |
| 0 | 0 | 0 | 0 | 0 |
| 0.015402915 | 0 | 0.010084085 | 0.136550917 | 0.011282995 |
| 0 | 0 | 0 | 0 | 0 |
| 0 | 0 | 0 | 0 | 0 |
| 0 | 0 | 0 | 0 | 0.001680842 |
| 0 | 0 | 0 | 0 | 0 |
| 0 | 0 | 0 | 0 | 0 |
| 0 | 0.003110708 | 0.001245731 | 0.003446042 | 0.003379346 |
| 0 | 0 | 0 | 0 | 0 |
| 0 | 0 | 0 | 0 | 0 |
| 0 | 0 | 0 | 0 | 0 |
| 0.052884719 | 0 | 0 | 0 | 0 |
| 0 | 0 | 0 | 0 | 0 |
| 0 | 0 | 0 | 0 | 0 |
| 0 | 0 | 0.007096519 | 0 | 0 |
| 0.00515415 | 0.001489458 | 0 | 0.005923154 | 0.003565783 |
| 0 | 0 | 0 | 0 | 0 |
| 0 | 0 | 0 | 0 | 0 |
| 0 | 0 | 0 | 0 | 0 |
| 0 | 0 | 0 | 0 | 0 |
| 0 | 0 | 0 | 0 | 0 |
| 0 | 0 | 0 | 0 | 0 |
| 0 | 0 | 0 | 0 | 0 |
| 0 | 0 | 0 | 0 | 0 |
| 0 | 0 | 0 | 0 | 0 |
| 0 | 0 | 0 | 0 | 0 |
| 0 | 0 | 0 | 0 | 0.003994134 |
| 0 | 0 | 0.001937477 | 0 | 0 |
| 0.000612164 | 0 | 0.005242473 | 0 | 0 |

| | | | | |
|---|---|---|---|---|
| 0 | 0 | 0 | 0 | 0 |
| 0 | 0 | 0 | 0 | 0 |
| 0 | 0 | 0 | 0 | 0 |
| 0 | 0 | 0 | 0 | 0 |
| 0 | 0 | 0 | 0 | 0 |
| 0 | 0 | 0 | 0 | 0 |
| 0 | 0.000755465 | 0 | 0.001802562 | 0 |
| 0 | 0 | 0 | 0 | 0 |
| 0 | 0 | 0 | 0 | 0 |
| 0 | 0 | 0 | 0 | 0 |
| 0 | 0 | 0 | 0 | 0 |
| 0 | 0 | 0 | 0 | 0 |
| 0 | 0 | 0 | 0 | 0 |
| 0 | 0 | 0 | 0 | 0 |
| 0 | 0 | 0 | 0 | 0 |
| 0 | 0 | 0 | 0 | 0 |
| 0 | 0 | 0 | 0 | 0 |
| 0 | 0 | 0 | 0 | 0 |
| 0.00083009 | 0.015652248 | 0.007781196 | 0.00300491 | 0 |
| 0 | 0.007628276 | 0 | 0 | 0 |
| 0.008877382 | 0 | 0.02061799 | 0 | 0.015248155 |
| 0 | 0 | 0 | 0 | 0 |
| 0 | 0 | 0 | 0 | 0 |
| 0 | 0 | 0 | 0 | 0 |
| 0 | 0 | 0.002111443 | 0.045288996 | 0 |
| 0 | 0 | 0 | 0 | 0 |
| 0 | 0 | 0 | 0 | 0 |
| 0 | 0 | 0 | 0 | 0 |
| 0 | 0 | 0 | 0 | 0 |
| 0 | 0 | 0 | 0 | 0 |
| 0.010128935 | 0.021074998 | 2.127613621 | 0.064253802 | 0.144601339 |
| 0 | 0 | 0 | 0 | 0 |
| 0.036384215 | 0.221772844 | 0 | 0 | 0 |
| 0 | 0 | 0 | 0 | 0 |
| 0 | 0 | 0 | 0 | 0 |
| 0 | 0 | 0 | 0 | 0 |
| 0 | 0 | 0 | 0 | 0 |
| 0 | 0 | 0 | 0 | 0 |
| 0 | 0 | 0 | 0 | 0 |
| 0 | 0.001364416 | 0 | 0 | 0 |
| 0 | 0 | 0 | 0 | 0 |
| 0 | 0.003182586 | 0 | 0.045799786 | 0.009075757 |
| 0 | 0 | 0 | 0 | 0 |
| 0 | 0 | 0 | 0 | 0 |
| 0 | 0 | 0 | 0 | 0 |
| 0 | 0 | 0 | 0 | 0 |
| 0 | 0 | 0 | 0 | 0 |
| 0 | 0 | 0 | 0 | 0 |

| | | | | |
|---|---|---|---|---|
| 0 | 0 | 0 | 0 | 0 |
| 0 | 0 | 0 | 0 | 0 |
| 0 | 0.002747429 | 0 | 0.198749299 | 0.002954546 |
| 0 | 0 | 0 | 0 | 0 |
| 0 | 0 | 2.401652605 | 0 | 0.124722076 |
| 0 | 0 | 0 | 0 | 0 |
| 0 | 0 | 0 | 0 | 0 |
| 0 | 0 | 0 | 0 | 0 |
| 0 | 0 | 0 | 0 | 0 |
| 0 | 0 | 0 | 0 | 0 |
| 0 | 0 | 0 | 0 | 0 |
| 0 | 0 | 0 | 0 | 0 |
| 0 | 0 | 0 | 0 | 0 |
| 0 | 0 | 0 | 0.100441422 | 0.009329449 |
| 0.006832758 | 0 | 0.00091239 | 0.04321737 | 0.005390167 |
| 0 | 0 | 0 | 0.002328045 | 0 |
| 0 | 0 | 0.001247504 | 0.023537208 | 0 |
| 0 | 0 | 0 | 0 | 0 |
| 0 | 0 | 0 | 0 | 0 |
| 0 | 0 | 0 | 0 | 0 |
| 0 | 0 | 0 | 0 | 0 |
| 0 | 0 | 0.003378419 | 0 | 0 |
| 0 | 0 | 0 | 0 | 0 |
| 0 | 0 | 0.052955129 | 0 | 0 |
| 0 | 0 | 0 | 0 | 0 |
| 0 | 0 | 0 | 0 | 0 |
| 0.256451358 | 0 | 0 | 0 | 0.13254327 |
| 0 | 0 | 0 | 0 | 0 |
| 0 | 0 | 0 | 0 | 0 |
| 0 | 0 | 0 | 0 | 0 |
| 0 | 0 | 0 | 0 | 0.001517339 |
| 0 | 0 | 0 | 0 | 0 |
| 0 | 0 | 0 | 0 | 0 |
| 0 | 0 | 0 | 0 | 0.000684895 |
| 0 | 0 | 0 | 0 | 0 |
| 0 | 0 | 0 | 0.003350418 | 0 |
| 0 | 0 | 0 | 0 | 0 |
| 0.004754535 | 0.000883271 | 0 | 0.059712804 | 0.001492632 |
| 0 | 0 | 0 | 0 | 0 |
| 0 | 0 | 0 | 0 | 0 |
| 0 | 0 | 0 | 0 | 0 |
| 0 | 0 | 0 | 0 | 0 |
| 0 | 0 | 0 | 0 | 0 |
| 0 | 0 | 0.0017575 | 0 | 0 |
| 0 | 0 | 0 | 0 | 0 |
| 0 | 0 | 0 | 0 | 0 |
| 0 | 0 | 0 | 0 | 0 |
| 0 | 0 | 0.004855687 | 0 | 0 |

| | | | | |
|---|---|---|---|---|
| 0.205557247 | 0 | 0 | 0 | 0 |
| 0 | 0 | 0 | 0 | 0 |
| 0.003397475 | 0 | 0 | 0.001171315 | 0 |
| 0 | 0 | 0 | 0 | 0 |
| 0.001424245 | 0 | 0.000988945 | 0.015614534 | 0.002086585 |
| 0 | 0 | 0 | 0 | 0 |
| 0 | 0 | 0 | 0 | 0 |
| 0 | 0.001384613 | 0 | 0.015279748 | 0.011991715 |
| 0 | 0 | 0 | 0 | 0 |
| 0.026338883 | 0.364280256 | 0.177172666 | 0.009790023 | 0.327693916 |
| 0 | 0 | 0 | 0 | 0 |
| 0 | 0.006850405 | 0 | 0.015691463 | 0.001620702 |
| 0 | 0 | 0.007158396 | 0.007935021 | 0.055496829 |
| 0 | 0 | 0.003539642 | 0.003012813 | 0.001867081 |
| 0 | 0 | 0.002568226 | 0 | 0 |
| 0 | 0 | 0 | 0 | 0 |
| 0 | 0 | 0 | 0 | 0 |
| 0 | 0 | 0 | 0 | 0 |
| 1.813134368 | 0 | 0.016402728 | 0 | 0.093896204 |
| 0 | 0 | 0 | 0 | 0 |
| 0 | 0 | 0 | 0 | 0 |
| 0 | 0 | 0 | 0 | 0 |
| 0 | 0 | 0 | 0 | 0 |
| 0 | 0 | 0 | 0 | 0 |
| 0 | 0 | 0 | 0 | 0 |
| 0 | 0.002420934 | 0.000969501 | 0 | 0.001022779 |
| 0 | 0.317253446 | 0 | 0 | 0 |
| 0 | 0 | 0 | 0 | 0 |
| 0 | 0 | 0 | 0 | 0.0058642 |
| 0 | 0 | 0.000997494 | 0 | 0 |
| 0 | 0 | 0 | 0 | 0.000877998 |
| 0 | 0 | 0 | 0 | 0 |
| 0 | 0 | 0 | 0 | 0 |
| 0 | 0 | 0.014707975 | 0 | 0.046281219 |
| 0 | 0 | 0 | 0 | 0 |
| 0.00424759 | 0 | 0.014746884 | 0 | 0.003733751 |
| 0.03830334 | 0 | 0 | 0.122450727 | 0.12962374 |
| 0 | 0 | 0 | 0 | 0 |
| 0 | 0 | 0 | 0 | 0 |
| 0.039061795 | 0 | 0.0024472 | 0.004556489 | 0.003657387 |
| 0 | 0 | 0 | 0 | 0.081704137 |
| 0 | 0 | 0.070365796 | 0 | 0.008408238 |
| 0 | 0 | 0 | 0 | 0 |
| 0 | 0 | 0.000937277 | 0 | 0 |
| 0 | 0 | 0.219502527 | 0 | 0 |
| 0 | 0 | 0 | 0 | 0.004789158 |
| 0.002430871 | 0.006322309 | 0 | 0.002873375 | 0.006613916 |
| 0 | 0 | 0 | 0 | 0 |

| | | | | |
|---|---|---|---|---|
| 0 | 0 | 0 | 0 | 0 |
| 0 | 0.001009375 | 0 | 0.007626595 | 0.003127179 |
| 0 | 0 | 0 | 0 | 0 |
| 0 | 0 | 0 | 0 | 0 |
| 0 | 0 | 0 | 0 | 0 |
| 0.034189382 | 0 | 0.085653565 | 0 | 0.00681211 |
| 0 | 0 | 0 | 0 | 0 |
| 0 | 0 | 0 | 0 | 0 |
| 0 | 0 | 0 | 0 | 0.066515747 |
| 0 | 0 | 0 | 0 | 0 |
| 0.004124008 | 0.007425616 | 0 | 0.012796149 | 0.009062797 |
| 0 | 0 | 0 | 0 | 0 |
| 0 | 0 | 0 | 0 | 0 |
| 0 | 0 | 0 | 0.003099784 | 0.001975863 |
| 0 | 0 | 0.024846545 | 0 | 0 |
| 0 | 0 | 0 | 0 | 0 |
| 0 | 0 | 0 | 0 | 0 |
| 0 | 0 | 0 | 0 | 0 |
| 0 | 0 | 2.037309296 | 0 | 0 |
| 0 | 0 | 0 | 0 | 0 |
| 0 | 0 | 0 | 0 | 0.386279234 |
| 0 | 0.827237768 | 0 | 0.359225815 | 0.352215969 |
| 0 | 0 | 0 | 0 | 0 |
| 0 | 0 | 0 | 0 | 0 |
| 0 | 0 | 0 | 0 | 0 |
| 0 | 0 | 0.001289089 | 0 | 0 |
| 0.04874382 | 0 | 0 | 0.003360987 | 0.001904319 |
| 0 | 0 | 0 | 0 | 0 |
| 0 | 0 | 0 | 0 | 0 |
| 0.761752653 | 0.077313163 | 0.198274059 | 1.662970157 | 0.688330683 |
| 0 | 0 | 0 | 0.003364731 | 0 |
| 0 | 0 | 0 | 0 | 0 |
| 0 | 0 | 0 | 0 | 0 |
| 0 | 0 | 0.002199584 | 0 | 0 |
| 0 | 0 | 0 | 0 | 0.001982648 |
| 0 | 0 | 0 | 0 | 0 |
| 0 | 3.697293713 | 0 | 0.087354965 | 0 |
| 0 | 0.000794501 | 0 | 0 | 0 |
| 0 | 0 | 0 | 0 | 0.337125183 |
| 0 | 0 | 0.134201019 | 0 | 0 |
| 0 | 0 | 0 | 0.015799831 | 0.015160831 |
| 8.03926728 | 0.087262603 | 2.963539528 | 4.491225604 | 7.347761559 |
| 0 | 0 | 0 | 0 | 0 |
| 0 | 0 | 0 | 0 | 0 |
| 0 | 0 | 0 | 0 | 0.003868357 |
| 0 | 0 | 0 | 0 | 0 |
| 0 | 0 | 0.001436521 | 0 | 0.002652063 |
| 0 | 0 | 0 | 0 | 0 |

| | | | | |
|---|---|---|---|---|
| 0 | 0.001106479 | 0 | 0.008580297 | 0 |
| 0.003262055 | 0 | 0.00353915 | 0 | 0 |
| 0 | 0 | 0.001509417 | 0.002248329 | 5.350031346 |
| 0 | 0 | 0 | 0 | 0 |
| 0 | 0 | 0 | 0 | 0 |
| 0 | 0 | 0 | 0 | 0.081023291 |
| 0 | 0 | 0 | 0 | 0 |
| 0 | 0 | 0 | 0.015164912 | 0 |
| 0 | 0 | 0 | 0 | 0 |
| 0 | 0 | 0 | 0 | 0.000917332 |
| 0 | 0 | 0 | 0 | 0 |
| 0 | 0 | 0 | 0 | 0 |
| 0 | 0 | 0 | 0 | 0 |
| 0.161936903 | 0 | 0.009219948 | 0.002686977 | 0 |
| 0 | 0 | 0 | 0 | 0 |
| 0.005639523 | 0.0023468 | 0 | 0 | 0.013384706 |
| 0 | 0.01105503 | 0.000520842 | 0.323513948 | 0.004395719 |
| 0.010024749 | 0 | 0 | 0 | 0.001557683 |
| 0 | 0 | 0.133242342 | 0 | 0 |
| 0 | 0 | 0 | 0 | 0 |
| 0.011028942 | 0 | 0.138502364 | 0 | 0.096716663 |
| 0.004350378 | 0 | 0 | 0.004499513 | 0.001912052 |
| 0 | 0 | 0 | 0 | 0 |
| 0 | 0 | 0 | 0 | 0.025676868 |
| 0 | 0 | 0 | 0 | 0 |
| 0.001317497 | 0 | 0 | 0 | 0.062248788 |
| 0 | 0 | 0 | 0 | 0.003952784 |
| 0.004017525 | 0 | 0.001255333 | 0 | 0 |
| 0 | 0 | 0 | 0 | 0 |
| 0 | 0 | 0 | 0 | 0 |
| 0.06396997 | 0 | 0.24891117 | 0 | 0.011847495 |
| 0 | 0 | 0 | 0.075562567 | 0.006590732 |
| 0.033238585 | 0.015447801 | 0.006424247 | 0.028353016 | 0.004267182 |
| 0 | 0.009791898 | 0 | 0 | 0 |
| 0 | 0 | 0 | 0 | 0.009756522 |
| 0 | 0 | 0 | 0 | 0 |
| 0 | 0 | 0 | 0.002563308 | 0 |
| 0 | 0.002940528 | 0 | 0 | 0.008518586 |
| 0 | 0.045560971 | 0 | 0.468345902 | 0.333417117 |
| 0 | 0.000858637 | 0 | 0.002253606 | 0.000580401 |
| 0 | 0 | 0 | 0 | 0 |
| 0 | 0 | 0 | 0 | 0 |
| 0 | 0 | 0.091324032 | 0 | 0 |
| 0 | 0 | 0.588580448 | 0 | 0.020919184 |
| 0.171724811 | 0 | 0.15958583 | 0.560395489 | 0 |
| 0 | 0 | 0 | 0.001365754 | 0 |
| 0 | 0 | 0 | 0 | 0 |
| 0 | 0 | 0 | 0 | 0 |

| | | | | |
|---|---|---|---|---|
| 0 | 0 | 0 | 0 | 0 |
| 0.017166633 | 0.021751434 | 0.04538733 | 1.595912139 | 0.273747191 |
| 0 | 0 | 0 | 0 | 0 |
| 0 | 0 | 0 | 0 | 0 |
| 0 | 0 | 0 | 0 | 0.0016607 |
| 0 | 0 | 0 | 0 | 0 |
| 0.01221644 | 0 | 0 | 0 | 0 |
| 0.001775009 | 0.003077678 | 0 | 0 | 0 |
| 0 | 0 | 0.001888562 | 0.02250464 | 0 |
| 0 | 0 | 0 | 0 | 0 |
| 0 | 0 | 0.006813687 | 0 | 0 |
| 0.00146165 | 0 | 0.002029836 | 0.07143048 | 0.015257367 |
| 0 | 0 | 0 | 0.02711972 | 0 |
| 0 | 0 | 0 | 0 | 0 |
| 0 | 0 | 0 | 0.000873373 | 0.001030935 |
| 0 | 0 | 0 | 0 | 0 |
| 0.017211386 | 0 | 0.009810506 | 0.059515153 | 0.000564526 |
| 0.00833685 | 0 | 0.004862604 | 0.063807566 | 0.081588677 |
| 0 | 0 | 0 | 0 | 0 |
| 0.002598487 | 0 | 0.049618187 | 0.00358342 | 0.006027599 |
| 0.001059659 | 0.015158018 | 0 | 0.233992712 | 0.004463292 |
| 0 | 0 | 0 | 0 | 0 |
| 0 | 0 | 0 | 0 | 0 |
| 0 | 0 | 0 | 0 | 0 |
| 0 | 0 | 0 | 0 | 0.038147914 |
| 0 | 0 | 0 | 0 | 0 |
| 0 | 0 | 0 | 0.070316452 | 0.168749495 |
| 0.187803963 | 0.018307654 | 0.132359539 | 0.054166447 | 0.108076599 |
| 0 | 0 | 0 | 0 | 0 |
| 0 | 0 | 0 | 0 | 0 |
| 0 | 0 | 0 | 0 | 0 |
| 0 | 0 | 0 | 0 | 0 |
| 0 | 0 | 0 | 0 | 0 |
| 0 | 0 | 0 | 0 | 0 |
| 0.0110414 | 0.14613723 | 0 | 0 | 0 |
| 4.792961257 | 0.006610051 | 0.062537678 | 1.272092757 | 0.052709719 |
| 0 | 0 | 0 | 0 | 0 |
| 0 | 0 | 0 | 0 | 0 |
| 0 | 0.001806763 | 0 | 0.002155496 | 0 |
| 0 | 0 | 0 | 0 | 0 |
| 0.161725564 | 0 | 0.174649859 | 0.585218365 | 0 |
| 0 | 0 | 0 | 0 | 0 |
| 0.005063542 | 0.002012002 | 0.002050968 | 0 | 0.000618194 |
| 0 | 0 | 0.004526105 | 0 | 0.008505176 |
| 0 | 0 | 0.041623767 | 0 | 0 |
| 0 | 0.38407724 | 0 | 0 | 0 |
| 0 | 0 | 0 | 0 | 0 |
| 0 | 0 | 0.002514909 | 0 | 0.000723577 |

| | | | | |
|---|---|---|---|---|
| 0.007663367 | 0 | 0.002455926 | 0 | 0 |
| 0 | 0 | 0 | 0 | 0 |
| 0 | 0 | 0 | 0 | 0 |
| 0 | 0 | 0 | 0.074465861 | 0.011160002 |
| 0 | 0 | 0 | 0 | 0 |
| 0 | 0 | 0 | 0 | 0 |
| 0 | 0 | 0 | 0 | 0 |
| 0 | 0 | 0 | 0.013187121 | 0.002093381 |
| 0 | 5.106909126 | 0 | 0.689155777 | 0 |
| 0 | 0 | 0 | 0 | 0 |
| 0 | 0 | 0 | 0 | 0 |
| 0 | 0.014491596 | 0 | 0.009740109 | 0.003966566 |
| 0 | 0 | 0 | 0 | 0 |
| 0 | 0 | 0 | 0 | 0.025494956 |
| 0.001554232 | 0.07613018 | 0 | 0.030743678 | 0.01963932 |
| 0 | 0 | 0 | 0 | 0 |
| 0 | 0 | 0 | 0 | 0 |
| 0 | 7.292673258 | 0 | 0.023878052 | 0 |
| 0 | 0 | 0 | 0 | 0 |
| 0 | 0 | 0 | 0 | 0 |
| 0 | 0 | 0 | 0 | 0 |
| 0 | 0 | 0 | 0 | 0 |
| 0 | 0 | 0 | 0 | 0 |
| 0.137173979 | 0.020117869 | 0.821894265 | 0 | 0.536001471 |
| 0.014768438 | 0 | 0 | 0 | 0.002906389 |
| 0 | 0 | 0 | 0.25023031 | 0 |
| 0 | 0 | 0 | 0 | 0 |
| 0 | 0 | 0 | 1.884129401 | 6.801636772 |
| 0 | 0 | 0 | 0 | 0 |
| 0 | 0 | 0 | 0 | 0 |
| 0 | 0 | 0 | 0.004024345 | 0.00050298 |
| 0 | 0 | 0 | 0 | 0 |
| 0.00661939 | 0 | 0 | 0 | 0.000893211 |
| 0 | 0 | 0 | 0 | 0 |
| 0 | 0 | 0 | 0 | 0 |
| 0 | 0.035162939 | 0 | 0 | 0 |
| 0.068674129 | 0.093433705 | 0 | 3.577858078 | 0 |
| 0 | 0 | 0 | 0 | 0 |
| 0 | 0 | 0.41217145 | 0 | 0.001077181 |
| 0 | 0 | 0 | 0 | 0.004320464 |
| 0.025521597 | 0 | 0.109777947 | 0 | 0.08437641 |
| 0 | 0 | 0 | 0 | 0 |
| 28.83675068 | 10.53134384 | 0.136771676 | 5.977277835 | 0.719256427 |
| 0 | 0 | 0 | 0 | 0 |
| 0 | 0 | 0 | 0 | 0 |
| 0 | 0 | 0 | 0 | 0 |
| 0 | 0 | 0 | 0 | 0 |
| 0 | 0 | 0.001131134 | 0 | 0.001988826 |

| | | | | |
|---|---|---|---|---|
| 0 | 0 | 0 | 0 | 0 |
| 0 | 1.673466335 | 0 | 0.011768023 | 0 |
| 0.291758958 | 0 | 0.093612966 | 0.006191344 | 0.141654007 |
| 0 | 0 | 0 | 0 | 0 |
| 0 | 0 | 0 | 0 | 0 |
| 0 | 0.010842656 | 0.003274378 | 0.005725528 | 0 |
| 0 | 0 | 0 | 0 | 0 |
| 0 | 0 | 0 | 0 | 0.000913207 |
| 0 | 0 | 0 | 0 | 0 |
| 0 | 0 | 0 | 0 | 0 |
| 0.007410536 | 0 | 0 | 0 | 0.015162057 |
| 4.079293454 | 0.768862951 | 0.366585449 | 1.660423119 | 2.528031545 |
| 0 | 0 | 0 | 0 | 0 |
| 0 | 0.030364506 | 0.004404387 | 0.077014453 | 0.013535249 |
| 1.792528152 | 0.005087139 | 0.215794827 | 0.051474401 | 0.062724184 |
| 0 | 0 | 0 | 0 | 0 |
| 0 | 0 | 0 | 0 | 0.001939806 |
| 0 | 0 | 0 | 0 | 0 |
| 0 | 0 | 0 | 0 | 0 |
| 0 | 0 | 0 | 0 | 0.002775454 |
| 0 | 0 | 0 | 0.0035443 | 0.002510229 |
| 21.02304216 | 0.464665964 | 2.881422104 | 0 | 5.592197664 |
| 2.301307527 | 7.852770176 | 5.97628733 | 0.698827569 | 6.17134509 |
| 0 | 0 | 0 | 0 | 0 |
| 0 | 0 | 0 | 0 | 0 |
| 0 | 0.152857506 | 0 | 5.18495452 | 0 |
| 0 | 0 | 0 | 0 | 0 |
| 0.004020881 | 0 | 0 | 0 | 0 |
| 0 | 0 | 0 | 0 | 0 |
| 0 | 0 | 0 | 0 | 0 |
| 0 | 0 | 0 | 0 | 0 |
| 0 | 0.091504457 | 0 | 0.272288124 | 0 |
| 0.327789877 | 0 | 0.007106984 | 0.054016279 | 0.002114693 |
| 0.008805818 | 0.000694016 | 0 | 0.005381816 | 0.010115506 |
| 0.003637513 | 0 | 0.066872515 | 0 | 0.003172099 |
| 0.004521747 | 0.394280972 | 0.023961184 | 0.021414619 | 0 |
| 0 | 0.002989561 | 0 | 0 | 0.001010407 |
| 0 | 0 | 0 | 0 | 0 |
| 0 | 2.987073147 | 0.014696526 | 0.152948928 | 0 |
| 0 | 0 | 0 | 0 | 0 |
| 0 | 0 | 0 | 0 | 0 |
| 0 | 0 | 0 | 0 | 0 |
| 0 | 0 | 0 | 0 | 0 |
| 0 | 0 | 0 | 0 | 0 |
| 0.01004168 | 0.419751294 | 0.319796846 | 0.005333309 | 0.011729464 |
| 0 | 0 | 0 | 0 | 0 |
| 0 | 0 | 0 | 0 | 0 |
| 0 | 0.014190997 | 0 | 0.339278695 | 0.098083318 |

| | | | | |
|---|---|---|---|---|
| 0 | 0 | 0 | 0 | 0 |
| 0 | 0 | 0 | 0 | 0 |
| 0 | 0 | 0 | 0 | 0 |
| 0 | 0 | 0 | 0 | 0.002202723 |
| 0 | 0 | 0 | 0 | 0 |
| 0 | 1.718145999 | 0 | 0.00739533 | 0 |
| 0 | 0.007903668 | 0 | 0 | 0 |
| 0 | 0 | 0 | 0 | 0.070629809 |
| 0 | 0 | 0 | 0 | 0 |
| 0 | 0 | 0 | 0 | 0 |
| 0 | 0.000670025 | 0.035239597 | 0 | 0 |
| 0 | 0 | 0 | 0 | 0 |
| 0 | 0.010950968 | 0.005481857 | 0 | 0 |
| 0 | 0 | 0 | 0 | 0 |
| 0 | 0 | 0 | 0 | 0 |
| 0 | 0 | 0 | 0 | 0 |
| 0 | 0 | 0 | 0 | 1.543167115 |
| 0 | 0.014316361 | 0 | 0.002439948 | 0.003770354 |
| 0 | 0 | 0.140714416 | 0 | 0 |
| 0.002127497 | 0 | 0.010072218 | 0 | 0 |
| 0 | 0 | 0 | 0 | 0.000418708 |
| 0 | 0 | 0 | 0 | 0 |
| 0 | 0 | 0 | 0.052937152 | 0.006701545 |
| 0 | 0 | 0 | 0 | 0 |
| 0 | 0 | 0 | 0 | 0 |
| 0 | 0 | 0 | 0 | 0 |
| 0 | 0 | 0 | 0 | 0 |
| 0 | 0 | 0 | 0 | 0 |
| 0 | 0 | 0 | 0 | 0 |
| 0 | 0 | 0.011512794 | 0.043943581 | 0 |
| 0 | 0 | 0 | 0.010912666 | 0 |
| 0 | 0 | 0 | 0 | 0 |
| 0 | 0.021368798 | 0 | 0 | 0 |
| 0 | 0 | 0 | 0 | 0 |
| 0 | 0 | 0 | 0.001906812 | 0 |
| 0 | 0 | 0 | 0 | 0 |
| 0 | 0.137707014 | 0 | 0 | 0 |
| 0.005451964 | 0.003025001 | 0.022865336 | 0 | 0.012300568 |
| 0 | 0 | 0 | 0 | 0 |
| 0 | 0 | 0 | 0 | 0.000630265 |
| 0 | 0 | 0 | 0.0057139 | 0.014388751 |
| 0 | 0 | 0 | 0 | 0 |
| 0 | 0 | 0 | 0 | 0 |
| 0 | 0.007344085 | 0 | 0 | 0 |
| 0 | 0.265986743 | 0 | 0.047381236 | 0 |
| 0 | 0 | 0 | 0 | 0 |
| 0 | 0 | 0 | 0 | 0 |
| 0 | 0 | 0 | 0 | 0 |

| | | | | |
|---|---|---|---|---|
| 0 | 0 | 0 | 0 | 0 |
| 0 | 0 | 0 | 0 | 0 |
| 0 | 0 | 0 | 0 | 0 |
| 0 | 0.019558429 | 0 | 0.019968094 | 0 |
| 0 | 0 | 0 | 0 | 0 |
| 0 | 0 | 0 | 0.901636296 | 0 |
| 0 | 0 | 0 | 0 | 0 |
| 0 | 0 | 0 | 0 | 0 |
| 0.185272328 | 0 | 0.551760135 | 0.023952953 | 0.218996762 |
| 0 | 0 | 0 | 0 | 0 |
| 0 | 0 | 0 | 0 | 0 |
| 0 | 0 | 0 | 0 | 0 |
| 0 | 0 | 0 | 0 | 0 |
| 0 | 0 | 0 | 0 | 0 |
| 0 | 0.035602388 | 0 | 0.723402509 | 0.234165358 |
| 0 | 0 | 0 | 0 | 1.088794336 |
| 0 | 0 | 0 | 0 | 0 |
| 0 | 0 | 0 | 0 | 0 |
| 0 | 0 | 0 | 0 | 0 |
| 0 | 0 | 0 | 0 | 0 |
| 0 | 0 | 0 | 0 | 0 |
| 0 | 0 | 0 | 0 | 0 |
| 0 | 0 | 0 | 0 | 0 |
| 0 | 0 | 0.017064834 | 0 | 0 |
| 0 | 0.091989959 | 0.021857693 | 0.036947624 | 0 |
| 0.003667076 | 0.012138612 | 0.452776341 | 0.881650446 | 0.393653136 |
| 0 | 0 | 0 | 0.037553291 | 0 |
| 0 | 0 | 0 | 0 | 0 |
| 0 | 0.217417015 | 0 | 0.012153524 | 0 |
| 0 | 0 | 0 | 0 | 0 |
| 0 | 0 | 0 | 0 | 0 |
| 0 | 0 | 0 | 0 | 0.008009012 |
| 0 | 0 | 0 | 0 | 0 |
| 0 | 0 | 0 | 0 | 0 |
| 0 | 0.037236228 | 0.004322268 | 0.009979166 | 0.027130794 |
| 0 | 0.005149859 | 0.026428516 | 0 | 0.00128929 |
| 0 | 0 | 0 | 0.087589973 | 0.017795587 |
| 2.527996912 | 0.116732355 | 0 | 4.354424438 | 2.463265642 |
| 0 | 0 | 0 | 0 | 0 |
| 0 | 0 | 0 | 0 | 0 |
| 0 | 0.000661288 | 0 | 0 | 0 |
| 0 | 0 | 0 | 0 | 0 |
| 2.995929685 | 0.017202468 | 11.19294791 | 0.07556124 | 0.582947866 |
| 0 | 0 | 0 | 0 | 0 |
| 0 | 0 | 0 | 0 | 0 |
| 0 | 0.000542094 | 0 | 0.000862302 | 0 |
| 0 | 0 | 0 | 0 | 0 |
| 0.028224618 | 0 | 0 | 0 | 0.468475511 |

| | | | | |
|---|---|---|---|---|
| 0 | 0 | 0.24724099 | 0 | 0 |
| 0 | 0 | 0 | 0 | 0 |
| 0 | 0 | 0 | 0 | 0 |
| 0 | 0 | 0 | 0 | 0 |
| 0 | 0 | 0 | 0 | 0 |
| 0 | 0.112375195 | 0 | 0 | 0 |
| 0 | 0 | 0 | 0 | 0 |
| 0 | 0 | 0 | 0 | 0 |
| 0 | 0 | 0 | 0 | 0 |
| 0 | 0 | 0 | 0 | 0 |
| 0 | 0 | 0 | 0 | 0 |
| 0.219704663 | 5.664677317 | 0 | 0.138788233 | 0.094630677 |
| 0 | 0.015527115 | 0 | 0.17381765 | 0.137536858 |
| 0 | 0 | 0 | 0 | 0 |
| 0.009408606 | 0.031391177 | 0.022370591 | 0.360347114 | 0.011695555 |
| 0 | 0 | 0 | 0 | 0 |
| 0 | 0 | 0 | 0 | 0 |
| 0 | 0 | 0 | 0 | 0 |
| 0 | 0 | 0 | 0 | 0 |
| 0 | 0 | 0 | 0 | 0 |
| 0 | 0 | 0 | 0 | 0 |
| 0 | 0 | 0 | 0 | 0 |
| 0 | 0.000842642 | 0.083181167 | 0.15355718 | 0.233175646 |
| 0 | 0 | 0 | 0 | 0 |
| 0 | 0 | 0 | 0 | 0 |
| 0 | 0 | 0 | 0 | 0 |
| 0 | 0.008900654 | 0 | 0 | 0 |
| 0 | 0 | 0 | 0 | 0 |
| 0 | 0.008015105 | 0 | 0 | 0 |
| 0 | 0 | 0 | 0 | 0 |
| 0 | 0 | 0 | 0 | 0 |
| 0.006289476 | 0.001282975 | 0.003853403 | 0.340560301 | 0.00514921 |
| 0 | 0 | 0 | 0 | 0 |
| 0.005175246 | 0 | 0.152082408 | 0 | 0.004197166 |
| 0 | 0.008302721 | 0.029093344 | 0 | 0.000701535 |
| 0 | 0.336616078 | 0 | 0.014046006 | 0.003391364 |
| 0 | 0 | 0 | 0 | 0 |
| 0 | 0.414234342 | 0 | 0 | 0 |
| 0 | 0 | 0 | 0.00113797 | 0 |
| 0 | 0 | 0 | 0 | 0 |
| 0 | 0 | 0 | 0 | 0 |
| 0 | 0 | 0 | 0 | 0 |
| 0 | 0 | 0 | 0 | 0 |
| 0 | 0 | 0 | 0 | 0.001856121 |
| 0.001650997 | 0 | 0.107924841 | 0 | 0.001727706 |
| 0 | 0 | 0 | 0 | 0 |
| 0 | 0 | 0 | 0.177744258 | 0 |
| 0 | 0 | 0 | 0 | 0.002614561 |

| | | | | |
|---|---|---|---|---|
| 0 | 0 | 0 | 0 | 0 |
| 0 | 0 | 0 | 0 | 0 |
| 0.002683284 | 0 | 0 | 0 | 0 |
| 0 | 0 | 0 | 0 | 0 |
| 0 | 0 | 0 | 0 | 0 |
| 0 | 0 | 0 | 0 | 0.003535641 |
| 0 | 1.796915949 | 0 | 0 | 0 |
| 0 | 0 | 0.031992679 | 0 | 0.008606461 |
| 0 | 0 | 0 | 0 | 0 |
| 0 | 0 | 0 | 0.003048083 | 0 |
| 0 | 0 | 0 | 0 | 0 |
| 0 | 0 | 0.003619137 | 0 | 0 |
| 0 | 0 | 0 | 0 | 0 |
| 0 | 0.036724556 | 0 | 0 | 0 |
| 0 | 0 | 0 | 0 | 0 |
| 0 | 0 | 0 | 0 | 0 |
| 0 | 0 | 0 | 0 | 0 |
| 0 | 0 | 0 | 0 | 0 |
| 0 | 0.044516455 | 0 | 0 | 0.318668864 |
| 0 | 0 | 0 | 0 | 0 |
| 0 | 0.013642457 | 0 | 0 | 0 |
| 0 | 0 | 0 | 0 | 0 |
| 0 | 0 | 0 | 0 | 0 |
| 0 | 0 | 0 | 0 | 0 |
| 0.00128847 | 0 | 0 | 0 | 0 |
| 0.004391475 | 0 | 0 | 0.002271009 | 0.001608429 |
| 0 | 0 | 0 | 0 | 0 |
| 0 | 0 | 0 | 0 | 0.031486639 |
| 0 | 0.048708268 | 0 | 0 | 0 |
| 0 | 0.055144613 | 2.242240867 | 0.216605074 | 0.031795426 |
| 0 | 0 | 0 | 0 | 0 |
| 0 | 0 | 0 | 0 | 0 |
| 0 | 0 | 0 | 0 | 0 |
| 0 | 0 | 0 | 0 | 0 |
| 0 | 0 | 0.011343891 | 0 | 0 |
| 0 | 0 | 0 | 0 | 0 |
| 0 | 0 | 0 | 0.00239619 | 0 |
| 0 | 0 | 0.003263322 | 0 | 0 |
| 0 | 0 | 0 | 0 | 0 |
| 0 | 0 | 0 | 0 | 0.000447712 |
| 3.974365545 | 0.130118246 | 1.049895439 | 30.47594938 | 3.855643281 |
| 0.010894027 | 0 | 0.044192185 | 0.436071376 | 0.040320651 |
| 0 | 0 | 0 | 0 | 0 |
| 0 | 0 | 0 | 0 | 0 |
| 0 | 0 | 0 | 0 | 0 |
| 0 | 0 | 0 | 0 | 0 |
| 0 | 0 | 0 | 0 | 0 |
| 0 | 0.010060332 | 0.011798676 | 0 | 0 |

| | | | | |
|---|---|---|---|---|
| 0 | 0 | 0 | 0 | 0 |
| 0.001918286 | 0 | 0.000739995 | 0.000881799 | 0.006245286 |
| 0 | 0 | 0 | 0 | 0 |
| 0 | 0 | 0 | 0 | 0 |
| 1.984633822 | 0.104088566 | 11.34852135 | 0.76433425 | 0.394990014 |
| 0 | 0 | 0 | 0 | 0 |
| 0 | 0 | 0 | 0 | 0 |
| 0 | 0 | 0 | 0 | 0 |
| 0.579958134 | 0 | 0.010344651 | 0.091351719 | 0.219509972 |
| 0 | 0 | 0 | 0 | 0 |
| 0 | 0 | 0 | 0 | 0 |
| 0 | 0 | 0 | 0 | 0 |
| 0 | 0.001795488 | 0 | 0.162366933 | 0.084501915 |
| 0.002800755 | 0.001765896 | 0.002651926 | 0.108233776 | 0.003170683 |
| 0.039440464 | 0.709674843 | 0 | 0.122932552 | 0 |
| 0.087334626 | 0.005434054 | 0.0474401 | 0.190597525 | 0.330433666 |
| 0 | 0 | 0 | 0 | 0.000968377 |
| 0 | 0 | 0 | 0 | 0 |
| 0 | 0.004441593 | 0 | 0 | 0 |
| 0 | 0 | 0 | 0 | 0 |
| 0 | 0 | 0 | 0 | 0 |
| 0 | 0 | 0 | 0 | 0 |
| 0 | 0 | 0 | 0 | 0 |
| 0 | 0 | 0 | 0 | 0 |
| 0 | 0 | 0 | 0 | 0 |
| 0 | 0 | 0 | 0 | 0 |
| 0.004023288 | 0.006643767 | 0 | 0.004755669 | 0.009964182 |
| 0 | 0 | 0.030772437 | 0 | 0.000475076 |
| 0 | 0 | 0 | 0 | 0 |
| 0 | 0 | 0 | 0 | 0 |
| 0 | 0 | 0 | 0 | 0 |
| 0 | 0 | 0 | 0 | 0 |
| 0 | 0 | 0 | 1.399816357 | 0.207738987 |
| 0 | 0 | 0 | 0 | 0 |
| 0 | 2.562759241 | 0 | 0 | 0 |
| 0 | 0 | 0 | 0.024571853 | 0 |
| 0 | 0 | 0 | 0 | 0 |
| 0 | 0 | 0 | 0 | 0.001383852 |
| 0 | 0 | 0 | 0 | 0 |
| 0 | 0 | 0 | 0 | 0 |
| 0 | 0.198881783 | 0 | 0.745385748 | 0 |
| 0 | 0 | 0 | 0 | 0 |
| 0 | 0 | 0 | 0 | 0 |
| 0 | 0 | 0 | 0 | 0 |
| 0 | 0 | 0 | 0 | 0 |
| 0 | 0 | 0 | 0 | 0 |
| 0 | 0 | 0 | 0 | 0 |
| 0 | 0 | 0 | 0 | 0 |

| | | | | |
|---|---|---|---|---|
| 0 | 0.001846853 | 0 | 0.07222006 | 0 |
| 0 | 0 | 0 | 0 | 0 |
| 0 | 0 | 0 | 0 | 0 |
| 0 | 0 | 0 | 0 | 0 |
| 0 | 0.001218827 | 0 | 0 | 0 |
| 0 | 0 | 0 | 0 | 0.003957519 |
| 0.003516693 | 0 | 0 | 0.020686847 | 0.004508104 |
| 0 | 0 | 0 | 0 | 0 |
| 0 | 0 | 0.007863762 | 0 | 0 |
| 0 | 0 | 0 | 0 | 0.003040544 |
| 0 | 0.12085066 | 0 | 0 | 0 |
| 0 | 0 | 0 | 0 | 0 |
| 0 | 0 | 0 | 0 | 0 |
| 0 | 0 | 0 | 0 | 0 |
| 0 | 0 | 0 | 0 | 0 |
| 0 | 0 | 0 | 0 | 0 |
| 0 | 0 | 0 | 0 | 0 |
| 0 | 0 | 0 | 0 | 0 |
| 0 | 0 | 0 | 0 | 0 |
| 0 | 0 | 0 | 0 | 0 |
| 0 | 0.001269894 | 0.014820561 | 0 | 0.045985352 |
| 0 | 0 | 0 | 0 | 0 |
| 0 | 0.002801377 | 0 | 0.00668417 | 0.009298975 |
| 0.004104738 | 0 | 0.000427528 | 0.002971816 | 0.001503408 |
| 0 | 0 | 0 | 0 | 0 |
| 0 | 0 | 0 | 0 | 0 |
| 0 | 0 | 0 | 0 | 0 |
| 0 | 0 | 0 | 0 | 0 |
| 0 | 0.085651458 | 0 | 0 | 0 |
| 0 | 0.001792576 | 0 | 0.032689573 | 0.644799529 |
| 0 | 0 | 0 | 0 | 0 |
| 0 | 0 | 0 | 0 | 0 |
| 0 | 0 | 0 | 0 | 0 |
| 0 | 0 | 0 | 0 | 0 |
| 0 | 0 | 0 | 0 | 0 |
| 0 | 0 | 0 | 0 | 0 |
| 0 | 0 | 0 | 0.052590383 | 0 |
| 0 | 0 | 0 | 0 | 0 |
| 0 | 0 | 0 | 0 | 0 |
| 0 | 0.031576959 | 0 | 0 | 0 |
| 0 | 0 | 0 | 0 | 0 |
| 0 | 0 | 0 | 0 | 0 |
| 0 | 0 | 0 | 0 | 0 |
| 0 | 0.003611624 | 0 | 0 | 0 |
| 0 | 0 | 0 | 0 | 0 |
| 0 | 0 | 0 | 0 | 0 |
| 0 | 0 | 0 | 0 | 0 |
| 0 | 0.116289173 | 0 | 0 | 0 |

| | | | | |
|---|---|---|---|---|
| 0 | 0 | 0.328618393 | 0 | 0 |
| 0 | 3.476136519 | 0 | 0 | 0 |
| 0 | 0 | 0 | 0 | 0 |
| 0 | 0 | 0.012476785 | 0 | 0 |
| 0 | 0 | 0 | 0 | 0 |
| 0 | 0.014253471 | 0 | 0.020751386 | 0 |
| 0 | 0 | 0.002004035 | 0 | 0 |
| 0.556910312 | 0.02024821 | 1.437052409 | 0.591949606 | 0.617240004 |
| 0.038130948 | 4.350067618 | 0.915374692 | 0.528829197 | 0.146007184 |
| 0.014238106 | 0.010251533 | 0.067529405 | 0.016722974 | 0.004596159 |
| 0 | 0 | 0 | 0 | 0 |
| 0 | 0 | 0 | 0 | 0 |
| 0 | 0 | 0 | 0 | 0 |
| 0 | 0 | 0 | 0 | 0 |
| 0 | 0 | 0 | 0 | 0 |
| 0 | 0 | 0 | 0 | 0 |
| 0.073500398 | 0 | 0 | 0 | 0 |
| 0 | 0 | 0 | 0 | 0 |
| 0 | 0 | 0 | 0 | 0 |
| 0 | 0 | 0 | 0 | 0 |
| 0 | 0 | 0 | 0 | 0 |
| 0 | 0 | 0 | 0 | 0 |
| 0 | 0 | 0 | 0 | 0 |
| 0 | 0 | 0 | 0 | 0 |
| 0 | 0.001658801 | 0 | 0 | 0 |
| 0 | 0 | 0 | 0 | 0 |
| 0 | 0 | 0 | 0 | 0 |
| 0 | 0 | 0 | 0 | 0.00152478 |
| 0 | 0 | 0 | 0 | 0 |
| 0 | 0 | 0 | 0 | 0 |
| 0 | 0 | 0 | 0 | 0.01745026 |
| 0 | 0 | 0 | 0 | 0 |
| 0 | 0 | 0 | 0 | 0 |
| 0 | 0 | 0 | 0 | 0 |
| 0 | 0.005331111 | 0 | 0 | 0 |
| 0 | 0 | 0 | 0 | 0 |
| 0 | 0 | 0 | 0 | 0 |
| 0 | 0 | 0.303615965 | 0 | 0 |
| 0 | 0 | 0 | 0 | 0 |
| 0.315933028 | 0 | 0.000612345 | 0 | 0.043120279 |
| 0 | 0 | 0 | 0 | 0 |
| 0 | 0.054621683 | 0 | 0.115272936 | 0.223264178 |
| 0 | 0 | 0 | 0 | 0 |
| 0 | 0 | 0 | 0 | 0 |
| 0 | 0 | 0 | 0 | 0 |
| 0 | 0.004766873 | 0 | 0 | 0 |
| 0 | 0 | 0 | 0 | 0 |
| 0 | 0 | 0 | 0 | 0 |

| | | | | |
|---|---|---|---|---|
| 0 | 0 | 0 | 0 | 0 |
| 0 | 0 | 0 | 0.126577687 | 0.027595201 |
| 0 | 0 | 0 | 0 | 0 |
| 0 | 0 | 0 | 0 | 0 |
| 0 | 1.527142099 | 0 | 0.065716508 | 0.255988314 |
| 0 | 0 | 0 | 0 | 0 |
| 0 | 0 | 0 | 0 | 0 |
| 0.017136563 | 0 | 0.052862881 | 0.056658748 | 0.051034871 |
| 0 | 0.03331493 | 0 | 0 | 0 |
| 0 | 0 | 0 | 0 | 0 |
| 0 | 0 | 0 | 0 | 0 |
| 0 | 0.185401756 | 0 | 0 | 0 |
| 0 | 0 | 0 | 0 | 0 |
| 0 | 0 | 0 | 0 | 0 |
| 0 | 0 | 0 | 0 | 0.006229235 |
| 0 | 0 | 0 | 0 | 0 |
| 0 | 0 | 0 | 0 | 0 |
| 0 | 0 | 0 | 0 | 0 |
| 0 | 0 | 0 | 0 | 0 |
| 0 | 0 | 0 | 0 | 0 |
| 0 | 0 | 0 | 0 | 0 |
| 0 | 0 | 0 | 0 | 0 |
| 0 | 0 | 0 | 0 | 0 |
| 0 | 0 | 0.006414915 | 0 | 0 |
| 0 | 1.479030692 | 0 | 0.036243183 | 0 |
| 0 | 0 | 0 | 0 | 0 |
| 0.008672051 | 0 | 0.188521353 | 0.002759796 | 0.001140189 |
| 0 | 0 | 0 | 0 | 0 |
| 0 | 0 | 0 | 0 | 0 |
| 0 | 0 | 0 | 0 | 0 |
| 0 | 0 | 0 | 0 | 0 |
| 0 | 0 | 0 | 0 | 0 |
| 0 | 0 | 0 | 0 | 0 |
| 0 | 0 | 0 | 0 | 0 |
| 0 | 0 | 0 | 0 | 0 |
| 0 | 0 | 0 | 0 | 0 |
| 0 | 0 | 0 | 0 | 0 |
| 0 | 0 | 0 | 0 | 0.052619864 |
| 0 | 0 | 0 | 0 | 0 |
| 0 | 0 | 0 | 0 | 0 |
| 0 | 0 | 0 | 0 | 0 |
| 0 | 0.369282489 | 0 | 0.083568398 | 0 |
| 0 | 0 | 0 | 0 | 0 |
| 0 | 0 | 0 | 0.025606256 | 0 |
| 0 | 0 | 0 | 0 | 0 |
| 0 | 0 | 0 | 0 | 0 |
| 0 | 0 | 0 | 0 | 0 |
| 0 | 0 | 0 | 0.043528201 | 0 |

| | | | | |
|---|---|---|---|---|
| 0 | 0.011602424 | 0 | 0.035686065 | 0.002144498 |
| 0 | 0 | 0 | 0 | 0 |
| 0 | 0 | 0 | 0 | 0.00257944 |
| 0 | 0 | 0 | 0 | 0 |
| 0 | 0 | 0 | 0 | 0 |
| 0 | 0.000630097 | 0 | 0 | 0 |
| 0 | 0 | 0 | 0 | 0 |
| 0 | 0 | 0 | 0 | 0 |
| 0 | 0 | 0 | 0 | 0 |
| 0 | 0 | 0 | 0 | 0 |
| 0 | 0 | 0 | 0 | 0 |
| 0 | 0 | 0 | 0 | 0 |
| 0 | 0 | 0 | 0 | 0 |
| 0 | 0 | 0 | 0 | 0 |
| 0 | 0 | 0 | 0 | 0 |
| 0 | 0 | 0 | 0 | 0 |
| 0 | 0 | 0 | 0 | 0 |
| 0 | 0 | 0 | 0 | 0.025175698 |
| 0 | 0 | 0 | 0 | 0 |
| 0 | 0 | 0 | 0 | 0 |
| 0.000726309 | 0 | 0.003530265 | 0.019531402 | 0.001064077 |
| 0 | 0 | 0 | 0 | 0 |
| 0 | 0.006096759 | 0.007643087 | 0.001897441 | 0.000895901 |
| 0 | 0 | 0 | 0 | 0.102291935 |
| 0 | 0 | 0 | 0 | 0 |
| 0 | 0.344360296 | 0 | 0.079741654 | 0 |
| 0 | 0 | 0 | 0 | 0 |
| 0 | 0 | 0 | 0 | 0 |
| 0 | 0 | 0 | 0 | 0 |
| 0 | 0 | 0 | 0 | 0 |
| 0 | 0 | 0 | 0 | 0 |
| 0 | 0 | 0 | 0 | 0 |
| 0.030924898 | 0.003723645 | 0.006561243 | 0.021323353 | 0.007341318 |
| 0 | 0 | 0 | 0 | 0 |
| 0 | 0.015099549 | 0 | 1.225746159 | 0.147283356 |
| 0 | 0 | 0.054884912 | 0 | 0.013802005 |
| 0 | 0 | 0 | 0 | 0 |
| 0 | 0.021476547 | 0 | 0.010381265 | 0 |
| 0.006378567 | 0 | 0.001291807 | 0.020616344 | 0 |
| 0 | 0 | 0 | 0 | 0 |
| 0 | 0 | 0 | 0 | 0 |
| 0 | 0 | 0 | 0 | 0 |
| 0 | 0.503815877 | 0 | 0.080998458 | 0 |
| 0 | 0 | 0 | 0 | 0 |
| 0 | 0 | 0 | 0 | 0 |
| 0.001234023 | 0 | 0 | 0.003828978 | 0.00072316 |
| 0 | 0 | 0 | 0 | 0 |
| 0 | 0 | 0 | 0 | 0 |

| | | | | |
|---|---|---|---|---|
| 0 | 0 | 0 | 0 | 0 |
| 0 | 0 | 0 | 0 | 0 |
| 0 | 0 | 0 | 0 | 0.001806606 |
| 0 | 0 | 0 | 0 | 0 |
| 0 | 0 | 0 | 0 | 0 |
| 0 | 0 | 0 | 0 | 0 |
| 0 | 0 | 0 | 0 | 0 |
| 0 | 0 | 0 | 0 | 0 |
| 0 | 0 | 0 | 0 | 0 |
| 0 | 0.094323941 | 0 | 0 | 0 |
| 0 | 0 | 0 | 0 | 0 |
| 0 | 0 | 0 | 0 | 0 |
| 0 | 0 | 0 | 0 | 0 |
| 0 | 0 | 0 | 0 | 0 |
| 0 | 0 | 0 | 0 | 0 |
| 0 | 0 | 0.00086834 | 0 | 0 |
| 0 | 0 | 0 | 0 | 0 |
| 0 | 0 | 0 | 0 | 0 |
| 0 | 0 | 0 | 0 | 0 |
| 0 | 0 | 0 | 1.791107203 | 0 |
| 0 | 0 | 0 | 0.018912434 | 0 |
| 0 | 0 | 0 | 0 | 0 |
| 0 | 0 | 0 | 0 | 0 |
| 0 | 0 | 0 | 0 | 0 |
| 0 | 0 | 0 | 0 | 0 |
| 0 | 0 | 0 | 0 | 0.284389026 |
| 0 | 0 | 0 | 0 | 0 |
| 0 | 0 | 0 | 0 | 0 |
| 0 | 0 | 0 | 0 | 0 |
| 0 | 0.001797842 | 0 | 0 | 0.011393098 |
| 0 | 0.01304142 | 0 | 0 | 0.05450084 |
| 0 | 0 | 0 | 0 | 0 |
| 0 | 0 | 0 | 0 | 0 |
| 0 | 0 | 0 | 0 | 0 |
| 0 | 0 | 0 | 0 | 0 |
| 0 | 0 | 0 | 0 | 0 |
| 0 | 0 | 0 | 0 | 0 |
| 0 | 0 | 0 | 0 | 0 |
| 0 | 0.049656051 | 0 | 0 | 0 |
| 0 | 0 | 0 | 0 | 0 |
| 0 | 0 | 0 | 0 | 0 |
| 0 | 0 | 0 | 0 | 0 |
| 0 | 0 | 0 | 0 | 0 |
| 0 | 0 | 0.336686423 | 0 | 0 |
| 0 | 0 | 0 | 0 | 0 |
| 0 | 0 | 0 | 0 | 0 |
| 0.221215377 | 0 | 0.073601992 | 0 | 0.001875526 |
| 0 | 0 | 0 | 0 | 0 |

| | | | | |
|---|---|---|---|---|
| 0 | 0 | 0 | 0 | 0 |
| 0 | 0 | 0 | 0 | 0 |
| 0 | 0 | 0 | 0 | 0 |
| 0 | 0 | 0 | 0.009967975 | 0 |
| 0.055853249 | 0.008788889 | 0.10359716 | 0.023146746 | 0.008126715 |
| 0 | 0 | 0 | 0 | 0 |
| 0 | 0 | 0 | 0 | 0 |
| 0 | 0 | 0 | 0 | 0 |
| 0 | 0 | 0 | 0 | 0 |
| 0 | 0 | 0 | 0 | 0 |
| 0 | 0 | 0 | 0 | 0 |
| 0 | 0 | 0.028765916 | 0 | 0 |
| 0 | 0.010130074 | 0.007954403 | 0 | 0 |
| 0 | 0 | 0 | 0 | 0 |
| 0 | 0 | 0 | 0 | 0.00103772 |
| 0.298616516 | 0 | 12.71696163 | 0 | 0.001966574 |
| 0 | 0 | 0 | 0.003168758 | 0.001496169 |
| 0 | 0 | 0 | 0 | 0 |
| 0.006233637 | 0 | 0.031834817 | 0.004783504 | 0.06643208 |
| 0 | 0 | 0 | 0 | 0 |
| 0 | 0 | 0 | 0 | 0 |
| 0 | 0 | 0 | 0 | 0 |
| 0 | 0 | 0 | 0 | 0 |
| 0 | 0 | 0 | 0 | 0 |
| 0 | 0 | 0 | 0 | 0 |
| 0 | 0 | 0 | 0 | 0 |
| 0 | 0.000729616 | 0 | 0.002176107 | 0.000924729 |
| 0 | 0 | 0 | 0 | 0 |
| 0 | 0 | 0 | 0 | 0 |
| 0 | 0 | 0 | 0 | 0 |
| 0 | 0 | 0.010267452 | 0 | 0.001921753 |
| 0 | 0 | 0 | 0 | 0 |
| 0 | 0 | 0 | 0 | 0 |
| 0 | 0 | 0 | 0 | 0 |
| 0 | 0 | 0 | 0 | 0 |
| 0 | 0 | 0 | 0 | 0 |
| 0 | 0 | 0 | 0 | 0 |
| 0 | 0 | 0.042583589 | 0 | 0.00196388 |
| 0 | 0 | 0 | 0.28349314 | 0 |
| 0 | 0 | 0 | 0 | 0 |
| 0 | 0 | 0 | 0 | 0 |
| 0 | 0 | 0 | 0 | 0 |
| 0 | 0 | 0 | 0 | 0 |
| 0.227848865 | 0 | 1.783272423 | 0 | 0.001820116 |
| 0 | 0 | 0 | 0 | 0 |
| 0 | 0 | 0 | 0 | 0 |
| 0 | 0 | 0 | 0 | 0 |
| 0 | 0 | 0 | 0 | 0 |

| | | | | |
|---|---|---|---|---|
| 0 | 0 | 0 | 0 | 0 |
| 0 | 0.000552342 | 0 | 0.001098255 | 0 |
| 0 | 0 | 0 | 0 | 0 |
| 0.001301999 | 0 | 0.001808125 | 0 | 0 |
| 0 | 0.028057207 | 0 | 0.002747757 | 0 |
| 0 | 0 | 0 | 0 | 0.002739934 |
| 0 | 0 | 0 | 0.002874971 | 0 |
| 0.004073104 | 0 | 0 | 0 | 0 |
| 0 | 0 | 0 | 0 | 0.002169452 |
| 0 | 0 | 0 | 0 | 0 |
| 0 | 0 | 0 | 0 | 0.001891426 |
| 0 | 0 | 0 | 0 | 0 |
| 0 | 0.032805531 | 0 | 0.027491705 | 0.007571993 |
| 0 | 0 | 0 | 0 | 0 |
| 0 | 0 | 0 | 0 | 0 |
| 0 | 0 | 0 | 0 | 0 |
| 0 | 0 | 0 | 0 | 0 |
| 0 | 0 | 0 | 0 | 0 |
| 0 | 0 | 0 | 0 | 0 |
| 0 | 0 | 0 | 0 | 0 |
| 0 | 0 | 0 | 0 | 0 |
| 0 | 0 | 0 | 0 | 0 |
| 0.013611265 | 0 | 0.050462564 | 0.005530592 | 0.001068275 |
| 0 | 0 | 0 | 0.072085829 | 0.016808021 |
| 0 | 0 | 0 | 0 | 0 |
| 0 | 0.009365349 | 0 | 0 | 0 |
| 0 | 0 | 0 | 0 | 0 |
| 0 | 0 | 0 | 0 | 0 |
| 0 | 0 | 0 | 0 | 0 |
| 0 | 0 | 0 | 0 | 0 |
| 0 | 0.001879603 | 0.069626248 | 0.00448479 | 0.000595561 |
| 0 | 0 | 0 | 0 | 0 |
| 0 | 0 | 0 | 0 | 0 |
| 0 | 0 | 0 | 0 | 0 |
| 0 | 0 | 0 | 0 | 0 |
| 0 | 0 | 0 | 0 | 0 |
| 0 | 0 | 0 | 0 | 0 |
| 0 | 0 | 0 | 0 | 0 |
| 0 | 0.000993922 | 0 | 0.004031596 | 0 |
| 0 | 0 | 0 | 0 | 0 |
| 0 | 0 | 0 | 0 | 0 |
| 0 | 0 | 0 | 0 | 0 |
| 0 | 0 | 0 | 0 | 0 |
| 0 | 0 | 0 | 0 | 0 |
| 0 | 0 | 0 | 0 | 0 |
| 0.002017394 | 0.044948619 | 0 | 0.064891765 | 0.007536719 |
| 0 | 0 | 0 | 0 | 0 |
| 0 | 0 | 0 | 0 | 0 |

| | | | | |
|---|---|---|---|---|
| 0 | 0 | 0 | 0 | 0 |
| 0 | 0 | 0 | 0 | 0 |
| 0 | 0.122706252 | 0 | 0.176139351 | 0 |
| 0 | 0 | 0 | 0 | 0 |
| 0 | 0 | 0 | 0 | 0 |
| 0 | 0 | 0 | 0 | 0 |
| 0 | 0 | 0 | 0 | 0 |
| 0 | 0 | 0 | 0 | 0 |
| 0 | 0 | 0 | 0 | 0 |
| 0 | 0 | 0 | 0 | 0 |
| 0 | 0.002257743 | 0.003616592 | 0 | 0 |
| 0 | 0 | 0 | 0 | 0 |
| 0 | 0 | 0 | 0 | 0 |
| 0 | 0 | 0 | 0 | 0 |
| 0 | 0 | 0 | 0 | 0 |
| 0 | 0 | 0 | 0 | 0 |
| 0 | 0 | 0 | 0 | 0 |
| 0 | 0 | 0 | 0 | 0 |
| 0 | 0 | 0 | 0 | 0 |
| 0 | 0.195261644 | 0 | 0.005613256 | 0 |
| 0 | 0 | 0 | 0 | 0 |
| 0 | 0 | 0 | 0 | 0 |
| 0 | 0 | 0 | 0 | 0 |
| 0 | 0 | 0 | 0 | 0 |
| 0 | 0 | 0 | 0 | 0 |
| 0 | 0 | 0 | 0 | 0 |
| 0 | 0 | 0 | 0 | 0 |
| 0 | 0.01278079 | 0 | 0.006776745 | 0 |
| 0 | 0 | 0 | 0 | 0 |
| 0 | 0 | 0 | 0 | 0 |
| 0 | 0.008819275 | 0 | 0.007671948 | 0.001707707 |
| 0 | 0 | 0 | 0 | 0 |
| 0.042817242 | 0.022186832 | 0 | 0.016288755 | 0.02667796 |
| 0.001508779 | 0 | 0.002095285 | 0.002340751 | 0 |
| 0 | 0 | 0 | 0 | 0 |
| 0 | 0 | 0 | 0 | 0 |
| 0 | 0 | 0 | 0 | 0 |
| 0 | 0 | 0 | 0 | 0 |
| 0 | 0 | 0 | 0 | 0 |
| 0 | 0 | 0 | 0 | 0 |
| 0 | 0 | 0 | 0 | 0 |
| 0.101627178 | 0 | 0.539649674 | 0 | 0.115077212 |
| 0.008138941 | 0.263211303 | 0 | 0.03520234 | 0.007587954 |
| 0 | 0 | 0 | 0 | 0 |
| 0 | 0 | 0 | 0 | 0 |
| 0 | 0.628163362 | 0 | 0.005897951 | 0 |
| 0 | 0 | 0 | 0 | 0 |
| 0 | 0 | 0 | 0 | 0 |

| | | | | |
|---|---|---|---|---|
| 0 | 0 | 0 | 0 | 0 |
| 0 | 0.015232262 | 0 | 0 | 0.009652829 |
| 0 | 0 | 0 | 0.034650264 | 0.002210887 |
| 0 | 0 | 0 | 0 | 0 |
| 0 | 0 | 0 | 0 | 0.035482265 |
| 0 | 0 | 0 | 0 | 0.094507581 |
| 0 | 0 | 0 | 0 | 0 |
| 0 | 0 | 0 | 0 | 0 |
| 0 | 0.186075691 | 0 | 0 | 0 |
| 0 | 0 | 0 | 0 | 0 |
| 0 | 0 | 0 | 0 | 0 |
| 0 | 0 | 0 | 0 | 0 |
| 0 | 0 | 0 | 0 | 0 |
| 0 | 0 | 0.255594378 | 0 | 0.011149848 |
| 0 | 0 | 0 | 0 | 0 |
| 0 | 0 | 0 | 0 | 0 |
| 0 | 0 | 0 | 0 | 0 |
| 0 | 0 | 0 | 0 | 0 |
| 0 | 0 | 0 | 0 | 0 |
| 0 | 0 | 0 | 0 | 0 |
| 0 | 0 | 0 | 0 | 0 |
| 0 | 0 | 0 | 0 | 0 |
| 0 | 0 | 0 | 0 | 0 |
| 0 | 0 | 0 | 0 | 0 |
| 0 | 0.084964034 | 0 | 0 | 0 |
| 0 | 0 | 0 | 0.010611005 | 0 |
| 0 | 0 | 0 | 0 | 0 |
| 0.000728187 | 0 | 0.796531374 | 0 | 0.004800724 |
| 0 | 0 | 0 | 0 | 0 |
| 0 | 0 | 0 | 0 | 0 |
| 0 | 0.009078509 | 0 | 0 | 0 |
| 0 | 0 | 0 | 0 | 0 |
| 0 | 0 | 0 | 0 | 0 |
| 0 | 0 | 0 | 0 | 0 |
| 0 | 0 | 0 | 0 | 0 |
| 0 | 0 | 0 | 0 | 0 |
| 0 | 0 | 0.047919547 | 0 | 0 |
| 0.095646698 | 0 | 0 | 0 | 0.050905466 |
| 0 | 0.001772627 | 0 | 5.18917894 | 0 |
| 0 | 0 | 0 | 0 | 0 |
| 0 | 0 | 0 | 0 | 0 |
| 0 | 0.034359557 | 0 | 0.001965345 | 0 |
| 0 | 0 | 0.255635047 | 0 | 0 |
| 0 | 0 | 0 | 0 | 0 |
| 0 | 0 | 0 | 0 | 0 |
| 0 | 0 | 0 | 0 | 0 |
| 0.034192012 | 0 | 0.051484869 | 0 | 0.001688524 |
| 0 | 0 | 0 | 0 | 0 |

| | | | | |
|---|---|---|---|---|
| 0 | 0 | 0 | 0 | 0 |
| 0.023753454 | 0 | 0.003702636 | 0.035598158 | 0.013671397 |
| 0 | 0 | 0 | 0 | 0 |
| 0 | 0 | 0 | 0 | 0 |
| 0 | 0 | 0 | 0 | 0 |
| 0 | 0 | 0 | 0 | 0 |
| 0 | 0 | 0 | 0 | 0 |
| 0 | 0 | 0 | 0 | 0 |
| 0 | 0 | 0 | 0 | 0 |
| 0 | 0 | 0 | 0 | 0 |
| 0 | 0 | 0 | 0 | 0 |
| 0 | 0 | 0 | 0 | 0 |
| 0 | 0.005108959 | 0 | 0.016430178 | 0 |
| 0 | 0 | 0 | 0 | 0 |
| 0.004230997 | 0 | 0.009233259 | 0.011252672 | 0.003099304 |
| 0 | 0 | 0 | 0 | 0 |
| 0 | 0 | 0 | 0 | 0 |
| 0 | 0 | 0 | 0 | 0 |
| 0 | 0 | 0 | 0 | 0 |
| 0 | 0 | 0 | 0 | 0 |
| 0 | 0 | 0 | 0 | 0 |
| 0 | 0 | 0 | 0 | 0 |
| 0 | 0.039544053 | 0.028979936 | 0.00212295 | 0.012529721 |
| 0 | 0 | 0 | 0 | 0 |
| 0 | 0 | 0 | 0 | 0 |
| 0 | 0 | 0 | 0 | 0 |
| 0 | 0 | 0 | 0 | 0 |
| 0 | 0 | 0 | 0 | 0 |
| 0 | 0 | 0 | 0 | 0 |
| 0 | 0 | 0 | 0 | 0 |
| 0 | 0 | 0 | 0 | 0 |
| 0 | 0 | 0 | 0 | 0 |
| 0 | 0 | 0 | 0 | 0 |
| 0 | 0 | 0 | 0 | 0 |
| 0 | 9.540634653 | 13.09222945 | 0 | 0 |
| 0 | 0 | 0 | 0 | 0 |
| 0 | 0 | 0 | 0 | 0 |
| 0 | 0 | 0 | 0 | 0 |
| 0 | 0 | 0 | 0 | 0.006296239 |
| 0 | 0 | 0 | 0 | 0 |
| 0.060325643 | 0 | 0.01623852 | 0.169040088 | 0.022192305 |
| 0 | 0 | 0 | 0 | 0 |
| 0 | 0 | 0 | 0 | 0 |
| 0 | 0 | 0 | 0 | 0 |
| 0 | 0 | 0 | 0 | 0 |
| 0.547986035 | 0.001582703 | 0.081973818 | 0.228156199 | 0.269911497 |
| 0 | 0 | 0 | 0 | 0 |
| 0 | 0 | 0 | 0 | 0 |

| | | | | |
|---|---|---|---|---|
| 0 | 0 | 0 | 0 | 0 |
| 0 | 0 | 0 | 0 | 0 |
| 0 | 0 | 0 | 0 | 0 |
| 0 | 0 | 0 | 0 | 0 |
| 0.00115382 | 0 | 0 | 0 | 0 |
| 0 | 0 | 0 | 0 | 0 |
| 0 | 0 | 0 | 0 | 0 |
| 0 | 0 | 0 | 0 | 0 |
| 0 | 0 | 0 | 0 | 0.00226482 |
| 0 | 0.003095773 | 0 | 0.007078833 | 0 |
| 0 | 0.00897857 | 0 | 0 | 0 |
| 0 | 0 | 0 | 0 | 0 |
| 0.000872561 | 0 | 0.001615669 | 0 | 0 |
| 0 | 0 | 0 | 0 | 0 |
| 0 | 0 | 0 | 0 | 0 |
| 0 | 0 | 0 | 0 | 0 |
| 0 | 0 | 0 | 0 | 0 |
| 0 | 0 | 0 | 0 | 0 |
| 0 | 0 | 0 | 0 | 0 |
| 0 | 0.496451438 | 0.170769535 | 0.106834252 | 0.141278758 |
| 0 | 0 | 0 | 0 | 0 |
| 0 | 0 | 0 | 0 | 0 |
| 0 | 0 | 0 | 0 | 0 |
| 0 | 0 | 0 | 0 | 0 |
| 0 | 0 | 0 | 0 | 0 |
| 0 | 0 | 0 | 0 | 0 |
| 0 | 0 | 0 | 0 | 0 |
| 0 | 0 | 0 | 0 | 0 |
| 0 | 0 | 0 | 0 | 0 |
| 0 | 0 | 0.023443956 | 0.00772833 | 0.004459925 |
| 0 | 0 | 0 | 0 | 0 |
| 0 | 0 | 0 | 0 | 0 |
| 0 | 0 | 0 | 0 | 0 |
| 0 | 0 | 0 | 0 | 0 |
| 0 | 0 | 0 | 0 | 0 |
| 0 | 0 | 0 | 0 | 0 |
| 0 | 0 | 0 | 0 | 0 |
| 0 | 0 | 0 | 0 | 0 |
| 0 | 0 | 0 | 0 | 0 |
| 0 | 0.415167424 | 0 | 0 | 0 |
| 0 | 0 | 0 | 0 | 0 |
| 0 | 0 | 0 | 0 | 0 |
| 0 | 0 | 0 | 0 | 0 |
| 0 | 0 | 0 | 0 | 0 |
| 0 | 0 | 0.010467755 | 0 | 0.004809052 |
| 0 | 0 | 0 | 0 | 0 |
| 0 | 0 | 0 | 0 | 0 |
| 0 | 0 | 0 | 0 | 0 |

| | | | | |
|---|---|---|---|---|
| 0 | 0 | 0 | 0 | 0 |
| 0 | 0 | 0 | 0 | 0 |
| 0 | 0 | 0 | 0 | 0 |
| 0 | 0 | 0 | 0 | 0 |
| 0 | 0 | 0 | 0 | 0 |
| 0.001089125 | 0.022818504 | 0 | 0.004130359 | 0 |
| 0 | 0 | 0 | 0 | 0 |
| 0 | 0 | 0 | 0 | 0 |
| 0 | 0 | 0 | 0 | 0 |
| 0 | 0 | 0 | 0 | 0 |
| 0 | 0 | 0 | 0 | 0 |
| 0 | 0 | 0 | 0 | 0 |
| 0 | 0 | 0 | 0 | 0 |
| 0 | 0 | 0 | 0 | 0 |
| 0 | 0 | 0 | 0 | 0 |
| 0 | 0 | 0 | 0 | 0 |
| 0 | 0 | 0 | 0 | 0 |
| 0 | 0 | 0 | 0 | 0 |
| 0 | 0 | 0 | 0 | 0 |
| 0 | 0 | 0 | 0 | 0 |
| 0 | 0 | 0 | 0 | 0 |
| 0 | 0 | 0 | 0 | 0 |
| 0 | 0 | 0 | 0 | 0 |
| 0 | 0 | 0 | 0 | 0 |
| 0 | 0 | 0 | 0 | 0 |
| 0 | 0 | 0 | 0.0023493 | 0 |
| 0 | 0 | 0 | 0 | 0 |
| 0 | 0 | 0 | 0 | 0 |
| 0 | 0 | 0 | 0 | 0 |
| 0 | 0 | 0 | 0 | 0 |
| 0 | 0 | 0 | 0 | 0 |
| 0 | 0 | 0 | 0 | 0 |
| 0 | 0 | 0 | 0 | 0 |
| 0 | 0 | 0 | 0 | 0 |
| 0 | 0 | 0 | 0 | 0 |
| 0 | 0 | 0 | 0 | 0 |
| 0 | 0 | 0 | 0 | 0 |
| 0 | 0 | 0 | 0 | 0 |
| 0 | 0.001999264 | 0 | 0 | 0 |
| 0 | 0 | 0 | 0 | 0 |
| 0 | 0 | 0 | 0 | 0 |
| 0 | 0 | 0 | 0 | 0 |
| 0 | 0 | 0 | 0 | 0 |
| 0 | 0 | 0 | 0 | 0 |
| 0 | 0 | 0 | 0 | 0 |
| 0 | 0 | 0 | 0 | 0 |
| 0 | 0 | 0 | 0 | 0 |
| 0 | 0 | 0 | 0 | 0 |

| | | | | |
|---|---|---|---|---|
| 0 | 0 | 0 | 0 | 0 |
| 0 | 0 | 0.084386086 | 0 | 0.018459722 |
| 0 | 0 | 0 | 0.005404146 | 0 |
| 0 | 0 | 0 | 0 | 0 |
| 0 | 0 | 0 | 0 | 0 |
| 0 | 0 | 0 | 0 | 0 |
| 0 | 0 | 0 | 0 | 0 |
| 0 | 0 | 0 | 0 | 0 |
| 0 | 0 | 0 | 0 | 0 |
| 0 | 0 | 0 | 0 | 0 |
| 0 | 0.001845626 | 0.00073911 | 0.014862555 | 0 |
| 0 | 0 | 0 | 0 | 0 |
| 0 | 0.155141178 | 0.199111238 | 0.21954863 | 0.013783404 |
| 0 | 0 | 0 | 0 | 0 |
| 0 | 0 | 0 | 0 | 0 |
| 0 | 0 | 0 | 0 | 0 |
| 0 | 0 | 0 | 0 | 0 |
| 0 | 0 | 0 | 0 | 0 |
| 0 | 0 | 0 | 0 | 0 |
| 0 | 0 | 0 | 0 | 0 |
| 0 | 0 | 0 | 0 | 0.004352569 |
| 0 | 0 | 0 | 0 | 0 |
| 0 | 0 | 0 | 0 | 0 |
| 0 | 0.166038066 | 0 | 3.075422955 | 1.687615703 |
| 0 | 0 | 0 | 0 | 0 |
| 0 | 0 | 0 | 0 | 0 |
| 0 | 0 | 0 | 0 | 0 |
| 0 | 0 | 0 | 0 | 0 |
| 0.002707073 | 0 | 0 | 0 | 0 |
| 0 | 0 | 0 | 0 | 0 |
| 0 | 0 | 0 | 0 | 0 |
| 0 | 0 | 0 | 0 | 0 |
| 0 | 0.00147782 | 0 | 0 | 0 |
| 0 | 0 | 0 | 0 | 0 |

| PHL018-0241_V1 | PHL018-0208_V1 | PHL018-0231_V1 | PHL018-0084_V2 | PHL018-0053_V2 |
|---|---|---|---|---|
| REF | REF | EYCF | EYCF | CTRL |
| 1.280773725 | 0.00676566 | 0 | 0.054501573 | 0 |
| 0.566350267 | 0.139109401 | 4.8764886 | 0.031031746 | 7.057208353 |
| 0 | 0.096275662 | 8.745479499 | 0 | 0 |
| 0 | 0 | 0 | 0 | 0 |
| 0 | 0 | 0 | 0 | 0 |
| 0 | 0 | 0 | 0 | 0 |
| 0 | 0 | 0 | 0 | 0 |
| 0 | 0 | 0 | 0 | 0 |
| 0 | 0 | 0 | 0 | 0 |
| 0 | 0 | 0 | 0 | 0 |
| 0 | 0 | 0 | 0 | 0 |
| 0 | 0 | 0 | 0 | 0 |
| 0 | 0 | 0 | 0 | 0 |
| 1.352454179 | 4.555911 | 1.602791265 | 6.13556448 | 9.585200833 |
| 0 | 0 | 0 | 0 | 0 |
| 0 | 0 | 0 | 0 | 0 |
| 0 | 0 | 0 | 0.006771099 | 0 |
| 0 | 0 | 0 | 0 | 0 |
| 0 | 0 | 0.017092708 | 0 | 0 |
| 0 | 0 | 0 | 0 | 0 |
| 0 | 0 | 0 | 0 | 0 |
| 0 | 0 | 0 | 0 | 0 |
| 0 | 0 | 0 | 0 | 0 |
| 0 | 0 | 0 | 0 | 0 |
| 0 | 0 | 0 | 0.009131472 | 0 |
| 0 | 0 | 0 | 0 | 0 |
| 0 | 0 | 0 | 0 | 0 |
| 0 | 0 | 0 | 0 | 0 |
| 0 | 0 | 0 | 0 | 0 |
| 0 | 0 | 0 | 0 | 0 |
| 0 | 0 | 0 | 0 | 0 |
| 0 | 0 | 0 | 0 | 0 |
| 0 | 0 | 0 | 0 | 0 |
| 0 | 0 | 0 | 0 | 0 |
| 0 | 0 | 0 | 0 | 0 |
| 0.083137303 | 1.265651188 | 12.04963559 | 4.823869794 | 10.0781726 |
| 0 | 0 | 0 | 0 | 0 |
| 0 | 0 | 0 | 0 | 0 |
| 0 | 0 | 0 | 0 | 0 |
| 0 | 0 | 0 | 0 | 0 |
| 0 | 0 | 0 | 0 | 0 |
| 0.001579348 | 0.320164237 | 0.162106541 | 0.001039154 | 0.008520886 |
| 0.172768505 | 0.257842141 | 0.256305537 | 0.348714118 | 0.15399405 |
| 0.183008217 | 0.009301 | 0.050855552 | 0 | 0.054566855 |
| 0.001555925 | 0 | 0.000848728 | 0 | 0.001089018 |
| 0 | 0 | 0 | 0 | 0 |

| | | | | |
|---|---|---|---|---|
| 0 | 0 | 0 | 0 | 0 |
| 0.05410414 | 0.003764488 | 0.138785826 | 0.113696335 | 0.291295301 |
| 0.044735239 | 0 | 0.077211182 | 0.270250926 | 0.006021334 |
| 0.724900272 | 0.319961193 | 0.001093418 | 0.364301033 | 0 |
| 0 | 0.000615841 | 0 | 0.036189063 | 0 |
| 0 | 0 | 0 | 0.000945892 | 0 |
| 0.160887601 | 0.952716793 | 1.177881595 | 0.001408834 | 0.451680535 |
| 0 | 0 | 0 | 0 | 0 |
| 0 | 0 | 0 | 0 | 0 |
| 0 | 0 | 0 | 0 | 0 |
| 0 | 0.00239885 | 0 | 0 | 0 |
| 0.066110682 | 0.001005017 | 0.000572415 | 0.719008377 | 0 |
| 0 | 0.004112342 | 0 | 0 | 0.000901601 |
| 0.020446161 | 0.004929464 | 0.019517749 | 0.269897465 | 0.00708077 |
| 0 | 0 | 0.002231541 | 0 | 0.00230089 |
| 0 | 0 | 0 | 0 | 0 |
| 0.444968662 | 0.288486999 | 0 | 0 | 0.017114949 |
| 0 | 0 | 0 | 0 | 0 |
| 0 | 0 | 0 | 0 | 0 |
| 0 | 0 | 0 | 0 | 0 |
| 1.472535964 | 0.016524867 | 0.11220574 | 0.315406446 | 0.561164506 |
| 0 | 0.021923708 | 0.005186829 | 0.01051074 | 0.001663828 |
| 0 | 0 | 0 | 0.558013629 | 0 |
| 0 | 0 | 0 | 0 | 0 |
| 0 | 0 | 0 | 0 | 0 |
| 0 | 0 | 0 | 0.007345593 | 0 |
| 0 | 0 | 0 | 0 | 0 |
| 0 | 0 | 0 | 0 | 0 |
| 0 | 0.00798254 | 0.003244 | 0.000640299 | 0.016082127 |
| 0 | 0 | 0 | 0 | 0 |
| 0.643113639 | 0.004968396 | 0.0494995 | 0 | 0.209268347 |
| 0 | 0 | 0 | 0 | 0 |
| 0 | 0 | 0 | 0 | 0 |
| 0 | 0 | 0 | 0 | 0 |
| 0 | 0 | 0 | 0 | 0 |
| 0 | 0 | 0 | 0 | 0 |
| 0 | 0 | 0 | 0 | 0 |
| 31.31456062 | 0.015541884 | 0 | 0.006069227 | 0.009415308 |
| 0 | 0 | 0 | 0 | 0 |
| 0 | 0 | 0 | 0 | 0 |
| 0 | 0 | 0 | 0 | 0 |
| 0 | 0 | 0 | 0 | 0 |
| 0 | 0 | 0 | 0 | 0 |
| 0 | 0 | 0 | 0 | 0 |
| 0 | 0 | 0 | 0.000611067 | 0 |
| 0 | 0 | 0 | 0 | 0 |
| 0.001044979 | 0 | 0 | 0 | 0 |
| 0 | 0 | 0 | 0 | 0 |

| | | | | |
|---|---|---|---|---|
| 0 | 0 | 0 | 0 | 0 |
| 0 | 0 | 0 | 0 | 0 |
| 0 | 0 | 0 | 0 | 0 |
| 0 | 0 | 0 | 0 | 0 |
| 0 | 0 | 0 | 0 | 0 |
| 0 | 0.00310928 | 0 | 0 | 0 |
| 0 | 0 | 0 | 0 | 0 |
| 0 | 0 | 0 | 0.075997672 | 0 |
| 0 | 0.134481229 | 0.001114106 | 0 | 0.149743142 |
| 0 | 0 | 0 | 0 | 0 |
| 0 | 0 | 0 | 0 | 0 |
| 0 | 0.045798148 | 0 | 0 | 0 |
| 0 | 0 | 0 | 0 | 0 |
| 0 | 0 | 0 | 0 | 0 |
| 0 | 0 | 0 | 0 | 0 |
| 0 | 0 | 0 | 0 | 0 |
| 0 | 0 | 0 | 0 | 0 |
| 0 | 0 | 0 | 0 | 0 |
| 0 | 0 | 0 | 0 | 0 |
| 0 | 0 | 0 | 0 | 0 |
| 0 | 0 | 0 | 0 | 0 |
| 0 | 0 | 0 | 0 | 0 |
| 0 | 0 | 0 | 0.000705533 | 0 |
| 0 | 0 | 0 | 0 | 0 |
| 0 | 0 | 0 | 0 | 0 |
| 0 | 0 | 0 | 0 | 0 |
| 0 | 0 | 0 | 0 | 0 |
| 0 | 0.001316622 | 0 | 0.006946756 | 0 |
| 0 | 0 | 0 | 0 | 0 |
| 0 | 0 | 0 | 0 | 0 |
| 0 | 0 | 0 | 0.079089982 | 0.0014882 |
| 0 | 0 | 0 | 0.001408963 | 0 |
| 0 | 0 | 0 | 0 | 0 |
| 0 | 0 | 0 | 0 | 0 |
| 0 | 0 | 0 | 0 | 0 |
| 0 | 0 | 0 | 0 | 0 |
| 0 | 0 | 0 | 0 | 0 |
| 0 | 0 | 0 | 0 | 0 |
| 0 | 0 | 0 | 0 | 0 |
| 0 | 0 | 0 | 0 | 0 |
| 0 | 0 | 0 | 0 | 0 |
| 0 | 0 | 0 | 0 | 0 |
| 0 | 0 | 0 | 0.001085692 | 0 |
| 0 | 0 | 0 | 0 | 0 |
| 0 | 0 | 0 | 0 | 0 |
| 0 | 0 | 0 | 0 | 0 |
| 0 | 0 | 0 | 0 | 0 |
| 0.010848517 | 0 | 0 | 0 | 0 |

| | | | | |
|---|---|---|---|---|
| 0 | 0 | 0 | 0 | 0 |
| 0 | 0 | 0 | 0 | 0 |
| 0 | 0 | 0 | 0.985554739 | 0 |
| 0 | 0 | 0 | 0 | 0 |
| 0 | 0 | 0 | 0.001362603 | 0 |
| 0 | 0 | 0 | 0 | 0 |
| 0 | 0 | 0 | 0 | 0 |
| 0 | 0 | 0 | 0 | 0 |
| 0 | 0 | 0 | 0 | 0 |
| 0 | 0 | 0 | 0 | 0 |
| 0 | 0 | 0 | 0 | 0 |
| 0 | 0 | 0 | 0 | 0 |
| 0 | 0 | 0 | 0 | 0 |
| 0 | 0 | 0 | 0 | 0 |
| 0 | 0 | 0 | 0 | 0 |
| 0 | 0 | 0 | 0 | 0 |
| 0 | 0 | 0 | 0 | 0 |
| 0 | 0 | 0 | 0 | 0 |
| 0 | 0 | 0 | 0 | 0 |
| 0 | 0 | 0 | 0 | 0 |
| 0 | 0 | 0 | 0.001464546 | 0 |
| 0 | 0 | 0 | 0 | 0 |
| 0 | 0 | 0 | 0 | 0 |
| 0 | 0 | 0 | 0 | 0 |
| 0 | 0 | 0 | 0 | 0 |
| 0 | 0 | 0 | 0 | 0 |
| 0 | 0 | 0 | 0.015645226 | 0 |
| 0 | 0 | 0 | 0 | 0 |
| 0 | 0 | 0 | 0 | 0 |
| 0 | 0 | 0 | 0 | 0 |
| 0 | 0 | 0 | 0 | 0 |
| 0.543556834 | 0.003286489 | 0.008360904 | 0 | 0.001401048 |
| 0 | 0 | 0 | 0 | 0 |
| 0 | 0 | 0 | 0 | 0 |
| 0 | 0 | 0 | 0 | 0 |
| 0 | 0 | 0 | 0 | 0 |
| 0 | 0 | 0 | 0 | 0 |
| 0 | 0 | 0 | 0 | 0 |
| 0 | 0 | 0 | 0 | 0 |
| 0 | 0 | 0 | 0 | 0 |
| 0 | 0 | 0 | 0 | 0 |
| 0 | 0 | 0 | 0 | 0 |
| 0 | 0 | 0 | 0 | 0 |
| 0 | 0 | 0 | 0 | 0 |
| 0 | 0 | 0 | 0 | 0 |
| 0 | 0 | 0 | 0 | 0 |
| 0 | 0 | 0 | 0 | 0 |
| 0 | 0 | 0 | 0 | 0 |

| | | | | |
|---|---|---|---|---|
| 0 | 0 | 0 | 2.326824227 | 0 |
| 0 | 0 | 0 | 0 | 0 |
| 0 | 0 | 0 | 0 | 0 |
| 0 | 0 | 0 | 0 | 0 |
| 0 | 0 | 0 | 0 | 0 |
| 0 | 0 | 0 | 0 | 0 |
| 0 | 0 | 0 | 0 | 0 |
| 0 | 0 | 0 | 0 | 0 |
| 0 | 0 | 0 | 0 | 0 |
| 0 | 0 | 0 | 0 | 0 |
| 0 | 0 | 0 | 0 | 0 |
| 0 | 0 | 0 | 0 | 0 |
| 0 | 0.000524141 | 0 | 0.00086421 | 0 |
| 0 | 0 | 0 | 0 | 0 |
| 0 | 0 | 0 | 0 | 0 |
| 0 | 0 | 0 | 0 | 0 |
| 0 | 0 | 0 | 0 | 0 |
| 0 | 0 | 0 | 0 | 0 |
| 0 | 0 | 0 | 0 | 0 |
| 0 | 0 | 0 | 0 | 0 |
| 0.040521635 | 0.068601277 | 0.052245334 | 0.009372015 | 0.116383515 |
| 0 | 0 | 0 | 0 | 0 |
| 0.001077431 | 0.000859905 | 0 | 0 | 0 |
| 0 | 0 | 0 | 0 | 0.002987436 |
| 0 | 0 | 0 | 0 | 0 |
| 0 | 0 | 0 | 0 | 0 |
| 0.03075001 | 0.002473205 | 0.007671627 | 0 | 0.014390006 |
| 0 | 0 | 0 | 0 | 0 |
| 0 | 0 | 0 | 0 | 0 |
| 0 | 0 | 0 | 0 | 0.000892501 |
| 0.003089016 | 0.360512228 | 0.308355055 | 0 | 0.00852348 |
| 0 | 0 | 0 | 0 | 0 |
| 0 | 0 | 0 | 0 | 0 |
| 0 | 0 | 0 | 0 | 0 |
| 0.003195822 | 0.016578955 | 0.002179077 | 0.046890998 | 0 |
| 0 | 0 | 0 | 0 | 0 |
| 0 | 0 | 0 | 0 | 0 |
| 0 | 0 | 0 | 0 | 0 |
| 0 | 0.013391192 | 0 | 0 | 0 |
| 0 | 0 | 0 | 0 | 0 |
| 0 | 0 | 0 | 0 | 0 |
| 0 | 0 | 0 | 0 | 0 |
| 0 | 0 | 0 | 0 | 0 |
| 0 | 0 | 0 | 0 | 0 |
| 0 | 0 | 0 | 0 | 0 |
| 0 | 0 | 0 | 0 | 0 |
| 0.001441761 | 0 | 0 | 0 | 0 |
| 0.002277429 | 0.003483796 | 0 | 0.004045858 | 0 |

| | | | | |
|---|---|---|---|---|
| 0 | 0 | 0 | 0 | 0 |
| 0 | 0 | 0 | 0 | 0 |
| 0 | 0.00304501 | 0.00208117 | 0 | 0.003944887 |
| 0 | 0.001732836 | 0 | 0 | 0.000633186 |
| 0 | 0 | 0 | 0 | 0 |
| 0 | 0 | 0 | 0 | 0 |
| 0 | 0.001078074 | 0 | 0 | 0 |
| 0 | 0.00696768 | 0 | 0 | 0 |
| 0.001308948 | 0 | 0 | 0.005610368 | 0 |
| 0 | 0 | 0 | 0 | 0 |
| 0 | 0 | 0 | 0 | 0 |
| 0 | 0 | 0 | 0 | 0 |
| 0 | 0 | 0 | 0 | 0 |
| 0 | 0 | 0 | 0 | 0 |
| 0 | 0 | 0 | 0 | 0 |
| 0 | 0 | 0 | 0 | 0 |
| 0 | 0 | 0 | 0 | 0 |
| 0 | 0 | 0 | 0 | 0 |
| 0 | 0 | 0 | 0.001066751 | 0 |
| 0 | 0 | 0 | 0 | 0 |
| 0 | 0.015906074 | 0 | 0 | 0.057789395 |
| 0 | 0 | 0 | 0 | 0 |
| 0 | 0 | 0 | 0 | 0 |
| 0 | 0 | 0 | 0 | 0 |
| 0.009615846 | 0.004965837 | 0.00555381 | 0 | 0.002474372 |
| 0 | 0 | 0 | 0 | 0 |
| 0 | 0 | 0 | 0 | 0 |
| 0 | 0 | 0 | 0 | 0 |
| 0 | 0 | 0 | 0 | 0 |
| 0 | 0 | 0 | 0 | 0 |
| 0.155335976 | 6.844842329 | 1.263570423 | 2.139867902 | 0.004029463 |
| 0 | 0 | 0 | 0 | 0 |
| 0 | 0 | 0 | 0.001644219 | 0 |
| 0 | 0 | 0 | 0 | 0 |
| 0 | 0 | 0 | 0 | 0 |
| 0 | 0 | 0 | 0 | 0 |
| 0 | 0 | 0 | 0 | 0 |
| 0 | 0 | 0 | 0 | 0 |
| 0 | 0 | 0 | 0 | 0 |
| 0 | 0 | 0 | 0 | 0 |
| 0 | 0 | 0 | 0 | 0 |
| 0.012803709 | 0 | 0 | 0.006907989 | 0.010641816 |
| 0 | 0 | 0 | 0 | 0 |
| 0 | 0 | 0 | 0 | 0 |
| 0 | 0 | 0 | 0 | 0 |
| 0 | 0 | 0 | 0 | 0 |
| 0 | 0 | 0 | 0 | 0 |
| 0 | 0 | 0 | 0 | 0 |

| | | | | |
|---|---|---|---|---|
| 0 | 0 | 0 | 0 | 0 |
| 0 | 0 | 0 | 0 | 0 |
| 0.143086752 | 0 | 0.057295977 | 0 | 0.053919065 |
| 0 | 0 | 0 | 0 | 0 |
| 0 | 0.002598249 | 0 | 0 | 0 |
| 0 | 0 | 0 | 0 | 0 |
| 0 | 0 | 0 | 0 | 0 |
| 0 | 0 | 0 | 0 | 0 |
| 0 | 0 | 0 | 0 | 0 |
| 0 | 0 | 0 | 0 | 0.006003439 |
| 0 | 0 | 0 | 0 | 0 |
| 0 | 0 | 0 | 0 | 0 |
| 0 | 0 | 0 | 0 | 0 |
| 0.012141582 | 0.030488936 | 0 | 0 | 0.00155453 |
| 0.003666324 | 0.00760791 | 0.001599929 | 0 | 0.002993808 |
| 0 | 0 | 0 | 0 | 0 |
| 0.010582867 | 0.00155589 | 0.001671061 | 0 | 0.00268021 |
| 0 | 0 | 0 | 0 | 0 |
| 0 | 0 | 0 | 0 | 0 |
| 0 | 0.00679281 | 0 | 0 | 0 |
| 0 | 0 | 0 | 0 | 0 |
| 0.002514029 | 0.00220711 | 0 | 0.002778955 | 0 |
| 0 | 0 | 0 | 0 | 0 |
| 0 | 0 | 0 | 0 | 0 |
| 0 | 0.012225542 | 0.000879556 | 0 | 0.032093798 |
| 0 | 0 | 0 | 0 | 0 |
| 0 | 0 | 0 | 0 | 0 |
| 0 | 0 | 0 | 0 | 0 |
| 0 | 0 | 0 | 0 | 0 |
| 0 | 0 | 0.003173886 | 0.002584147 | 0.005408747 |
| 0 | 0.000922551 | 0 | 0 | 0 |
| 0 | 0 | 0 | 0 | 0 |
| 0 | 0 | 0 | 0 | 0 |
| 0 | 0 | 0 | 0 | 0 |
| 0 | 0 | 0 | 0 | 0 |
| 0 | 0.310991825 | 0 | 0 | 0 |
| 0 | 0 | 0 | 0 | 0 |
| 0.006001372 | 0 | 0.00051689 | 0 | 0 |
| 0 | 0 | 0 | 0 | 0 |
| 0 | 0 | 0 | 0 | 0 |
| 0 | 0 | 0 | 0 | 0 |
| 0 | 0 | 0 | 0 | 0.001298366 |
| 0 | 0 | 0 | 0 | 0 |
| 0.003923496 | 0 | 0 | 0 | 0 |
| 0 | 0 | 0 | 0 | 0 |
| 0 | 0.036820728 | 0 | 0 | 0.006373166 |
| 0 | 0 | 0 | 0 | 0.004493236 |
| 0 | 0.022290255 | 0.000834778 | 0 | 0.001338898 |

| | | | | |
|---|---|---|---|---|
| 0 | 0.072668939 | 0 | 4.534870243 | 0 |
| 0 | 0 | 0.063516942 | 0.007101648 | 0 |
| 0.009743022 | 0 | 0 | 0 | 0 |
| 0 | 0.001962537 | 0 | 0 | 0 |
| 0.033646142 | 0.031504962 | 0.001011602 | 0 | 0 |
| 0 | 0.025858237 | 0 | 0 | 0.011338461 |
| 0 | 0 | 0 | 0 | 0 |
| 0.163768905 | 0.001778301 | 0.012559271 | 0.007916621 | 0.038338155 |
| 0 | 0 | 0 | 0 | 0.00262885 |
| 0.006124269 | 1.379282273 | 0.149077588 | 0.023270633 | 0.064966928 |
| 0 | 0 | 0.0007551 | 0 | 0 |
| 0 | 0.105108965 | 0.007376296 | 0.006731035 | 0 |
| 0.039022453 | 0.00854238 | 0.001824516 | 0 | 0.003316514 |
| 0.007111805 | 0.41813064 | 0 | 0 | 0 |
| 0 | 0 | 0.003752946 | 0.001448586 | 0 |
| 0 | 0.001335784 | 0.002008528 | 0 | 0.004100054 |
| 0 | 0.009908153 | 0 | 0 | 0.000638909 |
| 0 | 0.002317778 | 0 | 0 | 0 |
| 0 | 0.248077516 | 0.368969904 | 1.417348726 | 1.030432537 |
| 0 | 0.140093267 | 0.2003178 | 0 | 0 |
| 0 | 0 | 0 | 0 | 0 |
| 0 | 0 | 0 | 0 | 0 |
| 0 | 0.002387296 | 0.006526571 | 0 | 0 |
| 0 | 0 | 0 | 0.209209131 | 0 |
| 0 | 0 | 0 | 0 | 0 |
| 0.001558327 | 0 | 0 | 0 | 0 |
| 3.790778242 | 0.001385912 | 0 | 0 | 0 |
| 0.556541105 | 0 | 0.001884543 | 0.04786015 | 0.062925309 |
| 0 | 0 | 1.85377449 | 0 | 0 |
| 0 | 0 | 0 | 0 | 0 |
| 0 | 0 | 0 | 0.001760363 | 0 |
| 0 | 0 | 0 | 0 | 0 |
| 0 | 0 | 0 | 0 | 0 |
| 0 | 0 | 0.003335082 | 0 | 0.034472156 |
| 0 | 0 | 0 | 0 | 0 |
| 0 | 0.302055079 | 0.003878929 | 5.31219926 | 0 |
| 0.001177682 | 0.000939917 | 2.148198015 | 0.063229578 | 3.410198729 |
| 0 | 0 | 0.001106222 | 0 | 0.013011285 |
| 0 | 0 | 0 | 0 | 0 |
| 0 | 0 | 0.394860061 | 0 | 0 |
| 0 | 0 | 0 | 0 | 0 |
| 0 | 0 | 0 | 0.715273308 | 3.129237933 |
| 0 | 0 | 0 | 0.011288609 | 0 |
| 0 | 0 | 0 | 0 | 0.095925369 |
| 0 | 0 | 0 | 0 | 0.05716268 |
| 0 | 0 | 0 | 0.003150701 | 0 |
| 0.002583871 | 0 | 0 | 0.004590252 | 0.009325048 |
| 0 | 0 | 0 | 0.096985611 | 0 |

| | | | | |
|---|---|---|---|---|
| 0 | 0 | 0 | 0 | 0.001068322 |
| 0.002165742 | 0.033417551 | 0.004331697 | 0 | 0.002842198 |
| 0 | 0 | 0 | 0 | 0 |
| 0 | 0 | 0 | 0 | 0 |
| 0 | 0 | 0 | 0.005293147 | 0 |
| 0 | 0.00913633 | 0 | 0.049008473 | 0.002670765 |
| 0 | 0 | 0 | 0.025460651 | 0 |
| 0 | 0 | 0 | 0 | 0 |
| 0 | 0 | 0 | 0.001709773 | 0 |
| 0 | 0 | 0 | 0 | 0 |
| 0.005310869 | 0.028728573 | 0.013197353 | 0 | 0.021941603 |
| 0 | 0 | 0 | 0 | 0 |
| 0 | 0 | 0 | 0 | 0 |
| 0.013101098 | 0.005339274 | 0.001824614 | 0 | 0.00390199 |
| 0 | 0 | 0 | 0 | 0.043429241 |
| 0 | 0 | 0 | 0 | 0 |
| 0 | 0 | 0 | 0 | 0 |
| 0 | 0 | 0.071617643 | 0.000642539 | 0.491508711 |
| 0 | 0 | 0 | 0 | 0.0007787 |
| 0 | 0 | 0 | 0.021247391 | 0 |
| 0 | 0 | 0 | 0 | 0 |
| 0.095176343 | 0 | 0 | 0.297104763 | 0 |
| 0 | 0 | 0 | 0.184206938 | 0 |
| 0 | 0 | 0.055745266 | 0 | 0 |
| 0 | 0 | 0 | 0 | 0 |
| 0 | 0 | 0 | 0 | 0 |
| 0 | 0 | 0.059515801 | 1.46877904 | 0.05764452 |
| 0 | 0 | 0 | 0 | 0 |
| 0 | 0 | 0 | 0 | 0 |
| 3.850499273 | 0.003260941 | 0.031648272 | 0.230659331 | 1.305118498 |
| 0 | 0 | 0 | 0.0013438 | 0.003308988 |
| 0 | 0 | 0.001448259 | 0 | 0 |
| 0 | 0 | 0 | 0 | 0 |
| 0 | 0.001496361 | 0 | 0.001903281 | 0 |
| 0.003020802 | 0 | 0 | 0 | 0.006166737 |
| 0 | 0 | 0 | 0 | 0 |
| 0 | 0 | 0 | 0 | 0 |
| 0 | 0 | 0.005889264 | 0 | 0 |
| 0 | 0 | 0.020606747 | 0 | 0.609753719 |
| 0 | 0 | 0 | 0 | 0.092629627 |
| 0 | 0.102969521 | 0 | 0.005970852 | 0.012831442 |
| 0 | 0.368389255 | 1.210104554 | 3.399025515 | 0 |
| 0 | 0 | 0.003265363 | 0 | 0 |
| 0 | 0 | 0 | 0 | 0 |
| 0 | 0 | 0 | 0.228392385 | 0 |
| 0 | 0 | 0.001358811 | 0 | 0 |
| 0.0012026 | 0.031673523 | 0.001180793 | 0 | 0.033668798 |
| 0.000675385 | 0 | 0 | 0 | 0 |

| | | | | |
|---|---|---|---|---|
| 0.001483807 | 0 | 0 | 0 | 0 |
| 0 | 0 | 0 | 0 | 0 |
| 0.418108367 | 0.596714201 | 0 | 0 | 0.050235764 |
| 0 | 0 | 0 | 0 | 0 |
| 0 | 0 | 0 | 0.116513029 | 0 |
| 0 | 0.152393782 | 0 | 0.154921402 | 0 |
| 0 | 0 | 0 | 0 | 0 |
| 0 | 0 | 0 | 0 | 0 |
| 0 | 0.235088908 | 0 | 0 | 0.085612834 |
| 0 | 0 | 0 | 0 | 0 |
| 0 | 0 | 0 | 0 | 0 |
| 0 | 0 | 0 | 0 | 0 |
| 0 | 0 | 0 | 0 | 0 |
| 0.001610835 | 0.013927538 | 0.000732234 | 0.002331717 | 0.011744275 |
| 0 | 0 | 0 | 0 | 0 |
| 0.242326681 | 0 | 0.007438965 | 0.037437851 | 0.000734236 |
| 0.006511366 | 0.013363116 | 0.022960036 | 0 | 0.006510591 |
| 0 | 0 | 0 | 0 | 0.12928093 |
| 0 | 0 | 0 | 0.01631039 | 0 |
| 0 | 0 | 0.011334532 | 0 | 0 |
| 0.036634707 | 0 | 0.024619732 | 0.068032002 | 0 |
| 0 | 0 | 0.001029984 | 0.002129785 | 0.016991881 |
| 0 | 0 | 0 | 0 | 0 |
| 0 | 0.972350003 | 0.153050162 | 0 | 0 |
| 0 | 0 | 0 | 0 | 0 |
| 0 | 0.055255587 | 0 | 0.000725623 | 0.002412156 |
| 0 | 0.011545341 | 0 | 0.006293562 | 0.004907491 |
| 0 | 0 | 0 | 0 | 0 |
| 0 | 0 | 0 | 0 | 0 |
| 0 | 0 | 0 | 0.011767547 | 0 |
| 0 | 0.019710652 | 0 | 0.028894686 | 0 |
| 0.0833467 | 0.00614438 | 0 | 0 | 0.006442703 |
| 0.12525072 | 0.103524522 | 0.002260013 | 0.079768268 | 0.022306632 |
| 0 | 0.18700014 | 0 | 0 | 0.002797163 |
| 0.152564225 | 0 | 0.000889113 | 0 | 0 |
| 0 | 0 | 0 | 0 | 0 |
| 0.003169435 | 0.001609713 | 0.002357547 | 0 | 0.001260421 |
| 0 | 0 | 0.000737484 | 0.3850013 | 0.004534255 |
| 0 | 0.017100374 | 0 | 0.038195794 | 0 |
| 0.002947703 | 0 | 0 | 0 | 0 |
| 0 | 0 | 0 | 0 | 0 |
| 0 | 0 | 0 | 0 | 0 |
| 0 | 0 | 0 | 0.011440878 | 0 |
| 0 | 0.025409898 | 0.205236159 | 0 | 0 |
| 0.004474817 | 0 | 0.481614119 | 1.657033789 | 0.221649171 |
| 0 | 0 | 0 | 0 | 0 |
| 0 | 0 | 0 | 0.000593103 | 0 |
| 0 | 0 | 0 | 0 | 0 |

| | | | | |
|---|---|---|---|---|
| 0.003675009 | 0 | 0.002915852 | 0 | 0.02104527 |
| 7.469732939 | 0.308766628 | 0 | 0.00193942 | 0.033847419 |
| 0 | 0 | 0 | 0 | 0 |
| 0 | 0 | 0 | 0 | 0 |
| 0 | 0 | 0 | 0.00428099 | 0 |
| 0 | 0 | 0 | 0 | 0 |
| 0 | 0.413284734 | 0.284791451 | 0 | 0 |
| 0 | 0.015999248 | 0.001157822 | 0.004096617 | 0 |
| 0 | 0 | 0 | 0.006213828 | 0 |
| 0 | 0.004629954 | 0 | 0 | 0 |
| 0.002966161 | 0 | 0 | 0 | 0 |
| 0.819062811 | 0.011663523 | 0.091210567 | 0 | 0.192677727 |
| 0 | 0 | 0 | 0 | 0 |
| 0 | 0 | 0 | 0 | 0 |
| 0.001570753 | 0 | 0.000714014 | 0 | 0 |
| 0 | 0 | 0.000790281 | 0 | 0 |
| 0.062358899 | 0.028221545 | 0.028541838 | 0 | 0.001672262 |
| 0.009614806 | 1.285706779 | 0.770124586 | 0.447649656 | 0.040431649 |
| 0 | 0 | 0 | 0 | 0 |
| 0 | 0.046613887 | 0.003515491 | 0 | 0.005462283 |
| 0.09781678 | 0.011012148 | 0.035347483 | 0.011477785 | 0.036861696 |
| 0 | 0 | 0 | 0 | 0 |
| 0 | 0 | 0 | 0 | 0 |
| 0 | 0 | 0 | 0 | 0 |
| 0.123780304 | 0.003304011 | 0.00203237 | 0 | 0.025353314 |
| 0 | 0 | 0.112157101 | 0 | 0 |
| 0 | 0 | 0 | 0.01036905 | 0 |
| 0.030115747 | 0.590021483 | 0.107279226 | 0.439791176 | 0.041927912 |
| 0 | 0 | 0 | 0 | 0 |
| 0 | 0 | 0 | 0.004738755 | 0 |
| 0 | 0 | 0 | 0 | 0.006637675 |
| 0 | 0 | 0 | 0 | 0 |
| 0 | 0 | 0 | 0.032129586 | 0 |
| 0 | 0 | 0 | 0 | 0 |
| 0.052487554 | 0 | 0 | 0.989380083 | 0 |
| 0.123212273 | 0.003891016 | 0.041583162 | 0.027645184 | 1.09155071 |
| 0 | 0 | 0.000895759 | 0.023824396 | 0 |
| 0 | 0 | 0 | 0 | 0 |
| 0 | 0.151862581 | 0.002995733 | 0.001785482 | 0.06641995 |
| 0 | 0 | 0 | 0 | 0 |
| 0.006892402 | 0 | 0.442314867 | 0.772755554 | 0.164941883 |
| 0 | 0 | 0 | 0 | 0 |
| 0.001177365 | 0.000939664 | 0.065079396 | 0.015028478 | 0 |
| 0 | 0 | 0.035756899 | 0 | 0 |
| 0 | 0 | 0.082412136 | 0 | 0 |
| 0 | 0 | 0 | 0 | 0 |
| 0 | 0 | 0 | 0 | 0 |
| 0 | 0 | 0 | 0 | 0 |

| | | | | |
|---|---|---|---|---|
| 0 | 0.060823179 | 0 | 0 | 0.001439033 |
| 0 | 0 | 0 | 0 | 0 |
| 0 | 0 | 0 | 0 | 0 |
| 0 | 0 | 0 | 0 | 0 |
| 0 | 0 | 0 | 0 | 0 |
| 0 | 0 | 0 | 0 | 0 |
| 0 | 0 | 0 | 0 | 0 |
| 0 | 0 | 0 | 0 | 0 |
| 0 | 0.134533652 | 0 | 0.108149692 | 0 |
| 0 | 0.004238581 | 0 | 0 | 0 |
| 0 | 0 | 0 | 0 | 0 |
| 0.004817314 | 0 | 0 | 0.002420435 | 0 |
| 0 | 0 | 0 | 0 | 0 |
| 0 | 0 | 0 | 0 | 0 |
| 0.043005079 | 0 | 0 | 0.034810946 | 0 |
| 0 | 0 | 0 | 0 | 0 |
| 0.001414194 | 0.010158103 | 0 | 0 | 0 |
| 0 | 0 | 0 | 0 | 0 |
| 0 | 0 | 0 | 0 | 0 |
| 0 | 0 | 0 | 0 | 0 |
| 0 | 0 | 0 | 0 | 0 |
| 0 | 0 | 0 | 0 | 0 |
| 0 | 0 | 0 | 0 | 0 |
| 0.00121838 | 0.022781884 | 0.139186946 | 0.216995134 | 0.502826387 |
| 0 | 0.004744995 | 0 | 0.002266146 | 0 |
| 0 | 0 | 0 | 1.037629488 | 0 |
| 0 | 0 | 0 | 0.032163769 | 0 |
| 0 | 0 | 0 | 0.15448242 | 0.502774651 |
| 0 | 0 | 0 | 0 | 0 |
| 0 | 0 | 0 | 0 | 0 |
| 0 | 0 | 0 | 0 | 0 |
| 0 | 0 | 0 | 0 | 0 |
| 0 | 0.000387912 | 0 | 0.000767513 | 0 |
| 0 | 0 | 0 | 0 | 0 |
| 0 | 0 | 0 | 0.004965864 | 0 |
| 0 | 0 | 0 | 0 | 0 |
| 0 | 0 | 0 | 0.000736212 | 0 |
| 0 | 0 | 0 | 0.021226489 | 0 |
| 0 | 0 | 0 | 0.011698484 | 0 |
| 0 | 0 | 0 | 0 | 0 |
| 0.038651301 | 0.071251886 | 0.079177945 | 0.030351531 | 0.767289252 |
| 0 | 0 | 0 | 0.015125041 | 0 |
| 0.307181241 | 0.067173595 | 0.020206366 | 0.225426056 | 0.20591795 |
| 0 | 0 | 0 | 0.010547164 | 0 |
| 0 | 0 | 0 | 0.001263924 | 0 |
| 0 | 0 | 0 | 0 | 0 |
| 0 | 0 | 0 | 0 | 0 |
| 0.011110789 | 0 | 0 | 0 | 0 |

| | | | | |
|---|---|---|---|---|
| 0 | 0 | 0 | 0 | 0 |
| 0 | 0 | 0 | 0 | 0 |
| 0.303067803 | 0.746120348 | 0.505724665 | 0.321090881 | 0.599982234 |
| 0 | 0 | 0 | 0 | 0 |
| 0 | 0 | 0 | 0 | 0 |
| 0 | 0 | 0 | 0.001806736 | 0 |
| 0.011963007 | 0 | 0 | 0.066311054 | 0 |
| 0 | 0 | 0 | 0.00411965 | 0 |
| 0 | 0 | 0 | 0 | 0 |
| 0 | 0 | 0 | 0 | 0 |
| 0.141887082 | 0.006449241 | 0.026317489 | 0.1983475 | 0.004574541 |
| 1.834841557 | 6.132196137 | 2.87346317 | 2.618525929 | 0.940932166 |
| 0 | 0 | 0 | 0 | 0 |
| 0.03437096 | 0 | 0.003357979 | 0 | 0.005385852 |
| 0.015968397 | 1.064245556 | 0.357900762 | 0.401539314 | 0.196827478 |
| 0 | 0 | 0 | 0 | 0 |
| 0 | 0 | 0 | 0.001555704 | 0 |
| 0.027609947 | 0 | 0 | 0.000531698 | 0 |
| 0 | 0 | 0 | 0 | 0 |
| 0 | 0.038281084 | 0.268473954 | 0.080224602 | 0 |
| 0.000597599 | 2.559940196 | 0.073671318 | 0.006920297 | 3.129180453 |
| 0.232983194 | 0.084931876 | 1.797092759 | 0 | 0.464966497 |
| 0.273192377 | 36.07577362 | 31.19020764 | 1.793941522 | 6.448133949 |
| 0 | 0 | 0 | 0 | 0 |
| 0 | 0 | 0 | 0 | 0 |
| 0 | 0 | 0.017577943 | 0.007350252 | 0.015871733 |
| 0 | 0 | 0 | 0.001504909 | 0 |
| 0 | 0 | 0 | 0 | 0 |
| 0 | 0 | 0 | 0.10979085 | 0 |
| 0 | 0 | 0 | 0.092697047 | 0 |
| 0 | 0 | 0 | 0 | 0 |
| 0 | 0 | 0 | 0.107799303 | 0 |
| 3.51001418 | 0.194030707 | 0.004660129 | 1.50362365 | 0.006833706 |
| 0 | 0 | 0 | 0.06731047 | 0 |
| 0 | 0.001928655 | 0.010984796 | 0.138774624 | 0.006201705 |
| 0.092079474 | 0.015545808 | 0.011953211 | 0.015554022 | 0.002130191 |
| 0.004703955 | 0 | 0 | 0 | 0 |
| 0 | 0 | 0 | 0 | 0 |
| 0 | 0.007442074 | 0 | 0 | 0 |
| 0 | 0 | 0 | 0 | 0 |
| 0 | 0 | 0 | 0.014003877 | 0 |
| 0 | 0 | 0 | 0 | 0 |
| 0 | 0 | 0 | 0 | 0 |
| 0 | 0 | 0 | 0 | 0 |
| 0.11636503 | 0.060638605 | 0.00468145 | 0.01554531 | 0.004195962 |
| 0 | 0 | 0 | 0 | 0 |
| 0 | 0 | 0 | 0 | 0 |
| 0.005176261 | 0.001215062 | 0 | 0.966203239 | 0.00612705 |

| | | | | |
|---|---|---|---|---|
| 0 | 0 | 0 | 0 | 0 |
| 0 | 0 | 0 | 0 | 0 |
| 0 | 0 | 0 | 0 | 0 |
| 0 | 0 | 0 | 0 | 0 |
| 0 | 0 | 0 | 0 | 0 |
| 0 | 0 | 0 | 0 | 0 |
| 0 | 0 | 0 | 0 | 0 |
| 0.103930868 | 0.050084767 | 0.01274228 | 0.021413568 | 0 |
| 0 | 0 | 0 | 0 | 0 |
| 0 | 0 | 0 | 0 | 0 |
| 0.003114851 | 0 | 0.004182383 | 0.043700774 | 0 |
| 0 | 0 | 0 | 0 | 0 |
| 0 | 0 | 0 | 0.077686387 | 0 |
| 0 | 0 | 0.071378947 | 0 | 0 |
| 0 | 0 | 0 | 0 | 0 |
| 0 | 0 | 0 | 0 | 0 |
| 0 | 0 | 0 | 0 | 0 |
| 0.001595718 | 0 | 0 | 0.000944932 | 0 |
| 0 | 0 | 0 | 0 | 0 |
| 0 | 0 | 0 | 0.06959409 | 0.00141642 |
| 0 | 0 | 0 | 0 | 0 |
| 0 | 0 | 0 | 0 | 0 |
| 0 | 0 | 0 | 0 | 0 |
| 0 | 0.003580984 | 0 | 0 | 0 |
| 0 | 0 | 0 | 0 | 0 |
| 0 | 0 | 0 | 0 | 0 |
| 0 | 0 | 0 | 0 | 0 |
| 0 | 0 | 0 | 0 | 0 |
| 0 | 0 | 0 | 0 | 0 |
| 1.531248045 | 0 | 0 | 0 | 0 |
| 0 | 0 | 0 | 0.000596004 | 0 |
| 0 | 0 | 0 | 0 | 0 |
| 0 | 0 | 0 | 0 | 0 |
| 0 | 0 | 0 | 0 | 0 |
| 0 | 0 | 0 | 0 | 0 |
| 0 | 0 | 0 | 0 | 0 |
| 0 | 0 | 0 | 0 | 0 |
| 0.021499865 | 0 | 0.007744757 | 0.06613984 | 0.005146171 |
| 0 | 0 | 0 | 0 | 0 |
| 0 | 0 | 0 | 0.008529736 | 0 |
| 0 | 0 | 0 | 0.132375133 | 0 |
| 0 | 0 | 0 | 0 | 0 |
| 0 | 0 | 0 | 0 | 0 |
| 0.022257713 | 0 | 0 | 0 | 0 |
| 0.062874667 | 0 | 0 | 0 | 0 |
| 0 | 0 | 0 | 0 | 0 |
| 0 | 0 | 0 | 0.023433328 | 0 |
| 0 | 0 | 0 | 0 | 0 |

| | | | | |
|---|---|---|---|---|
| 0 | 0 | 0 | 0 | 0 |
| 0 | 0 | 0 | 0 | 0 |
| 0 | 0 | 0 | 0 | 0 |
| 0 | 0 | 0 | 0 | 0 |
| 0 | 0 | 0 | 0 | 0 |
| 0 | 0 | 0 | 0 | 0 |
| 0 | 0 | 0 | 0 | 0 |
| 0 | 0 | 0 | 0 | 0 |
| 0.011469133 | 0.076800901 | 0.287785104 | 0.015460659 | 0.001713173 |
| 0 | 0 | 0 | 0 | 0 |
| 0 | 0 | 0 | 0 | 0 |
| 0 | 0 | 0 | 0 | 0 |
| 0 | 0 | 0 | 0.003994331 | 0 |
| 0 | 0 | 0 | 0 | 0 |
| 0 | 0.14247019 | 0 | 0.028569696 | 0 |
| 0 | 0.810800834 | 0.426319956 | 0.367862694 | 0.046779523 |
| 0 | 0 | 0.002792751 | 0 | 0 |
| 0 | 0 | 0 | 0 | 0 |
| 0 | 0 | 0 | 0 | 0 |
| 0 | 0 | 0 | 0 | 0 |
| 0 | 0.000512139 | 0 | 0.013510732 | 0 |
| 0 | 0 | 0 | 0 | 0 |
| 0 | 0 | 0 | 0 | 0 |
| 0 | 0 | 0 | 0.020847852 | 0 |
| 0.187178432 | 0.019690917 | 0.010587062 | 0.053504952 | 0.005180508 |
| 0.002170412 | 0 | 0 | 2.432177072 | 0 |
| 0 | 0 | 0 | 0 | 0 |
| 0 | 0 | 0 | 0.001451054 | 0 |
| 0 | 0 | 0 | 0 | 0 |
| 0 | 0 | 0 | 0 | 0 |
| 0 | 0 | 0 | 0 | 0 |
| 0 | 0 | 0 | 0.009199814 | 0 |
| 0 | 0 | 0 | 0 | 0 |
| 0.001229802 | 0 | 0 | 0.002635566 | 0 |
| 0.011000048 | 0 | 0.004421289 | 0 | 0.003039125 |
| 0.002046237 | 0 | 0 | 0.036674566 | 0 |
| 0 | 0 | 0 | 0 | 0.04397008 |
| 4.63774617 | 0.2755041 | 0 | 0.015689099 | 0 |
| 0 | 0 | 0 | 0 | 0 |
| 0 | 0 | 0 | 0 | 0 |
| 0 | 0 | 0 | 0 | 0 |
| 0 | 0 | 0 | 0.019786957 | 0 |
| 0 | 0.012051084 | 0 | 0.033778927 | 0 |
| 0 | 0 | 0 | 0 | 0 |
| 0 | 0 | 0 | 0 | 0 |
| 0 | 0 | 0 | 0 | 0 |
| 0 | 0 | 0 | 0 | 0 |
| 0 | 1.056426637 | 0.316206065 | 0.005608376 | 1.359907311 |

| | | | | |
|---|---|---|---|---|
| 0 | 0 | 0 | 0.006000507 | 0 |
| 0 | 0 | 0 | 0 | 0 |
| 0 | 0 | 0 | 0 | 0 |
| 0 | 0 | 0 | 0 | 0 |
| 0.004127196 | 0 | 0.057033149 | 0.004163837 | 0 |
| 0 | 0 | 0 | 0 | 0 |
| 0 | 0 | 0 | 0 | 0 |
| 0 | 0 | 0 | 0 | 0 |
| 0 | 0 | 0 | 0 | 0 |
| 0 | 0 | 0 | 0.079951499 | 0 |
| 0 | 0 | 0 | 0 | 0 |
| 9.731172877 | 0.036635313 | 0 | 0.151819003 | 0.19487842 |
| 0 | 0.271875193 | 0 | 0 | 0 |
| 0 | 0 | 0 | 0 | 0 |
| 0.025191373 | 1.280185686 | 0.135533833 | 0.184663066 | 0.026911789 |
| 0 | 0 | 0 | 0 | 0 |
| 0 | 0 | 0 | 0 | 0 |
| 0 | 0 | 0 | 0 | 0 |
| 0 | 0.001298487 | 0 | 0.006606388 | 0 |
| 0 | 0 | 0 | 0 | 0 |
| 0 | 0 | 0 | 0.288059484 | 0 |
| 0 | 0 | 0 | 0 | 0 |
| 0 | 0 | 0 | 0 | 0.369549792 |
| 0 | 0 | 0 | 0 | 0 |
| 0 | 0 | 0 | 0 | 0 |
| 0 | 0 | 0 | 0 | 0 |
| 0 | 0 | 0 | 0 | 0 |
| 0 | 0 | 0 | 0 | 0 |
| 0 | 0 | 0 | 0 | 0 |
| 0 | 0 | 0 | 0 | 0 |
| 0 | 0 | 0 | 0 | 0 |
| 1.413211345 | 0.023617939 | 0.013514339 | 0.000815055 | 0.039738611 |
| 0 | 0 | 0 | 0 | 0 |
| 0.033349674 | 0 | 0 | 0.005564894 | 0.001654394 |
| 0.003340228 | 0 | 0 | 0.030065203 | 0 |
| 0.00143532 | 0 | 0 | 0.002266532 | 0.001004604 |
| 0 | 0 | 0 | 0 | 0 |
| 0 | 0 | 0 | 0 | 0 |
| 0 | 0 | 0 | 0 | 0 |
| 0 | 0 | 0 | 0 | 0 |
| 0 | 0 | 0 | 0 | 0 |
| 0 | 0 | 0 | 0 | 0 |
| 0 | 0 | 0 | 0 | 0 |
| 0 | 0 | 0 | 0 | 0 |
| 0.558061855 | 0.168948246 | 0.009572715 | 0.023728431 | 0 |
| 0 | 0 | 0 | 0 | 0 |
| 0 | 0 | 0 | 0 | 0 |
| 0 | 0 | 0 | 0.005242125 | 0 |

| | | | | |
|---|---|---|---|---|
| 0 | 0.001287303 | 0 | 0 | 0 |
| 0 | 0 | 0 | 0.001502362 | 0 |
| 0 | 0 | 0.005940349 | 0.018988032 | 0 |
| 0 | 0 | 0 | 0 | 0 |
| 0 | 0 | 0.003639725 | 0 | 0 |
| 0 | 0 | 0 | 0 | 0 |
| 0 | 0 | 0 | 0 | 0 |
| 0 | 0 | 0 | 0 | 0 |
| 0 | 0 | 0 | 0 | 0 |
| 0 | 0 | 0 | 0 | 0 |
| 0 | 0 | 0 | 0 | 0 |
| 0 | 0 | 0 | 0.001640366 | 0 |
| 0 | 0 | 0 | 0 | 0 |
| 0 | 0 | 0 | 0 | 0 |
| 0 | 0 | 0 | 0 | 0 |
| 0 | 0 | 0 | 0 | 0 |
| 0 | 0 | 0 | 0 | 0 |
| 0 | 0 | 0 | 0 | 0 |
| 0 | 13.40866773 | 0 | 0.104317804 | 0 |
| 0 | 0 | 0 | 0 | 0 |
| 0 | 0 | 0 | 0 | 0 |
| 0 | 0 | 0 | 0 | 0 |
| 0 | 0 | 0 | 0 | 0 |
| 0 | 0 | 0 | 0 | 0 |
| 0 | 0.002869284 | 0 | 0 | 0 |
| 0.016337563 | 0.007497496 | 0.021722599 | 0.118997232 | 0.014114995 |
| 0 | 0 | 0 | 0 | 0 |
| 0 | 0.414947567 | 0.039907333 | 0 | 0.145502816 |
| 0 | 0 | 0 | 0 | 0 |
| 0 | 0 | 0 | 0 | 0 |
| 0 | 0 | 0 | 0 | 0 |
| 0 | 0 | 0 | 0 | 0 |
| 0 | 0 | 0 | 0 | 0 |
| 0 | 0 | 0 | 0 | 0 |
| 0.000642028 | 0.000854012 | 0.00186781 | 0.137036734 | 0 |
| 0 | 0 | 0 | 0.002886957 | 0 |
| 0 | 0 | 0 | 0.010862195 | 0 |
| 0.000971352 | 0.00251954 | 0 | 0.000766937 | 0 |
| 0 | 0 | 0 | 0 | 0 |
| 0 | 0 | 0 | 0 | 0 |
| 2.874329655 | 0.07937734 | 1.403092357 | 0.007487047 | 18.78105263 |
| 0 | 0.000993092 | 0.07999527 | 0.387928535 | 1.123633466 |
| 0 | 0 | 0 | 0 | 0 |
| 0 | 0 | 0 | 0.16487202 | 0 |
| 0 | 0 | 0 | 0 | 0 |
| 0 | 0 | 0 | 0 | 0 |
| 0 | 0 | 0 | 0 | 0 |
| 0.034498616 | 0.003999292 | 0.000630783 | 0.00152171 | 0 |

| | | | | |
|---|---|---|---|---|
| 0 | 0 | 0 | 0 | 0 |
| 0 | 0 | 0.003568466 | 0.011425991 | 0.002601569 |
| 0 | 0 | 0 | 0 | 0 |
| 0 | 0 | 0 | 0 | 0 |
| 0 | 0.149737723 | 0 | 0.138447853 | 0 |
| 0 | 0 | 0 | 0 | 0 |
| 0 | 0 | 0 | 0 | 0 |
| 0 | 0 | 0 | 0.018254746 | 0 |
| 0.224272969 | 1.32310095 | 1.968077091 | 9.704024859 | 0.59453465 |
| 0 | 0 | 0 | 0.013282992 | 0 |
| 0 | 0 | 0.000366511 | 0 | 0 |
| 0 | 0 | 0 | 0 | 0 |
| 0 | 0 | 0 | 0 | 0 |
| 0.196078385 | 0 | 0.006717112 | 0 | 0.017196253 |
| 0 | 0 | 0 | 0 | 0 |
| 0 | 0.006203663 | 0.000954002 | 0 | 0 |
| 0 | 0.011448479 | 0.012072362 | 0.021357272 | 0.079602725 |
| 0 | 0 | 0 | 0 | 0 |
| 0 | 0 | 0 | 0 | 0 |
| 0 | 0 | 0 | 0 | 0 |
| 0 | 0 | 0 | 0 | 0 |
| 0 | 0 | 0 | 0 | 0 |
| 0 | 0 | 0 | 0 | 0 |
| 0 | 0 | 0 | 0 | 0 |
| 0 | 0 | 0 | 0 | 0 |
| 0 | 0 | 0 | 0 | 0 |
| 0 | 0 | 0 | 0 | 0 |
| 0 | 0.000641887 | 0.000987097 | 0.003016297 | 0 |
| 0 | 0 | 0 | 0 | 0 |
| 0 | 0 | 0 | 0.000937367 | 0 |
| 0 | 0 | 0 | 0 | 0 |
| 0 | 0 | 0 | 0 | 0 |
| 0 | 0 | 0 | 0 | 0 |
| 0 | 0 | 0 | 0.004493204 | 0 |
| 0 | 0 | 0 | 0 | 0 |
| 0.001162954 | 0 | 0.002156858 | 0 | 0.001627942 |
| 0 | 0.270946853 | 0 | 0 | 0 |
| 0 | 0 | 0 | 0 | 0 |
| 0 | 0 | 0 | 0 | 0 |
| 0 | 0 | 0 | 0 | 0 |
| 0.086313789 | 0.114358035 | 12.70809585 | 5.254942522 | 10.95673657 |
| 0 | 0 | 0 | 0 | 0 |
| 0 | 0 | 0 | 0 | 0 |
| 0 | 0 | 0 | 0 | 0 |
| 0 | 0 | 0 | 0 | 0 |
| 0 | 0 | 0 | 0 | 0 |
| 0 | 0 | 0 | 0 | 0 |
| 0 | 0 | 0 | 0 | 0 |

| | | | | |
|---|---|---|---|---|
| 0 | 0 | 0 | 0 | 0 |
| 0 | 0 | 0 | 0.000497659 | 0 |
| 0 | 0 | 0 | 0 | 0 |
| 0 | 0 | 0 | 0 | 0 |
| 0 | 0.038786548 | 0 | 0 | 0 |
| 0 | 0 | 0 | 0 | 0 |
| 0 | 0.263327361 | 0.006021504 | 0.121537279 | 0.000715398 |
| 0 | 0 | 0 | 0 | 0 |
| 0.001587192 | 0 | 0 | 0.002278505 | 0 |
| 0 | 0 | 0 | 0.05012439 | 0.000562927 |
| 0 | 0 | 0 | 0.000587187 | 0 |
| 0 | 0 | 0 | 0.030347449 | 0 |
| 0 | 0 | 0 | 0 | 0 |
| 0 | 0 | 0 | 0 | 0.009887969 |
| 0 | 0 | 0 | 0 | 0 |
| 0 | 0 | 0 | 0 | 0 |
| 0 | 0 | 0 | 0 | 0 |
| 0 | 0.05176167 | 0 | 0.065638218 | 0 |
| 0 | 0 | 0 | 0 | 0 |
| 0 | 0 | 0 | 0 | 0 |
| 0 | 0.149271954 | 0.148416008 | 0 | 0.073047831 |
| 0 | 0 | 0 | 0 | 0 |
| 0 | 0 | 0 | 0 | 0 |
| 0.003626819 | 0.001371122 | 0.00312373 | 0.044912973 | 0.000835023 |
| 0 | 0 | 0 | 0 | 0 |
| 0 | 0 | 0 | 0.002311523 | 0 |
| 0 | 0 | 0 | 0 | 0 |
| 0 | 0 | 0 | 0 | 0 |
| 0 | 0 | 0 | 0 | 0 |
| 6.315735754 | 0 | 0.117489707 | 0 | 1.553920314 |
| 0 | 0 | 0 | 0 | 0 |
| 0 | 0 | 0 | 0 | 0 |
| 0 | 0 | 0 | 0 | 0 |
| 0 | 2.356018914 | 0 | 2.098520764 | 0 |
| 0 | 0 | 0 | 0 | 0 |
| 0 | 0 | 0 | 0 | 0 |
| 0.09052296 | 0.111134586 | 0.008989769 | 0.010749236 | 0.007313814 |
| 0 | 0 | 0 | 0 | 0 |
| 0 | 0 | 0 | 0 | 0 |
| 0 | 0 | 0 | 0 | 0 |
| 0 | 0 | 0 | 0 | 0 |
| 0 | 0 | 0 | 0 | 0 |
| 0 | 0 | 0 | 0 | 0 |
| 0 | 0 | 0 | 0.013676494 | 0 |
| 0 | 0 | 0 | 0 | 0 |
| 0 | 0 | 0 | 0 | 0 |
| 0 | 0 | 0 | 0 | 0 |
| 0 | 0 | 0 | 0 | 0 |

| | | | | |
|---|---|---|---|---|
| 0 | 0.001475376 | 0.00605024 | 0.134766877 | 0.001976733 |
| 0 | 0 | 0 | 0 | 0 |
| 0 | 0 | 0 | 0 | 0 |
| 0 | 0 | 0 | 0 | 0 |
| 0 | 0 | 0 | 0 | 0 |
| 0 | 0 | 0 | 0 | 0 |
| 0 | 0.000378066 | 0 | 0 | 0 |
| 0.966759375 | 0.002356476 | 0.263904676 | 0.1635191 | 0.148534045 |
| 0.216011367 | 0.332192014 | 0.353314636 | 0.493840552 | 0.169806628 |
| 0.101450815 | 0.000537402 | 0.003917843 | 0.745011594 | 0.002945539 |
| 0 | 0 | 0 | 0.003535296 | 0 |
| 0 | 0 | 0 | 0 | 0 |
| 0 | 0 | 0 | 0.041898156 | 0 |
| 0 | 0.009524693 | 0.533024971 | 0 | 0 |
| 0 | 0 | 0 | 0.001859153 | 0 |
| 0 | 0 | 0 | 0 | 0 |
| 0 | 0.012204024 | 0 | 0 | 0.004197088 |
| 0 | 0 | 0 | 0 | 0 |
| 0 | 0 | 0 | 0 | 0 |
| 0 | 0 | 0 | 0.013078629 | 0 |
| 0 | 0 | 0 | 0 | 0 |
| 0 | 0 | 0 | 0 | 0 |
| 0 | 0 | 0 | 0 | 0 |
| 0 | 0 | 0 | 0 | 0 |
| 0 | 0 | 0 | 0 | 0 |
| 0 | 0 | 0 | 0.000538512 | 0 |
| 0 | 0 | 0 | 0 | 0 |
| 0 | 0 | 0 | 0 | 0 |
| 0 | 0 | 0 | 0 | 0 |
| 0 | 0 | 0 | 0 | 0 |
| 0 | 0 | 0 | 0.013661711 | 0 |
| 0 | 0 | 0 | 0 | 0 |
| 0 | 0 | 0 | 0 | 0 |
| 0 | 0 | 0 | 0 | 0 |
| 0.002735312 | 0 | 0 | 0.042211997 | 0 |
| 0 | 0 | 0 | 0 | 0 |
| 0 | 0 | 0 | 0 | 0 |
| 0 | 0 | 0 | 0 | 0 |
| 0 | 0 | 0 | 0 | 0 |
| 0.198081015 | 0.000654616 | 0.016330462 | 0 | 1.279120913 |
| 0 | 0 | 0 | 0 | 0 |
| 0 | 0 | 0 | 0.186237768 | 0 |
| 0 | 0 | 0 | 0 | 0 |
| 0 | 0 | 0 | 0 | 0 |
| 0 | 0 | 0 | 0 | 0 |
| 0 | 0 | 0 | 0 | 0 |
| 0 | 0 | 0 | 0 | 0 |
| 0 | 0 | 0 | 0 | 0 |

| | | | | |
|---|---|---|---|---|
| 0 | 0 | 0 | 0 | 0 |
| 0 | 0.010208864 | 0 | 0 | 0 |
| 0 | 0 | 0 | 0 | 0 |
| 0 | 0 | 0 | 0 | 0 |
| 0.108600556 | 0 | 0 | 0.059756355 | 0.048527525 |
| 0 | 0 | 0 | 0 | 0 |
| 0 | 0 | 0.005640343 | 0.001064884 | 0 |
| 0.029786235 | 0.147848914 | 0.029360123 | 0.001031488 | 0.001828762 |
| 0 | 0 | 0 | 0 | 0 |
| 0 | 0 | 0 | 0 | 0 |
| 0 | 0 | 0 | 0 | 0 |
| 0 | 0 | 0 | 0 | 0 |
| 0 | 0 | 0 | 0 | 0 |
| 0 | 0 | 0 | 0 | 0 |
| 0 | 0.005871956 | 0 | 0 | 0.033954744 |
| 0 | 0.0034363 | 0 | 0 | 0 |
| 0 | 0 | 0 | 0 | 0 |
| 0 | 0 | 0 | 0 | 0 |
| 0 | 0 | 0 | 0 | 0 |
| 0 | 0 | 0 | 0 | 0 |
| 0 | 0 | 0 | 0 | 0 |
| 0 | 0 | 0 | 0 | 0 |
| 0 | 0 | 0 | 0 | 0 |
| 0 | 0 | 0.000616718 | 0.013390013 | 0 |
| 0.192732619 | 0 | 0 | 0.065761736 | 0 |
| 0 | 0 | 0 | 0 | 0 |
| 0.076933777 | 0.524222609 | 0 | 0.041312155 | 0.027502618 |
| 0 | 0 | 0 | 0 | 0 |
| 0 | 0 | 0 | 0.001213546 | 0 |
| 0 | 0 | 0 | 0 | 0 |
| 0 | 0 | 0 | 0 | 0 |
| 0 | 0 | 0 | 0 | 0 |
| 0 | 0 | 0 | 0 | 0 |
| 0 | 0 | 0 | 0 | 0 |
| 0 | 0 | 0 | 0 | 0 |
| 0 | 0 | 0 | 0.018794777 | 0 |
| 0 | 0 | 0 | 0 | 0 |
| 0 | 0 | 0 | 0.001004776 | 0 |
| 0 | 0 | 0 | 0 | 0 |
| 0 | 0 | 0 | 0 | 0 |
| 0 | 0 | 0 | 0 | 0 |
| 0 | 0 | 0 | 0 | 0 |
| 0 | 0 | 0 | 0 | 0 |
| 0 | 0 | 0 | 0 | 0 |
| 0 | 0 | 0 | 0 | 0 |
| 0 | 0 | 0 | 0 | 0 |
| 0 | 0 | 0 | 0 | 0 |
| 0 | 0 | 0 | 0 | 0 |

| | | | | |
|---|---|---|---|---|
| 0 | 0 | 0.009335889 | 0 | 0 |
| 0 | 0 | 0 | 0.014827158 | 0 |
| 0 | 0 | 0 | 0 | 0 |
| 0 | 0 | 0 | 0 | 0 |
| 0 | 0 | 0 | 0 | 0 |
| 0.015096826 | 0 | 0.019665759 | 0 | 0 |
| 0 | 0 | 0 | 0 | 0 |
| 0 | 0 | 0 | 0 | 0 |
| 0 | 0 | 0 | 0 | 0 |
| 0 | 0 | 0 | 0 | 0 |
| 0 | 0 | 0 | 0 | 0 |
| 0 | 0 | 0 | 0 | 0 |
| 0 | 0 | 0 | 0 | 0 |
| 0 | 0 | 0 | 0 | 0 |
| 0 | 0 | 0 | 0 | 0 |
| 0 | 0 | 0 | 0 | 0 |
| 0 | 0 | 0 | 0 | 0 |
| 0 | 0 | 0 | 0 | 0 |
| 0 | 0 | 0 | 0.003905728 | 0 |
| 0 | 0 | 0 | 0 | 0 |
| 0.010808327 | 0 | 0.008475127 | 0.550606896 | 0 |
| 0 | 0 | 0 | 0 | 0 |
| 0.165507819 | 0.000907855 | 0 | 0.017738049 | 0 |
| 0 | 0 | 0 | 0 | 0 |
| 0 | 0 | 0 | 0 | 0 |
| 0 | 0 | 0 | 0 | 0 |
| 0 | 0 | 0 | 0 | 0 |
| 0 | 0 | 0 | 0 | 0 |
| 0 | 0 | 0.001074296 | 0 | 0 |
| 0 | 0 | 0 | 0 | 0 |
| 0 | 0 | 0 | 0 | 0 |
| 0 | 0 | 0 | 0 | 0 |
| 0.82612011 | 0.004463565 | 0.006682502 | 0.008831489 | 0.030290127 |
| 0 | 0 | 0 | 0 | 0 |
| 0 | 0.319410996 | 0 | 0 | 0 |
| 0 | 0.022173194 | 0.001632043 | 0 | 0.001308815 |
| 0 | 0 | 0 | 0 | 0 |
| 0 | 0 | 0 | 0 | 0 |
| 0.009393174 | 0.071613774 | 0.008899234 | 0.230690381 | 0.012975867 |
| 0 | 0 | 0 | 0 | 0 |
| 0 | 0 | 0 | 0 | 0 |
| 0 | 0 | 0 | 0 | 0 |
| 0 | 0.024913773 | 0 | 0.011410583 | 0 |
| 0 | 0 | 0 | 0 | 0 |
| 0 | 0 | 0 | 0 | 0 |
| 0 | 0 | 0 | 0 | 0 |
| 0 | 0 | 0 | 0 | 0 |
| 0 | 0 | 0 | 0 | 0 |

| | | | | |
|---|---|---|---|---|
| 0 | 0 | 0 | 0 | 0 |
| 0 | 0 | 0 | 0 | 0 |
| 0 | 0 | 0 | 0 | 0 |
| 0 | 0 | 0 | 0 | 0 |
| 0 | 0 | 0 | 0 | 0 |
| 0 | 0 | 0 | 0 | 0 |
| 0 | 0 | 0 | 0 | 0 |
| 0 | 0 | 0 | 0 | 1.320996261 |
| 0 | 0 | 0 | 0.024451882 | 0 |
| 0.011585069 | 0 | 0 | 0 | 0.001654813 |
| 0 | 0 | 0 | 0 | 0 |
| 0 | 0 | 0 | 0 | 0 |
| 0 | 0 | 0 | 0 | 0 |
| 0 | 0 | 0 | 0 | 0 |
| 0 | 0 | 0 | 0.025495125 | 0 |
| 0 | 0.100440127 | 0.074357769 | 0.012489383 | 0.025439888 |
| 0 | 0 | 0 | 0 | 0 |
| 0 | 0 | 0 | 0 | 0 |
| 0 | 0 | 0 | 0 | 0 |
| 0.930162415 | 0 | 0 | 0 | 0 |
| 0 | 0 | 0.022794817 | 0 | 0.012966237 |
| 0 | 0 | 0 | 0 | 0 |
| 0 | 0 | 0 | 0 | 0 |
| 0 | 0 | 0 | 0 | 0 |
| 0 | 0 | 0 | 0 | 0 |
| 0 | 0 | 0 | 0 | 0 |
| 0 | 0 | 0 | 0 | 0 |
| 0 | 0 | 0 | 0 | 0 |
| 0 | 0 | 0 | 0 | 0 |
| 0 | 0 | 0 | 0 | 0 |
| 0 | 0.497806993 | 0 | 0.003284153 | 0 |
| 0 | 0 | 0 | 0 | 0 |
| 0 | 0 | 0 | 0 | 0 |
| 0 | 0 | 0 | 0 | 0 |
| 0 | 0 | 0 | 0 | 0 |
| 0 | 0 | 0 | 0 | 0 |
| 0 | 0 | 0 | 0.001656139 | 0 |
| 0 | 0 | 0 | 0 | 0 |
| 0 | 0 | 0 | 0 | 0 |
| 0 | 0 | 0 | 0 | 0 |
| 0 | 0 | 0 | 0 | 0 |
| 0 | 0 | 0 | 0.121762232 | 0 |
| 0 | 0 | 0 | 0.002098969 | 0 |
| 0 | 0 | 0 | 0.000980915 | 0 |
| 0 | 0 | 0 | 0 | 0 |
| 0 | 0 | 0 | 0 | 0 |
| 0.356484175 | 0.177321436 | 0.005845355 | 0.649981583 | 0 |
| 0 | 0 | 0 | 0.003667268 | 0 |

| | | | | |
|---|---|---|---|---|
| 0 | 0 | 0 | 0 | 0 |
| 0 | 0 | 0 | 0 | 0 |
| 0 | 0 | 0 | 0 | 0 |
| 0.009603921 | 0 | 0 | 0 | 0 |
| 0 | 0 | 0 | 0 | 0 |
| 0 | 0 | 0 | 0 | 0 |
| 0 | 0 | 0 | 0 | 0 |
| 0 | 0 | 0 | 0 | 0 |
| 0 | 0 | 0 | 0 | 0 |
| 0 | 0 | 0 | 0 | 0 |
| 0 | 0 | 0 | 0 | 0 |
| 0.10250167 | 0.046127492 | 0.000943107 | 0.007020508 | 0 |
| 0.005327296 | 0 | 0.00197604 | 0 | 0 |
| 0 | 0 | 0 | 0 | 0 |
| 0 | 0.007811638 | 0 | 0 | 0 |
| 0 | 0 | 0 | 0 | 0 |
| 0 | 0 | 0 | 0.038997141 | 0 |
| 0 | 0 | 0 | 0 | 0 |
| 0 | 0.06015378 | 0 | 0.38053479 | 0.003599778 |
| 0 | 0.26408213 | 0.059275944 | 0.010989974 | 1.325246608 |
| 0 | 0 | 0 | 0 | 0 |
| 0 | 0 | 0 | 0 | 0 |
| 0 | 0 | 0 | 0 | 0 |
| 0 | 0 | 0 | 0 | 0 |
| 0 | 0 | 0 | 0 | 0 |
| 0 | 0 | 0 | 0 | 0 |
| 0 | 0.000624712 | 0.014090045 | 0.577229661 | 0.000856022 |
| 0 | 0 | 0 | 0 | 0 |
| 0 | 0 | 0 | 0 | 0 |
| 0 | 0 | 0 | 0 | 0 |
| 0.001330918 | 0 | 0 | 0.011909467 | 0 |
| 0 | 0 | 0 | 0 | 0 |
| 0 | 0 | 0 | 0 | 0 |
| 0 | 0 | 0 | 0 | 0 |
| 0 | 0 | 0 | 0 | 0 |
| 0 | 0 | 0 | 0 | 0 |
| 0 | 0 | 0 | 0 | 0 |
| 0.011064932 | 0.011567367 | 0 | 0.018334112 | 0 |
| 0 | 0 | 0 | 0 | 0 |
| 0 | 0 | 0 | 0.002537413 | 0 |
| 0 | 0 | 0.001951951 | 0.001351254 | 0 |
| 0 | 0 | 0 | 0 | 0 |
| 0 | 0 | 0 | 0 | 0 |
| 0 | 0 | 0 | 0.007481032 | 0.142136654 |
| 0 | 0 | 0 | 0 | 0 |
| 0 | 0 | 0 | 0 | 0 |
| 0 | 0 | 0 | 0.079229757 | 0 |
| 0 | 0 | 0 | 0 | 0 |

| | | | | |
|---|---|---|---|---|
| 0 | 0 | 0 | 0 | 0 |
| 0 | 0 | 0 | 0 | 0 |
| 0 | 0 | 0 | 0 | 0 |
| 0 | 0 | 0.002113769 | 0.043407688 | 0 |
| 0.004717189 | 0 | 0.002695669 | 0 | 0.001768735 |
| 0 | 0 | 0 | 0 | 0 |
| 0 | 0 | 0 | 0 | 0 |
| 0 | 0 | 0 | 0 | 0 |
| 0 | 0 | 0 | 0.001398116 | 0 |
| 0 | 0 | 0 | 0 | 0 |
| 0 | 0 | 0 | 0.00805855 | 0 |
| 0 | 0 | 0 | 0 | 0 |
| 0.019743103 | 0.000959128 | 0 | 0.000677752 | 0.003154229 |
| 0 | 0 | 0 | 0 | 0 |
| 0 | 0 | 0 | 0 | 0 |
| 0 | 0 | 0 | 0 | 0 |
| 0 | 0 | 0 | 0 | 0 |
| 0 | 0 | 0 | 0 | 0 |
| 0 | 0 | 0 | 0 | 0 |
| 0 | 0 | 0 | 0 | 0 |
| 0 | 0 | 0 | 0 | 0 |
| 0 | 0 | 0 | 0 | 0 |
| 0 | 0 | 0.002589563 | 0.071395455 | 0.003955612 |
| 0.040547643 | 0 | 0 | 0 | 0 |
| 0 | 0 | 0 | 0 | 0 |
| 0 | 0 | 0 | 0.000622188 | 0 |
| 0 | 0 | 0 | 0 | 0 |
| 0 | 0 | 0 | 0 | 0 |
| 0 | 0 | 0 | 0 | 0 |
| 0 | 0 | 0 | 0 | 0 |
| 0 | 0 | 0 | 0 | 0 |
| 0 | 0 | 0 | 0.002770178 | 0.000852665 |
| 0 | 0 | 0 | 0 | 0 |
| 0 | 0 | 0 | 0 | 0 |
| 0 | 0 | 0 | 0 | 0 |
| 0 | 0 | 0.001293582 | 0 | 0 |
| 0 | 0.032914054 | 0 | 0 | 0 |
| 0 | 0 | 0 | 0.203040708 | 0 |
| 0 | 0.773573327 | 0.001046957 | 0 | 0.002798685 |
| 0 | 0 | 0 | 0 | 0 |
| 0 | 0 | 0 | 0 | 0 |
| 0 | 0 | 0 | 0 | 0 |
| 0 | 0 | 0 | 0 | 0 |
| 0 | 0 | 0 | 0 | 0 |
| 0 | 0 | 0 | 0.421894719 | 0 |
| 0.119146508 | 0.000599003 | 0 | 0.110368994 | 0 |
| 0 | 0 | 0.105614102 | 0 | 0 |
| 0 | 0 | 0 | 0 | 0 |

| | | | | |
|---|---|---|---|---|
| 0 | 0 | 0 | 0 | 0 |
| 0 | 0 | 0 | 0 | 0 |
| 0.004811439 | 0 | 0 | 0 | 0 |
| 0 | 0 | 0 | 0 | 0 |
| 0 | 0 | 0 | 0.001499943 | 0 |
| 0 | 0 | 0 | 0 | 0 |
| 0 | 0 | 0 | 0 | 0 |
| 0 | 0 | 0 | 0 | 0 |
| 0 | 0 | 0.003659585 | 0 | 0 |
| 0 | 0 | 0 | 0 | 0 |
| 0.004325245 | 0.001380804 | 0 | 0 | 0 |
| 0 | 0 | 0 | 0.006118373 | 0 |
| 0 | 0 | 0 | 0 | 0 |
| 0 | 0 | 0 | 0 | 0 |
| 0 | 0 | 0 | 0 | 0 |
| 0 | 0 | 0 | 0 | 0 |
| 0 | 0.003044582 | 0 | 0 | 0 |
| 0 | 0 | 0 | 0 | 0 |
| 0 | 0 | 0 | 0 | 0 |
| 0.000688322 | 0.000732473 | 0 | 0.000724626 | 0 |
| 0 | 0 | 0 | 0 | 0 |
| 0 | 0 | 0 | 0 | 0 |
| 0 | 0 | 0 | 0 | 0 |
| 0 | 0 | 0 | 0 | 0 |
| 0 | 0 | 0 | 0 | 0 |
| 0 | 0 | 0 | 0 | 0 |
| 0 | 0 | 0 | 0 | 0 |
| 0 | 0.008308704 | 0 | 0 | 0 |
| 0 | 0 | 0 | 0.000884927 | 0 |
| 0 | 0 | 0 | 0 | 0 |
| 0.0088701 | 0.00062927 | 0 | 0 | 0.002586805 |
| 0 | 0 | 0 | 0 | 0 |
| 0.047604681 | 0.049866641 | 0.040074789 | 0.073366124 | 0.026631419 |
| 0.021049105 | 0.067757738 | 0 | 0 | 0 |
| 0 | 0 | 0 | 0 | 0 |
| 0 | 0 | 0 | 0 | 0 |
| 0 | 0 | 0 | 0 | 0 |
| 0 | 0 | 0 | 0 | 0 |
| 0 | 0 | 0 | 0 | 0 |
| 0 | 0 | 0 | 0 | 0 |
| 0 | 0 | 0 | 0 | 0 |
| 0 | 0.01532796 | 0.083809468 | 0.361699876 | 0.24182736 |
| 0.433772862 | 0.001098457 | 0 | 0.009599087 | 0.010435915 |
| 0 | 0 | 0 | 0 | 0 |
| 0 | 0 | 0 | 0 | 0 |
| 0 | 0 | 0 | 0 | 0 |
| 0 | 0 | 0 | 0.00102956 | 0 |
| 0 | 0 | 0 | 0 | 0 |

| | | | | |
|---|---|---|---|---|
| 0 | 0 | 0 | 0.02898363 | 0 |
| 0 | 0 | 0 | 0.013851153 | 0 |
| 0.113127193 | 0.004032697 | 0 | 0 | 0 |
| 0 | 0 | 0 | 0 | 0 |
| 0 | 0 | 0 | 0.388797963 | 0 |
| 0 | 0 | 0 | 0 | 0.012788822 |
| 0 | 0 | 0 | 0 | 0 |
| 0 | 0 | 0 | 0 | 0 |
| 0 | 0 | 0 | 0 | 0 |
| 0 | 0 | 0 | 0 | 0 |
| 0 | 0 | 0 | 0 | 0 |
| 0 | 0 | 0 | 0 | 0 |
| 0 | 0 | 0 | 0 | 0 |
| 0 | 0 | 0 | 0.47746873 | 0 |
| 0 | 0 | 0 | 0 | 0 |
| 0 | 0 | 0 | 0 | 0 |
| 0 | 0 | 0 | 0 | 0 |
| 0 | 0 | 0 | 0 | 0 |
| 0 | 0 | 0 | 0 | 0 |
| 0 | 0 | 0 | 0 | 0 |
| 0 | 0 | 0 | 0 | 0 |
| 0 | 0 | 0 | 0.095737725 | 0 |
| 0 | 0 | 0 | 0 | 0 |
| 0 | 0 | 0 | 0 | 0 |
| 0 | 0 | 0 | 0 | 0 |
| 0.009309172 | 0 | 0.001904242 | 0 | 0 |
| 0 | 0 | 0 | 0 | 0 |
| 0 | 0 | 0 | 0.398203234 | 0 |
| 0 | 0 | 0 | 0 | 0 |
| 0 | 0 | 0 | 0 | 0 |
| 0 | 0 | 0 | 0 | 0 |
| 0 | 0 | 0 | 0.043991337 | 0 |
| 0 | 0 | 0 | 0 | 0 |
| 0 | 0 | 0 | 0 | 0 |
| 0 | 0 | 0 | 0 | 0 |
| 0 | 0 | 0 | 0 | 0 |
| 0 | 0 | 0 | 0.000979974 | 0 |
| 0 | 0.000924457 | 0.085045354 | 0 | 1.191964071 |
| 0 | 0 | 1.429225908 | 1.734064454 | 0 |
| 0 | 0 | 0 | 0 | 0 |
| 0 | 0 | 0 | 0 | 0 |
| 0 | 0 | 0 | 0 | 0 |
| 0 | 0.001864133 | 0.00382223 | 0.105117186 | 0.000817395 |
| 0 | 0 | 0 | 0 | 0 |
| 0 | 0 | 0 | 0 | 0 |
| 0 | 0 | 0 | 0 | 0 |
| 0.003215837 | 0 | 0.006626899 | 0.008745738 | 0.002188295 |
| 0 | 0 | 0 | 0 | 0 |

| | | | | |
|---|---|---|---|---|
| 0 | 0 | 0 | 0 | 0 |
| 0.016682046 | 0.011514861 | 0.045375752 | 0.408847924 | 0.005719683 |
| 0 | 0 | 0 | 0 | 0 |
| 0 | 0 | 0 | 0 | 0 |
| 0 | 0 | 0 | 0 | 0 |
| 0 | 0 | 0 | 0 | 0 |
| 0 | 0 | 0 | 0 | 0 |
| 0 | 0 | 0 | 0 | 0 |
| 0 | 0 | 0 | 0 | 0 |
| 0 | 0 | 0 | 0 | 0 |
| 0 | 0 | 0 | 0 | 0 |
| 0 | 0 | 0 | 0 | 0 |
| 0 | 0 | 0 | 0 | 0 |
| 0 | 0 | 0 | 0 | 0 |
| 0.014054052 | 0 | 0 | 0.025299947 | 0.005164249 |
| 0 | 0 | 0 | 0 | 0 |
| 0 | 0 | 0 | 0 | 0 |
| 0 | 0 | 0 | 0 | 0 |
| 0 | 0 | 0 | 0.002455113 | 0 |
| 0 | 0 | 0 | 0 | 0 |
| 0 | 0 | 0 | 0 | 0 |
| 0 | 0.087023426 | 0.008131143 | 0 | 0.573585145 |
| 0 | 0.013035492 | 0 | 0 | 0 |
| 0 | 0 | 0 | 0 | 0 |
| 0 | 0 | 0 | 0 | 0 |
| 0 | 0 | 0 | 0 | 0 |
| 0 | 0 | 0 | 0 | 0 |
| 0 | 0 | 0 | 0 | 0 |
| 0 | 0 | 0 | 0.000881894 | 0 |
| 0 | 0 | 0 | 0 | 0 |
| 0 | 0 | 0 | 0 | 0 |
| 0 | 0 | 0 | 0 | 0 |
| 0 | 0 | 0 | 0 | 0 |
| 0 | 0 | 0 | 0 | 0 |
| 0.078197301 | 0 | 0 | 9.659725423 | 0 |
| 0 | 0 | 0 | 0 | 0 |
| 0 | 0 | 0 | 0 | 0 |
| 0 | 0 | 0 | 0 | 0 |
| 0 | 0.114404503 | 0.003909596 | 0 | 0.012058825 |
| 0 | 0 | 0 | 0 | 0 |
| 0.011369748 | 0 | 0.095052162 | 0 | 0.341020926 |
| 0 | 0 | 0 | 0 | 0 |
| 0 | 0 | 0 | 0 | 0 |
| 0 | 0 | 0 | 0 | 0 |
| 0 | 0 | 0 | 0 | 0 |
| 1.173280795 | 0.434097405 | 0.4464274 | 0.473016989 | 1.569705811 |
| 0 | 0 | 0 | 0 | 0 |
| 0 | 0 | 0 | 0 | 0 |

| | | | | |
|---|---|---|---|---|
| 0 | 0 | 0 | 0 | 0 |
| 0 | 0 | 0 | 0 | 0 |
| 0 | 0 | 0 | 0.006751516 | 0 |
| 0 | 0 | 0 | 0 | 0 |
| 0 | 0 | 0 | 0 | 0 |
| 0 | 0 | 0 | 0 | 0 |
| 0 | 0 | 0 | 0 | 0 |
| 0 | 0.032712032 | 0 | 0 | 0.004549324 |
| 0 | 0 | 0 | 0 | 0 |
| 0 | 0.058535487 | 0.002264558 | 0 | 0.000726423 |
| 0 | 0 | 0 | 0.00161324 | 0 |
| 0 | 0 | 0 | 0 | 0 |
| 0.000811545 | 0 | 0.003541463 | 0.020290764 | 0.00923023 |
| 0 | 0 | 0 | 0 | 0 |
| 0 | 0 | 0 | 0 | 0 |
| 0 | 0 | 0 | 0 | 0 |
| 0 | 0 | 0 | 0 | 0 |
| 0 | 0 | 0 | 0 | 0 |
| 0 | 0 | 0 | 0 | 0 |
| 0.177693843 | 0.356852763 | 0.083794707 | 0.001901075 | 0.13987511 |
| 0 | 0 | 0 | 0 | 0 |
| 0 | 0 | 0 | 0 | 0 |
| 0 | 0 | 0 | 0 | 0 |
| 0 | 0 | 0 | 0 | 0 |
| 0 | 0 | 0 | 0 | 0 |
| 0 | 0 | 0 | 0 | 0 |
| 0 | 0 | 0 | 0 | 0 |
| 0 | 0 | 0 | 0 | 0 |
| 0 | 0 | 0 | 0 | 0 |
| 0 | 0 | 0 | 0.468846958 | 0.002477133 |
| 0 | 0 | 0 | 0 | 0 |
| 0 | 0 | 0 | 0 | 0 |
| 0 | 0 | 0 | 0 | 0 |
| 0 | 0 | 0 | 0 | 0 |
| 0 | 0 | 0 | 0 | 0 |
| 0 | 0 | 0 | 0 | 0 |
| 0 | 0 | 0 | 0.524963069 | 0 |
| 0 | 0 | 0 | 0 | 0 |
| 0 | 0 | 0 | 0 | 0 |
| 0 | 0 | 0 | 0 | 0 |
| 0 | 0 | 0 | 0 | 0 |
| 0 | 0 | 0 | 0 | 0 |
| 0 | 0 | 0 | 0 | 0 |
| 0 | 0 | 0 | 0 | 0 |
| 0.013342681 | 0 | 0 | 0.001428448 | 0 |
| 0 | 0 | 0 | 0 | 0 |
| 0 | 0 | 0 | 0.043414243 | 0 |
| 0 | 0 | 0 | 0 | 0 |

| | | | | |
|---|---|---|---|---|
| 0 | 0 | 0 | 0 | 0 |
| 0 | 0 | 0 | 0 | 0 |
| 0 | 0 | 0 | 0 | 0 |
| 0 | 0 | 0 | 0 | 0 |
| 0 | 0 | 0 | 0 | 0 |
| 0.001350621 | 0.005524443 | 0.001749754 | 0.029858966 | 0.00088624 |
| 0 | 0 | 0 | 0 | 0 |
| 0 | 0 | 0 | 0 | 0 |
| 0 | 0 | 0 | 0 | 0 |
| 0 | 0 | 0 | 0 | 0 |
| 0 | 0 | 0 | 0 | 0 |
| 0 | 0 | 0 | 0 | 0 |
| 0 | 0 | 0 | 0 | 0 |
| 0 | 0 | 0 | 0 | 0 |
| 0 | 0 | 0 | 0 | 0 |
| 0 | 0 | 0 | 0 | 0 |
| 0 | 0 | 0 | 0 | 0 |
| 0 | 0 | 0 | 0 | 0 |
| 0 | 0 | 0 | 0 | 0 |
| 0 | 0 | 0 | 0 | 0 |
| 0 | 0 | 0 | 0 | 0 |
| 0 | 0 | 0 | 0 | 0 |
| 0 | 0 | 0 | 0 | 0 |
| 0 | 0 | 0 | 0 | 0 |
| 0 | 0 | 0 | 0 | 0 |
| 0 | 0 | 0 | 0 | 0.000924152 |
| 0 | 0 | 0 | 0 | 0 |
| 0 | 0 | 0 | 0 | 0 |
| 0 | 0 | 0 | 0 | 0 |
| 0 | 0 | 0 | 0 | 0 |
| 0 | 0 | 0 | 0 | 0 |
| 0 | 0 | 0 | 0 | 0 |
| 0 | 0 | 0 | 0 | 0 |
| 0 | 0 | 0 | 0 | 0 |
| 0 | 0 | 0 | 0 | 0 |
| 0 | 0 | 0 | 0 | 0 |
| 0 | 0 | 0 | 0 | 0 |
| 0 | 0 | 0 | 0 | 0 |
| 0 | 0 | 0 | 0 | 0 |
| 0 | 0 | 0 | 0 | 0 |
| 0 | 0 | 0 | 0 | 0 |
| 0 | 0 | 0 | 0 | 0 |
| 0 | 0 | 0 | 0 | 0 |
| 0 | 0 | 0 | 0 | 0 |
| 0 | 0 | 0 | 0 | 0 |
| 0 | 0 | 0 | 0 | 0 |
| 0 | 0 | 0 | 0 | 0 |
| 0 | 0 | 0 | 0 | 0 |

| | | | | |
|---|---|---|---|---|
| 0 | 0 | 0 | 0 | 0 |
| 0 | 0 | 0 | 0.178340512 | 0 |
| 0 | 0 | 0 | 0 | 0 |
| 0 | 0 | 0 | 0 | 0 |
| 0 | 0 | 0 | 0 | 0 |
| 0 | 0 | 0 | 0 | 0 |
| 0 | 0 | 0 | 0 | 0 |
| 0 | 0 | 0 | 0 | 0 |
| 0 | 0 | 0 | 0 | 0 |
| 0 | 0 | 0 | 0 | 0 |
| 0.003712525 | 0 | 0 | 0 | 0 |
| 0 | 0 | 0 | 0 | 0 |
| 0 | 0 | 0 | 0 | 0 |
| 0 | 0 | 0 | 0 | 0 |
| 0 | 0 | 0 | 0 | 0 |
| 0 | 0 | 0 | 0 | 0 |
| 0 | 0 | 0 | 0 | 0 |
| 0 | 0 | 0 | 0 | 0 |
| 0 | 0 | 0 | 0 | 0 |
| 0 | 0 | 0 | 0 | 0 |
| 0.035921498 | 0 | 0 | 0 | 0.001318639 |
| 0 | 0 | 0 | 0 | 0 |
| 0 | 0 | 0 | 0 | 0 |
| 0.01415035 | 0.00830404 | 0 | 0.631739205 | 0 |
| 0 | 0 | 0 | 0 | 0 |
| 0 | 0 | 0 | 0 | 0 |
| 0 | 0 | 0 | 0 | 0 |
| 0 | 0 | 0 | 1.134644191 | 0 |
| 0 | 0 | 0 | 0 | 0 |
| 0 | 0 | 0 | 0 | 0 |
| 0 | 0 | 0.004303664 | 0 | 0 |
| 0 | 0 | 0 | 0 | 0 |
| 0 | 0 | 0 | 0 | 0 |
| 0 | 0 | 0 | 0 | 0 |

| PHL018-0056_V3 | PHL018-0255_V1 | PHL018-0061_V2 | PHL018-0013_V3 | PHL018-0249_V1 |
|---|---|---|---|---|
| REF | REF | REF | EYCF | REF |
| 0.962587992 | 0.069117394 | 0.23969291 | 0.136067405 | 0.020030577 |
| 6.054554103 | 0.099828388 | 0.070050171 | 0.025367294 | 0.331169733 |
| 0.001312353 | 2.20906641 | 0 | 0.961812368 | 0 |
| 0 | 0 | 0 | 0 | 0 |
| 0 | 0 | 0 | 0 | 0 |
| 0 | 0 | 0 | 0.03169018 | 0 |
| 0 | 0 | 0 | 0 | 0 |
| 0 | 0 | 0 | 0 | 0 |
| 0 | 0 | 0 | 0 | 0 |
| 0 | 0 | 0 | 0 | 0 |
| 0 | 0 | 0 | 0 | 0 |
| 0 | 0 | 0 | 0 | 0 |
| 0 | 0 | 0 | 0 | 0 |
| 2.409821407 | 0.174785658 | 0.013175468 | 0.082338493 | 3.700442572 |
| 0 | 0 | 0 | 0 | 0 |
| 0 | 0 | 0 | 0 | 0 |
| 0 | 0 | 0 | 0 | 0 |
| 0 | 0 | 0 | 0 | 0 |
| 0 | 0 | 0 | 0 | 0 |
| 0 | 0 | 0 | 0 | 0 |
| 0 | 0 | 0 | 0 | 0 |
| 0 | 0 | 0 | 0 | 0 |
| 0 | 0 | 0 | 0 | 0 |
| 0 | 0 | 0 | 0 | 0 |
| 0.328554678 | 0.028504621 | 0.001869785 | 0.194275088 | 0 |
| 0 | 0 | 0 | 0 | 0 |
| 0 | 0 | 0 | 0 | 0 |
| 0 | 0 | 0 | 0 | 0 |
| 0 | 0 | 0 | 0 | 0 |
| 0 | 0 | 0 | 0 | 0 |
| 0 | 0 | 0 | 0 | 0 |
| 0 | 0 | 0 | 0 | 0 |
| 0 | 0 | 0 | 0 | 0 |
| 0 | 0 | 0 | 0 | 0 |
| 0 | 0 | 0 | 0 | 0 |
| 0.000748173 | 0.348720317 | 0.466998421 | 2.978648184 | 1.81195333 |
| 0 | 0 | 0 | 0 | 0 |
| 0 | 0 | 0 | 0 | 0 |
| 0 | 0 | 0 | 0 | 0 |
| 0 | 0 | 0 | 0 | 0 |
| 0 | 0 | 0 | 0 | 0 |
| 0.005922056 | 0.00197674 | 0.023852639 | 0 | 0.016376706 |
| 2.167038057 | 0.194479682 | 12.63685868 | 0.005188783 | 4.22689936 |
| 0.038357268 | 0.001620681 | 0 | 0.000981137 | 0 |
| 0.004200643 | 0.00382403 | 0 | 0 | 0.000944419 |
| 0 | 0 | 0 | 0 | 0 |

| | | | | |
|---|---|---|---|---|
| 0.017920246 | 0.001652009 | 0.000480869 | 0 | 0.020195795 |
| 0.023096355 | 0 | 0.006836951 | 0 | 0.003368231 |
| 0.016000099 | 0.007517726 | 0.00091178 | 0.034119893 | 0.000522183 |
| 0 | 0.165460845 | 0.503498436 | 0.700162234 | 0.384227132 |
| 0.014235324 | 0.001791087 | 0.01140842 | 0 | 0.000585455 |
| 0 | 0.015773362 | 0 | 0 | 0 |
| 0 | 0.000609083 | 0.004964202 | 0 | 0 |
| 0 | 0 | 0 | 0 | 0 |
| 0 | 0 | 0 | 0 | 0 |
| 0 | 0 | 0 | 0 | 0 |
| 0 | 0 | 0 | 0 | 0 |
| 0 | 0.372102977 | 0.449765196 | 0.53323293 | 0 |
| 0 | 0 | 0 | 0 | 0 |
| 0.249645711 | 0.010468951 | 0.016421664 | 0.003181452 | 0.006140591 |
| 0 | 0 | 0.00394867 | 0 | 0 |
| 0 | 0 | 0 | 0 | 0 |
| 0.006568864 | 0 | 0.113954328 | 0.005309572 | 0.285993854 |
| 0 | 0 | 0 | 0 | 0 |
| 0 | 0 | 0 | 0 | 0 |
| 0 | 0 | 0 | 0 | 0 |
| 0.052911627 | 0 | 0.017173748 | 0.046919249 | 0.013660449 |
| 0.015688061 | 0.005193285 | 0.031577099 | 0.005901127 | 0.000801614 |
| 0 | 0.560967762 | 0 | 0.011544423 | 0.145834677 |
| 0 | 0 | 0 | 0 | 0 |
| 0 | 0 | 0 | 0 | 0 |
| 0 | 0 | 0 | 0 | 0 |
| 0 | 0 | 0 | 0 | 0 |
| 0 | 0 | 0 | 0 | 0 |
| 0.020981821 | 0.000922738 | 0 | 0 | 0 |
| 0 | 0 | 0 | 0 | 0 |
| 1.065231594 | 0 | 0.003616098 | 0 | 0 |
| 0 | 0 | 0 | 0 | 0 |
| 0 | 0 | 0 | 0 | 0 |
| 0 | 0 | 0 | 0 | 0 |
| 0.004653684 | 0 | 0 | 0 | 0 |
| 0 | 0 | 0 | 0 | 0 |
| 0 | 0 | 0 | 0 | 0 |
| 0 | 0 | 5.05211262 | 0 | 15.12652496 |
| 0 | 0 | 0 | 0 | 0 |
| 0 | 0 | 0 | 0 | 0 |
| 0 | 0 | 0 | 0 | 0 |
| 0 | 0 | 0 | 0 | 0 |
| 0 | 0 | 0 | 0 | 0 |
| 0 | 0 | 0 | 0 | 0 |
| 0 | 0 | 0 | 0 | 0 |
| 0 | 0 | 0 | 0 | 0 |
| 0 | 0 | 0 | 0 | 0 |
| 0 | 0 | 0 | 0 | 0 |

| | | | | |
|---|---|---|---|---|
| 0 | 0 | 0 | 0 | 0 |
| 0.006989489 | 0 | 0 | 0 | 0 |
| 0 | 0 | 0 | 0 | 0 |
| 0 | 0 | 0 | 0 | 0 |
| 0.083062596 | 0 | 0 | 0.057016583 | 0 |
| 0.002921617 | 0 | 0 | 0 | 0 |
| 0 | 0 | 0 | 0 | 0 |
| 0 | 0 | 0 | 0.004110868 | 0 |
| 0 | 0.000976056 | 0.00519519 | 0.007030391 | 0 |
| 0 | 0 | 0 | 0 | 0 |
| 0 | 0 | 0 | 0 | 0 |
| 0 | 0 | 0 | 0 | 0 |
| 0 | 0 | 0 | 0 | 0 |
| 0 | 0 | 0 | 0 | 0 |
| 0 | 0 | 0 | 0 | 0 |
| 0 | 0 | 0 | 0.005285037 | 0 |
| 0 | 0 | 0 | 0 | 0 |
| 0 | 0 | 0 | 0 | 0 |
| 0 | 0.432349243 | 0.031032388 | 0.019461843 | 0.533013721 |
| 0 | 0 | 0 | 0 | 0 |
| 0 | 0 | 0 | 0 | 0 |
| 0 | 0 | 0 | 0 | 0 |
| 0 | 0 | 0 | 0 | 0 |
| 0 | 0 | 0 | 0 | 0 |
| 0 | 0 | 0 | 0 | 0 |
| 0 | 0 | 0 | 0 | 0 |
| 0 | 0 | 0 | 0 | 0 |
| 0 | 0 | 0 | 0 | 0 |
| 0 | 0 | 0 | 0 | 0 |
| 0 | 0 | 0 | 0 | 0 |
| 0 | 0.031999573 | 0.168261705 | 0.065670935 | 0.010754981 |
| 0.002676523 | 0.011235234 | 2.501084528 | 0 | 0.003610532 |
| 0 | 0 | 0 | 0 | 0 |
| 0 | 0 | 0 | 0 | 0 |
| 0 | 0 | 0 | 0 | 0 |
| 0 | 0 | 0 | 0 | 0 |
| 0 | 0 | 0 | 0 | 0 |
| 0 | 0 | 0 | 0 | 0 |
| 0 | 0 | 0 | 0 | 0.518999588 |
| 0 | 0 | 0 | 0 | 0 |
| 0 | 0 | 0 | 0 | 0 |
| 0 | 0 | 0 | 0 | 0 |
| 0 | 0 | 0.025868162 | 0.00044653 | 0.000695534 |
| 0 | 0 | 0 | 0 | 0 |
| 0 | 0 | 0 | 0 | 0 |
| 0 | 0 | 0 | 0 | 0 |
| 0 | 0 | 0 | 0 | 0 |
| 0 | 0 | 0 | 0 | 0 |

| | | | | |
|---|---|---|---|---|
| 0.004256218 | 0 | 0 | 0 | 0 |
| 0 | 0 | 0 | 0 | 0 |
| 0 | 0 | 0 | 0 | 0 |
| 0 | 0 | 0 | 0 | 0 |
| 0 | 0.05714235 | 0.249119065 | 0.132034848 | 0.007987338 |
| 0 | 0 | 0 | 0 | 0 |
| 0 | 0 | 0 | 0 | 0 |
| 0 | 0 | 0 | 0 | 0 |
| 0 | 0 | 0 | 0 | 0 |
| 0 | 0 | 0 | 0 | 0 |
| 0 | 0 | 0 | 0 | 0 |
| 0 | 0 | 0 | 0 | 0 |
| 0 | 0 | 0 | 0 | 0 |
| 0 | 0 | 0 | 0 | 0 |
| 0 | 0 | 0 | 0 | 0 |
| 0 | 0 | 8.447248341 | 0 | 2.678735856 |
| 0 | 0 | 0 | 0 | 0 |
| 0 | 0 | 0 | 0 | 0 |
| 0 | 0 | 0 | 0 | 0 |
| 0 | 0 | 0 | 0 | 0 |
| 0 | 0 | 0 | 0 | 0 |
| 0 | 0 | 0 | 0 | 0 |
| 0 | 0 | 0.004817216 | 0 | 0 |
| 0 | 0 | 0 | 0 | 0 |
| 0 | 0.006825141 | 0.001478454 | 0 | 0 |
| 0 | 0 | 0 | 0 | 0 |
| 0 | 0.007646176 | 0.012326738 | 0 | 0 |
| 0 | 0 | 0 | 0 | 0 |
| 0 | 0 | 0 | 0 | 0 |
| 0 | 0 | 0 | 0 | 0 |
| 0 | 0 | 0 | 0 | 0 |
| 0 | 0 | 1.515867022 | 0 | 0 |
| 0 | 0 | 0 | 0 | 0 |
| 0 | 0 | 0 | 0 | 0 |
| 0 | 0 | 0 | 0 | 0 |
| 0 | 0 | 0 | 0 | 0 |
| 0 | 0 | 0 | 0 | 0 |
| 0 | 0 | 0 | 0 | 0 |
| 0 | 0 | 0 | 0 | 0 |
| 0 | 0 | 0 | 0 | 0 |
| 0 | 0 | 0 | 0 | 0 |
| 0 | 0 | 0 | 0 | 0 |
| 0 | 0 | 0 | 0 | 0 |
| 0 | 0 | 0 | 0 | 0 |
| 0 | 0 | 0 | 0 | 0 |
| 0 | 0 | 0 | 0 | 0 |
| 0 | 0 | 0 | 0 | 0 |
| 0 | 0 | 0 | 0 | 0 |

| | | | | |
|---|---|---|---|---|
| 0 | 0 | 0 | 0 | 0.048820349 |
| 0 | 0 | 0 | 0 | 0 |
| 0 | 0 | 0 | 0 | 0 |
| 0 | 0 | 0 | 0 | 0 |
| 0 | 0 | 0 | 0 | 0 |
| 0 | 0 | 0 | 0 | 0 |
| 0 | 0 | 0 | 0 | 0 |
| 0 | 0 | 0 | 0 | 0 |
| 0 | 0 | 0 | 0 | 0 |
| 0 | 0 | 0 | 0 | 0 |
| 0 | 0 | 0 | 0 | 0 |
| 0 | 0 | 0 | 0.048603699 | 0 |
| 0.001477519 | 0.0005978 | 0.001044052 | 0 | 0 |
| 0 | 0 | 0 | 0 | 0 |
| 0 | 0.005351735 | 0 | 0 | 0 |
| 0 | 0 | 0 | 0 | 0 |
| 0 | 0 | 0 | 0 | 0 |
| 0 | 0 | 0 | 0 | 0 |
| 0 | 0 | 0 | 0 | 0 |
| 0 | 0 | 0 | 0 | 0 |
| 0.029928225 | 0.002794357 | 0.013339529 | 0.026051628 | 0.013353842 |
| 0 | 0 | 0 | 0 | 0 |
| 0.000606004 | 0 | 0 | 0 | 0 |
| 0 | 0 | 0.001357117 | 0 | 0 |
| 0 | 0 | 0 | 0 | 0 |
| 0 | 0 | 0 | 0 | 0 |
| 0.002413314 | 0.001952841 | 0 | 0.000464445 | 0.004702349 |
| 0 | 0 | 0 | 0 | 0 |
| 0 | 0 | 0 | 0 | 0 |
| 0.0117623 | 0 | 0 | 0 | 0 |
| 0.025660423 | 0 | 0 | 0 | 0 |
| 0 | 0 | 0 | 0 | 0 |
| 0 | 0 | 0 | 0 | 0 |
| 0 | 0.266577806 | 0.061639278 | 0.022582021 | 0.00835083 |
| 0.03145622 | 0.028545073 | 0.007832637 | 0.006918603 | 0.00181857 |
| 0 | 0 | 0 | 0 | 0 |
| 0 | 0 | 0 | 0 | 0 |
| 0 | 0 | 0 | 0 | 0 |
| 0 | 0 | 0 | 0 | 0 |
| 0 | 0 | 0 | 0 | 0 |
| 0 | 0 | 0 | 0 | 0 |
| 0 | 0 | 0 | 0 | 0 |
| 0.001556915 | 0 | 0 | 0 | 0 |
| 0 | 0 | 0 | 0 | 0 |
| 0 | 0 | 0 | 0 | 0 |
| 0 | 0.072878637 | 0.04897779 | 0 | 0.00153909 |
| 0.001946213 | 0 | 0 | 0 | 0 |
| 0.005764256 | 0 | 0.000905148 | 0.001602375 | 0 |

| | | | | |
|---|---|---|---|---|
| 0 | 0 | 0 | 0 | 0 |
| 0 | 0 | 0 | 0 | 0 |
| 0 | 0 | 0 | 0 | 0 |
| 0 | 0 | 0 | 0 | 0 |
| 0 | 0 | 0 | 0 | 0 |
| 0 | 0 | 0 | 0 | 0 |
| 0.004558526 | 0 | 0 | 0 | 0 |
| 0 | 0 | 0 | 0 | 0 |
| 0 | 0 | 0.019620195 | 0 | 0 |
| 0 | 0 | 0 | 0 | 0 |
| 0 | 0.000801535 | 0 | 0 | 0 |
| 0 | 0 | 0 | 0 | 0 |
| 0 | 0 | 0 | 0 | 0 |
| 0 | 0 | 0 | 0 | 0 |
| 0 | 0 | 0 | 0 | 0 |
| 0 | 0 | 0 | 0 | 0 |
| 0 | 0 | 0 | 0 | 0 |
| 0 | 0 | 0 | 0 | 0 |
| 0.011724421 | 0.002240066 | 0.069193172 | 0 | 0.008933007 |
| 0 | 0.060551306 | 0.194977927 | 0.074698074 | 0 |
| 0.489696661 | 0 | 0 | 0.001109461 | 0 |
| 0 | 0 | 0 | 0 | 0 |
| 0 | 0 | 0 | 0 | 0 |
| 0 | 0 | 0 | 0 | 0 |
| 0.015270937 | 0.001544645 | 0.002248089 | 0.00110209 | 0 |
| 0 | 0 | 0 | 0 | 0 |
| 0 | 0 | 0 | 0 | 0 |
| 0 | 0 | 0 | 0 | 0 |
| 0 | 0 | 0 | 0 | 0 |
| 0 | 0 | 0 | 0 | 0 |
| 0.40999896 | 0.025440067 | 0.261259849 | 0.079267239 | 0.013818898 |
| 0 | 0 | 0 | 0 | 0 |
| 0 | 0 | 0 | 0 | 0 |
| 0 | 0 | 0 | 0 | 0 |
| 0 | 0 | 0 | 0 | 0 |
| 0 | 0 | 0 | 0 | 0 |
| 0 | 0 | 0 | 0 | 0 |
| 0 | 0 | 0 | 0 | 0 |
| 0 | 0 | 0 | 0 | 0 |
| 0.001411368 | 0.000713795 | 0 | 0 | 0 |
| 0 | 0 | 0 | 0 | 0 |
| 0.066253596 | 0.053466371 | 0.00491912 | 0.001940303 | 0.001133362 |
| 0 | 0 | 0 | 0 | 0 |
| 0 | 0 | 0 | 0 | 0 |
| 0 | 0 | 0 | 0 | 0 |
| 0 | 0 | 0 | 0 | 0 |
| 0 | 0 | 0 | 0 | 0 |
| 0 | 0 | 0 | 0 | 0 |

| | | | | |
|---|---|---|---|---|
| 0 | 0 | 0 | 0 | 0 |
| 0 | 0 | 0 | 0 | 0 |
| 0.041897568 | 0.001097588 | 0 | 0 | 0 |
| 0 | 0 | 0 | 0 | 0 |
| 0 | 0 | 0 | 0 | 0.049930289 |
| 0 | 0 | 0 | 0 | 0 |
| 0 | 0 | 0 | 0 | 0 |
| 0 | 0 | 0 | 0 | 0 |
| 0 | 0 | 0 | 0 | 0 |
| 0 | 0.001505769 | 0 | 0 | 0 |
| 0 | 0 | 0 | 0 | 0 |
| 0 | 0 | 0 | 0 | 0 |
| 0 | 0 | 0 | 0 | 0 |
| 0.0013325 | 0 | 0.013417486 | 0 | 0 |
| 0.00494912 | 0.003671068 | 0.001165724 | 0.001904925 | 0 |
| 0 | 0.060563317 | 0.012272604 | 0 | 0.002581126 |
| 0.001879693 | 0 | 0 | 0 | 0 |
| 0 | 0 | 0.001202059 | 0 | 0 |
| 0 | 0 | 0 | 0 | 0 |
| 0 | 0 | 0 | 0 | 0 |
| 0 | 0 | 0 | 0 | 0 |
| 0.002828043 | 0.000858164 | 0 | 0.002449168 | 0 |
| 0 | 0 | 0 | 0 | 0 |
| 0 | 0 | 0 | 0 | 0 |
| 0 | 0 | 0 | 0 | 0 |
| 0 | 0 | 0 | 0 | 0 |
| 0 | 0.012212744 | 0 | 0 | 0 |
| 0 | 0 | 0 | 0 | 0 |
| 0 | 0 | 0 | 0 | 0 |
| 0.004908949 | 0 | 0.001156262 | 0 | 0 |
| 0 | 0 | 0 | 0 | 0 |
| 0 | 0 | 0 | 0 | 0 |
| 0 | 0 | 0 | 0 | 0 |
| 0 | 0 | 0 | 0 | 0 |
| 0 | 0 | 0 | 0 | 0 |
| 0.002824307 | 0 | 0 | 0 | 0 |
| 0 | 0 | 0 | 0 | 0 |
| 0.00568503 | 0.025229748 | 0.001757518 | 0 | 0.001677571 |
| 0 | 0 | 0 | 0 | 0 |
| 0 | 0 | 0 | 0 | 0 |
| 0 | 0 | 0 | 0 | 0 |
| 0 | 0 | 0 | 0 | 0 |
| 0 | 0 | 0 | 0 | 0 |
| 0.018536953 | 0 | 0 | 0 | 0 |
| 0 | 0 | 0 | 0.003013151 | 0 |
| 0 | 0 | 0 | 0 | 0 |
| 0 | 0 | 0 | 0 | 0 |
| 0.027113615 | 0 | 0 | 0 | 0 |

| | | | | |
|---|---|---|---|---|
| 0 | 0 | 0 | 0.047340773 | 0.001339567 |
| 0 | 0 | 0 | 0 | 0 |
| 0 | 0 | 0 | 0 | 0 |
| 0 | 0 | 0 | 0 | 0 |
| 0.00655647 | 0 | 0.001052947 | 0.004817776 | 0.002814139 |
| 0 | 0 | 0 | 0 | 0 |
| 0 | 0 | 0 | 0 | 0 |
| 0.00167097 | 0.001267631 | 0.001771124 | 0.009406206 | 0.043109232 |
| 0 | 0 | 0 | 0 | 0 |
| 0 | 0.044162238 | 0.007302144 | 0.01591004 | 0 |
| 0 | 0 | 0 | 0 | 0 |
| 0.003637551 | 0 | 0.017525352 | 0 | 0.004460839 |
| 0.01956532 | 0 | 0 | 0.001158573 | 0 |
| 0.010666806 | 0 | 0 | 0 | 0 |
| 0 | 0 | 0 | 0 | 0 |
| 0 | 0 | 0 | 0 | 0 |
| 0 | 0 | 0 | 0 | 0 |
| 0 | 0 | 0 | 0 | 0 |
| 2.34833301 | 0 | 0 | 0 | 0 |
| 0 | 0 | 0 | 0 | 0 |
| 0 | 0 | 0.005759853 | 0 | 0 |
| 0 | 0 | 0 | 0 | 0 |
| 0 | 0 | 0 | 0 | 0 |
| 0 | 0.052877118 | 0.264086709 | 0 | 0.006467035 |
| 0 | 0 | 0 | 0 | 0 |
| 0.001460808 | 0 | 0 | 0 | 0.004138211 |
| 0 | 0 | 17.05163477 | 0 | 3.652984767 |
| 0.498159573 | 0 | 0.002496539 | 0 | 0 |
| 0.008375679 | 0 | 0 | 0 | 0 |
| 0.045690832 | 0 | 0 | 0.000867754 | 0 |
| 0.004389075 | 0 | 0 | 0 | 0 |
| 0 | 0 | 0 | 0 | 0 |
| 0 | 0 | 0 | 0 | 0 |
| 0.004967219 | 0 | 0 | 0 | 0 |
| 0 | 0 | 0 | 0 | 0 |
| 0 | 0 | 0 | 0 | 0 |
| 2.126935414 | 0 | 0 | 0 | 0 |
| 0 | 0 | 0 | 0 | 0 |
| 0 | 0 | 0 | 0 | 0 |
| 2.544887043 | 0 | 0 | 0 | 0 |
| 0 | 0 | 0 | 0 | 0 |
| 0 | 0 | 0 | 0 | 0 |
| 0.00214443 | 0 | 0 | 0 | 0 |
| 0 | 0 | 0 | 0 | 0 |
| 0 | 0 | 0 | 0 | 0 |
| 0 | 0.000518913 | 0 | 0 | 0 |
| 0.003633261 | 0 | 0 | 0 | 0 |
| 0 | 0 | 0 | 0 | 0 |

| | | | | |
|---|---|---|---|---|
| 0 | 0 | 0 | 0 | 0 |
| 0 | 0 | 0.001434598 | 0 | 0 |
| 0 | 0 | 0 | 0 | 0 |
| 0 | 0 | 0 | 0 | 0 |
| 0 | 0 | 0 | 0 | 0 |
| 0.017169799 | 0 | 0.151051 | 0.006278252 | 0.001158072 |
| 0 | 0 | 0 | 0.001285987 | 0 |
| 0 | 0 | 0 | 0 | 0 |
| 0 | 0 | 0 | 0 | 0 |
| 0 | 0 | 0 | 0 | 0 |
| 0.013607952 | 0.001007148 | 0 | 0.001916241 | 0.00335792 |
| 0 | 0 | 0 | 0.000897977 | 0 |
| 0 | 0 | 0 | 0 | 0 |
| 0 | 0.001141803 | 0.000664716 | 0 | 0.002326422 |
| 0 | 0 | 0 | 0 | 0 |
| 0 | 0 | 0 | 0 | 0 |
| 0 | 0 | 0 | 0 | 0 |
| 0 | 0 | 0 | 0 | 0 |
| 0 | 0 | 0 | 0 | 0 |
| 0 | 0.000570554 | 0 | 0.068715903 | 0 |
| 0 | 0 | 0 | 0 | 0 |
| 0 | 0.088644011 | 0.32842003 | 0.15514876 | 0.238159225 |
| 0 | 0 | 0 | 0.102561779 | 0 |
| 0 | 0 | 0 | 0 | 0 |
| 0 | 0 | 0 | 0 | 0 |
| 0 | 0 | 0 | 0 | 0 |
| 0.151973741 | 0 | 0 | 0 | 0 |
| 0 | 0 | 0 | 0 | 0 |
| 0 | 0 | 0 | 0 | 0 |
| 0.184766682 | 0.103208058 | 0.116595313 | 0.110390609 | 0.149112085 |
| 0.0053182 | 0 | 0.001653508 | 0 | 0.002726142 |
| 0 | 0 | 0 | 0 | 0.004431747 |
| 0.075016274 | 0 | 0 | 0 | 0 |
| 0 | 0 | 0 | 0 | 0 |
| 0 | 0 | 0 | 0 | 0 |
| 0 | 0 | 0 | 0 | 0 |
| 0 | 0.200240465 | 0.159782857 | 0 | 1.087317773 |
| 0 | 0 | 0.000903363 | 0 | 0.002155676 |
| 0 | 0 | 0 | 0 | 0 |
| 0 | 0 | 0 | 0 | 0 |
| 0.002577834 | 0 | 0 | 0.003125471 | 0 |
| 0 | 0.418186534 | 0.980234427 | 0 | 0.378474885 |
| 0 | 0 | 0.000620654 | 0 | 0 |
| 0 | 0 | 0 | 0 | 0 |
| 0.022391072 | 0 | 0 | 0 | 0 |
| 0 | 0 | 0 | 0 | 0 |
| 0 | 0 | 0.00191186 | 0 | 0 |
| 0 | 0 | 0 | 0 | 0 |

| | | | | |
|---|---|---|---|---|
| 0 | 0.001823392 | 0 | 0 | 0 |
| 0.037968551 | 0 | 0.00195946 | 0 | 0.000431614 |
| 0.0027292 | 0 | 0 | 0.015231874 | 0 |
| 0 | 0 | 0 | 0 | 0 |
| 0 | 0 | 0.014543873 | 0 | 0 |
| 0.098223755 | 0 | 0 | 0 | 0.000761496 |
| 0 | 0 | 0 | 0 | 0 |
| 0 | 0 | 0 | 0 | 0 |
| 0.041069506 | 0 | 0.00276388 | 0 | 0 |
| 0 | 0 | 0 | 0 | 0 |
| 0 | 0 | 0 | 0 | 0 |
| 0 | 0 | 0 | 0 | 0 |
| 0 | 0 | 0 | 0 | 0 |
| 0 | 0.000733145 | 0 | 0.002266732 | 0 |
| 0 | 0 | 0 | 0 | 0 |
| 1.787323011 | 0.037097883 | 0.009339253 | 0.049872067 | 0.002228606 |
| 0.027729102 | 0.009843191 | 0.001109099 | 0.006645441 | 0.076046174 |
| 0.091216661 | 0 | 0 | 0 | 0 |
| 0.002431874 | 0 | 0 | 0 | 0 |
| 0 | 0 | 0 | 0 | 0 |
| 0 | 0 | 0 | 0 | 0 |
| 0.002124059 | 0 | 0 | 0 | 0 |
| 0 | 0 | 0 | 0 | 0 |
| 0 | 0 | 0 | 0.056018317 | 0 |
| 0 | 0.001928909 | 0 | 0 | 0 |
| 0.007925931 | 0 | 0 | 0.005769812 | 0 |
| 0 | 0 | 0 | 0.000958686 | 0 |
| 0.02080638 | 0.001020389 | 0 | 0 | 0 |
| 0 | 0 | 0 | 0 | 0 |
| 0.004546598 | 0 | 0 | 0 | 0 |
| 0.024245672 | 0.002452434 | 0.042653035 | 0.022601429 | 0 |
| 0.005271485 | 0 | 0 | 0 | 0 |
| 0.191803842 | 0.006527382 | 0.015200008 | 0.018007937 | 0.051505461 |
| 0 | 0.001870875 | 0.001815258 | 0 | 0.001039611 |
| 0 | 0 | 0 | 0 | 0 |
| 0 | 0 | 0 | 0 | 0 |
| 0.001620597 | 0 | 0 | 0 | 0 |
| 0.066157345 | 0.000769169 | 0 | 0.00248787 | 0 |
| 0 | 0.036112012 | 0.019426395 | 0.348976795 | 0.173361692 |
| 0 | 0 | 0 | 0 | 0 |
| 0.004102247 | 0 | 0 | 0 | 0 |
| 0 | 0 | 0 | 0 | 0 |
| 0 | 0 | 0 | 0 | 0 |
| 0.058173658 | 0 | 0.077320243 | 0.38247732 | 0 |
| 0.609857472 | 0 | 0 | 0 | 0 |
| 0 | 0 | 0 | 0 | 0 |
| 0 | 0 | 0 | 0 | 0 |
| 0 | 0 | 0 | 0 | 0 |

| | | | | |
|---|---|---|---|---|
| 0.002254928 | 0 | 0 | 0 | 0 |
| 0.479751954 | 0.053033379 | 0.016108217 | 0.169701336 | 0.008386624 |
| 0 | 0 | 0 | 0 | 0 |
| 0 | 0 | 0 | 0 | 0 |
| 0 | 0 | 0 | 0 | 0 |
| 0 | 0 | 0 | 0 | 0 |
| 0.020130657 | 0 | 0 | 0 | 0 |
| 0 | 0 | 0 | 0 | 0 |
| 0 | 0 | 0 | 0.000912736 | 0 |
| 0.018071345 | 0 | 0 | 0 | 0 |
| 0.032853164 | 0 | 0 | 0 | 0.001558045 |
| 0.011469309 | 0.002784271 | 0 | 0.002648735 | 0 |
| 0 | 0.113597728 | 0.824982638 | 0 | 0 |
| 0 | 0 | 0 | 0 | 0 |
| 0 | 0 | 0 | 0 | 0 |
| 0 | 0 | 0 | 0 | 0 |
| 0.0266078 | 0.000815564 | 0.001709249 | 0.00062069 | 0 |
| 0.041867394 | 0 | 0 | 0 | 0.00141194 |
| 0 | 0 | 0 | 0 | 0 |
| 0.421164163 | 0 | 0 | 0.004839661 | 0 |
| 0.013581095 | 0.003364209 | 0.002546074 | 0.129937923 | 0.129177508 |
| 0 | 0 | 0 | 0 | 0 |
| 0 | 0 | 0 | 0 | 0 |
| 0 | 0 | 0 | 0 | 0 |
| 0 | 0 | 0 | 0 | 0 |
| 0 | 0 | 0.001796017 | 0 | 0 |
| 0.049790814 | 0.00863367 | 0.002126471 | 0.045173638 | 0 |
| 0.28692646 | 0.025387168 | 0.014363152 | 0.03197501 | 0.029406512 |
| 0 | 0 | 0 | 0 | 0 |
| 0.002842715 | 0 | 0 | 0 | 0 |
| 0 | 0.001342839 | 0 | 0 | 0 |
| 0 | 0 | 0 | 0 | 0 |
| 0.003576249 | 0 | 0 | 0 | 0 |
| 0 | 0 | 0 | 0 | 0 |
| 0 | 1.052719175 | 0 | 0 | 0.19773366 |
| 0.098716603 | 0.035099205 | 0.066232614 | 0.101611043 | 0.057503103 |
| 0 | 0 | 0 | 0 | 0 |
| 0 | 0 | 0 | 0 | 0 |
| 0.010175323 | 0.005513717 | 0.005392604 | 0.006923813 | 0.002451096 |
| 0 | 0 | 0 | 0 | 0 |
| 0.612966528 | 0 | 0 | 0 | 0 |
| 0 | 0 | 0 | 0 | 0 |
| 0.001765899 | 0.000937752 | 0.002651636 | 0 | 0 |
| 0 | 0 | 0.005421397 | 0 | 0 |
| 0 | 0 | 0 | 0 | 0 |
| 0 | 0.219028135 | 0.108916519 | 0.330410737 | 0.106486853 |
| 0 | 0 | 0 | 0 | 0 |
| 0.003100395 | 0 | 0 | 0 | 0 |

| | | | | |
|---|---|---|---|---|
| 0 | 0 | 0 | 0 | 0 |
| 0 | 0.013509967 | 0.008481869 | 0 | 0 |
| 0 | 0 | 0 | 0 | 0 |
| 0.960228159 | 3.074188275 | 0.152054166 | 0.314525587 | 0.061731096 |
| 0 | 0 | 0 | 0 | 0 |
| 0.002675226 | 0 | 0 | 0 | 0 |
| 0 | 0 | 0 | 0 | 0 |
| 0 | 0 | 0.001950233 | 0.003983627 | 0 |
| 0 | 4.775904779 | 0.183231795 | 0.088640307 | 0.594162389 |
| 0.001148873 | 0.092733741 | 0.110407814 | 0.319845516 | 0.021387061 |
| 0 | 0 | 0 | 0 | 0 |
| 0 | 0.02152657 | 0.000696221 | 0.001137704 | 0 |
| 0 | 0 | 0 | 0 | 0 |
| 0 | 0 | 0 | 0 | 0 |
| 1.110426526 | 0.10707612 | 0.303491938 | 0.143993996 | 0.017000061 |
| 0 | 0 | 0 | 0 | 0 |
| 0 | 0.012229295 | 0.020234214 | 0.030921895 | 0.027361199 |
| 0 | 2.928921009 | 0.451832606 | 0 | 0 |
| 0 | 0 | 0 | 0 | 0 |
| 0 | 0.000667493 | 0.013989245 | 0.017399008 | 0 |
| 0 | 0 | 0.005687703 | 0 | 0 |
| 0 | 0 | 0 | 0 | 0 |
| 0 | 0.001651861 | 0 | 0 | 0 |
| 0.511806821 | 0 | 0.424606277 | 0.465395893 | 0.06048339 |
| 0.01937188 | 0 | 0.002933283 | 0 | 0 |
| 0 | 0.089287254 | 0.123273475 | 0.316435382 | 0 |
| 0 | 0.001498787 | 0.332146353 | 0 | 0.101422383 |
| 0 | 1.357468606 | 0.564852198 | 0.511214983 | 7.036146793 |
| 0 | 0 | 0 | 0 | 0 |
| 0 | 0 | 0 | 0 | 0 |
| 0 | 0.008429131 | 0.007276089 | 0.059726278 | 0.004199369 |
| 0 | 0 | 0 | 0 | 0 |
| 0.029342218 | 0 | 0.041467894 | 0.09459495 | 0.001966786 |
| 0 | 0 | 0 | 0 | 0 |
| 0 | 0.0021469 | 0 | 0 | 0 |
| 0 | 0.129877516 | 0.339312039 | 0 | 0.033140924 |
| 0 | 0.914280863 | 0.249592743 | 0.105372129 | 3.090918253 |
| 0 | 0 | 0 | 0 | 0 |
| 0 | 0 | 0 | 0.003405006 | 0 |
| 0.011460061 | 0 | 0 | 0.002035845 | 0 |
| 0.001181496 | 0.036282477 | 0.141427732 | 0.304234307 | 0 |
| 0 | 0.026405234 | 0.018272563 | 0 | 0 |
| 0.032882499 | 0.568409249 | 0 | 0.007855779 | 0.049328362 |
| 0 | 0.003622911 | 0.005672821 | 0 | 0 |
| 0 | 0 | 0 | 0 | 0 |
| 0 | 0 | 0 | 0 | 0 |
| 0 | 0 | 0 | 0 | 0 |
| 0.720088721 | 0.000689576 | 0.001405061 | 0.001476018 | 0 |

| | | | | |
|---|---|---|---|---|
| 0 | 0 | 0 | 0 | 0 |
| 0 | 0.086223678 | 0.032915577 | 0 | 0.076625727 |
| 0.464820028 | 0.002313628 | 0.010198028 | 0 | 0 |
| 0 | 0.002322284 | 0.00998362 | 0 | 0.021044267 |
| 0 | 0 | 0 | 0 | 0.00134644 |
| 0 | 0.065482958 | 0 | 0.00396312 | 0.042102626 |
| 0 | 0 | 0 | 0 | 0 |
| 0 | 0 | 0 | 0 | 0 |
| 0 | 0 | 0 | 0 | 0 |
| 0 | 0 | 0 | 0 | 0 |
| 0.811147701 | 0.001135786 | 0.060453768 | 0.159759211 | 0.005770381 |
| 3.033647436 | 0.713260252 | 0.6119147 | 0.857109439 | 1.009232517 |
| 0 | 0 | 0 | 0 | 0 |
| 0.092043556 | 0.064616487 | 0.001223328 | 0.006996699 | 0.019266732 |
| 0.291727059 | 0.005795803 | 0.009318952 | 0.008139223 | 0 |
| 0 | 10.23244621 | 1.297496838 | 0.126943498 | 0 |
| 0 | 0 | 0 | 0.001812877 | 0 |
| 0 | 0 | 0 | 0.001457866 | 0 |
| 0 | 0.197161957 | 0.027554067 | 0.135493754 | 0.004195199 |
| 0 | 0.00334139 | 0.059135118 | 0 | 0.017676243 |
| 0.243799937 | 0.068948834 | 0.115747363 | 0.211008451 | 0 |
| 0.000944447 | 0 | 0 | 0 | 0.465178004 |
| 0.001693198 | 0.365823885 | 0.158231339 | 0.369522408 | 0.977007641 |
| 0 | 0 | 0 | 0 | 0 |
| 0 | 0.004931626 | 0.013967081 | 0 | 0 |
| 0 | 0 | 0.010815189 | 2.145593879 | 2.833633797 |
| 0 | 0 | 0 | 0.709778305 | 0 |
| 0 | 0 | 0 | 0 | 0 |
| 0 | 0 | 0 | 0.14343489 | 0 |
| 0 | 0 | 0 | 0 | 0 |
| 0 | 0.011549814 | 0.110376374 | 0 | 0 |
| 0 | 0.057774838 | 0.210327028 | 0.216063287 | 0.06463687 |
| 3.092572631 | 0.024496127 | 0.003880475 | 0 | 0.005555936 |
| 0.055278175 | 0.02605061 | 0.017015184 | 0.367264313 | 0.000988594 |
| 0.111997004 | 0 | 0.000768348 | 0.00146483 | 0.004278151 |
| 0.115781265 | 0.143556617 | 0.004222655 | 0.009919177 | 0.280628611 |
| 0.032470583 | 0.003503336 | 0.009007852 | 0.005693527 | 0 |
| 0 | 0 | 0 | 0 | 0 |
| 0.136652945 | 4.527778842 | 0.115916052 | 0.056018513 | 0.552822925 |
| 0 | 0 | 0 | 0 | 0 |
| 0 | 0.052692522 | 0.031915069 | 0 | 0.049983468 |
| 0 | 0 | 0 | 0 | 0 |
| 0 | 0 | 0 | 0.027027913 | 0 |
| 0 | 0 | 0 | 0 | 0 |
| 2.37294571 | 0.034809921 | 0.024879871 | 0.033935085 | 0.016661988 |
| 0 | 0.004661814 | 0 | 0 | 0.061727517 |
| 0 | 0 | 0 | 0 | 0 |
| 0 | 0.155696624 | 0 | 0.003757318 | 0.032343074 |

| | | | | |
|---|---|---|---|---|
| 0 | 0 | 0 | 0 | 0 |
| 0 | 0 | 0 | 0 | 0 |
| 0 | 0 | 0 | 0 | 0 |
| 0.002662076 | 0.003818699 | 0.009576457 | 0 | 0 |
| 0 | 0 | 0 | 0 | 0 |
| 0 | 0.277335676 | 0.151788498 | 0 | 0.389240203 |
| 0 | 0 | 0 | 0 | 0.053075368 |
| 1.520083254 | 0.434371414 | 0.003147169 | 0.170741998 | 0.000600802 |
| 0 | 0.27483781 | 0 | 0 | 0 |
| 0 | 0 | 0 | 0 | 0 |
| 0.110507976 | 0.00212651 | 0.007618313 | 0.003890371 | 0 |
| 0 | 0 | 0 | 0 | 0 |
| 0.027139544 | 0 | 0 | 0 | 0.01050555 |
| 0 | 0 | 0.015927266 | 0 | 0.295678109 |
| 0 | 0 | 0 | 0 | 0 |
| 0 | 0 | 0 | 0 | 0 |
| 0 | 0.020438666 | 0.047594558 | 0.099764826 | 0.07503988 |
| 0.001346274 | 0.011847198 | 0.004598002 | 0 | 0.014679931 |
| 0 | 0 | 0 | 0 | 0 |
| 0.010319985 | 0 | 0 | 0.001168288 | 0 |
| 0 | 0 | 0.028594736 | 0 | 0 |
| 0 | 0 | 0 | 0.009549669 | 0 |
| 0.003363007 | 0.026061945 | 0.031075807 | 0.00298714 | 0 |
| 0 | 0 | 0 | 0 | 0 |
| 0 | 0 | 0 | 0 | 0 |
| 0 | 0 | 0 | 0 | 0 |
| 0 | 0 | 0 | 0 | 0 |
| 0 | 0 | 0 | 0 | 0 |
| 0 | 0 | 0 | 0 | 0 |
| 0 | 0.065634624 | 0.038305829 | 1.050361929 | 0.101828931 |
| 0 | 0 | 0.004500202 | 0 | 0 |
| 0 | 0 | 0 | 0 | 0 |
| 0 | 0 | 0 | 0 | 0 |
| 0 | 0 | 0 | 0 | 0 |
| 0.056151431 | 0.033167596 | 0 | 0 | 0.001590092 |
| 0 | 0 | 0 | 0 | 0 |
| 0 | 0.040413502 | 0.040644185 | 0.021953336 | 0.023433426 |
| 0.034452652 | 0 | 0 | 0.000395192 | 0 |
| 0 | 0 | 0 | 0 | 0 |
| 0 | 0.002458449 | 0 | 0 | 0 |
| 0 | 0.001818886 | 0 | 1.040183695 | 0 |
| 0 | 0 | 0 | 0 | 0 |
| 0 | 0 | 0 | 0 | 0.018736456 |
| 0 | 0.041955296 | 0.074840186 | 0 | 0.00224171 |
| 0 | 0.059905699 | 0.002196844 | 0.102760645 | 0.018697469 |
| 0 | 0.007276315 | 0.015045113 | 0 | 0 |
| 0 | 0.005920695 | 0.00382979 | 0.002252992 | 0 |
| 0 | 0 | 0 | 0 | 0 |

| | | | | |
|---|---|---|---|---|
| 0 | 0 | 0 | 0 | 0 |
| 0 | 0 | 0 | 0 | 0 |
| 0 | 0.032047163 | 0.213059285 | 0 | 0.035259868 |
| 0 | 1.008933966 | 0.1918055 | 0.836255205 | 0.03964704 |
| 0 | 0 | 0 | 0 | 0 |
| 0 | 0.37191677 | 0 | 0 | 0 |
| 0 | 0 | 0 | 0 | 0 |
| 0 | 0 | 0 | 0 | 0 |
| 5.794115191 | 0.002673652 | 0.092500683 | 0.002906858 | 0.02292221 |
| 0 | 0.002460663 | 0.000895317 | 0 | 0 |
| 0 | 0 | 0.026710735 | 0 | 0 |
| 0 | 0 | 0 | 0 | 0 |
| 0 | 0.000334234 | 0 | 0.010916751 | 0 |
| 0.034751565 | 0 | 0 | 0 | 0 |
| 0 | 0.278047791 | 0.25183204 | 0.502130045 | 0.433623444 |
| 0 | 0 | 0 | 0 | 0 |
| 0 | 0 | 0 | 0 | 0 |
| 0 | 0 | 0 | 0 | 0 |
| 0 | 0 | 0 | 0 | 0 |
| 0 | 0 | 0 | 0 | 0 |
| 0 | 0.037821222 | 0.005440772 | 0.035285512 | 0.031484254 |
| 0 | 0 | 0 | 0 | 0 |
| 0 | 0 | 0 | 0 | 0 |
| 0 | 0 | 0 | 0 | 0 |
| 0.20204682 | 0.062658191 | 0.397458694 | 0 | 4.452460166 |
| 0 | 7.900007173 | 0.409618908 | 1.091844139 | 1.534772655 |
| 0 | 1.592601148 | 0.007586069 | 0.017482238 | 0.052357925 |
| 0 | 0 | 0 | 0 | 0 |
| 1.327114935 | 0.223837066 | 0.000914454 | 0.007845188 | 0.119799605 |
| 0 | 0 | 0 | 0 | 0 |
| 0 | 0 | 0 | 0 | 0 |
| 0 | 0.014607793 | 0 | 0 | 0 |
| 0 | 0 | 0 | 0 | 0 |
| 0.055929287 | 0.004437811 | 0.019690693 | 0.005362804 | 0 |
| 0.005535741 | 0.001580997 | 0 | 0.000564013 | 0.007467511 |
| 0.010588381 | 0 | 0.000975915 | 0 | 0.002018298 |
| 0 | 0 | 0 | 0 | 0 |
| 0.051983098 | 0.452718598 | 0.087423493 | 1.195401128 | 0.387431207 |
| 0 | 0.662320064 | 0.001437959 | 2.071337727 | 0 |
| 0 | 0 | 0 | 0 | 0 |
| 0 | 0 | 0 | 0 | 0 |
| 0 | 0 | 0 | 0 | 0 |
| 0.528126226 | 0.357361001 | 0.205597489 | 1.552542682 | 0.560306421 |
| 0 | 0 | 0.000835069 | 0 | 0 |
| 0 | 0 | 0 | 0 | 0 |
| 0 | 0 | 0 | 0.001007221 | 0 |
| 0 | 0 | 0 | 0 | 0.002149235 |
| 0 | 0.179636978 | 0.112188182 | 1.063263415 | 0.033507966 |

| | | | | |
|---|---|---|---|---|
| 0 | 0.000471674 | 0 | 0.000897427 | 0 |
| 0 | 0 | 0 | 0 | 0 |
| 0 | 0 | 0 | 0 | 0 |
| 0 | 0 | 0 | 0 | 0 |
| 0.021923878 | 0 | 0.00109355 | 0.006552278 | 0 |
| 0.954527414 | 0 | 0.03931471 | 0 | 0 |
| 0 | 0 | 0 | 0 | 0 |
| 0 | 0 | 0 | 0.049843136 | 0 |
| 0 | 0 | 0 | 0 | 0 |
| 0 | 0.033937088 | 0 | 0.004782971 | 0 |
| 0 | 0.001916434 | 0.001255137 | 0 | 0 |
| 0 | 0.87572619 | 0.237420713 | 1.025708729 | 0.066766462 |
| 1.454406997 | 0.019332773 | 0.026481952 | 0.015146078 | 0.006319331 |
| 0 | 0 | 0 | 0 | 0 |
| 0.015511272 | 0.015930926 | 0.038362288 | 0.000861103 | 0.031587401 |
| 0 | 0 | 0 | 0 | 0 |
| 0.001452544 | 0 | 0 | 0.000359413 | 0 |
| 0 | 0 | 0 | 0.025526115 | 0 |
| 0.027452603 | 0.0019041 | 0.025495431 | 0 | 0 |
| 0 | 0.002287288 | 0.001024288 | 0.007532115 | 0.009385872 |
| 0 | 0.038359558 | 0.063935459 | 0.059112428 | 0 |
| 0 | 0 | 0 | 0 | 0 |
| 0 | 4.32582524 | 0.02658728 | 1.383686985 | 0.203708784 |
| 0 | 0 | 0 | 0.002446776 | 0 |
| 0 | 0 | 0 | 0 | 0 |
| 0 | 0 | 0 | 0 | 0 |
| 0 | 0 | 0 | 0 | 0 |
| 0 | 0 | 0 | 0 | 0 |
| 0 | 0.001956782 | 0.003797222 | 0.000930764 | 0 |
| 0 | 0 | 0 | 0 | 0 |
| 0 | 0 | 0 | 0 | 0 |
| 0.053416735 | 0.011510882 | 0.001641113 | 0.013855785 | 0 |
| 0 | 0 | 0 | 0 | 0.00370431 |
| 0.143679402 | 0.013965836 | 0.063266779 | 0.132189997 | 0 |
| 0.008516861 | 0 | 0.00106204 | 0 | 0.001182684 |
| 0 | 0.229294985 | 0.013234619 | 0.005593155 | 0.070350486 |
| 0 | 0.005397914 | 0 | 0 | 0 |
| 0 | 0.047536356 | 0 | 0 | 0 |
| 0 | 0.002969122 | 0 | 0.000996913 | 0 |
| 0 | 0 | 0 | 0 | 0 |
| 0.007683244 | 0.069037267 | 0.033329087 | 0.117429228 | 0.038146803 |
| 0 | 0.002782403 | 0 | 0 | 0 |
| 0 | 0 | 0 | 0 | 0 |
| 0 | 0 | 0 | 0 | 0.002200718 |
| 0.047872135 | 0.059604413 | 0.111945112 | 0.045875217 | 0.006324626 |
| 0 | 0 | 0 | 0 | 0 |
| 0 | 0.074216675 | 0 | 0.027450395 | 0 |
| 0.011436316 | 0.003257858 | 0 | 0 | 0 |

| | | | | |
|---|---|---|---|---|
| 0 | 0 | 0 | 0 | 0 |
| 0 | 0 | 0 | 0 | 0 |
| 0.032922774 | 0 | 0 | 0.002357588 | 0 |
| 0 | 0 | 0 | 0 | 0 |
| 0 | 0 | 0 | 0 | 0 |
| 0 | 0 | 0 | 0 | 0 |
| 0 | 0.432384833 | 0 | 0.261464758 | 0.313788787 |
| 0.260791206 | 0.181823948 | 0.007323576 | 0.00055663 | 0.112659826 |
| 0 | 0 | 0 | 0 | 0 |
| 0.001660258 | 0 | 0.000670389 | 0 | 0 |
| 0 | 0 | 0 | 0 | 0 |
| 0.003895131 | 0 | 0 | 0 | 0 |
| 0 | 0 | 0 | 0 | 0 |
| 0 | 0.117178224 | 0.00072494 | 0 | 0.117218614 |
| 0 | 0 | 0 | 0 | 0 |
| 0 | 0.002832589 | 0 | 0 | 0 |
| 0 | 0 | 0 | 0 | 0 |
| 0 | 0 | 0 | 0 | 0 |
| 0 | 1.561034936 | 0 | 0.904220954 | 0.302757247 |
| 0 | 0 | 0 | 0.000418148 | 0 |
| 0 | 0 | 0 | 0 | 0.000938417 |
| 0 | 0 | 0 | 0 | 0 |
| 0 | 0 | 0 | 0 | 0 |
| 0 | 0 | 0 | 0 | 0 |
| 0 | 0.001309005 | 0.000571541 | 0.000466981 | 0 |
| 0.18722797 | 0 | 0 | 0 | 0 |
| 0 | 0 | 0 | 0 | 0 |
| 0.002023718 | 0.582842767 | 0.196546947 | 1.117508972 | 0.076286153 |
| 0 | 0.090991487 | 0 | 0 | 0 |
| 0 | 0.252237526 | 0.347218862 | 0.202382141 | 0.14877397 |
| 0 | 0 | 0 | 0 | 0 |
| 0 | 0 | 0 | 0 | 0 |
| 0 | 0.052018452 | 0.005536546 | 0.002193295 | 0 |
| 0 | 0 | 0 | 0 | 0 |
| 0.003611104 | 0 | 0 | 0 | 0.104894193 |
| 0 | 0.007635572 | 0 | 0 | 0 |
| 0 | 0 | 0 | 0 | 0 |
| 0.033326704 | 0 | 0 | 0.001734868 | 0 |
| 0 | 0 | 0 | 0 | 0 |
| 0 | 0 | 0.000451855 | 0 | 0 |
| 3.223176861 | 0.319021482 | 0.538674347 | 1.171286122 | 1.696613087 |
| 3.078606712 | 0.002063049 | 0.097354172 | 0.451402812 | 0.005259943 |
| 0 | 0 | 0.029022708 | 0 | 0.250905555 |
| 0 | 0.005856755 | 0.004804419 | 0 | 0 |
| 0 | 0 | 0 | 0 | 0 |
| 0 | 0 | 0 | 0 | 0 |
| 0 | 0.003762637 | 0 | 0.002326659 | 0 |
| 0.076748119 | 0.048420199 | 0.012102654 | 0.024408454 | 0.001316064 |

| | | | | |
|---|---|---|---|---|
| 0 | 0 | 0 | 0 | 0 |
| 0.033449905 | 0.001218037 | 0.011030386 | 0.06476089 | 0 |
| 0.003125408 | 0 | 0.005889311 | 0 | 0 |
| 0 | 0 | 0 | 0 | 0 |
| 1.791289206 | 0.255432178 | 0.768610667 | 1.942928756 | 1.906799521 |
| 0 | 0 | 0 | 0 | 0.005552379 |
| 0 | 0 | 0 | 0 | 0 |
| 0 | 0 | 0 | 0.000819046 | 0 |
| 2.808544169 | 0 | 0.000944068 | 0 | 0 |
| 0 | 0 | 0 | 0 | 0 |
| 0 | 0 | 0 | 0 | 0 |
| 0 | 0 | 6.562143555 | 0 | 0 |
| 0 | 0.035505911 | 0.218338931 | 0.106586796 | 0.022506787 |
| 0.012520252 | 0.00080835 | 0 | 0.005229201 | 0.005030874 |
| 0 | 10.66276149 | 0.012298103 | 0 | 0.188197545 |
| 0.199141276 | 0.094590324 | 0.294257082 | 1.495176598 | 0.230742403 |
| 0 | 0.058898038 | 0.016126076 | 0.011179566 | 0 |
| 0 | 0 | 0 | 0 | 0 |
| 0 | 0 | 0 | 0 | 0 |
| 0 | 0.000724142 | 0 | 0 | 0 |
| 0 | 0 | 0 | 0.0031034 | 0 |
| 0 | 0 | 0 | 0 | 0 |
| 0 | 0 | 0 | 0 | 0 |
| 0 | 0 | 0 | 0 | 0 |
| 0 | 0.000751402 | 0.001603942 | 0 | 0 |
| 0 | 0 | 0 | 2.02088011 | 0 |
| 0 | 0.053039001 | 0.004815734 | 0.046522369 | 0.121271062 |
| 0.005202127 | 0.000686337 | 0 | 0.003264632 | 0 |
| 0 | 0 | 0 | 0 | 0 |
| 0 | 0.000945591 | 0 | 0 | 0 |
| 0 | 0 | 0 | 0 | 0 |
| 0 | 0 | 0 | 0 | 0 |
| 0 | 0 | 0 | 0 | 0.004275454 |
| 0 | 0.014389286 | 0.013235523 | 0.018890578 | 0.001439255 |
| 0 | 1.954766838 | 0.002774144 | 0 | 0.007754716 |
| 0 | 0.000952739 | 0.011092995 | 0.007401944 | 0 |
| 0 | 0 | 0 | 0 | 0 |
| 0 | 0.08126892 | 0 | 0.000570574 | 0 |
| 0 | 0 | 0 | 0 | 0 |
| 0 | 0 | 0 | 0 | 0 |
| 0 | 0.342084587 | 0.259946706 | 0.314146535 | 0.211379277 |
| 0.001907415 | 0 | 0 | 0 | 0 |
| 0 | 0.023460935 | 0 | 0.00433532 | 0.007690778 |
| 0 | 0 | 0 | 0 | 0.015683868 |
| 0 | 0 | 0 | 0 | 0 |
| 0 | 0 | 0 | 0 | 0 |
| 0 | 0 | 0 | 0 | 0 |
| 0 | 0 | 0 | 0 | 0 |

| | | | | |
|---|---|---|---|---|
| 0 | 1.535112443 | 0.032023603 | 0.110093577 | 0.083349072 |
| 0 | 0 | 0 | 0 | 0 |
| 0 | 0 | 0 | 0.024648586 | 0 |
| 0.001024868 | 0 | 0 | 0 | 0 |
| 0.007354484 | 0.013687782 | 0.046598537 | 0.09199951 | 0.03108558 |
| 0 | 0 | 0 | 0 | 0.004815729 |
| 0.048750942 | 0.144305195 | 0.00909964 | 0.216409315 | 0 |
| 0 | 0 | 0 | 0.000770018 | 0 |
| 0 | 0 | 0 | 0.004404449 | 0 |
| 0 | 0 | 0 | 0 | 0 |
| 0 | 0.010492851 | 0 | 0.002898019 | 0 |
| 0 | 0 | 0.001705244 | 0 | 0.000406919 |
| 0 | 0 | 0 | 0 | 0 |
| 0 | 0 | 0 | 0 | 0 |
| 0 | 0 | 0 | 0 | 0.001676406 |
| 0 | 0 | 0 | 0 | 0 |
| 0 | 0 | 0 | 0 | 0 |
| 0 | 0 | 0 | 0 | 0 |
| 0 | 0 | 0 | 0 | 0 |
| 0 | 0 | 0 | 0 | 0 |
| 0.023206807 | 0.108554006 | 0.048731425 | 0.131709905 | 0.008416946 |
| 0 | 0 | 0 | 0 | 0 |
| 0 | 0.017586491 | 0.055115686 | 0.056186231 | 0.001954497 |
| 0.016534036 | 0.029712369 | 0.001213856 | 0.002603446 | 0.001158639 |
| 0 | 0 | 0 | 0 | 0 |
| 0.012919854 | 0.000922471 | 0.006623357 | 0 | 0 |
| 0 | 0 | 0 | 0 | 0 |
| 0 | 0 | 0 | 0 | 0.002274042 |
| 0 | 0 | 0 | 0 | 0 |
| 0 | 0 | 0 | 0.00124899 | 0.001042221 |
| 0 | 0 | 0 | 0 | 0 |
| 0 | 0 | 0 | 0 | 0 |
| 0 | 0 | 0 | 0 | 0 |
| 0 | 0 | 0 | 0 | 0 |
| 0 | 0.093353763 | 0 | 0 | 0.022232123 |
| 0 | 0 | 0 | 0 | 0 |
| 0.080604007 | 0.022991577 | 0.003797121 | 0 | 0.006705131 |
| 0 | 0 | 0 | 0 | 0 |
| 0 | 0.143947317 | 0 | 0 | 0 |
| 0 | 0 | 0 | 0 | 0 |
| 0 | 0 | 0 | 0 | 0 |
| 0 | 0 | 0 | 0 | 0 |
| 0 | 0 | 0 | 0 | 0 |
| 0 | 0.030497494 | 0.030436319 | 0 | 0 |
| 0 | 0.009259486 | 0 | 0 | 0 |
| 0 | 0 | 0 | 0 | 0 |
| 0 | 0 | 0 | 0 | 0 |
| 0 | 0 | 0 | 0 | 0.00984564 |

| | | | | |
|---|---|---|---|---|
| 0.051756248 | 0 | 0.002285766 | 0.0004002 | 0 |
| 0 | 0.223290021 | 0 | 0 | 0 |
| 0 | 0 | 0.01358203 | 0 | 0.00129642 |
| 0 | 0 | 0 | 0 | 0 |
| 0 | 0 | 0 | 0 | 0 |
| 0 | 0 | 0 | 0 | 0 |
| 0 | 0 | 0 | 0 | 0 |
| 0 | 0.28940095 | 0.320025501 | 0.245727213 | 0.225940705 |
| 2.75778084 | 0.223407607 | 18.09636469 | 0.007119308 | 5.612996748 |
| 0.690037317 | 0 | 0.031578773 | 0 | 0 |
| 0 | 0.016722713 | 0.001465536 | 0 | 0 |
| 0 | 0 | 0 | 0.000637996 | 0 |
| 0 | 0.01950727 | 0.007570943 | 0 | 0 |
| 0 | 0 | 0 | 0 | 0 |
| 0 | 0 | 0 | 0 | 0 |
| 0 | 0 | 0 | 0 | 0 |
| 0 | 0 | 0 | 0 | 0 |
| 0 | 0 | 0 | 0.119268453 | 0.397018299 |
| 0 | 0 | 0 | 0 | 0 |
| 0 | 0.020711269 | 0.010651883 | 0.100207577 | 0.001553344 |
| 0 | 0 | 0 | 0 | 0 |
| 0 | 0 | 0 | 0 | 0 |
| 0 | 0 | 0 | 0.197081771 | 0 |
| 0 | 0 | 0 | 0 | 0 |
| 0.014013044 | 0.013143257 | 0.026876771 | 0 | 0.008469563 |
| 0 | 0 | 0 | 0 | 0 |
| 0 | 0 | 0 | 0 | 0 |
| 0 | 0 | 0 | 0 | 0 |
| 0 | 0 | 0 | 0 | 0 |
| 0 | 0 | 0 | 0 | 0 |
| 0.00451001 | 0 | 0 | 0 | 0 |
| 0 | 0 | 0 | 0 | 0 |
| 0 | 0 | 0 | 0 | 0 |
| 0 | 0 | 0 | 0 | 0.000619587 |
| 0 | 0.037857261 | 0 | 0 | 0.007735423 |
| 0 | 0 | 0.001215222 | 0 | 0 |
| 0 | 0 | 0 | 0 | 0 |
| 0 | 0 | 0 | 0 | 0 |
| 0 | 0.007003074 | 0.001358977 | 0.000634492 | 0 |
| 0 | 0 | 0.028523883 | 0 | 0 |
| 0 | 0 | 0 | 0 | 0 |
| 2.568191761 | 0.206237669 | 0.907878799 | 0.335927508 | 0.115886506 |
| 0 | 0 | 0 | 0 | 0 |
| 0 | 0 | 0 | 0 | 0 |
| 0.009440663 | 0 | 0 | 0 | 0 |
| 0.051569095 | 0 | 0 | 0 | 0 |
| 0 | 0 | 0 | 0 | 0 |
| 0 | 0 | 0 | 0 | 0 |

| | | | | |
|---|---|---|---|---|
| 0 | 0 | 0 | 0 | 0 |
| 0.000625611 | 0 | 0 | 0.002769186 | 0.006610755 |
| 0 | 0 | 0 | 0 | 0 |
| 0 | 0 | 0 | 0 | 0 |
| 0 | 1.334903336 | 0.359310064 | 0.149313363 | 0.194294707 |
| 0 | 0 | 0.025042071 | 0 | 0 |
| 0 | 0 | 0 | 0 | 0 |
| 0.473502352 | 0.005529713 | 0.114021823 | 0.00526053 | 0.023590857 |
| 0 | 0.013892085 | 0.001658966 | 0 | 0 |
| 0 | 0 | 0 | 0.018978656 | 0.12691253 |
| 0 | 0 | 0 | 0 | 0 |
| 0 | 0 | 1.929211156 | 0 | 0 |
| 0 | 0 | 0 | 0 | 0 |
| 0 | 0 | 0 | 0 | 0 |
| 0 | 0 | 0 | 0 | 0 |
| 0 | 0.009591757 | 0 | 0.124215515 | 0 |
| 0 | 0 | 0 | 0 | 0 |
| 0 | 0 | 0 | 0 | 0 |
| 0 | 0 | 0 | 0 | 0 |
| 0 | 0 | 0 | 0 | 0 |
| 0 | 0 | 0 | 0 | 0 |
| 0 | 0 | 0 | 0 | 0 |
| 0 | 0 | 0 | 0 | 0 |
| 0.031286534 | 0 | 0 | 0.00088114 | 0 |
| 0 | 0.080210768 | 0.031503646 | 0.191418703 | 0.080137357 |
| 0 | 0 | 0 | 0 | 0 |
| 0.005816074 | 0.001976661 | 0.017096701 | 0.001208853 | 0 |
| 0 | 0 | 0 | 0 | 0 |
| 0.004149541 | 0 | 0 | 0 | 0 |
| 0 | 0 | 0 | 0 | 0 |
| 0 | 0 | 0 | 0 | 0 |
| 0 | 0 | 0 | 0 | 0 |
| 0 | 0 | 0 | 0 | 0 |
| 0 | 0 | 0 | 0 | 0 |
| 0 | 0 | 0 | 0 | 0 |
| 0 | 0 | 0 | 0 | 0 |
| 0 | 0 | 0 | 0 | 0 |
| 0 | 0.037358068 | 0.008935428 | 0.006061 | 0.002574783 |
| 0 | 0 | 0 | 0.064733115 | 0 |
| 0 | 0 | 0 | 0 | 0 |
| 0 | 0 | 0 | 0 | 0 |
| 0 | 1.412933329 | 0.014112038 | 0 | 0.02288072 |
| 0 | 0 | 0 | 0 | 0 |
| 0 | 0 | 0 | 0 | 0 |
| 0 | 0 | 0 | 0 | 0 |
| 0 | 0 | 0 | 0 | 0 |
| 0 | 0 | 0 | 0 | 0 |
| 0 | 0.006144776 | 0 | 0 | 0 |

| | | | | |
|---|---|---|---|---|
| 0 | 0.048729201 | 0.038030593 | 0.148671118 | 0.044564194 |
| 0 | 0 | 0 | 0 | 0 |
| 0 | 0 | 0 | 0 | 0 |
| 0.000870699 | 0 | 0 | 0 | 0 |
| 0 | 0 | 0 | 0 | 0 |
| 0.522401812 | 0.128448025 | 0.290692775 | 0 | 0.001196726 |
| 0 | 0 | 0 | 0 | 0 |
| 0 | 0 | 0 | 0 | 0 |
| 0 | 0 | 0 | 0 | 0 |
| 0 | 0 | 0 | 0 | 0 |
| 0 | 0 | 0 | 0 | 0 |
| 0.039158369 | 0 | 0 | 0.000541514 | 0 |
| 0 | 0 | 0 | 0 | 0 |
| 0 | 0 | 0 | 0 | 0 |
| 0 | 0 | 0 | 0 | 0 |
| 0 | 0 | 0 | 0 | 0 |
| 0 | 0 | 0 | 0 | 0 |
| 1.654987494 | 0 | 0 | 0 | 0 |
| 0 | 0 | 0 | 0.00235601 | 0 |
| 0 | 0 | 0 | 0 | 0 |
| 0.006839067 | 0 | 0 | 0 | 0 |
| 0 | 0 | 0 | 0 | 0 |
| 0.042866317 | 0.090859741 | 0.104660161 | 0.031028576 | 0 |
| 0 | 0.03198445 | 0.003310253 | 0.921952269 | 1.112600147 |
| 0 | 0.001569433 | 0 | 0 | 0 |
| 0 | 0 | 0 | 0 | 0 |
| 0 | 0 | 0 | 0 | 0 |
| 0 | 0 | 0 | 0 | 0 |
| 0 | 0 | 0 | 0 | 0 |
| 0 | 0 | 0 | 0 | 0 |
| 0 | 0 | 0 | 0 | 0 |
| 0 | 0 | 0.012728292 | 0 | 0.026251167 |
| 0.087178668 | 0.00127271 | 0.014818502 | 0 | 0 |
| 0 | 0 | 0 | 0 | 0 |
| 1.732998312 | 0.042122032 | 0.04090139 | 0.021969735 | 0.006145318 |
| 0.075726895 | 0 | 0 | 0 | 0.001297179 |
| 0 | 0 | 0 | 0 | 0 |
| 0 | 0 | 0 | 0 | 0 |
| 0.101215265 | 0.001518803 | 0 | 0.002087032 | 0 |
| 0 | 0 | 0 | 0 | 0 |
| 0 | 0 | 0 | 0 | 0 |
| 0 | 0 | 0.004489562 | 0.597809766 | 0.002597175 |
| 0 | 0.458586057 | 0.048477693 | 0.011263228 | 0.068860514 |
| 0 | 0 | 0 | 0 | 0 |
| 0 | 0 | 0 | 0 | 0 |
| 0 | 0.033379489 | 0.11933081 | 0.00059633 | 0.002089956 |
| 0 | 0 | 0 | 0 | 0 |
| 0 | 0 | 0 | 0 | 0 |

| | | | | |
|---|---|---|---|---|
| 0 | 0 | 0 | 0 | 0 |
| 0 | 0 | 0 | 0 | 0 |
| 0 | 0 | 0 | 0 | 0 |
| 0 | 0 | 0 | 0 | 0 |
| 0 | 0 | 0 | 0 | 0 |
| 0 | 0 | 0 | 0 | 0 |
| 0 | 0 | 0 | 2.106562357 | 0 |
| 0 | 0 | 0 | 0 | 0 |
| 0.069190816 | 0.017325218 | 0.145137336 | 0 | 0.000815873 |
| 0 | 0.180134477 | 0.001691414 | 0 | 0 |
| 0 | 0.480146839 | 0 | 0 | 0 |
| 0 | 0 | 0 | 0 | 0 |
| 0 | 0 | 0 | 0 | 0 |
| 0 | 0 | 0.111370901 | 0 | 0.013423102 |
| 0 | 0 | 0 | 0 | 0 |
| 0.182126828 | 0 | 0 | 0 | 0 |
| 0 | 0 | 0 | 0 | 0 |
| 0 | 0 | 0 | 0 | 0 |
| 0 | 0 | 0 | 0 | 0 |
| 0 | 0.145326476 | 0.4176777 | 0.585136419 | 0.764847295 |
| 0 | 0 | 0.014870425 | 0.138699115 | 0.01198194 |
| 0 | 0 | 0 | 0 | 0 |
| 0 | 0 | 0 | 0 | 0 |
| 0 | 0.003807697 | 0.002216701 | 0 | 0 |
| 0.006719592 | 0 | 0 | 0 | 0 |
| 0 | 0 | 0 | 0 | 0 |
| 0 | 0 | 0 | 0 | 0 |
| 0 | 0 | 0 | 0 | 0 |
| 0 | 0 | 0 | 0 | 0 |
| 0 | 2.052003337 | 0.033920647 | 0.013622671 | 0.342617678 |
| 0.077416718 | 0.269382945 | 0.389244222 | 0.299738374 | 0.211016399 |
| 0 | 0 | 0 | 0 | 0 |
| 0 | 0 | 0 | 0.001555651 | 0 |
| 0 | 0 | 0 | 0 | 0 |
| 0 | 0 | 0 | 0 | 0 |
| 0 | 0 | 0 | 0 | 0 |
| 0 | 0 | 0 | 0 | 0 |
| 0 | 0 | 0 | 0 | 0 |
| 0 | 0.00261854 | 0.000846899 | 0 | 0.038640295 |
| 0 | 0 | 0 | 0 | 0 |
| 0 | 0 | 0 | 0 | 0 |
| 0 | 0.009720993 | 0.001342862 | 0 | 0.00531022 |
| 0 | 0.000837646 | 0 | 0 | 0 |
| 0 | 0 | 0 | 0 | 0 |
| 0 | 0 | 0 | 0 | 0 |
| 0 | 0 | 0 | 0 | 0 |
| 0 | 0 | 0 | 0 | 0 |
| 0 | 0.005284918 | 0.002461348 | 0 | 0 |

| | | | | |
|---|---|---|---|---|
| 0 | 0 | 0 | 0 | 0 |
| 0 | 0 | 0 | 0 | 0 |
| 0 | 0 | 0 | 0 | 0 |
| 0 | 0 | 0.002544674 | 0 | 0.000647712 |
| 2.316430707 | 0.012752692 | 0.086261535 | 0.516241211 | 0.000944858 |
| 0 | 0 | 0.08581964 | 0 | 0 |
| 0 | 0 | 0 | 0 | 0 |
| 0 | 0 | 0 | 0 | 0 |
| 0 | 0 | 0 | 0 | 0 |
| 0 | 0 | 0 | 0 | 0 |
| 0 | 0.013107395 | 0.064995125 | 0 | 0 |
| 0.012502913 | 0.011972127 | 0.02081103 | 0.010426853 | 0.001124398 |
| 0.01126628 | 0.001309301 | 0 | 0.010794896 | 0 |
| 0 | 0 | 0 | 0 | 0 |
| 0 | 0 | 0 | 0.000733476 | 0 |
| 0 | 0.036846721 | 0 | 0 | 0.016758929 |
| 0 | 0 | 0.032012305 | 0 | 0 |
| 0 | 0 | 0 | 0 | 0 |
| 0 | 0 | 0.033895064 | 0.001821986 | 0.001986602 |
| 0.033513072 | 0.03084185 | 0.231376744 | 0.192378445 | 0 |
| 0 | 0 | 0 | 0 | 0 |
| 0 | 0.160641255 | 0 | 0 | 0.023717458 |
| 0 | 0 | 0 | 0 | 0 |
| 0 | 0 | 0 | 0 | 0 |
| 0 | 0 | 0 | 0 | 0 |
| 0 | 0 | 0 | 0 | 0 |
| 0.008364847 | 0.001202351 | 0.000777738 | 0.001525095 | 0 |
| 0 | 0 | 0 | 0 | 0 |
| 0 | 0 | 0 | 0 | 0 |
| 0 | 0 | 0 | 0 | 0 |
| 0.002744787 | 0 | 0 | 0 | 0 |
| 0 | 0 | 0 | 0 | 0 |
| 0 | 0 | 0 | 0 | 0 |
| 0 | 0 | 0 | 0 | 0 |
| 0 | 0 | 0 | 0 | 0 |
| 0 | 0 | 0 | 0 | 0 |
| 0 | 0 | 0 | 0 | 0 |
| 0.186179029 | 0.007660428 | 0.013626614 | 0.016396927 | 0.001537162 |
| 0 | 0.207697216 | 0.001788003 | 0.006817531 | 0.216462512 |
| 0.004449389 | 0 | 0 | 0 | 0 |
| 0.031502828 | 0.007010278 | 0.013727424 | 0.000303137 | 0 |
| 0 | 0 | 0 | 0 | 0 |
| 0 | 0 | 0 | 0 | 0 |
| 0.094106328 | 0 | 0.001285871 | 0 | 0 |
| 0 | 0 | 0 | 0 | 0 |
| 0 | 0.061073529 | 0.002956054 | 0.003622895 | 0 |
| 0 | 0 | 0 | 0 | 0 |
| 0 | 0 | 0 | 0 | 0 |

| | | | | |
|---|---|---|---|---|
| 0 | 0 | 0 | 0 | 0 |
| 0 | 0.080099174 | 0.00989138 | 0.059010099 | 0.020381512 |
| 0 | 0 | 0 | 0 | 0 |
| 0.005721274 | 0 | 0 | 0 | 0 |
| 0.001010741 | 0.001380183 | 0 | 0 | 0.000681726 |
| 0.000978344 | 0 | 0.001382644 | 0 | 0 |
| 0 | 0.018612608 | 0 | 0 | 0.008381138 |
| 0 | 0.00375008 | 0 | 0 | 0 |
| 0 | 0 | 0.030653376 | 0 | 0 |
| 0 | 0.006878314 | 0.084535248 | 0 | 0 |
| 0 | 0.005055165 | 0 | 0 | 0 |
| 0 | 0 | 0 | 0 | 0 |
| 0.346655318 | 0.096928881 | 0.019514548 | 0.005574997 | 0.147842547 |
| 0.00245356 | 0 | 0.005393879 | 0 | 0 |
| 0 | 0 | 0 | 0 | 0 |
| 0 | 0 | 0 | 0 | 0 |
| 0 | 0 | 0 | 0 | 0 |
| 0 | 0 | 0 | 0 | 0 |
| 0 | 0 | 0 | 0 | 0 |
| 0 | 0 | 0 | 0 | 0 |
| 0 | 0 | 0 | 0 | 0 |
| 0 | 0 | 0 | 0 | 0 |
| 0.099939171 | 0 | 0 | 0 | 0.002401272 |
| 0 | 0.003642354 | 0.014631067 | 0.012993924 | 0.004452774 |
| 0 | 0 | 0 | 0 | 0 |
| 0 | 0.018425906 | 0 | 0 | 0 |
| 0 | 0.001916868 | 0.004292038 | 0 | 0 |
| 0 | 0 | 0 | 0 | 0 |
| 0 | 0 | 0 | 0 | 0 |
| 0 | 0 | 0 | 0 | 0 |
| 0 | 0.006022803 | 0.009617155 | 0.01522441 | 0.005928546 |
| 0 | 0 | 0.031607268 | 0 | 0 |
| 0 | 0 | 0 | 0 | 0 |
| 0 | 0 | 0 | 0 | 0 |
| 0 | 0 | 0 | 0.562156315 | 0.008177043 |
| 0 | 0 | 0 | 0 | 0 |
| 0 | 0 | 0 | 0 | 0 |
| 0 | 0 | 0 | 0 | 0 |
| 0.004318121 | 0 | 0 | 0 | 0 |
| 0 | 0 | 0 | 0 | 0 |
| 0 | 0 | 0 | 0 | 0 |
| 0 | 0 | 0 | 0 | 0 |
| 0 | 0.580023158 | 0.021785042 | 0 | 0 |
| 0 | 0 | 0 | 0 | 0 |
| 0 | 0 | 0 | 0 | 0 |
| 0.046012934 | 0.431685644 | 0.017151824 | 0.118205139 | 0.451998841 |
| 0 | 0 | 0 | 0 | 0 |
| 0 | 0 | 0 | 0 | 0 |

| | | | | |
|---|---|---|---|---|
| 0 | 0 | 0 | 0 | 0 |
| 0 | 0 | 0 | 0 | 0 |
| 0 | 0 | 0 | 0 | 0.053444379 |
| 0 | 0 | 0.03692942 | 0 | 0.000644809 |
| 0 | 0 | 0 | 0 | 0 |
| 0 | 0 | 0 | 0 | 0.000655328 |
| 0 | 0 | 0 | 0 | 0 |
| 0 | 0 | 0 | 0 | 0 |
| 0 | 0 | 0 | 0.006663976 | 0 |
| 0 | 0 | 0 | 0 | 0 |
| 0.024327451 | 0.000944911 | 0 | 0.000561821 | 0 |
| 0 | 0 | 0 | 0.00050328 | 0 |
| 0 | 0 | 0 | 0 | 0 |
| 0 | 0 | 0 | 0 | 0 |
| 0 | 0 | 0 | 0 | 0 |
| 0 | 0 | 0 | 0 | 0 |
| 0 | 0 | 0 | 0 | 0 |
| 0 | 0 | 0 | 0 | 0 |
| 0 | 0 | 0 | 0 | 0 |
| 0.000774297 | 0.000469918 | 0 | 0 | 0 |
| 0 | 0 | 0 | 0 | 0 |
| 0 | 0 | 0 | 0 | 0 |
| 0 | 0 | 0 | 0 | 0 |
| 0 | 0 | 0 | 0 | 0 |
| 0 | 0 | 0 | 0 | 0 |
| 0 | 0 | 0 | 0 | 0 |
| 0 | 0 | 0 | 0 | 0 |
| 0 | 0 | 0 | 0.017645302 | 0.020228496 |
| 0 | 0 | 0 | 0 | 0 |
| 0 | 0 | 0 | 0 | 0 |
| 0.006652013 | 0.004709927 | 0.001253461 | 0 | 0.020489086 |
| 0 | 0 | 0 | 0 | 0 |
| 0.023943361 | 0.015312373 | 0.004548113 | 0.015756118 | 0.012850027 |
| 0.00157855 | 0.000958015 | 0 | 0 | 0 |
| 0 | 0 | 0 | 0 | 0 |
| 0 | 0 | 0 | 0 | 0 |
| 0 | 0 | 0 | 0 | 0 |
| 0 | 0 | 0 | 0 | 0 |
| 0 | 0 | 0 | 0 | 0 |
| 0 | 0 | 0 | 0 | 0 |
| 0 | 0 | 0 | 0 | 0 |
| 0.749158883 | 0.121217365 | 0.001197341 | 0.030571734 | 0 |
| 0 | 0.067182824 | 0.01531634 | 0.053483809 | 0.044380994 |
| 0 | 0 | 0 | 0 | 0 |
| 0 | 0 | 0 | 0 | 0 |
| 0 | 0.149586074 | 0.016599883 | 0.008826743 | 0.025653449 |
| 0.001222371 | 0 | 0 | 0.000423444 | 0 |
| 0 | 0 | 0 | 0 | 0 |

| | | | | |
|---|---|---|---|---|
| 0 | 0 | 0 | 0 | 0 |
| 0 | 0.00180469 | 0.011843401 | 0.00795989 | 0.005652331 |
| 0.006315501 | 0.000574928 | 0 | 0.004922466 | 0 |
| 0 | 0 | 0 | 0 | 0 |
| 0 | 0.016066827 | 0.005701191 | 1.023929612 | 0.001190405 |
| 0 | 0 | 0.002081064 | 0.000850174 | 0 |
| 0 | 0 | 0 | 0 | 0 |
| 0 | 0 | 0 | 0 | 0 |
| 0 | 0 | 0 | 0 | 0 |
| 0 | 0 | 0 | 0 | 0 |
| 0 | 0 | 0 | 0 | 0 |
| 0 | 0 | 0 | 0 | 0 |
| 0 | 0 | 0 | 0 | 0 |
| 0.010882102 | 0 | 0 | 0 | 0 |
| 0 | 0 | 0 | 0 | 0 |
| 0 | 0 | 0 | 0 | 0 |
| 0 | 0 | 0.133235331 | 0 | 0.02905824 |
| 0 | 0 | 0 | 0 | 0 |
| 0 | 0 | 0 | 0 | 0 |
| 0 | 0 | 0 | 0 | 0 |
| 0 | 0 | 0 | 0 | 0 |
| 0 | 0 | 0 | 0 | 0 |
| 0 | 0 | 0 | 0 | 0 |
| 0 | 0 | 0 | 0 | 0 |
| 0 | 0.019900497 | 0 | 0.006160333 | 0.011233866 |
| 0.000785395 | 0 | 0 | 0 | 0 |
| 0 | 0 | 0 | 0 | 0 |
| 0 | 0.075674577 | 0.005742397 | 0 | 0.047446498 |
| 0 | 0 | 0 | 0 | 0 |
| 0 | 0 | 0 | 0 | 0 |
| 0 | 0 | 0 | 0 | 0 |
| 0.002525398 | 0 | 0.000648913 | 0 | 0 |
| 0 | 0 | 0 | 0 | 0 |
| 0 | 0 | 0 | 0 | 0 |
| 0 | 0 | 0 | 0 | 0 |
| 0 | 0 | 0 | 0 | 0 |
| 0 | 0 | 0 | 0 | 0 |
| 0 | 0 | 0 | 0.043030729 | 0 |
| 0 | 0.654338485 | 0.006046537 | 0 | 0.371780026 |
| 0 | 0 | 0 | 0 | 0 |
| 0 | 0 | 0 | 0 | 0 |
| 0.001136043 | 0.056191002 | 0.002207581 | 0 | 0.004789003 |
| 0.045454574 | 0 | 0.001670948 | 0.001213564 | 0 |
| 0 | 0 | 0 | 0 | 0 |
| 0 | 0 | 0 | 0 | 0 |
| 0 | 0.000733802 | 0 | 0.000523561 | 0 |
| 0.008440859 | 0 | 0 | 0.002552709 | 0 |
| 0 | 0 | 0 | 0 | 0 |

| | | | | |
|---|---|---|---|---|
| 0 | 0 | 0 | 0 | 0 |
| 0.078613135 | 0.020161321 | 0.01200597 | 0.006442097 | 0 |
| 0 | 0.054015761 | 0 | 0.010068939 | 0.013588123 |
| 0 | 0 | 0 | 0 | 0 |
| 0 | 0 | 0 | 0 | 0 |
| 0 | 0 | 0 | 0 | 0 |
| 0 | 0 | 0 | 0 | 0 |
| 0 | 0 | 0 | 0 | 0 |
| 0 | 0 | 0 | 0 | 0 |
| 0 | 0 | 0 | 0 | 0 |
| 0 | 0 | 0 | 0 | 0 |
| 0 | 0 | 0 | 0 | 0 |
| 0 | 0.822882669 | 0.038263547 | 0.012226808 | 0.019527144 |
| 0 | 0 | 0 | 0 | 0 |
| 0.038891303 | 0.031278741 | 0.002681129 | 0 | 0 |
| 0 | 0 | 0 | 0 | 0 |
| 0 | 0 | 0 | 0 | 0 |
| 0 | 0 | 0 | 0 | 0 |
| 0 | 0 | 0.011122585 | 0 | 0 |
| 0 | 0 | 0 | 0 | 0 |
| 0 | 0 | 0 | 0 | 0 |
| 0.007763075 | 0.019410882 | 0.03423004 | 0.035856258 | 0 |
| 0 | 0 | 0.062553892 | 0 | 0.086263355 |
| 0 | 0 | 0 | 0 | 0 |
| 0 | 0 | 0 | 0 | 0 |
| 0 | 0 | 0 | 0 | 0 |
| 0 | 0 | 0 | 0 | 0 |
| 0 | 0 | 0 | 0 | 0 |
| 0 | 0 | 0 | 0 | 0 |
| 0 | 0 | 0 | 0 | 0 |
| 0 | 0 | 0 | 0 | 0 |
| 0 | 0 | 0 | 0 | 0 |
| 0 | 0 | 0 | 0 | 0 |
| 0 | 0 | 0 | 0 | 0 |
| 14.45747847 | 7.572331619 | 0 | 45.37632605 | 17.86037193 |
| 0 | 0 | 0 | 0 | 0 |
| 0 | 0 | 0 | 0 | 0 |
| 0 | 0 | 0 | 0 | 0 |
| 0.006822085 | 0.009033374 | 0.024760645 | 0.001790341 | 0 |
| 0 | 0 | 0 | 0 | 0 |
| 0.051715625 | 0.0566973 | 0.323390827 | 0.072719055 | 0.032068238 |
| 0 | 0 | 0 | 0 | 0 |
| 0 | 0 | 0 | 0 | 0 |
| 0 | 0 | 0 | 0 | 0 |
| 0 | 0 | 0 | 0 | 0 |
| 1.50223709 | 0.073801661 | 0.827574931 | 0.004411039 | 0.002147132 |
| 0 | 0 | 0 | 0 | 0 |
| 0 | 0 | 0 | 0 | 0 |

| | | | | |
|---|---|---|---|---|
| 0 | 0 | 0 | 0 | 0 |
| 0 | 0 | 0 | 0 | 0 |
| 0 | 0 | 0 | 0 | 0 |
| 0 | 0 | 0 | 0 | 0 |
| 0.003138659 | 0 | 0 | 0 | 0 |
| 0 | 0 | 0 | 0 | 0 |
| 0 | 0 | 0 | 0 | 0 |
| 0 | 0 | 0 | 0 | 0 |
| 0.196243434 | 0 | 0 | 0 | 0 |
| 0 | 0.003401064 | 0.008139895 | 0 | 0.003149847 |
| 0 | 0.001992727 | 0.076102046 | 0.009668182 | 0.010851748 |
| 0 | 0 | 0 | 0 | 0 |
| 0.009433419 | 0.004617019 | 0 | 0.001581216 | 0.000615742 |
| 0 | 0 | 0 | 0 | 0 |
| 0 | 0 | 0 | 0 | 0 |
| 0 | 0 | 0 | 0 | 0 |
| 0 | 0 | 0 | 0 | 0 |
| 0 | 0 | 0 | 0 | 0 |
| 0 | 0 | 0 | 0 | 0 |
| 0.865642613 | 0.159940692 | 0.013780132 | 0.001876525 | 0.147974553 |
| 0 | 0 | 0 | 0 | 0 |
| 0 | 0 | 0 | 0 | 0 |
| 0 | 0 | 0 | 0 | 0 |
| 0 | 0 | 0 | 0 | 0 |
| 0 | 0 | 0 | 0 | 0 |
| 0 | 0 | 0 | 0 | 0 |
| 0 | 0 | 0 | 0 | 0 |
| 0 | 0 | 0 | 0 | 0 |
| 0 | 0 | 0 | 0 | 0 |
| 0.189072911 | 0 | 0.001636798 | 0 | 0 |
| 0 | 0 | 0 | 0 | 0 |
| 0 | 0 | 0 | 0.10862241 | 0 |
| 0 | 0 | 0 | 0 | 0 |
| 0 | 0 | 0 | 0 | 0 |
| 0 | 0 | 0 | 0 | 0 |
| 0.019283635 | 0 | 0 | 0 | 0 |
| 0 | 0 | 0 | 0 | 0 |
| 0 | 0 | 0 | 0 | 0 |
| 0 | 0 | 0 | 0 | 0 |
| 0 | 0 | 0 | 0.352724878 | 0.014322575 |
| 0 | 0 | 0 | 0 | 0 |
| 0 | 0 | 0 | 0 | 0 |
| 0 | 0.00778838 | 0 | 0.764842215 | 0.012148374 |
| 0 | 0 | 0 | 0 | 0 |
| 0.075809424 | 0.006947583 | 0.003235702 | 0.001468751 | 0 |
| 0 | 0.005168374 | 0 | 0 | 0 |
| 0 | 0 | 0 | 0 | 0 |
| 0 | 0 | 0 | 0 | 0 |

| | | | | |
|---|---|---|---|---|
| 0 | 0 | 0 | 0 | 0 |
| 0 | 0 | 0 | 0 | 0 |
| 0 | 0 | 0 | 0 | 0 |
| 0 | 0 | 0 | 0 | 0 |
| 0 | 0.006781593 | 0 | 0 | 0 |
| 0.005697451 | 0.24250426 | 0.014090866 | 0.012170924 | 0.002689979 |
| 0 | 0 | 0 | 0 | 0 |
| 0 | 0 | 0 | 0 | 0 |
| 0 | 0 | 0 | 0 | 0 |
| 0 | 0 | 0 | 0 | 0 |
| 0 | 0 | 0 | 0 | 0 |
| 0 | 0 | 0 | 0 | 0 |
| 0 | 0 | 0 | 0 | 0 |
| 0 | 0 | 0 | 0 | 0 |
| 0 | 0 | 0 | 0 | 0 |
| 0 | 0 | 0 | 0 | 0 |
| 0 | 0.008108862 | 0 | 0.298190943 | 0.014760852 |
| 0.048181443 | 0 | 0 | 0 | 0 |
| 0 | 0 | 0 | 0 | 0 |
| 0 | 0 | 0 | 0 | 0 |
| 0.000880817 | 0 | 0 | 0 | 0 |
| 0 | 0 | 0 | 0 | 0 |
| 0 | 0 | 0 | 0 | 0 |
| 0 | 0 | 0 | 0 | 0 |
| 0 | 0 | 0 | 0 | 0 |
| 0 | 0 | 0 | 0 | 0 |
| 0 | 0 | 0 | 0 | 0 |
| 0 | 0 | 0 | 0.588795316 | 0 |
| 0 | 0 | 0 | 0 | 0 |
| 0 | 0 | 0 | 0 | 0 |
| 0 | 0 | 0 | 0 | 0 |
| 0 | 0 | 0 | 0 | 0 |
| 0 | 0 | 0 | 0 | 0 |
| 0 | 0 | 0 | 0 | 0 |
| 0 | 0 | 0 | 0 | 0 |
| 0 | 0 | 0 | 0 | 0 |
| 0 | 0 | 0 | 0 | 0 |
| 0.025749921 | 0 | 0 | 0 | 0 |
| 0 | 0.098594962 | 0.010987145 | 0.831312238 | 0.325288831 |
| 0 | 0 | 0 | 0 | 0 |
| 0 | 0 | 0 | 0 | 0 |
| 0 | 0 | 0 | 0 | 0 |
| 0 | 0 | 0 | 0 | 0 |
| 0 | 0 | 0 | 0 | 0 |
| 0 | 0 | 0 | 0 | 0 |
| 0 | 0 | 0 | 0 | 0 |
| 0 | 0 | 0 | 0 | 0 |
| 0 | 0 | 0 | 0 | 0 |

| | | | | |
|---|---|---|---|---|
| 0 | 0 | 0 | 0 | 0 |
| 0 | 0.000774248 | 0.00285468 | 0.005646948 | 0.003585295 |
| 0.003644433 | 0 | 0 | 0 | 0 |
| 0 | 0 | 0 | 0 | 0 |
| 0 | 0 | 0 | 0 | 0 |
| 0 | 0 | 0 | 0 | 0 |
| 0 | 0 | 0 | 0 | 0 |
| 0 | 0 | 0 | 0 | 0 |
| 0 | 0 | 0 | 0 | 0 |
| 0 | 0.053963865 | 0 | 0 | 0 |
| 0.00334099 | 0.005238053 | 0 | 0 | 0.001502291 |
| 0 | 0 | 0 | 0 | 0 |
| 0 | 0.020286092 | 0.093319288 | 0.20044474 | 0 |
| 0 | 0 | 0 | 0.002591371 | 0 |
| 0 | 0 | 0 | 0 | 0 |
| 0 | 0 | 0 | 0 | 0 |
| 0 | 0 | 0 | 0 | 0 |
| 0 | 0 | 0 | 0 | 0 |
| 0 | 0 | 0 | 0 | 0 |
| 0 | 0 | 0 | 0 | 0 |
| 0.083642296 | 0 | 0.016772684 | 0 | 0 |
| 0 | 0 | 0.004941142 | 0 | 0 |
| 0 | 0 | 0 | 0 | 0 |
| 0 | 0.983376839 | 0.593162147 | 0.446805129 | 1.124781535 |
| 0.004343745 | 0 | 0 | 0 | 0 |
| 0 | 0 | 0 | 0 | 0 |
| 0 | 0 | 0 | 0 | 0 |
| 0 | 0 | 0 | 0 | 0 |
| 0.126642371 | 0 | 0 | 0 | 0 |
| 0 | 0 | 0 | 0 | 0 |
| 0 | 0.002693138 | 0 | 0 | 0 |
| 0 | 0 | 0 | 0 | 0 |
| 0 | 0.025796503 | 0 | 0 | 0.011628098 |
| 0 | 0 | 0 | 0 | 0 |

| PHL018-0011_V3 | PHL018-0063_V2 | PHL018-0078_V2 | PHL018-0098_V2 | PHL018-0239_V1 |
|---|---|---|---|---|
| CTRL | EYCF | CTRL | CTRL | REF |
| 0.075641342 | 0.141060743 | 0.008398554 | 0.062140208 | 0.109443957 |
| 1.312394481 | 0.070392687 | 1.13040426 | 2.766522256 | 1.069292312 |
| 11.97875047 | 8.548873543 | 0 | 0 | 0.001250191 |
| 0 | 0 | 0 | 0 | 0 |
| 0 | 0 | 0 | 0 | 0 |
| 0 | 0 | 0 | 0 | 0 |
| 0 | 0 | 0 | 0 | 0 |
| 0 | 0 | 0 | 0 | 0 |
| 0 | 0 | 0 | 0 | 0 |
| 0 | 0 | 0 | 0 | 0 |
| 0 | 0 | 0 | 0 | 0 |
| 0 | 0 | 0 | 0 | 0 |
| 0 | 0 | 0 | 0 | 0 |
| 26.02259811 | 1.750423831 | 0.599108149 | 3.144708307 | 0.31584346 |
| 0 | 0 | 0 | 0 | 0 |
| 0 | 0 | 0 | 0 | 0 |
| 0 | 0 | 0 | 0 | 0 |
| 0 | 0 | 0 | 0 | 0 |
| 0 | 0 | 0 | 0 | 0 |
| 0 | 0 | 0 | 0 | 0 |
| 0 | 0 | 0 | 0 | 0 |
| 0 | 0 | 0 | 0 | 0 |
| 0 | 0 | 0 | 0 | 0 |
| 0 | 0 | 0 | 0 | 0 |
| 0.017491045 | 0 | 0.001469662 | 0 | 0.084549878 |
| 0 | 0 | 0 | 0 | 0 |
| 0 | 0 | 0 | 0 | 0 |
| 0 | 0 | 0 | 0 | 0 |
| 0 | 0 | 0 | 0 | 0 |
| 0 | 0.001959418 | 0 | 0 | 0 |
| 0 | 0 | 0 | 0 | 0 |
| 0 | 0 | 0 | 0 | 0 |
| 0 | 0 | 0 | 0 | 0 |
| 0 | 0 | 0 | 0 | 0 |
| 0 | 0 | 0 | 0 | 0 |
| 0 | 0.549841896 | 1.633293625 | 2.761103036 | 2.216998681 |
| 0 | 0 | 0 | 0 | 0 |
| 0 | 0 | 0 | 0 | 0 |
| 0 | 0 | 0 | 0 | 0 |
| 0 | 0 | 0 | 0 | 0 |
| 0 | 0 | 0 | 0 | 0 |
| 0.001806731 | 0.077400762 | 0.032316165 | 0 | 0 |
| 0.279327022 | 0.310863528 | 13.71642452 | 0.008804754 | 0.019620111 |
| 0 | 0.026065957 | 0 | 0.018266053 | 0.081329387 |
| 0 | 0.017513555 | 0.011082153 | 0 | 0.006947348 |
| 0 | 0 | 0 | 0 | 0 |

| | | | | |
|---|---|---|---|---|
| 0.00259519 | 0.013771789 | 0.001700847 | 0 | 0.013325793 |
| 0.001666368 | 0 | 0 | 0.009050996 | 0.009910971 |
| 0.499358558 | 0.002176066 | 0.003654987 | 0.003842974 | 0.008194745 |
| 2.176197959 | 0.090458361 | 0.691485484 | 1.547493044 | 1.015124155 |
| 0.008871343 | 0 | 0.00390502 | 0.017824104 | 0.003583202 |
| 0 | 0 | 0 | 0 | 0 |
| 0.019391695 | 0.294686256 | 0.013935326 | 0.065548697 | 0 |
| 0 | 0 | 0 | 0 | 0 |
| 0 | 0 | 0 | 0 | 0 |
| 0 | 0 | 0 | 0 | 0 |
| 0 | 0 | 0.002816851 | 0.000722719 | 0 |
| 0.701367157 | 0.039815109 | 0.369581029 | 0 | 0.099146426 |
| 0 | 0 | 0 | 0 | 0 |
| 0.116218308 | 0.000538724 | 0.056287436 | 0.130128533 | 0.188293815 |
| 0 | 0 | 0 | 0 | 0 |
| 0 | 0 | 0 | 0 | 0 |
| 0.002004063 | 4.051777791 | 0.055273585 | 0.003346467 | 0 |
| 0 | 0 | 0 | 0 | 0 |
| 0 | 0 | 0 | 0 | 0 |
| 0 | 0 | 0 | 0 | 0 |
| 0.071637394 | 0.019924278 | 0.060322152 | 1.57050259 | 0.04783285 |
| 0.05819577 | 0 | 0.004752724 | 0.014531246 | 0.007925362 |
| 0 | 0 | 0.106392563 | 0 | 3.49515579 |
| 0 | 0 | 0 | 0 | 0 |
| 0 | 0 | 0 | 0 | 0 |
| 0 | 0 | 0 | 0 | 0 |
| 0 | 0 | 0.006384284 | 0 | 0 |
| 0 | 0 | 0 | 0 | 0 |
| 0.001159645 | 0.003076921 | 0.000760013 | 0 | 0 |
| 0 | 0 | 0 | 0 | 0 |
| 0.000616283 | 0 | 0 | 0.195686197 | 0 |
| 0 | 0 | 0 | 0 | 0 |
| 0 | 0 | 0 | 0 | 0 |
| 0.001005746 | 0 | 0 | 0 | 0 |
| 0 | 0 | 0 | 0 | 0 |
| 0 | 0 | 0.005294711 | 0 | 0 |
| 0 | 0 | 0 | 0 | 0 |
| 0 | 0.088468189 | 3.167826348 | 0 | 0.001372905 |
| 0 | 0 | 0 | 0 | 0 |
| 0 | 0 | 0 | 0 | 0 |
| 0 | 0 | 0 | 0 | 0 |
| 0 | 0 | 0 | 0 | 0 |
| 0 | 0 | 0 | 0 | 0 |
| 0 | 0 | 0 | 0 | 0 |
| 0 | 0 | 0 | 0 | 0 |
| 0 | 0 | 0 | 0.016173981 | 0 |
| 0 | 0 | 0.021300449 | 0 | 0 |
| 0 | 0.002835012 | 0.003601342 | 0 | 0 |

| | | | | |
|---|---|---|---|---|
| 0 | 0 | 0 | 0 | 0 |
| 0 | 0 | 0 | 0 | 0 |
| 0 | 0 | 0 | 0 | 0 |
| 0 | 0 | 0 | 0 | 0 |
| 0 | 0 | 0 | 0.013147456 | 0 |
| 0.012478785 | 0 | 0 | 0 | 0 |
| 0 | 0 | 0 | 0 | 0 |
| 0 | 0 | 0 | 0 | 0 |
| 0.000700944 | 0 | 0 | 0.021068382 | 0.021157526 |
| 0 | 0 | 0 | 0 | 0 |
| 0 | 0 | 0 | 0 | 0 |
| 0 | 0 | 0.020222489 | 0 | 0 |
| 0 | 0 | 0 | 0 | 0 |
| 0 | 0 | 0 | 0 | 0 |
| 0 | 0 | 0 | 0 | 0 |
| 0 | 0 | 0 | 0.001939046 | 0 |
| 0 | 0 | 0 | 0 | 0 |
| 0 | 0 | 0 | 0 | 0 |
| 0 | 0.14558912 | 0.225260024 | 0.119632262 | 0 |
| 0 | 0 | 0 | 0 | 0 |
| 0 | 0 | 0 | 0 | 0 |
| 0 | 0 | 0 | 0 | 0 |
| 0 | 0 | 0 | 0 | 0 |
| 0 | 0 | 0 | 0 | 0 |
| 0 | 0 | 0 | 0 | 0 |
| 0 | 0 | 0 | 0 | 0 |
| 0 | 0 | 0 | 0 | 0 |
| 0 | 0 | 0 | 0 | 0 |
| 0 | 0 | 0 | 0 | 0 |
| 0 | 0 | 0 | 0 | 0 |
| 0.356424013 | 0.013385851 | 0.002834028 | 0 | 0.101943522 |
| 0.000510355 | 0.005767629 | 0.006875392 | 0.003321439 | 0.007082625 |
| 0 | 0 | 0 | 0 | 0 |
| 0 | 0 | 0 | 0 | 0 |
| 0 | 0 | 0 | 0 | 0 |
| 0 | 0 | 0 | 0 | 0 |
| 0 | 0 | 0 | 0 | 0 |
| 0 | 0 | 0 | 0 | 0 |
| 0 | 0 | 0.027324908 | 0 | 0 |
| 0 | 0 | 0 | 0 | 0 |
| 0 | 0 | 0 | 0 | 0 |
| 0 | 0 | 0 | 0 | 0 |
| 0 | 0 | 0 | 0 | 0.001227959 |
| 0 | 0 | 0 | 0 | 0 |
| 0 | 0 | 0 | 0 | 0 |
| 0 | 0 | 0 | 0 | 0 |
| 0 | 0 | 0 | 0 | 0 |
| 0 | 0 | 0 | 0.001594108 | 0 |

| | | | | |
|---|---|---|---|---|
| 0.038764307 | 0 | 0 | 0 | 0 |
| 0 | 0 | 0 | 0 | 0.114519186 |
| 0 | 0 | 0 | 0 | 0 |
| 0 | 0 | 0 | 0 | 0 |
| 0.049602999 | 0 | 0.01342413 | 0 | 0.010171629 |
| 0 | 0 | 0 | 0 | 0 |
| 0 | 0 | 0 | 0 | 0 |
| 0 | 0 | 0 | 0 | 0 |
| 0 | 0 | 0 | 0 | 0 |
| 0 | 0 | 0 | 0 | 0 |
| 0 | 0 | 0 | 0 | 0 |
| 0 | 0 | 0 | 0 | 0 |
| 0 | 0 | 0 | 0 | 0 |
| 0 | 0 | 0 | 0 | 0 |
| 0 | 0 | 0 | 0 | 0 |
| 0 | 0 | 0 | 0 | 5.332712196 |
| 0 | 0 | 0 | 0 | 0 |
| 0 | 0 | 0 | 0 | 0 |
| 0 | 0 | 0 | 0 | 0 |
| 0 | 0 | 0 | 0 | 0 |
| 0 | 0 | 0 | 0 | 0 |
| 0 | 0 | 0 | 0 | 0 |
| 0 | 0 | 0 | 0 | 0 |
| 0 | 0 | 0 | 0 | 0 |
| 0 | 0 | 0 | 0 | 0 |
| 0 | 0 | 0 | 0 | 0 |
| 0 | 0 | 0 | 0 | 0 |
| 0 | 0 | 0 | 0 | 0 |
| 0 | 0 | 0 | 0 | 0 |
| 0 | 0 | 0 | 0 | 0 |
| 0 | 0 | 0 | 0.003666081 | 0 |
| 0.001373958 | 0.004339956 | 0.015222244 | 0.000806784 | 0 |
| 0 | 0 | 0 | 0 | 0 |
| 0 | 0 | 0 | 0 | 0 |
| 0 | 0 | 0 | 0 | 0 |
| 0 | 0 | 0 | 0 | 0 |
| 0 | 0 | 0 | 0 | 0 |
| 0 | 0 | 0 | 0 | 0 |
| 0 | 0 | 0 | 0 | 0 |
| 0 | 0 | 0 | 0 | 0 |
| 0 | 0 | 0 | 0 | 0 |
| 0 | 0 | 0 | 0 | 0 |
| 0 | 0 | 0 | 0 | 0 |
| 0 | 0 | 0 | 0 | 0 |
| 0 | 0 | 0 | 0 | 0 |
| 0 | 0 | 0 | 0 | 0 |
| 0 | 0 | 0 | 0 | 0 |
| 0 | 0 | 0.004468553 | 0 | 0 |

| | | | | |
|---|---|---|---|---|
| 0 | 0 | 0 | 0 | 0 |
| 0 | 0 | 0 | 0 | 0 |
| 0 | 0 | 0 | 0 | 0 |
| 0 | 0 | 0 | 0 | 0 |
| 0 | 0 | 0 | 0 | 0 |
| 0 | 0 | 0 | 0 | 0 |
| 0 | 0 | 0 | 0 | 0 |
| 0 | 0 | 0 | 0 | 0 |
| 0 | 0 | 0 | 0 | 0 |
| 0 | 0 | 0 | 0 | 0 |
| 0 | 0 | 0 | 0 | 0 |
| 0 | 0 | 0 | 0 | 0.000991546 |
| 0 | 0.003737621 | 0 | 0 | 0.000586472 |
| 0 | 0 | 0 | 0 | 0 |
| 0 | 0 | 0 | 0 | 0 |
| 0 | 0 | 0 | 0 | 0 |
| 0 | 0 | 0 | 0 | 0 |
| 0 | 0 | 0 | 0 | 0 |
| 0 | 0 | 0 | 0 | 0 |
| 0 | 0 | 0 | 0 | 0 |
| 0.094115995 | 0.004141306 | 0.014960227 | 0.04871742 | 0.04678666 |
| 0 | 0 | 0 | 0 | 0 |
| 0 | 0 | 0 | 0 | 0 |
| 0.001464837 | 0 | 0 | 0 | 0 |
| 0 | 0 | 0 | 0 | 0 |
| 0 | 0 | 0 | 0 | 0 |
| 0.001227111 | 0 | 0.000670191 | 0.003152431 | 0.00319306 |
| 0 | 0 | 0 | 0 | 0 |
| 0 | 0 | 0 | 0 | 0 |
| 0 | 0 | 0 | 0 | 0 |
| 0 | 0 | 0 | 0 | 0 |
| 0 | 0 | 0 | 0 | 0 |
| 0 | 0 | 0 | 0 | 0 |
| 0 | 0.035432726 | 0 | 0.017863837 | 0 |
| 0.004569917 | 0.055406775 | 0.011730623 | 0.010096448 | 0.025685349 |
| 0 | 0 | 0 | 0 | 0 |
| 0 | 0 | 0 | 0 | 0 |
| 0 | 0 | 0 | 0 | 0 |
| 0 | 0 | 0 | 0 | 0 |
| 0 | 0 | 0 | 0 | 0 |
| 0 | 0 | 0 | 0 | 0 |
| 0 | 0 | 0 | 0 | 0 |
| 0 | 0 | 0 | 0 | 0 |
| 0 | 0 | 0 | 0 | 0 |
| 0 | 0 | 0 | 0 | 0 |
| 0 | 0 | 0 | 0.041134347 | 0.010642562 |
| 0.006679806 | 0 | 0.001080949 | 0.003304953 | 0 |
| 0.000651329 | 0.000840093 | 0 | 0.000836628 | 0.001016893 |

| | | | | |
|---|---|---|---|---|
| 0 | 0 | 0 | 0 | 0 |
| 0 | 0 | 0 | 0 | 0 |
| 0 | 0 | 0 | 0 | 0 |
| 0 | 0 | 0 | 0 | 0 |
| 0 | 0 | 0 | 0 | 0 |
| 0 | 0 | 0 | 0 | 0 |
| 0 | 0 | 0.001519114 | 0 | 0 |
| 0 | 0.006900851 | 0 | 0 | 0 |
| 0 | 0 | 0.000700981 | 0 | 0.010854209 |
| 0 | 0 | 0 | 0 | 0 |
| 0 | 0 | 0 | 0 | 0 |
| 0 | 0 | 0 | 0 | 0 |
| 0 | 0 | 0 | 0 | 0 |
| 0 | 0 | 0 | 0 | 0 |
| 0.000770198 | 0 | 0 | 0 | 0 |
| 0 | 0 | 0 | 0 | 0 |
| 0 | 0 | 0 | 0 | 0 |
| 0 | 0 | 0.078585344 | 0 | 0 |
| 0 | 0.001220527 | 0.059511312 | 0 | 0 |
| 0 | 0 | 0 | 0 | 0 |
| 0 | 0 | 0 | 0 | 0.025425185 |
| 0 | 0 | 0 | 0 | 0 |
| 0 | 0 | 0 | 0 | 0 |
| 0 | 0 | 0 | 0 | 0 |
| 0 | 0.003934566 | 0 | 0.002493488 | 0.027276753 |
| 0 | 0 | 0 | 0 | 0 |
| 0 | 0 | 0 | 0 | 0 |
| 0 | 0 | 0 | 0 | 0 |
| 0 | 0 | 0 | 0 | 0 |
| 0 | 0 | 0 | 0 | 0 |
| 0.696012006 | 0.046863744 | 0.100648502 | 0.07585928 | 0.175921158 |
| 0 | 0 | 0 | 0 | 0 |
| 0 | 0.128559196 | 0 | 0 | 0 |
| 0 | 0 | 0 | 0 | 0 |
| 0 | 0 | 0 | 0 | 0 |
| 0 | 0 | 0 | 0 | 0 |
| 0 | 0 | 0 | 0 | 0 |
| 0 | 0 | 0 | 0 | 0 |
| 0 | 0 | 0 | 0 | 0 |
| 0 | 0.004892609 | 0 | 0 | 0 |
| 0 | 0 | 0 | 0 | 0 |
| 0.01629489 | 0 | 0.010799412 | 0.091624632 | 0.004192446 |
| 0 | 0 | 0 | 0 | 0 |
| 0 | 0 | 0 | 0 | 0 |
| 0 | 0 | 0 | 0 | 0 |
| 0 | 0 | 0 | 0 | 0 |
| 0 | 0 | 0 | 0 | 0 |
| 0 | 0 | 0 | 0 | 0 |

| | | | | |
|---|---|---|---|---|
| 0 | 0 | 0 | 0 | 0 |
| 0 | 0 | 0 | 0 | 0 |
| 0 | 0.018808178 | 0 | 0.019608077 | 0.055035931 |
| 0 | 0 | 0 | 0 | 0 |
| 0 | 0 | 0 | 0.665280957 | 0 |
| 0 | 0 | 0 | 0 | 0 |
| 0 | 0 | 0 | 0 | 0 |
| 0 | 0 | 0 | 0 | 0 |
| 0 | 0 | 0 | 0 | 0 |
| 0 | 0 | 0 | 0 | 0 |
| 0 | 0 | 0 | 0 | 0 |
| 0 | 0 | 0 | 0 | 0 |
| 0 | 0 | 0 | 0 | 0 |
| 0.020580376 | 0 | 0 | 0.006266159 | 0.017982937 |
| 0.00650097 | 0 | 0.001374398 | 0.002370451 | 0.007639553 |
| 0 | 0 | 0 | 0.004152896 | 0 |
| 0.001194721 | 0 | 0 | 0.002946455 | 0.004725347 |
| 0.000556059 | 0 | 0 | 0.014666027 | 0 |
| 0 | 0 | 0 | 0 | 0 |
| 0 | 0 | 0 | 0 | 0 |
| 0 | 0 | 0 | 0 | 0 |
| 0.001078492 | 0.000794888 | 0 | 0.001329904 | 0 |
| 0 | 0 | 0 | 0 | 0 |
| 0 | 0 | 0 | 0 | 0 |
| 0.003112739 | 0 | 0 | 0.024167462 | 0 |
| 0 | 0 | 0 | 0 | 0 |
| 0 | 0.000837944 | 0 | 0 | 0 |
| 0 | 0 | 0 | 0 | 0 |
| 0 | 0 | 0 | 0 | 0 |
| 0.001560049 | 0 | 0 | 0.022122758 | 0.00162376 |
| 0 | 0 | 0.008305346 | 0.001019126 | 0.001032261 |
| 0 | 0 | 0 | 0 | 0 |
| 0 | 0 | 0 | 0 | 0 |
| 0 | 0 | 0 | 0 | 0 |
| 0 | 0 | 0 | 0 | 0 |
| 0.015078948 | 0.001587675 | 0.012706083 | 0 | 0 |
| 0 | 0 | 0 | 0 | 0 |
| 0.011924124 | 0.020373361 | 0.000888055 | 0.003898735 | 0.128624024 |
| 0 | 0 | 0 | 0 | 0 |
| 0 | 0 | 0 | 0 | 0 |
| 0 | 0 | 0 | 0 | 0 |
| 0 | 0 | 0 | 0 | 0 |
| 0 | 0 | 0 | 0 | 0 |
| 0 | 0 | 0 | 0 | 0 |
| 0 | 0 | 0 | 0 | 0 |
| 0 | 0 | 0 | 0.003853442 | 0.003469428 |
| 0 | 0 | 0 | 0.004226086 | 0.001223015 |
| 0.001641262 | 0 | 0 | 0 | 0.005124867 |

| | | | | |
|---|---|---|---|---|
| 0.616740137 | 0 | 0 | 0 | 0 |
| 0 | 0 | 0 | 0.015860053 | 0 |
| 0 | 0.0009991 | 0 | 0 | 0.000705462 |
| 0 | 0 | 0 | 0 | 0 |
| 0.009319493 | 0.001507786 | 0.000993147 | 0.004437965 | 0.006151276 |
| 0.002594487 | 0 | 0 | 0.00571303 | 0.005400884 |
| 0 | 0 | 0 | 0 | 0 |
| 0.095903892 | 0 | 0 | 0.024555731 | 0.011275403 |
| 0 | 0 | 0 | 0 | 0 |
| 0.122495453 | 0.128284948 | 0 | 0.17852074 | 0.021876334 |
| 0 | 0 | 0 | 0 | 0 |
| 0.23229353 | 0.018217646 | 0.020111537 | 0 | 0 |
| 0.023723299 | 0.075862013 | 0 | 0.004128522 | 0.043808613 |
| 0 | 0 | 0.032372968 | 0 | 0.003326699 |
| 0.001770886 | 0 | 0 | 0.007481207 | 0 |
| 0 | 0 | 0 | 0 | 0 |
| 0 | 0 | 0 | 0 | 0 |
| 0 | 0 | 0 | 0 | 0 |
| 0 | 0 | 0 | 0.560796008 | 0 |
| 0 | 0 | 0 | 0 | 0 |
| 0 | 0 | 0 | 0 | 0 |
| 0 | 0 | 0 | 0 | 0 |
| 0 | 0 | 0.000862549 | 0 | 0 |
| 0 | 0 | 0 | 0 | 0 |
| 0 | 0 | 0 | 0 | 0 |
| 0 | 0.001149664 | 0.004381287 | 0.004121721 | 0.000927743 |
| 0 | 0 | 0.18682644 | 0.005205378 | 0 |
| 0 | 0 | 0.00647557 | 0.032121089 | 0 |
| 0 | 0 | 0 | 0 | 0 |
| 0 | 0 | 0 | 0 | 0 |
| 0 | 0 | 0 | 0 | 0 |
| 0 | 0 | 0 | 0 | 0 |
| 0 | 0 | 0 | 0 | 0 |
| 0.106662579 | 0 | 0 | 0 | 0 |
| 0 | 0 | 0 | 0 | 0 |
| 0 | 0 | 0 | 0 | 0 |
| 0 | 0 | 0.000827773 | 0 | 0.009728149 |
| 0 | 0 | 0 | 0 | 0 |
| 0 | 0 | 0 | 0 | 0 |
| 0.245615408 | 0 | 0 | 0 | 0 |
| 0 | 0 | 0 | 0 | 0 |
| 0 | 0 | 0 | 0 | 0 |
| 0 | 0 | 0 | 0 | 0 |
| 0 | 0 | 0 | 0 | 0 |
| 0 | 0 | 0 | 0 | 0 |
| 0.000815176 | 0 | 0 | 0 | 0 |
| 0.002216908 | 0.003267882 | 0 | 0.004271405 | 0.001442152 |
| 0 | 0 | 0 | 0 | 0 |

| | | | | |
|---|---|---|---|---|
| 0 | 0 | 0 | 0 | 0 |
| 0.001858162 | 0.16228959 | 0.001014842 | 0.003818875 | 0 |
| 0 | 0 | 0 | 0 | 0 |
| 0 | 0 | 0 | 0 | 0 |
| 0 | 0 | 0 | 0 | 0 |
| 0.138595373 | 0 | 0 | 0 | 0.023398912 |
| 0 | 0 | 0 | 0 | 0 |
| 0 | 0.0020806 | 0 | 0 | 0 |
| 0 | 0 | 0 | 0 | 0 |
| 0 | 0 | 0.004354014 | 0 | 0 |
| 0.01417614 | 0 | 0.002488613 | 0.013266672 | 0.038205104 |
| 0.001898039 | 0 | 0 | 0 | 0 |
| 0 | 0 | 0 | 0 | 0 |
| 0.006218134 | 0 | 0 | 0.000983034 | 0.003236037 |
| 0.09510047 | 0 | 0.003171877 | 0.010195202 | 0.003274288 |
| 0 | 0 | 0 | 0 | 0 |
| 0 | 0 | 0 | 0 | 0 |
| 0 | 0 | 0 | 0 | 0 |
| 0 | 0 | 0 | 0 | 0 |
| 0.070556739 | 0 | 0 | 0 | 0 |
| 0 | 0 | 0.105269851 | 0 | 0 |
| 0 | 0.006576602 | 0.304806044 | 0.145319036 | 0.162952468 |
| 0 | 0 | 0 | 0 | 0 |
| 0 | 0 | 0 | 0 | 0 |
| 0 | 0 | 0 | 0 | 0 |
| 0 | 0 | 0 | 0 | 0 |
| 0 | 0 | 0 | 0 | 0 |
| 0 | 0 | 0 | 0 | 0 |
| 0 | 0 | 0 | 0 | 0 |
| 0.211385873 | 2.98085078 | 0.071222386 | 0.185158492 | 0.334953683 |
| 0.003082757 | 0.001674181 | 0.001417818 | 0.005502012 | 0 |
| 0 | 0 | 0 | 0 | 0 |
| 0 | 0 | 0 | 0 | 0 |
| 0 | 0 | 0 | 0 | 0.004544551 |
| 0.020734291 | 0 | 0 | 0 | 0 |
| 0 | 0 | 0 | 0 | 0 |
| 0 | 1.988671022 | 0.138279882 | 0 | 0 |
| 0 | 0.003772962 | 0.002662682 | 0 | 0 |
| 0 | 0 | 0 | 0 | 0 |
| 0.448816281 | 0 | 0 | 0 | 0.027392508 |
| 0.068618538 | 0 | 0.052402353 | 0.07172429 | 0.015962248 |
| 0.075248147 | 0.017448276 | 0.272071766 | 0 | 1.543719324 |
| 0 | 0.002962523 | 0.003170946 | 0 | 0 |
| 0 | 0 | 0 | 0 | 0 |
| 0 | 0 | 0 | 0.014358452 | 0 |
| 0 | 0 | 0 | 0 | 0 |
| 0.009492618 | 0 | 0.002930329 | 0.000848223 | 0 |
| 0 | 0 | 0 | 0 | 0 |

| | | | | |
|---|---|---|---|---|
| 0 | 0.012948577 | 0.007509188 | 0 | 0.014840746 |
| 0 | 0 | 0 | 0 | 0 |
| 0.003816259 | 9.334652271 | 0.04016946 | 0.003565062 | 0.003611009 |
| 0 | 0 | 0 | 0 | 0 |
| 0 | 0 | 0.022738877 | 0 | 0 |
| 0.212694017 | 0 | 0.001175745 | 0 | 0 |
| 0 | 0 | 0 | 0 | 0 |
| 0 | 0 | 0 | 0 | 0 |
| 0.115920992 | 0 | 0.006284517 | 0 | 0 |
| 0 | 0 | 0 | 0 | 0 |
| 0 | 0 | 0 | 0 | 0 |
| 0 | 0 | 0 | 0 | 0 |
| 0 | 0 | 0 | 0 | 0 |
| 0 | 0 | 0 | 0.007574406 | 0.018940317 |
| 0 | 0 | 0 | 0 | 0 |
| 0.288555697 | 0 | 0.006881912 | 0 | 0.013490003 |
| 0.001197133 | 0.152643047 | 0.454839688 | 0.039365346 | 0.14204649 |
| 0 | 0 | 0 | 0 | 0.001261557 |
| 0.033592873 | 0 | 0 | 0 | 0 |
| 0.502374711 | 0 | 0 | 0.464285727 | 0 |
| 0 | 0 | 0 | 0.034883112 | 0.031406841 |
| 0 | 0 | 0 | 0.00285386 | 0.001445321 |
| 0 | 0 | 0 | 0 | 0 |
| 0 | 0 | 0 | 0 | 0 |
| 0 | 0 | 0.013239558 | 0 | 0 |
| 0 | 0 | 0.001722581 | 0 | 0 |
| 0.007852142 | 0 | 0.003596787 | 0.005465974 | 0.002109113 |
| 0.059149183 | 0 | 0 | 0 | 0 |
| 0 | 0 | 0 | 0 | 0 |
| 0 | 0 | 0 | 0 | 0 |
| 0.014061994 | 0.001277778 | 0.007364394 | 0 | 0 |
| 0.007179694 | 0.003598351 | 0 | 0.006197365 | 0.004184826 |
| 0.019687794 | 0.229348296 | 0.1012533 | 0.030627597 | 0.029030075 |
| 0 | 0 | 0.003424327 | 0 | 0 |
| 0.000839083 | 0.000824579 | 0.003360635 | 0 | 0.009897969 |
| 0 | 0 | 0 | 0 | 0 |
| 0 | 0 | 0 | 0 | 0.002058446 |
| 0.028419468 | 0.001282419 | 0.037378063 | 0.068141955 | 0.073447141 |
| 0 | 0 | 0.103694985 | 0.549595168 | 0.58552894 |
| 0 | 0 | 0 | 0.002274 | 0 |
| 0 | 0 | 0 | 0 | 0 |
| 0 | 0 | 0 | 0 | 0 |
| 0 | 0 | 0.000947438 | 0 | 0.002106522 |
| 0.619215191 | 0.00255833 | 0 | 0.067305675 | 0 |
| 0 | 0 | 0 | 0.002427843 | 0 |
| 0 | 0 | 0 | 0 | 0 |
| 0 | 0 | 0 | 0 | 0 |
| 0 | 0 | 0 | 0 | 0 |

| | | | | |
|---|---|---|---|---|
| 0 | 0 | 0 | 0 | 0 |
| 0.459696269 | 0.246197419 | 0.011222383 | 0.222249387 | 0.537147318 |
| 0 | 0.00047839 | 0.005435567 | 0 | 0 |
| 0 | 0 | 0 | 0 | 0 |
| 0 | 0 | 0 | 0 | 0 |
| 0 | 0 | 0 | 0 | 0 |
| 0 | 0 | 0 | 0 | 0 |
| 0.002832855 | 0.010588719 | 0 | 0 | 0 |
| 0.002652696 | 0 | 0 | 0 | 0 |
| 0 | 0 | 0 | 0 | 0 |
| 0.01213724 | 0.001009985 | 0.002779819 | 0.014483804 | 0.007742751 |
| 0.002041151 | 0.002149145 | 0 | 0.229224138 | 0.061914253 |
| 0.328670249 | 0.022245304 | 0 | 0 | 0 |
| 0 | 0 | 0 | 0 | 0 |
| 0 | 0 | 0 | 0 | 0 |
| 0 | 0 | 0 | 0 | 0 |
| 0.001844919 | 0.00710103 | 0 | 0.024013836 | 0.047579854 |
| 0.226190652 | 0 | 0.003197373 | 0.012305241 | 0.01218686 |
| 0 | 0 | 0 | 0 | 0 |
| 0 | 0 | 0 | 0.024503832 | 0 |
| 0.003170962 | 0.013711062 | 0.069504218 | 0.017812934 | 0.011661624 |
| 0 | 0.025010191 | 0 | 0 | 0 |
| 0 | 0 | 0 | 0 | 0 |
| 0 | 0 | 0 | 0 | 0.004093297 |
| 0.013141924 | 0.000785357 | 0 | 0 | 0 |
| 0 | 0 | 0 | 0 | 0 |
| 0.174439629 | 0 | 0.036923156 | 0 | 0.056684303 |
| 0.214348801 | 0.0415657 | 0.037304498 | 0.070419736 | 0.062440644 |
| 0 | 0 | 0 | 0 | 0 |
| 0 | 0 | 0 | 0 | 0 |
| 0 | 0 | 0 | 0.016103044 | 0 |
| 0 | 0 | 0 | 0 | 0 |
| 0.004091477 | 0 | 0 | 0 | 0 |
| 0 | 0 | 0 | 0 | 0 |
| 0.145868546 | 0 | 0.002994183 | 0 | 0 |
| 0.146401923 | 0.100896767 | 0.044444166 | 0.203975696 | 0.279826535 |
| 0.007572978 | 0 | 0 | 0 | 0 |
| 0 | 0 | 0 | 0 | 0 |
| 0.105880158 | 0.006332885 | 0.014229643 | 0.047850047 | 0.006923822 |
| 0 | 0 | 0 | 0 | 0 |
| 0 | 0 | 0 | 0 | 0 |
| 0 | 0 | 0 | 0 | 0 |
| 0.003198825 | 0 | 0 | 0.003806111 | 0.013668314 |
| 0 | 0.001557445 | 0 | 0.014364849 | 0.013704058 |
| 0.148203664 | 0 | 0 | 0 | 0.017114871 |
| 0 | 0 | 0.480083046 | 0.679791247 | 0.070970388 |
| 0 | 0 | 0 | 0 | 0 |
| 0.007094134 | 0 | 0 | 0.001619977 | 0 |

| | | | | |
|---|---|---|---|---|
| 0 | 0 | 0 | 0 | 0 |
| 0 | 0 | 0 | 0 | 0 |
| 0 | 0 | 0 | 0 | 0 |
| 0.067845488 | 0 | 0.328131984 | 0 | 0.956584311 |
| 0 | 0 | 0 | 0 | 0 |
| 0 | 0 | 0 | 0 | 0 |
| 0 | 0 | 0 | 0 | 0 |
| 0.006490513 | 0.001034322 | 0.016287002 | 0.026858727 | 0.10060332 |
| 0.046585273 | 3.858944442 | 0.151962441 | 0 | 0.289824534 |
| 0 | 0.002195839 | 0.19832049 | 0.03241584 | 0.118018642 |
| 0 | 0 | 0 | 0 | 0 |
| 0.014278175 | 0.013154411 | 0.00082085 | 0.009073583 | 0.001564345 |
| 0 | 0 | 0 | 0 | 0 |
| 2.556079688 | 0 | 0 | 0 | 0.58280388 |
| 0.15286096 | 0.095981526 | 0.038101942 | 0.0202323 | 0.071483667 |
| 0 | 0 | 0 | 0 | 0 |
| 0 | 0 | 0 | 0 | 0 |
| 0 | 5.752689212 | 0 | 0 | 0 |
| 0.01078756 | 0 | 0 | 0 | 0 |
| 0 | 0 | 0 | 0 | 0 |
| 0 | 0 | 0 | 0 | 0 |
| 0 | 0 | 0 | 0 | 0 |
| 0 | 0 | 0 | 0 | 0 |
| 1.384633868 | 0.017500375 | 0.586904735 | 0.130590912 | 0.825507943 |
| 0 | 0 | 0 | 0 | 0 |
| 0.027010452 | 0 | 0 | 0 | 0 |
| 0 | 0 | 0 | 0.001075316 | 0.256827405 |
| 0 | 0.243277448 | 1.667726197 | 4.32525411 | 0 |
| 0 | 0 | 0 | 0 | 0 |
| 0 | 0 | 0 | 0 | 0 |
| 0 | 0 | 0.029725745 | 0.02083274 | 0.014637797 |
| 0.438639563 | 0 | 0.001127805 | 0 | 0.024538158 |
| 0.03850418 | 0 | 0.005162404 | 0 | 0 |
| 0 | 0 | 0 | 0 | 0 |
| 0 | 0 | 0 | 0 | 0 |
| 0.184748352 | 0.050791242 | 0 | 0 | 0 |
| 0 | 0 | 0.08673049 | 0.776667485 | 1.462607977 |
| 0 | 0 | 0 | 0 | 0 |
| 0.123993561 | 0 | 0 | 0.012862095 | 0 |
| 0 | 0 | 0 | 0 | 0 |
| 0.001982512 | 0.002258192 | 0.137608504 | 0.011297306 | 0.919747237 |
| 0 | 0 | 0 | 0 | 0.168300219 |
| 1.220267011 | 3.189582597 | 2.921342601 | 12.45097494 | 1.239946343 |
| 0 | 0.002430046 | 0.000514485 | 0.012422788 | 0 |
| 0 | 0 | 0 | 0 | 0 |
| 0 | 0 | 0 | 0 | 0 |
| 0 | 0 | 0 | 0 | 0 |
| 0.028381829 | 0.000958096 | 0 | 0 | 0.054346247 |

| | | | | |
|---|---|---|---|---|
| 0 | 0 | 0 | 0 | 0 |
| 0 | 0.200174197 | 0.021344259 | 0 | 0 |
| 0.306340469 | 0 | 0.002722316 | 0.06295877 | 0.091007645 |
| 0 | 0 | 0 | 0 | 0 |
| 0 | 0 | 0 | 0 | 0 |
| 0.000818045 | 0.055348831 | 0.002859381 | 0.002858103 | 0 |
| 0 | 0 | 0 | 0.021740243 | 0 |
| 0 | 0 | 0 | 0 | 0 |
| 0 | 0 | 0 | 0 | 0 |
| 0 | 0 | 0 | 0 | 0 |
| 0.188007999 | 0.002855536 | 0.004009247 | 0.047565617 | 0.014007891 |
| 8.106267121 | 2.320297372 | 1.555051254 | 1.778865461 | 1.182336734 |
| 0.020695094 | 0 | 0 | 0 | 0 |
| 0.000660214 | 0.200317734 | 0.200481694 | 0.006558184 | 0.251735733 |
| 1.601404221 | 0 | 0.026710077 | 0.125460099 | 0.094405265 |
| 0 | 0 | 0 | 0 | 0 |
| 0 | 0 | 0 | 0 | 0 |
| 0.002118508 | 0 | 0 | 0 | 0 |
| 0 | 0 | 0 | 0 | 0 |
| 0.007894629 | 0 | 0 | 0 | 0.01276263 |
| 0.722433677 | 0.002015461 | 0.011574503 | 1.24904864 | 0.185894035 |
| 0.669917673 | 3.408756916 | 0 | 0.001480438 | 0.585187205 |
| 4.657520381 | 4.404098487 | 0.209831521 | 3.249307431 | 0.566901161 |
| 0 | 0 | 0 | 0 | 0 |
| 0 | 0 | 0 | 0 | 0 |
| 0 | 0.263852615 | 1.239327169 | 0.005471753 | 0 |
| 0.296402478 | 0 | 0.002232846 | 0 | 0 |
| 0 | 0 | 0.001362967 | 0 | 0 |
| 0 | 0 | 0.018880509 | 0 | 0 |
| 0.0357222 | 0 | 0 | 0 | 0 |
| 0.003393154 | 0 | 0 | 0 | 0 |
| 1.106871255 | 0.01213003 | 0 | 0.378032996 | 0.144429156 |
| 0.000837697 | 0 | 0 | 0 | 0.022669588 |
| 0 | 0.002118712 | 0.000697776 | 0.00919011 | 0.02775944 |
| 0.004976013 | 0 | 0.00286865 | 0.011078869 | 0 |
| 0.010255106 | 1.751305089 | 0.004356228 | 0.015221689 | 0.202606639 |
| 0.006237295 | 0.002839392 | 0.002003837 | 0.001131073 | 0.003818834 |
| 0 | 0 | 0 | 0 | 0 |
| 0 | 4.081890534 | 0.069910854 | 0.104362689 | 0.504251403 |
| 0 | 0 | 0 | 0 | 0 |
| 0 | 0 | 0 | 0 | 0 |
| 0 | 0 | 0 | 0 | 0 |
| 0 | 0 | 0 | 0 | 0 |
| 0 | 0 | 0 | 0 | 0 |
| 0.044613435 | 0.003671259 | 0.003075295 | 0.018026247 | 0.045308317 |
| 0 | 0 | 0 | 0 | 0 |
| 0 | 0 | 0 | 0 | 0 |
| 0.00679231 | 0.205060706 | 0 | 0.032482645 | 0.439408982 |

| | | | | |
|---|---|---|---|---|
| 0 | 0 | 0 | 0 | 0 |
| 0 | 0 | 0 | 0 | 0 |
| 0 | 0 | 0 | 0 | 0 |
| 0.007752435 | 0.003401086 | 0.002016201 | 0.005500583 | 0.003073918 |
| 0 | 0 | 0 | 0 | 0 |
| 0 | 0.011654811 | 0 | 0 | 0 |
| 0 | 0 | 0.973443981 | 0 | 0 |
| 0 | 0.000500738 | 0.000556581 | 0.006108746 | 0.006010692 |
| 0 | 0 | 0 | 0 | 0 |
| 0 | 0 | 0 | 0 | 0 |
| 0.001027876 | 0.003636392 | 0 | 0.001689984 | 0.007916915 |
| 0 | 0 | 0 | 0 | 0 |
| 0.001439978 | 0 | 0.002162735 | 0 | 0 |
| 0 | 0 | 0 | 0 | 0 |
| 0 | 0 | 0 | 0 | 0 |
| 0 | 0 | 0 | 0 | 0 |
| 0 | 0 | 0 | 0 | 0.174892485 |
| 0 | 0.032794815 | 0.002990942 | 0.005803353 | 0.002315634 |
| 0 | 0 | 0 | 0 | 0 |
| 0.00354975 | 0 | 0.023433096 | 0 | 0.084496577 |
| 0 | 0 | 0 | 0 | 0 |
| 0 | 0 | 0 | 0 | 0 |
| 0 | 0 | 0.035776555 | 0.001216518 | 0.019099055 |
| 0 | 0 | 0 | 0 | 0 |
| 0 | 0 | 0 | 0 | 0 |
| 0 | 0 | 0 | 0 | 0 |
| 0 | 0 | 0 | 0 | 0 |
| 0 | 0 | 0 | 0 | 0 |
| 0 | 0 | 0 | 0 | 0 |
| 0 | 0 | 0 | 0 | 0.006885576 |
| 0 | 0 | 0 | 0 | 0 |
| 0 | 0 | 0 | 0 | 0 |
| 0 | 0 | 0 | 0 | 0 |
| 0 | 0 | 0 | 0 | 0 |
| 0 | 0.000602392 | 0 | 0.008062759 | 0 |
| 0 | 0 | 0 | 0 | 0 |
| 0.754627725 | 0.014973452 | 0.15356861 | 0 | 0 |
| 0 | 0.00820872 | 0.001900867 | 0.040950961 | 0.001086779 |
| 0 | 0 | 0 | 0 | 0 |
| 0.004119521 | 0 | 0.001687419 | 0 | 0 |
| 0.040166063 | 0.002767838 | 0.00214018 | 0 | 0.358753233 |
| 0 | 0 | 0 | 0 | 0 |
| 0 | 0 | 0 | 0 | 0 |
| 0.002196961 | 0 | 0 | 0 | 0 |
| 0 | 0.445947583 | 0.071443522 | 0.287930739 | 0.272514264 |
| 0 | 0 | 0 | 0 | 0 |
| 0 | 0 | 0 | 0 | 0 |
| 0 | 0 | 0 | 0 | 0 |

| | | | | |
|---|---|---|---|---|
| 0 | 0 | 0 | 0 | 0 |
| 0 | 0 | 0 | 0 | 0 |
| 0 | 0.005194732 | 0 | 0 | 0 |
| 0 | 0 | 0.002268966 | 0 | 0.149001561 |
| 0 | 0 | 0 | 0 | 0 |
| 0 | 0 | 0 | 0 | 0 |
| 0 | 0 | 0 | 0 | 0 |
| 0.031653193 | 0 | 0 | 0 | 0 |
| 0.230166556 | 0.004245448 | 0.002883774 | 0.068316444 | 0.063701142 |
| 0 | 0 | 0 | 0 | 0 |
| 0 | 0 | 0 | 0 | 0.000714483 |
| 0 | 0 | 0 | 0 | 0 |
| 0 | 0 | 0 | 0 | 0 |
| 0 | 0 | 0 | 0 | 0 |
| 1.859800588 | 0 | 1.365682682 | 6.749450118 | 0 |
| 0.002219759 | 0 | 0 | 0 | 0.854082076 |
| 0 | 0 | 0 | 0 | 0 |
| 0 | 0 | 0 | 0 | 0 |
| 0 | 0 | 0 | 0 | 0 |
| 0 | 0 | 0 | 0 | 0 |
| 0 | 0 | 0.001202759 | 0 | 0.001719129 |
| 0 | 0 | 0 | 0 | 0 |
| 0 | 0 | 0 | 0 | 0 |
| 0 | 0 | 0 | 0 | 0 |
| 0.00705603 | 0.010401096 | 0.376735504 | 0 | 0.024676476 |
| 0.076880965 | 0.007058496 | 5.269089431 | 0.780183602 | 6.946570543 |
| 0 | 0 | 0 | 0.147355643 | 0 |
| 0 | 0 | 0 | 0 | 0 |
| 0 | 0.670738954 | 0.061454501 | 0 | 0 |
| 0 | 0 | 0 | 0 | 0 |
| 0 | 0 | 0 | 0 | 0 |
| 0.003271768 | 0 | 0 | 0 | 0 |
| 0 | 0 | 0 | 0 | 0 |
| 0 | 0 | 0.000823244 | 0.04352533 | 0.001568907 |
| 0.00347709 | 0.118252153 | 0.100105776 | 0.007656512 | 0.004394609 |
| 0.003511258 | 0.00168215 | 0 | 0.006494671 | 0.002192792 |
| 1.232359635 | 0 | 0 | 0 | 0 |
| 3.002358058 | 0.013880513 | 3.963764712 | 3.772325458 | 0.698243641 |
| 0 | 6.03123268 | 0.867300624 | 0 | 0 |
| 0 | 0 | 0 | 0 | 0 |
| 0 | 0.117538998 | 0 | 0 | 0 |
| 0.002189987 | 0 | 0 | 0 | 0 |
| 0.112757629 | 0.02015773 | 0.194445059 | 1.162389408 | 5.971753768 |
| 0 | 0.870005431 | 0 | 0 | 0 |
| 0 | 0 | 0 | 0 | 0 |
| 0 | 0.00257432 | 0.000726707 | 0 | 0.002423628 |
| 0 | 0 | 0 | 0 | 0 |
| 0.31690865 | 0.004296495 | 0.077278574 | 3.106234897 | 7.248719211 |

| | | | | |
|---|---|---|---|---|
| 0.097214892 | 0 | 0 | 0.009949142 | 0 |
| 0 | 0 | 0 | 0 | 0 |
| 0 | 0 | 0 | 0 | 0 |
| 0 | 0 | 0 | 0 | 0 |
| 0.00196725 | 0 | 0.001719073 | 0 | 0 |
| 0 | 0.954905426 | 0.016968281 | 0 | 0 |
| 0 | 0 | 0 | 0 | 0 |
| 0 | 0 | 0 | 0 | 0 |
| 0 | 0 | 0 | 0 | 0 |
| 0.020140385 | 0 | 0 | 0 | 0 |
| 0 | 0 | 0 | 0.000618732 | 0 |
| 0 | 0.544559967 | 0.048966357 | 0.047184385 | 2.607619932 |
| 0.337052224 | 0.006495778 | 0.050216102 | 0 | 0.418005269 |
| 0 | 0 | 0 | 0 | 0 |
| 0.002730149 | 0.123080677 | 0.047217572 | 0.034834795 | 0 |
| 0 | 0 | 0 | 0 | 0 |
| 0.00142441 | 0 | 0 | 0 | 0 |
| 0 | 0 | 0 | 0 | 0 |
| 0 | 0 | 0 | 0 | 0 |
| 0 | 0 | 0.003139877 | 0 | 0.000920593 |
| 0 | 0 | 0 | 0 | 0.063408474 |
| 0 | 0 | 0 | 0 | 0 |
| 0 | 0 | 0.17134745 | 0.050610401 | 0.811524615 |
| 0 | 0 | 0 | 0 | 0 |
| 0 | 0 | 0 | 0 | 0 |
| 0 | 0 | 0 | 0 | 0 |
| 0 | 0.02879786 | 0 | 0 | 0 |
| 0 | 0 | 0 | 0 | 0 |
| 0 | 0 | 0 | 0 | 0.000853201 |
| 0 | 0 | 0 | 0 | 0 |
| 0 | 0 | 0 | 0 | 0 |
| 0.006199813 | 0.006527836 | 0 | 0.047326649 | 0.200043161 |
| 0 | 0 | 0 | 0 | 0 |
| 0.051474503 | 0.007283334 | 0.011277419 | 0.014246772 | 0.063555115 |
| 0.00324796 | 0.002112232 | 0 | 0.005497201 | 0.000795436 |
| 0 | 0.851004311 | 0.011568281 | 0 | 0.012561311 |
| 0 | 0 | 0 | 0 | 0.004413023 |
| 0 | 0.008348439 | 0 | 0 | 0 |
| 0 | 0.026045969 | 0 | 0 | 0 |
| 0 | 0 | 0 | 0 | 0 |
| 0 | 0 | 0.070944866 | 0 | 0 |
| 0 | 0 | 0 | 0 | 0 |
| 0 | 0 | 0 | 0 | 0 |
| 0 | 0 | 0 | 0 | 0 |
| 0.085447225 | 0.001387174 | 0.00143908 | 0.00870315 | 0.029384398 |
| 0 | 0 | 0 | 0 | 0 |
| 0 | 0.153478453 | 0.416691549 | 0 | 0 |
| 0.003026217 | 0 | 0 | 0 | 0.002779238 |

| | | | | |
|---|---|---|---|---|
| 0 | 0 | 0 | 0 | 0 |
| 0 | 0 | 0 | 0 | 0 |
| 0 | 0 | 0 | 0 | 0 |
| 0 | 0 | 0 | 0 | 0 |
| 0.066329525 | 0 | 0 | 0 | 0 |
| 0 | 0 | 0 | 0 | 0 |
| 0 | 0.115286784 | 0.033001373 | 0 | 0 |
| 0.00882405 | 0 | 0.006626521 | 0.373280696 | 0.776656895 |
| 0 | 0 | 0 | 0 | 0 |
| 0 | 0.003866565 | 0 | 0 | 0 |
| 0 | 0 | 0 | 0 | 0 |
| 0 | 0 | 0 | 0 | 0 |
| 0 | 0 | 0 | 0 | 0 |
| 0 | 0.147870239 | 0 | 0 | 0 |
| 0 | 0 | 0 | 0 | 0 |
| 0 | 0 | 0 | 0 | 0 |
| 0 | 0 | 0 | 0 | 0 |
| 0 | 0 | 0 | 0 | 0 |
| 0 | 0.595573433 | 0.582530335 | 0 | 0 |
| 0 | 0 | 0 | 0 | 0 |
| 0 | 0 | 0 | 0 | 0 |
| 0 | 0 | 0 | 0 | 0 |
| 0 | 0 | 0 | 0 | 0 |
| 0 | 0 | 0 | 0 | 0 |
| 0.003290168 | 0.001060925 | 0.002021555 | 0 | 0 |
| 0.00455562 | 0 | 0 | 0.007742202 | 0.007659614 |
| 0 | 0 | 0 | 0 | 0 |
| 0.011833622 | 0.007836985 | 0.151177469 | 3.043742861 | 0.235270484 |
| 0 | 0 | 0 | 0 | 0 |
| 0 | 0.059997906 | 1.798421065 | 0.108909852 | 0.204345821 |
| 0 | 0.313266244 | 0 | 0 | 0 |
| 0 | 0 | 0 | 0 | 0 |
| 0 | 0 | 0 | 0 | 0 |
| 0 | 0 | 0 | 0 | 0 |
| 0.013771177 | 0.000405994 | 0.001203389 | 0 | 0.000573343 |
| 0 | 0 | 0 | 0 | 0 |
| 0 | 0 | 0 | 0 | 0 |
| 0.0012501 | 0 | 0 | 0.001712795 | 0.002602305 |
| 0 | 0 | 0 | 0 | 0 |
| 0 | 0 | 0 | 0 | 0 |
| 3.844673306 | 1.926249087 | 2.127063441 | 6.199902903 | 3.867573153 |
| 0.122620589 | 0 | 0.005997286 | 1.985977767 | 0.922130007 |
| 0 | 0 | 0.055139894 | 0 | 1.36068194 |
| 0.064403857 | 0 | 0 | 0 | 0 |
| 0 | 0 | 0 | 0 | 0 |
| 0 | 0 | 0 | 0 | 0 |
| 0 | 0 | 0 | 0 | 0 |
| 0.017362602 | 0 | 0 | 0 | 0.002237445 |

| | | | | |
|---|---|---|---|---|
| 0 | 0 | 0 | 0.192545293 | 0 |
| 0.002211098 | 0 | 0.001114707 | 0 | 0.001947336 |
| 0 | 0 | 0 | 0 | 0 |
| 0 | 0 | 0 | 0 | 0 |
| 0.984398036 | 0.246750952 | 2.445383418 | 5.637232739 | 5.926889613 |
| 0 | 0 | 0 | 0 | 0 |
| 0 | 0 | 0 | 0 | 0 |
| 0 | 0 | 0 | 0 | 0.014077368 |
| 0.496933443 | 0 | 0.030980277 | 0.476789262 | 0.050025768 |
| 0 | 0 | 0 | 0 | 0 |
| 0 | 0 | 0 | 0 | 0 |
| 0 | 0 | 0 | 0 | 0 |
| 0.095689046 | 0 | 0.093510062 | 0 | 0.346438822 |
| 0.012190668 | 0.136421662 | 0.005770249 | 0.021713582 | 0.005921309 |
| 0 | 0.173895659 | 0 | 0 | 0 |
| 0.40547821 | 0 | 0.225277893 | 0.051398101 | 1.029410326 |
| 0 | 0 | 0.003456869 | 0.002348717 | 0.636653833 |
| 0 | 0 | 0 | 0 | 0 |
| 0 | 0 | 0 | 0 | 0 |
| 0 | 0 | 0 | 0 | 0 |
| 0 | 0 | 0 | 0 | 0 |
| 0 | 0 | 0 | 0 | 0 |
| 0 | 0 | 0 | 0 | 0 |
| 0 | 0 | 0 | 0 | 0 |
| 0 | 0 | 0 | 0 | 0 |
| 0 | 0 | 0 | 0 | 0 |
| 0 | 0 | 0.032396812 | 0.006126918 | 0.035803177 |
| 0.00086255 | 0.004958695 | 0 | 0.000531811 | 0.00071822 |
| 0 | 0 | 0 | 0 | 0 |
| 0 | 0 | 0 | 0 | 0 |
| 0 | 0 | 0 | 0 | 0 |
| 0 | 0 | 0 | 0 | 0 |
| 0 | 0 | 0 | 0 | 2.008615586 |
| 0 | 0 | 0.010073981 | 0 | 0 |
| 0 | 0.349484007 | 0.071255384 | 0 | 0.083480995 |
| 0 | 0 | 0 | 0.000615195 | 0 |
| 0.34858902 | 0 | 0.002228007 | 0 | 0 |
| 0 | 0 | 0 | 0 | 0 |
| 0 | 0 | 0 | 0 | 0 |
| 0 | 0 | 0 | 0 | 0 |
| 0 | 0.326228261 | 0.216511125 | 2.94349915 | 2.37522755 |
| 0 | 0.003455018 | 0 | 0 | 0 |
| 0 | 0.017822591 | 0 | 0 | 0 |
| 0 | 0.071272 | 0 | 0 | 0 |
| 0 | 0 | 0 | 0 | 0 |
| 0 | 0 | 0 | 0 | 0 |
| 0 | 0 | 0 | 0 | 0 |
| 0 | 0 | 0 | 0 | 0 |

| | | | | |
|---|---|---|---|---|
| 0 | 0 | 0.036311877 | 0 | 0.408919545 |
| 0 | 0 | 0 | 0 | 0 |
| 0.394362715 | 0 | 0.040152074 | 0 | 0 |
| 0 | 0 | 0 | 0.0016065 | 0 |
| 0 | 0 | 0.1235641 | 0.002882074 | 0.006616897 |
| 0 | 0 | 0 | 0 | 0 |
| 0.001929305 | 0.005946408 | 0.003065301 | 0.081284288 | 0.814739006 |
| 0 | 0 | 0 | 0 | 0 |
| 0.005075718 | 0 | 0 | 0 | 0 |
| 0.229097528 | 0 | 0 | 0 | 0 |
| 0 | 0 | 0 | 0.002404159 | 0.009740577 |
| 0.006748855 | 0 | 0 | 0 | 0 |
| 0 | 0 | 0 | 0 | 0 |
| 0 | 0 | 0 | 0 | 0 |
| 0 | 0 | 0 | 0 | 0 |
| 0.001784355 | 0 | 0 | 0 | 0 |
| 0 | 0.013739453 | 0 | 0 | 0.006258964 |
| 0.001854824 | 0 | 0 | 0 | 0 |
| 0 | 0 | 0 | 0 | 0 |
| 0 | 0 | 0 | 0 | 0 |
| 0.052933405 | 0.0092304 | 0.279248394 | 0.473588902 | 0.313190645 |
| 0 | 0 | 0 | 0 | 0 |
| 0.002762714 | 0.001085984 | 0.006639014 | 0 | 0.00364235 |
| 0.00409439 | 0.003500547 | 0.00107336 | 0.007909862 | 0.008352735 |
| 0 | 0 | 0 | 0 | 0 |
| 0 | 0 | 0 | 0 | 0 |
| 0 | 0 | 0 | 0 | 0 |
| 0 | 0 | 0 | 0 | 0 |
| 0 | 0 | 0 | 0 | 0 |
| 0 | 0.293773795 | 0 | 1.085369373 | 1.653575676 |
| 0 | 0 | 0 | 0 | 0 |
| 0 | 0 | 0 | 0 | 0 |
| 0 | 0 | 0 | 0 | 0 |
| 0 | 0 | 0 | 1.184053743 | 0 |
| 0 | 0 | 0 | 0 | 0 |
| 0 | 0 | 0 | 0 | 0 |
| 0.003073885 | 0 | 0 | 0 | 0.029647948 |
| 0 | 0 | 0 | 0 | 0 |
| 0 | 0.00389701 | 0 | 0 | 0 |
| 0 | 0 | 0 | 0 | 0 |
| 0 | 0 | 0 | 0 | 0 |
| 0 | 0 | 0 | 0 | 0 |
| 0 | 0 | 0 | 0 | 0 |
| 0.000977743 | 0 | 0 | 0 | 0 |
| 0 | 0 | 0 | 0 | 0 |
| 0 | 0 | 0 | 0 | 0 |
| 0 | 0 | 0 | 0 | 0 |
| 0 | 0 | 0 | 0 | 0 |

| | | | | |
|---|---|---|---|---|
| 0.000528685 | 0 | 0.00173246 | 0 | 0.002384529 |
| 0 | 0.39985506 | 0.015700329 | 0 | 0 |
| 0 | 0 | 0 | 0 | 0.001307896 |
| 0 | 0 | 0 | 0 | 0 |
| 0 | 0 | 0 | 0 | 0 |
| 0 | 0.219779587 | 0.002428929 | 0 | 0 |
| 0.003928805 | 0 | 0 | 0 | 0 |
| 0.098435004 | 0.003600834 | 0.137366361 | 1.17625564 | 3.403425725 |
| 0.361538265 | 0.394093464 | 17.70905271 | 0.011085752 | 0.022169344 |
| 0.007125178 | 0.035483193 | 0.055742288 | 0.072030634 | 0 |
| 0 | 0 | 0 | 0 | 0 |
| 0 | 0 | 0 | 0 | 0 |
| 0.011173796 | 0 | 0 | 0 | 0.096859798 |
| 0 | 8.906128112 | 0 | 0 | 0 |
| 0 | 0 | 0 | 0 | 0 |
| 0 | 0 | 0 | 0 | 0 |
| 0 | 0 | 0 | 0 | 0 |
| 0 | 0 | 0 | 0 | 0 |
| 0 | 0 | 0 | 0 | 0 |
| 0.023633476 | 0 | 0.006104904 | 0 | 0 |
| 0 | 0 | 0 | 0 | 0 |
| 0 | 0 | 0 | 0 | 0 |
| 0 | 0 | 0 | 0 | 0 |
| 0 | 0 | 0 | 0 | 0 |
| 0 | 0 | 0 | 0 | 0.019648308 |
| 0 | 0 | 0 | 0 | 0 |
| 0 | 0 | 0 | 0 | 0 |
| 0 | 0 | 0 | 0 | 0 |
| 0 | 0 | 0 | 0 | 0 |
| 0 | 0 | 0 | 0 | 0 |
| 0 | 0 | 0 | 0 | 0 |
| 0 | 0 | 0 | 0 | 0 |
| 0 | 0 | 0 | 0 | 0 |
| 0 | 0 | 0 | 0 | 0 |
| 0.196921348 | 0 | 0 | 0 | 0 |
| 0 | 0 | 0 | 0.020988325 | 0 |
| 0 | 0 | 0 | 0 | 0 |
| 0 | 0 | 0 | 0 | 0 |
| 0 | 0.061314812 | 0 | 0.001076659 | 0 |
| 0 | 0 | 0.000576513 | 0.000542358 | 0 |
| 0 | 0 | 0 | 0 | 0 |
| 0.728194638 | 0.158657255 | 0.143340784 | 0 | 0.775502392 |
| 0 | 0 | 0 | 0 | 0 |
| 0.00824353 | 0 | 0 | 0 | 0 |
| 0.001885849 | 0 | 0 | 0 | 0 |
| 0.002820651 | 0.030259853 | 0.010954731 | 0 | 0 |
| 0 | 0 | 0 | 0 | 0 |
| 0 | 0 | 0 | 0 | 0 |

| | | | | |
|---|---|---|---|---|
| 0 | 0 | 0 | 0 | 0 |
| 0.010974713 | 0.000820599 | 0 | 0.0045764 | 0.000827747 |
| 0 | 0 | 0 | 0 | 0 |
| 0 | 0 | 0 | 0 | 0 |
| 0.02134844 | 0.895231381 | 0.040565451 | 0.079066565 | 0.515695713 |
| 0 | 0 | 0 | 0 | 0 |
| 0 | 0 | 0 | 0.04934758 | 0 |
| 0.013898865 | 0 | 0.000734605 | 0.004146503 | 0.007699899 |
| 0.005371936 | 0.062029193 | 0.001711443 | 0 | 0 |
| 0 | 0 | 0 | 0 | 0 |
| 0 | 0 | 0 | 0 | 0 |
| 0 | 0.322158882 | 0 | 0 | 0 |
| 0 | 0 | 0 | 0 | 0 |
| 0 | 0 | 0 | 0 | 0 |
| 0.079063541 | 0 | 0 | 0 | 0 |
| 0.082436416 | 0 | 0.001274235 | 0 | 0 |
| 0 | 0 | 0 | 0 | 0 |
| 0 | 0 | 0 | 0 | 0 |
| 0 | 0 | 0 | 0 | 0 |
| 0 | 0 | 0 | 0 | 0 |
| 0 | 0 | 0 | 0 | 0 |
| 0 | 0 | 0 | 0 | 0 |
| 0 | 0 | 0 | 0 | 0 |
| 0.000582016 | 0.003145754 | 0 | 0 | 0.008279053 |
| 0 | 0.12429391 | 0.086164371 | 0 | 0.105460086 |
| 0 | 0 | 0 | 0 | 0 |
| 0.031761721 | 0 | 0 | 0 | 0.005725272 |
| 0.003923605 | 0 | 0 | 0 | 0 |
| 0 | 0 | 0 | 0 | 0 |
| 0 | 0 | 0 | 0 | 0 |
| 0 | 0 | 0 | 0 | 0 |
| 0 | 0 | 0 | 0 | 0 |
| 0 | 0 | 0 | 0 | 0 |
| 0 | 0 | 0 | 0 | 0 |
| 0 | 0 | 0 | 0 | 0 |
| 0 | 0 | 0 | 0 | 0 |
| 0 | 0 | 0 | 0 | 0 |
| 0.176151697 | 0.002011831 | 0 | 0 | 0.016667773 |
| 0 | 0 | 0 | 0 | 0 |
| 0 | 0 | 0 | 0 | 0 |
| 0 | 0 | 0 | 0 | 0 |
| 0 | 3.121010492 | 0 | 0 | 0 |
| 0 | 0 | 0 | 0 | 0 |
| 0 | 0.167851082 | 0 | 0 | 0 |
| 0 | 0 | 0 | 0 | 0 |
| 0 | 0 | 0 | 0 | 0 |
| 0 | 0 | 0 | 0 | 0 |
| 0 | 0 | 0 | 0 | 0 |

| | | | | |
|---|---|---|---|---|
| 0.009344542 | 0 | 0.004556747 | 0.017147147 | 0.032131067 |
| 0 | 0 | 0 | 0 | 0 |
| 0 | 0 | 0 | 0 | 0 |
| 0 | 0 | 0 | 0 | 0 |
| 0 | 0 | 0 | 0 | 0 |
| 0 | 0.001282386 | 0.024706914 | 0.018078025 | 0 |
| 0 | 0 | 0 | 0 | 0 |
| 0 | 0 | 0 | 0 | 0 |
| 0 | 0 | 0 | 0 | 0 |
| 0 | 0 | 0 | 0 | 0 |
| 0 | 0 | 0 | 0 | 0 |
| 0.001073051 | 0 | 0 | 0 | 0 |
| 0.00139902 | 0 | 0 | 0 | 0 |
| 0 | 0 | 0 | 0 | 0 |
| 0 | 0 | 0 | 0 | 0 |
| 0 | 0 | 0 | 0 | 0 |
| 0 | 0 | 0 | 0 | 0 |
| 0 | 0 | 0 | 0 | 0 |
| 0 | 0 | 0 | 0 | 0 |
| 0 | 0 | 0 | 0 | 0 |
| 0.005795826 | 0 | 0.010340347 | 0.034741936 | 0.048260162 |
| 0 | 0 | 0 | 0 | 0 |
| 0.160789552 | 0.000539489 | 0.002132099 | 0 | 0.000761863 |
| 0 | 0 | 0.310517787 | 0 | 0.218951277 |
| 0 | 0 | 0 | 0 | 0 |
| 0 | 0.14141018 | 0 | 0 | 0 |
| 0 | 0 | 0 | 0 | 0 |
| 0 | 0 | 0 | 0 | 0 |
| 0 | 0.005604312 | 0 | 0 | 0 |
| 0 | 0 | 0 | 0 | 0 |
| 0 | 0 | 0 | 0 | 0 |
| 0 | 0 | 0 | 0 | 0 |
| 0.005026908 | 0.44359102 | 0 | 0.050247438 | 0.016885738 |
| 0 | 0 | 0 | 0 | 0 |
| 0.301064165 | 0.005121816 | 0.058716072 | 0 | 3.042116102 |
| 0.055190857 | 0 | 0 | 0.092890099 | 0 |
| 0 | 0 | 0 | 0 | 0 |
| 0 | 0.090684613 | 0 | 0 | 0 |
| 0 | 0 | 0 | 0.017216887 | 0.009050508 |
| 0 | 0 | 0 | 0 | 0 |
| 0 | 0 | 0 | 0 | 0 |
| 0 | 0 | 0.782917119 | 0 | 0 |
| 0.002975862 | 0.40576367 | 0.020045096 | 0 | 0.022197976 |
| 0 | 0 | 0 | 0 | 0 |
| 0 | 0 | 0 | 0 | 0 |
| 0 | 0 | 0.001505878 | 0 | 0.006149666 |
| 0 | 0 | 0 | 0 | 0 |
| 0 | 0 | 0 | 0 | 0 |

| | | | | |
|---|---|---|---|---|
| 0 | 0 | 0 | 0 | 0 |
| 0 | 0 | 0 | 0 | 0 |
| 0 | 0 | 0 | 0 | 0 |
| 0 | 0 | 0 | 0 | 0 |
| 0 | 0 | 0 | 0 | 0 |
| 0 | 0 | 0 | 0 | 0 |
| 0 | 0 | 0 | 0 | 0 |
| 0.038823666 | 0 | 0.177321065 | 0.497122921 | 0 |
| 0.00147616 | 0 | 0.001410868 | 0 | 0.022470501 |
| 0 | 0.072362485 | 0.004653113 | 0 | 0.054473158 |
| 0 | 0 | 0 | 0 | 0 |
| 0 | 0 | 0 | 0 | 0 |
| 0 | 0 | 0 | 0 | 0 |
| 0 | 0 | 0 | 0 | 0.022699587 |
| 0.005081896 | 0 | 0 | 0 | 0 |
| 0 | 0 | 0 | 0.017432775 | 0.001246409 |
| 0 | 0 | 0 | 0 | 0 |
| 0 | 0 | 0 | 0 | 0 |
| 0 | 0 | 0 | 0 | 0 |
| 0.315648192 | 0 | 1.096594844 | 0 | 0 |
| 0 | 0.016439066 | 0 | 0.001499424 | 0.001952678 |
| 0 | 0 | 0 | 0 | 0 |
| 0 | 0 | 0 | 0 | 0 |
| 0 | 0 | 0 | 0.001912302 | 0 |
| 0 | 0 | 0 | 0 | 0 |
| 0 | 0 | 0 | 0 | 0 |
| 0 | 0 | 0 | 0 | 0 |
| 0 | 0 | 0 | 0 | 0 |
| 0 | 0 | 0 | 0 | 0 |
| 0 | 0 | 0.043607893 | 2.14878486 | 0 |
| 0 | 0.004543251 | 0.083633706 | 0 | 0.373307389 |
| 0 | 0 | 0 | 0 | 0 |
| 0 | 0 | 0 | 0 | 0 |
| 0 | 0 | 0 | 0 | 0 |
| 0 | 0 | 0 | 0 | 0 |
| 0 | 0 | 0 | 0 | 0 |
| 0 | 0 | 0 | 0 | 0 |
| 0 | 0 | 0 | 0 | 0 |
| 0 | 0.062388293 | 0 | 0 | 0 |
| 0 | 0 | 0 | 0 | 0 |
| 0 | 0 | 0.002222159 | 0 | 0 |
| 0 | 0 | 0 | 0 | 0.529749692 |
| 0 | 0 | 0 | 0 | 0 |
| 0.003410948 | 0 | 0 | 0.013801221 | 0 |
| 0 | 0 | 0 | 0 | 0 |
| 0 | 0 | 0 | 0 | 0 |
| 0.001225876 | 0 | 0 | 0.078311589 | 0 |
| 0.000498134 | 0 | 0 | 0 | 0 |

| | | | | |
|---|---|---|---|---|
| 0 | 0 | 0 | 0 | 0 |
| 0 | 0 | 0 | 0 | 0 |
| 0 | 0 | 0 | 0 | 0 |
| 0 | 0 | 0 | 0 | 0 |
| 0.248034858 | 0.002699972 | 0.010170308 | 0.067288139 | 0.016045903 |
| 0 | 0 | 0 | 0 | 0 |
| 0 | 0 | 0 | 0 | 0 |
| 0 | 0 | 0 | 0 | 0 |
| 0 | 0 | 0 | 0 | 0 |
| 0 | 0 | 0 | 0 | 0 |
| 0 | 0 | 0.001058423 | 0 | 0.005042753 |
| 0.018648435 | 0 | 0 | 0.002177617 | 0.003529093 |
| 0.019928298 | 0.005390045 | 0.001397934 | 0 | 0.004376783 |
| 0 | 0 | 0 | 0 | 0 |
| 0.023901008 | 0 | 0 | 0 | 0.005210743 |
| 0 | 0 | 0.025740695 | 0.8148671 | 0 |
| 0 | 0.010330949 | 0.01620091 | 0 | 0.050892967 |
| 0 | 0 | 0 | 0 | 0 |
| 0.010269601 | 0.001655733 | 0.001226921 | 0.190118679 | 0 |
| 0.129061339 | 0.000617681 | 0.142118948 | 1.109423769 | 0.073708162 |
| 0 | 0 | 0 | 0 | 0 |
| 0 | 0 | 0 | 0 | 0 |
| 0 | 0 | 0 | 0 | 0 |
| 0 | 0 | 0 | 0 | 0 |
| 0 | 0 | 0 | 0 | 0 |
| 0 | 0 | 0 | 0 | 0 |
| 0.055572947 | 0 | 0.002384096 | 0.002242852 | 0.015902311 |
| 0 | 0 | 0 | 0 | 0 |
| 0 | 0 | 0 | 0 | 0 |
| 0 | 0 | 0 | 0 | 0 |
| 0.024550865 | 0 | 0 | 0 | 0 |
| 0 | 0 | 0 | 0 | 0 |
| 0 | 0 | 0 | 0 | 0 |
| 0 | 0 | 0 | 0 | 0 |
| 0 | 0 | 0 | 0 | 0 |
| 0 | 0 | 0 | 0 | 0 |
| 0 | 0 | 0 | 0 | 0 |
| 0.03195694 | 0.000689849 | 0 | 0.016350691 | 0.051911018 |
| 0 | 0 | 0 | 0.114859149 | 0.921174408 |
| 0.008484016 | 0 | 0 | 0 | 0 |
| 0.001334867 | 0 | 0.001603894 | 0.001646043 | 0 |
| 0 | 0 | 0 | 0 | 0 |
| 0 | 0 | 0 | 0 | 0 |
| 0 | 0 | 0 | 0.002852471 | 0.021875631 |
| 0 | 0 | 0 | 0 | 0 |
| 0 | 0 | 0 | 0 | 0.001844996 |
| 0 | 0 | 0 | 0 | 0 |
| 0 | 0 | 0 | 0 | 0 |

| | | | | |
|---|---|---|---|---|
| 0 | 0 | 0 | 0 | 0 |
| 0 | 0 | 0.000740446 | 0 | 0 |
| 0 | 0 | 0 | 0 | 0 |
| 0 | 0 | 0 | 0 | 0.002162808 |
| 0 | 0 | 0.008210137 | 0.000594133 | 0 |
| 0 | 0 | 0 | 0 | 0 |
| 0 | 0 | 0.025330209 | 0.013468872 | 0.002728493 |
| 0 | 0 | 0 | 0 | 0 |
| 0.019750592 | 0 | 0 | 0 | 0.018448751 |
| 0 | 0 | 0 | 0 | 0 |
| 0 | 0 | 0 | 0 | 0.039138838 |
| 0 | 0 | 0 | 0 | 0 |
| 0.007954048 | 0.034523138 | 0.013675975 | 0.019828398 | 0.028669458 |
| 0 | 0 | 0 | 0 | 0 |
| 0 | 0 | 0 | 0 | 0 |
| 0 | 0 | 0 | 0 | 0 |
| 0 | 0 | 0 | 0.053542202 | 0 |
| 0 | 0 | 0 | 0 | 0 |
| 0 | 0 | 0 | 0 | 0 |
| 0 | 0 | 0 | 0 | 0 |
| 0 | 0 | 0 | 0 | 0 |
| 0 | 0 | 0 | 0 | 0 |
| 0.004073086 | 0 | 0.007626963 | 0.0392638 | 0.005046935 |
| 0.003433129 | 0 | 0.008250073 | 0 | 0 |
| 0 | 0 | 0 | 0 | 0 |
| 0 | 0 | 0 | 0 | 0 |
| 0 | 0 | 0 | 0.028563322 | 0 |
| 0 | 0 | 0 | 0 | 0 |
| 0 | 0 | 0 | 0 | 0 |
| 0 | 0 | 0 | 0 | 0 |
| 0 | 0.00175331 | 0.037323244 | 0 | 0.006077494 |
| 0 | 0 | 0 | 0 | 0 |
| 0 | 0.000984173 | 0 | 0 | 0 |
| 0 | 0 | 0 | 0 | 0 |
| 0 | 0 | 0 | 0 | 0 |
| 0 | 0 | 0 | 0 | 0 |
| 0 | 0 | 0 | 0 | 0 |
| 0 | 0 | 0 | 0 | 0 |
| 0.026530861 | 0.005394262 | 0.020985415 | 0 | 0 |
| 0 | 0 | 0 | 0 | 0 |
| 0 | 0 | 0 | 0 | 0 |
| 0 | 0 | 0 | 0 | 0 |
| 0 | 0 | 0 | 0 | 0 |
| 0 | 0 | 0 | 0 | 0 |
| 0 | 0 | 0 | 0 | 0.000882031 |
| 0.072604078 | 0.361293764 | 0.444359676 | 0.035732228 | 0.508709289 |
| 0 | 0 | 0 | 0 | 0 |
| 0 | 0 | 0 | 0 | 0 |

| | | | | |
|---|---|---|---|---|
| 0 | 0 | 0 | 0 | 0 |
| 0 | 0 | 0 | 0 | 0 |
| 0 | 0.039310001 | 0 | 0 | 0 |
| 0 | 0 | 0.192213609 | 0 | 0 |
| 0 | 0 | 0 | 0 | 0 |
| 0 | 0 | 0 | 0 | 0 |
| 0 | 0 | 0 | 0 | 0 |
| 0 | 0 | 0 | 0 | 0 |
| 0 | 0 | 0 | 0 | 0 |
| 0 | 0 | 0 | 0 | 0 |
| 0.002375022 | 0 | 0 | 0 | 0 |
| 0 | 0.000816708 | 0 | 0.000683206 | 0 |
| 0 | 0 | 0 | 0 | 0 |
| 0 | 0 | 0 | 0 | 0 |
| 0 | 0 | 0 | 0 | 0 |
| 0 | 0 | 0 | 0 | 0 |
| 0 | 0 | 0 | 0 | 0 |
| 0 | 0 | 0 | 0 | 0 |
| 0 | 0 | 0 | 0 | 0 |
| 0 | 0.006093761 | 0 | 0 | 0.001024474 |
| 0 | 0 | 0 | 0 | 0 |
| 0 | 0 | 0 | 0 | 0 |
| 0 | 0 | 0 | 0 | 0 |
| 0 | 0 | 0 | 0 | 0 |
| 0 | 0 | 0 | 0 | 0 |
| 0 | 0 | 0 | 0 | 0 |
| 0 | 0 | 0 | 0 | 0 |
| 0 | 0 | 0.018085216 | 0 | 0 |
| 0 | 0 | 0 | 0 | 0 |
| 0 | 0 | 0 | 0 | 0 |
| 0.003551503 | 0.064068523 | 0.004987715 | 0.00260679 | 0.006336929 |
| 0 | 0 | 0 | 0 | 0 |
| 0.197542791 | 0.034734667 | 0.013941885 | 0.05246364 | 0.009606042 |
| 0 | 0 | 0 | 0 | 0.009398617 |
| 0 | 0 | 0 | 0 | 0 |
| 0 | 0 | 0 | 0 | 0 |
| 0 | 0 | 0 | 0 | 0 |
| 0 | 0 | 0 | 0 | 0 |
| 0 | 0 | 0 | 0 | 0 |
| 0 | 0 | 0 | 0 | 0 |
| 0 | 0 | 0 | 0 | 0 |
| 0.458794554 | 0 | 0.000705838 | 0.006142194 | 0.078691748 |
| 0 | 0.039745411 | 0.007739184 | 0.013550159 | 0.127005593 |
| 0 | 0 | 0 | 0 | 0 |
| 0 | 0 | 0 | 0 | 0 |
| 0.002844045 | 0.293462898 | 0.013668916 | 0 | 0.021017364 |
| 0.001118782 | 0 | 0 | 0 | 0 |
| 0 | 0 | 0 | 0 | 0 |

| | | | | |
|---|---|---|---|---|
| 0 | 0 | 0 | 0 | 0 |
| 0 | 0 | 0.013738253 | 0.61189344 | 0.005365129 |
| 0.004094376 | 0.004970334 | 0 | 0.035143828 | 0.010277941 |
| 0 | 0 | 0 | 0 | 0 |
| 0.418260496 | 0.006094593 | 0.007561971 | 0.191089106 | 0 |
| 0 | 0 | 0 | 0 | 0 |
| 0 | 0 | 0 | 0 | 0 |
| 0 | 0 | 0 | 0 | 0 |
| 0 | 0 | 0 | 0 | 0 |
| 0 | 0 | 0 | 0 | 0 |
| 0 | 0 | 0 | 0 | 0 |
| 0 | 0 | 0 | 0 | 0 |
| 0 | 0 | 0 | 0 | 0 |
| 0 | 0 | 0 | 0 | 0 |
| 0 | 0 | 0 | 0 | 0 |
| 0 | 0 | 0 | 0 | 0 |
| 0 | 0 | 0 | 0 | 0 |
| 0 | 0 | 0 | 0 | 0 |
| 0 | 0 | 0 | 0 | 0 |
| 0 | 0 | 0 | 0 | 0 |
| 0 | 0 | 0 | 0 | 0 |
| 0 | 0 | 0 | 0 | 0 |
| 0 | 0 | 0 | 0 | 0 |
| 0 | 0 | 0 | 0 | 0 |
| 0 | 0.009655456 | 1.314753305 | 0 | 0 |
| 0 | 0.001030184 | 0 | 0.000615561 | 0.001870483 |
| 0 | 0.000932249 | 0 | 0 | 0 |
| 0 | 0.000856555 | 0 | 0 | 0.511872037 |
| 0 | 0 | 0 | 0 | 0 |
| 0 | 0 | 0 | 0 | 0 |
| 0 | 0.004773162 | 0 | 0 | 0 |
| 0 | 0 | 0 | 0 | 0 |
| 0 | 0 | 0 | 0 | 0 |
| 0 | 0 | 0 | 0 | 0 |
| 0 | 0 | 0 | 0 | 0 |
| 0 | 0 | 0 | 0 | 0 |
| 0 | 0 | 0 | 0 | 0 |
| 0 | 0 | 0 | 0.001225478 | 0.04923715 |
| 0 | 0.930114715 | 0.549916806 | 0 | 1.238969149 |
| 0 | 0 | 0 | 0 | 0 |
| 0 | 0 | 0 | 0 | 0 |
| 0 | 0.057955047 | 0.001183071 | 0.000890385 | 0.006313023 |
| 0.001001988 | 0 | 0 | 0 | 0.002085816 |
| 0 | 0 | 0 | 0 | 0 |
| 0 | 0 | 0 | 0 | 0 |
| 0 | 0.001189467 | 0 | 0 | 0 |
| 0 | 0.015816673 | 0.011720372 | 0 | 0 |
| 0 | 0 | 0 | 0 | 0 |

| | | | | |
|---|---|---|---|---|
| 0 | 0 | 0 | 0 | 0 |
| 0.02262978 | 0.065575406 | 0.007605747 | 0.052471111 | 0.021540792 |
| 0 | 0.03820082 | 0 | 0 | 0 |
| 0 | 0 | 0 | 0 | 0 |
| 0 | 0 | 0 | 0 | 0 |
| 0 | 0 | 0 | 0 | 0 |
| 0 | 0 | 0 | 0 | 0 |
| 0 | 0 | 0 | 0 | 0 |
| 0 | 0 | 0 | 0 | 0 |
| 0 | 0 | 0 | 0 | 0 |
| 0 | 0 | 0 | 0 | 0 |
| 0 | 0 | 0 | 0 | 0 |
| 0 | 0.000753468 | 0.15990565 | 0 | 0.115767908 |
| 0 | 0 | 0 | 0 | 0 |
| 0.007717176 | 0.012975377 | 0.00658557 | 0.013877726 | 0.006024252 |
| 0 | 0 | 0 | 0 | 0 |
| 0 | 0 | 0 | 0 | 0 |
| 0 | 0 | 0 | 0 | 0 |
| 0 | 0 | 0 | 0 | 0 |
| 0 | 0 | 0 | 0 | 0 |
| 0 | 0 | 0 | 0 | 0 |
| 0.007815716 | 0 | 0.00258702 | 0.054759466 | 0.168121254 |
| 0 | 0.001475482 | 0.005963739 | 0 | 0 |
| 0 | 0 | 0 | 0 | 0.008125001 |
| 0 | 0 | 0 | 0 | 0 |
| 0 | 0 | 0 | 0 | 0 |
| 0 | 0 | 0 | 0 | 0 |
| 0 | 0 | 0 | 0 | 0 |
| 0 | 0 | 0 | 0 | 0 |
| 0 | 0 | 0 | 0 | 0 |
| 0 | 0 | 0 | 0 | 0 |
| 0 | 0 | 0 | 0 | 0 |
| 0 | 0 | 0 | 0 | 0 |
| 0 | 0 | 0 | 0 | 0 |
| 0 | 0 | 15.30829022 | 2.501616346 | 0.034915806 |
| 0 | 0 | 0 | 0 | 0 |
| 0 | 0 | 0 | 0 | 0 |
| 0 | 0 | 0 | 0 | 0 |
| 0.00141908 | 0.008890258 | 1.284234112 | 0.192001433 | 0.115454744 |
| 0 | 0 | 0 | 0 | 0 |
| 0.057893862 | 0.03157257 | 0.02802852 | 0.040750538 | 1.028361976 |
| 0 | 0 | 0 | 0 | 0 |
| 0 | 0 | 0 | 0 | 0 |
| 0 | 0 | 0 | 0 | 0 |
| 0 | 0 | 0 | 0 | 0 |
| 0.063856527 | 0.001610574 | 0.096007031 | 0.178392854 | 0.05231224 |
| 0 | 0 | 0 | 0 | 0 |
| 0 | 0 | 0 | 0 | 0 |

| | | | | |
|---|---|---|---|---|
| 0 | 0 | 0 | 0 | 0 |
| 0 | 0 | 0 | 0 | 0 |
| 0.020316744 | 0 | 0 | 0 | 0 |
| 0 | 0 | 0 | 0 | 0 |
| 0 | 0 | 0 | 0 | 0 |
| 0 | 0 | 0 | 0 | 0 |
| 0 | 0 | 0 | 0 | 0 |
| 0.002361897 | 0 | 0 | 0.007011544 | 0.005463009 |
| 0.10822826 | 0.001212282 | 0 | 0 | 0 |
| 0 | 0.149113646 | 0.018675254 | 0 | 0 |
| 0.312542497 | 0 | 0 | 0 | 0 |
| 0 | 0 | 0 | 0 | 0 |
| 0.001624675 | 0 | 0 | 0.008586038 | 0.005073074 |
| 0 | 0 | 0 | 0 | 0 |
| 0 | 0 | 0 | 0 | 0 |
| 0 | 0 | 0 | 0 | 0 |
| 0 | 0 | 0 | 0 | 0 |
| 0 | 0 | 0 | 0 | 0 |
| 0 | 0 | 0 | 0 | 0 |
| 0.107835908 | 3.030096729 | 0.130426447 | 0.147748815 | 0.3979997 |
| 0 | 0 | 0 | 0 | 0 |
| 0 | 0 | 0 | 0 | 0 |
| 0 | 0 | 0 | 0 | 0 |
| 0 | 0 | 0 | 0 | 0 |
| 0 | 0 | 0 | 0 | 0 |
| 0 | 0.008217121 | 0 | 0 | 0 |
| 0 | 0 | 0 | 0 | 0 |
| 0 | 0 | 0 | 0 | 0 |
| 0 | 0 | 0 | 0 | 0 |
| 0 | 0 | 0.001929799 | 0 | 0 |
| 0 | 0 | 0 | 0 | 0 |
| 0 | 0 | 0 | 0 | 0 |
| 0 | 0 | 0 | 0 | 0 |
| 0 | 0 | 0 | 0 | 0 |
| 0 | 0 | 0 | 0 | 0 |
| 0.003743823 | 0 | 0 | 0 | 0 |
| 0 | 0 | 0 | 0 | 0 |
| 0 | 0 | 0 | 0 | 0 |
| 0 | 0 | 0 | 0 | 0 |
| 0 | 0 | 0 | 0 | 0 |
| 0 | 0 | 0 | 0 | 0 |
| 0 | 0 | 0 | 0 | 0 |
| 0 | 0 | 0.021570579 | 0 | 0.01036646 |
| 0 | 0 | 0 | 0 | 0 |
| 0 | 0.00357517 | 0 | 0.001395687 | 0.002625396 |
| 0 | 0 | 0 | 0 | 0 |
| 0 | 0 | 0 | 0 | 0 |
| 0 | 0 | 0 | 0 | 0 |

| | | | | |
|---|---|---|---|---|
| 0 | 0 | 0 | 0 | 0 |
| 0 | 0 | 0 | 0 | 0 |
| 0 | 0 | 0 | 0 | 0 |
| 0 | 0 | 0 | 0 | 0 |
| 0 | 0 | 0 | 0 | 0 |
| 0.004345519 | 0.000747321 | 0.000791107 | 0.00133963 | 0.002412258 |
| 0 | 0 | 0 | 0 | 0 |
| 0 | 0 | 0 | 0 | 0 |
| 0 | 0 | 0 | 0 | 0 |
| 0 | 0 | 0 | 0 | 0 |
| 0 | 0 | 0 | 0 | 0 |
| 0 | 0 | 0 | 0 | 0 |
| 0 | 0 | 0 | 0 | 0 |
| 0 | 0 | 0 | 0 | 0 |
| 0 | 0 | 0 | 0 | 0 |
| 0 | 0 | 0 | 0 | 0 |
| 0.102083328 | 0 | 0 | 0 | 0 |
| 0 | 0 | 0 | 0 | 0.00055703 |
| 0 | 0 | 0 | 0 | 0 |
| 0 | 0 | 0 | 0 | 0 |
| 0 | 0 | 0 | 0 | 0 |
| 0 | 0 | 0 | 0 | 0 |
| 0 | 0 | 0 | 0 | 0 |
| 0 | 0 | 0 | 0 | 0 |
| 0 | 0 | 0 | 0 | 0 |
| 0 | 0.086668204 | 0.00098994 | 0 | 0 |
| 0 | 0 | 0 | 0 | 0 |
| 2.051488252 | 0 | 0 | 0 | 0 |
| 0 | 0 | 0 | 0 | 0 |
| 0 | 0 | 0 | 0 | 0 |
| 0 | 0 | 0 | 0 | 0 |
| 0 | 0 | 0 | 0 | 0 |
| 0 | 0 | 0 | 0 | 0 |
| 0 | 0 | 0 | 0 | 0 |
| 0 | 0 | 0 | 0 | 0 |
| 0 | 0 | 0 | 0 | 0 |
| 0 | 0 | 0 | 0 | 0 |
| 0 | 0 | 0 | 0 | 0 |
| 0 | 0 | 1.003930138 | 0 | 0.00127692 |
| 0 | 0 | 0 | 0 | 0 |
| 0 | 0 | 0 | 0 | 0 |
| 0 | 0 | 0 | 0 | 0 |
| 0 | 0 | 0 | 0 | 0 |
| 0 | 0 | 0 | 0 | 0 |
| 0 | 0.01141233 | 0.001757235 | 0 | 0 |
| 0 | 0 | 0 | 0 | 0 |
| 0 | 0 | 0 | 0 | 0 |
| 0 | 0 | 0 | 0 | 0 |

| | | | | |
|---|---|---|---|---|
| 0 | 0 | 0 | 0 | 0 |
| 0.005351675 | 0 | 0.001239992 | 0 | 0 |
| 0 | 0 | 0.00151812 | 0 | 0 |
| 0 | 0 | 0 | 0 | 0 |
| 0 | 0 | 0 | 0 | 0 |
| 0 | 0 | 0 | 0 | 0 |
| 0 | 0 | 0 | 0 | 0 |
| 0 | 0 | 0 | 0 | 0 |
| 0 | 0 | 0 | 0 | 0 |
| 0 | 0 | 0 | 0 | 0 |
| 0.000637054 | 0.041788323 | 0.045694732 | 0.001745689 | 0.01458755 |
| 0 | 0 | 0 | 0 | 0 |
| 0.542733998 | 0 | 0.087301837 | 0 | 0.631644482 |
| 0 | 0 | 0 | 0 | 0 |
| 0 | 0 | 0 | 0 | 0 |
| 0 | 0 | 0 | 0 | 0 |
| 0 | 0 | 0 | 0 | 0 |
| 0 | 0 | 0 | 0 | 0 |
| 0 | 0 | 0 | 0 | 0 |
| 0 | 0 | 0 | 0 | 0 |
| 0 | 0 | 0 | 0.004429423 | 0 |
| 0 | 0 | 0 | 0 | 0 |
| 0 | 0.001112812 | 0 | 0 | 0 |
| 1.135156321 | 0.051583394 | 0.611974061 | 5.953674685 | 3.045772982 |
| 0 | 0 | 0 | 0 | 0 |
| 0 | 0 | 0 | 0 | 0 |
| 0 | 0 | 0 | 0 | 0 |
| 0 | 0 | 0 | 0 | 0 |
| 0 | 0 | 0 | 0 | 0 |
| 0 | 0 | 0 | 0 | 0 |
| 0 | 0 | 0 | 0 | 0.006605261 |
| 0 | 0 | 0 | 0 | 0 |
| 0 | 0.043110193 | 0 | 0 | 0 |
| 0 | 0 | 0 | 0 | 0 |

| PHL018-0020_V3 | PHL018-0023_V3 | PHL018-0074_V2 | PHL018-0250_V1 | PHL018-0032_V3 |
|---|---|---|---|---|
| EYCF | EYCF | CTRL | REF | EYCF |
| 0.141235152 | 0.024076939 | 0.010742551 | 0.199310178 | 0.02189519 |
| 2.103285564 | 4.414497564 | 5.169660037 | 0.298156914 | 6.456326992 |
| 3.005420248 | 13.32070614 | 5.776239803 | 0 | 0 |
| 0 | 0 | 0 | 0 | 0 |
| 0 | 0 | 0 | 0 | 0 |
| 0 | 0 | 0 | 0 | 0 |
| 0 | 0 | 0 | 0 | 0 |
| 0 | 0 | 0 | 0 | 0 |
| 0 | 0 | 0 | 0 | 0 |
| 0 | 0 | 0 | 0 | 0 |
| 0 | 0 | 0 | 0 | 0 |
| 0 | 0 | 0 | 0 | 0 |
| 0 | 0 | 0 | 0 | 0 |
| 6.539803596 | 13.8274268 | 4.902405444 | 0 | 4.750044162 |
| 0 | 0 | 0 | 0 | 0 |
| 0 | 0 | 0 | 0 | 0 |
| 0 | 0 | 0 | 0 | 0 |
| 0 | 0 | 0 | 0 | 0 |
| 0 | 0 | 0 | 0 | 0 |
| 0 | 0 | 0 | 0 | 0 |
| 0 | 0 | 0 | 0 | 0 |
| 0 | 0 | 0 | 0 | 0 |
| 0 | 0 | 0 | 0 | 0 |
| 0 | 0 | 0 | 0 | 0 |
| 0 | 0 | 0 | 0 | 0 |
| 0 | 0 | 0 | 0 | 0 |
| 0 | 0 | 0 | 0 | 0 |
| 0 | 0 | 0 | 0 | 0 |
| 0 | 0 | 0 | 0 | 0 |
| 0 | 0 | 0 | 0 | 0 |
| 0 | 0 | 0 | 0 | 0 |
| 0 | 0 | 0 | 0 | 0 |
| 0 | 0 | 0 | 0 | 0 |
| 0 | 0 | 0 | 0 | 0 |
| 0 | 0 | 0 | 0 | 0 |
| 0 | 1.169581554 | 0 | 0 | 6.254472152 |
| 0 | 0 | 0 | 0 | 0 |
| 0 | 0 | 0 | 0 | 0 |
| 0 | 0 | 0 | 0 | 0 |
| 0 | 0 | 0 | 0 | 0 |
| 0 | 0 | 0 | 0 | 0 |
| 0.001094424 | 0 | 0 | 0.009726346 | 0.545017749 |
| 0 | 0.03884664 | 0 | 0.977385406 | 0.272277304 |
| 0.079858971 | 0.004290669 | 0.015953279 | 0.223714119 | 0.012660314 |
| 0 | 0 | 0.0010617 | 0.003418477 | 0 |
| 0 | 0 | 0 | 0 | 0 |

| | | | | |
|---|---|---|---|---|
| 0 | 0 | 0 | 0 | 0 |
| 0 | 0.002398382 | 0.003470967 | 0.203659433 | 0.001600984 |
| 0 | 0 | 0.003913526 | 0.018786692 | 0.011472509 |
| 0.200968432 | 0.459171072 | 0.419455459 | 0.17761703 | 0.735691647 |
| 0.00336865 | 0 | 0 | 0 | 0.000742075 |
| 0.034511282 | 0 | 0 | 0 | 0 |
| 0 | 0 | 0 | 0.324019661 | 0.018589965 |
| 0 | 0 | 0 | 0 | 0 |
| 0 | 0 | 0 | 0 | 0 |
| 0 | 0 | 0 | 0 | 0 |
| 0 | 0 | 0 | 0 | 0 |
| 0.254802041 | 0 | 0.008831301 | 0 | 0.652539333 |
| 0 | 0.003563203 | 0 | 0 | 0 |
| 0.10626268 | 0 | 0.011626357 | 0.08401541 | 0.048338421 |
| 0 | 0 | 0 | 0.006420088 | 0 |
| 0 | 0 | 0 | 0 | 0 |
| 0.038846635 | 0 | 0.412740843 | 0.011663416 | 0 |
| 0 | 0 | 0 | 0 | 0 |
| 0 | 0 | 0 | 0 | 0 |
| 0 | 0 | 0 | 0 | 0 |
| 0.01918235 | 0.021399526 | 0.118759628 | 0.043602563 | 0.100859036 |
| 0 | 0.001826552 | 0.006007743 | 0 | 0.00304818 |
| 0 | 0 | 0 | 0 | 0 |
| 0 | 0 | 0 | 0 | 0 |
| 0 | 0 | 0 | 0 | 0 |
| 0 | 0 | 0 | 0 | 0 |
| 0 | 0 | 0 | 0.01411029 | 0 |
| 0 | 0 | 0 | 0 | 0 |
| 0 | 0 | 0 | 0 | 0.007279075 |
| 0 | 0 | 0 | 0 | 0 |
| 0 | 0 | 0.228457559 | 0.045192339 | 0 |
| 0 | 0 | 0 | 0 | 0 |
| 0 | 0 | 0 | 0 | 0 |
| 0 | 0 | 0.005110339 | 0 | 0 |
| 0 | 0 | 0 | 0 | 0 |
| 0 | 0 | 0 | 0.023549687 | 0 |
| 0 | 0 | 0 | 0 | 0 |
| 2.677197271 | 0 | 0.011073231 | 0 | 0 |
| 0 | 0 | 0 | 0 | 0 |
| 0 | 0 | 0 | 0 | 0 |
| 0 | 0 | 0 | 0 | 0 |
| 0 | 0 | 0 | 0 | 0 |
| 0 | 0 | 0 | 0 | 0 |
| 0 | 0 | 0 | 0 | 0 |
| 0 | 0 | 0 | 0 | 0 |
| 0 | 0 | 0 | 0 | 0 |
| 0 | 0 | 0 | 0.002608969 | 0 |
| 0 | 0 | 0 | 0 | 0 |

| | | | | |
|---|---|---|---|---|
| 0 | 0 | 0 | 0 | 0 |
| 0 | 0 | 0 | 0 | 0 |
| 0 | 0 | 0 | 0 | 0 |
| 0 | 0 | 0 | 0 | 0 |
| 0 | 0 | 0.026259375 | 0 | 0 |
| 0 | 0 | 0 | 0 | 0 |
| 0 | 0 | 0 | 0 | 0 |
| 0 | 0 | 0.00124111 | 0 | 0 |
| 0 | 0 | 0 | 0.065882551 | 0 |
| 0 | 0 | 0 | 0 | 0 |
| 0 | 0 | 0 | 0 | 0 |
| 0 | 0.001639774 | 0 | 0 | 0.023488135 |
| 0 | 0 | 0 | 0 | 0 |
| 0 | 0 | 0 | 0 | 0 |
| 0 | 0 | 0 | 0 | 0 |
| 0 | 0 | 0 | 0 | 0 |
| 0 | 0 | 0 | 0 | 0 |
| 0 | 0 | 0 | 0 | 0 |
| 0 | 0 | 0 | 0.014330746 | 0 |
| 0 | 0 | 0 | 0 | 0 |
| 0 | 0 | 0 | 0 | 0 |
| 0 | 0 | 0 | 0 | 0 |
| 0 | 0 | 0 | 0 | 0 |
| 0 | 0 | 0 | 0 | 0 |
| 0 | 0 | 0 | 0 | 0 |
| 0 | 0 | 0 | 0 | 0 |
| 0 | 0 | 0 | 0 | 0 |
| 0 | 0 | 0 | 0 | 0 |
| 0 | 0 | 0 | 0 | 0 |
| 0 | 0 | 0 | 0 | 0 |
| 0.006875898 | 0 | 0 | 0 | 0.053256168 |
| 0.008656099 | 0 | 0 | 0 | 0.020593852 |
| 0 | 0 | 0 | 0 | 0 |
| 0 | 0 | 0 | 0 | 0 |
| 0 | 0 | 0 | 0 | 0 |
| 0 | 0 | 0 | 0 | 0 |
| 0 | 0 | 0 | 0 | 0 |
| 0 | 0 | 0 | 0 | 0 |
| 0 | 0 | 0 | 0 | 0 |
| 0 | 0 | 0 | 0 | 0 |
| 0 | 0 | 0 | 0 | 0 |
| 0 | 0 | 0 | 0 | 0 |
| 0.002382162 | 0 | 0 | 0 | 0.003526405 |
| 0 | 0 | 0 | 0 | 0 |
| 0 | 0 | 0 | 0 | 0 |
| 0 | 0 | 0 | 0 | 0 |
| 0 | 0 | 0 | 0.002639445 | 0 |
| 0 | 0 | 0 | 0 | 0 |

| | | | | |
|---|---|---|---|---|
| 0 | 0 | 0 | 0 | 0 |
| 0 | 0 | 0 | 0 | 0 |
| 0 | 0 | 0 | 0 | 0 |
| 0 | 0 | 0 | 0 | 0 |
| 0 | 0 | 0 | 0 | 0 |
| 0.045577233 | 0 | 0 | 0 | 0 |
| 0 | 0 | 0 | 0 | 0 |
| 0 | 0 | 0 | 0 | 0 |
| 0 | 0 | 0 | 0 | 0 |
| 0 | 0 | 0 | 0 | 0 |
| 0 | 0 | 0 | 0 | 0 |
| 0 | 0 | 0 | 0 | 0 |
| 0 | 0 | 0 | 0 | 0 |
| 0 | 0 | 0 | 0 | 0 |
| 0 | 0 | 0 | 0 | 0 |
| 0 | 0 | 0 | 0.268992844 | 0.066108025 |
| 0 | 0 | 0 | 0 | 0 |
| 0 | 0 | 0 | 0 | 0 |
| 0 | 0 | 0 | 0 | 0 |
| 0 | 0 | 0 | 0 | 0 |
| 0 | 0 | 0 | 0 | 0 |
| 0 | 0 | 0 | 0 | 0 |
| 0 | 0 | 0 | 0 | 0 |
| 0 | 0 | 0 | 0 | 0 |
| 0 | 0 | 0 | 0 | 0 |
| 0 | 0 | 0 | 0 | 0 |
| 0 | 0 | 0 | 0 | 0 |
| 0 | 0 | 0 | 0 | 0 |
| 0 | 0 | 0 | 0 | 0 |
| 0 | 0 | 0 | 0 | 0 |
| 0 | 0 | 0 | 0 | 0 |
| 0 | 0 | 0 | 0 | 0 |
| 0 | 0 | 0 | 0 | 0 |
| 0 | 0 | 0 | 0 | 0 |
| 0 | 0 | 0 | 0 | 0 |
| 0 | 0 | 0 | 0 | 0 |
| 0 | 0 | 0 | 0 | 0 |
| 0 | 0 | 0 | 0 | 0 |
| 0 | 0 | 0 | 0 | 0 |
| 0 | 0 | 0 | 0 | 0 |
| 0 | 0 | 0 | 0 | 0 |
| 0 | 0 | 0 | 0 | 0 |
| 0 | 0 | 0 | 0 | 0 |
| 0 | 0 | 0 | 0 | 0 |
| 0 | 0 | 0 | 0 | 0 |
| 0 | 0 | 0 | 0 | 0 |
| 0 | 0 | 0 | 0 | 0 |
| 0 | 0 | 0 | 0 | 0 |

| | | | | |
|---|---|---|---|---|
| 0 | 0 | 0 | 0 | 0 |
| 0 | 0 | 0 | 0 | 0 |
| 0 | 0 | 0 | 0 | 0 |
| 0 | 0 | 0 | 0 | 0 |
| 0 | 0 | 0 | 0 | 0 |
| 0.000758597 | 0 | 0 | 0 | 0 |
| 0 | 0 | 0 | 0 | 0 |
| 0 | 0 | 0 | 0 | 0 |
| 0 | 0 | 0 | 0 | 0 |
| 0 | 0 | 0 | 0 | 0 |
| 0 | 0 | 0 | 0 | 0 |
| 0 | 0 | 0 | 0 | 0 |
| 0 | 0 | 0 | 0 | 0 |
| 0 | 0 | 0 | 0 | 0 |
| 0 | 0 | 0 | 0 | 0 |
| 0 | 0 | 0 | 0 | 0 |
| 0 | 0 | 0 | 0 | 0 |
| 0 | 0 | 0 | 0 | 0 |
| 0 | 0 | 0 | 0 | 0 |
| 0 | 0 | 0 | 0 | 0 |
| 0.032334365 | 0.118173666 | 0.105201532 | 0.179856337 | 0.040150582 |
| 0 | 0 | 0 | 0 | 0 |
| 0 | 0 | 0 | 0.001075996 | 0 |
| 0 | 0 | 0 | 0 | 0.009851525 |
| 0 | 0 | 0 | 0 | 0 |
| 0 | 0 | 0 | 0 | 0 |
| 0.004955466 | 0.006799735 | 0 | 0.015473559 | 0.010545163 |
| 0 | 0 | 0 | 0 | 0 |
| 0 | 0 | 0 | 0 | 0 |
| 0 | 0 | 0 | 0 | 0 |
| 0 | 0 | 0.433453949 | 0.196009938 | 0 |
| 0 | 0 | 0 | 0 | 0 |
| 0 | 0 | 0 | 0 | 0 |
| 0 | 0 | 0 | 0 | 0 |
| 0.017993421 | 0.016575108 | 0.024532866 | 0.030053921 | 0.002305068 |
| 0 | 0 | 0 | 0 | 0 |
| 0 | 0 | 0 | 0 | 0 |
| 0 | 0 | 0 | 0 | 0 |
| 0 | 0 | 0 | 0 | 0 |
| 0 | 0 | 0 | 0 | 0 |
| 0 | 0 | 0 | 0 | 0 |
| 0 | 0 | 0 | 0 | 0 |
| 0 | 0 | 0 | 0 | 0 |
| 0 | 0 | 0 | 0 | 0 |
| 0 | 0 | 0 | 0 | 0 |
| 0 | 0 | 0 | 0 | 0 |
| 0.001498622 | 0 | 0 | 0 | 0 |
| 0 | 0 | 0 | 0.002274397 | 0 |

| | | | | |
|---|---|---|---|---|
| 0 | 0 | 0 | 0 | 0 |
| 0 | 0 | 0 | 0 | 0 |
| 0 | 0.003897631 | 0 | 0.002424679 | 0 |
| 0 | 0 | 0 | 0 | 0 |
| 0 | 0 | 0 | 0 | 0 |
| 0 | 0 | 0 | 0 | 0 |
| 0 | 0 | 0 | 0 | 0 |
| 0 | 0 | 0 | 0 | 0 |
| 0 | 0.000646557 | 0 | 0 | 0 |
| 0 | 0 | 0 | 0 | 0 |
| 0 | 0 | 0 | 0 | 0 |
| 0 | 0 | 0 | 0 | 0 |
| 0 | 0 | 0 | 0 | 0 |
| 0 | 0 | 0 | 0 | 0 |
| 0 | 0 | 0 | 0 | 0 |
| 0 | 0 | 0 | 0 | 0 |
| 0 | 0 | 0 | 0 | 0 |
| 0 | 0 | 0 | 0 | 0 |
| 0 | 0 | 0 | 0.003084062 | 0.002041806 |
| 0 | 0 | 0 | 0 | 0 |
| 0 | 0 | 0.00194275 | 0 | 0.016154582 |
| 0 | 0 | 0 | 0 | 0 |
| 0 | 0 | 0 | 0 | 0 |
| 0 | 0 | 0 | 0 | 0 |
| 0.010583031 | 0 | 0 | 0.027114471 | 0 |
| 0 | 0 | 0 | 0 | 0 |
| 0 | 0 | 0 | 0 | 0 |
| 0 | 0 | 0 | 0 | 0 |
| 0 | 0 | 0 | 0 | 0 |
| 0 | 0 | 0 | 0 | 0 |
| 0.120117476 | 0.001447707 | 0.029284318 | 0.005435792 | 0.391183792 |
| 0 | 0 | 0 | 0 | 0 |
| 0.034362855 | 0.038477347 | 0 | 0 | 0.027286821 |
| 0 | 0 | 0.007084628 | 0 | 0 |
| 0 | 0 | 0 | 0 | 0 |
| 0 | 0 | 0 | 0 | 0 |
| 0 | 0 | 0 | 0 | 0 |
| 0 | 0 | 0 | 0 | 0 |
| 0 | 0 | 0 | 0 | 0 |
| 0 | 0 | 0 | 0.001252985 | 0 |
| 0 | 0 | 0 | 0 | 0 |
| 0.050572958 | 0.014203577 | 0 | 0.003622887 | 0.009645446 |
| 0 | 0 | 0 | 0 | 0 |
| 0 | 0 | 0 | 0 | 0 |
| 0 | 0 | 0 | 0 | 0 |
| 0 | 0 | 0 | 0 | 0 |
| 0 | 0 | 0 | 0 | 0 |
| 0 | 0 | 0 | 0 | 0 |

| | | | | |
|---|---|---|---|---|
| 0 | 0 | 0 | 0 | 0 |
| 0 | 0 | 0 | 0 | 0 |
| 0.088290994 | 0.006022184 | 0.019198203 | 0.181162432 | 0.010822993 |
| 0 | 0 | 0 | 0 | 0 |
| 0 | 0 | 0 | 0 | 0 |
| 0 | 0 | 0 | 0 | 0 |
| 0 | 0 | 0 | 0 | 0 |
| 0 | 0 | 0 | 0 | 0 |
| 0 | 0 | 0 | 0 | 0 |
| 0 | 0 | 0 | 0 | 0 |
| 0 | 0 | 0 | 0 | 0 |
| 0 | 0 | 0 | 0 | 0 |
| 0 | 0 | 0 | 0 | 0 |
| 0.006464124 | 0 | 0.003367854 | 0.201695908 | 0 |
| 0.006097472 | 0.002957945 | 0.002779723 | 0.046866458 | 0.000940242 |
| 0 | 0 | 0 | 0 | 0 |
| 0 | 0.004092523 | 0.002217071 | 0.007509393 | 0 |
| 0 | 0 | 0 | 0 | 0 |
| 0 | 0 | 0 | 0 | 0 |
| 0 | 0 | 0 | 0 | 0 |
| 0 | 0 | 0 | 0 | 0 |
| 0 | 0 | 0 | 0.001757477 | 0 |
| 0 | 0 | 0 | 0 | 0 |
| 0 | 0 | 0 | 0 | 0 |
| 0 | 0 | 0 | 0.005233449 | 0 |
| 0 | 0 | 0 | 0 | 0 |
| 0 | 0 | 0 | 0 | 0 |
| 0 | 0 | 0 | 0 | 0 |
| 0 | 0 | 0 | 0 | 0 |
| 0 | 0 | 0 | 0.023969373 | 0 |
| 0 | 0 | 0 | 0.232416159 | 0 |
| 0 | 0 | 0 | 0 | 0 |
| 0 | 0 | 0 | 0 | 0 |
| 0 | 0 | 0 | 0 | 0 |
| 0 | 0 | 0 | 0 | 0 |
| 0 | 0 | 0 | 0 | 0 |
| 0 | 0 | 0 | 0 | 0 |
| 0.001641595 | 0 | 0 | 0.132800693 | 0.002126351 |
| 0 | 0 | 0 | 0 | 0 |
| 0 | 0 | 0 | 0 | 0 |
| 0 | 0 | 0 | 0 | 0 |
| 0 | 0 | 0 | 0 | 0 |
| 0 | 0 | 0 | 0 | 0 |
| 0 | 0 | 0 | 0 | 0 |
| 0 | 0 | 0 | 0 | 0 |
| 0 | 0 | 0 | 0.035403955 | 0 |
| 0 | 0 | 0 | 0.00205156 | 0 |
| 0.00119303 | 0 | 0 | 0 | 0 |

| | | | | |
|---|---|---|---|---|
| 0.016778743 | 0 | 0 | 0 | 0 |
| 0 | 0 | 0.001935237 | 0 | 0 |
| 0 | 0 | 0 | 0 | 0 |
| 0 | 0 | 0 | 0 | 0 |
| 0.001652273 | 0 | 0 | 0.013493451 | 0.002547836 |
| 0 | 0 | 0 | 0.034081987 | 0 |
| 0 | 0 | 0 | 0 | 0 |
| 0.060216628 | 0.135144205 | 0.016048643 | 0.003708636 | 0.090712363 |
| 0 | 0 | 0 | 0 | 0 |
| 0.638967509 | 15.1892261 | 0.121015476 | 0.906713841 | 1.553043749 |
| 0 | 0 | 0 | 0 | 0 |
| 0 | 0 | 0 | 0 | 0 |
| 0.000695339 | 0.002313025 | 0.007607809 | 0.050995669 | 0.002358898 |
| 0 | 0 | 0 | 0 | 0.00325687 |
| 0 | 0 | 0 | 0 | 0.004411041 |
| 0 | 0 | 0 | 0 | 0.011589044 |
| 0 | 0 | 0 | 0 | 0 |
| 0 | 0.001004138 | 0 | 0 | 0 |
| 0 | 0 | 0.218735152 | 0.084528786 | 0 |
| 0 | 0.023209082 | 0 | 0 | 0 |
| 0 | 0 | 0 | 0 | 0 |
| 0 | 0 | 0 | 0 | 0 |
| 0 | 0 | 0 | 0 | 0 |
| 0 | 0 | 0 | 0 | 0 |
| 0 | 0 | 0 | 0 | 0 |
| 0.001619786 | 0.000673521 | 0 | 0.002334378 | 0.000999096 |
| 0 | 0 | 0 | 0 | 0 |
| 0.039175428 | 0 | 0.017502153 | 0 | 0 |
| 0 | 0 | 0.033568427 | 0 | 0 |
| 0 | 0 | 0 | 0 | 0 |
| 0 | 0.008431787 | 0 | 0.004174858 | 0.00241219 |
| 0 | 0 | 0 | 0 | 0 |
| 0 | 0 | 0 | 0 | 0 |
| 0 | 0 | 0.038242931 | 0.00644509 | 0 |
| 0 | 0 | 0 | 0 | 0 |
| 0 | 0 | 0 | 0 | 0 |
| 0 | 0.014506595 | 0.043863301 | 0 | 0 |
| 0 | 0 | 0 | 0 | 0 |
| 0 | 0 | 0 | 0 | 0 |
| 0 | 0.000944496 | 0 | 0.865855999 | 0 |
| 0 | 0 | 0 | 0 | 0 |
| 0 | 0 | 0 | 0 | 0 |
| 0 | 0 | 0 | 0 | 0 |
| 0 | 0 | 0 | 0 | 0 |
| 0 | 0 | 0 | 0 | 0 |
| 0 | 0 | 0 | 0 | 0 |
| 0.002350054 | 0.000837577 | 0 | 0 | 0.00248491 |
| 0 | 0 | 0 | 0 | 0 |

| | | | | |
|---|---|---|---|---|
| 0 | 0 | 0 | 0 | 0 |
| 0.001500771 | 0 | 0 | 0.002162858 | 0.197171313 |
| 0 | 0.154526219 | 0 | 0 | 0 |
| 0 | 0 | 0 | 4.602804519 | 0 |
| 0 | 0 | 0 | 0 | 0 |
| 0 | 0 | 0.051207462 | 0 | 0.045259501 |
| 0 | 0 | 0 | 0 | 0.004896591 |
| 0 | 0 | 0 | 0 | 0 |
| 0 | 0 | 0 | 0 | 0 |
| 0 | 0 | 0 | 0 | 0 |
| 0.002760162 | 0.017598054 | 0 | 0.028581572 | 0.013336161 |
| 0.002874334 | 0 | 0 | 0 | 0 |
| 0 | 0 | 0 | 0 | 0 |
| 0.001158961 | 0 | 0.0069742 | 0.009186395 | 0.003752993 |
| 0 | 0.009264443 | 0.021169911 | 3.99120821 | 0 |
| 0 | 0 | 0 | 0 | 0 |
| 0 | 0 | 0 | 0 | 0 |
| 0 | 0 | 0 | 0 | 0 |
| 0 | 0 | 0 | 0 | 0 |
| 0 | 0 | 0.003421613 | 0 | 0 |
| 0 | 0 | 0 | 0 | 0 |
| 0.867111585 | 0 | 0 | 0 | 0.324602192 |
| 0 | 0 | 0 | 0 | 0.002543865 |
| 0 | 0 | 0 | 0 | 0 |
| 0 | 0 | 0 | 0 | 0 |
| 0 | 0 | 0 | 0 | 0 |
| 0 | 0.027431968 | 0.272915093 | 0 | 0 |
| 0 | 0 | 0 | 0 | 0 |
| 0 | 0 | 0 | 0 | 0 |
| 0.921595782 | 0.377203312 | 1.439079653 | 1.026221384 | 0.609443533 |
| 0.006093538 | 0.005721361 | 0.003656113 | 0 | 0 |
| 0 | 0 | 0 | 0 | 0 |
| 0 | 0 | 0 | 0 | 0 |
| 0 | 0 | 0 | 0 | 0 |
| 0 | 0 | 0 | 0 | 0.005810215 |
| 0 | 0 | 0 | 0 | 0 |
| 0 | 0 | 0 | 0 | 0 |
| 0 | 0 | 0 | 0 | 0 |
| 0 | 0.001799892 | 0 | 0 | 0.117000856 |
| 0 | 0.040485486 | 0.104278601 | 0.003623285 | 0 |
| 0.002143782 | 0 | 0.096688522 | 0.020596952 | 0.003746519 |
| 0.629535838 | 0 | 3.677925229 | 5.279042892 | 5.649213063 |
| 0 | 0 | 0 | 0 | 0 |
| 0 | 0 | 0 | 0 | 0 |
| 0 | 0 | 0 | 0 | 0 |
| 0 | 0 | 0 | 0 | 0 |
| 0.013000307 | 0.004781912 | 0 | 0.004563796 | 0.006245324 |
| 0 | 0 | 0 | 0 | 0 |

| | | | | |
|---|---|---|---|---|
| 0 | 0 | 0 | 0 | 0 |
| 0.001576802 | 0 | 0 | 0 | 0 |
| 0 | 0 | 0.848450577 | 0.02180637 | 0 |
| 0 | 0 | 0 | 0 | 0 |
| 0 | 0 | 0 | 0 | 0 |
| 0.251940247 | 0 | 0 | 0 | 0.030886687 |
| 0 | 0 | 0 | 0 | 0 |
| 0 | 0 | 0 | 0 | 0 |
| 0 | 0 | 0 | 0 | 0 |
| 0 | 0 | 0.007947583 | 0 | 0 |
| 0 | 0 | 0 | 0 | 0 |
| 0 | 0 | 0 | 0 | 0 |
| 0 | 0 | 0 | 0 | 0 |
| 0.002232486 | 0.001856574 | 0 | 0.015818787 | 0.021687977 |
| 0 | 0 | 0 | 0 | 0 |
| 0.01264871 | 0.003264482 | 0.004772117 | 0.000838109 | 0.015334611 |
| 0.001450322 | 0.013066219 | 0.010314319 | 0.103114175 | 0.004025562 |
| 0 | 0 | 0 | 0 | 0.00081515 |
| 0 | 0 | 0 | 0 | 0.00900922 |
| 0 | 0.360841463 | 0 | 0 | 0 |
| 0.003655636 | 0 | 0 | 0 | 0.014092648 |
| 0 | 0.00186537 | 0 | 0.001616312 | 0.00077824 |
| 0.000948681 | 0 | 0 | 0 | 0 |
| 0.001867802 | 0.003550391 | 0 | 0 | 0 |
| 0 | 0 | 0.012358626 | 0 | 0 |
| 0 | 0 | 0.006619453 | 0 | 0 |
| 0 | 0 | 0.003357464 | 0.002653465 | 0 |
| 0 | 0 | 0.000849899 | 0 | 0 |
| 0 | 0 | 0 | 0 | 0 |
| 0 | 0 | 0 | 0 | 0 |
| 0.002100335 | 0 | 0.001532005 | 0 | 0.001943254 |
| 0 | 0 | 0.002284032 | 0.039779317 | 0.003219059 |
| 0.442250083 | 0.00303042 | 0.019572365 | 0 | 2.500410353 |
| 0 | 0 | 0 | 0 | 0 |
| 0 | 0 | 0.010380702 | 0 | 0 |
| 0 | 0 | 0 | 0 | 0 |
| 0 | 0.003735966 | 0 | 0.001726481 | 0 |
| 0.001639528 | 0.00077912 | 0.005637756 | 0.012826787 | 0.02232024 |
| 0 | 0 | 0 | 0 | 0.090954217 |
| 0 | 0 | 0 | 0 | 0.006378318 |
| 0.003344624 | 0 | 0 | 0 | 0 |
| 0 | 0 | 0 | 0 | 0 |
| 0.001050819 | 0 | 0.00344915 | 0 | 0.001296306 |
| 0.002011198 | 0 | 0.002600569 | 0 | 0.055654337 |
| 0 | 0 | 0 | 0 | 0.785211305 |
| 0 | 0 | 0.001955423 | 0 | 0 |
| 0 | 0 | 0 | 0 | 0 |
| 0 | 0 | 0 | 0 | 0 |

| | | | | |
|---|---|---|---|---|
| 0 | 0 | 0.001899755 | 0 | 0 |
| 0.090000879 | 0.011412731 | 0.04085511 | 5.524313847 | 0.511430309 |
| 0 | 0 | 0 | 0 | 0 |
| 0 | 0 | 0 | 0 | 0 |
| 0 | 0 | 0 | 0 | 0 |
| 0 | 0 | 0 | 0 | 0 |
| 0 | 0 | 0.441279864 | 0.183254806 | 0 |
| 0 | 0 | 0 | 0 | 0 |
| 0 | 0 | 0 | 0 | 0 |
| 0 | 0 | 0 | 0 | 0 |
| 0.008063606 | 0 | 0 | 0 | 0.006143973 |
| 0.002119583 | 0.148359042 | 1.008407584 | 0.053286982 | 0.012093216 |
| 0 | 0 | 0 | 0 | 0 |
| 0 | 0 | 0 | 0 | 0.004814258 |
| 0 | 0.047748715 | 0.002084091 | 0.000784331 | 0 |
| 0 | 0 | 0 | 0 | 0 |
| 0.011175563 | 0 | 0.007608128 | 0.064900515 | 0.001148862 |
| 0.014829812 | 0.008847392 | 0.220455391 | 0.014557876 | 0.017300026 |
| 0 | 0 | 0.023616374 | 0 | 0 |
| 0 | 0.000522276 | 0.010765046 | 0.05490857 | 0 |
| 0.005890466 | 0.00532458 | 0 | 0.05807058 | 0.04810152 |
| 0 | 0 | 0 | 0 | 0 |
| 0 | 0 | 0 | 0 | 0 |
| 0 | 0 | 0 | 0 | 0 |
| 0 | 0 | 0 | 0.004134297 | 0.012341898 |
| 0 | 0 | 0 | 0 | 0 |
| 0.006066975 | 0 | 0.053380319 | 0 | 0.057067879 |
| 0.114053549 | 0.127672506 | 0.146230901 | 0.111672177 | 0.010325702 |
| 0 | 0 | 0 | 0 | 0 |
| 0 | 0 | 0 | 0.00094639 | 0 |
| 0 | 0.00995869 | 0 | 0 | 0.010809255 |
| 0 | 0 | 0 | 0 | 0 |
| 0 | 0 | 0 | 0 | 0 |
| 0 | 0 | 0 | 0 | 0 |
| 0.070450475 | 0 | 0 | 0 | 0.030286389 |
| 0.515511355 | 0.969286515 | 0.039315666 | 0.173506817 | 0.294102131 |
| 0 | 0 | 0 | 0 | 0 |
| 0 | 0 | 0 | 0 | 0 |
| 0 | 0 | 0.006613155 | 0.093560738 | 0.001242723 |
| 0 | 0 | 0 | 0 | 0 |
| 0 | 0.001401858 | 0 | 0 | 0.719769195 |
| 0 | 0 | 0 | 0 | 0 |
| 0.006118998 | 0.013400149 | 0.00312428 | 0.004899157 | 0 |
| 0.002756963 | 0.021453474 | 0.022623251 | 0 | 0.005101546 |
| 0 | 0 | 0 | 0 | 0.003111758 |
| 0 | 0 | 0 | 0.025412974 | 0 |
| 0 | 0 | 0 | 0 | 0 |
| 0 | 0.002117733 | 0 | 0.00122332 | 0 |

| | | | | |
|---|---|---|---|---|
| 0 | 0 | 0 | 0.061871824 | 0 |
| 0 | 0 | 0 | 0 | 0 |
| 0 | 0 | 0 | 0 | 0 |
| 0 | 0 | 0 | 0.102479179 | 0 |
| 0 | 0 | 0 | 0 | 0 |
| 0 | 0 | 0 | 0 | 0 |
| 0 | 0 | 0 | 0 | 0 |
| 0.004250396 | 0 | 0 | 0 | 0.013960426 |
| 0.715478481 | 0 | 0.314361222 | 0.337175869 | 0.065271213 |
| 0.007855733 | 0.005120431 | 0 | 0 | 0 |
| 0 | 0 | 0 | 0 | 0 |
| 0.017525561 | 0 | 0 | 0 | 0.016846595 |
| 0 | 0 | 0 | 0 | 0 |
| 0 | 0 | 0 | 0 | 0 |
| 0 | 0.001405749 | 0.015823238 | 0.053414086 | 0.024675788 |
| 0 | 0 | 0 | 0 | 0 |
| 0.047038981 | 0 | 0 | 0 | 0 |
| 0 | 0 | 0 | 0 | 0 |
| 0 | 0 | 0 | 0 | 0 |
| 0 | 0 | 0 | 0 | 0 |
| 0 | 0 | 0 | 0 | 0 |
| 0 | 0 | 0 | 0 | 0 |
| 0 | 0 | 0 | 0 | 0 |
| 0.010855126 | 0.006620028 | 0 | 0 | 0.19366802 |
| 0.004475019 | 0 | 0 | 0 | 0 |
| 0 | 0 | 0 | 0.007385128 | 0 |
| 0 | 0 | 0 | 0 | 0 |
| 0 | 0.170166696 | 0 | 0 | 0 |
| 0 | 0 | 0 | 0 | 0 |
| 0 | 0 | 0 | 0 | 0 |
| 0 | 0 | 0 | 0 | 0 |
| 0 | 0 | 0 | 0 | 0 |
| 0.002189241 | 0 | 0 | 0 | 0.005920723 |
| 0 | 0 | 0 | 0 | 0 |
| 0 | 0 | 0 | 0 | 0 |
| 0 | 0 | 0 | 0 | 0.030203161 |
| 0.147562415 | 0 | 0 | 0 | 0 |
| 0 | 0 | 0 | 0 | 0 |
| 0 | 0 | 0.002333122 | 0 | 0 |
| 0 | 0 | 0 | 0 | 0.001959481 |
| 0 | 0.063553111 | 0 | 0.363761462 | 0.027676219 |
| 0 | 0 | 0 | 0 | 0 |
| 31.90080132 | 0.487934448 | 23.97566738 | 0 | 7.8790631 |
| 0 | 0 | 0 | 0 | 0 |
| 0 | 0 | 0 | 0 | 0 |
| 0 | 0 | 0 | 0 | 0 |
| 0 | 0 | 0 | 0 | 0 |
| 0 | 0 | 0 | 0.00151309 | 0 |

| | | | | |
|---|---|---|---|---|
| 0 | 0 | 0 | 0 | 0 |
| 0 | 0 | 0 | 0.193215243 | 0 |
| 0.046296938 | 0.066749594 | 0.033310619 | 0.144321524 | 0.888582056 |
| 0 | 0 | 0 | 0 | 0 |
| 0 | 0 | 0 | 0 | 0 |
| 0.018631895 | 0 | 0.001951797 | 0 | 0.010819872 |
| 0 | 0 | 0 | 0 | 0 |
| 0 | 0 | 0.00131864 | 0 | 0 |
| 0 | 0 | 0 | 0 | 0 |
| 0 | 0 | 0 | 0 | 0 |
| 0.095357489 | 0 | 0.012973932 | 0.290991497 | 0.011428216 |
| 3.061240362 | 0.757446635 | 2.436507165 | 1.940399412 | 0.224417313 |
| 0 | 0 | 0 | 0 | 0 |
| 0.0050657 | 0.001330334 | 0.011376622 | 0.055330159 | 0.009127009 |
| 0.006933414 | 0.340889767 | 0.160914107 | 0.621332595 | 0.108664062 |
| 0 | 0 | 0 | 0 | 0 |
| 0 | 0 | 0.001493875 | 0 | 0 |
| 0 | 0 | 0 | 0 | 0 |
| 0 | 0 | 0 | 0 | 0 |
| 0 | 0 | 0.001781184 | 0 | 0.000941384 |
| 0.013044528 | 0.011881203 | 0.231526982 | 2.914788637 | 0.086973033 |
| 0 | 7.654251615 | 1.559466797 | 4.587010965 | 0 |
| 7.750807061 | 12.29094719 | 3.753128282 | 9.181095455 | 7.700935514 |
| 0 | 0 | 0 | 0 | 0 |
| 0 | 0 | 0 | 0 | 0 |
| 0 | 0.001238021 | 0 | 0 | 0.012166626 |
| 0.260278707 | 0 | 0 | 0 | 0 |
| 0 | 0 | 0 | 0 | 0 |
| 0 | 0 | 0 | 0 | 0 |
| 0 | 0 | 0 | 0 | 0 |
| 0 | 0 | 0 | 0 | 0 |
| 0.034403906 | 0 | 0 | 0 | 0.172718713 |
| 0 | 0 | 0 | 0.571872609 | 0.495774067 |
| 0.024958738 | 0 | 0 | 0 | 0.006802326 |
| 0.001172193 | 0 | 0.001099297 | 0.005309301 | 0.029592071 |
| 0.170255125 | 0.00880136 | 0.009062205 | 0 | 0.417007477 |
| 0 | 0 | 0.001215829 | 0.002775908 | 0.000616881 |
| 0 | 0 | 0 | 0 | 0 |
| 0 | 0 | 0 | 0 | 0 |
| 0 | 0 | 0 | 0 | 0 |
| 0 | 0 | 0 | 0 | 0 |
| 0 | 0 | 0 | 0 | 0 |
| 0 | 0 | 0 | 0 | 0 |
| 0 | 0 | 0 | 0 | 0 |
| 0.009970206 | 0 | 0.002870671 | 0.21653895 | 0 |
| 0 | 0 | 0 | 0 | 0 |
| 0 | 0 | 0 | 0 | 0 |
| 0.239902872 | 0.006580058 | 0.031165323 | 0.288572393 | 0.041873881 |

| | | | | |
|---|---|---|---|---|
| 0 | 0 | 0 | 0 | 0 |
| 0 | 0 | 0 | 0 | 0 |
| 0 | 0 | 0 | 0 | 0 |
| 0.008944806 | 0 | 0 | 0 | 0 |
| 0 | 0 | 0 | 0 | 0 |
| 0.001974999 | 0 | 0 | 0 | 0 |
| 0 | 0 | 0 | 0 | 0 |
| 0 | 0.003080209 | 0 | 0.507495766 | 0.009138321 |
| 0 | 0 | 0 | 0 | 0 |
| 0 | 0 | 0 | 0 | 0 |
| 0 | 0 | 0 | 0 | 0.001152136 |
| 0 | 0 | 0 | 0 | 0 |
| 0 | 0 | 0 | 0 | 0 |
| 1.094076755 | 0 | 0 | 0 | 0 |
| 0 | 0 | 0 | 0 | 0 |
| 0 | 0 | 0 | 0 | 0 |
| 0 | 0 | 0 | 0 | 0 |
| 0.013683878 | 0 | 0 | 0 | 0.010934049 |
| 0 | 0 | 0 | 0 | 0 |
| 0 | 0 | 0 | 0 | 0 |
| 0 | 0 | 0 | 0 | 0 |
| 0 | 0 | 0 | 0 | 0 |
| 0 | 0 | 0 | 0 | 0 |
| 0 | 0 | 0 | 0 | 0 |
| 0 | 0 | 0 | 0 | 0 |
| 0 | 0 | 0 | 0 | 0 |
| 0 | 0 | 0 | 0 | 0 |
| 0 | 0 | 0 | 0 | 0 |
| 0 | 0 | 0 | 0 | 0 |
| 0.013023638 | 5.516774597 | 0.120571462 | 0 | 0.045881234 |
| 0 | 0 | 0 | 0 | 0 |
| 0 | 0 | 0 | 0 | 0 |
| 0 | 0 | 0 | 0 | 0 |
| 0 | 0 | 0 | 0 | 0 |
| 0 | 0 | 0 | 0 | 0 |
| 0 | 0 | 0 | 0 | 0 |
| 0.026752693 | 0 | 0 | 0 | 0 |
| 0.002108283 | 0.006311824 | 0 | 0 | 0.003120971 |
| 0 | 0 | 0 | 0 | 0 |
| 0 | 0 | 0.001137602 | 0 | 0 |
| 0 | 0 | 0 | 0.007221912 | 0 |
| 0 | 0 | 0 | 0 | 0 |
| 0 | 0 | 0 | 0 | 0 |
| 0.365664264 | 0 | 0 | 0.005114424 | 0 |
| 0.100066408 | 0 | 0 | 0 | 0 |
| 0 | 0 | 0 | 0 | 0 |
| 0 | 0 | 0 | 0 | 0 |
| 0 | 0 | 0 | 0 | 0 |

| | | | | |
|---|---|---|---|---|
| 0 | 0 | 0 | 0 | 0 |
| 0 | 0 | 0 | 0 | 0 |
| 0 | 0 | 0 | 0 | 0 |
| 0 | 0 | 0 | 0 | 0 |
| 0 | 0 | 0 | 0 | 0 |
| 0 | 0 | 0.362843401 | 0 | 0 |
| 0 | 0 | 0 | 0 | 0 |
| 0 | 0 | 0 | 0 | 0 |
| 0.048558155 | 0.079554565 | 0.063626581 | 0.441672038 | 0.016141277 |
| 0 | 0 | 0 | 0 | 0 |
| 0 | 0 | 0 | 0 | 0 |
| 0 | 0 | 0 | 0 | 0 |
| 0 | 0 | 0 | 0 | 0 |
| 0 | 0 | 0 | 0 | 0 |
| 0.154834266 | 0 | 0 | 0 | 0 |
| 0 | 0 | 0 | 0 | 0 |
| 0 | 0 | 0 | 0 | 0 |
| 0 | 0 | 0 | 0 | 0 |
| 0 | 0 | 0 | 0 | 0 |
| 0 | 0 | 0 | 0 | 0 |
| 0 | 0 | 0 | 0 | 0 |
| 0 | 0 | 0 | 0 | 0 |
| 0 | 0 | 0 | 0 | 0 |
| 0 | 0 | 0 | 0 | 0 |
| 0 | 0.000853075 | 0 | 0.057162878 | 0.066435887 |
| 0.474084494 | 0.052531844 | 0 | 0.139031032 | 0.271645419 |
| 0 | 0 | 0 | 0.010508412 | 0 |
| 0 | 0 | 0 | 0 | 0 |
| 0 | 0 | 0 | 0 | 0 |
| 0 | 0 | 0 | 0 | 0 |
| 0 | 0 | 0 | 0 | 0 |
| 0.005505188 | 0 | 0 | 0 | 0 |
| 0 | 0 | 0 | 0 | 0 |
| 0 | 0 | 0 | 0 | 0.001351658 |
| 0 | 0.004754305 | 0 | 0.045676136 | 0.002227106 |
| 0 | 0 | 0 | 0.004087024 | 0.001967867 |
| 0.106454659 | 0.001095211 | 0 | 0 | 0.112302389 |
| 1.348783991 | 0.828072002 | 0 | 4.688103708 | 1.343458875 |
| 0 | 0.004021041 | 0 | 0 | 0 |
| 0 | 0 | 0 | 0 | 0 |
| 0 | 0 | 0 | 0 | 0 |
| 0 | 0 | 0 | 0 | 0 |
| 0.619374934 | 0 | 0 | 0 | 0.1860875 |
| 0 | 0 | 0 | 0 | 0 |
| 0 | 0 | 0 | 0 | 0 |
| 0.019344067 | 0 | 0 | 0 | 0 |
| 0 | 0 | 0 | 0 | 0 |
| 0.321977811 | 0 | 0.115553683 | 9.105584384 | 0.028512623 |

| | | | | |
|---|---|---|---|---|
| 0 | 0 | 0.003404835 | 0 | 0 |
| 0 | 0 | 0 | 0 | 0 |
| 0 | 0 | 0 | 0 | 0 |
| 0 | 0 | 0 | 0 | 0 |
| 0.001668321 | 0 | 0 | 0.005724584 | 0.022050708 |
| 0 | 0 | 0 | 0 | 0 |
| 0 | 0 | 0 | 0 | 0 |
| 0 | 0 | 0 | 0 | 0 |
| 0 | 0 | 0 | 0 | 0 |
| 0 | 0 | 0 | 0 | 0 |
| 0 | 0 | 0 | 0 | 0 |
| 0.496700093 | 0.026330797 | 0 | 4.315450958 | 0.04963737 |
| 1.318800352 | 0 | 0.000947212 | 0 | 0 |
| 0 | 0 | 0 | 0 | 0 |
| 0.051818527 | 0.002292191 | 0.002111 | 0.676613171 | 0.062479023 |
| 0 | 0 | 0 | 0 | 0 |
| 0.000958704 | 0 | 0 | 0 | 0 |
| 0 | 0 | 0.002617459 | 0 | 0 |
| 0.001932713 | 0 | 0 | 0 | 0 |
| 0 | 0 | 0 | 0 | 0 |
| 0 | 0 | 0 | 0 | 0 |
| 0 | 0 | 0 | 0 | 0 |
| 0.728229028 | 0 | 1.817654884 | 0.107658171 | 0 |
| 0 | 0 | 0 | 0 | 0 |
| 0 | 0 | 0 | 0 | 0 |
| 0 | 0 | 0 | 0 | 0 |
| 0 | 0 | 0 | 0 | 0 |
| 0 | 0 | 0 | 0 | 0 |
| 0 | 0 | 0 | 0 | 0 |
| 0 | 0 | 0 | 0 | 0 |
| 0 | 0 | 0 | 0 | 0 |
| 0.004649698 | 0.003271882 | 0.028567164 | 0.256011803 | 0.040372175 |
| 0 | 0 | 0 | 0 | 0 |
| 0 | 0 | 0.012121138 | 0 | 0.016366845 |
| 0 | 0.000769958 | 0 | 0.004447706 | 0.005996292 |
| 0.01491927 | 0.006699846 | 0.002611743 | 0 | 0.001932487 |
| 0 | 0 | 0 | 0 | 0 |
| 0 | 0 | 0 | 0 | 0 |
| 0.001861427 | 0 | 0 | 0 | 0 |
| 0 | 0 | 0 | 0 | 0 |
| 0 | 0 | 0 | 0 | 0 |
| 0 | 0 | 0 | 0 | 0 |
| 0 | 0 | 0 | 0 | 0 |
| 0 | 0 | 0 | 0 | 0 |
| 0.057231869 | 0.001706594 | 0 | 0.169342524 | 0.004430211 |
| 0 | 0 | 0 | 0 | 0 |
| 0.05928073 | 0 | 0 | 3.007952206 | 0 |
| 0.000862647 | 0 | 0 | 0 | 0 |

| | | | | |
|---|---|---|---|---|
| 0 | 0 | 0 | 0 | 0.001163378 |
| 0.003213984 | 0 | 0 | 0 | 0 |
| 0 | 0 | 0.004644353 | 0 | 0.010909398 |
| 0 | 0 | 0 | 0 | 0 |
| 0 | 0 | 0 | 0 | 0 |
| 0.006897848 | 0 | 0 | 0 | 0 |
| 0 | 0 | 0 | 0 | 0 |
| 0 | 0 | 0.037282337 | 0 | 0 |
| 0 | 0 | 0 | 0 | 0 |
| 0 | 0 | 0.00119893 | 0 | 0 |
| 0 | 0 | 0 | 0 | 0 |
| 0 | 0 | 0 | 0 | 0 |
| 0 | 0 | 0 | 0 | 0 |
| 0 | 0 | 0 | 0 | 0 |
| 0 | 0 | 0 | 0 | 0 |
| 0 | 0 | 0 | 0 | 0 |
| 0 | 0 | 0 | 0 | 0 |
| 0 | 0 | 0 | 0 | 0 |
| 0 | 0 | 0 | 0 | 0 |
| 0 | 0 | 0 | 0 | 0 |
| 0.008142189 | 0 | 0 | 0 | 0 |
| 0 | 0 | 0 | 0 | 0 |
| 0 | 0 | 0 | 0 | 0 |
| 0 | 0 | 0 | 0 | 0 |
| 0 | 0 | 0 | 0 | 0 |
| 0.032690155 | 0.00335408 | 0.01021908 | 0.003875004 | 0.009034311 |
| 0 | 0 | 0 | 0 | 0 |
| 0.039477015 | 0 | 0.396687959 | 0.013574441 | 0.015378771 |
| 0 | 0 | 0 | 0 | 0 |
| 0.32506262 | 0 | 0.204380439 | 0 | 0.308265928 |
| 0 | 0 | 0 | 0 | 0 |
| 0 | 0 | 0 | 0 | 0 |
| 0 | 0 | 0 | 0 | 0 |
| 0 | 0 | 0 | 0 | 0 |
| 0 | 0 | 0 | 0 | 0.003087195 |
| 0 | 0 | 0 | 0 | 0 |
| 0 | 0 | 0 | 0 | 0 |
| 0 | 0 | 0 | 0.004122749 | 0 |
| 0 | 0 | 0 | 0 | 0 |
| 0 | 0 | 0 | 0 | 0 |
| 15.22085314 | 16.7207288 | 19.61952537 | 6.415354302 | 14.43073999 |
| 0.042127365 | 0.014136463 | 3.229686598 | 4.937414735 | 0.230441275 |
| 0 | 0 | 0 | 0 | 0 |
| 0 | 0 | 0 | 0 | 0 |
| 0 | 0 | 0 | 0 | 0 |
| 0 | 0 | 0 | 0 | 0 |
| 0 | 0 | 0 | 0 | 0 |
| 0.046076072 | 0 | 0 | 0.005389246 | 0 |

| | | | | |
|---|---|---|---|---|
| 0 | 0 | 0 | 0 | 0 |
| 0 | 0 | 0 | 0 | 0.004003566 |
| 0 | 0 | 0 | 0 | 0 |
| 0 | 0 | 0 | 0 | 0 |
| 3.397206949 | 0 | 1.181737686 | 0 | 1.659440267 |
| 0 | 0 | 0 | 0 | 0 |
| 0 | 0 | 0 | 0 | 0 |
| 0 | 0 | 0 | 0 | 0 |
| 0.075099814 | 0.091029285 | 0.699439469 | 0.292570168 | 0.192268573 |
| 0 | 0 | 0 | 0 | 0 |
| 0 | 0.012777698 | 0 | 0 | 0 |
| 0 | 0 | 0 | 0 | 0 |
| 0.028431129 | 0 | 0 | 0 | 0 |
| 0.029045601 | 0.067961029 | 0.021006604 | 0.035474146 | 0.022318519 |
| 0.698776253 | 0 | 0 | 0 | 0 |
| 0.062212372 | 0 | 0 | 0 | 0.010278498 |
| 0 | 0 | 0 | 0.371642838 | 0.000985368 |
| 0 | 0 | 0 | 0 | 0 |
| 0 | 0 | 0 | 0 | 0 |
| 0 | 0 | 0 | 0 | 0 |
| 0 | 0 | 0 | 0 | 0 |
| 0 | 0 | 0 | 0 | 0 |
| 0 | 0 | 0 | 0 | 0 |
| 0 | 0 | 0 | 0 | 0 |
| 0 | 0 | 0 | 0 | 0 |
| 0 | 0 | 0 | 0 | 0 |
| 0.006482548 | 0 | 0 | 0.024023363 | 0 |
| 0 | 0 | 0 | 0 | 0 |
| 0 | 0 | 0 | 0 | 0 |
| 0 | 0 | 0 | 0 | 0 |
| 0 | 0 | 0 | 0 | 0 |
| 0 | 0 | 0 | 0 | 0 |
| 0 | 0 | 0 | 0 | 0 |
| 0 | 0 | 0 | 0 | 0 |
| 0.224913094 | 0 | 0 | 0 | 0.004075537 |
| 0 | 0 | 0 | 0.000696843 | 0.056368067 |
| 0 | 0 | 0 | 0 | 0 |
| 0 | 0 | 0 | 0.016450445 | 0 |
| 0 | 0 | 0 | 0 | 0 |
| 0 | 0 | 0 | 0 | 0 |
| 0 | 1.222093142 | 0 | 0 | 6.703651544 |
| 0 | 0 | 0 | 0 | 0 |
| 0 | 0 | 0 | 0 | 0 |
| 0 | 0 | 0 | 0 | 0 |
| 0 | 0 | 0 | 0 | 0 |
| 0 | 0 | 0 | 0 | 0 |
| 0 | 0 | 0 | 0 | 0 |
| 0 | 0 | 0 | 0 | 0 |

| | | | | |
|---|---|---|---|---|
| 0 | 0 | 0.009513924 | 0 | 0 |
| 0 | 0 | 0 | 0 | 0 |
| 0 | 0 | 0 | 0 | 0 |
| 0 | 0 | 0 | 0 | 0 |
| 0.055724889 | 0 | 0 | 0 | 0 |
| 0 | 0 | 0 | 0 | 0 |
| 0.000849944 | 0 | 0 | 0.379721827 | 0.001376161 |
| 0 | 0 | 0 | 0 | 0 |
| 0 | 0 | 0 | 0.011960136 | 0 |
| 0 | 0 | 0 | 0 | 0 |
| 0.003092089 | 0 | 0 | 0 | 0 |
| 0 | 0 | 0 | 0 | 0 |
| 0 | 0 | 0 | 0 | 0 |
| 0 | 0 | 0 | 0 | 0 |
| 0 | 0 | 0 | 0 | 0 |
| 0 | 0 | 0 | 0 | 0 |
| 0 | 0 | 0 | 0 | 0 |
| 0 | 0 | 0 | 0 | 0 |
| 0 | 0 | 0 | 0 | 0 |
| 0 | 0 | 0 | 0 | 0 |
| 0.010114925 | 0 | 0.268704063 | 0.186200352 | 0.006738079 |
| 0 | 0 | 0 | 0 | 0 |
| 0.009148516 | 0 | 0.001464809 | 0 | 0.00061934 |
| 0.001388895 | 0 | 0.00151961 | 0 | 0.001835744 |
| 0 | 0 | 0 | 0 | 0 |
| 0.0014045 | 0 | 0 | 0 | 0 |
| 0 | 0 | 0 | 0 | 0 |
| 0 | 0 | 0 | 0 | 0 |
| 0 | 0 | 0 | 0 | 0.007773183 |
| 0 | 0.540028658 | 0.79203376 | 0.016187403 | 0.006076753 |
| 0 | 0 | 0 | 0 | 0 |
| 0 | 0 | 0 | 0 | 0 |
| 0 | 0 | 0 | 0 | 0 |
| 0 | 0.027489456 | 0 | 0 | 7.019639206 |
| 0 | 0 | 0 | 0 | 0 |
| 0 | 0 | 0 | 0 | 0 |
| 0 | 0 | 0 | 0.050329579 | 0.002067293 |
| 0 | 0 | 0 | 0 | 0 |
| 0 | 0 | 0 | 0 | 0 |
| 0 | 0 | 0 | 0 | 0 |
| 0 | 0 | 0 | 0 | 0 |
| 0 | 0 | 0.008448867 | 0 | 0 |
| 0 | 0 | 0 | 0 | 0 |
| 0 | 0 | 0 | 0 | 0 |
| 0 | 0 | 0 | 0 | 0 |
| 0 | 0 | 0 | 0 | 0 |
| 0 | 0 | 0 | 0 | 0 |
| 0 | 0 | 0 | 0 | 0 |

| | | | | |
|---|---|---|---|---|
| 0.0040565 | 0 | 0.003503903 | 0.004717893 | 0.006518577 |
| 0 | 0 | 0 | 0 | 0 |
| 0 | 0 | 0 | 0 | 0 |
| 0 | 0 | 0 | 0 | 0 |
| 0 | 0 | 0 | 0 | 0 |
| 0 | 0 | 0 | 0 | 0 |
| 0 | 0 | 0 | 0 | 0 |
| 0.151478434 | 0.658482142 | 0 | 0 | 1.553003153 |
| 0 | 0.060197441 | 0 | 1.322032252 | 0.368657916 |
| 0.15491168 | 0 | 0.00918928 | 0.012328241 | 0.137066316 |
| 0 | 0 | 0 | 0 | 0 |
| 0 | 0 | 0 | 0 | 0 |
| 0 | 0 | 0 | 0 | 0 |
| 0 | 0 | 0 | 0 | 0 |
| 0 | 0 | 0 | 0 | 0 |
| 0 | 0 | 0 | 0 | 0 |
| 0 | 0 | 0.021140988 | 0 | 0 |
| 0 | 0 | 0 | 0 | 0 |
| 0 | 0 | 0 | 0 | 0 |
| 0 | 0 | 0.000634998 | 0 | 0 |
| 0 | 0 | 0 | 0 | 0 |
| 0 | 0 | 0 | 0 | 0 |
| 0 | 0 | 0 | 0 | 0 |
| 0 | 0 | 0 | 0 | 0 |
| 0 | 0 | 0 | 0 | 0 |
| 0 | 0 | 0 | 0 | 0 |
| 0 | 0 | 0 | 0 | 0 |
| 0 | 0 | 0 | 0 | 0 |
| 0 | 0 | 0 | 0 | 0 |
| 0 | 0 | 0 | 0 | 0 |
| 0 | 0 | 0 | 0 | 0 |
| 0 | 0 | 0 | 0 | 0 |
| 0 | 0 | 0 | 0 | 0 |
| 0 | 0 | 0 | 0 | 0 |
| 0.003360134 | 0 | 0 | 0 | 0.000956564 |
| 0 | 0 | 0 | 0 | 0 |
| 0 | 0 | 0 | 0 | 0 |
| 0 | 0 | 0.050363435 | 0 | 0 |
| 0 | 0 | 0 | 0 | 0 |
| 0 | 0 | 0 | 0.696660741 | 0 |
| 0 | 0 | 0 | 0 | 0 |
| 0.004617724 | 0 | 2.17418159 | 2.508580372 | 0.083006049 |
| 0 | 0 | 0 | 0 | 0 |
| 0 | 0 | 0 | 0 | 0 |
| 0 | 0 | 0 | 0 | 0 |
| 0 | 0 | 0 | 0 | 0 |
| 0 | 0 | 0 | 0 | 0 |
| 0 | 0 | 0 | 0 | 0 |

| | | | | |
|---|---|---|---|---|
| 0 | 0 | 0 | 0 | 0 |
| 0 | 0 | 0.032467568 | 1.138024727 | 0 |
| 0 | 0 | 0 | 0 | 0 |
| 0 | 0 | 0 | 0 | 0 |
| 0.475781665 | 0.151008154 | 0.060427378 | 0 | 0.023929254 |
| 0 | 0 | 0 | 0 | 0 |
| 0 | 0 | 0.013293305 | 0 | 0 |
| 0.003802225 | 0.007453276 | 0.019314611 | 0.036009007 | 0.001256379 |
| 0.011117971 | 0 | 0 | 0 | 0 |
| 0 | 0 | 0 | 0 | 0 |
| 0 | 0 | 0 | 0 | 0 |
| 0 | 0 | 0 | 0 | 0 |
| 0 | 0 | 0 | 0 | 0 |
| 0 | 0 | 0 | 0 | 0 |
| 0 | 0 | 0 | 0 | 0.001592004 |
| 0.000942183 | 0 | 0 | 0.004752444 | 0.020049526 |
| 0 | 0 | 0 | 0 | 0 |
| 0 | 0 | 0 | 0 | 0 |
| 0 | 0 | 0 | 0 | 0 |
| 0 | 0 | 0 | 0 | 0 |
| 0 | 0 | 0 | 0 | 0 |
| 0 | 0 | 0 | 0 | 0 |
| 0 | 0 | 0 | 0 | 0 |
| 0.00141022 | 0 | 0 | 0 | 0.000869834 |
| 0 | 0 | 0 | 0 | 0 |
| 0 | 0 | 0 | 0 | 0 |
| 0.007523867 | 0 | 0 | 0.022718942 | 0.012728991 |
| 0 | 0 | 0 | 0 | 0 |
| 0 | 0 | 0 | 0 | 0 |
| 0 | 0 | 0 | 0 | 0 |
| 0 | 0 | 0 | 0 | 0 |
| 0 | 0 | 0 | 0 | 0 |
| 0 | 0 | 0 | 0 | 0 |
| 0 | 0 | 0 | 0 | 0 |
| 0 | 0 | 0 | 0 | 0 |
| 0 | 0 | 0 | 0 | 0 |
| 0 | 0 | 0 | 0 | 0 |
| 0 | 0 | 0 | 0 | 0 |
| 0 | 0 | 0 | 0 | 0 |
| 0 | 0 | 0 | 0 | 0 |
| 0 | 0 | 0 | 0 | 0 |
| 0 | 0 | 0 | 0 | 0.027364516 |
| 0 | 0 | 0 | 0 | 0 |
| 0 | 0 | 0 | 0 | 0 |
| 0 | 0 | 0 | 0 | 0 |
| 0 | 0 | 0 | 0 | 0 |
| 0 | 0 | 0 | 0 | 0 |
| 0 | 0 | 0 | 0 | 0 |

| | | | | |
|---|---|---|---|---|
| 0.072777074 | 0.003530489 | 0.106610998 | 0 | 0.10305869 |
| 0 | 0 | 0 | 0 | 0 |
| 0 | 0 | 0 | 0 | 0 |
| 0 | 0 | 0 | 0 | 0 |
| 0 | 0 | 0 | 0 | 0 |
| 0 | 0.017334979 | 0 | 0.323586975 | 0 |
| 0 | 0 | 0 | 0 | 0 |
| 0 | 0 | 0 | 0 | 0 |
| 0 | 0 | 0 | 0 | 0 |
| 0 | 0 | 0 | 0 | 0 |
| 0 | 0 | 0 | 0 | 0 |
| 0.001083331 | 0 | 0 | 0 | 0 |
| 0 | 0 | 0 | 0 | 0 |
| 0 | 0 | 0 | 0 | 0 |
| 0 | 0 | 0 | 0 | 0 |
| 0 | 0 | 0 | 0 | 0 |
| 0 | 0 | 0 | 0 | 0 |
| 0 | 0 | 0 | 0 | 0 |
| 0 | 0 | 0 | 0 | 0 |
| 0 | 0 | 0 | 0 | 0 |
| 0.203158934 | 0 | 0.008706789 | 0.103441869 | 0.095065219 |
| 0 | 0 | 0 | 0 | 0 |
| 0.029263731 | 0.002458204 | 0.004527781 | 0.003691987 | 0.009298531 |
| 0 | 0 | 0 | 0 | 0 |
| 0 | 0 | 0 | 0 | 0 |
| 0 | 0 | 0 | 0 | 0 |
| 0 | 0 | 0 | 0 | 0 |
| 0 | 0 | 0 | 0 | 0 |
| 0 | 0 | 0.035556568 | 0 | 0 |
| 0 | 0 | 0 | 0 | 0 |
| 0 | 0 | 0 | 0 | 0 |
| 0 | 0 | 0 | 0 | 0 |
| 0 | 0 | 0 | 0.147875932 | 0.034576026 |
| 0 | 0 | 0 | 0 | 0 |
| 1.203660578 | 0 | 0.034677779 | 0 | 0 |
| 0 | 0 | 0.081905963 | 0.121225336 | 0.012742521 |
| 0 | 0 | 0 | 0 | 0 |
| 0 | 0 | 0 | 0 | 0 |
| 0.019527262 | 0 | 0.001967835 | 0.001728017 | 0.003565824 |
| 0 | 0 | 0 | 0 | 0 |
| 0 | 0 | 0 | 0 | 0 |
| 0 | 0 | 0 | 0 | 0 |
| 0.075710154 | 0.000749546 | 0.032871269 | 0.034927 | 0.006671228 |
| 0 | 0 | 0 | 0 | 0 |
| 0 | 0 | 0 | 0 | 0 |
| 0.001670194 | 0 | 0.025061269 | 0 | 0.014349053 |
| 0 | 0 | 0 | 0 | 0 |
| 0 | 0 | 0 | 0 | 0 |

| | | | | |
|---|---|---|---|---|
| 0 | 0 | 0 | 0 | 0 |
| 0 | 0 | 0 | 0 | 0 |
| 0 | 0 | 0 | 0 | 0 |
| 0 | 0 | 0 | 0 | 0 |
| 0 | 0 | 0 | 0 | 0 |
| 0 | 0 | 0 | 0 | 0 |
| 0 | 0 | 0 | 0 | 0 |
| 0 | 0.003643054 | 0.091066481 | 0 | 0 |
| 0.005141542 | 0 | 0 | 0 | 0 |
| 0 | 0 | 0 | 0.003069497 | 0 |
| 0 | 0 | 0 | 0 | 0 |
| 0 | 0 | 0 | 0 | 0 |
| 0.002993287 | 0 | 0 | 0 | 0 |
| 0 | 0 | 0 | 0 | 0 |
| 0 | 0 | 0 | 0 | 0 |
| 0 | 0.009651894 | 0.002645518 | 0 | 0 |
| 0 | 0 | 0 | 0 | 0 |
| 0 | 0 | 0 | 0 | 0 |
| 0 | 0 | 0 | 0 | 0 |
| 0.00303658 | 0 | 0 | 0.421088782 | 0 |
| 0 | 0.001050075 | 0 | 0 | 0.006542229 |
| 0 | 0 | 0 | 0 | 0 |
| 0 | 0 | 0 | 0 | 0 |
| 0 | 0 | 0 | 0 | 0 |
| 0 | 0 | 0 | 0 | 0 |
| 0 | 0 | 0 | 0 | 0 |
| 0 | 0 | 0 | 0 | 0 |
| 0 | 0 | 0 | 0 | 0 |
| 0 | 0 | 0 | 0 | 0 |
| 0.080693925 | 0 | 0 | 0 | 0 |
| 0 | 0 | 0.858575922 | 0 | 0.637120442 |
| 0 | 0 | 0 | 0 | 0 |
| 0 | 0 | 0 | 0 | 0 |
| 0 | 0 | 0 | 0 | 0 |
| 0 | 0 | 0 | 0 | 0 |
| 0 | 0 | 0 | 0 | 0 |
| 0 | 0 | 0 | 0 | 0 |
| 0 | 0 | 0 | 0 | 0 |
| 0 | 0 | 0 | 0 | 0 |
| 0 | 0 | 0 | 0 | 0 |
| 0 | 0 | 0 | 0 | 0 |
| 0 | 0 | 0 | 0 | 0 |
| 0 | 0 | 0 | 0 | 0 |
| 0 | 0 | 0 | 0 | 0 |
| 0 | 0 | 0 | 0 | 0 |
| 0 | 0 | 0 | 0 | 0 |
| 0 | 0.378755331 | 0 | 0.297744732 | 0.003206169 |
| 0 | 0 | 0 | 0 | 0 |

| | | | | |
|---|---|---|---|---|
| 0 | 0 | 0 | 0 | 0 |
| 0 | 0 | 0 | 0 | 0 |
| 0 | 0 | 0 | 0 | 0 |
| 0 | 0 | 0 | 0 | 0 |
| 0.049743201 | 0.003844545 | 0.44470501 | 0 | 0 |
| 0 | 0 | 0 | 0 | 0 |
| 0 | 0 | 0 | 0 | 0 |
| 0 | 0 | 0 | 0 | 0 |
| 0 | 0 | 0 | 0 | 0 |
| 0 | 0 | 0 | 0 | 0 |
| 0 | 0 | 0 | 0 | 0 |
| 0.010526056 | 0 | 0 | 0.02885958 | 0.002612856 |
| 0 | 0 | 0 | 0.005107394 | 0.001844375 |
| 0 | 0 | 0 | 0 | 0 |
| 0 | 0 | 0.011987469 | 0 | 0 |
| 0.051109468 | 0 | 0.005897773 | 0 | 0 |
| 0 | 0 | 0.000864168 | 0 | 0 |
| 0 | 0 | 0 | 0 | 0 |
| 0.01535758 | 0 | 0 | 0 | 0.069066566 |
| 0 | 0 | 0 | 2.304579463 | 0 |
| 0 | 0 | 0 | 0 | 0 |
| 0 | 0 | 0 | 0 | 0 |
| 0 | 0 | 0 | 0 | 0 |
| 0 | 0 | 0 | 0 | 0 |
| 0 | 0 | 0 | 0 | 0 |
| 0 | 0 | 0 | 0 | 0 |
| 0.009559945 | 0 | 0 | 0.000977124 | 0.000564572 |
| 0.001542983 | 0 | 0 | 0 | 0 |
| 0 | 0 | 0 | 0 | 0 |
| 0 | 0 | 0 | 0 | 0 |
| 0 | 0 | 0 | 0 | 0 |
| 0 | 0 | 0 | 0 | 0 |
| 0 | 0 | 0 | 0 | 0 |
| 0 | 0 | 0 | 0 | 0 |
| 0 | 0 | 0 | 0 | 0 |
| 0 | 0 | 0 | 0 | 0 |
| 0 | 0 | 0 | 0 | 0 |
| 0.010367388 | 0 | 0.001240653 | 0.171355886 | 0.003447134 |
| 0 | 0 | 0 | 0 | 0 |
| 0 | 0 | 0 | 0 | 0 |
| 0 | 0.00067244 | 0.002654082 | 0.012274663 | 0 |
| 0 | 0 | 0 | 0 | 0 |
| 0 | 0 | 0 | 0 | 0 |
| 0.000960845 | 0.007391262 | 0 | 0 | 0 |
| 0 | 0 | 0 | 0 | 0 |
| 0 | 0 | 0 | 0 | 0 |
| 0 | 0 | 0 | 0 | 0 |
| 0 | 0 | 0 | 0 | 0 |

| | | | | |
|---|---|---|---|---|
| 0 | 0 | 0 | 0 | 0 |
| 0 | 0 | 0 | 0 | 0 |
| 0 | 0 | 0 | 0 | 0 |
| 0.00201394 | 0 | 0 | 0.012577149 | 0 |
| 0 | 0.000776688 | 0 | 0.020189605 | 0.0017282 |
| 0.00120535 | 0 | 0 | 0.00144759 | 0.001951607 |
| 0.001710079 | 0 | 0 | 0 | 0 |
| 0 | 0 | 0 | 0 | 0 |
| 0 | 0 | 0 | 0 | 0 |
| 0 | 0 | 0 | 0 | 0 |
| 0 | 0 | 0 | 0 | 0 |
| 0 | 0 | 0 | 0 | 0 |
| 0.01909415 | 0 | 0 | 0.008058176 | 0.049200979 |
| 0 | 0 | 0 | 0 | 0 |
| 0 | 0 | 0 | 0 | 0 |
| 0 | 0 | 0 | 0 | 0 |
| 0 | 0 | 0 | 0 | 0 |
| 0 | 0 | 0 | 0 | 0 |
| 0 | 0 | 0 | 0 | 0 |
| 0 | 0 | 0 | 0 | 0 |
| 0 | 0 | 0 | 0 | 0 |
| 0 | 0 | 0 | 0 | 0 |
| 0.00093991 | 0.00136788 | 0 | 0.00270913 | 0 |
| 0.016082331 | 0.016833226 | 0 | 0.028238992 | 0 |
| 0 | 0 | 0 | 0 | 0 |
| 0 | 0 | 0 | 0 | 0 |
| 0 | 0 | 0 | 0 | 0 |
| 0 | 0 | 0 | 0 | 0 |
| 0 | 0 | 0 | 0 | 0 |
| 0 | 0 | 0 | 0 | 0 |
| 0.010741955 | 0 | 0.001719941 | 0 | 0.007999343 |
| 0 | 0 | 0 | 0 | 0 |
| 0 | 0 | 0 | 0 | 0 |
| 0 | 0 | 0 | 0 | 0 |
| 0 | 0 | 0 | 0 | 0 |
| 0 | 0 | 0 | 0 | 0 |
| 0 | 0 | 0 | 0.007220788 | 0.002575369 |
| 0 | 0 | 0 | 0 | 0 |
| 0 | 0 | 0 | 0 | 0 |
| 0 | 0 | 0 | 0 | 0 |
| 0 | 0 | 0 | 0 | 0 |
| 0 | 0 | 0 | 0 | 0 |
| 0 | 0 | 0 | 0 | 0 |
| 0 | 0 | 0 | 0 | 0 |
| 0.496642133 | 0 | 0 | 0 | 0 |
| 0.109608159 | 0.012164418 | 0 | 0.22017437 | 0.114041935 |
| 0 | 0 | 0 | 0 | 0 |
| 0 | 0 | 0 | 0 | 0 |

| | | | | |
|---|---|---|---|---|
| 0 | 0 | 0 | 0 | 0 |
| 0 | 0 | 0 | 0 | 0 |
| 0 | 0 | 0 | 0 | 0 |
| 0 | 0 | 0 | 0 | 0 |
| 0 | 0 | 0 | 0 | 0 |
| 0 | 0 | 0 | 0 | 0 |
| 0 | 0 | 0 | 0 | 0 |
| 0 | 0 | 0 | 0 | 0 |
| 0 | 0 | 0 | 0 | 0 |
| 0 | 0 | 0 | 0 | 0 |
| 0 | 0 | 0 | 0.001555015 | 0 |
| 0 | 0 | 0 | 0.001289801 | 0.001242057 |
| 0 | 0 | 0 | 0 | 0 |
| 0 | 0 | 0 | 0 | 0 |
| 0 | 0 | 0 | 0 | 0 |
| 0 | 0 | 0 | 0 | 0 |
| 0 | 0 | 0 | 0 | 0 |
| 0 | 0 | 0 | 0 | 0 |
| 0 | 0 | 0 | 0 | 0 |
| 0 | 0 | 0 | 0 | 0 |
| 0 | 0 | 0 | 0 | 0 |
| 0 | 0 | 0 | 0 | 0 |
| 0 | 0 | 0 | 0 | 0 |
| 0 | 0 | 0 | 0 | 0 |
| 0 | 0 | 0 | 0 | 0 |
| 0 | 0 | 0 | 0 | 0 |
| 0 | 0 | 0 | 0 | 0 |
| 0 | 0 | 0 | 0 | 0 |
| 0 | 0 | 0 | 0 | 0 |
| 0 | 0 | 0 | 0 | 0 |
| 0.004917295 | 0 | 0.002017532 | 0.003740166 | 0.007772125 |
| 0 | 0 | 0 | 0 | 0 |
| 0.071368498 | 0.039171842 | 0.02290503 | 0.121595992 | 0.434483593 |
| 0 | 0 | 0 | 0.003970648 | 0 |
| 0 | 0 | 0 | 0 | 0 |
| 0 | 0 | 0 | 0 | 0 |
| 0 | 0 | 0 | 0 | 0 |
| 0 | 0 | 0 | 0 | 0 |
| 0 | 0 | 0 | 0 | 0 |
| 0 | 0 | 0 | 0 | 0 |
| 0 | 0 | 0 | 0 | 0 |
| 0.079068477 | 0.003580702 | 0 | 0.012410471 | 0.037482909 |
| 0.020743836 | 0.001586293 | 0 | 0.186015092 | 0 |
| 0 | 0 | 0 | 0 | 0 |
| 0 | 0 | 0 | 0 | 0 |
| 0.088550637 | 0.002005765 | 0.009801551 | 0 | 0.001275145 |
| 0.004969801 | 0 | 0 | 0 | 0 |
| 0 | 0 | 0 | 0 | 0 |

| | | | | |
|---|---|---|---|---|
| 0 | 0 | 0 | 0 | 0 |
| 0.025728594 | 0 | 0 | 0 | 0.007857759 |
| 0.001167135 | 0 | 0 | 0.078775214 | 0.005669192 |
| 0 | 0 | 0 | 0 | 0 |
| 0 | 0.001162477 | 0.062450893 | 0 | 0.001077756 |
| 0 | 0.005846333 | 0.059548727 | 0.004793397 | 0 |
| 0 | 0 | 0 | 0 | 0 |
| 0 | 0 | 0 | 0 | 0 |
| 0 | 0 | 0 | 0 | 0 |
| 0 | 0 | 0 | 0 | 0 |
| 0 | 0 | 0 | 0 | 0 |
| 0 | 0 | 0 | 0 | 0 |
| 0 | 0 | 0 | 0 | 0 |
| 0 | 0 | 0 | 0 | 0 |
| 0 | 0 | 0 | 0 | 0 |
| 0 | 0 | 0 | 0 | 0 |
| 0 | 0 | 0 | 0 | 0 |
| 0 | 0 | 0 | 0 | 0 |
| 0 | 0 | 0 | 0 | 0 |
| 0 | 0 | 0 | 0 | 0 |
| 0 | 0 | 0 | 0 | 0 |
| 0 | 0 | 0 | 0 | 0 |
| 0 | 0 | 0 | 0 | 0 |
| 0 | 0 | 0 | 0 | 0.337922626 |
| 0.001923766 | 0 | 0 | 0 | 0 |
| 0.001935263 | 0.010461088 | 0 | 0 | 0 |
| 0 | 0 | 0 | 0 | 0 |
| 0.004458523 | 0 | 0 | 0 | 0 |
| 0 | 0 | 0 | 0 | 0 |
| 0 | 0 | 0 | 0 | 0 |
| 0 | 0 | 0 | 0 | 0 |
| 0.001484972 | 0 | 0 | 0 | 0 |
| 0 | 0 | 0 | 0 | 0 |
| 0 | 0 | 0 | 0 | 0 |
| 0 | 0 | 0 | 0 | 0 |
| 0 | 0 | 0 | 0 | 0 |
| 0.001474424 | 0 | 0 | 0 | 0 |
| 0 | 0 | 0 | 0 | 0.002450689 |
| 0 | 0 | 0 | 0 | 0 |
| 0 | 0 | 0 | 0 | 0 |
| 0 | 0 | 0 | 0 | 0 |
| 0 | 0 | 0 | 0 | 0 |
| 0.004612838 | 0 | 0.003453193 | 0.005364939 | 0.003593979 |
| 0 | 0 | 0 | 0 | 0 |
| 0 | 0 | 0 | 0 | 0 |
| 0 | 0 | 0 | 0 | 0 |
| 0 | 0 | 0 | 0 | 0 |
| 0 | 0 | 0 | 0 | 0 |

| | | | | |
|---|---|---|---|---|
| 0 | 0 | 0 | 0 | 0 |
| 0.042412623 | 0.00974339 | 0.028457872 | 0.022963552 | 0.013007946 |
| 0 | 0 | 0 | 0 | 0 |
| 0 | 0 | 0 | 0 | 0 |
| 0 | 0 | 0 | 0 | 0 |
| 0 | 0 | 0 | 0 | 0 |
| 0 | 0 | 0 | 0 | 0 |
| 0 | 0 | 0 | 0 | 0 |
| 0 | 0 | 0 | 0 | 0 |
| 0 | 0 | 0 | 0 | 0 |
| 0 | 0 | 0 | 0 | 0 |
| 0 | 0 | 0 | 0 | 0 |
| 0.032696547 | 0 | 0 | 0 | 0 |
| 0 | 0 | 0 | 0 | 0 |
| 0.029508824 | 0 | 0.006712939 | 0 | 0.008650106 |
| 0 | 0 | 0 | 0 | 0 |
| 0 | 0 | 0 | 0 | 0 |
| 0 | 0 | 0 | 0 | 0 |
| 0 | 0 | 0 | 0 | 0 |
| 0 | 0 | 0 | 0 | 0 |
| 0 | 0 | 0 | 0 | 0 |
| 0 | 0 | 0 | 0.096210986 | 0 |
| 0 | 0 | 0 | 0 | 0.008159724 |
| 0 | 0 | 0.000970055 | 0 | 0 |
| 0 | 0 | 0 | 0 | 0 |
| 0 | 0 | 0 | 0 | 0 |
| 0 | 0 | 0 | 0 | 0 |
| 0 | 0 | 0 | 0 | 0 |
| 0 | 0 | 0 | 0 | 0 |
| 0 | 0 | 0 | 0 | 0 |
| 0 | 0 | 0 | 0 | 0 |
| 0 | 0 | 0 | 0 | 0 |
| 0 | 0 | 0 | 0 | 0 |
| 0 | 0 | 0 | 0 | 0 |
| 0 | 0 | 0 | 0 | 6.333873464 |
| 0 | 0 | 0 | 0 | 0 |
| 0 | 0 | 0 | 0 | 0 |
| 0 | 0 | 0 | 0 | 0 |
| 0.00171921 | 0 | 0 | 0.36807078 | 0.009543794 |
| 0 | 0 | 0 | 0 | 0 |
| 0.137450318 | 0.009187216 | 0.003935346 | 0.156002433 | 0.01640144 |
| 0 | 0 | 0 | 0 | 0 |
| 0 | 0 | 0 | 0 | 0 |
| 0 | 0 | 0 | 0 | 0 |
| 0 | 0 | 0 | 0 | 0 |
| 0.045593543 | 0.087819223 | 0.083355505 | 1.308786322 | 0.501305185 |
| 0 | 0 | 0 | 0 | 0 |
| 0 | 0 | 0 | 0 | 0 |

| | | | | |
|---|---|---|---|---|
| 0 | 0 | 0 | 0 | 0 |
| 0 | 0 | 0 | 0 | 0 |
| 0 | 0 | 0 | 0 | 0 |
| 0 | 0 | 0 | 0 | 0 |
| 0 | 0 | 0 | 0 | 0 |
| 0 | 0 | 0 | 0 | 0 |
| 0 | 0 | 0 | 0 | 0 |
| 0 | 0 | 0 | 0.038488716 | 0 |
| 0 | 0.00149143 | 0 | 0 | 0.001290553 |
| 0.004315215 | 0 | 0 | 0.054450234 | 0.003193989 |
| 0 | 0 | 0.024564685 | 0 | 0 |
| 0 | 0 | 0 | 0 | 0 |
| 0 | 0 | 0.001538238 | 0 | 0.001300773 |
| 0 | 0 | 0 | 0 | 0 |
| 0 | 0 | 0 | 0 | 0 |
| 0 | 0 | 0 | 0 | 0 |
| 0 | 0 | 0 | 0 | 0 |
| 0 | 0 | 0 | 0 | 0 |
| 0 | 0 | 0 | 0 | 0 |
| 0.567119917 | 0.098238222 | 0.379525628 | 0.097144605 | 0.177603405 |
| 0 | 0 | 0 | 0 | 0 |
| 0 | 0 | 0 | 0 | 0 |
| 0 | 0 | 0 | 0 | 0 |
| 0 | 0 | 0 | 0 | 0 |
| 0 | 0 | 0 | 0 | 0 |
| 0 | 0 | 0 | 0 | 0 |
| 0 | 0 | 0 | 0 | 0 |
| 0 | 0 | 0 | 0 | 0 |
| 0 | 0 | 0 | 0 | 0 |
| 0 | 0 | 0.037469001 | 0.058093833 | 0.156443411 |
| 0 | 0 | 0 | 0 | 0 |
| 0 | 0 | 0 | 0 | 0 |
| 0 | 0 | 0 | 0 | 0 |
| 0 | 0 | 0 | 0 | 0 |
| 0 | 0 | 0 | 0 | 0 |
| 0 | 0 | 0 | 0 | 0 |
| 0 | 0 | 0 | 0 | 0 |
| 0 | 0 | 0 | 0 | 0 |
| 0 | 0 | 0 | 0 | 0 |
| 0 | 0 | 0 | 0 | 0 |
| 0 | 0 | 0 | 0 | 0 |
| 0 | 0 | 0 | 0 | 0 |
| 0 | 0 | 0 | 0 | 0 |
| 0 | 0 | 0 | 0 | 0 |
| 0.003996124 | 0 | 0 | 0 | 0 |
| 0 | 0 | 0 | 0 | 0 |
| 0 | 0 | 0 | 0 | 0 |
| 0 | 0 | 0 | 0 | 0 |

| | | | | |
|---|---|---|---|---|
| 0 | 0 | 0 | 0 | 0 |
| 0 | 0 | 0 | 0 | 0 |
| 0 | 0 | 0 | 0 | 0 |
| 0 | 0 | 0 | 0 | 0 |
| 0 | 0 | 0 | 0 | 0 |
| 0 | 0 | 0 | 0.035743797 | 0 |
| 0 | 0 | 0 | 0 | 0 |
| 0 | 0 | 0 | 0 | 0 |
| 0 | 0 | 0 | 0 | 0 |
| 0 | 0 | 0 | 0 | 0 |
| 0 | 0 | 0 | 0 | 0 |
| 0 | 0 | 0 | 0 | 0 |
| 0 | 0 | 0 | 0 | 0 |
| 0 | 0 | 0 | 0 | 0 |
| 0 | 0 | 0 | 0 | 0 |
| 0 | 0 | 0 | 0 | 0 |
| 0 | 0 | 0 | 0 | 0 |
| 0 | 0 | 0 | 0 | 0 |
| 0 | 0 | 0 | 0 | 0 |
| 0 | 0 | 0 | 0 | 0 |
| 0.001220843 | 0 | 0 | 0 | 0 |
| 0 | 0 | 0 | 0 | 0 |
| 0 | 0 | 0 | 0 | 0 |
| 0 | 0 | 0 | 0 | 0 |
| 0 | 0 | 0 | 0 | 0 |
| 0 | 0 | 0 | 0 | 0 |
| 0 | 0 | 0 | 0 | 0 |
| 0 | 0 | 0 | 0 | 0 |
| 0 | 0 | 0 | 0 | 0 |
| 0 | 0 | 0 | 0 | 0 |
| 0 | 0 | 0 | 0 | 0 |
| 0 | 0 | 0 | 0 | 0 |
| 0 | 0 | 0 | 0 | 0 |
| 0 | 0 | 0 | 0 | 0 |
| 0 | 0 | 0 | 0 | 0 |
| 0 | 0 | 0 | 0 | 0 |
| 0 | 0 | 0 | 0 | 0 |
| 0 | 0 | 0 | 0 | 0 |
| 1.33864773 | 0 | 0 | 0 | 0 |
| 0 | 0 | 0 | 0 | 0 |
| 0 | 0 | 0 | 0 | 0 |
| 0 | 0 | 0 | 0 | 0 |
| 0 | 0 | 0 | 0 | 0 |
| 0 | 0 | 0 | 0 | 0 |
| 0 | 0 | 0 | 0 | 0 |
| 0 | 0 | 0 | 0 | 0 |
| 0 | 0 | 0 | 0 | 0 |
| 0 | 0 | 0 | 0 | 0 |

| | | | | |
|---|---|---|---|---|
| 0 | 0 | 0 | 0 | 0 |
| 0.023380017 | 0 | 0.000859846 | 0 | 0 |
| 0 | 0 | 0 | 0 | 0 |
| 0 | 0 | 0 | 0 | 0 |
| 0 | 0 | 0 | 0 | 0 |
| 0 | 0 | 0 | 0 | 0 |
| 0 | 0 | 0 | 0 | 0 |
| 0 | 0 | 0 | 0 | 0 |
| 0 | 0 | 0 | 0 | 0 |
| 0 | 0 | 0 | 0 | 0 |
| 0 | 0 | 0 | 0.002471721 | 0 |
| 0 | 0 | 0 | 0 | 0 |
| 0.125061486 | 0 | 0 | 0 | 0 |
| 0 | 0 | 0 | 0 | 0 |
| 0 | 0 | 0 | 0 | 0 |
| 0 | 0 | 0 | 0 | 0 |
| 0 | 0 | 0 | 0 | 0 |
| 0 | 0 | 0 | 0 | 0 |
| 0 | 0 | 0 | 0 | 0 |
| 0 | 0 | 0 | 0 | 0 |
| 0.010705359 | 0.002388546 | 0 | 0 | 0.019084703 |
| 0 | 0 | 0 | 0 | 0 |
| 0 | 0 | 0 | 0 | 0 |
| 1.177106566 | 0 | 6.6458137 | 1.185799706 | 2.452313852 |
| 0 | 0 | 0 | 0 | 0 |
| 0 | 0 | 0 | 0 | 0 |
| 0 | 0 | 0 | 0 | 0 |
| 0 | 0 | 0 | 0 | 0 |
| 0 | 0 | 0 | 0 | 0 |
| 0 | 0 | 0 | 0 | 0 |
| 0 | 0 | 0 | 0 | 0 |
| 0 | 0 | 0 | 0 | 0 |
| 0.001647953 | 0 | 0 | 0 | 0 |
| 0 | 0 | 0 | 0 | 0 |

| PHL018-0004_V3 | PHL018-0115_V2 | PHL018-0052_V2 | PHL018-0052_V3 | PHL018-0060_V3 |
|---|---|---|---|---|
| CTRL | EYCF | EYCF | EYCF | CTRL |
| 0.012506613 | 0.672846464 | 0.087158427 | 0.263343243 | 0.005899917 |
| 1.862318548 | 8.489266613 | 1.846378318 | 1.826745136 | 0.020557931 |
| 6.618097711 | 2.38920984 | 6.747872454 | 5.827401616 | 0.965089237 |
| 0 | 0 | 0 | 0 | 0 |
| 0 | 0 | 0 | 0 | 0 |
| 0 | 0 | 0 | 0 | 0 |
| 0 | 0 | 0 | 0 | 0 |
| 0 | 0 | 0 | 0 | 0 |
| 0 | 0 | 0 | 0 | 0 |
| 0 | 0 | 0 | 0 | 0 |
| 0 | 0 | 0 | 0 | 0 |
| 0 | 0 | 0 | 0 | 0 |
| 0 | 0 | 0 | 0 | 0 |
| 5.890400462 | 0.385875418 | 28.88625569 | 7.378073355 | 0.168832023 |
| 0 | 0 | 0 | 0 | 0 |
| 0 | 0 | 0 | 0 | 0 |
| 0 | 0 | 0 | 0 | 0 |
| 0 | 0 | 0 | 0 | 0 |
| 0 | 0 | 0 | 0 | 0 |
| 0 | 0 | 0 | 0 | 0 |
| 0 | 0 | 0 | 0 | 0 |
| 0 | 0 | 0 | 0 | 0 |
| 0 | 0 | 0 | 0 | 0 |
| 0 | 0 | 0 | 0 | 0 |
| 0 | 0 | 0 | 0 | 0.098500778 |
| 0 | 0 | 0 | 0 | 0 |
| 0 | 0 | 0 | 0 | 0 |
| 0 | 0 | 0 | 0 | 0 |
| 0 | 0 | 0 | 0 | 0 |
| 0 | 0 | 0 | 0 | 0.001960495 |
| 0 | 0 | 0 | 0 | 0 |
| 0 | 0 | 0 | 0 | 0 |
| 0 | 0 | 0 | 0 | 0 |
| 0 | 0 | 0 | 0 | 0 |
| 0 | 0 | 0 | 0 | 0 |
| 4.981411513 | 0 | 5.5638658 | 0.89358681 | 0.419537501 |
| 0 | 0 | 0 | 0 | 0 |
| 0 | 0 | 0 | 0 | 0 |
| 0 | 0 | 0 | 0 | 0 |
| 0 | 0 | 0 | 0 | 0 |
| 0 | 0 | 0 | 0 | 0 |
| 0 | 0.209604904 | 0.009841344 | 0.284959921 | 0.004579981 |
| 0.095915802 | 0.011581031 | 0.019602229 | 0.719182545 | 2.160559637 |
| 0.046581103 | 0.257496885 | 0.010690999 | 0 | 0.009462637 |
| 0 | 0.002004818 | 0 | 0 | 0 |
| 0 | 0 | 0 | 0 | 0 |

| | | | | |
|---|---|---|---|---|
| 0.010204362 | 0 | 0 | 0 | 0.003508633 |
| 0.029513578 | 0 | 0.002334028 | 0 | 0 |
| 0.000947685 | 0 | 0.031735442 | 0.008998355 | 0 |
| 1.902814004 | 1.038054735 | 0 | 0 | 0.160308841 |
| 0 | 0 | 0 | 0 | 0 |
| 0 | 0 | 0 | 0 | 0.043714284 |
| 0.057739352 | 0.102694305 | 0.001945768 | 0.34877431 | 0.001655817 |
| 0 | 0 | 0 | 0 | 0 |
| 0 | 0 | 0 | 0 | 0.004476815 |
| 0 | 0 | 0 | 0 | 0 |
| 0 | 0 | 0 | 0 | 0.010079109 |
| 0.024925717 | 0.213906421 | 0 | 0.010043126 | 0.029944141 |
| 0 | 0 | 0 | 0 | 0 |
| 0.029033756 | 0.004116405 | 0.014134049 | 0.023557958 | 0 |
| 0 | 0 | 0 | 0.002521551 | 0 |
| 0 | 0 | 0 | 0 | 0 |
| 0 | 0.002257262 | 0.022378244 | 0 | 0.472052807 |
| 0 | 0 | 0 | 0 | 0 |
| 0 | 0 | 0 | 0 | 0 |
| 0 | 0 | 0 | 0 | 0 |
| 0.150842052 | 0.181530489 | 0.085194529 | 0.009004035 | 0 |
| 0.003273328 | 0.054181227 | 0.000987523 | 0.000994578 | 0 |
| 0.018846618 | 0 | 0 | 0 | 0.293774966 |
| 0 | 0 | 0 | 0 | 0 |
| 0 | 0 | 0 | 0 | 0 |
| 0 | 0.002839153 | 0 | 0 | 0 |
| 0 | 0 | 0 | 0 | 0 |
| 0 | 0 | 0 | 0 | 0 |
| 0.001395844 | 0.002786471 | 0 | 0.002968823 | 0 |
| 0 | 0 | 0 | 0 | 0 |
| 0.532865657 | 0 | 0.001342767 | 0 | 0 |
| 0 | 0 | 0 | 0 | 0 |
| 0 | 0 | 0 | 0 | 0 |
| 0 | 0 | 0 | 0 | 0 |
| 0 | 0 | 0 | 0 | 0 |
| 0 | 0 | 0 | 0 | 0 |
| 0 | 0 | 0 | 0 | 0 |
| 0 | 0 | 0 | 0 | 26.53683926 |
| 0 | 0 | 0 | 0 | 0 |
| 0 | 0 | 0 | 0 | 0 |
| 0 | 0 | 0 | 0 | 0 |
| 0 | 0 | 0 | 0 | 0 |
| 0 | 0 | 0 | 0 | 0 |
| 0 | 0 | 0 | 0 | 0 |
| 0 | 0 | 0 | 0 | 0 |
| 0 | 0 | 0 | 0 | 0 |
| 0 | 0 | 0.004395294 | 0 | 0 |
| 0 | 0 | 0 | 0 | 0 |

| | | | | |
|---|---|---|---|---|
| 0 | 0 | 0 | 0 | 0 |
| 0 | 0 | 0 | 0 | 0 |
| 0 | 0 | 0 | 0 | 0 |
| 0 | 0 | 0 | 0 | 0 |
| 0 | 0 | 0.333620617 | 0.000754726 | 0 |
| 0 | 0 | 0 | 0 | 0 |
| 0 | 0 | 0 | 0 | 0 |
| 0 | 0 | 0 | 0 | 0 |
| 0 | 0 | 0 | 0 | 0 |
| 0 | 0 | 0 | 0 | 0 |
| 0 | 0 | 0 | 0 | 0 |
| 0 | 0.001833196 | 0.088432519 | 0.083893041 | 0 |
| 0 | 0 | 0 | 0 | 0 |
| 0 | 0 | 0 | 0 | 0 |
| 0 | 0 | 0 | 0 | 0 |
| 0 | 0 | 0 | 0 | 0 |
| 0 | 0 | 0 | 0 | 0 |
| 0 | 0 | 0 | 0 | 0 |
| 0.015691333 | 2.031981886 | 0.04576087 | 0 | 0.295944696 |
| 0 | 0 | 0 | 0 | 0 |
| 0 | 0 | 0 | 0 | 0 |
| 0 | 0 | 0 | 0 | 0 |
| 0 | 0 | 0 | 0 | 0 |
| 0 | 0.00128389 | 0 | 0 | 0 |
| 0 | 0 | 0 | 0 | 0 |
| 0 | 0 | 0 | 0 | 0 |
| 0 | 0 | 0 | 0 | 0 |
| 0 | 0 | 0 | 0 | 0 |
| 0 | 0 | 0 | 0 | 0 |
| 0 | 0 | 0 | 0 | 0 |
| 0.017371653 | 0.140272044 | 0.006182984 | 0.015271356 | 0.015127151 |
| 0 | 0.006387048 | 0.001297298 | 0.000466631 | 0 |
| 0 | 0 | 0 | 0 | 0 |
| 0 | 0 | 0 | 0 | 0 |
| 0 | 0 | 0 | 0 | 0 |
| 0 | 0 | 0 | 0 | 0 |
| 0 | 0 | 0 | 0 | 0 |
| 0 | 0 | 0 | 0 | 0 |
| 0 | 0.30022084 | 0 | 0 | 0.278976649 |
| 0 | 0 | 0 | 0 | 0 |
| 0 | 0 | 0 | 0 | 0 |
| 0 | 0 | 0 | 0 | 0.012202401 |
| 0 | 0 | 0 | 0.002157404 | 0 |
| 0 | 0 | 0 | 0 | 0 |
| 0 | 0 | 0 | 0 | 0 |
| 0 | 0 | 0 | 0 | 0 |
| 0 | 0 | 0 | 0 | 0 |
| 0 | 0 | 0 | 0 | 0 |

| | | | | |
|---|---|---|---|---|
| 0 | 0 | 0 | 0 | 0 |
| 0 | 0 | 0 | 0 | 0 |
| 0 | 0 | 0 | 0 | 0 |
| 0 | 0 | 0 | 0 | 0 |
| 0 | 0.00466973 | 0.006452291 | 0 | 0 |
| 0 | 0 | 0 | 0 | 0 |
| 0 | 0 | 0 | 0 | 0 |
| 0 | 0 | 0 | 0 | 0 |
| 0 | 0 | 0 | 0 | 0 |
| 0 | 0 | 0 | 0 | 0 |
| 0 | 0 | 0 | 0 | 0 |
| 0 | 0 | 0 | 0 | 0 |
| 0 | 0 | 0 | 0 | 0 |
| 0 | 0 | 0 | 0 | 0 |
| 0 | 0 | 0 | 0 | 0 |
| 0 | 0 | 0 | 0 | 0.589240223 |
| 0 | 0 | 0 | 0 | 0 |
| 0 | 0 | 0 | 0 | 0 |
| 0 | 0 | 0 | 0 | 0 |
| 0 | 0 | 0 | 0 | 0 |
| 0 | 0 | 0 | 0 | 0 |
| 0 | 0 | 0 | 0 | 0 |
| 0 | 0 | 0 | 0 | 0 |
| 0 | 0 | 0 | 0 | 0 |
| 0 | 0.020071236 | 0 | 0 | 0 |
| 0 | 0 | 0 | 0 | 0 |
| 0 | 0 | 0 | 0 | 0.021585903 |
| 0 | 0 | 0 | 0 | 0 |
| 0 | 0 | 0 | 0 | 0 |
| 0 | 0 | 0 | 0 | 0.251891083 |
| 0 | 0 | 0 | 0 | 0 |
| 0 | 0 | 0 | 0 | 0.000477658 |
| 0 | 0 | 0 | 0 | 0 |
| 0 | 0 | 0 | 0 | 0 |
| 0 | 0 | 0 | 0 | 0 |
| 0 | 0 | 0 | 0 | 0 |
| 0 | 0 | 0 | 0 | 0 |
| 0 | 0 | 0 | 0 | 0 |
| 0 | 0 | 0 | 0 | 0 |
| 0 | 0 | 0 | 0 | 0 |
| 0 | 0 | 0 | 0 | 0 |
| 0 | 0 | 0 | 0 | 0 |
| 0 | 0 | 0 | 0 | 0 |
| 0 | 0 | 0 | 0 | 0 |
| 0 | 0 | 0 | 0 | 0 |
| 0 | 0 | 0 | 0 | 0 |
| 0 | 0 | 0 | 0 | 0 |
| 0.008689728 | 0 | 0 | 0 | 0 |

| | | | | |
|---|---|---|---|---|
| 0 | 0 | 0 | 0 | 0.886293453 |
| 0 | 0 | 0 | 0 | 0 |
| 0 | 0 | 0 | 0 | 0 |
| 0 | 0 | 0 | 0 | 0 |
| 0 | 0 | 0 | 0 | 0 |
| 0 | 0 | 0 | 0 | 0 |
| 0 | 0 | 0 | 0 | 0 |
| 0 | 0 | 0 | 0 | 0 |
| 0 | 0 | 0 | 0 | 0 |
| 0 | 0 | 0 | 0 | 0 |
| 0 | 0 | 0 | 0 | 0 |
| 0 | 0 | 0 | 0 | 0.028901535 |
| 0 | 0.001692402 | 0 | 0 | 0 |
| 0 | 0 | 0 | 0 | 0 |
| 0 | 0 | 0 | 0 | 0 |
| 0 | 0 | 0 | 0.001386274 | 0 |
| 0 | 0 | 0 | 0 | 0 |
| 0 | 0 | 0 | 0 | 0 |
| 0 | 0 | 0 | 0 | 0 |
| 0 | 0 | 0 | 0 | 0 |
| 0.049879549 | 0.040609603 | 0.022572074 | 0.048484922 | 0.007691525 |
| 0 | 0 | 0 | 0 | 0 |
| 0 | 0 | 0 | 0 | 0 |
| 0 | 0 | 0 | 0 | 0 |
| 0 | 0 | 0 | 0 | 0 |
| 0 | 0 | 0 | 0 | 0 |
| 0.009600833 | 0.008907171 | 0.001114021 | 0 | 0.001659023 |
| 0 | 0 | 0 | 0 | 0 |
| 0 | 0 | 0 | 0 | 0 |
| 0 | 0 | 0 | 0 | 0 |
| 0 | 0 | 0.011771188 | 0 | 0 |
| 0 | 0 | 0 | 0 | 0 |
| 0 | 0 | 0 | 0 | 0 |
| 0 | 0.003223126 | 0 | 0 | 0.004352391 |
| 0.006600873 | 0.134859084 | 0.03036889 | 0.13078373 | 0.002965639 |
| 0 | 0 | 0 | 0 | 0 |
| 0 | 0 | 0 | 0 | 0 |
| 0 | 0 | 0 | 0 | 0 |
| 0 | 0 | 0 | 0 | 0 |
| 0 | 0 | 0 | 0 | 0 |
| 0 | 0 | 0 | 0 | 0 |
| 0 | 0 | 0 | 0 | 0 |
| 0 | 0 | 0 | 0 | 0 |
| 0 | 0 | 0 | 0 | 0 |
| 0 | 0 | 0 | 0 | 0 |
| 0.026710188 | 0 | 0 | 0.004575029 | 0 |
| 0 | 0 | 0 | 0 | 0 |
| 0 | 0.00293448 | 0.002306082 | 0.027840901 | 0 |

| | | | | |
|---|---|---|---|---|
| 0 | 0 | 0 | 0 | 0 |
| 0 | 0 | 0 | 0 | 0 |
| 0 | 0 | 0 | 0 | 0 |
| 0.000996558 | 0 | 0 | 0 | 0 |
| 0 | 0 | 0 | 0 | 0 |
| 0 | 0 | 0 | 0 | 0 |
| 0 | 0 | 0 | 0 | 0 |
| 0 | 0 | 0.015300162 | 0.003852366 | 0 |
| 0 | 0 | 0 | 0 | 0 |
| 0 | 0 | 0 | 0 | 0 |
| 0 | 0 | 0 | 0 | 0 |
| 0 | 0 | 0 | 0 | 0 |
| 0 | 0 | 0 | 0 | 0 |
| 0 | 0 | 0 | 0 | 0 |
| 0 | 0 | 0 | 0 | 0.000624773 |
| 0 | 0 | 0 | 0 | 0 |
| 0 | 0 | 0 | 0 | 0 |
| 0 | 0 | 0 | 0 | 0 |
| 0.000597987 | 0 | 0 | 0.006510709 | 0.01289585 |
| 0 | 0 | 0 | 0 | 0.141469056 |
| 0 | 0.050992332 | 0.033796773 | 0.042212746 | 0 |
| 0 | 0 | 0 | 0 | 0 |
| 0 | 0 | 0 | 0 | 0 |
| 0 | 0 | 0 | 0 | 0 |
| 0.005257386 | 0.007288277 | 0 | 0 | 0 |
| 0 | 0 | 0 | 0 | 0 |
| 0 | 0 | 0 | 0 | 0 |
| 0 | 0 | 0 | 0 | 0 |
| 0 | 0 | 0 | 0.00508147 | 0 |
| 0 | 0 | 0 | 0 | 0 |
| 0.052969146 | 0.796019321 | 0.108795424 | 0.342742989 | 0.06935408 |
| 0 | 0 | 0 | 0 | 0 |
| 0 | 0.04058431 | 0.01362518 | 0.000816817 | 0 |
| 0 | 0 | 0.016455399 | 0 | 0.00301609 |
| 0 | 0 | 0 | 0 | 0 |
| 0 | 0 | 0 | 0 | 0 |
| 0 | 0 | 0 | 0 | 0 |
| 0 | 0 | 0 | 0 | 0 |
| 0 | 0 | 0 | 0 | 0 |
| 0 | 0 | 0 | 0 | 0 |
| 0 | 0 | 0 | 0 | 0 |
| 0.049365245 | 0 | 0.002792417 | 0 | 0.001336669 |
| 0 | 0 | 0 | 0 | 0 |
| 0 | 0 | 0 | 0 | 0 |
| 0 | 0 | 0 | 0 | 0 |
| 0 | 0 | 0 | 0 | 0 |
| 0 | 0 | 0 | 0 | 0 |
| 0 | 0 | 0 | 0 | 0 |

| | | | | |
|---|---|---|---|---|
| 0 | 0 | 0 | 0 | 0 |
| 0 | 0 | 0 | 0 | 0 |
| 0.036804299 | 0.257562868 | 0.015528073 | 0 | 0.012308327 |
| 0 | 0 | 0 | 0 | 0 |
| 0 | 0 | 0 | 0 | 0.008253891 |
| 0 | 0 | 0 | 0 | 0 |
| 0 | 0 | 0 | 0 | 0 |
| 0 | 0 | 0 | 0 | 0 |
| 0 | 0 | 0 | 0 | 0 |
| 0.001771628 | 0 | 0 | 0 | 0 |
| 0 | 0 | 0 | 0 | 0 |
| 0 | 0 | 0 | 0 | 0 |
| 0 | 0 | 0 | 0 | 0 |
| 0.00152915 | 0 | 0.00996464 | 0.003484662 | 0.002061049 |
| 0.004291185 | 0 | 0.005711462 | 0 | 0.00068045 |
| 0 | 0 | 0 | 0 | 0.016391521 |
| 0.005177034 | 0.001435377 | 0 | 0 | 0 |
| 0 | 0 | 0 | 0 | 0 |
| 0 | 0 | 0 | 0 | 0 |
| 0 | 0 | 0 | 0 | 0 |
| 0 | 0 | 0 | 0 | 0 |
| 0 | 0.001943604 | 0.003524757 | 0.02701897 | 0 |
| 0 | 0 | 0 | 0 | 0 |
| 0 | 0 | 0 | 0.003974543 | 0 |
| 0 | 0 | 0 | 0 | 0 |
| 0 | 0 | 0 | 0 | 0 |
| 0 | 0 | 0 | 0 | 0.123119705 |
| 0 | 0 | 0 | 0 | 0 |
| 0 | 0 | 0 | 0 | 0 |
| 0 | 0 | 0 | 0 | 0 |
| 0 | 0 | 0 | 0 | 0 |
| 0 | 0 | 0 | 0 | 0 |
| 0 | 0 | 0 | 0 | 0 |
| 0 | 0 | 0 | 0 | 0 |
| 0 | 0 | 0 | 0 | 0 |
| 0 | 0 | 0 | 0 | 0.042636628 |
| 0 | 0 | 0 | 0 | 0 |
| 0 | 0.021570443 | 0.004723725 | 0.001238925 | 0 |
| 0 | 0 | 0 | 0 | 0 |
| 0 | 0 | 0 | 0 | 0 |
| 0 | 0 | 0 | 0 | 0 |
| 0 | 0 | 0 | 0 | 0 |
| 0 | 0 | 0 | 0 | 0 |
| 0 | 0 | 0 | 0 | 0 |
| 0 | 0 | 0 | 0 | 0 |
| 0 | 0 | 0 | 0 | 0 |
| 0 | 0 | 0 | 0 | 0 |
| 0 | 0 | 0 | 0 | 0 |

| | | | | |
|---|---|---|---|---|
| 0 | 0.00129995 | 0 | 0 | 0 |
| 0 | 0 | 0 | 0 | 0 |
| 0.004351116 | 0 | 0.001476763 | 0 | 0.002199227 |
| 0 | 0 | 0 | 0 | 0 |
| 0.013132927 | 0.004779097 | 0 | 0 | 0.004056497 |
| 0 | 0 | 0 | 0 | 0 |
| 0 | 0 | 0 | 0 | 0 |
| 0.019942745 | 0.058855116 | 0.002429733 | 0.004078485 | 0 |
| 0 | 0 | 0 | 0 | 0 |
| 0 | 0.006904759 | 0.365639281 | 0.295766154 | 0.04129171 |
| 0 | 0 | 0 | 0 | 0 |
| 0 | 0.016666313 | 0.001648616 | 0.001614272 | 0.002864347 |
| 0.005987405 | 0.012067354 | 0.002501068 | 0 | 0 |
| 0 | 0.001309086 | 0 | 0 | 0 |
| 0 | 0 | 0 | 0 | 0 |
| 0 | 0 | 0 | 0 | 0 |
| 0 | 0 | 0 | 0 | 0 |
| 0 | 0 | 0 | 0 | 0 |
| 0.217972561 | 0 | 0.163276957 | 0 | 0 |
| 0 | 0 | 0 | 0 | 0 |
| 0 | 0 | 0 | 0 | 0 |
| 0 | 0 | 0 | 0 | 0 |
| 0 | 0 | 0.012903894 | 0 | 0 |
| 0 | 0 | 0 | 0 | 0.060095813 |
| 0 | 0 | 0 | 0 | 0 |
| 0 | 0 | 0.001213794 | 0.002648675 | 0 |
| 0 | 0 | 0 | 0 | 1.904946933 |
| 0.001297425 | 0 | 0 | 0 | 0 |
| 0 | 0 | 0 | 0.008135584 | 0 |
| 0 | 0.034775811 | 0 | 0 | 0 |
| 0 | 0 | 0 | 0 | 0 |
| 0 | 0 | 0 | 0 | 0 |
| 0 | 0 | 0 | 0 | 0 |
| 0.004560224 | 0 | 0 | 0.014655303 | 0 |
| 0 | 0 | 0 | 0 | 0 |
| 0.011967671 | 0 | 0 | 0 | 0 |
| 0.118278691 | 0 | 0 | 0 | 0 |
| 0 | 0 | 0.006318425 | 0 | 0 |
| 0 | 0 | 0.002593182 | 0 | 0 |
| 0 | 0 | 0 | 0 | 0 |
| 0.016296473 | 0 | 0 | 0 | 0 |
| 0 | 0 | 0 | 0 | 0 |
| 0 | 0 | 0 | 0 | 0 |
| 0 | 0 | 0 | 0 | 0 |
| 0 | 0 | 0 | 0 | 0 |
| 0 | 0 | 0 | 0 | 0.00224828 |
| 0 | 0.014981991 | 0.023547406 | 0.010388255 | 0.001123952 |
| 0 | 0 | 0 | 0 | 0 |

| | | | | |
|---|---|---|---|---|
| 0 | 0 | 0 | 0 | 0 |
| 0.022366369 | 0 | 0 | 0.046721569 | 0 |
| 0 | 0 | 0 | 0 | 0 |
| 0 | 0 | 0 | 0 | 0 |
| 0 | 0 | 0 | 0 | 0 |
| 0 | 0.00229447 | 0 | 0 | 0 |
| 0 | 0.042643569 | 0 | 0 | 0 |
| 0 | 0 | 0 | 0 | 0 |
| 0 | 0 | 0 | 0 | 0.233390646 |
| 0 | 0 | 0 | 0 | 0 |
| 0.006094131 | 0 | 0 | 0 | 0 |
| 0 | 0 | 2.408400148 | 1.161434171 | 0 |
| 0 | 0 | 0 | 0 | 0 |
| 0.0060453 | 0.003950834 | 0.008076798 | 0 | 0.000776009 |
| 0.079226575 | 0 | 0 | 0 | 0 |
| 0 | 0 | 0 | 0 | 0 |
| 0 | 0 | 0 | 0 | 0 |
| 0.030255705 | 0 | 0 | 0 | 0 |
| 0.001531975 | 0.047402443 | 0.002218454 | 0.001861919 | 0 |
| 0.001726177 | 0.048888716 | 0 | 0 | 0 |
| 0 | 0 | 0 | 0 | 0 |
| 0 | 0.185577161 | 0.902565075 | 0.922165538 | 0.029391638 |
| 0 | 0.002385824 | 0.010136913 | 0.068062204 | 0 |
| 0 | 0 | 0 | 0 | 0 |
| 0 | 0 | 0 | 0 | 0 |
| 0 | 0 | 0 | 0 | 0 |
| 0.247206063 | 0 | 0 | 0 | 0 |
| 0 | 0 | 0 | 0 | 0 |
| 0 | 0 | 0 | 0 | 0 |
| 0.770919552 | 0.874455383 | 0.30666556 | 6.0764828 | 0.021896233 |
| 0 | 0.000730998 | 0 | 0 | 0.003685223 |
| 0 | 0 | 0 | 0 | 0 |
| 0 | 0 | 0 | 0 | 0 |
| 0 | 0.001725573 | 0.004506271 | 0 | 0 |
| 0.005719427 | 0 | 0 | 0 | 0 |
| 0 | 0 | 0 | 0 | 0 |
| 0 | 0 | 0 | 0.789854293 | 3.263050672 |
| 0 | 0.00219652 | 0 | 0 | 0 |
| 0.136394714 | 0.402440221 | 0.27358269 | 0.638906424 | 0 |
| 0.009537517 | 0 | 0 | 0 | 0 |
| 0.081884933 | 0.019488093 | 0.003641294 | 0.00251678 | 0 |
| 2.224681423 | 5.152609426 | 0 | 0 | 0.057336624 |
| 0 | 0 | 0 | 0 | 0 |
| 0 | 0 | 0 | 0 | 0 |
| 0.004271467 | 0.009992538 | 0 | 0.002433479 | 0 |
| 0 | 0 | 0.000873129 | 0 | 0 |
| 0.00149036 | 0 | 0.000899247 | 0 | 0 |
| 0 | 0 | 0 | 0 | 0 |

| | | | | |
|---|---|---|---|---|
| 0 | 0.003059029 | 0.00305118 | 0 | 0 |
| 0 | 0 | 0 | 0 | 0 |
| 0.004175967 | 0 | 0.044220319 | 0 | 0.00168856 |
| 0.06265348 | 0 | 0 | 0 | 0 |
| 0 | 0 | 0 | 0 | 0.001384154 |
| 0 | 0 | 0 | 0 | 0 |
| 0 | 0 | 0 | 0 | 0 |
| 0 | 0 | 0.001554433 | 0 | 0 |
| 0.001282464 | 0.098885142 | 0 | 0 | 0 |
| 0 | 0 | 0 | 0 | 0 |
| 0 | 0 | 0 | 0 | 0 |
| 0 | 0 | 0 | 0 | 0 |
| 0 | 0 | 0 | 0 | 0 |
| 0 | 0.001383713 | 0 | 0 | 0.004484438 |
| 0 | 0 | 0 | 0 | 0 |
| 0.02123411 | 0.269525999 | 0.00372598 | 0 | 0.005986873 |
| 0.000720484 | 0.049740305 | 0.066077769 | 0.006384989 | 0.001456644 |
| 0 | 0.014926107 | 0.001056341 | 0.001773146 | 0 |
| 0 | 0 | 0 | 0.001695911 | 0 |
| 0 | 0 | 0 | 0 | 0 |
| 0 | 0 | 0.006574483 | 0.003908495 | 0 |
| 0 | 0.004518375 | 0.027229776 | 0 | 0 |
| 0.11624931 | 0 | 0 | 0 | 0 |
| 0.534192196 | 0 | 0 | 0 | 0 |
| 0 | 0.09717489 | 0 | 0 | 0 |
| 0.00094911 | 0 | 0.108234598 | 0.188408574 | 0 |
| 0.005487958 | 0.001901979 | 0 | 0 | 0 |
| 0 | 0.027684048 | 0 | 0 | 0 |
| 0 | 0 | 0 | 0 | 0 |
| 0 | 0 | 0 | 0 | 0 |
| 0.003709846 | 0.041368469 | 0.011961612 | 0.04508843 | 0 |
| 0.007259339 | 0 | 0.005318702 | 0 | 0 |
| 0.004607915 | 0.035726723 | 2.284813399 | 5.445821725 | 0.008428847 |
| 0 | 0.017655131 | 0 | 0 | 0 |
| 0 | 0 | 0 | 0 | 0.020192671 |
| 0 | 0 | 0 | 0 | 0 |
| 0 | 0.021532947 | 0 | 0 | 0 |
| 0.003025198 | 0.078972681 | 0.008003372 | 0.019267537 | 0.001725088 |
| 0 | 0 | 0 | 0 | 0.011300642 |
| 0 | 0 | 0 | 0 | 0 |
| 0 | 0 | 0.000601514 | 0 | 0 |
| 0 | 0 | 0 | 0 | 0 |
| 0.002436096 | 0.014328736 | 0 | 0 | 0 |
| 0.018710663 | 0 | 0.038636587 | 0 | 0.006121109 |
| 1.738319316 | 0 | 0 | 0 | 0 |
| 0 | 0 | 0.008265154 | 0.000578069 | 0.01572771 |
| 0 | 0 | 0 | 0 | 0 |
| 0 | 0 | 0 | 0 | 0 |

| | | | | |
|---|---|---|---|---|
| 0 | 0 | 0 | 0 | 0 |
| 1.431994486 | 0.018990052 | 0.361607064 | 0.000963467 | 0.039533686 |
| 0 | 0 | 0 | 0 | 0 |
| 0 | 0.002635668 | 0 | 0 | 0 |
| 0 | 0 | 0 | 0 | 0 |
| 0 | 0 | 0 | 0 | 0 |
| 0 | 0 | 0.012213133 | 0 | 0 |
| 0 | 0 | 0 | 0.001480088 | 0.001313126 |
| 0 | 0.031870355 | 0 | 0 | 0 |
| 0 | 0 | 0 | 0 | 0 |
| 0.000706906 | 0.004997888 | 0.001279589 | 0.001073941 | 0 |
| 0.015092367 | 0.057804327 | 0.003494303 | 0 | 0.005913398 |
| 0 | 0 | 0 | 0 | 0.028839206 |
| 0 | 0 | 0 | 0 | 0 |
| 0 | 0.001349283 | 0.010521869 | 0.003080534 | 0 |
| 0 | 0 | 0 | 0 | 0 |
| 0.019739524 | 0.032016771 | 0 | 0 | 0.002494285 |
| 0.189622251 | 0.075531291 | 0.057399978 | 0 | 0 |
| 0 | 0 | 0 | 0 | 0 |
| 0.111067292 | 0.009342109 | 0 | 0 | 0 |
| 0.061832652 | 0.007936867 | 0.019805645 | 0 | 0.00617337 |
| 0 | 0 | 0 | 0 | 0 |
| 0 | 0 | 0 | 0 | 0 |
| 0 | 0 | 0 | 0 | 0 |
| 0.075673247 | 0 | 0 | 0.010067463 | 0 |
| 0 | 0 | 0 | 0 | 0 |
| 0 | 0.150101062 | 0 | 0 | 0.017772487 |
| 0.107368048 | 0.066136031 | 0.082488498 | 0.035745637 | 0.01931962 |
| 0 | 0 | 0 | 0 | 0 |
| 0 | 0 | 0 | 0.001486814 | 0 |
| 0.012188041 | 0 | 0.004202266 | 0.009919422 | 0 |
| 0 | 0 | 0 | 0 | 0 |
| 0.000984968 | 0 | 0 | 0.002743359 | 0 |
| 0 | 0 | 0 | 0 | 0 |
| 0.054441393 | 0.048517532 | 0 | 0 | 0.001429447 |
| 1.185489419 | 0.63047011 | 0.200503624 | 0.233297282 | 0.046578236 |
| 0 | 0.027771345 | 0 | 0 | 0 |
| 0 | 0 | 0 | 0 | 0 |
| 0 | 0.007908866 | 0.007246917 | 0.004054827 | 0.011466779 |
| 0 | 0 | 0 | 0 | 0 |
| 1.648031417 | 0 | 0 | 0 | 0 |
| 0 | 0 | 0 | 0 | 0 |
| 0.032626814 | 0.0169403 | 0.006052596 | 0 | 0 |
| 0 | 0 | 0 | 0 | 0.001977844 |
| 0 | 0 | 0 | 0 | 0 |
| 0 | 0 | 0 | 0 | 0.066073832 |
| 0 | 0 | 0 | 0 | 0 |
| 0 | 0.006708013 | 0 | 0 | 0 |

| | | | | |
|---|---|---|---|---|
| 0.029726325 | 0 | 0 | 0 | 0 |
| 0 | 0 | 0 | 0 | 0 |
| 0 | 0 | 0 | 0 | 0 |
| 0.009913021 | 0.374282738 | 0 | 0 | 0.821711395 |
| 0 | 0 | 0 | 0 | 0 |
| 0 | 0 | 0 | 0 | 0 |
| 0 | 0 | 0 | 0 | 0 |
| 0 | 0.00263443 | 0 | 0 | 0 |
| 0.115970953 | 0 | 0.172551356 | 0.329523679 | 9.520151665 |
| 0 | 0 | 0 | 0 | 0.021608595 |
| 0 | 0 | 0 | 0 | 0 |
| 0.019900026 | 0 | 0 | 0 | 0.000914388 |
| 0 | 0 | 0 | 0 | 0 |
| 0 | 0 | 0 | 0 | 0 |
| 0.005038431 | 0.797191756 | 0 | 0.49980786 | 0.032068591 |
| 0 | 0 | 0 | 0 | 0 |
| 0 | 0 | 0 | 0 | 0 |
| 0 | 0.035267944 | 0 | 0 | 0.728819706 |
| 0 | 0 | 0 | 0 | 0 |
| 0 | 0 | 0 | 0 | 0.001360955 |
| 0 | 0 | 0 | 0 | 0 |
| 0 | 0 | 0 | 0 | 0.031976814 |
| 0 | 0 | 0 | 0 | 0 |
| 0.193233249 | 0.205431187 | 0.072232943 | 0.00163849 | 0 |
| 0.000846887 | 0.014528661 | 0 | 0 | 0 |
| 0 | 0 | 0 | 0 | 0.094259178 |
| 0.001007665 | 0 | 0 | 0 | 0.013411929 |
| 3.682982358 | 0 | 1.028272932 | 1.535348719 | 0.698829295 |
| 0 | 0 | 0 | 0 | 0 |
| 0 | 0 | 0 | 0 | 0 |
| 0 | 0 | 0 | 0 | 0.006518899 |
| 0 | 0.063057561 | 0 | 0 | 0.0047461 |
| 0.000669268 | 0.093939879 | 0 | 0.001398047 | 0 |
| 0 | 0 | 0 | 0 | 0 |
| 0 | 0 | 0 | 0 | 0.002188661 |
| 0 | 0.298384432 | 0 | 0 | 0.034294369 |
| 0 | 0 | 0.585021015 | 0 | 1.90306846 |
| 0 | 0 | 0 | 0 | 0 |
| 0 | 0 | 0 | 0 | 0 |
| 0 | 0 | 0.014796085 | 0.009959115 | 0 |
| 0.048810983 | 0.362691472 | 0.005497586 | 0.0026366 | 0.025584755 |
| 0 | 0 | 0 | 0 | 0.007110645 |
| 0.087702027 | 1.30073308 | 1.899603887 | 7.006670575 | 0.005787644 |
| 0.00170083 | 0.01367552 | 0 | 0 | 0 |
| 0 | 0 | 0 | 0 | 0 |
| 0 | 0 | 0 | 0 | 0 |
| 0 | 0 | 0 | 0 | 0 |
| 0 | 0 | 0.002832312 | 0.018818881 | 0 |

| | | | | |
|---|---|---|---|---|
| 0 | 0 | 0 | 0 | 0.009377681 |
| 0 | 0.003260641 | 0.007525924 | 0.004511719 | 0.03410363 |
| 0.042748446 | 0.226442712 | 0.006108955 | 0.009114958 | 0 |
| 0 | 0 | 0 | 0 | 0.00946983 |
| 0 | 0 | 0 | 0 | 0 |
| 0 | 0 | 0 | 0 | 0.016324212 |
| 0.004790564 | 0 | 0 | 0 | 0 |
| 0 | 0 | 0 | 0 | 0 |
| 0 | 0 | 0 | 0 | 0 |
| 0 | 0 | 0 | 0 | 0 |
| 0.01202689 | 0.065840518 | 0.095744423 | 0.315088981 | 0.001488703 |
| 1.965836687 | 2.914439897 | 6.978262056 | 2.311179544 | 0.362231214 |
| 0 | 0 | 0 | 0 | 0 |
| 0.005033026 | 0.051227696 | 0.005753946 | 0.00140852 | 0.00142815 |
| 0.092058004 | 0.095844742 | 0.00453433 | 0.003329902 | 0 |
| 0 | 0 | 0 | 0 | 1.587186784 |
| 0 | 0 | 0 | 0 | 0 |
| 0 | 0 | 0 | 0 | 0 |
| 0 | 0 | 0 | 0 | 0.006216407 |
| 0.011120093 | 0.027748272 | 0 | 0 | 0.019484531 |
| 0.105740344 | 0.025667 | 0.047664661 | 0 | 0.091085897 |
| 0 | 0 | 6.704087984 | 35.81959916 | 0.001899074 |
| 0.11425311 | 0.779173346 | 5.343010313 | 5.875574064 | 1.296385218 |
| 0 | 0 | 0 | 0 | 0 |
| 0 | 0 | 0 | 0 | 0.032747054 |
| 0.008381506 | 0.120874086 | 0.059124566 | 0 | 0.069775122 |
| 0 | 0.055872048 | 0.204876652 | 0.095880944 | 0 |
| 0 | 0 | 0 | 0 | 0 |
| 0 | 0 | 0 | 0 | 0 |
| 0 | 0 | 0 | 0 | 0 |
| 0 | 0 | 0 | 0 | 0.007034106 |
| 0 | 0.19724535 | 0.030409544 | 0 | 0.049998433 |
| 0 | 0.955170787 | 0.684217445 | 0 | 0 |
| 0.009995995 | 0.032138454 | 0 | 0.019128601 | 0 |
| 0.001996513 | 0.001453069 | 0 | 0 | 0 |
| 0 | 0.033939287 | 0.014689112 | 0.000520917 | 0.017869994 |
| 0 | 0.024795293 | 0 | 0.017611963 | 0.001190494 |
| 0 | 0 | 0 | 0 | 0 |
| 0.001069988 | 0.097120198 | 0.014041904 | 0.009143676 | 4.717526581 |
| 0 | 0 | 0 | 0 | 0 |
| 0 | 0 | 0 | 0 | 0.040966376 |
| 0 | 0 | 0 | 0 | 0 |
| 0 | 0 | 0 | 0 | 0 |
| 0 | 0 | 0 | 0 | 0 |
| 0.000782045 | 0.085538881 | 0.014155998 | 0.113859007 | 0.008608244 |
| 0 | 0 | 0 | 0 | 0.009912348 |
| 0 | 0 | 0 | 0 | 0 |
| 0.085845722 | 0.555541841 | 0.010814802 | 0.00477722 | 0 |

| | | | | |
|---|---|---|---|---|
| 0 | 0 | 0 | 0 | 0 |
| 0 | 0 | 0 | 0 | 0 |
| 0 | 0 | 0 | 0 | 0 |
| 0.001777423 | 0.007207283 | 0 | 0 | 0.004042711 |
| 0 | 0 | 0 | 0 | 0 |
| 0 | 0 | 0 | 0 | 0.009521324 |
| 0 | 0 | 0 | 0 | 0.009835552 |
| 0.010426635 | 0.272422365 | 0.142106572 | 0.109950319 | 0.004408919 |
| 0 | 0 | 0 | 0 | 0 |
| 0 | 0 | 0 | 0 | 0 |
| 0 | 0.00957066 | 0.001119777 | 0.002819442 | 0 |
| 0 | 0 | 0 | 0 | 0 |
| 0 | 0.076770339 | 0 | 0 | 0.06964728 |
| 0 | 0 | 0 | 0 | 0.017787594 |
| 0 | 0 | 0 | 0 | 0 |
| 0 | 0 | 0 | 0 | 0 |
| 0 | 0 | 0 | 0 | 0.000620126 |
| 0 | 0.000771034 | 0 | 0 | 0.000971765 |
| 0 | 0 | 0 | 0 | 0 |
| 0.008173997 | 0 | 0 | 0 | 0.000500783 |
| 0.001281067 | 0.019865097 | 0 | 0 | 0.025283428 |
| 0 | 0 | 0 | 0 | 0.176994661 |
| 0 | 0 | 0 | 0 | 0 |
| 0 | 0 | 0 | 0 | 0 |
| 0 | 0 | 0 | 0 | 0 |
| 0 | 0 | 0 | 0 | 0 |
| 0 | 0 | 0 | 0 | 0 |
| 0 | 0 | 0 | 0 | 0 |
| 0 | 0 | 0 | 0 | 0 |
| 0.35807956 | 0.253962475 | 0 | 0 | 0.578389103 |
| 0 | 0 | 0 | 0 | 0 |
| 0 | 0 | 0 | 0 | 0 |
| 0 | 0 | 0 | 0 | 0.002175705 |
| 0 | 0 | 0 | 0 | 0 |
| 0 | 0 | 0 | 0 | 0 |
| 0 | 0 | 0 | 0 | 0 |
| 0 | 0 | 0 | 0 | 0.029854802 |
| 0.011101828 | 0.003136154 | 0.001327075 | 0 | 0 |
| 0 | 0 | 0 | 0 | 0.006512188 |
| 0 | 0 | 0 | 0 | 0 |
| 0.00845093 | 0.010053503 | 0.025969754 | 0.012390915 | 0.004503219 |
| 0 | 0 | 0 | 0 | 0 |
| 0 | 0 | 0 | 0 | 0.003501042 |
| 0.062449535 | 0 | 0 | 0 | 0.042771413 |
| 0 | 0 | 0.712108625 | 0.123941045 | 0.026448066 |
| 0 | 0 | 0 | 0 | 0 |
| 0 | 0.004966452 | 0 | 0 | 0.000536521 |
| 0 | 0 | 0 | 0 | 0.212736818 |

| | | | | |
|---|---|---|---|---|
| 0 | 0 | 0 | 0 | 0 |
| 0 | 0 | 0 | 0 | 0 |
| 0 | 0 | 0.020182967 | 0 | 0.011598044 |
| 0 | 0 | 0.018292136 | 0 | 0.908502648 |
| 0 | 0 | 0 | 0 | 0 |
| 0 | 0 | 0 | 0 | 0.091783027 |
| 0 | 0 | 0 | 0 | 0 |
| 0 | 0 | 0 | 0 | 0 |
| 0.010400111 | 0.266004724 | 0.082361639 | 0.034672232 | 0.00116814 |
| 0 | 0 | 0 | 0 | 0 |
| 0 | 0 | 0 | 0 | 0 |
| 0 | 0 | 0 | 0 | 0 |
| 0 | 0 | 0 | 0 | 0 |
| 0 | 0 | 0 | 0 | 0 |
| 4.691259165 | 0.651439577 | 0 | 0 | 0.031479609 |
| 0 | 0.004444818 | 0.571708686 | 0.204311439 | 0 |
| 0 | 0 | 0 | 0 | 0 |
| 0 | 0 | 0 | 0 | 0 |
| 0 | 0 | 0 | 0 | 0 |
| 0 | 0 | 0 | 0 | 0 |
| 0 | 0 | 0 | 0 | 0.015928922 |
| 0 | 0 | 0 | 0 | 0 |
| 0 | 0 | 0 | 0 | 0 |
| 0 | 0 | 0 | 0 | 0 |
| 0.181754874 | 0.001695469 | 0 | 0.028386654 | 0.079903514 |
| 2.849855584 | 3.342321454 | 0 | 0 | 1.383460595 |
| 0 | 0 | 0 | 0 | 1.418184712 |
| 0 | 0 | 0 | 0.001121331 | 0 |
| 0.052572551 | 0.050769614 | 0.0330651 | 0.023915579 | 0.073861874 |
| 0 | 0 | 0 | 0 | 0.030994757 |
| 0 | 0 | 0 | 0 | 0 |
| 0 | 0.100181021 | 0 | 0.001661924 | 0.026392655 |
| 0 | 0 | 0 | 0 | 0 |
| 0 | 0.007243913 | 0 | 0 | 0 |
| 0.00388636 | 0.00186495 | 0.002976263 | 0.001816682 | 0 |
| 0.00105661 | 0.011337325 | 0 | 0.000802609 | 0 |
| 0.305091717 | 0.005713859 | 0.349155979 | 0 | 0 |
| 0 | 1.741342514 | 0 | 0 | 0.063519978 |
| 0 | 0 | 0 | 0 | 2.41357389 |
| 0 | 0 | 0 | 0 | 0 |
| 0 | 0 | 0 | 0 | 0 |
| 0 | 0 | 0 | 0 | 0 |
| 0.248057926 | 0.043595617 | 0 | 0 | 0.082320591 |
| 0 | 0 | 0 | 0 | 1.079051427 |
| 0 | 0 | 0 | 0 | 0 |
| 0 | 0.025977489 | 0.005617033 | 0.00288941 | 0 |
| 0 | 0 | 0 | 0 | 0 |
| 2.60928292 | 1.172316916 | 0.033773774 | 0.041186315 | 0.015690826 |

| | | | | |
|---|---|---|---|---|
| 0 | 0.007418517 | 0 | 0 | 0 |
| 0 | 0 | 0 | 0 | 0 |
| 0 | 0 | 0 | 0 | 0.027502751 |
| 0 | 0 | 0 | 0 | 0 |
| 0 | 0.005317915 | 0.001500197 | 0.002877932 | 0 |
| 0.001140147 | 0 | 0.019606202 | 0.105833066 | 0 |
| 0 | 0 | 0 | 0 | 0.037095743 |
| 0 | 0 | 0 | 0 | 0 |
| 0 | 0 | 0 | 0 | 0 |
| 0 | 0.005041107 | 0 | 0 | 0.295998561 |
| 0 | 0 | 0 | 0 | 0 |
| 0.402181016 | 0 | 2.227295923 | 0.095748688 | 0.169159338 |
| 0 | 0 | 0 | 0 | 0 |
| 0 | 0 | 0 | 0 | 0 |
| 0.001369263 | 0.734603436 | 1.361462024 | 0.033283287 | 0.044108578 |
| 0 | 0 | 0 | 0 | 0 |
| 0 | 0.002139165 | 0 | 0 | 0 |
| 0 | 0 | 0 | 0 | 0 |
| 0 | 0.002096345 | 0 | 0 | 0 |
| 0 | 0 | 0 | 0 | 0.004484193 |
| 0 | 0 | 0 | 0 | 0.028257907 |
| 0 | 0 | 0 | 0 | 0 |
| 0.193953261 | 0.002329612 | 0.084707016 | 0 | 0.005977063 |
| 0 | 0 | 0 | 0 | 0 |
| 0 | 0 | 0 | 0 | 0 |
| 0 | 0 | 0 | 0 | 0 |
| 0 | 0 | 0 | 0 | 0 |
| 0 | 0 | 0 | 0 | 0 |
| 0 | 0 | 0 | 0 | 0.014462629 |
| 0 | 0 | 0 | 0 | 0 |
| 0 | 0 | 0 | 0 | 0 |
| 0.092394156 | 0.050544455 | 0.004502753 | 0 | 0.015566561 |
| 0 | 0 | 0 | 0 | 0 |
| 0.009764304 | 0.2188432 | 0.003374247 | 0.005259355 | 0.002033929 |
| 0.001379828 | 0.012337851 | 0 | 0.000698752 | 0 |
| 0.002964608 | 0.054002976 | 0.011269264 | 0.004729071 | 0.179212352 |
| 0 | 0 | 0 | 0 | 0 |
| 0 | 0 | 0 | 0 | 0.021933435 |
| 0 | 0.003626002 | 0 | 0 | 0.000534155 |
| 0 | 0 | 0 | 0 | 0 |
| 0 | 0 | 0 | 0 | 0.074737681 |
| 0 | 0 | 0 | 0 | 0.303359027 |
| 0 | 0 | 0 | 0 | 0 |
| 0.018721777 | 0 | 0 | 0 | 0 |
| 0.002491992 | 0.191637802 | 0.003485634 | 0.023919826 | 0.000916038 |
| 0 | 0 | 0 | 0 | 0 |
| 0 | 0 | 0 | 0 | 0.087221344 |
| 0.000857083 | 0 | 0.000581786 | 0.00195314 | 0 |

| | | | | |
|---|---|---|---|---|
| 0 | 0 | 0 | 0 | 0 |
| 0 | 0 | 0 | 0 | 0 |
| 0 | 0.002923325 | 0 | 0 | 0 |
| 0 | 0 | 0 | 0 | 0 |
| 0 | 0 | 0 | 0 | 0 |
| 0 | 0 | 0 | 0 | 0.000679208 |
| 0.09130259 | 0 | 0 | 0 | 0.513450399 |
| 0 | 0 | 0 | 0 | 0.001491237 |
| 0 | 0 | 0 | 0 | 0 |
| 0 | 0 | 0 | 0 | 0.001614178 |
| 0 | 0 | 0 | 0 | 0 |
| 0 | 0 | 0 | 0.002173072 | 0 |
| 0 | 0 | 0 | 0 | 0 |
| 0 | 0 | 0 | 0 | 0.112475469 |
| 0 | 0 | 0 | 0 | 0 |
| 0 | 0 | 0 | 0 | 0 |
| 0 | 0 | 0 | 0 | 0.179395804 |
| 0 | 0 | 0 | 0 | 0.001845249 |
| 0.208300729 | 0 | 0 | 0 | 6.168700706 |
| 0 | 0 | 0 | 0 | 0 |
| 0.015540687 | 0 | 0 | 0 | 0 |
| 0 | 0 | 0 | 0 | 0 |
| 0 | 0 | 0 | 0 | 0 |
| 0 | 0 | 0 | 0 | 0 |
| 0 | 0 | 0 | 0 | 0 |
| 0.008225279 | 0.012104347 | 0 | 0.002883685 | 0 |
| 0 | 0 | 0 | 0 | 0 |
| 0.037364489 | 0.001802918 | 0.002428862 | 0 | 0.007373344 |
| 0 | 0 | 0 | 0 | 0.015165762 |
| 1.073607369 | 0.268475475 | 0.366902347 | 0 | 0.399181846 |
| 0 | 0 | 0 | 0 | 0 |
| 0 | 0 | 0 | 0 | 0 |
| 0 | 0 | 0 | 0 | 0.002937968 |
| 0 | 0 | 0 | 0 | 0 |
| 0.001105075 | 0.001103009 | 0 | 0.000503654 | 0.000446839 |
| 0 | 0 | 0 | 0 | 0 |
| 0 | 0 | 0 | 0 | 0.270748634 |
| 0.000752362 | 0.002816083 | 0 | 0.002857498 | 0 |
| 0 | 0 | 0 | 0 | 0 |
| 0 | 0 | 0 | 0 | 0 |
| 10.48626066 | 20.77749447 | 7.709826407 | 4.852719547 | 0.286089344 |
| 0.65665636 | 0.494503497 | 0 | 0 | 0.21390571 |
| 0 | 0 | 0 | 0 | 0 |
| 0 | 0 | 0 | 0 | 0.007463351 |
| 0 | 0 | 0 | 0 | 0 |
| 0 | 0 | 0 | 0 | 0 |
| 0 | 0 | 0 | 0 | 0.00134254 |
| 0 | 0.001738333 | 0 | 0.004233355 | 0.00067068 |

| | | | | |
|---|---|---|---|---|
| 0 | 0 | 0 | 0 | 0 |
| 0.001228366 | 0.045466764 | 0.006670494 | 0 | 0 |
| 0 | 0.001789976 | 0 | 0 | 0 |
| 0 | 0 | 0 | 0 | 0 |
| 5.32423627 | 0.046256852 | 0 | 0 | 0.281870745 |
| 0 | 0 | 0 | 0 | 0 |
| 0 | 0 | 0 | 0 | 0 |
| 0 | 0 | 0 | 0 | 0.010678732 |
| 0.546225816 | 0.348914787 | 0.094174321 | 0.002795101 | 0 |
| 0 | 0 | 0 | 0 | 0.00133814 |
| 0 | 0 | 0.00774561 | 0 | 0 |
| 0 | 0 | 0 | 0 | 0 |
| 0 | 0 | 0 | 0 | 0.023728865 |
| 0.036439667 | 0.080249325 | 0.01150987 | 0.074679848 | 0 |
| 0 | 0.241794081 | 0.708593284 | 0.491744137 | 0.195384905 |
| 2.847324694 | 0.097651278 | 0.07501443 | 0 | 0.069651619 |
| 0 | 0.071291646 | 0.007278467 | 0 | 0 |
| 0 | 0 | 0 | 0 | 0 |
| 0 | 0 | 0 | 0 | 0 |
| 0 | 0 | 0 | 0 | 0 |
| 0 | 0 | 0 | 0 | 0.000831415 |
| 0 | 0 | 0 | 0 | 0 |
| 0 | 0 | 0 | 0 | 0 |
| 0 | 0 | 0 | 0 | 0 |
| 0 | 0 | 0 | 0 | 0.010085909 |
| 0 | 0 | 0 | 0 | 0 |
| 0 | 0 | 0 | 0 | 0.011037371 |
| 0.00290706 | 0.017357945 | 0 | 0 | 0 |
| 0 | 0 | 0 | 0 | 0 |
| 0 | 0 | 0 | 0 | 0 |
| 0 | 0 | 0 | 0 | 0 |
| 0 | 0 | 0 | 0 | 0 |
| 0 | 0 | 0 | 0 | 0 |
| 0 | 0 | 0 | 0 | 0.00469414 |
| 0 | 0 | 0.13514243 | 0.190668364 | 0.082584759 |
| 0 | 0.031467935 | 0 | 0 | 0.007770176 |
| 0 | 0 | 0 | 0 | 0 |
| 0 | 0 | 0 | 0 | 0.001375734 |
| 0 | 0 | 0 | 0 | 0 |
| 0 | 0 | 0 | 0 | 0 |
| 5.262451257 | 0 | 3.266206307 | 0.079778113 | 0.040841501 |
| 1.046396497 | 0 | 0 | 0 | 0 |
| 0 | 0 | 0 | 0 | 0 |
| 0.021856222 | 0 | 0 | 0 | 0 |
| 0 | 0 | 0 | 0 | 0 |
| 0 | 0 | 0 | 0 | 0 |
| 0 | 0 | 0 | 0 | 0 |
| 0 | 0 | 0 | 0 | 0 |

| | | | | |
|---|---|---|---|---|
| 0 | 0 | 0 | 0 | 0.534579267 |
| 0 | 0 | 0 | 0 | 0 |
| 0 | 0 | 0 | 0 | 0 |
| 0 | 0 | 0 | 0 | 0 |
| 0 | 0 | 0 | 0 | 0.014560734 |
| 0 | 0 | 0 | 0 | 0 |
| 0.039056362 | 0.013433456 | 0.003821457 | 0 | 0.009674692 |
| 0 | 0 | 0 | 0 | 0 |
| 0 | 0 | 0 | 0 | 0 |
| 0 | 0 | 0.00521213 | 0.000504747 | 0.008657631 |
| 0 | 0 | 0 | 0 | 0.050551755 |
| 0 | 0 | 0 | 0 | 0 |
| 0 | 0 | 0 | 0 | 0 |
| 0 | 0 | 0 | 0 | 0 |
| 0 | 0 | 0 | 0 | 0 |
| 0 | 0 | 0 | 0 | 0 |
| 0 | 0 | 0 | 0 | 0 |
| 0 | 0 | 0 | 0 | 0 |
| 0 | 0 | 0 | 0 | 0 |
| 0 | 0 | 0 | 0 | 0 |
| 0.035575884 | 0 | 0.003820152 | 0 | 0 |
| 0 | 0 | 0 | 0 | 0 |
| 0 | 0.012170453 | 0.010433712 | 0.009430481 | 0.008217268 |
| 0.00571688 | 0.023611839 | 0.001605767 | 0 | 0.005579799 |
| 0 | 0 | 0 | 0 | 0 |
| 0 | 0 | 0 | 0 | 0 |
| 0 | 0 | 0 | 0 | 0 |
| 0 | 0 | 0 | 0 | 0 |
| 0 | 0 | 0 | 0 | 0.012731241 |
| 0 | 1.196837798 | 0.38492241 | 0.021120682 | 0.000573617 |
| 0 | 0 | 0 | 0 | 0.003750925 |
| 0 | 0 | 0 | 0 | 0 |
| 0 | 0 | 0 | 0 | 0 |
| 0 | 0 | 0 | 0 | 0 |
| 0 | 0 | 0 | 0 | 0.026199988 |
| 0 | 0 | 0 | 0 | 0 |
| 0 | 0 | 0 | 0 | 0 |
| 0 | 0 | 0 | 0 | 0 |
| 0 | 0 | 0 | 0 | 0 |
| 0 | 0 | 0 | 0 | 0.09954803 |
| 0 | 0 | 0 | 0 | 0 |
| 0 | 0 | 0 | 0 | 0 |
| 0 | 0 | 0 | 0 | 0 |
| 0 | 0 | 0 | 0 | 0.001744888 |
| 0 | 0.000873802 | 0 | 0 | 0.016833957 |
| 0 | 1.268273981 | 0 | 0 | 0 |
| 0 | 0 | 0 | 0 | 0 |
| 0 | 0 | 0 | 0 | 0.057198838 |

| | | | | |
|---|---|---|---|---|
| 0.004878828 | 0.024613203 | 0.009215266 | 0.003705994 | 0 |
| 0 | 0 | 0 | 0 | 0.078363398 |
| 0 | 0 | 0 | 0 | 0 |
| 0 | 0 | 0 | 0 | 0 |
| 0 | 0 | 0 | 0 | 0 |
| 0 | 0 | 0 | 0 | 0 |
| 0 | 0 | 0 | 0 | 0 |
| 3.312555688 | 0.065218571 | 0.041001978 | 0.000992666 | 0.127112597 |
| 0.145170053 | 0.014539722 | 0.019889022 | 0.98536864 | 2.797439335 |
| 0.036160169 | 0 | 0 | 0 | 0 |
| 0 | 0 | 0.002215664 | 0 | 0.003849549 |
| 0 | 0 | 0 | 0 | 0 |
| 0 | 0 | 0 | 0 | 0.019616159 |
| 0 | 0 | 0.003569501 | 0.003557553 | 0.273762495 |
| 0 | 0 | 0 | 0 | 0 |
| 0 | 0 | 0 | 0 | 0 |
| 0 | 0 | 0 | 0 | 0 |
| 0 | 0 | 0 | 0 | 0 |
| 0 | 0 | 0 | 0 | 0 |
| 0 | 0.01798608 | 0 | 0 | 0 |
| 0 | 0 | 0 | 0 | 0 |
| 0 | 0 | 0 | 0 | 0 |
| 0 | 0 | 0 | 0 | 0 |
| 0 | 0 | 0 | 0 | 0 |
| 0 | 0 | 0 | 0 | 0.009120274 |
| 0 | 0 | 0 | 0 | 0 |
| 0 | 0 | 0 | 0 | 0 |
| 0 | 0 | 0 | 0 | 0.023205378 |
| 0 | 0 | 0 | 0 | 0 |
| 0 | 0 | 0 | 0 | 0 |
| 0 | 0.178904053 | 0 | 0 | 0 |
| 0 | 0 | 0 | 0 | 0 |
| 0 | 0 | 0 | 0 | 0 |
| 0 | 0 | 0 | 0 | 0 |
| 0.004108875 | 0.005767306 | 0 | 0 | 0 |
| 0 | 0 | 0 | 0 | 0 |
| 0 | 0 | 0 | 0 | 0 |
| 0 | 0 | 0 | 0 | 0 |
| 0 | 0.005664585 | 0 | 0 | 0 |
| 0.100800207 | 0 | 0 | 0 | 0 |
| 0 | 0 | 0 | 0 | 0 |
| 0 | 0.302837733 | 0 | 0.170434732 | 0.100707703 |
| 0 | 0 | 0 | 0 | 0 |
| 0 | 0 | 0 | 0 | 0 |
| 0 | 0 | 0 | 0 | 0 |
| 0 | 0 | 0 | 0.000716387 | 0 |
| 0 | 0 | 0 | 0 | 0 |
| 0 | 0 | 0 | 0 | 0 |

| | | | | |
|---|---|---|---|---|
| 0 | 0 | 0 | 0 | 0 |
| 0 | 0.001433193 | 0.277239341 | 0.176403001 | 0.251980156 |
| 0 | 0 | 0 | 0 | 0 |
| 0 | 0 | 0 | 0 | 0 |
| 0.119382764 | 1.544599957 | 0.352976076 | 0.117319938 | 2.633167886 |
| 0 | 0 | 0.351586281 | 0 | 0 |
| 0 | 0 | 0 | 0 | 0 |
| 0.013761624 | 0.085512722 | 0.008303427 | 0.003689447 | 0.002545867 |
| 0 | 0.001680734 | 0 | 0 | 0.001452545 |
| 0 | 0 | 0 | 0 | 0.069078605 |
| 0 | 0 | 0 | 0 | 0 |
| 0 | 0.138077408 | 0 | 0 | 0.074717724 |
| 0 | 0 | 0 | 0 | 0 |
| 0 | 0 | 0 | 0 | 0 |
| 0.002849323 | 0 | 0 | 0 | 0 |
| 0.004446504 | 0 | 0 | 0 | 0 |
| 0 | 0 | 0 | 0 | 0 |
| 0 | 0 | 0 | 0 | 0 |
| 0 | 0 | 0 | 0 | 0 |
| 0 | 0 | 0 | 0 | 0 |
| 0 | 0 | 0 | 0 | 0 |
| 0 | 0 | 0 | 0 | 0 |
| 0 | 0 | 0 | 0 | 0 |
| 0 | 0.007575235 | 0 | 0.001596456 | 0 |
| 0 | 0 | 0.288834204 | 0.66301046 | 0 |
| 0 | 0 | 0 | 0 | 0 |
| 0 | 0 | 0.004252697 | 0.003082519 | 0.012378542 |
| 0 | 0.001964144 | 0 | 0.001024987 | 0 |
| 0 | 0 | 0 | 0 | 0 |
| 0 | 0 | 0 | 0 | 0 |
| 0 | 0 | 0 | 0 | 0 |
| 0 | 0.001024502 | 0 | 0.000623741 | 0 |
| 0 | 0 | 0 | 0 | 0 |
| 0 | 0 | 0 | 0 | 0 |
| 0 | 0 | 0 | 0 | 0 |
| 0 | 0 | 0 | 0 | 0 |
| 0 | 0 | 0 | 0 | 0 |
| 0 | 0 | 0 | 0 | 0.018451918 |
| 0 | 0 | 0 | 0 | 0 |
| 0 | 0 | 0 | 0 | 0 |
| 0 | 0 | 0 | 0 | 0 |
| 0 | 0 | 0 | 0 | 0.168246213 |
| 0 | 0 | 0 | 0 | 0 |
| 0 | 0 | 0 | 0 | 0 |
| 0 | 0 | 0 | 0 | 0 |
| 0 | 0 | 0 | 0 | 0 |
| 0 | 0 | 0 | 0 | 0 |
| 0 | 0 | 0 | 0 | 0.000437045 |

| | | | | |
|---|---|---|---|---|
| 0 | 0.222281331 | 0 | 0 | 0.025718381 |
| 0 | 0 | 0 | 0 | 0.005550223 |
| 0 | 0 | 0 | 0 | 0 |
| 0 | 0 | 0 | 0 | 0 |
| 0 | 0 | 0 | 0 | 0 |
| 0 | 0.054582692 | 0.000631829 | 0.001944382 | 0 |
| 0 | 0 | 0 | 0 | 0 |
| 0 | 0 | 0 | 0 | 0 |
| 0 | 0 | 0 | 0 | 0 |
| 0 | 0 | 0 | 0 | 0 |
| 0 | 0 | 0 | 0 | 0 |
| 0.003229032 | 0 | 0 | 0 | 0 |
| 0 | 0 | 0 | 0 | 0.00479165 |
| 0 | 0 | 0 | 0 | 0 |
| 0 | 0 | 0 | 0 | 0 |
| 0 | 0 | 0 | 0 | 0 |
| 0 | 0 | 0 | 0 | 0 |
| 0 | 0 | 0 | 0 | 0 |
| 0 | 0 | 0 | 0 | 0 |
| 0 | 0 | 0 | 0 | 0.001399294 |
| 0.008604144 | 0 | 0 | 0 | 0 |
| 0 | 0 | 0 | 0 | 0 |
| 0 | 0.002931378 | 0 | 0.004015559 | 0.001187528 |
| 0 | 0 | 0 | 0 | 0.446167157 |
| 0 | 0 | 0 | 0 | 0.01439966 |
| 0 | 0 | 0 | 0 | 0.013045571 |
| 0 | 0 | 0 | 0 | 0 |
| 0 | 0 | 0 | 0 | 0 |
| 0 | 0 | 0 | 0 | 0 |
| 0 | 0 | 0 | 0 | 0 |
| 0 | 0 | 0 | 0 | 0 |
| 0 | 0 | 0 | 0 | 0 |
| 0.062708295 | 0.36511467 | 0.005476357 | 0.139140518 | 0 |
| 0 | 0 | 0 | 0 | 0 |
| 0 | 0 | 0 | 0 | 0 |
| 0 | 0 | 0.074907054 | 0.005197128 | 0 |
| 0 | 0 | 0 | 0 | 0 |
| 0 | 0 | 0 | 0 | 0 |
| 0.007147852 | 0.002866537 | 0.004158809 | 0 | 0 |
| 0 | 0 | 0 | 0 | 0 |
| 0 | 0 | 0 | 0 | 0 |
| 0 | 0 | 0 | 0 | 0 |
| 0.014029468 | 0.021973178 | 0.019181415 | 0.030610214 | 0.937829295 |
| 0 | 0 | 0 | 0 | 0 |
| 0 | 0 | 0 | 0 | 0 |
| 0 | 0.048506515 | 0 | 0 | 0.016455236 |
| 0 | 0 | 0 | 0 | 0 |
| 0 | 0 | 0 | 0 | 0 |

| | | | | |
|---|---|---|---|---|
| 0 | 0 | 0 | 0 | 0 |
| 0 | 0 | 0 | 0 | 0 |
| 0 | 0 | 0 | 0 | 0 |
| 0 | 0 | 0 | 0 | 0 |
| 0 | 0 | 0 | 0 | 0 |
| 0 | 0 | 0 | 0 | 0 |
| 0 | 0 | 0 | 0 | 0 |
| 0.029321637 | 0 | 0 | 0 | 0 |
| 0 | 0.012747079 | 0 | 0 | 0.006136876 |
| 0.003418372 | 0 | 0.003093843 | 0 | 0.04722602 |
| 0 | 0 | 0 | 0 | 0 |
| 0 | 0 | 0 | 0 | 0 |
| 0 | 0 | 0 | 0 | 0 |
| 0 | 0 | 0 | 0 | 0 |
| 0 | 0.011571012 | 0 | 0 | 0 |
| 0.001201179 | 0.02397867 | 0 | 0 | 0 |
| 0 | 0 | 0 | 0 | 0 |
| 0 | 0 | 0 | 0 | 0 |
| 0 | 0 | 0 | 0 | 0 |
| 8.547949045 | 0 | 0 | 0 | 0.060454167 |
| 0.007025457 | 0 | 0.005677217 | 0 | 0 |
| 0 | 0 | 0 | 0 | 0 |
| 0 | 0 | 0 | 0 | 0 |
| 0 | 0 | 0 | 0 | 0 |
| 0 | 0.001620391 | 0 | 0 | 0 |
| 0 | 0 | 0 | 0 | 0.28497193 |
| 0 | 0 | 0 | 0 | 0 |
| 0 | 0 | 0 | 0 | 0.091578139 |
| 0 | 0 | 0 | 0 | 0 |
| 0 | 0 | 0 | 0 | 0.012584738 |
| 0 | 0.009744595 | 0.242106135 | 0.009789025 | 0.19922339 |
| 0 | 0 | 0 | 0 | 0 |
| 0 | 0 | 0 | 0 | 0.000500119 |
| 0 | 0 | 0 | 0 | 0 |
| 0 | 0 | 0 | 0 | 0 |
| 0 | 0 | 0 | 0 | 0 |
| 0 | 0 | 0 | 0 | 0 |
| 0 | 0 | 0 | 0 | 0 |
| 0 | 0 | 0 | 0.007689398 | 0.025656627 |
| 0 | 0 | 0 | 0 | 0 |
| 0 | 0 | 0 | 0 | 0 |
| 0 | 0 | 0 | 0 | 0.010078054 |
| 0 | 0 | 0 | 0 | 0.00042697 |
| 0 | 0 | 0 | 0 | 0 |
| 0 | 0 | 0 | 0 | 0 |
| 0 | 0 | 0 | 0 | 0 |
| 0.169690016 | 0 | 0.125312543 | 0 | 0 |
| 0 | 0 | 0 | 0 | 0 |

| | | | | |
|---|---|---|---|---|
| 0 | 0 | 0 | 0 | 0 |
| 0 | 0 | 0 | 0 | 0 |
| 0 | 0 | 0 | 0 | 0 |
| 0 | 0 | 0 | 0 | 0 |
| 0.077716205 | 0.04149037 | 0 | 0 | 0.011267324 |
| 0 | 0 | 0 | 0 | 0 |
| 0 | 0 | 0 | 0 | 0 |
| 0.005629064 | 0 | 0 | 0 | 0 |
| 0 | 0 | 0 | 0 | 0 |
| 0 | 0 | 0 | 0 | 0 |
| 0 | 0 | 0 | 0 | 0.002515268 |
| 0 | 0.043600319 | 0.003232054 | 0.006975307 | 0 |
| 0 | 0.00439312 | 0.01473894 | 0.028919509 | 0.004745847 |
| 0 | 0 | 0 | 0 | 0 |
| 0 | 0.028133376 | 0 | 0 | 0.001048008 |
| 0 | 0 | 0.050985707 | 0 | 0 |
| 0.002354215 | 0.024966786 | 0 | 0 | 0.004627444 |
| 0 | 0 | 0 | 0 | 0 |
| 0.056978038 | 0.325083507 | 0 | 0.003374447 | 0 |
| 0.00807007 | 0.001887887 | 0 | 0 | 0.018695117 |
| 0 | 0 | 0 | 0 | 0 |
| 0 | 0 | 0 | 0 | 0.008940822 |
| 0 | 0 | 0 | 0 | 0 |
| 0 | 0 | 0 | 0 | 0 |
| 0 | 0 | 0 | 0 | 0 |
| 0 | 0 | 0 | 0 | 0 |
| 0.018390295 | 0.059757601 | 0.028350319 | 0.082895375 | 0.000953353 |
| 0 | 0 | 0 | 0 | 0 |
| 0 | 0 | 0 | 0 | 0 |
| 0 | 0 | 0 | 0 | 0 |
| 0 | 0.002858159 | 0 | 0 | 0 |
| 0 | 0 | 0 | 0 | 0 |
| 0 | 0 | 0 | 0 | 0 |
| 0 | 0 | 0 | 0 | 0 |
| 0 | 0 | 0 | 0 | 0 |
| 0 | 0 | 0 | 0 | 0 |
| 0 | 0 | 0 | 0 | 0 |
| 0.003540804 | 0.1006038 | 0.001893658 | 0.002567365 | 0 |
| 0 | 0 | 0 | 0 | 0.091583254 |
| 0 | 0 | 0 | 0 | 0 |
| 0 | 0.005212197 | 0 | 0 | 0 |
| 0 | 0 | 0 | 0 | 0.00375423 |
| 0 | 0 | 0 | 0 | 0 |
| 0.001670634 | 0.005062087 | 0.101089946 | 0.157721776 | 0 |
| 0 | 0 | 0 | 0 | 0 |
| 0 | 0 | 0 | 0 | 0 |
| 0 | 0 | 0 | 0 | 0 |
| 0 | 0 | 0 | 0 | 0 |

| | | | | |
|---|---|---|---|---|
| 0 | 0 | 0 | 0 | 0 |
| 0 | 0 | 0 | 0 | 0.000824821 |
| 0 | 0 | 0 | 0 | 0.02020222 |
| 0.001500713 | 0.003120641 | 0.00067912 | 0 | 0 |
| 0 | 0 | 0 | 0 | 0 |
| 0 | 0 | 0 | 0.001364531 | 0 |
| 0.007038917 | 0 | 0 | 0 | 0.000981448 |
| 0 | 0 | 0.00548835 | 0.003566167 | 0 |
| 0 | 0 | 0 | 0 | 0 |
| 0 | 0 | 0 | 0 | 0.052396051 |
| 0 | 0 | 0 | 0 | 0.012953388 |
| 0 | 0 | 0 | 0 | 0 |
| 0.042729062 | 0 | 0 | 0 | 0.081488977 |
| 0 | 0 | 0 | 0 | 0.001518021 |
| 0 | 0 | 0 | 0 | 0 |
| 0 | 0 | 0 | 0 | 0 |
| 0 | 0 | 0 | 0 | 0 |
| 0 | 0 | 0 | 0 | 0 |
| 0 | 0 | 0 | 0 | 0 |
| 0 | 0 | 0 | 0 | 0 |
| 0 | 0 | 0 | 0 | 0 |
| 0 | 0 | 0 | 0 | 0 |
| 0.008638092 | 0 | 0.016903827 | 0.001064037 | 0 |
| 0 | 0.037809502 | 0 | 0 | 0.459137805 |
| 0 | 0 | 0 | 0 | 0 |
| 0 | 0 | 0 | 0 | 0.001919566 |
| 0 | 0 | 0 | 0 | 0.002254794 |
| 0 | 0 | 0 | 0 | 0 |
| 0 | 0 | 0 | 0 | 0 |
| 0 | 0 | 0 | 0 | 0 |
| 0.080175182 | 0.008119453 | 0.00918825 | 0 | 0.007894233 |
| 0 | 0 | 0.016579102 | 0 | 0 |
| 0 | 0 | 0 | 0 | 0 |
| 0 | 0 | 0 | 0 | 0 |
| 0 | 0 | 0 | 0 | 0.013972066 |
| 0.00190484 | 0 | 0 | 0 | 0 |
| 0 | 0.003105477 | 0 | 0 | 0 |
| 0 | 0 | 0 | 0 | 0 |
| 0 | 0.003297418 | 0.000996653 | 0.000836478 | 0.074657141 |
| 0 | 0 | 0 | 0 | 0 |
| 0 | 0 | 0 | 0 | 0 |
| 0 | 0 | 0 | 0 | 0 |
| 0 | 0 | 0 | 0 | 0 |
| 0 | 0 | 0 | 0 | 0 |
| 0 | 0 | 0 | 0 | 0 |
| 0.033135418 | 0.280447671 | 0.098211719 | 0.002502272 | 0.085274207 |
| 0 | 0 | 0.000834931 | 0 | 0 |
| 0 | 0 | 0 | 0 | 0.017627455 |

| | | | | |
|---|---|---|---|---|
| 0 | 0 | 0 | 0 | 0 |
| 0 | 0 | 0 | 0 | 0 |
| 0 | 0 | 0 | 0 | 0.081037726 |
| 0 | 0 | 0 | 0 | 0.011435329 |
| 0 | 0 | 0 | 0 | 0 |
| 0 | 0 | 0 | 0 | 0 |
| 0 | 0 | 0 | 0 | 0 |
| 0 | 0 | 0 | 0 | 0 |
| 0 | 0 | 0 | 0 | 0 |
| 0 | 0 | 0 | 0 | 0 |
| 0 | 0 | 0 | 0 | 0 |
| 0 | 0 | 0 | 0 | 0 |
| 0 | 0 | 0 | 0 | 0 |
| 0 | 0 | 0 | 0 | 0 |
| 0 | 0 | 0 | 0 | 0 |
| 0 | 0 | 0 | 0 | 0 |
| 0 | 0 | 0.005942689 | 0 | 0 |
| 0 | 0 | 0 | 0 | 0 |
| 0 | 0 | 0 | 0 | 0 |
| 0.00165866 | 0.001478178 | 0 | 0.002879841 | 0 |
| 0 | 0 | 0 | 0 | 0 |
| 0 | 0 | 0 | 0 | 0 |
| 0 | 0 | 0 | 0 | 0 |
| 0 | 0 | 0 | 0 | 0 |
| 0 | 0 | 0 | 0 | 0 |
| 0 | 0 | 0 | 0 | 0 |
| 0 | 0 | 0 | 0 | 0.003058952 |
| 0 | 0 | 0.017325224 | 0.007768388 | 0.000706879 |
| 0 | 0 | 0 | 0 | 0 |
| 0 | 0 | 0 | 0 | 0 |
| 0.004682013 | 0.005333614 | 0.007001117 | 0.001700931 | 0.00137187 |
| 0 | 0 | 0 | 0 | 0 |
| 0.04372671 | 0.027130663 | 0.193170929 | 0.361596133 | 0.005415794 |
| 0.010144465 | 0 | 0 | 0 | 0.002441627 |
| 0 | 0 | 0 | 0 | 0 |
| 0 | 0 | 0 | 0 | 0 |
| 0 | 0 | 0 | 0 | 0 |
| 0 | 0 | 0 | 0 | 0 |
| 0 | 0 | 0 | 0 | 0 |
| 0 | 0 | 0 | 0 | 0 |
| 0 | 0 | 0 | 0 | 0 |
| 0.090808951 | 0.251101692 | 0.174055206 | 0.002806433 | 0 |
| 0.016108997 | 0.004137949 | 0.105059329 | 0.011156823 | 0.035122906 |
| 0 | 0 | 0 | 0 | 0 |
| 0 | 0 | 0 | 0 | 0 |
| 0.001711663 | 0 | 0.025406283 | 0.032764845 | 2.29781915 |
| 0 | 0.003640382 | 0 | 0 | 0 |
| 0 | 0 | 0 | 0 | 0 |

| | | | | |
|---|---|---|---|---|
| 0 | 0 | 0 | 0 | 0 |
| 0 | 0.004025404 | 0.021563477 | 0.034499193 | 0.013547582 |
| 0.015364788 | 0 | 0.003673311 | 0 | 0.000781482 |
| 0 | 0 | 0 | 0 | 0 |
| 0.025461978 | 0.004620798 | 0.007960897 | 0.009670555 | 0.007643701 |
| 0 | 0 | 0 | 0 | 0 |
| 0 | 0 | 0 | 0 | 0 |
| 0 | 0 | 0 | 0 | 0 |
| 0 | 0 | 0 | 0 | 0 |
| 0 | 0 | 0 | 0 | 0 |
| 0 | 0 | 0 | 0 | 0 |
| 0 | 0 | 0 | 0 | 0 |
| 0 | 0 | 0 | 0 | 0 |
| 0 | 0 | 0 | 0 | 0 |
| 0 | 0 | 0 | 0 | 0 |
| 0 | 0 | 0 | 0 | 0 |
| 0 | 0 | 0 | 0 | 0.109088442 |
| 0 | 0 | 0 | 0 | 0 |
| 0 | 0 | 0 | 0 | 0 |
| 0 | 0 | 0 | 0 | 0 |
| 0 | 0 | 0 | 0 | 0 |
| 0 | 0.001163783 | 0 | 0 | 0 |
| 0 | 0 | 0 | 0 | 0 |
| 0 | 0 | 0 | 0 | 0 |
| 0.003822716 | 0 | 0 | 0 | 0.0367109 |
| 0.003124519 | 0.327161394 | 0 | 0 | 0.0006479 |
| 0 | 0 | 0 | 0 | 0 |
| 0 | 0 | 0 | 0 | 0.009741526 |
| 0 | 0 | 0 | 0 | 0 |
| 0 | 0 | 0 | 0 | 0 |
| 0 | 0 | 0 | 0 | 0.00152517 |
| 0 | 0 | 0 | 0 | 0 |
| 0 | 0 | 0 | 0 | 0 |
| 0 | 0 | 0 | 0 | 0 |
| 0 | 0 | 0 | 0 | 0 |
| 0 | 0 | 0 | 0 | 0 |
| 0 | 0 | 0 | 0 | 0 |
| 0.004067179 | 0 | 0 | 0 | 0 |
| 3.400253144 | 0.010618182 | 0.0017775 | 0 | 1.075012709 |
| 0 | 0 | 0 | 0 | 0 |
| 0 | 0 | 0 | 0 | 0 |
| 0 | 0 | 0 | 0 | 0.040590833 |
| 0.001688505 | 0.01835827 | 0.007641021 | 0.002198745 | 0 |
| 0 | 0 | 0 | 0 | 0 |
| 0 | 0 | 0 | 0 | 0 |
| 0 | 0 | 0 | 0 | 0 |
| 0.001107035 | 0.02992617 | 0 | 0 | 0 |
| 0 | 0 | 0 | 0 | 0 |

| | | | | |
|---|---|---|---|---|
| 0 | 0 | 0 | 0 | 0 |
| 0.016762501 | 0.140878779 | 0.046777634 | 0.030771227 | 0.001412073 |
| 0 | 0 | 0 | 0 | 0.000744142 |
| 0 | 0 | 0 | 0 | 0 |
| 0 | 0 | 0 | 0 | 0 |
| 0 | 0 | 0 | 0 | 0 |
| 0 | 0 | 0 | 0 | 0 |
| 0 | 0 | 0 | 0 | 0 |
| 0 | 0 | 0 | 0 | 0 |
| 0 | 0.008541406 | 0 | 0 | 0 |
| 0 | 0 | 0 | 0 | 0 |
| 0 | 0 | 0 | 0 | 0 |
| 0 | 0.012896282 | 0 | 0 | 0.079941618 |
| 0 | 0 | 0 | 0 | 0 |
| 0.012482131 | 0.023541339 | 0.005254474 | 0 | 0 |
| 0 | 0 | 0 | 0 | 0 |
| 0 | 0 | 0 | 0 | 0 |
| 0 | 0 | 0 | 0 | 0 |
| 0 | 0 | 0 | 0 | 0 |
| 0 | 0 | 0 | 0 | 0 |
| 0 | 0 | 0 | 0 | 0 |
| 0 | 0 | 0 | 0 | 0 |
| 0 | 0 | 0.035687469 | 0.000998401 | 0 |
| 0 | 0 | 0.002126037 | 0 | 0 |
| 0 | 0 | 0 | 0 | 0 |
| 0 | 0 | 0 | 0 | 0 |
| 0.004715857 | 0 | 0 | 0 | 0 |
| 0 | 0 | 0 | 0 | 0 |
| 0 | 0 | 0 | 0 | 0 |
| 0 | 0 | 0 | 0 | 0 |
| 0 | 0 | 0 | 0 | 0 |
| 0 | 0 | 0 | 0 | 0 |
| 0 | 0 | 0 | 0 | 0 |
| 0 | 0 | 0 | 0 | 0 |
| 0 | 10.33250293 | 1.780637818 | 0.470498076 | 2.976736383 |
| 0 | 0 | 0 | 0 | 0 |
| 0 | 0.001877186 | 0 | 0 | 0 |
| 0 | 0 | 0 | 0 | 0 |
| 0 | 0.001420774 | 0 | 0 | 0.011127652 |
| 0 | 0 | 0 | 0 | 0 |
| 0.196549734 | 0.467525499 | 0.032805679 | 0.088223112 | 0.030964382 |
| 0 | 0 | 0 | 0 | 0 |
| 0 | 0 | 0 | 0 | 0 |
| 0 | 0 | 0 | 0 | 0 |
| 0 | 0 | 0 | 0 | 0 |
| 0.037700848 | 3.636874232 | 0.032005582 | 0.012209935 | 0.001772605 |
| 0 | 0 | 0 | 0 | 0 |
| 0 | 0 | 0 | 0 | 0 |

| | | | | |
|---|---|---|---|---|
| 0 | 0 | 0 | 0 | 0 |
| 0 | 0 | 0 | 0 | 0 |
| 0 | 0 | 0 | 0 | 0 |
| 0 | 0 | 0 | 0 | 0.010066379 |
| 0 | 0.00165929 | 0 | 0 | 0 |
| 0 | 0 | 0 | 0 | 0 |
| 0 | 0 | 0 | 0 | 0 |
| 0 | 0 | 0 | 0 | 0 |
| 0 | 0 | 0 | 0 | 0 |
| 0 | 0.006062439 | 0.147970773 | 0.033869969 | 0.004430339 |
| 0 | 0 | 0 | 0 | 0.009751148 |
| 0 | 0 | 0 | 0 | 0 |
| 0.001955592 | 0.004182723 | 0 | 0 | 0.003012371 |
| 0 | 0 | 0 | 0 | 0 |
| 0 | 0 | 0 | 0 | 0 |
| 0 | 0 | 0 | 0 | 0 |
| 0 | 0 | 0 | 0 | 0 |
| 0 | 0 | 0 | 0 | 0 |
| 0 | 0 | 0 | 0 | 0 |
| 0.11537793 | 1.176442218 | 0.579960117 | 1.158611341 | 0.262089536 |
| 0 | 0 | 0 | 0 | 0 |
| 0 | 0 | 0 | 0 | 0 |
| 0 | 0 | 0 | 0 | 0 |
| 0 | 0 | 0 | 0 | 0 |
| 0 | 0 | 0 | 0 | 0 |
| 0 | 0 | 0 | 0 | 0.008708846 |
| 0 | 0 | 0 | 0 | 0 |
| 0 | 0 | 0 | 0 | 0 |
| 0 | 0 | 0 | 0 | 0 |
| 0 | 0 | 0.035847119 | 0 | 0 |
| 0 | 0 | 0 | 0 | 0 |
| 0 | 0 | 0 | 0 | 0 |
| 0 | 0 | 0 | 0 | 0 |
| 0 | 0 | 0 | 0 | 0 |
| 0 | 0 | 0 | 0 | 0 |
| 0 | 0 | 0 | 0 | 0 |
| 0 | 0 | 0 | 0 | 0 |
| 0 | 0 | 0 | 0 | 0 |
| 0 | 0 | 0 | 0 | 0 |
| 0 | 0 | 0 | 0 | 0 |
| 0 | 0 | 0 | 0 | 0 |
| 0 | 0 | 0 | 0 | 0 |
| 0 | 0 | 0 | 0 | 0.001253665 |
| 0 | 0 | 0 | 0 | 0 |
| 0 | 0.022145778 | 0.004650305 | 0.005854408 | 0 |
| 0 | 0 | 0 | 0 | 0.005948769 |
| 0 | 0 | 0 | 0 | 0 |
| 0 | 0 | 0 | 0 | 0 |

| | | | | |
|---|---|---|---|---|
| 0 | 0 | 0 | 0 | 0 |
| 0 | 0 | 0 | 0 | 0 |
| 0 | 0 | 0 | 0 | 0 |
| 0 | 0 | 0 | 0 | 0 |
| 0 | 0 | 0 | 0 | 0 |
| 0.002440958 | 0.054601365 | 0 | 0 | 0.00340752 |
| 0 | 0 | 0 | 0 | 0.005772144 |
| 0 | 0 | 0 | 0 | 0 |
| 0 | 0 | 0 | 0 | 0 |
| 0 | 0 | 0 | 0 | 0.004833968 |
| 0 | 0 | 0 | 0 | 0 |
| 0 | 0 | 0 | 0 | 0 |
| 0 | 0 | 0 | 0 | 0 |
| 0 | 0 | 0 | 0 | 0 |
| 0 | 0 | 0 | 0 | 0 |
| 0 | 0 | 0 | 0 | 0 |
| 0 | 0 | 0 | 0 | 0.017103302 |
| 0 | 0 | 0 | 0 | 0 |
| 0 | 0 | 0 | 0 | 0 |
| 0 | 0 | 0 | 0 | 0 |
| 0 | 0 | 0 | 0 | 0 |
| 0 | 0 | 0 | 0 | 0 |
| 0 | 0 | 0 | 0 | 0 |
| 0 | 0 | 0 | 0 | 0 |
| 0 | 0 | 0 | 0 | 0 |
| 0 | 0 | 0.000987312 | 0 | 0 |
| 0 | 0 | 0 | 0 | 0 |
| 0 | 1.054811665 | 0 | 0 | 0 |
| 0 | 0 | 0 | 0 | 0 |
| 0.008510401 | 0 | 0 | 0 | 0 |
| 0 | 0 | 0 | 0 | 0 |
| 0 | 0 | 0 | 0 | 0 |
| 0 | 0 | 0 | 0 | 0 |
| 0 | 0 | 0 | 0 | 0 |
| 0 | 0 | 0 | 0 | 0 |
| 0 | 0 | 0 | 0 | 0 |
| 0 | 0 | 0 | 0 | 0 |
| 0 | 0 | 0 | 0 | 0 |
| 0 | 0 | 0 | 0 | 0.089565894 |
| 0 | 0 | 0 | 0 | 0 |
| 0 | 0 | 0 | 0 | 0 |
| 0 | 0 | 0 | 0 | 0 |
| 0 | 0 | 0 | 0 | 0 |
| 0 | 0 | 0 | 0 | 0 |
| 0 | 0 | 0 | 0 | 0 |
| 0 | 0 | 0 | 0 | 0 |
| 0 | 0 | 0 | 0 | 0 |
| 0 | 0 | 0 | 0 | 0 |

| | | | | |
|---|---|---|---|---|
| 0 | 0 | 0 | 0 | 0 |
| 0 | 0 | 0 | 0 | 0.008419296 |
| 0 | 0 | 0 | 0 | 0 |
| 0 | 0 | 0 | 0 | 0 |
| 0 | 0 | 0 | 0 | 0 |
| 0 | 0.00233784 | 0 | 0 | 0 |
| 0 | 0 | 0 | 0 | 0 |
| 0 | 0 | 0 | 0 | 0 |
| 0 | 0 | 0 | 0 | 0 |
| 0 | 0 | 0 | 0 | 0 |
| 0 | 0.004464701 | 0 | 0 | 0.000689025 |
| 0 | 0 | 0 | 0 | 0 |
| 0 | 0.421864771 | 0 | 0 | 0.00697815 |
| 0 | 0 | 0 | 0 | 0 |
| 0 | 0 | 0 | 0 | 0 |
| 0 | 0 | 0 | 0 | 0 |
| 0 | 0 | 0 | 0 | 0 |
| 0 | 0 | 0 | 0 | 0 |
| 0 | 0 | 0 | 0 | 0 |
| 0 | 0 | 0 | 0 | 0 |
| 0.001037689 | 0.009386478 | 0.003756696 | 0.005517653 | 0 |
| 0 | 0.012014343 | 0 | 0 | 0 |
| 0 | 0 | 0 | 0 | 0 |
| 2.401351197 | 6.594107656 | 0 | 0.080642355 | 0.27894043 |
| 0 | 0 | 0 | 0 | 0 |
| 0 | 0 | 0 | 0 | 0 |
| 0 | 0 | 0 | 0 | 0 |
| 0 | 0 | 0 | 0 | 0 |
| 0 | 0 | 0 | 0 | 0 |
| 0 | 0 | 0 | 0 | 0 |
| 0 | 0 | 0 | 0 | 0.000857976 |
| 0 | 0 | 0 | 0 | 0 |
| 0.001637324 | 0 | 0 | 0 | 0 |
| 0 | 0 | 0 | 0 | 0 |

| PHL018-0263_V1 | PHL018-0233_V1 | PHL018-0080_V2 | PHL018-0024_V3 | PHL018-0253_V1 |
|---|---|---|---|---|
| REF | REF | CTRL | EYCF | REF |
| 0.062998175 | 0.073284629 | 0 | 0.120919577 | 0.66265759 |
| 0.204649623 | 0.12193644 | 1.868074331 | 0.420934241 | 1.10608701 |
| 7.20893639 | 0 | 0 | 1.663642814 | 0 |
| 0 | 0 | 0 | 0 | 0 |
| 0 | 0 | 0 | 0 | 0 |
| 0 | 0 | 0 | 0 | 0 |
| 0 | 0 | 0 | 0 | 0 |
| 0 | 0 | 0 | 0 | 0 |
| 0 | 0 | 0 | 0 | 0 |
| 0 | 0 | 0 | 0 | 0 |
| 0 | 0 | 0 | 0 | 0 |
| 0 | 0 | 0 | 0 | 0 |
| 0 | 0 | 0 | 0 | 0 |
| 4.321911908 | 0.038619667 | 2.380214082 | 3.747853502 | 1.025350406 |
| 0 | 0 | 0 | 0 | 0 |
| 0 | 0 | 0 | 0 | 0 |
| 0 | 0 | 0 | 0 | 0 |
| 0 | 0 | 0 | 0 | 0 |
| 0 | 0 | 0 | 0 | 0 |
| 0 | 0 | 0 | 0 | 0 |
| 0 | 0 | 0 | 0 | 0 |
| 0 | 0 | 0 | 0 | 0 |
| 0 | 0 | 0 | 0 | 0 |
| 0 | 0 | 0 | 0 | 0 |
| 0 | 0 | 0 | 0 | 0 |
| 0 | 0 | 0 | 0 | 0 |
| 0 | 0 | 0 | 0 | 0 |
| 0 | 0.021686591 | 0 | 0 | 0 |
| 0 | 0 | 0 | 0 | 0 |
| 0 | 0 | 0 | 0 | 0 |
| 0 | 0 | 0 | 0 | 0 |
| 0 | 0 | 0 | 0 | 0 |
| 0 | 0 | 0 | 0 | 0 |
| 0 | 0 | 0 | 0 | 0 |
| 0 | 0 | 0 | 0 | 0 |
| 2.166828924 | 0.074013453 | 18.33607364 | 0.071017163 | 2.365185956 |
| 0 | 0 | 0 | 0 | 0 |
| 0 | 0 | 0 | 0 | 0 |
| 0 | 0 | 0 | 0 | 0 |
| 0 | 0 | 0 | 0 | 0 |
| 0 | 0 | 0 | 0 | 0 |
| 0.19928894 | 0 | 0.006898281 | 0.001645248 | 0.023096542 |
| 4.213550263 | 5.967211539 | 0.031360968 | 0 | 1.863475386 |
| 0.044030328 | 0 | 0.049146371 | 0.046705637 | 0.078364025 |
| 0.022266152 | 0.002630072 | 0.002249701 | 0 | 0 |
| 0 | 0 | 0 | 0 | 0 |

| | | | | |
|---|---|---|---|---|
| 0 | 0 | 0.002368975 | 0 | 0 |
| 0.002146251 | 0 | 0 | 0 | 0.021755425 |
| 0.007763864 | 0 | 0 | 0.02437636 | 0.020129612 |
| 0.521026168 | 0.209152066 | 0.686862573 | 0.768957715 | 0.636686595 |
| 0 | 0 | 0 | 0.03026399 | 0.013541224 |
| 0 | 0 | 0 | 0 | 0 |
| 0.128536823 | 0 | 0.001074983 | 0.00162644 | 0.017488729 |
| 0 | 0 | 0 | 0 | 0 |
| 0 | 0 | 0 | 0 | 0 |
| 0 | 0 | 0 | 0 | 0 |
| 0.007926205 | 0 | 0 | 0 | 0 |
| 0.093958657 | 0.343766666 | 0.18340186 | 0.013937819 | 0.600459827 |
| 0 | 0 | 0 | 0 | 0 |
| 0.013797807 | 0 | 0.006736352 | 0.003328016 | 0 |
| 0.001412738 | 0 | 0 | 0 | 0 |
| 0 | 0 | 0 | 0 | 0 |
| 0.859659532 | 0.310437378 | 0 | 0 | 0.005450883 |
| 0 | 0 | 0 | 0 | 0 |
| 0 | 0 | 0 | 0 | 0 |
| 0 | 0 | 0 | 0 | 0 |
| 0.042216813 | 0.101763416 | 0.037828412 | 0.016704266 | 0.020797761 |
| 0.003882021 | 0.001607315 | 0.000763811 | 0.004457466 | 0.04339349 |
| 0 | 0.010015505 | 0 | 0 | 0.030529904 |
| 0 | 0 | 0 | 0 | 0 |
| 0.005415638 | 0 | 0 | 0 | 0 |
| 0 | 0 | 0 | 0 | 0 |
| 0 | 0 | 0.000684013 | 0 | 0 |
| 0 | 0 | 0 | 0 | 0 |
| 0.04208224 | 0 | 0 | 0 | 0.00504662 |
| 0 | 0 | 0 | 0 | 0 |
| 0.013427787 | 0 | 0 | 0.000935334 | 0 |
| 0 | 0 | 0 | 0 | 0 |
| 0 | 0 | 0 | 0 | 0 |
| 0 | 0 | 0 | 0 | 0.000547109 |
| 0 | 0 | 0 | 0 | 0 |
| 0 | 0 | 0 | 0 | 0 |
| 0 | 0 | 0 | 0 | 0 |
| 0 | 34.9308777 | 0.010188207 | 0 | 0 |
| 0 | 0 | 0 | 0 | 0 |
| 0 | 0 | 0 | 0 | 0 |
| 0 | 0 | 0 | 0 | 0 |
| 0 | 0 | 0 | 0 | 0 |
| 0 | 0 | 0 | 0 | 0 |
| 0 | 0 | 0 | 0 | 0 |
| 0 | 0 | 0 | 0 | 0 |
| 0 | 0 | 0 | 0 | 0 |
| 0 | 0 | 0 | 0 | 0 |
| 0.007637768 | 0 | 0 | 0.001834753 | 0 |

| | | | | |
|---|---|---|---|---|
| 0 | 0 | 0 | 0 | 0 |
| 0 | 0 | 0 | 0 | 0 |
| 0 | 0 | 0 | 0 | 0 |
| 0 | 0 | 0 | 0 | 0 |
| 0 | 0 | 0 | 0 | 0 |
| 0.000753396 | 0 | 0 | 0 | 0 |
| 0 | 0 | 0 | 0 | 0 |
| 0 | 0 | 0 | 0 | 0 |
| 0.020143804 | 0 | 0 | 0 | 0 |
| 0 | 0 | 0 | 0 | 0 |
| 0 | 0 | 0 | 0 | 0 |
| 0.016966718 | 0 | 0 | 0.003519977 | 0 |
| 0 | 0 | 0 | 0 | 0 |
| 0 | 0 | 0 | 0 | 0 |
| 0 | 0 | 0 | 0 | 0 |
| 0 | 0 | 0 | 0 | 0 |
| 0 | 0 | 0 | 0 | 0 |
| 0 | 0 | 0 | 0 | 0 |
| 0 | 0.10201958 | 0.014302634 | 0.377067895 | 0.405393107 |
| 0 | 0 | 0 | 0 | 0 |
| 0 | 0 | 0 | 0 | 0 |
| 0 | 0 | 0 | 0 | 0 |
| 0 | 0 | 0 | 0 | 0 |
| 0 | 0 | 0 | 0 | 0 |
| 0 | 0 | 0 | 0 | 0 |
| 0 | 0 | 0 | 0 | 0 |
| 0 | 0 | 0 | 0 | 0 |
| 0 | 0 | 0 | 0 | 0 |
| 0 | 0 | 0 | 0 | 0 |
| 0 | 0 | 0 | 0 | 0 |
| 0.011330493 | 0 | 0 | 0 | 1.18979556 |
| 0.128836208 | 0.002010963 | 0.000537541 | 0 | 0 |
| 0 | 0 | 0 | 0 | 0 |
| 0 | 0 | 0 | 0 | 0 |
| 0 | 0 | 0 | 0 | 0 |
| 0 | 0 | 0 | 0 | 0 |
| 0 | 0 | 0 | 0 | 0 |
| 0 | 0 | 0 | 0 | 0 |
| 0.310606408 | 0 | 0 | 0 | 0 |
| 0 | 0 | 0 | 0 | 0 |
| 0 | 0 | 0 | 0 | 0 |
| 0 | 0 | 0 | 0 | 0 |
| 0.004210374 | 0 | 0.00103552 | 0 | 0.004064605 |
| 0 | 0 | 0 | 0 | 0 |
| 0 | 0 | 0 | 0 | 0 |
| 0 | 0 | 0 | 0 | 0 |
| 0 | 0 | 0 | 0 | 0 |
| 0 | 0 | 0 | 0 | 0 |

| | | | | |
|---|---|---|---|---|
| 0 | 0 | 0 | 0 | 0 |
| 0 | 0 | 0 | 0 | 0 |
| 0 | 0 | 0 | 0 | 0 |
| 0 | 0 | 0 | 0 | 0 |
| 0 | 0 | 0.006706113 | 0.000404504 | 0.02158656 |
| 0 | 0 | 0 | 0 | 0 |
| 0 | 0 | 0 | 0 | 0 |
| 0 | 0 | 0 | 0 | 0 |
| 0 | 0 | 0 | 0 | 0 |
| 0 | 0 | 0 | 0 | 0 |
| 0 | 0 | 0 | 0 | 0 |
| 0 | 0 | 0 | 0 | 0 |
| 0 | 0 | 0 | 0 | 0 |
| 0 | 0 | 0 | 0 | 0 |
| 0 | 0 | 0 | 0 | 0 |
| 0 | 0 | 0 | 2.608980126 | 0 |
| 0 | 0 | 0 | 0 | 0 |
| 0 | 0 | 0 | 0 | 0 |
| 0 | 0 | 0 | 0 | 0 |
| 0 | 0 | 0 | 0 | 0 |
| 0 | 0 | 0 | 0 | 0 |
| 0 | 0 | 0 | 0 | 0 |
| 0 | 0 | 0 | 0 | 0 |
| 0 | 0 | 0 | 0 | 0 |
| 0 | 0 | 0 | 0.008900695 | 0 |
| 0 | 0 | 0 | 0 | 0 |
| 0 | 0 | 0 | 0 | 0 |
| 0 | 0 | 0 | 0 | 0 |
| 0 | 0 | 0 | 0 | 0 |
| 0 | 0 | 0 | 0 | 0 |
| 0 | 0 | 0 | 0 | 0 |
| 0.005530104 | 0 | 0 | 0 | 0.001281274 |
| 0 | 0 | 0 | 0 | 0 |
| 0 | 0 | 0 | 0 | 0 |
| 0 | 0 | 0 | 0 | 0 |
| 0 | 0 | 0 | 0 | 0 |
| 0 | 0 | 0 | 0 | 0 |
| 0 | 0 | 0 | 0 | 0 |
| 0 | 0 | 0 | 0 | 0 |
| 0 | 0 | 0 | 0 | 0 |
| 0 | 0 | 0 | 0 | 0 |
| 0 | 0 | 0 | 0 | 0 |
| 0 | 0 | 0 | 0 | 0 |
| 0 | 0 | 0 | 0 | 0 |
| 0 | 0 | 0 | 0 | 0 |
| 0 | 0 | 0 | 0 | 0 |
| 0 | 0 | 0 | 0 | 0 |
| 0 | 0 | 0 | 0 | 0 |

| | | | | |
|---|---|---|---|---|
| 0 | 0 | 0 | 0.003427664 | 0 |
| 0 | 0 | 0 | 0 | 0 |
| 0 | 0 | 0 | 0 | 0 |
| 0 | 0 | 0 | 0 | 0 |
| 0 | 0 | 0 | 0 | 0 |
| 0 | 0 | 0 | 0 | 0 |
| 0 | 0 | 0 | 0 | 0 |
| 0 | 0 | 0 | 0 | 0 |
| 0 | 0 | 0 | 0 | 0 |
| 0 | 0 | 0 | 0 | 0 |
| 0 | 0 | 0 | 0 | 0 |
| 0 | 0 | 0 | 0 | 0 |
| 0 | 0 | 0 | 0 | 0 |
| 0 | 0 | 0 | 0 | 0 |
| 0.01492996 | 0.001003852 | 0.004293354 | 0 | 0 |
| 0 | 0 | 0 | 0 | 0 |
| 0 | 0 | 0 | 0 | 0 |
| 0 | 0 | 0 | 0 | 0 |
| 0 | 0 | 0 | 0 | 0 |
| 0 | 0 | 0 | 0 | 0 |
| 0.047097023 | 0.006572858 | 0.042167009 | 0.014070815 | 0.017957341 |
| 0 | 0 | 0 | 0 | 0 |
| 0 | 0 | 0 | 0 | 0 |
| 0.019397461 | 0 | 0 | 0 | 0 |
| 0 | 0 | 0 | 0 | 0 |
| 0 | 0 | 0 | 0 | 0 |
| 0.016134233 | 0.002820541 | 0.001292477 | 0.006145883 | 0.00556273 |
| 0 | 0 | 0 | 0 | 0 |
| 0 | 0 | 0 | 0 | 0 |
| 0 | 0 | 0 | 0 | 0 |
| 0 | 0 | 0 | 0 | 0 |
| 0 | 0 | 0 | 0 | 0 |
| 0 | 0 | 0 | 0 | 0 |
| 0.004595569 | 0 | 0 | 0 | 0.028953766 |
| 0.002575112 | 0.003376304 | 0.018290736 | 0.012692467 | 0.073087167 |
| 0 | 0 | 0 | 0 | 0 |
| 0 | 0 | 0 | 0 | 0 |
| 0 | 0 | 0 | 0 | 0 |
| 0.003244762 | 0 | 0 | 0 | 0 |
| 0 | 0 | 0 | 0 | 0 |
| 0 | 0 | 0 | 0 | 0 |
| 0.0108014 | 0 | 0 | 0 | 0 |
| 0.005888388 | 0 | 0 | 0 | 0 |
| 0 | 0 | 0 | 0 | 0 |
| 0.009397763 | 0 | 0 | 0 | 0 |
| 0 | 0 | 0 | 0 | 0 |
| 0 | 0 | 0 | 0 | 0 |
| 0.001284566 | 0 | 0 | 0 | 0.001594405 |

| | | | | |
|---|---|---|---|---|
| 0 | 0 | 0 | 0 | 0 |
| 0 | 0 | 0 | 0 | 0 |
| 0 | 0 | 0 | 0 | 0 |
| 0 | 0 | 0 | 0 | 0 |
| 0 | 0 | 0 | 0 | 0 |
| 0 | 0 | 0 | 0 | 0 |
| 0.00156734 | 0 | 0 | 0 | 0 |
| 0.004484569 | 0 | 0 | 0 | 0 |
| 0 | 0 | 0 | 0 | 0 |
| 0.001886095 | 0 | 0 | 0 | 0 |
| 0.000681143 | 0 | 0 | 0 | 0 |
| 0 | 0 | 0 | 0 | 0 |
| 0 | 0 | 0 | 0 | 0 |
| 0 | 0 | 0 | 0 | 0 |
| 0.004340003 | 0 | 0 | 0 | 0 |
| 0 | 0 | 0 | 0 | 0.00070333 |
| 0 | 0 | 0 | 0 | 0 |
| 0 | 0.028044052 | 0 | 0 | 0 |
| 0.016609882 | 0.030505092 | 0 | 0 | 0.007206663 |
| 0 | 0 | 0 | 0 | 0.589657228 |
| 0 | 0.026950743 | 0 | 0.002446872 | 0 |
| 0 | 0 | 0 | 0 | 0 |
| 0 | 0 | 0 | 0 | 0 |
| 0 | 0 | 0 | 0 | 0 |
| 0.026252735 | 0 | 0.003066945 | 0 | 0 |
| 0.005041984 | 0 | 0 | 0 | 0 |
| 0 | 0 | 0 | 0 | 0 |
| 0 | 0 | 0 | 0 | 0 |
| 0.027924065 | 0 | 0 | 0 | 0 |
| 0 | 0 | 0 | 0 | 0 |
| 0.121454643 | 0.07891361 | 0.093835247 | 0.038273555 | 0.10357058 |
| 0 | 0 | 0 | 0 | 0 |
| 0 | 0 | 0 | 0.025218719 | 0 |
| 0 | 0 | 0 | 0 | 0 |
| 0 | 0 | 0 | 0 | 0 |
| 0 | 0 | 0 | 0 | 0 |
| 0 | 0 | 0 | 0 | 0 |
| 0 | 0 | 0 | 0 | 0 |
| 0 | 0 | 0 | 0 | 0 |
| 0.012738212 | 0 | 0.002078653 | 0 | 0 |
| 0 | 0 | 0 | 0 | 0 |
| 0.003507746 | 0.031562507 | 0.002121258 | 0 | 0.005577441 |
| 0 | 0.000756632 | 0 | 0 | 0 |
| 0 | 0 | 0 | 0 | 0 |
| 0 | 0 | 0 | 0 | 0 |
| 0 | 0 | 0 | 0 | 0 |
| 0 | 0 | 0 | 0 | 0 |
| 0 | 0 | 0 | 0 | 0 |

| | | | | |
|---|---|---|---|---|
| 0 | 0 | 0 | 0 | 0 |
| 0 | 0 | 0 | 0 | 0 |
| 0.022800038 | 0.001132341 | 0.047944582 | 0.0385204 | 0.070284067 |
| 0 | 0 | 0 | 0 | 0 |
| 0 | 0 | 0 | 0 | 0 |
| 0 | 0 | 0 | 0 | 0 |
| 0 | 0 | 0 | 0 | 0 |
| 0 | 0 | 0 | 0 | 0 |
| 0 | 0 | 0 | 0 | 0 |
| 0 | 0.001864134 | 0 | 0 | 0 |
| 0 | 0 | 0 | 0 | 0 |
| 0 | 0 | 0 | 0 | 0 |
| 0 | 0 | 0 | 0 | 0 |
| 0.004295141 | 0 | 0 | 0 | 0.000829287 |
| 0.018670735 | 0 | 0.001546158 | 0.000954826 | 0 |
| 0 | 0.021011272 | 0 | 0 | 0.021495864 |
| 0.022620061 | 0 | 0.002768399 | 0 | 0 |
| 0 | 0 | 0 | 0 | 0 |
| 0 | 0 | 0 | 0 | 0 |
| 0 | 0 | 0 | 0 | 0 |
| 0 | 0 | 0 | 0 | 0 |
| 0.002187799 | 0 | 0 | 0 | 0.001173364 |
| 0 | 0 | 0 | 0 | 0 |
| 0.001507372 | 0 | 0 | 0 | 0 |
| 0.005067123 | 0 | 0 | 0 | 0 |
| 0 | 0 | 0 | 0 | 0 |
| 0 | 0 | 0 | 0 | 0 |
| 0 | 0 | 0 | 0 | 0 |
| 0 | 0 | 0 | 0 | 0 |
| 0.05415105 | 0 | 0 | 0 | 0 |
| 0.024961898 | 0 | 0 | 0 | 0 |
| 0 | 0 | 0 | 0 | 0 |
| 0.006037046 | 0 | 0 | 0 | 0 |
| 0.002242245 | 0 | 0 | 0 | 0 |
| 0 | 0 | 0 | 0 | 0 |
| 0.013109454 | 0 | 0 | 0 | 0.022967545 |
| 0 | 0 | 0 | 0 | 0 |
| 5.202454749 | 0.001067839 | 0.009704922 | 0 | 0.005601998 |
| 0 | 0 | 0 | 0 | 0 |
| 0 | 0 | 0 | 0 | 0 |
| 0 | 0 | 0 | 0 | 0 |
| 0 | 0 | 0 | 0 | 0 |
| 0 | 0 | 0 | 0 | 0 |
| 0 | 0 | 0 | 0 | 0 |
| 0 | 0 | 0 | 0 | 0 |
| 0.374718588 | 0 | 0 | 0 | 0 |
| 0.023836298 | 0 | 0 | 0 | 0 |
| 0 | 0 | 0 | 0 | 0 |

| | | | | |
|---|---|---|---|---|
| 0.268023511 | 0 | 0 | 0.007882342 | 0.02236642 |
| 0 | 0 | 0 | 0 | 0 |
| 0.003564632 | 0 | 0 | 0 | 0 |
| 0 | 0 | 0 | 0 | 0 |
| 0.069677921 | 0.001791309 | 0.002633543 | 0.002690851 | 0.011623095 |
| 0.131995255 | 0 | 0 | 0 | 0 |
| 0 | 0 | 0 | 0 | 0 |
| 0.012567695 | 0.006591148 | 0.017115091 | 0.011605601 | 0.005199679 |
| 0.00830111 | 0 | 0 | 0 | 0 |
| 0.049964639 | 0.00097052 | 0.24143813 | 0.086725548 | 0.071458953 |
| 0 | 0 | 0 | 0 | 0 |
| 0.346496646 | 0 | 0.014610984 | 0 | 0.000823217 |
| 0.032557285 | 0.000753851 | 0.004836196 | 0.006097606 | 0.003746635 |
| 0.005894239 | 0 | 0 | 0.003704276 | 0.055637243 |
| 0 | 0 | 0 | 0 | 0.001712591 |
| 0 | 0 | 0 | 0 | 0 |
| 0.131102602 | 0 | 0 | 0 | 0 |
| 0 | 0 | 0 | 0.005672383 | 0 |
| 0.613643443 | 0 | 0 | 0.131134263 | 0 |
| 0 | 0 | 0 | 0 | 0 |
| 0 | 0 | 0 | 0 | 0 |
| 0 | 0 | 0 | 0 | 0 |
| 0.000889931 | 0 | 0 | 0 | 0 |
| 0 | 0 | 0 | 0 | 0 |
| 0 | 0 | 0 | 0 | 0 |
| 0.00251132 | 0 | 0.001173528 | 0 | 0 |
| 0.022835382 | 5.477576579 | 0 | 0 | 0.00842889 |
| 0 | 0 | 0 | 0 | 0 |
| 0.004628214 | 0 | 0 | 0 | 0 |
| 0 | 0 | 0 | 0 | 0 |
| 0.001347392 | 0 | 0 | 0 | 0 |
| 0 | 0 | 0 | 0 | 0 |
| 0 | 0 | 0 | 0 | 0 |
| 0.024632591 | 0 | 0 | 0 | 0 |
| 0 | 0 | 0 | 0 | 0 |
| 0 | 0 | 0 | 0.002675016 | 0 |
| 0.007686465 | 0 | 0.00186244 | 0.001150146 | 0 |
| 0 | 0 | 0 | 0 | 0 |
| 0 | 0 | 0 | 0 | 0 |
| 0 | 0 | 0 | 0 | 0 |
| 0 | 0 | 0 | 0 | 0 |
| 0 | 0 | 0 | 0 | 0 |
| 0 | 0 | 0 | 0 | 0 |
| 0 | 0 | 0 | 0 | 0 |
| 0 | 0 | 0 | 0 | 0 |
| 0 | 0 | 0 | 0 | 0 |
| 0 | 0 | 0.002626875 | 0 | 0.007537254 |
| 0 | 0 | 0 | 0 | 0 |

| | | | | |
|---|---|---|---|---|
| 0 | 0 | 0 | 0 | 0 |
| 0.009772558 | 0 | 0 | 0.001692082 | 0 |
| 0.014475291 | 0 | 0 | 0 | 0 |
| 0 | 0 | 0 | 0 | 0 |
| 0 | 0 | 0 | 0 | 0 |
| 0.003935617 | 0 | 0 | 0 | 0.097833533 |
| 0 | 0 | 0 | 0.003757034 | 0.071027431 |
| 0 | 0 | 0 | 0 | 0 |
| 0 | 0 | 0 | 0 | 0 |
| 0.000962622 | 0 | 0 | 0 | 0 |
| 0.060196364 | 0 | 0 | 0.016366892 | 0.002478723 |
| 0 | 0 | 0 | 0 | 0 |
| 0 | 0 | 0 | 0 | 0 |
| 0.006199154 | 0 | 0 | 0 | 0.002341773 |
| 0 | 0 | 0.006881659 | 0.008373589 | 0 |
| 0 | 0 | 0 | 0 | 0 |
| 0 | 0 | 0 | 0 | 0 |
| 0 | 0 | 0 | 0.005086521 | 0 |
| 0 | 0.004513508 | 0 | 0 | 0 |
| 0.009535495 | 0 | 0 | 0 | 0 |
| 0 | 0 | 0 | 0 | 0 |
| 0.450697316 | 0.061529439 | 0.188821645 | 0.342010244 | 0.239463249 |
| 0 | 0 | 0 | 0 | 0 |
| 0 | 0 | 0 | 0 | 0 |
| 0 | 0 | 0 | 0 | 0 |
| 0 | 0 | 0 | 0 | 0 |
| 0.003799122 | 0 | 0 | 0.036364759 | 0 |
| 0 | 0 | 0 | 0 | 0 |
| 0 | 0 | 0 | 0 | 0 |
| 0.134719628 | 0.282132249 | 0.162830792 | 1.341063205 | 0.873011482 |
| 0 | 0 | 0.005810372 | 0.001329738 | 0 |
| 0.021821265 | 0 | 0 | 0 | 0 |
| 0 | 0 | 0 | 0 | 0 |
| 0 | 0 | 0 | 0 | 0 |
| 0.000973634 | 0 | 0 | 0.001180068 | 0 |
| 0 | 0 | 0 | 0 | 0 |
| 0.008086545 | 0.060838177 | 0 | 0.987343035 | 0.963554698 |
| 0.02802157 | 0 | 0 | 0 | 0 |
| 0 | 0 | 0 | 0 | 0 |
| 0 | 0 | 0 | 0.031696291 | 0 |
| 0.033901999 | 0 | 0 | 0.011279621 | 0 |
| 0.857182113 | 7.039427175 | 2.707189696 | 1.32826024 | 5.478313007 |
| 0.016609722 | 0 | 0 | 0 | 0 |
| 0 | 0 | 0 | 0 | 0 |
| 0 | 0 | 0 | 0 | 0 |
| 0 | 0 | 0 | 0 | 0 |
| 0.006976977 | 0 | 0 | 0 | 0 |
| 0 | 0 | 0 | 0 | 0 |

| | | | | |
|---|---|---|---|---|
| 0.01692989 | 0 | 0 | 0 | 0 |
| 0 | 0 | 0 | 0 | 0 |
| 1.961585073 | 0.021188925 | 0 | 0 | 0.010568631 |
| 0 | 0 | 0 | 0 | 0 |
| 0 | 0 | 0 | 0 | 0 |
| 0.001455685 | 0 | 0 | 0 | 0.006558007 |
| 0.002446745 | 0 | 0 | 0 | 0 |
| 0 | 0 | 0 | 0 | 0 |
| 0.015129396 | 0 | 0 | 0.001358311 | 0 |
| 0.034630745 | 0 | 0 | 0 | 0 |
| 0 | 0 | 0 | 0 | 0 |
| 0 | 0 | 0 | 0 | 0 |
| 0 | 0 | 0 | 0 | 0 |
| 0 | 0 | 0 | 0.020765856 | 0 |
| 0 | 0 | 0 | 0 | 0 |
| 0.001014341 | 0.005142404 | 0.00151679 | 0.041144119 | 0.136112011 |
| 0.036651895 | 0.019064909 | 0.013239477 | 0.001090135 | 0.006946347 |
| 0 | 0 | 0 | 0 | 0 |
| 0 | 0 | 0 | 0 | 0 |
| 0 | 0 | 0 | 0 | 0 |
| 0 | 0 | 0.006886083 | 0 | 0 |
| 0 | 0 | 0 | 0.000843 | 0 |
| 0 | 0 | 0 | 0 | 0 |
| 0 | 0 | 0 | 0 | 0 |
| 0 | 0 | 0 | 0.011207198 | 0 |
| 0 | 0 | 0 | 0 | 0 |
| 0.002569134 | 0 | 0 | 0.000691967 | 0 |
| 0 | 0 | 0 | 0.058825462 | 0 |
| 0 | 0 | 0 | 0 | 0 |
| 0 | 0 | 0 | 0 | 0 |
| 0.009986188 | 0 | 0 | 0.006665697 | 0 |
| 0.019419366 | 0 | 0.007864635 | 0 | 0.008436168 |
| 0.165176096 | 0.00754214 | 0.122691212 | 0.422058042 | 0.030642014 |
| 0.0940671 | 0 | 0 | 0 | 0 |
| 0 | 0 | 0 | 0 | 0 |
| 0 | 0 | 0 | 0 | 0 |
| 0.001671611 | 0 | 0 | 0 | 0 |
| 0.007190025 | 0 | 0.010385038 | 0.02640753 | 0.005468742 |
| 0.043053737 | 0.049799623 | 0 | 0.137452296 | 0.084220079 |
| 0 | 0 | 0.000832434 | 0 | 0 |
| 0 | 0 | 0 | 0 | 0 |
| 0 | 0 | 0 | 0 | 0 |
| 0.050830807 | 0 | 0 | 0.001316414 | 0.001572788 |
| 0.018879084 | 0 | 0.026863716 | 0.027485776 | 0 |
| 0 | 0 | 0 | 0 | 0 |
| 0 | 0 | 0.001775771 | 0 | 0 |
| 0 | 0 | 0 | 0 | 0 |
| 0 | 0 | 0 | 0 | 0 |

| | | | | |
|---|---|---|---|---|
| 0 | 0 | 0 | 0 | 0 |
| 4.202160017 | 0.010899256 | 0.4167588 | 0.397498939 | 0.65948919 |
| 0 | 0 | 0 | 0 | 0 |
| 0 | 0 | 0 | 0 | 0 |
| 0 | 0 | 0 | 0 | 0 |
| 0 | 0 | 0 | 0 | 0 |
| 0 | 0 | 0 | 0 | 0 |
| 0 | 0 | 0 | 0 | 0 |
| 0.002989547 | 0 | 0 | 0 | 0 |
| 0 | 0 | 0 | 0.001743239 | 0 |
| 0.000661862 | 0 | 0.00061857 | 0.000891325 | 0.005324561 |
| 0.123232756 | 0.003734156 | 0.011670782 | 0.093466854 | 0.010786275 |
| 0 | 0.438498918 | 0 | 0 | 0 |
| 0 | 0 | 0 | 0 | 0 |
| 0.000759404 | 0 | 0 | 0 | 0 |
| 0 | 0 | 0 | 0 | 0 |
| 0.050824761 | 0 | 0.01597739 | 0.001306685 | 0.0066907 |
| 0.012595716 | 0 | 0.012052121 | 0.143939533 | 0 |
| 0 | 0 | 0 | 0 | 0 |
| 0.018889595 | 0 | 0 | 0.002360273 | 0 |
| 0.099346744 | 0.058099978 | 0.144059969 | 0.146109828 | 0.068998001 |
| 0 | 0 | 0 | 0 | 0 |
| 0 | 0 | 0 | 0 | 0 |
| 0 | 0 | 0 | 0 | 0 |
| 0.004403193 | 0 | 0 | 0 | 0 |
| 0 | 0 | 0 | 0 | 0 |
| 0 | 0.0065775 | 0 | 0.010070538 | 0.198184021 |
| 0.185100635 | 0.005975971 | 0.031474847 | 0.094730885 | 0.070156852 |
| 0 | 0.003668247 | 0 | 0 | 0 |
| 0 | 0 | 0 | 0 | 0.001179451 |
| 0.026083257 | 0 | 0.001269644 | 0 | 0 |
| 0.00052427 | 0 | 0 | 0 | 0 |
| 0 | 0 | 0 | 0 | 0 |
| 0 | 0 | 0 | 0 | 0 |
| 0 | 0 | 0.002268493 | 0 | 0 |
| 0.027427352 | 0.432653343 | 0.471413282 | 0.181449283 | 0.167554691 |
| 0 | 0 | 0 | 0 | 0 |
| 0 | 0 | 0 | 0 | 0 |
| 0.183361084 | 0 | 0.003795193 | 0.002019199 | 0.007237323 |
| 0 | 0 | 0 | 0 | 0 |
| 0 | 0 | 0 | 0 | 0 |
| 0 | 0 | 0 | 0 | 0 |
| 0 | 0 | 0 | 0.005519215 | 0.002014861 |
| 0 | 0 | 0 | 0 | 0.00442116 |
| 0 | 0 | 0 | 0.008361917 | 0 |
| 0 | 0.541153273 | 0.206786189 | 0 | 0 |
| 0 | 0 | 0 | 0 | 0 |
| 0 | 0 | 0 | 0 | 0 |

| | | | | |
|---|---|---|---|---|
| 0 | 0 | 0 | 0 | 0 |
| 0 | 0 | 0 | 0 | 0 |
| 0 | 0.010943197 | 0 | 0.000562002 | 0 |
| 0 | 0.048096807 | 0.128668494 | 0.278374151 | 0.423681627 |
| 0 | 0 | 0 | 0 | 0 |
| 0 | 0 | 0 | 0 | 0 |
| 0 | 0 | 0 | 0 | 0.001323146 |
| 0 | 0 | 0 | 0 | 0.008969963 |
| 0 | 0.662280124 | 0.38160607 | 0.139219824 | 0.495313244 |
| 0 | 0.038325507 | 0 | 0.030162173 | 0.007054726 |
| 0 | 0 | 0 | 0 | 0 |
| 0.001058637 | 0 | 0.006529987 | 0.001368633 | 0.001226382 |
| 0 | 0 | 0 | 0 | 0 |
| 0 | 0 | 0 | 0 | 0.076053031 |
| 0.022538615 | 0 | 0.002286057 | 0.022729164 | 0.256039491 |
| 0 | 0 | 0 | 0 | 0 |
| 0 | 0 | 0 | 0 | 0.642220538 |
| 0 | 1.024657916 | 0.00341171 | 0.057702553 | 0 |
| 0 | 0 | 0 | 0 | 0 |
| 0 | 0 | 0 | 0 | 0 |
| 0 | 0 | 0 | 0 | 0 |
| 0 | 0 | 0 | 0 | 0 |
| 0 | 0 | 0 | 0 | 0 |
| 0.110740191 | 0 | 0.003303088 | 0.065138081 | 0.574499029 |
| 0 | 0 | 0 | 0 | 0 |
| 0.314005531 | 0.092144053 | 0 | 1.808075611 | 0.062322976 |
| 0 | 0 | 0 | 0 | 0 |
| 7.038499208 | 0.295476592 | 0 | 6.985878267 | 2.08771767 |
| 0 | 0 | 0 | 0 | 0 |
| 0 | 0 | 0 | 0 | 0 |
| 0 | 0.000539753 | 0 | 0 | 0.059413927 |
| 0.000775739 | 0 | 0 | 0 | 0 |
| 0.008302747 | 0 | 0 | 0 | 0.000907392 |
| 0 | 0 | 0 | 0 | 0 |
| 0 | 0 | 0 | 0 | 0 |
| 0 | 0.022424847 | 0.006361273 | 0.00973032 | 0 |
| 0 | 0.190714515 | 0.088054579 | 0.026226285 | 3.505697 |
| 0 | 0 | 0 | 0 | 0 |
| 0 | 0 | 0 | 0.002137121 | 0 |
| 0 | 0 | 0 | 0.011755572 | 0 |
| 0.418415957 | 0.006391425 | 0.032081094 | 0.02084322 | 0.149022649 |
| 0 | 0 | 0 | 0 | 0 |
| 4.242865968 | 0.009584124 | 19.06703082 | 18.69314848 | 0.408585734 |
| 0 | 0 | 0 | 0.001286728 | 0 |
| 0 | 0 | 0 | 0 | 0 |
| 0 | 0 | 0 | 0 | 0 |
| 0 | 0 | 0 | 0 | 0 |
| 0.022707529 | 0.002561075 | 0 | 0 | 0.421220164 |

| | | | | |
|---|---|---|---|---|
| 0 | 0 | 0 | 0 | 0 |
| 0.076742847 | 0.027415243 | 0.098333479 | 0 | 0.029849103 |
| 0.021533669 | 0.00777442 | 0 | 0 | 0.004519162 |
| 0 | 0.001548064 | 0 | 0 | 0 |
| 0 | 0 | 0 | 0 | 0 |
| 0 | 0 | 0.002929512 | 0 | 0.018156108 |
| 0 | 0 | 0 | 0 | 0 |
| 0 | 0.019762759 | 0 | 0 | 0 |
| 0.004580083 | 0 | 0 | 0 | 0 |
| 0 | 0 | 0 | 0 | 0 |
| 0.013558613 | 0.009039199 | 0.000644326 | 0.004642194 | 0.292177451 |
| 2.051774525 | 0.211108116 | 3.278064412 | 2.49263346 | 0.67617515 |
| 0 | 0 | 0 | 0 | 0 |
| 0.180804453 | 0.014091237 | 0.022947651 | 0 | 0.007661772 |
| 0.039480288 | 0 | 0.000547989 | 0.021793556 | 0.019811434 |
| 0 | 3.016132276 | 0 | 0 | 0 |
| 0 | 0 | 0 | 0 | 0 |
| 0 | 0 | 0 | 0 | 0 |
| 0 | 0 | 0 | 0 | 0 |
| 0.003596709 | 0 | 0 | 0.00642423 | 0.132856805 |
| 2.716023322 | 0 | 0.005220391 | 0.081552061 | 0.102826814 |
| 0.493379728 | 0.000532196 | 1.023504449 | 0.923425663 | 0 |
| 2.11690028 | 0.015901968 | 9.415080456 | 10.6927658 | 0.921697017 |
| 0 | 0 | 0 | 0 | 0 |
| 0 | 0 | 0 | 0 | 0 |
| 0.00161565 | 1.441268022 | 0.012511189 | 2.925560562 | 1.731504813 |
| 0.05736289 | 0 | 0 | 0 | 0 |
| 0 | 0 | 0.007322281 | 0.002759493 | 0 |
| 0 | 0 | 0 | 0 | 0 |
| 0 | 0 | 0 | 0 | 0 |
| 0 | 0 | 0 | 0 | 0.047581108 |
| 0 | 0.209488617 | 0.098229092 | 0.075375165 | 0.266652882 |
| 0.145622967 | 0 | 0 | 0 | 0.023923883 |
| 0.004139582 | 0.009753815 | 0 | 0.043337613 | 0.229177047 |
| 0.089414656 | 0 | 0 | 0.000629342 | 0 |
| 0.049653768 | 0.206344984 | 0.460058981 | 0.032857756 | 0.00206615 |
| 0.00082698 | 0 | 0 | 0 | 0.011176888 |
| 0 | 0 | 0 | 0 | 0 |
| 0.030805634 | 0.012259344 | 0.065539711 | 0.010118476 | 0.220262764 |
| 0 | 0 | 0 | 0 | 0 |
| 0 | 0 | 0 | 0 | 0 |
| 0 | 0 | 0 | 0 | 0 |
| 0 | 0 | 0 | 0 | 0 |
| 0 | 0 | 0 | 0 | 0 |
| 0.00219664 | 0.008960242 | 0 | 0.029187579 | 0.538865179 |
| 0 | 0 | 0 | 0 | 0 |
| 0 | 0 | 0 | 0 | 0 |
| 0.132192962 | 0.007205667 | 0.32056003 | 0.003092612 | 0.009379369 |

| | | | | |
|---|---|---|---|---|
| 0 | 0 | 0 | 0 | 0 |
| 0 | 0 | 0 | 0 | 0 |
| 0 | 0 | 0 | 0 | 0 |
| 0 | 0.00090914 | 0 | 0 | 0.002409826 |
| 0 | 0 | 0 | 0 | 0 |
| 0 | 0 | 0 | 0 | 0.156610554 |
| 0 | 1.011061764 | 0 | 0 | 0 |
| 0.006891004 | 0 | 0 | 0 | 0.200866351 |
| 0 | 0 | 0 | 0 | 0 |
| 0 | 0 | 0 | 0 | 0 |
| 0.004865281 | 0.017415714 | 0 | 0.002059214 | 0.004920497 |
| 0 | 0 | 0 | 0 | 0 |
| 0 | 0.051952373 | 0 | 0 | 0 |
| 0 | 0.099363533 | 0 | 0 | 0 |
| 0 | 0 | 0 | 0 | 0 |
| 0 | 0 | 0 | 0 | 0 |
| 0.0249848 | 0 | 0.321915673 | 0 | 0 |
| 0 | 0.000842918 | 0 | 0.000779204 | 0 |
| 0.003247179 | 0 | 0 | 0 | 0 |
| 0.001565418 | 0 | 0 | 0 | 0 |
| 0 | 0 | 0 | 0 | 0 |
| 0 | 0 | 0 | 0 | 0 |
| 0 | 0.190088736 | 0 | 0 | 0.002146649 |
| 0 | 0 | 0 | 0 | 0 |
| 0 | 0 | 0 | 0 | 0 |
| 0 | 0 | 0 | 0 | 0 |
| 0 | 0 | 0 | 0 | 0 |
| 0 | 0 | 0 | 0 | 0 |
| 0 | 0 | 0 | 0 | 0 |
| 0.003727732 | 0 | 0.713764789 | 6.999293012 | 0.00607276 |
| 0 | 0 | 0 | 0 | 0.011274006 |
| 0 | 0 | 0 | 0 | 0 |
| 0 | 0 | 0 | 0 | 0 |
| 0 | 0 | 0 | 0 | 0 |
| 0 | 0 | 0 | 0 | 0 |
| 0 | 0 | 0 | 0 | 0 |
| 0 | 0.03785007 | 0 | 0.104699138 | 0.059101164 |
| 0 | 0 | 0 | 0 | 0 |
| 0 | 0 | 0 | 0 | 0 |
| 0 | 0 | 0 | 0 | 0 |
| 0.003312184 | 0 | 0 | 0 | 0 |
| 0 | 0 | 0 | 0 | 0 |
| 0 | 0 | 0 | 0 | 0.003841869 |
| 0 | 0 | 0 | 0 | 0.008273838 |
| 0.134952335 | 0.053918987 | 0.286591412 | 0 | 0.122325701 |
| 0 | 0 | 0 | 0 | 0 |
| 0 | 0 | 0 | 0 | 0.030042695 |
| 0 | 0 | 0 | 0 | 0 |

| | | | | |
|---|---|---|---|---|
| 0 | 0 | 0 | 0 | 0 |
| 0 | 0 | 0 | 0 | 0 |
| 0 | 0 | 0 | 0 | 0 |
| 0 | 0 | 0 | 0.064313177 | 0.664425012 |
| 0 | 0 | 0 | 0 | 0 |
| 0 | 0.235449345 | 0 | 0 | 0 |
| 0 | 0 | 0 | 0 | 0 |
| 0 | 0 | 0 | 0 | 0 |
| 0.241271661 | 0.016313409 | 0.021487299 | 0.007649439 | 0.003394552 |
| 0 | 0 | 0 | 0 | 0 |
| 0 | 0 | 0 | 0 | 0 |
| 0 | 0 | 0 | 0 | 0 |
| 0 | 0 | 0 | 0 | 0 |
| 0 | 0 | 0 | 0 | 0 |
| 2.178381774 | 0 | 0 | 0.880947714 | 0.087255989 |
| 0.047322573 | 0 | 0 | 0.00050534 | 0.006641315 |
| 0 | 0 | 0 | 0 | 0 |
| 0 | 0 | 0 | 0 | 0 |
| 0 | 0 | 0 | 0 | 0 |
| 0 | 0 | 0 | 0 | 0 |
| 0 | 0 | 0 | 0 | 0.855157581 |
| 0 | 0 | 0 | 0 | 0 |
| 0 | 0 | 0 | 0 | 0 |
| 0 | 0 | 0 | 0 | 0 |
| 0.587814415 | 0.611111044 | 0.00148638 | 0 | 0.060799656 |
| 0.177484542 | 0.460693281 | 1.407138963 | 4.548677065 | 0.448319794 |
| 0 | 0.634942797 | 0 | 0 | 0.068526882 |
| 0 | 0 | 0 | 0 | 0.003431016 |
| 0 | 0.002430782 | 0.048342173 | 0 | 0 |
| 0 | 0 | 0 | 0 | 0 |
| 0 | 0 | 0 | 0 | 0 |
| 0 | 0 | 0 | 0 | 0 |
| 0 | 0 | 0 | 0 | 0 |
| 0.000849379 | 0 | 0 | 0.001372625 | 0.06133396 |
| 0.052061735 | 0 | 0 | 0.001809322 | 0.004593591 |
| 0.000989283 | 0.000518831 | 0 | 0 | 0.000764026 |
| 0 | 0 | 0 | 0 | 0 |
| 1.579092588 | 0.123223798 | 0 | 0.702954267 | 1.014340347 |
| 0.61671344 | 0.00065526 | 0 | 0 | 0 |
| 0 | 0 | 0 | 0 | 0 |
| 0 | 0 | 0 | 0 | 0 |
| 0.001121855 | 0 | 0 | 0 | 0 |
| 0.572112702 | 0.326136339 | 0.004548413 | 0.144843672 | 0.072300843 |
| 0 | 1.384777042 | 0 | 0.037099748 | 0 |
| 0 | 0 | 0 | 0 | 0 |
| 0 | 0 | 0 | 0.003332077 | 0.019905022 |
| 0 | 0 | 0 | 0 | 0 |
| 4.510077282 | 0 | 0.001395443 | 0.47244746 | 0.963728729 |

| | | | | |
|---|---|---|---|---|
| 0 | 0 | 0 | 0.001439447 | 0 |
| 0 | 0 | 0 | 0 | 0 |
| 0 | 0 | 0 | 0 | 0 |
| 0 | 0 | 0 | 0 | 0 |
| 0 | 0 | 0 | 0 | 0.003210453 |
| 0 | 0 | 0.071632945 | 0 | 0 |
| 0 | 0 | 0 | 0 | 0 |
| 0 | 0 | 0 | 0 | 0 |
| 0 | 0 | 0 | 0 | 0 |
| 0 | 0 | 0 | 0 | 0 |
| 0 | 0 | 0 | 0 | 0.001310165 |
| 0 | 0.186471407 | 0.206834258 | 0.599929829 | 1.019665352 |
| 0 | 0 | 0.0842984 | 0 | 0.135796182 |
| 0 | 0 | 0 | 0 | 0 |
| 1.134070117 | 0 | 0.052718992 | 0.057181049 | 0.442576296 |
| 0 | 0 | 0 | 0 | 0 |
| 0 | 0 | 0 | 0 | 0.032888292 |
| 0 | 0 | 0 | 0 | 0 |
| 0 | 0.002357266 | 0 | 0 | 0 |
| 0 | 0 | 0 | 0 | 0.002886829 |
| 0.095077172 | 0 | 0 | 0 | 0 |
| 0 | 0 | 0 | 0 | 0 |
| 0.0120189 | 0.47924422 | 0.065354196 | 0.347140774 | 0.879702379 |
| 0 | 0 | 0 | 0 | 0 |
| 0 | 0 | 0 | 0 | 0 |
| 0 | 0 | 0 | 0 | 0 |
| 0 | 0 | 0 | 0 | 0 |
| 0 | 0 | 0 | 0 | 0 |
| 0 | 0 | 0 | 0 | 0 |
| 0 | 0 | 0 | 0 | 0 |
| 0 | 0 | 0 | 0 | 0 |
| 0.439854049 | 0 | 0.034205132 | 0.021238522 | 0.050107116 |
| 0 | 0 | 0 | 0 | 0 |
| 0 | 0 | 0 | 0.00349203 | 0.286270834 |
| 0.00086127 | 0 | 0 | 0 | 0 |
| 0.002775705 | 0.00485241 | 0.020753185 | 0.008074133 | 0.019025204 |
| 0 | 0 | 0 | 0 | 0 |
| 0 | 0.009178403 | 0.016163801 | 0 | 0.019478992 |
| 0 | 0 | 0 | 0.000599632 | 0 |
| 0 | 0 | 0 | 0 | 0 |
| 0 | 0.53333173 | 0 | 0 | 0 |
| 0 | 0 | 0 | 0 | 0.102517654 |
| 0 | 0 | 0 | 0 | 0 |
| 0 | 0 | 0 | 0 | 0.024875285 |
| 0.008908596 | 0.009270094 | 0 | 0.011311598 | 0.115692731 |
| 0 | 0 | 0 | 0 | 0 |
| 0 | 0 | 0 | 0.387037811 | 0.872625562 |
| 0 | 0 | 0 | 0 | 0 |

| | | | | |
|---|---|---|---|---|
| 0 | 0 | 0 | 0 | 0 |
| 0 | 0 | 0 | 0 | 0 |
| 0.004606859 | 0 | 0 | 0 | 0 |
| 0 | 0 | 0 | 0 | 0 |
| 0 | 0 | 0 | 0 | 0 |
| 0.01453186 | 0 | 0 | 0 | 0 |
| 0 | 0.050447296 | 0.074251147 | 0 | 0 |
| 0 | 0.003440744 | 0 | 0.003348071 | 0.00460013 |
| 0 | 0 | 0 | 0 | 0 |
| 0 | 0 | 0.00743092 | 0 | 0.009840659 |
| 0 | 0 | 0 | 0 | 0 |
| 0 | 0 | 0.004797986 | 0 | 0 |
| 0 | 0 | 0 | 0 | 0 |
| 0 | 0.047481759 | 0 | 0.167875478 | 0.469414322 |
| 0 | 0 | 0 | 0 | 0 |
| 0 | 0 | 0 | 0 | 0 |
| 0 | 0 | 0 | 0 | 0 |
| 0 | 0 | 0 | 0 | 0 |
| 0 | 0.025611655 | 0 | 1.929477959 | 0.604654887 |
| 0 | 0 | 0 | 0 | 0 |
| 0 | 0.002439133 | 0 | 0 | 0 |
| 0 | 0 | 0 | 0 | 0 |
| 0 | 0 | 0 | 0 | 0 |
| 0 | 0 | 0 | 0 | 0 |
| 0 | 0 | 0 | 0 | 0 |
| 0.002567056 | 0 | 0 | 0.006382224 | 0 |
| 0 | 0 | 0 | 0 | 0 |
| 0.930062371 | 0 | 0.013367175 | 0.005466062 | 0.22222652 |
| 0 | 0 | 0 | 0 | 0 |
| 0.23834769 | 0.063837413 | 1.884276547 | 1.134893069 | 1.119915134 |
| 0 | 0 | 0 | 0 | 0 |
| 0 | 0 | 0 | 0 | 0 |
| 0 | 0 | 0 | 0 | 0.018716994 |
| 0 | 0 | 0 | 0 | 0 |
| 0.00413864 | 0.000904382 | 0 | 0 | 0.005793264 |
| 0 | 0 | 0 | 0 | 0 |
| 0 | 0.494935736 | 0 | 0 | 0 |
| 0.000939229 | 0 | 0 | 0 | 0.002040101 |
| 0 | 0 | 0 | 0 | 0 |
| 0 | 0 | 0 | 0 | 0 |
| 1.902887651 | 3.868392232 | 19.59053188 | 7.824105931 | 3.369355768 |
| 0.190099215 | 0.096302465 | 0 | 0.76398866 | 1.196349659 |
| 0 | 0 | 0.318826784 | 0 | 0 |
| 0 | 0 | 0 | 0 | 0 |
| 0 | 0 | 0 | 0 | 0 |
| 0 | 0 | 0 | 0 | 0 |
| 0 | 0 | 0 | 0 | 0 |
| 0 | 0.000814456 | 0 | 0.003764467 | 0.028244942 |

| | | | | |
|---|---|---|---|---|
| 0 | 0 | 0 | 0 | 0 |
| 0.001150095 | 0 | 0 | 0.002942769 | 0.007031756 |
| 0 | 0 | 0 | 0 | 0.025934823 |
| 0 | 0 | 0 | 0 | 0 |
| 0.315028044 | 1.338537979 | 0.033093977 | 0.997894888 | 1.334025024 |
| 0 | 0 | 0 | 0 | 0 |
| 0 | 0 | 0 | 0 | 0 |
| 0 | 0 | 0 | 0 | 0 |
| 0.115605551 | 0 | 0.014489327 | 0.145220328 | 0 |
| 0 | 0 | 0 | 0 | 0 |
| 0 | 0 | 0 | 0 | 0 |
| 0 | 0 | 0 | 0 | 0 |
| 0 | 0 | 0 | 0.151096808 | 0.159126476 |
| 0.001144891 | 0.005403959 | 0.015408064 | 0.020166971 | 0.010610431 |
| 1.264592078 | 0.800602016 | 0.001804047 | 0 | 0 |
| 0.018601887 | 0.144530215 | 0.029502073 | 0.210580585 | 0.172685715 |
| 0.00079258 | 0 | 0.003148138 | 0 | 0.069436471 |
| 0 | 0 | 0 | 0 | 0 |
| 0 | 0 | 0 | 0 | 0 |
| 0 | 0 | 0 | 0 | 0 |
| 0 | 0 | 0 | 0 | 0 |
| 0 | 0 | 0 | 0 | 0 |
| 0 | 0 | 0 | 0 | 0 |
| 0 | 0 | 0 | 0 | 0 |
| 0 | 0 | 0 | 0 | 0 |
| 0 | 0 | 0 | 0 | 0 |
| 0.101137349 | 0.007981849 | 0 | 0.002645131 | 0.067197404 |
| 0 | 0 | 0 | 0.000785456 | 0 |
| 0 | 0 | 0 | 0 | 0 |
| 0 | 0 | 0 | 0 | 0 |
| 0 | 0 | 0 | 0 | 0 |
| 0 | 0 | 0 | 0 | 0 |
| 0 | 0 | 0 | 0.905923307 | 0 |
| 0 | 0 | 0 | 0.009551068 | 0.023412538 |
| 0 | 0.322145768 | 0.034917562 | 0 | 0 |
| 0.002248989 | 0 | 0 | 0 | 0 |
| 0.002298738 | 0 | 0.007519327 | 0 | 0.078779915 |
| 0 | 0 | 0 | 0 | 0 |
| 0 | 0 | 0 | 0 | 0 |
| 0 | 0 | 0 | 0 | 0 |
| 2.239078945 | 0.010834253 | 1.776244821 | 0.014466575 | 0.356315528 |
| 0 | 0.007324761 | 0 | 0 | 0 |
| 0 | 0 | 0 | 0 | 0 |
| 0 | 0 | 0.008450602 | 0 | 0 |
| 0 | 0 | 0 | 0 | 0 |
| 0 | 0 | 0 | 0 | 0 |
| 0 | 0 | 0 | 0 | 0 |
| 0 | 0 | 0 | 0 | 0 |

| | | | | |
|---|---|---|---|---|
| 0 | 0.09786984 | 0.036406715 | 0 | 0.716569912 |
| 0 | 0 | 0 | 0 | 0 |
| 0 | 0 | 0 | 0 | 0 |
| 0.001541646 | 0 | 0 | 0 | 0 |
| 0 | 0.026891615 | 0 | 0.243651763 | 0.032751666 |
| 0 | 0 | 0 | 0 | 0 |
| 0.073728411 | 0 | 0.012192462 | 0.015492369 | 0.049422359 |
| 0 | 0 | 0 | 0 | 0 |
| 0 | 0 | 0 | 0 | 0 |
| 0.001659048 | 0 | 0 | 0.003351344 | 0 |
| 0 | 0 | 0 | 0 | 0.012727022 |
| 0 | 0 | 0 | 0 | 0.083270937 |
| 0 | 0 | 0 | 0 | 0 |
| 0 | 0 | 0 | 0 | 0 |
| 0 | 0 | 0 | 0 | 0 |
| 0 | 0 | 0 | 0 | 0 |
| 0 | 0 | 0 | 1.652632156 | 0 |
| 0.002090358 | 0 | 0 | 0 | 0 |
| 0 | 0 | 0 | 0 | 0 |
| 0 | 0 | 0 | 0 | 0 |
| 0.074521845 | 0 | 0.056808378 | 0.024785409 | 0.186757992 |
| 0 | 0 | 0 | 0 | 0 |
| 0.002283259 | 0.001088599 | 0 | 0.00201263 | 0.004608802 |
| 0 | 0 | 0 | 0.002833614 | 0.014789136 |
| 0 | 0 | 0 | 0 | 0 |
| 0 | 0 | 0 | 0 | 0 |
| 0 | 0 | 0 | 0 | 0 |
| 0 | 0 | 0 | 0 | 0 |
| 0 | 0 | 0.145892625 | 0.075780074 | 0 |
| 0.031080247 | 0 | 0.477170331 | 0.070617862 | 0 |
| 0 | 0 | 0 | 0 | 0 |
| 0 | 0 | 0 | 0 | 0 |
| 0 | 0 | 0 | 0 | 0 |
| 0 | 0 | 0 | 0.560311723 | 0 |
| 0 | 0 | 0 | 0 | 0 |
| 0 | 0 | 0 | 0 | 0 |
| 0.176444296 | 0 | 0 | 0.001679488 | 0.031882177 |
| 0 | 0 | 0 | 0 | 0 |
| 0 | 0.002418245 | 0 | 0 | 0 |
| 0 | 0 | 0 | 0 | 0 |
| 0 | 0 | 0 | 0 | 0 |
| 0 | 0 | 0 | 0 | 0 |
| 0 | 0 | 0 | 0 | 0 |
| 0 | 0 | 0 | 0 | 0 |
| 0 | 0 | 0 | 0 | 0 |
| 0 | 0 | 0 | 0 | 0 |
| 0 | 0 | 0 | 0 | 0 |
| 0 | 0.002693767 | 0 | 0 | 0 |

| | | | | |
|---|---|---|---|---|
| 0.052630746 | 0 | 0 | 0.000641909 | 0.001342114 |
| 0 | 0.007079553 | 0.068126433 | 0 | 0 |
| 0 | 0.031977343 | 0 | 0 | 0 |
| 0 | 0 | 0 | 0 | 0 |
| 0 | 0 | 0 | 0 | 0 |
| 0 | 0 | 0 | 0 | 0 |
| 0 | 0 | 0 | 0 | 0 |
| 0.281007382 | 0.162917931 | 0.103487952 | 0.35195718 | 1.671374873 |
| 5.608565979 | 7.945265973 | 0.035836294 | 0 | 2.383824126 |
| 0.031685801 | 0 | 0 | 0 | 0.009637565 |
| 0.002546762 | 0 | 0 | 0 | 0.326254854 |
| 0 | 0 | 0 | 0 | 0 |
| 0 | 0 | 0 | 0 | 0 |
| 0.028387011 | 0 | 0 | 0 | 0 |
| 0 | 0 | 0 | 0 | 0 |
| 0 | 0 | 0 | 0 | 0 |
| 0.028411393 | 0 | 0 | 0 | 0.081443628 |
| 0 | 0.082130849 | 0 | 0 | 0 |
| 0 | 0 | 0 | 0 | 0 |
| 0.001079779 | 0 | 0 | 0.002617437 | 0 |
| 0 | 0 | 0 | 0 | 0 |
| 0 | 0 | 0 | 0 | 0 |
| 0 | 0 | 0 | 0 | 0 |
| 0 | 0 | 0 | 0 | 0 |
| 0 | 0 | 0 | 0.051064854 | 0 |
| 0 | 0 | 0 | 0 | 0 |
| 0 | 0 | 0 | 0 | 0 |
| 0 | 0 | 0 | 0 | 0 |
| 0 | 0 | 0 | 0 | 0 |
| 0 | 0 | 0 | 0 | 0 |
| 0.018363069 | 0 | 0 | 0 | 0.001772731 |
| 0 | 0 | 0 | 0 | 0 |
| 0 | 0 | 0 | 0 | 0 |
| 0 | 0 | 0 | 0 | 0 |
| 0 | 0 | 0 | 0 | 0 |
| 0 | 0 | 0 | 0 | 0 |
| 0 | 0 | 0 | 0 | 0 |
| 0 | 0 | 0 | 0 | 0 |
| 0 | 0 | 0 | 0 | 0.001595878 |
| 0 | 0 | 0 | 0 | 0 |
| 0 | 0 | 0 | 0 | 0 |
| 0.469038459 | 0.018237225 | 0.001338209 | 0.504108835 | 0.429300711 |
| 0 | 0 | 0 | 0 | 0 |
| 0 | 0 | 0 | 0 | 0 |
| 0 | 0 | 0 | 0 | 0 |
| 0 | 0 | 0 | 0 | 0 |
| 0 | 0 | 0 | 0 | 0 |
| 0 | 0 | 0 | 0 | 0 |

| | | | | |
|---|---|---|---|---|
| 0 | 0 | 0 | 0 | 0 |
| 0 | 0 | 0.001675264 | 0 | 0 |
| 0 | 0 | 0 | 0 | 0 |
| 0 | 0 | 0 | 0 | 0 |
| 0.255380032 | 0.091567892 | 0.168173921 | 0.185898337 | 0.868328149 |
| 0 | 0 | 0 | 0 | 0 |
| 0 | 0 | 0 | 0 | 0 |
| 0.020211357 | 0.000662492 | 0.015111477 | 0.003266221 | 0.003414532 |
| 0 | 0 | 0.00424357 | 0 | 0.001217601 |
| 0 | 0 | 0 | 0 | 0 |
| 0 | 0 | 0 | 0 | 0 |
| 0 | 0 | 0.016580898 | 0 | 0 |
| 0 | 0 | 0 | 0 | 0 |
| 0 | 0 | 0 | 0 | 0 |
| 0.03112393 | 0 | 0 | 0 | 0 |
| 0.007230778 | 0 | 0 | 0 | 0.922687044 |
| 0 | 0 | 0 | 0 | 0 |
| 0 | 0 | 0 | 0 | 0 |
| 0 | 0 | 0 | 0 | 0 |
| 0 | 0 | 0 | 0 | 0 |
| 0 | 0 | 0 | 0 | 0 |
| 0 | 0 | 0 | 0 | 0 |
| 0 | 0 | 0 | 0 | 0 |
| 0.000655922 | 0 | 0 | 0.001766653 | 0.00189964 |
| 0 | 0.050160359 | 0 | 0.136748875 | 0.067793696 |
| 0 | 0 | 0 | 0.002576226 | 0 |
| 1.420199996 | 0 | 0.000747567 | 0 | 0.455980636 |
| 0 | 0 | 0 | 0.000850695 | 0 |
| 0 | 0 | 0.00861154 | 0 | 0 |
| 0 | 0 | 0 | 0 | 0 |
| 0 | 0 | 0 | 0 | 0 |
| 0 | 0 | 0 | 0 | 0 |
| 0 | 0 | 0 | 0 | 0 |
| 0 | 0 | 0 | 0 | 0 |
| 0 | 0 | 0 | 0 | 0 |
| 0 | 0 | 0 | 0 | 0 |
| 0 | 0 | 0 | 0 | 0 |
| 0 | 0 | 0 | 0 | 0.716103008 |
| 0 | 0 | 0 | 0 | 0 |
| 0 | 0.000768937 | 0 | 0 | 0 |
| 0 | 0 | 0 | 0 | 0 |
| 0 | 0.349635254 | 0.016263346 | 0 | 0 |
| 0 | 0 | 0 | 0 | 0 |
| 0 | 0 | 0 | 0 | 0 |
| 0 | 0 | 0 | 0 | 0 |
| 0 | 0 | 0 | 0 | 0 |
| 0 | 0 | 0 | 0 | 0 |
| 0 | 0 | 0 | 0 | 0 |

| | | | | |
|---|---|---|---|---|
| 0 | 0.403710813 | 0 | 0.065947394 | 0.040666194 |
| 0 | 0 | 0 | 0 | 0 |
| 0 | 0 | 0 | 0 | 0 |
| 0 | 0 | 0 | 0 | 0 |
| 0 | 0 | 0 | 0 | 0 |
| 0 | 0.014663933 | 0 | 0.007569969 | 0 |
| 0 | 0 | 0 | 0 | 0 |
| 0 | 0 | 0 | 0 | 0 |
| 0 | 0 | 0 | 0 | 0 |
| 0 | 0 | 0 | 0 | 0 |
| 0 | 0 | 0 | 0 | 0 |
| 0 | 0 | 0 | 0 | 0.024321743 |
| 0 | 0 | 0 | 0 | 0 |
| 0 | 0 | 0 | 0 | 0 |
| 0 | 0 | 0 | 0 | 0 |
| 0 | 0 | 0 | 0 | 0 |
| 0 | 0 | 0 | 0 | 0 |
| 0 | 0 | 0 | 0 | 0 |
| 0 | 0 | 0 | 0 | 0 |
| 0 | 0 | 0 | 0 | 0 |
| 0.028957668 | 0 | 0 | 0.029027923 | 0 |
| 0 | 0 | 0 | 0 | 0 |
| 0.001924812 | 0.006008754 | 0.001027949 | 0.034438326 | 0.046719759 |
| 0 | 0.002419791 | 0 | 0 | 0.318759034 |
| 0 | 0 | 0 | 0 | 0 |
| 0 | 0.014184282 | 0 | 0 | 0 |
| 0 | 0 | 0 | 0 | 0 |
| 0 | 0 | 0 | 0 | 0 |
| 0 | 0 | 0 | 0.10117071 | 0 |
| 0 | 0 | 0 | 0 | 0 |
| 0 | 0 | 0 | 0 | 0 |
| 0 | 0 | 0 | 0 | 0 |
| 0.001287556 | 0 | 0.04067282 | 0 | 0.08949449 |
| 0 | 0 | 0 | 0 | 0 |
| 0 | 0 | 0.5534746 | 0 | 0.830543609 |
| 0.018804417 | 0 | 0 | 0 | 0 |
| 0 | 0 | 0 | 0 | 0 |
| 0 | 0 | 0 | 0 | 0 |
| 0.009560564 | 0 | 0 | 0 | 0.002076652 |
| 0 | 0 | 0 | 0 | 0 |
| 0 | 0 | 0 | 0 | 0 |
| 0 | 0 | 0 | 0 | 0 |
| 0 | 0.087455382 | 0.031343828 | 0.013323595 | 0.475393139 |
| 0 | 0 | 0 | 0 | 0 |
| 0 | 0 | 0 | 0 | 0 |
| 0 | 0.006014237 | 0 | 0.010581245 | 0.013499041 |
| 0 | 0 | 0 | 0 | 0 |
| 0 | 0 | 0 | 0 | 0 |

| | | | | |
|---|---|---|---|---|
| 0 | 0 | 0 | 0 | 0 |
| 0 | 0 | 0 | 0 | 0 |
| 0 | 0 | 0 | 0 | 0 |
| 0 | 0 | 0 | 0 | 0 |
| 0 | 0 | 0 | 0 | 0.003369604 |
| 0 | 0 | 0 | 0 | 0 |
| 0 | 0 | 0 | 0 | 0 |
| 0.009651543 | 0 | 0 | 0 | 0 |
| 0.001247707 | 0 | 0 | 0 | 0.52516614 |
| 0 | 0.004396166 | 0.009400942 | 0 | 0 |
| 0 | 0 | 0 | 0 | 0 |
| 0 | 0 | 0 | 0 | 0 |
| 0 | 0 | 0 | 0 | 0 |
| 0 | 0.226694686 | 0 | 0 | 0 |
| 0 | 0 | 0 | 0 | 0 |
| 0 | 0.010026929 | 0 | 0 | 0 |
| 0 | 0 | 0 | 0 | 0 |
| 0 | 0 | 0 | 0 | 0 |
| 0 | 0 | 0 | 0 | 0 |
| 0 | 0.108145573 | 0.038191822 | 0 | 0.872807701 |
| 0.003053978 | 1.123833923 | 0 | 0 | 0 |
| 0 | 0 | 0 | 0 | 0 |
| 0 | 0 | 0 | 0 | 0 |
| 0 | 0 | 0 | 0 | 0 |
| 0 | 0 | 0 | 0 | 0.00997803 |
| 0 | 0 | 0 | 0 | 0 |
| 0 | 0 | 0 | 0 | 0 |
| 0 | 0 | 0 | 0 | 0 |
| 0 | 0 | 0 | 0 | 0 |
| 0 | 0.017727772 | 0 | 0 | 0.063238958 |
| 0.213345319 | 0.088207501 | 0.138565326 | 0.187158238 | 0.298084628 |
| 0 | 0 | 0 | 0 | 0 |
| 0 | 0 | 0 | 0 | 0 |
| 0 | 0 | 0 | 0 | 0 |
| 0 | 0 | 0 | 0 | 0 |
| 0 | 0 | 0 | 0 | 0 |
| 0 | 0 | 0 | 0 | 0 |
| 0 | 0 | 0 | 0 | 0 |
| 0 | 0 | 0 | 0.033629678 | 0.011536576 |
| 0 | 0 | 0 | 0 | 0 |
| 0 | 0 | 0 | 0 | 0 |
| 0 | 0 | 0 | 0 | 5.570927114 |
| 0 | 0 | 0 | 0 | 0 |
| 0 | 0 | 0 | 0.002523692 | 0 |
| 0 | 0 | 0 | 0 | 0 |
| 0 | 0 | 0 | 0 | 0 |
| 0.0016118 | 0 | 0 | 0.023442493 | 0 |
| 0 | 0 | 0 | 0 | 0 |

| | | | | |
|---|---|---|---|---|
| 0 | 0 | 0 | 0 | 0 |
| 0 | 0 | 0 | 0 | 0 |
| 0 | 0 | 0 | 0 | 0 |
| 0 | 0 | 0 | 0 | 0 |
| 0.038876364 | 0.217269731 | 0 | 0.160816509 | 0.10113283 |
| 0 | 0 | 0 | 0 | 0 |
| 0 | 0 | 0 | 0 | 0.031916112 |
| 0 | 0 | 0 | 0 | 0 |
| 0 | 0 | 0 | 0 | 0 |
| 0 | 0 | 0 | 0 | 0 |
| 0 | 0 | 0 | 0.002470644 | 0 |
| 0.050152994 | 0.002713783 | 0 | 0.004631364 | 0.039194413 |
| 0 | 0 | 0 | 0.010322225 | 0.154355085 |
| 0 | 0 | 0 | 0 | 0 |
| 0 | 0 | 0 | 0.005735312 | 0.00245979 |
| 0 | 0 | 0.00329786 | 0 | 0.839074776 |
| 0 | 0 | 0 | 0.002523132 | 0 |
| 0 | 0 | 0 | 0 | 0 |
| 0.068357042 | 0 | 0.001014061 | 0.00467586 | 0.001920356 |
| 1.570672152 | 0 | 0 | 0 | 0.009345764 |
| 0 | 0 | 0 | 0 | 0 |
| 0 | 0 | 0 | 0 | 0 |
| 0 | 0 | 0 | 0 | 0 |
| 0 | 0 | 0 | 0 | 0 |
| 0 | 0 | 0 | 0 | 0 |
| 0 | 0 | 0 | 0 | 0 |
| 0.002081354 | 0 | 0 | 0.023544749 | 0.001461305 |
| 0.000956898 | 0 | 0 | 0 | 0 |
| 0 | 0 | 0 | 0 | 0 |
| 0 | 0 | 0 | 0 | 0 |
| 0 | 0 | 0 | 0.000866533 | 0 |
| 0.004170019 | 0 | 0 | 0 | 0 |
| 0 | 0 | 0 | 0 | 0 |
| 0 | 0 | 0 | 0 | 0 |
| 0 | 0 | 0 | 0 | 0 |
| 0 | 0 | 0 | 0 | 0 |
| 0 | 0 | 0 | 0 | 0 |
| 0.013562122 | 0.017386543 | 0 | 0.002435203 | 0.041896263 |
| 0 | 0.003960691 | 0 | 0 | 0 |
| 0 | 0 | 0 | 0 | 0 |
| 0 | 0.000657476 | 0 | 0.000364668 | 0 |
| 0 | 0 | 0 | 0 | 0 |
| 0 | 0 | 0 | 0 | 0 |
| 0 | 0 | 0 | 0 | 0 |
| 0 | 0 | 0 | 0 | 0 |
| 0 | 0 | 0 | 0 | 0.004821336 |
| 0 | 0 | 0 | 0 | 0 |
| 0 | 0 | 0 | 0 | 0 |

| | | | | |
|---|---|---|---|---|
| 0 | 0 | 0 | 0 | 0 |
| 0 | 0 | 0.000535487 | 0.227932773 | 0.379998489 |
| 0 | 0.079676061 | 0 | 0 | 0 |
| 0 | 0 | 0 | 0 | 0 |
| 0.003475191 | 0 | 0.000608978 | 0.000702003 | 0 |
| 0.008970136 | 0 | 0 | 0 | 0.001082447 |
| 0 | 0.000993203 | 0 | 0 | 0 |
| 0.009890105 | 0 | 0 | 0 | 0 |
| 0 | 0 | 0 | 0 | 0 |
| 0 | 0 | 0 | 0 | 0 |
| 0 | 0 | 0 | 0 | 0 |
| 0 | 0 | 0 | 0 | 0 |
| 0.054448528 | 0.078208732 | 0.131406596 | 0.00737726 | 0.013140861 |
| 0 | 0 | 0 | 0 | 0 |
| 0 | 0 | 0 | 0 | 0 |
| 0 | 0 | 0 | 0 | 0 |
| 0 | 0 | 0 | 0 | 0 |
| 0 | 0 | 0 | 0 | 0 |
| 0 | 0 | 0 | 0 | 0 |
| 0 | 0 | 0 | 0 | 0 |
| 0 | 0 | 0 | 0 | 0 |
| 0 | 0 | 0 | 0 | 0 |
| 0.003934546 | 0 | 0 | 0.000706484 | 0.001266109 |
| 0.010059615 | 0.005861979 | 0 | 0.008336751 | 0.018426639 |
| 0 | 0 | 0 | 0 | 0 |
| 0 | 0 | 0 | 0 | 0 |
| 0 | 0.012595537 | 0 | 0 | 0 |
| 0 | 0 | 0 | 0 | 0 |
| 0 | 0 | 0 | 0 | 0 |
| 0 | 0 | 0 | 0 | 0 |
| 0.004630723 | 0.00404765 | 0.052617311 | 0.017329976 | 0.027057691 |
| 0 | 0 | 0 | 0 | 0.098433194 |
| 0 | 0 | 0 | 0 | 0 |
| 0 | 0 | 0 | 0 | 0 |
| 0 | 0 | 0 | 0 | 0.826506991 |
| 0 | 0 | 0 | 0 | 0 |
| 0 | 0 | 0 | 0 | 0 |
| 0 | 0 | 0 | 0 | 0 |
| 0.031343304 | 0 | 0.000770873 | 0 | 0.035035758 |
| 0 | 0 | 0 | 0 | 0 |
| 0 | 0 | 0 | 0 | 0 |
| 0 | 0 | 0 | 0 | 0 |
| 0 | 0 | 0 | 0 | 0 |
| 0 | 0 | 0 | 0 | 0 |
| 0 | 0 | 0 | 0 | 0 |
| 0.053883869 | 0.060895854 | 0.251796811 | 0.353003387 | 0.187230973 |
| 0 | 0 | 0 | 0 | 0 |
| 0 | 0 | 0 | 0 | 0 |

| | | | | |
|---|---|---|---|---|
| 0 | 0 | 0 | 0 | 0 |
| 0 | 0 | 0 | 0 | 0 |
| 0 | 0.039689323 | 0.181399231 | 0.13051772 | 0.214861463 |
| 0 | 0 | 0 | 0 | 0 |
| 0 | 0 | 0 | 0 | 0 |
| 0 | 0 | 0 | 0 | 0 |
| 0 | 0 | 0 | 0 | 0 |
| 0 | 0 | 0 | 0 | 0 |
| 0 | 0 | 0 | 0 | 0 |
| 0.002353345 | 0 | 0 | 0 | 0 |
| 0 | 0 | 0 | 0 | 0.004198909 |
| 0 | 0 | 0 | 0 | 0 |
| 0 | 0 | 0 | 0 | 0 |
| 0 | 0 | 0 | 0 | 0 |
| 0 | 0 | 0 | 0 | 0 |
| 0 | 0 | 0 | 0 | 0 |
| 0.012295326 | 0 | 0 | 0 | 0 |
| 0 | 0 | 0 | 0 | 0 |
| 0 | 0 | 0 | 0 | 0 |
| 0 | 0 | 0 | 0 | 0 |
| 0 | 0 | 0 | 0 | 0 |
| 0 | 0 | 0 | 0 | 0 |
| 0 | 0 | 0 | 0 | 0 |
| 0 | 0 | 0 | 0 | 0 |
| 0 | 0 | 0 | 0 | 0 |
| 0 | 0 | 0 | 0 | 0 |
| 0 | 0 | 0 | 0 | 0 |
| 0 | 0.003004452 | 0 | 0.014878678 | 0.01516913 |
| 0 | 0 | 0 | 0 | 0 |
| 0 | 0 | 0 | 0 | 0 |
| 0 | 0.001166173 | 0 | 0.000616014 | 0.001287972 |
| 0 | 0 | 0 | 0 | 0 |
| 0.032309773 | 0.004062137 | 0.043846847 | 0.265002107 | 0.024995659 |
| 0.020126853 | 0.000988348 | 0.00253623 | 0.001827283 | 0.021394868 |
| 0 | 0 | 0 | 0 | 0 |
| 0 | 0 | 0 | 0 | 0 |
| 0 | 0 | 0 | 0 | 0 |
| 0 | 0 | 0 | 0 | 0 |
| 0 | 0 | 0 | 0 | 0 |
| 0 | 0 | 0 | 0 | 0 |
| 0 | 0 | 0 | 0 | 0 |
| 0.036048148 | 0 | 0 | 0.004266147 | 0.15712777 |
| 0 | 0.017060919 | 0.018449179 | 0.03136344 | 0.072373561 |
| 0 | 0 | 0 | 0 | 0 |
| 0 | 0 | 0 | 0 | 0 |
| 0.006730907 | 0.208720338 | 0.009286188 | 0.005179697 | 0.040224934 |
| 0 | 0 | 0 | 0 | 0.053556682 |
| 0 | 0 | 0 | 0 | 0 |

| | | | | |
|---|---|---|---|---|
| 0 | 0 | 0 | 0 | 0 |
| 0.013942024 | 0.031278757 | 0.004994866 | 0.007322498 | 0.00919721 |
| 0.038814387 | 0 | 0.006595557 | 0.007895493 | 0.007336897 |
| 0 | 0 | 0 | 0 | 0 |
| 0 | 0.009092867 | 0 | 0.109272302 | 6.393343347 |
| 0 | 0 | 0 | 0 | 0 |
| 0 | 0 | 0 | 0 | 0 |
| 0 | 0 | 0 | 0 | 0 |
| 0 | 0.057592368 | 0 | 0 | 0 |
| 0 | 0 | 0 | 0 | 0 |
| 0 | 0 | 0 | 0 | 0 |
| 0 | 0 | 0 | 0 | 0 |
| 0 | 0 | 0 | 0 | 0 |
| 0.005931727 | 0 | 0 | 0.003318184 | 0 |
| 0 | 0 | 0 | 0 | 0 |
| 0 | 0 | 0 | 0 | 0 |
| 0 | 0 | 0 | 0 | 0 |
| 0 | 0 | 0 | 0 | 0 |
| 0 | 0 | 0 | 0 | 0 |
| 0 | 0 | 0 | 0 | 0 |
| 0 | 0 | 0 | 0 | 0 |
| 0 | 0 | 0 | 0 | 0 |
| 0 | 0 | 0 | 0 | 0 |
| 0 | 0 | 0 | 0 | 0 |
| 0 | 0.296618175 | 0.00459941 | 0 | 0 |
| 0.002025295 | 0 | 0.002523762 | 0.000909151 | 0.002824144 |
| 0 | 0 | 0 | 0 | 0 |
| 0 | 0.016599941 | 0 | 0.019754735 | 0.037510322 |
| 0 | 0 | 0 | 0 | 0 |
| 0 | 0 | 0 | 0 | 0 |
| 0 | 0 | 0 | 0 | 0 |
| 0 | 0 | 0 | 0 | 0 |
| 0 | 0 | 0 | 0 | 0 |
| 0 | 0 | 0 | 0 | 0 |
| 0 | 0 | 0 | 0 | 0 |
| 0 | 0 | 0 | 0 | 0 |
| 0 | 0 | 0 | 0 | 0 |
| 0.001120006 | 0 | 0 | 0 | 0 |
| 0 | 4.122308252 | 1.307613021 | 0 | 14.11793768 |
| 0 | 0 | 0 | 0 | 0 |
| 0 | 0 | 0 | 0 | 0 |
| 0 | 0 | 0 | 0 | 0.001885391 |
| 0.045394834 | 0 | 0.001055363 | 0 | 0.001962233 |
| 0 | 0 | 0 | 0.007262629 | 0 |
| 0 | 0 | 0 | 0 | 0 |
| 0 | 0 | 0 | 0 | 0 |
| 0.008637464 | 0.027783584 | 0 | 0.002791684 | 0.049363465 |
| 0 | 0 | 0 | 0 | 0 |

| | | | | |
|---|---|---|---|---|
| 0 | 0 | 0 | 0 | 0 |
| 0.003269667 | 0.003429565 | 0.017112484 | 0.047202736 | 0.070704858 |
| 0 | 0 | 0 | 0 | 0 |
| 0 | 0 | 0 | 0 | 0 |
| 0 | 0 | 0 | 0 | 0 |
| 0 | 0 | 0 | 0 | 0 |
| 0 | 0 | 0 | 0 | 0 |
| 0 | 0 | 0 | 0 | 0 |
| 0 | 0 | 0 | 0 | 0 |
| 0.002060261 | 0 | 0 | 0 | 0 |
| 0 | 0 | 0 | 0 | 0 |
| 0 | 0 | 0 | 0 | 0 |
| 0 | 0 | 0.001292101 | 0 | 0.162606818 |
| 0 | 0 | 0 | 0 | 0 |
| 0 | 0 | 0.004064131 | 0.009662712 | 0 |
| 0 | 0 | 0 | 0 | 0 |
| 0 | 0 | 0 | 0 | 0 |
| 0 | 0 | 0 | 0 | 0 |
| 0 | 0 | 0 | 0.001943538 | 0 |
| 0 | 0 | 0 | 0 | 0 |
| 0 | 0 | 0 | 0 | 0 |
| 0.803947394 | 0 | 0.00124728 | 0 | 0.000773022 |
| 0.00184592 | 0 | 0 | 0.001822985 | 0 |
| 0 | 0 | 0 | 0 | 0 |
| 0 | 0 | 0 | 0 | 0 |
| 0 | 0 | 0 | 0 | 0 |
| 0 | 0 | 0 | 0 | 0 |
| 0 | 0 | 0 | 0 | 0 |
| 0 | 0 | 0 | 0 | 0.000625571 |
| 0 | 0 | 0 | 0 | 0 |
| 0 | 0 | 0 | 0 | 0 |
| 0 | 0 | 0 | 0 | 0 |
| 0 | 0 | 0 | 0 | 0 |
| 0 | 0 | 0 | 0 | 0 |
| 3.36688227 | 4.410921947 | 0 | 0 | 0.080480007 |
| 0 | 0 | 0 | 0 | 0 |
| 0 | 0 | 0 | 0 | 0 |
| 0 | 0 | 0 | 0 | 0 |
| 0.745264657 | 0 | 0.002491121 | 0 | 0.00180123 |
| 0 | 0 | 0 | 0 | 0 |
| 0.004301899 | 0.072279975 | 0.270491308 | 0.144833509 | 0.278940572 |
| 0 | 0 | 0 | 0 | 0 |
| 0 | 0 | 0 | 0 | 0 |
| 0 | 0 | 0 | 0 | 0 |
| 0 | 0 | 0 | 0 | 0 |
| 0.052811038 | 0.001674247 | 0.792264609 | 0.009286173 | 0.008453081 |
| 0 | 0 | 0 | 0 | 0 |
| 0 | 0 | 0 | 0 | 0 |

| | | | | |
|---|---|---|---|---|
| 0 | 0 | 0 | 0 | 0 |
| 0 | 0 | 0 | 0 | 0 |
| 0 | 0 | 0 | 0 | 0 |
| 0.003714839 | 0 | 0 | 0 | 0 |
| 0 | 0 | 0 | 0 | 0 |
| 0 | 0 | 0 | 0 | 0 |
| 0 | 0 | 0 | 0 | 0 |
| 0.130133462 | 0 | 0 | 0 | 0 |
| 0 | 0 | 0 | 0.00389427 | 0 |
| 0.036662959 | 0 | 0.001500651 | 0 | 0 |
| 0 | 0 | 0.002373976 | 0 | 0.041414612 |
| 0 | 0 | 0 | 0 | 0 |
| 0 | 0 | 0 | 0.000634056 | 0 |
| 0 | 0 | 0 | 0 | 0 |
| 0 | 0 | 0 | 0 | 0 |
| 0 | 0 | 0 | 0 | 0 |
| 0 | 0.000529306 | 0 | 0 | 0 |
| 0 | 0 | 0 | 0 | 0 |
| 0 | 0 | 0 | 0 | 0 |
| 0.964552072 | 0.007529509 | 1.369055046 | 0.997402697 | 0.080462114 |
| 0 | 0 | 0 | 0 | 0 |
| 0 | 0 | 0 | 0 | 0 |
| 0 | 0 | 0 | 0 | 0 |
| 0 | 0 | 0 | 0 | 0 |
| 0 | 0 | 0 | 0 | 0 |
| 0 | 0.016473819 | 0 | 0 | 0 |
| 0 | 0 | 0 | 0 | 0 |
| 0 | 0 | 0 | 0 | 0 |
| 0 | 0 | 0 | 0 | 0 |
| 0 | 0 | 0 | 0.009451766 | 0 |
| 0 | 0 | 0 | 0 | 0 |
| 0 | 0 | 0 | 0 | 0 |
| 0 | 0 | 0 | 0 | 0 |
| 0 | 0 | 0 | 0 | 0 |
| 0 | 0 | 0 | 0 | 0 |
| 0 | 0 | 0 | 0 | 0 |
| 0 | 0 | 0 | 0 | 0 |
| 0 | 0 | 0 | 0 | 0 |
| 0 | 0 | 0 | 0 | 0 |
| 0 | 0 | 0 | 0 | 0 |
| 0 | 0 | 0 | 0 | 0 |
| 0 | 0 | 0 | 0 | 0 |
| 0.002902874 | 0 | 0 | 0 | 0.028584196 |
| 0 | 0 | 0 | 0 | 0 |
| 0 | 0.002293619 | 0 | 0.001766875 | 0.026176114 |
| 0 | 0 | 0 | 0 | 0 |
| 0 | 0 | 0 | 0 | 0 |
| 0 | 0 | 0 | 0 | 0 |

| | | | | |
|---|---|---|---|---|
| 0 | 0 | 0 | 0 | 0 |
| 0 | 0 | 0 | 0 | 0 |
| 0 | 0 | 0 | 0 | 0 |
| 0 | 0 | 0 | 0 | 0 |
| 0 | 0 | 0 | 0 | 0 |
| 0.014039017 | 0.027111031 | 0 | 0.001055233 | 0.017492783 |
| 0 | 0 | 0 | 0 | 0 |
| 0 | 0 | 0 | 0 | 0.000938593 |
| 0 | 0 | 0 | 0 | 0 |
| 0 | 0 | 0 | 0 | 0 |
| 0 | 0 | 0 | 0 | 0 |
| 0 | 0 | 0 | 0 | 0 |
| 0 | 0 | 0 | 0 | 0 |
| 0 | 0 | 0 | 0 | 0 |
| 0 | 0 | 0 | 0 | 0 |
| 0 | 0 | 0 | 0 | 0 |
| 0 | 0 | 0 | 0 | 0.266092992 |
| 0.008242811 | 0 | 0 | 0 | 0 |
| 0 | 0 | 0 | 0 | 0 |
| 0 | 0 | 0 | 0 | 0 |
| 0 | 0 | 0 | 0 | 0 |
| 0 | 0 | 0 | 0 | 0 |
| 0 | 0 | 0 | 0 | 0 |
| 0 | 0 | 0 | 0 | 0 |
| 0 | 0 | 0 | 0 | 0 |
| 0.00153205 | 0 | 0 | 0 | 0 |
| 0 | 0 | 0 | 0 | 0 |
| 0 | 0 | 0 | 0.002316246 | 0 |
| 0 | 0 | 0 | 0 | 0 |
| 0 | 0 | 0 | 0 | 0 |
| 0 | 0 | 0 | 0 | 0 |
| 0 | 0 | 0 | 0 | 0 |
| 0 | 0 | 0 | 0 | 0 |
| 0 | 0 | 0 | 0 | 0 |
| 0 | 0 | 0 | 0 | 0 |
| 0 | 0 | 0 | 0 | 0 |
| 0 | 0 | 0 | 0 | 0 |
| 0 | 0 | 0 | 0 | 0 |
| 0 | 0 | 0.00689157 | 1.346186044 | 0 |
| 0 | 0 | 0 | 0 | 0 |
| 0 | 0 | 0 | 0 | 0 |
| 0 | 0 | 0 | 0 | 0 |
| 0 | 0 | 0 | 0 | 0 |
| 0 | 0 | 0 | 0 | 0 |
| 0 | 0 | 0 | 0 | 0 |
| 0 | 0 | 0 | 0 | 0 |
| 0 | 0 | 0 | 0 | 0 |
| 0 | 0 | 0 | 0 | 0 |

| | | | | |
|---|---|---|---|---|
| 0 | 0 | 0 | 0 | 0 |
| 0 | 0 | 0 | 0.0097467 | 0 |
| 0 | 0 | 0 | 0 | 0 |
| 0 | 0 | 0 | 0 | 0 |
| 0 | 0 | 0 | 0 | 0 |
| 0 | 0 | 0 | 0 | 0 |
| 0 | 0 | 0 | 0 | 0 |
| 0 | 0 | 0 | 0 | 0 |
| 0 | 0 | 0 | 0 | 0 |
| 0 | 0 | 0 | 0 | 0 |
| 0.434838269 | 0 | 0.002012968 | 0 | 0.000693093 |
| 0 | 0 | 0 | 0 | 0 |
| 0 | 0.016485926 | 0 | 0 | 0.247591688 |
| 0 | 0 | 0 | 0 | 0 |
| 0 | 0 | 0 | 0 | 0 |
| 0 | 0 | 0 | 0 | 0 |
| 0 | 0 | 0 | 0 | 0 |
| 0 | 0 | 0 | 0 | 0 |
| 0 | 0 | 0 | 0 | 0 |
| 0 | 0 | 0 | 0 | 0 |
| 0 | 0 | 0 | 0 | 0.004455171 |
| 0 | 0 | 0 | 0 | 0 |
| 0 | 0 | 0 | 0 | 0 |
| 4.492652409 | 0.650390339 | 3.088353039 | 0.05040049 | 3.063832134 |
| 0 | 0 | 0 | 0 | 0 |
| 0 | 0 | 0 | 0 | 0 |
| 0 | 0 | 0 | 0 | 0 |
| 0 | 0 | 0.003389374 | 0 | 0 |
| 0 | 0 | 0 | 0 | 0 |
| 0 | 0 | 0 | 0 | 0 |
| 0 | 0 | 0 | 0 | 0 |
| 0 | 0 | 0 | 0 | 0 |
| 0 | 0 | 0 | 0 | 0 |
| 0 | 0 | 0 | 0 | 0 |

| PHL018-0045_V3 | PHL018-0059_V3 | PHL018-0118_V2 | PHL018-0225_V1 | PHL018-0012_V3 |
|---|---|---|---|---|
| EYCF | CTRL | EYCF | CTRL | EYCF |
| 0.407326007 | 0.062026929 | 0.251855657 | 0.03119785 | 0.019439522 |
| 0.317317095 | 15.47103419 | 1.557366013 | 0.780260091 | 3.001134954 |
| 1.486131544 | 0.001978596 | 8.797902518 | 0 | 2.148194174 |
| 0 | 0 | 0 | 0 | 0 |
| 0 | 0 | 0 | 0 | 0 |
| 0 | 0 | 0 | 0 | 0 |
| 0 | 0 | 0 | 0 | 0 |
| 0 | 0 | 0 | 0 | 0 |
| 0 | 0 | 0 | 0 | 0 |
| 0 | 0 | 0 | 0 | 0 |
| 0 | 0 | 0 | 0 | 0 |
| 0 | 0 | 0 | 0 | 0 |
| 0 | 0 | 0 | 0 | 0 |
| 0.047388155 | 2.018707367 | 8.36742032 | 0.626047408 | 8.771779084 |
| 0 | 0 | 0 | 0 | 0 |
| 0 | 0 | 0 | 0 | 0 |
| 0 | 0 | 0 | 0 | 0 |
| 0 | 0 | 0 | 0 | 0 |
| 0.015630999 | 0 | 0 | 0 | 0 |
| 0 | 0 | 0 | 0 | 0 |
| 0 | 0 | 0 | 0 | 0 |
| 0 | 0 | 0 | 0 | 0 |
| 0 | 0 | 0 | 0 | 0 |
| 0 | 0 | 0 | 0 | 0 |
| 0 | 0 | 0 | 0.046937791 | 0 |
| 0 | 0 | 0 | 0 | 0 |
| 0 | 0 | 0 | 0 | 0 |
| 0 | 0 | 0 | 0 | 0 |
| 0 | 0 | 0 | 0 | 0 |
| 0 | 0 | 0 | 0 | 0 |
| 0 | 0 | 0 | 0 | 0 |
| 0 | 0 | 0 | 0 | 0 |
| 0 | 0 | 0 | 0 | 0 |
| 0 | 0 | 0 | 0 | 0 |
| 0 | 0 | 0 | 0 | 0 |
| 0 | 0 | 0.005059736 | 0.867868083 | 1.647619811 |
| 0 | 0 | 0 | 0 | 0 |
| 0 | 0 | 0 | 0 | 0 |
| 0 | 0 | 0 | 0 | 0 |
| 0 | 0 | 0 | 0 | 0 |
| 0 | 0 | 0 | 0 | 0 |
| 0.003067598 | 1.219411483 | 0.007509308 | 0 | 0.016412465 |
| 0.061101162 | 0 | 0.06677332 | 17.46871311 | 0.182001687 |
| 0 | 0.081254997 | 0.016623088 | 0 | 0.00171781 |
| 0 | 0.002638828 | 0 | 0.002569436 | 0 |
| 0 | 0 | 0 | 0 | 0 |

| | | | | |
|---|---|---|---|---|
| 0 | 0 | 0 | 0.007992108 | 0 |
| 0.007959288 | 0.001176409 | 0 | 0 | 0.042145442 |
| 0.11989803 | 0 | 0.097842954 | 0 | 0.004260812 |
| 0.326063634 | 0.393272557 | 0.450329031 | 0.284828338 | 0.433816617 |
| 0.002075187 | 0 | 0 | 0.046765204 | 0.005425413 |
| 0 | 0 | 0 | 0 | 0 |
| 0.044254938 | 0.41332972 | 0 | 0.004419945 | 0 |
| 0 | 0 | 0 | 0 | 0 |
| 0 | 0 | 0 | 0 | 0 |
| 0 | 0 | 0 | 0 | 0 |
| 0 | 0 | 0.001559023 | 0 | 0 |
| 0 | 0 | 2.230136399 | 0.069663651 | 0.019072204 |
| 0 | 0 | 0.00055123 | 0 | 0 |
| 0.08228983 | 0.028355191 | 0.156151762 | 0.063484375 | 0.063149122 |
| 0 | 0 | 0 | 0 | 0.002325935 |
| 0 | 0 | 0 | 0 | 0 |
| 0 | 0.07799152 | 0.001665895 | 0.595711937 | 0 |
| 0 | 0 | 0 | 0 | 0 |
| 0 | 0 | 0 | 0 | 0 |
| 0 | 0 | 0 | 0 | 0 |
| 0.055481734 | 0.033588764 | 0.149274341 | 0 | 0.122859309 |
| 0.01591172 | 0.001075111 | 0.035113866 | 0.006281033 | 0.001401615 |
| 0 | 0 | 0 | 0 | 0 |
| 0 | 0 | 0 | 0 | 0 |
| 0 | 0 | 0 | 0 | 0 |
| 0 | 0 | 0 | 0 | 0 |
| 0 | 0 | 0 | 0 | 0.000669431 |
| 0 | 0 | 0 | 0 | 0 |
| 0 | 0.01031533 | 0.001156759 | 0 | 0.000896535 |
| 0 | 0 | 0 | 0 | 0 |
| 0.162911533 | 0 | 0.009733517 | 0 | 0.032716601 |
| 0 | 0 | 0 | 0 | 0 |
| 0 | 0 | 0 | 0 | 0 |
| 0 | 0 | 0 | 0 | 0 |
| 0 | 0 | 0 | 0 | 0 |
| 0 | 0.004790853 | 0 | 0 | 0 |
| 0 | 0 | 0 | 0 | 0 |
| 0 | 5.700578237 | 0 | 25.29601344 | 0 |
| 0 | 0 | 0 | 0 | 0 |
| 0 | 0 | 0 | 0 | 0 |
| 0 | 0 | 0 | 0 | 0 |
| 0 | 0 | 0 | 0 | 0 |
| 0 | 0 | 0 | 0 | 0 |
| 0 | 0 | 0 | 0 | 0 |
| 0 | 0 | 0 | 0 | 0 |
| 0 | 0 | 0 | 0 | 0 |
| 0 | 0.000664602 | 0 | 0.002372791 | 0 |
| 0.926474446 | 0 | 0 | 0.000528823 | 0 |

| | | | | |
|---|---|---|---|---|
| 0 | 0 | 0 | 0 | 0 |
| 0.015914387 | 0 | 0.007754984 | 0 | 0 |
| 0 | 0 | 0 | 0 | 0 |
| 0 | 0 | 0 | 0 | 0 |
| 0 | 0.052009639 | 0.563507738 | 0 | 0 |
| 0 | 0 | 0 | 0 | 0 |
| 0 | 0 | 0 | 0 | 0 |
| 0 | 0 | 0 | 0 | 0 |
| 0 | 0 | 0 | 0 | 0.000812862 |
| 0 | 0 | 0 | 0 | 0 |
| 0 | 0 | 0 | 0 | 0 |
| 0 | 0.011461425 | 0 | 0 | 0 |
| 0 | 0 | 0 | 0 | 0 |
| 0 | 0 | 0 | 0 | 0 |
| 0 | 0 | 0 | 0 | 0 |
| 0.002563073 | 0 | 0 | 0 | 0 |
| 0 | 0 | 0 | 0 | 0 |
| 0 | 0 | 0 | 0 | 0 |
| 0 | 0.004473742 | 0 | 0 | 0.287513523 |
| 0 | 0 | 0 | 0 | 0 |
| 0 | 0 | 0 | 0 | 0 |
| 0 | 0 | 0 | 0 | 0 |
| 0 | 0 | 0 | 0 | 0 |
| 0 | 0 | 0 | 0 | 0 |
| 0 | 0 | 0 | 0 | 0 |
| 0 | 0 | 0 | 0 | 0 |
| 0 | 0 | 0 | 0 | 0 |
| 0.002218297 | 0 | 0.002549209 | 0 | 0 |
| 0 | 0 | 0 | 0 | 0 |
| 0 | 0 | 0 | 0 | 0 |
| 0.036596915 | 0.000961626 | 0.739084094 | 0.050094093 | 0.007521992 |
| 0.002843952 | 0.001008829 | 0.000848474 | 0.004256635 | 0 |
| 0 | 0 | 0 | 0 | 0 |
| 0 | 0 | 0 | 0 | 0 |
| 0 | 0 | 0 | 0 | 0 |
| 0 | 0 | 0 | 0 | 0 |
| 0 | 0 | 0 | 0 | 0 |
| 0 | 0 | 0 | 0 | 0 |
| 0 | 0 | 0 | 0.004814886 | 0 |
| 0 | 0 | 0 | 0 | 0 |
| 0 | 0 | 0 | 0 | 0 |
| 0 | 0 | 0 | 0 | 0 |
| 0 | 0.001166047 | 0 | 0 | 0.000608067 |
| 0 | 0 | 0 | 0 | 0 |
| 0 | 0 | 0 | 0 | 0 |
| 0 | 0 | 0 | 0 | 0 |
| 0 | 0 | 0 | 0 | 0 |
| 0 | 0 | 0 | 0 | 0 |

| | | | | |
|---|---|---|---|---|
| 0 | 0 | 0 | 0 | 0 |
| 0 | 0 | 0 | 0 | 0 |
| 0 | 0 | 0 | 0 | 0 |
| 0 | 0 | 0 | 0 | 0 |
| 0 | 0 | 0 | 0.038189163 | 0 |
| 0 | 0 | 0 | 0 | 0 |
| 0 | 0 | 0 | 0 | 0 |
| 0 | 0 | 0 | 0 | 0 |
| 0 | 0 | 0 | 0 | 0 |
| 0 | 0 | 0 | 0 | 0 |
| 0 | 0 | 0 | 0 | 0 |
| 0 | 0 | 0 | 0 | 0 |
| 0 | 0 | 0 | 0 | 0 |
| 0 | 0 | 0 | 0 | 0 |
| 0 | 0 | 0 | 0 | 0 |
| 0 | 0 | 0 | 0 | 3.850905894 |
| 0 | 0 | 0 | 0 | 0 |
| 0 | 0 | 0 | 0 | 0 |
| 0 | 0 | 0 | 0 | 0 |
| 0 | 0 | 0 | 0 | 0 |
| 0 | 0 | 0 | 0 | 0 |
| 0 | 0 | 0 | 0 | 0 |
| 0 | 0 | 0 | 0 | 0 |
| 0 | 0 | 0 | 0 | 0 |
| 0 | 0 | 0 | 0.009598473 | 0 |
| 0 | 0 | 0 | 0 | 0 |
| 0 | 0 | 0 | 0.00978121 | 0 |
| 0 | 0 | 0 | 0 | 0 |
| 0 | 0 | 0 | 0 | 0 |
| 0 | 0 | 0 | 0 | 0 |
| 0 | 0 | 0 | 0 | 0 |
| 0 | 0 | 0.218470259 | 0 | 0 |
| 0 | 0 | 0 | 0 | 0 |
| 0 | 0 | 0 | 0 | 0 |
| 0 | 0 | 0 | 0 | 0 |
| 0 | 0 | 0 | 0 | 0 |
| 0 | 0 | 0 | 0 | 0 |
| 0 | 0 | 0 | 0 | 0 |
| 0 | 0 | 0 | 0 | 0 |
| 0 | 0 | 0 | 0 | 0 |
| 0 | 0 | 0 | 0 | 0 |
| 0 | 0 | 0 | 0 | 0 |
| 0 | 0 | 0 | 0 | 0 |
| 0 | 0 | 0 | 0 | 0 |
| 0 | 0 | 0 | 0 | 0 |
| 0 | 0 | 0 | 0 | 0 |
| 0 | 0 | 0 | 0 | 0 |
| 0 | 0 | 0 | 0 | 0 |

| | | | | |
|---|---|---|---|---|
| 0 | 0 | 0 | 0 | 0 |
| 0 | 0 | 0 | 0 | 0 |
| 0 | 0 | 0 | 0 | 0 |
| 0 | 0 | 0 | 0 | 0 |
| 0 | 0 | 0 | 0 | 0 |
| 0 | 0 | 0 | 0 | 0 |
| 0 | 0 | 0 | 0 | 0 |
| 0 | 0 | 0 | 0 | 0 |
| 0 | 0 | 0 | 0 | 0.007714811 |
| 0 | 0 | 0 | 0 | 0 |
| 0 | 0 | 0 | 0 | 0 |
| 0 | 0 | 0 | 0 | 0 |
| 0.000981215 | 0 | 0.002341911 | 0 | 0 |
| 0 | 0 | 0 | 0 | 0 |
| 0 | 0 | 0 | 0 | 0 |
| 0 | 0 | 0 | 0 | 0 |
| 0 | 0 | 0 | 0 | 0 |
| 0 | 0 | 0 | 0 | 0 |
| 0 | 0 | 0 | 0 | 0 |
| 0 | 0 | 0 | 0 | 0 |
| 0.092465697 | 0.020131314 | 0.046123481 | 0.015546359 | 0.008416523 |
| 0 | 0 | 0 | 0 | 0 |
| 0.487763311 | 0 | 0 | 0 | 0 |
| 0.006887366 | 0 | 0 | 0 | 0 |
| 0 | 0 | 0 | 0 | 0 |
| 0 | 0 | 0 | 0 | 0 |
| 0 | 0.004244898 | 0.007990365 | 0.001377757 | 0 |
| 0 | 0 | 0 | 0 | 0 |
| 0 | 0 | 0 | 0 | 0 |
| 0 | 0 | 0 | 0 | 0 |
| 0 | 0.047351853 | 0 | 0 | 0 |
| 0 | 0 | 0 | 0 | 0 |
| 0 | 0 | 0 | 0 | 0 |
| 0 | 0 | 0 | 0 | 0.057575761 |
| 0.000716228 | 0.006323417 | 0.152331245 | 0.001319386 | 0 |
| 0 | 0 | 0 | 0 | 0 |
| 0 | 0 | 0 | 0 | 0 |
| 0 | 0 | 0 | 0 | 0 |
| 0 | 0.00853693 | 0 | 0 | 0 |
| 0 | 0 | 0 | 0 | 0 |
| 0 | 0 | 0 | 0 | 0 |
| 0 | 0 | 0 | 0 | 0 |
| 0 | 0 | 0 | 0 | 0 |
| 0 | 0 | 0 | 0 | 0 |
| 0 | 0 | 0 | 0 | 0 |
| 0 | 0 | 0 | 0 | 0 |
| 0 | 0 | 0.005141341 | 0.001904725 | 0 |
| 0.000850672 | 0 | 0 | 0 | 0 |

| | | | | |
|---|---|---|---|---|
| 0 | 0 | 0 | 0 | 0 |
| 0 | 0 | 0 | 0 | 0 |
| 0 | 0 | 0 | 0 | 0 |
| 0 | 0 | 0.001032329 | 0 | 0 |
| 0 | 0 | 0 | 0 | 0 |
| 0 | 0 | 0 | 0 | 0 |
| 0 | 0 | 0 | 0 | 0 |
| 0 | 0 | 0 | 0 | 0 |
| 0 | 0 | 0 | 0 | 0 |
| 0 | 0 | 0 | 0 | 0 |
| 0 | 0 | 0 | 0 | 0 |
| 0 | 0 | 0 | 0 | 0 |
| 0 | 0 | 0 | 0 | 0 |
| 0 | 0 | 0 | 0 | 0 |
| 0 | 0 | 0 | 0 | 0 |
| 0 | 0 | 0 | 0 | 0 |
| 0 | 0 | 0 | 0 | 0 |
| 0 | 0 | 0 | 0.104492796 | 0 |
| 0 | 0 | 0 | 0.083030689 | 0.001152242 |
| 0 | 0 | 0 | 0 | 0 |
| 0.214393418 | 0.021728948 | 0.022945392 | 0 | 0 |
| 0 | 0 | 0 | 0 | 0 |
| 0 | 0 | 0 | 0 | 0 |
| 0 | 0 | 0 | 0 | 0 |
| 0 | 0 | 0.005647807 | 0 | 0.003001566 |
| 0 | 0 | 0 | 0 | 0 |
| 0 | 0 | 0 | 0 | 0 |
| 0 | 0 | 0 | 0 | 0 |
| 0.003387322 | 0.04417228 | 0 | 0 | 0 |
| 0 | 0 | 0 | 0 | 0 |
| 1.655067956 | 0.074915721 | 0.752061433 | 0.128951402 | 0.002499539 |
| 0 | 0 | 0 | 0 | 0 |
| 0.028702518 | 0 | 0.126986006 | 0.013755792 | 0.016921227 |
| 0 | 0.002296782 | 0 | 0 | 0 |
| 0 | 0 | 0 | 0 | 0 |
| 0 | 0 | 0 | 0 | 0 |
| 0 | 0 | 0 | 0 | 0 |
| 0 | 0 | 0 | 0 | 0 |
| 0 | 0 | 0 | 0 | 0 |
| 0 | 0 | 0 | 0 | 0 |
| 0 | 0 | 0 | 0 | 0 |
| 0 | 0.001809576 | 0.004261434 | 0.011629139 | 0.000566192 |
| 0 | 0 | 0 | 0 | 0 |
| 0 | 0 | 0 | 0 | 0 |
| 0 | 0 | 0 | 0 | 0 |
| 0 | 0 | 0 | 0 | 0 |
| 0 | 0 | 0 | 0 | 0 |
| 0 | 0 | 0 | 0 | 0 |

| | | | | |
|---|---|---|---|---|
| 0 | 0 | 0 | 0 | 0 |
| 0 | 0 | 0 | 0 | 0 |
| 0 | 0.092706611 | 0.019301576 | 0.000884988 | 0 |
| 0 | 0 | 0 | 0 | 0 |
| 0 | 0 | 0 | 0 | 0 |
| 0 | 0 | 0 | 0.000775958 | 0 |
| 0 | 0 | 0 | 0 | 0 |
| 0 | 0 | 0 | 0 | 0 |
| 0 | 0 | 0 | 0 | 0 |
| 0 | 0.001039078 | 0 | 0 | 0 |
| 0 | 0 | 0 | 0 | 0 |
| 0 | 0 | 0 | 0 | 0 |
| 0 | 0 | 0 | 0 | 0 |
| 0 | 0.018331865 | 0.027034267 | 0 | 0 |
| 0.000876452 | 0.003109018 | 0.007147212 | 0 | 0.000648513 |
| 0 | 0 | 0.001306445 | 0 | 0 |
| 0 | 0.001416979 | 0.005958737 | 0 | 0.001108384 |
| 0 | 0.01373968 | 0 | 0 | 0 |
| 0 | 0 | 0 | 0 | 0 |
| 0 | 0 | 0 | 0 | 0 |
| 0 | 0 | 0 | 0 | 0 |
| 0 | 0 | 0 | 0 | 0 |
| 0 | 0 | 0 | 0 | 0 |
| 0.018516977 | 0 | 0 | 0 | 0 |
| 0 | 0 | 0 | 0 | 0 |
| 0 | 0 | 0 | 0 | 0 |
| 0 | 0 | 0 | 0 | 0 |
| 0 | 0 | 0 | 0 | 0 |
| 0 | 0 | 0 | 0 | 0 |
| 0 | 0 | 0 | 0 | 0 |
| 0 | 0 | 0 | 0 | 0 |
| 0 | 0 | 0 | 0 | 0 |
| 0 | 0 | 0 | 0 | 0 |
| 0 | 0.003687075 | 0 | 0.005744188 | 0 |
| 0 | 0 | 0 | 0 | 0 |
| 0 | 0 | 0.001074387 | 0 | 0.001665388 |
| 0 | 0 | 0 | 0 | 0 |
| 0 | 0 | 0.025906456 | 0 | 0.002304671 |
| 0 | 0 | 0 | 0 | 0 |
| 0 | 0 | 0 | 0 | 0 |
| 0 | 0 | 0 | 0 | 0 |
| 0 | 0 | 0 | 0 | 0 |
| 0 | 0 | 0 | 0 | 0 |
| 0 | 0 | 0 | 0 | 0.002342259 |
| 0 | 0 | 0 | 0 | 0 |
| 0 | 0 | 0 | 0 | 0 |
| 0 | 0 | 0 | 0 | 0 |
| 0.007545396 | 0 | 0.006002981 | 0 | 0 |

| | | | | |
|---|---|---|---|---|
| 0 | 0 | 0 | 0 | 0 |
| 0 | 0 | 0 | 0 | 0 |
| 0 | 0 | 0.011268249 | 0 | 0 |
| 0 | 0 | 0 | 0 | 0 |
| 0.002849976 | 0 | 0.011714843 | 0 | 0.001054392 |
| 0 | 0 | 0 | 0 | 0 |
| 0 | 0 | 0 | 0 | 0 |
| 0.011318793 | 0.016375274 | 0.030458178 | 0.007359074 | 0 |
| 0 | 0 | 0 | 0 | 0 |
| 12.61391551 | 0.08374258 | 0.242961515 | 0.060049163 | 0.479353043 |
| 0 | 0 | 0 | 0 | 0 |
| 0.001317647 | 0.206157189 | 0.005451138 | 0.003883646 | 0.028469032 |
| 0.002998445 | 0.002269082 | 0.004293916 | 0 | 0.002662369 |
| 0.353683505 | 0 | 0 | 0 | 0 |
| 0 | 0 | 0 | 0 | 0 |
| 0.000900233 | 0.001135423 | 0 | 0 | 0 |
| 0 | 0 | 0.00069444 | 0 | 0 |
| 0.002082711 | 0 | 0 | 0 | 0 |
| 0 | 0.003970555 | 0 | 0 | 0.003105833 |
| 0 | 0 | 0.011324746 | 0 | 0 |
| 0 | 0 | 0 | 0 | 0 |
| 0 | 0 | 0 | 0 | 0 |
| 0 | 0.038633086 | 0 | 0 | 0 |
| 0 | 0 | 0 | 0 | 0 |
| 0 | 0 | 0 | 0 | 0 |
| 0 | 0.080608501 | 0 | 0 | 0 |
| 0 | 0 | 0 | 3.478726933 | 0 |
| 0 | 0 | 0 | 0 | 0.015416454 |
| 0.373545491 | 0 | 0 | 0 | 0 |
| 0 | 0 | 0 | 0 | 0.000886255 |
| 0.013741057 | 0 | 0 | 0 | 0 |
| 0 | 0 | 0 | 0 | 0 |
| 0.02514357 | 0 | 0 | 0 | 0.049247187 |
| 0 | 0.004032501 | 0 | 0 | 0 |
| 0 | 0 | 0 | 0 | 0 |
| 0 | 0 | 0 | 0 | 0 |
| 0.00448689 | 0 | 0 | 0 | 0.01347524 |
| 0 | 0 | 0 | 0 | 0 |
| 0 | 0 | 0 | 0 | 0 |
| 0.263976064 | 0 | 0 | 0 | 0 |
| 0 | 0 | 0 | 0 | 0 |
| 0.002734392 | 0 | 0 | 0 | 0 |
| 0 | 0 | 0 | 0 | 0 |
| 0.001500597 | 0.019695236 | 0 | 0 | 0 |
| 0 | 0 | 0 | 0 | 0 |
| 0 | 0 | 0 | 0.008943316 | 0 |
| 0 | 0 | 0.006219533 | 0.004000287 | 0 |
| 0.018133745 | 0 | 0 | 0 | 0 |

| | | | | |
|---|---|---|---|---|
| 0 | 0 | 0.001088602 | 0 | 0 |
| 0.003559398 | 0.101621579 | 0 | 0.00089412 | 0 |
| 0 | 0 | 0 | 0 | 0 |
| 0 | 0 | 0 | 0 | 0 |
| 0.015167558 | 0 | 0 | 0 | 0 |
| 0.007981684 | 0.004745841 | 0 | 0.002310521 | 0.080320115 |
| 0 | 0 | 0 | 0 | 0 |
| 0 | 0.002232057 | 0 | 0.007606766 | 0 |
| 0 | 0 | 0 | 0 | 2.517183383 |
| 0 | 0 | 0 | 0 | 0 |
| 0 | 0 | 0.003787728 | 0 | 0.000587129 |
| 0 | 0 | 0 | 0 | 0 |
| 0 | 0 | 0 | 0 | 0 |
| 0 | 0 | 0.003578455 | 0 | 0.00110938 |
| 0.093802731 | 0 | 0.047874407 | 0 | 0 |
| 0.003312751 | 0 | 0.014394237 | 0 | 0 |
| 0 | 0 | 0 | 0 | 0.001291463 |
| 0 | 0.00161022 | 0 | 0 | 0 |
| 0 | 0 | 0.051629253 | 0 | 0 |
| 0 | 0 | 0 | 0 | 0 |
| 0.000807719 | 0 | 0 | 0.005207736 | 0 |
| 0 | 0 | 0 | 0.14884601 | 0 |
| 0 | 0 | 0 | 0 | 0 |
| 0 | 0 | 0 | 0 | 0 |
| 0 | 0 | 0 | 0 | 0 |
| 0 | 0 | 0 | 0 | 0 |
| 0.089681224 | 0 | 0 | 0 | 0 |
| 0 | 0 | 0 | 0 | 0 |
| 0 | 0 | 0 | 0 | 0 |
| 0.00842438 | 0.464625367 | 6.445557389 | 0.151477178 | 0.575917061 |
| 0 | 0.000962171 | 0 | 0.003747479 | 0 |
| 0 | 0 | 0 | 0 | 0 |
| 0 | 0 | 0 | 0 | 0 |
| 0.002641181 | 0 | 0 | 0 | 0 |
| 0.002708013 | 0 | 0 | 0 | 0 |
| 0 | 0 | 0 | 0 | 0 |
| 0 | 0 | 0 | 0.61704473 | 0 |
| 0 | 0 | 0.001621066 | 0 | 0.001696131 |
| 0 | 0 | 0.42026529 | 0 | 0 |
| 0.595492573 | 0 | 0 | 0 | 0 |
| 0.005135793 | 0.004469504 | 0.010623454 | 0 | 0 |
| 0 | 4.847069432 | 1.839974444 | 0.056305928 | 1.20024015 |
| 0 | 0 | 0 | 0 | 0 |
| 0.277821219 | 0 | 0 | 0 | 0 |
| 0 | 0 | 0 | 0 | 0 |
| 0.044661938 | 0 | 0 | 0 | 0 |
| 0.001724923 | 0.002447512 | 0 | 0 | 0.000797702 |
| 0 | 0 | 0 | 0 | 0 |

| | | | | |
|---|---|---|---|---|
| 0 | 0.009814414 | 0 | 0.000980136 | 0 |
| 0 | 0.004663141 | 0 | 0 | 0 |
| 0 | 0.005829209 | 0.005767818 | 0.2500744 | 0 |
| 0 | 0 | 0 | 0 | 0 |
| 0 | 0 | 0 | 0 | 0 |
| 0 | 0 | 0 | 0 | 0 |
| 0 | 0 | 0 | 0 | 0 |
| 0 | 0.027076843 | 0 | 0 | 0 |
| 0.049872676 | 0 | 0.02054743 | 0.008407933 | 0 |
| 0 | 0 | 0.132493902 | 0 | 0 |
| 0 | 0 | 0 | 0 | 0 |
| 0 | 0 | 0 | 0 | 0 |
| 0 | 0 | 0 | 0 | 0 |
| 0 | 0 | 0 | 0 | 0.003027388 |
| 0 | 0 | 0 | 0 | 0 |
| 0 | 0.005515353 | 0 | 0.004677374 | 0 |
| 0.002293164 | 0.131729466 | 0.004312218 | 0.236369397 | 0.000462759 |
| 0.067373082 | 0.00119795 | 0.020755198 | 0 | 0 |
| 0 | 0 | 0 | 0 | 0 |
| 0 | 0 | 0 | 0 | 0 |
| 0.000788193 | 0.031811566 | 0 | 0 | 0.112753476 |
| 0 | 0 | 0.011158192 | 0.006904519 | 0.001431403 |
| 0 | 0 | 0 | 0 | 0 |
| 0 | 0 | 0 | 0.009326217 | 0 |
| 0.001948349 | 0 | 0 | 0.054016187 | 0 |
| 0.737634067 | 0 | 0 | 0 | 0 |
| 0 | 0 | 0.001052768 | 0 | 0 |
| 0 | 0 | 0.000799484 | 0 | 0 |
| 0 | 0 | 0 | 0 | 0 |
| 0.000679361 | 0 | 0.038554735 | 0 | 0.001172919 |
| 0.064003338 | 0 | 0.067412818 | 0.01668433 | 0 |
| 0 | 0.009366889 | 0.014323645 | 0 | 0.004884622 |
| 0.102567666 | 0.028377188 | 0.460284055 | 0.187627421 | 0.00443942 |
| 0.001228324 | 0.006584226 | 0.001302977 | 0 | 0.002726624 |
| 0 | 0.076297148 | 0.00302248 | 0.001345848 | 0 |
| 0 | 0 | 0 | 0 | 0 |
| 0.002324663 | 0.024188937 | 0.001438469 | 0 | 0 |
| 0.013735959 | 0.051591465 | 0.041944542 | 0.021396299 | 0.038412634 |
| 0 | 0.942529465 | 0 | 0.048098643 | 0 |
| 0.004294034 | 0 | 0.010905738 | 0 | 0 |
| 0 | 0 | 0 | 0 | 0 |
| 0 | 0 | 0 | 0 | 0 |
| 0 | 0 | 0 | 0 | 0 |
| 0.00551899 | 0.006563088 | 0.033495594 | 0 | 0.000583381 |
| 0.757242279 | 0 | 0 | 0 | 0 |
| 0 | 0.016246796 | 0.032584189 | 0.001014074 | 0 |
| 0 | 0 | 0 | 0 | 0 |
| 0 | 0 | 0 | 0 | 0 |

| | | | | |
|---|---|---|---|---|
| 0 | 0 | 0 | 0 | 0 |
| 0 | 0.049470311 | 0.698996133 | 0 | 0.192464093 |
| 0 | 0 | 0 | 0 | 0 |
| 0 | 0 | 0 | 0 | 0.010607901 |
| 0 | 0 | 0 | 0 | 0 |
| 0 | 0 | 0 | 0 | 0 |
| 0 | 0.04648911 | 0 | 0 | 0 |
| 0 | 0 | 0 | 0 | 0.00046931 |
| 0.010834647 | 0 | 0 | 0 | 0.001118635 |
| 0 | 0.005676175 | 0 | 0 | 0 |
| 0.002045407 | 0.002708768 | 0.000813644 | 0.006405446 | 0 |
| 0.063066518 | 0.195398952 | 0.173308327 | 0 | 0.002930642 |
| 0 | 0 | 0 | 0 | 0 |
| 0 | 0 | 0.04212934 | 0 | 0 |
| 0 | 0.065045423 | 0 | 0.003674724 | 0 |
| 0 | 0 | 0 | 0 | 0 |
| 0 | 0.005065127 | 0.024026487 | 0 | 0.011727593 |
| 1.704662885 | 0.005786206 | 0.01061777 | 0 | 0.03723717 |
| 0 | 0 | 0 | 0 | 0 |
| 0.063010068 | 0.024592974 | 0 | 0 | 0 |
| 0 | 0 | 0.086808655 | 0.007934262 | 0.015853087 |
| 0 | 0 | 0.001093314 | 0.016454859 | 0 |
| 0 | 0 | 0 | 0 | 0 |
| 0 | 0 | 0 | 0 | 0 |
| 0 | 0.41564679 | 0 | 0 | 0 |
| 0 | 0 | 0 | 0 | 0 |
| 0.153483961 | 0 | 0.09973396 | 0.006024241 | 0.040651987 |
| 0.175835259 | 0.012435871 | 0.247097796 | 0.03719147 | 0.013722781 |
| 0 | 0 | 0 | 0 | 0 |
| 0 | 0 | 0.001351736 | 0 | 0 |
| 0 | 0.000714841 | 0 | 0 | 0 |
| 0 | 0 | 0 | 0 | 0 |
| 0 | 0 | 0 | 0 | 0 |
| 0 | 0 | 0 | 0 | 0 |
| 0 | 0 | 0 | 0.004145437 | 0 |
| 0.023441723 | 0.195436145 | 0.10557689 | 0.128816334 | 18.41271201 |
| 0 | 0 | 0 | 0 | 0 |
| 0 | 0 | 0 | 0 | 0 |
| 0 | 0.062185992 | 0.00345604 | 0.002667432 | 0.008142864 |
| 0 | 0 | 0 | 0 | 0 |
| 0.367452453 | 0 | 0 | 0 | 0 |
| 0 | 0 | 0 | 0 | 0 |
| 0.022868187 | 0 | 0.063817254 | 0.001944287 | 0.010673163 |
| 0.003566576 | 0.000642623 | 0.014052397 | 0 | 0 |
| 0 | 0 | 0 | 0 | 0.001816078 |
| 0 | 0 | 0 | 0.005656893 | 0 |
| 0 | 0 | 0.01861251 | 0 | 0 |
| 0.004117924 | 0 | 0 | 0 | 0 |

| | | | | |
|---|---|---|---|---|
| 0 | 0 | 0.015445554 | 0 | 0 |
| 0 | 0 | 0 | 0 | 0 |
| 0 | 0 | 0 | 0 | 0 |
| 0 | 0 | 0 | 0.003585425 | 0.002122342 |
| 0 | 0 | 0 | 0 | 0 |
| 0 | 0 | 0 | 0 | 0 |
| 0 | 0 | 0 | 0 | 0 |
| 0 | 0 | 0 | 0 | 0 |
| 0 | 0 | 0.137647733 | 0.248004044 | 0 |
| 0 | 0 | 0.001019758 | 0 | 0 |
| 0 | 0 | 0 | 0 | 0 |
| 0 | 0.000742736 | 0 | 0.001265608 | 0.042266322 |
| 0 | 0 | 0 | 0 | 0 |
| 0 | 0 | 0 | 0 | 0 |
| 0.391352454 | 0 | 0 | 0.102200367 | 0.033320141 |
| 0 | 0 | 0 | 0 | 0 |
| 0 | 0 | 0 | 0.006227687 | 0 |
| 0 | 0 | 0 | 0.708205682 | 0 |
| 0 | 0 | 0 | 0 | 0 |
| 0 | 0 | 0 | 0 | 0 |
| 0 | 0 | 0 | 0 | 0 |
| 0 | 0 | 0 | 0.06294775 | 0 |
| 0 | 0 | 0 | 0 | 0 |
| 0.089562227 | 0.016973635 | 0.548308867 | 0 | 0.071003377 |
| 0 | 0 | 0 | 0.012864986 | 0.00040796 |
| 0 | 0 | 0.062535989 | 0.086530553 | 0 |
| 0 | 0 | 0 | 0.013293223 | 0 |
| 0 | 0 | 0 | 0.636052871 | 0 |
| 0 | 0 | 0 | 0 | 0 |
| 0 | 0 | 0 | 0 | 0 |
| 0 | 0 | 0 | 0 | 0 |
| 0 | 0 | 0 | 0.108141743 | 0 |
| 0 | 0 | 0.021838674 | 0 | 0.018913989 |
| 0 | 0 | 0 | 0 | 0 |
| 0 | 0 | 0 | 0.001785145 | 0 |
| 0 | 0 | 0 | 0 | 0 |
| 0 | 0 | 0 | 0.157446092 | 0 |
| 0 | 0 | 0 | 0 | 0 |
| 0 | 0 | 0 | 0 | 0.002570403 |
| 0 | 0 | 0 | 0 | 0 |
| 0.008097353 | 0.013181668 | 0.012434766 | 0 | 0.023965913 |
| 0 | 0 | 0.218728306 | 0 | 0.715084368 |
| 0.798336294 | 19.26069709 | 3.243644804 | 2.796048627 | 0.115835976 |
| 0 | 0 | 0 | 0.002719701 | 0.019299496 |
| 0 | 0 | 0 | 0 | 0 |
| 0 | 0 | 0 | 0 | 0 |
| 0 | 0 | 0 | 0 | 0 |
| 0.002942821 | 0.010278407 | 0.001080579 | 0 | 0 |

| | | | | |
|---|---|---|---|---|
| 0 | 0 | 0 | 0 | 0 |
| 0 | 0 | 0 | 0.020514784 | 0 |
| 1.757499517 | 0.201576236 | 0.002589643 | 0.002998098 | 0.031310451 |
| 0 | 0 | 0 | 0 | 0 |
| 0 | 0 | 0 | 0 | 0 |
| 0 | 0 | 0.018224189 | 0.001102167 | 0.009992555 |
| 0 | 0 | 0 | 0 | 0 |
| 0 | 0.004260681 | 0 | 0 | 0 |
| 0 | 0 | 0 | 0 | 0 |
| 0 | 0 | 0 | 0 | 0 |
| 0.014914026 | 0.002418478 | 0.038308063 | 0.105380901 | 0.005832967 |
| 0.389892336 | 1.240115996 | 4.343438498 | 0.847331953 | 0.482048243 |
| 0 | 0 | 0 | 0 | 0 |
| 0 | 0.048287221 | 0.004207537 | 0.010165932 | 0 |
| 0.462608254 | 0.377436507 | 0.104084159 | 0.011182222 | 0.425827744 |
| 0 | 0 | 0 | 0 | 0 |
| 0 | 0 | 0 | 0 | 0 |
| 0 | 0 | 0 | 0 | 0 |
| 0 | 0 | 0 | 0 | 0 |
| 0.0528265 | 0 | 0.274507196 | 0.002909703 | 0 |
| 0.001428589 | 0 | 0.034665056 | 0 | 0.01109909 |
| 5.51462698 | 12.55750373 | 8.775572715 | 0.030329047 | 0.287502028 |
| 53.40134409 | 2.38270611 | 3.763181299 | 1.349089845 | 8.035245619 |
| 0 | 0 | 0 | 0 | 0 |
| 0 | 0 | 0 | 0 | 0 |
| 0 | 0 | 1.676376483 | 0.071742231 | 0.003641677 |
| 0 | 0 | 0.027697322 | 0 | 0 |
| 0 | 0 | 0 | 0.002401667 | 0 |
| 0 | 0 | 0 | 0.029516941 | 0.005023767 |
| 0 | 0 | 0 | 0 | 0 |
| 0 | 0 | 0 | 0 | 0 |
| 0.068614968 | 0.107963549 | 0 | 0.122205273 | 0.167280044 |
| 2.618128938 | 0.006623573 | 0.046829065 | 0 | 0.00097145 |
| 0 | 0 | 0 | 0.023514972 | 0.088526303 |
| 0.011120419 | 0 | 0 | 0 | 0 |
| 0 | 0.013138927 | 0.112556933 | 0.052270256 | 0.188224766 |
| 0.018400942 | 0 | 0 | 0.005649494 | 0.035598643 |
| 0 | 0 | 0 | 0 | 0 |
| 0 | 0.007248308 | 0.04692208 | 0.057744839 | 0 |
| 0 | 0 | 0 | 0 | 0 |
| 0 | 0 | 0 | 0.004827964 | 0 |
| 0 | 0 | 0 | 0 | 0 |
| 0 | 0 | 0 | 0 | 0 |
| 0 | 0 | 0 | 0 | 0 |
| 0.059964716 | 0.010274369 | 0.057068183 | 0.13714076 | 0.005860152 |
| 0 | 0 | 0 | 0 | 0 |
| 0 | 0 | 0 | 0 | 0 |
| 0.019925924 | 0 | 0.486439134 | 0.058327632 | 0 |

| | | | | |
|---|---|---|---|---|
| 0 | 0 | 0 | 0 | 0 |
| 0 | 0 | 0 | 0 | 0 |
| 0 | 0 | 0 | 0 | 0 |
| 0 | 0 | 0.002301529 | 0.002664541 | 0 |
| 0 | 0 | 0 | 0 | 0 |
| 0 | 0.000966744 | 0.038892248 | 0.047379881 | 0.002772744 |
| 0 | 0 | 0 | 0.002917165 | 0 |
| 0 | 0 | 0 | 0.001471116 | 0 |
| 0 | 0 | 0 | 0 | 0 |
| 0 | 0 | 0 | 0 | 0 |
| 0.004081087 | 0 | 0.004784815 | 0.001978395 | 0.001907192 |
| 0 | 0 | 0 | 0 | 0 |
| 0 | 0 | 0.006882717 | 0.016109828 | 0 |
| 0 | 0 | 0 | 0 | 0 |
| 0 | 0 | 0 | 0 | 0 |
| 0 | 0 | 0 | 0 | 0 |
| 0 | 0 | 0 | 0.00294281 | 0 |
| 0 | 0 | 0 | 0.008070147 | 0 |
| 0.001881568 | 0 | 0.000748471 | 0 | 0 |
| 0 | 0 | 0 | 0 | 0 |
| 0 | 0 | 0 | 0 | 0 |
| 0 | 0 | 0 | 0 | 0 |
| 0 | 0 | 0 | 0.004367325 | 0 |
| 0 | 0 | 0.380163947 | 0 | 0 |
| 0 | 0 | 0 | 0 | 0 |
| 0 | 0 | 0 | 0 | 0 |
| 0 | 0 | 0 | 0 | 0 |
| 0 | 0 | 0 | 0 | 0 |
| 0 | 0 | 0 | 0 | 0 |
| 0 | 0 | 1.059783524 | 0 | 0 |
| 0 | 0 | 0.050606578 | 0.013244755 | 0 |
| 0 | 0 | 0 | 0 | 0 |
| 0 | 0 | 0 | 0.000688321 | 0 |
| 0 | 0 | 0 | 0 | 0 |
| 0 | 0 | 0 | 0 | 0 |
| 0 | 0 | 0 | 0 | 0 |
| 0 | 0 | 0 | 0.030035318 | 0 |
| 0.000909135 | 0 | 0.003037822 | 0.006029086 | 0.001749011 |
| 0 | 0 | 0 | 0 | 0 |
| 0 | 0 | 0 | 0 | 0 |
| 0 | 0 | 0 | 0.03645477 | 0 |
| 0 | 0 | 0 | 0 | 0 |
| 0 | 0 | 0 | 0 | 0 |
| 0 | 0.181895661 | 0 | 0.051230871 | 0 |
| 0 | 0 | 0 | 0.010649278 | 0 |
| 0 | 0 | 0 | 0 | 0 |
| 0 | 0 | 0 | 0 | 0 |
| 0 | 0 | 0 | 0 | 0 |

| | | | | |
|---|---|---|---|---|
| 0 | 0 | 0 | 0 | 0 |
| 0 | 0 | 0 | 0 | 0 |
| 0 | 0 | 0 | 0.013759636 | 0 |
| 0 | 0 | 0 | 0 | 0 |
| 0 | 0 | 0 | 0 | 0 |
| 0 | 0 | 0 | 0.333581009 | 0 |
| 0 | 0 | 0 | 0 | 0 |
| 0 | 0 | 0 | 0 | 0 |
| 1.203193063 | 0.013046734 | 0.05985235 | 0 | 0.013359754 |
| 0 | 0 | 0 | 0 | 0 |
| 0 | 0 | 0 | 0 | 0 |
| 0 | 0 | 0 | 0 | 0 |
| 0 | 0 | 0 | 0 | 0 |
| 0 | 0 | 0 | 0 | 0 |
| 0.003964614 | 0 | 0.171759171 | 0.293129612 | 0 |
| 0.208157378 | 0 | 0.000768831 | 0 | 0.010868775 |
| 0 | 0 | 0 | 0 | 0 |
| 0 | 0 | 0 | 0 | 0 |
| 0 | 0 | 0 | 0 | 0 |
| 0 | 0 | 0 | 0 | 0 |
| 0 | 0 | 0 | 0.059342256 | 0 |
| 0 | 0 | 0 | 0 | 0 |
| 0 | 0 | 0 | 0 | 0 |
| 0 | 0 | 0 | 0 | 0 |
| 0.094072474 | 0 | 0.002815386 | 0.568773447 | 0.379238615 |
| 0 | 0 | 0.069215229 | 0.487194063 | 0.61617653 |
| 0 | 0 | 0.022862927 | 0.007880087 | 0 |
| 0 | 0 | 0 | 0 | 0 |
| 0 | 0 | 0 | 0.008549065 | 0 |
| 0 | 0 | 0 | 0 | 0 |
| 0 | 0 | 0 | 0 | 0 |
| 0 | 0 | 0 | 0.026763546 | 0 |
| 0 | 0 | 0 | 0 | 0 |
| 0.019529316 | 0 | 0.000375899 | 0.027706998 | 0.013634591 |
| 0 | 0.021356132 | 0.001651635 | 0.004541336 | 0 |
| 0.028249123 | 0.000520558 | 0.016199145 | 0 | 0.000407189 |
| 0.014954758 | 0.11084267 | 0 | 0 | 0 |
| 0 | 0.78412587 | 0.424497765 | 0.750914542 | 0 |
| 0 | 0 | 7.588548564 | 0 | 0 |
| 0 | 0 | 0 | 0 | 0 |
| 0 | 0 | 0 | 0 | 0 |
| 0 | 0 | 0 | 0 | 0 |
| 0 | 0.342871876 | 0 | 0.165311596 | 0.865250958 |
| 0 | 0 | 0 | 0.138095613 | 0 |
| 0 | 0 | 0 | 0 | 0 |
| 0.001042695 | 0 | 0 | 0.001280521 | 0 |
| 0 | 0 | 0 | 0 | 0 |
| 0 | 0 | 0.01563859 | 0.001020012 | 0.00512136 |

| | | | | |
|---|---|---|---|---|
| 0 | 0 | 0 | 0.036700098 | 0.000916561 |
| 0 | 0 | 0 | 0 | 0 |
| 0 | 0 | 0 | 0 | 0 |
| 0 | 0 | 0 | 0 | 0 |
| 0 | 0 | 0.001144706 | 0.003029156 | 0 |
| 0 | 0 | 0 | 0 | 0 |
| 0 | 0 | 0 | 0 | 0 |
| 0 | 0 | 0 | 0 | 0 |
| 0 | 0 | 0 | 0 | 0 |
| 0 | 0 | 0 | 0 | 0 |
| 0 | 0 | 0 | 0 | 0 |
| 0 | 0 | 0.816337581 | 1.14525795 | 7.209147681 |
| 0 | 0 | 0.044551222 | 0 | 0.098522835 |
| 0 | 0 | 0 | 0 | 0 |
| 0.04136241 | 1.336143978 | 0.185717552 | 0.02364673 | 0.311681625 |
| 0 | 0 | 0 | 0 | 0 |
| 0 | 0 | 0 | 0 | 0 |
| 0 | 0 | 0 | 0 | 0 |
| 0.019793853 | 0 | 0 | 0.153527958 | 0 |
| 0 | 0 | 0.003308515 | 0 | 0 |
| 0 | 0 | 0 | 0 | 0 |
| 0 | 0 | 0 | 0 | 0 |
| 0 | 0.203528018 | 0 | 0.008770498 | 0 |
| 0 | 0 | 0 | 0 | 0 |
| 0 | 0 | 0 | 0 | 0 |
| 0 | 0 | 0 | 0 | 0 |
| 0 | 0 | 0 | 0 | 0 |
| 0 | 0 | 0 | 0 | 0 |
| 0 | 0 | 0 | 0 | 0 |
| 0 | 0 | 0 | 0 | 0 |
| 0 | 0 | 0 | 0 | 0 |
| 0.002776216 | 0.048145835 | 0.168843394 | 0 | 0.012325241 |
| 0 | 0 | 0 | 0 | 0 |
| 0.02203709 | 0 | 0 | 0.019162083 | 0 |
| 0.02814701 | 0 | 0.010481947 | 0.001654803 | 0.001772496 |
| 0 | 0 | 0.023339775 | 0.002844323 | 0.006093233 |
| 0 | 0 | 0 | 0 | 0 |
| 0 | 0 | 0 | 0 | 0 |
| 0 | 0 | 0 | 0.000633708 | 0 |
| 0 | 0 | 0 | 0 | 0 |
| 0 | 0 | 0 | 0 | 0 |
| 0 | 0 | 0 | 0.005136336 | 0 |
| 0 | 0 | 0 | 0 | 0 |
| 0 | 0 | 0 | 0 | 0.343187563 |
| 0.025367828 | 0.009859014 | 0.031446659 | 0.047093143 | 0.013532169 |
| 0 | 0 | 0 | 0 | 0 |
| 0 | 0 | 0 | 0.09677182 | 0 |
| 0.001301969 | 0 | 0 | 0 | 0.000825743 |

| | | | | |
|---|---|---|---|---|
| 0 | 0 | 0 | 0 | 0 |
| 0 | 0 | 0 | 0 | 0 |
| 0.002440622 | 0 | 0.022329715 | 0 | 0 |
| 0 | 0 | 0 | 0 | 0 |
| 0 | 0 | 0 | 0 | 0.001526195 |
| 0 | 0 | 0 | 0 | 0 |
| 0 | 0 | 0 | 0.348907694 | 0 |
| 0 | 0.01399049 | 0 | 0.001769167 | 0.014354579 |
| 0 | 0 | 0 | 0 | 0 |
| 0 | 0 | 0 | 0 | 0 |
| 0 | 0 | 0 | 0 | 0 |
| 0.003931849 | 0 | 0 | 0 | 0 |
| 0 | 0 | 0 | 0 | 0 |
| 0 | 0 | 0 | 0.110244607 | 0 |
| 0 | 0 | 0 | 0 | 0 |
| 0 | 0 | 0 | 0 | 0 |
| 0 | 0 | 0 | 0 | 0 |
| 0 | 0 | 0 | 0 | 0 |
| 0 | 0 | 7.952479614 | 0 | 0 |
| 0 | 0 | 0 | 0 | 0 |
| 0 | 0 | 0 | 0 | 0 |
| 0 | 0 | 0 | 0 | 0 |
| 0 | 0 | 0 | 0 | 0 |
| 0 | 0 | 0 | 0 | 0 |
| 0 | 0 | 0 | 0 | 0 |
| 0.003478398 | 0.000692707 | 0.01441935 | 0 | 0.020861102 |
| 0 | 0 | 0 | 0 | 0 |
| 0 | 0 | 0.002138436 | 0 | 0.00079554 |
| 0 | 0 | 0 | 0.170490041 | 0 |
| 0 | 0.415535836 | 1.206614429 | 0.066513122 | 0.288758941 |
| 0 | 0 | 0 | 0 | 0 |
| 0 | 0 | 0 | 0 | 0 |
| 0 | 0 | 0 | 0 | 0 |
| 0 | 0 | 0 | 0 | 0 |
| 0.003645146 | 0 | 0.001068425 | 0.002120476 | 0 |
| 0 | 0 | 0 | 0 | 0 |
| 0 | 0 | 0 | 0 | 0 |
| 0.00130616 | 0 | 0.004503014 | 0 | 0.010792219 |
| 0 | 0 | 0 | 0 | 0 |
| 0 | 0 | 0 | 0.000938736 | 0 |
| 0.749842488 | 6.628174868 | 4.693377147 | 2.266795927 | 15.29625829 |
| 0 | 0.050497241 | 0.068713644 | 0 | 1.427061681 |
| 0 | 0 | 0 | 0 | 0 |
| 0 | 0 | 0 | 0 | 0.020951182 |
| 0 | 0 | 0 | 0 | 0 |
| 0 | 0 | 0 | 0 | 0 |
| 0 | 0 | 0 | 0.000910147 | 0 |
| 0.001382188 | 0 | 0.001099643 | 0.008911602 | 0.001406244 |

| | | | | |
|---|---|---|---|---|
| 0 | 0 | 0 | 0 | 0 |
| 0.002310253 | 0 | 0.010745203 | 0.000818421 | 0 |
| 0 | 0 | 0 | 0 | 0 |
| 0 | 0 | 0 | 0 | 0 |
| 0 | 4.897592194 | 0 | 0.784696304 | 3.080217607 |
| 0 | 0 | 0 | 0 | 0 |
| 0 | 0 | 0 | 0 | 0 |
| 0 | 0 | 0 | 0 | 0 |
| 1.510097285 | 0.029374971 | 0.01411758 | 0.000980658 | 0.714537132 |
| 0 | 0 | 0.00891315 | 0 | 0 |
| 0 | 0 | 0 | 0 | 0 |
| 0 | 0 | 0 | 0 | 0 |
| 0 | 0 | 0.077481694 | 0.007952357 | 0 |
| 0 | 0.007630893 | 0.352649362 | 0.004106178 | 0.018535343 |
| 0 | 0 | 0 | 0 | 0 |
| 0 | 0.534564712 | 0.076503159 | 0.021340167 | 0 |
| 0 | 0 | 0.000584604 | 0 | 0.000951494 |
| 0 | 0 | 0 | 0 | 0 |
| 0 | 0 | 0 | 0 | 0 |
| 0 | 0 | 0 | 0 | 0 |
| 0 | 0 | 0 | 0 | 0 |
| 0 | 0 | 0 | 0 | 0 |
| 0 | 0 | 0 | 0 | 0 |
| 0 | 0 | 0 | 0 | 0 |
| 0 | 0 | 0 | 0 | 0 |
| 0 | 0 | 0 | 0 | 0 |
| 0 | 0 | 0.049562884 | 0 | 0.001536531 |
| 0.005948117 | 0 | 0.002581208 | 0 | 0.002133912 |
| 0 | 0 | 0 | 0 | 0 |
| 0 | 0 | 0 | 0 | 0 |
| 0 | 0 | 0 | 0 | 0 |
| 0 | 0 | 0 | 0 | 0 |
| 0 | 0 | 0 | 0 | 0 |
| 0 | 0 | 0 | 0.007309329 | 0 |
| 0 | 0 | 0 | 0 | 0 |
| 0.039407859 | 0 | 0 | 0.071442186 | 0.003394178 |
| 0 | 0 | 0.001865094 | 0 | 0 |
| 0 | 0 | 0 | 0 | 0 |
| 0 | 0 | 0 | 0 | 0 |
| 0 | 0 | 0 | 0 | 0 |
| 0 | 0 | 0.006704474 | 0.213571108 | 1.759353557 |
| 0 | 0 | 0 | 0 | 0 |
| 0 | 0 | 0 | 0 | 0 |
| 0 | 0 | 0 | 0 | 0 |
| 0 | 0 | 0 | 0 | 0 |
| 0 | 0 | 0 | 0 | 0 |
| 0 | 0 | 0 | 0 | 0 |
| 0 | 0 | 0 | 0 | 0 |

| | | | | |
|---|---|---|---|---|
| 0 | 0 | 0 | 0 | 0 |
| 0 | 0 | 0 | 0 | 0 |
| 0 | 0 | 0 | 0 | 0 |
| 0 | 0 | 0 | 0 | 0 |
| 0 | 0 | 0 | 0.001259599 | 0 |
| 0 | 0 | 0 | 0 | 0 |
| 0 | 0 | 0.010643082 | 0 | 0.055594831 |
| 0 | 0 | 0 | 0 | 0 |
| 0 | 0 | 0 | 0 | 0 |
| 0 | 0 | 0 | 0 | 0 |
| 0 | 0.001051076 | 0 | 0 | 0 |
| 0 | 0 | 0 | 0 | 0 |
| 0 | 0 | 0 | 0 | 0 |
| 0 | 0 | 0 | 0 | 0 |
| 0 | 0 | 0 | 0 | 0 |
| 0 | 0 | 0 | 0 | 0 |
| 0 | 0 | 0.062733442 | 0 | 1.234779692 |
| 0 | 0 | 0 | 0 | 0 |
| 0 | 0 | 0 | 0 | 0 |
| 0 | 0 | 0 | 0 | 0 |
| 0 | 0 | 0.036367061 | 0.014462897 | 0 |
| 0 | 0 | 0 | 0 | 0 |
| 0 | 0 | 0 | 0.010812267 | 0 |
| 0.002909039 | 0.000647478 | 0.003403498 | 0.000630451 | 0.002785571 |
| 0 | 0 | 0 | 0 | 0 |
| 0.006460259 | 0 | 0 | 0.072147686 | 0 |
| 0 | 0 | 0 | 0 | 0 |
| 0 | 0 | 0 | 0 | 0 |
| 0 | 0 | 0 | 0 | 0 |
| 0.04383815 | 3.294167295 | 0.025863722 | 0.005897885 | 0.002186776 |
| 0 | 0 | 0 | 0 | 0 |
| 0 | 0 | 0 | 0 | 0 |
| 0 | 0 | 0 | 0 | 0 |
| 0 | 0 | 0 | 0.07978413 | 0.007572274 |
| 0 | 0 | 0 | 0 | 0 |
| 0 | 0 | 0 | 0 | 0 |
| 0.53807197 | 0 | 0.043438312 | 0 | 0.000792152 |
| 0 | 0 | 0 | 0 | 0 |
| 0 | 0 | 0 | 0 | 0 |
| 0 | 0 | 0 | 0 | 0 |
| 0 | 0 | 0 | 0 | 0 |
| 0 | 0 | 0 | 0.000791506 | 0 |
| 0 | 0 | 0 | 0 | 0 |
| 0 | 0 | 0 | 0.005833897 | 0 |
| 0 | 0 | 0 | 0 | 0 |
| 0 | 0 | 0 | 0 | 0 |
| 0 | 0 | 0 | 0 | 0 |
| 0 | 0 | 0 | 0 | 0 |

| | | | | |
|---|---|---|---|---|
| 0.009390689 | 0 | 0.026807927 | 0.005087908 | 0.012670715 |
| 0 | 0 | 0 | 0 | 0 |
| 0 | 0 | 0 | 0 | 0 |
| 0 | 0 | 0 | 0 | 0 |
| 0 | 0 | 0 | 0 | 0 |
| 0 | 0 | 0 | 0 | 0 |
| 0 | 0 | 0 | 0 | 0 |
| 0 | 2.99289769 | 0.093858065 | 0.119632621 | 0.01916521 |
| 0.077457761 | 0 | 0.089216822 | 23.72524453 | 0.268389036 |
| 0.089738811 | 0.012371514 | 0.102129454 | 0 | 0.117764077 |
| 0 | 0 | 0 | 0.001087384 | 0 |
| 0 | 0 | 0 | 0 | 0 |
| 0 | 0 | 0 | 0 | 0 |
| 0 | 0 | 0.549668812 | 0 | 0 |
| 0 | 0 | 0 | 0 | 0 |
| 0 | 0 | 0 | 0 | 0 |
| 0 | 0 | 0.035283275 | 0 | 0 |
| 0 | 0 | 0 | 0 | 0 |
| 0 | 0 | 0 | 0 | 0 |
| 0.003336932 | 0 | 0 | 0 | 0 |
| 0 | 0 | 0 | 0 | 0 |
| 0 | 0 | 0 | 0 | 0 |
| 0 | 0 | 0 | 0 | 0 |
| 0 | 0 | 0 | 0 | 0 |
| 0 | 0 | 0 | 0 | 0 |
| 0 | 0 | 0 | 0 | 0 |
| 0 | 0 | 0 | 0 | 0 |
| 0 | 0 | 0 | 0 | 0 |
| 0 | 0 | 0 | 0 | 0 |
| 0 | 0 | 0 | 0 | 0 |
| 0 | 0 | 0 | 0 | 0 |
| 0 | 0 | 0 | 0 | 0 |
| 0 | 0 | 0 | 0 | 0 |
| 0 | 0 | 0 | 0 | 0 |
| 0 | 0 | 0 | 0.001231926 | 0 |
| 0 | 0 | 0 | 0.001081989 | 0 |
| 0 | 0 | 0 | 0 | 0 |
| 0 | 0.159809326 | 0 | 0 | 0.034760004 |
| 0 | 0 | 0 | 0.001411648 | 0 |
| 0 | 0 | 0 | 0 | 0 |
| 0 | 0 | 0 | 0 | 0 |
| 0 | 0.232132698 | 0 | 0.06690022 | 0.422091635 |
| 0 | 0 | 0 | 0 | 0 |
| 0 | 0 | 0 | 0 | 0 |
| 0 | 0 | 0 | 0 | 0 |
| 0 | 0 | 0.051192427 | 0 | 0 |
| 0 | 0 | 0 | 0 | 0 |
| 0 | 0 | 0 | 0 | 0 |

| | | | | |
|---|---|---|---|---|
| 0 | 0 | 0 | 0 | 0 |
| 0 | 0.037099799 | 0.013089275 | 0 | 0 |
| 0 | 0 | 0 | 0 | 0 |
| 0 | 0 | 0 | 0 | 0 |
| 0 | 0 | 0.870667145 | 0.037666025 | 0.274563109 |
| 0 | 0 | 0 | 0 | 0 |
| 0 | 0 | 0 | 0 | 0 |
| 0.082916768 | 0.005317584 | 0.052550045 | 0 | 0.006065939 |
| 0 | 0 | 0 | 0.002369488 | 0 |
| 0 | 0 | 0 | 0 | 0 |
| 0 | 0 | 0 | 0 | 0 |
| 0 | 0 | 0 | 0 | 0 |
| 0 | 0 | 0 | 0 | 0 |
| 0 | 0 | 0 | 0 | 0 |
| 0 | 0 | 0 | 0 | 0 |
| 0 | 0 | 0.03070741 | 0.004116405 | 0.001202501 |
| 0 | 0 | 0 | 0 | 0 |
| 0 | 0 | 0 | 0 | 0 |
| 0 | 0 | 0 | 0 | 0 |
| 0 | 0 | 0 | 0 | 0 |
| 0 | 0 | 0 | 0 | 0 |
| 0 | 0 | 0 | 0 | 0 |
| 0 | 0 | 0 | 0 | 0 |
| 0.001418936 | 0 | 0 | 0.000746819 | 0.000899926 |
| 0 | 0 | 0 | 0.21628171 | 0 |
| 0 | 0 | 0 | 0 | 0 |
| 0 | 0.008593351 | 0.023894742 | 0 | 0 |
| 0 | 0 | 0 | 0 | 0.001011129 |
| 0.00716481 | 0 | 0 | 0 | 0 |
| 0 | 0 | 0 | 0 | 0 |
| 0 | 0 | 0 | 0 | 0 |
| 0 | 0 | 0 | 0 | 0 |
| 0 | 0 | 0 | 0 | 0 |
| 0 | 0 | 0 | 0 | 0 |
| 0 | 0 | 0 | 0 | 0 |
| 0 | 0 | 0 | 0 | 0 |
| 0 | 0 | 0 | 0 | 0 |
| 0 | 0 | 0 | 0.004903563 | 0 |
| 0 | 0 | 0 | 0 | 0 |
| 0 | 0 | 0 | 0 | 0 |
| 0 | 0 | 0 | 0 | 0 |
| 0 | 0 | 0 | 0.190366021 | 0 |
| 0 | 0 | 0 | 0 | 0 |
| 0 | 0 | 0 | 0 | 0 |
| 0 | 0 | 0 | 0 | 0 |
| 0 | 0 | 0 | 0 | 0 |
| 0 | 0 | 0 | 0 | 0 |
| 0 | 0 | 0 | 0 | 0 |

| | | | | |
|---|---|---|---|---|
| 0 | 0 | 0 | 0.007547621 | 0 |
| 0 | 0 | 0 | 0 | 0 |
| 0 | 0 | 0 | 0 | 0 |
| 0 | 0 | 0 | 0 | 0 |
| 0 | 0 | 0 | 0 | 0 |
| 0.265622549 | 0 | 0 | 0.293773131 | 0.343911504 |
| 0 | 0 | 0 | 0 | 0 |
| 0 | 0 | 0 | 0 | 0 |
| 0 | 0 | 0 | 0 | 0 |
| 0 | 0 | 0 | 0 | 0 |
| 0 | 0 | 0 | 0 | 0 |
| 0 | 0 | 0 | 0 | 0 |
| 0 | 0 | 0 | 0 | 0 |
| 0 | 0 | 0 | 0 | 0 |
| 0 | 0 | 0 | 0 | 0 |
| 0 | 0 | 0 | 0 | 0 |
| 0 | 0 | 0 | 0 | 0 |
| 0 | 0 | 0 | 0 | 0 |
| 0 | 0 | 0 | 0 | 0 |
| 0 | 0 | 0 | 0 | 0 |
| 0.004037156 | 0 | 0 | 0 | 0.030618952 |
| 0 | 0 | 0 | 0 | 0 |
| 0 | 0 | 0.001216914 | 0.066685739 | 0.026785707 |
| 0 | 0 | 0.651934556 | 0 | 0 |
| 0 | 0 | 0 | 0 | 0 |
| 0 | 0 | 0 | 0.024295207 | 0 |
| 0 | 0 | 0 | 0 | 0 |
| 0 | 0 | 0 | 0 | 0 |
| 0 | 0 | 0 | 0 | 0 |
| 0 | 0 | 0 | 0 | 0 |
| 0 | 0 | 0 | 0 | 0 |
| 0 | 0 | 0 | 0 | 0 |
| 0.005491078 | 0.00609758 | 1.308301266 | 0 | 0.007242767 |
| 0 | 0 | 0.216726707 | 0 | 0 |
| 0 | 0 | 0.328076359 | 0 | 0.74282914 |
| 0 | 0 | 0.003048365 | 0 | 0 |
| 0 | 0 | 0 | 0 | 0 |
| 0 | 0 | 0 | 0 | 0 |
| 0.001772748 | 0 | 0 | 0 | 0.004918915 |
| 0 | 0 | 0 | 0 | 0 |
| 0 | 0 | 0 | 0 | 0 |
| 0 | 0.069520528 | 0 | 0 | 0 |
| 0 | 0 | 0.012986986 | 0.024581644 | 0 |
| 0 | 0 | 0 | 0 | 0 |
| 0 | 0 | 0 | 0 | 0 |
| 0 | 0 | 0 | 0 | 0 |
| 0 | 0 | 0 | 0 | 0 |
| 0 | 0 | 0 | 0 | 0 |

| | | | | |
|---|---|---|---|---|
| 0 | 0 | 0 | 0 | 0 |
| 0 | 0 | 0 | 0 | 0 |
| 0 | 0 | 0 | 0 | 0 |
| 0 | 0 | 0 | 0 | 0 |
| 0 | 0 | 0 | 0 | 0 |
| 0 | 0 | 0 | 0 | 0 |
| 0 | 0 | 0 | 0 | 0 |
| 0 | 0 | 0 | 0 | 0 |
| 0 | 0 | 0 | 0 | 0 |
| 0 | 0 | 0 | 0.004099595 | 0 |
| 0 | 0 | 0 | 0 | 0 |
| 0.006392706 | 0 | 0 | 0 | 0 |
| 0 | 0 | 0 | 0 | 0 |
| 0 | 0 | 0 | 0 | 0 |
| 0.016614501 | 0 | 0 | 0 | 0 |
| 0.779292121 | 0.008284962 | 0.001161342 | 0 | 0.031323056 |
| 0 | 0 | 0 | 0 | 0 |
| 0 | 0 | 0 | 0 | 0 |
| 0 | 0 | 0 | 0 | 0 |
| 0 | 0.004624288 | 0.281276027 | 0 | 0 |
| 0 | 0 | 0 | 0.027483209 | 0.004028905 |
| 0 | 0 | 0 | 0 | 0 |
| 0 | 0 | 0 | 0 | 0 |
| 0 | 0 | 0 | 0 | 0 |
| 0 | 0 | 0 | 0 | 0 |
| 0 | 0 | 0 | 0 | 0 |
| 0 | 0 | 0 | 0 | 0 |
| 0 | 0 | 0 | 0 | 0 |
| 0 | 0 | 0 | 0 | 0 |
| 0 | 0 | 0 | 0.007763717 | 0 |
| 0 | 0 | 0.045712025 | 0.036530237 | 0.098698717 |
| 0 | 0 | 0 | 0 | 0 |
| 0 | 0 | 0 | 0 | 0 |
| 0 | 0 | 0 | 0 | 0 |
| 0 | 0 | 0 | 0 | 0 |
| 0 | 0 | 0 | 0 | 0 |
| 0 | 0 | 0 | 0 | 0 |
| 0 | 0 | 0 | 0 | 0 |
| 0 | 0 | 0 | 0 | 0 |
| 0 | 0 | 0 | 0 | 0 |
| 0 | 0 | 0 | 0 | 0 |
| 0 | 0 | 0 | 0.12255266 | 0 |
| 0 | 0 | 0 | 0 | 0 |
| 0 | 0 | 0 | 0.00184646 | 0.001648153 |
| 0 | 0 | 0 | 0 | 0 |
| 0 | 0 | 0 | 0 | 0 |
| 0.011100694 | 0 | 0 | 0 | 0.066815629 |
| 0 | 0 | 0 | 0 | 0 |

| | | | | |
|---|---|---|---|---|
| 0 | 0 | 0 | 0 | 0 |
| 0 | 0 | 0 | 0 | 0 |
| 0 | 0 | 0 | 0 | 0 |
| 0 | 0 | 0 | 0 | 0 |
| 0.004784451 | 0.06426647 | 0 | 0.024678051 | 1.61195315 |
| 0 | 0 | 0 | 0.001456631 | 0 |
| 0 | 0 | 0 | 0.000684404 | 0 |
| 0 | 0 | 0 | 0 | 0 |
| 0 | 0 | 0 | 0 | 0 |
| 0 | 0 | 0 | 0 | 0 |
| 0 | 0 | 0 | 0 | 0 |
| 0.009299519 | 0.00167558 | 0.005108508 | 0.011216688 | 0.003276665 |
| 0 | 0 | 0.002431644 | 0.002815178 | 0.009752931 |
| 0 | 0 | 0 | 0 | 0 |
| 0 | 0.002713432 | 0 | 0 | 0.00049941 |
| 0 | 0.978065185 | 0 | 0 | 0 |
| 0 | 0 | 0 | 0.001254831 | 0.001638094 |
| 0 | 0 | 0 | 0 | 0 |
| 0.169502548 | 0.081676352 | 0.034813722 | 0.002316366 | 0.244141196 |
| 0 | 0 | 0.002264098 | 0 | 0 |
| 0 | 0 | 0 | 0 | 0 |
| 0 | 0 | 0 | 0 | 0 |
| 0 | 0 | 0 | 0 | 0 |
| 0 | 0 | 0 | 0 | 0 |
| 0 | 0 | 0 | 0 | 0 |
| 0 | 0 | 0 | 0 | 0 |
| 0.020524508 | 0.00132752 | 0.020794958 | 0 | 0.002596019 |
| 0 | 0 | 0 | 0 | 0 |
| 0 | 0 | 0 | 0 | 0 |
| 0 | 0 | 0 | 0 | 0 |
| 0.002187365 | 0 | 0.003480456 | 0 | 0.000735681 |
| 0 | 0 | 0.007005964 | 0 | 0 |
| 0 | 0 | 0 | 0 | 0 |
| 0 | 0 | 0 | 0 | 0 |
| 0 | 0 | 0 | 0 | 0 |
| 0 | 0 | 0 | 0 | 0 |
| 0 | 0 | 0 | 0 | 0 |
| 0.024588465 | 0.000925083 | 0.015116197 | 0.001286795 | 0.080528036 |
| 0 | 0 | 0 | 0 | 0 |
| 0 | 0 | 0 | 0 | 0 |
| 0.020641975 | 0 | 0 | 0.02248114 | 0.022291238 |
| 0 | 0 | 0 | 0 | 0 |
| 0 | 0 | 0 | 0 | 0 |
| 0.002693185 | 0.003331469 | 0.001813011 | 0 | 0.006591468 |
| 0 | 0 | 0 | 0 | 0 |
| 0 | 0 | 0 | 0 | 0 |
| 0 | 0 | 0 | 0 | 0 |
| 0 | 0 | 0 | 0 | 0 |

| | | | | |
|---|---|---|---|---|
| 0 | 0 | 0 | 0 | 0 |
| 0 | 0 | 0.043811195 | 0 | 0 |
| 0 | 0 | 0 | 0 | 0 |
| 0.001302678 | 0 | 0.002245506 | 0 | 0 |
| 0 | 0.000952415 | 0.00176226 | 0 | 0 |
| 0.001299429 | 0 | 0.001033802 | 0.00119686 | 0 |
| 0 | 0 | 0 | 0.001552488 | 0 |
| 0.010867276 | 0 | 0 | 0 | 0 |
| 0 | 0 | 0 | 0 | 0 |
| 0 | 0 | 0 | 0 | 0 |
| 0 | 0.003224404 | 0 | 0.007270684 | 0 |
| 0 | 0 | 0 | 0 | 0 |
| 0 | 0.001455828 | 0 | 0.01913685 | 0.013551388 |
| 0 | 0 | 0 | 0.006603457 | 0 |
| 0 | 0 | 0 | 0 | 0 |
| 0 | 0 | 0 | 0 | 0 |
| 0 | 0 | 0 | 0 | 0 |
| 0 | 0 | 0 | 0 | 0 |
| 0 | 0 | 0 | 0 | 0 |
| 0 | 0 | 0 | 0 | 0 |
| 0 | 0 | 0 | 0 | 0 |
| 0 | 0 | 0 | 0 | 0 |
| 0.094842185 | 0.003067183 | 0.001773508 | 0 | 0.087870725 |
| 0 | 0.024883249 | 0 | 0 | 0 |
| 0 | 0 | 0 | 0 | 0 |
| 0 | 0 | 0 | 0.003253324 | 0 |
| 0 | 0 | 0 | 0 | 0 |
| 0 | 0 | 0 | 0 | 0 |
| 0 | 0 | 0 | 0 | 0 |
| 0 | 0 | 0 | 0 | 0 |
| 0 | 0.013038352 | 0.035054867 | 0 | 0.007858101 |
| 0 | 0.002479337 | 0.010704241 | 0 | 0 |
| 0 | 0 | 0 | 0 | 0 |
| 0 | 0 | 0 | 0 | 0 |
| 0 | 0 | 0 | 0 | 0 |
| 0.005905274 | 0 | 0 | 0 | 0.004194713 |
| 0.023046188 | 0 | 0.000763964 | 0 | 0.121321973 |
| 0 | 0 | 0 | 0 | 0 |
| 0 | 0 | 0.002585643 | 0 | 0.002970606 |
| 0 | 0 | 0 | 0 | 0 |
| 0 | 0 | 0 | 0 | 0 |
| 0 | 0 | 0 | 0 | 0 |
| 0 | 0 | 0 | 0 | 0 |
| 0 | 0 | 0 | 0 | 0 |
| 0 | 0 | 0 | 0 | 0 |
| 0.000672815 | 0.567389511 | 0.231776128 | 0.100547526 | 0.08948613 |
| 0.024623874 | 0 | 0 | 0 | 0 |
| 0 | 0 | 0 | 0 | 0 |

| | | | | |
|---|---|---|---|---|
| 0 | 0 | 0 | 0 | 0 |
| 0 | 0 | 0 | 0 | 0 |
| 0 | 0 | 0 | 0.042270335 | 0 |
| 0 | 0 | 0 | 0 | 0 |
| 0 | 0 | 0 | 0 | 0 |
| 0 | 0 | 0 | 0 | 0 |
| 0 | 0 | 0 | 0 | 0 |
| 0 | 0 | 0 | 0 | 0 |
| 0 | 0 | 0 | 0 | 0 |
| 0 | 0 | 0 | 0 | 0 |
| 0.005428341 | 0 | 0.000987129 | 0.003999888 | 0 |
| 0 | 0 | 0 | 0 | 0 |
| 0 | 0 | 0 | 0 | 0 |
| 0 | 0 | 0 | 0 | 0 |
| 0 | 0 | 0 | 0 | 0 |
| 0 | 0 | 0 | 0 | 0 |
| 0 | 0 | 0 | 0 | 0 |
| 0 | 0 | 0 | 0 | 0 |
| 0 | 0 | 0 | 0 | 0 |
| 0 | 0.002140206 | 0.000654551 | 0.000757791 | 0 |
| 0 | 0 | 0 | 0 | 0 |
| 0 | 0 | 0 | 0 | 0 |
| 0 | 0 | 0 | 0 | 0 |
| 0 | 0 | 0 | 0 | 0 |
| 0 | 0 | 0 | 0 | 0 |
| 0 | 0 | 0 | 0 | 0 |
| 0 | 0 | 0 | 0 | 0 |
| 0 | 0 | 0.001629836 | 0.006499337 | 0 |
| 0 | 0 | 0 | 0 | 0 |
| 0 | 0 | 0 | 0 | 0 |
| 0 | 0.000835754 | 0.006185601 | 0.001790308 | 0.015297523 |
| 0 | 0 | 0 | 0 | 0 |
| 0.022363572 | 0.016884879 | 0.063333177 | 0.02248808 | 0.159706027 |
| 0 | 0 | 0.012843846 | 0 | 0.001396217 |
| 0 | 0 | 0 | 0 | 0 |
| 0 | 0 | 0 | 0 | 0 |
| 0 | 0 | 0 | 0 | 0 |
| 0 | 0 | 0 | 0 | 0 |
| 0 | 0 | 0 | 0 | 0 |
| 0 | 0 | 0 | 0 | 0 |
| 0 | 0 | 0 | 0 | 0 |
| 0.127269027 | 0.082707552 | 0.06969532 | 0 | 0.231304061 |
| 0 | 0 | 0.037300866 | 0.0609882 | 0.31876756 |
| 0 | 0 | 0 | 0 | 0 |
| 0 | 0 | 0 | 0 | 0 |
| 0 | 0 | 0.02245931 | 0.01231661 | 0 |
| 0 | 0 | 0 | 0 | 0 |
| 0 | 0 | 0 | 0 | 0 |

| | | | | |
|---|---|---|---|---|
| 0 | 0 | 0 | 0 | 0 |
| 0 | 0 | 0 | 0 | 0 |
| 0 | 0.010473866 | 0.08008205 | 0 | 0.001117204 |
| 0 | 0 | 0 | 0 | 0 |
| 0 | 0.001330458 | 0 | 0.013324849 | 0.010407058 |
| 0.039507459 | 0 | 0.000466803 | 0 | 0 |
| 0 | 0 | 0 | 0 | 0 |
| 0 | 0 | 0 | 0 | 0 |
| 0 | 0 | 0 | 0 | 0 |
| 0 | 0 | 0 | 0 | 0 |
| 0 | 0 | 0 | 0 | 0 |
| 0 | 0 | 0 | 0 | 0 |
| 0 | 0 | 0 | 0 | 0 |
| 0.685309045 | 0 | 0 | 0 | 0 |
| 0 | 0 | 0 | 0 | 0 |
| 0 | 0 | 0 | 0 | 0 |
| 0 | 0 | 0 | 0 | 0 |
| 0 | 0 | 0 | 0 | 0 |
| 0 | 0 | 0 | 0 | 0 |
| 0 | 0 | 0 | 0 | 0 |
| 0 | 0 | 0 | 0 | 0 |
| 0 | 0 | 0 | 0 | 0 |
| 0 | 0 | 0 | 0 | 0 |
| 0 | 0 | 0 | 0 | 0 |
| 0 | 0 | 0.000824986 | 0.015663767 | 0 |
| 0.00166905 | 0.022695578 | 0.00049795 | 0 | 0 |
| 0 | 0 | 0 | 0 | 0 |
| 0 | 0 | 0 | 0 | 0.001796962 |
| 0 | 0 | 0 | 0 | 0 |
| 0 | 0 | 0 | 0 | 0 |
| 0 | 0 | 0 | 0 | 0 |
| 0 | 0 | 0.052255135 | 0 | 0 |
| 0 | 0 | 0 | 0 | 0 |
| 0 | 0 | 0 | 0 | 0 |
| 0 | 0 | 0 | 0 | 0.013016162 |
| 0 | 0 | 0 | 0 | 0 |
| 0 | 0 | 0 | 0 | 0 |
| 0.006437922 | 0 | 0 | 0 | 0 |
| 0.610882056 | 0.715648454 | 4.575696479 | 0 | 0 |
| 0 | 0 | 0 | 0 | 0 |
| 0 | 0 | 0 | 0 | 0 |
| 0 | 0 | 0.001260464 | 0 | 0 |
| 0.010887981 | 0 | 0.017824307 | 0.002507137 | 0.008985927 |
| 0 | 0 | 0 | 0 | 0 |
| 0 | 0 | 0 | 0 | 0 |
| 0 | 0 | 0 | 0 | 0.000534724 |
| 0.026906621 | 0 | 0.028287055 | 0 | 0.002607135 |
| 0 | 0 | 0 | 0 | 0 |

| | | | | |
|---|---|---|---|---|
| 0 | 0 | 0 | 0 | 0 |
| 0.009296164 | 0.004205645 | 0.122192457 | 0.006887132 | 0.006429916 |
| 0 | 0 | 0 | 0 | 0 |
| 0 | 0 | 0 | 0 | 0 |
| 0 | 0 | 0 | 0 | 0 |
| 0 | 0 | 0 | 0 | 0 |
| 0 | 0 | 0 | 0 | 0 |
| 0 | 0 | 0 | 0 | 0 |
| 0 | 0 | 0 | 0 | 0 |
| 0 | 0 | 0 | 0 | 0 |
| 0 | 0 | 0 | 0 | 0 |
| 0 | 0 | 0 | 0 | 0 |
| 0 | 0 | 0 | 0.046436564 | 0 |
| 0 | 0 | 0 | 0 | 0 |
| 0.009575104 | 0 | 0 | 0 | 0.001678005 |
| 0 | 0 | 0 | 0 | 0 |
| 0 | 0 | 0 | 0 | 0 |
| 0 | 0 | 0 | 0 | 0 |
| 0 | 0 | 0.002661232 | 0.002567482 | 0 |
| 0 | 0 | 0 | 0 | 0 |
| 0 | 0 | 0 | 0 | 0 |
| 0 | 0 | 0.000984376 | 0 | 0 |
| 0.004753833 | 0 | 0.006051301 | 0 | 0 |
| 0 | 0 | 0 | 0 | 0 |
| 0 | 0 | 0 | 0 | 0 |
| 0 | 0 | 0 | 0 | 0 |
| 0 | 0 | 0 | 0 | 0 |
| 0 | 0 | 0 | 0 | 0 |
| 0 | 0 | 0 | 0 | 0 |
| 0 | 0 | 0 | 0 | 0 |
| 0 | 0 | 0 | 0 | 0 |
| 0 | 0 | 0 | 0 | 0 |
| 0 | 0 | 0 | 0 | 0 |
| 0 | 0 | 0 | 0 | 0 |
| 0 | 0 | 0.002681277 | 4.485546588 | 0 |
| 0 | 0 | 0 | 0 | 0 |
| 0 | 0 | 0 | 0 | 0 |
| 0 | 0 | 0 | 0 | 0 |
| 0 | 0 | 0.039713963 | 0.077616152 | 0.006765495 |
| 0 | 0 | 0 | 0 | 0 |
| 0.018390749 | 0.011318246 | 0.045833135 | 0.090818035 | 0.124274332 |
| 0 | 0 | 0 | 0 | 0 |
| 0 | 0 | 0 | 0 | 0 |
| 0 | 0 | 0 | 0 | 0 |
| 0 | 0 | 0 | 0 | 0 |
| 0.215635338 | 0.155983403 | 0.306781718 | 0.011916863 | 0.306533603 |
| 0 | 0 | 0 | 0 | 0 |
| 0 | 0 | 0 | 0 | 0 |

| | | | | |
|---|---|---|---|---|
| 0 | 0 | 0.00359527 | 0 | 0 |
| 0 | 0 | 0 | 0 | 0 |
| 0 | 0 | 0 | 0 | 0 |
| 0 | 0 | 0 | 0.033588299 | 0 |
| 0 | 0 | 0 | 0.002658247 | 0 |
| 0 | 0 | 0 | 0 | 0 |
| 0 | 0 | 0 | 0 | 0 |
| 0 | 0 | 0 | 0 | 0 |
| 0 | 0 | 0 | 0 | 0 |
| 0 | 0.028163449 | 0.01440948 | 0 | 0.002570156 |
| 0 | 0 | 0 | 0.004820218 | 0 |
| 0 | 0 | 0 | 0 | 0 |
| 0 | 0 | 0 | 0 | 0 |
| 0 | 0 | 0 | 0 | 0 |
| 0 | 0 | 0 | 0 | 0 |
| 0 | 0 | 0 | 0 | 0 |
| 0 | 0 | 0 | 0 | 0 |
| 0 | 0 | 0 | 0 | 0 |
| 0 | 0 | 0 | 0 | 0 |
| 0.115046012 | 0.10739749 | 1.608360483 | 0.355072117 | 0.346892636 |
| 0 | 0 | 0 | 0 | 0 |
| 0 | 0 | 0 | 0 | 0 |
| 0 | 0 | 0 | 0 | 0 |
| 0 | 0 | 0 | 0 | 0 |
| 0 | 0 | 0 | 0 | 0 |
| 0 | 0 | 0 | 0.002384299 | 0 |
| 0 | 0 | 0 | 0 | 0 |
| 0 | 0 | 0 | 0 | 0 |
| 0 | 0 | 0 | 0 | 0 |
| 0.014998298 | 0 | 0.001652175 | 0 | 0.030219884 |
| 0 | 0 | 0 | 0 | 0 |
| 0 | 0 | 0 | 0 | 0 |
| 0 | 0 | 0 | 0 | 0 |
| 0 | 0 | 0 | 0 | 0 |
| 0 | 0 | 0 | 0 | 0 |
| 0 | 0 | 0 | 0 | 0 |
| 0 | 0 | 0 | 0 | 0 |
| 0 | 0 | 0 | 0 | 0 |
| 0 | 0 | 0 | 0 | 0 |
| 0 | 0 | 0 | 0 | 0 |
| 0 | 0 | 0 | 0 | 0 |
| 0 | 0 | 0 | 0 | 0 |
| 0 | 0 | 0 | 0 | 0 |
| 0 | 0 | 0 | 0 | 0 |
| 0.00304096 | 0 | 0.007903153 | 0.005041665 | 0 |
| 0 | 0 | 0 | 0 | 0 |
| 0 | 0 | 0 | 0 | 0 |
| 0.003652917 | 0 | 0 | 0 | 0 |

| | | | | |
|---|---|---|---|---|
| 0 | 0 | 0 | 0 | 0 |
| 0 | 0 | 0 | 0 | 0 |
| 0 | 0 | 0 | 0 | 0 |
| 0 | 0 | 0 | 0 | 0 |
| 0 | 0 | 0 | 0.002438438 | 0 |
| 0.009080779 | 0 | 0.009873488 | 0.004878998 | 0.001455815 |
| 0 | 0 | 0 | 0 | 0 |
| 0 | 0 | 0 | 0 | 0 |
| 0 | 0 | 0 | 0 | 0 |
| 0 | 0 | 0 | 0 | 0 |
| 0 | 0 | 0 | 0 | 0 |
| 0 | 0 | 0 | 0 | 0 |
| 0 | 0 | 0 | 0 | 0 |
| 0 | 0 | 0 | 0 | 0 |
| 0 | 0 | 0 | 0 | 0 |
| 0 | 0 | 0 | 0 | 0 |
| 0 | 0 | 0 | 0 | 0 |
| 0 | 0 | 0 | 0 | 0 |
| 0 | 0 | 0 | 0 | 0 |
| 0 | 0 | 0 | 0 | 0 |
| 0 | 0 | 0 | 0 | 0 |
| 0 | 0 | 0 | 0 | 0 |
| 0 | 0 | 0 | 0 | 0 |
| 0 | 0 | 0 | 0 | 0 |
| 0 | 0 | 0 | 0 | 0 |
| 0 | 0.002687203 | 0 | 0 | 0 |
| 0 | 0 | 0 | 0 | 0 |
| 0 | 0 | 0.016069289 | 0 | 0 |
| 0 | 0 | 0 | 0 | 0 |
| 0 | 0 | 0.012407537 | 0 | 0 |
| 0 | 0 | 0 | 0 | 0 |
| 0 | 0 | 0 | 0 | 0 |
| 0 | 0 | 0 | 0 | 0 |
| 0 | 0 | 0 | 0 | 0 |
| 0 | 0 | 0 | 0 | 0 |
| 0 | 0 | 0 | 0 | 0 |
| 0 | 0 | 0 | 0 | 0 |
| 0 | 0 | 0 | 0 | 0 |
| 0 | 0.071539784 | 0 | 0.003148408 | 3.855046394 |
| 0 | 0 | 0 | 0 | 0 |
| 0 | 0 | 0 | 0 | 0 |
| 0 | 0 | 0 | 0 | 0 |
| 0 | 0 | 0 | 0 | 0 |
| 0 | 0 | 0 | 0 | 0 |
| 0 | 0 | 0 | 0 | 0 |
| 0 | 0 | 0 | 0 | 0 |
| 0 | 0 | 0 | 0 | 0 |
| 0 | 0 | 0 | 0 | 0 |

| | | | | |
|---|---|---|---|---|
| 0 | 0 | 0 | 0 | 0 |
| 0 | 0.026607211 | 0.107980324 | 0.002497112 | 0 |
| 0 | 0 | 0 | 0 | 0 |
| 0 | 0 | 0 | 0 | 0 |
| 0 | 0 | 0 | 0 | 0 |
| 0 | 0 | 0.000554581 | 0 | 0 |
| 0 | 0 | 0 | 0 | 0 |
| 0 | 0 | 0 | 0 | 0 |
| 0 | 0 | 0 | 0 | 0 |
| 0 | 0 | 0 | 0.000635715 | 0 |
| 0 | 0.001049399 | 0 | 0 | 0 |
| 0 | 0 | 0 | 0 | 0 |
| 0 | 1.364694271 | 0.101671832 | 0.177916918 | 0.341479476 |
| 0 | 0 | 0 | 0 | 0 |
| 0 | 0 | 0 | 0 | 0 |
| 0 | 0 | 0 | 0 | 0 |
| 0 | 0 | 0 | 0 | 0 |
| 0 | 0 | 0 | 0 | 0 |
| 0 | 0 | 0 | 0 | 0 |
| 0 | 0 | 0 | 0 | 0 |
| 0.053144551 | 0 | 0 | 0.000622241 | 0.005498593 |
| 0 | 0 | 0 | 0 | 0 |
| 0 | 0 | 0 | 0 | 0 |
| 0 | 2.585870827 | 0.740876924 | 0.141409049 | 0 |
| 0 | 0 | 0 | 0 | 0 |
| 0 | 0 | 0 | 0 | 0 |
| 0 | 0 | 0 | 0 | 0 |
| 0 | 0 | 0 | 0 | 0 |
| 0 | 0 | 0 | 0 | 0 |
| 0 | 0 | 0 | 0 | 0 |
| 0 | 0 | 0.013539804 | 0 | 0 |
| 0 | 0 | 0 | 0 | 0 |
| 0 | 0 | 0 | 0 | 0 |
| 0 | 0 | 0 | 0 | 0 |

| PHL018-0230_V1 REF | PHL018-0014_V3 CTRL | PHL018-0246_V1 REF | PHL018-0100_V2 EYCF | PHL018-0173_V2 CTRL |
|---|---|---|---|---|
| 0.232357286 | 0.081658939 | 0.123619404 | 0.156896512 | 0 |
| 0 | 9.761997892 | 0.00913389 | 0.094307924 | 8.926578693 |
| 0 | 3.137903172 | 0 | 0.675975296 | 14.50660328 |
| 0 | 0 | 0 | 0 | 0 |
| 0 | 0 | 0 | 0 | 0 |
| 0 | 0 | 0 | 0 | 0 |
| 0 | 0 | 0 | 0 | 0 |
| 0 | 0 | 0 | 0 | 0 |
| 0 | 0 | 0 | 0 | 0 |
| 0 | 0 | 0 | 0 | 0 |
| 0 | 0 | 0 | 0 | 0 |
| 0 | 0 | 0 | 0 | 0 |
| 0 | 0 | 0 | 0 | 0 |
| 40.17244681 | 0.252539702 | 0.681212743 | 0.522242462 | 21.22202704 |
| 0 | 0 | 0 | 0 | 0 |
| 0 | 0 | 0 | 0 | 0 |
| 0 | 0 | 0 | 0 | 0 |
| 0 | 0 | 0 | 0 | 0 |
| 0 | 0 | 0 | 0 | 0 |
| 0 | 0 | 0 | 0 | 0 |
| 0 | 0 | 0 | 0 | 0 |
| 0 | 0 | 0 | 0 | 0 |
| 0 | 0 | 0 | 0 | 0 |
| 0 | 0 | 0 | 0 | 0 |
| 0 | 0 | 0.042689931 | 0.089298483 | 0 |
| 0 | 0 | 0 | 0 | 0 |
| 0 | 0 | 0 | 0 | 0 |
| 0 | 0 | 0 | 0 | 0 |
| 0 | 0 | 0 | 0 | 0 |
| 0 | 0 | 0 | 0 | 0 |
| 0 | 0 | 0 | 0 | 0 |
| 0 | 0 | 0 | 0 | 0 |
| 0 | 0 | 0 | 0 | 0 |
| 0 | 0 | 0 | 0 | 0 |
| 0 | 0 | 0 | 0 | 0 |
| 0 | 3.846065555 | 0.912039169 | 1.072172219 | 0 |
| 0 | 0 | 0 | 0 | 0 |
| 0 | 0 | 0 | 0 | 0 |
| 0 | 0 | 0 | 0 | 0 |
| 0 | 0 | 0 | 0 | 0 |
| 0 | 0 | 0 | 0 | 0 |
| 0.002252664 | 0.110376026 | 0.073740081 | 0.017083901 | 0.002481971 |
| 0.3089559 | 0.127016209 | 3.168688489 | 0.136729821 | 0.005778292 |
| 0.006279476 | 0.018776983 | 0.014079191 | 0.000933778 | 0 |
| 0.001864174 | 0.005264599 | 0.015225899 | 0 | 0 |
| 0 | 0 | 0 | 0 | 0 |

| | | | | |
|---|---|---|---|---|
| 0 | 0.004142557 | 0 | 0.000747865 | 0 |
| 0.018758266 | 0.002346997 | 0 | 0.004287528 | 0.013318679 |
| 0 | 0.011920711 | 0.006162646 | 0 | 0 |
| 0.189946148 | 1.710131234 | 0.727977999 | 0.364608868 | 0.460019271 |
| 0 | 0 | 0 | 0 | 0 |
| 0 | 0 | 0 | 0.008357156 | 0 |
| 0 | 0.001078115 | 0.002282492 | 0 | 0.010451687 |
| 0 | 0 | 0 | 0 | 0 |
| 0 | 0 | 0 | 0.007340256 | 0 |
| 0 | 0 | 0 | 0 | 0 |
| 0 | 0 | 0 | 0.004897996 | 0 |
| 0 | 0 | 0.125844764 | 0.048907276 | 0 |
| 0 | 0 | 0 | 0 | 0 |
| 0.019684937 | 0.044010391 | 0.006742928 | 0.018693901 | 0.043085367 |
| 0 | 0 | 0 | 0 | 0 |
| 0 | 0 | 0 | 0 | 0 |
| 0 | 0.053629854 | 0.017087099 | 0.16982637 | 0 |
| 0 | 0 | 0 | 0 | 0 |
| 0 | 0 | 0 | 0 | 0 |
| 0 | 0 | 0 | 0 | 0 |
| 1.572587788 | 0.047321315 | 0.004534217 | 0.001112884 | 0.062311579 |
| 0 | 0.003064145 | 0.00060817 | 0.152031483 | 0.006701754 |
| 0 | 0 | 0.041382754 | 0.079254195 | 0 |
| 0 | 0 | 0 | 0 | 0 |
| 0 | 0 | 0 | 0 | 0 |
| 0 | 0 | 0 | 0 | 0 |
| 0.281777851 | 0 | 0 | 0 | 0 |
| 0 | 0 | 0 | 0 | 0 |
| 0.002313385 | 0.012086444 | 0.020747333 | 0.002005075 | 0 |
| 0.001699623 | 0 | 0 | 0 | 0 |
| 0 | 0 | 0 | 0 | 0 |
| 0 | 0 | 0 | 0 | 0 |
| 0 | 0 | 0 | 0 | 0 |
| 0 | 0 | 0 | 0 | 0 |
| 0 | 0 | 0 | 0 | 0 |
| 0 | 0.00385882 | 0.001885283 | 0 | 0 |
| 0 | 0 | 0 | 0 | 0 |
| 0 | 3.744084354 | 0 | 8.37813635 | 0 |
| 0 | 0 | 0 | 0 | 0 |
| 0 | 0 | 0 | 0 | 0 |
| 0 | 0 | 0 | 0 | 0 |
| 0 | 0 | 0 | 0 | 0 |
| 0 | 0 | 0 | 0 | 0 |
| 0 | 0 | 0 | 0 | 0 |
| 0 | 0 | 0 | 0.011959595 | 0 |
| 0 | 0.277364303 | 0 | 0 | 0 |
| 0 | 0 | 0.00250635 | 0 | 0 |
| 0.001827005 | 0 | 0 | 0 | 0 |

| | | | | |
|---|---|---|---|---|
| 0 | 0 | 0 | 0 | 0 |
| 0.002088979 | 0 | 0 | 0 | 0.002092385 |
| 0 | 0 | 0 | 0 | 0 |
| 0 | 0 | 0 | 0 | 0 |
| 0 | 0.483931725 | 0.496578147 | 0.006838804 | 0 |
| 0 | 0 | 0 | 0 | 0.001113152 |
| 0 | 0 | 0 | 0 | 0 |
| 0 | 0 | 0 | 0 | 0 |
| 0.00052437 | 0 | 0.105028078 | 0 | 0 |
| 0 | 0 | 0 | 0 | 0 |
| 0 | 0 | 0 | 0 | 0 |
| 0 | 0.013180983 | 0.002502408 | 0.002638244 | 0 |
| 0 | 0 | 0 | 0 | 0 |
| 0 | 0 | 0 | 0 | 0 |
| 0 | 0 | 0 | 0 | 0 |
| 0 | 0 | 0 | 0.001223497 | 0 |
| 0 | 0 | 0 | 0 | 0 |
| 0 | 0 | 0 | 0 | 0 |
| 0 | 0.050491975 | 0.207113438 | 0.350100032 | 0.006331169 |
| 0 | 0 | 0 | 0 | 0 |
| 0 | 0 | 0 | 0 | 0 |
| 0 | 0 | 0 | 0 | 0 |
| 0 | 0 | 0 | 0 | 0 |
| 0 | 0 | 0 | 0 | 0 |
| 0 | 0 | 0 | 0 | 0 |
| 0 | 0 | 0 | 0 | 0 |
| 0 | 0 | 0 | 0 | 0 |
| 0 | 0 | 0 | 0 | 0 |
| 0 | 0 | 0 | 0 | 0 |
| 0 | 0 | 0 | 0 | 0 |
| 0 | 0 | 0 | 0.014486221 | 0 |
| 0 | 0.004073253 | 0.001712027 | 0.008211449 | 0 |
| 0 | 0 | 0 | 0 | 0 |
| 0 | 0 | 0 | 0 | 0 |
| 0 | 0 | 0 | 0 | 0 |
| 0 | 0 | 0 | 0 | 0 |
| 0 | 0 | 0 | 0 | 0 |
| 0 | 0 | 0 | 0 | 0 |
| 0 | 0.017769926 | 0 | 0.00517159 | 0 |
| 0 | 0 | 0 | 0 | 0 |
| 0 | 0 | 0 | 0 | 0 |
| 0 | 0 | 0 | 0 | 0 |
| 0 | 0.001384717 | 0 | 0 | 0.004911222 |
| 0 | 0 | 0 | 0 | 0 |
| 0 | 0 | 0 | 0 | 0 |
| 0 | 0 | 0 | 0 | 0 |
| 0 | 0 | 0 | 0 | 0 |
| 0 | 0 | 0 | 0 | 0 |

| | | | | |
|---|---|---|---|---|
| 0 | 0 | 0 | 0 | 0 |
| 0 | 0 | 0 | 0 | 0 |
| 0 | 0 | 0 | 0 | 0 |
| 0 | 0 | 0 | 0 | 0 |
| 0 | 0 | 0.001821264 | 0.083738841 | 0 |
| 0 | 0 | 0 | 0 | 0 |
| 0 | 0 | 0 | 0 | 0 |
| 0 | 0 | 0 | 0 | 0 |
| 0 | 0 | 0 | 0 | 0 |
| 0 | 0 | 0 | 0 | 0 |
| 0 | 0 | 0 | 0 | 0 |
| 0 | 0 | 0 | 0 | 0 |
| 0 | 0 | 0 | 0 | 0 |
| 0 | 0 | 0 | 0 | 0 |
| 0 | 0 | 0 | 0 | 0 |
| 0 | 0 | 8.899976832 | 1.11340781 | 0 |
| 0 | 0 | 0 | 0 | 0 |
| 0 | 0 | 0 | 0 | 0 |
| 0 | 0 | 0 | 0 | 0 |
| 0 | 0 | 0 | 0 | 0 |
| 0 | 0.001120749 | 0 | 0 | 0 |
| 0 | 0 | 0 | 0 | 0 |
| 0 | 0 | 0 | 0.002497302 | 0 |
| 0 | 0 | 0 | 0 | 0 |
| 0 | 0 | 0 | 0 | 0 |
| 0 | 0 | 0 | 0 | 0 |
| 0 | 0 | 0 | 0.025295045 | 0 |
| 0 | 0 | 0 | 0 | 0 |
| 0 | 0 | 0 | 0 | 0 |
| 0 | 0 | 0 | 0 | 0 |
| 0 | 0 | 0 | 0 | 0 |
| 0 | 0 | 0 | 0 | 0 |
| 0 | 0 | 0 | 0 | 0 |
| 0 | 0 | 0 | 0 | 0 |
| 0 | 0 | 0 | 0 | 0 |
| 0 | 0 | 0 | 0 | 0 |
| 0 | 0 | 0 | 0 | 0 |
| 0 | 0 | 0 | 0.268257843 | 0 |
| 0 | 0 | 0 | 0 | 0 |
| 0 | 0 | 0 | 0 | 0 |
| 0 | 0 | 0 | 0 | 0 |
| 0 | 0 | 0 | 0 | 0 |
| 0 | 0 | 0 | 0 | 0 |
| 0 | 0 | 0 | 0 | 0 |
| 0 | 0 | 0 | 0 | 0 |
| 0 | 0 | 0 | 0 | 0 |
| 0 | 0 | 0 | 0 | 0 |
| 0 | 0 | 0 | 0 | 0 |

| | | | | |
|---|---|---|---|---|
| 0 | 0 | 0 | 0 | 0 |
| 0 | 0 | 0 | 0 | 0 |
| 0 | 0 | 0 | 0 | 0 |
| 0 | 0 | 0 | 0 | 0 |
| 0 | 0 | 0 | 0 | 0 |
| 0 | 0 | 0 | 0 | 0 |
| 0 | 0 | 0 | 0 | 0 |
| 0 | 0 | 0 | 0 | 0 |
| 0 | 0 | 0 | 0 | 0 |
| 0 | 0 | 0 | 0 | 0 |
| 0 | 0 | 0 | 0 | 0 |
| 0 | 0 | 0 | 0.008784831 | 0 |
| 0.001124053 | 0 | 0.001260118 | 0 | 0 |
| 0 | 0 | 0 | 0 | 0 |
| 0 | 0 | 0 | 0 | 0 |
| 0 | 0 | 0 | 0 | 0 |
| 0 | 0 | 0 | 0 | 0 |
| 0 | 0 | 0 | 0 | 0 |
| 0 | 0 | 0 | 0 | 0 |
| 0 | 0 | 0 | 0 | 0 |
| 0.112091186 | 0.036107149 | 0.024346554 | 0.003542019 | 0.050523276 |
| 0 | 0 | 0 | 0 | 0 |
| 0 | 0 | 0 | 0 | 0 |
| 0 | 0 | 0 | 0 | 0.007683319 |
| 0 | 0 | 0 | 0 | 0 |
| 0 | 0 | 0 | 0 | 0 |
| 0 | 0.002880541 | 0.004345134 | 0.000442026 | 0.000612991 |
| 0 | 0 | 0 | 0 | 0 |
| 0 | 0 | 0 | 0 | 0 |
| 0.186607203 | 0 | 0 | 0 | 0.006339687 |
| 0.001220106 | 0.019238451 | 0 | 0 | 0.015887245 |
| 0 | 0 | 0 | 0 | 0 |
| 0 | 0 | 0 | 0 | 0 |
| 0 | 0 | 0 | 0 | 0.001929793 |
| 0 | 0.02317141 | 0.001533016 | 0.004444636 | 0.002054571 |
| 0 | 0 | 0 | 0 | 0 |
| 0 | 0 | 0 | 0 | 0 |
| 0 | 0 | 0 | 0 | 0 |
| 0 | 0 | 0 | 0 | 0 |
| 0 | 0 | 0 | 0 | 0 |
| 0 | 0 | 0 | 0 | 0 |
| 0 | 0 | 0 | 0 | 0 |
| 0 | 0 | 0 | 0 | 0 |
| 0 | 0 | 0 | 0 | 0 |
| 0 | 0 | 0 | 0 | 0 |
| 0 | 0 | 0 | 0 | 0 |
| 0 | 0 | 0 | 0 | 0 |
| 0.125873733 | 0 | 0 | 0 | 0 |

| | | | | |
|---|---|---|---|---|
| 0 | 0 | 0 | 0 | 0 |
| 0.019972733 | 0 | 0 | 0 | 0 |
| 0 | 0 | 0 | 0 | 0 |
| 0 | 0 | 0 | 0 | 0 |
| 0 | 0 | 0 | 0 | 0 |
| 0 | 0 | 0 | 0 | 0 |
| 0 | 0.348337551 | 0 | 0 | 0 |
| 0.01050653 | 0.019121574 | 0.001046962 | 0 | 0 |
| 0 | 0 | 0 | 0 | 0 |
| 0 | 0 | 0 | 0 | 0 |
| 0 | 0 | 0 | 0 | 0 |
| 0 | 0 | 0 | 0 | 0 |
| 0 | 0 | 0 | 0 | 0 |
| 0 | 0 | 0 | 0 | 0 |
| 0 | 0 | 0 | 0.006658519 | 0 |
| 0 | 0 | 0 | 0 | 0 |
| 0 | 0 | 0 | 0 | 0 |
| 0 | 0 | 0.018261141 | 0 | 0 |
| 0.002808022 | 0 | 0.031664453 | 0 | 0 |
| 0 | 0 | 0 | 0.041425018 | 0 |
| 0.018761358 | 0.012041773 | 0 | 0 | 0.014643076 |
| 0 | 0 | 0 | 0 | 0 |
| 0 | 0 | 0 | 0 | 0 |
| 0 | 0 | 0 | 0 | 0 |
| 0.003388491 | 0.012303528 | 0 | 0.007342246 | 0.003878875 |
| 0 | 0 | 0.001316489 | 0 | 0 |
| 0 | 0 | 0 | 0 | 0 |
| 0 | 0 | 0 | 0 | 0 |
| 0 | 0.060011782 | 0 | 0 | 0 |
| 0 | 0 | 0 | 0.000599572 | 0 |
| 0.568828822 | 0.019479484 | 0.071300167 | 0.06366552 | 0.129025122 |
| 0 | 0 | 0 | 0 | 0 |
| 0 | 0 | 0 | 0.000965403 | 0 |
| 0.002086122 | 0.0016365 | 0 | 0.006529246 | 0 |
| 0 | 0 | 0 | 0 | 0 |
| 0 | 0 | 0 | 0 | 0 |
| 0 | 0 | 0 | 0 | 0 |
| 0 | 0 | 0 | 0 | 0 |
| 0 | 0 | 0 | 0 | 0 |
| 0 | 0 | 0.003811719 | 0 | 0 |
| 0 | 0 | 0 | 0 | 0 |
| 0 | 0.056731698 | 0.007779688 | 0.017543136 | 0 |
| 0 | 0 | 0 | 0 | 0.004757782 |
| 0 | 0 | 0 | 0 | 0 |
| 0 | 0 | 0 | 0 | 0 |
| 0 | 0 | 0 | 0 | 0 |
| 0 | 0 | 0 | 0 | 0 |
| 0 | 0 | 0 | 0 | 0 |

| | | | | |
|---|---|---|---|---|
| 0 | 0 | 0 | 0 | 0 |
| 0 | 0 | 0 | 0 | 0 |
| 0.003898314 | 0.015542396 | 0.004884339 | 0 | 0 |
| 0 | 0 | 0 | 0 | 0 |
| 0 | 0 | 0 | 0 | 0 |
| 0 | 0 | 0 | 0 | 0 |
| 0 | 0 | 0 | 0 | 0 |
| 0 | 0 | 0 | 0 | 0 |
| 0 | 0 | 0 | 0 | 0 |
| 0 | 0 | 0 | 0 | 0 |
| 0 | 0 | 0 | 0 | 0 |
| 0 | 0 | 0 | 0 | 0 |
| 0 | 0 | 0 | 0 | 0 |
| 0.002280886 | 0.003936431 | 0.005398077 | 0.002013516 | 0 |
| 0.003555971 | 0.006793382 | 0.00140697 | 0 | 0.003352253 |
| 0 | 0 | 0.0143103 | 0.004529069 | 0 |
| 0.002621695 | 0.014134733 | 0.002939047 | 0.000860718 | 0.004535766 |
| 0 | 0 | 0 | 0 | 0 |
| 0 | 0 | 0 | 0 | 0 |
| 0 | 0 | 0 | 0 | 0 |
| 0 | 0 | 0 | 0 | 0 |
| 0.114889732 | 0 | 0 | 0 | 0 |
| 0 | 0 | 0 | 0 | 0 |
| 0.163763488 | 0 | 0 | 0 | 0 |
| 0 | 0 | 0.001160216 | 0 | 0 |
| 0 | 0 | 0 | 0 | 0 |
| 0 | 0 | 0 | 0.99748939 | 0 |
| 0 | 0 | 0 | 0 | 0 |
| 0 | 0 | 0 | 0 | 0 |
| 0.004979447 | 0 | 0.002093325 | 0 | 0 |
| 0 | 0 | 0 | 0 | 0.001651412 |
| 0 | 0 | 0 | 0 | 0 |
| 0 | 0 | 0 | 0 | 0 |
| 0 | 0 | 0 | 0 | 0 |
| 0 | 0 | 0 | 0 | 0 |
| 0 | 0.002427195 | 0.004817483 | 0.009621856 | 0 |
| 0 | 0 | 0 | 0 | 0 |
| 0 | 0.007633883 | 3.861255961 | 0.008981023 | 0 |
| 0 | 0 | 0 | 0 | 0 |
| 0 | 0 | 0 | 0 | 0 |
| 0 | 0.003856248 | 0 | 0 | 0 |
| 0 | 0 | 0 | 0 | 0 |
| 0 | 0 | 0 | 0 | 0 |
| 0 | 0 | 0 | 0 | 0.002690546 |
| 0 | 0 | 0 | 0 | 0 |
| 0.003740415 | 0 | 0 | 0 | 0 |
| 0.002051065 | 0 | 0 | 0 | 0 |
| 0 | 0 | 0 | 0.003783747 | 0.00688696 |

| | | | | |
|---|---|---|---|---|
| 0 | 0 | 0 | 0.182249367 | 0.06853572 |
| 0 | 0 | 0 | 0 | 0 |
| 0 | 0.012728303 | 0.002021045 | 0.001302126 | 0 |
| 0 | 0 | 0 | 0 | 0 |
| 0 | 0 | 0.0330422 | 0.001146306 | 0.003860626 |
| 0 | 0 | 0.019479079 | 0 | 0 |
| 0 | 0 | 0 | 0 | 0 |
| 0.037183343 | 0 | 0.002493934 | 0 | 0.005093192 |
| 0 | 0 | 0 | 0 | 0 |
| 0 | 0.010175963 | 0 | 0.191641835 | 0.379616306 |
| 0 | 0 | 0 | 0 | 0 |
| 0.005534686 | 0.057903761 | 0.011563213 | 0.01090242 | 0 |
| 0.001144979 | 0.004580828 | 0.018184003 | 0 | 0.001529128 |
| 0 | 0.009821783 | 0 | 0 | 0 |
| 0 | 0 | 0 | 0 | 0.000589752 |
| 0 | 0.008899108 | 0 | 0 | 0 |
| 0 | 0 | 0 | 0 | 0.001043299 |
| 0 | 0 | 0 | 0 | 0 |
| 0 | 0 | 0 | 0 | 0.02284675 |
| 0 | 0 | 0 | 0 | 0.075232132 |
| 0 | 0 | 0 | 0 | 0 |
| 0.649338664 | 0.256510664 | 0 | 0 | 0 |
| 0 | 0 | 0 | 0 | 0 |
| 0 | 0 | 0 | 0.25369164 | 0 |
| 0 | 0 | 0 | 0 | 0 |
| 0 | 0.069989166 | 0 | 0 | 0 |
| 0 | 0 | 0 | 0 | 0 |
| 0 | 0 | 0 | 0 | 0.009153553 |
| 0 | 0 | 0 | 0 | 0.084338355 |
| 0 | 0.00904162 | 0 | 0 | 0 |
| 0 | 0 | 0 | 0 | 0.002388946 |
| 0 | 0 | 0 | 0 | 0 |
| 0 | 0 | 0 | 0 | 0 |
| 0 | 0 | 0 | 0 | 0 |
| 0 | 0 | 0 | 0 | 0 |
| 0 | 0 | 0 | 0.000651177 | 0 |
| 0 | 0.028818525 | 0 | 0 | 0 |
| 0 | 0 | 0 | 0 | 0 |
| 0 | 0 | 0 | 0 | 0 |
| 0 | 0 | 0 | 0 | 1.699462756 |
| 0 | 0 | 0 | 0 | 0 |
| 0 | 0 | 0 | 0 | 0.290223499 |
| 0 | 0 | 0 | 0 | 0 |
| 0 | 0 | 0 | 0 | 0.018294614 |
| 0 | 0.031644311 | 0 | 0 | 0 |
| 0 | 0 | 0 | 0.014329647 | 0 |
| 0.004975343 | 0.002732105 | 0 | 0 | 0.002768586 |
| 0 | 0 | 0 | 0 | 0.001256485 |

| | | | | |
|---|---|---|---|---|
| 0 | 0 | 0 | 0 | 0.005233511 |
| 0 | 0 | 0 | 0 | 0.003712906 |
| 0 | 0.001773581 | 0 | 0 | 0 |
| 0 | 0 | 0 | 0 | 0 |
| 0 | 0 | 0 | 0 | 0 |
| 0 | 0.010451909 | 0 | 0 | 0 |
| 0 | 0 | 0 | 0 | 0 |
| 0 | 0 | 0 | 0 | 0 |
| 0 | 0 | 0 | 0.456245646 | 0 |
| 0 | 0 | 0 | 0 | 0 |
| 0 | 0 | 0.007642772 | 0 | 0.015680576 |
| 0.084720984 | 0 | 0 | 0 | 0.013511089 |
| 0 | 0 | 0 | 0 | 0 |
| 0.002862601 | 0.009768467 | 0 | 0.000516895 | 0.000955756 |
| 0 | 0 | 0 | 0 | 0.052946175 |
| 0 | 0 | 0 | 0 | 0.009015629 |
| 0 | 0 | 0 | 0.000601733 | 0 |
| 0 | 0.018684789 | 0 | 0 | 0.008603472 |
| 0 | 0 | 0 | 0 | 0.037892765 |
| 0 | 0 | 0 | 0.046492198 | 0 |
| 0 | 0 | 0 | 0 | 0 |
| 0 | 0.362938918 | 0 | 0.335448815 | 0 |
| 0 | 0 | 0 | 0 | 0 |
| 0.00840939 | 0 | 0 | 0 | 0 |
| 0 | 0 | 0 | 0 | 0 |
| 0 | 0 | 0 | 0 | 0 |
| 0 | 0 | 0 | 0 | 0 |
| 0 | 0 | 0 | 0 | 0 |
| 0 | 0 | 0 | 0 | 0 |
| 0.40176446 | 0.599896896 | 0.252442042 | 0.785945444 | 0.796083177 |
| 0 | 0 | 0 | 0.002337814 | 0 |
| 0 | 0 | 0 | 0 | 0 |
| 0 | 0.005982348 | 0 | 0 | 0 |
| 0 | 0 | 0 | 0 | 0 |
| 0 | 0 | 0 | 0 | 0 |
| 0 | 0 | 0 | 0 | 0 |
| 0 | 0 | 0.006590709 | 0.071448358 | 0 |
| 0 | 0.001716667 | 0 | 0 | 0 |
| 0 | 0 | 0 | 0 | 0 |
| 0 | 0 | 0 | 0 | 0 |
| 0.001176685 | 0 | 0 | 0 | 0 |
| 0 | 0 | 0.200050038 | 0.005482044 | 0 |
| 0 | 0.000943547 | 0 | 0 | 0 |
| 0 | 0 | 0 | 0 | 0 |
| 0 | 0.003123826 | 0 | 0 | 0.001994289 |
| 0 | 0 | 0 | 0 | 0 |
| 0.019142734 | 0.002906497 | 0 | 0 | 0.007009829 |
| 0 | 0 | 0 | 0 | 0 |

| | | | | |
|---|---|---|---|---|
| 0.001523801 | 0.961441302 | 0.017367259 | 0 | 0 |
| 0 | 0 | 0 | 0 | 0 |
| 0 | 0.006840927 | 0.003103504 | 0 | 0.001386454 |
| 0 | 0 | 0 | 0 | 0 |
| 0 | 0 | 1.361367558 | 0.08949315 | 0 |
| 0 | 0 | 0 | 0 | 0 |
| 0 | 0 | 0 | 0 | 0 |
| 0 | 0.00160773 | 0 | 0 | 0 |
| 0 | 0 | 0 | 0.000921104 | 0 |
| 0 | 0 | 0 | 0 | 0 |
| 0 | 0 | 0 | 0 | 0 |
| 0.045399762 | 0 | 0 | 0 | 0 |
| 0 | 0 | 0 | 0 | 0 |
| 0 | 0 | 0 | 0 | 0 |
| 0 | 0 | 0 | 0 | 0 |
| 0 | 0.012169677 | 0.001006429 | 0.03566342 | 0.003596884 |
| 0.179311692 | 2.230151286 | 0.093257642 | 0.002012395 | 0.002591401 |
| 0 | 0 | 0 | 0 | 0.001210943 |
| 0 | 0.002612424 | 0 | 0 | 0 |
| 0 | 0.022486337 | 0 | 0 | 0 |
| 0 | 0.006374906 | 0 | 0 | 0 |
| 0.009002994 | 0.004889478 | 0 | 0 | 0.00393078 |
| 0 | 0 | 0 | 0 | 0 |
| 0 | 0.001861245 | 0.126587526 | 0 | 0 |
| 0 | 0 | 0.005838354 | 0 | 0 |
| 0 | 0 | 0 | 0 | 0.067486512 |
| 0 | 0 | 0 | 0 | 0 |
| 0 | 0 | 0 | 0 | 0 |
| 0 | 0 | 0 | 0 | 0 |
| 0 | 0 | 0 | 0 | 0 |
| 0 | 0.006782749 | 0.002800171 | 0.005689866 | 0.008852832 |
| 0.001145826 | 0.004449388 | 0.013487529 | 0 | 0 |
| 0 | 0.070478611 | 0.162358956 | 0.024821571 | 0.012293554 |
| 0.001563483 | 0.008278912 | 0.013583734 | 0.000846946 | 0 |
| 0 | 0.023242211 | 0.014073866 | 0.001813512 | 0 |
| 0 | 0 | 0 | 0 | 0 |
| 0.006904257 | 0.029595571 | 0 | 0 | 0 |
| 0.528181851 | 0.026957244 | 0.015132609 | 0.008774721 | 0.013713784 |
| 0 | 0 | 0.01557759 | 0.044387718 | 0 |
| 0 | 0 | 0.000883747 | 0 | 0 |
| 0 | 0 | 0 | 0 | 0 |
| 0 | 0 | 0 | 0 | 0.004080631 |
| 0 | 0.003257739 | 0.001293188 | 0 | 0 |
| 0 | 0.025507214 | 0.001687552 | 0 | 0 |
| 0 | 0.409303652 | 0 | 0 | 0 |
| 0 | 0 | 0 | 0 | 0 |
| 0 | 0 | 0 | 0 | 0 |
| 0 | 0 | 0 | 0 | 0 |

| | | | | |
|---|---|---|---|---|
| 0 | 0 | 0 | 0 | 0 |
| 0.006306373 | 0.09572755 | 1.280213609 | 0.033972127 | 0.263193955 |
| 0 | 0 | 0 | 0 | 0 |
| 0 | 0 | 0 | 0.001724146 | 0 |
| 0 | 0 | 0 | 0 | 0 |
| 0 | 0 | 0 | 0 | 0 |
| 0 | 0.007208409 | 0 | 0 | 0.012174435 |
| 0 | 0 | 0 | 0 | 0 |
| 0.003127023 | 0 | 0 | 0 | 0 |
| 0 | 0.002696253 | 0 | 0 | 0 |
| 0 | 0.00248149 | 0.001094499 | 0.001128268 | 0 |
| 0.047117789 | 0.965542429 | 0.068472078 | 0.002941029 | 0.00670047 |
| 0 | 0 | 0.119261891 | 0.176888551 | 0 |
| 0 | 0 | 0 | 0 | 0 |
| 0.005825052 | 0.043657052 | 0 | 0 | 0 |
| 0 | 0 | 0 | 0 | 0 |
| 0.002044691 | 0.002021038 | 0.009168789 | 0.010337774 | 0.016179395 |
| 0.008493732 | 0.002623587 | 0 | 0 | 0.136121288 |
| 0 | 0 | 0 | 0 | 0 |
| 0 | 0.019469946 | 0 | 0 | 0.004488557 |
| 0 | 0.027987833 | 0.122446582 | 0.007919491 | 0.020909098 |
| 0 | 0 | 0 | 0 | 0 |
| 0 | 0 | 0 | 0 | 0 |
| 0 | 0 | 0 | 0 | 0 |
| 0 | 0 | 0 | 0 | 0 |
| 0 | 0 | 0 | 0 | 0 |
| 0 | 0.016457679 | 0 | 0.016836471 | 0.28706033 |
| 0 | 0.015822838 | 0.008542181 | 0.034316478 | 0.057690748 |
| 0 | 0 | 0.008817739 | 0 | 0 |
| 0.005136304 | 0 | 0 | 0 | 0 |
| 0 | 0 | 0.001078326 | 0 | 0 |
| 0 | 0 | 0 | 0 | 0 |
| 0 | 0 | 0 | 0 | 0.002180114 |
| 0 | 0 | 0 | 0 | 0 |
| 0 | 0 | 0.010727997 | 0 | 0 |
| 0.785902452 | 1.132224961 | 0.076537757 | 0.114239494 | 0.926992546 |
| 0 | 0 | 0 | 0 | 0 |
| 0 | 0 | 0 | 0 | 0 |
| 0.015205712 | 0.032792324 | 0.061986681 | 0.005191809 | 0.003323019 |
| 0 | 0 | 0 | 0 | 0 |
| 0 | 0.372352105 | 0 | 0 | 0 |
| 0 | 0 | 0 | 0 | 0 |
| 0.145427875 | 0.035331728 | 0 | 0 | 0.000841022 |
| 0.013781373 | 0 | 0.006906873 | 0 | 0 |
| 0 | 0 | 0 | 0 | 0 |
| 0 | 0 | 0 | 0.188265818 | 0 |
| 0 | 0 | 0 | 0 | 0 |
| 0.011007223 | 0 | 0 | 0 | 0 |

| | | | | |
|---|---|---|---|---|
| 0 | 0 | 0 | 0 | 0 |
| 0 | 0 | 0.035737098 | 0.035256759 | 0.03475742 |
| 0 | 0 | 0 | 0 | 0 |
| 0 | 0.025132052 | 2.490808697 | 0.448520984 | 0 |
| 0 | 0 | 0 | 0 | 0 |
| 0 | 0 | 0 | 0 | 0 |
| 0 | 0 | 0 | 0 | 0 |
| 0 | 0 | 0 | 0.002780316 | 0 |
| 0 | 0 | 7.231466665 | 1.6289586 | 0 |
| 0 | 0 | 0 | 0.093430525 | 0 |
| 0 | 0 | 0 | 0 | 0 |
| 0 | 0.001852247 | 0.003361215 | 0.008662301 | 0 |
| 0 | 0 | 0 | 0 | 0 |
| 0 | 0 | 0 | 0 | 0 |
| 0 | 0.029696158 | 0.013348349 | 0.037192731 | 0 |
| 0 | 0 | 0 | 0 | 0 |
| 0 | 0 | 0 | 0.042978451 | 0 |
| 0 | 0 | 0 | 0.321599624 | 0 |
| 0 | 0 | 0 | 0 | 0 |
| 0 | 0 | 0 | 0 | 0 |
| 0 | 0 | 0 | 0 | 0 |
| 0 | 0 | 0 | 0 | 0 |
| 0 | 0 | 0 | 0.003988257 | 0 |
| 0.919497693 | 0.249242195 | 0.023544952 | 0.04819852 | 0 |
| 0 | 0.005945747 | 0 | 0.023316615 | 0 |
| 0 | 0 | 0 | 0.156718763 | 0 |
| 0 | 0 | 0 | 0.01914882 | 0 |
| 0 | 0 | 4.008725466 | 3.848164853 | 0 |
| 0 | 0 | 0 | 0 | 0 |
| 0 | 0 | 0 | 0 | 0 |
| 0 | 0.06688313 | 0 | 0.027105844 | 0.003284539 |
| 0 | 0 | 0 | 0.020083858 | 0 |
| 0 | 0 | 0 | 0.00060086 | 0 |
| 0 | 0 | 0 | 0 | 0 |
| 0 | 0 | 0 | 0 | 0 |
| 0 | 0 | 0.471871563 | 0.000557833 | 0 |
| 0 | 0.907900716 | 5.056323917 | 1.013953596 | 0 |
| 0 | 0.00137658 | 0 | 0 | 0.000976473 |
| 0 | 0 | 0.001968201 | 0.026066083 | 0.000781574 |
| 0 | 0 | 0 | 0.001743819 | 0 |
| 0.368347705 | 0.143167343 | 0.059451437 | 0.004154964 | 0.021787575 |
| 0 | 0 | 0 | 0.089652463 | 0 |
| 1.44704896 | 0.788086388 | 0.015230901 | 0.008099634 | 0.422726091 |
| 0 | 0 | 0.001580033 | 0.138220309 | 0 |
| 0 | 0 | 0 | 0 | 0 |
| 0 | 0 | 0 | 0.087391084 | 0 |
| 0 | 0 | 0 | 0 | 0 |
| 0.407139111 | 0.142809345 | 0 | 0.027471092 | 0 |

| | | | | |
|---|---|---|---|---|
| 0 | 0 | 0 | 0 | 0 |
| 0 | 0 | 0.171931556 | 0.116460603 | 0 |
| 0.892029501 | 0.187212143 | 0.029261735 | 0 | 0.024899691 |
| 0 | 0.002529592 | 0 | 0.016173834 | 0 |
| 0 | 0 | 0 | 0 | 0 |
| 0 | 0 | 0.0557987 | 0.026756364 | 0 |
| 0 | 0 | 0 | 0 | 0 |
| 0 | 0 | 0 | 0.00955596 | 0 |
| 0 | 0 | 0 | 0 | 0 |
| 0 | 0 | 0 | 0 | 0 |
| 0.4102458 | 0.016083298 | 0.052899377 | 0.258113864 | 0.013853346 |
| 0 | 0.813752396 | 0.911676767 | 1.017199421 | 1.381555452 |
| 0 | 0 | 0 | 0 | 0 |
| 0 | 0.015188032 | 0.185300202 | 0.001268376 | 0.001978821 |
| 1.263988277 | 0.070835471 | 0.11111761 | 0.001749175 | 0.843801751 |
| 0 | 0 | 0 | 0.083653424 | 0 |
| 0 | 0 | 0 | 0 | 0 |
| 0 | 0 | 0 | 0 | 0 |
| 0 | 0 | 0 | 0.051528838 | 0 |
| 0 | 0 | 0 | 0.004235416 | 0.004195408 |
| 0.004773277 | 0.051032082 | 2.307459443 | 0.059594374 | 0.101426758 |
| 0 | 0.380208496 | 0 | 0.028802198 | 19.78631551 |
| 0 | 0.169613086 | 0.211916175 | 0.266555482 | 0.757046033 |
| 0 | 0 | 0 | 0 | 0 |
| 0 | 0 | 0.045328118 | 0.022808206 | 0 |
| 0 | 0.004615226 | 2.909302859 | 0.001180363 | 0 |
| 10.52587771 | 0 | 0 | 0.017377608 | 0.00081691 |
| 0 | 0 | 0 | 0 | 0 |
| 0 | 0 | 0.001743876 | 0 | 0.000779056 |
| 0 | 0 | 0 | 0 | 0 |
| 0 | 0 | 0 | 0.011407868 | 0 |
| 0 | 0 | 0.034460655 | 0.071838232 | 0 |
| 2.995914553 | 0 | 0.000936708 | 0 | 0.057329452 |
| 0 | 0.007422777 | 0.021965095 | 0.080883567 | 0 |
| 0 | 0 | 0.006336946 | 0 | 0 |
| 0 | 0.009094252 | 0.151834195 | 0.019291218 | 0.02466551 |
| 0 | 0.001550281 | 0 | 0.00264327 | 0 |
| 0 | 0 | 0 | 0 | 0 |
| 0 | 0.055088576 | 0 | 1.207021765 | 0 |
| 0 | 0 | 0 | 0 | 0 |
| 0 | 0 | 0.05366605 | 0.011686016 | 0 |
| 0 | 0 | 0 | 0 | 0 |
| 0 | 0 | 0.008733686 | 1.060415848 | 0 |
| 0 | 0 | 0 | 0 | 0 |
| 0 | 0.001525141 | 0.046980468 | 0.084253192 | 0.006058385 |
| 0 | 0 | 0 | 0 | 0 |
| 0 | 0 | 0 | 0 | 0 |
| 0 | 0.081684341 | 0.047031323 | 0.013927399 | 0.047241435 |

| | | | | |
|---|---|---|---|---|
| 0 | 0 | 0 | 0 | 0 |
| 0 | 0 | 0 | 0 | 0 |
| 0 | 0 | 0 | 0 | 0 |
| 0 | 0 | 0 | 0.001196811 | 0 |
| 0 | 0 | 0 | 0 | 0 |
| 0 | 0 | 0.001822896 | 0.048387835 | 0 |
| 0 | 0 | 0 | 0 | 0 |
| 0 | 0.086120456 | 0.004938018 | 0.098993076 | 0 |
| 0 | 0 | 0 | 0 | 0 |
| 0 | 0 | 0 | 0 | 0 |
| 1.115892061 | 0 | 0.002758476 | 0.00088862 | 0 |
| 0 | 0 | 0 | 0 | 0 |
| 0 | 0 | 0 | 0.002852874 | 0 |
| 0 | 0 | 0 | 0.12574896 | 0 |
| 0 | 0 | 0 | 0 | 0 |
| 0 | 0 | 0 | 0 | 0 |
| 0 | 0 | 0 | 0.016302188 | 0 |
| 0 | 0 | 0.017414127 | 0.001602805 | 0 |
| 0 | 0 | 0 | 0.004411045 | 0 |
| 0 | 0.00108688 | 0 | 0 | 0 |
| 0 | 0 | 0.000850059 | 0.025850457 | 0 |
| 0 | 0 | 0 | 0 | 0 |
| 0 | 0 | 0.05493663 | 0.013219731 | 0 |
| 0 | 0 | 0 | 0 | 0 |
| 0 | 0 | 0 | 0 | 0 |
| 0 | 0 | 0 | 0 | 0 |
| 0 | 0 | 0 | 0 | 0 |
| 0 | 0 | 0 | 0 | 0 |
| 0 | 0 | 0 | 0 | 0 |
| 0 | 0 | 0.469035904 | 0.1633833 | 0 |
| 0 | 0 | 0 | 0 | 0.001617645 |
| 0 | 0 | 0 | 0 | 0 |
| 0 | 0 | 0 | 0.010047949 | 0 |
| 0 | 0 | 0 | 0 | 0 |
| 0 | 0 | 0.008956407 | 0 | 0 |
| 0 | 0 | 0 | 0 | 0 |
| 0 | 0 | 0.079180165 | 0.069044535 | 0 |
| 0.011282698 | 0.00759819 | 0 | 0 | 0.010084053 |
| 0 | 0 | 0 | 0.079224923 | 0 |
| 0 | 0.005076875 | 0 | 0 | 0 |
| 0 | 0 | 0.004929525 | 0 | 0 |
| 0 | 0 | 0 | 0 | 0 |
| 0 | 0 | 0 | 0.001272012 | 0 |
| 0 | 0 | 0 | 0.007670324 | 0 |
| 0 | 0 | 0 | 0.134813715 | 0 |
| 0 | 0 | 0 | 0 | 0 |
| 0 | 0 | 0.058611568 | 0.004169357 | 0 |
| 0 | 0 | 0 | 0 | 0 |

| | | | | |
|---|---|---|---|---|
| 0 | 0 | 0 | 0 | 0 |
| 0 | 0 | 0 | 0.042396667 | 0 |
| 0 | 0 | 0 | 0.026113978 | 0 |
| 0 | 0.012921694 | 0 | 0.419145259 | 0 |
| 0 | 0 | 0 | 0.013904644 | 0 |
| 0 | 0 | 0 | 0 | 0 |
| 0 | 0 | 0 | 0 | 0 |
| 0 | 0 | 0 | 0 | 0 |
| 2.85287926 | 0.025014799 | 0 | 0.003976909 | 0.733745417 |
| 0 | 0 | 0 | 0 | 0 |
| 0 | 0 | 0.001151375 | 0.001359989 | 0 |
| 0 | 0 | 0 | 0 | 0 |
| 0 | 0 | 0 | 0 | 0 |
| 0 | 0 | 0 | 0 | 0 |
| 0 | 0 | 1.209237164 | 0.170480634 | 0 |
| 1.475518226 | 0 | 0 | 0.002265519 | 0.045832831 |
| 0 | 0 | 0 | 0 | 0 |
| 0 | 0 | 0 | 0 | 0 |
| 0 | 0 | 0 | 0 | 0 |
| 0 | 0 | 0 | 0 | 0 |
| 0 | 0 | 0 | 0.028954818 | 0 |
| 0 | 0 | 0 | 0 | 0 |
| 0 | 0 | 0 | 0 | 0 |
| 0 | 0 | 0 | 0 | 0 |
| 0.080796985 | 0.155033915 | 0.44657461 | 0.006506757 | 0 |
| 0 | 0 | 0.713919744 | 1.2158453 | 0 |
| 0 | 0 | 0.035919708 | 0.059898169 | 0 |
| 0 | 0 | 0 | 0 | 0 |
| 0 | 0 | 0 | 0 | 0 |
| 0 | 0 | 0 | 3.308160184 | 0 |
| 0 | 0 | 0 | 0 | 0 |
| 0 | 0 | 0 | 0.007202228 | 0 |
| 0 | 0 | 0 | 0 | 0 |
| 0 | 0.010190531 | 0 | 0.037465122 | 0 |
| 0 | 0.137824287 | 0.012497338 | 0 | 0.00099254 |
| 0.140968314 | 0.000989087 | 0 | 0 | 0.002806423 |
| 0 | 0 | 0 | 0 | 0 |
| 0 | 2.937036549 | 0.471052498 | 0.283496618 | 0 |
| 0 | 0 | 0.001735544 | 17.87047012 | 0 |
| 0 | 0 | 0 | 0 | 0 |
| 0 | 0 | 0 | 0 | 0 |
| 0 | 0 | 0 | 0.001290879 | 0 |
| 0.028955181 | 2.542706729 | 4.722013948 | 0.093333167 | 0.221472808 |
| 0 | 0 | 0 | 0 | 0 |
| 0 | 0 | 0 | 0 | 0 |
| 0 | 0.012415723 | 0 | 0.000838777 | 0 |
| 0 | 0 | 0 | 0 | 0 |
| 0.006343179 | 0.416867249 | 3.506024514 | 0.000572688 | 0.00132365 |

| | | | | |
|---|---|---|---|---|
| 0 | 0 | 0 | 0.015658658 | 0.001184457 |
| 0 | 0 | 0 | 0 | 0 |
| 0 | 0 | 0 | 0 | 0 |
| 0 | 0 | 0 | 0 | 0 |
| 0.006867843 | 0 | 0 | 0.005243912 | 0 |
| 0 | 0 | 0 | 0.001228332 | 0 |
| 0 | 0 | 0 | 0 | 0 |
| 0 | 0 | 0 | 0 | 0 |
| 0 | 0 | 0 | 0 | 0 |
| 0 | 0 | 0 | 0.00867744 | 0 |
| 0 | 0 | 0 | 0 | 0 |
| 0 | 2.884178869 | 0.282901895 | 1.288435363 | 0.321125532 |
| 0 | 0 | 0 | 0.22480513 | 0.297475452 |
| 0 | 0 | 0 | 0 | 0 |
| 0.080334339 | 2.457134408 | 0.059021504 | 0.068513407 | 0.011137855 |
| 0 | 0 | 0 | 0 | 0 |
| 0 | 0 | 0 | 0.006955309 | 0 |
| 0 | 0 | 0 | 0 | 0 |
| 0 | 0 | 0.00334475 | 0.003878941 | 0 |
| 0 | 0.085644342 | 0 | 0.000477903 | 0 |
| 0 | 0 | 0.120734789 | 0.003925054 | 0 |
| 0 | 0 | 0 | 0 | 0 |
| 0 | 3.913560715 | 0.651113498 | 0.971852518 | 0.08000822 |
| 0 | 0 | 0 | 0 | 0 |
| 0 | 0 | 0 | 0 | 0 |
| 0 | 0 | 0 | 0 | 0 |
| 0 | 0 | 0 | 0 | 0 |
| 0 | 0 | 0 | 0 | 0 |
| 0 | 0 | 0 | 0.00679141 | 0 |
| 0 | 0 | 0 | 0 | 0 |
| 0 | 0 | 0 | 0 | 0 |
| 0.012662533 | 0.029522921 | 0.301732894 | 0.011485429 | 0.029495764 |
| 0 | 0 | 0 | 0.113406249 | 0 |
| 0 | 0.031783364 | 0.011544681 | 0.068111263 | 0 |
| 0.13625755 | 0.003229123 | 0 | 0 | 0.003244972 |
| 0 | 0.044749363 | 0.638389982 | 0.034955527 | 0 |
| 0 | 0 | 0 | 0.00256582 | 0 |
| 0 | 0 | 0.09826235 | 0.129118381 | 0 |
| 0 | 0 | 0 | 0.02229663 | 0 |
| 0 | 0 | 0 | 0 | 0 |
| 0 | 0 | 0.488543042 | 0 | 0 |
| 0 | 0 | 0 | 0 | 0 |
| 0 | 0 | 0 | 0 | 0 |
| 0 | 0 | 0 | 0 | 0 |
| 0.06476704 | 0.019616229 | 0.025465057 | 0.059389598 | 0 |
| 0 | 0 | 0 | 0 | 0 |
| 0 | 0 | 0 | 0.061833868 | 0 |
| 0 | 0 | 0 | 0 | 0 |

| | | | | |
|---|---|---|---|---|
| 0 | 0 | 0 | 0 | 0 |
| 0 | 0 | 0 | 0 | 0 |
| 0 | 0 | 0 | 0 | 0.001944769 |
| 0 | 0 | 0 | 0 | 0 |
| 0 | 0 | 0 | 0 | 0 |
| 0 | 0 | 0 | 0 | 0 |
| 0 | 0 | 0.608821253 | 0 | 0 |
| 0 | 0.697276077 | 0.021995188 | 0.005562502 | 0.014509558 |
| 0 | 0 | 0 | 0 | 0 |
| 0 | 0 | 0 | 0.007167959 | 0 |
| 0 | 0 | 0 | 0 | 0.380540397 |
| 0.068155945 | 0 | 0 | 0 | 0 |
| 0 | 0 | 0 | 0 | 0 |
| 0 | 0 | 0.578002608 | 0.296632729 | 0 |
| 0 | 0 | 0 | 0 | 0 |
| 0 | 0 | 0 | 0 | 0 |
| 0 | 0 | 0.003136892 | 0.006568404 | 0 |
| 0 | 0 | 0 | 0 | 0 |
| 0 | 0 | 0.07535599 | 1.725345153 | 0.023897573 |
| 0 | 0 | 0 | 0 | 0 |
| 0 | 0 | 0 | 0.000382253 | 0.017139872 |
| 0 | 0 | 0 | 0 | 0 |
| 0 | 0 | 0 | 0 | 0 |
| 0 | 0 | 0 | 0 | 0 |
| 0 | 0.008688824 | 0.069671935 | 0 | 0 |
| 0.151700399 | 0.001974267 | 0.012539243 | 0 | 0.003676155 |
| 0 | 0 | 0 | 0 | 0 |
| 0.004960888 | 1.415252057 | 1.472810927 | 0 | 0.022617412 |
| 0 | 0 | 0.212708787 | 0 | 0 |
| 0.007164445 | 0 | 0.054080042 | 0.379821863 | 0 |
| 0 | 0 | 0 | 0 | 0 |
| 0 | 0 | 0 | 0 | 0 |
| 0 | 0 | 0 | 0.000913249 | 0 |
| 0 | 0 | 0 | 0 | 0 |
| 0.176005417 | 0 | 0 | 0 | 0.002201362 |
| 0 | 0 | 0 | 0 | 0 |
| 0 | 0 | 0 | 0.582448837 | 0 |
| 0.01433955 | 0.004401762 | 0 | 0 | 0 |
| 0 | 0 | 0 | 0 | 0 |
| 0 | 0 | 0 | 0 | 0 |
| 0.473132643 | 22.13288265 | 2.400043573 | 3.392772974 | 15.83448266 |
| 0 | 0.296648883 | 0.224600281 | 0.231398149 | 0.33765833 |
| 0 | 0 | 0 | 0 | 0 |
| 0 | 0 | 0 | 0.18776419 | 0 |
| 0 | 0 | 0 | 0 | 0 |
| 0 | 0 | 0 | 0 | 0 |
| 0 | 0 | 0 | 0.007835399 | 0 |
| 0 | 0 | 0.001109415 | 0.003335629 | 0 |

| | | | | |
|---|---|---|---|---|
| 0 | 0 | 0 | 0 | 0 |
| 0 | 0.001197778 | 0 | 0.020463283 | 0.001359424 |
| 0 | 0 | 0 | 0.004293406 | 0 |
| 0 | 0 | 0 | 0 | 0 |
| 0 | 3.370488829 | 3.578296395 | 1.074718462 | 2.375845984 |
| 0 | 0 | 0 | 0 | 0 |
| 0 | 0 | 0 | 0 | 0 |
| 0 | 0 | 0.006452737 | 0.089903592 | 0 |
| 0.697425159 | 0.110511524 | 0 | 0 | 0.49749444 |
| 0 | 0 | 0 | 0 | 0 |
| 0 | 0 | 0 | 0 | 0.0011519 |
| 0 | 0 | 0 | 0 | 0 |
| 0 | 0.050743666 | 0 | 0.053340347 | 0.174525548 |
| 0 | 0.077837138 | 0.009542073 | 0.013466609 | 0.033696494 |
| 0 | 0 | 2.16327555 | 0 | 0 |
| 0 | 0.926387415 | 0.121926714 | 0.139333679 | 0.001665729 |
| 0 | 0.049774081 | 0.016907567 | 0.000506664 | 0 |
| 0 | 0 | 0 | 0 | 0 |
| 0 | 0 | 0 | 0 | 0 |
| 0 | 0 | 0.010854672 | 0 | 0 |
| 0 | 0 | 0 | 0 | 0 |
| 0 | 0 | 0 | 0 | 0 |
| 0 | 0 | 0 | 0 | 0 |
| 0 | 0 | 0 | 0 | 0 |
| 0 | 0 | 0 | 0 | 0 |
| 0 | 0 | 0 | 0 | 0 |
| 0 | 0 | 0.035728853 | 0 | 0 |
| 0.030112376 | 0 | 0 | 0 | 0 |
| 0 | 0 | 0 | 0 | 0 |
| 0 | 0 | 0 | 0 | 0 |
| 0 | 0 | 0 | 0 | 0 |
| 0 | 0 | 0 | 0 | 0 |
| 0 | 0 | 0 | 0 | 0 |
| 0 | 0 | 0.045092887 | 0.005471791 | 0 |
| 0 | 0 | 1.169496557 | 0.004468534 | 0 |
| 1.15807352 | 0 | 0 | 0.00287537 | 0 |
| 0 | 0 | 0 | 0 | 0 |
| 0 | 0 | 0.074170601 | 0 | 0 |
| 0 | 0 | 0 | 0 | 0 |
| 0 | 0 | 0 | 0 | 0 |
| 0 | 4.040640658 | 0.952706192 | 1.122506305 | 0 |
| 0 | 0 | 0 | 0.061371734 | 0 |
| 0 | 0 | 0 | 0.008710565 | 0 |
| 0 | 0 | 0 | 0 | 0 |
| 0 | 0 | 0 | 0 | 0 |
| 0 | 0 | 0 | 0 | 0 |
| 0 | 0 | 0 | 0.02839565 | 0 |
| 0 | 0 | 0 | 0 | 0 |

| | | | | |
|---|---|---|---|---|
| 0 | 0.019686319 | 0.983590927 | 0.046538392 | 0 |
| 0 | 0 | 0 | 0 | 0 |
| 0 | 0 | 0 | 0 | 0 |
| 0 | 0 | 0 | 0 | 0 |
| 0 | 0 | 0 | 0.018589395 | 0 |
| 0 | 0 | 0 | 0 | 0 |
| 0.041461917 | 0.171685923 | 0.42440708 | 0.008592336 | 0.00070092 |
| 0 | 0 | 0 | 0 | 0 |
| 0 | 0 | 0 | 0 | 0 |
| 0 | 0 | 0 | 0.00503766 | 0.021877534 |
| 0 | 0 | 0 | 0.006435644 | 0 |
| 0 | 0 | 0 | 0 | 0 |
| 0 | 0 | 0 | 0 | 0 |
| 0 | 0 | 0 | 0 | 0 |
| 0 | 0 | 0 | 0 | 0 |
| 0 | 0 | 0 | 0 | 0 |
| 0 | 0 | 0 | 0 | 0.149895971 |
| 0 | 0 | 0 | 0 | 0 |
| 0 | 0 | 0 | 0 | 0 |
| 0 | 0 | 0 | 0 | 0 |
| 0 | 0.041392811 | 0.018485148 | 0 | 0 |
| 0 | 0 | 0 | 0 | 0 |
| 0 | 0.003631737 | 0.003501151 | 0.086115944 | 0 |
| 0 | 0.00069201 | 0 | 0.000589948 | 0 |
| 0 | 0 | 0 | 0 | 0 |
| 0 | 0 | 0.000864223 | 0.000974408 | 0 |
| 0 | 0 | 0 | 0 | 0 |
| 0 | 0 | 0 | 0 | 0 |
| 0 | 0 | 0.115296966 | 0 | 0 |
| 0.207133097 | 4.277997897 | 0.008170707 | 0 | 0 |
| 0 | 0 | 0 | 0.155349198 | 0 |
| 0 | 0 | 0 | 0 | 0 |
| 0 | 0 | 0 | 0 | 0 |
| 0 | 0 | 0 | 0 | 0 |
| 0 | 0 | 0 | 0 | 0 |
| 0 | 0 | 0 | 0 | 0 |
| 0.396339234 | 0 | 0.005728667 | 0 | 0.082713833 |
| 0 | 0 | 0 | 0 | 0 |
| 0 | 0 | 0 | 0 | 0 |
| 0 | 0 | 0 | 0 | 0 |
| 0 | 0 | 0 | 0 | 0 |
| 0 | 0 | 0 | 0 | 0 |
| 0 | 0 | 0 | 0 | 0 |
| 0 | 0 | 0 | 0.004930796 | 0 |
| 0 | 0 | 0 | 0 | 0 |
| 0 | 0 | 0 | 0 | 0 |
| 0 | 0 | 0 | 0 | 0 |
| 0 | 0 | 0 | 0 | 0 |

| | | | | |
|---|---|---|---|---|
| 0 | 0.003474926 | 0.00078823 | 0 | 0 |
| 0 | 0 | 0 | 0 | 0 |
| 0 | 0 | 0 | 0 | 0 |
| 0 | 0 | 0 | 0 | 0 |
| 0 | 0 | 0 | 0 | 0 |
| 0 | 0 | 0 | 0 | 0 |
| 0 | 0 | 0 | 0 | 0 |
| 0 | 3.851616466 | 0.181290832 | 0.609564028 | 0.64104683 |
| 0.434010508 | 0.137918852 | 3.816902021 | 0.193106381 | 0.008567236 |
| 0 | 0.024139356 | 0 | 0 | 0 |
| 0 | 0 | 0 | 0.298907894 | 0 |
| 0 | 0 | 0 | 0.001943041 | 0 |
| 0 | 0 | 0 | 0.012495494 | 0 |
| 0 | 0 | 0 | 0.05650536 | 0 |
| 0 | 0 | 0 | 0 | 0 |
| 0 | 0 | 0 | 0 | 0 |
| 0.000769773 | 0 | 0 | 0 | 0.103574779 |
| 0 | 0 | 0 | 0 | 0 |
| 0 | 0 | 0 | 0 | 0 |
| 0 | 0 | 0 | 0 | 0 |
| 0 | 0 | 0 | 0 | 0 |
| 0 | 0 | 0 | 0 | 0 |
| 0 | 0 | 0 | 0.024719523 | 0 |
| 0 | 0 | 0 | 0 | 0 |
| 0 | 0 | 0 | 0.016499885 | 0 |
| 0 | 0 | 0 | 0 | 0 |
| 0 | 0 | 0 | 0 | 0 |
| 0 | 0 | 0 | 0.274169882 | 0 |
| 0 | 0 | 0 | 0 | 0 |
| 0 | 0 | 0 | 0 | 0 |
| 0 | 0 | 0 | 0.064823946 | 0 |
| 0 | 0 | 0 | 0 | 0 |
| 0 | 0 | 0 | 0 | 0 |
| 0 | 0 | 0 | 0.003735242 | 0 |
| 0 | 0 | 0 | 0 | 0 |
| 0 | 0 | 0 | 0 | 0 |
| 0 | 0 | 0 | 0 | 0 |
| 0 | 0 | 0 | 0 | 0 |
| 0 | 0 | 0 | 0.04347833 | 0 |
| 0.002807731 | 0.000743371 | 0.003344327 | 0 | 0 |
| 0 | 0 | 0 | 0 | 0 |
| 0 | 0 | 0.061495927 | 0.066950015 | 0 |
| 0 | 0 | 0 | 0 | 0 |
| 0 | 0 | 0 | 0 | 0 |
| 0 | 0 | 0 | 0 | 0 |
| 0 | 0 | 0 | 0.000338682 | 0 |
| 0 | 0 | 0 | 0 | 0 |
| 0 | 0 | 0 | 0 | 0 |

| | | | | |
|---|---|---|---|---|
| 0 | 0 | 0 | 0 | 0 |
| 0 | 0.013889207 | 0 | 0.005385625 | 0 |
| 0 | 0 | 0 | 0 | 0 |
| 0 | 0 | 0 | 0 | 0 |
| 0 | 0.514605186 | 1.240084972 | 1.397670272 | 0 |
| 0 | 0 | 0 | 0.005417345 | 0 |
| 0 | 0 | 0 | 0 | 0 |
| 0.180448807 | 0.014839771 | 0 | 0 | 0.125422667 |
| 0 | 0 | 0 | 0.001935063 | 0 |
| 0 | 0 | 0 | 0 | 0 |
| 0 | 0 | 0 | 0 | 0 |
| 0 | 0 | 0 | 1.17865087 | 0 |
| 0 | 0 | 0 | 0 | 0 |
| 0 | 0 | 0 | 0 | 0 |
| 0 | 0 | 0 | 0 | 0 |
| 0 | 0 | 0.001304433 | 0 | 0 |
| 0 | 0 | 0 | 0 | 0 |
| 0 | 0 | 0 | 0 | 0 |
| 0 | 0 | 0 | 0 | 0 |
| 0 | 0 | 0 | 0 | 0 |
| 0 | 0 | 0 | 0 | 0 |
| 0 | 0 | 0 | 0 | 0 |
| 0 | 0 | 0 | 0 | 0 |
| 0.121525068 | 0 | 0.000867742 | 0 | 0 |
| 0 | 0 | 0.358422831 | 0.119131406 | 0 |
| 0 | 0 | 0 | 0 | 0 |
| 0.155041492 | 0.199932165 | 0 | 0.112877104 | 0.100161265 |
| 0 | 0 | 0 | 0 | 0 |
| 0.010522833 | 0 | 0 | 0 | 0 |
| 0 | 0 | 0 | 0 | 0 |
| 0 | 0 | 0 | 0 | 0 |
| 0 | 0 | 0 | 0 | 0 |
| 0 | 0 | 0 | 0 | 0 |
| 0 | 0 | 0 | 0 | 0 |
| 0 | 0 | 0 | 0 | 0 |
| 0 | 0 | 0 | 0 | 0 |
| 0 | 0 | 0 | 0 | 0 |
| 0 | 0 | 0.257406298 | 0.073285397 | 0 |
| 0 | 0 | 0 | 0 | 0 |
| 0 | 0 | 0 | 0 | 0 |
| 0 | 0 | 0 | 0 | 0 |
| 0 | 0 | 0 | 0 | 0 |
| 0 | 0 | 0 | 0 | 0 |
| 0 | 0 | 0 | 0.023148126 | 0 |
| 0 | 0 | 0 | 0 | 0 |
| 0 | 0 | 0 | 0 | 0 |
| 0 | 0 | 0 | 0 | 0 |
| 0 | 0 | 0 | 0.061715881 | 0 |

| | | | | |
|---|---|---|---|---|
| 0 | 0 | 0 | 0.081987668 | 0 |
| 0 | 0 | 0 | 0 | 0 |
| 0 | 0 | 0.000554482 | 0 | 0 |
| 0 | 0 | 0 | 0 | 0 |
| 0 | 0 | 0 | 0 | 0 |
| 0 | 0 | 0.003891145 | 0 | 0 |
| 0 | 0 | 0 | 0 | 0 |
| 0 | 0 | 0 | 0 | 0 |
| 0 | 0 | 0 | 0 | 0 |
| 0 | 0 | 0 | 0 | 0 |
| 0 | 0 | 0 | 0 | 0 |
| 0 | 0 | 0 | 0.004123 | 0 |
| 0 | 0 | 0 | 0.006831333 | 0 |
| 0 | 0 | 0 | 0 | 0 |
| 0 | 0 | 0 | 0 | 0 |
| 0 | 0 | 0 | 0 | 0 |
| 0 | 0 | 0 | 0 | 0 |
| 0 | 0.176820269 | 0 | 0 | 0 |
| 0 | 0 | 0 | 0 | 0 |
| 0 | 0 | 0 | 0 | 0 |
| 0 | 0.04734619 | 0 | 0 | 0 |
| 0 | 0 | 0 | 0 | 0 |
| 0 | 0 | 0.007366367 | 0.019863008 | 0 |
| 0 | 0.005661446 | 0 | 0.014157611 | 0 |
| 0 | 0 | 0 | 0 | 0 |
| 0 | 0 | 0 | 0 | 0 |
| 0 | 0 | 0 | 0 | 0 |
| 0 | 0 | 0 | 0 | 0 |
| 0 | 0 | 0 | 0 | 0 |
| 0 | 0 | 0 | 0 | 0 |
| 0 | 0 | 0 | 0 | 0 |
| 0 | 0 | 0 | 0 | 0 |
| 0.085011945 | 0.004183727 | 0.002299524 | 0.010864666 | 0.228285666 |
| 0 | 0 | 0 | 0 | 0 |
| 0 | 0 | 0.006856378 | 0.209534425 | 0.26219337 |
| 0.004023613 | 0 | 0 | 0 | 0.017769399 |
| 0 | 0 | 0 | 0 | 0 |
| 0 | 0 | 0 | 0 | 0 |
| 0 | 0.002090958 | 0 | 0.000611167 | 0 |
| 0 | 0 | 0 | 0 | 0 |
| 0 | 0 | 0 | 0 | 0 |
| 0 | 0 | 0 | 0 | 0 |
| 0 | 0 | 0.613108592 | 0.20992469 | 0 |
| 0 | 0 | 0 | 0 | 0 |
| 0 | 0 | 0 | 0 | 0 |
| 0 | 0 | 0.001761789 | 0.016175059 | 0 |
| 0 | 0 | 0 | 0 | 0 |
| 0 | 0 | 0 | 0 | 0 |

| | | | | |
|---|---|---|---|---|
| 0 | 0 | 0 | 0 | 0 |
| 0 | 0 | 0 | 0 | 0 |
| 0 | 0 | 0 | 0 | 0 |
| 0 | 0 | 0 | 0 | 0 |
| 0 | 0 | 0 | 0.002380051 | 0 |
| 0 | 0 | 0 | 0 | 0 |
| 0 | 0 | 0 | 0 | 0 |
| 0 | 0.003752633 | 0 | 0 | 0 |
| 0 | 0 | 0 | 0.098238578 | 0 |
| 0 | 0 | 0.001360968 | 0.039458207 | 0.000810662 |
| 0 | 0 | 0 | 0 | 0 |
| 0 | 0 | 0 | 0 | 0 |
| 0 | 0 | 0 | 0 | 0 |
| 0 | 0 | 0 | 0 | 0 |
| 0 | 0 | 0 | 0 | 0 |
| 5.632054683 | 0 | 0 | 0 | 0.160916146 |
| 0 | 0 | 0 | 0 | 0 |
| 0 | 0 | 0 | 0 | 0 |
| 0 | 0 | 0 | 0 | 0 |
| 0 | 0 | 1.108008805 | 0.274353079 | 0 |
| 0 | 0.008807803 | 0.00489487 | 0 | 0 |
| 0 | 0 | 0 | 0 | 0 |
| 0 | 0 | 0 | 0 | 0 |
| 0 | 0 | 0 | 0 | 0 |
| 0 | 0 | 0 | 0.003497978 | 0 |
| 0 | 0 | 0 | 0 | 0 |
| 0 | 0 | 0 | 0 | 0 |
| 0 | 0 | 0 | 0 | 0 |
| 0 | 0 | 0 | 0 | 0 |
| 0 | 0 | 0.132073641 | 0.0104317 | 0 |
| 0 | 0 | 0.219118234 | 0.187102638 | 0 |
| 0 | 0 | 0 | 0 | 0 |
| 0 | 0 | 0 | 0 | 0 |
| 0 | 0 | 0 | 0 | 0 |
| 0 | 0 | 0 | 0 | 0 |
| 0 | 0 | 0 | 0 | 0 |
| 0 | 0 | 0 | 0 | 0 |
| 0 | 0 | 0 | 0 | 0 |
| 0 | 0 | 0.1289971 | 0 | 0 |
| 0 | 0 | 0 | 0 | 0 |
| 0 | 0 | 0 | 0.002748056 | 0 |
| 0 | 0 | 0 | 0 | 0 |
| 0 | 0 | 0 | 0 | 0 |
| 0 | 0 | 0 | 0 | 0 |
| 0 | 0 | 0 | 0 | 0 |
| 0 | 0 | 0 | 0 | 0 |
| 0 | 0.002302115 | 0.002741535 | 0.000441581 | 0.300675778 |
| 0 | 0 | 0 | 0.001196242 | 0 |

| | | | | |
|---|---|---|---|---|
| 0 | 0 | 0 | 0 | 0 |
| 0 | 0 | 0 | 0 | 0 |
| 0 | 0 | 0 | 0 | 0 |
| 0 | 0 | 0 | 0 | 0 |
| 0 | 0.052885443 | 0.017409115 | 0.031999744 | 0 |
| 0 | 0 | 0 | 0 | 0 |
| 0 | 0 | 0 | 0 | 0 |
| 0 | 0 | 0 | 0 | 0 |
| 0 | 0 | 0 | 0 | 0 |
| 0 | 0 | 0 | 0 | 0 |
| 0 | 0 | 0 | 0.005584681 | 0 |
| 0.015218963 | 0.012834231 | 0.007345791 | 0.068396189 | 0 |
| 0 | 0 | 0 | 0.001712305 | 0 |
| 0 | 0 | 0 | 0 | 0 |
| 0 | 0 | 0 | 0 | 0.001290758 |
| 0 | 0 | 0 | 0.004036569 | 0 |
| 0 | 0 | 0.002733767 | 0.023366858 | 0 |
| 0 | 0 | 0 | 0 | 0 |
| 0 | 0 | 0.000717713 | 0.000809219 | 0.124725017 |
| 0.094041773 | 0.009738029 | 0.75133148 | 0.007094266 | 0 |
| 0 | 0 | 0 | 0 | 0 |
| 0 | 0 | 0 | 0.020513134 | 0 |
| 0 | 0 | 0 | 0 | 0 |
| 0 | 0 | 0 | 0 | 0 |
| 0 | 0 | 0 | 0 | 0 |
| 0 | 0 | 0 | 0 | 0 |
| 0.143351433 | 0 | 0 | 0 | 0 |
| 0 | 0 | 0 | 0 | 0 |
| 0 | 0 | 0 | 0 | 0 |
| 0 | 0 | 0 | 0 | 0 |
| 0 | 0 | 0 | 0.002742213 | 0 |
| 0 | 0 | 0 | 0.002073337 | 0 |
| 0 | 0 | 0 | 0 | 0 |
| 0 | 0 | 0 | 0 | 0 |
| 0 | 0 | 0 | 0.003542126 | 0 |
| 0 | 0 | 0 | 0 | 0 |
| 0 | 0 | 0 | 0 | 0 |
| 0.094159648 | 0.007156366 | 0.040069991 | 0.002504579 | 0.005744287 |
| 0 | 0 | 0.240800104 | 0.047968201 | 0 |
| 0 | 0 | 0 | 0 | 0 |
| 0 | 0 | 0 | 0 | 0 |
| 0 | 0 | 0 | 0 | 0 |
| 0 | 0 | 0 | 0 | 0 |
| 0 | 0 | 0 | 0 | 0.002179036 |
| 0 | 0 | 0 | 0 | 0 |
| 0 | 0 | 0 | 0.001021636 | 0 |
| 0 | 0 | 0 | 0 | 0 |
| 0 | 0 | 0 | 0 | 0 |

| | | | | |
|---|---|---|---|---|
| 0 | 0 | 0 | 0 | 0 |
| 0 | 0 | 0 | 0.06067907 | 0 |
| 0 | 0 | 0 | 0.336455492 | 0 |
| 0.06549594 | 0.000878006 | 0 | 0 | 0 |
| 0.000768942 | 0 | 0 | 0 | 0 |
| 0.018607383 | 0 | 0 | 0 | 0.001491018 |
| 0 | 0 | 0 | 0.004939342 | 0 |
| 0 | 0.001373353 | 0 | 0 | 0.001136548 |
| 0 | 0 | 0 | 0 | 0 |
| 0 | 0 | 0 | 0.015742068 | 0 |
| 0 | 0 | 0 | 0.01249384 | 0 |
| 0 | 0 | 0 | 0 | 0 |
| 0 | 0.001244765 | 0.019105993 | 0.109619784 | 0 |
| 0 | 0 | 0 | 0.001198393 | 0 |
| 0 | 0 | 0 | 0 | 0 |
| 0 | 0 | 1.784629328 | 0 | 0 |
| 0 | 0 | 0 | 0 | 0 |
| 0 | 0 | 0 | 0 | 0 |
| 0 | 0 | 0 | 0 | 0 |
| 0 | 0 | 0 | 0 | 0 |
| 0 | 0 | 0 | 0 | 0 |
| 0 | 0 | 0 | 0 | 0 |
| 0 | 0.012019838 | 0 | 0 | 0 |
| 0 | 0.003223596 | 0.006654101 | 0.003132901 | 0 |
| 0 | 0 | 0 | 0 | 0 |
| 0 | 0 | 0 | 0 | 0 |
| 0 | 0 | 0.001036055 | 0.005740612 | 0 |
| 0 | 0 | 0 | 0 | 0 |
| 0 | 0 | 0 | 0 | 0 |
| 0 | 0 | 0 | 0 | 0 |
| 0 | 0 | 0.001450928 | 0.007322669 | 0 |
| 0 | 0 | 0 | 0 | 0 |
| 0 | 0 | 0 | 0 | 0 |
| 0 | 0 | 0 | 0 | 0 |
| 0 | 0 | 0.005879389 | 0.000914343 | 0 |
| 0.142739823 | 0 | 0 | 0 | 0 |
| 0 | 0 | 0 | 0 | 0 |
| 0 | 0 | 0 | 0 | 0 |
| 0 | 0.00489642 | 0.00593333 | 0.017531906 | 0 |
| 0 | 0 | 0 | 0 | 0 |
| 0 | 0 | 0 | 0 | 0 |
| 0 | 0 | 0 | 0 | 0 |
| 0 | 0 | 0 | 0 | 0 |
| 0 | 0 | 0 | 0 | 0 |
| 0 | 0 | 0 | 0 | 0 |
| 0 | 0.221977448 | 0.467851388 | 0.046739459 | 0.050181014 |
| 0.00095557 | 0 | 0 | 0 | 0.021248249 |
| 0 | 0 | 0 | 0 | 0 |

| | | | | |
|---|---|---|---|---|
| 0 | 0 | 0 | 0 | 0 |
| 0 | 0 | 0 | 0 | 0 |
| 0 | 0 | 0 | 0.034379839 | 0 |
| 0 | 0 | 0 | 0.038522768 | 0 |
| 0 | 0 | 0 | 0 | 0.001628429 |
| 0 | 0 | 0 | 0.035449668 | 0 |
| 0 | 0 | 0 | 0 | 0.000557449 |
| 0 | 0 | 0 | 0 | 0 |
| 0 | 0 | 0 | 0.000365902 | 0 |
| 0 | 0 | 0 | 0 | 0 |
| 0 | 0 | 0 | 0.002245748 | 0 |
| 0.005747441 | 0 | 0 | 0 | 0.001107079 |
| 0 | 0 | 0 | 0 | 0 |
| 0 | 0 | 0 | 0 | 0 |
| 0 | 0 | 0 | 0 | 0 |
| 0 | 0 | 0 | 0 | 0 |
| 0 | 0 | 0 | 0 | 0 |
| 0 | 0 | 0 | 0 | 0 |
| 0 | 0 | 0 | 0 | 0 |
| 0 | 0.003049873 | 0 | 0 | 0 |
| 0 | 0 | 0 | 0 | 0 |
| 0 | 0 | 0 | 0 | 0 |
| 0 | 0 | 0 | 0 | 0 |
| 0 | 0 | 0 | 0 | 0 |
| 0 | 0 | 0 | 0 | 0.000680537 |
| 0 | 0 | 0 | 0 | 0 |
| 0 | 0 | 0 | 0 | 0 |
| 0 | 0 | 0.007551688 | 0 | 0 |
| 0 | 0 | 0 | 0 | 0 |
| 0 | 0 | 0 | 0 | 0 |
| 0 | 0.00547851 | 0.00397127 | 0.00146207 | 0.002703418 |
| 0 | 0 | 0 | 0 | 0 |
| 0 | 0.0486763 | 0.001976164 | 0.016410256 | 0.037471143 |
| 0 | 0 | 0.019745487 | 0.004481505 | 0.002205266 |
| 0 | 0 | 0 | 0 | 0 |
| 0 | 0 | 0 | 0 | 0 |
| 0 | 0 | 0 | 0 | 0 |
| 0 | 0 | 0 | 0 | 0 |
| 0 | 0 | 0 | 0 | 0 |
| 0 | 0 | 0 | 0 | 0 |
| 0 | 0 | 0 | 0 | 0 |
| 0.386887692 | 0.402730823 | 0 | 0.017690401 | 0.000484196 |
| 0 | 0.13111445 | 0.033671024 | 0.072454071 | 0.01789326 |
| 0 | 0 | 0 | 0 | 0 |
| 0 | 0 | 0 | 0 | 0 |
| 0 | 0.010414814 | 0.291941827 | 0.039544669 | 0 |
| 0 | 0 | 0 | 0.009537768 | 0 |
| 0 | 0 | 0 | 0 | 0 |

| | | | | |
|---|---|---|---|---|
| 0 | 0 | 0 | 0 | 0 |
| 0 | 0.004646419 | 0 | 0.398985317 | 0 |
| 0 | 0.112960022 | 0.021275663 | 0 | 0.003609362 |
| 0 | 0 | 0 | 0 | 0 |
| 0 | 0 | 0.009676477 | 0.02382924 | 0 |
| 0 | 0 | 0 | 0.00067428 | 0.010098814 |
| 0 | 0 | 0 | 0 | 0 |
| 0 | 0 | 0 | 0 | 0 |
| 0 | 0 | 0.157294294 | 0.021483963 | 0 |
| 0 | 0 | 0 | 0 | 0 |
| 0 | 0 | 0 | 0 | 0 |
| 0 | 0 | 0 | 0 | 0 |
| 0 | 0 | 0 | 0 | 0 |
| 0 | 0 | 0 | 0 | 0 |
| 0 | 0 | 0 | 0 | 0 |
| 0 | 0 | 0 | 0 | 0 |
| 0 | 0 | 0.592201108 | 0 | 0 |
| 0 | 0 | 0 | 0 | 0 |
| 0 | 0 | 0 | 0 | 0 |
| 0 | 0 | 0 | 0 | 0 |
| 0 | 0 | 0 | 0 | 0 |
| 0 | 0 | 0 | 0 | 0 |
| 0 | 0 | 0 | 0.027075216 | 0 |
| 0 | 0 | 0 | 0 | 0 |
| 0 | 0 | 0.029075595 | 0.003145988 | 0 |
| 0.054373014 | 0.341138247 | 0 | 0 | 0 |
| 0 | 0 | 0 | 0 | 0 |
| 0 | 0.011457932 | 0.156159249 | 0.000697717 | 0 |
| 0 | 0 | 0 | 0 | 0 |
| 0 | 0 | 0 | 0 | 0 |
| 0 | 0 | 0 | 0 | 0 |
| 0 | 0 | 0 | 0 | 0.040936848 |
| 0 | 0 | 0 | 0 | 0 |
| 0 | 0 | 0 | 0 | 0 |
| 0 | 0 | 0 | 0 | 0 |
| 0 | 0 | 0 | 0 | 0 |
| 0 | 0 | 0 | 0 | 0 |
| 0 | 0.001119784 | 0 | 0 | 0 |
| 0 | 0.668733953 | 0.177378676 | 0.222033892 | 0 |
| 0 | 0 | 0 | 0 | 0 |
| 0 | 0 | 0 | 0 | 0 |
| 0 | 0 | 0 | 0.012172638 | 0 |
| 0 | 0.004233753 | 0.001120417 | 0 | 0 |
| 0 | 0 | 0 | 0 | 0 |
| 0 | 0 | 0 | 0 | 0 |
| 0 | 0 | 0 | 0 | 0 |
| 0.003363674 | 0.002158938 | 0 | 0.001987765 | 0 |
| 0 | 0 | 0 | 0 | 0 |

| | | | | |
|---|---|---|---|---|
| 0 | 0 | 0 | 0 | 0 |
| 0.346299414 | 0.01225882 | 0.003892998 | 0.002647538 | 0.020483336 |
| 0 | 0 | 0 | 0.01652227 | 0 |
| 0 | 0 | 0 | 0 | 0 |
| 0 | 0 | 0 | 0 | 0 |
| 0 | 0 | 0 | 0 | 0 |
| 0 | 0 | 0 | 0 | 0 |
| 0 | 0 | 0 | 0 | 0 |
| 0 | 0 | 0 | 0 | 0 |
| 0 | 0 | 0 | 0 | 0 |
| 0 | 0 | 0 | 0 | 0 |
| 0 | 0 | 0 | 0 | 0 |
| 0 | 0 | 0.367628416 | 0.081898286 | 0 |
| 0 | 0 | 0 | 0 | 0 |
| 0 | 0.002377651 | 0.009438297 | 0.01250936 | 0 |
| 0 | 0 | 0 | 0 | 0 |
| 0 | 0 | 0 | 0 | 0 |
| 0 | 0 | 0 | 0 | 0 |
| 0 | 0 | 0 | 0.003267445 | 0 |
| 0 | 0 | 0 | 0 | 0 |
| 0 | 0 | 0 | 0 | 0 |
| 0.012993027 | 0.005670814 | 1.727240375 | 0.00341255 | 0 |
| 0.000544586 | 0 | 0 | 0 | 0 |
| 0.015329339 | 0 | 0 | 0 | 0 |
| 0 | 0 | 0 | 0 | 0 |
| 0 | 0 | 0 | 0 | 0 |
| 0 | 0 | 0 | 0 | 0.00090329 |
| 0 | 0 | 0 | 0 | 0 |
| 0 | 0 | 0 | 0 | 0 |
| 0 | 0 | 0 | 0 | 0 |
| 0 | 0 | 0 | 0 | 0 |
| 0 | 0 | 0 | 0 | 0 |
| 0 | 0 | 0 | 0 | 0 |
| 0 | 0 | 0 | 0 | 0 |
| 0 | 0 | 0 | 20.51361107 | 0.297767737 |
| 0 | 0 | 0 | 0 | 0 |
| 0 | 0 | 0 | 0 | 0 |
| 0 | 0 | 0 | 0 | 0 |
| 0.009200532 | 0.025650033 | 0.157622765 | 0.140062377 | 0.023629573 |
| 0 | 0 | 0 | 0 | 0 |
| 9.992041133 | 0.266724567 | 0.202035011 | 0.078528473 | 0.317593691 |
| 0 | 0 | 0 | 0 | 0 |
| 0 | 0 | 0 | 0 | 0 |
| 0 | 0 | 0 | 0 | 0 |
| 0 | 0 | 0 | 0 | 0 |
| 4.912667778 | 0.085835107 | 0.114029181 | 0.00104953 | 0.574422714 |
| 0 | 0 | 0 | 0 | 0 |
| 0 | 0 | 0 | 0 | 0 |

| | | | | |
|---|---|---|---|---|
| 0 | 0 | 0 | 0 | 0 |
| 0 | 0 | 0 | 0 | 0 |
| 0 | 0 | 0 | 0 | 0 |
| 0 | 0 | 0 | 0.004470102 | 0 |
| 0 | 0 | 0 | 0.001591979 | 0 |
| 0 | 0 | 0 | 0 | 0 |
| 0 | 0 | 0 | 0 | 0 |
| 0.001308824 | 0 | 0.012618394 | 0 | 0 |
| 0 | 0 | 0.002574898 | 0 | 0 |
| 0.498105357 | 0.120067436 | 0.0509809 | 0.002566106 | 0 |
| 0 | 0 | 0 | 0 | 0 |
| 0 | 0 | 0 | 0 | 0 |
| 0 | 0 | 0 | 0.001839312 | 0 |
| 0 | 0 | 0 | 0 | 0 |
| 0 | 0 | 0 | 0 | 0 |
| 0 | 0 | 0 | 0 | 0 |
| 0 | 0 | 0 | 0 | 0 |
| 0 | 0 | 0 | 0 | 0 |
| 0 | 0 | 0 | 0 | 0 |
| 0.663088929 | 0.138206413 | 0.380686324 | 0.185291891 | 0.152730221 |
| 0 | 0 | 0 | 0.000852278 | 0 |
| 0 | 0 | 0 | 0 | 0 |
| 0 | 0 | 0 | 0 | 0 |
| 0 | 0 | 0 | 0 | 0 |
| 0 | 0 | 0 | 0 | 0 |
| 0 | 0 | 0 | 0 | 0 |
| 0 | 0 | 0 | 0 | 0 |
| 0 | 0 | 0 | 0 | 0 |
| 0 | 0 | 0 | 0 | 0 |
| 0 | 0.014618968 | 0 | 0 | 0 |
| 0 | 0 | 0 | 0 | 0 |
| 0 | 0 | 0 | 0 | 0 |
| 0 | 0 | 0 | 0 | 0 |
| 0 | 0 | 0 | 0 | 0 |
| 0 | 0 | 0 | 0 | 0 |
| 0 | 0 | 0 | 0 | 0.002938869 |
| 0 | 0 | 0 | 0 | 0 |
| 0 | 0 | 0 | 0 | 0 |
| 0 | 0 | 0 | 0 | 0 |
| 0 | 0 | 0 | 0 | 0 |
| 0 | 0 | 0 | 0 | 0 |
| 0 | 0 | 0 | 0 | 0 |
| 0 | 0 | 0 | 0.00618562 | 0 |
| 0 | 0 | 0 | 0 | 0 |
| 0.005612532 | 0.008471715 | 0.008027621 | 0.0096452 | 0 |
| 0 | 0 | 0 | 0.003471851 | 0 |
| 0 | 0 | 0 | 0 | 0 |
| 0.000968676 | 0 | 0 | 0 | 0.043661508 |

| | | | | |
|---|---|---|---|---|
| 0 | 0 | 0 | 0 | 0 |
| 0 | 0 | 0 | 0.000372846 | 0 |
| 0 | 0 | 0 | 0 | 0 |
| 0 | 0 | 0 | 0 | 0 |
| 0 | 0 | 0 | 0 | 0 |
| 0.002600673 | 0.003060225 | 0.008098556 | 0.010331176 | 0 |
| 0 | 0 | 0 | 0 | 0 |
| 0 | 0 | 0 | 0 | 0 |
| 0 | 0 | 0 | 0 | 0 |
| 0 | 0 | 0 | 0 | 0 |
| 0 | 0 | 0 | 0 | 0 |
| 0 | 0 | 0 | 0 | 0 |
| 0 | 0 | 0 | 0 | 0 |
| 0 | 0 | 0.093396089 | 0 | 0 |
| 0 | 0 | 0 | 0 | 0 |
| 0 | 0 | 0 | 0 | 0 |
| 0 | 0 | 0 | 0.01620253 | 0 |
| 0 | 0 | 0 | 0 | 0 |
| 0 | 0 | 0 | 0 | 0 |
| 0 | 0 | 0 | 0 | 0 |
| 0 | 0 | 0 | 0 | 0 |
| 0 | 0 | 0 | 0 | 0 |
| 0 | 0 | 0 | 0 | 0 |
| 0 | 0 | 0 | 0 | 0 |
| 0 | 0 | 0 | 0 | 0 |
| 0.003841895 | 0.007020501 | 0 | 0 | 0 |
| 0 | 0 | 0 | 0 | 0 |
| 0 | 0 | 0 | 0.657129801 | 0 |
| 0 | 0 | 0 | 0 | 0 |
| 0 | 0 | 0 | 0 | 0 |
| 0 | 0 | 0 | 0 | 0 |
| 0 | 0 | 0 | 0 | 0 |
| 0 | 0 | 0 | 0 | 0 |
| 0 | 0 | 0 | 0 | 0 |
| 0 | 0 | 0 | 0 | 0 |
| 0 | 0 | 0 | 0 | 0 |
| 0 | 0 | 0 | 0 | 0 |
| 0 | 0 | 0 | 0 | 0 |
| 0 | 0 | 0 | 1.486326428 | 0 |
| 0 | 0 | 0 | 0 | 0 |
| 0 | 0 | 0 | 0 | 0 |
| 0 | 0 | 0 | 0 | 0 |
| 0 | 0 | 0 | 0 | 0 |
| 0 | 0 | 0 | 0 | 0 |
| 0 | 0 | 0 | 0 | 0 |
| 0 | 0 | 0 | 0 | 0 |
| 0 | 0 | 0 | 0 | 0 |
| 0 | 0 | 0 | 0 | 0 |

| | | | | |
|---|---|---|---|---|
| 0 | 0 | 0 | 0 | 0 |
| 0 | 0 | 0 | 0.051874378 | 0.001296182 |
| 0 | 0 | 0 | 0 | 0 |
| 0 | 0 | 0 | 0 | 0 |
| 0 | 0 | 0 | 0 | 0 |
| 0 | 0 | 0 | 0 | 0 |
| 0 | 0 | 0 | 0 | 0 |
| 0 | 0 | 0 | 0 | 0 |
| 0 | 0 | 0 | 0 | 0 |
| 0 | 0 | 0.002708376 | 0 | 0 |
| 0 | 0 | 0.017096399 | 0 | 0 |
| 0 | 0 | 0 | 0 | 0 |
| 0 | 0 | 0.157229638 | 0.183445101 | 0 |
| 0 | 0 | 0 | 0 | 0 |
| 0 | 0 | 0 | 0 | 0 |
| 0 | 0 | 0 | 0 | 0 |
| 0 | 0 | 0 | 0 | 0 |
| 0 | 0 | 0 | 0 | 0 |
| 0 | 0 | 0 | 0 | 0 |
| 0 | 0 | 0 | 0 | 0 |
| 0 | 0.208542081 | 0.001204987 | 0 | 0 |
| 0 | 0 | 0 | 0 | 0 |
| 0 | 0 | 0 | 0 | 0 |
| 0 | 1.615911451 | 2.902797395 | 0.582805142 | 0 |
| 0 | 0 | 0 | 0 | 0 |
| 0 | 0 | 0 | 0 | 0 |
| 0 | 0 | 0 | 0 | 0 |
| 0 | 0 | 0.050652989 | 0 | 0 |
| 0 | 0 | 0 | 0 | 0 |
| 0 | 0 | 0 | 0 | 0 |
| 0 | 0 | 0 | 0 | 0.004226839 |
| 0 | 0 | 0 | 0 | 0 |
| 0 | 0 | 0 | 0 | 0 |
| 0 | 0 | 0 | 0 | 0 |

| PHL018-0090_V2 | PHL018-0043_V3 | PHL018-0259_V1 | PHL018-0107_V2 | PHL018-0232_V1 |
|---|---|---|---|---|
| CTRL | EYCF | REF | EYCF | REF |
| 0.995340362 | 1.015118403 | 0.022544457 | 0.038155764 | 0.087047144 |
| 0.409503878 | 0.393038628 | 1.157779547 | 3.089843312 | 0.876555166 |
| 0.031280271 | 0.019574364 | 3.108616478 | 4.770358185 | 0 |
| 0 | 0 | 0 | 0 | 0 |
| 0 | 0 | 0 | 0 | 0 |
| 0 | 0 | 0 | 0 | 0 |
| 0 | 0 | 0 | 0 | 0 |
| 0 | 0 | 0 | 0 | 0 |
| 0 | 0 | 0 | 0 | 0 |
| 0 | 0 | 0 | 0 | 0 |
| 0 | 0 | 0 | 0 | 0 |
| 0 | 0 | 0 | 0 | 0 |
| 0 | 0 | 0 | 0 | 0 |
| 2.796317424 | 2.734491688 | 1.527864592 | 19.91818168 | 5.532108869 |
| 0 | 0 | 0 | 0 | 0 |
| 0 | 0 | 0 | 0 | 0 |
| 0 | 0 | 0 | 0 | 0 |
| 0 | 0 | 0 | 0 | 0 |
| 0 | 0 | 0 | 0 | 0 |
| 0 | 0 | 0 | 0 | 0 |
| 0 | 0 | 0 | 0 | 0 |
| 0 | 0 | 0 | 0 | 0 |
| 0 | 0 | 0 | 0 | 0 |
| 0 | 0 | 0 | 0 | 0 |
| 0 | 0 | 0 | 0 | 0 |
| 0 | 0 | 0 | 0 | 0 |
| 0 | 0 | 0 | 0 | 0 |
| 0 | 0 | 0 | 0 | 0 |
| 0 | 0 | 0 | 0 | 0 |
| 0 | 0 | 0 | 0 | 0 |
| 0 | 0 | 0 | 0 | 0 |
| 0 | 0 | 0 | 0 | 0 |
| 0 | 0 | 0 | 0 | 0 |
| 0 | 0 | 0 | 0 | 0 |
| 0 | 0 | 0 | 0 | 0 |
| 1.614485706 | 1.623965122 | 1.668184252 | 0 | 1.059980114 |
| 0 | 0 | 0 | 0 | 0 |
| 0 | 0 | 0 | 0 | 0 |
| 0 | 0 | 0 | 0 | 0 |
| 0 | 0 | 0 | 0 | 0 |
| 0 | 0 | 0 | 0 | 0 |
| 0 | 0 | 0.027954272 | 0.130325842 | 0.008238612 |
| 0.036903538 | 0.03672556 | 0.00951395 | 1.163549266 | 0.00404184 |
| 0.206605087 | 0.213787337 | 0.004718624 | 0.008094772 | 0.314672952 |
| 0 | 0 | 0.000583193 | 0.003209825 | 0.015785048 |
| 0 | 0 | 0 | 0 | 0 |

| | | | | |
|---|---|---|---|---|
| 0 | 0 | 0.002393466 | 0 | 0 |
| 0 | 0 | 0 | 0.031221017 | 0.073964303 |
| 0.000788599 | 0 | 0 | 0 | 0.002971158 |
| 3.386027086 | 3.450366607 | 0.600607595 | 0.014011907 | 0.546158493 |
| 0 | 0 | 0.001735333 | 0 | 0.008494487 |
| 0 | 0 | 0 | 0 | 0 |
| 0 | 0 | 1.509830237 | 0 | 0.001283851 |
| 0 | 0 | 0.018700858 | 0 | 0 |
| 0 | 0 | 0 | 0 | 0 |
| 0 | 0 | 0 | 0 | 0 |
| 0.002113357 | 0 | 0 | 0 | 0 |
| 0.055911849 | 0.051132487 | 0.020584166 | 0 | 0 |
| 0 | 0 | 0.000804712 | 0 | 0 |
| 0.193279392 | 0.198745157 | 0.001729643 | 0.006790306 | 0.066430531 |
| 0 | 0 | 0 | 0 | 0 |
| 0 | 0 | 0 | 0 | 0 |
| 0.00401463 | 0.004362021 | 0 | 0 | 0.016386158 |
| 0 | 0 | 0 | 0 | 0 |
| 0 | 0 | 0 | 0 | 0 |
| 0 | 0 | 0 | 0 | 0 |
| 0.275797037 | 0.267170982 | 0.018417591 | 0.024480309 | 0.37114334 |
| 0.024151395 | 0.025740403 | 0.008712163 | 0 | 0 |
| 0 | 0 | 0 | 0 | 0 |
| 0 | 0 | 0 | 0 | 0 |
| 0 | 0 | 0 | 0 | 0 |
| 0 | 0 | 0 | 0 | 0 |
| 0 | 0 | 0 | 0 | 0 |
| 0 | 0 | 0 | 0 | 0 |
| 0 | 0 | 0 | 0.002281343 | 0.011086416 |
| 0 | 0 | 0 | 0 | 0 |
| 0 | 0 | 0 | 0.002918737 | 0 |
| 0 | 0 | 0 | 0 | 0 |
| 0 | 0 | 0 | 0 | 0 |
| 0 | 0 | 0 | 0 | 0 |
| 0 | 0 | 0 | 0 | 0 |
| 0 | 0 | 0.001227595 | 0 | 0 |
| 0 | 0 | 0 | 0 | 0 |
| 0 | 0 | 0 | 0 | 0 |
| 0 | 0 | 0 | 0 | 0 |
| 0 | 0 | 0 | 0 | 0 |
| 0 | 0 | 0 | 0 | 0 |
| 0 | 0 | 0 | 0 | 0 |
| 0 | 0 | 0 | 0 | 0 |
| 0 | 0 | 0 | 0 | 0 |
| 0 | 0 | 0 | 0 | 0 |
| 0 | 0 | 0 | 0 | 0 |
| 0 | 0 | 0 | 0 | 0 |
| 0 | 0 | 0.002533939 | 0 | 0 |

| | | | | |
|---|---|---|---|---|
| 0 | 0 | 0 | 0 | 0 |
| 0 | 0 | 0 | 0 | 0 |
| 0 | 0 | 0 | 0 | 0 |
| 0 | 0 | 0 | 0 | 0 |
| 0 | 0 | 0.021035453 | 0 | 0 |
| 0 | 0 | 0 | 0 | 0 |
| 0 | 0 | 0 | 0 | 0 |
| 0 | 0 | 0 | 0 | 0 |
| 0.008600501 | 0.008449851 | 0 | 0 | 0 |
| 0 | 0 | 0 | 0 | 0 |
| 0 | 0 | 0 | 0 | 0 |
| 0 | 0 | 0.013331729 | 0 | 0.006142031 |
| 0 | 0 | 0 | 0 | 0 |
| 0 | 0 | 0 | 0 | 0 |
| 0 | 0 | 0 | 0 | 0 |
| 0 | 0 | 0 | 0 | 0.000949464 |
| 0 | 0 | 0 | 0 | 0 |
| 0 | 0 | 0 | 0 | 0 |
| 0 | 0 | 0.146198424 | 0 | 0 |
| 0 | 0 | 0.001471229 | 0 | 0 |
| 0 | 0 | 0 | 0 | 0 |
| 0 | 0 | 0 | 0 | 0 |
| 0 | 0 | 0 | 0 | 0 |
| 0 | 0 | 0 | 0 | 0 |
| 0 | 0 | 0 | 0 | 0 |
| 0 | 0 | 0 | 0 | 0 |
| 0 | 0 | 0 | 0 | 0 |
| 0 | 0 | 0 | 0 | 0 |
| 0 | 0 | 0 | 0 | 0 |
| 0 | 0 | 0 | 0 | 0 |
| 0.026149859 | 0.027801763 | 0.005195025 | 0 | 0 |
| 0.00238552 | 0 | 0.003592067 | 0 | 0.013909663 |
| 0 | 0 | 0 | 0 | 0 |
| 0 | 0 | 0 | 0 | 0 |
| 0 | 0 | 0 | 0 | 0 |
| 0 | 0 | 0 | 0 | 0 |
| 0 | 0 | 0 | 0 | 0 |
| 0 | 0 | 0 | 0 | 0 |
| 0 | 0 | 0.096455925 | 0 | 0 |
| 0 | 0 | 0 | 0 | 0 |
| 0 | 0 | 0 | 0 | 0 |
| 0 | 0 | 0 | 0 | 0 |
| 0.001181693 | 0.002370363 | 0 | 0 | 0 |
| 0 | 0 | 0 | 0 | 0 |
| 0 | 0 | 0.001526366 | 0 | 0 |
| 0 | 0 | 0 | 0 | 0 |
| 0 | 0 | 0 | 0 | 0 |
| 0 | 0 | 0 | 0 | 0 |

| | | | | |
|---|---|---|---|---|
| 0 | 0 | 0 | 0 | 0 |
| 0 | 0 | 0 | 0 | 0 |
| 0 | 0 | 0 | 0 | 0 |
| 0 | 0 | 0 | 0 | 0 |
| 0.000741545 | 0.001586632 | 0.004959257 | 0 | 0 |
| 0 | 0 | 0 | 0 | 0 |
| 0 | 0 | 0 | 0 | 0 |
| 0 | 0 | 0 | 0 | 0 |
| 0 | 0 | 0 | 0 | 0 |
| 0 | 0 | 0 | 0 | 0 |
| 0 | 0 | 0 | 0 | 0 |
| 0 | 0 | 0 | 0 | 0 |
| 0 | 0 | 0 | 0 | 0 |
| 0 | 0 | 0 | 0 | 0 |
| 0 | 0 | 0 | 0 | 0 |
| 0 | 0 | 0 | 0 | 0 |
| 0 | 0 | 0 | 0 | 0 |
| 0 | 0 | 0 | 0 | 0 |
| 0 | 0 | 0 | 0 | 0 |
| 0 | 0 | 0 | 0 | 0 |
| 0 | 0 | 0 | 0 | 0 |
| 0 | 0 | 0 | 0 | 0 |
| 0 | 0 | 0 | 0 | 0 |
| 0 | 0 | 0 | 0 | 0 |
| 0 | 0 | 0 | 0.009592585 | 0 |
| 0 | 0 | 0 | 0 | 0 |
| 0 | 0 | 0 | 0 | 0 |
| 0 | 0 | 0 | 0 | 0 |
| 0 | 0 | 0 | 0 | 0 |
| 0 | 0 | 0 | 0 | 0 |
| 0 | 0 | 0 | 0 | 0 |
| 0 | 0 | 0 | 0 | 0 |
| 0 | 0 | 0 | 0 | 0 |
| 0 | 0 | 0 | 0 | 0 |
| 0 | 0 | 0 | 0 | 0 |
| 0 | 0 | 0 | 0 | 0 |
| 0 | 0 | 0 | 0 | 0 |
| 0 | 0 | 0 | 0 | 0 |
| 0 | 0 | 0 | 0 | 0 |
| 0 | 0 | 0 | 0 | 0 |
| 0 | 0 | 0 | 0 | 0 |
| 0 | 0 | 0 | 0 | 0 |
| 0 | 0 | 0 | 0 | 0 |
| 0 | 0 | 0 | 0 | 0 |
| 0 | 0 | 0 | 0 | 0 |
| 0 | 0 | 0 | 0 | 0 |
| 0 | 0 | 0 | 0 | 0 |
| 0.002892401 | 0.003545593 | 0 | 0 | 0 |

| | | | | |
|---|---|---|---|---|
| 0 | 0 | 0 | 0 | 0 |
| 0 | 0 | 0 | 0 | 0 |
| 0 | 0 | 0 | 0 | 0 |
| 0 | 0 | 0 | 0 | 0 |
| 0 | 0 | 0 | 0 | 0 |
| 0 | 0 | 0 | 0 | 0 |
| 0 | 0 | 0 | 0 | 0 |
| 0 | 0 | 0 | 0 | 0 |
| 0 | 0 | 0 | 0 | 0 |
| 0 | 0 | 0 | 0 | 0 |
| 0 | 0 | 0 | 0 | 0 |
| 0 | 0 | 0 | 0 | 0 |
| 0 | 0.001509445 | 0 | 0 | 0 |
| 0 | 0 | 0 | 0 | 0 |
| 0 | 0 | 0 | 0 | 0 |
| 0 | 0 | 0 | 0 | 0 |
| 0 | 0 | 0 | 0 | 0 |
| 0 | 0 | 0 | 0 | 0 |
| 0 | 0 | 0 | 0 | 0 |
| 0 | 0 | 0 | 0 | 0 |
| 0.0517071 | 0.055975593 | 0.015853166 | 0.003070518 | 0.046825989 |
| 0 | 0 | 0 | 0 | 0 |
| 0.002006146 | 0 | 0 | 0 | 0 |
| 0 | 0 | 0 | 0 | 0.009673885 |
| 0 | 0 | 0 | 0 | 0 |
| 0 | 0 | 0 | 0 | 0 |
| 0.002048503 | 0.0027394 | 0.000893469 | 0.022352467 | 0.095188965 |
| 0 | 0 | 0 | 0 | 0 |
| 0 | 0 | 0 | 0 | 0 |
| 0 | 0 | 0 | 0 | 0 |
| 0 | 0 | 0 | 0 | 0 |
| 0 | 0 | 0 | 0 | 0 |
| 0 | 0 | 0 | 0 | 0 |
| 0.012898028 | 0.011498758 | 0.01328258 | 0 | 0 |
| 0.010527865 | 0.01254831 | 0.00831848 | 0 | 0.023856615 |
| 0 | 0 | 0 | 0 | 0 |
| 0 | 0 | 0 | 0 | 0 |
| 0 | 0 | 0 | 0 | 0 |
| 0 | 0 | 0 | 0 | 0 |
| 0 | 0 | 0 | 0 | 0 |
| 0 | 0 | 0 | 0 | 0 |
| 0 | 0 | 0 | 0 | 0 |
| 0 | 0 | 0 | 0 | 0 |
| 0 | 0 | 0 | 0 | 0 |
| 0 | 0 | 0 | 0 | 0 |
| 0 | 0 | 0 | 0.000713311 | 0 |
| 0.001982414 | 0.001656888 | 0 | 0 | 0 |
| 0 | 0 | 0 | 0 | 0 |

| | | | | |
|---|---|---|---|---|
| 0 | 0 | 0 | 0 | 0 |
| 0 | 0 | 0 | 0 | 0 |
| 0 | 0 | 0 | 0 | 0.004492059 |
| 0 | 0 | 0 | 0 | 0 |
| 0 | 0 | 0 | 0 | 0 |
| 0 | 0 | 0 | 0 | 0 |
| 0 | 0 | 0 | 0 | 0 |
| 0 | 0 | 0.002726902 | 0 | 0.001177786 |
| 0 | 0 | 0 | 0 | 0 |
| 0 | 0 | 0 | 0 | 0 |
| 0 | 0 | 0 | 0 | 0 |
| 0 | 0 | 0 | 0 | 0 |
| 0 | 0 | 0 | 0 | 0 |
| 0 | 0 | 0 | 0 | 0 |
| 0 | 0.001375511 | 0 | 0 | 0 |
| 0 | 0 | 0 | 0 | 0 |
| 0 | 0 | 0 | 0 | 0 |
| 0 | 0 | 0 | 0 | 0 |
| 0 | 0 | 0 | 0.006696586 | 0 |
| 0 | 0 | 0.03385226 | 0 | 0 |
| 0.001223362 | 0.002617543 | 0.002667888 | 0.049924685 | 0 |
| 0 | 0 | 0 | 0 | 0 |
| 0 | 0 | 0 | 0 | 0 |
| 0 | 0 | 0 | 0 | 0 |
| 0.003402653 | 0.008450491 | 0 | 0.010077716 | 0.023808527 |
| 0 | 0 | 0 | 0 | 0.001184793 |
| 0 | 0 | 0 | 0 | 0 |
| 0 | 0 | 0 | 0 | 0 |
| 0 | 0 | 0 | 0.007339526 | 0.003495511 |
| 0 | 0 | 0 | 0 | 0 |
| 0.060808729 | 0.052206894 | 0.010593164 | 0.121907283 | 0.117038092 |
| 0 | 0 | 0 | 0 | 0 |
| 0.003579213 | 0.003589779 | 0 | 0.078109962 | 0 |
| 0 | 0 | 0.005752794 | 0 | 0 |
| 0 | 0 | 0 | 0 | 0 |
| 0 | 0 | 0 | 0 | 0 |
| 0 | 0 | 0 | 0 | 0 |
| 0 | 0 | 0 | 0 | 0 |
| 0 | 0 | 0 | 0 | 0 |
| 0 | 0 | 0 | 0.000980422 | 0.020537326 |
| 0 | 0 | 0 | 0 | 0 |
| 0.205758826 | 0.209431682 | 0 | 0 | 0.011976149 |
| 0 | 0 | 0 | 0 | 0 |
| 0 | 0 | 0 | 0 | 0.001247784 |
| 0 | 0 | 0 | 0 | 0 |
| 0 | 0 | 0 | 0 | 0 |
| 0 | 0 | 0 | 0 | 0 |
| 0 | 0 | 0 | 0 | 0 |

| | | | | |
|---|---|---|---|---|
| 0 | 0 | 0 | 0 | 0 |
| 0 | 0 | 0 | 0 | 0 |
| 0.200566043 | 0.195230381 | 0.0070304 | 0.003517678 | 0.361201825 |
| 0 | 0 | 0 | 0 | 0 |
| 0 | 0 | 0 | 0 | 0 |
| 0 | 0 | 0 | 0 | 0 |
| 0 | 0 | 0 | 0 | 0 |
| 0 | 0 | 0 | 0 | 0 |
| 0 | 0 | 0 | 0 | 0 |
| 0 | 0 | 0 | 0 | 0 |
| 0 | 0 | 0 | 0 | 0 |
| 0 | 0 | 0 | 0 | 0 |
| 0 | 0 | 0 | 0 | 0 |
| 0 | 0 | 0 | 0 | 0 |
| 0.002310539 | 0.001404461 | 0 | 0.001833578 | 0.012926026 |
| 0 | 0 | 0 | 0 | 0 |
| 0.002153988 | 0.002560411 | 0.001391817 | 0.006789894 | 0.020739495 |
| 0 | 0 | 0 | 0 | 0 |
| 0 | 0 | 0 | 0 | 0 |
| 0 | 0 | 0 | 0 | 0 |
| 0 | 0 | 0 | 0 | 0 |
| 0 | 0 | 0 | 0.001178719 | 0 |
| 0 | 0 | 0 | 0 | 0 |
| 0 | 0 | 0 | 0 | 0 |
| 0 | 0 | 0 | 0 | 0 |
| 0 | 0 | 0 | 0 | 0 |
| 0 | 0 | 0 | 0 | 0 |
| 0 | 0 | 0 | 0 | 0 |
| 0 | 0 | 0 | 0 | 0 |
| 0.001562581 | 0 | 0 | 0 | 0 |
| 0 | 0 | 0 | 0 | 0 |
| 0 | 0 | 0 | 0 | 0 |
| 0 | 0 | 0 | 0 | 0 |
| 0 | 0 | 0 | 0 | 0 |
| 0 | 0 | 0 | 0 | 0 |
| 0 | 0 | 0 | 0 | 0.005961398 |
| 0 | 0 | 0 | 0 | 0 |
| 0 | 0 | 0 | 0.074935459 | 0.172836009 |
| 0 | 0 | 0 | 0 | 0 |
| 0 | 0 | 0 | 0 | 0 |
| 0 | 0 | 0 | 0 | 0 |
| 0 | 0 | 0 | 0 | 0 |
| 0 | 0 | 0 | 0 | 0 |
| 0 | 0 | 0 | 0 | 0 |
| 0 | 0 | 0 | 0 | 0 |
| 0 | 0 | 0 | 0 | 0 |
| 0 | 0 | 0 | 0 | 0 |
| 0 | 0 | 0 | 0.006098877 | 0.001651659 |

| | | | | |
|---|---|---|---|---|
| 0 | 0 | 0 | 0 | 0 |
| 0 | 0 | 0 | 0 | 0 |
| 0.003620704 | 0.001815696 | 0 | 0.000987693 | 0 |
| 0 | 0 | 0 | 0 | 0 |
| 0.002276736 | 0.003044609 | 0 | 0.007949713 | 0.042317753 |
| 0 | 0 | 0 | 0 | 0 |
| 0 | 0 | 0 | 0 | 0 |
| 0.065741734 | 0.068709803 | 0 | 0.024375908 | 0.009218283 |
| 0 | 0 | 0 | 0 | 0 |
| 0.046709335 | 0.046627282 | 0.002152035 | 0.879008249 | 0.033874854 |
| 0 | 0 | 0 | 0 | 0 |
| 0 | 0 | 0.00073457 | 0.017090792 | 0.001903624 |
| 0.003832548 | 0.005637664 | 0.017969615 | 0.001881867 | 0.083028068 |
| 0 | 0 | 0 | 0 | 0 |
| 0 | 0 | 0 | 0 | 0 |
| 0 | 0 | 0 | 0 | 0 |
| 0 | 0 | 0 | 0 | 0 |
| 0 | 0 | 0 | 0 | 0 |
| 0.173953377 | 0.175087769 | 0 | 0.007092587 | 0.035374164 |
| 0 | 0 | 0 | 0 | 0 |
| 0 | 0 | 0 | 0 | 0 |
| 0 | 0 | 0 | 0 | 0 |
| 0 | 0 | 0.051027318 | 0 | 0 |
| 0 | 0 | 0 | 0 | 0 |
| 0 | 0 | 0 | 0 | 0 |
| 0 | 0 | 0.028717935 | 0 | 0.350666296 |
| 0 | 0 | 0 | 0 | 0 |
| 0.02888008 | 0.031401671 | 0 | 0.080401858 | 0.033219182 |
| 0 | 0 | 0 | 0 | 0 |
| 0 | 0 | 0.002503995 | 0 | 0 |
| 0 | 0 | 0 | 0 | 0.011129103 |
| 0 | 0 | 0 | 0 | 0 |
| 0 | 0 | 0 | 0 | 0 |
| 0 | 0 | 0 | 0 | 0 |
| 0 | 0 | 0 | 0 | 0 |
| 0 | 0 | 0 | 0 | 0 |
| 0.050856363 | 0.048722619 | 0 | 0 | 0 |
| 0 | 0 | 0 | 0 | 0 |
| 0 | 0 | 0 | 0 | 0 |
| 0.023474618 | 0.017579457 | 0 | 0.108093556 | 0 |
| 0 | 0 | 0 | 0 | 0.035323072 |
| 0.047969426 | 0.056085721 | 0 | 0 | 0 |
| 0 | 0 | 0 | 0 | 0 |
| 0 | 0 | 0 | 0 | 0 |
| 0 | 0 | 0 | 0 | 0 |
| 0 | 0 | 0 | 0 | 0 |
| 0 | 0 | 0 | 0 | 0.002091517 |
| 0 | 0 | 0 | 0 | 0 |

| | | | | |
|---|---|---|---|---|
| 0 | 0 | 0 | 0 | 0 |
| 0 | 0 | 0.285470832 | 0 | 0 |
| 0 | 0 | 0 | 0 | 0 |
| 0 | 0 | 0 | 0 | 0 |
| 0 | 0 | 0 | 0 | 0 |
| 0.051811666 | 0.045313729 | 0 | 0 | 0.005491076 |
| 0 | 0 | 0.00123695 | 0 | 0 |
| 0 | 0 | 0 | 0 | 0 |
| 0 | 0 | 0 | 0 | 0 |
| 0 | 0 | 0 | 0 | 0 |
| 0.005324679 | 0.00712053 | 0 | 0.001383353 | 0.146480642 |
| 0 | 0 | 0 | 0 | 0 |
| 0 | 0 | 0 | 0 | 0 |
| 0.002874565 | 0.003523729 | 0.013234148 | 0.001829691 | 0.005716007 |
| 0 | 0 | 0 | 0.006611852 | 0.014002333 |
| 0 | 0 | 0 | 0 | 0 |
| 0 | 0 | 0 | 0 | 0 |
| 0 | 0 | 0 | 0 | 0 |
| 0 | 0 | 0 | 0 | 0 |
| 0 | 0 | 0.000417666 | 0 | 0 |
| 0 | 0 | 0.009381087 | 0 | 0 |
| 0 | 0 | 0.433129038 | 0 | 0 |
| 0 | 0 | 0 | 0 | 0 |
| 0 | 0 | 0 | 0 | 0 |
| 0 | 0 | 0 | 0 | 0 |
| 0 | 0 | 0 | 0 | 0 |
| 0 | 0 | 0 | 0 | 0 |
| 0 | 0 | 0 | 0 | 0 |
| 0 | 0 | 0 | 0 | 0 |
| 0.98771712 | 1.025966509 | 10.05122513 | 0.250187068 | 1.140353806 |
| 0 | 0 | 0.002717122 | 0 | 0.001428684 |
| 0 | 0 | 0 | 0 | 0 |
| 0 | 0 | 0 | 0 | 0 |
| 0.0046035 | 0.004309284 | 0 | 0 | 0 |
| 0.000865331 | 0 | 0 | 0.007081614 | 0 |
| 0 | 0 | 0 | 0 | 0 |
| 0 | 0 | 0.52151236 | 0 | 0 |
| 0 | 0 | 0 | 0 | 0.005826134 |
| 0 | 0 | 0.002044048 | 0.448241032 | 0 |
| 0 | 0 | 0 | 0 | 0 |
| 0 | 0 | 0.019039842 | 0 | 0 |
| 10.35384127 | 10.41084932 | 0 | 0 | 3.06047389 |
| 0 | 0 | 0 | 0 | 0.004775314 |
| 0 | 0 | 0 | 0.098770292 | 0 |
| 0 | 0 | 0 | 0 | 0 |
| 0 | 0 | 0 | 0 | 0 |
| 0.001033481 | 0 | 0 | 0.001127694 | 0.008047154 |
| 0 | 0 | 0 | 0 | 0 |

| | | | | |
|---|---|---|---|---|
| 0 | 0 | 0.015943306 | 0.004452432 | 0.01889682 |
| 0 | 0 | 0 | 0 | 0.004707012 |
| 0 | 0 | 0.019955746 | 0 | 0.024875539 |
| 0 | 0 | 0 | 0 | 0 |
| 0 | 0 | 0 | 0 | 0 |
| 0.014015735 | 0.014417549 | 0 | 0 | 0 |
| 0 | 0 | 0 | 0 | 0 |
| 0 | 0 | 0.022648188 | 0 | 0 |
| 0 | 0 | 0 | 0 | 0 |
| 0 | 0 | 0 | 0 | 0 |
| 0 | 0 | 0 | 0 | 0 |
| 0 | 0 | 0 | 0 | 0 |
| 0 | 0 | 0 | 0 | 0 |
| 0.008997991 | 0.00894742 | 0 | 0 | 0.495479047 |
| 0 | 0 | 0 | 0 | 0 |
| 0.435609419 | 0.446780947 | 0.118090943 | 0 | 0.000679313 |
| 0.000999229 | 0 | 0.028764169 | 0.011339329 | 0.01104323 |
| 0.005098904 | 0.006493912 | 0 | 0.123196763 | 0 |
| 0 | 0 | 0 | 0 | 0 |
| 0 | 0 | 0 | 0 | 0 |
| 0.010074506 | 0.004715315 | 0.012999109 | 0 | 0.016131777 |
| 0 | 0 | 0 | 0 | 0.002329013 |
| 0 | 0 | 0 | 0 | 0 |
| 0.190381884 | 0.180249857 | 0 | 0.064030263 | 1.36282438 |
| 0.095815722 | 0.090166561 | 0 | 0.001426613 | 0.00539505 |
| 0 | 0 | 0.002679213 | 0.002585348 | 0 |
| 0 | 0 | 0.000553278 | 0 | 0 |
| 0 | 0 | 0 | 0 | 0 |
| 0 | 0 | 0 | 0 | 0 |
| 0 | 0 | 0 | 0 | 0 |
| 0.087210078 | 0.077662905 | 0 | 0.002105315 | 0.006057822 |
| 0.039120919 | 0.041159758 | 0.079041179 | 0.007846916 | 0.09031469 |
| 0.084082794 | 0.096692231 | 0.43661521 | 0.017333473 | 0.138618774 |
| 0 | 0 | 0 | 0 | 0.002464693 |
| 0 | 0 | 0.000814591 | 0 | 0 |
| 0 | 0 | 0 | 0 | 0 |
| 0.001485667 | 0 | 0.005399867 | 0.001080735 | 0.00248776 |
| 0.058480321 | 0.057228712 | 0.012246543 | 0 | 0 |
| 0 | 0 | 0.209976525 | 0 | 0.009213698 |
| 0.001741556 | 0.001058604 | 0 | 0 | 0 |
| 0 | 0 | 0 | 0 | 0 |
| 0 | 0 | 0 | 0 | 0 |
| 0.054733165 | 0.045697208 | 0 | 0 | 0 |
| 0.060465031 | 0.056600623 | 0 | 0 | 0 |
| 0 | 0 | 0 | 0 | 0 |
| 0 | 0 | 0.006905034 | 0 | 0 |
| 0 | 0 | 0 | 0 | 0 |
| 0 | 0 | 0 | 0 | 0 |

| | | | | |
|---|---|---|---|---|
| 0 | 0 | 0 | 0 | 0.024519065 |
| 2.490873906 | 2.529631281 | 0.040471907 | 0.481110711 | 0.042085425 |
| 0 | 0 | 0 | 0 | 0 |
| 0 | 0 | 0 | 0 | 0 |
| 0 | 0 | 0 | 0 | 0 |
| 0 | 0 | 0 | 0 | 0 |
| 0 | 0 | 0 | 0 | 0 |
| 0 | 0 | 0 | 0.00176921 | 0 |
| 0 | 0 | 0.010535175 | 0 | 0.001516762 |
| 0 | 0 | 0 | 0.003347588 | 0 |
| 0.013333414 | 0.009177395 | 0 | 0 | 0.01354389 |
| 0.021028764 | 0.020309699 | 0.206366396 | 0.029000899 | 0.361298138 |
| 0 | 0 | 0 | 0 | 0 |
| 0 | 0 | 0 | 0 | 0 |
| 0 | 0 | 0.010466688 | 0 | 0.00282544 |
| 0 | 0 | 0 | 0 | 0 |
| 0.020943014 | 0.02608444 | 0 | 0.04772077 | 0.111912086 |
| 0.011461155 | 0.009267278 | 0 | 0.026175276 | 0.031800341 |
| 0 | 0 | 0 | 0 | 0 |
| 0 | 0 | 0 | 0.001321978 | 0 |
| 0.19818911 | 0.198774168 | 0.060182769 | 0.005545031 | 0.689532438 |
| 0 | 0 | 0 | 0 | 0.002316269 |
| 0 | 0 | 0 | 0 | 0 |
| 0 | 0 | 0 | 0 | 0 |
| 0 | 0 | 0.000775844 | 0 | 0 |
| 0 | 0 | 0 | 0 | 0 |
| 0 | 0 | 0.001671195 | 0 | 0.039896511 |
| 0.103747541 | 0.10924521 | 0.040407809 | 0.130885917 | 0.286026087 |
| 0 | 0 | 0 | 0 | 0 |
| 0 | 0 | 0 | 0 | 0 |
| 0 | 0 | 0 | 0 | 0.242310353 |
| 0 | 0 | 0 | 0 | 0 |
| 0 | 0 | 0 | 0 | 0 |
| 0 | 0 | 0 | 0 | 0 |
| 0.063148851 | 0.053762857 | 0 | 0 | 0.00197037 |
| 0.245287898 | 0.251105409 | 0.226482696 | 12.96308533 | 0.061227774 |
| 0 | 0 | 0 | 0 | 0 |
| 0 | 0 | 0 | 0 | 0 |
| 0.001943361 | 0 | 0.002623555 | 0.00878501 | 0.003835274 |
| 0 | 0 | 0 | 0 | 0 |
| 0 | 0 | 0 | 0 | 0 |
| 0 | 0 | 0 | 0 | 0 |
| 0 | 0 | 0 | 0.00680819 | 0.022237127 |
| 0 | 0 | 0.00106517 | 0 | 0.002862604 |
| 0 | 0 | 0 | 0 | 0 |
| 0 | 0 | 0.089877447 | 0 | 0 |
| 0 | 0 | 0 | 0 | 0 |
| 0 | 0 | 0 | 0 | 0.008593329 |

| | | | | |
|---|---|---|---|---|
| 0 | 0 | 0 | 0 | 0 |
| 0 | 0 | 0 | 0 | 0 |
| 0 | 0 | 0.001011062 | 0 | 0 |
| 0.010016575 | 0.011556349 | 0.009137355 | 0 | 0 |
| 0 | 0 | 0 | 0 | 0 |
| 0 | 0 | 0 | 0 | 0 |
| 0 | 0 | 0 | 0 | 0 |
| 0 | 0 | 0.001021793 | 0 | 0 |
| 2.281292436 | 2.291430391 | 0.069996687 | 0 | 0.253313847 |
| 0.639303448 | 0.649993412 | 0 | 0 | 0 |
| 0 | 0 | 0 | 0 | 0 |
| 0.00639797 | 0.007045961 | 0.003009383 | 0 | 0.001890608 |
| 0 | 0 | 0 | 0 | 0 |
| 0 | 0 | 0.61351228 | 0 | 0 |
| 0.220501838 | 0.221360419 | 0.010610679 | 0 | 0.009555825 |
| 0 | 0 | 0 | 0 | 0 |
| 0 | 0 | 0.2132057 | 0 | 0 |
| 0 | 0 | 0 | 0 | 0 |
| 0 | 0 | 0 | 0 | 0 |
| 0 | 0 | 0 | 0 | 0 |
| 0 | 0 | 0 | 0 | 0 |
| 0 | 0 | 0.000507998 | 0 | 0 |
| 0 | 0 | 0 | 0 | 0 |
| 0 | 0 | 0.447323262 | 0.000979278 | 0.006950494 |
| 0 | 0 | 0.001024564 | 0 | 0 |
| 0 | 0 | 0.086849465 | 0 | 0 |
| 0 | 0 | 0 | 0 | 0 |
| 2.901743733 | 2.972385309 | 1.175407183 | 0 | 1.907948295 |
| 0 | 0 | 0 | 0 | 0 |
| 0 | 0 | 0 | 0 | 0 |
| 0 | 0 | 0.012101484 | 0 | 0 |
| 0 | 0 | 0.001879425 | 0 | 0 |
| 0.303242582 | 0.300693286 | 0 | 0 | 0.001923416 |
| 0 | 0 | 0 | 0 | 0 |
| 0 | 0 | 0 | 0 | 0 |
| 0 | 0 | 0.00045102 | 0 | 0 |
| 0.491804006 | 0.488098903 | 0.51492537 | 0 | 0 |
| 0 | 0 | 0 | 0 | 0 |
| 0 | 0 | 0 | 0 | 0 |
| 0 | 0 | 0 | 0 | 0 |
| 0.240092732 | 0.243094838 | 0.00091858 | 0 | 0.006347952 |
| 0 | 0 | 0 | 0 | 0 |
| 5.221187347 | 5.170717547 | 5.311867415 | 0.585783885 | 4.107284434 |
| 0 | 0 | 0 | 0 | 0 |
| 0 | 0 | 0 | 0 | 0 |
| 0 | 0 | 0 | 0 | 0 |
| 0 | 0 | 0 | 0 | 0 |
| 0.314008799 | 0.314282817 | 0 | 0.001894315 | 0 |

| | | | | |
|---|---|---|---|---|
| 0 | 0 | 0 | 0 | 0 |
| 0.069985558 | 0.063714914 | 0.034634879 | 0 | 0 |
| 0.00416051 | 0.002225489 | 0 | 0.494609678 | 0.013585266 |
| 0 | 0 | 0 | 0 | 0 |
| 0 | 0 | 0 | 0 | 0 |
| 0.003605238 | 0.003725453 | 0.002739874 | 0 | 0 |
| 0 | 0 | 0 | 0 | 0 |
| 0 | 0 | 0 | 0 | 0 |
| 0 | 0 | 0 | 0 | 0 |
| 0 | 0 | 0 | 0 | 0 |
| 0.223851578 | 0.213040956 | 0.001187766 | 0.007800213 | 0.014620853 |
| 2.672623203 | 2.622906939 | 0.519946841 | 1.20710851 | 4.359990083 |
| 0 | 0 | 0 | 0 | 0 |
| 0 | 0 | 0 | 0.084664128 | 0.071699472 |
| 0.272023897 | 0.248320114 | 0.002020351 | 0.00530716 | 0.008071705 |
| 0 | 0 | 0 | 0 | 0 |
| 0 | 0 | 0 | 0 | 0 |
| 0 | 0 | 0 | 0 | 0 |
| 0 | 0 | 0 | 0 | 0 |
| 0 | 0 | 0.000978407 | 0.006058161 | 0 |
| 0.036291573 | 0.029988872 | 0 | 0 | 0.015694267 |
| 0.086400889 | 0.081771373 | 0.377892385 | 3.163110939 | 0.075888462 |
| 2.305375386 | 2.234881168 | 0.075458738 | 14.15954764 | 0.71559482 |
| 0 | 0 | 0 | 0 | 0 |
| 0 | 0 | 0 | 0 | 0 |
| 2.832476129 | 2.879516498 | 1.362630488 | 0 | 17.57706536 |
| 0 | 0 | 0 | 0 | 0 |
| 0 | 0 | 0.001038313 | 0 | 0 |
| 0 | 0 | 0.001608651 | 0 | 0 |
| 0 | 0 | 0 | 0 | 0 |
| 0.015293971 | 0.014897252 | 0.000411764 | 0 | 0 |
| 0.575828316 | 0.58417388 | 0.113134471 | 0 | 0 |
| 0.001678112 | 0.001683066 | 0 | 0 | 0 |
| 0.070702908 | 0.078505479 | 0.004651216 | 0.000698174 | 0 |
| 0.010245069 | 0.011293587 | 0 | 0 | 0 |
| 0.010461965 | 0.008139907 | 0.002350666 | 0.097344686 | 0 |
| 0.012678581 | 0.009583079 | 0 | 0 | 0 |
| 0 | 0 | 0 | 0 | 0 |
| 0.166054337 | 0.161037698 | 0.046601012 | 0.009472505 | 0.018450306 |
| 0 | 0 | 0 | 0 | 0 |
| 0 | 0 | 0.001339334 | 0 | 0 |
| 0 | 0 | 0 | 0 | 0 |
| 0 | 0 | 0 | 0 | 0 |
| 0 | 0 | 0 | 0 | 0 |
| 0.313885442 | 0.302846134 | 0.000788432 | 0.000710089 | 0.01607322 |
| 0 | 0 | 0 | 0 | 0 |
| 0 | 0 | 0 | 0 | 0 |
| 0.048408327 | 0.044789603 | 0.000760851 | 4.313356981 | 0.118303909 |

| | | | | |
|---|---|---|---|---|
| 0 | 0 | 0 | 0 | 0 |
| 0 | 0 | 0 | 0 | 0 |
| 0 | 0 | 0 | 0 | 0 |
| 0 | 0 | 0.000806373 | 0 | 0 |
| 0 | 0 | 0 | 0 | 0 |
| 0.005878297 | 0.005677292 | 0 | 0 | 0 |
| 0 | 0 | 0 | 0 | 0 |
| 0 | 0 | 0.006678079 | 0 | 0 |
| 0 | 0 | 0 | 0 | 0 |
| 0 | 0 | 0 | 0 | 0 |
| 0 | 0.001101422 | 0 | 0 | 0 |
| 0 | 0 | 0 | 0 | 0 |
| 0.008293308 | 0.011572577 | 0.00445595 | 0 | 0 |
| 0 | 0 | 0 | 0 | 0 |
| 0 | 0 | 0 | 0 | 0 |
| 0 | 0 | 0 | 0 | 0 |
| 0 | 0 | 0.013136138 | 0 | 0 |
| 0.000685658 | 0.000916909 | 0.000996848 | 0 | 0 |
| 0 | 0 | 0.236406687 | 0 | 0 |
| 0 | 0 | 0 | 0 | 0 |
| 0.019188314 | 0.023012489 | 0 | 0 | 0.000956279 |
| 0 | 0 | 0 | 0 | 0 |
| 0.003162057 | 0.003964239 | 0 | 0 | 0 |
| 0 | 0 | 0 | 0 | 0 |
| 0 | 0 | 0 | 0 | 0 |
| 0 | 0 | 0 | 0 | 0 |
| 0 | 0 | 0 | 0 | 0 |
| 0 | 0 | 0 | 0 | 0 |
| 0 | 0 | 0 | 0 | 0 |
| 0.065019065 | 0.067841595 | 1.702865427 | 0.004066983 | 0 |
| 0 | 0 | 0 | 0 | 0 |
| 0 | 0 | 0 | 0 | 0 |
| 0 | 0 | 0 | 0 | 0 |
| 0 | 0 | 0 | 0 | 0 |
| 0 | 0 | 0.000357057 | 0 | 0 |
| 0 | 0 | 0 | 0 | 0 |
| 0.042909292 | 0.038845747 | 0.093254345 | 0 | 0 |
| 0 | 0 | 0 | 0 | 0 |
| 0 | 0 | 0 | 0 | 0 |
| 0 | 0 | 0.000499909 | 0 | 0 |
| 0 | 0 | 0 | 0 | 0 |
| 0 | 0 | 0.01024681 | 0 | 0 |
| 0 | 0 | 0 | 0 | 0 |
| 0 | 0 | 0 | 0 | 0 |
| 0 | 0 | 0.094669808 | 0.152039215 | 0 |
| 0 | 0 | 0 | 0 | 0 |
| 0 | 0 | 0.017697826 | 0 | 0 |
| 0 | 0 | 0 | 0 | 0 |

| | | | | |
|---|---|---|---|---|
| 0 | 0 | 0 | 0 | 0 |
| 0 | 0 | 0 | 0 | 0 |
| 0 | 0 | 0.006304788 | 0 | 0 |
| 0.210660136 | 0.195689455 | 0.089612202 | 0 | 0 |
| 0 | 0 | 0 | 0 | 0 |
| 1.272269007 | 1.240090999 | 0 | 0 | 0 |
| 0 | 0 | 0 | 0 | 0 |
| 0 | 0 | 0 | 0 | 0 |
| 0 | 0 | 0.017300334 | 0.864052707 | 0.013887814 |
| 0.001742311 | 0.002071057 | 0 | 0 | 0 |
| 0 | 0 | 0.000624761 | 0 | 0 |
| 0 | 0 | 0 | 0 | 0 |
| 0 | 0 | 0 | 0 | 0 |
| 0 | 0 | 0 | 0 | 0 |
| 0 | 0 | 17.54900538 | 0 | 0 |
| 0 | 0 | 0.001077483 | 0.004852091 | 0 |
| 0 | 0 | 0 | 0 | 0 |
| 0 | 0 | 0 | 0 | 0 |
| 0 | 0 | 0 | 0 | 0 |
| 0 | 0 | 0 | 0 | 0 |
| 0 | 0 | 0 | 0 | 0 |
| 0 | 0 | 0 | 0 | 0 |
| 0 | 0 | 0 | 0 | 0 |
| 0 | 0 | 0 | 0 | 0 |
| 0 | 0 | 0 | 1.10987637 | 0 |
| 2.309905354 | 2.242341776 | 1.816658045 | 0.0037797 | 2.163090934 |
| 0 | 0 | 0.004586075 | 0 | 0 |
| 0 | 0 | 0 | 0 | 0 |
| 0.094662266 | 0.099486353 | 0.038448851 | 0 | 0 |
| 0 | 0 | 0 | 0 | 0 |
| 0 | 0 | 0 | 0 | 0 |
| 0 | 0 | 0 | 0 | 0 |
| 0 | 0 | 0.001492718 | 0 | 0 |
| 0.007850936 | 0.008479813 | 0.000548756 | 0 | 0 |
| 0 | 0 | 0.001265848 | 0.01112922 | 0.175580632 |
| 0 | 0 | 0 | 0.002110661 | 0.015459067 |
| 0.818067135 | 0.830917084 | 0 | 0 | 0 |
| 1.462611967 | 1.484056418 | 0.012846324 | 0.517826338 | 2.137606775 |
| 0 | 0 | 0.134965372 | 0 | 0 |
| 0 | 0 | 0 | 0 | 0 |
| 0 | 0 | 0 | 0 | 0 |
| 0 | 0 | 0 | 0 | 0 |
| 0.303738158 | 0.323594137 | 0.160705809 | 0 | 0 |
| 0 | 0 | 0 | 0 | 0 |
| 0 | 0 | 0 | 0 | 0 |
| 0 | 0 | 0.002300932 | 0 | 0 |
| 0 | 0 | 0 | 0 | 0 |
| 0.35325244 | 0.32013455 | 0 | 0 | 0.979613921 |

| | | | | |
|---|---|---|---|---|
| 0 | 0 | 0.000431603 | 0 | 0 |
| 0 | 0 | 0 | 0 | 0 |
| 0 | 0 | 0 | 0 | 0 |
| 0 | 0 | 0 | 0 | 0 |
| 0.004334973 | 0.003425516 | 0 | 0 | 0 |
| 0.041365641 | 0.039330897 | 0 | 0 | 0 |
| 0 | 0 | 0 | 0 | 0 |
| 0 | 0 | 0 | 0 | 0 |
| 0 | 0 | 0 | 0 | 0 |
| 0 | 0 | 0.002444062 | 0 | 0 |
| 0.00090464 | 0 | 0 | 0 | 0 |
| 0.47409754 | 0.460181995 | 0.073604909 | 7.688937357 | 0.258437058 |
| 0 | 0 | 0.176383266 | 0 | 0 |
| 0 | 0 | 0 | 0 | 0 |
| 0 | 0 | 0.441963714 | 0.307338253 | 0.084426655 |
| 0 | 0 | 0 | 0 | 0 |
| 0.001743771 | 0.002119901 | 0 | 0 | 0 |
| 0 | 0 | 0 | 0 | 0 |
| 0 | 0 | 0.001451948 | 0 | 0 |
| 0 | 0 | 0 | 0 | 0 |
| 0 | 0 | 0.016107865 | 0 | 0 |
| 0 | 0 | 0 | 0 | 0 |
| 0.25267699 | 0.252708664 | 0.141040992 | 0 | 0 |
| 0 | 0 | 0 | 0 | 0 |
| 0 | 0 | 0 | 0 | 0 |
| 0 | 0 | 0 | 0 | 0 |
| 0 | 0 | 0 | 0 | 0 |
| 0 | 0 | 0 | 0 | 0 |
| 0 | 0 | 0 | 0 | 0 |
| 0 | 0 | 0 | 0 | 0 |
| 0 | 0 | 0 | 0 | 0 |
| 0.08279832 | 0.095696874 | 0.009888099 | 0.120354173 | 0.453819284 |
| 0 | 0 | 2.405132787 | 0 | 0 |
| 0.027625653 | 0.032399155 | 0.00902073 | 0 | 0 |
| 0 | 0 | 0 | 0.002296928 | 0.012737671 |
| 0.001233473 | 0 | 0.061330603 | 0.001615101 | 0.005886559 |
| 0 | 0 | 0 | 0 | 0 |
| 0 | 0 | 0.002660422 | 0 | 0 |
| 0 | 0 | 0 | 0 | 0 |
| 0 | 0 | 0 | 0 | 0 |
| 0.712017297 | 0.693351351 | 0 | 0 | 0 |
| 0 | 0 | 0 | 0 | 0 |
| 0 | 0 | 0 | 0 | 0 |
| 0 | 0 | 0 | 0 | 0 |
| 0.420200322 | 0.416209779 | 0.000685185 | 0 | 0 |
| 0 | 0 | 0 | 0 | 0 |
| 0 | 0 | 4.145605085 | 0.044825564 | 0 |
| 0 | 0 | 0.000388837 | 0 | 0 |

| | | | | |
|---|---|---|---|---|
| 0 | 0 | 0 | 0 | 0 |
| 0 | 0 | 0 | 0 | 0 |
| 0.024956399 | 0.022944232 | 0 | 0 | 0.000734584 |
| 0 | 0 | 0 | 0 | 0 |
| 0 | 0 | 0 | 0 | 0 |
| 0 | 0 | 0 | 0 | 0 |
| 0 | 0 | 0.063809736 | 0 | 0 |
| 0 | 0 | 0 | 0 | 0 |
| 0 | 0 | 0 | 0 | 0 |
| 0 | 0 | 0 | 0 | 0 |
| 0 | 0 | 0 | 0 | 0 |
| 0 | 0 | 0 | 0 | 0 |
| 0 | 0 | 0 | 0 | 0 |
| 0.547842141 | 0.536620321 | 0 | 0.06448199 | 0 |
| 0 | 0 | 0 | 0 | 0 |
| 0 | 0 | 0 | 0 | 0 |
| 0 | 0 | 5.069160637 | 0 | 0 |
| 0 | 0 | 0 | 0 | 0 |
| 0.086666874 | 0.072714195 | 1.605606314 | 0 | 0.36028152 |
| 0 | 0 | 0 | 0 | 0 |
| 0 | 0 | 0 | 0 | 0 |
| 0 | 0 | 0 | 0 | 0 |
| 0 | 0 | 0 | 0 | 0 |
| 0 | 0 | 0 | 0 | 0 |
| 0 | 0 | 0 | 0 | 0 |
| 0.001404004 | 0.000938766 | 0 | 0.038874358 | 0.016310164 |
| 0 | 0 | 0 | 0 | 0 |
| 0.051018685 | 0.053064452 | 0 | 0 | 0.012081172 |
| 0 | 0 | 0 | 0 | 0 |
| 0 | 0 | 0.223719389 | 0.131011732 | 0 |
| 0 | 0 | 0 | 0 | 0 |
| 0 | 0 | 0 | 0 | 0 |
| 0 | 0 | 0.035336819 | 0 | 0 |
| 0 | 0 | 0 | 0 | 0 |
| 0.001287396 | 0.002705363 | 0 | 0.001404756 | 0 |
| 0 | 0 | 0 | 0 | 0 |
| 0 | 0 | 0 | 0 | 0 |
| 0.003756387 | 0.003348867 | 0 | 0 | 0.001572524 |
| 0 | 0 | 0 | 0 | 0 |
| 0 | 0 | 0 | 0 | 0 |
| 24.78930124 | 24.66360018 | 6.963625143 | 9.562986673 | 15.89333695 |
| 0.053771755 | 0.043307818 | 0.03464636 | 0 | 0.034532841 |
| 0.041955795 | 0.044357465 | 0 | 0 | 0.220223264 |
| 0 | 0 | 0 | 0 | 0 |
| 0 | 0 | 0.062586977 | 0 | 0 |
| 0 | 0 | 0 | 0 | 0 |
| 0 | 0 | 0 | 0 | 0 |
| 0.013415743 | 0.013510719 | 0 | 0 | 0 |

| | | | | |
|---|---|---|---|---|
| 0 | 0 | 0 | 0 | 0 |
| 0.056559681 | 0.05535974 | 0 | 0 | 0 |
| 0 | 0 | 0.001301741 | 0 | 0 |
| 0 | 0 | 0 | 0 | 0 |
| 1.410934778 | 1.466151908 | 1.629812518 | 0 | 0 |
| 0 | 0 | 0 | 0 | 0 |
| 0 | 0 | 0 | 0 | 0 |
| 0 | 0 | 0 | 0 | 0 |
| 0.037934378 | 0.040946756 | 0.010510866 | 0.189328983 | 1.438203888 |
| 0 | 0 | 0 | 0 | 0 |
| 0 | 0 | 0 | 0.066365625 | 0 |
| 0 | 0 | 0 | 0 | 0 |
| 0.591453869 | 0.609857484 | 0 | 0 | 0 |
| 0.028287546 | 0.034290191 | 0.500909061 | 0.009770617 | 0.053416447 |
| 0.065835635 | 0.068539265 | 0.052690318 | 0 | 0 |
| 0.3027652 | 0.313540865 | 0.42864458 | 0 | 0 |
| 0.024126275 | 0.024962867 | 0 | 0 | 0 |
| 0 | 0 | 0 | 0 | 0 |
| 0 | 0 | 0 | 0 | 0 |
| 0 | 0 | 0 | 0 | 0 |
| 0 | 0 | 0 | 0 | 0 |
| 0 | 0 | 0 | 0 | 0 |
| 0 | 0 | 0 | 0 | 0 |
| 0 | 0 | 0 | 0 | 0 |
| 0 | 0 | 0 | 0 | 0 |
| 0 | 0 | 0 | 0 | 0 |
| 0 | 0 | 0.000556572 | 0 | 0 |
| 0 | 0 | 0 | 0.001319791 | 0 |
| 0 | 0 | 0 | 0 | 0 |
| 0 | 0 | 0 | 0 | 0 |
| 0 | 0 | 0 | 0 | 0 |
| 0 | 0 | 0 | 0 | 0 |
| 0 | 0 | 0 | 0 | 0 |
| 0 | 0 | 0.001185016 | 0 | 0 |
| 0.138177342 | 0.142094638 | 0.01759734 | 0 | 0 |
| 0 | 0 | 0 | 0 | 0 |
| 0 | 0 | 0 | 0 | 0 |
| 0 | 0 | 0 | 0 | 0 |
| 0 | 0 | 0 | 0 | 0 |
| 0 | 0 | 0 | 0 | 0 |
| 0.924380173 | 0.91794263 | 1.674912111 | 0 | 1.099421924 |
| 0 | 0 | 0 | 0 | 0 |
| 0 | 0 | 0 | 0 | 0 |
| 0 | 0 | 0.099771187 | 0 | 0 |
| 0 | 0 | 0 | 0 | 0 |
| 0 | 0 | 0 | 0 | 0 |
| 0 | 0 | 0 | 0 | 0 |
| 0 | 0 | 0 | 0 | 0 |

| | | | | |
|---|---|---|---|---|
| 0 | 0 | 0.002888059 | 0 | 0 |
| 0 | 0 | 0 | 0 | 0 |
| 0 | 0 | 0 | 0 | 0 |
| 0 | 0 | 0 | 0 | 0 |
| 0.07041806 | 0.087906749 | 0 | 0 | 0 |
| 0 | 0 | 0 | 0 | 0 |
| 0.151432698 | 0.157400494 | 0 | 0 | 0.041919321 |
| 0 | 0 | 0 | 0 | 0 |
| 0 | 0 | 0 | 0 | 0 |
| 0 | 0 | 0 | 0 | 0 |
| 0 | 0 | 0.000619447 | 0 | 0 |
| 0 | 0 | 0 | 0 | 0 |
| 0 | 0 | 0 | 0 | 0 |
| 0 | 0 | 0 | 0 | 0 |
| 0 | 0 | 0 | 0 | 0 |
| 0 | 0 | 0 | 0 | 0 |
| 0 | 0 | 0.007443268 | 0 | 0 |
| 0 | 0 | 0 | 0 | 0 |
| 0 | 0 | 0 | 0 | 0 |
| 0 | 0 | 0 | 0 | 0 |
| 0.032614399 | 0.03086546 | 0 | 0 | 3.794556123 |
| 0 | 0 | 0 | 0 | 0 |
| 0.002951677 | 0.002713692 | 0.004291315 | 0 | 0 |
| 0 | 0 | 0.001192463 | 0 | 0.001442114 |
| 0 | 0 | 0 | 0 | 0 |
| 0 | 0 | 0.000562735 | 0 | 0 |
| 0 | 0 | 0 | 0 | 0 |
| 0 | 0 | 0 | 0 | 0 |
| 0 | 0 | 0 | 0 | 0 |
| 0.004013598 | 0.003472934 | 4.758601117 | 0.005409948 | 12.32693229 |
| 0 | 0 | 0 | 0 | 0 |
| 0 | 0 | 0 | 0 | 0 |
| 0 | 0 | 0 | 0 | 0 |
| 0 | 0 | 0 | 0.007263946 | 0 |
| 0 | 0 | 0 | 0 | 0 |
| 0 | 0 | 0 | 0 | 0 |
| 0.001026291 | 0.001372428 | 0 | 0 | 0 |
| 0 | 0 | 0 | 0 | 0 |
| 0 | 0 | 0 | 0 | 0 |
| 0 | 0 | 0 | 0 | 0 |
| 0 | 0 | 0 | 0 | 0 |
| 0 | 0 | 0 | 0 | 0 |
| 0 | 0 | 0 | 0 | 0 |
| 0 | 0 | 0 | 0 | 0 |
| 0 | 0 | 0.007766776 | 0 | 0 |
| 0 | 0 | 0 | 0 | 0 |
| 0 | 0 | 0 | 0 | 0 |
| 0.033454942 | 0.035319686 | 0 | 0 | 0 |

| | | | | |
|---|---|---|---|---|
| 0.042892986 | 0.048389681 | 0 | 0 | 0 |
| 0 | 0 | 0.018618834 | 0 | 0 |
| 0 | 0 | 0 | 0 | 0 |
| 0 | 0 | 0 | 0 | 0 |
| 0 | 0 | 0 | 0 | 0 |
| 0 | 0 | 0 | 0 | 0 |
| 0 | 0 | 0 | 0 | 0 |
| 0.635791107 | 0.629669859 | 0.794181152 | 0 | 1.281934269 |
| 0.041005735 | 0.039644738 | 0.014702704 | 1.435129005 | 0.003479614 |
| 0.039541401 | 0.041786125 | 0 | 0 | 0.030764521 |
| 0 | 0 | 0 | 0 | 0 |
| 0 | 0 | 0 | 0 | 0 |
| 0 | 0 | 0 | 0 | 0 |
| 0 | 0 | 0.006560623 | 0.460164219 | 0 |
| 0 | 0 | 0 | 0 | 0 |
| 0 | 0 | 0 | 0 | 0 |
| 0 | 0 | 0 | 0.003936132 | 0 |
| 0 | 0 | 0 | 0 | 0 |
| 0 | 0 | 0 | 0 | 0 |
| 0 | 0 | 0.001162684 | 0 | 0 |
| 0 | 0 | 0 | 0 | 0 |
| 0 | 0 | 0 | 0 | 0 |
| 0 | 0 | 0 | 0 | 0 |
| 0 | 0 | 0 | 0 | 0 |
| 0 | 0 | 0.063771677 | 0 | 0.102492801 |
| 0 | 0 | 0 | 0 | 0 |
| 0 | 0 | 0 | 0 | 0 |
| 0 | 0 | 0 | 0 | 0 |
| 0 | 0 | 0 | 0 | 0 |
| 0 | 0 | 0 | 0 | 0 |
| 0 | 0 | 0 | 0 | 0 |
| 0 | 0 | 0 | 0 | 0 |
| 0 | 0 | 0 | 0 | 0 |
| 0 | 0 | 0 | 0 | 0 |
| 0.003846518 | 0.002857684 | 0 | 0 | 0 |
| 0 | 0 | 0 | 0 | 0 |
| 0 | 0 | 0 | 0 | 0 |
| 0 | 0 | 0 | 0 | 0 |
| 0.001259336 | 0 | 0 | 0 | 0 |
| 0 | 0 | 0 | 0.001730526 | 0.350107595 |
| 0 | 0 | 0 | 0 | 0 |
| 0 | 0 | 0.189906133 | 0 | 0.95961801 |
| 0 | 0 | 0 | 0 | 0 |
| 0 | 0 | 0 | 0 | 0 |
| 0 | 0 | 0 | 0 | 0 |
| 0 | 0 | 0 | 0 | 0 |
| 0 | 0 | 0 | 0 | 0 |
| 0 | 0 | 0 | 0 | 0 |

| | | | | |
|---|---|---|---|---|
| 0 | 0 | 0 | 0 | 0 |
| 0.000477936 | 0 | 0 | 0 | 0 |
| 0 | 0 | 0 | 0 | 0 |
| 0 | 0 | 0 | 0 | 0 |
| 0.030738183 | 0.032765039 | 0.052460882 | 0.094787821 | 0.188541627 |
| 0 | 0 | 1.643698465 | 0 | 0 |
| 0 | 0 | 0 | 0 | 0 |
| 0.001347234 | 0 | 0.011588885 | 0.077177592 | 0.007049854 |
| 0.00403549 | 0.001499038 | 0 | 0 | 0 |
| 0 | 0 | 0 | 0 | 0 |
| 0 | 0 | 0 | 0 | 0 |
| 0 | 0 | 0.031703698 | 0 | 0 |
| 0 | 0 | 0 | 0 | 0 |
| 0 | 0 | 0 | 0 | 0 |
| 0 | 0 | 0 | 0 | 0 |
| 0 | 0 | 0.000424688 | 0 | 0 |
| 0 | 0 | 0 | 0 | 0 |
| 0 | 0 | 0 | 0 | 0 |
| 0 | 0 | 0 | 0 | 0 |
| 0 | 0 | 0 | 0 | 0 |
| 0 | 0 | 0 | 0 | 0 |
| 0 | 0 | 0 | 0 | 0 |
| 0 | 0 | 0 | 0 | 0 |
| 0 | 0 | 0 | 0 | 0 |
| 0.279217941 | 0.256864299 | 0.06447326 | 0.558407614 | 0 |
| 0 | 0 | 0 | 0 | 0 |
| 0.001421822 | 0.004040387 | 0 | 0 | 0.001785638 |
| 0 | 0 | 0 | 0 | 0 |
| 0 | 0 | 0 | 0 | 0 |
| 0 | 0 | 0 | 0 | 0 |
| 0 | 0 | 0 | 0 | 0 |
| 0 | 0 | 0 | 0 | 0 |
| 0 | 0 | 0 | 0 | 0 |
| 0 | 0 | 0 | 0 | 0 |
| 0 | 0 | 0 | 0 | 0 |
| 0 | 0 | 0 | 0 | 0 |
| 0 | 0 | 0 | 0 | 0 |
| 0 | 0 | 0.001059979 | 0 | 0 |
| 0 | 0 | 0 | 0 | 0 |
| 0 | 0 | 0 | 0 | 0 |
| 0 | 0 | 0 | 0 | 0 |
| 0 | 0 | 0 | 0 | 0 |
| 0 | 0 | 0 | 0 | 0 |
| 0 | 0 | 0 | 0 | 0 |
| 0 | 0 | 0 | 0 | 0 |
| 0 | 0 | 0 | 0 | 0 |
| 0 | 0 | 0 | 0 | 0 |
| 0 | 0 | 0 | 0 | 0 |

| | | | | |
|---|---|---|---|---|
| 0.016880899 | 0.020339229 | 0.003766408 | 0 | 0 |
| 0 | 0 | 0 | 0 | 0 |
| 0 | 0 | 0 | 0 | 0 |
| 0 | 0 | 0 | 0 | 0 |
| 0 | 0 | 0 | 0 | 0 |
| 0.07590633 | 0.071210414 | 0 | 0.003591945 | 0 |
| 0 | 0 | 0 | 0 | 0 |
| 0 | 0 | 0 | 0 | 0 |
| 0 | 0 | 0 | 0 | 0 |
| 0 | 0 | 0 | 0 | 0 |
| 0 | 0 | 0 | 0 | 0 |
| 0.001074792 | 0.001676835 | 0 | 0 | 0 |
| 0 | 0 | 0 | 0 | 0 |
| 0 | 0 | 0 | 0 | 0 |
| 0 | 0 | 0.003815248 | 0 | 0 |
| 0 | 0 | 0 | 0 | 0 |
| 0 | 0 | 0 | 0 | 0 |
| 0 | 0 | 0 | 0 | 0 |
| 0 | 0 | 0 | 0 | 0 |
| 0 | 0 | 0 | 0 | 0 |
| 0.00174157 | 0.002328948 | 0 | 0 | 0.09963924 |
| 0 | 0 | 0 | 0 | 0 |
| 0.065495492 | 0.065035215 | 0.001954163 | 0 | 0 |
| 0 | 0 | 0.043575624 | 0 | 0 |
| 0 | 0 | 0 | 0 | 0 |
| 0 | 0 | 0.021921293 | 0 | 0 |
| 0 | 0 | 0 | 0 | 0 |
| 0 | 0 | 0 | 0 | 0 |
| 0 | 0 | 0 | 0 | 0 |
| 0 | 0 | 0 | 0 | 0 |
| 0 | 0 | 0 | 0 | 0 |
| 0 | 0 | 0 | 0 | 0 |
| 0 | 0 | 1.526773738 | 0.134604653 | 0.027593193 |
| 0 | 0 | 0 | 0 | 0 |
| 0 | 0 | 0.174115667 | 0 | 0 |
| 0 | 0 | 0 | 0 | 0.000691949 |
| 0 | 0 | 0 | 0 | 0 |
| 0 | 0 | 0 | 0 | 0 |
| 0.002973979 | 0.001704433 | 0 | 0.007417348 | 0.003201396 |
| 0 | 0 | 0 | 0 | 0 |
| 0 | 0 | 0.170598377 | 0 | 0 |
| 0 | 0 | 0 | 0 | 0 |
| 0.231251933 | 0.250452823 | 0.009930928 | 0 | 0.025891787 |
| 0 | 0 | 0 | 0 | 0 |
| 0 | 0 | 0 | 0 | 0 |
| 0.200816237 | 0.188614894 | 0.06940454 | 0 | 0 |
| 0 | 0 | 0 | 0 | 0 |
| 0 | 0 | 0 | 0 | 0 |

| | | | | |
|---|---|---|---|---|
| 0 | 0 | 0 | 0 | 0 |
| 0 | 0 | 0 | 0 | 0 |
| 0 | 0 | 0 | 0 | 0 |
| 0 | 0 | 0 | 0 | 0 |
| 0 | 0 | 0 | 0 | 0 |
| 0 | 0 | 0 | 0 | 0 |
| 0 | 0 | 0 | 0 | 0 |
| 0 | 0 | 0 | 0 | 0.733033813 |
| 0 | 0 | 0.031169305 | 0 | 0 |
| 0.003860444 | 0.004890745 | 0.001476982 | 0 | 0 |
| 0 | 0 | 0 | 0 | 0 |
| 0 | 0 | 0 | 0 | 0 |
| 0 | 0 | 0 | 0 | 0 |
| 0 | 0 | 0 | 0 | 0 |
| 0 | 0 | 0 | 0 | 0 |
| 0 | 0 | 0 | 0 | 0 |
| 0 | 0 | 0 | 0 | 0 |
| 0 | 0 | 0 | 0 | 0 |
| 0 | 0 | 0 | 0 | 0 |
| 0 | 0 | 0 | 0 | 0 |
| 0.004593371 | 0.006980198 | 0.008044077 | 0 | 0.023074898 |
| 0 | 0 | 0 | 0 | 0 |
| 0 | 0 | 0 | 0 | 0 |
| 0 | 0 | 0 | 0 | 0 |
| 0.002161447 | 0 | 0 | 0 | 0 |
| 0 | 0 | 0 | 0 | 0 |
| 0 | 0 | 0 | 0 | 0 |
| 0 | 0 | 0.000859225 | 0 | 0 |
| 0 | 0 | 0 | 0 | 0 |
| 0 | 0 | 0 | 0 | 0 |
| 0.105774165 | 0.106901208 | 0.018897749 | 0.025352657 | 0.019589263 |
| 0 | 0 | 0 | 0 | 0 |
| 0 | 0 | 0 | 0 | 0 |
| 0 | 0 | 0 | 0 | 0 |
| 0 | 0 | 0 | 0 | 0 |
| 0 | 0 | 0 | 0 | 0 |
| 0 | 0 | 0 | 0 | 0 |
| 0 | 0 | 0 | 0 | 0 |
| 0 | 0 | 0.006788898 | 0 | 0 |
| 0 | 0 | 0 | 0 | 0 |
| 0 | 0 | 0 | 0 | 0 |
| 0 | 0 | 0.01567958 | 0 | 0 |
| 0.001230151 | 0.000940025 | 0 | 0 | 0 |
| 0 | 0 | 0 | 0 | 0 |
| 0 | 0 | 0 | 0 | 0 |
| 0 | 0 | 0 | 0 | 0 |
| 0.043998513 | 0.036397376 | 0 | 0.271309275 | 0 |
| 0 | 0 | 0 | 0 | 0 |

| | | | | |
|---|---|---|---|---|
| 0 | 0 | 0 | 0 | 0 |
| 0 | 0 | 0 | 0 | 0 |
| 0 | 0 | 0 | 0 | 0 |
| 0 | 0 | 0 | 0 | 0 |
| 0.060083622 | 0.061129893 | 0.090687031 | 0 | 0 |
| 0 | 0 | 0 | 0 | 0 |
| 0 | 0 | 0 | 0 | 0 |
| 0 | 0 | 0 | 0 | 0 |
| 0 | 0 | 0 | 0 | 0 |
| 0 | 0 | 0 | 0 | 0 |
| 0 | 0 | 0 | 0 | 0 |
| 0.08002124 | 0.084869962 | 0 | 0 | 0 |
| 0 | 0 | 0 | 0 | 0 |
| 0 | 0 | 0 | 0 | 0 |
| 0 | 0 | 0 | 0 | 0 |
| 0 | 0 | 0 | 0 | 0 |
| 0 | 0 | 0.002610787 | 0 | 0 |
| 0 | 0 | 0 | 0 | 0 |
| 0 | 0 | 0.013786413 | 0 | 0 |
| 0.015109572 | 0.016276707 | 0 | 0 | 0 |
| 0 | 0 | 0 | 0 | 0 |
| 0 | 0 | 0 | 0 | 0 |
| 0 | 0 | 0 | 0 | 0 |
| 0 | 0 | 0 | 0 | 0 |
| 0 | 0 | 0 | 0 | 0 |
| 0 | 0 | 0 | 0 | 0 |
| 0.00538133 | 0.00449768 | 0.001833675 | 0 | 0.001267182 |
| 0 | 0 | 0 | 0 | 0 |
| 0 | 0 | 0 | 0 | 0 |
| 0 | 0 | 0 | 0 | 0 |
| 0 | 0 | 0 | 0 | 0 |
| 0 | 0 | 0 | 0 | 0 |
| 0 | 0 | 0 | 0 | 0 |
| 0 | 0 | 0 | 0 | 0 |
| 0 | 0 | 0 | 0 | 0 |
| 0 | 0 | 0 | 0 | 0 |
| 0 | 0 | 0 | 0 | 0 |
| 0.061205111 | 0.075042001 | 0.001168273 | 0.001753645 | 0.002691161 |
| 0 | 0 | 0.036769205 | 0 | 0 |
| 0 | 0 | 0 | 0 | 0 |
| 0.00441221 | 0.002503163 | 0 | 0 | 0 |
| 0 | 0 | 0 | 0 | 0 |
| 0 | 0 | 0 | 0 | 0 |
| 0 | 0 | 0 | 0 | 0 |
| 0 | 0 | 0 | 0 | 0 |
| 0 | 0 | 0 | 0 | 0 |
| 0 | 0 | 0 | 0 | 0 |
| 0 | 0 | 0 | 0 | 0 |

| | | | | |
|---|---|---|---|---|
| 0 | 0 | 0 | 0 | 0 |
| 0 | 0 | 0 | 0 | 0 |
| 0 | 0 | 0 | 0 | 0 |
| 0 | 0 | 0 | 0 | 0 |
| 0 | 0 | 0 | 0.00252764 | 0.022546352 |
| 0 | 0 | 0 | 0 | 0.001251537 |
| 0 | 0 | 0.001027755 | 0 | 0 |
| 0 | 0 | 0 | 0 | 0 |
| 0 | 0 | 0 | 0 | 0 |
| 0 | 0 | 0 | 0 | 0 |
| 0 | 0 | 0.001750266 | 0 | 0 |
| 0 | 0 | 0 | 0 | 0 |
| 0.024343477 | 0.027029507 | 0.007614624 | 0 | 0.011487846 |
| 0 | 0 | 0 | 0 | 0 |
| 0 | 0 | 0 | 0 | 0 |
| 0 | 0 | 0 | 0 | 0 |
| 0 | 0 | 0 | 0 | 0 |
| 0 | 0 | 0 | 0 | 0 |
| 0 | 0 | 0 | 0 | 0 |
| 0 | 0 | 0 | 0 | 0 |
| 0 | 0 | 0 | 0 | 0 |
| 0 | 0 | 0 | 0 | 0 |
| 0.016901595 | 0.022601985 | 0 | 0.000635943 | 0.006831479 |
| 0.017193503 | 0.019313569 | 0.188976424 | 0.004002321 | 0.04261015 |
| 0 | 0 | 0 | 0 | 0 |
| 0 | 0 | 0 | 0 | 0 |
| 0 | 0 | 0 | 0 | 0.008158608 |
| 0 | 0 | 0 | 0 | 0 |
| 0 | 0 | 0 | 0 | 0 |
| 0 | 0 | 0 | 0 | 0 |
| 0 | 0 | 0.008187973 | 0.005672593 | 0 |
| 0 | 0 | 0.000608828 | 0 | 0 |
| 0 | 0 | 0.001250038 | 0 | 0 |
| 0 | 0 | 0 | 0 | 0 |
| 0 | 0 | 0 | 0 | 0 |
| 0 | 0 | 0 | 0 | 0 |
| 0 | 0 | 0 | 0 | 0 |
| 0 | 0 | 0 | 0 | 0 |
| 0 | 0 | 0 | 0.000799901 | 0.020714683 |
| 0 | 0 | 0 | 0 | 0 |
| 0 | 0 | 0 | 0 | 0 |
| 0 | 0 | 0 | 0 | 0 |
| 0 | 0 | 0 | 0 | 0 |
| 0 | 0 | 0 | 0 | 0 |
| 0 | 0 | 0 | 0 | 0 |
| 0.015963344 | 0.017034707 | 0.741848525 | 0.010204823 | 0.020048049 |
| 0 | 0 | 0 | 0 | 0 |
| 0 | 0 | 0 | 0 | 0 |

| | | | | |
|---|---|---|---|---|
| 0 | 0 | 0 | 0 | 0 |
| 0 | 0 | 0 | 0 | 0 |
| 0.053256412 | 0.057284179 | 0.104238128 | 0 | 0.413972585 |
| 0 | 0 | 0.003008467 | 0 | 0 |
| 0 | 0 | 0 | 0 | 0 |
| 0 | 0 | 0 | 0 | 0 |
| 0 | 0 | 0 | 0 | 0 |
| 0 | 0 | 0 | 0 | 0 |
| 0 | 0 | 0 | 0 | 0 |
| 0 | 0 | 0 | 0 | 0 |
| 0.002824915 | 0.002385899 | 0 | 0 | 0 |
| 0 | 0 | 0 | 0 | 0 |
| 0 | 0 | 0 | 0 | 0 |
| 0 | 0 | 0 | 0 | 0 |
| 0 | 0 | 0 | 0 | 0 |
| 0 | 0 | 0 | 0 | 0 |
| 0 | 0 | 0 | 0 | 0 |
| 0 | 0 | 0 | 0 | 0 |
| 0 | 0 | 0 | 0 | 0 |
| 0 | 0 | 0 | 0 | 0.001733396 |
| 0 | 0 | 0 | 0 | 0 |
| 0 | 0 | 0 | 0 | 0 |
| 0 | 0 | 0 | 0 | 0 |
| 0 | 0 | 0 | 0 | 0 |
| 0 | 0 | 0 | 0 | 0 |
| 0 | 0 | 0 | 0 | 0 |
| 0 | 0 | 0 | 0 | 0 |
| 0.016147344 | 0.013422892 | 0.002855179 | 0 | 0.080842482 |
| 0 | 0 | 0 | 0 | 0 |
| 0 | 0 | 0 | 0 | 0 |
| 0.006098171 | 0.007475322 | 0.000369411 | 0 | 0 |
| 0 | 0 | 0 | 0 | 0 |
| 0.053694531 | 0.060757274 | 0.039460988 | 0.02146131 | 0.054836361 |
| 0.004019779 | 0.002418987 | 0 | 0.004386226 | 0.005048362 |
| 0 | 0 | 0 | 0 | 0 |
| 0 | 0 | 0 | 0 | 0 |
| 0 | 0 | 0 | 0 | 0 |
| 0 | 0 | 0 | 0 | 0 |
| 0 | 0 | 0 | 0 | 0 |
| 0 | 0 | 0 | 0 | 0 |
| 0 | 0 | 0 | 0 | 0 |
| 0 | 0 | 0 | 0.001235922 | 0.00060964 |
| 0.030555094 | 0.026097927 | 0.006735057 | 0.312540549 | 0.010893134 |
| 0 | 0 | 0 | 0 | 0 |
| 0 | 0 | 0 | 0 | 0 |
| 0.009115713 | 0.011809221 | 0.002070771 | 0 | 0.006439638 |
| 0.001680896 | 0.001998054 | 0 | 0 | 0 |
| 0 | 0 | 0 | 0 | 0 |

| | | | | |
|---|---|---|---|---|
| 0 | 0 | 0 | 0 | 0 |
| 0 | 0 | 0.009457862 | 0 | 0 |
| 0.080814233 | 0.079681756 | 0.019991223 | 0.006580697 | 0.038173488 |
| 0 | 0 | 0 | 0 | 0 |
| 0 | 0 | 0.003360427 | 0 | 0 |
| 0 | 0 | 0 | 0 | 0 |
| 0 | 0 | 0 | 0 | 0 |
| 0 | 0 | 0 | 0 | 0 |
| 0 | 0 | 0 | 0 | 0 |
| 0 | 0 | 0 | 0 | 0 |
| 0 | 0 | 0 | 0 | 0 |
| 0 | 0 | 0 | 0 | 0 |
| 0 | 0 | 0 | 0 | 0 |
| 0 | 0 | 0 | 0 | 0 |
| 0 | 0 | 0 | 0 | 0 |
| 0 | 0 | 0 | 0 | 0 |
| 0 | 0 | 0 | 0 | 0 |
| 0 | 0 | 0 | 0 | 0 |
| 0 | 0 | 0 | 0 | 0 |
| 0 | 0 | 0 | 0 | 0 |
| 0 | 0 | 0 | 0 | 0 |
| 0 | 0 | 0 | 0 | 0 |
| 0 | 0 | 0 | 0 | 0 |
| 0 | 0 | 0 | 0 | 0 |
| 0.002186941 | 0 | 0.001734271 | 0 | 0 |
| 0.008000039 | 0.0077562 | 0.130847703 | 0 | 0.010047092 |
| 0 | 0 | 0 | 0 | 0 |
| 1.810675992 | 1.864860884 | 0.008743849 | 0 | 0 |
| 0 | 0 | 0 | 0 | 0 |
| 0 | 0 | 0 | 0 | 0 |
| 0 | 0 | 0 | 0 | 0 |
| 0 | 0 | 0 | 0 | 0 |
| 0 | 0 | 0 | 0 | 0 |
| 0 | 0 | 0 | 0 | 0 |
| 0 | 0 | 0 | 0 | 0 |
| 0 | 0 | 0 | 0 | 0 |
| 0 | 0 | 0 | 0 | 0.018562413 |
| 0 | 0 | 0 | 0 | 0 |
| 0 | 0 | 1.056654949 | 0.086784732 | 0.705356307 |
| 0 | 0 | 0 | 0 | 0 |
| 0 | 0 | 0 | 0 | 0 |
| 0.002169702 | 0 | 0.000630887 | 0 | 0 |
| 0.041951068 | 0.041336752 | 0 | 0 | 0.000756251 |
| 0 | 0 | 0 | 0 | 0 |
| 0 | 0 | 0 | 0 | 0 |
| 0 | 0 | 0.000671462 | 0 | 0 |
| 0.007369599 | 0.004516939 | 0 | 0 | 0 |
| 0 | 0 | 0 | 0 | 0 |

| | | | | |
|---|---|---|---|---|
| 0 | 0 | 0 | 0 | 0 |
| 0.012979963 | 0.007253967 | 0.006196448 | 0 | 0.041118155 |
| 0 | 0 | 0 | 0 | 0 |
| 0 | 0 | 0 | 0 | 0 |
| 0 | 0 | 0 | 0 | 0 |
| 0 | 0 | 0 | 0 | 0 |
| 0 | 0 | 0 | 0 | 0 |
| 0 | 0 | 0 | 0 | 0 |
| 0 | 0 | 0 | 0 | 0 |
| 0 | 0 | 0 | 0 | 0 |
| 0 | 0 | 0 | 0 | 0 |
| 0 | 0 | 0 | 0 | 0 |
| 0.062461139 | 0.069012776 | 0.004763778 | 0 | 0.00102877 |
| 0 | 0 | 0 | 0 | 0 |
| 0.017150271 | 0.014212949 | 0.001931509 | 0 | 0.003033618 |
| 0 | 0 | 0 | 0 | 0 |
| 0 | 0 | 0 | 0 | 0 |
| 0 | 0 | 0 | 0 | 0 |
| 0 | 0 | 0 | 0 | 0 |
| 0 | 0 | 0 | 0 | 0 |
| 0 | 0 | 0 | 0 | 0 |
| 0.048868188 | 0.054881253 | 0 | 0 | 0 |
| 0.007291496 | 0.009263159 | 0 | 0 | 0.001144656 |
| 0 | 0 | 0 | 0 | 0 |
| 0 | 0 | 0 | 0 | 0 |
| 0 | 0 | 0 | 0 | 0 |
| 0 | 0 | 0 | 0 | 0 |
| 0 | 0 | 0 | 0 | 0 |
| 0 | 0 | 0 | 0 | 0 |
| 0 | 0 | 0 | 0 | 0 |
| 0 | 0 | 0 | 0 | 0 |
| 0 | 0 | 0 | 0 | 0 |
| 0 | 0 | 0 | 0 | 0 |
| 0 | 0 | 0 | 0 | 0 |
| 3.188580055 | 3.196840687 | 2.767069034 | 0 | 0 |
| 0 | 0 | 0 | 0 | 0 |
| 0 | 0 | 0 | 0 | 0 |
| 0 | 0 | 0 | 0 | 0 |
| 0.002132074 | 0.001900771 | 0 | 0 | 0.001785086 |
| 0 | 0 | 0 | 0 | 0 |
| 0.549715958 | 0.568949756 | 0.462910646 | 0.083438291 | 0 |
| 0 | 0 | 0 | 0 | 0 |
| 0 | 0 | 0 | 0 | 0 |
| 0 | 0 | 0 | 0 | 0 |
| 0 | 0 | 0 | 0 | 0 |
| 0.008998196 | 0.009106063 | 0.002474989 | 0.825813846 | 2.163923216 |
| 0 | 0 | 0 | 0 | 0 |
| 0 | 0 | 0 | 0 | 0 |

| | | | | |
|---|---|---|---|---|
| 0.316893792 | 0.316598215 | 0 | 0 | 0 |
| 0 | 0 | 0 | 0 | 0 |
| 0 | 0 | 0 | 0 | 0 |
| 0 | 0 | 0.002400037 | 0 | 0 |
| 0 | 0 | 0 | 0 | 0 |
| 0 | 0 | 0 | 0 | 0 |
| 0 | 0 | 0 | 0 | 0 |
| 0 | 0 | 0 | 0 | 0 |
| 0 | 0 | 0.003952068 | 0 | 0 |
| 0 | 0 | 0.03993896 | 0.013754918 | 0.031065252 |
| 0 | 0 | 0 | 0 | 0 |
| 0 | 0 | 0 | 0 | 0 |
| 0.019295265 | 0.016165714 | 0 | 0 | 0.004087417 |
| 0 | 0 | 0 | 0 | 0 |
| 0 | 0 | 0 | 0 | 0 |
| 0 | 0 | 0 | 0 | 0 |
| 0 | 0 | 0 | 0 | 0 |
| 0 | 0 | 0 | 0 | 0 |
| 0 | 0 | 0 | 0 | 0 |
| 0.403695927 | 0.423634366 | 0.01098023 | 1.445896641 | 0.067044793 |
| 0 | 0 | 0 | 0 | 0 |
| 0 | 0 | 0 | 0 | 0 |
| 0 | 0 | 0 | 0 | 0 |
| 0 | 0 | 0 | 0 | 0 |
| 0 | 0 | 0 | 0 | 0 |
| 0 | 0 | 0 | 0 | 0 |
| 0 | 0 | 0 | 0 | 0 |
| 0 | 0 | 0 | 0 | 0 |
| 0 | 0 | 0 | 0 | 0 |
| 0 | 0 | 0 | 0 | 0 |
| 0 | 0 | 0 | 0 | 0 |
| 0 | 0 | 0 | 0 | 0 |
| 0 | 0 | 0 | 0 | 0 |
| 0 | 0 | 0 | 0 | 0 |
| 0 | 0 | 0 | 0 | 0 |
| 0 | 0 | 0 | 0 | 0 |
| 0 | 0 | 0 | 0 | 0 |
| 0 | 0 | 0 | 0 | 0 |
| 0 | 0 | 0 | 0 | 0 |
| 0 | 0 | 0.018804334 | 0 | 0 |
| 0 | 0 | 0 | 0 | 0 |
| 0 | 0 | 0 | 0 | 0 |
| 0 | 0 | 0 | 0 | 0 |
| 0 | 0 | 0 | 0 | 0 |
| 0.015936246 | 0.021830835 | 0 | 0 | 0.001708514 |
| 0 | 0 | 0 | 0 | 0 |
| 0 | 0 | 0 | 0 | 0 |
| 0 | 0 | 0 | 0 | 0 |

| | | | | |
|---|---|---|---|---|
| 0 | 0 | 0 | 0 | 0 |
| 0 | 0 | 0 | 0 | 0 |
| 0 | 0 | 0 | 0 | 0 |
| 0 | 0 | 0 | 0 | 0 |
| 0 | 0 | 0 | 0 | 0 |
| 0.002466457 | 0.000776075 | 0.001371069 | 0.005699227 | 0.00674178 |
| 0 | 0 | 0 | 0 | 0 |
| 0 | 0 | 0 | 0 | 0 |
| 0 | 0 | 0 | 0 | 0 |
| 0 | 0 | 0 | 0 | 0 |
| 0 | 0 | 0 | 0 | 0 |
| 0 | 0 | 0 | 0 | 0 |
| 0 | 0 | 0 | 0 | 0 |
| 0 | 0 | 0 | 0 | 0 |
| 0 | 0 | 0 | 0 | 0 |
| 0 | 0 | 0 | 0 | 0 |
| 0 | 0 | 0.007036182 | 0 | 0 |
| 0 | 0 | 0 | 0 | 0 |
| 0 | 0 | 0 | 0 | 0 |
| 0 | 0 | 0 | 0 | 0 |
| 0 | 0 | 0 | 0 | 0 |
| 0 | 0 | 0 | 0 | 0 |
| 0 | 0 | 0 | 0 | 0 |
| 0 | 0 | 0 | 0 | 0 |
| 0 | 0 | 0 | 0 | 0 |
| 0 | 0 | 0.003299358 | 0 | 0 |
| 0 | 0 | 0 | 0 | 0 |
| 0 | 0 | 0 | 0 | 0 |
| 0 | 0 | 0 | 0 | 0 |
| 0 | 0 | 0 | 0 | 0 |
| 0 | 0 | 0 | 0 | 0 |
| 0 | 0 | 0 | 0 | 0 |
| 0 | 0 | 0 | 0 | 0 |
| 0 | 0 | 0 | 0 | 0 |
| 0 | 0 | 0 | 0 | 0 |
| 0 | 0 | 0 | 0 | 0 |
| 0 | 0 | 0 | 0 | 0 |
| 0 | 0 | 0 | 0 | 0 |
| 0 | 0 | 1.248920323 | 0 | 0 |
| 0 | 0 | 0 | 0 | 0 |
| 0 | 0 | 0 | 0 | 0 |
| 0 | 0 | 0 | 0 | 0 |
| 0 | 0 | 0 | 0 | 0 |
| 0 | 0 | 0 | 0 | 0 |
| 0 | 0 | 0 | 0 | 0 |
| 0 | 0 | 0 | 0 | 0 |
| 0 | 0 | 0 | 0 | 0 |
| 0 | 0 | 0 | 0 | 0 |

| | | | | |
|---|---|---|---|---|
| 0 | 0 | 0 | 0 | 0 |
| 0 | 0 | 0 | 0 | 0 |
| 0 | 0 | 0 | 0 | 0 |
| 0 | 0 | 0 | 0 | 0 |
| 0 | 0 | 0 | 0 | 0 |
| 0 | 0 | 0 | 0 | 0 |
| 0 | 0 | 0 | 0 | 0 |
| 0 | 0 | 0 | 0 | 0 |
| 0 | 0 | 0 | 0 | 0 |
| 0 | 0 | 0 | 0 | 0 |
| 0 | 0 | 0.000773073 | 0.093066286 | 0.037396888 |
| 0 | 0 | 0 | 0 | 0 |
| 0 | 0 | 0.915265707 | 0 | 0 |
| 0 | 0 | 0 | 0 | 0 |
| 0 | 0 | 0 | 0 | 0 |
| 0 | 0 | 0 | 0 | 0 |
| 0 | 0 | 0 | 0 | 0 |
| 0 | 0 | 0 | 0 | 0 |
| 0 | 0 | 0 | 0 | 0 |
| 0 | 0 | 0 | 0 | 0 |
| 0 | 0 | 0 | 0 | 0.023044466 |
| 0 | 0 | 0 | 0 | 0 |
| 0 | 0 | 0 | 0 | 0 |
| 3.410101205 | 3.420705963 | 0 | 3.456760836 | 2.251050959 |
| 0 | 0 | 0 | 0 | 0 |
| 0 | 0 | 0 | 0 | 0 |
| 0 | 0 | 0 | 0 | 0 |
| 0 | 0 | 0 | 0 | 0 |
| 0 | 0 | 0 | 0 | 0 |
| 0 | 0 | 0 | 0 | 0 |
| 0 | 0 | 0 | 0 | 0 |
| 0 | 0 | 0 | 0 | 0 |
| 0 | 0.00121466 | 0 | 0 | 0 |
| 0 | 0 | 0 | 0 | 0 |

| PHL018-0081_V2 | PHL018-0226_V1 | PHL018-0227_V1 | PHL018-0057_V3 | PHL018-0035_V3 |
|---|---|---|---|---|
| EYCF | REF | REF | REF | CTRL |
| 0.472677985 | 0.56019522 | 0.039768227 | 2.4557838 | 0.005306179 |
| 1.169777072 | 0 | 0.909507407 | 2.522438291 | 2.342994599 |
| 1.917713938 | 0 | 0.047350304 | 6.060829594 | 0 |
| 0 | 0 | 0 | 0 | 0 |
| 0 | 0 | 0 | 0 | 0 |
| 0 | 0 | 0 | 0 | 0 |
| 0 | 0 | 0 | 0 | 0 |
| 0 | 0 | 0 | 0 | 0 |
| 0 | 0 | 0 | 0 | 0 |
| 0 | 0 | 0 | 0 | 0 |
| 0 | 0 | 0 | 0 | 0 |
| 0 | 0 | 0 | 0 | 0 |
| 0 | 0 | 0 | 0 | 0 |
| 4.815010719 | 0 | 1.513464863 | 1.17030236 | 0.3962533 |
| 0.005138662 | 0 | 0 | 0 | 0 |
| 0 | 0 | 0 | 0 | 0 |
| 0 | 0 | 0 | 0.0007443 | 0 |
| 0 | 0 | 0 | 0 | 0 |
| 0 | 0 | 0 | 0 | 0 |
| 0 | 0.060077806 | 0 | 0 | 0 |
| 0 | 0 | 0 | 0 | 0 |
| 0 | 0 | 0 | 0 | 0 |
| 0 | 0 | 0 | 0 | 0 |
| 0 | 0 | 0 | 0 | 0 |
| 0.084003833 | 0 | 0 | 0 | 0 |
| 0 | 0 | 0 | 0 | 0 |
| 0 | 0.000875351 | 0 | 0 | 0 |
| 0 | 0 | 0 | 0 | 0 |
| 0 | 0 | 0 | 0 | 0 |
| 0 | 0 | 0 | 0 | 0 |
| 0 | 0 | 0 | 0 | 0 |
| 0 | 0 | 0 | 0 | 0 |
| 0 | 0 | 0 | 0 | 0 |
| 0 | 0 | 0 | 0 | 0 |
| 0 | 0 | 0 | 0 | 0 |
| 1.483143766 | 0 | 32.13807342 | 0 | 0.016513704 |
| 0 | 0 | 0 | 0 | 0 |
| 0 | 0 | 0 | 0 | 0 |
| 0 | 0 | 0 | 0 | 0 |
| 0 | 0 | 0 | 0 | 0 |
| 0 | 0 | 0 | 0 | 0 |
| 0 | 0.002155096 | 0 | 0 | 0.612423906 |
| 0.116110831 | 0.042414716 | 0.032011686 | 5.986308665 | 0.04321022 |
| 0.005254793 | 0.00905542 | 0.07166589 | 0.009964569 | 0.082503254 |
| 0 | 0.008598694 | 0 | 0 | 0 |
| 0 | 0 | 0 | 0 | 0 |

| | | | | |
|---|---|---|---|---|
| 0 | 0 | 0 | 0 | 0 |
| 0 | 0.097963655 | 0.08113167 | 0.022700017 | 0.026137573 |
| 0 | 0.01502606 | 0 | 0 | 0 |
| 0.399556396 | 0 | 0.364424884 | 0.005756878 | 0.49164181 |
| 0 | 0.001105569 | 0 | 0 | 0 |
| 0 | 0 | 0 | 0 | 0 |
| 0.001086169 | 0 | 0 | 0 | 0.022151602 |
| 0 | 0 | 0 | 0 | 0 |
| 0 | 0 | 0 | 0 | 0 |
| 0 | 0 | 0 | 0 | 0 |
| 0 | 0 | 0 | 0 | 0 |
| 0.118046727 | 0 | 0 | 0 | 0.011264919 |
| 0 | 0 | 0 | 0 | 0.001975455 |
| 0.0941235 | 0.179604726 | 0.019253154 | 0 | 0.003266175 |
| 0 | 0 | 0 | 0.001183521 | 0 |
| 0 | 0 | 0 | 0 | 0 |
| 0 | 0 | 0.000807981 | 0.000946048 | 0 |
| 0 | 0 | 0 | 0 | 0 |
| 0 | 0 | 0 | 0 | 0 |
| 0.000811423 | 0 | 0 | 0 | 0 |
| 0.145764414 | 0.0260266 | 0.121860331 | 0.049769188 | 0.230253786 |
| 0.010804625 | 0.020327659 | 0 | 0 | 0.002546086 |
| 0 | 0 | 0 | 0 | 0 |
| 0 | 0 | 0 | 0 | 0 |
| 0 | 0 | 0 | 0 | 0 |
| 0 | 0 | 0 | 0 | 0 |
| 0.008600735 | 0.035633272 | 0 | 0.023299167 | 0 |
| 0 | 0 | 0 | 0 | 0 |
| 0 | 0 | 0 | 0 | 0.019246976 |
| 0 | 0 | 0 | 0 | 0 |
| 0 | 0 | 0.081718182 | 0.246316978 | 0 |
| 0 | 0 | 0 | 0 | 0 |
| 0 | 0 | 0 | 0 | 0 |
| 0 | 0 | 0 | 0 | 0 |
| 0 | 0 | 0 | 0 | 0 |
| 0 | 0 | 0 | 0 | 0 |
| 0 | 0 | 0 | 0 | 0 |
| 0.001559731 | 0 | 0 | 0 | 0 |
| 0 | 0 | 0 | 0 | 0 |
| 0 | 0 | 0 | 0 | 0 |
| 0 | 0 | 0 | 0 | 0 |
| 0 | 0 | 0 | 0 | 0 |
| 0 | 0 | 0 | 0 | 0 |
| 0 | 0 | 0 | 0 | 0 |
| 0 | 0 | 0 | 0 | 0 |
| 0 | 0 | 0 | 0 | 0 |
| 0 | 0 | 0 | 0 | 0 |
| 0 | 0 | 0 | 0.001556401 | 0 |

| | | | | |
|---|---|---|---|---|
| 0 | 0 | 0 | 0 | 0 |
| 0 | 0 | 0 | 0 | 0 |
| 0 | 0 | 0 | 0 | 0 |
| 0 | 0 | 0 | 0 | 0 |
| 0.008784629 | 0.081968223 | 0 | 0 | 0 |
| 0 | 0 | 0 | 0 | 0 |
| 0 | 0 | 0 | 0 | 0 |
| 0 | 0 | 0.001610659 | 0 | 0 |
| 0 | 0.001045121 | 0 | 0 | 0.080541552 |
| 0 | 0 | 0 | 0 | 0 |
| 0 | 0 | 0 | 0 | 0 |
| 0 | 0 | 0 | 0 | 0.016883235 |
| 0 | 0 | 0 | 0 | 0 |
| 0 | 0 | 0 | 0 | 0 |
| 0 | 0 | 0 | 0 | 0 |
| 0 | 0 | 0 | 0 | 0 |
| 0 | 0 | 0 | 0 | 0 |
| 0 | 0 | 0 | 0 | 0 |
| 0 | 0 | 0 | 0.775390643 | 0.002542741 |
| 0 | 0 | 0 | 0 | 0 |
| 0 | 0 | 0 | 0 | 0 |
| 0 | 0 | 0 | 0 | 0 |
| 0 | 0 | 0 | 0 | 0 |
| 0 | 0 | 0 | 0 | 0 |
| 0 | 0 | 0 | 0 | 0 |
| 0 | 0 | 0 | 0 | 0 |
| 0 | 0 | 0 | 0 | 0.029178713 |
| 0 | 0 | 0 | 0 | 0 |
| 0 | 0 | 0 | 0 | 0 |
| 0 | 0 | 0 | 0 | 0 |
| 0.372759132 | 0 | 0 | 0 | 0 |
| 0.006034827 | 0.034090488 | 0 | 0 | 0 |
| 0 | 0 | 0 | 0 | 0 |
| 0.001837089 | 0 | 0 | 0 | 0 |
| 0 | 0 | 0 | 0 | 0 |
| 0 | 0 | 0 | 0 | 0 |
| 0 | 0 | 0 | 0 | 0 |
| 0 | 0 | 0 | 0 | 0 |
| 0 | 0 | 0 | 0 | 0 |
| 0 | 0 | 0 | 0 | 0 |
| 0 | 0 | 0 | 0 | 0 |
| 0 | 0 | 0 | 0 | 0 |
| 0.001953086 | 0 | 0 | 0.009096563 | 0 |
| 0 | 0 | 0 | 0 | 0 |
| 0 | 0 | 0 | 0 | 0 |
| 0 | 0 | 0 | 0 | 0 |
| 0 | 0.001229472 | 0 | 0 | 0 |
| 0 | 0 | 0 | 0 | 0 |

| | | | | |
|---|---|---|---|---|
| 0 | 0.00933884 | 0 | 0 | 0 |
| 0 | 0 | 0 | 0 | 0 |
| 0 | 0 | 0 | 0 | 0 |
| 0 | 0 | 0 | 0 | 0 |
| 0 | 0 | 0 | 0 | 0.001512332 |
| 0 | 0 | 0 | 0 | 0 |
| 0 | 0 | 0 | 0 | 0 |
| 0 | 0 | 0 | 0 | 0 |
| 0 | 0 | 0 | 0 | 0 |
| 0 | 0 | 0 | 0 | 0 |
| 0 | 0 | 0 | 0 | 0 |
| 0 | 0 | 0 | 0 | 0 |
| 0 | 0 | 0 | 0 | 0 |
| 0 | 0 | 0 | 0 | 0 |
| 0 | 0 | 0 | 0 | 0 |
| 0 | 0 | 0 | 0 | 0 |
| 0 | 0 | 0 | 0 | 0 |
| 0 | 0 | 0 | 0 | 0 |
| 0 | 0 | 0 | 0 | 0 |
| 0 | 0 | 0 | 0 | 0 |
| 0 | 0 | 0 | 0 | 0 |
| 0 | 0 | 0 | 0 | 0 |
| 0 | 0 | 0 | 0 | 0 |
| 0 | 0 | 0 | 0 | 0 |
| 0.030569456 | 0 | 0 | 0 | 0.164773838 |
| 0 | 0 | 0 | 0 | 0 |
| 0 | 0 | 0 | 0 | 0.001179646 |
| 0 | 0 | 0 | 0 | 0 |
| 0 | 0 | 0 | 0 | 0 |
| 0 | 0 | 0 | 0 | 0 |
| 0 | 0 | 0 | 0 | 0 |
| 0 | 0 | 0 | 15.93697497 | 0 |
| 0 | 0 | 0 | 0 | 0 |
| 0 | 0 | 0 | 0 | 0 |
| 0 | 0 | 0 | 0 | 0 |
| 0 | 0 | 0 | 0 | 0 |
| 0 | 0 | 0 | 0 | 0 |
| 0 | 0 | 0 | 0 | 0 |
| 0 | 0 | 0 | 0 | 0 |
| 0 | 0 | 0 | 0 | 0 |
| 0 | 0 | 0 | 0 | 0 |
| 0 | 0 | 0 | 0 | 0 |
| 0 | 0 | 0 | 0 | 0 |
| 0 | 0 | 0 | 0 | 0 |
| 0 | 0 | 0 | 0 | 0 |
| 0 | 0 | 0 | 0 | 0 |
| 0 | 0 | 0 | 0 | 0 |
| 0 | 0 | 0 | 0.206523274 | 0 |

| | | | | |
|---|---|---|---|---|
| 0.021720655 | 0 | 0 | 0 | 0 |
| 0 | 0 | 0 | 0 | 0 |
| 0 | 0 | 0 | 0 | 0 |
| 0 | 0 | 0 | 0 | 0 |
| 0 | 0 | 0 | 0 | 0 |
| 0 | 0 | 0 | 0 | 0 |
| 0 | 0 | 0 | 0 | 0 |
| 0 | 0 | 0 | 0 | 0 |
| 0 | 0 | 0 | 0 | 0 |
| 0 | 0 | 0 | 0 | 0 |
| 0 | 0 | 0 | 0 | 0 |
| 0 | 0 | 0 | 0 | 0 |
| 0 | 0.064522092 | 0 | 0 | 0 |
| 0 | 0 | 0 | 0 | 0 |
| 0 | 0 | 0 | 0 | 0 |
| 0 | 0 | 0 | 0 | 0 |
| 0 | 0 | 0 | 0 | 0 |
| 0 | 0 | 0 | 0 | 0 |
| 0 | 0 | 0 | 0 | 0 |
| 0 | 0 | 0 | 0 | 0 |
| 0.110625597 | 0.011728968 | 0.025296581 | 0.033487448 | 0.138990262 |
| 0 | 0 | 0 | 0 | 0 |
| 0 | 0.001653982 | 0 | 0 | 0 |
| 0 | 0.012667774 | 0 | 0 | 0 |
| 0 | 0 | 0 | 0 | 0 |
| 0 | 0 | 0 | 0 | 0 |
| 0 | 0.000975811 | 0.006596477 | 0.015447367 | 0.026633402 |
| 0 | 0 | 0 | 0 | 0 |
| 0 | 0 | 0 | 0 | 0 |
| 0.002173409 | 0.001827009 | 0 | 0.000619759 | 0 |
| 0 | 0 | 0.005260454 | 0.28179075 | 0.0278504 |
| 0 | 0 | 0 | 0 | 0 |
| 0 | 0 | 0 | 0 | 0 |
| 0 | 0 | 0 | 0.059370063 | 0 |
| 0 | 0.009811918 | 0.003684919 | 0 | 0.004084137 |
| 0 | 0 | 0 | 0 | 0 |
| 0 | 0 | 0 | 0 | 0 |
| 0 | 0 | 0 | 0 | 0 |
| 0 | 0 | 0 | 0 | 0 |
| 0 | 0 | 0 | 0 | 0 |
| 0 | 0 | 0 | 0 | 0 |
| 0 | 0 | 0 | 0 | 0 |
| 0 | 0 | 0 | 0 | 0 |
| 0 | 0 | 0 | 0 | 0 |
| 0 | 0 | 0 | 0 | 0 |
| 0 | 0 | 0 | 0 | 0 |
| 0.001755271 | 0.002655924 | 0 | 0 | 0 |
| 0 | 0.064289742 | 0 | 0.001537349 | 0.002286795 |

| | | | | |
|---|---|---|---|---|
| 0 | 0 | 0 | 0 | 0 |
| 0 | 0 | 0 | 0 | 0 |
| 0 | 0 | 0 | 0 | 0 |
| 0 | 0 | 0 | 0 | 0 |
| 0 | 0 | 0 | 0 | 0 |
| 0 | 0 | 0 | 0 | 0 |
| 0 | 0 | 0 | 0 | 0.091146278 |
| 0 | 0 | 0 | 0 | 0.002390771 |
| 0 | 0 | 0 | 0 | 0.000819323 |
| 0 | 0 | 0 | 0 | 0.001869596 |
| 0 | 0 | 0 | 0 | 0 |
| 0 | 0 | 0 | 0 | 0 |
| 0 | 0 | 0 | 0 | 0 |
| 0 | 0 | 0 | 0 | 0 |
| 0 | 0 | 0 | 0 | 0 |
| 0 | 0 | 0 | 0 | 0 |
| 0 | 0 | 0 | 0 | 0 |
| 0 | 0 | 0 | 0.024586294 | 0 |
| 0 | 0 | 0 | 0.027204452 | 0 |
| 0 | 0 | 0.002448305 | 0 | 0 |
| 0.003119588 | 0.038170259 | 0.001313134 | 0.000922513 | 0.006861158 |
| 0 | 0 | 0 | 0 | 0 |
| 0 | 0 | 0 | 0 | 0 |
| 0 | 0 | 0 | 0 | 0 |
| 0 | 0.0028944 | 0.003652346 | 0.004123728 | 0.004956778 |
| 0 | 0 | 0 | 0 | 0 |
| 0 | 0 | 0 | 0 | 0 |
| 0 | 0 | 0 | 0 | 0 |
| 0 | 0 | 0 | 0 | 0.117666848 |
| 0 | 0 | 0 | 0 | 0 |
| 0.734026133 | 0.06941657 | 0 | 0.715970763 | 0.018574797 |
| 0 | 0 | 0 | 0 | 0 |
| 0.012993354 | 0 | 0 | 0 | 0 |
| 0 | 0 | 0 | 0 | 0 |
| 0 | 0 | 0 | 0 | 0 |
| 0 | 0 | 0 | 0 | 0 |
| 0 | 0 | 0 | 0 | 0 |
| 0 | 0 | 0 | 0 | 0 |
| 0 | 0 | 0 | 0 | 0 |
| 0 | 0 | 0 | 0 | 0 |
| 0 | 0 | 0 | 0 | 0 |
| 0.016886855 | 0.005459772 | 0.013385319 | 0.031287584 | 0 |
| 0 | 0.011727234 | 0 | 0 | 0 |
| 0 | 0 | 0 | 0 | 0 |
| 0 | 0 | 0 | 0 | 0 |
| 0 | 0 | 0 | 0 | 0 |
| 0 | 0 | 0 | 0 | 0 |
| 0 | 0 | 0 | 0 | 0 |

| | | | | |
|---|---|---|---|---|
| 0 | 0 | 0 | 0 | 0 |
| 0 | 0 | 0 | 0 | 0 |
| 0.006850596 | 0.006032966 | 0.066982711 | 0.002821697 | 0.059583378 |
| 0 | 0 | 0 | 0 | 0 |
| 0 | 0 | 0 | 0 | 0 |
| 0 | 0 | 0 | 0 | 0 |
| 0 | 0 | 0 | 0 | 0 |
| 0 | 0 | 0 | 0 | 0 |
| 0 | 0 | 0 | 0 | 0 |
| 0 | 0 | 0.000678176 | 0 | 0 |
| 0 | 0 | 0 | 0 | 0 |
| 0 | 0 | 0 | 0 | 0 |
| 0 | 0 | 0 | 0 | 0 |
| 0 | 0 | 0.000819498 | 0.002878602 | 0 |
| 0 | 0.000750431 | 0.004283793 | 0.006929734 | 0.016332037 |
| 0 | 0 | 0 | 0 | 0 |
| 0 | 0.001425082 | 0.002312052 | 0.002030351 | 0.00884685 |
| 0 | 0 | 0 | 0 | 0 |
| 0 | 0 | 0 | 0 | 0 |
| 0 | 0 | 0 | 0 | 0 |
| 0 | 0 | 0 | 0 | 0 |
| 0 | 0.049656665 | 0 | 0.000610943 | 0.000826157 |
| 0 | 0 | 0 | 0 | 0 |
| 0 | 0.00279199 | 0 | 0 | 0 |
| 0 | 0 | 0 | 0 | 0.010155987 |
| 0 | 0 | 0 | 0 | 0 |
| 0 | 0 | 0 | 0 | 0 |
| 0 | 0 | 0 | 0 | 0 |
| 0 | 0 | 0 | 0 | 0 |
| 0 | 0.010420762 | 0 | 0 | 0.004381825 |
| 0 | 0 | 0 | 0 | 0.027012138 |
| 0 | 0 | 0 | 0 | 0 |
| 0 | 0 | 0 | 0 | 0 |
| 0 | 0 | 0 | 0 | 0 |
| 0 | 0 | 0 | 0 | 0 |
| 0 | 0 | 0 | 0 | 0 |
| 0 | 0 | 0 | 0 | 0 |
| 0.021150007 | 0.221430535 | 0 | 0.004605517 | 0.003459934 |
| 0 | 0 | 0 | 0 | 0 |
| 0 | 0 | 0 | 0 | 0 |
| 0 | 0 | 0 | 0 | 0 |
| 0 | 0 | 0 | 0 | 0 |
| 0 | 0 | 0 | 0 | 0 |
| 0 | 0 | 0 | 0 | 0 |
| 0 | 0 | 0 | 0 | 0 |
| 0 | 0 | 0 | 0 | 0.012235067 |
| 0 | 0 | 0 | 0 | 0.002250265 |
| 0 | 0.011746332 | 0.004587855 | 0 | 0 |

| | | | | |
|---|---|---|---|---|
| 0 | 0 | 0 | 0 | 0 |
| 0 | 0 | 0 | 0.411556416 | 0 |
| 0 | 0 | 0.003157698 | 0.010451941 | 0.002307559 |
| 0 | 0 | 0 | 0 | 0 |
| 0 | 0.010031902 | 0.0075758 | 0.004077501 | 0.001160814 |
| 0 | 0.004863167 | 0 | 0 | 0.026026161 |
| 0 | 0 | 0 | 0 | 0 |
| 0 | 0.001900259 | 0.183951106 | 0.008422864 | 0.039051256 |
| 0 | 0 | 0 | 0 | 0 |
| 0.285657494 | 0 | 0.004236925 | 0.013332501 | 0.010901306 |
| 0 | 0 | 0 | 0.001121331 | 0 |
| 0 | 0 | 0 | 0 | 0.073096633 |
| 0 | 0.002510263 | 0.002262583 | 0.007225126 | 0.010258804 |
| 0 | 0.042630031 | 0 | 0 | 0.053418777 |
| 0.00195443 | 0 | 0 | 0.000557316 | 0 |
| 0 | 0 | 0 | 0 | 0 |
| 0 | 0 | 0 | 0 | 0 |
| 0 | 0 | 0 | 0 | 0 |
| 0.006577451 | 0 | 0 | 0.027133594 | 0.015386895 |
| 0 | 0 | 0 | 0 | 0 |
| 0 | 0 | 0 | 0 | 0 |
| 0 | 0 | 0 | 0 | 0 |
| 0 | 0 | 0 | 0 | 0 |
| 0 | 0 | 0 | 0 | 0 |
| 0 | 0 | 0 | 0 | 0 |
| 0.000948592 | 0 | 0.002395756 | 0 | 0.001706984 |
| 0 | 0.002132583 | 0 | 3.188575923 | 0 |
| 0.002294223 | 0 | 0.01477539 | 0.016028135 | 0 |
| 0 | 0 | 0 | 0 | 0 |
| 0.004229255 | 0 | 0 | 0 | 0 |
| 0 | 0 | 0 | 0.005869639 | 0.003052812 |
| 0 | 0 | 0 | 0 | 0 |
| 0 | 0 | 0 | 0 | 0 |
| 0.013232899 | 0 | 0.119688587 | 0 | 0 |
| 0 | 0 | 0 | 0.286251087 | 0 |
| 0 | 0 | 0 | 0 | 0 |
| 0 | 0 | 0.092609983 | 0.007393308 | 0.009997715 |
| 0 | 0 | 0 | 0 | 0 |
| 0 | 0 | 0 | 0 | 0 |
| 0 | 0 | 0 | 0.023454827 | 0 |
| 0 | 0 | 0 | 0 | 0 |
| 0 | 0 | 0 | 0 | 0.016037789 |
| 0 | 0 | 0 | 0 | 0 |
| 0 | 0.847990581 | 0 | 0.008474708 | 0.005042425 |
| 0 | 0 | 0 | 0.255596669 | 0 |
| 0 | 0 | 0 | 0 | 0 |
| 0.049938487 | 0.000991635 | 0 | 0 | 0 |
| 0 | 0 | 0 | 0 | 0 |

| | | | | |
|---|---|---|---|---|
| 0 | 0 | 0 | 0 | 0 |
| 0 | 0 | 0 | 0 | 0.020164923 |
| 0 | 0 | 0 | 0 | 0 |
| 0 | 0 | 0 | 0 | 0 |
| 0 | 0 | 0 | 0 | 0 |
| 0 | 0.00598792 | 0 | 0 | 0.012818195 |
| 0.048212578 | 0.02634346 | 0 | 0 | 0 |
| 0 | 0 | 0 | 0 | 0.009370596 |
| 0.027116937 | 0 | 0 | 0 | 0 |
| 0 | 0 | 0 | 0 | 0 |
| 0 | 0 | 0 | 0.04995147 | 0 |
| 0 | 0.807685363 | 0 | 0 | 0 |
| 0 | 0 | 0 | 0 | 0 |
| 0.012556323 | 0.001996907 | 0.006942388 | 0.000677392 | 0.010076152 |
| 0 | 0.014432223 | 0.003902472 | 0 | 0 |
| 0 | 0 | 0 | 0 | 0 |
| 0 | 0 | 0 | 0 | 0 |
| 0 | 0 | 0.001501348 | 0.009228965 | 0 |
| 0 | 0 | 0.001642023 | 0.03388604 | 0 |
| 0 | 0 | 0 | 0 | 0 |
| 0 | 0 | 0 | 0 | 0 |
| 1.451474138 | 0 | 0.270649366 | 0 | 0.443259455 |
| 0 | 0 | 0 | 0 | 0 |
| 0 | 0 | 0 | 0 | 0 |
| 0 | 0 | 0 | 0 | 0 |
| 0 | 0 | 0 | 0 | 0 |
| 0 | 0 | 0 | 0.015913564 | 0.019201895 |
| 0 | 0 | 0 | 0 | 0 |
| 0 | 0 | 0 | 0 | 0 |
| 0.141270343 | 1.856166414 | 0.923762529 | 0.493147631 | 1.386097841 |
| 0.00391389 | 0.011612081 | 0 | 0 | 0 |
| 0 | 0 | 0 | 0 | 0 |
| 0 | 0 | 0 | 0 | 0 |
| 0 | 0 | 0 | 0 | 0 |
| 0 | 0.007986431 | 0 | 0 | 0 |
| 0 | 0 | 0 | 0 | 0 |
| 0.543628638 | 0 | 0.406109352 | 0 | 0 |
| 0 | 0 | 0 | 0 | 0 |
| 0 | 0 | 0 | 0 | 0 |
| 0 | 0.078311677 | 0 | 0 | 0 |
| 0 | 0.0009381 | 0 | 0.0018563 | 0.002008168 |
| 0 | 0 | 5.295837722 | 1.07156812 | 0 |
| 0 | 0 | 0 | 0 | 0 |
| 0 | 0 | 0 | 1.269541284 | 0 |
| 0 | 0 | 0.000953814 | 0 | 0 |
| 0 | 0 | 0 | 0 | 0 |
| 0 | 0.00295381 | 0 | 0.00097416 | 0 |
| 0 | 0 | 0 | 0 | 0 |

| | | | | |
|---|---|---|---|---|
| 0 | 0.00242967 | 0 | 0 | 0 |
| 0 | 0 | 0 | 0 | 0 |
| 0 | 0 | 0 | 0.001310199 | 0.005315211 |
| 0 | 0 | 0.006190465 | 0.001522141 | 0 |
| 0 | 0 | 0 | 0.363603323 | 0 |
| 0 | 0 | 0.445309313 | 0.255078638 | 0 |
| 0 | 0 | 0 | 0 | 0 |
| 0 | 0 | 0 | 0 | 0 |
| 0.013908972 | 0 | 0 | 0 | 0 |
| 0 | 0 | 0 | 0.191761514 | 0 |
| 0 | 0.032740199 | 0 | 0 | 0 |
| 0 | 0 | 0 | 0 | 0 |
| 0 | 0 | 0 | 0 | 0.010792912 |
| 0 | 0 | 0.002228834 | 0.016963028 | 0 |
| 0 | 0 | 0 | 0 | 0 |
| 0 | 0 | 0.000774131 | 0 | 0.004366604 |
| 0.078423236 | 0.20967179 | 0 | 0.000941874 | 0.023180692 |
| 0 | 0.002313217 | 0 | 0 | 0 |
| 0 | 0.009587321 | 0 | 0 | 0 |
| 0 | 0 | 0 | 0 | 0 |
| 0 | 0 | 0.01703172 | 0.031337587 | 0.059391644 |
| 0 | 0 | 0 | 0.000655515 | 0 |
| 0 | 0 | 0 | 0 | 0 |
| 0 | 0 | 0 | 0 | 0 |
| 0.065620706 | 0 | 0 | 0 | 0 |
| 0.010815722 | 0.165535267 | 0 | 0 | 0 |
| 0 | 0 | 0 | 0.001913148 | 0 |
| 0 | 0 | 0 | 0 | 0 |
| 0 | 0 | 0 | 0 | 0 |
| 0 | 0 | 0 | 0 | 0 |
| 0.009020113 | 0.024585607 | 0 | 0 | 0.001721536 |
| 0 | 0.001027686 | 0.003705148 | 0 | 0.020166135 |
| 0.038024599 | 0 | 0.552688963 | 0.011101075 | 0.015709816 |
| 0 | 0 | 0 | 0 | 0.643391482 |
| 0.002778151 | 0.002668989 | 0 | 0 | 0 |
| 0 | 0 | 0 | 0 | 0 |
| 0 | 0.002359007 | 0.005847189 | 0 | 0.049236224 |
| 0 | 0.185638307 | 0 | 0.025919044 | 0.002961924 |
| 0 | 0 | 0.14082008 | 0 | 0 |
| 0 | 0 | 0 | 0 | 0 |
| 0 | 0 | 0 | 0 | 0 |
| 0 | 0 | 0 | 0 | 0 |
| 0 | 0.643875642 | 0 | 0 | 0 |
| 0 | 0 | 0 | 0 | 0 |
| 0 | 0 | 0.063820951 | 0 | 0 |
| 0 | 0 | 0 | 0 | 0 |
| 0 | 0 | 0 | 0 | 0 |
| 0 | 0 | 0 | 0.000741707 | 0 |

| | | | | |
|---|---|---|---|---|
| 0 | 0 | 0.00616355 | 0 | 0 |
| 0.00971903 | 0.054361918 | 0.331658266 | 0.312887508 | 0.020516318 |
| 0 | 0 | 0 | 0 | 0 |
| 0 | 0 | 0 | 0 | 0 |
| 0 | 0 | 0 | 0.000732013 | 0 |
| 0 | 0 | 0 | 0 | 0 |
| 0 | 0 | 0 | 0.267372413 | 0.023647545 |
| 0.002584121 | 0 | 0 | 0 | 0 |
| 0 | 0 | 0 | 0 | 0 |
| 0 | 0 | 0 | 0 | 0 |
| 0.011944576 | 0.027086904 | 0 | 0 | 0.001249661 |
| 0.023584459 | 0.003478165 | 0.369928196 | 0.005781338 | 1.328671521 |
| 0 | 0 | 0 | 0 | 0 |
| 0 | 0 | 0 | 0 | 0 |
| 0.002868469 | 0 | 0.017145508 | 0.00063619 | 0.197868543 |
| 0 | 0 | 0 | 0.002347149 | 0 |
| 0.001454383 | 0.006357426 | 0.008815627 | 0.018192597 | 0.002355436 |
| 0.020767892 | 0.002221912 | 0.016879754 | 0.033414841 | 0.050961361 |
| 0 | 0 | 0 | 0 | 0 |
| 0 | 0 | 0.05777658 | 0.056773326 | 0.010589326 |
| 0.134385299 | 0.001765106 | 0.171822453 | 0.005987603 | 0.003778521 |
| 0 | 0 | 0 | 0 | 0 |
| 0 | 0 | 0 | 0 | 0 |
| 0 | 0 | 0 | 0 | 0 |
| 0 | 0 | 0 | 0 | 0 |
| 0 | 0 | 0 | 0 | 0.001485004 |
| 0.005921652 | 0.160286838 | 0.06842221 | 0 | 0.015717553 |
| 0.145382301 | 0 | 0.207986055 | 0.023970835 | 0.244402876 |
| 0 | 0 | 0 | 0 | 0 |
| 0.002307436 | 0.002909511 | 0 | 0.022517441 | 0 |
| 0 | 0 | 0 | 0 | 0.003385786 |
| 0 | 0 | 0 | 0 | 0 |
| 0 | 0 | 0 | 0 | 0 |
| 0 | 0 | 0 | 0 | 0 |
| 0.542602307 | 0 | 0 | 0 | 0 |
| 0.51260598 | 0.274872818 | 0.327474096 | 0.31306504 | 1.656976599 |
| 0 | 0 | 0 | 0 | 0 |
| 0 | 0 | 0 | 0 | 0 |
| 0 | 0.008926642 | 0 | 0 | 0.274250201 |
| 0 | 0 | 0 | 0 | 0 |
| 0 | 0 | 0.072553546 | 0 | 0 |
| 0 | 0 | 0 | 0 | 0 |
| 0 | 0.33697835 | 0.001086042 | 0.009060326 | 0.016980796 |
| 0 | 0 | 0 | 0.015960486 | 0.028431209 |
| 0 | 0 | 0 | 0 | 0 |
| 0 | 0 | 0 | 0 | 0.151808123 |
| 0 | 0 | 0 | 0 | 0 |
| 0 | 0 | 0 | 0 | 0 |

| | | | | |
|---|---|---|---|---|
| 0 | 0 | 0.025792807 | 0.050185763 | 0 |
| 0 | 0 | 0 | 0 | 0 |
| 0 | 0 | 0 | 0 | 0.004727646 |
| 0 | 0.005379548 | 0 | 0.632219105 | 0 |
| 0 | 0 | 0 | 0 | 0 |
| 0 | 0 | 0 | 0 | 0 |
| 0 | 0.042069106 | 0 | 0 | 0 |
| 0 | 0 | 0.003017558 | 0.00248428 | 0 |
| 2.657049229 | 0 | 0 | 0 | 0.259261845 |
| 0 | 0 | 0.017145994 | 0.00231645 | 0.002684962 |
| 0 | 0 | 0 | 0 | 0 |
| 0.002399251 | 0 | 0 | 0 | 0.001199287 |
| 0 | 0 | 0 | 0 | 0 |
| 0 | 0 | 0 | 0 | 0 |
| 1.962488937 | 0.046046024 | 0 | 0 | 0 |
| 0 | 0 | 0 | 0 | 0 |
| 0 | 0 | 0 | 0 | 0.279354631 |
| 0 | 0 | 0 | 0 | 0 |
| 0 | 0 | 0 | 0 | 0 |
| 0 | 0 | 0 | 0 | 0 |
| 0 | 0 | 0 | 0 | 0 |
| 0.032422913 | 0 | 0 | 0 | 0 |
| 0 | 0 | 0 | 0 | 0 |
| 0 | 0 | 0.051216582 | 0 | 0.028217418 |
| 0 | 0.001258861 | 0 | 0 | 0 |
| 0 | 0 | 0 | 0.001711503 | 1.142159546 |
| 0 | 0 | 0 | 0 | 0 |
| 1.007246824 | 0 | 3.225956648 | 0 | 0 |
| 0 | 0 | 0 | 0 | 0 |
| 0 | 0.012595392 | 0 | 0 | 0 |
| 0.052092071 | 0 | 0 | 0 | 0 |
| 1.164499476 | 0 | 0 | 0 | 0.010985049 |
| 0 | 0 | 0 | 0.001968572 | 0 |
| 0 | 0 | 0 | 0 | 0 |
| 0.004067818 | 0 | 0 | 0 | 0 |
| 0.328295354 | 0 | 0 | 0 | 0.041519628 |
| 0 | 0 | 0.006665864 | 0 | 0 |
| 0 | 0 | 0 | 0 | 0 |
| 0 | 0.10054495 | 0 | 0 | 0 |
| 0.178792059 | 0.011067686 | 0 | 0 | 0 |
| 0.001278696 | 0.025797495 | 0.058517868 | 0.481209963 | 0.958827993 |
| 0 | 0 | 0 | 0 | 0.326348147 |
| 16.79769902 | 0 | 0.0197581 | 0.131457621 | 3.741911047 |
| 0.000891129 | 0 | 0 | 0 | 0.000601342 |
| 0 | 0.00982967 | 0 | 0 | 0 |
| 0 | 0 | 0 | 0 | 0 |
| 0 | 0 | 0 | 0 | 0 |
| 0.0055337 | 0.679669765 | 0.001164655 | 0.017795917 | 0 |

| | | | | |
|---|---|---|---|---|
| 0 | 0 | 0 | 0 | 0 |
| 0.023526334 | 0 | 0 | 0 | 0 |
| 0.024165746 | 1.896650868 | 0.018086551 | 0.00941208 | 0.007689611 |
| 0 | 0 | 0 | 0 | 0 |
| 0 | 0 | 0 | 0 | 0 |
| 0.003018036 | 0 | 0.001055399 | 0 | 0 |
| 0 | 0 | 0 | 0 | 0 |
| 0 | 0.001186628 | 0 | 0 | 0 |
| 0 | 0 | 0 | 0 | 0 |
| 0 | 0 | 0 | 0 | 0 |
| 0.105033065 | 0.042078924 | 0.00460387 | 0.002310248 | 0 |
| 2.932903697 | 0 | 2.124012355 | 0.822978759 | 1.978100293 |
| 0 | 0 | 0 | 0 | 0 |
| 0.008431435 | 0.052500913 | 0 | 0.006856614 | 0.004776466 |
| 0.008120804 | 0.42158815 | 0.083158058 | 0.172413594 | 0.084137331 |
| 0 | 0 | 0 | 0 | 0 |
| 0 | 0.001176279 | 0 | 0 | 0 |
| 0 | 0 | 0 | 0 | 0 |
| 0 | 0 | 0 | 0 | 0 |
| 0 | 0 | 0 | 0.143996442 | 0 |
| 0 | 0.423831608 | 0.005879956 | 0.004034018 | 7.632506067 |
| 0.047018669 | 0 | 0.003678667 | 0.015132146 | 0 |
| 7.059687625 | 0 | 0.678600195 | 1.085152735 | 0.417141485 |
| 0 | 0 | 0 | 0 | 0 |
| 0 | 0 | 0 | 0 | 0 |
| 3.619503667 | 0 | 0.016734134 | 0 | 0 |
| 0 | 42.14526527 | 0 | 0 | 0 |
| 0 | 0 | 0 | 0 | 0 |
| 0 | 0 | 0 | 0 | 0 |
| 0 | 0 | 0 | 0 | 0 |
| 0.006687212 | 0 | 0 | 0 | 0.000962687 |
| 0.263296353 | 0 | 0 | 0 | 0.062005972 |
| 0 | 0 | 0.003827667 | 0.019179206 | 0.003743251 |
| 0.541378455 | 0 | 0 | 0 | 0.001019471 |
| 0 | 0.901365979 | 0.003715159 | 0.007308008 | 0.000882354 |
| 0.02398338 | 0.387360744 | 0.03858927 | 0.024922592 | 0.010425787 |
| 0.089026127 | 0.001094109 | 0 | 0 | 0 |
| 0 | 0 | 0 | 0 | 0 |
| 0.029326765 | 0 | 0.019830957 | 0 | 0.002269819 |
| 0.340510513 | 0 | 0 | 0 | 0 |
| 0 | 0 | 0 | 0 | 0 |
| 0 | 0 | 0 | 0 | 0 |
| 0 | 0 | 0 | 0 | 0 |
| 0 | 0 | 0 | 0 | 0 |
| 0.028272244 | 0.913447968 | 0.010477781 | 0.068702059 | 0.00165899 |
| 0 | 0 | 0 | 0 | 0 |
| 0 | 0 | 0 | 0 | 0 |
| 0.003706962 | 3.354215424 | 0.003370418 | 0.059935268 | 0 |

| | | | | |
|---|---|---|---|---|
| 0 | 0 | 0 | 0 | 0 |
| 0 | 0 | 0 | 0 | 0 |
| 0 | 0 | 0 | 0 | 0 |
| 0 | 0.00110086 | 0 | 0 | 0.001178292 |
| 0 | 0 | 0 | 0 | 0 |
| 0 | 0 | 0 | 0 | 0 |
| 0 | 0 | 0 | 0 | 0 |
| 0 | 0.460302757 | 0 | 0 | 0.000650545 |
| 0 | 0 | 0 | 0 | 0 |
| 0 | 0 | 0 | 0 | 0 |
| 0 | 0.823426083 | 0 | 0.023484873 | 0 |
| 0 | 0 | 0 | 0 | 0 |
| 0.008939388 | 0 | 0 | 0 | 0 |
| 0 | 0 | 0 | 0 | 0 |
| 0 | 0 | 0 | 0 | 0 |
| 0 | 0 | 0 | 0 | 0 |
| 0.360140636 | 0 | 0 | 0 | 0 |
| 0.002185545 | 0 | 0 | 0 | 0 |
| 0 | 0 | 0 | 0 | 0 |
| 0 | 0 | 0 | 0.119485672 | 0 |
| 0 | 0 | 0 | 0 | 0 |
| 0 | 0 | 0 | 0 | 0 |
| 0 | 0 | 0.003181963 | 0 | 0 |
| 0 | 0 | 0 | 0 | 0 |
| 0 | 0 | 0 | 0 | 0 |
| 0 | 0 | 0 | 0 | 0 |
| 0 | 0 | 0 | 0 | 0 |
| 0 | 0 | 0 | 0 | 0 |
| 0 | 0 | 0 | 0 | 0 |
| 0.064536226 | 0 | 0.036450971 | 0.014833803 | 0.109797951 |
| 0 | 0 | 0 | 0 | 0 |
| 0 | 0 | 0 | 0 | 0 |
| 0 | 0 | 0 | 0 | 0 |
| 0 | 0 | 0 | 0 | 0 |
| 0 | 0 | 0 | 0 | 0 |
| 0 | 0 | 0 | 0 | 0 |
| 0.129723831 | 0 | 0 | 0 | 0 |
| 0.012099762 | 0.006850051 | 0.310766908 | 0.131933272 | 0.005554447 |
| 0 | 0 | 0 | 0 | 0 |
| 0 | 0 | 0 | 0 | 0.00219144 |
| 0 | 0.009348426 | 0 | 0 | 0.001459205 |
| 0 | 0 | 0 | 0 | 0 |
| 0 | 0 | 0 | 0 | 0 |
| 0 | 0 | 0 | 0.00366977 | 0 |
| 0.393668849 | 0 | 0 | 0 | 0 |
| 0 | 0 | 0 | 0 | 0 |
| 0.004223314 | 0 | 0 | 0 | 0 |
| 0 | 0 | 0 | 0 | 0 |

| | | | | |
|---|---|---|---|---|
| 0 | 0 | 0 | 0 | 0 |
| 0 | 0 | 0 | 0 | 0 |
| 0.08372362 | 0 | 0 | 0 | 0 |
| 0 | 0 | 0 | 0 | 0 |
| 0 | 0 | 0 | 0 | 0 |
| 0.581046176 | 0 | 0 | 0 | 0 |
| 0 | 0 | 0 | 0 | 0 |
| 0 | 0 | 0 | 0 | 0 |
| 0.035759128 | 0.014314189 | 0.121019356 | 0.058915422 | 0 |
| 0 | 0 | 0 | 0 | 0 |
| 0.003896212 | 0 | 0 | 0 | 0 |
| 0 | 0 | 0 | 0 | 0.015131507 |
| 0 | 0.010354779 | 0 | 0 | 0 |
| 0 | 0.016504011 | 0 | 0 | 0 |
| 0.804207918 | 0 | 3.223924282 | 0 | 3.290475077 |
| 0 | 0 | 0.001193258 | 0 | 0 |
| 0 | 0.00733551 | 0 | 0 | 0 |
| 0 | 0 | 0 | 0 | 0 |
| 0 | 0 | 0 | 0 | 0 |
| 0 | 0 | 0 | 0 | 0 |
| 0 | 0 | 0 | 0 | 0.001171512 |
| 0 | 0 | 0 | 0 | 0 |
| 0 | 0 | 0 | 0 | 0 |
| 0 | 0 | 0 | 0 | 0 |
| 0.010813299 | 0.033666156 | 0 | 3.37220993 | 0.012972299 |
| 0 | 0 | 1.054228359 | 9.670218923 | 0.478208917 |
| 0 | 0 | 0.011512052 | 0 | 0 |
| 0 | 0 | 0 | 0 | 0 |
| 0 | 0 | 0 | 0 | 0 |
| 0 | 0 | 0 | 0 | 0 |
| 0 | 0 | 0 | 0 | 0 |
| 0 | 0 | 0 | 0 | 0 |
| 0 | 0 | 0 | 0 | 0 |
| 0 | 0 | 0.000486176 | 0 | 0 |
| 0.002114526 | 0.004443777 | 0.000801064 | 0.003048339 | 0.036148204 |
| 0.004982379 | 0.03685688 | 0.00924884 | 0.001823297 | 0 |
| 0.117744417 | 0 | 0 | 0 | 0 |
| 1.232649461 | 0 | 12.84832301 | 0.231336261 | 3.065512581 |
| 0 | 0 | 0 | 0 | 0 |
| 0 | 0 | 0 | 0 | 0 |
| 0 | 0 | 0 | 0 | 0 |
| 0 | 0 | 0 | 0 | 0 |
| 0.218553174 | 0 | 1.031730936 | 1.565837433 | 0.042268925 |
| 1.385662435 | 0 | 0 | 0 | 0 |
| 0.308487828 | 0 | 0 | 0 | 0 |
| 0.06513847 | 0 | 0 | 0 | 0 |
| 0 | 0 | 0 | 0 | 0 |
| 0 | 5.933167058 | 1.754858117 | 0.100067936 | 1.394118089 |

| | | | | |
|---|---|---|---|---|
| 0 | 0.069764185 | 0 | 0 | 0 |
| 0 | 0 | 0 | 0 | 0 |
| 0 | 0 | 0 | 0 | 0 |
| 0 | 0 | 0 | 0 | 0 |
| 0.002512326 | 0.012906108 | 0 | 0 | 0 |
| 0 | 0 | 0 | 0 | 0 |
| 0.522231499 | 0 | 0 | 0 | 0 |
| 0 | 0 | 0 | 0 | 0 |
| 0 | 0 | 0 | 0 | 0 |
| 0 | 0 | 0 | 0 | 0 |
| 0 | 0 | 0 | 0 | 0 |
| 0.49111208 | 0 | 0.117465623 | 0 | 0.006951383 |
| 0 | 0.253442083 | 0 | 0.694689447 | 1.326540314 |
| 0 | 0 | 0 | 0 | 0 |
| 0.07037779 | 0.01058383 | 0.002201436 | 0 | 5.251671833 |
| 0 | 0 | 0 | 0 | 0 |
| 0 | 0.087784636 | 0 | 0 | 0 |
| 0 | 0 | 0 | 0 | 0 |
| 0 | 0.01308251 | 0.000857578 | 0 | 0 |
| 0 | 0 | 0 | 0 | 0 |
| 0 | 0 | 0 | 0 | 0 |
| 0 | 0 | 0 | 0 | 0 |
| 0 | 0 | 0.037732984 | 0.195701402 | 0.082437054 |
| 0 | 0 | 0 | 0 | 0 |
| 0 | 0 | 0 | 0 | 0 |
| 0 | 0 | 0 | 0 | 0 |
| 0.882390812 | 0 | 0 | 0 | 0 |
| 0 | 0 | 0 | 0 | 0 |
| 0.00523425 | 0 | 0 | 0 | 0 |
| 0 | 0 | 0 | 0 | 0 |
| 0 | 0 | 0 | 0 | 0 |
| 0 | 0.021129229 | 0.080304188 | 0.028988428 | 0.07236933 |
| 0 | 0 | 0 | 0 | 0 |
| 0.192334086 | 0.007213129 | 0 | 0 | 0 |
| 0.002711039 | 0.039653731 | 0.006778506 | 0.002344969 | 0 |
| 0.189421276 | 0 | 0.005825545 | 0.012091797 | 0 |
| 0 | 0 | 0 | 0 | 0 |
| 0 | 0 | 0 | 0 | 0 |
| 0 | 0 | 0.00094394 | 0 | 0 |
| 0.003054727 | 0 | 0 | 0 | 0 |
| 0 | 0 | 0 | 0.34994373 | 0 |
| 0 | 0 | 0 | 0 | 0 |
| 0 | 0 | 0 | 0 | 0 |
| 0 | 0 | 0.025417755 | 0 | 0 |
| 0.054481153 | 0.583025001 | 0.005463416 | 0.063970044 | 0 |
| 0 | 0 | 0 | 0 | 0 |
| 0.057523717 | 0 | 0 | 0 | 0.014152235 |
| 0.003788928 | 0 | 0 | 0 | 0 |

| | | | | |
|---|---|---|---|---|
| 0 | 0 | 0 | 0 | 0 |
| 0 | 0 | 0 | 0 | 0 |
| 0 | 0 | 0 | 0.006616103 | 0 |
| 0 | 0 | 0 | 0 | 0 |
| 0 | 0 | 0 | 0 | 0 |
| 0 | 0 | 0 | 0 | 0 |
| 0 | 0 | 0.197888026 | 0 | 0 |
| 0 | 0 | 0 | 0.001388509 | 0.010326982 |
| 0 | 0 | 0 | 0 | 0 |
| 0 | 0 | 0 | 0 | 0 |
| 0 | 0 | 0 | 0 | 0 |
| 0 | 0.050556698 | 0 | 0 | 0 |
| 0 | 0 | 0 | 0 | 0 |
| 0.419922933 | 0 | 0 | 0 | 0 |
| 0 | 0 | 0 | 0 | 0 |
| 0 | 0 | 0 | 0 | 0 |
| 0 | 0 | 0 | 0 | 0 |
| 0 | 0 | 0 | 0 | 0 |
| 0.127500931 | 0 | 1.698630575 | 5.410541883 | 0 |
| 0 | 0 | 0 | 0 | 0 |
| 0.00175674 | 0 | 0 | 0 | 0 |
| 0 | 0 | 0 | 0 | 0 |
| 0.001601567 | 0 | 0 | 0 | 0 |
| 0 | 0 | 0 | 0 | 0 |
| 0 | 0 | 0 | 0 | 0 |
| 0 | 0.067716154 | 0 | 0.008436773 | 0 |
| 0 | 0 | 0 | 0 | 0 |
| 0 | 0.74974867 | 0 | 0.001781121 | 7.052009924 |
| 0 | 0 | 0 | 0 | 0 |
| 0.839376192 | 0 | 0.020936083 | 0.799757542 | 1.519621391 |
| 0 | 0 | 0 | 0 | 0 |
| 0 | 0 | 0 | 0 | 0 |
| 0 | 0 | 0 | 0 | 0 |
| 0 | 0 | 0 | 0 | 0 |
| 0.00182382 | 0.241140548 | 0 | 0.005374073 | 0 |
| 0 | 0 | 0 | 0 | 0.001413612 |
| 0 | 0 | 0 | 0 | 0 |
| 0 | 0.006958646 | 0 | 0.004327612 | 0 |
| 0 | 0 | 0 | 0 | 0 |
| 0 | 0 | 0 | 0 | 0 |
| 14.67047865 | 15.43707299 | 5.085505519 | 1.062881235 | 20.45673241 |
| 0 | 0 | 0.166430062 | 0.20994083 | 0.00331017 |
| 0 | 0 | 0 | 0 | 0 |
| 0 | 0 | 0 | 0 | 0 |
| 0 | 0 | 0 | 0 | 0 |
| 0 | 0 | 0 | 0 | 0 |
| 0 | 0 | 0 | 0 | 0 |
| 0.00164247 | 0.048126963 | 0 | 0.001405077 | 0 |

| | | | | |
|---|---|---|---|---|
| 0 | 0 | 0 | 0 | 0 |
| 0.011343225 | 0 | 0 | 0 | 0 |
| 0 | 0 | 0 | 0 | 0 |
| 0 | 0 | 0 | 0 | 0 |
| 2.418103869 | 0 | 2.866529716 | 1.125217254 | 1.796467292 |
| 0 | 0 | 0 | 0 | 0 |
| 0 | 0 | 0 | 0 | 0 |
| 0 | 0 | 0 | 0 | 0.009209389 |
| 0.154233206 | 0.02917155 | 0.040718067 | 0.100536435 | 0.36687515 |
| 0 | 0 | 0 | 0 | 0 |
| 0 | 0 | 0 | 0 | 0 |
| 0 | 0 | 0 | 0 | 0 |
| 0 | 0 | 0 | 0 | 0.036291602 |
| 0.047281813 | 0.032717729 | 0.030800216 | 0.005179312 | 0.020233215 |
| 1.647525193 | 0 | 0 | 0 | 0 |
| 0 | 0 | 0.579160975 | 1.594261938 | 0.025543431 |
| 0 | 0.000629158 | 0 | 0 | 0.035915302 |
| 0 | 0 | 0 | 0 | 0 |
| 0 | 0 | 0 | 0 | 0 |
| 0 | 0 | 0 | 0 | 0 |
| 0 | 0 | 0 | 0 | 0 |
| 0.001198538 | 0 | 0 | 0 | 0 |
| 0 | 0 | 0 | 0 | 0 |
| 0 | 0 | 0 | 0 | 0 |
| 0 | 0 | 0 | 0 | 0 |
| 0 | 0 | 0 | 0 | 0 |
| 0.002820155 | 0 | 0.000986201 | 0.001732084 | 0 |
| 0.000979148 | 0.00082309 | 0 | 0.007329236 | 0 |
| 0 | 0 | 0 | 0 | 0 |
| 0 | 0 | 0 | 0 | 0 |
| 0 | 0 | 0 | 0 | 0 |
| 0.00184025 | 0 | 0 | 0 | 0 |
| 0 | 0 | 0 | 0 | 0 |
| 0 | 0 | 0 | 0 | 0.00138526 |
| 0.116237351 | 0 | 0 | 0 | 0 |
| 0 | 0 | 0 | 0.023927941 | 0 |
| 0 | 0 | 0 | 0 | 0 |
| 0 | 0 | 0 | 0.007472283 | 0 |
| 0 | 0 | 0 | 0 | 0 |
| 0 | 0 | 0 | 0 | 0 |
| 1.593580332 | 0 | 2.796116491 | 0 | 0.016226013 |
| 0 | 0 | 0 | 0 | 0 |
| 0 | 0 | 0 | 0 | 0 |
| 0 | 0 | 0 | 0 | 0 |
| 0 | 0 | 0 | 0 | 0 |
| 0 | 0 | 0 | 0 | 0 |
| 0 | 0 | 0 | 0 | 0 |
| 0 | 0 | 0 | 0 | 0 |

| | | | | |
|---|---|---|---|---|
| 0 | 0 | 0 | 0 | 0 |
| 0 | 0 | 0 | 0 | 0 |
| 0 | 0 | 0 | 0 | 0 |
| 0 | 0 | 0.011555531 | 0.005350556 | 0 |
| 0 | 0 | 0.132073522 | 0.021713466 | 0.019813634 |
| 0 | 0 | 0 | 0 | 0 |
| 0 | 0.053348411 | 0.002074241 | 0 | 0.574367156 |
| 0 | 0 | 0 | 0 | 0 |
| 0 | 0.000590672 | 0 | 0 | 0 |
| 0 | 0 | 0 | 0.073836656 | 0 |
| 0 | 0 | 0 | 0 | 0 |
| 0 | 0 | 0 | 0 | 0 |
| 0 | 0 | 0 | 0 | 0 |
| 0 | 0 | 0 | 0 | 0 |
| 0 | 0 | 0 | 0 | 0 |
| 0 | 0 | 0 | 0 | 0 |
| 0 | 0 | 0 | 0 | 0 |
| 0 | 0 | 0 | 0 | 0.003875044 |
| 0 | 0 | 0 | 0 | 0 |
| 0 | 0 | 0 | 0 | 0 |
| 0 | 0.095805129 | 0.216329624 | 0.002049491 | 0.088047018 |
| 0 | 0 | 0 | 0 | 0 |
| 0.02639612 | 0 | 0 | 0 | 0 |
| 0.000697179 | 0.016019029 | 0.001408632 | 0.031698236 | 0 |
| 0 | 0 | 0 | 0 | 0 |
| 0 | 0.002304738 | 0 | 0 | 0 |
| 0 | 0 | 0 | 0 | 0 |
| 0 | 0 | 0 | 0 | 0 |
| 0 | 0 | 0 | 0 | 0 |
| 0.409389009 | 0.001405801 | 0.766085567 | 0 | 5.279929904 |
| 0 | 0 | 0 | 0 | 0 |
| 0 | 0 | 0 | 0 | 0 |
| 0 | 0 | 0 | 0 | 0 |
| 0 | 0 | 0 | 0 | 0 |
| 0 | 0 | 0 | 0 | 0 |
| 0 | 0 | 0 | 0 | 0 |
| 0 | 0 | 0 | 0.01992809 | 0 |
| 0 | 0 | 0 | 0 | 0 |
| 0.003483391 | 0 | 0 | 0 | 0 |
| 0 | 0 | 0 | 0 | 0 |
| 0 | 0 | 0 | 0 | 0 |
| 0 | 0 | 0 | 0 | 0 |
| 0 | 0 | 0 | 0 | 0 |
| 0 | 0 | 0 | 0 | 0 |
| 0 | 0 | 0 | 0 | 0 |
| 0 | 0 | 0 | 0 | 0 |
| 0 | 0 | 0 | 0 | 0 |
| 0 | 0 | 0 | 0 | 0 |

| | | | | |
|---|---|---|---|---|
| 0.011002782 | 2.621081625 | 0 | 0.016471875 | 0 |
| 0 | 0 | 0 | 0 | 0 |
| 0 | 0 | 0 | 0 | 0 |
| 0 | 0 | 0 | 0 | 0 |
| 0 | 0 | 0 | 0 | 0 |
| 0 | 0 | 0 | 0 | 0 |
| 0 | 0.001777577 | 0 | 0 | 0 |
| 0.141987399 | 0 | 2.273200603 | 0 | 0.890572088 |
| 0.171527647 | 0.054112257 | 0.03895905 | 8.076448456 | 0.031784488 |
| 0 | 0 | 0 | 0 | 0 |
| 0.798442968 | 0 | 0 | 0 | 0 |
| 0 | 0 | 0 | 0 | 0 |
| 0 | 0 | 0 | 0 | 0 |
| 0 | 0 | 0 | 0 | 0 |
| 0 | 0 | 0 | 0 | 0 |
| 0 | 0 | 0 | 0 | 0 |
| 0 | 0 | 0 | 0 | 0 |
| 0 | 0 | 0 | 0 | 0 |
| 0 | 0 | 0 | 0 | 0 |
| 0 | 0 | 0 | 0 | 0 |
| 0 | 0 | 0 | 0 | 0 |
| 0 | 0 | 0 | 0 | 0 |
| 0 | 0 | 0 | 0 | 0 |
| 0 | 0 | 0 | 0 | 0 |
| 0 | 0 | 0 | 0 | 0.006911327 |
| 0 | 0 | 0 | 0 | 0 |
| 0 | 0 | 0 | 0 | 0 |
| 0 | 0 | 0 | 0 | 0 |
| 0 | 0 | 0 | 0.00206242 | 0 |
| 0 | 0 | 0 | 0 | 0 |
| 0 | 0 | 0 | 0 | 0 |
| 0 | 0 | 0 | 0 | 0 |
| 0 | 0 | 0 | 0 | 0 |
| 0 | 0 | 0 | 0 | 0 |
| 0.026035358 | 0 | 0 | 0 | 0 |
| 0 | 0 | 0 | 0 | 0 |
| 0 | 0 | 0 | 0 | 0 |
| 0 | 0 | 0 | 0 | 0 |
| 0 | 0 | 0 | 0 | 0 |
| 0 | 0 | 0.307932401 | 0.090525544 | 0.154309753 |
| 0 | 0 | 0 | 0 | 0 |
| 0.694095665 | 0.006495026 | 0 | 0 | 0 |
| 0 | 0 | 0 | 0 | 0 |
| 0 | 0 | 0 | 0 | 0 |
| 0 | 0 | 0 | 0 | 0 |
| 0 | 0 | 0.001516274 | 0 | 0 |
| 0 | 0 | 0 | 0 | 0 |
| 0 | 0 | 0 | 0 | 0 |

| | | | | |
|---|---|---|---|---|
| 0 | 0 | 0 | 0 | 0 |
| 0.004739553 | 0.000758887 | 0 | 0 | 0.000812266 |
| 0 | 0 | 0 | 0 | 0 |
| 0 | 0 | 0 | 0 | 0 |
| 1.519691361 | 0 | 0.043513566 | 0.163555216 | 0.007665716 |
| 0 | 0 | 0 | 0 | 0 |
| 0 | 0 | 0 | 0.026948736 | 0 |
| 0.003817182 | 0.023531165 | 0.010604708 | 0.079157356 | 0.001717248 |
| 0.002223267 | 0 | 0 | 0 | 0 |
| 0 | 0 | 0 | 0 | 0 |
| 0 | 0 | 0 | 0 | 0 |
| 0.585776359 | 0 | 0 | 0 | 0 |
| 0.10703705 | 0 | 0 | 0 | 0 |
| 0 | 0 | 0 | 0 | 0 |
| 0 | 0 | 0.011537344 | 0.017730359 | 0 |
| 0 | 0 | 0.002090314 | 0 | 0 |
| 0 | 0 | 0 | 0 | 0 |
| 0 | 0 | 0 | 0 | 0 |
| 0 | 0 | 0 | 0 | 0 |
| 0 | 0 | 0 | 0 | 0 |
| 0 | 0 | 0 | 0 | 0 |
| 0 | 0 | 0 | 0 | 0 |
| 0 | 0 | 0 | 0 | 0 |
| 0.001101152 | 0.083077028 | 0 | 0.012455304 | 0 |
| 0.975553091 | 0 | 0 | 0 | 0 |
| 0 | 0 | 0 | 0 | 0 |
| 0.002517822 | 0.020530326 | 0.008012327 | 0.170039206 | 0 |
| 0 | 0 | 0 | 0 | 0 |
| 0 | 0.013213025 | 0 | 0 | 0 |
| 0 | 0 | 0 | 0 | 0 |
| 0 | 0 | 0 | 0 | 0 |
| 0 | 0 | 0 | 0 | 0 |
| 0 | 0 | 0 | 0 | 0 |
| 0 | 0 | 0 | 0 | 0 |
| 0 | 0 | 0 | 0 | 0 |
| 0 | 0 | 0 | 0 | 0 |
| 0 | 0 | 0 | 0 | 0 |
| 0.228005954 | 0 | 0 | 0 | 0 |
| 0 | 0 | 0 | 0 | 0 |
| 0 | 0 | 0 | 0 | 0 |
| 0 | 0 | 0 | 0 | 0 |
| 0.001480413 | 0 | 0 | 0 | 0 |
| 0 | 0 | 0 | 0 | 0 |
| 0 | 0 | 0 | 0 | 0 |
| 0 | 0 | 0 | 0 | 0 |
| 0 | 0 | 0 | 0 | 0 |
| 0 | 0 | 0 | 0 | 0 |
| 0 | 0 | 0 | 0 | 0 |

| | | | | |
|---|---|---|---|---|
| 0 | 0 | 0 | 0 | 0.001704331 |
| 0 | 0 | 0 | 0 | 0 |
| 0 | 0 | 0 | 0 | 0 |
| 0 | 0 | 0 | 0 | 0 |
| 0 | 0 | 0 | 0 | 0 |
| 0 | 1.925978019 | 0 | 0.013324245 | 0 |
| 0 | 0 | 0 | 0 | 0 |
| 0 | 0 | 0 | 0 | 0 |
| 0 | 0 | 0 | 0 | 0 |
| 0 | 0 | 0 | 0 | 0 |
| 0 | 0 | 0 | 0 | 0 |
| 0 | 0.079996972 | 0 | 0 | 0 |
| 0 | 0 | 0 | 0 | 0 |
| 0 | 0 | 0 | 0 | 0 |
| 0.17820043 | 0 | 0 | 0 | 0 |
| 0 | 0 | 0 | 0 | 0 |
| 0 | 0 | 0 | 0 | 0 |
| 0 | 0 | 0 | 0 | 0 |
| 0 | 0.009112578 | 0 | 0 | 0 |
| 0 | 0 | 0 | 0 | 0 |
| 0.012610315 | 0 | 0.009969967 | 0.013862442 | 0 |
| 0 | 0 | 0 | 0 | 0 |
| 0.001038646 | 0.032886978 | 0 | 0.008523269 | 0 |
| 0 | 0 | 0 | 0 | 0 |
| 0 | 0 | 0 | 0 | 0 |
| 0 | 0 | 0 | 0 | 0 |
| 0 | 0 | 0 | 0 | 0 |
| 0 | 0 | 0 | 0 | 0 |
| 0 | 0 | 0 | 0 | 0 |
| 0 | 0 | 0 | 0 | 0 |
| 0 | 0 | 0 | 0 | 0 |
| 0 | 0 | 0 | 0 | 0 |
| 0.003242293 | 0.035704478 | 0.028251044 | 0.002588759 | 0.07818206 |
| 0 | 0 | 0 | 0 | 0 |
| 0 | 0.245747987 | 0.002856657 | 0.599748768 | 1.477475262 |
| 0 | 0.010060779 | 0.027007202 | 0 | 0 |
| 0 | 0 | 0 | 0 | 0 |
| 0 | 0 | 0 | 0 | 0 |
| 0 | 0 | 0.009804659 | 0.004805615 | 0 |
| 0 | 0 | 0 | 0 | 0 |
| 0 | 0 | 0 | 0 | 0 |
| 0 | 0 | 0 | 0 | 0 |
| 0.156238647 | 0 | 0.00453251 | 0.001638934 | 0.035460393 |
| 0 | 0 | 0 | 0 | 0 |
| 0 | 0 | 0 | 0 | 0 |
| 0.039962891 | 0 | 0.003176117 | 0 | 0 |
| 0 | 0 | 0 | 0 | 0 |
| 0 | 0 | 0 | 0 | 0 |

| | | | | |
|---|---|---|---|---|
| 0 | 0 | 0 | 0 | 0 |
| 0 | 0 | 0 | 0 | 0 |
| 0 | 0 | 0 | 0 | 0 |
| 0 | 0 | 0 | 0 | 0 |
| 0.005566724 | 0 | 0 | 0 | 0 |
| 0 | 0 | 0 | 0 | 0 |
| 0 | 0 | 0 | 0 | 0 |
| 0 | 0.054027696 | 0.174736095 | 0 | 0 |
| 0 | 0 | 0 | 0 | 0 |
| 0 | 0 | 0 | 0 | 0 |
| 0 | 0 | 0 | 0 | 0 |
| 0 | 0 | 0 | 0 | 0 |
| 0 | 0 | 0 | 0 | 0 |
| 0 | 0 | 0 | 0 | 0 |
| 0 | 0 | 0 | 0 | 0 |
| 0 | 0.104749394 | 0.004720704 | 0.014886234 | 0 |
| 0 | 0 | 0 | 0 | 0 |
| 0 | 0 | 0 | 0 | 0 |
| 0 | 0 | 0 | 0 | 0 |
| 0 | 0 | 2.176113025 | 2.280928145 | 0 |
| 0.014197763 | 0 | 0 | 0 | 0 |
| 0 | 0 | 0 | 0 | 0 |
| 0 | 0 | 0 | 0 | 0 |
| 0 | 0 | 0 | 0 | 0 |
| 0 | 0.041827974 | 0 | 0 | 0 |
| 0 | 0 | 1.104689523 | 0 | 0 |
| 0 | 0 | 0 | 0 | 0 |
| 0 | 0 | 0 | 0.00129984 | 0 |
| 0 | 0 | 0 | 0 | 0 |
| 0 | 0 | 0 | 0.073431559 | 0.002905023 |
| 0 | 0 | 1.203551907 | 0.978802946 | 0.09257548 |
| 0 | 0 | 0 | 0 | 0 |
| 0 | 0 | 0 | 0 | 0 |
| 0 | 0 | 0 | 0 | 0 |
| 0 | 0 | 0 | 0 | 0 |
| 0 | 0 | 0 | 0 | 0 |
| 0 | 0 | 0 | 0 | 0 |
| 0 | 0 | 0 | 0 | 0 |
| 0 | 0 | 0.00255526 | 0 | 0 |
| 0 | 0 | 0 | 0 | 0 |
| 0 | 0 | 0 | 0 | 0 |
| 0.509775426 | 0 | 0 | 0 | 0 |
| 0 | 0 | 0 | 0 | 0 |
| 0 | 0.00406863 | 0 | 0 | 0 |
| 0 | 0 | 0 | 0 | 0 |
| 0 | 0 | 0 | 0 | 0 |
| 0.002609227 | 0 | 0.006589842 | 0.150074521 | 0 |
| 0 | 0 | 0 | 0 | 0 |

| | | | | |
|---|---|---|---|---|
| 0 | 0 | 0 | 0 | 0 |
| 0 | 0 | 0 | 0 | 0 |
| 0 | 0 | 0 | 0 | 0 |
| 0.001039314 | 0 | 0 | 0 | 0 |
| 0.888008701 | 0 | 0.006564138 | 0.021328146 | 0.001948738 |
| 0 | 0 | 0 | 0 | 0 |
| 0 | 0 | 0 | 0 | 0 |
| 0 | 0 | 0 | 0 | 0 |
| 0 | 0 | 0 | 0 | 0 |
| 0 | 0 | 0 | 0 | 0 |
| 0 | 0 | 0 | 0 | 0 |
| 0.021349715 | 0.101615251 | 0.001594836 | 0.012604694 | 0 |
| 0 | 0.003271207 | 0 | 0 | 0 |
| 0 | 0 | 0 | 0 | 0 |
| 0 | 0 | 0 | 0 | 0 |
| 0 | 0 | 0 | 0 | 0.027140825 |
| 0.020583399 | 0 | 0 | 0 | 0 |
| 0 | 0 | 0 | 0 | 0 |
| 0.006375374 | 0.190445103 | 0 | 0.15043708 | 0.00122919 |
| 0 | 0.101714527 | 0.003604075 | 0 | 1.190603306 |
| 0 | 0 | 0 | 0 | 0 |
| 0 | 0 | 0 | 0 | 0 |
| 0 | 0 | 0 | 0 | 0 |
| 0 | 0 | 0 | 0 | 0 |
| 0 | 0 | 0 | 0 | 0 |
| 0 | 0 | 0 | 0 | 0 |
| 0.014770702 | 0.032242925 | 0.003429631 | 0.228418592 | 0 |
| 0 | 0 | 0 | 0 | 0 |
| 0 | 0 | 0 | 0 | 0 |
| 0 | 0 | 0 | 0 | 0 |
| 0 | 0.093529242 | 0 | 0 | 0 |
| 0 | 0 | 0 | 0.030664564 | 0 |
| 0 | 0 | 0 | 0 | 0 |
| 0 | 0 | 0 | 0 | 0 |
| 0 | 0 | 0 | 0 | 0 |
| 0 | 0 | 0 | 0 | 0 |
| 0 | 0 | 0 | 0 | 0 |
| 0.02219868 | 0.159811535 | 0.004312677 | 0.120559848 | 0.002902086 |
| 0 | 0 | 0 | 0 | 0 |
| 0.00842701 | 0 | 0 | 0 | 0 |
| 0 | 0.091235911 | 0 | 0.003780867 | 0 |
| 0.005837438 | 0 | 0 | 0 | 0 |
| 0 | 0 | 0 | 0 | 0 |
| 0 | 0.003311101 | 0 | 0.002807984 | 0.11720505 |
| 0 | 0 | 0 | 0 | 0 |
| 0 | 0 | 0 | 0 | 0 |
| 0 | 0 | 0 | 0 | 0 |
| 0 | 0 | 0 | 0 | 0 |

| | | | | |
|---|---|---|---|---|
| 0 | 0 | 0 | 0 | 0 |
| 0 | 0 | 0.248702068 | 0 | 0.825856433 |
| 0 | 0 | 0 | 0 | 0 |
| 0.00176913 | 0.024538204 | 0.001117025 | 0.027073573 | 0 |
| 0.002461259 | 0.394715603 | 0 | 0 | 0 |
| 0.001764718 | 0.044800335 | 0 | 0 | 0 |
| 0 | 0 | 0 | 0 | 0 |
| 0 | 0 | 0 | 0 | 0 |
| 0 | 0 | 0 | 0 | 0 |
| 0 | 0 | 0 | 0 | 0 |
| 0 | 0 | 0 | 0 | 0 |
| 0 | 0 | 0 | 0 | 0 |
| 0 | 0.005095251 | 0.011085387 | 0.00625805 | 0.001880566 |
| 0 | 0 | 0 | 0 | 0 |
| 0 | 0 | 0 | 0 | 0 |
| 0 | 0 | 0 | 0 | 0 |
| 0 | 0 | 0 | 0 | 0.004251436 |
| 0 | 0 | 0 | 0 | 0 |
| 0 | 0 | 0 | 0 | 0 |
| 0 | 0 | 0 | 0 | 0 |
| 0 | 0 | 0 | 0 | 0 |
| 0 | 0 | 0 | 0 | 0 |
| 0 | 0 | 0.004796125 | 0.125620462 | 0 |
| 0.011691639 | 0 | 0.011811322 | 0.002593058 | 0.029513065 |
| 0 | 0 | 0 | 0 | 0 |
| 0 | 0 | 0 | 0 | 0 |
| 0 | 0 | 0 | 0 | 0 |
| 0 | 0 | 0 | 0 | 0 |
| 0 | 0 | 0 | 0 | 0 |
| 0 | 0 | 0 | 0 | 0 |
| 0.023321956 | 0 | 0 | 0.015313366 | 0.040311064 |
| 0 | 0 | 0.000539397 | 0 | 0 |
| 0 | 0 | 0 | 0 | 0 |
| 0 | 0 | 0 | 0 | 0 |
| 0 | 0 | 0 | 0 | 0 |
| 0 | 0 | 0 | 0.001067212 | 0 |
| 0 | 0 | 0 | 0 | 0.024640511 |
| 0 | 0 | 0 | 0 | 0 |
| 0 | 0 | 0 | 0 | 0.003737639 |
| 0 | 0 | 0 | 0 | 0 |
| 0 | 0 | 0 | 0 | 0 |
| 0 | 0 | 0 | 0 | 0 |
| 0 | 0 | 0 | 0 | 0 |
| 0 | 0 | 0 | 0 | 0 |
| 0.26845158 | 0 | 0.021182725 | 0 | 0 |
| 0.238712287 | 0.003648473 | 0.230021185 | 0 | 0.055287988 |
| 0 | 0 | 0 | 0 | 0 |
| 0 | 0 | 0 | 0 | 0 |

| | | | | |
|---|---|---|---|---|
| 0.000784624 | 0 | 0 | 0 | 0 |
| 0 | 0 | 0 | 0 | 0 |
| 0.318657947 | 0 | 0.073852219 | 0 | 0 |
| 0 | 0 | 0 | 0 | 0 |
| 0 | 0 | 0 | 0 | 0 |
| 0 | 0 | 0 | 0 | 0 |
| 0 | 0.00155294 | 0 | 0 | 0 |
| 0 | 0 | 0 | 0 | 0 |
| 0 | 0 | 0 | 0 | 0 |
| 0 | 0 | 0 | 0 | 0.010130838 |
| 0 | 0.041963247 | 0 | 0 | 0 |
| 0.000943417 | 0.007666188 | 0.000953074 | 0.00397552 | 0 |
| 0 | 0 | 0 | 0 | 0 |
| 0 | 0 | 0 | 0 | 0 |
| 0 | 0 | 0 | 0 | 0 |
| 0 | 0 | 0 | 0 | 0 |
| 0 | 0 | 0 | 0 | 0 |
| 0 | 0 | 0 | 0 | 0 |
| 0 | 0 | 0 | 0 | 0 |
| 0.004189987 | 0.001643685 | 0 | 0 | 0 |
| 0 | 0 | 0 | 0 | 0 |
| 0 | 0 | 0 | 0 | 0 |
| 0 | 0 | 0 | 0 | 0 |
| 0 | 0 | 0 | 0 | 0 |
| 0 | 0 | 0 | 0 | 0 |
| 0 | 0 | 0 | 0.001731478 | 0 |
| 0 | 0 | 0 | 0 | 0 |
| 0.010201244 | 0 | 0 | 0 | 0 |
| 0 | 0 | 0 | 0 | 0 |
| 0 | 0 | 0 | 0 | 0 |
| 0 | 0.002219007 | 0 | 0.027942353 | 0.00215917 |
| 0 | 0 | 0 | 0 | 0 |
| 0.022569515 | 0 | 0.077268534 | 0.00630339 | 0.0298335 |
| 0 | 0 | 0.00302033 | 0.000568357 | 0 |
| 0 | 0 | 0 | 0 | 0 |
| 0 | 0 | 0 | 0 | 0 |
| 0 | 0 | 0 | 0 | 0 |
| 0 | 0 | 0 | 0 | 0 |
| 0 | 0 | 0 | 0 | 0 |
| 0 | 0 | 0 | 0 | 0 |
| 0 | 0 | 0 | 0 | 0 |
| 0.018109273 | 0 | 0.002431568 | 0.031114449 | 0.00515625 |
| 0.024296348 | 0 | 0.005501479 | 0 | 0.001758895 |
| 0 | 0 | 0 | 0 | 0 |
| 0 | 0 | 0 | 0 | 0 |
| 0.060130768 | 0 | 0.00886733 | 0 | 0.002541725 |
| 0 | 0.188831756 | 0 | 0 | 0 |
| 0 | 0 | 0 | 0 | 0 |

| | | | | |
|---|---|---|---|---|
| 0 | 0 | 0 | 0 | 0 |
| 0.055003281 | 0 | 0.004433484 | 0 | 0.017110556 |
| 0 | 0.007469393 | 0.003366205 | 0.003638236 | 0.001537457 |
| 0 | 0 | 0 | 0 | 0 |
| 0 | 0.18281803 | 0 | 0 | 0 |
| 0 | 1.076431776 | 0 | 0.001590563 | 0 |
| 0 | 0 | 0 | 0 | 0 |
| 0 | 0 | 0 | 0 | 0 |
| 0 | 0 | 0 | 0 | 0 |
| 0 | 0 | 0 | 0 | 0 |
| 0 | 0 | 0 | 0 | 0 |
| 0 | 0 | 0 | 0 | 0 |
| 0 | 0 | 0 | 0 | 0 |
| 0 | 0 | 0 | 0 | 0 |
| 0 | 0 | 0 | 0 | 0 |
| 0 | 0 | 0 | 0 | 0 |
| 0 | 0 | 0 | 0 | 0 |
| 0 | 0 | 0 | 0 | 0 |
| 0 | 0 | 0 | 0 | 0 |
| 0 | 0 | 0 | 0 | 0 |
| 0 | 0 | 0 | 0 | 0 |
| 0 | 0 | 0 | 0.008596712 | 0 |
| 0 | 0 | 0 | 0 | 0 |
| 0 | 0 | 0 | 0 | 0 |
| 0.001126611 | 0 | 0.007042267 | 0 | 0 |
| 0.001700017 | 0 | 0.039285962 | 0 | 1.501796128 |
| 0 | 0 | 0 | 0 | 0 |
| 0.034080893 | 0 | 0.002082446 | 0 | 0 |
| 0 | 0 | 0 | 0 | 0 |
| 0 | 0 | 0 | 0 | 0 |
| 0 | 0 | 0 | 0 | 0 |
| 0 | 0 | 0 | 0 | 0 |
| 0 | 0 | 0 | 0 | 0 |
| 0 | 0 | 0 | 0 | 0 |
| 0 | 0 | 0 | 0 | 0 |
| 0 | 0 | 0 | 0 | 0 |
| 0 | 0 | 0 | 0 | 0 |
| 0 | 0 | 0.001495853 | 0 | 0 |
| 0 | 0 | 0 | 0 | 0.124523837 |
| 0 | 0 | 0 | 0 | 0 |
| 0 | 0 | 0 | 0 | 0 |
| 0 | 0 | 0 | 0 | 0 |
| 0.006824609 | 2.071019043 | 0 | 0.013811709 | 0 |
| 0 | 0 | 0 | 0 | 0 |
| 0 | 0 | 0 | 0 | 0 |
| 0 | 0 | 0 | 0.000870678 | 0 |
| 0 | 0 | 0 | 0.037337923 | 0 |
| 0 | 0 | 0 | 0 | 0 |

| | | | | |
|---|---|---|---|---|
| 0 | 0 | 0 | 0 | 0 |
| 0.08370826 | 0.006690608 | 0.025577492 | 0.195393574 | 0.004691828 |
| 0 | 0 | 0 | 0 | 0 |
| 0 | 0 | 0 | 0 | 0 |
| 0 | 0 | 0 | 0 | 0 |
| 0 | 0 | 0 | 0 | 0 |
| 0 | 0 | 0 | 0 | 0 |
| 0 | 0 | 0 | 0 | 0 |
| 0 | 0 | 0 | 0 | 0 |
| 0 | 0 | 0 | 0 | 0 |
| 0 | 0 | 0 | 0 | 0 |
| 0 | 0 | 0 | 0 | 0 |
| 0 | 0 | 0 | 0.001930358 | 0.002088286 |
| 0 | 0 | 0 | 0 | 0 |
| 0.013688077 | 0.03567001 | 0.001296393 | 0 | 0 |
| 0 | 0 | 0 | 0 | 0 |
| 0 | 0 | 0 | 0 | 0 |
| 0 | 0 | 0 | 0 | 0 |
| 0 | 0 | 0 | 0 | 0 |
| 0 | 0 | 0 | 0 | 0 |
| 0 | 0 | 0 | 0 | 0 |
| 0 | 0.020905456 | 0 | 0 | 0.850284313 |
| 0.088318676 | 0 | 0 | 0.018728844 | 0 |
| 0 | 0.107371625 | 0 | 0 | 0 |
| 0 | 0 | 0 | 0 | 0 |
| 0.888318597 | 0 | 0 | 0 | 0 |
| 0 | 0 | 0 | 0 | 0 |
| 0 | 0 | 0 | 0 | 0 |
| 0 | 0 | 0 | 0 | 0 |
| 0 | 0 | 0 | 0 | 0 |
| 0 | 0 | 0 | 0 | 0 |
| 0 | 0 | 0 | 0 | 0 |
| 0 | 0 | 0 | 0 | 0 |
| 0 | 0 | 0 | 0 | 0 |
| 0 | 0 | 0 | 2.190013346 | 0 |
| 0 | 0 | 0 | 0 | 0 |
| 0 | 0 | 0 | 0 | 0 |
| 0 | 0 | 0 | 0 | 0 |
| 0 | 0.123002834 | 0 | 0.000893198 | 3.413054889 |
| 0 | 0 | 0 | 0 | 0 |
| 0.005717386 | 0.079680774 | 0.978789141 | 5.080717786 | 0.005414943 |
| 0 | 0 | 0 | 0 | 0 |
| 0 | 0 | 0 | 0 | 0 |
| 0 | 0 | 0 | 0 | 0 |
| 0 | 0 | 0 | 0 | 0 |
| 0.248748661 | 3.624255542 | 0.103633087 | 0.043783951 | 0.013019457 |
| 0 | 0 | 0 | 0 | 0 |
| 0 | 0 | 0 | 0 | 0 |

| | | | | |
|---|---|---|---|---|
| 0 | 0 | 0 | 0 | 0 |
| 0 | 0 | 0 | 0 | 0 |
| 0 | 0 | 0 | 0 | 0 |
| 0 | 0 | 0 | 0 | 0 |
| 0 | 0.01054329 | 0 | 0 | 0 |
| 0 | 0 | 0 | 0 | 0 |
| 0 | 0 | 0 | 0 | 0 |
| 0 | 0.001878204 | 0 | 0 | 0.022783551 |
| 0 | 0 | 0 | 0 | 0 |
| 0 | 0 | 0 | 0 | 0.01546156 |
| 0 | 0 | 0 | 0 | 0 |
| 0 | 0 | 0 | 0 | 0 |
| 0 | 0.001937937 | 0.006238308 | 0.008984302 | 0.001481605 |
| 0 | 0 | 0 | 0 | 0 |
| 0 | 0 | 0 | 0 | 0 |
| 0 | 0 | 0 | 0 | 0 |
| 0 | 0 | 0 | 0 | 0 |
| 0 | 0 | 0 | 0 | 0 |
| 0 | 0 | 0 | 0 | 0 |
| 0.428562612 | 1.192645507 | 0.013770286 | 0.783477925 | 0.004219946 |
| 0 | 0 | 0 | 0 | 0 |
| 0 | 0 | 0 | 0 | 0 |
| 0 | 0 | 0 | 0 | 0 |
| 0 | 0 | 0 | 0 | 0 |
| 0 | 0 | 0 | 0 | 0 |
| 0.019042528 | 0 | 0 | 0 | 0 |
| 0 | 0 | 0 | 0 | 0 |
| 0 | 0 | 0 | 0 | 0 |
| 0 | 0 | 0 | 0 | 0 |
| 0 | 0 | 0 | 0 | 0 |
| 0 | 0 | 0 | 0 | 0 |
| 0 | 0 | 0 | 0 | 0 |
| 0 | 0 | 0 | 0 | 0 |
| 0 | 0 | 0 | 0 | 0 |
| 0 | 0 | 0 | 0 | 0 |
| 0 | 0 | 0 | 0 | 0.008535349 |
| 0 | 0 | 0 | 0 | 0 |
| 0 | 0 | 0 | 0 | 0 |
| 0 | 0 | 0 | 0 | 0 |
| 0 | 0 | 0 | 0 | 0 |
| 0 | 0 | 0 | 0 | 0 |
| 0 | 0 | 0 | 0 | 0 |
| 0 | 0 | 0 | 0.028696196 | 0 |
| 0 | 0 | 0 | 0 | 0 |
| 0.017345329 | 0.031938916 | 0 | 0.021799522 | 0 |
| 0 | 0 | 0 | 0 | 0 |
| 0 | 0 | 0 | 0 | 0 |
| 0 | 0 | 0 | 0 | 0 |

| | | | | |
|---|---|---|---|---|
| 0 | 0 | 0 | 0 | 0 |
| 0 | 0 | 0 | 0 | 0 |
| 0 | 0 | 0 | 0 | 0 |
| 0 | 0 | 0 | 0 | 0 |
| 0 | 0 | 0 | 0 | 0 |
| 0.003288625 | 0.013476842 | 0 | 0.011487664 | 0 |
| 0 | 0 | 0 | 0 | 0 |
| 0 | 0.005145232 | 0 | 0 | 0 |
| 0 | 0 | 0 | 0 | 0 |
| 0 | 0 | 0 | 0 | 0 |
| 0 | 0 | 0 | 0 | 0 |
| 0 | 0 | 0 | 0 | 0 |
| 0 | 0 | 0 | 0 | 0 |
| 0 | 0 | 0 | 0 | 0 |
| 0 | 0 | 0 | 0 | 0 |
| 0 | 0 | 0 | 0 | 0 |
| 0.001495905 | 0 | 0 | 0 | 0 |
| 0 | 0 | 0 | 0 | 0 |
| 0 | 0 | 0 | 0 | 0 |
| 0 | 0 | 0 | 0 | 0 |
| 0 | 0.043072313 | 0 | 0 | 0 |
| 0 | 0 | 0 | 0 | 0 |
| 0 | 0 | 0 | 0 | 0 |
| 0 | 0 | 0 | 0 | 0 |
| 0 | 0 | 0 | 0 | 0 |
| 0 | 0 | 0 | 0 | 0 |
| 0 | 0 | 0 | 0 | 0 |
| 0 | 0 | 0 | 0 | 0 |
| 0 | 0 | 0 | 0 | 0 |
| 0 | 0 | 0 | 0 | 0 |
| 0 | 0 | 0 | 0 | 0 |
| 0 | 0 | 0 | 0 | 0 |
| 0 | 0 | 0 | 0 | 0 |
| 0 | 0 | 0 | 0 | 0 |
| 0 | 0 | 0 | 0 | 0 |
| 0 | 0 | 0 | 0 | 0 |
| 0 | 0 | 0 | 0 | 0 |
| 0 | 0 | 0 | 0 | 0 |
| 0 | 0 | 0.093207983 | 0 | 0 |
| 0 | 0 | 0 | 0 | 0 |
| 0 | 0 | 0 | 0 | 0 |
| 0 | 0 | 0 | 0 | 0 |
| 0 | 0 | 0 | 0 | 0 |
| 0 | 0 | 0 | 0 | 0 |
| 0 | 0 | 0 | 0 | 0 |
| 0 | 0 | 0 | 0 | 0 |
| 0 | 0 | 0 | 0 | 0 |
| 0 | 0 | 0 | 0 | 0 |

| | | | | |
|---|---|---|---|---|
| 0 | 0 | 0 | 0 | 0 |
| 0 | 0 | 0 | 0 | 0 |
| 0.017467419 | 0 | 0 | 0 | 0 |
| 0 | 0 | 0 | 0 | 0 |
| 0 | 0 | 0 | 0 | 0 |
| 0 | 0 | 0 | 0 | 0 |
| 0 | 0 | 0 | 0 | 0 |
| 0 | 0 | 0 | 0 | 0 |
| 0 | 0 | 0 | 0 | 0 |
| 0 | 0 | 0 | 0 | 0 |
| 0.001205283 | 0.014437864 | 0 | 0 | 0.000813337 |
| 0 | 0 | 0 | 0 | 0 |
| 0 | 0 | 0 | 0 | 1.005428199 |
| 0 | 0 | 0 | 0 | 0 |
| 0 | 0 | 0 | 0 | 0 |
| 0 | 0 | 0 | 0 | 0 |
| 0 | 0 | 0 | 0 | 0 |
| 0 | 0 | 0 | 0 | 0 |
| 0 | 0 | 0 | 0 | 0 |
| 0 | 0 | 0 | 0 | 0 |
| 0 | 0.04267529 | 0.003707436 | 0.002848757 | 0 |
| 0 | 0 | 0 | 0 | 0 |
| 0 | 0 | 0 | 0 | 0 |
| 3.300047813 | 0 | 3.008661809 | 1.069038685 | 6.080648383 |
| 0 | 0 | 0 | 0 | 0 |
| 0 | 0 | 0 | 0 | 0 |
| 0 | 0 | 0 | 0 | 0 |
| 0 | 0 | 0 | 0 | 0 |
| 0 | 0 | 0 | 0 | 0 |
| 0 | 0 | 0 | 0 | 0 |
| 0 | 0 | 0 | 0 | 0 |
| 0 | 0 | 0 | 0 | 0 |
| 0 | 0 | 0 | 0 | 0 |
| 0 | 0 | 0 | 0 | 0 |

| PHL018-0094_V2 | PHL018-0053_V3 | PHL018-0017_V3 | PHL018-0257_V1 | PHL018-0242_V1 |
|---|---|---|---|---|
| REF | CTRL | EYCF | REF | REF |
| 0.152808747 | 0 | 0.03740675 | 0.014982231 | 3.731851996 |
| 1.253173324 | 5.697791029 | 0.280354957 | 0.022371179 | 0.541932916 |
| 0 | 0 | 0 | 0 | 0 |
| 0 | 0 | 0 | 0 | 0 |
| 0 | 0 | 0 | 0 | 0 |
| 0 | 0 | 0 | 0 | 0 |
| 0 | 0 | 0 | 0 | 0 |
| 0 | 0 | 0 | 0 | 0 |
| 0 | 0 | 0 | 0 | 0 |
| 0 | 0 | 0 | 0 | 0 |
| 0 | 0 | 0 | 0 | 0 |
| 0 | 0 | 0 | 0 | 0 |
| 0 | 0 | 0 | 0 | 0 |
| 0.076108881 | 15.58751381 | 4.540237188 | 0.123502899 | 0.48461873 |
| 0 | 0 | 0 | 0 | 0 |
| 0 | 0 | 0 | 0 | 0 |
| 0 | 0 | 0 | 0 | 0 |
| 0 | 0 | 0 | 0 | 0 |
| 0 | 0 | 0 | 0 | 0 |
| 0 | 0 | 0 | 0 | 0 |
| 0 | 0 | 0 | 0 | 0 |
| 0 | 0 | 0 | 0 | 0 |
| 0 | 0 | 0 | 0 | 0 |
| 0 | 0 | 0 | 0 | 0 |
| 0 | 0 | 0 | 0 | 0.000937048 |
| 0 | 0 | 0 | 0 | 0 |
| 0 | 0 | 0 | 0 | 0.000490647 |
| 0 | 0 | 0 | 0 | 0 |
| 0 | 0 | 0 | 0 | 0 |
| 0 | 0 | 0 | 0 | 0 |
| 0 | 0 | 0 | 0 | 0 |
| 0 | 0 | 0 | 0 | 0 |
| 0 | 0 | 0 | 0 | 0 |
| 0 | 0 | 0 | 0 | 0 |
| 0 | 0 | 0 | 0 | 0 |
| 0 | 1.66706427 | 2.222215426 | 0.410400407 | 0.716631715 |
| 0 | 0 | 0 | 0 | 0 |
| 0 | 0 | 0 | 0 | 0 |
| 0 | 0 | 0 | 0 | 0 |
| 0 | 0 | 0 | 0 | 0 |
| 0 | 0 | 0 | 0 | 0 |
| 0 | 0.003220576 | 0 | 0.40746561 | 0.008304755 |
| 0.052221264 | 0.001527339 | 0.093707711 | 3.411362513 | 0.510092798 |
| 0.00350067 | 0.002200394 | 0.032167951 | 0 | 0 |
| 0.003453702 | 0 | 0.001955948 | 0.009030521 | 0.002380098 |
| 0 | 0 | 0 | 0 | 0 |

| | | | | |
|---|---|---|---|---|
| 0 | 0 | 0 | 0 | 0.049161757 |
| 0.006632492 | 0.010184133 | 0.00124568 | 0 | 0 |
| 0 | 0.003742491 | 0.007209809 | 0 | 0 |
| 2.859756517 | 0.230574311 | 0.279637954 | 0.098109961 | 0.335200565 |
| 0.025164821 | 0 | 0.004295759 | 0 | 0.000885269 |
| 0 | 0 | 0 | 0 | 0 |
| 0.000761674 | 0.250152538 | 0.016823116 | 0 | 0 |
| 0 | 0 | 0 | 0 | 0 |
| 0 | 0 | 0 | 0 | 0 |
| 0 | 0 | 0 | 0 | 0 |
| 0 | 0 | 0 | 0 | 0.001306188 |
| 0 | 0 | 0.823725402 | 0.441242969 | 0 |
| 0 | 0 | 0 | 0 | 0 |
| 0.094551127 | 0.007817515 | 0 | 0 | 0.044186182 |
| 0 | 0 | 0 | 0 | 0 |
| 0 | 0 | 0 | 0 | 0 |
| 0 | 0.001786164 | 0.000786514 | 0.539597996 | 0 |
| 0 | 0 | 0 | 0 | 0 |
| 0 | 0 | 0 | 0 | 0 |
| 0 | 0 | 0 | 0 | 0 |
| 0.187526056 | 3.774913003 | 0.012327057 | 0 | 0.005809503 |
| 0.004509956 | 0.003016221 | 0.003984459 | 0 | 0.001212124 |
| 0.414589903 | 0 | 0.672270641 | 0 | 0.056647422 |
| 0 | 0 | 0 | 0 | 0 |
| 0 | 0 | 0 | 0 | 0 |
| 0 | 0 | 0 | 0 | 0 |
| 0 | 0 | 0 | 0 | 0 |
| 0 | 0 | 0 | 0 | 0 |
| 0 | 0.001378078 | 0 | 0.038603826 | 0 |
| 0 | 0 | 0 | 0 | 0 |
| 0.265121227 | 0.02929467 | 0 | 0 | 0.001831293 |
| 0 | 0 | 0 | 0 | 0 |
| 0 | 0 | 0 | 0 | 0 |
| 0.00100077 | 0 | 0 | 0 | 0.011767569 |
| 0 | 0 | 0 | 0 | 0 |
| 0 | 0 | 0 | 0 | 0 |
| 0 | 0 | 0 | 0 | 0 |
| 0 | 0 | 0 | 17.80228731 | 0 |
| 0 | 0 | 0 | 0 | 0 |
| 0 | 0 | 0 | 0 | 0 |
| 0 | 0 | 0 | 0 | 0 |
| 0 | 0 | 0 | 0 | 0 |
| 0 | 0 | 0 | 0 | 0 |
| 0 | 0 | 0 | 0 | 0 |
| 0.449961476 | 0 | 0 | 0 | 0 |
| 0 | 0 | 0 | 0 | 0 |
| 0.04750618 | 0 | 0 | 0 | 0 |
| 0 | 0 | 0 | 0.029020208 | 0 |

| | | | | |
|---|---|---|---|---|
| 0 | 0 | 0 | 0 | 0 |
| 0.010419745 | 0 | 0 | 0 | 0 |
| 0 | 0 | 0 | 0 | 0 |
| 0 | 0 | 0 | 0 | 0 |
| 0.045174856 | 0 | 0.003887446 | 0 | 0 |
| 0 | 0 | 0 | 0 | 0 |
| 0 | 0 | 0 | 0 | 0 |
| 0 | 0 | 0 | 0 | 0 |
| 0.065562761 | 0.006663804 | 0.000733579 | 0 | 0.694180444 |
| 0 | 0 | 0 | 0 | 0 |
| 0 | 0 | 0 | 0 | 0 |
| 0 | 0.022242566 | 0 | 0.061387735 | 0 |
| 0 | 0 | 0 | 0 | 0 |
| 0 | 0 | 0 | 0 | 0 |
| 0 | 0 | 0 | 0 | 0 |
| 0 | 0 | 0.001579865 | 0 | 0 |
| 0 | 0 | 0 | 0 | 0 |
| 0 | 0 | 0 | 0 | 0 |
| 0.487336205 | 0 | 0.007958449 | 1.198202085 | 0 |
| 0 | 0 | 0 | 0 | 0 |
| 0 | 0 | 0 | 0 | 0 |
| 0 | 0 | 0 | 0 | 0 |
| 0.006675207 | 0 | 0 | 0 | 0 |
| 0 | 0 | 0 | 0 | 0 |
| 0 | 0 | 0 | 0 | 0 |
| 0 | 0 | 0 | 0 | 0 |
| 0 | 0 | 0 | 0 | 0 |
| 0.00422517 | 0 | 0 | 0 | 0 |
| 0 | 0 | 0 | 0 | 0 |
| 0 | 0 | 0 | 0 | 0 |
| 0.503914882 | 0 | 0.034790191 | 0.003897056 | 0.009576889 |
| 0 | 0 | 0.024391307 | 0.002574147 | 0.019146152 |
| 0 | 0 | 0 | 0 | 0 |
| 0 | 0 | 0 | 0 | 0 |
| 0 | 0 | 0 | 0 | 0 |
| 0 | 0 | 0 | 0 | 0 |
| 0 | 0 | 0 | 0 | 0 |
| 0 | 0 | 0 | 0 | 0 |
| 0 | 0 | 0.347631142 | 0.024589704 | 0 |
| 0 | 0 | 0 | 0 | 0 |
| 0 | 0 | 0 | 0 | 0 |
| 0 | 0 | 0 | 0 | 0 |
| 0.001173941 | 0 | 0 | 0 | 0 |
| 0 | 0 | 0 | 0 | 0 |
| 0.004171949 | 0 | 0 | 0 | 0 |
| 0 | 0 | 0 | 0 | 0 |
| 0 | 0 | 0 | 0.010091875 | 0 |
| 0 | 0 | 0 | 0 | 0 |

| | | | | |
|---|---|---|---|---|
| 0 | 0 | 0 | 0 | 0.005958039 |
| 0 | 0 | 0 | 0 | 0 |
| 0 | 0 | 0 | 0 | 0 |
| 0 | 0 | 0 | 0 | 0 |
| 0 | 0 | 0.083369864 | 0 | 0.000989974 |
| 0 | 0 | 0 | 0 | 0 |
| 0 | 0 | 0 | 0 | 0 |
| 0 | 0 | 0 | 0 | 0 |
| 0 | 0 | 0 | 0 | 0 |
| 0 | 0 | 0 | 0 | 0 |
| 0 | 0 | 0 | 0 | 0 |
| 0 | 0 | 0 | 0 | 0 |
| 0 | 0 | 0 | 0 | 0 |
| 0 | 0 | 0 | 0 | 0 |
| 0 | 0 | 0 | 0 | 0 |
| 0 | 0 | 0 | 0 | 0 |
| 0 | 0 | 0 | 0 | 0 |
| 0 | 0 | 0 | 0 | 0 |
| 0 | 0 | 0 | 0 | 0 |
| 0 | 0 | 0 | 0 | 0 |
| 0 | 0 | 0 | 0 | 0 |
| 0 | 0 | 0 | 0 | 0 |
| 0 | 0 | 0 | 0 | 0 |
| 0 | 0 | 0 | 0 | 0 |
| 0 | 0 | 0.018579911 | 0 | 0 |
| 0 | 0 | 0 | 0 | 0 |
| 0 | 0 | 0 | 0 | 0.078249324 |
| 0 | 0 | 0 | 0 | 0 |
| 0 | 0 | 0 | 0 | 0 |
| 0 | 0 | 0 | 0 | 0 |
| 0.00687809 | 0 | 0 | 0 | 0 |
| 0 | 0 | 0.133316465 | 0 | 0.003280771 |
| 0 | 0 | 0 | 0 | 0 |
| 0 | 0 | 0 | 0 | 0 |
| 0 | 0 | 0 | 0 | 0 |
| 0 | 0 | 0 | 0 | 0 |
| 0 | 0 | 0 | 0 | 0 |
| 0 | 0 | 0 | 0 | 0 |
| 0 | 0 | 0 | 0 | 0 |
| 0 | 0 | 0 | 0 | 0 |
| 0 | 0 | 0 | 0 | 0 |
| 0 | 0 | 0 | 0 | 0 |
| 0 | 0 | 0 | 0 | 0 |
| 0 | 0 | 0 | 0 | 0 |
| 0 | 0 | 0 | 0 | 0 |
| 0 | 0 | 0 | 0 | 0 |
| 0 | 0 | 0 | 0 | 0 |
| 0.051961149 | 0 | 0 | 0 | 0 |

| | | | | |
|---|---|---|---|---|
| 0 | 0 | 0 | 0 | 0 |
| 0 | 0 | 0 | 0 | 0 |
| 0 | 0 | 0 | 0 | 0 |
| 0 | 0 | 0 | 0 | 0 |
| 0 | 0 | 0 | 0 | 0 |
| 0 | 0 | 0 | 0 | 0 |
| 0 | 0 | 0 | 0 | 0 |
| 0 | 0 | 0 | 0 | 0 |
| 0 | 0 | 0 | 0 | 0 |
| 0 | 0 | 0 | 0 | 0 |
| 0 | 0 | 0 | 0 | 0 |
| 0.334808054 | 0 | 0 | 0 | 0 |
| 0.002055803 | 0 | 0 | 0.00167177 | 0 |
| 0 | 0 | 0 | 0 | 0 |
| 0 | 0 | 0 | 0.11410685 | 0 |
| 0 | 0 | 0 | 0 | 0 |
| 0 | 0 | 0 | 0 | 0 |
| 0 | 0 | 0 | 0 | 0 |
| 0 | 0 | 0 | 0 | 0 |
| 0 | 0 | 0 | 0 | 0 |
| 0.058007302 | 0.063851169 | 0.033812724 | 0.006251616 | 0.013696386 |
| 0 | 0 | 0 | 0.153133438 | 0 |
| 0 | 0 | 0 | 0 | 0.000515046 |
| 0 | 0 | 0 | 0 | 0 |
| 0 | 0 | 0 | 0 | 0 |
| 0 | 0 | 0 | 0 | 0 |
| 0.003866624 | 0.003645629 | 0.003424649 | 0.002548555 | 0.002278986 |
| 0 | 0 | 0 | 0 | 0 |
| 0 | 0 | 0 | 0 | 0 |
| 0 | 0 | 0 | 0 | 0 |
| 0 | 0.070985561 | 0 | 0 | 0 |
| 0 | 0 | 0 | 0 | 0 |
| 0 | 0 | 0 | 0 | 0 |
| 0 | 0 | 0.031670189 | 0.00095515 | 1.049645734 |
| 0.032285875 | 0.00190075 | 0.026783048 | 0.004677779 | 0.004073874 |
| 0 | 0 | 0 | 0 | 0 |
| 0 | 0 | 0 | 0 | 0 |
| 0 | 0 | 0 | 0 | 0 |
| 0 | 0 | 0.001158925 | 0.016756148 | 0 |
| 0 | 0 | 0 | 0 | 0 |
| 0 | 0 | 0 | 0.003758005 | 0 |
| 0 | 0 | 0 | 0.002234288 | 0 |
| 0 | 0 | 0.002485533 | 0.18306589 | 0 |
| 0 | 0 | 0 | 0 | 0 |
| 0 | 0 | 0 | 0 | 0 |
| 0 | 0 | 0 | 0.056156048 | 0 |
| 0 | 0 | 0 | 0 | 0 |
| 0.027058454 | 0 | 0 | 0.003188577 | 0.001088682 |

| | | | | |
|---|---|---|---|---|
| 0 | 0 | 0 | 0 | 0 |
| 0 | 0 | 0 | 0 | 0 |
| 0 | 0.004597312 | 0 | 0 | 0 |
| 0 | 0 | 0 | 0 | 0 |
| 0 | 0 | 0 | 0 | 0 |
| 0 | 0 | 0 | 0 | 0 |
| 0 | 0 | 0 | 0.030328096 | 0 |
| 0 | 0 | 0 | 0.04250282 | 0 |
| 0 | 0 | 0 | 0 | 0 |
| 0 | 0 | 0 | 0 | 0 |
| 0 | 0 | 0 | 0.006276257 | 0 |
| 0 | 0 | 0 | 0 | 0 |
| 0 | 0 | 0 | 0 | 0 |
| 0 | 0 | 0 | 0 | 0 |
| 0 | 0 | 0 | 0 | 0.001144327 |
| 0 | 0 | 0 | 0 | 0 |
| 0 | 0 | 0 | 0 | 0 |
| 0 | 0 | 0 | 0.023964727 | 0 |
| 0 | 0 | 0 | 0.017392993 | 0.003506078 |
| 0 | 0 | 0 | 0.045943961 | 0 |
| 0.002916808 | 0.002612594 | 0 | 0 | 0 |
| 0 | 0 | 0 | 0 | 0 |
| 0 | 0 | 0 | 0 | 0 |
| 0 | 0 | 0 | 0 | 0 |
| 0.002897428 | 0 | 0.004063209 | 0 | 0.002163141 |
| 0 | 0 | 0 | 0 | 0 |
| 0 | 0 | 0 | 0 | 0 |
| 0 | 0 | 0 | 0 | 0 |
| 0 | 0 | 0 | 0.048437141 | 0 |
| 0 | 0 | 0 | 0 | 0 |
| 0.652533466 | 0.005763119 | 0.427463635 | 0.010551704 | 0.136702053 |
| 0 | 0 | 0 | 0 | 0 |
| 0 | 0 | 0.024308628 | 0 | 0 |
| 0.00508713 | 0 | 0.00194562 | 0 | 0 |
| 0 | 0 | 0 | 0 | 0 |
| 0 | 0 | 0 | 0 | 0 |
| 0 | 0 | 0 | 0 | 0 |
| 0 | 0 | 0 | 0 | 0 |
| 0 | 0 | 0 | 0 | 0 |
| 0 | 0 | 0 | 0 | 0 |
| 0 | 0 | 0 | 0 | 0 |
| 0.04572788 | 0.0089206 | 0.008047748 | 0.029496712 | 0 |
| 0.001987155 | 0 | 0 | 0 | 0 |
| 0 | 0 | 0 | 0 | 0 |
| 0 | 0 | 0 | 0 | 0 |
| 0 | 0 | 0 | 0 | 0 |
| 0 | 0 | 0 | 0 | 0 |
| 0 | 0 | 0 | 0 | 0 |

| | | | | |
|---|---|---|---|---|
| 0 | 0 | 0 | 0 | 0 |
| 0 | 0 | 0 | 0 | 0 |
| 0 | 0 | 0.037774471 | 0 | 0 |
| 0 | 0 | 0 | 0 | 0 |
| 1.13079645 | 0 | 0 | 0 | 0 |
| 0 | 0 | 0 | 0 | 0 |
| 0 | 0 | 0 | 0 | 0 |
| 0 | 0 | 0 | 0 | 0 |
| 0 | 0 | 0 | 0 | 0 |
| 0 | 0.000999474 | 0 | 0 | 0 |
| 0 | 0 | 0 | 0 | 0 |
| 0 | 0 | 0 | 0 | 0 |
| 0 | 0 | 0 | 0 | 0 |
| 0 | 0 | 0.001063633 | 0 | 0.001132498 |
| 0.012937612 | 0.009469976 | 0.00263367 | 0.000746638 | 0.03061238 |
| 0 | 0 | 0.016213228 | 0.001598749 | 0 |
| 0.001426572 | 0 | 0 | 0 | 0.00159756 |
| 0 | 0 | 0 | 0 | 0.002271966 |
| 0 | 0 | 0 | 0 | 0 |
| 0 | 0 | 0 | 0 | 0 |
| 0 | 0 | 0 | 0 | 0 |
| 0.03305322 | 0 | 0 | 0.002687873 | 0.001442141 |
| 0 | 0 | 0 | 0 | 0 |
| 0 | 0 | 0 | 0 | 0 |
| 0.001376595 | 0.012741212 | 0 | 0 | 0 |
| 0 | 0 | 0 | 0 | 0 |
| 0 | 0 | 1.045476089 | 1.372206829 | 0 |
| 0 | 0 | 0 | 0 | 0 |
| 0 | 0 | 0 | 0.016083289 | 0 |
| 0.017075638 | 0.054504759 | 0 | 0 | 0 |
| 0 | 0 | 0 | 0 | 0 |
| 0 | 0 | 0 | 0 | 0 |
| 0 | 0 | 0.001868741 | 0.176417631 | 0 |
| 0 | 0 | 0.001561673 | 0 | 0 |
| 0 | 0 | 0 | 0 | 0 |
| 0 | 0 | 0.003156201 | 0.105838992 | 0.007201178 |
| 0 | 0 | 0 | 0 | 0 |
| 0.004314592 | 0 | 0.000850861 | 0.018814906 | 0.056772887 |
| 0 | 0 | 0 | 0 | 0 |
| 0 | 0 | 0 | 0 | 0 |
| 0 | 0 | 0 | 0 | 0 |
| 0 | 0 | 0 | 0 | 0 |
| 0 | 0 | 0 | 0 | 0 |
| 0 | 0 | 0 | 0 | 0 |
| 0 | 0 | 0 | 0 | 0 |
| 0 | 0 | 0 | 0 | 0 |
| 0.000876911 | 0 | 0 | 0 | 0 |
| 0 | 0 | 0 | 0 | 0 |

| | | | | |
|---|---|---|---|---|
| 0.025408867 | 0 | 0 | 0.005393231 | 13.45649619 |
| 0 | 0 | 0 | 0 | 0 |
| 0.000899238 | 0 | 0 | 0 | 0 |
| 0 | 0 | 0 | 0.002816816 | 0 |
| 0.15742134 | 0.03700648 | 0 | 0.000809286 | 0.018490157 |
| 0.00258165 | 0 | 0 | 0 | 0.00206506 |
| 0 | 0 | 0 | 0 | 0 |
| 0.008243071 | 0.032183808 | 0.002334165 | 0 | 0 |
| 0 | 0 | 0 | 0 | 0 |
| 0.005882062 | 0.140105234 | 0.097953439 | 0 | 0.06989624 |
| 0 | 0 | 0 | 0 | 0 |
| 0 | 0.001498636 | 0 | 0.718840328 | 0.001967365 |
| 0.011612592 | 0.00363766 | 0.001401571 | 0 | 0 |
| 0.001734741 | 0 | 0 | 0 | 0 |
| 0.01488016 | 0 | 0 | 0 | 0 |
| 0 | 0.001023887 | 0 | 0 | 0 |
| 0 | 0 | 0 | 0.000929484 | 0 |
| 0 | 0 | 0 | 0 | 0 |
| 0 | 0.312882099 | 0 | 0 | 0 |
| 0 | 0 | 0 | 0 | 0 |
| 0 | 0 | 0 | 0 | 0 |
| 0 | 0 | 0 | 0 | 0 |
| 0 | 0 | 0 | 0.050606329 | 0 |
| 0 | 0 | 0 | 0 | 0 |
| 0 | 0 | 0 | 0 | 0 |
| 0 | 0 | 0.003498156 | 0.003371839 | 0 |
| 0 | 0 | 0 | 3.609452711 | 0 |
| 0 | 0.004162963 | 0.066837846 | 0 | 0 |
| 0 | 0 | 0 | 0 | 0 |
| 0 | 0 | 0 | 0 | 0 |
| 0.004044835 | 0 | 0 | 0 | 0 |
| 0 | 0 | 0 | 0 | 0 |
| 0 | 0 | 0 | 0 | 0 |
| 0 | 0.066147454 | 0 | 0 | 0 |
| 0 | 0 | 0 | 0 | 3.512962256 |
| 0 | 0 | 0 | 0 | 0 |
| 0.011562422 | 0.727656532 | 0.012425292 | 0 | 0 |
| 0 | 0.03204939 | 0 | 0 | 0 |
| 0 | 0 | 0 | 0 | 0 |
| 0.00186565 | 0 | 0 | 0 | 0 |
| 0 | 0 | 0 | 0 | 0 |
| 0.107809121 | 0.37568594 | 0 | 0 | 0 |
| 0 | 0 | 0 | 0 | 0 |
| 0 | 0.016000471 | 0 | 0 | 0 |
| 0 | 0.122394624 | 0 | 0 | 0 |
| 0 | 0 | 0 | 0 | 0 |
| 0.004136136 | 0 | 0.013630733 | 0 | 0 |
| 0 | 0 | 0 | 0 | 0 |

| | | | | |
|---|---|---|---|---|
| 0.009903583 | 0.004357522 | 0 | 0 | 0 |
| 0 | 0 | 0 | 0 | 0 |
| 0 | 0 | 0 | 0 | 0.001964476 |
| 0 | 0 | 0 | 0 | 0 |
| 0 | 0 | 0 | 0 | 0 |
| 0.014116736 | 0.025418497 | 0 | 0 | 0.000972846 |
| 0 | 0 | 0 | 0 | 0 |
| 0 | 0 | 0 | 0 | 0 |
| 0 | 0 | 0.020277793 | 0 | 0 |
| 0 | 0 | 0 | 0 | 0 |
| 0.011083287 | 0.039408511 | 0.00688822 | 0.002703864 | 0.016642987 |
| 0.003541215 | 0 | 0 | 0 | 0 |
| 0 | 0 | 0 | 0 | 0 |
| 0.008567126 | 0.00227366 | 0 | 0.000851492 | 0.001865495 |
| 0 | 0.066140386 | 0 | 0 | 0 |
| 0 | 0 | 0 | 0 | 0 |
| 0 | 0 | 0 | 0 | 0 |
| 0.001621122 | 0.078825238 | 0 | 0 | 0 |
| 0 | 0.012099807 | 0 | 0 | 0 |
| 0.049801767 | 0 | 0 | 0 | 0 |
| 0 | 0 | 0 | 0 | 0 |
| 1.568335376 | 0 | 0.418534068 | 0.212998157 | 0 |
| 0 | 0 | 0 | 0 | 0 |
| 0 | 0 | 0 | 0 | 0 |
| 0 | 0 | 0 | 0 | 0 |
| 0 | 0 | 0 | 0 | 0 |
| 0 | 0.060398606 | 0.035008659 | 0 | 0 |
| 0 | 0 | 0 | 0 | 0 |
| 0 | 0 | 0 | 0 | 0 |
| 0.234408769 | 0.69237032 | 0.504458026 | 0.028082436 | 0.211722613 |
| 0.00403619 | 0.001349685 | 0.006112965 | 0.000722087 | 0.00253118 |
| 0 | 0.000812107 | 0 | 0.187347862 | 0 |
| 0 | 0 | 0 | 0 | 0 |
| 0 | 0.001092352 | 0 | 0 | 0 |
| 0.001934222 | 0.001283325 | 0 | 0 | 0 |
| 0 | 0 | 0 | 0 | 0 |
| 0 | 0 | 0 | 0.068429285 | 0 |
| 0 | 0.003476198 | 0 | 0.87180227 | 0 |
| 0 | 0.787935425 | 0 | 0 | 0 |
| 0 | 0.077683033 | 0 | 0 | 0 |
| 0.063583631 | 0.0221967 | 0 | 0 | 0.039217362 |
| 3.092793112 | 0 | 1.440181331 | 0.371913695 | 2.242668368 |
| 0 | 0.002122948 | 0 | 0.675792124 | 0 |
| 0 | 0.021422527 | 0 | 0 | 0 |
| 0.007503606 | 0.003689963 | 0 | 0 | 0 |
| 0 | 0 | 0 | 0 | 0 |
| 0 | 0.012016363 | 0.001079846 | 0 | 0.001149761 |
| 0 | 0 | 0 | 0 | 0 |

| | | | | |
|---|---|---|---|---|
| 0 | 0 | 0 | 0.007478799 | 0.002269779 |
| 0.003399619 | 0 | 0 | 0 | 0.002900643 |
| 0 | 0.002885971 | 0.002541598 | 0.16737019 | 0 |
| 0.003437653 | 0 | 0 | 0 | 0 |
| 0.568114821 | 0 | 0.030536837 | 0 | 0 |
| 0.00771164 | 0 | 0 | 0 | 0 |
| 0 | 0 | 0 | 0 | 0 |
| 0 | 0 | 0 | 0.013716354 | 0 |
| 0 | 0.107875187 | 0 | 0 | 0.001662151 |
| 0 | 0 | 0 | 0 | 0.053728596 |
| 0 | 0 | 0 | 0 | 0 |
| 0 | 0 | 0 | 0 | 0 |
| 0 | 0 | 0 | 0 | 0 |
| 0.014669072 | 0.010128083 | 0 | 0.00184524 | 0.002310088 |
| 0 | 0 | 0 | 0 | 0 |
| 0.155833676 | 0.002994835 | 0.045213806 | 0.004646519 | 0.004412941 |
| 0.022632981 | 0.000948418 | 0.035706787 | 0.062512557 | 0.02734673 |
| 0.000964849 | 0.006625672 | 0 | 0 | 0 |
| 0.001230429 | 0 | 0 | 0 | 0 |
| 0 | 0 | 0 | 0 | 0 |
| 0.028774207 | 0 | 0 | 0 | 0.029140851 |
| 0.007138996 | 0.001375144 | 0 | 0 | 0 |
| 0 | 0 | 0 | 0 | 0 |
| 0 | 0 | 0 | 0 | 0 |
| 0 | 0 | 0 | 0 | 0 |
| 0 | 0 | 0 | 0 | 0 |
| 0.004284709 | 0.009030181 | 0 | 0 | 0.003387014 |
| 0.005263592 | 0.002857343 | 0 | 0 | 0.04608413 |
| 0 | 0 | 0 | 0 | 0 |
| 0.00215662 | 0 | 0 | 0 | 0 |
| 0.002300124 | 0 | 0.005846362 | 0 | 0.003005117 |
| 0.009716073 | 0.005801812 | 0.001803354 | 0 | 0.004800279 |
| 0.108563388 | 0.05686583 | 0.032337519 | 0.044783291 | 0 |
| 0 | 0 | 0 | 0.101151406 | 0 |
| 0 | 0 | 0.0017563 | 0.089125173 | 0 |
| 0 | 0 | 0 | 0 | 0 |
| 0 | 0 | 0 | 0 | 0 |
| 0.079642499 | 0.006662625 | 0.015984144 | 0.013078135 | 0.004739479 |
| 0.247517087 | 0 | 0.027956835 | 0 | 0.217170346 |
| 0 | 0 | 0 | 0 | 0 |
| 0.007508682 | 0 | 0 | 0 | 0 |
| 0 | 0 | 0 | 0 | 0 |
| 0 | 0.003779428 | 0 | 0 | 0.002577412 |
| 0 | 0 | 0 | 0 | 0.008913028 |
| 0 | 1.960111535 | 0 | 0 | 0 |
| 0 | 0 | 0 | 0 | 0 |
| 0 | 0 | 0 | 0 | 0 |
| 0 | 0 | 0 | 0 | 0 |

| | | | | |
|---|---|---|---|---|
| 0 | 0.006472096 | 0 | 0 | 0 |
| 0.916416509 | 0.23854993 | 0.178378915 | 0.020868882 | 0.507257639 |
| 0 | 0 | 0 | 0 | 0 |
| 0 | 0 | 0 | 0.001420111 | 0.000711145 |
| 0 | 0 | 0 | 0 | 0 |
| 0 | 0 | 0 | 0 | 0 |
| 0 | 0.062847346 | 0 | 0 | 0 |
| 0 | 0 | 0 | 0 | 0 |
| 0 | 0 | 0.003533349 | 0.010088494 | 0 |
| 0 | 0 | 0 | 0 | 0 |
| 0.006622975 | 0 | 0 | 0 | 0.002399553 |
| 0.020020369 | 0.037423946 | 0.014037784 | 0 | 0.009097968 |
| 0 | 0 | 0.514311985 | 0 | 0 |
| 0 | 0 | 0 | 0 | 0 |
| 0 | 0 | 0.002585771 | 0 | 0 |
| 0 | 0 | 0 | 0 | 0 |
| 0.004691466 | 0.012180177 | 0.007508731 | 0 | 0.003197954 |
| 0.041572038 | 0.103407551 | 0.216670114 | 0 | 0.002965275 |
| 0 | 0 | 0 | 0 | 0 |
| 0.087345937 | 0.00369619 | 0 | 0 | 0.012708266 |
| 0.033866596 | 0.013314485 | 0.101991588 | 0.024837466 | 0.019551818 |
| 0 | 0 | 0 | 0 | 0 |
| 0 | 0 | 0 | 0 | 0 |
| 0 | 0 | 0 | 0 | 0 |
| 0 | 0.002110455 | 0 | 0 | 0 |
| 0.002700576 | 0 | 0 | 0 | 0 |
| 0 | 0 | 0 | 0 | 0 |
| 0.075567546 | 0.111875627 | 0.051261684 | 0.022198789 | 0.045817679 |
| 0 | 0 | 0 | 0 | 0 |
| 0.001348405 | 0 | 0 | 0 | 0 |
| 0 | 0 | 0.000756933 | 0.001931296 | 0 |
| 0 | 0 | 0 | 0 | 0 |
| 0 | 0 | 0 | 0 | 0 |
| 0 | 0 | 0 | 0 | 0 |
| 0 | 0 | 0.049588896 | 0.960089628 | 0 |
| 0.233084126 | 0.009941628 | 0.372897382 | 0.019631237 | 0.063984136 |
| 0 | 0 | 0 | 0 | 0 |
| 0 | 0 | 0 | 0 | 0 |
| 0.045507303 | 0.018445398 | 0.027847495 | 0.318501208 | 0.043858005 |
| 0 | 0 | 0 | 0 | 0 |
| 0 | 1.642763972 | 0 | 0 | 0 |
| 0 | 0 | 0 | 0 | 0 |
| 0.084265702 | 0 | 0.062726436 | 0 | 0.008442261 |
| 0.023452798 | 0 | 0 | 0 | 0.010686662 |
| 0.003187401 | 0 | 0 | 0 | 0 |
| 0 | 0 | 1.571687049 | 0.036773626 | 0 |
| 0 | 0 | 0 | 0 | 0 |
| 0.001307229 | 0 | 0 | 0 | 0 |

| | | | | |
|---|---|---|---|---|
| 0.002574416 | 0 | 0 | 0 | 0 |
| 0 | 0 | 0.11236325 | 0 | 0 |
| 0 | 0 | 0 | 0.003433312 | 0 |
| 6.651667713 | 0 | 0.043095096 | 0.087441223 | 1.821427495 |
| 0 | 0 | 0 | 0 | 0 |
| 0 | 0 | 0 | 0 | 0.001705266 |
| 0 | 0 | 0.001272787 | 0 | 0 |
| 0 | 0 | 0 | 0 | 0 |
| 2.132530854 | 0 | 1.380631259 | 3.026006795 | 0 |
| 0.094516607 | 0 | 0.004585288 | 0.09406192 | 0.037299721 |
| 0 | 0 | 0 | 0 | 0 |
| 0.050474126 | 0 | 0.009241036 | 0.001337774 | 0.002512174 |
| 0 | 0 | 0 | 0 | 0 |
| 0 | 0 | 0 | 0 | 0.30397461 |
| 0.137758073 | 0 | 0.181719436 | 0.006761598 | 0 |
| 0 | 0 | 0 | 0 | 0 |
| 0.067007705 | 0 | 0.001904762 | 0.093239076 | 0.011717831 |
| 0 | 0 | 0 | 9.810619275 | 0 |
| 0 | 0 | 0 | 0 | 0 |
| 0 | 0 | 0 | 0.006272009 | 0 |
| 0 | 0 | 0 | 0 | 0 |
| 0 | 0 | 0 | 0 | 0 |
| 0 | 0 | 0 | 0 | 0 |
| 0.334192666 | 0.331857708 | 0 | 0.021932043 | 0 |
| 0.008576212 | 0 | 0 | 0 | 0.312624863 |
| 0 | 0 | 0.159173671 | 0 | 0 |
| 0 | 0 | 0 | 0.003166844 | 0 |
| 0 | 0.608567406 | 3.78936812 | 0.367589517 | 0 |
| 0 | 0 | 0 | 0 | 0 |
| 0 | 0.006629328 | 0 | 0 | 0 |
| 0 | 0 | 0 | 0.010729491 | 0.084665055 |
| 3.378570566 | 0 | 0 | 0 | 0 |
| 0.03693051 | 0 | 0 | 0 | 0.011617126 |
| 0 | 0 | 0 | 0 | 0 |
| 0.009564435 | 0 | 0 | 0 | 0.084934921 |
| 0 | 0 | 0.185732728 | 0.011716359 | 0 |
| 0 | 0 | 3.235170419 | 1.074887707 | 0.982174659 |
| 0 | 0 | 0 | 0 | 0 |
| 0 | 0 | 0.00716377 | 0 | 0 |
| 0 | 0 | 0.002392474 | 0 | 0.014385176 |
| 0.170728511 | 0.363029275 | 0.018673342 | 0 | 0.215693353 |
| 0 | 0 | 0 | 0 | 0 |
| 0.003227045 | 0.391562964 | 15.02810674 | 0.004426183 | 0.015659956 |
| 0 | 0 | 0.004765058 | 0.001397462 | 0 |
| 0 | 0 | 0 | 0 | 0 |
| 0 | 0 | 0 | 0 | 0 |
| 0 | 0 | 0 | 0 | 0 |
| 0.002586999 | 0 | 0.012924318 | 0 | 0.030902126 |

| | | | | |
|---|---|---|---|---|
| 0 | 0 | 0 | 0 | 0 |
| 0 | 0 | 0.069613674 | 0.266865458 | 0 |
| 0.006199827 | 0.643431263 | 0.002173582 | 0 | 0.004860052 |
| 0 | 0 | 0 | 0 | 0 |
| 0 | 0 | 0 | 0 | 0 |
| 0 | 0 | 0.001369811 | 0.055920613 | 0.095167111 |
| 0 | 0 | 0 | 0 | 0 |
| 0 | 0 | 0 | 0 | 0.127851427 |
| 0 | 0 | 0 | 0 | 0.165195765 |
| 0 | 0 | 0 | 0 | 0.096457873 |
| 0.060668398 | 0.00266555 | 0.038199891 | 0 | 1.443328559 |
| 4.55863522 | 2.338156876 | 1.11490355 | 0.653537567 | 1.446081615 |
| 0 | 0 | 0 | 0 | 0 |
| 0.015547767 | 0.006015065 | 0.012437159 | 0.127520016 | 0.038255832 |
| 0.310620087 | 0.65557675 | 0 | 0 | 0.323694425 |
| 0 | 0 | 3.997096811 | 0.079373634 | 0 |
| 0 | 0 | 0 | 0 | 0.002825668 |
| 0 | 0 | 0 | 0 | 0 |
| 0 | 0 | 0 | 0 | 0 |
| 0.060170319 | 0 | 0.026368706 | 0 | 0.008797131 |
| 1.821212349 | 0.79352008 | 0.031480534 | 0 | 1.904854882 |
| 0 | 1.821196597 | 0.083869141 | 0 | 0 |
| 1.795835601 | 12.34342468 | 5.765041839 | 0.016380129 | 2.71046474 |
| 0 | 0 | 0 | 0 | 0 |
| 0 | 0 | 0 | 0 | 0.007092969 |
| 0 | 0.005110923 | 1.853148404 | 0.68176783 | 0 |
| 0.69340467 | 0 | 0 | 0 | 0 |
| 0 | 0 | 0 | 0.013962359 | 0 |
| 0.001551831 | 0 | 0 | 0 | 0 |
| 0 | 0 | 0 | 0 | 0 |
| 0 | 0 | 0 | 0 | 0 |
| 0.008348766 | 0 | 0.146911548 | 0.024845785 | 0 |
| 0 | 1.277458805 | 0.001972573 | 0 | 0.006767592 |
| 0.213895575 | 0 | 0.054654038 | 0 | 0.211237412 |
| 0.007289551 | 0.004763495 | 0 | 0 | 0.001386216 |
| 0.038171892 | 0.001354086 | 0.316213384 | 0.191349102 | 0.066660143 |
| 0.00492864 | 0 | 0.004799777 | 0 | 0 |
| 0 | 0 | 0 | 0 | 0 |
| 4.501821139 | 0 | 0.342356591 | 0.485219633 | 0 |
| 0 | 0 | 0 | 0 | 0 |
| 0 | 0 | 0 | 0 | 0 |
| 0 | 0 | 0 | 0 | 0 |
| 0 | 0 | 0 | 0 | 0 |
| 0 | 0 | 0 | 0 | 0 |
| 0.070683528 | 0.003088364 | 0.110606832 | 0.006168535 | 0.094841912 |
| 0 | 0 | 0 | 0 | 0 |
| 0 | 0 | 0 | 0 | 0 |
| 0.007018654 | 0.031976442 | 6.968842229 | 0 | 0.223279484 |

| | | | | |
|---|---|---|---|---|
| 0 | 0 | 0 | 0 | 0 |
| 0 | 0 | 0 | 0 | 0 |
| 0 | 0 | 0 | 0 | 0 |
| 0 | 0 | 0.003477163 | 0 | 0.000617049 |
| 0.003618695 | 0 | 0 | 0 | 0.038378886 |
| 0 | 0 | 0.570866337 | 0.057751265 | 0 |
| 0 | 0 | 0 | 0 | 0 |
| 0.001183058 | 0 | 0 | 0.004988956 | 0 |
| 0 | 0 | 0 | 0 | 0 |
| 0 | 0 | 0 | 0 | 0 |
| 0.009409674 | 0.000977191 | 0.007960424 | 0.005855371 | 0 |
| 0 | 0 | 0 | 0 | 0 |
| 0 | 0 | 0.11509876 | 0.251695559 | 0 |
| 0 | 0 | 0 | 0 | 0 |
| 0 | 0 | 0 | 0 | 0 |
| 0 | 0 | 0 | 0 | 0 |
| 0.288148217 | 0 | 0.012160945 | 0 | 0.256921637 |
| 0.010898558 | 0 | 0.005731348 | 0.012490811 | 0.015256062 |
| 1.054176044 | 0 | 0 | 0.019234603 | 0.004615359 |
| 0.08692252 | 0 | 0 | 0 | 0.002235747 |
| 0 | 0 | 0 | 0 | 0 |
| 0 | 0 | 0 | 0 | 0 |
| 0 | 0 | 0.05554711 | 0.002985575 | 0 |
| 0 | 0 | 0 | 0 | 0 |
| 0 | 0 | 0 | 0 | 0 |
| 0 | 0 | 0 | 0 | 0 |
| 0 | 0 | 0 | 0 | 0 |
| 0 | 0 | 0 | 0 | 0 |
| 0 | 0 | 0 | 0 | 0 |
| 1.056931654 | 0 | 0.105149408 | 0.011592605 | 0.113339498 |
| 0 | 0 | 0 | 0 | 0 |
| 0 | 0 | 0 | 0 | 0 |
| 0 | 0 | 0 | 0 | 0.430180397 |
| 0 | 0 | 0 | 0 | 0 |
| 0 | 0 | 0 | 0 | 0 |
| 0 | 0 | 0 | 0 | 0 |
| 0 | 0 | 0.101686144 | 0.004455195 | 0 |
| 0.035325055 | 0.005997269 | 0.018030411 | 0 | 0.000581751 |
| 0 | 0 | 0 | 0 | 0.03139448 |
| 0.012980607 | 0 | 0 | 0 | 0 |
| 0 | 0 | 0.008373055 | 0 | 0.344598551 |
| 0 | 0 | 0 | 0 | 0 |
| 0 | 0 | 0 | 0 | 0 |
| 0.011603099 | 0 | 0.004775363 | 0.027376882 | 0 |
| 0 | 0 | 0.141659007 | 0.04713661 | 0 |
| 0 | 0 | 0 | 0 | 0 |
| 0.000493599 | 0 | 0 | 0 | 0 |
| 0 | 0 | 0 | 0 | 0 |

| | | | | |
|---|---|---|---|---|
| 0 | 0 | 0 | 0 | 0 |
| 0 | 0 | 0 | 0 | 1.350609331 |
| 0 | 0 | 0 | 0 | 0 |
| 0 | 0 | 0.101631523 | 0.050382911 | 0 |
| 0 | 0 | 0 | 0 | 0 |
| 0 | 0 | 0.351328602 | 0 | 0 |
| 0 | 0 | 0 | 0 | 0 |
| 0 | 0 | 0 | 0 | 0.026040461 |
| 0.01886673 | 0.518235685 | 0 | 0 | 0.004546549 |
| 0 | 0 | 0.000809101 | 0 | 0 |
| 0 | 0 | 0.001077615 | 0 | 0 |
| 0 | 0 | 0 | 0.016019238 | 0 |
| 0 | 0 | 0 | 0 | 0 |
| 0 | 0 | 0 | 0 | 0.007801837 |
| 0.064358213 | 14.50133993 | 0.00884477 | 0.037612005 | 0.085525593 |
| 0.009571366 | 0.013629035 | 0 | 0 | 0 |
| 0 | 0 | 0 | 0 | 0.200035768 |
| 0 | 0 | 0 | 0 | 0 |
| 0 | 0 | 0 | 0 | 0 |
| 0 | 0 | 0 | 0 | 0 |
| 0.246708346 | 0 | 0.010947655 | 0 | 0.001840494 |
| 0 | 0 | 0 | 0 | 0 |
| 0 | 0 | 0 | 0 | 0 |
| 0.029271525 | 0 | 0 | 0 | 0 |
| 0.002808448 | 0 | 0.003544585 | 0.479578793 | 0.018555888 |
| 2.570843193 | 0 | 3.089793047 | 0.087847319 | 0.112349075 |
| 0 | 0 | 0.623811363 | 0.013642083 | 0 |
| 0 | 0 | 0 | 0 | 0 |
| 1.655667081 | 0 | 0 | 3.842128103 | 0 |
| 0 | 0 | 0 | 0 | 0.215535451 |
| 0 | 0 | 0 | 0 | 0 |
| 0 | 0 | 0.147616677 | 0 | 0 |
| 0 | 0 | 0 | 0 | 0 |
| 0.016798784 | 0 | 0.006783378 | 0 | 0.003695267 |
| 0 | 0.001180583 | 0.00441876 | 0.144355937 | 0.011346999 |
| 0.012577989 | 0.000625897 | 0 | 0.008282134 | 0.014085575 |
| 0 | 0.055548114 | 0.086462815 | 0 | 0 |
| 0.922585256 | 0 | 1.459971734 | 0.158983342 | 0.891787265 |
| 0 | 0 | 0 | 0.422345438 | 0 |
| 0 | 0 | 0 | 0 | 0 |
| 0 | 0 | 0 | 0 | 0 |
| 0 | 0 | 0 | 0 | 0.090514548 |
| 0.244929656 | 0 | 0.184041491 | 0.175744417 | 0.333902621 |
| 0 | 0 | 0.074899479 | 0 | 0 |
| 0 | 0 | 0 | 0 | 0 |
| 0.008440547 | 0 | 0.005744234 | 0 | 0 |
| 0 | 0 | 0 | 0 | 0 |
| 0.569511631 | 0.510743136 | 0.078201613 | 0.004717007 | 0.922703665 |

| | | | | |
|---|---|---|---|---|
| 0 | 0 | 0.004342609 | 0 | 0 |
| 0 | 0 | 0 | 0 | 0 |
| 0 | 0 | 0 | 0 | 0 |
| 0 | 0 | 0 | 0 | 0 |
| 0.076147392 | 0 | 0.001235306 | 0 | 0.023675141 |
| 0.006786213 | 0 | 0.038066591 | 0.0023607 | 0 |
| 0 | 0 | 0 | 0 | 0 |
| 0 | 0 | 0 | 0 | 0 |
| 0 | 0 | 0 | 0 | 0 |
| 0 | 0 | 0 | 0 | 0 |
| 0.001048491 | 0 | 0 | 0 | 0.001006424 |
| 1.714011848 | 0.392101558 | 0.602494077 | 0.198052353 | 6.505022143 |
| 0.746852938 | 0 | 0 | 0.006148545 | 0.120239807 |
| 0 | 0 | 0 | 0 | 0 |
| 0 | 0.038121766 | 0.001666736 | 1.702066254 | 0.934225941 |
| 0 | 0 | 0 | 0 | 0 |
| 0.00267724 | 0 | 0 | 0 | 0.071249713 |
| 0 | 0 | 0 | 0 | 0 |
| 0.000595282 | 0 | 0 | 0.004259907 | 0.007555157 |
| 0 | 0 | 0 | 0.004920372 | 0 |
| 0.092653653 | 0 | 0.054945018 | 0.016458458 | 0 |
| 0 | 0 | 0 | 0 | 0 |
| 1.20531798 | 0 | 0.315760266 | 0.184672232 | 0.112788289 |
| 0 | 0 | 0 | 0 | 0 |
| 0 | 0 | 0 | 0 | 0 |
| 0 | 0 | 0 | 0 | 0 |
| 0 | 0 | 0 | 0 | 0 |
| 0 | 0 | 0 | 0 | 0 |
| 0 | 0 | 0.005361813 | 0 | 0 |
| 0 | 0 | 0 | 0 | 0 |
| 0 | 0 | 0 | 0 | 0 |
| 0.090774466 | 0.040697343 | 0.013594889 | 0 | 0.046386084 |
| 0 | 0 | 0 | 0 | 0 |
| 0.043588219 | 0.002103278 | 0.049240348 | 0 | 0.110937869 |
| 0.012737448 | 0 | 0.001999518 | 0.006632235 | 0.011709374 |
| 0.486966697 | 0.003804938 | 0.011599297 | 0.111804494 | 0 |
| 0 | 0 | 0 | 0 | 0 |
| 0 | 0 | 0.007074411 | 0.200015491 | 0 |
| 0.010702074 | 0 | 0.002067436 | 0.001758337 | 0 |
| 0 | 0 | 0 | 0 | 0 |
| 0 | 0 | 0 | 0.094467924 | 0.090694816 |
| 0 | 0 | 0 | 0 | 0 |
| 0 | 0 | 0 | 0 | 0 |
| 0 | 0 | 0 | 0 | 0.017678471 |
| 0.011049435 | 0 | 0.092577119 | 0.001005141 | 0.051592495 |
| 0 | 0 | 0 | 0 | 0 |
| 0 | 0 | 0.220750086 | 0 | 0.311449032 |
| 0.044460132 | 0 | 0.010246545 | 0 | 0.001190177 |

| | | | | |
|---|---|---|---|---|
| 0 | 0 | 0 | 0 | 0 |
| 0 | 0 | 0 | 0 | 0.000909594 |
| 0.058107868 | 0 | 0 | 0 | 0.001301452 |
| 0 | 0 | 0 | 0 | 0 |
| 0 | 0 | 0 | 0 | 0 |
| 0 | 0 | 0 | 0 | 0.004105297 |
| 0 | 0 | 0 | 4.08603566 | 0 |
| 0 | 0.099181655 | 0 | 0 | 0.059816209 |
| 0 | 0 | 0 | 0 | 0 |
| 0 | 0 | 0.008330193 | 0 | 0 |
| 0 | 0 | 0 | 0 | 0.121363319 |
| 0 | 0 | 0 | 0.002820625 | 0 |
| 0 | 0 | 0 | 0 | 0 |
| 0 | 0 | 0.036523518 | 0.067279818 | 0 |
| 0 | 0 | 0 | 0 | 0 |
| 0 | 0 | 0 | 0 | 0 |
| 0 | 0 | 2.503126987 | 0 | 0.429827663 |
| 0 | 0 | 0 | 0 | 0 |
| 1.37943641 | 0 | 0 | 0.245489401 | 8.654676768 |
| 0 | 0 | 0 | 0 | 0 |
| 0 | 0 | 0 | 0 | 0.003744539 |
| 0 | 0 | 0 | 0 | 0 |
| 0 | 0 | 0 | 0 | 0 |
| 0 | 0 | 0 | 0 | 0 |
| 0.003273889 | 0 | 0 | 0.000732136 | 0 |
| 0.015691433 | 0.010202792 | 0.043826395 | 0 | 0.002538206 |
| 0 | 0 | 0 | 0 | 0 |
| 2.188286509 | 0.027513836 | 0.010589716 | 0 | 1.656711629 |
| 0 | 0 | 0 | 0 | 0 |
| 0 | 0 | 0.535555433 | 0.008524366 | 0.144780788 |
| 0 | 0 | 0 | 0 | 0 |
| 0 | 0 | 0 | 0 | 0 |
| 0 | 0 | 0 | 0.001611872 | 0 |
| 0 | 0 | 0 | 0 | 0 |
| 0.242635206 | 0 | 0 | 0 | 0.006956616 |
| 0 | 0 | 0 | 0 | 0 |
| 0 | 0 | 0 | 0.550625131 | 0 |
| 0 | 0 | 0 | 0 | 0.003250356 |
| 0 | 0 | 0 | 0 | 0 |
| 0 | 0 | 0 | 0 | 0 |
| 1.059020089 | 14.47331529 | 8.945157177 | 0.392565053 | 1.99008229 |
| 0.206239924 | 0.041141676 | 0 | 0 | 0.619801215 |
| 0.42856179 | 0 | 0.172542153 | 0 | 0 |
| 0 | 0 | 0 | 0.004764691 | 0 |
| 0 | 0 | 0 | 0 | 0 |
| 0 | 0 | 0 | 0 | 0 |
| 0 | 0 | 0 | 0 | 0 |
| 0.005594359 | 0 | 0.008652851 | 0 | 0.018978944 |

| | | | | |
|---|---|---|---|---|
| 0 | 0 | 0 | 0 | 0 |
| 0.043833926 | 0.001819097 | 0.004094079 | 0 | 0.018384241 |
| 0 | 0 | 0 | 0 | 0.016823232 |
| 0 | 0 | 0 | 0 | 0 |
| 0.879264621 | 0 | 1.240164334 | 0.17813357 | 2.997075849 |
| 0 | 0 | 0 | 0 | 0 |
| 0 | 0 | 0 | 0 | 0 |
| 0.006639333 | 0 | 0 | 0 | 0 |
| 0.047994136 | 3.599529954 | 0.251858982 | 0 | 0.001892488 |
| 0 | 0 | 0 | 0 | 0 |
| 0 | 0 | 0 | 0 | 0 |
| 0 | 0 | 0 | 0 | 0 |
| 0.135012118 | 0 | 0.030614038 | 0.021918133 | 0.015101039 |
| 0.001213035 | 0.010140849 | 0.029131344 | 0.000904232 | 0.003848881 |
| 0 | 0 | 0.239529961 | 0.44650298 | 0 |
| 0.316871551 | 0 | 0.032629412 | 0.022111787 | 0.590799855 |
| 0.013296123 | 0.00710384 | 0.003496092 | 0 | 1.637288508 |
| 0 | 0 | 0 | 0 | 0 |
| 0 | 0 | 0 | 0 | 0 |
| 0 | 0 | 0 | 0 | 0 |
| 0 | 0 | 0 | 0 | 0 |
| 0 | 0 | 0 | 0 | 0 |
| 0 | 0 | 0 | 0 | 0 |
| 0 | 0 | 0 | 0 | 0 |
| 0 | 0 | 0 | 0 | 0 |
| 0 | 0 | 0 | 0 | 0 |
| 0.072563785 | 0 | 0.066879896 | 0.084640774 | 1.287402503 |
| 0.006007975 | 0.000615013 | 0.00198596 | 0 | 0 |
| 0 | 0 | 0 | 0 | 0 |
| 0 | 0 | 0 | 0 | 0 |
| 0 | 0 | 0 | 0 | 0 |
| 0 | 0 | 0 | 0 | 0 |
| 0 | 0 | 0 | 0 | 0.217776874 |
| 0 | 0 | 0.008516521 | 0.002092488 | 0 |
| 1.730464971 | 0 | 0.581087183 | 1.15118868 | 0 |
| 0.14396387 | 0 | 0 | 0 | 0 |
| 0 | 0 | 0 | 0 | 0.005454968 |
| 0.095441154 | 0 | 0 | 0 | 0 |
| 0 | 0 | 0 | 0 | 0 |
| 0 | 0 | 0 | 0 | 0 |
| 0 | 1.777811003 | 2.078011945 | 0.050470318 | 0.767748225 |
| 0.168083792 | 0 | 0 | 0.071651713 | 0 |
| 0 | 0 | 0 | 0 | 0 |
| 0 | 0 | 0 | 2.570707976 | 0 |
| 0 | 0 | 0 | 0 | 0 |
| 0 | 0 | 0 | 0 | 0 |
| 0 | 0 | 0 | 0 | 0 |
| 0 | 0 | 0 | 0 | 0 |

| | | | | |
|---|---|---|---|---|
| 0 | 0 | 0.307584459 | 0.034969391 | 0 |
| 0 | 0 | 0 | 0 | 0.001807824 |
| 0 | 0 | 0 | 0.002061024 | 0 |
| 0.002916796 | 0 | 0 | 0 | 0 |
| 0.094701252 | 0 | 0.001369789 | 0.052591156 | 0.017293361 |
| 0 | 0 | 0.000712404 | 0 | 0.253095873 |
| 0.539975974 | 0.474591548 | 0.017805067 | 0 | 0.880660853 |
| 0 | 0 | 0 | 0 | 0 |
| 0 | 0 | 0 | 0 | 0 |
| 0.155088786 | 0 | 0 | 0 | 0.024811084 |
| 0 | 0 | 0.004229275 | 0 | 0 |
| 0 | 0 | 0 | 0 | 0.045976826 |
| 0 | 0 | 0 | 0 | 0 |
| 0 | 0 | 0 | 0 | 0 |
| 0 | 0 | 0 | 0 | 0 |
| 0 | 0 | 0 | 0 | 0 |
| 0 | 0 | 0 | 0 | 0 |
| 0 | 0 | 0 | 0 | 0 |
| 0 | 0 | 0 | 0 | 0 |
| 0 | 0 | 0 | 0 | 0 |
| 0.158845308 | 0.016073274 | 0.067805665 | 0.053803777 | 0.01116429 |
| 0 | 0 | 0 | 0 | 0.005235235 |
| 0.072758066 | 0 | 0.015613254 | 0.001639375 | 0.003078543 |
| 0.048237728 | 0.001751622 | 0.011826657 | 0 | 0 |
| 0 | 0 | 0 | 0.000697106 | 0 |
| 0 | 0 | 0 | 0.000859908 | 0.001076534 |
| 0 | 0 | 0 | 0.103678855 | 0 |
| 0 | 0 | 0 | 0 | 0 |
| 0 | 0 | 0 | 0 | 0 |
| 0 | 1.646770149 | 0.067098582 | 0 | 0.003151892 |
| 0 | 0 | 0 | 0 | 0 |
| 0 | 0 | 0 | 0 | 0 |
| 0 | 0 | 0 | 0 | 0 |
| 0 | 0 | 0 | 0 | 0 |
| 0 | 0 | 0 | 0 | 0 |
| 0 | 0 | 0 | 0 | 0 |
| 0 | 0.000974104 | 0.004932737 | 0 | 0.007763984 |
| 0 | 0 | 0 | 0 | 0 |
| 0 | 0 | 0 | 0 | 0 |
| 0 | 0 | 0 | 0.352458204 | 0 |
| 0 | 0 | 0 | 0 | 0 |
| 0 | 0 | 0 | 0 | 0 |
| 0 | 0 | 0 | 0 | 0 |
| 0 | 0 | 0 | 0 | 0 |
| 0 | 0 | 0 | 0 | 1.142834028 |
| 0 | 0 | 0 | 2.259049542 | 0 |
| 0 | 0 | 0 | 0 | 0 |
| 0 | 0 | 0 | 0 | 0 |

| | | | | |
|---|---|---|---|---|
| 0.006312832 | 0.004607308 | 0 | 0.004235193 | 0.003141992 |
| 0 | 0 | 0 | 0.752004032 | 0 |
| 0 | 0 | 0 | 0.002982581 | 0 |
| 0 | 0 | 0 | 0 | 0 |
| 0 | 0 | 0 | 0 | 0.002666428 |
| 0 | 0 | 0 | 0.081545635 | 0 |
| 0 | 0 | 0 | 0 | 0 |
| 1.89972273 | 0.004945694 | 0.598602945 | 0.028829514 | 1.781824098 |
| 0.066059587 | 0.001972326 | 0.112613893 | 4.478381929 | 0.625725754 |
| 0.127235412 | 0.002288457 | 0.103792245 | 0 | 0 |
| 0.008319902 | 0 | 0 | 0 | 0 |
| 0 | 0 | 0 | 0 | 0 |
| 0 | 0 | 0 | 0 | 0.041627781 |
| 0 | 0 | 0 | 0.640154552 | 0 |
| 0 | 0 | 0 | 0 | 0 |
| 0 | 0 | 0 | 0 | 0 |
| 0 | 0 | 0.008076693 | 0 | 0.002006578 |
| 0 | 0 | 0 | 0.218022509 | 0 |
| 0 | 0 | 0 | 0 | 0 |
| 0 | 0 | 0 | 0 | 0.015658778 |
| 0 | 0 | 0 | 0 | 0 |
| 0 | 0 | 0 | 0 | 0 |
| 0 | 0 | 0 | 0 | 0 |
| 0 | 0 | 0 | 0 | 0 |
| 0 | 0 | 0.005374873 | 0.000741289 | 0.103011652 |
| 0 | 0 | 0 | 0 | 0 |
| 0 | 0 | 0 | 0 | 0 |
| 0 | 0 | 0 | 0 | 0 |
| 0 | 0 | 0 | 0 | 0 |
| 0 | 0 | 0 | 0 | 0 |
| 0 | 0 | 0 | 0 | 0 |
| 0 | 0 | 0 | 0 | 0 |
| 0 | 0 | 0 | 0 | 0 |
| 0 | 0 | 0 | 0 | 0 |
| 0 | 0 | 0.004019086 | 0.050703272 | 0 |
| 0 | 0 | 0.000588321 | 0 | 0 |
| 0 | 0 | 0 | 0 | 0.004965006 |
| 0 | 0 | 0 | 0 | 0 |
| 0 | 0 | 0.002193057 | 0.003357317 | 0 |
| 0 | 0 | 0 | 0 | 0 |
| 0 | 0 | 0 | 0 | 0 |
| 0.100958826 | 0 | 0.278663672 | 0.790649755 | 0.571770403 |
| 0 | 0 | 0 | 0 | 0 |
| 0 | 0 | 0 | 0 | 0 |
| 0 | 0 | 0.001076538 | 0 | 0 |
| 0 | 0 | 0 | 0 | 0.026716377 |
| 0 | 0 | 0 | 0 | 0 |
| 0 | 0 | 0 | 0 | 0 |

| | | | | |
|---|---|---|---|---|
| 0 | 0 | 0 | 0 | 0 |
| 0.039250182 | 0 | 0.009654632 | 0 | 0 |
| 0 | 0 | 0 | 0 | 0 |
| 0 | 0 | 0 | 0 | 0 |
| 0.872283102 | 0.066331131 | 0.551810741 | 1.916630794 | 0 |
| 0 | 0 | 0 | 0 | 0.763864383 |
| 0 | 0 | 0 | 0 | 0 |
| 0.014722363 | 0.080186805 | 0.001407676 | 0 | 0.00574546 |
| 0.040758352 | 0 | 0.003045276 | 0 | 0 |
| 0 | 0 | 0.001055596 | 0 | 0 |
| 0 | 0 | 0 | 0 | 0 |
| 0 | 0 | 0 | 0.0602221 | 0 |
| 0 | 0 | 0 | 0 | 0 |
| 0 | 0 | 0 | 0 | 0 |
| 0 | 0.027755498 | 0 | 0 | 0.024267606 |
| 0.013929376 | 0 | 0 | 0 | 0.206252635 |
| 0 | 0 | 0 | 0 | 0 |
| 0 | 0 | 0 | 0 | 0 |
| 0 | 0 | 0 | 0 | 0 |
| 0 | 0 | 0 | 0 | 0 |
| 0 | 0 | 0 | 0 | 0 |
| 0 | 0 | 0 | 0 | 0 |
| 0 | 0 | 0 | 0 | 0 |
| 0.004246997 | 0.001613839 | 0.004872909 | 0.00345364 | 0.001080918 |
| 1.078457434 | 0 | 0.086710466 | 0.049010699 | 0 |
| 0 | 0 | 0 | 0 | 0 |
| 0.001235933 | 0.038798737 | 0 | 0 | 0.153236422 |
| 0.008180212 | 0 | 0.000586612 | 0 | 0 |
| 0 | 0 | 0 | 0.002347543 | 0 |
| 0 | 0 | 0 | 0 | 0 |
| 0 | 0 | 0 | 0 | 0 |
| 0 | 0 | 0 | 0 | 0 |
| 0 | 0 | 0 | 0 | 0 |
| 0 | 0 | 0 | 0 | 0 |
| 0 | 0 | 0 | 0 | 0 |
| 0 | 0 | 0 | 0 | 0.001767636 |
| 0 | 0 | 0 | 0 | 0 |
| 2.543192009 | 0 | 0.269292695 | 0 | 0 |
| 0 | 0 | 0.15943116 | 0 | 0 |
| 0 | 0 | 0 | 0 | 0 |
| 0 | 0 | 0 | 0 | 0 |
| 0 | 0 | 0.300920507 | 2.232982645 | 0 |
| 0 | 0 | 0 | 0 | 0 |
| 0 | 0 | 0 | 0.02144947 | 0 |
| 0 | 0 | 0 | 0 | 0 |
| 0 | 0 | 0 | 0 | 0 |
| 0 | 0 | 0 | 0 | 0 |
| 0 | 0 | 0 | 0 | 0 |

| | | | | |
|---|---|---|---|---|
| 0.02573639 | 0 | 0.458594038 | 0.018565798 | 0.298995979 |
| 0 | 0 | 0 | 0 | 0 |
| 0 | 0 | 0 | 0 | 0 |
| 0 | 0 | 0 | 0 | 0.012580191 |
| 0 | 0 | 0 | 0 | 0 |
| 0.056941036 | 0 | 0 | 0 | 0 |
| 0 | 0 | 0 | 0 | 0 |
| 0 | 0 | 0 | 0 | 0 |
| 0 | 0 | 0 | 0 | 0 |
| 0 | 0 | 0 | 0 | 0 |
| 0 | 0 | 0 | 0 | 0 |
| 0.003203226 | 0 | 0 | 0 | 0.050220267 |
| 0 | 0 | 0 | 0 | 0 |
| 0 | 0 | 0 | 0 | 0 |
| 0 | 0 | 0 | 0.002953882 | 0 |
| 0 | 0 | 0 | 0 | 0 |
| 0 | 0 | 0 | 0 | 0 |
| 0 | 0 | 0 | 0 | 0 |
| 0 | 0 | 0 | 0 | 0 |
| 0 | 0 | 0 | 0 | 0 |
| 0.024798748 | 0 | 0 | 0 | 0 |
| 0 | 0 | 0 | 0 | 0 |
| 0.059724628 | 0 | 0.101884706 | 0 | 0.147632356 |
| 0 | 0 | 0 | 0.525542963 | 0 |
| 0 | 0 | 0 | 0 | 0 |
| 0 | 0 | 0 | 1.231880683 | 0 |
| 0 | 0 | 0 | 0 | 0 |
| 0 | 0 | 0 | 0 | 0 |
| 0 | 0 | 0 | 0 | 0 |
| 0 | 0 | 0 | 0 | 0 |
| 0 | 0 | 0 | 0 | 0 |
| 0 | 0 | 0 | 0 | 0 |
| 0.003183114 | 0.007603002 | 0.001434806 | 0 | 0 |
| 0.111929489 | 0 | 0 | 0.013029656 | 0 |
| 0.670737988 | 0 | 0 | 0.008368503 | 0.116154437 |
| 0.002006962 | 0.013291661 | 0.001918953 | 0 | 0 |
| 0 | 0 | 0 | 0 | 0 |
| 0 | 0 | 0 | 0.086005137 | 0 |
| 0.022791629 | 0.001008125 | 0 | 0 | 0.019851543 |
| 0 | 0 | 0 | 0 | 0 |
| 0 | 0 | 0 | 0 | 0 |
| 0 | 0 | 0 | 0 | 0 |
| 0.232202583 | 0 | 0.966766197 | 2.793803734 | 0 |
| 0 | 0 | 0 | 0 | 0 |
| 0 | 0 | 0 | 0 | 0 |
| 0.010386486 | 0 | 0.228582028 | 0.001051796 | 0 |
| 0 | 0 | 0 | 0 | 0 |
| 0 | 0 | 0 | 0 | 0 |

| | | | | |
|---|---|---|---|---|
| 0 | 0 | 0 | 0 | 0 |
| 0 | 0 | 0 | 0 | 0 |
| 0 | 0 | 0 | 0 | 0 |
| 0 | 0 | 0 | 0 | 0 |
| 0 | 0 | 0 | 0 | 0 |
| 0 | 0 | 0 | 0 | 0 |
| 0 | 0 | 0 | 0 | 0 |
| 0 | 2.438453488 | 0 | 0 | 0 |
| 0.01505578 | 0 | 0.003282888 | 0.00476294 | 0.403003524 |
| 0.022405174 | 0 | 0.003609043 | 0.005416694 | 0 |
| 0 | 0 | 0 | 0 | 0 |
| 0 | 0 | 0 | 0 | 0 |
| 0 | 0 | 0 | 0 | 0 |
| 0 | 0 | 0 | 0.020033086 | 0 |
| 0 | 0 | 0 | 0 | 0 |
| 0 | 0.012807615 | 0 | 0 | 0 |
| 0 | 0 | 0 | 0 | 0 |
| 0 | 0 | 0 | 0 | 0 |
| 0 | 0 | 0 | 0 | 0 |
| 1.501097259 | 0 | 0.254797164 | 0.000892393 | 0 |
| 0 | 0.002477156 | 0.004799445 | 0.002597564 | 0.000929125 |
| 0 | 0 | 0 | 0 | 0 |
| 0 | 0 | 0 | 0 | 0 |
| 0 | 0 | 0 | 0.004732605 | 0 |
| 0 | 0 | 0 | 0 | 0.029456812 |
| 0 | 0 | 0 | 0 | 0 |
| 0 | 0 | 0 | 0 | 0 |
| 0 | 0 | 0 | 0.157206263 | 0 |
| 0 | 0 | 0 | 0 | 0 |
| 0 | 0 | 0.682142273 | 0 | 0 |
| 3.442308213 | 0 | 0.05042136 | 0.09471786 | 1.693907392 |
| 0 | 0 | 0 | 0 | 0 |
| 0 | 0 | 0 | 0 | 0 |
| 0 | 0 | 0 | 0 | 0 |
| 0 | 0 | 0 | 0 | 0 |
| 0 | 0 | 0 | 0 | 0 |
| 0 | 0 | 0 | 0 | 0 |
| 0 | 0 | 0 | 0 | 0 |
| 0 | 0 | 0 | 0 | 0 |
| 0 | 0 | 0 | 0 | 0 |
| 0 | 0 | 0 | 0 | 0 |
| 0 | 0 | 0 | 0 | 0 |
| 0 | 0 | 0 | 0.000937003 | 0.008993389 |
| 0 | 0 | 0 | 0 | 0 |
| 0 | 0 | 0 | 0 | 0 |
| 0 | 0 | 0 | 0 | 0 |
| 0.067699506 | 0.001699582 | 0 | 0 | 0 |
| 0 | 0 | 0 | 0.076262035 | 0 |

| | | | | |
|---|---|---|---|---|
| 0 | 0 | 0 | 0 | 0 |
| 0 | 0 | 0 | 0 | 0 |
| 0 | 0 | 0 | 0 | 0 |
| 0 | 0 | 0 | 0 | 0 |
| 0.037818529 | 0 | 0.326355857 | 0 | 0.000510258 |
| 0 | 0 | 0 | 0 | 0 |
| 0 | 0 | 0.004279589 | 0 | 0 |
| 0 | 0 | 0 | 0 | 0 |
| 0 | 0 | 0 | 0 | 0 |
| 0 | 0 | 0 | 0 | 0 |
| 0 | 0 | 0 | 0 | 0 |
| 0.005060773 | 0 | 0.018851345 | 0 | 0.018891177 |
| 0 | 0 | 0 | 0 | 0.212285697 |
| 0 | 0 | 0 | 0 | 0 |
| 0 | 0 | 0.016394188 | 0 | 0 |
| 0 | 0 | 0 | 0.001466807 | 0 |
| 0.035193199 | 0 | 0 | 0 | 0 |
| 0.003312033 | 0 | 0 | 0 | 0 |
| 0.169727733 | 0.001334813 | 0.001679335 | 0 | 0.014483314 |
| 1.679920199 | 0.029130644 | 0.004823939 | 0.033008238 | 2.232407667 |
| 0 | 0 | 0 | 0 | 0 |
| 0 | 0 | 0.114966428 | 0 | 0 |
| 0 | 0 | 0 | 0 | 0 |
| 0 | 0 | 0 | 0 | 0 |
| 0 | 0 | 0 | 0 | 0 |
| 0 | 0 | 0 | 0 | 0 |
| 0.004176585 | 0.000598557 | 0.002459955 | 0 | 0.002806312 |
| 0 | 0 | 0 | 0 | 0 |
| 0 | 0 | 0 | 0 | 0 |
| 0 | 0 | 0 | 0 | 0 |
| 0.00549188 | 0 | 0 | 0 | 0.00063622 |
| 0 | 0 | 0 | 0 | 0 |
| 0 | 0 | 0 | 0 | 0 |
| 0 | 0 | 0 | 0 | 0 |
| 0 | 0 | 0 | 0 | 0 |
| 0 | 0 | 0 | 0 | 0 |
| 0 | 0 | 0 | 0 | 0 |
| 0.010910058 | 0 | 0.007836443 | 0.005593695 | 0.082097171 |
| 0.164992241 | 0 | 0.742268794 | 0.282865178 | 0.043190206 |
| 0.026592426 | 0 | 0 | 0 | 0 |
| 0.004516093 | 0 | 0 | 0 | 0.006098615 |
| 0 | 0 | 0 | 0 | 0 |
| 0 | 0 | 0 | 0 | 0 |
| 0.001972955 | 0.085767245 | 0 | 0 | 0 |
| 0 | 0 | 0 | 0 | 0 |
| 0 | 0 | 0 | 0 | 0 |
| 0 | 0 | 0 | 0 | 0 |
| 0 | 0 | 0 | 0 | 0 |

| | | | | |
|---|---|---|---|---|
| 0 | 0 | 0 | 0 | 0 |
| 0 | 0 | 0.00124151 | 0 | 0.068360659 |
| 0 | 0 | 0 | 0 | 0 |
| 0.011785696 | 0 | 0.006306612 | 0 | 0.001620846 |
| 0 | 0 | 0.001008497 | 0 | 0 |
| 0 | 0 | 0 | 0 | 0 |
| 0.030498995 | 0 | 0.050649007 | 0 | 0.005392828 |
| 0.003072478 | 0 | 0 | 0 | 0.011952053 |
| 0 | 0 | 0 | 0 | 0 |
| 0 | 0 | 0 | 0 | 0 |
| 0.068512638 | 0 | 0.036838161 | 0.001375487 | 0.036353254 |
| 0 | 0 | 0 | 0 | 0 |
| 0.172657591 | 0 | 0.100817493 | 0.241500316 | 0.62798422 |
| 0 | 0 | 0 | 0 | 0 |
| 0 | 0 | 0 | 0 | 0 |
| 0 | 0 | 0 | 0 | 0 |
| 0 | 0 | 0 | 0 | 0 |
| 0 | 0 | 0 | 0 | 0 |
| 0 | 0 | 0 | 0 | 0 |
| 0 | 0 | 0 | 0 | 0 |
| 0 | 0 | 0 | 0 | 0 |
| 0 | 0 | 0 | 0 | 0 |
| 0.04805621 | 0.001843924 | 0.066782677 | 0 | 0 |
| 0.00706003 | 0 | 0.034732064 | 0.005296706 | 0.082632943 |
| 0 | 0 | 0 | 0 | 0 |
| 0 | 0 | 0 | 0 | 0 |
| 0 | 0.005725557 | 0 | 0 | 0 |
| 0 | 0 | 0.001330494 | 0 | 0 |
| 0 | 0 | 0 | 0 | 0 |
| 0 | 0 | 0 | 0 | 0 |
| 0 | 0 | 0.012674449 | 0.000577474 | 0.006024579 |
| 0 | 0.002186101 | 0 | 0 | 0 |
| 0 | 0 | 0 | 0 | 0 |
| 0 | 0 | 0 | 0 | 0 |
| 0 | 0 | 0 | 0 | 0 |
| 0 | 0 | 0 | 0 | 0 |
| 0 | 0 | 0 | 0 | 0.000768081 |
| 0 | 0 | 0 | 0 | 0 |
| 0 | 0 | 0.004978751 | 0.214976452 | 0.012437196 |
| 0 | 0 | 0 | 0 | 0 |
| 0 | 0 | 0 | 0 | 0 |
| 0 | 0 | 0 | 0 | 0 |
| 0 | 0 | 0 | 0 | 0 |
| 0 | 0 | 0 | 0 | 0 |
| 0 | 0 | 0 | 0 | 0.136451289 |
| 0.293785081 | 0 | 0.072951808 | 0.201036895 | 0.226029086 |
| 0 | 0 | 0 | 0 | 0 |
| 0 | 0 | 0 | 0 | 0 |

| | | | | |
|---|---|---|---|---|
| 0 | 0 | 0 | 0 | 0 |
| 0 | 0 | 0 | 0 | 0 |
| 0.210806707 | 0 | 0.055127235 | 0.029248159 | 0 |
| 0 | 0 | 0.023401956 | 0 | 0 |
| 0 | 0 | 0 | 0 | 0 |
| 0 | 0 | 0 | 0 | 0 |
| 0.009623493 | 0 | 0 | 0 | 0.435223218 |
| 0 | 0 | 0 | 0 | 0 |
| 0 | 0 | 0 | 0 | 0 |
| 0 | 0 | 0 | 0.01208133 | 0 |
| 0.002954089 | 0 | 0.00170885 | 0 | 0.000992449 |
| 0.000882093 | 0 | 0 | 0 | 0 |
| 0 | 0 | 0 | 0 | 0 |
| 0 | 0 | 0 | 0 | 0 |
| 0 | 0 | 0 | 0 | 0 |
| 0 | 0 | 0 | 0 | 0 |
| 0 | 0 | 0.003653725 | 0.220629796 | 0 |
| 0 | 0 | 0 | 0 | 0 |
| 0 | 0 | 0 | 0 | 0 |
| 0.00803114 | 0 | 0 | 0 | 0.003948473 |
| 0 | 0 | 0 | 0 | 0 |
| 0 | 0 | 0 | 0 | 0 |
| 0 | 0 | 0 | 0 | 0 |
| 0 | 0 | 0 | 0 | 0 |
| 0 | 0 | 0 | 0 | 0 |
| 0 | 0 | 0 | 0 | 0 |
| 0 | 0 | 0 | 0 | 0 |
| 0 | 0 | 0.0072959 | 0.002908641 | 0 |
| 0.007272109 | 0 | 0 | 0 | 0 |
| 0 | 0 | 0 | 0 | 0 |
| 0 | 0.004421446 | 0.000884966 | 0.026945085 | 0.030340874 |
| 0 | 0 | 0 | 0 | 0 |
| 0.08616834 | 0.075372775 | 0.020345206 | 0.004719115 | 0.015535701 |
| 0.181899842 | 0.002146146 | 0 | 0 | 0.002236028 |
| 0 | 0 | 0 | 0.003988831 | 0 |
| 0 | 0 | 0 | 0 | 0 |
| 0 | 0 | 0 | 0 | 0 |
| 0 | 0 | 0 | 0 | 0 |
| 0 | 0 | 0 | 0 | 0 |
| 0 | 0 | 0 | 0 | 0 |
| 0 | 0 | 0 | 0 | 0 |
| 0.085035745 | 0.013630351 | 0 | 0 | 0.013861172 |
| 0.085580555 | 0.017307256 | 0.081359546 | 0.026977523 | 0.304181992 |
| 0 | 0 | 0 | 0 | 0 |
| 0 | 0 | 0 | 0 | 0 |
| 0.115179931 | 0 | 0.052088063 | 0.349593841 | 0 |
| 0.005937316 | 0 | 0 | 0.000829847 | 0.140874772 |
| 0 | 0 | 0 | 0 | 0 |

| | | | | |
|---|---|---|---|---|
| 0 | 0 | 0 | 0 | 0 |
| 0.028928225 | 0 | 0.012083937 | 0.012663083 | 0 |
| 0.009346508 | 0.001144847 | 0.004032947 | 0 | 0.009930014 |
| 0 | 0 | 0 | 0 | 0 |
| 0 | 0 | 0.030792341 | 0.001027005 | 1.231510428 |
| 0.002048872 | 0.0235793 | 0 | 0 | 0 |
| 0 | 0 | 0 | 0 | 0 |
| 0 | 0 | 0 | 0 | 0 |
| 0 | 0 | 0 | 0 | 0 |
| 0 | 0 | 0 | 0 | 0 |
| 0 | 0 | 0 | 0 | 0 |
| 0 | 0 | 0 | 0 | 0 |
| 0 | 0 | 0 | 0 | 0 |
| 0 | 0 | 0 | 0 | 0 |
| 0 | 0 | 0 | 0 | 0 |
| 0 | 0 | 0 | 0 | 0 |
| 0.02260524 | 0 | 0 | 0 | 0 |
| 0 | 0 | 0 | 0 | 0 |
| 0 | 0 | 0 | 0 | 0 |
| 0 | 0 | 0 | 0 | 0 |
| 0 | 0 | 0 | 0 | 0 |
| 0 | 0 | 0 | 0 | 0 |
| 0 | 0 | 0 | 0.000727199 | 0 |
| 0 | 0 | 0 | 0 | 0 |
| 0.012245535 | 0 | 0.007893824 | 5.201826153 | 0 |
| 0 | 0 | 0.001044868 | 0 | 0 |
| 0 | 0 | 0 | 0 | 0 |
| 0 | 0 | 0.114126727 | 0.033101687 | 0.433399389 |
| 0 | 0 | 0 | 0 | 0 |
| 0 | 0 | 0 | 0 | 0 |
| 0 | 0 | 0 | 0.021012023 | 0 |
| 0 | 0 | 0 | 0 | 0 |
| 0 | 0 | 0 | 0 | 0 |
| 0 | 0 | 0 | 0 | 0 |
| 0 | 0 | 0 | 0 | 0 |
| 0 | 0 | 0 | 0 | 0 |
| 0 | 0 | 0 | 0 | 0 |
| 0.024524499 | 0.315328019 | 0 | 0 | 0 |
| 0 | 0 | 0.555534389 | 0.480222756 | 2.944987406 |
| 0 | 0 | 0 | 0 | 0 |
| 0 | 0 | 0 | 0 | 0 |
| 0.054964472 | 0 | 0.00430738 | 0.0057843 | 0 |
| 0.002991092 | 0.005715478 | 0 | 0.003210692 | 0.00133984 |
| 0 | 0 | 0 | 0 | 0.086270932 |
| 0 | 0 | 0 | 0 | 0 |
| 0 | 0 | 0 | 0 | 0.010790072 |
| 0.017387983 | 0 | 0.017967189 | 0.001637244 | 0.001366462 |
| 0 | 0 | 0 | 0 | 0 |

| | | | | |
|---|---|---|---|---|
| 0 | 0 | 0 | 0 | 0 |
| 0.076214145 | 0.012411864 | 0.021254332 | 0.00103295 | 0.002801864 |
| 0 | 0 | 0 | 0 | 0 |
| 0 | 0 | 0 | 0 | 0 |
| 0 | 0 | 0 | 0 | 0 |
| 0 | 0 | 0 | 0 | 0 |
| 0 | 0 | 0 | 0 | 0 |
| 0 | 0 | 0 | 0 | 0 |
| 0 | 0 | 0 | 0 | 0 |
| 0 | 0 | 0 | 0 | 0 |
| 0 | 0 | 0 | 0 | 0 |
| 0 | 0 | 0 | 0 | 0 |
| 0.020751618 | 0 | 0 | 0 | 0.04898393 |
| 0 | 0 | 0 | 0 | 0 |
| 0.074630224 | 0 | 0.038363277 | 0 | 0 |
| 0 | 0 | 0 | 0 | 0 |
| 0 | 0 | 0 | 0 | 0 |
| 0 | 0 | 0 | 0 | 0 |
| 0.001061869 | 0 | 0 | 0 | 0.011594156 |
| 0 | 0 | 0 | 0 | 0 |
| 0 | 0 | 0 | 0 | 0 |
| 0.551698958 | 0.006754836 | 0.000991468 | 0 | 0.738962089 |
| 0.013581872 | 0 | 0.053330316 | 0 | 0 |
| 0 | 0 | 0 | 0 | 0.197077643 |
| 0 | 0 | 0 | 0 | 0 |
| 0 | 0 | 0 | 0 | 0 |
| 0 | 0 | 0 | 0 | 0 |
| 0 | 0 | 0 | 0 | 0 |
| 0 | 0 | 0 | 0 | 0 |
| 0 | 0 | 0 | 0 | 0 |
| 0 | 0 | 0 | 0 | 0 |
| 0 | 0 | 0 | 0 | 0 |
| 0 | 0 | 0 | 0 | 0 |
| 0 | 0 | 0 | 0 | 0 |
| 11.36627206 | 0 | 0.759091413 | 0.153504828 | 0.108308313 |
| 0 | 0 | 0 | 0 | 0 |
| 0 | 0 | 0 | 0 | 0 |
| 0 | 0 | 0 | 0 | 0 |
| 0.13038007 | 0.040192065 | 0.001980201 | 0 | 0.091188678 |
| 0 | 0 | 0 | 0 | 0 |
| 0.013505037 | 0 | 0.069910775 | 0.043414153 | 0.022685608 |
| 0 | 0 | 0 | 0 | 0 |
| 0 | 0 | 0 | 0 | 0 |
| 0 | 0 | 0 | 0 | 0 |
| 0 | 0 | 0 | 0 | 0 |
| 0.12853077 | 0.814245427 | 0.10240432 | 0 | 0.019750665 |
| 0 | 0 | 0 | 0 | 0 |
| 0 | 0 | 0 | 0 | 0 |

| | | | | |
|---|---|---|---|---|
| 0 | 0 | 0 | 0 | 0 |
| 0 | 0 | 0 | 0 | 0 |
| 0 | 0 | 0 | 0 | 0 |
| 0 | 0 | 0 | 0 | 0 |
| 0.001099408 | 0 | 0 | 0 | 0.073050149 |
| 0 | 0 | 0 | 0 | 0 |
| 0 | 0 | 0 | 0 | 0 |
| 0.001827942 | 0.001247462 | 0.001098606 | 0 | 0.003216773 |
| 0 | 0 | 0.012910401 | 0 | 0 |
| 0 | 0.004515001 | 0 | 1.227367302 | 0.01825776 |
| 0 | 0 | 0.062116436 | 0.005126913 | 0 |
| 0 | 0 | 0 | 0 | 0 |
| 0.022863862 | 0 | 0 | 0 | 0 |
| 0 | 0 | 0 | 0 | 0 |
| 0 | 0 | 0 | 0 | 0 |
| 0 | 0 | 0 | 0 | 0 |
| 0 | 0 | 0 | 0 | 0 |
| 0 | 0 | 0 | 0 | 0 |
| 0 | 0 | 0 | 0 | 0 |
| 0.181304012 | 0.145578207 | 1.56248063 | 0.077412059 | 0.24147807 |
| 0 | 0 | 0 | 0 | 0 |
| 0 | 0 | 0 | 0 | 0 |
| 0 | 0 | 0 | 0 | 0 |
| 0 | 0 | 0 | 0 | 0 |
| 0 | 0 | 0 | 0 | 0 |
| 0 | 0 | 0.001296439 | 0 | 0 |
| 0 | 0 | 0 | 0 | 0 |
| 0 | 0 | 0 | 0 | 0 |
| 0 | 0 | 0 | 0 | 0 |
| 0.098666437 | 0.001574625 | 0 | 0 | 0 |
| 0 | 0 | 0 | 0 | 0 |
| 0 | 0 | 0 | 0 | 0 |
| 0 | 0 | 0 | 0 | 0 |
| 0 | 0 | 0 | 0 | 0 |
| 0 | 0 | 0 | 0 | 0 |
| 0.002660928 | 0 | 0 | 0 | 0.001489932 |
| 0 | 0 | 0 | 0 | 0 |
| 0 | 0 | 0 | 0 | 0 |
| 0 | 0 | 0 | 0 | 0 |
| 0 | 0 | 0 | 0 | 0 |
| 0 | 0 | 0 | 0 | 0 |
| 0 | 0 | 0 | 0 | 0 |
| 0.022725665 | 0 | 0.000898572 | 0.004126829 | 0 |
| 0 | 0 | 0 | 0 | 0 |
| 0.003282183 | 0 | 0.0083256 | 0 | 0.051458159 |
| 0 | 0 | 0 | 0.039537438 | 0 |
| 0 | 0 | 0 | 0 | 0 |
| 0 | 0 | 0 | 0 | 0 |

| | | | | |
|---|---|---|---|---|
| 0 | 0 | 0 | 0 | 0 |
| 0 | 0 | 0 | 0 | 0 |
| 0 | 0 | 0 | 0.00317728 | 0 |
| 0 | 0 | 0 | 0 | 0 |
| 0 | 0 | 0 | 0 | 0 |
| 0.01816088 | 0 | 0.050784233 | 0.007993661 | 0.007424848 |
| 0 | 0 | 0 | 0 | 0 |
| 0 | 0 | 0 | 0 | 0.004614369 |
| 0 | 0 | 0 | 0 | 0 |
| 0 | 0 | 0 | 0 | 0 |
| 0 | 0 | 0 | 0 | 0 |
| 0 | 0 | 0 | 0 | 0 |
| 0 | 0 | 0 | 0 | 0 |
| 0 | 0 | 0 | 0 | 0 |
| 0 | 0 | 0 | 0 | 0 |
| 0 | 0 | 0 | 0 | 0 |
| 0 | 0 | 0 | 0 | 0 |
| 0 | 0 | 0 | 0 | 0 |
| 0 | 0 | 0 | 0 | 0 |
| 0 | 0 | 0 | 0 | 0 |
| 0 | 0 | 0 | 0 | 0 |
| 0 | 0 | 0 | 0 | 0 |
| 0 | 0 | 0 | 0 | 0 |
| 0 | 0 | 0 | 0 | 0 |
| 0 | 0 | 0 | 0 | 0 |
| 0.000676349 | 0 | 0 | 0.010083409 | 0 |
| 0 | 0 | 0 | 0 | 0 |
| 0 | 0 | 0 | 0 | 0 |
| 0 | 0 | 0 | 0 | 0 |
| 0.142269878 | 0.001867129 | 0 | 0 | 0 |
| 0 | 0 | 0 | 0 | 0 |
| 0 | 0 | 0 | 0 | 0 |
| 0 | 0 | 0 | 0 | 0 |
| 0 | 0 | 0 | 0 | 0 |
| 0 | 0 | 0 | 0 | 0 |
| 0 | 0 | 0 | 0 | 0 |
| 0 | 0 | 0 | 0 | 0 |
| 0 | 0 | 0 | 0 | 0 |
| 0 | 0 | 0.274976568 | 0.147780973 | 1.015046382 |
| 0 | 0 | 0 | 0 | 0 |
| 0 | 0 | 0 | 0 | 0 |
| 0 | 0 | 0 | 0 | 0 |
| 0 | 0 | 0 | 0 | 0 |
| 0 | 0 | 0 | 0 | 0 |
| 0 | 0 | 0 | 0 | 0 |
| 0 | 0 | 0 | 0 | 0 |
| 0 | 0 | 0 | 0 | 0 |
| 0 | 0 | 0 | 0 | 0 |

| | | | | |
|---|---|---|---|---|
| 0 | 0 | 0 | 0 | 0 |
| 0 | 0 | 0.004243063 | 0 | 0.041382866 |
| 0 | 0 | 0 | 0 | 0 |
| 0 | 0 | 0 | 0 | 0 |
| 0 | 0 | 0 | 0 | 0 |
| 0 | 0 | 0 | 0.004156812 | 0 |
| 0 | 0 | 0 | 0 | 0 |
| 0.425937829 | 0 | 0 | 0 | 0 |
| 0 | 0 | 0 | 0 | 0 |
| 0 | 0 | 0 | 0 | 0 |
| 0.001056503 | 0 | 0.000888953 | 0.011151688 | 0.001183135 |
| 0 | 0 | 0 | 0 | 0 |
| 0 | 0 | 0 | 0.003898066 | 0.092895371 |
| 0 | 0 | 0 | 0 | 0 |
| 0 | 0 | 0 | 0 | 0 |
| 0 | 0 | 0 | 0 | 0 |
| 0 | 0 | 0 | 0 | 0 |
| 0 | 0 | 0 | 0 | 0 |
| 0 | 0 | 0 | 0 | 0 |
| 0 | 0 | 0.001664347 | 0.002673746 | 0 |
| 0.027450558 | 0.006659129 | 0.005413401 | 0.003261203 | 0 |
| 0 | 0 | 0 | 0 | 0 |
| 0 | 0 | 0 | 0 | 0 |
| 0.977643673 | 0 | 0.434245135 | 0.087245114 | 0.702493283 |
| 0 | 0 | 0 | 0 | 0 |
| 0 | 0 | 0 | 0 | 0 |
| 0 | 0 | 0 | 0 | 0 |
| 0 | 0 | 0 | 0 | 0 |
| 0 | 0 | 0 | 0 | 0 |
| 0 | 0 | 0 | 0 | 0 |
| 0.023153889 | 0 | 0 | 0 | 0 |
| 0 | 0 | 0 | 0 | 0 |
| 0 | 0 | 0.001898127 | 0 | 0 |
| 0 | 0 | 0 | 0 | 0 |

| PHL018-0034_V3 | PHL018-0054_V2 | PHL018-0236_V1 | PHL018-0009_V3 | PHL018-0111_V2 |
|---|---|---|---|---|
| CTRL | CTRL | EYCF | EYCF | EYCF |
| 0.029490996 | 0.006836642 | 0.00716298 | 0.151417376 | 2.119648988 |
| 0.09565281 | 5.252191898 | 2.094012294 | 3.41340299 | 0.859236901 |
| 0.104882531 | 6.271163174 | 20.05370321 | 4.06753337 | 1.932943991 |
| 0 | 0 | 0 | 0 | 0 |
| 0 | 0 | 0 | 0 | 0 |
| 0 | 0 | 0 | 0 | 0 |
| 0 | 0 | 0 | 0 | 0 |
| 0 | 0 | 0 | 0 | 0 |
| 0 | 0 | 0 | 0 | 0 |
| 0 | 0 | 0 | 0 | 0 |
| 0 | 0 | 0 | 0 | 0 |
| 0 | 0 | 0 | 0 | 0 |
| 0 | 0 | 0 | 0 | 0 |
| 0.280320465 | 32.28802011 | 14.24105721 | 22.30738169 | 3.346899062 |
| 0 | 0 | 0 | 0 | 0 |
| 0 | 0 | 0 | 0 | 0 |
| 0 | 0 | 0 | 0 | 0 |
| 0 | 0 | 0 | 0 | 0 |
| 0 | 0 | 0 | 0 | 0 |
| 0 | 0 | 0 | 0 | 0 |
| 0 | 0 | 0 | 0 | 0 |
| 0 | 0 | 0 | 0 | 0 |
| 0 | 0 | 0 | 0 | 0 |
| 0 | 0 | 0 | 0 | 0 |
| 0.128850714 | 0.135663066 | 0 | 0 | 0 |
| 0 | 0 | 0 | 0 | 0 |
| 0 | 0 | 0 | 0 | 0 |
| 0 | 0 | 0 | 0 | 0 |
| 0 | 0 | 0 | 0 | 0 |
| 0.002320963 | 0 | 0 | 0 | 0 |
| 0 | 0 | 0 | 0 | 0 |
| 0 | 0 | 0 | 0 | 0 |
| 0 | 0 | 0 | 0 | 0 |
| 0 | 0 | 0 | 0 | 0 |
| 0 | 0 | 0 | 0 | 0 |
| 0.061498444 | 1.829152144 | 0 | 0 | 0 |
| 0 | 0 | 0 | 0 | 0 |
| 0 | 0 | 0 | 0 | 0 |
| 0 | 0 | 0 | 0 | 0 |
| 0 | 0 | 0 | 0 | 0 |
| 0 | 0 | 0 | 0 | 0 |
| 0.001927851 | 1.108720471 | 0 | 2.3208484 | 0.207619731 |
| 0.291652695 | 0.001566058 | 0.010020672 | 0.044851192 | 3.902584647 |
| 0.023906552 | 0.020756812 | 0 | 0 | 0.001825985 |
| 0 | 0.001301298 | 0 | 0 | 0.001462745 |
| 0 | 0 | 0 | 0 | 0 |

| | | | | |
|---|---|---|---|---|
| 0.082336561 | 0 | 0 | 0 | 0 |
| 0 | 0 | 0 | 0 | 0.008086071 |
| 0 | 0.002158518 | 0 | 0 | 0 |
| 0.290114813 | 0.555328718 | 0.383148204 | 0.193798629 | 0.242409636 |
| 0.025572494 | 0.009357584 | 0 | 0.007762672 | 0 |
| 0 | 0 | 0 | 0 | 0 |
| 0 | 0.004974431 | 1.610540417 | 0.148520231 | 0 |
| 0 | 0 | 0 | 0 | 0 |
| 0 | 0 | 0 | 0 | 0 |
| 0 | 0 | 0 | 0 | 0 |
| 0 | 0 | 0 | 0 | 0 |
| 0.665188888 | 0.234111956 | 0 | 0.105945597 | 0.261825512 |
| 0 | 0 | 0 | 0 | 0 |
| 0.036266892 | 0.012468858 | 0.003449307 | 0.081878492 | 0.041847237 |
| 0 | 0.058348335 | 0 | 0 | 0 |
| 0 | 0 | 0 | 0 | 0 |
| 0.051321876 | 0 | 0.144737353 | 0 | 0 |
| 0 | 0 | 0 | 0 | 0 |
| 0 | 0 | 0 | 0 | 0 |
| 0 | 0 | 0 | 0 | 0 |
| 0.001442261 | 0.035527449 | 0 | 0.037195665 | 0.156144002 |
| 0.003482081 | 0.049482954 | 0.001934267 | 0.004744987 | 0.030393512 |
| 0 | 0.000991088 | 0 | 0 | 0 |
| 0 | 0 | 0 | 0 | 0 |
| 0 | 0 | 0 | 0 | 0 |
| 0 | 0 | 0 | 0 | 0 |
| 0 | 0 | 0 | 0 | 0 |
| 0 | 0 | 0 | 0 | 0 |
| 0 | 0.013847533 | 0 | 0.034235988 | 0.001778919 |
| 0 | 0 | 0 | 0 | 0 |
| 0 | 0 | 0 | 0 | 0.271461956 |
| 0 | 0 | 0 | 0 | 0 |
| 0 | 0 | 0 | 0 | 0 |
| 0 | 0 | 0 | 0 | 0 |
| 0 | 0 | 0 | 0 | 0 |
| 0 | 0 | 0 | 0 | 0 |
| 0 | 0 | 0 | 0 | 0 |
| 0.602394191 | 0 | 0.006314813 | 0 | 0.004576806 |
| 0 | 0 | 0 | 0 | 0 |
| 0 | 0 | 0 | 0 | 0 |
| 0 | 0 | 0 | 0 | 0 |
| 0 | 0 | 0 | 0 | 0 |
| 0 | 0 | 0 | 0 | 0 |
| 0 | 0 | 0 | 0 | 0 |
| 0 | 0 | 0 | 0 | 0 |
| 0 | 0 | 0 | 0 | 0 |
| 0 | 0 | 0.000551864 | 0 | 0 |
| 0 | 0 | 0.012326716 | 0.003068142 | 0 |

| | | | | |
|---|---|---|---|---|
| 0 | 0 | 0 | 0 | 0 |
| 0 | 0 | 0 | 0 | 0 |
| 0 | 0 | 0 | 0 | 0 |
| 0 | 0 | 0 | 0 | 0 |
| 0 | 0.026653561 | 0 | 0 | 0 |
| 0 | 0.001900656 | 0 | 0 | 0 |
| 0 | 0 | 0 | 0 | 0 |
| 0 | 0 | 0 | 0 | 0 |
| 0 | 0 | 0 | 0.082371777 | 0 |
| 0 | 0 | 0 | 0 | 0 |
| 0 | 0 | 0 | 0 | 0 |
| 0 | 0.015121655 | 0 | 0.000851956 | 0.002006291 |
| 0 | 0 | 0 | 0 | 0 |
| 0 | 0 | 0 | 0 | 0 |
| 0 | 0 | 0 | 0 | 0 |
| 0 | 0 | 0 | 0 | 0 |
| 0 | 0 | 0 | 0 | 0 |
| 0 | 0 | 0 | 0 | 0 |
| 0.045980363 | 0.007280323 | 0 | 0.227649718 | 0.397770826 |
| 0 | 0 | 0 | 0 | 0 |
| 0 | 0 | 0 | 0 | 0 |
| 0 | 0 | 0 | 0 | 0 |
| 0 | 0 | 0 | 0 | 0 |
| 0 | 0 | 0 | 0 | 0 |
| 0 | 0 | 0 | 0 | 0 |
| 0 | 0 | 0 | 0 | 0 |
| 0 | 0 | 0 | 0 | 0 |
| 0 | 0 | 0 | 0 | 0 |
| 0 | 0 | 0 | 0 | 0 |
| 0 | 0 | 0 | 0 | 0 |
| 0.010381748 | 0 | 0 | 0.010525424 | 0.224589483 |
| 0.031403415 | 0.015753811 | 0 | 0 | 0 |
| 0 | 0 | 0 | 0 | 0 |
| 0 | 0 | 0 | 0 | 0 |
| 0 | 0 | 0 | 0 | 0 |
| 0 | 0 | 0 | 0 | 0 |
| 0 | 0 | 0 | 0 | 0 |
| 0 | 0 | 0 | 0 | 0 |
| 0.030870776 | 0 | 0 | 0 | 0 |
| 0 | 0 | 0 | 0 | 0 |
| 0 | 0 | 0 | 0 | 0 |
| 0 | 0 | 0 | 0 | 0 |
| 0 | 0.000958364 | 0 | 0 | 0 |
| 0 | 0 | 0 | 0 | 0 |
| 0 | 0 | 0 | 0 | 0 |
| 0 | 0 | 0 | 0 | 0 |
| 0 | 0 | 0 | 0 | 0 |
| 0 | 0 | 0 | 0 | 0 |

| | | | | |
|---|---|---|---|---|
| 0 | 0 | 0 | 0 | 0 |
| 0 | 0 | 0 | 0 | 0 |
| 0 | 0 | 0 | 0 | 0 |
| 0 | 0 | 0 | 0 | 0 |
| 0.1519911 | 0.195755546 | 0 | 0.013951287 | 0 |
| 0 | 0 | 0 | 0 | 0 |
| 0 | 0 | 0 | 0 | 0 |
| 0 | 0 | 0 | 0 | 0 |
| 0 | 0 | 0 | 0 | 0 |
| 0 | 0 | 0 | 0 | 0 |
| 0 | 0 | 0 | 0 | 0 |
| 0 | 0 | 0 | 0 | 0 |
| 0 | 0 | 0 | 0 | 0 |
| 0 | 0 | 0 | 0 | 0 |
| 0 | 0 | 0 | 0 | 0 |
| 0 | 0 | 0 | 0 | 0 |
| 0 | 0 | 0 | 0 | 0 |
| 0 | 0 | 0 | 0 | 0 |
| 0 | 0 | 0 | 0 | 0 |
| 0 | 0 | 0 | 0 | 0 |
| 0 | 0 | 0 | 0.04109754 | 0 |
| 0 | 0 | 0 | 0 | 0 |
| 0 | 0 | 0 | 0 | 0 |
| 0 | 0 | 0 | 0 | 0 |
| 0 | 0 | 0 | 0 | 0 |
| 0 | 0 | 0 | 0 | 0 |
| 0.080253144 | 0 | 0 | 0 | 0 |
| 0 | 0 | 0 | 0 | 0 |
| 0 | 0 | 0 | 0 | 0 |
| 0 | 0 | 0 | 0 | 0 |
| 0 | 0 | 0 | 0 | 0.06303518 |
| 0 | 0 | 0 | 0 | 0 |
| 0 | 0 | 0 | 0 | 0 |
| 0 | 0 | 0 | 0 | 0 |
| 0 | 0 | 0 | 0 | 0 |
| 0 | 0 | 0 | 0 | 0 |
| 0 | 0 | 0 | 0 | 0 |
| 0 | 0 | 0 | 0 | 0 |
| 0 | 0 | 0 | 0 | 0 |
| 0 | 0 | 0 | 0 | 0 |
| 0 | 0 | 0 | 0 | 0 |
| 0 | 0 | 0 | 0 | 0 |
| 0 | 0 | 0 | 0 | 0 |
| 0 | 0 | 0 | 0 | 0 |
| 0 | 0 | 0 | 0 | 0 |
| 0 | 0 | 0 | 0 | 0 |
| 0 | 0 | 0 | 0 | 0 |
| 0 | 0 | 0 | 0 | 0 |

| | | | | |
|---|---|---|---|---|
| 0 | 0 | 0 | 0 | 0 |
| 0 | 0 | 0 | 0 | 0 |
| 0 | 0 | 0 | 0 | 0 |
| 0 | 0 | 0 | 0 | 0 |
| 0 | 0 | 0 | 0 | 0 |
| 0 | 0 | 0 | 0 | 0 |
| 0 | 0 | 0 | 0 | 0 |
| 0 | 0 | 0 | 0 | 0 |
| 0 | 0 | 0 | 0 | 0 |
| 0 | 0 | 0 | 0 | 0 |
| 0 | 0 | 0 | 0 | 0 |
| 0 | 0 | 0 | 0 | 0 |
| 0.00260535 | 0.000915428 | 0 | 0.000983154 | 0.001234802 |
| 0 | 0 | 0 | 0 | 0 |
| 0 | 0 | 0 | 0 | 0 |
| 0 | 0 | 0 | 0 | 0 |
| 0 | 0 | 0 | 0 | 0 |
| 0 | 0 | 0 | 0 | 0 |
| 0 | 0 | 0 | 0 | 0 |
| 0 | 0 | 0 | 0 | 0 |
| 0.005246101 | 0.0975629 | 0.023921773 | 0.01286783 | 0.009619926 |
| 0 | 0 | 0 | 0 | 0 |
| 0 | 0 | 0 | 0 | 0 |
| 0 | 0 | 0 | 0 | 0 |
| 0 | 0 | 0 | 0 | 0 |
| 0 | 0 | 0 | 0 | 0 |
| 0.001309374 | 0.016696615 | 0.006210417 | 0.00214112 | 0 |
| 0 | 0 | 0 | 0 | 0 |
| 0 | 0 | 0.00727411 | 0 | 0 |
| 0 | 0 | 0 | 0 | 0 |
| 0 | 0 | 0.335470723 | 0 | 0.004691114 |
| 0 | 0 | 0 | 0 | 0 |
| 0 | 0 | 0 | 0 | 0 |
| 0.07923625 | 0 | 0 | 0 | 0 |
| 0.017798406 | 0.033688747 | 0.013314451 | 0.004305855 | 0.061841257 |
| 0 | 0 | 0 | 0 | 0 |
| 0 | 0 | 0 | 0 | 0 |
| 0 | 0 | 0 | 0 | 0 |
| 0 | 0 | 0 | 0 | 0 |
| 0 | 0 | 0 | 0 | 0 |
| 0 | 0 | 0 | 0 | 0 |
| 0 | 0 | 0 | 0 | 0 |
| 0 | 0 | 0 | 0 | 0 |
| 0 | 0 | 0 | 0 | 0 |
| 0 | 0 | 0 | 0 | 0 |
| 0.086819339 | 0 | 0 | 0 | 0 |
| 0 | 0 | 0 | 0 | 0 |
| 0.002258728 | 0.001388864 | 0 | 0 | 0.002497881 |

| | | | | |
|---|---|---|---|---|
| 0 | 0 | 0 | 0 | 0 |
| 0 | 0 | 0 | 0 | 0 |
| 0 | 0 | 0 | 0 | 0 |
| 0 | 0 | 0 | 0 | 0 |
| 0 | 0 | 0 | 0 | 0 |
| 0 | 0 | 0 | 0 | 0 |
| 0 | 0 | 0 | 0.007765203 | 0 |
| 0 | 0.002281735 | 0 | 0.000980218 | 0.000769446 |
| 0 | 0 | 0 | 0 | 0 |
| 0 | 0 | 0 | 0 | 0 |
| 0 | 0 | 0 | 0.004429217 | 0 |
| 0 | 0 | 0 | 0 | 0 |
| 0 | 0 | 0 | 0 | 0 |
| 0 | 0 | 0 | 0 | 0 |
| 0 | 0 | 0 | 0 | 0 |
| 0 | 0 | 0 | 0 | 0 |
| 0 | 0 | 0 | 0 | 0 |
| 0 | 0 | 0 | 0 | 0 |
| 0 | 0 | 0 | 0 | 0.017056489 |
| 0 | 0 | 0 | 0 | 0 |
| 0 | 0 | 0.001603826 | 0.069304051 | 0.005353204 |
| 0 | 0 | 0 | 0 | 0 |
| 0 | 0 | 0 | 0 | 0 |
| 0 | 0 | 0 | 0 | 0 |
| 0 | 0.004730714 | 0.002788047 | 0.002032283 | 0 |
| 0 | 0 | 0 | 0 | 0 |
| 0 | 0 | 0 | 0 | 0 |
| 0 | 0 | 0 | 0 | 0 |
| 0 | 0.020033488 | 0 | 0.064486274 | 0 |
| 0 | 0 | 0 | 0 | 0 |
| 0.086054052 | 0.100894423 | 0.070462743 | 0.094960947 | 0.088367881 |
| 0 | 0 | 0 | 0 | 0 |
| 0 | 0 | 0 | 0.013795075 | 0.446855663 |
| 0 | 0 | 0 | 0 | 0 |
| 0 | 0 | 0 | 0 | 0 |
| 0 | 0 | 0 | 0 | 0 |
| 0 | 0 | 0 | 0 | 0 |
| 0 | 0 | 0 | 0 | 0 |
| 0 | 0 | 0 | 0 | 0 |
| 0 | 0 | 0 | 0 | 0 |
| 0 | 0 | 0 | 0 | 0 |
| 0.029304386 | 0 | 0.003305755 | 0.002491801 | 0.112545297 |
| 0 | 0 | 0 | 0 | 0 |
| 0 | 0 | 0 | 0 | 0 |
| 0 | 0 | 0 | 0 | 0 |
| 0 | 0 | 0 | 0 | 0 |
| 0 | 0 | 0 | 0 | 0 |
| 0 | 0 | 0 | 0 | 0 |

| | | | | |
|---|---|---|---|---|
| 0 | 0 | 0 | 0 | 0 |
| 0 | 0 | 0 | 0 | 0 |
| 0.016435761 | 0.01680767 | 0 | 0 | 0 |
| 0 | 0 | 0 | 0 | 0 |
| 0 | 0 | 0 | 0 | 5.257370942 |
| 0 | 0 | 0 | 0 | 0 |
| 0 | 0 | 0 | 0 | 0 |
| 0 | 0 | 0 | 0 | 0 |
| 0 | 0 | 0 | 0 | 0 |
| 0 | 0.00640507 | 0 | 0 | 0 |
| 0 | 0 | 0 | 0 | 0 |
| 0 | 0 | 0 | 0 | 0 |
| 0 | 0 | 0 | 0 | 0 |
| 0 | 0.002476735 | 0.007298321 | 0.000797991 | 0 |
| 0.001342603 | 0.00102211 | 0.001721088 | 0.001317275 | 0.002527623 |
| 0 | 0 | 0 | 0 | 0 |
| 0 | 0.004658411 | 0.00078441 | 0 | 0.000785454 |
| 0.002966683 | 0 | 0 | 0 | 0 |
| 0 | 0 | 0 | 0 | 0 |
| 0 | 0 | 0 | 0 | 0 |
| 0 | 0 | 0 | 0 | 0 |
| 0.003682537 | 0.000788478 | 0 | 0 | 0.000945388 |
| 0 | 0 | 0 | 0 | 0 |
| 0 | 0 | 0 | 0.007001343 | 0.001831959 |
| 0 | 0 | 0 | 0 | 0 |
| 0 | 0 | 0 | 0 | 0 |
| 3.103582911 | 0 | 0 | 0 | 0 |
| 0 | 0 | 0 | 0 | 0 |
| 0 | 0 | 0 | 0 | 0 |
| 0 | 0 | 0 | 0 | 0 |
| 0.004585728 | 0 | 0 | 0 | 0.016300486 |
| 0 | 0 | 0 | 0 | 0 |
| 0 | 0 | 0 | 0 | 0 |
| 0 | 0 | 0.00346985 | 0.000781097 | 0 |
| 0 | 0 | 0 | 0 | 0 |
| 0 | 0 | 0 | 0 | 0 |
| 0 | 0 | 0 | 0 | 0 |
| 0.008385941 | 0 | 0 | 0.00113486 | 0.002078619 |
| 0 | 0 | 0 | 0 | 0 |
| 0 | 0 | 0 | 0 | 0 |
| 0 | 0 | 0 | 0 | 0 |
| 0 | 0 | 0 | 0 | 0 |
| 0 | 0 | 0 | 0 | 0 |
| 0 | 0 | 0 | 0 | 0 |
| 0 | 0 | 0 | 0 | 0 |
| 0 | 0 | 0 | 0 | 0 |
| 0 | 0 | 0 | 0 | 0 |
| 0 | 0 | 0.002693981 | 0 | 0 |

| | | | | |
|---|---|---|---|---|
| 0 | 5.47295938 | 0 | 0 | 0 |
| 0 | 0.019568746 | 0 | 0 | 0 |
| 0 | 0 | 0 | 0 | 0 |
| 0 | 0 | 0 | 0 | 0 |
| 0.001212714 | 0.006370265 | 0.005596497 | 0.019275342 | 0.001494386 |
| 0 | 0 | 0 | 0 | 0.002436732 |
| 0 | 0 | 0 | 0 | 0 |
| 0 | 0.056681822 | 0.005752797 | 0 | 0 |
| 0 | 0 | 0 | 0 | 0 |
| 0.004905905 | 0.056822546 | 0.074658203 | 0.508496474 | 0 |
| 0 | 0 | 0 | 0 | 0 |
| 0 | 0.003687907 | 0 | 0.021388047 | 0 |
| 0.002857994 | 0.002098056 | 0.014916386 | 0.001001456 | 0.002725209 |
| 0 | 0 | 0 | 0 | 0 |
| 0 | 0 | 0 | 0 | 0 |
| 0 | 0 | 0.025456136 | 0 | 0.009440749 |
| 0 | 0 | 0 | 0 | 0 |
| 0 | 0 | 0 | 0 | 0 |
| 0 | 0 | 0.536400712 | 0 | 0.233299949 |
| 0 | 0 | 0.00081316 | 0 | 0 |
| 0 | 0 | 0 | 0 | 0 |
| 0 | 0 | 0 | 0 | 0 |
| 0 | 0 | 0 | 0.005580217 | 0 |
| 0.213062912 | 0 | 0 | 0 | 0 |
| 0 | 0 | 0 | 0 | 0 |
| 0 | 0.001357611 | 0.000731528 | 0 | 0.002441672 |
| 0.002225664 | 0 | 0 | 0 | 0 |
| 0 | 0 | 0 | 0 | 0.462091335 |
| 0 | 0 | 0 | 0 | 0 |
| 0.002446394 | 0 | 0 | 0 | 0 |
| 0 | 0 | 0 | 0 | 0 |
| 0 | 0 | 0 | 0.001944761 | 0 |
| 0 | 0 | 0 | 0 | 0 |
| 0 | 0.002130608 | 0.008849512 | 0 | 0 |
| 0 | 0 | 0 | 0 | 0 |
| 0 | 0 | 0.03542135 | 0 | 0 |
| 0 | 0 | 0 | 0 | 0.234993477 |
| 0 | 0 | 0 | 0 | 0 |
| 0 | 0 | 0 | 0 | 0 |
| 0 | 0 | 0 | 0 | 0 |
| 0 | 0 | 0 | 0 | 0 |
| 0 | 0 | 0 | 0 | 0 |
| 0 | 0 | 0 | 0 | 0 |
| 0 | 0 | 0 | 0 | 0 |
| 0 | 0 | 0 | 0 | 0 |
| 0.001565683 | 0 | 0 | 0 | 0 |
| 0.00887072 | 0 | 0.003638851 | 0.000870337 | 0.000910924 |
| 0 | 0 | 0 | 0 | 0 |

| | | | | |
|---|---|---|---|---|
| 0 | 0 | 0 | 0 | 0 |
| 0 | 0.013584888 | 0.001270835 | 0.034043194 | 0 |
| 0 | 0 | 0 | 0 | 0 |
| 0 | 0 | 0 | 0 | 0 |
| 0 | 0 | 0 | 0 | 0 |
| 0 | 0 | 0.003403418 | 0.001827989 | 0 |
| 0.00543823 | 0 | 0 | 0 | 0 |
| 0 | 0 | 0 | 0 | 0 |
| 0 | 0 | 0 | 0 | 0 |
| 0 | 0 | 0 | 0 | 0 |
| 0.00189081 | 0.001850727 | 0.013296476 | 0.007685579 | 0.024409321 |
| 0 | 0 | 0 | 0 | 0 |
| 0 | 0 | 0 | 0 | 0 |
| 0 | 0 | 0.001766509 | 0.002503775 | 0.00078616 |
| 0 | 0 | 0.083014321 | 0 | 0 |
| 0 | 0 | 0 | 0 | 0 |
| 0 | 0 | 0 | 0 | 0 |
| 0 | 0 | 0 | 0 | 0.005355426 |
| 0 | 0 | 0 | 0 | 0 |
| 0 | 0.01904676 | 0 | 0 | 0 |
| 0 | 0 | 0 | 0.004855888 | 0 |
| 0 | 0 | 0.33930719 | 0.110117714 | 0 |
| 0 | 0 | 0 | 0 | 0 |
| 0 | 0 | 0 | 0 | 0 |
| 0 | 0 | 0 | 0 | 0 |
| 0 | 0 | 0 | 0 | 0 |
| 0 | 0 | 0 | 0 | 0.015059169 |
| 0 | 0 | 0 | 0 | 0 |
| 0 | 0 | 0 | 0 | 0 |
| 0.081170522 | 0.294733125 | 8.790495366 | 0.415078164 | 0.120612237 |
| 0.01956339 | 0.009489605 | 0.001198436 | 0.001019168 | 0 |
| 0 | 0 | 0 | 0.093961081 | 0 |
| 0 | 0 | 0 | 0 | 0 |
| 0.000735623 | 0 | 0 | 0 | 0 |
| 0 | 0 | 0 | 0 | 0 |
| 0 | 0 | 0 | 0 | 0 |
| 0.088869031 | 0 | 0 | 0 | 0 |
| 0 | 0 | 0 | 0.097231738 | 0 |
| 0 | 0.301644665 | 0.722407966 | 0 | 0 |
| 0 | 0 | 0 | 0 | 0 |
| 0.008182021 | 0.100620515 | 0.001129538 | 0.021407108 | 0.010556384 |
| 0.533900989 | 1.893531452 | 2.59335526 | 0 | 2.665501508 |
| 0 | 0 | 0 | 0.012156612 | 0 |
| 0 | 0 | 0 | 0 | 0 |
| 0 | 0 | 0 | 0 | 0.003159305 |
| 0 | 0 | 0 | 0 | 0 |
| 0 | 0.003017384 | 0.008976162 | 0.000864165 | 0 |
| 0 | 0 | 0 | 0 | 0 |

| | | | | |
|---|---|---|---|---|
| 0 | 0 | 0 | 0 | 0.006416751 |
| 0 | 0 | 0 | 0 | 0.005704507 |
| 0.003331714 | 0 | 0.357050268 | 0 | 0 |
| 0 | 0 | 0 | 0 | 0 |
| 0 | 0 | 0 | 0 | 0 |
| 0 | 0 | 0.067138023 | 0 | 0 |
| 0 | 0 | 0 | 0 | 0 |
| 0 | 0 | 0 | 0.007170198 | 0.001563451 |
| 0 | 0 | 0 | 0.037924577 | 0.051834539 |
| 0 | 0 | 0 | 0 | 0 |
| 0 | 0.007334399 | 0 | 0 | 0 |
| 0 | 0 | 0 | 0 | 0 |
| 0 | 0 | 0 | 0 | 0 |
| 0.002703643 | 0.001122685 | 0.000756177 | 0.020015363 | 0.0045431 |
| 0 | 0 | 0 | 0 | 0 |
| 0.20360196 | 0.040797193 | 0 | 0.067089532 | 0 |
| 0.004045056 | 0.029903175 | 0 | 0 | 0.070614937 |
| 0 | 0 | 0 | 0.003299175 | 0 |
| 0 | 0 | 0 | 0.005609724 | 0 |
| 0 | 0 | 6.025534641 | 0 | 0 |
| 0 | 0.006127888 | 0.055513485 | 0 | 0 |
| 0 | 0 | 0 | 0 | 0.006339757 |
| 0 | 0 | 0 | 0 | 0 |
| 0.914902862 | 0.064676852 | 0 | 0 | 0.130077585 |
| 0.003780496 | 0 | 0 | 0 | 0 |
| 0.001682731 | 0 | 0.008844107 | 0.223158269 | 0.010655855 |
| 0 | 0.004012278 | 0 | 0.001591059 | 0 |
| 0 | 0 | 0 | 0 | 0 |
| 0 | 0 | 0 | 0.00182857 | 0 |
| 0 | 0 | 0 | 0 | 0.001662374 |
| 0.003699785 | 0.125104364 | 0 | 0 | 0.023850943 |
| 0.003064403 | 0.00944824 | 0.002121266 | 0 | 0.020139514 |
| 0 | 0 | 0.019075244 | 7.008419484 | 0 |
| 0 | 0.03772147 | 0 | 0.138254361 | 0 |
| 0 | 0.002385629 | 0 | 0 | 0.010726421 |
| 0 | 0 | 0 | 0 | 0 |
| 0 | 0.000903667 | 0 | 0 | 0 |
| 0.017122085 | 0.024028131 | 0.005711994 | 0.012548705 | 0.380247292 |
| 0.076470137 | 0 | 0 | 0.284322696 | 0 |
| 0 | 0.00077041 | 0.005967399 | 0 | 0 |
| 0 | 0 | 0 | 0 | 0 |
| 0 | 0 | 0 | 0 | 0 |
| 0 | 0 | 0 | 0 | 0 |
| 0.019163155 | 0.009930133 | 0 | 0 | 0.479723385 |
| 0 | 0 | 0.077561364 | 0 | 0 |
| 0 | 0 | 0 | 0 | 0.005772989 |
| 0 | 0 | 0 | 0 | 0 |
| 0 | 0 | 0 | 0 | 0 |

| | | | | |
|---|---|---|---|---|
| 0 | 0 | 0 | 0 | 0 |
| 0.093324053 | 0.344426438 | 0.056429043 | 0.007998014 | 1.02414685 |
| 0 | 0 | 0 | 0 | 0 |
| 0 | 0 | 0 | 0 | 0 |
| 0 | 0 | 0 | 0 | 0 |
| 0 | 0 | 0 | 0 | 0 |
| 0 | 0 | 0.008600694 | 0 | 0 |
| 0 | 0 | 0 | 0.001906547 | 0 |
| 0 | 0.001763061 | 0 | 0.007321524 | 0 |
| 0 | 0 | 0 | 0.001202483 | 0 |
| 0.002506626 | 0 | 0.000481988 | 0.001434614 | 0 |
| 0.004355972 | 0.007461352 | 0.00311105 | 0 | 0.017253368 |
| 0 | 0 | 0 | 0 | 0.521432667 |
| 0 | 0 | 0 | 0 | 0 |
| 0 | 0.002189499 | 0 | 0 | 0 |
| 0 | 0 | 0 | 0 | 0 |
| 0.00809313 | 0.017734313 | 0.005383583 | 0.001716854 | 0.002246146 |
| 0 | 0.002269734 | 0.103300755 | 0 | 0.36156009 |
| 0 | 0 | 0 | 0 | 0 |
| 0 | 0 | 0 | 0 | 0 |
| 0.231659513 | 0.019061307 | 0.002949411 | 0.002434451 | 0.028722649 |
| 0 | 0 | 0.0047495 | 0 | 0 |
| 0 | 0 | 0 | 0 | 0 |
| 0 | 0 | 0 | 0 | 0 |
| 0 | 0 | 0.006704582 | 0 | 0 |
| 0 | 0 | 0 | 0 | 0 |
| 0.025381874 | 0.077037595 | 0.001027488 | 0 | 0 |
| 0.012946322 | 0.102939425 | 0.001290795 | 0.085386251 | 0.052192923 |
| 0 | 0 | 0 | 0 | 0 |
| 0 | 0 | 0 | 0 | 0 |
| 0 | 0.002937609 | 0.000593582 | 0 | 0 |
| 0 | 0 | 0 | 0 | 0 |
| 0 | 0 | 0 | 0 | 0 |
| 0 | 0 | 0 | 0 | 0 |
| 0 | 0 | 0 | 0 | 0 |
| 0.066949904 | 0.307663051 | 0.111102205 | 0.226122107 | 0.205416842 |
| 0 | 0 | 0 | 0 | 0 |
| 0 | 0 | 0 | 0 | 0 |
| 0.001478772 | 0.005403715 | 0.002957203 | 0.010736472 | 0.003340773 |
| 0 | 0 | 0 | 0 | 0 |
| 0 | 0 | 0.065661832 | 0 | 0 |
| 0 | 0 | 0 | 0 | 0 |
| 0.04706718 | 0.004513161 | 0.011330196 | 0.00370139 | 0.003505045 |
| 0 | 0.003168998 | 0 | 0 | 0.001424865 |
| 0 | 0 | 0 | 0 | 0 |
| 0 | 0 | 0 | 0.569491573 | 0 |
| 0 | 0.197945823 | 0 | 0 | 0 |
| 0 | 0 | 0.002156365 | 0 | 0 |

| | | | | |
|---|---|---|---|---|
| 0 | 0 | 0 | 0 | 0.021132797 |
| 0 | 0 | 0 | 0 | 0 |
| 0 | 0 | 0 | 0 | 0.002874033 |
| 0.926053848 | 0.211928665 | 0 | 0.00061585 | 0 |
| 0 | 0 | 0 | 0 | 0 |
| 0 | 0 | 0 | 0 | 0 |
| 0 | 0 | 0 | 0 | 0 |
| 0.040056317 | 0 | 0 | 0 | 0 |
| 17.17239801 | 0.396829099 | 0 | 0 | 0 |
| 0.033847098 | 0 | 0 | 0 | 0 |
| 0 | 0 | 0 | 0 | 0 |
| 0.056531196 | 0.003891609 | 0 | 0.002163518 | 0 |
| 0 | 0 | 0 | 0 | 0 |
| 0 | 0 | 0 | 6.26416102 | 0 |
| 0.02431739 | 0.099552345 | 0 | 0.063136122 | 0.026330217 |
| 0 | 0 | 0 | 0 | 0 |
| 0.096670186 | 0.084271727 | 0 | 0 | 0 |
| 8.098998917 | 0 | 0 | 0 | 0 |
| 0 | 0 | 0 | 0 | 0 |
| 0 | 0 | 0 | 0 | 0 |
| 0 | 0 | 0 | 0 | 0 |
| 0 | 0 | 0 | 0 | 0 |
| 0 | 0 | 0 | 0 | 0 |
| 0.112657705 | 0.23982704 | 0.055029364 | 0.108030953 | 0.199302107 |
| 0.0028153 | 0 | 0 | 0 | 0 |
| 0.005416061 | 0 | 0 | 0.931068773 | 0 |
| 0.004913 | 0 | 0 | 0 | 0 |
| 0.536821188 | 1.070736553 | 0.233493092 | 0 | 0 |
| 0 | 0 | 0 | 0 | 0 |
| 0 | 0 | 0 | 0 | 0 |
| 0.265825036 | 0.020251825 | 0 | 0.001912098 | 0 |
| 0 | 0 | 0 | 0 | 0 |
| 0 | 0.00643625 | 0 | 0.006985186 | 0 |
| 0 | 0 | 0 | 0 | 0 |
| 0 | 0.003904032 | 0 | 0 | 0 |
| 0 | 0 | 0 | 0 | 0 |
| 2.586965297 | 0.970905372 | 0 | 0 | 0 |
| 0 | 0 | 0 | 0.002251081 | 0 |
| 0 | 0.153705992 | 0 | 0 | 0 |
| 0.006600433 | 0 | 0 | 0 | 0.026816932 |
| 0.067693317 | 0.021741003 | 0 | 0.044151724 | 0.180892982 |
| 0.168026922 | 0 | 0 | 0 | 0 |
| 0.003691202 | 0.612771822 | 4.688529114 | 0.46808749 | 0.130059861 |
| 0 | 0 | 0 | 0 | 0 |
| 0 | 0 | 0 | 0 | 0 |
| 0 | 0 | 0 | 0 | 0 |
| 0 | 0 | 0 | 0 | 0 |
| 0.028897419 | 0 | 0 | 0.000907272 | 0.04011981 |

| | | | | |
|---|---|---|---|---|
| 0 | 0 | 0 | 0 | 0 |
| 0.241360137 | 0 | 0 | 0 | 0 |
| 0.003324173 | 0.014930886 | 0.047044414 | 0.019786199 | 0.110371556 |
| 0 | 0 | 0 | 0 | 0 |
| 0 | 0 | 0 | 0 | 0 |
| 0.082749499 | 0.004983888 | 0 | 0.000685134 | 0 |
| 0 | 0 | 0 | 0 | 0 |
| 0 | 0 | 0 | 0 | 0 |
| 0.067120873 | 0 | 0 | 0 | 0 |
| 0 | 0.037585312 | 0 | 0 | 0 |
| 0.145563162 | 0.009193235 | 0.001673524 | 0.014089578 | 0.536240637 |
| 0.518466253 | 0.208724326 | 3.003949533 | 0.529185966 | 0.237627903 |
| 0 | 0 | 0 | 0 | 0 |
| 0.005166143 | 0.004022316 | 0 | 0 | 0.092114963 |
| 0.102148063 | 0.354799505 | 0.263738265 | 0.139619498 | 0.006650928 |
| 0 | 0 | 0 | 0 | 0 |
| 0 | 0 | 0 | 0 | 0 |
| 0 | 0 | 0 | 0 | 0 |
| 0 | 0 | 0 | 0 | 0 |
| 0.003046932 | 0.001228023 | 0 | 0 | 0.01527619 |
| 0.015683542 | 0.057685507 | 0.094819805 | 0.136699752 | 0.143449018 |
| 0 | 0.002413751 | 3.638314588 | 1.348952842 | 6.44107462 |
| 0.192919808 | 5.495743469 | 5.320847346 | 16.06009848 | 0.409303323 |
| 0 | 0 | 0 | 0 | 0 |
| 0.00589833 | 0 | 0 | 0 | 0 |
| 4.634149017 | 0.003992753 | 0 | 0 | 0 |
| 0 | 0 | 0 | 0 | 0 |
| 0 | 0 | 0 | 0 | 0 |
| 0.040076975 | 0.138087645 | 0 | 0 | 0 |
| 0 | 0 | 0 | 0 | 0 |
| 0.005430937 | 0 | 0 | 0 | 0 |
| 0.037704837 | 0.098892234 | 0 | 0.189528482 | 0.065415067 |
| 0 | 0 | 0 | 0.003727215 | 0.032126098 |
| 0.039023503 | 0.029773058 | 0 | 0.016050578 | 0.167465681 |
| 0 | 0 | 0.002155357 | 0 | 0.006663996 |
| 0.061197444 | 0.027074064 | 0.001091016 | 0.011929061 | 0.032669998 |
| 0.00097874 | 0.003576503 | 0 | 0.004417269 | 0.026935512 |
| 0 | 0 | 0 | 0 | 0 |
| 6.936300027 | 0 | 0 | 0.011167528 | 0 |
| 0 | 0 | 0 | 0 | 0 |
| 0.030690854 | 0 | 0 | 0 | 0 |
| 0 | 0 | 0 | 0 | 0 |
| 0 | 0 | 0 | 0 | 0 |
| 0 | 0 | 0 | 0 | 0 |
| 0.008781364 | 0.065972001 | 0 | 0.001133645 | 0.818217517 |
| 0 | 0 | 0 | 0 | 0 |
| 0 | 0 | 0 | 0 | 0 |
| 3.694909762 | 0.311317985 | 0.022726714 | 0 | 0.547312174 |

| | | | | |
|---|---|---|---|---|
| 0 | 0 | 0 | 0 | 0 |
| 0 | 0 | 0 | 0 | 0 |
| 0 | 0 | 0 | 0 | 0 |
| 0.001969558 | 0.019117407 | 0 | 0.004444525 | 0 |
| 0 | 0 | 0 | 0 | 0 |
| 0.337463259 | 0 | 0 | 0 | 0 |
| 0.280533483 | 0 | 0 | 0 | 0 |
| 0.176341662 | 0 | 0.002230328 | 0.078416843 | 0.452242611 |
| 0 | 0 | 0 | 0 | 0 |
| 0 | 0 | 0 | 0 | 0 |
| 0.003948419 | 0.001502946 | 0 | 0 | 0.00810917 |
| 0 | 0 | 0 | 0 | 0 |
| 0.206852801 | 0 | 0 | 0 | 0.063310224 |
| 0.099768702 | 1.08051183 | 0 | 0 | 0 |
| 0 | 0 | 0 | 0 | 0 |
| 0 | 0 | 0 | 0 | 0 |
| 0.06150513 | 0 | 0 | 0.231775339 | 0 |
| 0.038347972 | 0.001251168 | 0.001123621 | 0.00609159 | 0 |
| 0 | 0 | 0 | 0 | 0.009274047 |
| 0 | 0.002632808 | 0 | 0 | 0 |
| 0 | 0.004816778 | 0 | 0 | 0 |
| 0 | 0 | 0 | 0 | 0 |
| 0.033054083 | 0.098330901 | 0 | 0 | 0 |
| 0 | 0 | 0 | 0 | 0 |
| 0 | 0 | 0 | 0 | 0 |
| 0 | 0 | 0 | 0 | 0 |
| 0 | 0 | 0 | 0 | 0 |
| 0 | 0 | 0 | 0 | 0 |
| 0 | 0 | 0 | 0 | 0 |
| 0.003308858 | 0.025441895 | 0 | 0 | 0 |
| 0 | 0 | 0 | 0 | 0 |
| 0 | 0 | 0 | 0 | 0 |
| 0.029000949 | 0 | 0 | 0 | 0 |
| 0 | 0 | 0 | 0 | 0 |
| 0 | 0.004182742 | 0 | 0 | 0 |
| 0 | 0 | 0 | 0 | 0 |
| 0.084197443 | 0 | 0 | 0 | 0 |
| 0 | 0 | 0.002713607 | 0 | 0.01201297 |
| 0 | 0 | 0 | 0 | 0 |
| 0 | 0 | 0 | 0 | 0 |
| 0.012544017 | 0.506926268 | 0 | 0 | 0 |
| 0 | 0 | 0 | 0 | 0 |
| 0.09398983 | 0 | 0 | 0 | 0 |
| 0 | 0 | 0 | 0 | 0 |
| 0 | 0 | 0 | 0 | 0 |
| 0 | 0 | 0 | 0 | 0 |
| 0.051872144 | 0 | 0 | 0 | 0 |
| 0 | 0 | 0 | 0 | 0 |

| | | | | |
|---|---|---|---|---|
| 0 | 0 | 0 | 0 | 0 |
| 0 | 0 | 0 | 0 | 0 |
| 0 | 0 | 0 | 0 | 0 |
| 0.967676599 | 0.452218029 | 0 | 0.06034687 | 0 |
| 0 | 0 | 0 | 0 | 0 |
| 0 | 0 | 0 | 0 | 0 |
| 0 | 0 | 0 | 0 | 0 |
| 0 | 0 | 0 | 0 | 0 |
| 0.001792675 | 0.008773363 | 0.088835547 | 0.001256326 | 0.158578388 |
| 0 | 0 | 0 | 0 | 0 |
| 0 | 0 | 0 | 0 | 0 |
| 0 | 0 | 0 | 0 | 0 |
| 0 | 0 | 0 | 0 | 0 |
| 0 | 0 | 0 | 0 | 0 |
| 0.160296814 | 0.011979063 | 0 | 0 | 0 |
| 0 | 0 | 0.011993291 | 0 | 1.340374757 |
| 0 | 0 | 0 | 0 | 0 |
| 0 | 0 | 0 | 0 | 0 |
| 0 | 0 | 0 | 0 | 0 |
| 0 | 0 | 0 | 0 | 0 |
| 0.32134467 | 0 | 0 | 0 | 0 |
| 0 | 0 | 0 | 0 | 0 |
| 0 | 0 | 0 | 0 | 0 |
| 0 | 0 | 0 | 0 | 0 |
| 0.039452241 | 0 | 0 | 0 | 0.128033564 |
| 5.335108676 | 1.261580661 | 0 | 0.002506525 | 3.178118369 |
| 0 | 0 | 0 | 0.066380386 | 0 |
| 0 | 0.000853918 | 0 | 0 | 0 |
| 0 | 0 | 0 | 0 | 0 |
| 0 | 0 | 0 | 0 | 0 |
| 0 | 0 | 0 | 0 | 0 |
| 0.029286463 | 0 | 0 | 0 | 0 |
| 0 | 0 | 0 | 0 | 0 |
| 0.002090919 | 0.001285681 | 0 | 0.005523197 | 0.00396395 |
| 0.00079504 | 0.004842047 | 0 | 0 | 0.130082482 |
| 0.002622653 | 0 | 0 | 0 | 0.001923693 |
| 0 | 0 | 0 | 0 | 0 |
| 0.251086802 | 0.219089571 | 0 | 0.950325723 | 0.37057431 |
| 0.164904743 | 0 | 0 | 0 | 0 |
| 0 | 0 | 0 | 0 | 0 |
| 0 | 0 | 0 | 0 | 0 |
| 0 | 0.026442141 | 0 | 0 | 0 |
| 0.497138347 | 0.312205495 | 0 | 0.281805568 | 0 |
| 0 | 0 | 0 | 0 | 0 |
| 0 | 0 | 0 | 0 | 0 |
| 0.000709895 | 0.010133178 | 0.000682513 | 0.001567132 | 0 |
| 0 | 0.0248757 | 0 | 0 | 0 |
| 0.661604927 | 0.358382754 | 0 | 2.378732888 | 0.041518536 |

| | | | | |
|---|---|---|---|---|
| 0 | 0.114602966 | 0 | 0 | 0 |
| 0.027237942 | 0 | 0 | 0 | 0 |
| 0 | 0 | 0 | 0 | 0 |
| 0 | 0 | 0 | 0 | 0 |
| 0.00272887 | 0 | 0 | 0 | 0.009484518 |
| 0.006872867 | 0 | 0 | 0 | 0 |
| 0 | 0 | 0 | 0 | 0 |
| 0 | 0 | 0 | 0 | 0 |
| 0 | 0 | 0 | 0 | 0 |
| 0 | 0 | 0 | 0 | 0 |
| 0.000963722 | 0 | 0 | 0 | 0 |
| 0.677029038 | 1.059284548 | 0 | 1.621500871 | 9.224764219 |
| 0.137504839 | 0 | 0 | 0.136407019 | 0 |
| 0 | 0 | 0 | 0 | 0 |
| 0.000728293 | 0.567748875 | 0.044626173 | 1.480317548 | 0.142594976 |
| 0 | 0 | 0 | 0 | 0 |
| 0 | 0 | 0 | 0 | 0 |
| 0 | 0 | 0 | 0 | 0 |
| 0 | 0 | 0 | 0 | 0 |
| 0 | 0 | 0 | 0 | 0 |
| 0.033118855 | 0 | 0 | 0.337903666 | 0 |
| 0 | 0 | 0 | 0 | 0 |
| 0.888022293 | 0.010395804 | 0 | 0.053185607 | 1.318557406 |
| 0 | 0 | 0 | 0 | 0 |
| 0 | 0 | 0 | 0 | 0 |
| 0 | 0 | 0 | 0 | 0 |
| 0 | 0 | 0 | 0 | 0 |
| 0 | 0 | 0 | 0 | 0 |
| 0.00349871 | 0 | 0 | 0.003432706 | 0 |
| 0 | 0 | 0 | 0 | 0 |
| 0 | 0 | 0 | 0 | 0 |
| 0.028666894 | 0.049283446 | 0.004361325 | 0.004018012 | 0.070197574 |
| 0 | 0 | 0 | 0 | 0 |
| 0.045954659 | 0.028574923 | 0 | 0.038448085 | 0.015514617 |
| 0.003057968 | 0 | 0 | 0 | 0.00209346 |
| 0.49302422 | 0.002400859 | 0 | 0 | 0 |
| 0.002352535 | 0 | 0 | 0 | 0 |
| 0.124177894 | 0 | 0 | 0 | 0 |
| 0 | 0 | 0 | 0 | 0 |
| 0 | 0 | 0 | 0 | 0 |
| 0.166398198 | 0.305453483 | 0 | 0 | 0 |
| 0.024743646 | 0 | 0 | 0 | 0 |
| 0 | 0 | 0 | 0 | 0 |
| 0 | 0 | 0 | 0 | 0 |
| 0.041571186 | 0.013301215 | 0 | 0.001970383 | 0.034439882 |
| 0 | 0 | 0 | 0 | 0 |
| 0.032780431 | 0 | 1.406589113 | 0.435347768 | 2.07749039 |
| 0 | 0.003687408 | 0 | 0 | 0.004096127 |

| | | | | |
|---|---|---|---|---|
| 0 | 0 | 0 | 0 | 0.07922931 |
| 0 | 0 | 0 | 0 | 0 |
| 0 | 0 | 0 | 0.002445443 | 0 |
| 0 | 0 | 0 | 0 | 0 |
| 0 | 0 | 0 | 0 | 0 |
| 0 | 0 | 0 | 0 | 0 |
| 0 | 0 | 0 | 0 | 0 |
| 0.050608841 | 0 | 0 | 0 | 3.352845931 |
| 0 | 0 | 0 | 0 | 0 |
| 0 | 0 | 0 | 0 | 0 |
| 0 | 0 | 0 | 0 | 0 |
| 0 | 0 | 0 | 0 | 0 |
| 0 | 0 | 0 | 0 | 0 |
| 0 | 0 | 0 | 0 | 0 |
| 0 | 0 | 0 | 0 | 0 |
| 0 | 0 | 0 | 0 | 0 |
| 0 | 0 | 0 | 0 | 0 |
| 0 | 0 | 0 | 0 | 0 |
| 0.00599238 | 0.307423887 | 0 | 0 | 0 |
| 0 | 0 | 0 | 0 | 0 |
| 0.02623193 | 0.001293028 | 0 | 0 | 0 |
| 0 | 0 | 0 | 0 | 0 |
| 0 | 0 | 0 | 0 | 0 |
| 0 | 0 | 0 | 0 | 0 |
| 0.001535946 | 0 | 0 | 0 | 0.06489237 |
| 0.002991397 | 0.002134988 | 0.004745421 | 0 | 0.004991724 |
| 0 | 0 | 0 | 0 | 0 |
| 0.06368363 | 0.092157017 | 0 | 0.005027305 | 0.040590597 |
| 0 | 0 | 0 | 0 | 0 |
| 0.479782085 | 0.178542295 | 0 | 0.025733957 | 0 |
| 0 | 0 | 0 | 0 | 0 |
| 0 | 0 | 0 | 0 | 0 |
| 0 | 0 | 0 | 0 | 0 |
| 0 | 0 | 0 | 0 | 0 |
| 0.006269599 | 0 | 0 | 0.001730059 | 0 |
| 0 | 0 | 0 | 0 | 0 |
| 0 | 0 | 0 | 0 | 0 |
| 0.00088927 | 0 | 0 | 0 | 0.001826361 |
| 0.087462499 | 0 | 0 | 0 | 0 |
| 0 | 0 | 0 | 0 | 0 |
| 1.531338787 | 10.20648686 | 5.533489994 | 9.51782609 | 3.473627513 |
| 0 | 0.852241367 | 0.044184372 | 0.423916684 | 1.650241595 |
| 0 | 0.146357716 | 0 | 0 | 0 |
| 0 | 0 | 0 | 0 | 0 |
| 0 | 0 | 0 | 0 | 0 |
| 0 | 0 | 0 | 0 | 0 |
| 0.004541106 | 0 | 0 | 0 | 0 |
| 0.010057275 | 0.001611895 | 0 | 0.001731148 | 0.099834395 |

| | | | | |
|---|---|---|---|---|
| 0 | 0 | 0 | 0 | 0 |
| 0.007259465 | 0.023211537 | 0 | 0.001780628 | 0 |
| 0 | 0 | 0 | 0 | 0 |
| 0 | 0 | 0 | 0 | 0 |
| 2.014238986 | 1.10800751 | 0 | 1.273148347 | 0 |
| 0 | 0 | 0 | 0 | 0 |
| 0 | 0 | 0 | 0 | 0 |
| 0 | 0 | 0 | 0 | 0 |
| 0 | 0.008484553 | 0.150812657 | 0.007112013 | 1.106873368 |
| 0 | 0 | 0 | 0 | 0 |
| 0 | 0 | 0 | 0 | 0 |
| 0 | 0 | 0 | 0 | 0 |
| 0.029978682 | 0.05074642 | 0 | 0 | 0 |
| 0.002601582 | 0.023023981 | 0.410202213 | 0.025099606 | 0.004897811 |
| 0 | 0 | 0 | 0 | 0 |
| 0.120440508 | 0.156834809 | 0 | 0.151506864 | 0.121852055 |
| 0.006378585 | 0 | 0 | 0.027978068 | 0 |
| 0 | 0 | 0 | 0 | 0 |
| 0 | 0 | 0 | 0 | 0 |
| 0 | 0 | 0 | 0 | 0 |
| 0 | 0 | 0 | 0 | 0 |
| 0 | 0 | 0 | 0 | 0 |
| 0 | 0 | 0 | 0 | 0 |
| 0 | 0 | 0 | 0 | 0 |
| 0.014946445 | 0 | 0 | 0 | 0 |
| 0 | 0 | 0 | 0 | 0 |
| 0.028384734 | 0 | 0 | 0.000960319 | 0 |
| 0 | 0 | 0 | 0.001625419 | 0.002646338 |
| 0 | 0 | 0 | 0 | 0 |
| 0 | 0 | 0 | 0 | 0 |
| 0 | 0 | 0 | 0 | 0 |
| 0 | 0 | 0 | 0 | 0 |
| 0 | 0 | 0 | 0 | 0 |
| 0.029136872 | 0.029746851 | 0 | 0 | 0 |
| 4.065275152 | 0 | 0 | 0 | 0 |
| 0 | 0 | 0 | 0.005431897 | 0.006778512 |
| 0 | 0 | 0 | 0 | 0 |
| 0.041219791 | 0 | 0 | 0 | 0 |
| 0 | 0 | 0 | 0 | 0 |
| 0 | 0 | 0 | 0 | 0 |
| 0.073804158 | 1.901203144 | 0 | 0 | 0 |
| 0.400330532 | 0 | 0 | 0 | 0 |
| 0 | 0 | 0 | 0 | 0 |
| 0 | 0 | 0 | 0 | 0 |
| 0 | 0 | 0 | 0 | 0 |
| 0 | 0 | 0 | 0 | 0 |
| 0 | 0 | 0 | 0 | 0 |
| 0 | 0 | 0 | 0 | 0 |

| | | | | |
|---|---|---|---|---|
| 0.310580156 | 0.343384857 | 0 | 0 | 0 |
| 0 | 0 | 0 | 0 | 0 |
| 0 | 0 | 0 | 0 | 0 |
| 0 | 0 | 0 | 0 | 0.001284763 |
| 0.000798053 | 0 | 0 | 0 | 0 |
| 0 | 0 | 0 | 0 | 0 |
| 0.039114199 | 0.006197655 | 0 | 0.712073188 | 0.01556669 |
| 0 | 0 | 0 | 0 | 0 |
| 0.00092468 | 0 | 0 | 0 | 0 |
| 0.00098175 | 0 | 0 | 0 | 0 |
| 0.014297796 | 0 | 0 | 0 | 0 |
| 0 | 0 | 0 | 0 | 0 |
| 0 | 0 | 0 | 0 | 0 |
| 0 | 0 | 0 | 0 | 0 |
| 0 | 0 | 0 | 0 | 0 |
| 0 | 0 | 0 | 0 | 0 |
| 0 | 0 | 0 | 0 | 0 |
| 0 | 0 | 0 | 0 | 0 |
| 0.361746965 | 0 | 0 | 0 | 0 |
| 0 | 0 | 0 | 0 | 0 |
| 0.066103497 | 0.002848524 | 0 | 0.093001688 | 0 |
| 0 | 0 | 0 | 0 | 0 |
| 0.076842488 | 0.011894379 | 0 | 0.000771282 | 0 |
| 0.060814677 | 0.004589847 | 0.000940879 | 0 | 0.024585149 |
| 0 | 0 | 0 | 0 | 0 |
| 0 | 0 | 0 | 0 | 0 |
| 0 | 0 | 0 | 0 | 0 |
| 0 | 0 | 0 | 0 | 0 |
| 0 | 0 | 0 | 0 | 0 |
| 0 | 0.124937042 | 0.027469814 | 0.056016406 | 0 |
| 0.099831084 | 0 | 0 | 0 | 0 |
| 0 | 0 | 0 | 0 | 0 |
| 0 | 0 | 0 | 0 | 0 |
| 0.063266763 | 0 | 0.003445036 | 2.317766504 | 0 |
| 0 | 0 | 0 | 0 | 0 |
| 0 | 0 | 0 | 0 | 0 |
| 0 | 0 | 0 | 0.007079767 | 0.043336796 |
| 0 | 0 | 0 | 0 | 0 |
| 0 | 0 | 0 | 0 | 0 |
| 0.849294185 | 0 | 0 | 0 | 0 |
| 0 | 0 | 0 | 0 | 0 |
| 0 | 0 | 0 | 0 | 0 |
| 0 | 0 | 0 | 0 | 0 |
| 0 | 0 | 0 | 0 | 0 |
| 0.044286987 | 0 | 0 | 0 | 0 |
| 0 | 0 | 0 | 0 | 0 |
| 0 | 0 | 0 | 0 | 0 |
| 1.06356858 | 0 | 0 | 0 | 0 |

| | | | | |
|---|---|---|---|---|
| 0.006581484 | 0.013098659 | 0 | 0.001291465 | 0.033985334 |
| 0.460470256 | 0 | 0 | 0 | 0 |
| 0 | 0 | 0 | 0 | 0 |
| 0 | 0 | 0 | 0 | 0 |
| 0 | 0 | 0 | 0 | 0 |
| 0 | 0 | 0 | 0 | 0 |
| 0 | 0 | 0 | 0 | 0.003711125 |
| 0.386347148 | 0.433466348 | 0 | 1.056287311 | 0 |
| 0.390792866 | 0.001348217 | 0.01089699 | 0.053574613 | 5.137495564 |
| 0 | 0.025811181 | 0.001738494 | 0 | 0.049586668 |
| 0 | 0 | 0 | 0 | 0 |
| 0 | 0 | 0 | 0 | 0 |
| 0.030430003 | 0 | 0 | 0 | 0 |
| 0 | 0.02420414 | 0.031932795 | 0 | 0 |
| 0 | 0 | 0 | 0 | 0 |
| 0 | 0 | 0 | 0 | 0 |
| 0 | 0 | 0 | 0 | 0.085407166 |
| 0 | 0 | 0 | 0 | 0 |
| 0 | 0 | 0 | 0 | 0 |
| 0.014483208 | 0.009534176 | 0 | 0.023739024 | 0 |
| 0 | 0 | 0 | 0 | 0 |
| 0 | 0 | 0 | 0 | 0 |
| 0 | 0 | 0 | 0 | 0 |
| 0 | 0 | 0 | 0 | 0 |
| 0.01081199 | 0.008625697 | 0 | 0 | 0 |
| 0 | 0 | 0 | 0 | 0 |
| 0 | 0 | 0 | 0 | 0 |
| 0 | 0 | 0 | 0 | 0 |
| 0 | 0 | 0 | 0 | 0 |
| 0 | 0 | 0 | 0 | 0 |
| 0 | 0 | 0 | 0 | 0 |
| 0 | 0 | 0 | 0 | 0 |
| 0 | 0 | 0 | 0 | 0 |
| 0 | 0 | 0 | 0 | 0 |
| 0 | 0 | 0 | 0 | 0 |
| 0 | 0 | 0 | 0 | 0 |
| 0 | 0 | 0 | 0 | 0 |
| 0 | 0.056070646 | 0.024253394 | 0 | 0 |
| 0 | 0 | 0 | 0 | 0 |
| 0 | 0 | 0 | 0 | 0.017156776 |
| 0 | 0 | 0 | 0 | 0 |
| 0.031762184 | 0.137340734 | 0.189776576 | 0.266405508 | 2.465506935 |
| 0 | 0 | 0 | 0 | 0 |
| 0 | 0 | 0 | 0 | 0 |
| 0 | 0 | 0 | 0 | 0 |
| 0 | 0 | 0 | 0 | 0 |
| 0 | 0 | 0 | 0 | 0 |
| 0 | 0.009598365 | 0 | 0 | 0 |

| | | | | |
|---|---|---|---|---|
| 0 | 0 | 0 | 0 | 0 |
| 0 | 0.058722955 | 0 | 0 | 0.005054115 |
| 0 | 0 | 0 | 0 | 0 |
| 0 | 0 | 0 | 0 | 0 |
| 2.814941642 | 0.108386728 | 0.027193669 | 0.015365528 | 0 |
| 0 | 0 | 0 | 0 | 0 |
| 0 | 0.017624918 | 0 | 0 | 0 |
| 0.001913628 | 0.015023515 | 0.00588741 | 0.002816291 | 0.052811582 |
| 0.011702958 | 0.001909145 | 0 | 0 | 0 |
| 0 | 0 | 0 | 0 | 0 |
| 0 | 0 | 0 | 0 | 0 |
| 0 | 0 | 0 | 0 | 0 |
| 0 | 0 | 0 | 0 | 0 |
| 0 | 0 | 0 | 0 | 0 |
| 0 | 0 | 0 | 0 | 0.003112534 |
| 0.015559458 | 0.020373841 | 0 | 0.020558113 | 0 |
| 0 | 0 | 0 | 0 | 0 |
| 0 | 0 | 0 | 0 | 0 |
| 0 | 0 | 0 | 0 | 0 |
| 0 | 0 | 0 | 0 | 0 |
| 0 | 0 | 0 | 0 | 0 |
| 0 | 0 | 0 | 0 | 0 |
| 0 | 0 | 0 | 0 | 0 |
| 0.006831362 | 0 | 0 | 0 | 0.007015044 |
| 0 | 0 | 0 | 0 | 0 |
| 0 | 0 | 0 | 0 | 0 |
| 0.177785584 | 0 | 0.006261899 | 0 | 0.003888518 |
| 0.000996818 | 0 | 0 | 0 | 0 |
| 0 | 0 | 0 | 0 | 0 |
| 0 | 0 | 0 | 0 | 0 |
| 0 | 0 | 0 | 0 | 0 |
| 0 | 0 | 0 | 0 | 0 |
| 0 | 0 | 0 | 0 | 0 |
| 0 | 0 | 0 | 0 | 0 |
| 0 | 0 | 0 | 0 | 0 |
| 0 | 0 | 0 | 0 | 0 |
| 0 | 0 | 0 | 0 | 0 |
| 0.031067865 | 0.190469678 | 0 | 0.015812415 | 0 |
| 0.001146339 | 0 | 0 | 0 | 0 |
| 0 | 0 | 0 | 0 | 0 |
| 0 | 0 | 0 | 0 | 0 |
| 1.986688277 | 0 | 0 | 0 | 0 |
| 0 | 0 | 0 | 0 | 0 |
| 0 | 0 | 0 | 0 | 0 |
| 0 | 0 | 0 | 0 | 0 |
| 0 | 0 | 0 | 0 | 0 |
| 0 | 0 | 0 | 0 | 0 |
| 0 | 0 | 0 | 0 | 0 |

| | | | | |
|---|---|---|---|---|
| 0 | 0 | 0 | 0.106039414 | 0 |
| 0 | 0 | 0 | 0 | 0 |
| 0 | 0 | 0 | 0 | 0 |
| 0 | 0 | 0 | 0 | 0 |
| 0 | 0 | 0 | 0 | 0 |
| 0 | 0 | 0 | 0.025906356 | 0 |
| 0 | 0 | 0 | 0 | 0 |
| 0 | 0 | 0 | 0 | 0 |
| 0 | 0 | 0 | 0 | 0 |
| 0 | 0 | 0 | 0 | 0 |
| 0 | 0 | 0 | 0 | 0 |
| 0 | 0 | 0 | 0 | 0.009602146 |
| 0 | 0 | 0 | 0 | 0 |
| 0 | 0 | 0 | 0 | 0 |
| 0.019196543 | 0 | 0 | 0 | 0 |
| 0 | 0 | 0 | 0 | 0 |
| 0 | 0 | 0 | 0 | 0 |
| 0 | 0 | 0 | 0 | 0 |
| 0 | 0 | 0 | 0 | 0 |
| 0 | 0 | 0 | 0 | 0 |
| 0 | 0.010122406 | 0 | 0 | 0 |
| 0 | 0 | 0 | 0 | 0 |
| 0.017182894 | 0.001189196 | 0 | 0.015581558 | 1.040514935 |
| 0.014296982 | 0 | 0 | 0 | 0 |
| 0 | 0 | 0 | 0 | 0 |
| 0 | 0 | 0 | 0 | 0 |
| 0 | 0 | 0 | 0 | 0 |
| 0 | 0 | 0 | 0 | 0 |
| 0 | 0 | 0 | 0 | 0 |
| 0 | 0 | 0 | 0 | 0 |
| 0 | 0 | 0 | 0 | 0 |
| 0 | 0 | 0 | 0 | 0 |
| 0 | 0 | 1.717751676 | 0 | 0.328234364 |
| 0 | 0 | 0 | 0 | 0 |
| 0.109699403 | 0 | 0 | 0.129455447 | 0 |
| 0.001173902 | 0 | 0.000902898 | 0 | 0 |
| 0 | 0 | 0 | 0 | 0 |
| 0 | 0 | 0 | 0 | 0 |
| 0.005883815 | 0.002325784 | 0.015142987 | 0 | 0.031372022 |
| 0 | 0 | 0 | 0 | 0 |
| 0 | 0 | 0 | 0 | 0 |
| 0 | 0 | 0 | 0 | 0 |
| 1.787197323 | 0.039886565 | 0 | 0 | 0 |
| 0 | 0 | 0 | 0 | 0 |
| 0 | 0 | 0 | 0 | 0 |
| 0.024377255 | 0.002639737 | 0 | 0 | 0 |
| 0 | 0 | 0 | 0 | 0 |
| 0 | 0 | 0 | 0 | 0 |

| | | | | |
|---|---|---|---|---|
| 0 | 0 | 0 | 0 | 0 |
| 0 | 0 | 0 | 0 | 0 |
| 0 | 0 | 0 | 0 | 0 |
| 0 | 0 | 0 | 0 | 0 |
| 0 | 0 | 0 | 0 | 0 |
| 0 | 0 | 0 | 0 | 0 |
| 0 | 0 | 0 | 0 | 0 |
| 0 | 0.04452345 | 0.353613977 | 0 | 0 |
| 0.008269375 | 0 | 0 | 0 | 0 |
| 0.113853168 | 0.003954764 | 0 | 0 | 0 |
| 0 | 0 | 0 | 0 | 0 |
| 0 | 0 | 0 | 0 | 0 |
| 0 | 0 | 0 | 0 | 0 |
| 0 | 0 | 0 | 0 | 0 |
| 0 | 0 | 0 | 0 | 0 |
| 0 | 0.004377432 | 0.023587116 | 0 | 0.4520321 |
| 0 | 0 | 0 | 0 | 0 |
| 0 | 0 | 0 | 0 | 0 |
| 0 | 0 | 0 | 0 | 0 |
| 0.320048474 | 0 | 0 | 0 | 0 |
| 0.007784893 | 0.003047945 | 0 | 0 | 0 |
| 0 | 0 | 0 | 0 | 0 |
| 0 | 0 | 0 | 0 | 0 |
| 0 | 0 | 0 | 0 | 0 |
| 0 | 0 | 0 | 0 | 0 |
| 0 | 0 | 0 | 0 | 0 |
| 0 | 0 | 0 | 0 | 0 |
| 0 | 0 | 0 | 0 | 0 |
| 0 | 0 | 0 | 0 | 0 |
| 0.031342322 | 0 | 0 | 0.072763031 | 0 |
| 0.102989118 | 0.085586156 | 0 | 0.164900617 | 0 |
| 0 | 0 | 0 | 0 | 0 |
| 0 | 0 | 0 | 0 | 0 |
| 0 | 0 | 0 | 0 | 0 |
| 0 | 0 | 0 | 0 | 0 |
| 0 | 0 | 0 | 0 | 0 |
| 0 | 0 | 0 | 0 | 0 |
| 0 | 0 | 0 | 0 | 0 |
| 0 | 0 | 0 | 0 | 0 |
| 0 | 0 | 0 | 0 | 0 |
| 0 | 0 | 0 | 0 | 0 |
| 0.002872293 | 0 | 0 | 0 | 0 |
| 0.017598031 | 0 | 0 | 0 | 0 |
| 0 | 0.00545501 | 0 | 0 | 0 |
| 0 | 0 | 0 | 0 | 0 |
| 0 | 0 | 0 | 0 | 0 |
| 0 | 0 | 0.236093819 | 0 | 0.120890499 |
| 0.00088588 | 0 | 0 | 0 | 0 |

| | | | | |
|---|---|---|---|---|
| 0 | 0 | 0 | 0 | 0 |
| 0 | 0 | 0 | 0 | 0 |
| 0 | 0 | 0 | 0 | 0 |
| 0 | 0 | 0 | 0 | 0 |
| 0.007980597 | 0.000929931 | 0 | 0 | 0.13513685 |
| 0 | 0 | 0 | 0 | 0 |
| 0 | 0 | 0 | 0 | 0 |
| 0 | 0 | 0 | 0 | 0 |
| 0 | 0 | 0 | 0 | 0 |
| 0 | 0 | 0 | 0 | 0 |
| 0 | 0.082648214 | 0 | 0 | 0 |
| 0.014019463 | 0.003615006 | 0 | 0 | 0.057353477 |
| 0.022239679 | 0 | 0 | 0 | 0 |
| 0 | 0 | 0 | 0 | 0 |
| 0 | 0 | 0 | 0 | 0.018226144 |
| 0.492881315 | 0.332120752 | 0 | 0 | 0 |
| 0.017739167 | 0 | 0 | 0.00136506 | 0 |
| 0 | 0 | 0 | 0 | 0 |
| 0 | 0.03226107 | 0.058076285 | 0.123136334 | 0.442195864 |
| 0.001341364 | 0.001787042 | 0 | 0.014476648 | 0.009642015 |
| 0 | 0 | 0 | 0 | 0 |
| 0 | 0 | 0 | 0 | 0 |
| 0 | 0 | 0 | 0 | 0 |
| 0 | 0 | 0 | 0 | 0 |
| 0 | 0 | 0 | 0 | 0 |
| 0 | 0 | 0 | 0 | 0 |
| 0.004478735 | 0 | 0 | 0.004921552 | 0.045255734 |
| 0 | 0 | 0 | 0 | 0 |
| 0 | 0 | 0 | 0 | 0 |
| 0 | 0 | 0 | 0 | 0 |
| 0.002030753 | 0 | 0 | 0 | 0.000834142 |
| 0 | 0 | 0 | 0 | 0 |
| 0 | 0 | 0 | 0 | 0 |
| 0 | 0 | 0 | 0 | 0 |
| 0 | 0 | 0 | 0 | 0 |
| 0 | 0 | 0 | 0 | 0 |
| 0 | 0 | 0 | 0 | 0 |
| 0.025253438 | 0.015477902 | 0 | 0.001539815 | 0.004981375 |
| 0.409457963 | 0.072507321 | 0 | 0.016668739 | 0 |
| 0 | 0 | 0 | 0 | 0 |
| 0 | 0 | 0 | 0.001816729 | 0 |
| 0 | 0 | 0 | 0 | 0 |
| 0 | 0 | 0 | 0 | 0 |
| 0 | 0.001449585 | 0 | 0.001868196 | 0.012818181 |
| 0 | 0 | 0 | 0 | 0 |
| 0 | 0 | 0 | 0 | 0 |
| 0 | 0 | 0 | 0 | 0 |
| 0 | 0 | 0 | 0 | 0 |

| | | | | |
|---|---|---|---|---|
| 0 | 0 | 0 | 0 | 0 |
| 0.057503654 | 0.009911761 | 0 | 0 | 0 |
| 0 | 0 | 0 | 0 | 0 |
| 0.002438974 | 0 | 0 | 0.000652626 | 0.00068306 |
| 0 | 0.001409008 | 0.000474514 | 0 | 0.027452842 |
| 0 | 0.00181846 | 0 | 0 | 0 |
| 0.010758357 | 0 | 0 | 0 | 0 |
| 0 | 0 | 0 | 0 | 0 |
| 0 | 0 | 0 | 0 | 0 |
| 0 | 0 | 0 | 0 | 0 |
| 0.018687901 | 0 | 0 | 0 | 0 |
| 0 | 0 | 0 | 0 | 0 |
| 0.206837987 | 0.007538153 | 0 | 0.000771033 | 0 |
| 0.001996814 | 0 | 0 | 0 | 0 |
| 0 | 0 | 0 | 0 | 0 |
| 0 | 0 | 0 | 0 | 0 |
| 0 | 0 | 0 | 0 | 0 |
| 0 | 0 | 0 | 0 | 0 |
| 0 | 0 | 0 | 0 | 0 |
| 0 | 0 | 0 | 0 | 0 |
| 0 | 0 | 0 | 0 | 0 |
| 0 | 0 | 0 | 0 | 0 |
| 0 | 0.006144673 | 0 | 0.016853533 | 0.022314985 |
| 0.003907499 | 0.003067699 | 0 | 0.001916891 | 0 |
| 0 | 0 | 0 | 0 | 0 |
| 0.001983936 | 0 | 0 | 0 | 0 |
| 0 | 0 | 0 | 0 | 0 |
| 0.001865228 | 0 | 0 | 0 | 0 |
| 0 | 0 | 0 | 0 | 0 |
| 0 | 0 | 0 | 0 | 0 |
| 0.017306873 | 0.012648523 | 0 | 0 | 0 |
| 0 | 0 | 0 | 0 | 0 |
| 0 | 0 | 0 | 0 | 0 |
| 0 | 0 | 0 | 0 | 0 |
| 0.001934626 | 0 | 0 | 0 | 0 |
| 0 | 0.008264014 | 0 | 0 | 0 |
| 0 | 0 | 0 | 0 | 0 |
| 0 | 0 | 0 | 0 | 0 |
| 0 | 0 | 0 | 0 | 0 |
| 0 | 0 | 0 | 0 | 0 |
| 0 | 0 | 0 | 0 | 0 |
| 0.000889785 | 0 | 0 | 0 | 0 |
| 0 | 0 | 0 | 0 | 0 |
| 0 | 0 | 0 | 0 | 0 |
| 0 | 0 | 0 | 0 | 0 |
| 0.412607801 | 0.71931082 | 0.164358239 | 0.111065333 | 0.325626128 |
| 0 | 0 | 0 | 0 | 0 |
| 0 | 0 | 0 | 0 | 0 |

| | | | | |
|---|---|---|---|---|
| 0 | 0 | 0 | 0 | 0 |
| 0 | 0 | 0 | 0 | 0 |
| 0 | 0 | 0 | 0 | 0 |
| 0 | 0 | 0 | 0 | 0 |
| 0 | 0 | 0 | 0 | 0 |
| 0 | 0 | 0 | 0 | 0 |
| 0 | 0 | 0 | 0 | 0 |
| 0 | 0 | 0 | 0 | 0 |
| 0 | 0 | 0 | 0 | 0.015025814 |
| 0 | 0 | 0 | 0 | 0 |
| 0 | 0.001266097 | 0 | 0 | 0.028463529 |
| 0 | 0 | 0 | 0 | 0 |
| 0 | 0 | 0 | 0 | 0 |
| 0 | 0 | 0 | 0 | 0 |
| 0 | 0 | 0 | 0 | 0 |
| 0 | 0 | 0 | 0 | 0 |
| 0 | 0 | 0 | 0 | 0 |
| 0 | 0 | 0 | 0 | 0 |
| 0 | 0 | 0 | 0 | 0 |
| 0 | 0.013912226 | 0 | 0 | 0 |
| 0 | 0 | 0 | 0 | 0 |
| 0 | 0 | 0 | 0 | 0 |
| 0 | 0 | 0 | 0 | 0 |
| 0 | 0 | 0 | 0 | 0 |
| 0 | 0 | 0 | 0 | 0 |
| 0 | 0 | 0 | 0 | 0 |
| 0 | 0 | 0 | 0 | 0 |
| 0.013250125 | 0 | 0 | 0 | 0 |
| 0 | 0 | 0 | 0 | 0 |
| 0 | 0 | 0 | 0 | 0 |
| 0.009383745 | 0.001030348 | 0 | 0.002124627 | 0.028908168 |
| 0 | 0 | 0 | 0 | 0 |
| 0.005866799 | 0 | 11.26489138 | 0.395322128 | 0.001721299 |
| 0.000856462 | 0 | 0 | 0 | 0.007915413 |
| 0 | 0 | 0 | 0 | 0 |
| 0 | 0 | 0 | 0 | 0 |
| 0 | 0 | 0 | 0 | 0 |
| 0 | 0 | 0 | 0 | 0 |
| 0 | 0 | 0 | 0 | 0 |
| 0 | 0 | 0 | 0 | 0 |
| 0 | 0 | 0 | 0 | 0 |
| 0.414567215 | 0 | 0 | 0 | 0.139308534 |
| 0.049980966 | 0.062111026 | 0 | 0.091482776 | 0.383858186 |
| 0 | 0 | 0 | 0 | 0 |
| 0 | 0 | 0 | 0 | 0 |
| 0.342011214 | 0 | 0 | 0 | 0 |
| 0 | 0 | 0 | 0 | 0 |
| 0 | 0 | 0 | 0 | 0 |

| | | | | |
|---|---|---|---|---|
| 0 | 0 | 0 | 0 | 0 |
| 0 | 0 | 0.007445537 | 0 | 0 |
| 0.001541949 | 0.002641208 | 0.000988315 | 0 | 0.026917957 |
| 0 | 0 | 0 | 0 | 0 |
| 0.026470202 | 0.867451518 | 0 | 0 | 0 |
| 0.01917465 | 0.061811141 | 0 | 0 | 1.142032761 |
| 0 | 0 | 0 | 0 | 0 |
| 0 | 0 | 0 | 0 | 0 |
| 0 | 0 | 0 | 0 | 0 |
| 0 | 0 | 0 | 0 | 0 |
| 0 | 0 | 0 | 0 | 0 |
| 0 | 0 | 0 | 0 | 0 |
| 0 | 0 | 0 | 0 | 0 |
| 0 | 0 | 0.029240802 | 0 | 0 |
| 0 | 0 | 0 | 0 | 0 |
| 0 | 0 | 0 | 0 | 0 |
| 0.000865735 | 0 | 0 | 0 | 0 |
| 0 | 0 | 0 | 0 | 0 |
| 0 | 0 | 0 | 0 | 0 |
| 0 | 0 | 0 | 0 | 0 |
| 0 | 0 | 0 | 0 | 0 |
| 0 | 0 | 0 | 0 | 0 |
| 0 | 0 | 0 | 0 | 0 |
| 0 | 0 | 0 | 0 | 0 |
| 0.027321832 | 0 | 0 | 0 | 0 |
| 0.000639189 | 0.001459824 | 0 | 0 | 0 |
| 0 | 0 | 0 | 0 | 0 |
| 0.018394384 | 0 | 0 | 0 | 0 |
| 0 | 0 | 0 | 0 | 0 |
| 0 | 0 | 0 | 0 | 0 |
| 0 | 0 | 0 | 0 | 0 |
| 0 | 0 | 0 | 0 | 0 |
| 0 | 0 | 0 | 0 | 0 |
| 0 | 0 | 0 | 0 | 0 |
| 0 | 0 | 0 | 0 | 0 |
| 0 | 0 | 0 | 0 | 0 |
| 0 | 0 | 0 | 0 | 0 |
| 0 | 0 | 0 | 0 | 0 |
| 2.425405793 | 0 | 0 | 1.874225855 | 0 |
| 0 | 0 | 0 | 0 | 0 |
| 0 | 0 | 0 | 0 | 0 |
| 0.08228599 | 0 | 0.020089125 | 0 | 0 |
| 0.002565983 | 0.012209103 | 0 | 0.000629394 | 0.01998191 |
| 0 | 0 | 0 | 0 | 0 |
| 0 | 0 | 0 | 0 | 0 |
| 0.001968048 | 0 | 0 | 0 | 0 |
| 0 | 0 | 0.001258013 | 0.002246648 | 0 |
| 0 | 0 | 0 | 0 | 0 |

| | | | | |
|---|---|---|---|---|
| 0 | 0 | 0 | 0 | 0 |
| 0.018161725 | 0.05726931 | 0.010159236 | 0.004859755 | 0.061036557 |
| 0 | 0 | 0 | 0 | 0 |
| 0 | 0 | 0 | 0 | 0 |
| 0 | 0 | 0 | 0 | 0 |
| 0 | 0 | 0 | 0 | 0 |
| 0 | 0 | 0 | 0 | 0 |
| 0 | 0 | 0 | 0 | 0 |
| 0 | 0 | 0 | 0 | 0 |
| 0 | 0 | 0 | 0 | 0 |
| 0 | 0 | 0 | 0 | 0 |
| 0 | 0 | 0 | 0 | 0 |
| 0.873534147 | 0 | 0 | 0.022689261 | 0 |
| 0 | 0 | 0 | 0 | 0 |
| 0.046319183 | 0.001763116 | 0.003760526 | 0.00176732 | 0.013476642 |
| 0 | 0 | 0 | 0 | 0 |
| 0 | 0 | 0 | 0 | 0 |
| 0 | 0 | 0 | 0 | 0 |
| 0 | 0 | 0 | 0.006982547 | 0 |
| 0 | 0 | 0 | 0 | 0 |
| 0 | 0 | 0 | 0 | 0 |
| 0 | 0.003174452 | 0 | 0.123726862 | 0 |
| 0 | 0 | 0 | 0.015623377 | 0 |
| 0 | 0.000891728 | 0 | 0 | 0 |
| 0 | 0 | 0 | 0 | 0 |
| 0 | 0 | 0 | 0 | 0 |
| 0 | 0 | 0 | 0 | 0 |
| 0 | 0 | 0 | 0 | 0 |
| 0 | 0 | 0 | 0 | 0 |
| 0 | 0 | 0 | 0 | 0 |
| 0 | 0 | 0 | 0 | 0 |
| 0 | 0 | 0 | 0 | 0 |
| 0 | 0 | 0 | 0 | 0 |
| 0 | 0 | 0 | 0 | 0 |
| 0 | 0 | 0.006000392 | 0 | 18.78501951 |
| 0 | 0 | 0 | 0 | 0 |
| 0 | 0 | 0 | 0 | 0 |
| 0 | 0 | 0 | 0 | 0 |
| 0.021451339 | 0 | 0.001358752 | 0.007428231 | 0.394951305 |
| 0 | 0 | 0 | 0 | 0 |
| 0.168806303 | 0.21602169 | 0.072575921 | 0.197369603 | 0.039502253 |
| 0 | 0 | 0 | 0 | 0 |
| 0 | 0 | 0 | 0 | 0 |
| 0 | 0 | 0 | 0 | 0 |
| 0 | 0 | 0 | 0 | 0 |
| 0.073059822 | 0.001183398 | 1.206763143 | 0.001779329 | 0.428596045 |
| 0 | 0 | 0 | 0 | 0 |
| 0 | 0 | 0 | 0 | 0 |

| | | | | |
|---|---|---|---|---|
| 0 | 0 | 0 | 0 | 0 |
| 0 | 0 | 0 | 0 | 0 |
| 0 | 0 | 0 | 0 | 0 |
| 0 | 0 | 0 | 0 | 0 |
| 0 | 0 | 0 | 0 | 0 |
| 0 | 0 | 0 | 0 | 0 |
| 0 | 0 | 0 | 0 | 0 |
| 0 | 0 | 0.001076899 | 0 | 0.002300443 |
| 0 | 0 | 0 | 0 | 0 |
| 0 | 0.064812434 | 0 | 0.006960746 | 0.007285346 |
| 0 | 0.042110926 | 0 | 0 | 0 |
| 0 | 0 | 0 | 0 | 0 |
| 0.005200771 | 0 | 0 | 0 | 0 |
| 0 | 0 | 0 | 0 | 0 |
| 0 | 0 | 0 | 0 | 0 |
| 0 | 0 | 0 | 0 | 0 |
| 0 | 0 | 0 | 0 | 0 |
| 0 | 0 | 0 | 0 | 0 |
| 0 | 0 | 0 | 0 | 0 |
| 1.735674436 | 1.091197116 | 0.057814118 | 1.174360301 | 0.870120401 |
| 0 | 0 | 0 | 0 | 0 |
| 0 | 0 | 0 | 0 | 0 |
| 0 | 0 | 0 | 0 | 0 |
| 0 | 0 | 0 | 0 | 0 |
| 0 | 0 | 0 | 0 | 0 |
| 0 | 0 | 0 | 0 | 0 |
| 0 | 0 | 0 | 0 | 0 |
| 0 | 0 | 0 | 0 | 0 |
| 0 | 0 | 0 | 0 | 0 |
| 0 | 0 | 0.002537417 | 0 | 0 |
| 0 | 0 | 0 | 0 | 0 |
| 0 | 0 | 0 | 0 | 0 |
| 0 | 0 | 0 | 0 | 0 |
| 0 | 0 | 0 | 0 | 0 |
| 0 | 0 | 0 | 0 | 0 |
| 0.001141372 | 0 | 0 | 0.002799599 | 0 |
| 0 | 0 | 0 | 0 | 0 |
| 0 | 0 | 0 | 0 | 0 |
| 0 | 0 | 0 | 0 | 0 |
| 0 | 0 | 0 | 0 | 0 |
| 0 | 0 | 0 | 0 | 0 |
| 0 | 0 | 0 | 0 | 0 |
| 0 | 0 | 0 | 0 | 0 |
| 0 | 0 | 0 | 0 | 0 |
| 0 | 0 | 0 | 0 | 0.040394725 |
| 0 | 0 | 0 | 0 | 0 |
| 0 | 0 | 0 | 0 | 0 |
| 0 | 0 | 0 | 0 | 0 |

| | | | | |
|---|---|---|---|---|
| 0 | 0 | 0 | 0 | 0 |
| 0 | 0 | 0 | 0 | 0 |
| 0 | 0 | 0 | 0 | 0 |
| 0 | 0 | 0 | 0 | 0 |
| 0.009585644 | 0 | 0 | 0 | 0 |
| 0.235705766 | 0 | 0 | 0.000454936 | 0.056661973 |
| 0 | 0 | 0 | 0 | 0 |
| 0 | 0 | 0 | 0 | 0 |
| 0 | 0 | 0 | 0 | 0 |
| 0 | 0 | 0 | 0 | 0 |
| 0 | 0 | 0 | 0 | 0 |
| 0 | 0 | 0 | 0 | 0 |
| 0 | 0 | 0 | 0 | 0 |
| 0 | 0 | 0 | 0 | 0 |
| 0 | 0 | 0 | 0 | 0 |
| 0 | 0 | 0 | 0 | 0 |
| 0.024185126 | 0 | 0 | 0.056838918 | 0 |
| 0 | 0 | 0 | 0 | 0 |
| 0 | 0 | 0 | 0 | 0 |
| 0 | 0 | 0 | 0 | 0 |
| 0 | 0 | 0 | 0 | 0 |
| 0 | 0 | 0 | 0 | 0 |
| 0 | 0 | 0 | 0 | 0 |
| 0 | 0 | 0 | 0 | 0 |
| 0 | 0 | 0 | 0 | 0 |
| 0 | 0 | 0 | 0.000711596 | 0 |
| 0 | 0 | 0 | 0 | 0 |
| 0 | 0 | 0 | 0 | 0 |
| 0 | 0 | 0 | 0 | 0 |
| 0 | 0 | 0 | 0 | 0 |
| 0 | 0 | 0 | 0 | 0 |
| 0 | 0 | 0 | 0 | 0 |
| 0 | 0 | 0 | 0 | 0 |
| 0 | 0 | 0 | 0 | 0 |
| 0 | 0 | 0 | 0 | 0 |
| 0 | 0 | 0 | 0 | 0 |
| 0 | 0 | 0 | 0 | 0 |
| 0 | 0 | 0 | 0 | 0 |
| 0 | 0.612395454 | 0 | 0.466438869 | 0 |
| 0 | 0 | 0 | 0 | 0 |
| 0 | 0 | 0 | 0 | 0 |
| 0 | 0 | 0 | 0 | 0 |
| 0 | 0 | 0 | 0 | 0 |
| 0 | 0 | 0 | 0 | 0 |
| 0 | 0 | 0 | 0 | 0 |
| 0 | 0 | 0 | 0 | 0 |
| 0 | 0 | 0 | 0 | 0 |
| 0 | 0 | 0 | 0 | 0 |

| | | | | |
|---|---|---|---|---|
| 0 | 0 | 0 | 0 | 0 |
| 0.057969631 | 0 | 0 | 0 | 0 |
| 0 | 0 | 0 | 0 | 0 |
| 0 | 0 | 0 | 0 | 0 |
| 0 | 0 | 0 | 0 | 0 |
| 0.002372943 | 0 | 0 | 0 | 0 |
| 0 | 0 | 0 | 0 | 0 |
| 0 | 0 | 0 | 0 | 0 |
| 0 | 0 | 0 | 0 | 0 |
| 0.035007841 | 0 | 0 | 0 | 0 |
| 0.002945631 | 0 | 0 | 0 | 0.002094115 |
| 0 | 0 | 0 | 0 | 0 |
| 0.033879231 | 0.273102046 | 0 | 0.012772055 | 0 |
| 0 | 0 | 0 | 0 | 0 |
| 0 | 0 | 0 | 0 | 0 |
| 0 | 0 | 0 | 0 | 0 |
| 0 | 0 | 0 | 0 | 0 |
| 0 | 0 | 0 | 0 | 0 |
| 0 | 0 | 0 | 0 | 0 |
| 0.002639645 | 0 | 0 | 0 | 0.000774463 |
| 0.041624958 | 0 | 0 | 0.000676901 | 0.006848514 |
| 0 | 0 | 0 | 0 | 0 |
| 0 | 0 | 0 | 0 | 0 |
| 1.200967657 | 2.836616625 | 0 | 0.357568088 | 0 |
| 0 | 0 | 0 | 0 | 0 |
| 0 | 0 | 0 | 0 | 0 |
| 0 | 0 | 0 | 0 | 0 |
| 0 | 0 | 0 | 0 | 0 |
| 0 | 0 | 0 | 0 | 0 |
| 0 | 0 | 0 | 0 | 0 |
| 0 | 0 | 0 | 0 | 0 |
| 0 | 0 | 0 | 0 | 0 |
| 0 | 0 | 0 | 0 | 0 |
| 0 | 0 | 0 | 0 | 0 |

| PHL018-0024_V1 | PHL018-0019_V3 | PHL018-0014_V1 | PHL018-0093_V2 | PHL018-0008_V3 |
|---|---|---|---|---|
| EYCF | CTRL | CTRL | CTRL | CTRL |
| 0.064626105 | 0.029245444 | 0.068813854 | 0.225546222 | 0.420643212 |
| 0.644104153 | 0.087268701 | 1.314577306 | 1.99580469 | 0.78476302 |
| 0 | 0 | 0 | 0 | 2.694661336 |
| 0 | 0 | 0 | 0 | 0 |
| 0 | 0 | 0 | 0 | 0 |
| 0 | 0 | 0 | 0 | 0 |
| 0 | 0 | 0 | 0 | 0 |
| 0 | 0 | 0 | 0 | 0 |
| 0 | 0 | 0 | 0 | 0 |
| 0 | 0 | 0 | 0 | 0 |
| 0 | 0 | 0 | 0 | 0 |
| 0 | 0 | 0 | 0 | 0 |
| 0 | 0 | 0 | 0 | 0 |
| 0.522289059 | 4.819552729 | 0.203684563 | 14.15305378 | 2.270273182 |
| 0 | 0 | 0 | 0 | 0 |
| 0 | 0 | 0 | 0 | 0 |
| 0 | 0 | 0 | 0 | 0 |
| 0 | 0 | 0 | 0 | 0 |
| 0 | 0 | 0 | 0 | 0 |
| 0 | 0 | 0 | 0 | 0 |
| 0 | 0 | 0 | 0 | 0 |
| 0.027362899 | 0 | 0 | 0 | 0 |
| 0 | 0 | 0 | 0 | 0 |
| 0 | 0 | 0 | 0 | 0 |
| 0 | 0 | 0.041082168 | 0 | 0 |
| 0 | 0 | 0 | 0 | 0 |
| 0 | 0 | 0 | 0 | 0 |
| 0 | 0 | 0 | 0 | 0 |
| 0 | 0 | 0 | 0 | 0 |
| 0 | 0 | 0 | 0 | 0 |
| 0 | 0 | 0 | 0 | 0 |
| 0 | 0 | 0 | 0 | 0 |
| 0 | 0 | 0 | 0 | 0 |
| 0 | 0 | 0 | 0 | 0 |
| 0 | 0 | 0 | 0 | 0 |
| 0.144308132 | 10.11511569 | 5.182144894 | 0 | 2.891828439 |
| 0 | 0 | 0 | 0 | 0 |
| 0 | 0 | 0 | 0 | 0 |
| 0 | 0 | 0 | 0 | 0 |
| 0 | 0 | 0 | 0 | 0 |
| 0 | 0 | 0 | 0 | 0 |
| 0.048985573 | 0.01726829 | 0.020773952 | 0 | 0.106424175 |
| 0.043704189 | 0 | 0.089105148 | 0 | 1.007177209 |
| 0.045113572 | 0.011080058 | 0.11316857 | 0 | 0.01299124 |
| 0 | 0 | 0.003547415 | 0 | 0 |
| 0 | 0 | 0 | 0 | 0 |

| | | | | |
|---|---|---|---|---|
| 0 | 0.008244096 | 0 | 0 | 0 |
| 0.002357726 | 0.031391812 | 0 | 0.007684015 | 0.024429976 |
| 0.006628136 | 0.001744933 | 0.001609367 | 0 | 0.011542616 |
| 1.08642671 | 0.284832038 | 0.833491115 | 0.311211946 | 1.568320162 |
| 0.003278499 | 0.02875857 | 0.002571232 | 0 | 0.001525216 |
| 0 | 0 | 0 | 0 | 0 |
| 0.003032522 | 0 | 2.366062046 | 0 | 1.554800729 |
| 0 | 0 | 0 | 0 | 0 |
| 0 | 0 | 0 | 0 | 0 |
| 0 | 0 | 0 | 0 | 0 |
| 0 | 0 | 0 | 0 | 0.051800371 |
| 0.094165621 | 0.740500878 | 0.65149975 | 0 | 0.001885655 |
| 0 | 0 | 0 | 0 | 0 |
| 0.002714746 | 0.009112271 | 0.134842767 | 0 | 0.050326574 |
| 0 | 0 | 0 | 0 | 0 |
| 0 | 0 | 0 | 0 | 0 |
| 0 | 0 | 0.00128016 | 0 | 5.059005725 |
| 0 | 0 | 0 | 0 | 0 |
| 0 | 0 | 0 | 0 | 0 |
| 0 | 0 | 0 | 0 | 0 |
| 0.113981041 | 1.28170876 | 0.267212988 | 0.768968335 | 0.394095883 |
| 0.029986332 | 0.02933161 | 0.014638161 | 0.042670587 | 0.00550565 |
| 0 | 0 | 0 | 0 | 0 |
| 0 | 0 | 0 | 0 | 0 |
| 0 | 0 | 0 | 0 | 0 |
| 0 | 0 | 0 | 0 | 0 |
| 0 | 0.001919067 | 0 | 0 | 0 |
| 0 | 0 | 0 | 0 | 0 |
| 0 | 0 | 0.003792697 | 0 | 0.002185857 |
| 0 | 0 | 0 | 0 | 0 |
| 0.007324561 | 0 | 0 | 0 | 0.002581452 |
| 0 | 0 | 0 | 0 | 0 |
| 0 | 0 | 0 | 0 | 0 |
| 0 | 0 | 0 | 0 | 0 |
| 0 | 0 | 0 | 0 | 0 |
| 0 | 0 | 0 | 0 | 0 |
| 0 | 0 | 0 | 0 | 0 |
| 0 | 0 | 0.003819687 | 0 | 0.11303462 |
| 0 | 0 | 0 | 0 | 0 |
| 0 | 0 | 0 | 0 | 0 |
| 0 | 0 | 0 | 0 | 0 |
| 0 | 0 | 0 | 0 | 0 |
| 0 | 0 | 0 | 0 | 0 |
| 0 | 0 | 0 | 0 | 0 |
| 0 | 0 | 0 | 0 | 0 |
| 0 | 0 | 0 | 0 | 0 |
| 0 | 0 | 0 | 0 | 0.006337421 |
| 0.561653334 | 0 | 0 | 0 | 0.052555921 |

| | | | | |
|---|---|---|---|---|
| 0 | 0 | 0 | 0 | 0 |
| 0 | 0 | 0 | 0 | 0 |
| 0 | 0 | 0 | 0 | 0 |
| 0 | 0 | 0 | 0 | 0 |
| 0 | 0 | 0 | 0 | 0.03241482 |
| 0 | 0 | 0 | 0 | 0 |
| 0 | 0 | 0 | 0 | 0 |
| 0 | 0 | 0 | 2.001784853 | 0.015884172 |
| 0.003124039 | 0 | 0.005193911 | 0 | 0.001835049 |
| 0 | 0 | 0 | 0 | 0 |
| 0 | 0 | 0 | 0 | 0 |
| 0.051784752 | 0 | 0.015594899 | 0 | 0 |
| 0 | 0 | 0 | 0 | 0 |
| 0 | 0 | 0 | 0 | 0 |
| 0 | 0 | 0 | 0 | 0 |
| 0 | 0 | 0 | 0 | 0 |
| 0 | 0 | 0 | 0 | 0 |
| 0 | 0 | 0 | 0 | 0 |
| 0.010580056 | 0 | 0.001110299 | 0 | 0.002085606 |
| 0 | 0 | 0 | 0 | 0 |
| 0 | 0 | 0 | 0 | 0 |
| 0 | 0 | 0 | 0 | 0 |
| 0 | 0 | 0 | 0 | 0 |
| 0 | 0 | 0.005766808 | 0 | 0 |
| 0 | 0 | 0 | 0 | 0 |
| 0 | 0 | 0 | 0 | 0 |
| 0 | 0 | 0 | 0 | 0 |
| 0 | 0 | 0.001928807 | 0 | 0 |
| 0 | 0 | 0 | 0 | 0 |
| 0 | 0 | 0 | 0 | 0 |
| 0.022324131 | 0 | 0.025191608 | 0 | 0.003905632 |
| 0 | 0.000754063 | 0.043293549 | 0 | 0.015320518 |
| 0 | 0 | 0 | 0 | 0 |
| 0 | 0 | 0 | 0 | 0 |
| 0 | 0 | 0 | 0 | 0 |
| 0 | 0 | 0 | 0 | 0 |
| 0 | 0 | 0 | 0 | 0 |
| 0 | 0 | 0 | 0 | 0 |
| 0 | 0 | 0 | 0 | 0 |
| 0 | 0 | 0 | 0 | 0 |
| 0 | 0 | 0 | 0 | 0 |
| 0 | 0 | 0 | 0 | 0 |
| 0 | 0 | 0.000803863 | 0 | 0 |
| 0 | 0 | 0 | 0 | 0 |
| 0 | 0 | 0 | 0 | 0 |
| 0 | 0 | 0 | 0 | 0 |
| 0 | 0 | 0.00653146 | 0 | 0 |
| 0 | 0 | 0 | 0 | 0 |

| | | | | |
|---|---|---|---|---|
| 0 | 0 | 0 | 0 | 0 |
| 0 | 0 | 0 | 0 | 0 |
| 0 | 0 | 0 | 0 | 0 |
| 0 | 0 | 0 | 0 | 0 |
| 0.007479177 | 0 | 0.196885372 | 0 | 0 |
| 0 | 0 | 0 | 0 | 0 |
| 0 | 0 | 0 | 0 | 0 |
| 0 | 0 | 0 | 0 | 0 |
| 0 | 0 | 0 | 0 | 0 |
| 0 | 0 | 0 | 0 | 0 |
| 0 | 0 | 0 | 0 | 0 |
| 0 | 0 | 0 | 0 | 0 |
| 0 | 0 | 0 | 0 | 0 |
| 0 | 0 | 0.001780136 | 0 | 0 |
| 0 | 0 | 0 | 0 | 0 |
| 0 | 0 | 0 | 0 | 0 |
| 0 | 0 | 0 | 0 | 0 |
| 0 | 0 | 0 | 0 | 0 |
| 0 | 0 | 0 | 0 | 0 |
| 0 | 0 | 0 | 0 | 0 |
| 0 | 0 | 0 | 0 | 0 |
| 0 | 0 | 0 | 0 | 0 |
| 0 | 0 | 0.00265712 | 0 | 0 |
| 0 | 0 | 0 | 0 | 0 |
| 0 | 0 | 0 | 0 | 0 |
| 0 | 0 | 0 | 0 | 0 |
| 0 | 0 | 0 | 0 | 0 |
| 0 | 0 | 0 | 0 | 0 |
| 0 | 0 | 0 | 0 | 0 |
| 0 | 0 | 0 | 0 | 0 |
| 0 | 0 | 0 | 0 | 0 |
| 0.088840725 | 0 | 0.001203649 | 0 | 0.111403511 |
| 0 | 0 | 0 | 0 | 0 |
| 0 | 0 | 0 | 0 | 0 |
| 0 | 0 | 0 | 0 | 0 |
| 0 | 0 | 0 | 0 | 0 |
| 0 | 0 | 0 | 0 | 0 |
| 0 | 0 | 0 | 0 | 0 |
| 0 | 0 | 0 | 0 | 0 |
| 0 | 0 | 0 | 0 | 0 |
| 0 | 0 | 0 | 0 | 0 |
| 0 | 0 | 0 | 0 | 0 |
| 0 | 0 | 0 | 0 | 0 |
| 0 | 0 | 0 | 0 | 0 |
| 0 | 0 | 0 | 0 | 0 |
| 0 | 0 | 0 | 0 | 0 |
| 0 | 0 | 0 | 0 | 0 |
| 0.005243439 | 0 | 0 | 0.359576517 | 0 |

| | | | | |
|---|---|---|---|---|
| 1.700981429 | 0 | 0 | 0 | 0 |
| 0 | 0 | 0 | 0 | 0 |
| 0 | 0 | 0 | 0 | 0 |
| 0 | 0 | 0 | 0 | 0 |
| 0 | 0 | 0 | 0 | 0 |
| 0 | 0 | 0 | 0 | 0 |
| 0 | 0 | 0 | 0 | 0 |
| 0 | 0 | 0 | 0 | 0 |
| 0 | 0 | 0 | 0 | 0 |
| 0 | 0 | 0 | 0 | 0 |
| 0 | 0 | 0 | 0 | 0 |
| 0 | 0 | 0 | 0 | 0 |
| 0.001674194 | 0 | 0 | 0.001768256 | 0 |
| 0 | 0 | 0 | 0 | 0 |
| 0 | 0 | 0 | 0 | 0 |
| 0 | 0 | 0 | 0 | 0 |
| 0 | 0 | 0 | 0 | 0 |
| 0 | 0.000750927 | 0 | 0 | 0 |
| 0 | 0 | 0 | 0 | 0 |
| 0 | 0 | 0 | 0 | 0 |
| 0.11790951 | 0.179012409 | 0.109830535 | 0.015114118 | 0.147100051 |
| 0 | 0 | 0 | 0 | 0 |
| 0 | 0 | 0 | 0 | 0 |
| 0 | 0 | 0 | 0 | 0 |
| 0 | 0 | 0 | 0 | 0 |
| 0 | 0 | 0 | 0 | 0 |
| 0.038486332 | 0.001586453 | 0.050166835 | 0 | 0.002570028 |
| 0 | 0 | 0 | 0 | 0 |
| 0 | 0 | 0 | 0 | 0 |
| 0 | 0 | 0.002683637 | 0 | 0 |
| 0 | 0.022139936 | 0.015002345 | 0.212500278 | 0.002561882 |
| 0 | 0 | 0 | 0 | 0 |
| 0 | 0 | 0 | 0 | 0 |
| 0 | 0 | 0.003509626 | 0 | 0.006967127 |
| 0.004752456 | 0.008862229 | 0.006071901 | 0.106023358 | 0.028952618 |
| 0 | 0 | 0 | 0 | 0 |
| 0 | 0 | 0 | 0 | 0 |
| 0 | 0 | 0 | 0 | 0 |
| 0 | 0 | 0.003169003 | 0 | 0 |
| 0 | 0 | 0 | 0 | 0 |
| 0 | 0 | 0 | 0 | 0 |
| 0 | 0 | 0 | 0 | 0 |
| 0 | 0 | 0 | 0 | 0 |
| 0 | 0 | 0 | 0 | 0 |
| 0 | 0 | 0 | 0 | 0 |
| 0 | 0 | 0 | 0 | 0 |
| 0 | 0 | 0 | 0 | 0 |
| 0.000645091 | 0 | 0 | 0 | 0 |

| | | | | |
|---|---|---|---|---|
| 0 | 0 | 0 | 0 | 0 |
| 0 | 0 | 0 | 0 | 0 |
| 0 | 0 | 0 | 0 | 0 |
| 0 | 0 | 0 | 0 | 0 |
| 0 | 0 | 0 | 0 | 0 |
| 0 | 0 | 0 | 0 | 0 |
| 0 | 0 | 0.001658308 | 0.010287558 | 0 |
| 0.002781991 | 0 | 0.005023961 | 0 | 0 |
| 0 | 0 | 0 | 0 | 0.01092041 |
| 0 | 0 | 0 | 0 | 0 |
| 0 | 0 | 0 | 0 | 0 |
| 0 | 0 | 0 | 0 | 0 |
| 0 | 0 | 0 | 0 | 0 |
| 0 | 0 | 0 | 0 | 0.18099822 |
| 0 | 0 | 0 | 0 | 0.033068177 |
| 0 | 0 | 0 | 0 | 0 |
| 0 | 0 | 0 | 0 | 0 |
| 0 | 0 | 0 | 0 | 0 |
| 0 | 0 | 0 | 0 | 0.005375818 |
| 0 | 0 | 0 | 0 | 0 |
| 0.010161328 | 0 | 0 | 0 | 0 |
| 0 | 0 | 0 | 0 | 0 |
| 0 | 0 | 0 | 0 | 0 |
| 0 | 0 | 0 | 0 | 0 |
| 0.001922631 | 0.009680207 | 0.009920156 | 0 | 0.003049235 |
| 0 | 0 | 0 | 0 | 0 |
| 0 | 0 | 0 | 0 | 0 |
| 0 | 0 | 0 | 0 | 0 |
| 0.012155515 | 0 | 0.029584215 | 0 | 0 |
| 0 | 0 | 0 | 0 | 0 |
| 0.568910532 | 0.02965705 | 0.500247077 | 1.572401594 | 0.068841496 |
| 0 | 0 | 0 | 0 | 0 |
| 0 | 0 | 0 | 0 | 0 |
| 0 | 0 | 0 | 0 | 0.004866967 |
| 0 | 0 | 0 | 0 | 0 |
| 0 | 0 | 0 | 0 | 0 |
| 0 | 0 | 0 | 0 | 0 |
| 0 | 0 | 0 | 0 | 0 |
| 0 | 0 | 0 | 0 | 0 |
| 0 | 0 | 0 | 0 | 0 |
| 0 | 0 | 0 | 0 | 0 |
| 0.002176021 | 0.019274436 | 0.021893758 | 0.011080978 | 0.006327025 |
| 0 | 0 | 0 | 0 | 0 |
| 0 | 0 | 0 | 0 | 0 |
| 0 | 0 | 0 | 0 | 0 |
| 0 | 0 | 0 | 0 | 0 |
| 0 | 0 | 0 | 0 | 0 |
| 0 | 0 | 0 | 0 | 0 |

| | | | | |
|---|---|---|---|---|
| 0 | 0 | 0 | 0 | 0 |
| 0 | 0 | 0 | 0 | 0 |
| 0.041668381 | 0.010445171 | 0.126882532 | 0 | 0.010352082 |
| 0 | 0 | 0 | 0 | 0 |
| 0 | 0 | 0 | 0 | 0 |
| 0 | 0 | 0 | 0 | 0 |
| 0 | 0 | 0 | 0 | 0 |
| 0 | 0 | 0 | 0 | 0 |
| 0 | 0 | 0 | 0 | 0 |
| 0 | 0 | 0 | 0 | 0 |
| 0 | 0 | 0 | 0 | 0 |
| 0 | 0 | 0 | 0 | 0 |
| 0 | 0 | 0 | 0 | 0 |
| 0.000754936 | 0 | 0.005453315 | 0 | 0.000798204 |
| 0.005192506 | 0.004182979 | 0.004072331 | 0 | 0.002196042 |
| 0 | 0 | 0 | 0 | 0 |
| 0.002603214 | 0.006619624 | 0.003174775 | 0 | 0.001251097 |
| 0 | 0 | 0 | 0 | 0 |
| 0 | 0 | 0 | 0 | 0 |
| 0 | 0 | 0 | 0 | 0 |
| 0 | 0 | 0 | 0 | 0 |
| 0 | 0 | 0 | 0 | 0 |
| 0 | 0 | 0 | 0 | 0 |
| 0 | 0 | 0 | 0 | 0 |
| 0 | 0 | 0.019795265 | 0 | 0.00833014 |
| 0 | 0 | 0 | 0 | 0 |
| 0 | 0 | 0 | 0 | 0.01388984 |
| 0 | 0 | 0 | 0 | 0 |
| 0 | 0 | 0 | 0 | 0 |
| 0 | 0 | 0.003826684 | 0 | 0.00751485 |
| 0 | 0 | 0 | 0 | 0 |
| 0 | 0 | 0 | 0 | 0 |
| 0 | 0 | 0 | 0 | 0 |
| 0 | 0 | 0 | 0 | 0.00104174 |
| 0 | 0 | 0 | 0 | 0 |
| 0.029442357 | 0 | 0.206513696 | 0.008691574 | 0.007218503 |
| 0 | 0 | 0 | 0 | 0 |
| 0.001610444 | 0 | 0.012464055 | 0.002915869 | 0.003405487 |
| 0 | 0 | 0 | 0 | 0 |
| 0 | 0 | 0 | 0 | 0 |
| 0 | 0 | 0 | 0 | 0 |
| 0 | 0 | 0 | 0 | 0 |
| 0 | 0 | 0 | 0 | 0 |
| 0 | 0 | 0 | 0 | 0 |
| 0 | 0 | 0 | 0 | 0 |
| 0 | 0 | 0 | 0 | 0.004363236 |
| 0 | 0 | 0 | 0 | 0.009228559 |
| 0 | 0.043286406 | 0.00335491 | 0 | 0 |

| | | | | |
|---|---|---|---|---|
| 0.094304088 | 0 | 0 | 0 | 0.033085248 |
| 0 | 0 | 0.004476549 | 0 | 0.002402525 |
| 0 | 0 | 0 | 0 | 0 |
| 0 | 0 | 0 | 0 | 0 |
| 0.000675383 | 0 | 0.005807931 | 0.00152856 | 0 |
| 0.004772185 | 0 | 0 | 0 | 0.005433825 |
| 0 | 0 | 0 | 0 | 0 |
| 0.083309428 | 0 | 0.078480147 | 0 | 0.072068864 |
| 0 | 0 | 0 | 0 | 0.00655634 |
| 0.054643802 | 0.060410981 | 0 | 0 | 0.008941465 |
| 0 | 0 | 0 | 0.001198028 | 0 |
| 0.100920487 | 0 | 0.062382946 | 0 | 0.002112962 |
| 0.077688679 | 0.002332076 | 0.004497325 | 0.002058484 | 0.097968496 |
| 0.002302226 | 0 | 0 | 0 | 0 |
| 0 | 0 | 0.001809938 | 0 | 0 |
| 0 | 0 | 0 | 0 | 0.01353377 |
| 0 | 0 | 0 | 0 | 0 |
| 0 | 0 | 0 | 0 | 0 |
| 0.02846399 | 0.003081992 | 0 | 0.056524331 | 0.102906701 |
| 0.011285161 | 0 | 0 | 0 | 0 |
| 0 | 0 | 0 | 0 | 0 |
| 0 | 0 | 0 | 0 | 0 |
| 0 | 0 | 0.011702529 | 0 | 0 |
| 0 | 0 | 0 | 0 | 0.005327365 |
| 0 | 0 | 0 | 0 | 0 |
| 0 | 0.004691746 | 0.000910998 | 0 | 0.001166752 |
| 0 | 0 | 0 | 0 | 0 |
| 0 | 0 | 0 | 0.979626745 | 0 |
| 0 | 0 | 0 | 0 | 0 |
| 0 | 0 | 0.020854969 | 0 | 0 |
| 0 | 0 | 0 | 0 | 0 |
| 0 | 0 | 0 | 0 | 0 |
| 0 | 0 | 0 | 0 | 0 |
| 0 | 0 | 0 | 0 | 0 |
| 0 | 0 | 0 | 0 | 0 |
| 0.021485073 | 0 | 0 | 0 | 0.005915743 |
| 0 | 0.069981252 | 0 | 0.129781439 | 0.028304338 |
| 0 | 0 | 0 | 0 | 0 |
| 0 | 0 | 0 | 0 | 0 |
| 0 | 0 | 0 | 0 | 0 |
| 0 | 0 | 0 | 0 | 0 |
| 0 | 0 | 0 | 0.343880837 | 0.003288627 |
| 0 | 0 | 0 | 0 | 0 |
| 0 | 0 | 0 | 0.011348142 | 0 |
| 0 | 0 | 0 | 0 | 0 |
| 0 | 0 | 0 | 0 | 0 |
| 0 | 0.00307082 | 0.009912854 | 0 | 0 |
| 0.015694518 | 0 | 0.019537706 | 0 | 0 |

| | | | | |
|---|---|---|---|---|
| 0 | 0 | 0 | 0 | 0 |
| 0 | 0 | 0.077231634 | 0 | 0.01264798 |
| 0 | 0 | 0 | 0 | 0 |
| 0 | 0 | 0 | 0 | 0 |
| 0 | 0 | 0.010077757 | 0 | 0 |
| 0.008214461 | 0.103034168 | 0.03479947 | 0.010861278 | 0.009599497 |
| 0 | 0 | 0 | 0 | 0 |
| 0 | 0 | 0 | 0 | 0 |
| 0 | 0 | 0 | 0 | 0 |
| 0 | 0 | 0 | 0 | 0 |
| 0.005265138 | 0 | 0.02742514 | 0 | 0 |
| 0 | 0 | 0 | 0 | 0 |
| 0 | 0 | 0 | 0 | 0 |
| 0 | 0 | 0.001466598 | 0 | 0 |
| 0.022289152 | 0 | 0.001978578 | 0 | 0 |
| 0 | 0 | 0 | 0 | 0 |
| 0 | 0 | 0 | 0 | 0 |
| 0.013369653 | 0 | 0 | 0 | 0.001949781 |
| 0 | 0 | 0 | 0 | 0.001332797 |
| 0 | 0 | 0 | 0 | 0.005706649 |
| 0 | 0 | 0.002106932 | 0 | 0.399907797 |
| 0 | 0.22352929 | 0 | 0 | 0.288534217 |
| 0 | 0 | 0 | 0 | 0 |
| 0 | 0 | 0 | 0 | 0 |
| 0 | 0 | 0 | 0 | 0 |
| 0 | 0 | 0 | 0 | 0 |
| 0.017207484 | 0.171550925 | 0 | 0.002790077 | 0.847229081 |
| 0 | 0 | 0 | 0 | 0 |
| 0 | 0 | 0 | 0 | 0 |
| 0.153809395 | 0.102944034 | 0.206373609 | 1.110334256 | 0.093610166 |
| 0 | 0.004494924 | 0 | 0 | 0.000849532 |
| 0 | 0 | 0 | 0 | 0 |
| 0 | 0 | 0 | 0 | 0 |
| 0 | 0 | 0.057543078 | 0 | 0 |
| 0.002139136 | 0 | 0 | 0 | 0.000904695 |
| 0 | 0 | 0 | 0 | 0 |
| 9.691202358 | 0.164706894 | 0 | 0 | 0 |
| 0 | 0 | 0 | 0 | 0 |
| 0 | 0 | 0 | 0 | 0 |
| 0.09249108 | 0 | 0 | 0 | 0 |
| 0.038556865 | 0.021134133 | 0.104494253 | 0.092552522 | 0 |
| 2.183235797 | 0.632608321 | 0.032491269 | 4.862366404 | 4.782294694 |
| 0 | 0 | 0 | 0 | 0 |
| 0.028660823 | 0 | 0 | 0 | 2.086220007 |
| 0.003954014 | 0 | 0 | 0 | 0 |
| 0 | 0 | 0 | 0 | 0 |
| 0.008992933 | 0.002972817 | 0.001898208 | 0 | 0.00172879 |
| 0 | 0 | 0 | 0 | 0.004854478 |

| | | | | |
|---|---|---|---|---|
| 0 | 0.022572046 | 0.0026023 | 0 | 0.001066519 |
| 0 | 0 | 0 | 0 | 0 |
| 0.012713567 | 0 | 0 | 0 | 11.43533587 |
| 0 | 0 | 0 | 0 | 0 |
| 0.083485622 | 0 | 0 | 0 | 0 |
| 0 | 0 | 0 | 0 | 0.194776366 |
| 0 | 0 | 0 | 0 | 0 |
| 0 | 0 | 0 | 0 | 0 |
| 0.007386721 | 0 | 0 | 0 | 0 |
| 0 | 0 | 0 | 0 | 0 |
| 0 | 0 | 0 | 0 | 0 |
| 0 | 0 | 0 | 0 | 0 |
| 0 | 0 | 0 | 0 | 0 |
| 0 | 0 | 0.16809236 | 0.002788197 | 0 |
| 0 | 0 | 0 | 0 | 0 |
| 0.004278858 | 0 | 0.124002104 | 0.03776808 | 0.037135267 |
| 0.000790445 | 0.095293857 | 0.089725664 | 0.001073384 | 0 |
| 0.00072027 | 0 | 0 | 0 | 0 |
| 0.001224704 | 0 | 0 | 0 | 0 |
| 0 | 0 | 0 | 0 | 0.00334373 |
| 0 | 0.043747768 | 0.003854995 | 0.009807578 | 0.00105328 |
| 0.001375312 | 0.005642205 | 0 | 0.001556337 | 0 |
| 0 | 0 | 0 | 0 | 0 |
| 0 | 0 | 0.233883293 | 0 | 0 |
| 0.050788852 | 0 | 0 | 0 | 0 |
| 0 | 0 | 0 | 0 | 0 |
| 0.001756083 | 0 | 0.008025298 | 0.006132014 | 0 |
| 0.386582186 | 0 | 0 | 0 | 0 |
| 0 | 0 | 0 | 0 | 0 |
| 0 | 0 | 0 | 0 | 0 |
| 0.055327689 | 0 | 0.031894227 | 0.014508291 | 0.001815477 |
| 0.003413247 | 0.024186676 | 0.000880563 | 0.001158754 | 0.114882398 |
| 0.532979058 | 0.006363272 | 0.04359756 | 0.022433579 | 0.016607977 |
| 0.007762307 | 0 | 0.007609689 | 0 | 0.005745033 |
| 0 | 0.001239768 | 0 | 0 | 0.0038076 |
| 0 | 0 | 0 | 0 | 0 |
| 0.000734527 | 0.04273536 | 0.001515969 | 0 | 0.025110885 |
| 0.017233028 | 0.00492746 | 0.021537641 | 0.061364159 | 0.024699185 |
| 0.11080754 | 0.023076823 | 1.118508697 | 0 | 0.582933914 |
| 0 | 0 | 0 | 0 | 0.001158678 |
| 0 | 0 | 0 | 0 | 0 |
| 0 | 0 | 0 | 0 | 0 |
| 0 | 0 | 0.001773 | 0.010110255 | 0 |
| 0.195360232 | 0 | 0.09362715 | 0.78998182 | 0.002212546 |
| 0 | 0.802464739 | 0 | 0.006752459 | 0 |
| 0.001878541 | 0 | 0 | 0 | 0 |
| 0 | 0 | 0 | 0 | 0 |
| 0 | 0 | 0 | 0 | 0 |

| | | | | |
|---|---|---|---|---|
| 0 | 0.185821122 | 0 | 0.018890472 | 0 |
| 2.605218222 | 0.004962736 | 0.005384912 | 0.040390997 | 0.028690167 |
| 0 | 0 | 0 | 0 | 0 |
| 0 | 0 | 0 | 0 | 0 |
| 0 | 0 | 0 | 0 | 0 |
| 0 | 0 | 0 | 0 | 0 |
| 0 | 0 | 0.001743611 | 0.005900042 | 0 |
| 0 | 0.004260363 | 0.004963416 | 0 | 0 |
| 0.001671912 | 0 | 0 | 0 | 0 |
| 0.015168041 | 0.017819478 | 0.001956555 | 0 | 0 |
| 0 | 0.003037057 | 0.004201657 | 0.005265777 | 0 |
| 0.424536338 | 0.855318068 | 0.132024277 | 0 | 0.199700422 |
| 0 | 0 | 0 | 0 | 0.022105028 |
| 0 | 0 | 0 | 0 | 0 |
| 0.120796389 | 0.009458318 | 0.002754786 | 0 | 0.013642244 |
| 0 | 0 | 0 | 0 | 0 |
| 0.010760468 | 0.002498758 | 0.00984704 | 0.001378507 | 0.001931975 |
| 0.026092243 | 0.185482539 | 0.06554594 | 0.1453068 | 0.020899754 |
| 0 | 0 | 0 | 0 | 0 |
| 0.008385899 | 0 | 0 | 0.115967898 | 0.001266646 |
| 0.081864758 | 0.043806444 | 0.015556262 | 0 | 0.013946476 |
| 0 | 0 | 0 | 0 | 0 |
| 0 | 0 | 0 | 0 | 0 |
| 0 | 0 | 0 | 0 | 0 |
| 0 | 0 | 1.192519914 | 0 | 0.004463368 |
| 0 | 0 | 0 | 0 | 0 |
| 0.043398894 | 0 | 0 | 0 | 0.052441404 |
| 0.107038065 | 0.064486545 | 0.016074732 | 0.155626694 | 0.045880853 |
| 0 | 0 | 0 | 0 | 0.002752599 |
| 0 | 0 | 0 | 0.001093528 | 0.001135242 |
| 0 | 0.000801476 | 0.001478417 | 0 | 0 |
| 0 | 0 | 0 | 0 | 0 |
| 0 | 0 | 0 | 0 | 0 |
| 0 | 0 | 0 | 0 | 0 |
| 0 | 0 | 0 | 0 | 0.012097109 |
| 0.107714105 | 0.33331833 | 0.186552176 | 0.049097736 | 0.041015449 |
| 0 | 0 | 0 | 0 | 0.009405732 |
| 0 | 0 | 0 | 0 | 0 |
| 0.047217192 | 0.003071481 | 0.035410679 | 0 | 0.009288068 |
| 0 | 0 | 0 | 0 | 0 |
| 0 | 0.762853225 | 0 | 0 | 0 |
| 0 | 0 | 0 | 0 | 0 |
| 0.001333974 | 0 | 0.011012602 | 0.005887277 | 0 |
| 0.00563468 | 0.002701896 | 0 | 0 | 0.000851089 |
| 0.008460187 | 0 | 0 | 0.097077895 | 0 |
| 0 | 0 | 0 | 0 | 0 |
| 0 | 0 | 0 | 0 | 0 |
| 0 | 0 | 0 | 0 | 0 |

| | | | | |
|---|---|---|---|---|
| 0 | 0 | 0 | 0 | 0 |
| 0 | 0 | 0 | 0 | 0 |
| 0 | 0 | 0 | 0 | 0 |
| 1.923428267 | 0 | 0.056114608 | 0 | 1.622579879 |
| 0 | 0 | 0 | 0 | 0 |
| 0 | 0 | 0 | 0 | 0 |
| 0 | 0 | 0 | 0 | 0 |
| 0 | 0 | 0 | 0 | 0 |
| 0.012163945 | 0.225499724 | 0.296929335 | 0 | 0 |
| 0.080885246 | 0 | 0.945198506 | 0 | 0.171225616 |
| 0 | 0 | 0 | 0 | 0 |
| 0.000558214 | 0 | 0.004032287 | 0 | 0.000786943 |
| 0 | 0 | 0 | 0 | 0 |
| 0 | 0 | 0 | 0 | 0.201368163 |
| 0.028406118 | 0.185712879 | 0.008873209 | 0.025229533 | 0.056172009 |
| 0 | 0 | 0 | 0 | 0 |
| 0 | 0 | 0 | 0 | 0 |
| 0 | 0 | 0 | 0 | 0 |
| 0 | 0 | 0 | 0 | 0 |
| 0 | 0 | 0 | 0 | 0 |
| 0 | 0 | 0 | 0 | 0 |
| 0 | 0 | 0 | 0 | 0 |
| 0 | 0 | 0 | 0 | 0 |
| 0.507461075 | 0.007612302 | 0.153543889 | 0.418163798 | 0 |
| 0 | 0 | 0.016898743 | 0.17387816 | 0 |
| 0.509719755 | 0 | 0 | 0 | 0.161926826 |
| 0 | 0 | 0 | 0 | 0 |
| 3.933424273 | 19.68746339 | 1.042324633 | 0 | 1.115141168 |
| 0 | 0 | 0 | 0 | 0 |
| 0 | 0 | 0 | 0 | 0 |
| 0.001989824 | 0 | 0 | 0 | 0 |
| 0 | 0 | 0 | 0 | 0.013695432 |
| 0 | 0 | 0.010797289 | 0 | 0.004512467 |
| 0 | 0 | 0 | 0 | 0 |
| 0 | 0 | 0 | 0 | 0 |
| 0.036043627 | 0.033120288 | 0 | 0 | 0 |
| 0.090327855 | 0 | 0.617124098 | 0 | 1.757624218 |
| 0 | 0 | 0.00439527 | 0 | 0 |
| 0 | 0 | 0 | 0 | 0 |
| 0.01185346 | 0 | 0 | 0 | 0.000985727 |
| 0.471245903 | 0 | 0.125442099 | 0.000969588 | 0.421062854 |
| 0.007286505 | 0 | 0 | 0 | 0 |
| 2.33685901 | 2.628756289 | 1.194584664 | 0.438503825 | 0.181579335 |
| 0.001088491 | 0 | 0 | 0 | 0 |
| 0 | 0 | 0 | 0 | 0 |
| 0 | 0 | 0 | 0 | 0 |
| 0 | 0 | 0 | 0 | 0 |
| 0.001502061 | 0 | 0 | 0 | 0.063299092 |

| | | | | |
|---|---|---|---|---|
| 0 | 0 | 0 | 0 | 0 |
| 0 | 0.000656174 | 0 | 0.157685439 | 0 |
| 0.00287979 | 2.956033213 | 0.016344668 | 0.362848793 | 0.000652466 |
| 0 | 0 | 0 | 0 | 0 |
| 0 | 0 | 0 | 0 | 0.024514164 |
| 0 | 0 | 0.001170518 | 0 | 0.001713292 |
| 0 | 0 | 0 | 0 | 0 |
| 0 | 0 | 0 | 0 | 0.000520878 |
| 0 | 0 | 0 | 0 | 0 |
| 0 | 0 | 0 | 0 | 0 |
| 0.004847046 | 0 | 0.052102486 | 0.686726663 | 0.078580975 |
| 3.629685733 | 2.423283531 | 0.351085837 | 5.598428424 | 1.139528268 |
| 0 | 0 | 0 | 0 | 0 |
| 0.000653891 | 0 | 0.02744079 | 0.002071887 | 0 |
| 0.624189576 | 0.084366936 | 0.033677336 | 0.18053285 | 0.002542514 |
| 0 | 0 | 0 | 0 | 0 |
| 0 | 0 | 0 | 0 | 0 |
| 0 | 0 | 0 | 0 | 0 |
| 0 | 0 | 0 | 0 | 0 |
| 0.120778877 | 0 | 0.009098772 | 0 | 0.000879484 |
| 0.512218369 | 0 | 0.133454162 | 0.007125767 | 0.193649859 |
| 0.899472631 | 2.058269708 | 0.004785473 | 0 | 0 |
| 4.98287964 | 0.899079272 | 0.019138586 | 0.055146345 | 0.139293304 |
| 0 | 0 | 0 | 0 | 0 |
| 0 | 0 | 0 | 0 | 0 |
| 0.584582413 | 0.017248111 | 0.014024233 | 0 | 0.00471822 |
| 0 | 0 | 0 | 0 | 0 |
| 0.009003952 | 0 | 0 | 0 | 0 |
| 0 | 0 | 0 | 0 | 0 |
| 0 | 0 | 0 | 0 | 0 |
| 0 | 0 | 0 | 0 | 0 |
| 0 | 0 | 0.086907376 | 0 | 0.11084115 |
| 0.016178638 | 0 | 0.07106209 | 0.602760546 | 0.060089919 |
| 0.009490357 | 0 | 0 | 0.00923599 | 0.012208375 |
| 0.010130507 | 0.001378536 | 0.014550879 | 0.003346229 | 0.007671478 |
| 0.007711711 | 0.00294626 | 0.007569793 | 0.038823452 | 0.024063342 |
| 0 | 0.006708539 | 0 | 0 | 0.030160685 |
| 0 | 0 | 0 | 0 | 0 |
| 0.047542436 | 0.035462255 | 0.049969258 | 0.026900099 | 0 |
| 0 | 0 | 0 | 0 | 0 |
| 0 | 0 | 0 | 0 | 0.000525347 |
| 0 | 0 | 0 | 0 | 0 |
| 0 | 0 | 0 | 0 | 0 |
| 0 | 0 | 0 | 0 | 0 |
| 0.009652314 | 0 | 0.001328075 | 0.206804969 | 0 |
| 0 | 0 | 0 | 0 | 0 |
| 0 | 0 | 0 | 0 | 0 |
| 0.00413985 | 0.56451491 | 0.04699264 | 0.11419095 | 0.022706303 |

| | | | | |
|---|---|---|---|---|
| 0 | 0 | 0 | 0 | 0 |
| 0 | 0 | 0 | 0 | 0 |
| 0 | 0 | 0 | 0 | 0 |
| 0.041864395 | 0 | 0 | 0 | 0.002706083 |
| 0 | 0 | 0 | 0 | 0 |
| 0 | 0 | 0 | 0 | 0.003243565 |
| 0 | 0 | 0 | 0 | 0.083588923 |
| 0 | 0 | 0.010762839 | 0 | 0.455863611 |
| 0 | 0 | 0 | 0 | 0 |
| 0 | 0 | 0 | 0 | 0 |
| 0.002646882 | 0 | 0.001260652 | 0.018248154 | 0 |
| 0 | 0 | 0 | 0 | 0 |
| 0 | 0.004388187 | 0 | 0 | 0 |
| 0 | 0 | 0 | 0 | 0 |
| 0 | 0 | 0 | 0 | 0 |
| 0 | 0 | 0 | 0 | 0 |
| 0 | 0 | 0 | 0 | 0.066925327 |
| 0.001016986 | 0 | 0 | 0 | 0.000896061 |
| 0 | 0 | 0 | 0 | 0 |
| 0 | 0 | 0.059152655 | 0 | 0.00468697 |
| 0 | 0 | 0.003574069 | 0 | 0.002706669 |
| 0 | 0 | 0 | 0 | 0 |
| 0 | 0 | 0.011091269 | 0 | 0.033885472 |
| 0 | 0 | 0 | 0 | 0 |
| 0 | 0 | 0 | 0 | 0 |
| 0 | 0 | 0 | 0 | 0.003821055 |
| 0 | 0 | 0 | 0 | 0 |
| 0 | 0 | 0 | 0 | 0 |
| 0 | 0 | 0 | 0 | 0 |
| 3.260874418 | 0 | 0.002112905 | 0 | 0.06018328 |
| 0 | 0.000897118 | 0 | 0.090807771 | 0 |
| 0 | 0 | 0 | 0 | 0 |
| 0 | 0 | 0 | 0 | 0 |
| 0 | 0 | 0 | 0 | 0 |
| 0 | 0 | 0 | 0 | 0 |
| 0 | 0 | 0 | 0 | 0 |
| 0.122476828 | 0 | 0.067106107 | 0 | 0.050719416 |
| 0.031885921 | 0.004242541 | 0.001067164 | 0.073960189 | 0 |
| 0 | 0 | 0 | 0 | 0 |
| 0.001020017 | 0 | 0 | 0 | 0 |
| 0 | 0 | 0 | 0 | 0.013336533 |
| 0 | 0 | 0 | 0 | 0 |
| 0 | 0 | 0 | 0 | 0 |
| 0 | 0 | 0 | 0 | 0 |
| 0 | 0.390425285 | 0.068262103 | 0 | 0.029176503 |
| 0 | 0 | 0 | 0 | 0 |
| 0 | 0 | 0 | 0 | 0 |
| 0 | 0 | 0 | 0 | 0 |

| | | | | |
|---|---|---|---|---|
| 0 | 0 | 0 | 0 | 0 |
| 0 | 0 | 0 | 0 | 0 |
| 0 | 0 | 0 | 0 | 0 |
| 0 | 0 | 0.716699585 | 0 | 0.03262861 |
| 0 | 0 | 0 | 0 | 0 |
| 0 | 0 | 0 | 0 | 1.874302131 |
| 0 | 0 | 0 | 0 | 0 |
| 0 | 0 | 0 | 0 | 0 |
| 0.082009318 | 2.406427959 | 0.010793177 | 0.058748824 | 0.004021312 |
| 0 | 0 | 0 | 0 | 0 |
| 0 | 0 | 0 | 0 | 0 |
| 0 | 0 | 0 | 0 | 0 |
| 0 | 0 | 0 | 0 | 0.004838538 |
| 0 | 0 | 0 | 0 | 0 |
| 0.969764228 | 0 | 1.84422761 | 0 | 0.00379289 |
| 0 | 2.972687135 | 0.462816767 | 0.172161055 | 0 |
| 0 | 0 | 0 | 0 | 0 |
| 0 | 0 | 0 | 0 | 0 |
| 0 | 0 | 0 | 0 | 0 |
| 0 | 0 | 0 | 0 | 0 |
| 0 | 0 | 0.017443697 | 0 | 0 |
| 0 | 0 | 0 | 0 | 0 |
| 0 | 0 | 0 | 0 | 0 |
| 0 | 0 | 0 | 0 | 0 |
| 0.01705182 | 0.001251057 | 0.003173117 | 0 | 0.044333893 |
| 0.547240169 | 0 | 13.5648882 | 0.021109462 | 1.096757429 |
| 0 | 0 | 0 | 0 | 0 |
| 0 | 0 | 0.003044084 | 0 | 0.001100804 |
| 0 | 0.015312941 | 0.061032662 | 0 | 0 |
| 0 | 0 | 0 | 0 | 0 |
| 0 | 0 | 0 | 0 | 0 |
| 0 | 0 | 0.030095257 | 0 | 0 |
| 0 | 0 | 0 | 0 | 0 |
| 0 | 0 | 0.002310884 | 0.0040546 | 0.001262781 |
| 0.001475909 | 0.006054903 | 0.002030724 | 0 | 0.001820581 |
| 0.000521644 | 0.013618444 | 0.021891002 | 0.006611424 | 0 |
| 0 | 0.151380119 | 0.148692619 | 0 | 0.024664276 |
| 1.359473531 | 0 | 2.463565388 | 0 | 1.171459951 |
| 0 | 0 | 0 | 0 | 0 |
| 0 | 0 | 0 | 0 | 0 |
| 0 | 0 | 0 | 0 | 0 |
| 0 | 0 | 0 | 0 | 0 |
| 0.122370236 | 0 | 4.166689756 | 0.042914484 | 0.321323739 |
| 0.084808366 | 0 | 0 | 0 | 0 |
| 0 | 0 | 0 | 0 | 0 |
| 0 | 0.005529328 | 0.002379883 | 0 | 0 |
| 0 | 0 | 0 | 0 | 0 |
| 0.101300295 | 0 | 0.155178148 | 0.017105995 | 0.088237709 |

| | | | | |
|---|---|---|---|---|
| 0 | 0 | 0 | 0 | 0.004138299 |
| 0 | 0 | 0 | 0 | 0 |
| 0 | 0 | 0 | 0 | 0 |
| 0 | 0 | 0 | 0 | 0 |
| 0 | 0 | 0.001206379 | 0.007672938 | 0.00185407 |
| 0 | 0 | 0.001161725 | 0 | 0.001190295 |
| 0 | 0 | 0 | 0 | 0 |
| 0 | 0 | 0 | 0 | 0 |
| 0 | 0 | 0 | 0 | 0 |
| 0 | 0 | 0 | 0 | 0 |
| 0 | 0 | 0 | 0 | 0 |
| 2.204492093 | 0 | 0.499348785 | 0 | 1.846577792 |
| 0.008728969 | 0.020060928 | 0 | 0 | 0.578200387 |
| 0 | 0 | 0 | 0 | 0 |
| 0.196716077 | 0.060508031 | 1.262867979 | 0.026621257 | 0.048840845 |
| 0 | 0 | 0 | 0 | 0 |
| 0 | 0 | 0 | 0 | 0 |
| 0 | 0 | 0 | 0 | 0 |
| 0 | 0 | 0.010190566 | 0 | 0 |
| 0 | 0 | 0.002259942 | 0 | 0 |
| 0 | 0 | 0 | 0 | 0.071575112 |
| 0 | 0 | 0 | 0 | 0 |
| 0.061070547 | 0 | 2.848424092 | 0 | 0.458086486 |
| 0 | 0 | 0 | 0 | 0 |
| 0 | 0 | 0 | 0 | 0 |
| 0 | 0 | 0 | 0 | 0 |
| 0 | 0 | 0 | 0 | 0 |
| 0 | 0 | 0 | 0 | 0 |
| 0 | 0 | 0 | 0 | 0 |
| 0 | 0 | 0 | 0 | 0 |
| 0 | 0 | 0 | 0 | 0 |
| 0.096492536 | 0.008833234 | 0.031984398 | 0.003970667 | 0.010511444 |
| 0 | 0 | 0 | 0 | 0 |
| 0.004090207 | 0 | 0.023214562 | 0 | 0.017761873 |
| 0.000756906 | 0.013973406 | 0.015816841 | 0.004111363 | 0 |
| 0.012440738 | 0 | 0.127121752 | 0 | 0.053130863 |
| 0 | 0 | 0 | 0 | 0 |
| 0 | 0 | 0 | 0 | 0 |
| 0 | 0 | 0 | 0 | 0 |
| 0 | 0 | 0 | 0 | 0 |
| 0 | 0 | 0 | 0 | 0.225521101 |
| 0 | 0 | 0 | 0 | 0 |
| 0 | 0 | 0 | 0 | 0 |
| 0.037125537 | 0 | 0 | 0 | 0.027037559 |
| 0.002982515 | 0.001459946 | 0.007886767 | 0.029110129 | 0.067405046 |
| 0 | 0 | 0 | 0 | 0 |
| 0.297366328 | 0 | 0 | 0 | 0 |
| 0.002820925 | 0 | 0.000545815 | 0 | 0.000932063 |

| | | | | |
|---|---|---|---|---|
| 0 | 0 | 0 | 0 | 0 |
| 0 | 0 | 0 | 0 | 0 |
| 0.00115675 | 0 | 0 | 0 | 0 |
| 0 | 0 | 0 | 0 | 0 |
| 0 | 0 | 0 | 0.003717062 | 0 |
| 0 | 0 | 0 | 0 | 0 |
| 0 | 0 | 0 | 0 | 0 |
| 0 | 0.000611146 | 0.16421438 | 0 | 0.106264937 |
| 0 | 0 | 0 | 0 | 0 |
| 0 | 0 | 0 | 0 | 0 |
| 0 | 0 | 0.447737953 | 0 | 0 |
| 0 | 0 | 0.000607276 | 0.00079913 | 0 |
| 0 | 0 | 0 | 0 | 0 |
| 0 | 0 | 0 | 0 | 0.088823428 |
| 0 | 0 | 0 | 0 | 0 |
| 0 | 0 | 0 | 0 | 0 |
| 0 | 0 | 0 | 0 | 0 |
| 0 | 0 | 0 | 0 | 0 |
| 25.34119546 | 0 | 0 | 0 | 0 |
| 0 | 0 | 0 | 0 | 0 |
| 0 | 0 | 0 | 0 | 0 |
| 0 | 0 | 0 | 0 | 0 |
| 0 | 0 | 0 | 0 | 0 |
| 0 | 0 | 0 | 0 | 0 |
| 0 | 0.000683622 | 0 | 0 | 0 |
| 0.006767983 | 0.01689235 | 0.0008954 | 0.065041032 | 0.002018325 |
| 0 | 0 | 0 | 0 | 0 |
| 0.619816255 | 0 | 0.018229509 | 0.128939088 | 1.383056493 |
| 0 | 0 | 0 | 0 | 0 |
| 4.019218268 | 0 | 0.55988332 | 0 | 0.331788952 |
| 0 | 0 | 0 | 0 | 0 |
| 0 | 0 | 0 | 0 | 0 |
| 0 | 0 | 0 | 0 | 0 |
| 0 | 0 | 0 | 0 | 0 |
| 0.008910999 | 0.002238201 | 0 | 0.003210387 | 0 |
| 0 | 0 | 0 | 0 | 0 |
| 0 | 0 | 0 | 0 | 0 |
| 0 | 0.003001467 | 0 | 0.001120876 | 0.00305454 |
| 0 | 0 | 0 | 0 | 0 |
| 0 | 0 | 0 | 0 | 0 |
| 2.153721957 | 4.979617827 | 8.589304306 | 17.0974614 | 1.449749549 |
| 2.005070529 | 0 | 0.792319762 | 2.46884434 | 0.804617922 |
| 0 | 0.454457883 | 0.218631546 | 0 | 0 |
| 0 | 0 | 0 | 0 | 0 |
| 0 | 0 | 0 | 0 | 0 |
| 0 | 0 | 0 | 0 | 0 |
| 0 | 0 | 0 | 0 | 0 |
| 0.003111714 | 0.001282686 | 0 | 0.012899049 | 0.001904769 |

| | | | | |
|---|---|---|---|---|
| 0 | 0 | 0 | 0 | 0 |
| 0.002863743 | 0 | 0.005562726 | 0 | 0.027072755 |
| 0 | 0 | 0.005278772 | 0 | 0.001664182 |
| 0 | 0 | 0 | 0 | 0 |
| 1.206601957 | 0 | 3.370771703 | 0 | 2.182935578 |
| 0 | 0 | 0 | 0 | 0 |
| 0 | 0 | 0 | 0 | 0 |
| 0 | 0 | 0 | 0 | 0 |
| 0.036500947 | 5.012906102 | 0.109354707 | 0.596624736 | 0.185139387 |
| 0 | 0 | 0 | 0 | 0 |
| 0 | 0 | 0 | 0 | 0 |
| 0 | 0 | 0 | 0 | 0 |
| 0.160736918 | 0 | 0.130398918 | 0 | 0.170295412 |
| 0.004829565 | 0.005178462 | 0.026164946 | 0.015302714 | 0.004042537 |
| 0 | 0.05765293 | 0 | 0 | 0 |
| 0.486636384 | 0 | 0.720974665 | 0.154051122 | 0.546652333 |
| 0.005572319 | 0 | 0.128123283 | 0 | 0.036823044 |
| 0 | 0 | 0 | 0 | 0 |
| 0 | 0 | 0 | 0 | 0 |
| 0 | 0 | 0 | 0 | 0 |
| 0 | 0 | 0 | 0 | 0 |
| 0 | 0 | 0 | 0 | 0 |
| 0 | 0 | 0 | 0 | 0 |
| 0 | 0 | 0 | 0 | 0 |
| 0 | 0 | 0 | 0 | 0 |
| 0 | 0 | 0 | 0 | 0 |
| 0.014990334 | 0 | 0.346413125 | 0 | 0.011366798 |
| 0 | 0 | 0.006170995 | 0.004176292 | 0 |
| 0 | 0 | 0 | 0 | 0 |
| 0 | 0 | 0 | 0 | 0 |
| 0 | 0 | 0 | 0 | 0 |
| 0 | 0 | 0 | 0 | 0 |
| 0 | 0 | 0 | 0 | 0 |
| 0.007522389 | 0 | 0 | 0 | 0 |
| 0 | 0 | 0 | 0 | 0 |
| 0.000592941 | 0 | 0 | 0 | 0.117234821 |
| 0.006060569 | 0 | 0 | 0 | 2.986517938 |
| 0 | 0 | 0 | 0 | 0 |
| 0 | 0 | 0 | 0 | 0 |
| 0 | 0 | 0 | 0 | 0 |
| 0.013313665 | 10.94676803 | 5.415352222 | 0 | 3.008065994 |
| 0 | 0 | 0.120272966 | 0 | 0.067032125 |
| 0 | 0 | 0 | 0 | 0 |
| 0 | 0 | 0 | 0 | 0 |
| 0 | 0 | 0 | 0 | 0 |
| 0 | 0 | 0 | 0 | 0 |
| 0 | 0 | 0 | 0 | 0.617756817 |
| 0 | 0 | 0 | 0 | 0 |

| | | | | |
|---|---|---|---|---|
| 0 | 0 | 0 | 0 | 0.044899464 |
| 0 | 0 | 0 | 0 | 0 |
| 0 | 0 | 0 | 0 | 0 |
| 0.000774193 | 0 | 0 | 0 | 0 |
| 0.007407514 | 0 | 0.125553957 | 0 | 1.301884718 |
| 0 | 0 | 0 | 0 | 0 |
| 0.366705073 | 0 | 0.019359983 | 0 | 0.054181921 |
| 0 | 0 | 0 | 0 | 0 |
| 0 | 0 | 0 | 0 | 0 |
| 0.153091412 | 0 | 0.024449357 | 0 | 0.017342741 |
| 0 | 0 | 0.001739048 | 0 | 0.001559089 |
| 0 | 0 | 0 | 0 | 0.017507659 |
| 0 | 0 | 0 | 0 | 0 |
| 0 | 0 | 0 | 0 | 0 |
| 0 | 0 | 0 | 0 | 0 |
| 0 | 0 | 0 | 0 | 0 |
| 0.092910358 | 0 | 0 | 0 | 0 |
| 0.001002033 | 0.001121135 | 0.014304122 | 0 | 0 |
| 0 | 0 | 0 | 0 | 0 |
| 0 | 0 | 0 | 0 | 0 |
| 0.014222944 | 0 | 0.082601695 | 0.011229095 | 0.207386004 |
| 0 | 0 | 0 | 0 | 0 |
| 0.022072451 | 0 | 0.019953707 | 0 | 0.031245243 |
| 0.00162207 | 0.004174204 | 0.001004323 | 0.027093087 | 0.003601577 |
| 0 | 0 | 0 | 0 | 0 |
| 0 | 0 | 0.006121857 | 0 | 0 |
| 0 | 0 | 0 | 0 | 0 |
| 0 | 0 | 0 | 0 | 0 |
| 0.170445071 | 0 | 0 | 0 | 0 |
| 0.781370396 | 2.794867276 | 0.368350501 | 0 | 0.72445772 |
| 0 | 0 | 0 | 0 | 0 |
| 0 | 0 | 0 | 0 | 0 |
| 0 | 0 | 0 | 0 | 0 |
| 0.389068615 | 0.004070164 | 0 | 18.35013567 | 2.276990123 |
| 0 | 0 | 0 | 0 | 0 |
| 0 | 0 | 0 | 0 | 0 |
| 0.016642967 | 0 | 0.073724546 | 0.015709973 | 0.022103336 |
| 0 | 0 | 0 | 0 | 0 |
| 0 | 0 | 0 | 0 | 0 |
| 0 | 0 | 0 | 0 | 0 |
| 0 | 0 | 0 | 0 | 0 |
| 0 | 0 | 0 | 0 | 0 |
| 0 | 0 | 0 | 0 | 0.157389533 |
| 0 | 0 | 0 | 0 | 0 |
| 0 | 0 | 0 | 0 | 0 |
| 0 | 0 | 0 | 0 | 0 |
| 0 | 0 | 0 | 0 | 0 |
| 0 | 0 | 0 | 0 | 0 |

| | | | | |
|---|---|---|---|---|
| 0.00314173 | 0 | 0.017110884 | 0.067549921 | 0.00092272 |
| 0 | 0 | 0 | 0 | 0 |
| 0 | 0 | 0 | 0 | 0.091233885 |
| 0 | 0 | 0 | 0 | 0 |
| 0 | 0 | 0 | 0 | 0 |
| 0 | 0 | 0 | 0 | 0 |
| 0 | 0 | 0 | 0 | 0 |
| 0.619362472 | 0 | 0.768228092 | 0 | 6.310040215 |
| 0.05205384 | 0.002145722 | 0.127505304 | 0 | 1.38693789 |
| 0.002670192 | 0.028168556 | 0.068296192 | 0.571092899 | 0.032467141 |
| 0 | 0 | 0 | 0 | 0 |
| 0 | 0 | 0 | 0 | 0 |
| 0 | 0 | 0 | 0 | 0.00611249 |
| 0 | 0 | 0 | 0 | 0 |
| 0 | 0 | 0 | 0.002264288 | 0.002820796 |
| 0 | 0 | 0 | 0 | 0 |
| 0 | 0 | 0 | 0 | 0 |
| 0 | 0 | 0 | 0 | 0 |
| 0 | 0 | 0 | 0 | 0 |
| 0 | 0 | 0.011750907 | 0 | 0 |
| 0 | 0 | 0 | 0 | 0 |
| 0 | 0 | 0 | 0 | 0 |
| 0 | 0 | 0 | 0 | 0 |
| 0 | 0 | 0 | 0 | 0 |
| 0.225046354 | 0 | 0.078451449 | 0 | 0.029361509 |
| 0 | 0 | 0 | 0 | 0 |
| 0 | 0 | 0 | 0 | 0 |
| 0 | 0 | 0 | 0 | 0.03246911 |
| 0 | 0 | 0 | 0 | 0 |
| 0 | 0 | 0 | 0 | 0 |
| 0 | 0 | 0 | 0 | 0.082470449 |
| 0 | 0 | 0 | 0 | 0 |
| 0 | 0 | 0 | 0 | 0 |
| 0 | 0 | 0 | 0 | 0 |
| 0 | 0 | 0 | 0 | 0 |
| 0 | 0 | 0 | 0 | 0.012362154 |
| 0 | 0 | 0 | 0 | 0 |
| 0 | 0 | 0 | 0 | 0.009062873 |
| 0.002075423 | 0 | 0 | 0 | 0 |
| 0.001742461 | 0 | 0 | 0 | 0.051769398 |
| 0 | 0 | 0 | 0 | 0 |
| 0 | 0.03821526 | 0.378062638 | 0 | 0.62532622 |
| 0 | 0 | 0 | 0 | 0 |
| 0 | 0 | 0 | 0 | 0 |
| 0 | 0 | 0 | 0 | 0 |
| 0 | 0 | 0 | 0 | 0 |
| 0 | 0 | 0 | 0 | 0 |
| 0 | 0 | 0 | 0 | 0 |

| | | | | |
|---|---|---|---|---|
| 0 | 0 | 0 | 0 | 0 |
| 0 | 0.014276631 | 0.016418689 | 0 | 0.006828921 |
| 0 | 0 | 0 | 0 | 0 |
| 0 | 0 | 0 | 0 | 0 |
| 0.096249135 | 0.011335728 | 0.713670233 | 0 | 1.838765026 |
| 0 | 0 | 0 | 0 | 0 |
| 0 | 0 | 0 | 0 | 0 |
| 0.016208058 | 0.249163966 | 0.005957088 | 0.029547306 | 0.001173767 |
| 0 | 0 | 0 | 0 | 0.003281496 |
| 0 | 0 | 0 | 0 | 0 |
| 0 | 0 | 0 | 0 | 0 |
| 0 | 0.022269928 | 0 | 0 | 0 |
| 0 | 0 | 0 | 0 | 0 |
| 0 | 0 | 0 | 0 | 0 |
| 0 | 0 | 0 | 0 | 0.008262904 |
| 0 | 0 | 0 | 0 | 0.002646798 |
| 0 | 0 | 0 | 0 | 0 |
| 0 | 0 | 0 | 0 | 0 |
| 0 | 0 | 0 | 0 | 0 |
| 0 | 0 | 0 | 0 | 0 |
| 0 | 0 | 0 | 0 | 0 |
| 0 | 0 | 0 | 0 | 0 |
| 0 | 0 | 0 | 0 | 0 |
| 0.000960736 | 0 | 0 | 0.004174818 | 0 |
| 0.276950464 | 0.307573771 | 0.241338005 | 0 | 0.146592461 |
| 0 | 0 | 0 | 0 | 0 |
| 0 | 0 | 0.072541079 | 0.041524501 | 0.000557438 |
| 0 | 0 | 0 | 0 | 0.000782619 |
| 0 | 0 | 0 | 0 | 0 |
| 0 | 0 | 0 | 0 | 0 |
| 0 | 0 | 0 | 0 | 0 |
| 0 | 0 | 0 | 0 | 0 |
| 0 | 0 | 0 | 0 | 0 |
| 0 | 0 | 0 | 0 | 0 |
| 0 | 0 | 0 | 0 | 0 |
| 0 | 0 | 0 | 0 | 0 |
| 0 | 0 | 0 | 0 | 0 |
| 0 | 0 | 0.01022933 | 0 | 0.032967177 |
| 0 | 0 | 0 | 0 | 0 |
| 0 | 0 | 0 | 0 | 0 |
| 0 | 0 | 0 | 0 | 0 |
| 0 | 0 | 0 | 0 | 0 |
| 0 | 0 | 0 | 0 | 0 |
| 0 | 0 | 0 | 0 | 0 |
| 0 | 0 | 0 | 0 | 0 |
| 0 | 0 | 0 | 0 | 0 |
| 0 | 0 | 0 | 0 | 0 |
| 0 | 0 | 0 | 0 | 0 |

| | | | | |
|---|---|---|---|---|
| 0.153534577 | 0 | 0 | 0 | 0.787381795 |
| 0 | 0 | 0 | 0 | 0 |
| 0.008676511 | 0 | 0 | 0 | 0 |
| 0 | 0 | 0.000678738 | 0.005359008 | 0 |
| 0 | 0 | 0 | 0 | 0 |
| 0.002489153 | 0 | 0 | 0 | 0 |
| 0 | 0 | 0 | 0 | 0 |
| 0 | 0 | 0 | 0 | 0 |
| 0 | 0 | 0 | 0 | 0 |
| 0 | 0 | 0 | 0 | 0 |
| 0 | 0 | 0 | 0 | 0 |
| 0 | 0 | 0 | 0 | 0 |
| 0 | 0 | 0 | 0 | 0 |
| 0 | 0 | 0 | 0 | 0 |
| 0 | 0 | 0 | 0 | 0 |
| 0 | 0 | 0 | 0 | 0 |
| 0 | 0 | 0 | 0 | 0 |
| 0 | 0.002357952 | 0 | 0 | 0 |
| 0 | 0 | 0 | 0 | 0.000592596 |
| 0 | 0 | 0 | 0 | 0 |
| 0.089548939 | 0.001070436 | 0.020930182 | 2.990965042 | 0.008901659 |
| 0 | 0 | 0 | 0 | 0 |
| 0.052438787 | 0.008111297 | 0.002244336 | 0.134542796 | 0.014052684 |
| 0 | 0 | 0.247408982 | 0 | 0 |
| 0 | 0 | 0 | 0 | 0 |
| 0 | 0 | 0 | 0 | 0 |
| 0 | 0 | 0 | 0 | 0 |
| 0 | 0 | 0 | 0 | 0 |
| 0 | 0 | 0 | 0 | 0 |
| 0 | 0 | 0 | 0 | 0 |
| 0 | 0 | 0 | 0 | 0 |
| 0 | 0 | 0 | 0 | 0 |
| 0.009052296 | 1.906392135 | 0.006071901 | 0.0218193 | 0.002392778 |
| 0 | 0 | 0 | 0 | 0 |
| 0.012955578 | 0.017892873 | 0 | 0 | 0.667140506 |
| 0 | 0 | 0 | 0.107273901 | 0 |
| 0 | 0 | 0 | 0 | 0 |
| 0 | 0 | 0 | 0 | 0 |
| 0.001050258 | 0 | 0 | 0.046779298 | 0.000888362 |
| 0 | 0 | 0 | 0 | 0 |
| 0 | 0 | 0 | 0 | 0 |
| 0 | 0 | 0 | 0 | 0 |
| 0.002456134 | 0.221769433 | 0.025852628 | 0 | 0 |
| 0 | 0 | 0 | 0 | 0 |
| 0 | 0 | 0 | 0 | 0 |
| 0 | 0 | 0.126409443 | 0 | 0.015055455 |
| 0 | 0 | 0 | 0 | 0 |
| 0 | 0 | 0 | 0 | 0 |

| | | | | |
|---|---|---|---|---|
| 0 | 0 | 0 | 0 | 0 |
| 0 | 0 | 0 | 0 | 0 |
| 0 | 0 | 0 | 0 | 0 |
| 0 | 0 | 0 | 0 | 0 |
| 0 | 0 | 0 | 0 | 0 |
| 0 | 0 | 0 | 0 | 0 |
| 0 | 0 | 0 | 0 | 0 |
| 0 | 0 | 0.110481036 | 0.04760839 | 0 |
| 0.000548258 | 0 | 0.150682657 | 0 | 0.046181236 |
| 0.011050008 | 0.01910711 | 0 | 0 | 0.000849696 |
| 0 | 0 | 0 | 0 | 0 |
| 0 | 0 | 0 | 0 | 0 |
| 0 | 0 | 0 | 0 | 0 |
| 0 | 0 | 0 | 0 | 0.549939864 |
| 0.009417001 | 0 | 0 | 0 | 0 |
| 0 | 0.080284001 | 0 | 0 | 0 |
| 0 | 0 | 0 | 0 | 0 |
| 0 | 0 | 0 | 0 | 0 |
| 0 | 0 | 0 | 0 | 0 |
| 0 | 0 | 0 | 0 | 0 |
| 0.004542007 | 0.017324575 | 0.007030588 | 0 | 0.006548625 |
| 0 | 0 | 0 | 0 | 0 |
| 0 | 0 | 0 | 0 | 0 |
| 0 | 0 | 0 | 0 | 0 |
| 0 | 0 | 0 | 0 | 0 |
| 0 | 0 | 0 | 0 | 0 |
| 0 | 0 | 0 | 0 | 0 |
| 0 | 0 | 0 | 0 | 0 |
| 0 | 0 | 0 | 0 | 0 |
| 0 | 0 | 0 | 0 | 0 |
| 0.709164904 | 3.847745224 | 0.070296435 | 6.837067555 | 2.584583459 |
| 0 | 0 | 0 | 0 | 0 |
| 0 | 0 | 0 | 0 | 0 |
| 0 | 0 | 0 | 0 | 0 |
| 0 | 0 | 0 | 0 | 0 |
| 0 | 0 | 0 | 0 | 0 |
| 0 | 0 | 0 | 0 | 0 |
| 0 | 0 | 0 | 0 | 0 |
| 0.384237092 | 0.003646721 | 0 | 0 | 0 |
| 0 | 0 | 0 | 0 | 0 |
| 0 | 0 | 0 | 0 | 0 |
| 0 | 0 | 0 | 0 | 0 |
| 0 | 0 | 0 | 0 | 0 |
| 0 | 0 | 0 | 0 | 0 |
| 0 | 0 | 0 | 0 | 0 |
| 0 | 0 | 0 | 0 | 0 |
| 0.005261253 | 0.001584857 | 0 | 0.575408392 | 0.001497675 |
| 0 | 0 | 0 | 0 | 0 |

| | | | | |
|---|---|---|---|---|
| 0 | 0 | 0 | 0 | 0 |
| 0 | 0 | 0 | 0 | 0 |
| 0 | 0 | 0 | 0 | 0 |
| 0 | 0 | 0 | 0 | 0 |
| 0.018292169 | 0 | 0.299681371 | 0 | 0.019180715 |
| 0 | 0 | 0 | 0 | 0 |
| 0 | 0 | 0 | 0 | 0 |
| 0 | 0 | 0 | 0 | 0 |
| 0 | 0 | 0 | 0 | 0 |
| 0 | 0 | 0 | 0 | 0 |
| 0.014203534 | 0 | 0 | 0 | 0.049720386 |
| 0.001679075 | 0 | 0.002382459 | 0.0114005 | 0.011761421 |
| 0.004526934 | 0 | 0 | 0 | 0.005169299 |
| 0 | 0 | 0 | 0 | 0 |
| 0.005438184 | 0 | 0 | 0 | 0 |
| 0.070998479 | 0 | 0 | 1.62837159 | 1.170537423 |
| 0.003389947 | 0 | 0 | 0 | 0.004437625 |
| 0 | 0 | 0 | 0 | 0 |
| 0.04720093 | 0.001600342 | 0 | 0.067117872 | 0.001008205 |
| 0.384305791 | 0 | 0.001498948 | 0 | 0.110578369 |
| 0 | 0 | 0 | 0 | 0 |
| 0 | 0 | 0 | 0 | 0 |
| 0 | 0 | 0 | 0 | 0 |
| 0 | 0 | 0 | 0 | 0 |
| 0 | 0 | 0 | 0 | 0 |
| 0 | 0 | 0 | 0 | 0 |
| 0.033090932 | 0 | 0.002402351 | 0.021451774 | 0.000879082 |
| 0 | 0 | 0 | 0 | 0 |
| 0 | 0 | 0 | 0 | 0 |
| 0 | 0 | 0 | 0 | 0 |
| 0.00094247 | 0 | 0.000583541 | 0 | 0 |
| 0.016084044 | 0 | 0 | 0.002488252 | 0 |
| 0 | 0 | 0 | 0 | 0 |
| 0 | 0 | 0 | 0 | 0 |
| 0 | 0 | 0 | 0 | 0 |
| 0 | 0 | 0 | 0 | 0 |
| 0 | 0 | 0 | 0 | 0 |
| 0.022248312 | 0.041932464 | 0.003689811 | 0.024637239 | 0.075050962 |
| 0 | 0 | 0 | 0 | 0 |
| 0 | 0 | 0 | 0 | 0 |
| 0 | 0 | 0 | 0.001615789 | 0 |
| 0 | 0 | 0 | 0 | 0 |
| 0 | 0 | 0 | 0 | 0 |
| 0 | 0.084591882 | 0 | 0 | 0 |
| 0 | 0 | 0 | 0 | 0 |
| 0 | 0 | 0 | 0 | 0.001299374 |
| 0 | 0 | 0 | 0 | 0 |
| 0 | 0 | 0 | 0 | 0 |

| | | | | |
|---|---|---|---|---|
| 0 | 0 | 0 | 0 | 0 |
| 0 | 0 | 0 | 0 | 0 |
| 0 | 0 | 0 | 0 | 0 |
| 0.000617414 | 0.001842134 | 0.000637131 | 0.004471555 | 0 |
| 0.000763522 | 0.012600552 | 0.000590929 | 0 | 0 |
| 0 | 0 | 0 | 0.001003591 | 0 |
| 0 | 0 | 0 | 0 | 0 |
| 0 | 0 | 0.132877758 | 0 | 0 |
| 0 | 0 | 0 | 0 | 0 |
| 0 | 0 | 0 | 0 | 0 |
| 0 | 0 | 0 | 0 | 0 |
| 0 | 0 | 0 | 0 | 0 |
| 0.022758286 | 0.00587616 | 0.033421078 | 0 | 0.005861412 |
| 0 | 0 | 0 | 0 | 0 |
| 0 | 0 | 0 | 0 | 0 |
| 0 | 0 | 0 | 0 | 0 |
| 0 | 0 | 0 | 0 | 0 |
| 0 | 0 | 0 | 0 | 0 |
| 0 | 0 | 0 | 0 | 0 |
| 0 | 0 | 0 | 0 | 0 |
| 0 | 0 | 0 | 0 | 0 |
| 0 | 0 | 0 | 0 | 0 |
| 0.004802471 | 0.009457004 | 0.019625126 | 0.104605082 | 0.008530563 |
| 0.016094509 | 0 | 0.071580274 | 0 | 0.016058238 |
| 0 | 0 | 0 | 0 | 0 |
| 0 | 0 | 0 | 0 | 0 |
| 0 | 0 | 0 | 0 | 0 |
| 0 | 0 | 0 | 0 | 0 |
| 0 | 0 | 0 | 0 | 0 |
| 0 | 0 | 0 | 0 | 0 |
| 0.064899372 | 0 | 0.0156931 | 0 | 0.011096797 |
| 0 | 0 | 0 | 0 | 0 |
| 0 | 0 | 0 | 0 | 0 |
| 0 | 0 | 0 | 0 | 0 |
| 0 | 0 | 0 | 0 | 0 |
| 0 | 0 | 0 | 0 | 0 |
| 0 | 0 | 0 | 0 | 0 |
| 0 | 0 | 0 | 0 | 0 |
| 0.066869826 | 0 | 0.464523785 | 0.019194735 | 0.022226215 |
| 0 | 0 | 0 | 0 | 0 |
| 0 | 0 | 0 | 0 | 0 |
| 0 | 0 | 0 | 0 | 0 |
| 0 | 0 | 0 | 0 | 0 |
| 0 | 0 | 0 | 0 | 0 |
| 0 | 0 | 0 | 0 | 0 |
| 0.550715495 | 0.007135775 | 0.024680205 | 0.678126015 | 0.389927885 |
| 0 | 0 | 0 | 0 | 0 |
| 0 | 0 | 0 | 0 | 0 |

| | | | | |
|---|---|---|---|---|
| 0 | 0 | 0 | 0 | 0 |
| 0 | 0 | 0 | 0 | 0 |
| 0.413600594 | 0.093320847 | 0 | 0 | 0.070722781 |
| 0 | 0 | 0 | 0 | 0 |
| 0 | 0 | 0 | 0 | 0 |
| 0 | 0 | 0 | 0 | 0 |
| 0 | 0 | 0 | 0 | 0 |
| 0 | 0 | 0 | 0 | 0 |
| 0 | 0 | 0 | 0 | 0 |
| 0 | 0 | 0 | 0 | 0 |
| 0 | 0 | 0 | 0 | 0.001088103 |
| 0 | 0 | 0.002038562 | 0.000894198 | 0 |
| 0 | 0 | 0 | 0 | 0 |
| 0 | 0 | 0 | 0 | 0 |
| 0 | 0 | 0 | 0 | 0 |
| 0 | 0 | 0 | 0 | 0 |
| 0 | 0 | 0 | 0 | 0 |
| 0 | 0 | 0 | 0 | 0 |
| 0 | 0 | 0 | 0 | 0 |
| 0 | 0.001090722 | 0.000804786 | 0 | 0 |
| 0 | 0 | 0 | 0 | 0 |
| 0 | 0 | 0 | 0 | 0 |
| 0 | 0 | 0 | 0 | 0 |
| 0 | 0 | 0 | 0 | 0 |
| 0 | 0 | 0 | 0 | 0 |
| 0 | 0 | 0 | 0 | 0 |
| 0 | 0 | 0 | 0 | 0 |
| 0.002373441 | 0 | 0 | 0 | 0 |
| 0 | 0 | 0 | 0 | 0 |
| 0 | 0 | 0 | 0 | 0 |
| 0.000669998 | 0 | 0.002074183 | 0.002956926 | 0.00301069 |
| 0 | 0 | 0 | 0 | 0 |
| 0.313313592 | 0.045478502 | 0.011640301 | 0.170080002 | 0.020768674 |
| 0.002981122 | 0 | 0 | 0 | 0 |
| 0 | 0 | 0 | 0 | 0 |
| 0 | 0 | 0 | 0 | 0 |
| 0 | 0 | 0 | 0 | 0 |
| 0 | 0 | 0 | 0 | 0 |
| 0 | 0 | 0 | 0 | 0 |
| 0 | 0 | 0 | 0 | 0 |
| 0 | 0 | 0 | 0 | 0 |
| 0.116480113 | 0 | 0.056302454 | 0 | 0.154790819 |
| 0.104285037 | 0 | 0.025344942 | 0 | 0.086354692 |
| 0 | 0 | 0 | 0 | 0 |
| 0 | 0 | 0 | 0 | 0 |
| 0 | 0.001890969 | 0.004941496 | 0 | 0 |
| 0 | 0 | 0 | 0 | 0 |
| 0 | 0 | 0 | 0 | 0 |

| | | | | |
|---|---|---|---|---|
| 0 | 0 | 0 | 0 | 0 |
| 0 | 0.005255103 | 0 | 0 | 0.004534195 |
| 0.0324413 | 0.004270272 | 0.00073847 | 0 | 0 |
| 0 | 0 | 0 | 0 | 0 |
| 0.045139467 | 0 | 0 | 0 | 0.043900379 |
| 0.000926972 | 0.001244584 | 0 | 0 | 0 |
| 0 | 0 | 0 | 0 | 0 |
| 0 | 0 | 0 | 0 | 0 |
| 0 | 0 | 0 | 0 | 0 |
| 0 | 0 | 0 | 0 | 0 |
| 0 | 0 | 0 | 0 | 0 |
| 0 | 0 | 0 | 0 | 0 |
| 0 | 0 | 0 | 0 | 0 |
| 0.036891669 | 0 | 0 | 0 | 0 |
| 0 | 0 | 0 | 0 | 0 |
| 0 | 0 | 0 | 0 | 0 |
| 0 | 0 | 0 | 0 | 0 |
| 0 | 0 | 0 | 0 | 0 |
| 0 | 0 | 0 | 0 | 0 |
| 0 | 0 | 0 | 0 | 0 |
| 0 | 0 | 0 | 0 | 0 |
| 0 | 0 | 0 | 0 | 0 |
| 0 | 0 | 0 | 0 | 0 |
| 0 | 0 | 0 | 0 | 0 |
| 0 | 0 | 0 | 0 | 0 |
| 0.003361997 | 0.263755084 | 0.004897926 | 0 | 0.145951155 |
| 0 | 0 | 0 | 0 | 0 |
| 0 | 0 | 0 | 0 | 0.001217 |
| 0 | 0 | 0 | 0 | 0 |
| 0 | 0 | 0 | 0 | 0 |
| 0 | 0 | 0 | 0 | 0 |
| 0 | 0 | 0 | 0.029909288 | 0 |
| 0 | 0 | 0 | 0 | 0 |
| 0 | 0 | 0 | 0 | 0 |
| 0 | 0 | 0 | 0 | 0 |
| 0 | 0 | 0 | 0 | 0 |
| 0 | 0 | 0 | 0 | 0 |
| 0 | 0 | 0.062568575 | 0 | 0.007701201 |
| 0 | 1.031202864 | 0.137299015 | 0 | 0 |
| 0 | 0 | 0 | 0 | 0 |
| 0 | 0 | 0 | 0 | 0 |
| 0.00107272 | 0 | 0 | 1.564787166 | 0 |
| 0.002183261 | 0 | 0.012084191 | 0.051209474 | 0.000629561 |
| 0.07070639 | 0 | 0 | 0 | 0 |
| 0 | 0 | 0 | 0 | 0 |
| 0 | 0 | 0 | 0 | 0 |
| 0.005769026 | 0 | 0.02412638 | 0 | 0.001284141 |
| 0 | 0 | 0 | 0 | 0 |

| | | | | |
|---|---|---|---|---|
| 0 | 0 | 0 | 0 | 0 |
| 0.003831289 | 0.026791753 | 0.00454669 | 0.034337809 | 0.027545938 |
| 0 | 0 | 0 | 0 | 0 |
| 0 | 0 | 0 | 0 | 0 |
| 0 | 0 | 0 | 0 | 0 |
| 0 | 0 | 0 | 0 | 0 |
| 0 | 0 | 0 | 0 | 0 |
| 0 | 0 | 0 | 0 | 0 |
| 0 | 0 | 0 | 0 | 0 |
| 0 | 0 | 0 | 0 | 0 |
| 0 | 0 | 0 | 0 | 0 |
| 0 | 0 | 0 | 0 | 0 |
| 0 | 0 | 0.020687789 | 0 | 0.028262071 |
| 0 | 0 | 0 | 0 | 0 |
| 0.030095354 | 0.002672422 | 0.009119755 | 0 | 0.005808453 |
| 0 | 0 | 0 | 0 | 0 |
| 0 | 0 | 0 | 0 | 0 |
| 0.001474033 | 0 | 0 | 0 | 0 |
| 0.016118163 | 0 | 0 | 0 | 0 |
| 0.001908848 | 0 | 0 | 0 | 0 |
| 0 | 0 | 0 | 0 | 0 |
| 0.017592865 | 0 | 0.144027297 | 0 | 0.024305529 |
| 0.003965479 | 0 | 0.006510188 | 0 | 0 |
| 0 | 0 | 0.008975643 | 0 | 0 |
| 0 | 0 | 0 | 0 | 0 |
| 0 | 0 | 0 | 0 | 0 |
| 0 | 0 | 0 | 0 | 0 |
| 0 | 0 | 0 | 0 | 0 |
| 0 | 0 | 0 | 0 | 0 |
| 0 | 0 | 0 | 0 | 0 |
| 0 | 0 | 0 | 0 | 0 |
| 0 | 0 | 0 | 0 | 0 |
| 0 | 0 | 0 | 0 | 0 |
| 0 | 0 | 0 | 0 | 0 |
| 0 | 0 | 19.27399815 | 0 | 0 |
| 0 | 0 | 0 | 0 | 0 |
| 0 | 0 | 0 | 0 | 0 |
| 0 | 0 | 0 | 0 | 0 |
| 0.003747969 | 0 | 0.03239167 | 0 | 0.018823193 |
| 0 | 0 | 0 | 0 | 0 |
| 0.022473677 | 0.19904439 | 0.03966377 | 0.253842337 | 0.036641902 |
| 0 | 0 | 0 | 0 | 0 |
| 0 | 0 | 0 | 0 | 0 |
| 0 | 0 | 0 | 0 | 0 |
| 0 | 0 | 0 | 0 | 0 |
| 0.229894153 | 3.966993843 | 0.188349446 | 2.892606777 | 0.096109326 |
| 0 | 0 | 0 | 0 | 0 |
| 0 | 0 | 0 | 0 | 0 |

| | | | | |
|---|---|---|---|---|
| 0 | 0 | 0 | 0 | 0 |
| 0 | 0 | 0.00121256 | 0 | 0 |
| 0 | 0 | 0 | 0 | 0 |
| 0 | 0 | 0 | 0 | 0 |
| 0 | 0 | 0.001693859 | 0.012135623 | 0 |
| 0 | 0 | 0 | 0 | 0 |
| 0 | 0 | 0 | 0 | 0 |
| 0.004418632 | 0 | 0 | 0 | 0.00384743 |
| 0.008145266 | 0 | 0 | 0 | 0.012573663 |
| 0.025870348 | 0.001052557 | 0.047082971 | 0.002554955 | 0 |
| 0 | 0 | 0 | 0 | 0 |
| 0 | 0 | 0 | 0 | 0 |
| 0.002758482 | 0.001800376 | 0.000948859 | 0.005306664 | 0 |
| 0 | 0 | 0 | 0 | 0 |
| 0 | 0 | 0 | 0 | 0 |
| 0 | 0 | 0 | 0 | 0 |
| 0 | 0 | 0 | 0 | 0 |
| 0 | 0 | 0 | 0 | 0 |
| 0 | 0 | 0 | 0 | 0 |
| 0.405933652 | 0.289129835 | 0.135761585 | 0.040009173 | 0.535849824 |
| 0 | 0 | 0 | 0 | 0 |
| 0 | 0 | 0 | 0 | 0 |
| 0 | 0 | 0 | 0 | 0 |
| 0 | 0 | 0 | 0 | 0 |
| 0 | 0 | 0 | 0 | 0 |
| 0 | 0 | 0 | 0 | 0 |
| 0 | 0 | 0 | 0 | 0 |
| 0 | 0 | 0 | 0 | 0 |
| 0 | 0 | 0 | 0 | 0 |
| 0 | 0 | 0.030922227 | 0 | 0.00531899 |
| 0 | 0 | 0 | 0 | 0 |
| 0 | 0 | 0 | 0 | 0 |
| 0 | 0 | 0 | 0 | 0 |
| 0 | 0 | 0 | 0 | 0 |
| 0 | 0 | 0 | 0 | 0 |
| 0 | 0 | 0 | 0 | 0 |
| 0 | 0 | 0 | 0 | 0 |
| 0 | 0 | 0 | 0 | 0 |
| 0 | 0 | 0 | 0 | 0 |
| 0 | 0 | 0 | 0 | 0.005959061 |
| 0 | 0 | 0 | 0 | 0 |
| 0 | 0 | 0 | 0 | 0 |
| 0.033164552 | 0 | 0 | 0 | 0.017622572 |
| 0 | 0 | 0 | 0 | 0 |
| 0 | 0 | 0.0023797 | 0.013569864 | 0.012191115 |
| 0 | 0 | 0 | 0 | 0 |
| 0 | 0 | 0 | 0 | 0 |
| 0 | 0 | 0 | 0 | 0 |

| | | | | |
|---|---|---|---|---|
| 0 | 0 | 0 | 0 | 0 |
| 0 | 0 | 0 | 0 | 0 |
| 0 | 0 | 0 | 0 | 0 |
| 0 | 0 | 0 | 0 | 0 |
| 0 | 0 | 0 | 0 | 0 |
| 0.06125884 | 0.004494435 | 0.0072542 | 0.044028412 | 0.001365171 |
| 0 | 0 | 0 | 0 | 0 |
| 0 | 0 | 0 | 0 | 0 |
| 0 | 0 | 0 | 0 | 0 |
| 0 | 0 | 0 | 0 | 0 |
| 0 | 0 | 0 | 0 | 0 |
| 0 | 0 | 0 | 0 | 0 |
| 0 | 0 | 0 | 0 | 0 |
| 0 | 0 | 0 | 0 | 0.005042345 |
| 0 | 0 | 0 | 0 | 0 |
| 0 | 0 | 0 | 0 | 0 |
| 0 | 0 | 0.008978871 | 0 | 0 |
| 0 | 0 | 0 | 0.00191941 | 0 |
| 0 | 0 | 0 | 0 | 0 |
| 0 | 0 | 0 | 0 | 0 |
| 0 | 0 | 0 | 0 | 0 |
| 0 | 0 | 0 | 0 | 0 |
| 0 | 0 | 0 | 0 | 0 |
| 0 | 0 | 0.018625763 | 0 | 0 |
| 0 | 0 | 0 | 0 | 0 |
| 0 | 0 | 0.000926268 | 0 | 0 |
| 0 | 0 | 0 | 0 | 0 |
| 0.015619204 | 0 | 0.055286509 | 0 | 0 |
| 0 | 0 | 0 | 0 | 0 |
| 0 | 0 | 0.037736926 | 0 | 0 |
| 0 | 0 | 0 | 0 | 0 |
| 0 | 0 | 0 | 0 | 0 |
| 0 | 0 | 0 | 0 | 0 |
| 0 | 0 | 0 | 0 | 0 |
| 0 | 0 | 0 | 0 | 0 |
| 0 | 0 | 0 | 0 | 0 |
| 0 | 0 | 0 | 0 | 0 |
| 0 | 0 | 0 | 0 | 0 |
| 0 | 0 | 0 | 0 | 0 |
| 0 | 0 | 0 | 0 | 0 |
| 0 | 0 | 0 | 0 | 0 |
| 0 | 0 | 0 | 0 | 0 |
| 0 | 0 | 0 | 0 | 0 |
| 0 | 0 | 0 | 0 | 0 |
| 0 | 0 | 0 | 0 | 0.088022838 |
| 0 | 0 | 0 | 0 | 0 |
| 0 | 0 | 0 | 0 | 0 |
| 0 | 0 | 0 | 0 | 0 |

| | | | | |
|---|---|---|---|---|
| 0 | 0 | 0 | 0 | 0 |
| 0.000642475 | 0 | 0.040111062 | 0 | 0 |
| 0 | 0 | 0 | 0 | 0 |
| 0 | 0 | 0 | 0 | 0 |
| 0 | 0 | 0 | 0 | 0 |
| 0 | 0 | 0 | 0 | 0 |
| 0 | 0 | 0 | 0 | 0.000905558 |
| 0 | 0 | 0 | 0 | 0 |
| 0 | 0 | 0 | 0 | 0 |
| 0 | 0 | 0 | 0 | 0 |
| 0 | 0 | 0.003255512 | 0 | 0 |
| 0 | 0 | 0 | 0 | 0 |
| 0 | 0 | 0 | 0 | 0.522974677 |
| 0 | 0 | 0 | 0 | 0 |
| 0 | 0 | 0 | 0 | 0 |
| 0 | 0 | 0 | 0 | 0 |
| 0 | 0 | 0 | 0 | 0 |
| 0 | 0 | 0 | 0 | 0 |
| 0 | 0 | 0 | 0 | 0 |
| 0 | 0 | 0 | 0 | 0 |
| 0.003415354 | 0.072843663 | 0.010793552 | 0.035363829 | 0.02369784 |
| 0 | 0 | 0 | 0 | 0 |
| 0 | 0 | 0 | 0 | 0 |
| 0.578410519 | 1.885689914 | 1.92184796 | 1.134835845 | 3.462023117 |
| 0 | 0 | 0 | 0 | 0 |
| 0 | 0 | 0 | 0 | 0 |
| 0 | 0 | 0 | 0 | 0 |
| 0 | 0 | 0 | 0 | 0 |
| 0 | 0 | 0 | 0 | 0 |
| 0 | 0 | 0 | 0 | 0 |
| 0.006494826 | 0 | 0 | 0 | 0 |
| 0 | 0 | 0 | 0 | 0 |
| 0 | 0 | 0 | 0 | 0 |
| 0 | 0 | 0 | 0 | 0 |

| PHL018-0016_V1 | PHL018-0013_V1 | PHL018-0111_V3 | PHL018-0019_V1 | PHL018-0054_V3 |
|---|---|---|---|---|
| CTRL | EYCF | EYCF | CTRL | CTRL |
| 0.108495653 | 1.865747242 | 0.626841976 | 0.004827902 | 0.059529279 |
| 0.207110349 | 0.101030447 | 0 | 3.18585813 | 3.423068236 |
| 0.016731769 | 0 | 12.10059255 | 0 | 0 |
| 0 | 0 | 0 | 0 | 0 |
| 0 | 0 | 0 | 0 | 0 |
| 0 | 0 | 0 | 0 | 0 |
| 4.603652268 | 0 | 0 | 0 | 0 |
| 0 | 0 | 0 | 0 | 0 |
| 0 | 0 | 0 | 0 | 0 |
| 0 | 0 | 0 | 0 | 0 |
| 0 | 0 | 0 | 0 | 0 |
| 0 | 0 | 0 | 0 | 0 |
| 0 | 0 | 0 | 0 | 0 |
| 0.567185289 | 0.045556846 | 18.29107201 | 0.533230741 | 13.44070461 |
| 0 | 0 | 0 | 0 | 0 |
| 0 | 0 | 0 | 0 | 0 |
| 0 | 0 | 0 | 0 | 0 |
| 0 | 0 | 0 | 0 | 0 |
| 0 | 0 | 0 | 0 | 0 |
| 0 | 0 | 0 | 0 | 0 |
| 0 | 0.008455999 | 0 | 0 | 0 |
| 0 | 0 | 0 | 0 | 0 |
| 0 | 0 | 0 | 0 | 0 |
| 0 | 0 | 0 | 0 | 0 |
| 0 | 0.073485816 | 0 | 0 | 0.10819444 |
| 0 | 0 | 0 | 0 | 0 |
| 0 | 0 | 0 | 0 | 0 |
| 0 | 0 | 0 | 0 | 0 |
| 0 | 0 | 0 | 0 | 0 |
| 0 | 0 | 0 | 0 | 0 |
| 0 | 0 | 0 | 0 | 0 |
| 0 | 0 | 0 | 0 | 0 |
| 0 | 0 | 0 | 0 | 0 |
| 0 | 0 | 0 | 0 | 0 |
| 0 | 0 | 0 | 0 | 0 |
| 0 | 5.208077002 | 0 | 8.919387798 | 2.162442061 |
| 0 | 0 | 0 | 0 | 0 |
| 0 | 0 | 0 | 0 | 0 |
| 0 | 0 | 0 | 0 | 0 |
| 0 | 0 | 0 | 0 | 0 |
| 0 | 0 | 0 | 0 | 0 |
| 0.143406513 | 0.000746155 | 0.430626496 | 0.007995302 | 0.006971535 |
| 1.375771628 | 0.097134414 | 0 | 0.015640859 | 0.007141409 |
| 0.070947279 | 0.030128899 | 0 | 0.02294304 | 0.056472025 |
| 0 | 0 | 0 | 0 | 0 |
| 0 | 0 | 0 | 0 | 0 |

| | | | | |
|---|---|---|---|---|
| 0.160002556 | 0 | 0 | 0.330178144 | 0 |
| 0.001033575 | 0 | 0.00310125 | 0.026336259 | 0.001175757 |
| 0 | 0.573406895 | 0 | 0.00087103 | 0.008749421 |
| 0.332795448 | 0.279972299 | 0.176278195 | 0.774907195 | 1.375112093 |
| 0 | 0 | 0 | 0 | 0.02027314 |
| 0 | 0 | 0 | 0 | 0 |
| 0.39909436 | 0.00105375 | 0 | 0.006210209 | 0.774091216 |
| 0 | 0 | 0 | 0 | 0 |
| 0 | 0 | 0 | 0 | 0 |
| 0 | 0 | 0 | 0 | 0 |
| 0 | 0.004647323 | 0 | 0 | 0 |
| 2.836837775 | 0.241837656 | 0.02687075 | 0.70880247 | 0.335070357 |
| 0.002639229 | 0 | 0 | 0 | 0.002456418 |
| 0 | 0.015696983 | 0.027268375 | 0.053774996 | 0.034747451 |
| 0 | 0 | 0 | 0 | 0 |
| 0 | 0 | 0 | 0 | 0 |
| 0.77848833 | 0.005172813 | 0.001483415 | 0.028268504 | 0 |
| 0 | 0 | 0 | 0 | 0 |
| 0 | 0 | 0 | 0 | 0 |
| 0 | 0 | 0 | 0 | 0 |
| 0.20833473 | 0.019138705 | 0.03681824 | 0.185851657 | 0.069858344 |
| 0.005706815 | 0.003893365 | 0.004723679 | 0.000802283 | 0.001119286 |
| 0 | 0.013436503 | 0 | 0 | 0.00125541 |
| 0 | 0 | 0 | 0 | 0 |
| 0 | 0 | 0 | 0 | 0 |
| 0 | 0 | 0 | 0 | 0 |
| 0 | 0 | 0 | 0 | 0 |
| 0 | 0 | 0 | 0 | 0 |
| 0.006797173 | 0 | 0.004669553 | 0 | 0 |
| 0 | 0 | 0 | 0 | 0 |
| 0 | 0.002545161 | 0.015327468 | 0 | 0 |
| 0 | 0 | 0 | 0 | 0 |
| 0 | 0 | 0 | 0 | 0 |
| 0 | 0 | 0 | 0 | 0 |
| 0 | 0 | 0 | 0 | 0 |
| 0 | 0 | 0 | 0 | 0 |
| 0 | 0 | 0 | 0 | 0 |
| 0.014794725 | 0.000726325 | 0 | 0 | 0 |
| 0 | 0 | 0 | 0 | 0 |
| 0 | 0 | 0 | 0 | 0 |
| 0 | 0 | 0 | 0 | 0 |
| 0 | 0 | 0 | 0 | 0 |
| 0 | 0 | 0 | 0 | 0 |
| 0 | 0 | 0 | 0 | 0 |
| 0 | 0 | 0 | 0 | 0 |
| 0.032778567 | 0 | 0 | 0 | 0 |
| 0 | 0 | 0 | 0 | 0 |
| 0 | 0 | 0 | 0 | 0 |

| | | | | |
|---|---|---|---|---|
| 0 | 0 | 0 | 0 | 0 |
| 0 | 0 | 0.010823332 | 0 | 0 |
| 0 | 0 | 0 | 0 | 0 |
| 0 | 0 | 0 | 0 | 0 |
| 0 | 1.234730085 | 0 | 0 | 0.00738943 |
| 0 | 0 | 0 | 0 | 0 |
| 0 | 0 | 0 | 0 | 0 |
| 0 | 0 | 0 | 0 | 0 |
| 0 | 0 | 0 | 0 | 0 |
| 0 | 0 | 0 | 0 | 0 |
| 0 | 0 | 0 | 0 | 0 |
| 0.006404065 | 0 | 0 | 0.001125381 | 0.000753623 |
| 0 | 0 | 0 | 0 | 0 |
| 0 | 0 | 0 | 0 | 0 |
| 0 | 0 | 0 | 0 | 0 |
| 0 | 0 | 0 | 0 | 0.001164987 |
| 0 | 0 | 0 | 0 | 0 |
| 0 | 0 | 0 | 0 | 0 |
| 0.593908245 | 1.880118698 | 0 | 0 | 0.017437928 |
| 0 | 0 | 0 | 0 | 0 |
| 0 | 0 | 0 | 0 | 0 |
| 0 | 0 | 0 | 0 | 0 |
| 0 | 0 | 0 | 0 | 0 |
| 0 | 0 | 0 | 0 | 0 |
| 0 | 0 | 0 | 0 | 0 |
| 0 | 0 | 0 | 0 | 0 |
| 0 | 0 | 0 | 0 | 0 |
| 0 | 0 | 0 | 0 | 0 |
| 0 | 0 | 0 | 0 | 0 |
| 0 | 0 | 0 | 0 | 0 |
| 0.05896489 | 0 | 0 | 0 | 0 |
| 0 | 0.070115943 | 0 | 0 | 0.005461462 |
| 0 | 0 | 0 | 0 | 0 |
| 0 | 0 | 0 | 0 | 0 |
| 0 | 0 | 0 | 0 | 0 |
| 0 | 0 | 0 | 0 | 0 |
| 0 | 0 | 0 | 0 | 0 |
| 0 | 0 | 0 | 0 | 0 |
| 0 | 0 | 0 | 0 | 0 |
| 0 | 0 | 0 | 0 | 0 |
| 0 | 0 | 0 | 0 | 0 |
| 0 | 0 | 0 | 0 | 0 |
| 0 | 0 | 0 | 0 | 0.030106184 |
| 0 | 0 | 0 | 0 | 0 |
| 0 | 0 | 0 | 0 | 0 |
| 0 | 0 | 0 | 0 | 0 |
| 0 | 0 | 0 | 0 | 0 |
| 0 | 0 | 0 | 0 | 0 |

| | | | | |
|---|---|---|---|---|
| 0 | 0 | 0 | 0 | 0 |
| 0 | 0 | 0 | 0 | 0 |
| 0 | 0 | 0 | 0 | 0 |
| 0 | 0 | 0 | 0 | 0 |
| 0 | 0 | 0 | 0 | 0.050278303 |
| 0 | 0 | 0 | 0 | 0 |
| 0 | 0 | 0 | 0 | 0 |
| 0 | 0 | 0 | 0 | 0 |
| 0 | 0 | 0 | 0 | 0 |
| 0 | 0 | 0 | 0 | 0 |
| 0 | 0 | 0 | 0 | 0 |
| 0 | 0 | 0 | 0 | 0 |
| 0 | 0 | 0 | 0 | 0 |
| 0 | 0 | 0 | 0 | 0 |
| 0 | 0 | 0 | 0 | 0 |
| 0.006096931 | 0 | 0 | 0 | 0 |
| 0 | 0 | 0 | 0 | 0 |
| 0 | 0 | 0 | 0 | 0 |
| 0 | 0 | 0 | 0 | 0 |
| 0 | 0 | 0 | 0 | 0 |
| 0 | 0 | 0 | 0 | 0 |
| 0 | 0 | 0 | 0 | 0 |
| 0 | 0 | 0.001539499 | 0 | 0 |
| 0 | 0 | 0 | 0 | 0 |
| 0 | 0 | 0 | 0.001765474 | 0 |
| 0 | 0 | 0 | 0 | 0 |
| 0 | 0 | 0 | 0.000613324 | 0 |
| 0 | 0 | 0 | 0 | 0 |
| 0 | 0 | 0 | 0 | 0 |
| 0 | 0 | 0 | 0 | 0 |
| 0 | 0 | 0 | 0 | 0 |
| 0.007456806 | 0 | 0 | 0.006731602 | 0 |
| 0 | 0 | 0 | 0 | 0 |
| 0 | 0 | 0 | 0 | 0 |
| 0 | 0 | 0 | 0 | 0 |
| 0 | 0 | 0 | 0 | 0 |
| 0 | 0 | 0 | 0 | 0 |
| 0 | 0 | 0.658416037 | 0 | 0 |
| 0.000517497 | 0 | 0 | 0 | 0 |
| 0 | 0 | 0 | 0 | 0 |
| 0 | 0 | 0 | 0 | 0 |
| 0 | 0 | 0 | 0 | 0 |
| 0 | 0 | 0 | 0 | 0 |
| 0 | 0 | 0 | 0 | 0 |
| 0 | 0 | 0 | 0 | 0 |
| 0 | 0 | 0 | 0 | 0 |
| 0 | 0 | 0 | 0 | 0 |
| 0 | 0 | 0 | 0 | 0 |

| | | | | |
|---|---|---|---|---|
| 0 | 0 | 0 | 0 | 0 |
| 0 | 0 | 0 | 0 | 0 |
| 0 | 0 | 0 | 0 | 0 |
| 0 | 0 | 0 | 0 | 0 |
| 0 | 0 | 0 | 0 | 0 |
| 0 | 0 | 0 | 0 | 0 |
| 0 | 0 | 0 | 0 | 0 |
| 0 | 0 | 0 | 0 | 0 |
| 0 | 0 | 0 | 0 | 0 |
| 0 | 0 | 0 | 0 | 0 |
| 0 | 0 | 0 | 0 | 0 |
| 0 | 0 | 0 | 0 | 0 |
| 0.006523824 | 0.005274571 | 0 | 0 | 0.000927657 |
| 0 | 0 | 0 | 0 | 0 |
| 0.025883484 | 0.018517637 | 0 | 0 | 0 |
| 0 | 0 | 0 | 0 | 0 |
| 0.00172771 | 0 | 0 | 0 | 0 |
| 0 | 0 | 0 | 0 | 0 |
| 0 | 0 | 0 | 0 | 0 |
| 0 | 0 | 0 | 0 | 0 |
| 0.09110369 | 0.018564106 | 0.014140958 | 0.071875634 | 0.146564955 |
| 0 | 0 | 0 | 0 | 0 |
| 0 | 0 | 0 | 0 | 0 |
| 0 | 0 | 0 | 0 | 0.008139289 |
| 0 | 0 | 0 | 0 | 0 |
| 0 | 0 | 0 | 0 | 0 |
| 0.002885926 | 0 | 0 | 0.001583842 | 0.004545584 |
| 0 | 0 | 0 | 0 | 0 |
| 0 | 0 | 0 | 0 | 0 |
| 0 | 0 | 0 | 0 | 0 |
| 0 | 0 | 0.001690141 | 0 | 0 |
| 0 | 0 | 0 | 0 | 0 |
| 0 | 0 | 0 | 0 | 0 |
| 0.001397747 | 0 | 0 | 0 | 0 |
| 0.031498612 | 0.053033467 | 0.00378859 | 0 | 0.010721281 |
| 0 | 0 | 0 | 0 | 0 |
| 0 | 0 | 0 | 0 | 0 |
| 0 | 0 | 0 | 0 | 0 |
| 0 | 0 | 0 | 0 | 0 |
| 0 | 0 | 0 | 0 | 0 |
| 0 | 0 | 0 | 0 | 0 |
| 0 | 0 | 0 | 0 | 0 |
| 0 | 0 | 0 | 0 | 0 |
| 0 | 0 | 0 | 0 | 0 |
| 0 | 0 | 0 | 0 | 0 |
| 0 | 0 | 0 | 0 | 0 |
| 0 | 0 | 0.009376081 | 0 | 0 |
| 0.003888416 | 0.004438333 | 0 | 0 | 0 |

| | | | | |
|---|---|---|---|---|
| 0 | 0 | 0 | 0 | 0 |
| 0 | 0 | 0 | 0 | 0 |
| 0 | 0 | 0 | 0 | 0.014697944 |
| 0 | 0 | 0 | 0 | 0 |
| 0 | 0 | 0 | 0 | 0 |
| 0 | 0 | 0 | 0 | 0 |
| 0 | 0.001531615 | 0 | 0 | 0 |
| 0.002540758 | 0 | 0 | 0 | 0 |
| 0 | 0 | 0 | 0 | 0 |
| 0 | 0 | 0 | 0 | 0 |
| 0 | 0 | 0 | 0 | 0 |
| 0 | 0 | 0 | 0 | 0 |
| 0 | 0 | 0 | 0 | 0 |
| 0 | 0 | 0 | 0 | 0 |
| 0 | 0.003604917 | 0 | 0 | 0 |
| 0 | 0 | 0 | 0 | 0 |
| 0 | 0 | 0 | 0 | 0 |
| 0 | 0 | 0 | 0 | 0 |
| 0.009167234 | 0 | 0 | 0 | 0 |
| 0.047170536 | 0 | 0 | 0 | 0 |
| 0.003712087 | 0.000807061 | 0 | 0 | 0.002111366 |
| 0 | 0 | 0 | 0 | 0 |
| 0 | 0 | 0 | 0 | 0 |
| 0 | 0 | 0 | 0 | 0 |
| 0.013169367 | 0.001603396 | 0 | 0.00644285 | 0 |
| 0 | 0 | 0 | 0 | 0 |
| 0 | 0 | 0 | 0 | 0 |
| 0 | 0 | 0 | 0 | 0 |
| 0.049014101 | 0 | 0.015906561 | 0 | 0 |
| 0 | 0 | 0 | 0 | 0 |
| 0.601853892 | 0.054892394 | 0.007232613 | 0.039146466 | 0.092705628 |
| 0 | 0 | 0 | 0 | 0 |
| 0 | 0 | 0.064274836 | 0 | 0 |
| 0 | 0 | 0 | 0 | 0 |
| 0 | 0 | 0 | 0 | 0 |
| 0 | 0 | 0 | 0 | 0 |
| 0 | 0 | 0 | 0 | 0 |
| 0 | 0 | 0 | 0 | 0 |
| 0 | 0 | 0 | 0 | 0 |
| 0 | 0 | 0 | 0 | 0 |
| 0 | 0 | 0 | 0 | 0 |
| 0.009340469 | 0.006200271 | 0.0065051 | 0.003038323 | 0 |
| 0 | 0 | 0 | 0 | 0 |
| 0 | 0 | 0 | 0 | 0 |
| 0 | 0 | 0 | 0 | 0 |
| 0 | 0 | 0 | 0 | 0 |
| 0 | 0 | 0 | 0 | 0 |
| 0 | 0 | 0 | 0 | 0 |

| | | | | |
|---|---|---|---|---|
| 0 | 0 | 0 | 0 | 0 |
| 0 | 0 | 0 | 0 | 0 |
| 0.064881042 | 0.017279925 | 0 | 0.02289069 | 0.045419236 |
| 0 | 0 | 0 | 0 | 0 |
| 0 | 0 | 0 | 0 | 0 |
| 0 | 0 | 0 | 0 | 0 |
| 0 | 0 | 0 | 0 | 0 |
| 0 | 0 | 0 | 0 | 0 |
| 0 | 0 | 0 | 0 | 0 |
| 0.001141148 | 0 | 0 | 0 | 0.007529141 |
| 0 | 0 | 0 | 0 | 0 |
| 0 | 0 | 0 | 0 | 0 |
| 0 | 0 | 0 | 0 | 0 |
| 0.005791579 | 0 | 0 | 0.000843278 | 0 |
| 0.008649868 | 0.005542834 | 0 | 0.008584194 | 0.002589413 |
| 0 | 0.001483584 | 0 | 0 | 0 |
| 0.003890427 | 0.004341949 | 0 | 0.014274855 | 0 |
| 0 | 0 | 0 | 0 | 0 |
| 0 | 0 | 0 | 0 | 0 |
| 0 | 0 | 0 | 0 | 0 |
| 0 | 0 | 0 | 0 | 0 |
| 0.003043686 | 0.004275863 | 0 | 0 | 0 |
| 0 | 0 | 0 | 0 | 0 |
| 0 | 0 | 0 | 0 | 0 |
| 0 | 0 | 0 | 0 | 0 |
| 0 | 0 | 0 | 0 | 0 |
| 0 | 0 | 0 | 0 | 0 |
| 0 | 0 | 0 | 0 | 0 |
| 0 | 0 | 0 | 0 | 0 |
| 0 | 0 | 0 | 0 | 0 |
| 0 | 0 | 0 | 0 | 0.002449182 |
| 0 | 0 | 0 | 0 | 0 |
| 0 | 0 | 0 | 0 | 0 |
| 0 | 0 | 0 | 0 | 0 |
| 0 | 0 | 0 | 0 | 0 |
| 0.009352816 | 0 | 0 | 0.002383166 | 0 |
| 0 | 0 | 0 | 0 | 0 |
| 0 | 0.031785393 | 0.001925734 | 0.000899449 | 0.001673126 |
| 0 | 0 | 0 | 0 | 0 |
| 0 | 0.001683874 | 0 | 0 | 0 |
| 0 | 0 | 0 | 0 | 0.001931835 |
| 0 | 0 | 0 | 0 | 0 |
| 0 | 0 | 0 | 0 | 0 |
| 0 | 0 | 0 | 0 | 0 |
| 0 | 0 | 0 | 0 | 0 |
| 0 | 0 | 0 | 0 | 0 |
| 0 | 0 | 0 | 0 | 0 |
| 0 | 0 | 0 | 0 | 0 |

| | | | | |
|---|---|---|---|---|
| 0 | 0.02984965 | 0.392633545 | 0 | 10.254736 |
| 0 | 0 | 0 | 0 | 0.036312401 |
| 0 | 0.002425926 | 0 | 0 | 0 |
| 0 | 0 | 0 | 0 | 0 |
| 0.000986911 | 0.001126484 | 0.002422826 | 0 | 0.020488779 |
| 0 | 0 | 0 | 0 | 0 |
| 0 | 0 | 0 | 0 | 0 |
| 0 | 0 | 0 | 0 | 0.080650746 |
| 0 | 0 | 0 | 0 | 0 |
| 0 | 0.057777156 | 0.050562178 | 0.003269904 | 0.019058723 |
| 0 | 0 | 0 | 0 | 0 |
| 0.068442665 | 0.005624795 | 0.014338047 | 0.000837105 | 0 |
| 0.02616576 | 0.003160449 | 0 | 0.004444818 | 0 |
| 0 | 0 | 0 | 0 | 0 |
| 0 | 0 | 0 | 0 | 0 |
| 0 | 0 | 0 | 0 | 0 |
| 0.001360188 | 0 | 0 | 0 | 0 |
| 0 | 0 | 0 | 0 | 0 |
| 0 | 0.002041962 | 0.049773999 | 0 | 0 |
| 0 | 0.016367593 | 0 | 0 | 0 |
| 0 | 0 | 0 | 0 | 0 |
| 0 | 0 | 0 | 0 | 0 |
| 0 | 0 | 0 | 0 | 0 |
| 0 | 0 | 0 | 0 | 0 |
| 0 | 0 | 0 | 0 | 0 |
| 0 | 0 | 0 | 0 | 0 |
| 0 | 0 | 0 | 0 | 0 |
| 0 | 0.003116046 | 0.012446473 | 0.031600764 | 0 |
| 0 | 0 | 0.044854127 | 0 | 0 |
| 0 | 0 | 0 | 0 | 0 |
| 0 | 0 | 0 | 0 | 0 |
| 0 | 0 | 0 | 0 | 0 |
| 0 | 0 | 0 | 0 | 0 |
| 0 | 0 | 0.062471344 | 0 | 0 |
| 0 | 0 | 0 | 0 | 0 |
| 0 | 0 | 0 | 0 | 0.01841509 |
| 0 | 0 | 0.002991688 | 0 | 0 |
| 0 | 0 | 0 | 0 | 0 |
| 0 | 0 | 0 | 0 | 0 |
| 0 | 0 | 0 | 0 | 0 |
| 0 | 0 | 0.005938671 | 0 | 0 |
| 0 | 0 | 0 | 0 | 0 |
| 0 | 0 | 0 | 0 | 0 |
| 0 | 0 | 0 | 0 | 0 |
| 0 | 0 | 0 | 0 | 0 |
| 0 | 0 | 0 | 0 | 0 |
| 0.003308718 | 0 | 0 | 0 | 0 |
| 0 | 0 | 0 | 0 | 0 |

| | | | | |
|---|---|---|---|---|
| 0 | 0 | 0 | 0 | 0 |
| 0.021850186 | 0 | 0 | 0 | 0 |
| 0 | 0 | 0 | 0 | 0 |
| 0 | 0 | 0 | 0 | 0 |
| 0 | 0 | 0 | 0 | 0 |
| 0 | 0.011177177 | 0 | 0.003621993 | 0 |
| 0 | 0 | 0 | 0 | 0 |
| 0 | 0 | 0 | 0 | 0 |
| 0 | 0 | 0 | 0 | 0 |
| 0 | 0 | 0 | 0 | 0 |
| 0 | 0 | 0.005696326 | 0 | 0.010627562 |
| 0 | 0.002155625 | 0 | 0 | 0 |
| 0 | 0 | 0 | 0 | 0 |
| 0.1118854 | 0.002765542 | 0.001982696 | 0 | 0.00324836 |
| 0 | 0 | 0 | 0 | 0 |
| 0 | 0 | 0.09936277 | 0 | 0 |
| 0 | 0 | 0 | 0 | 0 |
| 0.002021022 | 0 | 0 | 0 | 0 |
| 0 | 0 | 0 | 0 | 0 |
| 0 | 0 | 0 | 0 | 0.006020563 |
| 0 | 0 | 0 | 0 | 0 |
| 0.203102124 | 0.131769951 | 0 | 0.274315029 | 0 |
| 0 | 0.02399421 | 0 | 0 | 0 |
| 0 | 0 | 0 | 0 | 0 |
| 0 | 0 | 0 | 0 | 0 |
| 0 | 0 | 0 | 0 | 0 |
| 0 | 0.001070921 | 0.001842657 | 0 | 0 |
| 0 | 0 | 0 | 0 | 0 |
| 0 | 0 | 0 | 0 | 0 |
| 2.110698656 | 0.105396321 | 1.326480268 | 0.614708516 | 0.449304527 |
| 0.001232802 | 0.000536057 | 0 | 0.011308567 | 0.003205461 |
| 0.002225333 | 0 | 0 | 0 | 0 |
| 0 | 0 | 0 | 0 | 0 |
| 0 | 0 | 0 | 0 | 0 |
| 0 | 0 | 0.002046347 | 0 | 0 |
| 0 | 0 | 0 | 0 | 0 |
| 0.345754366 | 0 | 0 | 0.059537108 | 0 |
| 0.010319258 | 0 | 0.001732196 | 0.00188779 | 0 |
| 0.152638371 | 0 | 0.928226162 | 0 | 0.521852575 |
| 0 | 0 | 0 | 0 | 0 |
| 0.03414644 | 0 | 0.007917102 | 0 | 0.246657619 |
| 0 | 0 | 0 | 3.092920683 | 2.670491127 |
| 0.00557491 | 0 | 0.002115738 | 0 | 0 |
| 0 | 0 | 0 | 0 | 0 |
| 0 | 0 | 0.001050695 | 0 | 0 |
| 0 | 0 | 0 | 0 | 0 |
| 0 | 0.000511346 | 0 | 0 | 0.001019232 |
| 0 | 0 | 0 | 0 | 0 |

| | | | | |
|---|---|---|---|---|
| 0 | 0 | 0 | 0.017464545 | 0 |
| 0 | 0 | 0 | 0 | 0 |
| 1.753722433 | 0.009742929 | 0.003697933 | 0.003454371 | 0.017135232 |
| 0 | 0 | 0 | 0 | 0 |
| 0.019447464 | 0.035375566 | 0 | 0 | 0 |
| 0 | 0 | 0 | 0 | 0 |
| 0 | 0 | 0 | 0 | 0 |
| 0.052039127 | 0 | 0 | 0.002209992 | 0 |
| 0 | 0 | 0 | 0 | 0 |
| 0 | 0 | 0 | 0 | 0 |
| 0 | 0 | 0 | 0 | 0.014864768 |
| 0 | 0 | 0 | 0 | 0 |
| 0 | 0 | 0 | 0 | 0 |
| 0 | 0 | 0.011184894 | 0 | 0 |
| 0 | 0 | 0 | 0 | 0 |
| 0.27667429 | 0.008920985 | 0.028567498 | 0.003385528 | 0.048232598 |
| 0.019274823 | 0.01071198 | 0.00590748 | 0.045471524 | 0.008622711 |
| 0 | 0 | 0 | 0 | 0 |
| 0 | 0 | 0 | 0 | 0 |
| 0 | 0 | 0 | 0 | 0 |
| 0 | 0.004361585 | 0.011614365 | 0.001390948 | 0.006830729 |
| 0.006280279 | 0 | 0 | 0.001536251 | 0 |
| 0 | 0 | 0 | 0 | 0 |
| 0 | 0.049845575 | 0.274408555 | 0 | 0.666831155 |
| 0 | 0 | 0 | 0 | 0 |
| 0.018829416 | 0.003256418 | 0 | 0 | 0.008438041 |
| 0.003849142 | 0 | 0 | 0 | 0.019703881 |
| 0 | 0 | 0 | 0 | 0 |
| 0 | 0 | 0 | 0 | 0 |
| 0 | 0 | 0 | 0 | 0 |
| 0.013172676 | 0.011773923 | 0.095133041 | 0 | 0.048997741 |
| 0.055175798 | 0.001660464 | 0 | 0.015568377 | 0.01702127 |
| 1.450495277 | 0.003613682 | 0 | 0.022688571 | 0 |
| 0.085495997 | 0 | 0.0027846 | 0 | 0.002903196 |
| 0 | 0.03049461 | 0.006956236 | 0 | 0 |
| 0 | 0 | 0 | 0 | 0 |
| 0.019588374 | 0 | 0 | 0.021606007 | 0 |
| 0.010072778 | 0.005429289 | 0.096165474 | 0.011977549 | 0.022678117 |
| 0 | 0.092883495 | 0 | 0.395950339 | 0 |
| 0 | 0 | 0 | 0 | 0.002146931 |
| 0 | 0 | 0 | 0 | 0 |
| 0 | 0 | 0 | 0 | 0 |
| 0 | 0.020298732 | 0.003081757 | 0 | 0.002856007 |
| 0.017528037 | 0 | 0.051833361 | 0 | 0.007826619 |
| 0 | 0 | 0.005946057 | 0 | 0 |
| 0 | 0.002262905 | 0.001247955 | 0 | 0 |
| 0 | 0 | 0 | 0 | 0 |
| 0 | 0 | 0 | 0 | 0 |

| | | | | |
|---|---|---|---|---|
| 0 | 0 | 0 | 0 | 0 |
| 0.025163303 | 0.073980947 | 0.056470991 | 0.002040116 | 0.760509858 |
| 0 | 0 | 0 | 0 | 0 |
| 0 | 0 | 0 | 0 | 0 |
| 0 | 0 | 0 | 0 | 0 |
| 0 | 0 | 0 | 0 | 0 |
| 0 | 0 | 0.002020448 | 0 | 0 |
| 0 | 0 | 0 | 0 | 0 |
| 0.001832324 | 0 | 0.033415811 | 0 | 0 |
| 0 | 0 | 0 | 0 | 0 |
| 0 | 0 | 0.069321945 | 0.00346521 | 0.001450322 |
| 0.353540879 | 0.083814992 | 0 | 0.837135429 | 0.011881623 |
| 0 | 0 | 0 | 0 | 0 |
| 0 | 0 | 0 | 0 | 0 |
| 0.011215041 | 0 | 0 | 0.004721376 | 0 |
| 0 | 0 | 0 | 0 | 0 |
| 0.038938775 | 0.007619259 | 0.002427762 | 0.001814288 | 0.008099712 |
| 0.002021916 | 0 | 0.028048976 | 0.004121593 | 0.011828863 |
| 0 | 0 | 0 | 0 | 0 |
| 0.001125366 | 0.005708979 | 0.058938195 | 0 | 0.007040959 |
| 0.002294611 | 0.325819294 | 0 | 0.741612933 | 0.02871291 |
| 0 | 0 | 0 | 0.001164072 | 0 |
| 0 | 0 | 0 | 0 | 0 |
| 0 | 0 | 0 | 0 | 0 |
| 0.005397525 | 0 | 0 | 0 | 0 |
| 0 | 0 | 0.002295904 | 0 | 0 |
| 0.102146817 | 0.009134403 | 0 | 0 | 0.029371662 |
| 0.021705616 | 0.033776099 | 0 | 0.042008841 | 0.13816143 |
| 0 | 0 | 0 | 0 | 0 |
| 0.001176722 | 0.002462422 | 0 | 0 | 0 |
| 0 | 0 | 0 | 0 | 0.000893056 |
| 0 | 0 | 0 | 0 | 0 |
| 0 | 0 | 0 | 0 | 0 |
| 0 | 0 | 0 | 0 | 0 |
| 0 | 0 | 0.045217992 | 0.045272344 | 0 |
| 0.129606004 | 0.05433724 | 0.664727574 | 0.329824894 | 0.678952105 |
| 0 | 0 | 0 | 0 | 0 |
| 0 | 0 | 0 | 0 | 0 |
| 0.029784879 | 0.025640983 | 0.003282634 | 0.001226571 | 0.007529371 |
| 0 | 0 | 0 | 0 | 0 |
| 0 | 0 | 0.004943688 | 0 | 0 |
| 0 | 0 | 0 | 0 | 0 |
| 0.014619628 | 0.00125154 | 0.00717812 | 0 | 0.03617187 |
| 0.001058623 | 0.005236134 | 0 | 0.002517619 | 0.020672964 |
| 0 | 0.002469408 | 0.034143224 | 0 | 0 |
| 0 | 0.175200964 | 0 | 0.000929007 | 0 |
| 0 | 0 | 0 | 0 | 0.300591328 |
| 0.000855591 | 0 | 0 | 0 | 0 |

| | | | | |
|---|---|---|---|---|
| 0 | 0 | 0.001114221 | 0 | 0 |
| 0 | 0 | 0 | 0 | 0 |
| 0 | 0 | 0 | 0 | 0 |
| 0.034693003 | 4.182965245 | 2.242401694 | 0 | 0.035591558 |
| 0 | 0 | 0 | 0 | 0 |
| 0 | 0 | 0 | 0 | 0 |
| 0 | 0 | 0 | 0 | 0 |
| 0 | 0 | 0 | 0 | 0.026208819 |
| 3.08291329 | 2.57383775 | 0 | 0.352568722 | 0.62829634 |
| 0 | 0 | 0 | 0.129709068 | 0 |
| 0 | 0 | 0 | 0 | 0 |
| 0 | 0.014121015 | 0 | 0.006235362 | 0.031084845 |
| 0 | 0 | 0 | 0 | 0 |
| 0 | 0 | 0 | 0 | 0 |
| 0.020529999 | 0.12779552 | 0.977343924 | 0.003258777 | 0.024694155 |
| 0 | 0 | 0 | 0 | 0 |
| 0.168140668 | 0.034742682 | 0 | 0 | 0.315122003 |
| 11.77182615 | 0 | 0 | 0.031580078 | 0 |
| 0 | 0 | 0 | 0 | 0 |
| 0 | 0.001247187 | 0 | 0 | 0 |
| 0 | 0 | 0 | 0 | 0 |
| 0 | 0 | 0 | 0.000771877 | 0 |
| 0 | 0 | 0 | 0 | 0 |
| 0.1439408 | 0.147152414 | 0.093029043 | 0 | 0.331908845 |
| 0.001718326 | 0 | 0 | 0 | 0 |
| 0 | 0 | 0 | 0 | 0 |
| 0 | 0 | 0 | 0 | 0 |
| 3.461789751 | 0 | 0 | 1.17526464 | 3.966933362 |
| 0 | 0 | 0 | 0 | 0 |
| 0 | 0 | 0 | 0 | 0 |
| 0.00198249 | 0.214821114 | 0 | 0 | 0 |
| 0 | 0 | 0 | 0.088145526 | 0 |
| 0.002866759 | 0 | 0 | 0 | 0.015104125 |
| 0 | 0 | 0 | 0 | 0 |
| 0 | 0 | 0 | 0 | 0.001457543 |
| 0 | 0 | 0.346452238 | 0.001541928 | 0 |
| 0.035820342 | 0.512110263 | 0 | 3.723489925 | 0.169165474 |
| 0 | 0 | 0 | 0 | 0 |
| 0.019530219 | 0 | 0.004864077 | 0 | 0.023665776 |
| 0 | 0 | 0.047937171 | 0 | 0 |
| 0.00762952 | 0.046147724 | 0.083885521 | 0 | 0.008679058 |
| 0 | 0 | 0 | 0 | 0 |
| 2.545815706 | 0.116791446 | 0.675994365 | 0.775225789 | 0.278977353 |
| 0.029823181 | 0 | 0 | 0 | 0 |
| 0 | 0 | 0 | 0 | 0 |
| 0 | 0 | 0 | 0 | 0 |
| 0 | 0 | 0 | 0 | 0 |
| 0 | 0.014852946 | 0.003592265 | 0 | 0 |

| | | | | |
|---|---|---|---|---|
| 0 | 0 | 0 | 0 | 0 |
| 0.024638157 | 0 | 0 | 0.040397455 | 0 |
| 0.007664798 | 0.002916263 | 0.111424793 | 0.001608391 | 0.216184583 |
| 0 | 0.030039954 | 0 | 0 | 0 |
| 0 | 0 | 0 | 0 | 0 |
| 0.147043839 | 0.148244682 | 0 | 0.001810041 | 0.01212113 |
| 0 | 0 | 0 | 0 | 0 |
| 0 | 0 | 0 | 0 | 0 |
| 0 | 0 | 0 | 0 | 0 |
| 0 | 0 | 0 | 0 | 0 |
| 0.112218067 | 0.102066554 | 0.108191889 | 0 | 0.065464152 |
| 0.3665066 | 0.815819062 | 0.291304076 | 0.909083271 | 0.299191181 |
| 0 | 0 | 0 | 0 | 0 |
| 0.001433258 | 0.001999502 | 0.003648909 | 0 | 0.003260842 |
| 0.063249893 | 0.003437857 | 0.203132154 | 0.00383727 | 0.371102991 |
| 0 | 0.063133704 | 0 | 0 | 0 |
| 0 | 0 | 0 | 0 | 0 |
| 0 | 0 | 0 | 0 | 0 |
| 0.003168834 | 0.036019148 | 0 | 0 | 0 |
| 0.005469713 | 0 | 0.020688952 | 0 | 0.002696262 |
| 0.053427843 | 0.696373557 | 0.013966453 | 0.020042547 | 0.192884231 |
| 0.02169756 | 0 | 9.519745051 | 0 | 0 |
| 0.038899251 | 1.12508098 | 4.389187923 | 0.044290628 | 2.248794286 |
| 0 | 0 | 0 | 0 | 0 |
| 0 | 0 | 0 | 0 | 0 |
| 0 | 5.669722976 | 0 | 13.52065541 | 0.004046095 |
| 0 | 0.854675767 | 0 | 0 | 0 |
| 0 | 0 | 0 | 0 | 0 |
| 0 | 0 | 0 | 0 | 0.008505068 |
| 0 | 0 | 0 | 0 | 0 |
| 0 | 0 | 0 | 0 | 0 |
| 0.185296869 | 0.081695642 | 0.102234637 | 0.105318902 | 0.28749091 |
| 0 | 0.011589564 | 0 | 0 | 0 |
| 0 | 0 | 0.022696732 | 0 | 0.06566589 |
| 0 | 0.0028542 | 0.001309605 | 0 | 0.005461523 |
| 0.830937954 | 0.155134393 | 0.150242958 | 0.173333155 | 0.037753485 |
| 0 | 0.005909454 | 0.060834043 | 0 | 0 |
| 0 | 0 | 0 | 0 | 0 |
| 0 | 0.840143607 | 0 | 0.411324378 | 0 |
| 0 | 0 | 0 | 0 | 0 |
| 0 | 0 | 0 | 0.000925022 | 0 |
| 0 | 0 | 0 | 0 | 0 |
| 0 | 0 | 0 | 0 | 0 |
| 0 | 0 | 0 | 0 | 0 |
| 0.030324774 | 0.022538988 | 0.009746604 | 0.00551071 | 0.043588391 |
| 0 | 0 | 0 | 0 | 0 |
| 0 | 0 | 0 | 0 | 0 |
| 0.467316131 | 0.011652076 | 0.027536291 | 2.106076928 | 0.742242054 |

| | | | | |
|---|---|---|---|---|
| 0 | 0 | 0 | 0 | 0 |
| 0 | 0 | 0 | 0 | 0 |
| 0 | 0 | 0 | 0 | 0 |
| 0.013824434 | 0.00792789 | 0 | 0 | 0.010256192 |
| 0 | 0 | 0 | 0 | 0 |
| 0 | 0 | 0 | 0 | 0 |
| 0 | 0 | 0 | 0 | 0 |
| 0 | 0 | 0.120090249 | 0 | 0.000838894 |
| 0 | 0 | 0 | 0 | 0 |
| 0 | 0 | 0 | 0 | 0 |
| 0.007140526 | 0.004414785 | 0.007060575 | 0 | 0.000761512 |
| 0 | 0 | 0 | 0 | 0 |
| 0 | 0 | 0.247070255 | 0.01174895 | 0 |
| 0 | 0 | 0 | 0.91641978 | 9.261020812 |
| 0 | 0 | 0 | 0 | 0 |
| 0 | 0 | 0 | 0 | 0 |
| 0 | 0 | 0 | 0.023680964 | 0 |
| 0.003715203 | 0.006784999 | 0 | 0.007005291 | 0.005705474 |
| 0 | 0.022311309 | 0 | 0 | 0 |
| 0 | 0 | 0.002010638 | 0 | 0.001905701 |
| 0 | 0 | 0.009363467 | 0 | 0.010137728 |
| 0 | 0 | 0 | 0 | 0 |
| 0.014991772 | 0.002444568 | 0 | 0.027940668 | 1.396972907 |
| 0.007312496 | 0.17329262 | 0 | 0 | 0 |
| 0 | 0 | 0 | 0 | 0 |
| 0 | 0 | 0 | 0 | 0 |
| 0 | 0 | 0 | 0 | 0 |
| 0 | 0 | 0 | 0 | 0 |
| 0 | 0 | 0 | 0 | 0 |
| 0.281393349 | 0.310090269 | 2.424382149 | 0 | 0.02782391 |
| 0 | 0 | 0 | 0 | 0 |
| 0 | 0 | 0 | 0 | 0 |
| 0 | 0 | 0 | 0 | 0 |
| 0 | 0 | 0 | 0 | 0 |
| 0 | 0 | 0 | 0.000723372 | 0.007669887 |
| 0 | 0 | 0 | 0 | 0 |
| 0 | 0 | 0 | 0.178666076 | 0 |
| 0 | 0 | 0.0026793 | 0.000770102 | 0.001289267 |
| 0 | 0 | 0 | 0 | 0 |
| 0 | 0 | 0 | 0 | 0 |
| 0 | 0 | 0 | 0 | 0.207051986 |
| 0 | 0 | 0 | 0 | 0 |
| 0 | 0 | 0 | 0 | 0 |
| 0.010639381 | 0 | 0 | 0.037019067 | 0 |
| 0 | 0.053370726 | 0 | 0.059024866 | 0 |
| 0 | 0 | 0 | 0 | 0 |
| 0 | 0.004780137 | 0 | 0.007317255 | 0.002245869 |
| 0.175391179 | 0 | 0 | 0 | 0 |

| | | | | |
|---|---|---|---|---|
| 0 | 0 | 0 | 0 | 0 |
| 0 | 0 | 0 | 0 | 0 |
| 0 | 0 | 0 | 0 | 0 |
| 1.83797229 | 0.653263601 | 0 | 0.101306863 | 0.186809134 |
| 0 | 0 | 0 | 0 | 0 |
| 0 | 0 | 0 | 1.852980102 | 0 |
| 0 | 0 | 0 | 0 | 0.13035586 |
| 0 | 0 | 0 | 0 | 0 |
| 0.015630911 | 0.002378868 | 0.171969042 | 0.006638118 | 0.028449828 |
| 0 | 0 | 0 | 0.003421214 | 0 |
| 0 | 0 | 0 | 0 | 0 |
| 0 | 0 | 0 | 0 | 0 |
| 0 | 0 | 0 | 0 | 0 |
| 0 | 0 | 0 | 0 | 0 |
| 1.38331747 | 1.716212683 | 1.864344258 | 0.157420773 | 0.016185464 |
| 0 | 0 | 0.174384947 | 0 | 0 |
| 0 | 0 | 0 | 0 | 0 |
| 0 | 0 | 0 | 0 | 0 |
| 0 | 0 | 0 | 0 | 0 |
| 0 | 0 | 0 | 0 | 0 |
| 0 | 0 | 0 | 0 | 0 |
| 0 | 0 | 0 | 0 | 0 |
| 0 | 0 | 0 | 0 | 0 |
| 0 | 0 | 0 | 0 | 0 |
| 0.068930647 | 0.036600371 | 0 | 0 | 0 |
| 9.211880844 | 3.674739534 | 0 | 11.28612915 | 0.032190806 |
| 0 | 0.121911576 | 0 | 0 | 0 |
| 0 | 0 | 0 | 0 | 0.006489941 |
| 0.141154249 | 0 | 0 | 0.064426901 | 0 |
| 0 | 0 | 0 | 0 | 0 |
| 0 | 0 | 0 | 0 | 0 |
| 0 | 0 | 0 | 0 | 0 |
| 0 | 0 | 0 | 0 | 0 |
| 0 | 0.003859588 | 0.008390407 | 0 | 0.001488979 |
| 0.00134793 | 0.002256555 | 0 | 0.00137385 | 0.003066709 |
| 0 | 0.009280645 | 0.004574322 | 0.007963389 | 0.001300675 |
| 0 | 0 | 0 | 0 | 0 |
| 0.195814126 | 0.518241685 | 0.299816402 | 5.103752064 | 0.632759315 |
| 0 | 5.237103061 | 0 | 0 | 0 |
| 0 | 0 | 0 | 0 | 0 |
| 0 | 0 | 0 | 0 | 0 |
| 0 | 0 | 0.002357872 | 0 | 0.054819942 |
| 0.530454976 | 1.887463165 | 0.444665776 | 0 | 0.59809365 |
| 0 | 0 | 0 | 0 | 0 |
| 0 | 0 | 0 | 0 | 0 |
| 0.093878734 | 0.00123641 | 0 | 0 | 0.001026855 |
| 0 | 0 | 0 | 0 | 0.042813636 |
| 0.001725693 | 0.01335052 | 0.012552614 | 0.120189931 | 0.004253349 |

| | | | | |
|---|---|---|---|---|
| 0.013511939 | 0 | 0.003042162 | 0 | 0.015370678 |
| 0 | 0 | 0 | 0 | 0 |
| 0 | 0 | 0 | 0 | 0 |
| 0 | 0 | 0 | 0 | 0 |
| 0.002775954 | 0.043222169 | 0 | 0 | 0 |
| 0 | 0 | 0 | 0 | 0 |
| 0 | 0 | 0 | 0 | 0 |
| 0 | 0.001691444 | 0 | 0 | 0 |
| 0 | 0 | 0 | 0 | 0 |
| 0.00514385 | 0 | 0 | 0 | 0 |
| 0 | 0 | 0 | 0 | 0 |
| 2.035399815 | 0.719702549 | 0.170666271 | 0.650150758 | 0.936287321 |
| 0.010859241 | 0.063942489 | 0 | 1.183754985 | 0 |
| 0 | 0 | 0 | 0 | 0 |
| 1.534072861 | 0.027624083 | 0.326572058 | 0.039265491 | 0.046633499 |
| 0 | 0 | 0 | 0 | 0 |
| 0 | 0.001437972 | 0 | 0 | 0 |
| 0 | 0 | 0 | 0 | 0 |
| 0 | 0 | 0.001180855 | 0 | 0 |
| 0 | 0.008218683 | 0 | 0 | 0 |
| 0 | 0.112364763 | 0.133154517 | 0.34100688 | 0 |
| 0.002575957 | 0 | 0 | 0 | 0 |
| 0.767379654 | 1.527590454 | 0.088413017 | 0.066500851 | 0.05171574 |
| 0 | 0 | 0 | 0 | 0 |
| 0 | 0 | 0 | 0 | 0 |
| 0 | 0 | 0 | 0 | 0 |
| 0 | 0 | 0 | 0 | 0 |
| 0 | 0 | 0 | 0 | 0 |
| 0 | 0 | 0 | 0.001813758 | 0 |
| 0 | 0 | 0 | 0 | 0 |
| 0 | 0 | 0 | 0 | 0 |
| 0.053836646 | 0.074130733 | 0.011888019 | 0.015351063 | 0.077282284 |
| 0 | 0 | 0 | 0 | 0 |
| 0.006243682 | 0.083692954 | 0 | 0.000979037 | 0.052267705 |
| 0 | 0.004892027 | 0.004299189 | 0.005072876 | 0.002123191 |
| 0.631993124 | 0.07750788 | 0.002625241 | 0.024523321 | 0.003345284 |
| 0 | 0 | 0 | 0 | 0 |
| 0 | 0 | 0 | 0.000808474 | 0 |
| 0 | 0.002900757 | 0 | 0 | 0 |
| 0 | 0 | 0 | 0 | 0 |
| 0.120679437 | 0 | 0 | 0 | 1.438128134 |
| 0 | 0 | 0 | 0.000814431 | 0 |
| 0 | 0 | 0 | 0 | 0 |
| 0 | 0 | 0 | 0 | 0 |
| 0.021654961 | 0.138977678 | 0.018055038 | 0.001041102 | 0.033232495 |
| 0 | 0 | 0 | 0 | 0 |
| 0.319881231 | 0 | 0 | 0.096881178 | 0 |
| 0 | 0.000735168 | 0 | 0 | 0.007473341 |

| | | | | |
|---|---|---|---|---|
| 0 | 0 | 0 | 0 | 0 |
| 0 | 0 | 0 | 0 | 0 |
| 0 | 0.00385873 | 0 | 0 | 0 |
| 0 | 0 | 0 | 0 | 0 |
| 0 | 0.001467491 | 0.001168985 | 0 | 0 |
| 0 | 0 | 0 | 0 | 0 |
| 0 | 0 | 0 | 0.037217649 | 0 |
| 0.003192685 | 0 | 0.017853027 | 0.001220279 | 0.000680978 |
| 0 | 0 | 0 | 0 | 0 |
| 0 | 0 | 0 | 0 | 0 |
| 0 | 0 | 0 | 0 | 0 |
| 0 | 0 | 0 | 0 | 0 |
| 0 | 0 | 0 | 0 | 0 |
| 0 | 0 | 0 | 0.215033917 | 0 |
| 0 | 0 | 0 | 0 | 0 |
| 0 | 0 | 0 | 0 | 0 |
| 0 | 0 | 0 | 0 | 0 |
| 0 | 0 | 0 | 0 | 0 |
| 1.34400505 | 2.444014758 | 0 | 0 | 0.126615627 |
| 0 | 0 | 0 | 0 | 0 |
| 0 | 0 | 0 | 0 | 0.005022826 |
| 0 | 0 | 0 | 0 | 0 |
| 0 | 0 | 0 | 0 | 0 |
| 0 | 0 | 0 | 0 | 0 |
| 0 | 0 | 0 | 0 | 0 |
| 0.011411292 | 0 | 0.041087587 | 0.015507631 | 0.00216351 |
| 0 | 0 | 0 | 0 | 0 |
| 0.000744626 | 0.543107922 | 0.091198193 | 0 | 0.001270588 |
| 0 | 0 | 0 | 0.039760651 | 0 |
| 0.939043969 | 0.511797516 | 0 | 2.093967723 | 0.471042307 |
| 0 | 0 | 0 | 0 | 0 |
| 0 | 0 | 0 | 0 | 0 |
| 0.04009928 | 0 | 0 | 0 | 0 |
| 0 | 0 | 0 | 0 | 0 |
| 0.005181942 | 0.002426583 | 0.000869843 | 0.000812552 | 0.00272067 |
| 0 | 0 | 0 | 0 | 0 |
| 0 | 0 | 0 | 0.002202635 | 0 |
| 0 | 0.027878796 | 0.001480526 | 0 | 0.0028299 |
| 0 | 0 | 0 | 0 | 0 |
| 0 | 0 | 0 | 0 | 0 |
| 1.319321654 | 1.692347382 | 31.48643771 | 4.820709843 | 14.30302985 |
| 0.020196523 | 1.163977922 | 0.097176423 | 0 | 2.473747015 |
| 0 | 0 | 0 | 0.062184601 | 0.229653058 |
| 0 | 0 | 0 | 0 | 0 |
| 0 | 0 | 0 | 0 | 0 |
| 0.038747943 | 0 | 0 | 0 | 0 |
| 0 | 0 | 0 | 0 | 0 |
| 0.003410268 | 0.055451969 | 0.002741731 | 0 | 0 |

| | | | | |
|---|---|---|---|---|
| 0 | 0 | 0 | 0 | 0 |
| 0.004430832 | 0.00084291 | 0 | 0 | 0.013020904 |
| 0.001293741 | 0 | 0 | 0 | 0 |
| 0 | 0 | 0 | 0 | 0 |
| 2.421460724 | 2.042685288 | 1.094724167 | 0 | 3.72772933 |
| 0 | 0 | 0 | 0 | 0 |
| 0 | 0 | 0 | 0 | 0 |
| 0 | 0 | 0 | 0 | 0 |
| 0.007005142 | 0.001262501 | 0.091719152 | 0.142233238 | 0.029568399 |
| 0 | 0 | 0 | 0 | 0 |
| 0 | 0 | 0 | 0 | 0 |
| 0 | 0 | 0 | 0 | 0 |
| 0 | 0.069481128 | 0 | 0.053754188 | 0.064892659 |
| 0.089759432 | 0.007551865 | 0.059916563 | 0.018207176 | 0.030105255 |
| 0 | 0 | 0 | 0.114168634 | 0 |
| 0.066236055 | 0.161397567 | 0.033172982 | 0.089459611 | 0.321977476 |
| 0.002671794 | 0.000726108 | 0 | 0 | 0.016499238 |
| 0 | 0 | 0 | 0 | 0 |
| 0 | 0 | 0 | 0 | 0 |
| 0 | 0 | 0 | 0 | 0 |
| 0 | 0 | 0 | 0 | 0 |
| 0 | 0 | 0 | 0 | 0 |
| 0 | 0 | 0 | 0 | 0 |
| 0 | 0 | 0 | 0 | 0 |
| 0 | 0 | 0 | 0 | 0 |
| 0 | 0 | 0 | 0 | 0 |
| 0.002157289 | 0.083342203 | 0 | 0.008118544 | 0 |
| 0.011984048 | 0.007836863 | 0.002247388 | 0 | 0.000852038 |
| 0 | 0 | 0 | 0 | 0 |
| 0 | 0 | 0 | 0 | 0 |
| 0 | 0 | 0 | 0 | 0 |
| 0 | 0 | 0 | 0 | 0.00320271 |
| 0 | 0 | 0 | 0 | 0 |
| 0 | 0.018521371 | 0 | 0 | 0.092665677 |
| 0 | 0 | 0 | 0.241117326 | 0 |
| 0 | 0 | 0 | 0.025830734 | 0 |
| 0 | 0.000673902 | 0 | 0 | 0 |
| 0 | 0.007470998 | 0 | 0.003544069 | 0 |
| 0 | 0 | 0 | 0 | 0 |
| 0 | 0 | 0 | 0 | 0 |
| 0 | 0.53900292 | 0 | 1.241375139 | 2.110691963 |
| 0 | 0 | 0 | 0 | 0 |
| 0 | 0 | 0 | 0.002112035 | 0 |
| 0 | 0 | 0 | 0 | 0 |
| 0 | 0 | 0 | 0 | 0 |
| 0 | 0 | 0 | 0 | 0 |
| 0 | 0 | 0 | 0 | 0 |
| 0 | 0 | 0 | 0 | 0 |

| | | | | |
|---|---|---|---|---|
| 0 | 0.231208998 | 0 | 0.008149614 | 0.079529871 |
| 0 | 0 | 0 | 0 | 0 |
| 0 | 0.004064187 | 0 | 0 | 0 |
| 0 | 0 | 0 | 0 | 0.002654278 |
| 0 | 0 | 0 | 0.016548678 | 0 |
| 0 | 0 | 0 | 0 | 0 |
| 0.000951892 | 0.053601302 | 0.001246323 | 0 | 0.083161863 |
| 0 | 0 | 0 | 0 | 0 |
| 0 | 0 | 0 | 0 | 0 |
| 0 | 0.004255734 | 0 | 0 | 0.00204493 |
| 0 | 0 | 0 | 0 | 0 |
| 0 | 0 | 0 | 0 | 0.024620991 |
| 0 | 0 | 0 | 0 | 0 |
| 0 | 0 | 0 | 0 | 0 |
| 0 | 0 | 0 | 0 | 0 |
| 0 | 0 | 0 | 0 | 0 |
| 0 | 0 | 0 | 0 | 0 |
| 0 | 0 | 0 | 0 | 0 |
| 0 | 0 | 0 | 0 | 0 |
| 0 | 0 | 0 | 0 | 0 |
| 0.001993759 | 0.039859682 | 0.409959687 | 0.006465765 | 0 |
| 0 | 0 | 0 | 0 | 0 |
| 0.070771335 | 0.021602224 | 0 | 0 | 0.014327488 |
| 0 | 0.001758563 | 0.000581891 | 0.001630696 | 0.013549049 |
| 0 | 0 | 0 | 0 | 0 |
| 0 | 0 | 0 | 0 | 0 |
| 0 | 0 | 0 | 0.014435076 | 0 |
| 0 | 0 | 0 | 0 | 0 |
| 0 | 0 | 0 | 0 | 0 |
| 2.830755354 | 0.035239095 | 0 | 0.826910122 | 0.09284451 |
| 0 | 0 | 0 | 0 | 0 |
| 0 | 0 | 0 | 0 | 0 |
| 0 | 0 | 0 | 0 | 0 |
| 0 | 0 | 1.469047935 | 0 | 0 |
| 0 | 0 | 0 | 0 | 0 |
| 0 | 0 | 0 | 0 | 0 |
| 0 | 0.020650077 | 0 | 0 | 0 |
| 0 | 0 | 0 | 0 | 0 |
| 0 | 0 | 0 | 0 | 0 |
| 0 | 0 | 0 | 0 | 0 |
| 0 | 0 | 0 | 0 | 0 |
| 0.026558783 | 0 | 0 | 0 | 0 |
| 0 | 0 | 0 | 0 | 0 |
| 0 | 0 | 0 | 0 | 0 |
| 0 | 0 | 0 | 0 | 0 |
| 0 | 0 | 0 | 0.576502307 | 0 |
| 0 | 0 | 0 | 0 | 0 |
| 0 | 0 | 0 | 0 | 0 |

| | | | | |
|---|---|---|---|---|
| 0 | 0.015574891 | 0.008139762 | 0 | 0.004569618 |
| 0 | 0 | 0 | 0 | 0 |
| 0 | 0 | 0 | 0 | 0 |
| 0 | 0 | 0 | 0 | 0 |
| 0 | 0 | 0 | 0 | 0 |
| 0 | 0 | 0 | 0.001476055 | 0 |
| 0 | 0.000783296 | 0 | 0 | 0 |
| 0.131200819 | 0.360639595 | 0 | 0.737282113 | 1.329166618 |
| 1.682320451 | 0.141427789 | 0 | 0.020197794 | 0.004440244 |
| 0 | 0.0497856 | 0.031245382 | 0.318718223 | 0.065390021 |
| 0 | 0 | 0 | 0 | 0 |
| 0 | 0 | 0 | 0 | 0 |
| 0 | 0 | 0 | 0 | 0 |
| 0.098693624 | 0 | 0 | 0 | 0 |
| 0 | 0 | 0 | 0 | 0 |
| 0 | 0 | 0 | 0 | 0 |
| 0.010052186 | 0.023585061 | 0.013405177 | 0 | 0 |
| 0 | 0.145044704 | 0 | 0 | 0 |
| 0 | 0 | 0 | 0 | 0 |
| 0.015946417 | 0 | 0 | 0 | 0.007886979 |
| 0 | 0 | 0 | 0 | 0 |
| 0 | 0 | 0 | 0 | 0 |
| 0 | 0 | 0 | 0 | 0 |
| 0 | 0 | 0 | 0 | 0 |
| 0.001958644 | 0 | 0 | 0.077548876 | 0.011140405 |
| 0 | 0 | 0 | 0 | 0 |
| 0 | 0 | 0 | 0 | 0 |
| 0 | 0.235826527 | 0 | 0 | 0 |
| 0 | 0 | 0 | 0 | 0 |
| 0 | 0 | 0 | 0 | 0 |
| 0 | 0 | 0 | 0 | 0 |
| 0 | 0 | 0 | 0 | 0 |
| 0 | 0 | 0 | 0 | 0 |
| 0 | 0 | 0 | 0 | 0 |
| 0 | 0 | 0.011370378 | 0 | 0 |
| 0 | 0 | 0 | 0 | 0 |
| 0 | 0 | 0 | 0 | 0 |
| 0 | 0 | 0 | 0 | 0.032058837 |
| 0 | 0.005884784 | 0 | 0 | 0 |
| 0 | 0 | 0.006875873 | 0.095566441 | 0 |
| 0 | 0 | 0 | 0 | 0 |
| 0.873951857 | 0.458147503 | 0 | 0.284937929 | 0.484088869 |
| 0 | 0 | 0 | 0 | 0 |
| 0 | 0 | 0 | 0 | 0 |
| 0 | 0 | 0 | 0 | 0 |
| 0 | 0 | 0 | 0 | 0 |
| 0 | 0 | 0 | 0 | 0 |
| 0 | 0 | 0 | 0 | 0.003444836 |

| | | | | |
|---|---|---|---|---|
| 0 | 0 | 0 | 0 | 0 |
| 0 | 0.001182366 | 0 | 0 | 0.022781738 |
| 0 | 0 | 0 | 0 | 0 |
| 0 | 0 | 0 | 0 | 0 |
| 2.526300781 | 2.797069102 | 0.177774703 | 0.044284192 | 0.127133701 |
| 0 | 0 | 0 | 0 | 0 |
| 0 | 0 | 0 | 0 | 0.027385916 |
| 0.01532984 | 0.004629063 | 0.022301712 | 0.002976118 | 0.020206695 |
| 0.012147839 | 0 | 0 | 0 | 0.007462222 |
| 0 | 0 | 0 | 0 | 0 |
| 0 | 0 | 0 | 0 | 0 |
| 0 | 0 | 0 | 0.002223328 | 0 |
| 0 | 0 | 0 | 0 | 0 |
| 0 | 0 | 0 | 0 | 0 |
| 0 | 0 | 0.001495202 | 0 | 0 |
| 0.144983775 | 0 | 0.016809213 | 0 | 0.107551396 |
| 0 | 0 | 0 | 0 | 0 |
| 0 | 0 | 0 | 0 | 0 |
| 0.004593566 | 0 | 0 | 0 | 0 |
| 0 | 0 | 0 | 0 | 0 |
| 0 | 0 | 0 | 0 | 0 |
| 0 | 0 | 0 | 0 | 0 |
| 0 | 0 | 0 | 0 | 0 |
| 0.002948159 | 0.000961457 | 0.007812024 | 0 | 0.001437307 |
| 0 | 0.247361151 | 0.327900622 | 0.087842713 | 0 |
| 0 | 0 | 0 | 0 | 0 |
| 0.003274231 | 1.202234329 | 0 | 0 | 0 |
| 0 | 0 | 0 | 0 | 0 |
| 0 | 0 | 0 | 0 | 0 |
| 0 | 0 | 0 | 0 | 0 |
| 0 | 0 | 0 | 0 | 0 |
| 0 | 0 | 0 | 0 | 0.000842342 |
| 0 | 0 | 0 | 0 | 0 |
| 0 | 0 | 0 | 0 | 0 |
| 0 | 0 | 0 | 0 | 0 |
| 0 | 0 | 0 | 0 | 0 |
| 0 | 0 | 0 | 0 | 0 |
| 0 | 0.012024496 | 0 | 0.020333595 | 0.122684278 |
| 0 | 0.169130372 | 0 | 0 | 0 |
| 0 | 0 | 0 | 0 | 0 |
| 0 | 0 | 0 | 0 | 0 |
| 0 | 0 | 0 | 0.057942058 | 0 |
| 0 | 0 | 0 | 0 | 0 |
| 0 | 0 | 0 | 0 | 0 |
| 0 | 0 | 0 | 0 | 0 |
| 0 | 0 | 0 | 0 | 0 |
| 0 | 0 | 0 | 0 | 0 |
| 0 | 0 | 0 | 0 | 0 |

| | | | | |
|---|---|---|---|---|
| 0 | 0.113569739 | 0 | 1.432006865 | 0 |
| 0 | 0 | 0 | 0 | 0 |
| 0 | 0 | 0 | 0 | 0 |
| 0 | 0 | 0 | 0 | 0 |
| 0 | 0 | 0 | 0 | 0 |
| 0 | 0 | 0 | 0 | 0 |
| 0 | 0 | 0 | 0 | 0 |
| 0 | 0 | 0 | 0 | 0 |
| 0 | 0 | 0 | 0 | 0 |
| 0 | 0 | 0 | 0 | 0 |
| 0 | 0 | 0 | 0 | 0 |
| 0 | 0 | 0 | 0 | 0 |
| 0 | 0 | 0 | 0 | 0 |
| 0 | 0 | 0 | 0 | 0 |
| 0 | 0 | 0 | 0 | 0 |
| 0 | 0 | 0 | 0 | 0 |
| 0 | 0 | 0 | 0 | 0 |
| 0 | 0 | 0 | 0 | 0 |
| 0 | 0 | 0 | 0 | 0 |
| 0 | 0 | 0 | 0 | 0 |
| 0 | 0 | 0.01670276 | 0.080578053 | 0.00811069 |
| 0 | 0 | 0 | 0 | 0 |
| 0.009269363 | 0.349653043 | 0.068195497 | 0 | 0 |
| 0.117318887 | 1.305905739 | 0 | 0 | 0 |
| 0 | 0 | 0 | 0 | 0 |
| 0.120164378 | 0 | 0 | 0 | 0 |
| 0 | 0 | 0 | 0 | 0 |
| 0 | 0 | 0 | 0 | 0 |
| 0 | 0 | 0 | 0 | 0 |
| 0 | 0 | 0 | 0 | 0 |
| 0 | 0 | 0 | 0 | 0 |
| 0 | 0 | 0 | 0.433179931 | 0 |
| 0.237603705 | 0 | 0.245717118 | 0.034884996 | 0 |
| 0 | 0 | 0 | 0 | 0 |
| 0.051691801 | 0.310964897 | 0 | 1.092608464 | 0 |
| 0.000597078 | 0.022565883 | 0 | 0 | 0.001358428 |
| 0 | 0 | 0 | 0 | 0 |
| 0 | 0 | 0 | 0.002828006 | 0 |
| 0.012200875 | 0.137336873 | 0.001004701 | 0.100657069 | 0.001571237 |
| 0 | 0 | 0 | 0 | 0 |
| 0 | 0 | 0 | 0 | 0 |
| 0 | 0.086722845 | 0 | 0 | 0 |
| 0.52920559 | 0.281028925 | 0 | 0.330872074 | 0.062772604 |
| 0 | 0 | 0 | 0 | 0 |
| 0 | 0 | 0 | 0 | 0 |
| 0.022873891 | 0.019195272 | 0 | 0.011112156 | 0.036477318 |
| 0 | 0 | 0 | 0 | 0 |
| 0 | 0 | 0 | 0 | 0 |

| | | | | |
|---|---|---|---|---|
| 0 | 0 | 0 | 0 | 0 |
| 0 | 0 | 0 | 0 | 0 |
| 0 | 0 | 0 | 0 | 0 |
| 0 | 0 | 0 | 0 | 0 |
| 0 | 0 | 0 | 0 | 0 |
| 0 | 0 | 0 | 0 | 0 |
| 0 | 3.946242644 | 0 | 0 | 0 |
| 0 | 0 | 0.023666114 | 0 | 0.047703166 |
| 0.036652531 | 0 | 0 | 0 | 0.022100393 |
| 0.000880742 | 0.016755016 | 0 | 0.049372306 | 0.011020896 |
| 0 | 0 | 0 | 0 | 0 |
| 0 | 0 | 0 | 0 | 0 |
| 0 | 0 | 0 | 0 | 0 |
| 0 | 0 | 0 | 0 | 0 |
| 0 | 0 | 0 | 0 | 0.001821761 |
| 0 | 0 | 0.036637743 | 0 | 0 |
| 0 | 0 | 0 | 0 | 0 |
| 0 | 0 | 0 | 0 | 0 |
| 0 | 0 | 0 | 0 | 0 |
| 0.567889496 | 0.209373807 | 0 | 6.688939013 | 0 |
| 0 | 0 | 0 | 0.040818707 | 0.004118219 |
| 0 | 0 | 0 | 0 | 0 |
| 0 | 0 | 0 | 0 | 0 |
| 0 | 0 | 0 | 0 | 0 |
| 0 | 0 | 0 | 0 | 0 |
| 0 | 0 | 0 | 0 | 0 |
| 0 | 0 | 0 | 0 | 0 |
| 0 | 0 | 0 | 0 | 0 |
| 0 | 0 | 0 | 0 | 0 |
| 0.010103801 | 0.025799141 | 0 | 0 | 0 |
| 0.439137256 | 0.795743614 | 0 | 0.062632703 | 0.578664381 |
| 0 | 0 | 0 | 0 | 0 |
| 0 | 0 | 0 | 0 | 0 |
| 0 | 0 | 0 | 0 | 0 |
| 0 | 0 | 0 | 0 | 0 |
| 0 | 0 | 0 | 0 | 0 |
| 0 | 0 | 0 | 0 | 0 |
| 0 | 0 | 0 | 0 | 0 |
| 0 | 0 | 0 | 0.001618098 | 0 |
| 0 | 0 | 0 | 0 | 0 |
| 0 | 0 | 0 | 0 | 0 |
| 0 | 0 | 0 | 0.031842087 | 0 |
| 0 | 0 | 0 | 0 | 0 |
| 0 | 0 | 0 | 0 | 0 |
| 0 | 0 | 0 | 0 | 0 |
| 0 | 0 | 0 | 0 | 0 |
| 0 | 0.025144733 | 0.010888778 | 0 | 0 |
| 0 | 0 | 0 | 0 | 0 |

| | | | | |
|---|---|---|---|---|
| 0 | 0 | 0 | 0 | 0 |
| 0 | 0 | 0 | 0 | 0 |
| 0 | 0 | 0 | 0 | 0 |
| 0 | 0 | 0 | 0 | 0 |
| 0.152922178 | 0.299929524 | 0.066704787 | 0.001688654 | 0.010554367 |
| 0 | 0 | 0 | 0 | 0 |
| 0 | 0 | 0 | 0 | 0 |
| 0 | 0 | 0 | 0 | 0 |
| 0 | 0 | 0 | 0 | 0 |
| 0 | 0 | 0 | 0 | 0 |
| 0 | 0 | 0 | 0 | 0.064130379 |
| 0.003910372 | 0.07036411 | 0.006525358 | 0 | 0.005233287 |
| 0 | 0.026577969 | 0 | 0 | 0 |
| 0 | 0 | 0 | 0.026826385 | 0 |
| 0 | 0 | 0 | 0.038591313 | 0.005583382 |
| 0 | 0 | 0 | 0 | 0.040696221 |
| 0 | 0 | 0 | 0 | 0 |
| 0 | 0 | 0 | 0 | 0 |
| 0.057303207 | 0.000662688 | 0.013682876 | 0 | 0.021398442 |
| 0 | 0.01124849 | 0.00521082 | 0 | 0.00336313 |
| 0 | 0 | 0 | 0 | 0 |
| 0 | 0 | 0 | 0 | 0 |
| 0 | 0 | 0 | 0 | 0 |
| 0 | 0 | 0 | 0 | 0 |
| 0 | 0 | 0 | 0 | 0 |
| 0 | 0 | 0 | 0 | 0 |
| 0 | 0.009429958 | 0.00298262 | 0 | 0.00352427 |
| 0 | 0 | 0 | 0 | 0 |
| 0 | 0 | 0 | 0 | 0 |
| 0 | 0 | 0 | 0 | 0 |
| 0 | 0 | 0.000901588 | 0 | 0 |
| 0 | 0.00424476 | 0 | 0 | 0 |
| 0 | 0 | 0 | 0 | 0 |
| 0 | 0 | 0 | 0 | 0 |
| 0 | 0 | 0 | 0 | 0 |
| 0 | 0 | 0 | 0 | 0 |
| 0 | 0 | 0 | 0 | 0 |
| 0.012336604 | 0.064608339 | 0.00190029 | 0 | 0.035992215 |
| 0 | 0.086876455 | 0 | 0 | 0.058582207 |
| 0 | 0 | 0 | 0 | 0 |
| 0 | 0.025028185 | 0 | 0 | 0 |
| 0 | 0 | 0 | 0 | 0 |
| 0 | 0 | 0 | 0 | 0 |
| 0.001936971 | 0 | 0 | 0 | 0.000979301 |
| 0 | 0 | 0 | 0 | 0 |
| 0 | 0.002928244 | 0 | 0 | 0 |
| 0 | 0 | 0 | 0 | 0 |
| 0 | 0 | 0 | 0 | 0 |

| | | | | |
|---|---|---|---|---|
| 0 | 0 | 0 | 0 | 0 |
| 0.000919746 | 0.374435977 | 0 | 0 | 0.006696119 |
| 0 | 0 | 0 | 0 | 0 |
| 0.000902202 | 0.008581632 | 0 | 0 | 0 |
| 0 | 0 | 0 | 0 | 0 |
| 0 | 0 | 0 | 0.001375886 | 0 |
| 0 | 0 | 0 | 0.008031189 | 0 |
| 0.009525603 | 0 | 0 | 0 | 0 |
| 0 | 0 | 0 | 0 | 0 |
| 0 | 0 | 0 | 0.00185954 | 0 |
| 0 | 0 | 0 | 0 | 0 |
| 0 | 0 | 0 | 0 | 0 |
| 0.064592968 | 0.278244149 | 0 | 0.80224239 | 0.024007845 |
| 0 | 0 | 0 | 0 | 0 |
| 0 | 0 | 0 | 0 | 0 |
| 0 | 0 | 0 | 0 | 0 |
| 0 | 0 | 0 | 0 | 0 |
| 0 | 0 | 0 | 0 | 0 |
| 0 | 0 | 0 | 0 | 0 |
| 0 | 0 | 0 | 0 | 0 |
| 0 | 0 | 0 | 0 | 0 |
| 0 | 0 | 0 | 0 | 0 |
| 0 | 0.047580183 | 0.025956968 | 0.004506142 | 0.006705745 |
| 0.004471784 | 0 | 0.009758267 | 0.110146189 | 0.007347795 |
| 0.005702493 | 0 | 0 | 0 | 0 |
| 0 | 0 | 0 | 0 | 0 |
| 0.034194316 | 0 | 0 | 0 | 0 |
| 0 | 0 | 0.000941024 | 0 | 0 |
| 0.301156024 | 0 | 0 | 0 | 0 |
| 0 | 0 | 0 | 0 | 0 |
| 0.022300284 | 0.036439546 | 0 | 0.051439549 | 0.031509693 |
| 0 | 0 | 0 | 0 | 0 |
| 0 | 0 | 0 | 0 | 0 |
| 0 | 0 | 0 | 0 | 0 |
| 0 | 0 | 0 | 0 | 0 |
| 0 | 0 | 0 | 0 | 0.001954031 |
| 0 | 0.000569329 | 0 | 0 | 0 |
| 0 | 0 | 0 | 0 | 0 |
| 0.011320552 | 0.003173716 | 0 | 0.003643654 | 0 |
| 0 | 0 | 0 | 0 | 0 |
| 0 | 0 | 0 | 0 | 0 |
| 0 | 0 | 0 | 0 | 0 |
| 0 | 0 | 0.010940479 | 0 | 0 |
| 0 | 0 | 0 | 0 | 0 |
| 0 | 0 | 0 | 0 | 0 |
| 0.721329313 | 0.555543832 | 0.051096429 | 0.230284291 | 0.350048823 |
| 0 | 0 | 0 | 0 | 0 |
| 0 | 0 | 0 | 0 | 0 |

| | | | | |
|---|---|---|---|---|
| 0 | 0 | 0 | 0 | 0 |
| 0 | 0 | 0 | 0 | 0 |
| 0 | 0 | 0 | 0.130982208 | 0 |
| 0 | 0 | 0 | 0 | 0 |
| 0 | 0 | 0 | 0 | 0 |
| 0 | 0 | 0 | 0 | 0 |
| 0 | 0 | 0 | 0 | 0 |
| 0 | 0 | 0 | 0 | 0 |
| 0 | 0.003076366 | 0 | 0 | 0 |
| 0.00181329 | 0 | 0 | 0 | 0 |
| 0 | 0.001716489 | 0 | 0 | 0 |
| 0 | 0.003294931 | 0 | 0 | 0 |
| 0 | 0 | 0 | 0 | 0 |
| 0 | 0 | 0 | 0 | 0 |
| 0 | 0 | 0 | 0 | 0 |
| 0 | 0 | 0 | 0 | 0 |
| 0 | 0 | 0 | 0 | 0 |
| 0 | 0 | 0 | 0 | 0 |
| 0 | 0 | 0 | 0 | 0 |
| 0.001923089 | 0 | 0 | 0 | 0.102089598 |
| 0 | 0 | 0 | 0 | 0 |
| 0 | 0 | 0 | 0 | 0 |
| 0 | 0 | 0 | 0 | 0 |
| 0 | 0 | 0 | 0 | 0 |
| 0 | 0 | 0 | 0 | 0 |
| 0 | 0 | 0 | 0 | 0 |
| 0 | 0 | 0 | 0 | 0 |
| 0 | 0.034548709 | 0 | 0.109180403 | 0 |
| 0 | 0 | 0 | 0 | 0 |
| 0 | 0 | 0 | 0 | 0 |
| 0.002753554 | 0.001815941 | 0 | 0.004116204 | 0.015035234 |
| 0 | 0 | 0 | 0 | 0 |
| 0.108703318 | 0.010055885 | 0.007209351 | 0.007820725 | 0.004121895 |
| 0.003920577 | 0.007789889 | 0 | 0 | 0 |
| 0 | 0 | 0 | 0 | 0 |
| 0 | 0 | 0 | 0 | 0 |
| 0 | 0 | 0 | 0 | 0 |
| 0 | 0 | 0 | 0 | 0 |
| 0 | 0 | 0 | 0 | 0 |
| 0 | 0 | 0 | 0 | 0 |
| 0 | 0 | 0 | 0 | 0 |
| 0 | 0.020548781 | 0.011862136 | 0.000893617 | 0 |
| 0.086732763 | 0.051531456 | 0.007924976 | 0.053998379 | 0.041312497 |
| 0 | 0 | 0 | 0 | 0 |
| 0 | 0 | 0 | 0 | 0 |
| 0.033031604 | 0.067889051 | 0.007747036 | 0.01132714 | 0.004565251 |
| 0 | 0.002464221 | 0 | 0 | 0 |
| 0 | 0 | 0 | 0 | 0 |

| | | | | |
|---|---|---|---|---|
| 0 | 0 | 0 | 0 | 0 |
| 0 | 0.005960603 | 0 | 0.007755628 | 0 |
| 0.024835441 | 0.002983977 | 0 | 0.002664527 | 0.005947763 |
| 0 | 0 | 0 | 0 | 0 |
| 0.412670739 | 0 | 0.066958575 | 0 | 0.288976787 |
| 0.044496904 | 0.00108228 | 0 | 0 | 0.253321242 |
| 0 | 0 | 0 | 0 | 0 |
| 0 | 0 | 0 | 0 | 0 |
| 0 | 0 | 0 | 0 | 0 |
| 0 | 0 | 0 | 0 | 0 |
| 0 | 0 | 0 | 0 | 0 |
| 0 | 0 | 0 | 0 | 0 |
| 0 | 0 | 0 | 0 | 0 |
| 0 | 0 | 0 | 0 | 0 |
| 0 | 0 | 0 | 0 | 0 |
| 0 | 0 | 0 | 0 | 0 |
| 0 | 0 | 0 | 0 | 0 |
| 0 | 0 | 0 | 0 | 0 |
| 0 | 0 | 0 | 0 | 0 |
| 0 | 0 | 0 | 0 | 0 |
| 0 | 0 | 0 | 0 | 0 |
| 0 | 0 | 0 | 0 | 0 |
| 0 | 0.007872853 | 0 | 0 | 0 |
| 0 | 0 | 0 | 0 | 0 |
| 0 | 0.003115008 | 0 | 0.00241554 | 0 |
| 0.147165875 | 0.000824638 | 0 | 0.462579004 | 0.001725881 |
| 0 | 0 | 0 | 0 | 0 |
| 0.004415135 | 0.002239797 | 0 | 0 | 0 |
| 0 | 0 | 0 | 0 | 0 |
| 0 | 0 | 0 | 0 | 0 |
| 0 | 0 | 0 | 0 | 0 |
| 0 | 0 | 0 | 0.001356056 | 0 |
| 0 | 0 | 0 | 0 | 0 |
| 0 | 0 | 0 | 0 | 0 |
| 0 | 0 | 0 | 0 | 0 |
| 0 | 0 | 0 | 0 | 0 |
| 0 | 0 | 0 | 0 | 0 |
| 0 | 0.003940118 | 0.001883188 | 0 | 0 |
| 0 | 0.331377503 | 0 | 0.052548315 | 0 |
| 0 | 0 | 0 | 0 | 0 |
| 0 | 0 | 0 | 0 | 0 |
| 0 | 0.009303893 | 0 | 0.006710197 | 0 |
| 0 | 0.012910782 | 0.004984058 | 0 | 0.004453996 |
| 0 | 0 | 0 | 0 | 0 |
| 0 | 0 | 0 | 0 | 0 |
| 0 | 0 | 0 | 0 | 0 |
| 0.009982954 | 0.005570786 | 0 | 0.004409132 | 0.004921036 |
| 0 | 0 | 0 | 0 | 0 |

| | | | | |
|---|---|---|---|---|
| 0 | 0 | 0 | 0 | 0 |
| 0.053745715 | 0.048566127 | 0.004352305 | 0.00663342 | 0.018389503 |
| 0 | 0 | 0 | 0.003298376 | 0 |
| 0 | 0 | 0 | 0 | 0 |
| 0 | 0 | 0 | 0 | 0 |
| 0 | 0 | 0 | 0 | 0 |
| 0 | 0 | 0 | 0 | 0 |
| 0 | 0 | 0 | 0 | 0 |
| 0 | 0 | 0 | 0 | 0 |
| 0 | 0 | 0 | 0 | 0 |
| 0 | 0 | 0 | 0 | 0 |
| 0 | 0 | 0 | 0 | 0 |
| 4.133661185 | 0.028540264 | 0.008959399 | 0 | 0 |
| 0 | 0 | 0 | 0 | 0 |
| 0 | 0.009162873 | 0.002856141 | 0 | 0.005062233 |
| 0 | 0 | 0 | 0 | 0 |
| 0 | 0 | 0 | 0 | 0 |
| 0 | 0 | 0 | 0 | 0 |
| 0 | 0.000661077 | 0.021169534 | 0 | 0 |
| 0 | 0 | 0 | 0 | 0 |
| 0 | 0 | 0 | 0 | 0 |
| 0 | 0.003521217 | 0.003646443 | 0 | 0.002047093 |
| 0 | 0 | 0 | 0.006644276 | 0 |
| 0 | 0 | 0 | 0 | 0 |
| 0 | 0 | 0 | 0 | 0 |
| 0 | 0 | 0 | 0 | 0 |
| 0 | 0 | 0 | 0 | 0 |
| 0 | 0 | 0 | 0 | 0 |
| 0 | 0 | 0 | 0 | 0 |
| 0 | 0 | 0 | 0 | 0 |
| 0 | 0 | 0 | 0 | 0 |
| 0 | 0 | 0 | 0 | 0 |
| 0 | 0 | 0 | 0 | 0 |
| 0 | 0 | 0 | 0 | 0 |
| 5.292153778 | 21.80827157 | 0 | 2.977086914 | 0 |
| 0 | 0 | 0 | 0 | 0 |
| 0 | 0 | 0 | 0 | 0 |
| 0 | 0 | 0 | 0 | 0 |
| 0.000770169 | 0.228563456 | 0 | 0.002616597 | 0.054611224 |
| 0 | 0 | 0 | 0 | 0 |
| 0.113942171 | 0.056401566 | 0.195147132 | 0.148509726 | 0.303223757 |
| 0 | 0 | 0 | 0 | 0 |
| 0 | 0 | 0 | 0 | 0 |
| 0 | 0 | 0 | 0 | 0 |
| 0 | 0 | 0 | 0 | 0 |
| 0.001581285 | 0.052543168 | 0.081090492 | 1.050554713 | 0.00479683 |
| 0 | 0 | 0 | 0 | 0 |
| 0 | 0 | 0 | 0 | 0 |

| | | | | |
|---|---|---|---|---|
| 0 | 0.009466872 | 0 | 0 | 0 |
| 0 | 0 | 0 | 0 | 0 |
| 0 | 0 | 0 | 0 | 0 |
| 0 | 0 | 0 | 0 | 0 |
| 0 | 0 | 0 | 0 | 0 |
| 0 | 0 | 0 | 0 | 0 |
| 0 | 0 | 0 | 0 | 0 |
| 0 | 0 | 0 | 0 | 0 |
| 0 | 0 | 0 | 0.005459054 | 0 |
| 0.024743991 | 0.001765213 | 0.012936542 | 0.003940594 | 0.004984514 |
| 0 | 0.010549457 | 0 | 0 | 0.11163143 |
| 0 | 0 | 0 | 0 | 0 |
| 0 | 0 | 0.001099511 | 0 | 0 |
| 0 | 0 | 0 | 0 | 0 |
| 0 | 0 | 0 | 0 | 0 |
| 0 | 0 | 0 | 0 | 0 |
| 0 | 0 | 0 | 0 | 0 |
| 0 | 0 | 0 | 0 | 0 |
| 0 | 0 | 0 | 0 | 0 |
| 0.418421466 | 0.737809095 | 0.152178944 | 0.221461374 | 0.35226652 |
| 0 | 0 | 0 | 0 | 0 |
| 0 | 0 | 0 | 0 | 0 |
| 0 | 0 | 0 | 0 | 0 |
| 0 | 0 | 0 | 0 | 0 |
| 0 | 0 | 0 | 0 | 0 |
| 0 | 0 | 0 | 0 | 0 |
| 0 | 0 | 0 | 0 | 0 |
| 0 | 0 | 0 | 0 | 0 |
| 0.00165834 | 0 | 0 | 0 | 0 |
| 0 | 0 | 0.013600404 | 0 | 0.00299954 |
| 0 | 0 | 0 | 0 | 0 |
| 0.001465459 | 0 | 0 | 0 | 0 |
| 0 | 0 | 0 | 0 | 0 |
| 0 | 0 | 0 | 0 | 0 |
| 0 | 0 | 0 | 0 | 0 |
| 0 | 0 | 0 | 0.006508655 | 0 |
| 0 | 0 | 0 | 0 | 0 |
| 0 | 0 | 0 | 0 | 0 |
| 0 | 0 | 0 | 0 | 0 |
| 0 | 0 | 0 | 0 | 0 |
| 0 | 0 | 0 | 0 | 0 |
| 0 | 0 | 0 | 0 | 0 |
| 0 | 0.003262213 | 0 | 0 | 0 |
| 0 | 0 | 0 | 0 | 0 |
| 0.004422794 | 0.025481825 | 0.006664035 | 0 | 0.003114556 |
| 0 | 0 | 0 | 0 | 0 |
| 0 | 0 | 0 | 0 | 0 |
| 0 | 0 | 0 | 0 | 0 |

| | | | | |
|---|---|---|---|---|
| 0 | 0 | 0 | 0 | 0 |
| 0 | 0 | 0 | 0 | 0 |
| 0 | 0 | 0 | 0 | 0 |
| 0 | 0 | 0 | 0 | 0 |
| 0 | 0 | 0 | 0 | 0 |
| 0.028615492 | 0.012682115 | 0.011836976 | 0.018108404 | 0 |
| 0 | 0 | 0 | 0 | 0 |
| 0 | 0 | 0 | 0 | 0 |
| 0 | 0 | 0 | 0 | 0 |
| 0 | 0 | 0 | 0 | 0 |
| 0 | 0 | 0 | 0 | 0 |
| 0 | 0 | 0 | 0 | 0 |
| 0 | 0 | 0 | 0 | 0 |
| 0 | 0 | 0 | 0 | 0 |
| 0 | 0 | 0 | 0 | 0 |
| 0 | 0 | 0 | 0 | 0 |
| 0 | 0 | 0 | 0 | 0 |
| 0 | 0 | 0 | 0 | 0 |
| 0 | 0 | 0 | 0 | 0 |
| 0 | 0 | 0 | 0 | 0 |
| 0 | 0 | 0 | 0 | 0 |
| 0 | 0 | 0 | 0 | 0 |
| 0 | 0 | 0 | 0 | 0 |
| 0 | 0 | 0 | 0 | 0 |
| 0 | 0 | 0 | 0 | 0 |
| 0.004918619 | 0 | 0 | 0 | 0 |
| 0 | 0 | 0 | 0 | 0 |
| 0.523473216 | 0 | 0 | 0 | 0 |
| 0 | 0 | 0 | 0 | 0 |
| 0 | 0 | 0 | 0 | 0 |
| 0 | 0 | 0 | 0 | 0 |
| 0 | 0 | 0 | 0 | 0 |
| 0 | 0 | 0 | 0 | 0 |
| 0 | 0 | 0 | 0 | 0 |
| 0 | 0 | 0 | 0 | 0 |
| 0 | 0 | 0 | 0 | 0 |
| 0 | 0 | 0 | 0 | 0 |
| 0 | 0 | 0 | 0 | 0 |
| 2.687266271 | 0.262111459 | 0 | 0 | 0.535746129 |
| 0 | 0 | 0 | 0 | 0 |
| 0 | 0 | 0 | 0 | 0 |
| 0 | 0 | 0 | 0 | 0 |
| 0 | 0 | 0 | 0 | 0 |
| 0 | 0 | 0 | 0 | 0 |
| 0 | 0 | 0 | 0 | 0 |
| 0 | 0 | 0 | 0 | 0 |
| 0 | 0 | 0 | 0 | 0 |
| 0 | 0 | 0 | 0 | 0 |

| | | | | |
|---|---|---|---|---|
| 0 | 0 | 0 | 0 | 0 |
| 0 | 0.030406543 | 0.003265096 | 0.039829977 | 0 |
| 0 | 0 | 0 | 0 | 0 |
| 0 | 0 | 0 | 0 | 0 |
| 0 | 0 | 0 | 0 | 0 |
| 0 | 0 | 0.00526755 | 0 | 0 |
| 0 | 0 | 0 | 0 | 0 |
| 0 | 0 | 0 | 0 | 0 |
| 0 | 0 | 0 | 0 | 0 |
| 0 | 0 | 0 | 0 | 0 |
| 0 | 0 | 0 | 0 | 0.002359839 |
| 0 | 0 | 0 | 0 | 0 |
| 0.159416241 | 0 | 0 | 0 | 1.001555127 |
| 0 | 0 | 0 | 0 | 0 |
| 0 | 0 | 0 | 0 | 0 |
| 0 | 0 | 0 | 0 | 0 |
| 0 | 0 | 0 | 0 | 0 |
| 0 | 0 | 0 | 0 | 0 |
| 0 | 0 | 0 | 0 | 0 |
| 0 | 0 | 0.002929793 | 0 | 0 |
| 0 | 0.00373835 | 0 | 0.009299106 | 0 |
| 0 | 0 | 0 | 0 | 0 |
| 0 | 0 | 0 | 0 | 0 |
| 2.919535121 | 1.32220655 | 0 | 2.85321179 | 3.268465742 |
| 0 | 0 | 0 | 0 | 0 |
| 0 | 0 | 0 | 0 | 0 |
| 0 | 0 | 0 | 0 | 0 |
| 0 | 0 | 0 | 0 | 0 |
| 0 | 0 | 0 | 0 | 0 |
| 0 | 0 | 0 | 0 | 0 |
| 0 | 0 | 0 | 0 | 0 |
| 0 | 0 | 0 | 0 | 0 |
| 0 | 0 | 0 | 0 | 0.002799345 |
| 0 | 0 | 0 | 0 | 0 |

| PHL018-0068_V2 | PHL018-0274_V2 | PHL018-0186_V2 | PHL018-0242_V3 | PHL018-0026_V3 |
|---|---|---|---|---|
| REF | REF | REF | REF | EYCF |
| 0.064461069 | 0.326506078 | 0.437960208 | 0.001218356 | 0.130330436 |
| 2.503328993 | 13.76688767 | 0 | 1.362809562 | 1.262472822 |
| 23.49127568 | 0.860238762 | 0 | 0 | 0 |
| 0 | 0 | 0 | 0 | 0 |
| 0 | 0 | 0 | 0 | 0 |
| 0 | 0 | 0 | 0 | 0 |
| 0 | 0 | 0 | 0 | 0 |
| 0 | 0 | 0 | 0 | 0 |
| 0 | 0 | 0 | 0 | 0 |
| 0 | 0 | 0 | 0 | 0 |
| 0 | 0 | 0 | 0 | 0 |
| 0 | 0 | 0 | 0 | 0 |
| 0 | 0 | 0 | 0 | 0 |
| 21.03401696 | 1.913462613 | 0 | 0.709844673 | 0 |
| 0 | 0 | 0 | 0 | 0 |
| 0 | 0 | 0 | 0 | 0 |
| 0 | 0 | 0.011616649 | 0 | 0.046102173 |
| 0 | 0 | 0 | 0 | 0 |
| 0 | 0 | 0 | 0 | 0 |
| 0 | 0 | 0 | 0 | 0 |
| 0 | 0 | 0 | 0 | 0 |
| 0 | 0 | 0 | 0 | 0 |
| 0 | 0 | 0 | 0 | 0 |
| 0 | 0 | 0 | 0 | 0 |
| 0 | 0 | 0 | 0 | 0 |
| 0 | 0 | 0 | 0 | 0 |
| 0 | 0 | 0 | 0 | 0 |
| 0 | 0 | 0 | 0 | 0 |
| 0 | 0 | 0 | 0 | 0 |
| 0 | 0 | 0 | 0 | 0 |
| 0 | 0 | 0 | 0 | 0 |
| 0 | 0 | 0 | 0 | 0 |
| 0 | 0 | 0 | 0 | 0 |
| 0 | 0 | 0 | 0 | 0 |
| 0 | 0 | 0 | 0 | 0 |
| 0 | 28.33361207 | 0 | 6.307201068 | 0 |
| 0 | 0 | 0 | 0 | 0 |
| 0 | 0 | 0 | 0 | 0 |
| 0 | 0 | 0 | 0 | 0 |
| 0 | 0 | 0 | 0 | 0 |
| 0 | 0 | 0 | 0 | 0 |
| 0 | 0 | 0 | 0.615970256 | 0 |
| 0.020481854 | 0.008214465 | 0.335452728 | 10.1174546 | 0.313351975 |
| 0 | 0.011439868 | 0.056274342 | 0.001736953 | 0 |
| 0 | 0 | 0.00265561 | 0 | 0 |
| 0 | 0 | 0 | 0 | 0 |

| | | | | |
|---|---|---|---|---|
| 0 | 0.012716524 | 0 | 0 | 0 |
| 0.01482164 | 0.001521477 | 0.098920598 | 0 | 0.00363163 |
| 0 | 0.003983704 | 0 | 0 | 0.039536358 |
| 0 | 0.11408337 | 0 | 1.142704488 | 0 |
| 0.005888572 | 0.001551489 | 0 | 0 | 0 |
| 0 | 0 | 0 | 0 | 0 |
| 0.000851058 | 0 | 0 | 0.034706165 | 0 |
| 0 | 0 | 0 | 0.003137192 | 0 |
| 0 | 0.003165521 | 0 | 0 | 0 |
| 0 | 0 | 0 | 0 | 0 |
| 0 | 0 | 0 | 0 | 0 |
| 0 | 0.087275183 | 0 | 0.076223977 | 0 |
| 0 | 0 | 0 | 0.001036768 | 0 |
| 0.23567357 | 0.018686591 | 0.136316153 | 0.003642611 | 0.142327633 |
| 0 | 0 | 0 | 0.010583342 | 0 |
| 0 | 0 | 0 | 0 | 0 |
| 0 | 0 | 0 | 0.027905601 | 0 |
| 0 | 0 | 0 | 0 | 0 |
| 0 | 0 | 0 | 0 | 0 |
| 0 | 0 | 0 | 0 | 0 |
| 0.128166835 | 0.142335546 | 0.042466363 | 0.389893019 | 0.24809691 |
| 0.022777198 | 0 | 0.009216591 | 0.00085034 | 0.005685182 |
| 0 | 0.281805545 | 0 | 0.090606816 | 0 |
| 0 | 0 | 0 | 0 | 0 |
| 0 | 0 | 0 | 0 | 0 |
| 0 | 0 | 0 | 0 | 0 |
| 0 | 0 | 0 | 0 | 0.127326633 |
| 0 | 0 | 0 | 0 | 0 |
| 0 | 0 | 0 | 0.610817284 | 0 |
| 0 | 0 | 0 | 0 | 0 |
| 0.003882782 | 0 | 0 | 0.015898237 | 0.353262 |
| 0 | 0 | 0 | 0 | 0 |
| 0 | 0 | 0 | 0 | 0 |
| 0 | 0 | 0 | 0 | 0 |
| 0 | 0 | 0 | 0 | 0 |
| 0 | 0 | 0 | 0 | 0 |
| 0 | 0 | 0 | 0 | 0 |
| 0 | 0 | 0 | 0.001288909 | 0 |
| 0 | 0 | 0 | 0 | 0 |
| 0 | 0 | 0 | 0 | 0 |
| 0 | 0 | 0 | 0 | 0 |
| 0 | 0 | 0 | 0 | 0 |
| 0 | 0 | 0 | 0 | 0 |
| 0 | 0 | 0 | 0 | 0 |
| 0 | 0 | 0 | 0 | 0 |
| 0 | 0 | 0 | 0 | 0 |
| 0.215813161 | 0 | 0 | 0.004993725 | 0 |
| 0 | 0 | 0 | 0.019974517 | 0 |

| | | | | |
|---|---|---|---|---|
| 0 | 0 | 0 | 0 | 0 |
| 0 | 0 | 0 | 0 | 0 |
| 0 | 0 | 0 | 0 | 0 |
| 0 | 0 | 0 | 0 | 0 |
| 0 | 0 | 0 | 0.010647001 | 0 |
| 0 | 0 | 0 | 0.002351653 | 0 |
| 0 | 0 | 0 | 0 | 0 |
| 0 | 0.53959018 | 0 | 0 | 0 |
| 0 | 0 | 0.001936643 | 0.003082202 | 0 |
| 0 | 0 | 0 | 0 | 0 |
| 0 | 0 | 0 | 0 | 0 |
| 0 | 0 | 0 | 0.017891881 | 0 |
| 0 | 0 | 0 | 0 | 0 |
| 0 | 0 | 0 | 0 | 0 |
| 0 | 0 | 0 | 0 | 0 |
| 0 | 0 | 0 | 0 | 0 |
| 0 | 0 | 0 | 0 | 0 |
| 0 | 0 | 0 | 0 | 0 |
| 0.001006517 | 0 | 0.381185676 | 0.033332015 | 0 |
| 0 | 0 | 0 | 0 | 0 |
| 0 | 0 | 0 | 0 | 0 |
| 0 | 0 | 0 | 0 | 0 |
| 0 | 0 | 0 | 0 | 0 |
| 0 | 0 | 0 | 0 | 0 |
| 0 | 0 | 0 | 0 | 0 |
| 0 | 0 | 0 | 0 | 0 |
| 0 | 0 | 0 | 0 | 0 |
| 0 | 0 | 0 | 0 | 0 |
| 0 | 0 | 0 | 0 | 0 |
| 0 | 0 | 0 | 0 | 0 |
| 0 | 0.001209146 | 0.018996398 | 0 | 0 |
| 0 | 0.000543643 | 0 | 0 | 0 |
| 0 | 0 | 0 | 0 | 0 |
| 0 | 0 | 0 | 0 | 0 |
| 0 | 0 | 0 | 0 | 0 |
| 0 | 0 | 0 | 0 | 0 |
| 0 | 0 | 0 | 0 | 0 |
| 0 | 0 | 0 | 0 | 0 |
| 0 | 0 | 0 | 0 | 0 |
| 0 | 0 | 0 | 0 | 0 |
| 0 | 0 | 0 | 0 | 0 |
| 0 | 0 | 0 | 0 | 0 |
| 0 | 0 | 0.002390386 | 0 | 0.015331807 |
| 0 | 0 | 0 | 0 | 0 |
| 0 | 0 | 0 | 0 | 0 |
| 0 | 0 | 0 | 0 | 0 |
| 0 | 0 | 0 | 0 | 0 |
| 0 | 0 | 0 | 0 | 0 |

| | | | | |
|---|---|---|---|---|
| 0 | 0 | 0 | 0 | 0 |
| 0 | 0 | 0 | 0 | 0 |
| 0 | 0 | 0 | 0 | 0 |
| 0 | 0 | 0 | 0 | 0 |
| 0 | 0 | 0 | 0.011111931 | 0 |
| 0 | 0 | 0 | 0 | 0 |
| 0 | 0 | 0 | 0 | 0 |
| 0 | 0 | 0 | 0 | 0 |
| 0 | 0 | 0 | 0 | 0 |
| 0 | 0 | 0 | 0 | 0 |
| 0 | 0 | 0 | 0 | 0 |
| 0 | 0 | 0 | 0 | 0 |
| 0 | 0 | 0 | 0 | 0 |
| 0 | 0 | 0 | 0 | 0 |
| 0 | 0 | 0 | 0 | 0 |
| 0 | 0 | 0 | 0 | 0 |
| 0 | 0 | 0 | 0 | 0 |
| 0 | 0 | 0 | 0 | 0 |
| 0 | 0 | 0 | 0 | 0 |
| 0 | 0 | 0 | 0 | 0 |
| 0 | 0 | 0 | 0 | 0 |
| 0 | 0 | 0 | 0 | 0 |
| 0 | 0 | 0 | 0 | 0 |
| 0 | 0 | 0 | 0 | 0 |
| 0 | 0.02124865 | 0 | 0.008888326 | 0 |
| 0 | 0 | 0 | 0 | 0 |
| 0 | 0 | 0 | 0 | 0 |
| 0 | 0 | 0 | 0 | 0 |
| 0 | 0 | 0 | 0 | 0 |
| 0 | 0 | 0 | 0 | 0 |
| 0 | 0 | 0.875797611 | 0 | 0 |
| 0 | 0 | 0 | 0 | 0 |
| 0 | 0 | 0 | 0 | 0 |
| 0 | 0 | 0 | 0 | 0 |
| 0 | 0 | 0 | 0 | 0 |
| 0 | 0 | 0 | 0 | 0 |
| 0 | 0 | 0 | 0 | 0 |
| 0 | 0 | 0 | 0 | 0 |
| 0 | 0 | 0 | 0 | 0 |
| 0 | 0 | 0 | 0 | 0 |
| 0 | 0 | 0 | 0 | 0 |
| 0 | 0 | 0 | 0 | 0 |
| 0 | 0 | 0 | 0 | 0 |
| 0 | 0 | 0 | 0 | 0 |
| 0 | 0 | 0 | 0 | 0 |
| 0 | 0 | 0 | 0 | 0 |
| 0 | 0 | 0 | 0 | 0 |
| 0 | 0 | 0 | 0 | 0 |

| | | | | |
|---|---|---|---|---|
| 0 | 0 | 0 | 0 | 0 |
| 0 | 0 | 0 | 0 | 0 |
| 0 | 0 | 0 | 0 | 0 |
| 0 | 0 | 0 | 0 | 0 |
| 0 | 0 | 0 | 0 | 0 |
| 0 | 0 | 0 | 0 | 0 |
| 0 | 0 | 0 | 0 | 0 |
| 0 | 0 | 0 | 0 | 0 |
| 0 | 0 | 0 | 0 | 0 |
| 0 | 0 | 0 | 0 | 0 |
| 0 | 0 | 0 | 0 | 0 |
| 0 | 0 | 0 | 0 | 0 |
| 0.000835292 | 0.001000355 | 0 | 0 | 0 |
| 0 | 0 | 0 | 0 | 0 |
| 0 | 0 | 0 | 0 | 0 |
| 0 | 0 | 0 | 0 | 0 |
| 0 | 0 | 0 | 0 | 0 |
| 0 | 0 | 0 | 0 | 0 |
| 0 | 0 | 0 | 0 | 0 |
| 0 | 0 | 0 | 0 | 0 |
| 0.024988724 | 0.012344798 | 0.056664051 | 0.058474085 | 0.02113213 |
| 0 | 0 | 0 | 0 | 0 |
| 0 | 0 | 0 | 0 | 0 |
| 0.002442957 | 0 | 0 | 0 | 0 |
| 0 | 0 | 0 | 0 | 0 |
| 0 | 0 | 0 | 0 | 0 |
| 0.000909553 | 0.005010738 | 0.002486287 | 0.023502007 | 0.002773376 |
| 0 | 0 | 0 | 0 | 0 |
| 0 | 0 | 0 | 0 | 0 |
| 0.001702954 | 0 | 0.028776802 | 0 | 0.042838841 |
| 0.252960829 | 0 | 0.034697682 | 0 | 0 |
| 0 | 0 | 0 | 0 | 0 |
| 0 | 0 | 0 | 0 | 0 |
| 0 | 0 | 0.015417265 | 0 | 0 |
| 0.003048562 | 0.056712084 | 0.020959576 | 0.009913487 | 0.003292179 |
| 0 | 0 | 0 | 0 | 0 |
| 0 | 0 | 0 | 0 | 0 |
| 0 | 0 | 0 | 0 | 0 |
| 0 | 0 | 0 | 0.000779093 | 0 |
| 0 | 0 | 0 | 0.029114831 | 0 |
| 0 | 0 | 0 | 0.001414497 | 0 |
| 0 | 0 | 0 | 0 | 0 |
| 0 | 0 | 0 | 0.067950426 | 0 |
| 0 | 0 | 0 | 0 | 0 |
| 0 | 0 | 0 | 0 | 0 |
| 0 | 0 | 0 | 0.027295756 | 0 |
| 0 | 0.000790611 | 0.001913922 | 0 | 0 |
| 0.00289665 | 0.001907984 | 0.034191668 | 0 | 0.002346096 |

| | | | | |
|---|---|---|---|---|
| 0 | 0 | 0 | 0 | 0 |
| 0 | 0 | 0 | 0 | 0 |
| 0 | 0 | 0.003617684 | 0 | 0.002522134 |
| 0 | 0 | 0 | 0 | 0 |
| 0 | 0 | 0 | 0 | 0 |
| 0 | 0 | 0 | 0 | 0 |
| 0 | 0 | 0 | 0.001358972 | 0 |
| 0 | 0 | 0 | 0.00548947 | 0 |
| 0 | 0.037768963 | 0 | 0 | 0 |
| 0 | 0 | 0 | 0 | 0 |
| 0 | 0 | 0 | 0 | 0 |
| 0 | 0 | 0 | 0 | 0 |
| 0 | 0 | 0 | 0 | 0 |
| 0 | 0 | 0 | 0 | 0 |
| 0 | 0 | 0 | 0 | 0 |
| 0 | 0 | 0 | 0 | 0 |
| 0 | 0 | 0 | 0 | 0 |
| 0 | 0 | 0 | 0 | 0 |
| 0 | 0 | 0.001464114 | 0.055341383 | 0.001403508 |
| 0 | 0 | 0 | 0 | 0 |
| 0 | 0 | 0.086658636 | 0 | 0.21241459 |
| 0 | 0 | 0 | 0 | 0 |
| 0 | 0 | 0 | 0 | 0 |
| 0 | 0 | 0 | 0 | 0 |
| 0.004856165 | 0 | 0.03378953 | 0 | 0 |
| 0 | 0 | 0 | 0 | 0 |
| 0 | 0 | 0 | 0 | 0 |
| 0 | 0 | 0 | 0 | 0 |
| 0 | 0 | 0 | 0.055392839 | 0 |
| 0 | 0 | 0 | 0 | 0 |
| 0.930806886 | 0.182721564 | 0.286641016 | 0.047177996 | 0.987213964 |
| 0 | 0 | 0 | 0 | 0 |
| 0 | 0 | 0 | 0 | 0.060003829 |
| 0 | 0 | 0 | 0 | 0 |
| 0 | 0 | 0 | 0 | 0 |
| 0 | 0 | 0 | 0 | 0 |
| 0 | 0 | 0 | 0 | 0 |
| 0 | 0 | 0 | 0 | 0 |
| 0 | 0 | 0 | 0 | 0 |
| 0 | 0 | 0 | 0 | 0 |
| 0 | 0 | 0 | 0 | 0 |
| 0 | 0.012091899 | 0 | 0.018033796 | 0 |
| 0 | 0 | 0 | 0 | 0 |
| 0 | 0 | 0 | 0 | 0 |
| 0 | 0 | 0 | 0 | 0 |
| 0 | 0 | 0 | 0 | 0 |
| 0 | 0 | 0 | 0 | 0 |
| 0 | 0 | 0 | 0 | 0 |

| | | | | |
|---|---|---|---|---|
| 0 | 0 | 0 | 0 | 0 |
| 0 | 0 | 0 | 0 | 0 |
| 0 | 0.013714011 | 0.068346054 | 0.004852371 | 0 |
| 0 | 0 | 0 | 0 | 0 |
| 0 | 0 | 14.25354405 | 0 | 0 |
| 0 | 0 | 0 | 0 | 0 |
| 0 | 0 | 0 | 0 | 0 |
| 0 | 0 | 0 | 0 | 0 |
| 0 | 0 | 0 | 0 | 0 |
| 0 | 0 | 0 | 0 | 0 |
| 0 | 0 | 0 | 0 | 0 |
| 0 | 0 | 0 | 0 | 0 |
| 0 | 0 | 0 | 0 | 0 |
| 0 | 0 | 0.002246386 | 0.002085512 | 0 |
| 0.003963701 | 0.001340322 | 0.014369256 | 0.00786888 | 0.00071094 |
| 0 | 0 | 0 | 0.006845048 | 0 |
| 0.001859645 | 0 | 0.0047533 | 0.001681103 | 0.001822617 |
| 0.003915515 | 0.004343741 | 0 | 0 | 0 |
| 0 | 0 | 0 | 0 | 0 |
| 0 | 0 | 0 | 0 | 0 |
| 0 | 0 | 0 | 0 | 0 |
| 0.002638003 | 0.001378605 | 0.032181554 | 0 | 0.002010926 |
| 0 | 0 | 0 | 0 | 0 |
| 0 | 0 | 0.030302797 | 0 | 0 |
| 0 | 0 | 0 | 0.001216656 | 0 |
| 0 | 0 | 0 | 0 | 0 |
| 0.032865109 | 0 | 0 | 0 | 0 |
| 0 | 0 | 0 | 0 | 0 |
| 0 | 0 | 0 | 0 | 0 |
| 0 | 0 | 0 | 0.002926879 | 0 |
| 0 | 0 | 0 | 0 | 0 |
| 0 | 0 | 0 | 0 | 0 |
| 0 | 0 | 0 | 0.051821148 | 0 |
| 0 | 0 | 0 | 0 | 0 |
| 0 | 0 | 0 | 0 | 0 |
| 0 | 0 | 0 | 0 | 0 |
| 0 | 0 | 0 | 0 | 0 |
| 0 | 0 | 0.138070489 | 0.007944385 | 0 |
| 0 | 0 | 0 | 0 | 0 |
| 0 | 0 | 0 | 0 | 0 |
| 0 | 0 | 0 | 0 | 0 |
| 0 | 0 | 0 | 0 | 0 |
| 0 | 0 | 0 | 0 | 0 |
| 0.026198932 | 0 | 0 | 0 | 0.005991364 |
| 0 | 0 | 0 | 0 | 0 |
| 0 | 0 | 0 | 0 | 0 |
| 0 | 0 | 0 | 0 | 0 |
| 0.006569251 | 0 | 0.011608711 | 0 | 0.004451272 |

| | | | | |
|---|---|---|---|---|
| 0 | 0 | 0 | 0 | 0 |
| 0 | 0 | 0 | 0 | 0 |
| 0 | 0 | 0 | 0 | 0 |
| 0 | 0 | 0 | 0 | 0 |
| 0.020975967 | 0 | 0.015072518 | 0.01039488 | 0.006164739 |
| 0 | 0 | 0 | 0 | 0 |
| 0 | 0 | 0 | 0 | 0 |
| 0.003542462 | 0.003054595 | 0 | 0.035119153 | 0 |
| 0 | 0 | 0 | 0 | 0 |
| 0.03286163 | 0.10704687 | 0.02867242 | 0.177139499 | 2.333280162 |
| 0 | 0 | 0 | 0 | 0 |
| 0 | 0 | 0 | 0.041109172 | 0 |
| 0.003403367 | 0.003056934 | 0.014589103 | 0.042399588 | 0.001297178 |
| 0 | 0.004023668 | 0.00224782 | 0 | 0 |
| 0.004375357 | 0 | 0.001739659 | 0 | 0.006170297 |
| 0 | 0 | 0 | 0.003030896 | 0 |
| 0 | 0 | 0 | 0 | 0 |
| 0 | 0 | 0 | 0 | 0.001267054 |
| 0.008589499 | 0 | 0.19637505 | 0 | 0.20900252 |
| 0.202694841 | 0 | 0.429788896 | 0 | 0 |
| 0 | 0 | 0 | 0 | 0 |
| 0.030424327 | 0 | 0 | 0 | 0 |
| 0 | 0 | 0 | 0.007176073 | 0 |
| 0 | 0 | 0 | 0 | 0 |
| 0 | 0 | 0 | 0 | 0 |
| 0 | 0.026348061 | 0 | 0 | 0 |
| 0 | 0 | 0 | 0 | 0 |
| 0 | 0 | 0.006849587 | 0 | 0 |
| 0 | 0 | 0 | 0 | 0 |
| 0.001019629 | 0 | 0 | 0 | 0.002331761 |
| 0.126811848 | 0 | 0.400618787 | 0 | 0.27966709 |
| 0 | 0 | 0 | 0 | 0 |
| 0.108595318 | 0 | 0.268454045 | 0 | 0 |
| 0.069339424 | 0 | 0 | 0 | 0 |
| 0 | 0 | 0 | 0 | 0 |
| 0 | 0 | 0 | 0 | 0 |
| 0.003370247 | 0 | 0 | 0 | 0 |
| 0 | 0 | 0 | 0 | 0 |
| 0 | 0 | 0 | 0 | 0 |
| 0 | 0 | 0 | 0.003022967 | 0 |
| 0 | 0 | 0 | 0 | 0 |
| 0 | 0 | 0 | 0 | 0 |
| 0 | 0 | 0 | 0 | 0 |
| 0 | 0 | 0 | 0 | 0.031495562 |
| 0 | 0 | 0 | 0 | 0 |
| 0 | 0 | 0 | 0 | 0 |
| 0 | 0.001771129 | 0 | 0.00454917 | 0 |
| 0 | 0 | 0 | 0 | 0 |

| | | | | |
|---|---|---|---|---|
| 0 | 0 | 0 | 0 | 0 |
| 0 | 0 | 0 | 0.003631439 | 0 |
| 0 | 0 | 0 | 0 | 0 |
| 0 | 0 | 0 | 0 | 0 |
| 0 | 0 | 0 | 0 | 0 |
| 0.176176139 | 0.089976145 | 0.059338455 | 0 | 0 |
| 0 | 0 | 0 | 0 | 0 |
| 0 | 0 | 0 | 0 | 0 |
| 0 | 0 | 0 | 0 | 0 |
| 0 | 0 | 0 | 0 | 0 |
| 0.0208275 | 0 | 0 | 0.049868428 | 0.052779023 |
| 0 | 0 | 0.391472313 | 0 | 0 |
| 0 | 0 | 0 | 0 | 0 |
| 0.001595414 | 0.012483151 | 0.025638026 | 0.003926099 | 0 |
| 0 | 0 | 0 | 0 | 0 |
| 0 | 0 | 0 | 0 | 0 |
| 0 | 0 | 0 | 0 | 0 |
| 0 | 0 | 0.025207156 | 0 | 0 |
| 0 | 0 | 0.014065844 | 0 | 0 |
| 0 | 0.007332567 | 0 | 0 | 0 |
| 0 | 0 | 0 | 0.027798525 | 0 |
| 0 | 0.476980572 | 0 | 0.199115485 | 0 |
| 0 | 0 | 0 | 0 | 0 |
| 0 | 0 | 0 | 0 | 0 |
| 0 | 0 | 0 | 0 | 0 |
| 0 | 0 | 0 | 0 | 0 |
| 0.037191804 | 0 | 0 | 0 | 0 |
| 0 | 0 | 0 | 0 | 0 |
| 0 | 0 | 0 | 0 | 0 |
| 0.422496767 | 0.784186479 | 0.489314534 | 0.372831311 | 0 |
| 0 | 0.005703495 | 0 | 0 | 0 |
| 0 | 0 | 0 | 0.012019935 | 0 |
| 0 | 0 | 0 | 0 | 0 |
| 0 | 0 | 0 | 0 | 0 |
| 0.009605355 | 0 | 0 | 0 | 0 |
| 0 | 0 | 0 | 0 | 0 |
| 0 | 0 | 0 | 0 | 0 |
| 0 | 0 | 0 | 0.04087492 | 0 |
| 0 | 0 | 0 | 0 | 0 |
| 0 | 0 | 0 | 0 | 0 |
| 0 | 0 | 0.008691648 | 0 | 0.012831075 |
| 0 | 0.366557089 | 0 | 2.824322 | 0 |
| 0 | 0 | 0 | 0.025137279 | 0 |
| 0 | 0 | 0 | 0 | 0 |
| 0.001479558 | 0.002126322 | 0 | 0 | 0 |
| 0 | 0 | 0 | 0 | 0.173214339 |
| 0.004359305 | 0 | 0.005701568 | 0 | 0.041100993 |
| 0 | 0 | 0 | 0 | 0 |

| | | | | |
|---|---|---|---|---|
| 0 | 0.006780562 | 0.003376693 | 0.002089916 | 0 |
| 0 | 0 | 0 | 0 | 0 |
| 0 | 0 | 0 | 0.080141574 | 0 |
| 0 | 0 | 0.126504915 | 0 | 0 |
| 0 | 0 | 0 | 0 | 0 |
| 0.001196751 | 0 | 0.005234162 | 0 | 0 |
| 0 | 0 | 0 | 0 | 0 |
| 0 | 0 | 0 | 0.020412095 | 0 |
| 0.240472097 | 0 | 0 | 0 | 0.008668834 |
| 0 | 0.02049137 | 0 | 0 | 0 |
| 0 | 0 | 0 | 0 | 0 |
| 0.010025583 | 0 | 0 | 0 | 0 |
| 0 | 0 | 0 | 0 | 0 |
| 0.016134397 | 0 | 0 | 0.062933096 | 0.230364221 |
| 0 | 0 | 0 | 0 | 0 |
| 0.01501042 | 0.274395618 | 0 | 0.002110778 | 0 |
| 0 | 0.008713974 | 0.043218846 | 0.038603027 | 0.075081032 |
| 0 | 0 | 0.005358083 | 0 | 0.016025227 |
| 0.009165475 | 0 | 0.041680868 | 0 | 0.002445366 |
| 0 | 0 | 0 | 0 | 0 |
| 0 | 0.06321396 | 0 | 0.007371333 | 0 |
| 0 | 0.004437552 | 0 | 0.004342061 | 0 |
| 0 | 0 | 0 | 0.027546457 | 0 |
| 0 | 0 | 0 | 0 | 0 |
| 0 | 0 | 0 | 0 | 0 |
| 0 | 0 | 0.324170736 | 0 | 0.720410104 |
| 0 | 0 | 0.0069983 | 0 | 0.002790783 |
| 0 | 0 | 0 | 0 | 0 |
| 0 | 0 | 0 | 0 | 0 |
| 0 | 0 | 0 | 0 | 0 |
| 0 | 0.000820777 | 0.069614212 | 0 | 0.044406774 |
| 0.006386036 | 0.006118393 | 0.003491281 | 0.001010261 | 0 |
| 0.0082085 | 0.056798986 | 0 | 0.046492083 | 0 |
| 0 | 0 | 0 | 0.27156527 | 0 |
| 0 | 0 | 0 | 0 | 0 |
| 0 | 0 | 0 | 0 | 0 |
| 0 | 0.001316672 | 0.00601056 | 0 | 0 |
| 0.018915452 | 0.048395822 | 0.275408688 | 0.018589123 | 0.008356512 |
| 0.306423311 | 0 | 0 | 0.053889951 | 0 |
| 0 | 0 | 0 | 0.002224172 | 0.004019002 |
| 0 | 0 | 0 | 0 | 0 |
| 0 | 0 | 0 | 0 | 0 |
| 0 | 0.00246386 | 0 | 0 | 0.010536814 |
| 0 | 0.004662078 | 0 | 0 | 0.057430515 |
| 0.043671281 | 2.664295383 | 0.975249065 | 0 | 0 |
| 0 | 0 | 0 | 0 | 0 |
| 0 | 0 | 0 | 0 | 0 |
| 0 | 0 | 0 | 0 | 0 |

| | | | | |
|---|---|---|---|---|
| 0 | 0 | 0 | 0 | 0 |
| 0.059457555 | 0.00252557 | 0.731343322 | 0.518339072 | 0 |
| 0 | 0 | 0 | 0 | 0 |
| 0 | 0 | 0 | 0 | 0 |
| 0 | 0 | 0.007140547 | 0 | 0 |
| 0 | 0 | 0 | 0 | 0 |
| 0.012735375 | 0 | 0.005117504 | 0 | 0 |
| 0 | 0 | 0 | 0 | 0 |
| 0 | 0 | 0.005063777 | 0.001979426 | 0 |
| 0.001277044 | 0 | 0 | 0 | 0 |
| 0.001958873 | 0.00375355 | 0.01038474 | 0 | 0.002820548 |
| 0.005187194 | 0.202208515 | 0.171441041 | 0.007180303 | 0.03978865 |
| 0 | 0 | 0 | 0 | 0 |
| 0 | 0 | 0 | 0 | 0 |
| 0 | 0.059576418 | 0.00471643 | 0.001843648 | 0 |
| 0 | 0 | 0 | 0 | 0 |
| 0.007749063 | 0 | 0.035568093 | 0.002644076 | 0.010076718 |
| 0.089054872 | 0.004251952 | 0.098226419 | 0.00374441 | 3.590501185 |
| 0 | 0 | 0 | 0 | 0 |
| 0.018827584 | 0.101236508 | 0 | 0.001823568 | 0.039980847 |
| 0.002115328 | 0.081742472 | 0.022428338 | 0.174509405 | 0.000537499 |
| 0 | 0 | 0 | 0 | 0 |
| 0 | 0 | 0 | 0 | 0 |
| 0 | 0 | 0 | 0 | 0 |
| 0 | 0 | 0 | 0 | 0 |
| 0 | 0 | 0 | 0 | 0 |
| 0 | 0.030228681 | 0 | 0 | 0 |
| 0.016987014 | 0.063664265 | 0 | 0.319053321 | 0 |
| 0 | 0 | 0 | 0 | 0 |
| 0 | 0 | 0 | 0 | 0 |
| 0 | 0 | 0 | 0.002826957 | 0 |
| 0 | 0 | 0 | 0 | 0 |
| 0 | 0 | 0 | 0 | 0 |
| 0 | 0 | 0 | 0 | 0 |
| 0 | 0.053184731 | 0 | 0.030076211 | 0 |
| 1.033713949 | 0.21387567 | 0.324856578 | 0.221163866 | 0 |
| 0 | 0 | 0 | 0 | 0 |
| 0 | 0 | 0 | 0 | 0 |
| 0 | 0 | 0.019910891 | 0.026325867 | 0 |
| 0 | 0 | 0 | 0 | 0 |
| 0.032100521 | 2.686444951 | 0.965320129 | 0 | 0 |
| 0 | 0 | 0.036236296 | 0 | 0 |
| 0.080115427 | 0 | 0.008931086 | 0.00276383 | 0.022545003 |
| 0.004156655 | 0.001212049 | 0.010059928 | 0 | 0 |
| 0.064343432 | 0 | 0 | 0 | 0 |
| 0 | 0 | 0 | 0.59020231 | 0 |
| 0 | 0 | 0 | 0 | 0 |
| 0.043818988 | 0 | 0.183518063 | 0 | 0.075490379 |

| | | | | |
|---|---|---|---|---|
| 0 | 0 | 0 | 0 | 0 |
| 0 | 0 | 0 | 0 | 0 |
| 0 | 0 | 0 | 0 | 0 |
| 0.018531017 | 0.008146122 | 12.85781869 | 0.022302988 | 0 |
| 0 | 0 | 0 | 0 | 0 |
| 0 | 0 | 0 | 0 | 0 |
| 0 | 0.062021514 | 0 | 0 | 0 |
| 0 | 0 | 0 | 0 | 0 |
| 0 | 0 | 0 | 0.159407017 | 0 |
| 0 | 0 | 0 | 0 | 0 |
| 0 | 0 | 0 | 0 | 0 |
| 0 | 0.004802991 | 0 | 0 | 0 |
| 0 | 0 | 0 | 0 | 0 |
| 0 | 0 | 0 | 0 | 0 |
| 0.004309185 | 0.489154586 | 0.174076238 | 0.004544709 | 0 |
| 0 | 0 | 0 | 0 | 0 |
| 0 | 0 | 0 | 0.007350937 | 0 |
| 0 | 0.046796558 | 0 | 0.031097861 | 0 |
| 0 | 0 | 0 | 0 | 0 |
| 0 | 0 | 0 | 0 | 0 |
| 0 | 0 | 0 | 0 | 0 |
| 0 | 0 | 0 | 0 | 0 |
| 0 | 0 | 0 | 0 | 0 |
| 0.668618851 | 0 | 0.974557676 | 0.019612185 | 0.385520708 |
| 0 | 0.053399819 | 0 | 0 | 0 |
| 0 | 0 | 0 | 0.687435635 | 0 |
| 0 | 0 | 0 | 0.000736227 | 0 |
| 0 | 0.179367719 | 0 | 2.239839731 | 0 |
| 0 | 0 | 0 | 0 | 0 |
| 0 | 0 | 0 | 0 | 0 |
| 0 | 0 | 0 | 0 | 0 |
| 0 | 0 | 0 | 0 | 0 |
| 0 | 0.000444212 | 0 | 0 | 0 |
| 0 | 0 | 0 | 0 | 0 |
| 0 | 0 | 0 | 0 | 0 |
| 0 | 0.036291441 | 0 | 0.013074319 | 0 |
| 0 | 0 | 0 | 0.738742969 | 0 |
| 0 | 0 | 0 | 0 | 0 |
| 0 | 0.011666466 | 0 | 0 | 0.117188656 |
| 0 | 0.001145951 | 0 | 0 | 0 |
| 0.019837815 | 0 | 0.435411049 | 0.382092083 | 0 |
| 0 | 0 | 0 | 0 | 0 |
| 0.524515192 | 0.121142077 | 0 | 2.406930416 | 0 |
| 0 | 0 | 0 | 0 | 0 |
| 0 | 0 | 0 | 0 | 0 |
| 0 | 0 | 0 | 0 | 0 |
| 0 | 0 | 0 | 0 | 0 |
| 0.014452927 | 0.003000225 | 0.002154953 | 0 | 0.000918111 |

| | | | | |
|---|---|---|---|---|
| 0 | 0 | 0 | 0 | 0 |
| 0 | 0 | 0 | 0.004397451 | 0 |
| 0.216596109 | 0.354860878 | 0.894705906 | 0.002922403 | 2.600911168 |
| 0 | 0 | 0.002232993 | 0 | 0 |
| 0 | 0.439327218 | 0 | 0 | 0 |
| 0 | 0.004357009 | 0 | 0.00364508 | 0 |
| 0 | 0 | 0 | 0 | 0 |
| 0 | 0 | 0 | 0 | 0 |
| 0 | 0 | 0 | 0 | 0 |
| 0 | 0 | 0 | 0 | 0 |
| 0.067561165 | 0.026282833 | 1.027403676 | 0.004782135 | 0.014862783 |
| 2.590849635 | 1.545737825 | 0 | 2.801985322 | 0 |
| 0 | 0 | 0 | 0 | 0 |
| 0.000734041 | 0.001172128 | 0.039157571 | 0.044901354 | 0 |
| 0.931501581 | 0.042674111 | 0.009774789 | 0 | 3.568602528 |
| 0 | 0 | 0 | 0 | 0 |
| 0 | 0 | 0 | 0 | 0 |
| 0 | 0 | 0 | 0 | 0 |
| 0 | 0 | 0 | 0 | 0 |
| 0 | 0 | 0 | 0 | 0 |
| 0.001900208 | 0 | 0.070830959 | 0.222651813 | 0 |
| 2.581875531 | 0.074238787 | 0.782958905 | 0 | 31.34433749 |
| 19.44083584 | 1.21367359 | 2.843762186 | 2.938143222 | 40.94995703 |
| 0 | 0 | 0 | 0 | 0 |
| 0 | 0 | 0 | 0 | 0 |
| 0 | 1.785193255 | 0 | 2.570776924 | 0 |
| 0 | 0 | 0.038856922 | 0 | 0 |
| 0 | 0 | 0 | 0 | 0 |
| 0 | 0.026857171 | 0 | 0 | 0 |
| 0 | 0 | 0 | 0 | 0 |
| 0 | 0 | 0 | 0 | 0 |
| 0 | 0 | 0 | 0 | 0 |
| 0 | 0.020969889 | 2.182089483 | 0.025539162 | 0.004259851 |
| 0 | 0.026028171 | 0 | 0 | 0 |
| 0.004303011 | 0.000883429 | 0.235858963 | 0.004214041 | 0.00866896 |
| 0 | 0.018004364 | 0.586616546 | 0.015588017 | 0 |
| 0 | 0.656986553 | 0.006082257 | 0 | 0 |
| 0 | 0 | 0 | 0 | 0 |
| 0 | 0 | 0 | 0.179284223 | 0 |
| 0 | 0 | 0 | 0 | 0 |
| 0 | 0 | 0 | 0.000784345 | 0 |
| 0 | 0 | 0 | 0 | 0 |
| 0 | 0 | 0 | 0 | 0 |
| 0 | 0 | 0 | 0 | 0 |
| 0.038766784 | 0.20854908 | 0.43704417 | 0.015998727 | 0 |
| 0 | 0 | 0.475991874 | 0 | 0 |
| 0 | 0 | 0 | 0 | 0 |
| 0 | 0.078753885 | 0.441446165 | 0.037372351 | 0 |

| | | | | |
|---|---|---|---|---|
| 0 | 0 | 0 | 0 | 0 |
| 0 | 0 | 0 | 0 | 0 |
| 0 | 0 | 0 | 0 | 0 |
| 0 | 0.002556075 | 0 | 0 | 0 |
| 0 | 0 | 0 | 0 | 0 |
| 0 | 0.009203692 | 0 | 0.024276997 | 0 |
| 0 | 0 | 0 | 0 | 0 |
| 0 | 0.234843331 | 0 | 0 | 0.039155008 |
| 0 | 0 | 0 | 0 | 0 |
| 0 | 0 | 0 | 0 | 0 |
| 0.001828505 | 0.002408823 | 0.573210555 | 0 | 0.020385105 |
| 0 | 0 | 0 | 0 | 0 |
| 0 | 0 | 0 | 0 | 0 |
| 0 | 0 | 0 | 0 | 0 |
| 0 | 0 | 0 | 0 | 0 |
| 0 | 0 | 0 | 0 | 0 |
| 0 | 0.041042327 | 0 | 0.07691194 | 0 |
| 0 | 0.007656563 | 0 | 0.001605386 | 0 |
| 0 | 0 | 0 | 0 | 0 |
| 0 | 0 | 0.032748943 | 0.008686756 | 0 |
| 0 | 0.000485875 | 0 | 0 | 0 |
| 0 | 0 | 0 | 0 | 0 |
| 0.003071206 | 0 | 0.001308347 | 0.022673472 | 0 |
| 0 | 0 | 0 | 0 | 0 |
| 0 | 0 | 0 | 0 | 0 |
| 0 | 0 | 0 | 0 | 0 |
| 0 | 0 | 0 | 0 | 0 |
| 0 | 0 | 0 | 0 | 0 |
| 0 | 0 | 0 | 0 | 0 |
| 0 | 0 | 0 | 0.20289847 | 0 |
| 0 | 0 | 0 | 0 | 0 |
| 0 | 0 | 0 | 0 | 0 |
| 0 | 0 | 0 | 0 | 0 |
| 0 | 0 | 0 | 0 | 0 |
| 0 | 0 | 0 | 0.002108433 | 0 |
| 0 | 0 | 0 | 0 | 0 |
| 0 | 0 | 0 | 0.010184802 | 0 |
| 0.001547862 | 0 | 0.021732581 | 0 | 0.003687254 |
| 0 | 0 | 0 | 0 | 0 |
| 0 | 0 | 0 | 0 | 0 |
| 0 | 0.015653378 | 0 | 0.001914567 | 0 |
| 0 | 0 | 0 | 0 | 0 |
| 0 | 0 | 0 | 0 | 0 |
| 0 | 0.536279108 | 0 | 0 | 0 |
| 0 | 0.007156658 | 0 | 0.041011877 | 0 |
| 0 | 0 | 0 | 0 | 0 |
| 0 | 0 | 0 | 0 | 0 |
| 0 | 0 | 0 | 0 | 0 |

| | | | | |
|---|---|---|---|---|
| 0 | 0 | 0 | 0 | 0 |
| 0 | 0 | 0 | 0 | 0 |
| 0 | 0 | 0 | 0.010153628 | 0 |
| 0 | 0 | 0 | 0.121786619 | 0 |
| 0 | 0 | 0 | 0 | 0 |
| 0 | 0.61650432 | 0 | 0 | 0 |
| 0 | 0 | 0 | 0 | 0 |
| 0 | 0 | 0 | 0 | 0 |
| 0.644697525 | 0.025054809 | 2.468384316 | 0.003095728 | 0.644617567 |
| 0 | 0 | 0 | 0 | 0 |
| 0 | 0 | 0 | 0 | 0 |
| 0 | 0 | 0 | 0 | 0 |
| 0 | 0.005369328 | 0 | 0 | 0 |
| 0 | 0 | 0 | 0 | 0 |
| 3.211244123 | 1.078649293 | 0 | 0.137525151 | 0 |
| 0.292044107 | 0 | 0 | 0 | 0.341772741 |
| 0 | 0 | 0 | 0 | 0 |
| 0 | 0 | 0 | 0 | 0 |
| 0 | 0 | 0 | 0 | 0 |
| 0 | 0 | 0 | 0 | 0 |
| 0 | 0 | 0 | 0 | 0 |
| 0 | 0 | 0 | 0 | 0 |
| 0 | 0 | 0 | 0 | 0 |
| 0 | 0 | 0.112292503 | 0 | 0 |
| 0 | 0.002705854 | 0.07080638 | 0.518934947 | 0.027508984 |
| 0.333334571 | 11.67110574 | 1.995082937 | 0.153161255 | 0 |
| 0 | 0 | 0 | 0.161746917 | 0 |
| 0 | 0 | 0 | 0 | 0 |
| 0 | 0.391127417 | 0 | 0.222797898 | 0 |
| 0 | 0 | 0 | 0 | 0 |
| 0 | 0 | 0 | 0 | 0 |
| 0 | 0 | 0 | 0 | 0 |
| 0 | 0 | 0 | 0 | 0 |
| 0 | 0.002247933 | 0.012493993 | 0 | 0 |
| 0.002485226 | 0 | 0 | 0.002912288 | 0 |
| 0.001171169 | 0.001309098 | 0.069073092 | 0 | 0.00282711 |
| 0 | 0 | 0 | 0 | 0 |
| 0 | 2.036323536 | 0 | 1.133452556 | 0 |
| 0 | 0 | 0 | 0 | 0 |
| 0 | 0 | 0 | 0 | 0 |
| 0 | 0 | 0 | 0 | 0 |
| 0 | 0 | 0 | 0 | 0 |
| 0 | 0.093278671 | 0 | 2.016473496 | 0.309266998 |
| 0 | 0 | 0 | 0 | 0 |
| 0 | 0 | 0 | 0 | 0 |
| 0 | 0.007441237 | 0 | 0 | 0 |
| 0.016032787 | 0 | 0 | 0 | 0 |
| 0.032995628 | 0 | 0.012299241 | 0.005281999 | 0 |

| | | | | |
|---|---|---|---|---|
| 0 | 0.007366771 | 0 | 0.004170486 | 0.084904741 |
| 0 | 0 | 0 | 0 | 0 |
| 0 | 0 | 0 | 0 | 0 |
| 0 | 0 | 0 | 0 | 0 |
| 0.00131234 | 0.011316045 | 0.016305979 | 0 | 0 |
| 0 | 0 | 0 | 0 | 0 |
| 0 | 0 | 0 | 0 | 0 |
| 0 | 0 | 0 | 0 | 0 |
| 0 | 0 | 0 | 0 | 0 |
| 0 | 0 | 0 | 0 | 0 |
| 0 | 0 | 0 | 0 | 0 |
| 0 | 0 | 0 | 0.309433096 | 0 |
| 0 | 0.409022504 | 0 | 0.108372459 | 0 |
| 0 | 0 | 0 | 0 | 0 |
| 0 | 0 | 0 | 1.130875978 | 0 |
| 0 | 0 | 0 | 0 | 0 |
| 0 | 0 | 0 | 0 | 0 |
| 0 | 0 | 0 | 0 | 0 |
| 0 | 0 | 0 | 0 | 0 |
| 0 | 0 | 0.002688086 | 0.012186208 | 0 |
| 0 | 0 | 0 | 0.178877584 | 0 |
| 0 | 0 | 0 | 0 | 0 |
| 0.001293513 | 0.211920497 | 0.084217601 | 0.406307829 | 0 |
| 0 | 0 | 0 | 0 | 0 |
| 0 | 0 | 0 | 0 | 0 |
| 0 | 0 | 0 | 0 | 0 |
| 0 | 0 | 0 | 0 | 0 |
| 0 | 0 | 0 | 0 | 0 |
| 0 | 0 | 0 | 0 | 0 |
| 0 | 0 | 0 | 0 | 0 |
| 0 | 0 | 0 | 0 | 0 |
| 0.011488486 | 0.007547653 | 0.037195488 | 0.015924418 | 0.005254542 |
| 0 | 0 | 0 | 0 | 0 |
| 0 | 0.012568925 | 0 | 0 | 0 |
| 0.000637263 | 0.001424627 | 0.068834381 | 0 | 0.002590824 |
| 0 | 0.008133146 | 0 | 0.00548726 | 0 |
| 0 | 0 | 0 | 0 | 0 |
| 0 | 0 | 0 | 0.008997475 | 0 |
| 0 | 0 | 0 | 0.001544271 | 0 |
| 0 | 0 | 0 | 0 | 0 |
| 0 | 0 | 0 | 0.018133148 | 0 |
| 0 | 0 | 0 | 0 | 0 |
| 0 | 0 | 0 | 0 | 0 |
| 0 | 0 | 0.430251286 | 0 | 0 |
| 0.024901444 | 0.126706997 | 0.324067652 | 0.001324158 | 0 |
| 0 | 0 | 0 | 0 | 0 |
| 0 | 0 | 0 | 0.057005634 | 0 |
| 0 | 0 | 0 | 0 | 0 |

| | | | | |
|---|---|---|---|---|
| 0 | 0 | 0 | 0 | 0 |
| 0 | 0 | 0 | 0 | 0 |
| 0.00519415 | 0 | 0 | 0 | 0.002309681 |
| 0 | 0 | 0 | 0 | 0 |
| 0.002414325 | 0 | 0 | 0 | 0 |
| 0 | 0 | 0 | 0 | 0 |
| 0 | 0 | 0 | 0.080150289 | 0 |
| 0 | 0 | 0.1507498 | 0.022202935 | 0 |
| 0 | 0 | 0 | 0 | 0 |
| 0 | 0 | 0 | 0 | 0 |
| 0 | 0 | 0 | 0 | 0 |
| 0.013212303 | 0 | 0.001969974 | 0.000928963 | 0.016786034 |
| 0 | 0 | 0 | 0 | 0 |
| 0 | 0 | 0 | 0.410319631 | 0 |
| 0 | 0 | 0 | 0 | 0 |
| 0 | 0 | 0 | 0 | 0 |
| 0 | 0 | 0 | 0 | 0 |
| 0 | 0 | 0 | 0 | 0 |
| 0 | 0 | 0 | 4.004991898 | 0 |
| 0 | 0 | 0 | 0 | 0 |
| 0 | 0 | 0 | 0 | 0 |
| 0 | 0 | 0 | 0 | 0 |
| 0 | 0 | 0 | 0 | 0 |
| 0 | 0 | 0 | 0 | 0 |
| 0 | 0 | 0 | 0.001446758 | 0 |
| 0.018506881 | 0.003546249 | 0.056543557 | 0 | 0 |
| 0 | 0 | 0 | 0 | 0 |
| 0 | 0 | 0.681386649 | 0.002212115 | 0 |
| 0 | 0 | 0 | 0 | 0 |
| 0 | 0.087904108 | 0 | 0 | 0 |
| 0 | 0 | 0 | 0 | 0 |
| 0 | 0 | 0 | 0 | 0 |
| 0 | 0 | 0 | 0 | 0 |
| 0 | 0 | 0 | 0 | 0 |
| 0 | 0 | 0.268064154 | 0 | 0.007625392 |
| 0 | 0 | 0 | 0 | 0 |
| 0 | 0 | 0 | 0 | 0 |
| 0 | 0 | 0.002072346 | 0 | 0 |
| 0 | 0 | 0 | 0 | 0 |
| 0 | 0 | 0 | 0 | 0 |
| 7.844048836 | 8.490586264 | 7.407500939 | 5.879776671 | 0 |
| 1.037753561 | 0.002274452 | 0.975240878 | 0.113739393 | 0 |
| 0 | 0 | 0 | 0.35872397 | 0 |
| 0 | 0 | 0 | 0 | 0 |
| 0 | 0 | 0 | 0 | 0 |
| 0 | 0 | 0 | 0 | 0 |
| 0 | 0 | 0 | 0 | 0 |
| 0.000735395 | 0.012506197 | 0.01809201 | 0 | 0 |

| | | | | |
|---|---|---|---|---|
| 0 | 0 | 0 | 0 | 0 |
| 0 | 0.021741391 | 0 | 0 | 0.000432456 |
| 0 | 0 | 0 | 0 | 0 |
| 0 | 0 | 0 | 0 | 0 |
| 0 | 0.357778933 | 0.1264108 | 1.671551245 | 0.997760321 |
| 0 | 0 | 0 | 0 | 0 |
| 0 | 0 | 0 | 0 | 0 |
| 0 | 0 | 0 | 0 | 0 |
| 0.357823268 | 0.056987009 | 1.481655177 | 0.003584618 | 3.399132845 |
| 0 | 0 | 0 | 0 | 0 |
| 0 | 0 | 0 | 0 | 0.04576418 |
| 0 | 0 | 0 | 0 | 0 |
| 0 | 0.006865777 | 0.015890188 | 0.167240284 | 0 |
| 0.007454623 | 0.007142194 | 0.023128081 | 0.037404371 | 0 |
| 0 | 0 | 0 | 0.080587916 | 0 |
| 0 | 0.000532803 | 8.060250893 | 0.129422082 | 0 |
| 0 | 0 | 0 | 0 | 0 |
| 0 | 0 | 0 | 0 | 0 |
| 0 | 0 | 0 | 0 | 0 |
| 0 | 0 | 0 | 0 | 0 |
| 0 | 0 | 0 | 0 | 0 |
| 0 | 0 | 0 | 0 | 0 |
| 0 | 0 | 0 | 0 | 0 |
| 0 | 0 | 0 | 0 | 0 |
| 0 | 0 | 0 | 0 | 0 |
| 0 | 0 | 0 | 0 | 0 |
| 0.007648981 | 0.008305527 | 0 | 0.012907275 | 0 |
| 0 | 0 | 0.010676485 | 0.001820552 | 0.000877247 |
| 0 | 0 | 0 | 0 | 0 |
| 0 | 0 | 0 | 0 | 0 |
| 0 | 0 | 0 | 0 | 0 |
| 0 | 0 | 0 | 0 | 0 |
| 0 | 0 | 0 | 0 | 0 |
| 0 | 0.002118947 | 0 | 0 | 0 |
| 0 | 0 | 0 | 0 | 0 |
| 0.002218733 | 0 | 0 | 0.004446002 | 0 |
| 0 | 0 | 0 | 0.001435058 | 0 |
| 0 | 0 | 0.004581638 | 0 | 0 |
| 0 | 0 | 0 | 0 | 0 |
| 0 | 0 | 0 | 0 | 0 |
| 0 | 2.717262066 | 0 | 0.824805726 | 0 |
| 0 | 0 | 0 | 0.006444496 | 0 |
| 0 | 0 | 0 | 0 | 0 |
| 0 | 0 | 0 | 0 | 0 |
| 0 | 0 | 0 | 0 | 0 |
| 0 | 0 | 0 | 0 | 0 |
| 0 | 0 | 0 | 0 | 0 |
| 0 | 0 | 0 | 0 | 0 |

| | | | | |
|---|---|---|---|---|
| 0.002310039 | 0 | 0 | 0.140862264 | 0 |
| 0 | 0 | 0 | 0 | 0 |
| 0 | 0 | 0 | 0 | 0 |
| 0 | 0.004787823 | 0 | 0 | 0 |
| 0 | 0 | 0 | 0 | 0 |
| 0 | 0 | 0 | 0 | 0 |
| 0 | 0 | 0.109323472 | 0.254198572 | 0 |
| 0 | 0.000902986 | 0 | 0 | 0 |
| 0 | 0 | 0 | 0 | 0 |
| 0 | 0 | 0 | 0 | 0 |
| 0 | 0 | 0 | 0 | 0 |
| 0 | 0 | 0.82502072 | 0 | 0 |
| 0 | 0 | 0 | 0 | 0 |
| 0 | 0 | 0 | 0 | 0 |
| 0 | 0 | 0 | 0 | 0 |
| 0 | 0 | 0 | 0 | 0 |
| 0 | 0 | 0 | 0 | 0 |
| 0 | 0 | 0 | 0 | 0 |
| 0 | 0 | 0 | 0 | 0 |
| 0 | 0 | 0 | 0 | 0 |
| 0.000928274 | 0 | 0.086919574 | 0.072642523 | 0 |
| 0 | 0 | 0 | 0 | 0 |
| 0 | 0.003727689 | 0 | 0.006911004 | 0 |
| 0 | 0.003489158 | 0 | 0.005377169 | 0.003608933 |
| 0 | 0 | 0 | 0 | 0 |
| 0 | 0 | 0 | 0 | 0 |
| 0 | 0 | 0 | 0 | 0 |
| 0 | 0 | 0 | 0 | 0 |
| 0 | 0 | 0 | 0 | 0 |
| 0.006289664 | 0.545107279 | 0.002083996 | 0 | 0.016980715 |
| 0 | 0 | 0 | 0 | 0 |
| 0 | 0 | 0 | 0 | 0 |
| 0 | 0 | 0 | 0 | 0 |
| 0 | 0.001117863 | 0 | 0 | 0 |
| 0 | 0 | 0 | 0 | 0 |
| 0 | 0 | 0 | 0 | 0 |
| 0.001139205 | 0 | 0 | 0.030997921 | 0.003473623 |
| 0 | 0 | 0 | 0 | 0 |
| 0 | 0 | 0 | 0 | 0 |
| 0 | 0 | 0 | 0.040198626 | 0 |
| 0 | 0 | 0 | 0 | 0 |
| 0 | 0 | 0 | 0 | 0 |
| 0 | 0 | 0 | 0 | 0 |
| 0 | 0 | 0 | 0 | 0 |
| 0 | 0 | 0 | 0 | 0 |
| 0 | 0 | 0 | 0 | 0 |
| 0 | 0 | 0 | 0 | 0 |
| 0 | 0 | 0 | 0.049701117 | 0 |

| | | | | |
|---|---|---|---|---|
| 0.004898366 | 0.016144168 | 0.096990961 | 0 | 0.048691112 |
| 0 | 0 | 0 | 0 | 0 |
| 0 | 0 | 0 | 0 | 0 |
| 0 | 0 | 0 | 0 | 0 |
| 0 | 0 | 0 | 0 | 0 |
| 0 | 0 | 0 | 0 | 0 |
| 0 | 0 | 0 | 0 | 0 |
| 1.560759182 | 1.804515876 | 1.952560735 | 0.398467306 | 0 |
| 0.029832241 | 0.006482504 | 0.443274555 | 12.51698994 | 0.502174243 |
| 0.06251904 | 0.091284156 | 0 | 0.019871155 | 0 |
| 0 | 0 | 0 | 0 | 0 |
| 0 | 0 | 0 | 0 | 0 |
| 0 | 0 | 0 | 0 | 0 |
| 0 | 0 | 0 | 0 | 0 |
| 0 | 0 | 0 | 0 | 0 |
| 0 | 0 | 0 | 0 | 0 |
| 0.070644934 | 0 | 0.377548545 | 0 | 0 |
| 0 | 0 | 0 | 0 | 0 |
| 0 | 0 | 0 | 0 | 0 |
| 0 | 0 | 0 | 0 | 0 |
| 0 | 0 | 0 | 0 | 0 |
| 0 | 0 | 0 | 0 | 0 |
| 0 | 0 | 0 | 0 | 0 |
| 0 | 0 | 0 | 0 | 0 |
| 0 | 0 | 0 | 0 | 0 |
| 0 | 0 | 0 | 0 | 0 |
| 0 | 0 | 0 | 0 | 0 |
| 0 | 0 | 0 | 0 | 0 |
| 0 | 0 | 0 | 0 | 0 |
| 0 | 0 | 0 | 0 | 0 |
| 0 | 0 | 0 | 0 | 0 |
| 0 | 0 | 0 | 0 | 0 |
| 0 | 0 | 0 | 0 | 0 |
| 0 | 0 | 0 | 0 | 0 |
| 0 | 0.00363624 | 0 | 0 | 0 |
| 0 | 0 | 0 | 0 | 0 |
| 0 | 0 | 0 | 0 | 0 |
| 0 | 0 | 0 | 0 | 0 |
| 0 | 0 | 0 | 0.004177164 | 0 |
| 0 | 0.002811093 | 0.12676995 | 0 | 0 |
| 0 | 0 | 0 | 0 | 0 |
| 0 | 0.10498513 | 0 | 0 | 0 |
| 0 | 0 | 0 | 0 | 0 |
| 0 | 0 | 0 | 0 | 0 |
| 0 | 0 | 0 | 0 | 0 |
| 0.002090709 | 0 | 0 | 0 | 0 |
| 0 | 0 | 0 | 0 | 0 |
| 0 | 0 | 0 | 0 | 0 |

| | | | | |
|---|---|---|---|---|
| 0 | 0 | 0 | 0 | 0 |
| 0 | 0 | 0.031288892 | 0.013614866 | 0.012267095 |
| 0 | 0 | 0 | 0 | 0 |
| 0 | 0 | 0 | 0 | 0 |
| 0 | 0.357458438 | 0 | 0.165061223 | 0 |
| 0 | 0 | 0 | 0 | 0 |
| 0 | 0 | 0 | 0 | 0 |
| 0.056827421 | 0.035103174 | 0.195722543 | 0.002628658 | 0.034769225 |
| 0 | 0 | 0 | 0 | 0 |
| 0 | 0 | 0 | 0 | 0 |
| 0 | 0 | 0 | 0 | 0 |
| 0 | 0 | 0 | 0 | 0 |
| 0 | 0 | 0 | 0 | 0 |
| 0 | 0 | 0 | 0 | 0 |
| 0 | 0 | 0 | 0 | 0 |
| 0 | 0.029202079 | 0 | 0 | 0 |
| 0 | 0 | 0 | 0 | 0 |
| 0 | 0 | 0 | 0 | 0 |
| 0 | 0 | 0 | 0 | 0 |
| 0 | 0 | 0 | 0 | 0 |
| 0 | 0 | 0 | 0 | 0 |
| 0 | 0 | 0 | 0 | 0 |
| 0 | 0 | 0 | 0 | 0 |
| 0.002372693 | 0 | 0.037092458 | 0 | 0.012331927 |
| 0 | 0 | 0 | 0.062744365 | 0 |
| 0 | 0.006215576 | 0 | 0 | 0 |
| 0 | 0.020980478 | 0.763220349 | 0.004577413 | 0.087825397 |
| 0 | 0 | 0 | 0 | 0 |
| 0.010556448 | 0 | 0 | 0 | 0.006258845 |
| 0 | 0 | 0 | 0 | 0 |
| 0 | 0 | 0 | 0 | 0 |
| 0 | 0 | 0 | 0 | 0 |
| 0 | 0 | 0 | 0 | 0 |
| 0 | 0 | 0 | 0 | 0 |
| 0 | 0 | 0 | 0 | 0 |
| 0 | 0 | 0 | 0 | 0 |
| 0 | 0 | 0 | 0 | 0 |
| 0 | 0.000581533 | 0 | 0 | 0 |
| 0 | 0 | 0 | 0 | 0 |
| 0 | 0.001829291 | 0 | 0 | 0 |
| 0 | 0 | 0 | 0 | 0 |
| 0 | 0 | 0 | 0 | 0 |
| 0 | 0 | 0 | 0 | 0 |
| 0 | 0 | 0 | 0 | 0 |
| 0 | 0 | 0 | 0 | 0 |
| 0 | 0 | 0 | 0 | 0 |
| 0 | 0 | 0 | 0 | 0 |
| 0 | 0 | 0 | 0 | 0 |

| | | | | |
|---|---|---|---|---|
| 0.009276314 | 0 | 3.743807775 | 0.117791222 | 0 |
| 0 | 0 | 0 | 0 | 0 |
| 0 | 0 | 0 | 0 | 0 |
| 0 | 0.001414817 | 0 | 0 | 0 |
| 0 | 0 | 0 | 0 | 0 |
| 0 | 0 | 0 | 0 | 0 |
| 0 | 0 | 0 | 0 | 0 |
| 0 | 0 | 0 | 0 | 0 |
| 0 | 0 | 0 | 0 | 0 |
| 0 | 0 | 0 | 0 | 0 |
| 0 | 0 | 0 | 0 | 0 |
| 0.000795362 | 0 | 0 | 0 | 0 |
| 0 | 0 | 0 | 0 | 0 |
| 0 | 0 | 0 | 0 | 0 |
| 0 | 0 | 0 | 0 | 0 |
| 0 | 0 | 0 | 0 | 0 |
| 0 | 0 | 0 | 0 | 0 |
| 0 | 0 | 2.481466358 | 0 | 0 |
| 0 | 0.002863317 | 0 | 0 | 0 |
| 0 | 0 | 0 | 0 | 0 |
| 0.003221965 | 0 | 0.389017012 | 0.00996766 | 0 |
| 0 | 0 | 0 | 0 | 0 |
| 0 | 0.068094982 | 0.005662625 | 0.001430503 | 0 |
| 0 | 0 | 0.1475844 | 0 | 0 |
| 0 | 0 | 0 | 0 | 0 |
| 0 | 0 | 0 | 0 | 0 |
| 0 | 0 | 0 | 0 | 0 |
| 0 | 0 | 0 | 0 | 0 |
| 0 | 0 | 0 | 0 | 0 |
| 0 | 0 | 0 | 0 | 0 |
| 0 | 0 | 0 | 0 | 0 |
| 0 | 0 | 0 | 0 | 0 |
| 0.052841736 | 0.009492581 | 0.063131253 | 0.028668733 | 0.012394087 |
| 0 | 0 | 0 | 0 | 0 |
| 0 | 0.364680497 | 0 | 0.056800363 | 0 |
| 0.003669514 | 0 | 0.116315783 | 0 | 0 |
| 0 | 0 | 0 | 0 | 0 |
| 0 | 0 | 0 | 0.07113841 | 0 |
| 0.002357986 | 0.005196069 | 0.061409149 | 0.00422767 | 0 |
| 0 | 0 | 0 | 0 | 0 |
| 0 | 0 | 0 | 0 | 0 |
| 0 | 0 | 0 | 0 | 0 |
| 0 | 0.008717397 | 0 | 0.018901283 | 0 |
| 0 | 0 | 0 | 0 | 0 |
| 0 | 0 | 0 | 0 | 0 |
| 0 | 0 | 0.117534868 | 0.048958618 | 0 |
| 0 | 0 | 0 | 0 | 0 |
| 0 | 0 | 0 | 0 | 0 |

| | | | | |
|---|---|---|---|---|
| 0 | 0 | 0 | 0 | 0 |
| 0 | 0 | 0 | 0 | 0 |
| 0 | 0 | 0 | 0 | 0 |
| 0 | 0 | 0 | 0 | 0 |
| 0 | 0 | 0 | 0 | 0 |
| 0 | 0 | 0 | 0 | 0 |
| 0 | 0 | 0 | 0 | 0 |
| 0 | 0.017231644 | 0 | 0 | 0 |
| 0 | 0 | 0 | 0 | 0 |
| 0 | 0 | 0 | 0.020694032 | 0 |
| 0 | 0 | 0 | 0 | 0 |
| 0 | 0 | 0 | 0 | 0 |
| 0 | 0 | 0 | 0 | 0 |
| 0 | 0 | 0 | 0 | 0 |
| 0 | 0 | 0 | 0 | 0 |
| 0.076556184 | 0 | 0.001544009 | 0 | 0.007950233 |
| 0 | 0 | 0 | 0 | 0 |
| 0 | 0 | 0 | 0 | 0 |
| 0 | 0 | 0 | 0 | 0 |
| 0 | 0.007831869 | 0 | 0 | 0 |
| 0 | 0 | 0 | 0.039108494 | 0 |
| 0 | 0 | 0 | 0 | 0 |
| 0 | 0 | 0 | 0 | 0 |
| 0 | 0 | 0 | 0.002805599 | 0 |
| 0 | 0 | 0 | 0 | 0 |
| 0 | 0 | 0 | 0 | 0 |
| 0 | 0 | 0 | 0 | 0 |
| 0 | 0 | 0 | 0 | 0 |
| 0 | 0 | 0 | 0 | 0 |
| 0 | 0 | 0 | 0 | 0 |
| 0 | 0.237422873 | 0 | 0.170811535 | 0 |
| 0 | 0 | 0 | 0 | 0 |
| 0 | 0 | 0 | 0.013253823 | 0 |
| 0 | 0 | 0 | 0 | 0 |
| 0 | 0 | 0 | 0 | 0 |
| 0 | 0 | 0 | 0 | 0 |
| 0 | 0 | 0 | 0 | 0 |
| 0 | 0 | 0 | 0 | 0 |
| 0 | 0 | 0 | 0 | 0 |
| 0 | 0 | 0 | 0 | 0 |
| 0 | 0 | 0 | 0 | 0 |
| 0 | 0 | 0 | 0 | 0 |
| 0 | 0 | 0 | 0 | 0 |
| 0 | 0 | 0 | 0 | 0.00325225 |
| 0 | 0 | 0 | 0 | 0 |
| 0 | 0 | 0 | 0 | 0 |
| 0.19149541 | 0 | 0.937516235 | 0 | 0.001558455 |
| 0 | 0.005836875 | 0 | 0 | 0 |

| | | | | |
|---|---|---|---|---|
| 0 | 0 | 0 | 0 | 0 |
| 0 | 0 | 0 | 0 | 0 |
| 0 | 0 | 0 | 0 | 0 |
| 0 | 0 | 0 | 0 | 0 |
| 0 | 0.42111482 | 0 | 0.016108248 | 0 |
| 0 | 0 | 0 | 0 | 0 |
| 0 | 0 | 0 | 0 | 0 |
| 0 | 0 | 0 | 0 | 0 |
| 0 | 0 | 0 | 0 | 0 |
| 0 | 0 | 0 | 0 | 0 |
| 0 | 0 | 0 | 0.019997391 | 0 |
| 0.005183437 | 0.022573616 | 0.064404844 | 0 | 0 |
| 0 | 0.003700315 | 0.002222606 | 0 | 0.002014389 |
| 0 | 0 | 0 | 0 | 0 |
| 0 | 0 | 0 | 0 | 0 |
| 0 | 0.158865387 | 0 | 0 | 0 |
| 0 | 0.001041707 | 0 | 0 | 0 |
| 0 | 0 | 0 | 0 | 0 |
| 0.056376246 | 0 | 0.291896952 | 0.00301051 | 0 |
| 0 | 0 | 0.021533991 | 0.155758285 | 0 |
| 0 | 0 | 0 | 0 | 0 |
| 0 | 0 | 0 | 0 | 0 |
| 0 | 0 | 0 | 0 | 0 |
| 0 | 0 | 0 | 0 | 0 |
| 0 | 0 | 0 | 0 | 0 |
| 0 | 0 | 0 | 0 | 0 |
| 0.006160058 | 0.00464997 | 0.034697897 | 0.001181226 | 0.00426887 |
| 0 | 0 | 0 | 0 | 0 |
| 0 | 0 | 0 | 0 | 0 |
| 0 | 0 | 0 | 0 | 0 |
| 0 | 0.001013651 | 0 | 0 | 0.00419379 |
| 0 | 0 | 0 | 0 | 0 |
| 0 | 0 | 0 | 0 | 0 |
| 0 | 0 | 0 | 0 | 0 |
| 0 | 0 | 0 | 0 | 0 |
| 0 | 0 | 0 | 0 | 0 |
| 0 | 0 | 0 | 0 | 0 |
| 0.002675933 | 0.026065123 | 0.043445093 | 0.003919708 | 0 |
| 0 | 0 | 0 | 0 | 0 |
| 0 | 0 | 0 | 0 | 0 |
| 0 | 0.001279741 | 0.014752451 | 0 | 0 |
| 0 | 0 | 0 | 0 | 0 |
| 0 | 0 | 0 | 0 | 0 |
| 2.840476383 | 0 | 0.161935421 | 0 | 0 |
| 0 | 0 | 0 | 0 | 0 |
| 0 | 0 | 0 | 0 | 0 |
| 0 | 0 | 0 | 0 | 0 |
| 0 | 0 | 0 | 0 | 0 |

| | | | | |
|---|---|---|---|---|
| 0 | 0 | 0 | 0 | 0 |
| 0 | 0 | 0 | 0 | 0 |
| 0 | 0 | 0 | 0 | 0 |
| 0 | 0 | 0.008956229 | 0 | 0 |
| 0 | 0 | 0 | 0.000903956 | 0 |
| 0 | 0 | 0.002474001 | 0 | 0 |
| 0 | 0 | 0 | 0 | 0 |
| 0 | 0 | 0 | 0 | 0 |
| 0 | 0 | 0 | 0 | 0 |
| 0 | 0 | 0 | 0.005631222 | 0 |
| 0 | 0 | 0 | 0 | 0 |
| 0 | 0 | 0 | 0 | 0 |
| 0 | 0.008472852 | 0.093602709 | 0.020208159 | 0 |
| 0 | 0 | 0 | 0 | 0 |
| 0 | 0 | 0 | 0 | 0 |
| 0 | 0 | 0 | 0 | 0 |
| 0 | 0 | 0 | 0 | 0 |
| 0 | 0 | 0 | 0 | 0 |
| 0 | 0 | 0 | 0 | 0 |
| 0 | 0 | 0 | 0 | 0 |
| 0 | 0 | 0 | 0 | 0 |
| 0 | 0 | 0 | 0 | 0 |
| 0.018976773 | 0.001239644 | 0.05251658 | 0.005458357 | 0.013972671 |
| 0 | 0 | 0 | 0.019054769 | 0 |
| 0 | 0 | 0 | 0 | 0 |
| 0 | 0 | 0 | 0 | 0 |
| 0 | 0.074812851 | 0 | 0 | 0 |
| 0 | 0 | 0 | 0 | 0 |
| 0 | 0 | 0 | 0 | 0 |
| 0 | 0 | 0 | 0 | 0 |
| 0 | 0.003225127 | 0 | 0 | 0 |
| 0 | 0 | 0 | 0 | 0 |
| 0 | 0 | 0 | 0 | 0 |
| 0 | 0 | 0 | 0 | 0 |
| 0 | 0 | 0.003005622 | 0 | 0 |
| 0.346113074 | 0 | 0 | 0 | 0 |
| 0.01966992 | 0 | 0 | 0 | 0 |
| 0 | 0 | 0 | 0 | 0 |
| 0 | 0 | 0 | 0.003647361 | 0 |
| 0 | 0 | 0 | 0 | 0 |
| 0 | 0 | 0 | 0 | 0 |
| 0 | 0 | 0 | 0 | 0 |
| 0 | 0 | 0 | 15.19739582 | 0 |
| 0 | 0 | 0 | 0 | 0 |
| 0 | 0 | 0 | 0 | 0 |
| 0 | 0.39595863 | 0 | 0.060028575 | 0 |
| 0.18064715 | 0 | 0 | 0 | 0.665631714 |
| 0 | 0 | 0 | 0 | 0 |

| | | | | |
|---|---|---|---|---|
| 0 | 0 | 0 | 0 | 0 |
| 0 | 0 | 0 | 0 | 0 |
| 0 | 0 | 0 | 0 | 0 |
| 0 | 0 | 0 | 0 | 0 |
| 0 | 0 | 0 | 0 | 0 |
| 0 | 0 | 0 | 0 | 0 |
| 0 | 0 | 0 | 0 | 0 |
| 0 | 0 | 0 | 0 | 0 |
| 0.000941143 | 0 | 0 | 0 | 0 |
| 0 | 0 | 0 | 0 | 0 |
| 0.001155264 | 0.002213691 | 0.002788834 | 0 | 0 |
| 0 | 0 | 0.000734777 | 0 | 0 |
| 0 | 0 | 0 | 0 | 0 |
| 0 | 0 | 0 | 0 | 0 |
| 0 | 0 | 0 | 0 | 0 |
| 0 | 0 | 0 | 0 | 0 |
| 0 | 0 | 0 | 0.005884729 | 0 |
| 0 | 0 | 0 | 0 | 0 |
| 0 | 0 | 0 | 0 | 0 |
| 0 | 0 | 0.003698473 | 0 | 0 |
| 0 | 0 | 0 | 0 | 0 |
| 0 | 0 | 0 | 0 | 0 |
| 0 | 0 | 0 | 0 | 0 |
| 0 | 0 | 0 | 0 | 0 |
| 0 | 0 | 0 | 0 | 0 |
| 0 | 0 | 0 | 0 | 0 |
| 0 | 0 | 0 | 0 | 0 |
| 0 | 0.160355785 | 0 | 0.029121683 | 0 |
| 0 | 0 | 0 | 0 | 0 |
| 0 | 0 | 0 | 0 | 0 |
| 0 | 0.0014412 | 0.004859509 | 0.016261222 | 0 |
| 0 | 0 | 0 | 0 | 0 |
| 0.053925679 | 0.052293128 | 0 | 0.086575995 | 0 |
| 0.004015836 | 0 | 0.010201216 | 0 | 0.00510206 |
| 0 | 0 | 0 | 0 | 0 |
| 0 | 0 | 0 | 0 | 0 |
| 0 | 0 | 0 | 0 | 0 |
| 0 | 0 | 0 | 0 | 0 |
| 0 | 0 | 0 | 0 | 0 |
| 0 | 0 | 0 | 0 | 0 |
| 0 | 0 | 0 | 0 | 0 |
| 0.010417497 | 0.156940908 | 0.407597905 | 0 | 0.184152866 |
| 0 | 0 | 0 | 0.022616232 | 0 |
| 0 | 0 | 0 | 0 | 0 |
| 0 | 0 | 0 | 0 | 0 |
| 0 | 0.006665006 | 0 | 0.04602163 | 0 |
| 0 | 0 | 0 | 0 | 0 |
| 0 | 0 | 0 | 0 | 0 |

| | | | | |
|---|---|---|---|---|
| 0 | 0 | 0 | 0 | 0 |
| 0 | 0.007906798 | 0 | 0.189548017 | 0 |
| 0 | 0 | 0.008251381 | 0.005365854 | 0 |
| 0 | 0 | 0 | 0 | 0 |
| 0 | 0 | 0 | 0 | 0 |
| 0.011654664 | 0 | 0 | 0 | 0 |
| 0 | 0 | 0 | 0 | 0 |
| 0 | 0 | 0 | 0 | 0 |
| 0 | 0 | 0 | 0 | 0 |
| 0 | 0 | 0 | 0 | 0 |
| 0 | 0 | 0 | 0 | 0 |
| 0 | 0 | 0 | 0 | 0 |
| 0 | 0 | 0 | 0 | 0 |
| 0 | 0 | 0 | 0 | 0 |
| 0 | 0 | 0 | 0 | 0 |
| 0 | 0 | 0 | 0 | 0 |
| 0 | 0 | 0 | 0 | 0 |
| 0 | 0 | 0 | 0 | 0 |
| 0 | 0 | 0 | 0 | 0 |
| 0 | 0 | 0 | 0 | 0 |
| 0 | 0 | 0 | 0 | 0 |
| 0 | 0 | 0.138740611 | 0 | 0.627223219 |
| 0 | 0 | 0 | 0 | 0 |
| 0 | 0 | 0 | 0 | 0 |
| 0 | 0 | 0 | 0.003025728 | 0 |
| 0 | 0.008082634 | 0.003089456 | 0 | 0 |
| 0 | 0 | 0 | 0 | 0 |
| 0 | 0 | 0 | 0.00340685 | 0 |
| 0 | 0 | 0 | 0 | 0 |
| 0 | 0 | 0 | 0 | 0 |
| 0 | 0 | 0 | 0 | 0 |
| 0 | 0 | 0 | 0 | 0.002374512 |
| 0 | 0 | 0 | 0 | 0 |
| 0 | 0 | 0 | 0 | 0 |
| 0 | 0 | 0 | 0 | 0 |
| 0 | 0 | 0 | 0 | 0 |
| 0 | 0 | 0 | 0 | 0 |
| 0 | 0 | 0 | 0 | 0 |
| 0 | 1.380391057 | 0 | 0 | 0 |
| 0 | 0 | 0 | 0 | 0 |
| 0 | 0 | 0 | 0 | 0 |
| 0 | 0 | 0 | 0 | 0 |
| 0.005347358 | 0.0074714 | 0.070649539 | 0 | 0.038214777 |
| 0 | 0 | 0 | 0 | 0 |
| 0 | 0 | 0 | 0 | 0 |
| 0 | 0 | 0 | 0 | 0 |
| 0.003749355 | 0.000979695 | 0.009825452 | 0 | 0.001818791 |
| 0 | 0 | 0 | 0 | 0 |

| | | | | |
|---|---|---|---|---|
| 0 | 0 | 0 | 0 | 0 |
| 0.022149664 | 0.049653848 | 0.014321727 | 0.007937961 | 0.012950241 |
| 0 | 0 | 0 | 0 | 0 |
| 0 | 0 | 0 | 0 | 0 |
| 0 | 0 | 0 | 0 | 0 |
| 0 | 0 | 0 | 0 | 0 |
| 0 | 0 | 0 | 0 | 0 |
| 0 | 0 | 0 | 0 | 0 |
| 0 | 0 | 0 | 0 | 0 |
| 0 | 0 | 0 | 0 | 0 |
| 0 | 0 | 0 | 0 | 0 |
| 0 | 0 | 0 | 0 | 0 |
| 0 | 0 | 0.045418003 | 0.029728568 | 0 |
| 0 | 0 | 0 | 0 | 0 |
| 0.003485678 | 0.001541349 | 0 | 0.002262272 | 0 |
| 0 | 0 | 0 | 0 | 0 |
| 0 | 0 | 0 | 0 | 0 |
| 0 | 0 | 0 | 0 | 0 |
| 0 | 0 | 0 | 0 | 0 |
| 0 | 0 | 0 | 0 | 0 |
| 0 | 0 | 0 | 0 | 0 |
| 0.012639547 | 0 | 0.019369243 | 0.008053771 | 0 |
| 0 | 0 | 0 | 0 | 0 |
| 0 | 0 | 0.048797181 | 0.035469753 | 0 |
| 0 | 0 | 0 | 0 | 0 |
| 0 | 0 | 0 | 0 | 0 |
| 0 | 0 | 0 | 0 | 0 |
| 0 | 0 | 0 | 0 | 0 |
| 0 | 0.005308291 | 0 | 0 | 0 |
| 0 | 0 | 0 | 0 | 0 |
| 0 | 0 | 0 | 0 | 0 |
| 0 | 0 | 0 | 0 | 0 |
| 0 | 0 | 0 | 0 | 0 |
| 0 | 0 | 0 | 0 | 0 |
| 0 | 0 | 3.34803129 | 0 | 0 |
| 0 | 0 | 0 | 0 | 0 |
| 0 | 0 | 0 | 0 | 0 |
| 0 | 0 | 0 | 0 | 0 |
| 0 | 0 | 0.001045542 | 0.029951992 | 0 |
| 0 | 0 | 0 | 0 | 0 |
| 0.041497124 | 0 | 0.13827638 | 0.076091779 | 0 |
| 0 | 0 | 0 | 0 | 0 |
| 0 | 0 | 0 | 0 | 0 |
| 0 | 0 | 0 | 0 | 0 |
| 0 | 0 | 0 | 0 | 0 |
| 0.60253072 | 0.173545546 | 2.837900101 | 0.025623503 | 0.473708468 |
| 0 | 0 | 0 | 0 | 0 |
| 0 | 0 | 0 | 0 | 0 |

| | | | | |
|---|---|---|---|---|
| 0 | 0 | 0 | 0 | 0 |
| 0 | 0 | 0 | 0 | 0 |
| 0 | 0 | 0 | 0 | 0 |
| 0 | 0 | 0 | 0 | 0 |
| 0 | 0.004119291 | 0 | 0 | 0 |
| 0 | 0 | 0 | 0 | 0 |
| 0 | 0 | 0 | 0 | 0 |
| 0 | 0 | 0 | 0 | 0 |
| 0 | 0 | 0 | 0 | 0 |
| 0 | 0 | 0 | 0.125020688 | 0 |
| 0 | 0 | 0 | 0 | 0 |
| 0 | 0 | 0 | 0 | 0 |
| 0 | 0 | 0.001539026 | 0.000816462 | 0 |
| 0 | 0 | 0 | 0 | 0 |
| 0 | 0 | 0 | 0 | 0 |
| 0 | 0 | 0 | 0 | 0 |
| 0 | 0 | 0 | 0 | 0 |
| 0 | 0 | 0 | 0 | 0 |
| 0 | 0 | 0 | 0 | 0 |
| 0.098763578 | 0.018704807 | 0 | 0.046993855 | 0.103300208 |
| 0 | 0 | 0 | 0 | 0 |
| 0 | 0 | 0 | 0 | 0 |
| 0 | 0 | 0 | 0 | 0 |
| 0 | 0 | 0 | 0 | 0 |
| 0 | 0 | 0 | 0 | 0 |
| 0 | 0 | 0 | 0 | 0 |
| 0 | 0 | 0 | 0 | 0 |
| 0 | 0 | 0 | 0 | 0 |
| 0 | 0 | 0 | 0 | 0 |
| 0.010557986 | 0 | 0 | 0.000776864 | 0 |
| 0 | 0 | 0 | 0 | 0 |
| 0 | 0 | 0 | 0 | 0 |
| 0 | 0 | 0 | 0 | 0 |
| 0 | 0.003747561 | 0 | 0 | 0 |
| 0 | 0 | 0 | 0 | 0 |
| 0 | 0.002278865 | 0 | 0 | 0 |
| 0 | 0 | 0 | 0 | 0 |
| 0 | 0 | 0 | 0 | 0 |
| 0 | 0 | 0 | 0 | 0 |
| 0 | 0 | 0 | 0 | 0 |
| 0 | 0 | 0 | 0 | 0 |
| 0 | 0 | 0 | 0 | 0 |
| 0 | 0 | 0 | 0 | 0 |
| 0 | 0 | 0 | 0 | 0 |
| 0.002372991 | 0.012607797 | 0.011150579 | 0 | 0 |
| 0 | 0 | 0 | 0 | 0 |
| 0 | 0 | 0 | 0 | 0 |
| 0 | 0 | 0 | 0 | 0.007407722 |

| | | | | |
|---|---|---|---|---|
| 0 | 0 | 0 | 0 | 0 |
| 0 | 0 | 0 | 0 | 0 |
| 0 | 0 | 0 | 0 | 0 |
| 0 | 0 | 0 | 0 | 0 |
| 0 | 0 | 0 | 0 | 0 |
| 0 | 0.008177823 | 0.006723506 | 0.020721762 | 0.000982125 |
| 0 | 0 | 0 | 0 | 0 |
| 0 | 0 | 0 | 0 | 0 |
| 0 | 0 | 0 | 0 | 0 |
| 0 | 0 | 0 | 0 | 0 |
| 0 | 0 | 0 | 0 | 0 |
| 0 | 0 | 0 | 0 | 0 |
| 0 | 0 | 0 | 0 | 0 |
| 0 | 0 | 0 | 0 | 0 |
| 0 | 0 | 0 | 0 | 0 |
| 0 | 0 | 0 | 0 | 0 |
| 0 | 0 | 0 | 0 | 0 |
| 0 | 0 | 0 | 0 | 0 |
| 0 | 0 | 0 | 0 | 0 |
| 0 | 0 | 0 | 0 | 0 |
| 0 | 0 | 0 | 0 | 0 |
| 0 | 0 | 0 | 0 | 0 |
| 0 | 0 | 0 | 0 | 0 |
| 0 | 0.029849395 | 0 | 0 | 0 |
| 0 | 0 | 0 | 0 | 0 |
| 0 | 0 | 0 | 0.002125398 | 0 |
| 0 | 0 | 0 | 0 | 0 |
| 0 | 0 | 0 | 0 | 0 |
| 0 | 0 | 0 | 0 | 0 |
| 0 | 0 | 0 | 0 | 0 |
| 0 | 0 | 0 | 0 | 0 |
| 0 | 0 | 0 | 0 | 0 |
| 0 | 0 | 0 | 0 | 0 |
| 0 | 0 | 0 | 0 | 0 |
| 0 | 0 | 0 | 0 | 0 |
| 0 | 0 | 0 | 0 | 0 |
| 0 | 0 | 0 | 0 | 0 |
| 0 | 0 | 0 | 0 | 0 |
| 0 | 0 | 4.333468609 | 0 | 0 |
| 0 | 0 | 0 | 0 | 0 |
| 0 | 0 | 0 | 0 | 0 |
| 0 | 0 | 0 | 0 | 0 |
| 0 | 0 | 0 | 0 | 0 |
| 0 | 0 | 0 | 0 | 0 |
| 0 | 0 | 0 | 0.004244376 | 0 |
| 0 | 0 | 0 | 0 | 0 |
| 0 | 0 | 0 | 0 | 0 |
| 0 | 0 | 0 | 0 | 0 |

| | | | | |
|---|---|---|---|---|
| 0 | 0 | 0 | 0 | 0 |
| 0 | 0.001036498 | 0 | 0 | 0 |
| 0 | 0 | 0 | 0 | 0 |
| 0 | 0 | 0 | 0 | 0 |
| 0 | 0 | 0 | 0 | 0 |
| 0 | 0 | 0 | 0 | 0 |
| 0 | 0 | 0 | 0 | 0 |
| 0 | 0 | 0 | 0 | 0 |
| 0 | 0 | 0 | 0 | 0 |
| 0 | 0 | 0 | 0 | 0 |
| 0 | 0 | 0.004928337 | 0.00423302 | 0 |
| 0 | 0 | 0 | 0 | 0 |
| 0 | 0.271739593 | 0 | 0 | 0 |
| 0 | 0 | 0 | 0 | 0 |
| 0 | 0 | 0 | 0 | 0 |
| 0 | 0 | 0 | 0 | 0 |
| 0 | 0 | 0 | 0 | 0 |
| 0 | 0 | 0 | 0 | 0 |
| 0 | 0 | 0 | 0 | 0 |
| 0 | 0 | 0 | 0 | 0 |
| 0 | 0 | 0 | 0.017943207 | 0 |
| 0 | 0 | 0 | 0 | 0 |
| 0 | 0 | 0 | 0 | 0 |
| 0 | 0.499235964 | 0 | 4.154214555 | 0 |
| 0 | 0 | 0 | 0 | 0 |
| 0 | 0 | 0 | 0 | 0 |
| 0 | 0 | 0 | 0 | 0 |
| 0 | 0 | 0 | 0 | 0 |
| 0 | 0 | 0 | 0 | 0 |
| 0 | 0 | 0 | 0 | 0 |
| 0 | 0 | 0 | 0 | 0 |
| 0 | 0 | 0 | 0 | 0 |
| 0 | 0 | 0 | 0 | 0 |
| 0 | 0 | 0 | 0 | 0 |

| PHL018-0018_V1 | PHL018-0010_V1 | PHL018-0235_V2 | PHL018-0139_V2 | PHL018-0017_V1 |
|---|---|---|---|---|
| CTRL | CTRL | EYCF | CTRL | EYCF |
| 0.05027702 | 0.067402363 | 0.410235362 | 0.019312761 | 0.298077447 |
| 0.085267051 | 2.35944805 | 0.396750839 | 0.058210441 | 1.335366247 |
| 0 | 13.35917996 | 14.4474223 | 0 | 0 |
| 0 | 0 | 0 | 0 | 0 |
| 0 | 0 | 0 | 0 | 0 |
| 0 | 0 | 0 | 0 | 0 |
| 0 | 0 | 0 | 0 | 0 |
| 0 | 0 | 0 | 0 | 0 |
| 0 | 0 | 0 | 0 | 0 |
| 0 | 0 | 0 | 0 | 0 |
| 0 | 0 | 0 | 0 | 0 |
| 0 | 0 | 0 | 0 | 0 |
| 0 | 0 | 0 | 0 | 0 |
| 0.840878845 | 20.49290142 | 1.457807747 | 0.303968175 | 3.262085247 |
| 0 | 0 | 0 | 0 | 0 |
| 0 | 0 | 0 | 0 | 0 |
| 0 | 0 | 0 | 0 | 0 |
| 0 | 0 | 0 | 0 | 0 |
| 0 | 0 | 0 | 0 | 0 |
| 0 | 0.027316721 | 0 | 0 | 0 |
| 0 | 0 | 0 | 0.003624572 | 0 |
| 0 | 0 | 0 | 0 | 0 |
| 0 | 0 | 0 | 0 | 0 |
| 0 | 0 | 0 | 0 | 0 |
| 0 | 0 | 0 | 0.015034917 | 0 |
| 0 | 0 | 0 | 0 | 0 |
| 0 | 0 | 0 | 0 | 0 |
| 0 | 0 | 0 | 0 | 0 |
| 0 | 0 | 0 | 0 | 0 |
| 0 | 0 | 0 | 0 | 0 |
| 0 | 0 | 0 | 0 | 0 |
| 0 | 0 | 0 | 0 | 0 |
| 0 | 0 | 0 | 0 | 0 |
| 0 | 0 | 0 | 0 | 0 |
| 0 | 0 | 0 | 0 | 0 |
| 0.625270185 | 0 | 0.281256514 | 0.097817128 | 0.549626544 |
| 0 | 0 | 0 | 0 | 0 |
| 0 | 0 | 0 | 0 | 0 |
| 0 | 0 | 0 | 0 | 0 |
| 0 | 0 | 0 | 0 | 0 |
| 0 | 0 | 0 | 0 | 0 |
| 0.007460153 | 0.002401713 | 0.047981315 | 0 | 0.003742896 |
| 0.012621811 | 0.017540563 | 0.477393639 | 7.175885957 | 0.444116464 |
| 0.00595109 | 0 | 0.008184565 | 0 | 0.033448937 |
| 0.004052163 | 0 | 0 | 0 | 0 |
| 0 | 0 | 0 | 0 | 0 |

| | | | | |
|---|---|---|---|---|
| 0.057768608 | 0 | 0 | 0 | 0 |
| 0 | 0.042277245 | 0.005940091 | 0 | 0 |
| 0 | 0.002511833 | 0.00210492 | 0 | 0.013863892 |
| 0.332347298 | 0 | 1.423978943 | 0.431898172 | 1.462710953 |
| 0 | 0 | 0 | 0.000937566 | 0.038950866 |
| 0 | 0 | 0 | 0.02417394 | 0 |
| 0 | 1.441106024 | 0 | 0 | 0.000845739 |
| 0.078663398 | 0 | 0.066886951 | 0 | 0 |
| 0 | 0 | 0 | 0 | 0 |
| 0 | 0 | 0 | 0 | 0 |
| 0 | 0 | 0 | 0.011205138 | 0 |
| 0.621180013 | 0 | 0.821875297 | 0.083515384 | 0.427351453 |
| 0 | 0 | 0 | 0 | 0 |
| 0.053672982 | 0.03750526 | 0 | 0 | 0.023167708 |
| 0 | 0 | 0 | 0 | 0 |
| 0 | 0 | 0 | 0 | 0 |
| 0 | 0 | 0.005715097 | 0.674936974 | 0.000622754 |
| 0 | 0 | 0 | 0 | 0 |
| 0 | 0 | 0 | 0 | 0 |
| 0 | 0 | 0 | 0 | 0 |
| 0.04568295 | 0.216433221 | 0.084938254 | 0 | 0.007200333 |
| 0.004208256 | 0.1629148 | 0.021843659 | 0.002727926 | 0.023886786 |
| 0 | 0 | 0 | 0.008279153 | 0.414346397 |
| 0 | 0 | 0 | 0 | 0 |
| 0 | 0.039818016 | 0 | 0 | 0 |
| 0 | 0 | 0 | 0 | 0 |
| 0 | 0 | 0 | 0 | 0 |
| 0 | 0 | 0 | 0 | 0 |
| 0 | 0 | 0.001488161 | 0 | 0 |
| 0 | 0 | 0 | 0 | 0 |
| 0 | 0 | 0 | 0 | 0 |
| 0 | 0 | 0 | 0 | 0 |
| 0 | 0 | 0 | 0 | 0 |
| 0 | 0.004456504 | 0 | 0 | 0 |
| 0 | 0 | 0 | 0 | 0 |
| 0 | 0 | 0 | 0 | 0 |
| 0 | 0 | 0 | 0 | 0 |
| 0 | 0 | 0 | 32.70673229 | 0 |
| 0 | 0 | 0 | 0 | 0 |
| 0 | 0 | 0 | 0 | 0 |
| 0 | 0 | 0 | 0 | 0 |
| 0 | 0 | 0 | 0 | 0 |
| 0 | 0 | 0 | 0 | 0 |
| 0 | 0 | 0 | 0 | 0 |
| 0.191629874 | 0 | 0 | 0 | 0 |
| 0 | 0 | 0 | 0 | 0 |
| 0 | 0 | 0 | 0 | 0 |
| 0.238585796 | 0 | 0 | 0 | 0.000910694 |

| | | | | |
|---|---|---|---|---|
| 0 | 0 | 0 | 0 | 0 |
| 0 | 0 | 0 | 0 | 0 |
| 0 | 0 | 0 | 0 | 0 |
| 0 | 0 | 0 | 0 | 0 |
| 0 | 0 | 0.000882738 | 0 | 0.000513007 |
| 0 | 0.001184873 | 0 | 0 | 0.000553962 |
| 0 | 0 | 0 | 0 | 0 |
| 0 | 0 | 0 | 0 | 0 |
| 0 | 0 | 0 | 0.005428611 | 0 |
| 0 | 0 | 0 | 0 | 0 |
| 0 | 0 | 0 | 0 | 0 |
| 0 | 0 | 0 | 0 | 0 |
| 0 | 0 | 0 | 0 | 0 |
| 0 | 0 | 0 | 0 | 0 |
| 0 | 0 | 0 | 0 | 0 |
| 0 | 0 | 0 | 0 | 0.004378228 |
| 0 | 0 | 0 | 0 | 0 |
| 0 | 0 | 0 | 0 | 0 |
| 0.94981638 | 0 | 0 | 0.078364897 | 0 |
| 0.00045099 | 0 | 0 | 0 | 0 |
| 0 | 0 | 0 | 0.005485016 | 0 |
| 0 | 0 | 0 | 0 | 0 |
| 0 | 0 | 0 | 0 | 0 |
| 0 | 0 | 0 | 0 | 0 |
| 0 | 0 | 0 | 0 | 0 |
| 0 | 0 | 0 | 0 | 0 |
| 0 | 0 | 0 | 0 | 0 |
| 0 | 0 | 0 | 0 | 0 |
| 0 | 0 | 0 | 0 | 0 |
| 0 | 0 | 0 | 0 | 0 |
| 0.053565281 | 0 | 0.011676507 | 0.014783379 | 0.13894215 |
| 0 | 0.002713686 | 0 | 0.001505733 | 0.002255511 |
| 0 | 0 | 0 | 0 | 0 |
| 0 | 0 | 0 | 0 | 0 |
| 0 | 0 | 0 | 0 | 0 |
| 0 | 0 | 0 | 0 | 0 |
| 0 | 0 | 0 | 0 | 0 |
| 0 | 0 | 0 | 0 | 0 |
| 0 | 0 | 0 | 0 | 0.053175572 |
| 0 | 0 | 0 | 0 | 0 |
| 0 | 0 | 0 | 0 | 0 |
| 0 | 0 | 0 | 0.004318193 | 0 |
| 0 | 0.001254637 | 0 | 0 | 0 |
| 0 | 0 | 0 | 0 | 0 |
| 0 | 0 | 0 | 0 | 0 |
| 0 | 0 | 0 | 0 | 0 |
| 0 | 0 | 0 | 0 | 0 |
| 0 | 0 | 0 | 0 | 0 |

| | | | | |
|---|---|---|---|---|
| 0 | 0 | 0 | 0 | 0 |
| 0 | 0 | 0 | 0 | 0 |
| 0 | 0 | 0 | 0 | 0 |
| 0 | 0 | 0 | 0 | 0 |
| 0.115668092 | 0 | 0 | 0.004980168 | 0.452755464 |
| 0 | 0 | 0 | 0 | 0 |
| 0 | 0 | 0 | 0 | 0 |
| 0 | 0 | 0 | 0 | 0 |
| 0 | 0 | 0 | 0 | 0 |
| 0 | 0 | 0 | 0 | 0 |
| 0 | 0 | 0 | 0 | 0 |
| 0 | 0 | 0 | 0 | 0 |
| 0 | 0 | 0 | 0 | 0 |
| 0 | 0 | 0 | 0 | 0 |
| 0 | 0 | 0 | 0 | 0 |
| 0 | 0 | 0 | 0 | 0 |
| 0 | 0 | 0 | 0 | 0 |
| 0 | 0 | 0 | 0 | 0 |
| 0 | 0 | 0 | 0 | 0 |
| 0 | 0 | 0 | 0 | 0 |
| 0 | 0 | 0 | 0 | 0 |
| 0 | 0 | 0 | 0.004460639 | 0 |
| 0 | 0 | 0 | 0 | 0.001795275 |
| 0 | 0 | 0 | 0 | 0 |
| 0 | 0 | 0.027589509 | 0.06673889 | 0.041324172 |
| 0 | 0 | 0 | 0 | 0 |
| 0 | 0 | 0 | 0.004252637 | 0 |
| 0 | 0 | 0 | 0 | 0 |
| 0 | 0 | 0 | 0 | 0 |
| 0 | 0 | 0 | 0 | 0 |
| 0 | 0 | 0 | 0.010854758 | 0.001662941 |
| 0 | 0 | 0 | 0 | 0.009921548 |
| 0 | 0 | 0 | 0 | 0 |
| 0 | 0 | 0 | 0 | 0 |
| 0 | 0 | 0 | 0 | 0 |
| 0 | 0 | 0 | 0 | 0 |
| 0 | 0 | 0 | 0 | 0 |
| 0.002945585 | 0 | 0 | 0.008135102 | 0 |
| 0 | 0 | 0 | 0 | 0 |
| 0 | 0 | 0 | 0 | 0 |
| 0 | 0 | 0 | 0 | 0 |
| 0 | 0 | 0 | 0 | 0 |
| 0 | 0 | 0 | 0 | 0 |
| 0 | 0 | 0 | 0 | 0 |
| 0 | 0 | 0 | 0 | 0 |
| 0 | 0 | 0 | 0 | 0 |
| 0 | 0 | 0 | 0 | 0 |
| 0 | 0 | 0 | 0 | 0 |

| | | | | |
|---|---|---|---|---|
| 0.161079534 | 0 | 0 | 0 | 0 |
| 0 | 0 | 0 | 0 | 0 |
| 0 | 0.010422047 | 0 | 0 | 0 |
| 0 | 0 | 0 | 0 | 0 |
| 0 | 0 | 0 | 0 | 0 |
| 0 | 0 | 0 | 0 | 0 |
| 0 | 0 | 0 | 0 | 0 |
| 0 | 0 | 0 | 0 | 0 |
| 0 | 0 | 0 | 0 | 0 |
| 0 | 0 | 0 | 0 | 0 |
| 0 | 0 | 0 | 0 | 0 |
| 0 | 0 | 0 | 0.001180469 | 0 |
| 0.002347541 | 0.00179764 | 0 | 0 | 0 |
| 0 | 0 | 0 | 0 | 0 |
| 0 | 0.004118115 | 0 | 0 | 0 |
| 0 | 0 | 0 | 0 | 0 |
| 0 | 0 | 0 | 0 | 0 |
| 0 | 0 | 0 | 0 | 0 |
| 0 | 0 | 0 | 0 | 0 |
| 0 | 0 | 0 | 0 | 0 |
| 0.006897532 | 0.02390155 | 0.09639215 | 0.025799065 | 0.145503305 |
| 0 | 0 | 0 | 0 | 0 |
| 0 | 0 | 0.002086993 | 0 | 0 |
| 0 | 0 | 0 | 0 | 0.001517307 |
| 0 | 0 | 0 | 0 | 0 |
| 0 | 0 | 0 | 0 | 0 |
| 0.001278124 | 0 | 0.0016039 | 0.002353277 | 0.005253737 |
| 0 | 0 | 0 | 0 | 0 |
| 0 | 0 | 0 | 0 | 0 |
| 0 | 0.561958219 | 0 | 0 | 0 |
| 0 | 0.005203341 | 0 | 0 | 0 |
| 0 | 0 | 0 | 0 | 0 |
| 0 | 0 | 0 | 0 | 0 |
| 0.059589623 | 0 | 0 | 0 | 0 |
| 0.01876759 | 0.085533922 | 0.013032285 | 0.001213464 | 0.02404674 |
| 0 | 0 | 0 | 0 | 0 |
| 0 | 0 | 0 | 0 | 0 |
| 0 | 0 | 0 | 0 | 0 |
| 0 | 0 | 0 | 0 | 0 |
| 0 | 0 | 0 | 0 | 0 |
| 0 | 0 | 0 | 0 | 0 |
| 0.001629841 | 0 | 0 | 0 | 0 |
| 0 | 0 | 0 | 0 | 0 |
| 0 | 0 | 0 | 0 | 0 |
| 0 | 0 | 0 | 0 | 0 |
| 0 | 0 | 0 | 0.004813956 | 0 |
| 0 | 0.00263098 | 0.000705525 | 0 | 0 |
| 0.019479947 | 0 | 0.006036653 | 0 | 0.003373299 |

| | | | | |
|---|---|---|---|---|
| 0 | 0 | 0 | 0 | 0 |
| 0 | 0 | 0 | 0 | 0 |
| 0 | 0 | 0.014214739 | 0 | 0 |
| 0 | 0 | 0.000590259 | 0 | 0 |
| 0 | 0 | 0 | 0 | 0 |
| 0 | 0 | 0 | 0 | 0 |
| 0 | 0 | 0.00082626 | 0 | 0 |
| 0 | 0 | 0 | 0 | 0 |
| 0 | 0 | 0 | 0 | 0 |
| 0 | 0 | 0 | 0 | 0 |
| 0 | 0 | 0 | 0 | 0 |
| 0 | 0 | 0 | 0 | 0 |
| 0 | 0 | 0 | 0 | 0 |
| 0 | 0 | 0 | 0 | 0 |
| 0 | 0 | 0 | 0.001590656 | 0 |
| 0 | 0 | 0 | 0 | 0 |
| 0 | 0 | 0 | 0 | 0 |
| 0 | 0 | 0 | 0 | 0 |
| 0 | 0 | 0.003541867 | 0.03444481 | 0.003018948 |
| 0 | 0 | 0 | 0.029226203 | 0 |
| 0.006978677 | 0.177166871 | 0.039881224 | 0 | 0 |
| 0 | 0 | 0 | 0 | 0 |
| 0 | 0 | 0 | 0 | 0 |
| 0 | 0 | 0 | 0 | 0 |
| 0 | 0 | 0 | 0 | 0.010455939 |
| 0 | 0 | 0.000839368 | 0 | 0 |
| 0 | 0 | 0 | 0 | 0 |
| 0 | 0 | 0 | 0 | 0 |
| 0 | 0 | 0 | 0 | 0 |
| 0 | 0 | 0 | 0 | 0 |
| 0.140793282 | 1.738598489 | 0.034446857 | 0.094434069 | 0.118625363 |
| 0 | 0 | 0 | 0 | 0 |
| 0 | 0 | 0 | 0 | 0 |
| 0 | 0 | 0 | 0.00370232 | 0.002695917 |
| 0 | 0 | 0 | 0 | 0 |
| 0 | 0 | 0 | 0 | 0 |
| 0 | 0 | 0 | 0 | 0 |
| 0 | 0 | 0 | 0 | 0 |
| 0 | 0 | 0 | 0 | 0 |
| 0.001281393 | 0 | 0.000383728 | 0 | 0 |
| 0 | 0 | 0 | 0 | 0 |
| 0.004413342 | 0 | 0.002349558 | 0 | 0.030950366 |
| 0 | 0 | 0 | 0 | 0 |
| 0 | 0 | 0 | 0 | 0 |
| 0 | 0 | 0 | 0 | 0 |
| 0 | 0 | 0 | 0 | 0 |
| 0 | 0 | 0 | 0 | 0 |
| 0 | 0 | 0 | 0 | 0 |

| | | | | |
|---|---|---|---|---|
| 0 | 0 | 0 | 0 | 0 |
| 0 | 0 | 0 | 0 | 0 |
| 0.00759415 | 0 | 0.00983419 | 0 | 0.032195558 |
| 0 | 0 | 0 | 0 | 0 |
| 0 | 0 | 0 | 0.487564597 | 0.05736833 |
| 0 | 0 | 0 | 0 | 0 |
| 0 | 0 | 0 | 0 | 0 |
| 0 | 0 | 0 | 0 | 0 |
| 0 | 0 | 0 | 0 | 0 |
| 0 | 0 | 0 | 0 | 0 |
| 0 | 0 | 0 | 0 | 0 |
| 0 | 0 | 0 | 0.004627824 | 0 |
| 0 | 0 | 0 | 0.002125544 | 0 |
| 0 | 0.001351001 | 0 | 0.002023988 | 0.044424746 |
| 0.001497783 | 0.001561104 | 0.000897057 | 0.002041768 | 0.009905252 |
| 0.220491474 | 0 | 0 | 0.022853451 | 0.006697819 |
| 0 | 0 | 0.001192469 | 0 | 0.003960052 |
| 0 | 0 | 0 | 0 | 0.038244891 |
| 0 | 0 | 0 | 0 | 0 |
| 0 | 0 | 0 | 0 | 0 |
| 0 | 0 | 0 | 0 | 0 |
| 0.016368483 | 0 | 0.008611671 | 0 | 0.002144878 |
| 0 | 0 | 0 | 0 | 0 |
| 0 | 0 | 0 | 0 | 0 |
| 0 | 0 | 0 | 0 | 0 |
| 0 | 0 | 0 | 0 | 0 |
| 3.713738189 | 0 | 0 | 0 | 2.123510355 |
| 0 | 0 | 0 | 0 | 0 |
| 0 | 0 | 0 | 0 | 0 |
| 0 | 0 | 0 | 0 | 0 |
| 0 | 0 | 0 | 0 | 0 |
| 0 | 0 | 0 | 0 | 0 |
| 0 | 0 | 0 | 0 | 0 |
| 0 | 0 | 0 | 0 | 0 |
| 0 | 0 | 0 | 0 | 0 |
| 0 | 0 | 0.010443215 | 0 | 0.002142044 |
| 0 | 0 | 0 | 0 | 0 |
| 0.111052643 | 0 | 0.071873543 | 0.000719602 | 0 |
| 0 | 0 | 0 | 0 | 0 |
| 0 | 0 | 0 | 0 | 0 |
| 0 | 0 | 0 | 0 | 0 |
| 0 | 0 | 0 | 0 | 0 |
| 0 | 0 | 0 | 0 | 0 |
| 0 | 0 | 0 | 0 | 0 |
| 0 | 0 | 0 | 0 | 0 |
| 0 | 0 | 0 | 0 | 0 |
| 0 | 0 | 0 | 0 | 0 |
| 0 | 0.016057694 | 0 | 0 | 0 |

| | | | | |
|---|---|---|---|---|
| 0 | 0 | 0.128978085 | 0 | 0 |
| 0 | 0 | 0 | 0 | 0 |
| 0 | 0 | 0 | 0 | 0 |
| 0 | 0 | 0 | 0 | 0 |
| 0.007914353 | 0.002900729 | 0 | 0.02977607 | 0 |
| 0 | 0 | 0 | 0 | 0 |
| 0 | 0 | 0 | 0 | 0 |
| 0 | 0 | 0 | 0.001974081 | 0.00184817 |
| 0 | 0 | 0 | 0 | 0 |
| 0 | 0.195227228 | 0.005385069 | 0.010754992 | 0.030207051 |
| 0 | 0 | 0 | 0 | 0 |
| 0.001351049 | 0 | 0.032007354 | 0 | 0.002717034 |
| 0.002562047 | 0.004476035 | 0.002864343 | 0 | 0.510961179 |
| 0 | 0 | 0 | 0 | 0 |
| 0.00122967 | 0 | 0 | 0 | 0 |
| 0 | 0 | 0 | 0 | 0 |
| 0 | 0 | 0 | 0 | 0 |
| 0 | 0.001589851 | 0 | 0 | 0 |
| 0 | 0 | 0 | 0 | 0 |
| 0 | 0.705699122 | 0 | 0 | 0 |
| 0 | 0 | 0 | 0 | 0 |
| 0 | 0 | 0 | 0 | 0 |
| 0 | 0 | 0 | 0 | 0 |
| 0 | 0 | 0 | 0.037711194 | 0 |
| 0 | 0 | 0 | 0 | 0 |
| 0 | 0 | 0.002383021 | 0 | 0.001661885 |
| 0 | 0 | 0 | 1.436073394 | 0 |
| 0 | 0.16477666 | 0 | 0 | 0 |
| 0 | 0 | 0 | 0 | 0 |
| 0.000818745 | 0 | 0.003922925 | 0 | 0 |
| 0 | 0 | 0 | 0 | 0 |
| 0 | 0 | 0 | 0 | 0 |
| 0 | 0 | 0 | 0 | 0 |
| 0 | 0.067872297 | 0 | 0 | 0 |
| 0 | 0 | 0 | 0 | 0 |
| 0 | 0.210266753 | 0 | 0 | 0 |
| 0 | 0.911478321 | 0 | 0 | 0 |
| 0 | 0 | 0 | 0 | 0 |
| 0 | 0 | 0 | 0 | 0 |
| 0 | 0 | 0 | 0 | 0 |
| 0 | 0 | 0 | 0 | 0 |
| 0 | 1.577453318 | 0 | 0 | 0 |
| 0 | 0 | 0 | 0 | 0 |
| 0 | 0 | 0 | 0 | 0 |
| 0 | 0 | 0 | 0 | 0 |
| 0 | 0 | 0 | 0.008513943 | 0 |
| 0.002474002 | 0 | 0 | 0 | 0.016533483 |
| 0 | 0 | 0 | 0 | 0 |

| | | | | |
|---|---|---|---|---|
| 0 | 0 | 0 | 0 | 0 |
| 0 | 0 | 0 | 0 | 0 |
| 0 | 0 | 0 | 0 | 0 |
| 0 | 0 | 0 | 0.008183258 | 0 |
| 0 | 0.035244483 | 0 | 0 | 0 |
| 0.006235444 | 0.11141244 | 0 | 0 | 0 |
| 0 | 0.003355205 | 0 | 0 | 0.004625518 |
| 0 | 0 | 0 | 0 | 0 |
| 0 | 0 | 0 | 0.702899042 | 0.00637832 |
| 0 | 0 | 0 | 0 | 0 |
| 0.000678006 | 0.003499699 | 0 | 0.000672184 | 0 |
| 0 | 0.020941639 | 0 | 0 | 0 |
| 0 | 1.884869564 | 0 | 0 | 0 |
| 0 | 0 | 0.225579105 | 0.00211682 | 0.00396361 |
| 0 | 0.076430025 | 0 | 0 | 0 |
| 0 | 0 | 0 | 0 | 0 |
| 0 | 0 | 0 | 0 | 0 |
| 0 | 0.039601267 | 0 | 0 | 0.002121478 |
| 0 | 0 | 0.001451817 | 0 | 0 |
| 0.04084195 | 0.001830088 | 0 | 0 | 0 |
| 0 | 0 | 0 | 0 | 0 |
| 0.175016643 | 0 | 0.424876622 | 0.214917866 | 0.357607028 |
| 0.057542907 | 0 | 0 | 0 | 0 |
| 0 | 0 | 0 | 0 | 0 |
| 0 | 0 | 0 | 0 | 0 |
| 0 | 0 | 0 | 0 | 0 |
| 0 | 0.008548722 | 0 | 0 | 0 |
| 0 | 0 | 0 | 0 | 0 |
| 0 | 0 | 0 | 0 | 0 |
| 0.077616262 | 3.927449688 | 3.776127622 | 0.099910643 | 0.565295713 |
| 0.001883092 | 0 | 0.01283988 | 0 | 0.034822478 |
| 0 | 0 | 0 | 0 | 0 |
| 0 | 0 | 0 | 0 | 0 |
| 0.000820647 | 0.00073315 | 0 | 0 | 0 |
| 0 | 0.041343563 | 0 | 0 | 0 |
| 0 | 0 | 0 | 0 | 0 |
| 0 | 0 | 17.12487076 | 1.01389243 | 0 |
| 0 | 0 | 0 | 0 | 0 |
| 0 | 0 | 0.01473805 | 0 | 0 |
| 0 | 0 | 0 | 0 | 0 |
| 0.048096003 | 0.012127226 | 0 | 0 | 0.005865334 |
| 0.415608486 | 0 | 0 | 1.026133447 | 0.499679288 |
| 0 | 0 | 0 | 0 | 0 |
| 0 | 0 | 0 | 0 | 0 |
| 0 | 0.001651052 | 0 | 0 | 0 |
| 0 | 0 | 0 | 0 | 0 |
| 0 | 0.00087782 | 0.000735615 | 0 | 0 |
| 0 | 0 | 0 | 0 | 0 |

| | | | | |
|---|---|---|---|---|
| 0.004773599 | 0 | 0.058451066 | 0 | 0 |
| 0 | 0 | 0 | 0 | 0 |
| 0 | 0 | 0.001731393 | 0.002149515 | 0.019836658 |
| 0 | 0 | 0 | 0 | 0 |
| 0.088965063 | 0 | 0 | 0 | 0.095301197 |
| 0 | 0.02060432 | 0 | 0 | 0 |
| 0 | 0 | 0 | 0 | 0 |
| 0 | 0 | 0 | 0 | 0 |
| 0 | 0.008157989 | 0 | 0 | 0 |
| 0 | 0 | 0 | 0 | 0 |
| 0 | 0 | 0 | 0 | 0 |
| 0 | 0 | 0 | 0 | 0 |
| 0 | 0 | 0 | 0 | 0 |
| 0.032080692 | 0 | 0 | 0.001427158 | 0 |
| 0 | 0 | 0 | 0 | 0 |
| 0.183850387 | 0.053600841 | 0 | 0 | 0.022076593 |
| 0.007303268 | 0.003182729 | 0.067282855 | 0.000529797 | 0.025296277 |
| 0 | 0.021139008 | 0.001036948 | 0 | 0 |
| 0 | 0.034409364 | 0 | 0 | 0 |
| 0 | 0 | 0 | 0 | 0 |
| 0.006061299 | 0 | 0.053064441 | 0 | 0.011460313 |
| 0.003719156 | 0.00073836 | 0 | 0 | 0 |
| 0 | 0 | 0 | 0 | 0 |
| 0 | 0 | 0 | 0 | 0 |
| 0 | 0.136631826 | 0 | 0 | 0 |
| 0 | 0 | 0 | 0 | 0 |
| 0.003392033 | 0.001346833 | 0 | 0 | 0 |
| 0 | 0 | 0 | 0 | 0 |
| 0 | 0 | 0 | 0 | 0 |
| 0 | 0 | 0 | 0 | 0 |
| 0.007050993 | 0.092593376 | 0.12057877 | 0.015515389 | 0 |
| 0.002820337 | 0 | 0.012284839 | 0 | 0.009519193 |
| 0.025876934 | 0 | 1.484522426 | 0.017667889 | 0.017447351 |
| 0.002798805 | 0.007084459 | 0.005866943 | 0 | 0 |
| 0 | 0 | 0 | 0.001980482 | 0.001854163 |
| 0 | 0 | 0 | 0 | 0 |
| 0 | 0 | 0.002291193 | 0 | 0.001433964 |
| 0.031240615 | 0.06455735 | 0.009372594 | 0 | 0 |
| 0 | 0 | 0 | 0.005084738 | 0.072834475 |
| 0 | 0.022861093 | 0 | 0 | 0 |
| 0 | 0 | 0.001049727 | 0 | 0 |
| 0 | 0 | 0 | 0 | 0 |
| 0.003355601 | 0.001537347 | 0 | 0 | 0.004791691 |
| 0.000808414 | 0 | 0.125133069 | 0 | 0 |
| 0 | 0.782348045 | 0 | 0 | 0 |
| 0 | 0 | 0 | 0.000746131 | 0.001397082 |
| 0 | 0 | 0 | 0 | 0 |
| 0 | 0 | 0 | 0 | 0 |

| | | | | |
|---|---|---|---|---|
| 0 | 0.109434546 | 0 | 0 | 0 |
| 0.16674936 | 0.031937274 | 0.010705395 | 0.165084401 | 1.436182538 |
| 0 | 0 | 0 | 0 | 0 |
| 0 | 0 | 0 | 0 | 0 |
| 0 | 0 | 0 | 0 | 0 |
| 0 | 0 | 0 | 0 | 0 |
| 0 | 0 | 0 | 0 | 0 |
| 0 | 0 | 0.013993309 | 0 | 0.004191897 |
| 0 | 0 | 0 | 0 | 0.006994185 |
| 0 | 0 | 0 | 0 | 0 |
| 0.001747715 | 0.003955481 | 0.011863135 | 0 | 0 |
| 0.006508178 | 0 | 0.151758211 | 0 | 0.135312924 |
| 0 | 0 | 0 | 0.065229244 | 0.795121787 |
| 0 | 0 | 0 | 0 | 0 |
| 0 | 0 | 0.010889172 | 0 | 0.00409478 |
| 0 | 0 | 0 | 0 | 0 |
| 0.00549033 | 0.015913845 | 0 | 0.000907198 | 0.056565712 |
| 0 | 7.11836454 | 0 | 0 | 0.008599923 |
| 0 | 0 | 0 | 0 | 0 |
| 0 | 0.137819848 | 0 | 0 | 0 |
| 0.005473185 | 0.002248108 | 0.001205708 | 0.100852283 | 0.432508886 |
| 0 | 0 | 0 | 0 | 0 |
| 0 | 0 | 0 | 0 | 0 |
| 0 | 0 | 0 | 0 | 0 |
| 0 | 0.004154965 | 0 | 0 | 0 |
| 0.001730712 | 0 | 0 | 0 | 0 |
| 0.005402307 | 0.187948875 | 0 | 0.006464042 | 0 |
| 0.017050378 | 0 | 0.542870881 | 0.096849834 | 0.038354305 |
| 0 | 0 | 0.001405505 | 0 | 0 |
| 0.001693732 | 0 | 0.000772888 | 0 | 0 |
| 0 | 0 | 0 | 0 | 0 |
| 0 | 0 | 0 | 0 | 0 |
| 0 | 0 | 0 | 0 | 0 |
| 0 | 0 | 0 | 0 | 0 |
| 0 | 0 | 0 | 0 | 0.141155837 |
| 0.087588714 | 0.048527145 | 0.129046224 | 0.091548702 | 0.480855729 |
| 0 | 0 | 0 | 0 | 0 |
| 0 | 0.003071135 | 0 | 0 | 0 |
| 0 | 0 | 0.011066022 | 0.004415918 | 0.004823305 |
| 0 | 0 | 0 | 0 | 0 |
| 0 | 0.298900977 | 0 | 0 | 0 |
| 0 | 0 | 0.006531526 | 0 | 0 |
| 0 | 0.00608742 | 0.001440358 | 0 | 0.085381259 |
| 0 | 0.004148675 | 0 | 0 | 0.00444497 |
| 0 | 0 | 0 | 0 | 0 |
| 0 | 0 | 0 | 0.084433208 | 2.024071236 |
| 0 | 0 | 0 | 0 | 0 |
| 0 | 0.023471041 | 0 | 0 | 0 |

| | | | | |
|---|---|---|---|---|
| 0 | 0.005002505 | 0 | 0 | 0 |
| 0 | 0 | 0 | 0 | 0.094071803 |
| 0 | 0 | 0 | 0.022806225 | 0 |
| 1.034955533 | 0.339064917 | 0.233880012 | 0.186400205 | 0.055245638 |
| 0 | 0 | 0 | 0 | 0 |
| 0 | 0 | 0 | 0 | 0 |
| 0 | 0 | 0 | 0 | 0.003401263 |
| 0.00109627 | 0 | 0.034017194 | 0 | 0 |
| 0.857514366 | 0 | 7.833156829 | 0.311872097 | 0.562553319 |
| 0.011891014 | 0 | 0.000874608 | 0.18769189 | 0.076532822 |
| 0 | 0 | 0 | 0 | 0 |
| 0.001677264 | 0 | 0 | 0 | 0.020394103 |
| 0 | 0 | 0 | 0 | 0 |
| 0 | 0 | 0 | 0.034790308 | 0 |
| 0.124152224 | 0.007913677 | 0.113291109 | 0.041028731 | 0.302798318 |
| 0 | 0 | 0 | 0 | 0 |
| 0.042607787 | 0 | 0 | 0.000894956 | 0.016589908 |
| 4.003218452 | 0 | 0 | 0 | 0 |
| 0 | 0.208281268 | 0 | 0 | 0 |
| 0 | 0 | 0 | 0.001039487 | 0 |
| 0 | 0 | 0 | 0 | 0 |
| 0 | 0 | 0 | 0.002007 | 0 |
| 0 | 0 | 0 | 0 | 0 |
| 0.162653627 | 0.855194063 | 0.068032226 | 0 | 0 |
| 0.012719819 | 0 | 0 | 0 | 0 |
| 0.082053278 | 0 | 0 | 0.65399886 | 0.12317814 |
| 0 | 0 | 0 | 0 | 0 |
| 0 | 0 | 1.180472664 | 0.396525948 | 1.44866882 |
| 0 | 0 | 0 | 0 | 0 |
| 0 | 0 | 0 | 0 | 0 |
| 0.275080008 | 0 | 0.382955994 | 0 | 0.009988947 |
| 0 | 0 | 0 | 0.026502354 | 0 |
| 0.004095288 | 0.008573804 | 0.133093368 | 0 | 0.015435017 |
| 0 | 0 | 0 | 0 | 0 |
| 0 | 0 | 0 | 0.003283662 | 0 |
| 1.022399671 | 0 | 0.197295408 | 0.024398176 | 0.174908209 |
| 0.023391068 | 0 | 0 | 0.528418401 | 1.70788478 |
| 0 | 0.001870897 | 0 | 0 | 0 |
| 0.0162381 | 0.013726862 | 0 | 0 | 0.023499146 |
| 0 | 0 | 0.040074047 | 0 | 0.008691674 |
| 0 | 0.045424375 | 0.449721354 | 0.126500893 | 0.022252748 |
| 0 | 0 | 0 | 0.056250217 | 0 |
| 0.098055954 | 0.249927964 | 0.525469074 | 0.820960743 | 10.3156987 |
| 0.005046048 | 0 | 0.001678997 | 0 | 0.007415758 |
| 0 | 0.007546445 | 0 | 0 | 0 |
| 0 | 0 | 0 | 0 | 0 |
| 0 | 0 | 0 | 0 | 0 |
| 0 | 0.047693245 | 0.004942781 | 0 | 0.487251446 |

| | | | | |
|---|---|---|---|---|
| 0 | 0 | 0 | 0.000660147 | 0 |
| 0.484591791 | 0 | 0.163374955 | 0.006988085 | 0.112365221 |
| 0.049506931 | 0.880373758 | 0.004442075 | 0 | 0.007400397 |
| 0.017036491 | 0 | 0 | 0 | 0 |
| 0 | 0 | 0 | 0.065192253 | 0 |
| 0.061347767 | 0 | 0 | 0 | 0.006507625 |
| 0 | 0 | 0 | 0 | 0 |
| 0 | 0 | 0 | 0 | 0 |
| 0 | 0 | 0 | 0 | 0 |
| 0 | 0.013396015 | 0 | 0 | 0 |
| 0.008192196 | 0.031009794 | 0.010031059 | 0.001985342 | 0.090907955 |
| 0.854352401 | 0 | 0.490848078 | 1.314665544 | 1.387015819 |
| 0 | 0 | 0 | 0 | 0 |
| 0.053048109 | 0 | 0.002667258 | 0 | 0.002735453 |
| 0.347746881 | 0.218732456 | 0.110782562 | 0 | 0.053603622 |
| 1.868206501 | 0 | 0 | 0.507034843 | 1.144944588 |
| 0 | 0 | 0 | 0 | 0 |
| 0 | 0.001024059 | 0 | 0 | 0 |
| 0 | 0 | 0 | 0.131782429 | 0 |
| 0.0101973 | 0.005716119 | 0.025148117 | 0.007433633 | 0 |
| 0.00701885 | 0.005270856 | 0.007798265 | 0.151273539 | 0.032715377 |
| 0.009807329 | 0.512869072 | 0.380419212 | 0 | 0.066854511 |
| 0.042659223 | 3.992718104 | 0.484291522 | 0.578384409 | 2.339606708 |
| 0 | 0 | 0 | 0 | 0 |
| 0 | 0 | 0 | 0.005633989 | 0 |
| 0 | 0 | 0.000876062 | 2.932240921 | 9.128659982 |
| 0 | 3.63121323 | 0 | 0.092817192 | 0 |
| 0 | 0 | 0.005083406 | 0.002682184 | 0 |
| 0 | 0.003455211 | 0 | 0 | 0 |
| 0 | 0 | 0 | 0 | 0 |
| 0 | 0 | 0 | 0.003503867 | 0.016089498 |
| 0.091760299 | 0 | 0.505228242 | 0.172376277 | 0.413816549 |
| 0 | 0.014476303 | 0 | 0.002965815 | 0.0295019 |
| 0.105222729 | 0 | 0.042924422 | 0.006643637 | 0.05187655 |
| 0.01048915 | 0.029986555 | 0.000492721 | 0 | 0 |
| 0.069683602 | 0.011107761 | 0.220556447 | 0.010589712 | 0.109529177 |
| 0.004585828 | 0 | 0.005885484 | 0.002435599 | 0.038612266 |
| 0 | 0 | 0 | 0 | 0 |
| 5.603116562 | 0 | 0.131503581 | 0.009835002 | 0.002394003 |
| 0 | 0 | 0 | 0 | 0 |
| 0 | 0 | 0 | 0.002812234 | 0 |
| 0 | 0 | 0 | 0 | 0 |
| 0 | 0 | 0 | 0.001810594 | 0 |
| 0 | 0 | 0 | 0 | 0 |
| 0.00773394 | 0 | 0.101750406 | 0.022810911 | 0.003589241 |
| 0.238959605 | 0 | 0 | 0.002963199 | 1.16086418 |
| 0 | 0 | 0 | 1.016812153 | 0 |
| 0.755902131 | 0.133353011 | 0.419621403 | 0 | 0.429713368 |

| | | | | |
|---|---|---|---|---|
| 0 | 0 | 0 | 0 | 0 |
| 0 | 0 | 0 | 0.055033337 | 0 |
| 0 | 0 | 0 | 0 | 0 |
| 0.017467756 | 0 | 0.005263821 | 0 | 0.003976824 |
| 0 | 0.021612162 | 0 | 0 | 0 |
| 0 | 0 | 0 | 0.03737161 | 1.142174941 |
| 0 | 0 | 0 | 0.506366015 | 0 |
| 0 | 0 | 0 | 0.003006695 | 0 |
| 0 | 0 | 0 | 0 | 0 |
| 0 | 0 | 0 | 0 | 0 |
| 0.013397879 | 0.00787029 | 0.013190632 | 0 | 0.010221068 |
| 0 | 0 | 0 | 0 | 0 |
| 0.295361336 | 0 | 0 | 0.004938837 | 0.027444655 |
| 0 | 0 | 0 | 0.031674849 | 0 |
| 0 | 0 | 0 | 0 | 0 |
| 0 | 0 | 0 | 0 | 0 |
| 0 | 0 | 0 | 0.017592624 | 0.081212385 |
| 0 | 0 | 0.0012201 | 0 | 0.003261709 |
| 0 | 0 | 0 | 0.02980624 | 0 |
| 0 | 0 | 0 | 0 | 0 |
| 0.012487983 | 0 | 0.028941805 | 0 | 0 |
| 0 | 0 | 0 | 0.034006153 | 0 |
| 0.017967517 | 0 | 0 | 0.000523919 | 0.010954549 |
| 0 | 0 | 0 | 0 | 0 |
| 0 | 0 | 0 | 0 | 0 |
| 0 | 0 | 0 | 0 | 0 |
| 0 | 0 | 0 | 0 | 0 |
| 0 | 0 | 0 | 0 | 0 |
| 0 | 0 | 0 | 0 | 0 |
| 0.05130906 | 0 | 0.110540247 | 1.546548676 | 0.057731253 |
| 0 | 0 | 0 | 0.002579601 | 0 |
| 0 | 0 | 0 | 0 | 0 |
| 0 | 0 | 0 | 0.038458485 | 0 |
| 0 | 0 | 0 | 0 | 0 |
| 0.000729681 | 0 | 0 | 0 | 0 |
| 0 | 0 | 0 | 0 | 0 |
| 0 | 0 | 0 | 0.025408908 | 0 |
| 0 | 0.006662339 | 0 | 0 | 0 |
| 0 | 0 | 0 | 0.014077242 | 0 |
| 0 | 0.000730151 | 0 | 0 | 0 |
| 0.021136572 | 0.029344608 | 0 | 0.002890356 | 0.002164803 |
| 0 | 0 | 0 | 0.004003821 | 0 |
| 0 | 0 | 0 | 0.006251069 | 0 |
| 0.003922787 | 0 | 0.001686783 | 0.020492586 | 0.007282098 |
| 0 | 0 | 0.372480617 | 0.039702058 | 0.224819862 |
| 0 | 0 | 0 | 0.006140167 | 0 |
| 0.037643455 | 0 | 0 | 0 | 0 |
| 0.597374535 | 0 | 0 | 0.002057836 | 0 |

| | | | | |
|---|---|---|---|---|
| 0 | 0 | 0 | 0 | 0 |
| 0 | 0 | 0 | 0 | 0 |
| 1.0515511 | 0 | 0 | 0.389096511 | 0 |
| 0.433642602 | 0 | 0 | 0.036618202 | 0.105860137 |
| 0 | 0 | 0 | 0 | 0 |
| 0 | 0 | 0 | 0.065970097 | 1.535130254 |
| 0 | 0.297036056 | 0 | 0 | 0 |
| 0 | 0 | 0 | 0 | 0 |
| 0.009642253 | 0.0474738 | 0 | 0.00870973 | 0.025854799 |
| 0 | 0 | 0 | 0 | 0.001601597 |
| 0 | 0 | 0.019820558 | 0.000759479 | 0.01592725 |
| 0 | 0 | 0 | 0 | 0 |
| 0 | 0 | 0 | 0 | 0 |
| 0 | 0 | 0 | 0 | 0 |
| 0 | 0 | 0.017952789 | 0.248224446 | 0.103047212 |
| 0 | 0.548840673 | 0 | 0 | 0 |
| 0 | 0 | 0 | 0 | 0 |
| 0 | 0 | 0 | 0 | 0 |
| 0 | 0 | 0 | 0 | 0 |
| 0 | 0 | 0 | 0 | 0 |
| 2.005916279 | 0 | 0 | 0.012345065 | 0.096719465 |
| 0 | 0 | 0 | 0.012386568 | 0 |
| 0 | 0 | 0 | 0 | 0 |
| 0 | 0 | 0 | 0 | 0 |
| 0.005795496 | 0 | 0.057951569 | 0.33550088 | 0.164652104 |
| 0.83481204 | 0 | 0.756739096 | 0.726395898 | 0.980968812 |
| 0.003936279 | 0 | 0 | 0.138259304 | 0.314903133 |
| 0 | 0 | 0 | 0 | 0 |
| 0.606047836 | 0 | 0 | 0.01288615 | 0 |
| 0 | 0 | 0 | 0.019389768 | 0.037150427 |
| 0 | 0 | 0 | 0 | 0 |
| 0 | 0 | 0 | 0.055343381 | 0 |
| 0 | 0 | 0 | 0 | 0 |
| 0.00053829 | 0 | 0.003116479 | 0.001734421 | 0.001498889 |
| 0.001552131 | 0 | 0.000708272 | 0 | 0.00493939 |
| 0.002351075 | 0.012135665 | 0.025033125 | 0 | 0.007419543 |
| 0 | 0.013679702 | 0.003100019 | 0 | 0.108846145 |
| 0.034158037 | 1.287895941 | 0.013992029 | 0.415676527 | 1.042643675 |
| 0 | 0 | 0 | 0.205477525 | 0 |
| 0 | 0 | 0 | 0 | 0 |
| 0 | 0 | 0 | 0 | 0 |
| 0 | 0 | 0.000567755 | 0 | 0 |
| 0.153785333 | 0 | 0.016590983 | 0.032276683 | 1.312893057 |
| 4.054589371 | 0 | 0 | 1.984597241 | 0.324009958 |
| 0.0293818 | 0 | 0 | 0.035082992 | 0 |
| 0.005494126 | 0.005660071 | 0 | 0 | 0.004410407 |
| 0 | 0 | 0 | 0 | 0 |
| 0.643212558 | 0.004226799 | 0.002833652 | 0.066372256 | 0 |

| | | | | |
|---|---|---|---|---|
| 0.008643829 | 0.009876044 | 0 | 0 | 0.013590017 |
| 2.610517163 | 0 | 0 | 0.008002013 | 0 |
| 0 | 0 | 0 | 0.042717071 | 0 |
| 0 | 0 | 0 | 0 | 0 |
| 0 | 0 | 0.000561012 | 0 | 0.001630173 |
| 0.006934343 | 0 | 0.269987807 | 0.027834561 | 0.145837424 |
| 0 | 0 | 0 | 0.018297525 | 0 |
| 0 | 0 | 0 | 0 | 0 |
| 0 | 0 | 0 | 0 | 0 |
| 0 | 0 | 0 | 0 | 0 |
| 0 | 0 | 0 | 0 | 0 |
| 0.130435013 | 0 | 0.664429117 | 0.02377469 | 0.191000209 |
| 0 | 0 | 0.07421206 | 0.015812453 | 0.467605948 |
| 0 | 0 | 0 | 0 | 0 |
| 0.020717993 | 0.009435983 | 0.401613271 | 0 | 0.116134175 |
| 0 | 0 | 0 | 0 | 0 |
| 0 | 0 | 0 | 0 | 0 |
| 0 | 0 | 0 | 0 | 0 |
| 0.00872356 | 0 | 0.010236231 | 0.002824051 | 0.027595988 |
| 0.449500021 | 0 | 0 | 0.001174283 | 0 |
| 0.049082213 | 0 | 0 | 0.02528022 | 0.080306337 |
| 0 | 0 | 0 | 0 | 0 |
| 0.577500855 | 0 | 0 | 0.013558402 | 0.058808417 |
| 0 | 0 | 0 | 0 | 0 |
| 0 | 0 | 0 | 0 | 0 |
| 0 | 0 | 0 | 0 | 0 |
| 0.163443767 | 0 | 0 | 0 | 0 |
| 0 | 0 | 0 | 0 | 0 |
| 0.022503779 | 0 | 0 | 0 | 0.001018904 |
| 0 | 0 | 0 | 0 | 0 |
| 0 | 0 | 0 | 0 | 0 |
| 0.020031583 | 0.003139617 | 0.005472489 | 0.003658344 | 0.165868224 |
| 0 | 0 | 0 | 0 | 0 |
| 0.025281938 | 0.02556653 | 0 | 0.058484662 | 0.174529624 |
| 0.001705706 | 0.013003465 | 0.026833679 | 0 | 0.009024242 |
| 1.255549857 | 0.019120347 | 0.823883414 | 0.045997598 | 0.013470186 |
| 0 | 0 | 0 | 0 | 0 |
| 0.115478345 | 0 | 0 | 0 | 0 |
| 0 | 0 | 0 | 0 | 0 |
| 0 | 0 | 0 | 0 | 0 |
| 0 | 0 | 0 | 0.131544036 | 0 |
| 0 | 0 | 0 | 0.001791856 | 0 |
| 0 | 0 | 0 | 0 | 0 |
| 0 | 0 | 0 | 0.008397737 | 0 |
| 0.002184378 | 0.00220167 | 0.481579359 | 0.004331244 | 0.099035305 |
| 0 | 0 | 0 | 0 | 0 |
| 0 | 0 | 0 | 0.345779585 | 0.077552295 |
| 0.007310571 | 0.003028925 | 0 | 0 | 0 |

| | | | | |
|---|---|---|---|---|
| 0 | 0 | 0 | 0 | 0 |
| 0 | 0 | 0 | 0.030344837 | 0 |
| 0 | 0.16291457 | 0 | 0 | 0 |
| 0 | 0 | 0 | 0 | 0 |
| 0 | 0 | 0 | 0 | 0 |
| 0 | 0 | 0 | 0 | 0 |
| 0.242183231 | 0 | 0.001069346 | 0.297190005 | 0 |
| 0 | 0 | 0 | 0.001301709 | 0 |
| 0 | 0 | 0 | 0 | 0 |
| 0.016957834 | 0 | 0 | 0 | 0.004347213 |
| 0 | 0 | 0 | 0 | 0 |
| 0.001414569 | 0 | 0.008895793 | 0 | 0 |
| 0 | 0 | 0 | 0 | 0 |
| 0 | 0 | 1.402352119 | 0.284374229 | 0 |
| 0 | 0 | 0 | 0 | 0 |
| 0 | 0 | 0 | 0 | 0 |
| 0 | 0 | 0 | 0.455012754 | 0 |
| 0 | 0 | 0 | 0.00632786 | 0 |
| 0 | 0 | 4.231416447 | 1.295324745 | 0 |
| 0 | 0 | 0 | 0 | 0 |
| 0 | 0 | 0 | 0.001095799 | 0 |
| 0 | 0 | 0 | 0 | 0 |
| 0 | 0 | 0 | 0 | 0 |
| 0 | 0 | 0 | 0 | 0 |
| 0.013401806 | 0.00065605 | 0 | 0 | 0 |
| 0.002659289 | 0.010062024 | 0.024484009 | 0 | 0 |
| 0 | 0 | 0 | 0 | 0 |
| 0.022048218 | 0.003282907 | 0.000978162 | 0.508826651 | 0.003552899 |
| 0 | 0 | 0 | 0.029561198 | 0 |
| 0.503600508 | 0 | 0 | 1.022428689 | 0.767545271 |
| 0 | 0 | 0 | 0 | 0 |
| 0 | 0 | 0 | 0 | 0 |
| 0 | 0 | 0 | 0 | 0 |
| 0 | 0 | 0 | 0 | 0 |
| 0.003114625 | 0.008005917 | 0.054719029 | 0 | 0.008520637 |
| 0 | 0 | 0 | 0 | 0 |
| 1.29768394 | 0 | 0 | 0 | 0 |
| 0 | 0 | 0.009506614 | 0 | 0.008632524 |
| 0 | 0 | 0 | 0 | 0 |
| 0.013933612 | 0 | 0 | 0 | 0 |
| 1.391891374 | 2.630037833 | 2.771098093 | 0.545779642 | 10.72532575 |
| 0 | 0.062605444 | 0.000326198 | 0.162529077 | 0.006571809 |
| 3.468279142 | 0 | 0 | 0 | 0.141512393 |
| 0.017174788 | 0 | 0 | 0.062630914 | 0 |
| 0 | 0 | 0 | 0 | 0 |
| 0 | 0 | 0 | 0 | 0 |
| 0 | 0 | 0 | 0.003348315 | 0 |
| 0.002066788 | 0 | 0.009549109 | 0.001317241 | 0.006029099 |

| | | | | |
|---|---|---|---|---|
| 0 | 0 | 0 | 0 | 0 |
| 0.006073892 | 0.009586454 | 0 | 0.000752717 | 0 |
| 0.010286246 | 0 | 0.000566499 | 0.000879131 | 0.000658447 |
| 0 | 0 | 0 | 0 | 0 |
| 1.793068383 | 0 | 0.477918785 | 0.207413362 | 8.105198896 |
| 0 | 0 | 0 | 0.014665211 | 0 |
| 0 | 0 | 0 | 0 | 0 |
| 0 | 0 | 0 | 0 | 0 |
| 0 | 8.599916908 | 0.00339027 | 0 | 0.003377609 |
| 0 | 0 | 0 | 0 | 0 |
| 0 | 0 | 0 | 0 | 0 |
| 0 | 0 | 0 | 0 | 0 |
| 0.076281038 | 0 | 0 | 0.04812567 | 0.048342886 |
| 0.003809237 | 0.186035943 | 0.19946307 | 0.001258843 | 0.035019812 |
| 0.213125645 | 0 | 0 | 0 | 0 |
| 0.218659393 | 0 | 0 | 0.030399637 | 0.14796794 |
| 0 | 0 | 0 | 0.043106474 | 0 |
| 0 | 0 | 0 | 0 | 0 |
| 0 | 0 | 0 | 0 | 0 |
| 0 | 0 | 0 | 0.020137615 | 0 |
| 0 | 0 | 0 | 0 | 0 |
| 0 | 0 | 0 | 0 | 0 |
| 0 | 0 | 0 | 0 | 0 |
| 0 | 0 | 0 | 0 | 0 |
| 0 | 0 | 0 | 0.009333402 | 0.019954166 |
| 0 | 0 | 0 | 0 | 0 |
| 0.013239444 | 0 | 0 | 0.003518246 | 0.039146082 |
| 0.002926241 | 0 | 0.001844836 | 0 | 0.001715417 |
| 0 | 0.006969453 | 0 | 0 | 0 |
| 0 | 0 | 0 | 0 | 0 |
| 0 | 0 | 0 | 0 | 0 |
| 0 | 0 | 0 | 0 | 0 |
| 0 | 0 | 0 | 0.000987525 | 0 |
| 0.027284444 | 0 | 0 | 0.005442045 | 0.029820389 |
| 0.000763792 | 0 | 0.612682034 | 0.007950958 | 0.244760203 |
| 0.011402286 | 0 | 0.003841366 | 0 | 0.033734475 |
| 0.001638915 | 0 | 0 | 0 | 0 |
| 0 | 0 | 0 | 0.003836163 | 0.015927454 |
| 0 | 0 | 0 | 0 | 0 |
| 0 | 0 | 0 | 0.041817027 | 0 |
| 0.668349804 | 0 | 0.304933247 | 0.117249488 | 0.1085559 |
| 0.139550848 | 0 | 0.00069146 | 0 | 0.001071588 |
| 0 | 0 | 0 | 0.026847914 | 0.009140182 |
| 0.87180409 | 0 | 0.149022643 | 0.011212779 | 0 |
| 0 | 0 | 0 | 0 | 0 |
| 0 | 0 | 0 | 0 | 0 |
| 0 | 0 | 0 | 0 | 0 |
| 0 | 0 | 0 | 0 | 0 |

| | | | | |
|---|---|---|---|---|
| 0.02276498 | 0 | 0 | 0.157652199 | 0.071372376 |
| 0 | 0 | 0 | 0 | 0 |
| 0 | 0 | 0 | 0 | 0 |
| 0 | 0 | 0 | 0 | 0 |
| 0.00317167 | 0 | 0.001599652 | 0.058420322 | 0.010226103 |
| 1.869759595 | 0 | 0 | 0 | 0 |
| 0.002818533 | 0 | 0.006877382 | 0.184581122 | 0.007993649 |
| 0 | 0 | 0 | 0 | 0 |
| 0 | 0.001184878 | 0 | 0 | 0 |
| 0 | 0.523666273 | 0 | 0 | 0 |
| 0.018418859 | 0 | 0 | 0.000753019 | 0 |
| 0 | 0 | 0 | 0 | 0 |
| 0 | 0 | 0 | 0 | 0 |
| 0 | 0 | 0 | 0 | 0 |
| 0 | 0 | 0 | 0 | 0 |
| 0 | 0 | 0 | 0 | 0 |
| 0 | 0 | 0 | 0 | 0.001494908 |
| 0 | 0 | 0 | 0 | 0 |
| 0 | 0 | 0 | 0 | 0 |
| 0 | 0 | 0 | 0 | 0 |
| 0.066637774 | 0.006392791 | 0.003214302 | 0.058823501 | 0 |
| 0 | 0 | 0 | 0 | 0 |
| 0 | 0 | 0 | 0.00407551 | 0.042276466 |
| 0.005409951 | 0.003657514 | 0 | 0.000869757 | 0.00339284 |
| 0 | 0 | 0 | 0 | 0 |
| 0.002760011 | 0 | 0.001928541 | 0 | 0.006244351 |
| 0 | 0 | 0 | 0 | 0 |
| 0 | 0 | 0 | 0 | 0 |
| 0.100363401 | 0 | 0 | 0 | 0 |
| 0.002104375 | 0 | 1.359339324 | 0 | 0.2718906 |
| 0 | 0 | 0 | 0.02633005 | 0 |
| 0 | 0 | 0 | 0 | 0 |
| 0 | 0 | 0 | 0.037880557 | 0 |
| 0 | 0.002232002 | 0 | 0 | 0 |
| 0 | 0 | 0 | 0.015452778 | 0 |
| 0 | 0 | 0 | 0.393842736 | 0 |
| 0 | 0.018741844 | 0 | 0.002539345 | 0.029887067 |
| 0 | 0 | 0 | 0 | 0 |
| 0.058668701 | 0 | 0 | 0.006685626 | 0 |
| 1.28279235 | 0 | 0 | 0 | 0 |
| 0 | 0 | 0 | 0 | 0 |
| 0 | 0 | 0 | 0 | 0 |
| 0 | 0 | 0 | 0 | 0 |
| 0 | 0 | 0 | 0.065425023 | 0.053149955 |
| 0 | 0 | 0 | 0.067292352 | 0 |
| 0 | 0 | 0 | 0 | 0.012372815 |
| 0 | 0 | 0 | 0.001268305 | 0 |
| 0.086493614 | 0 | 0 | 0 | 0 |

| | | | | |
|---|---|---|---|---|
| 0.007079958 | 0.006371947 | 0 | 0 | 0.010076254 |
| 0.277089253 | 0 | 0.335902129 | 0.018787385 | 0 |
| 0 | 0 | 0 | 0.000556105 | 0 |
| 0 | 0 | 0 | 0 | 0 |
| 0 | 0 | 0 | 0 | 0 |
| 0 | 0 | 0 | 0 | 0 |
| 0 | 0 | 0 | 0 | 0 |
| 0.124712607 | 0 | 0.00361211 | 0.536047912 | 1.184845972 |
| 0.016052195 | 0.018238445 | 0.600310065 | 9.633821121 | 0.593909542 |
| 0 | 0.049354695 | 0.01825988 | 0 | 0 |
| 0 | 0 | 0 | 0.023802071 | 0 |
| 0 | 0 | 0 | 0 | 0 |
| 0 | 0 | 0 | 0.045759878 | 0 |
| 0 | 0.080360109 | 0.0112419 | 0 | 0 |
| 0 | 0 | 0 | 0 | 0 |
| 0 | 0 | 0 | 0 | 0 |
| 0 | 0.227061835 | 0 | 0 | 0 |
| 0 | 0 | 0 | 0 | 0 |
| 0 | 0 | 0 | 0 | 0 |
| 0.166799265 | 0 | 0 | 0.009611076 | 0 |
| 0 | 0 | 0 | 0 | 0 |
| 0 | 0 | 0 | 0.001605907 | 0 |
| 0 | 0 | 0 | 0.071415246 | 0 |
| 0 | 0 | 0 | 0 | 0 |
| 0 | 0 | 0 | 0.038699967 | 0.025419634 |
| 0 | 0 | 0 | 0.008546144 | 0 |
| 0 | 0 | 0 | 0 | 0 |
| 0 | 0 | 0 | 0 | 0 |
| 0 | 0.025342596 | 0 | 0 | 0 |
| 0 | 0 | 0 | 0 | 0 |
| 0 | 0.034655715 | 0 | 0 | 0 |
| 0 | 0 | 0 | 0 | 0 |
| 0 | 0 | 0 | 0.009135256 | 0 |
| 0 | 0 | 0 | 0 | 0 |
| 0 | 0 | 0 | 0 | 0.013789862 |
| 0 | 0 | 0 | 0 | 0.013043163 |
| 0 | 0 | 0 | 0 | 0 |
| 0 | 0 | 0 | 0 | 0 |
| 0 | 0 | 0.001493958 | 0 | 0.000868221 |
| 0 | 0 | 0 | 0 | 0 |
| 0 | 0 | 0 | 0 | 0 |
| 0.231720781 | 0 | 0 | 0.343150814 | 0.110396885 |
| 0 | 0 | 0 | 0 | 0 |
| 0 | 0 | 0 | 0 | 0 |
| 0 | 0 | 0 | 0 | 0 |
| 0 | 0 | 0 | 0 | 0 |
| 0 | 0 | 0 | 0 | 0 |
| 0 | 0 | 0 | 0 | 0 |

| | | | | |
|---|---|---|---|---|
| 0 | 0 | 0 | 0 | 0 |
| 0 | 0 | 0 | 0.105585081 | 0.023724156 |
| 0 | 0 | 0 | 0 | 0 |
| 0 | 0 | 0 | 0 | 0 |
| 2.44615097 | 0.185171486 | 0.728663845 | 0.266311509 | 0.695089612 |
| 0.086728313 | 0 | 0 | 0 | 0 |
| 0 | 0 | 0 | 0 | 0 |
| 0.004403041 | 0.012396767 | 0.001118763 | 0.00753995 | 0.02879344 |
| 0 | 0 | 0 | 0 | 0.002967658 |
| 0 | 0 | 0 | 0 | 0.143878963 |
| 0 | 0 | 0 | 0 | 0 |
| 0 | 0 | 0 | 0 | 0 |
| 0 | 0 | 0 | 0 | 0 |
| 0 | 0 | 0 | 0 | 0 |
| 0 | 0.024166762 | 0 | 0 | 0 |
| 0 | 0.005996098 | 0 | 0 | 0 |
| 0 | 0 | 0 | 0 | 0 |
| 0 | 0 | 0 | 0 | 0 |
| 0 | 0 | 0 | 0 | 0 |
| 0 | 0 | 0 | 0 | 0 |
| 0 | 0 | 0 | 0.004341767 | 0 |
| 0 | 0 | 0 | 0 | 0 |
| 0 | 0 | 0 | 0 | 0 |
| 0.012817033 | 0.00247578 | 0 | 0 | 0.001286108 |
| 0.152253025 | 0 | 12.80334373 | 0.163473591 | 0.070229939 |
| 0 | 0 | 0 | 0 | 0 |
| 0.005069246 | 0.116426971 | 0.001518045 | 0.218932884 | 0 |
| 0 | 0 | 0 | 0 | 0 |
| 0.000941851 | 0 | 0.001128193 | 0 | 0 |
| 0 | 0 | 0 | 0 | 0 |
| 0 | 0 | 0 | 0 | 0 |
| 0 | 0 | 0 | 0 | 0 |
| 0 | 0 | 0 | 0 | 0 |
| 0 | 0 | 0 | 0 | 0 |
| 0 | 0 | 0 | 0 | 0 |
| 0 | 0 | 0 | 0 | 0 |
| 0 | 0 | 0 | 0 | 0 |
| 0.117135661 | 0 | 0 | 0.039139474 | 0 |
| 0 | 0 | 0 | 0 | 0 |
| 0 | 0 | 0 | 0 | 0 |
| 0 | 0 | 0 | 0 | 0 |
| 0 | 0 | 0.715097556 | 0.002031556 | 0.199189098 |
| 0 | 0 | 0 | 0 | 0 |
| 0 | 0 | 0.057268259 | 0 | 0 |
| 0 | 0 | 0 | 0 | 0 |
| 0 | 0 | 0 | 0 | 0 |
| 0 | 0 | 0 | 0 | 0 |
| 0 | 0 | 0 | 0.001271662 | 0 |

| | | | | |
|---|---|---|---|---|
| 0 | 0 | 0 | 0.002363128 | 0.151687861 |
| 0 | 0 | 0 | 0 | 0 |
| 0.006197841 | 0 | 0 | 0 | 0 |
| 0 | 0 | 0 | 0 | 0 |
| 0 | 0 | 0 | 0 | 0 |
| 0 | 0 | 0.053201922 | 0.418373164 | 0 |
| 0 | 0 | 0 | 0 | 0 |
| 0 | 0 | 0 | 0 | 0 |
| 0 | 0 | 0 | 0 | 0 |
| 0 | 0 | 0 | 0 | 0 |
| 0 | 0 | 0 | 0 | 0 |
| 0 | 0 | 0 | 0 | 0 |
| 0 | 0 | 0 | 0.003577273 | 0 |
| 0 | 0 | 0 | 0 | 0 |
| 0 | 0 | 0 | 0 | 0 |
| 0 | 0 | 0 | 0.08977468 | 0 |
| 0 | 0 | 0 | 0 | 0 |
| 0 | 0.760317638 | 0 | 0 | 0 |
| 0 | 0 | 0 | 0 | 0 |
| 0 | 0 | 0 | 0 | 0 |
| 0 | 0.043761394 | 0.025343362 | 0 | 0 |
| 0 | 0 | 0 | 0 | 0 |
| 0.009148799 | 0 | 0.094107009 | 0.006262787 | 0.171047599 |
| 0.042465393 | 0 | 0 | 0.016800775 | 0.00259069 |
| 0.049500557 | 0 | 0 | 0.010862566 | 0 |
| 0 | 0 | 0.002133128 | 0.007944802 | 0 |
| 0 | 0 | 0 | 0 | 0 |
| 0 | 0 | 0 | 0 | 0 |
| 0 | 0 | 0 | 0 | 0 |
| 0 | 0 | 0 | 0 | 0 |
| 0 | 0 | 0 | 0 | 0 |
| 0 | 0 | 0 | 0 | 0.009961493 |
| 0.00183596 | 0.586587749 | 0.76923061 | 0.002224683 | 0 |
| 0 | 0.267431972 | 0 | 0 | 0 |
| 0 | 0 | 0.099524343 | 0.015738843 | 0.435274926 |
| 0 | 0.16944855 | 0 | 0.000649169 | 0.001367469 |
| 0 | 0 | 0 | 0 | 0 |
| 0 | 0 | 0 | 0 | 0 |
| 0 | 0.063827752 | 0 | 0 | 0 |
| 0 | 0 | 0 | 0 | 0 |
| 0 | 0 | 0 | 0 | 0 |
| 0 | 0 | 0 | 0.000909824 | 0 |
| 0.133281654 | 0 | 0.781105141 | 0.031388081 | 0.39825338 |
| 0 | 0 | 0 | 0 | 0 |
| 0 | 0 | 0 | 0 | 0 |
| 0.015824564 | 0 | 0 | 0.001045912 | 0.06283213 |
| 0 | 0 | 0 | 0 | 0 |
| 0 | 0 | 0 | 0 | 0 |

| | | | | |
|---|---|---|---|---|
| 0 | 0 | 0 | 0 | 0 |
| 0 | 0 | 0 | 0 | 0 |
| 0 | 0 | 0 | 0 | 0 |
| 0 | 0 | 0 | 0 | 0 |
| 0 | 0 | 0 | 0 | 0 |
| 0 | 0 | 0 | 0 | 0 |
| 0 | 0 | 0 | 0 | 0 |
| 0 | 3.762991914 | 0.002035213 | 0 | 0 |
| 0.220580153 | 0 | 0.00355189 | 0.026947882 | 0.024923284 |
| 0.070448445 | 0 | 0.000867727 | 0 | 0.039166029 |
| 0 | 0 | 0 | 0 | 0 |
| 0 | 0 | 0 | 0 | 0 |
| 0 | 0 | 0 | 0 | 0 |
| 0 | 0 | 0 | 0.003860662 | 0 |
| 0 | 0 | 0 | 0 | 0 |
| 0 | 0.017828814 | 0 | 0 | 0 |
| 0 | 0 | 0 | 0 | 0 |
| 0 | 0 | 0 | 0 | 0 |
| 0 | 0 | 0 | 0 | 0 |
| 0.202438723 | 0 | 0.001191307 | 0.201735789 | 0.39504569 |
| 0 | 0 | 0 | 0 | 0 |
| 0 | 0 | 0 | 0 | 0 |
| 0 | 0 | 0 | 0 | 0 |
| 0 | 0 | 0 | 0 | 0 |
| 0 | 0 | 0 | 0 | 0 |
| 0 | 0 | 0 | 0 | 0 |
| 0 | 0 | 0 | 0 | 0 |
| 0 | 0 | 0.001261986 | 0.005918832 | 0 |
| 0 | 0 | 0 | 0 | 0 |
| 0 | 0 | 0 | 5.594989071 | 0.076106202 |
| 0.263287771 | 0 | 0.003006907 | 0.49075384 | 0.238866943 |
| 0 | 0 | 0 | 0 | 0 |
| 0 | 0 | 0 | 0 | 0 |
| 0 | 0 | 0 | 0 | 0 |
| 0 | 0 | 0 | 0 | 0 |
| 0 | 0 | 0 | 0 | 0 |
| 0 | 0 | 0 | 0 | 0 |
| 0 | 0 | 0 | 0.002069438 | 0 |
| 0 | 0 | 0.601399831 | 0.009870985 | 0 |
| 0 | 0 | 0 | 0 | 0 |
| 0 | 0 | 0 | 0 | 0.001264345 |
| 0 | 0 | 0 | 0.038670973 | 0 |
| 0.014724024 | 0 | 0 | 0.004969394 | 0 |
| 0 | 0 | 0 | 0 | 0 |
| 0 | 0 | 0 | 0 | 0 |
| 0 | 0 | 0 | 0 | 0 |
| 0 | 0.075177672 | 0 | 0 | 0 |
| 0 | 0 | 0 | 0 | 0 |

| | | | | |
|---|---|---|---|---|
| 0 | 0 | 0 | 0 | 0 |
| 0 | 0 | 0 | 0 | 0 |
| 0 | 0 | 0 | 0 | 0 |
| 0 | 0 | 0 | 0 | 0 |
| 0.041289952 | 0 | 0 | 0.001891405 | 0 |
| 0.035539017 | 0 | 0 | 0.006028608 | 0 |
| 0 | 0 | 0 | 0 | 0 |
| 0 | 0 | 0 | 0 | 0 |
| 0 | 0 | 0 | 0 | 0 |
| 0 | 0 | 0 | 0 | 0 |
| 0.009342871 | 0 | 0 | 0.00308755 | 0.022322006 |
| 0.017784021 | 0.004281843 | 0.084907946 | 0.009378361 | 0.018262796 |
| 0.003101271 | 0 | 0.020464241 | 0 | 0 |
| 0 | 0 | 0 | 0 | 0 |
| 0 | 0.035721959 | 0 | 0.006146391 | 0 |
| 0 | 0 | 0 | 0.083431982 | 0.043015409 |
| 0 | 0.004853204 | 0 | 0 | 0 |
| 0 | 0 | 0 | 0 | 0 |
| 0.021488558 | 1.577091964 | 0.005834398 | 0 | 0.010770425 |
| 0.00093525 | 0 | 0 | 0.276496942 | 0.001041696 |
| 0 | 0 | 0 | 0 | 0 |
| 0 | 0 | 0 | 0 | 0.032727952 |
| 0 | 0 | 0 | 0 | 0 |
| 0 | 0 | 0 | 0 | 0 |
| 0 | 0 | 0 | 0 | 0 |
| 0 | 0 | 0 | 0 | 0 |
| 0 | 0.042494217 | 0 | 0 | 0 |
| 0 | 0 | 0 | 0 | 0 |
| 0 | 0 | 0 | 0 | 0 |
| 0 | 0 | 0 | 0 | 0 |
| 0.002888474 | 0.111113511 | 0 | 0 | 0 |
| 0 | 0 | 0 | 0 | 0 |
| 0 | 0 | 0 | 0 | 0 |
| 0 | 0 | 0 | 0 | 0 |
| 0 | 0 | 0 | 0.005128885 | 0 |
| 0 | 0 | 0 | 0 | 0 |
| 0 | 0 | 0 | 0 | 0 |
| 0.0527633 | 0.027017075 | 0.022592689 | 0.000710094 | 0.174178295 |
| 0 | 0 | 0 | 0.024057077 | 0.317449676 |
| 0 | 0 | 0 | 0 | 0 |
| 0 | 0 | 0.001427518 | 0.029773908 | 0 |
| 0 | 0 | 0 | 0.001972321 | 0 |
| 0 | 0 | 0 | 0 | 0 |
| 0 | 0.031206888 | 0.314099792 | 0 | 0 |
| 0 | 0 | 0 | 0 | 0 |
| 0 | 0 | 0 | 0 | 0 |
| 0 | 0 | 0 | 0 | 0 |
| 0.023487359 | 0 | 0 | 0 | 0 |

| | | | | |
|---|---|---|---|---|
| 0 | 0 | 0 | 0 | 0 |
| 0.001664269 | 0 | 0 | 0.005699926 | 0.110799952 |
| 0 | 0 | 0 | 0.001213392 | 0 |
| 0.006492992 | 0.001325877 | 0.002518464 | 0 | 0.002238474 |
| 0 | 0 | 0 | 0 | 0 |
| 0 | 0.002380626 | 0 | 0 | 0 |
| 0 | 0 | 0 | 0.000535445 | 0.022892371 |
| 0.004642818 | 0 | 0.059321419 | 0 | 0 |
| 0 | 0 | 0 | 0 | 0.055534001 |
| 0 | 0 | 0 | 0.001062658 | 0 |
| 0 | 0 | 0 | 0.024164329 | 0.083520425 |
| 0 | 0 | 0 | 0 | 0 |
| 0.06693521 | 0 | 0.00430557 | 0.002868233 | 0.112416043 |
| 0 | 0 | 0 | 0.00110424 | 0 |
| 0 | 0.001355512 | 0 | 0 | 0 |
| 0 | 0 | 0 | 0 | 0 |
| 0 | 0 | 0 | 0 | 0 |
| 0 | 0 | 0 | 0 | 0 |
| 0 | 0 | 0 | 0 | 0 |
| 0 | 0 | 0 | 0 | 0 |
| 0 | 0 | 0 | 0 | 0 |
| 0 | 0 | 0 | 0 | 0 |
| 0 | 0.001031316 | 0 | 0 | 0.004500248 |
| 0.000613003 | 0.028234125 | 0 | 0.374367468 | 0.003793177 |
| 0 | 0 | 0 | 0 | 0 |
| 0.033047724 | 0 | 0 | 0.008677213 | 0 |
| 0 | 0 | 0 | 0.00328037 | 0.00261047 |
| 0 | 0 | 0 | 0 | 0 |
| 1.014714041 | 0 | 0 | 0 | 0 |
| 0 | 0 | 0 | 0 | 0 |
| 0.005985238 | 0 | 0 | 0.021629827 | 0.018458177 |
| 0 | 0 | 0 | 0.00074011 | 0 |
| 0 | 0 | 0 | 0 | 0 |
| 0 | 0 | 0 | 0 | 0 |
| 0.626750566 | 0 | 0 | 0.012677725 | 0 |
| 0 | 0 | 0 | 0 | 0 |
| 0 | 0 | 0 | 0 | 0 |
| 0 | 0 | 0 | 0 | 0 |
| 0.00065341 | 0 | 0.027785309 | 0 | 0.002729167 |
| 0 | 0 | 0 | 0 | 0 |
| 0 | 0 | 0 | 0 | 0 |
| 0 | 0 | 0 | 0 | 0 |
| 0 | 0 | 0 | 0 | 0 |
| 0 | 0 | 0 | 0 | 0 |
| 0 | 0 | 0 | 5.289256477 | 0 |
| 0.388052629 | 0 | 0.28693054 | 0.03889953 | 0.087377574 |
| 0 | 0 | 0 | 0 | 0 |
| 0 | 0 | 0 | 0 | 0 |

| | | | | |
|---|---|---|---|---|
| 0 | 0 | 0 | 0 | 0 |
| 0 | 0 | 0 | 0 | 0 |
| 0 | 0 | 0.403321606 | 0 | 0.080730577 |
| 0 | 0 | 0 | 0.003872312 | 0.643294384 |
| 0 | 0 | 0 | 0 | 0 |
| 0 | 0 | 0 | 0 | 0 |
| 0 | 0 | 0 | 0 | 0 |
| 0 | 0 | 0 | 0 | 0 |
| 0 | 0 | 0.02667439 | 0 | 0 |
| 0 | 0 | 0 | 0 | 0 |
| 0.005963518 | 0 | 0.002010729 | 0.003021849 | 0.002460095 |
| 0.001384997 | 0.000942727 | 0 | 0 | 0 |
| 0.08084518 | 0 | 0 | 0 | 0 |
| 0 | 0 | 0 | 0 | 0 |
| 0 | 0 | 0 | 0 | 0 |
| 0 | 0 | 0 | 0 | 0 |
| 0.00077921 | 0 | 0 | 0 | 0 |
| 0 | 0 | 0 | 0 | 0 |
| 0 | 0 | 0 | 0 | 0 |
| 0 | 0 | 0 | 0 | 0.002283756 |
| 0 | 0 | 0 | 0 | 0 |
| 0 | 0 | 0 | 0 | 0 |
| 0 | 0 | 0 | 0 | 0 |
| 0 | 0 | 0 | 0 | 0 |
| 0 | 0 | 0 | 0 | 0 |
| 0 | 0 | 0 | 0 | 0 |
| 0 | 0 | 0 | 0 | 0 |
| 0 | 0 | 0 | 0.010991001 | 0.041159892 |
| 0 | 0 | 0 | 0 | 0 |
| 0 | 0 | 0 | 0 | 0 |
| 0.001207884 | 0.004316399 | 0.017482889 | 0.001047824 | 0.003923965 |
| 0 | 0 | 0 | 0 | 0 |
| 0.014200074 | 0 | 0.113116735 | 0.040496395 | 0.028638488 |
| 0.003762095 | 0.026247658 | 0 | 0.000888046 | 0.006152391 |
| 0 | 0 | 0 | 0.003966516 | 0 |
| 0 | 0 | 0 | 0 | 0 |
| 0 | 0 | 0 | 0 | 0 |
| 0 | 0 | 0 | 0 | 0 |
| 0 | 0 | 0 | 0 | 0 |
| 0 | 0 | 0 | 0 | 0 |
| 0 | 0 | 0 | 0 | 0 |
| 0 | 0.005841122 | 0 | 0 | 0 |
| 0.015110839 | 0 | 0.028483678 | 0.007838942 | 0.054145207 |
| 0 | 0 | 0 | 0 | 0 |
| 0 | 0 | 0 | 0 | 0 |
| 0.246292166 | 0 | 0.074414031 | 0.008558795 | 0.029694856 |
| 0 | 0 | 0 | 0 | 0 |
| 0 | 0 | 0 | 0 | 0 |

| | | | | |
|---|---|---|---|---|
| 0 | 0 | 0 | 0 | 0 |
| 0.004233767 | 0 | 0.01308275 | 0 | 0.007175964 |
| 0.001290127 | 0 | 0 | 0.0008527 | 0.030934618 |
| 0 | 0 | 0 | 0 | 0 |
| 0.109019379 | 0.036197477 | 0.048533671 | 0.01736141 | 0.04732801 |
| 0 | 0 | 0 | 0 | 0 |
| 0 | 0 | 0 | 0 | 0 |
| 0 | 0 | 0 | 0 | 0 |
| 0 | 0 | 0 | 0 | 0 |
| 0 | 0 | 0 | 0 | 0 |
| 0 | 0 | 0 | 0.002705251 | 0 |
| 0 | 0 | 0 | 0 | 0 |
| 0 | 0 | 0 | 0 | 0 |
| 0 | 0.188586908 | 0 | 0 | 0 |
| 0 | 0 | 0 | 0 | 0 |
| 0 | 0 | 0 | 0 | 0 |
| 0.024808936 | 0 | 0 | 0.007540353 | 0 |
| 0 | 0 | 0 | 0 | 0 |
| 0 | 0 | 0 | 0 | 0 |
| 0 | 0 | 0 | 0 | 0 |
| 0 | 0 | 0 | 0 | 0 |
| 0 | 0 | 0 | 0 | 0 |
| 0 | 0 | 0 | 0.003133569 | 0 |
| 0 | 0 | 0 | 0 | 0 |
| 0.009037592 | 0 | 0.007641634 | 0.005973327 | 0.002302727 |
| 0 | 0 | 0.220369342 | 0 | 0.003474733 |
| 0 | 0 | 0 | 0 | 0 |
| 0.671813447 | 0 | 0 | 0.001714401 | 0.016050534 |
| 0 | 0 | 0 | 0 | 0 |
| 0 | 0 | 0 | 0 | 0 |
| 0.007087316 | 0 | 0.005957555 | 0 | 0 |
| 0 | 0 | 0 | 0 | 0 |
| 0 | 0 | 0 | 0 | 0 |
| 0 | 0 | 0 | 0 | 0 |
| 0 | 0 | 0 | 0 | 0 |
| 0 | 0 | 0 | 0 | 0 |
| 0 | 0 | 0 | 0 | 0 |
| 0 | 0 | 0 | 0 | 0 |
| 0 | 0 | 0.888072515 | 0.068196541 | 6.760252348 |
| 0 | 0 | 0 | 0 | 0 |
| 0 | 0 | 0 | 0 | 0 |
| 0.064206119 | 0 | 0.002316538 | 0 | 0 |
| 0.005725124 | 0.005754051 | 0 | 0 | 0.00763879 |
| 0 | 0 | 0 | 0 | 0 |
| 0 | 0 | 0 | 0.26609182 | 0 |
| 0 | 0 | 0 | 0 | 0 |
| 0.024085379 | 0.000978064 | 0.019452289 | 0 | 0.051824221 |
| 0 | 0 | 0 | 0 | 0 |

| | | | | |
|---|---|---|---|---|
| 0 | 0 | 0 | 0 | 0 |
| 0.013468879 | 0.07857335 | 0.084529292 | 0 | 0.013463189 |
| 0 | 0 | 0 | 0.045266576 | 0.021474754 |
| 0 | 0 | 0 | 0 | 0 |
| 0 | 0 | 0 | 0 | 0 |
| 0 | 0 | 0 | 0 | 0 |
| 0 | 0 | 0 | 0 | 0 |
| 0 | 0 | 0 | 0 | 0 |
| 0 | 0 | 0 | 0 | 0 |
| 0 | 0 | 0 | 0 | 0 |
| 0 | 0 | 0 | 0 | 0 |
| 0 | 0 | 0 | 0 | 0 |
| 0 | 0 | 0.001894966 | 0.063700927 | 0.020500546 |
| 0 | 0 | 0 | 0 | 0 |
| 0.018946696 | 0.003334028 | 0.00240707 | 0 | 0 |
| 0 | 0 | 0 | 0 | 0 |
| 0 | 0 | 0 | 0 | 0 |
| 0 | 0 | 0 | 0 | 0 |
| 0.000952725 | 0 | 0 | 0.001180681 | 0 |
| 0 | 0 | 0 | 0 | 0 |
| 0 | 0 | 0 | 0 | 0 |
| 0 | 0 | 0 | 0.051568816 | 0 |
| 0.000649912 | 0.009096362 | 0 | 0 | 0 |
| 0.004355745 | 0 | 0 | 0 | 0 |
| 0 | 0 | 0 | 0 | 0 |
| 0 | 0 | 0 | 0 | 0 |
| 0 | 0 | 0 | 0 | 0 |
| 0 | 0 | 0 | 0 | 0 |
| 0 | 0 | 0 | 0 | 0 |
| 0 | 0 | 0 | 0 | 0 |
| 0 | 0 | 0 | 0 | 0 |
| 0 | 0 | 0 | 0 | 0 |
| 0 | 0 | 0 | 0 | 0 |
| 0 | 0 | 0 | 0 | 0 |
| 26.17688429 | 0 | 0 | 0.001522654 | 0 |
| 0 | 0 | 0 | 0 | 0 |
| 0.000929949 | 0.004430922 | 0 | 0 | 0 |
| 0 | 0 | 0 | 0 | 0 |
| 0.006334599 | 0 | 0.002023434 | 0.019049963 | 0.000783952 |
| 0 | 0 | 0 | 0 | 0 |
| 0.113974778 | 0.001127606 | 0.041123595 | 0.012388281 | 0.10684339 |
| 0 | 0 | 0 | 0 | 0 |
| 0 | 0 | 0 | 0 | 0 |
| 0 | 0 | 0 | 0 | 0 |
| 0 | 0 | 0 | 0 | 0 |
| 0.005635926 | 3.209505431 | 0.051238163 | 0.265192927 | 0.006438337 |
| 0 | 0 | 0 | 0.193188835 | 0 |
| 0 | 0 | 0 | 0 | 0 |

| | | | | |
|---|---|---|---|---|
| 0 | 0 | 0 | 0 | 0 |
| 0 | 0 | 0 | 0 | 0 |
| 0 | 0 | 0 | 0 | 0 |
| 0 | 0 | 0 | 0.020422903 | 0 |
| 0.00180841 | 0 | 0 | 0 | 0 |
| 0 | 0 | 0 | 0 | 0 |
| 0 | 0 | 0 | 0 | 0 |
| 0 | 0 | 0 | 0.003948795 | 0 |
| 0 | 0 | 0 | 0 | 0.005451914 |
| 0.001059989 | 0 | 0.001015764 | 0 | 0.000787088 |
| 0 | 0 | 0 | 0.064060388 | 0 |
| 0 | 0 | 0 | 0 | 0 |
| 0.003522571 | 0.000740469 | 0 | 0 | 0 |
| 0 | 0 | 0 | 0 | 0 |
| 0 | 0 | 0 | 0 | 0 |
| 0 | 0 | 0 | 0 | 0 |
| 0 | 0 | 0 | 0.049778361 | 0 |
| 0 | 0 | 0 | 0 | 0 |
| 0 | 0 | 0 | 0 | 0 |
| 0.073403769 | 0.070081118 | 0.575130276 | 0.013713597 | 0.14739076 |
| 0 | 0 | 0 | 0 | 0 |
| 0 | 0 | 0 | 0 | 0 |
| 0 | 0 | 0 | 0 | 0 |
| 0 | 0 | 0 | 0 | 0 |
| 0 | 0 | 0 | 0 | 0 |
| 0.093267459 | 0 | 0 | 0.024121727 | 0.004106035 |
| 0 | 0 | 0 | 0 | 0 |
| 0 | 0 | 0 | 0 | 0 |
| 0 | 0 | 0 | 0 | 0 |
| 0 | 0 | 0 | 0 | 0 |
| 0 | 0 | 0 | 0 | 0 |
| 0 | 0 | 0 | 0 | 0 |
| 0 | 0 | 0 | 0 | 0 |
| 0 | 0 | 0 | 0 | 0 |
| 0 | 0 | 0 | 0 | 0 |
| 0 | 0 | 0 | 0 | 0.004431915 |
| 0 | 0 | 0 | 0.138531857 | 0 |
| 0 | 0 | 0 | 0.032750141 | 0 |
| 0 | 0 | 0 | 0 | 0 |
| 0 | 0 | 0 | 0.033901238 | 0 |
| 0 | 0 | 0 | 0 | 0 |
| 0 | 0 | 0 | 0 | 0 |
| 0.003679363 | 0 | 0 | 0.001215924 | 0 |
| 0 | 0 | 0 | 0 | 0 |
| 0.012125744 | 0.000825364 | 0.005671581 | 0.003949963 | 0.005948999 |
| 0 | 0 | 0 | 0.011127242 | 0.010591149 |
| 0 | 0 | 0 | 0 | 0 |
| 0 | 0 | 0 | 0 | 0 |

| | | | | |
|---|---|---|---|---|
| 0 | 0 | 0 | 0 | 0 |
| 0 | 0 | 0 | 0 | 0 |
| 0 | 0 | 0 | 0.002106337 | 0 |
| 0 | 0 | 0 | 0 | 0 |
| 0 | 0 | 0 | 0 | 0 |
| 0.086255864 | 0.011861198 | 0.001755584 | 0.003589845 | 0.007802036 |
| 0 | 0 | 0 | 0.000921683 | 0 |
| 0 | 0 | 0 | 0 | 0 |
| 0 | 0 | 0 | 0 | 0 |
| 0 | 0 | 0 | 0 | 0 |
| 0 | 0 | 0 | 0 | 0 |
| 0 | 0 | 0 | 0 | 0 |
| 0 | 0 | 0 | 0 | 0 |
| 0.044145723 | 0 | 0 | 0 | 0 |
| 0 | 0 | 0 | 0 | 0 |
| 0 | 0 | 0 | 0 | 0 |
| 0.111529738 | 0 | 0 | 0.009175462 | 0.035525706 |
| 0 | 0 | 0 | 0 | 0 |
| 0 | 0 | 0 | 0 | 0 |
| 0 | 0 | 0 | 0.003254933 | 0 |
| 0 | 0 | 0 | 0 | 0 |
| 0 | 0 | 0 | 0 | 0 |
| 0 | 0 | 0 | 0 | 0 |
| 0 | 0 | 0 | 0 | 0 |
| 0 | 0 | 0 | 0 | 0 |
| 0 | 0.001927575 | 0 | 0 | 0 |
| 0 | 0 | 0 | 0 | 0 |
| 0.004390317 | 0 | 0 | 0.057184412 | 0 |
| 0 | 0 | 0 | 0 | 0 |
| 0 | 0 | 0 | 0 | 0 |
| 0 | 0 | 0 | 0 | 0 |
| 0 | 0 | 0 | 0 | 0 |
| 0 | 0 | 0 | 0 | 0 |
| 0 | 0 | 0 | 0 | 0 |
| 0 | 0 | 0 | 0 | 0 |
| 0 | 0 | 0 | 0 | 0 |
| 0 | 0 | 0 | 0 | 0 |
| 0 | 0 | 0 | 0 | 0 |
| 3.804614457 | 0 | 1.0992845 | 0.445387184 | 0.006777399 |
| 0 | 0 | 0 | 0 | 0 |
| 0 | 0 | 0 | 0 | 0 |
| 0 | 0 | 0 | 0 | 0 |
| 0 | 0 | 0 | 0 | 0 |
| 0 | 0 | 0 | 0 | 0 |
| 0 | 0 | 0 | 0 | 0 |
| 0 | 0 | 0 | 0 | 0 |
| 0 | 0 | 0 | 0 | 0 |
| 0 | 0 | 0 | 0 | 0 |

| | | | | |
|---|---|---|---|---|
| 0 | 0 | 0 | 0 | 0 |
| 0 | 0 | 0 | 0.00043062 | 0 |
| 0 | 0 | 0 | 0 | 0 |
| 0 | 0 | 0 | 0 | 0 |
| 0 | 0 | 0 | 0 | 0 |
| 0.007743092 | 0 | 0 | 0 | 0 |
| 0 | 0 | 0 | 0 | 0 |
| 0 | 0 | 0 | 0 | 0 |
| 0 | 0 | 0 | 0 | 0 |
| 0 | 0 | 0 | 0.005067219 | 0 |
| 0.007583285 | 0 | 0.012717055 | 0 | 0 |
| 0 | 0 | 0 | 0.000807758 | 0 |
| 0.114921102 | 0 | 0 | 0.082093634 | 0 |
| 0 | 0 | 0 | 0 | 0 |
| 0 | 0 | 0 | 0 | 0 |
| 0 | 0 | 0 | 0 | 0 |
| 0 | 0 | 0 | 0 | 0 |
| 0 | 0 | 0 | 0 | 0 |
| 0 | 0 | 0 | 0 | 0 |
| 0.001577539 | 0 | 0 | 0 | 0 |
| 0.000769658 | 0.010313955 | 0 | 0 | 0 |
| 0 | 0 | 0 | 0 | 0 |
| 0 | 0 | 0 | 0 | 0 |
| 1.062693432 | 0 | 2.623532848 | 0.435681528 | 3.289181887 |
| 0 | 0 | 0 | 0 | 0 |
| 0 | 0 | 0 | 0 | 0 |
| 0 | 0 | 0 | 0 | 0 |
| 0 | 0 | 0 | 0 | 0 |
| 0 | 0 | 0 | 0 | 0 |
| 0 | 0 | 0 | 0 | 0 |
| 0 | 1.879304851 | 0 | 0 | 0 |
| 0 | 0 | 0 | 0 | 0 |
| 0 | 0 | 0 | 0 | 0.005448084 |
| 0 | 0 | 0 | 0 | 0 |

| PHL018-0011_V1 | PHL018-0003_V3 | PHL018-0005_V3 | PHL018-0050_V2 | PHL018-0253_V2 |
|---|---|---|---|---|
| CTRL | CTRL | EYCF | EYCF | REF |
| 0.107748806 | 0.1236639 | 1.062242076 | 0.13062384 | 0.003877013 |
| 1.156941535 | 3.462859792 | 0.722364721 | 0.891117908 | 0.084231129 |
| 5.713159401 | 0 | 9.985277611 | 0.037532889 | 0 |
| 0 | 0 | 0 | 0 | 0 |
| 0 | 0 | 0 | 0 | 0 |
| 0 | 0 | 0 | 0 | 0 |
| 0 | 0 | 0 | 0 | 0 |
| 0 | 0 | 0 | 0 | 0 |
| 0 | 0 | 0 | 0 | 0 |
| 0 | 0 | 0 | 0 | 0 |
| 0 | 0 | 0 | 0 | 0 |
| 0 | 0 | 0 | 0 | 0 |
| 0 | 0 | 0 | 0 | 0 |
| 1.456137144 | 1.06115304 | 6.328559663 | 7.251654923 | 0.236375229 |
| 0 | 0 | 0 | 0 | 0 |
| 0 | 0 | 0 | 0 | 0 |
| 0 | 0 | 0 | 0 | 0 |
| 0 | 0 | 0 | 0 | 0 |
| 0 | 0 | 0 | 0 | 0 |
| 0 | 0 | 0 | 0 | 0 |
| 0 | 0 | 0 | 0 | 0 |
| 0 | 0 | 0 | 0 | 0 |
| 0 | 0 | 0 | 0 | 0 |
| 0 | 0 | 0 | 0 | 0 |
| 0.095384081 | 0.041633578 | 0 | 0 | 0.147010577 |
| 0 | 0 | 0 | 0 | 0 |
| 0 | 0 | 0 | 0 | 0 |
| 0 | 0 | 0 | 0 | 0 |
| 0 | 0 | 0 | 0 | 0 |
| 0 | 0 | 0 | 0 | 0 |
| 0 | 0 | 0 | 0 | 0 |
| 0 | 0 | 0 | 0 | 0 |
| 0 | 0 | 0 | 0 | 0 |
| 0 | 0 | 0 | 0 | 0 |
| 0 | 0 | 0 | 0 | 0 |
| 0 | 0.658524913 | 0 | 0 | 0.577664997 |
| 0 | 0 | 0 | 0 | 0 |
| 0 | 0 | 0 | 0 | 0 |
| 0 | 0 | 0 | 0 | 0 |
| 0 | 0 | 0 | 0 | 0 |
| 0 | 0 | 0 | 0 | 0 |
| 0.0098074 | 0.029192731 | 0.003094998 | 0.115958014 | 0.003433222 |
| 0.001674396 | 0.295174663 | 0.37536133 | 0.309290496 | 0.06275912 |
| 0.037791991 | 0.124413781 | 0.008289212 | 0.004465464 | 0.007037039 |
| 0 | 0.027023337 | 0 | 0 | 0.001475915 |
| 0 | 0 | 0 | 0 | 0 |

| | | | | |
|---|---|---|---|---|
| 0.003305833 | 0 | 0 | 0 | 0 |
| 0.001654029 | 0 | 0.004784769 | 0.013704328 | 0 |
| 0 | 0 | 0 | 0 | 0 |
| 3.684142816 | 0.607627891 | 0.682070563 | 0.700471848 | 0.336638236 |
| 0.045539686 | 0 | 0 | 0 | 0 |
| 0 | 0 | 0 | 0 | 0.034091976 |
| 0.007977834 | 0.031737129 | 0 | 0.003771456 | 0 |
| 0 | 0.08526184 | 0 | 0 | 0 |
| 0 | 0 | 0 | 0 | 0 |
| 0 | 0 | 0 | 0 | 0 |
| 0 | 0 | 0.014778858 | 0 | 0 |
| 3.575933723 | 0.596393638 | 0.001316114 | 0.022693748 | 0.011944963 |
| 0 | 0.002156405 | 0 | 0 | 0.001832871 |
| 0.276151224 | 0 | 0 | 0.021181511 | 0.009217559 |
| 0 | 0 | 0 | 0.003705789 | 0 |
| 0 | 0 | 0 | 0 | 0 |
| 0 | 0.635709487 | 0.324765061 | 0.18474935 | 0.006058499 |
| 0 | 0 | 0 | 0 | 0 |
| 0 | 0 | 0 | 0 | 0 |
| 0 | 0 | 0 | 0 | 0 |
| 0.091399257 | 0.101251065 | 0.021698842 | 0.314554419 | 0 |
| 0.05967685 | 0.009285407 | 0.001159445 | 0.001269352 | 0.00551207 |
| 0 | 0.94475909 | 0 | 0 | 0.004215292 |
| 0 | 0 | 0 | 0 | 0 |
| 0 | 0 | 0 | 0 | 0 |
| 0 | 0 | 0 | 0 | 0 |
| 0 | 0 | 0 | 0.009935935 | 0 |
| 0 | 0 | 0 | 0 | 0 |
| 0 | 0.001131307 | 0.012501804 | 0 | 0 |
| 0 | 0 | 0 | 0 | 0 |
| 0.003033108 | 0 | 0.108668262 | 0 | 0 |
| 0 | 0 | 0 | 0 | 0 |
| 0 | 0 | 0 | 0 | 0 |
| 0.006696918 | 0 | 0 | 0 | 0 |
| 0 | 0 | 0 | 0 | 0 |
| 0 | 0.00796703 | 0 | 0 | 0 |
| 0 | 0 | 0 | 0 | 0 |
| 0 | 0.017425467 | 0.008435691 | 0.003249471 | 0 |
| 0 | 0 | 0 | 0 | 0 |
| 0 | 0 | 0 | 0 | 0 |
| 0 | 0 | 0 | 0 | 0 |
| 0 | 0 | 0 | 0 | 0 |
| 0 | 0 | 0 | 0 | 0 |
| 0 | 0 | 0 | 0 | 0 |
| 0 | 0 | 0 | 0 | 0.989824949 |
| 0 | 0 | 0 | 0 | 0 |
| 0 | 0 | 0 | 0 | 0 |
| 0 | 0 | 0 | 0.001852437 | 0 |

| | | | | |
|---|---|---|---|---|
| 0 | 0 | 0 | 0 | 0 |
| 0 | 0 | 0.001913404 | 0.001036811 | 0 |
| 0 | 0 | 0 | 0 | 0 |
| 0 | 0 | 0 | 0 | 0 |
| 0 | 0 | 0.016214075 | 0 | 0 |
| 0 | 0 | 0 | 0 | 0 |
| 0 | 0 | 0 | 0 | 0 |
| 0 | 0 | 0 | 0 | 0 |
| 0.030743628 | 0 | 0 | 0.002181206 | 0.001210874 |
| 0 | 0 | 0 | 0 | 0 |
| 0 | 0 | 0 | 0 | 0 |
| 0 | 0.002232832 | 0 | 0 | 0.00042174 |
| 0 | 0 | 0 | 0 | 0 |
| 0 | 0 | 0 | 0 | 0 |
| 0 | 0 | 0 | 0 | 0 |
| 0 | 0 | 0 | 0 | 0 |
| 0 | 0 | 0 | 0 | 0 |
| 0 | 0 | 0 | 0 | 0 |
| 0.069977074 | 0.218582779 | 0 | 0 | 0.006505706 |
| 0.051478538 | 0 | 0 | 0 | 0.001512593 |
| 0 | 0 | 0 | 0 | 0 |
| 0 | 0 | 0 | 0 | 0 |
| 0 | 0 | 0 | 0 | 0.003443497 |
| 0 | 0 | 0 | 0 | 0 |
| 0 | 0 | 0 | 0 | 0 |
| 0 | 0 | 0 | 0 | 0 |
| 0 | 0 | 0 | 0.000998862 | 0 |
| 0 | 0 | 0 | 0 | 0 |
| 0 | 0 | 0 | 0 | 0 |
| 0 | 0 | 0 | 0 | 0 |
| 0.202947423 | 0 | 0 | 0 | 0.048293888 |
| 0 | 0.017218455 | 0 | 0.001058752 | 0 |
| 0 | 0 | 0 | 0 | 0 |
| 0 | 0 | 0 | 0 | 0 |
| 0 | 0 | 0 | 0 | 0 |
| 0 | 0 | 0 | 0 | 0 |
| 0 | 0 | 0 | 0 | 0 |
| 0 | 0 | 0 | 0 | 0 |
| 0 | 0 | 0 | 0 | 0 |
| 0 | 0 | 0 | 0 | 0 |
| 0 | 0 | 0 | 0 | 0 |
| 0 | 0 | 0 | 0 | 0 |
| 0 | 0 | 0.014910523 | 0 | 0 |
| 0 | 0 | 0 | 0 | 0 |
| 0 | 0 | 0 | 0 | 0 |
| 0 | 0 | 0 | 0 | 0 |
| 0 | 0 | 0 | 0 | 0 |
| 0 | 0 | 0 | 0 | 0 |

| | | | | |
|---|---|---|---|---|
| 0 | 0 | 0 | 0 | 0 |
| 0 | 0 | 0 | 0 | 0 |
| 0 | 0 | 0 | 0 | 0 |
| 0 | 0 | 0 | 0 | 0 |
| 0 | 0.245565475 | 0 | 0 | 0.001534723 |
| 0 | 0 | 0 | 0 | 0 |
| 0 | 0 | 0 | 0 | 0 |
| 0 | 0 | 0 | 0 | 0 |
| 0 | 0 | 0 | 0 | 0 |
| 0 | 0 | 0 | 0 | 0 |
| 0 | 0 | 0 | 0 | 0 |
| 0 | 0 | 0 | 0 | 0 |
| 0 | 0 | 0 | 0 | 0 |
| 0 | 0 | 0 | 0 | 0 |
| 0 | 0 | 0 | 0 | 0 |
| 0 | 0.003669445 | 0 | 0 | 0 |
| 0 | 0 | 0 | 0 | 0 |
| 0 | 0 | 0 | 0 | 0 |
| 0 | 0 | 0 | 0 | 0 |
| 0 | 0 | 0 | 0 | 0 |
| 0 | 0 | 0 | 0 | 0 |
| 0 | 0 | 0 | 0 | 0.016854694 |
| 0.001505314 | 0 | 0 | 0 | 0.00568874 |
| 0 | 0 | 0 | 0 | 0 |
| 0 | 0.007054288 | 0 | 0 | 0 |
| 0 | 0 | 0 | 0 | 0 |
| 0.011876807 | 0 | 0 | 0 | 0.23840076 |
| 0 | 0 | 0 | 0 | 0 |
| 0 | 0 | 0 | 0 | 0 |
| 0 | 0 | 0 | 0 | 0 |
| 0.02246459 | 0 | 0 | 0 | 0 |
| 0 | 0 | 0 | 0.003053927 | 0 |
| 0 | 0 | 0 | 0 | 0 |
| 0 | 0 | 0 | 0 | 0 |
| 0 | 0 | 0 | 0 | 0 |
| 0 | 0 | 0 | 0 | 0 |
| 0 | 0 | 0 | 0 | 0 |
| 0 | 0 | 0 | 0 | 0 |
| 0 | 0 | 0 | 0 | 0.011420529 |
| 0 | 0 | 0 | 0 | 0 |
| 0 | 0 | 0 | 0 | 0 |
| 0 | 0 | 0 | 0 | 0 |
| 0 | 0 | 0 | 0 | 0 |
| 0 | 0 | 0 | 0 | 0 |
| 0 | 0 | 0 | 0 | 0 |
| 0 | 0 | 0 | 0 | 0 |
| 0 | 0 | 0 | 0 | 0 |
| 0 | 0 | 0 | 0 | 0 |

| | | | | |
|---|---|---|---|---|
| 0 | 0 | 0 | 0 | 0 |
| 0 | 0 | 0 | 0 | 0 |
| 0 | 0 | 0 | 0 | 0 |
| 0 | 0 | 0 | 0 | 0 |
| 0 | 0 | 0 | 0 | 0 |
| 0 | 0 | 0 | 0 | 0 |
| 0 | 0 | 0 | 0 | 0 |
| 0 | 0 | 0 | 0 | 0 |
| 0 | 0 | 0 | 0 | 0 |
| 0 | 0 | 0 | 0 | 0 |
| 0 | 0 | 0 | 0 | 0 |
| 0 | 0 | 0 | 0 | 0.104533477 |
| 0 | 0.004580766 | 0 | 0 | 0 |
| 0 | 0 | 0 | 0 | 0 |
| 0 | 0 | 0 | 0 | 0 |
| 0 | 0 | 0 | 0 | 0 |
| 0 | 0 | 0 | 0 | 0 |
| 0 | 0 | 0 | 0 | 0 |
| 0 | 0 | 0 | 0 | 0 |
| 0 | 0 | 0 | 0 | 0 |
| 0.085096783 | 0.035972714 | 0.092403134 | 0.01830445 | 0.033487549 |
| 0 | 0 | 0 | 0 | 0 |
| 0 | 0 | 0 | 0 | 0 |
| 0 | 0 | 0 | 0.022506581 | 0 |
| 0 | 0 | 0 | 0 | 0 |
| 0 | 0 | 0 | 0 | 0 |
| 0.000532886 | 0 | 0 | 0.000795528 | 0.004522286 |
| 0 | 0 | 0 | 0 | 0 |
| 0 | 0 | 0 | 0 | 0 |
| 0 | 0 | 0 | 0 | 0 |
| 0.003895441 | 0 | 0 | 0.122123035 | 0 |
| 0 | 0 | 0 | 0 | 0 |
| 0 | 0 | 0 | 0 | 0 |
| 0.038957801 | 0.032452997 | 0 | 0 | 0.033960882 |
| 0.019249981 | 0.023405765 | 0.021710868 | 0.010132261 | 0.000473669 |
| 0 | 0 | 0 | 0 | 0 |
| 0 | 0 | 0 | 0 | 0 |
| 0 | 0 | 0 | 0 | 0 |
| 0 | 0 | 0 | 0 | 0 |
| 0 | 0 | 0 | 0 | 0 |
| 0 | 0 | 0 | 0 | 0 |
| 0 | 0 | 0 | 0 | 0 |
| 0 | 0 | 0 | 0 | 0 |
| 0 | 0 | 0 | 0 | 0 |
| 0 | 0 | 0 | 0 | 0 |
| 0 | 0 | 0 | 0 | 0 |
| 0.018049271 | 0.002111852 | 0 | 0.001154794 | 0 |
| 0.002121349 | 0.001389965 | 0 | 0 | 0 |

| | | | | |
|---|---|---|---|---|
| 0 | 0 | 0 | 0 | 0 |
| 0 | 0 | 0 | 0 | 0 |
| 0 | 0 | 0.00081564 | 0.00092603 | 0 |
| 0 | 0 | 0 | 0 | 0 |
| 0 | 0 | 0 | 0 | 0 |
| 0 | 0 | 0 | 0 | 0 |
| 0 | 0 | 0 | 0 | 0 |
| 0 | 0 | 0 | 0 | 0 |
| 0 | 0 | 0 | 0 | 0 |
| 0 | 0 | 0 | 0 | 0 |
| 0 | 0 | 0 | 0 | 0 |
| 0 | 0 | 0 | 0 | 0 |
| 0 | 0 | 0 | 0 | 0 |
| 0 | 0 | 0 | 0 | 0 |
| 0 | 0 | 0.010555019 | 0 | 0 |
| 0 | 0 | 0 | 0 | 0 |
| 0 | 0 | 0 | 0 | 0 |
| 0 | 0 | 0.004234127 | 0 | 0 |
| 0 | 0.001817467 | 0.001059064 | 0.002447741 | 0.000801 |
| 0 | 0 | 0 | 0 | 0.083923099 |
| 0 | 0 | 0.003347631 | 0.055870335 | 0 |
| 0 | 0 | 0 | 0 | 0 |
| 0 | 0 | 0 | 0 | 0 |
| 0 | 0 | 0 | 0 | 0 |
| 0 | 0.007101695 | 0.00177354 | 0 | 0 |
| 0 | 0 | 0 | 0 | 0 |
| 0 | 0 | 0 | 0 | 0 |
| 0 | 0 | 0 | 0 | 0 |
| 0 | 0 | 0.010789784 | 0 | 0 |
| 0 | 0 | 0 | 0 | 0 |
| 1.251122069 | 0.141276528 | 0.247792147 | 1.805487117 | 0.08824503 |
| 0 | 0 | 0 | 0 | 0 |
| 0 | 0 | 0.057132998 | 0.042567951 | 0 |
| 0 | 0 | 0.003821574 | 0 | 0 |
| 0 | 0 | 0 | 0 | 0 |
| 0 | 0 | 0 | 0 | 0 |
| 0 | 0 | 0 | 0 | 0 |
| 0 | 0 | 0 | 0 | 0 |
| 0 | 0 | 0 | 0 | 0 |
| 0 | 0.024722579 | 0 | 0 | 0 |
| 0 | 0 | 0 | 0 | 0 |
| 0 | 0.027685245 | 0.056036601 | 0 | 0.00215073 |
| 0 | 0 | 0 | 0 | 0 |
| 0 | 0 | 0 | 0 | 0 |
| 0 | 0 | 0 | 0 | 0 |
| 0 | 0 | 0 | 0 | 0 |
| 0 | 0 | 0 | 0 | 0 |
| 0 | 0 | 0 | 0 | 0 |

| | | | | |
|---|---|---|---|---|
| 0 | 0 | 0 | 0 | 0 |
| 0 | 0 | 0 | 0 | 0 |
| 0.041132259 | 0.079058657 | 0.009031691 | 0.003219295 | 0.006513202 |
| 0 | 0 | 0 | 0 | 0 |
| 1.78981067 | 0 | 0 | 0.003569847 | 0.001102895 |
| 0 | 0 | 0 | 0 | 0 |
| 0 | 0 | 0 | 0 | 0 |
| 0 | 0 | 0 | 0 | 0 |
| 0 | 0 | 0 | 0 | 0 |
| 0 | 0.001794841 | 0 | 0 | 0.000726454 |
| 0 | 0 | 0 | 0 | 0 |
| 0 | 0 | 0 | 0 | 0 |
| 0 | 0 | 0 | 0 | 0 |
| 0.005075472 | 0 | 0.00510689 | 0.009092417 | 0 |
| 0.002003501 | 0.003324491 | 0.002107535 | 0 | 0.001449079 |
| 0 | 0.088708861 | 0 | 0 | 0.001861717 |
| 0.001660225 | 0.004953485 | 0.004366085 | 0 | 0 |
| 0 | 0 | 0.020998444 | 0 | 0 |
| 0 | 0 | 0 | 0 | 0 |
| 0 | 0 | 0 | 0 | 0 |
| 0 | 0 | 0 | 0 | 0 |
| 0.001873387 | 0.002630343 | 0 | 0 | 0 |
| 0 | 0 | 0 | 0 | 0 |
| 0 | 0 | 0 | 0 | 0 |
| 0 | 0 | 0 | 0.005112191 | 0 |
| 0 | 0 | 0 | 0 | 0 |
| 0 | 0 | 0 | 0 | 0 |
| 0 | 0 | 0 | 0 | 0 |
| 0 | 0 | 0 | 0 | 0 |
| 0.009213561 | 0 | 0 | 0.000970916 | 0 |
| 0 | 0 | 0 | 0 | 0 |
| 0 | 0 | 0 | 0 | 0 |
| 0 | 0 | 0 | 0 | 0 |
| 0 | 0 | 0 | 0.001934766 | 0 |
| 0 | 0 | 0 | 0 | 0 |
| 0.005238553 | 0 | 0 | 0 | 0 |
| 0 | 0 | 0 | 0 | 0 |
| 0.001647601 | 0.025777079 | 0 | 0.00084331 | 0.0117975 |
| 0 | 0 | 0 | 0 | 0 |
| 0 | 0 | 0 | 0 | 0 |
| 0 | 0 | 0 | 0 | 0 |
| 0 | 0 | 0 | 0 | 0 |
| 0 | 0 | 0 | 0 | 0 |
| 0 | 0 | 0 | 0.006547015 | 0 |
| 0 | 0 | 0 | 0 | 0 |
| 0 | 0 | 0 | 0.003241438 | 0 |
| 0 | 0 | 0 | 0 | 0 |
| 0 | 0 | 0 | 0 | 0 |

| | | | | |
|---|---|---|---|---|
| 1.933420792 | 0 | 0 | 0 | 1.142720493 |
| 0 | 0 | 0 | 0 | 0 |
| 0.003139562 | 0.012948873 | 0.000825647 | 0 | 0 |
| 0 | 0 | 0 | 0 | 0 |
| 0.007896756 | 0.004157813 | 0 | 0 | 0.019162135 |
| 0 | 0 | 0 | 0.009803599 | 0 |
| 0 | 0 | 0 | 0 | 0 |
| 0.035974385 | 0.008159326 | 0.02707187 | 0 | 0.02685995 |
| 0 | 0 | 0 | 0 | 0 |
| 0.042213673 | 0 | 0.311439761 | 0.625425802 | 0.032450606 |
| 0 | 0 | 0 | 0 | 0 |
| 0.001971516 | 0.018146579 | 0.004839075 | 0.007063716 | 0 |
| 0.013957671 | 0.016331148 | 0.008564757 | 0.007739448 | 0.001454192 |
| 0 | 0 | 0 | 0 | 0 |
| 0 | 0 | 0 | 0 | 0.001631567 |
| 0 | 0 | 0 | 0 | 0 |
| 0 | 0 | 0 | 0 | 0 |
| 0 | 0 | 0 | 0.002132228 | 0 |
| 0 | 0 | 0.027103735 | 0.226462602 | 0 |
| 0 | 0 | 0 | 0 | 0 |
| 0 | 0 | 0 | 0 | 0 |
| 0 | 0 | 0 | 0 | 0 |
| 0 | 0 | 0 | 0 | 0 |
| 0 | 0.004071204 | 0 | 0 | 0.026254986 |
| 0 | 0 | 0 | 0 | 0 |
| 0 | 0.052988808 | 0 | 0 | 0 |
| 0 | 0 | 0 | 0 | 0 |
| 0 | 0 | 0.013786817 | 0.086788862 | 0 |
| 0 | 0 | 0.105264028 | 0 | 0 |
| 0 | 0 | 0 | 0 | 0 |
| 0 | 0 | 0 | 0.019532082 | 0 |
| 0 | 0 | 0 | 0 | 0 |
| 0 | 0 | 0 | 0 | 0 |
| 0.155157138 | 0 | 0.001597526 | 0.00385419 | 0 |
| 0 | 0 | 0 | 0 | 0 |
| 0 | 0 | 0 | 0.003515834 | 0 |
| 0.045634014 | 0.014477987 | 0.018232142 | 0 | 0 |
| 0 | 0 | 0 | 0 | 0 |
| 0 | 0 | 0 | 0 | 0 |
| 0.065340078 | 0 | 0 | 0 | 0 |
| 0 | 0 | 0 | 0 | 0 |
| 0.105692088 | 0 | 0 | 0 | 0 |
| 0 | 0 | 0 | 0 | 0 |
| 0 | 0 | 0 | 0 | 0 |
| 0 | 0 | 0 | 0 | 0 |
| 0.042055059 | 0 | 0 | 0 | 0.010139105 |
| 0.062817182 | 0.001689635 | 0.010127056 | 0.014228359 | 0 |
| 0 | 0 | 0 | 0 | 0.001042831 |

| | | | | |
|---|---|---|---|---|
| 0 | 0 | 0 | 0 | 0 |
| 0 | 0.00642017 | 0.00848827 | 0 | 0 |
| 0 | 0 | 0 | 0 | 0 |
| 0 | 0 | 0.010899157 | 0 | 0 |
| 0.001427315 | 0 | 0 | 0 | 0 |
| 0.43125754 | 0 | 0.007976283 | 0.005093889 | 0 |
| 0 | 0 | 0 | 0 | 0.004527909 |
| 0 | 0 | 0 | 0 | 0 |
| 0 | 0 | 0 | 0 | 1.568107652 |
| 0 | 0 | 0 | 0 | 0 |
| 0.009893792 | 0.004013087 | 0 | 0.019496578 | 0.01206965 |
| 0.069854617 | 0 | 0 | 0.023440782 | 0 |
| 0 | 0 | 0 | 0 | 0 |
| 0.005608294 | 0.002333146 | 0.001311003 | 0 | 0.003305159 |
| 0 | 0 | 0 | 0 | 0 |
| 0 | 0 | 0 | 0 | 0 |
| 0 | 0 | 0 | 0 | 0 |
| 0.019607566 | 0 | 0 | 0 | 0 |
| 0 | 0 | 0 | 0 | 0 |
| 0.051440329 | 0 | 0 | 0.074636756 | 0 |
| 0 | 0.007855842 | 0 | 0.001202796 | 0 |
| 0 | 0.256863078 | 0 | 0 | 0.146670995 |
| 0 | 0 | 0 | 0 | 0.013800327 |
| 0 | 0 | 0 | 0 | 0 |
| 0 | 0 | 0 | 0 | 0 |
| 0 | 0 | 0 | 0 | 0 |
| 0.002026959 | 0 | 0.013030177 | 0.014121257 | 0 |
| 0.007016161 | 0 | 0 | 0 | 0 |
| 0 | 0 | 0 | 0 | 0 |
| 0.551388102 | 0.601475606 | 0.018896276 | 1.570030614 | 0.169079012 |
| 0 | 0.015234974 | 0 | 0.001514678 | 0.001569601 |
| 0 | 0 | 0 | 0 | 0 |
| 0 | 0 | 0 | 0 | 0 |
| 0 | 0 | 0 | 0 | 0 |
| 0.015381975 | 0 | 0.00453849 | 0.006048862 | 0 |
| 0 | 0 | 0 | 0 | 0 |
| 0 | 1.326257256 | 0 | 0 | 1.002611356 |
| 0 | 0.002378098 | 0 | 0 | 0 |
| 0 | 0 | 0 | 0 | 0 |
| 0 | 0 | 0 | 0 | 0 |
| 0.037738845 | 0 | 0.010957499 | 0.014989783 | 0 |
| 0.332060071 | 4.264131685 | 0 | 0.138784672 | 1.130829292 |
| 0 | 0 | 0 | 0 | 0 |
| 0 | 0 | 0.002817197 | 0 | 0.199796406 |
| 0 | 0 | 0.001013166 | 0 | 0 |
| 0 | 0 | 0 | 0.007482072 | 0 |
| 0 | 0.001006592 | 0.023755506 | 0.018462021 | 0 |
| 0 | 0 | 0 | 0 | 0 |

| | | | | |
|---|---|---|---|---|
| 0 | 0.017077026 | 0 | 0 | 0.001231567 |
| 0 | 0 | 0 | 0 | 0 |
| 0.007231639 | 1.380471823 | 0.759447191 | 0.4302162 | 0 |
| 0 | 0 | 0 | 0 | 0 |
| 0 | 0 | 0 | 0 | 0 |
| 0.801926687 | 0 | 0.010030765 | 0 | 0 |
| 0 | 0 | 0 | 0 | 0 |
| 0 | 0 | 0 | 0 | 0 |
| 0.014805854 | 0 | 0 | 0 | 0.008098443 |
| 0 | 0 | 0 | 0 | 0 |
| 0 | 0 | 0 | 0 | 0 |
| 0 | 0 | 0 | 0 | 0 |
| 0 | 0 | 0 | 0 | 0 |
| 0 | 0 | 0 | 0.001971164 | 0 |
| 0 | 0 | 0 | 0 | 0 |
| 0.010943959 | 0.032927038 | 0.001973526 | 0.009802738 | 0.013558082 |
| 0.005025384 | 0.058880517 | 0.011484103 | 0.004501344 | 0.023024122 |
| 0 | 0 | 0 | 0.01074933 | 0 |
| 0.027565163 | 0 | 0 | 0 | 0 |
| 0 | 0 | 0 | 0 | 0 |
| 0 | 0.009812391 | 0.002756807 | 0.138890063 | 0 |
| 0 | 0.012135655 | 0.003805989 | 0.024306169 | 0 |
| 0 | 0 | 0 | 0 | 0 |
| 0 | 0 | 0 | 0 | 0 |
| 0 | 0 | 0 | 0 | 0.044196203 |
| 0 | 0 | 0.017049334 | 0 | 0 |
| 0.002419911 | 0 | 0.001851322 | 0.002758721 | 0 |
| 0.204375451 | 0 | 0 | 0 | 0.000664583 |
| 0 | 0 | 0 | 0 | 0 |
| 0 | 0 | 0 | 0 | 0 |
| 0.004349884 | 0 | 0.023406757 | 0 | 0 |
| 0.016961144 | 0 | 0.001574283 | 0.002457604 | 0.003770467 |
| 0.005224774 | 0.013671422 | 5.136083949 | 0.010636232 | 0.017761026 |
| 0 | 0.003345058 | 0 | 0.000914565 | 0 |
| 0 | 0.003751817 | 0 | 0.009356328 | 0.001932677 |
| 0 | 0 | 0 | 0 | 0 |
| 0 | 0.004220456 | 0.001806848 | 0.000769271 | 0 |
| 0.412892908 | 0 | 0.002826078 | 0.022560208 | 0.002003851 |
| 0 | 0.866575347 | 0 | 0 | 0.09001073 |
| 0 | 0.015805919 | 0 | 0 | 0 |
| 0 | 0 | 0 | 0 | 0 |
| 0 | 0 | 0 | 0 | 0 |
| 0 | 0 | 0 | 0.006747751 | 0 |
| 0.033948465 | 0 | 0.031023313 | 0 | 0 |
| 0.32421731 | 0 | 0 | 0 | 0 |
| 0 | 0 | 0.00577623 | 0 | 0 |
| 0 | 0 | 0 | 0 | 0 |
| 0 | 0 | 0 | 0 | 0 |

| | | | | |
|---|---|---|---|---|
| 0.000746868 | 0 | 0 | 0 | 0 |
| 1.113340338 | 0.869297865 | 0.018773088 | 0.296959468 | 0.36133638 |
| 0 | 0 | 0 | 0 | 0 |
| 0 | 0 | 0 | 0 | 0 |
| 0 | 0 | 0 | 0 | 0 |
| 0 | 0 | 0 | 0 | 0 |
| 0 | 0 | 0 | 0.121721334 | 0 |
| 0 | 0.014558401 | 0 | 0 | 0 |
| 0 | 0 | 0 | 0 | 0 |
| 0.001995173 | 0 | 0 | 0 | 0 |
| 0.080591066 | 0.009787627 | 0 | 0.015077061 | 0 |
| 0.012915984 | 0.011734275 | 0.05381384 | 0.002268456 | 0.008261074 |
| 0.655988211 | 0 | 0 | 0 | 0.007549004 |
| 0 | 0 | 0 | 0 | 0 |
| 0 | 0 | 0 | 0 | 0 |
| 0 | 0 | 0 | 0 | 0 |
| 0.003204702 | 0.040746254 | 0.003183826 | 0.021528899 | 0 |
| 0.204540489 | 0 | 0.020665229 | 0.111582923 | 0 |
| 0 | 0 | 0 | 0 | 0 |
| 0 | 0 | 0.003946741 | 0 | 0 |
| 0.011383393 | 0.05748703 | 0.000965685 | 0.005756003 | 0.054047684 |
| 0 | 0.001603887 | 0 | 0.001841766 | 0 |
| 0 | 0 | 0.61062369 | 0 | 0 |
| 0 | 0 | 0 | 0 | 0 |
| 0 | 0.002598805 | 0 | 0 | 0 |
| 0 | 0 | 0 | 0 | 0 |
| 0.099311543 | 0 | 0 | 0 | 0.079156549 |
| 0.06049433 | 0.012603634 | 0.008416317 | 0.096835744 | 0.073591331 |
| 0 | 0 | 0 | 0 | 0 |
| 0 | 0 | 0 | 0 | 0 |
| 0 | 0.00235195 | 0 | 0 | 0 |
| 0 | 0 | 0 | 0 | 0 |
| 0.019425981 | 0 | 0 | 0 | 0 |
| 0 | 0 | 0 | 0 | 0 |
| 0.17634552 | 0.262880259 | 0 | 0 | 0.006900033 |
| 0.148620548 | 0.115928799 | 0.091370788 | 0.070998723 | 0.06437656 |
| 0 | 0 | 0 | 0 | 0 |
| 0 | 0 | 0 | 0 | 0 |
| 0.009388498 | 0.003717998 | 0.001519386 | 0.00733134 | 0.009384755 |
| 0 | 0 | 0 | 0 | 0 |
| 0.080665919 | 0 | 0 | 0 | 0 |
| 0 | 0 | 0 | 0 | 0 |
| 0.00541027 | 0.006364494 | 0.000922898 | 0.001309755 | 0.002326712 |
| 0.000564704 | 0.001387535 | 0.000594027 | 0.005311085 | 0 |
| 0.42361942 | 0 | 0.102624435 | 0 | 0 |
| 0 | 0.376834638 | 0 | 0 | 0.496575897 |
| 0 | 0 | 0 | 0 | 0 |
| 0.002966602 | 0 | 0 | 0.013490648 | 0 |

| | | | | |
|---|---|---|---|---|
| 0.03472716 | 0 | 0 | 0 | 0 |
| 0 | 0.613017825 | 0 | 0 | 0 |
| 0 | 0 | 0 | 0 | 0 |
| 0.314722021 | 0.162121211 | 0 | 0 | 0.036583417 |
| 0 | 0 | 0 | 0 | 0 |
| 0 | 0 | 0 | 0 | 0.02432127 |
| 0 | 0 | 0 | 0 | 0 |
| 0.01234078 | 0.036579576 | 0 | 0 | 0.002181853 |
| 0.231727087 | 1.991915328 | 0.118397615 | 0 | 0.228550737 |
| 0 | 0.133997198 | 0 | 0 | 0.150888651 |
| 0 | 0 | 0 | 0 | 0 |
| 0.005384599 | 0.004811092 | 0.003432849 | 0 | 0.012722159 |
| 0 | 0 | 0 | 0 | 0 |
| 2.512739158 | 0 | 0 | 0 | 0.707147043 |
| 0.069613024 | 0 | 0.145039132 | 0 | 0.020568211 |
| 0 | 0 | 0 | 0 | 0 |
| 0 | 2.640387427 | 0 | 0 | 0.004052362 |
| 0 | 0 | 0 | 0 | 0.164930968 |
| 0.089319914 | 0 | 0 | 0 | 0 |
| 0 | 0.00184133 | 0 | 0 | 0 |
| 0.137427708 | 0 | 0 | 0 | 0 |
| 0.009543937 | 0 | 0 | 0 | 0 |
| 0 | 0 | 0 | 0 | 0.004662074 |
| 1.77871335 | 0 | 0.117743757 | 0.063780382 | 0.122486402 |
| 0 | 0 | 0 | 0.019430998 | 0 |
| 0 | 0.498270032 | 0 | 0 | 0.84978107 |
| 0 | 0 | 0 | 0 | 0 |
| 0 | 7.300782248 | 0 | 0 | 0.651684932 |
| 0 | 0 | 0 | 0 | 0 |
| 0 | 0 | 0 | 0 | 0 |
| 0 | 0.053453736 | 0 | 0 | 0.003786155 |
| 3.476236344 | 0 | 0 | 0 | 0 |
| 0.042013382 | 0 | 0 | 0.005406938 | 0 |
| 0 | 0 | 0 | 0 | 0 |
| 0 | 0 | 0 | 0 | 0 |
| 0.767588208 | 0.067228806 | 0 | 0 | 0.01209189 |
| 0 | 1.887254036 | 0 | 0 | 0.125486742 |
| 0 | 0 | 0 | 0 | 0 |
| 0.097159707 | 0 | 0 | 0 | 0 |
| 0 | 0 | 0.028134271 | 0 | 0.000650442 |
| 0.269235244 | 0.001318681 | 0.004939807 | 0.178205853 | 0.173480201 |
| 0 | 0 | 0 | 0 | 0.012655008 |
| 0.070792868 | 1.31431148 | 30.76413841 | 0.975044437 | 0.003286746 |
| 0 | 0 | 0 | 0 | 0 |
| 0 | 0 | 0 | 0 | 0 |
| 0 | 0 | 0 | 0 | 0 |
| 0 | 0 | 0 | 0 | 0 |
| 0.112525399 | 0 | 0.005740274 | 0.211695667 | 0.002994158 |

| | | | | |
|---|---|---|---|---|
| 0 | 0 | 0 | 0 | 0.130259833 |
| 0 | 0 | 0 | 0 | 0.000409164 |
| 0.01118367 | 0.002026131 | 0.377599139 | 0.378724327 | 0.002296191 |
| 0 | 0 | 0 | 0 | 0.040799064 |
| 0 | 0.006962646 | 0 | 0 | 0 |
| 0.001420978 | 0.137464821 | 0 | 0 | 0.001695797 |
| 0 | 0 | 0 | 0 | 0 |
| 0 | 0 | 0 | 0.050691596 | 0 |
| 0 | 0 | 0 | 0 | 0 |
| 0 | 0 | 0 | 0 | 0 |
| 0.062340224 | 0.005719243 | 0.025895588 | 0.052666776 | 0 |
| 3.360170267 | 0.414208736 | 0.327214395 | 1.590739449 | 1.052017014 |
| 0.015129298 | 0 | 0 | 0 | 0 |
| 0.017011165 | 0.408186121 | 0 | 0.00359531 | 0.03953782 |
| 0.391017396 | 0.010151205 | 0.148462325 | 0.58567155 | 0.002037209 |
| 0.820211048 | 3.222865542 | 0 | 0 | 0.325270222 |
| 0 | 0 | 0 | 0 | 0 |
| 0 | 0 | 0 | 0 | 0 |
| 0 | 0 | 0 | 0 | 0 |
| 0.027280717 | 0 | 0.000460385 | 0.032537671 | 0.006731898 |
| 0.261548318 | 0 | 0.000468438 | 0.147318992 | 0.4224354 |
| 0 | 0 | 26.36981831 | 23.77363912 | 0.021527829 |
| 1.038067377 | 0.0149609 | 4.184706191 | 8.140313118 | 0.524712951 |
| 0 | 0 | 0 | 0 | 0.000759671 |
| 0 | 0 | 0 | 0 | 0.105098528 |
| 0 | 1.704258899 | 0 | 0 | 1.184775674 |
| 14.47973747 | 0 | 0 | 11.48196713 | 0 |
| 0 | 0 | 0 | 0 | 0 |
| 0 | 0 | 0 | 0 | 0 |
| 0.077284188 | 0 | 0 | 0 | 0 |
| 0.004747985 | 0.003678039 | 0 | 0 | 0.005991899 |
| 0.334647518 | 0.321900437 | 0 | 0.014058671 | 0.151073371 |
| 0.016551927 | 0.001787682 | 0.008227367 | 0.118770241 | 0.004630762 |
| 0 | 0.056282571 | 0 | 0 | 0.000993182 |
| 0.009243788 | 0 | 0.00088398 | 0.008172328 | 0 |
| 0.009566535 | 0.523846748 | 0.013012874 | 0.102827402 | 0.005905199 |
| 0.003505261 | 0 | 0.005698517 | 0 | 0.000887342 |
| 0 | 0 | 0 | 0 | 0 |
| 0 | 0 | 0.010151793 | 0 | 0.846585481 |
| 0 | 0 | 0 | 0 | 0 |
| 0.006680957 | 0 | 0 | 0 | 0.01247958 |
| 0 | 0 | 0 | 0 | 0 |
| 0 | 0 | 0 | 0 | 0 |
| 0 | 0 | 0 | 0 | 0 |
| 0.012038121 | 0.026409715 | 0.002967941 | 0.20605262 | 0.010026489 |
| 0 | 0 | 0 | 0 | 0 |
| 0 | 0 | 0 | 0 | 0 |
| 0.024504632 | 0.007008613 | 0.00620559 | 0 | 0.002707765 |

| | | | | |
|---|---|---|---|---|
| 0 | 0 | 0 | 0 | 0 |
| 0 | 0 | 0 | 0 | 0 |
| 0 | 0 | 0 | 0 | 0 |
| 0.051781147 | 0.020708181 | 0 | 0 | 0.002423361 |
| 0 | 0 | 0 | 0 | 0 |
| 0 | 0 | 0 | 0 | 0.066123912 |
| 0 | 0.011828617 | 0 | 0 | 0.297149293 |
| 0 | 0 | 0.300112557 | 0 | 0.000352094 |
| 0 | 0 | 0 | 0 | 0 |
| 0 | 0 | 0 | 0 | 0 |
| 0.001606918 | 0 | 0.001878176 | 0.024468959 | 0 |
| 0 | 0 | 0 | 0 | 0 |
| 0.005002603 | 0.003292473 | 0 | 0 | 0 |
| 0.182678161 | 0.088777013 | 0 | 0 | 0.193583217 |
| 0 | 0 | 0 | 0 | 0 |
| 0 | 0 | 0 | 0 | 0 |
| 0 | 0.187938534 | 0 | 0 | 0.079236799 |
| 0 | 0.027756203 | 0.001563539 | 0.005857996 | 0 |
| 0 | 0 | 0 | 0 | 0.311586273 |
| 0.009919321 | 0.010539579 | 0.001410053 | 0 | 0 |
| 0 | 0.00222489 | 0 | 0 | 0 |
| 0 | 0 | 0 | 0 | 0 |
| 0.069403411 | 0 | 0 | 0 | 0.013906611 |
| 0 | 0 | 0 | 0 | 0 |
| 0 | 0 | 0 | 0 | 0 |
| 0 | 0 | 0 | 0 | 0 |
| 0 | 0 | 0 | 0 | 0 |
| 0 | 0 | 0 | 0 | 0 |
| 0 | 0 | 0 | 0 | 0 |
| 0 | 0 | 0 | 0 | 1.586786048 |
| 0 | 0.014610994 | 0.003254401 | 0 | 0 |
| 0 | 0 | 0 | 0 | 0 |
| 0 | 0 | 0 | 0 | 0 |
| 0 | 0 | 0 | 0 | 0 |
| 0 | 0 | 0 | 0 | 0 |
| 0 | 0 | 0 | 0 | 0 |
| 0.534965929 | 0.112381225 | 0 | 0 | 0.038960862 |
| 0.004383137 | 0 | 0.008267528 | 0.055100356 | 0 |
| 0 | 0 | 0 | 0 | 0 |
| 0.005664993 | 0 | 0 | 0 | 0 |
| 0.102953491 | 0 | 0 | 0 | 0 |
| 0 | 0 | 0 | 0 | 0 |
| 0 | 0 | 0 | 0 | 0.012064802 |
| 0.007045316 | 0.009479863 | 0 | 0 | 0.016465454 |
| 0 | 0.457834807 | 0 | 0 | 0.214370596 |
| 0 | 0 | 0 | 0 | 0 |
| 0 | 0.010686809 | 0 | 0 | 0.002856423 |
| 0 | 0 | 0 | 0 | 0 |

| | | | | |
|---|---|---|---|---|
| 0 | 0 | 0 | 0 | 0 |
| 0 | 0 | 0 | 0 | 0 |
| 0 | 0 | 0 | 0 | 1.617196783 |
| 0.012328097 | 0.566564901 | 0 | 0 | 0.155018137 |
| 0 | 0 | 0 | 0 | 0 |
| 0 | 0.672183019 | 0 | 0 | 0 |
| 0 | 0 | 0 | 0 | 0 |
| 0.704468678 | 0 | 0 | 0 | 0 |
| 0.100890266 | 0.001463386 | 0.082666686 | 0.077112877 | 0.001824282 |
| 0 | 0 | 0 | 0 | 0 |
| 0 | 0 | 0 | 0 | 0.006166341 |
| 0 | 0 | 0 | 0 | 0 |
| 0 | 0 | 0 | 0 | 0 |
| 0.000723917 | 0 | 0 | 0 | 0 |
| 2.908934413 | 0.002103256 | 0.640234635 | 0 | 0.403065855 |
| 0.001285272 | 0 | 0.180662458 | 0 | 0 |
| 0 | 0 | 0 | 0 | 0 |
| 0 | 0 | 0 | 0 | 0 |
| 0 | 0 | 0 | 0 | 0 |
| 0 | 0 | 0 | 0 | 6.525278608 |
| 0 | 0.028869386 | 0 | 0 | 0.068858573 |
| 0 | 0 | 0 | 0 | 0 |
| 0 | 0 | 0 | 0 | 0 |
| 0 | 0 | 0 | 0 | 0 |
| 0 | 0.020994983 | 0.003610057 | 0.011198087 | 0.007996131 |
| 6.489431131 | 6.787830142 | 0 | 0 | 1.074437424 |
| 0 | 1.102205638 | 0 | 0 | 0.013637278 |
| 0.000760825 | 0 | 0 | 0 | 0 |
| 0 | 0 | 0 | 0 | 0.018074047 |
| 0 | 0 | 0 | 0 | 0.011617067 |
| 0 | 0 | 0 | 0 | 0 |
| 0.034572789 | 0 | 0 | 0 | 0.041573217 |
| 0 | 0 | 0 | 0 | 0 |
| 0 | 0 | 0.00123943 | 0 | 0 |
| 0.00194138 | 0.373133142 | 0.001361459 | 0.002318581 | 0.002745892 |
| 0.002287201 | 0.003853635 | 0.015237789 | 0.06432902 | 0 |
| 0.187252701 | 0.00265123 | 0 | 0 | 0.434040554 |
| 1.159091194 | 0.187594543 | 0 | 0.14651094 | 0.120682624 |
| 0 | 0 | 0 | 0 | 0 |
| 0 | 0 | 0 | 0 | 0 |
| 0 | 0 | 0 | 0 | 0 |
| 0.011239385 | 0 | 0 | 0 | 0.004952529 |
| 0.275252447 | 0.498812968 | 0.038357624 | 0.891199016 | 0.127482347 |
| 0 | 0.009452753 | 0 | 0 | 0.842939498 |
| 0 | 0 | 0 | 0 | 0 |
| 0.011556458 | 0 | 0.000911741 | 0 | 0.006780818 |
| 0 | 0 | 0 | 0 | 0 |
| 1.256078372 | 0.001615616 | 0 | 0.115436775 | 0.221362905 |

| | | | | |
|---|---|---|---|---|
| 0.08288866 | 0 | 0 | 0 | 0 |
| 0 | 0 | 0 | 0 | 2.139444012 |
| 0 | 0 | 0 | 0 | 0.079726279 |
| 0 | 0 | 0 | 0 | 0 |
| 0 | 0 | 0.035407176 | 0.000765214 | 0 |
| 0 | 0 | 0 | 0 | 0.005890704 |
| 0 | 0 | 0 | 0 | 2.232307114 |
| 0 | 0 | 0 | 0 | 0 |
| 0 | 0 | 0 | 0 | 0 |
| 0 | 0 | 0 | 0 | 1.874555707 |
| 0.00169959 | 0 | 0 | 0 | 0.001456211 |
| 0 | 0.733234047 | 0.223019976 | 2.559381883 | 0.447336827 |
| 0.323769096 | 0 | 0 | 0 | 0.003291898 |
| 0 | 0 | 0 | 0 | 0 |
| 0.048609429 | 0.252193725 | 0.398259543 | 0.065133765 | 0.010690294 |
| 0 | 0 | 0 | 0 | 0 |
| 0.006460566 | 0 | 0 | 0 | 0 |
| 0 | 0 | 0.02053898 | 0.005553959 | 0 |
| 0 | 0 | 0 | 0 | 0 |
| 0 | 0 | 0 | 0 | 0.336905822 |
| 0 | 0 | 0.044496422 | 0 | 0.024185162 |
| 0 | 0 | 0 | 0 | 0.060169969 |
| 0.009936113 | 1.349215236 | 0 | 0 | 0.120453526 |
| 0 | 0 | 0 | 0 | 0 |
| 0 | 0 | 0 | 0 | 0 |
| 0 | 0 | 0 | 0 | 0 |
| 0 | 0 | 0 | 0 | 0 |
| 0 | 0 | 0 | 0 | 0 |
| 0 | 0.057228018 | 0 | 0 | 0.00042482 |
| 0 | 0 | 0 | 0 | 11.49322303 |
| 0 | 0 | 0 | 0 | 0 |
| 0.051282546 | 0.063003201 | 0.016183636 | 0.00734958 | 0.029784326 |
| 0 | 1.194231267 | 0 | 0 | 0 |
| 0.104400995 | 0.023021943 | 0.00121276 | 0 | 0.000407664 |
| 0.001825302 | 0.003960736 | 0.015884321 | 0.061979917 | 0 |
| 0.004492158 | 0.659858464 | 0 | 0 | 0.163211103 |
| 0 | 0 | 0 | 0 | 0.001406273 |
| 0 | 0 | 0 | 0 | 0.002524821 |
| 0 | 0 | 0 | 0 | 0 |
| 0.006170105 | 0 | 0 | 0 | 0 |
| 0 | 0 | 0 | 0 | 0.176370051 |
| 0 | 0 | 0 | 0 | 0.009029156 |
| 0 | 0 | 0 | 0 | 0 |
| 0 | 0 | 0 | 0 | 0.018055941 |
| 0.0127502 | 0.008032957 | 0.041440475 | 0.042314888 | 0.000650261 |
| 0 | 0 | 0 | 0 | 0 |
| 0 | 0 | 0 | 0 | 0.195210622 |
| 0.012368624 | 0 | 0 | 0 | 0.000615031 |

| | | | | |
|---|---|---|---|---|
| 0 | 0 | 0 | 0 | 0 |
| 0 | 0 | 0 | 0 | 0 |
| 0 | 0 | 0.00444604 | 0.004997294 | 0 |
| 0 | 0 | 0 | 0 | 0 |
| 0.100815017 | 0 | 0.004869641 | 0 | 0 |
| 0 | 0 | 0 | 0 | 0 |
| 0 | 0 | 0 | 0 | 0 |
| 0.014050432 | 0 | 0 | 0 | 0.058814356 |
| 0 | 0 | 0 | 0 | 0 |
| 0 | 0 | 0 | 0 | 0.001500015 |
| 0 | 0 | 0 | 0 | 0 |
| 0 | 0 | 0 | 0 | 0 |
| 0 | 0 | 0 | 0 | 0 |
| 0 | 0 | 0 | 0 | 0.703548017 |
| 0 | 0 | 0 | 0 | 0 |
| 0 | 0 | 0 | 0 | 0 |
| 0 | 0 | 0 | 0 | 4.325523309 |
| 0 | 0 | 0 | 0 | 0.007522417 |
| 0 | 0 | 0 | 7.203353551 | 1.472746382 |
| 0 | 0 | 0 | 0 | 0 |
| 0 | 0 | 0 | 0 | 0.000855479 |
| 0 | 0 | 0 | 0 | 0 |
| 0 | 0 | 0 | 0 | 0 |
| 0 | 0 | 0 | 0 | 0 |
| 0 | 0 | 0 | 0 | 0 |
| 0.00136961 | 0 | 0.001920972 | 0.029170319 | 0 |
| 0 | 0 | 0 | 0 | 0 |
| 0.078051308 | 0.009202092 | 0 | 0.313003979 | 3.187365987 |
| 0 | 0 | 0 | 0 | 0.032735812 |
| 0 | 0.110168878 | 0 | 0 | 2.105848017 |
| 0 | 0 | 0 | 0 | 0 |
| 0 | 0 | 0 | 0 | 0 |
| 0 | 0 | 0 | 0 | 0.002127265 |
| 0 | 0 | 0 | 0 | 0 |
| 0.005262645 | 0.001791287 | 0 | 0.138987547 | 0.000507511 |
| 0 | 0 | 0 | 0 | 0 |
| 0 | 0 | 0 | 0 | 0.985129127 |
| 0.000723825 | 0 | 0.00418776 | 0.001296692 | 0 |
| 0 | 0 | 0 | 0 | 0 |
| 0 | 0 | 0 | 0 | 0 |
| 3.854681494 | 5.405965884 | 1.716699526 | 1.492837679 | 1.456976249 |
| 0.607862987 | 0.042962011 | 0 | 0.043306537 | 0.235537742 |
| 0 | 0.549876719 | 0 | 0 | 0.021522701 |
| 0.282731553 | 0.004283864 | 0 | 0 | 0 |
| 0 | 0 | 0 | 0 | 0 |
| 0 | 0 | 0 | 0 | 1.387773935 |
| 0 | 0 | 0 | 0 | 0.003920993 |
| 0.001005319 | 0 | 0.000906447 | 0.00475971 | 0 |

| | | | | |
|---|---|---|---|---|
| 0 | 0 | 0 | 0 | 0 |
| 0.028214888 | 0 | 0 | 0.002381718 | 0.000587639 |
| 0 | 0 | 0 | 0 | 0.002882581 |
| 0 | 0 | 0 | 0 | 0 |
| 3.414638476 | 8.909589562 | 0.599859932 | 0.646126472 | 0.408770866 |
| 0 | 0 | 0 | 0 | 0 |
| 0 | 0 | 0 | 0 | 0 |
| 0 | 0 | 0 | 0 | 0.004610565 |
| 0.230401459 | 0.013668886 | 0.029791385 | 0.182857693 | 0 |
| 0 | 0 | 0 | 0 | 0 |
| 0 | 0 | 0.015976073 | 0 | 0 |
| 0 | 0 | 0 | 0 | 0 |
| 0.283916243 | 0.216649353 | 0.022044593 | 0 | 0.047506455 |
| 0.004058676 | 0.011892763 | 0.001670646 | 0.082824876 | 0.003088692 |
| 0 | 2.477968015 | 0 | 0 | 0.009024516 |
| 1.433137343 | 0.345025398 | 0.026809097 | 0 | 0.273271452 |
| 0.006718905 | 0 | 0 | 0.00341947 | 0.722623517 |
| 0 | 0 | 0 | 0 | 0 |
| 0 | 0 | 0 | 0 | 0 |
| 0.232643296 | 0 | 0 | 0 | 0 |
| 0 | 0 | 0 | 0 | 0 |
| 0 | 0 | 0 | 0 | 0 |
| 0 | 0 | 0 | 0 | 0 |
| 0 | 0 | 0 | 0 | 0 |
| 0 | 0 | 0 | 0 | 0 |
| 0 | 0 | 0 | 0 | 0 |
| 0.031734657 | 0.147840723 | 0 | 0 | 0.00612716 |
| 0 | 0 | 0 | 0.005636593 | 0 |
| 0 | 0 | 0 | 0 | 0 |
| 0 | 0 | 0 | 0 | 0 |
| 0 | 0 | 0 | 0 | 0 |
| 0.00087607 | 0 | 0 | 0 | 0.000497859 |
| 0 | 0 | 0 | 0 | 0 |
| 0 | 0.07100798 | 0 | 0 | 0.017369065 |
| 0 | 0.762372123 | 0 | 0 | 0.016552613 |
| 0 | 0 | 0.018049714 | 0 | 0 |
| 0 | 0.00099494 | 0 | 0 | 0.001973219 |
| 0 | 0 | 0 | 0 | 0 |
| 0 | 0 | 0 | 0 | 0 |
| 0 | 0 | 0 | 0 | 0 |
| 0 | 0.652715346 | 0 | 0 | 0.078193573 |
| 0 | 0 | 0 | 0 | 0 |
| 0 | 0 | 0 | 0 | 0.075778055 |
| 0 | 0 | 0 | 0 | 0.011817495 |
| 0 | 0 | 0 | 0.017612846 | 0 |
| 0 | 0 | 0 | 0 | 0 |
| 0 | 0 | 0 | 0 | 0 |
| 0 | 0 | 0 | 0 | 0 |

| | | | | |
|---|---|---|---|---|
| 0 | 0 | 0 | 0 | 0.011224453 |
| 0 | 0 | 0 | 0 | 0 |
| 0.027149597 | 0 | 0 | 0 | 0 |
| 0.00458261 | 0 | 0 | 0 | 0 |
| 0 | 0.011628601 | 0 | 0 | 0.312538487 |
| 0 | 0 | 0 | 0 | 0 |
| 0.363004935 | 0.001711053 | 0 | 0.00395694 | 0.147615211 |
| 0 | 0 | 0 | 0 | 0 |
| 0.003010592 | 0 | 0 | 0 | 0 |
| 0.094293847 | 0 | 0.002353665 | 0 | 0 |
| 0 | 0 | 0 | 0 | 0.008083267 |
| 0.097780836 | 0 | 0 | 0 | 0.013460362 |
| 0 | 0 | 0 | 0 | 0 |
| 0 | 0 | 0 | 0 | 0 |
| 0 | 0 | 0 | 0 | 0 |
| 0 | 0 | 0 | 0 | 0 |
| 0 | 0 | 0 | 15.22240122 | 0 |
| 0 | 0 | 0 | 0 | 0 |
| 0 | 0 | 0 | 0 | 0 |
| 0 | 0 | 0 | 0 | 0 |
| 0.017113265 | 0 | 0.001373026 | 0.036373228 | 0.775381562 |
| 0 | 0 | 0 | 0 | 0 |
| 0.032312932 | 0.050310426 | 0 | 0 | 0.011199613 |
| 0.009103523 | 0.016177056 | 0.000748143 | 0.010829823 | 0.001584356 |
| 0 | 0 | 0 | 0 | 0 |
| 0 | 0 | 0 | 0 | 0 |
| 0 | 0 | 0 | 0 | 1.320760496 |
| 0 | 0 | 0 | 0 | 0 |
| 0 | 0 | 0 | 0 | 0 |
| 0 | 0 | 0.135024898 | 0 | 0 |
| 0 | 0 | 0 | 0 | 0.121842497 |
| 0 | 0 | 0 | 0 | 0 |
| 0 | 0 | 0 | 0 | 0.011285793 |
| 0 | 0 | 0 | 0.011973796 | 0 |
| 0 | 0 | 0 | 0 | 0.024368922 |
| 0 | 0 | 0 | 0 | 0.750175068 |
| 0.020111995 | 0.004248255 | 0.00711452 | 0.130726464 | 0.003681675 |
| 0 | 0 | 0 | 0 | 0 |
| 0 | 0.305116705 | 0 | 0 | 0.053759804 |
| 0 | 1.410470245 | 0 | 0 | 0.041712631 |
| 0 | 0 | 0 | 0 | 0 |
| 0 | 0 | 0 | 0 | 0 |
| 0 | 0 | 0 | 0 | 0 |
| 0.034477136 | 0 | 0 | 0 | 0.027565168 |
| 0 | 0 | 0 | 0 | 0 |
| 0 | 0 | 0 | 0 | 0 |
| 0 | 0 | 0 | 0 | 0 |
| 0 | 0 | 0 | 0 | 0 |

| | | | | |
|---|---|---|---|---|
| 0.013469106 | 0 | 0 | 0.138605417 | 0 |
| 0 | 1.575266242 | 0 | 0 | 0.002194293 |
| 0 | 0 | 0 | 0 | 0 |
| 0 | 0 | 0 | 0 | 0 |
| 0 | 0 | 0 | 0 | 0 |
| 0 | 0 | 0 | 0 | 0.012599676 |
| 0 | 0 | 0 | 0 | 0 |
| 2.704026068 | 0.381955708 | 0 | 0.190177553 | 0.04751272 |
| 0.002162228 | 0.384883772 | 0.537793901 | 0.46826405 | 0.089645224 |
| 0.153036623 | 0.000704499 | 0.091839615 | 0.155055644 | 0 |
| 0 | 0 | 0 | 0 | 0.513113844 |
| 0 | 0 | 0 | 0 | 0 |
| 0 | 0 | 0 | 0 | 0.0617684 |
| 0 | 0 | 0 | 0.141150907 | 0 |
| 0 | 0 | 0 | 0 | 0 |
| 0 | 0 | 0 | 0 | 0.041700204 |
| 0 | 0 | 0 | 0 | 0 |
| 0 | 0 | 0 | 0 | 0 |
| 0 | 0 | 0 | 0 | 0 |
| 0.054089227 | 0 | 0 | 0 | 0 |
| 0 | 0 | 0 | 0 | 0 |
| 0 | 0 | 0 | 0 | 0 |
| 0 | 0.002535872 | 0 | 0 | 0.013076123 |
| 0 | 0 | 0 | 0 | 0 |
| 0 | 0 | 0 | 0 | 0.019662209 |
| 0 | 0 | 0 | 0 | 0 |
| 0 | 0 | 0 | 0 | 0 |
| 0 | 0 | 0 | 0 | 0 |
| 0 | 0 | 0 | 0 | 0 |
| 0 | 0 | 0 | 0 | 0 |
| 0 | 0 | 0 | 0 | 0 |
| 0 | 0 | 0 | 0 | 0 |
| 0 | 0 | 0 | 0 | 0 |
| 0 | 0 | 0 | 0 | 0.00455089 |
| 0.206422285 | 0.013268644 | 0 | 0 | 0 |
| 0 | 0.077691519 | 0 | 0 | 0 |
| 0 | 0 | 0 | 0 | 0 |
| 0 | 0 | 0 | 0 | 0 |
| 0 | 0 | 0 | 0 | 0 |
| 0 | 0 | 0 | 0.00068433 | 0 |
| 0 | 0 | 0 | 0 | 0 |
| 0.75746093 | 0 | 0.015753452 | 0 | 0.032358016 |
| 0 | 0 | 0 | 0 | 0 |
| 0.003049493 | 0 | 0 | 0 | 0 |
| 0.004615898 | 0 | 0 | 0 | 0 |
| 0.000408299 | 0 | 0 | 0 | 0.000974529 |
| 0 | 0 | 0 | 0 | 0 |
| 0 | 0 | 0 | 0 | 0 |

| | | | | |
|---|---|---|---|---|
| 0 | 0 | 0 | 0 | 0 |
| 0.002762831 | 0 | 0.032986443 | 0.014105944 | 0 |
| 0 | 0 | 0 | 0 | 0 |
| 0 | 0 | 0 | 0 | 0 |
| 0.182518549 | 5.17902573 | 0.005485601 | 0 | 0.486959425 |
| 0 | 0 | 0 | 0 | 0.047044737 |
| 0 | 0 | 0 | 0 | 0 |
| 0.01304496 | 0.003007084 | 0.011674225 | 0.020055742 | 0.002168659 |
| 0.006415253 | 0 | 0 | 0 | 0.01159996 |
| 0 | 0 | 0 | 0 | 0 |
| 0 | 0 | 0 | 0 | 0 |
| 0 | 0 | 0 | 0 | 0 |
| 0 | 0 | 0 | 0 | 0 |
| 0 | 0 | 0 | 0 | 0 |
| 0.007264316 | 0 | 0 | 0 | 0 |
| 0.04694381 | 0 | 0 | 0.012100309 | 0.000671736 |
| 0 | 0 | 0 | 0 | 0 |
| 0 | 0 | 0 | 0 | 0 |
| 0 | 0 | 0 | 0 | 0 |
| 0 | 0 | 0 | 0 | 0 |
| 0 | 0 | 0 | 0 | 0 |
| 0 | 0 | 0 | 0 | 0 |
| 0 | 0 | 0 | 0 | 0 |
| 0.001179483 | 0 | 0 | 0.003924099 | 0 |
| 0 | 0.131556371 | 0 | 0 | 0.161495672 |
| 0 | 0 | 0 | 0 | 0 |
| 0.052089236 | 0.001298277 | 0 | 0.451528096 | 0.010176647 |
| 0.003407733 | 0 | 0 | 0.002761677 | 0.000645523 |
| 0 | 0 | 0 | 0 | 0 |
| 0 | 0 | 0 | 0 | 0 |
| 0 | 0 | 0 | 0 | 0 |
| 0 | 0 | 0 | 0 | 0 |
| 0 | 0 | 0 | 0 | 0 |
| 0 | 0 | 0 | 0 | 0 |
| 0 | 0 | 0 | 0 | 0 |
| 0 | 0 | 0 | 0 | 0 |
| 0 | 0 | 0 | 0 | 0 |
| 0.374260044 | 0.012426956 | 0 | 0 | 0.64217436 |
| 0 | 0 | 0 | 0 | 0 |
| 0 | 0 | 0 | 0 | 0 |
| 0 | 0 | 0 | 0 | 0 |
| 0 | 1.527214861 | 0 | 0 | 0.004613861 |
| 0 | 0 | 0 | 0 | 0 |
| 0 | 0 | 0 | 0 | 0 |
| 0 | 0 | 0 | 0 | 0 |
| 0 | 0 | 0 | 0 | 0 |
| 0 | 0 | 0 | 0 | 0 |
| 0 | 0 | 0 | 0 | 0 |

| | | | | |
|---|---|---|---|---|
| 0.230434452 | 0.289764552 | 0 | 0 | 0.036551473 |
| 0 | 0 | 0 | 0 | 0 |
| 0 | 0 | 0 | 0 | 0 |
| 0 | 0 | 0 | 0 | 0 |
| 0 | 0 | 0 | 0 | 0 |
| 0 | 0 | 0.916332853 | 0.07083728 | 0 |
| 0 | 0 | 0 | 0 | 0 |
| 0.002772467 | 0 | 0 | 0 | 0 |
| 0 | 0 | 0 | 0 | 0 |
| 0 | 0 | 0 | 0 | 0 |
| 0 | 0 | 0 | 0 | 0 |
| 0.006523772 | 0 | 0 | 0 | 0 |
| 0.008640557 | 0 | 0 | 0 | 0.030720136 |
| 0 | 0 | 0 | 0 | 0 |
| 0 | 0 | 0 | 0 | 0 |
| 0 | 0 | 0 | 0 | 0 |
| 0 | 0 | 0 | 0 | 0 |
| 0.077002956 | 0.2320903 | 0 | 0 | 0 |
| 0 | 0 | 0 | 0 | 0 |
| 0 | 0 | 0 | 0 | 0 |
| 0.003355864 | 0 | 0.000706024 | 0.076049763 | 0 |
| 0 | 0 | 0 | 0 | 0 |
| 0.014198011 | 0.001487674 | 0.010476973 | 0.004365696 | 0 |
| 0 | 0 | 0 | 0 | 2.247960698 |
| 0.003045422 | 0 | 0 | 0 | 0.047247344 |
| 0 | 0 | 0 | 0 | 0 |
| 0 | 0 | 0 | 0 | 0 |
| 0 | 0 | 0 | 0 | 0 |
| 0 | 0 | 0 | 0.001150224 | 0 |
| 0 | 0 | 0 | 0 | 0 |
| 0 | 0 | 0 | 0 | 0 |
| 0.025356037 | 0 | 0 | 0 | 0.014748539 |
| 0.002182988 | 0 | 0.055738478 | 0.332409273 | 0.001578897 |
| 0 | 0 | 0 | 0 | 0 |
| 0.28917297 | 0 | 0 | 0 | 0.004660644 |
| 0.122941422 | 0 | 0.031326207 | 0 | 0 |
| 0 | 0 | 0 | 0 | 0 |
| 0 | 0.365335484 | 0 | 0 | 0.011186828 |
| 0.012893891 | 0 | 0 | 0.015839087 | 0 |
| 0 | 0 | 0 | 0 | 0 |
| 0 | 0 | 0 | 0 | 0 |
| 0 | 0 | 0 | 0 | 0.00821907 |
| 0.018738337 | 0.226807821 | 0.011101793 | 0 | 0.032558225 |
| 0 | 0 | 0 | 0 | 0 |
| 0 | 0 | 0 | 0 | 0 |
| 0.001197361 | 0 | 0 | 0 | 0.013744972 |
| 0 | 0 | 0 | 0 | 0 |
| 0 | 0 | 0 | 0 | 0 |

| | | | | |
|---|---|---|---|---|
| 0 | 0 | 0 | 0 | 0 |
| 0 | 0 | 0 | 0 | 0 |
| 0 | 0 | 0 | 0 | 0 |
| 0 | 0 | 0 | 0 | 0 |
| 0 | 0 | 0 | 0 | 0.003281524 |
| 0 | 0 | 0 | 0 | 0 |
| 0 | 0 | 0 | 0 | 0 |
| 0 | 0 | 0 | 0.034940701 | 0 |
| 0 | 0 | 0 | 0 | 0.063751308 |
| 0 | 0 | 0 | 0 | 0 |
| 0 | 0 | 0 | 0 | 0 |
| 0 | 0 | 0 | 0 | 0 |
| 0 | 0 | 0 | 0 | 0 |
| 0 | 0 | 0 | 0 | 0 |
| 0 | 0 | 0 | 0 | 0 |
| 0 | 0 | 0.01440517 | 0 | 0 |
| 0 | 0 | 0 | 0 | 0 |
| 0 | 0 | 0 | 0 | 0 |
| 0 | 0 | 0 | 0 | 0 |
| 0.773823263 | 1.194693542 | 0 | 0 | 1.650097468 |
| 0 | 0.195223078 | 0 | 0 | 0 |
| 0 | 0 | 0 | 0 | 0 |
| 0 | 0 | 0 | 0 | 0 |
| 0 | 0 | 0 | 0 | 0 |
| 0.001639943 | 0 | 0 | 0 | 0 |
| 0 | 0 | 0 | 0 | 0 |
| 0 | 0 | 0 | 0 | 0 |
| 0 | 0 | 0 | 0 | 0 |
| 0 | 0 | 0 | 0 | 0 |
| 0 | 0 | 0 | 0 | 0.013150017 |
| 0.945219123 | 0.351319819 | 0 | 0 | 0.062546677 |
| 0 | 0 | 0 | 0 | 0 |
| 0 | 0 | 0 | 0 | 0.049702209 |
| 0 | 0 | 0 | 0 | 0 |
| 0 | 0 | 0 | 0 | 0 |
| 0 | 0 | 0 | 0 | 0 |
| 0 | 0 | 0 | 0 | 0 |
| 0 | 0 | 0 | 0 | 0 |
| 0 | 0.019619203 | 0 | 0 | 0.01023279 |
| 0 | 0 | 0 | 0 | 0 |
| 0 | 0 | 0 | 0 | 0 |
| 0 | 0 | 0 | 0 | 0.639999942 |
| 0 | 0 | 0 | 0 | 0.005334388 |
| 0.003147634 | 0 | 0 | 0 | 0 |
| 0 | 0 | 0 | 0 | 0 |
| 0 | 0 | 0 | 0 | 0 |
| 0 | 0 | 0.230343553 | 0.399430355 | 0 |
| 0.002018986 | 0 | 0 | 0 | 0 |

| | | | | |
|---|---|---|---|---|
| 0 | 0 | 0 | 0.026319434 | 0 |
| 0 | 0 | 0 | 0 | 0 |
| 0 | 0 | 0 | 0 | 0 |
| 0 | 0 | 0 | 0 | 0 |
| 0.003579344 | 0.167706316 | 0.063171781 | 0 | 0.10283468 |
| 0 | 0 | 0 | 0 | 0 |
| 0 | 0 | 0 | 0 | 0.040172966 |
| 0 | 0 | 0 | 0 | 0 |
| 0 | 0 | 0 | 0 | 0 |
| 0 | 0 | 0 | 0 | 0 |
| 0 | 0 | 0 | 0 | 0.032674555 |
| 0.006441812 | 0.003359451 | 0.0087124 | 0.024728867 | 0.001171454 |
| 0.007621935 | 0 | 0 | 0 | 0 |
| 0 | 0 | 0 | 0 | 0 |
| 0.005189631 | 0 | 0 | 0 | 0 |
| 0 | 0 | 0 | 0 | 0 |
| 0 | 0 | 0.001042357 | 0 | 0 |
| 0 | 0 | 0 | 0 | 0 |
| 0.009616189 | 0 | 0.085322223 | 0.00112349 | 0.011309656 |
| 0.048221601 | 0.003321422 | 0 | 0.355487525 | 0.021716126 |
| 0 | 0 | 0 | 0 | 0 |
| 0 | 0.644258739 | 0 | 0 | 0 |
| 0 | 0 | 0 | 0 | 0 |
| 0 | 0 | 0 | 0 | 0 |
| 0 | 0 | 0 | 0 | 0 |
| 0 | 0 | 0 | 0 | 0 |
| 0.048703727 | 0 | 0.013651834 | 0.018285916 | 0.000464057 |
| 0 | 0 | 0 | 0 | 0 |
| 0 | 0 | 0 | 0 | 0 |
| 0 | 0 | 0 | 0 | 0 |
| 0.001322353 | 0 | 0 | 0 | 0 |
| 0 | 0 | 0 | 0 | 0 |
| 0 | 0 | 0 | 0 | 0 |
| 0 | 0 | 0 | 0 | 0 |
| 0 | 0 | 0 | 0 | 0.067341136 |
| 0 | 0 | 0 | 0 | 0 |
| 0 | 0 | 0 | 0 | 0 |
| 0.02345843 | 0.010761591 | 0.33020139 | 0.035887204 | 0.002771815 |
| 0 | 0.873327009 | 0 | 0 | 0.001555832 |
| 0.011789673 | 0 | 0 | 0 | 0.015535419 |
| 0 | 0 | 0.033294024 | 0.009865405 | 0 |
| 0.005210572 | 0 | 0 | 0 | 0.000829107 |
| 0 | 0 | 0 | 0 | 0 |
| 0 | 0 | 0.14147767 | 0.070029398 | 0 |
| 0 | 0 | 0 | 0 | 0 |
| 0 | 0 | 0 | 0.069363839 | 0.013595979 |
| 0 | 0 | 0 | 0 | 0 |
| 0 | 0 | 0 | 0 | 0.010880797 |

| | | | | |
|---|---|---|---|---|
| 0 | 0 | 0 | 0 | 0 |
| 0 | 0 | 0 | 0 | 0.002342038 |
| 0 | 0 | 0 | 0 | 0 |
| 0.00090237 | 0 | 0 | 0.000646618 | 0 |
| 0 | 0.004700414 | 0 | 0 | 0 |
| 0 | 0.003033168 | 0.006362896 | 0.007159557 | 0 |
| 0 | 0 | 0 | 0 | 0.007524288 |
| 0 | 0 | 0.004602741 | 0 | 0 |
| 0.20584563 | 0 | 0 | 0 | 0 |
| 0 | 0 | 0 | 0 | 0.017255804 |
| 0 | 0 | 0 | 0 | 0 |
| 0 | 0 | 0 | 0 | 0 |
| 0 | 0.535273807 | 0 | 0 | 0.006208689 |
| 0 | 0 | 0 | 0 | 0 |
| 0.000790748 | 0 | 0 | 0 | 0 |
| 0 | 0 | 0 | 0 | 0 |
| 0 | 0 | 0 | 0 | 0 |
| 0 | 0 | 0 | 0 | 0 |
| 0 | 0 | 0 | 0 | 0 |
| 0 | 0 | 0 | 0 | 0 |
| 0 | 0 | 0 | 0 | 0 |
| 0 | 0 | 0 | 0 | 0 |
| 0.014487151 | 0 | 0.009037327 | 0.009506005 | 0 |
| 0 | 0.00502385 | 0 | 0 | 0.133163936 |
| 0 | 0 | 0 | 0 | 0 |
| 0 | 0 | 0 | 0 | 0.013782014 |
| 0 | 0 | 0 | 0 | 0.002304856 |
| 0 | 0 | 0 | 0 | 0 |
| 0 | 0 | 0 | 0 | 0 |
| 0 | 0 | 0 | 0 | 0 |
| 0 | 0.006856713 | 0 | 0 | 0.039450901 |
| 0 | 0 | 0 | 0 | 0.000462237 |
| 0 | 0 | 0 | 0 | 0 |
| 0 | 0 | 0 | 0 | 0 |
| 0 | 0 | 0 | 0 | 0.050238538 |
| 0 | 0 | 0 | 0.01248706 | 0 |
| 0 | 0 | 0 | 0 | 0 |
| 0 | 0 | 0 | 0 | 0 |
| 0.004608522 | 0.002008127 | 0.001170165 | 0 | 0.00050573 |
| 0 | 0 | 0 | 0 | 0 |
| 0 | 0 | 0 | 0 | 0 |
| 0 | 0 | 0 | 0 | 0 |
| 0 | 0 | 0 | 0 | 13.38447884 |
| 0 | 0 | 0 | 0 | 0 |
| 0 | 0 | 0 | 0 | 0.000585566 |
| 0.164706241 | 0.25736675 | 0.021620582 | 0 | 0.431054129 |
| 0 | 0 | 0.136540023 | 0 | 0 |
| 0 | 0 | 0 | 0 | 0 |

| | | | | |
|---|---|---|---|---|
| 0 | 0 | 0 | 0 | 0 |
| 0 | 0 | 0 | 0 | 0 |
| 0 | 0.011880347 | 0 | 0 | 0.052714421 |
| 0 | 0 | 0 | 0 | 0 |
| 0 | 0 | 0 | 0 | 0 |
| 0 | 0 | 0 | 0 | 0 |
| 0 | 0 | 0 | 0.022282166 | 0 |
| 0 | 0 | 0 | 0 | 0 |
| 0 | 0 | 0 | 0 | 0.006083155 |
| 0 | 0 | 0 | 0 | 0 |
| 0 | 0 | 0 | 0.000808349 | 0 |
| 0 | 0 | 0 | 0.001206868 | 0 |
| 0 | 0 | 0 | 0 | 0 |
| 0 | 0 | 0 | 0 | 0 |
| 0 | 0 | 0 | 0 | 0 |
| 0 | 0 | 0 | 0 | 0 |
| 0 | 0 | 0 | 0 | 0 |
| 0 | 0 | 0 | 0 | 0 |
| 0 | 0 | 0 | 0 | 0 |
| 0 | 0 | 0.008452999 | 0 | 0 |
| 0 | 0 | 0 | 0 | 0 |
| 0 | 0 | 0 | 0 | 0 |
| 0 | 0 | 0 | 0 | 0 |
| 0 | 0 | 0 | 0 | 0 |
| 0 | 0 | 0 | 0 | 0 |
| 0 | 0 | 0 | 0 | 0 |
| 0 | 0 | 0 | 0 | 0 |
| 0 | 0.006907228 | 0 | 0 | 0.005268765 |
| 0 | 0 | 0 | 0 | 0 |
| 0 | 0 | 0 | 0 | 0 |
| 0.009547431 | 0.023098097 | 0.003244724 | 0.047890296 | 0.003505821 |
| 0 | 0 | 0 | 0 | 0 |
| 0.075381672 | 0.007183726 | 0.355934791 | 0.036662974 | 0.01736796 |
| 0.007842614 | 0.001957597 | 0 | 0.004683198 | 0.001525233 |
| 0 | 0 | 0 | 0 | 0 |
| 0 | 0 | 0 | 0 | 0 |
| 0 | 0 | 0 | 0 | 0 |
| 0 | 0 | 0 | 0 | 0 |
| 0 | 0 | 0 | 0 | 0 |
| 0 | 0 | 0 | 0 | 0 |
| 0 | 0 | 0 | 0 | 0 |
| 0.236137153 | 0 | 0.326475506 | 0.089481478 | 0 |
| 0 | 0.078000316 | 0.010788586 | 0.114678708 | 0.023799148 |
| 0 | 0 | 0 | 0 | 0 |
| 0 | 0 | 0 | 0 | 0 |
| 0.009880451 | 0.343696729 | 0 | 0 | 0.020438332 |
| 0.010364646 | 0 | 0 | 0 | 0 |
| 0 | 0 | 0 | 0 | 0 |

| | | | | |
|---|---|---|---|---|
| 0 | 0 | 0 | 0 | 0 |
| 0 | 0.004274358 | 0 | 0 | 0.006126336 |
| 0.006693744 | 0.008811004 | 0 | 0.003559965 | 0.007156208 |
| 0 | 0 | 0 | 0 | 0 |
| 0.527114391 | 0.022043508 | 0 | 0.040392859 | 0.003189146 |
| 0.002763793 | 0 | 0.000684072 | 0.218725509 | 0 |
| 0 | 0 | 0 | 0 | 0 |
| 0 | 0 | 0 | 0 | 0 |
| 0 | 0 | 0 | 0 | 0 |
| 0 | 0 | 0 | 0 | 0 |
| 0 | 0 | 0 | 0 | 0.000703987 |
| 0 | 0 | 0 | 0 | 0 |
| 0 | 0 | 0 | 0 | 0.006964856 |
| 0 | 0 | 0 | 0 | 0 |
| 0 | 0 | 0 | 0 | 0 |
| 0 | 0 | 0 | 0 | 0 |
| 0 | 0 | 0 | 0 | 0.555590038 |
| 0 | 0 | 0 | 0 | 0 |
| 0 | 0 | 0 | 0 | 0 |
| 0 | 0 | 0 | 0 | 0 |
| 0 | 0 | 0 | 0 | 0 |
| 0 | 0 | 0 | 0 | 0 |
| 0 | 0 | 0 | 0 | 0 |
| 0 | 0 | 0 | 0 | 0 |
| 0 | 0.007261506 | 0 | 0 | 0 |
| 0.002601354 | 0.001947974 | 0 | 0 | 0 |
| 0 | 0 | 0 | 0 | 0 |
| 0.011944113 | 0.385478982 | 0 | 0 | 0.019540864 |
| 0 | 0 | 0 | 0 | 0 |
| 0 | 0 | 0 | 0 | 0 |
| 0 | 0 | 0 | 0 | 0 |
| 0 | 0 | 0 | 0 | 0 |
| 0 | 0 | 0 | 0 | 0 |
| 0 | 0 | 0 | 0 | 0 |
| 0 | 0 | 0 | 0 | 0 |
| 0 | 0 | 0 | 0 | 0 |
| 0 | 0 | 0 | 0 | 0 |
| 0.067325164 | 0 | 0 | 0 | 0 |
| 2.789945286 | 0 | 0 | 0 | 0.706029424 |
| 0 | 0 | 0 | 0 | 0 |
| 0 | 0 | 0 | 0 | 0 |
| 0 | 0 | 0 | 0 | 0.013321759 |
| 0.010442971 | 0 | 0 | 0.103673419 | 0 |
| 0 | 0 | 0 | 0 | 0 |
| 0 | 0 | 0 | 0 | 1.53035479 |
| 0 | 0 | 0 | 0 | 0.001115166 |
| 0.008520374 | 0 | 0.002800877 | 0.00190797 | 0 |
| 0 | 0 | 0 | 0 | 0 |

| | | | | |
|---|---|---|---|---|
| 0.049839802 | 0 | 0 | 0 | 0 |
| 0.0540917 | 0.020991803 | 0.012546378 | 0.061541916 | 0 |
| 0 | 0 | 0 | 0 | 0.204745185 |
| 0 | 0 | 0 | 0 | 0 |
| 0 | 0 | 0 | 0 | 0 |
| 0 | 0 | 0 | 0 | 0 |
| 0 | 0 | 0 | 0 | 0 |
| 0 | 0 | 0 | 0 | 0 |
| 0 | 0 | 0 | 0 | 0 |
| 0 | 0 | 0 | 0 | 0 |
| 0 | 0 | 0 | 0 | 0 |
| 0 | 0 | 0 | 0 | 0 |
| 0 | 0.726043267 | 0 | 0 | 0.01610595 |
| 0 | 0 | 0 | 0 | 0 |
| 0.016965776 | 0 | 0 | 0.019699308 | 0 |
| 0 | 0 | 0 | 0 | 0 |
| 0 | 0 | 0 | 0 | 0 |
| 0 | 0 | 0 | 0 | 0 |
| 0 | 0 | 0 | 0 | 0 |
| 0 | 0 | 0 | 0 | 0 |
| 0 | 0 | 0 | 0 | 0 |
| 0.071583735 | 0 | 0 | 0.010686514 | 0.002618487 |
| 0 | 0.011318474 | 0.01163901 | 0.002973206 | 0.001383314 |
| 0 | 0 | 0 | 0.002087546 | 0.002359901 |
| 0 | 0 | 0 | 0 | 0 |
| 0 | 0 | 0 | 0 | 0 |
| 0 | 0 | 0 | 0 | 0 |
| 0 | 0 | 0 | 0 | 0 |
| 0 | 0 | 0 | 0 | 0 |
| 0 | 0 | 0 | 0 | 0.009612351 |
| 0 | 0 | 0 | 0 | 0 |
| 0 | 0 | 0 | 0 | 0 |
| 0 | 0 | 0 | 0 | 0 |
| 0 | 0 | 0 | 0 | 0 |
| 0.020544095 | 0 | 0 | 0 | 0 |
| 0 | 0 | 0 | 0 | 0 |
| 0 | 0 | 0 | 0 | 0 |
| 0 | 0 | 0 | 0 | 0 |
| 0.0012325 | 0.050761384 | 0.005402081 | 0.040847147 | 0.004249174 |
| 0 | 0 | 0 | 0 | 0 |
| 0.002026016 | 0.024825979 | 0.027318355 | 0.022106903 | 0.054804612 |
| 0 | 0 | 0 | 0 | 0 |
| 0 | 0 | 0 | 0 | 0 |
| 0 | 0 | 0 | 0 | 0 |
| 0 | 0 | 0 | 0 | 0 |
| 0.039644933 | 0.001184336 | 0.931231257 | 0.726082829 | 0 |
| 0 | 0 | 0 | 0 | 0 |
| 0 | 0 | 0 | 0 | 0 |

| | | | | |
|---|---|---|---|---|
| 0 | 0 | 0 | 0 | 0 |
| 0 | 0 | 0 | 0 | 0 |
| 0.053753474 | 0 | 0 | 0 | 0 |
| 0 | 0 | 0 | 0 | 0 |
| 0.001919214 | 0 | 0 | 0.001575824 | 0 |
| 0 | 0 | 0 | 0 | 0 |
| 0 | 0 | 0 | 0 | 0 |
| 0 | 0 | 0 | 0.01674116 | 0 |
| 0.15314043 | 0 | 0 | 0 | 0 |
| 0 | 0.02142822 | 0.003037265 | 0.009421337 | 0 |
| 0.07330012 | 0.072072862 | 0 | 0 | 0.015920546 |
| 0 | 0 | 0 | 0 | 0 |
| 0.010885353 | 0 | 0 | 0.002527841 | 0 |
| 0 | 0 | 0 | 0 | 0 |
| 0 | 0 | 0 | 0 | 0 |
| 0 | 0 | 0 | 0 | 0 |
| 0 | 0 | 0 | 0 | 0 |
| 0 | 0 | 0 | 0 | 0 |
| 0 | 0 | 0 | 0 | 0 |
| 0.266260713 | 0.210854866 | 0.199118184 | 0.546418076 | 0.001998465 |
| 0 | 0 | 0 | 0 | 0.035354822 |
| 0 | 0 | 0 | 0 | 0 |
| 0 | 0 | 0 | 0 | 0 |
| 0 | 0 | 0 | 0 | 0 |
| 0 | 0 | 0 | 0 | 0 |
| 0 | 0 | 0 | 0 | 0.014979667 |
| 0 | 0 | 0 | 0 | 0 |
| 0 | 0 | 0 | 0 | 0 |
| 0 | 0 | 0 | 0 | 0 |
| 0 | 0 | 0 | 0 | 0 |
| 0 | 0 | 0 | 0 | 0.001531005 |
| 0 | 0 | 0 | 0 | 0 |
| 0 | 0 | 0 | 0 | 0 |
| 0 | 0 | 0 | 0 | 0 |
| 0 | 0 | 0 | 0 | 0 |
| 0.002787076 | 0 | 0 | 0 | 0 |
| 0 | 0 | 0 | 0 | 0 |
| 0 | 0 | 0 | 0 | 0 |
| 0 | 0 | 0 | 0 | 0 |
| 0 | 0 | 0 | 0 | 0 |
| 0 | 0 | 0 | 0 | 0 |
| 0 | 0 | 0 | 0 | 0 |
| 0.010738208 | 0 | 0 | 0 | 0.069414553 |
| 0 | 0 | 0 | 0 | 0 |
| 0.002359263 | 0.032652192 | 0 | 0.009660535 | 0 |
| 0 | 0 | 0 | 0 | 0 |
| 0 | 0 | 0 | 0 | 0 |
| 0 | 0 | 0.021116813 | 0 | 0 |

| | | | | |
|---|---|---|---|---|
| 0 | 0 | 0 | 0 | 0 |
| 0 | 0 | 0 | 0 | 0 |
| 0 | 0 | 0 | 0 | 0 |
| 0 | 0 | 0 | 0 | 0 |
| 0 | 0 | 0 | 0 | 0 |
| 0.024657939 | 0.10280368 | 0.010587074 | 0.022875452 | 0.01461332 |
| 0 | 0 | 0 | 0 | 0 |
| 0 | 0 | 0 | 0 | 0 |
| 0 | 0 | 0 | 0 | 0 |
| 0 | 0 | 0 | 0 | 0 |
| 0 | 0 | 0 | 0 | 0 |
| 0 | 0 | 0 | 0 | 0 |
| 0 | 0 | 0 | 0 | 0 |
| 0 | 0 | 0 | 0 | 0 |
| 0 | 0 | 0 | 0 | 0 |
| 0 | 0 | 0 | 0 | 0 |
| 0.068670703 | 0 | 0 | 0 | 0.086929532 |
| 0.001549366 | 0 | 0 | 0 | 0 |
| 0 | 0 | 0 | 0 | 0 |
| 0 | 0 | 0 | 0 | 0 |
| 0 | 0 | 0 | 0 | 0.000696327 |
| 0 | 0 | 0 | 0 | 0 |
| 0 | 0 | 0 | 0 | 0 |
| 0 | 0 | 0 | 0 | 0 |
| 0 | 0 | 0 | 0 | 0 |
| 0 | 0 | 0 | 0 | 0 |
| 0 | 0 | 0 | 0 | 0 |
| 0.004418318 | 0 | 0 | 0 | 0.003398045 |
| 0 | 0 | 0 | 0 | 0 |
| 0 | 0 | 0 | 0 | 0 |
| 0 | 0 | 0 | 0 | 0 |
| 0 | 0 | 0 | 0 | 0 |
| 0 | 0 | 0 | 0 | 0 |
| 0 | 0 | 0 | 0 | 0 |
| 0 | 0 | 0 | 0 | 0 |
| 0 | 0 | 0 | 0 | 0 |
| 0 | 0 | 0 | 0 | 0 |
| 0 | 0 | 0 | 0 | 0 |
| 0 | 0 | 0 | 0 | 0.458902602 |
| 0 | 0 | 0 | 0 | 0 |
| 0 | 0 | 0 | 0 | 0.006736855 |
| 0 | 0 | 0 | 0 | 0 |
| 0 | 0 | 0 | 0 | 0 |
| 0 | 0 | 0 | 0 | 0 |
| 0 | 0.035557251 | 0 | 0 | 0 |
| 0 | 0 | 0 | 0 | 0 |
| 0 | 0 | 0 | 0 | 0 |
| 0 | 0 | 0 | 0 | 0 |

| | | | | |
|---|---|---|---|---|
| 0 | 0 | 0 | 0 | 0 |
| 0.070284022 | 0.002175373 | 0 | 0 | 0.134024001 |
| 0 | 0 | 0 | 0 | 0 |
| 0 | 0 | 0 | 0 | 0 |
| 0 | 0 | 0 | 0 | 0 |
| 0 | 0 | 0 | 0 | 0 |
| 0 | 0 | 0 | 0 | 0 |
| 0 | 0 | 0 | 0 | 0 |
| 0 | 0 | 0 | 0 | 0 |
| 0 | 0 | 0 | 0 | 0 |
| 0.000553295 | 0.043245082 | 0 | 0 | 0.005135688 |
| 0 | 0 | 0 | 0 | 0 |
| 0.646206016 | 0.168988265 | 0 | 0.041370399 | 0.20296856 |
| 0 | 0 | 0 | 0 | 0 |
| 0 | 0 | 0 | 0 | 0 |
| 0 | 0 | 0 | 0 | 0 |
| 0 | 0 | 0 | 0 | 0 |
| 0 | 0 | 0 | 0 | 0 |
| 0 | 0 | 0 | 0 | 0 |
| 0 | 0 | 0 | 0 | 0 |
| 0 | 0 | 0.094712199 | 0.023641097 | 0 |
| 0 | 0 | 0 | 0 | 0 |
| 0 | 0 | 0 | 0 | 0 |
| 0.705858573 | 1.521434703 | 0 | 0.215843714 | 0.494221601 |
| 0 | 0 | 0 | 0 | 0 |
| 0 | 0 | 0 | 0 | 0 |
| 0 | 0 | 0 | 0 | 0 |
| 0 | 0 | 0 | 0 | 0 |
| 0 | 0 | 0 | 0 | 0 |
| 0 | 0 | 0 | 0 | 0 |
| 0.007900127 | 0 | 0 | 0 | 0 |
| 0 | 0 | 0 | 0 | 0 |
| 0 | 0 | 0 | 0 | 0 |
| 0 | 0 | 0 | 0 | 0 |

| PHL018-0138_V2 | PHL018-0012_V1 | PHL018-0006_V3 | PHL018-0109_V2 | PHL018-0199_V2 |
|---|---|---|---|---|
| EYCF | EYCF | EYCF | REF | CTRL |
| 0 | 0 | 0 | 0.010559662 | 0.063380655 |
| 0.108605697 | 36.12438425 | 3.268846967 | 1.074779333 | 1.816154914 |
| 0 | 0 | 4.098461158 | 0 | 6.052957816 |
| 0 | 0 | 0 | 0 | 0 |
| 0 | 0 | 0 | 0 | 0 |
| 0 | 0 | 0 | 0 | 0 |
| 0 | 0 | 0 | 0 | 0 |
| 0 | 0 | 0 | 0 | 0 |
| 0 | 0 | 0 | 0 | 0 |
| 0 | 0 | 0 | 0 | 0 |
| 0 | 0 | 0 | 0 | 0 |
| 0 | 0 | 0 | 0 | 0 |
| 0 | 0 | 0 | 0 | 0 |
| 0.61497973 | 9.262498767 | 3.06285306 | 0.583610301 | 2.213670556 |
| 0 | 0 | 0 | 0 | 0 |
| 0 | 0 | 0 | 0 | 0 |
| 0 | 0 | 0 | 0 | 0 |
| 0 | 0 | 0 | 0 | 0 |
| 0 | 0 | 0 | 0 | 0 |
| 0 | 0 | 0 | 0 | 0 |
| 0 | 0 | 0 | 0 | 0 |
| 0 | 0 | 0 | 0 | 0 |
| 0 | 0 | 0 | 0 | 0 |
| 0 | 0 | 0 | 0 | 0 |
| 0 | 0 | 0 | 0.23196912 | 0 |
| 0 | 0 | 0 | 0 | 0 |
| 0 | 0 | 0 | 0 | 0 |
| 0 | 0 | 0 | 0 | 0 |
| 0 | 0 | 0 | 0 | 0 |
| 0 | 0 | 0 | 0 | 0 |
| 0 | 0 | 0 | 0 | 0 |
| 0 | 0 | 0 | 0 | 0 |
| 0 | 0 | 0 | 0 | 0 |
| 0 | 0 | 0 | 0 | 0 |
| 0 | 0 | 0 | 0 | 0 |
| 1.7832696 | 0 | 0 | 0 | 4.154910719 |
| 0 | 0 | 0 | 0 | 0 |
| 0 | 0 | 0 | 0 | 0 |
| 0 | 0 | 0 | 0 | 0 |
| 0 | 0 | 0 | 0 | 0 |
| 0 | 0 | 0 | 0 | 0 |
| 0 | 0.006274566 | 0.009572725 | 0 | 0.022224917 |
| 11.34172875 | 0.053333251 | 0.075045782 | 0.145133037 | 0.80836811 |
| 0 | 0.01167375 | 0.029151366 | 0.038980173 | 0.043163388 |
| 0 | 0 | 0.006235845 | 0 | 0 |
| 0 | 0 | 0 | 0 | 0 |

| | | | | |
|---|---|---|---|---|
| 0 | 0 | 0 | 0 | 0.000966018 |
| 0 | 0.113706964 | 0.000617774 | 0 | 0 |
| 0 | 0 | 0.016686068 | 0 | 0 |
| 0.074855708 | 0.006682779 | 0.569658566 | 1.542424685 | 0.546259058 |
| 0 | 0 | 0 | 0.002669969 | 0.009241277 |
| 0 | 0 | 0 | 0.002250311 | 0 |
| 0.004964187 | 0 | 0 | 0 | 0.001602737 |
| 0 | 0 | 0 | 0 | 0 |
| 0 | 0 | 0 | 0 | 0 |
| 0 | 0 | 0 | 0 | 0 |
| 0 | 0 | 0 | 0.057295497 | 0 |
| 0.205479283 | 0.299016911 | 0 | 0.013362183 | 0.034812497 |
| 0 | 0 | 0 | 0 | 0 |
| 0.009428995 | 0.167927822 | 0 | 0.006190395 | 0.003698562 |
| 0 | 0 | 0 | 0 | 0 |
| 0 | 0 | 0 | 0 | 0 |
| 0.716024133 | 0.016596626 | 0.643379832 | 0 | 0.001223874 |
| 0 | 0 | 0 | 0 | 0 |
| 0 | 0 | 0 | 0 | 0 |
| 0 | 0 | 0 | 0 | 0 |
| 0 | 0.266584661 | 0.001670328 | 0.002180447 | 0.036252302 |
| 0.00230036 | 0 | 0 | 0.130583854 | 0.012653301 |
| 0 | 0 | 0 | 0.075938752 | 0.530786508 |
| 0 | 0 | 0 | 0 | 0 |
| 0 | 0 | 0 | 0 | 0 |
| 0 | 0 | 0 | 0 | 0 |
| 0 | 0 | 0 | 0 | 0 |
| 0 | 0 | 0 | 0 | 0 |
| 0 | 0.016604917 | 0 | 0.000561213 | 0.000971234 |
| 0 | 0 | 0 | 0 | 0 |
| 0 | 2.884434826 | 0 | 0 | 0 |
| 0 | 0 | 0 | 0 | 0 |
| 0 | 0 | 0 | 0 | 0 |
| 0 | 0 | 0 | 0 | 0 |
| 0 | 0 | 0 | 0 | 0 |
| 0 | 0 | 0 | 0 | 0 |
| 0 | 0 | 0 | 0 | 0 |
| 37.30153548 | 0 | 0 | 0 | 0 |
| 0 | 0 | 0 | 0 | 0 |
| 0 | 0 | 0 | 0 | 0 |
| 0 | 0 | 0 | 0 | 0 |
| 0 | 0 | 0 | 0 | 0 |
| 0 | 0 | 0 | 0 | 0 |
| 0 | 0 | 0 | 0 | 0 |
| 0 | 0 | 0 | 1.029943046 | 0 |
| 0 | 0 | 0 | 0 | 0 |
| 0.012324124 | 0 | 0 | 0.003254234 | 0 |
| 0 | 0.123113114 | 0 | 0.001181921 | 0 |

| | | | | |
|---|---|---|---|---|
| 0 | 0 | 0 | 0 | 0 |
| 0 | 0 | 0 | 0 | 0 |
| 0 | 0 | 0 | 0 | 0 |
| 0 | 0 | 0 | 0 | 0 |
| 0 | 0.000882054 | 0.009282553 | 0.030959415 | 0.074174194 |
| 0 | 0 | 0 | 0 | 0 |
| 0 | 0 | 0 | 0 | 0 |
| 0 | 0 | 0 | 0 | 0 |
| 0 | 0.001310775 | 0 | 0.017667907 | 0 |
| 0 | 0 | 0 | 0 | 0 |
| 0 | 0 | 0 | 0 | 0 |
| 0.000860471 | 0 | 0.008623435 | 0 | 0.001135941 |
| 0 | 0 | 0 | 0 | 0 |
| 0 | 0 | 0 | 0 | 0 |
| 0 | 0 | 0 | 0 | 0 |
| 0 | 0 | 0 | 0 | 0 |
| 0 | 0 | 0 | 0 | 0 |
| 0 | 0 | 0 | 0 | 0 |
| 0 | 0 | 0 | 0.593062005 | 0.010109342 |
| 0 | 0 | 0 | 0 | 0 |
| 0 | 0 | 0 | 0 | 0 |
| 0 | 0 | 0 | 0 | 0 |
| 0 | 0 | 0 | 0 | 0 |
| 0 | 0 | 0 | 0 | 0 |
| 0 | 0 | 0 | 0 | 0 |
| 0 | 0 | 0 | 0 | 0 |
| 0.003909964 | 0 | 0 | 0 | 0 |
| 0 | 0 | 0 | 0.001902731 | 0 |
| 0 | 0 | 0 | 0 | 0 |
| 0 | 0 | 0 | 0 | 0 |
| 0.003977914 | 0 | 0 | 0.002877488 | 0.281583424 |
| 0 | 0 | 0 | 0.009605069 | 0.008192517 |
| 0 | 0 | 0 | 0 | 0 |
| 0 | 0 | 0 | 0 | 0 |
| 0 | 0 | 0 | 0 | 0 |
| 0 | 0 | 0 | 0 | 0 |
| 0 | 0 | 0 | 0 | 0 |
| 0 | 0 | 0 | 0 | 0 |
| 0.015824592 | 0 | 0 | 0 | 0 |
| 0 | 0 | 0 | 0 | 0 |
| 0 | 0 | 0 | 0 | 0 |
| 0 | 0 | 0 | 0 | 0 |
| 0 | 0 | 0 | 0.015542717 | 0 |
| 0 | 0 | 0 | 0 | 0 |
| 0 | 0 | 0 | 0.001014533 | 0 |
| 0 | 0 | 0 | 0 | 0 |
| 0 | 0 | 0 | 0 | 0 |
| 0 | 0 | 0 | 0 | 0 |

| | | | | |
|---|---|---|---|---|
| 0 | 0 | 0 | 0 | 0 |
| 0 | 0 | 0 | 0 | 0 |
| 0 | 0 | 0 | 0 | 0 |
| 0 | 0 | 0 | 0 | 0 |
| 0.00250502 | 0 | 0 | 0 | 0.060765615 |
| 0 | 0 | 0 | 0 | 0 |
| 0 | 0 | 0 | 0 | 0 |
| 0 | 0 | 0 | 0 | 0 |
| 0 | 0 | 0 | 0 | 0 |
| 0 | 0 | 0 | 0 | 0 |
| 0 | 0 | 0 | 0 | 0 |
| 0 | 0 | 0 | 0 | 0 |
| 0 | 0 | 0 | 0 | 0 |
| 0 | 0 | 0 | 0 | 0 |
| 0 | 0 | 0 | 0 | 0 |
| 0 | 0 | 0 | 0 | 0 |
| 0 | 0 | 0 | 0 | 0 |
| 0 | 0 | 0 | 0 | 0 |
| 0 | 0 | 0 | 0 | 0 |
| 0 | 0 | 0 | 0 | 0 |
| 0 | 0 | 0 | 0 | 0 |
| 0 | 0 | 0 | 0 | 0 |
| 0 | 0 | 0 | 0 | 0 |
| 0 | 0 | 0 | 0 | 0 |
| 0.01653614 | 0 | 0 | 0 | 0 |
| 0 | 0 | 0 | 0 | 0 |
| 0 | 0 | 0 | 0.001043368 | 0 |
| 0 | 0 | 0 | 0 | 0 |
| 0 | 0 | 0 | 0 | 0 |
| 0 | 0 | 0 | 0 | 0 |
| 0 | 0 | 0 | 0 | 0 |
| 0 | 0 | 0 | 0 | 0 |
| 0 | 0 | 0 | 0 | 0 |
| 0 | 0 | 0 | 0 | 0 |
| 0 | 0 | 0 | 0 | 0 |
| 0 | 0 | 0 | 0 | 0 |
| 0 | 0 | 0 | 0 | 0 |
| 0 | 0 | 0 | 0 | 0 |
| 0 | 0 | 0 | 0 | 0 |
| 0 | 0 | 0 | 0 | 0 |
| 0 | 0 | 0 | 0 | 0 |
| 0 | 0 | 0 | 0 | 0 |
| 0 | 0 | 0 | 0 | 0 |
| 0 | 0 | 0 | 0 | 0 |
| 0 | 0 | 0 | 0 | 0 |
| 0 | 0 | 0 | 0 | 0 |
| 0 | 0 | 0 | 0 | 0 |
| 0 | 0 | 0 | 0 | 0 |

| | | | | |
|---|---|---|---|---|
| 0 | 0 | 0 | 9.379638901 | 0 |
| 0 | 0 | 0 | 0 | 0 |
| 0 | 0 | 0 | 0 | 0 |
| 0 | 0 | 0 | 0 | 0 |
| 0 | 0 | 0 | 0 | 0 |
| 0 | 0 | 0 | 0 | 0 |
| 0 | 0 | 0 | 0 | 0 |
| 0 | 0 | 0 | 0 | 0 |
| 0 | 0 | 0 | 0 | 0 |
| 0 | 0 | 0 | 0 | 0 |
| 0 | 0 | 0 | 0 | 0 |
| 0 | 0 | 0 | 0.075609437 | 0 |
| 0 | 0 | 0 | 0 | 0 |
| 0 | 0 | 0 | 0 | 0 |
| 0 | 0 | 0 | 0.001808305 | 0 |
| 0 | 0 | 0 | 0 | 0 |
| 0 | 0 | 0 | 0 | 0 |
| 0 | 0 | 0 | 0 | 0 |
| 0 | 0 | 0 | 0 | 0 |
| 0 | 0 | 0 | 0 | 0 |
| 0.001188247 | 0.055914145 | 0.018227025 | 0.126615771 | 0.02352975 |
| 0 | 0 | 0 | 0 | 0 |
| 0 | 0 | 0 | 0 | 0 |
| 0 | 0 | 0.000950356 | 0.002953798 | 0.000766775 |
| 0 | 0 | 0 | 0 | 0 |
| 0 | 0 | 0 | 0 | 0 |
| 0 | 0.001092721 | 0.001592248 | 0 | 0.011562051 |
| 0 | 0 | 0 | 0 | 0 |
| 0 | 0 | 0 | 0 | 0 |
| 0 | 0 | 0 | 0 | 0 |
| 0 | 0.266867989 | 0.005643383 | 0 | 0 |
| 0 | 0 | 0 | 0 | 0 |
| 0 | 0 | 0 | 0 | 0 |
| 0 | 0 | 0 | 0.044142779 | 0.006441007 |
| 0 | 0.008789973 | 0 | 0 | 0 |
| 0 | 0 | 0 | 0 | 0 |
| 0 | 0 | 0 | 0 | 0 |
| 0 | 0 | 0 | 0 | 0 |
| 0 | 0 | 0 | 0 | 0 |
| 0 | 0 | 0 | 0 | 0 |
| 0 | 0 | 0 | 0 | 0 |
| 0 | 0 | 0 | 0 | 0 |
| 0 | 0 | 0.009039502 | 0 | 0 |
| 0 | 0 | 0 | 0 | 0 |
| 0 | 0 | 0 | 0 | 0 |
| 0 | 0 | 0 | 0 | 0 |
| 0 | 0 | 0 | 0 | 0 |
| 0 | 0 | 0.000845138 | 0 | 0 |

| | | | | |
|---|---|---|---|---|
| 0 | 0 | 0 | 0 | 0 |
| 0 | 0 | 0 | 0 | 0 |
| 0 | 0.001907962 | 0 | 0.000960115 | 0 |
| 0.00105051 | 0 | 0 | 0 | 0 |
| 0 | 0 | 0 | 0 | 0 |
| 0 | 0 | 0 | 0 | 0 |
| 0 | 0 | 0 | 0 | 0 |
| 0 | 0 | 0.001822352 | 0 | 0 |
| 0 | 0 | 0 | 0 | 0 |
| 0 | 0 | 0 | 0 | 0 |
| 0 | 0 | 0 | 0 | 0 |
| 0 | 0 | 0 | 0 | 0 |
| 0 | 0 | 0 | 0 | 0 |
| 0 | 0 | 0 | 0 | 0 |
| 0 | 0 | 0 | 0.067300138 | 0 |
| 0 | 0 | 0 | 0 | 0 |
| 0 | 0 | 0 | 0 | 0 |
| 0.066831343 | 0 | 0 | 0 | 0 |
| 0.067938909 | 0 | 0 | 0 | 0.004045239 |
| 0 | 0 | 0 | 0.101941677 | 0 |
| 0 | 0.00287131 | 0 | 0.027584199 | 0 |
| 0 | 0 | 0 | 0 | 0 |
| 0 | 0 | 0 | 0 | 0 |
| 0 | 0 | 0 | 0 | 0 |
| 0 | 0.009853167 | 0.002938661 | 0 | 0.00440328 |
| 0 | 0 | 0 | 0 | 0 |
| 0 | 0 | 0 | 0 | 0 |
| 0 | 0 | 0 | 0 | 0 |
| 0 | 0 | 0.004807554 | 0 | 0.004202114 |
| 0 | 0 | 0 | 0 | 0 |
| 0.062294114 | 0.044336742 | 0.007302621 | 0.091706553 | 0.289834645 |
| 0 | 0 | 0 | 0 | 0 |
| 0 | 0 | 0.026467925 | 0 | 0 |
| 0 | 0 | 0.001407143 | 0.000937186 | 0 |
| 0 | 0 | 0 | 0 | 0 |
| 0 | 0 | 0 | 0 | 0 |
| 0 | 0 | 0 | 0 | 0 |
| 0 | 0 | 0 | 0 | 0 |
| 0 | 0 | 0 | 0 | 0 |
| 0 | 0 | 0 | 0 | 0 |
| 0 | 0 | 0 | 0 | 0 |
| 0.001238995 | 0 | 0.001267031 | 0 | 0.018145427 |
| 0 | 0 | 0 | 0 | 0 |
| 0 | 0 | 0 | 0 | 0 |
| 0 | 0 | 0 | 0 | 0 |
| 0 | 0 | 0 | 0 | 0 |
| 0 | 0 | 0 | 0 | 0 |
| 0 | 0 | 0 | 0 | 0 |

| | | | | |
|---|---|---|---|---|
| 0 | 0 | 0 | 0 | 0 |
| 0 | 0 | 0 | 0 | 0 |
| 0 | 0.014985515 | 0.029234003 | 0.04153696 | 0.032572308 |
| 0 | 0 | 0 | 0 | 0 |
| 0 | 0.022598581 | 0 | 0 | 0 |
| 0 | 0 | 0 | 0 | 0 |
| 0 | 0 | 0 | 0 | 0 |
| 0 | 0 | 0 | 0 | 0 |
| 0 | 0 | 0 | 0 | 0 |
| 0 | 0 | 0 | 0 | 0 |
| 0 | 0 | 0 | 0 | 0 |
| 0 | 0 | 0 | 0 | 0 |
| 0 | 0 | 0 | 0 | 0 |
| 0 | 0.005430065 | 0.002857245 | 0.012296207 | 0 |
| 0 | 0.00537817 | 0.003265314 | 0.003044673 | 0.018002758 |
| 0 | 0 | 0 | 0.004889586 | 0.00423095 |
| 0 | 0.007404597 | 0.005580798 | 0 | 0.001834454 |
| 0 | 0 | 0 | 0 | 0 |
| 0 | 0 | 0 | 0 | 0 |
| 0 | 0 | 0 | 0 | 0 |
| 0 | 0 | 0 | 0 | 0 |
| 0 | 0 | 0 | 0 | 0 |
| 0 | 0 | 0 | 0 | 0 |
| 0 | 0 | 0 | 0 | 0 |
| 0 | 0 | 0 | 0.001115866 | 0 |
| 0 | 0 | 0 | 0 | 0 |
| 0 | 0 | 0 | 2.551755657 | 7.272108997 |
| 0 | 0 | 0 | 0 | 0 |
| 0 | 0 | 0 | 0 | 0 |
| 0 | 0 | 0 | 0 | 0 |
| 0 | 0 | 0 | 0.006132879 | 0.001153646 |
| 0 | 0 | 0 | 0 | 0 |
| 0 | 0 | 0.004633856 | 0 | 0 |
| 0 | 0 | 0 | 0 | 0 |
| 0 | 0 | 0 | 0 | 0 |
| 0 | 0 | 0.034286782 | 0 | 0 |
| 0 | 0 | 0 | 0 | 0 |
| 0 | 0.003185468 | 0.001875425 | 0 | 0.007944024 |
| 0 | 0 | 0 | 0 | 0 |
| 0 | 0 | 0 | 0 | 0 |
| 0 | 0 | 0 | 0 | 0 |
| 0 | 0 | 0 | 0 | 0 |
| 0 | 0 | 0 | 0 | 0 |
| 0 | 0.021582817 | 0 | 0 | 0 |
| 0 | 0 | 0 | 0 | 0 |
| 0 | 0 | 0 | 0 | 0 |
| 0 | 0 | 0 | 0 | 0 |
| 0 | 0 | 0 | 0 | 0 |

| | | | | |
|---|---|---|---|---|
| 0 | 0 | 0 | 1.63584168 | 0 |
| 0 | 0 | 0 | 0 | 0 |
| 0 | 0 | 0 | 0 | 0 |
| 0 | 0 | 0 | 0 | 0 |
| 0 | 0.002186038 | 0 | 0 | 0.001586448 |
| 0 | 0 | 0 | 0.001195818 | 0 |
| 0 | 0 | 0 | 0 | 0 |
| 0.002156144 | 0.018385281 | 0.00771727 | 0 | 0.019346679 |
| 0 | 0 | 0 | 0 | 0 |
| 0.216404718 | 0 | 0.070168792 | 0.050328777 | 0.026817262 |
| 0 | 0 | 0 | 0 | 0 |
| 0 | 0.003773221 | 0.054108447 | 0 | 0.0110901 |
| 0 | 0.002044373 | 0.001654967 | 0.010339031 | 0 |
| 0 | 0 | 0 | 0 | 0 |
| 0 | 0 | 0 | 0 | 0 |
| 0 | 0.002455155 | 0 | 0 | 0 |
| 0 | 0 | 0 | 0 | 0 |
| 0 | 0 | 0 | 0 | 0 |
| 0 | 6.796191268 | 0 | 0 | 0 |
| 0 | 0 | 0 | 0.00179814 | 0 |
| 0 | 0 | 0 | 0 | 0 |
| 0 | 0 | 0 | 0 | 0 |
| 0 | 0 | 0 | 0 | 0 |
| 0 | 0 | 0 | 0.080015567 | 0 |
| 0 | 0 | 0 | 0 | 0 |
| 0 | 0 | 0.004626275 | 0 | 0.000933153 |
| 3.094258039 | 0 | 0 | 0 | 0.002703599 |
| 0 | 5.206137185 | 0 | 0 | 0.001128442 |
| 0 | 0 | 0 | 0 | 0 |
| 0 | 0 | 0 | 0 | 0 |
| 0 | 0 | 0 | 0 | 0 |
| 0 | 0 | 0 | 0 | 0 |
| 0 | 0 | 0 | 0 | 0 |
| 0 | 0.005294034 | 0 | 0 | 0 |
| 0 | 0 | 0 | 0 | 0 |
| 0 | 0.00386342 | 0 | 0 | 0 |
| 0 | 1.211717613 | 0.00677396 | 0 | 0 |
| 0 | 0 | 0 | 0 | 0 |
| 0 | 0 | 0 | 0 | 0 |
| 0 | 0 | 0 | 0 | 0.001472158 |
| 0 | 0 | 0 | 0 | 0 |
| 0 | 0 | 0 | 0 | 0 |
| 0 | 0 | 0 | 0 | 0 |
| 0 | 0 | 0 | 0 | 0 |
| 0 | 0 | 0 | 0 | 0 |
| 0 | 0 | 0 | 0.077060311 | 0 |
| 0 | 0 | 0 | 0 | 0.003094533 |
| 0 | 0 | 0 | 0 | 0 |

| | | | | |
|---|---|---|---|---|
| 0 | 0 | 0 | 0 | 0 |
| 0 | 0.00430212 | 0 | 0 | 0.001513031 |
| 0 | 0 | 0 | 0 | 0 |
| 0 | 0 | 0 | 0 | 0 |
| 0 | 0 | 0 | 0.013255351 | 0 |
| 0 | 0 | 0 | 0.016372296 | 0 |
| 0 | 0 | 0 | 0.000783015 | 0 |
| 0.001910328 | 0 | 0 | 0 | 0 |
| 0 | 0 | 0 | 0 | 0 |
| 0 | 0 | 0 | 0 | 0 |
| 0 | 0.001082024 | 0 | 0 | 0.00406364 |
| 0 | 0 | 0 | 0 | 0 |
| 0 | 0 | 0 | 0 | 0 |
| 0 | 0 | 0 | 0.003086444 | 0.001836102 |
| 0 | 0 | 0.043330766 | 0 | 0 |
| 0 | 0 | 0 | 0 | 0 |
| 0 | 0 | 0 | 0.401655154 | 0 |
| 0 | 0.023875345 | 0 | 0 | 0 |
| 0 | 0 | 0 | 0 | 0 |
| 0 | 0 | 0 | 0.152917253 | 0 |
| 0.239335956 | 0 | 0 | 0 | 0 |
| 0.16148113 | 0 | 0 | 0.448408357 | 1.248189669 |
| 0 | 0 | 0 | 0.234860161 | 0 |
| 0 | 0 | 0 | 0 | 0 |
| 0 | 0 | 0 | 0 | 0 |
| 0 | 0 | 0 | 0 | 0 |
| 0 | 0.008589948 | 0.046657484 | 0 | 0 |
| 0 | 0 | 0 | 0 | 0 |
| 0 | 0 | 0 | 0 | 0 |
| 0.096967127 | 0.13485527 | 0.083695794 | 0.153535962 | 0.447007323 |
| 0 | 0 | 0 | 0.000392608 | 0.001245649 |
| 0 | 0 | 0 | 0 | 0 |
| 0 | 0.008009568 | 0 | 0 | 0 |
| 0 | 0 | 0 | 0 | 0 |
| 0 | 0.140553616 | 0 | 0 | 0 |
| 0 | 0 | 0 | 0 | 0 |
| 1.779273052 | 0 | 0 | 0 | 0.308706204 |
| 0 | 0 | 0 | 0 | 0 |
| 0 | 0 | 0 | 0 | 0 |
| 0 | 0 | 0.205872018 | 0 | 0 |
| 0 | 0 | 0 | 0.035364908 | 0 |
| 1.165337299 | 0 | 0 | 1.779500445 | 0.673418451 |
| 0 | 0 | 0 | 0 | 0 |
| 0 | 0 | 0 | 0 | 0 |
| 0 | 0 | 0 | 0 | 0 |
| 0 | 0 | 0 | 0 | 0 |
| 0 | 0.002205134 | 0.004284255 | 0.002996072 | 0.000864165 |
| 0 | 0 | 0 | 0 | 0 |

| | | | | |
|---|---|---|---|---|
| 0 | 0 | 0.001982269 | 0 | 0.001599353 |
| 0 | 0 | 0 | 0.025063982 | 0 |
| 0.002054288 | 0.052642962 | 0.035413032 | 0.019868047 | 0 |
| 0 | 1.37945419 | 0 | 0 | 0 |
| 0.009670684 | 0 | 0 | 0 | 0.244614463 |
| 0 | 0.002760491 | 0 | 0 | 0 |
| 0 | 0 | 0 | 0 | 0 |
| 0 | 0 | 0.009133746 | 0 | 0 |
| 0.004145808 | 0 | 0 | 0.007046909 | 0.00386682 |
| 0 | 0 | 0 | 0 | 0 |
| 0 | 0 | 0 | 0 | 0 |
| 0 | 0 | 0 | 0 | 0 |
| 0 | 0 | 0 | 0 | 0 |
| 0 | 0 | 0 | 0 | 0 |
| 0 | 0 | 0 | 0 | 0 |
| 0 | 0 | 0 | 0.002903874 | 0.320622682 |
| 0.005400812 | 0.005543335 | 0.05902731 | 0.001126525 | 0.019913356 |
| 0 | 0.007253007 | 0 | 0.001759736 | 0 |
| 0 | 0 | 0 | 0.004654459 | 0 |
| 0 | 0 | 0 | 0 | 0 |
| 0 | 0.178416262 | 0.004785399 | 0 | 0 |
| 0 | 0 | 0 | 0 | 0 |
| 0 | 0 | 0 | 0 | 0 |
| 0 | 0 | 0 | 0 | 0 |
| 0.007255898 | 0 | 0 | 0 | 0 |
| 0 | 0 | 0 | 0 | 0 |
| 0 | 0 | 0 | 0.002655959 | 0 |
| 0 | 0 | 0 | 0 | 0 |
| 0 | 0.026663169 | 0 | 0 | 0 |
| 0 | 0 | 0 | 0 | 0 |
| 0 | 0 | 0 | 0.03775565 | 0 |
| 0 | 0 | 0.010434007 | 0.023627499 | 0.001803956 |
| 0.118658828 | 0.004967298 | 0 | 0.535065555 | 0.05171914 |
| 0 | 0.00167497 | 0 | 0 | 0.007384507 |
| 0 | 0 | 0.002661413 | 0.023687797 | 0.001952096 |
| 0 | 0 | 0 | 0 | 0 |
| 0 | 0 | 0.001069232 | 0.00059819 | 0.000862688 |
| 0.000981224 | 0.00661082 | 0 | 0.003274685 | 0.032653626 |
| 0 | 0 | 0 | 0.022589032 | 0.221724626 |
| 0 | 0 | 0 | 0 | 0 |
| 0 | 0 | 0 | 0.004750458 | 0 |
| 0 | 0 | 0 | 0 | 0.022013378 |
| 0 | 0 | 0 | 0 | 0 |
| 0 | 0 | 0 | 0.541325957 | 0 |
| 0 | 0.542489682 | 0 | 0 | 0.005771442 |
| 0 | 0 | 0.009115144 | 0 | 0 |
| 0 | 0 | 0 | 0 | 0 |
| 0 | 0 | 0 | 0 | 0 |

| | | | | |
|---|---|---|---|---|
| 0 | 0 | 0 | 0 | 0 |
| 0.000668521 | 0.329357974 | 0.105965479 | 0.066295153 | 0.275058192 |
| 0 | 0 | 0 | 0 | 0 |
| 0 | 0 | 0 | 0 | 0 |
| 0 | 0 | 0 | 0 | 0 |
| 0 | 0 | 0 | 0 | 0 |
| 0 | 0.01197962 | 0 | 0 | 0 |
| 0 | 0 | 0 | 0 | 0 |
| 0 | 0 | 0 | 0 | 0 |
| 0 | 0 | 0 | 0 | 0 |
| 0.00281511 | 0 | 0 | 0 | 0 |
| 0 | 0.006854985 | 0.002270155 | 0.089844613 | 0.003663254 |
| 0 | 0 | 0 | 0 | 0.09421217 |
| 0 | 0 | 0 | 0 | 0 |
| 0 | 0 | 0 | 0 | 0 |
| 0 | 0 | 0 | 0 | 0 |
| 0 | 0.016647736 | 0.011171494 | 0.004133581 | 0.018742329 |
| 0 | 0.156395837 | 0.007596361 | 0 | 0 |
| 0 | 0 | 0 | 0 | 0 |
| 0 | 0 | 0 | 0 | 0 |
| 0.079754046 | 0.008358113 | 0.003657343 | 0.003239701 | 0.041164361 |
| 0 | 0 | 0 | 0 | 0 |
| 0 | 0 | 0 | 0 | 0 |
| 0 | 0 | 0 | 0 | 0 |
| 0.002403617 | 0 | 0 | 0 | 0 |
| 0 | 0 | 0 | 0 | 0 |
| 0 | 0 | 0 | 0.16089717 | 0.128884173 |
| 0.020336424 | 0.020888089 | 0.106315336 | 0.343969971 | 0.095971338 |
| 0 | 0 | 0 | 0 | 0 |
| 0 | 0 | 0 | 0 | 0.002648194 |
| 0 | 0.00231858 | 0 | 0 | 0 |
| 0 | 0 | 0 | 0 | 0 |
| 0 | 0 | 0 | 0 | 0 |
| 0 | 0 | 0 | 0 | 0 |
| 0 | 0 | 0 | 0 | 0.248847207 |
| 0.039243164 | 3.81068504 | 0.102630526 | 0.166240554 | 0.260889748 |
| 0 | 0 | 0 | 0 | 0 |
| 0 | 0 | 0 | 0 | 0 |
| 0 | 0 | 0.022298214 | 0.010507532 | 0.007351099 |
| 0 | 0 | 0 | 0 | 0 |
| 0 | 0.389553031 | 0 | 0 | 0.007283451 |
| 0 | 0 | 0 | 0 | 0 |
| 0 | 0.002338768 | 0.032622771 | 0.015209207 | 0 |
| 0 | 0 | 0.002390372 | 0 | 0.002495864 |
| 0 | 0 | 0 | 0 | 0 |
| 0.022020012 | 0 | 0 | 0.786631227 | 1.015789798 |
| 0 | 0 | 0 | 0 | 0 |
| 0 | 0 | 0 | 0.001483492 | 0 |

| | | | | |
|---|---|---|---|---|
| 0 | 0 | 0 | 0 | 0 |
| 0 | 0 | 0 | 0 | 0.053673548 |
| 0 | 0 | 0 | 0 | 0 |
| 0 | 0 | 0 | 0.200230492 | 0.604627681 |
| 0 | 0 | 0 | 0 | 0 |
| 0 | 0 | 0 | 0.003428719 | 0 |
| 0 | 0 | 0 | 0 | 0 |
| 0 | 0 | 0 | 0 | 0.035137966 |
| 0.00823952 | 0 | 0 | 0.222888732 | 0.345679504 |
| 0 | 0 | 0 | 0.530085358 | 0.017858016 |
| 0 | 0 | 0 | 0.056464941 | 0 |
| 0 | 0.008030191 | 0 | 0.003182219 | 0.004195914 |
| 0 | 0 | 0 | 0 | 0 |
| 0 | 0 | 0 | 4.257055241 | 0 |
| 0.002229393 | 0.006792197 | 0 | 0 | 0.326338601 |
| 0 | 0 | 0 | 0.324600517 | 0 |
| 0.004447603 | 0 | 0 | 0.014190754 | 0.027522421 |
| 0.005137504 | 0 | 0 | 0 | 0 |
| 0 | 0 | 0 | 0 | 0 |
| 0 | 0 | 0 | 0.001217918 | 0 |
| 0 | 0 | 0 | 0.009903542 | 0 |
| 0 | 0 | 0 | 0.002110326 | 0 |
| 0 | 0 | 0 | 0 | 0 |
| 0 | 0.056489998 | 0 | 0.297274953 | 0 |
| 0 | 0 | 0 | 0.001276873 | 0 |
| 0.131261534 | 0 | 0 | 0.076178943 | 0 |
| 0 | 0 | 0 | 0 | 0 |
| 0.700203435 | 0 | 0 | 1.569373048 | 0.445290957 |
| 0 | 0.001132496 | 0 | 0 | 0 |
| 0 | 0 | 0 | 0 | 0 |
| 0.000463491 | 0 | 0 | 0 | 0.013384689 |
| 0.003639118 | 0 | 0 | 0.446103339 | 2.115862111 |
| 0 | 0 | 0 | 0 | 0.179383487 |
| 0 | 0 | 0 | 0 | 0 |
| 0 | 0 | 0 | 0 | 0 |
| 0 | 0 | 0 | 0 | 0.70772778 |
| 0.018779217 | 0 | 0 | 0.379101815 | 1.022495571 |
| 0 | 0 | 0 | 0 | 0 |
| 0 | 0 | 0 | 0 | 0 |
| 0 | 0 | 0 | 0 | 0 |
| 0 | 0.016947772 | 0.007327519 | 0.174153267 | 0.101385455 |
| 0 | 0 | 0 | 0.100247389 | 0 |
| 2.185881322 | 0.108816699 | 0.061718834 | 0.545140535 | 6.048017244 |
| 0 | 0 | 0 | 0 | 0 |
| 0 | 0 | 0 | 0 | 0 |
| 0 | 0 | 0 | 0 | 0 |
| 0 | 0 | 0 | 0 | 0 |
| 0 | 0 | 0 | 0.004019283 | 0 |

| | | | | |
|---|---|---|---|---|
| 0 | 0 | 0 | 0 | 0 |
| 0.003172289 | 0 | 0 | 0 | 0.104007668 |
| 0.001229783 | 0.297388331 | 0.041141843 | 0.007873391 | 0.076100778 |
| 0 | 0 | 0 | 0 | 0.00172357 |
| 0 | 0.004880966 | 0 | 0 | 0 |
| 0 | 0 | 0 | 0.00290323 | 0.002283781 |
| 0 | 0 | 0 | 0 | 0 |
| 0 | 0 | 0 | 0 | 0 |
| 0 | 0 | 0 | 0.015392283 | 0 |
| 0 | 0 | 0 | 0 | 0 |
| 0.00051747 | 0 | 0.007584898 | 0.014473705 | 0.068740071 |
| 0.952225628 | 1.170825145 | 1.289920473 | 5.528735999 | 2.24447435 |
| 0 | 0 | 0 | 0 | 0 |
| 0.00390932 | 0.003292294 | 0.077100053 | 0 | 0.030258501 |
| 0.003374093 | 0.228126599 | 0.053106816 | 0.094280664 | 0.015251027 |
| 0 | 0 | 0 | 0.008590859 | 0 |
| 0 | 0 | 0 | 0 | 0 |
| 0 | 0 | 0 | 0 | 0 |
| 0 | 0 | 0 | 0 | 0 |
| 0 | 0 | 0 | 0.102831765 | 0 |
| 0 | 0.010409919 | 0.005470202 | 0.092223901 | 0.017415503 |
| 0.564854184 | 1.395927361 | 29.54058938 | 0 | 0.001885329 |
| 9.534309689 | 0 | 41.22973434 | 2.186937358 | 2.550778252 |
| 0 | 0 | 0 | 0 | 0 |
| 0 | 0 | 0 | 0 | 0 |
| 1.365308316 | 0 | 0 | 0 | 0.812318562 |
| 0 | 0 | 0 | 0.004781492 | 0 |
| 0 | 0 | 0 | 0.002731793 | 0.002488227 |
| 0 | 0 | 0 | 0 | 0 |
| 0 | 0 | 0 | 0 | 0 |
| 0 | 0 | 0 | 0.027520813 | 0.034470675 |
| 0.073566465 | 0 | 0 | 0.093833928 | 0.361827159 |
| 0 | 2.591669989 | 0 | 0.002027028 | 0.004384959 |
| 0 | 0 | 0 | 0 | 0.214007739 |
| 0 | 0.00413565 | 0 | 0.000445955 | 0 |
| 0.004176744 | 0.015625858 | 0.076554055 | 0.003523107 | 0.000662726 |
| 0 | 0.007057106 | 0 | 0 | 0.012547606 |
| 0 | 0 | 0 | 0 | 0 |
| 0.018046648 | 0 | 0 | 0.37750021 | 0.044825225 |
| 0 | 0 | 0 | 0 | 0 |
| 0 | 0 | 0 | 0.015241603 | 0.264470914 |
| 0 | 0 | 0 | 0 | 0 |
| 0 | 0 | 0 | 0.090598228 | 0 |
| 0 | 0 | 0 | 0 | 0 |
| 0.000915981 | 0.010414004 | 0 | 0.006113897 | 0.030683996 |
| 0 | 0 | 0 | 0 | 0 |
| 0 | 0 | 0 | 0 | 0 |
| 0.005082649 | 0 | 0 | 0.030132268 | 0 |

| | | | | |
|---|---|---|---|---|
| 0 | 0 | 0 | 0 | 0 |
| 0 | 0 | 0 | 0 | 0 |
| 0 | 0 | 0 | 0 | 0 |
| 0 | 0 | 0 | 0.000595526 | 0.015974526 |
| 0 | 0 | 0 | 0 | 0 |
| 0.093358014 | 0 | 0 | 0 | 0.079759104 |
| 0.001367526 | 0 | 0 | 0.484100503 | 0 |
| 0.001005723 | 0 | 0 | 0.013699786 | 0.049669936 |
| 0 | 0 | 0 | 0 | 0 |
| 0 | 0 | 0 | 0 | 0 |
| 0.000521687 | 0.007029551 | 0 | 0.008290724 | 0 |
| 0 | 0 | 0 | 0 | 0 |
| 0 | 0 | 0 | 0 | 0 |
| 0 | 0 | 0 | 0 | 0.010162082 |
| 0 | 0 | 0 | 0 | 0 |
| 0 | 0 | 0 | 0 | 0 |
| 0 | 0 | 0 | 0.067210661 | 0.2290009 |
| 0 | 0 | 0 | 0 | 0.000358329 |
| 0 | 0 | 0 | 0.134647946 | 0 |
| 0 | 0 | 0 | 0 | 0 |
| 0 | 0 | 0 | 0 | 0 |
| 0 | 0 | 0 | 0 | 0 |
| 0.00116832 | 0 | 0 | 0.027214054 | 0.000688546 |
| 0 | 0 | 0 | 0 | 0 |
| 0 | 0 | 0 | 0 | 0 |
| 0 | 0 | 0 | 0 | 0 |
| 0 | 0 | 0 | 0 | 0 |
| 0 | 0 | 0 | 0 | 0 |
| 0 | 0 | 0 | 0 | 0 |
| 0 | 0 | 0.052755142 | 1.372162579 | 0.502548278 |
| 0 | 0 | 0 | 0 | 0 |
| 0 | 0 | 0 | 0 | 0 |
| 0.006050159 | 0 | 0 | 0.015143616 | 0 |
| 0 | 0 | 0 | 0 | 0 |
| 0 | 0 | 0 | 0 | 0.001254962 |
| 0 | 0 | 0 | 0 | 0 |
| 0.090782014 | 0 | 0.009551055 | 0.185238131 | 0.514765371 |
| 0 | 0.001301702 | 0.002709662 | 0 | 0 |
| 0 | 0 | 0 | 0 | 0 |
| 0 | 0 | 0 | 0 | 0 |
| 0.001795501 | 0 | 0 | 0 | 0 |
| 0 | 0 | 0 | 3.758893524 | 0 |
| 0 | 0 | 0 | 0 | 0 |
| 0.006147033 | 0 | 0 | 0 | 1.526975304 |
| 0.006184574 | 0 | 0 | 0 | 0.199009453 |
| 0 | 0 | 0 | 0 | 0 |
| 0 | 0 | 0 | 0.001867182 | 0 |
| 0 | 0 | 0 | 0 | 0 |

| | | | | |
|---|---|---|---|---|
| 0 | 0 | 0 | 0 | 0 |
| 0 | 0 | 0 | 0 | 0 |
| 0 | 0 | 0 | 0 | 1.560079711 |
| 0.021671138 | 0 | 0 | 0.093686289 | 0.134708249 |
| 0 | 0 | 0 | 0.118747023 | 0 |
| 0.036461217 | 0 | 0 | 0 | 0.811631709 |
| 0 | 0 | 0 | 0 | 0 |
| 0 | 0 | 0 | 0 | 0 |
| 0 | 0.732467887 | 0.01930823 | 0.009872916 | 0 |
| 0 | 0 | 0 | 0 | 0 |
| 0 | 0 | 0 | 0.010519953 | 0.060366576 |
| 0 | 0 | 0 | 0 | 0 |
| 0 | 0 | 0 | 0 | 0 |
| 0 | 0 | 0 | 0 | 0 |
| 0.009045584 | 0 | 0 | 4.961865574 | 0.255560784 |
| 0 | 0 | 0 | 0.282738854 | 0 |
| 0 | 0 | 0 | 0 | 0 |
| 0 | 0 | 0 | 0 | 0 |
| 0 | 0 | 0 | 0.000839407 | 0 |
| 0 | 0 | 0 | 0 | 0 |
| 0.008693381 | 0 | 0 | 0.065574943 | 0.69409538 |
| 0 | 0 | 0 | 0 | 0 |
| 0 | 0 | 0 | 0 | 0 |
| 0 | 0 | 0 | 0.00177274 | 0 |
| 0.304164115 | 0.003920763 | 0.002441498 | 0.004097742 | 0.127450675 |
| 0.064358386 | 0 | 0.002761456 | 4.02214815 | 4.757386113 |
| 0 | 0 | 0 | 0.023369796 | 0.172032198 |
| 0 | 0 | 0 | 0 | 0 |
| 0.001669868 | 0 | 0 | 0 | 0.07595038 |
| 0 | 0 | 0 | 0.030682118 | 0 |
| 0 | 0 | 0 | 0 | 0 |
| 0 | 0 | 0 | 0.018912605 | 0.026006465 |
| 0 | 0 | 0 | 0 | 0 |
| 0 | 0 | 0 | 0 | 0 |
| 0.005882528 | 0.001592377 | 0.082715039 | 0 | 0.014560778 |
| 0 | 0.001125617 | 0 | 0 | 0 |
| 0 | 0 | 0 | 0 | 0.13327813 |
| 0.145556318 | 0 | 0.264088709 | 3.126818438 | 0.782149253 |
| 0 | 0 | 5.288787139 | 0 | 0 |
| 0 | 0 | 0 | 0 | 0 |
| 0 | 0 | 0 | 0.374927649 | 0 |
| 0 | 0 | 0 | 0.004817501 | 0 |
| 0.088684484 | 0 | 0.096502139 | 0.054569355 | 0.105153847 |
| 0.056878948 | 0 | 0 | 0 | 0 |
| 0 | 0 | 0 | 0 | 0 |
| 0 | 0 | 0 | 0 | 0.000464336 |
| 0 | 0 | 0 | 0 | 0 |
| 0 | 0 | 0.008480873 | 0.427235513 | 0.415550136 |

| | | | | |
|---|---|---|---|---|
| 0.001838563 | 0 | 0 | 0.046218658 | 0 |
| 0 | 0 | 0 | 1.707356355 | 0 |
| 0 | 0 | 0 | 0 | 0 |
| 0 | 0 | 0 | 0 | 0 |
| 0 | 0 | 0 | 0.000634704 | 0.00137302 |
| 0 | 0 | 0 | 0 | 0 |
| 0 | 0 | 0 | 0.585494158 | 0 |
| 0 | 0 | 0 | 0.002795267 | 0 |
| 0 | 0 | 0 | 0 | 0 |
| 0 | 0 | 0 | 0 | 0 |
| 0.000763994 | 0 | 0 | 0.001092735 | 0 |
| 0.454582225 | 6.894492318 | 0 | 1.631355659 | 5.224625833 |
| 0 | 0.241044658 | 0.204403504 | 0.025553126 | 0.092600514 |
| 0 | 0 | 0 | 0 | 0 |
| 0.001347167 | 0.096273571 | 0.117100296 | 0 | 0.023659678 |
| 0 | 0 | 0 | 0 | 0 |
| 0 | 0.002705946 | 0 | 0.000893598 | 0 |
| 0 | 0 | 0 | 0 | 0 |
| 0 | 0 | 0 | 0.008524772 | 0 |
| 0 | 0 | 0 | 0.004458779 | 0 |
| 0 | 0 | 0 | 0.008256212 | 0.109465825 |
| 0 | 0 | 0 | 0 | 0 |
| 0.010497413 | 0 | 0 | 0.034095128 | 0.060493563 |
| 0 | 0 | 0 | 0.043635418 | 0 |
| 0 | 0 | 0 | 0 | 0 |
| 0 | 0 | 0 | 0 | 0 |
| 0 | 0 | 0 | 0 | 0 |
| 0 | 0 | 0 | 0 | 0 |
| 0 | 0 | 0 | 0.001818244 | 0 |
| 0 | 0 | 0 | 2.743876769 | 0 |
| 0 | 0 | 0 | 0 | 0 |
| 0 | 0.020505928 | 0.024772334 | 0.011430156 | 0.015247844 |
| 0 | 0 | 0 | 0 | 0.091246612 |
| 0.001871441 | 0 | 0 | 0.011896477 | 0.085028443 |
| 0 | 0.001224954 | 0 | 0 | 0 |
| 0 | 0 | 0 | 0.065755955 | 0.108468158 |
| 0 | 0 | 0 | 0 | 0 |
| 0 | 0 | 0 | 0.018419869 | 0 |
| 0 | 0 | 0 | 0 | 0.006127783 |
| 0 | 0 | 0 | 0 | 0 |
| 0 | 0 | 0 | 0.819949737 | 0 |
| 0 | 0 | 0 | 0.003711119 | 0 |
| 0 | 0 | 0 | 0 | 0 |
| 0 | 0.075622909 | 0 | 0.028603666 | 0 |
| 0.000636825 | 0.019709471 | 0.003093368 | 0.001821692 | 0.061870049 |
| 0 | 0 | 0 | 0.006273453 | 0 |
| 0.027594232 | 0 | 0 | 0 | 0.230218213 |
| 0 | 0 | 0 | 0 | 0 |

| | | | | |
|---|---|---|---|---|
| 0 | 0 | 0 | 0 | 0 |
| 0 | 0 | 0 | 0 | 0 |
| 0 | 0 | 0 | 0 | 0 |
| 0 | 0 | 0 | 0 | 0 |
| 0 | 0.120872918 | 0 | 0.001592279 | 0 |
| 0 | 0 | 0 | 0 | 0 |
| 0.006343868 | 0 | 0 | 0 | 0 |
| 0 | 0 | 0 | 0.020017606 | 0 |
| 0 | 0 | 0 | 0 | 0 |
| 0 | 0 | 0 | 0 | 0.01035309 |
| 0 | 0 | 0 | 0 | 0 |
| 0 | 0 | 0 | 0 | 0 |
| 0 | 0 | 0 | 0 | 0 |
| 0.233650596 | 0 | 0 | 0 | 0.347771954 |
| 0 | 0 | 0 | 0 | 0 |
| 0 | 0 | 0 | 0 | 0 |
| 0 | 0 | 0 | 0 | 0 |
| 0 | 0 | 0 | 0.060154941 | 0 |
| 0.01900192 | 0 | 0 | 6.079885835 | 0 |
| 0 | 0 | 0 | 0 | 0 |
| 0 | 0 | 0 | 0.000713275 | 0 |
| 0 | 0 | 0 | 0 | 0 |
| 0 | 0 | 0 | 0 | 0 |
| 0 | 0 | 0 | 0 | 0 |
| 0 | 0 | 0 | 0 | 0 |
| 0 | 0.00786376 | 0.030616915 | 0 | 0 |
| 0 | 0 | 0 | 0 | 0 |
| 0 | 0 | 0.00741779 | 0.394382342 | 0.431271056 |
| 0 | 0 | 0 | 0 | 0 |
| 0.01107157 | 0 | 0 | 0.675265173 | 1.271658738 |
| 0 | 0 | 0 | 0 | 0 |
| 0 | 0 | 0 | 0 | 0 |
| 0 | 0 | 0 | 0.002483119 | 0 |
| 0 | 0 | 0 | 0 | 0 |
| 0 | 0 | 0 | 0 | 0.000640763 |
| 0 | 0 | 0 | 0 | 0 |
| 0 | 0 | 0 | 0 | 0 |
| 0 | 0 | 0 | 0.001008316 | 0.00232665 |
| 0 | 0 | 0 | 0 | 0 |
| 0 | 0 | 0 | 0 | 0 |
| 0.505115694 | 3.033336713 | 3.336923648 | 1.664839294 | 4.248325086 |
| 0 | 2.319749475 | 0.010422463 | 0.0451466 | 0 |
| 0 | 0 | 0 | 0 | 0 |
| 0 | 0 | 0 | 0 | 0 |
| 0 | 0 | 0 | 0 | 0 |
| 0 | 0 | 0 | 0 | 0 |
| 0 | 0 | 0 | 0.001334931 | 0.01518149 |
| 0 | 0 | 0 | 0.002000638 | 0.001615738 |

| | | | | |
|---|---|---|---|---|
| 0 | 0 | 0 | 0 | 0 |
| 0 | 0 | 0 | 0.007270967 | 0.057811079 |
| 0 | 0 | 0 | 0 | 0 |
| 0 | 0 | 0 | 0 | 0 |
| 0.223886477 | 0 | 0.487227266 | 0.401995445 | 0.98142247 |
| 0 | 0 | 0 | 0 | 0 |
| 0 | 0 | 0 | 0 | 0 |
| 0 | 0 | 0 | 0.001454549 | 0 |
| 0 | 1.333883854 | 0.070811998 | 0 | 0 |
| 0 | 0 | 0 | 0.000997917 | 0 |
| 0 | 0 | 0.002825842 | 0 | 0 |
| 0 | 0 | 0 | 0 | 0 |
| 0.006989913 | 0 | 0 | 0.161028308 | 0.165405617 |
| 0.002234281 | 0.004342232 | 0.000703027 | 0.006063587 | 0.016307656 |
| 0 | 0 | 0 | 0.045590464 | 0.449483469 |
| 0.007051672 | 0 | 0 | 0.517342042 | 0.178154412 |
| 0 | 0.004509034 | 0.001977173 | 0.170317842 | 0.000613554 |
| 0 | 0 | 0 | 0 | 0 |
| 0 | 0 | 0 | 0 | 0 |
| 0 | 0 | 0 | 0 | 0 |
| 0 | 0 | 0 | 0.011573837 | 0 |
| 0 | 0 | 0 | 0 | 0 |
| 0 | 0 | 0 | 0 | 0 |
| 0 | 0 | 0 | 0 | 0 |
| 0 | 0 | 0 | 0 | 0 |
| 0 | 0 | 0 | 0 | 0 |
| 0.000775934 | 0 | 0 | 0.004439252 | 0.002774255 |
| 0 | 0.000921703 | 0 | 0 | 0 |
| 0 | 0 | 0 | 0 | 0 |
| 0 | 0 | 0 | 0.000821589 | 0 |
| 0 | 0 | 0 | 0 | 0 |
| 0 | 0 | 0 | 0.001278512 | 0 |
| 0 | 0.003974079 | 0 | 0.497954263 | 0 |
| 0 | 0 | 0 | 0.019253584 | 0.004796087 |
| 0 | 0 | 0 | 0 | 0.156441382 |
| 0 | 0.002772169 | 0 | 0 | 0 |
| 0 | 0 | 0 | 0 | 0 |
| 0 | 0 | 0 | 0.009271573 | 0 |
| 0 | 0 | 0 | 0 | 0 |
| 0 | 0 | 0 | 0 | 0 |
| 0.30308646 | 0 | 0 | 0 | 0.3896081 |
| 0 | 0 | 0 | 0 | 0 |
| 0 | 0 | 0 | 0 | 0 |
| 0.00285761 | 0 | 0 | 0 | 0 |
| 0 | 0 | 0 | 0 | 0 |
| 0 | 0 | 0 | 0.000983694 | 0 |
| 0 | 0 | 0 | 0.456083923 | 0.004081008 |
| 0 | 0 | 0 | 0 | 0 |

| | | | | |
|---|---|---|---|---|
| 0 | 0 | 0 | 0.07587035 | 0.053785934 |
| 0 | 0 | 0 | 0 | 0 |
| 0 | 0 | 0 | 0 | 0 |
| 0 | 0 | 0 | 0 | 0 |
| 0 | 0 | 0 | 0.087321705 | 0.007307982 |
| 0 | 0 | 0 | 0 | 0.165966487 |
| 0 | 0.001499357 | 0.014868603 | 0.539843899 | 0.012608484 |
| 0 | 0 | 0 | 0 | 0 |
| 0 | 0 | 0 | 0 | 0 |
| 0 | 0 | 0 | 0 | 0 |
| 0 | 0 | 0 | 0.006175935 | 0.030876592 |
| 0 | 0 | 0 | 0.004577525 | 0 |
| 0 | 0 | 0 | 0 | 0 |
| 0 | 0 | 0 | 0 | 0 |
| 0 | 0 | 0 | 0 | 0 |
| 0 | 0 | 0 | 0 | 0 |
| 0 | 0 | 0 | 0 | 0 |
| 0 | 0.016036399 | 0 | 0 | 0 |
| 0 | 0 | 0 | 0 | 0 |
| 0 | 0 | 0 | 0 | 0 |
| 0 | 0.005353017 | 0.191824709 | 0.091721113 | 0.002563957 |
| 0 | 0 | 0 | 0 | 0 |
| 0 | 0 | 0 | 0.00237693 | 0.120577142 |
| 0 | 0.001050043 | 0 | 0 | 0.001143054 |
| 0 | 0 | 0 | 0 | 0 |
| 0 | 0 | 0 | 0.002181866 | 0 |
| 0 | 0 | 0 | 0 | 0 |
| 0 | 0 | 0 | 0 | 0 |
| 0 | 0 | 0 | 0 | 0 |
| 0.003588983 | 0 | 0.006116992 | 2.570069275 | 0.005264391 |
| 0 | 0 | 0 | 0.15590007 | 0 |
| 0 | 0 | 0 | 0 | 0 |
| 0 | 0 | 0 | 0.024517737 | 0 |
| 0 | 0 | 0.005446711 | 0 | 0 |
| 0 | 0 | 0 | 0.001408189 | 0 |
| 0 | 0 | 0 | 0 | 0 |
| 0 | 0.010073051 | 0 | 0.00297523 | 0.00686523 |
| 0 | 0 | 0 | 0 | 0 |
| 0 | 0 | 0 | 0 | 0 |
| 0 | 0 | 0 | 0 | 0.097093723 |
| 0 | 0 | 0 | 0 | 0 |
| 0 | 0 | 0 | 0 | 0 |
| 0 | 0 | 0 | 0 | 0 |
| 0.005458671 | 0 | 0 | 0 | 0 |
| 0 | 0 | 0 | 0.004536616 | 0 |
| 0 | 0 | 0 | 0 | 0 |
| 0 | 0 | 0 | 0 | 0 |
| 0 | 0 | 0 | 0 | 0 |

| | | | | |
|---|---|---|---|---|
| 0 | 0.002448082 | 0 | 0 | 0 |
| 0 | 0 | 0 | 0 | 0 |
| 0 | 0 | 0 | 0 | 0 |
| 0 | 0 | 0 | 0 | 0 |
| 0 | 0 | 0 | 0.007479007 | 0 |
| 0 | 0 | 0 | 0 | 0 |
| 0 | 0 | 0 | 0 | 0 |
| 0.185970933 | 0 | 0.296616826 | 0.366624881 | 0.565020344 |
| 14.97955506 | 0.084833494 | 0.096670943 | 0.193664511 | 0.975540768 |
| 0 | 0.02716281 | 0 | 0 | 0.00107523 |
| 0 | 0 | 0 | 0 | 0 |
| 0 | 0 | 0 | 0 | 0 |
| 0 | 0 | 0 | 0 | 0.008031422 |
| 0 | 0 | 0 | 0 | 0 |
| 0 | 0 | 0 | 0 | 0 |
| 0 | 0 | 0 | 0 | 0 |
| 0 | 0.414530722 | 0.000890112 | 0 | 0 |
| 0 | 0 | 0 | 0 | 0 |
| 0 | 0 | 0 | 0 | 0 |
| 0 | 0 | 0 | 0 | 0.117246277 |
| 0 | 0 | 0 | 0 | 0 |
| 0 | 0 | 0 | 0 | 0 |
| 0 | 0 | 0 | 0 | 0.017126216 |
| 0 | 0 | 0 | 0 | 0 |
| 0.000587071 | 0 | 0 | 0.082177043 | 0.080795339 |
| 0 | 0 | 0 | 0.072097779 | 0 |
| 0 | 0 | 0 | 0 | 0 |
| 0 | 0 | 0 | 0 | 0 |
| 0 | 0 | 0 | 0 | 0 |
| 0 | 0 | 0 | 0 | 0 |
| 0 | 0 | 0 | 0 | 0 |
| 0 | 0 | 0 | 0 | 0 |
| 0 | 0 | 0 | 0 | 0 |
| 0 | 0 | 0 | 0 | 0 |
| 0 | 0 | 0 | 0 | 0.013103618 |
| 0 | 0 | 0 | 0.006196763 | 0 |
| 0 | 0 | 0 | 0 | 0 |
| 0 | 0 | 0 | 0 | 0 |
| 0.001418059 | 0 | 0 | 0 | 0.008628904 |
| 0 | 0.008835828 | 0 | 0 | 0.000491157 |
| 0 | 0 | 0 | 0 | 0 |
| 0.003531283 | 0.116564901 | 0 | 0 | 0.280973613 |
| 0 | 0 | 0 | 0 | 0 |
| 0 | 0 | 0 | 0 | 0 |
| 0 | 0 | 0 | 0 | 0 |
| 0 | 0 | 0 | 0 | 0 |
| 0 | 0 | 0 | 0 | 0 |
| 0 | 0 | 0 | 0.000794214 | 0 |

| | | | | |
|---|---|---|---|---|
| 0 | 0 | 0 | 0 | 0 |
| 0.00080726 | 0 | 0.006879391 | 0.054908486 | 0.000777069 |
| 0 | 0 | 0 | 0 | 0 |
| 0 | 0 | 0 | 0 | 0 |
| 0.003197878 | 0 | 0 | 0.071926829 | 0.129753364 |
| 0 | 0 | 0 | 0 | 0.086807945 |
| 0 | 0 | 0 | 0 | 0 |
| 0 | 0.080009376 | 0.001163523 | 0.01084902 | 0 |
| 0 | 0 | 0 | 0 | 0.027182305 |
| 0 | 0 | 0 | 0 | 0.070999974 |
| 0 | 0 | 0 | 0 | 0 |
| 0 | 0 | 0 | 0 | 0 |
| 0 | 0 | 0 | 0 | 0 |
| 0 | 0 | 0 | 0 | 0 |
| 0 | 0.008600337 | 0 | 0 | 0 |
| 0 | 0 | 0 | 0 | 0.102994128 |
| 0 | 0 | 0 | 0 | 0 |
| 0 | 0 | 0 | 0 | 0 |
| 0 | 0 | 0 | 0 | 0 |
| 0 | 0 | 0 | 0 | 0 |
| 0 | 0 | 0 | 0 | 0 |
| 0 | 0 | 0 | 0.034118375 | 0 |
| 0 | 0 | 0 | 0.002149775 | 0 |
| 0 | 0.004353509 | 0 | 0 | 0 |
| 0.163093966 | 0 | 0 | 0 | 0.098907037 |
| 0 | 0 | 0 | 0 | 0 |
| 0 | 0 | 0.200614577 | 0.02461677 | 0.006068265 |
| 0 | 0 | 0 | 0 | 0.00078241 |
| 0 | 0 | 0 | 0 | 0 |
| 0 | 0 | 0 | 0 | 0 |
| 0 | 0 | 0 | 0 | 0 |
| 0 | 0 | 0 | 0.006327803 | 0 |
| 0 | 0 | 0 | 0 | 0 |
| 0 | 0 | 0 | 0 | 0 |
| 0 | 0 | 0 | 0 | 0 |
| 0 | 0 | 0 | 0 | 0 |
| 0 | 0 | 0 | 0 | 0 |
| 0.003694374 | 0 | 0 | 0 | 0.079760627 |
| 0 | 0 | 0 | 0.01195815 | 0 |
| 0 | 0 | 0 | 0 | 0 |
| 0 | 0 | 0 | 0 | 0 |
| 0.003353595 | 0 | 0 | 0 | 0 |
| 0 | 0 | 0 | 0 | 0 |
| 0 | 0 | 0 | 0 | 0 |
| 0 | 0 | 0 | 0 | 0 |
| 0 | 0 | 0 | 0 | 0 |
| 0 | 0 | 0 | 0 | 0 |
| 0 | 0 | 0 | 0 | 0 |

| | | | | |
|---|---|---|---|---|
| 0.000423457 | 0 | 0 | 0.018035427 | 0.080359371 |
| 0 | 0 | 0 | 0.015666177 | 0 |
| 0 | 0 | 0 | 0 | 0 |
| 0 | 0 | 0 | 0 | 0 |
| 0 | 0 | 0 | 0 | 0 |
| 0 | 0.157829382 | 0 | 0 | 0 |
| 0 | 0 | 0 | 0 | 0 |
| 0 | 0 | 0 | 0 | 0 |
| 0 | 0 | 0 | 0 | 0 |
| 0 | 0 | 0 | 0.042636372 | 0 |
| 0 | 0 | 0 | 0 | 0 |
| 0 | 0.001911067 | 0 | 0.00086551 | 0 |
| 0 | 0 | 0 | 0 | 0 |
| 0 | 0 | 0 | 0 | 0 |
| 0 | 0 | 0 | 0 | 0 |
| 0 | 0 | 0 | 0 | 0 |
| 0 | 0 | 0 | 0 | 0 |
| 0 | 0 | 0 | 0 | 0.477377243 |
| 0 | 0 | 0 | 0 | 0 |
| 0 | 0 | 0 | 0 | 0 |
| 0 | 0.19591473 | 0 | 0 | 0.00121354 |
| 0 | 0 | 0 | 0 | 0 |
| 0.001031954 | 0 | 0 | 0.019089554 | 0.003065225 |
| 0 | 0 | 0 | 0.39158345 | 0 |
| 0 | 0 | 0 | 0.008838302 | 0 |
| 0.003005497 | 0 | 0 | 0 | 0 |
| 0 | 0 | 0 | 0 | 0 |
| 0 | 0 | 0 | 0 | 0 |
| 0 | 0 | 0 | 0 | 0 |
| 0 | 0 | 0 | 0 | 0 |
| 0 | 0 | 0 | 0 | 0 |
| 0 | 0 | 0 | 0 | 0.352727068 |
| 0 | 0.067145629 | 0 | 0 | 0 |
| 0 | 0 | 0 | 0 | 0 |
| 0 | 1.938565085 | 0.199965388 | 0.030663442 | 0.090868466 |
| 0 | 0 | 0.000793061 | 0 | 0 |
| 0 | 0 | 0 | 0 | 0 |
| 0 | 0 | 0 | 0 | 0 |
| 0 | 0.037620151 | 0 | 0 | 0 |
| 0 | 0 | 0 | 0 | 0 |
| 0 | 0 | 0 | 0 | 0 |
| 0 | 0 | 0 | 0 | 0 |
| 0.001258638 | 0 | 0 | 0.023002814 | 0.034962325 |
| 0 | 0 | 0 | 0 | 0 |
| 0 | 0 | 0 | 0 | 0 |
| 0 | 0 | 0 | 0.023192702 | 0.062130196 |
| 0 | 0 | 0 | 0 | 0 |
| 0 | 0 | 0 | 0 | 0 |

| | | | | |
|---|---|---|---|---|
| 0 | 0 | 0 | 0 | 0 |
| 0 | 0 | 0 | 0 | 0 |
| 0 | 0 | 0 | 0 | 0 |
| 0 | 0 | 0 | 0 | 0 |
| 0 | 0 | 0 | 0 | 0 |
| 0 | 0 | 0 | 0 | 0 |
| 0 | 0 | 0 | 0 | 0 |
| 0 | 0 | 0 | 0 | 0 |
| 0 | 0 | 0 | 0 | 0 |
| 0 | 0 | 0 | 0.006708338 | 0.00084947 |
| 0 | 0 | 0 | 0 | 0 |
| 0 | 0 | 0 | 0 | 0 |
| 0 | 0 | 0 | 0 | 0 |
| 0 | 0 | 0 | 0 | 0 |
| 0 | 0 | 0 | 0.049320172 | 0.021281121 |
| 0 | 0.203247587 | 0.002071776 | 0 | 0 |
| 0 | 0 | 0 | 0 | 0 |
| 0 | 0 | 0 | 0 | 0 |
| 0 | 0 | 0 | 0 | 0 |
| 0 | 0 | 0 | 0.09690627 | 0.013645035 |
| 0.033326108 | 0 | 0 | 0 | 0.001745836 |
| 0 | 0 | 0 | 0 | 0 |
| 0 | 0 | 0 | 0 | 0 |
| 0 | 0 | 0 | 0 | 0 |
| 0 | 0 | 0 | 0 | 0 |
| 0 | 0 | 0 | 0 | 0 |
| 0 | 0 | 0 | 0 | 0 |
| 0 | 0 | 0 | 0 | 0 |
| 0 | 0 | 0 | 0 | 0 |
| 0 | 0 | 0 | 0.049052578 | 0.093263741 |
| 0.051594141 | 0 | 0 | 0.116318238 | 0.192808646 |
| 0 | 0 | 0 | 0 | 0 |
| 0 | 0 | 0 | 0 | 0 |
| 0 | 0 | 0 | 0 | 0 |
| 0 | 0 | 0 | 0 | 0 |
| 0 | 0 | 0 | 0 | 0 |
| 0 | 0 | 0 | 0 | 0 |
| 0 | 0 | 0 | 0 | 0 |
| 0.003711614 | 0 | 0 | 0 | 0 |
| 0 | 0 | 0 | 0 | 0 |
| 0 | 0 | 0 | 0 | 0.00159735 |
| 0 | 0 | 0 | 0.105887892 | 0 |
| 0 | 0 | 0 | 0.002122751 | 0 |
| 0 | 0 | 0 | 0 | 0 |
| 0 | 0 | 0 | 0 | 0 |
| 0 | 0 | 0 | 0 | 0 |
| 0 | 0 | 0.220923164 | 0 | 0 |
| 0 | 0 | 0 | 0 | 0 |

| | | | | |
|---|---|---|---|---|
| 0 | 0 | 0 | 0 | 0 |
| 0 | 0 | 0 | 0 | 0 |
| 0 | 0 | 0 | 0 | 0 |
| 0 | 0 | 0 | 0 | 0 |
| 0 | 0 | 0 | 0.126931571 | 0 |
| 0 | 0 | 0 | 0.013795497 | 0 |
| 0 | 0 | 0 | 0 | 0 |
| 0 | 0 | 0 | 0 | 0 |
| 0 | 0 | 0 | 0 | 0 |
| 0 | 0 | 0 | 0 | 0 |
| 0 | 0 | 0 | 0.036108366 | 0 |
| 0 | 0.004076045 | 0.005866048 | 0.002905764 | 0.010353214 |
| 0 | 0 | 0.000474461 | 0 | 0.003445283 |
| 0 | 0 | 0 | 0 | 0 |
| 0 | 0.0032788 | 0 | 0 | 0 |
| 0 | 0 | 0 | 0.139300353 | 0 |
| 0.002481668 | 0 | 0 | 0 | 0.016608231 |
| 0 | 0 | 0 | 0 | 0 |
| 0 | 0.003086405 | 0 | 0.006859637 | 0 |
| 0 | 0.001119418 | 0 | 0.178962918 | 0.002047203 |
| 0 | 0 | 0 | 0 | 0 |
| 0 | 0 | 0 | 0 | 0 |
| 0 | 0 | 0 | 0 | 0 |
| 0 | 0 | 0 | 0 | 0 |
| 0 | 0 | 0 | 0 | 0 |
| 0 | 0 | 0 | 0 | 0 |
| 0 | 0.00197349 | 0 | 0.001083371 | 0 |
| 0 | 0 | 0 | 0 | 0 |
| 0 | 0 | 0 | 0 | 0 |
| 0 | 0 | 0 | 0 | 0 |
| 0 | 0 | 0 | 0 | 0.001992442 |
| 0 | 0 | 0 | 0 | 0 |
| 0 | 0 | 0 | 0 | 0 |
| 0 | 0 | 0 | 0 | 0 |
| 0 | 0 | 0 | 0 | 0 |
| 0 | 0 | 0 | 0 | 0 |
| 0 | 0 | 0 | 0 | 0 |
| 0.00056553 | 0 | 0.005089274 | 0.010137871 | 0.01735792 |
| 0 | 0 | 0 | 0.00843187 | 0.0347967 |
| 0 | 0 | 0 | 0 | 0 |
| 0 | 0.00984224 | 0 | 0.000753681 | 0 |
| 0 | 0 | 0 | 0 | 0 |
| 0 | 0 | 0 | 0 | 0 |
| 0 | 0 | 0 | 0.011034992 | 0 |
| 0 | 0 | 0 | 0 | 0 |
| 0 | 0 | 0 | 0 | 0 |
| 0 | 0 | 0 | 0 | 0 |
| 0 | 0 | 0 | 0.011771626 | 0 |

| | | | | |
|---|---|---|---|---|
| 0 | 0 | 0 | 0 | 0 |
| 0 | 0 | 0 | 0.014489249 | 0 |
| 0 | 0 | 0 | 0 | 0 |
| 0 | 0 | 0 | 0 | 0.000580112 |
| 0 | 0 | 0.002167306 | 0 | 0.000807067 |
| 0 | 0 | 0 | 0.001203744 | 0 |
| 0 | 0 | 0 | 0 | 0.003799944 |
| 0 | 0 | 0 | 0.573349836 | 0 |
| 0 | 0 | 0 | 0 | 0.41801831 |
| 0 | 0 | 0 | 0.000774708 | 0 |
| 0.023384301 | 0 | 0 | 0.027837362 | 0.037869094 |
| 0 | 0 | 0 | 0 | 0 |
| 0.014079623 | 0 | 0 | 0.004989941 | 0.03906569 |
| 0 | 0 | 0 | 0 | 0 |
| 0 | 0 | 0 | 0 | 0 |
| 0 | 0 | 0 | 0 | 0 |
| 0 | 0 | 0 | 0 | 0 |
| 0 | 0 | 0 | 0 | 0 |
| 0 | 0 | 0 | 0 | 0 |
| 0 | 0 | 0 | 0 | 0 |
| 0 | 0 | 0 | 0 | 0 |
| 0 | 0 | 0 | 0 | 0 |
| 0 | 0.002694352 | 0 | 0.00484973 | 0 |
| 0 | 0 | 0.002771806 | 0.010153441 | 0.041692396 |
| 0 | 0 | 0 | 0 | 0 |
| 0 | 0 | 0 | 0.002317691 | 0.02866677 |
| 0 | 0 | 0 | 0.019281328 | 0 |
| 0 | 0 | 0 | 0 | 0 |
| 0 | 0 | 0 | 0 | 0 |
| 0 | 0 | 0 | 0 | 0 |
| 0 | 0 | 0 | 0.014826817 | 0.042715083 |
| 0 | 0 | 0 | 0 | 0.02241029 |
| 0 | 0 | 0 | 0 | 0 |
| 0 | 0 | 0 | 0 | 0 |
| 0 | 0 | 0 | 0.007458282 | 0 |
| 0 | 0.004348384 | 0 | 0 | 0 |
| 0 | 0.003682769 | 0 | 0 | 0 |
| 0 | 0 | 0 | 0 | 0 |
| 0 | 0 | 0.064577229 | 0.00059033 | 0.000383109 |
| 0 | 0 | 0 | 0 | 0 |
| 0 | 0 | 0 | 0 | 0 |
| 0 | 0 | 0 | 0 | 0 |
| 0 | 0 | 0 | 0 | 0 |
| 0 | 0 | 0 | 0 | 0 |
| 0.016128766 | 0 | 0 | 0 | 0 |
| 0.030503566 | 0.072078422 | 0.054449906 | 0.094139983 | 0.047527206 |
| 0 | 0 | 0 | 0 | 0 |
| 0 | 0 | 0 | 0 | 0 |

| | | | | |
|---|---|---|---|---|
| 0 | 0 | 0 | 0 | 0 |
| 0 | 0 | 0 | 0 | 0 |
| 0.146243525 | 0 | 0 | 0 | 0 |
| 0 | 0 | 0 | 0 | 0.076336211 |
| 0 | 0 | 0 | 0 | 0 |
| 0 | 0 | 0 | 0 | 0 |
| 0 | 0 | 0 | 0 | 0 |
| 0 | 0 | 0 | 0 | 0 |
| 0 | 0.003075428 | 0 | 0.000819319 | 0 |
| 0 | 0 | 0 | 0 | 0 |
| 0 | 0 | 0 | 0 | 0 |
| 0 | 0 | 0 | 0 | 0 |
| 0 | 0 | 0 | 0 | 0 |
| 0 | 0 | 0 | 0 | 0 |
| 0 | 0 | 0 | 0 | 0 |
| 0 | 0 | 0 | 0 | 0 |
| 0.002768609 | 0 | 0 | 0 | 0 |
| 0 | 0 | 0 | 0 | 0 |
| 0 | 0 | 0 | 0 | 0 |
| 0 | 0 | 0 | 0.004128311 | 0 |
| 0 | 0 | 0 | 0 | 0 |
| 0 | 0 | 0 | 0 | 0 |
| 0 | 0 | 0 | 0 | 0 |
| 0 | 0 | 0 | 0 | 0 |
| 0 | 0 | 0 | 0 | 0 |
| 0 | 0 | 0 | 0 | 0 |
| 0 | 0 | 0 | 0 | 0 |
| 0.016585395 | 0 | 0 | 0 | 0.008210628 |
| 0 | 0 | 0 | 0 | 0 |
| 0 | 0 | 0 | 0 | 0 |
| 0 | 0.000903586 | 0 | 0.000954866 | 0.005193534 |
| 0 | 0 | 0 | 0 | 0 |
| 0.024417358 | 0.14667198 | 0.038389066 | 0.076024939 | 0.06222093 |
| 0 | 0 | 0 | 0 | 0.000420152 |
| 0.000824086 | 0 | 0 | 0 | 0 |
| 0 | 0 | 0 | 0 | 0 |
| 0 | 0 | 0 | 0 | 0 |
| 0 | 0 | 0 | 0 | 0 |
| 0 | 0 | 0 | 0 | 0 |
| 0 | 0 | 0 | 0 | 0 |
| 0 | 0 | 0 | 0 | 0 |
| 0 | 0 | 0 | 0.074141918 | 0.372526691 |
| 0.018146478 | 0.311254915 | 0 | 0.076038734 | 0.248487038 |
| 0 | 0 | 0 | 0 | 0 |
| 0 | 0 | 0 | 0 | 0 |
| 0 | 0 | 0 | 0.002064575 | 0.011314328 |
| 0 | 0.003785773 | 0 | 0.000751997 | 0 |
| 0 | 0 | 0 | 0.000961442 | 0 |

| | | | | |
|---|---|---|---|---|
| 0 | 0 | 0 | 0 | 0 |
| 0 | 0 | 0 | 0.011641432 | 0.051373884 |
| 0 | 0.004117803 | 0.003125115 | 0.025642739 | 0.002857625 |
| 0 | 0 | 0 | 0 | 0 |
| 0 | 0 | 0 | 0 | 0 |
| 0 | 0.031003786 | 0 | 0.001962778 | 0 |
| 0 | 0 | 0 | 0 | 0 |
| 0 | 0 | 0 | 0 | 0 |
| 0 | 0 | 0 | 0 | 0 |
| 0 | 0 | 0 | 0 | 0 |
| 0 | 0 | 0 | 0 | 0 |
| 0 | 0 | 0 | 0 | 0 |
| 0 | 0 | 0 | 0 | 0 |
| 0 | 0 | 0 | 0.053883471 | 0 |
| 0 | 0 | 0 | 0 | 0 |
| 0 | 0 | 0 | 0 | 0 |
| 0 | 0 | 0 | 0 | 0 |
| 0 | 0 | 0 | 0 | 0 |
| 0 | 0 | 0 | 0 | 0 |
| 0 | 0 | 0 | 0 | 0 |
| 0 | 0 | 0 | 0.000545578 | 0 |
| 0 | 0.000621056 | 0 | 0 | 0 |
| 0 | 0 | 0 | 0 | 0 |
| 0 | 0 | 0 | 0 | 0 |
| 0 | 0 | 0 | 0 | 0.001108275 |
| 0 | 0 | 0.001381827 | 0.001771627 | 0.001254261 |
| 0 | 0 | 0 | 0 | 0 |
| 0 | 0 | 0 | 0.037651645 | 0.179392235 |
| 0 | 0 | 0 | 0 | 0 |
| 0 | 0 | 0 | 0 | 0 |
| 0 | 0 | 0 | 0 | 0 |
| 0 | 0 | 0.000454418 | 0 | 0 |
| 0 | 0 | 0 | 0 | 0 |
| 0 | 0 | 0 | 0 | 0 |
| 0 | 0 | 0 | 0 | 0 |
| 0 | 0 | 0 | 0 | 0 |
| 0 | 0 | 0 | 0 | 0 |
| 0 | 0 | 0 | 0 | 0 |
| 0 | 0 | 0 | 0.376680971 | 14.25702477 |
| 0 | 0 | 0 | 0 | 0 |
| 0 | 0 | 0 | 0 | 0 |
| 0 | 0 | 0 | 0.004193328 | 0 |
| 0 | 0 | 0 | 0 | 0 |
| 0 | 0 | 0 | 0 | 0 |
| 0 | 0 | 0 | 0 | 0 |
| 0 | 0 | 0 | 0 | 0 |
| 0 | 0 | 0 | 0 | 0.011340226 |
| 0 | 0 | 0 | 0 | 0 |

| | | | | |
|---|---|---|---|---|
| 0 | 0 | 0 | 0 | 0 |
| 0 | 0.014674374 | 0.012213868 | 0 | 0.001079946 |
| 0 | 0 | 0 | 0 | 0 |
| 0 | 0 | 0 | 0 | 0 |
| 0 | 0 | 0 | 0 | 0 |
| 0 | 0 | 0 | 0 | 0 |
| 0 | 0 | 0 | 0 | 0 |
| 0 | 0 | 0 | 0 | 0 |
| 0 | 0 | 0 | 0 | 0 |
| 0 | 0 | 0 | 0 | 0 |
| 0 | 0 | 0 | 0 | 0 |
| 0 | 0 | 0 | 0 | 0 |
| 0 | 0 | 0 | 0.004287764 | 0.28411526 |
| 0 | 0 | 0 | 0 | 0 |
| 0 | 0.00566941 | 0 | 0.001750663 | 0 |
| 0 | 0 | 0 | 0 | 0 |
| 0 | 0 | 0 | 0 | 0 |
| 0 | 0 | 0 | 0 | 0 |
| 0 | 0 | 0 | 0.004518936 | 0 |
| 0 | 0 | 0 | 0 | 0 |
| 0 | 0 | 0 | 0 | 0 |
| 0 | 0.002024658 | 0 | 0.104303552 | 0.001983597 |
| 0 | 0 | 0 | 0.030386062 | 0.043186578 |
| 0 | 0 | 0 | 0 | 0 |
| 0 | 0 | 0 | 0 | 0 |
| 0 | 0 | 0 | 0 | 0 |
| 0 | 0 | 0 | 0 | 0 |
| 0 | 0 | 0 | 0 | 0 |
| 0 | 0 | 0 | 0 | 0 |
| 0 | 0 | 0 | 0 | 0 |
| 0 | 0 | 0 | 0 | 0 |
| 0 | 0 | 0 | 0 | 0 |
| 0 | 0 | 0 | 0 | 0 |
| 0 | 0 | 0 | 0 | 0 |
| 6.176588973 | 0 | 0 | 7.769885439 | 0.074291083 |
| 0 | 0 | 0 | 0.300094709 | 0 |
| 0 | 0 | 0 | 0 | 0 |
| 0 | 0 | 0 | 0 | 0 |
| 0.001800595 | 0 | 0.00122756 | 0.108738166 | 0.08864357 |
| 0 | 0 | 0 | 0 | 0 |
| 0.008610517 | 0.026739831 | 0.023847844 | 0.104682222 | 0.281660601 |
| 0 | 0 | 0 | 0 | 0 |
| 0 | 0 | 0 | 0 | 0 |
| 0 | 0 | 0 | 0 | 0 |
| 0 | 0 | 0 | 0 | 0 |
| 0.000821538 | 4.614346324 | 0.021633295 | 0.00058752 | 0.003728118 |
| 0 | 0 | 0 | 0.027010961 | 0 |
| 0 | 0 | 0 | 0.008320342 | 0 |

| | | | | |
|---|---|---|---|---|
| 0 | 0 | 0 | 0 | 0 |
| 0 | 0 | 0 | 0 | 0 |
| 0 | 0 | 0 | 0 | 0 |
| 0.002296251 | 0 | 0 | 0 | 0 |
| 0 | 0 | 0 | 0 | 0 |
| 0 | 0 | 0 | 0 | 0 |
| 0 | 0 | 0 | 0 | 0 |
| 0 | 0 | 0 | 0.000846701 | 0 |
| 0 | 0 | 0 | 0 | 0 |
| 0 | 0 | 0.019719515 | 0 | 0.001325857 |
| 0.00656711 | 0 | 0 | 0 | 0 |
| 0 | 0 | 0 | 0 | 0 |
| 0 | 0.001488081 | 0 | 0 | 0.002429837 |
| 0 | 0 | 0 | 0 | 0 |
| 0 | 0 | 0 | 0 | 0 |
| 0 | 0 | 0 | 0 | 0 |
| 0 | 0 | 0 | 0 | 0 |
| 0 | 0 | 0 | 0 | 0 |
| 0 | 0 | 0 | 0 | 0 |
| 0.006465658 | 0.085650795 | 0.03877828 | 0.009164467 | 0.023213264 |
| 0 | 0 | 0 | 0 | 0 |
| 0 | 0 | 0 | 0 | 0 |
| 0 | 0 | 0 | 0 | 0 |
| 0 | 0 | 0 | 0 | 0 |
| 0 | 0 | 0 | 0 | 0 |
| 0.000523934 | 0 | 0 | 0 | 0 |
| 0 | 0 | 0 | 0 | 0 |
| 0 | 0 | 0 | 0 | 0 |
| 0 | 0 | 0 | 0 | 0 |
| 0 | 0.01439507 | 0 | 0 | 0 |
| 0 | 0 | 0 | 0 | 0 |
| 0 | 0 | 0 | 0.00108897 | 0 |
| 0 | 0 | 0 | 0 | 0 |
| 0 | 0 | 0 | 0 | 0 |
| 0 | 0 | 0 | 0 | 0 |
| 0 | 0 | 0.002313253 | 0 | 0 |
| 0 | 0 | 0 | 0 | 0 |
| 0 | 0 | 0 | 0 | 0 |
| 0 | 0 | 0 | 0 | 0 |
| 0 | 0 | 0 | 0 | 0.328424978 |
| 0 | 0 | 0 | 0 | 0 |
| 0 | 0 | 0 | 0 | 0 |
| 0 | 0 | 0 | 0.064682612 | 0 |
| 0 | 0 | 0 | 0 | 0 |
| 0 | 0 | 0 | 0 | 0.001760471 |
| 0 | 0 | 0 | 0 | 0 |
| 0 | 0 | 0 | 0 | 0 |
| 0 | 0 | 0 | 0 | 0 |

| | | | | |
|---|---|---|---|---|
| 0 | 0 | 0 | 0 | 0 |
| 0 | 0 | 0 | 0 | 0 |
| 0.001677519 | 0 | 0 | 0 | 0 |
| 0 | 0 | 0 | 0 | 0 |
| 0 | 0 | 0 | 0 | 0 |
| 0 | 0 | 0 | 0.003154538 | 0 |
| 0 | 0 | 0 | 0 | 0 |
| 0 | 0 | 0 | 0 | 0 |
| 0 | 0 | 0 | 0 | 0 |
| 0 | 0 | 0 | 0 | 0 |
| 0 | 0 | 0 | 0 | 0 |
| 0 | 0 | 0 | 0 | 0 |
| 0 | 0 | 0 | 0 | 0 |
| 0 | 0 | 0 | 0 | 0 |
| 0 | 0 | 0 | 0 | 0 |
| 0 | 0 | 0 | 0 | 0 |
| 0 | 0 | 0 | 0.292934687 | 0 |
| 0 | 0.00800626 | 0 | 0 | 0 |
| 0 | 0 | 0 | 0 | 0 |
| 0 | 0 | 0 | 0 | 0 |
| 0 | 0 | 0 | 0 | 0 |
| 0 | 0 | 0 | 0 | 0 |
| 0 | 0 | 0 | 0 | 0 |
| 0 | 0 | 0 | 0 | 0 |
| 0 | 0 | 0 | 0 | 0 |
| 0 | 0 | 0.002743896 | 0 | 0 |
| 0 | 0 | 0 | 0 | 0 |
| 0 | 0 | 0 | 0.037202831 | 0 |
| 0 | 0 | 0 | 0 | 0 |
| 0 | 0 | 0 | 0.031048695 | 0 |
| 0 | 0 | 0 | 0 | 0 |
| 0 | 0 | 0 | 0 | 0 |
| 0 | 0 | 0 | 0 | 0 |
| 0 | 0 | 0 | 0 | 0 |
| 0 | 0 | 0 | 0 | 0 |
| 0 | 0 | 0 | 0 | 0 |
| 0 | 0 | 0 | 0 | 0 |
| 0 | 0 | 0 | 0 | 0 |
| 0.192505066 | 0 | 0 | 0.125176349 | 0 |
| 0 | 0 | 0 | 0 | 0 |
| 0 | 0 | 0 | 0 | 0 |
| 0 | 0 | 0 | 0 | 0 |
| 0 | 0 | 0 | 0 | 0 |
| 0 | 0 | 0 | 0 | 0 |
| 0 | 0 | 0 | 0 | 0 |
| 0 | 0 | 0 | 0 | 0 |
| 0 | 0 | 0 | 0 | 0 |
| 0 | 0 | 0 | 0 | 0 |

| | | | | |
|---|---|---|---|---|
| 0 | 0 | 0 | 0 | 0 |
| 0 | 0 | 0 | 0.010202862 | 0 |
| 0 | 0 | 0 | 0 | 0 |
| 0 | 0 | 0 | 0 | 0 |
| 0 | 0 | 0 | 0 | 0 |
| 0 | 0 | 0 | 0 | 0 |
| 0 | 0 | 0 | 0 | 0 |
| 0 | 0 | 0 | 0 | 0 |
| 0 | 0 | 0 | 0 | 0 |
| 0 | 0 | 0 | 0 | 0.003534633 |
| 0 | 0 | 0.009184614 | 0 | 0.001926707 |
| 0 | 0 | 0 | 0 | 0 |
| 0.000396914 | 0 | 0 | 0.331412788 | 0 |
| 0 | 0 | 0 | 0 | 0 |
| 0 | 0 | 0 | 0 | 0 |
| 0 | 0 | 0 | 0 | 0 |
| 0 | 0 | 0 | 0 | 0 |
| 0 | 0 | 0 | 0 | 0 |
| 0 | 0 | 0 | 0 | 0 |
| 0 | 0 | 0 | 0 | 0 |
| 0 | 0 | 0.002237239 | 0 | 0.00180507 |
| 0 | 0 | 0 | 0 | 0 |
| 0 | 0 | 0 | 0 | 0 |
| 0.001208212 | 0 | 2.900303965 | 0.158984893 | 0.761241955 |
| 0 | 0 | 0 | 0 | 0 |
| 0 | 0 | 0 | 0 | 0 |
| 0 | 0 | 0 | 0 | 0 |
| 0 | 0 | 0 | 0 | 0 |
| 0 | 0 | 0 | 0 | 0 |
| 0 | 0 | 0 | 0 | 0 |
| 0 | 0 | 0 | 0 | 0 |
| 0 | 0 | 0 | 0 | 0 |
| 0 | 0 | 0 | 0 | 0.00063292 |
| 0 | 0 | 0 | 0 | 0 |

| PHL018-0162_V3 REF | PHL018-0007_V3 EYCF | PHL018-0254_V2 REF | PHL018-0020_V1 EYCF | PHL018-0022_V1 CTRL |
|---|---|---|---|---|
| 0.063505085 | 0.044720885 | 1.955664155 | 0.372176419 | 0.436803633 |
| 0.174987194 | 5.319147292 | 0 | 6.955510862 | 0.890818016 |
| 0 | 4.670267392 | 0 | 0 | 0 |
| 0 | 0 | 0 | 0 | 0 |
| 0 | 0 | 0 | 0 | 0 |
| 0 | 0 | 0 | 0 | 0 |
| 0 | 0 | 0 | 0 | 0 |
| 0 | 0 | 0 | 0 | 0 |
| 0 | 0 | 0 | 0 | 0 |
| 0 | 0 | 0 | 0 | 0 |
| 0 | 0 | 0 | 0 | 0 |
| 0 | 0 | 0 | 0 | 0 |
| 0 | 0 | 0 | 0 | 0 |
| 0.62431038 | 3.167454081 | 0 | 0 | 0.341779741 |
| 0 | 0 | 0 | 0 | 0 |
| 0 | 0 | 0 | 0 | 0 |
| 0 | 0 | 0 | 0 | 0 |
| 0 | 0 | 0 | 0 | 0 |
| 0 | 0 | 0 | 0 | 0 |
| 0 | 0 | 0 | 0 | 0 |
| 0 | 0 | 0 | 0 | 0 |
| 0 | 0 | 0 | 0 | 0 |
| 0 | 0 | 0 | 0 | 0 |
| 0 | 0 | 0 | 0 | 0 |
| 0 | 0.022209872 | 0 | 0 | 0 |
| 0 | 0 | 0 | 0 | 0 |
| 0 | 0 | 0 | 0 | 0 |
| 0 | 0 | 0 | 0 | 0 |
| 0 | 0 | 0 | 0 | 0 |
| 0 | 0 | 0 | 0 | 0 |
| 0 | 0 | 0 | 0 | 0 |
| 0 | 0 | 0 | 0 | 0 |
| 0 | 0 | 0 | 0 | 0 |
| 0 | 0 | 0 | 0 | 0 |
| 0 | 0 | 0 | 0 | 0 |
| 10.34108483 | 0 | 0 | 0 | 0 |
| 0 | 0 | 0 | 0 | 0 |
| 0 | 0 | 0 | 0 | 0 |
| 0 | 0 | 0 | 0 | 0 |
| 0 | 0 | 0 | 0 | 0 |
| 0 | 0 | 0 | 0 | 0 |
| 0.026678728 | 0 | 0.050581962 | 0 | 0.045156823 |
| 7.259076374 | 0.004609862 | 1.167047993 | 0.036216223 | 5.908268838 |
| 0.056585125 | 0.002535768 | 0.338027147 | 0.13752791 | 0.134696683 |
| 0.007084998 | 0 | 0.012975069 | 0.003424428 | 0 |
| 0 | 0 | 0 | 0 | 0 |

| | | | | |
|---|---|---|---|---|
| 0 | 0 | 0 | 0 | 0 |
| 0.001018885 | 0 | 0.044263083 | 1.047271701 | 0 |
| 0 | 0.006931455 | 0 | 0 | 0.060121849 |
| 1.643505549 | 0.473348367 | 0 | 0.435700501 | 0.291407682 |
| 0 | 0 | 0 | 0.010189627 | 0 |
| 0 | 0 | 0 | 0 | 0 |
| 0.789574176 | 0.000998371 | 0.002426057 | 0 | 0 |
| 0 | 0 | 0 | 0 | 0 |
| 0 | 0 | 0 | 0 | 0 |
| 0 | 0 | 0 | 0 | 0 |
| 0 | 0 | 0 | 0 | 0 |
| 2.167541897 | 1.348382241 | 0 | 0.340346095 | 0 |
| 0 | 0 | 0 | 0 | 0 |
| 0.101674778 | 0 | 0.006563758 | 0.049431662 | 0.028825961 |
| 0.014593694 | 0 | 0 | 0 | 0 |
| 0 | 0 | 0 | 0 | 0 |
| 0.007076487 | 0 | 0.001058614 | 0.083830688 | 0 |
| 0 | 0 | 0 | 0 | 0 |
| 0 | 0 | 0 | 0 | 0 |
| 0 | 0 | 0 | 0 | 0 |
| 0.10776951 | 0.642017551 | 0.182576606 | 0.219968366 | 0.097976961 |
| 0.003569412 | 0.005142973 | 0 | 0.005284771 | 0.015665763 |
| 0 | 0 | 0 | 0 | 0 |
| 0 | 0 | 0 | 0 | 0 |
| 0 | 0 | 0 | 0 | 0 |
| 0 | 0 | 0 | 0 | 0 |
| 0 | 0 | 0.005488719 | 0 | 0 |
| 0 | 0 | 0 | 0 | 0 |
| 0 | 0 | 0.008085843 | 0 | 0.003961601 |
| 0 | 0 | 0 | 0 | 0 |
| 0 | 0.525685741 | 0.016711119 | 0 | 0 |
| 0 | 0 | 0 | 0 | 0 |
| 0 | 0 | 0 | 0 | 0 |
| 0 | 0 | 0 | 0 | 0 |
| 0 | 0 | 0 | 0 | 0 |
| 0 | 0 | 0 | 0 | 0 |
| 0 | 0 | 0 | 0 | 0 |
| 0 | 0 | 0 | 5.878242858 | 0 |
| 0 | 0 | 0 | 0 | 0 |
| 0 | 0 | 0 | 0 | 0 |
| 0 | 0 | 0 | 0 | 0 |
| 0 | 0 | 0 | 0 | 0 |
| 0 | 0 | 0 | 0 | 0 |
| 0 | 0 | 0 | 0 | 0 |
| 0 | 0 | 0 | 0 | 0 |
| 0 | 0 | 0 | 0 | 0 |
| 0 | 0.078274924 | 0 | 0.010454045 | 0.002026907 |
| 0.004233437 | 0 | 0 | 0 | 0 |

| | | | | |
|---|---|---|---|---|
| 0 | 0 | 0 | 0 | 0 |
| 0 | 0 | 0 | 0 | 0.045720722 |
| 0 | 0 | 0 | 0 | 0 |
| 0 | 0 | 0 | 0 | 0 |
| 0 | 0 | 0 | 0 | 0.021149254 |
| 0 | 0 | 0 | 0 | 0 |
| 0 | 0 | 0 | 0 | 0 |
| 0 | 0 | 0 | 0 | 0 |
| 0 | 0 | 0.003517493 | 0.102367237 | 0.051236938 |
| 0 | 0 | 0 | 0 | 0 |
| 0 | 0 | 0 | 0 | 0 |
| 0 | 0 | 0.114129794 | 0 | 0.016966533 |
| 0 | 0 | 0 | 0 | 0 |
| 0 | 0 | 0 | 0 | 0 |
| 0 | 0 | 0 | 0 | 0 |
| 0 | 0 | 0 | 0 | 0 |
| 0 | 0 | 0 | 0 | 0 |
| 0 | 0 | 0 | 0 | 0 |
| 0.300521625 | 0 | 0.771627316 | 0 | 0.230857408 |
| 0 | 0.05517952 | 0 | 0 | 0 |
| 0 | 0 | 0 | 0 | 0 |
| 0 | 0 | 0 | 0 | 0 |
| 0 | 0 | 0 | 0 | 0 |
| 0 | 0 | 0 | 0 | 0 |
| 0 | 0 | 0 | 0 | 0 |
| 0 | 0 | 0 | 0 | 0 |
| 0 | 0 | 0 | 0 | 0.003096207 |
| 0 | 0 | 0 | 0 | 0 |
| 0 | 0 | 0 | 0 | 0 |
| 0 | 0 | 0 | 0 | 0 |
| 0 | 0.109291995 | 0 | 0 | 0 |
| 0.007426829 | 0 | 0.013838779 | 0.00119015 | 0 |
| 0 | 0 | 0 | 0 | 0 |
| 0 | 0 | 0 | 0 | 0 |
| 0 | 0 | 0 | 0 | 0 |
| 0 | 0 | 0 | 0 | 0 |
| 0 | 0 | 0 | 0 | 0 |
| 0 | 0 | 0 | 0 | 0 |
| 0 | 0 | 0 | 0 | 0 |
| 0 | 0 | 0 | 0 | 0 |
| 0 | 0 | 0 | 0 | 0 |
| 0 | 0 | 0 | 0 | 0 |
| 0.001178229 | 0.049817147 | 0.003739194 | 0.030034494 | 0.000592703 |
| 0 | 0 | 0 | 0 | 0 |
| 0 | 0 | 0 | 0 | 0 |
| 0 | 0 | 0 | 0 | 0 |
| 0 | 0 | 0.250237151 | 0.00793212 | 0 |
| 0 | 0 | 0 | 0 | 0 |

| | | | | |
|---|---|---|---|---|
| 0 | 0 | 0.012709626 | 0 | 0 |
| 0 | 0 | 0 | 0 | 0 |
| 0 | 0 | 0 | 0 | 0 |
| 0 | 0 | 0 | 0 | 0 |
| 0.011153938 | 0.026505993 | 0 | 0 | 0 |
| 0 | 0 | 0 | 0.03190237 | 0 |
| 0 | 0 | 0 | 0 | 0 |
| 0 | 0 | 0 | 0 | 0 |
| 0 | 0 | 0 | 0 | 0 |
| 0 | 0 | 0 | 0 | 0 |
| 0 | 0 | 0 | 0 | 0 |
| 0 | 0 | 0 | 0 | 0 |
| 0 | 0 | 0 | 0 | 0 |
| 0 | 0 | 0 | 0 | 0 |
| 0 | 0 | 0 | 0 | 0 |
| 0 | 0 | 0 | 0 | 0 |
| 0 | 0 | 0 | 0 | 0 |
| 0 | 0 | 0 | 0 | 0 |
| 0 | 0 | 0 | 0 | 0 |
| 0 | 0 | 0 | 0 | 0 |
| 0 | 0 | 0 | 0 | 0 |
| 0 | 0 | 0 | 0 | 0 |
| 0 | 0 | 0 | 0 | 0 |
| 0 | 0 | 0 | 0 | 0 |
| 0 | 0 | 0 | 0 | 0 |
| 0 | 0 | 0 | 0 | 0 |
| 0 | 0 | 0 | 0 | 0 |
| 0 | 0 | 0 | 0 | 0 |
| 0 | 0 | 0 | 0 | 0 |
| 0 | 0 | 0 | 0 | 0 |
| 0.003626572 | 0 | 0 | 0 | 0 |
| 0 | 0 | 0 | 0 | 0 |
| 0 | 0 | 0 | 0 | 0 |
| 0 | 0 | 0 | 0 | 0 |
| 0 | 0 | 0 | 0 | 0 |
| 0 | 0 | 0 | 0 | 0 |
| 0 | 0 | 0 | 0 | 0 |
| 0 | 0 | 0 | 0 | 0 |
| 0 | 0 | 0 | 0 | 0 |
| 0 | 0 | 0 | 0 | 0 |
| 0 | 0 | 0 | 0 | 0 |
| 0 | 0 | 0 | 0 | 0 |
| 0 | 0 | 0 | 0 | 0 |
| 0 | 0 | 0 | 0 | 0 |
| 0 | 0 | 0 | 0 | 0 |
| 0 | 0 | 0 | 0 | 0 |
| 0 | 0 | 0 | 0 | 0 |
| 0 | 0 | 0 | 0.082774181 | 0 |

| | | | | |
|---|---|---|---|---|
| 0 | 0 | 0 | 0 | 0 |
| 0 | 0 | 0 | 0 | 0 |
| 0 | 0 | 0 | 0 | 0 |
| 0 | 0 | 0 | 0 | 0 |
| 0 | 0 | 0 | 0 | 0 |
| 0 | 0 | 0 | 0 | 0 |
| 0 | 0 | 0 | 0 | 0 |
| 0 | 0 | 0 | 0 | 0 |
| 0 | 0 | 0 | 0 | 0 |
| 0 | 0 | 0 | 0 | 0 |
| 0 | 0 | 0 | 0 | 0 |
| 0 | 0 | 0 | 0 | 0 |
| 0 | 0.001286088 | 0.002381115 | 0 | 0.000943582 |
| 0 | 0 | 0 | 0 | 0 |
| 0 | 0 | 0 | 0 | 0 |
| 0 | 0 | 0 | 0 | 0 |
| 0 | 0 | 0 | 0 | 0 |
| 0 | 0 | 0 | 0 | 0 |
| 0 | 0 | 0 | 0 | 0 |
| 0 | 0 | 0 | 0 | 0 |
| 0.031564607 | 0.067330925 | 0.035987908 | 0.065925634 | 0.012702753 |
| 0 | 0 | 0 | 0 | 0 |
| 0 | 0 | 0 | 0 | 0.001238431 |
| 0 | 0.007881007 | 0 | 0 | 0 |
| 0 | 0 | 0 | 0 | 0 |
| 0 | 0 | 0 | 0 | 0 |
| 0.045868672 | 0.001200366 | 0.048399032 | 0.008107952 | 0.004315374 |
| 0 | 0 | 0 | 0 | 0 |
| 0 | 0 | 0 | 0 | 0 |
| 0 | 0 | 0 | 0 | 0.000659565 |
| 0 | 0 | 0 | 0 | 0 |
| 0 | 0 | 0 | 0 | 0 |
| 0 | 0 | 0 | 0 | 0 |
| 0.061361613 | 0.00335906 | 0 | 0 | 0 |
| 0 | 0 | 0.02679523 | 0.13161489 | 0.02181065 |
| 0 | 0 | 0 | 0 | 0 |
| 0 | 0 | 0 | 0 | 0 |
| 0 | 0 | 0 | 0 | 0 |
| 0 | 0 | 0 | 0 | 0 |
| 0 | 0 | 0 | 0 | 0 |
| 0 | 0 | 0 | 0 | 0 |
| 0 | 0 | 0 | 0 | 0 |
| 0 | 0 | 0.02082525 | 0 | 0 |
| 0 | 0 | 0 | 0 | 0 |
| 0 | 0 | 0 | 0 | 0 |
| 0 | 0 | 0 | 0 | 0 |
| 0.001058894 | 0 | 0 | 0 | 0.005468768 |
| 0 | 0.001274267 | 0.011009733 | 0 | 0.007526016 |

| | | | | |
|---|---|---|---|---|
| 0 | 0 | 0 | 0 | 0 |
| 0 | 0 | 0 | 0 | 0 |
| 0 | 0 | 0 | 0 | 0 |
| 0 | 0.00121482 | 0 | 0 | 0 |
| 0 | 0 | 0 | 0 | 0 |
| 0 | 0 | 0 | 0 | 0 |
| 0 | 0 | 0.005387324 | 0 | 0 |
| 0 | 0 | 0.003956673 | 0 | 0.004939011 |
| 0 | 0 | 0 | 0 | 0 |
| 0 | 0 | 0 | 0 | 0 |
| 0 | 0 | 0.001915569 | 0 | 0 |
| 0 | 0 | 0 | 0 | 0 |
| 0 | 0 | 0 | 0 | 0 |
| 0 | 0 | 0 | 0 | 0 |
| 0 | 0 | 0 | 0 | 0 |
| 0 | 0 | 0 | 0 | 0 |
| 0 | 0 | 0 | 0 | 0 |
| 0 | 0 | 0 | 0 | 0 |
| 0.033454393 | 0 | 0.00454871 | 0 | 0.045257892 |
| 0 | 0.12214668 | 0 | 0 | 0 |
| 0 | 0.006451702 | 0 | 0 | 0 |
| 0 | 0 | 0 | 0 | 0 |
| 0 | 0 | 0 | 0 | 0 |
| 0 | 0 | 0 | 0 | 0 |
| 0.011632042 | 0 | 0.007690648 | 0.004526945 | 0.004388587 |
| 0 | 0 | 0 | 0 | 0 |
| 0 | 0 | 0 | 0 | 0 |
| 0 | 0 | 0 | 0 | 0 |
| 0 | 0 | 0 | 0 | 0 |
| 0 | 0 | 0 | 0 | 0 |
| 0.238626782 | 0.107529175 | 0.011195752 | 0.333626542 | 0.624435869 |
| 0 | 0 | 0 | 0 | 0 |
| 0 | 0 | 0 | 0 | 0 |
| 0 | 0 | 0 | 0 | 0 |
| 0 | 0 | 0 | 0 | 0 |
| 0 | 0 | 0 | 0 | 0 |
| 0 | 0 | 0 | 0 | 0 |
| 0 | 0 | 0 | 0 | 0 |
| 0 | 0 | 0 | 0 | 0 |
| 0 | 0 | 0.00322222 | 0 | 0 |
| 0 | 0 | 0 | 0 | 0 |
| 0.009560348 | 0.032058582 | 0.00560939 | 0.150931755 | 0.014900922 |
| 0 | 0 | 0 | 0 | 0 |
| 0 | 0 | 0 | 0 | 0 |
| 0 | 0 | 0 | 0 | 0 |
| 0 | 0 | 0 | 0 | 0 |
| 0 | 0 | 0 | 0 | 0 |
| 0 | 0 | 0 | 0 | 0 |

| | | | | |
|---|---|---|---|---|
| 0 | 0 | 0 | 0 | 0 |
| 0 | 0 | 0 | 0 | 0 |
| 0.063762084 | 0.001574211 | 0.312343335 | 0.166734407 | 0.157538296 |
| 0 | 0 | 0 | 0 | 0 |
| 0.731645034 | 0 | 0 | 0 | 0 |
| 0 | 0 | 0 | 0 | 0 |
| 0 | 0 | 0 | 0 | 0 |
| 0 | 0 | 0 | 0 | 0 |
| 0 | 0 | 0 | 0 | 0 |
| 0 | 0 | 0 | 0.000735502 | 0 |
| 0 | 0 | 0 | 0 | 0 |
| 0 | 0 | 0 | 0 | 0 |
| 0 | 0 | 0.004228416 | 0 | 0 |
| 0.001957462 | 0.001491246 | 0.001073704 | 0.003258823 | 0.003829361 |
| 0.008616684 | 0.002461657 | 0.010191322 | 0.004890421 | 0.010114035 |
| 0 | 0.005490686 | 0 | 0 | 0 |
| 0.00531805 | 0 | 0.018175458 | 0.003343318 | 0.002881009 |
| 0 | 0 | 0 | 0 | 0 |
| 0 | 0 | 0 | 0 | 0 |
| 0 | 0 | 0 | 0 | 0 |
| 0 | 0 | 0 | 0 | 0 |
| 0 | 0.001054987 | 0.010482411 | 0 | 0.006935277 |
| 0 | 0 | 0 | 0 | 0 |
| 0 | 0 | 0.016603392 | 0 | 0 |
| 0 | 0 | 0 | 0.008468751 | 0.002780078 |
| 0 | 0 | 0 | 0 | 0 |
| 0 | 0 | 0 | 0 | 0 |
| 0 | 0 | 0 | 0 | 0 |
| 0 | 0 | 0 | 0 | 0 |
| 0 | 0 | 0 | 0.094589179 | 0.005015971 |
| 0 | 0 | 0 | 0.001541859 | 0 |
| 0 | 0 | 0 | 0 | 0 |
| 0 | 0 | 0.015091472 | 0 | 0 |
| 0 | 0 | 0 | 0 | 0 |
| 0 | 0 | 0 | 0 | 0 |
| 0 | 0 | 0.008647943 | 0 | 0 |
| 0 | 0 | 0 | 0 | 0 |
| 0.045236696 | 0 | 0.058978981 | 0.086897305 | 0.022328235 |
| 0 | 0 | 0 | 0 | 0 |
| 0 | 0 | 0 | 0 | 0 |
| 0 | 0 | 0 | 0 | 0 |
| 0 | 0 | 0 | 0 | 0 |
| 0 | 0 | 0 | 0 | 0 |
| 0 | 0 | 0 | 0 | 0 |
| 0 | 0 | 0 | 0 | 0 |
| 0 | 0 | 0.003521524 | 0 | 0.002093251 |
| 0 | 0 | 0.002172415 | 0 | 0.014167604 |
| 0 | 0 | 0 | 0 | 0 |

| | | | | |
|---|---|---|---|---|
| 0 | 0 | 9.564074914 | 0 | 0 |
| 0 | 0.042845007 | 0 | 0 | 0 |
| 0.001547181 | 0 | 0.015275837 | 0 | 0.101244259 |
| 0 | 0 | 0 | 0 | 0 |
| 0.027629894 | 0.013563386 | 0.009605603 | 0.010336484 | 0.002512279 |
| 0 | 0 | 0 | 0.008211203 | 0 |
| 0 | 0 | 0 | 0 | 0 |
| 0.199101732 | 0.002181713 | 0.059916434 | 0.118882932 | 0.019208246 |
| 0 | 0 | 0 | 0 | 0 |
| 0.006184666 | 0.064892181 | 0.177291642 | 0.029102113 | 0.01022915 |
| 0 | 0 | 0 | 0 | 0 |
| 0 | 0 | 0.052492823 | 0 | 0.021034021 |
| 0.00294787 | 0.009357358 | 0.006467845 | 0.00758459 | 0.004613509 |
| 0 | 0 | 0 | 0 | 0.155607794 |
| 0 | 0 | 0 | 0 | 0.000988518 |
| 0.015980041 | 0 | 0 | 0 | 0 |
| 0 | 0 | 0 | 0 | 0 |
| 0 | 0.001706144 | 0 | 0 | 0 |
| 0.021162393 | 0 | 0.010447167 | 0 | 0.013351422 |
| 0 | 0.031014819 | 0 | 0 | 0 |
| 0 | 0 | 0 | 0 | 0 |
| 0 | 0 | 0 | 0 | 0 |
| 0 | 0 | 0 | 0 | 0 |
| 0 | 0 | 0 | 0 | 0 |
| 0 | 0 | 0 | 0 | 0 |
| 0.0101098 | 0 | 0.004943787 | 0.003637576 | 0 |
| 0 | 0 | 0 | 0 | 0 |
| 0.01245623 | 0 | 0 | 0.084834786 | 0.003790576 |
| 0 | 0.033476817 | 0 | 0 | 0 |
| 0 | 0 | 0 | 0 | 0 |
| 0 | 0 | 0.00075785 | 0 | 0.000720765 |
| 0 | 0 | 0 | 0 | 0 |
| 0 | 0 | 0 | 0 | 0 |
| 0 | 0 | 0.003694609 | 0 | 0 |
| 0 | 0 | 0 | 0 | 0 |
| 0 | 0 | 0 | 0 | 0 |
| 0.006703308 | 0 | 0 | 0 | 0.001522871 |
| 0 | 0 | 0 | 0 | 0 |
| 0 | 0 | 0 | 0 | 0 |
| 0 | 0 | 0 | 0 | 0 |
| 0 | 0 | 0 | 0 | 0 |
| 0 | 0 | 0 | 0 | 0 |
| 0 | 0 | 0 | 0 | 0 |
| 0 | 0 | 0.004931196 | 0.003139885 | 0 |
| 0 | 0 | 0 | 0 | 0 |
| 0 | 0 | 0 | 0 | 0 |
| 0.001897711 | 0.001897516 | 0 | 0.001292461 | 0 |
| 0 | 0 | 0 | 0 | 0 |

| | | | | |
|---|---|---|---|---|
| 0 | 0 | 0 | 0 | 0 |
| 0 | 0 | 0 | 0 | 0 |
| 0 | 0 | 0 | 0 | 0 |
| 0 | 0 | 0 | 0 | 0 |
| 0 | 0 | 0 | 0 | 0 |
| 0.001121008 | 0.022631364 | 0 | 0 | 0 |
| 0 | 0 | 0 | 0 | 0 |
| 0 | 0 | 0 | 0 | 0 |
| 0 | 0 | 0 | 0 | 0 |
| 0 | 0 | 0 | 0 | 0 |
| 0.068259477 | 0.013371914 | 0.037174144 | 0.001180673 | 0.011700233 |
| 0 | 0 | 0 | 0 | 0 |
| 0 | 0 | 0 | 0 | 0 |
| 0.002047242 | 0.002573412 | 0.003284629 | 0 | 0.004085097 |
| 0.262865855 | 0.003550678 | 0 | 0 | 0.032823932 |
| 0 | 0 | 0 | 0 | 0 |
| 0 | 0 | 0 | 0 | 0 |
| 0 | 0 | 0 | 0 | 0 |
| 0 | 0 | 0 | 0.004748861 | 0 |
| 0 | 0.005891862 | 0 | 0 | 0 |
| 0.017425972 | 0 | 0 | 0 | 0 |
| 0 | 0.007406054 | 0 | 0 | 0 |
| 0 | 0 | 0 | 0 | 0 |
| 0 | 0 | 0 | 0 | 0 |
| 0 | 0 | 0 | 0 | 0 |
| 0 | 0 | 0 | 0 | 0 |
| 0 | 0 | 0 | 0 | 0.109951521 |
| 0 | 0 | 0 | 0 | 0 |
| 0 | 0 | 0 | 0 | 0 |
| 0.376003739 | 0.589130726 | 0.098513672 | 0.839982559 | 0.811183867 |
| 0.004305562 | 0 | 0 | 0.004918796 | 0 |
| 0 | 0 | 0.022140489 | 0 | 0 |
| 0 | 0 | 0 | 0 | 0 |
| 0 | 0 | 0 | 0 | 0 |
| 0.074508436 | 0 | 0 | 0 | 0 |
| 0 | 0 | 0 | 0 | 0 |
| 0 | 0 | 0 | 0 | 0 |
| 0 | 0 | 0.01982994 | 0 | 0 |
| 0 | 0.289844318 | 0 | 0 | 0.599790586 |
| 0.992461864 | 0 | 0 | 0 | 0.009383094 |
| 0 | 0.270992264 | 0 | 0 | 0.040498283 |
| 0.745606689 | 0 | 0 | 2.410768501 | 0 |
| 0 | 0 | 0.002123241 | 0 | 0 |
| 0 | 0 | 0 | 0 | 0 |
| 0 | 0 | 0 | 0 | 0 |
| 0 | 0 | 0 | 0 | 0 |
| 0.001059893 | 0.003835409 | 0.003052196 | 0.003368651 | 0 |
| 0 | 0 | 0 | 0 | 0 |

| | | | | |
|---|---|---|---|---|
| 0.003487279 | 0 | 0.002689927 | 0.001484411 | 0 |
| 0 | 0.004137788 | 0 | 0.003035183 | 0 |
| 0.023461444 | 0.001018114 | 0.002199139 | 0 | 0 |
| 0 | 0 | 0 | 0 | 0 |
| 0 | 0 | 0 | 0 | 0 |
| 0 | 0 | 0 | 0 | 0 |
| 0 | 0 | 0 | 0 | 0 |
| 0 | 0 | 0.015677286 | 0 | 0 |
| 0 | 0 | 0 | 0.007702375 | 0 |
| 0 | 0 | 0 | 0 | 0 |
| 0 | 0 | 0 | 0 | 0.081284799 |
| 0 | 0 | 0 | 0 | 0 |
| 0 | 0 | 0 | 0 | 0 |
| 0.000591535 | 0 | 0.000730053 | 0.001074327 | 0.009720593 |
| 0 | 0 | 0 | 0 | 0 |
| 0.004930927 | 0.0364499 | 0 | 0.093192056 | 0.129682688 |
| 0.016567049 | 0.001171041 | 0.013490473 | 0.033965939 | 0.003007108 |
| 0 | 0.021104409 | 0 | 0 | 0 |
| 0.001058506 | 0.000604798 | 0.00348366 | 0 | 0 |
| 0 | 0 | 0 | 0 | 0 |
| 0 | 0 | 0 | 0 | 0 |
| 0 | 0 | 0 | 0 | 0 |
| 0 | 0 | 0 | 0 | 0 |
| 0 | 0 | 0 | 0.635602536 | 0 |
| 0 | 0 | 0 | 0 | 0 |
| 0 | 0 | 0 | 0 | 0 |
| 0 | 0.02006972 | 0 | 0 | 0.004326553 |
| 0 | 0 | 0 | 0 | 0 |
| 0 | 0 | 0 | 0 | 0 |
| 0 | 0 | 0 | 0 | 0 |
| 0 | 0.014132376 | 0 | 0 | 0.051677562 |
| 0.010571019 | 0.029778421 | 0.02700301 | 0.005692652 | 0 |
| 0.070221202 | 0.03209783 | 0.043621212 | 0.296515434 | 0.009890854 |
| 0 | 0 | 0.00455396 | 0 | 0.020474361 |
| 0.003591348 | 0 | 0 | 0 | 0.0039344 |
| 0 | 0 | 0 | 0 | 0 |
| 0 | 0 | 0 | 0 | 0.001987114 |
| 0.011915535 | 0.013616358 | 0 | 0.02772704 | 0.0101011 |
| 0 | 0 | 0 | 0 | 0 |
| 0.000541232 | 0.006030247 | 0 | 0 | 0 |
| 0 | 0 | 0 | 0.01416373 | 0 |
| 0 | 0 | 0 | 0 | 0 |
| 0 | 0.028847926 | 0 | 0 | 0.015396682 |
| 0.002971326 | 0.116774075 | 0 | 0 | 0 |
| 0 | 0 | 0 | 0 | 0 |
| 0 | 0 | 0 | 0 | 0 |
| 0 | 0 | 0 | 0 | 0 |
| 0 | 0 | 0 | 0 | 0 |

| | | | | |
|---|---|---|---|---|
| 0 | 0 | 0 | 0 | 0 |
| 0.375241889 | 0.736491329 | 1.477279552 | 0.777133917 | 0.485976135 |
| 0 | 0 | 0 | 0 | 0 |
| 0 | 0 | 0 | 0 | 0 |
| 0 | 0 | 0 | 0 | 0 |
| 0 | 0 | 0 | 0 | 0 |
| 0 | 0 | 0 | 0 | 0 |
| 0 | 0 | 0 | 0 | 0.001219872 |
| 0 | 0 | 0 | 0 | 0.001211523 |
| 0 | 0 | 0 | 0.003124977 | 0 |
| 0.012903319 | 0 | 0 | 0.005478218 | 0.017702618 |
| 0.080873176 | 0.00370809 | 0.020947982 | 0.009519954 | 0.039259843 |
| 0 | 0 | 0 | 0 | 0 |
| 0 | 0 | 0 | 0 | 0 |
| 0.002114992 | 0 | 0.001423774 | 0 | 0.000677051 |
| 0 | 0 | 0 | 0 | 0 |
| 0.025619468 | 0.009224068 | 0.048510869 | 0.010755909 | 0.008032785 |
| 0.006378172 | 0.020043627 | 0 | 0 | 0.037432642 |
| 0 | 0 | 0 | 0 | 0 |
| 0.001035413 | 0.018086174 | 0 | 0 | 0 |
| 0.087765518 | 0.013441352 | 0.063650354 | 0.01947272 | 0.01890412 |
| 0 | 0 | 0 | 0 | 0 |
| 0 | 0 | 0 | 0 | 0 |
| 0 | 0 | 0 | 0 | 0 |
| 0.023411563 | 0 | 0 | 0 | 0 |
| 0 | 0 | 0 | 0 | 0 |
| 0 | 0.337193183 | 0 | 0.629898715 | 0.067089657 |
| 0.013271017 | 0.026814048 | 0.048898567 | 0.105840978 | 0.069307108 |
| 0 | 0 | 0 | 0 | 0 |
| 0 | 0 | 0 | 0 | 0 |
| 0.006810345 | 0 | 0 | 0 | 0 |
| 0 | 0 | 0 | 0 | 0 |
| 0 | 0 | 0 | 0 | 0 |
| 0 | 0 | 0 | 0 | 0 |
| 0.027319669 | 0.001532234 | 0 | 0.384684849 | 0.041706905 |
| 0.166662717 | 0.683693461 | 0.340673326 | 0.358259082 | 0.100486685 |
| 0 | 0 | 0 | 0 | 0 |
| 0 | 0 | 0 | 0 | 0 |
| 0.029658124 | 0 | 0.035431767 | 0.016159243 | 0.02812803 |
| 0 | 0 | 0 | 0 | 0 |
| 0 | 0 | 0 | 0 | 0 |
| 0 | 0 | 0 | 0 | 0 |
| 0.005908854 | 0.130928418 | 0.006581045 | 0.005496608 | 0.016070421 |
| 0 | 0.005088158 | 0 | 0 | 0.000489969 |
| 0 | 0 | 0 | 0 | 0 |
| 0 | 0 | 0 | 0 | 0 |
| 0 | 0 | 0 | 0 | 0 |
| 0 | 0 | 0.001110329 | 0.00520816 | 0.001055996 |

| | | | | |
|---|---|---|---|---|
| 0 | 0 | 0 | 0 | 0 |
| 0 | 0 | 0 | 0 | 0 |
| 0 | 0 | 0 | 0 | 0 |
| 0 | 0.0199484 | 0.122637363 | 0.000685908 | 6.415581111 |
| 0 | 0 | 0 | 0 | 0 |
| 0 | 0 | 0 | 0 | 0 |
| 0 | 0 | 0 | 0 | 0 |
| 0 | 0 | 0 | 0.012681446 | 0 |
| 0.340592586 | 0 | 0 | 1.943123896 | 11.16060801 |
| 0 | 0 | 0 | 0.026564769 | 0 |
| 0 | 0 | 0 | 0 | 0 |
| 0.002251488 | 0.009556356 | 0 | 0.0041621 | 0.007361909 |
| 0 | 0 | 0 | 0 | 0 |
| 0 | 0.00138142 | 0 | 0 | 0 |
| 0.005971929 | 0.061874214 | 0 | 0 | 0 |
| 0 | 0 | 0 | 0 | 0 |
| 0 | 0 | 0 | 0.369254785 | 0 |
| 0 | 0 | 0 | 0 | 0 |
| 0 | 0 | 0 | 0 | 0 |
| 0 | 0 | 0 | 0 | 0 |
| 0 | 0 | 0 | 0 | 0 |
| 0 | 0 | 0 | 0 | 0 |
| 0 | 0 | 0 | 0 | 0 |
| 0.434761024 | 0.232559699 | 0.081723604 | 0.364269661 | 0.384121142 |
| 0 | 0.00378535 | 0 | 0.001845848 | 0 |
| 0 | 0 | 0 | 0 | 0 |
| 0 | 0 | 0 | 0 | 0 |
| 9.249220882 | 0 | 0 | 0 | 0 |
| 0 | 0 | 0 | 0 | 0 |
| 0 | 0 | 0 | 0 | 0 |
| 0.723095625 | 0 | 0 | 0 | 0 |
| 0 | 0 | 0 | 0 | 0 |
| 0 | 0 | 0 | 0.00275535 | 0 |
| 0 | 0 | 0 | 0 | 0 |
| 0 | 0 | 0 | 0 | 0 |
| 0.006628154 | 0.033326728 | 0 | 0 | 0 |
| 0 | 0.16003668 | 0 | 0.318090376 | 0 |
| 0 | 0 | 0 | 0 | 0 |
| 0 | 0 | 0 | 0 | 0 |
| 0 | 0 | 0 | 0 | 0 |
| 0.022216115 | 0.014809225 | 0.030655279 | 0.119993874 | 0.088370988 |
| 0 | 0.222789025 | 0 | 0 | 0 |
| 0.013140402 | 26.83095507 | 0 | 0 | 1.510451599 |
| 0 | 0 | 0 | 0 | 0 |
| 0 | 0 | 0 | 0 | 0 |
| 0 | 0 | 0 | 0 | 0 |
| 0 | 0 | 0 | 0 | 0 |
| 0 | 0 | 3.552818233 | 0.001010483 | 0.004571462 |

| | | | | |
|---|---|---|---|---|
| 0 | 0 | 0 | 0 | 0 |
| 0 | 0 | 0 | 0 | 0 |
| 0.005866901 | 1.297188589 | 0.147447648 | 0.227635504 | 0.135015414 |
| 0 | 0 | 0 | 0 | 0 |
| 0 | 0 | 0 | 0 | 0 |
| 0.002801041 | 0 | 0 | 0.019649165 | 0 |
| 0 | 0 | 0 | 0 | 0 |
| 0 | 0 | 0 | 0 | 0 |
| 0 | 0 | 0 | 0 | 0 |
| 0 | 0 | 0 | 0 | 0 |
| 0.007680341 | 0.004188848 | 0 | 0.114839774 | 0.286225637 |
| 3.380255399 | 3.272339151 | 2.965520751 | 2.413237182 | 0.856897044 |
| 0 | 0 | 0 | 0 | 0 |
| 0.067253425 | 0 | 0.097881827 | 0.044905712 | 0.001326727 |
| 0.515434233 | 0.054795322 | 0.71839643 | 0.360345077 | 0.092527822 |
| 0 | 0 | 0 | 0 | 0 |
| 0 | 0 | 0 | 0 | 0 |
| 0 | 0 | 0 | 0 | 0 |
| 0 | 0 | 0 | 0 | 0 |
| 0 | 0 | 0 | 0 | 0 |
| 0.955201175 | 0.09663255 | 0.70328187 | 0.235350664 | 0.458504619 |
| 0.052721413 | 3.186742174 | 0.351590139 | 0 | 0.015018388 |
| 0.63758615 | 1.959027279 | 6.704404426 | 0.314713163 | 1.682317905 |
| 0 | 0 | 0 | 0 | 0 |
| 0 | 0 | 0 | 0 | 0 |
| 0.005785324 | 0 | 0 | 0 | 0 |
| 0 | 0.001199762 | 0 | 0.001906793 | 0 |
| 0 | 0 | 0 | 0 | 0 |
| 0 | 0.033816498 | 0 | 0 | 0 |
| 0 | 0 | 0 | 0 | 0 |
| 0 | 0.008298 | 0 | 0 | 0 |
| 0.10358973 | 0.230123189 | 0 | 0.105920783 | 0 |
| 0.072604633 | 0 | 0.376567335 | 0.001220947 | 0.010100313 |
| 0.00615185 | 0.008748401 | 0 | 0 | 0 |
| 0.006862611 | 0.021363181 | 0.009053716 | 0.01595563 | 0.033331659 |
| 0.153461987 | 0.01318969 | 2.871862323 | 0.031000893 | 0.076135288 |
| 0 | 0.002512322 | 0 | 0 | 0 |
| 0 | 0 | 0 | 0 | 0 |
| 0 | 0.002391276 | 0 | 0 | 0 |
| 0 | 0 | 0 | 0 | 0 |
| 0 | 0 | 0 | 0 | 0 |
| 0 | 0 | 0 | 0 | 0 |
| 0 | 0 | 0 | 0 | 0 |
| 0 | 0 | 0 | 0 | 0 |
| 0.002966203 | 0.002118499 | 0.067724589 | 0.01944415 | 0.005440079 |
| 0 | 0 | 0 | 0 | 0 |
| 0 | 0 | 0 | 0 | 0 |
| 0.034215116 | 1.295888459 | 20.20445069 | 0.03929467 | 0.004724799 |

| | | | | |
|---|---|---|---|---|
| 0 | 0 | 0 | 0 | 0 |
| 0 | 0 | 0 | 0 | 0 |
| 0 | 0 | 0 | 0 | 0 |
| 0 | 0.006139 | 0 | 0.013774271 | 0 |
| 0 | 0 | 0 | 0 | 0 |
| 0 | 0 | 0 | 0 | 0 |
| 0 | 0 | 0 | 0 | 0.042233542 |
| 0 | 0.02807881 | 0.15360032 | 0 | 0.105637837 |
| 0 | 0 | 0 | 0 | 0 |
| 0 | 0 | 0 | 0 | 0 |
| 0.000703904 | 0.001809853 | 0.093606133 | 0 | 0.008881902 |
| 0 | 0 | 0 | 0 | 0 |
| 0.007704034 | 0.001760742 | 0 | 0 | 0 |
| 0 | 0 | 0 | 0.485792231 | 0 |
| 0 | 0 | 0 | 0 | 0 |
| 0 | 0 | 0 | 0 | 0 |
| 0 | 0 | 0 | 0 | 0 |
| 0 | 0 | 0 | 0.005587287 | 0 |
| 0 | 0 | 0 | 0 | 0 |
| 0 | 0 | 0 | 0 | 0 |
| 0 | 0 | 0 | 0 | 0 |
| 0 | 0 | 0 | 0 | 0 |
| 0 | 0 | 0 | 0 | 0 |
| 0 | 0 | 0 | 0 | 0 |
| 0 | 0 | 0 | 0 | 0 |
| 0 | 0 | 0 | 0 | 0 |
| 0 | 0 | 0 | 0 | 0 |
| 0 | 0 | 0 | 0 | 0 |
| 0 | 0 | 0 | 0 | 0 |
| 0 | 0.002224485 | 0 | 0.077859066 | 0 |
| 0 | 0 | 0 | 0 | 0 |
| 0 | 0 | 0 | 0 | 0 |
| 0 | 0 | 0 | 0 | 0 |
| 0 | 0 | 0 | 0 | 0 |
| 0 | 0 | 0 | 0 | 0 |
| 0 | 0 | 0 | 0 | 0 |
| 0 | 0 | 0 | 0.288722043 | 0 |
| 0.005213834 | 0 | 0 | 0.010348484 | 0.001049119 |
| 0 | 0 | 0 | 0 | 0 |
| 0 | 0 | 0 | 0 | 0 |
| 0 | 0 | 0 | 0 | 0 |
| 0 | 0 | 0 | 0 | 0 |
| 0 | 0 | 0 | 0 | 0 |
| 0 | 0 | 0 | 0.954712357 | 0 |
| 1.176946715 | 0.003672598 | 0 | 0 | 0 |
| 0 | 0 | 0 | 0 | 0 |
| 0 | 0 | 0 | 0 | 0 |
| 0 | 0 | 0 | 0 | 0 |

| | | | | |
|---|---|---|---|---|
| 0 | 0 | 0 | 0 | 0 |
| 0 | 0 | 0 | 0 | 0 |
| 0 | 0 | 0 | 0 | 0 |
| 0 | 0 | 0 | 0 | 0 |
| 0 | 0 | 0 | 0 | 0 |
| 0 | 0 | 0 | 0 | 0 |
| 0 | 0 | 0 | 0 | 0 |
| 0 | 0 | 0 | 0 | 0 |
| 0.036570093 | 0.020895053 | 0.084435304 | 0.087872515 | 0.079580062 |
| 0 | 0 | 0 | 0 | 0 |
| 0 | 0 | 0 | 0 | 0 |
| 0 | 0 | 0 | 0 | 0 |
| 0 | 0 | 0 | 0 | 0 |
| 0 | 0 | 0 | 0 | 0 |
| 0 | 3.051400184 | 0 | 0.051684426 | 3.541344168 |
| 0 | 0 | 0 | 0 | 0.451087858 |
| 0 | 0 | 0 | 0 | 0.199790285 |
| 0 | 0 | 0 | 0 | 0 |
| 0 | 0 | 0 | 0 | 0 |
| 0 | 0 | 0 | 0 | 0 |
| 0 | 0.049008913 | 0 | 0 | 0 |
| 0 | 0 | 0 | 0 | 0 |
| 0 | 0 | 0 | 0 | 0 |
| 0 | 0 | 0 | 0 | 0 |
| 0.257307515 | 0 | 0.054864884 | 0.001645471 | 0.588727722 |
| 2.138529601 | 1.499682995 | 0 | 6.403458899 | 12.06641108 |
| 0 | 0.243989145 | 0 | 0 | 0 |
| 0 | 0 | 0 | 0 | 0 |
| 0 | 0 | 0 | 0 | 0 |
| 0 | 0 | 0 | 0 | 0 |
| 0 | 0 | 0 | 0 | 0 |
| 0 | 0 | 0 | 0 | 0 |
| 0 | 0 | 0 | 0 | 0 |
| 0 | 0.001179597 | 0 | 0.003515149 | 0 |
| 0.04634755 | 0.000971802 | 0.044868358 | 0.0159276 | 0 |
| 0 | 0.001545629 | 0.088657791 | 0 | 0.010584022 |
| 0 | 0 | 0 | 0 | 0 |
| 0 | 0.012798734 | 0 | 3.776752055 | 0.610506658 |
| 0 | 0 | 0 | 0 | 0 |
| 0 | 0 | 0 | 0 | 0 |
| 0 | 0 | 0 | 0 | 0 |
| 0 | 0 | 0.034915095 | 0 | 0 |
| 7.75978967 | 0.534133975 | 0 | 1.397279754 | 0.581663912 |
| 0 | 0 | 0 | 0 | 0 |
| 0 | 0 | 0 | 0 | 0 |
| 0 | 0.01480559 | 0 | 0.001357539 | 0 |
| 0 | 0 | 0 | 0 | 0 |
| 3.311148949 | 0.085276365 | 0.005318789 | 0.808858122 | 0.001064951 |

| | | | | |
|---|---|---|---|---|
| 0 | 0 | 0 | 0 | 0 |
| 0 | 0 | 0 | 0 | 0 |
| 0 | 0 | 0 | 0 | 0 |
| 0 | 0 | 0 | 0 | 0 |
| 0 | 0 | 0 | 0.002064438 | 0.005929901 |
| 0.003405499 | 0.001853143 | 0 | 0 | 0 |
| 0 | 0 | 0 | 0 | 0 |
| 0 | 0 | 0 | 0 | 0 |
| 0 | 0 | 0 | 0 | 0 |
| 0 | 0 | 0 | 0 | 0 |
| 0 | 0 | 0 | 0 | 0 |
| 0 | 2.790329128 | 0 | 2.134817634 | 6.109509288 |
| 0 | 0 | 0 | 0 | 0 |
| 0 | 0 | 0 | 0 | 0 |
| 0.391067093 | 0 | 0.831163252 | 0.001856962 | 0.585667283 |
| 0 | 0 | 0 | 0 | 0 |
| 0 | 0 | 0 | 0.007012548 | 0 |
| 0 | 0 | 0 | 0 | 0 |
| 0 | 0 | 0 | 0.014416066 | 0 |
| 0 | 0 | 0 | 0 | 0 |
| 0 | 0.108526712 | 0 | 0 | 0 |
| 0 | 0 | 0 | 0 | 0 |
| 0 | 0.059368818 | 1.496854296 | 1.167309758 | 0.651698974 |
| 0 | 0 | 0 | 0 | 0 |
| 0 | 0.003177021 | 0 | 0 | 0 |
| 0 | 0 | 0 | 0 | 0 |
| 0 | 0 | 0 | 0 | 0 |
| 0 | 0 | 0 | 0 | 0 |
| 0 | 0 | 0 | 0 | 0 |
| 0 | 0 | 0 | 0 | 0 |
| 0 | 0 | 0 | 0 | 0 |
| 0.046247986 | 0.01732768 | 0.107605535 | 0.049226065 | 0.088991241 |
| 0 | 0 | 0 | 0 | 0 |
| 0 | 0.018755988 | 0 | 0 | 0.241261407 |
| 0 | 0.000934462 | 0.091233924 | 0 | 0.011134133 |
| 0 | 0.014937469 | 0 | 0.042502744 | 0.024499509 |
| 0 | 0 | 0 | 0 | 0 |
| 0 | 0 | 0 | 0 | 0 |
| 0 | 0 | 0 | 0.001023731 | 0 |
| 0 | 0 | 0 | 0 | 0 |
| 0 | 0.0149679 | 0 | 0 | 0 |
| 0 | 0 | 0 | 0 | 0 |
| 0 | 0 | 0 | 0 | 0 |
| 0 | 0 | 0 | 0 | 0 |
| 0 | 0.006075554 | 0.063031494 | 0.114554567 | 0.146936553 |
| 0 | 0 | 0 | 0 | 0 |
| 0.243185597 | 0 | 0 | 0.848249772 | 0 |
| 0 | 0 | 0 | 0 | 0 |

| | | | | |
|---|---|---|---|---|
| 0 | 0 | 0 | 0 | 0 |
| 0 | 0 | 0 | 0.027363769 | 0 |
| 0 | 0.032941526 | 0 | 0 | 0 |
| 0 | 0 | 0 | 0 | 0 |
| 0 | 0 | 0 | 0 | 0 |
| 0 | 0 | 0 | 0 | 0 |
| 0 | 0 | 0 | 0 | 0 |
| 0 | 0 | 0 | 0 | 0.082381235 |
| 0 | 0 | 0 | 0 | 0 |
| 0 | 0 | 0 | 0 | 0 |
| 0 | 0 | 0 | 0 | 0 |
| 0 | 0 | 0.033897139 | 0 | 0.007562095 |
| 0 | 0 | 0 | 0 | 0 |
| 0.615964191 | 0 | 0 | 0 | 0 |
| 0 | 0 | 0 | 0 | 0 |
| 0 | 0 | 0 | 0 | 0 |
| 0 | 0 | 0 | 0 | 0 |
| 0 | 0 | 0 | 0 | 0 |
| 0 | 0.63887385 | 9.186727351 | 0 | 0.03134827 |
| 0 | 0.005133384 | 0 | 0 | 0 |
| 0 | 0 | 0 | 0.004949331 | 0 |
| 0 | 0 | 0 | 0 | 0 |
| 0 | 0 | 0 | 0 | 0 |
| 0 | 0 | 0 | 0 | 0 |
| 0 | 0.001005768 | 0 | 0 | 0 |
| 0.013348935 | 0.005656112 | 0.024804769 | 0.013279671 | 0.023062807 |
| 0 | 0 | 0 | 0 | 0 |
| 2.613777635 | 0.315060575 | 0.001087116 | 1.092045111 | 0.008788313 |
| 0 | 0 | 0 | 0 | 0 |
| 0 | 0.044499174 | 0 | 1.077967257 | 0 |
| 0 | 0 | 0 | 0 | 0 |
| 0 | 0 | 0 | 0 | 0 |
| 0 | 0.00212573 | 0 | 0 | 0 |
| 0 | 0 | 0 | 0 | 0 |
| 0.002986385 | 0 | 0 | 0 | 0.017342208 |
| 0 | 0 | 0 | 0 | 0 |
| 0 | 0 | 0 | 0 | 0 |
| 0 | 0.001426663 | 0.01342702 | 0 | 0.016956855 |
| 0 | 0 | 0 | 0 | 0 |
| 0 | 0 | 0 | 0 | 0 |
| 4.79591257 | 7.463270988 | 1.135377541 | 11.80026859 | 5.258982602 |
| 0.095695996 | 1.351206059 | 2.157950326 | 3.416130946 | 0.916072781 |
| 0 | 0 | 0 | 0 | 0 |
| 0 | 0 | 0 | 0 | 0 |
| 0 | 0 | 0 | 0 | 0 |
| 0 | 0 | 0 | 0 | 0 |
| 0 | 0 | 0 | 0 | 0 |
| 0 | 0.000647017 | 0.058872387 | 0.053600656 | 0.10068502 |

| | | | | |
|---|---|---|---|---|
| 0 | 0 | 0 | 0 | 0 |
| 0 | 0 | 0 | 0 | 0.011279123 |
| 0 | 0 | 0 | 0 | 0 |
| 0 | 0 | 0 | 0 | 0 |
| 6.203632003 | 3.406940974 | 0 | 7.469797762 | 2.471645747 |
| 0 | 0 | 0 | 0 | 0 |
| 0 | 0 | 0 | 0 | 0 |
| 0 | 0 | 0 | 0 | 0 |
| 0.509122733 | 0.258169411 | 0 | 0.139608807 | 0.212963515 |
| 0 | 0 | 0 | 0 | 0 |
| 0 | 0 | 0 | 0 | 0 |
| 0 | 0 | 0 | 0 | 0 |
| 0.021927938 | 0.185752024 | 0 | 0.371334387 | 0 |
| 0.006261252 | 0.02583744 | 0.005580921 | 0.02132154 | 0.003062205 |
| 1.715811328 | 0 | 0 | 1.327597963 | 0 |
| 0 | 1.510725223 | 0.243426703 | 0.187988039 | 0.552486054 |
| 0.001655544 | 0 | 0.007429879 | 0.00533014 | 0 |
| 0 | 0 | 0 | 0 | 0 |
| 0 | 0 | 0 | 0 | 0 |
| 0 | 0 | 0 | 0 | 0 |
| 0 | 0 | 0 | 0 | 0 |
| 0 | 0 | 0 | 0 | 0 |
| 0 | 0 | 0 | 0 | 0 |
| 0 | 0 | 0 | 0 | 0 |
| 0 | 0 | 0 | 0 | 0 |
| 0 | 0 | 0 | 0 | 0 |
| 0.345102568 | 0.013160372 | 0.001938175 | 0.019608668 | 0 |
| 0.000738357 | 0.005062503 | 0.051030331 | 0 | 0 |
| 0 | 0 | 0 | 0 | 0 |
| 0 | 0 | 0 | 0 | 0 |
| 0 | 0 | 0 | 0 | 0 |
| 0 | 0 | 0 | 0 | 0 |
| 0 | 0 | 0 | 0 | 0 |
| 0.257430919 | 0.002299637 | 0 | 0 | 0 |
| 0.00091543 | 0 | 0 | 0 | 0 |
| 0 | 0 | 0 | 0.001396113 | 0 |
| 0 | 0 | 0 | 0 | 0 |
| 0 | 0 | 0 | 0 | 0 |
| 0 | 0 | 0 | 0 | 0 |
| 0 | 0 | 0 | 0 | 0 |
| 0.861973859 | 0 | 0 | 0 | 0 |
| 0.118584214 | 0.155750594 | 0 | 0 | 0 |
| 0 | 0 | 0 | 0 | 0 |
| 0.005783586 | 0 | 0 | 0 | 0 |
| 0 | 0 | 0 | 0 | 0 |
| 0 | 0 | 0 | 0 | 0 |
| 0 | 0 | 0 | 0 | 0 |
| 0 | 0 | 0 | 0 | 0 |

| | | | | |
|---|---|---|---|---|
| 0 | 0.011270698 | 0 | 0 | 0 |
| 0 | 0 | 0 | 0 | 0 |
| 0 | 0 | 0 | 0 | 0 |
| 0 | 0 | 0 | 0 | 0.023169549 |
| 0 | 0 | 0 | 0.198396129 | 0 |
| 0 | 0 | 0 | 0 | 0 |
| 0.003453174 | 0.002916668 | 0.063000404 | 0.14765399 | 0 |
| 0 | 0 | 0 | 0 | 0 |
| 0 | 0 | 0 | 0 | 0 |
| 0 | 0 | 0 | 0 | 0 |
| 0 | 0.000832221 | 0 | 0 | 0 |
| 0 | 0 | 0 | 0 | 0 |
| 0 | 0 | 0 | 0 | 0 |
| 0 | 0 | 0 | 0 | 0 |
| 0 | 0 | 0 | 0 | 0 |
| 0 | 0 | 0 | 0 | 0 |
| 0 | 0 | 0 | 0 | 0.156791721 |
| 0 | 0 | 0.000712568 | 0 | 0 |
| 0 | 0 | 0 | 0 | 0 |
| 0 | 0 | 0 | 0 | 0 |
| 2.275600782 | 0.00277683 | 0.025050427 | 1.718050245 | 0 |
| 0 | 0 | 0 | 0 | 0 |
| 0.005518171 | 0.008287669 | 0 | 0 | 0 |
| 0.008692072 | 0.00656845 | 0 | 0.004010221 | 0 |
| 0 | 0 | 0 | 0 | 0 |
| 0 | 0 | 0 | 0.003154106 | 0 |
| 0 | 0 | 0 | 0 | 0 |
| 0 | 0 | 0 | 0 | 0 |
| 0 | 0 | 0 | 0 | 0 |
| 0.341803575 | 0 | 0.005976518 | 0.001374202 | 0.34649087 |
| 0 | 0 | 0 | 0 | 0 |
| 0 | 0 | 0 | 0 | 0 |
| 0 | 0 | 0 | 0 | 0 |
| 0 | 0 | 0 | 0 | 0 |
| 0 | 0 | 0 | 0 | 0 |
| 0 | 0 | 0 | 0 | 0 |
| 0 | 0 | 0 | 0.000716832 | 0.009471548 |
| 0 | 0 | 0 | 0 | 0 |
| 0 | 0 | 0 | 0 | 0 |
| 0 | 0 | 0 | 0 | 0 |
| 0 | 0 | 0 | 0 | 0 |
| 0 | 0 | 0 | 0 | 0 |
| 0 | 0 | 0 | 0 | 0 |
| 0 | 0.012242347 | 0 | 0 | 0 |
| 0 | 0.114211218 | 0 | 0 | 0 |
| 0 | 0 | 0 | 0 | 0 |
| 0 | 0 | 0 | 0 | 0 |
| 0 | 0 | 0 | 0 | 0 |

| | | | | |
|---|---|---|---|---|
| 0 | 0.026202894 | 0 | 0.00143838 | 0.093492429 |
| 0 | 0 | 0 | 0 | 0 |
| 0 | 0 | 0 | 0 | 0 |
| 0 | 0 | 0 | 0 | 0 |
| 0 | 0 | 0 | 0 | 0 |
| 0 | 0 | 0 | 0 | 0 |
| 0 | 0 | 0 | 0 | 0 |
| 0.483112731 | 2.264575872 | 9.100378533 | 0.318586527 | 0.944138493 |
| 9.381571607 | 0.007305876 | 1.37994218 | 0.040639562 | 7.442858392 |
| 0 | 0.013186268 | 0 | 0.108677376 | 0.010642003 |
| 0 | 0 | 0 | 0 | 0 |
| 0 | 0 | 0 | 0 | 0 |
| 0 | 0.075859286 | 0 | 0 | 0 |
| 0 | 0.003004816 | 2.622138046 | 0 | 0 |
| 0 | 0 | 0 | 0 | 0 |
| 0 | 0 | 0 | 0 | 0 |
| 0.006826416 | 0 | 0 | 0 | 0.074181775 |
| 0 | 0 | 0 | 0 | 0 |
| 0 | 0 | 0 | 0 | 0 |
| 0 | 0.000937232 | 0 | 0 | 0 |
| 0 | 0 | 0 | 0 | 0 |
| 0 | 0 | 0 | 0 | 0 |
| 0 | 0 | 0 | 0 | 0 |
| 0 | 0 | 0 | 0 | 0 |
| 0 | 0.008010962 | 0 | 0 | 0 |
| 0 | 0 | 0 | 0 | 0 |
| 0 | 0 | 0 | 0 | 0 |
| 0 | 0 | 0 | 0 | 0 |
| 0 | 0 | 0 | 0 | 0 |
| 0 | 0 | 0 | 0 | 0 |
| 0 | 0.011688498 | 0 | 0 | 0 |
| 0 | 0 | 0 | 0 | 0 |
| 0 | 0 | 0 | 0 | 0 |
| 0 | 0 | 0 | 0 | 0 |
| 0.001461045 | 0 | 0 | 0.014179658 | 0.001714936 |
| 0 | 0 | 0 | 0 | 0 |
| 0 | 0 | 0 | 0 | 0 |
| 0 | 0 | 0 | 0 | 0 |
| 0 | 0 | 0 | 0 | 0 |
| 0.003012001 | 0 | 0.165606178 | 4.405916253 | 0 |
| 0 | 0 | 0 | 0 | 0 |
| 0.21829577 | 0.998353861 | 0 | 0.148144109 | 0 |
| 0 | 0 | 0 | 0 | 0 |
| 0 | 0 | 0 | 0 | 0 |
| 0 | 0 | 0 | 0 | 0 |
| 0 | 0.006438074 | 0 | 0 | 0 |
| 0 | 0 | 0 | 0 | 0 |
| 0 | 0.01589278 | 0 | 0 | 0 |

| | | | | |
|---|---|---|---|---|
| 0 | 0 | 0 | 0 | 0 |
| 0 | 0 | 0.003024634 | 0.002781859 | 0.00447475 |
| 0 | 0 | 0 | 0 | 0 |
| 0 | 0 | 0 | 0 | 0 |
| 0 | 0.671088723 | 0 | 0.777626705 | 1.107593871 |
| 0.207023676 | 0 | 0 | 0 | 0 |
| 0 | 0.003773154 | 0 | 0 | 0 |
| 0.020916864 | 0.005701523 | 0.005920847 | 0.006796116 | 0.034011932 |
| 0 | 0 | 0 | 0 | 0 |
| 0 | 0 | 0 | 0 | 0 |
| 0 | 0 | 0 | 0 | 0 |
| 0 | 0 | 0 | 0 | 0 |
| 0 | 0 | 0 | 0 | 0 |
| 0 | 0 | 0 | 0 | 0 |
| 0 | 0 | 0 | 0 | 0 |
| 0 | 0.021301056 | 0 | 0.015642355 | 0 |
| 0 | 0 | 0 | 0 | 0 |
| 0 | 0 | 0 | 0 | 0 |
| 0 | 0 | 0 | 0 | 0 |
| 0 | 0 | 0 | 0 | 0 |
| 0 | 0 | 0 | 0 | 0 |
| 0 | 0 | 0 | 0 | 0 |
| 0 | 0 | 0 | 0 | 0 |
| 0 | 0 | 0.114982692 | 0 | 0.002144237 |
| 0 | 0 | 0 | 0 | 0 |
| 0 | 0 | 0 | 0 | 0 |
| 0.033568019 | 0 | 4.138916032 | 0.020068852 | 0.001248003 |
| 0 | 0 | 0 | 0 | 0 |
| 0 | 0 | 0.015603574 | 0 | 0.008480013 |
| 0 | 0 | 0 | 0 | 0 |
| 0 | 0 | 0 | 0 | 0 |
| 0 | 0 | 0 | 0 | 0 |
| 0 | 0 | 0 | 0 | 0 |
| 0 | 0 | 0 | 0 | 0 |
| 0 | 0 | 0 | 0 | 0 |
| 0 | 0 | 0 | 0 | 0 |
| 0 | 0 | 0 | 0 | 0 |
| 0 | 0.090250434 | 0 | 0 | 0 |
| 0 | 0 | 0 | 0 | 0 |
| 0 | 0 | 0 | 0 | 0 |
| 0 | 0 | 0 | 0 | 0 |
| 0.481549803 | 0 | 0 | 0 | 0 |
| 0 | 0 | 0 | 0 | 0 |
| 0 | 0 | 0 | 0 | 0 |
| 0 | 0 | 0 | 0 | 0 |
| 0 | 0 | 0 | 0 | 0 |
| 0 | 0 | 0 | 0 | 0 |
| 0 | 0 | 0 | 0 | 0 |

| | | | | |
|---|---|---|---|---|
| 0.099417234 | 0 | 0 | 0.093197723 | 0 |
| 0 | 0 | 0 | 0 | 0 |
| 0 | 0 | 0 | 0 | 0 |
| 0 | 0 | 0 | 0 | 0 |
| 0 | 0 | 0 | 0 | 0 |
| 0 | 0 | 0.005923003 | 0 | 0.078670057 |
| 0 | 0 | 0 | 0 | 0 |
| 0 | 0 | 0 | 0 | 0 |
| 0 | 0 | 0 | 0 | 0 |
| 0 | 0 | 0 | 0 | 0 |
| 0 | 0 | 0 | 0 | 0 |
| 0 | 0 | 0 | 0.005213255 | 0 |
| 0 | 0.030107627 | 0 | 0 | 0 |
| 0 | 0 | 0 | 0 | 0 |
| 0 | 0 | 0 | 0 | 0 |
| 0 | 0 | 0 | 0 | 0 |
| 0 | 0 | 0 | 0 | 0 |
| 0 | 0 | 0 | 0 | 0 |
| 0 | 0 | 0 | 0 | 0 |
| 0 | 0 | 0 | 0 | 0 |
| 0.039194479 | 0.108665533 | 0.317584955 | 0.225039828 | 0.06425232 |
| 0 | 0 | 0 | 0 | 0 |
| 0.003759476 | 0.009546898 | 0.779489775 | 0.042674011 | 0.382440369 |
| 0 | 0 | 0 | 0 | 0 |
| 0 | 0 | 0 | 0 | 0 |
| 0 | 0.08396285 | 0 | 0 | 0 |
| 0 | 0 | 0 | 0 | 0 |
| 0 | 0 | 0 | 0 | 0 |
| 0 | 0 | 0.004686047 | 0 | 0 |
| 0 | 0 | 0 | 0 | 0 |
| 0 | 0 | 0 | 0 | 0 |
| 0 | 0 | 0 | 0 | 0 |
| 0.007823854 | 0 | 0.064694789 | 0.107370041 | 0 |
| 0 | 0 | 0 | 0 | 0 |
| 0 | 0 | 0 | 0.018112137 | 0 |
| 0.112067916 | 0.003407879 | 0.192937302 | 0.16077491 | 0.000921165 |
| 0 | 0 | 0 | 0 | 0 |
| 0 | 0 | 0 | 0 | 0 |
| 0.002360106 | 0.006431277 | 0.003808999 | 0.015331942 | 0.031964186 |
| 0 | 0 | 0 | 0 | 0 |
| 0 | 0 | 0 | 0 | 0 |
| 0 | 0 | 0 | 0 | 0 |
| 0.037786262 | 0 | 0 | 0.192288331 | 1.353249924 |
| 0 | 0 | 0 | 0 | 0 |
| 0 | 0 | 0 | 0 | 0 |
| 0 | 0.060107866 | 0 | 0.01928972 | 0.015039284 |
| 0 | 0 | 0 | 0 | 0 |
| 0 | 0 | 0 | 0 | 0 |

| | | | | |
|---|---|---|---|---|
| 0 | 0 | 0 | 0 | 0 |
| 0 | 0 | 0 | 0 | 0 |
| 0 | 0 | 0 | 0 | 0 |
| 0 | 0 | 0 | 0 | 0 |
| 0 | 0 | 0 | 0 | 0 |
| 0 | 0 | 0 | 0 | 0 |
| 0 | 0 | 0 | 0 | 0 |
| 0.026880216 | 0.204780654 | 0 | 0 | 0 |
| 0 | 0 | 0 | 0 | 0.018354619 |
| 0 | 0 | 0.001071533 | 0 | 0.002038197 |
| 0 | 0 | 0 | 0 | 0 |
| 0 | 0 | 0 | 0 | 0 |
| 0 | 0 | 0 | 0 | 0 |
| 0 | 0 | 0 | 0 | 0 |
| 0 | 0 | 0 | 0 | 0 |
| 0 | 0.034946897 | 0 | 0 | 0 |
| 0 | 0 | 0 | 0 | 0 |
| 0 | 0 | 0 | 0 | 0 |
| 0 | 0 | 0 | 0 | 0 |
| 0 | 0 | 0 | 0.010131517 | 0 |
| 0 | 0 | 0 | 0 | 0 |
| 0 | 0 | 0 | 0 | 0 |
| 0 | 0 | 0 | 0 | 0 |
| 0 | 0 | 0 | 0 | 0 |
| 0 | 0 | 0 | 0.001572607 | 0 |
| 0 | 0 | 0 | 0 | 0 |
| 0 | 0 | 0 | 0 | 0 |
| 0 | 0 | 0 | 0 | 0 |
| 0 | 0 | 0 | 0 | 0 |
| 0 | 3.090619378 | 0 | 0 | 0 |
| 0 | 0.05395146 | 0 | 0 | 0.113604076 |
| 0 | 0 | 0 | 0 | 0 |
| 0 | 0 | 0 | 0 | 0 |
| 0 | 0 | 0 | 0 | 0 |
| 0 | 0 | 0 | 0 | 0 |
| 0 | 0 | 0 | 0 | 0 |
| 0 | 0 | 0 | 0 | 0 |
| 0 | 0 | 0 | 0 | 0 |
| 0 | 0 | 0 | 0 | 0 |
| 0 | 0 | 0 | 0 | 0 |
| 0 | 0 | 0 | 0 | 0 |
| 0 | 0 | 0 | 0 | 0 |
| 0 | 0 | 0 | 0 | 0 |
| 0 | 0 | 0 | 0 | 0 |
| 0 | 0 | 0 | 0 | 0 |
| 0 | 0 | 0 | 0 | 0 |
| 0 | 0.194263615 | 0 | 0 | 0 |
| 0 | 0 | 0 | 0 | 0 |

| | | | | |
|---|---|---|---|---|
| 0 | 0 | 0 | 0 | 0 |
| 0 | 0 | 0 | 0 | 0 |
| 0 | 0 | 0 | 0 | 0 |
| 0 | 0 | 0 | 0 | 0 |
| 0.417588538 | 0.015229631 | 0 | 0.204137394 | 0.017023508 |
| 0 | 0 | 0 | 0 | 0 |
| 0 | 0 | 0 | 0 | 0 |
| 0 | 0 | 0 | 0 | 0 |
| 0 | 0 | 0 | 0 | 0 |
| 0 | 0 | 0 | 0 | 0 |
| 0 | 0 | 0 | 0 | 0 |
| 0 | 0.000621886 | 0.179328148 | 0.021497045 | 0.041520355 |
| 0 | 0 | 0 | 0 | 0.103056001 |
| 0 | 0 | 0 | 0 | 0 |
| 0 | 0 | 0.095201747 | 0 | 0.001298109 |
| 0 | 0 | 0 | 0.634093106 | 0 |
| 0 | 0.004942477 | 0 | 0 | 0 |
| 0 | 0 | 0 | 0 | 0 |
| 0 | 0.046304745 | 0.001356188 | 0.051078255 | 0.060944165 |
| 0.005918503 | 0 | 0.006861726 | 0.125323529 | 0.148623292 |
| 0 | 0 | 0 | 0 | 0 |
| 0 | 0 | 0 | 0 | 0 |
| 0 | 0 | 0 | 0 | 0 |
| 0 | 0 | 0 | 0 | 0 |
| 0 | 0 | 0 | 0 | 0 |
| 0 | 0 | 0 | 0 | 0 |
| 0.003880446 | 0.006240924 | 0 | 0 | 0.097337002 |
| 0 | 0 | 0 | 0 | 0 |
| 0 | 0 | 0 | 0 | 0 |
| 0 | 0 | 0 | 0 | 0 |
| 0 | 0.021781752 | 0.016688273 | 0 | 0.006692862 |
| 0 | 0 | 0 | 0 | 0 |
| 0 | 0 | 0 | 0 | 0 |
| 0 | 0 | 0 | 0 | 0 |
| 0 | 0 | 0 | 0 | 0 |
| 0 | 0 | 0 | 0 | 0 |
| 0 | 0 | 0 | 0 | 0 |
| 0 | 0.007847809 | 0.486080482 | 0.005768578 | 0.101433394 |
| 0 | 0.211440442 | 0 | 0 | 0 |
| 0 | 0 | 0 | 0 | 0 |
| 0 | 0 | 0.073614743 | 0 | 0.00308484 |
| 0 | 0 | 0 | 0 | 0 |
| 0 | 0 | 0 | 0 | 0 |
| 0.001527553 | 0.003297236 | 0.014453619 | 0 | 0 |
| 0 | 0 | 0 | 0 | 0 |
| 0 | 0 | 0 | 0 | 0 |
| 0 | 0.01456199 | 0 | 0 | 0 |
| 0 | 0 | 0 | 0 | 0 |

| | | | | |
|---|---|---|---|---|
| 0 | 0 | 0 | 0 | 0 |
| 0 | 0 | 0 | 0 | 1.337863588 |
| 0 | 0 | 0 | 0 | 0 |
| 0.001600882 | 0.001219595 | 0 | 0 | 0.00104393 |
| 0.004949309 | 0 | 0.002443309 | 0.008764059 | 0 |
| 0.001703349 | 0.013625395 | 0.008671642 | 0.00145011 | 0 |
| 0 | 0 | 0 | 0.001880987 | 0 |
| 0 | 0 | 0 | 0 | 0 |
| 0 | 0 | 0 | 0 | 0 |
| 0 | 0 | 0 | 0 | 0 |
| 0 | 0.016628208 | 0 | 0 | 0 |
| 0 | 0 | 0 | 0 | 0 |
| 0 | 0.025647458 | 0.321500014 | 0.039158775 | 0 |
| 0 | 0 | 0 | 0 | 0 |
| 0 | 0 | 0 | 0 | 0 |
| 0 | 0 | 0 | 0 | 0 |
| 0 | 0 | 0 | 0 | 0 |
| 0 | 0 | 0 | 0 | 0 |
| 0 | 0 | 0 | 0 | 0 |
| 0 | 0 | 0 | 0 | 0 |
| 0 | 0 | 0 | 0 | 0 |
| 0 | 0 | 0 | 0 | 0 |
| 0 | 0.002845938 | 0 | 0.026007695 | 0 |
| 0 | 0 | 0 | 0.048036483 | 0 |
| 0 | 0 | 0 | 0 | 0 |
| 0 | 0 | 0 | 0 | 0 |
| 0 | 0 | 0 | 0 | 0 |
| 0 | 0 | 0 | 0 | 0 |
| 0 | 0 | 0 | 0 | 0 |
| 0 | 0 | 0 | 0 | 0 |
| 0 | 0 | 0 | 0.035302479 | 0 |
| 0 | 0 | 0 | 0 | 0 |
| 0 | 0 | 0 | 0 | 0 |
| 0 | 0 | 0 | 0 | 0 |
| 0 | 0 | 0 | 0 | 0 |
| 0 | 0 | 0 | 0 | 0 |
| 0 | 0 | 0 | 0 | 0 |
| 0 | 0 | 0 | 0 | 0 |
| 0 | 0 | 0.010631689 | 0.001280072 | 0 |
| 0 | 0 | 0 | 0 | 0 |
| 0 | 0 | 0 | 0 | 0 |
| 0 | 0 | 0 | 0 | 0 |
| 0 | 0 | 0 | 0 | 0 |
| 0 | 0 | 0 | 0 | 0 |
| 0 | 0 | 0 | 4.866879619 | 0 |
| 0.501750248 | 0.062360491 | 0 | 0.192401358 | 0.178413085 |
| 0 | 0 | 0 | 0 | 0 |
| 0 | 0 | 0 | 0 | 0 |

| | | | | |
|---|---|---|---|---|
| 0 | 0 | 0 | 0 | 0 |
| 0 | 0 | 0 | 0 | 0 |
| 0.125377007 | 0 | 0 | 0 | 0 |
| 0 | 0 | 0 | 0 | 0 |
| 0 | 0 | 0 | 0 | 0 |
| 0 | 0 | 0 | 0 | 0 |
| 0 | 0 | 0 | 0 | 0 |
| 0 | 0 | 0 | 0 | 0 |
| 0 | 0 | 0.015381709 | 0 | 0 |
| 0 | 0 | 0 | 0 | 0 |
| 0 | 0 | 0 | 0 | 0 |
| 0 | 0 | 0 | 0 | 0 |
| 0 | 0 | 0 | 0 | 0 |
| 0 | 0 | 0 | 0 | 0 |
| 0 | 0 | 0 | 0 | 0 |
| 0 | 0 | 0 | 0 | 0 |
| 0 | 0 | 0 | 0 | 0 |
| 0 | 0 | 0 | 0 | 0 |
| 0 | 0 | 0 | 0 | 0 |
| 0 | 0 | 0.002287686 | 0 | 0 |
| 0 | 0 | 0 | 0 | 0 |
| 0 | 0 | 0 | 0 | 0 |
| 0 | 0 | 0 | 0 | 0 |
| 0 | 0 | 0 | 0 | 0 |
| 0 | 0 | 0 | 0 | 0 |
| 0 | 0 | 0 | 0 | 0 |
| 0 | 0 | 0 | 0 | 0 |
| 0 | 0 | 0 | 0 | 0 |
| 0 | 0 | 0 | 0 | 0 |
| 0 | 0 | 0 | 0 | 0 |
| 0.014042585 | 0 | 0 | 0.005521419 | 0.029737035 |
| 0 | 0 | 0 | 0 | 0 |
| 0.067068493 | 0.619089413 | 0.092938882 | 0.045334433 | 0.019532828 |
| 0.001030627 | 0.001962899 | 0.01717154 | 0.002339741 | 0.002217826 |
| 0 | 0 | 0 | 0 | 0 |
| 0 | 0 | 0 | 0 | 0 |
| 0 | 0 | 0 | 0 | 0 |
| 0 | 0 | 0 | 0 | 0 |
| 0 | 0 | 0 | 0 | 0 |
| 0 | 0 | 0 | 0 | 0 |
| 0 | 0 | 0 | 0 | 0 |
| 0.146031409 | 0.024652126 | 0 | 0.239600003 | 0.231140805 |
| 0 | 0.11474238 | 0 | 0.101660268 | 0.263599509 |
| 0 | 0 | 0 | 0 | 0 |
| 0 | 0 | 0 | 0 | 0 |
| 0.02093714 | 0 | 0 | 0.232131622 | 0.066296587 |
| 0 | 0 | 0 | 0.010001194 | 0 |
| 0 | 0 | 0 | 0 | 0 |

| | | | | |
|---|---|---|---|---|
| 0 | 0 | 0 | 0 | 0 |
| 0 | 0 | 0 | 0 | 0 |
| 0.004535696 | 0.014135786 | 0.012722284 | 0.034260877 | 0.003387924 |
| 0 | 0 | 0 | 0 | 0 |
| 0 | 0.049473564 | 0 | 0 | 1.941484517 |
| 0 | 0 | 0 | 0 | 0 |
| 0 | 0 | 0 | 0 | 0 |
| 0 | 0 | 0 | 0 | 0 |
| 0 | 0 | 0 | 0 | 0 |
| 0 | 0 | 0 | 0 | 0 |
| 0 | 0 | 0 | 0 | 0 |
| 0 | 0 | 0 | 0 | 0 |
| 0 | 0 | 0 | 0 | 0 |
| 0 | 0 | 0 | 0 | 0 |
| 0 | 0 | 0 | 0 | 0 |
| 0 | 0 | 0 | 0 | 0 |
| 0 | 0 | 0 | 0 | 0 |
| 0 | 0 | 0 | 0 | 0 |
| 0 | 0 | 0 | 0 | 0 |
| 0 | 0 | 0 | 0 | 0 |
| 0 | 0 | 0 | 0 | 0 |
| 0 | 0 | 0 | 0 | 0 |
| 0 | 0 | 0 | 0 | 0 |
| 0 | 0 | 0 | 0 | 0 |
| 0 | 0 | 0 | 0 | 0.025926878 |
| 0.020511226 | 0 | 0.00653952 | 0.00465648 | 0.019461055 |
| 0 | 0 | 0 | 0 | 0 |
| 0.110757765 | 0 | 0 | 0 | 0 |
| 0 | 0 | 0 | 0 | 0 |
| 0 | 0 | 0 | 0 | 0 |
| 0.001072975 | 0 | 0 | 0 | 0 |
| 0.046166967 | 0 | 0 | 0 | 0 |
| 0 | 0 | 0 | 0 | 0 |
| 0 | 0 | 0 | 0 | 0 |
| 0 | 0 | 0 | 0 | 0 |
| 0 | 0 | 0 | 0 | 0 |
| 0 | 0 | 0 | 0.007450052 | 0 |
| 0 | 0 | 0 | 0 | 0 |
| 0 | 0 | 0 | 0 | 0 |
| 0 | 0 | 0 | 0 | 0 |
| 0 | 0 | 0 | 0 | 0 |
| 0 | 0.101710795 | 0 | 0 | 0 |
| 0 | 0.023327562 | 0 | 0.002336643 | 0.065842605 |
| 0 | 0 | 0 | 0 | 0 |
| 0 | 0 | 0 | 0 | 0 |
| 0.001381485 | 0 | 0 | 0 | 0 |
| 0 | 0.003298762 | 0 | 0.001429842 | 0.013553386 |
| 0 | 0 | 0 | 0 | 0 |

| | | | | |
|---|---|---|---|---|
| 0 | 0 | 0 | 0 | 0 |
| 0.003145796 | 0.019298542 | 0.033920229 | 0.124264151 | 0.027207536 |
| 0 | 0 | 0 | 0 | 0 |
| 0 | 0 | 0 | 0 | 0 |
| 0 | 0 | 0 | 0 | 0 |
| 0 | 0 | 0 | 0 | 0 |
| 0 | 0 | 0 | 0 | 0 |
| 0 | 0 | 0 | 0 | 0 |
| 0 | 0 | 0 | 0 | 0 |
| 0 | 0 | 0 | 0 | 0 |
| 0 | 0 | 0 | 0 | 0 |
| 0 | 0 | 0 | 0 | 0 |
| 0 | 0 | 0 | 0.2176589 | 0.681424978 |
| 0 | 0 | 0 | 0 | 0 |
| 0.034681682 | 0 | 0 | 0.09363807 | 0.041919997 |
| 0 | 0 | 0 | 0 | 0 |
| 0 | 0 | 0 | 0 | 0 |
| 0 | 0 | 0 | 0 | 0 |
| 0 | 0 | 0 | 0 | 0 |
| 0 | 0 | 0 | 0 | 0 |
| 0 | 0 | 0 | 0 | 0 |
| 0 | 0 | 1.443232477 | 0.127860266 | 0.175145648 |
| 0 | 0 | 0 | 0 | 0 |
| 0 | 0 | 0.394567634 | 0 | 0 |
| 0 | 0 | 0 | 0 | 0 |
| 0 | 0 | 0 | 0 | 0 |
| 0 | 0 | 0 | 0 | 0 |
| 0 | 0 | 0 | 0 | 0 |
| 0 | 0 | 0 | 0 | 0 |
| 0 | 0 | 0 | 0 | 0 |
| 0 | 0 | 0 | 0 | 0 |
| 0 | 0 | 0 | 0 | 0 |
| 0 | 0 | 0 | 0 | 0 |
| 0 | 0 | 0 | 0 | 0 |
| 3.011110517 | 2.912725822 | 0 | 0 | 7.117337253 |
| 0 | 0 | 0 | 0 | 0 |
| 0 | 0 | 0 | 0 | 0 |
| 0 | 0 | 0 | 0 | 0 |
| 0.063572253 | 0.001850868 | 0.243871783 | 0.039987413 | 0.011406791 |
| 0 | 0 | 0 | 0 | 0 |
| 0.033038757 | 0.121976752 | 0.038534927 | 0.289551995 | 0.152989292 |
| 0 | 0 | 0 | 0 | 0 |
| 0 | 0 | 0 | 0 | 0 |
| 0 | 0 | 0 | 0 | 0 |
| 0 | 0 | 0 | 0 | 0 |
| 0.725574209 | 0.012587958 | 3.934876698 | 0.186566968 | 0.34568944 |
| 0 | 0 | 0 | 0 | 0 |
| 0 | 0 | 0 | 0 | 0 |

| | | | | |
|---|---|---|---|---|
| 0 | 0 | 0 | 0 | 0 |
| 0 | 0 | 0 | 0 | 0 |
| 0 | 0 | 0 | 0 | 0 |
| 0 | 0 | 0 | 0 | 0 |
| 0 | 0 | 0 | 0.000858858 | 0 |
| 0 | 0 | 0 | 0 | 0 |
| 0 | 0 | 0 | 0 | 0 |
| 0 | 0 | 0 | 0.001835987 | 0 |
| 0 | 0 | 0 | 0 | 0 |
| 0.006504614 | 0 | 0.053936627 | 0.101337452 | 0.099015667 |
| 0 | 0 | 0 | 0 | 0 |
| 0 | 0 | 0 | 0 | 0 |
| 0.005165641 | 0 | 0.002452024 | 0 | 0 |
| 0 | 0 | 0 | 0 | 0 |
| 0 | 0 | 0 | 0 | 0 |
| 0 | 0 | 0 | 0 | 0 |
| 0 | 0 | 0 | 0 | 0 |
| 0 | 0 | 0 | 0 | 0 |
| 0 | 0 | 0 | 0 | 0 |
| 0.883658972 | 0.013943502 | 0.457228349 | 0.575210777 | 1.195381952 |
| 0 | 0 | 0 | 0 | 0 |
| 0 | 0 | 0 | 0 | 0 |
| 0 | 0 | 0 | 0 | 0 |
| 0 | 0 | 0 | 0 | 0 |
| 0 | 0 | 0 | 0 | 0 |
| 0 | 0 | 0 | 0 | 0 |
| 0 | 0 | 0 | 0 | 0 |
| 0 | 0 | 0 | 0 | 0 |
| 0 | 0 | 0 | 0 | 0 |
| 0 | 0 | 0 | 0 | 0.098964343 |
| 0 | 0 | 0 | 0 | 0 |
| 0 | 0 | 0 | 0 | 0 |
| 0 | 0 | 0 | 0 | 0 |
| 0 | 0 | 0 | 0 | 0 |
| 0 | 0 | 0 | 0 | 0 |
| 0 | 0 | 0 | 0 | 0 |
| 0 | 0 | 0 | 0 | 0 |
| 0 | 0 | 0 | 0 | 0 |
| 0 | 0 | 0 | 0 | 0 |
| 0 | 0 | 0 | 0 | 0 |
| 0 | 0 | 0 | 0 | 0 |
| 0 | 0 | 0 | 0 | 0 |
| 0 | 0.008062879 | 0 | 0 | 0 |
| 0 | 0 | 0 | 0 | 0 |
| 0.000996556 | 0.003416413 | 0.043662018 | 0.004524783 | 0 |
| 0 | 0 | 0 | 0 | 0 |
| 0 | 0 | 0 | 0 | 0 |
| 0 | 0 | 0 | 0 | 0 |

| | | | | |
|---|---|---|---|---|
| 0 | 0 | 0 | 0 | 0 |
| 0 | 0 | 0 | 0 | 0 |
| 0 | 0 | 0 | 0 | 0 |
| 0 | 0 | 0 | 0 | 0 |
| 0 | 0 | 0 | 0 | 0 |
| 0 | 0.000708468 | 0 | 0 | 0 |
| 0 | 0 | 0 | 0 | 0 |
| 0 | 0 | 0 | 0 | 0 |
| 0 | 0 | 0 | 0 | 0 |
| 0 | 0 | 0 | 0 | 0 |
| 0 | 0 | 0 | 0 | 0 |
| 0 | 0 | 0 | 0 | 0 |
| 0 | 0 | 0 | 0 | 0 |
| 0 | 0 | 0 | 0 | 0 |
| 0 | 0 | 0 | 0 | 0 |
| 0 | 0 | 0 | 0 | 0 |
| 0 | 0.030077837 | 0 | 0 | 0 |
| 0 | 0.003141058 | 0 | 0.000624014 | 0 |
| 0 | 0 | 0 | 0 | 0 |
| 0 | 0 | 0 | 0 | 0 |
| 0 | 0 | 0 | 0 | 0 |
| 0 | 0 | 0 | 0 | 0 |
| 0 | 0 | 0 | 0 | 0 |
| 0 | 0 | 0 | 0.011424255 | 0 |
| 0 | 0 | 0 | 0 | 0 |
| 0 | 0 | 0.004547928 | 0.001585093 | 0.001138257 |
| 0 | 0 | 0 | 0 | 0 |
| 0 | 0.121421726 | 0 | 0 | 0 |
| 0 | 0 | 0 | 0 | 0 |
| 0 | 0 | 0 | 0 | 0 |
| 0 | 0 | 0 | 0 | 0 |
| 0 | 0 | 0 | 0 | 0 |
| 0 | 0 | 0 | 0 | 0 |
| 0 | 0 | 0 | 0 | 0 |
| 0 | 0 | 0 | 0 | 0 |
| 0 | 0 | 0 | 0 | 0 |
| 0 | 0 | 0 | 0 | 0 |
| 0 | 0 | 0 | 0 | 0.063008652 |
| 0 | 0 | 0 | 0.59722282 | 0 |
| 0 | 0 | 0 | 0 | 0 |
| 0 | 0 | 0 | 0 | 0 |
| 0 | 0 | 0 | 0 | 0 |
| 0 | 0 | 0 | 0 | 0 |
| 0 | 0 | 0 | 0 | 0 |
| 0 | 0 | 0 | 0 | 0 |
| 0 | 0 | 0 | 0 | 0 |
| 0 | 0 | 0 | 0 | 0 |
| 0 | 0 | 0 | 0 | 0 |

| | | | | |
|---|---|---|---|---|
| 0 | 0 | 0 | 0 | 0 |
| 0 | 0 | 0 | 0.343960254 | 0 |
| 0 | 0 | 0 | 0 | 0 |
| 0 | 0 | 0 | 0 | 0 |
| 0 | 0 | 0 | 0 | 0 |
| 0 | 0.001305234 | 0 | 0 | 0 |
| 0 | 0 | 0 | 0 | 0 |
| 0 | 0 | 0 | 0 | 0 |
| 0 | 0 | 0 | 0 | 0 |
| 0 | 0 | 0 | 0 | 0 |
| 0.015632768 | 0.002284954 | 0.004262504 | 0.011637311 | 0 |
| 0 | 0 | 0 | 0 | 0 |
| 0 | 0.551865902 | 0 | 0.377326961 | 0 |
| 0 | 0 | 0 | 0 | 0 |
| 0 | 0 | 0 | 0 | 0 |
| 0 | 0 | 0 | 0 | 0 |
| 0 | 0 | 0 | 0 | 0 |
| 0 | 0 | 0 | 0 | 0 |
| 0 | 0 | 0 | 0 | 0 |
| 0 | 0 | 0 | 0 | 0 |
| 0.000553476 | 0 | 0.026184805 | 0.039956938 | 0 |
| 0 | 0 | 0 | 0 | 0.007655403 |
| 0 | 0 | 0 | 0 | 0 |
| 0.624017792 | 2.783548783 | 0 | 4.28055713 | 0.948442873 |
| 0 | 0 | 0 | 0 | 0 |
| 0 | 0 | 0 | 0 | 0 |
| 0 | 0 | 0 | 0 | 0 |
| 0 | 0 | 0 | 0 | 0 |
| 0 | 0 | 0 | 0 | 0 |
| 0 | 0 | 0 | 0 | 0 |
| 0 | 0 | 0 | 0 | 0 |
| 0 | 0 | 0 | 0 | 0 |
| 0 | 0 | 0 | 0 | 0 |
| 0 | 0 | 0 | 0 | 0 |

| PHL018-0008_V1 | PHL018-0114_V2 | PHL018-0060_V2 | PHL018-0026_V1 | PHL018-0138_V3 |
|---|---|---|---|---|
| CTRL | EYCF | CTRL | EYCF | EYCF |
| 0.221190043 | 0.04895508 | 0.047537996 | 0.03693267 | 0.275994922 |
| 0.981826418 | 0.288155405 | 13.53364556 | 0.00233776 | 0.03424169 |
| 0 | 0 | 6.050857771 | 14.81137721 | 0.073713645 |
| 0 | 0 | 0 | 0 | 0 |
| 0 | 0 | 0 | 0 | 0 |
| 0 | 0 | 0 | 0 | 0 |
| 0 | 0 | 0 | 0 | 0 |
| 0 | 0 | 0 | 0 | 0 |
| 0 | 0 | 0 | 0 | 0 |
| 0 | 0 | 0 | 0 | 0 |
| 0 | 0 | 0 | 0 | 0 |
| 0 | 0 | 0 | 0 | 0 |
| 0 | 0 | 0 | 0 | 0 |
| 1.758704888 | 3.029174536 | 3.178077059 | 10.76620672 | 6.540699844 |
| 0 | 0 | 0 | 0 | 0 |
| 0 | 0 | 0 | 0 | 0 |
| 0 | 0 | 0 | 0 | 0 |
| 0 | 0 | 0 | 0 | 0 |
| 0 | 0 | 0 | 0 | 0 |
| 0 | 0 | 0 | 0 | 0 |
| 0 | 0 | 0 | 0 | 0 |
| 0 | 0 | 0 | 0 | 0 |
| 0 | 0 | 0 | 0 | 0 |
| 0 | 0 | 0 | 0 | 0 |
| 0 | 0.012520147 | 0 | 0 | 0 |
| 0 | 0 | 0 | 0 | 0 |
| 0 | 0 | 0 | 0 | 0 |
| 0 | 0 | 0 | 0 | 0 |
| 0 | 0 | 0 | 0 | 0 |
| 0 | 0 | 0 | 0 | 0 |
| 0 | 0 | 0 | 0 | 0 |
| 0 | 0 | 0 | 0 | 0 |
| 0 | 0 | 0 | 0 | 0 |
| 0 | 0 | 0 | 0 | 0 |
| 0 | 0 | 0 | 0 | 0 |
| 5.91631577 | 5.27798025 | 2.353930685 | 0 | 1.627901015 |
| 0 | 0 | 0 | 0 | 0 |
| 0 | 0 | 0 | 0 | 0 |
| 0 | 0 | 0 | 0 | 0 |
| 0 | 0 | 0 | 0 | 0 |
| 0 | 0 | 0 | 0 | 0 |
| 0.386692156 | 0.032347974 | 0.014728724 | 0 | 0.001524301 |
| 1.798786465 | 0.788236076 | 1.052694426 | 0.059404645 | 0.44787087 |
| 0.036485367 | 0.001221377 | 0.068367172 | 0.013036109 | 0 |
| 0.007206757 | 0 | 0.002143057 | 0 | 0 |
| 0 | 0 | 0 | 0 | 0 |

| | | | | |
|---|---|---|---|---|
| 0 | 0 | 0.001909496 | 0 | 0 |
| 0.089316523 | 0 | 0 | 0.01492032 | 0.107828706 |
| 0.008146533 | 0.003153123 | 0.007307048 | 0 | 0.090485125 |
| 0.32439388 | 1.315506796 | 1.209934548 | 0 | 0.111153563 |
| 0.001548748 | 0 | 0.002922708 | 0 | 0.013206596 |
| 0 | 0.011761321 | 0 | 0 | 0 |
| 0 | 0.561661865 | 0.052225251 | 0 | 0 |
| 0 | 0 | 0 | 0 | 0 |
| 0 | 0 | 0 | 0 | 0 |
| 0 | 0 | 0 | 0 | 0 |
| 0 | 0 | 0 | 0 | 0.006680893 |
| 0.008910942 | 0.007296361 | 0.059801784 | 0 | 0.106951734 |
| 0 | 0 | 0 | 0 | 0 |
| 0.028443728 | 0.010331624 | 0.039418861 | 0.079700981 | 0.005207457 |
| 0 | 0 | 0 | 0 | 0.14514961 |
| 0 | 0 | 0 | 0 | 0 |
| 2.431786915 | 0.190263854 | 0.001508067 | 0 | 0 |
| 0 | 0 | 0 | 0 | 0 |
| 0 | 0 | 0 | 0 | 0 |
| 0 | 0 | 0 | 0 | 0 |
| 0.078528498 | 0 | 0.018986291 | 0 | 0.147413866 |
| 0.000489362 | 0.001195871 | 0.010186444 | 0.004669713 | 0.004214759 |
| 0 | 0.113053172 | 0 | 0 | 0 |
| 0 | 0 | 0 | 0 | 0 |
| 0 | 0 | 0 | 0 | 0 |
| 0 | 0 | 0 | 0 | 0 |
| 0 | 0 | 0 | 0 | 0.465431616 |
| 0 | 0 | 0 | 0 | 0 |
| 0.006886397 | 0.000874208 | 0.001163519 | 0 | 0 |
| 0 | 0 | 0 | 0 | 0 |
| 0.129568549 | 0 | 0 | 0.205126056 | 0.110613395 |
| 0 | 0 | 0 | 0 | 0 |
| 0 | 0 | 0 | 0 | 0 |
| 0 | 0 | 0 | 0 | 0 |
| 0 | 0 | 0 | 0 | 0 |
| 0.001327365 | 0 | 0 | 0 | 0 |
| 0 | 0 | 0 | 0 | 0 |
| 0.045098659 | 0.646338909 | 0 | 0 | 0 |
| 0 | 0 | 0 | 0 | 0 |
| 0 | 0 | 0 | 0 | 0 |
| 0 | 0 | 0 | 0 | 0 |
| 0 | 0 | 0 | 0 | 0 |
| 0 | 0 | 0 | 0 | 0 |
| 0 | 0 | 0 | 0 | 0 |
| 0 | 0 | 0 | 0 | 0 |
| 0 | 0 | 0 | 0 | 0 |
| 0.002621748 | 0 | 0 | 0 | 0 |
| 0 | 0 | 0 | 0 | 0 |

| | | | | |
|---|---|---|---|---|
| 0 | 0 | 0 | 0 | 0 |
| 0 | 0 | 0 | 0 | 0 |
| 0 | 0 | 0 | 0 | 0 |
| 0 | 0 | 0 | 0 | 0 |
| 0.056816252 | 0 | 0 | 0 | 0 |
| 0 | 0 | 0 | 0 | 0 |
| 0 | 0 | 0 | 0 | 0 |
| 0 | 0 | 0 | 0 | 0 |
| 0 | 0 | 0 | 0 | 0.013141597 |
| 0 | 0 | 0 | 0 | 0 |
| 0 | 0 | 0 | 0 | 0 |
| 0.003294912 | 0.001533692 | 0.001020626 | 0 | 0 |
| 0 | 0 | 0 | 0 | 0 |
| 0 | 0 | 0 | 0 | 0 |
| 0 | 0 | 0 | 0 | 0 |
| 0 | 0 | 0 | 0 | 0 |
| 0 | 0 | 0 | 0 | 0 |
| 0 | 0 | 0 | 0 | 0 |
| 0.419321376 | 0.075582127 | 0 | 0 | 0 |
| 0 | 0 | 0 | 0 | 0 |
| 0 | 0 | 0 | 0 | 0 |
| 0 | 0 | 0 | 0 | 0 |
| 0 | 0 | 0 | 0 | 0 |
| 0 | 0 | 0 | 0 | 0 |
| 0 | 0 | 0 | 0 | 0 |
| 0 | 0 | 0 | 0 | 0 |
| 0 | 0 | 0 | 0 | 0 |
| 0 | 0 | 0 | 0 | 0 |
| 0 | 0 | 0 | 0 | 0 |
| 0 | 0 | 0 | 0 | 0 |
| 0.004231168 | 0.021851196 | 0.015619196 | 0 | 0 |
| 0.003214349 | 0.025649016 | 0.024578846 | 0 | 0.001020623 |
| 0 | 0 | 0 | 0 | 0 |
| 0 | 0 | 0 | 0 | 0 |
| 0 | 0 | 0 | 0 | 0 |
| 0 | 0 | 0 | 0 | 0 |
| 0 | 0 | 0 | 0 | 0 |
| 0 | 0 | 0 | 0 | 0 |
| 0 | 0 | 0.054781996 | 0 | 0 |
| 0 | 0 | 0 | 0 | 0 |
| 0 | 0 | 0 | 0 | 0 |
| 0 | 0 | 0 | 0 | 0 |
| 0 | 0 | 0 | 0 | 0 |
| 0 | 0 | 0 | 0 | 0 |
| 0 | 0 | 0 | 0 | 0 |
| 0 | 0 | 0 | 0 | 0 |
| 0 | 0 | 0 | 0 | 0 |
| 0 | 0 | 0 | 0 | 0 |

| | | | | |
|---|---|---|---|---|
| 0 | 0 | 0 | 0 | 0 |
| 0 | 0 | 0 | 0 | 0 |
| 0 | 0 | 0 | 0 | 0 |
| 0 | 0 | 0 | 0 | 0 |
| 0 | 0.120692183 | 0.048283047 | 0 | 0 |
| 0 | 0 | 0 | 0 | 0 |
| 0 | 0 | 0 | 0 | 0 |
| 0 | 0 | 0 | 0 | 0 |
| 0 | 0 | 0 | 0 | 0 |
| 0 | 0 | 0 | 0 | 0 |
| 0 | 0 | 0 | 0 | 0 |
| 0 | 0 | 0 | 0 | 0 |
| 0 | 0 | 0 | 0 | 0 |
| 0 | 0 | 0 | 0 | 0 |
| 0 | 0 | 0 | 0 | 0 |
| 0 | 19.95309181 | 0 | 0 | 0 |
| 0 | 0 | 0 | 0 | 0 |
| 0 | 0 | 0 | 0 | 0 |
| 0 | 0 | 0 | 0 | 0 |
| 0 | 0 | 0 | 0 | 0 |
| 0 | 0 | 0 | 0 | 0 |
| 0 | 0 | 0 | 0 | 0 |
| 0 | 0 | 0 | 0 | 0 |
| 0 | 0 | 0 | 0 | 0 |
| 0 | 0.032894849 | 0 | 0 | 0 |
| 0 | 0 | 0 | 0 | 0 |
| 0 | 0 | 0 | 0 | 0 |
| 0 | 0 | 0 | 0 | 0 |
| 0 | 0 | 0 | 0 | 0 |
| 0 | 0 | 0 | 0 | 0 |
| 0 | 0 | 0 | 0 | 0 |
| 0 | 0.010912577 | 0 | 0 | 0 |
| 0 | 0 | 0 | 0 | 0 |
| 0 | 0 | 0 | 0 | 0 |
| 0 | 0 | 0 | 0 | 0 |
| 0 | 0 | 0 | 0 | 0 |
| 0 | 0 | 0 | 0 | 0 |
| 0 | 0 | 0 | 0 | 0 |
| 0 | 0 | 0 | 0 | 0 |
| 0 | 0 | 0 | 0 | 0 |
| 0 | 0 | 0 | 0 | 0 |
| 0 | 0 | 0 | 0 | 0 |
| 0 | 0 | 0 | 0 | 0 |
| 0 | 0 | 0 | 0 | 0 |
| 0 | 0 | 0 | 0 | 0 |
| 0 | 0 | 0 | 0 | 0 |
| 0 | 0 | 0 | 0 | 0 |
| 0 | 0 | 0 | 0.011570274 | 0 |

| | | | | |
|---|---|---|---|---|
| 0 | 0 | 0 | 0 | 0 |
| 0 | 0 | 0 | 0 | 0 |
| 0 | 0 | 0 | 0 | 0 |
| 0 | 0 | 0 | 0 | 0 |
| 0 | 0 | 0 | 0 | 0 |
| 0 | 0 | 0 | 0 | 0 |
| 0 | 0 | 0 | 0 | 0 |
| 0 | 0 | 0 | 0 | 0 |
| 0 | 0 | 0 | 0 | 0 |
| 0 | 0 | 0 | 0 | 0 |
| 0 | 0 | 0 | 0 | 0 |
| 0 | 0 | 0 | 0 | 0 |
| 0.000844959 | 0 | 0 | 0 | 0 |
| 0 | 0 | 0 | 0 | 0 |
| 0 | 0 | 0 | 0 | 0 |
| 0 | 0 | 0 | 0 | 0 |
| 0 | 0 | 0 | 0 | 0 |
| 0 | 0 | 0.002266355 | 0 | 0 |
| 0 | 0 | 0 | 0 | 0 |
| 0 | 0 | 0 | 0 | 0 |
| 0.048976002 | 0.022502847 | 0.072232174 | 0.027739523 | 0.032656905 |
| 0 | 0 | 0 | 0 | 0 |
| 0 | 0 | 0 | 0 | 0.001386504 |
| 0 | 0 | 0 | 0 | 0.003570238 |
| 0 | 0 | 0 | 0 | 0 |
| 0 | 0 | 0 | 0 | 0 |
| 0 | 0.006745286 | 0.008823666 | 0.007199442 | 0 |
| 0 | 0 | 0 | 0 | 0 |
| 0 | 0 | 0 | 0 | 0 |
| 0 | 0 | 0 | 0.019726102 | 0.002625513 |
| 0 | 0 | 0 | 0 | 0.633269254 |
| 0 | 0 | 0 | 0 | 0 |
| 0 | 0 | 0 | 0 | 0 |
| 0 | 0.000808961 | 0 | 0 | 0 |
| 0.029399321 | 0.00495229 | 0.002521849 | 0.012359513 | 0.036499135 |
| 0 | 0 | 0 | 0 | 0 |
| 0 | 0 | 0 | 0 | 0 |
| 0 | 0 | 0 | 0 | 0 |
| 0 | 0 | 0 | 0 | 0 |
| 0 | 0 | 0 | 0 | 0 |
| 0 | 0 | 0 | 0 | 0 |
| 0.002874511 | 0 | 0 | 0 | 0 |
| 0 | 0 | 0 | 0 | 0 |
| 0 | 0 | 0 | 0 | 0 |
| 0 | 0 | 0 | 0 | 0 |
| 0 | 0.03526089 | 0 | 0 | 0 |
| 0 | 0 | 0 | 0.009133791 | 0 |
| 0 | 0 | 0 | 0 | 0.00061057 |

| | | | | |
|---|---|---|---|---|
| 0 | 0 | 0 | 0 | 0 |
| 0 | 0 | 0 | 0 | 0 |
| 0 | 0 | 0 | 0 | 0 |
| 0 | 0 | 0.001246035 | 0 | 0 |
| 0 | 0 | 0 | 0 | 0 |
| 0 | 0 | 0 | 0 | 0 |
| 0 | 0 | 0 | 0 | 0 |
| 0.002527308 | 0 | 0.006575975 | 0 | 0 |
| 0.005485396 | 0 | 0 | 0 | 0 |
| 0 | 0 | 0 | 0 | 0 |
| 0 | 0 | 0.00343634 | 0 | 0 |
| 0 | 0 | 0 | 0 | 0 |
| 0 | 0 | 0 | 0 | 0 |
| 0 | 0 | 0 | 0 | 0 |
| 0 | 0 | 0 | 0 | 0.008086395 |
| 0 | 0 | 0 | 0 | 0 |
| 0 | 0 | 0 | 0 | 0 |
| 0 | 0 | 0 | 0 | 0 |
| 0.013558857 | 0.003745155 | 0.004984575 | 0 | 0.002980538 |
| 0 | 0.003218832 | 0 | 0 | 0 |
| 0.024836027 | 0 | 0 | 0.002936238 | 0.17256642 |
| 0 | 0 | 0 | 0 | 0 |
| 0 | 0 | 0 | 0 | 0 |
| 0 | 0 | 0 | 0 | 0 |
| 0.003929895 | 0 | 0.007303902 | 0.008750186 | 0 |
| 0 | 0 | 0 | 0 | 0 |
| 0 | 0 | 0 | 0 | 0 |
| 0 | 0 | 0 | 0 | 0 |
| 0.166057433 | 0.005891701 | 0 | 0 | 0 |
| 0 | 0 | 0 | 0 | 0 |
| 0.081774618 | 0.033004985 | 0.043792474 | 3.864481954 | 0.340523259 |
| 0 | 0 | 0 | 0 | 0 |
| 0 | 0.00210457 | 0 | 0 | 0.015409052 |
| 0.002090871 | 0.002846738 | 0.003031069 | 0 | 0 |
| 0 | 0 | 0 | 0 | 0 |
| 0 | 0 | 0 | 0 | 0 |
| 0 | 0 | 0 | 0 | 0 |
| 0.060804937 | 0 | 0 | 0 | 0 |
| 0 | 0 | 0 | 0 | 0 |
| 0 | 0 | 0.001080064 | 0 | 0 |
| 0 | 0 | 0 | 0 | 0 |
| 0.034758967 | 0.013802295 | 0.004592505 | 0 | 0.01647659 |
| 0 | 0 | 0 | 0 | 0 |
| 0 | 0 | 0 | 0 | 0 |
| 0 | 0 | 0 | 0 | 0 |
| 0 | 0 | 0 | 0 | 0 |
| 0 | 0 | 0 | 0 | 0 |
| 0 | 0 | 0 | 0 | 0 |

| | | | | |
|---|---|---|---|---|
| 0 | 0 | 0 | 0 | 0 |
| 0 | 0 | 0 | 0 | 0 |
| 0.02771806 | 0.001083191 | 0.065509091 | 0.015593538 | 0 |
| 0 | 0 | 0 | 0 | 0 |
| 0 | 0 | 0 | 0 | 0 |
| 0 | 0 | 0 | 0 | 0 |
| 0 | 0 | 0 | 0 | 0 |
| 0 | 0 | 0 | 0 | 0 |
| 0 | 0 | 0 | 0 | 0 |
| 0 | 0 | 0 | 0 | 0 |
| 0 | 0 | 0 | 0 | 0 |
| 0 | 0 | 0 | 0 | 0 |
| 0 | 0 | 0 | 0 | 0 |
| 0.00114304 | 0.004549061 | 0 | 0.008944059 | 0 |
| 0.003773718 | 0 | 0.011362095 | 0 | 0 |
| 0 | 0 | 0 | 0 | 0 |
| 0.001074953 | 0 | 0.00503459 | 0.002051536 | 0 |
| 0 | 0 | 0 | 0 | 0 |
| 0 | 0 | 0 | 0 | 0 |
| 0 | 0 | 0 | 0 | 0 |
| 0 | 0 | 0 | 0 | 0 |
| 0 | 0 | 0 | 0 | 0.000485281 |
| 0 | 0 | 0 | 0 | 0 |
| 0 | 0 | 0 | 0 | 0 |
| 0 | 0 | 0 | 0 | 0 |
| 0 | 0 | 0 | 0 | 0 |
| 1.05652431 | 0 | 0 | 0 | 0 |
| 0 | 0 | 0 | 0 | 0 |
| 0 | 0 | 0 | 0 | 0 |
| 0 | 0 | 0 | 0 | 0.008657544 |
| 0 | 0 | 0 | 0 | 0 |
| 0 | 0 | 0 | 0 | 0 |
| 0 | 0 | 0 | 0 | 0 |
| 0 | 0 | 0 | 0 | 0 |
| 0 | 0 | 0 | 0 | 0 |
| 0 | 0 | 0 | 0 | 0 |
| 0 | 0 | 0 | 0 | 0 |
| 0 | 0.001787606 | 0.020121193 | 0 | 0 |
| 0 | 0 | 0 | 0 | 0 |
| 0 | 0 | 0 | 0 | 0 |
| 0 | 0 | 0 | 0 | 0 |
| 0 | 0 | 0 | 0 | 0 |
| 0 | 0 | 0 | 0 | 0 |
| 0 | 0 | 0 | 0 | 0 |
| 0 | 0 | 0 | 0 | 0 |
| 0 | 0 | 0 | 0 | 0.002499765 |
| 0 | 0 | 0 | 0 | 0.000660898 |
| 0 | 0 | 0 | 0.001972825 | 0 |

| | | | | |
|---|---|---|---|---|
| 0 | 0.007952833 | 0 | 0 | 0 |
| 0.005254508 | 0 | 0 | 0.095088523 | 0 |
| 0.001219672 | 0 | 0.00566705 | 0 | 0 |
| 0 | 0 | 0 | 0 | 0 |
| 0 | 0.001927751 | 0.002280639 | 0 | 0.003409283 |
| 0 | 0 | 0 | 0 | 0.00305753 |
| 0 | 0 | 0 | 0 | 0 |
| 0 | 0.042634187 | 0.013106954 | 0.015592908 | 0 |
| 0 | 0 | 0 | 0 | 0.00082858 |
| 0.007978071 | 0.028780212 | 0.111042497 | 0.472289698 | 10.62394377 |
| 0 | 0 | 0 | 0 | 0.003340086 |
| 0.281398743 | 0 | 0.022522452 | 0 | 0.001513185 |
| 0.048715012 | 0.010636661 | 0.008254106 | 0 | 0 |
| 0.000784299 | 0 | 0 | 0 | 0 |
| 0 | 0 | 0 | 0.024158314 | 0 |
| 0 | 0 | 0 | 0 | 0 |
| 0 | 0 | 0 | 0 | 0 |
| 0 | 0 | 0 | 0 | 0 |
| 0.074238056 | 0 | 0 | 0.041390385 | 1.920036774 |
| 0 | 0 | 0 | 0 | 0 |
| 0 | 0 | 0 | 0 | 0 |
| 0 | 0 | 0 | 0.116880695 | 0 |
| 0 | 0 | 0 | 0 | 0 |
| 0 | 0.001198473 | 0 | 0 | 0 |
| 0 | 0 | 0 | 0 | 0 |
| 0 | 0.003149745 | 0.001341478 | 0.001147937 | 0 |
| 0 | 2.526547207 | 0 | 0 | 0.003351949 |
| 0 | 0 | 0 | 0 | 0 |
| 0 | 0 | 0 | 0 | 0.035417514 |
| 0 | 0 | 0.005750885 | 0.002165318 | 0 |
| 0 | 0 | 0 | 0 | 0.004124379 |
| 0 | 0 | 0 | 0 | 0 |
| 0 | 0 | 0 | 0 | 0.01571133 |
| 0 | 0 | 0 | 0 | 0.038246679 |
| 0 | 0 | 0 | 0 | 0 |
| 0 | 0 | 0 | 0.021535285 | 0.001016766 |
| 0 | 0 | 0 | 0 | 0.117262832 |
| 0 | 0 | 0 | 0 | 0 |
| 0 | 0 | 0 | 0 | 0 |
| 0.008223978 | 0 | 0 | 0 | 3.80185553 |
| 0 | 0 | 0 | 0 | 0 |
| 0 | 0 | 0 | 0 | 0 |
| 0 | 0 | 0 | 0 | 0 |
| 0 | 0 | 0 | 0 | 0 |
| 0 | 0 | 0.003347896 | 0 | 0 |
| 0 | 0 | 0 | 0 | 0 |
| 0.000748001 | 0.001566783 | 0 | 0.005710201 | 0.001039088 |
| 0 | 0 | 0 | 0 | 0 |

| | | | | |
|---|---|---|---|---|
| 0 | 0 | 0 | 0 | 0 |
| 0.018669391 | 0.008171284 | 0 | 0 | 0 |
| 0 | 0 | 0 | 0 | 0 |
| 0 | 0 | 0 | 0 | 0 |
| 0 | 0 | 0 | 0.009171396 | 0 |
| 0.003534846 | 0 | 0.004379789 | 0 | 0.005565172 |
| 0 | 0 | 0 | 0 | 0 |
| 0 | 0 | 0 | 0 | 0 |
| 0 | 0 | 0 | 0 | 0 |
| 0 | 0 | 0 | 0 | 0 |
| 0 | 0.008587611 | 0.001015964 | 0.001086732 | 0 |
| 0 | 0 | 0.011188224 | 0.011611077 | 0 |
| 0 | 0 | 0 | 0 | 0 |
| 0 | 0.001802921 | 0 | 0.124229469 | 0 |
| 0 | 0 | 0 | 0 | 0.08111895 |
| 0 | 0 | 0 | 0 | 0 |
| 0 | 0 | 0 | 0 | 0 |
| 0.002303495 | 0 | 0 | 0 | 0.026530164 |
| 0 | 0 | 0 | 0 | 0 |
| 0.000903222 | 0 | 0 | 0 | 0 |
| 0 | 0 | 0 | 0 | 0 |
| 0.150838993 | 0.397742284 | 1.985903833 | 0 | 0.422794858 |
| 0 | 0 | 0.014801053 | 0 | 0 |
| 0 | 0 | 0 | 0.001206432 | 0 |
| 0 | 0 | 0 | 0 | 0 |
| 0 | 0 | 0 | 0 | 0 |
| 0 | 0 | 0 | 0 | 2.708152708 |
| 0 | 0 | 0 | 0 | 0 |
| 0 | 0 | 0 | 0 | 0 |
| 0.163069289 | 0.234209654 | 0.431218417 | 0.121295927 | 0.027078248 |
| 0.005401451 | 0.001528923 | 0 | 0 | 0 |
| 0 | 0.00214656 | 0 | 0 | 0 |
| 0 | 0 | 0 | 0 | 0 |
| 0 | 0 | 0 | 0 | 0 |
| 0.01574077 | 0 | 0 | 0 | 0.017655058 |
| 0 | 0 | 0 | 0 | 0 |
| 0 | 0 | 0.216494764 | 0 | 0.051120306 |
| 0 | 0.003215899 | 0.033018428 | 0 | 0 |
| 0 | 0 | 0.010242616 | 0 | 5.044752285 |
| 0 | 0 | 0 | 0 | 0 |
| 0 | 0.015007245 | 0.004537243 | 0.002869773 | 0.020200461 |
| 0 | 1.288614916 | 3.344203076 | 0 | 0.005960166 |
| 0 | 0.001893839 | 0.037640731 | 0 | 0 |
| 0 | 0 | 4.541064285 | 0 | 0 |
| 0 | 0 | 0 | 0 | 0 |
| 0 | 0 | 0 | 0.001376258 | 0 |
| 0 | 0.001555672 | 0 | 0.003720746 | 0.007737895 |
| 0 | 0 | 0 | 0 | 0 |

| | | | | |
|---|---|---|---|---|
| 0.049712659 | 0 | 0 | 0 | 0 |
| 0 | 0 | 0 | 0 | 0 |
| 5.261034799 | 0.41878801 | 0.003480915 | 0 | 0 |
| 0.010361387 | 0 | 0 | 0 | 0 |
| 0 | 0.036017385 | 0 | 0 | 0 |
| 0.081933533 | 0 | 0.033479442 | 0 | 0 |
| 0 | 0 | 0 | 0 | 0 |
| 0 | 0 | 0 | 0 | 0 |
| 0.003067652 | 0 | 0 | 0 | 0 |
| 0 | 0 | 0 | 0 | 0.248993803 |
| 0 | 0 | 0 | 0 | 0 |
| 0 | 0 | 0 | 0 | 0 |
| 0 | 0 | 0 | 0 | 0 |
| 0 | 0 | 0 | 0 | 0 |
| 0 | 0 | 0 | 0 | 0 |
| 0.025752313 | 0.100928311 | 0.051293538 | 0 | 0 |
| 0.327804367 | 0.002068157 | 0.019818663 | 0.017987168 | 0 |
| 0.083100048 | 0 | 0 | 0 | 0.025087483 |
| 0.003523191 | 0 | 0 | 0.061223502 | 0 |
| 0.00335179 | 0 | 0 | 0 | 0 |
| 0.006561159 | 0 | 0.004289006 | 0.026986845 | 0.041109365 |
| 0 | 0.001090434 | 0 | 0.009935324 | 0 |
| 0 | 0 | 0 | 0 | 0 |
| 0 | 0 | 0.177228342 | 0 | 0 |
| 0 | 0.001581458 | 0 | 0.002561638 | 0 |
| 0.001418919 | 0 | 0 | 0 | 0.712157649 |
| 0 | 0.001193428 | 0 | 0.000652424 | 0.001329686 |
| 0 | 0 | 0 | 0 | 0 |
| 0 | 0 | 0 | 0 | 0 |
| 0 | 0 | 0 | 0 | 0 |
| 0 | 0.015066141 | 0.007344391 | 0.029770016 | 0 |
| 0.054780348 | 0 | 0.030255436 | 0 | 0 |
| 0.021896644 | 0.020613629 | 0.094652445 | 0.043902443 | 0.055162194 |
| 0.007404276 | 0.020309672 | 0.002752241 | 0 | 0 |
| 0.021894043 | 0.001844938 | 0 | 0 | 0 |
| 0 | 0 | 0 | 0 | 0 |
| 0.002446706 | 0 | 0.001488215 | 0.017829038 | 0 |
| 0.0534023 | 0.007287163 | 0.015518037 | 0.012117248 | 0.023342597 |
| 0.606435903 | 0.066991002 | 0.069023195 | 0 | 0 |
| 0.002133314 | 0 | 0 | 0 | 0.006401166 |
| 0 | 0 | 0 | 0 | 0 |
| 0 | 0 | 0 | 0 | 0 |
| 0 | 0 | 0 | 0 | 0 |
| 0 | 0 | 0.076165141 | 0 | 0.104463957 |
| 0 | 0 | 0 | 0.001774963 | 0 |
| 0.00246504 | 0 | 0 | 0 | 0 |
| 0 | 0 | 0 | 0 | 0 |
| 0 | 0 | 0 | 0 | 0 |

| | | | | |
|---|---|---|---|---|
| 0 | 0 | 0 | 0 | 0 |
| 0.008058931 | 0.105006413 | 0.150924532 | 0.010178191 | 0.003951211 |
| 0 | 0 | 0 | 0 | 0 |
| 0 | 0.000451035 | 0 | 0 | 0 |
| 0 | 0 | 0 | 0 | 0 |
| 0 | 0 | 0 | 0 | 0 |
| 0 | 0 | 0 | 0 | 0.626715744 |
| 0.002366806 | 0 | 0.041416628 | 0 | 0 |
| 0 | 0 | 0.265187831 | 0 | 0 |
| 0 | 0 | 0 | 0 | 0 |
| 0 | 0 | 0 | 0.04050679 | 0 |
| 0.114914317 | 0.002472974 | 0.022527653 | 0.08562206 | 0 |
| 0.013494912 | 0.142032049 | 0 | 0 | 0 |
| 0 | 0 | 0 | 0 | 0 |
| 0.001010477 | 0 | 0 | 0.00250703 | 0 |
| 0 | 0 | 0 | 0 | 0.023038351 |
| 0.000737765 | 0.000772671 | 0.011312164 | 0.002464026 | 0 |
| 0.008380053 | 0 | 0.01121381 | 0.37744032 | 10.87379849 |
| 0 | 0 | 0 | 0 | 0 |
| 0.024720268 | 0 | 0.005201187 | 0.001483596 | 0.058184069 |
| 0.031193677 | 0.039641741 | 0.022482975 | 0 | 0.008877945 |
| 0 | 0 | 0 | 0 | 0 |
| 0 | 0 | 0 | 0 | 0 |
| 0 | 0 | 0 | 0 | 0 |
| 0 | 0 | 0 | 0 | 0 |
| 0 | 0 | 0 | 0 | 0 |
| 0.003567101 | 0.446928677 | 0.075159089 | 0 | 0 |
| 0.04973141 | 0.031335333 | 0.013526085 | 0.219724726 | 0.234719131 |
| 0 | 0 | 0 | 0 | 0 |
| 0.007315577 | 0 | 0 | 0.002326949 | 0 |
| 0 | 0 | 0.012094584 | 0 | 0 |
| 0 | 0 | 0 | 0 | 0 |
| 0 | 0 | 0 | 0 | 0 |
| 0 | 0 | 0 | 0 | 0 |
| 0.1500956 | 0.104798153 | 0 | 0 | 0 |
| 0.140963364 | 0.413157147 | 0.151469343 | 0.111659244 | 0.625387753 |
| 0.003010751 | 0 | 0 | 0 | 0 |
| 0 | 0 | 0 | 0 | 0 |
| 0.086038593 | 0 | 0.008899167 | 0 | 0.01309533 |
| 0 | 0 | 0 | 0 | 0 |
| 0 | 0 | 0 | 0.001194651 | 0 |
| 0 | 0 | 0 | 0 | 0 |
| 0 | 0.002062435 | 0 | 0.095403297 | 0.02701938 |
| 0 | 0 | 0 | 0.000697804 | 0 |
| 0.014410655 | 0 | 0 | 0 | 0.612890118 |
| 0 | 0.093214382 | 0 | 0 | 0 |
| 0 | 0 | 0 | 0 | 0 |
| 0 | 0 | 0 | 0 | 0.00295564 |

| | | | | |
|---|---|---|---|---|
| 0 | 0 | 0 | 0 | 0 |
| 0 | 0 | 0 | 0 | 0 |
| 0 | 0.000436175 | 0 | 0 | 0 |
| 0.194070955 | 0.135440565 | 0.329736534 | 0 | 0 |
| 0 | 0 | 0 | 0 | 0 |
| 0.00091794 | 0 | 0 | 0 | 0 |
| 0 | 0 | 0 | 0 | 0 |
| 0 | 0.001322413 | 0.001232036 | 0 | 0 |
| 0 | 0.358570001 | 0.049290376 | 0 | 0.014133205 |
| 0.005834285 | 0.051855218 | 0 | 0 | 0 |
| 0 | 0 | 0 | 0 | 0 |
| 0.002197483 | 0.003363664 | 0.001130989 | 0 | 0 |
| 0 | 0 | 0 | 0 | 0 |
| 0.002329487 | 0 | 0 | 0 | 0 |
| 0.137676496 | 0.011833265 | 0.012443938 | 0 | 0.081965949 |
| 0 | 0 | 0 | 0 | 0 |
| 0 | 0.032967088 | 0.129044613 | 0 | 0 |
| 0 | 0.636412535 | 0 | 0 | 0 |
| 0 | 0 | 0 | 0 | 0 |
| 0 | 0 | 0 | 0 | 0 |
| 0 | 0 | 0 | 0 | 0 |
| 0 | 0 | 0 | 0 | 0 |
| 0 | 0 | 0 | 0 | 0 |
| 0 | 0 | 0.378030604 | 0.079878904 | 0.731306436 |
| 0 | 0 | 0 | 0 | 0 |
| 0.049584068 | 0.105397584 | 0 | 0 | 0 |
| 0 | 0 | 0 | 0 | 0 |
| 0.762851222 | 2.823116189 | 0.943159505 | 0 | 0 |
| 0 | 0 | 0 | 0 | 0 |
| 0 | 0 | 0 | 0 | 0 |
| 0.002464994 | 0.022029845 | 0 | 0 | 0 |
| 0.016412201 | 0 | 0 | 0 | 0 |
| 0.000875486 | 0.003536648 | 0 | 0 | 0 |
| 0 | 0 | 0 | 0 | 0 |
| 0 | 0 | 0 | 0 | 0 |
| 0 | 0 | 0 | 0 | 0 |
| 0.511966589 | 0.194498372 | 0.374652698 | 0 | 0 |
| 0 | 0 | 0 | 0 | 0 |
| 0 | 0 | 0 | 0 | 0.052067043 |
| 0.01923777 | 0 | 0 | 0.007389195 | 0 |
| 0.026756549 | 0.003057001 | 0.002893286 | 0 | 0.042980848 |
| 0 | 0 | 0 | 0 | 0 |
| 0.37077782 | 8.907134172 | 3.351324033 | 0.128456618 | 15.86548609 |
| 0.001130107 | 0 | 0.005355914 | 0 | 0 |
| 0 | 0 | 0 | 0 | 0 |
| 0 | 0 | 0 | 0 | 0 |
| 0 | 0 | 0 | 0 | 0 |
| 0.008382248 | 0 | 0.00195641 | 0 | 0.00211221 |

| | | | | |
|---|---|---|---|---|
| 0 | 0 | 0 | 0 | 0 |
| 0 | 0.065098191 | 0.009505838 | 0 | 0 |
| 0.056994792 | 0 | 0.005209554 | 0.765161993 | 0.066973885 |
| 0 | 0 | 0 | 0 | 0 |
| 0 | 0 | 0 | 0 | 0 |
| 0.001472075 | 0.002620933 | 0.001969865 | 0 | 0 |
| 0 | 0 | 0 | 0 | 0 |
| 0.002983618 | 0 | 0 | 0 | 0 |
| 0 | 0 | 0 | 0 | 0 |
| 0 | 0 | 0 | 0 | 0 |
| 0.041281067 | 0.001345065 | 0.002455132 | 0.046745433 | 0.031654863 |
| 1.268737095 | 3.163170545 | 1.359535655 | 2.001362147 | 2.143924707 |
| 0 | 0 | 0 | 0 | 0 |
| 0 | 0.016590277 | 0.009715488 | 0 | 0 |
| 0.008894242 | 0.049680344 | 0.03549682 | 1.061952169 | 0.254522753 |
| 0 | 7.729331341 | 0 | 0 | 0 |
| 0 | 0 | 0 | 0 | 0 |
| 0 | 0 | 0 | 0 | 0 |
| 0 | 0 | 0 | 0 | 0 |
| 0.215514447 | 0 | 0.000842659 | 0 | 0.00151161 |
| 0.005382162 | 0.004348399 | 0.007716608 | 0.012326153 | 0.400278632 |
| 0 | 0 | 0.208693375 | 51.7312779 | 9.746751368 |
| 0.447936235 | 5.09653968 | 2.816609274 | 4.308501082 | 0.618136428 |
| 0 | 0 | 0 | 0 | 0 |
| 0 | 0 | 0 | 0 | 0 |
| 0.010503381 | 0 | 0.002465816 | 0 | 0.001382288 |
| 0 | 0 | 0.91391792 | 1.128690478 | 0 |
| 0 | 0 | 0 | 0 | 0 |
| 0 | 0.00342906 | 0 | 0 | 0 |
| 0 | 0 | 0 | 0 | 0 |
| 0 | 0.001421088 | 0 | 0 | 0 |
| 0.100607235 | 0.117378945 | 0.069774775 | 0 | 0 |
| 0 | 0.010656646 | 0 | 0 | 0.467552503 |
| 0.012070161 | 0.002708839 | 0 | 0 | 0 |
| 0.004352813 | 0 | 0.001386843 | 0 | 0.004457303 |
| 0.02323851 | 0.007874064 | 0.09717728 | 0 | 0.0014242 |
| 0.022283023 | 0 | 0.001472501 | 0 | 0.002338789 |
| 0 | 0 | 0 | 0 | 0 |
| 0 | 0 | 0.014716323 | 0 | 0.004833146 |
| 0 | 0 | 0 | 0 | 0 |
| 0.000752304 | 0.000630319 | 0.005872416 | 0 | 0 |
| 0 | 0 | 0 | 0 | 0 |
| 0 | 0 | 0 | 0 | 0 |
| 0 | 0 | 0 | 0 | 0 |
| 0.000779438 | 0 | 0 | 0 | 0.161276978 |
| 0 | 0.004574373 | 0 | 0 | 0 |
| 0 | 0 | 0 | 0 | 0 |
| 0.130219761 | 0.004677326 | 0.004193844 | 0 | 0.002546902 |

| | | | | |
|---|---|---|---|---|
| 0 | 0 | 0 | 0 | 0 |
| 0 | 0 | 0 | 0 | 0 |
| 0 | 0 | 0 | 0.00544677 | 0 |
| 0.005480571 | 0.002087229 | 0.001481587 | 0 | 0 |
| 0 | 0 | 0 | 0 | 0 |
| 0.052364385 | 0.082646981 | 0 | 0 | 0 |
| 0 | 0.799410395 | 0 | 0 | 0 |
| 0.082523735 | 0.001920628 | 0.032890284 | 0.009916389 | 0 |
| 0 | 0 | 0 | 0 | 0 |
| 0 | 0 | 0 | 0 | 0 |
| 0.000924834 | 0 | 0.001443833 | 0.012884756 | 0.001079179 |
| 0 | 0 | 0 | 0 | 0 |
| 0 | 0.005597313 | 0 | 0.001081798 | 0 |
| 0 | 0.613944291 | 0 | 0 | 0 |
| 0 | 0 | 0 | 0 | 0 |
| 0 | 0 | 0 | 0 | 0 |
| 0.307323792 | 0.012966789 | 0 | 0 | 0 |
| 0.001693787 | 0.002096459 | 0.002232208 | 0 | 0 |
| 0 | 0 | 0 | 0 | 0 |
| 0.020829756 | 0 | 0 | 0 | 0 |
| 0.008618379 | 0.003581805 | 0 | 0 | 0 |
| 0 | 0 | 0 | 0 | 0 |
| 0.003550573 | 0.009296413 | 0 | 0 | 0 |
| 0 | 0 | 0 | 0 | 0 |
| 0 | 0 | 0 | 0 | 0 |
| 0.003283084 | 0 | 0 | 0 | 0 |
| 0 | 0 | 0 | 0 | 0 |
| 0 | 0 | 0 | 0 | 0 |
| 0 | 0 | 0 | 0 | 0 |
| 0.039061521 | 0.124871926 | 1.447184084 | 0 | 0 |
| 0 | 0.003966944 | 0 | 0 | 0 |
| 0 | 0 | 0 | 0 | 0 |
| 0 | 0.058804966 | 0 | 0 | 0 |
| 0 | 0 | 0 | 0 | 0 |
| 0 | 0.000770177 | 0 | 0 | 0 |
| 0 | 0 | 0 | 0 | 0 |
| 0.085567661 | 0.057986961 | 0.050220526 | 0 | 0 |
| 0.00532364 | 0 | 0.006285755 | 0.004781226 | 0.005089745 |
| 0 | 0 | 0 | 0 | 0 |
| 0 | 0 | 0.004994381 | 0 | 0.000772345 |
| 0.006757794 | 0.001077016 | 0 | 0 | 0 |
| 0 | 0 | 0 | 0 | 0 |
| 0 | 0 | 0 | 0 | 0 |
| 0 | 0.295089867 | 0.004747721 | 0 | 0 |
| 0.109697841 | 0 | 0 | 0 | 0 |
| 0 | 0 | 0 | 0 | 0 |
| 0 | 0.002337212 | 0 | 0 | 0 |
| 0 | 0 | 0 | 0 | 0 |

| | | | | |
|---|---|---|---|---|
| 0 | 0 | 0 | 0 | 0 |
| 0 | 0 | 0 | 0 | 0 |
| 0 | 0 | 0 | 0 | 0 |
| 0.042363689 | 0.062311062 | 0.008162965 | 0 | 0 |
| 0 | 0 | 0 | 0 | 0 |
| 1.423506985 | 0.979332498 | 0 | 0 | 0 |
| 0 | 0 | 0 | 0 | 0 |
| 0 | 0 | 0 | 0 | 0 |
| 0.002375414 | 0 | 0 | 0.121372817 | 0.05435697 |
| 0 | 0 | 0 | 0 | 0 |
| 0 | 0.000808572 | 0 | 0 | 0 |
| 0 | 0 | 0 | 0 | 0 |
| 0.013731739 | 0 | 0 | 0 | 0 |
| 0 | 0 | 0 | 0 | 0 |
| 0 | 2.028829987 | 1.032251222 | 0 | 0 |
| 0 | 0 | 0 | 0 | 0.006381271 |
| 0 | 0 | 0 | 0 | 0 |
| 0 | 0 | 0 | 0 | 0 |
| 0 | 0 | 0 | 0 | 0 |
| 0 | 0 | 0 | 0 | 0 |
| 0 | 0.105663181 | 0 | 0 | 0 |
| 0 | 0 | 0 | 0 | 0 |
| 0 | 0 | 0 | 0 | 0 |
| 0 | 0 | 0 | 0 | 0 |
| 0.054598645 | 0.045213617 | 0.054654498 | 0 | 0.018414688 |
| 6.818050135 | 3.877027635 | 2.196958056 | 0 | 0 |
| 0 | 0.078939979 | 0 | 0 | 0 |
| 0 | 0 | 0 | 0 | 0 |
| 0 | 0.281357957 | 0 | 0 | 0 |
| 0 | 0.009921499 | 0 | 0 | 0 |
| 0 | 0 | 0 | 0 | 0 |
| 0 | 0 | 0 | 0 | 0 |
| 0.001967587 | 0 | 0 | 0 | 0 |
| 0.00298373 | 0 | 0 | 0 | 0 |
| 0.001787725 | 0.006319046 | 0.01520078 | 0.001279445 | 0 |
| 0 | 0 | 0 | 0.000904413 | 0.003686511 |
| 0.142799908 | 0 | 0.499898362 | 0 | 0 |
| 3.037780158 | 0.488873885 | 8.147465732 | 0 | 0 |
| 0 | 0.958822856 | 0 | 0 | 0 |
| 0 | 0 | 0 | 0 | 0 |
| 0 | 0 | 0 | 0 | 0 |
| 0 | 0 | 0 | 0 | 0 |
| 0.380991928 | 0.592988511 | 0.202197391 | 0 | 0 |
| 0 | 0 | 0.010128866 | 0 | 0 |
| 0 | 0 | 0 | 0 | 0 |
| 0 | 0 | 0 | 0.009996202 | 0 |
| 0 | 0 | 0 | 0 | 0 |
| 0.181669019 | 0.165545108 | 0.028181036 | 0 | 0 |

| | | | | |
|---|---|---|---|---|
| 0.004266796 | 0 | 0 | 0.001357189 | 0.042083391 |
| 0 | 0 | 0 | 0 | 0 |
| 0 | 0 | 0 | 0 | 0 |
| 0 | 0 | 0 | 0 | 0 |
| 0 | 0 | 0 | 0.001520143 | 0 |
| 0.005113559 | 0.123533591 | 0 | 0 | 0 |
| 0 | 0 | 0 | 0 | 0 |
| 0 | 0 | 0 | 0 | 0 |
| 0 | 0 | 0 | 0 | 0 |
| 0 | 0 | 0 | 0 | 0 |
| 0 | 0 | 0 | 0 | 0 |
| 1.883307304 | 0.018470539 | 11.6032557 | 0 | 0.071320091 |
| 0.274330259 | 0.049605855 | 0.618519238 | 0 | 0.000714538 |
| 0 | 0 | 0 | 0 | 0 |
| 6.729466821 | 0.069891221 | 0.521602311 | 0.072470549 | 0.015867655 |
| 0 | 0 | 0 | 0 | 0 |
| 0.000854411 | 0 | 0 | 0 | 0 |
| 0 | 0 | 0 | 0 | 0 |
| 0.004844423 | 0 | 0 | 0.000684854 | 0.003439599 |
| 0 | 0 | 0 | 0 | 0 |
| 0.082442941 | 0.009178874 | 0.297503461 | 0 | 0 |
| 0 | 0 | 0 | 0 | 0 |
| 1.114479512 | 0.292533386 | 0.045330269 | 0 | 0 |
| 0 | 0 | 0 | 0 | 0 |
| 0 | 0 | 0 | 0 | 0 |
| 0 | 0 | 0 | 0 | 0 |
| 0 | 0 | 0 | 0 | 0 |
| 0 | 0 | 0 | 0 | 0 |
| 0.002581422 | 0 | 0 | 0 | 0 |
| 0 | 0 | 0 | 0 | 0 |
| 0 | 0 | 0 | 0 | 0 |
| 0.005312663 | 0.009180644 | 0.041766356 | 0.002027831 | 0 |
| 0 | 0 | 0 | 0 | 0 |
| 0.022427271 | 0.010840798 | 0.014946389 | 0 | 0 |
| 0 | 0 | 0 | 0.000984227 | 0.002435765 |
| 0.056509131 | 0.014389545 | 0 | 0 | 0.009419987 |
| 0 | 0 | 0 | 0 | 0 |
| 0 | 0 | 0 | 0 | 0 |
| 0.002369901 | 0.001241015 | 0.001541602 | 0 | 0 |
| 0 | 0 | 0 | 0 | 0 |
| 0.002929248 | 0.130536697 | 0 | 0 | 0 |
| 0 | 0 | 0 | 0 | 0 |
| 0 | 0 | 0 | 0 | 0 |
| 0 | 0 | 0 | 0 | 0 |
| 0.022522507 | 0 | 0.009253081 | 0.003231876 | 0.222445186 |
| 0 | 0 | 0 | 0 | 0 |
| 0 | 0.312769326 | 0 | 0.894353804 | 0 |
| 0.000961004 | 0 | 0 | 0 | 0 |

| | | | | |
|---|---|---|---|---|
| 0 | 0 | 0 | 0 | 0 |
| 0 | 0 | 0 | 0 | 0 |
| 0 | 0 | 0 | 0 | 0.007298985 |
| 0 | 0 | 0 | 0 | 0 |
| 0.006039062 | 0 | 0 | 0 | 0.026055833 |
| 0 | 0 | 0 | 0 | 0 |
| 0 | 0.03977719 | 0 | 0 | 0 |
| 0 | 0.006929256 | 0 | 0 | 0 |
| 0 | 0 | 0 | 0 | 0 |
| 0 | 0 | 0 | 0 | 0 |
| 0 | 0 | 0 | 0 | 0 |
| 0 | 0 | 0 | 0 | 0 |
| 0 | 0 | 0 | 0 | 0 |
| 0.133204279 | 0 | 0 | 0 | 0 |
| 0 | 0 | 0 | 0 | 0 |
| 0 | 0 | 0 | 0 | 0 |
| 0.052164587 | 0 | 0 | 0 | 0 |
| 0 | 0 | 0 | 0 | 0 |
| 0 | 2.56563667 | 0.817860143 | 0 | 0.152864402 |
| 0 | 0 | 0 | 0 | 0 |
| 0 | 0 | 0 | 0 | 0 |
| 0 | 0 | 0 | 0 | 0 |
| 0 | 0 | 0 | 0 | 0 |
| 0 | 0 | 0 | 0 | 0 |
| 0 | 0 | 0 | 0 | 0 |
| 0.003941279 | 0 | 0 | 0 | 0.029565237 |
| 0 | 0 | 0 | 0 | 0 |
| 0.068821867 | 0.001454491 | 0 | 0 | 0.028938486 |
| 0 | 0 | 0 | 0 | 0 |
| 0.862408878 | 0.111754738 | 0.659495363 | 0 | 0 |
| 0 | 0 | 0 | 0 | 0 |
| 0 | 0 | 0 | 0 | 0 |
| 0 | 0.00324228 | 0 | 0 | 0 |
| 0 | 0 | 0 | 0 | 0 |
| 0 | 0 | 0 | 0.003625936 | 0.010465213 |
| 0 | 0 | 0 | 0 | 0 |
| 0 | 0 | 0 | 0 | 0 |
| 0.001499707 | 0 | 0 | 0 | 0 |
| 0 | 0 | 0 | 0 | 0 |
| 0 | 0 | 0 | 0 | 0 |
| 4.432849896 | 2.441017082 | 4.50505358 | 1.514841179 | 0.686477473 |
| 1.945014144 | 0.227359831 | 0.932978697 | 0 | 0.00457501 |
| 0 | 0.276303063 | 0 | 0 | 0 |
| 0.007676165 | 0.007724087 | 0 | 0 | 0 |
| 0 | 0 | 0 | 0 | 0 |
| 0 | 0 | 0 | 0 | 0 |
| 0 | 0 | 0 | 0 | 0 |
| 0.002380502 | 0 | 0 | 0 | 0.00731648 |

| | | | | |
|---|---|---|---|---|
| 0 | 0 | 0 | 0 | 0 |
| 0.054174002 | 0.003846589 | 0 | 0 | 0 |
| 0 | 0.004305408 | 0 | 0 | 0 |
| 0 | 0 | 0 | 0 | 0 |
| 2.65282615 | 2.231687328 | 0.561862915 | 0 | 0 |
| 0 | 0 | 0 | 0 | 0 |
| 0 | 0 | 0 | 0 | 0 |
| 0 | 0 | 0 | 0 | 0 |
| 0.068763726 | 0 | 0.03646581 | 0.594056088 | 10.03543557 |
| 0 | 0 | 0 | 0 | 0 |
| 0 | 0 | 0 | 0.028377494 | 0.082573822 |
| 0 | 0 | 0 | 0 | 0 |
| 0.019479506 | 0.105743418 | 0.009119994 | 0 | 0 |
| 0.010054534 | 0.005935234 | 0.016308516 | 0.00750114 | 0 |
| 0 | 0 | 0 | 0 | 0 |
| 0.195616069 | 0.152553821 | 0.440958317 | 0 | 0 |
| 0.00047458 | 0 | 0 | 0 | 0.020041664 |
| 0 | 0 | 0 | 0 | 0 |
| 0 | 0 | 0 | 0 | 0 |
| 0 | 0 | 0 | 0 | 0 |
| 0 | 0 | 0 | 0 | 0 |
| 0 | 0 | 0 | 0 | 0 |
| 0 | 0 | 0 | 0 | 0 |
| 0 | 0 | 0 | 0 | 0 |
| 0 | 0.012013038 | 0 | 0 | 0 |
| 0 | 0 | 0 | 0 | 0 |
| 0.173732853 | 0.009652289 | 0.00322124 | 0 | 0 |
| 0 | 0 | 0.001557775 | 0.00281417 | 0 |
| 0 | 0 | 0 | 0 | 0 |
| 0 | 0 | 0 | 0 | 0 |
| 0 | 0 | 0 | 0 | 0 |
| 0 | 0 | 0 | 0 | 0 |
| 0 | 0 | 0 | 0 | 0 |
| 0 | 0.047713794 | 0 | 0 | 0 |
| 0 | 0.34484413 | 0 | 0 | 0 |
| 0.003411506 | 0.004325112 | 0 | 0 | 0 |
| 0 | 0.001345456 | 0.099461749 | 0 | 0 |
| 0.021871807 | 0 | 0 | 0 | 0 |
| 0 | 0 | 0 | 0 | 0 |
| 0 | 0 | 0 | 0 | 0 |
| 6.136481755 | 0.846852891 | 2.430658304 | 0 | 1.715045917 |
| 0.251321902 | 0 | 0 | 0 | 0 |
| 0 | 0.001599069 | 0 | 0 | 0 |
| 0 | 0.041582536 | 0 | 0 | 0 |
| 0 | 0 | 0 | 0 | 0 |
| 0 | 0 | 0 | 0 | 0 |
| 0 | 0.051630542 | 0 | 0 | 0 |
| 0 | 0 | 0 | 0 | 0 |

| | | | | |
|---|---|---|---|---|
| 0.061520897 | 0.093265775 | 0 | 0 | 0 |
| 0 | 0 | 0 | 0 | 0 |
| 0 | 0 | 0 | 0 | 0 |
| 0 | 0 | 0 | 0 | 0 |
| 0.04323626 | 0.204385417 | 0 | 0 | 0 |
| 0 | 0 | 0 | 0 | 0 |
| 0 | 0.001322203 | 0 | 0 | 0.022228983 |
| 0 | 0 | 0 | 0 | 0 |
| 0 | 0 | 0 | 0 | 0 |
| 0.00115897 | 0 | 0 | 0 | 0 |
| 0.000956845 | 0.025052929 | 0 | 0 | 0 |
| 0.039745948 | 0 | 0.000615582 | 0 | 0 |
| 0 | 0 | 0 | 0 | 0 |
| 0 | 0 | 0 | 0 | 0 |
| 0 | 0 | 0 | 0 | 0 |
| 0 | 0 | 0 | 0 | 0 |
| 0 | 0 | 0 | 0 | 0 |
| 0 | 0 | 0 | 0 | 0 |
| 0 | 0 | 0 | 0 | 0 |
| 0 | 0 | 0 | 0 | 0 |
| 0.002554127 | 0.03147027 | 0.042220103 | 0 | 0.000375679 |
| 0 | 0 | 0 | 0 | 0 |
| 0.017731745 | 0.007115878 | 0 | 0 | 0 |
| 0.007662589 | 0.001234636 | 0.002136194 | 0.001265535 | 0 |
| 0 | 0 | 0 | 0 | 0 |
| 0.000869244 | 0 | 0 | 0 | 0.001159213 |
| 0 | 0 | 0 | 0 | 0 |
| 0 | 0 | 0 | 0 | 0 |
| 0 | 0 | 0.020690876 | 0 | 0 |
| 0.222898144 | 0.002665397 | 0.949306165 | 0.014773593 | 0 |
| 0 | 0.036456147 | 0 | 0 | 0 |
| 0 | 0 | 0 | 0 | 0 |
| 0 | 0 | 0 | 0 | 0 |
| 0.391533853 | 0.011701842 | 0 | 0 | 0 |
| 0 | 0.120426014 | 0 | 0 | 0 |
| 0 | 0 | 0 | 0 | 0 |
| 0 | 0.010427737 | 0 | 0 | 0.056862091 |
| 0 | 0 | 0 | 0 | 0 |
| 0 | 0 | 0 | 0 | 0 |
| 0 | 0 | 0 | 0 | 0 |
| 0 | 0 | 0 | 0 | 0 |
| 0 | 0 | 0 | 0 | 0 |
| 0 | 0 | 0 | 0 | 0 |
| 0 | 0.004422481 | 0 | 0 | 0 |
| 0 | 0 | 0.003502701 | 0 | 0 |
| 0 | 0 | 0 | 0 | 0 |
| 0 | 0 | 0 | 0 | 0 |
| 0 | 0 | 0 | 0 | 0 |

| | | | | |
|---|---|---|---|---|
| 0 | 0 | 0 | 0.007111418 | 0.000660801 |
| 0 | 0.037326188 | 0 | 0 | 0 |
| 0 | 0.243925402 | 0 | 0 | 0 |
| 0 | 0 | 0 | 0 | 0 |
| 0 | 0 | 0 | 0 | 0 |
| 0 | 0.004400369 | 0 | 0 | 0 |
| 0 | 0 | 0 | 0 | 0 |
| 1.527131681 | 0.306577487 | 0.545924109 | 0.041635902 | 0 |
| 2.309917347 | 1.019711783 | 1.421008991 | 0.087637014 | 0.593085799 |
| 0.002252477 | 0.001814654 | 0 | 0 | 0 |
| 0 | 0 | 0 | 0 | 0 |
| 0 | 0.000635371 | 0 | 0 | 0 |
| 0.010053649 | 0.026716205 | 0 | 0 | 0 |
| 0 | 0 | 0 | 0 | 0.011825299 |
| 0.002272175 | 0 | 0 | 0 | 0 |
| 0 | 0 | 0 | 0 | 0 |
| 0 | 0 | 0.027359153 | 0 | 0 |
| 0 | 0.672769883 | 0 | 0 | 0 |
| 0 | 0 | 0 | 0 | 0 |
| 0 | 0.004062838 | 0 | 0 | 0 |
| 0 | 0 | 0 | 0 | 0 |
| 0 | 0 | 0 | 0 | 0 |
| 0 | 0 | 0 | 0 | 0 |
| 0 | 0 | 0 | 0 | 0 |
| 0.016485409 | 0.007586312 | 0 | 0 | 0 |
| 0 | 0 | 0 | 0 | 0 |
| 0 | 0 | 0 | 0 | 0 |
| 0 | 0 | 0 | 0 | 0 |
| 0 | 0 | 0 | 0 | 0 |
| 0 | 0 | 0 | 0 | 0 |
| 0.040235078 | 0 | 0 | 0 | 0.118257244 |
| 0 | 0 | 0 | 0 | 0 |
| 0 | 0 | 0 | 0 | 0 |
| 0 | 0 | 0 | 0 | 0 |
| 0.008446305 | 0.005026099 | 0 | 0 | 0 |
| 0 | 0 | 0 | 0 | 0 |
| 0 | 0 | 0 | 0 | 0 |
| 0 | 0 | 0.151006475 | 0 | 0 |
| 0.009238581 | 0.001382242 | 0.002312741 | 0 | 0 |
| 0.330835326 | 0 | 0 | 0 | 0 |
| 0 | 0 | 0 | 0 | 0 |
| 0.007022278 | 0.034549187 | 0.181564063 | 0 | 0.060708024 |
| 0 | 0 | 0 | 0 | 0 |
| 0 | 0 | 0 | 0 | 0 |
| 0.002313814 | 0 | 0 | 0 | 0 |
| 0 | 0 | 0 | 0 | 0 |
| 0 | 0 | 0 | 0 | 0 |
| 0 | 0 | 0 | 0 | 0 |

| | | | | |
|---|---|---|---|---|
| 0 | 0 | 0 | 0 | 0 |
| 0.007012343 | 0 | 0.00063834 | 0 | 0 |
| 0 | 0 | 0 | 0 | 0 |
| 0 | 0 | 0 | 0 | 0 |
| 1.313368101 | 0.013201867 | 0.052879999 | 0 | 0.00533666 |
| 0 | 0.079476478 | 0 | 0 | 0 |
| 0 | 0 | 0 | 0.006789082 | 0 |
| 0.002420426 | 0 | 0.000899696 | 0.009431185 | 0.012776912 |
| 0.005034804 | 0.001265525 | 0 | 0 | 0 |
| 0 | 0 | 0 | 0 | 0 |
| 0 | 0 | 0 | 0 | 0 |
| 0 | 0 | 0 | 0 | 0 |
| 0 | 0 | 0 | 0 | 0 |
| 0 | 0 | 0 | 0 | 0 |
| 0.000851947 | 0 | 0 | 0 | 0 |
| 0.002624022 | 0 | 0.013265094 | 0.001836236 | 0 |
| 0 | 0 | 0 | 0 | 0 |
| 0 | 0 | 0 | 0 | 0 |
| 0 | 0 | 0 | 0 | 0 |
| 0 | 0 | 0 | 0 | 0 |
| 0 | 0 | 0 | 0 | 0 |
| 0 | 0 | 0 | 0 | 0 |
| 0 | 0 | 0 | 0 | 0 |
| 0.000698227 | 0 | 0 | 0.009327901 | 0 |
| 0.096445324 | 0 | 0.19829888 | 0 | 0.117537684 |
| 0 | 0 | 0 | 0 | 0 |
| 0.013570408 | 0 | 0.009613752 | 0.045704059 | 0.004923539 |
| 0 | 0 | 0 | 0 | 0 |
| 0 | 0 | 0 | 0 | 0 |
| 0 | 0 | 0 | 0 | 0 |
| 0 | 0 | 0 | 0 | 0 |
| 0 | 0 | 0 | 0 | 0 |
| 0 | 0 | 0 | 0 | 0 |
| 0 | 0 | 0 | 0 | 0 |
| 0 | 0 | 0 | 0 | 0 |
| 0 | 0 | 0 | 0 | 0 |
| 0 | 0 | 0 | 0 | 0 |
| 0.014899643 | 0.001371833 | 0 | 0 | 0 |
| 0 | 0 | 0.059284909 | 0 | 0 |
| 0 | 0 | 0 | 0 | 0 |
| 0 | 0 | 0 | 0 | 0 |
| 0 | 0 | 0 | 0 | 0 |
| 0 | 0 | 0 | 0 | 0 |
| 0 | 0 | 0 | 0 | 0 |
| 0 | 0 | 0 | 0 | 0 |
| 0 | 0 | 0 | 0 | 0 |
| 0 | 0 | 0 | 0 | 0 |
| 0 | 0 | 0 | 0 | 0 |

| | | | | |
|---|---|---|---|---|
| 0.114105498 | 0.00251588 | 0.078019667 | 0 | 0 |
| 0 | 0 | 0 | 0 | 0 |
| 0 | 0 | 0 | 0 | 0 |
| 0 | 0 | 0 | 0 | 0 |
| 0 | 0 | 0 | 0 | 0 |
| 0 | 0.001821745 | 0.016681471 | 0.003983653 | 0.104966563 |
| 0 | 0 | 0 | 0 | 0 |
| 0 | 0 | 0 | 0 | 0 |
| 0 | 0 | 0 | 0 | 0 |
| 0 | 0 | 0 | 0 | 0 |
| 0 | 0.046911202 | 0 | 0 | 0 |
| 0.001287307 | 0 | 0 | 0 | 0 |
| 0 | 0 | 0 | 0 | 0 |
| 0 | 0 | 0 | 0 | 0 |
| 0 | 0.019750905 | 0 | 0 | 0 |
| 0 | 0 | 0 | 0 | 0 |
| 0 | 0 | 0 | 0 | 0 |
| 0 | 0 | 0 | 0 | 0 |
| 0.005346218 | 0 | 0 | 0.000850266 | 0 |
| 0 | 0 | 0 | 0 | 0 |
| 0.010429616 | 0 | 0 | 0.014099235 | 0.004346504 |
| 0 | 0 | 0 | 0 | 0 |
| 0.021074954 | 0.000919671 | 0.002937662 | 0.009426868 | 0.021225361 |
| 0 | 0.002735624 | 0 | 0 | 0 |
| 0 | 0 | 0 | 0 | 0 |
| 0 | 0 | 0 | 0 | 0 |
| 0 | 0 | 0 | 0 | 0 |
| 0 | 0 | 0 | 0 | 0 |
| 0 | 0 | 0 | 0 | 0 |
| 0 | 0 | 0 | 0 | 0 |
| 0 | 0 | 0 | 0 | 0 |
| 0 | 0 | 0 | 0 | 0 |
| 0.0059621 | 0 | 0.022696645 | 0.001177096 | 0.003255791 |
| 0 | 0 | 0 | 0 | 0 |
| 0.308950275 | 0.052194594 | 0.517394434 | 0 | 0.025750774 |
| 0 | 0 | 0 | 0.002518855 | 0.000687536 |
| 0 | 0 | 0 | 0 | 0 |
| 0 | 0 | 0 | 0 | 0 |
| 0.001144933 | 0 | 0 | 0 | 0.023698269 |
| 0 | 0 | 0 | 0 | 0 |
| 0 | 0 | 0 | 0 | 0 |
| 0 | 0.018404059 | 0 | 0 | 0 |
| 0 | 0.031547516 | 0.003981069 | 0 | 0.00130183 |
| 0 | 0 | 0 | 0 | 0 |
| 0 | 0 | 0 | 0 | 0 |
| 0.003012442 | 0.004639667 | 0.012251428 | 0 | 0 |
| 0 | 0 | 0 | 0 | 0 |
| 0 | 0 | 0 | 0 | 0 |

| | | | | |
|---|---|---|---|---|
| 0 | 0 | 0 | 0 | 0 |
| 0 | 0 | 0 | 0 | 0 |
| 0 | 0 | 0 | 0 | 0 |
| 0 | 0 | 0 | 0 | 0 |
| 0 | 0 | 0 | 0 | 0 |
| 0 | 0 | 0 | 0 | 0 |
| 0 | 0 | 0 | 0 | 0 |
| 0 | 0 | 0 | 0 | 0 |
| 0.004482602 | 0.131625263 | 0 | 0 | 0 |
| 0.075744395 | 0.003058435 | 0 | 0 | 0 |
| 0 | 0 | 0 | 0 | 0 |
| 0 | 0 | 0 | 0 | 0 |
| 0 | 0 | 0 | 0 | 0 |
| 0 | 1.520614778 | 0 | 0 | 0 |
| 0 | 0 | 0 | 0 | 0 |
| 0 | 0 | 0 | 0.134345867 | 0.011525005 |
| 0 | 0 | 0 | 0 | 0 |
| 0 | 0 | 0 | 0 | 0 |
| 0 | 0 | 0 | 0 | 0 |
| 0 | 0.129432421 | 0.869967847 | 0 | 0 |
| 0.006001746 | 0 | 0 | 0 | 0.003439157 |
| 0 | 0 | 0 | 0 | 0 |
| 0 | 0 | 0 | 0 | 0 |
| 0 | 0.004008266 | 0 | 0 | 0 |
| 0 | 0 | 0 | 0 | 0 |
| 0 | 0 | 0 | 0 | 0 |
| 0 | 0 | 0 | 0 | 0 |
| 0 | 0 | 0 | 0 | 0 |
| 0 | 0 | 0 | 0 | 0 |
| 0 | 0.032552118 | 0.03569764 | 0 | 0 |
| 2.033583632 | 0.074901481 | 1.108382626 | 0 | 0 |
| 0 | 0 | 0 | 0 | 0 |
| 0 | 0 | 0 | 0 | 0 |
| 0 | 0 | 0 | 0 | 0 |
| 0 | 0 | 0 | 0 | 0 |
| 0 | 0 | 0 | 0 | 0 |
| 0 | 0 | 0 | 0 | 0 |
| 0 | 0 | 0 | 0 | 0 |
| 0 | 0 | 0 | 0 | 0.57817819 |
| 0 | 0 | 0 | 0 | 0 |
| 0 | 0.000958518 | 0 | 0 | 0 |
| 0 | 0 | 0 | 0 | 0 |
| 0 | 0.004463956 | 0 | 0 | 0 |
| 0 | 0 | 0 | 0 | 0.002078371 |
| 0 | 0 | 0 | 0 | 0 |
| 0 | 0 | 0 | 0 | 0 |
| 0 | 0 | 0 | 2.250456709 | 0.04949058 |
| 0 | 0 | 0 | 0 | 0 |

| | | | | |
|---|---|---|---|---|
| 0 | 0 | 0 | 0 | 0 |
| 0 | 0 | 0 | 0 | 0 |
| 0 | 0 | 0 | 0 | 0 |
| 0.000659016 | 0 | 0 | 0 | 0 |
| 0.201196288 | 0.008917661 | 0.024197173 | 0 | 0 |
| 0 | 0 | 0 | 0 | 0 |
| 0 | 0 | 0 | 0 | 0 |
| 0 | 0 | 0 | 0 | 0 |
| 0 | 0 | 0 | 0 | 0 |
| 0 | 0 | 0 | 0 | 0 |
| 0.088055925 | 0.008583057 | 0 | 0 | 0 |
| 0.004957424 | 0 | 0.001913597 | 0.003275026 | 0.046564188 |
| 0.010199019 | 0 | 0 | 0 | 0.002742195 |
| 0 | 0 | 0 | 0 | 0 |
| 0 | 0 | 0 | 0 | 0.003633293 |
| 2.765885866 | 0.15109568 | 0 | 0 | 0 |
| 0.048100536 | 0 | 0.002943566 | 0 | 0.00195568 |
| 0 | 0 | 0 | 0 | 0 |
| 0.011405746 | 0.001058453 | 0.008049924 | 0 | 0.073646095 |
| 0 | 0.00394862 | 0 | 0.003058066 | 0.068505688 |
| 0 | 0 | 0 | 0 | 0 |
| 0 | 0 | 0 | 0 | 0 |
| 0 | 0 | 0 | 0 | 0 |
| 0 | 0 | 0 | 0 | 0 |
| 0 | 0 | 0 | 0 | 0 |
| 0 | 0 | 0 | 0 | 0 |
| 0.001510631 | 0 | 0.00067382 | 0 | 0.034751152 |
| 0 | 0 | 0 | 0 | 0 |
| 0 | 0 | 0 | 0 | 0 |
| 0 | 0 | 0 | 0 | 0 |
| 0 | 0 | 0 | 0 | 0.004567218 |
| 0 | 0 | 0 | 0 | 0 |
| 0 | 0 | 0 | 0 | 0 |
| 0 | 0 | 0 | 0 | 0 |
| 0 | 0 | 0 | 0 | 0 |
| 0 | 0 | 0 | 0 | 0 |
| 0 | 0 | 0 | 0 | 0 |
| 0.023580138 | 0.000755994 | 0.001207419 | 0.002525646 | 0.001704672 |
| 0 | 0.017579917 | 0 | 0 | 0 |
| 0 | 0 | 0 | 0 | 0 |
| 0 | 0 | 0.001339326 | 0 | 0.004905224 |
| 0 | 0 | 0 | 0 | 0 |
| 0 | 0 | 0 | 0 | 0 |
| 0 | 0 | 0 | 0 | 0.001784338 |
| 0 | 0 | 0 | 0 | 0 |
| 0 | 0 | 0 | 0 | 0 |
| 0 | 0 | 0 | 0 | 0 |
| 0 | 0 | 0 | 0 | 0 |

| | | | | |
|---|---|---|---|---|
| 0 | 0 | 0 | 0 | 0 |
| 0 | 0 | 0 | 0 | 0 |
| 0 | 0 | 0 | 0 | 0 |
| 0.007852478 | 0 | 0 | 0.003746591 | 0.000779164 |
| 0 | 0 | 0.00077348 | 0 | 0 |
| 0 | 0 | 0 | 0 | 0.002984529 |
| 0 | 0.010831055 | 0 | 0 | 0 |
| 0 | 0 | 0 | 0 | 0 |
| 0 | 0 | 0 | 0 | 0 |
| 0 | 0 | 0 | 0 | 0 |
| 0 | 0.002912412 | 0.048754533 | 0 | 0 |
| 0 | 0 | 0 | 0 | 0 |
| 0.013253099 | 0.026094694 | 0.010049664 | 0 | 0 |
| 0 | 0 | 0 | 0 | 0 |
| 0 | 0 | 0 | 0 | 0 |
| 0 | 0 | 0 | 0 | 0 |
| 0 | 0 | 0 | 0 | 0 |
| 0 | 0 | 0 | 0 | 0 |
| 0 | 0 | 0 | 0 | 0 |
| 0 | 0 | 0 | 0 | 0 |
| 0 | 0 | 0 | 0 | 0 |
| 0 | 0 | 0 | 0 | 0 |
| 0.022512148 | 0 | 0.00408669 | 0.026144865 | 0.007563665 |
| 0.004324553 | 0.036448991 | 0 | 0 | 0 |
| 0 | 0 | 0 | 0 | 0 |
| 0 | 0 | 0 | 0 | 0 |
| 0 | 0.004889377 | 0 | 0 | 0 |
| 0 | 0 | 0 | 0 | 0 |
| 0 | 0 | 0 | 0 | 0 |
| 0 | 0 | 0 | 0 | 0 |
| 0.028603402 | 0.004279535 | 0.019745446 | 0 | 0 |
| 0 | 0 | 0 | 0 | 0 |
| 0 | 0 | 0 | 0 | 0 |
| 0 | 0 | 0 | 0 | 0 |
| 0 | 0 | 0 | 0 | 0 |
| 0 | 0 | 0 | 0 | 0 |
| 0 | 0 | 0 | 0 | 0 |
| 0 | 0 | 0 | 0 | 0 |
| 0 | 0.002069019 | 0.00073433 | 0 | 0.00137217 |
| 0 | 0 | 0 | 0 | 0 |
| 0 | 0 | 0 | 0 | 0 |
| 0 | 0 | 0 | 0 | 0 |
| 0 | 0 | 0 | 0 | 0 |
| 0 | 0 | 0 | 0 | 0 |
| 0 | 0 | 0 | 0 | 0 |
| 0.594449287 | 0.334345877 | 0.138424664 | 0 | 0 |
| 0 | 0 | 0 | 0.336579728 | 0.076730097 |
| 0 | 0 | 0 | 0 | 0 |

| | | | | |
|---|---|---|---|---|
| 0 | 0 | 0 | 0 | 0 |
| 0 | 0 | 0 | 0 | 0 |
| 0.058537078 | 0.060061898 | 0.463396216 | 0 | 2.413753546 |
| 0 | 0.013513024 | 0 | 0 | 0 |
| 0 | 0 | 0 | 0 | 0 |
| 0 | 0 | 0 | 0 | 0 |
| 0 | 0 | 0 | 0 | 0 |
| 0 | 0 | 0 | 0 | 0 |
| 0 | 0 | 0 | 0 | 0 |
| 0 | 0 | 0 | 0 | 0 |
| 0.000534233 | 0 | 0 | 0 | 0 |
| 0.00139582 | 0 | 0 | 0.000570837 | 0 |
| 0 | 0 | 0 | 0 | 0 |
| 0 | 0 | 0 | 0 | 0 |
| 0 | 0 | 0 | 0 | 0 |
| 0 | 0 | 0 | 0 | 0 |
| 0.000981624 | 0 | 0 | 0 | 0 |
| 0 | 0 | 0 | 0 | 0 |
| 0 | 0 | 0 | 0 | 0 |
| 0 | 0 | 0 | 0 | 0 |
| 0 | 0 | 0 | 0 | 0 |
| 0 | 0 | 0 | 0 | 0 |
| 0 | 0 | 0 | 0 | 0 |
| 0 | 0 | 0 | 0 | 0 |
| 0 | 0 | 0 | 0 | 0 |
| 0 | 0 | 0 | 0 | 0 |
| 0 | 0 | 0 | 0 | 0 |
| 0 | 0 | 0 | 0 | 0 |
| 0 | 0 | 0 | 0 | 0 |
| 0 | 0 | 0 | 0 | 0 |
| 0.003804135 | 0.003665394 | 0.003223997 | 0.003339668 | 0 |
| 0 | 0 | 0 | 0 | 0 |
| 0.018374669 | 0.111763515 | 0.055362757 | 0.036753798 | 0.027096033 |
| 0 | 0.000567269 | 0 | 0 | 0 |
| 0 | 0.001468839 | 0 | 0 | 0 |
| 0 | 0 | 0 | 0 | 0 |
| 0 | 0 | 0 | 0 | 0 |
| 0 | 0 | 0 | 0 | 0 |
| 0 | 0 | 0 | 0 | 0 |
| 0 | 0 | 0 | 0 | 0 |
| 0 | 0 | 0 | 0 | 0 |
| 0.192446884 | 0 | 0.014749914 | 0.012483172 | 0.117152369 |
| 0.078269576 | 0.006491061 | 0.47471218 | 0 | 0.002509119 |
| 0 | 0 | 0 | 0 | 0 |
| 0 | 0 | 0 | 0 | 0 |
| 0 | 0.041808153 | 0 | 0 | 0 |
| 0 | 0 | 0 | 0 | 0 |
| 0 | 0 | 0 | 0 | 0 |

| | | | | |
|---|---|---|---|---|
| 0 | 0 | 0 | 0 | 0 |
| 0.004266852 | 0.013017616 | 0.035995994 | 0 | 0.009277537 |
| 0.006501044 | 0.001588681 | 0.003866402 | 0 | 0 |
| 0 | 0 | 0 | 0 | 0 |
| 0.139285675 | 0.003443032 | 0 | 0.009576308 | 0.002019019 |
| 0 | 0 | 0 | 0.010285816 | 0 |
| 0 | 0 | 0 | 0 | 0 |
| 0 | 0 | 0 | 0 | 0 |
| 0 | 0 | 0 | 0 | 0 |
| 0 | 0 | 0 | 0 | 0 |
| 0 | 0 | 0 | 0 | 0 |
| 0 | 0 | 0 | 0 | 0 |
| 0 | 0 | 0 | 0 | 0 |
| 0 | 0 | 0.018889529 | 0 | 0 |
| 0 | 0 | 0 | 0 | 0 |
| 0 | 0 | 0 | 0 | 0 |
| 0 | 0.02542124 | 0 | 0 | 0 |
| 0 | 0 | 0 | 0 | 0 |
| 0 | 0 | 0 | 0 | 0 |
| 0 | 0 | 0 | 0 | 0 |
| 0 | 0 | 0 | 0 | 0 |
| 0 | 0 | 0 | 0.003659381 | 0 |
| 0 | 0.004875871 | 0 | 0 | 0 |
| 0 | 0 | 0 | 0 | 0 |
| 0 | 0.552149158 | 0 | 0 | 0 |
| 0.493884787 | 0 | 0.002604471 | 0.001200075 | 0 |
| 0 | 0 | 0 | 0 | 0 |
| 0 | 0.002555308 | 0 | 0 | 0 |
| 0 | 0 | 0 | 0 | 0 |
| 0 | 0 | 0 | 0 | 0 |
| 0 | 0 | 0 | 0 | 0 |
| 0 | 0 | 0 | 0.009772239 | 0.003676892 |
| 0 | 0 | 0 | 0 | 0 |
| 0 | 0 | 0 | 0 | 0 |
| 0 | 0 | 0 | 0 | 0 |
| 0 | 0 | 0 | 0 | 0 |
| 0 | 0 | 0 | 0 | 0 |
| 0.011087846 | 0 | 0 | 0 | 0 |
| 0.43258623 | 0 | 0 | 0 | 0 |
| 0 | 0 | 0 | 0 | 0 |
| 0 | 0 | 0 | 0 | 0 |
| 0 | 0 | 0 | 0 | 0 |
| 0.000540925 | 0 | 0 | 0.003957336 | 0.000601142 |
| 0 | 0 | 0 | 0 | 0 |
| 0 | 0 | 0 | 0 | 0 |
| 0 | 0 | 0 | 0 | 0 |
| 0 | 0 | 0.003691117 | 0.001316076 | 0.009426207 |
| 0 | 0 | 0 | 0 | 0 |

| | | | | |
|---|---|---|---|---|
| 0 | 0 | 0 | 0 | 0 |
| 0.055688745 | 0.003827487 | 0.012420029 | 0.014613682 | 0.03669777 |
| 0 | 0.002809434 | 0 | 0 | 0 |
| 0 | 0 | 0 | 0 | 0 |
| 0 | 0 | 0 | 0 | 0 |
| 0 | 0 | 0 | 0 | 0 |
| 0 | 0 | 0 | 0 | 0 |
| 0 | 0 | 0 | 0 | 0 |
| 0 | 0 | 0 | 0 | 0 |
| 0 | 0 | 0 | 0 | 0 |
| 0 | 0 | 0 | 0 | 0 |
| 0 | 0 | 0 | 0 | 0 |
| 1.093104019 | 0.055102563 | 0 | 0 | 0 |
| 0 | 0 | 0 | 0 | 0 |
| 0.010198335 | 0 | 0.005807214 | 0 | 0.00090428 |
| 0 | 0 | 0 | 0 | 0 |
| 0 | 0 | 0 | 0 | 0 |
| 0 | 0 | 0 | 0 | 0 |
| 0 | 0 | 0 | 0 | 0 |
| 0 | 0 | 0 | 0 | 0 |
| 0 | 0 | 0 | 0 | 0 |
| 0 | 0 | 0 | 0 | 0.020248104 |
| 0 | 0.042530846 | 0.0010956 | 0 | 0.004640407 |
| 0 | 0 | 0.000979035 | 0 | 0 |
| 0 | 0 | 0 | 0 | 0 |
| 0 | 0 | 0 | 0 | 0 |
| 0 | 0 | 0 | 0 | 0 |
| 0 | 0 | 0 | 0 | 0 |
| 0 | 0 | 0 | 0 | 0 |
| 0 | 0 | 0 | 0 | 0 |
| 0 | 0 | 0 | 0 | 0 |
| 0 | 0 | 0 | 0 | 0 |
| 0 | 0 | 0 | 0 | 0 |
| 0 | 0 | 0 | 0 | 0 |
| 11.50241127 | 0 | 0 | 0 | 0 |
| 0 | 0 | 0 | 0 | 0 |
| 0 | 0 | 0 | 0 | 0 |
| 0 | 0 | 0 | 0 | 0 |
| 0 | 0.275023991 | 0 | 0 | 0.081412836 |
| 0 | 0 | 0 | 0 | 0 |
| 0.061058037 | 0.06354727 | 0.156814256 | 0 | 0.004930106 |
| 0 | 0 | 0 | 0 | 0 |
| 0 | 0 | 0 | 0 | 0 |
| 0 | 0 | 0 | 0 | 0 |
| 0 | 0 | 0 | 0 | 0 |
| 0.457892792 | 0 | 0.005603064 | 0.057953125 | 0.152587251 |
| 0 | 0 | 0 | 0 | 0 |
| 0 | 0 | 0 | 0 | 0 |

| | | | | |
|---|---|---|---|---|
| 0 | 0 | 0 | 0 | 0 |
| 0 | 0 | 0 | 0 | 0 |
| 0 | 0 | 0 | 0 | 0 |
| 0 | 0 | 0 | 0 | 0 |
| 0 | 0 | 0 | 0 | 0 |
| 0 | 0 | 0 | 0 | 0 |
| 0 | 0 | 0 | 0 | 0 |
| 0.001652873 | 0 | 0 | 0 | 0.005313821 |
| 0.073551944 | 0 | 0 | 0 | 0 |
| 0.057045757 | 0.001118817 | 0.084103097 | 0 | 0 |
| 0 | 0.055693743 | 0.005527966 | 0 | 0 |
| 0 | 0 | 0 | 0 | 0 |
| 0.000626488 | 0 | 0 | 0 | 0 |
| 0 | 0 | 0 | 0 | 0 |
| 0 | 0 | 0 | 0 | 0 |
| 0 | 0 | 0 | 0 | 0 |
| 0 | 0 | 0 | 0 | 0 |
| 0 | 0 | 0 | 0 | 0 |
| 0 | 0 | 0 | 0 | 0 |
| 0.471745212 | 0.010122695 | 0.703689156 | 0 | 0.035632428 |
| 0 | 0 | 0 | 0 | 0 |
| 0 | 0 | 0 | 0 | 0 |
| 0 | 0 | 0 | 0 | 0 |
| 0 | 0 | 0 | 0 | 0 |
| 0 | 0 | 0 | 0 | 0 |
| 0 | 0 | 0 | 0 | 0 |
| 0 | 0 | 0 | 0 | 0 |
| 0 | 0 | 0 | 0 | 0 |
| 0 | 0 | 0 | 0 | 0 |
| 0 | 0 | 0.001551041 | 0 | 0 |
| 0 | 0 | 0 | 0 | 0 |
| 0 | 0 | 0 | 0 | 0 |
| 0 | 0 | 0 | 0 | 0 |
| 0 | 0 | 0 | 0 | 0 |
| 0 | 0 | 0 | 0 | 0 |
| 0 | 0 | 0.002683091 | 0 | 0 |
| 0 | 0 | 0 | 0 | 0 |
| 0 | 0 | 0 | 0 | 0 |
| 0 | 0 | 0 | 0 | 0 |
| 0 | 0.404030939 | 0 | 0 | 0 |
| 0 | 0 | 0 | 0 | 0 |
| 0 | 0 | 0 | 0 | 0 |
| 0.002781087 | 0 | 0 | 0 | 0 |
| 0 | 0 | 0 | 0 | 0 |
| 0.007681458 | 0 | 0.001168066 | 0 | 0.005820398 |
| 0 | 0.024877658 | 0 | 0 | 0 |
| 0 | 0 | 0 | 0 | 0 |
| 0 | 0 | 0 | 0.013674524 | 0 |

| | | | | |
|---|---|---|---|---|
| 0 | 0 | 0 | 0 | 0 |
| 0 | 0 | 0 | 0 | 0 |
| 0 | 0.001868742 | 0 | 0 | 0 |
| 0 | 0 | 0 | 0 | 0 |
| 0 | 0 | 0 | 0 | 0 |
| 0.008080441 | 0.003821891 | 0.020637493 | 0.001243664 | 0.001738061 |
| 0 | 0 | 0 | 0 | 0 |
| 0 | 0 | 0 | 0 | 0 |
| 0 | 0 | 0 | 0 | 0 |
| 0 | 0 | 0 | 0 | 0 |
| 0 | 0 | 0 | 0 | 0 |
| 0 | 0 | 0 | 0 | 0 |
| 0 | 0.009575271 | 0 | 0 | 0 |
| 0 | 0 | 0 | 0 | 0 |
| 0 | 0 | 0 | 0 | 0 |
| 0 | 0 | 0 | 0 | 0 |
| 0 | 0 | 0 | 0 | 0 |
| 0 | 0 | 0 | 0 | 0 |
| 0 | 0 | 0 | 0 | 0 |
| 0 | 0 | 0 | 0 | 0 |
| 0 | 0.007280218 | 0 | 0 | 0 |
| 0 | 0 | 0 | 0 | 0 |
| 0 | 0 | 0 | 0 | 0 |
| 0 | 0.007201494 | 0 | 0 | 0 |
| 0 | 0 | 0 | 0 | 0 |
| 0 | 0 | 0.00181862 | 0 | 0 |
| 0 | 0 | 0 | 0 | 0 |
| 0 | 0 | 0 | 0 | 0 |
| 0 | 0 | 0 | 0 | 0 |
| 0 | 0 | 0 | 0 | 0 |
| 0 | 0 | 0 | 0 | 0 |
| 0 | 0 | 0 | 0 | 0 |
| 0 | 0 | 0 | 0 | 0 |
| 0 | 0 | 0 | 0 | 0 |
| 0 | 0 | 0 | 0 | 0 |
| 0 | 0 | 0 | 0 | 0 |
| 0 | 0 | 0 | 0 | 0 |
| 0 | 0 | 0 | 0 | 0 |
| 0.252409656 | 0 | 0.653000765 | 0 | 0 |
| 0 | 0 | 0 | 0 | 0 |
| 0 | 0 | 0 | 0 | 0 |
| 0 | 0 | 0 | 0 | 0 |
| 0 | 0 | 0 | 0 | 0 |
| 0 | 0 | 0 | 0 | 0 |
| 0.001266531 | 0 | 0 | 0 | 0 |
| 0 | 0 | 0 | 0 | 0 |
| 0 | 0 | 0 | 0 | 0 |
| 0 | 0 | 0 | 0 | 0 |

| | | | | |
|---|---|---|---|---|
| 0 | 0 | 0 | 0 | 0 |
| 0.273735792 | 0.011308562 | 0.000976282 | 0 | 0 |
| 0 | 0 | 0 | 0 | 0 |
| 0 | 0 | 0 | 0 | 0 |
| 0 | 0 | 0 | 0 | 0 |
| 0 | 0 | 0 | 0 | 0 |
| 0 | 0 | 0 | 0 | 0 |
| 0 | 0 | 0 | 0 | 0 |
| 0 | 0 | 0 | 0 | 0 |
| 0 | 0 | 0 | 0 | 0 |
| 0 | 0 | 0.002343667 | 0 | 0 |
| 0 | 0 | 0 | 0 | 0 |
| 0.068112356 | 0.087547532 | 0.478950096 | 0 | 0 |
| 0 | 0 | 0 | 0 | 0 |
| 0 | 0 | 0 | 0 | 0 |
| 0 | 0 | 0 | 0 | 0 |
| 0 | 0 | 0 | 0 | 0 |
| 0 | 0 | 0 | 0 | 0 |
| 0 | 0 | 0 | 0 | 0 |
| 0 | 0 | 0 | 0 | 0 |
| 0 | 0 | 0.001081218 | 0 | 0 |
| 0 | 0 | 0 | 0 | 0 |
| 0 | 0 | 0 | 0 | 0 |
| 2.204967172 | 1.070188927 | 1.301782686 | 0 | 0 |
| 0 | 0 | 0 | 0 | 0 |
| 0 | 0 | 0 | 0 | 0 |
| 0 | 0 | 0 | 0 | 0 |
| 0 | 0 | 0.000993448 | 0 | 0 |
| 0 | 0 | 0 | 0 | 0 |
| 0 | 0 | 0 | 0 | 0 |
| 0 | 0 | 0 | 0 | 0 |
| 0 | 0 | 0 | 0 | 0 |
| 0 | 0 | 0 | 0 | 0 |
| 0 | 0 | 0 | 0 | 0 |

| PHL018-0187_V2 | PHL018-0264_V2 | PHL018-0112_V2 | PHL018-0025_V1 | PHL018-0015_V1 |
|---|---|---|---|---|
| CTRL | REF | CTRL | CTRL | CTRL |
| 3.991336379 | 0.00229368 | 0.053442633 | 0.019514605 | 0.040120809 |
| 0.753458119 | 0.174855614 | 0.121052758 | 0.22926343 | 0 |
| 0.162775075 | 8.355559309 | 0 | 10.71114525 | 0 |
| 0 | 0 | 0 | 0 | 0 |
| 0 | 0 | 0 | 0 | 0 |
| 0 | 0 | 0 | 0 | 0 |
| 0 | 0 | 0 | 0 | 0 |
| 0 | 0 | 0 | 0 | 0 |
| 0 | 0 | 0 | 0 | 0 |
| 0 | 0 | 0 | 0 | 0 |
| 0 | 0 | 0 | 0 | 0 |
| 0 | 0 | 0 | 0 | 0 |
| 0 | 0 | 0 | 0 | 0 |
| 5.401837052 | 0.392807354 | 0.628014371 | 13.96058661 | 24.67702285 |
| 0 | 0 | 0 | 0 | 0 |
| 0 | 0 | 0 | 0 | 0 |
| 0 | 0 | 0 | 0 | 0 |
| 0 | 0 | 0 | 0 | 0 |
| 0 | 0 | 0 | 0 | 0 |
| 0 | 0 | 0 | 0 | 0 |
| 0 | 0 | 0 | 0 | 0 |
| 0.002466407 | 0 | 0 | 0 | 0 |
| 0 | 0 | 0 | 0 | 0 |
| 0 | 0 | 0 | 0 | 0 |
| 0 | 0 | 0.313466876 | 0 | 0 |
| 0 | 0 | 0 | 0 | 0 |
| 0 | 0 | 0 | 0 | 0 |
| 0 | 0 | 0 | 0 | 0 |
| 0 | 0 | 0 | 0 | 0 |
| 0 | 0 | 0.178558241 | 0 | 0 |
| 0 | 0 | 0 | 0 | 0 |
| 0 | 0 | 0 | 0 | 0 |
| 0 | 0 | 0 | 0 | 0 |
| 0 | 0 | 0 | 0 | 0 |
| 0 | 0 | 0 | 0 | 0 |
| 0 | 6.378289809 | 0.22782455 | 0.17081064 | 0 |
| 0 | 0 | 0 | 0 | 0 |
| 0 | 0 | 0 | 0 | 0 |
| 0 | 0 | 0 | 0 | 0 |
| 0 | 0 | 0 | 0 | 0 |
| 0 | 0 | 0 | 0 | 0 |
| 0.033387746 | 0.03784446 | 0 | 0.418863596 | 0.032014746 |
| 0.012621253 | 0.362174205 | 2.154702498 | 0.00984836 | 1.9976092 |
| 0 | 0.021552212 | 0.006225825 | 1.561311571 | 0.010467039 |
| 0 | 0 | 0 | 0.006268124 | 0 |
| 0 | 0 | 0 | 0 | 0 |

| | | | | |
|---|---|---|---|---|
| 0 | 0 | 0.000786125 | 0 | 0 |
| 0.001275109 | 0.004060728 | 0 | 0 | 0.044925339 |
| 0.211071953 | 0.000790013 | 0.001430954 | 0 | 0 |
| 0.137176174 | 0.384046691 | 1.348910573 | 0.215951266 | 0 |
| 0.022837305 | 0 | 0.010107352 | 0 | 0 |
| 0 | 0 | 0.016279485 | 0 | 0 |
| 0.001640053 | 0.02841891 | 0.000695612 | 0.005241459 | 0 |
| 0 | 0 | 0 | 0 | 0 |
| 0 | 0 | 0.029898614 | 0 | 0 |
| 0 | 0 | 0 | 0 | 0 |
| 0 | 0.014114312 | 0 | 0.049993485 | 0 |
| 0.000964523 | 0.001626158 | 0.132055066 | 0.006693484 | 0 |
| 0 | 0 | 0 | 0 | 0 |
| 0.058140438 | 0.003667158 | 0.027233587 | 0 | 0 |
| 0 | 0.027924096 | 0 | 0 | 0 |
| 0 | 0 | 0 | 0 | 0 |
| 0.03810782 | 0.002765007 | 0 | 0.000980194 | 0 |
| 0 | 0 | 0 | 0 | 0 |
| 0 | 0 | 0 | 0 | 0 |
| 0 | 0 | 0 | 0 | 0 |
| 0 | 0.076338626 | 0.010352886 | 0 | 0.056591947 |
| 0.007186088 | 0.004365961 | 0.010379358 | 0 | 0 |
| 0 | 0 | 0.014043908 | 0 | 0 |
| 0 | 0 | 0 | 0 | 0 |
| 0 | 0 | 0 | 0 | 0 |
| 0 | 0 | 0 | 0 | 0 |
| 0 | 0 | 0 | 0 | 0 |
| 0 | 0 | 0 | 0 | 0 |
| 0 | 0.000698166 | 0 | 0.017923091 | 0.00322726 |
| 0 | 0 | 0 | 0 | 0 |
| 0 | 0.033599162 | 0.001493455 | 0 | 1.697946034 |
| 0 | 0 | 0 | 0 | 0 |
| 0 | 0 | 0 | 0 | 0 |
| 0.000538719 | 0 | 0 | 0 | 0 |
| 0 | 0 | 0 | 0 | 0 |
| 0 | 0 | 0 | 0.00103079 | 0 |
| 0 | 0 | 1.131974246 | 0 | 0 |
| 0.001413063 | 0 | 0.003596018 | 0 | 0 |
| 0 | 0 | 0 | 0 | 0 |
| 0 | 0 | 0 | 0 | 0 |
| 0 | 0 | 0 | 0 | 0 |
| 0 | 0 | 0.042756493 | 0 | 0 |
| 0 | 0 | 0 | 0 | 0 |
| 0 | 0 | 0 | 0 | 0 |
| 0 | 0 | 0 | 0 | 0 |
| 0 | 0 | 0 | 0 | 0 |
| 0 | 0 | 0 | 0 | 0 |
| 0 | 0 | 0 | 0 | 0 |

| | | | | |
|---|---|---|---|---|
| 0 | 0 | 0 | 0 | 0 |
| 0 | 0 | 0 | 0 | 0 |
| 0 | 0 | 0 | 0 | 0 |
| 0 | 0 | 0 | 0 | 0 |
| 0 | 0 | 0.003188017 | 0.000807456 | 0 |
| 0 | 0.001117986 | 0 | 0 | 0 |
| 0 | 0 | 0 | 0 | 0 |
| 0 | 0 | 0.00136141 | 0 | 0 |
| 0 | 0 | 0.003025664 | 0 | 0 |
| 0 | 0 | 0 | 0 | 0 |
| 0 | 0 | 0 | 0 | 0 |
| 0 | 0.006124234 | 0 | 0.003980237 | 0 |
| 0 | 0 | 0 | 0 | 0 |
| 0 | 0.00057238 | 0 | 0 | 0 |
| 0 | 0 | 0 | 0 | 0 |
| 0 | 0 | 0.00120035 | 0 | 0 |
| 0 | 0 | 0 | 0 | 0 |
| 0 | 0 | 0 | 0 | 0 |
| 0 | 0.09937681 | 0.010530258 | 0.109277647 | 0 |
| 0 | 0 | 0 | 0 | 0 |
| 0 | 0 | 0 | 0 | 0 |
| 0 | 0 | 0 | 0 | 0 |
| 0 | 0 | 0 | 0 | 0 |
| 0 | 0 | 0 | 0 | 0 |
| 0 | 0 | 0 | 0 | 0 |
| 0 | 0 | 0 | 0 | 0 |
| 0 | 0 | 0.001400158 | 0 | 0 |
| 0 | 0.001183527 | 0 | 0 | 0 |
| 0 | 0 | 0 | 0 | 0 |
| 0 | 0 | 0 | 0 | 0 |
| 0 | 0.009274625 | 0.039787495 | 0 | 0 |
| 0 | 0.006315893 | 0.001236757 | 0.00832054 | 0 |
| 0 | 0 | 0 | 0 | 0 |
| 0 | 0 | 0 | 0 | 0 |
| 0 | 0 | 0 | 0 | 0 |
| 0 | 0 | 0 | 0 | 0 |
| 0 | 0 | 0 | 0 | 0 |
| 0 | 0 | 0 | 0 | 0 |
| 0 | 0.617928275 | 0 | 0 | 0 |
| 0 | 0 | 0 | 0 | 0 |
| 0 | 0 | 0 | 0 | 0 |
| 0 | 0 | 0 | 0 | 0 |
| 0 | 0.000591906 | 0.001072122 | 0.001923446 | 0 |
| 0 | 0 | 0 | 0 | 0 |
| 0 | 0 | 0 | 0 | 0 |
| 0 | 0 | 0 | 0 | 0 |
| 0 | 0 | 0 | 0 | 0 |
| 0 | 0 | 0 | 0 | 0 |

| | | | | |
|---|---|---|---|---|
| 0 | 0 | 0 | 0 | 0 |
| 0 | 0 | 0 | 0 | 0 |
| 0 | 0 | 0 | 0 | 0 |
| 0 | 0 | 0 | 0 | 0 |
| 0 | 0 | 0.107511142 | 0 | 0 |
| 0 | 0 | 0 | 0 | 0 |
| 0 | 0 | 0 | 0 | 0 |
| 0 | 0 | 0 | 0 | 0 |
| 0 | 0 | 0 | 0 | 0 |
| 0 | 0 | 0 | 0 | 0 |
| 0 | 0 | 0 | 0 | 0 |
| 0 | 0 | 0 | 0 | 0 |
| 0 | 0 | 0 | 0 | 0 |
| 0 | 0 | 0 | 0 | 0 |
| 0 | 0 | 0 | 0 | 0 |
| 0 | 0 | 2.962562237 | 0 | 0 |
| 0 | 0 | 0 | 0 | 0 |
| 0 | 0 | 0 | 0 | 0 |
| 0 | 0 | 0 | 0 | 0 |
| 0 | 0 | 0 | 0 | 0 |
| 0 | 0 | 0 | 0 | 0 |
| 0 | 0 | 0 | 0 | 0 |
| 0.018567409 | 0 | 0.001575036 | 0 | 0 |
| 0 | 0 | 0 | 0 | 0 |
| 0 | 0 | 0 | 0 | 0 |
| 0 | 0 | 0 | 0 | 0 |
| 0 | 0 | 0.017968633 | 0 | 0 |
| 0 | 0 | 0 | 0 | 0 |
| 0 | 0 | 0 | 0 | 0 |
| 0 | 0 | 0 | 0 | 0 |
| 0 | 0 | 0.007092051 | 0 | 0 |
| 0.003574611 | 0.080355909 | 0 | 0.001344015 | 0 |
| 0 | 0 | 0 | 0 | 0 |
| 0 | 0 | 0 | 0 | 0 |
| 0 | 0 | 0 | 0 | 0 |
| 0 | 0 | 0 | 0 | 0 |
| 0 | 0 | 0 | 0 | 0 |
| 0 | 0 | 0.002569958 | 0 | 0 |
| 0 | 0 | 0 | 0 | 0 |
| 0 | 0 | 0 | 0 | 0 |
| 0 | 0 | 0 | 0 | 0 |
| 0 | 0 | 0 | 0 | 0 |
| 0 | 0 | 0 | 0 | 0 |
| 0 | 0 | 0 | 0 | 0 |
| 0 | 0 | 0 | 0 | 0 |
| 0 | 0 | 0 | 0 | 0 |
| 0 | 0 | 0 | 0 | 0 |
| 0 | 0 | 0 | 0 | 0.272275926 |

| | | | | |
|---|---|---|---|---|
| 0 | 0 | 1.376910644 | 0 | 0 |
| 0 | 0 | 0 | 0 | 0 |
| 0 | 0 | 0 | 0 | 0 |
| 0 | 0 | 0 | 0 | 0 |
| 0 | 0 | 0 | 0 | 0 |
| 0 | 0 | 0 | 0 | 0 |
| 0 | 0 | 0 | 0 | 0 |
| 0 | 0 | 0 | 0 | 0 |
| 0 | 0 | 0 | 0 | 0 |
| 0 | 0 | 0 | 0 | 0 |
| 0 | 0 | 0 | 0 | 0 |
| 0 | 0 | 0.112715547 | 0 | 0 |
| 0.003018134 | 0 | 0 | 0 | 0 |
| 0 | 0 | 0 | 0 | 0 |
| 0 | 0 | 0 | 0 | 0 |
| 0 | 0 | 0 | 0 | 0 |
| 0 | 0 | 0 | 0 | 0 |
| 0 | 0 | 0 | 0 | 0 |
| 0 | 0 | 0 | 0 | 0 |
| 0 | 0 | 0 | 0 | 0 |
| 0.005266973 | 0.082456852 | 0.014041868 | 0.201649635 | 0.040009102 |
| 0 | 0 | 0 | 0 | 0 |
| 0.000660206 | 0 | 0 | 0 | 0 |
| 0 | 0 | 0 | 0.001791141 | 0 |
| 0 | 0 | 0 | 0 | 0 |
| 0 | 0 | 0 | 0 | 0 |
| 0.004820139 | 0.003283482 | 0.001672702 | 0.009002759 | 0.012272708 |
| 0 | 0 | 0 | 0 | 0 |
| 0 | 0 | 0 | 0 | 0 |
| 0.01734625 | 0 | 0 | 0 | 0 |
| 0 | 0.001636537 | 0 | 0 | 0.020744279 |
| 0 | 0 | 0 | 0 | 0 |
| 0 | 0 | 0 | 0 | 0 |
| 0 | 0 | 0.044662899 | 0 | 0 |
| 0.005385244 | 0.000917108 | 0.006229355 | 0 | 0.011923078 |
| 0 | 0 | 0 | 0 | 0 |
| 0 | 0 | 0 | 0 | 0 |
| 0 | 0 | 0 | 0 | 0 |
| 0 | 0.037112512 | 0 | 0 | 0.001733497 |
| 0 | 0 | 0 | 0 | 0 |
| 0 | 0.000605212 | 0 | 0.000786674 | 0 |
| 0 | 0 | 0 | 0 | 0 |
| 0 | 0.001429846 | 0 | 0.00092928 | 0 |
| 0 | 0 | 0 | 0 | 0 |
| 0 | 0 | 0 | 0 | 0 |
| 0 | 0 | 0.18228016 | 0 | 0 |
| 0 | 0 | 0 | 0 | 0 |
| 0 | 0 | 0 | 0 | 0.004389966 |

| | | | | |
|---|---|---|---|---|
| 0 | 0 | 0 | 0 | 0 |
| 0 | 0 | 0 | 0 | 0 |
| 0 | 0 | 0 | 0 | 0.002588038 |
| 0 | 0 | 0 | 0 | 0 |
| 0 | 0 | 0 | 0 | 0 |
| 0 | 0 | 0 | 0 | 0 |
| 0 | 0 | 0 | 0.005668457 | 0 |
| 0 | 0.003288246 | 0 | 0.001373839 | 0 |
| 0.341682066 | 0 | 0 | 0 | 0 |
| 0 | 0 | 0 | 0 | 0 |
| 0 | 0 | 0 | 0 | 0 |
| 0 | 0 | 0 | 0 | 0 |
| 0 | 0 | 0 | 0 | 0 |
| 0 | 0 | 0 | 0 | 0 |
| 0 | 0.004379375 | 0 | 0.052487747 | 0 |
| 0 | 0 | 0 | 0 | 0 |
| 0 | 0 | 0 | 0 | 0 |
| 0 | 0 | 0 | 0 | 0.010364101 |
| 0 | 0.000830828 | 0.009932217 | 0 | 0.010513333 |
| 0 | 0 | 0.139858971 | 0 | 0 |
| 0.023290302 | 0 | 0.001331912 | 0 | 0.000637325 |
| 0 | 0 | 0 | 0 | 0 |
| 0 | 0 | 0 | 0 | 0 |
| 0 | 0 | 0 | 0 | 0 |
| 0 | 0.00194786 | 0 | 0.018989178 | 0.017726526 |
| 0 | 0 | 0 | 0 | 0 |
| 0 | 0 | 0 | 0 | 0 |
| 0 | 0 | 0 | 0 | 0 |
| 0 | 0 | 0 | 0.039641105 | 0 |
| 0 | 0 | 0 | 0 | 0 |
| 0.302868546 | 0.221322538 | 0.280095335 | 0 | 0.008903869 |
| 0 | 0 | 0 | 0 | 0 |
| 0 | 0 | 0 | 0 | 0 |
| 0 | 0 | 0.005701795 | 0 | 0 |
| 0 | 0 | 0 | 0 | 0 |
| 0 | 0 | 0 | 0 | 0 |
| 0 | 0 | 0 | 0 | 0 |
| 0 | 0 | 0 | 0 | 0 |
| 0 | 0 | 0 | 0 | 0 |
| 0.000768802 | 0 | 0 | 0 | 0 |
| 0 | 0 | 0 | 0 | 0 |
| 0 | 0.00440915 | 0.003993156 | 0.001790986 | 0 |
| 0 | 0 | 0 | 0 | 0 |
| 0 | 0 | 0 | 0 | 0 |
| 0 | 0 | 0 | 0 | 0 |
| 0 | 0 | 0 | 0 | 0 |
| 0 | 0 | 0 | 0 | 0 |
| 0 | 0 | 0 | 0 | 0 |

| | | | | |
|---|---|---|---|---|
| 0 | 0 | 0 | 0 | 0 |
| 0 | 0 | 0 | 0 | 0 |
| 0 | 0.019146764 | 0.001044597 | 1.701275614 | 0.010396742 |
| 0 | 0 | 0 | 0 | 0 |
| 0 | 0 | 1.18646623 | 0 | 0 |
| 0 | 0 | 0 | 0 | 0 |
| 0 | 0 | 0 | 0 | 0 |
| 0 | 0 | 0 | 0 | 0 |
| 0 | 0 | 0 | 0 | 0 |
| 0 | 0 | 0 | 0 | 0 |
| 0 | 0 | 0 | 0 | 0 |
| 0 | 0 | 0 | 0 | 0 |
| 0 | 0 | 0 | 0 | 0 |
| 0 | 0.007138526 | 0 | 0.006959163 | 0.001767735 |
| 0 | 0.016623629 | 0.000952862 | 0.045540747 | 0.005288997 |
| 0 | 0 | 0 | 0 | 0 |
| 0 | 0.001678328 | 0.000868561 | 0.02127006 | 0.007896581 |
| 0 | 0 | 0 | 0 | 0 |
| 0 | 0 | 0 | 0 | 0 |
| 0 | 0 | 0 | 0 | 0 |
| 0 | 0 | 0 | 0 | 0 |
| 0.001848594 | 0 | 0.000588047 | 0 | 0.004126945 |
| 0 | 0 | 0 | 0 | 0 |
| 0 | 0.004361826 | 0 | 0 | 0 |
| 0 | 0 | 0 | 0 | 0 |
| 0 | 0 | 0 | 0 | 0 |
| 0 | 0 | 3.985090937 | 0 | 0 |
| 0 | 0 | 0 | 0 | 0 |
| 0 | 0 | 0 | 0 | 0 |
| 0 | 0 | 0 | 0 | 0 |
| 0 | 0 | 0 | 0 | 0 |
| 0 | 0 | 0 | 0 | 0 |
| 0 | 0 | 0 | 0 | 0 |
| 0 | 0 | 0 | 0 | 0 |
| 0 | 0 | 0 | 0.003123924 | 0 |
| 0 | 0.008646 | 0 | 0.003792942 | 0.017235333 |
| 0 | 0 | 0 | 0 | 0 |
| 0.086515616 | 0.00842981 | 0.002462733 | 0.520119958 | 0 |
| 0 | 0 | 0 | 0 | 0 |
| 0 | 0 | 0 | 0 | 0 |
| 0 | 0 | 0 | 0 | 0 |
| 0 | 0 | 0 | 0 | 0 |
| 0 | 0 | 0 | 0 | 0 |
| 0 | 0 | 0 | 0 | 0.001463791 |
| 0 | 0 | 0 | 0 | 0 |
| 0 | 0 | 0 | 0 | 0 |
| 0 | 0 | 0 | 0 | 0 |
| 0 | 0 | 0 | 0 | 0.007993276 |

| | | | | |
|---|---|---|---|---|
| 2.90779105 | 0.183917482 | 0 | 0 | 0 |
| 0 | 0 | 0 | 0.044069234 | 0 |
| 0 | 0 | 0.000821244 | 0.154923586 | 0 |
| 0 | 0 | 0 | 0 | 0 |
| 0 | 0.014369155 | 0.003718129 | 0.003779967 | 0.003953642 |
| 0 | 0 | 0.003031374 | 0 | 0 |
| 0 | 0 | 0 | 0 | 0 |
| 0 | 0.04411955 | 0.01100264 | 0.013090304 | 0.062623457 |
| 0 | 0 | 0 | 0 | 0 |
| 0.102730265 | 0.036577698 | 0 | 0.060394819 | 1.107015673 |
| 0 | 0 | 0 | 0 | 0 |
| 0.001350989 | 0.033406707 | 0 | 0.096715091 | 0.001316096 |
| 0.002049547 | 0.014781767 | 0.007475933 | 0.015533245 | 0.154570983 |
| 0 | 0.01049596 | 0 | 0 | 0 |
| 0 | 0 | 0.00107286 | 0 | 0 |
| 0 | 0 | 0 | 0.001685637 | 0 |
| 0 | 0 | 0 | 0.001430102 | 0.002179608 |
| 0 | 0 | 0 | 0 | 0 |
| 0 | 0.183877796 | 0 | 0 | 0.08573179 |
| 0 | 0 | 0 | 0 | 0 |
| 0 | 0.003969822 | 0 | 0 | 0.01499165 |
| 0 | 0 | 0 | 0 | 0 |
| 0 | 0.002971339 | 0 | 0.003089793 | 0 |
| 0 | 0 | 0 | 0 | 0 |
| 0 | 0 | 0 | 0 | 0 |
| 0 | 0 | 0 | 0.003269687 | 0.006201448 |
| 0 | 0.003587939 | 0 | 0 | 0 |
| 0 | 0 | 0 | 0 | 0.604863358 |
| 0 | 0 | 0 | 0 | 0 |
| 0 | 0 | 0 | 0 | 0 |
| 0 | 0.007437893 | 0 | 0 | 0.45687239 |
| 0.000852875 | 0 | 0 | 0 | 0 |
| 0 | 0 | 0 | 0 | 0.00221594 |
| 0 | 0 | 0 | 0 | 0 |
| 0 | 0 | 0 | 0 | 0 |
| 0 | 0 | 0 | 0 | 0 |
| 0 | 0.036499741 | 0 | 0 | 0.004613422 |
| 0 | 0 | 0 | 0 | 0 |
| 0 | 0 | 0 | 0 | 0 |
| 0 | 0 | 0 | 0 | 0 |
| 0 | 0 | 0 | 0 | 0 |
| 0 | 0 | 0 | 0 | 0 |
| 0 | 0 | 0 | 0 | 0 |
| 0.027501931 | 0 | 0 | 0 | 0.045745587 |
| 0 | 0 | 0 | 0 | 0 |
| 0 | 0 | 0 | 0 | 0 |
| 0 | 0 | 0.002014611 | 0 | 0 |
| 0 | 0 | 0 | 0 | 0 |

| | | | | |
|---|---|---|---|---|
| 0 | 0 | 0 | 0 | 0 |
| 0 | 0.018645149 | 0 | 0.016661947 | 0 |
| 0 | 0.015162569 | 0 | 0 | 0 |
| 0 | 0 | 0.042822103 | 0 | 0 |
| 0 | 0 | 0 | 0 | 0 |
| 0.005611651 | 0.025842782 | 0.000991719 | 0 | 0 |
| 0 | 0 | 0 | 0 | 0 |
| 0 | 0 | 0 | 0 | 0 |
| 0 | 0 | 0.549936498 | 0 | 0 |
| 0 | 0 | 0 | 0 | 0 |
| 0.001627143 | 0.001524065 | 0.002070408 | 0.026248622 | 0.01673179 |
| 0 | 0 | 0 | 0 | 1.202317056 |
| 0 | 0 | 0 | 0 | 0.037263376 |
| 0 | 0.006959323 | 0.000652006 | 0.098725622 | 0.001871924 |
| 0 | 0 | 0 | 0.004970958 | 0 |
| 0 | 0 | 0 | 0 | 0 |
| 0 | 0 | 0 | 0 | 0 |
| 0 | 0 | 0 | 0 | 0 |
| 0 | 0 | 0.0023132 | 0 | 0 |
| 0 | 0.004460847 | 0 | 0 | 0 |
| 0 | 0 | 0 | 0 | 0 |
| 0 | 0.331552576 | 0.339095171 | 0 | 0 |
| 0 | 0 | 0.025573862 | 0 | 0 |
| 0 | 0 | 0 | 0 | 0 |
| 0 | 0 | 0 | 0 | 0 |
| 0 | 0 | 0 | 0 | 0 |
| 0 | 0.071294312 | 0 | 0 | 0.002931732 |
| 0 | 0 | 0 | 0 | 0 |
| 0 | 0 | 0 | 0 | 0 |
| 0.543414972 | 0.54029645 | 0.225227611 | 0.064468445 | 0.441454627 |
| 0 | 0 | 0 | 0.002698145 | 0 |
| 0 | 0 | 0 | 0 | 0 |
| 0 | 0 | 0 | 0 | 0.015862176 |
| 0 | 0 | 0 | 0 | 0 |
| 0 | 0.002600647 | 0.001962734 | 0 | 0 |
| 0 | 0 | 0 | 0 | 0 |
| 0 | 0 | 0.163481195 | 0 | 0 |
| 0 | 0.000978401 | 0 | 0.001430726 | 0 |
| 0 | 0 | 0 | 0 | 0 |
| 0 | 0 | 0 | 0 | 0 |
| 0.045285474 | 0.147176039 | 0.001429386 | 0 | 0 |
| 0.04383849 | 0.055314075 | 1.627175204 | 0 | 2.953847864 |
| 0 | 0.002688835 | 0 | 0.000655319 | 0 |
| 0 | 0 | 0 | 0 | 0.003919411 |
| 0 | 0 | 0 | 0 | 0 |
| 0 | 0 | 0 | 0 | 0 |
| 0.000884286 | 0 | 0 | 0 | 0.007717124 |
| 0 | 0 | 0 | 0 | 0 |

| | | | | |
|---|---|---|---|---|
| 0 | 0 | 0 | 0.003237862 | 0 |
| 0 | 0 | 0 | 0 | 0 |
| 0.08808419 | 0.010443563 | 0.001891647 | 0.183600204 | 0.007844719 |
| 0 | 0 | 0 | 0 | 0 |
| 0 | 0.001883385 | 0 | 0 | 0 |
| 0 | 0.007344454 | 0 | 0 | 0 |
| 0 | 0 | 0 | 0 | 0 |
| 0 | 0 | 0 | 0 | 0 |
| 0.019403789 | 0.023092406 | 0.001161871 | 0 | 0.001853199 |
| 0.049512519 | 0.161187772 | 0 | 0 | 0 |
| 0 | 0 | 0 | 0 | 0 |
| 0 | 0 | 0 | 0 | 0 |
| 0 | 0 | 0 | 0 | 0.016337702 |
| 0 | 0.002542446 | 0 | 0.015998003 | 0 |
| 0 | 0 | 0 | 0 | 0 |
| 0.001157049 | 0 | 0.009978598 | 0 | 0 |
| 0.001923702 | 0 | 0.002538415 | 0.035131296 | 0.023772257 |
| 0.002856607 | 0 | 0 | 0 | 0 |
| 0.003311732 | 0 | 0 | 0 | 0 |
| 0 | 0 | 0 | 0 | 0 |
| 0 | 0 | 0.001371054 | 0 | 0 |
| 0 | 0.002322265 | 0 | 0 | 0.000805097 |
| 0 | 0 | 0 | 0 | 0 |
| 0 | 0 | 0 | 0.134870091 | 0 |
| 0 | 0 | 0.015420561 | 0 | 0 |
| 0.071237462 | 0 | 0.005254722 | 0 | 0 |
| 0.005969714 | 0.007116504 | 0.001150906 | 0 | 0.004625989 |
| 0 | 0 | 0 | 0 | 0 |
| 0 | 0 | 0 | 0 | 0 |
| 0 | 0 | 0 | 0 | 0 |
| 0.051590349 | 0.003479195 | 0.002800835 | 0 | 0 |
| 0 | 0 | 0.002870795 | 0.230642742 | 0.001998089 |
| 0.930063735 | 0.168192755 | 0.055661659 | 0.010164332 | 0 |
| 0 | 0.002359249 | 0 | 0.127648358 | 0.001363199 |
| 0 | 0 | 0 | 0.39918087 | 0 |
| 0 | 0 | 0 | 0 | 0 |
| 0 | 0 | 0 | 0.002901871 | 0.001074967 |
| 0.008698659 | 0.001745916 | 0.014757794 | 0 | 0 |
| 0 | 0.125375555 | 0.204905958 | 0 | 0 |
| 0 | 0 | 0 | 0 | 0.002749347 |
| 0 | 0 | 0.000669017 | 0 | 0 |
| 0 | 0 | 0 | 0 | 0 |
| 0.13845113 | 0 | 0 | 0 | 0 |
| 0.017784317 | 0.070719506 | 0.002605777 | 0 | 0 |
| 0 | 0 | 0 | 0 | 0.00693476 |
| 0 | 0 | 0.001915143 | 0 | 0 |
| 0 | 0 | 0 | 0 | 0 |
| 0 | 0 | 0 | 0 | 0 |

| | | | | |
|---|---|---|---|---|
| 0 | 0.053210401 | 0.005991202 | 0 | 0 |
| 0.134052086 | 2.647598422 | 0.542234719 | 0.522090371 | 0.303564016 |
| 0 | 0 | 0 | 0 | 0 |
| 0 | 0 | 0 | 0 | 0 |
| 0 | 0 | 0 | 0 | 0.001123813 |
| 0 | 0 | 0 | 0 | 0 |
| 0 | 0 | 0 | 0 | 0 |
| 0 | 0 | 0 | 0.001781433 | 0 |
| 0 | 0 | 0 | 0 | 0 |
| 0.008613363 | 0.003457592 | 0.001739653 | 0 | 0 |
| 0.001887449 | 0 | 0.00213478 | 0 | 0 |
| 0.001874272 | 0.040377403 | 0.010334399 | 1.620723647 | 0.006339805 |
| 0 | 0 | 0 | 0 | 0 |
| 0.003987703 | 0 | 0 | 0 | 0 |
| 0 | 0 | 0 | 0.041966011 | 0 |
| 0 | 0 | 0 | 0 | 0 |
| 0 | 0.012547202 | 0.002794278 | 0.050932996 | 0.053482898 |
| 0.001425378 | 0.072628467 | 0 | 0 | 0.023605585 |
| 0 | 0 | 0 | 0 | 0 |
| 0 | 0.04820145 | 0 | 0 | 0 |
| 0.002264666 | 0.156544693 | 0.027855522 | 0.03846302 | 0.013972438 |
| 0 | 0 | 0 | 0 | 0 |
| 0 | 0 | 0 | 0 | 0 |
| 0 | 0 | 0 | 0 | 0 |
| 0 | 0.001069204 | 0 | 0 | 0 |
| 0 | 0 | 0 | 0 | 0 |
| 0 | 0 | 0.056127301 | 0 | 0 |
| 0.004573311 | 0.122420673 | 0.030627215 | 0.251437126 | 0 |
| 0 | 0 | 0 | 0 | 0 |
| 0 | 0 | 0 | 0 | 0 |
| 0 | 0 | 0.001533608 | 0.001414994 | 0.005451361 |
| 0 | 0 | 0 | 0 | 0 |
| 0 | 0 | 0 | 0 | 0 |
| 0 | 0 | 0 | 0 | 0.000443982 |
| 0 | 0 | 0.061385818 | 0 | 0 |
| 0.550358683 | 0.277016045 | 0.288201125 | 0.094110324 | 15.3600714 |
| 0 | 0 | 0 | 0 | 0 |
| 0 | 0 | 0 | 0 | 0 |
| 0.004157035 | 0.014184161 | 0.008312061 | 0.062360523 | 0.003133671 |
| 0 | 0 | 0 | 0 | 0 |
| 0 | 0 | 0 | 0 | 0 |
| 0 | 0 | 0 | 0 | 0 |
| 0 | 0.001182544 | 0 | 0 | 0 |
| 0.021766829 | 0 | 0 | 0 | 0.003251378 |
| 0 | 0 | 0 | 0 | 0 |
| 0 | 0.002696311 | 0.684805684 | 0 | 0 |
| 0 | 0 | 0 | 0 | 0 |
| 0 | 0.001581864 | 0 | 0 | 0.1048835 |

| | | | | |
|---|---|---|---|---|
| 0 | 0 | 0 | 0 | 0.002045497 |
| 0 | 0 | 0 | 0 | 0 |
| 0 | 0 | 0 | 0 | 0 |
| 0 | 0 | 0.032609154 | 0.010166031 | 0 |
| 0 | 0 | 0 | 0 | 0 |
| 0 | 0 | 0.004481071 | 0 | 0 |
| 0 | 0 | 0 | 0 | 0 |
| 0.018228605 | 0 | 0.059620066 | 0 | 0 |
| 0 | 0.236697356 | 0.646559967 | 0 | 0 |
| 0 | 0 | 0.063066925 | 0 | 0 |
| 0 | 0 | 0 | 0 | 0 |
| 0.001207577 | 0.002073643 | 0.001707272 | 0 | 0 |
| 0 | 0 | 0 | 0 | 0 |
| 0 | 0 | 0 | 0 | 0 |
| 0.656026199 | 0.000622246 | 0.063820682 | 0 | 0 |
| 0 | 0 | 0 | 0 | 0 |
| 0.01191521 | 0 | 0.002021486 | 0 | 0 |
| 0 | 0 | 0.269337868 | 0 | 0 |
| 0 | 0 | 0 | 0 | 0 |
| 0 | 0 | 0.005793633 | 0 | 0 |
| 0 | 0 | 0 | 0 | 0 |
| 0 | 0 | 0.061025454 | 0 | 0 |
| 0 | 0 | 0.047163257 | 0 | 0 |
| 0.178537785 | 0.052296088 | 0 | 0 | 0 |
| 3.415167359 | 0 | 0 | 0 | 0 |
| 0 | 1.158690018 | 1.22036607 | 0 | 0 |
| 0 | 0 | 0.00399399 | 0 | 0 |
| 0 | 6.71036613 | 2.569041018 | 1.745124384 | 0 |
| 0 | 0 | 0 | 0 | 0 |
| 0 | 0 | 0 | 0 | 0 |
| 0 | 0 | 0.042987767 | 0 | 0 |
| 0 | 0.014677062 | 0 | 0 | 0 |
| 0.009828256 | 0.000836877 | 0 | 0 | 0 |
| 0 | 0 | 0 | 0 | 0 |
| 0 | 0 | 0 | 0 | 0 |
| 0 | 0 | 0.132566747 | 0 | 0 |
| 0 | 0 | 1.495821486 | 0 | 0 |
| 0.000628225 | 0 | 0 | 0 | 0 |
| 0 | 0 | 0 | 0 | 0 |
| 0 | 0 | 0 | 0 | 0 |
| 0.001737681 | 0.026945838 | 0.076486294 | 0 | 0.012696004 |
| 0 | 0 | 0.319345533 | 0 | 0 |
| 0.158195086 | 0.739132415 | 0.038705944 | 12.31679052 | 0.457476551 |
| 0.007400543 | 0 | 0.00499365 | 0 | 0 |
| 0 | 0 | 0 | 0 | 0 |
| 0 | 0 | 2.803929857 | 0 | 0 |
| 0 | 0 | 0 | 0 | 0 |
| 0.029012391 | 0.001956563 | 0.000984425 | 0.007417675 | 0 |

| | | | | |
|---|---|---|---|---|
| 0 | 0 | 0 | 0 | 0 |
| 0 | 0.213700068 | 0.017757414 | 0.00617848 | 0 |
| 0.277004053 | 0.013129106 | 0 | 0.001422136 | 1.610608483 |
| 0 | 0 | 0.000612045 | 0 | 0 |
| 0 | 0 | 0 | 0 | 0 |
| 0.015073356 | 0 | 0.001932831 | 0 | 0 |
| 0 | 0 | 0 | 0 | 0 |
| 0 | 0 | 0 | 0 | 0 |
| 0 | 0 | 0 | 0 | 0 |
| 0 | 0 | 0 | 0 | 0 |
| 0.697725729 | 0 | 0.008894678 | 0 | 0 |
| 0.151747446 | 2.749584583 | 0.501592362 | 2.850748294 | 0 |
| 0 | 0 | 0 | 0 | 0 |
| 0.003536384 | 0.002649886 | 0.001399927 | 0.079436629 | 0.003062264 |
| 0.23706432 | 0.111719344 | 0.075332532 | 0 | 0.009803549 |
| 0 | 0 | 0.157140695 | 0 | 0 |
| 0 | 0 | 0 | 0 | 0 |
| 0 | 0 | 0 | 0 | 0 |
| 0 | 0 | 0.061018497 | 0 | 0 |
| 0.118583349 | 0 | 0.009616521 | 0 | 0 |
| 0.076898779 | 0.133765029 | 0.170844888 | 0.805328514 | 0 |
| 0.000964613 | 0.022045532 | 0 | 1.12478649 | 13.3169396 |
| 0.600430822 | 1.502525428 | 0.05750538 | 1.154637659 | 7.225617975 |
| 0 | 0 | 0 | 0 | 0 |
| 0 | 0 | 0.014308911 | 0 | 0 |
| 0 | 0.009453037 | 0.000558337 | 0 | 0 |
| 0 | 0.042048445 | 0 | 0 | 0.103998855 |
| 0 | 0 | 0 | 0 | 0 |
| 0 | 0 | 0 | 0 | 0 |
| 0 | 0 | 0 | 0 | 0 |
| 0 | 0 | 0.00376875 | 0 | 0 |
| 0 | 0 | 0.3523468 | 0 | 0 |
| 0 | 0.001681121 | 0.058997299 | 0 | 0.038247673 |
| 0.182035821 | 0.000961489 | 0.002757452 | 0 | 0 |
| 0.020878654 | 0.002219122 | 0 | 0 | 0.005770043 |
| 0.032958377 | 0.008194021 | 0.00845641 | 0.52090087 | 0 |
| 0.027120699 | 0 | 0.003500904 | 0 | 0 |
| 0 | 0 | 0 | 0 | 0 |
| 0 | 0.029880939 | 0.034533999 | 0 | 0 |
| 0 | 0 | 0 | 0 | 0 |
| 0 | 0 | 0.015956346 | 0 | 0 |
| 0 | 0 | 0 | 0 | 0 |
| 0 | 0 | 0.127760467 | 0 | 0 |
| 0 | 0 | 0.723238952 | 0 | 0 |
| 0.170757849 | 0 | 0.04132961 | 0.001059252 | 0.005462045 |
| 0 | 0 | 0.164894619 | 0 | 0 |
| 0 | 0 | 0 | 0 | 0 |
| 0 | 0.010485433 | 0.005222884 | 0.720393716 | 0 |

| | | | | |
|---|---|---|---|---|
| 0 | 0 | 0 | 0 | 0 |
| 0 | 0 | 0 | 0 | 0 |
| 0 | 0 | 0 | 0 | 0 |
| 0 | 0 | 0.00100643 | 0 | 0 |
| 0 | 0 | 0 | 0 | 0 |
| 0 | 0 | 0.127405104 | 0 | 0 |
| 0 | 0 | 0.357550152 | 0 | 0 |
| 0 | 0.001193005 | 0.002160895 | 0 | 0 |
| 0 | 0 | 0 | 0 | 0 |
| 0 | 0 | 0 | 0 | 0 |
| 0.0314928 | 0 | 0.003549507 | 0 | 0.031466058 |
| 0 | 0 | 0 | 0 | 0 |
| 0 | 0 | 0.005725025 | 0 | 0 |
| 0 | 0 | 0 | 0 | 0 |
| 0 | 0 | 0 | 0 | 0 |
| 0 | 0 | 0 | 0 | 0 |
| 0 | 0.206958958 | 0.010003825 | 0 | 0 |
| 0.004400062 | 0 | 0.001555202 | 0 | 0 |
| 0 | 0 | 0.037639146 | 0 | 0 |
| 0 | 0.001858375 | 0.005750391 | 0 | 0 |
| 0 | 0 | 0.004421357 | 0 | 0 |
| 0 | 0 | 0 | 0 | 0 |
| 0 | 0 | 0 | 0 | 0 |
| 0 | 0 | 0 | 0 | 0 |
| 0 | 0 | 0 | 0 | 0 |
| 0 | 0 | 0 | 0 | 0 |
| 0 | 0 | 0 | 0 | 0 |
| 0 | 0 | 0 | 0 | 0 |
| 0 | 0 | 0 | 0 | 0 |
| 0.063339893 | 0.003526453 | 2.330114076 | 0 | 0 |
| 0 | 0 | 0 | 0 | 0 |
| 0 | 0 | 0 | 0 | 0 |
| 0 | 0 | 0 | 0 | 0 |
| 0 | 0 | 0 | 0 | 0 |
| 0.008055321 | 0 | 0 | 0 | 0 |
| 0 | 0 | 0 | 0 | 0 |
| 0 | 0 | 0.093914972 | 0.008784452 | 0 |
| 0.009880685 | 0 | 0 | 0 | 0.000908065 |
| 0 | 0 | 0.047489074 | 0 | 0 |
| 0 | 0 | 0.000935902 | 0 | 0 |
| 0.665903749 | 0.001966019 | 0.00623185 | 0 | 0 |
| 0 | 0 | 0 | 0 | 0 |
| 0 | 0 | 0 | 0 | 0 |
| 0 | 0 | 3.412298731 | 0 | 0 |
| 0 | 0.053931325 | 0.191960205 | 0 | 0 |
| 0 | 0 | 0 | 0 | 0 |
| 0 | 0 | 0 | 0 | 0 |
| 0 | 0 | 0 | 0 | 0 |

| | | | | |
|---|---|---|---|---|
| 0 | 0 | 0 | 0 | 0 |
| 0 | 0 | 0.414994978 | 0 | 0 |
| 0 | 0 | 0.314393164 | 0 | 0 |
| 0 | 0 | 0.378163764 | 0 | 0 |
| 0 | 0 | 0.017743123 | 0 | 0 |
| 0 | 0 | 0.609451368 | 0 | 0 |
| 0 | 0 | 0 | 0 | 0 |
| 0 | 0 | 0 | 0 | 0 |
| 0.007713461 | 0.003130756 | 0.002181058 | 0.001408658 | 1.396187264 |
| 0 | 0 | 0 | 0 | 0 |
| 0 | 0 | 0.007641669 | 0 | 0 |
| 0 | 0 | 0 | 0 | 0 |
| 0 | 0 | 0 | 0 | 0 |
| 0 | 0 | 0 | 0 | 0 |
| 1.010876058 | 4.331641678 | 0.260496182 | 0.439000245 | 0 |
| 0 | 0.047702376 | 0 | 0 | 0.052605513 |
| 0 | 0 | 0 | 0 | 0 |
| 0 | 0 | 0 | 0 | 0 |
| 0 | 0 | 0 | 0 | 0 |
| 0 | 0 | 0.098994937 | 0 | 0 |
| 0 | 0 | 0.034355269 | 0 | 0 |
| 0 | 0 | 0 | 0 | 0 |
| 0 | 0 | 0 | 0 | 0 |
| 0.027710606 | 0 | 0 | 0 | 0 |
| 0 | 0.038232829 | 0.083358012 | 0.003865264 | 1.248405397 |
| 0 | 0.46659992 | 1.202700895 | 0.092380441 | 0 |
| 0 | 0 | 0.006487042 | 0 | 0 |
| 0 | 0 | 0.001910555 | 0 | 0 |
| 0 | 0 | 0 | 0 | 0 |
| 0 | 0 | 0 | 0 | 0 |
| 0 | 0 | 0 | 0 | 0 |
| 0 | 0 | 0.227285649 | 0 | 0 |
| 0 | 0 | 0 | 0 | 0 |
| 0.047798032 | 0 | 0 | 0 | 0 |
| 0.001064271 | 0.000996851 | 0.0006771 | 0.017978368 | 0.020087674 |
| 0.003761553 | 0 | 0.000797712 | 0 | 0 |
| 0 | 0 | 0.240050121 | 0 | 0 |
| 0.015503169 | 22.87086827 | 0.326073383 | 0 | 0 |
| 0 | 0.536636198 | 0 | 0 | 0 |
| 0 | 0 | 0 | 0 | 0 |
| 0 | 0 | 0 | 0 | 0 |
| 0.018555497 | 0.001398393 | 0 | 0 | 0 |
| 0 | 0.523347156 | 0.340543564 | 0 | 0 |
| 0 | 0 | 0.646271751 | 0 | 0 |
| 0 | 0 | 0.052610939 | 0 | 0 |
| 0.005701759 | 0 | 0.001058025 | 0 | 0 |
| 0 | 0 | 0 | 0 | 0 |
| 0.192742332 | 0.032437322 | 0.834111992 | 0.904864307 | 0 |

| | | | | |
|---|---|---|---|---|
| 0 | 0 | 0 | 0 | 0 |
| 0 | 0 | 0.101327203 | 0 | 0 |
| 0 | 0 | 0 | 0 | 0 |
| 0 | 0 | 0 | 0 | 0 |
| 0.013909383 | 0 | 0 | 0 | 0 |
| 0 | 0 | 0.007919185 | 0 | 0 |
| 0 | 0 | 0 | 0 | 0 |
| 0 | 0 | 0 | 0 | 0 |
| 0 | 0 | 0 | 0 | 0 |
| 0 | 0 | 0 | 0 | 0 |
| 0 | 0 | 0 | 0 | 0 |
| 10.57789249 | 0 | 0.528435504 | 5.183875784 | 0 |
| 0.004848404 | 0 | 0.189621873 | 0 | 0 |
| 0 | 0 | 0 | 0 | 0 |
| 0.018523504 | 0.187198008 | 0.001654012 | 1.459733066 | 0.279975754 |
| 0 | 0 | 0 | 0 | 0 |
| 0.039505065 | 0 | 0 | 0 | 0 |
| 0 | 0 | 0 | 0 | 0 |
| 0 | 0 | 0 | 0 | 0 |
| 0 | 0 | 0.007434805 | 0 | 0 |
| 0 | 0.100390446 | 0 | 0 | 0 |
| 0 | 0 | 0 | 0 | 0 |
| 0 | 0.015565262 | 0.13867638 | 0.00587999 | 0 |
| 0 | 0 | 0 | 0 | 0 |
| 0 | 0 | 0 | 0 | 0 |
| 0 | 0 | 0 | 0 | 0 |
| 0 | 0 | 0 | 0 | 0 |
| 0 | 0 | 0 | 0 | 0 |
| 0 | 0 | 0 | 0 | 0 |
| 0 | 0 | 0 | 0 | 0 |
| 0 | 0 | 0 | 0 | 0 |
| 0.005060387 | 0.161746237 | 0.045609129 | 0.477378483 | 0.030040283 |
| 0 | 0 | 2.139732241 | 0 | 0 |
| 0.137622459 | 0 | 0.012867126 | 0 | 0 |
| 0.004912215 | 0 | 0 | 0 | 0 |
| 0 | 0.6371191 | 0.049688056 | 0.474145082 | 0 |
| 0 | 0 | 0 | 0 | 0 |
| 0 | 0 | 0 | 0 | 0 |
| 0 | 0 | 0 | 0 | 0 |
| 0 | 0 | 0 | 0 | 0 |
| 0 | 0 | 0.009911092 | 0 | 0 |
| 0 | 0 | 0.002341449 | 0 | 0 |
| 0 | 0 | 0 | 0 | 0 |
| 0.001666248 | 0.002497109 | 0.062191561 | 0 | 0 |
| 0.681495876 | 0 | 0.05712578 | 0 | 0.022915483 |
| 0 | 0 | 0 | 0 | 0 |
| 0.748806689 | 0 | 0.137429481 | 0 | 0 |
| 0 | 0.000714485 | 0 | 0 | 0 |

| | | | | |
|---|---|---|---|---|
| 0 | 0 | 0 | 0 | 0 |
| 0 | 0 | 0 | 0 | 0 |
| 0 | 0.002734501 | 0 | 0 | 0 |
| 0 | 0 | 0 | 0 | 0 |
| 0 | 0 | 0 | 0 | 0 |
| 0 | 0.037303352 | 0 | 0 | 0 |
| 0 | 0 | 0.418648941 | 0 | 0 |
| 0 | 0 | 0.000501177 | 0 | 0 |
| 0 | 0 | 0 | 0 | 0 |
| 0 | 0 | 0 | 0 | 0 |
| 0 | 0 | 0 | 0 | 0 |
| 0 | 0 | 0 | 0 | 0 |
| 0 | 0 | 0 | 0 | 0 |
| 0 | 0 | 0.390666599 | 0 | 0 |
| 0 | 0 | 0 | 0 | 0 |
| 0 | 0 | 0 | 0 | 0 |
| 0 | 0 | 0.161245072 | 0 | 0 |
| 0 | 0 | 0.058324046 | 0 | 0 |
| 0.175584525 | 2.054724355 | 0.073716341 | 2.704638949 | 0 |
| 0 | 0 | 0 | 0 | 0 |
| 0 | 0 | 0 | 0 | 0 |
| 0 | 0 | 0 | 0 | 0 |
| 0 | 0 | 0 | 0 | 0 |
| 0 | 0 | 0 | 0 | 0 |
| 0 | 0 | 0 | 0 | 0 |
| 0.035288763 | 0 | 0 | 0.007541496 | 0.017981049 |
| 0 | 0 | 0 | 0 | 0 |
| 0.197506545 | 0.017552987 | 0.288014361 | 0.050832554 | 0 |
| 0 | 0 | 0 | 0 | 0 |
| 0.003419886 | 2.407394957 | 1.555817248 | 0.004371857 | 0 |
| 0 | 0 | 0 | 0 | 0 |
| 0 | 0 | 0 | 0 | 0 |
| 0 | 0 | 0.005759816 | 0 | 0 |
| 0 | 0 | 0 | 0 | 0 |
| 0.014162722 | 0 | 0 | 0 | 0 |
| 0 | 0 | 0 | 0 | 0 |
| 0 | 0 | 0 | 0 | 0 |
| 0.008258472 | 0 | 0 | 0 | 0 |
| 0 | 0 | 0 | 0 | 0 |
| 0 | 0 | 0.0171746 | 0 | 0 |
| 15.21479401 | 9.156811105 | 5.196227265 | 2.982982047 | 13.9372796 |
| 0.585571285 | 0.378301503 | 0.386821633 | 0 | 0 |
| 0 | 0 | 0 | 0 | 0 |
| 0 | 0 | 0.301755524 | 0 | 0 |
| 0 | 0 | 0 | 0 | 0 |
| 0 | 0 | 0 | 0 | 0 |
| 0 | 0 | 0 | 0 | 0 |
| 0.042337756 | 0 | 0.005559949 | 0 | 0 |

| | | | | |
|---|---|---|---|---|
| 0 | 0 | 0 | 0 | 0 |
| 0.00273313 | 0 | 0.098148115 | 0 | 0 |
| 0 | 0 | 0 | 0 | 0 |
| 0 | 0 | 0 | 0 | 0 |
| 0 | 0.294931281 | 7.335614259 | 0 | 0 |
| 0 | 0 | 0.107458488 | 0 | 0 |
| 0 | 0 | 0 | 0 | 0 |
| 0 | 0 | 0.002622046 | 0 | 0 |
| 0.004730441 | 0.511583868 | 0.002469377 | 0.044357377 | 1.983030572 |
| 0 | 0 | 0 | 0 | 0 |
| 0 | 0 | 0 | 0 | 0 |
| 0 | 0 | 0 | 0 | 0 |
| 0 | 0 | 0.055268165 | 0 | 0 |
| 0.042661562 | 0.012843964 | 0.006646949 | 0.011129995 | 0.017316543 |
| 0 | 0.03308426 | 0 | 0 | 0 |
| 0 | 0.284069028 | 0.269843174 | 0.021202057 | 0 |
| 0.001130111 | 0.00141136 | 0.012941782 | 0 | 0 |
| 0 | 0 | 0 | 0 | 0 |
| 0 | 0 | 0 | 0 | 0 |
| 0 | 0 | 0.050034521 | 0 | 0 |
| 0 | 0 | 0 | 0 | 0 |
| 0 | 0 | 0 | 0 | 0 |
| 0 | 0 | 0 | 0 | 0 |
| 0 | 0 | 0 | 0 | 0 |
| 0 | 0 | 0 | 0 | 0 |
| 0 | 0 | 0 | 0 | 0 |
| 0 | 0.002147662 | 0.050015163 | 0.027517193 | 0 |
| 0 | 0 | 0 | 0 | 0 |
| 0 | 0 | 0 | 0 | 0 |
| 0 | 0 | 0 | 0 | 0 |
| 0 | 0 | 1.869361006 | 0 | 0 |
| 0 | 0 | 0.000785697 | 0 | 0 |
| 0 | 0 | 0 | 0 | 0 |
| 0 | 0 | 0.015776142 | 0 | 0 |
| 0 | 0 | 0.183479894 | 0 | 0 |
| 0 | 0 | 0 | 0 | 0 |
| 0.001092562 | 0.001023349 | 0 | 0 | 0 |
| 0 | 0 | 0.002739905 | 0 | 0 |
| 0 | 0 | 0 | 0 | 0 |
| 0 | 0 | 0 | 0 | 0 |
| 0 | 6.749333615 | 0.337155132 | 0.176202724 | 0 |
| 0 | 0 | 0 | 0 | 0 |
| 0 | 0 | 0.016963034 | 0 | 0 |
| 0 | 0 | 0 | 0 | 0 |
| 0 | 0 | 0 | 0 | 0 |
| 0 | 0 | 0.000831215 | 0 | 0 |
| 0 | 0 | 0 | 0 | 0 |
| 0 | 0 | 0 | 0 | 0 |

| | | | | |
|---|---|---|---|---|
| 0 | 0 | 0.017164639 | 0 | 0 |
| 0 | 0 | 0 | 0 | 0 |
| 0 | 0 | 0.004208457 | 0 | 0 |
| 0 | 0.003921761 | 0 | 0 | 0 |
| 0 | 0 | 0.003228414 | 0 | 0 |
| 0 | 0 | 0 | 0 | 0 |
| 0.006200498 | 0.017071126 | 0.067739298 | 0 | 0 |
| 0 | 0 | 0 | 0 | 0 |
| 0 | 0 | 0 | 0 | 0 |
| 0.031146737 | 0.033605061 | 0 | 0 | 0 |
| 0 | 0 | 0.01410961 | 0 | 0 |
| 0 | 0 | 0.003205891 | 0 | 0 |
| 0 | 0 | 0 | 0 | 0 |
| 0 | 0 | 0 | 0 | 0 |
| 0 | 0 | 0 | 0 | 0 |
| 0 | 0 | 0 | 0 | 0 |
| 0 | 0 | 0 | 0 | 0.106163187 |
| 0 | 0.001691969 | 0 | 0 | 0 |
| 0 | 0 | 0 | 0 | 0 |
| 0 | 0 | 0 | 0 | 0 |
| 0.021824023 | 0.308633058 | 0.182549128 | 0 | 0 |
| 0 | 0 | 0 | 0 | 0 |
| 0 | 0.002217728 | 0.008201331 | 0 | 0 |
| 0.001052701 | 0 | 0.000744154 | 0 | 0 |
| 0 | 0 | 0 | 0 | 0 |
| 0 | 0 | 0 | 0 | 0 |
| 0 | 0 | 0 | 0 | 0 |
| 0 | 0 | 0 | 0 | 0 |
| 0 | 0 | 0 | 0 | 0 |
| 0 | 0.033585463 | 0 | 22.96615148 | 0.003689879 |
| 0 | 0 | 0.000840411 | 0 | 0 |
| 0 | 0 | 0 | 0 | 0 |
| 0 | 0 | 0 | 0 | 0 |
| 36.24431743 | 0 | 0.020503558 | 0 | 0 |
| 0 | 0 | 0 | 0 | 0 |
| 0 | 0 | 0 | 0 | 0 |
| 0 | 0.005346284 | 0.083242959 | 0 | 0 |
| 0 | 0 | 0 | 0 | 0 |
| 0 | 0 | 0 | 0 | 0 |
| 0 | 0 | 0 | 0 | 0 |
| 0 | 0 | 0 | 0 | 0 |
| 0 | 0 | 0 | 0 | 0 |
| 0 | 0 | 0 | 0 | 0 |
| 0 | 0 | 0.030565174 | 0 | 0 |
| 0 | 0 | 0.323945149 | 0 | 0 |
| 0 | 0 | 0 | 0 | 0 |
| 0 | 0 | 0.002474137 | 0 | 0 |
| 0 | 0 | 0.005502566 | 0 | 0 |

| | | | | |
|---|---|---|---|---|
| 0.15782889 | 0.001591478 | 0 | 0 | 0.001226097 |
| 0 | 0 | 0 | 0 | 0 |
| 0 | 0 | 0 | 0 | 0 |
| 0 | 0 | 0 | 0 | 0 |
| 0 | 0 | 0 | 0 | 0 |
| 0 | 0 | 0 | 0 | 0 |
| 0.0008708 | 0 | 0 | 0 | 0 |
| 0 | 0.060279688 | 0.424406704 | 0.285152313 | 0 |
| 0.019854452 | 0.461031463 | 2.848582592 | 0.005141182 | 2.546169884 |
| 0.016916568 | 0.004057846 | 0 | 0 | 0.027466081 |
| 0 | 0 | 0.003491106 | 0 | 0 |
| 0 | 0 | 0 | 0 | 0 |
| 0 | 0 | 0.114878486 | 0 | 0 |
| 0 | 0.011507129 | 0 | 0.005809321 | 0 |
| 0 | 0 | 0 | 0 | 0 |
| 0 | 0 | 0 | 0 | 0 |
| 0 | 0.050334466 | 0 | 0 | 0.030649828 |
| 0 | 0 | 0 | 0 | 0 |
| 0 | 0 | 0 | 0 | 0 |
| 0 | 0 | 0 | 0 | 0 |
| 0 | 0 | 0 | 0 | 0 |
| 0 | 0 | 0 | 0 | 0 |
| 0 | 0 | 0 | 0 | 0 |
| 0 | 0 | 0 | 0 | 0 |
| 0 | 0 | 0.119452807 | 0 | 0 |
| 0 | 0 | 0 | 0 | 0 |
| 0 | 0 | 0 | 0 | 0 |
| 0 | 0 | 0 | 0 | 0 |
| 0 | 0 | 0 | 0 | 0 |
| 0 | 0 | 0 | 0 | 0 |
| 0 | 0.00163497 | 0.000987142 | 0 | 0 |
| 0 | 0 | 0 | 0 | 0 |
| 0 | 0 | 0 | 0 | 0 |
| 0 | 0 | 0.003692873 | 0 | 0 |
| 0 | 0 | 0.003489853 | 0 | 0 |
| 0 | 0 | 0.001021702 | 0 | 0 |
| 0 | 0 | 0 | 0 | 0 |
| 0 | 0 | 0 | 0 | 0 |
| 0 | 0 | 0 | 0 | 0 |
| 0 | 0.008650096 | 0 | 0 | 0 |
| 0 | 0 | 0 | 0 | 0 |
| 0 | 0.279197135 | 0.920538894 | 0 | 0 |
| 0 | 0 | 0 | 0 | 0 |
| 0 | 0 | 0 | 0 | 0 |
| 0 | 0 | 0 | 0 | 0 |
| 0 | 0 | 0.007547381 | 0 | 0 |
| 0 | 0 | 0 | 0 | 0 |
| 0 | 0 | 0 | 0 | 0 |

| | | | | |
|---|---|---|---|---|
| 0 | 0 | 0 | 0 | 0 |
| 0 | 0 | 0.000722699 | 0.003422925 | 0 |
| 0 | 0 | 0 | 0 | 0 |
| 0 | 0 | 0 | 0 | 0 |
| 0 | 0.336941189 | 0.195822347 | 1.013052235 | 0 |
| 0 | 0 | 2.42806797 | 0 | 0 |
| 0 | 0 | 0 | 0.004905109 | 0 |
| 0.01561009 | 0.002474358 | 0.006519003 | 0 | 0.089486844 |
| 0 | 0 | 0 | 0 | 0 |
| 0 | 0 | 0.107241656 | 0 | 0 |
| 0 | 0 | 0 | 0 | 0 |
| 0 | 0 | 0.170756534 | 0 | 0 |
| 0 | 0 | 0 | 0 | 0 |
| 0 | 0 | 0 | 0 | 0 |
| 0 | 0 | 0 | 0 | 0 |
| 0.041032114 | 0.001365632 | 0.001060103 | 0 | 0 |
| 0 | 0 | 0 | 0 | 0 |
| 0 | 0 | 0 | 0 | 0 |
| 0 | 0 | 0 | 0 | 0 |
| 0 | 0 | 0 | 0 | 0 |
| 0 | 0 | 0 | 0 | 0 |
| 0 | 0 | 0.00150744 | 0 | 0 |
| 0 | 0 | 0 | 0 | 0 |
| 0.011430902 | 0 | 0 | 0 | 0.004049333 |
| 0 | 0 | 0.103056533 | 0.479698678 | 0 |
| 0 | 0 | 0 | 0 | 0 |
| 0.47274963 | 0.00249265 | 0 | 0.004860039 | 0.183481258 |
| 0 | 0 | 0 | 0 | 0 |
| 0 | 0 | 0 | 0 | 0 |
| 0 | 0 | 0 | 0 | 0 |
| 0 | 0 | 0 | 0 | 0 |
| 0 | 0 | 0 | 0 | 0 |
| 0 | 0 | 0 | 0 | 0 |
| 0 | 0 | 0 | 0 | 0 |
| 0 | 0 | 0 | 0 | 0 |
| 0 | 0 | 0 | 0 | 0 |
| 0 | 0 | 0 | 0 | 0 |
| 0 | 0 | 0.114435596 | 0 | 0 |
| 0 | 0 | 1.649141523 | 0 | 0 |
| 0 | 0 | 0 | 0 | 0 |
| 0 | 0 | 0 | 0 | 0 |
| 0 | 0 | 0 | 0 | 0 |
| 0 | 0 | 0 | 0 | 0 |
| 0 | 0 | 0.06767023 | 0 | 0 |
| 0 | 0 | 0 | 0 | 0 |
| 0 | 0 | 0 | 0 | 0 |
| 0 | 0 | 0 | 0 | 0 |
| 0 | 0 | 0 | 0 | 0 |

| | | | | |
|---|---|---|---|---|
| 0 | 0 | 0.043975578 | 0 | 0 |
| 0 | 0 | 0.002406875 | 0 | 0 |
| 0 | 0 | 0.000751039 | 0 | 0 |
| 0.067052469 | 0 | 0 | 0 | 0 |
| 0 | 0 | 0 | 0 | 0 |
| 0 | 0 | 0.070976186 | 0 | 0 |
| 0 | 0 | 0.000709024 | 0 | 0 |
| 0 | 0 | 0 | 0 | 0 |
| 0 | 0 | 0 | 0 | 0 |
| 0 | 0 | 0 | 0 | 0 |
| 0 | 0 | 0 | 0 | 0 |
| 0.00555612 | 0 | 0 | 0 | 0 |
| 0 | 0 | 0 | 0 | 0 |
| 0 | 0 | 0 | 0 | 0 |
| 0 | 0 | 0 | 0 | 0 |
| 0 | 0 | 0 | 0 | 0 |
| 0 | 0 | 0 | 0 | 0 |
| 0 | 0 | 0 | 0 | 0 |
| 0 | 0 | 0 | 0 | 0 |
| 0 | 0 | 0 | 0 | 0 |
| 0.00393235 | 0.071726242 | 0.000526695 | 0.000566952 | 0.583522693 |
| 0 | 0 | 0 | 0 | 0 |
| 0.012807752 | 0 | 0.033480576 | 0.001432037 | 0 |
| 0 | 0 | 0 | 0 | 0 |
| 0 | 0 | 0 | 0 | 0 |
| 0 | 0 | 0.024640879 | 0 | 0 |
| 0 | 0 | 0 | 0 | 0 |
| 0 | 0 | 0 | 0 | 0 |
| 0 | 0 | 0 | 0.364055615 | 0 |
| 0 | 0 | 0 | 0 | 0 |
| 0 | 0 | 0 | 0 | 0 |
| 0 | 0 | 0.203102874 | 0 | 0 |
| 0.026681436 | 0 | 0.002907032 | 0.002011646 | 0.015301284 |
| 0 | 0 | 0 | 0 | 0 |
| 0.040069307 | 0 | 0.505392077 | 0 | 0 |
| 0 | 0.008463319 | 0 | 0 | 0.003189257 |
| 0 | 0 | 0 | 0 | 0 |
| 0 | 0 | 0.011459957 | 0 | 0 |
| 0.002272009 | 0.001489656 | 0 | 0 | 0.164155161 |
| 0 | 0 | 0 | 0 | 0 |
| 0 | 0 | 0 | 0 | 0 |
| 0 | 0 | 0.128108178 | 0 | 0 |
| 0 | 0.019409232 | 0.063776619 | 0.011158865 | 0 |
| 0 | 0 | 0 | 0 | 0 |
| 0 | 0 | 0 | 0 | 0 |
| 0 | 0 | 0.011275374 | 0 | 0 |
| 0 | 0 | 0 | 0 | 0 |
| 0 | 0 | 0 | 0 | 0 |

| | | | | |
|---|---|---|---|---|
| 0 | 0 | 0 | 0 | 0 |
| 0 | 0 | 0 | 0 | 0 |
| 0 | 0 | 0 | 0 | 0 |
| 0 | 0 | 0 | 0 | 0 |
| 0 | 0 | 0 | 0 | 0 |
| 0 | 0 | 0 | 0 | 0 |
| 0 | 0 | 0 | 0 | 0 |
| 0 | 0.019860124 | 0 | 0.019733446 | 0 |
| 0.006523217 | 0.002777263 | 0.041585192 | 0 | 0 |
| 0 | 0.010380824 | 0 | 0.017461952 | 0 |
| 0 | 0 | 0 | 0 | 0 |
| 0 | 0 | 0 | 0 | 0 |
| 0 | 0 | 0 | 0 | 0 |
| 0 | 0 | 0 | 0 | 0 |
| 0 | 0 | 0 | 0 | 0 |
| 0.001496677 | 0.003204262 | 0 | 0 | 0.030549018 |
| 0 | 0 | 0 | 0 | 0 |
| 0 | 0 | 0 | 0 | 0 |
| 0 | 0 | 0 | 0 | 0 |
| 0 | 0.00251504 | 0.645666378 | 0.000490369 | 0 |
| 0 | 0.011085691 | 4.702404596 | 0 | 0 |
| 0 | 0 | 0 | 0 | 0 |
| 0 | 0 | 0 | 0 | 0 |
| 0 | 0 | 0.009180447 | 0 | 0 |
| 0 | 0 | 0 | 0 | 0 |
| 0 | 0 | 0.634272529 | 0 | 0 |
| 0 | 0 | 0 | 0 | 0 |
| 0 | 0 | 0 | 0 | 0 |
| 0 | 0 | 0 | 0 | 0 |
| 0 | 0 | 0.346630964 | 0 | 0 |
| 0.416897301 | 0.526563741 | 0.467006748 | 0 | 0 |
| 0 | 0 | 0 | 0 | 0 |
| 0 | 0 | 0.000560269 | 0 | 0 |
| 0 | 0 | 0 | 0 | 0 |
| 0 | 0 | 0 | 0 | 0 |
| 0 | 0 | 0 | 0 | 0 |
| 0 | 0 | 0 | 0 | 0 |
| 0 | 0 | 0 | 0 | 0 |
| 0 | 0 | 0.003488962 | 0 | 0 |
| 0 | 0 | 0 | 0 | 0 |
| 0 | 0 | 0 | 0 | 0 |
| 0 | 0 | 0 | 0 | 0 |
| 0 | 0 | 0.003188819 | 0 | 0 |
| 0 | 0 | 0 | 0 | 0 |
| 0 | 0 | 0 | 0 | 0 |
| 0 | 0 | 0 | 0 | 0 |
| 0 | 0.016605907 | 0 | 0.002997901 | 1.53769814 |
| 0 | 0 | 0 | 0 | 0 |

| | | | | |
|---|---|---|---|---|
| 0 | 0 | 0 | 0 | 0.001950675 |
| 0 | 0 | 0 | 0 | 0 |
| 0 | 0 | 0 | 0 | 0 |
| 0 | 0 | 0.00133121 | 0 | 0 |
| 0 | 0.037523876 | 0.025383647 | 0.000895862 | 0 |
| 0 | 0 | 0.014029785 | 0 | 0 |
| 0 | 0 | 0 | 0 | 0 |
| 0 | 0 | 0 | 0 | 0 |
| 0 | 0 | 0 | 0 | 0 |
| 0 | 0 | 0 | 0 | 0 |
| 0 | 0 | 0.032052298 | 0 | 0 |
| 0.143703668 | 0 | 0.010399131 | 0 | 0.008477668 |
| 0.021152996 | 0 | 0.001163015 | 0 | 0 |
| 0 | 0 | 0 | 0 | 0 |
| 0 | 0 | 0.039917834 | 0 | 0 |
| 0 | 0 | 0.14571213 | 0 | 0 |
| 0.005587371 | 0.001962531 | 0.008886851 | 0 | 0 |
| 0 | 0 | 0 | 0 | 0 |
| 0.298935584 | 0.279837321 | 0.012394702 | 0 | 0.011443169 |
| 0.010324706 | 0.082410732 | 0.015041326 | 0 | 0 |
| 0 | 0 | 0 | 0 | 0 |
| 0 | 0 | 0 | 0 | 0 |
| 0 | 0 | 0 | 0 | 0 |
| 0 | 0 | 0 | 0 | 0 |
| 0 | 0 | 0 | 0 | 0 |
| 0 | 0 | 0 | 0 | 0 |
| 0.035252987 | 0.001347745 | 0 | 0 | 0 |
| 0 | 0 | 0 | 0 | 0 |
| 0 | 0 | 0 | 0 | 0 |
| 0 | 0 | 0 | 0 | 0 |
| 0.013660176 | 0 | 0 | 0 | 0 |
| 0 | 0.000825068 | 0 | 0 | 0 |
| 0 | 0 | 0 | 0 | 0 |
| 0 | 0 | 0 | 0 | 0 |
| 0 | 0 | 0 | 0 | 0 |
| 0 | 0 | 0 | 0 | 0 |
| 0 | 0 | 0 | 0 | 0 |
| 0.156563985 | 0.002012523 | 0.005346426 | 0.000653985 | 0 |
| 0 | 0 | 0.001607664 | 1.250238392 | 0 |
| 0 | 0.001064128 | 0.001734713 | 0 | 0 |
| 0 | 0 | 0.002668728 | 0 | 0 |
| 0 | 0 | 0.001246151 | 0 | 0 |
| 0 | 0 | 0 | 0 | 0 |
| 0.004035797 | 0.006565495 | 0 | 0 | 0 |
| 0 | 0 | 0 | 0 | 0 |
| 0 | 0 | 0 | 0 | 0 |
| 0 | 0 | 0 | 0 | 0 |
| 0 | 0 | 0 | 0 | 0 |

| | | | | |
|---|---|---|---|---|
| 0 | 0 | 0 | 0 | 0 |
| 0 | 0 | 0.010010258 | 0 | 0 |
| 0 | 0 | 0 | 0 | 0 |
| 0 | 0 | 0 | 0 | 0 |
| 0 | 0 | 0 | 0 | 0 |
| 0 | 0 | 0 | 0.005109533 | 0 |
| 0 | 0 | 0.007513174 | 0 | 0 |
| 0 | 0.001956829 | 0 | 0 | 0 |
| 0 | 0 | 0 | 0 | 0 |
| 0 | 0 | 0.01789302 | 0 | 0 |
| 0 | 0 | 0 | 0 | 0 |
| 0 | 0 | 0 | 0 | 0 |
| 0.050968447 | 0.003990621 | 0.005354242 | 0 | 0 |
| 0 | 0 | 0 | 0 | 0 |
| 0 | 0 | 0 | 0 | 0 |
| 0 | 0 | 0.158599354 | 0 | 0 |
| 0 | 0 | 0 | 0 | 0 |
| 0 | 0 | 0 | 0 | 0 |
| 0 | 0 | 0 | 0 | 0 |
| 0 | 0 | 0 | 0 | 0 |
| 0 | 0 | 0 | 0 | 0 |
| 0 | 0 | 0 | 0 | 0 |
| 0.019739323 | 0.00447625 | 0.001233803 | 0 | 0.006241153 |
| 0 | 0.126771091 | 0.050749562 | 0 | 0 |
| 0 | 0 | 0 | 0 | 0 |
| 0 | 0 | 0.007219323 | 0 | 0 |
| 0 | 0 | 0 | 0 | 0 |
| 0 | 0 | 0 | 0 | 0 |
| 0 | 0 | 0 | 0 | 0 |
| 0 | 0 | 0 | 0 | 0 |
| 0 | 0.051645987 | 0.038519178 | 0 | 0 |
| 0 | 0 | 0 | 0 | 0 |
| 0 | 0 | 0 | 0 | 0 |
| 0 | 0 | 0 | 0 | 0 |
| 0 | 0.001455423 | 0.089466488 | 0 | 0 |
| 0 | 0 | 0 | 0 | 0.001769498 |
| 0 | 0 | 0 | 0 | 0 |
| 0 | 0 | 0 | 0 | 0 |
| 0 | 0.018726894 | 0 | 0.00501155 | 0.031029645 |
| 0 | 0 | 0 | 0 | 0 |
| 0 | 0 | 0 | 0 | 0 |
| 0 | 0 | 0 | 0 | 0 |
| 0 | 0 | 0 | 0 | 0 |
| 0 | 0 | 0 | 0 | 0 |
| 0 | 0 | 0 | 0 | 0 |
| 0.205572562 | 0.258940657 | 0.696508181 | 0.183459875 | 0 |
| 0 | 0.001345799 | 0 | 0 | 0 |
| 0 | 0 | 0 | 0 | 0 |

| | | | | |
|---|---|---|---|---|
| 0 | 0 | 0 | 0 | 0 |
| 0 | 0 | 0 | 0 | 0 |
| 0 | 0 | 0 | 0 | 0 |
| 0 | 0 | 0 | 0 | 0 |
| 0 | 0 | 0 | 0 | 0 |
| 0 | 0 | 0 | 0 | 0 |
| 0 | 0 | 0 | 0 | 0 |
| 0 | 0 | 0 | 0 | 0 |
| 0 | 0.000509628 | 0.002461577 | 0 | 0 |
| 0 | 0 | 0 | 0 | 0 |
| 0.008428066 | 0 | 0.003777019 | 0 | 0 |
| 0.00379868 | 0 | 0 | 0 | 0 |
| 0 | 0 | 0 | 0 | 0 |
| 0 | 0 | 0 | 0 | 0 |
| 0 | 0 | 0 | 0 | 0 |
| 0 | 0 | 0 | 0 | 0 |
| 0 | 0.001970502 | 0 | 0.001920991 | 0 |
| 0 | 0 | 0 | 0 | 0 |
| 0 | 0 | 0 | 0 | 0 |
| 0 | 0 | 0 | 0 | 0 |
| 0 | 0 | 0 | 0 | 0 |
| 0 | 0 | 0 | 0 | 0 |
| 0 | 0 | 0 | 0 | 0 |
| 0 | 0 | 0 | 0 | 0 |
| 0 | 0 | 0 | 0 | 0 |
| 0 | 0 | 0 | 0 | 0 |
| 0 | 0 | 0 | 0 | 0 |
| 0 | 0 | 0 | 0 | 0 |
| 0 | 0 | 0 | 0 | 0 |
| 0 | 0 | 0 | 0 | 0 |
| 0 | 0.00067879 | 0 | 0 | 0 |
| 0 | 0 | 0 | 0 | 0 |
| 0.056377059 | 0.038224973 | 0.006424055 | 0.033806995 | 0 |
| 0 | 0.003825638 | 0.003464696 | 0.009421932 | 0.001396101 |
| 0 | 0 | 0 | 0 | 0 |
| 0 | 0 | 0 | 0 | 0 |
| 0 | 0 | 0 | 0 | 0 |
| 0 | 0 | 0 | 0 | 0 |
| 0 | 0 | 0 | 0 | 0 |
| 0 | 0 | 0 | 0 | 0 |
| 0 | 0 | 0 | 0 | 0 |
| 0.336434555 | 0.01556157 | 0 | 0 | 0 |
| 0.457100217 | 0 | 0.033802833 | 0.271647328 | 0 |
| 0 | 0 | 0 | 0 | 0 |
| 0 | 0 | 0 | 0 | 0 |
| 0 | 0.009132059 | 0.038509361 | 0 | 0 |
| 0 | 0 | 0 | 0 | 0 |
| 0 | 0 | 0 | 0 | 0 |

| | | | | |
|---|---|---|---|---|
| 0 | 0 | 0 | 0 | 0 |
| 0 | 0.00889626 | 0.004309512 | 0 | 0 |
| 0.004128224 | 0.045675457 | 0.015320765 | 0.028036029 | 0.011310752 |
| 0 | 0 | 0 | 0 | 0 |
| 1.713193776 | 0 | 0.002271823 | 0.001128677 | 0 |
| 0.165838817 | 0 | 0.003572228 | 0.001831078 | 0 |
| 0 | 0 | 0 | 0 | 0 |
| 0 | 0 | 0 | 0 | 0 |
| 0 | 0 | 0 | 0 | 0 |
| 0 | 0 | 0 | 0 | 0 |
| 0 | 0 | 0 | 0 | 0 |
| 0 | 0 | 0 | 0 | 0 |
| 0 | 0 | 0 | 0 | 0 |
| 0 | 0.01198462 | 0 | 0 | 0 |
| 0 | 0 | 0 | 0 | 0.005799216 |
| 0 | 0 | 0 | 0 | 0 |
| 0 | 0 | 0 | 0 | 0 |
| 0 | 0 | 0 | 0 | 0 |
| 0 | 0 | 0 | 0 | 0 |
| 0 | 0 | 0 | 0 | 0 |
| 0 | 0 | 0 | 0 | 0 |
| 0 | 0 | 0 | 0 | 0 |
| 0 | 0 | 0 | 0 | 0 |
| 0 | 0 | 0 | 0 | 0 |
| 0 | 0 | 0.525982432 | 0 | 0 |
| 0 | 0 | 0 | 0.047268736 | 0 |
| 0 | 0 | 0 | 0 | 0 |
| 0 | 0.000971779 | 0.018129928 | 0 | 0 |
| 0 | 0 | 0 | 0 | 0 |
| 0 | 0 | 0 | 0 | 0 |
| 0 | 0 | 0 | 0 | 0 |
| 0 | 0 | 0 | 0 | 0 |
| 0 | 0 | 0 | 0 | 0 |
| 0 | 0 | 0 | 0 | 0 |
| 0 | 0 | 0 | 0 | 0 |
| 0 | 0 | 0 | 0 | 0 |
| 0 | 0 | 0 | 0 | 0 |
| 0 | 0.031711143 | 0 | 0 | 0 |
| 0.533894376 | 0 | 0.041516458 | 0 | 0 |
| 0 | 0 | 0 | 0 | 0 |
| 0 | 0 | 0 | 0 | 0 |
| 0 | 0 | 0 | 0 | 0 |
| 0.136108802 | 0.000804331 | 0 | 0 | 0 |
| 0 | 0 | 0 | 0 | 0 |
| 0 | 0 | 0 | 0 | 0 |
| 0 | 0 | 0 | 0 | 0 |
| 0.00262738 | 0 | 0.000835782 | 0 | 0 |
| 0 | 0 | 0 | 0 | 0 |

| | | | | |
|---|---|---|---|---|
| 0 | 0 | 0 | 0 | 0 |
| 0.017197962 | 0.000970391 | 0.006151855 | 0 | 0.011606531 |
| 0 | 0 | 0.033762464 | 0 | 0 |
| 0 | 0 | 0 | 0 | 0 |
| 0 | 0 | 0 | 0 | 0 |
| 0 | 0 | 0 | 0 | 0 |
| 0 | 0 | 0 | 0 | 0 |
| 0 | 0 | 0 | 0 | 0 |
| 0 | 0 | 0 | 0 | 0 |
| 0 | 0.000815275 | 0 | 0 | 0 |
| 0 | 0 | 0 | 0 | 0 |
| 0 | 0 | 0 | 0 | 0 |
| 0 | 0 | 0.017651151 | 0 | 0 |
| 0 | 0 | 0 | 0 | 0 |
| 0.001808468 | 0.007501574 | 0.001095777 | 0 | 0.001048666 |
| 0 | 0 | 0 | 0 | 0 |
| 0 | 0 | 0 | 0 | 0 |
| 0 | 0 | 0 | 0 | 0 |
| 0.012003805 | 0 | 0.005333735 | 0 | 0 |
| 0 | 0 | 0 | 0 | 0 |
| 0 | 0 | 0 | 0 | 0 |
| 0.004059562 | 0.024240258 | 0.011407074 | 0 | 0 |
| 0.012282801 | 0 | 0 | 0 | 0 |
| 0 | 0 | 0 | 0 | 0 |
| 0 | 0 | 0 | 0 | 0 |
| 0 | 0 | 0.006191421 | 0 | 0 |
| 0 | 0 | 0 | 0 | 0 |
| 0 | 0 | 0 | 0 | 0 |
| 0 | 0 | 0 | 0 | 0 |
| 0 | 0 | 0 | 0 | 0 |
| 0 | 0 | 0 | 0 | 0 |
| 0 | 0 | 0 | 0 | 0 |
| 0 | 0 | 0 | 0 | 0 |
| 0 | 0 | 0 | 0 | 0 |
| 0 | 1.78659714 | 6.800242176 | 0 | 0 |
| 0 | 0 | 0 | 0 | 0 |
| 0 | 0.001881358 | 0 | 0 | 0 |
| 0 | 0 | 0 | 0 | 0 |
| 0.001520237 | 0.001186609 | 0.024502135 | 0.002313589 | 0.000617072 |
| 0 | 0 | 0 | 0 | 0 |
| 0.282614651 | 0.062773181 | 0.382537443 | 0.034850567 | 0.147543038 |
| 0 | 0 | 0 | 0 | 0 |
| 0 | 0 | 0 | 0 | 0 |
| 0 | 0 | 0.576266244 | 0 | 0 |
| 0 | 0 | 0 | 0 | 0 |
| 0.207046089 | 0.027042994 | 0.174529802 | 0.081465618 | 0.353056881 |
| 0 | 0 | 0 | 0 | 0 |
| 0 | 0 | 0 | 0 | 0 |

| | | | | |
|---|---|---|---|---|
| 0 | 0 | 0 | 0 | 0 |
| 0 | 0 | 0.002515653 | 0 | 0 |
| 0 | 0 | 0 | 0 | 0 |
| 0 | 0 | 0.244219357 | 0 | 0 |
| 0.327274831 | 0 | 0 | 0 | 0 |
| 0 | 0 | 0 | 0 | 0 |
| 0 | 0 | 0 | 0 | 0 |
| 0 | 0 | 0.005246682 | 0 | 0.002282324 |
| 0 | 0.002310602 | 0.004334673 | 0 | 0 |
| 0 | 0.032404868 | 0 | 0.028803249 | 0 |
| 0 | 0 | 0 | 0 | 0 |
| 0 | 0 | 0 | 0 | 0 |
| 0.001740488 | 0.011877386 | 0.000632752 | 0 | 0 |
| 0 | 0 | 0 | 0 | 0 |
| 0 | 0 | 0 | 0 | 0 |
| 0 | 0 | 0 | 0 | 0 |
| 0 | 0 | 0 | 0 | 0 |
| 0 | 0 | 0 | 0 | 0 |
| 0 | 0 | 0 | 0 | 0 |
| 1.701195398 | 0.108826447 | 0.000563194 | 1.341812001 | 0 |
| 0 | 0 | 0 | 0 | 0 |
| 0 | 0 | 0 | 0 | 0 |
| 0 | 0 | 0 | 0 | 0 |
| 0 | 0 | 0 | 0 | 0 |
| 0 | 0 | 0 | 0 | 0 |
| 0 | 0 | 0.009756278 | 0 | 0 |
| 0 | 0 | 0 | 0 | 0 |
| 0 | 0 | 0 | 0 | 0 |
| 0 | 0 | 0 | 0 | 0 |
| 0 | 0 | 0 | 0 | 0 |
| 0 | 0 | 0 | 0 | 0 |
| 0 | 0 | 0.020331429 | 0 | 0 |
| 0 | 0 | 0 | 0 | 0 |
| 0 | 0 | 0 | 0 | 0 |
| 0 | 0 | 0 | 0 | 0 |
| 0 | 0 | 0.001458082 | 0 | 0 |
| 0 | 0 | 0 | 0 | 0 |
| 0 | 0 | 0 | 0 | 0 |
| 0 | 0 | 0 | 0 | 0 |
| 0 | 0 | 0 | 0 | 0 |
| 0 | 0 | 0 | 0 | 0 |
| 0 | 0 | 0 | 0 | 0 |
| 0 | 0.022744438 | 0.335194496 | 0 | 0 |
| 0 | 0 | 0 | 0 | 0 |
| 0.150075429 | 0 | 0.003173833 | 0.00360622 | 0.001012461 |
| 0 | 0 | 0 | 0 | 0 |
| 0 | 0 | 0 | 0 | 0 |
| 0 | 0 | 0 | 0 | 0 |

| | | | | |
|---|---|---|---|---|
| 0 | 0 | 0 | 0 | 0 |
| 0 | 0 | 0 | 0 | 0 |
| 0 | 0 | 0 | 0 | 0 |
| 0 | 0 | 0 | 0 | 0 |
| 0 | 0 | 0 | 0 | 0 |
| 0.015983138 | 0.007412639 | 0.001711226 | 0 | 0.00642465 |
| 0 | 0 | 0 | 0 | 0 |
| 0 | 0 | 0 | 0 | 0 |
| 0 | 0 | 0 | 0 | 0 |
| 0 | 0 | 0 | 0 | 0 |
| 0 | 0 | 0 | 0 | 0 |
| 0 | 0 | 0 | 0 | 0 |
| 0 | 0 | 0 | 0 | 0 |
| 0 | 0 | 0.013127232 | 0 | 0 |
| 0 | 0 | 0 | 0 | 0 |
| 0 | 0 | 0 | 0 | 0 |
| 0 | 0 | 0.163182338 | 0 | 0 |
| 0 | 0.019332127 | 0 | 0 | 0 |
| 0 | 0 | 0.002653131 | 0 | 0 |
| 0 | 0 | 0 | 0 | 0 |
| 0 | 0 | 0 | 0 | 0 |
| 0 | 0 | 0 | 0 | 0 |
| 0 | 0 | 0 | 0 | 0 |
| 0 | 0 | 0 | 0 | 0 |
| 0 | 0 | 0 | 0 | 0 |
| 0 | 0 | 0 | 0.003546128 | 0 |
| 0 | 0 | 0 | 0 | 0 |
| 0 | 0 | 0.020957425 | 0 | 0 |
| 0 | 0 | 0 | 0 | 0 |
| 0 | 0 | 0 | 0 | 0 |
| 0 | 0 | 0 | 0 | 0 |
| 0 | 0 | 0 | 0 | 0 |
| 0 | 0 | 0 | 0 | 0 |
| 0 | 0 | 0 | 0 | 0 |
| 0 | 0 | 0 | 0 | 0 |
| 0 | 0 | 0 | 0 | 0 |
| 0 | 0 | 0 | 0 | 0 |
| 0 | 0 | 0 | 0 | 0 |
| 0 | 0 | 0 | 0 | 0 |
| 0 | 0 | 0 | 0 | 0 |
| 0 | 0 | 0 | 0 | 0 |
| 0 | 0 | 0 | 0 | 0 |
| 0 | 0 | 0 | 0 | 0 |
| 0 | 0 | 0 | 0 | 0 |
| 0 | 0 | 0 | 0 | 0 |
| 0 | 0 | 0 | 0 | 0 |
| 0 | 0 | 0 | 0 | 0 |
| 0 | 0 | 0 | 0 | 0 |

| | | | | |
|---|---|---|---|---|
| 0 | 0 | 0 | 0 | 0 |
| 0 | 0 | 0.004863325 | 0.001903653 | 0 |
| 0 | 0 | 0.00396944 | 0 | 0 |
| 0 | 0 | 0 | 0 | 0 |
| 0 | 0 | 0 | 0 | 0 |
| 0 | 0 | 0 | 0 | 0 |
| 0.002593908 | 0 | 0 | 0 | 0 |
| 0 | 0 | 0 | 0 | 0 |
| 0 | 0 | 0 | 0 | 0 |
| 0 | 0 | 0 | 0 | 0 |
| 0 | 0 | 0 | 0.002077235 | 0 |
| 0 | 0 | 0 | 0 | 0 |
| 0 | 0 | 0.04775737 | 0 | 0 |
| 0 | 0 | 0 | 0 | 0 |
| 0 | 0 | 0 | 0 | 0 |
| 0 | 0 | 0 | 0 | 0 |
| 0 | 0 | 0 | 0 | 0 |
| 0 | 0 | 0 | 0 | 0 |
| 0 | 0 | 0 | 0 | 0 |
| 0 | 0 | 0 | 0 | 0 |
| 0.001385323 | 0 | 0.001370998 | 0 | 0.023804434 |
| 0 | 0 | 0 | 0 | 0 |
| 0 | 0 | 0 | 0 | 0 |
| 0.013197408 | 5.133012724 | 0.740174292 | 0.435748994 | 0 |
| 0 | 0 | 0 | 0 | 0 |
| 0 | 0 | 0.010404772 | 0 | 0 |
| 0 | 0 | 0 | 0 | 0 |
| 0 | 0 | 0.05106299 | 0 | 0 |
| 0 | 0 | 0 | 0 | 0 |
| 0 | 0 | 0 | 0 | 0 |
| 0 | 0 | 0.004229135 | 0 | 0 |
| 0 | 0 | 0 | 0 | 0 |
| 0 | 0 | 0 | 0.001108849 | 0.000788661 |
| 0 | 0 | 0 | 0 | 0 |

| PHL018-0009_V1 | PHL018-0055_V2 | PHL018-0003_V1 | PHL018-0130_V2 | PHL018-0189_V2 |
|---|---|---|---|---|
| EYCF | EYCF | CTRL | REF | EYCF |
| 0.099228151 | 1.566319314 | 0.417643892 | 0.202415032 | 0.115491228 |
| 2.231969728 | 0.110882609 | 0.365190601 | 0.723025603 | 0.791950631 |
| 4.667916553 | 0 | 0 | 0 | 0 |
| 0 | 0 | 0 | 0 | 0 |
| 0 | 0 | 0 | 0 | 0 |
| 0 | 0 | 0 | 0 | 0 |
| 0 | 0 | 0 | 0.110106838 | 0 |
| 0 | 0 | 0 | 0 | 0 |
| 0 | 0 | 0 | 0 | 0 |
| 0 | 0 | 0 | 0 | 0 |
| 0 | 0 | 0 | 0 | 0 |
| 0 | 0 | 0 | 0 | 0 |
| 0 | 0 | 0 | 0 | 0 |
| 1.212360122 | 0.681418937 | 0.68544311 | 0.179650378 | 0 |
| 0 | 0 | 0 | 0 | 0 |
| 0 | 0 | 0 | 0 | 0 |
| 0 | 0 | 0 | 0 | 0 |
| 0 | 0 | 0 | 0 | 0 |
| 0 | 0.015758634 | 0 | 0 | 0 |
| 0 | 0 | 0 | 0 | 0 |
| 0 | 0 | 0 | 0 | 0 |
| 0 | 0 | 0 | 0 | 0 |
| 0 | 0 | 0 | 0 | 0 |
| 0 | 0 | 0 | 0 | 0 |
| 0 | 0 | 0.362347697 | 0.026926962 | 0 |
| 0 | 0 | 0 | 0 | 0 |
| 0 | 0 | 0 | 0 | 0 |
| 0 | 0 | 0 | 0 | 0 |
| 0 | 0 | 0 | 0 | 0 |
| 0 | 0 | 0 | 0 | 0 |
| 0 | 0 | 0 | 0 | 0 |
| 0 | 0 | 0 | 0 | 0 |
| 0 | 0 | 0 | 0 | 0 |
| 0 | 0 | 0 | 0 | 0 |
| 0 | 0 | 0 | 0 | 0 |
| 0 | 3.594759573 | 2.199595994 | 0 | 0 |
| 0 | 0 | 0 | 0 | 0 |
| 0 | 0 | 0 | 0 | 0 |
| 0 | 0 | 0 | 0 | 0 |
| 0 | 0 | 0 | 0 | 0 |
| 0 | 0 | 0 | 0 | 0 |
| 0.089428451 | 0.00488008 | 0.014927981 | 0 | 0.036993819 |
| 0.054053107 | 1.768793253 | 0.342112268 | 0.848321931 | 13.97282659 |
| 0.132263919 | 0.041283655 | 0.117925281 | 0.013770742 | 0.163386623 |
| 0.002072992 | 0.001311192 | 0 | 0.001378919 | 0.005727098 |
| 0 | 0 | 0 | 0 | 0 |

| | | | | |
|---|---|---|---|---|
| 0 | 0 | 0.002163574 | 0 | 0 |
| 0 | 0.001402893 | 0 | 0 | 0.013230124 |
| 0.003056497 | 2.230221215 | 0 | 0.056114402 | 0 |
| 0.732806818 | 0.340913346 | 0.780670818 | 0.37321882 | 0.167534539 |
| 0 | 0 | 0.005085677 | 0 | 0 |
| 0 | 0 | 0.051152592 | 0 | 0 |
| 0.008914903 | 0.00100245 | 1.210442781 | 0 | 0 |
| 0 | 0 | 0.076951354 | 0 | 0 |
| 0 | 0 | 0 | 0.032230448 | 0 |
| 0 | 0 | 0 | 0 | 0 |
| 0 | 0 | 0 | 0 | 0 |
| 0.01817537 | 1.696299444 | 0.402591486 | 0 | 0.059869915 |
| 0 | 0 | 0 | 0 | 0 |
| 0.043036754 | 0 | 0.003264474 | 0.00943758 | 0 |
| 0 | 0 | 0 | 0 | 0 |
| 0 | 0 | 0 | 0 | 0 |
| 0 | 0.00639728 | 2.467875517 | 0.000776275 | 0.01563211 |
| 0 | 0 | 0 | 0 | 0 |
| 0 | 0 | 0 | 0 | 0 |
| 0 | 0 | 0 | 0 | 0 |
| 0.008245756 | 0.086862438 | 0.031045892 | 0.000598357 | 0.0165678 |
| 0.001173025 | 0.055023161 | 0.003562664 | 0.010674179 | 0 |
| 0.022366625 | 0 | 0 | 0 | 0 |
| 0 | 0 | 0 | 0 | 0 |
| 0 | 0 | 0 | 0 | 0 |
| 0 | 0 | 0 | 0 | 0 |
| 0 | 0.00233882 | 0 | 0 | 0 |
| 0 | 0 | 0 | 0 | 0 |
| 0.00480204 | 0 | 0.002486006 | 0 | 0.00293978 |
| 0 | 0 | 0 | 0 | 0 |
| 0 | 0.007263765 | 0.002018446 | 0 | 0 |
| 0 | 0 | 0 | 0 | 0 |
| 0 | 0 | 0 | 0 | 0 |
| 0 | 0 | 0 | 0 | 0 |
| 0 | 0 | 0 | 0 | 0 |
| 0 | 0 | 0 | 0 | 0 |
| 0 | 0 | 0 | 0 | 0 |
| 0 | 0 | 0.055375885 | 0.047686815 | 0 |
| 0 | 0 | 0 | 0 | 0 |
| 0 | 0 | 0 | 0 | 0 |
| 0 | 0 | 0 | 0 | 0 |
| 0 | 0 | 0 | 0 | 0 |
| 0 | 0 | 0 | 0 | 0 |
| 0 | 0 | 0 | 0 | 0 |
| 0 | 0 | 0 | 0 | 0 |
| 0 | 0 | 0 | 0 | 0 |
| 0 | 0.0033023 | 0.035237332 | 0 | 0 |
| 0.002133242 | 0 | 0.009653053 | 0 | 0 |

| | | | | |
|---|---|---|---|---|
| 0 | 0 | 0 | 0 | 0 |
| 0 | 0 | 0 | 0 | 0 |
| 0 | 0 | 0 | 0 | 0 |
| 0 | 0 | 0 | 0 | 0 |
| 0 | 0 | 0 | 0 | 0 |
| 0 | 0 | 0 | 0 | 0 |
| 0 | 0 | 0 | 0 | 0 |
| 0 | 0 | 0.021745254 | 0 | 0 |
| 0 | 0 | 0 | 0 | 0 |
| 0 | 0 | 0 | 0 | 0 |
| 0 | 0 | 0 | 0 | 0 |
| 0.004738834 | 0 | 0 | 0 | 0 |
| 0 | 0 | 0 | 0 | 0 |
| 0 | 0 | 0 | 0 | 0 |
| 0 | 0 | 0 | 0 | 0 |
| 0 | 0 | 0 | 0 | 0 |
| 0 | 0 | 0 | 0 | 0 |
| 0 | 0 | 0 | 0 | 0 |
| 0.017103659 | 2.220442193 | 0.401566956 | 0.912284922 | 0 |
| 0 | 0 | 0.118220166 | 0 | 0 |
| 0 | 0 | 0 | 0 | 0 |
| 0 | 0 | 0 | 0 | 0 |
| 0 | 0 | 0 | 0 | 0 |
| 0 | 0 | 0 | 0 | 0 |
| 0 | 0 | 0 | 0 | 0 |
| 0 | 0 | 0 | 0 | 0 |
| 0 | 0 | 0 | 0 | 0 |
| 0 | 0 | 0 | 0 | 0 |
| 0 | 0 | 0 | 0 | 0 |
| 0 | 0 | 0 | 0 | 0 |
| 0.003986975 | 0.095244765 | 0.027230956 | 0.105524437 | 0 |
| 0 | 0.001002542 | 0.028415689 | 0 | 0 |
| 0 | 0 | 0 | 0 | 0 |
| 0 | 0 | 0 | 0 | 0 |
| 0 | 0 | 0 | 0 | 0 |
| 0 | 0 | 0 | 0 | 0 |
| 0 | 0 | 0 | 0 | 0 |
| 0 | 0 | 0 | 0 | 0 |
| 0 | 0 | 0 | 0 | 0 |
| 0 | 0 | 0 | 0 | 0 |
| 0 | 0 | 0 | 0 | 0 |
| 0 | 0 | 0 | 0 | 0 |
| 0 | 0 | 0.000878183 | 0.075961591 | 0 |
| 0 | 0 | 0 | 0 | 0 |
| 0 | 0 | 0 | 0 | 0 |
| 0 | 0 | 0 | 0 | 0 |
| 0 | 0 | 0 | 0 | 0 |
| 0 | 0 | 0 | 0 | 0 |

| | | | | |
|---|---|---|---|---|
| 0 | 0 | 0 | 0 | 0 |
| 0 | 0 | 0.077566414 | 0 | 0 |
| 0 | 0 | 0 | 0 | 0 |
| 0 | 0 | 0 | 0 | 0 |
| 0.006514675 | 0 | 0.156860498 | 0.012388575 | 0.003753641 |
| 0 | 0 | 0 | 0 | 0 |
| 0 | 0 | 0 | 0 | 0 |
| 0 | 0 | 0 | 0 | 0 |
| 0 | 0 | 0 | 0 | 0 |
| 0 | 0 | 0 | 0 | 0 |
| 0 | 0 | 0 | 0 | 0 |
| 0 | 0 | 0 | 0 | 0 |
| 0 | 0 | 0 | 0 | 0 |
| 0 | 0 | 0 | 0.00209894 | 0 |
| 0 | 0 | 0 | 0 | 0 |
| 0.029593707 | 4.326308574 | 0 | 0 | 0 |
| 0 | 0 | 0 | 0 | 0 |
| 0 | 0 | 0 | 0 | 0 |
| 0 | 0 | 0 | 0 | 0 |
| 0 | 0 | 0 | 0 | 0 |
| 0 | 0 | 0 | 0 | 0 |
| 0 | 0 | 0 | 0 | 0 |
| 0 | 0 | 0.005999076 | 0.001790276 | 0 |
| 0 | 0 | 0 | 0 | 0 |
| 0 | 0 | 0.007661683 | 0 | 0 |
| 0 | 0 | 0 | 0 | 0 |
| 0 | 0 | 0.010233994 | 0.004390245 | 0 |
| 0 | 0 | 0 | 0 | 0 |
| 0 | 0 | 0 | 0 | 0 |
| 0 | 0.011956345 | 0 | 0 | 0 |
| 0 | 0 | 0 | 0.021189527 | 0 |
| 0 | 0 | 0 | 0 | 0 |
| 0 | 0 | 0 | 0 | 0 |
| 0 | 0 | 0 | 0 | 0 |
| 0 | 0 | 0 | 0 | 0 |
| 0 | 0 | 0 | 0 | 0 |
| 0 | 0 | 0 | 0 | 0 |
| 0 | 0 | 0 | 0 | 0 |
| 0 | 0 | 0 | 0 | 0 |
| 0 | 0 | 0 | 0 | 0 |
| 0 | 0 | 0 | 0 | 0 |
| 0 | 0 | 0 | 0 | 0 |
| 0 | 0 | 0 | 0 | 0 |
| 0 | 0 | 0 | 0 | 0 |
| 0 | 0 | 0 | 0 | 0 |
| 0 | 0 | 0 | 0 | 0 |
| 0 | 0 | 0 | 0 | 0 |
| 0 | 0 | 0 | 0 | 0 |

| | | | | |
|---|---|---|---|---|
| 0 | 0 | 0 | 0 | 0 |
| 0 | 0 | 0 | 0 | 0 |
| 0 | 0 | 0 | 0 | 0 |
| 0 | 0 | 0 | 0 | 0 |
| 0 | 0 | 0 | 0 | 0 |
| 0 | 0 | 0 | 0 | 0 |
| 0 | 0 | 0 | 0 | 0 |
| 0 | 0 | 0 | 0 | 0 |
| 0 | 0 | 0 | 0 | 0 |
| 0 | 0 | 0 | 0 | 0 |
| 0 | 0 | 0 | 0 | 0 |
| 0 | 0 | 0.003403729 | 0 | 0 |
| 0 | 0.011806563 | 0.000671071 | 0 | 0 |
| 0 | 0 | 0 | 0 | 0 |
| 0 | 0 | 0 | 0 | 0 |
| 0 | 0 | 0 | 0 | 0 |
| 0 | 0 | 0 | 0 | 0 |
| 0.002046108 | 0 | 0 | 0 | 0 |
| 0 | 0 | 0 | 0 | 0 |
| 0 | 0 | 0 | 0 | 0 |
| 0.034537661 | 0.020695721 | 0.019134814 | 0.015960796 | 0.044513147 |
| 0 | 0 | 0 | 0 | 0 |
| 0 | 0 | 0 | 0.00047743 | 0 |
| 0.002369463 | 0.00287753 | 0 | 0.007565409 | 0 |
| 0 | 0 | 0 | 0 | 0 |
| 0 | 0 | 0 | 0 | 0 |
| 0.026994998 | 0.003816688 | 0 | 0.004013833 | 0.025125764 |
| 0 | 0 | 0 | 0 | 0 |
| 0 | 0 | 0 | 0 | 0 |
| 0 | 0 | 0 | 0 | 0 |
| 0.376995826 | 0 | 0 | 0 | 0 |
| 0 | 0 | 0 | 0 | 0 |
| 0 | 0 | 0 | 0 | 0 |
| 0 | 0 | 0.030097698 | 0.001330125 | 0 |
| 0.008870543 | 0.021320754 | 0.003061507 | 0.024782254 | 0.032750046 |
| 0 | 0 | 0 | 0 | 0 |
| 0 | 0 | 0 | 0 | 0 |
| 0 | 0 | 0 | 0 | 0 |
| 0 | 0 | 0 | 0 | 0 |
| 0 | 0 | 0 | 0 | 0 |
| 0 | 0 | 0 | 0 | 0 |
| 0 | 0 | 0 | 0 | 0 |
| 0 | 0 | 0 | 0 | 0 |
| 0 | 0 | 0 | 0 | 0 |
| 0 | 0 | 0 | 0 | 0 |
| 0 | 0 | 0.005829777 | 0.321346946 | 0 |
| 0.002241036 | 0.000728991 | 0.00066296 | 0 | 0 |
| 0 | 0 | 0 | 0 | 0 |

| | | | | |
|---|---|---|---|---|
| 0 | 0 | 0 | 0 | 0 |
| 0 | 0 | 0 | 0 | 0 |
| 0.002310541 | 0 | 0 | 0 | 0 |
| 0 | 0 | 0 | 0 | 0 |
| 0 | 0 | 0 | 0 | 0 |
| 0 | 0 | 0 | 0 | 0 |
| 0 | 0 | 0 | 0 | 0 |
| 0 | 0 | 0 | 0 | 0 |
| 0 | 0 | 0 | 0.033806856 | 0 |
| 0 | 0 | 0 | 0 | 0 |
| 0 | 0 | 0 | 0 | 0 |
| 0 | 0 | 0 | 0 | 0 |
| 0 | 0 | 0 | 0 | 0 |
| 0 | 0 | 0 | 0 | 0 |
| 0 | 0 | 0 | 0 | 0 |
| 0 | 0 | 0 | 0 | 0 |
| 0 | 0 | 0 | 0 | 0 |
| 0 | 0 | 0 | 0 | 0.095968828 |
| 0 | 0.007807295 | 0.004585494 | 0 | 0.057158163 |
| 0 | 0 | 0.109333033 | 0.025090488 | 0 |
| 0.042358072 | 0 | 0 | 0 | 0.006668913 |
| 0 | 0 | 0 | 0 | 0 |
| 0 | 0 | 0 | 0 | 0 |
| 0 | 0 | 0 | 0 | 0 |
| 0.013188198 | 0.004766682 | 0.002167461 | 0.003007738 | 0.010883275 |
| 0 | 0 | 0 | 0 | 0 |
| 0 | 0 | 0 | 0 | 0 |
| 0 | 0 | 0 | 0 | 0 |
| 0.051241734 | 0 | 0 | 0 | 0 |
| 0 | 0 | 0 | 0 | 0 |
| 0.074145773 | 0.365893287 | 0.031285666 | 0.076253551 | 0.179508571 |
| 0 | 0 | 0 | 0 | 0 |
| 0 | 0 | 0 | 0 | 0.374113067 |
| 0 | 0.080342914 | 0 | 0 | 0 |
| 0 | 0 | 0 | 0 | 0 |
| 0 | 0 | 0 | 0 | 0 |
| 0 | 0 | 0 | 0 | 0 |
| 0 | 0 | 0 | 0.001297498 | 0 |
| 0 | 0 | 0 | 0 | 0 |
| 0 | 0 | 0 | 0 | 0 |
| 0 | 0 | 0 | 0 | 0 |
| 0.006396993 | 0 | 0.010957261 | 0.000756475 | 0.008925812 |
| 0 | 0 | 0 | 0 | 0 |
| 0 | 0 | 0 | 0 | 0 |
| 0 | 0 | 0 | 0 | 0 |
| 0 | 0 | 0 | 0 | 0 |
| 0 | 0 | 0 | 0 | 0 |
| 0 | 0 | 0 | 0 | 0 |

| | | | | |
|---|---|---|---|---|
| 0 | 0 | 0 | 0 | 0 |
| 0 | 0 | 0 | 0 | 0 |
| 0.134767119 | 0.041096715 | 0.106783504 | 0.001187349 | 0.131804026 |
| 0 | 0 | 0 | 0 | 0 |
| 0 | 0 | 0 | 0.772604422 | 0 |
| 0 | 0 | 0 | 0 | 0 |
| 0 | 0 | 0 | 0 | 0 |
| 0 | 0 | 0 | 0 | 0 |
| 0 | 0 | 0 | 0 | 0 |
| 0 | 0 | 0 | 0 | 0 |
| 0 | 0 | 0 | 0 | 0 |
| 0 | 0 | 0 | 0 | 0 |
| 0 | 0 | 0 | 0 | 0 |
| 0.003616693 | 0.003244235 | 0 | 0.01627171 | 0.003220534 |
| 0.004613347 | 0.001853786 | 0.00917864 | 0.014296629 | 0.007360974 |
| 0 | 0 | 0.014038494 | 0 | 0 |
| 0.01607869 | 0.000704074 | 0.005122402 | 0.005183095 | 0.004892516 |
| 0 | 0 | 0 | 0 | 0 |
| 0 | 0 | 0 | 0 | 0 |
| 0 | 0 | 0 | 0 | 0 |
| 0 | 0 | 0 | 0 | 0 |
| 0 | 0 | 0 | 0 | 0 |
| 0 | 0 | 0 | 0 | 0 |
| 0.019471793 | 0.002955874 | 0 | 0 | 0 |
| 0 | 0 | 0 | 0 | 0 |
| 0 | 0 | 0 | 0 | 0 |
| 0 | 0 | 0 | 0.870197232 | 6.132134193 |
| 0 | 0 | 0 | 0 | 0 |
| 0 | 0 | 0 | 0 | 0 |
| 0 | 0 | 0 | 0 | 0 |
| 0 | 0 | 0 | 0 | 0 |
| 0 | 0 | 0 | 0 | 0 |
| 0 | 0 | 0 | 0 | 0 |
| 0 | 0 | 0 | 0 | 0 |
| 0 | 0 | 0 | 0 | 0 |
| 0 | 0 | 0.001924149 | 0 | 0.003780609 |
| 0 | 0 | 0 | 0 | 0 |
| 0.012624873 | 0.090234826 | 0.006535869 | 0 | 0.042541813 |
| 0 | 0 | 0 | 0 | 0 |
| 0 | 0 | 0 | 0 | 0 |
| 0 | 0 | 0 | 0 | 0 |
| 0 | 0 | 0 | 0 | 0 |
| 0 | 0 | 0 | 0 | 0 |
| 0 | 0 | 0 | 0 | 0 |
| 0 | 0 | 0 | 0 | 0 |
| 0 | 0 | 0 | 0 | 0 |
| 0 | 0 | 0 | 0 | 0 |
| 0 | 0.00161205 | 0 | 0 | 0 |

| | | | | |
|---|---|---|---|---|
| 0 | 0 | 0.05373632 | 0.332832155 | 0 |
| 0 | 0 | 0 | 0 | 0 |
| 0 | 0.001109531 | 0 | 0 | 0.003744847 |
| 0 | 0 | 0 | 0 | 0 |
| 0.066476411 | 0 | 0 | 0.02324432 | 0.011081403 |
| 0 | 0 | 0 | 0 | 0 |
| 0 | 0 | 0 | 0 | 0 |
| 0.032984539 | 0.00281652 | 0.00711501 | 0 | 0.065549365 |
| 0 | 0 | 0 | 0 | 0 |
| 0 | 0.154184139 | 0 | 0.021710226 | 0.796348603 |
| 0 | 0.003208005 | 0 | 0 | 0 |
| 0.004569369 | 0 | 0.011039244 | 0 | 0 |
| 0.00643691 | 0.00413406 | 0.04836215 | 0.01324041 | 0.002051929 |
| 0 | 0 | 0 | 0.005102225 | 0.030596903 |
| 0 | 0 | 0 | 0 | 0 |
| 0 | 0 | 0 | 0 | 0 |
| 0 | 0 | 0 | 0 | 0 |
| 0 | 0 | 0 | 0 | 0 |
| 0 | 0 | 0 | 0 | 0 |
| 0 | 0 | 0 | 0 | 0 |
| 0 | 0 | 0 | 0 | 0 |
| 0 | 0 | 0 | 0 | 0 |
| 0.003831957 | 0 | 0 | 0 | 0 |
| 0 | 0 | 0.119142304 | 0 | 0 |
| 0 | 0 | 0 | 0 | 0 |
| 0 | 0 | 0 | 0 | 0.021292499 |
| 0 | 0 | 0 | 0 | 0 |
| 0 | 0.005028804 | 0 | 0 | 0 |
| 0 | 0 | 0 | 0 | 0 |
| 0 | 0.000900756 | 0 | 0 | 0 |
| 0 | 0 | 0 | 0 | 0 |
| 0 | 0 | 0 | 0 | 0 |
| 0 | 0 | 0 | 0 | 0 |
| 0 | 0 | 0 | 0 | 0 |
| 0 | 0 | 0 | 0 | 0 |
| 0 | 0 | 0 | 0 | 0 |
| 0 | 0 | 0 | 0 | 0 |
| 0 | 0 | 0 | 0 | 0 |
| 0 | 0 | 0 | 0 | 0 |
| 0 | 0.004143501 | 0 | 0 | 0 |
| 0 | 0 | 0 | 0 | 0 |
| 0 | 0 | 0 | 0 | 0 |
| 0 | 0 | 0 | 0 | 0 |
| 0 | 0 | 0 | 0.013166014 | 0 |
| 0 | 0 | 0 | 0 | 0 |
| 0 | 0 | 0.11548364 | 0 | 0 |
| 0.003585984 | 0.012248159 | 0.010148627 | 0.002289923 | 0.002431736 |
| 0 | 0.00790565 | 0 | 0 | 0 |

| | | | | |
|---|---|---|---|---|
| 0 | 0.044247824 | 0 | 0 | 0 |
| 0.002003794 | 0 | 0.016044498 | 0 | 0 |
| 0 | 0 | 0 | 0 | 0 |
| 0 | 0 | 0 | 0 | 0 |
| 0 | 0 | 0 | 0 | 0 |
| 0 | 0 | 0 | 0 | 0 |
| 0.003768689 | 0 | 0 | 0 | 0 |
| 0 | 0 | 0 | 0 | 0.0019267 |
| 0.001373867 | 0 | 0 | 0 | 0 |
| 0 | 0 | 0 | 0 | 0 |
| 0.018344618 | 0.005967355 | 0 | 0.005491139 | 0.035789387 |
| 0 | 0 | 0 | 0 | 0.001850638 |
| 0 | 0 | 0 | 0 | 0 |
| 0.004951735 | 0.000939609 | 0 | 0 | 0.000932745 |
| 0 | 0.004278222 | 0 | 0 | 0 |
| 0 | 0 | 0 | 0 | 0 |
| 0 | 0 | 0 | 0.000517648 | 0 |
| 0 | 0 | 0 | 0 | 0 |
| 0 | 0.019251344 | 0 | 0 | 0 |
| 0 | 0 | 0 | 0.033922051 | 0 |
| 0 | 0 | 0.050177584 | 0 | 0 |
| 0 | 0 | 0.09153853 | 0 | 0.214077626 |
| 0 | 0 | 0 | 0 | 0 |
| 0 | 0 | 0 | 0 | 0 |
| 0 | 0 | 0 | 0 | 0 |
| 0 | 0 | 0 | 0 | 0 |
| 0 | 0 | 0 | 0 | 0 |
| 0 | 0 | 0 | 0 | 0 |
| 0 | 0 | 0 | 0 | 0 |
| 1.047903569 | 0.460352728 | 0.307180349 | 0.852752077 | 0.683262746 |
| 0 | 0.018167335 | 0.007246386 | 0.000837971 | 0.005062348 |
| 0 | 0 | 0 | 0 | 0 |
| 0 | 0 | 0 | 0 | 0 |
| 0 | 0 | 0 | 0 | 0 |
| 0 | 0 | 0 | 0 | 0 |
| 0 | 0 | 0 | 0 | 0 |
| 0 | 0 | 0.213567194 | 0 | 0 |
| 0 | 0 | 0 | 0 | 0 |
| 0 | 0 | 0 | 0 | 0 |
| 0 | 0 | 0 | 0 | 0 |
| 0 | 0.000579346 | 0 | 0.019496672 | 0 |
| 0 | 0.685608441 | 0.748117654 | 0.289072665 | 0.154322038 |
| 0 | 0 | 0 | 0 | 0 |
| 0 | 0 | 0 | 0 | 0 |
| 0 | 0 | 0 | 0 | 0 |
| 0 | 0 | 0 | 0 | 0 |
| 0.010948704 | 0.019660772 | 0 | 0.026431577 | 0 |
| 0 | 0 | 0 | 0 | 0 |

| | | | | |
|---|---|---|---|---|
| 0.002306391 | 0 | 0 | 0 | 0.000744769 |
| 0 | 0 | 0 | 0.023526988 | 0 |
| 0 | 0.006474399 | 5.608743166 | 0 | 0.033488577 |
| 0 | 0 | 0 | 0 | 0 |
| 0 | 0.399382433 | 0 | 0 | 0 |
| 0 | 0 | 0 | 0 | 0 |
| 0 | 0 | 0 | 0 | 0 |
| 0.001846426 | 0 | 0 | 0 | 0 |
| 0.003584736 | 0 | 0 | 0 | 0 |
| 0 | 0 | 0 | 0 | 0 |
| 0 | 0.097904882 | 0 | 0 | 0 |
| 0 | 0 | 0 | 0 | 0 |
| 0 | 0 | 0 | 0 | 0 |
| 0.036365307 | 0.004072395 | 0 | 0.028075789 | 0 |
| 0 | 0 | 0 | 0 | 0 |
| 0 | 0.026520888 | 0.023475495 | 0.068425373 | 0 |
| 0.00774574 | 0.068589824 | 0.010336767 | 0.002679215 | 0.008754267 |
| 0.00125477 | 0 | 0 | 0 | 0.001654503 |
| 0 | 0 | 0 | 0.001064395 | 0 |
| 0 | 0 | 0 | 0 | 0 |
| 0 | 0 | 0 | 0 | 0 |
| 0 | 0.002045845 | 0 | 0.002390577 | 0.006092697 |
| 0 | 0 | 0 | 0 | 0 |
| 0 | 0 | 0 | 0 | 0 |
| 0 | 0 | 0 | 0 | 0 |
| 0.161217628 | 0 | 0 | 0 | 0 |
| 0 | 0 | 0 | 0.001831461 | 0 |
| 0 | 0 | 0 | 0.004139383 | 0 |
| 0 | 0 | 0 | 0 | 0 |
| 0 | 0 | 0 | 0 | 0 |
| 0 | 0.030272249 | 0 | 0.005173335 | 0 |
| 0.011520686 | 0.005077373 | 0.014365467 | 0.035597575 | 0.013720765 |
| 1.880341807 | 0.005371496 | 0.0153876 | 0.04236713 | 0.075184786 |
| 0.002028388 | 0 | 0.007350644 | 0 | 0 |
| 0 | 0.014834679 | 0.038224423 | 0 | 0 |
| 0 | 0 | 0 | 0 | 0 |
| 0.002559215 | 0 | 0 | 0 | 0.002169326 |
| 0.036025647 | 0.027533871 | 0.001899578 | 0.033549118 | 0 |
| 1.399634712 | 0 | 0.371642724 | 0 | 0 |
| 0 | 0 | 0 | 0 | 0 |
| 0 | 0 | 0 | 0 | 0 |
| 0 | 0 | 0 | 0 | 0 |
| 0 | 0 | 0 | 0 | 0.00902092 |
| 0.002538833 | 0.379917551 | 0 | 0.036477845 | 0.004708758 |
| 0.009936487 | 0 | 0 | 0 | 0 |
| 0 | 0 | 0 | 0 | 0 |
| 0 | 0.008862429 | 0 | 0 | 0 |
| 0 | 0 | 0 | 0 | 0 |

| | | | | |
|---|---|---|---|---|
| 0 | 0 | 0 | 0.001480423 | 0 |
| 0.548507138 | 0.341805488 | 0.794643585 | 3.261814651 | 3.919641805 |
| 0 | 0 | 0.011788749 | 0 | 0 |
| 0 | 0 | 0 | 0 | 0 |
| 0 | 0 | 0 | 0 | 0 |
| 0 | 0 | 0 | 0 | 0 |
| 0.028383872 | 0 | 0 | 0 | 0 |
| 0 | 0 | 0 | 0 | 0 |
| 0 | 0 | 0 | 0 | 0 |
| 0 | 0 | 0 | 0 | 0 |
| 0.001773279 | 0.006345164 | 0.007868765 | 0.001011047 | 0.001526984 |
| 0.112068662 | 0.054416588 | 0.011981209 | 0.087347055 | 0.328663275 |
| 0 | 0 | 0.217497146 | 0.690848796 | 0 |
| 0 | 0 | 0 | 0 | 0 |
| 0 | 0 | 0 | 0 | 0.002628035 |
| 0 | 0.00261621 | 0 | 0 | 0 |
| 0.043769285 | 0.00402685 | 0.01666257 | 0.012281065 | 0.037375991 |
| 0.002410485 | 0.006011519 | 0 | 0.005222549 | 0 |
| 0 | 0.003627972 | 0 | 0 | 0 |
| 0 | 0.003903566 | 0 | 0 | 0 |
| 0.012583697 | 0.021178762 | 0.066467321 | 0.076644338 | 0.084920639 |
| 0 | 0 | 0 | 0 | 0 |
| 0 | 0 | 0 | 0 | 0 |
| 0 | 0 | 0 | 0 | 0 |
| 0 | 0 | 1.386133489 | 0 | 0 |
| 0 | 0 | 0 | 0 | 0 |
| 0 | 0.012706655 | 0 | 0.012500866 | 0 |
| 0.02849374 | 0.043695684 | 0.027696031 | 0.15526445 | 0.075361134 |
| 0 | 0 | 0 | 0 | 0 |
| 0 | 0 | 0 | 0 | 0 |
| 0.007487463 | 0 | 0 | 0 | 0 |
| 0 | 0.00502609 | 0 | 0 | 0 |
| 0 | 0 | 0 | 0 | 0 |
| 0 | 0 | 0 | 0 | 0 |
| 0 | 0 | 0.560530424 | 0 | 0 |
| 0.770599109 | 0.061626948 | 0.153868609 | 0.41399957 | 0.32664669 |
| 0 | 0 | 0 | 0 | 0 |
| 0 | 0 | 0 | 0 | 0 |
| 0.013988351 | 0.010889599 | 0.007675005 | 0.008589061 | 0 |
| 0 | 0.008396656 | 0 | 0 | 0 |
| 0.011703684 | 0 | 0 | 0 | 0 |
| 0 | 0.000947195 | 0 | 0 | 0 |
| 0.01546842 | 0 | 0 | 0 | 0.023802426 |
| 0 | 0.000798273 | 0.00043558 | 0.001846914 | 0 |
| 0 | 0 | 0 | 0 | 0 |
| 4.270087364 | 0.001154693 | 0.836481342 | 0.215457959 | 0 |
| 0 | 0 | 0 | 0 | 0 |
| 0 | 0 | 0 | 0 | 0 |

| | | | | |
|---|---|---|---|---|
| 0 | 0.01178178 | 0 | 0.005830751 | 0 |
| 0 | 0 | 0 | 0 | 0 |
| 0 | 0 | 0 | 0 | 0 |
| 0 | 2.014931348 | 0.12085364 | 0.485699076 | 0 |
| 0 | 0 | 0 | 0 | 0 |
| 0 | 0 | 0 | 0.001405088 | 0 |
| 0 | 0 | 0 | 0.001099192 | 0 |
| 0 | 0 | 0.005484172 | 0 | 0 |
| 0 | 0 | 0.599017904 | 0 | 0 |
| 0 | 0.017901809 | 0.415929987 | 0.019188543 | 0 |
| 0 | 0 | 0 | 0 | 0 |
| 0.00437605 | 0 | 0.012250329 | 0 | 0 |
| 0 | 0 | 0 | 0 | 0 |
| 0 | 0 | 0.266581245 | 0.188659862 | 0 |
| 0.006018546 | 0 | 0.025618705 | 0.11465841 | 0.003023194 |
| 0 | 0 | 0 | 0 | 0 |
| 0 | 0.049855028 | 0.15263603 | 0.057234563 | 0 |
| 0 | 0 | 0 | 0 | 0 |
| 0 | 0 | 0 | 0 | 0 |
| 0 | 0.00675629 | 0.005844591 | 0 | 0 |
| 0 | 0 | 0 | 0 | 0 |
| 0 | 0 | 0.002804433 | 0.011891151 | 0 |
| 0 | 0 | 0.027196775 | 0.000536085 | 0 |
| 0.026088312 | 0 | 0 | 0.378383597 | 0.009901175 |
| 0 | 0 | 0 | 0 | 0 |
| 0.030964937 | 0 | 0.379711896 | 0.636645577 | 0 |
| 0 | 0 | 0.272563331 | 0.008214875 | 0 |
| 0 | 0 | 1.009154179 | 1.279214676 | 0 |
| 0 | 0 | 0 | 0 | 0 |
| 0 | 0 | 0 | 0 | 0 |
| 0.001890785 | 0 | 0.00570999 | 0.004150472 | 0.036506627 |
| 0 | 0 | 0.038581354 | 0 | 0 |
| 0 | 0.22814199 | 0.000744981 | 0.001005077 | 0 |
| 0 | 0 | 0 | 0 | 0 |
| 0 | 0 | 0.016267772 | 0 | 0 |
| 0 | 0 | 0.243593255 | 0 | 0 |
| 0.463276304 | 0.41017343 | 0.963814791 | 0.160933719 | 0 |
| 0 | 0 | 0 | 0 | 0 |
| 0 | 0 | 0 | 0 | 0 |
| 0 | 0 | 0.000600579 | 0.007639599 | 0 |
| 0.007696403 | 0.104619181 | 0.001448877 | 0.005398765 | 0 |
| 0.00202559 | 0 | 0.179527809 | 0.168154114 | 0 |
| 0.030496153 | 1.033546159 | 0.513127385 | 0.004689865 | 0 |
| 0 | 0 | 0.002243842 | 0 | 0 |
| 0 | 0 | 0 | 0 | 0 |
| 0 | 0 | 0 | 3.492602908 | 0 |
| 0 | 0 | 0 | 0 | 0 |
| 0.001682174 | 0.012980744 | 0.030576794 | 0.266087312 | 0 |

| | | | | |
|---|---|---|---|---|
| 0 | 0 | 0 | 0 | 0 |
| 0 | 0 | 1.270812617 | 0 | 0 |
| 0.443452378 | 0.058953672 | 0.000556544 | 0.041189527 | 0.01437762 |
| 0 | 0.03109128 | 0 | 0.000927581 | 0 |
| 0 | 0 | 0.017680189 | 0 | 0 |
| 0.000635154 | 0 | 0.017975375 | 0 | 0 |
| 0 | 0 | 0 | 0 | 0 |
| 0 | 0 | 0 | 0.027230808 | 0 |
| 0 | 0 | 0 | 0 | 0 |
| 0 | 0 | 0 | 0 | 0 |
| 0.023484759 | 0.337077828 | 0.041528479 | 0.477707084 | 0.051096932 |
| 0.253165432 | 0.754244505 | 0.492048915 | 0.935549324 | 1.865590606 |
| 0 | 0 | 0 | 0 | 0 |
| 0.03246529 | 0.014914665 | 0.003734936 | 0.002955155 | 0.009655896 |
| 0.490582383 | 0.89648935 | 0.001858909 | 0.254015698 | 0.028576833 |
| 0 | 0 | 0 | 0 | 0 |
| 0 | 0 | 0 | 0 | 0 |
| 0 | 0 | 0 | 0 | 0 |
| 0 | 0 | 0.097625375 | 0 | 0 |
| 0 | 0.017158048 | 0 | 0.005552097 | 0 |
| 0.422425176 | 0.14671612 | 0.049615214 | 0.237620563 | 0.052658577 |
| 0.014215398 | 1.338629169 | 0 | 0 | 4.79952198 |
| 0.198033138 | 3.967042119 | 0.002595477 | 2.087310289 | 8.781152139 |
| 0 | 0 | 0 | 0 | 0 |
| 0 | 0 | 0 | 0 | 0 |
| 0 | 0 | 2.33059323 | 0 | 0 |
| 0 | 0 | 0 | 0 | 0.282622256 |
| 0 | 0 | 0.00079612 | 0 | 0 |
| 0 | 0 | 0 | 0 | 0 |
| 0 | 0 | 0 | 0 | 0 |
| 0 | 0 | 0.012797066 | 0.002531323 | 0 |
| 0.201071248 | 0.079556813 | 0.118902974 | 0.108166037 | 0 |
| 0 | 0 | 0 | 0.109458781 | 0 |
| 0 | 0 | 0.023822814 | 0 | 0 |
| 0 | 0 | 0 | 0.0011422 | 0 |
| 0.038091561 | 0.29826034 | 0.01645447 | 0.005884924 | 0.024441993 |
| 0 | 0 | 0.000819319 | 0.004358313 | 0 |
| 0 | 0 | 0 | 0 | 0 |
| 0.020705722 | 0 | 0.635019713 | 0 | 0 |
| 0 | 0 | 0 | 0 | 0 |
| 0 | 0 | 0.019716974 | 0.019000563 | 0 |
| 0 | 0 | 0 | 0 | 0 |
| 0 | 0 | 0.037707067 | 0 | 0 |
| 0 | 0 | 0 | 0 | 0 |
| 0.001121009 | 0.657928995 | 0.027082718 | 0.012079958 | 0.024917057 |
| 0 | 0 | 0 | 0 | 0 |
| 0 | 0 | 0.306352782 | 0 | 0 |
| 0.029749352 | 1.598831007 | 0 | 0 | 0.01502839 |

| | | | | |
|---|---|---|---|---|
| 0 | 0 | 0 | 0 | 0 |
| 0 | 0 | 0 | 0 | 0 |
| 0 | 0 | 0 | 0 | 0 |
| 0.002627436 | 0 | 0.006594997 | 0 | 0 |
| 0 | 0 | 0 | 0 | 0 |
| 0 | 0 | 0.276816962 | 0.260668763 | 0 |
| 0 | 0 | 6.471885657 | 0 | 0 |
| 0.156272451 | 0 | 0.229544469 | 0.052983702 | 0.021197683 |
| 0 | 0 | 0 | 0 | 0 |
| 0 | 0 | 0 | 0 | 0 |
| 0 | 0.023422296 | 0.002387151 | 0.005945687 | 0 |
| 0 | 0 | 0 | 0 | 0 |
| 0 | 0 | 0.061096329 | 0 | 0 |
| 0 | 0 | 0.796006551 | 0 | 0 |
| 0 | 0 | 0 | 0 | 0 |
| 0 | 0 | 0 | 0 | 0 |
| 0.465150348 | 0 | 0.097511091 | 0.172774871 | 0 |
| 0.007086696 | 0.006891721 | 0.003821633 | 0.004242556 | 0 |
| 0 | 0 | 0.003056242 | 0.056361717 | 0 |
| 0 | 0 | 0.029915444 | 0.002072462 | 0 |
| 0 | 0 | 0.003259411 | 0 | 0 |
| 0 | 0 | 0 | 0 | 0 |
| 0 | 0.057751389 | 0.305076211 | 0 | 0 |
| 0 | 0 | 0 | 0 | 0 |
| 0 | 0 | 0 | 0 | 0 |
| 0 | 0 | 0 | 0 | 0 |
| 0 | 0 | 0 | 0 | 0 |
| 0 | 0 | 0 | 0 | 0 |
| 0 | 0 | 0 | 0 | 0 |
| 0 | 0 | 0.442814491 | 2.127076151 | 0.012497262 |
| 0 | 0.055024936 | 0.018222968 | 0 | 0 |
| 0 | 0 | 0 | 0 | 0 |
| 0 | 0 | 0.043443353 | 0 | 0 |
| 0 | 0 | 0 | 0 | 0 |
| 0 | 0 | 0 | 0 | 0 |
| 0 | 0 | 0 | 0 | 0 |
| 0 | 0 | 0.045892885 | 0.016879142 | 0 |
| 0 | 0.03435607 | 0 | 0 | 0.140153226 |
| 0 | 0 | 0 | 0 | 0 |
| 0.000666358 | 0 | 0 | 0 | 0 |
| 0 | 0 | 0.008458979 | 0.071678073 | 0 |
| 0 | 0 | 0 | 0 | 0 |
| 0 | 0 | 0.001226644 | 0 | 0 |
| 0 | 0 | 0.015095404 | 0.001396504 | 0 |
| 0 | 0.315193043 | 0.095312413 | 0 | 0 |
| 0 | 0 | 0 | 0 | 0 |
| 0 | 0 | 0.011224998 | 0 | 0 |
| 0 | 0 | 0 | 0 | 0 |

| | | | | |
|---|---|---|---|---|
| 0 | 0 | 0 | 0 | 0 |
| 0 | 0 | 0.058242913 | 0 | 0 |
| 0 | 0 | 0 | 0 | 0 |
| 0 | 0 | 0.753930568 | 0.085646697 | 0 |
| 0 | 0 | 0 | 0 | 0 |
| 0 | 0 | 0.99569422 | 0 | 0 |
| 0 | 0 | 0 | 0 | 0 |
| 0 | 0 | 0 | 0 | 0 |
| 0.012733804 | 0.022159101 | 0.002786973 | 0.005454055 | 0.011466635 |
| 0 | 0 | 0 | 0 | 0 |
| 0 | 0 | 0.000919741 | 0 | 0 |
| 0 | 0 | 0.001437678 | 0 | 0 |
| 0 | 0 | 0 | 0 | 0 |
| 0 | 0 | 0 | 0 | 0 |
| 0 | 0.193799863 | 1.745356412 | 1.172087105 | 0.342557992 |
| 0 | 0.003088689 | 0 | 0 | 0 |
| 0 | 0 | 0 | 0 | 0 |
| 0 | 0 | 0 | 0 | 0 |
| 0 | 0 | 0 | 0 | 0 |
| 0 | 0 | 0 | 0 | 0 |
| 0 | 0 | 0.099503195 | 0.036585845 | 0 |
| 0 | 0 | 0 | 0 | 0 |
| 0 | 0 | 0 | 0 | 0 |
| 0 | 0 | 0 | 0 | 0 |
| 0.001460936 | 0.054334685 | 0.047900472 | 0.054808951 | 0.495346642 |
| 0 | 0.189696041 | 9.607059853 | 18.81952647 | 3.166257458 |
| 0.016029756 | 0 | 0.051948109 | 0.088265673 | 0 |
| 0 | 0 | 0 | 0 | 0 |
| 0 | 0 | 0.17060064 | 0 | 0 |
| 0 | 0 | 0.431855962 | 0 | 0 |
| 0 | 0 | 0 | 0 | 0 |
| 0 | 0 | 0.103260186 | 0 | 0 |
| 0 | 0 | 0 | 0 | 0 |
| 0.028868638 | 0.070176692 | 0 | 0.001556995 | 0.002498499 |
| 0.00353534 | 0.004878862 | 0.00221847 | 0.006413588 | 0.029543638 |
| 0.001363122 | 0.071216971 | 0.002665922 | 0.086320304 | 0.000855892 |
| 0 | 0 | 0 | 0 | 0 |
| 0.810902043 | 0.660659573 | 0.73433295 | 1.921357365 | 6.027750324 |
| 0 | 10.21526119 | 0 | 0 | 0 |
| 0 | 0 | 0 | 0 | 0 |
| 0 | 0 | 0 | 0 | 0 |
| 0 | 0 | 0 | 0 | 0 |
| 7.468307407 | 0.03228266 | 0.220510476 | 0.07359999 | 2.226903138 |
| 0 | 0 | 0.030916591 | 0 | 0 |
| 0 | 0 | 0 | 0 | 0 |
| 0.008178784 | 0 | 0.001337096 | 0 | 0 |
| 0.021595908 | 0 | 0.002026177 | 0 | 0 |
| 0 | 0.035395047 | 0.067454907 | 0.85695735 | 0.34748935 |

| | | | | |
|---|---|---|---|---|
| 0 | 0 | 0 | 0 | 0 |
| 0 | 0 | 0 | 0 | 0 |
| 0 | 0 | 0.005871505 | 0 | 0 |
| 0 | 0 | 0 | 0 | 0 |
| 0 | 0.004057696 | 0 | 0.007315352 | 0 |
| 0 | 0 | 0 | 0 | 0 |
| 0 | 0 | 0.025320107 | 0.086060933 | 0 |
| 0 | 0 | 0 | 0.003742398 | 0 |
| 0 | 0 | 0 | 0 | 0 |
| 0 | 0 | 0 | 0 | 0 |
| 0 | 0 | 0 | 0 | 0 |
| 0.444352612 | 3.445787039 | 0.31288086 | 0.448987921 | 0 |
| 2.314895478 | 0 | 0 | 0.000738134 | 0.590149064 |
| 0 | 0 | 0 | 0 | 0 |
| 1.953424989 | 0.000670389 | 0.072753505 | 0 | 0.315664919 |
| 0 | 0 | 0 | 0 | 0 |
| 0 | 0 | 0 | 0 | 0 |
| 0 | 0 | 0 | 0 | 0 |
| 0 | 0.201544667 | 0 | 0.000617945 | 0 |
| 0 | 0.011727859 | 0 | 0.003426011 | 0 |
| 0.460827454 | 0.061295695 | 0.182272753 | 0 | 0 |
| 0 | 0 | 0 | 0 | 0 |
| 0.482016886 | 0.088750494 | 0.045725019 | 0.531967981 | 0.115137763 |
| 0 | 0 | 0 | 0 | 0 |
| 0 | 0 | 0 | 0 | 0 |
| 0 | 0 | 0 | 0 | 0 |
| 0 | 0 | 0 | 0 | 0 |
| 0 | 0 | 0 | 0 | 0 |
| 0.000795573 | 0 | 0.00109831 | 0 | 0 |
| 0 | 0 | 0 | 0 | 0 |
| 0 | 0 | 0 | 0 | 0 |
| 0.183379684 | 0.035086927 | 0.053532941 | 0.346121343 | 0.05382911 |
| 0 | 0 | 0.663651812 | 0 | 0 |
| 0.030919499 | 0.469077117 | 0.094065577 | 0.005941616 | 0.004170432 |
| 0 | 0.065116618 | 0.00307185 | 0.080321041 | 0.002794276 |
| 0.003505894 | 0.159661448 | 0.092399692 | 0 | 0 |
| 0.000658392 | 0 | 0.00075744 | 0 | 0 |
| 0 | 0 | 0.033780107 | 0 | 0 |
| 0 | 0 | 0.000588184 | 0 | 0 |
| 0 | 0 | 0 | 0 | 0 |
| 0 | 0 | 0.22682636 | 0.140398624 | 0 |
| 0 | 0 | 0.249052893 | 0 | 0 |
| 0 | 0 | 0 | 0 | 0 |
| 0 | 0.122979226 | 0 | 0.098645393 | 0 |
| 0.003653289 | 0.020334592 | 0.017315902 | 0.072514385 | 0.005872302 |
| 0 | 0 | 0 | 0 | 0 |
| 0 | 0 | 0.356026197 | 0.243393583 | 0 |
| 0 | 0 | 0 | 0 | 0 |

| | | | | |
|---|---|---|---|---|
| 0 | 0 | 0 | 0 | 0 |
| 0 | 0 | 0.017316801 | 0 | 0 |
| 0 | 0.000955957 | 0 | 0.002211738 | 0.000759179 |
| 0 | 0 | 0 | 0 | 0 |
| 0 | 0 | 0 | 0 | 0 |
| 0 | 0 | 0 | 0 | 0 |
| 0 | 0 | 1.22436271 | 0 | 0 |
| 0.014511331 | 0.00668081 | 0.009524024 | 0.551817614 | 0 |
| 0 | 0 | 0 | 0 | 0 |
| 0 | 0 | 0.000969518 | 0 | 0 |
| 0 | 0.421043062 | 0 | 0 | 0 |
| 0 | 0 | 0 | 0 | 0 |
| 0 | 0 | 0 | 0 | 0 |
| 0 | 0.210891381 | 0.160197025 | 0 | 0 |
| 0 | 0 | 0 | 0 | 0 |
| 0 | 0 | 0 | 0 | 0 |
| 0 | 0 | 0 | 0 | 0 |
| 0 | 0 | 0.062263001 | 0 | 0 |
| 0 | 7.361336756 | 0.011333165 | 3.690208085 | 0 |
| 0 | 0 | 0 | 0 | 0 |
| 0 | 0 | 0.012006456 | 0 | 0 |
| 0 | 0 | 0 | 0 | 0 |
| 0 | 0 | 0 | 0 | 0 |
| 0 | 0 | 0 | 0 | 0 |
| 0 | 0.000807902 | 0 | 0 | 0 |
| 0 | 0.038033571 | 0 | 0 | 0.008542018 |
| 0 | 0 | 0 | 0 | 0 |
| 0.002441248 | 0.017013382 | 0.011795738 | 0.315327492 | 0.002006625 |
| 0 | 0 | 0.105711041 | 0 | 0 |
| 1.02738612 | 0.30430103 | 0.924170331 | 0.661142111 | 1.039453258 |
| 0 | 0 | 0 | 0 | 0 |
| 0 | 0 | 0 | 0 | 0 |
| 0 | 0 | 0.089289798 | 0.000935277 | 0 |
| 0 | 0 | 0 | 0 | 0 |
| 0 | 0.021822065 | 0.001968144 | 0.110573636 | 0 |
| 0 | 0 | 0 | 0 | 0 |
| 0 | 0 | 0.457554289 | 0 | 0 |
| 0 | 0.014938848 | 0 | 0 | 0 |
| 0 | 0 | 0 | 0 | 0 |
| 0 | 0 | 0 | 0 | 0 |
| 30.20839674 | 0.824566778 | 3.837782852 | 7.610159499 | 5.244615113 |
| 0.542146553 | 0.140061775 | 0.004631824 | 1.148912064 | 0 |
| 0 | 0 | 0.146144966 | 0 | 0 |
| 0 | 0 | 0.063504698 | 0.02108066 | 0 |
| 0 | 0 | 0.093797398 | 0 | 0 |
| 0 | 0 | 0 | 0 | 0 |
| 0 | 0 | 0 | 0 | 0 |
| 0 | 0.020464292 | 0.002215555 | 0.000683217 | 0 |

| | | | | |
|---|---|---|---|---|
| 0 | 0 | 0 | 0 | 0 |
| 0 | 0 | 0.00167118 | 0.001581177 | 0 |
| 0 | 0 | 0 | 0 | 0 |
| 0 | 0 | 0.017874344 | 0 | 0 |
| 14.81762643 | 0.15278487 | 2.495356164 | 0.815374312 | 1.901819946 |
| 0 | 0 | 0 | 0 | 0 |
| 0 | 0 | 0 | 0 | 0 |
| 0 | 0 | 0.030605266 | 0 | 0 |
| 0.013625934 | 0.008840981 | 0 | 0.008069655 | 0 |
| 0 | 0 | 0 | 0 | 0 |
| 0 | 0 | 0 | 0 | 0 |
| 0 | 0 | 0 | 0 | 0 |
| 0 | 0.000973948 | 0.092853885 | 0.067430128 | 0 |
| 0.080717059 | 0.009778517 | 0.010344676 | 0.013641523 | 0.029715553 |
| 0 | 0 | 1.077842261 | 0 | 0 |
| 0.946935117 | 0.120690227 | 0.067314428 | 0.933074191 | 0 |
| 0.002957737 | 0 | 0 | 0 | 0 |
| 0 | 0 | 0 | 0 | 0 |
| 0 | 0 | 0.090962762 | 0 | 0 |
| 0 | 0 | 0 | 0 | 0 |
| 0 | 0 | 0 | 0 | 0 |
| 0 | 0 | 0 | 0 | 0 |
| 0 | 0 | 0 | 0 | 0 |
| 0 | 0 | 0 | 0 | 0 |
| 0 | 0 | 0 | 0 | 0 |
| 0 | 0 | 0 | 0 | 0 |
| 0.036995416 | 0.054208013 | 0.003823661 | 0.007895849 | 0 |
| 0 | 0.022366021 | 0 | 0 | 0 |
| 0 | 0 | 0 | 0 | 0 |
| 0 | 0 | 0 | 0 | 0 |
| 0 | 0 | 0 | 0 | 0 |
| 0 | 0 | 0 | 0 | 0 |
| 0 | 0 | 0 | 0 | 0 |
| 0 | 0 | 0.014699182 | 0.021481128 | 0 |
| 0 | 0 | 0.621282655 | 0 | 0 |
| 0 | 0.00215607 | 0 | 0 | 0.001556596 |
| 0 | 0 | 0 | 0.010323832 | 0 |
| 0 | 0.014251591 | 0 | 0.005450084 | 0 |
| 0 | 0 | 0 | 0 | 0 |
| 0 | 0 | 0 | 0 | 0 |
| 0 | 0.374009432 | 0.27594802 | 0 | 0 |
| 0 | 0 | 0 | 0 | 0 |
| 0 | 0 | 0.079577877 | 0 | 0 |
| 0 | 0 | 0 | 0 | 0 |
| 0 | 0 | 0 | 0 | 0 |
| 0 | 0 | 0 | 0 | 0 |
| 0 | 0 | 0 | 0 | 0 |
| 0 | 0 | 0 | 0 | 0 |

| | | | | |
|---|---|---|---|---|
| 0 | 0 | 0.138009882 | 0.145351864 | 0 |
| 0 | 0 | 0 | 0 | 0 |
| 0 | 0 | 0.021846498 | 0.01225791 | 0 |
| 0 | 0.001535534 | 0 | 0 | 0 |
| 0.003145474 | 0.138839956 | 0.114572898 | 0.053112645 | 0 |
| 0 | 0 | 0 | 0 | 0 |
| 0.077848994 | 0.014298384 | 0.002663317 | 0.001630512 | 0 |
| 0 | 0 | 0 | 0 | 0 |
| 0 | 0 | 0 | 0.00166238 | 0 |
| 0 | 0 | 0 | 0.003611419 | 0 |
| 0 | 0 | 0.005319519 | 0.00790905 | 0 |
| 0 | 0 | 0 | 0.161128206 | 0 |
| 0 | 0 | 0 | 0 | 0 |
| 0 | 0 | 0 | 0 | 0 |
| 0 | 0 | 0 | 0 | 0 |
| 0 | 0 | 0 | 0 | 0 |
| 0 | 0.010336125 | 0 | 0 | 0.173550975 |
| 0 | 0 | 0 | 0.000696696 | 0 |
| 0 | 0 | 0 | 0 | 0 |
| 0 | 0 | 0 | 0 | 0 |
| 0 | 0 | 0.268627242 | 0.004484532 | 0.522301444 |
| 0 | 0 | 0 | 0 | 0 |
| 0 | 0.119034065 | 0.041458255 | 0.021878411 | 0 |
| 0.000847735 | 0.058714144 | 0 | 0.002706714 | 0.006706791 |
| 0 | 0 | 0 | 0 | 0 |
| 0 | 0.087488315 | 0 | 0 | 0 |
| 0 | 0 | 0 | 0 | 0 |
| 0 | 0 | 0 | 0 | 0 |
| 0 | 0 | 0 | 0 | 0 |
| 2.502707112 | 0.048618405 | 0.141276213 | 0.016556285 | 0.88413633 |
| 0 | 0 | 0.030426723 | 0 | 0 |
| 0 | 0 | 0 | 0 | 0 |
| 0 | 0 | 0 | 0 | 0 |
| 0 | 0 | 0 | 1.946643669 | 0 |
| 0 | 0 | 0.60600735 | 0 | 0 |
| 0 | 0 | 0.645298964 | 0.199405742 | 0 |
| 0 | 0 | 0 | 0.153041051 | 0 |
| 0 | 0 | 0 | 0 | 0 |
| 0 | 0 | 1.015929265 | 0 | 0 |
| 0 | 0 | 0.151999454 | 0 | 0 |
| 0 | 0 | 0 | 0 | 0 |
| 0 | 0 | 0 | 0 | 0 |
| 0 | 0 | 0 | 0 | 0 |
| 0 | 0 | 0 | 0 | 0 |
| 0 | 0 | 0.04192408 | 0 | 0 |
| 0 | 0 | 0 | 0 | 0 |
| 0 | 0 | 0 | 0 | 0 |
| 0 | 0 | 0 | 0 | 0 |

| | | | | |
|---|---|---|---|---|
| 0 | 0 | 0 | 0 | 0.007732227 |
| 0 | 0 | 0.070763532 | 0 | 0 |
| 0 | 0 | 0 | 0 | 0 |
| 0 | 0 | 0 | 0 | 0 |
| 0 | 0 | 0 | 0 | 0 |
| 0 | 0 | 0 | 0 | 0 |
| 0 | 0 | 0 | 0 | 0 |
| 2.349031705 | 0.981403791 | 0.218520974 | 2.656498931 | 8.156785434 |
| 0.064074084 | 2.314829151 | 0.462788154 | 1.263486619 | 17.29803824 |
| 0 | 0.0501234 | 0.001720124 | 0 | 0 |
| 0 | 0 | 0.005651898 | 0.008169807 | 0 |
| 0 | 0 | 0 | 0 | 0 |
| 0 | 0 | 0.022345134 | 0 | 0 |
| 0 | 0.171773136 | 0 | 0 | 0 |
| 0 | 0 | 0 | 0.004347511 | 0 |
| 0 | 0 | 0 | 0 | 0 |
| 0 | 0.084896259 | 0 | 0.000797156 | 0 |
| 0 | 0 | 0 | 0 | 0 |
| 0 | 0 | 0 | 0 | 0 |
| 0.00392618 | 0.046582521 | 0.049780251 | 0 | 0 |
| 0 | 0 | 0 | 0 | 0 |
| 0 | 0 | 0 | 0 | 0 |
| 0 | 0 | 0.260235107 | 0 | 0 |
| 0 | 0 | 0 | 0 | 0 |
| 0 | 0.000818003 | 0.033847861 | 0.050755055 | 0 |
| 0 | 0 | 0.065337846 | 0 | 0 |
| 0 | 0 | 0 | 0 | 0 |
| 0 | 0 | 0 | 0 | 0 |
| 0 | 0 | 0 | 0 | 0 |
| 0 | 0 | 0 | 0 | 0 |
| 0 | 0 | 0 | 0 | 0 |
| 0 | 0 | 0 | 0 | 0 |
| 0 | 0 | 0 | 0 | 0 |
| 0 | 0 | 0.0033795 | 0.007381868 | 0 |
| 0 | 0 | 0.038876489 | 0 | 0 |
| 0 | 0 | 0 | 0 | 0 |
| 0 | 0 | 0 | 0 | 0 |
| 0 | 0 | 0 | 0 | 0 |
| 0 | 0 | 0.002058945 | 0 | 0 |
| 0 | 0 | 0 | 0 | 0.058322727 |
| 0 | 0 | 0 | 0 | 0 |
| 0.231352115 | 0 | 0.549930959 | 0.443770099 | 0 |
| 0 | 0 | 0 | 0 | 0 |
| 0 | 0 | 0 | 0 | 0 |
| 0 | 0 | 0 | 0 | 0 |
| 0 | 0 | 0 | 0 | 0 |
| 0 | 0 | 0 | 0 | 0 |
| 0 | 0 | 0 | 0 | 0 |

| | | | | |
|---|---|---|---|---|
| 0 | 0 | 0 | 0 | 0 |
| 0 | 0 | 0 | 0 | 0.028690102 |
| 0 | 0 | 0 | 0 | 0 |
| 0 | 0 | 0 | 0 | 0 |
| 0.103884989 | 6.198804835 | 0.438392397 | 0 | 0 |
| 0 | 0 | 0.042115733 | 0 | 0 |
| 0 | 0 | 0 | 0 | 0 |
| 0.03191031 | 0.023559826 | 0 | 0.005788964 | 0.004808688 |
| 0 | 0 | 0.003798735 | 0.000924811 | 0 |
| 0 | 0 | 0 | 0 | 0 |
| 0 | 0 | 0 | 0 | 0 |
| 0 | 0 | 0.19716149 | 0 | 0 |
| 0 | 0 | 0 | 0 | 0 |
| 0 | 0 | 0 | 0 | 0 |
| 0 | 0 | 0 | 0 | 0.000769352 |
| 0.004277134 | 0 | 0 | 0.11909161 | 0.003601829 |
| 0 | 0 | 0 | 0 | 0 |
| 0 | 0 | 0 | 0 | 0 |
| 0 | 0 | 0.005250175 | 0 | 0 |
| 0 | 0 | 0 | 0 | 0 |
| 0 | 0 | 0 | 0 | 0 |
| 0 | 0 | 0 | 0 | 0 |
| 0 | 0 | 0 | 0 | 0 |
| 0 | 0.008955955 | 0 | 0 | 0 |
| 0 | 0 | 0.003391994 | 0.048852904 | 0 |
| 0 | 0 | 0 | 0 | 0 |
| 0 | 5.011120218 | 0.775676672 | 5.174836448 | 0 |
| 0 | 0 | 0 | 0 | 0 |
| 0 | 0 | 0 | 0 | 0 |
| 0 | 0 | 0 | 0 | 0 |
| 0 | 0 | 0 | 0 | 0 |
| 0 | 0 | 0 | 0 | 0 |
| 0 | 0 | 0 | 0 | 0 |
| 0 | 0 | 0 | 0 | 0 |
| 0 | 0 | 0 | 0 | 0 |
| 0 | 0 | 0 | 0 | 0 |
| 0 | 0 | 0 | 0 | 0 |
| 0.001883875 | 0 | 0.722843661 | 0.009586385 | 0 |
| 0 | 0 | 0.190580431 | 0 | 0 |
| 0 | 0 | 0 | 0 | 0 |
| 0 | 0 | 0 | 0 | 0 |
| 0 | 0 | 0.592695692 | 0 | 0 |
| 0 | 0 | 0 | 0.004754167 | 0 |
| 0 | 0 | 0.032966638 | 0 | 0 |
| 0 | 0 | 0 | 0 | 0 |
| 0 | 0 | 0.00487846 | 0 | 0 |
| 0 | 0 | 0 | 0 | 0 |
| 0 | 0 | 0.024222954 | 0 | 0 |

| | | | | |
|---|---|---|---|---|
| 0.197795387 | 0.55184076 | 0.115514757 | 0.12444585 | 0 |
| 0 | 0 | 0 | 0 | 0 |
| 0 | 0 | 0 | 0.002902491 | 0 |
| 0 | 0 | 0 | 0 | 0 |
| 0 | 0 | 0 | 0 | 0 |
| 0.232159463 | 0 | 0 | 0 | 0.008482357 |
| 0 | 0 | 0 | 0 | 0 |
| 0 | 0 | 0 | 0 | 0 |
| 0 | 0 | 0 | 0 | 0 |
| 0 | 0 | 0 | 0 | 0 |
| 0 | 0 | 0 | 0 | 0 |
| 0 | 0 | 0 | 0 | 0 |
| 0 | 0 | 0 | 0 | 0 |
| 0 | 0 | 0 | 0 | 0 |
| 0 | 0 | 0.000936101 | 0 | 0 |
| 0 | 0 | 0 | 0 | 0 |
| 0 | 0 | 0 | 0 | 0 |
| 0 | 0 | 0 | 0 | 0 |
| 0 | 0 | 0 | 0.000438398 | 0 |
| 0 | 0 | 0 | 0 | 0 |
| 0 | 0 | 0 | 0 | 0 |
| 0 | 0 | 0 | 0 | 0 |
| 0.013260881 | 0.124377102 | 0.001307644 | 0.004284445 | 0 |
| 0 | 1.66280114 | 0.000797882 | 0.044980159 | 0 |
| 0 | 0 | 0 | 0 | 0 |
| 0 | 0 | 0 | 0 | 0 |
| 0 | 0 | 0 | 0 | 0 |
| 0 | 0 | 0 | 0 | 0 |
| 0 | 0 | 0 | 0 | 0 |
| 0 | 0 | 0 | 0.001200287 | 0 |
| 0 | 0 | 0 | 0 | 0 |
| 0.053017749 | 0 | 0.088532201 | 0 | 0 |
| 0.207866401 | 0.433821229 | 0 | 0.038707521 | 0.10938961 |
| 0 | 0 | 0 | 0 | 0 |
| 2.080467583 | 0 | 0 | 0.045179698 | 2.30352475 |
| 0 | 0.021611383 | 0 | 0.007765291 | 0 |
| 0 | 0 | 0 | 0 | 0 |
| 0 | 0 | 0.024355299 | 0 | 0 |
| 0 | 0 | 0 | 0 | 0 |
| 0 | 0 | 0 | 0 | 0 |
| 0 | 0 | 0 | 0 | 0 |
| 0 | 0 | 0.465515159 | 1.006867979 | 0 |
| 0 | 0.031421135 | 0.097465281 | 0 | 0 |
| 0 | 0 | 0 | 0 | 0 |
| 0 | 0 | 0 | 0 | 0 |
| 0 | 0.025147271 | 0.00844411 | 0.001627459 | 0 |
| 0 | 0 | 0 | 0 | 0 |
| 0 | 0 | 0 | 0 | 0 |

| | | | | |
|---|---|---|---|---|
| 0 | 0 | 0 | 0 | 0 |
| 0 | 0 | 0 | 0 | 0 |
| 0 | 0 | 0 | 0 | 0 |
| 0 | 0 | 0 | 0 | 0 |
| 0 | 0 | 0 | 0 | 0 |
| 0 | 0 | 0 | 0 | 0 |
| 0 | 0 | 0 | 0.70977995 | 0 |
| 0 | 0 | 0 | 0.005700824 | 0 |
| 0 | 0 | 0.011867022 | 0.01257942 | 0 |
| 0 | 0.064554288 | 0.126473813 | 0 | 0 |
| 0 | 0 | 0 | 0 | 0 |
| 0 | 0 | 0 | 0 | 0 |
| 0 | 0 | 0 | 0 | 0 |
| 0 | 0 | 0 | 0 | 0 |
| 0 | 0 | 0 | 0 | 0 |
| 0 | 0 | 0 | 0 | 0 |
| 0 | 0 | 0 | 0 | 0 |
| 0 | 0 | 0 | 0 | 0 |
| 0 | 0 | 0 | 0 | 0 |
| 0 | 0.19596402 | 2.268719011 | 0.692822436 | 0 |
| 0 | 0 | 0.253412808 | 0 | 0 |
| 0 | 0 | 0 | 0 | 0 |
| 0 | 0 | 0 | 0 | 0 |
| 0 | 0 | 0 | 0.013181084 | 0 |
| 0 | 0 | 0 | 0 | 0 |
| 0 | 0 | 0 | 0 | 0 |
| 0 | 0 | 0 | 0 | 0 |
| 0 | 0 | 0 | 0 | 0 |
| 0 | 0 | 0 | 0 | 0 |
| 0.376088355 | 0 | 0.01699873 | 0.389727392 | 0 |
| 0.560265243 | 0.822138114 | 0.404118473 | 1.054113382 | 0.018030824 |
| 0 | 0 | 0 | 0 | 0 |
| 0 | 0 | 0.002937094 | 0 | 0 |
| 0 | 0 | 0 | 0 | 0 |
| 0 | 0 | 0 | 0 | 0 |
| 0 | 0 | 0 | 0 | 0 |
| 0 | 0 | 0 | 0 | 0 |
| 0 | 0 | 0 | 0 | 0 |
| 0 | 0 | 0.002612878 | 0 | 0 |
| 0 | 0 | 0 | 0 | 0 |
| 0 | 0 | 0.004315782 | 0 | 0 |
| 0 | 0 | 0 | 0 | 0 |
| 0 | 0 | 0 | 0.001087378 | 0 |
| 0 | 0 | 0 | 0 | 0 |
| 0 | 0 | 0 | 0 | 0 |
| 0 | 0 | 0 | 0 | 0 |
| 0 | 0.062209641 | 0 | 0 | 0.077293569 |
| 0 | 0 | 0 | 0 | 0 |

| | | | | |
|---|---|---|---|---|
| 0 | 0 | 0 | 0 | 0 |
| 0 | 0 | 0 | 0 | 0 |
| 0 | 0 | 0 | 0 | 0 |
| 0 | 0 | 0.004743257 | 0 | 0 |
| 0 | 0.020539061 | 0.067079564 | 0.005991233 | 0.115488137 |
| 0 | 0 | 0 | 0 | 0 |
| 0 | 0 | 0 | 0 | 0 |
| 0 | 0 | 0 | 0 | 0 |
| 0 | 0 | 0 | 0 | 0 |
| 0 | 0 | 0 | 0 | 0 |
| 0 | 0 | 0.019214672 | 0.017615709 | 0 |
| 0.00191959 | 0.181083818 | 0.027257624 | 0.179492705 | 0.002066217 |
| 0 | 0.221235634 | 0 | 0.00245505 | 0 |
| 0 | 0 | 0 | 0 | 0 |
| 0 | 0 | 0.010674609 | 0.007339735 | 0.005038726 |
| 0.047981355 | 0.756534384 | 1.24780653 | 0.137197932 | 0.968725833 |
| 0 | 0.018409907 | 0.041055181 | 0.012626633 | 0 |
| 0 | 0 | 0 | 0 | 0 |
| 0.016404062 | 0.011662885 | 0 | 0.008287372 | 0.03301204 |
| 0 | 0.010083521 | 0 | 0.001731325 | 0 |
| 0 | 0 | 0 | 0 | 0 |
| 0 | 0 | 0.031340455 | 0.00721046 | 0 |
| 0 | 0 | 0 | 0 | 0 |
| 0 | 0 | 0 | 0 | 0 |
| 0 | 0 | 0 | 0 | 0 |
| 0 | 0 | 0 | 0 | 0 |
| 0 | 0 | 0.000899813 | 0 | 0 |
| 0 | 0 | 0 | 0 | 0 |
| 0 | 0 | 0 | 0 | 0 |
| 0 | 0 | 0 | 0 | 0 |
| 0 | 0 | 0 | 0 | 0.000556692 |
| 0 | 0.074906974 | 0 | 0 | 0 |
| 0 | 0 | 0 | 0 | 0 |
| 0 | 0 | 0 | 0 | 0 |
| 0 | 0 | 0 | 0 | 0 |
| 0 | 0 | 0 | 0 | 0 |
| 0 | 0 | 0 | 0 | 0 |
| 0.003460573 | 0.549883783 | 0.005852334 | 0.010911077 | 0.045890866 |
| 0.000832472 | 0 | 0.347073637 | 0 | 0 |
| 0 | 0 | 0 | 0 | 0 |
| 0.012955376 | 0.087188736 | 0 | 0 | 0 |
| 0 | 0 | 0.01209567 | 0 | 0 |
| 0 | 0 | 0 | 0 | 0 |
| 0 | 0 | 0 | 0 | 0 |
| 0 | 0 | 0 | 0 | 0 |
| 0 | 0 | 0 | 0 | 0 |
| 0 | 0 | 0 | 0 | 0 |
| 0 | 0 | 0 | 0 | 0 |

| | | | | |
|---|---|---|---|---|
| 0 | 0 | 0 | 0 | 0 |
| 0 | 0.352711796 | 0.137751244 | 1.421054177 | 0 |
| 0 | 0 | 0.002244972 | 0 | 0 |
| 0 | 0 | 0 | 0 | 0.002026053 |
| 0.001995178 | 0 | 0 | 0 | 0.002067045 |
| 0.001609355 | 0.001465828 | 0 | 0.019526213 | 0 |
| 0 | 0.022578835 | 0.008285517 | 0 | 0 |
| 0 | 0 | 0 | 0 | 0 |
| 0 | 0 | 0.007086926 | 0 | 0 |
| 0 | 0 | 0.013941367 | 0 | 0 |
| 0 | 0 | 0.062575787 | 0 | 0 |
| 0 | 0 | 0 | 0 | 0 |
| 0.01601122 | 0.312933199 | 0.036445171 | 0.000456446 | 0 |
| 0 | 0 | 0.001671566 | 0 | 0 |
| 0 | 0 | 0 | 0 | 0 |
| 0 | 0 | 0 | 0 | 0 |
| 0 | 0 | 0 | 0 | 0 |
| 0 | 0 | 0 | 0 | 0 |
| 0 | 0.024335373 | 0 | 0 | 0 |
| 0 | 0 | 0 | 0 | 0 |
| 0 | 0 | 0 | 0 | 0 |
| 0 | 0 | 0 | 0 | 0 |
| 0 | 0 | 0 | 0 | 0.016641911 |
| 0.002073231 | 0.012364093 | 0.045998873 | 0.054138673 | 0.005802145 |
| 0 | 0 | 0 | 0 | 0 |
| 0 | 0 | 0.016758834 | 0 | 0 |
| 0 | 0 | 0.0048002 | 0 | 0 |
| 0 | 0 | 0.000567787 | 0 | 0 |
| 0 | 0 | 0 | 0 | 0 |
| 0 | 0 | 0 | 0 | 0 |
| 0.033582213 | 0.011045398 | 0.048099714 | 0.015860211 | 0.029309504 |
| 0 | 0 | 0 | 0 | 0 |
| 0 | 0 | 0 | 0 | 0 |
| 0 | 0 | 0 | 0 | 0 |
| 0 | 0 | 0.067695383 | 0.087834081 | 0 |
| 0 | 0 | 0 | 0 | 0 |
| 0 | 0 | 0 | 0.003559925 | 0 |
| 0 | 0 | 0 | 0 | 0 |
| 0 | 0 | 0.004739659 | 0 | 0.002497628 |
| 0 | 0 | 0 | 0 | 0 |
| 0 | 0 | 0 | 0 | 0 |
| 0 | 0 | 0 | 0 | 0 |
| 0 | 0 | 0 | 0 | 0 |
| 0 | 0 | 0 | 0 | 0 |
| 0 | 0 | 7.714060872 | 0 | 0 |
| 0.244361722 | 0.449533434 | 0.055074304 | 0.088131988 | 0.054309633 |
| 0 | 0 | 0 | 0 | 0 |
| 0 | 0 | 0 | 0 | 0 |

| | | | | |
|---|---|---|---|---|
| 0 | 0 | 0 | 0 | 0 |
| 0 | 0 | 0 | 0 | 0 |
| 0 | 0 | 0.008407221 | 0 | 0 |
| 0 | 0 | 0.351707337 | 0.043684072 | 0 |
| 0 | 0 | 0 | 0 | 0 |
| 0 | 0 | 0 | 0 | 0 |
| 0 | 0 | 0 | 0 | 0 |
| 0 | 0 | 0 | 0 | 0 |
| 0 | 0 | 0.003175667 | 0.016787823 | 0 |
| 0 | 0 | 0 | 0 | 0 |
| 0 | 0.005102892 | 0 | 0.000919967 | 0 |
| 0 | 0.001741402 | 0 | 0 | 0.00086434 |
| 0 | 0 | 0 | 0 | 0 |
| 0 | 0 | 0 | 0 | 0 |
| 0 | 0 | 0 | 0 | 0 |
| 0 | 0 | 0 | 0 | 0 |
| 0 | 0 | 0.000974514 | 0 | 0 |
| 0 | 0 | 0 | 0 | 0 |
| 0 | 0 | 0 | 0 | 0 |
| 0 | 0 | 0 | 0.000813356 | 0 |
| 0 | 0 | 0 | 0 | 0 |
| 0 | 0 | 0 | 0 | 0 |
| 0 | 0 | 0 | 0 | 0 |
| 0 | 0 | 0 | 0 | 0 |
| 0 | 0 | 0 | 0 | 0 |
| 0 | 0 | 0 | 0 | 0 |
| 0 | 0 | 0 | 0 | 0 |
| 0 | 0.016262205 | 0.017513521 | 0 | 0 |
| 0 | 0 | 0 | 0 | 0 |
| 0 | 0 | 0 | 0 | 0 |
| 0.002626182 | 0.034550704 | 0.007704223 | 0 | 0.001648956 |
| 0 | 0 | 0 | 0 | 0 |
| 0.101896266 | 0.024108705 | 0.015435178 | 0.013589743 | 0.018188761 |
| 0.004414359 | 0.001182551 | 0 | 0.010363614 | 0 |
| 0 | 0 | 0 | 0 | 0 |
| 0 | 0 | 0 | 0 | 0 |
| 0 | 0 | 0 | 0 | 0 |
| 0 | 0 | 0 | 0 | 0 |
| 0 | 0 | 0 | 0 | 0 |
| 0 | 0 | 0 | 0 | 0 |
| 0 | 0 | 0 | 0 | 0 |
| 0.053725904 | 1.211937528 | 0 | 0.010679554 | 0.029139892 |
| 0.070801624 | 0.166244332 | 0.039906107 | 0.023378616 | 0 |
| 0 | 0 | 0 | 0 | 0 |
| 0 | 0 | 0 | 0 | 0 |
| 0 | 0.158387242 | 0.167158374 | 0 | 0 |
| 0 | 0 | 0 | 0 | 0 |
| 0 | 0 | 0 | 0 | 0 |

| | | | | |
|---|---|---|---|---|
| 0 | 0 | 0 | 0 | 0 |
| 0 | 0 | 0.051383859 | 0 | 0.003819677 |
| 0.005609989 | 0.00733333 | 0.012262513 | 0.126628031 | 0.0220741 |
| 0 | 0 | 0 | 0 | 0 |
| 0 | 0 | 0.005153172 | 0.612796084 | 0 |
| 0.00896249 | 0.015811584 | 0 | 0.1104042 | 0.0014601 |
| 0 | 0 | 0 | 0 | 0 |
| 0 | 0 | 0 | 0 | 0 |
| 0 | 0 | 0 | 0 | 0 |
| 0 | 0 | 0 | 0 | 0 |
| 0 | 0 | 0 | 0 | 0 |
| 0 | 0 | 0 | 0 | 0 |
| 0 | 0 | 0 | 0 | 0 |
| 0 | 0 | 0 | 0 | 0 |
| 0 | 0 | 0 | 0 | 0 |
| 0 | 0 | 0 | 0 | 0 |
| 0 | 0 | 0 | 0.188983957 | 0 |
| 0 | 0 | 0 | 0 | 0 |
| 0 | 0 | 0 | 0 | 0 |
| 0 | 0 | 0 | 0 | 0 |
| 0 | 0 | 0 | 0 | 0 |
| 0 | 0 | 0 | 0 | 0 |
| 0 | 0.003477304 | 0 | 0 | 0 |
| 0 | 0 | 0 | 0 | 0 |
| 0 | 0 | 0.040069541 | 0 | 0 |
| 0.006459793 | 0.000980615 | 0.003210452 | 0 | 0.004088495 |
| 0 | 0 | 0 | 0 | 0 |
| 0 | 0.029107665 | 0.027163186 | 0.312914185 | 0 |
| 0 | 0 | 0 | 0 | 0 |
| 0 | 0 | 0 | 0 | 0 |
| 0 | 0 | 0 | 0 | 0 |
| 0 | 0 | 0 | 0 | 0 |
| 0 | 0 | 0 | 0 | 0 |
| 0 | 0 | 0 | 0 | 0 |
| 0 | 0 | 0 | 0 | 0 |
| 0 | 0 | 0 | 0 | 0 |
| 0 | 0 | 0 | 0 | 0 |
| 0 | 0 | 0 | 0 | 0 |
| 0 | 0 | 0 | 0 | 0 |
| 0 | 0 | 0 | 0 | 0 |
| 0 | 0 | 0 | 0 | 0 |
| 0 | 0 | 0.008707114 | 0 | 0 |
| 0 | 0 | 0 | 0 | 0.007229538 |
| 0 | 0 | 0 | 0 | 0 |
| 0 | 0 | 0 | 0 | 0 |
| 0 | 0 | 0.001029678 | 0 | 0 |
| 0 | 0 | 0.002464545 | 0 | 0 |
| 0 | 0 | 0 | 0 | 0 |

| | | | | |
|---|---|---|---|---|
| 0 | 0 | 0 | 0 | 0 |
| 0.00775902 | 0.051293094 | 0.002245968 | 0.03156638 | 0.019990177 |
| 0 | 0 | 0.178221699 | 0 | 0 |
| 0 | 0 | 0 | 0 | 0 |
| 0 | 0 | 0 | 0 | 0 |
| 0 | 0 | 0 | 0 | 0 |
| 0 | 0 | 0 | 0 | 0 |
| 0 | 0 | 0 | 0 | 0 |
| 0 | 0 | 0 | 0 | 0 |
| 0 | 0 | 0 | 0 | 0 |
| 0 | 0 | 0 | 0 | 0 |
| 0 | 0 | 0 | 0 | 0 |
| 0 | 0 | 0.45474819 | 0 | 0 |
| 0 | 0 | 0 | 0 | 0 |
| 0.002184526 | 0 | 0 | 0.001992836 | 0.005878476 |
| 0 | 0 | 0 | 0 | 0 |
| 0 | 0 | 0 | 0 | 0 |
| 0 | 0 | 0 | 0 | 0 |
| 0 | 0 | 0 | 0 | 0 |
| 0 | 0 | 0 | 0 | 0 |
| 0 | 0 | 0 | 0 | 0 |
| 0.003677789 | 0.003256743 | 0 | 0.000978561 | 0 |
| 0 | 0.00554134 | 0 | 0.000751944 | 0 |
| 0 | 9.00592596 | 0 | 0 | 0 |
| 0 | 0 | 0 | 0 | 0 |
| 0 | 0 | 0 | 0 | 0 |
| 0 | 0 | 0 | 0 | 0 |
| 0 | 0 | 0 | 0 | 0 |
| 0 | 0 | 0 | 0 | 0 |
| 0 | 0 | 0 | 0 | 0 |
| 0 | 0 | 0 | 0 | 0 |
| 0 | 0 | 0 | 0 | 0 |
| 0 | 0 | 0 | 0 | 0 |
| 0 | 0 | 0 | 0 | 0 |
| 0 | 0 | 0 | 9.46452468 | 0.001887004 |
| 0 | 0 | 0 | 0 | 0 |
| 0 | 0 | 0 | 0 | 0 |
| 0 | 0 | 0 | 0 | 0 |
| 0 | 0.070852637 | 0.001901358 | 0.0329809 | 0.008763052 |
| 0 | 0 | 0 | 0 | 0 |
| 0.41136013 | 0.039575293 | 0.029739612 | 0.101858213 | 0.128197 |
| 0 | 0 | 0 | 0 | 0 |
| 0 | 0 | 0 | 0 | 0 |
| 0 | 0 | 0 | 0 | 0 |
| 0 | 0 | 0 | 0 | 0 |
| 0.411910561 | 0.213677217 | 0 | 0.456701792 | 0.106294841 |
| 0 | 0 | 0 | 0 | 0 |
| 0 | 0 | 0 | 0 | 0 |

| | | | | |
|---|---|---|---|---|
| 0 | 0 | 0 | 0 | 0 |
| 0 | 0 | 0 | 0 | 0 |
| 0 | 0 | 0 | 0 | 0 |
| 0 | 0 | 0.014894897 | 0 | 0 |
| 0 | 0 | 0 | 0 | 0 |
| 0 | 0 | 0 | 0 | 0 |
| 0 | 0 | 0 | 0 | 0 |
| 0 | 0 | 0 | 0 | 0.003326428 |
| 0 | 0 | 0 | 0 | 0 |
| 0.008911233 | 0 | 0.001272642 | 0 | 0.000694587 |
| 0 | 0 | 0.038475893 | 0 | 0 |
| 0 | 0 | 0 | 0 | 0 |
| 0 | 0.008890654 | 0 | 0.001198703 | 0 |
| 0 | 0 | 0 | 0 | 0 |
| 0 | 0 | 0 | 0 | 0 |
| 0 | 0 | 0 | 0 | 0 |
| 0 | 0 | 0.233738201 | 0 | 0 |
| 0 | 0 | 0 | 0 | 0 |
| 0 | 0 | 0 | 0 | 0 |
| 0.049990237 | 1.581038724 | 0.454673219 | 0.08151349 | 0.340606401 |
| 0 | 0 | 0 | 0.174436055 | 0 |
| 0 | 0 | 0 | 0 | 0 |
| 0 | 0 | 0 | 0 | 0 |
| 0 | 0 | 0 | 0 | 0 |
| 0 | 0 | 0 | 0 | 0 |
| 0 | 0 | 0.000737671 | 0 | 0 |
| 0 | 0 | 0 | 0 | 0 |
| 0 | 0 | 0 | 0 | 0 |
| 0 | 0 | 0 | 0 | 0 |
| 0 | 0 | 0 | 0.001596792 | 0 |
| 0 | 0 | 0 | 0 | 0 |
| 0 | 0.021217052 | 0 | 0.055384007 | 0 |
| 0 | 0 | 0 | 0 | 0 |
| 0 | 0 | 0 | 0 | 0 |
| 0 | 0 | 0 | 0 | 0 |
| 0.017994562 | 0 | 0 | 0 | 0 |
| 0 | 0 | 0 | 0 | 0 |
| 0 | 0 | 0 | 0 | 0 |
| 0 | 0 | 0 | 0 | 0 |
| 0 | 0 | 0 | 0 | 0 |
| 0 | 0 | 0 | 0 | 0 |
| 0 | 0 | 0 | 0 | 0 |
| 0 | 0 | 0.002760942 | 0.105005964 | 0 |
| 0 | 0 | 0 | 0 | 0 |
| 0.001757586 | 0.057935162 | 0.00727919 | 0.004609662 | 0 |
| 0 | 0.086640669 | 0.022458775 | 0 | 0 |
| 0 | 0 | 0 | 0 | 0 |
| 0 | 0 | 0 | 0 | 0 |

| | | | | |
|---|---|---|---|---|
| 0 | 0 | 0 | 0 | 0 |
| 0 | 0 | 0 | 0 | 0 |
| 0 | 0 | 0 | 0 | 0 |
| 0 | 0 | 0 | 0 | 0 |
| 0 | 0 | 0.000617254 | 0 | 0 |
| 0 | 0 | 0.016561366 | 0.00658334 | 0.0004237 |
| 0 | 0 | 0 | 0 | 0 |
| 0 | 0 | 0 | 0 | 0 |
| 0 | 0 | 0 | 0 | 0 |
| 0 | 0 | 0 | 0 | 0 |
| 0 | 0 | 0 | 0 | 0 |
| 0 | 0 | 0 | 0 | 0 |
| 0 | 0 | 0 | 0 | 0 |
| 0 | 0 | 0 | 0 | 0 |
| 0 | 0 | 0 | 0 | 0 |
| 0 | 0 | 0 | 0 | 0 |
| 0 | 0 | 0.121160696 | 0.013611725 | 0 |
| 0 | 0 | 0 | 0 | 0 |
| 0 | 0 | 0 | 0 | 0 |
| 0 | 0 | 0 | 0 | 0 |
| 0 | 0 | 0 | 0 | 0 |
| 0 | 0 | 0 | 0 | 0 |
| 0 | 0 | 0 | 0 | 0 |
| 0 | 0 | 0.040082919 | 0 | 0 |
| 0 | 0 | 0 | 0 | 0 |
| 0 | 0 | 0 | 0 | 0 |
| 0 | 0 | 0 | 0 | 0 |
| 0 | 0 | 0 | 0.001050945 | 0 |
| 0 | 0 | 0 | 0 | 0 |
| 0 | 0 | 0 | 0 | 0 |
| 0 | 0 | 0 | 0 | 0 |
| 0 | 0 | 0 | 0 | 0 |
| 0 | 0 | 0 | 0 | 0 |
| 0 | 0 | 0 | 0 | 0 |
| 0 | 0 | 0 | 0 | 0 |
| 0 | 0 | 0 | 0 | 0 |
| 0 | 0 | 0 | 0 | 0 |
| 0 | 0 | 0 | 0 | 0 |
| 0 | 0 | 1.436094168 | 0 | 0 |
| 0 | 0 | 0 | 0 | 0 |
| 0 | 0 | 0.033866702 | 0 | 0 |
| 0 | 0 | 0 | 0 | 0 |
| 0 | 0 | 0 | 0 | 0 |
| 0 | 0 | 0 | 0 | 0 |
| 0 | 0 | 0.008262634 | 0 | 0 |
| 0 | 0 | 0 | 0 | 0 |
| 0 | 0 | 0 | 0 | 0 |
| 0 | 0 | 0 | 0 | 0 |

| | | | | |
|---|---|---|---|---|
| 0 | 0 | 0 | 0 | 0 |
| 0 | 0 | 0 | 0.240046393 | 0 |
| 0 | 0 | 0 | 0 | 0 |
| 0 | 0 | 0 | 0 | 0 |
| 0 | 0 | 0 | 0 | 0 |
| 0 | 0 | 0.002781002 | 0 | 0 |
| 0 | 0 | 0 | 0.001250528 | 0 |
| 0 | 0 | 0 | 0 | 0 |
| 0 | 0 | 0 | 0 | 0 |
| 0 | 0 | 0 | 0 | 0 |
| 0.003022723 | 0.001042859 | 0.001517437 | 0 | 0 |
| 0 | 0 | 0 | 0 | 0 |
| 0.080959578 | 0 | 0.004330968 | 0.213257695 | 0 |
| 0 | 0 | 0 | 0 | 0 |
| 0 | 0 | 0 | 0 | 0 |
| 0 | 0 | 0 | 0 | 0 |
| 0 | 0 | 0 | 0 | 0 |
| 0 | 0 | 0 | 0 | 0 |
| 0 | 0 | 0 | 0 | 0 |
| 0 | 0 | 0.002367525 | 0 | 0 |
| 0.011713731 | 0 | 0.03137969 | 0 | 0.006934665 |
| 0 | 0 | 0 | 0 | 0 |
| 0 | 0 | 0 | 0 | 0 |
| 3.067739684 | 1.074619138 | 1.782326708 | 0.249889305 | 0.689534961 |
| 0 | 0 | 0 | 0 | 0 |
| 0 | 0 | 0 | 0 | 0 |
| 0 | 0 | 0 | 0 | 0 |
| 0 | 0 | 0 | 0 | 0 |
| 0 | 0 | 0 | 0 | 0 |
| 0 | 0 | 0 | 0 | 0 |
| 0 | 0 | 0 | 0 | 0 |
| 0 | 0 | 0 | 0 | 0 |
| 0.00264037 | 0 | 0.002025057 | 0 | 0 |
| 0 | 0 | 0 | 0 | 0 |

| PHL018-0023_V1 | PHL018-0004_V1 | PHL018-0057_V2 | PHL018-0029_V1 | PHL018-0188_V2 |
|---|---|---|---|---|
| EYCF | CTRL | REF | EYCF | CTRL |
| 0.103991558 | 0.35467679 | 2.481226671 | 0.04922078 | 0.004712992 |
| 5.556569401 | 1.396059576 | 0.912387912 | 0.271814669 | 0.112025801 |
| 12.98403287 | 1.796963099 | 3.487829338 | 0.138103721 | 11.04361099 |
| 0 | 0 | 0 | 0 | 0 |
| 0 | 0 | 0 | 0 | 0 |
| 0 | 0 | 0 | 0 | 0 |
| 0 | 0 | 0 | 0 | 0 |
| 0 | 0 | 0 | 0 | 0 |
| 0 | 0 | 0 | 0 | 0 |
| 0 | 0 | 0 | 0 | 0 |
| 0 | 0 | 0 | 0 | 0 |
| 0 | 0 | 0 | 0 | 0 |
| 0 | 0 | 0 | 0 | 0 |
| 2.220159364 | 0.940620489 | 1.488540988 | 1.095409067 | 15.00895778 |
| 0 | 0 | 0 | 0 | 0 |
| 0 | 0 | 0 | 0 | 0 |
| 0 | 0 | 0.003288645 | 0 | 0 |
| 0 | 0 | 0 | 0 | 0 |
| 0 | 0 | 0 | 0 | 0 |
| 0 | 0 | 0 | 0 | 0 |
| 0 | 0 | 0 | 0 | 0 |
| 0 | 0 | 0 | 0 | 0 |
| 0 | 0 | 0 | 0 | 0 |
| 0 | 0 | 0 | 0 | 0 |
| 0 | 0 | 0 | 0 | 0 |
| 0 | 0 | 0 | 0 | 0 |
| 0 | 0 | 0 | 0 | 0 |
| 0 | 0 | 0 | 0 | 0 |
| 0 | 0 | 0 | 0 | 0 |
| 0 | 0 | 0 | 0 | 0 |
| 0 | 0 | 0 | 0 | 0 |
| 0 | 0 | 0 | 0 | 0 |
| 0 | 0 | 0 | 0 | 0 |
| 0 | 0 | 0 | 0 | 0 |
| 0 | 0.151314742 | 2.260857211 | 0.657478293 | 0 |
| 0 | 0 | 0 | 0 | 0 |
| 0 | 0 | 0 | 0 | 0 |
| 0 | 0 | 0 | 0 | 0 |
| 0 | 0 | 0 | 0 | 0 |
| 0 | 0 | 0 | 0 | 0 |
| 0 | 0 | 0 | 0.037627921 | 0.461196472 |
| 0.02098128 | 13.07971512 | 0.771388182 | 0.128482652 | 0.0463552 |
| 0.004121882 | 0.007852963 | 0.002703465 | 0.1344069 | 0.022047014 |
| 0 | 0.000918115 | 0.001670659 | 0.038953414 | 0 |
| 0 | 0 | 0 | 0 | 0 |

| | | | | |
|---|---|---|---|---|
| 0 | 0.068815712 | 0 | 0.013400199 | 0 |
| 0.111920751 | 0.078586054 | 0.003475697 | 0 | 0.029919341 |
| 0 | 0.006768517 | 0.870975832 | 0.003899865 | 0.000930014 |
| 0 | 1.242307611 | 0.325455746 | 0.425238141 | 0.960944969 |
| 0.001650618 | 0 | 0.654260424 | 0.023730262 | 0 |
| 0 | 0 | 0 | 0.016001384 | 0 |
| 0 | 0.006141885 | 0.001064396 | 0.002068138 | 7.832721439 |
| 0 | 0 | 0 | 0 | 0 |
| 0 | 0 | 0 | 0 | 0 |
| 0 | 0 | 0 | 0 | 0 |
| 0 | 0 | 0 | 0 | 0.068899209 |
| 0 | 0.435512522 | 0.011830948 | 0.104113704 | 0.011571096 |
| 0.000984122 | 0 | 0 | 0 | 0.005848718 |
| 0.007972921 | 0.047128127 | 0.053169512 | 0.01300108 | 0.001036087 |
| 0 | 0 | 0 | 0 | 0 |
| 0 | 0 | 0 | 0 | 0 |
| 0 | 0.004020033 | 0 | 0.011280445 | 4.652644944 |
| 0 | 0 | 0 | 0 | 0 |
| 0 | 0 | 0 | 0 | 0 |
| 0 | 0 | 0 | 0 | 0 |
| 0.515295723 | 0.311415479 | 0.025977456 | 0 | 0.187580032 |
| 0.018726142 | 0.002285911 | 0.049347816 | 0.009469985 | 0.001370579 |
| 0 | 0 | 0.218428396 | 0.172877204 | 0 |
| 0 | 0 | 0 | 0 | 0 |
| 0.000713156 | 0 | 0.024222535 | 0 | 0 |
| 0 | 0 | 0 | 0 | 0 |
| 0 | 0 | 0.000677276 | 0 | 0 |
| 0 | 0 | 0 | 0 | 0 |
| 0 | 0 | 0 | 0.027153973 | 0.008986004 |
| 0 | 0 | 0.001066233 | 0 | 0 |
| 0 | 0.03626265 | 0 | 0 | 0 |
| 0 | 0 | 0.00183897 | 0 | 0 |
| 0 | 0 | 0 | 0 | 0 |
| 0 | 0 | 0 | 0 | 0 |
| 0 | 0 | 0 | 0.006957379 | 0 |
| 0 | 0 | 0 | 0 | 0 |
| 0 | 0 | 0 | 0 | 0 |
| 0 | 0 | 0 | 0 | 0.099510505 |
| 0 | 0 | 0 | 0 | 0 |
| 0 | 0 | 0 | 0 | 0 |
| 0 | 0 | 0 | 0 | 0 |
| 0 | 0 | 0 | 0 | 0 |
| 0 | 0 | 0 | 0 | 0 |
| 0 | 0 | 0 | 0 | 0 |
| 0 | 0 | 0 | 1.635141774 | 0 |
| 0 | 0 | 0 | 0 | 0.009685282 |
| 0 | 0 | 0 | 0 | 0 |
| 0 | 0 | 0 | 0 | 0 |

| | | | | |
|---|---|---|---|---|
| 0 | 0 | 0 | 0 | 0 |
| 0 | 0 | 0.002779826 | 0 | 0 |
| 0 | 0 | 0 | 0 | 0 |
| 0 | 0 | 0 | 0 | 0 |
| 0 | 0 | 0 | 0 | 0 |
| 0 | 0 | 0 | 0.001605496 | 0 |
| 0 | 0 | 0 | 0 | 0 |
| 0 | 0 | 0 | 0 | 0 |
| 0 | 0 | 0 | 0.00236728 | 0 |
| 0 | 0 | 0 | 0 | 0 |
| 0 | 0 | 0 | 0 | 0 |
| 0 | 0 | 0 | 0.013741826 | 0.048255812 |
| 0 | 0 | 0 | 0 | 0 |
| 0 | 0 | 0 | 0 | 0 |
| 0 | 0 | 0 | 0 | 0 |
| 0 | 0 | 0 | 0 | 0 |
| 0 | 0 | 0 | 0 | 0 |
| 0 | 0 | 0 | 0 | 0 |
| 0 | 0 | 2.383521643 | 0.489183313 | 0 |
| 0 | 0 | 0 | 0 | 0 |
| 0 | 0 | 0 | 0 | 0 |
| 0 | 0 | 0 | 0 | 0 |
| 0 | 0 | 0 | 0 | 0 |
| 0 | 0 | 0 | 0 | 0 |
| 0 | 0 | 0 | 0 | 0 |
| 0 | 0 | 0 | 0 | 0 |
| 0 | 0 | 0 | 0 | 0 |
| 0 | 0 | 0.001968144 | 0 | 0 |
| 0 | 0 | 0 | 0 | 0 |
| 0 | 0 | 0 | 0 | 0 |
| 0 | 0.00594798 | 0.018771686 | 0.03694832 | 0 |
| 0.000757399 | 0.006824944 | 0.002128987 | 0.006894428 | 0.00048228 |
| 0 | 0 | 0 | 0 | 0 |
| 0 | 0 | 0 | 0 | 0 |
| 0 | 0 | 0 | 0 | 0 |
| 0 | 0 | 0 | 0 | 0 |
| 0 | 0 | 0 | 0 | 0 |
| 0 | 0 | 0 | 0 | 0 |
| 0 | 0 | 0 | 0 | 0 |
| 0 | 0 | 0 | 0 | 0 |
| 0 | 0 | 0 | 0 | 0 |
| 0 | 0 | 0 | 0 | 0 |
| 0 | 0.001126937 | 0.028503951 | 0 | 0 |
| 0 | 0 | 0 | 0 | 0 |
| 0 | 0 | 0 | 0 | 0 |
| 0 | 0 | 0 | 0 | 0 |
| 0 | 0 | 0 | 0 | 0.009155899 |
| 0 | 0 | 0 | 0 | 0 |

| | | | | |
|---|---|---|---|---|
| 0 | 0 | 0 | 0 | 0 |
| 0 | 0 | 0 | 0 | 0 |
| 0 | 0 | 0 | 0 | 0 |
| 0 | 0 | 0 | 0 | 0 |
| 0 | 0 | 0.018376006 | 0.027070362 | 0 |
| 0 | 0 | 0 | 0 | 0 |
| 0 | 0 | 0 | 0 | 0 |
| 0 | 0 | 0 | 0 | 0 |
| 0 | 0 | 0 | 0 | 0 |
| 0 | 0 | 0 | 0 | 0 |
| 0 | 0 | 0 | 0 | 0 |
| 0 | 0 | 0 | 0 | 0 |
| 0 | 0 | 0 | 0 | 0 |
| 0 | 0 | 0 | 0 | 0 |
| 0 | 0 | 0 | 0 | 0 |
| 0 | 0 | 0 | 3.919027628 | 0 |
| 0 | 0 | 0 | 0 | 0 |
| 0 | 0 | 0 | 0 | 0 |
| 0 | 0 | 0 | 0 | 0 |
| 0 | 0 | 0 | 0 | 0 |
| 0 | 0 | 0 | 0 | 0 |
| 0 | 0 | 0 | 0 | 0 |
| 0 | 0 | 0.00090377 | 0.003512079 | 0 |
| 0 | 0 | 0 | 0 | 0 |
| 0 | 0 | 0 | 0 | 0 |
| 0 | 0 | 0 | 0 | 0 |
| 0 | 0 | 0 | 0.001997119 | 0 |
| 0 | 0 | 0 | 0 | 0 |
| 0 | 0 | 0 | 0 | 0 |
| 0 | 0 | 0 | 0 | 0 |
| 0 | 0 | 0 | 0 | 0 |
| 0 | 0 | 14.7467453 | 0.003005102 | 0 |
| 0 | 0 | 0 | 0 | 0 |
| 0 | 0 | 0 | 0 | 0 |
| 0 | 0 | 0 | 0 | 0 |
| 0 | 0 | 0 | 0 | 0 |
| 0 | 0 | 0 | 0 | 0 |
| 0 | 0 | 0 | 0 | 0 |
| 0 | 0 | 0 | 0 | 0 |
| 0 | 0 | 0 | 0 | 0 |
| 0 | 0 | 0 | 0 | 0 |
| 0 | 0 | 0 | 0 | 0 |
| 0 | 0 | 0 | 0 | 0 |
| 0 | 0 | 0 | 0 | 0 |
| 0 | 0 | 0 | 0 | 0 |
| 0 | 0 | 0 | 0 | 0 |
| 0 | 0 | 0 | 0 | 0 |
| 0 | 0 | 0 | 0 | 0 |

| | | | | |
|---|---|---|---|---|
| 0 | 0 | 0 | 1.601645575 | 0 |
| 0 | 0 | 0 | 0 | 0 |
| 0 | 0 | 0 | 0 | 0 |
| 0 | 0 | 0 | 0 | 0 |
| 0 | 0 | 0 | 0 | 0 |
| 0 | 0 | 0 | 0 | 0 |
| 0 | 0 | 0 | 0 | 0 |
| 0 | 0 | 0 | 0 | 0 |
| 0 | 0 | 0 | 0 | 0 |
| 0 | 0 | 0 | 0 | 0 |
| 0 | 0 | 0 | 0 | 0 |
| 0 | 0 | 0 | 0.010981816 | 0 |
| 0 | 0.001937606 | 0.000783509 | 0.001353218 | 0 |
| 0 | 0 | 0 | 0 | 0 |
| 0 | 0 | 0 | 0 | 0 |
| 0 | 0 | 0 | 0 | 0 |
| 0 | 0 | 0 | 0 | 0 |
| 0 | 0 | 0 | 0 | 0 |
| 0 | 0 | 0 | 0 | 0 |
| 0 | 0 | 0 | 0 | 0 |
| 0.072859887 | 0.012478735 | 0.019044652 | 0.078119688 | 0.031194937 |
| 0 | 0 | 0 | 0 | 0 |
| 0 | 0.001236213 | 0 | 0 | 0 |
| 0.003260867 | 0 | 0 | 0 | 0 |
| 0 | 0 | 0 | 0 | 0 |
| 0 | 0 | 0 | 0 | 0 |
| 0.00855922 | 0.001406578 | 0.008318367 | 0.00663086 | 0.001352876 |
| 0 | 0 | 0 | 0 | 0 |
| 0 | 0 | 0 | 0 | 0 |
| 0 | 0 | 0 | 0 | 0 |
| 0.125984471 | 0 | 0.157760607 | 0 | 0.002889831 |
| 0 | 0 | 0 | 0 | 0 |
| 0 | 0 | 0 | 0 | 0 |
| 0 | 0 | 0 | 0.012950232 | 0 |
| 0.063276521 | 0.080931388 | 0.169667756 | 0.003086771 | 0.028934148 |
| 0 | 0 | 0 | 0 | 0 |
| 0 | 0 | 0 | 0 | 0 |
| 0 | 0 | 0 | 0 | 0 |
| 0 | 0 | 0 | 0 | 0 |
| 0 | 0 | 0 | 0 | 0 |
| 0 | 0 | 0 | 0 | 0 |
| 0 | 0 | 0 | 0.006371028 | 0 |
| 0 | 0 | 0 | 0.002352792 | 0 |
| 0 | 0 | 0 | 0 | 0 |
| 0 | 0 | 0 | 0 | 0 |
| 0 | 0.01870279 | 0.058309468 | 0.052117246 | 0 |
| 0 | 0.003402997 | 0 | 0 | 0 |
| 0 | 0.001306528 | 0 | 0.002346364 | 0.0006155 |

| | | | | |
|---|---|---|---|---|
| 0 | 0 | 0 | 0 | 0 |
| 0 | 0 | 0 | 0 | 0 |
| 0.002649816 | 0 | 0 | 0 | 0.042182342 |
| 0 | 0 | 0 | 0.001677676 | 0 |
| 0 | 0 | 0 | 0 | 0 |
| 0 | 0 | 0 | 0 | 0 |
| 0 | 0 | 0 | 0 | 0 |
| 0 | 0 | 0 | 0.001686471 | 0.002654378 |
| 0.007142912 | 0 | 0 | 0 | 0 |
| 0 | 0 | 0 | 0.008101382 | 0 |
| 0 | 0 | 0 | 0 | 0.001332676 |
| 0 | 0 | 0 | 0.00885982 | 0 |
| 0 | 0 | 0 | 0 | 0 |
| 0 | 0 | 0 | 0 | 0 |
| 0 | 0 | 0 | 0 | 0.067930781 |
| 0 | 0 | 0 | 0 | 0 |
| 0 | 0 | 0 | 0 | 0 |
| 0 | 0.074118544 | 0.008976038 | 0 | 0 |
| 0 | 0.023347697 | 0.003626762 | 0 | 0 |
| 0 | 0 | 0 | 0 | 0 |
| 0.048722845 | 0.002800019 | 0.001019018 | 0 | 0.112880046 |
| 0 | 0 | 0 | 0 | 0 |
| 0 | 0 | 0 | 0 | 0 |
| 0 | 0 | 0 | 0 | 0 |
| 0.004753485 | 0 | 0 | 0.001748278 | 0.005044705 |
| 0 | 0 | 0 | 0 | 0 |
| 0 | 0 | 0 | 0 | 0 |
| 0 | 0 | 0 | 0 | 0 |
| 0 | 0 | 0 | 0.008967686 | 0.162152199 |
| 0 | 0 | 0 | 0 | 0 |
| 0.749624725 | 0.467153652 | 0.204179702 | 0.058234779 | 0.037511542 |
| 0 | 0 | 0 | 0 | 0 |
| 0 | 0 | 0 | 0 | 0 |
| 0 | 0 | 0 | 0 | 0 |
| 0 | 0 | 0 | 0 | 0 |
| 0 | 0 | 0 | 0 | 0 |
| 0 | 0 | 0 | 0 | 0 |
| 0 | 0 | 0 | 0 | 0 |
| 0 | 0 | 0 | 0 | 0 |
| 0 | 0 | 0 | 0 | 0 |
| 0 | 0 | 0 | 0 | 0 |
| 0 | 0.030010807 | 0.010310892 | 0 | 0 |
| 0 | 0 | 0 | 0 | 0 |
| 0 | 0 | 0 | 0 | 0 |
| 0 | 0 | 0 | 0 | 0 |
| 0 | 0 | 0 | 0 | 0 |
| 0 | 0 | 0 | 0 | 0 |
| 0 | 0 | 0 | 0 | 0 |

| | | | | |
|---|---|---|---|---|
| 0 | 0 | 0 | 0 | 0 |
| 0 | 0 | 0 | 0 | 0 |
| 0 | 0.006588026 | 0.000959039 | 0.156527897 | 0.023463156 |
| 0 | 0 | 0 | 0 | 0 |
| 0 | 0 | 0.066166275 | 0 | 0 |
| 0 | 0 | 0 | 0 | 0 |
| 0 | 0 | 0 | 0 | 0 |
| 0 | 0 | 0 | 0 | 0 |
| 0 | 0 | 0 | 0 | 0 |
| 0.002621166 | 0 | 0 | 0 | 0.16432202 |
| 0 | 0 | 0 | 0 | 0 |
| 0 | 0 | 0 | 0 | 0 |
| 0 | 0 | 0 | 0 | 0 |
| 0.001809932 | 0 | 0.001324889 | 0.006635921 | 0.004321837 |
| 0 | 0 | 0.039585497 | 0.002644109 | 0.00277442 |
| 0 | 0 | 0 | 0.023657971 | 0 |
| 0.004468069 | 0 | 0.002242747 | 0.002367142 | 0.008806206 |
| 0 | 0 | 0 | 0 | 0 |
| 0 | 0 | 0 | 0 | 0 |
| 0 | 0 | 0 | 0 | 0 |
| 0 | 0 | 0 | 0 | 0 |
| 0 | 0.00098898 | 0 | 0.001748335 | 0 |
| 0 | 0 | 0 | 0 | 0 |
| 0 | 0 | 0 | 0 | 0 |
| 0 | 0 | 0 | 0 | 0 |
| 0 | 0 | 0 | 0 | 0 |
| 2.496452232 | 0.18874481 | 0 | 0 | 0 |
| 0 | 0 | 0 | 0 | 0 |
| 0 | 0 | 0 | 0 | 0 |
| 0 | 0 | 0 | 0 | 0 |
| 0 | 0 | 0 | 0 | 0 |
| 0 | 0 | 0 | 0 | 0 |
| 0 | 0 | 0 | 0.003216254 | 0 |
| 0 | 0 | 0 | 0 | 0 |
| 0 | 0 | 0 | 0 | 0 |
| 0.00230175 | 0 | 0 | 0.017072242 | 0.004071277 |
| 0 | 0 | 0 | 0 | 0 |
| 0 | 0 | 0 | 0.204772054 | 0 |
| 0 | 0 | 0 | 0 | 0 |
| 0 | 0 | 0 | 0 | 0 |
| 0 | 0 | 0 | 0 | 0 |
| 0 | 0 | 0 | 0 | 0 |
| 0 | 0 | 0 | 0 | 0 |
| 0 | 0 | 0 | 0 | 0 |
| 0 | 0 | 0 | 0.004301866 | 0 |
| 0 | 0 | 0 | 0 | 0 |
| 0 | 0 | 0 | 0 | 0 |
| 0 | 0 | 0 | 0 | 0 |

| | | | | |
|---|---|---|---|---|
| 0 | 0 | 0.002031145 | 0.166826878 | 0 |
| 0 | 0 | 0 | 0 | 0 |
| 0 | 0 | 0.006126088 | 0.001627773 | 0 |
| 0 | 0 | 0 | 0 | 0 |
| 0.004452819 | 0.00182384 | 0 | 0.001228272 | 0.003222013 |
| 0 | 0 | 0 | 0 | 0 |
| 0 | 0 | 0 | 0 | 0 |
| 0.085396516 | 0.032504237 | 0.010633121 | 0.021234222 | 0.243582384 |
| 0 | 0 | 0 | 0 | 0 |
| 0.199434109 | 0.006775851 | 0.05068902 | 0.049392671 | 0.211652369 |
| 0 | 0 | 0 | 0 | 0 |
| 0 | 0 | 0 | 0.034299622 | 0.074125134 |
| 0.036796886 | 0 | 0.012969039 | 0.006547446 | 0.071413137 |
| 0 | 0.002997507 | 0.00545445 | 0 | 0.050247693 |
| 0 | 0 | 0 | 0 | 0 |
| 0 | 0 | 0 | 0.001293263 | 0.001899799 |
| 0 | 0 | 0 | 0.000752373 | 0 |
| 0.001553065 | 0 | 0 | 0 | 0 |
| 0.071818574 | 0.003896415 | 0 | 0 | 0.10191671 |
| 0.274821932 | 0 | 0 | 0 | 0 |
| 0 | 0 | 0 | 0 | 0 |
| 0 | 0 | 0 | 0 | 0 |
| 0 | 0 | 0 | 0 | 0 |
| 0 | 0 | 0 | 0.029923987 | 0 |
| 0 | 0 | 0 | 0 | 0 |
| 0 | 0 | 0.002788731 | 0.018262514 | 0 |
| 0 | 5.829675503 | 1.505312597 | 0 | 0 |
| 0 | 0.131892017 | 0 | 0 | 0 |
| 0 | 0 | 0.036166599 | 0 | 0 |
| 0 | 0.004730415 | 0 | 0 | 0.001516604 |
| 0.064300872 | 0 | 0 | 0 | 0 |
| 0 | 0 | 0 | 0 | 0 |
| 0 | 0 | 0 | 0 | 0 |
| 0 | 0.002672398 | 0 | 0 | 0 |
| 0 | 0 | 0 | 0 | 0 |
| 0 | 0.032894937 | 0.029928839 | 0 | 0 |
| 0.192540782 | 0.074993744 | 0.002897869 | 0 | 0 |
| 0 | 0 | 0 | 0 | 0 |
| 0 | 0 | 0 | 0 | 0 |
| 2.96003586 | 0 | 0 | 0 | 0 |
| 0 | 0.448037472 | 0 | 0 | 0 |
| 0 | 0 | 0.016648692 | 0 | 0 |
| 0 | 0 | 0 | 0 | 0 |
| 0 | 0 | 0 | 0 | 0 |
| 0 | 0 | 0 | 0 | 0 |
| 0 | 0 | 0 | 0 | 0 |
| 0 | 0.001270568 | 0.000867002 | 0 | 0 |
| 0 | 0 | 0 | 0 | 0 |

| | | | | |
|---|---|---|---|---|
| 0 | 0 | 0.00327782 | 0 | 0 |
| 0 | 0.001064961 | 0 | 0 | 0 |
| 1.119035233 | 0 | 0 | 0 | 0 |
| 0 | 0 | 0 | 0 | 0 |
| 0 | 0 | 0 | 0 | 0 |
| 0.095618964 | 0.037026866 | 0.024128033 | 0 | 0 |
| 0 | 0 | 0 | 0 | 0 |
| 0 | 0 | 0 | 0 | 0 |
| 0 | 0 | 0 | 0 | 0 |
| 0 | 0 | 0 | 0 | 0 |
| 0.001127052 | 0.002031179 | 0.031152507 | 0.011399213 | 0.00215298 |
| 0.027675173 | 0 | 0 | 0 | 0 |
| 0 | 0.016839018 | 0 | 0 | 0 |
| 0.001064782 | 0 | 0.002244762 | 0.082278241 | 0.004068055 |
| 0.044387558 | 0 | 0 | 0 | 0 |
| 0 | 0 | 0 | 0 | 0 |
| 0 | 0 | 0 | 0 | 0 |
| 0 | 0 | 0 | 0 | 0 |
| 0 | 0.043474657 | 0 | 0 | 0 |
| 0 | 0.065752704 | 0 | 0 | 0 |
| 0 | 0 | 0 | 0 | 0 |
| 0 | 0 | 0 | 0.208452746 | 0 |
| 0 | 0 | 0.025562061 | 0 | 0 |
| 0 | 0 | 0 | 0 | 0 |
| 0 | 0 | 0 | 0 | 0 |
| 0 | 0 | 0 | 0 | 0 |
| 0.002770813 | 0.001486183 | 0.038672224 | 0 | 0 |
| 0 | 0 | 0 | 0 | 0 |
| 0 | 0 | 0 | 0 | 0 |
| 0.127356975 | 0.206538964 | 0.599209437 | 0.172695443 | 10.97739917 |
| 0.01199134 | 0 | 0.00575315 | 0 | 0 |
| 0 | 0 | 0 | 0.002051531 | 0 |
| 0 | 0 | 0 | 0 | 0 |
| 0 | 0 | 0 | 0 | 0 |
| 0 | 0.006436822 | 0 | 0 | 0 |
| 0 | 0 | 0 | 0 | 0 |
| 0 | 0 | 0 | 0 | 0 |
| 0 | 0 | 0 | 0.009440126 | 0.001382144 |
| 1.422366716 | 0 | 0 | 0 | 0 |
| 0.09020952 | 0 | 0 | 0 | 0 |
| 0.019258065 | 0.014874425 | 0.019889726 | 0 | 0.028241466 |
| 0 | 1.769453837 | 1.228634355 | 0.039138469 | 0.19223349 |
| 0 | 0 | 0 | 0.005027771 | 0.000844089 |
| 0 | 0 | 0 | 0 | 0 |
| 0.000592475 | 0 | 0 | 0 | 0 |
| 0 | 0 | 0 | 0 | 0 |
| 0.009555104 | 0.002365421 | 0.026471242 | 0 | 0.006435314 |
| 0 | 0 | 0.002618735 | 0 | 0 |

| | | | | |
|---|---|---|---|---|
| 0 | 0 | 0 | 0.012611939 | 0 |
| 0.015645619 | 0.026294469 | 0 | 0 | 0 |
| 0.001853533 | 0 | 0.002532703 | 0.017497183 | 9.551201983 |
| 0 | 0 | 0 | 0 | 0 |
| 0 | 0 | 0.072426867 | 0.65726182 | 0 |
| 0.028945376 | 0 | 0.000898048 | 0 | 0 |
| 0 | 0 | 0 | 0 | 0 |
| 0 | 0 | 0 | 0.001285039 | 0 |
| 0.194297436 | 0 | 0 | 0 | 0.056383086 |
| 0 | 0.006586382 | 0 | 0 | 0 |
| 0 | 0 | 0 | 0 | 0 |
| 0 | 0 | 0 | 0 | 0 |
| 0 | 0 | 0 | 0 | 0 |
| 0.00348682 | 0 | 0.002642472 | 0 | 0.000870692 |
| 0 | 0 | 0 | 0 | 0 |
| 0.042636514 | 0.266999615 | 0.008072455 | 0.087543679 | 0 |
| 0.003553243 | 0.00434535 | 0.010820202 | 0.122549137 | 0.001319826 |
| 0 | 0 | 0.006573062 | 0 | 0.004352457 |
| 0.000550535 | 0 | 0 | 0 | 0.000779017 |
| 0 | 0 | 0 | 0 | 0 |
| 0 | 0 | 0 | 0.001585231 | 0 |
| 0 | 0.001061133 | 0.001930903 | 0.000833728 | 0.001312225 |
| 0 | 0.354548272 | 0 | 0 | 0 |
| 0 | 0 | 0 | 0 | 0.012904166 |
| 0 | 0 | 0 | 0 | 0 |
| 0 | 0.008435712 | 0 | 0 | 0.002483769 |
| 0.002255435 | 0.001451699 | 0.003434083 | 0 | 0.001436168 |
| 0 | 0.009921935 | 0.000501516 | 0 | 0 |
| 0 | 0 | 0 | 0 | 0 |
| 0 | 0 | 0 | 0 | 0 |
| 0.09674022 | 0.004416053 | 0.008236618 | 0.003469676 | 0 |
| 0.003068677 | 0 | 0 | 0.009828437 | 0.003799458 |
| 0.044070407 | 0.007835826 | 0.010266162 | 0.032999265 | 0.065116751 |
| 0 | 0 | 0 | 0.010234655 | 0.029995282 |
| 0 | 0 | 0 | 0 | 0 |
| 0 | 0 | 0 | 0 | 0 |
| 0.005062883 | 0 | 0 | 0.001335833 | 0 |
| 0.013427561 | 0.011301836 | 0.012298991 | 0.004526962 | 0.002740422 |
| 0 | 0 | 0.173894012 | 0.020695039 | 0.000542874 |
| 0 | 0 | 0 | 0 | 0 |
| 0 | 0 | 0 | 0 | 0 |
| 0 | 0 | 0 | 0 | 0 |
| 0 | 0.025187112 | 0.001206106 | 0 | 0 |
| 0 | 0.063833209 | 0.048476566 | 0.002990182 | 0.027524853 |
| 0 | 0.026743371 | 0 | 0 | 0 |
| 0 | 0 | 0 | 0 | 0 |
| 0 | 0 | 0 | 0 | 0 |
| 0 | 0 | 0 | 0 | 0 |

| | | | | |
|---|---|---|---|---|
| 0 | 0 | 0 | 0 | 0 |
| 0.034247772 | 0.077604856 | 0.330508184 | 0.900847027 | 0.0460548 |
| 0 | 0 | 0 | 0 | 0 |
| 0 | 0.000731524 | 0 | 0 | 0 |
| 0 | 0 | 0.004312473 | 0 | 0 |
| 0 | 0 | 0 | 0 | 0 |
| 0.114147946 | 0 | 0.141571121 | 0 | 0 |
| 0 | 0 | 0 | 0 | 0.001720943 |
| 0 | 0.001105694 | 0 | 0.007167104 | 0.019598414 |
| 0 | 0.003072043 | 0 | 0 | 0.016281303 |
| 0.002789023 | 0.01480991 | 0 | 0 | 0.000739973 |
| 0.074518288 | 0.001002717 | 0.010947642 | 0.190392552 | 0.053732776 |
| 0 | 0 | 0 | 1.051100515 | 0 |
| 0 | 0 | 0 | 0 | 0 |
| 0.015400268 | 0 | 0 | 0.028522504 | 0 |
| 0 | 0 | 0 | 0 | 0 |
| 0.001460255 | 0.005404343 | 0.014109765 | 0.004615404 | 0.009298303 |
| 0.015401854 | 0.06314025 | 0.03496772 | 0 | 0.090277141 |
| 0 | 0 | 0 | 0 | 0 |
| 0.032157574 | 0.033077278 | 0.001982288 | 0 | 0 |
| 0.004517674 | 0.002423148 | 0.014991659 | 0.071013918 | 0.001398381 |
| 0 | 0 | 0 | 0 | 0.00447444 |
| 0 | 0 | 0 | 0 | 0 |
| 0 | 0 | 0 | 0 | 0 |
| 0 | 0 | 0 | 0 | 0.021481511 |
| 0 | 0 | 0 | 0 | 0 |
| 0 | 0 | 0 | 0.010523539 | 0.04653487 |
| 1.583734562 | 0.136756329 | 0.036554288 | 0.042089224 | 0.763519245 |
| 0 | 0 | 0 | 0 | 0 |
| 0 | 0 | 0.000847943 | 0 | 0 |
| 0.008801578 | 0 | 0 | 0.001736991 | 0.000911298 |
| 0 | 0 | 0 | 0 | 0 |
| 0 | 0 | 0 | 0 | 0 |
| 0 | 0 | 0 | 0 | 0 |
| 0 | 0.535495776 | 0 | 0.007229764 | 0 |
| 0.379911845 | 0.319063674 | 0.214547946 | 0.23424863 | 0.582860365 |
| 0 | 0 | 0.001836704 | 0 | 0 |
| 0 | 0 | 0 | 0 | 0 |
| 0.005676531 | 0 | 0 | 0.030454113 | 0.021739869 |
| 0 | 0 | 0 | 0 | 0 |
| 0 | 0.020335223 | 0 | 0 | 0 |
| 0 | 0 | 0 | 0 | 0 |
| 0.080203742 | 0.025473645 | 0.002809299 | 0.010007273 | 0.041524561 |
| 0.02113185 | 0 | 0.008136976 | 0 | 0 |
| 0.048489039 | 0.012483888 | 0.016925991 | 0 | 0 |
| 0 | 0 | 0 | 0.437119788 | 0 |
| 0 | 0 | 0 | 0 | 0 |
| 0.014037533 | 0 | 0 | 0 | 0 |

| | | | | |
|---|---|---|---|---|
| 0 | 0 | 0 | 0 | 0 |
| 0 | 0 | 0 | 0 | 0 |
| 0 | 0 | 0 | 0 | 0 |
| 0 | 0 | 3.151086826 | 0.184718954 | 0 |
| 0 | 0 | 0 | 0 | 0 |
| 0 | 0 | 0 | 0 | 0 |
| 0 | 0 | 0 | 0 | 0 |
| 0 | 0 | 0 | 0 | 0 |
| 0 | 1.973073744 | 0 | 1.6215601 | 0.033016994 |
| 0.245780479 | 0 | 0.077494308 | 0.506486094 | 0 |
| 0 | 0 | 0 | 0 | 0 |
| 0 | 0.004306945 | 0.003526734 | 0.007783086 | 0 |
| 0 | 0 | 0 | 0 | 0 |
| 0 | 0 | 0 | 0 | 0 |
| 0.032440792 | 0.378511866 | 0 | 0.387870557 | 0 |
| 0 | 0 | 0 | 0 | 0 |
| 0 | 0 | 0.151636766 | 0.060465489 | 0 |
| 0 | 0 | 0 | 2.339602244 | 0 |
| 0 | 0 | 0 | 0 | 0 |
| 0 | 0 | 0 | 0.026895442 | 0 |
| 0 | 0 | 0 | 0 | 0 |
| 0 | 0 | 0 | 0.024662737 | 0 |
| 0 | 0 | 0 | 0 | 0 |
| 0 | 0.399694931 | 0.261488905 | 0.176453496 | 0.03711461 |
| 0 | 0.006250222 | 0 | 0.005386012 | 0 |
| 0 | 0 | 0 | 0.472311936 | 0 |
| 0 | 0 | 0.002400921 | 0.003958205 | 0 |
| 0 | 0 | 0 | 5.467225644 | 0 |
| 0 | 0 | 0 | 0 | 0 |
| 0 | 0 | 0 | 0 | 0 |
| 0 | 0.042498913 | 0 | 0 | 0 |
| 0 | 0 | 0 | 0.318636835 | 0 |
| 0.001361522 | 0.004780019 | 0.170046291 | 0.206309972 | 0 |
| 0 | 0 | 0 | 0 | 0 |
| 0 | 0 | 0 | 0 | 0 |
| 0 | 0 | 0 | 0.31631376 | 0 |
| 0 | 0 | 2.385138352 | 1.233128013 | 0 |
| 0 | 0 | 0 | 0 | 0 |
| 0 | 0 | 0 | 0.015323714 | 0 |
| 0 | 0 | 0 | 0.002179927 | 0 |
| 0.161284695 | 0 | 0.045486192 | 0.168645059 | 0 |
| 0 | 0 | 0 | 0.133219453 | 0 |
| 0.078162389 | 0.022304403 | 0.67486301 | 2.212174235 | 0.174483095 |
| 0.004613434 | 0.010257974 | 0 | 0.00381773 | 0 |
| 0 | 0 | 0 | 0 | 0 |
| 0 | 0 | 0 | 0 | 0 |
| 0 | 0 | 0 | 0 | 0 |
| 0.000578754 | 0.001986732 | 0.022142987 | 0.020877946 | 0 |

| | | | | |
|---|---|---|---|---|
| 0 | 0 | 0 | 0 | 0 |
| 0 | 0 | 0 | 0.127632218 | 0 |
| 0.127974185 | 0.094749222 | 0.012562654 | 0.008230002 | 0.512051983 |
| 0 | 0 | 0.025286203 | 0.01536623 | 0 |
| 0 | 0 | 0.028780401 | 0.11261288 | 0 |
| 0 | 0 | 0 | 0.046414378 | 0 |
| 0 | 0.028332401 | 0 | 0 | 0 |
| 0 | 0 | 0 | 0 | 0 |
| 0 | 0 | 0 | 0 | 0 |
| 0 | 0 | 0 | 0 | 0 |
| 0.00581031 | 0.393845184 | 1.06117496 | 0.122675199 | 0 |
| 3.894571339 | 1.59576764 | 1.339652918 | 0.717463119 | 5.100615439 |
| 0 | 0 | 0 | 0 | 0 |
| 0 | 0.010594812 | 0.02455775 | 0.208900395 | 0.001247793 |
| 0.777216396 | 0.20992069 | 0.199312133 | 0.062787336 | 0.087186722 |
| 0 | 0 | 0 | 0.695154969 | 0 |
| 0 | 0 | 0 | 0 | 0 |
| 0 | 0 | 0 | 0 | 0 |
| 0 | 0 | 0 | 0.083970342 | 0 |
| 0 | 0.17655639 | 0.041867094 | 0.366389417 | 0 |
| 0.16177184 | 0.016064226 | 0.007129626 | 0.414973656 | 0.04086146 |
| 5.508513865 | 0.993102511 | 0.006385552 | 0 | 4.002100912 |
| 21.49800808 | 0.270524399 | 1.508102782 | 0.703944679 | 5.829468509 |
| 0 | 0 | 0 | 0 | 0 |
| 0 | 0 | 0 | 0.018404749 | 0 |
| 0 | 0 | 0.005126059 | 1.494186526 | 0 |
| 3.68716762 | 0 | 0 | 0 | 0 |
| 0 | 0 | 0 | 0 | 0 |
| 0 | 0 | 0 | 0 | 0 |
| 0 | 0 | 0 | 0 | 0 |
| 0 | 0 | 0.001572759 | 0.034973023 | 0 |
| 0 | 0 | 0.103824345 | 0.11000281 | 0 |
| 0.026107112 | 0.041049059 | 0.197877359 | 0.001131652 | 0.17890518 |
| 0 | 0 | 0 | 0.06227785 | 0 |
| 0.018461805 | 0.030103209 | 0.00648683 | 0 | 0 |
| 0.025027186 | 0 | 0.155671807 | 0.130498591 | 0.024586379 |
| 0.001143456 | 0.022079381 | 0 | 0.014043365 | 0 |
| 0 | 0 | 0 | 0 | 0 |
| 0 | 0 | 0 | 0.111880373 | 0.007770278 |
| 0 | 0 | 0 | 0 | 0 |
| 0 | 0 | 0 | 0.150603641 | 0 |
| 0 | 0 | 0 | 0 | 0 |
| 0 | 0 | 0 | 0.140851678 | 0 |
| 0 | 0 | 0 | 0 | 0 |
| 0 | 0.031526981 | 0.265385597 | 0.043884976 | 0 |
| 0 | 0 | 0 | 0 | 0 |
| 0 | 0 | 0 | 0 | 0 |
| 0.07676478 | 0.003892851 | 0.402952043 | 0.15881182 | 0.060977374 |

| | | | | |
|---|---|---|---|---|
| 0 | 0 | 0 | 0 | 0 |
| 0 | 0 | 0 | 0 | 0 |
| 0 | 0 | 0 | 0 | 0 |
| 0 | 0.043373347 | 0 | 0.016789785 | 0 |
| 0 | 0 | 0 | 0 | 0 |
| 0 | 0 | 0 | 0.2760181 | 0 |
| 0 | 0 | 0 | 0 | 0 |
| 0.010133066 | 0.059574964 | 0 | 0.000764834 | 0.004654658 |
| 0 | 0 | 0 | 0.006988511 | 0 |
| 0 | 0 | 0 | 0 | 0 |
| 0.009701762 | 0.009661285 | 0.001500755 | 0.011849109 | 0.000971334 |
| 0 | 0 | 0 | 0 | 0 |
| 0 | 0 | 0 | 0.258722105 | 0.000680383 |
| 0 | 0 | 0 | 0.17631378 | 0 |
| 0 | 0 | 0 | 0 | 0 |
| 0 | 0 | 0 | 0 | 0 |
| 0 | 0 | 0.4542799 | 0.013218926 | 0 |
| 0 | 0 | 0.002855645 | 0.000770634 | 0 |
| 0 | 0 | 0 | 0.022705512 | 0 |
| 0 | 0.006191818 | 0.160152627 | 0 | 0 |
| 0.000406151 | 0 | 0 | 0 | 0 |
| 0 | 0 | 0 | 0 | 0 |
| 0 | 0 | 0.003703898 | 0.114970041 | 0 |
| 0 | 0 | 0 | 0 | 0 |
| 0 | 0 | 0 | 0 | 0 |
| 0 | 0 | 0 | 0 | 0 |
| 0 | 0 | 0 | 0 | 0 |
| 0 | 0 | 0 | 0 | 0 |
| 0 | 0 | 0 | 0 | 0 |
| 0 | 0.573222156 | 0 | 0.824607835 | 0 |
| 0.007497044 | 0 | 0.016548167 | 0 | 0 |
| 0 | 0 | 0 | 0 | 0 |
| 0 | 0 | 0.003170141 | 0 | 0 |
| 0 | 0 | 0 | 0 | 0 |
| 0 | 0 | 0 | 0 | 0 |
| 0 | 0 | 0 | 0 | 0 |
| 0 | 0 | 0 | 0.020285256 | 0 |
| 0.010382106 | 0.006981601 | 0.002722319 | 0 | 0.006084657 |
| 0 | 0 | 0 | 0 | 0 |
| 0 | 0 | 0 | 0 | 0 |
| 0 | 0 | 0 | 0.007352399 | 0.00154295 |
| 0 | 0 | 0 | 0 | 0 |
| 0 | 0 | 0 | 0 | 0 |
| 0 | 0 | 0.014452197 | 0.006544588 | 0 |
| 0 | 0 | 0 | 0.068514985 | 0 |
| 0 | 0 | 0 | 0 | 0 |
| 0 | 0 | 0 | 0.006701227 | 0 |
| 0 | 0 | 0 | 0 | 0 |

| | | | | |
|---|---|---|---|---|
| 0 | 0 | 0 | 0 | 0 |
| 0 | 0 | 0 | 0 | 0 |
| 0 | 0 | 0 | 0.644553091 | 0 |
| 0 | 0 | 1.003370819 | 0.273598623 | 0 |
| 0 | 0 | 0 | 0 | 0 |
| 0 | 0 | 0 | 0 | 0 |
| 0 | 0 | 0 | 0 | 0 |
| 0 | 0 | 0 | 0 | 0 |
| 0.134210712 | 0.011004336 | 0.034792005 | 0.004971488 | 0.031072185 |
| 0 | 0 | 0 | 0.012532749 | 0 |
| 0 | 0 | 0 | 0.017928397 | 0 |
| 0 | 0 | 0 | 0 | 0 |
| 0 | 0 | 0 | 0 | 0 |
| 0 | 0 | 0 | 0 | 0 |
| 0 | 0 | 0.894152471 | 2.005805561 | 0 |
| 0 | 0.086085678 | 0.548649866 | 0 | 0.105931443 |
| 0 | 0 | 0 | 0 | 0 |
| 0 | 0 | 0 | 0 | 0 |
| 0 | 0 | 0 | 0 | 0 |
| 0 | 0 | 0 | 0 | 0 |
| 0 | 0.008204033 | 0 | 0.038509988 | 0 |
| 0 | 0 | 0 | 0 | 0 |
| 0 | 0 | 0 | 0 | 0 |
| 0 | 0 | 0 | 0 | 0 |
| 0 | 13.73987898 | 0.019721333 | 0.055921396 | 0.005601023 |
| 2.053969982 | 6.700243178 | 5.467143867 | 10.70504483 | 0 |
| 0 | 0 | 0 | 0.790946245 | 0 |
| 0 | 0 | 0 | 0 | 0 |
| 0 | 0 | 0.545827776 | 0.191342735 | 0 |
| 0 | 0 | 0 | 0 | 0 |
| 0 | 0 | 0 | 0 | 0 |
| 0 | 0 | 0 | 0.055657518 | 0 |
| 0 | 0 | 0 | 0 | 0 |
| 0.003892385 | 0.009157354 | 0 | 0 | 0 |
| 0.002874982 | 0.000854062 | 0.009583643 | 0.419395746 | 0 |
| 0.001406952 | 0.072043705 | 0.014830604 | 0 | 0 |
| 0 | 0 | 0 | 0 | 0 |
| 0.986577579 | 0 | 1.51998656 | 0.795025039 | 0 |
| 0 | 0.90987905 | 0 | 0.08666526 | 0 |
| 0 | 0 | 0 | 0 | 0 |
| 0 | 0 | 0 | 0.02686105 | 0 |
| 0 | 0 | 0 | 0.00125511 | 0 |
| 0 | 0.012374875 | 1.158931247 | 0.117322906 | 0 |
| 0 | 0 | 0 | 0 | 0 |
| 0 | 0 | 0 | 0 | 0 |
| 0 | 0 | 0 | 0.003445222 | 0 |
| 0 | 0 | 0 | 0 | 0 |
| 0.040110286 | 0 | 0 | 4.594239926 | 0 |

| | | | | |
|---|---|---|---|---|
| 0 | 0 | 0 | 0.008897594 | 0 |
| 0 | 0 | 0 | 0.692558226 | 0 |
| 0 | 0 | 0 | 0 | 0 |
| 0 | 0 | 0 | 0 | 0 |
| 0 | 0.008794362 | 0.025235136 | 0.002657576 | 0 |
| 0 | 0.003040096 | 0 | 0.001194296 | 0 |
| 0 | 0 | 0 | 0 | 0 |
| 0 | 0 | 0 | 0 | 0 |
| 0 | 0 | 0 | 0 | 0 |
| 0 | 0 | 0 | 0.01209237 | 0 |
| 0 | 0 | 0 | 0 | 0 |
| 0 | 0 | 0.536031368 | 2.178327709 | 0 |
| 0.068084729 | 0 | 0.203453752 | 0.06950574 | 0 |
| 0 | 0 | 0 | 0 | 0 |
| 0.002633616 | 0 | 0 | 0.203259895 | 0.777429763 |
| 0 | 0 | 0 | 0 | 0 |
| 0 | 0 | 0 | 0 | 0 |
| 0 | 0 | 0.559200233 | 0 | 0 |
| 0.000532697 | 0 | 0 | 0.001975526 | 0 |
| 0 | 0 | 0.010838351 | 0.022502856 | 0 |
| 0 | 0 | 0 | 0.150173667 | 0 |
| 0 | 0 | 0 | 0 | 0 |
| 0.02641676 | 0.047563981 | 0.031344182 | 1.003599181 | 0 |
| 0 | 0 | 0 | 0 | 0 |
| 0 | 0 | 0 | 0 | 0 |
| 0 | 0 | 0 | 0 | 0 |
| 0 | 0 | 0 | 0 | 0 |
| 0 | 0 | 0 | 0 | 0 |
| 0 | 0 | 0 | 0 | 0 |
| 0 | 0 | 0 | 0 | 0 |
| 0 | 0 | 0 | 0 | 0 |
| 0.054153987 | 0.001692036 | 0.052957659 | 0.021802588 | 0.025945967 |
| 0 | 0 | 0 | 0 | 0 |
| 0 | 0.112088355 | 0.119516757 | 0.015275642 | 0 |
| 0.001190868 | 0.069860647 | 0.011158113 | 0 | 0 |
| 0.003070347 | 0.01242191 | 0.597713256 | 0.108689214 | 0.011404576 |
| 0 | 0 | 0 | 0.001527382 | 0 |
| 0 | 0 | 0 | 0 | 0 |
| 0 | 0 | 0 | 0 | 0 |
| 0 | 0 | 0 | 0 | 0 |
| 0 | 0 | 0 | 0.146161575 | 0 |
| 0 | 0 | 0 | 0 | 0 |
| 0 | 0 | 0 | 0 | 0 |
| 0 | 0.237654251 | 0.118088507 | 0 | 0 |
| 0.027652219 | 0.363730659 | 0.198834893 | 0.059156496 | 0 |
| 0 | 0 | 0 | 0 | 0 |
| 0 | 0 | 0 | 0.395331205 | 0 |
| 0 | 0 | 0.005198166 | 0.001282555 | 0 |

| | | | | |
|---|---|---|---|---|
| 0 | 0 | 0 | 0 | 0 |
| 0 | 0 | 0 | 0 | 0 |
| 0 | 0 | 0.029029853 | 0 | 0 |
| 0 | 0 | 0 | 0 | 0 |
| 0.001230466 | 0 | 0 | 0 | 0 |
| 0 | 0 | 0.006636472 | 0 | 0 |
| 0 | 0.011789338 | 0 | 0.343689919 | 0 |
| 0 | 0 | 0.003450963 | 0.010264867 | 0 |
| 0 | 0 | 0 | 0 | 0 |
| 0 | 0 | 0.012451557 | 0.009286422 | 0 |
| 0 | 0 | 0 | 0 | 0 |
| 0 | 0.012713352 | 0 | 0 | 0 |
| 0 | 0 | 0 | 0 | 0 |
| 0 | 0 | 0 | 0.317259367 | 0 |
| 0 | 0 | 0 | 0 | 0 |
| 0 | 0 | 0 | 0 | 0 |
| 0 | 0 | 0 | 0 | 0 |
| 0 | 0 | 0 | 0 | 0 |
| 0 | 3.241507139 | 1.137303822 | 5.547000994 | 0 |
| 0 | 0 | 0 | 0 | 0 |
| 0 | 0 | 0 | 0 | 0 |
| 0 | 0 | 0 | 0 | 0 |
| 0 | 0 | 0 | 0 | 0 |
| 0 | 0 | 0 | 0 | 0 |
| 0 | 0 | 0 | 0 | 0 |
| 0.006708826 | 0.084755295 | 0.03819104 | 0 | 0.026989161 |
| 0 | 0 | 0 | 0 | 0 |
| 0.13316467 | 0.003538517 | 0.004113746 | 0.446067622 | 0 |
| 0 | 0 | 0 | 0.139931551 | 0 |
| 0 | 0.251753712 | 0.937354333 | 0.179833019 | 0 |
| 0 | 0 | 0 | 0 | 0 |
| 0 | 0 | 0 | 0 | 0 |
| 0 | 0 | 0 | 0 | 0 |
| 0 | 0 | 0 | 0 | 0 |
| 0.002452477 | 0.003367519 | 0.001531937 | 0.004299501 | 0 |
| 0 | 0 | 0 | 0 | 0 |
| 0 | 0 | 0 | 0 | 0 |
| 0 | 0.012896372 | 0.009126072 | 0.000750565 | 0 |
| 0 | 0 | 0 | 0 | 0 |
| 0 | 0 | 0 | 0 | 0 |
| 5.218900528 | 7.806098253 | 5.032909681 | 2.365498535 | 7.034282304 |
| 0.078630283 | 1.469175312 | 0.171779822 | 0.03420122 | 0.013115244 |
| 0 | 0 | 0 | 0 | 0 |
| 0 | 0 | 0.007327258 | 0.547031461 | 0 |
| 0 | 0 | 0 | 0 | 0 |
| 0 | 0 | 0 | 0 | 0 |
| 0 | 0 | 0 | 0.011924277 | 0 |
| 0.004269996 | 0.004549011 | 0.00776031 | 0.005510134 | 0 |

| | | | | |
|---|---|---|---|---|
| 0 | 0 | 0 | 0 | 0 |
| 0 | 0.000974799 | 0.001951185 | 0.016390166 | 0 |
| 0 | 0 | 0 | 0 | 0 |
| 0 | 0 | 0 | 0 | 0 |
| 0 | 0.208996562 | 4.016911935 | 1.917523498 | 0 |
| 0 | 0 | 0 | 0 | 0 |
| 0 | 0 | 0 | 0 | 0 |
| 0 | 0 | 0 | 0.159323814 | 0 |
| 0.276290769 | 0.540799614 | 0.032589881 | 0 | 0.883504666 |
| 0 | 0 | 0.00129028 | 0 | 0 |
| 0 | 0 | 0 | 0 | 0.034865181 |
| 0 | 0 | 0 | 0 | 0 |
| 0 | 0 | 0.065150312 | 0.103741218 | 0 |
| 0.020262742 | 0.012343331 | 0.006144907 | 0.002561765 | 0.517635167 |
| 0 | 0 | 0 | 0 | 0 |
| 0 | 0 | 0.532762629 | 0.353313947 | 0 |
| 0 | 0 | 0 | 0.069512823 | 0 |
| 0 | 0 | 0 | 0 | 0 |
| 0 | 0 | 0 | 0 | 0 |
| 0 | 0 | 0 | 0 | 0 |
| 0 | 0 | 0 | 0 | 0 |
| 0 | 0 | 0 | 0 | 0 |
| 0 | 0 | 0 | 0 | 0 |
| 0 | 0 | 0 | 0 | 0 |
| 0 | 0 | 0 | 0.019418846 | 0 |
| 0 | 0 | 0 | 0 | 0 |
| 0 | 0 | 0.172354392 | 0.034834617 | 0 |
| 0.002611361 | 0.00079096 | 0.002158919 | 0 | 0.000489061 |
| 0 | 0 | 0 | 0 | 0 |
| 0 | 0 | 0 | 0 | 0 |
| 0 | 0 | 0 | 0 | 0 |
| 0 | 0 | 0 | 0 | 0 |
| 0 | 0 | 0 | 0 | 0 |
| 0 | 0 | 0.104858067 | 0 | 0 |
| 0 | 0 | 0 | 0.137725344 | 0 |
| 0.176805829 | 0.000686232 | 0.007908502 | 0 | 0 |
| 0 | 0 | 0 | 0.001102193 | 0 |
| 0 | 0 | 0 | 0 | 0 |
| 0 | 0 | 0 | 0 | 0 |
| 0 | 0 | 0 | 0 | 0 |
| 0 | 0.184060261 | 2.354433462 | 0.088266054 | 0 |
| 0 | 1.153593921 | 0 | 0.078030306 | 0 |
| 0 | 0 | 0 | 0 | 0 |
| 0 | 0.012584859 | 0 | 0 | 0 |
| 0 | 0 | 0 | 0 | 0 |
| 0 | 0 | 0 | 0 | 0 |
| 0 | 0 | 0 | 0 | 0 |
| 0 | 0 | 0 | 0 | 0 |

| | | | | |
|---|---|---|---|---|
| 0 | 0 | 0 | 0.594188985 | 0 |
| 0 | 0 | 0 | 0 | 0 |
| 0 | 0 | 0 | 0 | 0 |
| 0 | 0 | 0 | 0 | 0 |
| 0.03013523 | 0 | 0.007409973 | 0.033173641 | 0 |
| 0 | 0 | 0 | 0 | 0 |
| 0.020458953 | 0 | 0 | 0.167278089 | 0 |
| 0 | 0 | 0 | 0 | 0 |
| 0 | 0 | 0 | 0.000557466 | 0 |
| 0 | 0 | 0.037230068 | 0 | 0 |
| 0 | 0 | 0 | 0.06187084 | 0 |
| 0 | 0 | 0 | 0 | 0 |
| 0.00500993 | 0 | 0 | 0 | 0 |
| 0 | 0 | 0 | 0 | 0 |
| 0 | 0 | 0 | 0.008536407 | 0 |
| 0 | 0 | 0 | 0 | 0 |
| 0 | 0 | 0 | 0 | 0.130131414 |
| 0 | 0 | 0 | 0 | 0 |
| 0 | 0 | 0 | 0 | 0 |
| 0 | 0 | 0 | 0 | 0 |
| 0.002676375 | 0 | 0.001567307 | 0.169333881 | 0 |
| 0 | 0 | 0 | 0 | 0 |
| 0 | 0 | 0.003841632 | 0.006966729 | 0 |
| 0 | 0.005256389 | 0.029036159 | 0.000589989 | 0.003250095 |
| 0 | 0 | 0 | 0 | 0 |
| 0.001032295 | 0 | 0 | 0 | 0 |
| 0 | 0 | 0 | 0 | 0 |
| 0 | 0 | 0 | 0 | 0 |
| 0 | 0 | 0 | 0 | 0 |
| 1.57330982 | 0 | 0 | 0.911968689 | 0.533029285 |
| 0 | 0 | 0 | 0 | 0 |
| 0 | 0 | 0 | 0 | 0 |
| 0 | 0 | 0 | 0 | 0 |
| 0 | 0 | 0 | 0 | 0 |
| 0 | 0 | 0 | 0 | 0 |
| 0 | 0 | 0 | 1.474507717 | 0 |
| 0.024633953 | 0.005167724 | 0.18037648 | 0.001476459 | 0.145046033 |
| 0 | 0 | 0 | 0 | 0 |
| 0 | 0 | 0 | 0 | 0 |
| 0 | 0 | 0 | 0 | 0 |
| 0 | 0 | 0 | 0 | 0 |
| 0 | 0 | 0 | 0 | 0 |
| 0 | 0 | 0 | 0 | 0 |
| 0 | 0 | 0.000611811 | 0 | 0 |
| 0 | 0 | 0 | 0 | 0 |
| 0 | 0 | 0 | 0 | 0 |
| 0 | 0 | 0 | 0 | 0 |
| 0 | 0 | 0 | 0 | 0 |

| | | | | |
|---|---|---|---|---|
| 0.013024403 | 0.06241867 | 0.039881968 | 0.001110986 | 0 |
| 0 | 0 | 0 | 0 | 0 |
| 0 | 0 | 0 | 0.001508911 | 0 |
| 0 | 0 | 0 | 0 | 0 |
| 0 | 0 | 0 | 0 | 0 |
| 0 | 0 | 0 | 0 | 0 |
| 0 | 0 | 0 | 0 | 0 |
| 1.514384785 | 0 | 1.666485697 | 0.363241647 | 0 |
| 0.032512962 | 16.26551467 | 0.981129552 | 0.163175476 | 0.053848489 |
| 0.021434232 | 0.241706503 | 0.249032894 | 0 | 0 |
| 0 | 0 | 0 | 0.005087981 | 0 |
| 0 | 0 | 0 | 0 | 0 |
| 0 | 0 | 0 | 0.043557088 | 0 |
| 0 | 0 | 0 | 0 | 0 |
| 0 | 0 | 0 | 0 | 0 |
| 0 | 0 | 0 | 0 | 0 |
| 0.262733802 | 0 | 0.076191757 | 0 | 0 |
| 0 | 0 | 0 | 0 | 0 |
| 0 | 0 | 0 | 0 | 0 |
| 0 | 0 | 0.122570155 | 0 | 0 |
| 0 | 0 | 0 | 0 | 0 |
| 0 | 0 | 0 | 0 | 0 |
| 0 | 0 | 0 | 0 | 0 |
| 0 | 0 | 0 | 0 | 0 |
| 0 | 0 | 0.022148039 | 0.022126411 | 0 |
| 0 | 0 | 0 | 0 | 0 |
| 0 | 0 | 0 | 0 | 0 |
| 0 | 0 | 0 | 0 | 0 |
| 0 | 0 | 0 | 0 | 0 |
| 0 | 0 | 0 | 0 | 0 |
| 0 | 0 | 0 | 0 | 0 |
| 0 | 0 | 0 | 0 | 0 |
| 0 | 0 | 0 | 0 | 0 |
| 0 | 0 | 0 | 0 | 0 |
| 0 | 0.022254238 | 0 | 0 | 0 |
| 0 | 0 | 0 | 0 | 0 |
| 0 | 0 | 0 | 0 | 0 |
| 0 | 0 | 0 | 0 | 0 |
| 0 | 0 | 0 | 0.000566164 | 0 |
| 0.396025909 | 0.263369822 | 0 | 0 | 0 |
| 0 | 0 | 0 | 0 | 0 |
| 0 | 0 | 0 | 0 | 0 |
| 0 | 0 | 0 | 0 | 0 |
| 0 | 0 | 0 | 0 | 0 |
| 0 | 0 | 0 | 0 | 0 |
| 0 | 0 | 0 | 0 | 0 |
| 0 | 0 | 0 | 0 | 0 |
| 0 | 0 | 0 | 0 | 0 |

| | | | | |
|---|---|---|---|---|
| 0 | 0 | 0 | 0 | 0 |
| 0 | 0 | 0 | 0.02549758 | 0 |
| 0 | 0 | 0 | 0 | 0 |
| 0 | 0 | 0 | 0 | 0 |
| 0.072671629 | 0.340550555 | 0.400291843 | 0.504095739 | 0.009246454 |
| 0 | 0 | 0 | 0 | 0 |
| 0 | 0 | 0 | 0 | 0 |
| 0.035930567 | 0.006167073 | 0.064058866 | 0.013728765 | 0.009321117 |
| 0 | 0 | 0.002100889 | 0.027415325 | 0 |
| 0 | 0 | 0 | 0.031793404 | 0 |
| 0 | 0 | 0 | 0 | 0 |
| 0 | 0 | 0 | 0 | 0 |
| 0 | 0 | 0 | 0 | 0 |
| 0 | 0 | 0 | 0 | 0 |
| 0 | 0.008682804 | 0.037524458 | 0 | 0 |
| 0 | 0.19968318 | 0 | 0.042199295 | 0 |
| 0 | 0 | 0 | 0 | 0 |
| 0 | 0 | 0 | 0 | 0 |
| 0 | 0 | 0 | 0 | 0 |
| 0 | 0 | 0 | 0 | 0 |
| 0 | 0 | 0 | 0 | 0 |
| 0 | 0 | 0 | 0 | 0 |
| 0 | 0 | 0 | 0 | 0 |
| 0.001209248 | 0.005114715 | 0.000809309 | 0.008910836 | 0 |
| 0 | 0 | 0 | 0.263960646 | 0 |
| 0 | 0 | 0 | 0 | 0 |
| 0 | 0.13362773 | 0.119543111 | 0.567301915 | 0.000503037 |
| 0 | 0 | 0.000584559 | 0 | 0 |
| 0 | 0.003325468 | 0 | 0 | 0 |
| 0 | 0 | 0 | 0 | 0 |
| 0 | 0 | 0 | 0 | 0 |
| 0 | 0 | 0 | 0 | 0 |
| 0 | 0 | 0 | 0 | 0 |
| 0 | 0 | 0 | 0 | 0 |
| 0 | 0 | 0 | 0 | 0 |
| 0 | 0 | 0 | 0 | 0 |
| 0 | 0 | 0 | 0 | 0 |
| 0 | 0 | 0 | 0.312369969 | 0 |
| 0 | 0 | 0 | 0 | 0 |
| 0 | 0 | 0 | 0 | 0 |
| 0 | 0 | 0 | 0 | 0 |
| 0 | 0 | 0 | 0 | 0 |
| 0 | 0 | 0 | 0 | 0 |
| 0 | 0 | 0 | 0 | 0 |
| 0 | 0 | 0 | 0 | 0 |
| 0 | 0 | 0 | 0 | 0 |
| 0 | 0 | 0 | 0 | 0 |
| 0 | 0 | 0 | 0 | 0 |

| | | | | |
|---|---|---|---|---|
| 0 | 0 | 0.090840923 | 0.184094573 | 0 |
| 0 | 0 | 0 | 0.004174293 | 0 |
| 0 | 0 | 0 | 0 | 0 |
| 0 | 0 | 0 | 0 | 0 |
| 0 | 0 | 0 | 0 | 0 |
| 0.040625961 | 0 | 0.030645898 | 0 | 0 |
| 0 | 0 | 0 | 0 | 0 |
| 0 | 0 | 0 | 0 | 0 |
| 0 | 0 | 0 | 0 | 0 |
| 0 | 0 | 0 | 0 | 0 |
| 0 | 0 | 0 | 0 | 0 |
| 0 | 0.007584922 | 0 | 0 | 0 |
| 0 | 0 | 0 | 0 | 0 |
| 0 | 0 | 0 | 0 | 0 |
| 0 | 0 | 0 | 0 | 0 |
| 0 | 0 | 0 | 0 | 0 |
| 0 | 0 | 0 | 0 | 0 |
| 0 | 0 | 0 | 0 | 0 |
| 0 | 0 | 0 | 0 | 0 |
| 0 | 0 | 0 | 0 | 0 |
| 0 | 0.830434913 | 0.103762966 | 0 | 0.012049407 |
| 0 | 0 | 0 | 0 | 0 |
| 0.011297431 | 0.092292536 | 0.029008035 | 0.143709363 | 0 |
| 0 | 0 | 1.336100245 | 0.662076057 | 0 |
| 0 | 0 | 0 | 0 | 0 |
| 0 | 0 | 0 | 0.089933352 | 0 |
| 0 | 0 | 0 | 0.038129885 | 0 |
| 0 | 0 | 0 | 0 | 0 |
| 0 | 0 | 0 | 0 | 0 |
| 0 | 0 | 0 | 0 | 0 |
| 0 | 0 | 0 | 0 | 0 |
| 0 | 0 | 0 | 0 | 0 |
| 0.345477598 | 0.532994739 | 0.021446767 | 0.001234708 | 6.25409451 |
| 0 | 0 | 0 | 0 | 0 |
| 0.072070151 | 0 | 0.222961073 | 0.0999519 | 0 |
| 0.000816347 | 0 | 0.057175882 | 0 | 0.00086636 |
| 0 | 0 | 0 | 0 | 0 |
| 0 | 0 | 0 | 0 | 0 |
| 0 | 0.152667131 | 0.124082226 | 0 | 0.07495574 |
| 0 | 0 | 0 | 0 | 0 |
| 0 | 0 | 0 | 0 | 0 |
| 0 | 0 | 0 | 0 | 0 |
| 0 | 0.184483027 | 0 | 1.475160302 | 0.002577815 |
| 0 | 0 | 0 | 0 | 0 |
| 0 | 0 | 0 | 0 | 0 |
| 0 | 0 | 0 | 0.088330582 | 0 |
| 0 | 0 | 0 | 0 | 0 |
| 0 | 0 | 0 | 0 | 0 |

| | | | | |
|---|---|---|---|---|
| 0 | 0 | 0 | 0 | 0 |
| 0 | 0 | 0 | 0 | 0 |
| 0 | 0 | 0.00410494 | 0 | 0 |
| 0 | 0 | 0 | 0 | 0 |
| 0 | 0 | 0 | 0 | 0 |
| 0 | 0 | 0 | 0 | 0 |
| 0 | 0 | 0 | 0 | 0 |
| 1.595646736 | 0.002617748 | 0 | 0 | 0 |
| 0 | 0.012267376 | 0.001924352 | 0.021271021 | 0 |
| 0 | 0.003952834 | 0 | 0.006211454 | 0.00230032 |
| 0 | 0 | 0 | 0 | 0 |
| 0 | 0 | 0.001126021 | 0 | 0 |
| 0 | 0 | 0 | 0 | 0 |
| 0 | 0 | 0 | 0.01398458 | 0 |
| 0 | 0 | 0 | 0 | 0 |
| 0.022392377 | 0.01075236 | 0 | 0 | 0 |
| 0 | 0 | 0 | 0 | 0 |
| 0 | 0 | 0 | 0 | 0 |
| 0 | 0 | 0 | 0 | 0 |
| 0 | 0 | 0.937899025 | 1.235669438 | 0 |
| 0 | 0 | 0.006956578 | 0 | 0 |
| 0 | 0 | 0 | 0 | 0 |
| 0 | 0 | 0 | 0 | 0 |
| 0 | 0 | 0.004436063 | 0.004309671 | 0 |
| 0 | 0 | 0 | 0 | 0 |
| 0 | 0 | 0 | 0 | 0 |
| 0 | 0 | 0 | 0 | 0 |
| 0 | 0 | 0 | 0 | 0 |
| 0 | 0 | 0 | 0 | 0 |
| 0 | 0 | 0 | 0.045083035 | 0 |
| 0 | 0 | 0.28336765 | 0.351061408 | 0 |
| 0 | 0 | 0 | 0 | 0 |
| 0 | 0 | 0 | 0 | 0 |
| 0 | 0 | 0 | 0 | 0 |
| 0 | 0 | 0 | 0 | 0 |
| 0 | 0 | 0 | 0 | 0 |
| 0 | 0 | 0 | 0 | 0 |
| 0 | 0 | 0 | 0 | 0 |
| 0 | 0 | 0 | 0 | 0 |
| 0 | 0 | 0 | 0 | 0 |
| 0 | 0 | 0 | 0 | 0 |
| 0 | 0 | 0.001511626 | 0.998991879 | 0 |
| 0 | 0 | 0 | 0 | 0 |
| 0 | 0 | 0 | 0 | 0 |
| 0 | 0 | 0 | 0 | 0 |
| 0 | 0 | 0 | 0 | 0 |
| 0.07295308 | 0.008899368 | 0.015980768 | 0 | 0.355641621 |
| 0 | 0 | 0 | 0 | 0 |

| | | | | |
|---|---|---|---|---|
| 0 | 0 | 0 | 0 | 0 |
| 0 | 0 | 0 | 0 | 0 |
| 0 | 0 | 0 | 0 | 0 |
| 0 | 0 | 0 | 0 | 0 |
| 0.055724623 | 0.393135855 | 0.005253084 | 0.047975519 | 0 |
| 0 | 0 | 0 | 0.040212789 | 0 |
| 0 | 0 | 0 | 0 | 0 |
| 0 | 0 | 0 | 0 | 0 |
| 0 | 0 | 0 | 0 | 0 |
| 0 | 0 | 0 | 0 | 0 |
| 0 | 0 | 0 | 0.0586433 | 0 |
| 0.006981767 | 0.080888016 | 0.038012743 | 0.067561283 | 0 |
| 0 | 0 | 0.014109657 | 0 | 0 |
| 0 | 0 | 0 | 0 | 0 |
| 0 | 0 | 0 | 0 | 0.002289145 |
| 0 | 0 | 0 | 0 | 0 |
| 0 | 0.00261554 | 0 | 0 | 0 |
| 0 | 0 | 0 | 0 | 0 |
| 0.167171946 | 0.014530512 | 0.088860587 | 0 | 0.004397433 |
| 0 | 0.001200786 | 0.006336574 | 0.081703048 | 0 |
| 0 | 0 | 0 | 0 | 0 |
| 0 | 0 | 0 | 0.045330617 | 0 |
| 0 | 0 | 0 | 0 | 0 |
| 0 | 0 | 0 | 0 | 0 |
| 0 | 0 | 0 | 0 | 0 |
| 0 | 0 | 0 | 0 | 0 |
| 0.003587985 | 0.028482452 | 0.046750659 | 0.015423059 | 0.002221218 |
| 0 | 0 | 0.01129013 | 0 | 0 |
| 0 | 0 | 0 | 0 | 0 |
| 0 | 0 | 0 | 0 | 0 |
| 0.001016792 | 0.002399662 | 0.000595442 | 0.001028403 | 0 |
| 0 | 0 | 0.129272465 | 0 | 0 |
| 0 | 0 | 0 | 0 | 0 |
| 0 | 0 | 0 | 0 | 0 |
| 0 | 0 | 0 | 0 | 0 |
| 0 | 0 | 0 | 0 | 0 |
| 0 | 0 | 0 | 0 | 0 |
| 0.00488152 | 0.091500158 | 0.370788849 | 0.014932203 | 0 |
| 0 | 0 | 0 | 0.519112909 | 0 |
| 0 | 0 | 0 | 0 | 0 |
| 0.001664064 | 0.002601161 | 0.004733235 | 0 | 0 |
| 0 | 0 | 0 | 0 | 0 |
| 0 | 0 | 0 | 0 | 0 |
| 8.883266709 | 0 | 0.009305182 | 0.002499967 | 0.216973892 |
| 0 | 0 | 0 | 0 | 0 |
| 0 | 0 | 0 | 0 | 0 |
| 0 | 0 | 0 | 0 | 0 |
| 0 | 0 | 0 | 0.004496551 | 0 |

| | | | | |
|---|---|---|---|---|
| 0 | 0 | 0 | 0 | 0 |
| 0 | 0 | 0 | 0.018314898 | 0 |
| 0 | 0 | 0 | 0 | 0 |
| 0.001480227 | 0.012861998 | 0.001733666 | 0 | 0.00333819 |
| 0.001029665 | 0 | 0 | 0.001909275 | 0 |
| 0.008859212 | 0 | 0 | 0 | 0 |
| 0 | 0 | 0.003364783 | 0 | 0 |
| 0 | 0 | 0 | 0 | 0 |
| 0 | 0 | 0 | 0 | 0 |
| 0 | 0 | 0 | 0.017732705 | 0 |
| 0 | 0 | 0 | 0 | 0 |
| 0 | 0 | 0 | 0 | 0 |
| 0 | 0 | 0.450093891 | 0.064736495 | 0 |
| 0 | 0 | 0 | 0.002059884 | 0 |
| 0 | 0 | 0 | 0 | 0 |
| 0 | 0 | 0 | 0 | 0 |
| 0 | 0 | 0 | 0 | 0 |
| 0 | 0 | 0 | 0 | 0 |
| 0 | 0 | 0 | 0 | 0 |
| 0 | 0 | 0 | 0 | 0 |
| 0 | 0 | 0 | 0 | 0 |
| 0 | 0 | 0 | 0 | 0 |
| 0.019688517 | 0.018230489 | 0.014361626 | 0 | 0.010997225 |
| 0.016507749 | 0 | 0.010741191 | 0.045553954 | 0 |
| 0 | 0 | 0 | 0 | 0 |
| 0 | 0 | 0 | 0.043536748 | 0 |
| 0 | 0 | 0 | 0.005006702 | 0 |
| 0 | 0 | 0 | 0 | 0 |
| 0 | 0 | 0 | 0 | 0 |
| 0 | 0 | 0 | 0 | 0 |
| 0 | 0.022805806 | 0.025711255 | 0.013828358 | 0 |
| 0.003127174 | 0.015718926 | 0 | 0 | 0 |
| 0 | 0 | 0 | 0 | 0 |
| 0 | 0 | 0 | 0 | 0 |
| 0 | 0 | 0 | 3.236840347 | 0 |
| 0 | 0.00077741 | 0 | 0 | 0 |
| 0 | 0.019225582 | 0 | 0 | 0.051023692 |
| 0 | 0 | 0 | 0 | 0 |
| 0.001140474 | 0 | 0 | 0.028178272 | 0.006570442 |
| 0 | 0 | 0 | 0 | 0 |
| 0 | 0 | 0 | 0 | 0 |
| 0 | 0 | 0 | 0 | 0 |
| 0 | 0 | 0 | 0 | 0 |
| 0 | 0 | 0 | 0 | 0 |
| 0 | 0 | 0 | 0 | 0 |
| 0 | 0.487341171 | 0.346861288 | 0.303596007 | 0 |
| 0 | 0 | 0 | 0 | 0 |
| 0 | 0 | 0 | 0 | 0 |

| | | | | |
|---|---|---|---|---|
| 0 | 0 | 0 | 0 | 0 |
| 0 | 0 | 0 | 0 | 0 |
| 0 | 0 | 0 | 0.12566185 | 0 |
| 0 | 0 | 0 | 0 | 0 |
| 0 | 0 | 0 | 0 | 0 |
| 0 | 0 | 0.003864212 | 0 | 0 |
| 0 | 0 | 0 | 0 | 0 |
| 0 | 0 | 0 | 0 | 0 |
| 0 | 0 | 0.015890351 | 0 | 0 |
| 0 | 0 | 0 | 0.004701567 | 0.000789324 |
| 0 | 0.001701484 | 0.004024964 | 0 | 0 |
| 0 | 0.002794353 | 0 | 0 | 0 |
| 0 | 0 | 0 | 0 | 0 |
| 0 | 0 | 0 | 0 | 0 |
| 0 | 0 | 0 | 0 | 0 |
| 0 | 0 | 0 | 0 | 0 |
| 0 | 0 | 0 | 0.002161622 | 0 |
| 0 | 0 | 0 | 0 | 0 |
| 0 | 0 | 0 | 0 | 0 |
| 0 | 0.000902584 | 0 | 0 | 0.001302186 |
| 0 | 0 | 0 | 0 | 0 |
| 0 | 0 | 0 | 0 | 0 |
| 0 | 0 | 0 | 0 | 0 |
| 0 | 0 | 0 | 0 | 0 |
| 0 | 0 | 0 | 0 | 0 |
| 0 | 0 | 0 | 0 | 0 |
| 0 | 0 | 0 | 0 | 0 |
| 0 | 0 | 0 | 0.007455619 | 0 |
| 0 | 0 | 0 | 0 | 0 |
| 0 | 0 | 0 | 0 | 0 |
| 0 | 0.001744679 | 0.001940111 | 0.001827717 | 0.008789885 |
| 0 | 0 | 0 | 0 | 0 |
| 0.172402693 | 0.047042427 | 0.001638303 | 0.055353164 | 0.027834414 |
| 0 | 0 | 0 | 0.002168624 | 0.0007585 |
| 0 | 0 | 0 | 0.033516021 | 0 |
| 0 | 0 | 0 | 0 | 0 |
| 0 | 0 | 0 | 0 | 0 |
| 0 | 0 | 0 | 0 | 0 |
| 0 | 0 | 0 | 0 | 0 |
| 0 | 0 | 0 | 0 | 0 |
| 0 | 0 | 0 | 0 | 0 |
| 0.003740048 | 0.086291111 | 0.349758523 | 0 | 0.027326283 |
| 0 | 0 | 0.040229519 | 0.105285625 | 0 |
| 0 | 0 | 0 | 0 | 0 |
| 0 | 0 | 0 | 0 | 0 |
| 0 | 0.006519986 | 0.012753987 | 0.033297738 | 0 |
| 0 | 0 | 0 | 0 | 0 |
| 0 | 0 | 0 | 0 | 0 |

| | | | | |
|---|---|---|---|---|
| 0 | 0 | 0 | 0 | 0 |
| 0 | 0 | 0 | 0.002599679 | 0 |
| 0.002144583 | 0 | 0.006781779 | 0.019738545 | 0.000682791 |
| 0 | 0 | 0 | 0 | 0 |
| 0.076713099 | 0.001983857 | 6.542030822 | 0.059404126 | 0 |
| 0.038669466 | 0 | 0 | 0.001517238 | 0 |
| 0 | 0 | 0 | 0 | 0 |
| 0 | 0 | 0 | 0 | 0 |
| 0 | 0 | 0 | 0 | 0 |
| 0 | 0 | 0 | 0 | 0 |
| 0 | 0 | 0 | 0 | 0 |
| 0 | 0 | 0 | 0 | 0 |
| 0 | 0 | 0 | 0 | 0 |
| 0 | 0 | 0 | 0 | 0 |
| 0 | 0 | 0 | 0 | 0 |
| 0 | 0 | 0 | 0 | 0 |
| 0 | 0 | 0 | 0.034342976 | 0 |
| 0 | 0 | 0 | 0 | 0 |
| 0 | 0 | 0 | 0 | 0 |
| 0 | 0 | 0 | 0 | 0 |
| 0 | 0 | 0 | 0 | 0 |
| 0 | 0 | 0.006869457 | 0 | 0 |
| 0 | 0 | 0 | 0 | 0 |
| 0 | 0 | 0 | 0 | 0 |
| 0 | 0.003185286 | 0 | 0.010368192 | 0 |
| 0.016002003 | 0 | 0 | 0.002157963 | 0.00132085 |
| 0 | 0 | 0 | 0 | 0 |
| 0 | 0 | 0.020806237 | 0.11321263 | 0 |
| 0 | 0 | 0 | 0 | 0 |
| 0 | 0 | 0 | 0 | 0 |
| 0 | 0 | 0 | 0 | 0 |
| 0 | 0 | 0.001095697 | 0 | 0 |
| 0 | 0 | 0 | 0 | 0 |
| 0 | 0 | 0 | 0 | 0 |
| 0 | 0.001084179 | 0 | 0 | 0 |
| 0 | 0 | 0 | 0 | 0 |
| 0 | 0 | 0 | 0 | 0 |
| 0.355385868 | 0.010935886 | 0.016583017 | 0 | 0 |
| 0 | 0 | 1.105856691 | 0 | 0 |
| 0 | 0 | 0 | 0 | 0 |
| 0 | 0 | 0.006248479 | 0 | 0 |
| 0 | 0 | 0 | 0.000975441 | 0 |
| 0.010528063 | 0.056967143 | 0.030513107 | 0.000902397 | 0 |
| 0 | 0 | 0 | 0 | 0 |
| 0 | 0 | 0 | 0 | 0 |
| 0 | 0 | 0 | 0 | 0 |
| 0.009008371 | 0.073092358 | 0.02461842 | 0.000828294 | 0 |
| 0 | 0 | 0 | 0 | 0 |

| | | | | |
|---|---|---|---|---|
| 0 | 0 | 0 | 0 | 0 |
| 0.040128628 | 0.078261722 | 0.127079663 | 0.004180619 | 0.016267173 |
| 0 | 0 | 0 | 0 | 0 |
| 0 | 0 | 0 | 0 | 0 |
| 0 | 0 | 0 | 0 | 0 |
| 0 | 0 | 0 | 0 | 0 |
| 0 | 0 | 0 | 0 | 0 |
| 0 | 0 | 0 | 0 | 0 |
| 0 | 0 | 0 | 0 | 0 |
| 0 | 0 | 0 | 0 | 0 |
| 0 | 0 | 0 | 0 | 0 |
| 0 | 0 | 0 | 0 | 0 |
| 0 | 0 | 0.010661465 | 0.004603422 | 0 |
| 0 | 0 | 0 | 0 | 0 |
| 0 | 0.069660918 | 0 | 0 | 0.004330017 |
| 0 | 0 | 0 | 0 | 0 |
| 0 | 0 | 0 | 0 | 0 |
| 0 | 0 | 0 | 0 | 0 |
| 0 | 0 | 0.001391157 | 0 | 0 |
| 0 | 0 | 0 | 0 | 0 |
| 0 | 0 | 0 | 0 | 0 |
| 0 | 0.001085913 | 0.000740998 | 0.015997458 | 0 |
| 0.108899437 | 0 | 0.006073562 | 0.007703449 | 0 |
| 0 | 0 | 3.277286281 | 0 | 0 |
| 0 | 0 | 0 | 0 | 0 |
| 0 | 0 | 0 | 0 | 0 |
| 0 | 0 | 0 | 0 | 0 |
| 0 | 0 | 0 | 0 | 0 |
| 0 | 0 | 0 | 0 | 0 |
| 0 | 0 | 0 | 0 | 0 |
| 0 | 0 | 0 | 0 | 0 |
| 0 | 0 | 0 | 0 | 0 |
| 0 | 0 | 0 | 0 | 0 |
| 0 | 0 | 0 | 0 | 0 |
| 0 | 0.018733023 | 0.092620078 | 0.647418833 | 0 |
| 0 | 0 | 0 | 0 | 0 |
| 0 | 0 | 0 | 0 | 0 |
| 0 | 0 | 0 | 0 | 0 |
| 0 | 0 | 0 | 0.00681617 | 0 |
| 0 | 0 | 0 | 0 | 0 |
| 0 | 0.185956573 | 0.133139648 | 0.198053961 | 0.103406346 |
| 0 | 0 | 0 | 0 | 0 |
| 0 | 0 | 0 | 0 | 0 |
| 0 | 0 | 0 | 0 | 0 |
| 0 | 0 | 0 | 0 | 0 |
| 3.135532565 | 0.809883892 | 0.173199522 | 0.000656003 | 1.431961519 |
| 0 | 0 | 0 | 0 | 0 |
| 0 | 0 | 0 | 0 | 0 |

| | | | | |
|---|---|---|---|---|
| 0.004621472 | 0 | 0.038791332 | 0 | 0 |
| 0 | 0 | 0 | 0 | 0 |
| 0 | 0 | 0 | 0 | 0 |
| 0 | 0 | 0 | 0 | 0 |
| 0 | 0.00168862 | 0 | 0 | 0 |
| 0 | 0 | 0 | 0 | 0 |
| 0 | 0 | 0 | 0 | 0 |
| 0 | 0 | 0 | 0 | 0 |
| 0 | 0 | 0 | 0.003703307 | 0 |
| 0.000845771 | 0 | 0 | 0.064584893 | 0.070685002 |
| 0 | 0 | 0.009402385 | 0.045559691 | 0 |
| 0 | 0 | 0 | 0 | 0 |
| 0.001033338 | 0.00744915 | 0.002178472 | 0 | 0 |
| 0 | 0 | 0 | 0 | 0 |
| 0 | 0 | 0 | 0 | 0 |
| 0 | 0 | 0 | 0 | 0 |
| 0 | 0 | 0 | 0 | 0 |
| 0 | 0 | 0 | 0 | 0 |
| 0 | 0 | 0 | 0 | 0 |
| 0.071004214 | 1.016562334 | 0.136375906 | 0.02790738 | 0.020693139 |
| 0 | 0 | 0 | 0 | 0 |
| 0 | 0 | 0 | 0 | 0 |
| 0 | 0 | 0 | 0 | 0 |
| 0 | 0 | 0 | 0 | 0 |
| 0 | 0 | 0 | 0 | 0 |
| 0 | 0 | 0 | 0 | 0 |
| 0 | 0 | 0 | 0 | 0 |
| 0 | 0 | 0 | 0 | 0 |
| 0 | 0 | 0 | 0 | 0 |
| 0 | 0 | 0 | 0 | 0 |
| 0 | 0 | 0 | 0 | 0 |
| 0 | 0 | 0 | 0 | 0 |
| 0 | 0 | 0 | 0 | 0 |
| 0 | 0 | 0 | 0 | 0 |
| 0 | 0 | 0 | 0 | 0 |
| 0 | 0 | 0 | 0 | 0 |
| 0 | 0 | 0 | 0 | 0 |
| 0 | 0 | 0 | 0 | 0 |
| 0 | 0 | 0 | 0 | 0 |
| 0 | 0 | 0 | 0 | 0 |
| 0 | 0 | 0 | 0 | 0 |
| 0 | 0 | 0 | 0 | 0 |
| 0 | 0 | 0.001253597 | 0.124803704 | 0 |
| 0 | 0 | 0 | 0 | 0 |
| 0.002246028 | 0.005782579 | 0.018414096 | 0.004193862 | 0 |
| 0 | 0 | 0 | 0 | 0 |
| 0 | 0 | 0 | 0 | 0 |
| 0 | 0 | 0 | 0 | 0.026063445 |

| | | | | |
|---|---|---|---|---|
| 0 | 0 | 0 | 0 | 0 |
| 0 | 0 | 0 | 0 | 0 |
| 0 | 0 | 0 | 0.03196961 | 0 |
| 0 | 0 | 0 | 0 | 0 |
| 0 | 0 | 0 | 0 | 0 |
| 0.000515922 | 0.031214651 | 0.056044804 | 0.029221555 | 0 |
| 0 | 0 | 0 | 0 | 0 |
| 0 | 0 | 0 | 0 | 0 |
| 0 | 0 | 0 | 0 | 0 |
| 0 | 0 | 0 | 0 | 0 |
| 0 | 0 | 0 | 0 | 0 |
| 0 | 0 | 0 | 0 | 0 |
| 0 | 0 | 0 | 0 | 0 |
| 0 | 0 | 0 | 0 | 0 |
| 0 | 0 | 0 | 0 | 0 |
| 0 | 0 | 0 | 0 | 0 |
| 0 | 0 | 0 | 0.49386399 | 0 |
| 0 | 0.001226889 | 0 | 0 | 0 |
| 0 | 0 | 0 | 0 | 0 |
| 0 | 0 | 0 | 0 | 0 |
| 0 | 0 | 0 | 0.000453777 | 0 |
| 0 | 0 | 0 | 0 | 0 |
| 0 | 0 | 0 | 0 | 0 |
| 0 | 0 | 0 | 0 | 0 |
| 0 | 0 | 0 | 0 | 0 |
| 0 | 0 | 0 | 0.003264815 | 0.004496252 |
| 0 | 0 | 0 | 0 | 0 |
| 0 | 0 | 0 | 1.244804793 | 0 |
| 0 | 0 | 0 | 0 | 0 |
| 0 | 0 | 0.001638576 | 0 | 0 |
| 0 | 0 | 0 | 0 | 0 |
| 0 | 0 | 0 | 0 | 0 |
| 0 | 0 | 0 | 0 | 0 |
| 0 | 0 | 0 | 0 | 0 |
| 0 | 0 | 0 | 0 | 0 |
| 0 | 0 | 0 | 0 | 0 |
| 0 | 0 | 0 | 0 | 0 |
| 0 | 0 | 0.008444443 | 0 | 0 |
| 0 | 0 | 0.232432263 | 0 | 0 |
| 0 | 0 | 0 | 0 | 0 |
| 0 | 0 | 0 | 0 | 0 |
| 0 | 0 | 0 | 0 | 0 |
| 0 | 0 | 0 | 0 | 0 |
| 0 | 0 | 0 | 0 | 0 |
| 0 | 0 | 0 | 0 | 0 |
| 0 | 0 | 0 | 0 | 0 |
| 0 | 0 | 0 | 0 | 0 |
| 0 | 0 | 0 | 0 | 0 |

| | | | | |
|---|---|---|---|---|
| 0 | 0 | 0 | 0 | 0 |
| 0 | 0 | 0.229845638 | 0.001168425 | 0 |
| 0 | 0 | 0 | 0.003755063 | 0 |
| 0 | 0 | 0 | 0 | 0 |
| 0 | 0 | 0 | 0 | 0 |
| 0 | 0 | 0 | 0.004406208 | 0 |
| 0 | 0 | 0 | 0 | 0 |
| 0 | 0 | 0 | 0 | 0 |
| 0 | 0 | 0 | 0 | 0 |
| 0 | 0 | 0 | 0 | 0 |
| 0 | 0 | 0 | 0.048193718 | 0 |
| 0 | 0 | 0 | 0 | 0 |
| 0 | 0 | 0.000652467 | 0.066768374 | 0 |
| 0 | 0 | 0 | 0 | 0 |
| 0 | 0 | 0 | 0 | 0 |
| 0 | 0 | 0 | 0 | 0 |
| 0 | 0 | 0 | 0 | 0 |
| 0 | 0 | 0 | 0 | 0 |
| 0 | 0 | 0 | 0 | 0 |
| 0 | 0 | 0 | 0.001750512 | 0 |
| 0.00825216 | 0.003458626 | 0.016857671 | 0 | 0.005906373 |
| 0 | 0 | 0 | 0 | 0 |
| 0 | 0 | 0 | 0 | 0 |
| 0 | 0.285489135 | 0.457675927 | 0.846580827 | 0 |
| 0 | 0 | 0 | 0.002424285 | 0 |
| 0 | 0 | 0 | 0 | 0 |
| 0 | 0 | 0 | 0 | 0 |
| 0 | 0 | 0 | 0 | 0 |
| 0 | 0 | 0 | 0 | 0 |
| 0 | 0 | 0 | 0 | 0 |
| 0 | 0 | 0.002867938 | 0 | 0 |
| 0 | 0 | 0 | 0 | 0 |
| 0 | 0 | 0 | 0 | 0 |
| 0 | 0 | 0 | 0 | 0 |

| PHL018-0030_V1 | PHL018-0028_V1 | PHL018-0248_V2 | PHL018-0034_V1 | PHL018-0021_V1 |
|---|---|---|---|---|
| CTRL | CTRL | REF | CTRL | EYCF |
| 0.672799023 | 0.024413801 | 0.506863697 | 0.044836225 | 0.309738372 |
| 0.103082275 | 0.877863146 | 0.012545431 | 0.092225097 | 2.591521221 |
| 0 | 3.320101994 | 0.111456787 | 6.93784612 | 0.077748542 |
| 0 | 0 | 0 | 0 | 0 |
| 0 | 0 | 0 | 0 | 0 |
| 0 | 0 | 0 | 0 | 0 |
| 0 | 0 | 0 | 0 | 0 |
| 0 | 0 | 0 | 0 | 0 |
| 0 | 0 | 0 | 0 | 0 |
| 0 | 0 | 0 | 0 | 0 |
| 0 | 0 | 0 | 0 | 0 |
| 0 | 0 | 0 | 0 | 0 |
| 0 | 0 | 0 | 0 | 0 |
| 0.383628685 | 0.578454619 | 0.12209345 | 2.424410033 | 0.127981959 |
| 0 | 0 | 0 | 0 | 0 |
| 0 | 0 | 0 | 0 | 0 |
| 0 | 0 | 0 | 0 | 0 |
| 0 | 0 | 0 | 0 | 0 |
| 0 | 0 | 0 | 0 | 0.011884758 |
| 0 | 0 | 0 | 0 | 0 |
| 0 | 0 | 0 | 0 | 0 |
| 0 | 0 | 0 | 0 | 0 |
| 0 | 0 | 0 | 0 | 0 |
| 0 | 0 | 0 | 0 | 0 |
| 0 | 0 | 0 | 0.187952956 | 0 |
| 0 | 0 | 0 | 0 | 0 |
| 0 | 0 | 0 | 0 | 0 |
| 0 | 0 | 0 | 0 | 0 |
| 0 | 0 | 0 | 0 | 0 |
| 0 | 0 | 0 | 0 | 0 |
| 0 | 0 | 0 | 0 | 0 |
| 0 | 0 | 0 | 0 | 0 |
| 0 | 0 | 0 | 0 | 0 |
| 0 | 0 | 0 | 0 | 0 |
| 0 | 0 | 0 | 0 | 0 |
| 0.971793224 | 0.235884931 | 1.497577643 | 0.112568253 | 0 |
| 0 | 0 | 0 | 0 | 0 |
| 0 | 0 | 0 | 0 | 0 |
| 0 | 0 | 0 | 0 | 0 |
| 0 | 0 | 0 | 0 | 0 |
| 0 | 0 | 0 | 0 | 0 |
| 0 | 0.002211057 | 0.715401266 | 0.030230984 | 0.470689442 |
| 4.281727666 | 0.308282596 | 0.306672361 | 0.022825791 | 0.66054609 |
| 0.002380263 | 0.668718757 | 0.000909083 | 0.013371206 | 0.101929509 |
| 0.00343217 | 0.013360024 | 0.002883833 | 0 | 0.01276691 |
| 0 | 0 | 0 | 0 | 0 |

| | | | | |
|---|---|---|---|---|
| 0.089281078 | 0 | 0.006115925 | 0 | 0 |
| 0.000655751 | 0 | 0 | 0 | 0.141595438 |
| 0.044098829 | 0 | 0 | 0.025307544 | 0.023185674 |
| 0.082978084 | 0.302055765 | 0.078603535 | 0.299057911 | 0.563996747 |
| 0.002066849 | 0 | 0 | 0.055037875 | 0 |
| 0 | 0.073316843 | 0 | 0 | 0 |
| 0 | 0 | 0 | 0.033255924 | 0.01279608 |
| 0 | 0 | 0 | 0 | 0 |
| 0 | 0 | 0 | 0 | 0 |
| 0 | 0 | 0 | 0 | 0 |
| 0 | 0 | 0.001151007 | 0.001032289 | 0 |
| 0.261571031 | 2.820486972 | 0.065421642 | 0.579444196 | 0 |
| 0 | 0 | 0 | 0.001362631 | 0 |
| 0.013423094 | 0.001987793 | 0.007715563 | 0 | 0.110182923 |
| 0 | 0 | 0 | 0 | 0 |
| 0 | 0 | 0 | 0 | 0 |
| 0.072456457 | 0.000817516 | 0.026812023 | 0 | 0.621726355 |
| 0 | 0 | 0 | 0 | 0 |
| 0 | 0 | 0 | 0 | 0 |
| 0 | 0 | 0 | 0 | 0 |
| 0.024822134 | 0 | 0.008152974 | 0 | 0.630115112 |
| 0.004494645 | 0.001577721 | 0.002492276 | 0.011016423 | 0.003383083 |
| 1.455619491 | 0.001327199 | 0.141166613 | 0.034919704 | 0 |
| 0 | 0 | 0 | 0 | 0 |
| 0 | 0 | 0 | 0 | 0 |
| 0 | 0 | 0 | 0 | 0 |
| 0 | 0 | 0 | 0 | 0 |
| 0 | 0 | 0 | 0 | 0 |
| 0 | 0 | 0 | 0.002042491 | 0.020287228 |
| 0 | 0 | 0 | 0 | 0 |
| 0 | 0 | 0 | 0 | 0.022760852 |
| 0 | 0 | 0 | 0 | 0 |
| 0 | 0 | 0 | 0 | 0 |
| 0 | 0 | 0.000658383 | 0 | 0 |
| 0 | 0 | 0 | 0 | 0 |
| 0 | 0 | 0 | 0 | 0.002212169 |
| 0 | 0 | 0 | 0 | 0 |
| 0 | 0 | 0 | 0.002904037 | 0.024101272 |
| 0 | 0 | 0 | 0 | 0 |
| 0 | 0 | 0 | 0 | 0 |
| 0 | 0 | 0 | 0 | 0 |
| 0 | 0 | 0 | 0 | 0 |
| 0 | 0 | 0 | 0 | 0 |
| 0 | 0 | 0 | 0 | 0 |
| 0 | 0 | 0 | 0.161104994 | 0 |
| 0 | 0 | 0 | 0 | 0 |
| 0 | 0 | 0 | 0 | 0 |
| 0 | 0 | 0 | 0 | 0.001495379 |

| | | | | |
|---|---|---|---|---|
| 0 | 0 | 0 | 0 | 0 |
| 0 | 0 | 0 | 0 | 0 |
| 0 | 0 | 0 | 0 | 0 |
| 0 | 0 | 0 | 0 | 0 |
| 0 | 0.498350553 | 0 | 0.001272129 | 1.615904979 |
| 0 | 0 | 0.000875238 | 0.000588722 | 0 |
| 0 | 0 | 0 | 0 | 0 |
| 0.002235342 | 0 | 0 | 0 | 0 |
| 0 | 0 | 0 | 0 | 0.008992542 |
| 0 | 0 | 0 | 0 | 0 |
| 0 | 0 | 0 | 0 | 0 |
| 0 | 0 | 0 | 0 | 0.172974519 |
| 0 | 0 | 0 | 0 | 0 |
| 0 | 0 | 0 | 0 | 0 |
| 0 | 0 | 0 | 0 | 0 |
| 0 | 0 | 0 | 0 | 0 |
| 0 | 0 | 0 | 0 | 0 |
| 0 | 0 | 0 | 0 | 0 |
| 4.184503083 | 0 | 0.261878578 | 0.066330599 | 0 |
| 0 | 0 | 0 | 0 | 0 |
| 0 | 0 | 0 | 0 | 0 |
| 0 | 0 | 0 | 0 | 0 |
| 0 | 0 | 0 | 0.000506381 | 0 |
| 0 | 0 | 0 | 0 | 0 |
| 0 | 0 | 0 | 0 | 0 |
| 0 | 0 | 0 | 0 | 0 |
| 0 | 0 | 0 | 0 | 0 |
| 0 | 0 | 0 | 0 | 0 |
| 0 | 0 | 0 | 0 | 0 |
| 0 | 0 | 0 | 0 | 0 |
| 0.027843644 | 0.007937899 | 0.028661144 | 0.025704957 | 0 |
| 0.109968595 | 0 | 0.018040822 | 0.008239837 | 0.008531504 |
| 0 | 0 | 0 | 0 | 0 |
| 0 | 0 | 0 | 0 | 0 |
| 0 | 0 | 0 | 0 | 0 |
| 0 | 0 | 0 | 0 | 0 |
| 0 | 0 | 0 | 0 | 0 |
| 0 | 0 | 0 | 0 | 0 |
| 0 | 0 | 0.038234088 | 0 | 0 |
| 0 | 0 | 0 | 0 | 0 |
| 0 | 0 | 0 | 0 | 0 |
| 0 | 0 | 0 | 0 | 0 |
| 0 | 0 | 0 | 0.003290091 | 0 |
| 0 | 0 | 0 | 0 | 0 |
| 0 | 0 | 0 | 0 | 0 |
| 0 | 0 | 0 | 0 | 0 |
| 0 | 0 | 0 | 0 | 0 |
| 0 | 0 | 0 | 0 | 0 |

| | | | | |
|---|---|---|---|---|
| 0.004383502 | 0 | 0 | 0 | 0 |
| 0 | 0 | 0 | 0 | 0 |
| 0 | 0 | 0 | 0 | 0 |
| 0 | 0 | 0 | 0 | 0 |
| 0.053295973 | 0 | 0.006688099 | 0.030252148 | 0 |
| 0 | 0 | 0 | 0 | 0 |
| 0 | 0 | 0 | 0 | 0 |
| 0 | 0 | 0 | 0 | 0 |
| 0 | 0 | 0 | 0 | 0 |
| 0 | 0 | 0 | 0 | 0 |
| 0 | 0 | 0 | 0 | 0 |
| 0 | 0 | 0 | 0 | 0 |
| 0 | 0 | 0 | 0 | 0 |
| 0 | 0 | 0 | 0 | 0 |
| 0 | 0 | 0 | 0 | 0 |
| 0 | 0 | 0 | 0 | 0 |
| 0 | 0 | 0 | 0 | 0 |
| 0 | 0 | 0 | 0 | 0 |
| 0 | 0 | 0 | 0 | 0 |
| 0 | 0 | 0 | 0 | 0 |
| 0 | 0 | 0 | 0 | 0 |
| 0 | 0 | 0 | 0 | 0 |
| 0 | 0 | 0 | 0 | 0 |
| 0 | 0 | 0 | 0 | 0 |
| 0 | 0 | 0 | 0.095036913 | 0 |
| 0 | 0 | 0 | 0 | 0 |
| 0 | 0 | 0 | 0.049961006 | 0 |
| 0 | 0 | 0 | 0 | 0 |
| 0 | 0 | 0 | 0 | 0 |
| 0 | 0 | 0 | 0 | 0 |
| 0 | 0 | 0 | 0 | 0 |
| 0 | 0 | 0.177314799 | 0.11079969 | 0.036169192 |
| 0 | 0 | 0 | 0 | 0 |
| 0 | 0 | 0 | 0 | 0 |
| 0 | 0 | 0 | 0 | 0 |
| 0 | 0 | 0 | 0 | 0 |
| 0 | 0 | 0 | 0 | 0 |
| 0 | 0 | 0 | 0 | 0 |
| 0 | 0 | 0 | 0 | 0 |
| 0 | 0 | 0 | 0 | 0 |
| 0 | 0 | 0 | 0 | 0 |
| 0 | 0 | 0 | 0 | 0 |
| 0 | 0 | 0 | 0 | 0 |
| 0 | 0 | 0 | 0 | 0 |
| 0 | 0 | 0 | 0 | 0 |
| 0 | 0 | 0 | 0 | 0 |
| 0 | 0 | 0 | 0 | 0 |
| 0 | 0 | 0 | 0 | 0 |

| | | | | |
|---|---|---|---|---|
| 0 | 0 | 0 | 0 | 0 |
| 0 | 0 | 0 | 0 | 0 |
| 0 | 0 | 0 | 0 | 0 |
| 0 | 0 | 0 | 0 | 0 |
| 0 | 0 | 0 | 0 | 0 |
| 0 | 0 | 0 | 0 | 0 |
| 0 | 0 | 0 | 0 | 0 |
| 0 | 0 | 0 | 0 | 0 |
| 0 | 0 | 0 | 0 | 0 |
| 0 | 0 | 0 | 0 | 0 |
| 0 | 0 | 0 | 0 | 0 |
| 0 | 0 | 0 | 0 | 0 |
| 0.001379679 | 0 | 0 | 0.000661619 | 0.000876212 |
| 0 | 0 | 0 | 0 | 0 |
| 0.017217119 | 0.000665124 | 0 | 0 | 0 |
| 0 | 0 | 0 | 0 | 0 |
| 0 | 0 | 0 | 0 | 0 |
| 0 | 0 | 0 | 0 | 0 |
| 0 | 0 | 0 | 0 | 0 |
| 0 | 0 | 0 | 0 | 0 |
| 0 | 0.03208941 | 0 | 0.017009692 | 0.048330019 |
| 0 | 0 | 0 | 0 | 0 |
| 0 | 0 | 0 | 0 | 0 |
| 0 | 0 | 0 | 0 | 0 |
| 0 | 0 | 0 | 0 | 0 |
| 0 | 0 | 0 | 0 | 0 |
| 0.002253507 | 0 | 0.002008233 | 0.000900549 | 0.016696936 |
| 0 | 0 | 0 | 0 | 0 |
| 0 | 0 | 0 | 0 | 0 |
| 0 | 0 | 0 | 0 | 0 |
| 0 | 0 | 0 | 0 | 0.038518995 |
| 0 | 0 | 0 | 0 | 0 |
| 0 | 0 | 0 | 0 | 0 |
| 0.145435162 | 0.001634255 | 0.006111497 | 0.003024078 | 0 |
| 0.008182539 | 0.002485605 | 0 | 0.003219611 | 0.020253412 |
| 0 | 0.005982797 | 0 | 0 | 0 |
| 0 | 0 | 0 | 0 | 0 |
| 0 | 0 | 0 | 0 | 0 |
| 0 | 0 | 0 | 0 | 0 |
| 0 | 0 | 0 | 0 | 0 |
| 0 | 0 | 0 | 0 | 0 |
| 0 | 0 | 0 | 0 | 0 |
| 0 | 0 | 0 | 0 | 0 |
| 0 | 0 | 0 | 0 | 0 |
| 0 | 0 | 0 | 0 | 0 |
| 0 | 0 | 0.545377831 | 1.377139998 | 0 |
| 0 | 0 | 0 | 0 | 0 |
| 0.001046607 | 0.000708524 | 0.000639562 | 0 | 0.024498336 |

| | | | | |
|---|---|---|---|---|
| 0 | 0 | 0 | 0 | 0 |
| 0 | 0 | 0 | 0 | 0 |
| 0 | 0 | 0 | 0 | 0.002082425 |
| 0 | 0 | 0 | 0 | 0 |
| 0 | 0 | 0 | 0 | 0 |
| 0 | 0 | 0 | 0 | 0 |
| 0 | 0 | 0.003186427 | 0 | 0.002433005 |
| 0 | 0 | 0.005286443 | 0 | 0.015014918 |
| 0 | 0 | 0 | 0 | 0 |
| 0 | 0 | 0 | 0 | 0 |
| 0 | 0 | 0 | 0 | 0.001174831 |
| 0 | 0 | 0 | 0 | 0 |
| 0 | 0 | 0 | 0 | 0 |
| 0 | 0 | 0 | 0 | 0 |
| 0 | 0 | 0.001764659 | 0 | 0 |
| 0 | 0 | 0 | 0 | 0 |
| 0 | 0 | 0 | 0 | 0 |
| 0 | 0 | 0 | 0 | 0 |
| 0.021439913 | 0.001981551 | 0 | 0 | 0.001158821 |
| 0 | 0 | 0.036150091 | 0 | 0 |
| 0 | 0 | 0.001439175 | 0 | 0.187176957 |
| 0 | 0 | 0 | 0 | 0 |
| 0 | 0 | 0 | 0 | 0 |
| 0 | 0 | 0 | 0 | 0 |
| 0.001336848 | 0.0147818 | 0 | 0.002136931 | 0.033960419 |
| 0 | 0.001280719 | 0 | 0 | 0 |
| 0 | 0 | 0 | 0 | 0 |
| 0 | 0 | 0 | 0 | 0 |
| 0 | 0 | 0.010232253 | 0 | 0.137467841 |
| 0 | 0 | 0 | 0 | 0 |
| 0.018471759 | 0.003516992 | 0.043563597 | 0.03021228 | 0.732356876 |
| 0 | 0 | 0 | 0 | 0 |
| 0 | 0 | 0 | 0 | 0 |
| 0 | 0 | 0 | 0 | 0.001626155 |
| 0 | 0 | 0 | 0 | 0 |
| 0 | 0 | 0 | 0 | 0 |
| 0 | 0 | 0 | 0 | 0 |
| 0 | 0 | 0 | 0 | 0 |
| 0 | 0 | 0 | 0 | 0 |
| 0 | 0.004098483 | 0 | 0 | 0 |
| 0 | 0 | 0 | 0 | 0 |
| 0.037321543 | 0.008564138 | 0.022832154 | 0 | 0 |
| 0.000705274 | 0 | 0 | 0 | 0 |
| 0 | 0 | 0 | 0 | 0 |
| 0 | 0 | 0 | 0 | 0 |
| 0 | 0 | 0 | 0 | 0 |
| 0 | 0 | 0 | 0 | 0 |
| 0 | 0 | 0 | 0 | 0 |

| | | | | |
|---|---|---|---|---|
| 0 | 0 | 0 | 0 | 0 |
| 0 | 0 | 0 | 0 | 0 |
| 0.000844384 | 0.698239264 | 0 | 0.009920561 | 0.110200404 |
| 0 | 0 | 0 | 0 | 0 |
| 0 | 0 | 0 | 0 | 0 |
| 0 | 0 | 0 | 0 | 0.001567296 |
| 0 | 0 | 0 | 0 | 0 |
| 0 | 0 | 0 | 0 | 0 |
| 0 | 0 | 0 | 0 | 0 |
| 0 | 0 | 0 | 0.001851688 | 0 |
| 0 | 0 | 0 | 0 | 0 |
| 0 | 0 | 0 | 0 | 0 |
| 0 | 0 | 0 | 0 | 0 |
| 0 | 0.017136171 | 0 | 0.00671266 | 0.003259622 |
| 0.00096279 | 0.00250936 | 0.000823679 | 0.001846807 | 0.017854416 |
| 3.697458469 | 0.006350148 | 0.183867311 | 0.001384076 | 0 |
| 0.000658208 | 0.003119117 | 0 | 0 | 0.010311092 |
| 0 | 0 | 0 | 0 | 0 |
| 0 | 0 | 0 | 0 | 0 |
| 0 | 0 | 0 | 0 | 0 |
| 0 | 0 | 0 | 0 | 0 |
| 0 | 0 | 0.000635404 | 0.00132969 | 0.028427056 |
| 0 | 0 | 0 | 0 | 0 |
| 0 | 0 | 0 | 0 | 0 |
| 0 | 0 | 0 | 0 | 0 |
| 0 | 0 | 0 | 0 | 0 |
| 0 | 0 | 0 | 0 | 0 |
| 0 | 0 | 0 | 0 | 0 |
| 0 | 0 | 0 | 0 | 0 |
| 0 | 0 | 0 | 0 | 0 |
| 0 | 0 | 0 | 0 | 0 |
| 0 | 0 | 0 | 0 | 0 |
| 0 | 0 | 0 | 0 | 0 |
| 0 | 0 | 0 | 0 | 0 |
| 0 | 0 | 0 | 0 | 0 |
| 0 | 0 | 0.005076519 | 0 | 0.037685178 |
| 0 | 0 | 0 | 0 | 0 |
| 0.083858854 | 0.035376012 | 0.00691877 | 0.012648959 | 0 |
| 0 | 0 | 0 | 0 | 0 |
| 0 | 0 | 0 | 0 | 0 |
| 0 | 0 | 0 | 0 | 0.009579676 |
| 0 | 0 | 0 | 0 | 0 |
| 0 | 0 | 0 | 0 | 0 |
| 0 | 0 | 0 | 0 | 0 |
| 0 | 0 | 0 | 0 | 0 |
| 0 | 0 | 0 | 0 | 0 |
| 0 | 0 | 0 | 0 | 0 |
| 0 | 0 | 0.010958941 | 0 | 0.009188095 |

| | | | | |
|---|---|---|---|---|
| 0 | 0 | 0.027836229 | 0 | 0 |
| 0 | 0.135450056 | 0 | 0 | 0 |
| 0 | 0.007372994 | 0 | 0 | 0 |
| 0 | 0 | 0 | 0 | 0 |
| 0.00250458 | 0.002225383 | 0 | 0.023821028 | 0 |
| 0 | 0 | 0 | 0 | 0 |
| 0 | 0 | 0 | 0 | 0 |
| 0 | 0.107444758 | 0 | 0.001402959 | 0.039389631 |
| 0 | 0 | 0 | 0 | 0 |
| 0 | 0 | 0.006449453 | 0.145473616 | 0.012256527 |
| 0 | 0 | 0 | 0 | 0 |
| 0.000926366 | 0 | 0.010897124 | 0 | 0.134137016 |
| 0 | 0.020187332 | 0 | 0.006570874 | 0.016734817 |
| 0 | 0 | 0 | 0 | 0 |
| 0 | 0 | 0 | 0.020273981 | 0 |
| 0 | 0 | 0 | 0 | 0.007369043 |
| 0 | 0 | 0.002050784 | 0 | 0 |
| 0 | 0 | 0 | 0 | 0 |
| 0 | 0 | 0 | 0 | 0 |
| 0 | 0 | 0 | 0 | 0 |
| 0 | 0 | 0.001109948 | 0 | 0 |
| 0 | 0 | 0 | 0 | 0 |
| 0 | 0 | 0 | 0.000521561 | 0 |
| 0 | 0 | 0.172497296 | 0.543219182 | 0 |
| 0 | 0 | 0 | 0 | 0 |
| 0.005115281 | 0.021089065 | 0 | 0 | 0.107334821 |
| 0 | 0 | 0 | 0.001574483 | 0 |
| 0 | 0 | 0 | 0 | 2.958631777 |
| 0 | 0 | 0 | 0 | 0 |
| 0 | 0 | 0 | 0 | 0 |
| 0 | 0 | 0 | 0 | 0 |
| 0 | 0 | 0 | 0 | 0 |
| 0 | 0 | 0 | 0 | 0 |
| 0 | 0 | 0 | 0 | 0 |
| 0 | 0 | 0 | 0 | 0 |
| 0 | 0 | 0 | 0 | 0.01159589 |
| 0 | 0 | 0 | 0 | 0.265445977 |
| 0 | 0 | 0 | 0 | 0 |
| 0 | 0 | 0 | 0 | 0 |
| 0 | 0 | 0 | 0 | 0 |
| 0 | 0 | 0 | 0 | 0 |
| 0 | 0 | 0 | 0 | 0 |
| 0 | 0 | 0 | 0 | 0 |
| 0 | 0 | 0 | 0 | 0.035384719 |
| 0 | 0 | 0 | 0 | 0 |
| 0 | 0 | 0 | 0 | 0 |
| 0.003307853 | 0 | 0 | 0 | 0.002908751 |
| 0 | 0 | 0 | 0 | 0 |

| | | | | |
|---|---|---|---|---|
| 0 | 0 | 0 | 0 | 0 |
| 0 | 0 | 0 | 0 | 0.092104078 |
| 0 | 0 | 0 | 0 | 0 |
| 0 | 0.010381763 | 0.124755209 | 0.016284114 | 0 |
| 0 | 0 | 0 | 0.006030224 | 0 |
| 0.004609439 | 0 | 0 | 0.026909657 | 0 |
| 0 | 0 | 0 | 0 | 0 |
| 0 | 0 | 0 | 0 | 0.00131694 |
| 0 | 0 | 2.055809098 | 0.544855719 | 0 |
| 0 | 0 | 0 | 0 | 0 |
| 0 | 0 | 0.000745715 | 0.005350401 | 0 |
| 0 | 0 | 0 | 0 | 0 |
| 0 | 0 | 0 | 0 | 0 |
| 0 | 0.006504 | 0.000704514 | 0.00168493 | 0 |
| 0 | 0 | 0 | 0 | 0 |
| 0 | 0 | 0 | 0 | 0 |
| 0 | 0.16863259 | 0 | 0 | 0 |
| 0 | 0 | 0 | 0 | 0.038273403 |
| 0 | 0 | 0 | 0 | 0 |
| 0 | 0 | 0.145463981 | 0 | 0 |
| 0 | 0 | 0 | 0 | 0.012922967 |
| 0.193659477 | 0.114420109 | 0.082308066 | 0.297179363 | 0 |
| 0 | 0 | 0.003788452 | 0.320583035 | 0.008734767 |
| 0 | 0 | 0 | 0 | 0 |
| 0 | 0 | 0 | 0 | 0 |
| 0 | 0 | 0 | 0 | 0 |
| 0 | 0 | 0 | 0 | 0.007863244 |
| 0 | 0 | 0 | 0 | 0 |
| 0 | 0 | 0 | 0 | 0 |
| 0.117166979 | 0.578874239 | 0.052666181 | 0.18121822 | 0.076455119 |
| 0 | 0.039535526 | 0.010515051 | 0 | 0 |
| 0 | 0 | 0 | 0 | 0 |
| 0 | 0 | 0 | 0 | 0 |
| 0 | 0 | 0 | 0 | 0 |
| 0 | 0 | 0 | 0 | 0 |
| 0 | 0 | 0 | 0 | 0 |
| 0 | 0 | 1.744576736 | 1.401921455 | 0 |
| 0 | 0 | 0 | 0 | 0 |
| 0 | 0 | 0 | 0 | 0 |
| 0 | 0 | 0 | 0 | 0 |
| 0 | 0.006202515 | 0 | 0.025279821 | 0.021325798 |
| 0.147349008 | 0.747003427 | 0.111580121 | 0.616958349 | 0 |
| 0 | 0 | 0 | 0 | 0 |
| 0 | 0 | 0 | 0.002924432 | 0 |
| 0 | 0 | 0 | 0 | 0.00957091 |
| 0 | 0 | 0 | 0 | 0 |
| 0 | 0 | 0.001823696 | 0.001453862 | 0.002888123 |
| 0 | 0 | 0 | 0 | 0 |

| | | | | |
|---|---|---|---|---|
| 0 | 0.071182166 | 0.001750105 | 0 | 0.136005373 |
| 0 | 0 | 0 | 0 | 0 |
| 0.003504153 | 0.00301918 | 0.006131962 | 0.00366633 | 1.448553506 |
| 0 | 0 | 0 | 0 | 0 |
| 0.014884459 | 0.713141013 | 0.166880403 | 0 | 0 |
| 0 | 0 | 0 | 0 | 0 |
| 0 | 0 | 0 | 0 | 0 |
| 0 | 0 | 0.00140108 | 0 | 0.001664131 |
| 0 | 0 | 0 | 0.005817414 | 0 |
| 0 | 0 | 0 | 0 | 0 |
| 0 | 0 | 0 | 0 | 0 |
| 0 | 0 | 0 | 0 | 0 |
| 0 | 0 | 0 | 0 | 0.00219098 |
| 0 | 0 | 0 | 0.004259917 | 0 |
| 0 | 0 | 0 | 0 | 0 |
| 0.153717843 | 0.11298629 | 0.016438449 | 0.005865472 | 0 |
| 0.006595363 | 0.137170859 | 0.002742842 | 0.001581389 | 0.437709616 |
| 0 | 0 | 0 | 0 | 0.008764383 |
| 0 | 0 | 0 | 0 | 0 |
| 0 | 0 | 0 | 0 | 0 |
| 0 | 0 | 0 | 0 | 0.014076904 |
| 0.00467517 | 0 | 0.001818031 | 0 | 0 |
| 0 | 0 | 0 | 0 | 0 |
| 0 | 0 | 0 | 0 | 0 |
| 0 | 0.001014236 | 0 | 0 | 0 |
| 0 | 0 | 0 | 0 | 0.003678503 |
| 0 | 0 | 0 | 0.002007586 | 0.001181661 |
| 0 | 0 | 0 | 0.031903324 | 0 |
| 0 | 0 | 0 | 0 | 0 |
| 0 | 0 | 0 | 0 | 0.017188692 |
| 0 | 0.000838181 | 0.002837246 | 0.023749649 | 0 |
| 0 | 0.058732379 | 0 | 0.011633995 | 0.043207781 |
| 0.069548782 | 0.011600667 | 0.003490516 | 0.005297761 | 0.015945601 |
| 0 | 0 | 0.028474347 | 0.043827746 | 0.170015908 |
| 0 | 0.181031647 | 0 | 0 | 0.325552167 |
| 0 | 0 | 0 | 0 | 0 |
| 0 | 0.011025639 | 0 | 0 | 0.012109374 |
| 0.002307741 | 0 | 0.024299108 | 0.008002056 | 0.030214104 |
| 0.074112742 | 0.015310566 | 0.000846144 | 0.002150133 | 0 |
| 0 | 0 | 0 | 0 | 0 |
| 0 | 0 | 0 | 0 | 0 |
| 0 | 0 | 0 | 0 | 0 |
| 0 | 0.001258051 | 0.000851702 | 0.002291565 | 0.00134881 |
| 0 | 0 | 0 | 0.067383436 | 0.034146652 |
| 0 | 0 | 0 | 0 | 0.292775056 |
| 0 | 0 | 0 | 0 | 0.009831594 |
| 0 | 0 | 0 | 0 | 0 |
| 0 | 0 | 0 | 0 | 0 |

| | | | | |
|---|---|---|---|---|
| 0 | 0 | 0 | 0 | 0 |
| 0.124332272 | 0.008597073 | 0.122871429 | 1.587781214 | 0.514631866 |
| 0 | 0 | 0 | 0 | 0 |
| 0 | 0 | 0 | 0 | 0 |
| 0 | 0 | 0 | 0 | 0 |
| 0 | 0 | 0 | 0 | 0 |
| 0 | 0 | 0 | 0 | 0.006821753 |
| 0 | 0 | 0 | 0 | 0.00188796 |
| 0 | 0 | 0 | 0 | 0.003937578 |
| 0 | 0 | 0 | 0 | 0.018754463 |
| 0.001977271 | 0 | 0 | 0 | 0.006849452 |
| 0.00722913 | 0.225210791 | 0 | 0.011298865 | 0.145555 |
| 0 | 0 | 0 | 0.04892022 | 0 |
| 0 | 0 | 0 | 0 | 0 |
| 0 | 0.000733008 | 0 | 0 | 0.025672374 |
| 0 | 0 | 0 | 0 | 0 |
| 0.001882262 | 0.014271466 | 0.001006437 | 0.009387356 | 0.017213676 |
| 0 | 0 | 0 | 0 | 0.355669528 |
| 0 | 0 | 0 | 0 | 0 |
| 0 | 0 | 0 | 0 | 0.004836716 |
| 0.005629151 | 0.00344941 | 0.01079406 | 0.115237943 | 0.004931012 |
| 0 | 0.000858447 | 0 | 0 | 0 |
| 0 | 0 | 0 | 0 | 0 |
| 0 | 0 | 0 | 0 | 0 |
| 0 | 0 | 0 | 0 | 0 |
| 0 | 0 | 0 | 0 | 0 |
| 0 | 0.001134913 | 0.012498286 | 0.039875278 | 0 |
| 0.007427119 | 0.01735341 | 0.002647498 | 0.082115642 | 0.028301063 |
| 0 | 0 | 0 | 0 | 0 |
| 0.001244284 | 0 | 0 | 0 | 0.014856226 |
| 0 | 0 | 0 | 0 | 0 |
| 0 | 0 | 0 | 0 | 0 |
| 0 | 0 | 0 | 0 | 0 |
| 0 | 0 | 0 | 0 | 0 |
| 0 | 0 | 0.009228442 | 0 | 0 |
| 0.084148872 | 0.275908888 | 0.020163298 | 0.231068239 | 0.019070445 |
| 0 | 0 | 0 | 0 | 0 |
| 0 | 0 | 0 | 0 | 0 |
| 0.013488768 | 0 | 0.015241264 | 0.003173216 | 0.117668148 |
| 0 | 0 | 0 | 0 | 0 |
| 0 | 0 | 0 | 0 | 0.132258151 |
| 0 | 0 | 0 | 0 | 0 |
| 0.042048551 | 0.001282002 | 0.008100555 | 0.013788753 | 0.001963556 |
| 0 | 0 | 0.000798054 | 0.034355564 | 0 |
| 0 | 0 | 0 | 0 | 0 |
| 0 | 0.161317776 | 0.083444937 | 0.011684225 | 0 |
| 0 | 0 | 0 | 0 | 0 |
| 0 | 0 | 0 | 0 | 0 |

| | | | | |
|---|---|---|---|---|
| 0 | 0 | 0 | 0 | 0 |
| 0 | 0 | 0 | 0 | 0 |
| 0 | 0 | 0 | 0.001358766 | 0 |
| 0.680584218 | 3.927137014 | 2.161867281 | 0.053531779 | 0 |
| 0 | 0 | 0 | 0 | 0 |
| 0 | 0 | 0 | 0 | 0 |
| 0 | 0 | 0 | 0 | 0 |
| 0.028670977 | 0 | 0 | 0 | 0 |
| 0.032859267 | 4.747881848 | 14.88975173 | 0.614822884 | 0 |
| 0.001931031 | 0.010103982 | 0.017724822 | 0.110041764 | 0 |
| 0 | 0 | 0 | 0 | 0 |
| 0.030016051 | 0.005722304 | 0.003874005 | 0 | 0 |
| 0 | 0 | 0 | 0 | 0 |
| 0 | 0 | 0 | 0 | 0 |
| 0.261643757 | 0.385522471 | 0.064850155 | 0.07741192 | 0 |
| 0 | 0 | 0 | 0 | 0 |
| 0.106026858 | 0.01341894 | 0.01926141 | 0.008904503 | 0 |
| 0 | 6.383000787 | 0.637917099 | 0 | 0 |
| 0 | 0 | 0 | 0 | 0 |
| 0 | 0 | 0 | 0 | 0 |
| 0 | 0 | 0 | 0 | 0 |
| 0 | 0 | 0 | 0.13102702 | 0 |
| 0 | 0 | 0 | 0 | 0 |
| 0.032909249 | 0.065304241 | 0.032553392 | 0.21936265 | 0.357218138 |
| 0 | 0 | 0.002418039 | 0 | 0 |
| 0 | 0.605985101 | 0.211289568 | 0.005587515 | 0 |
| 0 | 0 | 0.013152426 | 0.061559589 | 0 |
| 0.735951937 | 1.987993357 | 0.184303852 | 1.030765022 | 0 |
| 0 | 0 | 0 | 0 | 0 |
| 0 | 0 | 0 | 0 | 0 |
| 0 | 0.004370972 | 0 | 0.020909807 | 0 |
| 0 | 0.018345005 | 0.101891283 | 0 | 0 |
| 0 | 0.401615054 | 0.000409479 | 0 | 0.00405298 |
| 0 | 0 | 0 | 0 | 0 |
| 0 | 0 | 0 | 0 | 0 |
| 0 | 0 | 0.147500542 | 0 | 0 |
| 0 | 1.502725387 | 2.58448494 | 0.020079196 | 0 |
| 0 | 0.000850557 | 0 | 0 | 0 |
| 0 | 0.102969603 | 0 | 0 | 0 |
| 0 | 0.002194218 | 0.000660217 | 0.080528322 | 0 |
| 0.004468195 | 0 | 0.018759048 | 0.015554439 | 0.005885556 |
| 0 | 0 | 0.015370312 | 0.244131941 | 0 |
| 1.290207624 | 0.290931917 | 0.135659582 | 0.03674771 | 0.001260794 |
| 0 | 0.138851584 | 0.002004156 | 0 | 0 |
| 0 | 0 | 0 | 0 | 0 |
| 0 | 0 | 0 | 0 | 0 |
| 0 | 0 | 0 | 0 | 0 |
| 0.004376601 | 0.008248803 | 0.024890676 | 0.003243569 | 0 |

| | | | | |
|---|---|---|---|---|
| 0 | 0 | 0 | 0 | 0 |
| 0 | 0.011164984 | 1.378785094 | 0 | 0 |
| 0.001144218 | 0 | 0.000611808 | 0 | 0.005086722 |
| 0 | 0.001953721 | 0 | 0 | 0 |
| 0 | 0 | 0 | 0 | 0 |
| 0.097197827 | 0.101268213 | 0.030202763 | 0 | 0 |
| 0 | 0 | 0 | 0 | 0 |
| 0 | 0 | 0 | 0 | 0 |
| 0 | 0 | 0 | 0 | 0 |
| 0 | 0 | 0 | 0 | 0 |
| 0.099984312 | 0.033494705 | 0.022625886 | 0.050461114 | 0.146736071 |
| 0.045539735 | 0.654777408 | 0.439376298 | 1.370096435 | 2.151720906 |
| 0 | 0 | 0 | 0 | 0 |
| 0.034958682 | 0.260942037 | 0.023986588 | 0.002519488 | 0.032596711 |
| 0.004777245 | 0.011507864 | 0.027246632 | 0.021381799 | 0.224107904 |
| 0 | 14.34545782 | 0 | 1.798148232 | 0 |
| 0 | 0 | 0 | 0 | 0 |
| 0.001414727 | 0 | 0 | 0 | 0 |
| 0 | 0 | 0 | 0 | 0 |
| 0.001388103 | 0 | 0.012700077 | 0.029436899 | 0 |
| 0.02008727 | 0 | 0.103378018 | 0.005869964 | 0.011162464 |
| 0 | 0.00803186 | 0 | 0 | 1.017183436 |
| 0.0755952 | 0.137672978 | 0.274541802 | 2.113666692 | 2.442637664 |
| 0 | 0 | 0 | 0 | 0 |
| 0 | 0 | 0 | 0 | 0 |
| 0 | 0 | 0.001407703 | 2.087287839 | 0 |
| 0 | 0 | 0 | 0 | 0 |
| 0 | 0 | 0 | 0.000784906 | 0 |
| 0 | 0 | 0 | 0.241607534 | 0 |
| 0 | 0 | 0 | 0 | 0 |
| 0 | 0.011688525 | 0 | 0 | 0 |
| 0 | 0.150596225 | 0.048164641 | 0.079288884 | 0 |
| 0 | 0.009568194 | 0.001233843 | 0 | 0.693179277 |
| 0.015543967 | 0 | 0.044222335 | 0.125030993 | 0 |
| 0.014849439 | 0 | 0.002443055 | 0 | 0.031757881 |
| 0.112648159 | 0.102879661 | 0.067691596 | 0.004850087 | 0.050056575 |
| 0.011454377 | 0.0223506 | 0.003962979 | 0.002584892 | 0 |
| 0 | 0 | 0 | 0 | 0 |
| 2.744572322 | 0.410002821 | 0.278663343 | 0.412360293 | 0 |
| 0 | 0 | 0 | 0 | 0 |
| 0 | 0 | 0.023645299 | 0.054783354 | 0 |
| 0 | 0 | 0 | 0 | 0 |
| 0 | 0 | 0 | 0 | 0 |
| 0 | 0 | 0 | 0 | 0 |
| 0.008153189 | 0.00848115 | 0.016587265 | 0.003623741 | 0.010861085 |
| 0 | 0 | 0 | 0 | 0 |
| 0 | 0 | 0 | 0.15579189 | 0 |
| 13.59769074 | 0.789460109 | 0.036939277 | 0.050614149 | 1.407642712 |

| | | | | |
|---|---|---|---|---|
| 0 | 0 | 0 | 0 | 0 |
| 0 | 0 | 0 | 0 | 0 |
| 0 | 0 | 0 | 0 | 0 |
| 0.024744969 | 0 | 0.001087481 | 0.005039131 | 0 |
| 0 | 0 | 0 | 0 | 0 |
| 0 | 0 | 0.121657854 | 0.319145982 | 0 |
| 0 | 0 | 0 | 0 | 0 |
| 0.447443168 | 0.390589818 | 0.004669839 | 0.14090232 | 0 |
| 0.012096212 | 0 | 0 | 0 | 0 |
| 0 | 0 | 0 | 0 | 0 |
| 0.004719066 | 0.003578035 | 0.011506081 | 0 | 0.052027967 |
| 0 | 0 | 0 | 0 | 0 |
| 0 | 0 | 0.124251838 | 0 | 0.058150282 |
| 0.119196658 | 0 | 0 | 0 | 0 |
| 0 | 0 | 0 | 0 | 0 |
| 0 | 0 | 0 | 0 | 0 |
| 0 | 0 | 0.005554865 | 0.074594173 | 0 |
| 0.076998884 | 0.001116987 | 0.006721789 | 0 | 0 |
| 0 | 0 | 0 | 0 | 0 |
| 0 | 0 | 0 | 0 | 0 |
| 0.007538779 | 0.000826963 | 0 | 0 | 0 |
| 0 | 0 | 0 | 0 | 0 |
| 0.002174062 | 0.006224397 | 0.000581231 | 0 | 0 |
| 0 | 0 | 0 | 0 | 0 |
| 0 | 0 | 0 | 0 | 0 |
| 0 | 0 | 0 | 0 | 0 |
| 0 | 0 | 0 | 0 | 0 |
| 0 | 0 | 0 | 0 | 0 |
| 0 | 0 | 0 | 0 | 0 |
| 0.049923822 | 0.058470125 | 0.134992696 | 0.023485602 | 0 |
| 0.03464048 | 0 | 0 | 0 | 0 |
| 0 | 0 | 0 | 0 | 0 |
| 0 | 0 | 0 | 0.104086699 | 0 |
| 0 | 0 | 0 | 0 | 0 |
| 0.022063942 | 0.001955992 | 0 | 0 | 0 |
| 0 | 0 | 0 | 0 | 0 |
| 0.082727667 | 0.083754789 | 0.01170903 | 0.032800449 | 0 |
| 0.000798954 | 0.000757217 | 0.001367031 | 0 | 0.157700869 |
| 0 | 0 | 0 | 0 | 0 |
| 0 | 0 | 0 | 0 | 0 |
| 0.152022307 | 0.338355271 | 0.01907888 | 0.062261155 | 0 |
| 0 | 0 | 0 | 0 | 0 |
| 0 | 0 | 0 | 0 | 0 |
| 0 | 0 | 0 | 0.132903231 | 0.067210892 |
| 0 | 0 | 0.018238938 | 0 | 0 |
| 0 | 0 | 0 | 0 | 0 |
| 0 | 0.00449677 | 0.003409636 | 0.001310553 | 0 |
| 0 | 0 | 0 | 0 | 0 |

| | | | | |
|---|---|---|---|---|
| 0 | 0 | 0 | 0 | 0 |
| 0 | 0 | 0 | 0 | 0 |
| 0 | 0.910626313 | 0 | 0 | 0 |
| 1.05269325 | 0 | 0.124591269 | 1.580984552 | 0 |
| 0 | 0 | 0 | 0 | 0 |
| 0 | 0 | 0 | 0 | 0 |
| 0 | 0 | 0 | 0 | 0 |
| 0 | 0 | 0 | 0 | 0 |
| 0.001101891 | 0.001044329 | 0.001178353 | 0.004861346 | 1.002384284 |
| 0.001064818 | 0 | 0 | 0 | 0 |
| 0 | 0.004107003 | 0.001348094 | 0 | 0 |
| 0 | 0 | 0 | 0 | 0 |
| 0 | 0 | 0.001787315 | 0.000616526 | 0 |
| 0 | 0 | 0 | 0 | 0 |
| 0.03230741 | 0.068669149 | 0.064187098 | 0.092197804 | 0 |
| 0 | 0 | 0 | 0 | 0 |
| 0 | 0 | 0 | 0 | 0 |
| 0 | 0 | 0 | 0 | 0 |
| 0 | 0 | 0 | 0 | 0 |
| 0 | 0 | 0 | 0.006043114 | 0 |
| 0 | 0 | 0.589807537 | 0 | 0 |
| 0 | 0 | 0 | 0 | 0 |
| 0 | 0 | 0 | 0 | 0 |
| 0 | 0 | 0 | 0 | 0 |
| 0.08863224 | 0.085967095 | 0.007759973 | 0.008202372 | 0.013825346 |
| 12.18993923 | 0.168132208 | 1.593167239 | 0.408810012 | 18.34077334 |
| 0.063618488 | 0.173119986 | 0.003092411 | 0.073958706 | 0 |
| 0 | 0 | 0 | 0.017126271 | 0 |
| 4.771974467 | 0 | 0.700250643 | 0 | 0 |
| 0 | 0 | 0 | 0 | 0 |
| 0 | 0 | 0 | 0 | 0 |
| 0 | 0 | 0 | 0 | 0 |
| 0 | 0 | 0 | 0 | 0 |
| 0.030726427 | 0.030006738 | 0 | 0.002256669 | 0.006328819 |
| 0.025541775 | 0.122928631 | 0.002194887 | 0 | 0.499090653 |
| 0.002095662 | 0 | 0.001896301 | 0 | 0.029689788 |
| 0 | 0 | 0 | 0 | 0 |
| 0.110855106 | 0.06900761 | 0.211789222 | 0.446230414 | 0 |
| 0 | 0 | 6.54472103 | 22.34013078 | 0 |
| 0 | 0 | 0.013994895 | 0 | 0 |
| 0 | 0 | 0 | 0 | 0 |
| 0 | 0 | 0 | 0 | 0 |
| 0.135257592 | 1.370650093 | 0.042639137 | 0.105004903 | 0 |
| 0 | 0 | 0 | 0 | 0 |
| 0 | 0 | 0 | 0 | 0 |
| 0 | 0.003075794 | 0 | 0.002197105 | 0 |
| 0 | 0 | 0 | 0 | 0 |
| 0 | 0 | 0.429307633 | 0.062304482 | 0.019778311 |

| | | | | |
|---|---|---|---|---|
| 0 | 0.078238926 | 0 | 0 | 0 |
| 0 | 0 | 0 | 0 | 0 |
| 0 | 0 | 0 | 0 | 0 |
| 0 | 0 | 0 | 0 | 0 |
| 0.001806362 | 0 | 0 | 0 | 0.002523821 |
| 0 | 0 | 0.005022554 | 0.001001003 | 0 |
| 0 | 0.00528125 | 0 | 0 | 0 |
| 0 | 0 | 0 | 0 | 0 |
| 0 | 0 | 0 | 0 | 0 |
| 0 | 0 | 0.01434766 | 0.02521394 | 0 |
| 0 | 0 | 0.014041901 | 0 | 0 |
| 0.391290413 | 2.495692927 | 0.125283244 | 0.680720241 | 0 |
| 0.038931933 | 0.263521173 | 0.017776084 | 0.004824738 | 1.220694888 |
| 0 | 0 | 0 | 0 | 0 |
| 0.004178114 | 0.001237454 | 0.037978324 | 0.040673006 | 6.510512819 |
| 0 | 0 | 0 | 0 | 0 |
| 0.005812964 | 0.007230953 | 0 | 0 | 0 |
| 0 | 0 | 0 | 0.012708091 | 0 |
| 0.002746577 | 0.003036946 | 0 | 0 | 0 |
| 0 | 0.038004514 | 0 | 0.009541694 | 0 |
| 0.068369004 | 0 | 0.008429904 | 0 | 0 |
| 0 | 0 | 0 | 0 | 0 |
| 0.351460519 | 0.006538825 | 0.264084195 | 0 | 0.000678441 |
| 0 | 0 | 0 | 0 | 0 |
| 0 | 0 | 0 | 0 | 0 |
| 0 | 0 | 0 | 0 | 0 |
| 0 | 0 | 0 | 0 | 0 |
| 0 | 0 | 0 | 0 | 0 |
| 0 | 0 | 0 | 0.00162426 | 0 |
| 0 | 0 | 0 | 0 | 0 |
| 0 | 0 | 0 | 0 | 0 |
| 0.041746994 | 0.059413457 | 0.004348428 | 0.05147894 | 0.053370013 |
| 0 | 0 | 0 | 0 | 0 |
| 0.084914035 | 0.158517599 | 0.020710211 | 0.198989707 | 0 |
| 0.002456037 | 0 | 0.001313232 | 0 | 0.030972967 |
| 0.071916374 | 0.101542646 | 1.195681958 | 0.30235931 | 0.007181279 |
| 0 | 0 | 0 | 0 | 0 |
| 0 | 0 | 0.046104124 | 0 | 0 |
| 0 | 0 | 0 | 0 | 0 |
| 0 | 0 | 0 | 0 | 0 |
| 0 | 0.002479048 | 0.027971963 | 0.136185813 | 0 |
| 0 | 0 | 0 | 0.130957283 | 0 |
| 0 | 0 | 0 | 0 | 0 |
| 0 | 0 | 0 | 0 | 0 |
| 0.101616595 | 0.244865215 | 0.002956949 | 0.006298408 | 0.106680415 |
| 0 | 0 | 0 | 0 | 0 |
| 0 | 0.062875237 | 0.127816387 | 1.697601299 | 0 |
| 0 | 0 | 0 | 0.001097373 | 0.000622843 |

| | | | | |
|---|---|---|---|---|
| 0 | 0 | 0 | 0 | 0 |
| 0 | 0 | 0 | 0 | 0 |
| 0 | 0 | 0 | 0 | 0 |
| 0 | 0 | 0 | 0 | 0 |
| 0 | 0 | 0 | 0 | 0 |
| 0 | 0 | 0 | 0 | 0 |
| 0 | 0 | 7.162502546 | 0 | 0 |
| 0.061443127 | 0.000999886 | 0.027257462 | 0.00145705 | 0 |
| 0 | 0 | 0 | 0 | 0 |
| 0 | 0 | 0 | 0 | 0 |
| 0 | 0 | 0 | 0 | 0 |
| 0 | 0 | 0 | 0 | 0 |
| 0 | 0 | 0 | 0 | 0 |
| 0 | 0 | 0.072839029 | 0 | 0 |
| 0 | 0 | 0 | 0 | 0 |
| 0 | 0 | 0 | 0 | 0 |
| 0 | 0 | 0 | 0 | 0 |
| 0 | 0 | 0 | 0 | 0 |
| 0 | 2.248354001 | 4.170751898 | 17.56351281 | 0 |
| 0 | 0 | 0 | 0 | 0 |
| 0 | 0 | 0.005036342 | 0 | 0 |
| 0 | 0 | 0 | 0 | 0 |
| 0 | 0 | 0 | 0 | 0 |
| 0 | 0 | 0 | 0 | 0 |
| 0.001699362 | 0 | 0.003028803 | 0.000543281 | 0 |
| 0.035234124 | 0.001906022 | 0 | 0 | 0.173904186 |
| 0 | 0 | 0 | 0 | 0 |
| 0.003149516 | 0 | 0.050184156 | 0.008608918 | 0 |
| 0 | 0 | 0 | 0 | 0 |
| 1.430741672 | 0.096539671 | 0.319890968 | 0.638172646 | 0 |
| 0 | 0 | 0 | 0 | 0 |
| 0 | 0 | 0 | 0 | 0 |
| 0 | 0 | 0.001066915 | 0 | 0 |
| 0 | 0 | 0 | 0 | 0 |
| 0.001180193 | 0.001597915 | 0 | 0.160569911 | 0.014562134 |
| 0 | 0 | 0 | 0 | 0 |
| 0.119089422 | 0 | 0 | 0 | 0 |
| 0.003443589 | 0.028557352 | 0.000613757 | 0 | 0.001214982 |
| 0 | 0 | 0 | 0 | 0 |
| 0 | 0 | 0 | 0 | 0 |
| 1.052818638 | 5.696094315 | 0.384209435 | 4.403072591 | 0.828392813 |
| 0.605393547 | 0.340770187 | 0 | 0.140623901 | 0 |
| 0 | 0 | 0 | 0 | 0 |
| 0 | 0 | 0 | 0 | 0 |
| 0 | 0 | 0 | 0 | 0 |
| 0 | 0.004165916 | 0 | 0 | 0 |
| 0 | 0.005915491 | 0 | 0.002706809 | 0 |
| 0.031581595 | 0.051984894 | 0 | 0.054899954 | 0.005399952 |

| | | | | |
|---|---|---|---|---|
| 0 | 0 | 0 | 0 | 0 |
| 0 | 0 | 0.001336092 | 0 | 0 |
| 0 | 0 | 0 | 0.003148938 | 0 |
| 0 | 0 | 0 | 0 | 0 |
| 1.349156424 | 0.909877167 | 0.135416966 | 0.585984568 | 0 |
| 0 | 0 | 0 | 0 | 0 |
| 0 | 0 | 0 | 0 | 0 |
| 0 | 0 | 0 | 0 | 0 |
| 0 | 0 | 0 | 0 | 3.877124995 |
| 0 | 0 | 0 | 0 | 0 |
| 0 | 0 | 0 | 0 | 0 |
| 0 | 0 | 0 | 0 | 0 |
| 0.021093258 | 0.064180882 | 0.014767494 | 0.064329628 | 0 |
| 0.001865613 | 0.020996827 | 0 | 0.011451473 | 0 |
| 0 | 0 | 2.562315759 | 0 | 0 |
| 0.052663242 | 0.046611264 | 0.061229019 | 0.251516467 | 0 |
| 0 | 0 | 0.013811385 | 0 | 0 |
| 0 | 0 | 0 | 0 | 0 |
| 0 | 0 | 0 | 0 | 0 |
| 0 | 0 | 0 | 0 | 0 |
| 0 | 0 | 0 | 0 | 0 |
| 0 | 0 | 0 | 0 | 0 |
| 0 | 0 | 0 | 0 | 0 |
| 0 | 0 | 0 | 0 | 0 |
| 0 | 0 | 0.001699974 | 0.101041684 | 0 |
| 0 | 0 | 0 | 0 | 0 |
| 0.074260264 | 0.052053958 | 0.003302631 | 0.000673179 | 0 |
| 0 | 0 | 0 | 0 | 0.001207179 |
| 0 | 0 | 0 | 0 | 0 |
| 0.001714707 | 0 | 0 | 0 | 0 |
| 0 | 0 | 0 | 0 | 0 |
| 0 | 0 | 0 | 0.001269011 | 0 |
| 0 | 0 | 0 | 0 | 0 |
| 0.02324659 | 0.038753064 | 0.00941117 | 0 | 0 |
| 9.307680873 | 0 | 2.825990187 | 0.028064952 | 0 |
| 0 | 0 | 0 | 0 | 0 |
| 0 | 0 | 0.000801149 | 0.009520344 | 0 |
| 0.032010238 | 0 | 0.002775524 | 0.014271694 | 0.084833149 |
| 0 | 0 | 0 | 0 | 0 |
| 0 | 0 | 0 | 0 | 0 |
| 0.106599028 | 0.263476504 | 0.349907871 | 0.127421071 | 0 |
| 0 | 0 | 0.187454238 | 0.053952048 | 0 |
| 0 | 0 | 0.002499426 | 0 | 0 |
| 0 | 0 | 0 | 0 | 0 |
| 0 | 0 | 0 | 0 | 0 |
| 0 | 0 | 0 | 0 | 0 |
| 0 | 0 | 0 | 0 | 0 |
| 0 | 0 | 0 | 0 | 0 |

| | | | | |
|---|---|---|---|---|
| 0 | 2.25460408 | 0.305839457 | 0.096809773 | 0 |
| 0 | 0 | 0 | 0 | 0 |
| 0 | 0 | 0 | 0 | 0 |
| 0 | 0 | 0 | 0 | 0.006381653 |
| 0 | 0.020543157 | 0.045532914 | 0.007903844 | 0 |
| 0 | 0 | 0 | 0 | 0 |
| 0.000805236 | 0 | 0.009816684 | 0.000463377 | 0.025978749 |
| 0 | 0 | 0 | 0 | 0 |
| 0 | 0.001750696 | 0 | 0.004797019 | 0 |
| 0 | 0 | 0.007950336 | 0 | 0 |
| 0 | 0 | 0.024644065 | 0 | 0 |
| 0 | 0 | 0 | 0 | 0 |
| 0 | 0 | 0 | 0 | 0 |
| 0 | 0 | 0 | 0 | 0 |
| 0 | 0 | 0 | 0 | 0 |
| 0 | 0 | 0 | 0 | 0 |
| 0 | 0 | 0 | 0 | 0 |
| 0 | 0 | 0 | 0 | 0 |
| 0 | 0 | 0 | 0.332110075 | 0 |
| 0 | 0 | 0 | 0 | 0 |
| 0.522994941 | 0 | 0.007378445 | 0.052939298 | 0.003310746 |
| 0 | 0 | 0.001908938 | 0.02011653 | 0 |
| 0.121088508 | 0.040471554 | 0.018808744 | 0.066664041 | 0 |
| 0.073686934 | 0.008373381 | 0.00048245 | 0.001298066 | 0.012606643 |
| 0 | 0.00255588 | 0 | 0 | 0 |
| 0.001064498 | 0 | 0 | 0 | 0 |
| 0 | 0 | 0 | 0 | 0 |
| 0 | 0 | 0 | 0 | 0 |
| 0 | 0 | 0 | 0 | 0 |
| 0 | 1.036921074 | 0 | 0.001245481 | 3.759914011 |
| 0 | 0.002515024 | 0 | 0 | 0 |
| 0 | 0 | 0 | 0 | 0 |
| 0 | 0 | 0 | 0 | 0 |
| 0.377245593 | 0.026636539 | 0 | 0.621967056 | 0 |
| 0 | 0 | 0 | 0 | 0 |
| 0 | 0 | 0 | 0 | 0 |
| 0 | 0 | 0.001609786 | 0 | 0.640766128 |
| 0 | 0 | 0 | 0 | 0 |
| 0 | 0 | 0 | 0 | 0 |
| 0 | 0 | 0.21319591 | 0 | 0 |
| 0 | 0 | 0 | 0 | 0 |
| 0 | 0 | 0 | 0 | 0 |
| 0 | 0 | 0 | 0 | 0 |
| 0 | 0 | 0.02688219 | 0 | 0 |
| 0 | 0 | 0 | 0.110336711 | 0 |
| 0 | 0 | 0 | 0 | 0 |
| 0 | 0 | 0 | 0 | 0 |
| 0 | 0 | 0 | 0 | 0 |

| | | | | |
|---|---|---|---|---|
| 0 | 0.004409176 | 0.000865222 | 0.036781485 | 0.004727265 |
| 1.462525638 | 0 | 3.628409019 | 0.084632728 | 0 |
| 0 | 0 | 0 | 0 | 0 |
| 0 | 0 | 0 | 0 | 0 |
| 0 | 0 | 0 | 0 | 0 |
| 0 | 0 | 0 | 0 | 0 |
| 0 | 0.002357959 | 0 | 0 | 0 |
| 0.483989957 | 0.385798503 | 0.062542477 | 0.205403903 | 0 |
| 5.13017585 | 0.37071759 | 0.439479834 | 0.02533478 | 0.827830932 |
| 0.001060939 | 0 | 0.003592769 | 0 | 1.133082345 |
| 0 | 0 | 0.000665692 | 0 | 0 |
| 0 | 0 | 0 | 0 | 0 |
| 0 | 0 | 0 | 0 | 0 |
| 0 | 1.507463091 | 0.027548568 | 0.215150452 | 0 |
| 0 | 0 | 0 | 0 | 0 |
| 0 | 0 | 0 | 0 | 0 |
| 0 | 0 | 0 | 0 | 0 |
| 0 | 0 | 0 | 0 | 0 |
| 0 | 0 | 0 | 0 | 0 |
| 0.010996952 | 0.020671245 | 0 | 0.433414686 | 0 |
| 0 | 0 | 0 | 0 | 0 |
| 0 | 0 | 0 | 0 | 0 |
| 0 | 0 | 0 | 0 | 0 |
| 0 | 0 | 0 | 0 | 0 |
| 0 | 0.001358936 | 0.02998522 | 0.014546379 | 0 |
| 0 | 0 | 0 | 0 | 0 |
| 0 | 0 | 0 | 0 | 0 |
| 0 | 0 | 0 | 0.754479018 | 0 |
| 0 | 0 | 0 | 0 | 0 |
| 0 | 0 | 0 | 0 | 0 |
| 0 | 0 | 0 | 0 | 0 |
| 0 | 0 | 0 | 0 | 0 |
| 0 | 0 | 0 | 0 | 0 |
| 0 | 0 | 0 | 0 | 0 |
| 0 | 0 | 0 | 0 | 0 |
| 0 | 0 | 0 | 0 | 0 |
| 0 | 0 | 0.001093786 | 0.021908324 | 0 |
| 0 | 0 | 0 | 0 | 0.024126274 |
| 0 | 0.001139751 | 0 | 0 | 0 |
| 0.002907772 | 0 | 0 | 0 | 0.142194289 |
| 0 | 0 | 0 | 0 | 0 |
| 0.060330972 | 0.038501262 | 0.021208759 | 0.21882408 | 0 |
| 0 | 0 | 0 | 0 | 0 |
| 0 | 0 | 0 | 0 | 0 |
| 0 | 0 | 0 | 0 | 0 |
| 0 | 0 | 0 | 0 | 0 |
| 0 | 0 | 0 | 0 | 0 |
| 0 | 0 | 0 | 0 | 0 |

| | | | | |
|---|---|---|---|---|
| 0 | 0 | 0 | 0 | 0 |
| 0 | 0.001903227 | 0 | 0 | 0.035431027 |
| 0 | 0 | 0 | 0 | 0 |
| 0 | 0 | 0 | 0 | 0 |
| 1.984548377 | 4.241358108 | 12.66018024 | 0.579880393 | 0.336907262 |
| 0 | 0 | 0 | 0 | 0 |
| 0 | 0.005664508 | 0 | 0 | 0 |
| 0.002058415 | 0.004145628 | 0.001761 | 0.003356152 | 0.138570194 |
| 0 | 0.005113199 | 0 | 0.002759642 | 0 |
| 0 | 0 | 0 | 0.024871228 | 0 |
| 0 | 0 | 0 | 0 | 0 |
| 0 | 0 | 1.273303458 | 0 | 0 |
| 0 | 0 | 0 | 0 | 0 |
| 0 | 0 | 0 | 0 | 0 |
| 0 | 0 | 0 | 0 | 0 |
| 0.001428198 | 0.266487961 | 0.003818256 | 0.098452376 | 0 |
| 0 | 0 | 0 | 0 | 0 |
| 0 | 0 | 0 | 0 | 0 |
| 0 | 0 | 0 | 0 | 0 |
| 0 | 0 | 0 | 0 | 0 |
| 0 | 0 | 0 | 0 | 0 |
| 0 | 0 | 0 | 0 | 0 |
| 0 | 0 | 0 | 0 | 0 |
| 0.002493944 | 0 | 0.0013335 | 0 | 0.00837185 |
| 0 | 0.03736573 | 0.154237415 | 0.484455362 | 0 |
| 0 | 0 | 0 | 0 | 0 |
| 0 | 0.020653258 | 0.893297383 | 0.223456062 | 0.535990474 |
| 0.00085779 | 0 | 0 | 0 | 0 |
| 0 | 0 | 0 | 0 | 0 |
| 0 | 0 | 0 | 0 | 0 |
| 0 | 0 | 0 | 0 | 0 |
| 0 | 0 | 0 | 0 | 0 |
| 0 | 0 | 0 | 0 | 0 |
| 0 | 0 | 0 | 0 | 0 |
| 0 | 0 | 0 | 0 | 0 |
| 0 | 0 | 0 | 0 | 0 |
| 0 | 0 | 0 | 0 | 0 |
| 0.110949348 | 0.086308142 | 0.001429497 | 0.42243704 | 0 |
| 0 | 0 | 0 | 0 | 0 |
| 0 | 0 | 0 | 0 | 0 |
| 0 | 0 | 0 | 0 | 0 |
| 5.453175731 | 0 | 0.185425393 | 0 | 0 |
| 0 | 0 | 0 | 0 | 0 |
| 0 | 0 | 0 | 0 | 0 |
| 0 | 0 | 0 | 0 | 0 |
| 0 | 0 | 0 | 0 | 0 |
| 0 | 0 | 0 | 0 | 0 |
| 0 | 0 | 0 | 0 | 0 |

| | | | | |
|---|---|---|---|---|
| 0.053473748 | 0.070611168 | 0.126329892 | 0.00867016 | 0 |
| 0 | 0 | 0 | 0 | 0 |
| 0 | 0 | 0 | 0 | 0 |
| 0 | 0.000722408 | 0 | 0 | 0 |
| 0 | 0 | 0 | 0 | 0 |
| 1.84792208 | 3.251885778 | 0 | 0 | 0 |
| 0 | 0 | 0 | 0 | 0 |
| 0 | 0 | 0 | 0 | 0 |
| 0 | 0 | 0 | 0 | 0 |
| 0 | 0 | 0 | 0 | 0 |
| 0 | 0 | 0 | 0 | 0 |
| 0.002299018 | 0.004669111 | 0 | 0 | 0 |
| 0 | 0 | 0 | 0 | 0 |
| 0 | 0 | 0 | 0 | 0 |
| 0 | 0 | 0 | 0 | 0 |
| 0 | 0 | 0 | 0 | 0 |
| 0 | 0 | 0 | 0 | 0 |
| 0 | 0 | 0 | 0 | 0 |
| 0 | 0 | 0.001944841 | 0 | 0 |
| 0 | 0 | 0 | 0 | 0 |
| 0 | 0 | 0 | 0 | 0.183185445 |
| 0 | 0 | 0 | 0 | 0 |
| 0.204992847 | 1.284876238 | 0.031145658 | 0.080576573 | 0.078824005 |
| 0 | 0.003886753 | 0.069730379 | 0.686740242 | 0 |
| 0 | 0 | 0 | 0 | 0 |
| 0 | 0 | 0.345210677 | 0 | 0 |
| 0 | 0 | 0 | 0 | 0 |
| 0 | 0 | 0 | 0 | 0 |
| 0 | 0.003216718 | 0.984537175 | 0 | 0 |
| 0 | 0 | 0 | 0 | 0 |
| 0 | 0 | 0 | 0 | 0 |
| 0.040548224 | 0 | 0 | 0 | 0 |
| 0.005455026 | 0 | 0 | 0 | 6.580226817 |
| 0 | 0 | 0 | 0 | 0 |
| 0.047856585 | 0.24968352 | 0.015252873 | 0.004499885 | 1.347539859 |
| 0.002862166 | 0 | 0 | 0 | 0.053676066 |
| 0 | 0 | 0 | 0 | 0 |
| 0 | 0 | 0 | 0 | 0 |
| 0.029600187 | 0 | 0.000833005 | 0 | 0.847668592 |
| 0 | 0 | 0 | 0 | 0 |
| 0 | 0 | 0 | 0 | 0 |
| 0 | 0 | 0 | 0 | 0 |
| 0.129109961 | 0.457521114 | 1.550658975 | 0.057655471 | 0 |
| 0 | 0 | 0.004470978 | 0 | 0 |
| 0 | 0 | 0 | 0 | 0 |
| 0.383197798 | 0.024637605 | 0.023399898 | 0 | 0 |
| 0 | 0 | 0 | 0 | 0 |
| 0 | 0 | 0 | 0 | 0 |

| | | | | |
|---|---|---|---|---|
| 0 | 0 | 0 | 0 | 0 |
| 0 | 0 | 0 | 0 | 0 |
| 0 | 0 | 0 | 0 | 0 |
| 0 | 0 | 0 | 0 | 0 |
| 0 | 0 | 0 | 0 | 0 |
| 0 | 0 | 0 | 0 | 0 |
| 0 | 0 | 0 | 0 | 0 |
| 0 | 0 | 0 | 0 | 0 |
| 0.005421739 | 0.007025307 | 0.017031507 | 0.060774329 | 0 |
| 0 | 0 | 0.00139431 | 0.003215563 | 0 |
| 0 | 0 | 0 | 0 | 0 |
| 0 | 0 | 0 | 0 | 0 |
| 0 | 0 | 0 | 0 | 0 |
| 0 | 0 | 0 | 0 | 0 |
| 0 | 0 | 0 | 0 | 0 |
| 0 | 0 | 0 | 0 | 0.015130225 |
| 0 | 0 | 0 | 0 | 0 |
| 0 | 0 | 0 | 0 | 0 |
| 0 | 0 | 0 | 0 | 0 |
| 0.358417651 | 0.282654267 | 0.174744175 | 0.003090265 | 5.817502094 |
| 0 | 0 | 0 | 0.005507202 | 0 |
| 0 | 0 | 0 | 0 | 0 |
| 0 | 0 | 0 | 0 | 0 |
| 0 | 0 | 0.025843635 | 0.059466925 | 0 |
| 0.001238413 | 0 | 0 | 0 | 0 |
| 0 | 0 | 0 | 0 | 0 |
| 0 | 0 | 0 | 0 | 0 |
| 0 | 0.349381474 | 0 | 0 | 0 |
| 0 | 0 | 0 | 0 | 0 |
| 0 | 0 | 0.015233717 | 0.010019145 | 0 |
| 0.056898783 | 0.172578652 | 0.228893224 | 0.428283314 | 0 |
| 0 | 0 | 0 | 0 | 0 |
| 0 | 0 | 0 | 0 | 0 |
| 0 | 0 | 0 | 0 | 0 |
| 0 | 0 | 0 | 0 | 0 |
| 0 | 0 | 0 | 0 | 0 |
| 0 | 0 | 0 | 0 | 0 |
| 0 | 0 | 0 | 0 | 0 |
| 0 | 0 | 0.031236648 | 0 | 0 |
| 0 | 0 | 0 | 0 | 0 |
| 0 | 0 | 0 | 0 | 0 |
| 0 | 0 | 0 | 0 | 0 |
| 0 | 0 | 0.000516844 | 0 | 0 |
| 0 | 0.002087139 | 0 | 0 | 0 |
| 0 | 0 | 0 | 0 | 0 |
| 0 | 0 | 0 | 0 | 0 |
| 0 | 0 | 0 | 0 | 4.360426868 |
| 0 | 0 | 0 | 0 | 0 |

| | | | | |
|---|---|---|---|---|
| 0 | 0 | 0 | 0 | 0 |
| 0 | 0 | 0 | 0 | 0 |
| 0 | 0 | 0 | 0 | 0 |
| 0 | 0 | 0 | 0 | 0 |
| 0.990045711 | 0.012453012 | 0.004346494 | 0.027556136 | 0 |
| 0 | 0 | 0 | 0 | 0 |
| 0 | 0 | 0 | 0 | 0 |
| 0 | 0 | 0 | 0 | 0 |
| 0 | 0 | 0 | 0 | 0 |
| 0 | 0 | 0 | 0 | 0 |
| 0 | 0 | 0.002093238 | 0 | 0 |
| 0.020431216 | 0.092900612 | 0.036206848 | 0.011197371 | 0.033118514 |
| 0.034246727 | 0 | 0 | 0.02044808 | 0.011301235 |
| 0 | 0 | 0 | 0 | 0 |
| 0.002224288 | 0 | 0 | 0 | 0.011112527 |
| 0.864316289 | 0.001971904 | 0 | 0 | 0 |
| 0 | 0 | 0.01104292 | 0 | 0.003041441 |
| 0 | 0 | 0 | 0 | 0 |
| 0.005009539 | 0 | 0.001418071 | 0 | 0 |
| 0.000961902 | 0 | 0.025304766 | 0 | 0.010995989 |
| 0 | 0 | 0 | 0 | 0 |
| 0 | 0 | 0 | 0 | 0 |
| 0 | 0 | 0 | 0 | 0 |
| 0 | 0 | 0 | 0 | 0 |
| 0 | 0 | 0 | 0 | 0 |
| 0 | 0 | 0 | 0 | 0 |
| 0.005087385 | 0 | 0 | 0 | 0.034463106 |
| 0 | 0 | 0 | 0 | 0 |
| 0 | 0 | 0 | 0 | 0 |
| 0 | 0 | 0 | 0 | 0 |
| 0 | 0 | 0.048775232 | 0 | 0.000665893 |
| 0 | 0 | 0 | 0 | 0.001678232 |
| 0 | 0 | 0 | 0 | 0 |
| 0 | 0 | 0 | 0 | 0 |
| 0 | 0 | 0 | 0 | 0 |
| 0 | 0 | 0 | 0 | 0 |
| 0 | 0 | 0 | 0 | 0 |
| 0.043953708 | 0.053817412 | 0.008534192 | 0.003532593 | 0.022612137 |
| 0 | 1.101832004 | 0.086540865 | 0.064584585 | 0 |
| 0 | 0 | 0 | 0 | 0 |
| 0.059446325 | 0.148112614 | 0 | 0.003644222 | 0 |
| 0 | 0 | 0 | 0 | 0 |
| 0 | 0 | 0 | 0 | 0 |
| 0 | 0 | 0 | 0 | 0 |
| 0 | 0 | 0 | 0 | 0 |
| 0 | 0 | 0 | 0 | 0 |
| 0 | 0 | 0 | 0 | 0 |
| 0 | 0 | 0 | 0 | 0 |

| | | | | |
|---|---|---|---|---|
| 0 | 0 | 0 | 0 | 0 |
| 0 | 0 | 0.001497658 | 0 | 0 |
| 0 | 0 | 0 | 0 | 0 |
| 0.003816011 | 0 | 0.001836364 | 0 | 0.011148042 |
| 0 | 0.001257899 | 0 | 0 | 0.069455346 |
| 0 | 0 | 0 | 0 | 0.004351398 |
| 0 | 0 | 0 | 0.000887914 | 0 |
| 0 | 0.002121409 | 0 | 0 | 0 |
| 0 | 0 | 0 | 0.028840388 | 0 |
| 0 | 0 | 0 | 0 | 0 |
| 0 | 0 | 0 | 0 | 0 |
| 0 | 0 | 0 | 0 | 0 |
| 0.554798544 | 0.184426609 | 0.108621587 | 0.003372663 | 0 |
| 0 | 0 | 0.011025307 | 0 | 0 |
| 0 | 0 | 0.003933621 | 0 | 0 |
| 0 | 0 | 0 | 0 | 0 |
| 0 | 0 | 0 | 0 | 0 |
| 0 | 0 | 0 | 0 | 0 |
| 0 | 0 | 0 | 0 | 0 |
| 0 | 0 | 0 | 0 | 0 |
| 0 | 0 | 0 | 0 | 0 |
| 0 | 0 | 0 | 0 | 0 |
| 0.013357055 | 0 | 0 | 0 | 0.03483624 |
| 0 | 0 | 0.006067984 | 0.006046795 | 0 |
| 0 | 0 | 0 | 0 | 0 |
| 0 | 0.006067682 | 0 | 0 | 0 |
| 0 | 0.00886956 | 0.002183527 | 0 | 0 |
| 0 | 0 | 0.001092293 | 0 | 0 |
| 0 | 0 | 0 | 0 | 0 |
| 0 | 0 | 0 | 0 | 0 |
| 0.107428581 | 0.003058261 | 0.007644721 | 0.01733101 | 0 |
| 0 | 0 | 0 | 0 | 0 |
| 0 | 0 | 0 | 0 | 0 |
| 0 | 0 | 0 | 0 | 0 |
| 0.018312805 | 0 | 0 | 0 | 0 |
| 0 | 0 | 0 | 0 | 0 |
| 0 | 0 | 0 | 0 | 0 |
| 0 | 0 | 0 | 0 | 0 |
| 0 | 0.000796154 | 0.008983283 | 0.002094748 | 0.081517906 |
| 0 | 0 | 0 | 0 | 0 |
| 0 | 0 | 0 | 0 | 0 |
| 0 | 0 | 0 | 0 | 0 |
| 0 | 0 | 0 | 0 | 0 |
| 0 | 0 | 0 | 0 | 0 |
| 0 | 0 | 0 | 0 | 0 |
| 0.283812097 | 0.465821911 | 0.085677249 | 0.262702982 | 0.975385601 |
| 0 | 0 | 0 | 0 | 0.432624859 |
| 0 | 0 | 0 | 0 | 0 |

| | | | | |
|---|---|---|---|---|
| 0 | 0 | 0 | 0 | 0 |
| 0 | 0 | 0 | 0 | 0 |
| 0 | 0 | 0.073774281 | 0 | 0 |
| 0 | 0 | 0 | 0 | 0 |
| 0 | 0 | 0 | 0 | 0 |
| 0 | 0 | 0.019646964 | 0 | 0 |
| 0 | 0 | 0 | 0 | 0 |
| 0 | 0 | 0 | 0 | 0 |
| 0 | 0 | 0 | 0 | 0 |
| 0 | 0 | 0 | 0 | 0.001461253 |
| 0.095136737 | 0 | 0.00145757 | 0.00039217 | 0.001558104 |
| 0 | 0 | 0 | 0 | 0.002067782 |
| 0 | 0 | 0 | 0 | 0 |
| 0 | 0 | 0 | 0 | 0 |
| 0 | 0 | 0 | 0 | 0 |
| 0 | 0 | 0 | 0 | 0 |
| 0 | 0 | 0 | 0 | 0 |
| 0 | 0 | 0 | 0 | 0 |
| 0 | 0 | 0 | 0 | 0 |
| 0.004699655 | 0 | 0 | 0 | 0 |
| 0 | 0 | 0 | 0 | 0 |
| 0 | 0 | 0 | 0 | 0 |
| 0 | 0 | 0 | 0 | 0 |
| 0 | 0 | 0 | 0 | 0 |
| 0 | 0 | 0 | 0 | 0 |
| 0 | 0 | 0 | 0 | 0 |
| 0 | 0.011850515 | 0 | 0 | 0 |
| 0 | 0 | 0.005134027 | 0.006331174 | 0 |
| 0 | 0 | 0 | 0 | 0 |
| 0 | 0 | 0 | 0 | 0 |
| 0.009317284 | 0.032378697 | 0.005812229 | 0.001191483 | 0.006903467 |
| 0 | 0 | 0 | 0 | 0 |
| 0.006671295 | 0.013243681 | 0.017739149 | 0.018157565 | 0.022214741 |
| 0.003316544 | 0.001964556 | 0 | 0.004594587 | 0.000702095 |
| 0 | 0.001271713 | 0 | 0 | 0 |
| 0 | 0 | 0 | 0 | 0 |
| 0 | 0 | 0 | 0 | 0 |
| 0 | 0 | 0 | 0 | 0 |
| 0 | 0 | 0 | 0 | 0 |
| 0 | 0 | 0 | 0 | 0 |
| 0 | 0 | 0 | 0 | 0 |
| 0.136617001 | 0.015113052 | 0 | 0.031583336 | 1.353732956 |
| 0.018613591 | 0.122460932 | 0.015267101 | 0.034837654 | 0 |
| 0 | 0 | 0 | 0 | 0 |
| 0 | 0 | 0 | 0 | 0 |
| 0.519417354 | 0.133651557 | 0.688390941 | 0.037318815 | 0 |
| 0.00462278 | 0.011358894 | 0 | 0 | 0 |
| 0 | 0 | 0 | 0 | 0 |

| | | | | |
|---|---|---|---|---|
| 0 | 0 | 0 | 0 | 0 |
| 0.007572895 | 0.002018613 | 0.001822137 | 0.246218987 | 0 |
| 0.002874931 | 0.004453911 | 0.00331092 | 0.014210808 | 0.013763892 |
| 0 | 0 | 0 | 0 | 0 |
| 0 | 0 | 0 | 0.770386448 | 0 |
| 0 | 0.001018123 | 0 | 0 | 0.107629047 |
| 0 | 0 | 0 | 0 | 0 |
| 0 | 0 | 0 | 0 | 0 |
| 0 | 0 | 0 | 0 | 0 |
| 0 | 0 | 0 | 0 | 0 |
| 0 | 0 | 0 | 0 | 0 |
| 0 | 0 | 0 | 0 | 0 |
| 0 | 0 | 0 | 0 | 0 |
| 0 | 0 | 0 | 0 | 0 |
| 0 | 0 | 0 | 0 | 0 |
| 0 | 0 | 0 | 0 | 0 |
| 0 | 0 | 0.155353674 | 0.322602725 | 0 |
| 0 | 0 | 0 | 0 | 0 |
| 0 | 0 | 0 | 0 | 0 |
| 0 | 0 | 0 | 0 | 0 |
| 0 | 0 | 0 | 0 | 0 |
| 0 | 0 | 0 | 0 | 0 |
| 0 | 0 | 0 | 0 | 0 |
| 0 | 0 | 0 | 0 | 0 |
| 0 | 0.008420901 | 0.263510907 | 0.002622023 | 0 |
| 0 | 0.068421451 | 0 | 0 | 0.01071253 |
| 0 | 0 | 0 | 0 | 0 |
| 0.009248341 | 0.000842809 | 0.004374464 | 0.002217501 | 0 |
| 0 | 0 | 0 | 0 | 0 |
| 0 | 0 | 0 | 0 | 0 |
| 0 | 0 | 0.003897545 | 0 | 0 |
| 0 | 0 | 0 | 0 | 0.005514019 |
| 0 | 0 | 0 | 0 | 0 |
| 0 | 0 | 0 | 0 | 0 |
| 0 | 0 | 0 | 0 | 0 |
| 0 | 0 | 0 | 0 | 0 |
| 0 | 0 | 0 | 0 | 0 |
| 0 | 0 | 0 | 0 | 0.235290763 |
| 0.002164037 | 0.046738884 | 0 | 0.797155162 | 0 |
| 0 | 0 | 0 | 0 | 0 |
| 0 | 0 | 0 | 0 | 0 |
| 0.020487 | 0.009896897 | 0.00212705 | 0.003433789 | 0 |
| 0 | 0.002615939 | 0.001377439 | 0.031943038 | 0.004908154 |
| 0 | 0 | 0 | 0.01815863 | 0 |
| 0 | 0 | 0 | 0 | 0 |
| 0.003175428 | 0 | 0 | 0 | 0 |
| 0 | 0 | 0.001505151 | 0 | 0.00214529 |
| 0 | 0 | 0 | 0 | 0 |

| | | | | |
|---|---|---|---|---|
| 0 | 0 | 0 | 0 | 0 |
| 0.008347137 | 0.001893611 | 0 | 0.012604665 | 0.017820828 |
| 0 | 0 | 0.004729079 | 0 | 0 |
| 0 | 0 | 0 | 0 | 0 |
| 0 | 0 | 0 | 0 | 0 |
| 0 | 0 | 0 | 0 | 0 |
| 0 | 0 | 0 | 0 | 0 |
| 0 | 0 | 0 | 0 | 0 |
| 0 | 0 | 0 | 0 | 0 |
| 0 | 0 | 0 | 0 | 0 |
| 0 | 0 | 0 | 0 | 0 |
| 0 | 0 | 0 | 0 | 0 |
| 1.196263433 | 0.233978203 | 0.011644359 | 0.016565281 | 0 |
| 0 | 0 | 0 | 0 | 0 |
| 0 | 0.001049354 | 0.048544948 | 0 | 0.109974374 |
| 0 | 0 | 0 | 0 | 0 |
| 0 | 0 | 0 | 0 | 0 |
| 0 | 0 | 0 | 0 | 0 |
| 0 | 0 | 0 | 0 | 0 |
| 0 | 0 | 0 | 0 | 0 |
| 0 | 0 | 0 | 0 | 0 |
| 0 | 0 | 0.00837219 | 0.001042869 | 0 |
| 0 | 0 | 0 | 0 | 0 |
| 0 | 0 | 0 | 0 | 0 |
| 0 | 0 | 0 | 0 | 0 |
| 0 | 0 | 0 | 0 | 0 |
| 0 | 0 | 0 | 0 | 0 |
| 0 | 0 | 0 | 0 | 0 |
| 0 | 0 | 0 | 0 | 0 |
| 0 | 0 | 0 | 0 | 0 |
| 0 | 0 | 0 | 0 | 0 |
| 0 | 0 | 0 | 0 | 0 |
| 0 | 0 | 0 | 0 | 0 |
| 0 | 0 | 0 | 0 | 0 |
| 0 | 6.995619505 | 6.972670988 | 0.181419793 | 0.043889275 |
| 0 | 0 | 0 | 0 | 0 |
| 0 | 0 | 0 | 0 | 0 |
| 0 | 0 | 0 | 0 | 0 |
| 0.001520193 | 0 | 0.010683046 | 0 | 0.038066322 |
| 0 | 0 | 0 | 0 | 0 |
| 0.042449401 | 1.104992139 | 0.243425536 | 0.089642966 | 0.013603131 |
| 0 | 0 | 0 | 0 | 0 |
| 0 | 0 | 0 | 0 | 0 |
| 0 | 0 | 0 | 0 | 0 |
| 0 | 0 | 0 | 0 | 0 |
| 0.049716393 | 0.01479079 | 0.042676061 | 0.013684679 | 4.755934527 |
| 0 | 0 | 0 | 0 | 0 |
| 0 | 0 | 0 | 0 | 0 |

| | | | | |
|---|---|---|---|---|
| 0 | 0 | 0 | 0 | 0 |
| 0 | 0 | 0 | 0 | 0 |
| 0 | 0 | 0 | 0 | 0 |
| 0 | 0 | 0 | 0 | 0 |
| 0 | 0 | 0 | 0 | 0 |
| 0 | 0 | 0 | 0 | 0 |
| 0 | 0 | 0 | 0 | 0 |
| 0 | 0 | 0 | 0 | 0 |
| 0 | 0 | 0.006460357 | 0 | 0 |
| 0 | 0.004907909 | 0.017021339 | 0 | 0.374431266 |
| 0 | 0 | 0 | 0.056239284 | 0 |
| 0 | 0 | 0 | 0 | 0 |
| 0.001278689 | 0 | 0.007292899 | 0.004087931 | 0.004060374 |
| 0 | 0 | 0 | 0 | 0 |
| 0 | 0 | 0 | 0 | 0 |
| 0 | 0 | 0 | 0 | 0 |
| 0 | 0 | 0 | 0 | 0 |
| 0 | 0 | 0 | 0 | 0 |
| 0 | 0 | 0 | 0 | 0 |
| 1.457746378 | 1.764296929 | 0.053755205 | 0.051849297 | 4.130824101 |
| 0 | 0 | 0 | 0 | 0 |
| 0 | 0 | 0 | 0 | 0 |
| 0 | 0 | 0 | 0 | 0 |
| 0 | 0 | 0 | 0 | 0 |
| 0 | 0 | 0 | 0 | 0 |
| 0 | 0 | 0 | 0 | 0 |
| 0 | 0 | 0 | 0 | 0 |
| 0 | 0 | 0 | 0 | 0 |
| 0 | 0 | 0 | 0 | 0 |
| 0 | 0 | 0 | 0 | 0 |
| 0 | 0 | 0 | 0 | 0 |
| 0 | 0 | 0 | 0 | 0 |
| 0 | 0 | 0 | 0 | 0 |
| 0 | 0 | 0 | 0 | 0 |
| 0 | 0 | 0 | 0 | 0 |
| 0 | 0.00319988 | 0 | 0 | 0.006861455 |
| 0 | 0 | 0 | 0 | 0 |
| 0 | 0 | 0 | 0 | 0 |
| 0 | 0 | 0 | 0 | 0 |
| 0 | 0 | 0 | 0 | 0 |
| 0 | 0 | 0 | 0 | 0 |
| 0 | 0 | 0 | 0 | 0 |
| 0.000788378 | 0.000933991 | 0.001854785 | 0.010283313 | 0 |
| 0 | 0 | 0 | 0 | 0 |
| 0.012293124 | 0 | 0.067445502 | 0 | 0.00633625 |
| 0 | 0 | 0 | 0 | 0 |
| 0 | 0 | 0 | 0 | 0 |
| 0 | 0 | 0 | 0 | 0 |

| | | | | |
|---|---|---|---|---|
| 0 | 0 | 0 | 0 | 0 |
| 0 | 0 | 0 | 0 | 0 |
| 0 | 0.001725811 | 0 | 0 | 0 |
| 0 | 0 | 0 | 0 | 0 |
| 0 | 0 | 0.004297462 | 0 | 0 |
| 0.021280684 | 0.049319486 | 0.175942945 | 0.175654571 | 0 |
| 0 | 0 | 0 | 0 | 0 |
| 0 | 0 | 0 | 0 | 0 |
| 0 | 0 | 0 | 0 | 0 |
| 0 | 0 | 0 | 0 | 0 |
| 0 | 0 | 0 | 0 | 0 |
| 0 | 0 | 0 | 0 | 0 |
| 0 | 0 | 0 | 0 | 0 |
| 0.066490093 | 0 | 0 | 0 | 0 |
| 0 | 0 | 0 | 0 | 0 |
| 0 | 0 | 0 | 0 | 0 |
| 0 | 0 | 0 | 0.061738588 | 0 |
| 0 | 0 | 0 | 0 | 0.005409452 |
| 0 | 0 | 0 | 0 | 0 |
| 0 | 0 | 0 | 0 | 0 |
| 0 | 0 | 0 | 0 | 0 |
| 0 | 0 | 0 | 0 | 0 |
| 0 | 0 | 0 | 0 | 0 |
| 0 | 0 | 0 | 0 | 0 |
| 0 | 0 | 0 | 0 | 0 |
| 0 | 0 | 0.006006884 | 0 | 0.047300213 |
| 0 | 0 | 0 | 0 | 0 |
| 0 | 0 | 1.316585129 | 3.645258609 | 0 |
| 0 | 0 | 0 | 0 | 0 |
| 0 | 0 | 0 | 0.009224429 | 0 |
| 0 | 0 | 0 | 0 | 0 |
| 0 | 0 | 0 | 0 | 0 |
| 0 | 0 | 0 | 0 | 0 |
| 0 | 0 | 0 | 0 | 0 |
| 0 | 0 | 0 | 0 | 0 |
| 0 | 0 | 0 | 0 | 0 |
| 0 | 0 | 0 | 0 | 0 |
| 0 | 0 | 0 | 0 | 0 |
| 0.020088978 | 0 | 0 | 0 | 0 |
| 0 | 0 | 0 | 0 | 0 |
| 0 | 0 | 0 | 0 | 0 |
| 0 | 0 | 0 | 0 | 0 |
| 0 | 0 | 0 | 0 | 0 |
| 0 | 0 | 0 | 0 | 0 |
| 0 | 0 | 0 | 0 | 0 |
| 0 | 0 | 0 | 0 | 0 |
| 0 | 0 | 0 | 0 | 0 |
| 0 | 0 | 0 | 0 | 0 |

| | | | | |
|---|---|---|---|---|
| 0 | 0 | 0 | 0 | 0 |
| 0.023229805 | 0 | 0.007643614 | 0.024136127 | 0 |
| 0 | 0 | 0 | 0 | 0 |
| 0 | 0 | 0 | 0 | 0 |
| 0 | 0 | 0 | 0 | 0 |
| 0 | 0 | 0.000873472 | 0 | 0 |
| 0 | 0 | 0 | 0 | 0 |
| 0 | 0 | 0 | 0 | 0 |
| 0 | 0 | 0 | 0 | 0 |
| 0 | 0 | 0 | 0 | 0 |
| 0.001559876 | 0.008546937 | 0.003336236 | 0 | 0.005696251 |
| 0 | 0 | 0 | 0 | 0 |
| 0.145770216 | 0.195833362 | 0 | 0.042837395 | 0 |
| 0 | 0 | 0 | 0 | 0 |
| 0 | 0 | 0 | 0 | 0 |
| 0 | 0 | 0 | 0 | 0 |
| 0 | 0 | 0 | 0 | 0 |
| 0 | 0 | 0 | 0 | 0 |
| 0 | 0 | 0 | 0 | 0 |
| 0 | 0 | 0.001214556 | 0 | 0 |
| 0 | 0 | 0.003386078 | 0 | 0.070381679 |
| 0 | 0 | 0 | 0 | 0 |
| 0 | 0 | 0 | 0 | 0 |
| 0.929857872 | 0.939819764 | 0.226155217 | 1.777190515 | 0 |
| 0 | 0 | 0 | 0 | 0 |
| 0 | 0 | 0 | 0 | 0 |
| 0 | 0 | 0 | 0 | 0 |
| 0 | 0 | 0 | 0 | 0 |
| 0 | 0 | 0 | 0 | 0 |
| 0 | 0 | 0 | 0 | 0 |
| 0 | 0 | 0.002908005 | 0 | 0.015542905 |
| 0 | 0 | 0 | 0 | 0 |
| 0.089096477 | 0.001972947 | 0 | 0 | 0 |
| 0 | 0 | 0 | 0 | 0 |

| PHL018-0236_V2 | PHL018-0270_V2 | PHL018-0031_V1 | PHL018-0002_V1 | PHL018-0176_V2 |
|---|---|---|---|---|
| EYCF | REF | CTRL | CTRL | REF |
| 0.019621469 | 0.063556803 | 0.768218383 | 0.659727742 | 0.135828505 |
| 0.259375352 | 2.764144366 | 0.492458397 | 5.555062784 | 1.195227277 |
| 12.94911487 | 0 | 6.791173506 | 3.855698681 | 6.699702213 |
| 0 | 0 | 0 | 0 | 0 |
| 0 | 0 | 0 | 0 | 0 |
| 0 | 0 | 0 | 0 | 0 |
| 0 | 0 | 0.039003004 | 0 | 0 |
| 0 | 0 | 0 | 0 | 0 |
| 0 | 0 | 0 | 0 | 0 |
| 0 | 0 | 0 | 0 | 0 |
| 0 | 0 | 0 | 0 | 0 |
| 0 | 0 | 0 | 0 | 0 |
| 0 | 0 | 0 | 0 | 0 |
| 3.574056606 | 4.602091562 | 16.93022118 | 1.569982994 | 0.969878494 |
| 0 | 0 | 0 | 0 | 0 |
| 0 | 0 | 0 | 0 | 0 |
| 0 | 0 | 0 | 0 | 0 |
| 0 | 0 | 0 | 0 | 0 |
| 0 | 0 | 0 | 0 | 0 |
| 0 | 0 | 0 | 0 | 0 |
| 0 | 0 | 0 | 0 | 0 |
| 0 | 0 | 0 | 0 | 0 |
| 0 | 0 | 0 | 0 | 0 |
| 0 | 0 | 0 | 0 | 0 |
| 0 | 0 | 0 | 0.544462652 | 0 |
| 0 | 0 | 0 | 0 | 0 |
| 0 | 0 | 0 | 0 | 0 |
| 0 | 0 | 0 | 0 | 0 |
| 0 | 0 | 0 | 0 | 0 |
| 0 | 0 | 0 | 0 | 0 |
| 0 | 0 | 0 | 0 | 0 |
| 0 | 0 | 0 | 0 | 0 |
| 0 | 0 | 0 | 0 | 0 |
| 0 | 0 | 0 | 0 | 0 |
| 0 | 0 | 0 | 0 | 0 |
| 0 | 4.353688365 | 0 | 0 | 2.473588122 |
| 0 | 0 | 0 | 0 | 0 |
| 0 | 0 | 0 | 0 | 0 |
| 0 | 0 | 0 | 0 | 0 |
| 0 | 0 | 0 | 0 | 0 |
| 0 | 0 | 0 | 0 | 0 |
| 0.009021871 | 0 | 0.059433009 | 0 | 0 |
| 0.008245966 | 0.005813602 | 0.145626342 | 0.010725725 | 0.025295539 |
| 0 | 0.022613827 | 0.075121843 | 0 | 0.009206543 |
| 0 | 0 | 0.012546764 | 0 | 0 |
| 0 | 0 | 0 | 0 | 0 |

| | | | | |
|---|---|---|---|---|
| 0 | 0 | 0.002845654 | 0 | 0 |
| 0.009509677 | 0 | 0.005406991 | 0 | 0.147461169 |
| 0 | 0 | 0.104521727 | 0.076001884 | 0 |
| 0.174539829 | 0.564354421 | 0.32279041 | 0.290409319 | 0.346907388 |
| 0 | 0 | 0.00674082 | 0.032639423 | 0.003428927 |
| 0 | 0 | 0.047612662 | 0 | 0 |
| 0.002223892 | 0 | 1.038779377 | 0 | 0.927711681 |
| 0 | 0 | 0 | 0 | 0 |
| 0 | 0 | 0 | 0 | 0 |
| 0 | 0 | 0 | 0 | 0 |
| 0 | 0 | 0 | 0 | 0 |
| 0 | 0.105447771 | 0.081517463 | 0.771415438 | 0.078900917 |
| 0 | 0 | 0 | 0 | 0 |
| 0.041011554 | 0.165185492 | 0.014025823 | 0.016363957 | 0.051675394 |
| 0 | 0 | 0 | 0 | 0 |
| 0 | 0 | 0 | 0 | 0 |
| 2.272189478 | 0.065111435 | 0 | 0 | 4.172641779 |
| 0 | 0 | 0 | 0 | 0 |
| 0 | 0 | 0 | 0 | 0 |
| 0 | 0 | 0 | 0 | 0 |
| 0 | 0.199140408 | 0.030398124 | 0.065448017 | 0.211019237 |
| 0.004082051 | 0.008137508 | 0.009371631 | 0.008379451 | 0.095025601 |
| 0 | 0 | 0 | 0 | 0 |
| 0 | 0 | 0 | 0 | 0 |
| 0 | 0 | 0 | 0 | 0 |
| 0 | 0 | 0 | 0 | 0 |
| 0 | 0 | 0 | 0 | 0 |
| 0 | 0 | 0 | 0 | 0 |
| 0 | 0 | 0 | 0 | 0 |
| 0 | 0 | 0 | 0 | 0 |
| 0 | 0 | 0 | 0.108597266 | 0.0046815 |
| 0 | 0 | 0 | 0 | 0 |
| 0 | 0 | 0 | 0 | 0 |
| 0 | 0 | 0 | 0 | 0 |
| 0 | 0 | 0 | 0 | 0 |
| 0 | 0 | 0.004621813 | 0 | 0 |
| 0 | 0 | 0 | 0 | 0 |
| 0.125504315 | 0 | 0 | 0 | 0.088357057 |
| 0 | 0 | 0 | 0 | 0 |
| 0 | 0 | 0 | 0 | 0 |
| 0 | 0 | 0 | 0 | 0 |
| 0 | 0 | 0 | 0 | 0 |
| 0 | 0 | 0 | 0 | 0 |
| 0 | 0 | 0 | 0 | 0 |
| 0 | 0 | 0 | 0.588884164 | 0 |
| 0 | 0 | 0 | 0 | 0 |
| 0 | 0 | 0.001228874 | 0.12043334 | 0.003947092 |
| 0 | 0 | 0.00473419 | 0.048679257 | 0.088417155 |

| | | | | |
|---|---|---|---|---|
| 0 | 0 | 0 | 0 | 0 |
| 0 | 0 | 0 | 0 | 0 |
| 0 | 0 | 0 | 0 | 0 |
| 0 | 0 | 0 | 0 | 0 |
| 0 | 0.005385236 | 5.067319311 | 0 | 0 |
| 0 | 0 | 0 | 0 | 0 |
| 0 | 0 | 0 | 0 | 0 |
| 0 | 0 | 0.003666741 | 0 | 0 |
| 0 | 0 | 0 | 0 | 0 |
| 0 | 0 | 0 | 0 | 0 |
| 0 | 0 | 0 | 0 | 0 |
| 0 | 0 | 0 | 0 | 0 |
| 0 | 0 | 0 | 0 | 0 |
| 0 | 0 | 0 | 0 | 0 |
| 0 | 0 | 0 | 0 | 0 |
| 0 | 0 | 0 | 0 | 0 |
| 0 | 0 | 0 | 0 | 0 |
| 0 | 0 | 0 | 0 | 0 |
| 0 | 0.014930665 | 0.24816377 | 0.833531805 | 0.012378361 |
| 0 | 0 | 0 | 0 | 0 |
| 0 | 0 | 0 | 0 | 0 |
| 0 | 0 | 0 | 0 | 0 |
| 0 | 0 | 0 | 0.003506649 | 0 |
| 0 | 0 | 0 | 0.000823202 | 0 |
| 0 | 0 | 0 | 0 | 0 |
| 0 | 0 | 0 | 0 | 0 |
| 0 | 0 | 0 | 0 | 0 |
| 0 | 0 | 0 | 0.003861559 | 0 |
| 0 | 0 | 0 | 0 | 0 |
| 0 | 0 | 0 | 0 | 0 |
| 0 | 0.081756629 | 0.087634116 | 0.008275669 | 0.031915128 |
| 0.000617805 | 0 | 0.032643879 | 0 | 0 |
| 0 | 0 | 0 | 0 | 0 |
| 0 | 0 | 0 | 0 | 0 |
| 0 | 0 | 0 | 0 | 0 |
| 0 | 0 | 0 | 0 | 0 |
| 0 | 0 | 0 | 0 | 0 |
| 0 | 0 | 0 | 0 | 0 |
| 0 | 0 | 0 | 0 | 0.178010967 |
| 0 | 0 | 0 | 0 | 0 |
| 0 | 0 | 0 | 0 | 0 |
| 0 | 0 | 0 | 0 | 0 |
| 0 | 0 | 0.001540047 | 0 | 0.007739921 |
| 0 | 0 | 0 | 0 | 0 |
| 0 | 0 | 0 | 0 | 0 |
| 0 | 0 | 0 | 0 | 0 |
| 0 | 0 | 0 | 0 | 0 |
| 0 | 0 | 0 | 0 | 0 |

| | | | | |
|---|---|---|---|---|
| 0 | 0 | 0 | 0 | 0 |
| 0 | 0 | 0 | 0 | 0 |
| 0 | 0 | 0 | 0 | 0 |
| 0 | 0 | 0 | 0 | 0 |
| 0 | 0 | 0 | 0 | 0.074082218 |
| 0 | 0 | 0 | 0 | 0 |
| 0 | 0 | 0 | 0 | 0 |
| 0 | 0 | 0 | 0 | 0 |
| 0 | 0 | 0 | 0 | 0 |
| 0 | 0 | 0 | 0 | 0 |
| 0 | 0 | 0 | 0 | 0 |
| 0 | 0 | 0 | 0 | 0 |
| 0 | 0 | 0 | 0 | 0 |
| 0 | 0 | 0 | 0.00111809 | 0 |
| 0 | 0 | 0 | 0 | 0 |
| 0 | 0 | 3.001129829 | 0 | 2.675437322 |
| 0 | 0 | 0 | 0 | 0 |
| 0 | 0 | 0 | 0 | 0 |
| 0 | 0 | 0 | 0 | 0 |
| 0 | 0 | 0 | 0 | 0 |
| 0 | 0 | 0 | 0 | 0 |
| 0 | 0 | 0 | 0 | 0 |
| 0 | 0 | 0 | 0 | 0 |
| 0 | 0 | 0 | 0 | 0 |
| 0 | 0 | 0 | 0 | 0 |
| 0 | 0 | 0 | 0 | 0 |
| 0 | 0 | 0.079748797 | 0.15391883 | 0 |
| 0 | 0 | 0 | 0 | 0 |
| 0 | 0 | 0 | 0 | 0 |
| 0 | 0 | 0 | 0 | 0 |
| 0 | 0.012801459 | 0 | 0.007300071 | 0 |
| 0 | 0 | 0 | 0 | 0 |
| 0 | 0 | 0 | 0 | 0 |
| 0 | 0 | 0 | 0 | 0 |
| 0 | 0 | 0 | 0 | 0 |
| 0 | 0 | 0 | 0 | 0 |
| 0 | 0 | 0 | 0 | 0 |
| 0 | 0 | 0 | 0 | 0 |
| 0 | 0 | 0 | 0 | 0 |
| 0 | 0 | 0 | 0 | 0 |
| 0 | 0 | 0 | 0 | 0 |
| 0 | 0 | 0 | 0 | 0 |
| 0 | 0 | 0 | 0 | 0 |
| 0 | 0 | 0 | 0 | 0 |
| 0 | 0 | 0 | 0 | 0 |
| 0 | 0 | 0 | 0 | 0 |
| 0 | 0 | 0 | 0 | 0 |
| 0 | 0 | 0 | 0 | 0 |

| | | | | |
|---|---|---|---|---|
| 0 | 0 | 0 | 0 | 0 |
| 0 | 0 | 0 | 0 | 0 |
| 0 | 0 | 0 | 0 | 0 |
| 0 | 0 | 0 | 0 | 0 |
| 0 | 0 | 0 | 0 | 0 |
| 0 | 0 | 0 | 0 | 0 |
| 0 | 0 | 0 | 0 | 0 |
| 0 | 0 | 0 | 0 | 0 |
| 0 | 0 | 0 | 0 | 0 |
| 0 | 0 | 0 | 0 | 0 |
| 0 | 0 | 0 | 0 | 0 |
| 0 | 0 | 0 | 0 | 0 |
| 0 | 0 | 0 | 0.001085131 | 0.000972784 |
| 0 | 0 | 0 | 0 | 0 |
| 0 | 0 | 0 | 0 | 0 |
| 0 | 0 | 0 | 0 | 0 |
| 0 | 0 | 0 | 0 | 0 |
| 0 | 0 | 0 | 0 | 0 |
| 0 | 0 | 0 | 0 | 0 |
| 0 | 0 | 0 | 0 | 0 |
| 0.038260463 | 0.015029657 | 0.008251535 | 0.015555158 | 0.018916279 |
| 0 | 0 | 0 | 0 | 0 |
| 0 | 0 | 0 | 0 | 0 |
| 0 | 0 | 0 | 0.001763141 | 0.001517375 |
| 0 | 0 | 0 | 0 | 0 |
| 0 | 0 | 0 | 0 | 0 |
| 0.004159305 | 0.002775316 | 0.00048055 | 0 | 0 |
| 0 | 0 | 0 | 0 | 0 |
| 0 | 0 | 0 | 0 | 0 |
| 0.002383916 | 0 | 0.002399288 | 0 | 0 |
| 0 | 0 | 0.004949943 | 0 | 0 |
| 0 | 0 | 0 | 0 | 0 |
| 0 | 0 | 0 | 0 | 0 |
| 0 | 0.006272815 | 0.088249451 | 0.001653278 | 0 |
| 0.001991544 | 0.008825038 | 0.02362311 | 0.015181523 | 0.000946765 |
| 0 | 0 | 0 | 0 | 0 |
| 0 | 0 | 0 | 0 | 0 |
| 0 | 0 | 0 | 0 | 0 |
| 0 | 0 | 0 | 0 | 0 |
| 0 | 0 | 0 | 0 | 0 |
| 0 | 0 | 0 | 0 | 0 |
| 0 | 0 | 0 | 0 | 0 |
| 0 | 0 | 0 | 0 | 0 |
| 0 | 0 | 0 | 0 | 0 |
| 0 | 0 | 0 | 0 | 0 |
| 0 | 0 | 0.172777713 | 0 | 0.047550062 |
| 0 | 0.004131965 | 0.000775077 | 0 | 0 |
| 0 | 0 | 0.001785474 | 0.002195111 | 0 |

| | | | | |
|---|---|---|---|---|
| 0 | 0 | 0 | 0 | 0 |
| 0 | 0 | 0 | 0 | 0 |
| 0 | 0 | 0 | 0 | 0 |
| 0 | 0 | 0 | 0.001395136 | 0.001072024 |
| 0 | 0 | 0 | 0 | 0 |
| 0 | 0 | 0 | 0 | 0 |
| 0 | 0 | 0 | 0 | 0 |
| 0 | 0 | 0.000549997 | 0 | 0 |
| 0 | 0 | 0 | 0 | 0 |
| 0 | 0 | 0 | 0 | 0 |
| 0 | 0 | 0 | 0 | 0 |
| 0 | 0 | 0 | 0 | 0 |
| 0 | 0 | 0 | 0 | 0 |
| 0 | 0 | 0 | 0 | 0 |
| 0 | 0 | 0 | 0 | 0 |
| 0 | 0 | 0 | 0 | 0 |
| 0 | 0 | 0 | 0 | 0 |
| 0 | 0 | 0 | 0 | 0 |
| 0 | 0 | 0 | 0 | 0 |
| 0 | 0.057446091 | 0 | 0.045317459 | 0 |
| 0.050033412 | 0.007904583 | 0.057396685 | 0.012701717 | 0.070597386 |
| 0 | 0 | 0 | 0 | 0 |
| 0 | 0 | 0 | 0 | 0 |
| 0 | 0 | 0 | 0 | 0 |
| 0 | 0 | 0 | 0.002336541 | 0 |
| 0 | 0 | 0 | 0 | 0 |
| 0 | 0 | 0 | 0 | 0 |
| 0 | 0 | 0 | 0 | 0 |
| 0 | 0 | 0 | 0 | 0 |
| 0 | 0 | 0.001303652 | 0 | 0 |
| 0.582501171 | 0.111182465 | 0.197935629 | 0.09408749 | 0.110518552 |
| 0 | 0 | 0 | 0 | 0 |
| 0.092300914 | 0 | 0 | 0 | 0 |
| 0 | 0.002183149 | 0.051690109 | 0 | 0 |
| 0 | 0 | 0 | 0 | 0 |
| 0 | 0 | 0 | 0 | 0 |
| 0 | 0 | 0 | 0 | 0 |
| 0 | 0 | 0 | 0 | 0 |
| 0 | 0 | 0 | 0 | 0 |
| 0 | 0 | 0 | 0 | 0 |
| 0 | 0 | 0 | 0 | 0 |
| 0.00239367 | 0 | 0.110417267 | 0.008991239 | 0.003413797 |
| 0 | 0 | 0 | 0 | 0 |
| 0 | 0 | 0 | 0 | 0 |
| 0 | 0 | 0 | 0 | 0 |
| 0 | 0 | 0 | 0 | 0 |
| 0 | 0 | 0 | 0 | 0 |
| 0 | 0 | 0 | 0 | 0 |

| | | | | |
|---|---|---|---|---|
| 0 | 0 | 0 | 0 | 0 |
| 0 | 0 | 0 | 0 | 0 |
| 0 | 0.006239431 | 0.089490368 | 0 | 0.005715444 |
| 0 | 0 | 0 | 0 | 0 |
| 0 | 2.297728549 | 0.001250096 | 1.191868301 | 0.005786691 |
| 0 | 0 | 0 | 0 | 0 |
| 0 | 0 | 0 | 0 | 0 |
| 0 | 0 | 0 | 0 | 0 |
| 0 | 0 | 0 | 0 | 0 |
| 0 | 0 | 0 | 0 | 0 |
| 0 | 0 | 0 | 0 | 0 |
| 0 | 0 | 0 | 0 | 0 |
| 0 | 0 | 0 | 0 | 0 |
| 0.008673542 | 0.001060877 | 0.034626005 | 0.016636622 | 0.00500064 |
| 0.004874126 | 0 | 0.013304126 | 0.002019317 | 0 |
| 0 | 0 | 0 | 0.017295562 | 0 |
| 0.001041306 | 0.001496527 | 0 | 0 | 0.002227629 |
| 0 | 0 | 0 | 0 | 0 |
| 0 | 0 | 0 | 0 | 0 |
| 0 | 0 | 0 | 0 | 0 |
| 0 | 0 | 0 | 0 | 0 |
| 0 | 0 | 0.000675759 | 0.001869294 | 0 |
| 0 | 0 | 0 | 0 | 0 |
| 0 | 0 | 0 | 0 | 0 |
| 0 | 0 | 0 | 0 | 0 |
| 0 | 0 | 0 | 0 | 0 |
| 0 | 0 | 0 | 0 | 0 |
| 0 | 0 | 0 | 0 | 0 |
| 0 | 0 | 0 | 0 | 0 |
| 0 | 0 | 0 | 0 | 0 |
| 0 | 0 | 0 | 0 | 0 |
| 0 | 0 | 0 | 0 | 0 |
| 0 | 0 | 0 | 0 | 0 |
| 0 | 0 | 0 | 0 | 0 |
| 0 | 0 | 0 | 0 | 0 |
| 0 | 0 | 0 | 0 | 0 |
| 0 | 0.001131541 | 0.003396086 | 0.000782858 | 0 |
| 0 | 0 | 0 | 0 | 0 |
| 0 | 0 | 0 | 0 | 0 |
| 0 | 0 | 0.004288823 | 0 | 0 |
| 0 | 0 | 0 | 0 | 0 |
| 0 | 0 | 0 | 0 | 0 |
| 0 | 0 | 0.002372889 | 0.009724321 | 0 |
| 0 | 0 | 0 | 0 | 0 |
| 0 | 0 | 0 | 0 | 0 |
| 0 | 0 | 0 | 0 | 0 |
| 0 | 0 | 0.001799661 | 0 | 0 |

| | | | | |
|---|---|---|---|---|
| 0 | 0 | 0 | 0.933310974 | 0 |
| 0 | 0 | 0 | 0.310289798 | 0 |
| 0 | 0.003537502 | 0 | 0 | 0 |
| 0 | 0 | 0 | 0 | 0 |
| 0.003797227 | 0.002609983 | 0 | 0.002626501 | 0 |
| 0 | 0 | 0 | 0 | 0 |
| 0 | 0 | 0 | 0 | 0 |
| 0.004396972 | 0 | 0.007985545 | 0 | 0 |
| 0 | 0 | 0 | 0 | 0 |
| 0.615405196 | 0.019539911 | 0.12372033 | 0.151472784 | 0 |
| 0 | 0 | 0 | 0 | 0 |
| 0 | 0 | 0.00059263 | 0 | 0 |
| 0.043910743 | 0.012381739 | 0.001198749 | 0 | 0.089973158 |
| 0 | 0 | 0 | 0.004196784 | 0 |
| 0 | 0 | 0 | 0 | 0.001019112 |
| 0 | 0 | 0 | 0 | 0.010709972 |
| 0 | 0 | 0 | 0 | 0 |
| 0 | 0 | 0 | 0.002879101 | 0 |
| 0.009651418 | 0 | 0.007261034 | 0 | 0.021447213 |
| 0.041559634 | 0 | 0 | 0.005962898 | 0 |
| 0 | 0 | 0 | 0 | 0 |
| 0 | 0 | 0 | 0 | 0 |
| 0 | 0 | 0 | 0 | 0 |
| 0 | 0 | 0 | 0 | 0 |
| 0 | 0 | 0 | 0 | 0 |
| 0.000809254 | 0 | 0 | 0 | 0 |
| 0 | 0 | 0 | 0 | 0 |
| 0 | 0 | 0 | 0 | 0.003349619 |
| 0.020216836 | 0 | 0 | 0.13532926 | 0 |
| 0.00166524 | 0 | 0.001256983 | 0 | 0 |
| 0 | 0 | 0 | 0 | 0 |
| 0 | 0 | 0 | 0 | 0 |
| 0 | 0 | 0 | 0 | 0 |
| 0 | 0 | 0 | 0.004209303 | 0 |
| 0 | 0 | 0 | 0 | 0 |
| 0 | 0 | 0 | 0 | 0 |
| 0 | 0 | 0.001780625 | 0 | 0 |
| 0 | 0 | 0 | 0 | 0 |
| 0 | 0 | 0 | 0 | 0 |
| 0 | 0.000978568 | 0 | 0 | 0.006312064 |
| 0 | 0 | 0 | 0 | 0 |
| 0 | 0 | 0 | 0 | 0 |
| 0 | 0 | 0 | 0 | 0 |
| 0.002868638 | 0.109251613 | 0 | 0 | 0.018596296 |
| 0 | 0 | 0 | 0 | 0 |
| 0 | 0 | 0.000893849 | 0 | 0 |
| 0.001006371 | 0 | 0.01758035 | 0 | 0 |
| 0 | 0 | 0 | 0.007023812 | 0 |

| | | | | |
|---|---|---|---|---|
| 0 | 0 | 0 | 0 | 0 |
| 0.019344661 | 0 | 0.057001342 | 0 | 0.475106687 |
| 0 | 0 | 0 | 0 | 0 |
| 0 | 0 | 0 | 0 | 0 |
| 0.014522504 | 0.005146328 | 0 | 0 | 0.00624187 |
| 0.027266775 | 0.017998598 | 0.004786498 | 0.021016625 | 0 |
| 0 | 0.002280013 | 0 | 0 | 0 |
| 0.000820144 | 0 | 0 | 0 | 0 |
| 0 | 0 | 0 | 0 | 0 |
| 0 | 0 | 0 | 0 | 0 |
| 0.00165479 | 0.012155263 | 0 | 0.002437575 | 0 |
| 0 | 0 | 0 | 0 | 0 |
| 0 | 0 | 0 | 0 | 0 |
| 0.000868534 | 0.004243969 | 0 | 0.001842318 | 0.080018973 |
| 0.018630576 | 0 | 0 | 0 | 0.002255986 |
| 0.003188576 | 0 | 0 | 0 | 0 |
| 0.000849314 | 0 | 0 | 0 | 0.001211271 |
| 0 | 0 | 0 | 0 | 0 |
| 0 | 0 | 0 | 0 | 0.013447562 |
| 0.00479139 | 0 | 0 | 0.09417719 | 0 |
| 0 | 0 | 0.00746748 | 0 | 1.264963246 |
| 0.399269968 | 0 | 0.218556387 | 0.006769557 | 0.341993879 |
| 0 | 0 | 0 | 0 | 0 |
| 0 | 0 | 0 | 0 | 0 |
| 0 | 0 | 0 | 0 | 0 |
| 0 | 0 | 0 | 0 | 0 |
| 0 | 0 | 0 | 0 | 0 |
| 0 | 0 | 0 | 0 | 0 |
| 0 | 0 | 0 | 0 | 0 |
| 0.988681377 | 0.878592288 | 0.075779648 | 0.059409962 | 0.11075471 |
| 0 | 0 | 0.000762468 | 0 | 0 |
| 0 | 0 | 0 | 0 | 0 |
| 0 | 0 | 0 | 0 | 0 |
| 0.001001464 | 0 | 0.00089993 | 0 | 0 |
| 0.005490556 | 0 | 0 | 0 | 0.002013558 |
| 0 | 0 | 0 | 0 | 0 |
| 0 | 0 | 0 | 0 | 0 |
| 0 | 0 | 0 | 0 | 0 |
| 0.416520223 | 0 | 0.051771092 | 0.46692192 | 0 |
| 0 | 0 | 0 | 0 | 0 |
| 0.003284537 | 0.026475387 | 0.010779508 | 0.321092262 | 0.010590662 |
| 0 | 3.15778619 | 1.402941713 | 0 | 6.490622306 |
| 0 | 0 | 0 | 0 | 0 |
| 0 | 0 | 0 | 0 | 0 |
| 0.003382944 | 0 | 0.009206741 | 0 | 0 |
| 0 | 0 | 0 | 0 | 0 |
| 0.012140694 | 0 | 0.003717413 | 0 | 0.001710101 |
| 0 | 0 | 0 | 0 | 0 |

| | | | | |
|---|---|---|---|---|
| 0 | 0 | 0.005783186 | 0 | 0.001318736 |
| 0 | 0 | 0 | 0.045744183 | 0 |
| 5.397276978 | 0.158619249 | 0 | 0 | 9.410616641 |
| 0 | 0 | 0 | 0 | 0 |
| 0 | 0.044885014 | 0 | 0 | 0 |
| 0.010007109 | 0 | 0.00067444 | 0 | 0 |
| 0 | 0 | 0 | 0 | 0 |
| 0 | 0 | 0 | 0 | 0 |
| 0 | 0.003558932 | 0 | 0 | 0 |
| 0.02739972 | 0 | 0.00802246 | 0 | 0 |
| 0 | 0 | 0 | 0 | 0 |
| 0 | 0 | 0 | 0 | 0 |
| 0 | 0 | 0 | 0 | 0 |
| 0.006190258 | 0.251263616 | 0 | 0 | 0 |
| 0 | 0 | 0 | 0 | 0 |
| 0 | 0.001127415 | 0.098127385 | 0.01300006 | 0.014730821 |
| 0.000579671 | 0 | 0.183461072 | 0.001536982 | 0 |
| 0 | 0 | 0.004366827 | 0.036880632 | 0.003515484 |
| 0.004340981 | 0.008820232 | 0.000807076 | 0.001786035 | 0 |
| 1.448917325 | 0.013887765 | 0 | 0 | 0 |
| 0 | 0.343848757 | 0.017725029 | 0.000726388 | 0.129194784 |
| 0 | 0 | 0.006706811 | 0.005125607 | 0 |
| 0 | 0 | 0 | 0 | 0 |
| 0 | 0.683045846 | 0.014835218 | 0 | 0 |
| 0 | 0.000583946 | 0 | 0 | 0 |
| 0.207130259 | 0.031276988 | 0.219444931 | 0 | 0 |
| 0 | 0.002115208 | 0.001388703 | 0.022926737 | 0 |
| 0 | 0 | 0 | 0 | 0 |
| 0 | 0 | 0 | 0 | 0 |
| 0 | 0 | 0 | 0 | 0 |
| 0.162577516 | 0.076006795 | 0.028816568 | 0.033016573 | 0 |
| 0 | 0 | 0.009896998 | 0.006083823 | 0.000892464 |
| 0.012710857 | 0.008848361 | 0.003640813 | 0.019747575 | 0.026908648 |
| 0 | 0 | 0 | 0 | 0 |
| 0 | 0.002043706 | 0.05914695 | 0 | 0.00144863 |
| 0 | 0 | 0 | 0.003719289 | 0 |
| 0 | 0 | 0.005227749 | 0 | 0.03200957 |
| 0.00365092 | 0.010897552 | 0.019078929 | 0.039093617 | 0.026835341 |
| 0 | 0 | 0.038132556 | 0 | 3.452503174 |
| 0.002985 | 0 | 0.000619007 | 0.093606004 | 0 |
| 0 | 0 | 0 | 0.003721698 | 0 |
| 0 | 0 | 0 | 0 | 0 |
| 0 | 0 | 0 | 0.003897629 | 0 |
| 0 | 0.031822041 | 0.051417884 | 0.420994612 | 0.018759664 |
| 0.008580234 | 0 | 0 | 0 | 1.83921086 |
| 0 | 0 | 0.120989159 | 0 | 0 |
| 0 | 0 | 0 | 0.002896135 | 0 |
| 0 | 0 | 0 | 0 | 0 |

| | | | | |
|---|---|---|---|---|
| 0 | 0 | 0.011449792 | 0 | 0 |
| 0 | 0.096245442 | 0.199313873 | 0.148902369 | 0.112701402 |
| 0 | 0 | 0 | 0 | 0 |
| 0 | 0 | 0 | 0 | 0.008924546 |
| 0 | 0 | 0 | 0 | 0 |
| 0 | 0 | 0 | 0 | 0 |
| 0 | 0 | 0.001717931 | 0 | 0 |
| 0 | 0 | 0 | 0.000779371 | 0 |
| 0 | 0 | 0 | 0.000928844 | 0 |
| 0.009176842 | 0 | 0 | 0 | 0 |
| 0.002843731 | 0.032695262 | 0.002913176 | 0.00263903 | 0.004055664 |
| 0.018002229 | 0.0140014 | 0.09866103 | 0 | 0.577220106 |
| 0 | 0 | 0 | 0 | 0 |
| 0 | 0 | 0 | 0 | 0 |
| 0 | 0 | 0 | 0 | 0.004188031 |
| 0 | 0 | 0 | 0 | 0 |
| 0.002233348 | 0.007917227 | 0.002408302 | 0.002960834 | 0.001486403 |
| 0.01488253 | 0.049443853 | 0.002501047 | 0.024598856 | 0.004686066 |
| 0 | 0 | 0 | 0.004602831 | 0 |
| 0 | 0.010461588 | 0 | 0.015806824 | 0.005011812 |
| 0.000767715 | 0.031775969 | 0.002317996 | 0.003867598 | 0.012481831 |
| 0 | 0 | 0 | 0 | 0.001907394 |
| 0 | 0 | 0 | 0 | 0 |
| 0 | 0 | 0 | 0 | 0 |
| 0 | 0 | 0.005841998 | 0 | 0.286614669 |
| 0 | 0 | 0 | 0 | 0 |
| 0.105027355 | 0 | 0.004085674 | 0.252558571 | 0.151516739 |
| 0.000652774 | 0.043389135 | 0.07443855 | 0.01968802 | 0.049806976 |
| 0 | 0 | 0 | 0 | 0 |
| 0 | 0 | 0 | 0 | 0 |
| 0 | 0.008304687 | 0 | 0 | 0 |
| 0 | 0 | 0 | 0 | 0 |
| 0 | 0 | 0 | 0 | 0 |
| 0 | 0 | 0 | 0 | 0 |
| 0 | 0 | 0 | 0.201167472 | 0 |
| 0.012704986 | 1.097930394 | 0.027996179 | 0.097399258 | 0.319849592 |
| 0 | 0 | 0 | 0 | 0 |
| 0.005593459 | 0 | 0 | 0 | 0 |
| 0 | 0.002893262 | 0 | 0 | 0 |
| 0 | 0 | 0 | 0 | 0 |
| 0.006316389 | 0 | 0 | 0 | 1.763102115 |
| 0 | 0 | 0 | 0 | 0 |
| 0.0022011 | 0.006326682 | 0.000659313 | 0.146390412 | 0.023020491 |
| 0 | 0 | 0.000509244 | 0.001408676 | 0 |
| 0.013959578 | 0 | 0 | 0 | 0 |
| 0 | 0 | 0 | 0 | 0.327757272 |
| 0 | 0 | 0 | 0 | 0 |
| 0.000954192 | 0 | 0.001852094 | 0.001518011 | 0.001360847 |

| | | | | |
|---|---|---|---|---|
| 0 | 0 | 0 | 0 | 0 |
| 0 | 0 | 0 | 0 | 0 |
| 0 | 0 | 0 | 0 | 0 |
| 0 | 0.529309845 | 0.013975621 | 0.142554122 | 0.051388875 |
| 0 | 0 | 0 | 0 | 0 |
| 0 | 0 | 0 | 0 | 0 |
| 0 | 0 | 0 | 0 | 0 |
| 0 | 0.013000864 | 0 | 0 | 0.001090265 |
| 0 | 0 | 0 | 0 | 0 |
| 0 | 0.116347377 | 0 | 0 | 0 |
| 0 | 0 | 0 | 0 | 0 |
| 0 | 0.005098812 | 0.009564391 | 0 | 0.001751481 |
| 0 | 0 | 0 | 0 | 0 |
| 0 | 0 | 0 | 4.165377809 | 0 |
| 0.097288997 | 0.027834581 | 0.371800276 | 2.153393743 | 0.178256906 |
| 0 | 0 | 0 | 0 | 0 |
| 0 | 0 | 0 | 0 | 0 |
| 0 | 0 | 0 | 0 | 0 |
| 0 | 0 | 0 | 0 | 0 |
| 0 | 0 | 0 | 0 | 0 |
| 0 | 0 | 0 | 0 | 0 |
| 0 | 0 | 0 | 0 | 0 |
| 0 | 0 | 0 | 0 | 0 |
| 0.438200842 | 1.54869529 | 0.342582957 | 0.64622321 | 2.41086034 |
| 0 | 0 | 0.082521135 | 0.156160933 | 0 |
| 0 | 0 | 0 | 0 | 3.347366027 |
| 0 | 0 | 0 | 0 | 0 |
| 0.021692691 | 0 | 0.581776001 | 0 | 0.877988193 |
| 0 | 0 | 0 | 0 | 0 |
| 0 | 0 | 0 | 0 | 0 |
| 0 | 0.005338216 | 0.000429149 | 0.001758691 | 0 |
| 0 | 0 | 0 | 0 | 0 |
| 0 | 0 | 0.057592859 | 0 | 0.002447827 |
| 0 | 0 | 0 | 0 | 0 |
| 0 | 0 | 0 | 0.124543729 | 0 |
| 0 | 0 | 0 | 0.512217991 | 0 |
| 0 | 0.278084747 | 0 | 0.045835427 | 0 |
| 0.002413621 | 0 | 0 | 0 | 0.025310665 |
| 0.012926519 | 0 | 0.037375289 | 0 | 0.002431055 |
| 0 | 0 | 0.020432486 | 0 | 0.016436917 |
| 0.011519567 | 0 | 0.105727958 | 0.116795522 | 0.004107235 |
| 0 | 0.016339453 | 0 | 0.557536393 | 0 |
| 11.21108717 | 0.721743537 | 0.437398335 | 12.07553216 | 5.646494151 |
| 0 | 0 | 0 | 0 | 0 |
| 0 | 0 | 0 | 0 | 0 |
| 0 | 0 | 0 | 0 | 0 |
| 0 | 0 | 0 | 0 | 0 |
| 0 | 0.107423494 | 0.006278497 | 0.004172411 | 0 |

| | | | | |
|---|---|---|---|---|
| 0 | 0 | 0 | 0 | 0 |
| 0 | 0 | 0 | 0 | 0 |
| 0.056870103 | 0.004119102 | 0.014151956 | 0.043197047 | 0.008605499 |
| 0 | 0 | 0 | 0 | 0 |
| 0 | 0 | 0 | 0 | 0 |
| 0 | 0.001537044 | 0 | 0 | 0.001355816 |
| 0 | 0 | 0 | 0 | 0 |
| 0 | 0 | 0.00038958 | 0 | 0 |
| 0 | 0 | 0 | 0 | 0 |
| 0.46837446 | 0 | 0 | 0 | 0 |
| 0.032435443 | 0 | 0.36381783 | 0.037699413 | 0.086391782 |
| 1.795948251 | 1.239625104 | 1.712743881 | 2.414262751 | 2.087972625 |
| 0 | 0 | 0 | 0 | 0 |
| 0.000799216 | 0 | 0.021717966 | 0 | 0 |
| 0.098880595 | 0.063903167 | 0.030425938 | 0.31795525 | 0.37078701 |
| 0 | 0 | 0 | 0 | 0 |
| 0 | 0 | 0.006178915 | 0 | 0 |
| 0 | 0 | 0 | 0 | 0 |
| 0 | 0 | 0 | 0 | 0 |
| 0.002318023 | 0.032612448 | 0.003946753 | 0.003073091 | 0.018791496 |
| 0 | 0.090807161 | 0.001204739 | 0.007405702 | 0.000885196 |
| 24.03024261 | 0 | 29.09048684 | 0.816226425 | 0.031899092 |
| 18.00733627 | 0.373072327 | 2.475576675 | 4.646766266 | 0.163204593 |
| 0 | 0 | 0 | 0 | 0 |
| 0 | 0 | 0 | 0 | 0 |
| 0 | 0.008551215 | 0 | 0 | 0.002970045 |
| 0 | 0 | 0 | 0 | 0 |
| 0 | 0 | 0 | 0 | 0 |
| 0 | 0 | 0 | 0.397452817 | 0 |
| 0 | 0 | 0 | 0 | 0 |
| 0 | 0 | 0.001624084 | 0 | 0 |
| 0 | 0.62839804 | 0 | 0.072712119 | 0.286809806 |
| 0.005932282 | 0.00131164 | 0.257356216 | 0 | 0.641261273 |
| 0 | 0 | 0.120204036 | 0.278341101 | 0.018197311 |
| 0.004216563 | 0.001009982 | 0 | 0.027950284 | 0.006299925 |
| 0.001793164 | 0.030726611 | 0.001933635 | 0.014080726 | 0.015540937 |
| 0.002131904 | 0.018191879 | 0.016236061 | 0.100511924 | 0.00817127 |
| 0 | 0 | 0 | 0 | 0 |
| 0 | 0.023893567 | 0 | 0.004707798 | 0.021178673 |
| 0 | 0 | 0 | 0 | 0 |
| 0 | 0 | 0 | 0 | 0 |
| 0 | 0 | 0 | 0 | 0 |
| 0 | 0 | 0 | 0 | 0 |
| 0 | 0 | 0 | 0 | 0 |
| 0.012426701 | 0 | 0.481558095 | 0.037328405 | 0.035221011 |
| 0 | 0 | 0 | 0 | 2.187035081 |
| 0 | 0 | 0 | 0 | 0 |
| 0.000948734 | 0 | 0.060360248 | 0.071693034 | 0.067653127 |

| | | | | |
|---|---|---|---|---|
| 0 | 0 | 0 | 0 | 0 |
| 0 | 0 | 0 | 0 | 0 |
| 0 | 0 | 0 | 0 | 0 |
| 0 | 0.004046176 | 0.001301117 | 0.006576253 | 0 |
| 0 | 0 | 0 | 0 | 0 |
| 0 | 0 | 0 | 0 | 0.0567398 |
| 0 | 0 | 0 | 0 | 0 |
| 0.01751785 | 0 | 0.847396016 | 0.192334596 | 0.01724217 |
| 0 | 0 | 0 | 0 | 0 |
| 0 | 0 | 0 | 0 | 0 |
| 0 | 0.001072945 | 0.004991342 | 0.001583612 | 0 |
| 0 | 0 | 0 | 0 | 0 |
| 0 | 0 | 0.014379731 | 0.014905692 | 0 |
| 0 | 0.082951059 | 0 | 0 | 0 |
| 0 | 0 | 0 | 0 | 0 |
| 0 | 0 | 0 | 0 | 0 |
| 0 | 0 | 0.065143904 | 0 | 0 |
| 0.000497203 | 0 | 0.004289217 | 0 | 0.004254594 |
| 0 | 0.018609733 | 0 | 0 | 0 |
| 0 | 0 | 0 | 0 | 0.006235026 |
| 0 | 0 | 0.007621262 | 0 | 0.001417452 |
| 0 | 0 | 0 | 0 | 0 |
| 0 | 0.014623133 | 0 | 0.014629321 | 0 |
| 0 | 0 | 0 | 0 | 0 |
| 0 | 0 | 0 | 0 | 0 |
| 0 | 0 | 0 | 0 | 0 |
| 0 | 0 | 0 | 0 | 0 |
| 0 | 0 | 0 | 0 | 0 |
| 0 | 0 | 0 | 0 | 0 |
| 0 | 0 | 0 | 0.001791585 | 0.07131072 |
| 0 | 0.004901369 | 0 | 0.009666348 | 0.003186692 |
| 0 | 0 | 0 | 0 | 0 |
| 0 | 0 | 0 | 0 | 0.004074882 |
| 0 | 0 | 0 | 0 | 0 |
| 0 | 0 | 0 | 0 | 0 |
| 0 | 0 | 0 | 0 | 0 |
| 0 | 0.22727879 | 0 | 0 | 0 |
| 0.002401545 | 0 | 0.003816363 | 0.004021666 | 0.022172547 |
| 0 | 0 | 0 | 0 | 0 |
| 0.000623356 | 0.001791728 | 0.007797373 | 0.005619561 | 0 |
| 0 | 0.000681743 | 0.052815217 | 0.016351043 | 0 |
| 0 | 0 | 0 | 0 | 0 |
| 0 | 0 | 0 | 0 | 0 |
| 0 | 0 | 0 | 0 | 2.377116547 |
| 0 | 0 | 0 | 0 | 0 |
| 0 | 0 | 0 | 0 | 0 |
| 0 | 0.003624622 | 0 | 0 | 0 |
| 0 | 0 | 0 | 0 | 0 |

| | | | | |
|---|---|---|---|---|
| 0 | 0 | 0 | 0 | 0 |
| 0 | 0 | 0 | 0 | 0 |
| 0 | 0 | 0 | 0 | 0 |
| 0 | 0.038668047 | 0 | 0 | 0 |
| 0 | 0 | 0 | 0 | 0 |
| 0 | 0 | 0 | 0 | 0 |
| 0 | 0 | 0 | 0 | 0 |
| 0 | 0 | 0 | 0 | 0 |
| 0.015166074 | 0.009019089 | 0.062972839 | 0.023572831 | 0.092982011 |
| 0 | 0 | 0 | 0 | 0 |
| 0 | 0.002507917 | 0 | 0 | 0 |
| 0 | 0 | 0 | 0 | 0 |
| 0 | 0 | 0 | 0 | 0 |
| 0 | 0 | 0 | 0 | 0 |
| 0 | 0 | 0 | 1.92502174 | 0.014828888 |
| 0.08115063 | 0 | 0.250643011 | 0 | 0 |
| 0 | 0 | 0.005848748 | 0 | 0 |
| 0 | 0 | 0 | 0 | 0 |
| 0 | 0 | 0 | 0 | 0 |
| 0 | 0 | 0 | 0 | 0 |
| 0 | 0.004597591 | 0 | 0.106205006 | 0 |
| 0 | 0 | 0 | 0 | 0 |
| 0.063002007 | 0 | 0 | 0 | 0 |
| 0 | 0.127415995 | 0 | 0 | 0 |
| 0 | 0.004419247 | 0.019895176 | 0 | 0.012864023 |
| 0 | 0.243254939 | 0.812796427 | 2.800581958 | 3.188842908 |
| 0 | 0 | 0 | 1.235245745 | 0 |
| 0 | 0 | 0 | 0 | 0 |
| 0 | 0 | 0 | 0 | 0.054444905 |
| 0 | 0 | 0 | 0 | 0 |
| 0 | 0 | 0 | 0 | 0 |
| 0 | 0.381746064 | 0.003416694 | 0.151570911 | 0.004707103 |
| 0 | 0.002853366 | 0 | 0 | 0 |
| 0.001532753 | 0 | 0.014403186 | 0.017127101 | 0.011554451 |
| 0 | 0 | 0.132859741 | 0 | 0 |
| 0.002550311 | 0 | 0.01251293 | 0.003719148 | 0.001273018 |
| 0.238971988 | 0 | 0 | 0 | 0.30797335 |
| 0 | 0.164925613 | 0 | 0 | 0.209252719 |
| 0 | 0 | 0 | 0 | 0 |
| 0 | 0 | 0 | 0 | 0 |
| 0 | 0 | 0 | 0 | 0 |
| 0 | 0 | 0 | 0.00440925 | 0 |
| 0 | 0.007512871 | 0.586820014 | 0.392952766 | 0 |
| 0 | 0 | 0 | 0 | 0 |
| 0 | 0 | 0 | 0 | 0 |
| 0.001932879 | 0.001215316 | 0.000521074 | 0.293725566 | 0 |
| 0 | 0 | 0 | 0 | 0 |
| 0 | 0 | 0.002905466 | 0.094659625 | 0.058463692 |

| | | | | |
|---|---|---|---|---|
| 0.010189582 | 0 | 0.0267731 | 0 | 0.001228069 |
| 0 | 0 | 0 | 0 | 0 |
| 0 | 0 | 0 | 0 | 0 |
| 0 | 0 | 0 | 0 | 0 |
| 0.000571544 | 0 | 0.003697904 | 0 | 0 |
| 0 | 0 | 0 | 0.001276924 | 0 |
| 0 | 0 | 0 | 0 | 0 |
| 0 | 0 | 0 | 0 | 0 |
| 0 | 0 | 0 | 0 | 0 |
| 0 | 0 | 0 | 0.002091454 | 0 |
| 0 | 0 | 0.001768466 | 0 | 0 |
| 0 | 7.176520464 | 1.453179051 | 1.315948359 | 0 |
| 0 | 0.338155336 | 0.014178791 | 0 | 0.152980532 |
| 0 | 0 | 0 | 0 | 0 |
| 0.029248844 | 0.005462224 | 0.164115502 | 0.000657225 | 0.000707017 |
| 0 | 0 | 0 | 0 | 0 |
| 0 | 0 | 0 | 0.004114757 | 0.002787052 |
| 0 | 0 | 0 | 0 | 0 |
| 0 | 0.00083263 | 0.002030409 | 0.023042228 | 0.000619698 |
| 0 | 0 | 0.002597768 | 0 | 0 |
| 0 | 0 | 0 | 0.111440611 | 0.028902002 |
| 0 | 0 | 0 | 0 | 0 |
| 0 | 2.421368443 | 0.054369092 | 0.029127067 | 0.130356476 |
| 0 | 0 | 0 | 0 | 0.018679703 |
| 0 | 0 | 0 | 0 | 0 |
| 0 | 0 | 0 | 0 | 0 |
| 0 | 0 | 0 | 0 | 0 |
| 0 | 0 | 0 | 0 | 0 |
| 0 | 0 | 0 | 0 | 0 |
| 0 | 0 | 0 | 0 | 0 |
| 0 | 0 | 0 | 0 | 0 |
| 0.00107216 | 0.069030956 | 0.00624321 | 0.00909698 | 0.029664344 |
| 0 | 3.734387883 | 0 | 0 | 0.022837488 |
| 0.003535176 | 0.020168547 | 0.044590146 | 0.125405158 | 0.038042566 |
| 0.001942764 | 0 | 0.0151136 | 0.00331148 | 0.002176998 |
| 0 | 0.015168549 | 0.074460787 | 0.03083094 | 0 |
| 0 | 0 | 0.001062643 | 0 | 0 |
| 0 | 0 | 0 | 0 | 0 |
| 0 | 0.006873593 | 0 | 0 | 0.007730518 |
| 0 | 0 | 0 | 0 | 0.004459971 |
| 0 | 0 | 0 | 0 | 0 |
| 0 | 0 | 0 | 0 | 0 |
| 0 | 0 | 0 | 0 | 0 |
| 0 | 0 | 0.000609102 | 0 | 0 |
| 0.002460622 | 0 | 0.134880156 | 0.075572906 | 0.018716174 |
| 0 | 0 | 0 | 0 | 0 |
| 0 | 0.081822877 | 0 | 0 | 0 |
| 0.000905064 | 0.000557454 | 0.005716363 | 0.003599634 | 0.002397166 |

| | | | | |
|---|---|---|---|---|
| 0 | 0 | 0 | 0 | 0 |
| 0 | 0 | 0 | 0 | 0 |
| 0.001413833 | 0 | 0 | 0.013682913 | 0 |
| 0 | 0 | 0 | 0 | 0 |
| 0.846390363 | 0 | 0 | 0 | 0 |
| 0 | 0 | 0.000393476 | 0 | 0 |
| 0 | 0 | 0 | 0 | 0 |
| 0 | 0.03012749 | 0.013822379 | 0.003186304 | 0.012758665 |
| 0 | 0 | 0 | 0 | 0 |
| 0 | 0 | 0 | 0 | 0 |
| 0 | 0 | 0 | 0 | 0 |
| 0 | 0 | 0 | 0 | 0 |
| 0 | 0 | 0 | 0 | 0 |
| 0 | 0 | 0 | 0 | 0 |
| 0 | 0 | 0 | 0 | 0 |
| 0 | 0 | 0 | 0 | 0 |
| 0 | 0.038800276 | 0 | 0 | 0 |
| 0 | 0 | 0 | 0 | 0 |
| 0 | 0 | 0 | 1.346100393 | 0 |
| 0 | 0 | 0 | 0.000797865 | 0 |
| 0 | 0.002400023 | 0 | 0 | 0 |
| 0 | 0 | 0 | 0 | 0 |
| 0 | 0 | 0 | 0 | 0 |
| 0 | 0 | 0 | 0 | 0 |
| 0 | 0 | 0 | 0.003762181 | 0 |
| 0.003054328 | 0 | 0.005901025 | 0.005905139 | 0.001633505 |
| 0 | 0 | 0 | 0 | 0 |
| 0 | 0 | 0.00644755 | 0.006605665 | 0 |
| 0 | 0 | 0 | 0 | 0 |
| 0 | 0.62847135 | 0.209775146 | 0.236385871 | 0.206345749 |
| 0 | 0 | 0 | 0 | 0 |
| 0 | 0 | 0 | 0 | 0 |
| 0 | 0.001890312 | 0 | 0.001918128 | 0 |
| 0 | 0 | 0 | 0 | 0 |
| 0 | 0 | 0.010210641 | 0.003005707 | 0.002092215 |
| 0 | 0 | 0 | 0 | 0 |
| 0 | 0 | 0 | 0 | 0 |
| 0 | 0.003914738 | 0.00489553 | 0.054168312 | 0.016186709 |
| 0 | 0 | 0 | 0 | 0 |
| 0 | 0 | 0.035620913 | 0 | 0 |
| 0.599036 | 27.61912682 | 1.580600708 | 0.759730831 | 1.482879272 |
| 0 | 2.130100246 | 0.649458698 | 0.857939888 | 0.566054017 |
| 0 | 0 | 0 | 0 | 0 |
| 0 | 0 | 0 | 0 | 0 |
| 0 | 0 | 0 | 0 | 0 |
| 0 | 0 | 0 | 0 | 0 |
| 0 | 0 | 0 | 0 | 0 |
| 0.001201035 | 0 | 0.027715591 | 0.004936 | 0.006166401 |

| | | | | |
|---|---|---|---|---|
| 0 | 0 | 0 | 0 | 0 |
| 0 | 0.006391514 | 0.03077241 | 0.045202422 | 0.000528554 |
| 0 | 0 | 0 | 0.01759796 | 0 |
| 0 | 0 | 0 | 0 | 0 |
| 0 | 0.060102584 | 0.383440223 | 2.384633072 | 0.000480993 |
| 0 | 0 | 0 | 0 | 0.090512573 |
| 0 | 0 | 0 | 0 | 0 |
| 0 | 0 | 0 | 0 | 0 |
| 0.016652828 | 0.011345939 | 0.053340015 | 0.490279223 | 0.104499465 |
| 0 | 0 | 0 | 0 | 0 |
| 0 | 0 | 0 | 0 | 0 |
| 0 | 0 | 0 | 0 | 0 |
| 0 | 0.099367124 | 0.044397921 | 0.335938555 | 0.055466708 |
| 0.046780606 | 0.088440034 | 0 | 0.001565144 | 0.003367441 |
| 0 | 0 | 0 | 0 | 0 |
| 0 | 0.540035494 | 0.300603563 | 1.070502038 | 0.741254567 |
| 0 | 0 | 0.002203283 | 0 | 0.002913998 |
| 0 | 0 | 0 | 0 | 0 |
| 0 | 0 | 0 | 0 | 0 |
| 0 | 0 | 0 | 0 | 0 |
| 0 | 0 | 0 | 0 | 0 |
| 0 | 0 | 0 | 0.007970616 | 0 |
| 0 | 0 | 0 | 0 | 0 |
| 0 | 0 | 0 | 0 | 0 |
| 0 | 0 | 0 | 0 | 0 |
| 0 | 0 | 0 | 0 | 0 |
| 0 | 0.007340912 | 0.009220006 | 0.003533091 | 0.022646202 |
| 0.001127683 | 0 | 0.007701474 | 0.001162787 | 0 |
| 0 | 0 | 0 | 0 | 0 |
| 0 | 0 | 0 | 0 | 0 |
| 0 | 0 | 0 | 0 | 0 |
| 0 | 0 | 0 | 0 | 0 |
| 0 | 0 | 0 | 2.415996003 | 0 |
| 0 | 0 | 0 | 0.005572165 | 0.000936612 |
| 0 | 0 | 0 | 0 | 0 |
| 0 | 0.022080658 | 0 | 0.014027504 | 0.001653725 |
| 0 | 0 | 0 | 0 | 0 |
| 0 | 0 | 0.023171624 | 0 | 0.001171189 |
| 0 | 0 | 0 | 0 | 0 |
| 0 | 0 | 0 | 0 | 0 |
| 0 | 4.635344741 | 0 | 0 | 2.630472111 |
| 0 | 0 | 0 | 0.068797688 | 0 |
| 0 | 0 | 0 | 0 | 0 |
| 0 | 0 | 0 | 0 | 0 |
| 0 | 0 | 0 | 0 | 0 |
| 0 | 0 | 0 | 0 | 0 |
| 0 | 0 | 0 | 0 | 0 |
| 0 | 0 | 0 | 0 | 0 |

| | | | | |
|---|---|---|---|---|
| 0 | 0.005126265 | 0 | 0.058018934 | 0 |
| 0 | 0 | 0 | 0.00295081 | 0 |
| 0 | 0 | 0 | 0 | 0 |
| 0 | 0 | 0 | 0 | 0.001012145 |
| 0 | 0.010149203 | 0 | 0 | 0 |
| 0 | 1.936981005 | 0 | 0 | 0 |
| 0.001656082 | 0 | 0.002884778 | 0 | 0.00090841 |
| 0 | 0 | 0 | 0.001795768 | 0 |
| 0 | 0 | 0 | 0 | 0 |
| 0 | 0 | 0.000720619 | 0.010985775 | 0 |
| 0 | 0 | 0 | 0.001228819 | 0 |
| 0 | 0 | 0 | 0 | 0 |
| 0 | 0 | 0 | 0 | 0 |
| 0 | 0 | 0 | 0 | 0 |
| 0 | 0 | 0 | 0 | 0 |
| 0 | 0 | 0 | 0 | 0 |
| 0 | 0 | 0 | 0 | 0 |
| 0.009063003 | 0 | 0 | 0.006332317 | 0 |
| 0 | 0 | 0 | 0 | 0 |
| 0 | 0 | 0 | 0 | 0 |
| 0 | 0 | 0 | 0.228482321 | 0 |
| 0 | 0 | 0 | 0 | 0 |
| 0 | 0 | 0.023802003 | 0 | 0.005151245 |
| 0.001070587 | 0 | 0.009235631 | 0.001892424 | 0.001017898 |
| 0 | 0 | 0 | 0 | 0 |
| 0 | 0 | 0 | 0.012279491 | 0 |
| 0 | 0 | 0 | 0 | 0 |
| 0 | 0 | 0 | 0 | 0 |
| 0 | 0 | 0 | 0 | 0 |
| 0.145180042 | 0 | 0.625289018 | 0 | 0.351110127 |
| 0 | 0 | 0 | 0 | 0 |
| 0 | 0 | 0 | 0 | 0 |
| 0 | 0 | 0 | 0 | 0 |
| 0.001778502 | 0 | 0 | 0 | 0.024821055 |
| 0 | 0 | 0 | 0 | 0 |
| 0 | 0 | 0 | 0 | 0 |
| 0 | 0.002994753 | 0.321325613 | 0 | 0.002971859 |
| 0 | 0 | 0 | 0 | 0 |
| 0 | 0 | 0 | 0 | 0 |
| 0 | 0 | 0 | 0 | 0 |
| 0 | 0 | 0 | 0 | 0 |
| 0 | 0 | 0 | 0 | 0 |
| 0 | 0 | 0 | 0 | 0 |
| 0 | 0 | 0 | 0 | 0 |
| 0 | 0 | 0 | 0.039778643 | 0 |
| 0 | 0 | 0 | 0 | 0 |
| 0 | 0 | 0 | 0 | 0 |
| 0 | 0 | 0 | 0 | 0 |

| | | | | |
|---|---|---|---|---|
| 0.002815975 | 0 | 0.003588674 | 0.003393859 | 0 |
| 0 | 0 | 0 | 0 | 0 |
| 0 | 0 | 0 | 0 | 0 |
| 0 | 0 | 0 | 0 | 0 |
| 0 | 0 | 0 | 0 | 0 |
| 0 | 0 | 0 | 0 | 0 |
| 0 | 0 | 0.000530539 | 0 | 0.00084201 |
| 0 | 0.394760353 | 0.413259682 | 0.698861768 | 3.779658902 |
| 0.013863028 | 0.006929893 | 0.176355067 | 0.015981513 | 0.025501892 |
| 0.036506047 | 0.05990276 | 0.126242126 | 0.037271597 | 0.028525531 |
| 0 | 0 | 0.005132789 | 0 | 0.002106768 |
| 0 | 0 | 0 | 0 | 0 |
| 0 | 0 | 0 | 0 | 0 |
| 0.010560522 | 0 | 0.03255949 | 0.014789242 | 0 |
| 0 | 0 | 0 | 0 | 0 |
| 0 | 0 | 0 | 0 | 0 |
| 0 | 0 | 0 | 0.005944945 | 0 |
| 0 | 0 | 0 | 0 | 0 |
| 0 | 0 | 0 | 0 | 0 |
| 0 | 0 | 0 | 0.095641025 | 0 |
| 0 | 0 | 0 | 0 | 0 |
| 0 | 0 | 0 | 0 | 0 |
| 0 | 0 | 0 | 0 | 0 |
| 0 | 0 | 0 | 0 | 0 |
| 0 | 0 | 0 | 0 | 0 |
| 0 | 0 | 0 | 0 | 0 |
| 0 | 0 | 0 | 0 | 0 |
| 0 | 0 | 0 | 0 | 0 |
| 0 | 0 | 0 | 0 | 0 |
| 0 | 0 | 0 | 0 | 0 |
| 0 | 0 | 0 | 0.045848868 | 0 |
| 0 | 0 | 0 | 0 | 0 |
| 0 | 0 | 0 | 0 | 0 |
| 0 | 0 | 0 | 0 | 0 |
| 0 | 0 | 0 | 0.046428804 | 0 |
| 0 | 0 | 0 | 0 | 0 |
| 0 | 0 | 0 | 0 | 0 |
| 0 | 0.022915418 | 0 | 0 | 0 |
| 0 | 0.015749083 | 0 | 0 | 0.013458034 |
| 0 | 0 | 0 | 0 | 0.537975722 |
| 0 | 0 | 0 | 0 | 0 |
| 0.037703607 | 0 | 0.188785868 | 0.897809249 | 0.027074021 |
| 0 | 0 | 0 | 0 | 0 |
| 0 | 0 | 0 | 0 | 0 |
| 0 | 0 | 0.021215642 | 0 | 0 |
| 0 | 0 | 0 | 0.047832607 | 0 |
| 0 | 0 | 0 | 0 | 0 |
| 0 | 0 | 0 | 0 | 0 |

| | | | | |
|---|---|---|---|---|
| 0 | 0 | 0 | 0 | 0 |
| 0 | 0 | 0.412964243 | 0 | 0 |
| 0 | 0 | 0 | 0 | 0 |
| 0 | 0 | 0 | 0 | 0 |
| 0.003391912 | 0.118386358 | 4.127889913 | 1.63190482 | 0 |
| 0 | 0 | 0 | 0.174755167 | 0 |
| 0 | 0 | 0 | 0.013928164 | 0 |
| 0.000814119 | 0.005148099 | 0.02563453 | 0.00885033 | 0.008359761 |
| 0 | 0 | 0 | 0 | 0 |
| 0 | 0 | 0 | 0 | 0 |
| 0 | 0 | 0 | 0 | 0 |
| 0 | 0 | 0 | 0 | 0 |
| 0 | 0 | 0 | 0 | 0 |
| 0 | 0 | 0 | 0 | 0 |
| 0 | 0 | 0 | 0 | 0 |
| 0.020052656 | 0 | 0 | 0.107648764 | 0.003222381 |
| 0 | 0 | 0 | 0 | 0 |
| 0 | 0 | 0 | 0 | 0 |
| 0 | 0 | 0 | 0 | 0 |
| 0 | 0 | 0 | 0 | 0 |
| 0 | 0 | 0 | 0 | 0 |
| 0 | 0 | 0 | 0 | 0 |
| 0 | 0 | 0 | 0 | 0 |
| 0 | 0 | 0 | 0.002988968 | 0 |
| 0 | 0 | 0 | 0 | 0 |
| 0 | 0 | 0 | 0 | 0 |
| 0.007861604 | 0 | 1.56471651 | 0 | 0.0141529 |
| 0 | 0 | 0 | 0.001799094 | 0 |
| 0 | 0 | 0 | 0 | 0 |
| 0 | 0 | 0 | 0 | 0 |
| 0 | 0 | 0 | 0 | 0 |
| 0 | 0 | 0 | 0 | 0 |
| 0 | 0 | 0 | 0 | 0 |
| 0 | 0 | 0 | 0.006449957 | 0 |
| 0 | 0 | 0 | 0 | 0 |
| 0 | 0 | 0 | 0 | 0 |
| 0 | 0 | 0 | 0 | 0 |
| 0 | 0 | 0.648211578 | 1.174367042 | 0.009990588 |
| 0 | 0 | 0 | 0 | 0.225686175 |
| 0 | 0.006518463 | 0 | 0 | 0 |
| 0 | 0 | 0 | 0 | 0 |
| 0 | 0 | 0 | 0 | 0 |
| 0 | 0 | 0 | 0 | 0 |
| 0 | 0 | 0 | 0 | 0 |
| 0 | 0 | 0 | 0 | 0 |
| 0 | 0 | 0 | 0 | 0 |
| 0 | 0 | 0 | 0 | 0 |
| 0 | 0.012747643 | 0 | 0 | 0 |

| | | | | |
|---|---|---|---|---|
| 0 | 0.956092261 | 0.005489138 | 0 | 0.015902358 |
| 0 | 0 | 0 | 0 | 0 |
| 0 | 0 | 0 | 0 | 0 |
| 0 | 0 | 0.001300331 | 0.003197326 | 0 |
| 0 | 0 | 0 | 0 | 0 |
| 0 | 0 | 0 | 0 | 0 |
| 0 | 0 | 0 | 0 | 0 |
| 0 | 0 | 0 | 0 | 0 |
| 0 | 0 | 0 | 0 | 0 |
| 0 | 0 | 0 | 0 | 0 |
| 0 | 0 | 0 | 0 | 0 |
| 0 | 0 | 0 | 0.003271981 | 0.003334613 |
| 0 | 0 | 0 | 0.00464014 | 0 |
| 0 | 0 | 0 | 0 | 0 |
| 0 | 0 | 0 | 0 | 0 |
| 0 | 0 | 0 | 0 | 0 |
| 0 | 0 | 0 | 0 | 0 |
| 0 | 0 | 0 | 0 | 4.113442405 |
| 0 | 0 | 0.00110805 | 0 | 0 |
| 0 | 0 | 0 | 0 | 0 |
| 0 | 0 | 0.124380064 | 0.010417658 | 0.00120074 |
| 0 | 0 | 0 | 0 | 0 |
| 0.003367102 | 0 | 0.085421027 | 0.057795693 | 0.019461076 |
| 0 | 0 | 0.3921332 | 0 | 0 |
| 0 | 0 | 0 | 0 | 0 |
| 0 | 0 | 0 | 0.056360769 | 0 |
| 0 | 0 | 0 | 0 | 0 |
| 0 | 0 | 0 | 0 | 0 |
| 0 | 0 | 0 | 0 | 0 |
| 0 | 0 | 0 | 0 | 0 |
| 0 | 0 | 0 | 0 | 0 |
| 0 | 0 | 0 | 0 | 0 |
| 0.213095176 | 0 | 0.013422221 | 0.015621567 | 0.0186986 |
| 0 | 0.063695316 | 0 | 0 | 0 |
| 0 | 2.361572515 | 0.017929129 | 0 | 0.943420307 |
| 0 | 0.164219417 | 0.011919652 | 0.00353117 | 0.000759739 |
| 0 | 0 | 0 | 0 | 0 |
| 0 | 0 | 0 | 0 | 0 |
| 0 | 0.005534549 | 0.008139292 | 0.012457287 | 0.00505286 |
| 0 | 0 | 0 | 0 | 0 |
| 0 | 0 | 0 | 0 | 0 |
| 0 | 0 | 0 | 0 | 0 |
| 0 | 0 | 0 | 0 | 0 |
| 0 | 0 | 0 | 0 | 0 |
| 0 | 0 | 0 | 0 | 0 |
| 0 | 0 | 0 | 0.824184657 | 0.001020048 |
| 0 | 0 | 0 | 0 | 0 |
| 0 | 0 | 0 | 0 | 0 |

| | | | | |
|---|---|---|---|---|
| 0 | 0 | 0 | 0 | 0 |
| 0 | 0 | 0 | 0 | 0 |
| 0 | 0 | 0 | 0 | 0 |
| 0 | 0 | 0 | 0 | 0 |
| 0 | 0 | 0 | 0 | 0 |
| 0 | 0 | 0 | 0 | 0 |
| 0 | 0 | 0 | 0 | 0 |
| 0 | 0 | 0 | 0.103355462 | 0.10527163 |
| 0 | 0.009245306 | 0 | 0.40226092 | 0 |
| 0 | 0 | 0 | 0 | 0 |
| 0 | 0 | 0 | 0 | 0 |
| 0 | 0 | 0 | 0 | 0 |
| 0 | 0 | 0 | 0.001081413 | 0 |
| 0 | 0 | 0 | 0 | 0 |
| 0 | 0 | 0 | 0 | 0 |
| 0.00275429 | 0 | 0 | 0 | 0.002274166 |
| 0 | 0 | 0 | 0 | 0 |
| 0 | 0 | 0 | 0 | 0 |
| 0 | 0 | 0 | 0 | 0 |
| 0 | 1.617963304 | 0 | 0 | 0.04777318 |
| 0 | 0 | 0 | 0 | 0.004534482 |
| 0 | 0 | 0 | 0 | 0 |
| 0 | 0 | 0 | 0 | 0 |
| 0 | 0 | 0.009786319 | 0 | 0.00220308 |
| 0 | 0 | 0 | 0 | 0.000838253 |
| 0 | 0 | 0 | 0 | 0 |
| 0 | 0 | 0 | 0 | 0 |
| 0 | 0 | 0 | 0 | 0 |
| 0 | 0 | 0 | 0 | 0 |
| 0 | 0.853760612 | 0 | 0.412904989 | 0 |
| 0 | 0.055539867 | 0 | 1.886745634 | 0.46629539 |
| 0 | 0 | 0 | 0 | 0 |
| 0 | 0 | 0 | 0 | 0 |
| 0 | 0 | 0 | 0 | 0 |
| 0 | 0 | 0 | 0 | 0 |
| 0 | 0 | 0 | 0 | 0 |
| 0 | 0 | 0 | 0 | 0 |
| 0 | 0 | 0 | 0 | 0 |
| 0 | 0 | 0 | 0 | 0 |
| 0 | 0 | 0 | 0 | 0 |
| 0 | 0 | 0 | 0 | 0 |
| 0 | 0 | 0 | 0 | 0 |
| 0 | 0 | 0 | 0 | 0 |
| 0 | 0 | 0.002170617 | 0 | 0 |
| 0 | 0 | 0 | 0 | 0 |
| 0 | 0 | 0 | 0 | 0 |
| 0.096161373 | 0 | 0 | 0.964792133 | 0.015238126 |
| 0 | 0 | 0.00039015 | 0 | 0 |

| | | | | |
|---|---|---|---|---|
| 0 | 0 | 0 | 0 | 0 |
| 0 | 0 | 0 | 0 | 0 |
| 0 | 0 | 0 | 0 | 0 |
| 0 | 0 | 0 | 0 | 0 |
| 0 | 1.049343459 | 0.21183998 | 0.038801743 | 0.135185212 |
| 0 | 0 | 0 | 0 | 0 |
| 0 | 0 | 0 | 0 | 0 |
| 0 | 0 | 0 | 0 | 0 |
| 0 | 0 | 0 | 0 | 0 |
| 0 | 0 | 0 | 0 | 0 |
| 0 | 0 | 0 | 0 | 0 |
| 0.000923509 | 0 | 0.070207732 | 0.012855478 | 0.00570738 |
| 0 | 0 | 0.003150295 | 0.009682653 | 0 |
| 0 | 0 | 0 | 0 | 0 |
| 0 | 0 | 0.00050594 | 0 | 0 |
| 0 | 1.35516399 | 0 | 0 | 1.075774919 |
| 0 | 0.002381868 | 0.001659515 | 0.043316177 | 0.009116971 |
| 0 | 0 | 0 | 0 | 0 |
| 0.384724153 | 0.002009979 | 0.039965531 | 0.080500633 | 0.096240062 |
| 0 | 0 | 0 | 0 | 0 |
| 0 | 0 | 0.033576966 | 0 | 0 |
| 0 | 0 | 0 | 0 | 0 |
| 0 | 0 | 0 | 0 | 0 |
| 0 | 0 | 0 | 0 | 0 |
| 0 | 0 | 0 | 0 | 0 |
| 0 | 0 | 0 | 0 | 0 |
| 0.020608788 | 0.001051534 | 0.005785927 | 0.051087009 | 0.004521812 |
| 0 | 0 | 0 | 0 | 0 |
| 0 | 0 | 0 | 0 | 0 |
| 0 | 0 | 0 | 0 | 0 |
| 0 | 0 | 0.001937784 | 0.016859817 | 0 |
| 0 | 0 | 0 | 0.028701437 | 0 |
| 0 | 0 | 0 | 0.01840158 | 0 |
| 0 | 0 | 0 | 0 | 0 |
| 0 | 0 | 0 | 0 | 0 |
| 0 | 0 | 0 | 0 | 0 |
| 0 | 0 | 0 | 0 | 0 |
| 0.00038847 | 0.016609281 | 0.090901671 | 0.105576946 | 0.03601176 |
| 0 | 0 | 0 | 0.080373751 | 0.036484362 |
| 0 | 0 | 0 | 0 | 0 |
| 0 | 0 | 0.025091936 | 0 | 0.040376125 |
| 0 | 0 | 0 | 0 | 0 |
| 0 | 0 | 0 | 0 | 0 |
| 0.014401297 | 0 | 0 | 0.002291081 | 0.00102694 |
| 0 | 0 | 0 | 0 | 0 |
| 0 | 0 | 0 | 0 | 0 |
| 0 | 0 | 0 | 0.598792731 | 0 |
| 0 | 0 | 0 | 0 | 0 |

| | | | | |
|---|---|---|---|---|
| 0 | 0 | 0 | 0 | 0 |
| 0 | 0.421726792 | 0 | 0 | 0 |
| 0 | 0 | 0 | 0 | 0 |
| 0 | 0 | 0.001139246 | 0.001000443 | 0.000645743 |
| 0 | 0 | 0.001358527 | 0 | 0 |
| 0 | 0 | 1.060167744 | 0 | 0 |
| 0 | 0 | 0 | 0 | 0 |
| 0.00271486 | 0 | 0 | 0 | 0 |
| 0 | 0.001233471 | 0.002115432 | 0 | 0 |
| 0 | 0 | 0 | 0 | 0 |
| 0 | 0 | 0 | 0.040342069 | 0 |
| 0 | 0 | 0 | 0 | 0 |
| 0 | 0.006150225 | 0.02111205 | 0.007800907 | 0.040433752 |
| 0 | 0 | 0 | 0 | 0 |
| 0 | 0 | 0 | 0 | 0 |
| 0 | 0 | 0 | 0 | 0 |
| 0 | 0 | 0 | 0 | 0 |
| 0 | 0 | 0 | 0 | 0 |
| 0 | 0 | 0 | 0 | 0 |
| 0 | 0 | 0 | 0 | 0 |
| 0 | 0 | 0 | 0 | 0 |
| 0 | 0 | 0 | 0 | 0 |
| 0.003944506 | 0 | 0.007443637 | 0.065367253 | 0.024109555 |
| 0 | 0 | 0 | 0 | 0.005215852 |
| 0 | 0 | 0 | 0 | 0 |
| 0 | 0 | 0 | 0 | 0 |
| 0 | 0.449153078 | 0 | 0 | 0 |
| 0 | 0 | 0 | 0.001071138 | 0 |
| 0 | 0 | 0 | 0 | 0 |
| 0 | 0 | 0 | 0 | 0 |
| 0 | 0.018510912 | 0.004234501 | 0.004789533 | 0.007840597 |
| 0 | 0 | 0 | 0 | 0 |
| 0 | 0 | 0 | 0 | 0 |
| 0 | 0 | 0 | 0 | 0 |
| 0 | 0 | 0 | 0 | 0 |
| 0 | 0 | 0 | 0 | 0 |
| 0 | 0 | 0 | 0 | 0 |
| 0 | 0.139012405 | 0 | 0 | 0 |
| 0.000797379 | 0 | 0 | 0 | 0 |
| 0 | 0 | 0 | 0 | 0 |
| 0 | 0 | 0 | 0 | 0 |
| 0 | 0 | 0 | 0 | 0 |
| 0 | 0 | 0 | 0 | 0 |
| 0 | 0 | 0 | 0 | 0 |
| 0 | 0 | 0 | 0 | 0 |
| 0.010757284 | 0.093936172 | 0 | 0.198418609 | 0.02351296 |
| 0 | 0 | 0 | 0 | 0 |
| 0 | 0 | 0 | 0 | 0 |

| | | | | |
|---|---|---|---|---|
| 0 | 0 | 0 | 0 | 0 |
| 0 | 0 | 0 | 0 | 0 |
| 0 | 0 | 0 | 0 | 0 |
| 0 | 0 | 0 | 0 | 0 |
| 0 | 0 | 0 | 0 | 0 |
| 0 | 0 | 0 | 0 | 0 |
| 0 | 0 | 0 | 0.004119071 | 0 |
| 0 | 0 | 0 | 0 | 0 |
| 0 | 0 | 0 | 0 | 0 |
| 0 | 0 | 0 | 0 | 0 |
| 0 | 0 | 0.004185381 | 0 | 0.002613968 |
| 0 | 0 | 0 | 0 | 0 |
| 0 | 0 | 0 | 0 | 0 |
| 0 | 0 | 0 | 0 | 0 |
| 0 | 0 | 0 | 0 | 0 |
| 0 | 0 | 0 | 0 | 0 |
| 0 | 0 | 0 | 0 | 0 |
| 0 | 0 | 0 | 0 | 0 |
| 0 | 0 | 0 | 0 | 0 |
| 0 | 0 | 0.002312722 | 0 | 0 |
| 0 | 0 | 0 | 0 | 0 |
| 0 | 0 | 0 | 0 | 0 |
| 0 | 0 | 0 | 0 | 0 |
| 0 | 0 | 0 | 0 | 0 |
| 0 | 0 | 0 | 0 | 0 |
| 0 | 0 | 0 | 0 | 0 |
| 0 | 0 | 0 | 0 | 0 |
| 0 | 0 | 0 | 0 | 0 |
| 0 | 0 | 0 | 0 | 0 |
| 0 | 0 | 0 | 0 | 0 |
| 0 | 0.002824551 | 0.011656292 | 0 | 0.009284799 |
| 0 | 0 | 0 | 0 | 0 |
| 6.548452036 | 0.005739285 | 0.023377155 | 1.013481796 | 0.051665501 |
| 0 | 0.021784015 | 0 | 0.003284769 | 0.000831442 |
| 0 | 0 | 0 | 0 | 0 |
| 0 | 0 | 0 | 0 | 0 |
| 0 | 0 | 0 | 0 | 0 |
| 0 | 0 | 0 | 0 | 0 |
| 0 | 0 | 0 | 0 | 0 |
| 0 | 0 | 0 | 0 | 0 |
| 0 | 0 | 0 | 0 | 0 |
| 0.001056021 | 0.028329948 | 0.266087235 | 0.019600106 | 0.063589766 |
| 0 | 0.292751529 | 0.071215505 | 0.053821281 | 0 |
| 0 | 0 | 0 | 0 | 0 |
| 0 | 0 | 0 | 0 | 0 |
| 0 | 0 | 0.094669537 | 0 | 0 |
| 0 | 0 | 0 | 0.004309168 | 0.00540824 |
| 0 | 0 | 0 | 0 | 0 |

| | | | | |
|---|---|---|---|---|
| 0 | 0 | 0 | 0 | 0 |
| 0.008685904 | 0 | 0.004521678 | 0 | 0.009611097 |
| 0 | 0.001508435 | 0.001509085 | 0 | 0 |
| 0 | 0 | 0 | 0 | 0 |
| 0.001396747 | 0 | 0.356057315 | 1.148437343 | 0.008366438 |
| 0 | 0 | 0.137519395 | 0 | 0.002132911 |
| 0 | 0 | 0 | 0 | 0 |
| 0 | 0 | 0 | 0 | 0 |
| 0 | 0 | 0 | 0 | 0 |
| 0 | 0 | 0 | 0 | 0 |
| 0 | 0 | 0 | 0 | 0 |
| 0 | 0 | 0 | 0 | 0 |
| 0 | 0 | 0 | 0 | 0 |
| 0.018673427 | 0 | 0.001109872 | 0 | 0 |
| 0 | 0 | 0 | 0 | 0 |
| 0 | 0 | 0 | 0 | 0 |
| 0 | 0 | 0 | 0 | 0 |
| 0 | 0 | 0 | 0 | 0 |
| 0 | 0 | 0 | 0 | 0 |
| 0.014176617 | 0 | 0 | 0 | 0 |
| 0 | 0 | 0 | 0 | 0 |
| 0 | 0 | 0 | 0 | 0 |
| 0 | 0 | 0 | 0 | 0 |
| 0 | 0 | 0 | 0 | 0 |
| 0 | 0 | 0 | 0 | 0 |
| 0 | 0.001250592 | 0.01595192 | 0 | 0.027302699 |
| 0 | 0 | 0 | 0 | 0 |
| 0 | 0 | 0 | 0 | 0 |
| 0 | 0 | 0 | 0 | 0 |
| 0 | 0 | 0 | 0 | 0 |
| 0 | 0 | 0 | 0 | 0 |
| 0 | 0 | 0.001920044 | 0 | 0 |
| 0 | 0 | 0 | 0 | 0 |
| 0 | 0 | 0 | 0 | 0 |
| 0 | 0 | 0 | 0 | 0 |
| 0 | 0 | 0 | 0 | 0 |
| 0 | 0 | 0.05778789 | 0 | 0.056257726 |
| 0 | 0.022407472 | 0.000934047 | 0 | 0.02182439 |
| 0 | 5.396933393 | 0 | 0 | 0 |
| 0 | 0 | 0 | 0 | 0 |
| 0 | 0 | 0 | 0 | 0 |
| 0.010226797 | 0 | 0 | 0.251345404 | 0 |
| 0.001309982 | 0 | 0.002354337 | 0.003087454 | 0.000622755 |
| 0 | 0 | 0 | 0 | 0 |
| 0 | 0 | 0 | 0 | 0 |
| 0 | 0 | 0.001805721 | 0 | 0 |
| 0.006234717 | 0 | 0.007443462 | 0.0023616 | 0 |
| 0 | 0 | 0 | 0 | 0 |

| | | | | |
|---|---|---|---|---|
| 0 | 0 | 0 | 0 | 0 |
| 0.001826283 | 0.007268312 | 0.024238141 | 0.082506117 | 0.028249925 |
| 0 | 0 | 0 | 0 | 0 |
| 0 | 0 | 0 | 0 | 0 |
| 0 | 0 | 0 | 0 | 0 |
| 0 | 0 | 0 | 0 | 0 |
| 0 | 0 | 0 | 0 | 0 |
| 0 | 0 | 0 | 0 | 0 |
| 0 | 0 | 0 | 0 | 0 |
| 0 | 0 | 0 | 0 | 0 |
| 0 | 0 | 0 | 0 | 0 |
| 0 | 0 | 0 | 0 | 0 |
| 0 | 0.005122166 | 0 | 0 | 0.028168338 |
| 0 | 0 | 0 | 0 | 0 |
| 0.001926777 | 0 | 0.014355121 | 0 | 0 |
| 0 | 0 | 0 | 0 | 0 |
| 0 | 0 | 0 | 0 | 0 |
| 0 | 0 | 0 | 0 | 0 |
| 0 | 0.027013044 | 0 | 0 | 0.000829068 |
| 0 | 0 | 0 | 0 | 0 |
| 0 | 0 | 0 | 0 | 0 |
| 0 | 0 | 0 | 0.00136834 | 0 |
| 0.008459851 | 0 | 0.008552411 | 0 | 0.024319008 |
| 0 | 0 | 0.078717649 | 0 | 0 |
| 0 | 0 | 0 | 0 | 0 |
| 0 | 0 | 0 | 0 | 0 |
| 0 | 0 | 0 | 0 | 0 |
| 0 | 0 | 0 | 0 | 0 |
| 0 | 0 | 0 | 0.004244786 | 0 |
| 0 | 0 | 0 | 0 | 0 |
| 0 | 0 | 0 | 0 | 0 |
| 0 | 0 | 0 | 0 | 0 |
| 0 | 0 | 0 | 0 | 0 |
| 0 | 0 | 0 | 0 | 0 |
| 3.069227061 | 0.083276897 | 0 | 12.54531162 | 0.265963533 |
| 0 | 0 | 0 | 0 | 0 |
| 0 | 0 | 0 | 0 | 0 |
| 0 | 0 | 0 | 0 | 0 |
| 0 | 0 | 0.00463106 | 0 | 0 |
| 0 | 0 | 0 | 0 | 0 |
| 0.005699043 | 0.125955958 | 0.010796129 | 0.01449827 | 0.018672072 |
| 0 | 0 | 0 | 0 | 0 |
| 0 | 0 | 0 | 0 | 0 |
| 0 | 0 | 0 | 0 | 0.001078968 |
| 0 | 0 | 0 | 0 | 0 |
| 0.452342328 | 0.025851533 | 0.299702669 | 0.073645812 | 1.201206243 |
| 0 | 0 | 0 | 0 | 0 |
| 0 | 0 | 0 | 0 | 0 |

| | | | | |
|---|---|---|---|---|
| 0.009738391 | 0 | 0 | 0 | 0 |
| 0.013212873 | 0 | 0 | 0 | 0 |
| 0 | 0 | 0 | 0 | 0 |
| 0 | 0 | 0 | 0 | 0.005824719 |
| 0 | 0 | 0.005913308 | 0.010816322 | 0 |
| 0 | 0 | 0 | 0 | 0 |
| 0 | 0 | 0 | 0.00577924 | 0 |
| 0 | 0 | 0 | 0 | 0 |
| 0 | 0 | 0.47806901 | 0 | 0 |
| 0 | 0 | 0.001859833 | 0 | 0 |
| 0 | 0 | 0 | 0 | 0 |
| 0 | 0 | 0 | 0 | 0 |
| 0 | 0 | 0.001090698 | 0.004693272 | 0 |
| 0 | 0 | 0 | 0 | 0 |
| 0 | 0 | 0 | 0 | 0 |
| 0 | 0 | 0 | 0 | 0 |
| 0 | 0 | 0 | 0 | 0 |
| 0 | 0 | 0 | 0 | 0 |
| 0 | 0 | 0 | 0 | 0 |
| 0.075923065 | 0.007978683 | 1.345688228 | 0.271129461 | 0.05264192 |
| 0 | 0 | 0 | 0 | 0 |
| 0 | 0 | 0 | 0 | 0 |
| 0 | 0 | 0 | 0 | 0 |
| 0 | 0 | 0 | 0 | 0 |
| 0 | 0 | 0 | 0 | 0 |
| 0 | 0 | 0 | 0 | 0 |
| 0 | 0 | 0 | 0 | 0 |
| 0 | 0 | 0 | 0 | 0 |
| 0 | 0 | 0 | 0 | 0 |
| 0.004812035 | 0 | 0.019545216 | 0 | 0 |
| 0 | 0 | 0 | 0 | 0 |
| 0 | 0 | 0 | 0 | 0 |
| 0 | 0 | 0 | 0 | 0 |
| 0 | 0 | 0 | 0 | 0 |
| 0 | 0 | 0 | 0 | 0 |
| 0 | 0.004466258 | 0 | 0 | 0 |
| 0 | 0 | 0 | 0 | 0 |
| 0 | 0 | 0 | 0 | 0 |
| 0 | 0 | 0 | 0 | 0 |
| 0 | 0 | 0 | 0 | 0 |
| 0 | 0 | 0 | 0 | 0 |
| 0 | 0 | 0 | 0 | 0 |
| 0 | 0 | 0 | 0 | 0 |
| 0 | 0 | 0 | 0 | 0 |
| 0.000563713 | 0.004050742 | 0.04999755 | 0.019430737 | 0.002813842 |
| 0 | 0 | 0 | 0 | 0 |
| 0 | 0 | 0 | 0 | 0 |
| 0 | 0 | 0 | 0 | 0 |

| | | | | |
|---|---|---|---|---|
| 0 | 0 | 0 | 0 | 0 |
| 0 | 0 | 0 | 0 | 0 |
| 0 | 0 | 0 | 0 | 0 |
| 0 | 0 | 0 | 0 | 0 |
| 0 | 0 | 0 | 0 | 0 |
| 0 | 0.002116825 | 0.038913418 | 0.001813225 | 0.007652341 |
| 0 | 0 | 0 | 0 | 0 |
| 0 | 0 | 0 | 0.001162995 | 0 |
| 0 | 0 | 0 | 0 | 0 |
| 0 | 0 | 0 | 0 | 0 |
| 0 | 0 | 0 | 0 | 0 |
| 0 | 0 | 0 | 0 | 0 |
| 0 | 0 | 0 | 0 | 0 |
| 0 | 0 | 0 | 0 | 0 |
| 0 | 0 | 0 | 0 | 0 |
| 0 | 0 | 0 | 0 | 0 |
| 0 | 0 | 0 | 0 | 0.009828249 |
| 0 | 0 | 0 | 0.001374205 | 0 |
| 0 | 0 | 0 | 0 | 0 |
| 0 | 0 | 0 | 0 | 0 |
| 0 | 0 | 0 | 0 | 0 |
| 0 | 0 | 0 | 0 | 0 |
| 0 | 0 | 0 | 0 | 0 |
| 0 | 0 | 0 | 0 | 0 |
| 0 | 0 | 0 | 0 | 0 |
| 0 | 0 | 0 | 0 | 0 |
| 0 | 0 | 0 | 0 | 0 |
| 0 | 0 | 0 | 0.394493945 | 0 |
| 0 | 0 | 0 | 0 | 0 |
| 0 | 0 | 0 | 0.0451352 | 0 |
| 0 | 0 | 0 | 0 | 0 |
| 0 | 0 | 0 | 0 | 0 |
| 0 | 0 | 0 | 0 | 0 |
| 0 | 0 | 0 | 0 | 0 |
| 0 | 0 | 0 | 0 | 0 |
| 0 | 0 | 0 | 0 | 0 |
| 0 | 0 | 0 | 0 | 0 |
| 0 | 0 | 0 | 0.071498563 | 0 |
| 0 | 0 | 0 | 0 | 0.630114623 |
| 0 | 0 | 0 | 0 | 0 |
| 0 | 0 | 0 | 0 | 0 |
| 0 | 0 | 0 | 0 | 0 |
| 0 | 0 | 0 | 0 | 0 |
| 0 | 0 | 0 | 0 | 0 |
| 0 | 0 | 0 | 0 | 0 |
| 0 | 0 | 0 | 0 | 0 |
| 0 | 0 | 0 | 0 | 0 |
| 0 | 0 | 0 | 0 | 0 |

| | | | | |
|---|---|---|---|---|
| 0 | 0 | 0 | 0 | 0 |
| 0 | 0 | 0.011304481 | 0 | 0 |
| 0 | 0 | 0 | 0 | 0 |
| 0 | 0 | 0 | 0 | 0 |
| 0 | 0 | 0 | 0 | 0 |
| 0 | 0 | 0 | 0.021842158 | 0 |
| 0 | 0 | 0 | 0 | 0 |
| 0 | 0 | 0 | 0 | 0 |
| 0 | 0 | 0 | 0 | 0 |
| 0 | 0 | 0 | 0 | 0 |
| 0 | 0 | 0.001663182 | 0 | 0 |
| 0 | 0 | 0 | 0 | 0 |
| 0 | 0.255673602 | 0.000612509 | 0.25121268 | 0.470174076 |
| 0 | 0 | 0 | 0 | 0 |
| 0 | 0 | 0 | 0 | 0 |
| 0 | 0 | 0 | 0 | 0 |
| 0 | 0 | 0 | 0.007828867 | 0 |
| 0 | 0 | 0 | 0 | 0 |
| 0 | 0 | 0 | 0 | 0 |
| 0 | 0 | 0 | 0 | 0 |
| 0 | 0 | 0.03443581 | 0 | 0.03750665 |
| 0 | 0 | 0 | 0 | 0 |
| 0 | 0 | 0 | 0 | 0 |
| 0 | 1.520577852 | 0.748823521 | 1.906922164 | 4.474890812 |
| 0 | 0 | 0 | 0 | 0 |
| 0 | 0 | 0 | 0 | 0 |
| 0 | 0 | 0 | 0 | 0 |
| 0 | 0 | 0 | 0 | 0 |
| 0 | 0 | 0 | 0 | 0 |
| 0 | 0 | 0 | 0 | 0 |
| 0 | 0 | 0 | 0 | 0 |
| 0 | 0 | 0 | 0 | 0 |
| 0 | 0 | 0 | 0 | 0 |
| 0 | 0 | 0 | 0 | 0 |

| PHL018-0199_V3 | PHL018-0243_V3 | PHL018-0269_V3 | PHL018-0073_V2 | PHL018-0241_V2 |
|---|---|---|---|---|
| CTRL | REF | REF | CTRL | REF |
| 0.104952947 | 0.077899896 | 0.045371866 | 0.27372041 | 0.061560867 |
| 5.600896097 | 9.14940569 | 1.089803372 | 0.018433556 | 9.440831662 |
| 2.333841528 | 0 | 0.107001813 | 1.295082277 | 2.869186889 |
| 0 | 0 | 0 | 0 | 0 |
| 0 | 0 | 0 | 0 | 0 |
| 0 | 0 | 0 | 0 | 0 |
| 0 | 0 | 0 | 0 | 0 |
| 0 | 0 | 0 | 0 | 0 |
| 0 | 0 | 0 | 0 | 0 |
| 0 | 0 | 0 | 0 | 0 |
| 0 | 0 | 0 | 0 | 0 |
| 0 | 0 | 0 | 0 | 0 |
| 0 | 0 | 0 | 0 | 0 |
| 6.709097343 | 2.085869423 | 3.237456616 | 0.152034249 | 3.441999872 |
| 0 | 0 | 0 | 0 | 0 |
| 0 | 0 | 0 | 0 | 0 |
| 0 | 0 | 0 | 0 | 0 |
| 0 | 0 | 0 | 0 | 0 |
| 0 | 0 | 0 | 0 | 0 |
| 0 | 0 | 0 | 0 | 0 |
| 0 | 0 | 0 | 0 | 0 |
| 0 | 0 | 0 | 0 | 0 |
| 0 | 0 | 0 | 0 | 0 |
| 0 | 0 | 0 | 0 | 0 |
| 0 | 0 | 0.006199603 | 0 | 0 |
| 0 | 0 | 0 | 0 | 0 |
| 0 | 0 | 0 | 0 | 0 |
| 0 | 0 | 0 | 0 | 0 |
| 0 | 0 | 0 | 0 | 0 |
| 0 | 0 | 0 | 0 | 0 |
| 0 | 0 | 0 | 0 | 0 |
| 0 | 0 | 0 | 0 | 0 |
| 0 | 0 | 0 | 0 | 0 |
| 0 | 0 | 0 | 0 | 0 |
| 0 | 0 | 0 | 0 | 0 |
| 7.481678571 | 2.905377651 | 6.490671426 | 0.110414156 | 0.00302362 |
| 0 | 0 | 0 | 0 | 0 |
| 0 | 0 | 0 | 0 | 0 |
| 0 | 0 | 0 | 0 | 0 |
| 0 | 0 | 0 | 0 | 0 |
| 0 | 0 | 0 | 0 | 0 |
| 0.001235327 | 0.003104921 | 0.002881736 | 0.075039163 | 0 |
| 0.185908539 | 0.04122972 | 0.040695673 | 8.351686939 | 0.077696617 |
| 0.006414492 | 0.004751879 | 0.049353453 | 0.130003511 | 0.104621611 |
| 0 | 0 | 0.001514132 | 0.083431069 | 0.004244058 |
| 0 | 0 | 0 | 0 | 0 |

| | | | | |
|---|---|---|---|---|
| 0.002050446 | 0.000951448 | 0.002698228 | 0 | 0 |
| 0 | 0.022691429 | 0 | 0 | 0.20261943 |
| 0 | 0 | 0 | 0 | 0 |
| 1.273538122 | 0.226557249 | 0.573892425 | 0.301024213 | 0.092181454 |
| 0.001448514 | 0.004854341 | 0.021776126 | 0 | 0 |
| 0 | 0 | 0 | 0 | 0 |
| 0.013026185 | 0.098455508 | 0 | 0.003784762 | 0 |
| 0 | 0 | 0 | 0 | 0 |
| 0 | 0 | 0 | 0 | 0 |
| 0 | 0 | 0 | 0 | 0 |
| 0 | 0 | 0.012962803 | 0.005795773 | 0 |
| 0.051217643 | 0.467398099 | 0.339034673 | 0.581210199 | 0 |
| 0 | 0 | 0 | 0 | 0 |
| 0.006330355 | 0.088766487 | 0.020985075 | 0.001807018 | 0.001848453 |
| 0 | 0 | 0 | 0 | 0 |
| 0 | 0 | 0 | 0 | 0 |
| 1.642015523 | 0.000688808 | 0 | 1.128874028 | 0.134303792 |
| 0 | 0 | 0 | 0 | 0 |
| 0 | 0 | 0 | 0 | 0 |
| 0 | 0 | 0 | 0 | 0 |
| 0 | 0.047253378 | 0.343024404 | 0.00801974 | 0.182768016 |
| 0.002809715 | 0.069623546 | 0.010795525 | 0.000717119 | 0 |
| 0.001112267 | 0 | 0 | 0 | 0 |
| 0 | 0 | 0 | 0 | 0 |
| 0 | 0 | 0 | 0 | 0 |
| 0 | 0 | 0 | 0 | 0 |
| 0 | 0 | 0 | 0 | 0.002815393 |
| 0 | 0 | 0 | 0 | 0 |
| 0 | 0.005952082 | 0 | 0.00298156 | 0 |
| 0 | 0 | 0 | 0 | 0 |
| 0 | 0.000753138 | 0 | 0 | 0.111322483 |
| 0 | 0 | 0 | 0 | 0 |
| 0 | 0 | 0 | 0 | 0 |
| 0 | 0 | 0 | 0 | 0 |
| 0 | 0 | 0 | 0 | 0 |
| 0 | 0 | 0 | 0 | 0 |
| 0 | 0 | 0 | 0 | 0 |
| 0.038880743 | 0 | 0 | 0.028478834 | 9.578567075 |
| 0 | 0 | 0 | 0 | 0 |
| 0 | 0 | 0 | 0 | 0 |
| 0 | 0 | 0 | 0 | 0 |
| 0 | 0 | 0 | 0 | 0 |
| 0 | 0 | 0 | 0 | 0 |
| 0 | 0 | 0 | 0 | 0 |
| 0 | 0 | 0 | 0 | 0 |
| 0 | 0 | 0 | 0 | 0 |
| 0 | 0.001232627 | 0 | 0 | 0.00194343 |
| 0 | 0 | 0 | 0 | 0 |

| | | | | |
|---|---|---|---|---|
| 0 | 0 | 0 | 0 | 0 |
| 0 | 0 | 0 | 0 | 0 |
| 0 | 0 | 0 | 0 | 0 |
| 0 | 0 | 0 | 0 | 0 |
| 0.002257545 | 0.002458823 | 0 | 0 | 0 |
| 0 | 0 | 0 | 0 | 0 |
| 0 | 0 | 0 | 0 | 0 |
| 0 | 0 | 0 | 0 | 0 |
| 0 | 0 | 0 | 0.00086644 | 0 |
| 0 | 0 | 0 | 0 | 0 |
| 0 | 0 | 0 | 0 | 0 |
| 0 | 0 | 0 | 0 | 0 |
| 0 | 0 | 0 | 0 | 0 |
| 0 | 0 | 0 | 0 | 0 |
| 0 | 0 | 0 | 0 | 0 |
| 0 | 0 | 0 | 0 | 0 |
| 0 | 0 | 0 | 0 | 0 |
| 0 | 0 | 0 | 0 | 0 |
| 0.022283134 | 0.190673893 | 0.046719151 | 0.046909616 | 0 |
| 0 | 0 | 0 | 0 | 0 |
| 0 | 0 | 0 | 0 | 0 |
| 0 | 0 | 0 | 0 | 0 |
| 0 | 0 | 0 | 0 | 0 |
| 0 | 0 | 0 | 0 | 0 |
| 0 | 0 | 0 | 0 | 0 |
| 0 | 0 | 0 | 0 | 0 |
| 0 | 0 | 0 | 0 | 0 |
| 0 | 0 | 0 | 0 | 0 |
| 0 | 0 | 0 | 0 | 0 |
| 0 | 0 | 0 | 0 | 0 |
| 0.024244328 | 0.503693754 | 0.411988744 | 0.039768212 | 0 |
| 0.026209851 | 0.001403295 | 0 | 0.002018725 | 0 |
| 0 | 0 | 0 | 0 | 0 |
| 0 | 0 | 0 | 0 | 0 |
| 0 | 0 | 0 | 0 | 0 |
| 0 | 0 | 0 | 0 | 0 |
| 0 | 0 | 0 | 0 | 0 |
| 0 | 0 | 0 | 0 | 0 |
| 0 | 0 | 0 | 0 | 0 |
| 0 | 0 | 0 | 0 | 0 |
| 0 | 0 | 0 | 0 | 0 |
| 0 | 0 | 0 | 0 | 0 |
| 0 | 0 | 0 | 0 | 0 |
| 0 | 0 | 0 | 0 | 0 |
| 0 | 0 | 0 | 0 | 0 |
| 0 | 0 | 0 | 0 | 0 |
| 0 | 0 | 0 | 0 | 0 |
| 0 | 0 | 0 | 0 | 0 |

| | | | | |
|---|---|---|---|---|
| 0 | 0 | 0 | 0 | 0 |
| 0 | 0 | 0 | 0 | 0 |
| 0 | 0 | 0 | 0 | 0 |
| 0 | 0 | 0 | 0 | 0 |
| 0.011743821 | 0.028906696 | 0 | 0 | 0 |
| 0 | 0 | 0.002308989 | 0 | 0 |
| 0 | 0 | 0 | 0 | 0 |
| 0 | 0 | 0 | 0 | 0 |
| 0 | 0 | 0 | 0 | 0 |
| 0 | 0 | 0 | 0 | 0 |
| 0 | 0 | 0 | 0 | 0 |
| 0 | 0 | 0 | 0 | 0 |
| 0 | 0 | 0 | 0 | 0 |
| 0 | 0 | 0 | 0 | 0 |
| 0 | 0 | 0 | 0 | 0 |
| 11.35386036 | 0 | 2.91894715 | 0 | 0 |
| 0 | 0 | 0 | 0 | 0 |
| 0 | 0 | 0 | 0 | 0 |
| 0 | 0 | 0 | 0 | 0 |
| 0 | 0 | 0 | 0 | 0 |
| 0 | 0 | 0 | 0 | 0 |
| 0 | 0 | 0 | 0 | 0 |
| 0 | 0 | 0 | 0 | 0 |
| 0 | 0 | 0 | 0 | 0 |
| 0 | 0 | 0.203624729 | 0 | 0 |
| 0 | 0 | 0 | 0 | 0 |
| 0.003537806 | 0 | 0 | 0 | 0 |
| 0 | 0 | 0 | 0 | 0 |
| 0 | 0 | 0 | 0 | 0 |
| 0 | 0 | 0 | 0.009192412 | 0 |
| 0 | 0.122212922 | 0 | 0 | 0 |
| 0 | 0 | 0 | 0 | 0 |
| 0 | 0 | 0 | 0 | 0 |
| 0 | 0 | 0 | 0 | 0 |
| 0 | 0 | 0 | 0 | 0 |
| 0 | 0 | 0 | 0 | 0 |
| 0 | 0 | 0 | 0 | 0 |
| 0 | 0 | 0 | 0 | 0 |
| 0 | 0 | 0 | 0.009429174 | 0 |
| 0 | 0 | 0 | 0 | 0 |
| 0 | 0 | 0 | 0 | 0 |
| 0 | 0 | 0 | 0 | 0 |
| 0 | 0 | 0 | 0 | 0 |
| 0 | 0 | 0 | 0 | 0 |
| 0 | 0 | 0 | 0 | 0 |
| 0 | 0 | 0 | 0 | 0 |
| 0 | 0 | 0 | 0 | 0 |
| 0 | 0 | 0 | 0 | 0 |

| | | | | |
|---|---|---|---|---|
| 0 | 0 | 0 | 0 | 0 |
| 0 | 0 | 0 | 0 | 0 |
| 0 | 0 | 0 | 0 | 0 |
| 0 | 0 | 0 | 0 | 0 |
| 0 | 0 | 0 | 0 | 0 |
| 0 | 0 | 0 | 0 | 0 |
| 0 | 0 | 0 | 0 | 0 |
| 0 | 0 | 0 | 0 | 0 |
| 0 | 0 | 0 | 0 | 0 |
| 0 | 0 | 0 | 0 | 0 |
| 0 | 0 | 0 | 0 | 0 |
| 0 | 0 | 0 | 0 | 0 |
| 0 | 0 | 0 | 0.001671593 | 0 |
| 0 | 0 | 0 | 0 | 0 |
| 0 | 0 | 0 | 0.000604636 | 0 |
| 0 | 0 | 0 | 0 | 0 |
| 0 | 0 | 0 | 0 | 0 |
| 0 | 0 | 0 | 0 | 0 |
| 0 | 0 | 0 | 0 | 0 |
| 0 | 0 | 0 | 0 | 0 |
| 0.029774099 | 0.089158785 | 0.126465027 | 0.023961988 | 0.03806124 |
| 0 | 0 | 0 | 0 | 0 |
| 0 | 0 | 0 | 0 | 0 |
| 0 | 0.001678243 | 0 | 0 | 0 |
| 0 | 0 | 0 | 0 | 0 |
| 0 | 0 | 0 | 0 | 0 |
| 0.007271501 | 0 | 0.020297313 | 0.005258367 | 0.021574877 |
| 0 | 0 | 0 | 0 | 0 |
| 0 | 0 | 0 | 0 | 0 |
| 0 | 0 | 0.001551128 | 0 | 0 |
| 0 | 0 | 0 | 0 | 0.004124532 |
| 0 | 0 | 0 | 0 | 0 |
| 0 | 0 | 0 | 0 | 0 |
| 0.002543538 | 0.005114426 | 0.011258425 | 0 | 0.006202844 |
| 0.005207692 | 0.010471382 | 0.008746819 | 0.002485513 | 0.016179585 |
| 0 | 0 | 0 | 0 | 0 |
| 0 | 0 | 0 | 0 | 0 |
| 0 | 0 | 0 | 0 | 0 |
| 0 | 0 | 0 | 0 | 0 |
| 0 | 0 | 0 | 0 | 0 |
| 0 | 0 | 0 | 0 | 0 |
| 0 | 0 | 0 | 0 | 0 |
| 0 | 0 | 0 | 0 | 0 |
| 0 | 0 | 0 | 0 | 0 |
| 0 | 0 | 0 | 0 | 0 |
| 0 | 0 | 0 | 0 | 0 |
| 0.000902167 | 0 | 0 | 0 | 0 |
| 0 | 0.000447731 | 0 | 0.017712439 | 0 |

| | | | | |
|---|---|---|---|---|
| 0 | 0 | 0 | 0 | 0 |
| 0 | 0 | 0 | 0 | 0 |
| 0 | 0 | 0 | 0 | 0 |
| 0 | 0 | 0 | 0 | 0 |
| 0 | 0 | 0 | 0 | 0 |
| 0 | 0 | 0 | 0 | 0 |
| 0 | 0 | 0 | 0 | 0 |
| 0 | 0 | 0 | 0 | 0 |
| 0 | 0.007940555 | 0 | 0 | 0 |
| 0 | 0 | 0 | 0 | 0 |
| 0 | 0 | 0 | 0 | 0 |
| 0 | 0 | 0 | 0 | 0 |
| 0 | 0 | 0 | 0 | 0 |
| 0 | 0 | 0 | 0 | 0 |
| 0 | 0 | 0 | 0.001523272 | 0 |
| 0 | 0 | 0 | 0.002770199 | 0 |
| 0 | 0 | 0 | 0 | 0 |
| 0 | 0 | 0 | 0.053904013 | 0 |
| 0 | 0 | 0 | 0.048800029 | 0 |
| 0 | 0.043831081 | 0 | 0 | 0 |
| 0 | 0 | 0 | 0 | 0 |
| 0 | 0 | 0 | 0 | 0 |
| 0 | 0 | 0 | 0 | 0 |
| 0 | 0 | 0 | 0 | 0 |
| 0.001769712 | 0.005782486 | 0.013073057 | 0.017756702 | 0.004909153 |
| 0 | 0 | 0 | 0.002095645 | 0 |
| 0 | 0 | 0 | 0 | 0 |
| 0 | 0 | 0 | 0 | 0 |
| 0 | 0 | 0 | 0.039386789 | 0 |
| 0 | 0 | 0 | 0 | 0 |
| 0.061404941 | 0.44992527 | 0.185389222 | 0.060035321 | 0.006229444 |
| 0 | 0 | 0 | 0 | 0 |
| 0 | 0 | 0 | 0 | 0 |
| 0 | 0.002768875 | 0 | 0 | 0 |
| 0 | 0 | 0 | 0 | 0 |
| 0 | 0 | 0 | 0 | 0 |
| 0 | 0 | 0 | 0 | 0 |
| 0 | 0 | 0 | 0 | 0 |
| 0 | 0 | 0 | 0 | 0 |
| 0 | 0 | 0 | 0.004790258 | 0 |
| 0 | 0 | 0 | 0 | 0 |
| 0.014688286 | 0.003020575 | 0.005451158 | 0.059566547 | 0.003174941 |
| 0 | 0.000844791 | 0 | 0 | 0 |
| 0 | 0 | 0 | 0 | 0 |
| 0 | 0 | 0 | 0 | 0 |
| 0 | 0 | 0 | 0 | 0 |
| 0 | 0 | 0 | 0 | 0 |
| 0 | 0 | 0 | 0 | 0 |

| | | | | |
|---|---|---|---|---|
| 0 | 0 | 0 | 0 | 0 |
| 0 | 0 | 0 | 0 | 0 |
| 0.003353375 | 0.001053564 | 0.040743013 | 0.143452963 | 0.082723219 |
| 0 | 0 | 0 | 0 | 0 |
| 0 | 12.00418062 | 0.072695868 | 0 | 0 |
| 0 | 0 | 0 | 0 | 0 |
| 0 | 0 | 0 | 0 | 0 |
| 0 | 0 | 0 | 0 | 0 |
| 0 | 0 | 0 | 0 | 0 |
| 0 | 0 | 0 | 0 | 0 |
| 0 | 0 | 0 | 0 | 0 |
| 0 | 0 | 0 | 0 | 0 |
| 0 | 0 | 0 | 0 | 0 |
| 0 | 0 | 0 | 0.001256271 | 0 |
| 0.001338263 | 0.005189625 | 0.021109741 | 0.005391815 | 0 |
| 0.032749429 | 0 | 0 | 0 | 0 |
| 0 | 0.001971037 | 0.006097857 | 0.007088641 | 0.003107651 |
| 0 | 0 | 0 | 0.001466341 | 0 |
| 0 | 0 | 0 | 0 | 0 |
| 0 | 0 | 0 | 0 | 0 |
| 0 | 0 | 0 | 0 | 0 |
| 0 | 0.000790793 | 0.001834875 | 0.012371439 | 0 |
| 0 | 0 | 0 | 0 | 0 |
| 0 | 0 | 0 | 0 | 0 |
| 0 | 0 | 0 | 0.004446197 | 0 |
| 0 | 0 | 0 | 0 | 0 |
| 0.963391226 | 0 | 0 | 0 | 0 |
| 0 | 0 | 0 | 0 | 0 |
| 0 | 0 | 0 | 0 | 0 |
| 0 | 0 | 0 | 0 | 0 |
| 0 | 0 | 0 | 0 | 0 |
| 0 | 0 | 0 | 0 | 0 |
| 0 | 0 | 0 | 0 | 0 |
| 0.000906906 | 0 | 0 | 0 | 0 |
| 0 | 0 | 0 | 0 | 0 |
| 0 | 0 | 0 | 0 | 0 |
| 0 | 0 | 0 | 0 | 0 |
| 0.000741177 | 0.000745162 | 0.008837099 | 3.078939564 | 0.002349731 |
| 0 | 0 | 0 | 0 | 0 |
| 0 | 0 | 0 | 0 | 0 |
| 0 | 0 | 0 | 0 | 0 |
| 0 | 0 | 0 | 0 | 0 |
| 0 | 0 | 0 | 0 | 0 |
| 0 | 0 | 0 | 0 | 0.003891644 |
| 0 | 0 | 0 | 0 | 0 |
| 0 | 0 | 0 | 0 | 0 |
| 0 | 0 | 0 | 0 | 0 |
| 0 | 0.003911173 | 0 | 0 | 0 |

| | | | | |
|---|---|---|---|---|
| 0 | 0 | 0.001534036 | 0 | 0 |
| 0 | 0.003249793 | 0.017446614 | 0 | 0 |
| 0 | 0 | 0 | 0.004468317 | 0 |
| 0 | 0 | 0 | 0 | 0 |
| 0.005802211 | 0.003541711 | 0.011279363 | 0.004046002 | 0.00525559 |
| 0 | 0 | 0 | 0.002199124 | 0 |
| 0 | 0 | 0 | 0 | 0 |
| 0.029918099 | 0.009052888 | 0.09666997 | 0.003150757 | 0.038215615 |
| 0 | 0 | 0 | 0 | 0 |
| 0.017364655 | 0 | 0.091724519 | 0.008768393 | 0.005220317 |
| 0 | 0 | 0 | 0 | 0 |
| 0 | 0 | 0 | 0.040405233 | 0 |
| 0 | 0.044714595 | 0.014104731 | 0.055432712 | 0.006358787 |
| 0 | 0 | 0.038311437 | 0 | 0 |
| 0 | 0 | 0 | 0 | 0 |
| 0 | 0 | 0 | 0 | 0 |
| 0 | 0 | 0 | 0 | 0 |
| 0 | 0 | 0 | 0 | 0 |
| 0 | 0 | 0 | 0 | 0 |
| 0 | 0 | 0 | 0 | 0 |
| 0 | 0.004662179 | 0 | 0 | 0 |
| 0 | 0 | 0 | 0.012506804 | 0 |
| 0 | 0 | 0 | 0 | 0 |
| 0 | 0 | 0 | 0 | 0 |
| 0 | 0 | 0 | 0 | 0 |
| 0 | 0 | 0.001579655 | 0.014543647 | 0.025117239 |
| 0 | 0 | 0.653974107 | 0.046851528 | 0 |
| 0 | 0.000740947 | 0 | 0 | 0 |
| 0 | 0 | 0 | 0.002256151 | 0 |
| 0 | 0 | 0 | 0 | 0 |
| 0 | 0 | 0 | 0 | 0 |
| 0 | 0 | 0 | 0 | 0 |
| 0 | 0 | 0 | 0 | 0 |
| 0 | 0 | 0 | 0 | 0 |
| 0 | 0 | 0 | 0 | 0 |
| 0 | 0 | 0 | 0 | 0 |
| 0 | 0.053019576 | 0 | 0 | 0.196755479 |
| 0 | 0 | 0 | 0 | 0 |
| 0 | 0 | 0 | 0 | 0 |
| 0 | 0 | 0.001661393 | 0 | 0 |
| 0 | 0 | 0.227497195 | 0 | 0 |
| 0 | 0.106019861 | 0 | 0 | 0 |
| 0 | 0 | 0 | 0 | 0 |
| 0 | 0.044426512 | 0 | 0 | 0 |
| 0 | 0.027320781 | 0 | 0 | 0 |
| 0 | 0 | 0 | 0 | 0 |
| 0.002273669 | 0.003301845 | 0.001178656 | 0 | 0 |
| 0 | 0 | 0 | 0 | 0 |

| | | | | |
|---|---|---|---|---|
| 0 | 0 | 0 | 0 | 0 |
| 0.004234973 | 0 | 0 | 0 | 0 |
| 0 | 0 | 0 | 0 | 0 |
| 0 | 0 | 0 | 0.00265433 | 0 |
| 0 | 0 | 0 | 0 | 0 |
| 0.022086404 | 0.325275444 | 0.032491696 | 0 | 0 |
| 0 | 0 | 0 | 0 | 0 |
| 0.002316164 | 0 | 0 | 0 | 0 |
| 0 | 0 | 0 | 0 | 0 |
| 0 | 0 | 0 | 0 | 0 |
| 0.011538969 | 0 | 0.019739708 | 0.007009275 | 0 |
| 0 | 0 | 0 | 0 | 0 |
| 0 | 0 | 0 | 0 | 0 |
| 0.001526201 | 0 | 0.0566254 | 0.020339033 | 0.003110443 |
| 0 | 0 | 0.013380206 | 0.02034015 | 0 |
| 0 | 0 | 0 | 0 | 0 |
| 0 | 0 | 0 | 0 | 0 |
| 0 | 0.000639954 | 0.01319896 | 0 | 0 |
| 0 | 0 | 0 | 0 | 0 |
| 0 | 0.100683405 | 0 | 0 | 0 |
| 0 | 0 | 0 | 0 | 0 |
| 0 | 0 | 0.218515317 | 0.256693752 | 0 |
| 0 | 0 | 0 | 0.004488551 | 0 |
| 0 | 0 | 0 | 0 | 0 |
| 0 | 0 | 0 | 0 | 0 |
| 0 | 0 | 0 | 0 | 0 |
| 0 | 0.04800972 | 0 | 0 | 0.00262309 |
| 0 | 0 | 0 | 0 | 0 |
| 0 | 0 | 0 | 0 | 0 |
| 1.225926056 | 0.916997912 | 0.858836442 | 1.233763023 | 3.750243131 |
| 0.000443746 | 0 | 0 | 0.012354397 | 0.001641258 |
| 0 | 0 | 0 | 0 | 0 |
| 0 | 0 | 0 | 0 | 0 |
| 0 | 0 | 0 | 0 | 0 |
| 0 | 0.003365291 | 0 | 0 | 0 |
| 0 | 0 | 0 | 0 | 0 |
| 0 | 0 | 0 | 1.397583468 | 0 |
| 0 | 0 | 0 | 0 | 0 |
| 0 | 0 | 0.028584979 | 0.013628297 | 0 |
| 0 | 0 | 0.001604846 | 0 | 0 |
| 0 | 0.066856813 | 0.03902583 | 0 | 0 |
| 9.175724092 | 7.059597046 | 6.951075638 | 0 | 3.30447048 |
| 0 | 0 | 0 | 0 | 0 |
| 0 | 0 | 0 | 0 | 0 |
| 0 | 0 | 0 | 0 | 0 |
| 0 | 0 | 0 | 0 | 0 |
| 0.00150503 | 0.001134841 | 0.001755446 | 0 | 0 |
| 0 | 0 | 0 | 0 | 0 |

| | | | | |
|---|---|---|---|---|
| 0 | 0 | 0.001082963 | 0.069492585 | 0.002943522 |
| 0 | 0 | 0 | 0 | 0 |
| 3.716294266 | 0.003179808 | 0 | 2.51886385 | 0 |
| 0.001679739 | 0.001477674 | 0 | 0 | 0 |
| 0 | 0.014857426 | 0.222566684 | 0 | 0 |
| 0 | 0 | 0.092175043 | 0 | 0 |
| 0 | 0 | 0 | 0 | 0 |
| 0 | 0 | 0 | 0 | 0 |
| 0 | 0.024608745 | 0 | 0 | 0 |
| 0 | 0 | 0 | 0 | 0 |
| 0 | 0 | 0 | 0 | 0 |
| 0 | 0 | 0 | 0 | 0 |
| 0 | 0 | 0 | 0 | 0 |
| 0 | 0 | 0.038536017 | 0 | 0 |
| 0 | 0 | 0 | 0 | 0 |
| 0.091242663 | 0.001154915 | 0.010208553 | 0 | 0 |
| 0 | 0.000548615 | 0.037905642 | 0.984350111 | 0.004324892 |
| 0 | 0 | 0 | 0 | 0 |
| 0 | 0 | 0 | 0 | 0 |
| 0 | 0 | 0 | 0 | 0 |
| 0 | 0.043789392 | 0.005704107 | 0 | 0 |
| 0 | 0 | 0.001640618 | 0.00091545 | 0.00200666 |
| 0 | 0 | 0 | 0 | 0 |
| 0 | 0.815544604 | 0.034973309 | 0 | 0 |
| 0.018529715 | 0 | 0 | 0 | 0 |
| 0 | 0 | 0 | 0 | 0 |
| 0 | 0.006036087 | 0 | 0 | 0 |
| 0 | 0.005436005 | 0 | 0 | 0 |
| 0 | 0 | 0 | 0 | 0 |
| 0 | 0 | 0 | 0 | 0 |
| 0.013170805 | 0.018538264 | 0.026764442 | 0.001523909 | 0 |
| 0.001832698 | 0.005264434 | 0.010179209 | 0.031807466 | 0.017430436 |
| 0.075781845 | 0.01428549 | 0.176006413 | 0.055973185 | 0.463111109 |
| 0 | 0.003591667 | 0 | 0.005425171 | 0 |
| 0.001529896 | 0 | 0 | 0 | 0 |
| 0 | 0 | 0 | 0 | 0 |
| 0 | 0 | 0.004205866 | 0.001711233 | 0 |
| 0.014276135 | 0.020554756 | 0.206944378 | 0.012809097 | 0.005028797 |
| 0.04503984 | 0.155064501 | 0.225203207 | 0 | 0 |
| 0 | 0 | 0 | 0 | 0 |
| 0 | 0 | 0 | 0 | 0 |
| 0.001034312 | 0 | 0.002573668 | 0 | 0 |
| 0 | 0 | 0 | 0 | 0 |
| 0 | 0.045370092 | 0.346413663 | 0.074172273 | 0 |
| 0 | 0 | 0.647749705 | 0 | 0 |
| 0 | 0.002897374 | 0 | 0 | 0 |
| 0 | 0 | 0 | 0 | 0 |
| 0 | 0 | 0 | 0 | 0 |

| | | | | |
|---|---|---|---|---|
| 0 | 0 | 0 | 0 | 0.013255178 |
| 0.072288477 | 0.246039854 | 0.689469918 | 2.726995211 | 1.759918141 |
| 0.008481401 | 0 | 0 | 0 | 0 |
| 0 | 0 | 0 | 0 | 0 |
| 0 | 0 | 0 | 0 | 0 |
| 0 | 0 | 0 | 0 | 0 |
| 0 | 0 | 0 | 0 | 0 |
| 0 | 0 | 0 | 0 | 0 |
| 0 | 0 | 0 | 0 | 0 |
| 0 | 0.002105504 | 0 | 0 | 0 |
| 0.003747784 | 0.005203337 | 0 | 0 | 0.00113157 |
| 0.005582431 | 0.008819556 | 0.241433744 | 0.071222811 | 0.018119179 |
| 0.076975746 | 0 | 0 | 0 | 0 |
| 0 | 0 | 0.004465559 | 0 | 0 |
| 0 | 0 | 0.012419563 | 0 | 0 |
| 0 | 0 | 0 | 0 | 0 |
| 0.012147173 | 0.011460944 | 0.038654008 | 0.053718738 | 0.020439814 |
| 0 | 0.009512092 | 0.037728002 | 0 | 0.00423001 |
| 0 | 0 | 0 | 0 | 0 |
| 0 | 0.012986815 | 0 | 0.002904873 | 0 |
| 0.265182707 | 0.018019321 | 0.027873452 | 0.063968516 | 0.061402891 |
| 0 | 0 | 0 | 0 | 0 |
| 0 | 0 | 0 | 0 | 0 |
| 0 | 0 | 0 | 0 | 0 |
| 0 | 0.005859836 | 0 | 0 | 0 |
| 0 | 0.003198237 | 0 | 0 | 0 |
| 0 | 0.876293705 | 0 | 0.008666291 | 0 |
| 0.173698299 | 0.061574317 | 0.235728711 | 0.100650515 | 0.031494698 |
| 0 | 0 | 0 | 0.001299673 | 0 |
| 0 | 0 | 0 | 0 | 0 |
| 0.000879143 | 0 | 0 | 0 | 0.00209034 |
| 0 | 0 | 0 | 0 | 0 |
| 0 | 0 | 0 | 0 | 0 |
| 0 | 0 | 0 | 0 | 0 |
| 0.065151629 | 0 | 0.01304702 | 0 | 0.006507681 |
| 0.206960475 | 0.380247645 | 0.251234946 | 0.045500539 | 1.380380181 |
| 0 | 0 | 0 | 0 | 0 |
| 0 | 0 | 0 | 0 | 0 |
| 0 | 0.0053349 | 0.00157188 | 0.02494237 | 0 |
| 0 | 0 | 0 | 0 | 0 |
| 0 | 0 | 0.635217527 | 0 | 0 |
| 0 | 0 | 0 | 0 | 0 |
| 0 | 0.000925857 | 0.009547837 | 0 | 0.024815905 |
| 0 | 0 | 0 | 0.017359995 | 0.000704685 |
| 0 | 0.020355817 | 0 | 0 | 0 |
| 0.229813449 | 0.011390825 | 0.188344213 | 0 | 0 |
| 0 | 0 | 0 | 0 | 0 |
| 0 | 0 | 0 | 0 | 0 |

| | | | | |
|---|---|---|---|---|
| 0 | 0 | 0 | 0 | 0 |
| 0 | 0 | 0 | 0 | 0 |
| 0 | 0 | 0.003281248 | 0 | 0 |
| 0.068107804 | 0.99458008 | 1.33998516 | 0 | 0 |
| 0 | 0 | 0 | 0 | 0 |
| 0 | 0 | 0 | 0 | 0 |
| 0 | 0.020482136 | 0 | 0 | 0 |
| 0.008315869 | 0 | 0 | 0 | 0 |
| 0.020076423 | 1.330615274 | 0.397786392 | 17.49514449 | 0 |
| 0.066783045 | 0 | 0.440285821 | 0 | 0 |
| 0 | 0 | 0 | 0 | 0 |
| 0 | 0.003616048 | 0 | 0.007617072 | 0 |
| 0 | 0 | 0 | 0 | 0 |
| 0.01180142 | 0 | 0 | 0 | 0 |
| 0.065578983 | 0.130158001 | 0.045718406 | 0 | 0 |
| 0 | 0 | 0 | 0 | 0 |
| 0.146380124 | 0.17719367 | 0.097481359 | 0.005599351 | 0.198717685 |
| 0 | 0 | 0 | 0 | 0 |
| 0 | 0 | 0 | 0 | 0 |
| 0 | 0 | 0 | 0 | 0 |
| 0 | 0 | 0 | 0 | 0 |
| 0 | 0 | 0 | 0 | 0 |
| 0 | 0 | 0 | 0 | 0 |
| 0 | 1.105385001 | 0.167261773 | 0.010780267 | 0 |
| 0 | 0 | 0 | 0 | 0 |
| 0 | 0 | 0.852766327 | 0 | 0 |
| 0 | 0 | 0 | 0 | 0 |
| 0.025115159 | 0.389844044 | 0.401902333 | 1.644506252 | 0 |
| 0 | 0 | 0 | 0 | 0 |
| 0 | 0 | 0 | 0 | 0 |
| 0.002830593 | 0.002845812 | 0.01035785 | 0.012101006 | 0 |
| 0 | 0 | 0.028545122 | 0 | 0 |
| 0.008363712 | 0.000509617 | 0.03547388 | 0.083849998 | 0 |
| 0 | 0 | 0 | 0 | 0 |
| 0 | 0 | 0 | 0 | 0 |
| 0.092612796 | 0 | 0.030518488 | 0 | 0 |
| 0.234843885 | 0 | 0.801056031 | 0 | 0 |
| 0 | 0 | 0 | 0 | 0 |
| 0 | 0.002509537 | 0 | 0 | 0 |
| 0 | 0 | 0 | 0 | 0 |
| 0 | 0.032046502 | 0.171456427 | 0.028159528 | 0.004167122 |
| 0 | 0.08220747 | 0 | 0 | 0 |
| 18.69627652 | 0.04399229 | 0.036477318 | 0.006200275 | 0.259321774 |
| 0 | 0.057560189 | 0 | 0 | 0 |
| 0 | 0 | 0 | 0 | 0 |
| 0 | 0 | 0 | 0 | 0 |
| 0 | 0 | 0 | 0 | 0 |
| 0 | 0.047856602 | 0 | 0.00064274 | 0 |

| | | | | |
|---|---|---|---|---|
| 0 | 0 | 0 | 0 | 0 |
| 0.014395209 | 0.094795557 | 0 | 0.125894221 | 0 |
| 0.00870958 | 0.00913712 | 0.010600417 | 0.002259176 | 0.482004308 |
| 0 | 0 | 0 | 0 | 0 |
| 0 | 0 | 0 | 0 | 0 |
| 0 | 0.001949423 | 0 | 0 | 0 |
| 0 | 0 | 0.024946436 | 0 | 0 |
| 0 | 0 | 0 | 0 | 0 |
| 0 | 0 | 0 | 0 | 0 |
| 0 | 0 | 0 | 0 | 0 |
| 0.056884489 | 0.140172364 | 0.00722759 | 0.083279972 | 0.016796287 |
| 1.847931998 | 1.611292262 | 2.256854136 | 2.22727234 | 1.669306277 |
| 0 | 0 | 0 | 0 | 0 |
| 0.004012553 | 0 | 0.029641176 | 2.128155246 | 0.075371146 |
| 0.14782563 | 0.303916806 | 0.101793376 | 0.003258441 | 0.376671799 |
| 0.025131345 | 0.628216112 | 0 | 0 | 0 |
| 0 | 0 | 0 | 0 | 0 |
| 0 | 0.004059952 | 0 | 0 | 0 |
| 0 | 0 | 0.007501985 | 0 | 0 |
| 0 | 0.033100006 | 0.344358056 | 0.002657657 | 0 |
| 0 | 0.315643256 | 0.095955307 | 0.097349412 | 0.011114023 |
| 0 | 0 | 0 | 0.007479506 | 0.005985485 |
| 0.899180913 | 0.904607126 | 4.441889005 | 0.474111406 | 0.504811365 |
| 0 | 0 | 0 | 0 | 0 |
| 0 | 0 | 0 | 0 | 0 |
| 5.559171576 | 0.003941906 | 0.013356639 | 1.048061449 | 1.639585506 |
| 0 | 0 | 0 | 0 | 0 |
| 0 | 0 | 0 | 0 | 0 |
| 0 | 0 | 0 | 0 | 0 |
| 0 | 0 | 0 | 0 | 0 |
| 0 | 0 | 0 | 0 | 0 |
| 0.252563343 | 0.564926681 | 0.564123438 | 0 | 0 |
| 0 | 0 | 0.017518098 | 0.009733518 | 0.046303234 |
| 0.04484237 | 0.018735984 | 0.018566615 | 0 | 0 |
| 0 | 0.009501583 | 0.000979844 | 0 | 0 |
| 0.003289478 | 0.006440266 | 0.006192732 | 0.099157465 | 0.005214264 |
| 0.003177591 | 0.004371661 | 0 | 0 | 0 |
| 0 | 0 | 0 | 0 | 0 |
| 0.025324696 | 0.001222121 | 0 | 1.968846435 | 0 |
| 0 | 0 | 0 | 0 | 0 |
| 0.010366604 | 0 | 0 | 0 | 0 |
| 0 | 0 | 0 | 0 | 0 |
| 0 | 0.040054814 | 0 | 0 | 0 |
| 0 | 0 | 0 | 0 | 0 |
| 0.019546125 | 0.002580462 | 0.007983253 | 0.007067365 | 0.001564809 |
| 0 | 0 | 0 | 0 | 0 |
| 0 | 0 | 0 | 0 | 0 |
| 0 | 0.024183617 | 0.072224881 | 2.386273414 | 0.007550351 |

| | | | | |
|---|---|---|---|---|
| 0 | 0 | 0 | 0 | 0 |
| 0 | 0 | 0 | 0 | 0 |
| 0 | 0 | 0 | 0 | 0 |
| 0.000673094 | 0.002368496 | 0 | 0 | 0 |
| 0 | 0 | 0 | 0 | 0 |
| 0.019023123 | 0 | 0 | 0 | 0 |
| 0 | 0 | 0 | 0 | 0 |
| 0.126196343 | 0.020237703 | 0.003130499 | 0 | 0.005399802 |
| 0 | 0 | 0 | 0 | 0 |
| 0 | 0 | 0 | 0 | 0 |
| 0 | 0.007159952 | 0 | 0.00223619 | 0 |
| 0 | 0 | 0 | 0 | 0 |
| 0 | 0 | 0 | 0 | 0 |
| 0.123941255 | 0.042929334 | 0 | 0 | 0 |
| 0 | 0 | 0 | 0 | 0 |
| 0 | 0 | 0 | 0 | 0 |
| 0.536633509 | 0 | 0 | 0 | 0 |
| 0 | 0.000627421 | 0 | 0 | 0 |
| 0 | 0.239987432 | 0 | 0 | 0 |
| 0 | 0.005092466 | 0.070896212 | 0 | 0 |
| 0 | 0 | 0 | 0 | 0 |
| 0 | 0 | 0 | 0 | 0 |
| 0 | 0.008318769 | 0 | 0 | 0 |
| 0 | 0 | 0 | 0 | 0 |
| 0 | 0 | 0 | 0 | 0 |
| 0 | 0 | 0 | 0 | 0 |
| 0 | 0 | 0 | 0 | 0 |
| 0 | 0 | 0 | 0 | 0 |
| 0 | 0 | 0 | 0 | 0 |
| 0.012250315 | 0 | 1.317188766 | 0 | 0.063333831 |
| 0 | 0 | 0 | 0 | 0 |
| 0 | 0 | 0 | 0 | 0 |
| 0 | 0 | 0 | 0 | 0 |
| 0 | 0 | 0 | 0 | 0 |
| 0.000745105 | 0 | 0 | 0 | 0 |
| 0 | 0 | 0 | 0 | 0 |
| 1.639092796 | 0.246113093 | 0.122935561 | 0 | 0 |
| 0 | 0.000957004 | 0.008635408 | 0.004302213 | 0.015591628 |
| 0 | 0 | 0 | 0 | 0 |
| 0 | 0.003303775 | 0 | 0 | 0 |
| 0 | 0.024393128 | 0.03148264 | 0.066037798 | 0 |
| 0 | 0 | 0 | 0 | 0 |
| 0 | 0 | 0 | 0 | 0 |
| 0.783270083 | 0.003252775 | 0.054149629 | 0 | 0 |
| 0 | 0.069184535 | 0 | 0 | 0 |
| 0 | 0 | 0 | 0 | 0 |
| 0 | 0.00712296 | 0 | 0 | 0 |
| 0 | 0 | 0 | 0 | 0 |

| | | | | |
|---|---|---|---|---|
| 0 | 0 | 0 | 0 | 0 |
| 0 | 0 | 0 | 0 | 0 |
| 0 | 0 | 0 | 0 | 0 |
| 0.011993405 | 0.12137215 | 0 | 0 | 0 |
| 0 | 0 | 0 | 0 | 0 |
| 0.070676874 | 0 | 0 | 0 | 0 |
| 0 | 0 | 0 | 0 | 0 |
| 0 | 0 | 0 | 0 | 0 |
| 0 | 0.013198683 | 0.023138788 | 0.002373388 | 0.00589611 |
| 0 | 0 | 0 | 0 | 0 |
| 0.0009387 | 0 | 0 | 0 | 0 |
| 0 | 0 | 0 | 0 | 0 |
| 0 | 0 | 0 | 0 | 0 |
| 0 | 0 | 0 | 0 | 0 |
| 0.190570829 | 0.245343311 | 0.201249444 | 0.593882675 | 0 |
| 0 | 0 | 0 | 0.04207243 | 0 |
| 0 | 0.008589019 | 0 | 0.016506608 | 0 |
| 0 | 0 | 0 | 0 | 0 |
| 0 | 0 | 0 | 0 | 0 |
| 0 | 0 | 0 | 0 | 0 |
| 0.080139218 | 0 | 0 | 0 | 0 |
| 0 | 0 | 0 | 0 | 0 |
| 0 | 0 | 0 | 0 | 0 |
| 0 | 0 | 0 | 0 | 0 |
| 0.014666359 | 0.004397695 | 0.002400931 | 0.372045179 | 0.038339046 |
| 0.9244872 | 1.357612843 | 3.978315122 | 0.08970579 | 0.942615277 |
| 0.140299203 | 0 | 0 | 0 | 0 |
| 0 | 0 | 0 | 0 | 0 |
| 0.009448225 | 0 | 0 | 0.393598471 | 0.07666646 |
| 0 | 0 | 0 | 0 | 0 |
| 0 | 0 | 0 | 0 | 0 |
| 0.026347182 | 0 | 0 | 0 | 0 |
| 0 | 0.008393083 | 0 | 0 | 0 |
| 0 | 0.000552624 | 0 | 0 | 0 |
| 0 | 0 | 0.024296573 | 0.252479472 | 0.020816555 |
| 0.001120361 | 0.000482736 | 0.00298691 | 0.001041668 | 0 |
| 0 | 0 | 0 | 0 | 0 |
| 0.889894003 | 9.015102845 | 12.82061812 | 0 | 0 |
| 0 | 0 | 0 | 0.55473561 | 0 |
| 0 | 0 | 0 | 0 | 0 |
| 0 | 0 | 0 | 0 | 0 |
| 0 | 0 | 0 | 0 | 0 |
| 0.082673611 | 0.767852927 | 0.629937032 | 0 | 0 |
| 0 | 0 | 0 | 0 | 0 |
| 0 | 0 | 0 | 0 | 0 |
| 0 | 0 | 0 | 0 | 0 |
| 0 | 0 | 0 | 0 | 0 |
| 0.062806312 | 1.912534184 | 0.438035736 | 0.076774262 | 0.152015001 |

| | | | | |
|---|---|---|---|---|
| 0 | 0.001267715 | 0 | 0 | 0 |
| 0 | 0 | 0 | 0 | 0 |
| 0 | 0 | 0 | 0 | 0 |
| 0 | 0 | 0 | 0 | 0 |
| 0.001076063 | 0.020014199 | 0 | 0 | 0 |
| 0 | 0 | 0 | 0 | 0 |
| 0 | 0 | 0 | 0 | 0 |
| 0 | 0.00173255 | 0 | 0 | 0 |
| 0 | 0 | 0 | 0 | 0 |
| 0 | 0 | 0 | 0 | 0 |
| 0.000548918 | 0 | 0 | 0 | 0 |
| 0.259766172 | 0.429338315 | 0.003087772 | 1.461096101 | 0 |
| 0 | 0.085363703 | 0.313060713 | 0 | 0 |
| 0 | 0 | 0 | 0 | 0 |
| 0.00705198 | 0.006255789 | 0.001290248 | 1.567008509 | 0 |
| 0 | 0 | 0 | 0 | 0 |
| 0 | 0 | 0 | 0 | 0.001600971 |
| 0 | 0 | 0 | 0 | 0 |
| 0 | 0.010235263 | 0 | 0.002169167 | 0 |
| 0 | 0 | 0.001567481 | 0 | 0 |
| 0 | 0 | 0 | 0 | 0 |
| 0 | 0 | 0 | 0 | 0 |
| 0.034823904 | 1.624729788 | 0.050308731 | 0.054072745 | 0.317753452 |
| 0 | 0 | 0 | 0 | 0 |
| 0 | 0 | 0 | 0 | 0 |
| 0 | 0 | 0 | 0 | 0 |
| 0 | 0 | 0 | 0 | 0 |
| 0 | 0 | 0 | 0 | 0 |
| 0 | 0 | 0 | 0 | 0 |
| 0 | 0 | 0 | 0 | 0 |
| 0 | 0 | 0 | 0 | 0 |
| 0.01399554 | 0.031929861 | 0.087062414 | 0.09488286 | 0.058448435 |
| 0.117980366 | 0 | 0 | 0 | 0 |
| 0.033883971 | 0.107740947 | 0 | 0.060236801 | 0 |
| 0.000522528 | 0.001050676 | 0.002979631 | 0.001133595 | 0 |
| 0.015268325 | 0.158018972 | 0.018856336 | 0.015587647 | 0 |
| 0 | 0 | 0 | 0 | 0 |
| 0 | 0 | 0 | 0 | 0 |
| 0.002801438 | 0 | 0 | 0 | 0 |
| 0 | 0 | 0 | 0 | 0 |
| 0 | 0.403875154 | 0 | 0 | 0 |
| 0 | 0 | 0 | 0 | 0 |
| 0 | 0 | 0 | 0 | 0 |
| 0 | 0 | 0.015215715 | 0 | 0 |
| 0.018873908 | 0.075211514 | 0.00613731 | 0.027173156 | 0.003535725 |
| 0 | 0 | 0 | 0 | 0 |
| 0.232570987 | 0 | 0 | 0 | 0 |
| 0 | 0.001142102 | 0.001261912 | 0.000880169 | 0 |

| | | | | |
|---|---|---|---|---|
| 0 | 0 | 0 | 0 | 0 |
| 0 | 0 | 0 | 0 | 0 |
| 0 | 0.022479882 | 0 | 0 | 0 |
| 0 | 0 | 0 | 0 | 0 |
| 0 | 0 | 0 | 0 | 0 |
| 0 | 0 | 0 | 0 | 0 |
| 0 | 0 | 0 | 0 | 0 |
| 0.001005553 | 0.000673973 | 0.041701798 | 0 | 0 |
| 0 | 0 | 0 | 0 | 0 |
| 0 | 0 | 0 | 0 | 0 |
| 0 | 0 | 0 | 0 | 0 |
| 0 | 0 | 0 | 0.001175136 | 0 |
| 0 | 0 | 0 | 0 | 0 |
| 0.349825569 | 0 | 0 | 0 | 0 |
| 0 | 0 | 0 | 0 | 0 |
| 0 | 0 | 0 | 0 | 0 |
| 0 | 0 | 0 | 0 | 0 |
| 0 | 0 | 0 | 0 | 0 |
| 0 | 0 | 0 | 1.844574972 | 0.063721279 |
| 0 | 0 | 0 | 0 | 0 |
| 0 | 0 | 0 | 0 | 0.002044648 |
| 0 | 0 | 0 | 0 | 0 |
| 0 | 0 | 0 | 0 | 0 |
| 0 | 0 | 0 | 0 | 0 |
| 0 | 0 | 0 | 0.003050243 | 0 |
| 0 | 0.000802971 | 0.00099367 | 0.03915865 | 0.00126601 |
| 0 | 0 | 0 | 0 | 0 |
| 0.069731739 | 0.089598181 | 1.011023315 | 0.345126363 | 0 |
| 0 | 0 | 0 | 0 | 0 |
| 1.754896704 | 0.854662215 | 0.485188033 | 0.163997747 | 0.156848064 |
| 0 | 0 | 0 | 0 | 0 |
| 0 | 0 | 0 | 0 | 0 |
| 0 | 0.000663915 | 0 | 0 | 0 |
| 0 | 0 | 0 | 0 | 0 |
| 0.001673928 | 0.001009757 | 0 | 0.001089447 | 0 |
| 0 | 0 | 0 | 0 | 0 |
| 0 | 0 | 0 | 0 | 0 |
| 0.003418953 | 0 | 0 | 0.002266373 | 0.0018065 |
| 0 | 0 | 0 | 0 | 0 |
| 0 | 0 | 0 | 0 | 0 |
| 3.923102132 | 7.054947139 | 6.898381906 | 0.853564085 | 35.79820902 |
| 0 | 1.073694788 | 0.727396849 | 0 | 0.02248022 |
| 0 | 0 | 0 | 0 | 0 |
| 0 | 0 | 0 | 0 | 0 |
| 0 | 0 | 0 | 0 | 0 |
| 0 | 0 | 0 | 0.862772783 | 0 |
| 0 | 0 | 0 | 0 | 0 |
| 0 | 0 | 0 | 0.016024951 | 0.000955825 |

| | | | | |
|---|---|---|---|---|
| 0 | 0 | 0 | 0 | 0 |
| 0.015195597 | 0.011224135 | 0.014468516 | 0 | 0 |
| 0 | 0 | 0 | 0 | 0 |
| 0 | 0 | 0 | 0 | 0 |
| 0.639061968 | 2.655676662 | 1.680055489 | 0.026479199 | 0 |
| 0.011288932 | 0 | 0 | 0 | 0 |
| 0 | 0 | 0 | 0 | 0 |
| 0 | 0 | 0 | 0 | 0 |
| 0 | 0.086236158 | 0.326747166 | 0.002855078 | 0.053256996 |
| 0 | 0 | 0 | 0 | 0 |
| 0 | 0 | 0 | 0 | 0 |
| 0 | 0 | 0 | 0 | 0 |
| 0.184865237 | 0.382018567 | 0.244620155 | 0 | 0 |
| 0.016485165 | 0.027188475 | 0.02189263 | 0.058065641 | 0.049326304 |
| 0.068900153 | 0 | 0 | 0 | 0 |
| 0.186650581 | 0.852389693 | 0.806058212 | 0.056221631 | 0 |
| 0 | 0 | 0 | 0 | 0 |
| 0 | 0 | 0 | 0 | 0 |
| 0 | 0 | 0 | 0 | 0 |
| 0 | 0 | 0 | 0 | 0 |
| 0 | 0 | 0 | 0 | 0 |
| 0 | 0 | 0 | 0 | 0 |
| 0 | 0 | 0 | 0 | 0 |
| 0 | 0 | 0 | 0 | 0 |
| 0 | 0 | 0 | 0 | 0 |
| 0 | 0 | 0 | 0 | 0 |
| 0.001114994 | 0.03601177 | 0.008670092 | 0.068183183 | 0 |
| 0 | 0 | 0 | 0.002047107 | 0 |
| 0 | 0 | 0 | 0 | 0 |
| 0 | 0 | 0 | 0 | 0 |
| 0 | 0 | 0 | 0 | 0 |
| 0.001182306 | 0 | 0 | 0 | 0 |
| 0 | 0 | 0 | 0 | 0 |
| 0 | 0.004143637 | 0 | 0 | 0 |
| 0.020278381 | 0 | 0 | 0 | 0 |
| 0 | 0.01646144 | 0 | 0 | 0 |
| 0 | 0 | 0.081840101 | 0.007664792 | 0 |
| 0 | 0.016753264 | 0 | 0 | 0 |
| 0 | 0 | 0 | 0 | 0 |
| 0 | 0 | 0 | 0 | 0 |
| 0.752886753 | 3.125855398 | 6.714775002 | 0.120603551 | 0.004238271 |
| 0 | 0 | 0 | 0 | 0 |
| 0 | 0 | 0 | 0 | 0 |
| 0 | 0.040058146 | 0 | 0 | 0 |
| 0 | 0 | 0 | 0 | 0 |
| 0 | 0 | 0 | 0 | 0 |
| 0 | 0 | 0 | 0 | 0 |
| 0 | 0 | 0 | 0 | 0 |

| | | | | |
|---|---|---|---|---|
| 0.003099493 | 0.453054615 | 0 | 0.079195787 | 0 |
| 0 | 0 | 0 | 0 | 0 |
| 0 | 0 | 0.012516049 | 0 | 0 |
| 0 | 0 | 0 | 0 | 0 |
| 0.016704947 | 0 | 0.053814105 | 0 | 0 |
| 0 | 0 | 0 | 0 | 0 |
| 0 | 0.069285315 | 0.006962646 | 0.019772344 | 0 |
| 0 | 0.006992586 | 0 | 0 | 0 |
| 0 | 0.001361603 | 0 | 0 | 0 |
| 0 | 0 | 0 | 0 | 0 |
| 0 | 0.007602719 | 0 | 0 | 0 |
| 0 | 0 | 0.039360812 | 0 | 0 |
| 0 | 0 | 0 | 0 | 0 |
| 0 | 0 | 0 | 0 | 0 |
| 0 | 0.019586415 | 0 | 0 | 0 |
| 0 | 0 | 0 | 0 | 0 |
| 0 | 0 | 0 | 0.047869627 | 0 |
| 0 | 0 | 0 | 0 | 0 |
| 0 | 0 | 0 | 0 | 0 |
| 0 | 0 | 0 | 0 | 0 |
| 0.08022472 | 0.17156627 | 0.22703732 | 0.007760926 | 0.009169532 |
| 0 | 0 | 0 | 0 | 0 |
| 0.034756984 | 0.003038596 | 0 | 0 | 0 |
| 0.00104514 | 0.001050759 | 0.000928789 | 0.001133685 | 0 |
| 0 | 0 | 0 | 0 | 0 |
| 0 | 0.003364789 | 0 | 0 | 0 |
| 0 | 0 | 0 | 0 | 0 |
| 0 | 0 | 0 | 0 | 0 |
| 0 | 0 | 0 | 0.15971314 | 0 |
| 0.005801917 | 0.002376453 | 2.11339507 | 1.837224782 | 0.521834205 |
| 0 | 0 | 0 | 0 | 0 |
| 0 | 0 | 0 | 0 | 0 |
| 0 | 0 | 0 | 0 | 0 |
| 0 | 0.084321327 | 0 | 0 | 0 |
| 0 | 0 | 0 | 0 | 0 |
| 0 | 0 | 0 | 0 | 0 |
| 0 | 0 | 0.020047194 | 0.013172096 | 0 |
| 0 | 0 | 0 | 0 | 0 |
| 0 | 0 | 0 | 0 | 0 |
| 0.050050104 | 0 | 0 | 0 | 0 |
| 0 | 0 | 0 | 0 | 0 |
| 0 | 0 | 0 | 0 | 0 |
| 0 | 0 | 0 | 0 | 0 |
| 0 | 0 | 0 | 0 | 0 |
| 0 | 0 | 0 | 0 | 0 |
| 0 | 0 | 0 | 0 | 0 |
| 0 | 0 | 0 | 0 | 0 |
| 0 | 0 | 0 | 0 | 0 |

| | | | | |
|---|---|---|---|---|
| 0 | 0.000484566 | 0.001249263 | 0.002265501 | 0 |
| 0 | 0 | 0 | 0 | 0 |
| 0 | 0.020922852 | 0.005938499 | 0.004556238 | 0 |
| 0 | 0 | 0 | 0 | 0 |
| 0 | 0 | 0 | 0 | 0 |
| 0 | 0 | 0 | 0 | 0 |
| 0 | 0 | 0 | 0 | 0 |
| 0.289999042 | 1.294929718 | 1.607749696 | 1.39945218 | 3.284489711 |
| 0.233263853 | 0.053495453 | 0.053336642 | 10.28790936 | 0.112725149 |
| 0 | 0.052244602 | 0 | 0 | 0 |
| 0 | 0 | 0 | 0 | 0 |
| 0 | 0 | 0 | 0 | 0 |
| 0 | 0 | 0 | 0 | 0 |
| 0 | 0 | 0 | 0.132642629 | 0 |
| 0 | 0.001697367 | 0 | 0 | 0 |
| 0 | 0 | 0 | 0 | 0 |
| 0 | 0 | 0 | 0 | 0 |
| 0 | 0 | 0 | 0 | 0 |
| 0 | 0 | 0 | 0 | 0 |
| 0.007715584 | 0.001463597 | 0.016300707 | 0 | 0 |
| 0 | 0 | 0 | 0 | 0 |
| 0 | 0 | 0 | 0 | 0 |
| 0 | 0 | 0 | 0 | 0 |
| 0 | 0 | 0 | 0 | 0 |
| 0 | 0.098977878 | 0.020860118 | 0 | 0 |
| 0 | 0 | 0 | 0 | 0 |
| 0 | 0 | 0 | 0 | 0 |
| 0 | 0 | 0 | 0 | 0 |
| 0 | 0 | 0 | 0 | 0 |
| 0 | 0 | 0 | 0 | 0 |
| 0 | 0 | 0.001796764 | 0 | 0 |
| 0 | 0 | 0 | 0 | 0 |
| 0 | 0 | 0 | 0 | 0 |
| 0 | 0.002440238 | 0 | 0 | 0 |
| 0.003500988 | 0 | 0 | 0 | 0 |
| 0 | 0 | 0 | 0 | 0 |
| 0 | 0 | 0 | 0 | 0 |
| 0 | 0 | 0 | 0 | 0 |
| 0.005540019 | 0 | 0 | 0 | 0 |
| 0 | 0 | 0 | 0 | 0.569486785 |
| 0 | 0 | 0 | 0 | 0 |
| 0.117813098 | 0.075526287 | 0.312401649 | 0 | 0.008393238 |
| 0 | 0 | 0 | 0 | 0 |
| 0 | 0 | 0 | 0 | 0 |
| 0 | 0 | 0 | 0 | 0 |
| 0 | 0 | 0 | 0 | 0 |
| 0 | 0 | 0 | 0 | 0 |
| 0 | 0 | 0 | 0 | 0 |

| | | | | |
|---|---|---|---|---|
| 0 | 0 | 0 | 0 | 0 |
| 0 | 0.027260976 | 0 | 0 | 0.002068615 |
| 0 | 0 | 0 | 0 | 0 |
| 0 | 0 | 0 | 0 | 0 |
| 0.017637669 | 0.109452307 | 0.669664839 | 0.947141556 | 0 |
| 0.028717679 | 0.01397036 | 0 | 0 | 0 |
| 0 | 0 | 0.001093737 | 0 | 0 |
| 0.001226213 | 0.00431482 | 0.020023328 | 0.003325248 | 0.015549696 |
| 0.002244602 | 0 | 0 | 0 | 0 |
| 0.007224799 | 0 | 0 | 0 | 0 |
| 0 | 0 | 0 | 0 | 0 |
| 0 | 0.447900889 | 0 | 0 | 0 |
| 0 | 0 | 0 | 0 | 0 |
| 0 | 0 | 0 | 0 | 0 |
| 0 | 0.010269578 | 0 | 0 | 0 |
| 0.008685131 | 0 | 0 | 0 | 0 |
| 0 | 0 | 0 | 0 | 0 |
| 0 | 0 | 0 | 0 | 0 |
| 0 | 0 | 0 | 0 | 0 |
| 0 | 0 | 0 | 0 | 0 |
| 0 | 0 | 0 | 0 | 0 |
| 0 | 0 | 0 | 0 | 0 |
| 0 | 0 | 0 | 0 | 0 |
| 0 | 0.006934797 | 0 | 0.003836971 | 0 |
| 0 | 0 | 0 | 0 | 0 |
| 0 | 0 | 0 | 0 | 0 |
| 0.001617626 | 0 | 0.001006281 | 0.387080785 | 0 |
| 0 | 0 | 0 | 0 | 0 |
| 0 | 0 | 0 | 0 | 0 |
| 0 | 0 | 0 | 0 | 0 |
| 0 | 0 | 0 | 0 | 0 |
| 0 | 0 | 0 | 0 | 0 |
| 0 | 0 | 0 | 0 | 0 |
| 0 | 0 | 0 | 0 | 0 |
| 0 | 0 | 0 | 0 | 0 |
| 0 | 0 | 0.001124506 | 0 | 0 |
| 0 | 0 | 0 | 0 | 0 |
| 0 | 0 | 0 | 0 | 0 |
| 0 | 0 | 0 | 0 | 0 |
| 0 | 0 | 0 | 0 | 0 |
| 0 | 0 | 0 | 0 | 0 |
| 0 | 0 | 0 | 0 | 0 |
| 0 | 0 | 0 | 0 | 0 |
| 0 | 0 | 0 | 0 | 0 |
| 0 | 0 | 0 | 0 | 0 |
| 0 | 0 | 0 | 0 | 0 |
| 0 | 0 | 0 | 0 | 0 |
| 0 | 0.032756176 | 0 | 0 | 0 |

| | | | | |
|---|---|---|---|---|
| 0.003042475 | 0.03181186 | 0 | 0 | 0 |
| 0 | 0 | 0 | 0 | 0 |
| 0 | 0.087868353 | 0 | 0 | 0 |
| 0 | 0 | 0 | 0 | 0 |
| 0 | 0 | 0 | 0 | 0 |
| 0 | 0 | 0 | 0 | 0.002793706 |
| 0 | 0 | 0 | 0 | 0 |
| 0 | 0 | 0 | 0 | 0 |
| 0 | 0 | 0 | 0 | 0 |
| 0 | 0 | 0 | 0 | 0 |
| 0 | 0 | 0 | 0 | 0 |
| 0 | 0 | 0 | 0 | 0 |
| 0 | 0 | 0 | 0 | 0 |
| 0 | 0 | 0 | 0 | 0 |
| 0 | 0 | 0 | 0 | 0 |
| 0 | 0 | 0 | 0 | 0 |
| 0 | 0 | 0 | 0 | 0 |
| 0 | 0 | 0 | 0 | 0 |
| 0 | 0 | 0 | 0 | 0 |
| 0 | 0 | 0.019157705 | 0 | 0 |
| 0 | 0.00088536 | 0.00684766 | 0 | 0.000837545 |
| 0 | 0 | 0 | 0 | 0 |
| 0 | 0 | 0 | 0.206051318 | 0.001410345 |
| 0 | 0 | 0.265317104 | 0 | 0 |
| 0 | 0 | 0 | 0 | 0 |
| 0 | 0 | 0 | 0 | 0 |
| 0 | 0 | 0 | 0 | 0 |
| 0 | 0 | 0 | 0 | 0 |
| 0 | 0 | 0 | 4.607674132 | 0 |
| 0 | 0 | 0 | 0 | 0 |
| 0 | 0 | 0 | 0 | 0 |
| 0.001565594 | 0 | 0.461913099 | 0 | 0 |
| 0.001041538 | 0 | 0 | 0.234768022 | 0.058444625 |
| 0 | 0 | 0 | 0 | 0 |
| 0.004099265 | 0.087858724 | 0.352659003 | 0 | 0 |
| 0 | 0.022519626 | 0 | 0 | 0.002384713 |
| 0 | 0 | 0 | 0 | 0 |
| 0 | 0 | 0 | 0 | 0 |
| 0 | 0 | 0 | 0.002306978 | 0 |
| 0 | 0 | 0 | 0 | 0 |
| 0 | 0 | 0 | 0 | 0 |
| 0 | 0 | 0.261076433 | 0 | 0 |
| 0.005199797 | 0.144558734 | 0.041136319 | 1.538093205 | 0 |
| 0 | 0 | 0 | 0 | 0 |
| 0 | 0 | 0 | 0 | 0 |
| 0 | 0.014621354 | 0.025967962 | 0 | 0 |
| 0 | 0 | 0 | 0 | 0 |
| 0 | 0 | 0 | 0 | 0 |

| | | | | |
|---|---|---|---|---|
| 0 | 0 | 0 | 0 | 0 |
| 0 | 0 | 0 | 0 | 0 |
| 0 | 0 | 0 | 0 | 0 |
| 0 | 0 | 0 | 0 | 0 |
| 0 | 0 | 0 | 0 | 0 |
| 0 | 0 | 0 | 0 | 0 |
| 0 | 0 | 0 | 0 | 0 |
| 0 | 0.017268607 | 0.008364426 | 0 | 0 |
| 0 | 0.001860338 | 0.01857419 | 0 | 0 |
| 0.000924648 | 0 | 0.010353567 | 0.008425076 | 0 |
| 0 | 0 | 0 | 0 | 0 |
| 0 | 0 | 0.004337219 | 0 | 0 |
| 0 | 0 | 0 | 0 | 0 |
| 0 | 0.112577839 | 0.033146478 | 0.036576782 | 0 |
| 0 | 0 | 0 | 0 | 0 |
| 0 | 0 | 0.001131863 | 0 | 0 |
| 0 | 0 | 0 | 0 | 0 |
| 0 | 0 | 0 | 0 | 0 |
| 0 | 0 | 0 | 0 | 0 |
| 0.010968087 | 1.837842644 | 1.968704121 | 0 | 0 |
| 0.01596287 | 0.003439006 | 0.013003654 | 0.002473609 | 0 |
| 0 | 0 | 0 | 0 | 0 |
| 0 | 0 | 0 | 0 | 0 |
| 0 | 0 | 0 | 0 | 0 |
| 0 | 0 | 0 | 0 | 0 |
| 0 | 0 | 0 | 0 | 0 |
| 0 | 0 | 0 | 0 | 0 |
| 0 | 0 | 0 | 0.000777974 | 0 |
| 0 | 0 | 0 | 0 | 0 |
| 0.082031217 | 0.473788873 | 0 | 0 | 0.331204065 |
| 0.506059346 | 1.287412984 | 0.423175306 | 0.04186779 | 0.044773476 |
| 0 | 0 | 0 | 0 | 0 |
| 0 | 0 | 0 | 0 | 0 |
| 0 | 0 | 0 | 0 | 0 |
| 0 | 0 | 0 | 0 | 0 |
| 0 | 0 | 0 | 0 | 0 |
| 0 | 0 | 0 | 0 | 0 |
| 0 | 0 | 0 | 0 | 0 |
| 0 | 0 | 0 | 0.014463337 | 0 |
| 0 | 0 | 0 | 0 | 0 |
| 0 | 0 | 0 | 0 | 0 |
| 0 | 0 | 0 | 0 | 0 |
| 0 | 0 | 0 | 0 | 0 |
| 0 | 0 | 0 | 0 | 0 |
| 0 | 0 | 0 | 0 | 0 |
| 0 | 0 | 0 | 0 | 0 |
| 0 | 0 | 0.248247129 | 0.007475533 | 0.890766082 |
| 0 | 0.00243501 | 0 | 0 | 0 |

| | | | | |
|---|---|---|---|---|
| 0 | 0 | 0 | 0 | 0 |
| 0 | 0 | 0 | 0 | 0 |
| 0 | 0 | 0 | 0 | 0 |
| 0 | 0 | 0 | 0 | 0 |
| 0.232521349 | 0.010072739 | 0.057780154 | 0 | 0.000882292 |
| 0 | 0 | 0 | 0 | 0 |
| 0 | 0 | 0 | 0 | 0 |
| 0 | 0 | 0 | 0 | 0 |
| 0 | 0 | 0 | 0 | 0 |
| 0 | 0 | 0 | 0 | 0 |
| 0 | 0 | 0 | 0 | 0 |
| 0.003477437 | 0.102553238 | 0 | 0.026404344 | 0 |
| 0 | 0.00134057 | 0 | 0 | 0 |
| 0 | 0 | 0 | 0 | 0 |
| 0 | 0 | 0 | 0 | 0.002800456 |
| 0.19380101 | 2.061861521 | 0.201164122 | 0 | 0 |
| 0.00505195 | 0.010158223 | 0 | 0 | 0 |
| 0 | 0 | 0 | 0 | 0 |
| 0.134143606 | 0 | 0.067340045 | 0.013487692 | 0 |
| 0 | 0.051272258 | 0.032080955 | 0.497039402 | 0 |
| 0 | 0 | 0 | 0 | 0 |
| 0 | 0 | 0 | 0 | 0 |
| 0 | 0 | 0 | 0 | 0 |
| 0 | 0 | 0 | 0 | 0 |
| 0 | 0 | 0 | 0 | 0 |
| 0 | 0 | 0 | 0 | 0 |
| 0 | 0.011387356 | 0.021423283 | 0.014610437 | 0 |
| 0 | 0 | 0 | 0 | 0 |
| 0 | 0 | 0 | 0 | 0 |
| 0 | 0 | 0 | 0 | 0 |
| 0 | 0 | 0 | 0 | 0 |
| 0 | 0 | 0 | 0 | 0 |
| 0 | 0 | 0 | 0 | 0 |
| 0 | 0 | 0 | 0 | 0 |
| 0 | 0 | 0 | 0 | 0 |
| 0 | 0 | 0 | 0 | 0 |
| 0 | 0 | 0 | 0 | 0 |
| 0.01572476 | 0.003431477 | 0.004360168 | 0.061881132 | 0.014298551 |
| 0.066417749 | 0.168779672 | 0 | 0 | 0 |
| 0 | 0 | 0.009622757 | 0 | 0 |
| 0 | 0 | 0 | 0 | 0 |
| 0 | 0 | 0 | 0 | 0 |
| 0 | 0 | 0 | 0 | 0 |
| 0.001446067 | 0.025260493 | 0.003092232 | 0 | 0 |
| 0 | 0 | 0 | 0 | 0 |
| 0 | 0 | 0 | 0 | 0.005379281 |
| 0 | 0.018707963 | 0 | 0 | 0 |
| 0 | 0 | 0 | 0 | 0 |

| | | | | |
|---|---|---|---|---|
| 0 | 0 | 0 | 0 | 0 |
| 0 | 0.036501604 | 0.366889664 | 0 | 2.747241415 |
| 0 | 0 | 0 | 0 | 0 |
| 0.001136613 | 0.00095227 | 0 | 0 | 0 |
| 0 | 0 | 0.002732429 | 0.026872317 | 0.006405628 |
| 0 | 0 | 0 | 0 | 0 |
| 0.005147319 | 0 | 0 | 0 | 0 |
| 0 | 0 | 0.001382447 | 0 | 0 |
| 0.044475324 | 0 | 0.002871994 | 0 | 0 |
| 0 | 0 | 0 | 0 | 0 |
| 0 | 0 | 0 | 0 | 0 |
| 0 | 0 | 0 | 0 | 0 |
| 0.005908458 | 0.004860184 | 0.005429701 | 0 | 0.003192852 |
| 0 | 0 | 0 | 0 | 0 |
| 0 | 0 | 0 | 0 | 0 |
| 0 | 0 | 0 | 0 | 0 |
| 0 | 0 | 0.035269924 | 0 | 0 |
| 0 | 0 | 0 | 0 | 0 |
| 0 | 0 | 0 | 0 | 0 |
| 0 | 0 | 0 | 0 | 0 |
| 0 | 0 | 0 | 0 | 0 |
| 0 | 0 | 0 | 0 | 0 |
| 0 | 0.012088402 | 0 | 0 | 0.0016817 |
| 0.015023061 | 0.028319684 | 0.018496171 | 0 | 0.027121005 |
| 0 | 0 | 0 | 0 | 0 |
| 0.000924556 | 0 | 0 | 0 | 0 |
| 0 | 0 | 0 | 0 | 0 |
| 0 | 0 | 0 | 0 | 0 |
| 0 | 0 | 0 | 0 | 0 |
| 0 | 0 | 0 | 0 | 0 |
| 0.0626948 | 0.053319416 | 0.005825578 | 0.006201839 | 0.003437661 |
| 0.006708239 | 0 | 0 | 0 | 0 |
| 0 | 0 | 0 | 0 | 0 |
| 0 | 0 | 0 | 0 | 0 |
| 0 | 0 | 0.046269795 | 0 | 0 |
| 0 | 0 | 0 | 0 | 0 |
| 0 | 0 | 0 | 0 | 0 |
| 0 | 0 | 0 | 0 | 0 |
| 0 | 0.001509321 | 0 | 0 | 0 |
| 0 | 0 | 0 | 0 | 0 |
| 0 | 0 | 0 | 0 | 0 |
| 0 | 0 | 0 | 0 | 0 |
| 0 | 0 | 0 | 0 | 0 |
| 0 | 0 | 0 | 0 | 0 |
| 0 | 0.013980694 | 0 | 0 | 0 |
| 0.013355277 | 0.040281242 | 0.305842158 | 0 | 1.506785377 |
| 0 | 0 | 0 | 0 | 0 |
| 0 | 0 | 0 | 0 | 0 |

| | | | | |
|---|---|---|---|---|
| 0 | 0 | 0 | 0 | 0 |
| 0 | 0 | 0 | 0 | 0 |
| 0 | 0 | 0 | 0 | 0 |
| 0.006204199 | 0 | 0 | 0 | 0 |
| 0 | 0 | 0.002888594 | 0 | 0 |
| 0 | 0 | 0 | 0.008940357 | 0 |
| 0 | 0 | 0 | 0 | 0 |
| 0 | 0 | 0 | 0 | 0 |
| 0 | 0 | 0 | 0 | 0 |
| 0 | 0 | 0 | 0 | 0 |
| 0 | 0.002312876 | 0 | 0.003669718 | 0 |
| 0 | 0 | 0 | 0 | 0 |
| 0 | 0 | 0 | 0 | 0 |
| 0 | 0 | 0 | 0 | 0 |
| 0 | 0 | 0 | 0 | 0 |
| 0 | 0 | 0 | 0 | 0 |
| 0 | 0 | 0 | 0 | 0 |
| 0 | 0 | 0 | 0 | 0 |
| 0 | 0 | 0 | 0 | 0 |
| 0 | 0 | 0 | 0 | 0 |
| 0 | 0 | 0 | 0 | 0 |
| 0 | 0 | 0 | 0 | 0 |
| 0 | 0 | 0 | 0 | 0 |
| 0 | 0 | 0 | 0 | 0 |
| 0 | 0 | 0 | 0 | 0 |
| 0 | 0 | 0 | 0 | 0 |
| 0 | 0 | 0 | 0 | 0 |
| 0.029195071 | 0 | 0 | 0.003446913 | 0.003462987 |
| 0 | 0 | 0 | 0 | 0 |
| 0 | 0 | 0 | 0 | 0 |
| 0.005242022 | 0.002790109 | 0 | 0.177106284 | 0.00171074 |
| 0 | 0 | 0 | 0 | 0 |
| 0.049766263 | 0.091608699 | 0.054566658 | 0.02951562 | 0.051361158 |
| 0 | 0.003126602 | 0.004551935 | 0.01646991 | 0 |
| 0 | 0 | 0 | 0 | 0 |
| 0 | 0 | 0 | 0 | 0 |
| 0 | 0 | 0 | 0 | 0 |
| 0 | 0 | 0 | 0 | 0 |
| 0 | 0 | 0 | 0 | 0 |
| 0 | 0 | 0 | 0 | 0 |
| 0 | 0 | 0 | 0 | 0 |
| 0.001767292 | 0.250527845 | 0.25720994 | 0.100643464 | 0.001167247 |
| 0.014891994 | 0.023991373 | 0 | 0.067144731 | 0 |
| 0 | 0 | 0 | 0 | 0 |
| 0 | 0 | 0 | 0 | 0 |
| 0.001814937 | 0.017204266 | 0.77821905 | 0.103216148 | 0 |
| 0 | 0 | 0.001057456 | 0 | 0.002155645 |
| 0 | 0 | 0 | 0 | 0 |

| | | | | |
|---|---|---|---|---|
| 0 | 0 | 0 | 0 | 0 |
| 0 | 0 | 0.004969512 | 0.00876739 | 0 |
| 0 | 0.004194191 | 0.03858561 | 0.033581731 | 0.029235543 |
| 0 | 0 | 0 | 0 | 0 |
| 0 | 1.1063574 | 0.213752758 | 0.906520965 | 0 |
| 0 | 0 | 0 | 0 | 0 |
| 0 | 0 | 0 | 0 | 0 |
| 0 | 0 | 0 | 0 | 0 |
| 0 | 0 | 0 | 0 | 0 |
| 0 | 0 | 0 | 0 | 0 |
| 0 | 0 | 0 | 0 | 0 |
| 0 | 0 | 0 | 0 | 0 |
| 0 | 0 | 0 | 0 | 0 |
| 0 | 0 | 0 | 0 | 0 |
| 0 | 0 | 0 | 0 | 0 |
| 0 | 0 | 0 | 0 | 0 |
| 0 | 0 | 0 | 0 | 0 |
| 0 | 0 | 0 | 0 | 0 |
| 0 | 0 | 0 | 0 | 0 |
| 0 | 0 | 0 | 0 | 0 |
| 0 | 0 | 0 | 0 | 0 |
| 0 | 0 | 0 | 0 | 0 |
| 0 | 0.004243296 | 0 | 0 | 0 |
| 0 | 0 | 0 | 0 | 0 |
| 0 | 0.001819265 | 0 | 0.021394974 | 0 |
| 0 | 0 | 0.003680267 | 0.004541516 | 0.002885499 |
| 0 | 0 | 0 | 0 | 0 |
| 0.195121256 | 0 | 0.001098458 | 0.001532323 | 0 |
| 0 | 0 | 0 | 0 | 0 |
| 0 | 0 | 0 | 0 | 0 |
| 0 | 0 | 0 | 0 | 0 |
| 0 | 0 | 0.013902544 | 0 | 0 |
| 0 | 0 | 0 | 0 | 0 |
| 0 | 0 | 0 | 0 | 0 |
| 0 | 0 | 0 | 0 | 0 |
| 0 | 0 | 0 | 0 | 0 |
| 0 | 0 | 0 | 0 | 0 |
| 0 | 0.011294836 | 0 | 0 | 0 |
| 0 | 2.864468069 | 0 | 0 | 0.237878429 |
| 0 | 0 | 0 | 0 | 0 |
| 0 | 0 | 0 | 0 | 0 |
| 0 | 0.153869795 | 0 | 0.021849486 | 0 |
| 0 | 0 | 0 | 0.001783529 | 0 |
| 0 | 0 | 0 | 0.38568398 | 0 |
| 0 | 0 | 0 | 0 | 0 |
| 0 | 0 | 0 | 0 | 0 |
| 0.019563812 | 0.003371828 | 0.00130394 | 0.00272845 | 0 |
| 0 | 0 | 0 | 0 | 0 |

| | | | | |
|---|---|---|---|---|
| 0 | 0 | 0 | 0 | 0 |
| 0.007934764 | 0.006027387 | 0.006032887 | 0.001147601 | 0.015372701 |
| 0 | 0 | 0 | 0 | 0 |
| 0 | 0 | 0 | 0 | 0 |
| 0 | 0 | 0 | 0 | 0 |
| 0 | 0 | 0 | 0 | 0 |
| 0 | 0 | 0 | 0 | 0 |
| 0 | 0 | 0 | 0 | 0 |
| 0 | 0 | 0 | 0 | 0 |
| 0 | 0 | 0 | 0 | 0 |
| 0 | 0 | 0 | 0 | 0 |
| 0 | 0 | 0 | 0 | 0 |
| 0.025536034 | 0.475237648 | 0.009565945 | 0 | 0 |
| 0 | 0 | 0 | 0 | 0 |
| 0 | 0.015030488 | 0.002051482 | 0 | 0 |
| 0 | 0 | 0 | 0 | 0 |
| 0 | 0 | 0 | 0 | 0 |
| 0 | 0 | 0 | 0 | 0 |
| 0 | 0.00195619 | 0 | 0 | 0 |
| 0 | 0 | 0 | 0 | 0 |
| 0 | 0 | 0 | 0 | 0 |
| 0 | 0 | 0 | 0.159729196 | 0 |
| 0.011779797 | 0 | 0.019093794 | 0 | 0 |
| 0 | 0 | 0 | 0 | 0 |
| 0 | 0 | 0 | 0 | 0 |
| 0 | 0 | 0 | 0 | 0 |
| 0 | 0 | 0 | 0 | 0 |
| 0 | 0 | 0 | 0 | 0 |
| 0 | 0.027931409 | 0 | 0 | 0 |
| 0 | 0 | 0 | 0 | 0 |
| 0 | 0 | 0 | 0 | 0 |
| 0 | 0 | 0 | 0 | 0 |
| 0 | 0 | 0 | 0 | 0 |
| 0 | 0 | 0 | 0 | 0 |
| 0 | 0.016161573 | 2.958814004 | 5.690007132 | 0 |
| 0 | 0 | 0 | 0 | 0 |
| 0 | 0 | 0 | 0 | 0 |
| 0 | 0 | 0 | 0 | 0 |
| 0 | 0 | 0.007712433 | 0.375617801 | 0 |
| 0 | 0 | 0 | 0 | 0 |
| 0.215948441 | 0.050341409 | 0.422429534 | 0.342453154 | 4.095735114 |
| 0 | 0 | 0 | 0 | 0 |
| 0 | 0 | 0 | 0 | 0 |
| 0 | 0 | 0 | 0 | 0 |
| 0 | 0 | 0 | 0 | 0 |
| 0 | 0.033380784 | 0.061618433 | 0.069869502 | 1.117160649 |
| 0 | 0 | 0 | 0 | 0 |
| 0 | 0 | 0 | 0 | 0 |

| | | | | |
|---|---|---|---|---|
| 0 | 0 | 0 | 0 | 0 |
| 0 | 0 | 0 | 0 | 0 |
| 0 | 0 | 0 | 0 | 0 |
| 0 | 0 | 0 | 0 | 0 |
| 0 | 0 | 0 | 0 | 0 |
| 0 | 0 | 0 | 0 | 0 |
| 0 | 0 | 0 | 0 | 0 |
| 0 | 0 | 0 | 0.002076125 | 0 |
| 0 | 0 | 0.009794272 | 0 | 0 |
| 0 | 0 | 0 | 0.012445435 | 0 |
| 0 | 0 | 0 | 0 | 0 |
| 0 | 0 | 0 | 0 | 0 |
| 0 | 0.001063639 | 0.002632489 | 0 | 0 |
| 0 | 0 | 0 | 0 | 0 |
| 0 | 0 | 0 | 0 | 0 |
| 0 | 0 | 0 | 0 | 0 |
| 0 | 0 | 0 | 0 | 0 |
| 0 | 0 | 0 | 0 | 0 |
| 0 | 0 | 0 | 0 | 0 |
| 0.011864801 | 0.109061411 | 0.032510523 | 1.057587177 | 0.100007076 |
| 0 | 0 | 0 | 0 | 0 |
| 0 | 0 | 0 | 0 | 0 |
| 0 | 0 | 0 | 0 | 0 |
| 0 | 0 | 0 | 0 | 0 |
| 0 | 0 | 0 | 0 | 0 |
| 0 | 0 | 0 | 0 | 0 |
| 0 | 0 | 0 | 0 | 0 |
| 0 | 0 | 0 | 0 | 0 |
| 0 | 0 | 0 | 0 | 0 |
| 0 | 0.00465544 | 0 | 0 | 0 |
| 0 | 0 | 0 | 0 | 0 |
| 0 | 0 | 0.00246103 | 0 | 0 |
| 0 | 0 | 0 | 0 | 0 |
| 0 | 0 | 0 | 0 | 0 |
| 0 | 0 | 0 | 0 | 0 |
| 0 | 0 | 0 | 0 | 0 |
| 0 | 0 | 0 | 0 | 0 |
| 0 | 0 | 0 | 0 | 0 |
| 0 | 0 | 0 | 0 | 0 |
| 0 | 0 | 0 | 0 | 0 |
| 0 | 0 | 0 | 0 | 0 |
| 0 | 0 | 0 | 0 | 0 |
| 0 | 0.161796037 | 0.04260545 | 0 | 0 |
| 0 | 0 | 0 | 0 | 0 |
| 0.00159198 | 0 | 0 | 0 | 0 |
| 0 | 0.114264607 | 0 | 0 | 0 |
| 0 | 0 | 0 | 0 | 0 |
| 0 | 0 | 0 | 0 | 0 |

| | | | | |
|---|---|---|---|---|
| 0 | 0 | 0 | 0 | 0 |
| 0 | 0 | 0 | 0 | 0 |
| 0 | 0 | 0 | 0 | 0 |
| 0 | 0 | 0 | 0 | 0 |
| 0 | 0 | 0 | 0 | 0 |
| 0.007394954 | 0.012877984 | 0.005955613 | 0.197528509 | 0 |
| 0 | 0 | 0 | 0 | 0 |
| 0 | 0 | 0 | 0 | 0 |
| 0 | 0 | 0 | 0 | 0 |
| 0 | 0 | 0 | 0 | 0 |
| 0 | 0 | 0 | 0 | 0 |
| 0 | 0 | 0 | 0 | 0 |
| 0 | 0 | 0 | 0 | 0 |
| 0 | 0 | 0 | 0 | 0 |
| 0 | 0 | 0 | 0 | 0 |
| 0 | 0 | 0 | 0 | 0 |
| 0 | 0 | 0 | 0 | 0 |
| 0 | 0 | 0 | 0 | 0 |
| 0 | 0 | 0 | 0 | 0 |
| 0 | 0 | 0 | 0 | 0 |
| 0 | 0 | 0 | 0 | 0 |
| 0 | 0 | 0 | 0 | 0 |
| 0 | 0 | 0 | 0 | 0 |
| 0 | 0.003771665 | 0 | 0 | 0 |
| 0 | 0 | 0 | 0 | 0 |
| 0 | 0 | 0 | 0 | 0 |
| 0 | 0 | 0 | 0 | 0 |
| 0 | 0 | 0 | 0 | 0 |
| 0 | 0 | 0 | 0 | 0 |
| 0 | 0 | 0 | 0 | 0 |
| 0 | 0 | 0 | 0 | 0 |
| 0 | 0 | 0 | 0 | 0 |
| 0 | 0 | 0 | 0 | 0 |
| 0 | 0 | 0 | 0 | 0 |
| 0 | 0 | 0 | 0 | 0 |
| 0 | 0 | 0 | 0 | 0 |
| 0 | 0 | 0 | 0 | 0 |
| 0 | 0 | 0 | 0 | 0 |
| 0 | 0 | 0.075082152 | 0 | 0 |
| 0 | 0 | 0 | 0 | 0 |
| 0 | 0 | 0 | 0 | 0 |
| 0 | 0 | 0 | 0 | 0 |
| 0 | 0 | 0 | 0 | 0 |
| 0 | 0 | 0 | 0 | 0 |
| 0 | 0 | 0 | 0 | 0 |
| 0 | 0 | 0 | 0 | 0 |
| 0 | 0 | 0 | 0 | 0 |
| 0 | 0 | 0 | 0 | 0 |

| | | | | |
|---|---|---|---|---|
| 0 | 0 | 0 | 0 | 0 |
| 0 | 0.066590076 | 0.003218933 | 0 | 0 |
| 0 | 0 | 0.005629905 | 0 | 0 |
| 0 | 0 | 0 | 0 | 0 |
| 0 | 0 | 0 | 0 | 0 |
| 0 | 0 | 0 | 0 | 0 |
| 0 | 0 | 0 | 0 | 0 |
| 0 | 0 | 0 | 0 | 0 |
| 0 | 0 | 0 | 0 | 0 |
| 0.0022081 | 0 | 0 | 0 | 0 |
| 0.000774358 | 0 | 0.002107469 | 0.044518032 | 0.003682386 |
| 0 | 0 | 0 | 0 | 0 |
| 0 | 0.270654513 | 0 | 0 | 0 |
| 0 | 0 | 0 | 0 | 0 |
| 0 | 0 | 0 | 0 | 0 |
| 0 | 0 | 0 | 0 | 0 |
| 0 | 0 | 0 | 0 | 0 |
| 0 | 0 | 0 | 0 | 0 |
| 0 | 0 | 0 | 0 | 0 |
| 0 | 0 | 0 | 0 | 0 |
| 0 | 0.001580305 | 0 | 0 | 0.008720599 |
| 0 | 0 | 0.01161229 | 0 | 0 |
| 0 | 0 | 0 | 0 | 0 |
| 0.549007805 | 1.158084033 | 1.856960001 | 0.979351128 | 3.330844955 |
| 0 | 0 | 0 | 0 | 0 |
| 0 | 0 | 0 | 0 | 0 |
| 0 | 0 | 0 | 0 | 0 |
| 0 | 0 | 0 | 0 | 0 |
| 0 | 0 | 0 | 0 | 0 |
| 0 | 0 | 0 | 0 | 0 |
| 0 | 0 | 0 | 0 | 0 |
| 0 | 0 | 0 | 0 | 0 |
| 0 | 0 | 0 | 0 | 0 |
| 0 | 0 | 0 | 0 | 0 |

| PHL018-0005_V1 | PHL018-0064_V2 | PHL018-0263_V3 | PHL018-0237_V2 | PHL018-0185_V2 |
|---|---|---|---|---|
| EYCF | CTRL | REF | CTRL | REF |
| 0.048470004 | 0.067828844 | 0.012354479 | 0.034029708 | 0 |
| 0.384757696 | 2.163026346 | 0.103594078 | 0.047461628 | 2.625351398 |
| 5.528516598 | 17.22541266 | 0.00876096 | 1.695829952 | 0 |
| 0 | 0 | 0 | 0 | 0 |
| 0 | 0 | 0 | 0 | 0 |
| 0 | 0 | 0 | 0 | 0 |
| 0 | 0 | 0 | 0 | 0 |
| 0 | 0 | 0 | 0 | 0 |
| 0 | 0 | 0 | 0 | 0 |
| 0 | 0 | 0 | 0 | 0 |
| 0 | 0 | 0 | 0 | 0 |
| 0 | 0 | 0 | 0 | 0 |
| 0 | 0 | 0 | 0 | 0 |
| 23.16253484 | 11.06576545 | 0.158845351 | 0.539935535 | 0.636324341 |
| 0 | 0 | 0 | 0 | 0 |
| 0 | 0 | 0 | 0 | 0 |
| 0 | 0 | 0 | 0 | 0 |
| 0 | 0 | 0 | 0 | 0 |
| 0 | 0 | 0 | 0 | 0 |
| 0 | 0 | 0 | 0 | 0 |
| 0 | 0 | 0 | 0 | 0 |
| 0 | 0 | 0 | 0 | 0 |
| 0 | 0 | 0 | 0 | 0 |
| 0 | 0 | 0 | 0 | 0 |
| 0 | 0 | 0 | 0 | 0 |
| 0 | 0 | 0 | 0 | 0 |
| 0 | 0 | 0 | 0 | 0 |
| 0 | 0 | 0 | 0 | 0 |
| 0 | 0 | 0 | 0 | 0 |
| 0 | 0 | 0 | 0 | 0 |
| 0 | 0 | 0 | 0 | 0 |
| 0 | 0 | 0 | 0 | 0 |
| 0 | 0 | 0 | 0 | 0 |
| 0 | 0 | 0 | 0 | 0 |
| 0 | 0 | 0 | 0 | 0 |
| 0 | 0.18714897 | 1.52650147 | 0.563709885 | 0 |
| 0 | 0 | 0 | 0 | 0 |
| 0 | 0 | 0 | 0 | 0 |
| 0 | 0 | 0 | 0 | 0 |
| 0 | 0 | 0 | 0 | 0 |
| 0 | 0 | 0 | 0 | 0 |
| 0.001089863 | 0.001231528 | 0.095665436 | 0 | 0 |
| 0.026015341 | 0.051395918 | 0.98962248 | 6.085382278 | 0.039435275 |
| 0.003276359 | 0.056687423 | 0.003822308 | 0 | 0.013585785 |
| 0 | 0 | 0 | 0 | 0.001780886 |
| 0 | 0 | 0 | 0 | 0 |

| | | | | |
|---|---|---|---|---|
| 0 | 0 | 0 | 0 | 0 |
| 0.007467145 | 0 | 0.001769088 | 0 | 0.022230091 |
| 0 | 0.011039949 | 0.000487582 | 0 | 0.057505128 |
| 0.098713655 | 0.516536701 | 0.394011587 | 0.168564575 | 0.423405826 |
| 0.001810426 | 0 | 0 | 0 | 0 |
| 0 | 0 | 0 | 0 | 0 |
| 0.222253115 | 0.008944524 | 0.005477844 | 0 | 0 |
| 0 | 0 | 0 | 0.052890397 | 0 |
| 0 | 0 | 0 | 0 | 0 |
| 0 | 0 | 0 | 0 | 0 |
| 0 | 0 | 0.002322959 | 0 | 0 |
| 0 | 0.192878229 | 0 | 0 | 0 |
| 0 | 0 | 0 | 0 | 0 |
| 0.017930586 | 0.002704666 | 0 | 0 | 0.057774501 |
| 0.005444461 | 0 | 0 | 0 | 0 |
| 0 | 0 | 0 | 0 | 0 |
| 0.006044492 | 0 | 0.047989191 | 0.358780238 | 0.007352152 |
| 0 | 0 | 0 | 0 | 0 |
| 0 | 0 | 0 | 0 | 0 |
| 0 | 0 | 0 | 0 | 0 |
| 0.038981343 | 0.575059412 | 0.221144647 | 0 | 0.144768013 |
| 0.009186375 | 0.010168606 | 0.002245493 | 0 | 0.002902272 |
| 0 | 0 | 0 | 0.229133132 | 0 |
| 0 | 0 | 0 | 0 | 0 |
| 0 | 0 | 0 | 0 | 0 |
| 0 | 0 | 0 | 0 | 0 |
| 0 | 0 | 0 | 0 | 0 |
| 0 | 0 | 0 | 0 | 0 |
| 0 | 0 | 0.008234888 | 0 | 0 |
| 0 | 0 | 0.001829099 | 0 | 0 |
| 0 | 0.128504011 | 0.042745779 | 0 | 0 |
| 0 | 0 | 0 | 0 | 0 |
| 0 | 0 | 0 | 0 | 0 |
| 0 | 0 | 0 | 0 | 0 |
| 0 | 0 | 0.002068828 | 0 | 0 |
| 0 | 0.00738796 | 0 | 0 | 0 |
| 0 | 0 | 0 | 0 | 0 |
| 0 | 0 | 0 | 46.12183967 | 4.273321654 |
| 0 | 0 | 0 | 0 | 0 |
| 0 | 0 | 0 | 0 | 0 |
| 0 | 0 | 0 | 0 | 0 |
| 0 | 0 | 0 | 0 | 0 |
| 0 | 0 | 0 | 0 | 0 |
| 0 | 0 | 0 | 0 | 0 |
| 0 | 0 | 0 | 0 | 0 |
| 0 | 0 | 0 | 0 | 0 |
| 0.001983054 | 0.009079678 | 0 | 0 | 0 |
| 0 | 0.481431525 | 0 | 0 | 0 |

| | | | | |
|---|---|---|---|---|
| 0 | 0 | 0 | 0 | 0 |
| 0 | 0 | 0 | 0 | 0 |
| 0 | 0 | 0 | 0 | 0 |
| 0 | 0 | 0 | 0 | 0 |
| 0.108382349 | 3.392943666 | 0 | 0 | 0.021473023 |
| 0 | 0 | 0 | 0 | 0 |
| 0 | 0 | 0 | 0 | 0 |
| 0 | 0 | 0 | 0 | 0 |
| 0 | 0.002184168 | 0.007668906 | 0 | 0 |
| 0 | 0 | 0 | 0 | 0 |
| 0 | 0 | 0 | 0 | 0 |
| 0 | 0.000950916 | 0.019318804 | 0 | 0 |
| 0 | 0 | 0 | 0 | 0 |
| 0 | 0 | 0 | 0 | 0 |
| 0 | 0 | 0 | 0 | 0 |
| 0 | 0 | 0 | 0 | 0 |
| 0 | 0 | 0 | 0 | 0 |
| 0 | 0 | 0 | 0 | 0 |
| 0 | 0 | 0.313208546 | 0.423265171 | 0 |
| 0 | 0 | 0 | 0 | 0 |
| 0 | 0 | 0 | 0 | 0 |
| 0 | 0 | 0 | 0 | 0 |
| 0 | 0 | 0 | 0 | 0 |
| 0 | 0 | 0 | 0 | 0 |
| 0 | 0 | 0 | 0 | 0 |
| 0 | 0 | 0 | 0 | 0 |
| 0 | 0 | 0 | 0 | 0 |
| 0 | 0 | 0 | 0 | 0 |
| 0 | 0 | 0 | 0 | 0 |
| 0 | 0 | 0.002680908 | 0 | 0 |
| 0 | 0.078572795 | 0 | 0.104208836 | 0 |
| 0 | 0.000397571 | 0.025003737 | 0 | 0.004085016 |
| 0 | 0 | 0 | 0 | 0 |
| 0 | 0 | 0 | 0 | 0 |
| 0 | 0 | 0 | 0 | 0 |
| 0 | 0 | 0 | 0 | 0 |
| 0 | 0 | 0 | 0 | 0 |
| 0 | 0 | 0 | 0 | 0 |
| 0 | 0 | 0.784751392 | 0 | 0 |
| 0 | 0 | 0 | 0 | 0 |
| 0 | 0 | 0 | 0 | 0 |
| 0 | 0 | 0 | 0 | 0 |
| 0 | 0.004901644 | 0.013963098 | 0 | 0.004459319 |
| 0 | 0 | 0 | 0 | 0 |
| 0 | 0 | 0 | 0 | 0 |
| 0 | 0 | 0 | 0 | 0 |
| 0 | 0 | 0 | 0 | 0 |
| 0 | 0 | 0 | 0 | 0 |

| | | | | |
|---|---|---|---|---|
| 0 | 0 | 0 | 0 | 0 |
| 0 | 0 | 0 | 0 | 0 |
| 0 | 0 | 0 | 0 | 0 |
| 0 | 0 | 0 | 0 | 0 |
| 0 | 0 | 0 | 0.00340773 | 0 |
| 0 | 0 | 0 | 0 | 0 |
| 0 | 0 | 0 | 0 | 0 |
| 0 | 0 | 0 | 0 | 0 |
| 0 | 0 | 0 | 0 | 0 |
| 0 | 0 | 0 | 0 | 0 |
| 0 | 0 | 0 | 0 | 0 |
| 0 | 0 | 0 | 0 | 0 |
| 0 | 0 | 0 | 0 | 0 |
| 0 | 0 | 0 | 0 | 0 |
| 0 | 0 | 0 | 0 | 0 |
| 0 | 0 | 0 | 0 | 0 |
| 0 | 0 | 0 | 0 | 0 |
| 0 | 0 | 0 | 0 | 0 |
| 0 | 0 | 0 | 0 | 0 |
| 0 | 0 | 0 | 0 | 0 |
| 0 | 0 | 0 | 0 | 0 |
| 0 | 0 | 0 | 0 | 0 |
| 0 | 0 | 0 | 0 | 0 |
| 0 | 0 | 0 | 0 | 0 |
| 0 | 0 | 0 | 0.017833639 | 0 |
| 0 | 0 | 0 | 0 | 0 |
| 0 | 0 | 0 | 0 | 0 |
| 0 | 0 | 0 | 0 | 0 |
| 0 | 0 | 0 | 0 | 0 |
| 0 | 0 | 0 | 0 | 0 |
| 0 | 0 | 0 | 0 | 0 |
| 0 | 0 | 0.011507152 | 0 | 0 |
| 0 | 0 | 0 | 0 | 0 |
| 0 | 0 | 0 | 0 | 0 |
| 0 | 0 | 0 | 0 | 0 |
| 0 | 0 | 0 | 0 | 0 |
| 0 | 0 | 0 | 0 | 0 |
| 0 | 0 | 0 | 0 | 0 |
| 0 | 0 | 0 | 0 | 0 |
| 0 | 0 | 0 | 0 | 0 |
| 0 | 0 | 0 | 0 | 0 |
| 0 | 0 | 0 | 0 | 0 |
| 0 | 0 | 0 | 0 | 0 |
| 0 | 0 | 0 | 0 | 0 |
| 0 | 0 | 0 | 0 | 0 |
| 0 | 0 | 0 | 0 | 0 |
| 0 | 0 | 0 | 0 | 0 |
| 0 | 0 | 0 | 0 | 0 |

| | | | | |
|---|---|---|---|---|
| 0 | 0 | 0 | 0 | 0 |
| 0 | 0 | 0 | 0 | 0 |
| 0 | 0 | 0 | 0 | 0 |
| 0 | 0 | 0 | 0 | 0 |
| 0 | 0 | 0 | 0 | 0 |
| 0 | 0 | 0 | 0 | 0 |
| 0 | 0 | 0 | 0 | 0 |
| 0 | 0 | 0 | 0 | 0 |
| 0 | 0 | 0 | 0 | 0 |
| 0 | 0 | 0 | 0 | 0 |
| 0 | 0 | 0 | 0 | 0 |
| 0 | 0 | 0 | 0 | 0 |
| 0.000453191 | 0 | 0 | 0 | 0 |
| 0 | 0 | 0 | 0 | 0 |
| 0 | 0 | 0 | 0 | 0 |
| 0 | 0 | 0 | 0 | 0 |
| 0 | 0 | 0 | 0 | 0 |
| 0 | 0 | 0 | 0 | 0 |
| 0 | 0 | 0 | 0 | 0 |
| 0 | 0 | 0 | 0 | 0 |
| 0.058467726 | 0.124748702 | 0.174275853 | 0.001554273 | 0.026703853 |
| 0 | 0 | 0 | 0 | 0 |
| 0 | 0 | 0 | 0 | 0.002260883 |
| 0 | 0 | 0.01360742 | 0.003889906 | 0 |
| 0 | 0 | 0 | 0 | 0 |
| 0 | 0 | 0 | 0 | 0 |
| 0 | 0.001911861 | 0.005404015 | 0 | 0.009276449 |
| 0 | 0 | 0 | 0 | 0 |
| 0 | 0 | 0 | 0 | 0 |
| 0.005455673 | 0.012784191 | 0 | 0 | 0 |
| 0.588824995 | 1.191760521 | 0.051848768 | 0 | 0.007615213 |
| 0 | 0 | 0 | 0 | 0 |
| 0 | 0 | 0 | 0 | 0 |
| 0 | 0 | 0.009924074 | 0.049583482 | 0 |
| 0.038777334 | 0.001246002 | 0.000754697 | 0.003438396 | 0.032616134 |
| 0 | 0 | 0 | 0 | 0 |
| 0 | 0 | 0 | 0 | 0 |
| 0 | 0 | 0 | 0 | 0 |
| 0 | 0 | 0 | 0 | 0 |
| 0 | 0 | 0 | 0 | 0 |
| 0 | 0 | 0 | 0 | 0 |
| 0 | 0 | 0 | 0 | 0 |
| 0 | 0 | 0 | 0 | 0 |
| 0 | 0 | 0 | 0 | 0 |
| 0 | 0 | 0 | 0 | 0 |
| 0 | 0 | 0 | 0.035048139 | 0 |
| 0 | 0 | 0.001225704 | 0 | 0.001650215 |
| 0.000785795 | 0 | 0.002151269 | 0 | 0 |

| | | | | |
|---|---|---|---|---|
| 0 | 0 | 0 | 0 | 0 |
| 0 | 0 | 0 | 0 | 0 |
| 0 | 0 | 0 | 0 | 0 |
| 0 | 0 | 0 | 0 | 0 |
| 0 | 0 | 0 | 0 | 0 |
| 0 | 0 | 0 | 0 | 0 |
| 0 | 0 | 0.00287091 | 0 | 0 |
| 0 | 0 | 0.006571537 | 0 | 0 |
| 0 | 0 | 0 | 0 | 0 |
| 0 | 0 | 0 | 0 | 0 |
| 0 | 0 | 0 | 0 | 0 |
| 0 | 0 | 0 | 0 | 0 |
| 0 | 0 | 0 | 0 | 0 |
| 0 | 0 | 0 | 0 | 0 |
| 0 | 0 | 0.004451787 | 0 | 0 |
| 0 | 0 | 0 | 0 | 0 |
| 0 | 0 | 0 | 0 | 0 |
| 0 | 0 | 0.007835768 | 0 | 0 |
| 0 | 0 | 0.004922609 | 0.032101955 | 0 |
| 0 | 0 | 0.089317152 | 0.175033946 | 0 |
| 0.024165903 | 0.005518503 | 0.002622154 | 0 | 0.005539876 |
| 0 | 0 | 0 | 0 | 0 |
| 0 | 0 | 0 | 0 | 0 |
| 0 | 0 | 0 | 0 | 0 |
| 0.004293637 | 0 | 0.004007279 | 0 | 0.005179358 |
| 0 | 0 | 0 | 0 | 0 |
| 0 | 0 | 0 | 0 | 0 |
| 0 | 0 | 0 | 0 | 0 |
| 0 | 0.012266721 | 0.00860446 | 0 | 0 |
| 0 | 0 | 0 | 0 | 0 |
| 0.449430194 | 2.107232929 | 0.154542112 | 0.015104185 | 0.497921633 |
| 0 | 0 | 0 | 0 | 0 |
| 0 | 0 | 0 | 0 | 0 |
| 0.01570004 | 0.078204284 | 0 | 0 | 0 |
| 0 | 0 | 0 | 0 | 0 |
| 0 | 0 | 0 | 0 | 0 |
| 0 | 0 | 0 | 0 | 0 |
| 0 | 0 | 0 | 0 | 0 |
| 0 | 0 | 0 | 0 | 0 |
| 0 | 0 | 0 | 0 | 0 |
| 0 | 0 | 0 | 0 | 0 |
| 0.007804669 | 0.008985547 | 0.004233051 | 0.007292939 | 0 |
| 0 | 0 | 0 | 0 | 0 |
| 0 | 0 | 0 | 0 | 0 |
| 0 | 0 | 0 | 0 | 0 |
| 0 | 0 | 0 | 0 | 0 |
| 0 | 0 | 0 | 0 | 0 |
| 0 | 0 | 0 | 0 | 0 |

| | | | | |
|---|---|---|---|---|
| 0 | 0 | 0 | 0 | 0 |
| 0 | 0 | 0 | 0 | 0 |
| 0.00425285 | 0.056414163 | 0.002657646 | 0 | 0.010120907 |
| 0 | 0 | 0 | 0 | 0 |
| 0 | 0.049890299 | 0 | 0 | 0 |
| 0 | 0 | 0 | 0 | 0 |
| 0 | 0 | 0 | 0 | 0 |
| 0 | 0 | 0 | 0 | 0 |
| 0 | 0 | 0 | 0 | 0 |
| 0 | 0 | 0 | 0 | 0 |
| 0 | 0 | 0 | 0 | 0 |
| 0 | 0 | 0 | 0 | 0 |
| 0 | 0 | 0 | 0 | 0 |
| 0.015530988 | 0.004552382 | 0.038602925 | 0 | 0.002033714 |
| 0.003879373 | 0.00098019 | 0.016103992 | 0 | 0.006434499 |
| 0 | 0 | 0.003522447 | 0.023848805 | 0 |
| 0.00230619 | 0.001675258 | 0.001578413 | 0 | 0.004143911 |
| 0 | 0 | 0 | 0 | 0 |
| 0 | 0 | 0 | 0 | 0 |
| 0 | 0 | 0 | 0 | 0 |
| 0 | 0 | 0 | 0 | 0 |
| 0.000693944 | 0 | 0.001424857 | 0 | 0 |
| 0 | 0 | 0 | 0 | 0 |
| 0 | 0 | 0 | 0 | 0 |
| 0 | 0 | 0 | 0 | 0 |
| 0 | 0 | 0 | 0 | 0 |
| 0 | 0 | 0 | 0 | 0 |
| 0 | 0 | 0 | 0 | 0 |
| 0 | 0 | 0 | 0 | 0 |
| 0 | 0.000972234 | 0 | 0 | 0 |
| 0 | 0 | 0 | 0 | 0 |
| 0 | 0 | 0 | 0 | 0 |
| 0 | 0 | 0 | 0 | 0 |
| 0 | 0 | 0 | 0 | 0 |
| 0 | 0 | 0 | 0 | 0 |
| 0 | 0 | 0.277835748 | 0 | 0 |
| 0 | 0 | 0 | 0 | 0 |
| 0 | 0 | 0.004251689 | 0 | 0.015907317 |
| 0 | 0 | 0 | 0 | 0 |
| 0 | 0 | 0 | 0 | 0 |
| 0 | 0 | 0 | 0 | 0 |
| 0 | 0 | 0 | 0 | 0 |
| 0 | 0 | 0 | 0 | 0 |
| 0 | 0 | 0 | 0 | 0 |
| 0 | 0 | 0 | 0 | 0 |
| 0 | 0 | 0 | 0 | 0 |
| 0 | 0 | 0 | 0 | 0 |
| 0 | 0 | 0 | 0 | 0 |

| | | | | |
|---|---|---|---|---|
| 0 | 0 | 0 | 0 | 0 |
| 0 | 0 | 0 | 0 | 0.175570216 |
| 0 | 0 | 0 | 0 | 0.001506986 |
| 0 | 0 | 0 | 0 | 0 |
| 0 | 0 | 0.010698364 | 0 | 0.000909704 |
| 0 | 0.025697208 | 0 | 0 | 0 |
| 0 | 0 | 0 | 0 | 0 |
| 0.007687908 | 0.005212326 | 0.16311561 | 0 | 0.024227858 |
| 0 | 0 | 0 | 0 | 0 |
| 0.011093731 | 0 | 0.046641626 | 0.006686019 | 0 |
| 0 | 0 | 0 | 0 | 0 |
| 0 | 0.001768327 | 0.064977976 | 0 | 0 |
| 0.052317806 | 0.000596151 | 0.057186835 | 0 | 0.002297028 |
| 0 | 0 | 0.003118996 | 0 | 0 |
| 0 | 0 | 0 | 0 | 0 |
| 0.00138596 | 0 | 0 | 0 | 0 |
| 0 | 0 | 0.00189699 | 0 | 0 |
| 0 | 0 | 0 | 0 | 0 |
| 0 | 2.256868065 | 0.001275841 | 0 | 0 |
| 0 | 0.008490331 | 0 | 0 | 0 |
| 0 | 0 | 0 | 0 | 0 |
| 0 | 0 | 0 | 0 | 0 |
| 0 | 0 | 0 | 0 | 0 |
| 0 | 0 | 0 | 0 | 0 |
| 0 | 0 | 0 | 0 | 0 |
| 0 | 0.004860544 | 0 | 0 | 0.005053632 |
| 0 | 0 | 0.000738137 | 4.166364875 | 0 |
| 0.29432516 | 0.171714219 | 0 | 0 | 0 |
| 0 | 0.034657737 | 0 | 0 | 0 |
| 0 | 0 | 0 | 0 | 0 |
| 0 | 0 | 0.001579538 | 0 | 0 |
| 0 | 0 | 0 | 0 | 0 |
| 0 | 0 | 0 | 0 | 0 |
| 0 | 0.183209115 | 0 | 0 | 0 |
| 0 | 0 | 0 | 0 | 0 |
| 0 | 0 | 0.005597582 | 0 | 0 |
| 0.007923679 | 0.0497861 | 0.018563897 | 0 | 0.038416723 |
| 0 | 0 | 0 | 0 | 0 |
| 0 | 0 | 0 | 0 | 0 |
| 0 | 0.2399011 | 0.011611269 | 0 | 0 |
| 0.018350445 | 0 | 0 | 0 | 0 |
| 0 | 0.489286262 | 0 | 0 | 0 |
| 0 | 0 | 0 | 0 | 0 |
| 0 | 0 | 0 | 0 | 0 |
| 0 | 0 | 0 | 0 | 0 |
| 0 | 0 | 0 | 0 | 0 |
| 0.003566111 | 0.005396848 | 0 | 0 | 0.001848409 |
| 0.003843773 | 0 | 0 | 0 | 0 |

| | | | | |
|---|---|---|---|---|
| 0 | 0 | 0.002595244 | 0 | 0 |
| 0.001743603 | 0.001447525 | 0 | 0 | 0 |
| 0 | 0 | 0 | 0 | 0 |
| 0 | 0.043678502 | 0 | 0 | 0 |
| 0 | 0.012162021 | 0 | 0 | 0 |
| 0.014394814 | 0 | 0 | 0 | 0.007861568 |
| 0 | 0 | 0 | 0 | 0.032376414 |
| 0 | 0.000879635 | 0 | 0 | 0.001129774 |
| 0 | 0 | 0 | 0.481398513 | 0 |
| 0 | 0 | 0 | 0 | 0 |
| 0 | 0 | 0.008779168 | 0 | 0 |
| 0.071558892 | 0 | 0 | 0 | 0 |
| 0 | 0.000572202 | 0 | 0 | 0 |
| 0 | 0 | 0.006516806 | 0 | 0 |
| 0.002335547 | 0.182476951 | 0 | 0 | 0.014204083 |
| 0 | 0 | 0 | 0 | 0 |
| 0 | 0 | 0 | 0 | 0 |
| 0 | 0.12473911 | 0.004420115 | 0 | 0 |
| 0 | 0.028356215 | 0 | 0 | 0 |
| 0 | 0 | 0 | 0 | 0 |
| 0 | 0.002408858 | 0 | 0 | 0 |
| 0 | 0 | 0.701518668 | 0.102201079 | 0 |
| 0 | 0 | 0 | 0 | 0 |
| 0 | 0 | 0 | 0 | 0 |
| 0.009768804 | 0 | 0 | 0 | 0 |
| 0 | 0 | 0 | 0 | 0 |
| 0.404196972 | 0.827821257 | 0.052673391 | 0 | 0.007610535 |
| 0 | 0 | 0 | 0 | 0 |
| 0 | 0 | 0 | 0 | 0 |
| 0.098965307 | 0.322515873 | 0.183519411 | 0.080989331 | 1.859878899 |
| 0.001043982 | 0.001263945 | 0 | 0.004164975 | 0 |
| 0 | 0 | 0.000940475 | 0 | 0 |
| 0 | 0 | 0 | 0 | 0 |
| 0.008871814 | 0 | 0 | 0 | 0 |
| 0 | 0 | 0 | 0 | 0 |
| 0 | 0 | 0 | 0 | 0 |
| 0 | 0 | 0 | 0.409215726 | 0 |
| 0 | 0 | 0.002012837 | 0 | 0 |
| 0 | 0.003777451 | 0 | 0 | 0 |
| 0 | 0.776641531 | 0 | 0 | 0 |
| 0.029887876 | 0.047174716 | 0.000974095 | 0 | 0 |
| 2.301194732 | 2.024265349 | 0.045937335 | 0.010178256 | 1.017379006 |
| 0 | 0 | 0 | 0 | 0 |
| 0 | 0 | 0 | 0 | 0 |
| 0 | 0.011057774 | 0 | 0 | 0 |
| 0 | 0 | 0 | 0 | 0 |
| 0.001991712 | 0.004340447 | 0.009542239 | 0.001461325 | 0.002477657 |
| 0 | 0 | 0 | 0 | 0 |

| | | | | |
|---|---|---|---|---|
| 0 | 0.001190085 | 0 | 0 | 0.002377676 |
| 0 | 0.012293608 | 0 | 0 | 0 |
| 0.017579356 | 0 | 0.008021163 | 0.03715859 | 0.003239766 |
| 0 | 0 | 0 | 0 | 0 |
| 0 | 0 | 0 | 0 | 0 |
| 0.004501828 | 0 | 0.002310871 | 0 | 0 |
| 0 | 0 | 0 | 0 | 0 |
| 0 | 0 | 0 | 0 | 0 |
| 0 | 0 | 0 | 0 | 0 |
| 0 | 0 | 0 | 0 | 0 |
| 0 | 0 | 0 | 0 | 0 |
| 0 | 0.001505255 | 0 | 0 | 0 |
| 0 | 0 | 0 | 0 | 0 |
| 0 | 0 | 0.001141201 | 0 | 0 |
| 0 | 0 | 0 | 0 | 0 |
| 0 | 0 | 0 | 0.029159218 | 0 |
| 0.004974734 | 0.017874396 | 0.008072744 | 0.000529835 | 0.047644682 |
| 0.000974856 | 0.001321886 | 0.003602964 | 0 | 0 |
| 0.012597674 | 0.00899063 | 0 | 0 | 0 |
| 0 | 0.093692917 | 0 | 0 | 0 |
| 0.03478606 | 0.078158286 | 0 | 0 | 0 |
| 0.004839718 | 0.001983192 | 0 | 0 | 0.004013685 |
| 0 | 0 | 0 | 0 | 0 |
| 0 | 0 | 0 | 0 | 0 |
| 0 | 0 | 0 | 0 | 0 |
| 0.101682398 | 0 | 0 | 0 | 0 |
| 0.005296842 | 0.008682038 | 0.002509815 | 0 | 0 |
| 0 | 0 | 0 | 0 | 0 |
| 0 | 0 | 0 | 0 | 0 |
| 0 | 0 | 0 | 0 | 0 |
| 0.022929943 | 0.054772108 | 0.014792483 | 0 | 0.003083726 |
| 0.006005601 | 0 | 0.025136564 | 0 | 0 |
| 4.399012481 | 0.041963658 | 0.156921867 | 0.007503345 | 0 |
| 0 | 0 | 0.049183337 | 0 | 0 |
| 0.014172338 | 0.115478771 | 0 | 0 | 0 |
| 0 | 0 | 0 | 0 | 0 |
| 0.008549718 | 0 | 0 | 0 | 0.002638314 |
| 0.068884187 | 0.016265406 | 0.00143673 | 0 | 0 |
| 0 | 0 | 0.031307572 | 0.011822872 | 0 |
| 0 | 0.006033638 | 0 | 0.000699583 | 0.079286438 |
| 0 | 0 | 0 | 0 | 0 |
| 0 | 0 | 0 | 0 | 0 |
| 0.00209288 | 0 | 0 | 0 | 0.002314229 |
| 0.059234826 | 0.450748969 | 0.095954404 | 0 | 0 |
| 0 | 0 | 0 | 0 | 0 |
| 0.036103927 | 0.048923379 | 0 | 0 | 0 |
| 0 | 0 | 0 | 0 | 0 |
| 0 | 0 | 0 | 0 | 0 |

| | | | | |
|---|---|---|---|---|
| 0 | 0.004912559 | 0.012544476 | 0 | 0 |
| 0 | 0.015186224 | 1.60772902 | 0.00303143 | 1.208939084 |
| 0 | 0 | 0 | 0 | 0 |
| 0 | 0 | 0 | 0 | 0 |
| 0 | 0 | 0 | 0 | 0 |
| 0 | 0 | 0 | 0 | 0 |
| 0.566883651 | 1.09128388 | 0 | 0 | 0 |
| 0 | 0.00063052 | 0 | 0 | 0 |
| 0 | 0 | 0 | 0.001494244 | 0 |
| 0 | 0 | 0 | 0 | 0 |
| 0.005353335 | 0 | 0.00161645 | 0 | 0.003917328 |
| 0.270175979 | 0.087681283 | 0.058749067 | 0 | 0.022950888 |
| 0 | 0 | 0 | 0.175377741 | 0 |
| 0 | 0 | 0 | 0 | 0 |
| 0.00234852 | 0.020997009 | 0 | 0 | 0 |
| 0 | 0 | 0 | 0 | 0 |
| 0.005111092 | 0 | 0.015910974 | 0 | 0.030628326 |
| 1.416873006 | 1.177253914 | 0.026807089 | 0 | 0.008874955 |
| 0 | 0 | 0 | 0 | 0 |
| 0.007921823 | 0.074977474 | 0.007419429 | 0 | 0 |
| 0 | 0.000823403 | 0.065982025 | 0.020583588 | 0.011844574 |
| 0 | 0 | 0 | 0 | 0 |
| 0.119324 | 0 | 0 | 0 | 0 |
| 0 | 0 | 0 | 0.005028279 | 0 |
| 0.060563409 | 0 | 0 | 0 | 0 |
| 0 | 0.001510167 | 0 | 0 | 0 |
| 0 | 0.288197468 | 0 | 0 | 0 |
| 0.005240683 | 0.085940272 | 0.111872694 | 0.004375463 | 0.01648564 |
| 0 | 0 | 0 | 0 | 0 |
| 0 | 0 | 0 | 0 | 0 |
| 0 | 0 | 0 | 0 | 0 |
| 0 | 0 | 0 | 0 | 0 |
| 0 | 0.003612271 | 0 | 0 | 0 |
| 0 | 0 | 0.000562055 | 0 | 0 |
| 0 | 0 | 0 | 0.001733142 | 0 |
| 0.061169265 | 0.113455976 | 0.167358341 | 0.051163271 | 1.431065683 |
| 0 | 0 | 0 | 0 | 0 |
| 0 | 0 | 0 | 0 | 0 |
| 0 | 0.0095005 | 0.052181933 | 0 | 0 |
| 0 | 0 | 0 | 0 | 0 |
| 0 | 0 | 0 | 0 | 0 |
| 0 | 0 | 0 | 0 | 0 |
| 0.002166577 | 0.030952197 | 0 | 0 | 0.011005339 |
| 0 | 0 | 0 | 0 | 0.008890681 |
| 0 | 0.235062948 | 0.009345648 | 0 | 0 |
| 0 | 0 | 0.753011572 | 0.363624242 | 0 |
| 0 | 0 | 0.000786747 | 0 | 0 |
| 0 | 0.003479582 | 0 | 0 | 0 |

| | | | | |
|---|---|---|---|---|
| 0 | 0.008061861 | 0.004272645 | 0 | 0 |
| 0 | 0 | 0 | 0 | 0 |
| 0 | 0 | 0 | 0 | 0 |
| 0 | 0 | 0 | 0.325095761 | 0 |
| 0 | 0 | 0 | 0 | 0 |
| 0 | 0 | 0 | 0 | 0 |
| 0 | 0 | 0 | 0 | 0 |
| 0 | 0 | 0 | 0 | 0 |
| 0 | 0 | 1.880137937 | 1.631449012 | 0 |
| 0 | 0 | 0 | 0.063757809 | 0 |
| 0 | 0 | 0 | 0 | 0 |
| 0.001813249 | 0 | 0.000930774 | 0.003658562 | 0 |
| 0 | 0 | 0 | 0 | 0 |
| 0 | 0 | 0 | 0 | 0 |
| 0.009476555 | 0.013043255 | 0.019713988 | 0.122683411 | 0 |
| 0 | 0 | 0 | 0 | 0 |
| 0 | 0 | 0 | 0.382173981 | 0.002967548 |
| 0 | 0 | 0 | 0 | 0 |
| 0 | 0 | 0 | 0 | 0 |
| 0 | 0 | 0 | 0 | 0 |
| 0 | 0 | 0 | 0 | 0 |
| 0 | 0 | 0 | 0 | 0 |
| 0 | 0 | 0 | 0 | 0 |
| 0.457138208 | 1.10760942 | 0.097414301 | 0 | 0 |
| 0 | 0.001096191 | 0 | 0 | 0 |
| 0 | 0 | 0.316285426 | 0.107349582 | 0 |
| 0 | 0 | 0 | 0.001667306 | 0 |
| 0 | 0 | 3.817410189 | 0.258444231 | 0 |
| 0 | 0 | 0 | 0 | 0 |
| 0 | 0 | 0 | 0 | 0 |
| 0 | 0 | 0 | 0.009538547 | 0 |
| 0 | 0 | 0 | 0 | 0 |
| 0 | 0.014293698 | 0 | 0 | 0 |
| 0 | 0 | 0 | 0 | 0 |
| 0 | 0 | 0 | 0 | 0 |
| 0 | 0 | 0 | 0.054557173 | 0 |
| 0 | 0 | 0.000825735 | 0.24572791 | 0 |
| 0 | 0 | 0 | 0 | 0 |
| 0 | 0 | 0 | 0 | 0 |
| 0 | 0 | 0 | 0 | 0 |
| 0.002609233 | 0.557947815 | 0.162956388 | 0 | 0.005770386 |
| 0 | 0 | 0 | 0 | 0 |
| 0.462384656 | 0.309736326 | 0.579139 | 0.600741782 | 6.876425802 |
| 0 | 0.007216075 | 0 | 0.05642365 | 0 |
| 0 | 0 | 0 | 0 | 0 |
| 0 | 0 | 0 | 0 | 0 |
| 0 | 0 | 0 | 0 | 0 |
| 0 | 0 | 0.004114632 | 0 | 0 |

| | | | | |
|---|---|---|---|---|
| 0 | 0 | 0 | 0 | 0 |
| 0 | 0 | 0.169336514 | 0.020441628 | 0 |
| 0.026392872 | 0.209843188 | 0.068425716 | 0 | 0.872756986 |
| 0 | 0 | 0 | 0 | 0 |
| 0 | 0 | 0 | 0 | 0 |
| 0 | 0 | 0 | 0.007385942 | 0 |
| 0 | 0 | 0 | 0 | 0.468065239 |
| 0 | 0 | 0 | 0 | 0 |
| 0 | 0 | 0 | 0 | 0 |
| 0 | 0 | 0 | 0 | 0 |
| 0.004428191 | 0.01779123 | 0.004882896 | 0.012815407 | 0.032371498 |
| 0.302338643 | 3.845598243 | 2.560658622 | 0.489390339 | 0.080567591 |
| 0 | 0 | 0 | 0 | 0 |
| 0 | 0 | 0.013447129 | 0.001558412 | 0.026129084 |
| 0.177564733 | 1.254128385 | 0.070168353 | 0 | 0.095550195 |
| 0 | 0 | 0 | 5.366306782 | 0 |
| 0 | 0 | 0 | 0 | 0 |
| 0 | 0 | 0 | 0 | 0 |
| 0 | 0 | 0 | 0 | 0 |
| 0.006349638 | 0.09172642 | 0 | 0 | 0 |
| 0.003848939 | 0.476108345 | 0.466837073 | 0 | 0.004788015 |
| 43.41332222 | 0.859966469 | 0.009665536 | 0 | 0.22294457 |
| 0.787067507 | 1.500998294 | 1.929574344 | 0.236628925 | 0.620644977 |
| 0 | 0 | 0 | 0 | 0 |
| 0 | 0 | 0 | 0 | 0 |
| 0 | 0 | 0.003043953 | 0.492005298 | 0 |
| 0.004439099 | 0 | 0 | 0 | 0 |
| 0 | 0 | 0 | 0 | 0 |
| 0 | 0 | 0 | 0 | 0 |
| 0 | 0 | 0 | 0 | 0 |
| 0 | 0 | 0 | 0.001835492 | 0 |
| 0.02868022 | 0 | 0 | 0.085837315 | 0 |
| 0.061480436 | 0.022350641 | 0.022307488 | 0 | 0 |
| 0 | 0 | 0 | 0 | 0 |
| 0.008449087 | 0.006783629 | 0.046224061 | 0 | 0.000553185 |
| 0.013899686 | 0.006213527 | 0.026187676 | 0.102878935 | 0.370394202 |
| 0.000885291 | 0.000571636 | 0 | 0.007307327 | 0 |
| 0 | 0 | 0 | 0 | 0 |
| 0.016980404 | 0 | 0.100192206 | 0.44988552 | 0.031425593 |
| 0 | 0 | 0 | 0 | 0 |
| 0 | 0 | 0 | 0 | 0 |
| 0 | 0 | 0 | 0 | 0 |
| 0 | 0 | 0 | 0 | 0 |
| 0 | 0 | 0 | 0 | 0 |
| 0.001916057 | 0.003374201 | 0 | 0.041982782 | 0.21321867 |
| 0 | 0 | 0 | 0 | 0 |
| 0 | 0 | 0 | 0 | 0 |
| 0.007774332 | 0.032765181 | 0.377499426 | 0.005549886 | 0.069701892 |

| | | | | |
|---|---|---|---|---|
| 0 | 0 | 0 | 0 | 0 |
| 0 | 0 | 0 | 0 | 0 |
| 0 | 0 | 0 | 0 | 0 |
| 0 | 0 | 0.00548688 | 0.001797257 | 0 |
| 0 | 0 | 0 | 0 | 0 |
| 0 | 0 | 0 | 0 | 0 |
| 0.003575752 | 0 | 0 | 0.001967654 | 0 |
| 0.00409827 | 0 | 0.06409327 | 0.137415899 | 0 |
| 0 | 0 | 0 | 0 | 0 |
| 0 | 0 | 0 | 0 | 0 |
| 0.002976189 | 0.008167395 | 0.006959672 | 0.000909849 | 0.004936439 |
| 0 | 0 | 0 | 0 | 0 |
| 0 | 0.004346803 | 0 | 0.014817582 | 0 |
| 0 | 0 | 0 | 0 | 0 |
| 0 | 0 | 0 | 0 | 0 |
| 0 | 0 | 0 | 0 | 0 |
| 0 | 0 | 0.196946709 | 0 | 0 |
| 0 | 0 | 0 | 0.002272286 | 0 |
| 0 | 0 | 0 | 0 | 0 |
| 0 | 0 | 0 | 0 | 0 |
| 0 | 0 | 0 | 0 | 0 |
| 0 | 0 | 0 | 0 | 0 |
| 0 | 0 | 0 | 0 | 0 |
| 0 | 0 | 0 | 0 | 0 |
| 0 | 0 | 0 | 0 | 0 |
| 0 | 0 | 0 | 0 | 0 |
| 0 | 0 | 0 | 0 | 0 |
| 0 | 0 | 0 | 0 | 0 |
| 0 | 0 | 0 | 0 | 0 |
| 0 | 0.033336482 | 0 | 0.479808808 | 0 |
| 0.006641561 | 0 | 0 | 0.002723109 | 0 |
| 0 | 0 | 0 | 0 | 0 |
| 0 | 0 | 0 | 0 | 0 |
| 0 | 0 | 0 | 0 | 0 |
| 0 | 0 | 0 | 0.003617339 | 0 |
| 0 | 0 | 0 | 0 | 0 |
| 0 | 0 | 0 | 0.012249283 | 0 |
| 0.006158524 | 0.018572479 | 0.001149558 | 0 | 0.002089391 |
| 0 | 0 | 0 | 0 | 0 |
| 0 | 0 | 0 | 0 | 0 |
| 0 | 0 | 0 | 0 | 0 |
| 0 | 0 | 0 | 0 | 0 |
| 0 | 0 | 0 | 0 | 0 |
| 0 | 0 | 0 | 0.017651826 | 0 |
| 0 | 0 | 0.095287708 | 0.024662534 | 0 |
| 0 | 0 | 0 | 0 | 0 |
| 0 | 0 | 0 | 0 | 0 |
| 0 | 0 | 0 | 0 | 0 |

| | | | | |
|---|---|---|---|---|
| 0 | 0 | 0 | 0 | 0 |
| 0 | 0 | 0 | 0 | 0 |
| 0 | 0 | 0 | 0 | 0 |
| 0 | 0 | 0 | 0.445272782 | 0 |
| 0 | 0 | 0 | 0 | 0 |
| 0 | 0 | 0 | 0.232766761 | 0 |
| 0 | 0 | 0 | 0 | 0 |
| 0 | 0 | 0 | 0 | 0 |
| 0.011582227 | 0.24324474 | 0.007927161 | 0 | 0.064035976 |
| 0 | 0 | 0 | 0 | 0 |
| 0 | 0 | 0 | 0 | 0 |
| 0 | 0 | 0 | 0 | 0 |
| 0 | 0 | 0 | 0 | 0 |
| 0 | 0 | 0 | 0 | 0 |
| 0 | 0.146452174 | 1.802425582 | 0.440607372 | 0 |
| 0.919008344 | 0.776272437 | 0 | 0 | 0 |
| 0 | 0 | 0 | 0 | 0 |
| 0 | 0 | 0 | 0 | 0 |
| 0 | 0 | 0 | 0 | 0 |
| 0 | 0 | 0 | 0 | 0 |
| 0 | 0 | 0 | 0 | 0 |
| 0 | 0 | 0 | 0 | 0 |
| 0 | 0 | 0 | 0 | 0 |
| 0 | 0 | 0 | 0 | 0 |
| 0 | 0.001319211 | 0.032161301 | 0.692286083 | 0.001882612 |
| 0.004707181 | 0 | 0.297861774 | 2.467334507 | 26.63576977 |
| 0 | 0 | 0.280329189 | 0.182872312 | 0 |
| 0 | 0 | 0 | 0 | 0 |
| 0 | 0 | 0 | 0.06530911 | 0.272567886 |
| 0 | 0 | 0 | 0 | 0 |
| 0 | 0 | 0 | 0 | 0 |
| 0 | 0 | 0 | 0 | 0 |
| 0.00054964 | 0 | 0 | 0 | 0 |
| 0 | 0 | 0 | 0 | 0 |
| 0 | 0.000967387 | 0 | 0.003297674 | 0.118946789 |
| 0 | 0.000957356 | 0.003769122 | 0.000932422 | 0.001171044 |
| 0 | 0.145089672 | 0.005535243 | 0 | 0 |
| 0.658826001 | 0 | 4.89831775 | 0.572408166 | 3.161381347 |
| 0 | 0 | 8.485291552 | 0 | 0 |
| 0 | 0 | 0 | 0 | 0 |
| 0 | 0 | 0 | 0 | 0 |
| 0 | 0 | 0 | 0 | 0 |
| 0.029136826 | 0 | 0.053090519 | 0.156723131 | 0 |
| 0 | 0 | 0 | 0 | 0 |
| 0 | 0 | 0 | 0 | 0 |
| 0 | 0.00116611 | 0 | 0 | 0 |
| 0 | 0 | 0 | 0 | 0 |
| 0.000639354 | 0.042315485 | 0.035225964 | 0.008209177 | 0.030753362 |

| | | | | |
|---|---|---|---|---|
| 0 | 0 | 0 | 0 | 0 |
| 0 | 0 | 0 | 0 | 0 |
| 0 | 0 | 0 | 0 | 0 |
| 0 | 0 | 0 | 0 | 0 |
| 0.005696121 | 0 | 0 | 0 | 0 |
| 0 | 0 | 0 | 0 | 0 |
| 0 | 0 | 0 | 0 | 0 |
| 0 | 0 | 0 | 0 | 0 |
| 0 | 0 | 0 | 0 | 0 |
| 0 | 0 | 0 | 0 | 0 |
| 0 | 0 | 0 | 0 | 0 |
| 0 | 0 | 0 | 1.821465808 | 0 |
| 0 | 0 | 0 | 0.004638655 | 0 |
| 0 | 0 | 0 | 0 | 0 |
| 0.008051467 | 0.178652131 | 0.225246683 | 0 | 0.003642141 |
| 0 | 0 | 0 | 0 | 0 |
| 0 | 0 | 0 | 0 | 0 |
| 0.014171858 | 0 | 0 | 0 | 0 |
| 0 | 0 | 0 | 0.0243592 | 0 |
| 0 | 0 | 0 | 0.004110288 | 0 |
| 0 | 0 | 0 | 0.024843632 | 0 |
| 0 | 0 | 0 | 0 | 0 |
| 0 | 0.026434116 | 0.348879561 | 0.170951215 | 0 |
| 0 | 0.018288666 | 0 | 0 | 0 |
| 0 | 0 | 0 | 0 | 0 |
| 0 | 0 | 0 | 0 | 0 |
| 0 | 0 | 0 | 0 | 0 |
| 0 | 0 | 0 | 0 | 0 |
| 0 | 0 | 0 | 0 | 0 |
| 0 | 0 | 0 | 0 | 0 |
| 0 | 0 | 0 | 0 | 0 |
| 0.016146761 | 0 | 0.238658048 | 0 | 0.06537893 |
| 0 | 0 | 0 | 0 | 0 |
| 0.016133287 | 0.004710783 | 0 | 0 | 0 |
| 0 | 0.000893008 | 0.002681913 | 0 | 0.001274389 |
| 0 | 0.002110526 | 0.536493363 | 0.010900693 | 0.080827844 |
| 0 | 0 | 0 | 0 | 0 |
| 0 | 0 | 0 | 0.034445553 | 0 |
| 0 | 0 | 0 | 0 | 0 |
| 0 | 0 | 0 | 0 | 0 |
| 0 | 0 | 0 | 0.021901577 | 0 |
| 0 | 0 | 0 | 0.000814539 | 0 |
| 0 | 0 | 0 | 0 | 0 |
| 0 | 0 | 0 | 0.002340522 | 0 |
| 0.000756884 | 0.004838365 | 0.006527179 | 0.054144473 | 0.429849375 |
| 0 | 0 | 0 | 0 | 0 |
| 0 | 0 | 0 | 0 | 0 |
| 0 | 0.003466839 | 0 | 0 | 0 |

| | | | | |
|---|---|---|---|---|
| 0 | 0 | 0 | 0 | 0 |
| 0 | 0 | 0 | 0 | 0 |
| 0.020666117 | 0 | 0.003857564 | 0 | 0 |
| 0 | 0 | 0 | 0 | 0 |
| 0 | 0.010457288 | 0 | 0 | 0 |
| 0 | 0 | 0.032633131 | 0 | 0 |
| 0 | 0 | 0 | 0.0128975 | 0 |
| 0 | 0.220541331 | 0.002428739 | 0.010577152 | 0 |
| 0 | 0 | 0 | 0 | 0 |
| 0 | 0 | 0 | 0.001281024 | 0.037164631 |
| 0 | 0 | 0 | 0 | 0 |
| 0 | 0 | 0 | 0 | 0 |
| 0 | 0 | 0 | 0 | 0 |
| 0 | 0 | 0 | 0.072587861 | 0 |
| 0 | 0 | 0 | 0 | 0 |
| 0 | 0 | 0 | 0 | 0 |
| 0 | 0 | 0 | 0 | 0 |
| 0 | 0 | 0 | 0 | 0 |
| 0 | 8.740925014 | 38.47996317 | 0.540923746 | 0 |
| 0 | 0 | 0 | 0 | 0 |
| 0 | 0 | 0 | 0.001095878 | 0 |
| 0 | 0 | 0 | 0 | 0 |
| 0 | 0 | 0 | 0 | 0 |
| 0 | 0 | 0 | 0 | 0 |
| 0 | 0 | 0 | 0 | 0 |
| 0.021420764 | 0.006551759 | 0.001302119 | 0 | 0.019634682 |
| 0 | 0 | 0 | 0 | 0 |
| 0 | 0.010688116 | 0.080152882 | 0 | 0 |
| 0 | 0 | 0 | 0 | 0 |
| 0 | 0 | 0.203101888 | 0.119247237 | 0 |
| 0 | 0 | 0 | 0 | 0 |
| 0 | 0 | 0 | 0 | 0 |
| 0 | 0 | 0.001046717 | 0.007373666 | 0 |
| 0 | 0 | 0 | 0 | 0 |
| 0.000886091 | 0.00171646 | 0.010309861 | 0 | 0.010287966 |
| 0 | 0 | 0 | 0 | 0 |
| 0 | 0 | 0 | 0 | 0 |
| 0 | 0 | 0 | 0.011987692 | 0.008338449 |
| 0 | 0 | 0 | 0 | 0 |
| 0 | 0 | 0 | 0 | 0 |
| 1.934035 | 2.894513254 | 3.494908613 | 0.739761619 | 28.15227166 |
| 0.193076443 | 0.057860057 | 0.376541319 | 0.003240017 | 0 |
| 0 | 0 | 0.064445483 | 0 | 0 |
| 0 | 0 | 0 | 0 | 0 |
| 0 | 0 | 0 | 0 | 0 |
| 0 | 0 | 0 | 0 | 0 |
| 0 | 0 | 0 | 0 | 0 |
| 0 | 0 | 0 | 0.003805638 | 0.009264996 |

| | | | | |
|---|---|---|---|---|
| 0 | 0 | 0 | 0 | 0.766406115 |
| 0.000957591 | 0 | 0 | 0 | 0 |
| 0 | 0 | 0 | 0 | 0 |
| 0 | 0 | 0 | 0 | 0 |
| 0.188476283 | 0 | 0.031951241 | 0.351422569 | 0 |
| 0 | 0 | 0 | 0 | 0 |
| 0 | 0 | 0 | 0 | 0 |
| 0 | 0 | 0 | 0 | 0 |
| 1.584115236 | 1.412059382 | 0.348345012 | 0 | 0.130275328 |
| 0 | 0 | 0 | 0 | 0 |
| 0 | 0 | 0 | 0 | 0 |
| 0 | 0 | 0 | 0 | 0 |
| 0.00824124 | 0 | 0 | 0.022967407 | 0 |
| 0.008170779 | 0.008546978 | 0.008052887 | 0.001798477 | 0.251261777 |
| 0 | 0 | 0.056925706 | 0 | 0 |
| 0.010459617 | 0.061693599 | 0.01362929 | 0.050915698 | 0 |
| 0 | 0 | 0.168696367 | 0 | 0 |
| 0 | 0 | 0 | 0 | 0 |
| 0 | 0 | 0 | 0 | 0 |
| 0 | 0 | 0 | 0 | 0 |
| 0 | 0 | 0 | 0 | 0 |
| 0 | 0 | 0 | 0 | 0 |
| 0 | 0 | 0 | 0 | 0 |
| 0 | 0 | 0 | 0 | 0 |
| 0 | 0 | 0 | 0.007383673 | 0 |
| 0 | 0 | 0 | 0 | 0 |
| 0 | 0 | 0.00416585 | 0.002571212 | 0 |
| 0 | 0.000806321 | 0 | 0 | 0 |
| 0 | 0 | 0 | 0 | 0 |
| 0 | 0 | 0 | 0 | 0 |
| 0 | 0 | 0 | 0 | 0 |
| 0 | 0 | 0 | 0 | 0 |
| 0 | 0 | 0 | 0.209864304 | 0 |
| 0 | 0 | 0 | 0 | 0 |
| 0 | 0 | 0 | 0.072510408 | 0 |
| 0 | 0 | 0.008074205 | 0 | 0 |
| 0 | 0 | 0 | 0 | 0 |
| 0 | 0 | 0.009024727 | 0.001000811 | 0 |
| 0 | 0 | 0 | 0 | 0 |
| 0 | 0 | 0 | 0 | 0 |
| 0 | 0.194393941 | 1.592907801 | 0.157065106 | 0 |
| 0 | 0 | 0 | 0 | 0 |
| 0 | 0 | 0 | 0 | 0 |
| 0 | 0 | 0 | 0 | 0 |
| 0 | 0 | 0 | 0 | 0 |
| 0 | 0 | 0 | 0 | 0 |
| 0 | 0 | 0 | 0 | 0 |
| 0 | 0 | 0 | 0 | 0 |

| | | | | |
|---|---|---|---|---|
| 0 | 0 | 0 | 0.540202664 | 0 |
| 0 | 0 | 0 | 0 | 0 |
| 0 | 0 | 0 | 0 | 0 |
| 0 | 0 | 0 | 0 | 0 |
| 0 | 0 | 0 | 0.002648138 | 0 |
| 0 | 0 | 0 | 0 | 0 |
| 0 | 0.050963538 | 0.466479991 | 0 | 0 |
| 0 | 0 | 0 | 0 | 0 |
| 0 | 0 | 0 | 0 | 0 |
| 0 | 0 | 0 | 0 | 0 |
| 0 | 0 | 0 | 0 | 0 |
| 0 | 0 | 0 | 0 | 0 |
| 0 | 0 | 0 | 0 | 0 |
| 0 | 0 | 0 | 0 | 0 |
| 0 | 0 | 0 | 0 | 0 |
| 0 | 0 | 0 | 0 | 0 |
| 0.013784186 | 0.039115426 | 0 | 0 | 0 |
| 0.00366177 | 0 | 0 | 0 | 0 |
| 0 | 0 | 0 | 0 | 0 |
| 0 | 0 | 0 | 0 | 0 |
| 0 | 0 | 0.012409297 | 0.018771671 | 0 |
| 0 | 0 | 0 | 0 | 0 |
| 0 | 0 | 0 | 0.009618899 | 0 |
| 0.000790347 | 0.003189567 | 0.002163731 | 0.001739639 | 0 |
| 0 | 0 | 0 | 0 | 0 |
| 0 | 0 | 0 | 0.007525404 | 0 |
| 0 | 0 | 0 | 0 | 0 |
| 0 | 0 | 0 | 0 | 0 |
| 0 | 0 | 0 | 0 | 0 |
| 0.311673426 | 0.460706097 | 0.01459742 | 0.003129685 | 0 |
| 0 | 0 | 0 | 0 | 0 |
| 0 | 0 | 0 | 0 | 0 |
| 0 | 0 | 0 | 0 | 0 |
| 0 | 0 | 0 | 0 | 0 |
| 0 | 0 | 0 | 0 | 0 |
| 0 | 0 | 0 | 0 | 0 |
| 0 | 0.041186859 | 0.007614529 | 0 | 0 |
| 0 | 0 | 0 | 0 | 0 |
| 0 | 0 | 0 | 0 | 0 |
| 0 | 0 | 0 | 0.011654408 | 0 |
| 0 | 0 | 0 | 0 | 0 |
| 0 | 0 | 0 | 0 | 0 |
| 0 | 0 | 0 | 0 | 0 |
| 0 | 0 | 0 | 0 | 0 |
| 0 | 0 | 0 | 0 | 0 |
| 0 | 0 | 0 | 0 | 0 |
| 0 | 0 | 0 | 0 | 0 |
| 0 | 0 | 0 | 0 | 0 |

| | | | | |
|---|---|---|---|---|
| 0.002409583 | 0.001235552 | 0.000582063 | 0 | 0.010109151 |
| 0 | 0 | 0 | 0 | 0 |
| 0 | 0 | 0 | 0 | 0 |
| 0 | 0 | 0 | 0 | 0 |
| 0 | 0 | 0 | 0 | 0 |
| 0 | 0 | 0 | 0 | 0 |
| 0 | 0 | 0 | 0 | 0 |
| 0 | 0 | 0.076050583 | 0.206458248 | 5.223942842 |
| 0.035819686 | 0.081023141 | 1.37913218 | 7.935475778 | 0.048341532 |
| 0.658264035 | 0.021752268 | 0.077120498 | 0 | 0 |
| 0 | 0 | 0 | 0 | 0 |
| 0 | 0 | 0 | 0 | 0 |
| 0 | 0 | 0 | 0 | 0 |
| 0 | 0 | 0.078460088 | 0 | 0 |
| 0 | 0 | 0 | 0 | 0 |
| 0 | 0 | 0 | 0 | 0 |
| 0.060208639 | 0.055309564 | 0 | 0 | 0.09917835 |
| 0 | 0 | 0 | 0 | 0 |
| 0 | 0 | 0 | 0 | 0 |
| 0 | 0 | 0 | 0 | 0 |
| 0 | 0 | 0 | 0 | 0 |
| 0 | 0 | 0 | 0 | 0 |
| 0 | 0 | 0 | 0 | 0 |
| 0 | 0 | 0 | 0 | 0 |
| 0 | 0 | 0 | 0 | 0 |
| 0 | 0 | 0 | 0 | 0 |
| 0 | 0 | 0 | 0 | 0 |
| 0 | 0 | 0 | 0 | 0 |
| 0 | 0 | 0 | 0 | 0 |
| 0 | 0 | 0 | 0 | 0 |
| 0 | 0 | 0.00254137 | 0 | 0 |
| 0 | 0 | 0 | 0 | 0 |
| 0 | 0 | 0 | 0 | 0 |
| 0 | 0 | 0 | 0 | 0 |
| 0 | 0 | 0 | 0 | 0 |
| 0 | 0 | 0 | 0 | 0 |
| 0 | 0 | 0 | 0 | 0 |
| 0 | 0 | 0 | 0 | 0 |
| 0 | 0 | 0 | 0 | 0 |
| 0 | 0 | 0.004793918 | 0 | 0 |
| 0 | 0 | 0 | 0 | 0 |
| 0.438388442 | 0 | 0 | 0 | 0 |
| 0 | 0 | 0 | 0 | 0 |
| 0 | 0 | 0 | 0 | 0 |
| 0 | 0 | 0 | 0 | 0 |
| 0 | 0 | 0 | 0 | 0 |
| 0 | 0 | 0 | 0 | 0 |
| 0 | 0 | 0 | 0 | 0 |

| | | | | |
|---|---|---|---|---|
| 0 | 0 | 0 | 0 | 0 |
| 0.018805228 | 16.7342759 | 0 | 0 | 0.003819328 |
| 0 | 0 | 0 | 0 | 0 |
| 0 | 0 | 0 | 0 | 0 |
| 0.047755498 | 0.078822254 | 0.280944438 | 0.449593127 | 0.577902119 |
| 0 | 0 | 0 | 0 | 0 |
| 0 | 0 | 0 | 0 | 0.01775269 |
| 0.020915239 | 0.053612809 | 0.006108516 | 0 | 0.011364273 |
| 0 | 0 | 0 | 0 | 0 |
| 0 | 0 | 0 | 0 | 0 |
| 0 | 0 | 0 | 0 | 0 |
| 0 | 0 | 0 | 0 | 0 |
| 0 | 0 | 0 | 0 | 0 |
| 0 | 0 | 0 | 0 | 0 |
| 0.002411623 | 0 | 0 | 0 | 0 |
| 0.004534908 | 0 | 0 | 0 | 0 |
| 0 | 0 | 0 | 0 | 0 |
| 0 | 0 | 0 | 0 | 0 |
| 0 | 0 | 0 | 0 | 0 |
| 0 | 0 | 0 | 0 | 0 |
| 0 | 0 | 0 | 0 | 0 |
| 0 | 0 | 0 | 0 | 0 |
| 0 | 0 | 0 | 0 | 0 |
| 0 | 0 | 0.003364084 | 0 | 0.004140986 |
| 0 | 0 | 0 | 0 | 0 |
| 0 | 0 | 0 | 0 | 0 |
| 0 | 0 | 0.029889267 | 0.012879337 | 0.270799242 |
| 0 | 0 | 0 | 0 | 0 |
| 0 | 0 | 0 | 0 | 0 |
| 0 | 0 | 0 | 0 | 0 |
| 0 | 0 | 0 | 0 | 0 |
| 0 | 0 | 0 | 0 | 0 |
| 0 | 0 | 0 | 0 | 0 |
| 0 | 0 | 0 | 0 | 0 |
| 0 | 0 | 0 | 0 | 0 |
| 0 | 0 | 0.002619677 | 0 | 0 |
| 0 | 0 | 0 | 0 | 0 |
| 0 | 0 | 0 | 0 | 0 |
| 0 | 0 | 0 | 0 | 0 |
| 0 | 0 | 0 | 0 | 0 |
| 0 | 0 | 0 | 0 | 0 |
| 0 | 0 | 0 | 0.052270177 | 0 |
| 0 | 0 | 0 | 0 | 0 |
| 0 | 0 | 0 | 0 | 0 |
| 0 | 0 | 0 | 0 | 0 |
| 0 | 0 | 0 | 0 | 0 |
| 0 | 0 | 0 | 0 | 0 |
| 0 | 0 | 0 | 0 | 0 |

| | | | | |
|---|---|---|---|---|
| 0 | 0 | 0 | 0.212106067 | 0 |
| 0 | 0 | 0 | 0 | 0 |
| 0 | 0 | 0 | 0 | 0 |
| 0 | 0 | 0 | 0 | 0 |
| 0 | 0 | 0 | 0 | 0 |
| 0.041205084 | 0.003162639 | 0 | 0 | 0 |
| 0 | 0 | 0 | 0 | 0 |
| 0 | 0 | 0 | 0 | 0 |
| 0 | 0 | 0 | 0 | 0 |
| 0 | 0 | 0 | 0 | 0 |
| 0 | 0 | 0 | 0 | 0 |
| 0 | 0 | 0 | 0 | 0 |
| 0 | 0 | 0 | 0 | 0 |
| 0 | 0 | 0 | 0 | 0 |
| 0 | 0 | 0 | 0 | 0 |
| 0 | 0 | 0 | 0 | 0 |
| 0 | 0 | 0 | 0 | 0 |
| 0 | 0 | 0 | 0 | 0 |
| 0 | 0 | 0 | 0 | 0 |
| 0 | 0 | 0 | 0 | 0 |
| 0.001087701 | 0.012642004 | 0.082314796 | 0 | 0 |
| 0 | 0 | 0 | 0 | 0 |
| 0.005887228 | 0 | 0.003223491 | 0.023109197 | 0.009764817 |
| 0 | 0 | 0 | 0.466699994 | 0 |
| 0 | 0 | 0 | 0 | 0 |
| 0 | 0 | 0 | 0 | 0 |
| 0 | 0 | 0 | 0 | 0 |
| 0 | 0 | 0 | 0 | 0 |
| 0 | 0 | 0 | 0 | 0 |
| 0 | 0 | 0 | 0 | 0 |
| 0 | 0 | 0 | 0 | 0 |
| 0 | 0 | 0 | 0 | 0 |
| 0.024993921 | 0.00551801 | 0.001698067 | 0.000606776 | 0.011888124 |
| 0 | 0 | 0 | 0 | 0 |
| 0 | 0 | 0 | 0.004342064 | 0 |
| 0.060907097 | 0.001571216 | 0 | 0 | 0 |
| 0 | 0 | 0 | 0 | 0 |
| 0 | 0 | 0 | 0 | 0 |
| 0.000682312 | 0.004791221 | 0.010857541 | 0 | 0 |
| 0 | 0 | 0 | 0 | 0 |
| 0 | 0 | 0 | 0 | 0 |
| 0 | 0 | 0 | 0 | 0 |
| 0 | 0 | 0.187364231 | 0.206342173 | 0 |
| 0 | 0 | 0 | 0 | 0 |
| 0 | 0 | 0 | 0 | 0 |
| 0 | 0.015188839 | 0.000487868 | 0.015689822 | 0 |
| 0 | 0 | 0.028060463 | 0 | 0 |
| 0 | 0 | 0 | 0 | 0 |

| | | | | |
|---|---|---|---|---|
| 0 | 0 | 0 | 0 | 0 |
| 0 | 0 | 0 | 0 | 0 |
| 0 | 0 | 0 | 0 | 0 |
| 0 | 0 | 0 | 0 | 0 |
| 0 | 0 | 0 | 0 | 0 |
| 0 | 0 | 0 | 0 | 0 |
| 0 | 0 | 0 | 0 | 0 |
| 0.148628746 | 0.046552024 | 0.004714349 | 0 | 0.714939763 |
| 0 | 0 | 0 | 0.002776649 | 0 |
| 0 | 0.001264189 | 0 | 0.003950306 | 0 |
| 0 | 0 | 0 | 0 | 0 |
| 0 | 0 | 0 | 0 | 0 |
| 0 | 0 | 0 | 0 | 0 |
| 0 | 0 | 0 | 0 | 0 |
| 0 | 0 | 0 | 0 | 0 |
| 0.03306817 | 0.03467154 | 0.005273632 | 0 | 0 |
| 0 | 0 | 0 | 0 | 0 |
| 0 | 0 | 0 | 0 | 0 |
| 0 | 0 | 0 | 0 | 0 |
| 0 | 0 | 0.059743869 | 0.308985746 | 0 |
| 0 | 0 | 0.002237598 | 0 | 0 |
| 0 | 0 | 0 | 0 | 0 |
| 0 | 0 | 0 | 0 | 0 |
| 0 | 0 | 0 | 0.000941294 | 0 |
| 0 | 0 | 0 | 0 | 0 |
| 0 | 0 | 0 | 0 | 0 |
| 0 | 0 | 0 | 0 | 0 |
| 0 | 0 | 0.004709686 | 0 | 0 |
| 0 | 0 | 0 | 0 | 0 |
| 0 | 0 | 0 | 0.037535775 | 0 |
| 0 | 0 | 0.11988133 | 0.047958936 | 0 |
| 0 | 0 | 0 | 0 | 0 |
| 0 | 0 | 0 | 0 | 0 |
| 0 | 0 | 0 | 0 | 0 |
| 0 | 0 | 0 | 0 | 0 |
| 0 | 0 | 0 | 0 | 0 |
| 0 | 0 | 0 | 0 | 0 |
| 0 | 0 | 0 | 0 | 0 |
| 0 | 0 | 0 | 0.003883947 | 0 |
| 0 | 0 | 0 | 0 | 0 |
| 0 | 0 | 0 | 0 | 0 |
| 0 | 0 | 0 | 0 | 0 |
| 0 | 0 | 0 | 0 | 0 |
| 0 | 0 | 0 | 0 | 0 |
| 0 | 0 | 0 | 0 | 0 |
| 0 | 0 | 0 | 0 | 0 |
| 0 | 0.325166839 | 0.031547943 | 0 | 0.050696585 |
| 0 | 0 | 0 | 0 | 0 |

| | | | | |
|---|---|---|---|---|
| 0 | 0 | 0 | 0 | 0 |
| 0 | 0 | 0 | 0 | 0 |
| 0 | 0 | 0 | 0 | 0 |
| 0 | 0 | 0 | 0 | 0 |
| 0 | 0.088108967 | 0.011091138 | 0.048234327 | 0.315821419 |
| 0 | 0 | 0 | 0 | 0 |
| 0 | 0 | 0 | 0 | 0 |
| 0 | 0 | 0 | 0 | 0 |
| 0 | 0 | 0 | 0 | 0 |
| 0 | 0 | 0 | 0 | 0 |
| 0 | 0 | 0 | 0.008405605 | 0 |
| 0 | 0 | 0.001924804 | 0.008816298 | 0.083680037 |
| 0 | 0 | 0 | 0 | 0.010975559 |
| 0 | 0 | 0 | 0 | 0 |
| 0 | 0.004654776 | 0 | 0 | 0 |
| 0 | 0 | 0 | 0.087084785 | 0.257434424 |
| 0.015075375 | 0 | 0 | 0 | 0 |
| 0 | 0 | 0 | 0 | 0 |
| 0.211657992 | 0.370255241 | 0 | 0 | 0 |
| 0 | 0.782612203 | 0.071795571 | 0 | 0 |
| 0 | 0 | 0 | 0 | 0 |
| 0 | 0 | 0 | 0.029106357 | 0 |
| 0 | 0 | 0 | 0 | 0 |
| 0 | 0 | 0 | 0 | 0 |
| 0 | 0 | 0 | 0 | 0 |
| 0 | 0 | 0 | 0 | 0 |
| 0.00175548 | 0.008632208 | 0.079437242 | 0 | 0 |
| 0 | 0 | 0 | 0 | 0 |
| 0 | 0 | 0 | 0 | 0 |
| 0 | 0 | 0 | 0 | 0 |
| 0 | 0 | 0 | 0 | 0 |
| 0 | 0.004804094 | 0.011881734 | 0 | 0 |
| 0 | 0 | 0 | 0 | 0 |
| 0 | 0 | 0 | 0 | 0 |
| 0 | 0 | 0 | 0 | 0 |
| 0 | 0 | 0 | 0 | 0 |
| 0 | 0 | 0 | 0 | 0 |
| 0.000967892 | 0.001562432 | 0.007949391 | 0.046514515 | 0.173025235 |
| 0 | 0 | 0 | 0.210977326 | 0 |
| 0 | 0 | 0 | 0 | 0 |
| 0.002898805 | 0 | 0 | 0 | 0 |
| 0 | 0 | 0 | 0 | 0 |
| 0 | 0 | 0 | 0 | 0 |
| 0.033967854 | 0.007414043 | 0 | 0 | 0 |
| 0 | 0 | 0 | 0 | 0 |
| 0 | 0 | 0 | 0 | 0 |
| 0 | 0 | 0 | 0 | 0 |
| 0 | 0 | 0 | 0 | 0 |

| | | | | |
|---|---|---|---|---|
| 0 | 0 | 0 | 0 | 0 |
| 0 | 0 | 0 | 0.009900587 | 0 |
| 0 | 0 | 0 | 0 | 0 |
| 0.001002774 | 0.001456866 | 0.001029484 | 0 | 0 |
| 0.000775047 | 0.000750678 | 0.003501045 | 0 | 0.007198953 |
| 0 | 0 | 0.000821534 | 0 | 0 |
| 0 | 0 | 0 | 0 | 0 |
| 0.001699221 | 0 | 0 | 0 | 0 |
| 0 | 0 | 0 | 0 | 0 |
| 0 | 0 | 0 | 0 | 0 |
| 0 | 0 | 0 | 0 | 0 |
| 0 | 0 | 0 | 0 | 0 |
| 0 | 0.002294917 | 0.010824767 | 0.08840011 | 0 |
| 0 | 0 | 0 | 0 | 0 |
| 0 | 0 | 0 | 0 | 0 |
| 0 | 0 | 0 | 0 | 0 |
| 0 | 0 | 0 | 0 | 0 |
| 0 | 0 | 0 | 0 | 0 |
| 0 | 0 | 0 | 0 | 0 |
| 0 | 0 | 0 | 0 | 0 |
| 0 | 0 | 0 | 0 | 0 |
| 0 | 0 | 0 | 0 | 0 |
| 0.005147963 | 0.002417502 | 0.014413878 | 0 | 0 |
| 0.016015311 | 0 | 0.02986002 | 0.026742468 | 0.032670299 |
| 0 | 0 | 0 | 0 | 0 |
| 0 | 0 | 0 | 0 | 0 |
| 0 | 0 | 0 | 0.003608668 | 0 |
| 0 | 0 | 0 | 0 | 0 |
| 0 | 0 | 0 | 0 | 0 |
| 0 | 0 | 0 | 0 | 0 |
| 0 | 0 | 0.004821423 | 0.003062852 | 0 |
| 0 | 0 | 0 | 0 | 0 |
| 0 | 0 | 0 | 0 | 0 |
| 0 | 0 | 0 | 0 | 0 |
| 0 | 0 | 0 | 0.002888805 | 0 |
| 0 | 0.056532239 | 0 | 0 | 0 |
| 0 | 0 | 0 | 0 | 0 |
| 0 | 0.075996114 | 0 | 0 | 0 |
| 0 | 0 | 0.585146935 | 0 | 0.002196829 |
| 0 | 0 | 0 | 0 | 0 |
| 0 | 0 | 0 | 0 | 0 |
| 0 | 0.00233857 | 0 | 0 | 0 |
| 0 | 0 | 0 | 0 | 0 |
| 0 | 0 | 0 | 0 | 0 |
| 0 | 0 | 0 | 0 | 0 |
| 0.045447375 | 0 | 0.079092543 | 0.151049397 | 0.360798051 |
| 0 | 0 | 0.047462997 | 0 | 0 |
| 0 | 0 | 0 | 0 | 0 |

| | | | | |
|---|---|---|---|---|
| 0 | 0 | 0 | 0.002936752 | 0 |
| 0 | 0 | 0 | 0 | 0 |
| 0 | 0 | 0 | 0.061242726 | 0 |
| 0 | 0 | 0 | 0 | 0 |
| 0 | 0 | 0 | 0 | 0 |
| 0 | 0 | 0 | 0 | 0 |
| 0 | 0 | 0 | 0 | 0 |
| 0 | 0 | 0 | 0 | 0 |
| 0 | 0 | 0.004053984 | 0 | 0 |
| 0 | 0 | 0 | 0 | 0 |
| 0 | 0 | 0 | 0 | 0.001980241 |
| 0 | 0 | 0 | 0 | 0 |
| 0 | 0 | 0 | 0 | 0 |
| 0 | 0 | 0 | 0 | 0 |
| 0 | 0 | 0 | 0 | 0 |
| 0 | 0 | 0 | 0 | 0 |
| 0 | 0 | 0 | 0 | 0 |
| 0 | 0 | 0 | 0 | 0 |
| 0 | 0 | 0 | 0 | 0 |
| 0.010133153 | 0 | 0 | 0.001219759 | 0.005777508 |
| 0 | 0 | 0 | 0 | 0 |
| 0 | 0 | 0 | 0 | 0 |
| 0 | 0 | 0 | 0 | 0 |
| 0 | 0 | 0 | 0 | 0 |
| 0 | 0 | 0 | 0 | 0 |
| 0 | 0 | 0 | 0 | 0 |
| 0 | 0 | 0 | 0 | 0 |
| 0 | 0 | 0 | 0.001542708 | 0 |
| 0 | 0 | 0 | 0 | 0 |
| 0 | 0 | 0 | 0 | 0 |
| 0.000952156 | 0.004742834 | 0.00404972 | 0.002993998 | 0.021207583 |
| 0 | 0 | 0 | 0 | 0 |
| 0.284916073 | 0.039772153 | 0.02496997 | 0.00608359 | 0.003929388 |
| 0 | 0 | 0.036452288 | 0 | 0 |
| 0 | 0 | 0 | 0 | 0 |
| 0 | 0 | 0 | 0 | 0 |
| 0 | 0 | 0 | 0 | 0 |
| 0 | 0 | 0 | 0 | 0 |
| 0 | 0 | 0 | 0 | 0 |
| 0 | 0 | 0 | 0 | 0 |
| 0 | 0 | 0 | 0 | 0 |
| 0.143445259 | 0.013465616 | 0.014806656 | 0.005290909 | 0.037067969 |
| 0 | 0 | 0.009425618 | 0.093377271 | 0 |
| 0 | 0 | 0 | 0 | 0 |
| 0 | 0 | 0 | 0 | 0 |
| 0 | 0 | 0.013855512 | 0.026433485 | 0.006449942 |
| 0 | 0 | 0 | 0 | 0 |
| 0 | 0 | 0 | 0 | 0 |

| | | | | |
|---|---|---|---|---|
| 0 | 0 | 0 | 0 | 0 |
| 0 | 0 | 0.021451684 | 0.004015208 | 0 |
| 0.001355983 | 0.001125726 | 0.027643164 | 0 | 0.021848327 |
| 0 | 0 | 0 | 0 | 0 |
| 0 | 0.071757227 | 0 | 0.050045332 | 0 |
| 0 | 0.064015233 | 0 | 0 | 0 |
| 0 | 0 | 0 | 0 | 0 |
| 0 | 0 | 0 | 0 | 0 |
| 0 | 0 | 0 | 0 | 0 |
| 0 | 0 | 0 | 0 | 0 |
| 0 | 0 | 0 | 0 | 0 |
| 0 | 0 | 0 | 0 | 0 |
| 0 | 0 | 0 | 0 | 0 |
| 0 | 0 | 0 | 0 | 0 |
| 0 | 0 | 0 | 0 | 0 |
| 0 | 0 | 0 | 0 | 0 |
| 0 | 0 | 0 | 0 | 0 |
| 0 | 0 | 0 | 0 | 0 |
| 0 | 0 | 0 | 0 | 0 |
| 0 | 0 | 0 | 0 | 0 |
| 0 | 0 | 0 | 0 | 0 |
| 0 | 0 | 0 | 0 | 0 |
| 0 | 0 | 0 | 0 | 0 |
| 0 | 0 | 0 | 0 | 0 |
| 0 | 0 | 0.003605761 | 0.129665718 | 0 |
| 0 | 0.002022152 | 0 | 0 | 0 |
| 0 | 0 | 0 | 0 | 0 |
| 0 | 0 | 0.000799686 | 0.005143576 | 0 |
| 0 | 0 | 0 | 0 | 0 |
| 0 | 0 | 0 | 0 | 0 |
| 0 | 0 | 0 | 0 | 0 |
| 0.001267529 | 0 | 0 | 0 | 0.019155013 |
| 0 | 0 | 0 | 0 | 0 |
| 0 | 0 | 0 | 0 | 0 |
| 0 | 0 | 0 | 0 | 0 |
| 0 | 0 | 0 | 0 | 0 |
| 0 | 0 | 0 | 0 | 0 |
| 0.017265274 | 0 | 0.009519072 | 0 | 0 |
| 0 | 0 | 0.083680082 | 1.44073612 | 0 |
| 0 | 0 | 0 | 0 | 0 |
| 0 | 0 | 0 | 0 | 0 |
| 0 | 0 | 0.001430935 | 0.004601886 | 0 |
| 0.001934154 | 0.002185562 | 0.001654728 | 0 | 0.006950824 |
| 0 | 0 | 0 | 0 | 0 |
| 0 | 0 | 0 | 0 | 0 |
| 0 | 0 | 0 | 0.001836694 | 0 |
| 0 | 0.006686964 | 0.018479294 | 0 | 0.010633401 |
| 0 | 0 | 0 | 0 | 0 |

| | | | | |
|---|---|---|---|---|
| 0 | 0 | 0 | 0 | 0 |
| 0.026134907 | 0.012204557 | 0.006228639 | 0.002396907 | 0.196616456 |
| 0 | 0 | 0 | 0 | 0 |
| 0 | 0 | 0 | 0 | 0 |
| 0 | 0 | 0 | 0 | 0 |
| 0 | 0 | 0 | 0 | 0 |
| 0 | 0 | 0 | 0 | 0 |
| 0 | 0 | 0 | 0 | 0 |
| 0 | 0 | 0 | 0 | 0 |
| 0 | 0 | 0 | 0 | 0 |
| 0 | 0 | 0 | 0 | 0 |
| 0 | 0 | 0 | 0 | 0 |
| 0 | 0 | 0.002870046 | 0.163245438 | 0 |
| 0 | 0 | 0 | 0 | 0 |
| 0 | 0.005448149 | 0.003982653 | 0 | 0.001608603 |
| 0 | 0 | 0 | 0 | 0 |
| 0 | 0 | 0 | 0 | 0 |
| 0 | 0 | 0 | 0 | 0 |
| 0 | 0 | 0 | 0 | 0 |
| 0 | 0 | 0 | 0 | 0 |
| 0 | 0 | 0 | 0 | 0 |
| 0 | 0.076383175 | 0.065318377 | 0 | 0 |
| 0.009514435 | 0.012901386 | 0.015478318 | 0 | 0 |
| 0 | 0 | 0 | 0 | 0 |
| 0 | 0 | 0 | 0 | 0 |
| 0 | 0 | 0 | 0 | 0 |
| 0 | 0 | 0 | 0 | 0 |
| 0 | 0 | 0 | 0 | 0 |
| 0 | 0 | 0 | 0 | 0 |
| 0 | 0 | 0 | 0 | 0 |
| 0 | 0 | 0 | 0 | 0 |
| 0 | 0 | 0 | 0 | 0 |
| 0 | 0 | 0 | 0 | 0 |
| 0 | 0 | 0 | 0 | 0 |
| 0 | 0.20487221 | 7.028761354 | 0 | 0 |
| 0 | 0 | 0 | 0 | 0 |
| 0 | 0 | 0 | 0 | 0 |
| 0 | 0 | 0 | 0 | 0 |
| 0 | 0.0184246 | 0.009764716 | 0.001046777 | 1.120724579 |
| 0 | 0 | 0 | 0 | 0 |
| 0.014327366 | 0.042787034 | 0.045702976 | 0.089727677 | 0.361552321 |
| 0 | 0 | 0 | 0 | 0 |
| 0 | 0 | 0 | 0 | 0 |
| 0 | 0 | 0 | 0 | 0 |
| 0 | 0 | 0 | 0 | 0 |
| 2.924183239 | 0.374882996 | 0.072174893 | 0 | 0.813005893 |
| 0 | 0 | 0 | 0 | 0 |
| 0 | 0 | 0 | 0 | 0 |

| | | | | |
|---|---|---|---|---|
| 0 | 0 | 0 | 0 | 0 |
| 0 | 0 | 0 | 0 | 0 |
| 0 | 0 | 0 | 0 | 0 |
| 0 | 0 | 0 | 0 | 0 |
| 0 | 0 | 0 | 0 | 0 |
| 0 | 0 | 0 | 0 | 0 |
| 0 | 0 | 0 | 0 | 0 |
| 0 | 0.02412365 | 0 | 0 | 0 |
| 0.000661458 | 0 | 0.00203723 | 0 | 0 |
| 0.000763952 | 0.002219795 | 0.220780947 | 0 | 0 |
| 0 | 0 | 0 | 0.007093705 | 0 |
| 0 | 0 | 0 | 0 | 0 |
| 0 | 0.00379691 | 0.001916472 | 0 | 0 |
| 0 | 0 | 0 | 0 | 0 |
| 0 | 0 | 0 | 0 | 0 |
| 0 | 0 | 0 | 0 | 0 |
| 0 | 0 | 0 | 0 | 0 |
| 0 | 0 | 0 | 0 | 0 |
| 0 | 0 | 0 | 0 | 0 |
| 0.078258409 | 0.074510321 | 0.790124538 | 0.023406233 | 1.453185638 |
| 0 | 0 | 0 | 0 | 0 |
| 0 | 0 | 0 | 0 | 0 |
| 0 | 0 | 0 | 0 | 0 |
| 0 | 0 | 0 | 0 | 0 |
| 0 | 0 | 0 | 0 | 0 |
| 0 | 0 | 0 | 0 | 0 |
| 0 | 0 | 0 | 0 | 0 |
| 0 | 0 | 0 | 0 | 0 |
| 0 | 0 | 0 | 0 | 0 |
| 0.001953831 | 0 | 0 | 0 | 0 |
| 0 | 0 | 0 | 0 | 0 |
| 0 | 0 | 0 | 0 | 0 |
| 0 | 0 | 0 | 0 | 0 |
| 0 | 0 | 0 | 0 | 0 |
| 0 | 0 | 0 | 0 | 0 |
| 0 | 0 | 0 | 0 | 0 |
| 0 | 0 | 0 | 0 | 0 |
| 0 | 0 | 0 | 0 | 0 |
| 0 | 0 | 0 | 0 | 0 |
| 0 | 0 | 0 | 0 | 0 |
| 0 | 0 | 0 | 0 | 0 |
| 0 | 0 | 0 | 0 | 0 |
| 0 | 0 | 0.008932925 | 0 | 0 |
| 0 | 0 | 0 | 0 | 0 |
| 0.001560577 | 0.001209207 | 0 | 0.002919749 | 0 |
| 0 | 0 | 0 | 0 | 0 |
| 0 | 0 | 0 | 0 | 0 |
| 0 | 0 | 0 | 0 | 0 |

| | | | | |
|---|---|---|---|---|
| 0 | 0 | 0 | 0 | 0 |
| 0 | 0 | 0 | 0 | 0 |
| 0 | 0 | 0 | 0 | 0 |
| 0 | 0 | 0 | 0 | 0 |
| 0 | 0 | 0 | 0 | 0 |
| 0.002912578 | 0.002256799 | 0.04915824 | 0.012565366 | 0 |
| 0 | 0 | 0 | 0 | 0 |
| 0 | 0 | 0 | 0 | 0 |
| 0 | 0 | 0 | 0 | 0 |
| 0 | 0 | 0 | 0 | 0 |
| 0 | 0 | 0 | 0 | 0 |
| 0 | 0 | 0 | 0 | 0 |
| 0 | 0 | 0 | 0 | 0 |
| 0 | 0 | 0 | 0 | 0 |
| 0 | 0 | 0 | 0 | 0 |
| 0 | 0 | 0 | 0 | 0 |
| 0 | 0 | 0 | 0.010731401 | 0 |
| 0 | 0 | 0 | 0 | 0 |
| 0 | 0 | 0 | 0 | 0 |
| 0 | 0 | 0 | 0 | 0 |
| 0 | 0 | 0 | 0 | 0 |
| 0 | 0 | 0 | 0 | 0 |
| 0 | 0 | 0 | 0 | 0 |
| 0 | 0 | 0 | 0 | 0 |
| 0 | 0 | 0 | 0 | 0 |
| 0 | 0 | 0.00374169 | 0 | 0 |
| 0 | 0 | 0 | 0 | 0 |
| 0 | 0 | 0 | 0 | 0 |
| 0 | 0 | 0 | 0 | 0 |
| 0 | 0 | 0.092761076 | 0 | 0 |
| 0 | 0 | 0 | 0 | 0 |
| 0 | 0 | 0 | 0 | 0 |
| 0 | 0 | 0 | 0 | 0 |
| 0 | 0 | 0 | 0 | 0 |
| 0 | 0 | 0 | 0 | 0 |
| 0 | 0 | 0 | 0 | 0 |
| 0 | 0 | 0 | 0 | 0 |
| 0 | 0 | 0 | 0 | 0 |
| 0 | 0 | 0 | 1.45191163 | 0 |
| 0 | 0 | 0 | 0 | 0 |
| 0 | 0 | 0 | 0 | 0 |
| 0 | 0 | 0 | 0 | 0 |
| 0 | 0 | 0 | 0 | 0 |
| 0 | 0 | 0 | 0 | 0 |
| 0 | 0 | 0 | 0 | 0 |
| 0 | 0 | 0 | 0 | 0 |
| 0 | 0 | 0 | 0 | 0 |
| 0 | 0 | 0 | 0 | 0 |

| | | | | |
|---|---|---|---|---|
| 0 | 0 | 0 | 0 | 0 |
| 0 | 0 | 0 | 0 | 0 |
| 0 | 0 | 0 | 0 | 0 |
| 0 | 0 | 0 | 0 | 0 |
| 0 | 0 | 0 | 0 | 0 |
| 0 | 0 | 0 | 0 | 0 |
| 0 | 0 | 0 | 0 | 0 |
| 0 | 0 | 0 | 0 | 0 |
| 0 | 0 | 0 | 0 | 0 |
| 0 | 0 | 0 | 0 | 0 |
| 0 | 0 | 0.003682209 | 0 | 0.004532579 |
| 0 | 0 | 0 | 0 | 0 |
| 0 | 0 | 0 | 0.169737062 | 0 |
| 0 | 0 | 0 | 0 | 0 |
| 0 | 0 | 0 | 0 | 0 |
| 0 | 0 | 0 | 0 | 0 |
| 0 | 0 | 0 | 0 | 0 |
| 0 | 0 | 0 | 0 | 0 |
| 0 | 0 | 0 | 0 | 0 |
| 0 | 0 | 0 | 0 | 0 |
| 0.005547055 | 0.016789502 | 0.00373722 | 0 | 0 |
| 0 | 0 | 0 | 0 | 0 |
| 0 | 0 | 0 | 0 | 0 |
| 1.154538468 | 1.810597697 | 1.573017915 | 0.417064472 | 2.306536636 |
| 0 | 0 | 0 | 0 | 0 |
| 0 | 0 | 0 | 0 | 0 |
| 0 | 0 | 0 | 0 | 0 |
| 0 | 0 | 0 | 0 | 0 |
| 0 | 0 | 0 | 0 | 0 |
| 0 | 0 | 0 | 0 | 0 |
| 0 | 0 | 0 | 0 | 0 |
| 0 | 0 | 0 | 0 | 0 |
| 0 | 0 | 0 | 0 | 0 |
| 0 | 0 | 0 | 0 | 0 |

| PHL018-0095_V2 | PHL018-0027_V1 | PHL018-0006_V1 | PHL018-0045_V2 | PHL018-0140_V2 |
|---|---|---|---|---|
| CTRL | EYCF | EYCF | EYCF | CTRL |
| 0.068307994 | 0.383877232 | 0.008804776 | 0.097384282 | 0.01874823 |
| 3.889068111 | 4.411332954 | 6.243075024 | 1.321128018 | 1.470644254 |
| 10.73897932 | 15.34303176 | 0 | 4.048177918 | 7.577410985 |
| 0 | 0 | 0 | 0 | 0 |
| 0 | 0 | 0 | 0 | 0 |
| 0 | 0 | 0 | 0 | 0 |
| 0 | 0 | 0 | 0 | 0 |
| 0 | 0 | 0 | 0 | 0 |
| 0 | 0 | 0 | 0 | 0 |
| 0 | 0 | 0 | 0 | 0 |
| 0 | 0 | 0 | 0 | 0 |
| 0 | 0 | 0 | 0 | 0 |
| 0 | 0 | 0 | 0 | 0 |
| 3.522492914 | 3.615227506 | 2.133175699 | 46.25759854 | 6.859366816 |
| 0 | 0 | 0 | 0 | 0 |
| 0 | 0 | 0 | 0 | 0 |
| 0 | 0 | 0 | 0.004698401 | 0 |
| 0 | 0 | 0 | 0 | 0 |
| 0 | 0 | 0 | 0 | 0 |
| 0 | 0 | 0 | 0 | 0 |
| 0 | 0 | 0 | 0 | 0 |
| 0 | 0 | 0 | 0 | 0 |
| 0 | 0 | 0 | 0 | 0 |
| 0 | 0 | 0 | 0 | 0 |
| 0 | 0 | 0.014251903 | 0 | 0 |
| 0 | 0 | 0 | 0 | 0 |
| 0 | 0 | 0 | 0 | 0 |
| 0 | 0 | 0 | 0 | 0 |
| 0 | 0 | 0 | 0 | 0 |
| 0 | 0 | 0 | 0 | 0 |
| 0 | 0 | 0 | 0 | 0 |
| 0 | 0 | 0 | 0 | 0 |
| 0 | 0 | 0 | 0 | 0 |
| 0 | 0 | 0 | 0 | 0 |
| 0 | 0 | 0 | 0 | 0 |
| 1.421869832 | 0 | 8.0888034 | 0 | 3.362507458 |
| 0 | 0 | 0 | 0 | 0 |
| 0 | 0 | 0 | 0 | 0 |
| 0 | 0 | 0 | 0 | 0 |
| 0 | 0 | 0 | 0 | 0 |
| 0 | 0 | 0 | 0 | 0 |
| 0.00449918 | 0.044064349 | 0.989668086 | 0.001062613 | 0 |
| 0.018017984 | 0.008065605 | 0.023824572 | 0.003023628 | 0.107842815 |
| 0 | 0.134368168 | 0.112776839 | 0.015681796 | 0.02474858 |
| 0.001181987 | 0.005788111 | 0.032240437 | 0 | 0 |
| 0 | 0 | 0 | 0 | 0 |

| | | | | |
|---|---|---|---|---|
| 0 | 0 | 0.005497935 | 0 | 0.001734354 |
| 0 | 0.005160764 | 0 | 0.005152314 | 0.015908947 |
| 0.061650373 | 0 | 0 | 0.081497894 | 0.003069287 |
| 0.218550072 | 0.328421678 | 0.974293193 | 0.19229484 | 4.496216368 |
| 0 | 0.049100462 | 0 | 0.034887854 | 0 |
| 0 | 0 | 0.001399846 | 0 | 0 |
| 0.129620029 | 0.106845422 | 0.685682575 | 0 | 0.064796934 |
| 0 | 0 | 0.003400763 | 0 | 0 |
| 0 | 0 | 0 | 0 | 0 |
| 0 | 0 | 0 | 0 | 0 |
| 0 | 0 | 0 | 0 | 0 |
| 1.441696046 | 0.682535516 | 0.246815651 | 0 | 0.034096069 |
| 0 | 0 | 0 | 0 | 0 |
| 0.056021462 | 0.074434335 | 0.016065564 | 0.176771491 | 0.029308753 |
| 0.001664878 | 0 | 0 | 0 | 0.000837746 |
| 0 | 0 | 0 | 0 | 0 |
| 1.062161929 | 1.041853213 | 0.919178121 | 0 | 0.001116089 |
| 0 | 0 | 0 | 0 | 0 |
| 0 | 0 | 0 | 0 | 0 |
| 0 | 0 | 0 | 0 | 0 |
| 0.091894933 | 0.136598278 | 0.031702829 | 0.06813948 | 0.32174728 |
| 0.124404279 | 0.07219373 | 0.001344265 | 0.043162698 | 0.008481118 |
| 0 | 0 | 0 | 0 | 0.004529792 |
| 0 | 0 | 0 | 0 | 0 |
| 0.033019012 | 0 | 0 | 0 | 0 |
| 0 | 0 | 0 | 0 | 0 |
| 0 | 0 | 0 | 0 | 0 |
| 0 | 0 | 0 | 0 | 0 |
| 0 | 0 | 0.004422095 | 0 | 0 |
| 0 | 0 | 0 | 0 | 0 |
| 0 | 0 | 0 | 0.052967666 | 0.508417004 |
| 0 | 0 | 0 | 0 | 0 |
| 0 | 0 | 0 | 0 | 0 |
| 0 | 0 | 0 | 0 | 0 |
| 0 | 0 | 0 | 0 | 0 |
| 0 | 0 | 0 | 0 | 0 |
| 0 | 0 | 0 | 0 | 0 |
| 1.269600579 | 0.022073751 | 0 | 0 | 0 |
| 0 | 0 | 0 | 0 | 0 |
| 0 | 0 | 0 | 0 | 0 |
| 0 | 0 | 0 | 0 | 0 |
| 0 | 0 | 0 | 0 | 0 |
| 0 | 0 | 0 | 0 | 0 |
| 0 | 0 | 0 | 0 | 0 |
| 0 | 0 | 0 | 0 | 0 |
| 0 | 0 | 0 | 0 | 0 |
| 0 | 0 | 0.000949697 | 0 | 0 |
| 0 | 1.422206608 | 0 | 0.315255218 | 0 |

| | | | | |
|---|---|---|---|---|
| 0 | 0 | 0 | 0 | 0 |
| 0 | 0 | 0 | 0.131506192 | 0 |
| 0 | 0 | 0 | 0 | 0 |
| 0 | 0 | 0 | 0 | 0 |
| 0 | 0.006121942 | 0.001967302 | 0 | 0 |
| 0 | 0 | 0 | 0 | 0 |
| 0 | 0 | 0 | 0 | 0 |
| 0 | 0.002136697 | 0 | 0 | 0 |
| 0 | 0 | 0 | 0 | 0 |
| 0 | 0 | 0 | 0 | 0 |
| 0 | 0 | 0 | 0 | 0 |
| 0 | 0 | 0.008404528 | 0 | 0 |
| 0 | 0 | 0 | 0 | 0 |
| 0 | 0 | 0 | 0 | 0 |
| 0 | 0 | 0 | 0 | 0 |
| 0 | 0 | 0 | 0 | 0 |
| 0 | 0 | 0 | 0 | 0 |
| 0 | 0 | 0 | 0 | 0 |
| 0 | 0.025616797 | 0.329871183 | 0 | 0.078246446 |
| 0 | 0 | 0 | 0 | 0 |
| 0 | 0 | 0 | 0 | 0 |
| 0 | 0 | 0 | 0 | 0 |
| 0 | 0 | 0 | 0 | 0 |
| 0 | 0 | 0 | 0 | 0 |
| 0 | 0 | 0 | 0 | 0 |
| 0 | 0 | 0 | 0 | 0 |
| 0 | 0 | 0 | 0 | 0 |
| 0 | 0 | 0 | 0.000665961 | 0 |
| 0 | 0 | 0 | 0 | 0 |
| 0 | 0 | 0 | 0 | 0 |
| 0 | 0.065498325 | 0.002576505 | 0 | 0.019867781 |
| 0.036150082 | 0.000776423 | 0.01729906 | 0 | 0 |
| 0 | 0 | 0 | 0 | 0 |
| 0 | 0 | 0 | 0 | 0 |
| 0 | 0 | 0 | 0 | 0 |
| 0 | 0 | 0 | 0 | 0 |
| 0 | 0 | 0 | 0 | 0 |
| 0 | 0 | 0 | 0 | 0 |
| 0 | 0 | 0 | 0 | 0 |
| 0 | 0 | 0 | 0 | 0 |
| 0 | 0 | 0 | 0 | 0 |
| 0 | 0 | 0 | 0 | 0 |
| 0.000652872 | 0 | 0 | 0 | 0 |
| 0 | 0 | 0 | 0 | 0 |
| 0 | 0 | 0 | 0 | 0 |
| 0 | 0 | 0 | 0 | 0 |
| 0 | 0 | 0 | 0 | 0 |
| 0 | 0 | 0 | 0 | 0 |

| | | | | |
|---|---|---|---|---|
| 0 | 0 | 0 | 0 | 0 |
| 0 | 0 | 0.001503812 | 0 | 0 |
| 0 | 0 | 0 | 0 | 0 |
| 0 | 0 | 0 | 0 | 0 |
| 0 | 0 | 0.215344666 | 0 | 0 |
| 0 | 0 | 0 | 0 | 0 |
| 0 | 0 | 0 | 0 | 0 |
| 0 | 0 | 0 | 0 | 0 |
| 0 | 0 | 0 | 0 | 0 |
| 0 | 0 | 0 | 0 | 0 |
| 0 | 0 | 0 | 0 | 0 |
| 0 | 0 | 0 | 0 | 0 |
| 0 | 0 | 0 | 0 | 0 |
| 0 | 0 | 0 | 0 | 0 |
| 0 | 0 | 0 | 0 | 0 |
| 0 | 0 | 0 | 0 | 0 |
| 0 | 0 | 0 | 0 | 0 |
| 0 | 0 | 0 | 0 | 0 |
| 0 | 0 | 0 | 0 | 0 |
| 0 | 0 | 0 | 0 | 0 |
| 0 | 0 | 0 | 0 | 0 |
| 0 | 0 | 0 | 0 | 0 |
| 0 | 0 | 0 | 0 | 0 |
| 0 | 0 | 0 | 0 | 0 |
| 0.018103437 | 0 | 0.007395345 | 0 | 0 |
| 0 | 0 | 0 | 0 | 0 |
| 0 | 0 | 0.000782975 | 0 | 0 |
| 0 | 0 | 0 | 0 | 0 |
| 0 | 0 | 0 | 0 | 0 |
| 0 | 0 | 0 | 0 | 0 |
| 0 | 0 | 0 | 0 | 0 |
| 0 | 0 | 0 | 0 | 0 |
| 0 | 0 | 0 | 0 | 0 |
| 0 | 0 | 0 | 0 | 0 |
| 0 | 0 | 0 | 0 | 0 |
| 0 | 0 | 0 | 0 | 0 |
| 0 | 0 | 0 | 0 | 0 |
| 0 | 0 | 0 | 0 | 0 |
| 0 | 0 | 0 | 0 | 0 |
| 0 | 0 | 0 | 0 | 0 |
| 0 | 0 | 0 | 0 | 0 |
| 0 | 0 | 0 | 0 | 0 |
| 0 | 0 | 0 | 0 | 0 |
| 0 | 0 | 0 | 0 | 0 |
| 0 | 0 | 0 | 0.003059222 | 0 |
| 0 | 0 | 0 | 0 | 0 |
| 0 | 0 | 0 | 0 | 0 |
| 0 | 0 | 0 | 0 | 0.005628722 |

| | | | | |
|---|---|---|---|---|
| 0 | 0 | 0 | 0 | 0 |
| 0 | 0 | 0 | 0 | 0 |
| 0 | 0 | 0 | 0 | 0 |
| 0 | 0 | 0 | 0 | 0 |
| 0 | 0 | 0 | 0 | 0 |
| 0 | 0 | 0 | 0 | 0 |
| 0 | 0 | 0 | 0 | 0 |
| 0 | 0 | 0 | 0 | 0 |
| 0 | 0 | 0 | 0 | 0 |
| 0 | 0 | 0 | 0 | 0 |
| 0 | 0 | 0 | 0 | 0 |
| 0 | 0 | 0 | 0 | 0 |
| 0.001455118 | 0 | 0 | 0.001590695 | 0 |
| 0 | 0 | 0 | 0 | 0 |
| 0.001127859 | 0 | 0 | 0 | 0 |
| 0 | 0 | 0 | 0 | 0 |
| 0 | 0 | 0 | 0 | 0 |
| 0 | 0 | 0 | 0 | 0 |
| 0 | 0 | 0 | 0 | 0 |
| 0 | 0 | 0 | 0 | 0 |
| 0.002332046 | 0.050888544 | 0.038686767 | 0.049570304 | 0.056325954 |
| 0 | 0 | 0 | 0 | 0 |
| 0.001193632 | 0 | 0 | 0 | 0 |
| 0.007295565 | 0 | 0 | 0.002067674 | 0 |
| 0 | 0 | 0 | 0 | 0 |
| 0 | 0 | 0 | 0 | 0 |
| 0.000905419 | 0.008167481 | 0.009315396 | 0 | 0.008200727 |
| 0 | 0 | 0 | 0 | 0 |
| 0 | 0 | 0 | 0 | 0 |
| 0 | 0 | 0 | 0.112013413 | 0 |
| 0 | 0 | 0 | 0 | 0 |
| 0 | 0 | 0 | 0 | 0 |
| 0 | 0 | 0 | 0 | 0 |
| 0.007601076 | 0 | 0.048195243 | 0 | 0 |
| 0.008092552 | 0.003910724 | 0.003146459 | 0 | 0.00381757 |
| 0 | 0 | 0 | 0 | 0 |
| 0 | 0 | 0 | 0 | 0 |
| 0 | 0 | 0 | 0 | 0 |
| 0 | 0 | 0 | 0 | 0 |
| 0 | 0 | 0 | 0 | 0 |
| 0 | 0 | 0 | 0 | 0 |
| 0 | 0 | 0 | 0 | 0 |
| 0 | 0 | 0 | 0 | 0 |
| 0 | 0 | 0 | 0 | 0 |
| 0 | 0 | 0 | 0 | 0 |
| 0 | 0 | 0 | 0 | 0.023262349 |
| 0 | 0 | 0 | 0 | 0 |
| 0.000901089 | 0 | 0 | 0.001225836 | 0 |

| | | | | |
|---|---|---|---|---|
| 0 | 0 | 0 | 0 | 0 |
| 0 | 0 | 0 | 0.002964042 | 0 |
| 0 | 0 | 0.003467408 | 0 | 0 |
| 0 | 0 | 0 | 0.000779098 | 0 |
| 0 | 0 | 0 | 0 | 0 |
| 0 | 0 | 0 | 0 | 0 |
| 0 | 0 | 0.000736573 | 0 | 0 |
| 0 | 0 | 0.007438334 | 0 | 0 |
| 0 | 0 | 0 | 0 | 0.001072186 |
| 0 | 0 | 0 | 0 | 0 |
| 0 | 0 | 0 | 0 | 0 |
| 0 | 0 | 0 | 0 | 0 |
| 0 | 0 | 0 | 0 | 0 |
| 0 | 0 | 0 | 0.254659524 | 0 |
| 0 | 0 | 0 | 0 | 0 |
| 0 | 0 | 0 | 0 | 0 |
| 0 | 0 | 0 | 0 | 0 |
| 0 | 0 | 0 | 0 | 0 |
| 0 | 0 | 0 | 0 | 0.001106694 |
| 0 | 0 | 0 | 0 | 0 |
| 0.040012865 | 0.008640325 | 0 | 0.018619473 | 0 |
| 0 | 0 | 0 | 0 | 0 |
| 0 | 0 | 0 | 0 | 0 |
| 0 | 0 | 0 | 0 | 0 |
| 0 | 0.014950885 | 0.008225 | 0 | 0.003243278 |
| 0 | 0 | 0 | 0 | 0 |
| 0 | 0 | 0 | 0 | 0 |
| 0 | 0 | 0 | 0 | 0 |
| 0 | 0 | 0.044151959 | 0 | 0 |
| 0 | 0 | 0 | 0 | 0 |
| 0.325425023 | 0.323809641 | 0.19634741 | 1.490785941 | 0.098229953 |
| 0 | 0 | 0 | 0 | 0 |
| 0 | 0 | 0 | 0.008616858 | 0 |
| 0 | 0 | 0 | 0 | 0 |
| 0 | 0 | 0 | 0 | 0 |
| 0 | 0 | 0 | 0 | 0 |
| 0 | 0 | 0 | 0 | 0 |
| 0 | 0 | 0 | 0 | 0 |
| 0 | 0 | 0 | 0 | 0 |
| 0 | 0 | 0.002660594 | 0 | 0 |
| 0 | 0 | 0 | 0 | 0 |
| 0.036069378 | 0.013996643 | 0.013575627 | 0 | 0.009992517 |
| 0 | 0 | 0 | 0 | 0 |
| 0 | 0 | 0.021434702 | 0 | 0 |
| 0 | 0 | 0 | 0 | 0 |
| 0 | 0 | 0 | 0 | 0 |
| 0 | 0 | 0 | 0 | 0 |
| 0 | 0 | 0 | 0 | 0 |

| | | | | |
|---|---|---|---|---|
| 0 | 0 | 0 | 0 | 0 |
| 0 | 0 | 0 | 0 | 0 |
| 0 | 0.1209271 | 0.112749986 | 0.019254256 | 0.024326301 |
| 0 | 0 | 0 | 0 | 0 |
| 0 | 0 | 0 | 0 | 0 |
| 0 | 0 | 0 | 0 | 0 |
| 0 | 0 | 0 | 0 | 0 |
| 0 | 0 | 0 | 0 | 0 |
| 0 | 0 | 0 | 0 | 0 |
| 0 | 0.000959644 | 0.001336332 | 0 | 0 |
| 0 | 0 | 0 | 0 | 0 |
| 0 | 0 | 0 | 0 | 0 |
| 0 | 0 | 0 | 0 | 0 |
| 0 | 0 | 0 | 0.001912758 | 0 |
| 0.003249389 | 0.003349903 | 0.005331239 | 0.004538852 | 0.003270107 |
| 0 | 0 | 0 | 0 | 0 |
| 0 | 0.002999004 | 0.010124115 | 0 | 0.001862997 |
| 0 | 0 | 0 | 0 | 0 |
| 0 | 0 | 0 | 0 | 0 |
| 0 | 0 | 0 | 0 | 0 |
| 0 | 0 | 0 | 0 | 0 |
| 0 | 0 | 0 | 0.001826801 | 0 |
| 0 | 0 | 0 | 0 | 0 |
| 0 | 0 | 0 | 0 | 0 |
| 0 | 0 | 0 | 0 | 0 |
| 0 | 0 | 0 | 0 | 0 |
| 0 | 0 | 4.476100271 | 0.000713242 | 0 |
| 0 | 0 | 0 | 0 | 0 |
| 0 | 0 | 0 | 0 | 0 |
| 0 | 0 | 0 | 0 | 0 |
| 0 | 0 | 0 | 0 | 0 |
| 0 | 0 | 0 | 0 | 0 |
| 0 | 0 | 0 | 0 | 0 |
| 0.006055571 | 0 | 0.001844054 | 0 | 0 |
| 0 | 0 | 0 | 0 | 0 |
| 0 | 0.00245788 | 0 | 0.001216259 | 0 |
| 0 | 0 | 0 | 0 | 0 |
| 0.004199128 | 0.005565882 | 0.129751597 | 0 | 0 |
| 0 | 0 | 0 | 0 | 0 |
| 0 | 0 | 0 | 0 | 0 |
| 0 | 0 | 0 | 0 | 0 |
| 0 | 0 | 0 | 0 | 0 |
| 0 | 0 | 0 | 0 | 0 |
| 0 | 0 | 0 | 0.002850981 | 0 |
| 0 | 0 | 0 | 0 | 0 |
| 0 | 0 | 0 | 0 | 0 |
| 0 | 0 | 0 | 0 | 0 |
| 0.020344788 | 0.002247226 | 0 | 0.002162259 | 0 |

| | | | | |
|---|---|---|---|---|
| 0 | 0 | 0 | 0 | 0 |
| 0 | 0 | 0 | 0 | 0 |
| 0 | 0 | 0 | 0 | 0.00201315 |
| 0 | 0 | 0 | 0 | 0 |
| 0 | 0.010114879 | 0.001926189 | 0.00641699 | 0.016456566 |
| 0 | 0 | 0 | 0 | 0 |
| 0 | 0 | 0 | 0 | 0 |
| 0.003878999 | 0.144327106 | 0.00607495 | 0.004197585 | 0.009936769 |
| 0 | 0 | 0 | 0 | 0 |
| 0.010177155 | 0.025480486 | 0 | 0.097965256 | 0 |
| 0 | 0 | 0 | 0.001397054 | 0 |
| 0.001395741 | 0.008057924 | 0.104995452 | 0 | 0 |
| 0 | 0.028814759 | 0.023913099 | 0 | 0.007458283 |
| 0 | 0 | 0 | 0.024334874 | 0 |
| 0 | 0 | 0 | 0.006295477 | 0 |
| 0.002225038 | 0 | 0 | 0 | 0 |
| 0 | 0 | 0 | 0 | 0 |
| 0 | 0 | 0 | 0.001875769 | 0 |
| 0 | 0 | 0 | 0 | 0.267098525 |
| 0 | 0.316262115 | 0 | 0 | 0 |
| 0 | 0 | 0 | 0 | 0 |
| 0 | 0 | 0 | 0 | 0 |
| 0 | 0.012839297 | 0.008364484 | 0 | 0 |
| 0 | 0 | 0.001010392 | 0 | 0 |
| 0 | 0 | 0 | 0 | 0 |
| 0 | 0 | 0.413305201 | 0 | 0 |
| 0 | 0 | 0 | 0 | 0 |
| 0 | 0.060570938 | 0 | 0 | 0.003001427 |
| 0 | 0 | 0 | 0.82507818 | 0 |
| 0 | 0.002092783 | 0 | 0 | 0 |
| 0 | 0 | 0 | 0 | 0 |
| 0 | 0 | 0 | 0 | 0 |
| 0 | 0 | 0 | 0 | 0 |
| 0 | 0 | 0 | 0 | 0.007465798 |
| 0 | 0 | 0 | 0 | 0 |
| 0 | 0 | 0 | 0 | 0.007047264 |
| 0 | 0 | 0 | 1.873864774 | 0.091442023 |
| 0 | 0.009264751 | 0 | 0 | 0 |
| 0 | 0 | 0 | 0 | 0 |
| 0 | 0 | 0 | 0.005293065 | 0 |
| 0 | 0 | 0 | 0 | 0.030718365 |
| 0 | 0 | 0 | 0.037679258 | 0 |
| 0 | 0 | 0 | 0.028089072 | 0 |
| 0 | 0 | 0 | 0 | 0 |
| 0 | 0 | 0 | 0.025538424 | 0 |
| 0 | 0 | 0.002417742 | 0 | 0 |
| 0.057353047 | 0.002845692 | 0.002054737 | 0.015646261 | 0 |
| 0 | 0.021237644 | 0 | 0 | 0 |

| | | | | |
|---|---|---|---|---|
| 0 | 0 | 0 | 0.001232351 | 0 |
| 0.058954648 | 0.023321902 | 0.017058316 | 0 | 0.010348348 |
| 0 | 0 | 0 | 0.006872883 | 0 |
| 0 | 0 | 0 | 0 | 0 |
| 0 | 0 | 0 | 0 | 0 |
| 0 | 0.022164232 | 0 | 0.007188607 | 0 |
| 0 | 0 | 0 | 0 | 0 |
| 0 | 0 | 0.00089706 | 0 | 0 |
| 0 | 0 | 0.011245909 | 0 | 0 |
| 0 | 0 | 0 | 0 | 0 |
| 0.001961215 | 0.029172859 | 0.010189527 | 0.025965586 | 0.00169176 |
| 0.002888451 | 0 | 0 | 0 | 0 |
| 0 | 0 | 0 | 0 | 0 |
| 0.003970409 | 0.002728817 | 0.001773312 | 0.001575384 | 0.003196579 |
| 0 | 0 | 0 | 0.030741536 | 0.088944986 |
| 0 | 0 | 0 | 0 | 0 |
| 0 | 0 | 0 | 0 | 0 |
| 0 | 0 | 0 | 0.001095074 | 0.026700925 |
| 0 | 0 | 0 | 0 | 0.005953966 |
| 0 | 0.035016669 | 0 | 0 | 0.008625506 |
| 0 | 0 | 0.001637718 | 0 | 0 |
| 0.137661983 | 0.385652923 | 0.148650634 | 0 | 0 |
| 0 | 0.035217428 | 0 | 0 | 0 |
| 0 | 0 | 0 | 0 | 0 |
| 0 | 0 | 0 | 0 | 0 |
| 0 | 0 | 0 | 0 | 0 |
| 0 | 0 | 0 | 0.046363621 | 0.322621024 |
| 0 | 0 | 0 | 0 | 0 |
| 0 | 0 | 0 | 0 | 0 |
| 0.192828968 | 1.269959997 | 0.57667425 | 0.380355442 | 0.691503894 |
| 0 | 0 | 0.002234238 | 0 | 0 |
| 0 | 0 | 0 | 0 | 0 |
| 0 | 0 | 0 | 0 | 0 |
| 0 | 0 | 0 | 0.001946248 | 0.003327475 |
| 0 | 0 | 0 | 0 | 0.011547198 |
| 0 | 0 | 0 | 0 | 0 |
| 0 | 0 | 0.325466852 | 0 | 0 |
| 0 | 0.001946974 | 0.03072712 | 0 | 0 |
| 0 | 0 | 0 | 0 | 0.13900454 |
| 0 | 0 | 0 | 0 | 0.005673556 |
| 0.054184194 | 0.314522494 | 0 | 0.003515372 | 0.270239367 |
| 3.753133841 | 7.721865856 | 4.039620594 | 4.912496411 | 4.426663638 |
| 0 | 0.001783554 | 0.033351867 | 0 | 0 |
| 0 | 0 | 0.006030038 | 1.766245957 | 0 |
| 0 | 0 | 0 | 0.002921964 | 0.001235187 |
| 0 | 0 | 0 | 0.054051314 | 0 |
| 0.001141972 | 0.001412756 | 0 | 0.003495444 | 0 |
| 0 | 0 | 0 | 0 | 0 |

| | | | | |
|---|---|---|---|---|
| 0.002818003 | 0.003776724 | 0.039646245 | 0 | 0 |
| 0 | 0 | 0 | 0 | 0.00054363 |
| 2.335717185 | 2.336714462 | 2.121887895 | 0 | 0 |
| 0 | 0 | 0 | 0 | 0.048741166 |
| 0 | 0.002725712 | 0 | 0 | 0 |
| 0 | 0 | 0 | 0.007292938 | 0 |
| 0 | 0 | 0 | 0 | 0 |
| 0 | 0 | 0 | 0 | 0 |
| 0 | 0 | 0 | 0 | 0.005696287 |
| 0 | 0 | 0 | 0.001410561 | 0 |
| 0 | 0 | 0 | 0 | 0 |
| 0 | 0 | 0 | 0 | 0 |
| 0 | 0 | 0 | 0 | 0 |
| 0.026768453 | 0.002102589 | 0.000975972 | 0.018207809 | 0 |
| 0 | 0 | 0 | 0 | 0 |
| 0.023309837 | 0.029371074 | 0.008199078 | 0 | 0.046315412 |
| 0.00485815 | 0.076492335 | 0.281933876 | 0 | 0.002222329 |
| 0.00214635 | 0.015212608 | 0 | 0.001140578 | 0 |
| 0 | 0.022574516 | 0 | 0 | 0 |
| 0 | 0 | 0 | 0 | 0 |
| 0 | 0 | 0.001331773 | 0.115709794 | 0 |
| 0 | 0.007393928 | 0 | 0 | 0 |
| 0 | 0 | 0 | 0 | 0.070066635 |
| 0 | 0 | 0 | 0 | 0.665300316 |
| 0.002063823 | 0 | 0 | 0 | 0 |
| 0.150103368 | 0 | 0 | 0 | 0 |
| 0.004205086 | 0.049709861 | 0 | 0 | 0.016363336 |
| 0 | 0.452124989 | 0 | 0.008899714 | 0.000714167 |
| 0 | 0 | 0 | 0 | 0 |
| 0 | 0 | 0 | 0.001223716 | 0 |
| 0.174821617 | 0.015385434 | 0 | 0.045498747 | 0.008796812 |
| 0 | 0.001638414 | 0.103125507 | 0 | 0.002878895 |
| 0.396756903 | 0.155063154 | 0.023750834 | 0 | 0.003654797 |
| 0 | 0.001788496 | 0.003320708 | 0.001475034 | 0 |
| 0 | 0.004148726 | 0.014220846 | 0 | 0 |
| 0 | 0 | 0 | 0 | 0 |
| 0 | 0 | 0 | 0 | 0 |
| 0.012410344 | 0.077427169 | 0.009829701 | 0.148817356 | 0.003660707 |
| 0 | 0 | 2.393292387 | 0 | 0 |
| 0.014695256 | 0 | 0 | 0 | 0 |
| 0 | 0 | 0 | 0 | 0 |
| 0 | 0.00647265 | 0 | 0 | 0 |
| 0 | 0.025071817 | 0 | 0 | 0.001932205 |
| 0.580098989 | 0.097636186 | 0 | 0.097566 | 0.058833617 |
| 0 | 0 | 0 | 0.087542927 | 1.244078479 |
| 0 | 0 | 0 | 0 | 0 |
| 0 | 0 | 0 | 0 | 0 |
| 0 | 0 | 0 | 0 | 0 |

| | | | | |
|---|---|---|---|---|
| 0.007931216 | 0 | 0 | 0 | 0 |
| 0.017785358 | 0.622205086 | 7.253246055 | 0.042390968 | 0.725926522 |
| 0 | 0 | 0.001145889 | 0 | 0 |
| 0 | 0 | 0 | 0 | 0 |
| 0 | 0 | 0 | 0 | 0 |
| 0 | 0 | 0 | 0 | 0 |
| 0 | 0 | 0 | 0 | 0 |
| 0.001567659 | 0 | 0.003215053 | 0 | 0 |
| 0 | 0.003026741 | 0 | 0 | 0 |
| 0 | 0 | 0 | 0 | 0 |
| 0 | 0.004243943 | 0 | 0 | 0.000654134 |
| 0 | 0.03027653 | 0.054361342 | 0.003567156 | 0.008769165 |
| 0 | 0 | 0.059626055 | 0 | 0 |
| 0 | 0 | 0 | 0 | 0 |
| 0 | 0 | 0 | 0 | 0 |
| 0 | 0 | 0 | 0.004511761 | 0 |
| 0.0020419 | 0.026196333 | 0.01715388 | 0.001543215 | 0.013471137 |
| 0 | 0.073781871 | 0 | 0.364601776 | 0.238302828 |
| 0 | 0 | 0 | 0 | 0 |
| 0 | 0 | 0 | 0.099677059 | 0.094307572 |
| 0.008890795 | 0.022432122 | 0.114201122 | 0 | 0.038615442 |
| 0 | 0 | 0.018947328 | 0 | 0 |
| 0 | 0 | 0 | 0 | 0 |
| 0 | 0 | 0 | 0 | 0 |
| 0.004324214 | 0.011752842 | 0.669319753 | 0 | 0.088222533 |
| 0 | 0 | 0 | 0 | 0 |
| 0.035333572 | 0 | 0 | 0 | 0 |
| 0.275530503 | 0.269953349 | 0.036175682 | 0 | 0.109863802 |
| 0 | 0 | 0 | 0 | 0 |
| 0.001799753 | 0 | 0 | 0 | 0 |
| 0.002401444 | 0 | 0.016343805 | 0.010889694 | 0.000805585 |
| 0 | 0 | 0 | 0 | 0 |
| 0 | 0 | 0 | 0.021817723 | 0 |
| 0 | 0 | 0 | 0 | 0 |
| 0 | 0 | 0.28911366 | 0 | 0.010904175 |
| 0.071967995 | 0.08287186 | 0.085393019 | 0.001789195 | 0.805166303 |
| 0 | 0 | 0 | 0 | 0 |
| 0 | 0 | 0 | 0 | 0 |
| 0.007975939 | 0.004743841 | 0.037286824 | 0 | 0.003704673 |
| 0 | 0 | 0 | 0 | 0 |
| 0 | 0 | 0 | 0.083471918 | 1.169712325 |
| 0 | 0 | 0 | 0.006049914 | 0 |
| 0.053291777 | 0.003649877 | 0 | 0.006337217 | 0.018564769 |
| 0 | 0 | 0 | 0 | 0 |
| 0 | 0 | 0 | 0.270687838 | 0 |
| 0 | 0 | 0.331421768 | 0 | 0 |
| 0 | 0 | 0 | 0 | 0 |
| 0 | 0 | 0 | 0.01310428 | 0 |

| | | | | |
|---|---|---|---|---|
| 0 | 0 | 0 | 0 | 0.019386044 |
| 0 | 0 | 0.0008816 | 0 | 0 |
| 0 | 0 | 0 | 0 | 0 |
| 0 | 0 | 0.076642859 | 0 | 0.003057668 |
| 0 | 0 | 0 | 0 | 0 |
| 0 | 0 | 0 | 0 | 0 |
| 0 | 0 | 0 | 0.008726371 | 0 |
| 0 | 0 | 0.003902088 | 0 | 0 |
| 0.13710051 | 0 | 0.453531622 | 0 | 0.052359605 |
| 0.017844647 | 0 | 0.165216481 | 0 | 0 |
| 0 | 0 | 0 | 0 | 0 |
| 0.004158592 | 0.002786699 | 0.003980059 | 0 | 0.004394362 |
| 0 | 0 | 0 | 0 | 0 |
| 0 | 0 | 0.006980766 | 0 | 0 |
| 0.134178725 | 0 | 0.005419186 | 0.017068966 | 0.007770511 |
| 0 | 0 | 0 | 0 | 0 |
| 0.01410043 | 0 | 0.027632791 | 0 | 0 |
| 0 | 0 | 0 | 0 | 0 |
| 0 | 0 | 0 | 0 | 0 |
| 0 | 0 | 0 | 0 | 0 |
| 0 | 0 | 0 | 0 | 0 |
| 0 | 0 | 0 | 0 | 0 |
| 0 | 0 | 0.00091624 | 0 | 0 |
| 1.167203229 | 0.709682664 | 0 | 0.324338878 | 0.581168208 |
| 0 | 0 | 0 | 0 | 0.002938747 |
| 0 | 0 | 0.045508695 | 0 | 0 |
| 0 | 0 | 0.023277369 | 0 | 0 |
| 0 | 0 | 3.117251316 | 0 | 3.825016075 |
| 0 | 0 | 0 | 0 | 0 |
| 0 | 0 | 0 | 0 | 0 |
| 0 | 0 | 0.012768678 | 0 | 0 |
| 0 | 0 | 0.129600901 | 0 | 0 |
| 0.005692291 | 0.015701855 | 0 | 0 | 0.001238613 |
| 0 | 0 | 0 | 0 | 0 |
| 0 | 0 | 0.00875256 | 0 | 0 |
| 0.440141571 | 0 | 0.005249169 | 0 | 0 |
| 0 | 0 | 0.446588388 | 0 | 0 |
| 0 | 0 | 0 | 0 | 0 |
| 0 | 0 | 0 | 0 | 0 |
| 0 | 0 | 0 | 0 | 0 |
| 0.03391012 | 0.048531402 | 0 | 0.001187197 | 0.041553818 |
| 0 | 0 | 0.030010965 | 0 | 0 |
| 24.15348492 | 4.375827361 | 3.304032824 | 0.872010493 | 0.091269253 |
| 0 | 0 | 0.001330445 | 0 | 0.002273355 |
| 0 | 0 | 0 | 0 | 0 |
| 0 | 0 | 0 | 0 | 0 |
| 0 | 0 | 0 | 0 | 0 |
| 0 | 0.002224842 | 0 | 0 | 0 |

| | | | | |
|---|---|---|---|---|
| 0 | 0 | 0 | 0 | 0 |
| 0 | 0 | 0.07338521 | 0 | 0 |
| 0.198831734 | 0.209010739 | 0.01891957 | 0.396548287 | 0.013417073 |
| 0 | 0 | 0 | 0 | 0 |
| 0 | 0 | 0.007459161 | 0 | 0 |
| 0.001629697 | 0.002613502 | 0.001213125 | 0 | 0 |
| 0 | 0 | 0 | 0 | 0.004666256 |
| 0 | 0 | 0 | 0 | 0 |
| 0 | 0 | 0 | 0 | 0 |
| 0 | 0 | 0 | 0 | 0 |
| 0.562858326 | 0.055839859 | 0.00107998 | 0.07349298 | 0.01748854 |
| 6.054233465 | 3.162099965 | 0.687796682 | 0 | 1.930178825 |
| 0 | 0 | 0 | 0 | 0 |
| 0.009986305 | 0.13685098 | 0.371113938 | 0 | 0.000980485 |
| 0.070250501 | 0.848107549 | 0.002204412 | 1.193787723 | 0.136760846 |
| 0 | 0 | 0 | 0 | 0 |
| 0 | 0 | 0 | 0 | 0 |
| 0 | 0 | 0 | 0 | 0 |
| 0 | 0 | 0.006764545 | 0 | 0 |
| 0.04889302 | 0.229411768 | 0 | 0.174977891 | 0.022076657 |
| 0.316081753 | 0.0323714 | 0 | 0.001125811 | 0.158572604 |
| 0.148288909 | 0.411162263 | 0 | 16.03376734 | 0 |
| 2.468416296 | 1.749806838 | 0.132006889 | 1.637871579 | 0.12945796 |
| 0 | 0 | 0 | 0 | 0 |
| 0 | 0 | 0 | 0 | 0 |
| 0.00158667 | 0 | 0.018439565 | 0 | 0.006387144 |
| 0.291172004 | 0 | 0 | 0 | 0 |
| 0 | 0 | 0 | 0 | 0 |
| 0 | 0 | 0 | 0 | 0 |
| 0 | 0 | 0 | 0 | 0 |
| 0 | 0 | 0.003394536 | 0 | 0 |
| 0 | 0 | 0.16134126 | 0.066959663 | 0 |
| 0 | 0 | 0.018190355 | 0.043356201 | 0 |
| 0 | 0.043915266 | 0.000845826 | 0 | 0.016542882 |
| 0.011932483 | 0.001261702 | 0 | 0.005072771 | 0.00076978 |
| 0.010929668 | 0.034886702 | 0.002615111 | 0 | 0.000846104 |
| 0.004669845 | 0 | 0 | 0 | 0.001021656 |
| 0 | 0 | 0 | 0 | 0 |
| 1.551269537 | 0 | 0.262236319 | 0 | 0 |
| 0 | 0 | 0 | 0 | 0 |
| 0 | 0 | 0.004782608 | 0 | 0 |
| 0 | 0 | 0 | 0 | 0 |
| 0 | 0 | 0 | 0 | 0 |
| 0 | 0 | 0 | 0 | 0 |
| 0.206855216 | 0.02372233 | 0.001376417 | 0.009170914 | 0.00844275 |
| 0 | 0 | 0 | 0 | 0 |
| 0 | 0 | 0 | 0 | 0 |
| 0.051755251 | 0.049183084 | 0 | 0 | 0.022405379 |

| | | | | |
|---|---|---|---|---|
| 0 | 0 | 0 | 0 | 0 |
| 0 | 0 | 0 | 0 | 0 |
| 0 | 0 | 0 | 0.003160471 | 0 |
| 0 | 0 | 0.000782077 | 0 | 0.001233552 |
| 0 | 0 | 0 | 0 | 0 |
| 0 | 0 | 0.275599326 | 0 | 0 |
| 0 | 0 | 0.777880396 | 0 | 0 |
| 0.023121951 | 0 | 0 | 0.01102841 | 0.012486005 |
| 0 | 0 | 0 | 0 | 0 |
| 0 | 0 | 0 | 0 | 0 |
| 0.00819088 | 0 | 0 | 0.020118974 | 0 |
| 0 | 0 | 0 | 0 | 0 |
| 0 | 0 | 0.014681043 | 0 | 0 |
| 0 | 0 | 1.203542717 | 0 | 0 |
| 0 | 0 | 0 | 0 | 0 |
| 0 | 0 | 0 | 0 | 0 |
| 0 | 0 | 0.143814757 | 0 | 0 |
| 0.009470445 | 0 | 0.007432372 | 0 | 0 |
| 0 | 0 | 0 | 0 | 0 |
| 0 | 0 | 0.01128027 | 0 | 0.002234759 |
| 0 | 0 | 0.001449456 | 0 | 0.00152413 |
| 0 | 0 | 0 | 0 | 0 |
| 0.023147741 | 0 | 0.003552997 | 0 | 0 |
| 0 | 0 | 0 | 0 | 0 |
| 0 | 0 | 0 | 0 | 0 |
| 0 | 0 | 0 | 0 | 0 |
| 0 | 0 | 0 | 0 | 0 |
| 0 | 0 | 0 | 0 | 0 |
| 0 | 0 | 0 | 0 | 0 |
| 0.014185864 | 0 | 0.216791636 | 0 | 0.34263145 |
| 0 | 0 | 0.001886583 | 0.004113848 | 0 |
| 0 | 0 | 0 | 0 | 0 |
| 0 | 0 | 0.004924504 | 0 | 0 |
| 0 | 0 | 0 | 0 | 0 |
| 0 | 0.002424664 | 0 | 0 | 0 |
| 0 | 0 | 0 | 0 | 0 |
| 0.008146694 | 0 | 0.131456537 | 0 | 0 |
| 0.008089344 | 0.012707918 | 0 | 0.001473842 | 0.004651954 |
| 0 | 0 | 0 | 0 | 0 |
| 0.011398402 | 0.004112846 | 0 | 0.008560774 | 0 |
| 0 | 0.108053744 | 0.000518855 | 0 | 0.01582198 |
| 0 | 0 | 0 | 0 | 0 |
| 0 | 0 | 0 | 0 | 0 |
| 0.005237141 | 0 | 0.010382631 | 0 | 0.080940306 |
| 0.134281302 | 0 | 0.120353364 | 0 | 0 |
| 0 | 0 | 0 | 0 | 0 |
| 0 | 0 | 0.002452084 | 0 | 0 |
| 0 | 0 | 0 | 0 | 0 |

| | | | | |
|---|---|---|---|---|
| 0 | 0 | 0 | 0 | 0 |
| 0 | 0 | 0 | 0 | 0 |
| 0 | 0 | 0 | 0 | 0 |
| 0 | 0 | 0.005317412 | 0 | 0 |
| 0 | 0 | 0 | 0 | 0 |
| 0 | 0 | 0.7783994 | 0 | 0 |
| 0 | 0 | 0 | 0 | 0 |
| 0 | 0 | 0 | 0 | 0 |
| 0.016203569 | 0.147604651 | 0 | 0.331325914 | 0.028336545 |
| 0 | 0 | 0 | 0 | 0 |
| 0 | 0 | 0.001999593 | 0 | 0 |
| 0 | 0 | 0 | 0 | 0 |
| 0 | 0 | 0 | 0 | 0 |
| 0 | 0 | 0 | 0 | 0.001114056 |
| 0.02061616 | 0 | 0.245542402 | 0 | 8.322478741 |
| 0 | 0 | 0 | 0.17250432 | 0 |
| 0 | 0.027566364 | 0 | 0 | 0 |
| 0 | 0 | 0 | 0 | 0 |
| 0 | 0 | 0 | 0 | 0 |
| 0 | 0 | 0 | 0 | 0 |
| 0 | 0 | 0.000777576 | 0 | 0 |
| 0 | 0 | 0 | 0 | 0 |
| 0 | 0 | 0 | 0 | 0 |
| 0 | 0.136960769 | 0 | 0 | 0 |
| 0 | 0 | 0.002989654 | 0.001062385 | 0.212512527 |
| 0 | 2.465614062 | 2.71924758 | 0 | 2.65370405 |
| 0 | 0 | 0.001334369 | 0 | 0 |
| 0 | 0 | 0 | 0 | 0 |
| 0 | 0 | 0.015160111 | 0 | 0.034355902 |
| 0 | 0 | 0.552055858 | 0 | 0 |
| 0 | 0 | 0 | 0 | 0 |
| 0 | 0 | 0.35368743 | 0 | 0 |
| 0 | 0 | 0 | 0 | 0 |
| 0.011344361 | 0.002407851 | 0 | 0 | 0.000503678 |
| 0.002473931 | 0.011335365 | 1.066790547 | 0.004674332 | 0.001659803 |
| 0.008743847 | 0 | 0 | 0.004130232 | 0 |
| 0 | 0 | 0 | 0.482149723 | 0.19291951 |
| 0.408535803 | 2.342237736 | 0.013649194 | 0 | 0 |
| 0 | 0 | 0 | 0 | 0 |
| 0 | 0 | 0 | 0 | 0 |
| 0 | 0 | 0 | 0 | 0 |
| 0 | 0 | 0 | 0 | 0 |
| 0.722227836 | 0.177108752 | 0.407109322 | 0 | 0.269355614 |
| 0 | 0 | 0.001336667 | 0 | 0 |
| 0 | 0 | 0 | 0 | 0 |
| 0.013806162 | 0 | 0.004756842 | 0 | 0 |
| 0 | 0 | 0 | 0 | 0 |
| 3.264332107 | 0.143609836 | 0 | 0 | 0.55957547 |

| | | | | |
|---|---|---|---|---|
| 0 | 0 | 0 | 0 | 0 |
| 0 | 0 | 0 | 0 | 0 |
| 0 | 0 | 0 | 0 | 0 |
| 0 | 0 | 0 | 0 | 0 |
| 0.012628295 | 0 | 0 | 0.002591727 | 0 |
| 0 | 0 | 0 | 0 | 0 |
| 0 | 0 | 0 | 0 | 0 |
| 0 | 0 | 0 | 0 | 0 |
| 0 | 0 | 0 | 0 | 0 |
| 0 | 0 | 0 | 0 | 0 |
| 0 | 0 | 0 | 0 | 0 |
| 1.604140705 | 2.858153834 | 0.028067855 | 0 | 0.416832905 |
| 0 | 0 | 0 | 0 | 0 |
| 0 | 0 | 0 | 0 | 0 |
| 0.071260489 | 0.178859497 | 1.406920854 | 0.001070476 | 0 |
| 0 | 0 | 0 | 0 | 0 |
| 0 | 0 | 0 | 0 | 0 |
| 0 | 0 | 0 | 0 | 0 |
| 0.001544936 | 0 | 0 | 0 | 0 |
| 0 | 0 | 0 | 0 | 0 |
| 0 | 0 | 0.006295648 | 0 | 0 |
| 0 | 0 | 0 | 0 | 0 |
| 0.006223568 | 0.46129412 | 0.56174858 | 0 | 0.187465495 |
| 0 | 0 | 0 | 0 | 0 |
| 0 | 0 | 0 | 0 | 0 |
| 0 | 0 | 0 | 0 | 0 |
| 0 | 0 | 0 | 0 | 0 |
| 0 | 0 | 0 | 0 | 0 |
| 0 | 0 | 0.001302446 | 0 | 0 |
| 0 | 0 | 0 | 0 | 0 |
| 0 | 0 | 0 | 0 | 0 |
| 0.000980252 | 0.075793066 | 0.212651603 | 0.006945459 | 0.066424468 |
| 0 | 0 | 0 | 0 | 0 |
| 0.140059079 | 0.083639377 | 0.007342857 | 0 | 0.008706829 |
| 0.00655512 | 0.000871986 | 0 | 0.003775571 | 0 |
| 0 | 0.003372286 | 0.026610687 | 0 | 0.001920308 |
| 0 | 0 | 0 | 0 | 0 |
| 0.207053271 | 0 | 0.004644475 | 0 | 0 |
| 0.006073279 | 0 | 0 | 0 | 0 |
| 0 | 0 | 0 | 0 | 0 |
| 0 | 0 | 0 | 0 | 0 |
| 0 | 0 | 0.013256307 | 0 | 0 |
| 0 | 0 | 0 | 0 | 0 |
| 0 | 0 | 0 | 0 | 0.044349895 |
| 0.22101143 | 0.008589933 | 0.000797448 | 0.007969963 | 0.00356376 |
| 0 | 0 | 0 | 0 | 0 |
| 0 | 0 | 0.204305807 | 0 | 0 |
| 0 | 0.001218681 | 0 | 0 | 0.001189652 |

| | | | | |
|---|---|---|---|---|
| 0 | 0 | 0 | 0 | 0 |
| 0 | 0 | 0.013834349 | 0 | 0 |
| 0 | 0.028207145 | 0 | 0.010624229 | 0 |
| 0 | 0 | 0 | 0 | 0 |
| 0 | 0 | 0 | 0.010031435 | 0 |
| 0 | 0 | 0 | 0 | 0.001119132 |
| 0 | 0 | 0.556530399 | 0 | 0.067780338 |
| 0 | 0 | 0.00077891 | 0 | 0 |
| 0 | 0 | 0 | 0 | 0 |
| 0 | 0 | 0.001532954 | 0 | 0 |
| 0 | 0 | 0 | 0 | 0 |
| 0 | 0 | 0 | 0.011555405 | 0 |
| 0 | 0 | 0 | 0 | 0 |
| 0 | 0 | 0.078243183 | 0 | 0 |
| 0 | 0 | 0 | 0 | 0 |
| 0 | 0 | 0 | 0 | 0 |
| 0 | 0 | 0 | 0 | 0 |
| 0 | 0 | 0 | 0 | 0 |
| 0 | 0 | 0.100907055 | 0 | 0.234220107 |
| 0 | 0 | 0 | 0 | 0 |
| 0 | 0 | 0.002060766 | 0 | 0.003348897 |
| 0 | 0 | 0 | 0 | 0 |
| 0 | 0 | 0 | 0 | 0 |
| 0 | 0 | 0 | 0 | 0 |
| 0.019345245 | 0 | 0.002831665 | 0 | 0 |
| 0.001163544 | 0.003998451 | 0 | 0.037263554 | 0.002732245 |
| 0 | 0 | 0 | 0 | 0 |
| 0.047453238 | 0.075925559 | 0 | 0 | 0.104298646 |
| 0 | 0 | 0.015573723 | 0 | 0 |
| 1.67966872 | 2.471773866 | 1.873343587 | 0 | 0.624735809 |
| 0 | 0 | 0 | 0 | 0 |
| 0 | 0 | 0 | 0 | 0 |
| 0 | 0 | 0 | 0 | 0 |
| 0 | 0 | 0 | 0 | 0 |
| 0.001016101 | 0 | 0 | 0.008639358 | 0 |
| 0 | 0 | 0 | 0 | 0 |
| 0 | 0 | 0.158621472 | 0 | 0 |
| 0.001383573 | 0 | 0 | 0 | 0 |
| 0 | 0 | 0 | 0 | 0 |
| 0 | 0 | 0 | 0 | 0 |
| 2.81234756 | 10.56327578 | 5.190218622 | 0 | 6.674020093 |
| 0.014516538 | 2.230915079 | 0 | 0 | 0.644838523 |
| 0 | 0 | 0.073760177 | 0 | 0 |
| 0 | 0 | 0.040223278 | 0 | 0 |
| 0 | 0 | 0.081700576 | 0 | 0 |
| 0 | 0 | 0 | 0 | 0 |
| 0 | 0 | 0 | 0 | 0 |
| 0.019948454 | 0 | 0 | 0 | 0 |

| | | | | |
|---|---|---|---|---|
| 0 | 0 | 0 | 0 | 0 |
| 0.034825201 | 0.001552538 | 0 | 0.002880971 | 0.001894445 |
| 0 | 0 | 0 | 0 | 0 |
| 0 | 0 | 0.004147147 | 0 | 0 |
| 0.561196244 | 5.919952915 | 4.028276153 | 0 | 4.939189288 |
| 0 | 0 | 0 | 0 | 0 |
| 0 | 0 | 0 | 0 | 0 |
| 0 | 0 | 0.000764077 | 0 | 0 |
| 0.10093817 | 0.338535699 | 0 | 0.346005883 | 0.661510827 |
| 0 | 0 | 0 | 0 | 0 |
| 0 | 0 | 0 | 0.004159035 | 0 |
| 0 | 0 | 0 | 0 | 0 |
| 0.111027252 | 0.213083473 | 0.023632947 | 0 | 0 |
| 0.011243551 | 0.00834577 | 0.011406505 | 0 | 0.022856781 |
| 0.029620044 | 0 | 0.16691552 | 0 | 0 |
| 0 | 0.299240476 | 0.270029922 | 0 | 2.180536933 |
| 0 | 0.002607934 | 0 | 0 | 0.000783327 |
| 0 | 0 | 0 | 0 | 0 |
| 0 | 0 | 0.00295263 | 0 | 0 |
| 0 | 0 | 0 | 0 | 0 |
| 0 | 0 | 0 | 0 | 0 |
| 0 | 0 | 0 | 0 | 0 |
| 0 | 0 | 0 | 0 | 0 |
| 0 | 0 | 0 | 0 | 0 |
| 0 | 0 | 0 | 0 | 0 |
| 0 | 0 | 0 | 0 | 0 |
| 0 | 0 | 0 | 0 | 0 |
| 0.000954644 | 0.000984174 | 0 | 0.001623364 | 0 |
| 0 | 0 | 0 | 0 | 0 |
| 0 | 0 | 0 | 0 | 0 |
| 0 | 0 | 0 | 0 | 0 |
| 0 | 0 | 0 | 0 | 0 |
| 0 | 0 | 0 | 0 | 0 |
| 0 | 0 | 0.005746557 | 0 | 0 |
| 0 | 0 | 0.184642207 | 0 | 0 |
| 0 | 0 | 0 | 0 | 0 |
| 0 | 0 | 0 | 0 | 0 |
| 0 | 0 | 0 | 0 | 0 |
| 0 | 0 | 0 | 0 | 0 |
| 0 | 0 | 0 | 0 | 0 |
| 1.467477387 | 0 | 0.972907759 | 0 | 3.540317508 |
| 0 | 0 | 0 | 0 | 0.235318799 |
| 0 | 0 | 0.021120568 | 0 | 0 |
| 0 | 0 | 0.004472962 | 0 | 0.005173743 |
| 0 | 0 | 0 | 0 | 0 |
| 0 | 0 | 0 | 0 | 0 |
| 0 | 0 | 0 | 0 | 0 |
| 0 | 0 | 0 | 0 | 0 |

| | | | | |
|---|---|---|---|---|
| 0 | 0 | 0.020807739 | 0 | 0 |
| 0 | 0 | 0 | 0 | 0 |
| 0 | 0 | 0.037710727 | 0 | 0 |
| 0 | 0 | 0 | 0 | 0 |
| 0.019038691 | 0 | 0.344620979 | 0 | 0 |
| 0 | 0 | 0 | 0 | 0 |
| 0 | 0.001200735 | 0 | 0 | 0.060755557 |
| 0 | 0 | 0 | 0 | 0 |
| 0 | 0 | 0 | 0 | 0 |
| 0 | 0 | 0 | 0.154460553 | 0 |
| 0.008238988 | 0 | 0.03109053 | 0 | 0 |
| 0 | 0 | 0 | 0 | 0 |
| 0 | 0 | 0 | 0 | 0 |
| 0 | 0 | 0 | 0 | 0 |
| 0 | 0 | 0 | 0 | 0 |
| 0 | 0 | 0 | 0 | 0 |
| 0 | 0 | 0 | 6.565850912 | 0 |
| 0 | 0 | 0 | 0 | 0 |
| 0 | 0 | 0 | 0 | 0 |
| 0 | 0 | 0 | 0 | 0 |
| 0.000924055 | 0.00762111 | 0.026885283 | 0 | 0.02715444 |
| 0 | 0 | 0.000858024 | 0 | 0 |
| 0 | 0 | 0.023801536 | 0 | 0 |
| 0.005981642 | 0.000934344 | 0.001387841 | 0.001695288 | 0.002918681 |
| 0 | 0 | 0 | 0 | 0 |
| 0.000641545 | 0 | 0 | 0 | 0 |
| 0 | 0 | 0 | 0 | 0 |
| 0 | 0 | 0 | 0 | 0 |
| 0 | 0 | 0 | 0 | 0 |
| 0 | 0 | 0.021472354 | 0.047023862 | 0.010501665 |
| 0 | 0 | 0.0009796 | 0 | 0 |
| 0 | 0 | 0 | 0 | 0 |
| 0 | 0 | 0 | 0 | 0 |
| 0 | 0 | 0 | 0 | 0 |
| 0 | 0 | 0.056403963 | 0 | 0 |
| 0 | 0 | 0.353674781 | 0 | 0 |
| 0 | 0 | 0.015411866 | 0 | 0 |
| 0 | 0 | 0 | 0 | 0 |
| 0 | 0 | 0.048806214 | 0 | 0 |
| 0 | 0 | 0.126043848 | 0 | 0 |
| 0 | 0 | 0 | 0 | 0 |
| 0 | 0 | 0 | 0 | 0 |
| 0 | 0 | 0 | 0 | 0 |
| 0 | 0 | 0 | 0 | 0 |
| 0 | 0 | 0.011093705 | 0 | 0 |
| 0 | 0 | 0 | 0 | 0 |
| 0 | 0 | 0.001669628 | 0 | 0 |
| 0 | 0 | 0 | 0 | 0 |

| | | | | |
|---|---|---|---|---|
| 0.008581945 | 0.022319602 | 0 | 0.002321697 | 0.004514619 |
| 0 | 0 | 0.051695018 | 0 | 0 |
| 0 | 0 | 0 | 0 | 0 |
| 0 | 0 | 0 | 0 | 0 |
| 0 | 0 | 0 | 0 | 0 |
| 0 | 0 | 0 | 0 | 0 |
| 0.001049585 | 0 | 0 | 0 | 0 |
| 0.615218115 | 1.967574691 | 0.488755097 | 0 | 2.506390857 |
| 0.025410547 | 0.008206155 | 0.025491676 | 0.002863341 | 0.155592078 |
| 1.080798759 | 0.237743642 | 0.004079664 | 0.005074038 | 0.056411436 |
| 0 | 0 | 0.00670238 | 0 | 0 |
| 0 | 0 | 0 | 0 | 0 |
| 0 | 0 | 0 | 0 | 0 |
| 0 | 0 | 0 | 0 | 0 |
| 0 | 0 | 0 | 0 | 0 |
| 0 | 0 | 0 | 0 | 0 |
| 0 | 0.019078737 | 0 | 0 | 0 |
| 0 | 0 | 0 | 0 | 0 |
| 0 | 0 | 0 | 0 | 0 |
| 0 | 0 | 0.000507443 | 0 | 0 |
| 0 | 0 | 0 | 0 | 0 |
| 0 | 0 | 0 | 0 | 0 |
| 0 | 0 | 0 | 0 | 0 |
| 0 | 0 | 0 | 0 | 0 |
| 0 | 0 | 0.011909356 | 0 | 0 |
| 0 | 0 | 0.005423723 | 0 | 0 |
| 0 | 0 | 0 | 0 | 0 |
| 0 | 0 | 0 | 0 | 0 |
| 0 | 0 | 0 | 0.002803144 | 0 |
| 0 | 0 | 0 | 0 | 0 |
| 0 | 0 | 0 | 0 | 0 |
| 0 | 0 | 0 | 0 | 0 |
| 0 | 0 | 0 | 0 | 0 |
| 0 | 0 | 0.004156055 | 0 | 0 |
| 0 | 0 | 0.018757271 | 0 | 0.001901072 |
| 0 | 0 | 0.240565031 | 0 | 0 |
| 0 | 0 | 0 | 0 | 0 |
| 0 | 0 | 0 | 0 | 0 |
| 0.00997268 | 0.000956387 | 0 | 0 | 0 |
| 0 | 0 | 0 | 0 | 0.055651691 |
| 0 | 0 | 0 | 0 | 0 |
| 0.33185918 | 0 | 0.04056668 | 0 | 0 |
| 0 | 0 | 0 | 0 | 0 |
| 0 | 0 | 0 | 0 | 0 |
| 0 | 0 | 0 | 0 | 0 |
| 0.075617194 | 0 | 0 | 0 | 0 |
| 0 | 0 | 0 | 0 | 0 |
| 0 | 0 | 0 | 0 | 0 |

| | | | | |
|---|---|---|---|---|
| 0 | 0 | 0 | 0 | 0 |
| 0 | 0 | 0 | 0 | 0 |
| 0 | 0 | 0 | 0 | 0 |
| 0 | 0 | 0 | 0 | 0 |
| 0.03396497 | 0.141584845 | 0.212000982 | 0 | 0.035915369 |
| 0 | 0 | 0.021957922 | 0 | 0 |
| 0 | 0 | 0 | 0 | 0 |
| 0.026299697 | 0.023020662 | 0 | 0.028900801 | 0.04219799 |
| 0 | 0 | 0.005690249 | 0 | 0 |
| 0 | 0 | 0 | 0 | 0 |
| 0 | 0 | 0 | 0 | 0 |
| 0 | 0 | 0.015078575 | 0 | 0 |
| 0 | 0 | 0 | 0 | 0 |
| 0 | 0 | 0 | 0 | 0 |
| 0 | 0 | 0 | 0.001039533 | 0.002988167 |
| 0.182261062 | 0.093394866 | 0 | 0 | 0.006713243 |
| 0 | 0 | 0 | 0 | 0 |
| 0 | 0 | 0 | 0 | 0 |
| 0 | 0 | 0 | 0 | 0 |
| 0 | 0 | 0 | 0 | 0 |
| 0 | 0 | 0 | 0 | 0 |
| 0 | 0 | 0 | 0 | 0 |
| 0 | 0 | 0 | 0 | 0 |
| 0.001503034 | 0 | 0 | 0.02391588 | 0 |
| 0.057120897 | 0 | 0.056131342 | 0 | 0 |
| 0 | 0 | 0 | 0 | 0 |
| 0.13098877 | 0.009920527 | 0.003195217 | 0.008515749 | 0.002569282 |
| 0 | 0 | 0 | 0 | 0 |
| 0 | 0 | 0 | 0 | 0 |
| 0 | 0 | 0 | 0 | 0 |
| 0 | 0 | 0 | 0 | 0 |
| 0 | 0 | 0 | 0 | 0 |
| 0 | 0 | 0 | 0 | 0 |
| 0 | 0 | 0 | 0 | 0 |
| 0 | 0 | 0 | 0 | 0 |
| 0 | 0 | 0 | 0 | 0 |
| 0 | 0 | 0 | 0 | 0 |
| 0.008660384 | 0 | 0.043948775 | 0 | 0 |
| 0 | 0 | 0.194821778 | 0 | 0 |
| 0 | 0 | 0 | 0 | 0 |
| 0 | 0 | 0 | 0 | 0 |
| 0.080135618 | 0 | 0.041773526 | 0 | 0 |
| 0 | 0 | 0 | 0 | 0 |
| 0 | 0 | 0.032424966 | 0 | 0 |
| 0 | 0 | 0 | 0 | 0 |
| 0 | 0 | 0.003018358 | 0 | 0 |
| 0 | 0 | 0 | 0 | 0 |
| 0 | 0 | 0.000570698 | 0 | 0 |

| | | | | |
|---|---|---|---|---|
| 0.00646391 | 0.061878677 | 0.014317114 | 0 | 0 |
| 0 | 0 | 0 | 0 | 0 |
| 0 | 0 | 0 | 0 | 0 |
| 0 | 0 | 0 | 0 | 0 |
| 0 | 0 | 0 | 0 | 0 |
| 0 | 0 | 0 | 0 | 0 |
| 0 | 0 | 0 | 0 | 0 |
| 0 | 0 | 0 | 0 | 0 |
| 0 | 0 | 0 | 0 | 0 |
| 0 | 0 | 0 | 0 | 0 |
| 0 | 0 | 0 | 0 | 0 |
| 0 | 0 | 0 | 0 | 0.00637436 |
| 0 | 0 | 0 | 0 | 0 |
| 0 | 0 | 0 | 0 | 0 |
| 0 | 0 | 0 | 0 | 0 |
| 0 | 0 | 0 | 0 | 0 |
| 0 | 0 | 0 | 0 | 0 |
| 0 | 0 | 0 | 0 | 0 |
| 0 | 0 | 0 | 0 | 0 |
| 0 | 0 | 0 | 0 | 0 |
| 0 | 0.032844907 | 0 | 0.001090805 | 0.004303695 |
| 0 | 0 | 0 | 0 | 0 |
| 0.12583909 | 0.000835182 | 0 | 0 | 0 |
| 0 | 0 | 0 | 0 | 0 |
| 0 | 0 | 0 | 0 | 0 |
| 0 | 0 | 0 | 0 | 0 |
| 0 | 0 | 0 | 0 | 0 |
| 0 | 0 | 0 | 0 | 0 |
| 0 | 0 | 0 | 0 | 0 |
| 0 | 0 | 0 | 0 | 0 |
| 0 | 0 | 0 | 0 | 0 |
| 0 | 0 | 0.006756596 | 0 | 0 |
| 0.051590019 | 0.044842972 | 0 | 0.054400198 | 0.039957228 |
| 0 | 0 | 0 | 0 | 0 |
| 0 | 0 | 0 | 0 | 0 |
| 0 | 0 | 0 | 0 | 0.001021147 |
| 0 | 0 | 0 | 0 | 0 |
| 0 | 0 | 0.009025643 | 0 | 0 |
| 0.001408362 | 0 | 0 | 0.002594482 | 0.002126011 |
| 0 | 0 | 0 | 0 | 0 |
| 0 | 0 | 0 | 0 | 0 |
| 0 | 0 | 0.848199493 | 0 | 0 |
| 0.011253111 | 0.002546605 | 0.05805294 | 0 | 0.008838914 |
| 0 | 0 | 0 | 0 | 0 |
| 0 | 0 | 0 | 0 | 0 |
| 0.009808816 | 0.000674149 | 0.002086159 | 0 | 0 |
| 0 | 0 | 0 | 0 | 0 |
| 0 | 0 | 0 | 0 | 0 |

| | | | | |
|---|---|---|---|---|
| 0 | 0 | 0 | 0 | 0 |
| 0 | 0 | 0 | 0 | 0 |
| 0 | 0.146997407 | 0 | 0.539070116 | 0 |
| 0 | 0 | 0 | 0 | 0 |
| 0 | 0 | 0 | 0 | 0 |
| 0 | 0 | 0 | 0 | 0 |
| 0 | 0 | 0 | 0 | 0 |
| 0 | 0.029532006 | 0 | 2.118618679 | 0.034975807 |
| 0 | 0 | 0.002149991 | 0 | 0.00061657 |
| 0.002469615 | 0.004629105 | 0.14761708 | 0 | 0.00112971 |
| 0 | 0 | 0 | 0 | 0 |
| 0 | 0.009034304 | 0 | 0 | 0 |
| 0 | 0 | 0 | 0 | 0 |
| 0 | 0 | 0 | 0 | 0 |
| 0 | 0.001683427 | 0 | 0 | 0 |
| 0 | 0.009336783 | 0 | 0.015588542 | 0.002445321 |
| 0 | 0 | 0 | 0 | 0 |
| 0 | 0 | 0 | 0 | 0 |
| 0 | 0 | 0 | 0 | 0 |
| 0.534103935 | 0 | 1.646802457 | 0 | 11.6783567 |
| 0 | 0 | 0.495482102 | 0 | 0.007894071 |
| 0 | 0 | 0 | 0 | 0 |
| 0 | 0 | 0 | 0 | 0 |
| 0 | 0 | 0 | 0 | 0.001184443 |
| 0 | 0 | 0 | 0 | 0 |
| 0 | 0 | 0 | 0 | 0 |
| 0 | 0 | 0 | 0 | 0 |
| 0 | 0 | 0 | 0 | 0 |
| 0 | 0 | 0 | 0 | 0 |
| 0 | 0 | 0.02300661 | 0 | 0 |
| 0 | 1.640819538 | 0.302761589 | 0 | 0 |
| 0 | 0 | 0 | 0 | 0 |
| 0 | 0 | 0 | 0 | 0 |
| 0 | 0 | 0 | 0 | 0 |
| 0 | 0 | 0 | 0 | 0 |
| 0 | 0 | 0 | 0 | 0 |
| 0 | 0 | 0 | 0 | 0 |
| 0 | 0 | 0 | 0 | 0 |
| 0 | 0 | 0.005422404 | 0 | 0 |
| 0 | 0 | 0 | 0 | 0 |
| 0 | 0 | 0 | 0 | 0 |
| 0 | 0 | 0 | 0 | 0 |
| 0 | 0 | 0.001115085 | 0 | 0 |
| 0 | 0 | 0 | 0 | 0 |
| 0 | 0 | 0 | 0 | 0 |
| 0 | 0 | 0 | 0 | 0 |
| 0 | 0 | 0 | 0.107283075 | 0.084655601 |
| 0 | 0.001705121 | 0 | 0 | 0 |

| | | | | |
|---|---|---|---|---|
| 0 | 0 | 0 | 0 | 0 |
| 0 | 0 | 0 | 0 | 0 |
| 0 | 0 | 0 | 0 | 0 |
| 0 | 0 | 0.002133566 | 0 | 0 |
| 0.048652939 | 0.054860223 | 0.031689536 | 0 | 0.08092519 |
| 0 | 0 | 0 | 0 | 0 |
| 0 | 0 | 0 | 0 | 0 |
| 0 | 0 | 0 | 0 | 0.001132358 |
| 0 | 0 | 0 | 0 | 0 |
| 0 | 0 | 0 | 0 | 0 |
| 0 | 0 | 0 | 0 | 0 |
| 0.052771505 | 0.00145077 | 0 | 0.001395917 | 0.000944142 |
| 0.001821141 | 0 | 0 | 0 | 0 |
| 0 | 0 | 0 | 0 | 0 |
| 0.00303851 | 0 | 0.009066315 | 0 | 0 |
| 0 | 0 | 0.137028029 | 0 | 0 |
| 0 | 0.000743876 | 0 | 0.003374251 | 0.003449245 |
| 0 | 0 | 0 | 0.004176275 | 0 |
| 0.197663288 | 0.103628141 | 0 | 0.244770067 | 0.033243288 |
| 0.018782687 | 0.016973112 | 0 | 0 | 0.003500452 |
| 0 | 0 | 0 | 0 | 0 |
| 0 | 0 | 0.037943836 | 0 | 0 |
| 0 | 0 | 0 | 0 | 0 |
| 0 | 0 | 0 | 0 | 0 |
| 0 | 0 | 0 | 0 | 0 |
| 0 | 0 | 0 | 0 | 0 |
| 0.014679787 | 0.013601332 | 0 | 0.002685876 | 0.007854203 |
| 0 | 0.026157675 | 0 | 0 | 0 |
| 0 | 0 | 0 | 0 | 0 |
| 0 | 0 | 0 | 0 | 0 |
| 0.002948916 | 0 | 0 | 0 | 0 |
| 0 | 0 | 0 | 0 | 0 |
| 0 | 0 | 0 | 0 | 0 |
| 0 | 0 | 0 | 0 | 0 |
| 0 | 0 | 0 | 0 | 0 |
| 0 | 0 | 0 | 0 | 0 |
| 0 | 0 | 0 | 0 | 0 |
| 0.010359113 | 0.021206537 | 0.000566536 | 0.0054105 | 0.004170064 |
| 0 | 0 | 0.922311515 | 0 | 0 |
| 0 | 0 | 0 | 0 | 0 |
| 0.001772869 | 0 | 0 | 0 | 0 |
| 0 | 0 | 0.002541941 | 0 | 0 |
| 0 | 0 | 0 | 0 | 0 |
| 0 | 0 | 0 | 0.004291418 | 0 |
| 0 | 0 | 0 | 0 | 0 |
| 0 | 0 | 0 | 0 | 0 |
| 0 | 0 | 0 | 0 | 0 |
| 0 | 0 | 0 | 0 | 0 |

| | | | | |
|---|---|---|---|---|
| 0 | 0 | 0 | 0 | 0 |
| 0 | 0 | 0 | 0 | 0 |
| 0 | 0 | 0 | 0 | 0 |
| 0.001609866 | 0 | 0 | 0.000977701 | 0 |
| 0 | 0.001539312 | 0.079208732 | 0.001632244 | 0 |
| 0.002752887 | 0.001135217 | 0 | 0 | 0 |
| 0 | 0 | 0.001922379 | 0 | 0.002246019 |
| 0 | 0 | 0 | 0 | 0 |
| 0 | 0 | 0 | 0 | 0 |
| 0 | 0 | 0.010173882 | 0 | 0 |
| 0 | 0 | 0.021825394 | 0 | 0 |
| 0 | 0 | 0 | 0 | 0 |
| 0.061134021 | 0.002016802 | 0.167259077 | 0 | 0.013453204 |
| 0 | 0 | 0 | 0 | 0 |
| 0 | 0 | 0 | 0 | 0 |
| 0 | 0 | 0 | 0 | 0 |
| 0 | 0 | 0 | 0 | 0 |
| 0 | 0 | 0 | 0 | 0 |
| 0 | 0 | 0 | 0 | 0 |
| 0 | 0 | 0 | 0 | 0 |
| 0 | 0 | 0 | 0 | 0 |
| 0 | 0 | 0 | 0 | 0 |
| 0.037137065 | 0 | 0 | 0.009490941 | 0.005184811 |
| 0.008865928 | 0.003394923 | 0.267408292 | 0 | 0 |
| 0 | 0 | 0 | 0 | 0 |
| 0.001122441 | 0 | 0.002148507 | 0 | 0 |
| 0 | 0 | 0.007262694 | 0 | 0 |
| 0 | 0 | 0 | 0 | 0 |
| 0 | 0 | 0 | 0 | 0 |
| 0 | 0 | 0 | 0 | 0 |
| 0.12494104 | 0.082415995 | 0.12828204 | 0 | 0.043116929 |
| 0 | 0 | 0 | 0 | 0 |
| 0 | 0 | 0 | 0 | 0 |
| 0 | 0 | 0 | 0 | 0 |
| 0 | 0 | 0.067986191 | 0 | 0 |
| 0 | 0 | 0 | 0 | 0.000755418 |
| 0 | 0 | 0 | 0 | 0 |
| 0 | 0 | 0 | 0.04375941 | 0 |
| 0 | 0.003340342 | 0.000775254 | 0.002582714 | 0 |
| 0 | 0 | 0 | 0 | 0 |
| 0 | 0 | 0 | 0 | 0 |
| 0 | 0 | 0 | 0 | 0 |
| 0 | 0 | 0 | 0 | 0 |
| 0 | 0 | 0 | 0 | 0 |
| 0 | 0 | 6.206713848 | 0 | 0 |
| 0.011759413 | 0.55803303 | 0.22833133 | 0 | 0.119957667 |
| 0 | 0 | 0 | 0 | 0 |
| 0 | 0 | 0 | 0 | 0 |

| | | | | |
|---|---|---|---|---|
| 0 | 0 | 0 | 0 | 0 |
| 0 | 0 | 0 | 0 | 0 |
| 0 | 0 | 0.016091633 | 0 | 0 |
| 0 | 0 | 0.00901092 | 0 | 0 |
| 0 | 0 | 0 | 0 | 0 |
| 0 | 0 | 0 | 0 | 0 |
| 0 | 0 | 0 | 0 | 0 |
| 0 | 0 | 0 | 0 | 0 |
| 0 | 0.003283874 | 0 | 0 | 0 |
| 0 | 0 | 0 | 0 | 0 |
| 0.008542957 | 0 | 0 | 0 | 0 |
| 0 | 0 | 0 | 0 | 0 |
| 0 | 0 | 0 | 0 | 0 |
| 0 | 0 | 0 | 0 | 0 |
| 0 | 0 | 0 | 0 | 0 |
| 0 | 0 | 0 | 0 | 0 |
| 0 | 0 | 0 | 0 | 0 |
| 0 | 0 | 0 | 0 | 0 |
| 0 | 0 | 0 | 0 | 0 |
| 0 | 0 | 0 | 0 | 0 |
| 0 | 0 | 0 | 0 | 0 |
| 0 | 0 | 0 | 0 | 0 |
| 0 | 0 | 0 | 0 | 0 |
| 0 | 0 | 0 | 0 | 0 |
| 0 | 0 | 0 | 0 | 0 |
| 0 | 0 | 0 | 0 | 0 |
| 0 | 0 | 0 | 0 | 0 |
| 0 | 0 | 0 | 0 | 0 |
| 0 | 0 | 0 | 0 | 0 |
| 0 | 0 | 0 | 0 | 0 |
| 0.001310232 | 0.004438216 | 0.002507965 | 0 | 0.000565108 |
| 0 | 0 | 0 | 0 | 0 |
| 0.029991593 | 0.015459661 | 0.016016012 | 0 | 0.042649627 |
| 0.002442969 | 0.002289579 | 0.001912982 | 0 | 0.008046146 |
| 0 | 0 | 0 | 0 | 0 |
| 0 | 0 | 0 | 0 | 0 |
| 0 | 0 | 0 | 0 | 0 |
| 0 | 0 | 0 | 0 | 0 |
| 0 | 0 | 0 | 0 | 0 |
| 0 | 0 | 0 | 0 | 0 |
| 0 | 0 | 0 | 0 | 0 |
| 0.116396023 | 0.058431474 | 0 | 0.170414759 | 0.018173506 |
| 0.066436204 | 0.131817628 | 0.013550694 | 0 | 0.014906444 |
| 0 | 0 | 0 | 0 | 0 |
| 0 | 0 | 0 | 0 | 0 |
| 0.002203392 | 0 | 0.043079729 | 0 | 0 |
| 0 | 0 | 0 | 0 | 0 |
| 0 | 0 | 0 | 0 | 0 |

| | | | | |
|---|---|---|---|---|
| 0 | 0 | 0 | 0 | 0 |
| 0.003651186 | 0.013880222 | 0.078406424 | 0 | 0 |
| 0 | 0.010992248 | 0.080361837 | 0 | 0.010462148 |
| 0 | 0 | 0 | 0 | 0 |
| 0 | 0.44739639 | 0.000814794 | 0 | 0.053548217 |
| 0 | 0 | 0 | 0 | 0 |
| 0 | 0 | 0 | 0 | 0 |
| 0 | 0 | 0 | 0 | 0 |
| 0 | 0 | 0 | 0 | 0 |
| 0 | 0 | 0 | 0 | 0 |
| 0 | 0 | 0 | 0 | 0 |
| 0 | 0 | 0 | 0 | 0 |
| 0 | 0 | 0 | 0 | 0 |
| 0 | 0 | 0 | 0 | 0 |
| 0 | 0 | 0 | 0 | 0 |
| 0 | 0 | 0 | 0 | 0 |
| 0 | 0 | 0 | 0 | 0 |
| 0 | 0 | 0 | 0 | 0 |
| 0 | 0 | 0 | 0 | 0 |
| 0 | 0 | 0 | 0 | 0 |
| 0 | 0 | 0 | 0 | 0 |
| 0 | 0 | 0 | 0 | 0 |
| 0 | 0 | 0 | 0 | 0 |
| 0 | 0 | 0 | 0 | 0 |
| 0 | 0 | 0.018081692 | 0 | 0.002874094 |
| 0 | 0.001594827 | 0.003595649 | 0 | 0 |
| 0 | 0 | 0 | 0 | 0 |
| 0 | 0.002210054 | 0 | 0 | 0 |
| 0 | 0 | 0 | 0 | 0 |
| 0 | 0 | 0 | 0 | 0 |
| 0 | 0 | 0 | 0 | 0 |
| 0.00271321 | 0 | 0 | 0 | 0 |
| 0 | 0 | 0 | 0 | 0 |
| 0 | 0 | 0 | 0 | 0 |
| 0 | 0 | 0 | 0 | 0 |
| 0 | 0 | 0 | 0 | 0 |
| 0 | 0 | 0 | 0 | 0 |
| 0 | 0 | 0 | 0.04488957 | 0.003099402 |
| 5.646015751 | 1.348067289 | 0.41018322 | 0 | 4.393923567 |
| 0 | 0 | 0 | 0 | 0 |
| 0 | 0 | 0 | 0 | 0 |
| 0.023255459 | 0 | 0.002294544 | 0 | 0 |
| 0.004879468 | 0.016005831 | 0 | 0.002451532 | 0.002901705 |
| 0 | 0 | 0 | 0 | 0 |
| 0 | 0 | 0 | 0 | 0 |
| 0 | 0 | 0 | 0 | 0 |
| 0.093986488 | 0.003847767 | 0 | 0.004615833 | 0.001707324 |
| 0 | 0 | 0 | 0 | 0 |

| | | | | |
|---|---|---|---|---|
| 0 | 0 | 0 | 0 | 0 |
| 0.017339506 | 0.005296554 | 0.001639022 | 0.029667691 | 0.004739522 |
| 0 | 0 | 0.039597121 | 0 | 0 |
| 0 | 0 | 0 | 0 | 0 |
| 0 | 0 | 0 | 0 | 0 |
| 0 | 0 | 0 | 0 | 0 |
| 0 | 0 | 0 | 0 | 0 |
| 0 | 0 | 0 | 0 | 0 |
| 0 | 0 | 0 | 0 | 0 |
| 0 | 0 | 0 | 0 | 0 |
| 0 | 0 | 0 | 0 | 0 |
| 0 | 0 | 0 | 0 | 0 |
| 0 | 0.091682421 | 0.56114323 | 0 | 0 |
| 0 | 0 | 0 | 0 | 0 |
| 0.019751377 | 0.019536845 | 0 | 0 | 0.00349196 |
| 0 | 0 | 0 | 0 | 0 |
| 0 | 0 | 0 | 0 | 0 |
| 0 | 0 | 0 | 0 | 0 |
| 0 | 0 | 0 | 0 | 0 |
| 0 | 0 | 0 | 0 | 0 |
| 0 | 0 | 0 | 0 | 0 |
| 0 | 0 | 0 | 0 | 0 |
| 0.068886764 | 0 | 0 | 0.052747858 | 0 |
| 0 | 0 | 0.003359184 | 0 | 0 |
| 0 | 0 | 0 | 0 | 0 |
| 0 | 0 | 0 | 0 | 0 |
| 0 | 0 | 0 | 0 | 0 |
| 0 | 0 | 0 | 0 | 0 |
| 0 | 0 | 0 | 0 | 0 |
| 0 | 0 | 0 | 0 | 0 |
| 0 | 0 | 0 | 0 | 0 |
| 0 | 0 | 0 | 0 | 0 |
| 0 | 0 | 0 | 0 | 0 |
| 0 | 0 | 0 | 0 | 0 |
| 0.174213726 | 0 | 0 | 0 | 0 |
| 0 | 0 | 0 | 0 | 0 |
| 0 | 0 | 0 | 0 | 0 |
| 0 | 0 | 0 | 0 | 0 |
| 0.011779457 | 0.001079452 | 0.001753695 | 0.00734465 | 0 |
| 0 | 0 | 0 | 0 | 0 |
| 0 | 0 | 0.019542926 | 0 | 0.127786377 |
| 0 | 0 | 0 | 0 | 0 |
| 0 | 0 | 0 | 0 | 0 |
| 0 | 0 | 0 | 0 | 0 |
| 0 | 0 | 0 | 0 | 0 |
| 3.06049995 | 0.428298642 | 0 | 0.533047562 | 0.027584628 |
| 0 | 0 | 0 | 0 | 0 |
| 0 | 0 | 0 | 0 | 0 |

| | | | | |
|---|---|---|---|---|
| 0 | 0 | 0 | 0.071632407 | 0 |
| 0 | 0 | 0 | 0 | 0 |
| 0 | 0 | 0 | 0 | 0 |
| 0 | 0 | 0.009447819 | 0 | 0 |
| 0 | 0 | 0 | 0 | 0 |
| 0 | 0 | 0 | 0 | 0 |
| 0 | 0 | 0 | 0 | 0 |
| 0 | 0 | 0 | 0 | 0 |
| 0 | 0 | 0 | 0 | 0 |
| 0.00394219 | 0.003793192 | 0.09331758 | 0 | 0 |
| 0 | 0 | 0.018038643 | 0 | 0 |
| 0 | 0 | 0 | 0 | 0 |
| 0.003853159 | 0.00079447 | 0.000983397 | 0 | 0 |
| 0 | 0 | 0 | 0 | 0 |
| 0 | 0 | 0 | 0 | 0 |
| 0 | 0 | 0 | 0 | 0 |
| 0 | 0 | 0.027509466 | 0 | 0 |
| 0 | 0 | 0 | 0 | 0 |
| 0 | 0 | 0 | 0 | 0 |
| 0.443102204 | 0.4388946 | 0.954287951 | 0.216366361 | 0.048780492 |
| 0 | 0 | 0 | 0 | 0 |
| 0 | 0 | 0 | 0 | 0 |
| 0 | 0 | 0 | 0 | 0 |
| 0 | 0 | 0 | 0 | 0 |
| 0 | 0 | 0 | 0 | 0 |
| 0 | 0 | 0 | 0 | 0 |
| 0 | 0 | 0 | 0 | 0 |
| 0 | 0 | 0 | 0 | 0 |
| 0 | 0 | 0 | 0 | 0 |
| 0 | 0.011087088 | 0 | 0 | 0 |
| 0 | 0 | 0 | 0 | 0 |
| 0 | 0 | 0 | 0 | 0 |
| 0 | 0 | 0 | 0 | 0 |
| 0 | 0 | 0 | 0 | 0 |
| 0 | 0 | 0 | 0 | 0 |
| 0 | 0 | 0 | 0 | 0 |
| 0 | 0 | 0 | 0 | 0 |
| 0 | 0 | 0 | 0 | 0 |
| 0 | 0 | 0 | 0 | 0 |
| 0 | 0 | 0 | 0 | 0 |
| 0 | 0 | 0 | 0 | 0 |
| 0 | 0 | 0 | 0 | 0 |
| 0 | 0 | 0 | 0 | 0 |
| 0 | 0 | 0 | 0 | 0 |
| 0.008804589 | 0.002878054 | 0 | 0 | 0.000864461 |
| 0 | 0 | 0 | 0 | 0 |
| 0 | 0 | 0 | 0 | 0 |
| 0 | 0 | 0 | 0 | 0 |

| | | | | |
|---|---|---|---|---|
| 0 | 0 | 0 | 0 | 0 |
| 0 | 0 | 0 | 0 | 0 |
| 0 | 0 | 0 | 0 | 0 |
| 0 | 0 | 0 | 0 | 0 |
| 0 | 0 | 0 | 0 | 0 |
| 0.012664987 | 0 | 0.001994639 | 0.00381663 | 0.003549445 |
| 0 | 0 | 0 | 0 | 0 |
| 0 | 0 | 0 | 0 | 0 |
| 0 | 0 | 0 | 0 | 0 |
| 0 | 0 | 0 | 0 | 0 |
| 0 | 0 | 0 | 0 | 0 |
| 0 | 0 | 0 | 0 | 0 |
| 0 | 0 | 0 | 0 | 0 |
| 0 | 0 | 0 | 0 | 0 |
| 0 | 0 | 0 | 0 | 0 |
| 0 | 0 | 0 | 0 | 0 |
| 0 | 0 | 0 | 0 | 0 |
| 0 | 0 | 0 | 0 | 0 |
| 0 | 0 | 0 | 0 | 0 |
| 0 | 0 | 0 | 0 | 0 |
| 0 | 0 | 0 | 0 | 0 |
| 0 | 0 | 0 | 0 | 0 |
| 0 | 0 | 0 | 0 | 0 |
| 0 | 0 | 0.01079349 | 0 | 0 |
| 0 | 0 | 0 | 0 | 0 |
| 0 | 0.01421851 | 0.005039918 | 0 | 0 |
| 0 | 0 | 0 | 0 | 0 |
| 0 | 0 | 0 | 0 | 0 |
| 0 | 0 | 0 | 0 | 0 |
| 0 | 0.015835724 | 0 | 0 | 0 |
| 0 | 0 | 0 | 0 | 0 |
| 0 | 0 | 0 | 0 | 0 |
| 0 | 0 | 0 | 0 | 0 |
| 0 | 0 | 0 | 0 | 0 |
| 0 | 0 | 0 | 0 | 0 |
| 0 | 0 | 0 | 0 | 0 |
| 0 | 0 | 0 | 0 | 0 |
| 0 | 0 | 0 | 0 | 0 |
| 0 | 0.720433376 | 0.122154207 | 0 | 0 |
| 0 | 0 | 0 | 0 | 0 |
| 0 | 0 | 0.002065437 | 0 | 0 |
| 0 | 0 | 0 | 0 | 0 |
| 0 | 0 | 0 | 0 | 0 |
| 0 | 0 | 0 | 0 | 0 |
| 0 | 0 | 0.003621131 | 0 | 0 |
| 0 | 0 | 0 | 0 | 0 |
| 0 | 0 | 0 | 0 | 0 |
| 0 | 0 | 0 | 0 | 0 |

| | | | | |
|---|---|---|---|---|
| 0 | 0 | 0 | 0 | 0 |
| 0 | 0.00795668 | 0 | 0 | 0 |
| 0 | 0 | 0 | 0 | 0 |
| 0 | 0 | 0 | 0 | 0 |
| 0 | 0 | 0 | 0 | 0 |
| 0 | 0 | 0 | 0 | 0 |
| 0 | 0 | 0 | 0 | 0 |
| 0 | 0 | 0 | 0 | 0 |
| 0 | 0 | 0 | 0 | 0 |
| 0 | 0 | 0 | 0 | 0 |
| 0 | 0.00242294 | 0.444694859 | 0 | 0 |
| 0 | 0 | 0 | 0 | 0 |
| 0 | 0.51182838 | 0.007621068 | 0 | 0 |
| 0 | 0 | 0 | 0 | 0 |
| 0 | 0 | 0 | 0 | 0 |
| 0 | 0 | 0 | 0 | 0 |
| 0 | 0 | 0 | 0 | 0 |
| 0 | 0 | 0 | 0 | 0 |
| 0 | 0 | 0 | 0 | 0 |
| 0 | 0 | 0.001871708 | 0 | 0 |
| 0.001192676 | 0 | 0 | 0 | 0 |
| 0 | 0 | 0 | 0 | 0 |
| 0 | 0 | 0 | 0 | 0 |
| 0.921222756 | 3.110319214 | 1.297735897 | 0 | 2.33205879 |
| 0 | 0 | 0 | 0 | 0 |
| 0 | 0 | 0 | 0 | 0 |
| 0 | 0 | 0 | 0 | 0 |
| 0 | 0 | 0 | 0 | 0 |
| 0 | 0.01073948 | 0 | 0 | 0 |
| 0 | 0 | 0 | 0 | 0 |
| 0 | 0 | 0 | 0 | 0 |
| 0 | 0 | 0 | 0 | 0 |
| 0 | 0 | 0.00864519 | 0 | 0 |
| 0 | 0 | 0 | 0 | 0 |

| PHL018-0243_V2 | PHL018-0115_V3 | PHL018-0216_V3 | PHL018-0230_V3 | PHL018-0277_V2 |
|---|---|---|---|---|
| REF | EYCF | CTRL | REF | REF |
| 0.002375916 | 0.541304829 | 0.00984782 | 0.121060391 | 0.009536649 |
| 2.913791971 | 10.5460983 | 0.481737296 | 2.561845506 | 0.292290984 |
| 0 | 4.915466678 | 0 | 0.919632055 | 3.061609822 |
| 0 | 0 | 0 | 0 | 0 |
| 0 | 0 | 0 | 0 | 0 |
| 0 | 0 | 0 | 0 | 0 |
| 0 | 0 | 0 | 0 | 0 |
| 0 | 0 | 0 | 0 | 0 |
| 0 | 0 | 0 | 0 | 0 |
| 0 | 0 | 0 | 0 | 0 |
| 0 | 0 | 0 | 0 | 0 |
| 0 | 0 | 0 | 0 | 0 |
| 0 | 0 | 0 | 0 | 0 |
| 1.165519394 | 0 | 0.737477737 | 0.844369646 | 2.531599393 |
| 0 | 0 | 0 | 0 | 0 |
| 0 | 0 | 0 | 0 | 0 |
| 0 | 0 | 0 | 0 | 0 |
| 0 | 0 | 0 | 0 | 0 |
| 0 | 0 | 0 | 0 | 0 |
| 0 | 0 | 0 | 0 | 0 |
| 0 | 0 | 0.001692406 | 0 | 0 |
| 0 | 0 | 0 | 0 | 0 |
| 0 | 0 | 0 | 0 | 0 |
| 0 | 0 | 0 | 0 | 0 |
| 0 | 0 | 0.000556055 | 0.151875249 | 0 |
| 0 | 0 | 0 | 0 | 0 |
| 0 | 0 | 0 | 0 | 0 |
| 0 | 0 | 0 | 0 | 0 |
| 0 | 0 | 0 | 0 | 0 |
| 0 | 0 | 0 | 0 | 0 |
| 0 | 0 | 0 | 0 | 0 |
| 0 | 0 | 0 | 0 | 0 |
| 0 | 0 | 0 | 0 | 0 |
| 0 | 0 | 0 | 0 | 0 |
| 0 | 0 | 0 | 0 | 0 |
| 0 | 3.964157141 | 5.940940544 | 0 | 2.380321955 |
| 0 | 0 | 0 | 0 | 0 |
| 0 | 0 | 0 | 0 | 0 |
| 0 | 0 | 0 | 0 | 0 |
| 0 | 0 | 0 | 0 | 0 |
| 0 | 0 | 0 | 0 | 0 |
| 0.006311855 | 0.058293331 | 0.003733436 | 0 | 0.013426529 |
| 0.024491119 | 0.001994084 | 1.742938427 | 0.002041553 | 0.171586012 |
| 0.009879425 | 0.004700979 | 0.184269384 | 0.001544134 | 0 |
| 0 | 0 | 0 | 0 | 0.023391614 |
| 0 | 0 | 0 | 0 | 0 |

| | | | | |
|---|---|---|---|---|
| 0 | 0 | 0.002097415 | 0 | 0.005413579 |
| 0.1418327 | 0.079375127 | 0 | 0.003403215 | 0 |
| 0.001250238 | 0.013659076 | 0.002820001 | 0.009379666 | 0 |
| 0.561297722 | 0.414142827 | 0.880491323 | 0.141932125 | 0.684570372 |
| 0 | 0.009188484 | 0.000729623 | 0.000946458 | 0.05910764 |
| 0 | 0 | 0.047253495 | 0 | 0 |
| 0 | 0.010364751 | 0.016872027 | 0 | 0.001862883 |
| 0 | 0 | 0 | 0 | 0 |
| 0 | 0 | 0 | 0 | 0 |
| 0 | 0 | 0 | 0 | 0 |
| 0 | 0 | 0 | 0 | 0.001222592 |
| 0 | 0.013715003 | 0.004167456 | 0.00707925 | 0.968358508 |
| 0.001404028 | 0 | 0 | 0.001580017 | 0.000691642 |
| 0 | 0.057527541 | 0 | 0.020463535 | 0.246243767 |
| 0 | 0 | 0 | 0 | 0 |
| 0 | 0 | 0 | 0 | 0 |
| 0.008592427 | 0 | 2.860190178 | 0 | 0.002090242 |
| 0 | 0 | 0 | 0 | 0 |
| 0 | 0 | 0 | 0 | 0 |
| 0 | 0 | 0 | 0 | 0 |
| 1.25617864 | 0.074352009 | 0.002340843 | 0.082353918 | 0.010875392 |
| 0 | 0.040811585 | 0 | 0.006047554 | 0.017018223 |
| 0 | 0 | 0.033615216 | 0 | 0 |
| 0 | 0 | 0 | 0 | 0 |
| 0 | 0 | 0 | 0 | 0 |
| 0 | 0 | 0 | 0 | 0 |
| 0 | 0 | 0 | 0 | 0 |
| 0 | 0 | 0 | 0 | 0 |
| 0.005303449 | 0 | 0 | 0 | 0.000725708 |
| 0 | 0 | 0 | 0 | 0 |
| 0 | 0 | 0 | 0.029123323 | 0 |
| 0 | 0 | 0 | 0.023318041 | 0 |
| 0 | 0 | 0 | 0 | 0 |
| 0 | 0 | 0 | 0 | 0 |
| 0 | 0 | 0 | 0 | 0 |
| 0.048994952 | 0 | 0.001161324 | 0 | 0 |
| 0 | 0 | 0 | 0 | 0 |
| 4.572058349 | 0 | 0.561123127 | 0 | 0 |
| 0 | 0 | 0 | 0 | 0 |
| 0 | 0 | 0 | 0 | 0 |
| 0 | 0 | 0 | 0 | 0 |
| 0 | 0 | 0 | 0 | 0 |
| 0 | 0 | 0 | 0 | 0 |
| 0 | 0 | 0 | 0 | 0 |
| 0 | 0 | 0 | 0.035202837 | 0 |
| 0 | 0 | 0 | 0 | 0 |
| 0 | 0 | 0 | 0 | 0 |
| 0 | 0.289560431 | 0 | 0 | 0 |

| | | | | |
|---|---|---|---|---|
| 0 | 0 | 0 | 0 | 0 |
| 0 | 0 | 0 | 0 | 0 |
| 0 | 0 | 0 | 0 | 0 |
| 0 | 0 | 0 | 0 | 0 |
| 2.074436242 | 0 | 0 | 0 | 0 |
| 0 | 0 | 0 | 0 | 0 |
| 0 | 0 | 0 | 0 | 0 |
| 0 | 0 | 0 | 0 | 0 |
| 0 | 0.003114284 | 0 | 0 | 0.000913858 |
| 0 | 0 | 0 | 0 | 0 |
| 0 | 0 | 0 | 0 | 0 |
| 0 | 0.001205207 | 0 | 0 | 0.004668273 |
| 0 | 0 | 0 | 0 | 0 |
| 0 | 0 | 0 | 0 | 0 |
| 0 | 0 | 0 | 0 | 0 |
| 0 | 0.003672903 | 0 | 0 | 0 |
| 0 | 0 | 0 | 0 | 0 |
| 0 | 0 | 0 | 0 | 0 |
| 0 | 0 | 0.08360713 | 0 | 0 |
| 0 | 0 | 0 | 0 | 0 |
| 0 | 0 | 0 | 0 | 0 |
| 0 | 0 | 0 | 0 | 0 |
| 0 | 0.001784263 | 0 | 0.001141711 | 0 |
| 0 | 0 | 0 | 0 | 0 |
| 0 | 0 | 0 | 0 | 0 |
| 0 | 0 | 0 | 0 | 0 |
| 0 | 0 | 0 | 0 | 0 |
| 0 | 0.003194193 | 0 | 0.003372425 | 0 |
| 0 | 0 | 0 | 0 | 0 |
| 0 | 0 | 0 | 0.164147009 | 0 |
| 0 | 0.126527705 | 0.000714847 | 0.002511412 | 0.030274543 |
| 0 | 0.002159521 | 0.006936915 | 0 | 0.126865098 |
| 0 | 0 | 0 | 0 | 0 |
| 0 | 0 | 0 | 0 | 0 |
| 0 | 0 | 0 | 0 | 0 |
| 0 | 0 | 0 | 0 | 0 |
| 0 | 0 | 0 | 0 | 0 |
| 0 | 0 | 0 | 0 | 0 |
| 0 | 0 | 0 | 0 | 0 |
| 0 | 0 | 0 | 0 | 0 |
| 0 | 0 | 0 | 0 | 0 |
| 0 | 0 | 0 | 0 | 0 |
| 0 | 0 | 0 | 0 | 0 |
| 0 | 0 | 0 | 0 | 0 |
| 0 | 0 | 0 | 0 | 0 |
| 0 | 0 | 0 | 0 | 0 |
| 0 | 0 | 0 | 0 | 0 |
| 0 | 0 | 0 | 0 | 0 |

| | | | | |
|---|---|---|---|---|
| 0 | 0 | 0 | 0 | 0 |
| 0 | 0 | 0 | 0 | 0 |
| 0 | 0 | 0 | 0 | 0 |
| 0 | 0 | 0 | 0 | 0 |
| 0 | 0 | 0.008485556 | 0.028047594 | 0.402923416 |
| 0 | 0 | 0 | 0 | 0 |
| 0 | 0 | 0 | 0 | 0 |
| 0 | 0 | 0 | 0 | 0 |
| 0 | 0 | 0 | 0 | 0 |
| 0 | 0 | 0 | 0 | 0 |
| 0 | 0 | 0 | 0 | 0 |
| 0 | 0 | 0 | 0 | 0 |
| 0 | 0 | 0 | 0 | 0 |
| 0 | 0 | 0 | 0 | 0 |
| 0 | 0 | 0 | 0 | 0 |
| 0 | 0 | 0.287851585 | 0 | 0 |
| 0 | 0 | 0 | 0 | 0 |
| 0 | 0 | 0 | 0 | 0 |
| 0 | 0 | 0 | 0 | 0 |
| 0 | 0 | 0 | 0 | 0 |
| 0 | 0 | 0 | 0 | 0 |
| 0 | 0 | 0 | 0 | 0 |
| 0 | 0 | 0 | 0 | 0 |
| 0 | 0 | 0 | 0 | 0 |
| 0 | 0 | 0.091452818 | 0 | 0 |
| 0 | 0 | 0 | 0 | 0 |
| 0 | 0 | 0 | 0.006164262 | 0 |
| 0 | 0 | 0 | 0 | 0 |
| 0 | 0 | 0 | 0 | 0 |
| 0 | 0 | 0 | 0 | 0 |
| 0 | 0 | 0 | 0.036127216 | 0 |
| 0 | 0 | 0 | 0 | 0.007779368 |
| 0 | 0 | 0 | 0 | 0 |
| 0 | 0 | 0 | 0 | 0 |
| 0 | 0 | 0 | 0 | 0 |
| 0 | 0 | 0 | 0 | 0 |
| 0 | 0 | 0 | 0 | 0 |
| 0 | 0 | 0 | 0 | 0 |
| 0 | 0 | 0 | 0 | 0 |
| 0 | 0 | 0 | 0 | 0 |
| 0 | 0 | 0 | 0 | 0 |
| 0 | 0 | 0 | 0 | 0 |
| 0 | 0 | 0 | 0 | 0 |
| 0 | 0 | 0 | 0 | 0 |
| 0 | 0 | 0 | 0 | 0 |
| 0 | 0 | 0 | 0 | 0 |
| 0 | 0 | 0 | 0 | 0 |
| 0 | 0 | 0 | 0 | 0 |

| | | | | |
|---|---|---|---|---|
| 0 | 0 | 0 | 0 | 0 |
| 0 | 0 | 0 | 0 | 0 |
| 0 | 0 | 0 | 0 | 0 |
| 0 | 0 | 0 | 0 | 0 |
| 0 | 0 | 0 | 0 | 0 |
| 0 | 0 | 0 | 0 | 0 |
| 0 | 0 | 0 | 0 | 0 |
| 0 | 0 | 0 | 0 | 0 |
| 0 | 0.011043429 | 0 | 0 | 0 |
| 0 | 0 | 0 | 0 | 0 |
| 0 | 0 | 0 | 0 | 0 |
| 0 | 0 | 0 | 0 | 0 |
| 0 | 0 | 0 | 0.002237578 | 0.001763074 |
| 0 | 0 | 0 | 0 | 0.001048763 |
| 0 | 0 | 0 | 0 | 0 |
| 0 | 0 | 0 | 0 | 0 |
| 0 | 0 | 0 | 0 | 0 |
| 0 | 0 | 0 | 0 | 0 |
| 0 | 0 | 0 | 0 | 0 |
| 0 | 0 | 0 | 0 | 0 |
| 0.214141401 | 0.012185068 | 0.027866021 | 0.010877707 | 0.037360624 |
| 0 | 0 | 0 | 0 | 0 |
| 0 | 0 | 0 | 0 | 0 |
| 0 | 0 | 0 | 0 | 0 |
| 0 | 0 | 0 | 0 | 0 |
| 0 | 0 | 0 | 0 | 0 |
| 0.005715916 | 0.001730804 | 0.000901584 | 0.001218254 | 0.002133132 |
| 0 | 0 | 0 | 0 | 0 |
| 0 | 0 | 0 | 0 | 0 |
| 0 | 0 | 0 | 0 | 0 |
| 0.01553945 | 0 | 0 | 0 | 0 |
| 0 | 0 | 0 | 0 | 0 |
| 0 | 0 | 0 | 0 | 0 |
| 0 | 0.104254591 | 0.002128748 | 0 | 0 |
| 0 | 0.046215922 | 0.005540064 | 0.00272216 | 0.019065736 |
| 0 | 0 | 0 | 0 | 0 |
| 0 | 0 | 0 | 0 | 0 |
| 0 | 0 | 0 | 0 | 0 |
| 0 | 0 | 0 | 0 | 0 |
| 0 | 0 | 0 | 0 | 0 |
| 0 | 0 | 0 | 0 | 0 |
| 0 | 0 | 0 | 0 | 0 |
| 0 | 0 | 0 | 0 | 0 |
| 0 | 0 | 0 | 0 | 0 |
| 0 | 0 | 0 | 0 | 0 |
| 0 | 0 | 0.001678328 | 0 | 2.0308285 |
| 0 | 0.001674964 | 0 | 0 | 0 |
| 0 | 0 | 0 | 0.006983585 | 0 |

| | | | | |
|---|---|---|---|---|
| 0 | 0 | 0 | 0 | 0 |
| 0 | 0 | 0 | 0 | 0 |
| 0 | 0 | 0 | 0 | 0 |
| 0 | 0 | 0 | 0 | 0 |
| 0 | 0 | 0 | 0 | 0 |
| 0 | 0 | 0 | 0 | 0 |
| 0 | 0 | 0.017115233 | 0 | 0.000725272 |
| 0 | 0 | 0.002321722 | 0 | 0.000732421 |
| 0 | 0 | 0 | 0 | 0 |
| 0 | 0 | 0 | 0 | 0 |
| 0 | 0 | 0 | 0 | 0 |
| 0 | 0 | 0 | 0 | 0 |
| 0 | 0 | 0 | 0 | 0 |
| 0 | 0 | 0 | 0 | 0 |
| 0 | 0 | 0 | 0 | 0 |
| 0 | 0 | 0 | 0 | 0 |
| 0 | 0 | 0 | 0 | 0 |
| 0 | 0 | 0 | 0 | 0 |
| 0 | 0 | 0.01168026 | 0 | 0 |
| 0 | 0 | 0.00836565 | 0 | 0 |
| 0.01799863 | 0.00764888 | 0 | 0.001455077 | 0.002038243 |
| 0 | 0 | 0 | 0 | 0 |
| 0 | 0 | 0 | 0 | 0 |
| 0 | 0 | 0 | 0 | 0 |
| 0.012946892 | 0.008624823 | 0.002139387 | 0 | 0 |
| 0 | 0 | 0 | 0 | 0 |
| 0 | 0 | 0 | 0 | 0 |
| 0 | 0 | 0 | 0 | 0 |
| 0 | 0.00764288 | 0 | 0 | 0.002656776 |
| 0 | 0 | 0 | 0 | 0 |
| 0.006160835 | 0.032833276 | 0 | 0.136307523 | 0.115776069 |
| 0 | 0 | 0 | 0 | 0 |
| 0 | 0.016065631 | 0 | 0 | 0.000698827 |
| 0 | 0 | 0.001792735 | 0 | 0 |
| 0 | 0 | 0 | 0 | 0 |
| 0 | 0 | 0 | 0 | 0 |
| 0 | 0 | 0 | 0 | 0 |
| 0 | 0 | 0 | 0 | 0 |
| 0 | 0 | 0 | 0 | 0 |
| 0 | 0 | 0.003163617 | 0 | 0.001122759 |
| 0 | 0 | 0 | 0 | 0 |
| 0 | 0.037806518 | 0.002219565 | 0.004144292 | 0.009166171 |
| 0 | 0 | 0 | 0 | 0 |
| 0 | 0 | 0 | 0 | 0 |
| 0 | 0 | 0 | 0 | 0 |
| 0 | 0 | 0 | 0 | 0 |
| 0 | 0 | 0 | 0 | 0 |
| 0 | 0 | 0 | 0 | 0 |

| | | | | |
|---|---|---|---|---|
| 0 | 0 | 0 | 0 | 0 |
| 0 | 0 | 0 | 0 | 0 |
| 0.003504669 | 0 | 0.195852158 | 0 | 0 |
| 0 | 0 | 0 | 0 | 0 |
| 0 | 0 | 0 | 4.873279313 | 0 |
| 0 | 0 | 0 | 0 | 0 |
| 0 | 0 | 0 | 0 | 0 |
| 0 | 0 | 0 | 0 | 0 |
| 0 | 0 | 0 | 0 | 0 |
| 0.001869784 | 0 | 0 | 0.002504944 | 0 |
| 0 | 0 | 0 | 0 | 0 |
| 0 | 0 | 0 | 0 | 0 |
| 0 | 0 | 0 | 0 | 0 |
| 0.002904969 | 0 | 0 | 0 | 0.000795014 |
| 0.019714191 | 0.001064837 | 0.003928975 | 0 | 0.001093633 |
| 0 | 0 | 0.003711605 | 0 | 0 |
| 0.006981604 | 0.002426573 | 0.002896697 | 0 | 0.000747658 |
| 0 | 0 | 0 | 0.000883274 | 0 |
| 0 | 0 | 0 | 0 | 0 |
| 0 | 0 | 0 | 0 | 0 |
| 0 | 0 | 0 | 0 | 0 |
| 0 | 0 | 0 | 0.007195161 | 0 |
| 0 | 0 | 0 | 0 | 0 |
| 0 | 0 | 0 | 0 | 0 |
| 0 | 0 | 0 | 0 | 0 |
| 0 | 0 | 0 | 0 | 0 |
| 0 | 0 | 1.968204355 | 0 | 0 |
| 0 | 0 | 0 | 0 | 0 |
| 0 | 0 | 0 | 0 | 0 |
| 0 | 0 | 0 | 0 | 0 |
| 0 | 0 | 0 | 0 | 0 |
| 0 | 0 | 0 | 0 | 0 |
| 0 | 0 | 0 | 0 | 0 |
| 0 | 0 | 0 | 0 | 0 |
| 0 | 0 | 0 | 0 | 0 |
| 0 | 0 | 0.037509726 | 0 | 0.028758635 |
| 0 | 0 | 0 | 0 | 0 |
| 0 | 0.00779773 | 0 | 0.006457125 | 0.01158892 |
| 0 | 0 | 0 | 0 | 0 |
| 0 | 0 | 0 | 0 | 0 |
| 0 | 0 | 0 | 0 | 0 |
| 0 | 0 | 0 | 0 | 0 |
| 0 | 0 | 0 | 0 | 0 |
| 0 | 0 | 0 | 0 | 0 |
| 0 | 0 | 0 | 0 | 0 |
| 0 | 0 | 0 | 0 | 0 |
| 0 | 0 | 0 | 0 | 0.001838337 |
| 0 | 0 | 0 | 0 | 0 |

| | | | | |
|---|---|---|---|---|
| 0 | 0.000732542 | 0 | 0 | 0 |
| 0.018547264 | 0 | 0 | 0.029568779 | 0 |
| 0 | 0 | 0 | 0 | 0 |
| 0 | 0 | 0 | 0 | 0 |
| 0.008374082 | 0.004809101 | 0.000751525 | 0.005957533 | 0.024419173 |
| 0 | 0 | 0 | 0 | 0 |
| 0 | 0 | 0 | 0 | 0 |
| 0.113738321 | 0.00512317 | 0.040381402 | 0 | 0 |
| 0 | 0 | 0 | 0 | 0 |
| 0 | 1.368499442 | 0 | 0.009781073 | 0.0678206 |
| 0 | 0 | 0 | 0 | 0 |
| 0.006408232 | 0.005122762 | 0.000833897 | 0 | 0.037355205 |
| 0.034998275 | 0.007285869 | 0.031416187 | 0.031681341 | 0.001596352 |
| 0 | 0 | 0 | 0 | 0 |
| 0 | 0.000999113 | 0 | 0 | 0 |
| 0 | 0 | 0 | 0 | 0 |
| 0 | 0 | 0 | 0 | 0 |
| 0 | 0 | 0 | 0.046307104 | 0 |
| 0.033368911 | 0.010460869 | 0 | 0.015830556 | 0 |
| 0 | 0 | 0 | 0.004426455 | 0 |
| 0 | 0 | 0 | 0 | 0 |
| 0 | 0 | 0 | 0 | 0 |
| 0 | 0 | 0 | 0 | 0 |
| 0 | 0 | 0.002102487 | 0.068561798 | 0.063507108 |
| 0 | 0 | 0 | 0 | 0 |
| 0 | 0 | 0.001473497 | 0 | 0 |
| 0 | 0 | 0.018061675 | 0 | 0.002486319 |
| 0 | 0.00615729 | 0 | 0 | 0 |
| 0 | 0.001544644 | 0 | 0.025277921 | 0 |
| 0 | 0 | 0 | 0 | 0 |
| 0 | 0 | 0 | 0 | 0 |
| 0 | 0 | 0 | 0 | 0 |
| 0 | 0 | 0 | 0 | 0 |
| 0 | 0.001479782 | 0 | 0.057286238 | 0 |
| 0 | 0 | 0 | 0 | 0 |
| 0 | 0 | 0 | 0 | 0 |
| 0.167512146 | 0.025439434 | 0 | 0.21938551 | 0 |
| 0 | 0 | 0 | 0 | 0 |
| 0 | 0 | 0 | 0 | 0 |
| 0 | 0 | 0 | 0 | 0 |
| 0 | 0 | 0 | 0 | 0 |
| 0 | 0 | 0 | 0 | 0 |
| 0 | 0.001730208 | 0 | 0 | 0 |
| 0 | 0.0194468 | 0 | 0 | 0 |
| 0 | 0 | 0 | 0 | 0 |
| 0 | 0 | 0.001257747 | 0.079828633 | 0 |
| 0 | 0.006800974 | 0.001221607 | 0 | 0.012139261 |
| 0 | 0 | 0 | 0.002097589 | 0 |

| | | | | |
|---|---|---|---|---|
| 0 | 0 | 0 | 0.001872188 | 0 |
| 0 | 0 | 0 | 0 | 0 |
| 0 | 0 | 0 | 0 | 0 |
| 0 | 0 | 0 | 0 | 0 |
| 0 | 0 | 0 | 0 | 0 |
| 0 | 0.116552021 | 0 | 0.002600222 | 0.017528754 |
| 0 | 0 | 0 | 0 | 0 |
| 0.001793084 | 0 | 0 | 0 | 0 |
| 0 | 0 | 0 | 0 | 0 |
| 0 | 0 | 0 | 0 | 0 |
| 0.001929532 | 0.010925859 | 0 | 0.00784113 | 0.004224499 |
| 0 | 0 | 0 | 0.001413255 | 0 |
| 0 | 0 | 0 | 0 | 0 |
| 0.006380237 | 0 | 0 | 0 | 0.005238312 |
| 0.025822611 | 0.002047885 | 0 | 0.004900884 | 0 |
| 0 | 0 | 0 | 0 | 0 |
| 0 | 0 | 0 | 0 | 0 |
| 0 | 0.00433321 | 0 | 0 | 0 |
| 0 | 0 | 0 | 0 | 0 |
| 0 | 0 | 0 | 0.010763396 | 0 |
| 0 | 0 | 0 | 0 | 0 |
| 0 | 0 | 0.806679505 | 0.009341838 | 0 |
| 0 | 0 | 0 | 0 | 0 |
| 0 | 0 | 0 | 0 | 0 |
| 0 | 0.076714457 | 0 | 0 | 0 |
| 0 | 0 | 0 | 0 | 0 |
| 0.006917866 | 0.002633412 | 0 | 0.002780353 | 0 |
| 0 | 0 | 0 | 0 | 0 |
| 0 | 0 | 0 | 0 | 0 |
| 1.713213914 | 0.211822604 | 0.700825261 | 0.407609066 | 0.406948183 |
| 0 | 0 | 0 | 0 | 0.007107556 |
| 0 | 0 | 0 | 0 | 0 |
| 0 | 0 | 0 | 0 | 0 |
| 0 | 0 | 0 | 0 | 0 |
| 0 | 0 | 0 | 0 | 0 |
| 0 | 0 | 0 | 0 | 0 |
| 0 | 0 | 0 | 0 | 0.316463865 |
| 0 | 0.002269251 | 0.002149207 | 0 | 0 |
| 0 | 0.046652175 | 0 | 0.014290122 | 0 |
| 0 | 0 | 0 | 0 | 0 |
| 0 | 0.216808294 | 0.001083427 | 0.072144316 | 0.042039273 |
| 7.801471498 | 0 | 0.230152966 | 0 | 0.777256125 |
| 0 | 0.002645721 | 0 | 0 | 0 |
| 0 | 0 | 0 | 0 | 0 |
| 0 | 0.004129365 | 0 | 0 | 0 |
| 0 | 0 | 0 | 0 | 0 |
| 0.001048622 | 0 | 0 | 0.002458462 | 0 |
| 0 | 0 | 0 | 0 | 0 |

| | | | | |
|---|---|---|---|---|
| 0 | 0 | 0 | 0.000909999 | 0.016730546 |
| 0 | 0 | 0 | 0.00949722 | 0 |
| 0.009696135 | 0 | 0.192703478 | 0.002893201 | 0.002532961 |
| 0 | 0 | 0 | 0 | 0 |
| 0 | 0 | 0.00639377 | 0 | 0.078841209 |
| 0 | 0 | 0 | 0 | 0 |
| 0 | 0 | 0 | 0 | 0 |
| 0 | 0 | 0.004088542 | 0 | 0.00327408 |
| 0.000812111 | 0.072494437 | 0 | 0 | 0.005778589 |
| 0 | 0 | 0 | 0 | 0 |
| 0 | 0 | 0 | 0 | 0 |
| 0 | 0 | 0 | 0 | 0 |
| 0 | 0 | 0 | 0 | 0.001306249 |
| 0.001170489 | 0 | 0 | 0 | 0.008648973 |
| 0 | 0 | 0 | 0 | 0 |
| 0 | 0.120957298 | 0.000793542 | 0.001715622 | 0.194321915 |
| 0.028895259 | 0.003377042 | 0.007696138 | 0.011409525 | 0.130271859 |
| 0 | 0 | 0 | 0.002599149 | 0 |
| 0 | 0.021103785 | 0 | 0.00270077 | 0.021280413 |
| 0 | 0 | 0 | 0 | 0 |
| 0 | 0 | 0 | 0 | 0 |
| 0 | 0.008813684 | 0 | 0 | 0 |
| 0 | 0.031146651 | 0 | 0.129361634 | 0 |
| 0 | 0 | 0 | 0 | 0 |
| 0 | 0.044975449 | 0 | 0 | 0.121751647 |
| 0 | 0 | 0 | 0 | 0.005482754 |
| 0 | 0.022293344 | 0 | 0.004526401 | 0.003170246 |
| 0.000610906 | 0.001356556 | 0 | 0 | 0 |
| 0 | 0 | 0 | 0.001486313 | 0 |
| 0 | 0 | 0 | 0 | 0 |
| 0 | 0.003586428 | 0 | 0.003442315 | 0.027926991 |
| 0.004013187 | 0 | 0.008545619 | 0.007527036 | 0.007787976 |
| 0 | 0.021983843 | 0.00662981 | 0.004234901 | 0.007130006 |
| 0 | 0 | 0 | 0 | 0.001226159 |
| 0.009948842 | 0 | 0.00154123 | 0 | 0 |
| 0 | 0 | 0 | 0 | 0 |
| 0.002826437 | 0 | 0 | 0 | 0 |
| 0 | 0.276830797 | 0 | 0.06748412 | 0.013510104 |
| 0 | 0 | 0.084996192 | 0 | 0.05362653 |
| 0.001004018 | 0.00989892 | 0 | 0.003012978 | 0 |
| 0 | 0 | 0 | 0.007235744 | 0 |
| 0 | 0 | 0 | 0 | 0 |
| 0 | 0.000734043 | 0 | 0.00275556 | 0.001809344 |
| 0 | 0 | 0 | 0.02319344 | 0.214546405 |
| 0 | 0 | 0 | 0 | 0 |
| 0 | 0 | 0 | 0 | 0 |
| 0 | 0 | 0 | 0.00435098 | 0 |
| 0 | 0 | 0 | 0 | 0 |

| | | | | |
|---|---|---|---|---|
| 0 | 0 | 0 | 0 | 0 |
| 2.779367296 | 0.354031161 | 0.560914267 | 0.092897201 | 0.214865952 |
| 0 | 0 | 0 | 0 | 0 |
| 0 | 0 | 0 | 0 | 0 |
| 0 | 0 | 0 | 0 | 0 |
| 0 | 0 | 0 | 0 | 0 |
| 0 | 0 | 0 | 0 | 0 |
| 0 | 0.010617042 | 0 | 0 | 0 |
| 0 | 0 | 0 | 0 | 0 |
| 0 | 0.007128319 | 0 | 0 | 0 |
| 0 | 0.006129788 | 0 | 0.001632532 | 0.010004828 |
| 0.052971691 | 0.005182172 | 0.009962138 | 0.001389544 | 0.007299172 |
| 0 | 0 | 0.005917985 | 0 | 0 |
| 0 | 0 | 0 | 0 | 0 |
| 0.000856023 | 0 | 0 | 0 | 0 |
| 0 | 0 | 0 | 0 | 0 |
| 0.079426498 | 0.032267343 | 0.010618092 | 0.002442138 | 0.001282837 |
| 0.011493847 | 0.03895985 | 0 | 0.004755349 | 0.003608148 |
| 0 | 0.003125926 | 0 | 0 | 0 |
| 0 | 0.010967554 | 0 | 0.00864605 | 0 |
| 0.034106269 | 0.028445417 | 0.052419886 | 0.005036921 | 0.371963325 |
| 0 | 0 | 0 | 0 | 0 |
| 0 | 0 | 0 | 0 | 0 |
| 0 | 0 | 0 | 0 | 0 |
| 0 | 0 | 0.019180818 | 0 | 0 |
| 0 | 0 | 0 | 0 | 0 |
| 0 | 0.259050499 | 0 | 0.038033446 | 0.136455712 |
| 0.142145392 | 0.036317964 | 0.010647679 | 0 | 0.029058933 |
| 0 | 0 | 0 | 0 | 0 |
| 0.001721492 | 0 | 0 | 0 | 0 |
| 0 | 0.000816109 | 0 | 0 | 0.000754361 |
| 0 | 0 | 0 | 0 | 0 |
| 0 | 0 | 0 | 0.012023362 | 0 |
| 0 | 0 | 0 | 0 | 0 |
| 0 | 0 | 0 | 0.034989039 | 0 |
| 1.105764402 | 0.342729197 | 1.006217879 | 0.323459294 | 0.715100499 |
| 0 | 0 | 0 | 0 | 0 |
| 0 | 0 | 0 | 0 | 0 |
| 0.058098634 | 0.001172834 | 0.00733122 | 0 | 0.013298246 |
| 0 | 0 | 0 | 0 | 0 |
| 0 | 0 | 0 | 0 | 0 |
| 0 | 0 | 0 | 0 | 0 |
| 0.000641636 | 0.003704467 | 0.006308552 | 0.415185942 | 0.012818733 |
| 0.018791129 | 0.004814644 | 0 | 0.001936494 | 0 |
| 0 | 0.020421769 | 0 | 0 | 0 |
| 0 | 0 | 1.529393671 | 0 | 0 |
| 0 | 0 | 0 | 0 | 0.004223524 |
| 0 | 0.002223571 | 0 | 0 | 0 |

| | | | | |
|---|---|---|---|---|
| 0.011652938 | 0 | 0 | 0.001120819 | 0 |
| 0 | 0.026483427 | 0.004586218 | 0 | 0 |
| 0 | 0 | 0.00357086 | 0 | 0 |
| 0 | 0.032690875 | 0.90028073 | 0.985341596 | 0.018815597 |
| 0 | 0 | 0 | 0 | 0 |
| 0 | 0 | 0.00168654 | 0 | 0 |
| 0 | 0 | 0 | 0 | 0 |
| 0 | 0 | 0.001546611 | 0 | 0.000731851 |
| 0 | 0 | 0.115750122 | 0 | 1.02941471 |
| 0 | 0 | 0.191596862 | 0 | 0.002924612 |
| 0 | 0 | 0 | 0 | 0 |
| 0 | 0.006836645 | 0.002070489 | 0 | 0.019594961 |
| 0 | 0 | 0 | 0 | 0 |
| 0 | 0 | 0 | 0.241705871 | 0 |
| 0 | 0.110864326 | 0.005125718 | 0.206488962 | 0.112541891 |
| 0 | 0 | 0 | 0 | 0 |
| 0.001284504 | 0 | 0.330736631 | 0 | 0.001687368 |
| 0 | 0 | 0.10865646 | 0 | 0.34861426 |
| 0 | 0 | 0 | 0 | 0 |
| 0 | 0 | 0 | 0 | 0 |
| 0 | 0 | 0 | 0 | 0 |
| 0 | 0 | 0 | 0 | 0 |
| 0 | 0 | 0 | 0 | 0 |
| 0 | 0.879222864 | 0 | 0.059421466 | 0.355592621 |
| 0 | 0 | 0 | 0.246431493 | 0.002201507 |
| 0 | 0 | 0.208174303 | 0 | 0 |
| 0 | 0 | 0 | 0 | 0.095610049 |
| 0 | 0 | 0.720446354 | 0 | 2.026409592 |
| 0 | 0 | 0 | 0 | 0 |
| 0 | 0 | 0 | 0 | 0 |
| 0 | 0 | 0.103260146 | 0 | 0.034289358 |
| 0 | 0 | 0 | 0 | 0 |
| 0 | 0 | 0 | 0 | 0 |
| 0 | 0 | 0 | 0 | 0 |
| 0 | 0 | 0 | 0.04901892 | 0 |
| 0 | 0 | 0 | 0.103868508 | 0 |
| 0 | 0 | 1.069916666 | 2.699613822 | 0.646462357 |
| 0 | 0 | 0 | 0 | 0 |
| 0 | 0.009930625 | 0 | 0 | 0 |
| 0 | 0 | 0 | 0 | 0.012061974 |
| 0.003892269 | 0.423101992 | 0.122353867 | 0.005582541 | 0.056017631 |
| 0 | 0 | 0 | 0.031701895 | 0 |
| 12.00767458 | 0 | 0.515392627 | 6.644655021 | 0.032697998 |
| 0 | 0 | 0 | 0 | 0 |
| 0 | 0 | 0 | 0 | 0 |
| 0 | 0 | 0 | 0 | 0 |
| 0 | 0 | 0 | 0 | 0 |
| 0 | 0.005133843 | 0 | 0 | 0.002937634 |

| | | | | |
|---|---|---|---|---|
| 0 | 0 | 0 | 0 | 0 |
| 0 | 0 | 0.050683979 | 0 | 0.022851212 |
| 0.564093131 | 0.151683587 | 0 | 0.259303873 | 0.007148445 |
| 0 | 0 | 0 | 0 | 0 |
| 0 | 0 | 0.070685691 | 0 | 0 |
| 0 | 0.002907648 | 0.015146078 | 0 | 0.008873516 |
| 0.123738504 | 0 | 0 | 0 | 0 |
| 0 | 0 | 0 | 0 | 0 |
| 0 | 0 | 0 | 0 | 0 |
| 0 | 0 | 0 | 0 | 0 |
| 0 | 0.055221834 | 0.003370936 | 0.000728789 | 0.09740835 |
| 0.73583507 | 1.787763837 | 0.69281386 | 0.712296012 | 1.253320632 |
| 0 | 0 | 0 | 0 | 0 |
| 0.005871023 | 0.006146008 | 0.008731323 | 0.001835258 | 0.194416203 |
| 0.231990515 | 0.103323522 | 0.000955641 | 0.030164722 | 1.435302136 |
| 0 | 0 | 8.505998284 | 0 | 0 |
| 0 | 0 | 0 | 0 | 0 |
| 0 | 0 | 0 | 0 | 0 |
| 0 | 0 | 0.058114135 | 0 | 0 |
| 0 | 0.004690977 | 0 | 0.000600331 | 0.011562803 |
| 2.142089462 | 0.002314201 | 0.00809931 | 0.010384158 | 0.055795012 |
| 0.011896319 | 1.262050469 | 0 | 0.023385161 | 0 |
| 0.851055134 | 20.23030453 | 0.099983647 | 0.459249331 | 0.945227939 |
| 0 | 0 | 0 | 0 | 0 |
| 0 | 0 | 0.000738429 | 0 | 0 |
| 0 | 0.00675945 | 0.005642676 | 0 | 7.703755958 |
| 0 | 0 | 0 | 0.268611567 | 0 |
| 0 | 0 | 0 | 0 | 0.005763542 |
| 0 | 0.041354275 | 0 | 0.116741117 | 0.290350044 |
| 0 | 0 | 0 | 0 | 0 |
| 0 | 0 | 0 | 0.03076713 | 0 |
| 0 | 0.10227518 | 0.020805237 | 0.058436197 | 0.305951209 |
| 0 | 0 | 0 | 0 | 0.023808863 |
| 0 | 0.109338918 | 0.000704019 | 0.017884386 | 0.141584279 |
| 0.000702376 | 0.005263891 | 0 | 0.011032886 | 0 |
| 0.006755153 | 0.091287257 | 0.005441687 | 0.043891809 | 0.024561414 |
| 0 | 0.004347012 | 0 | 0.012238854 | 0.003826773 |
| 0 | 0 | 0 | 0 | 0 |
| 0.017221374 | 0 | 0.314230015 | 0 | 0.898645276 |
| 0 | 0 | 0 | 0 | 0 |
| 0 | 0.000424558 | 0 | 0 | 0 |
| 0 | 0 | 0 | 0 | 0 |
| 0 | 0 | 0.028282906 | 0 | 0 |
| 0 | 0 | 0 | 0 | 0 |
| 0 | 0.080459699 | 0.003341491 | 0.024768804 | 0.004743545 |
| 0 | 0 | 0.300701127 | 0 | 0 |
| 0 | 0 | 0 | 0 | 0 |
| 1.260130436 | 0.845429644 | 0.002591197 | 0.104572813 | 0.363756458 |

| | | | | |
|---|---|---|---|---|
| 0 | 0 | 0 | 0 | 0 |
| 0 | 0 | 0 | 0 | 0 |
| 0 | 0 | 0 | 0 | 0 |
| 0 | 0.001093459 | 0.000813697 | 0 | 0 |
| 0 | 0 | 0 | 0 | 0 |
| 0 | 0 | 0.006108542 | 0 | 0.13347495 |
| 0 | 0 | 0.170819178 | 0 | 0.349670613 |
| 0 | 0.037084914 | 0.011979975 | 0.015378398 | 0.014349357 |
| 0 | 0 | 0 | 0 | 0 |
| 0 | 0 | 0 | 0 | 0 |
| 0 | 0.001739748 | 0 | 0.002204188 | 0.005574802 |
| 0 | 0 | 0 | 0 | 0 |
| 0 | 0 | 0 | 0.002144377 | 0.00525665 |
| 0 | 0.352883848 | 0.39091481 | 0 | 0.772852056 |
| 0 | 0 | 0 | 0 | 0 |
| 0 | 0 | 0 | 0 | 0 |
| 0 | 0 | 0.034711315 | 0 | 0.218950463 |
| 0 | 0.005069051 | 0 | 0 | 0.001427968 |
| 0 | 0 | 0 | 0 | 0 |
| 0 | 0 | 0 | 0.001103206 | 0 |
| 0 | 0 | 0 | 0 | 0 |
| 0 | 0 | 0 | 0 | 0 |
| 0 | 0 | 0.0013047 | 0.007757018 | 0.084992549 |
| 0 | 0 | 0 | 0 | 0 |
| 0 | 0 | 0 | 0 | 0 |
| 0 | 0 | 0 | 0 | 0 |
| 0 | 0 | 0 | 0 | 0 |
| 0 | 0 | 0 | 0 | 0 |
| 0 | 0 | 0 | 0 | 0 |
| 0 | 0 | 0.139662935 | 0 | 0.046574256 |
| 0 | 0 | 0 | 0.001350256 | 0 |
| 0 | 0 | 0 | 0 | 0 |
| 0 | 0 | 0.001576495 | 0 | 0 |
| 0 | 0 | 0 | 0 | 0 |
| 0 | 0.018813731 | 0 | 0 | 0 |
| 0 | 0 | 0 | 0 | 0 |
| 0 | 0 | 0.017522325 | 0 | 0.134199497 |
| 0.000884296 | 0.020029085 | 0 | 0.001451243 | 0.066249853 |
| 0 | 0 | 0 | 0 | 0 |
| 0 | 0.009732529 | 0 | 0.002044894 | 0 |
| 0 | 0 | 0.000539833 | 0.003306804 | 0.077826171 |
| 0 | 0 | 0 | 0 | 0 |
| 0 | 0 | 0 | 0 | 0 |
| 0 | 0.00185925 | 0.013968638 | 0 | 1.53649558 |
| 0 | 0 | 0 | 0 | 0 |
| 0 | 0 | 0 | 0 | 0 |
| 0 | 0 | 0.000728921 | 0 | 0 |
| 0 | 0 | 0 | 0 | 0 |

| | | | | |
|---|---|---|---|---|
| 0 | 0 | 0 | 0 | 0 |
| 0 | 0 | 0 | 0 | 0 |
| 0 | 0 | 0 | 0 | 0 |
| 0 | 0 | 0.022892696 | 0 | 0 |
| 0 | 0 | 0 | 0 | 0 |
| 0 | 0 | 0.053606533 | 0 | 0 |
| 0 | 0 | 0 | 0 | 0 |
| 0 | 0.009103581 | 0 | 0 | 0 |
| 0.241479102 | 0.049762838 | 0 | 0.031166317 | 0.048563574 |
| 0 | 0 | 0 | 0 | 0.00181429 |
| 0 | 0 | 0 | 0 | 0 |
| 0 | 0 | 0.006611512 | 0 | 0 |
| 0 | 0 | 0 | 0 | 0 |
| 0 | 0.000987533 | 0 | 0 | 0 |
| 0 | 2.439330497 | 1.866603517 | 0.641497611 | 0.058644823 |
| 0 | 0 | 0 | 0 | 0 |
| 0 | 0 | 0 | 0 | 0 |
| 0 | 0 | 0 | 0 | 0 |
| 0 | 0 | 0 | 0 | 0 |
| 0 | 0 | 0 | 0 | 0 |
| 0 | 0 | 0.000809015 | 0.035855995 | 0.42914442 |
| 0 | 0 | 0 | 0 | 0 |
| 0 | 0 | 0 | 0 | 0 |
| 0 | 0.001287681 | 0 | 0 | 0 |
| 0 | 0 | 0.064387947 | 0 | 0.066529485 |
| 3.364624693 | 0 | 5.908169582 | 27.56612707 | 1.158411239 |
| 0 | 0 | 0.171457396 | 0.028014159 | 0 |
| 0 | 0 | 0 | 0 | 0 |
| 0.003761715 | 0 | 0.301863579 | 0 | 0 |
| 0 | 0.009341454 | 0.08403222 | 0 | 0 |
| 0 | 0 | 0 | 0 | 0 |
| 0 | 0.014459501 | 0.002003198 | 0.0407102 | 0 |
| 0 | 0 | 0 | 0.000651308 | 0 |
| 0 | 0.01173592 | 0 | 0.005566868 | 0.002829904 |
| 0.011358503 | 0.001050929 | 0.009853809 | 0 | 0.032639515 |
| 0 | 0.002822941 | 0 | 0.00167324 | 0.003845363 |
| 0 | 2.846960278 | 0 | 0.000647526 | 0 |
| 1.019194731 | 2.255134977 | 1.187644526 | 15.00188743 | 0.13915332 |
| 0 | 0 | 0 | 0 | 0 |
| 0 | 0 | 0 | 0 | 0 |
| 0 | 0 | 0 | 0 | 0 |
| 0 | 0 | 0 | 0 | 0 |
| 0 | 2.110512923 | 2.900735371 | 0.022007675 | 0.514465777 |
| 0 | 0 | 0 | 0 | 0 |
| 0 | 0 | 0 | 0 | 0 |
| 0 | 0.000844542 | 0 | 0.17080363 | 0.014571996 |
| 0 | 0 | 0 | 0 | 0 |
| 0 | 0.001793942 | 0.534109664 | 1.262063111 | 0.001105473 |

| | | | | |
|---|---|---|---|---|
| 0 | 0.008695338 | 0 | 0 | 0 |
| 0 | 0 | 0 | 0 | 0 |
| 0 | 0 | 0.014488634 | 0 | 0 |
| 0 | 0 | 0 | 0 | 0 |
| 0 | 0.001331879 | 0 | 0 | 0.002462215 |
| 0 | 0 | 0.334259847 | 0.004965204 | 0.002766256 |
| 0 | 0 | 0 | 0 | 0 |
| 0 | 0 | 0 | 0 | 0 |
| 0 | 0 | 0 | 0 | 0 |
| 0 | 0 | 0 | 0 | 0 |
| 0 | 0 | 0 | 0 | 0 |
| 0 | 0.313944044 | 0.781071458 | 0.008864926 | 0.182291955 |
| 0 | 0.45316098 | 0.863412731 | 0 | 0.186083505 |
| 0 | 0 | 0 | 0 | 0 |
| 0.023122025 | 0.238364298 | 0.011032441 | 0 | 0.077833089 |
| 0 | 0 | 0 | 0 | 0 |
| 0 | 0.007366659 | 0 | 0.003582461 | 0 |
| 0 | 0 | 0 | 0 | 0.004752894 |
| 0 | 0 | 0 | 0.004276251 | 0 |
| 0 | 0.004303954 | 0 | 0 | 0 |
| 0 | 0.075308104 | 0.073507676 | 0.03186309 | 0 |
| 0 | 0 | 0 | 0 | 0 |
| 0 | 1.000004811 | 0.330588945 | 0.001386024 | 0.186264087 |
| 0 | 0 | 0 | 0 | 0 |
| 0 | 0 | 0 | 0.003095391 | 0 |
| 0 | 0 | 0 | 0 | 0 |
| 0 | 0 | 0 | 0 | 0 |
| 0 | 0 | 0 | 0 | 0 |
| 0 | 0 | 0 | 0 | 0.001068718 |
| 0 | 0 | 0 | 0.848336225 | 0 |
| 0 | 0 | 0 | 0 | 0 |
| 0.19240104 | 0.030731223 | 0.038067904 | 0 | 0.032947948 |
| 0 | 0 | 0 | 0 | 0 |
| 0 | 0.058041033 | 0.002600762 | 0.038832413 | 0.06830225 |
| 0 | 0.001293502 | 0 | 0.000910452 | 0.002191996 |
| 0.030037062 | 0.011672367 | 0.037458326 | 0.015257879 | 0.350135881 |
| 0 | 0 | 0 | 0 | 0 |
| 0 | 0 | 0 | 0 | 0.04516048 |
| 0 | 0 | 0 | 0 | 0 |
| 0 | 0 | 0 | 0 | 0 |
| 0 | 0 | 0.206486799 | 0 | 0.067572739 |
| 0 | 0 | 0 | 0 | 0.123618751 |
| 0 | 0 | 0 | 0 | 0 |
| 0 | 0.008950753 | 0 | 0.009820791 | 0 |
| 0 | 0.018476323 | 0.00248907 | 0.01390174 | 0.003140851 |
| 0 | 0 | 0 | 0 | 0 |
| 0 | 0 | 0.102443612 | 0 | 0.161366296 |
| 0 | 0.001958438 | 0 | 0.005089765 | 0 |

| | | | | |
|---|---|---|---|---|
| 0 | 0 | 0 | 0 | 0 |
| 0 | 0 | 0.004273126 | 0 | 0 |
| 0 | 0.005600955 | 0 | 0.003246618 | 0 |
| 0 | 0 | 0 | 0 | 0 |
| 0 | 0 | 0 | 0 | 0 |
| 0 | 0 | 0 | 0.026733314 | 0 |
| 0 | 0 | 0.101070766 | 0 | 0 |
| 0 | 0.165999426 | 0.07131538 | 0.015768653 | 0 |
| 0 | 0 | 0 | 0 | 0 |
| 0.000919241 | 0 | 0.012759465 | 0 | 0 |
| 0 | 0 | 0 | 0 | 0 |
| 0 | 0 | 0 | 0 | 0 |
| 0 | 0 | 0 | 0 | 0 |
| 0 | 0 | 0 | 0 | 0 |
| 0 | 0.254827547 | 0 | 0 | 0 |
| 0 | 0 | 0 | 0 | 0 |
| 0 | 0 | 0.004895178 | 0 | 0 |
| 0 | 0 | 0 | 0 | 0 |
| 0 | 0 | 0.662471589 | 0.473782325 | 0.673383719 |
| 0 | 0 | 0 | 0 | 0 |
| 0 | 0 | 0 | 0 | 0.000737872 |
| 0 | 0 | 0 | 0 | 0 |
| 0 | 0 | 0 | 0 | 0 |
| 0 | 0 | 0 | 0 | 0 |
| 0 | 0 | 0 | 0.003919705 | 0.10273511 |
| 0.003116255 | 0.020314689 | 0 | 0.082453031 | 0.011147803 |
| 0 | 0 | 0 | 0 | 0 |
| 0 | 0.158782038 | 0.024382158 | 0 | 0.020034197 |
| 0 | 0 | 0 | 0 | 0 |
| 0 | 0 | 0.180835692 | 0.185213035 | 1.771848498 |
| 0 | 0 | 0 | 0 | 0 |
| 0 | 0 | 0 | 0.001144422 | 0 |
| 0 | 0 | 0 | 0 | 0 |
| 0 | 0 | 0 | 0 | 0 |
| 0 | 0 | 0 | 0.000874994 | 0.010150115 |
| 0 | 0 | 0 | 0 | 0 |
| 0 | 0 | 0 | 0 | 0 |
| 0 | 0 | 0 | 0.001489293 | 0.003476953 |
| 0 | 0 | 0 | 0 | 0 |
| 0 | 0 | 0 | 0 | 0 |
| 26.05432064 | 4.188152727 | 9.082335579 | 2.763062602 | 17.34425153 |
| 0 | 1.06107832 | 0.073115663 | 0.665911781 | 0 |
| 0 | 0 | 0.018544433 | 0 | 0 |
| 0 | 0 | 0.003134107 | 0 | 0 |
| 0 | 0 | 0 | 0 | 0 |
| 0 | 0 | 0 | 0 | 0 |
| 0 | 0 | 0 | 0 | 0 |
| 0 | 0.005037829 | 0 | 0.002166973 | 0.004139254 |

| | | | | |
|---|---|---|---|---|
| 0.004990263 | 0 | 0 | 0 | 0 |
| 0 | 0.000431819 | 0 | 0.015805005 | 0.003193173 |
| 0 | 0 | 0 | 0.033368833 | 0 |
| 0 | 0 | 0 | 0 | 0 |
| 0 | 3.912327556 | 1.846184489 | 0.117459575 | 2.987685979 |
| 0 | 0 | 3.245121052 | 0 | 0 |
| 0 | 0 | 0 | 0 | 0 |
| 0 | 0 | 0 | 0.000644516 | 0 |
| 0.100324641 | 0.269201282 | 0 | 0.049975394 | 0.0081483 |
| 0 | 0 | 0 | 0 | 0 |
| 0 | 0 | 0 | 0 | 0 |
| 0 | 0 | 0 | 0 | 0 |
| 0 | 0.050210385 | 0.129863471 | 0.133489997 | 0.050505256 |
| 0.124925871 | 0.004470592 | 0.026870227 | 0.004114912 | 0.013774471 |
| 0 | 0 | 0 | 0 | 0 |
| 0 | 0.458284758 | 0.425371865 | 0.534307904 | 0.285189216 |
| 0 | 0 | 0.033715419 | 0 | 0.007151801 |
| 0 | 0 | 0 | 0 | 0 |
| 0 | 0 | 0 | 0 | 0 |
| 0 | 0 | 0 | 0 | 0 |
| 0 | 0 | 0 | 0 | 0 |
| 0 | 0 | 0 | 0 | 0 |
| 0 | 0 | 0 | 0 | 0 |
| 0 | 0 | 0 | 0 | 0 |
| 0 | 0 | 0 | 0 | 0 |
| 0 | 0 | 0 | 0 | 0 |
| 0 | 0.039331874 | 0.241275071 | 0.064839698 | 0 |
| 0 | 0.001751906 | 0 | 0.004521392 | 0 |
| 0 | 0 | 0 | 0 | 0 |
| 0 | 0 | 0 | 0 | 0 |
| 0 | 0 | 0 | 0 | 0 |
| 0 | 0 | 0 | 0 | 0.003607089 |
| 0 | 0 | 0 | 0 | 0 |
| 0 | 0 | 0.005978897 | 0 | 0 |
| 0 | 0 | 0.125164525 | 0 | 0.445934439 |
| 0 | 0 | 0 | 0.124338789 | 0.004370933 |
| 0 | 0 | 0.202989329 | 0 | 0.000638231 |
| 0 | 0 | 0.01661406 | 0 | 0 |
| 0 | 0 | 0 | 0 | 0 |
| 0 | 0 | 0 | 0 | 0 |
| 0 | 4.219283975 | 1.233385661 | 0 | 2.590981981 |
| 0 | 0 | 0.001603319 | 0.191804292 | 0.133697136 |
| 0 | 0 | 0.000935085 | 0 | 0 |
| 0 | 0 | 0.010703761 | 0 | 0 |
| 0 | 0 | 0 | 0 | 0 |
| 0 | 0 | 0 | 0 | 0 |
| 0 | 0 | 10.46955217 | 0 | 0 |
| 0 | 0 | 0 | 0 | 0 |

| | | | | |
|---|---|---|---|---|
| 0 | 0.094310177 | 0.091175685 | 0.054230489 | 0 |
| 0 | 0 | 0 | 0 | 0 |
| 0 | 0 | 0 | 0 | 0 |
| 0 | 0 | 0 | 0 | 0 |
| 0 | 0 | 0.253666944 | 0 | 0 |
| 0 | 0 | 0 | 0 | 0 |
| 0 | 0.070504567 | 0.30618026 | 0 | 0.032013381 |
| 0 | 0 | 0 | 0 | 0 |
| 0 | 0 | 0 | 0 | 0.002453311 |
| 0 | 0.109632459 | 0 | 0.035294928 | 0 |
| 0 | 0 | 0.00093761 | 0 | 0 |
| 0 | 0.001168249 | 0 | 0 | 0 |
| 0 | 0 | 0 | 0 | 0 |
| 0 | 0 | 0 | 0 | 0 |
| 0 | 0 | 0 | 0 | 0 |
| 0 | 0 | 0 | 0.001417181 | 0 |
| 0 | 0 | 0 | 0 | 0 |
| 0 | 0.000856203 | 0 | 0.020691057 | 0 |
| 0 | 0 | 0 | 0 | 0.087163551 |
| 0 | 0 | 0 | 0 | 0 |
| 0 | 0.008902799 | 0.241997086 | 0 | 0 |
| 0 | 0 | 0 | 0 | 0 |
| 0 | 0 | 0.001420877 | 0.007021502 | 0.083179806 |
| 0 | 0.001801806 | 0.001263459 | 0.001560898 | 0.002903912 |
| 0 | 0 | 0 | 0 | 0 |
| 0 | 0 | 0 | 0.00161132 | 0 |
| 0 | 0 | 0 | 0 | 0 |
| 0 | 0 | 0 | 0 | 0 |
| 0 | 0 | 0 | 0 | 0 |
| 0.005689382 | 0 | 0.005715013 | 0 | 0 |
| 0 | 0 | 0.001189074 | 0 | 0 |
| 0 | 0 | 0 | 0 | 0 |
| 0 | 0 | 0 | 0 | 0 |
| 0 | 1.809358983 | 0.076524461 | 0 | 0.497469427 |
| 0 | 0 | 0 | 0 | 0 |
| 0 | 0 | 0 | 0 | 0 |
| 0 | 0 | 0.000903379 | 0 | 0.020518847 |
| 0 | 0 | 0 | 0 | 0 |
| 0 | 0 | 0 | 0 | 0 |
| 0 | 0 | 0 | 0 | 0.127996453 |
| 0 | 0 | 0 | 0 | 0 |
| 0 | 0 | 0.000906412 | 0 | 0 |
| 0 | 0 | 0 | 0 | 0 |
| 0 | 0 | 0 | 0.004659295 | 0 |
| 0 | 0 | 0 | 0.575099079 | 0 |
| 0 | 0 | 0 | 0 | 0 |
| 0 | 0 | 0 | 0 | 0 |
| 0 | 0 | 0 | 0 | 0.06172881 |

| | | | | |
|---|---|---|---|---|
| 0 | 0.005070726 | 0 | 0.001679582 | 0.025916731 |
| 0 | 0 | 0.068889147 | 0 | 0.176064607 |
| 0 | 0 | 0.139408548 | 0 | 0 |
| 0 | 0 | 0 | 0 | 0 |
| 0 | 0 | 0 | 0 | 0 |
| 0 | 0 | 0.015684247 | 0 | 0 |
| 0 | 0 | 0 | 0 | 0 |
| 2.012967539 | 2.09928732 | 1.403504283 | 1.181317372 | 0.73754839 |
| 0.043222965 | 0.002575057 | 2.284274184 | 0.004284078 | 0.21349893 |
| 0 | 0.047261492 | 0 | 0.041295417 | 0.010645233 |
| 0 | 0 | 0.026648216 | 0 | 0 |
| 0 | 0 | 0.003715451 | 0 | 0 |
| 0 | 0 | 0 | 0 | 0.040007284 |
| 0 | 0 | 0 | 0 | 0 |
| 0 | 0 | 0 | 0 | 0 |
| 0 | 0 | 0 | 0 | 0 |
| 0.043464132 | 0.014803792 | 0 | 0 | 0 |
| 0 | 0 | 1.603857248 | 0 | 0 |
| 0 | 0 | 0 | 0 | 0 |
| 0 | 0 | 0.00158388 | 0.008560788 | 0.103262556 |
| 0 | 0 | 0 | 0 | 0 |
| 0 | 0 | 0 | 0 | 0 |
| 0 | 0 | 0 | 0 | 0 |
| 0 | 0 | 0 | 0 | 0 |
| 0 | 0 | 0.008872472 | 0.003637985 | 0.060949509 |
| 0 | 0 | 0 | 0 | 0 |
| 0 | 0 | 0 | 0 | 0 |
| 0 | 0 | 0 | 0 | 0 |
| 0 | 0 | 0 | 0 | 0 |
| 0 | 0 | 0 | 0 | 0 |
| 0 | 0 | 0 | 0.004960759 | 0 |
| 0 | 0 | 0 | 0 | 0 |
| 0 | 0 | 0.008921279 | 0 | 0 |
| 0 | 0 | 0.00262534 | 0 | 0 |
| 0 | 0 | 0 | 0.008133481 | 0 |
| 0 | 0 | 0.269496885 | 0 | 0 |
| 0 | 0 | 0 | 0 | 0 |
| 0 | 0 | 0 | 0 | 0 |
| 0 | 0 | 0 | 0 | 0 |
| 0 | 0.308941428 | 0 | 0 | 0 |
| 0 | 0 | 0 | 0 | 0 |
| 0 | 0.093851929 | 0.157425507 | 0.320457267 | 0.752504301 |
| 0 | 0 | 0 | 0 | 0 |
| 0 | 0 | 0 | 0 | 0 |
| 0 | 0 | 0 | 0 | 0 |
| 0 | 0 | 0 | 0.003173665 | 0 |
| 0 | 0 | 0 | 0 | 0 |
| 0 | 0 | 0 | 0 | 0 |

| | | | | |
|---|---|---|---|---|
| 0 | 0 | 0 | 0 | 0 |
| 0.014548135 | 0 | 0 | 0 | 0 |
| 0 | 0 | 0 | 0 | 0 |
| 0 | 0 | 0 | 0 | 0 |
| 0.150009991 | 0.79362246 | 0.063475844 | 0.695704796 | 0.426774076 |
| 0 | 0 | 0 | 0 | 0 |
| 0.001715037 | 0 | 0 | 0 | 0 |
| 0.026485084 | 0.057673548 | 0 | 0.008546199 | 0.027356391 |
| 0 | 0 | 0 | 0 | 0.003501839 |
| 0 | 0 | 0.060661511 | 0 | 0 |
| 0 | 0 | 0 | 0 | 0 |
| 0 | 0 | 0 | 0 | 0 |
| 0 | 0 | 0 | 0 | 0 |
| 0 | 0 | 0 | 0 | 0 |
| 0 | 0 | 0 | 0 | 0 |
| 0 | 0 | 0 | 0.043082566 | 0.020278627 |
| 0 | 0 | 0 | 0 | 0 |
| 0 | 0 | 0 | 0 | 0 |
| 0 | 0 | 0.011918996 | 0 | 0 |
| 0 | 0 | 0 | 0 | 0 |
| 0 | 0 | 0 | 0 | 0 |
| 0 | 0 | 0 | 0 | 0 |
| 0 | 0 | 0 | 0 | 0 |
| 0 | 0.00049255 | 0 | 0.002773512 | 0.00384461 |
| 0 | 0 | 0 | 0 | 0 |
| 0 | 0 | 0 | 0 | 0 |
| 0.006311605 | 0.076283419 | 0 | 0 | 0.009993786 |
| 0 | 0 | 0 | 0.001112939 | 0 |
| 0 | 0 | 0 | 0 | 0 |
| 0 | 0 | 0 | 0 | 0 |
| 0 | 0 | 0 | 0 | 0 |
| 0 | 0.003463938 | 0 | 0 | 0 |
| 0 | 0 | 0 | 0 | 0 |
| 0 | 0 | 0 | 0 | 0 |
| 0 | 0 | 0 | 0 | 0 |
| 0 | 0 | 0 | 0 | 0 |
| 0 | 0 | 0 | 0 | 0 |
| 0 | 0.000462007 | 0.005815985 | 0.076311415 | 0 |
| 0 | 0 | 0 | 0 | 0 |
| 0 | 0 | 0 | 0 | 0 |
| 0 | 0 | 0 | 0 | 0 |
| 0 | 0 | 0 | 0 | 0.29750392 |
| 0 | 0 | 0 | 0.011850855 | 0 |
| 0 | 0 | 0 | 0 | 0 |
| 0 | 0 | 0 | 0 | 0 |
| 0 | 0 | 0 | 0 | 0 |
| 0 | 0 | 0 | 0 | 0 |
| 0 | 0 | 0.114597928 | 0 | 0 |

| | | | | |
|---|---|---|---|---|
| 0 | 0.127518497 | 0.017286683 | 0 | 0 |
| 0 | 0 | 0 | 0 | 0 |
| 0 | 0 | 0 | 0 | 0 |
| 0 | 0 | 0 | 0.006131491 | 0 |
| 0 | 0 | 0 | 0 | 0 |
| 0 | 0 | 0.010226871 | 0.017503975 | 0 |
| 0 | 0 | 0 | 0 | 0 |
| 0 | 0.000476732 | 0 | 0 | 0 |
| 0 | 0 | 0 | 0 | 0 |
| 0 | 0 | 0 | 0 | 0 |
| 0 | 0 | 0.487509194 | 0 | 0 |
| 0 | 0.004843224 | 0 | 0.002556735 | 0 |
| 0 | 0 | 0 | 0.002407466 | 0 |
| 0 | 0 | 0 | 0 | 0 |
| 0 | 0 | 0.000577485 | 0 | 0 |
| 0 | 0 | 0 | 0 | 0 |
| 0 | 0 | 0 | 0 | 0 |
| 0 | 0 | 0 | 0 | 0 |
| 0 | 0 | 0 | 0 | 0 |
| 0 | 0 | 0 | 0 | 0 |
| 0 | 0.033026376 | 0 | 0.32360468 | 0.001612016 |
| 0 | 0 | 0 | 0 | 0 |
| 0 | 0.049969295 | 0.001344486 | 0.019184554 | 0.011451707 |
| 0 | 0.343386741 | 0.005414391 | 0 | 0 |
| 0 | 0 | 0 | 0 | 0 |
| 0 | 0 | 0 | 0.371154716 | 0 |
| 0 | 0 | 0 | 0 | 0 |
| 0 | 0 | 0 | 0 | 0 |
| 0 | 0 | 0 | 0 | 0 |
| 0 | 0 | 0 | 0 | 0 |
| 0 | 0 | 0 | 0 | 0 |
| 0 | 0 | 0 | 0 | 0 |
| 0.073729749 | 0.007348138 | 0 | 0.00136108 | 0.010247833 |
| 0 | 0 | 0 | 0 | 0 |
| 0 | 3.118310174 | 1.144734565 | 0 | 0.190793975 |
| 0 | 0 | 0 | 0.012669632 | 0.000764973 |
| 0 | 0 | 0 | 0 | 0 |
| 0 | 0 | 0 | 0 | 0 |
| 0 | 0.005204981 | 0 | 0.144270371 | 0.000663609 |
| 0 | 0 | 0 | 0 | 0 |
| 0 | 0 | 0 | 0 | 0 |
| 0 | 0 | 0.00129469 | 0 | 0 |
| 0 | 0 | 0.020771071 | 0 | 0.095184144 |
| 0 | 0 | 0 | 0 | 0 |
| 0 | 0 | 0 | 0 | 0 |
| 0 | 0.111170133 | 0.009333169 | 0.454476923 | 0.00944909 |
| 0 | 0 | 0 | 0 | 0 |
| 0 | 0 | 0 | 0 | 0 |

| | | | | |
|---|---|---|---|---|
| 0 | 0 | 0 | 0 | 0 |
| 0 | 0 | 0 | 0 | 0 |
| 0 | 0 | 0 | 0 | 0 |
| 0 | 0 | 0 | 0 | 0 |
| 0 | 0 | 0 | 0 | 0 |
| 0 | 0 | 0 | 0 | 0 |
| 0 | 0 | 0 | 0 | 0 |
| 1.362115625 | 0.104848641 | 0 | 0.313562359 | 0.012703742 |
| 0 | 0 | 0.004677191 | 0.749826096 | 0 |
| 0 | 0 | 0 | 0 | 0.004443079 |
| 0 | 0 | 0 | 0 | 0 |
| 0 | 0 | 0 | 0 | 0 |
| 0 | 0 | 0 | 0 | 0 |
| 0 | 0 | 0.829503982 | 0 | 0 |
| 0 | 0 | 0 | 0 | 0.003248651 |
| 0 | 0.080398532 | 0 | 0.014266333 | 0 |
| 0 | 0 | 0 | 0 | 0 |
| 0 | 0 | 0 | 0 | 0 |
| 0 | 0 | 0 | 0 | 0 |
| 0 | 1.10244929 | 0.747119674 | 0 | 0.035205298 |
| 0 | 0.003175357 | 0.000689191 | 0 | 0.01869771 |
| 0 | 0 | 0 | 0 | 0 |
| 0 | 0 | 0 | 0 | 0 |
| 0 | 0 | 0 | 0 | 0.098712456 |
| 0 | 0.002282789 | 0 | 0 | 0 |
| 0 | 0 | 0 | 0 | 0 |
| 0 | 0 | 0 | 0 | 0 |
| 0 | 0 | 0 | 0 | 0 |
| 0 | 0 | 0 | 0 | 0 |
| 0 | 0 | 0.108969406 | 1.354147389 | 0.27551108 |
| 0 | 0 | 0.241347053 | 0.130640093 | 0.275782892 |
| 0 | 0 | 0 | 0 | 0 |
| 0 | 0 | 0 | 0 | 0 |
| 0 | 0 | 0 | 0 | 0 |
| 0 | 0 | 0 | 0 | 0 |
| 0 | 0 | 0 | 0 | 0 |
| 0 | 0 | 0 | 0 | 0 |
| 0 | 0 | 0 | 0 | 0 |
| 0 | 0 | 0 | 0 | 0 |
| 0 | 0 | 0 | 0 | 0 |
| 0 | 0 | 0 | 0 | 0 |
| 0 | 0 | 0 | 0 | 0 |
| 0 | 0 | 0.000966808 | 0 | 0 |
| 0 | 0 | 0 | 0 | 0 |
| 0 | 0.225203068 | 0 | 0 | 0 |
| 0 | 0 | 0 | 0 | 0 |
| 0.622667705 | 0 | 0 | 0.020446077 | 0 |
| 0 | 0 | 0 | 0 | 0.002944146 |

| | | | | |
|---|---|---|---|---|
| 0 | 0 | 0 | 0 | 0 |
| 0 | 0 | 0 | 0 | 0 |
| 0 | 0 | 0 | 0 | 0 |
| 0 | 0 | 0 | 0 | 0 |
| 0.001551244 | 0.094038605 | 0.020186193 | 0.009455795 | 0.173051034 |
| 0 | 0 | 0 | 0 | 0 |
| 0 | 0 | 0 | 0 | 0 |
| 0 | 0 | 0 | 0 | 0 |
| 0 | 0 | 0 | 0 | 0 |
| 0 | 0 | 0 | 0 | 0 |
| 0 | 0 | 0.114478209 | 0 | 0 |
| 0 | 0.004842158 | 0 | 0.002272153 | 0.002431295 |
| 0 | 0.000773623 | 0 | 0.002178106 | 0.012394881 |
| 0 | 0 | 0 | 0 | 0 |
| 0 | 0 | 0 | 0 | 0 |
| 0.086251622 | 0 | 0.081363782 | 0 | 0.322618205 |
| 0 | 0.024000361 | 0 | 0.006019358 | 0 |
| 0 | 0 | 0 | 0.001586149 | 0 |
| 0 | 0.050108964 | 0 | 0.016249047 | 0 |
| 0 | 0.002659636 | 0.005079862 | 0 | 0.001966723 |
| 0 | 0 | 0 | 0 | 0 |
| 0 | 0 | 0 | 0 | 0 |
| 0 | 0 | 0 | 0 | 0 |
| 0 | 0 | 0 | 0 | 0 |
| 0 | 0 | 0 | 0 | 0 |
| 0 | 0 | 0 | 0 | 0 |
| 0 | 0.026712143 | 0 | 0.024202274 | 0.005603646 |
| 0 | 0 | 0 | 0.000758655 | 0 |
| 0 | 0 | 0 | 0 | 0 |
| 0 | 0 | 0 | 0 | 0 |
| 0 | 0 | 0 | 0.005668295 | 0 |
| 0 | 0.007480516 | 0 | 0.001224484 | 0 |
| 0 | 0 | 0 | 0.004117405 | 0 |
| 0 | 0 | 0 | 0 | 0 |
| 0 | 0 | 0 | 0 | 0 |
| 0 | 0 | 0 | 0 | 0 |
| 0 | 0 | 0 | 0 | 0 |
| 0.006454783 | 0.00724207 | 0.000884163 | 0.001433657 | 0.066104485 |
| 0 | 0 | 0.013116092 | 0.384348527 | 0.024825954 |
| 0 | 0.015257235 | 0 | 0.001010734 | 0 |
| 0 | 0.00225935 | 0 | 0.001113196 | 0 |
| 0 | 0 | 0 | 0 | 0 |
| 0 | 0 | 0 | 0 | 0 |
| 0 | 0 | 0 | 0 | 0 |
| 0 | 0 | 0 | 0 | 0 |
| 0 | 0 | 0 | 0 | 0 |
| 0 | 0 | 0 | 0.025413782 | 0 |
| 0 | 0 | 0 | 0 | 0 |

| | | | | |
|---|---|---|---|---|
| 0 | 0 | 0 | 0 | 0 |
| 0 | 0 | 0.191810608 | 0 | 0 |
| 0 | 0 | 0 | 0 | 0 |
| 0.001055906 | 0.001406824 | 0 | 0.002723093 | 0.000866921 |
| 0.000979336 | 0 | 0 | 0 | 0 |
| 0 | 0 | 0 | 0.006815448 | 0 |
| 0 | 0 | 0 | 0 | 0.006940572 |
| 0 | 0 | 0 | 0 | 0 |
| 0 | 0 | 0 | 0 | 0 |
| 0 | 0 | 0 | 0 | 0 |
| 0 | 0.010171708 | 0.003784637 | 0.005932172 | 0 |
| 0 | 0 | 0 | 0 | 0 |
| 0 | 0.01059568 | 0.005844008 | 0.018951936 | 0.056843588 |
| 0 | 0 | 0 | 0 | 0 |
| 0 | 0 | 0 | 0 | 0 |
| 0 | 0 | 0 | 0 | 0 |
| 0 | 0 | 0 | 0 | 0 |
| 0 | 0 | 0 | 0 | 0 |
| 0 | 0 | 0 | 0 | 0 |
| 0 | 0 | 0 | 0 | 0 |
| 0 | 0 | 0 | 0 | 0 |
| 0 | 0 | 0 | 0 | 0 |
| 0 | 0.003611124 | 0 | 0.016867956 | 0.037424953 |
| 0.013665563 | 0.053071933 | 0.025728407 | 0.030538823 | 0.124371802 |
| 0 | 0 | 0 | 0 | 0 |
| 0 | 0 | 0.000931406 | 0 | 0 |
| 0 | 0 | 0.011640838 | 0 | 0 |
| 0 | 0 | 0 | 0 | 0 |
| 0 | 0 | 0 | 0 | 0 |
| 0 | 0 | 0 | 0 | 0 |
| 0 | 0 | 0.007150105 | 0.003864595 | 0.062480205 |
| 0 | 0 | 0 | 0 | 0 |
| 0 | 0 | 0 | 0 | 0 |
| 0 | 0 | 0 | 0 | 0 |
| 0 | 0 | 0.001998163 | 0 | 0 |
| 0 | 0 | 0 | 0 | 0 |
| 0 | 0.002916642 | 0 | 0 | 0 |
| 0 | 0 | 0 | 0 | 0 |
| 0 | 0 | 0.085701038 | 0 | 0.049045793 |
| 0 | 0 | 0 | 0 | 0 |
| 0 | 0 | 0 | 0 | 0 |
| 0 | 0 | 0 | 0 | 0 |
| 0 | 0 | 0 | 0 | 0 |
| 0 | 0 | 0 | 0 | 0 |
| 0 | 0 | 0.578102639 | 0 | 0 |
| 0.075464309 | 0 | 1.782166314 | 0.046412526 | 0.290311853 |
| 0 | 0 | 0 | 0 | 0 |
| 0 | 0 | 0 | 0 | 0 |

| | | | | |
|---|---|---|---|---|
| 0 | 0 | 0 | 0 | 0 |
| 0 | 0 | 0 | 0 | 0 |
| 0 | 0 | 0.07643195 | 0 | 0 |
| 0 | 0 | 0.004017961 | 0 | 0 |
| 0 | 0 | 0 | 0 | 0 |
| 0 | 0 | 0 | 0 | 0 |
| 0 | 0 | 0 | 0 | 0 |
| 0 | 0 | 0 | 0 | 0 |
| 0 | 0 | 0 | 0 | 0 |
| 0 | 0 | 0 | 0 | 0 |
| 0 | 0.000753727 | 0 | 0 | 0 |
| 0 | 0 | 0 | 0 | 0 |
| 0 | 0 | 0 | 0 | 0 |
| 0 | 0 | 0 | 0 | 0 |
| 0 | 0 | 0 | 0 | 0 |
| 0 | 0 | 0 | 0 | 0 |
| 0 | 0 | 0 | 0 | 0 |
| 0 | 0 | 0 | 0 | 0 |
| 0 | 0 | 0 | 0 | 0 |
| 0 | 0 | 0 | 0 | 0.000821283 |
| 0 | 0 | 0 | 0 | 0 |
| 0 | 0 | 0 | 0 | 0 |
| 0 | 0 | 0 | 0 | 0 |
| 0 | 0 | 0 | 0 | 0 |
| 0 | 0 | 0 | 0 | 0 |
| 0 | 0 | 0 | 0.142287914 | 0 |
| 0 | 0 | 0 | 0 | 0 |
| 0 | 0 | 0 | 0 | 0.020449985 |
| 0 | 0 | 0 | 0 | 0 |
| 0 | 0 | 0 | 0 | 0 |
| 0 | 0.034492556 | 0 | 0 | 0.002116701 |
| 0 | 0 | 0 | 0 | 0 |
| 0.025444513 | 1.30999662 | 0.022723034 | 1.100672719 | 0.020071257 |
| 0.00509834 | 0.013245796 | 0.00088459 | 0.000956232 | 0.007743816 |
| 0 | 0 | 0 | 0 | 0 |
| 0 | 0 | 0 | 0 | 0 |
| 0 | 0 | 0 | 0 | 0 |
| 0 | 0 | 0 | 0 | 0 |
| 0 | 0 | 0 | 0 | 0 |
| 0 | 0 | 0 | 0 | 0 |
| 0 | 0 | 0 | 0 | 0 |
| 0 | 0.133023447 | 0 | 0.036951878 | 0.088796496 |
| 0 | 0.015156697 | 0.065721002 | 0 | 0.01211114 |
| 0 | 0 | 0 | 0 | 0 |
| 0 | 0 | 0 | 0 | 0 |
| 0 | 0 | 0.02100028 | 0 | 0.05962355 |
| 0 | 0.014359888 | 0 | 0.00155499 | 0 |
| 0 | 0 | 0 | 0 | 0 |

| | | | | |
|---|---|---|---|---|
| 0 | 0 | 0 | 0 | 0 |
| 0 | 0 | 0.027722372 | 0 | 0 |
| 0.040693144 | 0.003668823 | 0.020703667 | 0.003443146 | 0.00577766 |
| 0 | 0 | 0 | 0 | 0 |
| 0 | 0.726648686 | 0 | 0.021764393 | 0.132378307 |
| 0 | 0.003009853 | 0 | 0 | 0 |
| 0 | 0 | 0 | 0 | 0 |
| 0 | 0 | 0 | 0 | 0 |
| 0 | 0 | 0 | 0 | 0 |
| 0 | 0 | 0 | 0 | 0 |
| 0 | 0 | 0 | 0 | 0 |
| 0 | 0 | 0 | 0 | 0 |
| 0 | 0 | 0 | 0 | 0 |
| 0 | 0 | 0 | 0 | 0 |
| 0 | 0 | 0 | 0 | 0 |
| 0 | 0 | 0 | 0 | 0 |
| 0 | 0 | 0.45356657 | 0 | 0 |
| 0 | 0 | 0 | 0 | 0 |
| 0 | 0 | 0 | 0 | 0 |
| 0 | 0 | 0 | 0 | 0 |
| 0 | 0 | 0 | 0 | 0 |
| 0 | 0.000819762 | 0 | 0 | 0 |
| 0 | 0 | 0.025811364 | 0 | 0 |
| 0 | 0 | 0 | 0 | 0 |
| 0 | 0 | 0.773729273 | 0 | 0.001242158 |
| 0 | 0 | 0.009902714 | 0 | 0 |
| 0 | 0 | 0 | 0 | 0 |
| 0 | 0 | 0.019425423 | 0 | 0.25313468 |
| 0 | 0 | 0 | 0 | 0 |
| 0 | 0 | 0 | 0 | 0 |
| 0 | 0 | 0 | 0 | 0 |
| 0 | 0 | 0 | 0 | 0 |
| 0 | 0 | 0 | 0 | 0 |
| 0 | 0 | 0 | 0 | 0 |
| 0 | 0 | 0 | 0 | 0 |
| 0 | 0 | 0 | 0 | 0 |
| 0 | 0 | 0 | 0 | 0 |
| 0 | 0.171909097 | 0 | 0 | 0 |
| 0 | 0 | 0.081517371 | 0 | 17.41830887 |
| 0 | 0 | 0 | 0 | 0 |
| 0 | 0 | 0 | 0 | 0 |
| 0 | 0.120258346 | 0.003342241 | 0.009032321 | 0.008811419 |
| 0 | 0.002035116 | 0 | 0.001432449 | 0.021528564 |
| 0 | 0 | 0 | 0 | 0 |
| 0 | 0 | 0 | 0 | 0 |
| 0 | 0 | 0 | 0 | 0 |
| 0 | 0.007264436 | 0 | 0.004747961 | 0.005116037 |
| 0 | 0 | 0 | 0 | 0 |

| | | | | |
|---|---|---|---|---|
| 0 | 0 | 0 | 0 | 0 |
| 0.003194248 | 0.033555504 | 0.00682116 | 0.031568242 | 0.017752619 |
| 0 | 0 | 0.002021992 | 0 | 0 |
| 0 | 0 | 0 | 0 | 0 |
| 0 | 0 | 0 | 0 | 0 |
| 0 | 0 | 0 | 0 | 0 |
| 0 | 0 | 0 | 0 | 0 |
| 0 | 0 | 0 | 0 | 0 |
| 0 | 0 | 0 | 0 | 0 |
| 0 | 0 | 0 | 0 | 0 |
| 0 | 0 | 0 | 0 | 0 |
| 0 | 0 | 0 | 0 | 0 |
| 0 | 0.00069212 | 0.091258788 | 0 | 0.209839024 |
| 0 | 0.002664004 | 0 | 0 | 0 |
| 0 | 0.019592737 | 0 | 0 | 0.022637905 |
| 0 | 0 | 0 | 0 | 0 |
| 0 | 0 | 0 | 0 | 0 |
| 0 | 0 | 0 | 0 | 0 |
| 0 | 0.021900417 | 0 | 0 | 0 |
| 0 | 0 | 0 | 0 | 0 |
| 0 | 0 | 0 | 0 | 0 |
| 0.01233586 | 0.027258953 | 0.002349152 | 0 | 0.012598252 |
| 0 | 0.004620502 | 0 | 0 | 0 |
| 0 | 0 | 0 | 0 | 0 |
| 0 | 0 | 0 | 0 | 0 |
| 0 | 0 | 0 | 0 | 0 |
| 0 | 0 | 0 | 0 | 0 |
| 0 | 0 | 0 | 0 | 0 |
| 0 | 0 | 0 | 0.197511079 | 0 |
| 0 | 0 | 0 | 0 | 0 |
| 0 | 0 | 0 | 0 | 0 |
| 0 | 0 | 0 | 0 | 0 |
| 0 | 0 | 0 | 0 | 0 |
| 0 | 0 | 0 | 0 | 0 |
| 0 | 0 | 0.042184011 | 11.56510164 | 0 |
| 0 | 0 | 0 | 0 | 0 |
| 0 | 0.002468256 | 0 | 0 | 0 |
| 0 | 0 | 0 | 0 | 0 |
| 0.019229423 | 0.006004712 | 0.023459127 | 0 | 0.010854092 |
| 0 | 0 | 0 | 0 | 0 |
| 0.369619386 | 0.03445772 | 0.236050575 | 0.14299527 | 0.593217489 |
| 0 | 0 | 0 | 0 | 0 |
| 0 | 0 | 0 | 0 | 0 |
| 0 | 0 | 0 | 0 | 0 |
| 0 | 0 | 0 | 0 | 0 |
| 5.502999782 | 0.570406477 | 0.014984815 | 0.016776946 | 1.119327347 |
| 0 | 0 | 0 | 0 | 0 |
| 0 | 0 | 0 | 0 | 0 |

| | | | | |
|---|---|---|---|---|
| 0 | 0 | 0 | 0 | 0 |
| 0 | 0 | 0 | 0 | 0 |
| 0 | 0 | 0 | 0 | 0 |
| 0 | 0 | 0 | 0 | 0 |
| 0 | 0.001714233 | 0 | 0.022815527 | 0 |
| 0 | 0 | 0 | 0 | 0 |
| 0 | 0 | 0 | 0 | 0 |
| 0 | 0 | 0 | 0 | 0 |
| 0 | 0 | 0 | 0 | 0 |
| 0 | 0.000803831 | 0.007850985 | 0 | 0.023281033 |
| 0 | 0 | 0.004967991 | 0 | 0 |
| 0 | 0 | 0 | 0 | 0 |
| 0 | 0 | 0 | 0 | 0.005809851 |
| 0 | 0 | 0 | 0 | 0 |
| 0 | 0 | 0 | 0 | 0 |
| 0 | 0 | 0 | 0 | 0 |
| 0 | 0 | 0 | 0 | 0 |
| 0 | 0 | 0 | 0 | 0 |
| 0 | 0 | 0 | 0 | 0 |
| 0.029917721 | 0.289863056 | 0.016164604 | 0.009844377 | 0.045032337 |
| 0 | 0 | 0 | 0 | 0 |
| 0 | 0 | 0 | 0 | 0 |
| 0 | 0 | 0 | 0 | 0 |
| 0 | 0 | 0 | 0 | 0 |
| 0 | 0 | 0 | 0 | 0 |
| 0 | 0 | 0 | 0 | 0 |
| 0 | 0 | 0 | 0 | 0 |
| 0 | 0 | 0 | 0 | 0 |
| 0 | 0 | 0 | 0 | 0 |
| 0 | 0 | 0 | 0 | 0 |
| 0 | 0 | 0 | 0 | 0 |
| 0 | 0 | 0 | 0 | 0 |
| 0 | 0 | 0 | 0 | 0 |
| 0 | 0 | 0 | 0 | 0 |
| 0 | 0 | 0 | 0 | 0 |
| 0 | 0 | 0.006188998 | 0 | 0 |
| 0 | 0 | 0 | 0 | 0 |
| 0 | 0 | 0 | 0 | 0 |
| 0 | 0 | 0 | 0 | 0 |
| 0 | 0 | 1.019833028 | 0 | 0 |
| 0 | 0 | 0 | 0 | 0 |
| 0 | 0 | 0 | 0 | 0 |
| 0 | 0.015258919 | 0.000756995 | 0.006751004 | 0 |
| 0 | 0 | 0 | 0 | 0 |
| 0 | 0.016748796 | 0 | 0.010170821 | 0 |
| 0 | 0 | 0.136701903 | 0 | 0 |
| 0 | 0 | 0 | 0 | 0 |
| 0 | 0 | 0 | 0 | 0 |

| | | | | |
|---|---|---|---|---|
| 0 | 0 | 0 | 0 | 0 |
| 0 | 0 | 0 | 0 | 0 |
| 0 | 0 | 0 | 0 | 0 |
| 0 | 0 | 0 | 0 | 0 |
| 0 | 0 | 0 | 0 | 0 |
| 0 | 0.006251814 | 0.002075284 | 0.002933626 | 0.038072049 |
| 0 | 0 | 0 | 0 | 0 |
| 0 | 0 | 0 | 0 | 0 |
| 0 | 0 | 0 | 0 | 0 |
| 0 | 0 | 0 | 0 | 0 |
| 0 | 0 | 0 | 0 | 0 |
| 0 | 0 | 0 | 0 | 0 |
| 0 | 0 | 0.007673137 | 0 | 0 |
| 0 | 0 | 0 | 0 | 0 |
| 0 | 0 | 0 | 0 | 0 |
| 0 | 0 | 0 | 0 | 0 |
| 0 | 0 | 0.001355472 | 0.033700778 | 0 |
| 0 | 0 | 0 | 0.037404311 | 0 |
| 0 | 0 | 0 | 0 | 0 |
| 0 | 0 | 0 | 0 | 0 |
| 0 | 0 | 0 | 0 | 0 |
| 0 | 0 | 0 | 0 | 0 |
| 0 | 0 | 0 | 0 | 0 |
| 0 | 0 | 0.015549069 | 0 | 0 |
| 0 | 0 | 0 | 0.002547981 | 0 |
| 0 | 0 | 0 | 0 | 0.000945255 |
| 0 | 0 | 0 | 0 | 0 |
| 0 | 0 | 0 | 0 | 0 |
| 0 | 0 | 0 | 0 | 0 |
| 0 | 0.028587721 | 0 | 0.015598389 | 0 |
| 0 | 0 | 0 | 0 | 0 |
| 0 | 0 | 0 | 0 | 0 |
| 0 | 0 | 0 | 0 | 0 |
| 0 | 0 | 0 | 0 | 0 |
| 0 | 0 | 0 | 0 | 0 |
| 0 | 0 | 0 | 0 | 0 |
| 0 | 0 | 0 | 0 | 0 |
| 0 | 0 | 0 | 0.068345778 | 0 |
| 0 | 1.263185618 | 0.105460187 | 0 | 0 |
| 0 | 0 | 0 | 0 | 0 |
| 0 | 0 | 0 | 0 | 0 |
| 0 | 0 | 0 | 0 | 0 |
| 0 | 0 | 0 | 0 | 0 |
| 0 | 0 | 0 | 0 | 0 |
| 0 | 0 | 0 | 0 | 0 |
| 0 | 0 | 0 | 0 | 0 |
| 0 | 0 | 0 | 0 | 0 |
| 0 | 0 | 0 | 0 | 0 |

| | | | | |
|---|---|---|---|---|
| 0 | 0 | 0 | 0 | 0 |
| 0 | 0.441923486 | 0.001608541 | 0 | 0.015730554 |
| 0 | 0 | 0 | 0 | 0 |
| 0 | 0 | 0 | 0 | 0 |
| 0 | 0 | 0 | 0 | 0 |
| 0 | 0 | 0 | 0.001324686 | 0 |
| 0 | 0 | 0 | 0 | 0 |
| 0 | 0 | 0 | 0 | 0 |
| 0 | 0 | 0 | 0 | 0 |
| 0 | 0 | 0 | 0 | 0.008497401 |
| 0 | 0.001797092 | 0.000936114 | 0 | 0.024363124 |
| 0 | 0 | 0 | 0 | 0 |
| 0 | 0.630851346 | 0.361640327 | 0.193228721 | 0.012561271 |
| 0 | 0 | 0 | 0 | 0 |
| 0 | 0 | 0 | 0 | 0 |
| 0 | 0 | 0 | 0 | 0 |
| 0 | 0 | 0 | 0 | 0 |
| 0 | 0 | 0 | 0 | 0 |
| 0 | 0 | 0 | 0 | 0 |
| 0 | 0 | 0 | 0 | 0 |
| 0 | 0 | 0 | 0 | 0.102280296 |
| 0 | 0 | 0 | 0 | 0 |
| 0 | 0 | 0 | 0 | 0 |
| 12.90469195 | 3.699716415 | 1.265792495 | 1.217077884 | 3.524165843 |
| 0 | 0 | 0 | 0 | 0 |
| 0 | 0 | 0 | 0 | 0 |
| 0 | 0 | 0 | 0 | 0 |
| 0 | 0 | 0 | 0 | 0 |
| 0 | 0 | 0 | 0 | 0.000482806 |
| 0 | 0 | 0 | 0 | 0 |
| 0 | 0 | 0 | 0 | 0 |
| 0 | 0 | 0 | 0 | 0 |
| 0 | 0 | 0 | 0 | 0 |
| 0 | 0 | 0 | 0 | 0 |

| PHL018-0107_V3 | PHL018-0150_V2 | PHL018-0044_V2 | PHL018-0191_V2 | PHL018-0149_V3 |
|---|---|---|---|---|
| EYCF | EYCF | CTRL | EYCF | REF |
| 0.428306687 | 0.073486429 | 0.21809005 | 0.301729526 | 0 |
| 1.066168382 | 0.885377022 | 0.136235323 | 1.799280672 | 0.750415951 |
| 5.033485858 | 2.51720327 | 0.024241954 | 0.596366703 | 0 |
| 0 | 0 | 0 | 0 | 0 |
| 0 | 0 | 0 | 0 | 0 |
| 0 | 0 | 0.013956384 | 0 | 0 |
| 0 | 0 | 0 | 0 | 0 |
| 0 | 0 | 0 | 0 | 0 |
| 0 | 0 | 0 | 0 | 0 |
| 0 | 0 | 0 | 0 | 0 |
| 0 | 0 | 0 | 0 | 0 |
| 0 | 0 | 0 | 0 | 0 |
| 0 | 0 | 0 | 0 | 0 |
| 1.722141032 | 1.094475867 | 0.229394657 | 0.399790757 | 9.930427456 |
| 0 | 0 | 0 | 0 | 0 |
| 0 | 0 | 0 | 0 | 0 |
| 0 | 0 | 0 | 0 | 0 |
| 0 | 0 | 0 | 0 | 0 |
| 0 | 0 | 0 | 0 | 0 |
| 0 | 0 | 0 | 0 | 0 |
| 0 | 0 | 0 | 0 | 0 |
| 0 | 0 | 0 | 0 | 0 |
| 0 | 0 | 0 | 0 | 0 |
| 0 | 0 | 0 | 0 | 0 |
| 0 | 0 | 0.037628835 | 0.226882589 | 0 |
| 0 | 0 | 0 | 0 | 0 |
| 0 | 0 | 0 | 0 | 0 |
| 0 | 0 | 0 | 0 | 0 |
| 0 | 0 | 0 | 0 | 0 |
| 0 | 0 | 0 | 0.006743953 | 0 |
| 0 | 0 | 0 | 0 | 0 |
| 0 | 0 | 0 | 0 | 0 |
| 0 | 0 | 0 | 0 | 0 |
| 0 | 0 | 0 | 0 | 0 |
| 0 | 0 | 0 | 0 | 0 |
| 4.298826094 | 1.38785749 | 1.100142896 | 6.431892045 | 0 |
| 0 | 0 | 0 | 0 | 0 |
| 0 | 0 | 0 | 0 | 0 |
| 0 | 0 | 0 | 0 | 0 |
| 0 | 0 | 0 | 0 | 0 |
| 0 | 0 | 0 | 0 | 0 |
| 0.033491229 | 0.007384051 | 0.068761309 | 0.001955766 | 0.005919743 |
| 0.080566002 | 1.433358864 | 0.88910658 | 0.03153534 | 0.175813555 |
| 0.004775417 | 0.045086397 | 0.007687599 | 0.034207676 | 0.010465259 |
| 0 | 0 | 0.043108237 | 0 | 0 |
| 0 | 0 | 0 | 0 | 0 |

| | | | | |
|---|---|---|---|---|
| 0 | 0.001637488 | 0.063850529 | 0 | 0 |
| 0 | 0 | 0 | 0 | 0.005654836 |
| 0.009729674 | 0.000677422 | 0.105101675 | 0.020833174 | 1.059431228 |
| 2.385089958 | 0.450384349 | 0.285255128 | 0.724665755 | 0.123201135 |
| 0 | 0 | 0 | 0.01452413 | 0.063339798 |
| 0 | 0 | 0 | 0 | 0 |
| 0.003290272 | 7.091076808 | 0.15419493 | 0.002209609 | 0.00501606 |
| 0 | 0 | 0 | 0 | 0 |
| 0.066148834 | 0 | 0 | 0 | 0 |
| 0 | 0 | 0 | 0 | 0 |
| 0.00251927 | 0.078062897 | 0.001892609 | 0 | 0 |
| 0.065790818 | 0.528788168 | 0.432369754 | 0.653379078 | 0.545743988 |
| 0 | 0 | 0 | 0 | 0 |
| 0.184976579 | 0.010534155 | 0.027185903 | 0.117453137 | 0 |
| 0 | 0 | 0 | 0 | 0.001848271 |
| 0 | 0 | 0 | 0 | 0 |
| 0.051954965 | 1.530768386 | 0.014734253 | 0.000723125 | 0 |
| 0 | 0 | 0 | 0 | 0 |
| 0 | 0 | 0 | 0 | 0 |
| 0 | 0 | 0 | 0 | 0 |
| 0.029879737 | 0.044691572 | 0.004676509 | 0.226671244 | 0.051878688 |
| 0.105787566 | 0.004211702 | 0.005227127 | 0.068905551 | 0 |
| 0.014203409 | 0 | 0.408756653 | 0 | 0 |
| 0 | 0 | 0 | 0 | 0 |
| 0 | 0 | 0 | 0 | 0 |
| 0 | 0 | 0 | 0 | 0 |
| 0 | 0 | 0 | 0 | 0.013476166 |
| 0 | 0 | 0 | 0 | 0 |
| 0.013956983 | 0 | 0.002674803 | 0 | 0 |
| 0 | 0 | 0 | 0 | 0 |
| 0 | 0 | 0 | 0.007115933 | 0.003365409 |
| 0 | 0 | 0 | 0 | 0 |
| 0 | 0 | 0 | 0 | 0 |
| 0 | 0 | 0 | 0 | 0 |
| 0 | 0 | 0 | 0 | 0 |
| 0 | 0.002175997 | 0 | 0 | 0 |
| 0 | 0 | 0 | 0 | 0 |
| 0.002362404 | 0.073013065 | 0 | 0 | 0 |
| 0 | 0 | 0 | 0 | 0 |
| 0 | 0 | 0 | 0 | 0 |
| 0 | 0 | 0 | 0 | 0 |
| 0 | 0 | 0 | 0 | 0 |
| 0 | 0 | 0 | 0 | 0 |
| 0 | 0 | 0 | 0 | 0 |
| 0 | 0 | 0 | 0 | 0 |
| 0 | 0 | 0 | 0 | 0 |
| 0 | 0.34058283 | 0 | 0 | 0.006058816 |
| 0.004527132 | 0.144991387 | 0 | 0 | 0 |

| | | | | |
|---|---|---|---|---|
| 0 | 0 | 0 | 0 | 0 |
| 0 | 0 | 0 | 0 | 0 |
| 0 | 0 | 0 | 0 | 0 |
| 0 | 0 | 0 | 0 | 0 |
| 0.110434792 | 0.441226957 | 0.873673512 | 0 | 0.003718777 |
| 0 | 0 | 0.011821667 | 0 | 0 |
| 0 | 0 | 0 | 0 | 0 |
| 0 | 0 | 0 | 0 | 0 |
| 0.011110251 | 0.001206204 | 0.001010485 | 0.016861422 | 0 |
| 0 | 0 | 0 | 0 | 0 |
| 0 | 0 | 0 | 0 | 0 |
| 0.00065587 | 0 | 0.008446707 | 0 | 0 |
| 0 | 0 | 0 | 0 | 0 |
| 0 | 0 | 0 | 0 | 0 |
| 0 | 0 | 0 | 0 | 0 |
| 0 | 0 | 0 | 0 | 0 |
| 0 | 0 | 0 | 0 | 0 |
| 0 | 0 | 0 | 0 | 0 |
| 0.119462681 | 0.285396355 | 2.149702905 | 0.000522646 | 0 |
| 0 | 0 | 0 | 0 | 0 |
| 0 | 0 | 0 | 0 | 0 |
| 0 | 0 | 0 | 0 | 0 |
| 0 | 0 | 0 | 0.001475406 | 0 |
| 0 | 0 | 0 | 0 | 0 |
| 0 | 0 | 0 | 0 | 0 |
| 0 | 0 | 0 | 0 | 0 |
| 0 | 0 | 0 | 0 | 0 |
| 0 | 0 | 0 | 0 | 0 |
| 0 | 0 | 0 | 0 | 0 |
| 0 | 0 | 0 | 0 | 0 |
| 0.168679783 | 0.069901174 | 0.043574439 | 0.076923124 | 0 |
| 0.00621553 | 0.00073186 | 0.007259207 | 0.039776615 | 0 |
| 0 | 0 | 0 | 0 | 0 |
| 0 | 0 | 0 | 0 | 0 |
| 0 | 0 | 0 | 0 | 0 |
| 0 | 0 | 0 | 0 | 0 |
| 0 | 0 | 0 | 0 | 0 |
| 0 | 0 | 0 | 0 | 0 |
| 0.590469819 | 0 | 0 | 0 | 0 |
| 0 | 0 | 0 | 0 | 0 |
| 0 | 0 | 0 | 0 | 0 |
| 0 | 0 | 0 | 0 | 0 |
| 0 | 0.000845915 | 0 | 0 | 0.058466235 |
| 0 | 0 | 0 | 0 | 0 |
| 0 | 0 | 0 | 0 | 0 |
| 0 | 0 | 0 | 0 | 0 |
| 0 | 0.033168407 | 0 | 0 | 0 |
| 0 | 0 | 0 | 0 | 0 |

| | | | | |
|---|---|---|---|---|
| 0 | 0 | 0 | 0 | 0 |
| 0 | 0 | 0 | 0 | 0 |
| 0 | 0 | 0 | 0 | 0 |
| 0 | 0 | 0 | 0 | 0 |
| 0.014002133 | 0.008663223 | 0.111680537 | 0.101156129 | 0.007034616 |
| 0 | 0 | 0 | 0 | 0 |
| 0 | 0 | 0 | 0 | 0 |
| 0 | 0 | 0 | 0 | 0 |
| 0 | 0 | 0 | 0 | 0 |
| 0 | 0 | 0 | 0 | 0 |
| 0 | 0 | 0 | 0 | 0 |
| 0 | 0 | 0 | 0 | 0 |
| 0 | 0 | 0 | 0 | 0 |
| 0 | 0 | 0 | 0 | 0 |
| 0 | 0 | 0 | 0 | 0 |
| 0 | 0.010011042 | 0 | 1.905933203 | 0 |
| 0 | 0 | 0 | 0 | 0 |
| 0 | 0 | 0 | 0 | 0 |
| 0 | 0 | 0 | 0 | 0 |
| 0 | 0 | 0 | 0 | 0 |
| 0 | 0 | 0 | 0 | 0 |
| 0 | 0 | 0 | 0 | 0 |
| 0 | 0 | 0 | 0 | 0.03513754 |
| 0 | 0 | 0 | 0 | 0 |
| 0 | 0 | 0 | 0 | 0 |
| 0 | 0 | 0 | 0 | 0 |
| 0 | 0 | 0 | 0.050142678 | 0 |
| 0 | 0 | 0 | 0 | 0 |
| 0 | 0 | 0 | 0 | 0 |
| 0 | 0 | 0 | 0 | 0 |
| 0 | 0 | 0 | 0 | 0 |
| 0.002109229 | 0.437572777 | 0.001131832 | 0 | 0 |
| 0 | 0 | 0 | 0 | 0 |
| 0 | 0 | 0 | 0 | 0 |
| 0 | 0 | 0 | 0 | 0 |
| 0 | 0 | 0 | 0 | 0 |
| 0 | 0 | 0 | 0 | 0 |
| 0 | 0 | 0 | 0 | 0 |
| 0 | 0 | 0 | 0 | 0 |
| 0 | 0 | 0 | 0 | 0 |
| 0 | 0 | 0 | 0 | 0 |
| 0 | 0 | 0 | 0 | 0 |
| 0 | 0 | 0 | 0 | 0 |
| 0 | 0 | 0 | 0 | 0 |
| 0 | 0 | 0 | 0 | 0 |
| 0 | 0 | 0 | 0 | 0 |
| 0 | 0 | 0 | 0 | 0 |
| 0 | 0 | 0 | 0 | 0 |

| | | | | |
|---|---|---|---|---|
| 0 | 0 | 0 | 0 | 0 |
| 0 | 0 | 0 | 0 | 0 |
| 0 | 0 | 0 | 0 | 0 |
| 0 | 0 | 0 | 0 | 0 |
| 0 | 0 | 0 | 0 | 0 |
| 0 | 0 | 0 | 0 | 0 |
| 0 | 0 | 0 | 0 | 0 |
| 0 | 0 | 0 | 0 | 0 |
| 0 | 0 | 0 | 0 | 0 |
| 0 | 0 | 0 | 0 | 0 |
| 0 | 0 | 0 | 0 | 0 |
| 0 | 0 | 0 | 0 | 0 |
| 0.00080733 | 0 | 0.004982041 | 0.001265061 | 0 |
| 0 | 0 | 0 | 0 | 0 |
| 0 | 0 | 0.032437185 | 0 | 0 |
| 0 | 0 | 0 | 0 | 0 |
| 0 | 0 | 0 | 0 | 0 |
| 0 | 0 | 0 | 0 | 0 |
| 0 | 0 | 0 | 0 | 0 |
| 0 | 0 | 0 | 0 | 0 |
| 0.02377484 | 0.062244838 | 0.022680535 | 0.096641888 | 0.021574423 |
| 0 | 0 | 0 | 0 | 0 |
| 0 | 0 | 0 | 0 | 0 |
| 0 | 0 | 0 | 0 | 0 |
| 0 | 0 | 0 | 0 | 0 |
| 0 | 0 | 0 | 0 | 0 |
| 0 | 0.002463587 | 0.00235868 | 0.005510122 | 0.001172681 |
| 0 | 0 | 0 | 0 | 0 |
| 0 | 0 | 0 | 0 | 0 |
| 0 | 0 | 0 | 0 | 0.020970297 |
| 0 | 0 | 0 | 0 | 0 |
| 0 | 0 | 0 | 0 | 0 |
| 0 | 0 | 0 | 0 | 0 |
| 0 | 0 | 0.006435434 | 0.462166715 | 0 |
| 0.095270494 | 0.000983005 | 0.251662363 | 0.025723785 | 0 |
| 0 | 0 | 0 | 0 | 0 |
| 0 | 0 | 0 | 0 | 0 |
| 0 | 0 | 0 | 0 | 0 |
| 0 | 0 | 0 | 0 | 0 |
| 0 | 0 | 0 | 0 | 0 |
| 0 | 0 | 0.015767031 | 0 | 0.000823424 |
| 0 | 0 | 0 | 0 | 0 |
| 0 | 0 | 0.001095601 | 0 | 0 |
| 0 | 0 | 0 | 0 | 0 |
| 0 | 0 | 0 | 0 | 0 |
| 0.004909447 | 0 | 0 | 0.057566302 | 0 |
| 0 | 0 | 0 | 0.002856617 | 0.001215908 |
| 0 | 0.000560413 | 0.003192465 | 0.001566792 | 0 |

| | | | | |
|---|---|---|---|---|
| 0 | 0 | 0 | 0 | 0 |
| 0 | 0 | 0 | 0 | 0 |
| 0 | 0 | 0 | 0 | 0 |
| 0 | 0 | 0 | 0 | 0 |
| 0 | 0 | 0 | 0 | 0 |
| 0 | 0 | 0 | 0 | 0 |
| 0 | 0 | 0 | 0 | 0 |
| 0.000754612 | 0 | 0 | 0 | 0 |
| 0 | 0 | 0 | 0.009571143 | 0 |
| 0 | 0 | 0 | 0 | 0 |
| 0 | 0 | 0 | 0 | 0 |
| 0 | 0 | 0 | 0 | 0 |
| 0 | 0 | 0 | 0 | 0 |
| 0 | 0 | 0 | 0 | 0 |
| 0 | 0.077092807 | 0 | 0 | 0 |
| 0 | 0 | 0 | 0 | 0 |
| 0 | 0 | 0 | 0 | 0 |
| 0 | 0 | 0 | 0 | 0 |
| 0 | 0.007836621 | 0.001909833 | 0 | 0 |
| 0 | 0 | 0 | 0 | 0 |
| 0 | 0.010719101 | 0 | 0.022329296 | 0.01040479 |
| 0 | 0 | 0 | 0 | 0 |
| 0 | 0 | 0 | 0 | 0 |
| 0 | 0 | 0 | 0 | 0 |
| 0 | 0.006263459 | 0.002798479 | 0.009806307 | 0 |
| 0 | 0 | 0 | 0 | 0 |
| 0 | 0 | 0 | 0 | 0 |
| 0 | 0 | 0 | 0 | 0 |
| 0 | 0 | 0 | 0 | 0 |
| 0 | 0 | 0 | 0 | 0 |
| 0.728056557 | 0.303215574 | 0.235734911 | 0.296573974 | 0.703564421 |
| 0 | 0 | 0 | 0 | 0 |
| 0.029040039 | 0 | 0.014166505 | 0.001504292 | 0.049943082 |
| 0 | 0.041588673 | 0 | 0 | 0.029884994 |
| 0 | 0 | 0 | 0 | 0 |
| 0 | 0 | 0 | 0 | 0 |
| 0 | 0 | 0 | 0 | 0 |
| 0 | 0 | 0 | 0 | 0 |
| 0 | 0 | 0 | 0 | 0 |
| 0 | 0 | 0 | 0.001035791 | 0 |
| 0 | 0 | 0 | 0 | 0 |
| 0.011214623 | 0.011499839 | 0.021326491 | 0.018850198 | 0 |
| 0 | 0 | 0 | 0 | 0 |
| 0 | 0 | 0 | 0 | 0 |
| 0 | 0 | 0 | 0 | 0 |
| 0 | 0 | 0 | 0 | 0 |
| 0 | 0 | 0 | 0 | 0 |
| 0 | 0 | 0 | 0 | 0 |

| | | | | |
|---|---|---|---|---|
| 0 | 0 | 0 | 0 | 0 |
| 0 | 0 | 0 | 0 | 0 |
| 0.00617623 | 0.050243079 | 0.001855965 | 0.043578473 | 0.009039108 |
| 0 | 0 | 0 | 0 | 0 |
| 0 | 0 | 0 | 0 | 0 |
| 0 | 0 | 0 | 0 | 0 |
| 0 | 0 | 0 | 0 | 0 |
| 0 | 0 | 0 | 0 | 0 |
| 0 | 0 | 0 | 0 | 0 |
| 0 | 0.001085479 | 0.002667423 | 0 | 0 |
| 0 | 0 | 0 | 0 | 0 |
| 0 | 0 | 0 | 0 | 0 |
| 0 | 0 | 0 | 0 | 0 |
| 0.003003374 | 0 | 0.002051176 | 0.013201789 | 0.012487308 |
| 0.002253538 | 0 | 0.005320786 | 0.007264232 | 0.001717773 |
| 0.047771659 | 0 | 0.003884048 | 0.000864147 | 0 |
| 0.001027082 | 0.001027955 | 0.001928996 | 0.00321881 | 0 |
| 0.00517871 | 0 | 0 | 0 | 0 |
| 0 | 0 | 0 | 0 | 0 |
| 0 | 0 | 0 | 0 | 0 |
| 0 | 0 | 0 | 0 | 0 |
| 0 | 0 | 0.00273638 | 0.002490572 | 0 |
| 0 | 0 | 0 | 0 | 0 |
| 0 | 0 | 0 | 0 | 0 |
| 0.001858312 | 0 | 0 | 0.002995115 | 0 |
| 0 | 0 | 0 | 0 | 0 |
| 0 | 0 | 0 | 0 | 0 |
| 0 | 0 | 0 | 0 | 0 |
| 0 | 0 | 0 | 0 | 0 |
| 0.001676435 | 0 | 0.001439345 | 0.029421521 | 0.001789022 |
| 0 | 0 | 0 | 0 | 0 |
| 0 | 0 | 0 | 0 | 0 |
| 0 | 0 | 0 | 0 | 0 |
| 0.00293979 | 0 | 0 | 0 | 0 |
| 0.001961011 | 0 | 0 | 0 | 0 |
| 0 | 0 | 0 | 0 | 0 |
| 0 | 0 | 0 | 0 | 0 |
| 0 | 0 | 0.029379087 | 0.003911432 | 0 |
| 0 | 0 | 0 | 0 | 0 |
| 0 | 0 | 0.013845679 | 0 | 0 |
| 0 | 0 | 0 | 0 | 0 |
| 0 | 0 | 0 | 0 | 0 |
| 0 | 0 | 0 | 0 | 0 |
| 0 | 0 | 0 | 0 | 0.026195276 |
| 0 | 0 | 0 | 0 | 0 |
| 0 | 0 | 0 | 0.01042386 | 0 |
| 0 | 0 | 0 | 0.00423985 | 0 |
| 0.044445405 | 0 | 0 | 0 | 0.048129374 |

| | | | | |
|---|---|---|---|---|
| 0 | 0 | 0.012977688 | 0 | 0 |
| 0 | 0 | 0 | 0 | 0 |
| 0 | 0 | 0.001563352 | 0 | 0 |
| 0 | 0 | 0 | 0 | 0 |
| 0.002198363 | 0.005476132 | 0.002883618 | 0.002405868 | 0 |
| 0 | 0 | 0 | 0.001069905 | 0 |
| 0 | 0 | 0 | 0 | 0 |
| 0.002396714 | 0.014255438 | 0.001837284 | 0.045373489 | 0.009656522 |
| 0 | 0 | 0 | 0 | 0 |
| 0.343025031 | 0.014128258 | 0.034087116 | 0.016747799 | 2.052777432 |
| 0 | 0 | 0 | 0 | 0 |
| 0.039300122 | 0.00086805 | 0.006399364 | 0.002912255 | 0 |
| 0.001850309 | 0.107985317 | 0.017430791 | 0.02503589 | 0 |
| 0.00967938 | 0 | 0 | 0 | 0 |
| 0 | 0 | 0 | 0 | 0 |
| 0 | 0 | 0.004305881 | 0 | 0 |
| 0 | 0 | 0 | 0 | 0 |
| 0 | 0 | 0 | 0 | 0 |
| 0 | 0 | 0 | 0.0044602 | 0.986949327 |
| 0 | 0 | 0.031138131 | 0 | 0 |
| 0 | 0 | 0 | 0 | 0 |
| 0 | 0 | 0 | 0 | 0 |
| 0 | 0 | 0 | 0 | 0 |
| 0 | 0 | 0 | 0.031512018 | 0 |
| 0 | 0 | 0 | 0 | 0 |
| 0 | 0.000958654 | 0.003597899 | 0.001715319 | 0 |
| 0 | 0.000769149 | 0.003780166 | 0 | 0 |
| 0 | 0.003014125 | 0 | 0 | 0 |
| 0 | 0 | 0 | 0 | 0 |
| 0 | 0 | 0 | 0 | 0.006197403 |
| 0 | 0 | 0 | 0 | 0 |
| 0 | 0 | 0 | 0 | 0 |
| 0 | 0 | 0 | 0 | 0 |
| 0 | 0 | 0 | 0 | 0.011935765 |
| 0 | 0 | 0 | 0 | 0 |
| 0 | 0 | 0 | 0 | 0 |
| 0 | 0 | 0 | 0 | 0.002689909 |
| 0 | 0 | 0 | 0 | 0 |
| 0 | 0 | 0 | 0 | 0 |
| 0 | 0 | 0 | 0 | 0 |
| 0 | 0 | 0 | 0 | 0.436000776 |
| 0 | 0 | 0 | 0 | 0 |
| 0 | 0 | 0 | 0 | 0 |
| 0.001157504 | 0 | 0 | 0 | 0.101319076 |
| 0 | 0 | 0 | 0 | 0 |
| 0 | 0 | 0 | 0 | 0 |
| 0.008040255 | 0 | 0.007030998 | 0.00426627 | 0.001361941 |
| 0 | 0 | 0 | 0 | 0 |

| | | | | |
|---|---|---|---|---|
| 0 | 0 | 0 | 0 | 0 |
| 0.002662384 | 0.036772142 | 0 | 0 | 0 |
| 0 | 0 | 0 | 0 | 0 |
| 0 | 0 | 0 | 0 | 0 |
| 0 | 0 | 0 | 0 | 0 |
| 0 | 0.02948371 | 0 | 0 | 0 |
| 0 | 0 | 0 | 0.000700568 | 0 |
| 0.007583824 | 0 | 0 | 0 | 0 |
| 0 | 0 | 0 | 0.001532343 | 0 |
| 0 | 0 | 0 | 0 | 0 |
| 0.010609208 | 0 | 0 | 0.028742378 | 0 |
| 0 | 0 | 0 | 0 | 0 |
| 0 | 0 | 0 | 0 | 0 |
| 0.000771004 | 0.000617327 | 0 | 0.00276146 | 0 |
| 0 | 0.002290289 | 0 | 0 | 0.554609232 |
| 0 | 0 | 0 | 0 | 0 |
| 0 | 0 | 0 | 0 | 0 |
| 0 | 0 | 0 | 0 | 0.024021022 |
| 0 | 0 | 0 | 0 | 0 |
| 0 | 0 | 0.01157732 | 0 | 0 |
| 0 | 0.623240694 | 0 | 0 | 0 |
| 0 | 0.174023551 | 0.133468671 | 0.169635708 | 0 |
| 0 | 0 | 0.008113895 | 0 | 0 |
| 0 | 0 | 0 | 0 | 0 |
| 0 | 0 | 0 | 0 | 0 |
| 0 | 0 | 0 | 0 | 0 |
| 0 | 0 | 0 | 0 | 0 |
| 0 | 0 | 0 | 0 | 0 |
| 0 | 0 | 0 | 0 | 0.040900535 |
| 0.47427779 | 1.210719378 | 0.127352015 | 0.501242844 | 0.066331513 |
| 0.001743549 | 0 | 0.011975738 | 0.005620293 | 0 |
| 0.004406199 | 0.00078399 | 0 | 0 | 0 |
| 0 | 0 | 0 | 0 | 0 |
| 0 | 0 | 0 | 0 | 0 |
| 0 | 0 | 0 | 0 | 0 |
| 0 | 0 | 0 | 0 | 0 |
| 0 | 0 | 0 | 0.238442862 | 0 |
| 0.00681077 | 0.002307143 | 0 | 0 | 0 |
| 0.009873102 | 0 | 0 | 0 | 0 |
| 0.006994258 | 0 | 0 | 0 | 0.001390804 |
| 0.034650403 | 0 | 0 | 0 | 0.005153654 |
| 2.424768292 | 3.718520199 | 0 | 4.275454717 | 0.484927916 |
| 0.001439794 | 0 | 0 | 0 | 0 |
| 0 | 0 | 0 | 0 | 0 |
| 0 | 0 | 0 | 0 | 0 |
| 0 | 0 | 0 | 0 | 0 |
| 0.05166929 | 0 | 0.001665953 | 0 | 0 |
| 0 | 0 | 0 | 0 | 0 |

| | | | | |
|---|---|---|---|---|
| 0 | 0 | 0.015269479 | 0.000979975 | 0 |
| 0 | 0 | 0 | 0 | 0 |
| 0.102151364 | 3.569233055 | 0.007602132 | 0.005007341 | 0 |
| 0 | 0 | 0 | 0 | 0 |
| 0 | 0.251641662 | 0 | 0 | 0 |
| 0 | 0 | 0 | 0 | 0 |
| 0 | 0 | 0 | 0 | 0 |
| 0 | 0 | 0 | 0 | 0 |
| 0 | 0 | 0 | 0 | 0.000523641 |
| 0 | 0 | 0 | 0 | 0 |
| 0 | 0 | 0 | 0 | 0 |
| 0 | 0 | 0 | 0 | 0 |
| 0 | 0 | 0 | 0 | 0 |
| 0.005445626 | 0 | 0 | 0 | 0 |
| 0 | 0 | 0 | 0 | 0.001187821 |
| 0.104263741 | 0.12762355 | 0.144491396 | 0.082620009 | 0.015777173 |
| 0.007289845 | 0.002746746 | 0.283950576 | 0.024573764 | 0.003105231 |
| 0 | 0 | 0 | 0 | 0 |
| 0 | 0 | 0 | 0 | 0 |
| 0.122953995 | 0 | 0 | 0 | 0 |
| 0.069715236 | 0 | 0.004059983 | 0.015484967 | 0 |
| 0.001243506 | 0 | 0.007740413 | 0.001336139 | 0.003222758 |
| 0 | 0.017742359 | 0 | 0 | 0 |
| 0.133013386 | 0 | 0.01092234 | 0 | 0 |
| 0 | 0 | 0 | 0 | 0 |
| 0 | 0 | 0 | 0 | 0.052101249 |
| 0.011159859 | 0 | 0 | 0 | 0 |
| 0 | 0 | 0 | 0 | 0 |
| 0 | 0 | 0 | 0 | 0.006949174 |
| 0 | 0 | 0 | 0 | 0 |
| 0.017388084 | 0 | 0.00333236 | 0.056902735 | 0.026192768 |
| 0 | 0.057080188 | 0.022522234 | 0 | 0.002352422 |
| 0.008227559 | 8.023278879 | 0.005676409 | 0.014207872 | 0 |
| 0.004632137 | 0.002360184 | 0.007230976 | 0 | 0 |
| 0.001202243 | 0.01708636 | 0.006451346 | 0 | 0.007102229 |
| 0 | 0 | 0 | 0 | 0 |
| 0 | 0.0142511 | 0 | 0 | 0 |
| 0.088087213 | 0.011477708 | 0.006911892 | 0.004092531 | 0 |
| 0.112663343 | 0.02100718 | 0.08993895 | 0.302225341 | 0 |
| 0.002378029 | 0 | 0 | 0.022053593 | 0 |
| 0 | 0 | 0 | 0 | 0 |
| 0 | 0 | 0 | 0.055893993 | 0 |
| 0.001864164 | 0 | 0.002667541 | 0.023925099 | 0.023682843 |
| 0.014433727 | 0.716002147 | 0 | 0 | 0 |
| 0 | 0 | 0 | 0 | 0 |
| 0 | 0.002175937 | 0 | 0 | 0.029864346 |
| 0 | 0 | 0 | 0 | 0 |
| 0 | 0 | 0 | 0 | 0 |

| | | | | |
|---|---|---|---|---|
| 0 | 0 | 0 | 0 | 0 |
| 0.151452951 | 0.057799383 | 0.64196546 | 0.212417501 | 0.023952332 |
| 0 | 0 | 0 | 0 | 0 |
| 0 | 0 | 0 | 0 | 0 |
| 0 | 0 | 0 | 0 | 0 |
| 0 | 0 | 0 | 0 | 0.042221675 |
| 0 | 0 | 0 | 0 | 0 |
| 0 | 0 | 0 | 0 | 0 |
| 0 | 0 | 0 | 0.003480622 | 0 |
| 0 | 0 | 0 | 0 | 0 |
| 0 | 0.0018528 | 0.001806154 | 0.01262042 | 0.014912844 |
| 0.004386857 | 0.115660602 | 0.021186256 | 0.023287661 | 0.021496553 |
| 0.241960248 | 0 | 0 | 0 | 0 |
| 0 | 0 | 0 | 0 | 0 |
| 0 | 0.048508163 | 0 | 0 | 0.000735941 |
| 0 | 0 | 0 | 0 | 0 |
| 0.008370797 | 0.007760594 | 0.002836957 | 0.018146521 | 0 |
| 0.003145552 | 0.019232799 | 0 | 0.068877716 | 0.069751729 |
| 0 | 0 | 0 | 0 | 0 |
| 0 | 0.006690415 | 0.002789958 | 0 | 0 |
| 0.018173482 | 0.004183455 | 0.226003981 | 0.023798869 | 0 |
| 0 | 0.032818149 | 0.006067436 | 0 | 0 |
| 0 | 0 | 0 | 0 | 3.405777803 |
| 0 | 0 | 0 | 0 | 0 |
| 0 | 0.239291062 | 0.00204816 | 0 | 0 |
| 0 | 0 | 0 | 0 | 0.002858276 |
| 0 | 0.119030945 | 0 | 0 | 0.015382632 |
| 0.104063484 | 0.145957734 | 0.00984675 | 0.020538419 | 0 |
| 0 | 0 | 0 | 0 | 0 |
| 0 | 0 | 0.001875393 | 0 | 0 |
| 0 | 0 | 0 | 0 | 0 |
| 0 | 0 | 0 | 0 | 0 |
| 0 | 0 | 0 | 0.040418508 | 0 |
| 0 | 0 | 0 | 0 | 0 |
| 0 | 0.02190061 | 0 | 0.214773632 | 0 |
| 0.299528697 | 0.075252575 | 0.086833611 | 0.166482343 | 0.028033533 |
| 0 | 0 | 0 | 0 | 0 |
| 0 | 0 | 0 | 0 | 0 |
| 0.02353026 | 0.002384837 | 0.017261622 | 0.00426719 | 0.00227039 |
| 0 | 0 | 0 | 0 | 0 |
| 0 | 0 | 0 | 0 | 0 |
| 0 | 0 | 0 | 0 | 0 |
| 0 | 0 | 0 | 0 | 0.000689535 |
| 0.023057015 | 0 | 0.001312246 | 0.001720458 | 0.007722501 |
| 0 | 0 | 0 | 0 | 0 |
| 0.360227335 | 0 | 0.181486334 | 1.193731205 | 0 |
| 0 | 0 | 0 | 0 | 0 |
| 0 | 0 | 0 | 0 | 0 |

| | | | | |
|---|---|---|---|---|
| 0.001769239 | 0.001618965 | 0 | 0 | 0 |
| 0 | 0 | 0 | 0 | 0 |
| 0 | 0 | 0 | 0 | 0 |
| 0.143706912 | 0.015521732 | 1.689055613 | 0.002452782 | 0 |
| 0 | 0 | 0 | 0 | 0 |
| 0 | 0 | 0 | 0 | 0 |
| 0 | 0 | 0 | 0 | 0 |
| 0 | 0 | 0.004046168 | 0.08675832 | 0.00344856 |
| 0.361347349 | 0.052107409 | 1.251329561 | 1.03953029 | 0 |
| 0 | 0 | 0 | 0 | 0 |
| 0 | 0 | 0 | 0 | 0 |
| 0 | 0.00808233 | 0.088400738 | 0.011930916 | 0 |
| 0 | 0 | 0 | 0 | 0 |
| 0 | 0 | 0 | 0 | 0 |
| 0.199916815 | 0.033347797 | 0.220991086 | 0.394115222 | 0 |
| 0 | 0 | 0 | 0 | 0 |
| 0 | 0 | 0.062270515 | 0 | 0 |
| 0 | 0 | 0 | 2.615936369 | 0 |
| 0 | 0 | 0 | 0 | 0 |
| 0 | 0 | 0.025347299 | 0 | 0 |
| 0 | 0 | 0 | 0 | 0 |
| 0 | 0 | 0 | 0 | 0 |
| 0 | 0 | 0 | 0 | 0 |
| 0 | 0.2278126 | 0.095240853 | 0 | 0 |
| 0 | 0 | 0 | 0 | 0 |
| 0 | 0 | 0 | 0 | 0 |
| 0.436534001 | 0 | 0 | 0 | 0 |
| 6.868790825 | 11.5939957 | 2.559284868 | 0 | 0 |
| 0 | 0 | 0 | 0 | 0 |
| 0 | 0 | 0 | 0 | 0 |
| 0 | 0.043215717 | 0.134176853 | 0 | 0 |
| 0 | 0 | 0 | 0.001324642 | 0 |
| 0 | 0 | 0.013305856 | 0 | 0.068316976 |
| 0 | 0 | 0 | 0 | 0 |
| 0 | 0 | 0 | 0 | 0 |
| 0 | 0.049700006 | 0 | 0.205631642 | 0 |
| 1.393566763 | 0 | 0.611019316 | 0.892175966 | 0 |
| 0 | 0 | 0 | 0 | 0 |
| 0 | 0 | 0.006992109 | 0.015054647 | 0 |
| 0.070807476 | 0 | 0.000620342 | 0 | 0 |
| 0.334281522 | 0.042799602 | 0.031386055 | 0.087055781 | 0.277247751 |
| 0.056854657 | 0 | 0 | 0 | 0 |
| 9.316900931 | 5.157404953 | 0.024436025 | 1.317700324 | 0.355438536 |
| 0 | 0 | 0 | 0 | 0 |
| 0 | 0 | 0 | 0 | 0 |
| 0 | 0 | 0 | 0 | 0 |
| 0 | 0 | 0 | 0 | 0 |
| 0 | 0 | 0.002748517 | 0.003960896 | 0 |

| | | | | |
|---|---|---|---|---|
| 0 | 0 | 0 | 0 | 0 |
| 0 | 0.016473284 | 0 | 0 | 0 |
| 0.033031057 | 0 | 0 | 0.102518076 | 0.225922612 |
| 0 | 0 | 0 | 0 | 0 |
| 0 | 0 | 0 | 0 | 0 |
| 0.003340482 | 0.002815429 | 0.24510538 | 0.000629706 | 0 |
| 0 | 0 | 0 | 0 | 0 |
| 0 | 0 | 0 | 0 | 0 |
| 0 | 0 | 0 | 0 | 0 |
| 0 | 0 | 0 | 0 | 0 |
| 0.051494599 | 0.135798147 | 0.139455843 | 0.043165703 | 2.364143697 |
| 1.851786308 | 1.986733386 | 0.363239858 | 1.012721676 | 0 |
| 0 | 0 | 0 | 0 | 0 |
| 0 | 0.000946763 | 0.253805111 | 0.012493568 | 0 |
| 0.190462905 | 0.853521775 | 0.003000122 | 0.089109311 | 0.125009508 |
| 0 | 0 | 3.039800966 | 0 | 0 |
| 0.000756934 | 0 | 0.000487413 | 0 | 0 |
| 0.000620879 | 0 | 0.005774938 | 0 | 0 |
| 0 | 0 | 0.345525328 | 0 | 0 |
| 0.028158329 | 0 | 0 | 0.024081907 | 0 |
| 0.096421403 | 0.011028958 | 0.519771524 | 0.137645469 | 0.06467806 |
| 0.330532958 | 0.488396093 | 0 | 0.159937729 | 35.93794054 |
| 4.449187976 | 0.666695548 | 0.329897615 | 0.451693635 | 0.612222244 |
| 0 | 0 | 0 | 0 | 0 |
| 0 | 0 | 0 | 0 | 0 |
| 0.007482705 | 0 | 4.009620438 | 0.009261895 | 0 |
| 0 | 0 | 0.107973188 | 0 | 0 |
| 0 | 0.001993854 | 0 | 0 | 0.03840144 |
| 0 | 0 | 0 | 0 | 0 |
| 0 | 0 | 0 | 0 | 0 |
| 0 | 0 | 0 | 0.03682119 | 0 |
| 0.311233155 | 0.203106956 | 0.139377102 | 0.102623112 | 0 |
| 0.031956825 | 0.002883064 | 0 | 0 | 0 |
| 0 | 0.007832369 | 0.103510341 | 0.045792863 | 0 |
| 0.002228018 | 0.000475715 | 0.00398525 | 0 | 0 |
| 0.062651744 | 0.007026229 | 0.132944902 | 0.036363877 | 0.001711151 |
| 0.007688301 | 0.001894114 | 0.008674368 | 0.022417739 | 0 |
| 0 | 0 | 0 | 0 | 0 |
| 0.000716435 | 0.093693778 | 0.740186227 | 0 | 0 |
| 0 | 0 | 0 | 0 | 0 |
| 0.001257903 | 0 | 0 | 0.014320603 | 0 |
| 0 | 0 | 0 | 0 | 0 |
| 0 | 0 | 0.118816706 | 0 | 0 |
| 0 | 0 | 0 | 0 | 0 |
| 0.039563597 | 0.002795099 | 0.052201303 | 0.112528603 | 1.858239749 |
| 0 | 0 | 0 | 0 | 0 |
| 0 | 0 | 0 | 0 | 0 |
| 0.00112293 | 0 | 0.971350115 | 0.62918118 | 0 |

| | | | | |
|---|---|---|---|---|
| 0 | 0 | 0 | 0 | 0 |
| 0 | 0 | 0 | 0 | 0 |
| 0 | 0 | 0 | 0 | 0 |
| 0.000991762 | 0 | 0.008515016 | 0.008525126 | 0 |
| 0 | 0 | 0 | 0 | 0 |
| 0 | 0 | 0 | 0.245801502 | 0 |
| 0.062324349 | 0.824337166 | 0 | 0 | 0 |
| 0.124661183 | 0.013591036 | 0.684614241 | 0.025168283 | 0 |
| 0 | 0 | 0 | 0 | 0 |
| 0 | 0 | 0 | 0 | 0 |
| 0.003755503 | 0.009020871 | 0.024894117 | 0.004945174 | 0.299905205 |
| 0 | 0 | 0 | 0 | 0 |
| 0 | 0.003716931 | 0 | 0.007966991 | 0 |
| 0 | 0 | 0 | 0 | 0 |
| 0 | 0 | 0 | 0 | 0 |
| 0 | 0 | 0 | 0 | 0 |
| 0.257130018 | 0 | 0 | 0 | 0 |
| 0 | 0.000588994 | 0.025855388 | 0.007574809 | 0 |
| 0 | 0 | 0.001869164 | 0 | 0 |
| 0 | 0 | 0 | 0 | 0 |
| 0 | 0 | 0 | 0 | 0 |
| 0 | 0 | 0 | 0 | 0 |
| 0.000636085 | 0.382145257 | 0.127429549 | 0.00075941 | 0 |
| 0 | 0 | 0.000887942 | 0 | 0 |
| 0 | 0 | 0 | 0 | 0 |
| 0 | 0 | 0 | 0 | 0 |
| 0 | 0 | 0 | 0 | 0 |
| 0 | 0 | 0 | 0 | 0 |
| 0 | 0 | 0 | 0 | 0 |
| 0 | 0.209180729 | 0.030517817 | 0 | 0 |
| 0 | 0 | 0 | 0.022901865 | 0 |
| 0 | 0 | 0 | 0 | 0 |
| 0 | 0 | 0 | 0.001204363 | 0 |
| 0 | 0 | 0 | 0 | 0 |
| 0 | 0 | 0.001696679 | 0 | 0 |
| 0 | 0 | 0 | 0 | 0 |
| 0 | 0 | 0 | 0.092924177 | 0 |
| 0.00673221 | 0.001347587 | 0 | 0.017916833 | 0.017818291 |
| 0 | 0 | 0 | 0.007433134 | 0 |
| 0 | 0 | 0 | 0 | 0 |
| 0.591468097 | 0 | 0.152150065 | 0 | 0 |
| 0 | 0 | 0 | 0 | 0 |
| 0 | 0 | 0 | 0.0016989 | 0 |
| 0.005811354 | 0.003635184 | 0 | 0 | 0 |
| 0 | 0 | 0.086598456 | 0.346044578 | 0 |
| 0 | 0 | 0 | 0 | 0 |
| 0 | 0 | 0.001525575 | 0 | 0 |
| 0 | 0 | 0 | 0 | 0 |

| | | | | |
|---|---|---|---|---|
| 0 | 0 | 0 | 0 | 0 |
| 0 | 0 | 0 | 0 | 0 |
| 0 | 0 | 0 | 1.950058877 | 0 |
| 0.587252498 | 0.029936803 | 1.864598692 | 0.195875416 | 0 |
| 0 | 0 | 0 | 0 | 0 |
| 1.105228575 | 0 | 0 | 1.815528312 | 0 |
| 0 | 0 | 0 | 0 | 0 |
| 0 | 0 | 0 | 0 | 0 |
| 0.002837035 | 0.030975787 | 0.008303888 | 0.016627495 | 0.176149289 |
| 0 | 0.001330382 | 0 | 0 | 0 |
| 0 | 0 | 0 | 0.003137422 | 0 |
| 0 | 0 | 0 | 0 | 0 |
| 0 | 0 | 0.007912409 | 0 | 0 |
| 0 | 0 | 0 | 0 | 0 |
| 0.495235352 | 0.112487291 | 1.494036319 | 0 | 0.003673241 |
| 0 | 0 | 0 | 0 | 0.047426192 |
| 0 | 0 | 0 | 0 | 0 |
| 0 | 0 | 0 | 0 | 0 |
| 0 | 0 | 0 | 0 | 0 |
| 0 | 0 | 0 | 0 | 0 |
| 0 | 0 | 0 | 0.015892631 | 0 |
| 0 | 0 | 0 | 0 | 0 |
| 0 | 0 | 0 | 0 | 0 |
| 0 | 0 | 0 | 0 | 0 |
| 0.002729677 | 1.105912895 | 0.06900696 | 0.001629455 | 0.010403565 |
| 0.223550756 | 0.37881438 | 9.243318153 | 2.768076415 | 0 |
| 3.648684748 | 0 | 0.345771365 | 0.041818194 | 0 |
| 0 | 0 | 0 | 0 | 0 |
| 0.0042427 | 0.001698523 | 0.239335177 | 0.151008629 | 0 |
| 0 | 0 | 0 | 0 | 0 |
| 0 | 0 | 0 | 0 | 0 |
| 0 | 0 | 0 | 0.604966468 | 0 |
| 0 | 0 | 0 | 0 | 0 |
| 0.002429705 | 0 | 0.002607605 | 0.040030747 | 0 |
| 0 | 0 | 0.171001056 | 0.007647307 | 0.000813761 |
| 0 | 0 | 0.003847007 | 0.001858216 | 0.046449878 |
| 0 | 0 | 0 | 0 | 0 |
| 0.669128538 | 1.643946221 | 0.894393575 | 0.148594041 | 0 |
| 0 | 11.6735042 | 4.655556125 | 0 | 0 |
| 0 | 0 | 0 | 0 | 0 |
| 0 | 0 | 0 | 0 | 0 |
| 0 | 0 | 0 | 0 | 0 |
| 0.546553084 | 0.25142175 | 1.668407021 | 0.100648034 | 0.055613956 |
| 0 | 0 | 0 | 0 | 0 |
| 0 | 0 | 0 | 0 | 0 |
| 0.002323519 | 0 | 0.000959094 | 0.003200775 | 0 |
| 0 | 0 | 0 | 0 | 0 |
| 0.181950051 | 0.070904007 | 0.46449722 | 0.0163175 | 0.027781914 |

| | | | | |
|---|---|---|---|---|
| 0 | 0 | 0.010026676 | 0.010076606 | 0 |
| 0 | 0 | 0 | 0 | 0 |
| 0 | 0 | 0 | 0 | 0 |
| 0 | 0 | 0 | 0 | 0 |
| 0.003593824 | 0.002200444 | 0.012024637 | 0 | 0.044636337 |
| 0 | 0 | 0 | 0 | 0 |
| 0 | 0 | 0 | 0 | 0 |
| 0 | 0 | 0.005995142 | 0 | 0 |
| 0 | 0 | 0 | 0 | 0 |
| 0 | 0 | 0 | 0 | 0 |
| 0 | 0 | 0 | 0 | 0 |
| 2.282841405 | 0.82935352 | 0.228777175 | 1.924107843 | 0 |
| 0.011262683 | 0.006763355 | 0.0673044 | 0.79715186 | 0.039218058 |
| 0 | 0 | 0 | 0 | 0 |
| 0.275046455 | 0.037193427 | 0.183933043 | 0.007662031 | 0 |
| 0 | 0 | 0 | 0 | 0 |
| 0 | 0 | 0 | 0 | 0 |
| 0.261494475 | 0 | 0.00122627 | 0 | 0 |
| 0 | 0 | 0 | 0 | 0 |
| 0 | 0 | 0.012495596 | 0 | 0 |
| 0.012418924 | 0 | 0.063785055 | 0 | 0 |
| 0 | 0 | 0 | 0 | 0 |
| 0.374021923 | 0.088089794 | 2.533226685 | 1.592423031 | 0.001588303 |
| 0 | 0 | 0 | 0 | 0 |
| 0 | 0 | 0 | 0 | 0 |
| 0 | 0 | 0 | 0 | 0 |
| 0 | 0 | 0 | 0.096567281 | 0 |
| 0 | 0 | 0 | 0 | 0 |
| 0 | 0 | 0 | 0 | 0 |
| 0 | 0 | 0 | 0 | 0 |
| 0 | 0 | 0 | 0 | 0 |
| 0.018718012 | 0.053598079 | 0.072182475 | 0.034656911 | 0 |
| 0 | 0 | 0 | 0 | 0 |
| 0.042001134 | 0 | 0.010205977 | 0.005250966 | 0.336707647 |
| 0 | 0.000657551 | 0.005508563 | 0.002206041 | 0.04632364 |
| 0 | 0.033058834 | 0.049992488 | 0.014456262 | 0 |
| 0 | 0 | 0 | 0.004569214 | 0 |
| 0 | 0 | 0 | 0 | 0 |
| 0 | 0 | 0.014682181 | 0.032313806 | 0 |
| 0 | 0 | 0 | 0 | 0 |
| 0 | 0.536845662 | 0.028159895 | 0 | 0 |
| 0 | 0 | 0 | 0 | 0 |
| 0 | 0 | 0 | 0 | 0 |
| 0 | 0 | 0 | 0 | 0 |
| 0.058248267 | 0.010040175 | 0.062079007 | 0.017385375 | 0 |
| 0 | 0 | 0 | 0 | 0 |
| 0.355483801 | 0 | 0 | 0.012951576 | 0 |
| 0.003634571 | 0 | 0.000821198 | 0.009249443 | 0 |

| | | | | |
|---|---|---|---|---|
| 0 | 0 | 0 | 0 | 0 |
| 0 | 0 | 0 | 0 | 0 |
| 0.028866574 | 0 | 0.002020444 | 0 | 0 |
| 0 | 0 | 0 | 0 | 0 |
| 0 | 0 | 0 | 0 | 0 |
| 0 | 0 | 0 | 0 | 0 |
| 0 | 0.005899639 | 0.450990565 | 0.671722318 | 0 |
| 0.355785992 | 0.019139014 | 0.047491005 | 0.000707551 | 0.026502611 |
| 0 | 0 | 0 | 0 | 0 |
| 0.001166375 | 0.000778245 | 0 | 0 | 0 |
| 0 | 0 | 0 | 0 | 0 |
| 0 | 0 | 0 | 0 | 0.027741963 |
| 0 | 0 | 0 | 0 | 0 |
| 0 | 0 | 0 | 0.209610975 | 0 |
| 0 | 0 | 0 | 0 | 0 |
| 0 | 0 | 0 | 0 | 0 |
| 0 | 0 | 0 | 0 | 0 |
| 0 | 0 | 0 | 0 | 0 |
| 0 | 1.856561412 | 0.622485985 | 0.002801911 | 0 |
| 0 | 0 | 0 | 0 | 0 |
| 0 | 0 | 0 | 0 | 0.003042314 |
| 0 | 0 | 0 | 0 | 0 |
| 0 | 0 | 0 | 0 | 0 |
| 0 | 0 | 0 | 0 | 0 |
| 0 | 0 | 0 | 0 | 0 |
| 0.00094144 | 0.000603034 | 0 | 0.004889257 | 0 |
| 0 | 0 | 0 | 0 | 0 |
| 0.598779231 | 0.001032938 | 3.065553527 | 0.069638882 | 0.013064742 |
| 0 | 0.085740656 | 0 | 0 | 0 |
| 0.819543059 | 0.390995867 | 0.413818732 | 1.162789754 | 0 |
| 0 | 0 | 0 | 0 | 0 |
| 0 | 0 | 0 | 0 | 0 |
| 0 | 0 | 0 | 0 | 0 |
| 0 | 0 | 0 | 0 | 0 |
| 0 | 0 | 0 | 0.003533544 | 0.007670599 |
| 0 | 0 | 0 | 0 | 0 |
| 0 | 0 | 0 | 0 | 0 |
| 0 | 0 | 0.006728035 | 0.001804292 | 0 |
| 0 | 0 | 0 | 0 | 0 |
| 0 | 0 | 0 | 0 | 0 |
| 7.836025823 | 2.337585 | 1.86175789 | 4.907700211 | 1.871888764 |
| 0 | 0.0123351 | 0.759146167 | 1.171064915 | 0.042725065 |
| 0 | 0.05021078 | 0 | 0.93338705 | 0 |
| 0 | 0 | 0 | 0 | 0 |
| 0 | 0 | 0 | 0 | 0 |
| 0 | 0 | 0 | 0 | 0 |
| 0 | 0 | 0 | 0 | 0 |
| 0.001421556 | 0 | 0.143028768 | 0.002863972 | 0.106327217 |

| | | | | |
|---|---|---|---|---|
| 0 | 0 | 0 | 0 | 0 |
| 0.029609297 | 0.002195146 | 0 | 0.009328466 | 0 |
| 0 | 0 | 0 | 0 | 0 |
| 0 | 0 | 0 | 0 | 0 |
| 5.445536927 | 0.945938529 | 1.827355075 | 1.61530282 | 0.215532029 |
| 0 | 0 | 0 | 0 | 0 |
| 0 | 0 | 0 | 0 | 0 |
| 0 | 0 | 0 | 0 | 0 |
| 0.001095024 | 0.025133897 | 0 | 0.530121572 | 0.123394782 |
| 0 | 0 | 0 | 0 | 0 |
| 0 | 0 | 0 | 0 | 0.008342346 |
| 0 | 0 | 0 | 0 | 0 |
| 0.136393379 | 0.026590882 | 0.020203478 | 0.124513417 | 0 |
| 0.055893929 | 0.053843646 | 0.007732623 | 0.01798601 | 0 |
| 0 | 0.014921447 | 0 | 0 | 0 |
| 0.070949207 | 0.404108769 | 0.248237169 | 0.034491584 | 0 |
| 0.08294275 | 0.00151277 | 0.006488616 | 0.017763377 | 0 |
| 0 | 0 | 0 | 0 | 0 |
| 0 | 0 | 0 | 0 | 0 |
| 0 | 0 | 0 | 0 | 0 |
| 0 | 0 | 0 | 0 | 0 |
| 0 | 0 | 0 | 0 | 0 |
| 0 | 0 | 0 | 0 | 0 |
| 0 | 0 | 0 | 0 | 0 |
| 0 | 0 | 0 | 0 | 0 |
| 0 | 0 | 0.003371992 | 0 | 0 |
| 0.010185813 | 0.004077789 | 0.325656034 | 0.096206429 | 0.001878435 |
| 0 | 0.00089058 | 0.078586481 | 0 | 0.011587167 |
| 0 | 0 | 0 | 0 | 0 |
| 0 | 0 | 0 | 0 | 0 |
| 0 | 0 | 0 | 0 | 0 |
| 0 | 0 | 0 | 0 | 0 |
| 0 | 0 | 0 | 0 | 0 |
| 0.023902291 | 0 | 0.020438485 | 0 | 0 |
| 0 | 0.010673538 | 0 | 0.793150052 | 0 |
| 0 | 0.134614894 | 0.007134571 | 0.027266453 | 0.600562208 |
| 0 | 0 | 0 | 0 | 0 |
| 0 | 0 | 0 | 0 | 0 |
| 0 | 0 | 0 | 0 | 0 |
| 0 | 0 | 0 | 0 | 0 |
| 4.594119771 | 0.149307032 | 0.206893627 | 7.067381638 | 0 |
| 0 | 0 | 0 | 0 | 0 |
| 0 | 0 | 0 | 0 | 0 |
| 0 | 0 | 0 | 0.070089764 | 0 |
| 0 | 0 | 0 | 0 | 0 |
| 0 | 0 | 0 | 0 | 0 |
| 0 | 0 | 0 | 0 | 0 |
| 0 | 0 | 0 | 0 | 0 |

| | | | | |
|---|---|---|---|---|
| 0.048916633 | 0 | 0.58270871 | 0 | 0 |
| 0 | 0 | 0 | 0 | 0 |
| 0 | 0 | 0.002002844 | 0 | 0 |
| 0 | 0 | 0 | 0 | 0 |
| 0 | 0 | 0 | 0 | 0 |
| 0 | 0 | 0 | 0 | 0 |
| 0.173398139 | 0.006640004 | 0.005461458 | 0.20133504 | 0.000431001 |
| 0 | 0 | 0 | 0 | 0 |
| 0 | 0 | 0.00685317 | 0 | 0 |
| 0 | 0 | 0.002122207 | 0 | 0 |
| 0 | 0 | 0 | 0.02067001 | 0 |
| 0 | 0 | 0 | 0 | 0 |
| 0 | 0 | 0 | 0 | 0 |
| 0 | 0 | 0 | 0 | 0 |
| 0 | 0 | 0 | 0 | 0 |
| 0 | 0 | 0 | 0 | 0 |
| 0 | 0 | 0 | 0 | 0 |
| 0 | 0 | 0 | 0 | 0 |
| 0 | 0 | 0 | 0 | 0 |
| 0 | 0 | 0 | 0 | 0 |
| 0.237937284 | 0.039510337 | 0.067402379 | 0.012371775 | 0.012857238 |
| 0 | 0 | 0 | 0 | 0 |
| 0.005937647 | 0 | 0.09282453 | 0.024633733 | 0 |
| 0.032734865 | 0.000563659 | 0.010766156 | 0.009612797 | 0 |
| 0 | 0 | 0 | 0 | 0 |
| 0 | 0 | 0 | 0 | 0 |
| 0 | 0 | 0 | 0 | 0 |
| 0 | 0 | 0 | 0 | 0 |
| 0 | 0 | 0 | 0 | 0 |
| 0.013171417 | 5.88634203 | 0.007067905 | 0.022000027 | 0.065066871 |
| 0 | 0 | 0 | 0 | 0 |
| 0 | 0 | 0 | 0 | 0 |
| 0 | 0 | 0 | 0 | 0 |
| 0 | 0 | 0 | 0 | 10.76205056 |
| 0 | 0 | 0 | 0 | 0 |
| 0 | 0 | 0 | 0 | 0 |
| 0.030609739 | 0.004055382 | 0 | 0 | 0 |
| 0 | 0 | 0 | 0 | 0 |
| 0 | 0 | 0 | 0 | 0 |
| 0 | 0 | 0 | 0 | 0 |
| 0 | 0 | 0 | 0 | 0 |
| 0 | 0 | 0 | 0 | 0 |
| 0 | 0 | 0 | 0 | 0 |
| 0 | 0 | 0 | 0 | 0 |
| 0 | 0 | 0 | 0 | 0 |
| 0 | 0 | 0 | 0 | 0 |
| 0 | 0 | 0 | 0 | 0 |
| 0 | 0 | 0 | 0.79463132 | 0 |

| | | | | |
|---|---|---|---|---|
| 0.011741292 | 0.000909776 | 0.003861589 | 0.064944938 | 0.116348321 |
| 0 | 0 | 0 | 0 | 0 |
| 0.173966871 | 0.000900982 | 0 | 0 | 0 |
| 0 | 0 | 0 | 0 | 0 |
| 0 | 0 | 0 | 0 | 0 |
| 0 | 0 | 0 | 0 | 0 |
| 0 | 0 | 0.00187491 | 0 | 0 |
| 0.363225177 | 0.251904925 | 0.537150301 | 0.903277847 | 0 |
| 0.095715652 | 1.920462695 | 1.189618429 | 0.040989265 | 0.263742996 |
| 0 | 0.014746585 | 0 | 0.06615047 | 0 |
| 0 | 0 | 0 | 0.333119485 | 0 |
| 0 | 0 | 0 | 0 | 0 |
| 0 | 0 | 0 | 0 | 0 |
| 0 | 0.057264773 | 0.013225782 | 0 | 0.034219696 |
| 0 | 0 | 0 | 0 | 0 |
| 0 | 0 | 0 | 0 | 0 |
| 0.035660442 | 0 | 0 | 0 | 0 |
| 0 | 0 | 0.042200196 | 0 | 0 |
| 0 | 0 | 0 | 0 | 0 |
| 0.015443893 | 0 | 0.024309522 | 0 | 0 |
| 0 | 0 | 0 | 0 | 0 |
| 0 | 0 | 0 | 0 | 0 |
| 0 | 0 | 0 | 0 | 0 |
| 0 | 0 | 0 | 0 | 0 |
| 0 | 0 | 0 | 0 | 0 |
| 0 | 0 | 0 | 0 | 0 |
| 0 | 0 | 0 | 0 | 0 |
| 0 | 0 | 0.81691808 | 0 | 0 |
| 0 | 0 | 0 | 0 | 0 |
| 0 | 0 | 0 | 0 | 0 |
| 0.004474671 | 0 | 0 | 0 | 0 |
| 0 | 0 | 0 | 0 | 0 |
| 0 | 0 | 0 | 0 | 0 |
| 0 | 0 | 0 | 0 | 0 |
| 0 | 0.001652118 | 0 | 0.010264359 | 0 |
| 0 | 0 | 0 | 0 | 0 |
| 0 | 0 | 0 | 0 | 0 |
| 0 | 0 | 0 | 0 | 0 |
| 0 | 0 | 0.021508634 | 0.000806523 | 0 |
| 0 | 0 | 0 | 0 | 0.013834489 |
| 0 | 0 | 0 | 0 | 0 |
| 0.220791011 | 0.015304534 | 0.050238239 | 2.094697552 | 0 |
| 0 | 0 | 0 | 0 | 0 |
| 0 | 0 | 0 | 0 | 0 |
| 0 | 0 | 0 | 0 | 0 |
| 0.007577706 | 0 | 0 | 0 | 0 |
| 0 | 0 | 0 | 0 | 0 |
| 0 | 0 | 0 | 0 | 0 |

| | | | | |
|---|---|---|---|---|
| 0 | 0 | 0 | 0 | 0 |
| 0.065633218 | 0 | 0.00421899 | 0.00749913 | 0.02397238 |
| 0 | 0 | 0 | 0 | 0 |
| 0 | 0 | 0 | 0 | 0 |
| 0.018878627 | 0.044390534 | 1.444422251 | 0.417039771 | 0 |
| 0 | 0 | 0 | 0 | 0 |
| 0 | 0 | 0 | 0 | 0 |
| 0.003131695 | 0.036840752 | 0.018614731 | 0.012942245 | 0.017628207 |
| 0 | 0.001540696 | 0 | 0.007968784 | 0 |
| 0 | 0 | 0 | 0 | 0 |
| 0 | 0 | 0 | 0 | 0 |
| 0 | 0 | 0 | 0 | 0 |
| 0 | 0 | 0 | 0 | 0 |
| 0 | 0 | 0 | 0 | 0 |
| 0.001187094 | 0 | 0 | 0 | 0 |
| 0.04095036 | 0.002007437 | 0 | 0.002432021 | 0.956824845 |
| 0 | 0 | 0 | 0 | 0 |
| 0 | 0 | 0 | 0 | 0 |
| 0 | 0 | 0 | 0 | 0 |
| 0 | 0 | 0 | 0 | 0 |
| 0 | 0 | 0 | 0 | 0 |
| 0 | 0 | 0 | 0 | 0 |
| 0 | 0 | 0 | 0 | 0 |
| 0.003752618 | 0 | 0.004698609 | 0 | 0.013348791 |
| 0.367451037 | 1.049416053 | 0.139746301 | 0 | 0 |
| 0 | 0 | 0 | 0.00136109 | 0 |
| 0 | 0.019999989 | 0.1160301 | 0 | 0 |
| 0 | 0 | 0 | 0.000539334 | 0 |
| 0 | 0 | 0 | 0 | 0.026572611 |
| 0 | 0 | 0 | 0 | 0 |
| 0 | 0 | 0 | 0 | 0 |
| 0 | 0 | 0 | 0 | 0 |
| 0 | 0 | 0 | 0 | 0 |
| 0 | 0 | 0 | 0 | 0 |
| 0 | 0 | 0 | 0 | 0 |
| 0 | 0 | 0 | 0 | 0 |
| 0 | 0 | 0 | 0 | 0 |
| 0 | 0.002348606 | 0.002308558 | 0.024513777 | 0 |
| 0 | 0 | 0.008672972 | 0 | 0 |
| 0 | 0 | 0 | 0 | 0 |
| 0 | 0 | 0 | 0 | 0 |
| 0 | 0 | 0 | 0.327664778 | 0 |
| 0 | 0 | 0 | 0 | 0 |
| 0 | 0 | 0 | 0 | 0 |
| 0 | 0 | 0 | 0 | 0 |
| 0 | 0 | 0 | 0 | 0 |
| 0 | 0 | 0 | 0 | 0 |
| 0 | 0 | 0 | 0 | 0 |

| | | | | |
|---|---|---|---|---|
| 0.47769712 | 0 | 0.236899783 | 2.060000747 | 0 |
| 0 | 0 | 0 | 0 | 0 |
| 0 | 0 | 0 | 0 | 0 |
| 0 | 0 | 0 | 0 | 0 |
| 0 | 0 | 0 | 0 | 0 |
| 0.002492972 | 0.000499018 | 0 | 0.023066417 | 0 |
| 0 | 0 | 0 | 0 | 0 |
| 0 | 0 | 0 | 0 | 0 |
| 0 | 0 | 0 | 0 | 0 |
| 0 | 0 | 0 | 0 | 0 |
| 0 | 0 | 0 | 0 | 0 |
| 0 | 0 | 0.000618767 | 0 | 0 |
| 0 | 0 | 0 | 0.014957265 | 0 |
| 0 | 0 | 0 | 0 | 0 |
| 0 | 0 | 0 | 0 | 0 |
| 0 | 0 | 0 | 0 | 0 |
| 0 | 0 | 0 | 0 | 0 |
| 0 | 0 | 0 | 0 | 0 |
| 0 | 0 | 0.001794749 | 0 | 0 |
| 0 | 0 | 0 | 0 | 0 |
| 0 | 0.001828497 | 0 | 0.209688171 | 0 |
| 0 | 0 | 0 | 0 | 0 |
| 0.004719471 | 0 | 0.254095386 | 0.004225865 | 0.03617431 |
| 0 | 0 | 1.43271177 | 0 | 0 |
| 0 | 0 | 0 | 0 | 0 |
| 0 | 0 | 0 | 0 | 0 |
| 0 | 0 | 0 | 0 | 0 |
| 0 | 0 | 0 | 0 | 0 |
| 0 | 0.005997256 | 0 | 0 | 0 |
| 0 | 0 | 0 | 0 | 0 |
| 0 | 0 | 0 | 0 | 0 |
| 0 | 0 | 0 | 0 | 0 |
| 0.0098217 | 0.169273475 | 0.004743374 | 0.055185214 | 0.00598932 |
| 0 | 0 | 0 | 0 | 0 |
| 0.008785464 | 0.006155054 | 0.080851309 | 0.843889459 | 0.035158059 |
| 0 | 0.003786338 | 0 | 0 | 0.003754827 |
| 0 | 0 | 0 | 0 | 0 |
| 0 | 0 | 0 | 0 | 0 |
| 0 | 0 | 0.034976659 | 0 | 0 |
| 0 | 0 | 0 | 0 | 0 |
| 0 | 0 | 0 | 0 | 0 |
| 0 | 0 | 0.033871051 | 0 | 0 |
| 0.026115959 | 0.007041301 | 0.143572686 | 0.180633417 | 0 |
| 0 | 0 | 0 | 0 | 0 |
| 0 | 0 | 0 | 0 | 0 |
| 0 | 0.039313554 | 0.021123526 | 0.002463547 | 0 |
| 0 | 0 | 0 | 0 | 0 |
| 0 | 0 | 0 | 0 | 0 |

| | | | | |
|---|---|---|---|---|
| 0 | 0 | 0 | 0 | 0 |
| 0 | 0 | 0 | 0 | 0 |
| 0 | 0 | 0 | 0 | 0 |
| 0 | 0 | 0 | 0 | 0 |
| 0 | 0 | 0 | 0.002384098 | 0 |
| 0 | 0 | 0 | 0 | 0 |
| 0 | 0 | 0.003730185 | 0 | 0 |
| 0 | 0 | 0 | 0 | 0 |
| 0 | 0 | 0 | 0 | 0 |
| 0 | 0 | 0.003977086 | 0.01522456 | 0 |
| 0 | 0 | 0 | 0 | 0 |
| 0 | 0 | 0 | 0 | 0 |
| 0 | 0 | 0 | 0 | 0 |
| 1.136363332 | 0.001651295 | 0 | 0 | 0 |
| 0 | 0 | 0 | 0 | 0 |
| 0.00343158 | 0.000686899 | 0 | 0 | 0.006866324 |
| 0 | 0 | 0 | 0 | 0 |
| 0 | 0 | 0 | 0 | 0 |
| 0 | 0 | 0 | 0 | 0 |
| 1.593991414 | 0.128677277 | 0.275577657 | 0 | 0 |
| 0.008960104 | 0 | 0.016347457 | 0.009025846 | 0 |
| 0 | 0 | 0 | 0 | 0 |
| 0 | 0 | 0 | 0 | 0 |
| 0 | 0 | 0 | 0.002302236 | 0 |
| 0 | 0 | 0 | 0 | 0 |
| 0 | 0 | 0 | 0.129989319 | 0 |
| 0 | 0 | 0 | 0 | 0 |
| 0 | 0 | 0 | 0 | 0 |
| 0 | 0 | 0 | 0 | 0 |
| 0.017338275 | 0.007297166 | 0.031728616 | 0.113251996 | 0 |
| 0.119582311 | 0.253604421 | 0.259482154 | 0.056503287 | 0 |
| 0 | 0 | 0 | 0 | 0 |
| 0 | 0 | 0 | 0 | 0 |
| 0 | 0 | 0 | 0 | 0 |
| 0 | 0 | 0 | 0 | 0 |
| 0 | 0 | 0 | 0 | 0 |
| 0 | 0 | 0 | 0 | 0 |
| 0 | 0 | 0 | 0 | 0 |
| 0 | 0 | 0 | 0 | 0 |
| 0 | 0 | 0 | 0 | 0 |
| 0 | 0 | 0 | 0 | 0 |
| 0 | 0 | 0 | 0 | 0 |
| 0 | 0 | 0 | 0 | 0 |
| 0 | 0 | 0 | 0 | 0 |
| 0 | 0 | 0 | 0 | 0 |
| 0 | 0 | 0 | 0 | 0 |
| 0 | 0 | 0 | 1.210614155 | 0 |
| 0 | 0 | 0.001148981 | 0 | 0 |

| | | | | |
|---|---|---|---|---|
| 0 | 0 | 0 | 0 | 0 |
| 0 | 0 | 0 | 0 | 0 |
| 0 | 0 | 0 | 0 | 0 |
| 0 | 0 | 0 | 0.00176191 | 0 |
| 0.376599421 | 0.047410445 | 0.171984986 | 0.007784056 | 0.008126845 |
| 0 | 0 | 0 | 0 | 0 |
| 0 | 0 | 0 | 0 | 0 |
| 0 | 0 | 0 | 0 | 0 |
| 0 | 0 | 0 | 0 | 0 |
| 0 | 0 | 0 | 0 | 0 |
| 0 | 0.025511919 | 0 | 0 | 0 |
| 0.009108946 | 0.00145867 | 0.021506959 | 0.005097655 | 0 |
| 0 | 0 | 0.012440342 | 0.027267551 | 0.02126578 |
| 0 | 0 | 0 | 0 | 0 |
| 0 | 0 | 0 | 0 | 0 |
| 0.836029702 | 0.042395324 | 0 | 0 | 0 |
| 0.002452051 | 0 | 0 | 0 | 0 |
| 0 | 0 | 0 | 0 | 0 |
| 0 | 0.776749205 | 0.004811534 | 0.029027002 | 0.222788407 |
| 0.415844308 | 0.004687014 | 0.055333425 | 0.486657533 | 0 |
| 0 | 0 | 0 | 0 | 0.005995426 |
| 0 | 0 | 0 | 0 | 0 |
| 0 | 0 | 0 | 0 | 0 |
| 0 | 0 | 0 | 0 | 0 |
| 0 | 0 | 0 | 0.004485069 | 0 |
| 0 | 0 | 0 | 0 | 0 |
| 0.006314688 | 0.003611461 | 0.008906968 | 0.065750729 | 0 |
| 0 | 0 | 0 | 0 | 0 |
| 0 | 0 | 0 | 0 | 0 |
| 0 | 0 | 0 | 0 | 0 |
| 0.001636121 | 0 | 0.002633874 | 0.002563751 | 0 |
| 0 | 0 | 0 | 0 | 0 |
| 0 | 0 | 0 | 0 | 0 |
| 0 | 0 | 0 | 0 | 0 |
| 0 | 0 | 0 | 0 | 0 |
| 0 | 0 | 0 | 0.001197624 | 0 |
| 0 | 0 | 0 | 0 | 0 |
| 0.002298983 | 0.003451406 | 0.017888004 | 0.007719502 | 0 |
| 0.192873041 | 0 | 0.124827596 | 0 | 0 |
| 0 | 0 | 0 | 0 | 0 |
| 0 | 0 | 0 | 0.00051377 | 0 |
| 0 | 0 | 0 | 0 | 0 |
| 0 | 0 | 0 | 0 | 0 |
| 0 | 0.015865791 | 0 | 0.000572352 | 0 |
| 0 | 0 | 0 | 0 | 0 |
| 0 | 0 | 0.05601432 | 0 | 0 |
| 0 | 0 | 0 | 0 | 0 |
| 0 | 0 | 0 | 0 | 0 |

| | | | | |
|---|---|---|---|---|
| 0 | 0 | 0 | 0 | 0 |
| 0 | 0.000874877 | 0.298835691 | 0.000815322 | 0 |
| 0 | 0 | 0 | 0.249088229 | 0 |
| 0.001116485 | 0.001251526 | 0.016295961 | 0.000999713 | 0 |
| 0 | 0 | 0 | 0.002596209 | 0.001420799 |
| 0.001603729 | 0 | 0 | 0 | 0 |
| 0 | 0 | 0 | 0.070044483 | 0 |
| 0 | 0 | 0 | 0 | 0 |
| 0 | 0 | 0 | 0 | 0 |
| 0 | 0 | 0 | 0.051292036 | 0 |
| 0.028968663 | 0 | 0 | 0 | 0 |
| 0 | 0 | 0 | 0 | 0 |
| 0.007122876 | 0.005576409 | 0.544961011 | 0.059385334 | 0 |
| 0 | 0 | 0 | 0.009603447 | 0 |
| 0 | 0 | 0 | 0 | 0 |
| 0 | 0 | 0 | 0 | 0 |
| 0 | 0 | 0 | 0 | 0 |
| 0 | 0 | 0 | 0 | 0 |
| 0 | 0 | 0 | 0 | 0 |
| 0 | 0 | 0 | 0 | 0 |
| 0 | 0 | 0 | 0 | 0 |
| 0 | 0 | 0 | 0 | 0 |
| 0.01834152 | 0 | 0.060171818 | 0 | 0.0370177 |
| 0.011559663 | 0.008270955 | 0.01003042 | 0.002202265 | 0 |
| 0 | 0 | 0 | 0 | 0 |
| 0 | 0 | 0 | 0.003578067 | 0 |
| 0 | 0 | 0 | 0 | 0 |
| 0 | 0 | 0 | 0 | 0 |
| 0 | 0 | 0 | 0 | 0 |
| 0 | 0 | 0 | 0 | 0 |
| 0.016732402 | 0.009675829 | 0.036663281 | 0.032461808 | 0 |
| 0.008446467 | 0 | 0 | 0 | 0 |
| 0 | 0 | 0 | 0 | 0 |
| 0 | 0 | 0 | 0 | 0 |
| 0 | 0 | 0 | 0 | 0 |
| 0 | 0 | 0 | 0 | 0 |
| 0 | 0 | 0 | 0 | 0.001317418 |
| 0 | 0 | 0 | 0 | 0 |
| 0 | 0.000629725 | 0.001477128 | 0 | 0 |
| 0 | 0 | 0 | 0 | 0 |
| 0 | 0 | 0 | 0 | 0 |
| 0 | 0 | 0 | 0 | 0 |
| 0 | 0 | 0 | 0 | 0 |
| 0 | 0 | 0 | 0 | 0 |
| 0 | 0 | 0 | 0 | 0 |
| 0.059337194 | 0.1565202 | 0.55865447 | 0.165434641 | 0 |
| 0 | 0 | 0 | 0 | 0 |
| 0 | 0 | 0 | 0 | 0 |

| | | | | |
|---|---|---|---|---|
| 0 | 0 | 0 | 0 | 0 |
| 0 | 0 | 0 | 0 | 0 |
| 0 | 0.085387399 | 0 | 0 | 0 |
| 0.002611855 | 0 | 0 | 0.07039436 | 0 |
| 0 | 0 | 0 | 0 | 0 |
| 0 | 0 | 0 | 0 | 0 |
| 0 | 0 | 0 | 0 | 0 |
| 0 | 0 | 0 | 0 | 0 |
| 0 | 0 | 0.000585744 | 0 | 0 |
| 0 | 0 | 0 | 0 | 0 |
| 0 | 0.000383157 | 0.003423846 | 0 | 0.011915794 |
| 0 | 0 | 0.002811497 | 0 | 0 |
| 0 | 0 | 0 | 0 | 0 |
| 0 | 0 | 0 | 0 | 0 |
| 0 | 0 | 0 | 0 | 0 |
| 0 | 0 | 0 | 0 | 0 |
| 0 | 0 | 0.013337201 | 0 | 0 |
| 0 | 0 | 0 | 0 | 0 |
| 0 | 0 | 0 | 0 | 0 |
| 0 | 0.000508132 | 0 | 0 | 0 |
| 0 | 0 | 0 | 0 | 0 |
| 0 | 0 | 0 | 0 | 0 |
| 0 | 0 | 0 | 0 | 0 |
| 0 | 0 | 0 | 0 | 0 |
| 0 | 0 | 0 | 0 | 0 |
| 0 | 0 | 0 | 0 | 0 |
| 0 | 0 | 0 | 0 | 0 |
| 0 | 0.006935446 | 0.018592317 | 0 | 0 |
| 0 | 0 | 0 | 0 | 0 |
| 0 | 0 | 0 | 0 | 0 |
| 0.001999098 | 0.001309613 | 0.053636719 | 0.008136425 | 0.000692647 |
| 0 | 0 | 0 | 0 | 0 |
| 0.033973281 | 0.448913057 | 0.007020394 | 0.016249215 | 0 |
| 0 | 0.002589803 | 0 | 0.001158484 | 0 |
| 0 | 0 | 0 | 0 | 0 |
| 0 | 0 | 0 | 0 | 0 |
| 0 | 0 | 0 | 0 | 0 |
| 0 | 0 | 0 | 0 | 0 |
| 0 | 0 | 0 | 0 | 0 |
| 0 | 0 | 0 | 0 | 0 |
| 0 | 0 | 0 | 0 | 0 |
| 0.199465758 | 0 | 0.445467585 | 0.013678967 | 0 |
| 0.130702716 | 0.036238397 | 0.018839199 | 0.11513889 | 0 |
| 0 | 0 | 0 | 0 | 0 |
| 0 | 0 | 0 | 0 | 0 |
| 0 | 0.002447056 | 0.040999929 | 0.067046063 | 0 |
| 0 | 0 | 0 | 0 | 0 |
| 0 | 0 | 0 | 0 | 0 |

| | | | | |
|---|---|---|---|---|
| 0 | 0 | 0 | 0 | 0 |
| 0 | 0.005854343 | 0.007133696 | 0.008332528 | 0 |
| 0.002846951 | 0.032949092 | 0.013610421 | 0.001158723 | 0 |
| 0 | 0 | 0 | 0 | 0 |
| 0 | 0.002978294 | 0.012641513 | 0.038857746 | 1.386179692 |
| 0.003419586 | 0 | 0.001511162 | 0 | 1.302686876 |
| 0 | 0 | 0 | 0 | 0 |
| 0 | 0 | 0 | 0 | 0 |
| 0 | 0 | 0 | 0 | 0 |
| 0 | 0 | 0 | 0 | 0 |
| 0 | 0 | 0 | 0 | 0 |
| 0 | 0 | 0 | 0 | 0 |
| 0 | 0 | 0 | 0 | 0 |
| 0 | 0 | 0 | 0.039152255 | 0 |
| 0 | 0 | 0 | 0 | 0 |
| 0 | 0 | 0 | 0 | 0 |
| 0 | 0 | 0 | 0 | 0 |
| 0 | 0 | 0 | 0 | 0 |
| 0 | 0 | 0 | 0 | 0 |
| 0 | 0 | 0 | 0 | 0 |
| 0 | 0 | 0 | 0 | 0 |
| 0 | 0 | 0 | 0 | 0 |
| 0 | 0 | 0 | 0 | 0 |
| 0 | 0 | 0 | 0 | 0 |
| 0 | 0 | 0.012590398 | 0.031513368 | 0 |
| 0 | 0.001718048 | 0 | 0.001152789 | 0 |
| 0 | 0 | 0 | 0 | 0 |
| 6.088625334 | 0.032998025 | 0 | 0 | 0 |
| 0 | 0 | 0 | 0 | 0 |
| 0 | 0 | 0 | 0 | 0 |
| 0 | 0 | 0 | 0 | 0 |
| 0 | 0 | 0 | 0 | 0 |
| 0 | 0 | 0 | 0 | 0 |
| 0 | 0 | 0 | 0 | 0 |
| 0 | 0 | 0 | 0 | 0 |
| 0 | 0 | 0 | 0 | 0 |
| 0 | 0 | 0 | 0 | 0 |
| 0 | 0 | 0 | 0 | 0 |
| 1.59125699 | 0.920937192 | 0 | 0 | 0 |
| 0 | 0 | 0 | 0 | 0 |
| 0 | 0 | 0 | 0 | 0 |
| 0.037477467 | 0.001491054 | 0.006995046 | 0 | 0 |
| 0.007321834 | 0.000862125 | 0.003004502 | 0.049170366 | 0.090282691 |
| 0 | 0 | 0 | 0 | 0 |
| 0 | 0 | 0 | 0 | 0 |
| 0 | 0 | 0 | 0.005767795 | 0 |
| 0 | 0.00290154 | 0.016263831 | 0.01917399 | 0.00125559 |
| 0 | 0 | 0 | 0 | 0 |

| | | | | |
|---|---|---|---|---|
| 0 | 0 | 0 | 0 | 0 |
| 0.145492603 | 0.002995535 | 0.188489773 | 0.021402427 | 0.154629928 |
| 0 | 0 | 0 | 0 | 0 |
| 0 | 0 | 0 | 0 | 0 |
| 0 | 0 | 0 | 0 | 0 |
| 0 | 0 | 0 | 0 | 0 |
| 0 | 0 | 0 | 0 | 0 |
| 0 | 0 | 0 | 0 | 0 |
| 0 | 0 | 0 | 0 | 0 |
| 0 | 0 | 0 | 0 | 0 |
| 0 | 0 | 0 | 0 | 0 |
| 0 | 0 | 0 | 0 | 0 |
| 0.005053382 | 0.000703677 | 0.087009935 | 0 | 0 |
| 0 | 0 | 0 | 0 | 0 |
| 0.027211102 | 0.008299958 | 0.025031539 | 0.00719351 | 0 |
| 0 | 0 | 0 | 0 | 0 |
| 0 | 0 | 0 | 0 | 0 |
| 0 | 0 | 0 | 0 | 0 |
| 0 | 0 | 0.002153773 | 0 | 0 |
| 0 | 0 | 0 | 0 | 0 |
| 0 | 0 | 0 | 0 | 0 |
| 0.043775556 | 0 | 0.018846926 | 0.230852717 | 0 |
| 0.022881617 | 0 | 0 | 0.008930861 | 0 |
| 0 | 0 | 0 | 0 | 0 |
| 0 | 0 | 0 | 0 | 0 |
| 0 | 0 | 0 | 0 | 0 |
| 0 | 0 | 0 | 0 | 0 |
| 0 | 0 | 0 | 0 | 0 |
| 0 | 0 | 0 | 0 | 0 |
| 0 | 0 | 0 | 0 | 0 |
| 0 | 0 | 0 | 0 | 0 |
| 0 | 0 | 0 | 0 | 0 |
| 0 | 0 | 0 | 0 | 0 |
| 0 | 0 | 0 | 0 | 0 |
| 0 | 0.06864287 | 21.4251659 | 18.97172185 | 14.91113853 |
| 0 | 0 | 0 | 0 | 0 |
| 0 | 0 | 0 | 0 | 0 |
| 0 | 0 | 0 | 0 | 0 |
| 0.012962062 | 0.002645491 | 0.096832352 | 0.244644076 | 0 |
| 0 | 0 | 0 | 0 | 0 |
| 0.208060222 | 0.093604066 | 0.141117964 | 0.088762568 | 0.002431908 |
| 0 | 0 | 0 | 0 | 0 |
| 0 | 0 | 0 | 0 | 0 |
| 0 | 0 | 0 | 0 | 0 |
| 0 | 0 | 0 | 0 | 0 |
| 0.013567546 | 0.003133634 | 0.094086094 | 0.287125758 | 0.232494845 |
| 0 | 0 | 0 | 0 | 0 |
| 0 | 0 | 0 | 0 | 0 |

| | | | | |
|---|---|---|---|---|
| 0 | 0 | 0.003741046 | 0 | 0 |
| 0 | 0 | 0 | 0 | 0 |
| 0 | 0 | 0 | 0 | 0 |
| 0 | 0 | 0 | 0 | 0 |
| 0 | 0 | 0 | 0 | 0 |
| 0 | 0 | 0 | 0 | 0 |
| 0 | 0 | 0 | 0 | 0 |
| 0 | 0 | 0.00121064 | 0.001010065 | 0 |
| 0 | 0 | 0 | 0 | 0 |
| 0.014034605 | 0.005107832 | 0.0150622 | 0.00297032 | 0 |
| 0 | 0 | 0.012997015 | 0 | 0 |
| 0 | 0 | 0 | 0 | 0 |
| 0.000997647 | 0 | 0.003747426 | 0 | 0.002281387 |
| 0 | 0 | 0 | 0 | 0 |
| 0 | 0 | 0 | 0 | 0 |
| 0 | 0 | 0 | 0 | 0 |
| 0 | 0 | 0 | 0 | 0 |
| 0 | 0 | 0 | 0 | 0 |
| 0 | 0 | 0 | 0 | 0 |
| 0.581180386 | 0.183611846 | 0.182021577 | 0.680210969 | 0.438608626 |
| 0 | 0 | 0 | 0 | 0 |
| 0 | 0 | 0 | 0 | 0 |
| 0 | 0 | 0 | 0 | 0 |
| 0 | 0 | 0 | 0 | 0 |
| 0 | 0 | 0 | 0 | 0 |
| 0 | 0 | 0 | 0 | 0 |
| 0 | 0 | 0 | 0 | 0 |
| 0 | 0 | 0 | 0 | 0 |
| 0 | 0 | 0 | 0 | 0 |
| 0.037970434 | 0 | 0 | 0 | 0 |
| 0 | 0 | 0 | 0 | 0 |
| 0 | 0 | 0.021131317 | 0 | 0 |
| 0 | 0 | 0 | 0 | 0 |
| 0 | 0 | 0 | 0 | 0 |
| 0 | 0 | 0 | 0 | 0 |
| 0 | 0 | 0 | 0.003087729 | 0 |
| 0 | 0 | 0 | 0 | 0 |
| 0 | 0 | 0 | 0 | 0 |
| 0 | 0 | 0 | 0 | 0 |
| 0 | 0 | 0 | 0 | 0 |
| 0 | 0 | 0 | 0 | 0 |
| 0 | 0 | 0 | 0 | 0 |
| 0 | 0 | 0.001386289 | 0 | 0 |
| 0 | 0 | 0 | 0 | 0 |
| 0.006672159 | 0.002170295 | 0.009174628 | 0.009708274 | 0 |
| 0 | 0 | 0 | 0 | 0 |
| 0 | 0 | 0 | 0 | 0 |
| 0 | 0 | 0 | 0 | 0 |

| | | | | |
|---|---|---|---|---|
| 0 | 0 | 0 | 0 | 0 |
| 0 | 0 | 0 | 0 | 0 |
| 0 | 0 | 0 | 0 | 0 |
| 0 | 0 | 0 | 0 | 0 |
| 0 | 0 | 0 | 0 | 0 |
| 0.02272593 | 0.007727154 | 0.200798974 | 0.007665735 | 0.012932908 |
| 0 | 0 | 0 | 0 | 0 |
| 0 | 0 | 0 | 0 | 0 |
| 0 | 0 | 0 | 0 | 0 |
| 0 | 0 | 0 | 0 | 0 |
| 0 | 0 | 0 | 0 | 0 |
| 0 | 0 | 0 | 0 | 0 |
| 0 | 0 | 0 | 0 | 0 |
| 0 | 0 | 0 | 0 | 0 |
| 0 | 0 | 0 | 0 | 0 |
| 0 | 0 | 0 | 0 | 0 |
| 0 | 0 | 0 | 0.057143405 | 0 |
| 0 | 0 | 0 | 0 | 0 |
| 0 | 0 | 0 | 0 | 0 |
| 0 | 0 | 0 | 0 | 0 |
| 0 | 0 | 0 | 0 | 0 |
| 0 | 0 | 0 | 0 | 0 |
| 0 | 0 | 0 | 0 | 0 |
| 0 | 0 | 0 | 0 | 0 |
| 0 | 0 | 0 | 0 | 0 |
| 0 | 0 | 0.014894133 | 0 | 0 |
| 0 | 0 | 0 | 0 | 0 |
| 0 | 0 | 0.066687809 | 0 | 0 |
| 0 | 0 | 0 | 0 | 0 |
| 0 | 0 | 0 | 0 | 0 |
| 0 | 0 | 0 | 0 | 0 |
| 0 | 0 | 0 | 0 | 0 |
| 0 | 0 | 0 | 0 | 0 |
| 0 | 0 | 0 | 0 | 0 |
| 0 | 0 | 0 | 0 | 0 |
| 0 | 0 | 0 | 0 | 0 |
| 0 | 0 | 0 | 0 | 0 |
| 0 | 0 | 0 | 0 | 0 |
| 0 | 0.166252489 | 0.196195397 | 0 | 0 |
| 0 | 0 | 0 | 0 | 0 |
| 0 | 0 | 0 | 0 | 0 |
| 0 | 0 | 0 | 0 | 0 |
| 0 | 0 | 0 | 0 | 0 |
| 0 | 0 | 0 | 0 | 0 |
| 0 | 0 | 0 | 0 | 0 |
| 0 | 0 | 0 | 0 | 0 |
| 0 | 0 | 0 | 0 | 0 |
| 0 | 0 | 0 | 0 | 0 |

| | | | | |
|---|---|---|---|---|
| 0 | 0 | 0 | 0 | 0 |
| 0 | 0 | 0.07817874 | 0 | 0 |
| 0 | 0 | 0 | 0 | 0 |
| 0 | 0 | 0 | 0 | 0 |
| 0 | 0 | 0 | 0 | 0 |
| 0 | 0 | 0 | 0 | 0 |
| 0 | 0 | 0 | 0 | 0 |
| 0 | 0 | 0 | 0 | 0 |
| 0 | 0 | 0 | 0 | 0 |
| 0 | 0 | 0 | 0 | 0 |
| 0 | 0 | 0.011265473 | 0.002451924 | 0 |
| 0 | 0 | 0 | 0 | 0 |
| 0.186564471 | 0.139958207 | 0 | 0 | 0 |
| 0 | 0 | 0 | 0 | 0 |
| 0 | 0 | 0 | 0 | 0 |
| 0 | 0 | 0 | 0 | 0 |
| 0 | 0 | 0 | 0 | 0 |
| 0 | 0 | 0 | 0 | 0 |
| 0 | 0 | 0 | 0 | 0 |
| 0 | 0 | 0 | 0 | 0 |
| 0 | 0.003152475 | 0.163552726 | 0.005391869 | 0.029305736 |
| 0 | 0 | 0 | 0 | 0 |
| 0 | 0.001299094 | 0 | 0 | 0 |
| 1.054268558 | 0.756415936 | 1.572925944 | 1.792661659 | 0.193483647 |
| 0 | 0 | 0 | 0 | 0 |
| 0 | 0 | 0 | 0 | 0 |
| 0 | 0 | 0 | 0 | 0 |
| 0 | 0 | 0 | 0 | 0 |
| 0 | 0 | 0 | 0 | 0 |
| 0 | 0 | 0 | 0 | 0 |
| 0 | 0 | 0 | 0 | 0 |
| 0 | 0 | 0 | 0 | 0 |
| 0 | 0 | 0 | 0.002617722 | 0 |
| 0 | 0 | 0 | 0 | 0 |

| PHL018-0032_V1 | PHL018-0215_V3 | PHL018-0033_V1 | PHL018-0223_V3 | PHL018-0128_V2 |
|---|---|---|---|---|
| EYCF | CTRL | EYCF | REF | CTRL |
| 0.0601523 | 0 | 0.114420213 | 0.538883852 | 0.223411537 |
| 0.247373381 | 7.174583481 | 30.95392113 | 0.305613757 | 15.29459486 |
| 0 | 0 | 2.023594944 | 25.04819017 | 0 |
| 0 | 0 | 0 | 0 | 0 |
| 0 | 0 | 0 | 0 | 0 |
| 0 | 0 | 0 | 0 | 0 |
| 0 | 0 | 0 | 0 | 0 |
| 0 | 0 | 0 | 0 | 0 |
| 0 | 0 | 0 | 0 | 0 |
| 0 | 0 | 0 | 0 | 0 |
| 0 | 0 | 0 | 0 | 0 |
| 0 | 0 | 0 | 0 | 0 |
| 0 | 0 | 0 | 0 | 0 |
| 0.483923898 | 2.383812729 | 0.410397389 | 5.27748745 | 9.673641241 |
| 0 | 0 | 0 | 0 | 0 |
| 0 | 0 | 0 | 0 | 0 |
| 0 | 0 | 0 | 0 | 0 |
| 0 | 0 | 0 | 0 | 0 |
| 0 | 0 | 0 | 0 | 0 |
| 0 | 0 | 0 | 0 | 0 |
| 0 | 0 | 0 | 0 | 0 |
| 0 | 0 | 0 | 0 | 0 |
| 0 | 0 | 0 | 0 | 0 |
| 0 | 0 | 0 | 0 | 0 |
| 0.326552349 | 0 | 0 | 0 | 0 |
| 0 | 0 | 0 | 0 | 0 |
| 0 | 0 | 0 | 0 | 0 |
| 0 | 0 | 0 | 0 | 0 |
| 0 | 0 | 0 | 0 | 0 |
| 0.25517796 | 0 | 0 | 0 | 0 |
| 0 | 0 | 0 | 0 | 0 |
| 0 | 0 | 0 | 0 | 0 |
| 0 | 0 | 0 | 0 | 0 |
| 0 | 0 | 0 | 0 | 0 |
| 0 | 0 | 0 | 0 | 0 |
| 0.216295742 | 0 | 2.718465317 | 0 | 6.769616927 |
| 0 | 0 | 0 | 0 | 0 |
| 0 | 0 | 0 | 0 | 0 |
| 0 | 0 | 0 | 0 | 0 |
| 0 | 0 | 0 | 0 | 0 |
| 0 | 0 | 0 | 0 | 0 |
| 0.011051585 | 0.003441923 | 0.023201717 | 0.007805904 | 0.003911647 |
| 0.796832022 | 0.769591717 | 0.177783225 | 0.391201028 | 0.022790916 |
| 0.016125377 | 0.025843113 | 0.17013404 | 0 | 0.077656765 |
| 0 | 0.016918687 | 0.003698311 | 0.001745868 | 0 |
| 0 | 0 | 0 | 0 | 0 |

| | | | | |
|---|---|---|---|---|
| 0.006600924 | 0 | 0.268013674 | 0 | 0.010488224 |
| 0 | 0.002291385 | 0.107167699 | 0.019346833 | 0 |
| 0.004244576 | 0.105308112 | 0.012496294 | 0 | 0.001461071 |
| 0.673536724 | 0.832475717 | 0.211155673 | 0.304080769 | 0.583570931 |
| 0.020536439 | 0 | 0 | 0 | 0 |
| 0.035196873 | 0 | 0 | 0 | 0 |
| 0 | 0 | 0.006774177 | 0 | 0.000947003 |
| 0 | 0 | 0 | 0 | 0 |
| 0.018309568 | 0 | 0 | 0 | 0 |
| 0 | 0 | 0 | 0 | 0 |
| 0 | 0 | 0 | 0 | 0 |
| 0.503311478 | 0 | 0.00810642 | 0.006728462 | 0.034752859 |
| 0 | 0 | 0.001020614 | 0 | 0 |
| 0.028341838 | 0 | 0.05188945 | 0.377931345 | 0.073925311 |
| 0 | 0 | 0.002894013 | 0.049885158 | 0 |
| 0 | 0 | 0 | 0 | 0 |
| 0.001246641 | 13.50032075 | 0.044242463 | 0.003978214 | 0.001859519 |
| 0 | 0 | 0 | 0 | 0 |
| 0 | 0 | 0 | 0 | 0 |
| 0 | 0 | 0 | 0 | 0 |
| 0.00880841 | 0.017192254 | 0.266354663 | 0.122657088 | 0.042760904 |
| 0.017893789 | 0 | 0.003139093 | 0.027266546 | 0.008074523 |
| 0.278112467 | 0 | 0 | 0 | 0.153206115 |
| 0 | 0 | 0 | 0 | 0 |
| 0 | 0 | 0 | 0 | 0 |
| 0 | 0 | 0 | 0 | 0 |
| 0 | 0 | 0.03548284 | 0.016986373 | 0 |
| 0 | 0 | 0 | 0 | 0 |
| 0 | 0 | 0.017401833 | 0 | 0.186220632 |
| 0 | 0 | 0 | 0 | 0 |
| 0.001022301 | 0 | 0.384149379 | 0.056994495 | 0 |
| 0 | 0 | 0 | 0 | 0 |
| 0 | 0 | 0 | 0 | 0 |
| 0 | 0 | 0 | 0 | 0 |
| 0 | 0 | 0 | 0 | 0 |
| 0 | 0 | 0 | 0 | 0 |
| 0.014458795 | 0 | 0 | 0 | 0 |
| 0.014951633 | 0 | 0.011673206 | 0 | 0 |
| 0 | 0 | 0 | 0 | 0 |
| 0 | 0 | 0 | 0 | 0 |
| 0 | 0 | 0 | 0 | 0 |
| 0.103636481 | 0 | 0 | 0 | 0 |
| 0 | 0 | 0 | 0 | 0 |
| 0 | 0 | 0 | 0 | 0 |
| 0 | 0 | 0 | 0 | 0 |
| 0 | 0 | 0.060632438 | 0 | 0 |
| 0 | 0.000898956 | 0 | 0.001046917 | 0.007209844 |
| 0 | 0 | 0 | 0.001197741 | 0.168369604 |

| | | | | |
|---|---|---|---|---|
| 0 | 0 | 0 | 0 | 0 |
| 0 | 0 | 0 | 0 | 0 |
| 0 | 0 | 0 | 0 | 0 |
| 0 | 0 | 0 | 0 | 0 |
| 0.001540422 | 0 | 0 | 0.008096463 | 0 |
| 0 | 0 | 0 | 0 | 0 |
| 0 | 0 | 0 | 0 | 0 |
| 0 | 0 | 0 | 0 | 0.013437279 |
| 0 | 0 | 0.002022786 | 0.001309578 | 0 |
| 0 | 0 | 0 | 0 | 0 |
| 0 | 0 | 0 | 0 | 0 |
| 0 | 0 | 0.00164389 | 0 | 0 |
| 0 | 0 | 0 | 0 | 0 |
| 0 | 0 | 0 | 0 | 0 |
| 0 | 0 | 0 | 0 | 0 |
| 0.000939045 | 0 | 0 | 0 | 0 |
| 0 | 0 | 0 | 0 | 0 |
| 0 | 0 | 0 | 0 | 0 |
| 0.051658631 | 0 | 0 | 0.034841798 | 0 |
| 0 | 0 | 0 | 0 | 0 |
| 0 | 0 | 0 | 0 | 0 |
| 0 | 0 | 0 | 0 | 0 |
| 0 | 0 | 0 | 0 | 0 |
| 0 | 0 | 0 | 0 | 0 |
| 0 | 0 | 0 | 0 | 0 |
| 0 | 0 | 0 | 0 | 0 |
| 0 | 0 | 0 | 0 | 0 |
| 0 | 0 | 0 | 0 | 0 |
| 0 | 0 | 0 | 0 | 0 |
| 0 | 0 | 0 | 0 | 0 |
| 0.020846826 | 0 | 0 | 0 | 0.00060185 |
| 0.002539758 | 0 | 0.001767339 | 0 | 0.027570846 |
| 0 | 0 | 0 | 0 | 0 |
| 0 | 0 | 0 | 0 | 0 |
| 0 | 0 | 0 | 0 | 0 |
| 0 | 0 | 0 | 0 | 0 |
| 0 | 0 | 0 | 0 | 0 |
| 0 | 0 | 0 | 0 | 0 |
| 0 | 0 | 0 | 0 | 0 |
| 0 | 0 | 0 | 0 | 0 |
| 0 | 0 | 0 | 0 | 0 |
| 0 | 0 | 0 | 0 | 0 |
| 0.000978519 | 0 | 0 | 0 | 0.002189373 |
| 0 | 0 | 0 | 0 | 0 |
| 0 | 0 | 0 | 0 | 0 |
| 0 | 0 | 0 | 0 | 0 |
| 0 | 0 | 0 | 0 | 0 |
| 0 | 0 | 0 | 0 | 0 |

| | | | | |
|---|---|---|---|---|
| 0 | 0 | 0 | 0 | 0 |
| 0 | 0 | 0 | 0 | 0 |
| 0 | 0 | 0 | 0 | 0 |
| 0 | 0 | 0 | 0 | 0 |
| 0.131037777 | 0 | 0.003418375 | 0 | 0 |
| 0 | 0 | 0 | 0 | 0 |
| 0 | 0 | 0 | 0 | 0 |
| 0 | 0 | 0 | 0 | 0 |
| 0 | 0 | 0 | 0 | 0 |
| 0 | 0 | 0 | 0 | 0 |
| 0 | 0 | 0 | 0 | 0 |
| 0 | 0 | 0 | 0 | 0 |
| 0 | 0 | 0 | 0 | 0 |
| 0 | 0 | 0 | 0 | 0 |
| 0 | 0 | 0 | 0 | 0 |
| 4.038151163 | 0 | 0 | 0 | 0 |
| 0 | 0 | 0 | 0 | 0 |
| 0 | 0 | 0 | 0 | 0 |
| 0 | 0 | 0 | 0 | 0 |
| 0 | 0 | 0 | 0 | 0 |
| 0 | 0 | 0 | 0 | 0 |
| 0 | 0 | 0 | 0 | 0 |
| 0.003414125 | 0 | 0 | 0 | 0 |
| 0 | 0 | 0 | 0 | 0 |
| 0 | 0 | 0 | 0 | 0.022950961 |
| 0 | 0 | 0 | 0 | 0 |
| 0.021304506 | 0 | 0 | 0 | 0 |
| 0 | 0 | 0 | 0 | 0 |
| 0 | 0 | 0 | 0 | 0 |
| 0 | 0 | 0 | 0 | 0 |
| 0.0103566 | 0 | 0 | 0 | 0 |
| 0 | 0 | 0.003398546 | 0.000916776 | 0 |
| 0 | 0 | 0 | 0 | 0 |
| 0 | 0 | 0 | 0 | 0 |
| 0 | 0 | 0 | 0 | 0 |
| 0 | 0 | 0 | 0 | 0 |
| 0 | 0 | 0 | 0 | 0 |
| 0.021696672 | 0 | 0 | 0 | 0 |
| 0 | 0 | 0 | 0 | 0 |
| 0 | 0 | 0 | 0 | 0 |
| 0 | 0 | 0 | 0 | 0 |
| 0 | 0 | 0 | 0 | 0 |
| 0 | 0 | 0 | 0 | 0 |
| 0 | 0 | 0 | 0 | 0 |
| 0 | 0 | 0 | 0 | 0 |
| 0 | 0 | 0 | 0 | 0 |
| 0 | 0 | 0 | 0 | 0 |
| 0 | 0 | 0 | 0 | 0 |

| | | | | |
|---|---|---|---|---|
| 0.170876055 | 0 | 0 | 0 | 0 |
| 0 | 0 | 0 | 0 | 0 |
| 0 | 0 | 0 | 0 | 0 |
| 0 | 0 | 0 | 0 | 0 |
| 0 | 0 | 0 | 0 | 0 |
| 0 | 0 | 0 | 0 | 0 |
| 0 | 0 | 0 | 0 | 0 |
| 0 | 0 | 0 | 0 | 0 |
| 0 | 0 | 0 | 0 | 0 |
| 0 | 0 | 0 | 0 | 0 |
| 0 | 0 | 0 | 0 | 0 |
| 0.013274174 | 0 | 0 | 0 | 0 |
| 0 | 0 | 0.003252076 | 0.003509059 | 0.002556016 |
| 0 | 0 | 0 | 0 | 0 |
| 0 | 0 | 0 | 0 | 0 |
| 0 | 0 | 0 | 0 | 0 |
| 0 | 0 | 0 | 0 | 0 |
| 0 | 0 | 0 | 0 | 0 |
| 0 | 0 | 0 | 0 | 0 |
| 0 | 0 | 0 | 0 | 0 |
| 0.016019919 | 0.02591555 | 0.030403 | 0.039038531 | 0.043446683 |
| 0 | 0 | 0 | 0 | 0 |
| 0 | 0 | 0 | 0 | 0 |
| 0 | 0 | 0 | 0.006157715 | 0.062522488 |
| 0 | 0 | 0 | 0 | 0 |
| 0 | 0 | 0 | 0 | 0 |
| 0.00356222 | 0.001968601 | 0.002596879 | 0.002101566 | 0.002277209 |
| 0 | 0 | 0 | 0 | 0 |
| 0 | 0 | 0 | 0 | 0 |
| 0 | 0 | 0 | 0.001430821 | 0 |
| 0 | 0 | 0 | 0.05903821 | 0 |
| 0 | 0 | 0 | 0 | 0 |
| 0 | 0 | 0 | 0 | 0 |
| 0.008900345 | 0 | 0 | 0 | 0 |
| 0.027669423 | 0 | 0.049586409 | 0 | 0.016113175 |
| 0 | 0 | 0 | 0 | 0 |
| 0 | 0 | 0 | 0 | 0 |
| 0 | 0 | 0 | 0 | 0 |
| 0 | 0 | 0 | 0 | 0 |
| 0 | 0 | 0 | 0 | 0 |
| 0 | 0 | 0 | 0 | 0 |
| 0 | 0 | 0 | 0 | 0 |
| 0 | 0 | 0 | 0 | 0 |
| 0 | 0 | 0 | 0 | 0 |
| 0 | 0 | 0 | 0 | 0 |
| 0.308011836 | 0 | 0 | 0 | 0 |
| 0 | 0 | 0 | 0.003004425 | 0 |
| 0.001080437 | 0 | 0 | 0.015058913 | 0.006043518 |

| | | | | |
|---|---|---|---|---|
| 0 | 0 | 0 | 0 | 0 |
| 0 | 0 | 0 | 0 | 0 |
| 0 | 0 | 0 | 0 | 0 |
| 0 | 0 | 0 | 0 | 0 |
| 0 | 0 | 0 | 0 | 0 |
| 0 | 0 | 0 | 0 | 0 |
| 0 | 0 | 0 | 0 | 0.006882297 |
| 0 | 0 | 0 | 0 | 0 |
| 0 | 0 | 0 | 0 | 0.007343973 |
| 0 | 0 | 0 | 0 | 0 |
| 0 | 0 | 0 | 0 | 0 |
| 0 | 0 | 0 | 0 | 0 |
| 0 | 0 | 0 | 0 | 0 |
| 0 | 0 | 0 | 0.001786568 | 0 |
| 0 | 0 | 0 | 0 | 0 |
| 0 | 0 | 0 | 0 | 0 |
| 0 | 0 | 0 | 0 | 0 |
| 0 | 0 | 0 | 0 | 0 |
| 0.003571092 | 0.004781951 | 0.001911548 | 0.001701646 | 0 |
| 0.090345551 | 0 | 0 | 0 | 0 |
| 0 | 0 | 0 | 0.282272301 | 0 |
| 0 | 0 | 0 | 0 | 0 |
| 0 | 0 | 0 | 0 | 0 |
| 0 | 0 | 0 | 0 | 0 |
| 0 | 0.008953385 | 0.006162184 | 0 | 0 |
| 0 | 0 | 0 | 0 | 0 |
| 0 | 0 | 0 | 0 | 0 |
| 0 | 0 | 0 | 0 | 0 |
| 0 | 0 | 0.004544148 | 0 | 0.005443254 |
| 0 | 0 | 0 | 0.008810945 | 0 |
| 0.242382661 | 0.001008526 | 0.0120254 | 5.113698338 | 0.333766645 |
| 0 | 0 | 0 | 0 | 0 |
| 0 | 0.092976831 | 0 | 0.001251793 | 0 |
| 0.002698369 | 0 | 0 | 0 | 0 |
| 0 | 0 | 0 | 0 | 0 |
| 0 | 0 | 0 | 0 | 0 |
| 0 | 0 | 0 | 0 | 0 |
| 0 | 0 | 0 | 0.026049802 | 0 |
| 0 | 0 | 0 | 0 | 0 |
| 0 | 0 | 0 | 0 | 0 |
| 0 | 0 | 0 | 0 | 0 |
| 0.016400389 | 0.020119906 | 0.006551634 | 0 | 0.002491622 |
| 0 | 0 | 0 | 0 | 0 |
| 0 | 0 | 0 | 0 | 0 |
| 0 | 0 | 0 | 0 | 0 |
| 0 | 0 | 0 | 0 | 0 |
| 0 | 0 | 0 | 0 | 0 |
| 0 | 0 | 0 | 0 | 0 |

| | | | | |
|---|---|---|---|---|
| 0 | 0 | 0 | 0 | 0 |
| 0 | 0 | 0 | 0 | 0 |
| 0.001144077 | 0.028214376 | 0.174352006 | 0.000859039 | 0.103813992 |
| 0 | 0 | 0 | 0 | 0 |
| 1.829182149 | 0 | 0 | 0 | 0 |
| 0 | 0 | 0 | 0 | 0 |
| 0 | 0 | 0 | 0 | 0 |
| 0 | 0 | 0 | 0 | 0 |
| 0 | 0 | 0 | 0 | 0 |
| 0 | 0 | 0 | 0 | 0.002081041 |
| 0 | 0 | 0 | 0 | 0 |
| 0 | 0 | 0 | 0 | 0 |
| 0 | 0 | 0 | 0 | 0 |
| 0.001685882 | 0.002445645 | 0.004106037 | 0 | 0 |
| 0.001043606 | 0.019849175 | 0.003631067 | 0 | 0.002853889 |
| 0 | 0 | 0 | 0 | 0 |
| 0.001387278 | 0.004216608 | 0.011032734 | 0 | 0.000886841 |
| 0 | 0 | 0 | 0.001038889 | 0 |
| 0 | 0 | 0 | 0 | 0 |
| 0 | 0 | 0 | 0 | 0 |
| 0 | 0 | 0 | 0 | 0 |
| 0.001610121 | 0 | 0 | 0.013298692 | 0.005603952 |
| 0 | 0 | 0 | 0 | 0 |
| 0 | 0 | 0 | 0.04341856 | 0 |
| 0 | 0 | 0 | 0 | 0 |
| 0 | 0 | 0 | 0 | 0 |
| 1.12102735 | 0 | 0 | 0 | 0 |
| 0 | 0 | 0 | 0 | 0 |
| 0 | 0 | 0 | 0 | 0 |
| 0 | 0 | 0.003601595 | 0.003206113 | 0 |
| 0 | 0 | 0 | 0 | 0 |
| 0 | 0 | 0 | 0 | 0 |
| 0 | 0 | 0 | 0 | 0 |
| 0 | 0 | 0 | 0 | 0 |
| 0 | 0 | 0 | 0 | 0 |
| 0 | 0 | 0 | 0 | 0.010127088 |
| 0 | 0 | 0 | 0 | 0 |
| 0 | 0.015433916 | 0.01783098 | 0 | 0 |
| 0 | 0 | 0 | 0 | 0 |
| 0 | 0 | 0 | 0 | 0 |
| 0 | 0 | 0 | 0 | 0 |
| 0 | 0 | 0 | 0 | 0 |
| 0 | 0 | 0 | 0 | 0 |
| 0 | 0 | 0 | 0 | 0 |
| 0 | 0 | 0 | 0 | 0 |
| 0 | 0 | 0 | 0 | 0 |
| 0 | 0 | 0 | 0 | 0 |
| 0.001361278 | 0 | 0.006441446 | 0.013492066 | 0 |

| | | | | |
|---|---|---|---|---|
| 0 | 0 | 0 | 0 | 0 |
| 0 | 0 | 0 | 0 | 0 |
| 0.002061249 | 0 | 0 | 0 | 0 |
| 0 | 0 | 0 | 0 | 0 |
| 0.006221446 | 0 | 0.002623826 | 0.004459083 | 0 |
| 0.001844479 | 0 | 0 | 0.001731176 | 0 |
| 0 | 0 | 0 | 0 | 0 |
| 0.008456462 | 0.002555723 | 0.048180085 | 0.027382647 | 0 |
| 0 | 0 | 0 | 0 | 0 |
| 0 | 0.113535681 | 0.01023532 | 1.946834667 | 0.602125913 |
| 0 | 0 | 0 | 0.059634556 | 0 |
| 0 | 0 | 0.031152133 | 0 | 0.00811295 |
| 0.003014921 | 0.001688068 | 0.026500933 | 0.002859508 | 0.008757559 |
| 0 | 0 | 0 | 0 | 0.002516306 |
| 0 | 0 | 0 | 0 | 0 |
| 0 | 0.003685921 | 0.001657596 | 0 | 0 |
| 0 | 0 | 0 | 0 | 0 |
| 0 | 0 | 0 | 0 | 0 |
| 0 | 0.087748474 | 0 | 0.04618811 | 0 |
| 0 | 0 | 0 | 0 | 0 |
| 0 | 0 | 0 | 0 | 0 |
| 0 | 0 | 0 | 0 | 0 |
| 0 | 0 | 0.000911518 | 0 | 0 |
| 0.141352019 | 0 | 0 | 0 | 0 |
| 0 | 0 | 0 | 0 | 0 |
| 0 | 0.002144912 | 0.007202261 | 0 | 0.003308213 |
| 0 | 0 | 0 | 0 | 0 |
| 0 | 0 | 0.443253121 | 0.070735038 | 0 |
| 0 | 0 | 0 | 0 | 0 |
| 0 | 0 | 0 | 0 | 0 |
| 0 | 0 | 0 | 0 | 0 |
| 0 | 0 | 0 | 0 | 0 |
| 0 | 0 | 0 | 0 | 0 |
| 0 | 0 | 0 | 0 | 0 |
| 0 | 0 | 0 | 0 | 0 |
| 0 | 0 | 0 | 0.018364632 | 0 |
| 0 | 0.001215739 | 0.102347841 | 0.016990079 | 0 |
| 0 | 0 | 0 | 0 | 0 |
| 0 | 0 | 0 | 0 | 0 |
| 0 | 0 | 0 | 0.424074161 | 0 |
| 0 | 0 | 0 | 0 | 0 |
| 0 | 0 | 0 | 0 | 0.010036884 |
| 0 | 0 | 0 | 0 | 0 |
| 0 | 0 | 0.004144568 | 0.056012771 | 0 |
| 0 | 0 | 0 | 0 | 0 |
| 0 | 0 | 0 | 0 | 0 |
| 0.003447608 | 0.005779296 | 0.001279511 | 0.001035465 | 0 |
| 0 | 0 | 0 | 0 | 0 |

| | | | | |
|---|---|---|---|---|
| 0 | 0 | 0 | 0 | 0 |
| 0.001284314 | 0 | 0 | 0 | 0 |
| 0 | 0 | 0 | 0 | 0 |
| 0.001236819 | 0 | 0 | 1.472972075 | 0 |
| 0 | 0 | 0 | 0 | 0 |
| 0.000724109 | 0 | 0 | 0 | 0.086947852 |
| 0 | 0 | 0 | 0 | 0 |
| 0 | 0 | 0 | 0 | 0 |
| 0.873523844 | 0 | 0 | 0 | 0 |
| 0 | 0 | 0 | 0.003457626 | 0 |
| 0.004619145 | 0.027615495 | 0 | 0 | 0.000939548 |
| 0 | 0 | 0 | 0 | 0 |
| 0 | 0 | 0 | 0 | 0 |
| 0.002181968 | 0.001151018 | 0 | 0.001787288 | 0.00946814 |
| 0 | 0.315418701 | 0 | 0 | 0 |
| 0 | 0 | 0 | 0 | 0 |
| 0 | 0 | 0 | 0 | 0 |
| 0 | 0 | 0 | 0 | 0 |
| 0 | 0 | 0 | 0.00214008 | 0 |
| 0 | 0 | 0 | 0 | 0 |
| 0 | 0 | 0 | 0 | 0 |
| 0.349858573 | 0 | 0 | 0 | 2.339993475 |
| 0.005082341 | 0 | 0 | 0 | 0 |
| 0 | 0 | 0 | 0 | 0 |
| 0 | 0 | 0 | 0 | 0 |
| 0 | 0 | 0 | 0 | 0 |
| 0 | 0 | 0.003591946 | 0 | 0 |
| 0 | 0 | 0 | 0 | 0 |
| 0 | 0 | 0 | 0 | 0 |
| 0.254701631 | 0.069173176 | 6.16532736 | 0.085470653 | 2.262598327 |
| 0.00201858 | 0.004164661 | 0 | 0 | 0 |
| 0 | 0 | 0 | 0 | 0 |
| 0 | 0.105617437 | 0.001730449 | 0 | 0 |
| 0 | 0 | 0 | 0 | 0 |
| 0.001253964 | 0 | 0 | 0 | 0 |
| 0 | 0 | 0 | 0 | 0 |
| 0.143257046 | 0 | 0 | 0 | 0 |
| 0 | 0 | 0 | 0 | 0 |
| 0 | 0 | 0 | 0 | 0 |
| 0 | 1.450928956 | 0 | 0 | 0 |
| 0.000815371 | 0 | 0 | 0 | 0 |
| 1.487357767 | 0 | 0 | 0 | 4.988607303 |
| 0 | 0 | 0 | 0 | 0 |
| 0 | 0 | 0 | 0 | 0.001467257 |
| 0 | 0 | 0 | 0.022583412 | 0 |
| 0 | 0 | 0 | 0 | 0 |
| 0 | 0.002979509 | 0 | 0.008096463 | 0.003318941 |
| 0 | 0 | 0 | 0 | 0 |

| | | | | |
|---|---|---|---|---|
| 0 | 0.009803236 | 0.002645168 | 0 | 0 |
| 0 | 0 | 0 | 0 | 0 |
| 0.001150997 | 0.661204192 | 0.088103753 | 0.000972264 | 0 |
| 0.001337186 | 0 | 0 | 0 | 0 |
| 0 | 0 | 0 | 0 | 0 |
| 0 | 0 | 0 | 0 | 0 |
| 0 | 0 | 0.002506094 | 0.003549169 | 0 |
| 0 | 0 | 0 | 0 | 0 |
| 0 | 0 | 0 | 0 | 0 |
| 0 | 0 | 0 | 0 | 0 |
| 0 | 0 | 0 | 0 | 0 |
| 0 | 0 | 0 | 0 | 0 |
| 0 | 0 | 0 | 0 | 0 |
| 0 | 0.003510543 | 0 | 0 | 0 |
| 0 | 0 | 0 | 0 | 0 |
| 0.019259935 | 0 | 0.001454523 | 0 | 0.002004318 |
| 0.002647768 | 0.080981433 | 0.011745937 | 0.072130644 | 0.002715261 |
| 0 | 0 | 0 | 0.01517211 | 0 |
| 0 | 0 | 0 | 0.056408563 | 0 |
| 0 | 0 | 0 | 0 | 0 |
| 0.001877029 | 0 | 0 | 0.015268304 | 0.068440038 |
| 0 | 0.005754865 | 0.002404355 | 0.004107721 | 0 |
| 0 | 0 | 0 | 0 | 0 |
| 0 | 0 | 1.266858297 | 0 | 0 |
| 0.003711893 | 0 | 0 | 0 | 0 |
| 0.003051972 | 0 | 0 | 0.754508124 | 0 |
| 0 | 0 | 0 | 0 | 0 |
| 0 | 0 | 0 | 0 | 0 |
| 0 | 0 | 0 | 0 | 0 |
| 0 | 0.000869244 | 0 | 0 | 0 |
| 0.001278153 | 0 | 0 | 0.097530667 | 0.002144838 |
| 0.001429177 | 0.018659282 | 0.387862981 | 0 | 0.002487093 |
| 0.037194385 | 0.002193353 | 2.073434648 | 0.012516373 | 0.095060229 |
| 0 | 0 | 0.009046838 | 0 | 0.002424032 |
| 0 | 0 | 0.027442849 | 0 | 0.045329717 |
| 0 | 0 | 0 | 0 | 0 |
| 0 | 0.000991471 | 0.002853597 | 0 | 0.009786875 |
| 0.02822151 | 0 | 0.005579113 | 0.045872265 | 0.065535579 |
| 0.408696628 | 0 | 0 | 0 | 0 |
| 0.00065551 | 0 | 0 | 0 | 0 |
| 0 | 0 | 0 | 0 | 0 |
| 0 | 0 | 0 | 0 | 0 |
| 0 | 0 | 0.002336195 | 0 | 0 |
| 0.001752416 | 0 | 0 | 1.848672634 | 0.014189976 |
| 0 | 0 | 0 | 0 | 0 |
| 0.001573146 | 0.000676179 | 0 | 0 | 0 |
| 0 | 0 | 0 | 0 | 0 |
| 0 | 0 | 0 | 0 | 0 |

| | | | | |
|---|---|---|---|---|
| 0.003803921 | 0 | 0 | 0 | 0 |
| 0.465179085 | 0.032753009 | 1.76037676 | 0.008367049 | 0.075938051 |
| 0 | 0 | 0 | 0 | 0 |
| 0 | 0 | 0 | 0 | 0 |
| 0 | 0 | 0 | 0 | 0 |
| 0 | 0 | 0 | 0 | 0 |
| 0 | 0 | 0 | 0.004780965 | 0 |
| 0 | 0 | 0 | 0 | 0.001001346 |
| 0 | 0 | 0 | 0 | 0 |
| 0 | 0 | 0 | 0.006795485 | 0 |
| 0 | 0 | 0.008134995 | 0.021761714 | 0.009203221 |
| 0.013059898 | 0.007718536 | 0.156851794 | 0 | 0.740978251 |
| 0 | 0 | 0 | 0 | 0 |
| 0 | 0 | 0 | 0 | 0 |
| 0 | 0 | 0.002333472 | 0 | 0.011115311 |
| 0 | 0 | 0 | 0 | 0 |
| 0.004250537 | 0.027788524 | 0.023189288 | 0 | 0.001521647 |
| 0 | 0.004695938 | 0.076178766 | 0 | 0.00164609 |
| 0 | 0 | 0 | 0 | 0 |
| 0 | 0 | 0 | 0 | 0.01368173 |
| 0.030332961 | 0 | 0.105416845 | 0 | 0.021184232 |
| 0 | 0 | 0 | 0 | 0.028963869 |
| 0 | 0 | 0 | 0 | 0 |
| 0 | 0 | 0 | 0 | 0 |
| 0 | 0 | 0 | 0 | 0 |
| 0 | 0 | 0 | 0 | 0 |
| 0.042227724 | 0 | 0 | 0 | 0 |
| 0.037457407 | 0.106333145 | 0.022304766 | 0.782467762 | 0.123141485 |
| 0 | 0 | 0 | 0 | 0 |
| 0 | 0 | 0 | 0 | 0 |
| 0 | 0 | 0 | 0 | 0 |
| 0 | 0 | 0 | 0.00144855 | 0 |
| 0 | 0 | 0 | 0 | 0 |
| 0 | 0 | 0 | 0 | 0 |
| 0.401929198 | 0 | 0 | 0 | 0.0528068 |
| 0.318864507 | 0.07167998 | 0.04389475 | 0.102660119 | 0.253102425 |
| 0 | 0 | 0 | 0 | 0 |
| 0 | 0 | 0 | 0 | 0 |
| 0 | 0 | 0.022716222 | 0 | 0.00445782 |
| 0 | 0 | 0 | 0 | 0 |
| 0 | 0 | 0 | 0 | 0 |
| 0 | 0 | 0 | 0 | 0 |
| 0 | 0.003511189 | 0.00349813 | 0 | 0 |
| 0 | 0.002477298 | 0 | 0 | 0 |
| 0 | 0 | 0 | 0.0033913 | 0 |
| 0.776850295 | 0 | 0 | 0 | 0 |
| 0 | 0 | 0 | 0 | 0 |
| 0 | 0 | 0 | 0.014235787 | 0 |

| | | | | |
|---|---|---|---|---|
| 0 | 0 | 0 | 0.00483371 | 0 |
| 0 | 0 | 0 | 0 | 0 |
| 0 | 0 | 0 | 0 | 0 |
| 0.035617051 | 0 | 0 | 0.050922244 | 0 |
| 0 | 0 | 0 | 0 | 0 |
| 0 | 0 | 0 | 0 | 0 |
| 0 | 0 | 0 | 0 | 0.011660625 |
| 0.154951528 | 0 | 0.045560275 | 0 | 0 |
| 0.452945844 | 0 | 2.130442006 | 0 | 0 |
| 0.008866694 | 0 | 0 | 0 | 0 |
| 0 | 0 | 0 | 0 | 0 |
| 0.006388692 | 0 | 0.006722756 | 0 | 0.001626991 |
| 0 | 0 | 0 | 0 | 0 |
| 0 | 0 | 0 | 0 | 0 |
| 0.030345943 | 0 | 0.043307345 | 0 | 0.073075639 |
| 0 | 0 | 0 | 0 | 0 |
| 0.000670909 | 0 | 0 | 0 | 0 |
| 1.310889015 | 0 | 0 | 0 | 0.317084093 |
| 0 | 0 | 0 | 0 | 0 |
| 0.041189305 | 0 | 0 | 0 | 0 |
| 0 | 0 | 0 | 0 | 0 |
| 0.033418556 | 0 | 0 | 0 | 0 |
| 0.038167136 | 0 | 0 | 0 | 0 |
| 0 | 0 | 0 | 0 | 0 |
| 0 | 0 | 0 | 0 | 0 |
| 1.06525703 | 0 | 0 | 0 | 0 |
| 0.004166049 | 0 | 0 | 0 | 0 |
| 1.38122399 | 0 | 0 | 0 | 3.403833258 |
| 0 | 0 | 0 | 0 | 0 |
| 0 | 0 | 0 | 0 | 0 |
| 0.004090103 | 0 | 0 | 0 | 0 |
| 0 | 0 | 0 | 0 | 0 |
| 0 | 0 | 0 | 0 | 0 |
| 0 | 0 | 0 | 0 | 0 |
| 0 | 0 | 0 | 0 | 0 |
| 0.084258348 | 0 | 0 | 0 | 0.019733555 |
| 0.530995706 | 0 | 0 | 0 | 0 |
| 0 | 0 | 0 | 0 | 0 |
| 0 | 0 | 0 | 0 | 0.003387395 |
| 0 | 0 | 0 | 0 | 0 |
| 0.083710545 | 0.000433699 | 0.959070704 | 0.122061596 | 0 |
| 0.366375912 | 0 | 0 | 0 | 0 |
| 0.026539855 | 0.048559014 | 4.107442123 | 6.840302092 | 0.775700087 |
| 0.007422502 | 0 | 0 | 0 | 0 |
| 0 | 0 | 0 | 0 | 0 |
| 0.034489088 | 0 | 0 | 0 | 0 |
| 0 | 0 | 0 | 0 | 0 |
| 0 | 0 | 0 | 0.006678838 | 0 |

| | | | | |
|---|---|---|---|---|
| 0 | 0 | 0 | 0 | 0 |
| 0.062536461 | 0 | 0.003189742 | 0 | 0 |
| 0 | 0.548090421 | 1.345655601 | 0.250664442 | 0.025437564 |
| 0 | 0 | 0 | 0 | 0 |
| 0 | 0 | 0 | 0 | 0 |
| 0.004206663 | 0 | 0 | 0 | 0.017002561 |
| 0 | 0 | 0.029015424 | 0 | 0 |
| 0 | 0 | 0 | 0 | 0 |
| 0 | 0 | 0 | 0 | 0 |
| 0 | 0 | 0 | 0 | 0 |
| 0.002198658 | 0.079165276 | 0.006590676 | 0.092766727 | 0.018668314 |
| 0.376744358 | 1.108911665 | 0.12611794 | 6.725043701 | 2.915990666 |
| 0 | 0 | 0 | 0 | 0 |
| 0.006388527 | 0.169288055 | 0.03708883 | 0.001850223 | 0.00163359 |
| 0.024884343 | 0.030603968 | 0.016015025 | 1.627504276 | 0.016520752 |
| 0.540082178 | 0 | 0 | 0 | 0 |
| 0 | 0 | 0 | 0 | 0 |
| 0 | 0 | 0 | 0 | 0 |
| 0.001210679 | 0 | 0 | 0 | 0 |
| 0.007662413 | 0 | 0 | 0 | 0 |
| 0.019703778 | 0 | 0.663663986 | 0.020914857 | 0 |
| 0 | 30.7608309 | 0.027439675 | 7.043675203 | 0 |
| 0.005623376 | 31.50730954 | 0.586224666 | 2.486062265 | 10.29961026 |
| 0 | 0 | 0 | 0 | 0 |
| 0.020700956 | 0 | 0 | 0 | 0 |
| 0 | 0 | 0.4453882 | 0 | 0.449989331 |
| 0 | 0 | 0 | 0 | 0 |
| 0 | 0 | 0 | 0 | 0 |
| 0 | 0 | 0 | 0 | 0.003537267 |
| 0 | 0 | 0 | 0 | 0 |
| 0.009693748 | 0 | 0 | 0 | 0 |
| 0.341123627 | 0 | 0 | 0 | 0.132347121 |
| 0.172829915 | 0 | 0 | 0.058298985 | 0.019950123 |
| 0.003311462 | 0 | 0 | 0 | 0.004346718 |
| 0.004241795 | 0.001219595 | 0.000797766 | 0.00387363 | 0 |
| 0.032290087 | 0.28013803 | 0.240918749 | 0 | 0.095624409 |
| 0.000760775 | 0 | 0.00127056 | 0 | 0.053562069 |
| 0 | 0 | 0 | 0 | 0 |
| 0.076493452 | 0 | 0.089784582 | 0 | 0 |
| 0 | 0 | 0 | 0 | 0 |
| 0.018724202 | 0 | 0 | 0 | 0 |
| 0 | 0 | 0 | 0 | 0 |
| 0.638095406 | 0 | 0 | 0 | 0 |
| 0.531279324 | 0 | 0 | 0 | 0 |
| 0.037721305 | 0 | 0.001499948 | 0.009103928 | 0.051979027 |
| 0.271147028 | 0 | 0 | 0 | 0 |
| 0 | 0 | 0 | 0 | 0 |
| 0.157090934 | 0 | 0.961064948 | 0.218660341 | 0.026502447 |

| | | | | |
|---|---|---|---|---|
| 0 | 0 | 0 | 0 | 0 |
| 0 | 0 | 0 | 0 | 0 |
| 0 | 0 | 0 | 0 | 0 |
| 0.003521159 | 0 | 0 | 0 | 0 |
| 0 | 0 | 0 | 0 | 0 |
| 0.367099348 | 0 | 0 | 0 | 0.009680862 |
| 0.167943864 | 0 | 0 | 0 | 0 |
| 0.004226222 | 0 | 0 | 0.109161234 | 0.056525237 |
| 0 | 0 | 0 | 0 | 0 |
| 0 | 0 | 0 | 0 | 0 |
| 0.009889351 | 0 | 0.000949199 | 0.043976854 | 0.015768503 |
| 0 | 0 | 0 | 0 | 0 |
| 0.017467091 | 0 | 0 | 0 | 0 |
| 0.012512802 | 0 | 1.120605734 | 0 | 0 |
| 0 | 0 | 0 | 0 | 0 |
| 0 | 0 | 0 | 0 | 0 |
| 0.114637691 | 0 | 0 | 0 | 0.030453968 |
| 0.004258272 | 0 | 0 | 0 | 0.005293113 |
| 0.048542338 | 0 | 0 | 0 | 0 |
| 0.008448563 | 0 | 0 | 0 | 0 |
| 0.005800813 | 0 | 0 | 0 | 0 |
| 0 | 0 | 0 | 0 | 0 |
| 0.001800144 | 0 | 0 | 0 | 0 |
| 0 | 0 | 0 | 0 | 0 |
| 0 | 0 | 0 | 0 | 0 |
| 0 | 0 | 0 | 0 | 0 |
| 0 | 0 | 0 | 0 | 0 |
| 0 | 0 | 0 | 0 | 0 |
| 0 | 0 | 0 | 0 | 0 |
| 0.390083198 | 0 | 0 | 0 | 0 |
| 0 | 0 | 0 | 0 | 0 |
| 0 | 0 | 0 | 0 | 0 |
| 0 | 0 | 0 | 0 | 0 |
| 0 | 0 | 0 | 0 | 0 |
| 0 | 0 | 0 | 0 | 0 |
| 0 | 0 | 0 | 0 | 0 |
| 0.03956108 | 0 | 0 | 0 | 0 |
| 0 | 0 | 0.002410541 | 0.039828202 | 0 |
| 0.014698935 | 0 | 0 | 0 | 0 |
| 0.000569462 | 0 | 0 | 0 | 0 |
| 0.005416929 | 0 | 0 | 0 | 0.002424004 |
| 0 | 0 | 0 | 0 | 0 |
| 0.001138994 | 0 | 0 | 0 | 0 |
| 3.974595228 | 0 | 0 | 0 | 0.474605539 |
| 0.17241484 | 0 | 0 | 0 | 0.069428255 |
| 0 | 0 | 0 | 0 | 0 |
| 0.004662886 | 0 | 0 | 0 | 0 |
| 0.010798676 | 0 | 0 | 0 | 0 |

| | | | | |
|---|---|---|---|---|
| 0 | 0 | 0 | 0 | 0 |
| 0.298715629 | 0 | 0 | 0 | 0 |
| 2.053209288 | 0 | 0 | 0 | 0 |
| 0.450817224 | 0 | 0 | 0 | 0 |
| 0.000744554 | 0 | 0 | 0 | 0 |
| 1.176911109 | 0 | 0 | 0 | 0.281258914 |
| 0 | 0 | 0 | 0 | 0 |
| 0 | 0 | 0 | 0 | 0 |
| 0.029461462 | 0.005390457 | 0.793179516 | 1.690042443 | 0.027317403 |
| 0 | 0 | 0 | 0 | 0 |
| 0.022773906 | 0 | 0 | 0 | 0 |
| 0 | 0 | 0 | 0 | 0 |
| 0 | 0 | 0 | 0 | 0 |
| 0 | 0 | 0 | 0 | 0 |
| 0.29082593 | 0 | 0.574180661 | 0 | 0.211886848 |
| 0 | 0 | 0 | 0 | 0 |
| 0 | 0 | 0 | 0 | 0 |
| 0 | 0 | 0 | 0 | 0 |
| 0 | 0 | 0 | 0 | 0 |
| 0.240286861 | 0 | 0 | 0 | 0 |
| 0.019331041 | 0 | 0 | 0 | 0 |
| 0 | 0 | 0 | 0 | 0 |
| 0 | 0 | 0 | 0 | 0 |
| 0 | 0 | 0 | 0 | 0 |
| 0.041200459 | 0.037354964 | 0.009122313 | 0.013973788 | 0.006634405 |
| 6.687465587 | 0.001280313 | 0.099492999 | 0.001988057 | 8.794636987 |
| 0.023334471 | 0 | 0 | 0 | 0 |
| 0.003051568 | 0 | 0 | 0 | 0 |
| 0 | 0 | 0.041871429 | 0.285142194 | 0.087310793 |
| 0 | 0 | 0 | 0 | 0 |
| 0 | 0 | 0 | 0 | 0 |
| 0.086534816 | 0 | 0 | 0 | 0 |
| 0.002493921 | 0 | 0 | 0 | 0 |
| 0 | 0 | 0.002783943 | 0 | 0 |
| 0 | 0.139254418 | 0.041570225 | 0.005336218 | 0 |
| 0.003494725 | 0.000422472 | 0 | 0.026240411 | 0.003475208 |
| 0.049889355 | 0 | 0 | 0 | 0 |
| 0.076949895 | 0.012562158 | 0.56338825 | 0 | 0.607991784 |
| 0 | 0 | 0 | 8.440662725 | 0 |
| 0 | 0 | 0 | 0 | 0 |
| 0 | 0 | 0 | 0 | 0 |
| 0 | 0 | 0 | 0 | 0 |
| 0.03760788 | 0.091408304 | 3.653022345 | 0.100626481 | 0.207469009 |
| 0.443919034 | 0 | 0 | 0 | 0 |
| 0.093046768 | 0 | 0 | 0 | 0 |
| 0 | 0 | 0 | 0 | 0.001234621 |
| 0 | 0 | 0 | 0 | 0 |
| 0.42218155 | 0 | 0.017740156 | 0.011138669 | 0.026880958 |

| | | | | |
|---|---|---|---|---|
| 0 | 0 | 0 | 0 | 0.003422344 |
| 0.25230049 | 0 | 0 | 0 | 0 |
| 0 | 0 | 0 | 0 | 0 |
| 0 | 0 | 0 | 0 | 0 |
| 0 | 0 | 0 | 0.001286398 | 0.016063176 |
| 0.043209648 | 0 | 0 | 0 | 0 |
| 0 | 0 | 0 | 0 | 0 |
| 0 | 0 | 0 | 0 | 0 |
| 0 | 0 | 0 | 0 | 0 |
| 0 | 0 | 0 | 0 | 0 |
| 0.001123653 | 0 | 0 | 0 | 0 |
| 0.396798746 | 0 | 0.001256995 | 0 | 0 |
| 0.094436009 | 0 | 0 | 0 | 0.00648323 |
| 0 | 0 | 0 | 0 | 0 |
| 0.00584973 | 0.001277459 | 0.26603642 | 0.007226066 | 0.108929137 |
| 0 | 0 | 0 | 0 | 0 |
| 0 | 0 | 0 | 0 | 0 |
| 0 | 0 | 0 | 0 | 0 |
| 0 | 0 | 0 | 0 | 0 |
| 0.005318526 | 0 | 0 | 0 | 0 |
| 0 | 0 | 0 | 0 | 0 |
| 0 | 0 | 0 | 0 | 0 |
| 0.026857751 | 0 | 0 | 0 | 0.109149957 |
| 0 | 0 | 0 | 0 | 0 |
| 0 | 0 | 0 | 0 | 0 |
| 0 | 0 | 0 | 0 | 0 |
| 0 | 0 | 0 | 0 | 0 |
| 0 | 0 | 0 | 0 | 0 |
| 0 | 0 | 0 | 0 | 0 |
| 0 | 0 | 0 | 0 | 0 |
| 0 | 0 | 0 | 0 | 0 |
| 0.060481871 | 0.033864081 | 0.139382782 | 0.001378946 | 0.039446732 |
| 0.380344304 | 0 | 0 | 0 | 0 |
| 0.010398115 | 0 | 0 | 0.011435698 | 0.004379313 |
| 0.00237696 | 0 | 0 | 0.025700528 | 0.004963739 |
| 0.059445286 | 0 | 0.043782717 | 0 | 0.015235385 |
| 0 | 0 | 0 | 0 | 0 |
| 0 | 0 | 0 | 0 | 0 |
| 0 | 0 | 0 | 0.001230255 | 0 |
| 0 | 0 | 0 | 0 | 0 |
| 0.001350119 | 0 | 0 | 0 | 0 |
| 0.012669523 | 0 | 0 | 0 | 0 |
| 0 | 0 | 0 | 0 | 0 |
| 0.012384413 | 0 | 0 | 0 | 0 |
| 0.032469412 | 0.011171662 | 0.020204662 | 0 | 0.019093429 |
| 0 | 0 | 0 | 0 | 0 |
| 0.452734114 | 0 | 0 | 0 | 0 |
| 0.000590582 | 0 | 0 | 0 | 0 |

| | | | | |
|---|---|---|---|---|
| 0 | 0 | 0 | 0 | 0 |
| 0 | 0 | 0 | 0 | 0 |
| 0 | 0 | 0 | 0 | 0 |
| 0 | 0 | 0 | 0 | 0 |
| 0 | 0 | 0 | 0 | 0 |
| 0 | 0 | 0 | 0 | 0 |
| 0.098457024 | 0 | 0 | 0 | 0 |
| 0 | 0 | 0 | 0 | 0 |
| 0 | 0 | 0 | 0 | 0 |
| 0 | 0 | 0 | 0 | 0 |
| 0 | 0 | 0 | 0.055404918 | 0 |
| 0 | 0 | 0 | 0 | 0 |
| 0 | 0 | 0 | 0 | 0 |
| 0.241816472 | 0 | 0 | 0 | 0 |
| 0 | 0 | 0 | 0 | 0 |
| 0 | 0 | 0 | 0 | 0 |
| 0.492497234 | 0 | 0 | 0 | 0 |
| 0.043798508 | 0 | 0 | 0 | 0 |
| 0.012938557 | 0 | 0 | 0 | 0 |
| 0 | 0 | 0 | 0 | 0 |
| 0 | 0 | 0 | 0 | 0 |
| 0 | 0 | 0 | 0 | 0 |
| 0 | 0 | 0 | 0 | 0 |
| 0 | 0 | 0 | 0 | 0 |
| 0.00255834 | 0 | 0 | 0 | 0 |
| 0 | 0.07744078 | 0.055822563 | 0.152712177 | 0.015607546 |
| 0 | 0 | 0 | 0 | 0 |
| 0.014366764 | 0 | 0.486803471 | 0.002883759 | 0.000848705 |
| 0.000602193 | 0 | 0 | 0 | 0 |
| 0.890039547 | 0 | 0.138370557 | 0 | 0.152053326 |
| 0 | 0 | 0 | 0 | 0 |
| 0 | 0 | 0 | 0 | 0 |
| 0.001652185 | 0 | 0 | 0 | 0 |
| 0 | 0 | 0 | 0 | 0 |
| 0.001218341 | 0 | 0 | 0.136877027 | 0.002044468 |
| 0 | 0 | 0 | 0 | 0 |
| 0 | 0 | 0 | 0 | 0 |
| 0 | 0 | 0.000962011 | 0 | 0 |
| 0 | 0 | 0 | 0 | 0 |
| 0.009303967 | 0 | 0 | 0 | 0 |
| 5.732950063 | 2.43374879 | 4.015809882 | 1.359896282 | 9.36529393 |
| 0.091077021 | 0 | 0 | 0 | 0.001698157 |
| 0 | 0 | 0 | 0 | 0 |
| 0.211032496 | 0 | 0 | 0 | 0 |
| 0 | 0 | 0 | 0 | 0 |
| 0 | 0 | 0 | 0 | 0 |
| 0 | 0 | 0 | 0 | 0 |
| 0.001234346 | 0 | 0.011834332 | 0 | 0.004909811 |

| | | | | |
|---|---|---|---|---|
| 0 | 0 | 0 | 0 | 0 |
| 0.016081942 | 0 | 0 | 0 | 0.025671573 |
| 0.001482853 | 0 | 0 | 0 | 0 |
| 0 | 0 | 0 | 0 | 0 |
| 0.60657486 | 0.060090103 | 15.07023728 | 0.062028143 | 0.742590527 |
| 0.00072328 | 0 | 0 | 0 | 0 |
| 0 | 0 | 0 | 0 | 0 |
| 0 | 0 | 0 | 0 | 0 |
| 0.00253551 | 0.110753743 | 0.42835306 | 0.078373405 | 0.014917987 |
| 0 | 0 | 0 | 0 | 0 |
| 0 | 0.465493887 | 0 | 0.263695512 | 0 |
| 0 | 0 | 0 | 0 | 0 |
| 0.13433176 | 0 | 0 | 0 | 0.038847693 |
| 0.00977402 | 0 | 0.283315104 | 0.004365379 | 0.072644391 |
| 0 | 0 | 0 | 0 | 0 |
| 0.309065443 | 0 | 0 | 0 | 0.010932206 |
| 0.003353994 | 0 | 0.000811806 | 0 | 0 |
| 0 | 0 | 0 | 0 | 0 |
| 0 | 0 | 0 | 0 | 0 |
| 0.095232328 | 0 | 0 | 0 | 0 |
| 0 | 0 | 0 | 0 | 0 |
| 0 | 0 | 0 | 0 | 0 |
| 0 | 0 | 0 | 0 | 0 |
| 0 | 0 | 0 | 0 | 0 |
| 0 | 0 | 0 | 0 | 0 |
| 0 | 0 | 0 | 0 | 0 |
| 0.066063611 | 0 | 0 | 0 | 0.004350222 |
| 0 | 0 | 0.000796527 | 0.002094958 | 0 |
| 0 | 0 | 0 | 0 | 0 |
| 0 | 0 | 0 | 0 | 0 |
| 0.196228745 | 0 | 0 | 0 | 0 |
| 0 | 0 | 0 | 0 | 0 |
| 0.116289379 | 0 | 0 | 0 | 0 |
| 0.030747432 | 0 | 0 | 0 | 0 |
| 0.263431887 | 0 | 0 | 0 | 0 |
| 0 | 0 | 0 | 0.025912078 | 0 |
| 0 | 0 | 0.005415343 | 0 | 0 |
| 0 | 0 | 0 | 0.005104907 | 0 |
| 0 | 0 | 0 | 0 | 0 |
| 0 | 0 | 0 | 0 | 0 |
| 0.329207281 | 0 | 2.760871387 | 0 | 0.678221707 |
| 0 | 0 | 0 | 0 | 0 |
| 0.054363165 | 0 | 0 | 0 | 0 |
| 0 | 0 | 0 | 0 | 0 |
| 0 | 0 | 0 | 0 | 0 |
| 0.001137968 | 0 | 0 | 0 | 0 |
| 0 | 0 | 0 | 0 | 0 |
| 0 | 0 | 0 | 0 | 0 |

| | | | | |
|---|---|---|---|---|
| 0.04449164 | 0 | 0 | 0 | 0 |
| 0 | 0 | 0 | 0 | 0 |
| 0.021725856 | 0 | 0 | 0 | 0 |
| 0 | 0 | 0 | 0 | 0 |
| 0 | 0 | 0 | 0 | 0 |
| 0 | 0 | 0 | 0 | 0 |
| 0.012510527 | 0 | 0.004251616 | 0.013762754 | 0 |
| 0 | 0 | 0 | 0 | 0 |
| 0 | 0 | 0 | 0 | 0 |
| 0 | 0 | 0 | 0 | 0 |
| 0.007585531 | 0 | 0 | 0 | 0 |
| 0.01106791 | 0 | 0 | 0 | 0 |
| 0 | 0 | 0 | 0 | 0 |
| 0 | 0 | 0 | 0 | 0 |
| 0 | 0 | 0 | 0 | 0 |
| 0 | 0 | 0 | 0 | 0 |
| 0 | 0 | 0 | 0 | 0 |
| 0 | 0 | 0 | 0 | 0 |
| 0 | 0 | 0 | 0 | 0 |
| 0 | 0 | 0 | 0 | 0 |
| 0.00041549 | 0 | 0.004047775 | 0 | 0.001652679 |
| 0 | 0 | 0 | 0 | 0 |
| 0.022914212 | 0 | 0 | 0 | 0.000683587 |
| 0.003260089 | 0 | 0.000756198 | 0 | 0 |
| 0 | 0 | 0 | 0 | 0 |
| 0 | 0 | 0 | 0 | 0 |
| 0 | 0 | 0 | 0 | 0 |
| 0 | 0 | 0 | 0 | 0 |
| 0 | 0 | 0 | 0 | 0 |
| 0.003128021 | 0.010777042 | 0.550975525 | 0 | 0.361893272 |
| 0.049218326 | 0 | 0 | 0 | 0 |
| 0 | 0 | 0 | 0 | 0 |
| 0 | 0 | 0 | 0 | 0 |
| 0 | 0 | 0 | 0 | 1.119567694 |
| 0 | 0 | 0 | 0 | 0 |
| 0.055828538 | 0 | 0 | 0 | 0 |
| 0.217728118 | 0 | 0 | 0.043837902 | 0 |
| 0 | 0 | 0 | 0 | 0 |
| 0 | 0 | 0 | 0 | 0 |
| 0 | 0 | 0 | 0 | 0 |
| 0 | 0 | 0 | 0 | 0 |
| 0 | 0 | 0 | 0 | 0 |
| 0 | 0 | 0 | 0 | 0 |
| 0.023030967 | 0 | 0 | 0 | 0 |
| 0.042628225 | 0 | 0 | 0 | 0 |
| 0 | 0 | 0 | 0 | 0 |
| 0.003327772 | 0 | 0 | 0 | 0 |
| 0.00437414 | 0 | 0 | 0 | 0 |

| | | | | |
|---|---|---|---|---|
| 0 | 0 | 0.002441089 | 0 | 0 |
| 0 | 0 | 0 | 0 | 0 |
| 0 | 0 | 0 | 0 | 0 |
| 0 | 0 | 0 | 0 | 0 |
| 0 | 0 | 0 | 0 | 0 |
| 0 | 0 | 0 | 0 | 0 |
| 0 | 0 | 0 | 0 | 0 |
| 0.715576189 | 0.320764944 | 0 | 0.536507072 | 0.812391945 |
| 1.046424082 | 1.042403011 | 0.228622113 | 0.591999905 | 0.032853216 |
| 0 | 0 | 0.001556032 | 0.519168315 | 0.198695492 |
| 0.011433241 | 0 | 0 | 0 | 0 |
| 0 | 0 | 0 | 0 | 0 |
| 0.043018503 | 0 | 0 | 0 | 0 |
| 0 | 0 | 0.007091604 | 0.365573966 | 0 |
| 0 | 0 | 0 | 0 | 0 |
| 0 | 0 | 0 | 0 | 0 |
| 0 | 0 | 0 | 0 | 0 |
| 0 | 0 | 0 | 0 | 0 |
| 0 | 0 | 0 | 0 | 0 |
| 0 | 0 | 0 | 0 | 0 |
| 0 | 0 | 0 | 0 | 0 |
| 0 | 0 | 0 | 0 | 0 |
| 0 | 0 | 0 | 0 | 0 |
| 0 | 0 | 0 | 0 | 0 |
| 0.03350154 | 0 | 0 | 0 | 0 |
| 0 | 0 | 0 | 0 | 0 |
| 0 | 0 | 0 | 0 | 0 |
| 0 | 0 | 0 | 0 | 0 |
| 0 | 0 | 0 | 0 | 0 |
| 0 | 0 | 0 | 0 | 0 |
| 0.002102238 | 0 | 0 | 0 | 0 |
| 0 | 0 | 0 | 0 | 0 |
| 0 | 0 | 0 | 0 | 0 |
| 0.005113809 | 0 | 0 | 0 | 0 |
| 0.017518433 | 0 | 0 | 0 | 0.001055794 |
| 0.000777085 | 0 | 0 | 0 | 0 |
| 0 | 0 | 0 | 0 | 0 |
| 0 | 0.064340646 | 0 | 0 | 0 |
| 0.000695209 | 0 | 0 | 0.002544763 | 0.001296236 |
| 0 | 0 | 0.508311557 | 0.010682523 | 0 |
| 0 | 0 | 0 | 0 | 0 |
| 0.413837902 | 0 | 0 | 0 | 0.105314306 |
| 0 | 0 | 0 | 0 | 0 |
| 0 | 0 | 0 | 0 | 0 |
| 0 | 0 | 0 | 0 | 0 |
| 0.020145474 | 0 | 0 | 0 | 0 |
| 0 | 0 | 0 | 0 | 0 |
| 0 | 0 | 0 | 0 | 0 |

| | | | | |
|---|---|---|---|---|
| 0 | 0 | 0 | 0 | 0 |
| 0 | 0.001530979 | 0 | 0.009211993 | 0 |
| 0 | 0 | 0 | 0 | 0 |
| 0 | 0 | 0 | 0 | 0 |
| 0.305438804 | 0 | 0.525716234 | 0 | 0.83168914 |
| 0.173113136 | 0 | 0 | 0 | 0 |
| 0 | 0 | 0 | 0 | 0 |
| 0.058011114 | 0.011867945 | 0.098073885 | 0.193079617 | 0.044652059 |
| 0.000556942 | 0 | 0.002066979 | 0 | 0 |
| 0.057125881 | 0 | 0 | 0 | 0 |
| 0 | 0 | 0 | 0 | 0 |
| 0.21682671 | 0 | 0 | 0 | 0 |
| 0 | 0 | 0 | 0 | 0 |
| 0 | 0 | 0 | 0 | 0 |
| 0 | 0 | 0 | 0.00058968 | 0 |
| 0 | 0 | 0 | 0 | 0.017078101 |
| 0 | 0 | 0 | 0 | 0 |
| 0 | 0 | 0 | 0 | 0 |
| 0 | 0 | 0 | 0 | 0 |
| 0 | 0 | 0 | 0 | 0 |
| 0 | 0 | 0 | 0 | 0 |
| 0.002522359 | 0 | 0 | 0 | 0 |
| 0 | 0 | 0 | 0 | 0 |
| 0.001287278 | 0 | 0 | 0.012323673 | 0 |
| 0.050680259 | 0 | 0 | 0 | 0 |
| 0 | 0 | 0 | 0 | 0.001093765 |
| 0.010890597 | 0.962289817 | 0.064928711 | 0 | 0.054661094 |
| 0 | 0 | 0 | 0 | 0 |
| 0 | 0 | 0 | 0 | 0 |
| 0 | 0 | 0 | 0 | 0 |
| 0 | 0 | 0 | 0 | 0 |
| 0 | 0 | 0 | 0 | 0 |
| 0 | 0 | 0 | 0 | 0 |
| 0 | 0 | 0 | 0 | 0 |
| 0 | 0 | 0 | 0 | 0 |
| 0 | 0 | 0 | 0 | 0 |
| 0 | 0 | 0 | 0 | 0 |
| 0.17025095 | 0 | 0 | 0 | 0 |
| 1.106862776 | 0 | 0 | 0 | 0 |
| 0 | 0 | 0 | 0 | 0.001351994 |
| 0 | 0 | 0 | 0 | 0 |
| 0 | 0 | 0 | 0 | 0 |
| 0 | 0 | 0 | 0 | 0 |
| 0.042855433 | 0 | 0 | 0 | 0 |
| 0 | 0 | 0 | 0 | 0 |
| 0 | 0 | 0 | 0 | 0 |
| 0 | 0 | 0 | 0 | 0 |
| 0 | 0 | 0 | 0 | 0 |

| | | | | |
|---|---|---|---|---|
| 0.083870955 | 0 | 0 | 0 | 0 |
| 0 | 0 | 0 | 0 | 0 |
| 0 | 0 | 0 | 0 | 0 |
| 0 | 0 | 0 | 0 | 0 |
| 0 | 0 | 0 | 0 | 0 |
| 0.978748748 | 0 | 0 | 0.028714596 | 0 |
| 0.006039804 | 0 | 0 | 0 | 0 |
| 0 | 0 | 0 | 0 | 0 |
| 0 | 0 | 0 | 0 | 0 |
| 0 | 0 | 0 | 0 | 0 |
| 0 | 0 | 0 | 0 | 0 |
| 0 | 0 | 0 | 0 | 0 |
| 0 | 0 | 0 | 0 | 0 |
| 0 | 0 | 0 | 0 | 0 |
| 0 | 0 | 0 | 0 | 0 |
| 0 | 0 | 0 | 0 | 0 |
| 0 | 0 | 0 | 0 | 0 |
| 0 | 0 | 0 | 0 | 0 |
| 0 | 0 | 0 | 0 | 0 |
| 0 | 0 | 0 | 0 | 0 |
| 0 | 0 | 0.440217985 | 0 | 0 |
| 0 | 0 | 0 | 0 | 0 |
| 0.008701815 | 0 | 0.014363792 | 0 | 0.018715097 |
| 0 | 0 | 0 | 0 | 0 |
| 0 | 0 | 0 | 0 | 0 |
| 0.035362946 | 0 | 0.211974019 | 0 | 0 |
| 0 | 0 | 0 | 0 | 0 |
| 0 | 0 | 0 | 0 | 0 |
| 0 | 0 | 0 | 0.003354269 | 0 |
| 0 | 0 | 0 | 0 | 0 |
| 0 | 0 | 0 | 0 | 0 |
| 0.003866132 | 0 | 0 | 0 | 0 |
| 0.010802446 | 0.007148038 | 0.961659827 | 0.019210499 | 0.250743612 |
| 0 | 0 | 0 | 0 | 0 |
| 0.288864971 | 0 | 0 | 0 | 0.005057248 |
| 0 | 0 | 0 | 0.00513854 | 0.010434856 |
| 0 | 0 | 0 | 0 | 0 |
| 0.007208532 | 0 | 0 | 0 | 0 |
| 0.009674696 | 0 | 0.012485398 | 0.008915302 | 0.025188649 |
| 0 | 0 | 0 | 0 | 0 |
| 0 | 0 | 0 | 0 | 0 |
| 0.553682067 | 0 | 0 | 0 | 0 |
| 0.044016371 | 0 | 0.222708922 | 0 | 0 |
| 0 | 0 | 0 | 0 | 0 |
| 0 | 0 | 0 | 0 | 0 |
| 0.01437469 | 0 | 0 | 0 | 0 |
| 0 | 0 | 0 | 0 | 0 |
| 0 | 0 | 0 | 0 | 0 |

| | | | | |
|---|---|---|---|---|
| 0 | 0 | 0 | 0 | 0 |
| 0 | 0 | 0 | 0 | 0 |
| 0 | 0 | 0 | 0 | 0 |
| 0 | 0 | 0 | 0 | 0 |
| 0 | 0 | 0 | 0 | 0 |
| 0 | 0 | 0 | 0 | 0 |
| 0 | 0 | 0 | 0 | 0 |
| 0 | 0 | 0 | 0 | 0 |
| 0.038413701 | 0 | 0 | 0 | 0 |
| 0.00386969 | 0 | 0 | 0 | 0 |
| 0 | 0 | 0 | 0 | 0 |
| 0 | 0 | 0 | 0 | 0 |
| 0 | 0 | 0 | 0 | 0 |
| 0 | 0 | 0 | 0 | 0 |
| 0 | 0 | 0 | 0 | 0 |
| 0 | 0.05859368 | 0 | 0.013423819 | 0 |
| 0 | 0 | 0 | 0 | 0 |
| 0 | 0 | 0 | 0 | 0 |
| 0 | 0 | 0 | 0 | 0 |
| 0.161460482 | 0 | 0 | 0 | 0.008062392 |
| 4.409943407 | 0 | 0.004812394 | 0 | 0 |
| 0 | 0 | 0 | 0 | 0 |
| 0 | 0 | 0 | 0 | 0 |
| 0.016537387 | 0 | 0 | 0 | 0 |
| 0 | 0 | 0 | 0 | 0 |
| 0.285597215 | 0 | 0 | 0 | 0 |
| 0 | 0 | 0 | 0 | 0 |
| 0 | 0 | 0 | 0 | 0 |
| 0 | 0 | 0 | 0 | 0 |
| 0.157669378 | 0 | 0 | 0 | 0 |
| 0.155801612 | 0 | 0 | 0.01303199 | 0.646016141 |
| 0 | 0 | 0 | 0 | 0 |
| 0.001193161 | 0 | 0 | 0 | 0 |
| 0 | 0 | 0 | 0 | 0 |
| 0 | 0 | 0 | 0 | 0 |
| 0 | 0 | 0 | 0 | 0 |
| 0 | 0 | 0 | 0 | 0 |
| 0 | 0 | 0 | 0 | 0 |
| 0.001819632 | 0 | 0 | 0 | 0 |
| 0 | 0 | 0 | 0 | 0 |
| 0 | 0 | 0 | 0 | 0 |
| 0.02833729 | 0 | 0 | 0 | 0 |
| 0.001891771 | 0 | 0 | 0 | 0 |
| 0 | 0 | 0 | 0 | 0 |
| 0 | 0 | 0 | 0 | 0 |
| 0 | 0 | 0 | 0 | 0 |
| 0 | 0.199775857 | 0.424752141 | 7.47252114 | 0 |
| 0.001652629 | 0 | 0 | 0 | 0 |

| | | | | |
|---|---|---|---|---|
| 0 | 0 | 0 | 0 | 0 |
| 0 | 0 | 0 | 0 | 0 |
| 0 | 0 | 0 | 0 | 0 |
| 0.003796837 | 0 | 0 | 0 | 0 |
| 0.011647056 | 0 | 0 | 0 | 1.299388112 |
| 0.017951977 | 0 | 0 | 0 | 0 |
| 0 | 0 | 0 | 0 | 0 |
| 0 | 0 | 0 | 0 | 0 |
| 0 | 0 | 0 | 0 | 0 |
| 0 | 0 | 0 | 0 | 0 |
| 0.022342306 | 0 | 0 | 0 | 0 |
| 0.007909358 | 0.03484637 | 0.003913869 | 0 | 0.005767803 |
| 0 | 0.005572772 | 0.002743554 | 0 | 0 |
| 0 | 0 | 0 | 0 | 0 |
| 0.054917065 | 0 | 0 | 0 | 0 |
| 0.278009231 | 0 | 0 | 0 | 4.511584159 |
| 0.054749112 | 0 | 0 | 0 | 0.009678817 |
| 0 | 0 | 0 | 0 | 0 |
| 0.004258852 | 0 | 0.02833946 | 0.005845963 | 0 |
| 0.001042643 | 0 | 0.077633116 | 0.004892979 | 0 |
| 0 | 0 | 0 | 0 | 0 |
| 0 | 0 | 0 | 0 | 0 |
| 0 | 0 | 0 | 0 | 0 |
| 0 | 0 | 0 | 0 | 0 |
| 0 | 0 | 0 | 0 | 0 |
| 0 | 0 | 0 | 0 | 0 |
| 0.000696265 | 0 | 0 | 0.010978711 | 0 |
| 0 | 0 | 0 | 0 | 0 |
| 0 | 0 | 0 | 0 | 0 |
| 0 | 0 | 0 | 0 | 0 |
| 0.002525609 | 0 | 0 | 0 | 0.001648174 |
| 0 | 0 | 0 | 0.035909317 | 0 |
| 0 | 0 | 0 | 0 | 0 |
| 0 | 0 | 0 | 0 | 0 |
| 0 | 0 | 0 | 0 | 0 |
| 0 | 0 | 0 | 0 | 0 |
| 0 | 0 | 0 | 0 | 0 |
| 0.009537503 | 0 | 0.014817163 | 0.001748691 | 0.003143032 |
| 0.004535308 | 0 | 0 | 0 | 0 |
| 0.000527754 | 0 | 0 | 0 | 0 |
| 0.031775247 | 0 | 0 | 0.001621061 | 0.001486307 |
| 0.000568677 | 0 | 0 | 0 | 0 |
| 0 | 0 | 0 | 0 | 0 |
| 0 | 0 | 0 | 0.198556428 | 0 |
| 0 | 0 | 0 | 0 | 0 |
| 0 | 0 | 0 | 0 | 0 |
| 0 | 0 | 0 | 0 | 0 |
| 0.001114906 | 0 | 0 | 0 | 0 |

| | | | | |
|---|---|---|---|---|
| 0 | 0 | 0 | 0 | 0 |
| 0.131562823 | 0 | 0 | 0 | 0 |
| 0 | 0 | 0 | 0 | 0 |
| 0.000861734 | 0 | 0 | 0.005241023 | 0.001542459 |
| 0 | 0.000772956 | 0.002447147 | 0.003420679 | 0 |
| 0 | 0 | 0 | 0.005576482 | 0.016001567 |
| 0.004682988 | 0 | 0 | 0 | 0 |
| 0 | 0 | 0 | 0 | 0 |
| 0 | 0 | 0 | 0 | 0 |
| 0.012746037 | 0 | 0 | 0 | 0 |
| 0 | 0 | 0 | 0 | 0 |
| 0 | 0 | 0 | 0 | 0 |
| 0.057786257 | 0 | 0.040126658 | 0 | 0.013849562 |
| 0 | 0 | 0 | 0 | 0 |
| 0 | 0 | 0 | 0 | 0 |
| 0.037333163 | 0 | 0 | 0 | 0 |
| 0 | 0 | 0 | 0 | 0 |
| 0 | 0 | 0 | 0 | 0 |
| 0 | 0 | 0 | 0 | 0 |
| 0 | 0 | 0 | 0 | 0 |
| 0 | 0 | 0 | 0 | 0 |
| 0 | 0 | 0 | 0 | 0 |
| 0.009008681 | 0 | 0.000895552 | 0.018662049 | 0.00263952 |
| 0.018983132 | 0 | 0.010303625 | 0 | 0 |
| 0 | 0 | 0 | 0 | 0 |
| 0.003224422 | 0 | 0 | 0 | 0 |
| 0 | 0 | 0 | 0 | 0.117379843 |
| 0 | 0 | 0 | 0 | 0 |
| 0 | 0 | 0 | 0 | 0 |
| 0 | 0 | 0 | 0 | 0 |
| 0.02134486 | 0 | 0.001747439 | 0 | 0.004280804 |
| 0 | 0 | 0 | 0 | 0 |
| 0 | 0 | 0 | 0 | 0 |
| 0 | 0 | 0 | 0 | 0 |
| 0.0126318 | 0 | 0 | 0 | 0 |
| 0 | 0 | 0 | 0 | 0 |
| 0 | 0 | 0 | 0 | 0 |
| 0 | 0 | 0 | 0 | 0 |
| 0 | 0 | 0 | 0 | 0.030559418 |
| 0 | 0 | 0 | 0 | 0 |
| 0 | 0 | 0 | 0 | 0 |
| 0 | 0 | 0 | 0 | 0 |
| 0 | 0 | 0 | 0 | 0 |
| 0 | 0 | 0 | 0 | 0 |
| 0 | 0 | 0 | 0 | 0 |
| 0.225786022 | 0.004906955 | 0 | 0 | 0.217487894 |
| 0 | 0 | 0 | 0 | 0 |
| 0 | 0 | 0 | 0 | 0 |

| | | | | |
|---|---|---|---|---|
| 0 | 0 | 0 | 0 | 0 |
| 0 | 0 | 0 | 0 | 0 |
| 0 | 0 | 0 | 0 | 0 |
| 0 | 0 | 0 | 0 | 0 |
| 0 | 0 | 0 | 0 | 0 |
| 0 | 0 | 0 | 0 | 0 |
| 0 | 0 | 0 | 0 | 0 |
| 0 | 0 | 0 | 0 | 0 |
| 0 | 0.003123358 | 0 | 0 | 0 |
| 0 | 0 | 0 | 0 | 0 |
| 0.001231166 | 0 | 0.004454998 | 0 | 0 |
| 0.00055144 | 0 | 0 | 0.00124216 | 0 |
| 0 | 0 | 0 | 0 | 0 |
| 0 | 0 | 0 | 0 | 0 |
| 0 | 0 | 0 | 0 | 0 |
| 0 | 0 | 0 | 0 | 0 |
| 0 | 0 | 0 | 0 | 0 |
| 0 | 0 | 0 | 0 | 0 |
| 0 | 0 | 0 | 0 | 0 |
| 0 | 0 | 0 | 0 | 0 |
| 0 | 0 | 0 | 0 | 0 |
| 0 | 0 | 0 | 0 | 0 |
| 0 | 0 | 0 | 0 | 0 |
| 0 | 0 | 0 | 0 | 0 |
| 0 | 0 | 0 | 0 | 0 |
| 0 | 0 | 0 | 0 | 0 |
| 0 | 0 | 0 | 0 | 0 |
| 0 | 0 | 0.013579553 | 0 | 0.03827203 |
| 0 | 0 | 0 | 0 | 0 |
| 0 | 0 | 0.001470123 | 0 | 0 |
| 0.00448861 | 0 | 0.013274811 | 0.002685714 | 0.002719971 |
| 0 | 0 | 0 | 0 | 0 |
| 0.007654733 | 0.028509247 | 0.123760592 | 0.106025153 | 0.09523076 |
| 0.006157986 | 0 | 0.002547928 | 0 | 0 |
| 0 | 0 | 0 | 0 | 0 |
| 0 | 0 | 0 | 0 | 0 |
| 0 | 0 | 0 | 0 | 0 |
| 0 | 0 | 0 | 0 | 0 |
| 0 | 0 | 0 | 0 | 0 |
| 0 | 0 | 0 | 0 | 0 |
| 0 | 0 | 0 | 0 | 0 |
| 0 | 0 | 0.000932388 | 0.58160682 | 0 |
| 0.034932565 | 0 | 0 | 0 | 0 |
| 0 | 0 | 0 | 0 | 0 |
| 0 | 0 | 0 | 0 | 0 |
| 0.028070646 | 0 | 0.016414702 | 0 | 0 |
| 0 | 0 | 0 | 0 | 0 |
| 0 | 0 | 0 | 0 | 0 |

| | | | | |
|---|---|---|---|---|
| 0 | 0 | 0 | 0 | 0 |
| 0.00513035 | 0 | 0 | 0 | 0.062495796 |
| 0.014182937 | 0.003670595 | 0.023909133 | 0 | 0.00148983 |
| 0 | 0 | 0 | 0 | 0 |
| 0.001913982 | 0 | 0 | 0 | 0.003806585 |
| 0.003570862 | 0 | 0 | 0 | 0 |
| 0 | 0 | 0 | 0 | 0 |
| 0 | 0 | 0 | 0 | 0 |
| 0 | 0 | 0 | 0 | 0 |
| 0 | 0 | 0 | 0 | 0 |
| 0 | 0 | 0 | 0 | 0 |
| 0 | 0 | 0 | 0 | 0 |
| 0 | 0 | 0 | 0 | 0 |
| 0 | 0 | 0 | 0 | 0 |
| 0 | 0 | 0 | 0 | 0 |
| 0 | 0 | 0 | 0 | 0 |
| 0 | 0 | 0 | 0 | 0 |
| 0 | 0 | 0 | 0 | 0 |
| 0 | 0 | 0 | 0 | 0 |
| 0 | 0 | 0 | 0 | 0 |
| 0 | 0 | 0 | 0 | 0 |
| 0 | 0 | 0 | 0 | 0 |
| 0 | 0 | 0 | 0 | 0 |
| 0 | 0 | 0 | 0 | 0 |
| 0.032102861 | 0 | 0.002062097 | 0 | 0 |
| 0 | 0.001121166 | 0.026045512 | 0 | 0.006669905 |
| 0 | 0 | 0 | 0 | 0 |
| 0.922941725 | 0 | 0 | 0 | 0 |
| 0 | 0 | 0 | 0 | 0 |
| 0 | 0 | 0 | 0 | 0 |
| 0 | 0 | 0 | 0 | 0 |
| 0 | 0.000983196 | 0 | 0.000654299 | 0.001299803 |
| 0 | 0 | 0 | 0 | 0 |
| 0 | 0 | 0 | 0 | 0 |
| 0 | 0 | 0 | 0 | 0 |
| 0 | 0 | 0 | 0 | 0 |
| 0 | 0 | 0 | 0 | 0 |
| 0 | 0 | 0.005047563 | 0 | 0 |
| 1.521086731 | 0 | 0 | 0 | 0.488272868 |
| 0 | 0 | 0 | 0 | 0 |
| 0 | 0 | 0 | 0 | 0 |
| 0.001257659 | 0 | 0.002750522 | 0 | 0 |
| 0 | 0 | 0.000925293 | 0 | 0 |
| 0 | 0 | 0 | 0 | 0 |
| 0.000788572 | 0 | 0 | 0 | 0 |
| 0 | 0 | 0 | 0 | 0 |
| 0.00762814 | 0 | 0 | 0.029211002 | 0 |
| 0 | 0 | 0 | 0 | 0 |

| | | | | |
|---|---|---|---|---|
| 0 | 0 | 0 | 0 | 0 |
| 0.029357193 | 0.006205809 | 0.042643649 | 0.011202218 | 0.017707374 |
| 0.125922261 | 0 | 0 | 0 | 0 |
| 0 | 0 | 0 | 0 | 0 |
| 0 | 0 | 0 | 0 | 0 |
| 0 | 0 | 0 | 0 | 0 |
| 0 | 0 | 0 | 0 | 0 |
| 0 | 0 | 0 | 0 | 0 |
| 0 | 0 | 0 | 0 | 0 |
| 0 | 0 | 0 | 0 | 0 |
| 0 | 0 | 0 | 0 | 0 |
| 0 | 0 | 0 | 0 | 0 |
| 0.036629297 | 0 | 0 | 0 | 0 |
| 0 | 0 | 0 | 0 | 0 |
| 0.000600065 | 0 | 0.003340536 | 0.061276675 | 0 |
| 0 | 0 | 0 | 0 | 0 |
| 0 | 0 | 0 | 0 | 0 |
| 0 | 0 | 0 | 0 | 0 |
| 0.000663828 | 0 | 0 | 0 | 0 |
| 0 | 0 | 0 | 0 | 0 |
| 0 | 0 | 0 | 0 | 0 |
| 0.002357247 | 0 | 0.016129963 | 0.006194849 | 0 |
| 0 | 0 | 0 | 0.111355482 | 0 |
| 0 | 0 | 0 | 0.456901186 | 0 |
| 0 | 0 | 0 | 0 | 0 |
| 0.018082801 | 0 | 0 | 0 | 0 |
| 0 | 0 | 0 | 0 | 0 |
| 0 | 0 | 0 | 0 | 0 |
| 0 | 0 | 0 | 0 | 0.001422719 |
| 0 | 0 | 0 | 0 | 0 |
| 0 | 0 | 0 | 0 | 0 |
| 0 | 0 | 0 | 0 | 0 |
| 0 | 0 | 0 | 0 | 0 |
| 0 | 0 | 0 | 0 | 0 |
| 20.125473 | 0 | 0 | 0 | 0 |
| 0 | 0 | 0 | 0 | 0 |
| 0 | 0 | 0 | 0 | 0 |
| 0 | 0 | 0 | 0 | 0 |
| 0.000784665 | 0.012710841 | 0.045593036 | 0.003755972 | 0 |
| 0 | 0 | 0 | 0 | 0 |
| 0.417208864 | 0.016329958 | 0.023989571 | 0.007725958 | 0.007870804 |
| 0 | 0 | 0 | 0 | 0 |
| 0 | 0 | 0 | 0 | 0 |
| 2.933529534 | 0 | 0 | 0 | 0 |
| 0 | 0 | 0 | 0 | 0 |
| 1.23809045 | 0.136330035 | 0.660035029 | 1.441168032 | 0.085008814 |
| 0 | 0 | 0 | 0 | 0 |
| 0 | 0 | 0 | 0 | 0 |

| | | | | |
|---|---|---|---|---|
| 0 | 0 | 0 | 0 | 0 |
| 0.001312013 | 0 | 0 | 0 | 0 |
| 0 | 0 | 0 | 0 | 0 |
| 0.087164881 | 0 | 0 | 0 | 0 |
| 0 | 0 | 0 | 0 | 0 |
| 0 | 0 | 0 | 0 | 0 |
| 0 | 0 | 0 | 0 | 0 |
| 0.002394309 | 0 | 0.000908796 | 0.00392244 | 0 |
| 0.002046327 | 0 | 0 | 0 | 0 |
| 0 | 0 | 0.012334687 | 0 | 0.01263239 |
| 0 | 0 | 0 | 0 | 0 |
| 0 | 0 | 0 | 0 | 0 |
| 0.006737604 | 0 | 0 | 0.009106167 | 0 |
| 0 | 0 | 0 | 0 | 0 |
| 0 | 0 | 0 | 0 | 0 |
| 0 | 0 | 0 | 0 | 0 |
| 0 | 0 | 0 | 0 | 0 |
| 0 | 0 | 0 | 0 | 0 |
| 0 | 0 | 0 | 0 | 0 |
| 0.00976645 | 0.285841946 | 0.511262816 | 0.164611357 | 0.031691483 |
| 0 | 0 | 0 | 0 | 0 |
| 0 | 0 | 0 | 0 | 0 |
| 0 | 0 | 0 | 0 | 0 |
| 0 | 0 | 0 | 0 | 0 |
| 0 | 0 | 0 | 0 | 0 |
| 0.005479692 | 0 | 0 | 0 | 0 |
| 0 | 0 | 0 | 0 | 0 |
| 0 | 0 | 0 | 0 | 0 |
| 0 | 0 | 0 | 0 | 0 |
| 0 | 0 | 0.032119904 | 0 | 0 |
| 0 | 0 | 0 | 0 | 0 |
| 0.00127975 | 0 | 0 | 0 | 0 |
| 0 | 0 | 0 | 0 | 0 |
| 0 | 0 | 0 | 0 | 0 |
| 0 | 0 | 0 | 0 | 0 |
| 0.001552581 | 0 | 0 | 0 | 0 |
| 0 | 0 | 0 | 0 | 0 |
| 0 | 0 | 0 | 0 | 0 |
| 0 | 0 | 0 | 0 | 0 |
| 0 | 0 | 0 | 0 | 0 |
| 0 | 0 | 0 | 0 | 0 |
| 0 | 0 | 0 | 0 | 0 |
| 0.014527409 | 0 | 0 | 0 | 0 |
| 0 | 0 | 0 | 0 | 0 |
| 0.00305767 | 0 | 0.002239724 | 0 | 0.003120609 |
| 0 | 0 | 0 | 0 | 0 |
| 0 | 0 | 0 | 0 | 0 |
| 0 | 0.002960148 | 0 | 0.059693853 | 0 |

| | | | | |
|---|---|---|---|---|
| 0 | 0 | 0 | 0 | 0 |
| 0 | 0 | 0 | 0 | 0 |
| 0 | 0 | 0 | 0 | 0 |
| 0 | 0 | 0 | 0 | 0 |
| 0.001528395 | 0 | 0 | 0 | 0 |
| 0.001681967 | 0 | 0.000836021 | 0.053042543 | 0.026522234 |
| 0 | 0 | 0 | 0 | 0 |
| 0 | 0 | 0 | 0 | 0 |
| 0 | 0 | 0 | 0 | 0 |
| 0 | 0 | 0 | 0 | 0 |
| 0 | 0 | 0 | 0 | 0 |
| 0 | 0 | 0 | 0 | 0 |
| 0 | 0 | 0 | 0.195821535 | 0 |
| 0.005683367 | 0 | 0 | 0 | 0 |
| 0 | 0 | 0 | 0 | 0 |
| 0 | 0 | 0 | 0 | 0 |
| 0.160885436 | 0 | 0 | 0 | 0 |
| 0.000591838 | 0 | 0 | 0 | 0 |
| 0.006134462 | 0 | 0 | 0 | 0 |
| 0 | 0 | 0 | 0 | 0 |
| 0 | 0 | 0 | 0 | 0 |
| 0 | 0 | 0 | 0 | 0 |
| 0 | 0 | 0 | 0 | 0 |
| 0 | 0 | 0 | 0 | 0.023758124 |
| 0 | 0 | 0 | 0 | 0 |
| 0 | 0 | 0 | 0 | 0 |
| 0 | 0 | 0 | 0 | 0 |
| 0.014345794 | 0 | 0 | 0 | 0 |
| 0 | 0 | 0 | 0 | 0 |
| 0 | 0 | 0 | 0 | 0 |
| 0 | 0 | 0 | 0 | 0 |
| 0 | 0 | 0 | 0 | 0 |
| 0 | 0 | 0 | 0 | 0 |
| 0 | 0 | 0 | 0 | 0 |
| 0 | 0 | 0 | 0 | 0 |
| 0 | 0 | 0 | 0 | 0 |
| 0 | 0 | 0 | 0 | 0 |
| 0 | 0 | 0 | 0 | 0 |
| 0 | 0 | 0 | 0 | 0 |
| 0 | 0 | 0 | 0 | 0 |
| 0.000898479 | 0 | 0 | 0 | 0 |
| 0 | 0 | 0 | 0 | 0 |
| 0 | 0 | 0 | 0 | 0 |
| 0 | 0 | 0 | 0 | 0 |
| 0 | 0 | 0 | 0 | 0 |
| 0 | 0 | 0 | 0 | 0 |
| 0 | 0 | 0 | 0 | 0 |
| 0.000824109 | 0 | 0 | 0 | 0 |

| | | | | |
|---|---|---|---|---|
| 0 | 0 | 0 | 0 | 0 |
| 0.051516163 | 0 | 0 | 0 | 0 |
| 0 | 0 | 0 | 0 | 0 |
| 0 | 0 | 0 | 0 | 0 |
| 0 | 0 | 0 | 0 | 0 |
| 0 | 0 | 0 | 0 | 0 |
| 0 | 0 | 0 | 0 | 0 |
| 0 | 0 | 0 | 0 | 0 |
| 0 | 0 | 0 | 0 | 0 |
| 0 | 0 | 0 | 0 | 0 |
| 0 | 0.10748016 | 0.001715851 | 0 | 0 |
| 0 | 0 | 0 | 0 | 0 |
| 0.027372154 | 0 | 0 | 0 | 0.280190942 |
| 0 | 0 | 0 | 0 | 0 |
| 0 | 0 | 0 | 0 | 0 |
| 0 | 0 | 0 | 0 | 0 |
| 0 | 0 | 0 | 0 | 0 |
| 0 | 0 | 0 | 0 | 0 |
| 0 | 0 | 0 | 0 | 0 |
| 0 | 0 | 0 | 0 | 0 |
| 0.009831662 | 0.3575113 | 0.012439182 | 0 | 0 |
| 0 | 0 | 0 | 0 | 0 |
| 0 | 0 | 0 | 0 | 0 |
| 1.481122467 | 0 | 0 | 1.617966475 | 0.909818822 |
| 0 | 0 | 0 | 0 | 0 |
| 0.028675336 | 0 | 0 | 0 | 0 |
| 0 | 0 | 0 | 0 | 0 |
| 0.018272408 | 0 | 0 | 0 | 0 |
| 0 | 0 | 0 | 0 | 0 |
| 0 | 0 | 0 | 0 | 0 |
| 0 | 0 | 0 | 0 | 0 |
| 0 | 0 | 0 | 0 | 0 |
| 0 | 0 | 0 | 0 | 0 |
| 0 | 0 | 0 | 0 | 0 |

| PHL018-0037_V2 | PHL018-0190_V3 | PHL018-0245_V3 | PHL018-0046_V2 | PHL018-0194_V3 |
|---|---|---|---|---|
| CTRL | REF | REF | CTRL | EYCF |
| 0 | 0.233404187 | 0.167766833 | 0.776237137 | 0.242125262 |
| 0.014973145 | 0.047666744 | 1.314944101 | 0.414797135 | 0.158844119 |
| 12.74796779 | 0 | 1.391507269 | 19.66178984 | 0 |
| 0 | 0 | 0 | 0 | 0 |
| 0 | 0 | 0 | 0 | 0 |
| 0 | 0 | 0 | 0 | 0 |
| 0 | 0 | 0 | 0 | 0 |
| 0 | 0 | 0 | 0 | 0 |
| 0 | 0 | 0 | 0 | 0 |
| 0 | 0 | 0 | 0 | 0 |
| 0 | 0 | 0 | 0 | 0 |
| 0 | 0 | 0 | 0 | 0 |
| 0 | 0 | 0 | 0 | 0 |
| 0.597219228 | 1.11277325 | 1.894722888 | 5.59329827 | 0.103553571 |
| 0 | 0 | 0 | 0 | 0 |
| 0 | 0 | 0 | 0 | 0 |
| 0 | 0 | 0 | 0 | 0 |
| 0.000559729 | 0 | 0 | 0 | 0 |
| 0 | 0 | 0 | 0 | 0 |
| 0 | 0 | 0 | 0 | 0 |
| 0 | 0 | 0 | 0 | 0 |
| 0 | 0 | 0 | 0.004674865 | 0 |
| 0 | 0 | 0 | 0 | 0 |
| 0 | 0 | 0 | 0 | 0 |
| 0 | 0.116479495 | 0.164065569 | 0 | 0.364995263 |
| 0 | 0 | 0 | 0 | 0 |
| 0 | 0 | 0 | 0 | 0 |
| 0 | 0 | 0 | 0 | 0 |
| 0 | 0 | 0 | 0 | 0 |
| 0 | 0 | 0 | 0 | 0 |
| 0 | 0 | 0 | 0 | 0 |
| 0 | 0 | 0 | 0 | 0 |
| 0 | 0 | 0 | 0 | 0 |
| 0 | 0 | 0 | 0 | 0 |
| 0 | 0 | 0 | 0 | 0 |
| 0 | 0.991922322 | 0.619685589 | 1.02471492 | 0.347915368 |
| 0 | 0 | 0 | 0 | 0 |
| 0 | 0 | 0 | 0 | 0 |
| 0 | 0 | 0 | 0 | 0 |
| 0 | 0 | 0 | 0 | 0 |
| 0 | 0 | 0 | 0 | 0 |
| 0.009531485 | 0.007580261 | 0 | 0.010809315 | 0 |
| 0.854788325 | 2.905374589 | 0.449806433 | 0.16928023 | 8.052285712 |
| 0.000956894 | 0 | 0.057567125 | 0.003347978 | 0.005344949 |
| 0 | 0 | 0.00544313 | 0 | 0 |
| 0 | 0 | 0 | 0 | 0 |

| | | | | |
|---|---|---|---|---|
| 0 | 0 | 0 | 0 | 0.015890781 |
| 0.013942563 | 0 | 0.016258132 | 0.005823481 | 0 |
| 0 | 0 | 0.071628083 | 0 | 0.010674849 |
| 0 | 0.24805091 | 0.657571027 | 0.198719431 | 0.238514552 |
| 0.031590409 | 0.03490335 | 0.005938685 | 0.017181317 | 0 |
| 0 | 0 | 0 | 0 | 0.072956441 |
| 0.018899989 | 0 | 0 | 0 | 0.000892771 |
| 0 | 0.028473094 | 0 | 0 | 0 |
| 0 | 0.031278432 | 0 | 0 | 0 |
| 0 | 0 | 0 | 0 | 0 |
| 0 | 0 | 0 | 0 | 0 |
| 0.596651959 | 0.770525739 | 2.45446822 | 1.822529064 | 0.067573014 |
| 0.001142318 | 0 | 0 | 0.00094041 | 0 |
| 0.093332532 | 0.033509052 | 0.068174443 | 0.219428705 | 0.010709527 |
| 0 | 0 | 0 | 0 | 0 |
| 0 | 0 | 0 | 0 | 0 |
| 0 | 0.27459878 | 0.197644795 | 0.002397982 | 0.001972159 |
| 0 | 0 | 0 | 0 | 0 |
| 0 | 0 | 0 | 0 | 0 |
| 0 | 0 | 0 | 0 | 0 |
| 0.069352616 | 0.008377963 | 0.101563733 | 0.016224638 | 0.016214932 |
| 0.081198995 | 0.003562003 | 0.018849926 | 0.020825345 | 0.004757575 |
| 0 | 0.017312543 | 0.000829113 | 0 | 0.237104074 |
| 0 | 0 | 0 | 0 | 0 |
| 0 | 0 | 0 | 0 | 0 |
| 0.001447397 | 0 | 0 | 0 | 0 |
| 0.007271584 | 0 | 0 | 0.011281884 | 0 |
| 0 | 0 | 0 | 0 | 0 |
| 0.011186754 | 0 | 0 | 0.0027135 | 0 |
| 0.060760457 | 0 | 0 | 0 | 0 |
| 0 | 0 | 0 | 0 | 0 |
| 0.033413142 | 0 | 0 | 0 | 0 |
| 0 | 0 | 0 | 0 | 0 |
| 0 | 0 | 0 | 0 | 0.002052783 |
| 0 | 0 | 0 | 0 | 0 |
| 0 | 0 | 0 | 0 | 0 |
| 0 | 0 | 0 | 0 | 0 |
| 0 | 14.82690931 | 0 | 0 | 0 |
| 0 | 0 | 0 | 0 | 0 |
| 0 | 0 | 0 | 0 | 0 |
| 0 | 0 | 0 | 0 | 0 |
| 0 | 0 | 0 | 0 | 0 |
| 0 | 0 | 0 | 0 | 0 |
| 0 | 0 | 0 | 0 | 0 |
| 0 | 0 | 0 | 0 | 0.192114515 |
| 0 | 0 | 0 | 0 | 0 |
| 0 | 0 | 0 | 0 | 0 |
| 0 | 0.04963312 | 0 | 0.184882233 | 0 |

| | | | | |
|---|---|---|---|---|
| 0 | 0 | 0 | 0 | 0 |
| 0 | 0 | 0 | 0 | 0 |
| 0 | 0 | 0 | 0 | 0 |
| 0 | 0 | 0 | 0 | 0 |
| 0 | 0 | 0 | 0 | 0 |
| 0 | 0 | 0 | 0 | 0 |
| 0 | 0 | 0 | 0 | 0 |
| 0 | 0 | 0 | 0 | 0.001310459 |
| 0.001207464 | 0 | 0 | 0 | 0 |
| 0 | 0 | 0 | 0 | 0 |
| 0 | 0 | 0 | 0 | 0 |
| 0 | 0 | 0 | 0.00129832 | 0 |
| 0 | 0 | 0 | 0 | 0 |
| 0 | 0 | 0 | 0 | 0 |
| 0 | 0 | 0 | 0 | 0 |
| 0 | 0 | 0 | 0 | 0 |
| 0 | 0 | 0 | 0 | 0 |
| 0 | 0 | 0 | 0 | 0 |
| 0 | 0.00326088 | 0.037465764 | 0.020027691 | 0.535315985 |
| 0 | 0 | 0 | 0 | 0 |
| 0 | 0 | 0 | 0 | 0 |
| 0 | 0 | 0 | 0 | 0 |
| 0 | 0.00141218 | 0 | 0 | 0 |
| 0 | 0 | 0 | 0 | 0 |
| 0 | 0 | 0 | 0 | 0 |
| 0 | 0 | 0 | 0 | 0 |
| 0 | 0 | 0 | 0 | 0 |
| 0 | 0.001544943 | 0.00504973 | 0 | 0 |
| 0 | 0 | 0 | 0 | 0 |
| 0 | 0 | 0 | 0 | 0 |
| 0.598062886 | 0.025355333 | 0.158188274 | 0.084856758 | 0.172342974 |
| 0 | 0.049718237 | 0.004161838 | 0 | 0 |
| 0 | 0 | 0 | 0 | 0 |
| 0 | 0 | 0 | 0 | 0.001698734 |
| 0 | 0 | 0 | 0 | 0 |
| 0 | 0 | 0 | 0 | 0 |
| 0 | 0 | 0 | 0 | 0 |
| 0 | 0 | 0 | 0 | 0 |
| 0 | 0.014780555 | 0 | 0 | 0 |
| 0 | 0 | 0 | 0 | 0 |
| 0 | 0 | 0 | 0 | 0 |
| 0 | 0 | 0 | 0 | 0.011380244 |
| 0 | 0 | 0 | 0 | 0 |
| 0 | 0 | 0 | 0 | 0 |
| 0 | 0.001201318 | 0 | 0 | 0 |
| 0 | 0 | 0 | 0 | 0 |
| 0.000532669 | 0 | 0 | 0 | 0 |
| 0 | 0 | 0 | 0 | 0 |

| | | | | |
|---|---|---|---|---|
| 0 | 0 | 0 | 0 | 0 |
| 0 | 0 | 0 | 0 | 0 |
| 0 | 0 | 0 | 0 | 0 |
| 0 | 0 | 0 | 0 | 0 |
| 0 | 0.019879418 | 0.387446799 | 0 | 0.020334843 |
| 0 | 0 | 0 | 0 | 0 |
| 0 | 0 | 0 | 0 | 0 |
| 0 | 0 | 0 | 0 | 0 |
| 0 | 0 | 0 | 0 | 0 |
| 0 | 0 | 0 | 0 | 0 |
| 0 | 0 | 0 | 0 | 0 |
| 0 | 0 | 0 | 0 | 0 |
| 0 | 0 | 0 | 0 | 0 |
| 0 | 0 | 0.001775429 | 0 | 0 |
| 0 | 0 | 0 | 0 | 0 |
| 0.486906683 | 2.642880483 | 0 | 3.340249926 | 0 |
| 0 | 0 | 0 | 0 | 0 |
| 0 | 0 | 0 | 0 | 0 |
| 0 | 0 | 0 | 0 | 0 |
| 0 | 0.100995717 | 0 | 0 | 0 |
| 0 | 0 | 0 | 0 | 0 |
| 0 | 0 | 0 | 0 | 0 |
| 0 | 0 | 0.002650094 | 0 | 0 |
| 0 | 0 | 0 | 0 | 0 |
| 0 | 0 | 0 | 0 | 0 |
| 0 | 0 | 0 | 0 | 0 |
| 0 | 0.027986984 | 0 | 0 | 0.05479809 |
| 0 | 0 | 0 | 0 | 0 |
| 0 | 0 | 0 | 0 | 0 |
| 0 | 0 | 0 | 0 | 0 |
| 0 | 0 | 0 | 0 | 0 |
| 0 | 0 | 0 | 0 | 0.589423225 |
| 0 | 0 | 0 | 0 | 0 |
| 0 | 0 | 0 | 0 | 0 |
| 0 | 0 | 0 | 0 | 0 |
| 0 | 0 | 0 | 0 | 0 |
| 0 | 0 | 0 | 0 | 0 |
| 0 | 0 | 0 | 0 | 0 |
| 0 | 0 | 0 | 0 | 0 |
| 0 | 0 | 0 | 0 | 0 |
| 0 | 0 | 0 | 0 | 0 |
| 0 | 0 | 0 | 0 | 0 |
| 0 | 0 | 0 | 0 | 0 |
| 0 | 0 | 0 | 0 | 0 |
| 0 | 0 | 0 | 0 | 0 |
| 0 | 0 | 0 | 0 | 0 |
| 0 | 0.002868199 | 0 | 0 | 0 |
| 0 | 0 | 0 | 0 | 0 |

| | | | | |
|---|---|---|---|---|
| 0 | 0 | 0 | 0 | 0 |
| 0 | 0 | 0 | 0 | 0 |
| 0 | 0 | 0 | 0 | 0 |
| 0 | 0 | 0 | 0 | 0 |
| 0 | 0 | 0 | 0 | 0 |
| 0 | 0 | 0 | 0 | 0 |
| 0 | 0 | 0 | 0 | 0 |
| 0 | 0 | 0 | 0 | 0 |
| 0 | 0 | 0 | 0 | 0 |
| 0 | 0 | 0 | 0 | 0 |
| 0 | 0 | 0 | 0 | 0 |
| 0 | 0.00904657 | 0 | 0 | 0 |
| 0.020383291 | 0.000461276 | 0.01110437 | 0.00079907 | 0.002792993 |
| 0 | 0 | 0 | 0 | 0 |
| 0.016150137 | 0 | 0 | 0 | 0 |
| 0 | 0 | 0 | 0 | 0 |
| 0 | 0 | 0 | 0 | 0 |
| 0 | 0 | 0 | 0 | 0 |
| 0 | 0 | 0 | 0 | 0 |
| 0 | 0 | 0 | 0 | 0 |
| 0.037204432 | 0.003737392 | 0.039019188 | 0.083294356 | 0.003686277 |
| 0 | 0 | 0 | 0 | 0 |
| 0.000928911 | 0 | 0 | 0.00180256 | 0 |
| 0 | 0 | 0 | 0 | 0.001601685 |
| 0 | 0 | 0 | 0 | 0 |
| 0 | 0 | 0 | 0 | 0 |
| 0 | 0.000669714 | 0.007713602 | 0 | 0.0016101 |
| 0 | 0 | 0 | 0 | 0 |
| 0 | 0 | 0 | 0 | 0 |
| 0 | 0 | 0 | 0 | 0 |
| 0.067780044 | 0 | 0 | 0 | 0 |
| 0 | 0 | 0 | 0 | 0 |
| 0 | 0 | 0 | 0 | 0 |
| 0 | 0.167439317 | 0.014657697 | 0 | 0.007321676 |
| 0.034047506 | 0.033109165 | 0.064285118 | 0.008506059 | 0.004796976 |
| 0.003158058 | 0 | 0 | 0 | 0 |
| 0 | 0 | 0 | 0 | 0 |
| 0 | 0 | 0 | 0 | 0 |
| 0.030521265 | 0 | 0 | 0 | 0 |
| 0 | 0 | 0 | 0 | 0 |
| 0.001558504 | 0 | 0 | 0 | 0 |
| 0 | 0 | 0 | 0 | 0 |
| 0.005523071 | 0 | 0 | 0 | 0 |
| 0 | 0 | 0 | 0 | 0 |
| 0.026839369 | 0 | 0 | 0 | 0 |
| 0 | 0 | 0 | 0 | 0.043198514 |
| 0.004901047 | 0.001215201 | 0 | 0.00368392 | 0 |
| 0.03464164 | 0.000933115 | 0.000497949 | 0.006927597 | 0 |

| | | | | |
|---|---|---|---|---|
| 0 | 0 | 0 | 0 | 0 |
| 0.002519178 | 0 | 0 | 0 | 0 |
| 0 | 0 | 0 | 0 | 0 |
| 0 | 0 | 0 | 0 | 0 |
| 0 | 0 | 0 | 0 | 0 |
| 0 | 0 | 0 | 0 | 0 |
| 0 | 0 | 0 | 0.00936828 | 0 |
| 0 | 0 | 0 | 0.000995855 | 0 |
| 0 | 0 | 0 | 0 | 0 |
| 0 | 0 | 0 | 0 | 0 |
| 0 | 0 | 0 | 0 | 0 |
| 0 | 0 | 0 | 0 | 0 |
| 0 | 0 | 0 | 0 | 0 |
| 0 | 0 | 0 | 0 | 0 |
| 0 | 0 | 0 | 0 | 0 |
| 0 | 0 | 0 | 0 | 0 |
| 0 | 0 | 0 | 0 | 0 |
| 0 | 0 | 0 | 0 | 0.045751508 |
| 0.00499215 | 0.014370149 | 0 | 0.000880665 | 0.019700427 |
| 0 | 0.144945334 | 0 | 0 | 0.06922134 |
| 0.236697757 | 0 | 0 | 0.010652379 | 0 |
| 0 | 0 | 0 | 0 | 0 |
| 0 | 0 | 0 | 0 | 0 |
| 0 | 0 | 0 | 0 | 0 |
| 0 | 0 | 0.005441658 | 0 | 0 |
| 0 | 0 | 0 | 0 | 0 |
| 0 | 0 | 0 | 0 | 0 |
| 0 | 0 | 0 | 0 | 0 |
| 0.007180264 | 0 | 0 | 0.001724079 | 0 |
| 0 | 0 | 0 | 0 | 0 |
| 1.348877852 | 0.138513663 | 0.30429802 | 0.331647564 | 0.078872713 |
| 0 | 0 | 0 | 0 | 0 |
| 0 | 0.002559695 | 0 | 0 | 0 |
| 0.094873193 | 0 | 0 | 0 | 0 |
| 0 | 0 | 0 | 0 | 0 |
| 0 | 0 | 0 | 0 | 0 |
| 0 | 0 | 0 | 0 | 0 |
| 0 | 0 | 0 | 0 | 0 |
| 0 | 0 | 0 | 0 | 0 |
| 0 | 0 | 0.011521624 | 0 | 0.00078469 |
| 0 | 0 | 0 | 0 | 0 |
| 0 | 0.081387542 | 0.071853053 | 0 | 0.02674582 |
| 0.003175518 | 0 | 0.000626362 | 0 | 0 |
| 0 | 0 | 0 | 0 | 0 |
| 0 | 0 | 0 | 0 | 0 |
| 0 | 0 | 0 | 0 | 0 |
| 0 | 0 | 0 | 0 | 0 |
| 0 | 0 | 0 | 0 | 0 |

| | | | | |
|---|---|---|---|---|
| 0 | 0 | 0 | 0 | 0 |
| 0 | 0 | 0 | 0 | 0 |
| 0 | 0 | 0.050618804 | 0.002445215 | 0 |
| 0 | 0 | 0 | 0 | 0 |
| 0 | 0 | 0 | 0 | 0.247121091 |
| 0 | 0 | 0 | 0 | 0 |
| 0 | 0 | 0 | 0 | 0 |
| 0 | 0 | 0 | 0 | 0 |
| 0 | 0 | 0 | 0 | 0 |
| 0 | 0 | 0 | 0 | 0 |
| 0 | 0 | 0 | 0 | 0 |
| 0 | 0 | 0 | 0 | 0 |
| 0 | 0 | 0 | 0 | 0 |
| 0.002188413 | 0 | 0.008028845 | 0.008917935 | 0 |
| 0.002709372 | 0 | 0.001282599 | 0.001338289 | 0.002935043 |
| 0 | 0.008087347 | 0 | 0 | 0.02003249 |
| 0.001029029 | 0 | 0.002192109 | 0 | 0 |
| 0 | 0 | 0 | 0 | 0 |
| 0 | 0 | 0 | 0 | 0 |
| 0 | 0 | 0 | 0 | 0 |
| 0 | 0 | 0 | 0 | 0 |
| 0.035670506 | 0.000529743 | 0 | 0.009865013 | 0 |
| 0 | 0 | 0 | 0 | 0 |
| 0.715698426 | 0 | 0 | 0 | 0 |
| 0 | 0 | 0 | 0 | 0 |
| 0 | 0 | 0 | 0 | 0 |
| 0 | 0 | 0 | 0 | 0.552112631 |
| 0 | 0 | 0 | 0 | 0 |
| 0 | 0 | 0 | 0 | 0 |
| 0 | 0 | 0 | 0 | 0 |
| 0 | 0 | 0 | 0 | 0.007518589 |
| 0 | 0 | 0 | 0 | 0 |
| 0.002691438 | 0 | 0 | 0 | 0 |
| 0 | 0 | 0 | 0 | 0 |
| 0 | 0 | 0 | 0 | 0 |
| 0 | 0 | 0 | 0 | 0 |
| 0 | 0 | 0 | 0 | 0 |
| 0.000700255 | 0 | 0.014088575 | 0 | 0.041959 |
| 0 | 0 | 0 | 0 | 0 |
| 0 | 0 | 0 | 0 | 0 |
| 0.001684442 | 0 | 0 | 0 | 0 |
| 0 | 0 | 0 | 0 | 0 |
| 0 | 0 | 0 | 0 | 0 |
| 0 | 0 | 0 | 0 | 0 |
| 0 | 0 | 0 | 0 | 0 |
| 0 | 0 | 0 | 0 | 0 |
| 0 | 0 | 0 | 0 | 0 |
| 0.010460936 | 0 | 0.004684455 | 0.004887852 | 0 |

| | | | | |
|---|---|---|---|---|
| 0.183778706 | 0.107887551 | 0 | 5.87634445 | 0 |
| 0 | 0 | 0.048899225 | 0 | 0.069693529 |
| 0 | 0 | 0 | 0 | 0.000790509 |
| 0 | 0 | 0 | 0 | 0 |
| 0 | 0 | 0.016450908 | 0.006285848 | 0.000596496 |
| 0 | 0 | 0 | 0 | 0 |
| 0 | 0 | 0 | 0 | 0 |
| 0 | 0.015389285 | 0.009743479 | 0.04744384 | 0 |
| 0 | 0 | 0 | 0 | 0 |
| 0 | 0.022314436 | 0.009038534 | 0.155087278 | 0.003734501 |
| 0.039640058 | 0 | 0 | 0 | 0 |
| 0.002389632 | 0 | 0 | 0.00241436 | 0 |
| 0.00131827 | 0.000469863 | 0.001755169 | 0.003662756 | 0 |
| 0.371141476 | 0 | 0 | 0.092713022 | 0 |
| 0.002033716 | 0 | 0 | 0.003976342 | 0 |
| 0 | 0 | 0 | 0 | 0 |
| 0 | 0 | 0 | 0 | 0 |
| 0.013520387 | 0 | 0.001523919 | 0.000795044 | 0 |
| 0 | 0 | 0 | 0 | 0 |
| 0 | 0 | 0 | 0 | 0 |
| 0 | 0 | 0 | 0 | 0 |
| 0 | 0 | 0.004219239 | 0 | 0 |
| 0 | 0 | 0.001609874 | 0 | 0 |
| 0 | 0.069498624 | 0 | 0 | 0 |
| 0 | 0 | 0 | 0 | 0 |
| 0.000575793 | 0 | 0.002271472 | 0 | 0 |
| 0 | 1.073324106 | 0 | 0 | 0.024761854 |
| 0 | 0 | 0 | 0 | 0 |
| 0.000786039 | 0 | 0 | 0 | 0 |
| 0 | 0.003003054 | 0.001635941 | 0.006096332 | 0 |
| 0.00597262 | 0 | 0 | 0.04145488 | 0 |
| 0 | 0 | 0 | 0 | 0 |
| 0 | 0 | 0 | 0 | 0 |
| 0 | 0 | 0 | 0 | 0 |
| 0 | 0 | 0 | 0 | 0 |
| 0 | 0 | 0 | 0 | 0 |
| 0.011748968 | 0 | 0 | 0 | 0 |
| 0 | 0 | 0 | 0 | 0 |
| 0 | 0 | 0 | 0.003797656 | 0 |
| 0 | 0 | 0 | 0 | 0 |
| 0 | 0 | 0 | 0.662668695 | 0 |
| 0 | 0 | 0 | 0 | 0 |
| 0 | 0 | 0 | 0 | 0 |
| 0.000618506 | 0 | 0 | 0 | 0 |
| 0 | 0 | 0 | 0.116407174 | 0 |
| 0 | 0.020153716 | 0 | 0 | 0 |
| 0 | 0.008847467 | 0.032767154 | 0 | 0.001939214 |
| 0 | 0 | 0.001281976 | 0.048155015 | 0 |

| | | | | |
|---|---|---|---|---|
| 0 | 0 | 0 | 0 | 0 |
| 0 | 0.003802947 | 0 | 0 | 0 |
| 0 | 0 | 0 | 0 | 0 |
| 0 | 0 | 0 | 0 | 0 |
| 0.016513811 | 0 | 0 | 0.049815732 | 0 |
| 0.154152219 | 0.003216209 | 0.013349019 | 0 | 0 |
| 0 | 0 | 0 | 0 | 0 |
| 0 | 0 | 0 | 0 | 0 |
| 0 | 0 | 0 | 0 | 0.028325136 |
| 0 | 0 | 0 | 0 | 0 |
| 0 | 0.00435197 | 0.023481965 | 0.001884734 | 0 |
| 0 | 0 | 0 | 0 | 0 |
| 0 | 0 | 0 | 0 | 0 |
| 0 | 0.000783147 | 0.001950293 | 0 | 0 |
| 0 | 0 | 0 | 0 | 0 |
| 0 | 0 | 0 | 0 | 0 |
| 0.103162013 | 0 | 0 | 0 | 0 |
| 0 | 0 | 0 | 0 | 0 |
| 0 | 0 | 0 | 0.070392288 | 0 |
| 0.020380661 | 0 | 0 | 0 | 0.051557667 |
| 0 | 0 | 0 | 0 | 0 |
| 0 | 0.365462408 | 0 | 0 | 0 |
| 0.060792832 | 0 | 0 | 0.382114962 | 0 |
| 0 | 0 | 0 | 0 | 0 |
| 0 | 0 | 0 | 0 | 0 |
| 0 | 0 | 0 | 0 | 0 |
| 0.414088075 | 0 | 0 | 0 | 0 |
| 0 | 0 | 0 | 0 | 0 |
| 0 | 0 | 0 | 0 | 0 |
| 0 | 0.268902554 | 0.297306947 | 0.120805227 | 0.041090505 |
| 0 | 0.003984774 | 0 | 0 | 0.007947889 |
| 0 | 0 | 0 | 0 | 0 |
| 0 | 0 | 0 | 0 | 0 |
| 0 | 0 | 0.003045232 | 0.005866071 | 0 |
| 0 | 0 | 0 | 0.01056715 | 0 |
| 0 | 0 | 0 | 0 | 0 |
| 0 | 0.084545181 | 0 | 0 | 0.345884489 |
| 0.002939432 | 0 | 0 | 0 | 0 |
| 0 | 0 | 0 | 0 | 0 |
| 0 | 0 | 0 | 0 | 0 |
| 0.011346235 | 0 | 0.039482563 | 0.022585092 | 0.024938026 |
| 0 | 1.90508362 | 0.126346396 | 0 | 0.728188262 |
| 0.002308037 | 0 | 0 | 0 | 0 |
| 0 | 0 | 0 | 0 | 0 |
| 0 | 0 | 0.004521664 | 0 | 0 |
| 0 | 0 | 0 | 0 | 0 |
| 0.000710966 | 0 | 0 | 0.014266702 | 0 |
| 0 | 0 | 0 | 0 | 0 |

| | | | | |
|---|---|---|---|---|
| 0 | 0 | 0.003374044 | 0 | 0 |
| 0 | 0 | 0 | 0.237757301 | 0 |
| 0 | 0.008804477 | 0.012023951 | 0 | 0 |
| 0 | 0 | 0 | 0 | 0 |
| 0 | 0.744819569 | 0.465102566 | 0.015244936 | 0.013284105 |
| 0 | 0 | 0 | 0 | 0.001883113 |
| 0 | 0 | 0 | 0 | 0 |
| 0 | 0 | 0 | 0 | 0 |
| 0 | 0 | 0 | 0 | 0 |
| 0 | 0 | 0 | 0 | 0 |
| 0 | 0 | 0 | 0 | 0 |
| 0 | 0 | 0 | 0 | 0 |
| 0 | 0 | 0 | 0 | 0 |
| 0 | 0 | 0.063161451 | 0.00416493 | 0.003223849 |
| 0 | 0 | 0 | 0 | 0 |
| 0 | 0.080166278 | 0.064589493 | 0.076009645 | 0.074321791 |
| 0.006874037 | 0.001306705 | 0.047591557 | 0 | 0.00383965 |
| 0.557057546 | 0 | 0 | 0.001031325 | 0 |
| 0.014378232 | 0 | 0.001470553 | 0.005918415 | 0 |
| 0 | 0 | 0 | 0 | 0 |
| 0 | 0.002881936 | 0.003588481 | 0.50200255 | 0.005938837 |
| 0 | 0 | 0 | 0 | 0.002429343 |
| 0 | 0 | 0 | 0 | 0 |
| 0 | 0 | 0.276370547 | 0 | 0.261823194 |
| 0.000642445 | 0.023203606 | 0 | 0 | 0 |
| 0.018110665 | 0 | 0 | 0 | 0 |
| 0.003054326 | 0 | 0.003614744 | 0.002424661 | 0.001550967 |
| 0.003589688 | 0 | 0 | 0 | 0 |
| 0 | 0 | 0 | 0 | 0 |
| 0 | 0 | 0 | 0.08206532 | 0 |
| 0.055415522 | 0.005992449 | 0.081095578 | 0.151203717 | 0.001348007 |
| 0.004452453 | 0 | 0 | 0.009163666 | 0 |
| 0 | 0.010968755 | 0.039301285 | 0.017450098 | 0 |
| 0.002700171 | 0 | 0 | 0.00166718 | 0 |
| 0.001204522 | 0 | 0 | 0 | 0.002446593 |
| 0.001425792 | 0 | 0 | 0 | 0 |
| 0.002129252 | 0 | 0 | 0.001314674 | 0.000864976 |
| 0.198821731 | 0.01835918 | 0.057741665 | 0.003701291 | 0.018602415 |
| 0 | 0.085005368 | 0.572561661 | 0 | 0.051633459 |
| 0 | 0.008022856 | 0.001127877 | 0 | 0 |
| 0 | 0 | 0 | 0 | 0 |
| 0.005863226 | 0 | 0 | 0 | 0 |
| 0.01967309 | 0.001183454 | 0.00442078 | 0.052585104 | 0.001517448 |
| 0.025217541 | 0 | 0.195989406 | 0 | 0.032211277 |
| 0 | 0 | 0 | 0 | 0 |
| 0.070610315 | 0 | 0 | 0 | 0 |
| 0 | 0 | 0 | 0 | 0 |
| 0.000507484 | 0 | 0 | 0 | 0 |

| | | | | |
|---|---|---|---|---|
| 0.00222201 | 0 | 0 | 0 | 0 |
| 0.001588307 | 0.006038517 | 0.002416802 | 0.24320771 | 0.27076433 |
| 0 | 0 | 0 | 0 | 0 |
| 0 | 0 | 0 | 0.003563609 | 0.002372551 |
| 0.002448606 | 0 | 0 | 0 | 0 |
| 0.00143214 | 0 | 0 | 0 | 0 |
| 0.064443294 | 0 | 0 | 0 | 0 |
| 0 | 0 | 0.002268883 | 0 | 0 |
| 0 | 0.002566462 | 0 | 0 | 0.006117456 |
| 0 | 0 | 0 | 0 | 0 |
| 0.029001311 | 0.012660535 | 0.004589646 | 0.001873928 | 0.00410977 |
| 0.194141089 | 0 | 0.03596545 | 0.204382885 | 0.012498306 |
| 0 | 1.310914616 | 0.02585864 | 0 | 0.013119736 |
| 0 | 0 | 0 | 0 | 0 |
| 0 | 0 | 0 | 0.004300196 | 0 |
| 0.04098985 | 0 | 0 | 0 | 0 |
| 0 | 0 | 0.010447873 | 0 | 0.017393404 |
| 0.592026373 | 0 | 0.021429197 | 0.001981234 | 0 |
| 0 | 0 | 0 | 0 | 0 |
| 0 | 0 | 0 | 0.042271043 | 0 |
| 0 | 0.087741365 | 0.003447806 | 0.008319238 | 0.015902896 |
| 0 | 0 | 0 | 0 | 0 |
| 0 | 0 | 0 | 0 | 0 |
| 0 | 0 | 0 | 0 | 0 |
| 0 | 0 | 0 | 0 | 0 |
| 0 | 0 | 0 | 0 | 0 |
| 0.097049557 | 0 | 0 | 0.284295558 | 0.007300908 |
| 0 | 0.009196856 | 0.022445115 | 0.029394079 | 0.060927368 |
| 0.012263427 | 0 | 0 | 0 | 0.018202843 |
| 0 | 0 | 0.001105064 | 0 | 0 |
| 0 | 0 | 0 | 0 | 0 |
| 0.074392946 | 0 | 0 | 0.00230901 | 0 |
| 0 | 0 | 0.001946301 | 0 | 0 |
| 0 | 0 | 0 | 0 | 0 |
| 0 | 1.047644925 | 0 | 0 | 0 |
| 0 | 0.15471739 | 0.293817961 | 0.349851892 | 0.03891795 |
| 0 | 0 | 0 | 0 | 0 |
| 0 | 0 | 0 | 0 | 0 |
| 0.007161985 | 0 | 0 | 0.013266177 | 0.003879263 |
| 0 | 0 | 0 | 0 | 0 |
| 0 | 0 | 0 | 0 | 0 |
| 0 | 0 | 0 | 0 | 0 |
| 0.00116008 | 0.017228348 | 0.067617052 | 0.041902009 | 0 |
| 0.064122011 | 0 | 0 | 0.004667954 | 0 |
| 0 | 0 | 0.004571322 | 0 | 0 |
| 0 | 0.31608463 | 0 | 0 | 0 |
| 0 | 0 | 0 | 0 | 0 |
| 0.000905225 | 0 | 0 | 0.016208607 | 0 |

| | | | | |
|---|---|---|---|---|
| 0 | 0 | 0 | 0 | 0 |
| 0 | 0 | 0 | 0 | 0 |
| 0 | 0 | 0 | 0 | 0 |
| 0 | 0.00593941 | 0.125524239 | 0.3981373 | 0.475290829 |
| 0 | 0 | 0 | 0 | 0 |
| 0 | 0 | 0 | 0 | 0 |
| 0.002094995 | 0 | 0 | 0 | 0 |
| 0 | 0.158110711 | 0 | 0 | 0.001534459 |
| 0 | 0.665315473 | 2.019262779 | 0 | 3.801781633 |
| 0 | 0.02582451 | 0.220802739 | 0 | 0.057027441 |
| 0 | 0 | 0 | 0 | 0 |
| 0 | 0.004306395 | 0.00842626 | 0 | 0.00213639 |
| 0 | 0 | 0 | 0 | 0 |
| 0 | 0 | 0 | 0 | 0 |
| 0 | 0.062442669 | 0.294254035 | 0 | 0.153512637 |
| 0 | 0 | 0 | 0 | 0 |
| 0 | 0 | 0 | 0 | 0.202012722 |
| 0 | 0.42328254 | 0 | 0 | 0 |
| 0 | 0 | 0 | 0 | 0 |
| 0 | 0.056861827 | 0 | 0 | 0 |
| 0 | 0 | 0 | 0 | 0 |
| 0 | 0.01699223 | 0 | 0 | 0 |
| 0 | 0 | 0 | 0 | 0.001815926 |
| 0 | 0.077630421 | 0 | 0.735401624 | 0.111687785 |
| 0 | 0.008351764 | 0.038190575 | 0.648805723 | 0 |
| 0 | 0 | 0 | 0 | 0 |
| 0 | 0.006853317 | 0 | 0 | 0.001830721 |
| 0 | 0.416915562 | 4.913607407 | 0 | 0 |
| 0 | 0 | 0 | 0 | 0 |
| 0 | 0 | 0 | 0 | 0 |
| 0 | 0.09120698 | 0 | 0 | 0.07301226 |
| 0 | 0.268367322 | 0 | 0 | 2.010596525 |
| 0.002633975 | 0 | 0.106412128 | 0.202105759 | 0.143235753 |
| 0 | 0 | 0 | 0 | 0 |
| 0 | 0 | 0.145250733 | 0 | 0 |
| 0 | 0.619427211 | 0 | 0 | 0.595491888 |
| 0 | 0.055668814 | 0.68297413 | 0 | 1.204182109 |
| 0 | 0 | 0 | 0 | 0 |
| 0 | 0.012007921 | 0 | 0 | 0 |
| 0 | 0.001100858 | 0 | 0 | 0.008351625 |
| 0.093760741 | 0 | 0.001653441 | 0.144919569 | 0.002364789 |
| 0 | 0 | 0 | 0 | 0 |
| 0.011359484 | 2.102717813 | 9.224623933 | 1.940391899 | 0.018155873 |
| 0 | 0 | 0 | 0 | 0.067157324 |
| 0 | 0 | 0 | 0 | 0 |
| 0 | 0 | 0 | 0 | 0 |
| 0 | 0 | 0 | 0 | 0 |
| 0 | 0.001418913 | 0.015680161 | 0.005300038 | 0.14081079 |

| | | | | |
|---|---|---|---|---|
| 0 | 0 | 0 | 0 | 0 |
| 0 | 0.145102198 | 0 | 0 | 0.098068005 |
| 0.200172015 | 0 | 0.012914133 | 0.110228771 | 0 |
| 0 | 0.002573022 | 0.146918651 | 0 | 0.003338457 |
| 0 | 0 | 0 | 0 | 0 |
| 0 | 0.002276946 | 0.008839059 | 0 | 0.115493793 |
| 0 | 0 | 0 | 0 | 0 |
| 0 | 0 | 0 | 0 | 0 |
| 0 | 0 | 0 | 0 | 0 |
| 0 | 0 | 0 | 0 | 0 |
| 3.709897634 | 0.017361028 | 0.429228493 | 0.317301261 | 0.300554578 |
| 0 | 0.249002505 | 0.856085516 | 6.164617562 | 0.136553576 |
| 0 | 0 | 0 | 0 | 0 |
| 0 | 0.003963539 | 0.056979751 | 0 | 0.006160157 |
| 0.306961456 | 0.004401184 | 0.026163503 | 1.555149892 | 0.010770788 |
| 0 | 1.68385073 | 0 | 0 | 5.295732445 |
| 0 | 0 | 0 | 0 | 0 |
| 0 | 0 | 0 | 0 | 0 |
| 0 | 0 | 0 | 0 | 0 |
| 0.324217651 | 0.006462915 | 0 | 0.015185702 | 0.014546118 |
| 0.168109695 | 0.002378545 | 0 | 0.299392834 | 0.079833474 |
| 30.21958946 | 0 | 0.83230613 | 2.681758199 | 0 |
| 0 | 0.393550276 | 1.311475688 | 2.863207304 | 0.228473268 |
| 0 | 0 | 0 | 0 | 0 |
| 0 | 0 | 0 | 0 | 0.009524157 |
| 0 | 2.016607538 | 0 | 0 | 0 |
| 0 | 0 | 0 | 0.085445352 | 0 |
| 0 | 0 | 0 | 0.004360662 | 0 |
| 0.082834217 | 0 | 0 | 0 | 0 |
| 0 | 0 | 0 | 0 | 0 |
| 0 | 0.026234734 | 0 | 0 | 0.054910237 |
| 0.026822004 | 0.076083795 | 0.219572194 | 0 | 0.063513134 |
| 0.044734179 | 0 | 0 | 0.202475656 | 0.001648722 |
| 0.07565444 | 0.032292707 | 0 | 0 | 0.030174675 |
| 0.017619826 | 0.002715732 | 0.002677045 | 0.542337572 | 0.003385439 |
| 0.061666408 | 0.028606247 | 0.113053056 | 0.017775062 | 0.089871436 |
| 0 | 0.001952349 | 0.015520959 | 0 | 0.006739764 |
| 0 | 0 | 0 | 0 | 0 |
| 0 | 0.113342276 | 0.408890934 | 0 | 1.896712625 |
| 0 | 0.158532338 | 0 | 0 | 0 |
| 0 | 0.014511193 | 0 | 0 | 0.023111902 |
| 0 | 0 | 0 | 0 | 0 |
| 0 | 0 | 0 | 0 | 0 |
| 0 | 0 | 0 | 0 | 0 |
| 1.440793126 | 0.00797825 | 0.144156658 | 0.377536948 | 0.007388242 |
| 0 | 0.088798438 | 0 | 0 | 0 |
| 0 | 0 | 0 | 0 | 0 |
| 0 | 1.220102964 | 0.534459253 | 0 | 0.00205667 |

| | | | | |
|---|---|---|---|---|
| 0 | 0 | 0 | 0 | 0 |
| 0 | 0.040239211 | 0 | 0 | 0 |
| 0 | 0 | 0 | 0 | 0 |
| 0 | 0.004079898 | 0 | 0 | 0.000807304 |
| 0.160301412 | 0 | 0 | 0 | 0 |
| 0 | 0.395833587 | 0 | 0 | 0.169980499 |
| 0 | 0.212913247 | 0 | 0 | 0 |
| 0.096114392 | 0 | 0.078776487 | 0 | 0.27129447 |
| 0 | 0 | 0 | 0 | 0 |
| 0 | 0 | 0 | 0 | 0 |
| 0.067107482 | 0.002524403 | 0.002305083 | 0.053132365 | 0.002157894 |
| 0 | 0 | 0 | 0 | 0 |
| 0 | 0.032565272 | 0 | 0 | 0 |
| 0 | 0.231642483 | 0 | 0 | 0 |
| 0 | 0 | 0 | 0 | 0 |
| 0 | 0 | 0 | 0 | 0 |
| 0 | 0 | 0 | 0 | 0.044536112 |
| 0 | 0.003362413 | 0 | 0.005278654 | 0.004191594 |
| 0 | 0.295140713 | 0 | 0 | 0.00204781 |
| 0 | 0 | 0.143636166 | 0 | 0.013635446 |
| 0 | 0.002240429 | 0.119802158 | 0 | 0 |
| 0 | 0 | 0 | 0 | 0 |
| 0 | 0.327736066 | 0.025945276 | 0 | 0.002934083 |
| 0 | 0 | 0 | 1.455183074 | 0 |
| 0 | 0 | 0 | 0 | 0 |
| 0 | 0 | 0 | 0 | 0 |
| 0 | 0 | 0 | 0 | 0 |
| 0 | 0 | 0 | 0 | 0 |
| 0 | 0 | 0 | 0 | 0 |
| 0 | 0 | 0 | 0 | 0.051899934 |
| 0 | 0.026509984 | 0 | 0 | 0 |
| 0 | 0 | 0 | 0 | 0 |
| 0.00631442 | 0.001610193 | 0 | 0 | 0 |
| 0 | 0 | 0 | 0 | 0 |
| 0 | 0 | 0 | 0 | 0.000857926 |
| 0 | 0 | 0 | 0 | 0 |
| 0 | 0.085085498 | 0.053272808 | 0 | 0.00347693 |
| 0.009143185 | 0.004273903 | 0.004257367 | 0.00222111 | 0 |
| 0 | 0 | 0 | 0 | 0 |
| 0.001478415 | 0 | 0 | 0 | 0.001651605 |
| 0 | 0 | 0 | 0 | 0.00328496 |
| 0 | 0 | 0 | 0 | 0 |
| 0 | 0.000803011 | 0 | 0 | 0 |
| 0 | 0.920988512 | 0 | 0 | 0.040209244 |
| 0 | 0.068265154 | 0.051161908 | 0 | 0 |
| 0 | 0 | 0 | 0 | 0 |
| 0 | 0.011370587 | 0 | 0 | 0.004339165 |
| 0 | 0 | 0 | 0 | 0 |

| | | | | |
|---|---|---|---|---|
| 0 | 0 | 0 | 0 | 0 |
| 0 | 0 | 0 | 0 | 0 |
| 0 | 0.34735023 | 0 | 0 | 0.0618289 |
| 0 | 0.418042643 | 0.031231887 | 0 | 0.064504426 |
| 0 | 0 | 0 | 0 | 0 |
| 0 | 0 | 0 | 0 | 0 |
| 0 | 0 | 0 | 0 | 0 |
| 0 | 0 | 0 | 0 | 0 |
| 1.593198517 | 0.001964809 | 0.00880743 | 0.03165391 | 0.002939204 |
| 0 | 0 | 0 | 0 | 0.002535994 |
| 0 | 0 | 0 | 0 | 0.00255197 |
| 0 | 0 | 0 | 0 | 0.001279915 |
| 0 | 0.004470307 | 0 | 0 | 0 |
| 0 | 0 | 0 | 0 | 0 |
| 0 | 0.059443662 | 4.348020021 | 5.530354143 | 0.090824142 |
| 1.772975283 | 0 | 0.004148302 | 1.126372918 | 0 |
| 0 | 0 | 0 | 0 | 0 |
| 0 | 0 | 0 | 0 | 0 |
| 0 | 0 | 0 | 0 | 0 |
| 0 | 0 | 0 | 0 | 0 |
| 0 | 0.028094471 | 0.013656157 | 0 | 0.205159414 |
| 0 | 0.514585987 | 0 | 0 | 0 |
| 0 | 0 | 0 | 0 | 0 |
| 0 | 0 | 0 | 0 | 0 |
| 0.046674811 | 0.070241001 | 0.013234311 | 0.086455967 | 8.84991614 |
| 0 | 2.673247504 | 5.838593839 | 1.952890995 | 11.58341087 |
| 0 | 0.094189273 | 0 | 0 | 0.187695185 |
| 0 | 0 | 0.001428722 | 0 | 0 |
| 0 | 0.043835942 | 0.520172679 | 0 | 0 |
| 0 | 0.365348835 | 0 | 0 | 0 |
| 0 | 0 | 0 | 0 | 0 |
| 0 | 0.040622753 | 0.109316346 | 0 | 0.053740877 |
| 0 | 0 | 0 | 0 | 0 |
| 0.01739721 | 0.000370196 | 0.006760669 | 0.019238765 | 0.007120093 |
| 0 | 0.001829899 | 0.011898954 | 0.00158497 | 0.000869012 |
| 0.072734219 | 0.004311722 | 0.053330067 | 0.01157728 | 0.000614286 |
| 0 | 0.585712724 | 0 | 0 | 0 |
| 0 | 0.075626815 | 0.748694256 | 1.227362564 | 0.126935274 |
| 0 | 0 | 0 | 0 | 0.325260638 |
| 0 | 0 | 0 | 0 | 0 |
| 0 | 0 | 0 | 0 | 0 |
| 0.001543307 | 0 | 0 | 0.019057832 | 0 |
| 0 | 0.577420366 | 2.308570574 | 0.048227189 | 0.895255436 |
| 0.011645195 | 0.065967315 | 0 | 0 | 0 |
| 0 | 0 | 0 | 0 | 0.404295165 |
| 0 | 0.025870496 | 0.022210014 | 0 | 0 |
| 0 | 0 | 0 | 0 | 0 |
| 0.000912901 | 0.0010846 | 0 | 0 | 0.155062602 |

| | | | | |
|---|---|---|---|---|
| 0 | 0.011161266 | 0 | 0 | 0.01469145 |
| 0 | 0 | 0 | 0 | 0 |
| 0 | 0 | 0 | 0 | 0 |
| 0 | 0 | 0 | 0 | 0 |
| 0.081840364 | 0 | 0.005614882 | 0.0027201 | 0.004129988 |
| 0 | 0.045595615 | 0.010620986 | 0 | 0 |
| 0 | 0 | 0 | 0 | 0 |
| 0 | 0 | 0 | 0 | 0 |
| 0 | 0 | 0 | 0 | 0 |
| 0 | 0 | 0 | 0 | 0 |
| 0 | 0.000739381 | 0 | 0 | 0.000658368 |
| 0 | 0.79771178 | 2.441855861 | 0.182611917 | 0.190032596 |
| 0 | 0.024960163 | 0 | 0 | 0.092304425 |
| 0 | 0 | 0 | 0 | 0 |
| 0.320001657 | 0.015831451 | 0.001623403 | 0.129219615 | 0 |
| 0 | 0 | 0 | 0 | 0 |
| 0 | 0 | 0 | 0.006060061 | 0.000692214 |
| 0 | 0 | 0 | 0 | 0 |
| 0 | 0 | 0.048785369 | 0.022906624 | 0 |
| 0 | 0.051228002 | 0.080917382 | 0 | 0.001547363 |
| 0 | 0.005000759 | 0.06849439 | 0 | 0.08506107 |
| 0 | 0 | 0 | 0 | 0 |
| 0 | 0.499230987 | 0.326130355 | 0.012580448 | 0.205063579 |
| 0 | 0 | 0 | 0 | 0 |
| 0 | 0 | 0 | 0 | 0 |
| 0 | 0 | 0 | 0 | 0 |
| 0 | 0.004901174 | 0 | 0 | 0 |
| 0 | 0 | 0 | 0 | 0 |
| 0 | 0.001509896 | 0 | 0 | 0 |
| 0 | 0 | 0 | 0 | 0 |
| 0 | 0 | 0 | 0 | 0 |
| 0.004577124 | 0 | 0.036413946 | 0.011932322 | 0.059397089 |
| 0 | 0 | 0 | 0 | 0.012921039 |
| 0 | 0.027770984 | 0.147939763 | 0.297117874 | 0.006579834 |
| 0.06006417 | 0.003597379 | 0.050441099 | 0.013411819 | 0 |
| 0 | 1.610004426 | 0.688408136 | 0 | 0.235048914 |
| 0 | 0 | 0 | 0 | 0 |
| 0 | 0.019443074 | 0 | 0 | 0 |
| 0 | 0 | 0 | 0 | 0 |
| 0 | 0 | 0 | 0 | 0 |
| 0 | 0.103009987 | 0 | 0 | 0.004129326 |
| 0 | 0 | 0 | 0 | 0.059887221 |
| 0 | 0 | 0 | 0 | 0 |
| 0 | 0 | 0 | 0 | 0 |
| 0.152056809 | 0.00523863 | 0.037602894 | 0.240217932 | 0.158872036 |
| 0 | 0 | 0 | 0 | 0 |
| 0 | 0.398764038 | 0 | 0 | 0.185200609 |
| 0.003679792 | 0.005246266 | 0.013427832 | 0.006248087 | 0 |

| | | | | |
|---|---|---|---|---|
| 0 | 0 | 0 | 0 | 0 |
| 0 | 0 | 0 | 0 | 0 |
| 0.022466399 | 0 | 0 | 0.036231629 | 0.005278463 |
| 0 | 0 | 0 | 0 | 0 |
| 0 | 0 | 0 | 0 | 0 |
| 0.02250081 | 0 | 0 | 0 | 0 |
| 0 | 0.369773057 | 0 | 0 | 0 |
| 0 | 0.02588522 | 0 | 0 | 0.022191356 |
| 0 | 0 | 0 | 0 | 0 |
| 0 | 0 | 0 | 0 | 0 |
| 0.18461066 | 0 | 0.409134982 | 0 | 0 |
| 1.807397559 | 0 | 0.00121134 | 0.001263936 | 0 |
| 0 | 0 | 0 | 0 | 0 |
| 0 | 0.025238699 | 0 | 0 | 0 |
| 0 | 0 | 0 | 0 | 0 |
| 0 | 0 | 0 | 0 | 0 |
| 0 | 0 | 0 | 0 | 0 |
| 0 | 0.071163525 | 0 | 0 | 0 |
| 0 | 0 | 0 | 0 | 0.028019111 |
| 0 | 0 | 0 | 0.003084549 | 0 |
| 0 | 0 | 0 | 0 | 0 |
| 0 | 0 | 0 | 0 | 0 |
| 0 | 0 | 0 | 0 | 0 |
| 0 | 0 | 0 | 0 | 0 |
| 0 | 0 | 0 | 0 | 0.004317053 |
| 0.210527721 | 0 | 0.001607458 | 0.058517515 | 0.002452292 |
| 0 | 0 | 0 | 0 | 0 |
| 0.001624883 | 0.011512752 | 0.001398562 | 0.006566791 | 0.062557977 |
| 0 | 0.033879619 | 0 | 0 | 0.0225462 |
| 0 | 0.147264418 | 0.261954691 | 0 | 0.120913043 |
| 0 | 0 | 0 | 0 | 0 |
| 0 | 0 | 0 | 0 | 0 |
| 0 | 0.005633487 | 0 | 0 | 0.000792036 |
| 0 | 0 | 0 | 0 | 0 |
| 0.102165261 | 0.001803797 | 0 | 0.125379607 | 0.000963694 |
| 0 | 0.012689981 | 0 | 0 | 0 |
| 0 | 0 | 0 | 0 | 0 |
| 0.001794546 | 0.001876217 | 0.006796208 | 0.001551221 | 0.028978022 |
| 0 | 0 | 0 | 0 | 0 |
| 0 | 0 | 0 | 0 | 0 |
| 0 | 1.370959948 | 15.78407497 | 10.85919907 | 0.298407341 |
| 0.265087613 | 0 | 0.1837587 | 0.072347624 | 0 |
| 0 | 0 | 0.845025384 | 0 | 0 |
| 0 | 0.172962195 | 0 | 0 | 0 |
| 0 | 0 | 0 | 0 | 0 |
| 0 | 0 | 0 | 0 | 0 |
| 0 | 0 | 0 | 0 | 0 |
| 0.042727521 | 0 | 0.009607789 | 0.016708259 | 0.186881395 |

| | | | | |
|---|---|---|---|---|
| 0 | 0 | 0 | 0 | 0 |
| 0.017872505 | 0.010025219 | 0 | 0.003798971 | 0.006248733 |
| 0 | 0.030085342 | 0 | 0.020494642 | 0 |
| 0 | 0 | 0 | 0 | 0.00494684 |
| 0 | 1.187397824 | 1.473929015 | 0.203311829 | 0.661523265 |
| 0 | 0 | 0 | 0 | 0 |
| 0 | 0 | 0 | 0 | 0 |
| 0 | 0 | 0 | 0 | 0 |
| 1.468504492 | 0.003197779 | 0.020081908 | 0.085983065 | 0.00133704 |
| 0.085249866 | 0 | 0 | 0 | 0 |
| 0 | 0 | 0 | 0.004905196 | 0 |
| 0 | 0 | 0 | 0 | 0 |
| 0 | 0.041941315 | 0.052729188 | 0 | 0.021829241 |
| 0 | 0.006653236 | 0.033965905 | 0.00972458 | 0 |
| 0 | 0.030672783 | 0 | 0 | 0.596306503 |
| 0 | 0.173888182 | 0.331746879 | 0.16562767 | 0.039814809 |
| 0.001665786 | 0 | 0 | 0.000561008 | 0.001999341 |
| 0 | 0 | 0 | 0 | 0 |
| 0 | 0.029361031 | 0 | 0 | 0 |
| 0 | 0 | 0 | 0 | 0 |
| 0 | 0 | 0 | 0 | 0 |
| 0 | 0.013829438 | 0 | 0 | 0 |
| 0 | 0 | 0 | 0 | 0 |
| 0 | 0 | 0 | 0 | 0 |
| 0 | 0 | 0 | 0 | 0.01239049 |
| 0 | 0 | 0 | 0 | 0 |
| 0 | 0.019399208 | 0.156151594 | 0 | 0.022065694 |
| 0.017978789 | 0 | 0.010023331 | 0.022384945 | 0 |
| 0 | 0 | 0 | 0 | 0 |
| 0 | 0 | 0 | 0 | 0 |
| 0 | 0 | 0 | 0 | 0 |
| 0 | 0.000707796 | 0 | 0 | 0 |
| 0 | 0.226059559 | 0 | 0 | 0.036110271 |
| 0 | 0 | 0.02691299 | 0 | 0.008067412 |
| 0 | 1.401972306 | 0 | 0 | 0.306079508 |
| 0.002062584 | 0 | 0 | 0 | 0.029151713 |
| 0.004743472 | 0 | 0 | 0 | 0.119364668 |
| 0 | 0 | 0 | 0 | 0 |
| 0 | 0 | 0 | 0 | 0 |
| 0 | 0 | 0 | 0 | 0 |
| 0 | 0.132055064 | 0.623090614 | 1.087647529 | 0.373709295 |
| 0 | 0 | 0.63355313 | 0 | 0 |
| 0 | 0 | 0 | 0 | 0 |
| 0 | 0 | 0 | 0 | 0.002770346 |
| 0 | 0 | 0 | 0 | 0 |
| 0 | 0 | 0 | 0 | 0 |
| 0 | 0.018438105 | 0 | 0 | 0.001326057 |
| 0 | 0 | 0 | 0 | 0 |

| | | | | |
|---|---|---|---|---|
| 0 | 0.095096145 | 0.179059405 | 0 | 0.686664637 |
| 0 | 0 | 0 | 0 | 0 |
| 0 | 0 | 0 | 0 | 0 |
| 0 | 0 | 0 | 0 | 0 |
| 0 | 0.009031101 | 0.039834684 | 0 | 0.022243799 |
| 0 | 0.182410508 | 0 | 0 | 0 |
| 0.001510668 | 0 | 0 | 0.009327393 | 0.007977918 |
| 0 | 0 | 0 | 0 | 0 |
| 0 | 0 | 0 | 0 | 0 |
| 1.161283261 | 0.002862209 | 0.003001194 | 0.013504615 | 0 |
| 0 | 0.010795356 | 0.022547843 | 0 | 0.032372473 |
| 0 | 0.031893912 | 0 | 0.041763744 | 0.00375676 |
| 0 | 0 | 0 | 0 | 0 |
| 0 | 0 | 0 | 0 | 0 |
| 0 | 0 | 0 | 0 | 0 |
| 0 | 0.002556325 | 0 | 0 | 0 |
| 0 | 0 | 0 | 0.033898227 | 0 |
| 0 | 0 | 0.009969219 | 0 | 0 |
| 0 | 0 | 0 | 0 | 0 |
| 0 | 0 | 0 | 0 | 0 |
| 0 | 0.000956897 | 0.02178731 | 0 | 0.026583979 |
| 0 | 0 | 0 | 0 | 0 |
| 0 | 0.041313497 | 0.06214351 | 0 | 0.042371954 |
| 0.001551682 | 0.004022232 | 0.009181951 | 0 | 0.000859565 |
| 0 | 0 | 0 | 0 | 0 |
| 0 | 0 | 0.020483534 | 0.01068646 | 0 |
| 0 | 0.740911451 | 0 | 0 | 0 |
| 0 | 0 | 0 | 0 | 0 |
| 0 | 0.207665212 | 0 | 0 | 0.073911376 |
| 1.383996419 | 0 | 0.192938713 | 1.462361664 | 0 |
| 0 | 0.073563499 | 0 | 0 | 0 |
| 0 | 0 | 0 | 0 | 0 |
| 0 | 0 | 0 | 0 | 0 |
| 7.187459288 | 0 | 0 | 0 | 0.034423618 |
| 0 | 0.203524173 | 0 | 0 | 0 |
| 0 | 0 | 0 | 0 | 0.097549806 |
| 0.058423199 | 0 | 0 | 0.25261662 | 0.003585126 |
| 0 | 0 | 0 | 0 | 0 |
| 0 | 9.333064753 | 0 | 0 | 0 |
| 0 | 0.01342228 | 0 | 0 | 0 |
| 0 | 0 | 0 | 0 | 0 |
| 0 | 0 | 0 | 0 | 0 |
| 0 | 0 | 0 | 0 | 0 |
| 0 | 0.00960512 | 0 | 0 | 0.015736955 |
| 0 | 0 | 0 | 0 | 0 |
| 0 | 0 | 0 | 0 | 0 |
| 0 | 0 | 0 | 0 | 0.001504132 |
| 0 | 0.017749235 | 0 | 0 | 0 |

| | | | | |
|---|---|---|---|---|
| 0.465077668 | 0.001298419 | 0.019939849 | 0.139453945 | 0.001079076 |
| 0 | 0.108911852 | 0 | 0 | 0 |
| 0 | 0 | 0 | 0 | 0.008060637 |
| 0 | 0 | 0 | 0 | 0 |
| 0 | 0 | 0 | 0 | 0 |
| 0 | 0 | 0 | 0 | 1.290786143 |
| 0.002217058 | 0 | 0 | 0 | 0 |
| 0 | 0.332555164 | 1.008192292 | 0.718632813 | 0.026274641 |
| 1.08071866 | 3.8220499 | 0.599184678 | 0.235958223 | 9.809113293 |
| 0.020898997 | 0.147243981 | 0.17077973 | 0.322594242 | 0 |
| 0 | 0 | 0 | 0 | 0 |
| 0 | 0 | 0 | 0 | 0.000589801 |
| 0 | 0 | 0 | 0 | 0 |
| 0 | 0 | 0 | 0 | 0 |
| 0 | 0 | 0 | 0 | 0 |
| 0 | 0 | 0 | 0 | 0 |
| 0 | 0 | 0 | 0.123124134 | 0 |
| 0 | 0 | 0 | 0 | 0 |
| 0 | 0 | 0 | 0 | 0 |
| 0 | 0.03385808 | 0.188493995 | 0 | 0.025282644 |
| 0 | 0 | 0 | 0 | 0 |
| 0 | 0 | 0 | 0 | 0 |
| 0 | 0 | 0 | 0 | 0.058754689 |
| 0 | 0 | 0 | 0 | 0 |
| 0 | 0.0046589 | 0.126350613 | 0 | 0.010684729 |
| 0 | 0 | 0 | 0 | 0 |
| 0 | 0 | 0 | 0 | 0 |
| 0 | 0 | 0 | 0 | 0.183558889 |
| 0.13957367 | 0 | 0 | 0 | 0 |
| 0 | 0 | 0 | 0 | 0 |
| 0 | 0 | 0 | 0 | 0.075065657 |
| 0 | 0 | 0 | 0 | 0 |
| 0 | 0 | 0 | 0 | 0 |
| 0 | 0 | 0 | 0 | 0 |
| 0 | 0.036154148 | 0 | 0 | 0 |
| 0 | 0 | 0 | 0 | 0 |
| 0 | 0 | 0 | 0 | 0 |
| 0 | 0 | 0 | 0 | 0 |
| 0 | 0 | 0 | 0 | 0 |
| 0 | 0 | 0.070478757 | 0 | 0 |
| 0 | 0 | 0 | 0 | 0 |
| 0.318447984 | 0.087963995 | 0.453104546 | 0.79381943 | 0.224498787 |
| 0 | 0 | 0 | 0 | 0 |
| 0 | 0 | 0 | 0 | 0 |
| 0 | 0 | 0 | 0 | 0 |
| 0 | 0.000641422 | 0 | 0.016667056 | 0 |
| 0 | 0 | 0 | 0 | 0 |
| 0 | 0 | 0.000836427 | 0 | 0.001004872 |

| | | | | |
|---|---|---|---|---|
| 0 | 0 | 0 | 0 | 0 |
| 0.255084847 | 0 | 0.00924149 | 0 | 0.000834783 |
| 0 | 0 | 0 | 0 | 0 |
| 0 | 0 | 0 | 0 | 0 |
| 0 | 3.012751769 | 1.974857917 | 0 | 4.556333047 |
| 0 | 0.681304415 | 0 | 0 | 0 |
| 0 | 0 | 0.005111414 | 0 | 0.008435148 |
| 0.127436108 | 0 | 0.010283081 | 0.059131839 | 0.006078933 |
| 0 | 0.020705929 | 0 | 0 | 0.007831728 |
| 0 | 0 | 0 | 0 | 0 |
| 0 | 0 | 0 | 0 | 0 |
| 0 | 0.636555505 | 0 | 0 | 0 |
| 0 | 0 | 0 | 0 | 0.114422994 |
| 0 | 0 | 0 | 0 | 0 |
| 0 | 0 | 0 | 0.000839249 | 0 |
| 0.008540521 | 0 | 0 | 0.010753223 | 0.069389131 |
| 0 | 0 | 0 | 0 | 0 |
| 0 | 0 | 0 | 0 | 0 |
| 0 | 0 | 0 | 0 | 0 |
| 0 | 0 | 0 | 0 | 0 |
| 0 | 0 | 0 | 0 | 0 |
| 0 | 0 | 0 | 0 | 0 |
| 0 | 0 | 0 | 0 | 0 |
| 0.093074241 | 0 | 0.001779832 | 0.034872426 | 0.000848519 |
| 0 | 0.041402913 | 0.116303058 | 0 | 0 |
| 0 | 0 | 0 | 0.000940325 | 0 |
| 0.849588427 | 0.02120804 | 0 | 0.055296886 | 0.07822752 |
| 0 | 0.000764773 | 0 | 0 | 0 |
| 0.261012193 | 0 | 0 | 0 | 0 |
| 0 | 0 | 0 | 0 | 0 |
| 0 | 0 | 0 | 0 | 0 |
| 0 | 0 | 0 | 0 | 0 |
| 0 | 0 | 0 | 0 | 0 |
| 0 | 0 | 0 | 0 | 0 |
| 0 | 0 | 0 | 0 | 0 |
| 0.003586525 | 0 | 0 | 0 | 0 |
| 0 | 0 | 0 | 0 | 0 |
| 0 | 1.198787382 | 0.766098493 | 0 | 0.05571313 |
| 0 | 0 | 0 | 0 | 0 |
| 0 | 0 | 0 | 0 | 0 |
| 0 | 0 | 0 | 0 | 0 |
| 0 | 0.08267628 | 0 | 0 | 0 |
| 0 | 0 | 0 | 0 | 0 |
| 0 | 0 | 0 | 0 | 0.069885297 |
| 0 | 0 | 0 | 0 | 0 |
| 0 | 0 | 0 | 0 | 0 |
| 0 | 0 | 0 | 0 | 0 |
| 0 | 0 | 0 | 0 | 0.028434199 |

| | | | | |
|---|---|---|---|---|
| 0 | 0.134692556 | 0.001871052 | 0 | 0.067289756 |
| 0 | 0 | 0 | 0 | 0 |
| 0 | 0 | 0 | 0 | 0.005494275 |
| 0 | 0 | 0.001015414 | 0.01589255 | 0 |
| 0 | 0 | 0 | 0 | 0 |
| 0 | 0.767426153 | 0 | 0 | 0 |
| 0 | 0 | 0 | 0 | 0 |
| 0 | 0 | 0 | 0 | 0 |
| 0 | 0 | 0 | 0 | 0 |
| 0 | 0 | 0 | 0 | 0.059977498 |
| 0 | 0 | 0 | 0 | 0 |
| 0 | 0 | 0.000729209 | 0.004945664 | 0 |
| 0 | 0 | 0 | 0 | 0 |
| 0 | 0 | 0 | 0 | 0 |
| 0 | 0 | 0 | 0 | 0 |
| 0 | 0 | 0 | 0 | 0.001398431 |
| 0 | 0 | 0 | 0 | 0 |
| 0 | 0 | 0 | 0 | 0 |
| 0 | 0.001158167 | 0 | 0 | 0 |
| 0 | 0 | 0 | 0 | 0 |
| 0.049753795 | 0.020876838 | 0.045688318 | 0.013561887 | 0 |
| 0 | 0 | 0 | 0 | 0 |
| 0.025008077 | 0 | 0.043275764 | 0.004671184 | 0.15153355 |
| 0 | 0.007495381 | 8.742720995 | 0 | 0.156076662 |
| 0 | 0.039828764 | 0 | 0 | 0 |
| 0 | 0.083279902 | 0 | 0 | 0 |
| 0 | 0 | 0 | 0 | 0 |
| 0 | 0 | 0 | 0 | 0 |
| 0 | 0 | 0 | 0 | 0 |
| 0 | 0 | 0 | 0 | 0 |
| 0 | 0 | 0 | 0 | 0 |
| 0 | 0.001004201 | 0 | 0 | 0.002575214 |
| 0.024994412 | 0.008791699 | 0 | 0.002673333 | 0.004397228 |
| 0 | 0 | 0 | 0 | 0 |
| 0 | 0.028277415 | 0 | 0 | 0.077056916 |
| 0.039956007 | 0 | 0.00149525 | 0.001560173 | 0 |
| 0 | 0 | 0 | 0 | 0 |
| 0 | 0.060686719 | 0 | 0 | 0 |
| 0 | 0 | 0.024212898 | 0.00135344 | 0.029868237 |
| 0 | 0 | 0 | 0 | 0 |
| 0 | 0 | 0 | 0 | 0 |
| 0 | 0 | 0 | 0 | 0 |
| 0 | 0.122215044 | 0.210065918 | 0 | 0.434500696 |
| 0 | 0 | 0 | 0 | 0 |
| 0 | 0 | 0 | 0 | 0 |
| 0 | 0.773093721 | 1.110385002 | 0 | 0.044619495 |
| 0 | 0 | 0 | 0 | 0 |
| 0 | 0 | 0 | 0 | 0 |

| | | | | |
|---|---|---|---|---|
| 0 | 0 | 0 | 0 | 0 |
| 0 | 0 | 0 | 0 | 0 |
| 0 | 0 | 0 | 0 | 0 |
| 0 | 0 | 0 | 0 | 0 |
| 0 | 0 | 0 | 0 | 0 |
| 0 | 0 | 0 | 0 | 0 |
| 0 | 0 | 0 | 0 | 0 |
| 0 | 0 | 0 | 0 | 0 |
| 0 | 0.734740055 | 0 | 0.480830063 | 0.006617674 |
| 0 | 0.006808634 | 0 | 0 | 0 |
| 0 | 0 | 0 | 0 | 0 |
| 0.037663575 | 0 | 0 | 0 | 0 |
| 0 | 0 | 0 | 0 | 0 |
| 0 | 0 | 0 | 0 | 0.082565729 |
| 0 | 0 | 0 | 0.006800029 | 0 |
| 0.028020395 | 0 | 0 | 0.052433004 | 0 |
| 0 | 0 | 0 | 0 | 0 |
| 0 | 0 | 0 | 0 | 0 |
| 0 | 0 | 0 | 0 | 0 |
| 0 | 0.018193643 | 0 | 0 | 0.014909036 |
| 0 | 0 | 0.001699878 | 0 | 0 |
| 0 | 0 | 0 | 0 | 0 |
| 0 | 0 | 0 | 0 | 0 |
| 0 | 0 | 0 | 0 | 0.001395295 |
| 0 | 0 | 0 | 0 | 0 |
| 0 | 0 | 0 | 0 | 0 |
| 0 | 0 | 0 | 0 | 0 |
| 0 | 0 | 0 | 0 | 0 |
| 0 | 0 | 0 | 0 | 0 |
| 0 | 0.858723147 | 0 | 0 | 0.008866189 |
| 0 | 0.027223563 | 0.376017414 | 2.691541498 | 0.081590632 |
| 0 | 0 | 0 | 0 | 0 |
| 0 | 0.001009439 | 0.00104743 | 0 | 0 |
| 0 | 0.126885394 | 0 | 0 | 0 |
| 0 | 0 | 0 | 0 | 0 |
| 0 | 0 | 0 | 0 | 0 |
| 0 | 0 | 0 | 0 | 0 |
| 0 | 0 | 0 | 0 | 0 |
| 0 | 0 | 0 | 0 | 0 |
| 0 | 0 | 0 | 0 | 0 |
| 0 | 0 | 0 | 0 | 0 |
| 0 | 0.232062808 | 0 | 0 | 0.021735252 |
| 0 | 0 | 0 | 0 | 0.006752848 |
| 0 | 0 | 0 | 0 | 0 |
| 0 | 0 | 0 | 0 | 0 |
| 0 | 0 | 0 | 0 | 0 |
| 0 | 0 | 0 | 0.222306947 | 0 |
| 0 | 0 | 0 | 0 | 0.03064683 |

| | | | | |
|---|---|---|---|---|
| 0 | 0 | 0 | 0 | 0 |
| 0 | 0 | 0 | 0 | 0 |
| 0 | 0 | 0 | 0 | 0 |
| 0 | 0 | 0 | 0 | 0 |
| 0 | 1.01826393 | 0.582685564 | 0.151468758 | 0.085184008 |
| 0 | 0 | 0 | 0 | 0 |
| 0 | 0 | 0 | 0 | 0 |
| 0 | 0 | 0 | 0 | 0 |
| 0 | 0 | 0 | 0 | 0 |
| 0 | 0 | 0 | 0 | 0 |
| 0 | 0 | 0 | 0 | 0.007458903 |
| 0.059867964 | 0.004510553 | 0.010584708 | 0.04012008 | 0.048381382 |
| 0.003149449 | 0.003741799 | 0.002422759 | 0 | 0.030865908 |
| 0 | 0 | 0 | 0 | 0 |
| 0 | 0 | 0 | 0 | 0 |
| 0 | 0 | 0 | 0 | 0.272156884 |
| 0.008703736 | 0.018279754 | 0 | 0 | 0.004277129 |
| 0.104163326 | 0 | 0 | 0 | 0 |
| 0.329212481 | 0.00183907 | 0 | 0.921956135 | 0 |
| 0.013714816 | 0 | 0 | 0.003342634 | 0.001832709 |
| 0 | 0 | 0 | 0 | 0 |
| 0 | 0 | 0 | 0 | 0 |
| 0 | 0 | 0 | 0 | 0 |
| 0 | 0 | 0 | 0 | 0 |
| 0 | 0 | 0 | 0 | 0 |
| 0 | 0 | 0 | 0 | 0 |
| 0.085174904 | 0.009483774 | 0.010268565 | 0.070715186 | 0.000587453 |
| 0.18977806 | 0 | 0 | 0 | 0 |
| 0 | 0 | 0 | 0 | 0 |
| 0 | 0 | 0 | 0 | 0 |
| 0.016064384 | 0.000934814 | 0.003103989 | 0.003441185 | 0.005826707 |
| 0 | 0 | 0 | 0 | 0 |
| 0 | 0 | 0 | 0 | 0 |
| 0 | 0 | 0 | 0 | 0 |
| 0 | 0 | 0 | 0 | 0.003532789 |
| 0 | 0 | 0 | 0 | 0.003629163 |
| 0 | 0 | 0 | 0 | 0 |
| 0.071749066 | 0.055716234 | 0.097044374 | 0.018914767 | 0.124798608 |
| 0 | 0.006714687 | 0 | 0 | 0.476206844 |
| 0 | 0.016668994 | 0.012323652 | 0 | 0.00408171 |
| 0.012762095 | 0.0381382 | 0 | 0.000993844 | 0 |
| 0 | 0.006875904 | 0 | 0 | 0 |
| 0 | 0 | 0 | 0 | 0 |
| 0.023738158 | 0 | 0 | 0.028680867 | 0 |
| 0 | 0 | 0 | 0 | 0 |
| 0 | 0 | 0 | 0 | 0.000930339 |
| 0 | 0 | 0 | 0 | 0 |
| 0 | 0 | 0 | 0 | 0 |

| | | | | |
|---|---|---|---|---|
| 0 | 0 | 0 | 0 | 0 |
| 0 | 0.01096004 | 0 | 0 | 0.037504806 |
| 0 | 0.211362383 | 0 | 0 | 0 |
| 0.015928885 | 0.003061989 | 0.003600862 | 0.003536197 | 0.000545298 |
| 0 | 0 | 0 | 0 | 0 |
| 0 | 0 | 0 | 0.007407494 | 0 |
| 0 | 0.008749204 | 0 | 0 | 0.007761161 |
| 0.268191929 | 0 | 0.016897114 | 0.276734099 | 0.000995097 |
| 0 | 0.178685563 | 0 | 0 | 0.033224221 |
| 0 | 0.002752016 | 0 | 0 | 0.008821726 |
| 0 | 0.017430055 | 0 | 0 | 0.019825875 |
| 0 | 0 | 0 | 0 | 0 |
| 0 | 0.024719803 | 0.408400666 | 0.136613308 | 0.378937439 |
| 0 | 0.009702568 | 0 | 0 | 0.005820271 |
| 0 | 0 | 0 | 0 | 0 |
| 0 | 0 | 0 | 0 | 0 |
| 0 | 0 | 0 | 0 | 0 |
| 0 | 0 | 0 | 0 | 0 |
| 0 | 0 | 0 | 0 | 0 |
| 0 | 0 | 0 | 0 | 0 |
| 0 | 0 | 0 | 0 | 0 |
| 0 | 0 | 0 | 0 | 0 |
| 0 | 0 | 0.019573231 | 0.016503509 | 0.021207637 |
| 0 | 0.096869294 | 0.023097085 | 0.008520185 | 0.013814209 |
| 0 | 0 | 0 | 0 | 0 |
| 0 | 0.016604775 | 0 | 0 | 0.00591416 |
| 0 | 0.000455107 | 0 | 0 | 0 |
| 0 | 0 | 0 | 0 | 0 |
| 0 | 0 | 0 | 0 | 0 |
| 0 | 0 | 0 | 0 | 0 |
| 0 | 0.009914301 | 0.008156469 | 0 | 0.003215913 |
| 0 | 0 | 0 | 0 | 0 |
| 0 | 0 | 0 | 0 | 0 |
| 0 | 0 | 0 | 0 | 0 |
| 0 | 0.021967235 | 0 | 0 | 0.06232863 |
| 0 | 0 | 0 | 0.03246119 | 0 |
| 0.009696911 | 0 | 0 | 0.042765791 | 0 |
| 0 | 0 | 0 | 0 | 0 |
| 0 | 0 | 0 | 0.006811354 | 0 |
| 0 | 0 | 0 | 0 | 0 |
| 0 | 0 | 0 | 0 | 0 |
| 0 | 0 | 0 | 0 | 0 |
| 0 | 0 | 0 | 0 | 0 |
| 0 | 0 | 0 | 0 | 0 |
| 0 | 0 | 0 | 0 | 0 |
| 0 | 0.125068336 | 0.364629339 | 0.055819268 | 0.202639126 |
| 0.016667947 | 0 | 0 | 0 | 0 |
| 0 | 0 | 0 | 0 | 0 |

| | | | | |
|---|---|---|---|---|
| 0 | 0 | 0 | 0 | 0 |
| 0 | 0 | 0 | 0 | 0 |
| 0 | 0.061641524 | 0 | 0 | 0 |
| 0 | 0.010545646 | 0 | 0 | 0 |
| 0.004913572 | 0 | 0 | 0 | 0 |
| 0 | 0 | 0 | 0 | 0 |
| 0 | 0 | 0 | 0.022155601 | 0 |
| 0 | 0 | 0 | 0 | 0 |
| 0 | 0 | 0 | 0.011164099 | 0 |
| 0 | 0 | 0 | 0 | 0 |
| 0.018794288 | 0 | 0 | 0.004104904 | 0.001817832 |
| 0.004199446 | 0 | 0.00542179 | 0 | 0 |
| 0 | 0 | 0 | 0 | 0 |
| 0 | 0 | 0 | 0 | 0 |
| 0 | 0 | 0 | 0 | 0 |
| 0 | 0 | 0 | 0 | 0 |
| 0 | 0 | 0 | 0 | 0 |
| 0 | 0 | 0 | 0 | 0 |
| 0 | 0 | 0 | 0 | 0 |
| 0 | 0 | 0.009029901 | 0 | 0.011192802 |
| 0 | 0 | 0 | 0 | 0 |
| 0 | 0 | 0 | 0 | 0 |
| 0 | 0 | 0 | 0 | 0 |
| 0 | 0 | 0 | 0 | 0 |
| 0.787931214 | 0 | 0 | 0 | 0 |
| 0 | 0.00131368 | 0 | 0 | 0 |
| 0 | 0 | 0 | 0 | 0 |
| 0 | 0.008382233 | 0 | 0 | 0 |
| 0 | 0 | 0 | 0 | 0 |
| 0 | 0 | 0 | 0 | 0 |
| 0 | 0.000553796 | 0.010688276 | 0.003237781 | 0.001923155 |
| 0 | 0 | 0 | 0 | 0 |
| 0 | 0.003536573 | 0.012810505 | 0.069131068 | 0.001374141 |
| 0 | 0 | 0.001840914 | 0 | 0.002808446 |
| 0 | 0 | 0 | 0 | 0 |
| 0 | 0 | 0 | 0 | 0 |
| 0 | 0 | 0 | 0 | 0 |
| 0 | 0 | 0 | 0 | 0 |
| 0 | 0 | 0 | 0 | 0 |
| 0 | 0 | 0 | 0 | 0 |
| 0 | 0 | 0 | 0 | 0 |
| 0 | 0 | 0 | 0.246738216 | 0.227087813 |
| 0 | 0.048013203 | 0.109738277 | 0.006698534 | 0.016871428 |
| 0 | 0 | 0 | 0 | 0 |
| 0 | 0 | 0 | 0 | 0 |
| 0 | 1.009770272 | 0.054212715 | 0 | 0.248033114 |
| 0 | 0 | 0 | 0 | 0 |
| 0 | 0 | 0 | 0 | 0 |

| | | | | |
|---|---|---|---|---|
| 0 | 0 | 0 | 0 | 0 |
| 0 | 0.007426875 | 0 | 0 | 0 |
| 0.001244661 | 0 | 0 | 0.004098651 | 0.004213538 |
| 0 | 0 | 0 | 0 | 0 |
| 0.333254855 | 0.005037723 | 0 | 0 | 0.078388353 |
| 0.57396734 | 0 | 0 | 0.21322975 | 0 |
| 0 | 0 | 0 | 0 | 0 |
| 0 | 0 | 0 | 0 | 0 |
| 0 | 0.002730375 | 0 | 0 | 0 |
| 0 | 0 | 0 | 0 | 0 |
| 0 | 0 | 0 | 0 | 0 |
| 0 | 0 | 0 | 0 | 0 |
| 0 | 0 | 0 | 0 | 0 |
| 0 | 0 | 0 | 0 | 0 |
| 0 | 0 | 0 | 0 | 0 |
| 0 | 0 | 0 | 0 | 0 |
| 0 | 0.000830256 | 0 | 0 | 0.992357212 |
| 0 | 0 | 0 | 0 | 0 |
| 0 | 0 | 0 | 0 | 0 |
| 0 | 0 | 0 | 0 | 0 |
| 0.000728243 | 0 | 0 | 0 | 0 |
| 0 | 0 | 0 | 0 | 0 |
| 0 | 0 | 0 | 0 | 0 |
| 0 | 0 | 0 | 0 | 0 |
| 0 | 0.026973974 | 0.002225645 | 0 | 0.004514312 |
| 0.028721381 | 0 | 0.001831865 | 0 | 0 |
| 0 | 0 | 0 | 0 | 0 |
| 0 | 0.045667207 | 0 | 0 | 0.018976291 |
| 0 | 0 | 0 | 0 | 0 |
| 0 | 0 | 0 | 0 | 0 |
| 0 | 0 | 0 | 0 | 0 |
| 0.0007541 | 0 | 0 | 0 | 0 |
| 0 | 0 | 0 | 0 | 0 |
| 0 | 0 | 0 | 0 | 0 |
| 0 | 0 | 0 | 0 | 0 |
| 0 | 0 | 0 | 0 | 0 |
| 0.000524121 | 0 | 0 | 0 | 0 |
| 0 | 0 | 0 | 0.061307356 | 0 |
| 0 | 0 | 1.191582996 | 0 | 0.539337794 |
| 0 | 0 | 0 | 0 | 0 |
| 0.007193464 | 0 | 0 | 0 | 0 |
| 0 | 0.001064006 | 0.003753772 | 0 | 0.018758978 |
| 0.34779914 | 0.001312439 | 0.013277887 | 0.097833431 | 0.001051773 |
| 0 | 0 | 0 | 0 | 0 |
| 0 | 0 | 0 | 0 | 0 |
| 0 | 0.002642351 | 0 | 0 | 0 |
| 0 | 0.008533045 | 0.019687567 | 0.174447303 | 0.008313198 |
| 0 | 0 | 0 | 0 | 0 |

| | | | | |
|---|---|---|---|---|
| 0 | 0 | 0 | 0 | 0 |
| 0.027487759 | 0.032301398 | 0.054416002 | 0.0092574 | 0.003722161 |
| 0 | 0 | 0 | 0 | 0 |
| 0 | 0 | 0 | 0 | 0 |
| 0 | 0 | 0 | 0 | 0 |
| 0 | 0 | 0 | 0 | 0 |
| 0 | 0 | 0 | 0 | 0 |
| 0 | 0 | 0 | 0 | 0 |
| 0 | 0 | 0 | 0 | 0 |
| 0 | 0 | 0 | 0 | 0 |
| 0 | 0 | 0 | 0 | 0 |
| 0 | 0.020202918 | 0 | 0 | 0 |
| 0 | 0.009205878 | 0.025426628 | 0 | 0.05937751 |
| 0 | 0 | 0 | 0 | 0 |
| 0.046112888 | 0.028824247 | 0.013520545 | 0 | 0.021517243 |
| 0 | 0 | 0 | 0 | 0 |
| 0 | 0 | 0 | 0 | 0 |
| 0 | 0 | 0 | 0 | 0 |
| 0 | 0.037128792 | 0 | 0 | 0.01470225 |
| 0 | 0 | 0 | 0 | 0 |
| 0 | 0 | 0 | 0 | 0 |
| 0.024479187 | 0 | 0 | 0.004534291 | 0 |
| 0 | 0.006108478 | 0.036546159 | 0.006000621 | 0.003024712 |
| 0 | 0 | 0 | 0.01271359 | 0 |
| 0 | 0 | 0 | 0 | 0 |
| 0 | 0 | 0 | 0 | 0 |
| 0.003634079 | 0 | 0 | 0 | 0 |
| 0 | 0 | 0 | 0 | 0 |
| 0 | 0 | 0 | 0 | 0 |
| 0 | 0 | 0 | 0 | 0 |
| 0 | 0 | 0 | 0 | 0 |
| 0 | 0 | 0 | 0 | 0 |
| 0 | 0 | 0 | 0 | 0 |
| 0 | 0 | 0 | 0 | 0 |
| 10.48906722 | 5.489447428 | 0 | 0 | 14.41364785 |
| 0 | 0 | 0 | 0 | 0 |
| 0 | 0 | 0 | 0 | 0 |
| 0 | 0 | 0 | 0 | 0 |
| 0.001629695 | 0.014134325 | 0 | 0 | 0.083789302 |
| 0 | 0 | 0 | 0 | 0 |
| 0 | 0.023089695 | 0.051880505 | 0.247659093 | 0.312755876 |
| 0 | 0 | 0 | 0 | 0 |
| 0 | 0 | 0 | 0 | 0 |
| 0 | 0 | 0 | 0 | 0 |
| 0 | 0 | 0 | 0 | 0 |
| 2.934890739 | 0.136553542 | 0.055934668 | 0.055522635 | 0.002548642 |
| 0 | 0 | 0 | 0 | 0 |
| 0 | 0 | 0 | 0 | 0 |

| | | | | |
|---|---|---|---|---|
| 0 | 0 | 0 | 0 | 0 |
| 0 | 0 | 0.000806236 | 0 | 0 |
| 0 | 0 | 0 | 0 | 0 |
| 0 | 0 | 0 | 0 | 0 |
| 0 | 0.000753752 | 0.002627925 | 0.075797491 | 0 |
| 0 | 0 | 0 | 0 | 0 |
| 0 | 0 | 0 | 0 | 0 |
| 0 | 0 | 0 | 0 | 0 |
| 0 | 0 | 0 | 0 | 0 |
| 0 | 0 | 0 | 0.002020511 | 0 |
| 0 | 0.015171304 | 0 | 0 | 0 |
| 0 | 0 | 0 | 0 | 0 |
| 0.010195289 | 0.002090056 | 0.002365874 | 0.015305313 | 0.003248377 |
| 0 | 0 | 0 | 0 | 0 |
| 0 | 0 | 0 | 0 | 0 |
| 0 | 0 | 0 | 0 | 0 |
| 0 | 0 | 0 | 0 | 0 |
| 0 | 0 | 0 | 0 | 0 |
| 0 | 0 | 0 | 0 | 0 |
| 1.105135039 | 0.017419144 | 2.037145566 | 0.050975671 | 0.022226754 |
| 0 | 0 | 0 | 0 | 0 |
| 0 | 0 | 0 | 0 | 0 |
| 0 | 0 | 0 | 0 | 0 |
| 0 | 0 | 0 | 0 | 0 |
| 0 | 0 | 0 | 0 | 0 |
| 0 | 0.009465029 | 0 | 0 | 0 |
| 0 | 0 | 0 | 0 | 0 |
| 0 | 0 | 0 | 0 | 0 |
| 0 | 0 | 0 | 0 | 0 |
| 0 | 0 | 0.081490845 | 0.004462856 | 0.000965885 |
| 0 | 0 | 0 | 0 | 0 |
| 0.000996732 | 0 | 0 | 0 | 0 |
| 0 | 0 | 0 | 0 | 0 |
| 0 | 0 | 0 | 0 | 0 |
| 0 | 0.029828852 | 0 | 0 | 0 |
| 0 | 0 | 0 | 0 | 0 |
| 0 | 1.054964008 | 0 | 0 | 0 |
| 0 | 0 | 0 | 0 | 0 |
| 0 | 0 | 0 | 0 | 0 |
| 0 | 0.248336294 | 0 | 0 | 0 |
| 0 | 0 | 0 | 0 | 0 |
| 0 | 0 | 0 | 0 | 0 |
| 0 | 0.001546352 | 0 | 0.000913211 | 0.007510469 |
| 0 | 0 | 0 | 0 | 0 |
| 0 | 0.013342707 | 0.012064821 | 0.040448844 | 0.027495462 |
| 0 | 0 | 0 | 0 | 0.024193071 |
| 0 | 0 | 0 | 0 | 0 |
| 0.002342097 | 0 | 0 | 0 | 0 |

| | | | | |
|---|---|---|---|---|
| 0 | 0 | 0 | 0 | 0 |
| 0 | 0 | 0 | 0 | 0 |
| 0 | 0 | 0 | 0 | 0 |
| 0 | 0 | 0 | 0 | 0 |
| 0 | 0 | 0 | 0 | 0 |
| 0.152334051 | 0.034507229 | 0.091397462 | 0.026807216 | 0.092749061 |
| 0 | 0 | 0 | 0 | 0 |
| 0 | 0 | 0 | 0 | 0 |
| 0 | 0 | 0 | 0 | 0 |
| 0 | 0 | 0 | 0 | 0 |
| 0 | 0 | 0 | 0 | 0 |
| 0 | 0 | 0 | 0 | 0 |
| 0 | 0 | 0 | 0 | 0 |
| 0 | 0 | 0 | 0 | 0 |
| 0 | 0 | 0 | 0 | 0 |
| 0 | 0 | 0 | 0 | 0 |
| 0 | 0.021575786 | 0.113909222 | 0 | 0 |
| 0 | 0 | 0 | 0 | 0 |
| 0 | 0 | 0 | 0 | 0 |
| 0 | 0 | 0 | 0 | 0 |
| 0 | 0 | 0 | 0 | 0 |
| 0 | 0 | 0 | 0 | 0 |
| 0 | 0 | 0 | 0 | 0 |
| 0 | 0 | 0 | 0 | 0.020912019 |
| 0 | 0 | 0 | 0 | 0 |
| 0 | 0 | 0 | 0 | 0 |
| 0 | 0 | 0 | 0 | 0 |
| 0 | 0 | 0.290048413 | 0 | 0.945020209 |
| 0 | 0 | 0 | 0 | 0 |
| 0.009472936 | 0 | 0 | 0 | 0 |
| 0 | 0 | 0 | 0 | 0 |
| 0 | 0 | 0 | 0 | 0 |
| 0 | 0 | 0 | 0 | 0 |
| 0 | 0 | 0 | 0 | 0 |
| 0 | 0 | 0 | 0 | 0 |
| 0 | 0 | 0 | 0 | 0 |
| 0 | 0 | 0 | 0 | 0 |
| 0 | 0 | 0 | 0 | 0 |
| 0 | 1.255246187 | 0 | 0 | 0.038275483 |
| 0 | 0 | 0 | 0 | 0 |
| 0 | 0 | 0 | 0 | 0 |
| 0 | 0 | 0 | 0 | 0 |
| 0 | 0 | 0 | 0 | 0 |
| 0 | 0 | 0 | 0 | 0 |
| 0 | 0 | 0 | 0 | 0 |
| 0 | 0 | 0 | 0 | 0 |
| 0 | 0 | 0 | 0 | 0 |
| 0 | 0 | 0 | 0 | 0 |

| | | | | |
|---|---|---|---|---|
| 0 | 0 | 0 | 0 | 0 |
| 0 | 0 | 0.071413875 | 0.021905923 | 0.006951042 |
| 0 | 0 | 0 | 0 | 0 |
| 0 | 0 | 0 | 0 | 0 |
| 0 | 0 | 0 | 0 | 0 |
| 0 | 0 | 0 | 0 | 0 |
| 0 | 0 | 0 | 0 | 0 |
| 0 | 0 | 0 | 0 | 0 |
| 0 | 0 | 0 | 0 | 0 |
| 0 | 0 | 0 | 0 | 0 |
| 0 | 0 | 0.002813983 | 0 | 0 |
| 0 | 0 | 0 | 0 | 0 |
| 0 | 0.209733661 | 0 | 0 | 0.011082063 |
| 0 | 0 | 0 | 0 | 0 |
| 0 | 0 | 0 | 0 | 0 |
| 0 | 0 | 0 | 0 | 0 |
| 0 | 0 | 0 | 0 | 0 |
| 0 | 0 | 0 | 0 | 0 |
| 0 | 0 | 0 | 0 | 0 |
| 0 | 0 | 0 | 0 | 0 |
| 0.001485067 | 0 | 0.003075717 | 0 | 0 |
| 0.014929445 | 0 | 0 | 0 | 0 |
| 0 | 0 | 0 | 0 | 0.010377093 |
| 0 | 0.4051586 | 1.247208214 | 0 | 0.250037137 |
| 0 | 0 | 0 | 0 | 0 |
| 0 | 0 | 0 | 0 | 0 |
| 0 | 0 | 0 | 0 | 0 |
| 0 | 0 | 0 | 0 | 0 |
| 0 | 0 | 0 | 0 | 0 |
| 0 | 0 | 0 | 0 | 0 |
| 0 | 0 | 0.005822 | 0 | 0 |
| 0 | 0 | 0 | 0 | 0 |
| 0 | 0.000742383 | 0 | 0 | 0 |
| 0 | 0 | 0 | 0 | 0 |

| PHL018-0043_V2 | PHL018-0030_V2 | PHL018-0035_V2 | PHL018-0169_V3 | PHL018-0205_V3 |
|---|---|---|---|---|
| EYCF | CTRL | CTRL | REF | REF |
| 2.436034178 | 0 | 1.168426779 | 0.072760026 | 0.02975936 |
| 0.293690928 | 0.402633521 | 1.553055496 | 0.417778775 | 2.333095014 |
| 5.205311889 | 26.67134714 | 1.271071658 | 1.920319036 | 1.133699724 |
| 0 | 0 | 0 | 0 | 0 |
| 0 | 0 | 0 | 0 | 0 |
| 0 | 0 | 0 | 0 | 0 |
| 0 | 0 | 0 | 0 | 0 |
| 0 | 0 | 0 | 0 | 0 |
| 0 | 0 | 0 | 0 | 0 |
| 0 | 0 | 0 | 0 | 0 |
| 0 | 0 | 0 | 0 | 0 |
| 0 | 0 | 0 | 0 | 0 |
| 0 | 0 | 0 | 0 | 0 |
| 11.54540269 | 0.108719027 | 1.733028789 | 1.997498531 | 0.998519506 |
| 0 | 0 | 0 | 0 | 0 |
| 0 | 0 | 0 | 0 | 0 |
| 0 | 0 | 0 | 0 | 0 |
| 0 | 0 | 0 | 0 | 0 |
| 0 | 0 | 0 | 0 | 0 |
| 0 | 0 | 0 | 0 | 0 |
| 0 | 0 | 0 | 0 | 0 |
| 0 | 0 | 0 | 0 | 0 |
| 0 | 0 | 0 | 0 | 0 |
| 0 | 0 | 0 | 0 | 0 |
| 0 | 0 | 0 | 0.021519513 | 0 |
| 0 | 0 | 0 | 0 | 0 |
| 0 | 0 | 0 | 0 | 0 |
| 0 | 0 | 0 | 0 | 0 |
| 0 | 0 | 0 | 0 | 0 |
| 0 | 0 | 0 | 0 | 0 |
| 0 | 0 | 0 | 0 | 0 |
| 0 | 0 | 0 | 0 | 0 |
| 0 | 0 | 0 | 0 | 0 |
| 0 | 0 | 0 | 0 | 0 |
| 0 | 0 | 0 | 0 | 0 |
| 0 | 0 | 0.772463997 | 0.015282235 | 0.007013895 |
| 0 | 0 | 0 | 0 | 0 |
| 0 | 0 | 0 | 0 | 0 |
| 0 | 0 | 0 | 0 | 0 |
| 0 | 0 | 0 | 0 | 0 |
| 0 | 0 | 0 | 0 | 0 |
| 0.011027608 | 0.041174589 | 0.005273401 | 0.003360123 | 0.32477749 |
| 0.002762901 | 0.423559233 | 1.291253433 | 0.555394072 | 0.363705459 |
| 0.022347286 | 0.05634615 | 0.007436477 | 0.556302726 | 0.040171811 |
| 0.002705766 | 0 | 0.004987384 | 0.028071251 | 0 |
| 0 | 0 | 0 | 0 | 0 |

| | | | | |
|---|---|---|---|---|
| 0 | 0 | 0 | 0.00394699 | 0 |
| 0.027853409 | 0.132436483 | 0 | 0 | 0.000816549 |
| 0.361773544 | 0.003166358 | 0.229983409 | 0 | 0 |
| 0.106303754 | 0.102281469 | 0.148656981 | 0.485549743 | 0.137558361 |
| 0 | 0 | 0.000371008 | 0.003151999 | 0 |
| 0 | 0 | 0 | 0 | 0 |
| 0 | 0.001026149 | 0.002144825 | 0 | 0.130624846 |
| 0 | 0 | 0 | 0 | 0 |
| 0 | 0 | 0 | 0 | 0 |
| 0 | 0 | 0 | 0 | 0 |
| 0 | 0 | 0 | 0 | 0 |
| 0 | 0 | 0.062564513 | 0.198789492 | 0 |
| 0 | 0.008000655 | 0 | 0 | 0 |
| 0.003367048 | 0.165841369 | 0.000853365 | 0.005256247 | 0.042935573 |
| 0 | 0 | 0 | 0 | 0 |
| 0 | 0 | 0 | 0 | 0 |
| 0 | 0 | 0 | 0.688274065 | 10.76001731 |
| 0 | 0 | 0 | 0 | 0 |
| 0 | 0 | 0 | 0 | 0 |
| 0 | 0 | 0 | 0 | 0 |
| 0.005159003 | 0.369641608 | 0.000541046 | 0.000957627 | 0.040898944 |
| 0 | 0.005103786 | 0.003047938 | 0.00251753 | 0 |
| 0 | 0 | 0.418780636 | 0.117385407 | 0.572467693 |
| 0 | 0 | 0 | 0 | 0 |
| 0 | 0 | 0.00299218 | 0 | 0 |
| 0 | 0 | 0 | 0 | 0 |
| 0 | 0 | 0 | 0 | 0 |
| 0 | 0 | 0 | 0 | 0 |
| 0 | 0 | 0 | 0 | 0.006762127 |
| 0 | 0 | 0 | 0 | 0 |
| 0.026117978 | 0.018726399 | 0 | 0 | 0.001056961 |
| 0 | 0 | 0 | 0 | 0 |
| 0 | 0 | 0 | 0 | 0 |
| 0 | 0 | 0 | 0 | 0 |
| 0 | 0 | 0 | 0 | 0 |
| 0 | 0 | 0 | 0 | 0 |
| 0 | 0 | 0 | 0 | 0 |
| 0 | 0 | 0 | 0 | 0.225845731 |
| 0 | 0 | 0 | 0 | 0 |
| 0 | 0 | 0 | 0 | 0 |
| 0 | 0 | 0 | 0 | 0 |
| 0 | 0 | 0 | 0 | 0 |
| 0 | 0 | 0 | 0 | 0 |
| 0 | 0 | 0 | 0 | 0 |
| 0 | 0 | 0 | 0.000987947 | 0 |
| 0 | 0 | 0 | 0 | 0 |
| 0 | 0 | 0 | 0 | 0 |
| 0 | 0 | 0 | 0 | 0 |

| | | | | |
|---|---|---|---|---|
| 0 | 0 | 0 | 0 | 0 |
| 0 | 0 | 0 | 0 | 0 |
| 0 | 0 | 0 | 0 | 0 |
| 0 | 0 | 0 | 0 | 0 |
| 0 | 0.00193648 | 0 | 0 | 0 |
| 0 | 0 | 0 | 0 | 0 |
| 0 | 0 | 0 | 0 | 0 |
| 0 | 0 | 0 | 0 | 0 |
| 0 | 0 | 0 | 0.004519154 | 0 |
| 0 | 0 | 0 | 0 | 0 |
| 0 | 0 | 0 | 0 | 0 |
| 0 | 0 | 0 | 0 | 0.004535978 |
| 0 | 0 | 0 | 0 | 0 |
| 0 | 0 | 0 | 0 | 0 |
| 0 | 0 | 0 | 0 | 0 |
| 0 | 0 | 0 | 0 | 0 |
| 0 | 0 | 0 | 0 | 0 |
| 0 | 0 | 0 | 0 | 0 |
| 0.005838286 | 0.161650286 | 0.006933404 | 1.543014618 | 0.003105237 |
| 0 | 0 | 0 | 0 | 0 |
| 0 | 0 | 0 | 0 | 0 |
| 0 | 0 | 0 | 0 | 0 |
| 0 | 0 | 0 | 0.000760451 | 0 |
| 0 | 0 | 0 | 0 | 0 |
| 0 | 0 | 0 | 0 | 0 |
| 0 | 0 | 0 | 0 | 0 |
| 0 | 0 | 0 | 0 | 0 |
| 0 | 0 | 0 | 0 | 0 |
| 0 | 0 | 0 | 0 | 0 |
| 0 | 0 | 0 | 0 | 0 |
| 0 | 0 | 0.003634944 | 0.056455555 | 0 |
| 0.011598035 | 0 | 0.000794453 | 0 | 0.001750577 |
| 0 | 0 | 0 | 0 | 0 |
| 0 | 0 | 0 | 0 | 0 |
| 0 | 0 | 0 | 0 | 0 |
| 0 | 0 | 0 | 0 | 0 |
| 0 | 0 | 0 | 0 | 0 |
| 0 | 0 | 0 | 0 | 0 |
| 0 | 0 | 0 | 0 | 0 |
| 0 | 0 | 0 | 0 | 0 |
| 0 | 0 | 0 | 0 | 0 |
| 0 | 0 | 0 | 0 | 0 |
| 0 | 0 | 0 | 0 | 0 |
| 0 | 0 | 0 | 0 | 0 |
| 0 | 0 | 0 | 0 | 0 |
| 0 | 0 | 0 | 0 | 0 |
| 0 | 0 | 0 | 0 | 0 |
| 0 | 0 | 0 | 0 | 0 |

| | | | | |
|---|---|---|---|---|
| 0 | 0 | 0 | 0 | 0 |
| 0 | 0 | 0 | 0.002346941 | 0 |
| 0 | 0 | 0 | 0 | 0 |
| 0 | 0 | 0 | 0 | 0 |
| 0 | 0 | 0.012446659 | 0.114115449 | 0.001650653 |
| 0 | 0 | 0 | 0 | 0 |
| 0 | 0 | 0 | 0 | 0 |
| 0 | 0 | 0 | 0 | 0 |
| 0 | 0 | 0 | 0 | 0 |
| 0 | 0 | 0 | 0 | 0 |
| 0 | 0 | 0 | 0 | 0 |
| 0 | 0 | 0 | 0 | 0 |
| 0 | 0 | 0 | 0 | 0 |
| 0 | 0 | 0 | 0 | 0 |
| 0 | 0 | 0 | 0 | 0 |
| 0 | 0.001075699 | 0 | 1.49989292 | 0 |
| 0 | 0 | 0 | 0 | 0 |
| 0 | 0 | 0 | 0 | 0 |
| 0 | 0 | 0 | 0 | 0 |
| 0 | 0 | 0 | 0 | 0 |
| 0 | 0 | 0 | 0 | 0 |
| 0 | 0 | 0 | 0 | 0 |
| 0 | 0 | 0 | 0.002578686 | 0 |
| 0 | 0 | 0 | 0 | 0 |
| 0 | 0 | 0 | 0 | 0 |
| 0 | 0 | 0 | 0 | 0 |
| 0 | 0 | 0 | 0.008614813 | 0 |
| 0 | 0 | 0 | 0 | 0 |
| 0 | 0 | 0 | 0 | 0 |
| 0 | 0 | 0 | 0 | 0 |
| 0 | 0 | 0 | 0 | 0 |
| 0 | 0 | 1.424438131 | 0 | 0 |
| 0 | 0 | 0 | 0 | 0 |
| 0 | 0 | 0 | 0 | 0 |
| 0 | 0 | 0 | 0 | 0 |
| 0 | 0 | 0 | 0 | 0 |
| 0 | 0 | 0 | 0 | 0 |
| 0 | 0 | 0 | 0.013791563 | 0 |
| 0 | 0 | 0 | 0 | 0 |
| 0 | 0 | 0 | 0 | 0 |
| 0 | 0 | 0 | 0 | 0 |
| 0 | 0 | 0 | 0 | 0 |
| 0 | 0 | 0 | 0 | 0 |
| 0 | 0 | 0 | 0 | 0 |
| 0 | 0 | 0 | 0 | 0 |
| 0 | 0 | 0 | 0 | 0 |
| 0 | 0 | 0 | 0 | 0 |
| 0 | 0 | 0 | 0 | 0 |

| | | | | |
|---|---|---|---|---|
| 0 | 0 | 0 | 0 | 0 |
| 0 | 0 | 0 | 0 | 0 |
| 0 | 0 | 0 | 0 | 0 |
| 0 | 0 | 0 | 0 | 0 |
| 0 | 0 | 0 | 0 | 0 |
| 0 | 0 | 0 | 0 | 0 |
| 0 | 0 | 0 | 0 | 0 |
| 0 | 0 | 0 | 0 | 0 |
| 0 | 0 | 0 | 0 | 0 |
| 0 | 0 | 0 | 0 | 0 |
| 0 | 0 | 0 | 0 | 0 |
| 0 | 0 | 0 | 0.001511853 | 0 |
| 0 | 0.002014279 | 0.007894098 | 0 | 0 |
| 0 | 0 | 0 | 0 | 0 |
| 0 | 0 | 0 | 0 | 0 |
| 0 | 0 | 0 | 0 | 0 |
| 0 | 0 | 0 | 0 | 0 |
| 0 | 0 | 0 | 0 | 0 |
| 0 | 0 | 0 | 0 | 0 |
| 0 | 0 | 0 | 0 | 0 |
| 0.007117922 | 0.125697511 | 0.021648098 | 0.07175566 | 0.009335544 |
| 0 | 0 | 0 | 0 | 0 |
| 0 | 0 | 0 | 0 | 0 |
| 0 | 0 | 0 | 0 | 0 |
| 0 | 0 | 0 | 0 | 0 |
| 0 | 0 | 0 | 0 | 0 |
| 0.002637924 | 0.001096678 | 0.002865305 | 0.001420008 | 0.003682994 |
| 0 | 0 | 0 | 0 | 0 |
| 0 | 0 | 0 | 0 | 0 |
| 0 | 0.093865544 | 0 | 0 | 0 |
| 0 | 0.049740687 | 0.008949497 | 0 | 0 |
| 0 | 0 | 0 | 0 | 0 |
| 0 | 0 | 0 | 0 | 0 |
| 0 | 0 | 0.127087922 | 0.016391507 | 0 |
| 0.06610127 | 0 | 0.033506194 | 0.014051745 | 0 |
| 0 | 0 | 0 | 0 | 0 |
| 0 | 0 | 0 | 0 | 0 |
| 0 | 0 | 0 | 0 | 0 |
| 0 | 0 | 0 | 0 | 0 |
| 0 | 0 | 0 | 0 | 0 |
| 0 | 0 | 0 | 0 | 0 |
| 0 | 0 | 0 | 0 | 0 |
| 0 | 0 | 0 | 0 | 0 |
| 0 | 0 | 0 | 0 | 0 |
| 0 | 0 | 0 | 0 | 0 |
| 0 | 0 | 0.000812778 | 0.039920589 | 0 |
| 0 | 0 | 0 | 0 | 0 |
| 0 | 0.00065486 | 0 | 0 | 0 |

| | | | | |
|---|---|---|---|---|
| 0 | 0 | 0 | 0 | 0 |
| 0 | 0 | 0 | 0 | 0 |
| 0 | 0 | 0 | 0 | 0 |
| 0 | 0.001109884 | 0 | 0 | 0 |
| 0 | 0 | 0 | 0 | 0 |
| 0 | 0 | 0 | 0 | 0 |
| 0 | 0 | 0 | 0 | 0.000993833 |
| 0 | 0 | 0 | 0 | 0.007226129 |
| 0 | 0 | 0 | 0 | 0 |
| 0 | 0 | 0 | 0 | 0 |
| 0 | 0 | 0 | 0 | 0 |
| 0 | 0 | 0 | 0 | 0 |
| 0 | 0 | 0 | 0 | 0 |
| 0 | 0 | 0 | 0 | 0 |
| 0 | 0 | 0 | 0 | 0 |
| 0 | 0 | 0 | 0 | 0 |
| 0 | 0 | 0 | 0 | 0 |
| 0 | 0 | 0 | 0 | 0 |
| 0 | 0.001775969 | 0.006650807 | 0.003449358 | 0.003692167 |
| 0 | 0 | 0 | 0.01317602 | 0 |
| 0.12827955 | 0.047810174 | 0 | 0.001211465 | 0 |
| 0 | 0 | 0 | 0 | 0 |
| 0 | 0 | 0 | 0 | 0 |
| 0 | 0 | 0 | 0 | 0 |
| 0.004918244 | 0.027975039 | 0 | 0 | 0.002080821 |
| 0 | 0 | 0 | 0.002335556 | 0 |
| 0 | 0 | 0 | 0 | 0 |
| 0 | 0 | 0 | 0 | 0 |
| 0.004906895 | 0 | 0 | 0 | 0 |
| 0 | 0 | 0 | 0 | 0 |
| 0.002813169 | 1.577391139 | 0.023151081 | 0.045608447 | 0.037120288 |
| 0 | 0 | 0 | 0 | 0 |
| 0.054526945 | 0 | 0 | 0 | 0.140000654 |
| 0 | 0 | 0 | 0.003687946 | 0 |
| 0 | 0 | 0 | 0 | 0 |
| 0 | 0 | 0 | 0 | 0 |
| 0 | 0 | 0 | 0 | 0 |
| 0 | 0 | 0 | 0 | 0 |
| 0 | 0 | 0 | 0 | 0 |
| 0 | 0 | 0 | 0 | 0 |
| 0 | 0 | 0 | 0 | 0 |
| 0 | 0 | 0.003249093 | 0.029237995 | 0.001727044 |
| 0 | 0.002883084 | 0 | 0 | 0 |
| 0 | 0 | 0 | 0 | 0 |
| 0 | 0 | 0 | 0 | 0 |
| 0 | 0 | 0 | 0 | 0 |
| 0 | 0 | 0 | 0 | 0 |
| 0 | 0 | 0 | 0 | 0 |

| | | | | |
|---|---|---|---|---|
| 0 | 0 | 0 | 0 | 0 |
| 0 | 0 | 0 | 0 | 0 |
| 0.016520817 | 0.08074639 | 0.001288349 | 0.646013987 | 0.033514538 |
| 0 | 0 | 0 | 0 | 0 |
| 0 | 0 | 0 | 0 | 0 |
| 0 | 0 | 0 | 0 | 0 |
| 0 | 0 | 0 | 0 | 0 |
| 0 | 0 | 0 | 0 | 0 |
| 0 | 0 | 0 | 0 | 0 |
| 0 | 0 | 0 | 0 | 0 |
| 0 | 0 | 0 | 0 | 0 |
| 0 | 0 | 0 | 0 | 0 |
| 0 | 0 | 0 | 0 | 0 |
| 0 | 0 | 0 | 0.085181501 | 0 |
| 0.007148585 | 0.008996078 | 0.000587604 | 0.012064374 | 0.002157978 |
| 0 | 0 | 0 | 0.026278406 | 0 |
| 0.014088945 | 0.001921917 | 0 | 0.002133042 | 0.001229411 |
| 0 | 0 | 0.001731199 | 0 | 0 |
| 0 | 0 | 0 | 0 | 0 |
| 0 | 0 | 0 | 0 | 0 |
| 0 | 0 | 0 | 0 | 0 |
| 0 | 0 | 0 | 0 | 0.007583684 |
| 0 | 0 | 0 | 0 | 0 |
| 0 | 0 | 0 | 0 | 0 |
| 0 | 0 | 0 | 0 | 0 |
| 0 | 0 | 0 | 0 | 0 |
| 0 | 0 | 10.11542207 | 0 | 0 |
| 0 | 0 | 0 | 0 | 0 |
| 0 | 0 | 0 | 0 | 0 |
| 0 | 0.003346147 | 0 | 0 | 0 |
| 0 | 0 | 0 | 0 | 0 |
| 0 | 0 | 0 | 0 | 0 |
| 0 | 0 | 0 | 0 | 0 |
| 0 | 0 | 0 | 0 | 0 |
| 0 | 0 | 0 | 0 | 0 |
| 0 | 0 | 0 | 0 | 0.105292423 |
| 0 | 0 | 0 | 0 | 0 |
| 0 | 0 | 0.022780513 | 0.093274642 | 0.007436577 |
| 0 | 0 | 0 | 0 | 0 |
| 0 | 0 | 0 | 0 | 0 |
| 0 | 0 | 0 | 0 | 0 |
| 0 | 0 | 0 | 0 | 0 |
| 0 | 0 | 0 | 0 | 0 |
| 0 | 0 | 0 | 0 | 0 |
| 0 | 0 | 0 | 0 | 0 |
| 0 | 0 | 0 | 0 | 0 |
| 0 | 0 | 0 | 0 | 0 |
| 0 | 0 | 0 | 0 | 0 |

| | | | | |
|---|---|---|---|---|
| 0 | 0.630672168 | 0 | 0.23052707 | 0 |
| 0 | 0 | 0 | 0 | 0 |
| 0 | 0 | 0.007596587 | 0 | 0 |
| 0 | 0 | 0 | 0 | 0 |
| 0 | 0.005180171 | 0 | 0 | 0.003898412 |
| 0 | 0 | 0 | 0 | 0 |
| 0 | 0 | 0 | 0 | 0 |
| 0.009980459 | 0.011105289 | 0 | 0.006004594 | 0.010109292 |
| 0 | 0 | 0 | 0 | 0 |
| 8.308023735 | 0.401946904 | 0.008895212 | 0.014006831 | 0.02956989 |
| 0 | 0 | 0 | 0 | 0 |
| 0.022075114 | 0 | 0 | 1.724932283 | 0.026603077 |
| 0.001586345 | 0.00487297 | 0.011972239 | 0.013852768 | 0.214754517 |
| 0 | 0.005063695 | 0 | 0 | 0 |
| 0 | 0 | 0 | 0 | 0 |
| 0.002116771 | 0 | 0 | 0 | 0 |
| 0 | 0 | 0 | 0 | 0 |
| 0.012396049 | 0 | 0 | 0 | 0 |
| 0.036726871 | 0.370767924 | 0 | 0 | 0 |
| 0 | 0.177984008 | 0 | 0 | 0 |
| 0 | 0 | 0 | 0 | 0 |
| 0 | 0 | 0 | 0 | 0 |
| 0 | 0 | 0 | 0 | 0 |
| 0 | 0 | 0 | 0 | 0 |
| 0 | 0 | 0 | 0 | 0 |
| 0.003490077 | 0 | 0.087205818 | 0.051720227 | 0.002675233 |
| 0 | 0 | 0.550775332 | 0.00118223 | 0 |
| 0.002730895 | 0.401700127 | 0 | 0 | 0 |
| 0 | 0 | 0 | 0 | 0 |
| 0 | 0 | 0 | 0 | 0 |
| 0 | 0.009616433 | 0 | 0 | 0 |
| 0 | 0 | 0 | 0 | 0 |
| 0.007564085 | 0 | 0 | 0 | 0 |
| 0.022553407 | 0 | 0 | 0 | 0 |
| 0 | 0 | 0 | 0 | 0 |
| 0 | 0 | 0 | 0 | 0 |
| 0.004887269 | 0 | 0 | 0 | 0 |
| 0 | 0 | 0 | 0 | 0 |
| 0 | 0 | 0 | 0 | 0 |
| 0.007557258 | 7.992158 | 0 | 0 | 0 |
| 0 | 0 | 0 | 0 | 0 |
| 0.002652185 | 0.445904192 | 0 | 0 | 0 |
| 0 | 0 | 0 | 0 | 0 |
| 0 | 0.019734374 | 0 | 0 | 0 |
| 0 | 0 | 0 | 0 | 0 |
| 0 | 0 | 0 | 0.015686048 | 0 |
| 0.008425069 | 0 | 0 | 0.004947553 | 0 |
| 0 | 0 | 0 | 0 | 0 |

| | | | | |
|---|---|---|---|---|
| 0 | 0 | 0 | 0 | 0 |
| 0.009415604 | 0 | 0 | 0 | 0.007170433 |
| 0 | 0 | 0 | 0 | 0 |
| 0 | 0 | 0 | 0.064488775 | 0 |
| 0 | 0 | 0 | 0 | 0 |
| 0 | 0 | 0.000815422 | 0 | 0 |
| 0 | 0 | 0 | 0 | 0 |
| 0 | 0 | 0 | 0 | 0.001936596 |
| 0 | 0 | 0 | 0.678435938 | 0 |
| 0 | 0 | 0 | 0 | 0 |
| 0.019357529 | 0 | 0.000945752 | 0 | 0.063170195 |
| 0 | 0 | 0 | 0 | 0 |
| 0 | 0 | 0 | 0 | 0 |
| 0.001101688 | 0.003206074 | 0 | 0.006879312 | 0.001435602 |
| 0.106120415 | 5.142017498 | 0 | 0 | 0 |
| 0.001572874 | 0 | 0 | 0 | 0 |
| 0 | 0.000895749 | 0 | 0 | 0 |
| 0 | 0.023400207 | 0 | 0 | 0 |
| 0 | 0.036853624 | 0 | 0.001262409 | 0 |
| 0 | 0 | 0 | 0 | 0 |
| 0 | 0 | 0 | 0 | 0.00508235 |
| 0 | 0 | 0 | 0.358851992 | 0.527972437 |
| 0.003136348 | 0 | 0.001695775 | 0.00630303 | 0 |
| 0 | 0 | 0 | 0 | 0 |
| 0 | 0 | 0 | 0 | 0 |
| 0 | 0 | 0 | 0 | 0 |
| 0.006211512 | 0 | 0 | 0 | 0 |
| 0 | 0 | 0 | 0 | 0 |
| 0 | 0 | 0 | 0 | 0 |
| 0.207991884 | 0.614685778 | 0.249821812 | 0.325110171 | 4.426828668 |
| 0 | 0 | 0.014244943 | 0.0062764 | 0.003756633 |
| 0 | 0 | 0 | 0 | 0 |
| 0.001611317 | 0 | 0 | 0 | 0 |
| 0 | 0 | 0 | 0 | 0 |
| 0 | 0 | 0 | 0 | 0 |
| 0 | 0 | 0 | 0 | 0 |
| 0 | 0 | 0 | 0 | 0 |
| 0 | 0 | 0 | 0 | 0.039926117 |
| 0 | 0 | 0 | 0 | 0 |
| 0 | 0.003034885 | 0 | 0 | 0 |
| 0.00126799 | 0.016605171 | 0.006978254 | 0.01228618 | 0 |
| 0 | 0 | 0.360885285 | 0.142062518 | 3.512618053 |
| 0 | 0 | 0 | 0 | 0.03580894 |
| 0 | 0 | 0 | 0 | 0.004881675 |
| 0 | 0.007433134 | 0 | 0 | 0 |
| 0 | 0 | 0 | 0 | 0 |
| 0.002661694 | 0.007469279 | 0 | 0 | 0.001061766 |
| 0 | 0 | 0 | 0 | 0 |

| | | | | |
|---|---|---|---|---|
| 0.002814929 | 0 | 0.005469647 | 0.121475766 | 0 |
| 0 | 0 | 0 | 0 | 0 |
| 0 | 0 | 0 | 0.037508698 | 24.42933683 |
| 0 | 0 | 0 | 0 | 0 |
| 0 | 0 | 0.035380535 | 0.00101549 | 0 |
| 0.030741985 | 0 | 0 | 0 | 0 |
| 0 | 0 | 0 | 0 | 0 |
| 0 | 0 | 0 | 0 | 0.0048943 |
| 0 | 0 | 0 | 0 | 0 |
| 0 | 0 | 0 | 0 | 0 |
| 0 | 0 | 0 | 0 | 0 |
| 0 | 0 | 0 | 0 | 0 |
| 0 | 0 | 0 | 0 | 0 |
| 0 | 0 | 0.002796838 | 0 | 0.005135697 |
| 0 | 0 | 0 | 0 | 0 |
| 0 | 0.101834598 | 0.016308538 | 0.035352621 | 0 |
| 0.052940171 | 0.009361508 | 0.030189308 | 1.18405064 | 0.005303981 |
| 0.007593362 | 0.007474255 | 0 | 0 | 0 |
| 0 | 0 | 0 | 0 | 0 |
| 0 | 0 | 0 | 0 | 0 |
| 0.003243318 | 0 | 0 | 0 | 0 |
| 0.011727193 | 0 | 0.002810089 | 0.010100491 | 0.000992313 |
| 0 | 0 | 0 | 0 | 0 |
| 0 | 0 | 0 | 0 | 0 |
| 0 | 0 | 0 | 0 | 0 |
| 0 | 0.214931516 | 0 | 0.008081751 | 0 |
| 0 | 0.003056017 | 0 | 0.001004955 | 0 |
| 0 | 0 | 0 | 0 | 0 |
| 0 | 0 | 0 | 0 | 0 |
| 0 | 0 | 0 | 0 | 0 |
| 0 | 0.138412592 | 0.001799166 | 0.000955332 | 0 |
| 0.001852106 | 0 | 0.001341168 | 0.020794529 | 0 |
| 0.027711442 | 0 | 0.014812887 | 0.030105638 | 0.961854732 |
| 0.003610291 | 0 | 0 | 0 | 0.029571425 |
| 0 | 0 | 0 | 0.004438784 | 0 |
| 0 | 0 | 0 | 0 | 0 |
| 0 | 0.002319802 | 0 | 0.003432847 | 0 |
| 0 | 0.230920814 | 0.001263991 | 0.000958802 | 0.002321004 |
| 0 | 0 | 0.294169177 | 0.120657708 | 0 |
| 0 | 0 | 0 | 0 | 0 |
| 0 | 0 | 0 | 0 | 0 |
| 0 | 0.009429809 | 0 | 0 | 0 |
| 0 | 0 | 0 | 0 | 0.003966923 |
| 0.000695204 | 0 | 0.001127658 | 0 | 0.000517666 |
| 0 | 0 | 0 | 0 | 0 |
| 0 | 0 | 0 | 0 | 0 |
| 0 | 0 | 0 | 0 | 0 |
| 0 | 0 | 0 | 0 | 0 |

| | | | | |
|---|---|---|---|---|
| 0 | 0 | 0 | 0 | 0 |
| 0.049511672 | 0.861339018 | 0.370550742 | 1.60933349 | 0.154518655 |
| 0 | 0 | 0 | 0 | 0.000556599 |
| 0 | 0 | 0 | 0 | 0 |
| 0 | 0 | 0 | 0 | 0 |
| 0 | 0 | 0 | 0 | 0 |
| 0 | 0.04541298 | 0.00887749 | 0 | 0 |
| 0.000932112 | 0 | 0 | 0 | 0 |
| 0 | 0 | 0 | 0 | 0 |
| 0 | 0 | 0 | 0 | 0 |
| 0 | 0.001049724 | 0 | 0 | 0.006546997 |
| 0.009939869 | 0.064107327 | 0.009804529 | 0.07172805 | 0.003000596 |
| 0 | 0 | 0 | 0.110014304 | 0 |
| 0 | 0 | 0 | 0 | 0 |
| 0 | 0 | 0 | 0.011807822 | 0 |
| 0 | 0 | 0 | 0 | 0 |
| 0.009631787 | 0.091509424 | 0.005935314 | 0.003863214 | 0.00386729 |
| 0.115962813 | 0.256490562 | 0 | 0 | 0.006389443 |
| 0 | 0 | 0 | 0 | 0 |
| 0.01337255 | 0.200139853 | 0.001291131 | 0 | 0 |
| 0.007400911 | 0.071697998 | 0.01401862 | 0.29124755 | 0.148468017 |
| 0 | 0 | 0 | 0 | 0 |
| 0 | 0 | 0 | 0 | 0 |
| 0 | 0 | 0 | 0 | 0 |
| 0 | 0 | 0 | 0 | 0.000913751 |
| 0 | 0.001559267 | 0 | 0 | 0 |
| 0 | 3.253719003 | 0 | 0.015936388 | 0.002326117 |
| 0.022356217 | 0 | 0.007344934 | 0.046577832 | 0.018882002 |
| 0 | 0 | 0 | 0 | 0 |
| 0 | 0 | 0.000759404 | 0 | 0 |
| 0.00133268 | 0 | 0 | 0 | 0 |
| 0 | 0 | 0 | 0 | 0 |
| 0 | 0 | 0 | 0 | 0 |
| 0 | 0 | 0 | 0 | 0 |
| 0 | 0 | 0 | 0 | 0 |
| 7.317871614 | 1.084277176 | 0.066061155 | 0.198018449 | 0.314349198 |
| 0 | 0.005430163 | 0 | 0 | 0 |
| 0 | 0 | 0 | 0 | 0 |
| 0.010725126 | 0.023408767 | 0 | 0.508333144 | 0.021391587 |
| 0 | 0 | 0 | 0 | 0 |
| 0 | 0 | 0 | 0 | 0 |
| 0 | 0 | 0 | 0 | 0 |
| 0 | 0.004062526 | 0.001100733 | 0 | 0 |
| 0 | 0.003268577 | 0.000607278 | 0 | 0 |
| 0 | 0.004007876 | 0.001595646 | 0 | 0 |
| 0 | 0 | 0 | 0.221720614 | 0 |
| 0 | 0 | 0 | 0 | 0 |
| 0 | 0.004578948 | 0 | 0 | 0 |

| | | | | |
|---|---|---|---|---|
| 0 | 0.001891812 | 0 | 0 | 0 |
| 0 | 0.014281296 | 0 | 0 | 0 |
| 0 | 0 | 0 | 0.018976743 | 0 |
| 0 | 0 | 1.438410806 | 0.149565528 | 0.002017787 |
| 0 | 0 | 0 | 0 | 0 |
| 0 | 0 | 0 | 0.002867144 | 0 |
| 0 | 0 | 0 | 0 | 0 |
| 0 | 0 | 0 | 0.00817778 | 0 |
| 0 | 0 | 0 | 1.04470108 | 4.616021958 |
| 0 | 0 | 0 | 0.057885186 | 0 |
| 0 | 0 | 0 | 0 | 0 |
| 0 | 0 | 0.001930185 | 0.000745383 | 0.001772152 |
| 0 | 0 | 0 | 0 | 0 |
| 0 | 0 | 0 | 0.020777575 | 0 |
| 0 | 0 | 0 | 1.425475308 | 0.084685366 |
| 0 | 0 | 0 | 0 | 0 |
| 0 | 0 | 0.213998958 | 1.184645257 | 0 |
| 0 | 0 | 0 | 0 | 0.00069442 |
| 0 | 0 | 0 | 0 | 0 |
| 0 | 0 | 0 | 0.008819825 | 0 |
| 0 | 0 | 0 | 0 | 0 |
| 0 | 0 | 0 | 0.009688467 | 0 |
| 0 | 0 | 0 | 0 | 0 |
| 0 | 0 | 0.05216513 | 0.145308437 | 0 |
| 0 | 0.00377277 | 0 | 0 | 0 |
| 0 | 0 | 0 | 0.318797286 | 0 |
| 0 | 0 | 0 | 0.001660712 | 0 |
| 0 | 0 | 0 | 1.797358419 | 0 |
| 0 | 0 | 0 | 0 | 0 |
| 0 | 0 | 0 | 0 | 0 |
| 0 | 0 | 0.00272941 | 0.032427605 | 0.001566213 |
| 0 | 0 | 0 | 0.019970279 | 0 |
| 0 | 0 | 0.007400314 | 0.031435709 | 0 |
| 0 | 0 | 0 | 0 | 0 |
| 0 | 0 | 0 | 0.101528397 | 0 |
| 0 | 0 | 0 | 0 | 0.03651927 |
| 0 | 0 | 0.398263708 | 3.289527884 | 1.628473152 |
| 0 | 0.005765 | 0 | 0 | 0 |
| 0 | 0 | 0 | 0.010863211 | 0 |
| 0 | 0 | 0 | 0.014271798 | 0 |
| 0 | 0.001691251 | 0.001346667 | 0.166966885 | 0.004327434 |
| 0 | 0 | 0 | 0 | 0 |
| 0.448976542 | 0.000869513 | 0.159529912 | 1.537828428 | 6.360626254 |
| 0 | 0 | 0 | 0 | 0 |
| 0 | 0 | 0 | 0 | 0 |
| 0 | 0 | 0 | 0 | 0 |
| 0 | 0 | 0 | 0 | 0 |
| 0.016567198 | 0 | 0.00930838 | 0 | 0.003529978 |

| | | | | |
|---|---|---|---|---|
| 0 | 0 | 0 | 0 | 0 |
| 0 | 0 | 0 | 0 | 0 |
| 0.153266324 | 1.172701519 | 0 | 0.008858111 | 0.031701632 |
| 0 | 0 | 0.004613055 | 0.004898936 | 0 |
| 0 | 0 | 0.003910728 | 0.411532921 | 0 |
| 0 | 0.00416723 | 0 | 0.041868469 | 0.002104494 |
| 0 | 0 | 0 | 0 | 0 |
| 0 | 0 | 0 | 0 | 0.006113753 |
| 0 | 0 | 0 | 0 | 0 |
| 0 | 0 | 0 | 0 | 0 |
| 0.049972251 | 0.003280299 | 0.689447409 | 0.076251265 | 0.046862933 |
| 3.403687396 | 0.017418916 | 0.297421545 | 0.678177183 | 0.623510604 |
| 0 | 0 | 0 | 0 | 0 |
| 0.047038515 | 0.001770116 | 0.095579305 | 0.450540396 | 0.01943794 |
| 0.060395527 | 2.614543976 | 0.037741068 | 0.073107229 | 0.064543141 |
| 0 | 0 | 0 | 0.649193337 | 0.009312523 |
| 0 | 0 | 0 | 0 | 0 |
| 0 | 0 | 0 | 0 | 0 |
| 0 | 0 | 0 | 0.022079551 | 0 |
| 0 | 0.071830904 | 0.002823928 | 0.002165896 | 0 |
| 0 | 0.867198248 | 0 | 0.080861987 | 0.083392553 |
| 9.505652476 | 1.892215202 | 0.006223395 | 0 | 1.276043611 |
| 17.70766035 | 0.153646957 | 0.248475296 | 0.065648226 | 2.842784676 |
| 0 | 0 | 0 | 0 | 0 |
| 0 | 0 | 0 | 0.012793406 | 0 |
| 0 | 0 | 0 | 5.175961052 | 2.766583749 |
| 0 | 0 | 0 | 0 | 0.104123688 |
| 0 | 0 | 0 | 0 | 0 |
| 0 | 0.000522667 | 0 | 0 | 0 |
| 0 | 0 | 0 | 0 | 0 |
| 0 | 0 | 0 | 0.012153593 | 0 |
| 0 | 0 | 0.065856948 | 0.132514569 | 0.014528192 |
| 0 | 0.048288319 | 0 | 0.007062624 | 0.001077525 |
| 0 | 0 | 0 | 0.135278561 | 0 |
| 0.001146102 | 0.043915122 | 0.004002099 | 0 | 0.00284472 |
| 0.104628364 | 0.012983845 | 0.314486699 | 0.032022527 | 1.48489326 |
| 0 | 0 | 0 | 0.047127828 | 0.003303588 |
| 0 | 0 | 0 | 0 | 0 |
| 0 | 0 | 0 | 3.935307701 | 0.008575688 |
| 0 | 0 | 0 | 0 | 0 |
| 0 | 0 | 0 | 0.006966428 | 0 |
| 0 | 0 | 0 | 0 | 0 |
| 0 | 0 | 0 | 0 | 0 |
| 0 | 0 | 0 | 0 | 0 |
| 0.002394317 | 0.020322786 | 0.076258272 | 0.003866618 | 0.005757175 |
| 0 | 0 | 0 | 0 | 0 |
| 0 | 0 | 0 | 0.161921623 | 0 |
| 0.405310735 | 0.1498907 | 1.170716009 | 0 | 0.091625577 |

| | | | | |
|---|---|---|---|---|
| 0 | 0 | 0 | 0 | 0 |
| 0 | 0 | 0 | 0 | 0 |
| 0 | 0 | 0 | 0 | 0 |
| 0 | 0 | 0.000689598 | 0.003661669 | 0 |
| 0 | 0 | 0 | 0 | 0 |
| 0 | 0 | 0 | 0.890471822 | 0 |
| 0 | 0 | 0 | 0 | 0 |
| 0 | 0 | 0 | 0.084066501 | 0.035975626 |
| 0 | 0 | 0 | 0 | 0 |
| 0 | 0 | 0 | 0 | 0 |
| 0 | 0.01625959 | 0.001152044 | 0.000815626 | 0.008814346 |
| 0 | 0 | 0 | 0 | 0 |
| 0.017246499 | 0 | 0 | 0 | 0 |
| 0 | 0 | 0 | 0.076037686 | 0 |
| 0 | 0 | 0 | 0 | 0 |
| 0 | 0 | 0 | 0 | 0 |
| 0 | 0 | 0.198353306 | 0.021231387 | 0 |
| 0 | 0 | 0.06185877 | 0.002885708 | 0.014235219 |
| 0 | 0 | 0 | 0 | 0 |
| 0 | 0 | 0 | 0 | 0 |
| 0 | 0 | 0 | 0 | 0 |
| 0 | 0 | 0 | 0 | 0 |
| 0 | 0 | 0.000737145 | 0.012916645 | 0 |
| 0 | 0 | 0 | 0 | 0 |
| 0 | 0 | 0 | 0 | 0 |
| 0 | 0 | 0 | 0 | 0 |
| 0 | 0 | 0 | 0 | 0 |
| 0 | 0 | 0 | 0 | 0 |
| 0 | 0 | 0 | 0 | 0 |
| 0 | 0 | 0 | 1.659702963 | 0 |
| 0 | 0.006294592 | 0.000907363 | 0.00176659 | 0 |
| 0 | 0 | 0 | 0 | 0 |
| 0 | 0 | 0.00066803 | 0.045935529 | 0 |
| 0 | 0 | 0 | 0 | 0 |
| 0 | 0.010737506 | 0 | 0 | 0 |
| 0 | 0 | 0 | 0 | 0 |
| 0 | 0 | 0 | 0.043806213 | 0.092030325 |
| 0.000961966 | 0.008864967 | 0 | 0 | 0 |
| 0 | 0 | 0 | 0 | 0 |
| 0 | 0 | 0 | 0 | 0 |
| 0 | 0 | 0 | 0.015061472 | 0 |
| 0 | 0 | 0 | 0.028665737 | 0 |
| 0 | 0 | 0 | 0 | 0 |
| 0 | 0 | 0 | 0 | 0 |
| 0 | 0 | 0 | 0 | 0 |
| 0 | 0 | 0 | 0 | 0 |
| 0 | 0 | 0 | 0.007380397 | 0 |
| 0 | 0 | 0 | 0 | 0 |

| | | | | |
|---|---|---|---|---|
| 0 | 0 | 0 | 0 | 0 |
| 0 | 0 | 0 | 0 | 0 |
| 0 | 0 | 0 | 0 | 0 |
| 0 | 0 | 0.023766563 | 1.237762167 | 0 |
| 0 | 0 | 0 | 0 | 0 |
| 0 | 0 | 0 | 0 | 0.000641711 |
| 0 | 0 | 0 | 0 | 0 |
| 0 | 0 | 0 | 0 | 0 |
| 0.128027731 | 1.295341132 | 0.002017499 | 0.00166641 | 0.013789474 |
| 0 | 0 | 0 | 0 | 0 |
| 0 | 0 | 0 | 0.002042632 | 0 |
| 0 | 0 | 0 | 0 | 0 |
| 0 | 0 | 0 | 0 | 0 |
| 0 | 0 | 0 | 0 | 0 |
| 0 | 4.464273964 | 0.036567881 | 0.938689657 | 0.011388506 |
| 0 | 0.055050896 | 0.065825897 | 0 | 0 |
| 0 | 0 | 0 | 0 | 0 |
| 0 | 0 | 0 | 0 | 0 |
| 0 | 0 | 0 | 0 | 0 |
| 0 | 0 | 0 | 0.003828587 | 0 |
| 0 | 0 | 0 | 0.324559407 | 0 |
| 0 | 0 | 0 | 0 | 0 |
| 0 | 0 | 0 | 0 | 0 |
| 0 | 0 | 0 | 0 | 0 |
| 0 | 0.014377745 | 0.098064208 | 0.022675963 | 0.17813428 |
| 0 | 0 | 23.86025486 | 0.706365777 | 0 |
| 0 | 0 | 0 | 0.094337138 | 0 |
| 0 | 0 | 0 | 0 | 0 |
| 0 | 0 | 5.189342428 | 0.072203165 | 0 |
| 0 | 0 | 0 | 0 | 0 |
| 0 | 0 | 0 | 0 | 0 |
| 0 | 0 | 0 | 0 | 0 |
| 0 | 0 | 0 | 0 | 0 |
| 0 | 0.001414486 | 0 | 0.005382409 | 0 |
| 0.062238429 | 0 | 0.184417504 | 0.3631162 | 0.016825354 |
| 0 | 0.00894341 | 0.081332033 | 0 | 0.014302293 |
| 0.032715893 | 0 | 0 | 0.009883797 | 0 |
| 0 | 0 | 0.489185064 | 0.291604791 | 0.160817613 |
| 0 | 0 | 0 | 4.682928878 | 0.160286614 |
| 0 | 0 | 0 | 0.000831571 | 0 |
| 0 | 0 | 0 | 0.012678608 | 0 |
| 0 | 0.004804057 | 0 | 0.000789895 | 0 |
| 0 | 0 | 0.294211723 | 0.064487816 | 0.073857023 |
| 0 | 0 | 0 | 0 | 0 |
| 0 | 0 | 0 | 0.068622767 | 0 |
| 0 | 0 | 0 | 0 | 0 |
| 0 | 0 | 0 | 0 | 0 |
| 0.351045713 | 0.001420855 | 0 | 0.105391985 | 0.027721199 |

| | | | | |
|---|---|---|---|---|
| 0 | 0 | 0 | 0.005291658 | 0 |
| 0 | 0 | 0 | 0.124663229 | 0 |
| 0 | 0 | 0 | 0 | 0 |
| 0 | 0 | 0 | 0 | 0 |
| 0 | 0 | 0.014515559 | 0.001756153 | 0.00134958 |
| 0 | 0 | 0 | 0 | 0 |
| 0 | 0 | 0 | 0 | 0 |
| 0 | 0 | 0 | 0 | 0 |
| 0 | 0 | 0 | 0 | 0 |
| 0 | 0 | 0 | 0 | 0 |
| 0 | 0 | 0 | 0 | 0 |
| 0 | 0.004622706 | 6.485907473 | 1.409001789 | 3.536010208 |
| 0.024578799 | 0 | 0.079869759 | 0.037093747 | 0.288828096 |
| 0 | 0 | 0 | 0 | 0 |
| 0.01907429 | 0.023179576 | 0.002549961 | 0.285467021 | 0.854727433 |
| 0 | 0 | 0 | 0 | 0 |
| 0 | 0.000636503 | 0 | 0 | 0 |
| 0 | 0 | 0.038589368 | 0 | 0 |
| 0 | 0 | 0 | 0 | 0 |
| 0 | 0 | 0 | 0.078737032 | 0 |
| 0 | 0 | 0 | 0.066485025 | 0 |
| 0 | 0 | 0 | 0 | 0 |
| 0 | 0.020535159 | 0.181829807 | 2.324679124 | 0 |
| 0 | 0 | 0 | 0 | 0 |
| 0 | 0 | 0 | 0 | 0 |
| 0 | 0 | 0 | 0 | 0 |
| 0 | 0 | 0 | 0 | 0 |
| 0 | 0 | 0 | 0 | 0 |
| 0 | 0 | 0 | 0 | 0 |
| 0 | 0 | 0 | 0 | 0 |
| 0 | 0 | 0 | 0 | 0 |
| 0.00951983 | 0.234297563 | 0.047152266 | 0.015520134 | 0.011898889 |
| 0 | 0 | 0 | 0 | 0 |
| 0 | 0 | 0.042429178 | 0.033062484 | 0 |
| 0 | 0.009988794 | 0.078516609 | 0 | 0.01064938 |
| 0.013160892 | 0 | 8.004228359 | 0.044950402 | 0.116796509 |
| 0 | 0 | 0 | 0 | 0 |
| 0 | 0 | 0 | 0 | 0 |
| 0 | 0 | 0 | 0 | 0 |
| 0 | 0 | 0 | 0 | 0 |
| 0 | 0 | 0 | 0.004036486 | 0 |
| 0 | 0 | 0 | 0.006936767 | 0 |
| 0 | 0 | 0 | 0.164921335 | 0 |
| 0 | 0 | 0.03980457 | 0.046282495 | 0 |
| 0.004681745 | 0 | 0.382514736 | 0.007840643 | 0.007585588 |
| 0 | 0 | 0 | 0 | 0 |
| 0 | 0 | 0 | 0.118116645 | 0.03938041 |
| 0 | 0 | 0 | 0 | 0 |

| | | | | |
|---|---|---|---|---|
| 0 | 0 | 0 | 0 | 0 |
| 0 | 0 | 0 | 0 | 0 |
| 0 | 0 | 0 | 0 | 0 |
| 0 | 0 | 0 | 0 | 0 |
| 0.001273115 | 0 | 0 | 0 | 0 |
| 0 | 0 | 0 | 0 | 0 |
| 0 | 0 | 0 | 0 | 0 |
| 0 | 0 | 0.173075033 | 0.008387738 | 0 |
| 0 | 0 | 0 | 0 | 0 |
| 0 | 0 | 0 | 0.003050338 | 0 |
| 0 | 2.802545748 | 0 | 0 | 0 |
| 0 | 0.001062034 | 0.00092493 | 0 | 0 |
| 0 | 0 | 0 | 0 | 0 |
| 0 | 0 | 0 | 0 | 0 |
| 0 | 0 | 0 | 0 | 0 |
| 0 | 0 | 0 | 0 | 0 |
| 0 | 0 | 0 | 0.238371605 | 0 |
| 0 | 0 | 0 | 0 | 0 |
| 0 | 0 | 0.079261617 | 1.338113154 | 0.954598995 |
| 0 | 0 | 0 | 0 | 0 |
| 0 | 0 | 0.000825948 | 0 | 0 |
| 0 | 0 | 0 | 0 | 0 |
| 0 | 0 | 0 | 0 | 0 |
| 0 | 0 | 0 | 0 | 0 |
| 0 | 0 | 0.000768255 | 0.00326346 | 0 |
| 0.010008471 | 0.011039739 | 0.043531433 | 0 | 0.032003909 |
| 0 | 0 | 0 | 0 | 0 |
| 0 | 0.003678539 | 0.000961097 | 0.290547251 | 0.064856942 |
| 0 | 0 | 0 | 0 | 0 |
| 0 | 0 | 0.664009973 | 0.518500074 | 0.328775536 |
| 0 | 0 | 0 | 0 | 0 |
| 0 | 0 | 0 | 0 | 0 |
| 0 | 0 | 0 | 0 | 0 |
| 0 | 0 | 0 | 0 | 0 |
| 0 | 0.002215333 | 0.00120048 | 0 | 0.002204383 |
| 0 | 0 | 0 | 0 | 0 |
| 0 | 0 | 0 | 0 | 0 |
| 0 | 0 | 0.029384459 | 0 | 0.000535999 |
| 0 | 0 | 0 | 0 | 0 |
| 0 | 0 | 0 | 0 | 0 |
| 5.353771759 | 13.87755765 | 3.54445022 | 5.102761187 | 4.513635898 |
| 0 | 0 | 0.307259036 | 2.643382306 | 0 |
| 0 | 0 | 0 | 0 | 0.000460514 |
| 0 | 0 | 0 | 0 | 0 |
| 0 | 0 | 0 | 0 | 0 |
| 0 | 0 | 0 | 0 | 0 |
| 0 | 0 | 0 | 0.001876115 | 0 |
| 0.004722686 | 0.001108364 | 0.003243336 | 0.005904571 | 0.000708998 |

| | | | | |
|---|---|---|---|---|
| 0 | 0 | 0 | 0 | 0 |
| 0 | 0 | 0 | 0.0043863 | 0 |
| 0 | 0 | 0 | 0.003546658 | 0 |
| 0 | 0 | 0 | 0.080256413 | 0 |
| 0 | 0.001867424 | 5.080998331 | 1.404583452 | 3.475087234 |
| 0 | 0 | 0 | 0 | 0 |
| 0 | 0 | 0 | 0 | 0 |
| 0 | 0 | 0 | 0.019139071 | 0 |
| 0.129242641 | 0.566904655 | 0.000951749 | 0 | 0.005971137 |
| 0 | 0 | 0 | 0 | 0 |
| 0.016287438 | 0 | 0 | 0 | 0 |
| 0 | 0 | 0 | 0 | 0 |
| 0 | 0.00332324 | 0.038280932 | 0.063438795 | 0 |
| 0.007487522 | 0.038404503 | 0.003226059 | 0.015316169 | 0.245491539 |
| 0 | 0 | 0 | 0.529845557 | 0 |
| 0 | 0 | 0.303970145 | 0.206385334 | 0 |
| 0 | 0 | 0 | 0.075860387 | 0.001809239 |
| 0 | 0 | 0 | 0 | 0 |
| 0 | 0 | 0 | 0 | 0 |
| 0 | 0 | 0 | 0 | 0 |
| 0 | 0 | 0 | 0 | 0 |
| 0 | 0 | 0 | 0 | 0 |
| 0 | 0 | 0 | 0 | 0 |
| 0 | 0 | 0 | 0 | 0 |
| 0 | 0 | 0 | 0 | 0 |
| 0 | 0 | 0 | 0 | 0 |
| 0 | 0 | 0.041695159 | 0.001668048 | 0 |
| 0 | 0 | 0 | 0.001197767 | 0.005621432 |
| 0 | 0 | 0 | 0 | 0 |
| 0 | 0 | 0 | 0 | 0 |
| 0 | 0 | 0 | 0.034299307 | 0 |
| 0 | 0 | 0 | 0 | 0 |
| 0 | 0 | 0 | 0 | 0 |
| 0 | 0 | 0.029895543 | 0.022421069 | 0 |
| 0 | 0 | 0 | 0.399469417 | 0 |
| 0 | 0 | 0 | 0 | 0 |
| 0 | 0 | 0 | 0 | 0 |
| 0 | 0 | 0.023408349 | 0 | 0 |
| 0 | 0 | 0 | 0 | 0 |
| 0 | 0 | 0 | 0 | 0 |
| 0 | 0 | 0.805588507 | 0.023757504 | 0.014251895 |
| 0 | 0 | 0 | 0 | 0 |
| 0 | 0 | 0 | 0 | 0 |
| 0 | 0 | 0 | 0 | 0 |
| 0 | 0 | 0 | 0 | 0 |
| 0 | 0 | 0 | 0 | 0 |
| 0 | 0 | 0 | 0.000859224 | 0 |
| 0 | 0 | 0 | 0 | 0 |

| | | | | |
|---|---|---|---|---|
| 0 | 0 | 0 | 1.258985349 | 0 |
| 0 | 0 | 0 | 0 | 0 |
| 0 | 0 | 0 | 0 | 0 |
| 0 | 0 | 0 | 0 | 0 |
| 0 | 0 | 0 | 0.010756731 | 0 |
| 0 | 0 | 0 | 0.095876368 | 0 |
| 0 | 0 | 0 | 0.225636039 | 0.00406089 |
| 0 | 0 | 0 | 0 | 0 |
| 0 | 0 | 0 | 0 | 0 |
| 0 | 1.209729544 | 0 | 0 | 0 |
| 0 | 0 | 0 | 0.056749385 | 0 |
| 0 | 0 | 0 | 0 | 0 |
| 0 | 0 | 0 | 0 | 0 |
| 0 | 0 | 0 | 0 | 0 |
| 0 | 0 | 0 | 0 | 0 |
| 0 | 0 | 0 | 0 | 0 |
| 4.814660667 | 0.053607736 | 0 | 0.680557756 | 0 |
| 0 | 0 | 0 | 0 | 0 |
| 0 | 0 | 0 | 0 | 0 |
| 0 | 0 | 0 | 0 | 0 |
| 0 | 0.003133902 | 0 | 0.091922928 | 0.01703989 |
| 0 | 0 | 0 | 0 | 0 |
| 0 | 0.011110766 | 0 | 0.087689226 | 0 |
| 0.004224829 | 0 | 0.00703646 | 0.007959863 | 0 |
| 0 | 0 | 0 | 0 | 0 |
| 0 | 0 | 0 | 0 | 0 |
| 0 | 0 | 0 | 0 | 0 |
| 0 | 0 | 0 | 0 | 0 |
| 0 | 0 | 0 | 0 | 0 |
| 0.156790528 | 0 | 0.003082165 | 0.731320575 | 0.001617037 |
| 0 | 0 | 0 | 0 | 0 |
| 0 | 0 | 0 | 0 | 0 |
| 0 | 0 | 0 | 0 | 0 |
| 0.189014814 | 0.474327836 | 0.001796949 | 0.364834186 | 0 |
| 0 | 0 | 0 | 0 | 0 |
| 0 | 0 | 0 | 0.999268611 | 0 |
| 0 | 0.195322732 | 0.001339805 | 0.005081556 | 0 |
| 0 | 0 | 0 | 0 | 0 |
| 0 | 0 | 0 | 0 | 0 |
| 0 | 0 | 0 | 0 | 0 |
| 0 | 0 | 0 | 0 | 0 |
| 0 | 0 | 0 | 0 | 0 |
| 0 | 0 | 0 | 0 | 0 |
| 0 | 0 | 0 | 0 | 0 |
| 0 | 0 | 0 | 0 | 0 |
| 0 | 0 | 0 | 0 | 0 |
| 0 | 0 | 0 | 0 | 0 |
| 0 | 0 | 0 | 0 | 0 |

| | | | | |
|---|---|---|---|---|
| 0 | 0.00188996 | 0 | 0.001398383 | 0.003475785 |
| 0 | 0 | 0 | 0 | 0 |
| 0 | 0 | 0 | 0 | 0 |
| 0 | 0 | 0 | 0 | 0 |
| 0 | 0 | 0 | 0 | 0 |
| 0 | 0 | 0 | 0 | 0 |
| 0 | 0 | 0 | 0 | 0 |
| 0 | 0.141660994 | 0.572417665 | 0.301702337 | 0.142133158 |
| 0.005606647 | 0.45203198 | 1.712153985 | 0.75753968 | 0.445949623 |
| 0.014725632 | 0.022717651 | 0.02446133 | 0 | 0.148292639 |
| 0 | 0 | 0 | 0 | 0 |
| 0 | 0 | 0 | 0 | 0 |
| 0 | 0 | 0 | 0.037550185 | 0 |
| 16.09471352 | 0 | 0 | 0 | 0.60778164 |
| 0 | 0 | 0 | 0 | 0 |
| 0 | 0 | 0 | 0.009945074 | 0 |
| 0 | 0.013104455 | 0.002162411 | 0 | 0 |
| 0 | 0 | 0 | 0 | 0 |
| 0 | 0 | 0 | 0 | 0 |
| 0.001765424 | 0 | 0.024758318 | 0.043557091 | 0 |
| 0 | 0 | 0 | 0 | 0 |
| 0 | 0 | 0 | 0 | 0 |
| 0 | 0 | 0 | 0.1687235 | 0 |
| 0 | 0 | 0 | 0 | 0 |
| 0 | 0 | 0.01426076 | 0.012115624 | 0 |
| 0 | 0 | 0 | 0 | 0 |
| 0 | 0 | 0 | 0 | 0 |
| 0 | 0 | 0 | 0 | 0 |
| 0 | 0.007454134 | 0 | 0 | 0 |
| 0 | 0 | 0 | 0 | 0 |
| 0 | 0 | 0 | 0 | 0 |
| 0 | 0 | 0 | 0 | 0 |
| 0 | 0 | 0 | 0 | 0 |
| 0 | 0 | 0 | 0.001389899 | 0 |
| 0 | 0 | 0 | 0 | 0 |
| 0 | 0 | 0 | 0 | 0 |
| 0 | 0 | 0 | 0 | 0 |
| 0 | 0 | 0 | 0 | 0 |
| 0 | 0 | 0 | 0 | 0 |
| 0 | 0 | 0 | 0 | 0 |
| 0 | 0 | 0 | 0 | 0 |
| 0 | 0 | 0 | 0.154286409 | 0 |
| 0 | 0 | 0 | 0 | 0 |
| 0 | 0 | 0 | 0 | 0 |
| 0 | 0 | 0 | 0 | 0 |
| 0 | 0 | 0 | 0 | 0 |
| 0 | 0 | 0 | 0 | 0 |
| 0 | 0 | 0 | 0 | 0 |

| | | | | |
|---|---|---|---|---|
| 0 | 0 | 0 | 0 | 0 |
| 0.001465365 | 0 | 0 | 0.318207387 | 0.00040918 |
| 0 | 0 | 0 | 0 | 0 |
| 0 | 0 | 0 | 0 | 0 |
| 0.537601672 | 0 | 5.007919018 | 0.140064775 | 4.659112484 |
| 0 | 0 | 0 | 0 | 0 |
| 0 | 0 | 0 | 0 | 0 |
| 0.01363118 | 0.143348381 | 0.002303178 | 0.003112977 | 0.002114607 |
| 0 | 0 | 0.006689855 | 0.008436523 | 0 |
| 0 | 0 | 0 | 0 | 0 |
| 0 | 0 | 0 | 0 | 0 |
| 0 | 0 | 0 | 0 | 0 |
| 0 | 0 | 0 | 0 | 0 |
| 0 | 0 | 0 | 0 | 0 |
| 0 | 0 | 0 | 0 | 0 |
| 0 | 0 | 0 | 0.004628335 | 0.005335222 |
| 0 | 0 | 0 | 0 | 0 |
| 0 | 0 | 0 | 0 | 0 |
| 0 | 0 | 0 | 0 | 0 |
| 0 | 0 | 0 | 0 | 0 |
| 0 | 0 | 0 | 0 | 0 |
| 0 | 0 | 0 | 0 | 0 |
| 0 | 0 | 0 | 0 | 0 |
| 0 | 0 | 0.000906005 | 0 | 0.001663653 |
| 0 | 0 | 0 | 0 | 0.093399644 |
| 0 | 0 | 0 | 0 | 0 |
| 0.071602301 | 0.017364434 | 0.032317129 | 0.002815991 | 0 |
| 0 | 0.006512175 | 0 | 0 | 0 |
| 0 | 0 | 0 | 0 | 0 |
| 0 | 0 | 0 | 0 | 0 |
| 0 | 0 | 0 | 0 | 0 |
| 0 | 0 | 0 | 0 | 0 |
| 0 | 0 | 0 | 0 | 0 |
| 0 | 0 | 0 | 0 | 0 |
| 0 | 0 | 0 | 0 | 0 |
| 0 | 0 | 0 | 0 | 0 |
| 0 | 0 | 0 | 0 | 0 |
| 0 | 0 | 0 | 0.09512231 | 0 |
| 0 | 0 | 0 | 0 | 0 |
| 0 | 0 | 0 | 0 | 0 |
| 0 | 0 | 0 | 0 | 0 |
| 0 | 0 | 0 | 0 | 0 |
| 0 | 0 | 0 | 0 | 0 |
| 0 | 0 | 0 | 0 | 0 |
| 0 | 0 | 0 | 0 | 0 |
| 0 | 0 | 0 | 0 | 0 |
| 0 | 0 | 0 | 0 | 0 |
| 0 | 0 | 0 | 0 | 0 |

| | | | | |
|---|---|---|---|---|
| 0 | 0 | 0.186401044 | 0.441586833 | 0.012306023 |
| 0 | 0 | 0 | 0 | 0 |
| 0 | 0 | 0 | 0 | 0 |
| 0 | 0 | 0 | 0 | 0 |
| 0 | 0 | 0 | 0.001533049 | 0 |
| 0 | 0 | 0 | 0.207096031 | 0.078746381 |
| 0 | 0 | 0 | 0 | 0 |
| 0 | 0 | 0 | 0 | 0 |
| 0 | 0 | 0 | 0 | 0 |
| 0 | 0 | 0 | 0 | 0 |
| 0 | 0 | 0 | 0 | 0 |
| 0 | 0 | 0 | 0 | 0 |
| 0 | 0 | 0 | 0.015881042 | 0 |
| 0 | 0 | 0 | 0 | 0 |
| 0 | 0 | 0 | 0 | 0 |
| 0 | 0 | 0 | 0 | 0 |
| 0 | 0 | 0 | 0 | 0 |
| 0 | 0 | 0 | 0 | 0 |
| 0 | 0 | 0 | 0 | 0 |
| 0 | 0 | 0 | 0 | 0 |
| 0.005873675 | 0.010877527 | 0.038073581 | 0 | 0.003810406 |
| 0 | 0 | 0 | 0 | 0 |
| 0.062940808 | 0 | 0.008203933 | 0.04356174 | 0.003556891 |
| 0 | 0 | 0.188342387 | 0.007973995 | 0 |
| 0 | 0 | 0 | 0.002825861 | 0 |
| 0 | 0 | 0 | 0 | 0 |
| 0 | 0 | 0 | 0 | 0 |
| 0 | 0 | 0 | 0 | 0 |
| 0.023351472 | 0 | 0 | 0 | 0.001992384 |
| 0 | 0 | 0 | 0 | 0 |
| 0 | 0 | 0 | 0 | 0 |
| 0 | 0 | 0 | 0 | 0 |
| 0.571333905 | 0.065857238 | 0.005762212 | 0.000679923 | 0.493577913 |
| 0 | 0 | 0 | 0.247294549 | 0 |
| 0.030128546 | 0 | 0.09793115 | 0.218744963 | 0.274997365 |
| 0.018244261 | 0.088980734 | 0.009419124 | 0 | 0.002672999 |
| 0 | 0 | 0 | 0 | 0 |
| 0 | 0 | 0 | 0 | 0 |
| 0.007229517 | 0.021891886 | 0.029910921 | 0 | 0.002182409 |
| 0 | 0 | 0 | 0 | 0 |
| 0 | 0 | 0 | 0 | 0 |
| 0 | 0 | 0 | 0 | 0 |
| 0 | 0 | 0.04840887 | 0.090764946 | 0.299209141 |
| 0 | 0 | 0 | 0 | 0 |
| 0 | 0 | 0 | 0 | 0 |
| 0 | 0 | 0.003678951 | 0.020316107 | 0 |
| 0 | 0 | 0 | 0 | 0 |
| 0 | 0 | 0 | 0 | 0 |

| | | | | |
|---|---|---|---|---|
| 0 | 0 | 0 | 0 | 0 |
| 0 | 0 | 0 | 0 | 0 |
| 0 | 0 | 0.029551934 | 0 | 0 |
| 0 | 0 | 0 | 0 | 0 |
| 0 | 0 | 0 | 0 | 0 |
| 0 | 0 | 0 | 0 | 0 |
| 0 | 0 | 0 | 0 | 0 |
| 0 | 5.536724392 | 0 | 0 | 0 |
| 0 | 0 | 0 | 0.059116169 | 0 |
| 0 | 0.002719357 | 0 | 0.030985637 | 0.00104371 |
| 0 | 0 | 0 | 0 | 0 |
| 0 | 0 | 0 | 0 | 0 |
| 0 | 0 | 0 | 0 | 0 |
| 0 | 0 | 0 | 0 | 0 |
| 0 | 0 | 0 | 0 | 0 |
| 0.017836077 | 0.020601708 | 0 | 0 | 0 |
| 0 | 0 | 0 | 0 | 0 |
| 0 | 0 | 0 | 0 | 0 |
| 0 | 0 | 0 | 0 | 0 |
| 0 | 0 | 0 | 0.384520617 | 0 |
| 0 | 0 | 0 | 0 | 0 |
| 0 | 0 | 0 | 0 | 0 |
| 0 | 0 | 0 | 0 | 0 |
| 0 | 0 | 0 | 0.003427996 | 0 |
| 0 | 0 | 0 | 0 | 0 |
| 0 | 0 | 0 | 0 | 0 |
| 0 | 0 | 0 | 0 | 0 |
| 0 | 0 | 0 | 0 | 0 |
| 0 | 0 | 0 | 0 | 0 |
| 0 | 0 | 0.130410876 | 0.023184707 | 0 |
| 0 | 0 | 0.129541135 | 0.265001487 | 0.051329857 |
| 0 | 0 | 0 | 0 | 0 |
| 0 | 0 | 0 | 0 | 0 |
| 0 | 0 | 0 | 0 | 0 |
| 0 | 0 | 0 | 0 | 0 |
| 0 | 0 | 0 | 0 | 0 |
| 0 | 0 | 0 | 0 | 0 |
| 0 | 0 | 0 | 0 | 0 |
| 0 | 0 | 0 | 0 | 0 |
| 0 | 0 | 0 | 0 | 0 |
| 0 | 0 | 0 | 0.008575894 | 0 |
| 0 | 0 | 0.000966991 | 0.415285584 | 0 |
| 0 | 0 | 0 | 0.001566243 | 0 |
| 0 | 0 | 0 | 0 | 0 |
| 0 | 0 | 0 | 0 | 0 |
| 0 | 0 | 0 | 0 | 0 |
| 0.030493847 | 0.291149047 | 0.002671594 | 0 | 0 |
| 0 | 0 | 0 | 0 | 0 |

| | | | | |
|---|---|---|---|---|
| 0 | 0 | 0 | 0 | 0 |
| 0 | 0 | 0 | 0 | 0 |
| 0 | 0 | 0 | 0 | 0 |
| 0 | 0 | 0 | 0 | 0 |
| 0 | 0 | 0.004847139 | 0.038152253 | 0.032068209 |
| 0 | 0 | 0 | 0 | 0 |
| 0 | 0 | 0 | 0 | 0 |
| 0 | 0 | 0 | 0 | 0 |
| 0 | 0 | 0 | 0 | 0 |
| 0 | 0 | 0 | 0 | 0 |
| 0 | 0 | 0.005792171 | 0 | 0 |
| 0.00605234 | 0 | 0.061753487 | 0.002942714 | 0.001817226 |
| 0.027961062 | 0 | 0.004952094 | 0.031191226 | 0.005800891 |
| 0 | 0 | 0 | 0 | 0 |
| 0.001041488 | 0.006927735 | 0 | 0 | 0.000969397 |
| 0 | 0 | 0.062336367 | 0.338516384 | 0 |
| 0 | 0 | 0.00045669 | 0.047852551 | 0 |
| 0 | 0 | 0 | 0 | 0 |
| 0 | 1.055329137 | 0.003896674 | 0 | 0.001100811 |
| 0 | 0.008426039 | 0 | 0.070864729 | 0 |
| 0 | 0 | 0 | 0 | 0 |
| 0 | 0 | 0 | 0 | 0 |
| 0 | 0 | 0 | 0 | 0 |
| 0 | 0 | 0 | 0 | 0 |
| 0 | 0 | 0 | 0 | 0 |
| 0 | 0 | 0 | 0 | 0 |
| 0 | 0.008552751 | 0.009408792 | 0 | 0 |
| 0 | 0 | 0 | 0 | 0 |
| 0 | 0 | 0 | 0 | 0 |
| 0 | 0 | 0 | 0 | 0 |
| 0 | 0 | 0 | 0 | 0.000652809 |
| 0 | 0 | 0 | 0 | 0 |
| 0 | 0 | 0 | 0 | 0 |
| 0 | 0 | 0 | 0 | 0 |
| 0 | 0 | 0 | 0 | 0 |
| 0 | 0 | 0 | 0 | 0 |
| 0 | 0 | 0 | 0 | 0 |
| 0.004188399 | 0.10378453 | 1.205526495 | 0.021087481 | 0.005274416 |
| 0 | 0 | 0 | 0 | 0 |
| 0 | 0 | 0 | 0 | 0 |
| 0 | 0 | 0 | 0.010989463 | 0.005494505 |
| 0 | 0 | 0 | 0 | 0 |
| 0 | 0 | 0 | 0 | 0 |
| 0 | 0.017542925 | 0 | 0 | 0 |
| 0 | 0 | 0 | 0 | 0 |
| 0 | 0 | 0 | 0 | 0 |
| 0 | 0 | 0 | 0 | 0 |
| 0 | 0 | 0 | 0 | 0 |

| | | | | |
|---|---|---|---|---|
| 0 | 0 | 0 | 0 | 0 |
| 0 | 0 | 0.035138928 | 0.021179671 | 0 |
| 0 | 0 | 0 | 0 | 0 |
| 0 | 0.000835685 | 0.000776322 | 0 | 0 |
| 0 | 0 | 0.000540019 | 0.001911618 | 0.003966445 |
| 0 | 0 | 0 | 0.001233563 | 0 |
| 0 | 0 | 0 | 0.003000182 | 0 |
| 0 | 0.037907514 | 0 | 0 | 0 |
| 0 | 0 | 0 | 0.002511573 | 0 |
| 0 | 0 | 0 | 0 | 0 |
| 0 | 0 | 0 | 0 | 0 |
| 0 | 0 | 0 | 0 | 0 |
| 0 | 0 | 0.283956174 | 0.138212044 | 0 |
| 0 | 0 | 0 | 0 | 0 |
| 0 | 0 | 0 | 0 | 0 |
| 0 | 0 | 0 | 0 | 0 |
| 0 | 0 | 0 | 0 | 0 |
| 0 | 0 | 0 | 0 | 0 |
| 0 | 0 | 0 | 0 | 0 |
| 0 | 0 | 0 | 0 | 0 |
| 0 | 0 | 0 | 0 | 0 |
| 0 | 0 | 0 | 0 | 0 |
| 0.001429539 | 0 | 0.003623109 | 0 | 0.000997941 |
| 0 | 0 | 0.004702933 | 0.006129471 | 0 |
| 0 | 0 | 0 | 0 | 0 |
| 0 | 0 | 0 | 0.023974601 | 0 |
| 0 | 0 | 0 | 0 | 0 |
| 0 | 0 | 0 | 0 | 0 |
| 0 | 0 | 0 | 0 | 0 |
| 0 | 0 | 0 | 0 | 0 |
| 0 | 0 | 0.017774902 | 0.014586348 | 0.007788897 |
| 0 | 0 | 0 | 0 | 0 |
| 0 | 0 | 0 | 0 | 0 |
| 0 | 0 | 0 | 0 | 0 |
| 0 | 0 | 0 | 0.015465919 | 0 |
| 0.00145797 | 0 | 0 | 0 | 0 |
| 0 | 0 | 0 | 0 | 0 |
| 0 | 0 | 0 | 0 | 0 |
| 0 | 0 | 0 | 0 | 0.185773733 |
| 0 | 0 | 0 | 0 | 0 |
| 0 | 0 | 0 | 0 | 0 |
| 0 | 0 | 0 | 0 | 0 |
| 0 | 0 | 0 | 0 | 0 |
| 0 | 0 | 0 | 0 | 0 |
| 0 | 0 | 0 | 0 | 0 |
| 0.012755127 | 0 | 0.66158297 | 0.489253038 | 0.07979241 |
| 0 | 0 | 0 | 0 | 0 |
| 0 | 0 | 0 | 0 | 0 |

| | | | | |
|---|---|---|---|---|
| 0 | 0 | 0 | 0 | 0 |
| 0 | 0 | 0 | 0 | 0 |
| 0 | 0 | 0 | 0 | 0 |
| 0 | 0 | 0 | 0.152121394 | 0 |
| 0 | 0.001638764 | 0 | 0 | 0 |
| 0 | 0 | 0 | 0.000441023 | 0 |
| 0 | 0.002742603 | 0 | 0 | 0 |
| 0 | 0 | 0 | 0 | 0 |
| 0 | 0 | 0 | 0.005541459 | 0 |
| 0 | 0 | 0 | 0 | 0 |
| 0.000957299 | 0 | 0 | 0.010895311 | 0 |
| 0 | 0 | 0 | 0 | 0 |
| 0 | 0 | 0 | 0 | 0 |
| 0 | 0 | 0 | 0 | 0 |
| 0 | 0 | 0 | 0 | 0 |
| 0 | 0 | 0 | 0 | 0 |
| 0.001407188 | 0 | 0 | 0 | 0 |
| 0 | 0 | 0 | 0 | 0 |
| 0 | 0 | 0 | 0 | 0 |
| 0 | 0 | 0.004780441 | 0 | 0 |
| 0 | 0 | 0 | 0 | 0 |
| 0 | 0 | 0 | 0 | 0 |
| 0 | 0 | 0 | 0 | 0 |
| 0 | 0 | 0 | 0 | 0 |
| 0 | 0 | 0 | 0 | 0 |
| 0 | 0 | 0 | 0 | 0 |
| 0 | 0 | 0 | 0 | 0 |
| 0 | 0 | 0 | 0.018151176 | 0.003736323 |
| 0 | 0 | 0 | 0 | 0 |
| 0 | 0 | 0 | 0 | 0 |
| 0 | 0 | 0.027326636 | 0.001341971 | 0.002900495 |
| 0 | 0 | 0 | 0 | 0 |
| 0.02424208 | 0.117404525 | 0.002200856 | 0.010712385 | 0.072238674 |
| 0 | 0.007801045 | 0 | 0 | 0.000516224 |
| 0 | 0 | 0 | 0 | 0 |
| 0 | 0 | 0 | 0 | 0 |
| 0 | 0 | 0 | 0 | 0 |
| 0 | 0 | 0 | 0 | 0 |
| 0 | 0 | 0 | 0 | 0 |
| 0 | 0 | 0 | 0 | 0 |
| 0 | 0 | 0 | 0 | 0 |
| 0 | 0.107848977 | 0.118595887 | 0 | 0 |
| 0 | 0 | 0.269481445 | 0.082574964 | 0.170289616 |
| 0 | 0 | 0 | 0 | 0 |
| 0 | 0 | 0 | 0 | 0 |
| 0.012053028 | 0 | 0.192319006 | 0.031876836 | 0.109260644 |
| 0 | 0 | 0 | 0 | 0 |
| 0 | 0 | 0 | 0 | 0 |

| | | | | |
|---|---|---|---|---|
| 0 | 0 | 0 | 0 | 0 |
| 0 | 0 | 0 | 0.009612025 | 0.011315819 |
| 0.001331349 | 0.007425975 | 0.00337425 | 0.027233534 | 0.000619597 |
| 0 | 0 | 0 | 0 | 0 |
| 0 | 0 | 0.135247651 | 0 | 0 |
| 0.028455229 | 0.026097348 | 0 | 0.007055376 | 0 |
| 0 | 0 | 0 | 0 | 0 |
| 0 | 0 | 0 | 0 | 0 |
| 0 | 0 | 0 | 0 | 0 |
| 0 | 0 | 0 | 0 | 0 |
| 0 | 0 | 0 | 0 | 0 |
| 0 | 0 | 0 | 0 | 0 |
| 0 | 0 | 0 | 0 | 0 |
| 0.024618739 | 0 | 0 | 0 | 0 |
| 0 | 0 | 0 | 0 | 0 |
| 0 | 0 | 0 | 0 | 0 |
| 0 | 0 | 0 | 0.040841484 | 0 |
| 0 | 0 | 0 | 0 | 0 |
| 0 | 0 | 0 | 0 | 0 |
| 0 | 0 | 0 | 0 | 0 |
| 0 | 0 | 0 | 0 | 0 |
| 0 | 0 | 0 | 0 | 0 |
| 0 | 0 | 0 | 0 | 0 |
| 0 | 0 | 0 | 0 | 0 |
| 0 | 0 | 0 | 0.002559433 | 0.006468045 |
| 0 | 0.005085906 | 0 | 0.030104527 | 0 |
| 0 | 0 | 0 | 0 | 0 |
| 0 | 0 | 0.072906708 | 0.006147882 | 0 |
| 0 | 0 | 0 | 0 | 0 |
| 0 | 0 | 0 | 0 | 0 |
| 0 | 0 | 0 | 0 | 0 |
| 0 | 0 | 0 | 0 | 0.000450473 |
| 0 | 0 | 0 | 0 | 0 |
| 0 | 0 | 0 | 0 | 0 |
| 0 | 0 | 0 | 0 | 0 |
| 0 | 0 | 0 | 0 | 0 |
| 0 | 0 | 0 | 0 | 0 |
| 0 | 0 | 0 | 0 | 0 |
| 0.55618887 | 0 | 1.624004562 | 0.866729386 | 0 |
| 0 | 0 | 0 | 0 | 0 |
| 0 | 0 | 0 | 0 | 0 |
| 0 | 0 | 0 | 0.043616647 | 0 |
| 0 | 0.001074581 | 0 | 0.001391396 | 0.003093245 |
| 0 | 0 | 0 | 0 | 0 |
| 0 | 0 | 0 | 0 | 0 |
| 0 | 0 | 0 | 0 | 0 |
| 0.001694656 | 0 | 0.00200435 | 0.001824484 | 0 |
| 0 | 0 | 0 | 0 | 0 |

| | | | | |
|---|---|---|---|---|
| 0 | 0 | 0 | 0 | 0 |
| 0.040984928 | 0.018409398 | 0.035227047 | 0.039328612 | 0.009122344 |
| 0 | 0 | 0 | 0 | 0 |
| 0 | 0 | 0 | 0 | 0 |
| 0 | 0 | 0 | 0 | 0 |
| 0 | 0 | 0 | 0 | 0 |
| 0 | 0 | 0 | 0 | 0 |
| 0 | 0 | 0 | 0 | 0 |
| 0 | 0 | 0 | 0 | 0 |
| 0 | 0 | 0 | 0 | 0 |
| 0 | 0 | 0 | 0 | 0 |
| 0 | 0 | 0 | 0 | 0 |
| 0 | 0 | 0 | 2.787677278 | 0.006662523 |
| 0 | 0 | 0 | 0 | 0 |
| 0.001333096 | 0 | 0 | 0 | 0.001861231 |
| 0 | 0 | 0 | 0 | 0 |
| 0 | 0 | 0 | 0 | 0 |
| 0 | 0 | 0 | 0 | 0 |
| 0 | 0 | 0 | 0 | 0 |
| 0 | 0 | 0 | 0 | 0 |
| 0 | 0 | 0 | 0 | 0 |
| 0 | 0 | 0 | 0.025605992 | 0 |
| 0 | 0.009758863 | 0 | 0.027438244 | 0 |
| 0 | 0 | 0.794719166 | 0.002903565 | 0 |
| 0 | 0 | 0 | 0 | 0 |
| 0 | 0 | 0 | 0 | 0 |
| 0 | 0 | 0 | 0 | 0 |
| 0 | 0 | 0 | 0 | 0 |
| 0 | 0 | 0 | 0 | 0 |
| 0 | 0 | 0 | 0 | 0 |
| 0 | 0 | 0 | 0 | 0 |
| 0 | 0 | 0 | 0 | 0 |
| 0 | 0 | 0 | 0 | 0 |
| 0 | 0 | 0 | 0 | 0 |
| 0 | 0 | 0.071701697 | 9.885652568 | 0 |
| 0 | 0 | 0 | 0 | 0 |
| 0 | 0 | 0 | 0 | 0 |
| 0 | 0 | 0 | 0 | 0 |
| 0.006101202 | 0.014584788 | 0 | 0.045276528 | 0.002028172 |
| 0 | 0 | 0 | 0 | 0 |
| 0.042982773 | 0.232262289 | 0.008318908 | 0.294691971 | 0.066739847 |
| 0 | 0 | 0 | 0 | 0 |
| 0 | 0 | 0 | 0 | 0 |
| 0 | 0 | 0 | 0 | 0 |
| 0 | 0 | 0 | 0 | 0 |
| 1.322246351 | 4.596555667 | 0.030841509 | 0.026491255 | 0.119511778 |
| 0 | 0 | 0 | 0 | 0 |
| 0 | 0 | 0 | 0 | 0 |

| | | | | |
|---|---|---|---|---|
| 0 | 0 | 0 | 0 | 0 |
| 0 | 0 | 0 | 0 | 0 |
| 0 | 0 | 0 | 0 | 0 |
| 0 | 0 | 0 | 0.016732192 | 0 |
| 0 | 0 | 0 | 0 | 0 |
| 0 | 0 | 0 | 0 | 0 |
| 0 | 0 | 0 | 0 | 0 |
| 0 | 0 | 0 | 0 | 0 |
| 0 | 0 | 0 | 0 | 0 |
| 0.062131139 | 0 | 0 | 0 | 0.195483273 |
| 0 | 0 | 0.004210311 | 0.054896804 | 0 |
| 0 | 0 | 0 | 0 | 0 |
| 0 | 0.008400768 | 0 | 0 | 0 |
| 0 | 0 | 0 | 0 | 0 |
| 0 | 0 | 0 | 0 | 0 |
| 0 | 0 | 0 | 0 | 0 |
| 0 | 0 | 0 | 0 | 0 |
| 0 | 0 | 0 | 0 | 0 |
| 0 | 0 | 0 | 0 | 0 |
| 0.118191535 | 0.471345349 | 0.144903876 | 0.313710016 | 0.750054845 |
| 0 | 0 | 0 | 0 | 0 |
| 0 | 0 | 0 | 0 | 0 |
| 0 | 0 | 0 | 0 | 0 |
| 0 | 0 | 0 | 0 | 0 |
| 0 | 0 | 0 | 0 | 0 |
| 0 | 0 | 0 | 0 | 0 |
| 0 | 0 | 0 | 0 | 0 |
| 0 | 0 | 0 | 0 | 0 |
| 0 | 0 | 0 | 0 | 0 |
| 0 | 0 | 0 | 0 | 0 |
| 0 | 0 | 0 | 0 | 0 |
| 0 | 0 | 0 | 0 | 0 |
| 0 | 0 | 0 | 0 | 0 |
| 0 | 0 | 0 | 0 | 0 |
| 0 | 0 | 0 | 0 | 0 |
| 0 | 0 | 0 | 0 | 0.002981123 |
| 0 | 0 | 0 | 0 | 0 |
| 0 | 0 | 0 | 0 | 0 |
| 0 | 0 | 0 | 0.003366536 | 0 |
| 0 | 0 | 0 | 0 | 0 |
| 0 | 0 | 0 | 0 | 0 |
| 0 | 0 | 0 | 0 | 0 |
| 0 | 0 | 0.000962315 | 0.176627405 | 0 |
| 0 | 0 | 0 | 0 | 0 |
| 0.030210405 | 0 | 0.010329754 | 0 | 0 |
| 0 | 0 | 0 | 0.021821703 | 0 |
| 0 | 0 | 0 | 0 | 0 |
| 0 | 0 | 0 | 0 | 0 |

| | | | | |
|---|---|---|---|---|
| 0 | 0 | 0 | 0 | 0 |
| 0 | 0 | 0 | 0 | 0 |
| 0 | 0 | 0 | 0 | 0 |
| 0 | 0 | 0 | 0 | 0 |
| 0 | 0 | 0 | 0 | 0 |
| 0 | 0.00815561 | 0.007982144 | 0.010775612 | 0 |
| 0 | 0 | 0 | 0 | 0 |
| 0 | 0 | 0 | 0 | 0 |
| 0 | 0 | 0 | 0 | 0 |
| 0 | 0 | 0 | 0 | 0 |
| 0 | 0 | 0 | 0 | 0 |
| 0 | 0 | 0 | 0 | 0 |
| 0 | 0 | 0 | 0 | 0 |
| 0 | 0 | 0 | 0 | 0 |
| 0 | 0 | 0 | 0 | 0 |
| 0 | 0 | 0 | 0 | 0 |
| 0 | 0 | 0 | 0.025095825 | 0 |
| 0 | 0 | 0 | 0 | 0 |
| 0 | 0 | 0 | 0 | 0 |
| 0 | 0 | 0 | 0 | 0 |
| 0.001392615 | 0 | 0 | 0 | 0 |
| 0 | 0 | 0 | 0 | 0 |
| 0 | 0 | 0 | 0 | 0 |
| 0 | 0 | 0.001098137 | 0 | 0 |
| 0 | 0 | 0 | 0 | 0 |
| 0.004801019 | 0 | 0 | 0 | 0.002137203 |
| 0 | 0 | 0 | 0 | 0 |
| 0 | 0 | 0 | 0.013119291 | 0 |
| 0 | 0 | 0 | 0 | 0 |
| 0 | 0 | 0 | 0 | 0 |
| 0 | 0 | 0 | 0 | 0 |
| 0 | 0 | 0.001911622 | 0 | 0 |
| 0 | 0 | 0 | 0 | 0 |
| 0 | 0 | 0 | 0 | 0 |
| 0 | 0 | 0 | 0 | 0 |
| 0 | 0 | 0 | 0 | 0 |
| 0 | 0 | 0 | 0 | 0 |
| 0 | 0 | 0 | 0 | 0 |
| 0 | 0 | 0.041823479 | 0.101202144 | 0.038925223 |
| 0 | 0 | 0 | 0 | 0 |
| 0 | 0 | 0 | 0 | 0 |
| 0 | 0 | 0 | 0 | 0 |
| 0 | 0 | 0 | 0 | 0 |
| 0 | 0 | 0 | 0 | 0 |
| 0 | 0 | 0 | 0 | 0 |
| 0 | 0 | 0 | 0 | 0 |
| 0 | 0 | 0 | 0 | 0 |
| 0 | 0 | 0 | 0 | 0 |

| | | | | |
|---|---|---|---|---|
| 0 | 0 | 0 | 0 | 0 |
| 0 | 0.001956614 | 0 | 0.024932611 | 0 |
| 0 | 0 | 0 | 0 | 0 |
| 0 | 0 | 0 | 0 | 0 |
| 0 | 0 | 0 | 0 | 0 |
| 0 | 0 | 0 | 0 | 0 |
| 0 | 0 | 0 | 0 | 0 |
| 0 | 0 | 0 | 0 | 0 |
| 0 | 0 | 0 | 0 | 0 |
| 0 | 0 | 0 | 0 | 0 |
| 0.005086631 | 0 | 0.003966725 | 0.022115898 | 0.00072839 |
| 0 | 0 | 0 | 0 | 0 |
| 0 | 0 | 0 | 0.000620552 | 0 |
| 0 | 0 | 0 | 0 | 0 |
| 0 | 0 | 0 | 0 | 0 |
| 0 | 0 | 0 | 0 | 0 |
| 0 | 0 | 0 | 0 | 0 |
| 0 | 0 | 0 | 0 | 0 |
| 0 | 0 | 0 | 0 | 0 |
| 0 | 0 | 0 | 0 | 0 |
| 0.001191375 | 0 | 0 | 0 | 0 |
| 0 | 0 | 0 | 0 | 0 |
| 0 | 0 | 0 | 0 | 0 |
| 0 | 0 | 0.532953216 | 1.15091164 | 0.613944044 |
| 0 | 0 | 0 | 0 | 0 |
| 0 | 0 | 0 | 0 | 0 |
| 0 | 0 | 0 | 0 | 0 |
| 0 | 0 | 0 | 0 | 0 |
| 0 | 0 | 0 | 0 | 0 |
| 0 | 0 | 0 | 0 | 0 |
| 0 | 0.008507331 | 0 | 0 | 0 |
| 0 | 0 | 0 | 0 | 0 |
| 0 | 0 | 0 | 0.000674611 | 0 |
| 0 | 0 | 0 | 0 | 0 |

| PHL018-0175_V3 CTRL | PHL018-0237_V3 CTRL | PHL018-0177_V3 CTRL | PHL018-0253_V3 REF | PHL018-0029_V2 EYCF |
|---|---|---|---|---|
| 0 | 0.010406229 | 0.001719664 | 1.535039297 | 0.008964878 |
| 0 | 0.267079799 | 0.551821787 | 0.373444711 | 0.851963662 |
| 2.439297074 | 0 | 0 | 0.185071198 | 2.697983447 |
| 0 | 0 | 0 | 0 | 0 |
| 0 | 0 | 0 | 0 | 0 |
| 0 | 0 | 0 | 0 | 0 |
| 0 | 0 | 0 | 0 | 0 |
| 0 | 0 | 0 | 0 | 0 |
| 0 | 0 | 0 | 0 | 0 |
| 0 | 0 | 0 | 0 | 0 |
| 0 | 0 | 0 | 0 | 0 |
| 0 | 0 | 0 | 0 | 0 |
| 0 | 0 | 0 | 0 | 0 |
| 10.896454 | 0.452829239 | 1.196339406 | 0.029646746 | 0.190871076 |
| 0 | 0 | 0 | 0 | 0 |
| 0 | 0 | 0 | 0 | 0 |
| 0 | 0 | 0 | 0 | 0 |
| 0 | 0 | 0 | 0 | 0 |
| 0 | 0 | 0 | 0 | 0 |
| 0 | 0 | 0 | 0 | 0 |
| 0 | 0 | 0 | 0 | 0 |
| 0 | 0 | 0 | 0 | 0 |
| 0 | 0 | 0 | 0 | 0 |
| 0 | 0 | 0 | 0 | 0 |
| 0 | 0 | 0 | 0 | 0 |
| 0 | 0 | 0 | 0 | 0 |
| 0 | 0 | 0 | 0.00295327 | 0 |
| 0 | 0 | 0 | 0 | 0 |
| 0 | 0 | 0 | 0 | 0 |
| 0 | 0.051868601 | 0 | 0 | 0 |
| 0 | 0 | 0 | 0 | 0 |
| 0 | 0 | 0 | 0 | 0 |
| 0 | 0 | 0 | 0 | 0 |
| 0 | 0 | 0 | 0 | 0 |
| 0 | 0 | 0 | 0 | 0 |
| 0 | 3.660423387 | 2.536633561 | 0.116736188 | 4.996502873 |
| 0 | 0 | 0 | 0 | 0 |
| 0 | 0 | 0 | 0 | 0 |
| 0 | 0 | 0 | 0 | 0 |
| 0 | 0 | 0 | 0 | 0 |
| 0 | 0 | 0 | 0 | 0 |
| 0 | 0.001009767 | 0.001868915 | 0.024236307 | 0.001082549 |
| 0.705855022 | 0.655102078 | 0.014845884 | 1.205903907 | 0.258339037 |
| 0.08356825 | 0.01043133 | 0.028347139 | 0.008693457 | 0.004082757 |
| 0.188052273 | 0 | 0 | 0 | 0 |
| 0 | 0 | 0 | 0 | 0 |

| | | | | |
|---|---|---|---|---|
| 0 | 0 | 0.000715869 | 0 | 0 |
| 0 | 0 | 0.002708703 | 0.007185023 | 0.007759357 |
| 0 | 0 | 0.002239652 | 0.040925745 | 0.009623571 |
| 0 | 0.164276613 | 0.326206158 | 0.28103993 | 0 |
| 0 | 0.00627535 | 0 | 0 | 0.003681176 |
| 0 | 0 | 0.097058599 | 0 | 0 |
| 0 | 0.084649347 | 0.169182763 | 0 | 0 |
| 0 | 0 | 0 | 0 | 0 |
| 0 | 0 | 0 | 0 | 0 |
| 0 | 0 | 0 | 0 | 0 |
| 0 | 0 | 0 | 0.010220743 | 0 |
| 0 | 1.414096041 | 0.160561391 | 0.039453511 | 0.397735607 |
| 0 | 0.000988308 | 0 | 0.004694834 | 0.001907179 |
| 0 | 0.010294506 | 0.018524197 | 0.032274992 | 0.077605981 |
| 0 | 0 | 0 | 0.001121051 | 0 |
| 0 | 0 | 0 | 0 | 0 |
| 0.878360428 | 0.598332699 | 0.01865734 | 0 | 0.001200785 |
| 0.181158708 | 0 | 0 | 0 | 0 |
| 0 | 0 | 0 | 0 | 0 |
| 0 | 0 | 0 | 0 | 0 |
| 0 | 0.037469087 | 0.006990847 | 0.026549882 | 0.450012797 |
| 0 | 0.006160522 | 0.013690053 | 0.016821166 | 0.019466077 |
| 0 | 0.001454683 | 0.005258559 | 0 | 0 |
| 0 | 0 | 0 | 0 | 0 |
| 0 | 0 | 0 | 0 | 0 |
| 0 | 0 | 0 | 0 | 0 |
| 0.023499736 | 0 | 0 | 0 | 0 |
| 0 | 0 | 0 | 0 | 0 |
| 0 | 0 | 0 | 0.004407534 | 0 |
| 0 | 0 | 0 | 0 | 0 |
| 0 | 0 | 0.000849991 | 0.00321498 | 0 |
| 0 | 0 | 0 | 0 | 0 |
| 0 | 0 | 0 | 0 | 0 |
| 0 | 0 | 0 | 0 | 0 |
| 0 | 0 | 0 | 0 | 0 |
| 0 | 0 | 0 | 0 | 0 |
| 0 | 0 | 0 | 0 | 0 |
| 0 | 31.49155686 | 0.001364435 | 0.00786407 | 0.000843024 |
| 0 | 0 | 0 | 0 | 0 |
| 0 | 0 | 0 | 0 | 0 |
| 0 | 0 | 0 | 0 | 0 |
| 0 | 0 | 0 | 0 | 0 |
| 0 | 0 | 0 | 0 | 0 |
| 0 | 0 | 0 | 0 | 0 |
| 0 | 0 | 0 | 0 | 0.004933533 |
| 0 | 0 | 0 | 0 | 0 |
| 0 | 0 | 0 | 0 | 0.009669658 |
| 0 | 0 | 0 | 0 | 0.467443349 |

| | | | | |
|---|---|---|---|---|
| 0 | 0 | 0 | 0 | 0 |
| 0 | 0 | 0 | 0 | 0 |
| 0 | 0 | 0 | 0 | 0 |
| 0 | 0 | 0 | 0 | 0 |
| 0 | 0.323487567 | 0 | 0 | 0.19011903 |
| 0 | 0 | 0 | 0.003736519 | 0 |
| 0 | 0 | 0 | 0 | 0 |
| 0 | 0 | 0 | 0 | 0 |
| 0 | 0 | 0.004350414 | 0.000587674 | 0.001343964 |
| 0 | 0 | 0 | 0 | 0 |
| 0 | 0 | 0 | 0 | 0 |
| 0 | 0 | 0.00273582 | 0.002046833 | 0 |
| 0 | 0 | 0 | 0 | 0 |
| 0 | 0 | 0 | 0 | 0 |
| 0 | 0 | 0 | 0 | 0 |
| 0 | 0 | 0 | 0 | 0 |
| 0 | 0 | 0 | 0 | 0 |
| 0 | 0 | 0 | 0 | 0 |
| 0 | 0.105077004 | 0.036895856 | 0.404999218 | 2.328000422 |
| 0 | 0.000903446 | 0.016895493 | 0 | 0.000484282 |
| 0 | 0 | 0 | 0 | 0 |
| 0 | 0 | 0 | 0 | 0 |
| 0 | 0 | 0 | 0 | 0 |
| 0 | 0 | 0 | 0 | 0 |
| 0 | 0 | 0 | 0 | 0 |
| 0 | 0 | 0 | 0 | 0 |
| 0 | 0 | 0 | 0 | 0 |
| 0 | 0 | 0 | 0.001318522 | 0 |
| 0 | 0 | 0 | 0 | 0 |
| 0 | 0 | 0 | 0 | 0 |
| 0 | 0.132245717 | 0.01476107 | 0.012870317 | 0.134315782 |
| 0 | 0.002129739 | 0 | 0.004754255 | 0 |
| 0 | 0 | 0 | 0 | 0 |
| 0 | 0 | 0 | 0 | 0 |
| 0 | 0 | 0 | 0 | 0 |
| 0 | 0 | 0 | 0 | 0 |
| 0 | 0 | 0 | 0 | 0 |
| 0 | 0 | 0 | 0 | 0 |
| 0 | 0 | 0 | 0 | 0.911321089 |
| 0 | 0 | 0 | 0 | 0 |
| 0 | 0 | 0 | 0 | 0 |
| 0 | 0 | 0 | 0 | 0 |
| 0 | 0 | 0 | 0 | 0 |
| 0 | 0 | 0 | 0 | 0 |
| 0 | 0 | 0 | 0.002343445 | 0 |
| 0 | 0 | 0.00148438 | 0 | 0 |
| 0 | 0 | 0 | 0.051619664 | 0 |
| 0 | 0 | 0 | 0 | 0 |

| | | | | |
|---|---|---|---|---|
| 0 | 0 | 0 | 0 | 0 |
| 0 | 0 | 0 | 0 | 0 |
| 0 | 0 | 0 | 0 | 0 |
| 0 | 0 | 0 | 0 | 0 |
| 0 | 0.028335078 | 0.030786128 | 0 | 0 |
| 0 | 0 | 0 | 0 | 0 |
| 0 | 0 | 0 | 0 | 0 |
| 0 | 0 | 0 | 0 | 0 |
| 0 | 0 | 0 | 0 | 0 |
| 0 | 0 | 0 | 0 | 0 |
| 0 | 0 | 0 | 0 | 0 |
| 0 | 0 | 0 | 0 | 0 |
| 0 | 0 | 0 | 0 | 0 |
| 0 | 0 | 0 | 0 | 0 |
| 0 | 0 | 0 | 0 | 0 |
| 0 | 0 | 0 | 1.255135902 | 0 |
| 0 | 0 | 0 | 0 | 0 |
| 0 | 0 | 0 | 0 | 0 |
| 0 | 0 | 0 | 0 | 0 |
| 0 | 0 | 0 | 0 | 0 |
| 0 | 0 | 0 | 0 | 0 |
| 0 | 0 | 0 | 0 | 0 |
| 0 | 0 | 0 | 0 | 0 |
| 0 | 0 | 0 | 0 | 0 |
| 0 | 0.049231893 | 0 | 0 | 0 |
| 0 | 0 | 0 | 0 | 0 |
| 0 | 0 | 0 | 0 | 0 |
| 0 | 0 | 0 | 0 | 0 |
| 0 | 0 | 0 | 0 | 0 |
| 0 | 0 | 0 | 0 | 0 |
| 0 | 0 | 0 | 0 | 0 |
| 0 | 0 | 0.001015176 | 0 | 0 |
| 0 | 0 | 0 | 0 | 0 |
| 0 | 0 | 0 | 0 | 0 |
| 0 | 0 | 0 | 0 | 0 |
| 0 | 0 | 0 | 0 | 0 |
| 0 | 0 | 0 | 0 | 0 |
| 0 | 0 | 0 | 0 | 0 |
| 0 | 0 | 0 | 0 | 0 |
| 0 | 0 | 0 | 0 | 0 |
| 0 | 0 | 0 | 0 | 0 |
| 0 | 0 | 0 | 0 | 0 |
| 0 | 0 | 0 | 0 | 0 |
| 0 | 0 | 0 | 0 | 0 |
| 0 | 0 | 0 | 0 | 0 |
| 0 | 0 | 0 | 0 | 0 |
| 0 | 0 | 0.040642473 | 0 | 0 |
| 0 | 0 | 0 | 0 | 0 |

| | | | | |
|---|---|---|---|---|
| 0 | 0 | 0 | 0 | 0 |
| 0 | 0 | 0 | 0 | 0 |
| 0 | 0 | 0 | 0 | 0 |
| 0 | 0 | 0 | 0 | 0 |
| 0 | 0 | 0 | 0 | 0 |
| 0 | 0 | 0 | 0 | 0 |
| 0 | 0 | 0 | 0 | 0 |
| 0 | 0 | 0 | 0 | 0 |
| 0 | 0 | 0 | 0 | 0 |
| 0 | 0 | 0 | 0 | 0 |
| 0 | 0 | 0 | 0 | 0 |
| 0 | 0 | 0.003744629 | 0 | 0 |
| 0 | 0.000671816 | 0 | 0.007348564 | 0 |
| 0 | 0 | 0 | 0 | 0 |
| 0 | 0 | 0 | 0.407595528 | 0 |
| 0 | 0 | 0 | 0 | 0 |
| 0 | 0 | 0 | 0 | 0 |
| 0 | 0 | 0 | 0 | 0 |
| 0 | 0 | 0 | 0 | 0 |
| 0 | 0 | 0 | 0 | 0 |
| 0.003602182 | 0.079136474 | 0.021069418 | 0.008636603 | 0.039726901 |
| 0 | 0 | 0 | 0 | 0 |
| 0 | 0 | 0 | 0.001033375 | 0 |
| 0 | 0 | 0 | 0.001228124 | 0 |
| 0 | 0 | 0 | 0 | 0 |
| 0 | 0 | 0 | 0 | 0 |
| 0.009090579 | 0.004572145 | 0.007404511 | 0.005944255 | 0 |
| 0 | 0 | 0 | 0 | 0 |
| 0 | 0 | 0 | 0 | 0 |
| 0 | 0 | 0 | 0 | 0.00146839 |
| 0 | 0 | 0 | 0 | 0 |
| 0 | 0 | 0 | 0 | 0 |
| 0 | 0 | 0 | 0.001933567 | 0 |
| 0 | 0 | 0.006882171 | 0.020872705 | 0 |
| 0 | 0 | 0.003309041 | 0.010728011 | 0.006352582 |
| 0 | 0 | 0 | 0 | 0 |
| 0 | 0 | 0 | 0 | 0 |
| 0 | 0 | 0 | 0 | 0 |
| 0 | 0 | 0 | 0 | 0 |
| 0 | 0 | 0 | 0 | 0 |
| 0 | 0 | 0 | 0 | 0 |
| 0 | 0 | 0 | 0 | 0 |
| 0 | 0 | 0 | 0 | 0 |
| 0 | 0 | 0 | 0 | 0 |
| 0 | 0 | 0 | 0 | 0 |
| 0 | 0 | 0 | 0 | 0.063403398 |
| 0 | 0.001548623 | 0 | 0 | 0 |
| 0 | 0.000582436 | 0 | 0.021661032 | 0.002653771 |

| | | | | |
|---|---|---|---|---|
| 0 | 0 | 0 | 0 | 0 |
| 0 | 0 | 0 | 0 | 0 |
| 0 | 0 | 0 | 0 | 0 |
| 0 | 0 | 0 | 0 | 0 |
| 0 | 0 | 0 | 0 | 0 |
| 0 | 0 | 0 | 0 | 0 |
| 0 | 0 | 0 | 0 | 0 |
| 0 | 0 | 0 | 0 | 0 |
| 0 | 0 | 0 | 0 | 0.004152782 |
| 0 | 0 | 0 | 0 | 0 |
| 0 | 0 | 0 | 0 | 0 |
| 0 | 0 | 0 | 0 | 0 |
| 0 | 0 | 0 | 0 | 0 |
| 0.085621258 | 0 | 0 | 0 | 0 |
| 0 | 0 | 0 | 0 | 0 |
| 0 | 0 | 0 | 0 | 0 |
| 0 | 0 | 0 | 0 | 0 |
| 0 | 0 | 0 | 0 | 0 |
| 0.005378957 | 0.004294417 | 0 | 0.006294036 | 0.001111299 |
| 0 | 0 | 0.095598431 | 0 | 0 |
| 0 | 0.010703462 | 0 | 0 | 0.008196393 |
| 0 | 0 | 0 | 0 | 0 |
| 0 | 0 | 0 | 0 | 0 |
| 0 | 0 | 0 | 0 | 0 |
| 0.003318648 | 0 | 0.007530133 | 0.015190257 | 0 |
| 0.002214004 | 0 | 0 | 0 | 0 |
| 0 | 0 | 0 | 0 | 0 |
| 0 | 0 | 0 | 0 | 0 |
| 0 | 0 | 0 | 0.016826043 | 0 |
| 20.56320096 | 0 | 0 | 0 | 0 |
| 0 | 0.464023608 | 0.086024451 | 0.691110531 | 0.10727712 |
| 0.56862962 | 0 | 0 | 0 | 0 |
| 0 | 0 | 0 | 0 | 0 |
| 0 | 0.010390149 | 0 | 0 | 0 |
| 0 | 0 | 0 | 0 | 0 |
| 0 | 0 | 0 | 0 | 0 |
| 0 | 0 | 0 | 0 | 0 |
| 0 | 0 | 0 | 0.001685025 | 0 |
| 0 | 0 | 0 | 0 | 0 |
| 0.147836904 | 0 | 0 | 0 | 0 |
| 0 | 0 | 0 | 0 | 0 |
| 0 | 0.000818614 | 0.009280116 | 0.034793808 | 0.020185211 |
| 0 | 0 | 0 | 0.004293131 | 0.001472708 |
| 0 | 0 | 0 | 0 | 0 |
| 0 | 0 | 0 | 0 | 0 |
| 0 | 0 | 0 | 0 | 0 |
| 0 | 0 | 0 | 0 | 0 |
| 0 | 0 | 0 | 0 | 0 |

| | | | | |
|---|---|---|---|---|
| 0 | 0 | 0 | 0 | 0 |
| 0 | 0 | 0 | 0 | 0 |
| 0.090002857 | 0.006784167 | 0.033293464 | 0 | 0.001983589 |
| 0 | 0 | 0 | 0 | 0 |
| 0 | 0.002497722 | 0 | 0 | 0 |
| 0 | 0 | 0 | 0 | 0 |
| 0 | 0 | 0 | 0 | 0 |
| 0 | 0 | 0 | 0 | 0 |
| 0 | 0 | 0 | 0 | 0 |
| 0 | 0 | 0 | 0 | 0 |
| 0 | 0 | 0 | 0 | 0 |
| 0 | 0 | 0 | 0 | 0 |
| 0 | 0 | 0 | 0 | 0 |
| 0 | 0.000681612 | 0.023128467 | 0.001704163 | 0 |
| 0.001434043 | 0.002062798 | 0.010629393 | 0 | 0.001005219 |
| 0 | 0 | 0.005341712 | 0.021584739 | 0 |
| 0.002042455 | 0 | 0.001977345 | 0.003739524 | 0.000916285 |
| 0 | 0 | 0 | 0 | 0 |
| 0 | 0 | 0 | 0 | 0 |
| 0 | 0 | 0 | 0 | 0 |
| 0 | 0 | 0 | 0 | 0 |
| 0 | 0.0011573 | 0 | 0.025317921 | 0.002274646 |
| 0 | 0 | 0 | 0 | 0 |
| 0 | 0 | 0 | 0 | 0 |
| 0 | 0 | 0 | 0 | 0 |
| 0 | 0 | 0 | 0 | 0 |
| 0 | 0 | 1.400271832 | 0 | 0 |
| 0 | 0 | 0 | 0 | 0 |
| 0 | 0 | 0 | 0 | 0 |
| 0 | 0 | 0.07229566 | 0 | 0 |
| 0 | 0 | 0.001025893 | 0 | 0 |
| 0 | 0 | 0 | 0 | 0 |
| 0 | 0 | 0 | 0 | 0 |
| 0 | 0 | 0 | 0 | 0 |
| 0 | 0 | 0 | 0 | 0 |
| 0 | 0 | 0 | 0 | 0 |
| 0 | 0 | 0 | 0 | 0 |
| 0.18068587 | 0 | 0.003363961 | 0.020903285 | 0.000779416 |
| 5.785819836 | 0 | 0 | 0 | 0 |
| 0 | 0 | 0 | 0 | 0 |
| 0 | 0.0101431 | 0 | 0 | 0 |
| 0 | 0 | 0 | 0 | 0 |
| 0 | 0 | 0 | 0 | 0 |
| 0 | 0 | 0 | 0.008278743 | 0 |
| 0 | 0 | 0 | 0 | 0 |
| 0 | 0 | 0 | 0 | 0 |
| 0 | 0 | 0 | 0 | 0 |
| 0 | 0 | 0 | 0 | 0 |

| | | | | |
|---|---|---|---|---|
| 0 | 1.429463144 | 0.00111924 | 0 | 4.934402174 |
| 0 | 0 | 0 | 0 | 0 |
| 0 | 0 | 0 | 0.001262788 | 0 |
| 0 | 0 | 0 | 0 | 0 |
| 0.003691627 | 0 | 0.003762048 | 0.002032778 | 0 |
| 0 | 0 | 0 | 0 | 0 |
| 0 | 0 | 0 | 0 | 0 |
| 0.013617441 | 0.068094988 | 0.030980861 | 0.00213704 | 0 |
| 0 | 0 | 0 | 0 | 0 |
| 0.538693678 | 0 | 0.002377963 | 0 | 0.018890194 |
| 0 | 0 | 0 | 0 | 0 |
| 0 | 0.013306903 | 0.001565397 | 0.005075065 | 0.000967189 |
| 0.001308275 | 0.001368645 | 0.035622286 | 0.001283206 | 0.009537417 |
| 0 | 0.002598274 | 0 | 0.007470624 | 0.001949887 |
| 0 | 0.00140762 | 0 | 0 | 0 |
| 0 | 0 | 0.001188333 | 0 | 0.00165199 |
| 0 | 0 | 0 | 0 | 0 |
| 0 | 0 | 0 | 0 | 0 |
| 0 | 0 | 0 | 0 | 0 |
| 0 | 0 | 0 | 0 | 0 |
| 0 | 0 | 0 | 0 | 0 |
| 0 | 0 | 0 | 0 | 0 |
| 0 | 0 | 0 | 0 | 0 |
| 0 | 0 | 0 | 0 | 0 |
| 0 | 0 | 0 | 0 | 0 |
| 0.108380787 | 0 | 0 | 0 | 0 |
| 0 | 3.377890134 | 0 | 0 | 22.65801362 |
| 0 | 0 | 0 | 0 | 0 |
| 0 | 0 | 0 | 0 | 0 |
| 0 | 0.00205019 | 0 | 0 | 0.012308598 |
| 0 | 0 | 0 | 0 | 0 |
| 0 | 0 | 0 | 0 | 0 |
| 0 | 0 | 0 | 0 | 0 |
| 0 | 0 | 0 | 0 | 0 |
| 0 | 0 | 0 | 0 | 0 |
| 0 | 0 | 0 | 0.014650914 | 0 |
| 0 | 0 | 0 | 0 | 0 |
| 0 | 0 | 0 | 0 | 0 |
| 0 | 0 | 0 | 0 | 0 |
| 0 | 0 | 0 | 0.001047963 | 0 |
| 0 | 0 | 0 | 0 | 0 |
| 0 | 0 | 0 | 0 | 0.088313263 |
| 0 | 0 | 0 | 0 | 0 |
| 0 | 0 | 0 | 0.012041084 | 0 |
| 0 | 0 | 0 | 0 | 0 |
| 0 | 0 | 0 | 0 | 0 |
| 0 | 0 | 0.001433255 | 0 | 0.001328317 |
| 0 | 0 | 0 | 0 | 0 |

| | | | | |
|---|---|---|---|---|
| 0 | 0 | 0 | 0 | 0 |
| 0 | 0 | 0.013134457 | 0 | 0 |
| 0 | 0 | 0 | 0 | 0 |
| 1.954048159 | 0 | 0 | 0 | 0 |
| 0 | 0 | 0 | 0 | 0 |
| 0 | 0.035131365 | 0.006773173 | 0.068072364 | 0 |
| 0 | 0 | 0 | 0 | 0 |
| 0 | 0 | 0 | 0 | 0 |
| 0 | 0 | 0 | 0 | 0 |
| 0 | 0 | 0 | 0 | 0 |
| 0.002812996 | 0 | 0.014925906 | 0.016979028 | 0 |
| 0 | 0 | 0 | 0 | 0 |
| 0 | 0 | 0 | 0 | 0 |
| 0.001635432 | 0.001497036 | 0.004453025 | 0.00481228 | 0.000687831 |
| 0 | 0 | 0 | 0 | 0.228971431 |
| 0 | 0 | 0 | 0 | 0 |
| 0 | 0 | 0 | 0 | 0 |
| 0 | 0 | 0 | 0.00572382 | 0 |
| 0 | 0 | 0 | 0 | 0.006344787 |
| 0 | 0 | 0 | 0 | 0.025709228 |
| 0 | 0 | 0.110885997 | 0 | 0 |
| 0 | 0.09405807 | 0.290609891 | 0.203425614 | 0 |
| 0 | 0 | 0.002347615 | 0.028414545 | 0.009138068 |
| 0 | 0 | 0 | 0 | 0 |
| 0 | 0 | 0 | 0 | 0.047995562 |
| 0 | 0 | 0 | 0 | 0 |
| 0 | 0 | 0 | 0.017972371 | 0 |
| 0 | 0 | 0 | 0 | 0 |
| 0 | 0 | 0 | 0 | 0 |
| 0 | 0.099999273 | 1.16510554 | 0.541966508 | 0.659709872 |
| 0 | 0.00101562 | 0.00469937 | 0 | 0.002644287 |
| 0 | 0 | 0 | 0 | 0 |
| 0 | 0 | 0 | 0 | 0 |
| 0 | 0 | 0 | 0 | 0 |
| 0 | 0.005021562 | 0 | 0 | 0 |
| 0 | 0 | 0 | 0 | 0 |
| 0 | 1.019523031 | 0 | 0 | 0.001474051 |
| 0 | 0 | 0.002521566 | 0 | 0 |
| 0 | 0 | 0 | 0 | 0.003180681 |
| 0 | 0 | 0 | 0 | 1.065912324 |
| 0 | 0.011252357 | 0.041083109 | 0.030331001 | 0.025069271 |
| 0 | 0.019657427 | 1.062794897 | 2.856977632 | 3.742633836 |
| 0 | 0 | 0.001616942 | 0 | 0 |
| 0 | 0 | 0 | 0 | 0 |
| 0 | 0 | 0 | 0 | 0 |
| 0 | 0 | 0 | 0 | 0 |
| 0 | 0 | 0 | 0 | 0 |
| 0 | 0 | 0 | 0 | 0 |

| | | | | |
|---|---|---|---|---|
| 0.058110057 | 0 | 0 | 0 | 0 |
| 0 | 0 | 0 | 0.008184067 | 0 |
| 0.034696593 | 0 | 0.044500284 | 0 | 0 |
| 0 | 0 | 0 | 0 | 0 |
| 0 | 0 | 0 | 0.002861193 | 0 |
| 0 | 0 | 0 | 0 | 0 |
| 0 | 0 | 0 | 0 | 0 |
| 0 | 0 | 0 | 0 | 0 |
| 0 | 0.027058209 | 0 | 0.076464498 | 0.074564153 |
| 0 | 0 | 0 | 0.030440865 | 0 |
| 0 | 0 | 0 | 0 | 0 |
| 0 | 0.000959936 | 0 | 0 | 0 |
| 0 | 0.001119923 | 0 | 0 | 0 |
| 0.00196894 | 0 | 0.002859261 | 0.002574947 | 0 |
| 0 | 0 | 0 | 0 | 0 |
| 0 | 0.012877561 | 0.013406781 | 0.49139803 | 0.091808269 |
| 0.141043233 | 0.003389941 | 0.001857502 | 0.005352958 | 0.0095639 |
| 0 | 0 | 0 | 0 | 0 |
| 0 | 0 | 0 | 0.006450735 | 0 |
| 0 | 0 | 0 | 0 | 0.195656904 |
| 0 | 0 | 0.00546229 | 0 | 0 |
| 0 | 0 | 0 | 0 | 0 |
| 0 | 0 | 0.079381631 | 0 | 0 |
| 0 | 0 | 0 | 0 | 1.014138441 |
| 0 | 0 | 0 | 0 | 0 |
| 0 | 0.033609886 | 0 | 0.004113421 | 0.040316007 |
| 0 | 0 | 0.004192203 | 0.003680963 | 0.005342237 |
| 0 | 0 | 0.00116069 | 0 | 0 |
| 0 | 0 | 0 | 0 | 0 |
| 0 | 0 | 0 | 0 | 0 |
| 0 | 0.065284602 | 0.0009963 | 0.008828756 | 0.001292695 |
| 0 | 0.01129968 | 0.00712973 | 0 | 0.001376607 |
| 0.004674663 | 0.07873502 | 0.01725402 | 0.002751051 | 0 |
| 0 | 0 | 0 | 0.007008933 | 0 |
| 0 | 0.061276977 | 0 | 0 | 0.012602469 |
| 0 | 0 | 0 | 0 | 0 |
| 0 | 0 | 0 | 0 | 0 |
| 0 | 0.002593208 | 0 | 0.156036938 | 0.03243476 |
| 0 | 0.045463424 | 0.050511024 | 0.158781272 | 0 |
| 0 | 0 | 0 | 0 | 0.00136382 |
| 0 | 0 | 0 | 0 | 0 |
| 0 | 0 | 0 | 0 | 0 |
| 0 | 0.000775628 | 0 | 0.007110479 | 0.115860361 |
| 0 | 0.010121593 | 0 | 0.009278855 | 0.440556301 |
| 0 | 0 | 0.211097346 | 0 | 0 |
| 0 | 0.028268086 | 0 | 0 | 0 |
| 0 | 0 | 0 | 0 | 0 |
| 0 | 0 | 0 | 0 | 0 |

| | | | | |
|---|---|---|---|---|
| 0 | 0 | 0 | 0 | 0 |
| 0.415683715 | 0.090223949 | 0.725130739 | 0.08981875 | 0.001767859 |
| 0 | 0 | 0 | 0 | 0 |
| 0 | 0 | 0.000495115 | 0 | 0 |
| 0 | 0 | 0 | 0 | 0 |
| 0 | 0 | 0 | 0 | 0 |
| 0 | 0 | 0 | 0 | 0 |
| 0 | 0 | 0 | 0 | 0.000775944 |
| 0 | 0 | 0.001995633 | 0 | 0 |
| 0 | 0 | 0 | 0.003851972 | 0 |
| 0 | 0.002275722 | 0 | 0.001969529 | 0.001689059 |
| 0.003240232 | 0.026075014 | 0.008445609 | 0.007171192 | 0 |
| 0 | 0 | 0 | 0 | 0 |
| 0 | 0 | 0 | 0 | 0 |
| 0 | 0 | 0 | 0 | 0 |
| 0 | 0 | 0 | 0 | 0 |
| 0.018748838 | 0.00458271 | 0.029686483 | 0.078141333 | 0.000786084 |
| 0 | 0 | 0 | 0.044323456 | 0.12015744 |
| 0 | 0 | 0 | 0 | 0 |
| 0 | 0.002626815 | 0 | 0.02028215 | 0 |
| 0.013371718 | 0.021172165 | 0.759452147 | 0.003308411 | 0.012159761 |
| 0 | 0 | 0.003265245 | 0 | 0 |
| 0 | 0 | 0 | 0 | 0 |
| 0 | 0 | 0 | 0 | 0 |
| 0 | 0 | 0.006246001 | 0 | 0 |
| 0 | 0.001386819 | 0 | 0 | 0.001115084 |
| 0 | 0.086204188 | 0.157348388 | 0.007697407 | 0.033006301 |
| 0.00768225 | 0.077956426 | 0.037884031 | 0.003014016 | 0 |
| 0 | 0 | 0 | 0 | 0 |
| 0 | 0 | 0 | 0 | 0 |
| 0.008449137 | 0 | 0.00249384 | 0.007815601 | 0 |
| 0 | 0 | 0 | 0 | 0 |
| 0 | 0 | 0 | 0 | 0 |
| 0 | 0 | 0 | 0 | 0 |
| 0 | 0.004026992 | 0.017823129 | 0 | 0.104552546 |
| 0 | 0.076509627 | 0.624250464 | 0.263128762 | 0.299297608 |
| 0 | 0.000787437 | 0 | 0 | 0 |
| 0 | 0 | 0 | 0 | 0 |
| 0 | 0.008179235 | 0.006307682 | 0.008985471 | 0.000797162 |
| 0 | 0 | 0 | 0 | 0 |
| 0 | 0 | 0.215896537 | 0 | 0 |
| 0 | 0.001149807 | 0 | 0 | 0 |
| 0 | 0 | 0 | 0.008092766 | 0.002582444 |
| 0.003056745 | 0 | 0.012106236 | 0.002725609 | 0 |
| 0 | 0.005919822 | 0.006406095 | 0 | 0 |
| 0 | 0.05827026 | 0.306625538 | 0 | 0 |
| 0 | 0 | 0 | 0 | 0 |
| 0 | 0 | 0 | 0 | 0 |

| | | | | |
|---|---|---|---|---|
| 0 | 0 | 0 | 0.002629249 | 0 |
| 0 | 0 | 0.002582784 | 0 | 0 |
| 0 | 0.000551883 | 0 | 0 | 0 |
| 0 | 0.021216696 | 0.013387044 | 2.264534046 | 0.012594871 |
| 0 | 0 | 0 | 0 | 0 |
| 0 | 0.000456151 | 0 | 0 | 0 |
| 0 | 0 | 0 | 0 | 0.018071654 |
| 0 | 0.008941279 | 0.011613233 | 0 | 0 |
| 0.029141212 | 0.178749761 | 0.076882762 | 0 | 0 |
| 0 | 0.045760764 | 0.021754046 | 0 | 0 |
| 0 | 0 | 0 | 0 | 0 |
| 0 | 0.003023981 | 0.004081072 | 0 | 0.005043026 |
| 0 | 0 | 0 | 0 | 0 |
| 0 | 0 | 0 | 4.291049431 | 0.005866674 |
| 0.000795109 | 0.057255502 | 0.005933587 | 0 | 0.027049864 |
| 0 | 0 | 0 | 0 | 0 |
| 0 | 0 | 0.116097657 | 0 | 0 |
| 0 | 0 | 0 | 0 | 0 |
| 0 | 0 | 0 | 0 | 0 |
| 0 | 0 | 0 | 0 | 0 |
| 0 | 0 | 0 | 0 | 0 |
| 0 | 0.002339621 | 0 | 0 | 0 |
| 0 | 0 | 0 | 0 | 0 |
| 0 | 0.513992698 | 0 | 0.230707223 | 0.618757284 |
| 0 | 0 | 0 | 0.153959645 | 0.004384306 |
| 0 | 0.4438401 | 0.010125937 | 0 | 0 |
| 0 | 0 | 0 | 0 | 0.003812166 |
| 1.225846502 | 0.602987265 | 0 | 0 | 0.023942577 |
| 0 | 0 | 0.001461721 | 0 | 0 |
| 0 | 0 | 0 | 0 | 0 |
| 0 | 0.019925485 | 0.001322499 | 0.002654218 | 0 |
| 0 | 0 | 0.001780057 | 0 | 0 |
| 0 | 0.000497205 | 0 | 0.088882397 | 0.037313001 |
| 0 | 0 | 0 | 0 | 0 |
| 0 | 0 | 0 | 0 | 0 |
| 0 | 0.032127403 | 0.000961141 | 0 | 0.031919246 |
| 0 | 0.022892533 | 0.340121489 | 0 | 0 |
| 0 | 0 | 0 | 0 | 0 |
| 0 | 0 | 0 | 0 | 0.000937463 |
| 0 | 0.000961993 | 0 | 0 | 0.001675915 |
| 0 | 0.163098961 | 0.105333742 | 0.593133696 | 0.664172033 |
| 0 | 0.150192472 | 0.271375627 | 0 | 0 |
| 38.90880894 | 0.013920277 | 0.423140789 | 0 | 0.012229084 |
| 0 | 0 | 0 | 0 | 0 |
| 0 | 0 | 0 | 0 | 0.010094741 |
| 0 | 0 | 0 | 0 | 0 |
| 0 | 0 | 0 | 0 | 0 |
| 0 | 0 | 0 | 0.010414278 | 0.013708409 |

| | | | | |
|---|---|---|---|---|
| 0 | 0 | 0 | 0 | 0 |
| 0 | 0.082249713 | 0.009596099 | 0 | 0.00056767 |
| 0 | 0.002228646 | 0.006874771 | 0.083348621 | 0.798219703 |
| 0 | 0 | 0 | 0 | 0 |
| 0 | 0 | 0.035043194 | 0 | 0 |
| 0 | 0.001170428 | 0.005077199 | 0 | 0.002666429 |
| 0 | 0 | 0 | 0 | 0 |
| 0 | 0 | 0 | 0 | 0 |
| 0 | 0.03918563 | 0 | 0.774095465 | 0 |
| 0 | 0 | 0 | 0 | 0 |
| 0.003137427 | 0.001823446 | 0.006960737 | 0 | 0.014661578 |
| 0.04145507 | 1.440308558 | 0.765188109 | 0.277208744 | 0.499502322 |
| 0 | 0 | 0 | 0 | 0 |
| 0.395226053 | 0 | 0.003414687 | 0.009840459 | 0 |
| 0.000444721 | 0.007443888 | 0.120014331 | 0.039936494 | 0.210151361 |
| 0 | 0 | 0.003024396 | 0 | 0 |
| 0 | 0 | 0 | 0 | 0 |
| 0 | 0 | 0 | 0 | 0 |
| 0 | 0.069669951 | 0 | 0 | 0 |
| 0 | 0.02238039 | 0.005733815 | 0.008637406 | 0.005958125 |
| 0.001606994 | 0.24376645 | 0.213917837 | 0.050820507 | 0.087003013 |
| 4.864311117 | 0.009984302 | 0 | 0 | 3.802838499 |
| 0 | 0.059184803 | 0.203052485 | 0.066407676 | 1.706360792 |
| 0 | 0 | 0 | 0 | 0 |
| 0 | 0.004044329 | 0 | 0 | 0 |
| 0.00070024 | 0.002197658 | 14.80106794 | 0 | 0 |
| 0.003145155 | 0 | 0 | 0 | 0 |
| 0 | 0.001115804 | 0.000553171 | 0 | 0 |
| 0 | 0 | 0 | 0 | 0.018315066 |
| 0 | 0 | 0 | 0 | 0 |
| 0 | 0.012979838 | 0 | 0 | 0 |
| 0 | 0 | 0.422269203 | 0 | 0.222900483 |
| 0 | 0.000936363 | 0.002382953 | 0.08802506 | 0 |
| 0 | 0.015851899 | 0.00776434 | 0 | 0.002143448 |
| 0 | 0.007786944 | 0.001000859 | 0.027967211 | 0.014046201 |
| 0.008278894 | 0.014434868 | 0.003339575 | 0.337547677 | 0.015111184 |
| 0 | 0.002460686 | 0 | 0 | 0.001582829 |
| 0 | 0 | 0 | 0 | 0 |
| 0.017096431 | 0.08306747 | 0.002988453 | 0 | 0 |
| 0 | 0 | 0 | 0 | 0 |
| 0 | 0 | 0 | 0 | 0 |
| 0 | 0 | 0 | 0 | 0 |
| 0 | 0 | 0.002308385 | 0 | 0 |
| 0 | 0 | 0 | 0 | 0 |
| 0 | 0.014524701 | 0.000896096 | 0.184478192 | 0.020139293 |
| 0 | 0 | 0.436518219 | 0.028068326 | 0 |
| 0 | 0 | 0 | 0 | 0 |
| 0.325803094 | 0 | 0 | 0.027451344 | 0.385690464 |

| | | | | |
|---|---|---|---|---|
| 0 | 0 | 0 | 0 | 0 |
| 0 | 0 | 0 | 0 | 0 |
| 0 | 0 | 0 | 0 | 0 |
| 0 | 0 | 0.003436824 | 0.005364803 | 0.003008235 |
| 0 | 0 | 0 | 0 | 0 |
| 0 | 0.69375803 | 0 | 0.079090834 | 0 |
| 0 | 1.349700529 | 0 | 0 | 0 |
| 0 | 0.005467794 | 0.010014583 | 2.653797113 | 0.023936106 |
| 0 | 0 | 0 | 0 | 0 |
| 0 | 0 | 0 | 0 | 0 |
| 0 | 0.00459577 | 0 | 0.066643904 | 0.00945989 |
| 0 | 0 | 0 | 0 | 0 |
| 0 | 0 | 0 | 0 | 0 |
| 0 | 0 | 0 | 0 | 0 |
| 0 | 0 | 0 | 0 | 0 |
| 0 | 0 | 0 | 0 | 0 |
| 0 | 0.102131131 | 0.083190104 | 0 | 0 |
| 0 | 0.002142489 | 0.001770268 | 0 | 0.004265702 |
| 0 | 0 | 0.162621701 | 0.582785249 | 0 |
| 0 | 0.006210549 | 0.000957893 | 0 | 0 |
| 0 | 0 | 0 | 0 | 0.000728796 |
| 0 | 0 | 0 | 0 | 0 |
| 0 | 0.012703562 | 0 | 0.536635293 | 0.003593959 |
| 0 | 0 | 0 | 0 | 0 |
| 0 | 0 | 0 | 0 | 0 |
| 0 | 0 | 0 | 0 | 0 |
| 0 | 0 | 0 | 0 | 0 |
| 0 | 0 | 0 | 0 | 0 |
| 0 | 0 | 0 | 0 | 0 |
| 0 | 1.531420559 | 0.017749285 | 0.357972703 | 0.022593617 |
| 0 | 0 | 0 | 0 | 0 |
| 0 | 0 | 0 | 0 | 0 |
| 0 | 0 | 0 | 0.016790343 | 0 |
| 0 | 0 | 0 | 0 | 0 |
| 0 | 0 | 0 | 0 | 0 |
| 0 | 0 | 0 | 0 | 0 |
| 0 | 0.024485818 | 0.14938915 | 0 | 0 |
| 0 | 0 | 0.0028802 | 0.001750818 | 0.004671311 |
| 0 | 0 | 0 | 0 | 0 |
| 0 | 0 | 0 | 0 | 0.001810098 |
| 0 | 0 | 0 | 0 | 0.02833197 |
| 0 | 0 | 0 | 0 | 0 |
| 0 | 0 | 0 | 0 | 0 |
| 0 | 1.346946622 | 0.467975625 | 0 | 0 |
| 0 | 0.146307901 | 0.197452644 | 0 | 0 |
| 0 | 0 | 0 | 0 | 0 |
| 0 | 0.002957225 | 0.000684168 | 0 | 0.001902226 |
| 0 | 0 | 0 | 0 | 0 |

| | | | | |
|---|---|---|---|---|
| 0 | 0 | 0 | 0 | 0 |
| 0 | 0 | 0 | 0 | 0 |
| 0 | 0 | 0 | 0 | 0 |
| 0 | 0.008513577 | 0.022024339 | 0.460156279 | 0.019747989 |
| 0 | 0 | 0 | 0 | 0 |
| 0 | 0 | 1.577577205 | 0 | 0 |
| 0 | 0 | 0 | 0 | 0 |
| 0 | 0 | 0 | 0 | 0 |
| 0 | 0.010945644 | 0.00273094 | 0.023341808 | 0.066956175 |
| 0 | 0 | 0.001199571 | 0 | 0.000741162 |
| 0 | 0 | 0 | 0 | 0 |
| 0 | 0 | 0 | 0 | 0 |
| 0 | 0.004256988 | 0 | 0 | 0.004698055 |
| 0 | 0 | 0 | 0.00857211 | 0 |
| 0 | 0.750492559 | 0.152541771 | 0.420507806 | 0 |
| 0 | 0.002481209 | 0 | 0 | 1.211741824 |
| 0 | 0 | 0 | 0 | 0 |
| 0 | 0 | 0 | 0 | 0 |
| 0 | 0 | 0 | 0 | 0 |
| 0 | 0 | 0 | 0 | 0 |
| 0 | 0.008205406 | 0.012908844 | 0 | 0 |
| 0 | 0 | 0 | 0 | 0 |
| 0 | 0 | 0 | 0 | 0 |
| 0 | 0 | 0 | 0 | 0 |
| 0.417612272 | 0.089345193 | 0.008758658 | 0.037545596 | 0.005411585 |
| 0 | 1.337869657 | 0.225647605 | 6.670543758 | 0 |
| 0 | 0 | 0.012379268 | 1.017665598 | 0 |
| 0 | 0.005170067 | 0.005799367 | 0 | 0 |
| 0 | 0.00794372 | 0.02348322 | 0 | 0 |
| 0 | 0.051783043 | 0 | 0 | 0 |
| 0 | 0 | 0 | 0 | 0 |
| 0 | 0.262988066 | 0 | 0 | 0 |
| 0 | 0 | 0 | 0 | 0 |
| 0 | 0.000539165 | 0.001403306 | 0.186531906 | 0 |
| 2.401926895 | 0 | 0.00154147 | 0.001943468 | 0 |
| 0 | 0.00156993 | 0 | 0.003925133 | 0 |
| 0 | 0 | 0 | 0 | 0 |
| 0 | 0.320330927 | 0.533781194 | 0 | 0 |
| 0 | 0 | 0 | 0 | 0 |
| 0 | 0 | 0 | 0 | 0 |
| 0 | 0 | 0 | 0 | 0 |
| 0 | 0.001958345 | 0 | 0 | 0.027293485 |
| 0 | 0.423060522 | 0.126565153 | 5.47452616 | 0 |
| 0 | 2.036311555 | 0 | 0 | 0 |
| 0 | 0 | 0 | 0 | 0 |
| 0 | 0.00133858 | 0 | 0.001487428 | 0 |
| 0 | 0 | 0 | 0 | 0 |
| 0.00135897 | 0.021562138 | 4.615735297 | 0.042653731 | 0.014479441 |

| | | | | |
|---|---|---|---|---|
| 0 | 0 | 0 | 0 | 0.04944043 |
| 0 | 0 | 0 | 0 | 0 |
| 0 | 0 | 0 | 0 | 0 |
| 0 | 0 | 0 | 0 | 0 |
| 0 | 0 | 0 | 0 | 0.000565789 |
| 0 | 0.034050432 | 0.02136001 | 0 | 0 |
| 0 | 0.249000637 | 0 | 0 | 0 |
| 0 | 0 | 0 | 0 | 0 |
| 0 | 0 | 0 | 0 | 0 |
| 0 | 0.001412554 | 0 | 0 | 0 |
| 0 | 0.005922714 | 0 | 0 | 0 |
| 0 | 0.01898844 | 1.976316198 | 4.017100753 | 4.463075659 |
| 0 | 0.009585191 | 0.286806924 | 0 | 0 |
| 0 | 0 | 0 | 0 | 0 |
| 0 | 0.053099831 | 0.029888542 | 0.631500153 | 0.000872447 |
| 0 | 0 | 0 | 0 | 0 |
| 0 | 0 | 0 | 0.038923039 | 0 |
| 0 | 0 | 0 | 0 | 0 |
| 0 | 0 | 0 | 0.084932101 | 0 |
| 0 | 0.006129619 | 0 | 0 | 0 |
| 0 | 0.037498918 | 0.165313667 | 0.186586528 | 0.008230285 |
| 0 | 0 | 0 | 0 | 0 |
| 0 | 0.076292946 | 0.171492924 | 9.76127834 | 0 |
| 0 | 0 | 0 | 0 | 0 |
| 0 | 0 | 0 | 0 | 0 |
| 0 | 0 | 0 | 0 | 0 |
| 0 | 0 | 0 | 0 | 0 |
| 0 | 0 | 0 | 0 | 0 |
| 0 | 0.001466039 | 0 | 0 | 0 |
| 0 | 0 | 0 | 0 | 0 |
| 0 | 0 | 0 | 0 | 0 |
| 0.020188546 | 0.014520096 | 0.043365417 | 0.030692613 | 0 |
| 0 | 0 | 0.022349437 | 0 | 0 |
| 0 | 0.156554288 | 0.01601961 | 0 | 0.023471933 |
| 0 | 0.000854239 | 0 | 0.002776494 | 0 |
| 0.033684586 | 0.029072133 | 0.003821577 | 0.019272814 | 0.009444729 |
| 0 | 0 | 0 | 0 | 0 |
| 0 | 0 | 0.00264575 | 0 | 0 |
| 0 | 0.000588836 | 0.000908201 | 0 | 0 |
| 0 | 0 | 0 | 0 | 0 |
| 0 | 0 | 0.069373924 | 0 | 0 |
| 0 | 1.015584915 | 0 | 0 | 0 |
| 0 | 0 | 0 | 0 | 0 |
| 0 | 0 | 0 | 0 | 0 |
| 0 | 0.007741898 | 0.002725625 | 0.045024356 | 0.011547717 |
| 0 | 0 | 0 | 0 | 0 |
| 0 | 0 | 0.392478321 | 0 | 0 |
| 0 | 0.006048981 | 0.000920698 | 0.002387943 | 0.004778407 |

| | | | | |
|---|---|---|---|---|
| 0 | 0 | 0 | 0 | 0 |
| 0 | 0 | 0 | 0 | 0 |
| 0 | 0 | 0.010067778 | 0.004352005 | 0 |
| 0 | 0 | 0 | 0 | 0 |
| 0 | 0 | 0.003471462 | 0 | 0 |
| 0 | 0 | 0 | 0 | 0 |
| 0 | 0.257330636 | 0.531733609 | 0 | 0 |
| 0 | 0.028110615 | 0.001711454 | 0.447585717 | 0 |
| 0 | 0 | 0 | 0 | 0 |
| 0 | 0 | 0.004116776 | 0 | 0.007283867 |
| 0 | 0 | 0 | 0 | 0 |
| 0 | 0 | 0 | 0 | 0 |
| 0 | 0 | 0 | 0 | 0 |
| 0 | 0 | 0 | 0 | 0 |
| 0 | 0 | 0 | 0 | 0 |
| 0 | 0 | 0 | 0 | 0 |
| 0 | 16.51426379 | 14.70741262 | 0 | 0 |
| 0 | 0 | 0 | 0 | 0 |
| 0 | 3.181829943 | 0 | 0 | 0 |
| 0 | 0 | 0 | 0 | 0 |
| 0 | 0 | 0 | 0 | 0 |
| 0 | 0 | 0 | 0 | 0 |
| 0 | 0 | 0 | 0 | 0 |
| 0 | 0 | 0 | 0 | 0 |
| 0.023553673 | 0 | 0 | 0.001379244 | 0 |
| 0 | 0 | 0 | 0.048771389 | 0.00184774 |
| 0 | 0 | 0 | 0 | 0 |
| 0 | 0.091403406 | 0.007805796 | 0.001533735 | 0.028443874 |
| 0 | 0 | 1.615861803 | 0 | 0 |
| 0 | 0.374388951 | 0.6620907 | 0 | 1.123786795 |
| 0 | 0 | 0 | 0 | 0 |
| 0 | 0 | 0 | 0 | 0 |
| 0 | 0 | 0.000936619 | 0 | 0 |
| 0 | 0 | 0 | 0 | 0 |
| 0 | 0.000656776 | 0 | 0.011494483 | 0.153497154 |
| 0 | 0 | 0 | 0 | 0 |
| 0 | 0 | 0 | 0 | 0 |
| 0 | 0 | 0 | 0.20426069 | 0.000798965 |
| 0 | 0 | 0 | 0 | 0 |
| 0 | 0 | 0 | 0 | 0 |
| 0 | 2.343327682 | 9.782787959 | 4.020349256 | 9.230679895 |
| 0 | 0.012410699 | 0.410813043 | 6.341205937 | 0.621691087 |
| 0 | 0.033455359 | 0 | 0 | 0 |
| 0 | 0 | 0 | 0 | 0 |
| 0 | 0 | 0 | 0 | 0 |
| 0 | 0 | 0 | 0 | 0 |
| 0 | 0 | 0 | 0 | 0 |
| 0 | 0.001182941 | 0 | 0.038633444 | 0 |

| | | | | |
|---|---|---|---|---|
| 0 | 0 | 0 | 0 | 0 |
| 0 | 0.012471711 | 0.053133388 | 0.019393512 | 0.004402531 |
| 0 | 0.033751078 | 0 | 0.000666192 | 0 |
| 0 | 0 | 0 | 0 | 0 |
| 0 | 1.255083572 | 1.655983637 | 5.055748425 | 0 |
| 0 | 0 | 0 | 0 | 0 |
| 0 | 0 | 0 | 0 | 0 |
| 0 | 0 | 0 | 0 | 0 |
| 0 | 0.001063089 | 0.000702716 | 0.085813173 | 1.966821163 |
| 0 | 0 | 0 | 0 | 0 |
| 0 | 0 | 0 | 0 | 0 |
| 0 | 0 | 0 | 0 | 0 |
| 0 | 0.060444178 | 0.076929351 | 0 | 0 |
| 0 | 0.006359044 | 0.052752844 | 0.012264833 | 0.032723487 |
| 2.47044855 | 0 | 0.016167289 | 0 | 0 |
| 0 | 0.046814578 | 0.146767333 | 0.386735172 | 0 |
| 0 | 0.001886663 | 0.085478965 | 0 | 0.028148549 |
| 0 | 0 | 0 | 0 | 0 |
| 0 | 0 | 0 | 0 | 0 |
| 0 | 0 | 0 | 0 | 0 |
| 0 | 0 | 0 | 0 | 0 |
| 0 | 0 | 0 | 0 | 0 |
| 0 | 0 | 0 | 0 | 0 |
| 0 | 0 | 0 | 0 | 0 |
| 0 | 0 | 0 | 0 | 0.079515645 |
| 0 | 0 | 0 | 0 | 0 |
| 0 | 0.00300764 | 0.01154448 | 0 | 0 |
| 0 | 0 | 0 | 0.015813194 | 0.000992292 |
| 0 | 0 | 0 | 0 | 0 |
| 0 | 0 | 0 | 0 | 0 |
| 0 | 0 | 0 | 0 | 0 |
| 0 | 0 | 0 | 0 | 0 |
| 0 | 0 | 0 | 0 | 0 |
| 0 | 0 | 0.010475384 | 0 | 0 |
| 0.000731288 | 0.004972717 | 0.036283791 | 0 | 0 |
| 0 | 0 | 0 | 0 | 0.054524095 |
| 0 | 0 | 0 | 0.078209117 | 0 |
| 0 | 0 | 0.003313727 | 0.014745572 | 0 |
| 0 | 0 | 0 | 0 | 0 |
| 0 | 0 | 0 | 0 | 0 |
| 0 | 0.813901068 | 1.757853015 | 0.135241897 | 0.455496171 |
| 0 | 0 | 0 | 0 | 0 |
| 0 | 0 | 0.006801972 | 0 | 0 |
| 0 | 0.001132827 | 0 | 0 | 0 |
| 0 | 0 | 0 | 0 | 0 |
| 0 | 0 | 0 | 0 | 0 |
| 0 | 0 | 0 | 0 | 0 |
| 0 | 0 | 0 | 0 | 0 |

| | | | | |
|---|---|---|---|---|
| 0 | 0.001520131 | 0.031065913 | 0 | 0 |
| 0 | 0 | 0 | 0 | 0 |
| 0 | 0 | 0 | 0 | 0 |
| 0 | 0 | 0 | 0 | 0 |
| 0 | 0.028424212 | 0.012958738 | 0 | 0 |
| 0 | 0 | 0.002581846 | 0 | 0 |
| 0.001649137 | 0 | 0.365032947 | 0.532316158 | 0.106099073 |
| 0 | 0 | 0 | 0 | 0 |
| 0 | 0 | 0 | 0 | 0 |
| 0 | 0 | 0 | 0.004319783 | 0 |
| 0 | 0 | 0.017600883 | 0.010699235 | 0 |
| 0 | 0.037993095 | 0 | 0 | 0 |
| 0 | 0 | 0 | 0 | 0 |
| 0 | 0 | 0 | 0 | 0 |
| 0 | 0 | 0 | 0 | 0 |
| 0 | 0 | 0 | 0 | 0 |
| 0 | 0 | 0 | 0 | 0 |
| 0 | 0 | 0 | 0.682543322 | 0 |
| 0 | 0 | 0 | 0 | 0 |
| 0 | 0 | 0 | 0 | 0 |
| 0.002854673 | 0.376585123 | 1.578574699 | 0.177891455 | 0.092687926 |
| 0 | 0 | 0.029159092 | 0 | 0 |
| 0 | 0.013340657 | 0.00533456 | 0 | 0.003531415 |
| 0 | 0 | 0.000847062 | 0.003478519 | 0.00533829 |
| 0 | 0 | 0 | 0 | 0 |
| 0 | 0 | 0 | 0.036286858 | 0 |
| 0 | 0 | 0 | 0 | 0 |
| 0 | 0 | 0 | 0 | 0 |
| 0 | 0 | 0.003397979 | 0 | 0 |
| 0.011887445 | 0.016230017 | 0.002682065 | 0.00105398 | 0.127674226 |
| 0 | 0 | 0 | 0 | 0 |
| 0 | 0 | 0 | 0 | 0 |
| 0 | 0 | 0 | 0 | 0 |
| 0 | 0.025024387 | 0 | 0 | 0 |
| 0 | 0 | 0 | 0 | 0 |
| 0 | 0 | 0 | 0 | 0 |
| 0 | 0.004031499 | 0.00339166 | 0.078116435 | 0 |
| 0 | 0 | 0 | 0 | 0 |
| 0 | 0.007092211 | 0 | 0 | 0 |
| 0 | 0.03443221 | 0 | 0 | 0 |
| 0 | 0 | 0 | 0 | 0 |
| 0 | 0 | 0 | 0 | 0 |
| 0 | 0 | 0 | 0 | 0 |
| 0 | 0 | 0.002629627 | 0 | 0 |
| 0 | 0 | 0 | 0 | 0 |
| 0 | 0 | 0 | 0 | 0 |
| 0 | 0 | 0 | 0 | 0 |
| 0 | 0.001222659 | 0 | 0 | 0 |

| | | | | |
|---|---|---|---|---|
| 0 | 0.000630352 | 0 | 0.025610071 | 0.121641659 |
| 0 | 0.027499875 | 0 | 0 | 0 |
| 0 | 0 | 0.051776471 | 0.115184853 | 0 |
| 0 | 0 | 0 | 0 | 0 |
| 0 | 0 | 0 | 0 | 0 |
| 0 | 0 | 0 | 0 | 0 |
| 0 | 0 | 0 | 0 | 0 |
| 0 | 0.119737419 | 0.738216016 | 0.475144056 | 0.625880221 |
| 0.863505718 | 0.889499844 | 0.034336484 | 1.596512886 | 0.318224757 |
| 0 | 0 | 0.001195202 | 0.008395571 | 0.022153853 |
| 0 | 0 | 0 | 0 | 0 |
| 0 | 0 | 0 | 0 | 0 |
| 0 | 0 | 0 | 0 | 0 |
| 0 | 0 | 0 | 0.003204914 | 0.037497655 |
| 0 | 0 | 0 | 0 | 0 |
| 0 | 0 | 0.067904293 | 0 | 0 |
| 0 | 0 | 0 | 0.002300542 | 0 |
| 0 | 0 | 0 | 0 | 0 |
| 0 | 0 | 0 | 0 | 0 |
| 0 | 0 | 0 | 0 | 0.007042028 |
| 0 | 0 | 0 | 0 | 0 |
| 0 | 0 | 0 | 0 | 0 |
| 0 | 0 | 0 | 0 | 0 |
| 0 | 0 | 0 | 0 | 0 |
| 0 | 0.081300202 | 0.005599681 | 0.028395239 | 0 |
| 0 | 0 | 0 | 0 | 0 |
| 0 | 0 | 0 | 0 | 0 |
| 0 | 0 | 0 | 0 | 0 |
| 0 | 0 | 0 | 0 | 0 |
| 0 | 0 | 0 | 0 | 0 |
| 0 | 0 | 0 | 0 | 0.015967862 |
| 0 | 0 | 0 | 0 | 0 |
| 0 | 0 | 0 | 0 | 0 |
| 0 | 0 | 0 | 0 | 0 |
| 0 | 0 | 0 | 0 | 0.00695415 |
| 0 | 0 | 0 | 0 | 0 |
| 0 | 0 | 0.032070934 | 0 | 0 |
| 0 | 0 | 0 | 0 | 0 |
| 0 | 0 | 0.001083807 | 0 | 0 |
| 0 | 0 | 0.000727945 | 0.01337344 | 0 |
| 0 | 0 | 0 | 0 | 0 |
| 0 | 0.557358147 | 1.199166154 | 1.579765308 | 1.523583779 |
| 0 | 0 | 0 | 0 | 0 |
| 0 | 0 | 0 | 0 | 0 |
| 0 | 0 | 0 | 0 | 0 |
| 0 | 0.001821662 | 0 | 0 | 0.000450684 |
| 0 | 0 | 0 | 0 | 0 |
| 0 | 0 | 0 | 0 | 0 |

| | | | | |
|---|---|---|---|---|
| 0 | 0 | 0 | 0 | 0 |
| 0 | 0.169111611 | 0 | 0 | 0 |
| 0 | 0 | 0 | 0 | 0 |
| 0 | 0 | 0 | 0 | 0 |
| 1.337446762 | 0.104599151 | 0.061188871 | 0.485002298 | 0.467954325 |
| 0 | 0 | 1.128913817 | 0 | 0 |
| 0 | 0 | 0 | 0 | 0 |
| 0 | 0.003407878 | 0.006261052 | 0.004510781 | 0.007951762 |
| 0 | 0.000800621 | 0.007640634 | 0 | 0 |
| 0 | 0 | 0 | 0 | 0 |
| 0 | 0 | 0 | 0 | 0 |
| 0 | 0 | 0 | 0 | 0 |
| 0 | 0 | 0 | 0 | 0 |
| 0 | 0 | 0 | 0 | 0 |
| 0 | 0.003104622 | 0 | 0.019934649 | 0 |
| 0 | 0.029903994 | 0 | 0.133883044 | 0.006150936 |
| 0 | 0 | 0 | 0 | 0 |
| 0 | 0 | 0 | 0 | 0 |
| 0 | 0 | 0 | 0 | 0 |
| 0 | 0 | 0 | 0 | 0 |
| 0 | 0 | 0 | 0 | 0 |
| 0 | 0 | 0 | 0 | 0 |
| 0 | 0 | 0 | 0 | 0 |
| 0 | 0.003122723 | 0 | 0.033181532 | 0.001115934 |
| 0 | 0.208684789 | 0 | 0 | 0.105906009 |
| 0 | 0 | 0 | 0 | 0 |
| 0.000910035 | 0.042999934 | 0 | 2.663110919 | 0.000680432 |
| 0 | 0.000835379 | 0 | 0 | 0 |
| 0 | 0 | 0 | 0 | 0 |
| 0 | 0 | 0 | 0 | 0 |
| 0 | 0 | 0 | 0 | 0 |
| 0 | 0 | 0 | 0 | 0 |
| 0 | 0 | 0 | 0 | 0 |
| 0 | 0 | 0 | 0 | 0 |
| 0 | 0 | 0 | 0 | 0 |
| 0 | 0 | 0 | 0 | 0 |
| 0 | 0 | 0 | 0 | 0 |
| 0 | 0.393636039 | 0.003576523 | 0 | 0.047626444 |
| 0 | 0 | 0 | 0 | 0 |
| 0 | 0 | 0 | 0 | 0 |
| 0 | 0 | 0 | 0 | 0 |
| 0 | 0.005784264 | 0 | 0 | 0 |
| 0 | 0 | 0 | 0 | 0 |
| 0 | 0 | 0 | 0 | 0 |
| 0 | 0 | 0 | 0 | 0 |
| 0 | 0 | 0 | 0 | 0 |
| 0 | 0 | 0 | 0 | 0 |
| 0 | 0 | 0.127439599 | 0 | 0 |

| | | | | |
|---|---|---|---|---|
| 0 | 0.001492167 | 0.012082698 | 0 | 0 |
| 0 | 0 | 0 | 0 | 0 |
| 0 | 0 | 0 | 0 | 0 |
| 0 | 0 | 0 | 0.002598296 | 0 |
| 0 | 0 | 0 | 0 | 0 |
| 0 | 0 | 0 | 0 | 0.032619284 |
| 0 | 0 | 0 | 0 | 0 |
| 0 | 0 | 0.001359662 | 0 | 0 |
| 0 | 0 | 0 | 0 | 0 |
| 0 | 0 | 0 | 0 | 0 |
| 0 | 0 | 0 | 0 | 0 |
| 0 | 0 | 0 | 0.037985197 | 0.000857261 |
| 0 | 0 | 0 | 0 | 0 |
| 0 | 0 | 0 | 0 | 0 |
| 0 | 0 | 0 | 0 | 0 |
| 0 | 0 | 0 | 0 | 0 |
| 0 | 0 | 0 | 0 | 0 |
| 0 | 0 | 0.01858683 | 0 | 0 |
| 0 | 0.002024146 | 0 | 0 | 0.001220649 |
| 0 | 0 | 0 | 0 | 0 |
| 0 | 0 | 0 | 0.107551152 | 0 |
| 0 | 0 | 0 | 0 | 0 |
| 0.000834235 | 0 | 0.001009551 | 0.884526769 | 0 |
| 0 | 0.850488219 | 0 | 0 | 0 |
| 0 | 0 | 0 | 0 | 0 |
| 0 | 0 | 0 | 0 | 0 |
| 0 | 0 | 0 | 0 | 0 |
| 0 | 0 | 0 | 0 | 0 |
| 0 | 0 | 0 | 0 | 0 |
| 0 | 0 | 0 | 0 | 0 |
| 0 | 0 | 0 | 0 | 0 |
| 0 | 0 | 0.002537757 | 0 | 0 |
| 0 | 0 | 0 | 0 | 0.086964664 |
| 0 | 0 | 0 | 0 | 0 |
| 0 | 0.017363107 | 0.349059359 | 0 | 0 |
| 0 | 0 | 0 | 0 | 0 |
| 0 | 0 | 0 | 0 | 0 |
| 0 | 0.001219117 | 0 | 0 | 0 |
| 0 | 0 | 0 | 0.078236828 | 0.006302914 |
| 0 | 0 | 0 | 0 | 0 |
| 0 | 0 | 0 | 0 | 0 |
| 0 | 0 | 0 | 0 | 0 |
| 0.0250132 | 0.013748987 | 0.007182662 | 0.004989943 | 0 |
| 0 | 0 | 0 | 0 | 0 |
| 0 | 0 | 0 | 0 | 0 |
| 0 | 0.108838571 | 0.010389937 | 0.027740163 | 0.000755233 |
| 0 | 0 | 0 | 0 | 0 |
| 0 | 0 | 0 | 0 | 0 |

| | | | | |
|---|---|---|---|---|
| 0 | 0 | 0 | 0 | 0 |
| 0 | 0 | 0 | 0 | 0 |
| 0 | 0 | 0 | 0 | 0 |
| 0 | 0 | 0 | 0 | 0 |
| 0 | 0 | 0 | 0 | 0 |
| 0 | 0 | 0 | 0 | 0 |
| 0 | 0 | 0 | 0 | 0 |
| 0 | 0 | 0 | 0 | 0 |
| 0 | 0.734591889 | 0.006107864 | 0.014645137 | 0.006545144 |
| 0 | 0 | 0.001259002 | 0 | 0.013029515 |
| 0 | 0 | 0 | 0 | 0 |
| 0 | 0 | 0 | 0 | 0 |
| 0 | 0 | 0 | 0 | 0 |
| 0 | 0 | 0.285694136 | 0.685163609 | 0 |
| 0 | 0.005130736 | 0 | 0.014110632 | 0 |
| 0 | 0.00107084 | 0 | 0.00513151 | 0 |
| 0 | 0 | 0 | 0 | 0 |
| 0 | 0 | 0 | 0 | 0 |
| 0 | 0 | 0 | 0 | 0 |
| 0 | 0.031080085 | 3.205833141 | 0 | 0 |
| 0 | 0 | 0.106303478 | 0 | 0 |
| 0 | 0 | 0 | 0 | 0 |
| 0 | 0 | 0 | 0 | 0 |
| 0 | 0 | 0 | 0 | 0.006116767 |
| 0 | 0 | 0 | 0.077193716 | 0 |
| 0 | 0 | 0 | 0 | 0 |
| 0 | 0 | 0 | 0 | 0 |
| 0 | 0.046783104 | 0 | 0 | 0 |
| 0 | 0 | 0 | 0 | 0 |
| 0 | 0 | 4.07404394 | 0 | 0 |
| 0 | 1.165396618 | 0.215789981 | 0 | 0.216630482 |
| 0 | 0 | 0 | 0 | 0 |
| 0 | 0 | 0 | 0 | 0 |
| 0 | 0 | 0 | 0 | 0 |
| 0 | 0 | 0 | 0 | 0 |
| 0 | 0 | 0 | 0 | 0 |
| 0 | 0 | 0 | 0 | 0 |
| 0 | 0 | 0 | 0 | 0 |
| 0 | 0.008337793 | 0 | 0 | 0 |
| 0 | 0 | 0 | 0 | 0 |
| 0 | 0 | 0 | 0 | 0 |
| 0 | 0 | 0 | 0 | 0 |
| 0 | 0.044086954 | 0 | 0 | 0 |
| 0 | 0 | 0 | 0 | 0 |
| 0 | 0 | 0 | 0 | 0 |
| 0 | 0 | 0 | 0 | 0 |
| 0 | 0 | 0 | 0.009135808 | 0 |
| 0 | 0 | 0 | 0 | 0.000636736 |

| | | | | |
|---|---|---|---|---|
| 0 | 0 | 0 | 0 | 0 |
| 0 | 0 | 0 | 0 | 0 |
| 0 | 0 | 0 | 0 | 0 |
| 0 | 0.001473679 | 0 | 0 | 0 |
| 0 | 0.26356585 | 0.019104717 | 1.349840205 | 0 |
| 0 | 0 | 0 | 0 | 0 |
| 0 | 0 | 0 | 0 | 0 |
| 0 | 0 | 0 | 0 | 0 |
| 0 | 0 | 0 | 0 | 0 |
| 0 | 0 | 0 | 0 | 0 |
| 0 | 0 | 0.021650298 | 0 | 0 |
| 0 | 0.002652988 | 0 | 0.128395745 | 0.002031579 |
| 0 | 0 | 0 | 0.001021893 | 0 |
| 0 | 0 | 0 | 0 | 0 |
| 0 | 0.033503485 | 0 | 0 | 0 |
| 0 | 0.006483947 | 2.845572037 | 1.195571668 | 0 |
| 0 | 0.006267135 | 0 | 0.015669068 | 0.010937679 |
| 0 | 0 | 0 | 0 | 0 |
| 0 | 0.013631534 | 0.002821749 | 0.184578286 | 0.663478884 |
| 0 | 0.036159322 | 0.028607854 | 0 | 0.03615447 |
| 0 | 0 | 0 | 0 | 0 |
| 0 | 0 | 0.000570064 | 0 | 0 |
| 0 | 0 | 0.001054574 | 0 | 0 |
| 0 | 0 | 0 | 0 | 0 |
| 0 | 0 | 0 | 0 | 0 |
| 0 | 0 | 0 | 0 | 0 |
| 0 | 0.001201083 | 0 | 0.006005885 | 0.001770525 |
| 0 | 0 | 0 | 0 | 0 |
| 0 | 0 | 0 | 0 | 0 |
| 0 | 0 | 0 | 0 | 0 |
| 0 | 0.000850931 | 0 | 0.010850228 | 0.001642077 |
| 0 | 0 | 0 | 0.005974652 | 0 |
| 0 | 0 | 0 | 0 | 0 |
| 0 | 0 | 0 | 0 | 0 |
| 0 | 0 | 0 | 0 | 0 |
| 0 | 0 | 0 | 0 | 0 |
| 0 | 0 | 0 | 0 | 0 |
| 0 | 0.002869631 | 0.002627948 | 0.414036544 | 0.012818621 |
| 0 | 0.000862894 | 0.166029538 | 0 | 0 |
| 0 | 0 | 0 | 0.001185504 | 0 |
| 0 | 0 | 0 | 0.04909368 | 0.001535654 |
| 0 | 0 | 0 | 0 | 0 |
| 0 | 0 | 0 | 0 | 0 |
| 0 | 0 | 0 | 0.005097893 | 0.022810068 |
| 0 | 0 | 0 | 0 | 0 |
| 0 | 0 | 0 | 0 | 0 |
| 0 | 0 | 0 | 0 | 0 |
| 0 | 0 | 0 | 0 | 0 |

| | | | | |
|---|---|---|---|---|
| 0 | 0 | 0 | 0 | 0 |
| 0 | 0 | 0 | 0 | 0 |
| 0 | 0 | 0 | 0 | 0 |
| 0 | 0.001114893 | 0.000644839 | 0.010220656 | 0.00159367 |
| 0.009554867 | 0 | 0 | 0 | 0 |
| 0 | 0 | 0 | 0.00194635 | 0 |
| 0 | 0 | 0 | 0 | 0 |
| 0 | 0 | 0 | 0 | 0.045337368 |
| 0 | 0 | 0 | 0 | 0 |
| 0 | 0 | 0 | 0 | 0 |
| 0 | 0 | 0 | 0.011900459 | 0 |
| 0 | 0 | 0 | 0 | 0 |
| 0 | 0.008956759 | 0.004723371 | 0.201872214 | 0.01863985 |
| 0 | 0 | 0 | 0 | 0 |
| 0 | 0 | 0 | 0 | 0 |
| 0 | 0 | 0 | 0 | 0 |
| 0 | 0 | 0 | 0 | 0 |
| 0 | 0 | 0 | 0 | 0 |
| 0 | 0 | 0 | 0 | 0 |
| 0 | 0 | 0 | 0 | 0 |
| 0 | 0 | 0 | 0 | 0 |
| 0 | 0 | 0 | 0 | 0 |
| 0 | 0.002601615 | 0 | 0.023850009 | 0.011714367 |
| 0 | 0.002660662 | 0.013731652 | 0 | 0 |
| 0 | 0 | 0 | 0 | 0 |
| 0 | 0 | 0 | 0 | 0.00064817 |
| 0 | 0 | 0 | 0 | 0.036773923 |
| 0 | 0 | 0 | 0 | 0 |
| 0 | 0 | 0 | 0 | 0 |
| 0 | 0 | 0 | 0 | 0 |
| 0 | 0.018758436 | 0.020804449 | 0 | 0.087698421 |
| 0 | 0 | 0.001383932 | 0 | 0 |
| 0 | 0 | 0 | 0 | 0 |
| 0 | 0 | 0 | 0 | 0 |
| 0 | 0.088361078 | 0.006751735 | 0.05715546 | 0 |
| 0 | 0 | 0 | 0 | 0 |
| 0 | 0 | 0 | 0 | 0.01674033 |
| 0 | 0 | 0 | 0 | 0 |
| 0 | 0 | 0 | 0.003681692 | 0 |
| 0 | 0 | 0 | 0 | 0 |
| 0 | 0 | 0 | 0 | 0 |
| 0 | 0 | 0 | 0 | 0 |
| 0 | 0 | 1.572558557 | 0 | 0 |
| 0 | 0 | 0 | 0 | 0 |
| 0 | 0 | 0 | 0 | 0 |
| 0 | 0.0149715 | 0.008159742 | 0 | 0.344625359 |
| 0 | 0 | 0 | 0 | 0 |
| 0 | 0 | 0 | 0 | 0 |

| | | | | |
|---|---|---|---|---|
| 0 | 0 | 0 | 0 | 0 |
| 0 | 0 | 0 | 0 | 0 |
| 0 | 0.050203993 | 0 | 0 | 0 |
| 0 | 0 | 0 | 0 | 0 |
| 0 | 0 | 0 | 0.00113878 | 0 |
| 0 | 0 | 0 | 0.026011847 | 0 |
| 0 | 0 | 0 | 0.007900574 | 0 |
| 0 | 0 | 0 | 0 | 0 |
| 0 | 0 | 0 | 0 | 0 |
| 0 | 0 | 0 | 0 | 0 |
| 0 | 0 | 0 | 0.009458322 | 0 |
| 0 | 0 | 0 | 0.002477422 | 0 |
| 0 | 0 | 0 | 0 | 0 |
| 0 | 0 | 0 | 0 | 0 |
| 0 | 0 | 0 | 0 | 0 |
| 0 | 0 | 0 | 0 | 0 |
| 0 | 0 | 0 | 0 | 0 |
| 0 | 0 | 0 | 0 | 0 |
| 0 | 0 | 0 | 0 | 0 |
| 0.215383856 | 0 | 0 | 0.005061356 | 0 |
| 0 | 0 | 0 | 0 | 0 |
| 0 | 0 | 0 | 0 | 0 |
| 0 | 0 | 0 | 0 | 0 |
| 0 | 0 | 0 | 0 | 0 |
| 0 | 0 | 0 | 0 | 0 |
| 0 | 0 | 0.049560662 | 0 | 0 |
| 0 | 0 | 0 | 0 | 0 |
| 0 | 0.014610798 | 0.041464735 | 0 | 0 |
| 0 | 0 | 0 | 0 | 0 |
| 0 | 0 | 0 | 0 | 0 |
| 0 | 0 | 0.002449158 | 0 | 0.001783445 |
| 0 | 0 | 0 | 0 | 0 |
| 0 | 0.571911033 | 0.017062332 | 0.002853633 | 1.465345611 |
| 0.006860942 | 0 | 0.001037848 | 0.007178102 | 0 |
| 0 | 0 | 0 | 0 | 0 |
| 0 | 0 | 0 | 0 | 0 |
| 0 | 0 | 0 | 0 | 0 |
| 0 | 0 | 0 | 0 | 0 |
| 0 | 0 | 0 | 0 | 0 |
| 0 | 0 | 0 | 0 | 0 |
| 0 | 0 | 0 | 0 | 0 |
| 0 | 0 | 0.013702811 | 1.424486825 | 0.237109601 |
| 0 | 0.003343866 | 0.079804924 | 0.182827017 | 0.202074371 |
| 0 | 0 | 0 | 0 | 0 |
| 0 | 0 | 0 | 0 | 0 |
| 0.041327693 | 0.006612373 | 0.005001281 | 0.011127462 | 0.015268576 |
| 0 | 0 | 0 | 0.079833399 | 0 |
| 0 | 0 | 0 | 0 | 0 |

| | | | | |
|---|---|---|---|---|
| 0 | 0 | 0 | 0 | 0 |
| 0 | 0.002212511 | 0.01215703 | 0.00345732 | 0 |
| 0.004323285 | 0.002799817 | 0.01046367 | 0.002692343 | 0.009928447 |
| 0 | 0 | 0 | 0 | 0 |
| 0 | 0.021151453 | 0 | 4.46236114 | 0.167765638 |
| 0 | 0 | 0 | 0.017367847 | 0 |
| 0 | 0 | 0 | 0 | 0 |
| 0 | 0 | 0 | 0 | 0 |
| 0 | 0 | 0 | 0 | 0 |
| 0 | 0 | 0 | 0 | 0 |
| 0 | 0 | 0 | 0 | 0 |
| 0 | 0 | 0 | 0 | 0 |
| 0 | 0 | 0 | 0 | 0 |
| 0 | 0 | 0 | 0 | 0.325265529 |
| 0 | 0 | 0 | 0 | 0 |
| 0 | 0 | 0 | 0 | 0 |
| 0 | 0 | 0.001678538 | 0.002947679 | 0 |
| 0 | 0 | 0 | 0 | 0 |
| 0 | 0 | 0 | 0 | 0 |
| 0 | 0 | 0 | 0 | 0 |
| 0 | 0 | 0 | 0 | 0 |
| 0 | 0 | 0 | 0 | 0 |
| 0 | 0 | 0.00281705 | 0 | 0 |
| 0 | 0 | 0 | 0 | 0 |
| 0 | 0.008697286 | 0.000615967 | 0 | 0 |
| 0.004437691 | 0 | 0.001032747 | 0 | 0.061830745 |
| 0 | 0 | 0 | 0 | 0 |
| 0 | 0.055425901 | 0.08936059 | 0 | 0 |
| 0 | 0 | 0 | 0 | 0 |
| 0 | 0 | 0 | 0 | 0 |
| 0 | 0 | 0 | 0 | 0 |
| 0 | 0 | 0 | 0 | 0 |
| 0 | 0 | 0 | 0 | 0 |
| 0 | 0 | 0 | 0 | 0 |
| 0 | 0 | 0 | 0 | 0.002901647 |
| 0 | 0 | 0 | 0 | 0 |
| 0 | 0 | 0 | 0 | 0 |
| 0 | 0 | 0 | 0.012442629 | 0 |
| 0 | 1.762840524 | 2.739259074 | 0 | 6.50985953 |
| 0 | 0 | 0 | 0 | 0 |
| 0 | 0 | 0 | 0 | 0 |
| 0 | 0.001162237 | 0 | 0 | 0 |
| 0 | 0.00215041 | 0 | 0.027106255 | 0.09682714 |
| 0 | 0 | 0 | 0 | 0 |
| 0 | 0 | 0.012785322 | 0 | 0 |
| 0 | 0 | 0 | 0 | 0 |
| 0 | 0.005208702 | 0.000951361 | 0.003427047 | 0.003526823 |
| 0 | 0 | 1.28628478 | 0 | 0 |

| | | | | |
|---|---|---|---|---|
| 0 | 0 | 0 | 0 | 0 |
| 0 | 0 | 0.00260096 | 0.014270199 | 0.006860728 |
| 0 | 0 | 0.00948924 | 0 | 0 |
| 0 | 0 | 0 | 0 | 0 |
| 0 | 0 | 0 | 0 | 0 |
| 0 | 0 | 0 | 0 | 0 |
| 0 | 0 | 0 | 0 | 0 |
| 0 | 0 | 0 | 0 | 0 |
| 0 | 0 | 0 | 0 | 0 |
| 0 | 0 | 0 | 0 | 0 |
| 0 | 0 | 0 | 0 | 0 |
| 0 | 0 | 0 | 0 | 0 |
| 0 | 0.023036893 | 0.056257883 | 0 | 0 |
| 0 | 0 | 0 | 0 | 0 |
| 0 | 0.004097411 | 0.00124731 | 0.00808763 | 0.02265732 |
| 0 | 0 | 0 | 0 | 0 |
| 0 | 0 | 0 | 0 | 0 |
| 0 | 0 | 0 | 0 | 0 |
| 0 | 0 | 0 | 0 | 0.023786043 |
| 0 | 0 | 0 | 0 | 0 |
| 0 | 0 | 0 | 0 | 0 |
| 0 | 0 | 0.032093864 | 0.080221184 | 0.0088551 |
| 0 | 0.031734712 | 0 | 0 | 0.00610653 |
| 0 | 0 | 0.0012617 | 0 | 0 |
| 0 | 0 | 0 | 0 | 0 |
| 0 | 0 | 0 | 0 | 0 |
| 0 | 0 | 0 | 0 | 0 |
| 0 | 0 | 0 | 0 | 0 |
| 0 | 0 | 0 | 0 | 0 |
| 0 | 0.021598182 | 0.008064523 | 0 | 0 |
| 0 | 0 | 0 | 0 | 0 |
| 0 | 0 | 0 | 0 | 0 |
| 0 | 0 | 0 | 0 | 0 |
| 0 | 0 | 0 | 0 | 0 |
| 0 | 0 | 0.03536772 | 5.194939949 | 0.07503942 |
| 0 | 0 | 0 | 0 | 0 |
| 0 | 0 | 0 | 0 | 0 |
| 0 | 0 | 0 | 0 | 0 |
| 0.006671527 | 0.004864418 | 0.053089806 | 0.011633216 | 0.107021626 |
| 0 | 0 | 0 | 0 | 0 |
| 0 | 0.13862283 | 0.223312762 | 0.521298796 | 0.054891825 |
| 0 | 0 | 0 | 0 | 0 |
| 0 | 0 | 0 | 0 | 0 |
| 0 | 0 | 0 | 0 | 0 |
| 0 | 0 | 0 | 0 | 0 |
| 0.00373575 | 0.015415421 | 0.015822896 | 0.032570349 | 0.800955248 |
| 0 | 0 | 0 | 0 | 0 |
| 0 | 0 | 0 | 0 | 0 |

| | | | | |
|---|---|---|---|---|
| 0 | 0 | 0 | 0 | 0 |
| 0 | 0 | 0 | 0 | 0 |
| 0 | 0 | 0 | 0 | 0 |
| 0 | 0 | 0 | 0 | 0 |
| 0 | 0 | 0 | 0.019144162 | 0 |
| 0 | 0 | 0 | 0 | 0 |
| 0 | 0 | 0 | 0 | 0 |
| 0 | 0 | 0 | 0 | 0 |
| 0 | 0 | 0.001190992 | 0 | 0 |
| 0 | 0.006157928 | 0 | 0.007167117 | 0 |
| 0 | 0.117123405 | 0.108802701 | 0 | 0 |
| 0 | 0 | 0 | 0 | 0 |
| 0 | 0.001037734 | 0.001920677 | 0 | 0 |
| 0 | 0 | 0 | 0 | 0 |
| 0 | 0 | 0 | 0 | 0 |
| 0 | 0 | 0 | 0 | 0 |
| 0 | 0 | 0 | 0 | 0 |
| 0 | 0 | 0 | 0 | 0 |
| 0 | 0 | 0 | 0 | 0 |
| 0.006356985 | 0.002586238 | 0.008975075 | 0.587491551 | 0.657117925 |
| 0 | 0 | 0 | 0 | 0 |
| 0 | 0 | 0 | 0 | 0 |
| 0 | 0 | 0 | 0 | 0 |
| 0 | 0 | 0 | 0 | 0 |
| 0 | 0 | 0 | 0 | 0 |
| 0 | 0.017169897 | 0 | 0 | 0 |
| 0 | 0 | 0 | 0 | 0 |
| 0 | 0 | 0 | 0 | 0 |
| 0 | 0 | 0 | 0 | 0 |
| 0 | 0.000789923 | 0 | 0 | 0.014184891 |
| 0 | 0 | 0 | 0 | 0 |
| 0 | 0 | 0 | 0 | 0 |
| 0 | 0 | 0 | 0 | 0 |
| 0 | 0 | 0 | 0 | 0 |
| 0 | 0 | 0 | 0 | 0 |
| 0 | 0 | 0 | 0 | 0 |
| 0 | 0 | 0 | 0 | 0 |
| 0 | 0 | 0 | 0 | 0 |
| 0 | 0 | 0 | 0 | 0 |
| 0 | 0 | 0 | 0 | 0 |
| 0 | 0 | 0 | 0 | 0 |
| 0 | 0 | 0 | 0 | 0 |
| 0 | 0.005220901 | 0 | 0.025914595 | 0 |
| 0 | 0 | 0 | 0 | 0 |
| 0 | 0 | 0 | 0.039258904 | 0.002790187 |
| 0 | 0 | 0.083227367 | 0 | 0 |
| 0 | 0 | 0 | 0 | 0 |
| 0 | 0 | 0 | 0 | 0 |

| | | | | |
|---|---|---|---|---|
| 0 | 0 | 0 | 0 | 0 |
| 0 | 0 | 0 | 0 | 0 |
| 0 | 0 | 0 | 0 | 0 |
| 0 | 0 | 0 | 0 | 0 |
| 0 | 0 | 0 | 0 | 0 |
| 0 | 0.006476463 | 0.005244262 | 0.003886183 | 0.001527521 |
| 0 | 0 | 0 | 0 | 0 |
| 0 | 0 | 0 | 0.004050445 | 0 |
| 0 | 0 | 0 | 0 | 0 |
| 0 | 0 | 0 | 0 | 0 |
| 0 | 0 | 0 | 0 | 0 |
| 0 | 0 | 0 | 0 | 0 |
| 0.039407533 | 0 | 0 | 0 | 0 |
| 0 | 0 | 0 | 0 | 0 |
| 0 | 0 | 0 | 0 | 0 |
| 0 | 0 | 0 | 0 | 0 |
| 0 | 0.17298702 | 0 | 0 | 0 |
| 0 | 0 | 0 | 0 | 0 |
| 0 | 0 | 0 | 0 | 0 |
| 0 | 0 | 0 | 0 | 0 |
| 0 | 0 | 0.001216132 | 0 | 0 |
| 0 | 0 | 0 | 0 | 0 |
| 0 | 0 | 0 | 0 | 0 |
| 0 | 0 | 0 | 0 | 0 |
| 0 | 0 | 0 | 0 | 0 |
| 0 | 0 | 0 | 0 | 0 |
| 0 | 0 | 0 | 0 | 0 |
| 0 | 0.941812595 | 0.000526226 | 0.001137361 | 0 |
| 0 | 0 | 0 | 0 | 0 |
| 0 | 0 | 0 | 0.005561854 | 0 |
| 0 | 0 | 0 | 0 | 0 |
| 0 | 0 | 0 | 0 | 0 |
| 0 | 0 | 0 | 0 | 0 |
| 0 | 0 | 0 | 0 | 0 |
| 0 | 0 | 0 | 0 | 0 |
| 0 | 0 | 0 | 0 | 0 |
| 0 | 0 | 0 | 0 | 0 |
| 0 | 0 | 0 | 0 | 0 |
| 0 | 0.245008515 | 0 | 0 | 0 |
| 0 | 0 | 0 | 0 | 0 |
| 0 | 0 | 0 | 0 | 0 |
| 0 | 0 | 0 | 0 | 0 |
| 0 | 0 | 0 | 0 | 0 |
| 0 | 0 | 0 | 0 | 0 |
| 0 | 0 | 0 | 0 | 0 |
| 0 | 0 | 0 | 0 | 0 |
| 0 | 0 | 0 | 0 | 0 |
| 0 | 0 | 0 | 0 | 0 |

| | | | | |
|---|---|---|---|---|
| 0 | 0 | 0 | 0 | 0 |
| 0 | 0 | 0.035899255 | 0 | 0 |
| 0 | 0 | 0 | 0.000887799 | 0.013895044 |
| 0 | 0 | 0 | 0 | 0 |
| 0 | 0 | 0 | 0 | 0 |
| 0 | 0 | 0 | 0 | 0 |
| 0 | 0 | 0 | 0 | 0 |
| 0 | 0 | 0 | 0 | 0 |
| 0 | 0 | 0 | 0 | 0 |
| 0 | 0 | 0 | 0 | 0 |
| 0.040840709 | 0 | 0.00087864 | 0 | 0 |
| 0 | 0 | 0 | 0 | 0 |
| 0 | 0 | 0.422111832 | 0 | 2.894085776 |
| 0 | 0 | 0 | 0 | 0 |
| 0 | 0 | 0 | 0 | 0 |
| 0 | 0 | 0 | 0.007646757 | 0 |
| 0 | 0 | 0 | 0 | 0 |
| 0 | 0 | 0 | 0 | 0 |
| 0 | 0 | 0 | 0 | 0 |
| 0 | 0 | 0 | 0 | 0 |
| 0.001658034 | 0 | 0 | 0.001445567 | 0.002686041 |
| 0 | 0 | 0 | 0 | 0 |
| 0 | 0 | 0 | 0 | 0 |
| 0 | 0.005587927 | 0.98159946 | 0.374420486 | 1.297071104 |
| 0 | 0 | 0 | 0 | 0 |
| 0 | 0 | 0 | 0 | 0 |
| 0 | 0 | 0 | 0 | 0 |
| 0 | 0 | 0 | 0 | 0 |
| 0 | 0 | 0 | 0 | 0.000739622 |
| 0 | 0 | 0 | 0 | 0 |
| 0 | 0 | 0 | 0 | 0 |
| 0 | 0 | 0 | 0 | 0 |
| 0 | 0.001621844 | 0 | 0 | 0 |
| 0 | 0 | 0 | 0 | 0 |

| PHL018-0024_V2 | PHL018-0217_V3 | PHL018-0007_V1 | PHL018-0234_V3 | PHL018-0195_V3 |
|---|---|---|---|---|
| EYCF | REF | EYCF | CTRL | CTRL |
| 0.159619492 | 0.112740381 | 1.884733516 | 0.12018427 | 0.127150133 |
| 0.584978797 | 0.200802255 | 4.522863975 | 1.084944032 | 0.439704824 |
| 0 | 7.219909114 | 0 | 5.071459762 | 5.75063057 |
| 0 | 0 | 0 | 0 | 0 |
| 0 | 0 | 0 | 0 | 0 |
| 0 | 0 | 0 | 0 | 0 |
| 0 | 0 | 0 | 0 | 0 |
| 0 | 0 | 0 | 0 | 0 |
| 0 | 0 | 0 | 0 | 0 |
| 0 | 0 | 0 | 0 | 0 |
| 0 | 0 | 0 | 0 | 0 |
| 0 | 0 | 0 | 0 | 0 |
| 0 | 0 | 0 | 0 | 0 |
| 0.007957754 | 3.157421232 | 0 | 1.875242364 | 1.518753498 |
| 0 | 0 | 0 | 0 | 0 |
| 0 | 0 | 0 | 0 | 0 |
| 0 | 0 | 0 | 0 | 0 |
| 0 | 0 | 0 | 0 | 0 |
| 0 | 0 | 0 | 0 | 0 |
| 0 | 0 | 0 | 0 | 0 |
| 0 | 0 | 0 | 0 | 0 |
| 0 | 0 | 0 | 0 | 0 |
| 0 | 0 | 0 | 0 | 0 |
| 0 | 0 | 0 | 0 | 0 |
| 0 | 0 | 0 | 0 | 0 |
| 0 | 0 | 0 | 0 | 0 |
| 0 | 0 | 0 | 0 | 0 |
| 0 | 0 | 0 | 0 | 0 |
| 0 | 0 | 0 | 0 | 0 |
| 0 | 0 | 0 | 0 | 0 |
| 0 | 0 | 0 | 0 | 0 |
| 0 | 0 | 0 | 0 | 0 |
| 0 | 0 | 0 | 0 | 0 |
| 0 | 0 | 0 | 0 | 0 |
| 0 | 0 | 0 | 0 | 0 |
| 0.251059063 | 0 | 0 | 0 | 0 |
| 0 | 0 | 0 | 0 | 0 |
| 0 | 0 | 0 | 0 | 0 |
| 0 | 0 | 0 | 0 | 0 |
| 0 | 0 | 0 | 0 | 0 |
| 0 | 0 | 0 | 0 | 0 |
| 0.019872234 | 0.002211645 | 0.01711463 | 0 | 0 |
| 0.246361928 | 0.005034525 | 0.245216966 | 0.665861179 | 0.040899083 |
| 0.000766718 | 0.001994602 | 0.066970288 | 0.056184314 | 0.132862595 |
| 0.00856804 | 0 | 0.020846086 | 0 | 0 |
| 0 | 0 | 0 | 0 | 0 |

| | | | | |
|---|---|---|---|---|
| 0.068596326 | 0 | 0 | 0 | 0 |
| 0.002710751 | 0.000699368 | 0.232333423 | 0.003801742 | 0 |
| 0 | 0 | 0.04723439 | 0 | 0.000699184 |
| 0.282914995 | 0.811939893 | 0 | 0.063135766 | 0.165364346 |
| 0 | 0.006612971 | 0 | 0 | 0 |
| 0.063082593 | 0 | 0 | 0 | 0 |
| 0 | 0.013992719 | 0 | 0 | 0.004984994 |
| 0 | 0 | 0 | 0 | 0 |
| 0.022632743 | 0 | 0 | 0 | 0 |
| 0 | 0 | 0 | 0 | 0 |
| 0.000970757 | 0 | 0.008344646 | 0 | 0 |
| 0.080154995 | 0.157647603 | 0 | 0 | 0.351803696 |
| 0 | 0 | 0 | 0 | 0 |
| 0.041427213 | 0.174833191 | 0.13826837 | 0.006878808 | 0.012333058 |
| 0 | 0 | 0 | 0 | 0 |
| 0 | 0 | 0 | 0 | 0 |
| 0.003371234 | 0.055197035 | 0.008629042 | 0 | 0.374853006 |
| 0 | 0 | 0 | 0 | 0 |
| 0 | 0 | 0 | 0 | 0 |
| 0 | 0 | 0 | 0 | 0 |
| 0 | 0.013047486 | 0.424353512 | 0.117842373 | 0.067047468 |
| 0.008820809 | 0.027873912 | 0.020191943 | 0.017992333 | 0.008211005 |
| 0 | 0 | 0 | 0 | 0.001805804 |
| 0 | 0 | 0 | 0 | 0 |
| 0 | 0 | 0 | 0 | 0 |
| 0 | 0 | 0 | 0 | 0 |
| 0 | 0 | 0 | 0.008395842 | 0.000961199 |
| 0 | 0 | 0 | 0 | 0 |
| 0.000720279 | 0 | 0.00106521 | 0 | 0 |
| 0 | 0 | 0 | 0 | 0 |
| 0 | 0 | 2.392697685 | 0 | 0.02499303 |
| 0 | 0 | 0 | 0 | 0 |
| 0 | 0 | 0 | 0 | 0 |
| 0 | 0 | 0 | 0 | 0 |
| 0 | 0 | 0 | 0 | 0 |
| 0 | 0 | 0 | 0 | 0 |
| 0 | 0 | 0 | 0 | 0 |
| 0.002958512 | 0 | 0 | 0 | 50.65152552 |
| 0 | 0 | 0 | 0 | 0 |
| 0 | 0 | 0 | 0 | 0 |
| 0 | 0 | 0 | 0 | 0 |
| 0 | 0 | 0 | 0 | 0 |
| 0 | 0 | 0 | 0 | 0 |
| 0 | 0 | 0 | 0 | 0 |
| 0 | 0 | 0 | 0.0246177 | 0 |
| 0 | 0 | 0 | 0 | 0 |
| 0 | 0 | 0 | 0 | 0 |
| 0 | 0 | 0 | 0 | 0.331993558 |

| | | | | |
|---|---|---|---|---|
| 0 | 0 | 0 | 0 | 0 |
| 0 | 0 | 0 | 0 | 0 |
| 0 | 0 | 0 | 0 | 0 |
| 0 | 0 | 0 | 0 | 0 |
| 0.105530815 | 0 | 0 | 0.031308455 | 0 |
| 0 | 0 | 0 | 0 | 0 |
| 0 | 0 | 0 | 0 | 0 |
| 0.005621828 | 0 | 0 | 0 | 0 |
| 0 | 0 | 0.003420522 | 0 | 0 |
| 0 | 0 | 0 | 0 | 0 |
| 0 | 0 | 0 | 0 | 0 |
| 0.005475778 | 0 | 0 | 0 | 0 |
| 0 | 0 | 0 | 0 | 0 |
| 0 | 0 | 0 | 0 | 0 |
| 0 | 0 | 0 | 0 | 0 |
| 0 | 0 | 0 | 0 | 0 |
| 0 | 0 | 0 | 0 | 0 |
| 0 | 0 | 0 | 0 | 0 |
| 0.70904858 | 0 | 1.531740125 | 0.025817092 | 0.035855856 |
| 0 | 0 | 0 | 0 | 0 |
| 0 | 0 | 0 | 0 | 0 |
| 0 | 0 | 0 | 0 | 0 |
| 0 | 0 | 0 | 0 | 0 |
| 0 | 0 | 0 | 0 | 0 |
| 0 | 0 | 0 | 0 | 0 |
| 0 | 0 | 0 | 0 | 0 |
| 0 | 0 | 0 | 0 | 0 |
| 0 | 0 | 0 | 0.000807289 | 0 |
| 0 | 0 | 0 | 0 | 0 |
| 0 | 0 | 0 | 0 | 0 |
| 0.010911308 | 0.238517962 | 0 | 0.040510277 | 0 |
| 0.002817701 | 0.000666381 | 0.003711286 | 0 | 0.000453223 |
| 0 | 0 | 0 | 0 | 0 |
| 0 | 0 | 0 | 0 | 0 |
| 0 | 0 | 0 | 0 | 0 |
| 0 | 0 | 0 | 0 | 0 |
| 0 | 0 | 0 | 0 | 0 |
| 0 | 0 | 0 | 0 | 0 |
| 0 | 0 | 0 | 0 | 0 |
| 0 | 0 | 0 | 0 | 0 |
| 0 | 0 | 0 | 0 | 0 |
| 0 | 0 | 0 | 0 | 0 |
| 0 | 0.000577673 | 0 | 0 | 0 |
| 0 | 0 | 0 | 0 | 0 |
| 0.006510413 | 0 | 0 | 0 | 0 |
| 0 | 0 | 0 | 0 | 0 |
| 0 | 0 | 0 | 0.305468557 | 0 |
| 0 | 0 | 0 | 0 | 0 |

| | | | | |
|---|---|---|---|---|
| 0 | 0 | 0.006139232 | 0 | 0 |
| 0 | 0 | 0 | 0 | 0 |
| 0 | 0 | 0 | 0 | 0 |
| 0 | 0 | 0 | 0 | 0 |
| 0.3024227 | 0 | 0.004590361 | 0 | 0 |
| 0 | 0 | 0 | 0 | 0 |
| 0 | 0 | 0 | 0 | 0 |
| 0 | 0 | 0 | 0 | 0 |
| 0 | 0 | 0 | 0 | 0 |
| 0 | 0 | 0 | 0 | 0 |
| 0 | 0 | 0 | 0 | 0 |
| 0 | 0 | 0 | 0 | 0 |
| 0 | 0 | 0 | 0 | 0 |
| 0 | 0 | 0 | 0 | 0 |
| 0 | 0 | 0 | 0 | 0 |
| 1.448063694 | 0 | 0 | 0 | 0 |
| 0 | 0 | 0 | 0 | 0 |
| 0 | 0 | 0 | 0 | 0 |
| 0 | 0 | 0 | 0 | 0 |
| 0 | 0 | 0 | 0 | 0 |
| 0 | 0 | 0 | 0 | 0 |
| 0 | 0 | 0 | 0 | 0 |
| 0 | 0 | 0 | 0 | 0 |
| 0 | 0 | 0 | 0 | 0 |
| 0 | 0 | 0 | 0 | 0 |
| 0 | 0 | 0 | 0 | 0 |
| 0 | 0 | 0 | 0 | 0 |
| 0 | 0 | 0 | 0 | 0 |
| 0 | 0 | 0 | 0 | 0 |
| 0 | 0 | 0 | 0 | 0 |
| 0.009233033 | 0.001310157 | 0 | 0 | 0 |
| 0.076398956 | 0 | 0 | 0 | 0 |
| 0 | 0 | 0 | 0 | 0 |
| 0 | 0 | 0 | 0 | 0 |
| 0 | 0 | 0 | 0 | 0 |
| 0 | 0 | 0 | 0 | 0 |
| 0 | 0 | 0 | 0 | 0 |
| 0 | 0 | 0 | 0 | 0 |
| 0 | 0 | 0 | 0.008157773 | 0 |
| 0 | 0 | 0 | 0 | 0 |
| 0 | 0 | 0 | 0 | 0 |
| 0 | 0 | 0 | 0 | 0 |
| 0 | 0 | 0 | 0 | 0 |
| 0 | 0 | 0 | 0 | 0 |
| 0 | 0 | 0 | 0 | 0 |
| 0 | 0 | 0 | 0 | 0 |
| 0 | 0 | 0 | 0 | 0 |
| 0 | 0 | 0 | 0 | 0 |

| | | | | |
|---|---|---|---|---|
| 0 | 0 | 0 | 0 | 0 |
| 0 | 0 | 0 | 0 | 0 |
| 0 | 0 | 0 | 0 | 0 |
| 0 | 0 | 0 | 0 | 0 |
| 0 | 0 | 0 | 0 | 0 |
| 0 | 0 | 0 | 0 | 0 |
| 0 | 0 | 0 | 0 | 0 |
| 0 | 0 | 0 | 0 | 0 |
| 0 | 0 | 0 | 0 | 0 |
| 0 | 0 | 0 | 0 | 0 |
| 0 | 0 | 0 | 0 | 0 |
| 0 | 0 | 0 | 0 | 0 |
| 0.006416245 | 0 | 0 | 0.001071259 | 0.003335897 |
| 0 | 0 | 0 | 0 | 0 |
| 0 | 0 | 0 | 0 | 0 |
| 0 | 0 | 0 | 0 | 0 |
| 0 | 0 | 0 | 0 | 0 |
| 0 | 0 | 0 | 0 | 0 |
| 0 | 0 | 0 | 0 | 0 |
| 0 | 0 | 0 | 0 | 0 |
| 0.005089567 | 0.011348918 | 0.022177436 | 0.096143896 | 0.02089504 |
| 0 | 0 | 0 | 0 | 0 |
| 0 | 0 | 0 | 0 | 0 |
| 0 | 0.001075876 | 0 | 0.007101661 | 0.001300855 |
| 0 | 0 | 0 | 0 | 0 |
| 0 | 0 | 0 | 0 | 0 |
| 0 | 0.00260368 | 0.005635905 | 0 | 0.006356825 |
| 0 | 0 | 0 | 0 | 0 |
| 0 | 0 | 0 | 0 | 0 |
| 0 | 0.002785638 | 0.018591788 | 0.000998415 | 0.000777265 |
| 0 | 0 | 0.032069647 | 0 | 0.007966105 |
| 0 | 0 | 0 | 0 | 0 |
| 0 | 0 | 0 | 0 | 0 |
| 0.047989451 | 0 | 0 | 0.001224108 | 0 |
| 0.013009636 | 0.007384211 | 0.193621972 | 0.006255634 | 0.001217501 |
| 0 | 0 | 0 | 0 | 0 |
| 0 | 0 | 0 | 0 | 0 |
| 0 | 0 | 0 | 0 | 0 |
| 0 | 0 | 0 | 0 | 0 |
| 0 | 0 | 0 | 0 | 0 |
| 0 | 0 | 0.002770165 | 0 | 0 |
| 0 | 0 | 0 | 0 | 0 |
| 0 | 0 | 0 | 0 | 0 |
| 0 | 0 | 0 | 0 | 0 |
| 0 | 0 | 0 | 0.003504555 | 0 |
| 0 | 0 | 0 | 0 | 0 |
| 0 | 0 | 0.009942329 | 0 | 0 |
| 0.001348512 | 0 | 0.007291639 | 0.005572418 | 0.005494952 |

| | | | | |
|---|---|---|---|---|
| 0 | 0 | 0 | 0 | 0 |
| 0 | 0 | 0 | 0 | 0 |
| 0 | 0 | 0.002733517 | 0.001018392 | 0 |
| 0 | 0 | 0 | 0 | 0 |
| 0 | 0 | 0 | 0 | 0 |
| 0 | 0 | 0 | 0 | 0 |
| 0 | 0 | 0.00106457 | 0 | 0 |
| 0 | 0 | 0 | 0 | 0 |
| 0 | 0 | 0 | 0 | 0 |
| 0 | 0 | 0 | 0 | 0 |
| 0 | 0 | 0 | 0 | 0 |
| 0 | 0 | 0 | 0 | 0 |
| 0 | 0 | 0 | 0 | 0 |
| 0 | 0 | 0 | 0 | 0 |
| 0.001328847 | 0 | 0.014149536 | 0 | 0 |
| 0 | 0 | 0 | 0 | 0 |
| 0 | 0 | 0 | 0 | 0 |
| 0.002056107 | 0 | 0 | 0 | 0 |
| 0.001885713 | 0 | 0 | 0.00302246 | 0 |
| 0 | 0.016725546 | 0 | 0.157160846 | 0 |
| 0 | 0.024639382 | 0.036462299 | 0.747345288 | 0.002603165 |
| 0 | 0 | 0 | 0 | 0 |
| 0 | 0 | 0 | 0 | 0 |
| 0 | 0 | 0 | 0 | 0 |
| 0 | 0 | 0.008358469 | 0.01051843 | 0.003447825 |
| 0 | 0 | 0 | 0 | 0 |
| 0 | 0 | 0 | 0 | 0 |
| 0 | 0 | 0 | 0 | 0 |
| 0.00143831 | 0 | 0 | 0 | 0 |
| 0 | 0 | 0 | 0 | 0 |
| 0.050389319 | 0.39559097 | 0.296773976 | 0.177437228 | 0.396379487 |
| 0 | 0 | 0 | 0 | 0 |
| 0 | 0.048116939 | 0 | 0 | 0 |
| 0 | 0 | 0 | 0.001325428 | 0 |
| 0 | 0 | 0 | 0 | 0 |
| 0 | 0 | 0 | 0 | 0 |
| 0 | 0 | 0 | 0 | 0 |
| 0 | 0 | 0 | 0 | 0 |
| 0 | 0 | 0 | 0 | 0 |
| 0 | 0 | 0 | 0 | 0 |
| 0 | 0 | 0 | 0 | 0 |
| 0 | 0.003585942 | 0.07589071 | 0 | 0.031217807 |
| 0 | 0 | 0 | 0 | 0.00313769 |
| 0 | 0 | 0 | 0 | 0 |
| 0 | 0 | 0 | 0 | 0 |
| 0 | 0 | 0 | 0 | 0 |
| 0 | 0 | 0 | 0 | 0 |
| 0 | 0 | 0 | 0 | 0 |

| | | | | |
|---|---|---|---|---|
| 0 | 0 | 0 | 0 | 0 |
| 0 | 0 | 0 | 0 | 0 |
| 0 | 0.002026233 | 0.076023443 | 0.05979319 | 0.130051162 |
| 0 | 0 | 0 | 0 | 0 |
| 0.921763666 | 0.116055802 | 0 | 0 | 0 |
| 0 | 0 | 0 | 0 | 0 |
| 0 | 0 | 0 | 0 | 0 |
| 0 | 0 | 0 | 0 | 0 |
| 0 | 0 | 0 | 0 | 0 |
| 0 | 0 | 0.009174158 | 0 | 0 |
| 0 | 0 | 0 | 0 | 0 |
| 0 | 0 | 0 | 0 | 0 |
| 0 | 0 | 0 | 0 | 0 |
| 0 | 0 | 0 | 0.008115367 | 0 |
| 0 | 0 | 0.002648678 | 0.006219732 | 0.004842054 |
| 0.003718783 | 0 | 0.026810818 | 0 | 0 |
| 0 | 0 | 0.002743574 | 0.004361124 | 0 |
| 0 | 0 | 0 | 0 | 0 |
| 0 | 0 | 0 | 0 | 0 |
| 0 | 0 | 0 | 0 | 0 |
| 0 | 0 | 0 | 0 | 0 |
| 0.001786328 | 0 | 0.005448661 | 0.007381595 | 0.006512771 |
| 0 | 0 | 0 | 0 | 0 |
| 0 | 0 | 0.001919697 | 0 | 0 |
| 0 | 0 | 0 | 0 | 0 |
| 0 | 0 | 0 | 0 | 0 |
| 0.556883795 | 13.72613899 | 0 | 0 | 0 |
| 0 | 0 | 0 | 0 | 0 |
| 0 | 0 | 0.001317288 | 0 | 0 |
| 0 | 0 | 0 | 0 | 0 |
| 0 | 0 | 0.006073303 | 0 | 0 |
| 0 | 0 | 0 | 0 | 0 |
| 0.023105723 | 0 | 0 | 0 | 0 |
| 0 | 0.001948395 | 0 | 0 | 0 |
| 0 | 0 | 0 | 0 | 0 |
| 0 | 0 | 0.062824162 | 0 | 0 |
| 0 | 0 | 0 | 0 | 0 |
| 0.029737459 | 0 | 0.005289842 | 0.002473123 | 0 |
| 0 | 0 | 0 | 0 | 0 |
| 0 | 0 | 0 | 0 | 0 |
| 0 | 0 | 0 | 0 | 0 |
| 0 | 0 | 0 | 0 | 0 |
| 0 | 0 | 0 | 0 | 0 |
| 0 | 0 | 0.028215838 | 0 | 0 |
| 0 | 0 | 0 | 0 | 0 |
| 0 | 0 | 0 | 0 | 0 |
| 0 | 0 | 0 | 0 | 0 |
| 0 | 0 | 0 | 0 | 0.007287653 |

| | | | | |
|---|---|---|---|---|
| 0.174733482 | 0 | 0 | 0.179956157 | 0 |
| 0 | 0 | 0 | 0 | 0 |
| 0 | 0 | 0 | 0 | 0.001404451 |
| 0 | 0 | 0.004441145 | 0 | 0 |
| 0 | 0.006900523 | 0.007307797 | 0 | 0.006459489 |
| 0 | 0 | 0.001702289 | 0 | 0 |
| 0 | 0 | 0 | 0 | 0 |
| 0.001978997 | 0 | 0.010975163 | 0 | 0.086016758 |
| 0 | 0 | 0 | 0 | 0 |
| 0 | 0.095195131 | 0.463428798 | 0.195177922 | 0.005541209 |
| 0 | 0.000872318 | 0 | 0 | 0.002900508 |
| 0.015926909 | 0.000987983 | 0.014769563 | 0 | 0 |
| 0.004159076 | 0.001311485 | 0.009665539 | 0.058051956 | 0.003737798 |
| 0.000902366 | 0 | 0 | 0 | 0.001032137 |
| 0 | 0 | 0.002711124 | 0 | 0.000873691 |
| 0 | 0 | 0 | 0 | 0 |
| 0 | 0 | 0 | 0 | 0 |
| 0 | 0 | 0 | 0 | 0 |
| 0 | 0 | 0.161799504 | 0.02855349 | 0.003293164 |
| 0 | 0 | 0 | 0 | 0 |
| 0 | 0 | 0.220967217 | 0 | 0 |
| 0 | 0 | 0 | 0 | 0 |
| 0 | 0 | 0.001360035 | 0 | 0 |
| 0.061221699 | 0 | 0 | 0 | 0 |
| 0.005709317 | 0 | 0 | 0 | 0 |
| 0 | 0 | 0.002046889 | 0.003050334 | 0.001385231 |
| 0 | 0 | 0.002737105 | 0 | 2.951826634 |
| 0 | 0 | 0.303219049 | 0 | 0.005025386 |
| 0 | 0 | 0 | 0 | 0 |
| 0 | 0.001796174 | 0.012899195 | 0 | 0 |
| 0 | 0 | 0.210582052 | 0.001909351 | 0 |
| 0 | 0.009615527 | 0 | 0 | 0 |
| 0 | 0 | 0 | 0 | 0 |
| 0 | 0.002853927 | 0.005688528 | 0 | 0 |
| 0 | 0 | 0 | 0 | 0 |
| 0 | 0 | 0 | 0 | 0 |
| 0 | 0 | 0.357915246 | 0 | 0 |
| 0 | 0 | 0 | 0 | 0 |
| 0 | 0 | 0 | 0 | 0.003523147 |
| 0 | 0 | 0.148274313 | 0.001336738 | 0.003954464 |
| 0 | 0 | 0 | 0 | 0 |
| 0 | 0 | 0 | 0 | 0 |
| 0 | 0 | 0 | 0 | 0 |
| 0 | 0 | 0 | 0.020069226 | 0.007493079 |
| 0 | 0 | 0 | 0 | 0 |
| 0 | 0 | 0 | 0 | 0 |
| 0.004589885 | 0 | 0.023863762 | 0 | 0.002707011 |
| 0 | 0 | 0 | 0 | 0 |

| | | | | |
|---|---|---|---|---|
| 0 | 0 | 0 | 0 | 0 |
| 0 | 0.07187744 | 0 | 0 | 0 |
| 0 | 0 | 0 | 0 | 0 |
| 0.013275778 | 0 | 0 | 0 | 0 |
| 0 | 0 | 0 | 0 | 0 |
| 0 | 0.001496184 | 0.027566885 | 0.212376211 | 0 |
| 0 | 0 | 0 | 0 | 0 |
| 0 | 0 | 0 | 0 | 0 |
| 0.09232038 | 0.176488248 | 0 | 0 | 0 |
| 0 | 0 | 0 | 0 | 0 |
| 0 | 0.002974836 | 0.024706366 | 0 | 0 |
| 0 | 0 | 0 | 0 | 0.001896258 |
| 0 | 0 | 0 | 0 | 0 |
| 0 | 0.001171032 | 0.001922227 | 0 | 0.005097271 |
| 0 | 0.062087374 | 0.006668396 | 0 | 0 |
| 0 | 0 | 0 | 0 | 0 |
| 0 | 0 | 0 | 0 | 0 |
| 0 | 0 | 0 | 0 | 0 |
| 0 | 0.007229022 | 0.005260926 | 0 | 0.001130263 |
| 0.013954858 | 0 | 0 | 0.055947583 | 0 |
| 0 | 0.00656104 | 0 | 0 | 0 |
| 0.072217563 | 0 | 0 | 0 | 0 |
| 0 | 0 | 0 | 0.068087696 | 0 |
| 0 | 0 | 0 | 0 | 0 |
| 0 | 0 | 0 | 0 | 0 |
| 0 | 0 | 0 | 0 | 0 |
| 0 | 0.0060946 | 0.003275177 | 0 | 0 |
| 0 | 0 | 0 | 0 | 0 |
| 0 | 0 | 0 | 0 | 0 |
| 0.263099989 | 0.433927937 | 0.305506476 | 0.427482344 | 0.386651765 |
| 0.083476892 | 0.001271123 | 0.036514199 | 0 | 0 |
| 0 | 0 | 0 | 0 | 0 |
| 0 | 0 | 0.008032654 | 0 | 0 |
| 0 | 0 | 0 | 0.002097132 | 0 |
| 0 | 0.008460379 | 0 | 0 | 0 |
| 0 | 0 | 0.023692964 | 0 | 0 |
| 0 | 0 | 0 | 1.360782258 | 0 |
| 0 | 0 | 0 | 0 | 0 |
| 0 | 0 | 0 | 0 | 0 |
| 0 | 0.397398717 | 0 | 0 | 0 |
| 0.002035355 | 0.009049525 | 0 | 0.023101229 | 0 |
| 0 | 5.00643907 | 0 | 0 | 0 |
| 0 | 0 | 0 | 0 | 0 |
| 0 | 0.000929138 | 0 | 0.004058648 | 0 |
| 0 | 0.00152039 | 0.225121751 | 0 | 0 |
| 0 | 0 | 0 | 0.049065228 | 0 |
| 0 | 0.002222966 | 0.002132508 | 0.017419742 | 0.00073304 |
| 0 | 0 | 0 | 0 | 0 |

| | | | | |
|---|---|---|---|---|
| 0.006325862 | 0 | 0.011986382 | 0 | 0 |
| 0 | 0.002708481 | 0 | 0.000927934 | 0 |
| 0.005746314 | 0.114155375 | 0.006771956 | 0 | 0.028652855 |
| 0 | 0 | 0 | 0 | 0 |
| 0.257717385 | 0 | 0 | 0 | 0 |
| 0 | 0.003794746 | 0.609058528 | 0 | 0 |
| 0 | 0 | 0 | 0 | 0 |
| 0 | 0 | 0 | 0 | 0 |
| 0 | 0 | 0.062880547 | 0 | 0 |
| 0 | 0 | 0 | 0 | 0 |
| 0 | 0 | 0 | 0 | 0 |
| 0 | 0 | 0 | 0 | 0 |
| 0 | 0 | 0 | 0 | 0.001112194 |
| 0 | 0.032031477 | 0.003173788 | 0 | 0.000818232 |
| 0 | 0 | 0 | 0 | 0 |
| 0.587362904 | 0 | 0.054772429 | 0.003080131 | 0.012948543 |
| 0.210882671 | 0 | 0.174568526 | 0.002958804 | 0.001594679 |
| 0 | 0.000712175 | 0.100480205 | 0.053645942 | 0.003444396 |
| 0 | 0.017760456 | 0.001656447 | 0 | 0.002745305 |
| 0 | 0 | 0 | 0 | 0 |
| 0 | 0.009849892 | 0 | 0.011473645 | 0.006252553 |
| 0 | 0 | 0.050755472 | 0.001056018 | 0.005549234 |
| 0 | 0 | 0 | 0 | 0 |
| 0 | 0.343803925 | 0 | 0 | 0 |
| 0 | 0.204889619 | 0 | 0 | 0 |
| 0 | 0 | 0.001508945 | 0.011093461 | 0 |
| 0 | 0 | 0.004653357 | 0 | 0 |
| 0 | 0 | 0 | 0 | 0 |
| 0 | 0 | 0 | 0 | 0 |
| 0 | 0 | 0 | 0 | 0 |
| 0 | 0.043764696 | 0.06215132 | 0.081303191 | 0.017790868 |
| 0 | 0 | 0.010552028 | 0.01900097 | 0.003570539 |
| 0.0025476 | 0.062392498 | 0.005651413 | 0.021834559 | 0.005099453 |
| 0.004867946 | 0 | 0 | 0 | 0 |
| 0 | 0 | 0 | 0.013719365 | 0 |
| 0 | 0.002161453 | 0 | 0 | 0 |
| 0 | 0 | 0.001986937 | 0.004793987 | 0.002853969 |
| 0 | 0.010602262 | 0.027969786 | 0 | 0.010987992 |
| 0.127116421 | 0 | 0 | 0 | 0 |
| 0.010472365 | 0 | 0 | 0 | 0 |
| 0 | 0 | 0 | 0 | 0 |
| 0 | 0 | 0 | 0 | 0 |
| 0.001496507 | 0 | 0 | 0 | 0 |
| 0.036089116 | 0.006059487 | 0.02911182 | 0.245322159 | 0 |
| 0 | 0 | 0 | 0 | 0 |
| 0 | 0 | 0 | 0.000721208 | 0 |
| 0 | 0 | 0 | 0 | 0 |
| 0 | 0 | 0 | 0 | 0 |

| | | | | |
|---|---|---|---|---|
| 0 | 0 | 0.010861167 | 0 | 0 |
| 0.127355177 | 0 | 1.483660575 | 0 | 0.551410312 |
| 0 | 0 | 0 | 0 | 0 |
| 0 | 0.003281103 | 0 | 0 | 0 |
| 0 | 0 | 0 | 0 | 0 |
| 0 | 0 | 0 | 0 | 0.000984403 |
| 0 | 0 | 0 | 0 | 0 |
| 0 | 0 | 0 | 0 | 0 |
| 0 | 0 | 0 | 0 | 0 |
| 0 | 0.001499757 | 0.00307728 | 0 | 0.012240278 |
| 0.003850402 | 0.005367823 | 0.049675304 | 0.005806086 | 0.003824641 |
| 0.008150138 | 0 | 0.020423345 | 0.110432513 | 0.017349685 |
| 0.115777229 | 0.677274201 | 0 | 0 | 0 |
| 0 | 0 | 0 | 0 | 0 |
| 0 | 0 | 0 | 0.006405536 | 0 |
| 0 | 0 | 0 | 0 | 0 |
| 0.000848827 | 0 | 0.065433418 | 0.00210455 | 0.007099687 |
| 0 | 0.012508604 | 0.019554904 | 0.013356425 | 0.015360623 |
| 0 | 0 | 0 | 0 | 0 |
| 0 | 0.022675128 | 0 | 0 | 0 |
| 0.011817284 | 0 | 0.038836467 | 0.001205734 | 0.005444243 |
| 0 | 0.003091233 | 0 | 0 | 0 |
| 0 | 0 | 0 | 0.003970898 | 0 |
| 0 | 0 | 0 | 0 | 0 |
| 0 | 0 | 0 | 0 | 0 |
| 0 | 0 | 0 | 0 | 0 |
| 0.003672079 | 0.047406594 | 0.046239825 | 0 | 0.190489809 |
| 0.009071132 | 0.14126013 | 0.422061736 | 0.275269944 | 0.005387363 |
| 0 | 0 | 0 | 0 | 0 |
| 0.000841685 | 0.001061637 | 0.007779726 | 0 | 0.000722047 |
| 0 | 0 | 0 | 0 | 0 |
| 0 | 0.001518545 | 0 | 0 | 0 |
| 0 | 0 | 0 | 0 | 0 |
| 0 | 0 | 0.000586173 | 0 | 0 |
| 0.862035081 | 0 | 0 | 0 | 0 |
| 0.072825245 | 0.051017422 | 2.837172602 | 0.789476614 | 0.365724675 |
| 0 | 0 | 0 | 0 | 0 |
| 0 | 0 | 0 | 0 | 0 |
| 0.003443156 | 0.001085733 | 0.025778416 | 0 | 0 |
| 0 | 0 | 0 | 0 | 0 |
| 0 | 0 | 0 | 0 | 0 |
| 0 | 0 | 0 | 0 | 0 |
| 0.001742854 | 0.0018136 | 0 | 0.03418525 | 0.007027074 |
| 0 | 0 | 0 | 0 | 0 |
| 0 | 0 | 0 | 0.085991595 | 0 |
| 0.048506071 | 0 | 0 | 0 | 0 |
| 0 | 0 | 0 | 0 | 0 |
| 0 | 0 | 0.068784481 | 0.004495818 | 0.00093333 |

| | | | | |
|---|---|---|---|---|
| 0 | 0 | 0.02133473 | 0 | 0 |
| 0 | 0 | 0 | 0 | 0 |
| 0 | 0 | 0 | 0 | 0 |
| 0.955258631 | 0 | 0 | 0 | 0 |
| 0 | 0 | 0 | 0 | 0 |
| 0 | 0 | 0 | 0 | 0 |
| 0 | 0 | 0 | 0 | 0 |
| 0.09842394 | 0 | 0 | 0 | 0.021342122 |
| 17.01643339 | 0 | 0 | 4.058288582 | 0.000940234 |
| 0.005986886 | 0 | 0.006036763 | 0 | 0 |
| 0 | 0 | 0 | 0 | 0 |
| 0.053871994 | 0 | 0 | 0 | 0 |
| 0 | 0 | 0 | 0 | 0 |
| 0 | 0 | 0 | 0 | 0 |
| 0.098379433 | 0.396252545 | 0.704911914 | 0 | 0 |
| 0 | 0 | 0 | 0 | 0 |
| 0.001256059 | 0 | 0 | 0 | 0 |
| 1.691050839 | 0 | 0 | 0 | 0 |
| 0 | 0 | 0 | 0 | 0 |
| 0.076766232 | 0 | 0 | 0 | 0 |
| 0.368399883 | 0 | 0 | 0 | 0 |
| 0.018236953 | 0 | 0 | 0 | 0 |
| 0 | 0 | 0 | 0 | 0 |
| 0.167731813 | 0.144195043 | 0.325102162 | 0.570844451 | 0.099728462 |
| 0 | 0.004478564 | 0.076140349 | 0 | 0 |
| 0 | 0 | 0 | 0 | 0 |
| 0 | 0 | 0 | 0 | 0.011709125 |
| 1.694077187 | 0.101611314 | 0 | 6.828572367 | 0 |
| 0 | 0 | 0 | 0 | 0 |
| 0 | 0 | 0 | 0 | 0 |
| 0.028927917 | 0 | 0 | 0.007917115 | 0 |
| 0 | 0 | 0.051570159 | 1.834813961 | 0 |
| 0.037125473 | 0 | 0 | 0.088638367 | 0.023577689 |
| 0 | 0 | 0 | 0 | 0 |
| 0 | 0 | 0 | 0 | 0 |
| 0.41777141 | 0.024264537 | 0 | 0 | 0 |
| 0.394623947 | 0 | 0 | 0 | 0 |
| 0 | 0 | 0 | 0 | 0 |
| 0.005466355 | 0 | 0 | 0 | 0.018583752 |
| 0 | 0 | 0.015131455 | 0 | 0 |
| 0.034329317 | 0.032122286 | 0.058356908 | 0.125616361 | 0.059219488 |
| 0 | 0 | 0 | 0 | 0 |
| 0 | 30.38497567 | 0 | 9.963199079 | 0.203714997 |
| 0.067124446 | 0.005227539 | 0 | 0 | 0 |
| 0 | 0 | 0 | 0 | 0 |
| 0 | 0 | 0 | 0 | 0 |
| 0 | 0 | 0 | 0 | 0 |
| 0.217328678 | 0 | 0.753508847 | 0 | 0 |

| | | | | |
|---|---|---|---|---|
| 0 | 0 | 0 | 0 | 0 |
| 1.353868778 | 0 | 0 | 0 | 0 |
| 0 | 0.019321812 | 0.055563331 | 0.002606075 | 0.061971411 |
| 0.000697211 | 0 | 0.00876431 | 0 | 0 |
| 0 | 0 | 0 | 0 | 0 |
| 0.109750436 | 0.000801104 | 0.09111058 | 0 | 0.000581175 |
| 0 | 0 | 0 | 0 | 0 |
| 0 | 0 | 0 | 0 | 0 |
| 0 | 0 | 0 | 0 | 0 |
| 0 | 0 | 0 | 0 | 0 |
| 0.01583182 | 0.059108373 | 0.580653547 | 0.009536985 | 0.009597564 |
| 0.245340899 | 0.807926412 | 0.688818743 | 0.89539339 | 2.015289527 |
| 0 | 0 | 0 | 0 | 0 |
| 0.105024071 | 0 | 0.681498163 | 0 | 0.004885888 |
| 0.031956211 | 0.078123343 | 0.281367063 | 0.440766028 | 0.219465226 |
| 7.978553271 | 0 | 0 | 0 | 0.290062287 |
| 0 | 0 | 0 | 0 | 0 |
| 0 | 0 | 0 | 0 | 0 |
| 0.159795541 | 0 | 0 | 0 | 0 |
| 0.006781451 | 0.006744187 | 0 | 0.022993069 | 0 |
| 0.055731466 | 0.041507761 | 0.718039179 | 0.005653867 | 0.022766525 |
| 0 | 3.876482526 | 0 | 0.005135708 | 0.018871232 |
| 0 | 13.24531183 | 8.184432331 | 0.636037911 | 0.479958893 |
| 0 | 0 | 0 | 0 | 0 |
| 0 | 0 | 0 | 0 | 0 |
| 0.229818368 | 0 | 0 | 0 | 0 |
| 0 | 0.019417685 | 0 | 0 | 0 |
| 0 | 0 | 0 | 0.001016704 | 0 |
| 0 | 0 | 0 | 0 | 0 |
| 0 | 0 | 0 | 0 | 0 |
| 0.013660081 | 0 | 0 | 0 | 0 |
| 0.110908606 | 0.131148394 | 0 | 0.128840637 | 0 |
| 0 | 0.067678807 | 0.887305611 | 0 | 0 |
| 0.040834941 | 0.029558644 | 0.168151266 | 0.010384067 | 0 |
| 0.048793 | 0.003925445 | 0.069672836 | 0.02664695 | 0 |
| 0.040301797 | 0.02175926 | 0.220235715 | 0.238081637 | 0.010456295 |
| 0.00341833 | 0.018324386 | 0.025487235 | 0 | 0 |
| 0 | 0 | 0 | 0 | 0 |
| 0.299991507 | 0 | 0.778264158 | 0.040561033 | 0 |
| 0 | 0 | 0 | 0 | 0 |
| 0.031333097 | 0 | 0 | 0 | 0 |
| 0 | 0 | 0 | 0 | 0 |
| 0 | 0 | 0 | 0 | 0 |
| 0 | 0 | 0 | 0 | 0 |
| 0.046408012 | 0.021844774 | 3.317468608 | 0.001482284 | 0.007500717 |
| 0.001039702 | 0 | 0 | 0 | 0 |
| 0 | 0 | 0 | 0 | 0 |
| 0 | 0.003325825 | 0 | 0 | 0.054983483 |

| | | | | |
|---|---|---|---|---|
| 0 | 0 | 0 | 0 | 0 |
| 0 | 0 | 0 | 0 | 0 |
| 0 | 0 | 0 | 0 | 0 |
| 0.000764316 | 0 | 0 | 0 | 0 |
| 0 | 0 | 0 | 0 | 0 |
| 0.245039219 | 0 | 0 | 0 | 0 |
| 0.157105562 | 0 | 0.472184643 | 0.000691561 | 0 |
| 0.006681437 | 0.025781354 | 0.418320767 | 0.065100337 | 0.007240076 |
| 0 | 0 | 0 | 0 | 0 |
| 0 | 0 | 0 | 0 | 0 |
| 0.005958721 | 0 | 0.038238938 | 0.004690106 | 0.002008183 |
| 0 | 0 | 0 | 0 | 0 |
| 0.012856981 | 0.002291506 | 0.195306883 | 0 | 0 |
| 0.553511048 | 0 | 0 | 0 | 0 |
| 0 | 0 | 0 | 0 | 0 |
| 0 | 0 | 0 | 0 | 0 |
| 0.092274725 | 0 | 0 | 0 | 0 |
| 0.058817344 | 0 | 0 | 0 | 0.00121583 |
| 0.600272446 | 0 | 0 | 0.024856247 | 0 |
| 0.019971199 | 0 | 0 | 0.000704226 | 0 |
| 0.011017531 | 0 | 0 | 0 | 0 |
| 0 | 0 | 0 | 0 | 0 |
| 0.291265605 | 0 | 0.016991306 | 0 | 0 |
| 0 | 0 | 0 | 0 | 0 |
| 0 | 0 | 0 | 0 | 0 |
| 0 | 0 | 0 | 0 | 0 |
| 0 | 0 | 0 | 0 | 0 |
| 0 | 0 | 0 | 0 | 0 |
| 0 | 0 | 0 | 0 | 0 |
| 0.013482536 | 1.493274571 | 0 | 0 | 0 |
| 0.08581771 | 0 | 0 | 0 | 0.005607731 |
| 0 | 0 | 0 | 0 | 0 |
| 0.148082189 | 0 | 0 | 0 | 0 |
| 0 | 0 | 0 | 0 | 0 |
| 0.002030608 | 0 | 0 | 0 | 0 |
| 0 | 0 | 0 | 0 | 0 |
| 0.016458948 | 0.315581696 | 0 | 0 | 0 |
| 0 | 0.001022515 | 0.123485289 | 0.022034924 | 0.038635498 |
| 0.006742542 | 0 | 0 | 0 | 0 |
| 0 | 0 | 0 | 0 | 0 |
| 0.009465343 | 0.075790434 | 0 | 0.018997925 | 0 |
| 0 | 0 | 0 | 0 | 0 |
| 0.014012894 | 0 | 0 | 0 | 0 |
| 0.118438204 | 0 | 0 | 0 | 0 |
| 0 | 0 | 0 | 0 | 0 |
| 0 | 0 | 0 | 0 | 0 |
| 0.032180214 | 0 | 0 | 0 | 0 |
| 0 | 0 | 0 | 0 | 0 |

| | | | | |
|---|---|---|---|---|
| 0 | 0 | 0 | 0 | 0 |
| 0.029485404 | 0 | 0 | 0 | 0 |
| 0 | 0 | 0 | 0 | 0 |
| 0.266463069 | 0 | 0 | 0 | 0 |
| 0 | 0 | 0 | 0 | 0 |
| 0 | 0 | 0 | 0 | 0 |
| 0 | 0 | 0 | 0 | 0 |
| 0 | 0 | 0 | 0 | 0 |
| 0.00198764 | 0.046537247 | 0.822322983 | 0.118000191 | 0.066073157 |
| 0.001000399 | 0 | 0 | 0 | 0.003604442 |
| 0 | 0 | 0 | 0 | 0 |
| 0 | 0 | 0 | 0 | 0 |
| 0 | 0 | 0 | 0 | 0 |
| 0 | 0 | 0.106039591 | 0 | 0 |
| 0.360485059 | 0 | 0 | 0 | 1.760660027 |
| 0 | 0.001086895 | 0 | 1.163518339 | 0 |
| 0 | 0 | 0 | 0 | 0 |
| 0 | 0 | 0 | 0 | 0 |
| 0 | 0 | 0 | 0 | 0 |
| 0 | 0 | 0 | 0 | 0 |
| 0.058703665 | 0.057150702 | 0 | 0 | 0 |
| 0 | 0 | 0 | 0 | 0 |
| 0 | 0 | 0 | 0 | 0 |
| 0 | 0 | 0 | 0 | 0 |
| 0.011102689 | 0 | 0.018147986 | 0.107534721 | 0.002757102 |
| 5.879535756 | 0 | 0 | 1.667517908 | 2.198722642 |
| 0.229732988 | 0 | 1.259593733 | 0.000718501 | 0 |
| 0 | 0 | 0 | 0 | 0 |
| 0 | 0 | 2.901591596 | 0 | 0.040101883 |
| 0.419625887 | 0 | 0 | 0 | 0 |
| 0 | 0 | 0 | 0 | 0 |
| 0 | 0 | 0 | 0.026954743 | 0 |
| 0 | 0 | 0 | 0 | 0 |
| 0.007489957 | 0.002214201 | 0.073383723 | 0 | 0.008433235 |
| 0.016711883 | 0.000729661 | 0.029372781 | 0.00113326 | 0.002205604 |
| 0.010722819 | 0.00309469 | 0.017537676 | 0.034646607 | 0.002182733 |
| 0 | 2.659055696 | 0 | 0 | 0 |
| 0.137291452 | 0.909136835 | 11.16346687 | 2.502225546 | 0.243299381 |
| 0.001836262 | 0 | 0 | 0.035157423 | 0 |
| 0 | 0 | 0 | 0 | 0 |
| 0 | 0 | 0 | 0.00185158 | 0 |
| 0.000824391 | 0 | 0 | 0 | 0 |
| 0.100335269 | 0.05779564 | 0.221474701 | 0 | 0.65066413 |
| 0 | 0 | 0 | 0 | 0 |
| 0 | 0 | 0 | 0 | 0 |
| 0 | 0.00928413 | 0 | 0.002276761 | 0 |
| 0 | 0 | 0 | 0 | 0 |
| 0.021395463 | 0.031138383 | 0.892957968 | 0.001511314 | 0.007059337 |

| | | | | |
|---|---|---|---|---|
| 0.004090928 | 0 | 0 | 0.001352386 | 0.020530206 |
| 0 | 0 | 0 | 0 | 0 |
| 0 | 0 | 0 | 0 | 0 |
| 0 | 0 | 0 | 0 | 0 |
| 0 | 0.036025747 | 0.016941067 | 0 | 0 |
| 0 | 0 | 1.6031219 | 0 | 0 |
| 0 | 0 | 0 | 0 | 0 |
| 0 | 0 | 0 | 0 | 0 |
| 0 | 0 | 0 | 0 | 0 |
| 0 | 0 | 0 | 0 | 0 |
| 0.007479731 | 0 | 0 | 0 | 0 |
| 0.427990184 | 2.047482893 | 0 | 4.808321128 | 0.443712719 |
| 0.010603113 | 0 | 0.343518709 | 0 | 0 |
| 0 | 0 | 0 | 0 | 0 |
| 0.08973343 | 0.001559607 | 0.097178022 | 0 | 0 |
| 0 | 0 | 0 | 0 | 0 |
| 0 | 0 | 0.003271033 | 0 | 0.001827156 |
| 0 | 0 | 0 | 0 | 0 |
| 0 | 0 | 0.048541269 | 0 | 0.005312712 |
| 0.016480902 | 0 | 0.006093342 | 0 | 0 |
| 0 | 0 | 0 | 0.375153213 | 0 |
| 0 | 0 | 0 | 0 | 0 |
| 0.300891559 | 0 | 0 | 0.01636808 | 0 |
| 0 | 0 | 0 | 0 | 0 |
| 0 | 0 | 0 | 0 | 0 |
| 0 | 0 | 0 | 0 | 0 |
| 0 | 0 | 0 | 0 | 0 |
| 0 | 0 | 0 | 0 | 0 |
| 0 | 0 | 0 | 0 | 0 |
| 0 | 0 | 0 | 0 | 0 |
| 0 | 0 | 0 | 0 | 0 |
| 0.019254135 | 0 | 0.091186715 | 0.009766504 | 0.047192344 |
| 0 | 0 | 0 | 0 | 0 |
| 0.002595612 | 0 | 0.061135566 | 0.012618561 | 0 |
| 0.015229183 | 0.003554898 | 0.023034086 | 0.035306815 | 0.001018012 |
| 0.740671738 | 0 | 0 | 0.049729095 | 0.002843404 |
| 0 | 0 | 0 | 0 | 0 |
| 0 | 0 | 0 | 0 | 0 |
| 0 | 0 | 0 | 0 | 0 |
| 0 | 0 | 0 | 0 | 0 |
| 0 | 0 | 0.143356871 | 0 | 0 |
| 0 | 0 | 0 | 0 | 0 |
| 0 | 0 | 0 | 0 | 0 |
| 0 | 0 | 0 | 0.037082015 | 0 |
| 0.035070244 | 0.00258037 | 0.073907449 | 0.001288176 | 0.01169984 |
| 0 | 0 | 0 | 0 | 0 |
| 0.035572704 | 0 | 0 | 0 | 0 |
| 0.000921394 | 0.003660853 | 0 | 0.001218384 | 0 |

| | | | | |
|---|---|---|---|---|
| 0 | 0 | 0 | 0 | 0 |
| 0 | 0 | 0 | 0 | 0 |
| 0.031032206 | 0 | 0 | 0.006217383 | 0 |
| 0 | 0 | 0 | 0 | 0 |
| 0 | 0 | 0 | 0.601032579 | 0 |
| 0 | 0 | 0 | 0 | 0 |
| 0.108689659 | 0 | 0 | 0 | 0.018077207 |
| 0.047195716 | 0 | 1.707635763 | 0.002516459 | 0 |
| 0 | 0 | 0 | 0 | 0 |
| 0.023408461 | 0 | 0 | 0 | 0 |
| 0 | 0 | 0 | 0 | 0 |
| 0 | 0 | 0.017055838 | 0 | 0 |
| 0 | 0 | 0 | 0 | 0 |
| 0 | 0 | 0 | 0 | 0 |
| 0 | 0 | 0 | 0 | 0 |
| 0 | 0 | 0 | 0 | 0 |
| 0.048885116 | 0 | 0 | 0 | 0 |
| 0 | 0 | 0 | 0 | 0 |
| 0 | 0 | 0 | 0.660551462 | 0.065757508 |
| 0 | 0 | 0 | 0 | 0 |
| 0.018125703 | 0 | 0 | 0 | 0 |
| 0 | 0 | 0 | 0 | 0 |
| 0 | 0 | 0 | 0 | 0 |
| 0 | 0 | 0 | 0 | 0 |
| 0.002554486 | 0 | 0.006847248 | 0 | 0 |
| 0 | 0.003088578 | 0.240012909 | 0.009194197 | 0.005912859 |
| 0 | 0 | 0 | 0 | 0 |
| 0.894616989 | 0.001847452 | 0.882986795 | 0 | 0.002284544 |
| 0.760626861 | 0 | 0 | 0 | 0 |
| 0.848810928 | 0.05924181 | 0 | 1.44097052 | 0.010205857 |
| 0 | 0 | 0 | 0 | 0 |
| 0 | 0 | 0 | 0 | 0 |
| 0 | 0 | 0 | 0.002272331 | 0 |
| 0 | 0 | 0 | 0 | 0 |
| 0.025850735 | 0.001438506 | 0.006114033 | 0.016128069 | 0 |
| 0.116339305 | 0 | 0 | 0 | 0 |
| 0.287956028 | 0 | 0 | 0 | 0 |
| 0 | 0 | 0.020095225 | 0.010219835 | 0.003330472 |
| 0 | 0 | 0 | 0 | 0 |
| 0 | 0 | 0 | 0 | 0 |
| 1.528345423 | 2.861034212 | 2.150972195 | 9.952053214 | 17.90732986 |
| 0.030944794 | 0 | 0.47404067 | 1.022978496 | 0.52022523 |
| 3.193241413 | 0 | 0 | 0 | 0 |
| 0.004502448 | 0 | 0 | 0 | 0 |
| 0.070225301 | 0 | 0 | 0 | 0 |
| 0 | 0 | 0 | 0.876832956 | 0 |
| 0 | 0 | 0 | 0 | 0 |
| 0.008730113 | 0 | 0.11581781 | 0.002452171 | 0.000881082 |

| | | | | |
|---|---|---|---|---|
| 0 | 0 | 0 | 0 | 0 |
| 0.01373357 | 0.04680399 | 0.014646907 | 0 | 0 |
| 0 | 0 | 0 | 0 | 0 |
| 0 | 0 | 0 | 0 | 0 |
| 0.187306267 | 1.025551575 | 0.549862281 | 0 | 0.328924388 |
| 0 | 0 | 0 | 0 | 0 |
| 0 | 0 | 0 | 0 | 0 |
| 0 | 0 | 0 | 0 | 0 |
| 0 | 0.021621168 | 1.060432681 | 0.016467386 | 0.379165601 |
| 0 | 0 | 0 | 0 | 0 |
| 0 | 0 | 0.001103197 | 0 | 0 |
| 0 | 0 | 0 | 0 | 0 |
| 0.039349179 | 0 | 0.063759211 | 0 | 0 |
| 0.002103296 | 0.002984554 | 0.002099611 | 0.024567669 | 0.051062545 |
| 2.55769311 | 0 | 0 | 0 | 0 |
| 0.093373877 | 0.074616013 | 0.042690261 | 0.097567043 | 0 |
| 0.027119142 | 0.001721779 | 0.001615011 | 0 | 0.000780684 |
| 0 | 0 | 0 | 0 | 0 |
| 0 | 0 | 0 | 0 | 0 |
| 0 | 0 | 0 | 0 | 0 |
| 0 | 0 | 0 | 0 | 0 |
| 0 | 0 | 0 | 0 | 0 |
| 0 | 0 | 0 | 0 | 0 |
| 0 | 0 | 0 | 0 | 0 |
| 0 | 0 | 0 | 0 | 0 |
| 0 | 0 | 0 | 0 | 0 |
| 0.127243569 | 0 | 0 | 0.027729023 | 0 |
| 0 | 0 | 0.008913467 | 0.002558228 | 0 |
| 0 | 0 | 0 | 0 | 0 |
| 0.008084142 | 0 | 0 | 0 | 0 |
| 0 | 0 | 0 | 0 | 0 |
| 0 | 0 | 0 | 0 | 0 |
| 0 | 0 | 0 | 0 | 0 |
| 0.012542525 | 0 | 0 | 0 | 0 |
| 0.793754597 | 0 | 0 | 0 | 0 |
| 0.120644241 | 0.11471483 | 0.237153709 | 0 | 0 |
| 0 | 0 | 0.006323469 | 0 | 0 |
| 0.002926088 | 0 | 0 | 0 | 0 |
| 0 | 0 | 0 | 0 | 0 |
| 0 | 0 | 0 | 0 | 0 |
| 0.024036001 | 0 | 0 | 0 | 0 |
| 0 | 0 | 0.003526446 | 0 | 0 |
| 0 | 0 | 0 | 0 | 0 |
| 0 | 0 | 0 | 0 | 0 |
| 0 | 0 | 0 | 0 | 0 |
| 0 | 0.00089574 | 0 | 0 | 0 |
| 0 | 0 | 0 | 0 | 0 |
| 0 | 0 | 0 | 0 | 0 |

| | | | | |
|---|---|---|---|---|
| 0.989972678 | 0 | 0 | 0 | 0 |
| 0 | 0 | 0 | 0 | 0 |
| 0.001048701 | 0 | 0 | 0 | 0 |
| 0 | 0 | 0.003590119 | 0 | 0 |
| 0.001935603 | 0 | 0.039502977 | 0 | 0 |
| 0 | 0 | 0 | 0 | 0 |
| 0.015796164 | 0.000687037 | 0.021346863 | 0.011204128 | 0.0032709 |
| 0 | 0 | 0 | 0 | 0 |
| 0 | 0 | 0 | 0 | 0 |
| 0 | 0.018380745 | 0 | 0 | 0 |
| 0.038971402 | 0 | 0 | 0 | 0 |
| 0.05795547 | 0 | 0 | 0 | 0 |
| 0 | 0 | 0 | 0 | 0 |
| 0 | 0 | 0 | 0 | 0 |
| 0 | 0 | 0 | 0 | 0 |
| 0 | 0 | 0 | 0 | 0 |
| 0 | 0 | 0 | 7.478371786 | 0 |
| 0 | 0.001321028 | 0 | 0.003269948 | 0 |
| 0 | 0 | 0 | 0 | 0 |
| 0 | 0 | 0 | 0 | 0 |
| 0.05617953 | 0.010956132 | 0.027992612 | 0 | 0.007711053 |
| 0 | 0 | 0 | 0 | 0 |
| 0.011630509 | 0.005591371 | 0 | 0 | 0 |
| 0.015089102 | 0.001122688 | 0 | 0.004110113 | 0.002763392 |
| 0 | 0 | 0 | 0 | 0 |
| 0 | 0 | 0.02307288 | 0 | 0.000857943 |
| 0 | 0 | 0 | 0 | 0 |
| 0 | 0 | 0 | 0 | 0 |
| 0 | 0 | 0 | 0 | 0 |
| 0 | 0.031392908 | 0 | 1.083327697 | 0.138363635 |
| 0 | 0 | 0 | 0 | 0 |
| 0 | 0 | 0 | 0 | 0 |
| 0 | 0 | 0 | 0 | 0 |
| 0 | 0 | 0 | 1.34793027 | 0 |
| 0 | 0 | 0 | 0 | 0 |
| 0 | 0 | 0 | 0 | 0 |
| 0 | 0 | 0.022117919 | 0.000935057 | 0 |
| 0 | 0 | 0 | 0 | 0 |
| 0 | 0 | 0 | 0 | 0 |
| 0 | 0 | 0 | 0 | 0 |
| 0 | 0 | 0 | 0 | 0 |
| 0 | 0 | 0 | 0 | 0 |
| 0 | 0 | 0 | 0 | 0 |
| 0 | 0.002872487 | 0 | 0 | 0 |
| 0 | 0 | 0 | 0 | 0 |
| 0 | 0 | 0 | 0 | 0 |
| 0 | 0 | 0 | 0 | 0 |
| 0 | 0 | 0 | 0 | 0 |

| | | | | |
|---|---|---|---|---|
| 0.002554042 | 0.002243526 | 0.028530863 | 0.007840112 | 0 |
| 0 | 0 | 0 | 0 | 0 |
| 0 | 0 | 0 | 0 | 0 |
| 0 | 0 | 0 | 0 | 0 |
| 0 | 0 | 0 | 0 | 0 |
| 0 | 0 | 0 | 0 | 0 |
| 0 | 0 | 0.003888862 | 0 | 0 |
| 0.780171074 | 0.021618342 | 3.496878802 | 0.861146006 | 0.374569741 |
| 0.289217276 | 0.009481098 | 0.288392876 | 0.800220661 | 0.064360547 |
| 0 | 0.058272595 | 0.429401156 | 0 | 0.025310193 |
| 0 | 0 | 0 | 0 | 0 |
| 0 | 0 | 0 | 0 | 0 |
| 0 | 0 | 0 | 0 | 0 |
| 0 | 0 | 0 | 0 | 0 |
| 0 | 0 | 0 | 0 | 0 |
| 0.195641744 | 0 | 0 | 0 | 0 |
| 0 | 0 | 0 | 0 | 0.000913794 |
| 0 | 0 | 0 | 0 | 0 |
| 0 | 0 | 0 | 0 | 0 |
| 0.04595522 | 0 | 0.003300338 | 0 | 0 |
| 0 | 0 | 0 | 0 | 0 |
| 0 | 0 | 0 | 0 | 0 |
| 0.001076357 | 0 | 0 | 0 | 0 |
| 0 | 0 | 0 | 0 | 0 |
| 0.005603916 | 0 | 0 | 0 | 0 |
| 0 | 0 | 0 | 0 | 0 |
| 0 | 0 | 0 | 0 | 0 |
| 0 | 0 | 0 | 0 | 0 |
| 0 | 0 | 0 | 0 | 0 |
| 0 | 0 | 0 | 0 | 0 |
| 0.005247677 | 0 | 0 | 0 | 0 |
| 0 | 0 | 0 | 0 | 0 |
| 0 | 0 | 0 | 0 | 0 |
| 0.043517935 | 0 | 0 | 0 | 0 |
| 0.059190235 | 0 | 0 | 0 | 0 |
| 0 | 0 | 0 | 0 | 0 |
| 0 | 0 | 0 | 0 | 0 |
| 0 | 0 | 0 | 0 | 0 |
| 0 | 0 | 0 | 0 | 0 |
| 0 | 0 | 0.655102944 | 0 | 0 |
| 0 | 0 | 0 | 0 | 0 |
| 0.047536943 | 0 | 1.878972624 | 0.00124228 | 0 |
| 0 | 0 | 0 | 0 | 0 |
| 0 | 0 | 0 | 0 | 0 |
| 0 | 0 | 0.015944806 | 0 | 0 |
| 0 | 0 | 0 | 0 | 0 |
| 0 | 0 | 0 | 0 | 0 |
| 0 | 0.012053356 | 0 | 0 | 0 |

| | | | | |
|---|---|---|---|---|
| 0 | 0 | 0 | 0 | 0 |
| 0.012842892 | 0.00295944 | 0.002556773 | 0.034110159 | 0 |
| 0 | 0 | 0 | 0 | 0 |
| 0 | 0 | 0 | 0 | 0 |
| 0.81861714 | 0.022648536 | 0 | 0.052510612 | 0.093608008 |
| 0 | 0 | 0 | 0 | 0 |
| 0 | 0 | 0 | 0 | 0 |
| 0.001392397 | 0.018001694 | 0.097296997 | 0.03094246 | 0.10869762 |
| 0.005329318 | 0 | 0 | 0 | 0 |
| 0 | 0 | 0 | 0 | 0 |
| 0 | 0 | 0 | 0 | 0 |
| 0 | 0 | 0 | 0 | 0 |
| 0 | 0 | 0 | 0 | 0 |
| 0.000892732 | 0 | 0 | 0 | 0 |
| 0 | 0 | 0 | 0 | 0 |
| 0.041058926 | 0 | 0 | 0 | 0 |
| 0 | 0 | 0 | 0 | 0 |
| 0 | 0 | 0 | 0 | 0 |
| 0 | 0 | 0 | 0 | 0 |
| 0 | 0 | 0 | 0 | 0 |
| 0 | 0 | 0 | 0 | 0 |
| 0 | 0 | 0 | 0 | 0 |
| 0 | 0 | 0 | 0 | 0 |
| 0.002209176 | 0.00075995 | 0.035864044 | 0.003540913 | 0.001378298 |
| 0 | 0 | 0 | 9.788353768 | 0 |
| 0 | 0 | 0 | 0 | 0 |
| 0.187910393 | 0.011120975 | 0 | 0.24349971 | 0 |
| 0 | 0 | 0 | 0 | 0 |
| 0 | 0 | 0.014838702 | 0 | 0 |
| 0 | 0 | 0 | 0 | 0 |
| 0 | 0 | 0 | 0 | 0 |
| 0 | 0 | 0 | 0 | 0 |
| 0 | 0 | 0 | 0 | 0 |
| 0 | 0 | 0 | 0 | 0 |
| 0 | 0 | 0 | 0 | 0 |
| 0 | 0 | 0 | 0 | 0 |
| 0 | 0 | 0 | 0 | 0 |
| 0.074787007 | 0.2134913 | 0 | 0 | 0 |
| 0 | 0 | 0 | 0 | 0 |
| 0 | 0 | 0 | 0 | 0 |
| 0 | 0 | 0 | 0 | 0 |
| 0.419912993 | 0 | 0 | 0 | 0 |
| 0 | 0 | 0 | 0 | 0 |
| 0 | 0 | 0 | 0 | 0 |
| 0 | 0 | 0 | 0 | 0 |
| 0 | 0 | 0 | 0 | 0 |
| 0 | 0 | 0 | 0 | 0 |
| 0 | 0 | 0 | 0 | 0 |

| | | | | |
|---|---|---|---|---|
| 0.404385507 | 0 | 0 | 0 | 0.022820779 |
| 0 | 0 | 0 | 0 | 0 |
| 0 | 0 | 0 | 0 | 0 |
| 0 | 0 | 0 | 0 | 0 |
| 0 | 0 | 0 | 0 | 0 |
| 0 | 0 | 0.869261418 | 0.001102655 | 0.020258603 |
| 0 | 0 | 0 | 0 | 0 |
| 0 | 0 | 0 | 0 | 0 |
| 0 | 0 | 0 | 0 | 0 |
| 0 | 0 | 0 | 0 | 0 |
| 0 | 0 | 0 | 0 | 0 |
| 0 | 0 | 0.09630792 | 0.001428068 | 0.00238232 |
| 0 | 0 | 0 | 0 | 0 |
| 0 | 0 | 0 | 0 | 0 |
| 0 | 0 | 0 | 0 | 0 |
| 0 | 0 | 0 | 0 | 0 |
| 0 | 0 | 0 | 0 | 0 |
| 0 | 0 | 0 | 0 | 0 |
| 0 | 0 | 0 | 0 | 0 |
| 0 | 0 | 0 | 0 | 0 |
| 0 | 0.000945967 | 0.059893248 | 0 | 0.005318578 |
| 0 | 0 | 0 | 0 | 0 |
| 0.05859824 | 0 | 0.042302824 | 0.038965681 | 0.011700541 |
| 0.003467597 | 0 | 0.623586521 | 0 | 0 |
| 0 | 0 | 0 | 0 | 0 |
| 0 | 0 | 0 | 0 | 0 |
| 0 | 0 | 0 | 0 | 0 |
| 0 | 0 | 0 | 0 | 0 |
| 0 | 0 | 0 | 0 | 0 |
| 0 | 0 | 0 | 0 | 0 |
| 0 | 0 | 0 | 0 | 0 |
| 0 | 0 | 0 | 0 | 0 |
| 0.001655772 | 0 | 0.045125988 | 0.011729314 | 0.009942924 |
| 0 | 0 | 0 | 0 | 0 |
| 0.009944395 | 0 | 0.416322547 | 0 | 0 |
| 0 | 0 | 0.000842133 | 0 | 0 |
| 0 | 0 | 0 | 0 | 0 |
| 0 | 0 | 0 | 0 | 0 |
| 0.00483006 | 0 | 0.152196858 | 0 | 0.00150673 |
| 0 | 0 | 0 | 0 | 0 |
| 0 | 0 | 0 | 0 | 0 |
| 0 | 0 | 0 | 0 | 0 |
| 1.744909612 | 0 | 0 | 0.485434459 | 0.002862955 |
| 0 | 0 | 0 | 0 | 0 |
| 0 | 0 | 0 | 0 | 0 |
| 1.200639358 | 0 | 0.001582942 | 0.001347969 | 0 |
| 0 | 0 | 0 | 0 | 0 |
| 0 | 0 | 0 | 0 | 0 |

| | | | | |
|---|---|---|---|---|
| 0 | 0 | 0 | 0 | 0 |
| 0 | 0 | 0 | 0 | 0 |
| 0 | 0.128683856 | 0 | 0 | 0 |
| 0 | 0 | 0 | 0 | 0 |
| 0 | 0 | 0 | 0 | 0 |
| 0 | 0 | 0 | 0 | 0 |
| 0 | 0 | 0 | 0 | 0 |
| 0 | 0 | 0.34766146 | 0 | 0 |
| 0.014708161 | 0 | 0 | 0 | 0 |
| 0.014909461 | 0 | 0 | 0 | 0 |
| 0 | 0 | 0 | 0 | 0 |
| 0 | 0 | 0 | 0 | 0 |
| 0 | 0 | 0 | 0 | 0 |
| 0 | 0 | 0 | 0 | 0 |
| 0 | 0 | 0 | 0 | 0 |
| 0 | 0 | 0.114123495 | 0.000683013 | 0.009925525 |
| 0 | 0 | 0 | 0 | 0 |
| 0 | 0 | 0 | 0 | 0 |
| 0 | 0 | 0 | 0 | 0 |
| 0.48226783 | 0.011618273 | 0 | 0 | 0 |
| 0.228520307 | 0 | 0 | 0 | 0 |
| 0 | 0 | 0 | 0 | 0 |
| 0 | 0 | 0 | 0 | 0 |
| 0 | 0 | 0 | 0 | 0 |
| 0 | 0 | 0.022919339 | 0 | 0.000958187 |
| 0.033047158 | 0 | 0 | 0 | 0 |
| 0 | 0 | 0 | 0 | 0 |
| 0 | 0 | 0 | 0 | 0 |
| 0 | 0 | 0 | 0 | 0 |
| 0.023983086 | 0 | 0 | 0 | 0 |
| 0.070529025 | 0.375061332 | 0.414051036 | 0.309209404 | 0.014837864 |
| 0 | 0 | 0 | 0 | 0 |
| 0 | 0 | 0 | 0 | 0 |
| 0 | 0 | 0 | 0 | 0 |
| 0 | 0 | 0 | 0 | 0 |
| 0 | 0 | 0 | 0 | 0 |
| 0 | 0 | 0 | 0 | 0 |
| 0 | 0 | 0 | 0 | 0 |
| 0 | 0 | 0 | 0 | 0 |
| 0 | 0 | 0 | 0 | 0 |
| 0 | 0 | 0 | 0 | 0 |
| 0 | 0 | 0 | 0 | 0 |
| 0.001634643 | 0 | 0 | 0 | 0 |
| 0 | 0 | 0 | 0 | 0.000946596 |
| 0 | 0 | 0 | 0 | 0 |
| 0 | 0 | 0 | 0 | 0 |
| 0 | 0.026410776 | 2.809956783 | 0.166175385 | 0 |
| 0.001547005 | 0 | 0 | 0 | 0 |

| | | | | |
|---|---|---|---|---|
| 0 | 0 | 0 | 0 | 0 |
| 0 | 0 | 0 | 0 | 0 |
| 0 | 0 | 0 | 0 | 0 |
| 0 | 0 | 0 | 0 | 0 |
| 0.083587089 | 0.020478201 | 0 | 0.107778425 | 0.078483375 |
| 0 | 0 | 0 | 0 | 0 |
| 0 | 0 | 0 | 0 | 0 |
| 0 | 0 | 0 | 0 | 0 |
| 0 | 0 | 0 | 0 | 0 |
| 0 | 0 | 0 | 0 | 0 |
| 0 | 0 | 0 | 0 | 0 |
| 0.016672376 | 0.00186773 | 0.019952328 | 0.011603334 | 0.004892973 |
| 0 | 0 | 0.003988571 | 0 | 0.001136532 |
| 0 | 0 | 0 | 0 | 0 |
| 0 | 0 | 0 | 0.007921452 | 0 |
| 0.005117817 | 0 | 0 | 0.123505556 | 0.004244016 |
| 0.064790075 | 0.000638447 | 0 | 0 | 0 |
| 0 | 0 | 0 | 0 | 0 |
| 0.000996665 | 0.009271233 | 0.001105465 | 0 | 0 |
| 0.004771581 | 0.00225694 | 0.0512407 | 0.009081987 | 0 |
| 0 | 0 | 0 | 0 | 0 |
| 0 | 0 | 0 | 0 | 0 |
| 0 | 0 | 0 | 0 | 0 |
| 0 | 0 | 0 | 0 | 0 |
| 0 | 0 | 0 | 0 | 0 |
| 0.001716067 | 0 | 0 | 0 | 0 |
| 0.000695216 | 0.042090811 | 0 | 0.020301266 | 0 |
| 0 | 0 | 0 | 0 | 0 |
| 0 | 0 | 0 | 0 | 0 |
| 0 | 0 | 0 | 0 | 0 |
| 0 | 0.001863756 | 0.00961501 | 0 | 0 |
| 0 | 0 | 0 | 0.003048406 | 0 |
| 0 | 0 | 0 | 0 | 0 |
| 0 | 0 | 0 | 0 | 0 |
| 0 | 0 | 0 | 0 | 0 |
| 0 | 0 | 0 | 0 | 0 |
| 0 | 0 | 0 | 0 | 0 |
| 0.016056447 | 0.00759464 | 0.117755892 | 0.125530411 | 0.018286381 |
| 0.751860089 | 0 | 0 | 0 | 0 |
| 0 | 0.004361869 | 0 | 0.002177538 | 0 |
| 0.003731007 | 0 | 0.059315614 | 0.002893167 | 0.000355631 |
| 0 | 0 | 0 | 0 | 0 |
| 0 | 0 | 0 | 0 | 0 |
| 0 | 0 | 0.110643923 | 0 | 0 |
| 0 | 0 | 0 | 0 | 0 |
| 0 | 0 | 0 | 0 | 0 |
| 0 | 0 | 0 | 0 | 0 |
| 0 | 0 | 0 | 0 | 0 |

| | | | | |
|---|---|---|---|---|
| 0 | 0 | 0 | 0 | 0 |
| 0.232449393 | 0 | 0 | 0 | 0 |
| 0 | 0 | 0 | 0 | 0 |
| 0.005162617 | 0.001424441 | 0.009543649 | 0.000948148 | 0.000738132 |
| 0 | 0 | 0.007523844 | 0.125093567 | 0.002224969 |
| 0 | 0 | 0 | 0 | 0 |
| 0 | 0 | 0 | 0 | 0 |
| 0 | 0 | 0 | 0.102702466 | 0.000865925 |
| 0 | 0.038683165 | 0 | 0 | 0 |
| 0 | 0 | 0 | 0 | 0 |
| 0 | 0 | 0 | 0 | 0 |
| 0 | 0 | 0 | 0 | 0 |
| 0.573637351 | 0.000576987 | 0 | 0.100311475 | 0.016481804 |
| 0 | 0 | 0.002148709 | 0 | 0 |
| 0 | 0 | 0 | 0 | 0 |
| 0 | 0 | 0 | 0 | 0 |
| 0 | 0 | 0 | 0 | 0 |
| 0 | 0 | 0 | 0 | 0 |
| 0 | 0 | 0 | 0 | 0 |
| 0 | 0 | 0 | 0 | 0 |
| 0 | 0 | 0 | 0 | 0 |
| 0 | 0 | 0 | 0 | 0 |
| 0 | 0.006457955 | 0.058125237 | 0.013938839 | 0.011712589 |
| 0 | 0.000896542 | 0.095657777 | 0.018014816 | 0.008333405 |
| 0 | 0 | 0 | 0 | 0 |
| 0.00944872 | 0 | 0 | 0 | 0 |
| 0.003644797 | 0 | 0 | 0 | 0 |
| 0 | 0 | 0 | 0 | 0 |
| 0 | 0 | 0 | 0 | 0 |
| 0 | 0 | 0 | 0 | 0 |
| 0.064027653 | 0.002329603 | 0 | 0.103858321 | 0 |
| 0 | 0 | 0 | 0 | 0 |
| 0 | 0 | 0 | 0 | 0 |
| 0 | 0 | 0 | 0 | 0 |
| 0.00075076 | 0 | 0 | 0 | 0 |
| 0 | 0 | 0 | 0 | 0 |
| 0 | 0.021150281 | 0 | 0.002096758 | 0 |
| 0 | 0 | 0 | 0 | 0 |
| 0.00568236 | 0 | 0.128784563 | 0 | 0 |
| 0 | 0 | 0 | 0 | 0 |
| 0 | 0 | 0 | 0 | 0 |
| 0 | 0 | 0 | 0 | 0 |
| 0 | 0 | 0 | 0 | 0 |
| 0 | 0 | 0 | 0 | 0 |
| 0 | 0 | 0 | 0 | 0 |
| 0.33913535 | 0.003941272 | 0.480593417 | 0.138280607 | 0.259049298 |
| 0 | 0 | 0 | 0 | 0 |
| 0 | 0 | 0 | 0 | 0 |

| | | | | |
|---|---|---|---|---|
| 0.044076009 | 0 | 0 | 0 | 0 |
| 0 | 0 | 0 | 0 | 0 |
| 0 | 0 | 0 | 0 | 0 |
| 0 | 0 | 0 | 0 | 0 |
| 0 | 0 | 0 | 0 | 0 |
| 0 | 0 | 0 | 0 | 0 |
| 0 | 0 | 0 | 0 | 0 |
| 0 | 0 | 0 | 0 | 0 |
| 0 | 0 | 0 | 0 | 0 |
| 0 | 0 | 0 | 0 | 0 |
| 0 | 0 | 0.029485855 | 0 | 0 |
| 0.005506096 | 0 | 0.001781256 | 0 | 0 |
| 0 | 0 | 0 | 0 | 0 |
| 0 | 0 | 0 | 0 | 0 |
| 0 | 0 | 0 | 0 | 0 |
| 0 | 0 | 0 | 0 | 0 |
| 0.103904466 | 0 | 0 | 0 | 0 |
| 0 | 0 | 0 | 0 | 0 |
| 0 | 0 | 0 | 0 | 0 |
| 0.058078641 | 0.001735009 | 0.001808246 | 0 | 0 |
| 0 | 0 | 0 | 0 | 0 |
| 0 | 0 | 0 | 0 | 0 |
| 0 | 0 | 0 | 0 | 0 |
| 0 | 0 | 0 | 0 | 0 |
| 0 | 0 | 0 | 0 | 0 |
| 0 | 0 | 0 | 0 | 0 |
| 0 | 0 | 0 | 0 | 0 |
| 0.001353133 | 0 | 0 | 0 | 0 |
| 0 | 0 | 0 | 0 | 0 |
| 0 | 0 | 0 | 0 | 0 |
| 0.106794013 | 0.000662468 | 0.029127563 | 0.004822972 | 0.002102619 |
| 0 | 0 | 0 | 0 | 0 |
| 0.003862255 | 0.989950851 | 0.008342283 | 0.029567797 | 0.29226511 |
| 0 | 0 | 0.004377656 | 0 | 0.0007128 |
| 0 | 0 | 0 | 0 | 0 |
| 0 | 0 | 0 | 0 | 0 |
| 0 | 0 | 0 | 0 | 0 |
| 0 | 0 | 0 | 0 | 0 |
| 0 | 0 | 0 | 0 | 0 |
| 0 | 0 | 0 | 0 | 0 |
| 0 | 0 | 0 | 0 | 0 |
| 0 | 0.065812007 | 1.129261586 | 0.050124568 | 0.066423332 |
| 0.042173753 | 0.093669001 | 0 | 0.239773294 | 0.021672508 |
| 0 | 0 | 0 | 0 | 0 |
| 0 | 0 | 0 | 0 | 0 |
| 1.039581072 | 0 | 0 | 0.011030551 | 0.005556469 |
| 0 | 0 | 0.00706526 | 0.001276225 | 0.004305339 |
| 0 | 0 | 0 | 0 | 0 |

| | | | | |
|---|---|---|---|---|
| 0 | 0 | 0 | 0 | 0 |
| 0.00256131 | 0 | 0 | 0 | 0 |
| 0.002243911 | 0 | 0.019081299 | 0.01428644 | 0.007272061 |
| 0 | 0 | 0 | 0 | 0 |
| 0.025680382 | 0.046892007 | 0 | 0 | 0 |
| 0 | 0.064708727 | 0 | 0 | 0.171718203 |
| 0 | 0 | 0 | 0 | 0 |
| 0 | 0 | 0 | 0 | 0 |
| 0 | 0 | 0 | 0 | 0 |
| 0 | 0 | 0 | 0 | 0 |
| 0 | 0 | 0 | 0 | 0 |
| 0 | 0 | 0 | 0 | 0 |
| 0 | 0 | 0 | 0 | 0 |
| 0 | 0 | 0 | 0.04526139 | 0 |
| 0 | 0 | 0 | 0 | 0 |
| 0 | 0 | 0 | 0 | 0 |
| 0 | 0 | 0 | 0 | 0 |
| 0 | 0 | 0 | 0 | 0 |
| 0 | 0 | 0 | 0 | 0 |
| 0 | 0 | 0 | 0 | 0 |
| 0 | 0 | 0 | 0 | 0 |
| 0 | 0 | 0 | 0 | 0 |
| 0 | 0 | 0 | 0 | 0 |
| 0 | 0 | 0 | 0 | 0 |
| 0.017671067 | 0 | 0 | 0 | 0.15917236 |
| 0 | 0 | 0.002292704 | 0.157166088 | 0.005851694 |
| 0 | 0 | 0 | 0 | 0 |
| 0.754124039 | 0.047799979 | 0 | 0 | 0 |
| 0 | 0 | 0 | 0 | 0 |
| 0 | 0 | 0 | 0 | 0 |
| 0 | 0 | 0 | 0 | 0 |
| 0 | 0 | 0.003015848 | 0 | 0 |
| 0 | 0 | 0 | 0 | 0 |
| 0 | 0 | 0 | 0 | 0 |
| 0 | 0 | 0 | 0 | 0 |
| 0 | 0 | 0 | 0 | 0 |
| 0 | 0 | 0 | 0 | 0 |
| 0 | 0 | 0.219775262 | 0.004534656 | 0 |
| 0.111852738 | 1.116528132 | 0 | 2.096367707 | 2.0498051 |
| 0 | 0 | 0 | 0 | 0 |
| 0.00375901 | 0 | 0 | 0 | 0 |
| 0.00246668 | 0 | 0.010197643 | 0 | 0 |
| 0.001244709 | 0.001962476 | 0.018637924 | 0.007315169 | 0 |
| 0 | 0 | 0 | 0 | 0 |
| 0 | 0 | 0 | 0 | 0 |
| 0 | 0 | 0 | 0 | 0 |
| 0.002856243 | 0.002401764 | 0 | 0.006294812 | 0.001089001 |
| 0 | 0 | 0 | 0 | 0 |

| | | | | |
|---|---|---|---|---|
| 0 | 0 | 0 | 0 | 0 |
| 0.018821165 | 0.005303522 | 0.220305903 | 0.005957171 | 0.007557652 |
| 0 | 0 | 0 | 0 | 0 |
| 0 | 0 | 0 | 0 | 0 |
| 0 | 0 | 0 | 0 | 0 |
| 0 | 0 | 0 | 0 | 0 |
| 0 | 0 | 0 | 0 | 0 |
| 0 | 0 | 0 | 0 | 0 |
| 0 | 0 | 0 | 0 | 0 |
| 0 | 0 | 0 | 0 | 0 |
| 0 | 0 | 0 | 0 | 0 |
| 0 | 0 | 0 | 0 | 0 |
| 0.935095022 | 0 | 0 | 0 | 0 |
| 0 | 0 | 0 | 0 | 0 |
| 0.005991614 | 0.011099877 | 0.015229692 | 0 | 0 |
| 0 | 0 | 0 | 0 | 0 |
| 0 | 0 | 0 | 0 | 0 |
| 0 | 0 | 0 | 0 | 0 |
| 0 | 0.002873888 | 0 | 0 | 0 |
| 0 | 0 | 0 | 0 | 0 |
| 0 | 0 | 0 | 0 | 0 |
| 0.001961413 | 0 | 0.089468615 | 0 | 0 |
| 0 | 0.013366743 | 0 | 0.015362645 | 0.012767894 |
| 0 | 0 | 0 | 0 | 0 |
| 0 | 0 | 0 | 0 | 0 |
| 0 | 0 | 0 | 0 | 0 |
| 0 | 0 | 0 | 0 | 0 |
| 0 | 0 | 0 | 0 | 0 |
| 0 | 0 | 0 | 0 | 0 |
| 0 | 0 | 0 | 0 | 0 |
| 0 | 0 | 0 | 0 | 0 |
| 0 | 0 | 0 | 0 | 0 |
| 0 | 0 | 0 | 0 | 0 |
| 0 | 0 | 0 | 0 | 0 |
| 15.83690814 | 0.003579933 | 9.534821716 | 0 | 0 |
| 0 | 0 | 0 | 0 | 0 |
| 0 | 0 | 0 | 0 | 0 |
| 0 | 0 | 0 | 0 | 0 |
| 0.001469031 | 0 | 0.04426527 | 0 | 0 |
| 0 | 0 | 0 | 0 | 0 |
| 0.064980905 | 0.045272894 | 2.708150273 | 0.060719113 | 0.095857789 |
| 0 | 0 | 0 | 0 | 0 |
| 0 | 0 | 0 | 0 | 0 |
| 0 | 0 | 0 | 0 | 0 |
| 0 | 0 | 0 | 0 | 0 |
| 0.004021552 | 0.006657627 | 1.399501145 | 0.228499535 | 1.475704148 |
| 0 | 0 | 0 | 0 | 0 |
| 0 | 0 | 0 | 0 | 0 |

| | | | | |
|---|---|---|---|---|
| 0 | 0 | 0 | 0 | 0 |
| 0.000614079 | 0 | 0 | 0 | 0 |
| 0 | 0 | 0 | 0 | 0 |
| 0.024287501 | 0 | 0 | 0 | 0 |
| 0 | 0 | 0.008246058 | 0 | 0 |
| 0 | 0 | 0 | 0 | 0 |
| 0 | 0 | 0 | 0 | 0 |
| 0 | 0 | 0 | 0 | 0 |
| 0 | 0.003060415 | 0.293862639 | 0 | 0 |
| 0.001229089 | 0 | 0.005180393 | 0 | 0 |
| 0 | 0 | 0 | 0 | 0 |
| 0 | 0 | 0 | 0 | 0 |
| 0.012493841 | 0 | 0.009327312 | 0.006354209 | 0.003710058 |
| 0 | 0 | 0 | 0 | 0 |
| 0 | 0 | 0 | 0 | 0 |
| 0 | 0 | 0 | 0 | 0 |
| 0 | 0 | 0 | 0 | 0 |
| 0 | 0 | 0 | 0 | 0 |
| 0 | 0 | 0 | 0 | 0 |
| 0.557303132 | 0.064939631 | 0.441190182 | 5.427110498 | 0.07356598 |
| 0 | 0 | 0 | 0 | 0 |
| 0 | 0 | 0 | 0 | 0 |
| 0 | 0 | 0 | 0 | 0 |
| 0 | 0 | 0 | 0 | 0 |
| 0 | 0 | 0 | 0 | 0 |
| 0 | 0 | 0 | 0 | 0 |
| 0 | 0 | 0 | 0 | 0 |
| 0 | 0 | 0 | 0 | 0 |
| 0 | 0 | 0 | 0 | 0 |
| 0 | 0 | 0 | 0 | 0 |
| 0 | 0 | 0 | 0 | 0 |
| 0 | 0 | 0 | 0 | 0 |
| 0 | 0 | 0 | 0 | 0 |
| 0 | 0 | 0 | 0 | 0 |
| 0 | 0 | 0 | 0 | 0 |
| 0 | 0 | 0.007676217 | 0 | 0 |
| 0 | 0 | 0 | 0 | 0 |
| 0 | 0 | 0 | 0 | 0 |
| 0 | 0 | 0 | 0 | 0 |
| 0 | 0 | 0 | 0 | 0 |
| 0 | 0 | 0 | 0 | 0 |
| 0 | 0 | 0 | 0 | 0 |
| 0.009599243 | 0 | 0 | 0.001371194 | 0 |
| 0 | 0 | 0 | 0 | 0 |
| 0.008436095 | 0.001900115 | 0.096466355 | 0.000885339 | 0.002756943 |
| 0.004554938 | 0 | 0 | 0 | 0 |
| 0 | 0 | 0 | 0 | 0 |
| 0 | 0 | 0 | 0.015066299 | 0 |

| | | | | |
|---|---|---|---|---|
| 0 | 0 | 0 | 0 | 0 |
| 0 | 0 | 0 | 0 | 0 |
| 0 | 0 | 0 | 0 | 0 |
| 0 | 0 | 0 | 0 | 0 |
| 0.003815231 | 0 | 0 | 0 | 0 |
| 0.037037489 | 0.002411466 | 0.103616388 | 0.570060998 | 0.000514541 |
| 0 | 0 | 0 | 0 | 0 |
| 0 | 0 | 0 | 0 | 0 |
| 0 | 0 | 0 | 0 | 0 |
| 0 | 0 | 0 | 0 | 0 |
| 0 | 0 | 0 | 0 | 0 |
| 0 | 0 | 0 | 0 | 0 |
| 0 | 0 | 0 | 0 | 0 |
| 0 | 0 | 0 | 0 | 0 |
| 0 | 0 | 0 | 0 | 0 |
| 0 | 0 | 0 | 0 | 0 |
| 0.025100475 | 0 | 0 | 0.000601287 | 0 |
| 0 | 0 | 0.014338102 | 0 | 0 |
| 0 | 0 | 0 | 0 | 0 |
| 0.551574403 | 0 | 0 | 0 | 0 |
| 0 | 0.000822372 | 0 | 0 | 0.000596604 |
| 0 | 0 | 0 | 0 | 0 |
| 0 | 0 | 0 | 0 | 0 |
| 0 | 0 | 0 | 0 | 0 |
| 0 | 0 | 0 | 0 | 0 |
| 0 | 0 | 0 | 0 | 0 |
| 0 | 0 | 0 | 0 | 0 |
| 0 | 0 | 0 | 0 | 0 |
| 0 | 0 | 0 | 0 | 0 |
| 0 | 0 | 0 | 0 | 0 |
| 0 | 0 | 0 | 0 | 0 |
| 0 | 0 | 0.034310547 | 0 | 0 |
| 0 | 0 | 0 | 0 | 0 |
| 0 | 0 | 0 | 0 | 0 |
| 0 | 0 | 0 | 0 | 0 |
| 0 | 0 | 0 | 0 | 0 |
| 0 | 0 | 0 | 0 | 0 |
| 0 | 0 | 0 | 0 | 0 |
| 0 | 0 | 0 | 0 | 0 |
| 0 | 0 | 0 | 0 | 0 |
| 0 | 0 | 0 | 0 | 0 |
| 0 | 0 | 0 | 0 | 0 |
| 0 | 0 | 0 | 0 | 0 |
| 0 | 0 | 0 | 0 | 0 |
| 0 | 0 | 0 | 0 | 0 |
| 0 | 0 | 0 | 0 | 0 |
| 0 | 0 | 0 | 0 | 0 |

| | | | | |
|---|---|---|---|---|
| 0 | 0 | 0 | 0 | 0 |
| 0.094852415 | 0 | 0 | 0 | 0 |
| 0 | 0 | 0 | 0 | 0 |
| 0 | 0 | 0 | 0 | 0 |
| 0 | 0 | 0 | 0 | 0 |
| 0 | 0 | 0.003319485 | 0 | 0 |
| 0 | 0 | 0 | 0 | 0 |
| 0 | 0 | 0 | 0 | 0 |
| 0 | 0 | 0 | 0 | 0 |
| 0 | 0 | 0 | 0 | 0 |
| 0.025719648 | 0 | 0.013897921 | 0 | 0 |
| 0 | 0 | 0 | 0 | 0 |
| 0 | 0 | 0 | 0 | 0 |
| 0 | 0 | 0 | 0 | 0 |
| 0 | 0 | 0 | 0 | 0 |
| 0 | 0 | 0 | 0 | 0 |
| 0 | 0 | 0 | 0 | 0 |
| 0 | 0 | 0 | 0 | 0 |
| 0 | 0 | 0 | 0 | 0 |
| 0 | 0 | 0.001623109 | 0 | 0 |
| 0 | 0.000844242 | 0.033902821 | 0 | 0.004019336 |
| 0.00650698 | 0 | 0 | 0 | 0 |
| 0 | 0 | 0 | 0 | 0 |
| 0.968295073 | 0 | 0 | 0 | 0.25336795 |
| 0 | 0 | 0 | 0 | 0 |
| 0 | 0 | 0 | 0 | 0 |
| 0 | 0 | 0 | 0 | 0 |
| 0 | 0 | 0 | 0 | 0 |
| 0 | 0 | 0.019134164 | 0 | 0 |
| 0 | 0 | 0 | 0 | 0 |
| 0 | 0 | 0 | 0 | 0 |
| 0 | 0 | 0 | 0 | 0 |
| 0.017132415 | 0 | 0 | 0 | 0 |
| 0 | 0 | 0 | 0 | 0 |

| PHL018-0231_V2 | PHL018-0263_V2 | PHL018-0031_V2 | PHL018-0149_V2 | PHL018-0137_V3 |
|---|---|---|---|---|
| EYCF | REF | CTRL | REF | EYCF |
| 0.023223636 | 0.004919638 | 0.04340888 | 1.792657624 | 0.106959526 |
| 6.508209607 | 0.311016917 | 0.595022856 | 1.673840565 | 0.945229062 |
| 0 | 5.889190361 | 3.335125103 | 4.560947788 | 0 |
| 0 | 0 | 0 | 0 | 0 |
| 0 | 0 | 0 | 0 | 0 |
| 0 | 0 | 0 | 0 | 0 |
| 0 | 0 | 0 | 0 | 0 |
| 0 | 0 | 0 | 0 | 0 |
| 0 | 0 | 0 | 0 | 0 |
| 0 | 0 | 0 | 0 | 0 |
| 0 | 0 | 0 | 0 | 0 |
| 0 | 0 | 0 | 0 | 0 |
| 0 | 0 | 0 | 0 | 0 |
| 0.878825903 | 23.07146897 | 4.048628945 | 0 | 0.39878427 |
| 0 | 0 | 0 | 0 | 0 |
| 0 | 0 | 0 | 0 | 0 |
| 0 | 0 | 0 | 0.002042875 | 0 |
| 0 | 0 | 0 | 0 | 0 |
| 0 | 0 | 0 | 0 | 0 |
| 0 | 0 | 0 | 0 | 0 |
| 0 | 0 | 0 | 0 | 0 |
| 0 | 0 | 0 | 0 | 0 |
| 0 | 0 | 0 | 0 | 0 |
| 0 | 0 | 0 | 0 | 0 |
| 0 | 0.009885354 | 0 | 0 | 0.725862101 |
| 0 | 0 | 0 | 0 | 0 |
| 0 | 0 | 0 | 0 | 0 |
| 0 | 0 | 0 | 0 | 0 |
| 0 | 0 | 0 | 0 | 0 |
| 0 | 0 | 0 | 0 | 0 |
| 0 | 0 | 0 | 0 | 0 |
| 0 | 0 | 0 | 0 | 0 |
| 0 | 0 | 0 | 0 | 0 |
| 0 | 0 | 0 | 0 | 0 |
| 0 | 0 | 0 | 0 | 0 |
| 0 | 0.001708093 | 1.109779837 | 0 | 0.883126103 |
| 0 | 0 | 0 | 0 | 0 |
| 0 | 0 | 0 | 0 | 0 |
| 0 | 0 | 0 | 0 | 0 |
| 0 | 0 | 0 | 0 | 0 |
| 0 | 0 | 0 | 0 | 0 |
| 0.002046423 | 0.002396758 | 0.003071948 | 0.001950779 | 0 |
| 0.392299222 | 0.087959075 | 0.00228934 | 0.056618876 | 1.224224134 |
| 0.001864236 | 0.005290503 | 0.000719605 | 0.018233117 | 0.092943318 |
| 0.001344049 | 0 | 0.010376189 | 0 | 0 |
| 0 | 0 | 0 | 0 | 0 |

| | | | | |
|---|---|---|---|---|
| 0 | 0 | 0.004706718 | 0.030935185 | 0 |
| 0.507630973 | 0 | 0 | 0.017272552 | 0.004691377 |
| 0 | 0.000482048 | 0.062917318 | 0 | 0.001825411 |
| 0 | 0.209192748 | 0.18289386 | 0 | 0.465621607 |
| 0 | 0.000324275 | 0.000643231 | 0 | 0.001841936 |
| 0 | 0 | 0 | 0 | 0 |
| 0 | 0 | 0 | 0.002424372 | 0 |
| 0 | 0 | 0 | 0 | 0 |
| 0 | 0 | 0 | 0 | 0 |
| 0 | 0 | 0 | 0.154827294 | 0 |
| 0 | 0 | 0 | 0 | 0 |
| 0 | 0.100253524 | 1.013674983 | 0 | 0.051977569 |
| 0 | 0 | 0 | 0 | 0 |
| 0.012878487 | 0.008801298 | 0.004438536 | 0.292480765 | 0.031986965 |
| 0 | 0 | 0 | 0 | 0 |
| 0 | 0 | 0 | 0 | 0 |
| 0 | 0.016769206 | 0 | 0.063400607 | 0.005517643 |
| 0 | 0 | 0 | 0 | 0 |
| 0 | 0 | 0 | 0 | 0 |
| 0 | 0 | 0 | 0 | 0 |
| 0.214238171 | 0.01718184 | 0.002251279 | 0.070218642 | 0.049021731 |
| 0.00273796 | 0 | 0.004227464 | 0.035548125 | 0.028162412 |
| 0 | 0.063744007 | 0 | 0 | 0 |
| 0 | 0 | 0 | 0 | 0 |
| 0 | 0 | 0 | 0 | 0 |
| 0 | 0 | 0 | 0 | 0 |
| 0 | 0 | 0 | 0 | 0 |
| 0 | 0 | 0 | 0 | 0 |
| 0 | 0 | 0 | 0 | 0 |
| 0 | 0 | 0 | 0 | 0 |
| 7.458608104 | 0.00771425 | 0 | 0.002129336 | 0.207660423 |
| 0 | 0 | 0 | 0 | 0 |
| 0 | 0 | 0 | 0 | 0 |
| 0 | 0 | 0 | 0 | 0 |
| 0 | 0 | 0 | 0 | 0 |
| 0 | 0 | 0.000819053 | 0 | 0 |
| 0 | 0 | 0 | 0 | 0 |
| 0 | 0.001256265 | 0 | 0 | 0 |
| 0 | 0 | 0 | 0 | 0 |
| 0 | 0 | 0 | 0 | 0 |
| 0 | 0 | 0 | 0 | 0 |
| 0 | 0 | 0 | 0 | 0 |
| 0 | 0 | 0 | 0 | 0 |
| 0 | 0 | 0 | 0 | 0 |
| 0 | 0 | 0.003548357 | 0 | 0 |
| 0 | 0 | 0 | 0 | 0 |
| 0 | 0 | 0 | 0 | 0 |
| 0.449419401 | 0 | 0 | 0.328615357 | 0.060940702 |

| | | | | |
|---|---|---|---|---|
| 0 | 0 | 0 | 0 | 0 |
| 0 | 0 | 0 | 0 | 0 |
| 0 | 0 | 0 | 0 | 0 |
| 0 | 0 | 0 | 0 | 0 |
| 0 | 0 | 0.214934052 | 0 | 0 |
| 0 | 0 | 0 | 0 | 0 |
| 0 | 0 | 0 | 0 | 0 |
| 0 | 0 | 0 | 0 | 0 |
| 0.001764302 | 0 | 0.000681029 | 0.011806537 | 0.001422002 |
| 0 | 0 | 0 | 0 | 0 |
| 0 | 0 | 0 | 0 | 0 |
| 0 | 0.003321663 | 0 | 0 | 0 |
| 0 | 0 | 0 | 0 | 0 |
| 0 | 0 | 0 | 0 | 0 |
| 0 | 0 | 0 | 0 | 0 |
| 0 | 0 | 0 | 0 | 0 |
| 0 | 0 | 0 | 0 | 0 |
| 0 | 0 | 0 | 0 | 0 |
| 0.085129826 | 0.201755524 | 0.160080762 | 0.117764641 | 0 |
| 0 | 0 | 0 | 0 | 0 |
| 0 | 0 | 0 | 0 | 0 |
| 0 | 0 | 0 | 0 | 0 |
| 0 | 0 | 0 | 0 | 0 |
| 0 | 0 | 0 | 0 | 0 |
| 0 | 0 | 0 | 0 | 0 |
| 0 | 0 | 0 | 0 | 0 |
| 0 | 0 | 0 | 0 | 0 |
| 0 | 0 | 0 | 0.008761907 | 0.004102 |
| 0 | 0 | 0 | 0 | 0 |
| 0 | 0 | 0 | 0 | 0 |
| 0 | 0 | 0.005671841 | 0.642358947 | 0.888218343 |
| 0 | 0.002777526 | 0.001156994 | 0 | 0.001183261 |
| 0 | 0 | 0 | 0 | 0 |
| 0 | 0 | 0 | 0 | 0 |
| 0 | 0 | 0 | 0 | 0 |
| 0 | 0 | 0 | 0 | 0 |
| 0 | 0 | 0 | 0 | 0 |
| 0 | 0 | 0 | 0 | 0 |
| 0 | 2.077682195 | 0 | 0 | 0 |
| 0 | 0 | 0 | 0 | 0 |
| 0 | 0 | 0 | 0 | 0 |
| 0 | 0 | 0 | 0.002107071 | 0 |
| 0 | 0 | 0 | 0.000509536 | 0 |
| 0 | 0 | 0 | 0 | 0 |
| 0 | 0 | 0 | 0.002716182 | 0 |
| 0 | 0 | 0 | 0 | 0 |
| 0 | 0 | 0 | 0 | 0 |
| 0 | 0 | 0 | 0 | 0 |

| | | | | |
|---|---|---|---|---|
| 0 | 0 | 0 | 0 | 0 |
| 0 | 0 | 0 | 0 | 0 |
| 0 | 0 | 0 | 0 | 0 |
| 0 | 0 | 0 | 0 | 0 |
| 0 | 0.001571389 | 0 | 0.16588517 | 0 |
| 0 | 0 | 0 | 0 | 0 |
| 0 | 0 | 0 | 0 | 0 |
| 0 | 0 | 0 | 0 | 0 |
| 0 | 0 | 0 | 0 | 0 |
| 0 | 0 | 0 | 0 | 0 |
| 0 | 0 | 0 | 0 | 0 |
| 0 | 0 | 0 | 0 | 0 |
| 0 | 0 | 0 | 0 | 0 |
| 0 | 0 | 0 | 0 | 0 |
| 0 | 0 | 0 | 0 | 0 |
| 0 | 0 | 0 | 0 | 0 |
| 0 | 0 | 0 | 0 | 0 |
| 0 | 0 | 0 | 0 | 0 |
| 0 | 0 | 0 | 0 | 0 |
| 0 | 0 | 0 | 0 | 0 |
| 0 | 0 | 0 | 0 | 0 |
| 0 | 0 | 0 | 0.071043523 | 0 |
| 0 | 0 | 0 | 0 | 0 |
| 0 | 0 | 0 | 0 | 0 |
| 0 | 0 | 0 | 0 | 0 |
| 0 | 0 | 0 | 0 | 0 |
| 0 | 0 | 0 | 0 | 0 |
| 0 | 0 | 0 | 0 | 0 |
| 0 | 0 | 0 | 0 | 0 |
| 0 | 0 | 0 | 0 | 0 |
| 0 | 0 | 0 | 0 | 0.076253694 |
| 0 | 0.001121633 | 0 | 0 | 0.000682614 |
| 0 | 0 | 0 | 0 | 0 |
| 0 | 0 | 0 | 0 | 0 |
| 0 | 0 | 0 | 0 | 0 |
| 0 | 0 | 0 | 0 | 0 |
| 0 | 0 | 0 | 0 | 0 |
| 0 | 0 | 0 | 0 | 0 |
| 0 | 0 | 0 | 0 | 0 |
| 0 | 0 | 0 | 0 | 0 |
| 0 | 0 | 0 | 0 | 0 |
| 0 | 0 | 0 | 0 | 0 |
| 0 | 0 | 0 | 0 | 0 |
| 0 | 0 | 0 | 0 | 0 |
| 0 | 0 | 0 | 0 | 0 |
| 0 | 0 | 0 | 0 | 0 |
| 0 | 0 | 0 | 0 | 0 |
| 0 | 0 | 0 | 0 | 0 |

| | | | | |
|---|---|---|---|---|
| 0 | 0 | 0 | 0 | 0 |
| 0 | 0 | 0 | 0 | 0 |
| 0 | 0 | 0 | 0 | 0 |
| 0 | 0 | 0 | 0 | 0 |
| 0 | 0 | 0 | 0 | 0 |
| 0 | 0 | 0 | 0 | 0 |
| 0 | 0 | 0 | 0 | 0 |
| 0 | 0 | 0 | 0 | 0 |
| 0 | 0 | 0 | 0 | 0 |
| 0 | 0 | 0 | 0 | 0 |
| 0 | 0 | 0 | 0 | 0 |
| 0 | 0 | 0 | 0 | 0 |
| 0 | 0 | 0 | 0.001297888 | 0.002177316 |
| 0 | 0 | 0 | 0 | 0 |
| 0 | 0 | 0 | 0 | 0 |
| 0 | 0 | 0 | 0 | 0 |
| 0 | 0 | 0 | 0 | 0 |
| 0 | 0 | 0 | 0 | 0 |
| 0 | 0 | 0 | 0 | 0 |
| 0 | 0 | 0 | 0 | 0 |
| 0.007425015 | 0.011467426 | 0.004094418 | 0.040041196 | 0.028904513 |
| 0 | 0 | 0 | 0 | 0 |
| 0 | 0 | 0 | 0.001596985 | 0 |
| 0 | 0 | 0 | 0 | 0 |
| 0 | 0 | 0 | 0 | 0 |
| 0 | 0 | 0 | 0 | 0 |
| 0.002470945 | 0 | 0 | 0.000883297 | 0.011143175 |
| 0 | 0 | 0 | 0 | 0 |
| 0 | 0 | 0 | 0 | 0 |
| 0 | 0 | 0 | 0.023436653 | 0 |
| 0.25760052 | 0 | 0 | 0 | 0.056011941 |
| 0 | 0 | 0 | 0 | 0 |
| 0 | 0 | 0 | 0 | 0 |
| 0 | 0 | 0.00187669 | 0.041155254 | 0 |
| 0.008281904 | 0.001678797 | 0.01776031 | 0.065921823 | 0.075227266 |
| 0 | 0 | 0 | 0 | 0 |
| 0 | 0 | 0 | 0 | 0 |
| 0 | 0 | 0 | 0 | 0 |
| 0 | 0 | 0 | 0 | 0 |
| 0 | 0 | 0 | 0 | 0 |
| 0 | 0 | 0 | 0 | 0 |
| 0 | 0 | 0 | 0 | 0 |
| 0 | 0 | 0 | 0 | 0 |
| 0 | 0 | 0 | 0 | 0 |
| 0 | 0 | 0 | 0 | 0 |
| 0 | 0 | 0 | 1.596742891 | 0 |
| 0 | 0 | 0 | 0 | 0 |
| 0.001311535 | 0 | 0 | 0.019972543 | 0.00094382 |

| | | | | |
|---|---|---|---|---|
| 0 | 0 | 0 | 0 | 0 |
| 0 | 0 | 0 | 0 | 0 |
| 0 | 0 | 0 | 0 | 0 |
| 0 | 0 | 0 | 0 | 0 |
| 0 | 0 | 0 | 0 | 0 |
| 0 | 0 | 0 | 0 | 0 |
| 0 | 0 | 0 | 0 | 0 |
| 0 | 0 | 0 | 0 | 0 |
| 0 | 0 | 0.008417389 | 0 | 0 |
| 0 | 0 | 0 | 0 | 0 |
| 0 | 0 | 0 | 0 | 0 |
| 0 | 0 | 0 | 0 | 0 |
| 0 | 0 | 0 | 0 | 0 |
| 0 | 0 | 0 | 0 | 0 |
| 0 | 0 | 0 | 0 | 0 |
| 0 | 0 | 0 | 0 | 0 |
| 0 | 0 | 0 | 0 | 0 |
| 0 | 0 | 0 | 0 | 0 |
| 0.002167462 | 0 | 0 | 0 | 0.004415351 |
| 0 | 0 | 0 | 0 | 0.06137744 |
| 0 | 0.001595321 | 0 | 0.148755926 | 0.007645803 |
| 0 | 0 | 0 | 0 | 0 |
| 0 | 0 | 0 | 0 | 0 |
| 0 | 0 | 0 | 0 | 0 |
| 0.00342029 | 0 | 0 | 0.004191987 | 0.005625928 |
| 0 | 0 | 0 | 0 | 0 |
| 0 | 0 | 0 | 0 | 0 |
| 0 | 0 | 0 | 0 | 0 |
| 0 | 0 | 0 | 0 | 0 |
| 0 | 0 | 0 | 0 | 0 |
| 0.009222838 | 0.030938894 | 0.021639588 | 0.468550523 | 0.207595933 |
| 0 | 0 | 0 | 0 | 0 |
| 0 | 0.031724148 | 0.027992095 | 0 | 0 |
| 0 | 0 | 0.005870294 | 0 | 0 |
| 0 | 0 | 0 | 0 | 0 |
| 0 | 0 | 0 | 0 | 0 |
| 0 | 0 | 0 | 0 | 0 |
| 0 | 0 | 0 | 0 | 0 |
| 0 | 0 | 0 | 0 | 0 |
| 0 | 0 | 0.002266082 | 0 | 0 |
| 0 | 0 | 0 | 0 | 0 |
| 0 | 0.002092506 | 0.001956755 | 0 | 0.026955277 |
| 0 | 0 | 0 | 0 | 0 |
| 0 | 0 | 0 | 0 | 0 |
| 0 | 0 | 0 | 0 | 0 |
| 0 | 0 | 0 | 0 | 0 |
| 0 | 0 | 0 | 0 | 0 |
| 0 | 0 | 0 | 0 | 0 |

| | | | | |
|---|---|---|---|---|
| 0 | 0 | 0 | 0 | 0 |
| 0 | 0 | 0 | 0 | 0 |
| 0 | 0.004879617 | 0.001116831 | 0.021049674 | 0.100740826 |
| 0 | 0 | 0 | 0 | 0 |
| 0 | 0 | 0 | 0 | 5.128278919 |
| 0 | 0 | 0 | 0 | 0 |
| 0 | 0 | 0 | 0 | 0 |
| 0 | 0 | 0 | 0 | 0 |
| 0 | 0 | 0 | 0 | 0 |
| 0.00148187 | 0 | 0 | 0 | 0 |
| 0 | 0 | 0 | 0 | 0 |
| 0 | 0 | 0 | 0 | 0 |
| 0 | 0 | 0 | 0 | 0 |
| 0 | 0 | 0.005184058 | 0 | 0 |
| 0.005489581 | 0.001369569 | 0.001222503 | 0 | 0.003646586 |
| 0 | 0 | 0 | 0 | 0 |
| 0.001202863 | 0 | 0 | 0.001651167 | 0.010525892 |
| 0 | 0 | 0 | 0 | 0 |
| 0 | 0 | 0 | 0 | 0 |
| 0 | 0 | 0 | 0 | 0 |
| 0 | 0 | 0 | 0 | 0 |
| 0.002171683 | 0 | 0 | 0.020122194 | 0.001750346 |
| 0 | 0 | 0 | 0 | 0 |
| 0.03501269 | 0 | 0 | 0.0008665 | 0 |
| 0 | 0 | 0 | 0 | 0 |
| 0 | 0 | 0 | 0 | 0 |
| 0 | 0 | 0 | 0 | 0 |
| 0 | 0 | 0 | 0 | 0 |
| 0 | 0 | 0 | 0 | 0 |
| 0.00157068 | 0 | 0.001515726 | 0 | 0.001085096 |
| 0 | 0 | 0 | 0 | 0 |
| 0 | 0 | 0 | 0 | 0 |
| 0 | 0 | 0 | 0 | 0 |
| 0 | 0.000609079 | 0 | 0 | 0 |
| 0 | 0 | 0 | 0 | 0 |
| 0 | 0 | 0.008790317 | 0 | 0 |
| 0 | 0 | 0 | 0 | 0 |
| 0.004092743 | 0.002654803 | 0.066615703 | 0.016620215 | 0.018849669 |
| 0 | 0 | 0 | 0 | 0 |
| 0 | 0 | 0 | 0 | 0 |
| 0 | 0 | 0.025081379 | 0 | 0 |
| 0 | 0 | 0 | 0 | 0 |
| 0 | 0 | 0 | 0 | 0 |
| 0.004067061 | 0 | 0 | 0.019481812 | 0 |
| 0 | 0 | 0 | 0 | 0 |
| 0 | 0 | 0 | 0 | 0 |
| 0 | 0 | 0 | 0 | 0 |
| 0 | 0 | 0 | 0.014744013 | 0.006468975 |

| | | | | |
|---|---|---|---|---|
| 1.493312982 | 0 | 0 | 0 | 0 |
| 0 | 0 | 0 | 0 | 0 |
| 0.003412004 | 0 | 0 | 0 | 0 |
| 0 | 0 | 0 | 0 | 0 |
| 0.001373122 | 0.001298924 | 0.002650159 | 0.002356102 | 0.002371535 |
| 0 | 0 | 0 | 0 | 0 |
| 0 | 0 | 0 | 0 | 0 |
| 0.001283155 | 0.002080832 | 0 | 0.013210384 | 0.058728041 |
| 0 | 0 | 0 | 0 | 0 |
| 0.021822498 | 0.061393717 | 0.224310053 | 0.007375457 | 0.007994827 |
| 0 | 0 | 0 | 0 | 0 |
| 0 | 0 | 0 | 0.011110974 | 0 |
| 0.027737369 | 0 | 0 | 0.008593324 | 0.026614193 |
| 0 | 0 | 0 | 0.047435522 | 0 |
| 0 | 0 | 0 | 0.003569208 | 0 |
| 0 | 0 | 0 | 0 | 0 |
| 0 | 0 | 0 | 0 | 0 |
| 0 | 0 | 0 | 0 | 0 |
| 0.259482328 | 0 | 0 | 0.009342538 | 0.088484654 |
| 0 | 0 | 0.027676396 | 0.724684675 | 0.014357475 |
| 0 | 0 | 0 | 0 | 0 |
| 0 | 0 | 0 | 0.001787662 | 0 |
| 0 | 0 | 0 | 0 | 0 |
| 0 | 0 | 0 | 0 | 0 |
| 0 | 0 | 0 | 0 | 0 |
| 0 | 0 | 0.000866017 | 0 | 0.001291615 |
| 0 | 0 | 0 | 0 | 0 |
| 0 | 0 | 0 | 0 | 0.004373379 |
| 0 | 0 | 0 | 0 | 0 |
| 0 | 0 | 0 | 0 | 0.082126565 |
| 0 | 0 | 0 | 0.457079629 | 0.002771945 |
| 0 | 0 | 0 | 0 | 0 |
| 0 | 0 | 0 | 0 | 0 |
| 0 | 0.000713723 | 0 | 0.012838244 | 0 |
| 0 | 0 | 0 | 0 | 0 |
| 0.053464113 | 0.005534058 | 0 | 0 | 0.082253316 |
| 0.00686686 | 0.00412432 | 0 | 0 | 0.083458113 |
| 0 | 0 | 0 | 0 | 0 |
| 0 | 0 | 0 | 0 | 0 |
| 0.018877031 | 0 | 0 | 0.263983253 | 0 |
| 0 | 0 | 0 | 0 | 0 |
| 0 | 0 | 0 | 0 | 0.994178418 |
| 0 | 0 | 0 | 0 | 0 |
| 0 | 0 | 0 | 0.002636186 | 0.002289254 |
| 0 | 0 | 0 | 0 | 0 |
| 0 | 0 | 0 | 0 | 0 |
| 0 | 0 | 0.001076961 | 0.003829853 | 0 |
| 0 | 0 | 0 | 0 | 0 |

| | | | | |
|---|---|---|---|---|
| 0 | 0 | 0 | 0 | 0 |
| 0 | 0.004297923 | 0 | 0 | 0 |
| 0 | 0 | 0 | 0 | 0 |
| 0 | 0 | 0 | 0 | 0 |
| 0 | 0 | 0 | 0 | 0 |
| 0.014942846 | 0.000890886 | 0 | 0.16081572 | 0.022518719 |
| 0 | 0 | 0 | 0 | 0 |
| 0 | 0.008526658 | 0 | 0 | 0 |
| 0 | 0 | 0 | 0 | 0 |
| 0 | 0 | 0 | 0 | 0 |
| 0.031858796 | 0.006199669 | 0.001229765 | 0 | 0.008510334 |
| 0 | 0 | 0 | 18.52770574 | 0 |
| 0 | 0 | 0 | 0 | 0 |
| 0.001444732 | 0 | 0 | 0.003305302 | 0.001940722 |
| 0 | 0.163172687 | 0 | 0 | 0.003085755 |
| 0 | 0 | 0 | 0 | 0 |
| 0 | 0 | 0 | 0 | 0 |
| 0 | 0.000759992 | 0 | 0 | 0.005665909 |
| 0 | 0 | 0 | 0 | 0 |
| 0 | 0 | 0 | 0 | 0.010806011 |
| 0 | 0 | 0 | 0 | 0 |
| 0 | 0.071518973 | 0.106924108 | 0 | 0.179297254 |
| 0 | 0 | 0 | 0 | 0.148647691 |
| 0 | 0 | 0 | 0 | 0 |
| 0 | 0 | 0 | 0 | 0 |
| 0 | 0 | 0 | 0 | 0 |
| 0 | 0.262283328 | 0 | 0 | 0.060722843 |
| 0 | 0 | 0 | 0 | 0 |
| 0 | 0 | 0 | 0 | 0 |
| 1.194721582 | 0.36716613 | 0.716275663 | 0.309466757 | 0.290565032 |
| 0 | 0.001986801 | 0 | 0.005325664 | 0 |
| 0 | 0 | 0 | 0.000983824 | 0 |
| 0 | 0 | 0 | 0 | 0 |
| 0 | 0 | 0 | 0 | 0 |
| 0 | 0 | 0 | 0.005969973 | 0 |
| 0 | 0 | 0 | 0 | 0 |
| 0 | 0.043513876 | 0.69939277 | 0 | 0 |
| 0 | 0 | 0 | 0.001052807 | 0 |
| 0 | 0.420437238 | 0 | 0 | 0 |
| 0 | 0.006468458 | 0 | 0 | 0.003936637 |
| 0 | 0.000642027 | 0 | 0.005604469 | 0.000911706 |
| 11.92681072 | 4.099174806 | 0.302134143 | 0 | 2.252734844 |
| 0 | 0 | 0 | 0 | 0 |
| 0 | 0 | 0 | 0 | 13.19586135 |
| 0 | 0 | 0 | 0.014943222 | 0.002056903 |
| 0 | 0 | 0 | 0 | 0 |
| 0 | 0.001179244 | 0 | 0 | 0 |
| 0 | 0 | 0 | 0 | 0 |

| | | | | |
|---|---|---|---|---|
| 0.001025401 | 0 | 0.005442385 | 0.000879729 | 0 |
| 0.001146601 | 0 | 0 | 0 | 0 |
| 0 | 0.039084233 | 0 | 0.001498372 | 0.013674227 |
| 0.218900258 | 0.002068211 | 0 | 0 | 0.031238445 |
| 0 | 0 | 0 | 0.33339671 | 0 |
| 0.003251165 | 0.005272261 | 0 | 0.002975245 | 0.006239029 |
| 0 | 0 | 0 | 0 | 0 |
| 0 | 0 | 0.000831792 | 0 | 0 |
| 0 | 0 | 0 | 0 | 0 |
| 0 | 0 | 0 | 0 | 0 |
| 0 | 0 | 0 | 0 | 0 |
| 0 | 0 | 0 | 0 | 0 |
| 0 | 0 | 0 | 0 | 0 |
| 0.022959441 | 0.001880416 | 0.001342797 | 0 | 0.00186919 |
| 0 | 0 | 0 | 0 | 0 |
| 0.006524512 | 0 | 0.011193306 | 1.013479448 | 0.022328449 |
| 0.003013225 | 0.010750093 | 0.13589118 | 0.036364523 | 0.001387781 |
| 0 | 0 | 0 | 0.011097717 | 0 |
| 0.000829982 | 0 | 0 | 0.012639281 | 0.008839749 |
| 0 | 1.317449242 | 0 | 0 | 0.116290915 |
| 0.031646012 | 0.002052754 | 0 | 0.10382261 | 0.019821986 |
| 0 | 0 | 0.002473458 | 0.007396661 | 0 |
| 0 | 0 | 0 | 0 | 0.706549083 |
| 0 | 0 | 0 | 0 | 0 |
| 0 | 0.001978955 | 0 | 0 | 0 |
| 0 | 0.004076753 | 0 | 0.428637284 | 0 |
| 0 | 0 | 0 | 0.005907377 | 0 |
| 0.009360515 | 0 | 0 | 0 | 0 |
| 0 | 0 | 0 | 0 | 0 |
| 0.001527478 | 0 | 0 | 0 | 0 |
| 0.014545702 | 0.000786269 | 0.001122943 | 0.119468372 | 0.012281895 |
| 0 | 0 | 0 | 0 | 0.006325549 |
| 0.012388721 | 0.010938001 | 0.030818027 | 0.028579484 | 0.03328377 |
| 0 | 0 | 0 | 0 | 0 |
| 0 | 0 | 0.05027334 | 0 | 0.001945421 |
| 0 | 0 | 0 | 0 | 0 |
| 0.000746684 | 0 | 0 | 0.001708286 | 0.002865793 |
| 0 | 0.008838202 | 0.001314858 | 0.281219317 | 0.005602957 |
| 0 | 0 | 0 | 0 | 0.366113086 |
| 0 | 0 | 0 | 0.054204532 | 0 |
| 0 | 0 | 0 | 0 | 0 |
| 0 | 0 | 0 | 0 | 0 |
| 0 | 0 | 0 | 0.000749222 | 0.008379213 |
| 0 | 0 | 0.023754034 | 0 | 0.100247457 |
| 1.043080447 | 0.006750628 | 0 | 0.033928521 | 0.313262626 |
| 0 | 0 | 0.008599817 | 0 | 0 |
| 0 | 0 | 0 | 0 | 0 |
| 0 | 0 | 0 | 0 | 0 |

| | | | | |
|---|---|---|---|---|
| 0 | 0 | 0 | 0 | 0 |
| 0.086730667 | 0.018064758 | 0.035833222 | 0.321075392 | 1.278055365 |
| 0 | 0 | 0 | 0 | 0 |
| 0 | 0 | 0 | 0 | 0 |
| 0 | 0 | 0 | 0.007590122 | 0 |
| 0 | 0 | 0 | 0 | 0 |
| 0.02183825 | 0 | 0 | 0 | 0.044921496 |
| 0 | 0 | 0 | 0 | 0 |
| 0 | 0 | 0 | 0.011456765 | 0 |
| 0 | 0 | 0 | 0.006945008 | 0 |
| 0 | 0 | 0.0005706 | 0.005241969 | 0 |
| 0.00352296 | 0 | 0.004532933 | 0.079591848 | 0.12371911 |
| 0 | 0.157811934 | 0 | 0.002079265 | 0 |
| 0 | 0 | 0 | 0 | 0 |
| 0 | 0 | 0 | 0 | 0 |
| 0 | 0 | 0 | 0 | 0 |
| 0.035498716 | 0 | 0.001394168 | 0.004426692 | 0.078895377 |
| 0 | 0.353878451 | 0 | 0.031490587 | 0.016966688 |
| 0 | 0 | 0 | 0.002064484 | 0 |
| 0 | 0.00197487 | 0 | 0.088360716 | 0.02343676 |
| 0.010429061 | 0.04659516 | 0.010885738 | 0.00639105 | 0.033328618 |
| 0 | 0 | 0 | 0 | 0 |
| 0 | 0 | 0 | 0 | 0 |
| 0 | 0 | 0 | 0 | 0 |
| 0 | 0 | 0 | 0 | 0 |
| 0.00117107 | 0 | 0 | 0.006028223 | 0 |
| 0 | 0 | 0 | 0 | 0.026365987 |
| 0.095462878 | 0.050807726 | 0.026414374 | 0.26608145 | 0.143777332 |
| 0 | 0 | 0 | 0 | 0 |
| 0 | 0 | 0 | 0.007725431 | 0 |
| 0 | 0 | 0 | 0 | 0 |
| 0 | 0 | 0 | 0 | 0 |
| 0 | 0 | 0 | 0 | 0 |
| 0 | 0 | 0 | 0.001322915 | 0 |
| 0.011433594 | 0 | 0.028344834 | 0 | 0 |
| 11.89648403 | 0.558773583 | 0.014778431 | 0.076532594 | 0.660353561 |
| 0 | 0 | 0 | 0 | 0 |
| 0 | 0 | 0 | 0 | 0 |
| 0 | 0.001131354 | 0 | 0.006464269 | 0 |
| 0 | 0 | 0 | 0 | 0 |
| 0.596084015 | 0.007951222 | 0 | 0.03485467 | 0.191141738 |
| 0 | 0 | 0 | 0 | 0 |
| 0.005424207 | 0.001511844 | 0.003925821 | 0.008289269 | 0.000975856 |
| 0 | 0 | 0 | 0 | 0.00697206 |
| 0 | 0.13737351 | 0 | 0 | 0 |
| 0 | 0 | 0.059395204 | 0.960496457 | 0.612303902 |
| 0 | 0 | 0 | 0 | 0.006903343 |
| 0 | 0 | 0 | 0.228997767 | 0 |

| | | | | |
|---|---|---|---|---|
| 0.011840211 | 0 | 0 | 0 | 0.00163595 |
| 0 | 0 | 0 | 0 | 0 |
| 0 | 0 | 0 | 0 | 0 |
| 0 | 0.003041416 | 0 | 0.790811918 | 1.210439652 |
| 0 | 0 | 0 | 0 | 0 |
| 0 | 0 | 0 | 0 | 0 |
| 0 | 0 | 0 | 0 | 0 |
| 0 | 0.001575242 | 0 | 0 | 0 |
| 0 | 0.036301333 | 10.15384658 | 0.241620202 | 0.936726894 |
| 0 | 0 | 0 | 0 | 0 |
| 0 | 0 | 0 | 0 | 0 |
| 0 | 0.001533686 | 0.096742488 | 0 | 0 |
| 0 | 0 | 0 | 0 | 0 |
| 0 | 0 | 0 | 0 | 0 |
| 0.009833534 | 0.002910883 | 0.104836179 | 0.57971123 | 0.028216164 |
| 0 | 0 | 0 | 0 | 0 |
| 0 | 0 | 0 | 0 | 0 |
| 0 | 0.020658562 | 7.655006469 | 0 | 0 |
| 0 | 0 | 0 | 0 | 0 |
| 0 | 0 | 0 | 0 | 0 |
| 0 | 0 | 0 | 0 | 0 |
| 0 | 0 | 0 | 0 | 0 |
| 0 | 0 | 0 | 0 | 0 |
| 0.073221712 | 0.063149308 | 0.013783751 | 0.624325895 | 0.479707723 |
| 0.054721927 | 0 | 0 | 0 | 0.00407814 |
| 0 | 0 | 0 | 0 | 2.039481548 |
| 0 | 0 | 0 | 0.016631691 | 0 |
| 0 | 0 | 0.704433589 | 0 | 0.100585109 |
| 0 | 0 | 0 | 0 | 0 |
| 0 | 0 | 0 | 0 | 0 |
| 0.005700496 | 0 | 0 | 0 | 0 |
| 0 | 0 | 0 | 0 | 0 |
| 0.001259563 | 0.003858196 | 0 | 0 | 0.032389394 |
| 0 | 0 | 0 | 0 | 0 |
| 0 | 0.000688313 | 0 | 0 | 0 |
| 0 | 0 | 0 | 0 | 0 |
| 0.11800037 | 0 | 0 | 1.447202618 | 0.363083723 |
| 0 | 0 | 0 | 0 | 0 |
| 0 | 0 | 0 | 0.024495276 | 0.002040474 |
| 0 | 0 | 0 | 0 | 0 |
| 0.093087191 | 0.009563433 | 0.000700431 | 0.267455042 | 0.009610803 |
| 0 | 0 | 0 | 0 | 0 |
| 8.428327167 | 1.591502198 | 13.75032211 | 0 | 0.15188649 |
| 0 | 0 | 0 | 0 | 0.02966694 |
| 0 | 0 | 0 | 0 | 0 |
| 0 | 0 | 0 | 0 | 0 |
| 0 | 0 | 0 | 0 | 0 |
| 0 | 0 | 0 | 0.014035724 | 0.122314151 |

| | | | | |
|---|---|---|---|---|
| 0 | 0 | 0 | 0 | 0 |
| 0 | 0.005156182 | 0 | 0 | 0.04324555 |
| 0.541996818 | 0.017802417 | 0 | 0.068529912 | 0.014205058 |
| 0 | 0 | 0 | 0.012410899 | 0 |
| 0 | 0 | 0 | 0 | 0 |
| 0 | 0.00053425 | 0.009855554 | 0.009044645 | 0 |
| 0 | 0 | 0 | 0 | 0 |
| 0 | 0 | 0 | 0 | 0 |
| 0 | 0 | 0 | 0 | 0 |
| 0 | 0 | 0 | 0 | 0 |
| 0.029974203 | 0.001664649 | 0.059634032 | 1.404514078 | 0.009928265 |
| 1.897853224 | 2.091314296 | 0.600822898 | 1.239515389 | 3.396400309 |
| 0 | 0 | 0 | 0 | 0 |
| 0.001772569 | 0.00377277 | 0.097501403 | 0.003041503 | 0.00612285 |
| 0.877747037 | 0 | 0 | 0.014530725 | 0.010856342 |
| 0 | 0 | 0 | 0 | 0 |
| 0 | 0 | 0 | 0 | 0 |
| 0 | 0 | 0 | 0 | 0 |
| 0 | 0 | 0 | 0 | 0 |
| 0 | 0.000685616 | 0.000489595 | 0.05571467 | 0.018109042 |
| 0.002581109 | 0.03767097 | 0 | 0.098320578 | 0.111545688 |
| 0.357910988 | 8.128349942 | 0.480464498 | 0.654418347 | 0 |
| 1.111691615 | 7.70898439 | 2.708653504 | 1.123318103 | 0.111533096 |
| 0 | 0 | 0 | 0 | 0 |
| 0 | 0 | 0 | 0 | 0 |
| 0 | 1.564390666 | 0.193808362 | 0 | 0 |
| 0 | 0 | 0 | 1.69772591 | 0 |
| 0.00861456 | 0 | 0 | 0 | 0.000826573 |
| 0 | 0.016444707 | 0 | 0 | 0.021694931 |
| 0 | 0 | 0 | 0 | 0 |
| 0 | 0 | 0.007142955 | 0 | 0 |
| 0 | 0.055453939 | 0 | 0 | 0.116717571 |
| 0.75294994 | 0 | 0.000813896 | 0 | 0.03763026 |
| 0.013184815 | 0 | 0 | 0 | 0.022401383 |
| 0.008210704 | 0 | 0 | 0.035818277 | 0.045827236 |
| 0.003824069 | 0.043719259 | 0.138938827 | 0.046423435 | 0.00616429 |
| 0 | 0 | 0.006416558 | 0.016955305 | 0 |
| 0 | 0 | 0 | 0 | 0 |
| 0 | 0 | 1.693447289 | 0 | 0.05049425 |
| 0 | 0 | 0 | 0 | 0 |
| 0 | 0 | 0.00406184 | 0 | 0 |
| 0 | 0 | 0 | 0 | 0 |
| 0 | 0 | 0 | 0 | 0 |
| 0 | 0 | 0 | 0 | 0 |
| 0.022458723 | 0.001944771 | 0.221148212 | 0.642396537 | 0.035399425 |
| 0 | 0 | 0 | 0.117551626 | 0 |
| 0 | 0 | 0 | 0 | 0 |
| 0 | 0.068671625 | 0.976700665 | 0.831996966 | 0.002119927 |

| | | | | |
|---|---|---|---|---|
| 0 | 0 | 0 | 0 | 0 |
| 0 | 0 | 0 | 0 | 0 |
| 0 | 0 | 0 | 0 | 0 |
| 0 | 0.000452051 | 0.017933751 | 0 | 0.004707498 |
| 0 | 0 | 0 | 0 | 0 |
| 0 | 0.013308286 | 0 | 0 | 0 |
| 0 | 0.006763767 | 0 | 0 | 0 |
| 0 | 0.005130281 | 0.001485207 | 0 | 0 |
| 0 | 0 | 0 | 0 | 0 |
| 0 | 0 | 0 | 0 | 0 |
| 0.000827904 | 0 | 0.000599203 | 0.225871992 | 0.002621453 |
| 0 | 0 | 0 | 0 | 0 |
| 0 | 0 | 0 | 0 | 0 |
| 0 | 0 | 0 | 0 | 0 |
| 0 | 0 | 0 | 0 | 0 |
| 0 | 0 | 0 | 0 | 0 |
| 0 | 0.002745934 | 0 | 0 | 0 |
| 0 | 0.001536785 | 0.020285478 | 0 | 0 |
| 0 | 0 | 0 | 0.957260885 | 0.043213293 |
| 0.003107773 | 0 | 0 | 0.044526737 | 0.016281334 |
| 0 | 0 | 0 | 0 | 0 |
| 0 | 0 | 0 | 0 | 0 |
| 0 | 0 | 0.045050087 | 0.064763984 | 0 |
| 0 | 0 | 0 | 0 | 0 |
| 0 | 0 | 0 | 0 | 0 |
| 0 | 0 | 0 | 0 | 0 |
| 0 | 0 | 0 | 0 | 0 |
| 0 | 0 | 0 | 0 | 0 |
| 0 | 0 | 0 | 0 | 0 |
| 0 | 0.322680014 | 0 | 0 | 0 |
| 0 | 0 | 0 | 0 | 0 |
| 0 | 0 | 0 | 0 | 0 |
| 0 | 0.001459708 | 0 | 0 | 0 |
| 0 | 0 | 0 | 0 | 0 |
| 0 | 0 | 0 | 0 | 0 |
| 0 | 0 | 0 | 0 | 0 |
| 0 | 0.021031464 | 0.012872975 | 0 | 0 |
| 0.002978551 | 0 | 0.026714442 | 0.033821567 | 0.003833002 |
| 0 | 0 | 0 | 0 | 0 |
| 0 | 0 | 0.00116739 | 0 | 0 |
| 0 | 0 | 0.061392795 | 0 | 0.091611118 |
| 0 | 0 | 0 | 0 | 0 |
| 0 | 0 | 0 | 0 | 0 |
| 0 | 0 | 0 | 0 | 0 |
| 0 | 0.098239136 | 0.083706406 | 0 | 0.119803713 |
| 0 | 0 | 0 | 0 | 0 |
| 0 | 0 | 0 | 0 | 0 |
| 0 | 0 | 0 | 0 | 0 |

| | | | | |
|---|---|---|---|---|
| 0 | 0 | 0 | 0 | 0 |
| 0 | 0 | 0 | 0 | 0 |
| 0 | 0 | 0.065348423 | 0 | 0 |
| 0 | 0.010174467 | 0 | 0.284242136 | 0 |
| 0 | 0 | 0 | 0 | 0 |
| 0 | 0 | 0 | 0 | 0 |
| 0 | 0 | 0 | 0 | 0 |
| 0 | 0 | 0 | 0 | 0 |
| 0.054611165 | 0.134016114 | 0 | 0.145119613 | 0.024484145 |
| 0 | 0 | 0.001690066 | 0 | 0 |
| 0 | 0.000420288 | 0 | 0 | 0 |
| 0 | 0 | 0 | 0 | 0 |
| 0 | 0 | 0 | 0 | 0 |
| 0 | 0 | 0 | 0.000719967 | 0 |
| 0 | 0.004787216 | 0.076414169 | 0 | 0.067580614 |
| 0 | 0 | 0 | 0.008308688 | 0.113652546 |
| 0 | 0 | 0 | 0 | 0 |
| 0 | 0 | 0 | 0 | 0 |
| 0 | 0 | 0 | 0 | 0 |
| 0 | 0 | 0 | 0 | 0 |
| 0 | 0.001498166 | 0.013729527 | 0 | 0 |
| 0 | 0 | 0 | 0 | 0 |
| 0 | 0 | 0 | 0 | 0 |
| 0 | 0 | 0 | 0 | 0 |
| 0.014492389 | 0.006451427 | 0 | 0.00146277 | 0.048424015 |
| 1.526134514 | 1.645708415 | 0.069268862 | 1.087122262 | 22.32740269 |
| 0 | 0 | 0 | 0 | 0 |
| 0 | 0 | 0 | 0.003329368 | 0.005280642 |
| 0 | 0 | 0 | 0 | 0.010011995 |
| 0 | 0 | 0 | 0 | 0 |
| 0 | 0 | 0 | 0 | 0 |
| 0 | 0 | 0 | 0 | 0 |
| 0 | 0 | 0 | 0.00177087 | 0 |
| 0 | 0.000615264 | 0 | 0.128249174 | 0 |
| 0 | 0.004257792 | 0.04343528 | 0.001930789 | 0.001151666 |
| 0 | 0 | 0.000682299 | 0.050195552 | 0.001831698 |
| 0 | 0 | 0 | 0 | 0 |
| 4.644500116 | 0.29470022 | 0 | 0 | 7.375112477 |
| 0 | 0 | 1.561854927 | 0 | 0 |
| 0 | 0 | 0 | 0 | 0 |
| 0 | 0 | 0.05894191 | 0 | 0 |
| 0.006414285 | 0 | 0 | 0 | 0 |
| 8.676767803 | 0.0119469 | 0.068762613 | 0.043103831 | 0.040326891 |
| 0 | 0 | 0 | 0 | 0 |
| 0 | 0 | 0 | 0 | 0 |
| 0 | 0 | 0 | 0 | 0.030271418 |
| 0 | 0 | 0 | 0 | 0 |
| 0.004802029 | 0.025092169 | 0.07491662 | 0.001144399 | 0.094916136 |

| | | | | |
|---|---|---|---|---|
| 0 | 0 | 0 | 0.8470644 | 0.000687175 |
| 0 | 0 | 0 | 0 | 0 |
| 0 | 0 | 0 | 0 | 0 |
| 0 | 0 | 0 | 0 | 0 |
| 0 | 0.000802984 | 0.000764543 | 0.027868129 | 0 |
| 0 | 0 | 0 | 0 | 0 |
| 0 | 0 | 0 | 0 | 0 |
| 0 | 0 | 0 | 0 | 0 |
| 0 | 0 | 0 | 0 | 0 |
| 0 | 0 | 0 | 0 | 0 |
| 0 | 0 | 0 | 0 | 0 |
| 0 | 0 | 0.416326615 | 5.933552411 | 0.242795358 |
| 0 | 0.055613996 | 0 | 0 | 0.006903343 |
| 0 | 0 | 0 | 0 | 0 |
| 0.051539123 | 0.044946698 | 0.031167826 | 0.010022585 | 0.001846213 |
| 0 | 0 | 0 | 0 | 0 |
| 0 | 0 | 0 | 0.00259747 | 0 |
| 0 | 0 | 0 | 0 | 0 |
| 0 | 0 | 0.015887348 | 0 | 0 |
| 0 | 0 | 0 | 0.007639919 | 0 |
| 0 | 0.00161732 | 0 | 0 | 0 |
| 0 | 0 | 0 | 0 | 0 |
| 0.102883137 | 0 | 0.050489523 | 0.039695109 | 0.056195671 |
| 0 | 0 | 0 | 0 | 0 |
| 0 | 0 | 0 | 0 | 0 |
| 0 | 0 | 0 | 0 | 0 |
| 0 | 0 | 0 | 0 | 0 |
| 0 | 0 | 0 | 0 | 0 |
| 0 | 0 | 0 | 0 | 0 |
| 0 | 0 | 0 | 0 | 0 |
| 0 | 0 | 0 | 0 | 0 |
| 0.015753772 | 0.004579206 | 0.012907855 | 0.021421163 | 0.208085361 |
| 0 | 0 | 0 | 0 | 0 |
| 0.002821433 | 0.001565263 | 0.032529311 | 0 | 0.658957045 |
| 0 | 0 | 0.00204191 | 0.042413028 | 0.000885932 |
| 0 | 0.000804253 | 0.602071921 | 0.031273555 | 0.087939671 |
| 0 | 0 | 0 | 0 | 0 |
| 0 | 0 | 0 | 0 | 0 |
| 0 | 0.000895928 | 0.174659467 | 0 | 0 |
| 0 | 0 | 0 | 0 | 0 |
| 0 | 0 | 0 | 0 | 0 |
| 0 | 0 | 0 | 0 | 0 |
| 0 | 0 | 0 | 0 | 0 |
| 0 | 0 | 0 | 0.646092124 | 0 |
| 0.003031876 | 0.006453105 | 0.005668734 | 0.807334081 | 0.007854585 |
| 0 | 0 | 0 | 0 | 0 |
| 0 | 0 | 0 | 0 | 0 |
| 0 | 0 | 0.002421371 | 0.011686088 | 0.004539964 |

| | | | | |
|---|---|---|---|---|
| 0 | 0 | 0 | 0 | 0 |
| 0 | 0 | 0 | 0 | 0 |
| 0.007643315 | 0 | 0.001323878 | 0 | 0 |
| 0 | 0 | 0 | 0 | 0 |
| 0 | 0.006607372 | 0 | 0 | 0 |
| 0 | 0 | 0 | 0 | 0 |
| 0 | 0.036558769 | 5.138431748 | 0 | 0 |
| 0.018138534 | 0 | 0 | 0.011750677 | 0.015343975 |
| 0 | 0 | 0 | 0 | 0 |
| 0 | 0 | 0 | 0 | 0 |
| 0 | 0 | 0 | 0 | 0 |
| 0 | 0 | 0 | 0.019845144 | 0 |
| 0 | 0 | 0 | 0 | 0 |
| 0 | 0.029383884 | 0.019006246 | 0 | 0.394767692 |
| 0 | 0 | 0 | 0 | 0 |
| 0 | 0 | 0 | 0 | 0 |
| 0 | 0 | 0 | 0 | 0 |
| 0 | 0 | 0.001736529 | 0 | 0 |
| 0 | 0 | 0.004445909 | 1.354304094 | 0 |
| 0 | 0 | 0 | 0 | 0 |
| 0 | 0 | 0 | 0 | 0 |
| 0 | 0 | 0 | 0 | 0 |
| 0 | 0 | 0 | 0 | 0 |
| 0 | 0 | 0 | 0 | 0 |
| 0.010144801 | 0 | 0.001798137 | 0 | 0 |
| 0.003175383 | 0.002002532 | 0.001021428 | 0.016951051 | 0.008124802 |
| 0 | 0 | 0 | 0 | 0 |
| 0.002244716 | 0.00364015 | 0 | 0.067107532 | 0.027436351 |
| 0 | 0 | 0 | 0 | 0 |
| 0 | 0.034530304 | 0.139883353 | 0 | 0 |
| 0 | 0 | 0 | 0 | 0 |
| 0 | 0 | 0 | 0 | 0 |
| 0 | 0 | 0 | 0 | 0 |
| 0 | 0 | 0 | 0 | 0 |
| 0 | 0 | 0 | 0.316890888 | 0 |
| 0 | 0 | 0 | 0 | 0.195528402 |
| 0 | 0 | 0 | 0 | 0 |
| 0.002097698 | 0 | 0 | 0.005399074 | 0 |
| 0 | 0 | 0 | 0 | 0 |
| 0 | 0 | 0 | 0 | 0 |
| 9.685562251 | 1.630314098 | 0.572816425 | 1.91461956 | 10.16917981 |
| 0.776514718 | 0.176920735 | 0.00103726 | 1.524869962 | 0.021746588 |
| 0 | 0 | 4.687889764 | 0 | 0.087156512 |
| 0 | 0 | 0 | 0 | 0 |
| 0 | 0 | 0 | 0 | 0 |
| 0 | 0 | 0 | 0 | 0 |
| 0 | 0 | 0 | 0 | 0 |
| 0 | 0 | 0.080651325 | 0.023853189 | 0.002108613 |

| | | | | |
|---|---|---|---|---|
| 0 | 0 | 0 | 0 | 0 |
| 0 | 0.001666185 | 0.002148277 | 0.044221719 | 0.000985856 |
| 0 | 0 | 0 | 0 | 0.00483598 |
| 0 | 0 | 0 | 0 | 0 |
| 5.807164606 | 0.331554374 | 0.871012627 | 2.811089952 | 0.373391443 |
| 0 | 0 | 0 | 0 | 0 |
| 0 | 0 | 0 | 0 | 0 |
| 0 | 0 | 0 | 0 | 0 |
| 0.083614607 | 0.725801559 | 0 | 0.051051394 | 0.145100881 |
| 0 | 0 | 0 | 0 | 0 |
| 0 | 0 | 0 | 0 | 0 |
| 0 | 0 | 0 | 0 | 0 |
| 0 | 0.007690168 | 0.001445135 | 0.194430873 | 0 |
| 0.010228116 | 0.001990375 | 0.041257682 | 0.004036533 | 0.035801276 |
| 0.034058454 | 0 | 0 | 0 | 0 |
| 0.140668662 | 0.037157279 | 0.014255055 | 1.159466583 | 0 |
| 0 | 0.00100471 | 0 | 0.014731321 | 0.022827701 |
| 0 | 0 | 0 | 0 | 0 |
| 0 | 0 | 0 | 0 | 0 |
| 0 | 0 | 0 | 0 | 0 |
| 0 | 0 | 0 | 0 | 0 |
| 0 | 0 | 0 | 0 | 0 |
| 0 | 0 | 0 | 0 | 0 |
| 0 | 0 | 0 | 0 | 0 |
| 0 | 0 | 0 | 0 | 0 |
| 0 | 0 | 0 | 0 | 0 |
| 0.041251505 | 0 | 0.010546206 | 0.050841787 | 0.010102041 |
| 0 | 0 | 0 | 0.000894067 | 0 |
| 0 | 0 | 0 | 0 | 0 |
| 0 | 0 | 0 | 0 | 0 |
| 0 | 0 | 0 | 0 | 0 |
| 0 | 0 | 0 | 0 | 0 |
| 0 | 0 | 0 | 0 | 0 |
| 0 | 0 | 0 | 0 | 0 |
| 0 | 0 | 0.015793151 | 0 | 0.018099828 |
| 0 | 0 | 0 | 0.05378097 | 0 |
| 0 | 0 | 0 | 0.002290443 | 0 |
| 0.009106733 | 0 | 0 | 0 | 0 |
| 0 | 0 | 0 | 0 | 0 |
| 0 | 0 | 0 | 0 | 0 |
| 0 | 0 | 0.167411708 | 0 | 0.952839956 |
| 0 | 0 | 0 | 0 | 0 |
| 0 | 0 | 0 | 0 | 0 |
| 0 | 0 | 0 | 0 | 0 |
| 0 | 0 | 0 | 0 | 0 |
| 0 | 0 | 0 | 0 | 0 |
| 0 | 0.941773051 | 0 | 0 | 0 |
| 0 | 0 | 0 | 0 | 0 |

| | | | | |
|---|---|---|---|---|
| 0.016164394 | 0 | 0 | 0 | 0 |
| 0 | 0 | 0 | 0 | 0 |
| 0 | 0.035679075 | 0 | 0 | 0 |
| 0 | 0.001276251 | 0 | 0 | 0.009288196 |
| 0 | 0 | 0 | 0 | 0 |
| 0 | 0 | 0 | 0 | 0 |
| 0 | 0.003722707 | 0 | 0.086809504 | 0.006709664 |
| 0 | 0 | 0 | 0 | 0 |
| 0 | 0 | 0 | 0 | 0 |
| 0 | 0.000600881 | 0 | 0 | 0 |
| 0 | 0.012501392 | 0.016945123 | 0 | 0 |
| 0 | 0 | 0 | 0 | 0 |
| 0 | 0 | 0 | 0 | 0 |
| 0 | 0 | 0 | 0 | 0 |
| 0 | 0 | 0 | 0 | 0 |
| 0 | 0 | 0 | 0 | 0 |
| 0 | 0 | 0 | 0 | 0 |
| 0 | 0 | 0 | 0.013399925 | 0 |
| 0 | 0 | 0 | 0 | 0 |
| 0 | 0 | 0 | 0 | 0 |
| 0 | 0.006952133 | 0 | 0.001730837 | 0.042780071 |
| 0 | 0 | 0 | 0 | 0 |
| 0 | 0.003307842 | 0 | 0 | 0.01772154 |
| 0.000659566 | 0.001470683 | 0.00127298 | 0.007356259 | 0.001139145 |
| 0 | 0 | 0 | 0 | 0 |
| 0 | 0 | 0.010700537 | 0 | 0 |
| 0 | 0 | 0 | 0 | 0 |
| 0 | 0 | 0 | 0 | 0 |
| 0 | 0 | 0.005496627 | 0 | 0 |
| 0.001186589 | 0 | 0 | 0 | 0.396485054 |
| 0 | 0 | 0 | 0 | 0 |
| 0 | 0 | 0 | 0 | 0 |
| 0 | 0 | 0 | 0 | 0 |
| 0 | 0.01770496 | 0 | 0.004230176 | 0 |
| 0 | 0 | 0 | 0 | 0 |
| 0 | 0 | 0 | 0 | 0 |
| 0.047866733 | 0 | 0 | 0 | 0.028507315 |
| 0 | 0 | 0 | 0 | 0 |
| 0 | 0 | 0 | 0 | 0 |
| 0 | 0 | 0 | 0 | 0 |
| 0 | 0 | 0 | 0 | 0 |
| 0 | 0 | 0.136244331 | 0 | 0 |
| 0 | 0 | 0 | 0 | 0 |
| 0 | 0 | 0 | 0 | 0.009747696 |
| 0 | 0 | 0.001507491 | 0 | 0 |
| 0 | 0 | 0 | 0 | 0 |
| 0 | 0 | 0 | 0 | 0 |
| 0 | 0.014510376 | 0 | 0 | 0 |

| | | | | |
|---|---|---|---|---|
| 0 | 0 | 0 | 0.076568167 | 0.006741684 |
| 0 | 0 | 0 | 0 | 0.025187635 |
| 0 | 0.050093754 | 0 | 0 | 0 |
| 0 | 0 | 0 | 0 | 0 |
| 0 | 0 | 0 | 0 | 0 |
| 0 | 0 | 0 | 0 | 0 |
| 0 | 0 | 0 | 0 | 0 |
| 2.125855255 | 0.555264996 | 0.040611115 | 0.218661759 | 0.304156655 |
| 0.472339122 | 0.116973033 | 0.002150062 | 0.082911089 | 1.82589081 |
| 0.011439193 | 0 | 0.023761835 | 0 | 0.193326121 |
| 0 | 0 | 0 | 0 | 0 |
| 0 | 0 | 0 | 0 | 0 |
| 0 | 0 | 0 | 0 | 0 |
| 0 | 0.005349633 | 2.314771048 | 0 | 0 |
| 0 | 0 | 0 | 0 | 0 |
| 0 | 0 | 0 | 0 | 0 |
| 0.199688187 | 0 | 0 | 0.267534352 | 0 |
| 0 | 0 | 0.035714017 | 0 | 0 |
| 0 | 0 | 0 | 0 | 0 |
| 0 | 0.007169781 | 0.046079165 | 0 | 0 |
| 0 | 0 | 0 | 0 | 0 |
| 0 | 0 | 0 | 0 | 0 |
| 0 | 0 | 0 | 0 | 0 |
| 0 | 0 | 0 | 0 | 0 |
| 0 | 0.000793192 | 0 | 0.34710098 | 0 |
| 0 | 0 | 0 | 0 | 0 |
| 0 | 0 | 0 | 0 | 0 |
| 0 | 0 | 0 | 0 | 0 |
| 0 | 0 | 0 | 0 | 0 |
| 0 | 0 | 0 | 0 | 0 |
| 0 | 0 | 0 | 0 | 0.016370358 |
| 0 | 0 | 0 | 0 | 0 |
| 0 | 0 | 0 | 0 | 0 |
| 0 | 0 | 0 | 0 | 0 |
| 0 | 0 | 0 | 0 | 0 |
| 0 | 0 | 0 | 0 | 0 |
| 0 | 0 | 0 | 0 | 0 |
| 0.090747594 | 0 | 0 | 0 | 0 |
| 0 | 0.000684264 | 0.364029005 | 0 | 0 |
| 1.683974408 | 0 | 0 | 0.016108419 | 0.011829033 |
| 0 | 0 | 0 | 0 | 0 |
| 0 | 0.02667082 | 0.04514491 | 0 | 1.60858013 |
| 0 | 0 | 0 | 0 | 0 |
| 0 | 0 | 0 | 0 | 0 |
| 0 | 0 | 0 | 0 | 0 |
| 0 | 0 | 0 | 0 | 0.06249049 |
| 0 | 0 | 0 | 0 | 0 |
| 0 | 0 | 0.001275578 | 0 | 0 |

| | | | | |
|---|---|---|---|---|
| 0 | 0 | 0 | 0 | 0 |
| 0 | 0.005713129 | 0.003554301 | 0 | 0 |
| 0 | 0 | 0 | 0 | 0 |
| 0 | 0 | 0 | 0 | 0 |
| 0 | 0.034492591 | 5.811403413 | 1.498091316 | 0.277386829 |
| 0 | 0 | 0 | 0 | 0 |
| 0 | 0 | 0 | 0 | 0 |
| 0.007222491 | 0.008967286 | 0 | 0.025173028 | 0.006237026 |
| 0 | 0.000548174 | 0 | 0 | 0 |
| 0 | 0 | 0 | 0 | 0 |
| 0 | 0 | 0 | 0 | 0 |
| 0 | 0 | 0 | 0 | 0 |
| 0 | 0 | 0 | 0 | 0 |
| 0 | 0 | 0 | 0 | 0 |
| 0 | 0 | 0 | 0 | 0 |
| 0 | 0 | 0.023990497 | 0 | 0.03177979 |
| 0 | 0 | 0 | 0 | 0 |
| 0 | 0 | 0 | 0 | 0 |
| 0 | 0 | 0 | 0 | 0 |
| 0 | 0 | 0 | 0 | 0 |
| 0 | 0 | 0 | 0 | 0 |
| 0 | 0 | 0 | 0 | 0 |
| 0 | 0 | 0 | 0 | 0 |
| 0 | 0 | 0 | 0.044743413 | 0 |
| 0 | 0 | 0.213489869 | 0 | 0 |
| 0 | 0 | 0 | 0 | 0 |
| 0.005538116 | 0.001593386 | 0 | 0.049659285 | 0.01213617 |
| 0 | 0 | 0 | 0 | 0 |
| 0 | 0 | 0 | 0.008201378 | 0 |
| 0 | 0 | 0 | 0 | 0 |
| 0 | 0 | 0 | 0 | 0 |
| 0 | 0 | 0 | 0 | 0 |
| 0 | 0 | 0 | 0 | 0 |
| 0 | 0 | 0 | 0 | 0 |
| 0 | 0 | 0 | 0 | 0 |
| 0 | 0 | 0 | 0 | 0 |
| 0 | 0 | 0 | 0 | 0 |
| 0 | 0 | 0 | 0 | 0 |
| 0 | 0.015435591 | 0 | 0 | 0 |
| 0 | 0 | 0 | 0 | 0 |
| 0 | 0 | 0 | 0 | 0 |
| 0 | 0 | 9.651842166 | 0 | 0 |
| 0 | 0 | 0 | 0 | 0 |
| 0 | 0 | 0 | 0 | 0 |
| 0 | 0 | 0 | 0 | 0 |
| 0 | 0 | 0 | 0 | 0 |
| 0 | 0 | 0 | 0 | 0 |
| 0 | 0 | 0 | 0 | 0 |

| | | | | |
|---|---|---|---|---|
| 0 | 0.003269328 | 0.017671718 | 0.182621148 | 0.022632603 |
| 0 | 0 | 0 | 0 | 0 |
| 0 | 0 | 0 | 0 | 0 |
| 0.000668618 | 0 | 0 | 0 | 0 |
| 0 | 0 | 0 | 0 | 0 |
| 0 | 0.000473463 | 0.003568811 | 0 | 0 |
| 0 | 0 | 0 | 0 | 0 |
| 0 | 0 | 0 | 0 | 0 |
| 0 | 0 | 0 | 0 | 0 |
| 0 | 0 | 0 | 0 | 0.043266843 |
| 0 | 0 | 0 | 0 | 0 |
| 0 | 0 | 0 | 0.00308961 | 0 |
| 0 | 0 | 0 | 0 | 0 |
| 0 | 0 | 0 | 0 | 0 |
| 0 | 0 | 0 | 0 | 0 |
| 0 | 0 | 0 | 0 | 0 |
| 0 | 0 | 0 | 0 | 0 |
| 0 | 0 | 0 | 3.157255915 | 0 |
| 0 | 0 | 0 | 0 | 0 |
| 0 | 0 | 0 | 0 | 0 |
| 0.042986864 | 0.004100575 | 0 | 0.129997806 | 0.04143282 |
| 0 | 0 | 0 | 0 | 0 |
| 0 | 0 | 0.141286591 | 0.010959234 | 0.01187956 |
| 0 | 0 | 0 | 0.793178104 | 0 |
| 0 | 0 | 0 | 0 | 0 |
| 0 | 0 | 0 | 0 | 0 |
| 0 | 0 | 0 | 0 | 0 |
| 0 | 0 | 0 | 0 | 0 |
| 0 | 0.518319346 | 0.454966168 | 0 | 0 |
| 0 | 0 | 0 | 0 | 0 |
| 0 | 0 | 0 | 0 | 0 |
| 0 | 0 | 0 | 0 | 0 |
| 0.088570358 | 0 | 0.154514697 | 0.005131639 | 0.007151888 |
| 0 | 0 | 0 | 0 | 0 |
| 0 | 0.077586575 | 0 | 0 | 0.209216379 |
| 0 | 0 | 0 | 0.17881298 | 0.038898279 |
| 0 | 0 | 0 | 0 | 0 |
| 0 | 0 | 0 | 0 | 0 |
| 0.021566339 | 0.000692537 | 0 | 0.020517675 | 0.012047042 |
| 0 | 0 | 0 | 0 | 0 |
| 0 | 0 | 0 | 0 | 0 |
| 0 | 0.870194149 | 0 | 0 | 0 |
| 0 | 0.053243249 | 1.159657194 | 0.028275473 | 0.082507237 |
| 0 | 0 | 0 | 0 | 0 |
| 0 | 0 | 0 | 0 | 0 |
| 0.007733228 | 0.001768548 | 0.00631456 | 3.312375495 | 0.001598172 |
| 0 | 0 | 0 | 0 | 0 |
| 0 | 0 | 0 | 0 | 0 |

| | | | | |
|---|---|---|---|---|
| 0 | 0 | 0 | 0 | 0 |
| 0 | 0 | 0 | 0 | 0 |
| 0 | 0 | 0 | 0 | 0 |
| 0 | 0 | 0 | 0 | 0 |
| 0 | 0 | 0 | 0 | 0 |
| 0 | 0 | 0 | 0 | 0 |
| 0 | 0 | 0 | 0 | 0 |
| 0 | 0.000776808 | 0 | 0 | 0.044785876 |
| 0 | 0 | 0 | 0 | 0.143102835 |
| 0 | 0.002815177 | 0.061885847 | 0 | 0.004467985 |
| 0 | 0 | 0 | 0 | 0 |
| 0 | 0 | 0 | 0 | 0 |
| 0 | 0 | 0 | 0 | 0 |
| 0 | 0.343399008 | 0 | 0 | 0 |
| 0 | 0 | 0 | 0 | 0 |
| 0.001406608 | 0 | 0 | 0.084474623 | 0.004627372 |
| 0 | 0 | 0 | 0 | 0 |
| 0 | 0 | 0 | 0 | 0 |
| 0 | 0 | 0 | 0 | 0 |
| 0 | 0.213210031 | 0 | 0 | 0 |
| 0 | 0 | 0 | 0 | 0 |
| 0 | 0 | 0 | 0 | 0 |
| 0 | 0 | 0 | 0 | 0 |
| 0 | 0 | 0 | 0.070039165 | 0 |
| 0 | 0 | 0 | 0.004426989 | 0 |
| 0 | 0 | 0 | 0 | 0 |
| 0 | 0 | 0 | 0 | 0 |
| 0 | 0 | 0 | 0 | 0 |
| 0 | 0 | 0 | 0 | 0 |
| 0 | 0 | 0 | 0 | 0 |
| 0 | 0.18195268 | 0.037037746 | 1.046553259 | 0.078604852 |
| 0 | 0 | 0 | 0 | 0 |
| 0 | 0 | 0 | 0 | 0.00214414 |
| 0 | 0 | 0 | 0 | 0 |
| 0 | 0 | 0 | 0 | 0 |
| 0 | 0 | 0 | 0 | 0 |
| 0 | 0 | 0 | 0 | 0 |
| 0 | 0 | 0 | 0 | 0 |
| 0 | 0 | 0.007460449 | 0 | 0 |
| 0 | 0 | 0 | 0 | 0 |
| 0 | 0 | 0 | 0 | 0 |
| 0 | 0.069981469 | 0 | 0 | 0 |
| 0 | 0 | 0 | 0 | 0 |
| 0 | 0 | 0 | 0.011601125 | 0 |
| 0 | 0 | 0 | 0 | 0 |
| 0 | 0 | 0 | 0 | 0 |
| 0.012136591 | 0.182802949 | 0 | 0.541093037 | 0 |
| 0 | 0 | 0 | 0 | 0.00288732 |

| | | | | |
|---|---|---|---|---|
| 0 | 0 | 0 | 0 | 0 |
| 0 | 0 | 0 | 0 | 0 |
| 0 | 0 | 0 | 0 | 0 |
| 0 | 0.002242237 | 0 | 0 | 0 |
| 0.013369895 | 0 | 0 | 0.04443178 | 0.017517544 |
| 0 | 0 | 0 | 0 | 0 |
| 0 | 0 | 0 | 0 | 0 |
| 0 | 0 | 0 | 0 | 0 |
| 0 | 0 | 0 | 0 | 0 |
| 0 | 0 | 0 | 0.025615349 | 0 |
| 0 | 0 | 0 | 0 | 0 |
| 0.000853432 | 0.000518989 | 0.009265194 | 0.135821364 | 0.001473973 |
| 0 | 0 | 0.008348796 | 0.00631696 | 0.004662781 |
| 0 | 0 | 0 | 0 | 0 |
| 0 | 0 | 0 | 0 | 0.006739624 |
| 0.144015226 | 0 | 0 | 0.690091008 | 0 |
| 0 | 0.000931383 | 0.001741919 | 0 | 0.007557736 |
| 0 | 0 | 0 | 0 | 0 |
| 0.004038899 | 0 | 0 | 0.091479126 | 0.416119678 |
| 0 | 0.005473218 | 0 | 0.041262426 | 0 |
| 0 | 0 | 0 | 0 | 0 |
| 0 | 0 | 0 | 0 | 0 |
| 0 | 0 | 0 | 0 | 0 |
| 0 | 0 | 0 | 0 | 0 |
| 0 | 0 | 0 | 0 | 0 |
| 0 | 0 | 0 | 0 | 0 |
| 0 | 0.000411182 | 0.005219966 | 0.096876035 | 0.007979909 |
| 0 | 0 | 0 | 0 | 0 |
| 0 | 0 | 0 | 0 | 0 |
| 0 | 0 | 0 | 0 | 0 |
| 0 | 0 | 0 | 0.003452239 | 0 |
| 0 | 0 | 0 | 0.054511798 | 0.004766025 |
| 0 | 0 | 0 | 0 | 0 |
| 0 | 0 | 0 | 0 | 0 |
| 0 | 0 | 0 | 0 | 0 |
| 0 | 0 | 0 | 0 | 0 |
| 0 | 0 | 0 | 0 | 0 |
| 0.005384888 | 0.001091552 | 0.058720263 | 0.107874457 | 0.023870783 |
| 0 | 0 | 0 | 0 | 0.047355595 |
| 0 | 0 | 0 | 0.043054029 | 0 |
| 0 | 0 | 0 | 0.001729553 | 0 |
| 0 | 0 | 0 | 0 | 0 |
| 0 | 0 | 0 | 0 | 0 |
| 0.241150197 | 0.00097118 | 0 | 0.016099175 | 0.005976175 |
| 0 | 0 | 0 | 0 | 0 |
| 0 | 0 | 0 | 0 | 0 |
| 0 | 0 | 0 | 0 | 0 |
| 0 | 0 | 0 | 0 | 0 |

| | | | | |
|---|---|---|---|---|
| 0 | 0 | 0 | 0 | 0 |
| 0 | 0 | 0 | 0 | 0 |
| 0 | 0 | 0 | 0 | 0 |
| 0.002928949 | 0 | 0 | 0.018307923 | 0 |
| 0.001164237 | 0 | 0.002996009 | 0 | 0 |
| 0 | 0 | 0 | 0 | 0 |
| 0 | 0.001810781 | 0.005681674 | 0 | 0 |
| 0 | 0 | 0 | 0 | 0 |
| 0 | 0 | 0 | 0.015893707 | 0 |
| 0 | 0 | 0 | 0 | 0 |
| 0 | 0 | 0 | 0 | 0 |
| 0 | 0 | 0 | 0 | 0 |
| 0 | 0 | 0.107906617 | 0.039187687 | 0.010074538 |
| 0 | 0 | 0 | 0 | 0 |
| 0 | 0 | 0 | 0 | 0 |
| 0 | 0 | 0 | 0 | 0 |
| 0 | 0 | 0 | 0 | 0 |
| 0 | 0 | 0 | 0 | 0 |
| 0 | 0 | 0 | 0 | 0 |
| 0 | 0 | 0 | 0 | 0 |
| 0 | 0 | 0 | 0 | 0 |
| 0 | 0 | 0 | 0 | 0 |
| 0.010545019 | 0 | 0.001130675 | 0.022784925 | 0.014165224 |
| 0 | 0 | 0.00177898 | 0 | 0.008225063 |
| 0 | 0 | 0 | 0 | 0 |
| 0 | 0 | 0 | 0 | 0 |
| 0 | 0 | 0 | 0 | 0 |
| 0 | 0 | 0 | 0 | 0 |
| 0 | 0 | 0 | 0 | 0 |
| 0 | 0 | 0 | 0 | 0 |
| 0 | 0.001059268 | 0.002101163 | 0 | 0 |
| 0 | 0 | 0.006499367 | 0 | 0 |
| 0 | 0 | 0.001668052 | 0 | 0 |
| 0 | 0 | 0 | 0 | 0 |
| 0 | 0 | 0 | 1.459611799 | 0 |
| 0 | 0 | 0 | 0.007030004 | 0 |
| 0 | 0 | 0 | 0 | 0 |
| 0 | 0 | 0 | 0 | 0 |
| 0 | 0 | 0.008888646 | 0.002054618 | 0 |
| 0 | 0 | 0 | 0 | 0 |
| 0 | 0 | 0 | 0 | 0 |
| 0 | 0 | 0 | 0 | 0 |
| 0 | 0 | 0 | 0 | 0 |
| 0 | 0 | 0 | 0 | 0 |
| 0 | 0 | 0.078629612 | 0 | 0 |
| 0.025122656 | 0.034563485 | 0.09728752 | 0 | 0.090138486 |
| 0 | 0 | 0 | 0 | 0 |
| 0 | 0 | 0 | 0 | 0 |

| | | | | |
|---|---|---|---|---|
| 0 | 0 | 0 | 0 | 0 |
| 0 | 0 | 0 | 0 | 0 |
| 0 | 0.195459662 | 0.025829737 | 0 | 0.142226102 |
| 0 | 0 | 0 | 0 | 0 |
| 0 | 0 | 0 | 0 | 0 |
| 0 | 0 | 0 | 0 | 0 |
| 0 | 0 | 0 | 0 | 0 |
| 0 | 0 | 0 | 0 | 0 |
| 0 | 0.004284389 | 0 | 0.002339776 | 0 |
| 0 | 0 | 0 | 0 | 0 |
| 0 | 0 | 0.004038201 | 0.006410946 | 0 |
| 0 | 0 | 0 | 0.003828625 | 0 |
| 0 | 0 | 0 | 0 | 0 |
| 0 | 0 | 0 | 0 | 0 |
| 0 | 0 | 0 | 0 | 0 |
| 0 | 0 | 0 | 0 | 0 |
| 0 | 0 | 0 | 0 | 0 |
| 0 | 0 | 0 | 0 | 0 |
| 0 | 0 | 0 | 0 | 0 |
| 0 | 0 | 0.020465081 | 0.023295522 | 0 |
| 0 | 0 | 0 | 0 | 0 |
| 0 | 0 | 0 | 0 | 0 |
| 0 | 0 | 0 | 0 | 0 |
| 0 | 0 | 0 | 0 | 0 |
| 0 | 0 | 0 | 0 | 0 |
| 0 | 0 | 0 | 0 | 0 |
| 0 | 0 | 0 | 0 | 0 |
| 0 | 0.000889227 | 0.004444949 | 0 | 0 |
| 0 | 0 | 0 | 0 | 0 |
| 0 | 0 | 0 | 0 | 0 |
| 0 | 0.004279883 | 0.236448174 | 0.010079673 | 0.001176313 |
| 0 | 0 | 0 | 0 | 0 |
| 0.022340405 | 0.058670744 | 0.01373656 | 0.043252164 | 0.078534586 |
| 0.002222338 | 0 | 0 | 0.003119931 | 0.004885016 |
| 0 | 0 | 0 | 0 | 0 |
| 0 | 0 | 0 | 0 | 0 |
| 0 | 0 | 0 | 0 | 0 |
| 0 | 0 | 0 | 0 | 0 |
| 0 | 0 | 0 | 0 | 0 |
| 0 | 0 | 0 | 0 | 0 |
| 0 | 0 | 0 | 0 | 0 |
| 0.376855915 | 0.007912769 | 0 | 0.038094916 | 0.071538847 |
| 0 | 0.000803332 | 0.017783324 | 0.278802098 | 0.022359004 |
| 0 | 0 | 0 | 0 | 0 |
| 0 | 0 | 0 | 0 | 0 |
| 0 | 0.001160871 | 0.255323457 | 0.040534505 | 0.010880004 |
| 0 | 0 | 0 | 0.005637243 | 0 |
| 0 | 0 | 0 | 0 | 0 |

| | | | | |
|---|---|---|---|---|
| 0 | 0 | 0 | 0 | 0 |
| 0 | 0.00252479 | 0 | 0 | 0.006214537 |
| 0.001939895 | 0 | 0.001404017 | 0.003952728 | 0.02484892 |
| 0 | 0 | 0 | 0 | 0 |
| 0.001742527 | 0.005651548 | 0 | 0 | 0.259712091 |
| 0 | 0 | 0 | 0 | 0 |
| 0 | 0 | 0 | 0 | 0 |
| 0 | 0 | 0 | 0 | 0 |
| 0 | 0 | 0 | 0 | 0 |
| 0 | 0 | 0 | 0 | 0 |
| 0 | 0 | 0 | 0 | 0 |
| 0 | 0 | 0 | 0 | 0 |
| 0 | 0 | 0 | 0 | 0 |
| 0 | 0 | 0 | 0 | 0 |
| 0 | 0 | 0 | 0 | 0 |
| 0 | 0 | 0 | 0 | 0 |
| 0 | 0 | 0 | 0 | 0 |
| 0 | 0 | 0 | 0 | 0 |
| 0 | 0 | 0 | 0 | 0 |
| 0 | 0 | 0 | 0 | 0 |
| 0 | 0 | 0 | 0 | 0 |
| 0 | 0 | 0 | 0.016734276 | 0 |
| 0 | 0 | 0 | 0 | 0 |
| 0 | 0 | 0 | 0 | 0 |
| 0 | 0 | 0.164309253 | 0 | 0.002301012 |
| 0 | 0 | 0 | 0.001034865 | 0.005555432 |
| 0 | 0 | 0 | 0 | 0 |
| 0 | 0 | 0 | 2.114787866 | 0 |
| 0 | 0 | 0 | 0 | 0 |
| 0 | 0 | 0 | 0 | 0 |
| 0 | 0 | 0 | 0 | 0 |
| 0 | 0 | 0 | 0 | 0 |
| 0 | 0 | 0 | 0 | 0 |
| 0 | 0 | 0 | 0 | 0 |
| 0 | 0 | 0 | 0 | 0 |
| 0 | 0 | 0 | 0 | 0 |
| 0 | 0 | 0 | 0 | 0 |
| 0.271557472 | 0 | 0 | 0 | 0.07534578 |
| 0 | 0.009175826 | 0 | 6.173327246 | 0 |
| 0 | 0 | 0 | 0 | 0 |
| 0 | 0 | 0 | 0.006586779 | 0 |
| 0 | 0.042087217 | 0.016626671 | 0 | 0 |
| 0 | 0 | 0 | 0.051756891 | 0.006272417 |
| 0 | 0 | 0 | 0 | 0 |
| 0 | 0 | 0 | 0 | 0 |
| 0 | 0 | 0 | 0 | 0 |
| 0.019445466 | 0 | 0.004765744 | 0.066731888 | 0.121899293 |
| 0 | 0 | 0 | 0 | 0 |

| | | | | |
|---|---|---|---|---|
| 0 | 0 | 0 | 0.186506437 | 0 |
| 0.034078612 | 0.008526397 | 0.012402827 | 0.047113789 | 0.053812648 |
| 0 | 0 | 0 | 0 | 0 |
| 0 | 0 | 0 | 0 | 0 |
| 0 | 0 | 0 | 0 | 0 |
| 0 | 0 | 0 | 0 | 0 |
| 0 | 0 | 0 | 0 | 0 |
| 0 | 0 | 0 | 0 | 0 |
| 0 | 0 | 0 | 0 | 0 |
| 0 | 0 | 0 | 0 | 0 |
| 0 | 0 | 0 | 0 | 0 |
| 0 | 0 | 0 | 0 | 0 |
| 0 | 0.001502193 | 0 | 0.00264912 | 0.006399533 |
| 0 | 0 | 0 | 0 | 0 |
| 0.009469399 | 0.005512439 | 0.005623438 | 0.017081529 | 0.006989194 |
| 0 | 0 | 0 | 0 | 0 |
| 0 | 0 | 0 | 0 | 0 |
| 0 | 0 | 0 | 0 | 0 |
| 0 | 0 | 0 | 0.00368714 | 0 |
| 0 | 0 | 0 | 0 | 0 |
| 0 | 0 | 0 | 0 | 0 |
| 0.013353411 | 0.000580034 | 0 | 0.098606675 | 0.007687613 |
| 0 | 0 | 0 | 0.03191192 | 0 |
| 0 | 0 | 0 | 0.684190108 | 0 |
| 0 | 0 | 0 | 0 | 0 |
| 0 | 0 | 0 | 0 | 0 |
| 0 | 0 | 0 | 0 | 0 |
| 0 | 0 | 0 | 0 | 0 |
| 0 | 0 | 0 | 0 | 0 |
| 0 | 0.001123136 | 0 | 0 | 0 |
| 0 | 0 | 0 | 0 | 0 |
| 0 | 0 | 0 | 0 | 0 |
| 0 | 0 | 0 | 0 | 0 |
| 0 | 0 | 0 | 0 | 0 |
| 0 | 24.78624128 | 7.709437439 | 0 | 0 |
| 0 | 0 | 0 | 0 | 0 |
| 0 | 0 | 0 | 0 | 0 |
| 0 | 0 | 0 | 0 | 0 |
| 0.000714375 | 0.002316936 | 0.005055458 | 0.001634368 | 0.016450739 |
| 0 | 0 | 0 | 0 | 0 |
| 0.11273355 | 0.003462399 | 0.013598659 | 0.034254366 | 0.232562488 |
| 0 | 0 | 0 | 0 | 0 |
| 0 | 0 | 0 | 0 | 0 |
| 0 | 0 | 0 | 0 | 0 |
| 0 | 0 | 0 | 0 | 0 |
| 1.577223686 | 0.002973159 | 0.030903176 | 0.011744703 | 0.32087291 |
| 0 | 0 | 0 | 0 | 0 |
| 0 | 0 | 0 | 0 | 0 |

| | | | | |
|---|---|---|---|---|
| 0 | 0 | 0 | 0 | 0 |
| 0 | 0 | 0 | 0 | 0 |
| 0 | 0 | 0 | 0 | 0 |
| 0 | 0 | 0 | 0 | 0 |
| 0.004264214 | 0 | 0 | 0.000477186 | 0 |
| 0 | 0 | 0 | 0 | 0 |
| 0 | 0 | 0 | 0 | 0 |
| 0 | 0 | 0 | 0 | 0 |
| 0 | 0 | 0.006871721 | 0 | 0 |
| 0 | 0.001938503 | 0 | 0.00389715 | 0 |
| 0 | 0 | 0.003406472 | 0 | 0 |
| 0 | 0 | 0 | 0 | 0 |
| 0.00116839 | 0 | 0 | 0.005613464 | 0.002690587 |
| 0 | 0 | 0 | 0 | 0 |
| 0 | 0 | 0 | 0 | 0 |
| 0 | 0 | 0 | 0 | 0 |
| 0 | 0 | 0 | 0 | 0 |
| 0 | 0 | 0 | 0 | 0 |
| 0 | 0 | 0 | 0 | 0 |
| 0 | 0.052279557 | 1.004768515 | 0.364378795 | 0.138420175 |
| 0 | 0 | 0 | 0 | 0 |
| 0 | 0 | 0 | 0 | 0 |
| 0 | 0 | 0 | 0 | 0 |
| 0 | 0 | 0 | 0 | 0 |
| 0 | 0 | 0 | 0 | 0 |
| 0 | 0 | 0 | 0 | 0 |
| 0 | 0 | 0 | 0 | 0 |
| 0 | 0 | 0 | 0 | 0 |
| 0 | 0 | 0 | 0 | 0 |
| 0 | 0.009194427 | 0 | 0 | 0.137477021 |
| 0 | 0 | 0 | 0 | 0 |
| 0 | 0 | 0 | 0 | 0 |
| 0 | 0 | 0 | 0 | 0 |
| 0 | 0 | 0 | 0 | 0 |
| 0 | 0 | 0 | 0 | 0 |
| 0 | 0 | 0 | 0 | 0 |
| 0 | 0 | 0 | 0 | 0 |
| 0 | 0 | 0 | 0 | 0 |
| 0 | 0 | 0 | 0 | 0 |
| 0 | 0 | 0 | 0 | 0 |
| 0 | 0 | 0 | 0 | 0 |
| 0 | 0 | 0 | 0 | 0 |
| 0 | 0.002383116 | 0 | 0.095671638 | 0 |
| 0 | 0 | 0 | 0 | 0 |
| 0 | 0 | 0.037891893 | 0.027318701 | 0.003373944 |
| 0 | 0 | 0 | 0 | 0 |
| 0 | 0 | 0 | 0 | 0 |
| 0 | 0 | 0 | 0 | 0 |

| | | | | |
|---|---|---|---|---|
| 0 | 0 | 0 | 0 | 0 |
| 0 | 0 | 0 | 0 | 0 |
| 0 | 0 | 0 | 0 | 0 |
| 0 | 0 | 0 | 0 | 0 |
| 0 | 0 | 0 | 0 | 0 |
| 0.002916754 | 0 | 0.018014103 | 0.004128942 | 0.015616439 |
| 0 | 0 | 0 | 0 | 0 |
| 0 | 0 | 0 | 0 | 0 |
| 0 | 0 | 0 | 0 | 0 |
| 0 | 0 | 0 | 0 | 0 |
| 0 | 0 | 0 | 0 | 0 |
| 0 | 0 | 0 | 0 | 0 |
| 0 | 0 | 0 | 0 | 0 |
| 0 | 0 | 0 | 0 | 0 |
| 0 | 0 | 0 | 0 | 0 |
| 0 | 0 | 0 | 0 | 0 |
| 0 | 0 | 0.002389951 | 0 | 0 |
| 0 | 0 | 0 | 0 | 0 |
| 0 | 0 | 0 | 0 | 0 |
| 0 | 0 | 0 | 0 | 0 |
| 0 | 0 | 0 | 0.001595819 | 0 |
| 0 | 0 | 0 | 0 | 0 |
| 0 | 0 | 0 | 0 | 0 |
| 0 | 0 | 0 | 0 | 0 |
| 0 | 0 | 0 | 0 | 0 |
| 0 | 0 | 0 | 0 | 0 |
| 0 | 0 | 0 | 0 | 0 |
| 0 | 0.000692154 | 0 | 0 | 0 |
| 0 | 0 | 0 | 0 | 0 |
| 0 | 0 | 0 | 0 | 0 |
| 0 | 0 | 0 | 0 | 0 |
| 0 | 0 | 0 | 0 | 0 |
| 0 | 0 | 0 | 0 | 0 |
| 0 | 0 | 0 | 0 | 0 |
| 0 | 0 | 0 | 0 | 0 |
| 0 | 0 | 0 | 0 | 0 |
| 0 | 0 | 0 | 0 | 0 |
| 0 | 0 | 0 | 0 | 0 |
| 0 | 0.898859648 | 0.053042307 | 0.971042111 | 1.591908913 |
| 0 | 0 | 0 | 0 | 0 |
| 0 | 0 | 0 | 0 | 0 |
| 0 | 0 | 0 | 0 | 0 |
| 0 | 0 | 0 | 0 | 0 |
| 0 | 0 | 0 | 0 | 0 |
| 0 | 0 | 0 | 0 | 0 |
| 0 | 0 | 0 | 0 | 0 |
| 0 | 0 | 0 | 0 | 0 |
| 0 | 0 | 0 | 0 | 0 |

| | | | | |
|---|---|---|---|---|
| 0 | 0 | 0 | 0 | 0 |
| 0.000979662 | 0.001456283 | 0 | 0 | 0.034779726 |
| 0 | 0.001134589 | 0 | 0 | 0 |
| 0 | 0 | 0 | 0 | 0 |
| 0 | 0 | 0 | 0 | 0 |
| 0 | 0 | 0 | 0 | 0 |
| 0 | 0 | 0 | 0 | 0 |
| 0 | 0 | 0 | 0 | 0 |
| 0 | 0 | 0 | 0 | 0 |
| 0 | 0 | 0 | 0 | 0 |
| 0.000641395 | 0 | 0.027233985 | 0 | 0 |
| 0 | 0 | 0 | 0 | 0 |
| 0 | 0.002425987 | 0.033280048 | 0 | 0 |
| 0 | 0 | 0 | 0 | 0 |
| 0 | 0 | 0 | 0 | 0 |
| 0 | 0 | 0 | 0 | 0 |
| 0 | 0 | 0 | 0 | 0 |
| 0 | 0 | 0 | 0 | 0 |
| 0 | 0 | 0 | 0 | 0 |
| 0 | 0 | 0 | 0 | 0 |
| 0 | 0 | 0 | 0 | 0.001249234 |
| 0 | 0 | 0 | 0 | 0 |
| 0 | 0 | 0 | 0 | 0 |
| 0 | 1.735169302 | 0.417882537 | 2.148279205 | 1.843095037 |
| 0 | 0 | 0 | 0 | 0 |
| 0 | 0 | 0 | 0 | 0 |
| 0 | 0 | 0 | 0 | 0 |
| 0 | 0 | 0 | 0 | 0 |
| 0.011340651 | 0 | 0 | 0 | 0 |
| 0 | 0 | 0 | 0 | 0 |
| 0 | 0 | 0 | 0 | 0 |
| 0 | 0 | 0 | 0 | 0 |
| 0 | 0 | 0 | 0.002545771 | 0 |
| 0 | 0 | 0 | 0 | 0 |

| PHL018-0017_V2 | PHL018-0244_V3 | PHL018-0134_V3 | PHL018-0023_V2 | PHL018-0157_V3 |
|---|---|---|---|---|
| EYCF | REF | CTRL | EYCF | REF |
| 0.012485958 | 0.012354222 | 0.340851366 | 0.022615717 | 0.027835541 |
| 2.187543686 | 0.338407355 | 1.22080078 | 2.321420948 | 0.100682538 |
| 0 | 2.94426112 | 0 | 1.995789159 | 0 |
| 0 | 0 | 0 | 0 | 0 |
| 0 | 0 | 0 | 0 | 0 |
| 0 | 0 | 0 | 0 | 0 |
| 0 | 0 | 0 | 0 | 0 |
| 0 | 0 | 0 | 0 | 0 |
| 0 | 0 | 0 | 0 | 0 |
| 0 | 0 | 0 | 0 | 0 |
| 0 | 0 | 0 | 0 | 0 |
| 0 | 0 | 0 | 0 | 0 |
| 0 | 0 | 0 | 0 | 0 |
| 5.557506768 | 1.468138516 | 1.095614358 | 1.542026096 | 0.147810901 |
| 0 | 0 | 0 | 0 | 0 |
| 0 | 0 | 0 | 0 | 0 |
| 0 | 0 | 0 | 0 | 0 |
| 0 | 0 | 0 | 0 | 0 |
| 0 | 0 | 0 | 0 | 0 |
| 0 | 0 | 0 | 0 | 0 |
| 0 | 0 | 0 | 0 | 0 |
| 0 | 0 | 0 | 0 | 0 |
| 0 | 0 | 0 | 0 | 0 |
| 0 | 0 | 0 | 0 | 0 |
| 0 | 0.026770495 | 0 | 0 | 0 |
| 0 | 0 | 0 | 0 | 0 |
| 0 | 0 | 0 | 0 | 0 |
| 0 | 0 | 0 | 0 | 0 |
| 0 | 0 | 0 | 0 | 0 |
| 0 | 0 | 0 | 0 | 0 |
| 0 | 0 | 0 | 0 | 0 |
| 0 | 0 | 0 | 0 | 0 |
| 0 | 0 | 0 | 0 | 0 |
| 0 | 0 | 0 | 0 | 0 |
| 0 | 0 | 0 | 0 | 0 |
| 0 | 4.843255479 | 0 | 1.35065799 | 0.557457135 |
| 0 | 0 | 0 | 0 | 0 |
| 0 | 0 | 0 | 0 | 0 |
| 0 | 0 | 0 | 0 | 0 |
| 0 | 0 | 0 | 0 | 0 |
| 0 | 0 | 0 | 0 | 0 |
| 1.462945187 | 1.085593357 | 0 | 0.020266517 | 0 |
| 1.012562561 | 7.807473865 | 0.003740275 | 8.452602961 | 10.79924271 |
| 0.012276304 | 0.026277208 | 0.024046219 | 0.010959506 | 0.00780075 |
| 0.001936108 | 0.00281614 | 0.001748216 | 0.029056824 | 0.00721034 |
| 0 | 0 | 0 | 0 | 0 |

| | | | | |
|---|---|---|---|---|
| 0 | 0.010285739 | 0 | 0 | 0.003177218 |
| 0.06239206 | 0 | 0.003896837 | 0 | 0 |
| 0 | 0 | 0.010525345 | 0 | 0.003401983 |
| 0 | 0.257560588 | 0.396358147 | 0.404489571 | 0.662058814 |
| 0 | 0.003047135 | 0.069503687 | 0 | 0 |
| 0 | 0 | 0 | 0 | 0.017448178 |
| 3.854387023 | 0 | 0.017264027 | 0.024845631 | 0 |
| 0 | 0 | 0 | 0 | 0 |
| 0 | 0 | 0 | 0 | 0.028661963 |
| 0 | 0 | 0 | 0 | 0 |
| 0 | 0.000899191 | 0 | 0 | 0 |
| 0.404234067 | 0.082879111 | 0.584422789 | 0.000515712 | 0.643657555 |
| 0 | 0 | 0 | 0 | 0 |
| 0.132510871 | 0.010337971 | 0.154549409 | 0.023550432 | 0 |
| 0 | 0.112989381 | 0 | 0.007658991 | 0 |
| 0 | 0 | 0 | 0 | 0 |
| 2.485865054 | 0.000720624 | 0 | 0.015544701 | 16.6039981 |
| 0 | 0 | 0 | 0 | 0 |
| 0 | 0 | 0 | 0 | 0 |
| 0 | 0 | 0 | 0 | 0 |
| 1.347023024 | 0.064618546 | 0.039194672 | 0.007557835 | 0 |
| 0.009014957 | 0.003302979 | 0.014245152 | 0.002076899 | 0 |
| 0 | 0 | 0 | 0 | 0.00307524 |
| 0 | 0 | 0 | 0 | 0 |
| 0 | 0 | 0 | 0 | 0 |
| 0 | 0 | 0 | 0 | 0 |
| 0 | 0.000622717 | 0 | 0 | 0 |
| 0 | 0 | 0 | 0 | 0 |
| 0.021143352 | 0.047814483 | 0 | 0 | 0 |
| 0 | 0 | 0 | 0 | 0 |
| 0 | 0 | 0 | 0 | 0 |
| 0 | 0 | 0 | 0 | 0 |
| 0 | 0 | 0 | 0 | 0 |
| 0 | 0 | 0 | 0 | 0 |
| 0 | 0 | 0 | 0 | 0 |
| 0 | 0 | 0 | 0 | 0 |
| 0 | 0 | 0 | 0 | 0 |
| 0.061034674 | 0 | 0 | 0 | 0.341732853 |
| 0 | 0 | 0 | 0 | 0 |
| 0 | 0 | 0 | 0 | 0 |
| 0 | 0 | 0 | 0 | 0 |
| 0 | 0 | 0 | 0 | 0 |
| 0 | 0 | 0 | 0 | 0 |
| 0 | 0 | 0 | 0 | 0 |
| 0 | 0.13641716 | 0 | 0 | 0 |
| 0 | 0 | 0 | 0 | 0 |
| 0 | 0 | 0 | 0 | 0 |
| 0 | 0 | 0 | 0.001573755 | 0 |

| | | | | |
|---|---|---|---|---|
| 0 | 0 | 0 | 0 | 0 |
| 0 | 0 | 0 | 0 | 0 |
| 0 | 0 | 0 | 0 | 0 |
| 0 | 0 | 0 | 0 | 0 |
| 0 | 0.122089753 | 0 | 0 | 0.057067425 |
| 0 | 0.027350176 | 0 | 0 | 0 |
| 0 | 0 | 0 | 0 | 0 |
| 0 | 0.001077389 | 0 | 0 | 0 |
| 0 | 0.004200774 | 0 | 0 | 0 |
| 0 | 0 | 0 | 0 | 0 |
| 0 | 0 | 0 | 0 | 0 |
| 0 | 0.034334193 | 0 | 0 | 0 |
| 0 | 0 | 0 | 0 | 0 |
| 0 | 0 | 0 | 0 | 0 |
| 0 | 0 | 0 | 0 | 0 |
| 0 | 0 | 0 | 0 | 0 |
| 0 | 0 | 0 | 0 | 0 |
| 0 | 0 | 0 | 0 | 0 |
| 0 | 0.092188316 | 0.428373866 | 0 | 0.420703404 |
| 0 | 0 | 0 | 0 | 0 |
| 0 | 0 | 0 | 0 | 0 |
| 0 | 0 | 0 | 0 | 0 |
| 0 | 0 | 0 | 0 | 0 |
| 0 | 0.001147658 | 0 | 0 | 0 |
| 0 | 0 | 0 | 0 | 0 |
| 0 | 0 | 0 | 0 | 0 |
| 0 | 0 | 0 | 0 | 0 |
| 0 | 0 | 0 | 0 | 0 |
| 0 | 0 | 0 | 0 | 0 |
| 0 | 0 | 0 | 0 | 0 |
| 0 | 0.018969897 | 0.418345359 | 0 | 0.066739721 |
| 0 | 0 | 0 | 0.000974428 | 0.013415097 |
| 0 | 0 | 0 | 0 | 0 |
| 0 | 0 | 0 | 0 | 0 |
| 0 | 0 | 0 | 0 | 0 |
| 0 | 0 | 0 | 0 | 0 |
| 0 | 0 | 0 | 0 | 0 |
| 0 | 0 | 0 | 0 | 0 |
| 0 | 0 | 0 | 0 | 0.277514696 |
| 0 | 0 | 0 | 0 | 0 |
| 0 | 0 | 0 | 0 | 0 |
| 0 | 0 | 0 | 0 | 0 |
| 0 | 0 | 0 | 0 | 0.00233648 |
| 0 | 0 | 0 | 0 | 0 |
| 0 | 0 | 0.00709207 | 0 | 0 |
| 0 | 0 | 0 | 0 | 0 |
| 0 | 0 | 0 | 0 | 0 |
| 0 | 0 | 0 | 0 | 0 |

| | | | | |
|---|---|---|---|---|
| 0 | 0 | 0 | 0 | 0 |
| 0 | 0 | 0 | 0 | 0 |
| 0 | 0 | 0 | 0 | 0 |
| 0 | 0 | 0 | 0 | 0 |
| 0 | 0.008092894 | 0 | 0.004664711 | 0.008637281 |
| 0 | 0 | 0.020616744 | 0 | 0 |
| 0 | 0 | 0 | 0 | 0 |
| 0 | 0 | 0 | 0 | 0 |
| 0 | 0 | 0 | 0 | 0 |
| 0 | 0 | 0 | 0 | 0 |
| 0 | 0 | 0 | 0 | 0 |
| 0 | 0 | 0 | 0 | 0 |
| 0 | 0 | 0 | 0 | 0 |
| 0 | 0 | 0 | 0 | 0 |
| 0 | 0 | 0 | 0 | 0 |
| 0 | 0 | 0 | 0.173699587 | 0 |
| 0 | 0 | 0 | 0 | 0 |
| 0 | 0 | 0 | 0 | 0 |
| 0 | 0 | 0 | 0 | 0 |
| 0 | 0 | 0 | 0 | 0 |
| 0 | 0 | 0 | 0 | 0 |
| 0 | 0 | 0 | 0 | 0 |
| 0 | 0 | 0 | 0 | 0.002808396 |
| 0 | 0 | 0 | 0 | 0 |
| 0 | 0 | 0.038313412 | 0.007998101 | 0.013144374 |
| 0 | 0 | 0 | 0 | 0 |
| 0 | 0 | 0 | 0 | 0.010979196 |
| 0 | 0 | 0 | 0 | 0 |
| 0 | 0 | 0 | 0 | 0 |
| 0 | 0 | 0 | 0 | 0 |
| 0 | 0 | 0.013140207 | 0 | 0 |
| 0 | 0.002258513 | 0.016065154 | 0 | 0 |
| 0 | 0 | 0 | 0 | 0 |
| 0 | 0 | 0 | 0 | 0 |
| 0 | 0 | 0 | 0 | 0 |
| 0 | 0 | 0 | 0 | 0 |
| 0 | 0 | 0 | 0 | 0 |
| 0 | 0 | 0 | 0 | 0 |
| 0 | 0 | 0 | 0 | 0 |
| 0 | 0 | 0 | 0 | 0 |
| 0 | 0 | 0 | 0 | 0 |
| 0 | 0 | 0 | 0 | 0 |
| 0 | 0 | 0 | 0 | 0 |
| 0 | 0 | 0 | 0 | 0 |
| 0 | 0 | 0 | 0 | 0 |
| 0 | 0 | 0 | 0 | 0 |
| 0 | 0 | 0 | 0 | 0 |
| 0 | 0 | 0 | 0 | 0 |

| | | | | |
|---|---|---|---|---|
| 0 | 0 | 0 | 0 | 0 |
| 0 | 0 | 0 | 0 | 0 |
| 0 | 0 | 0 | 0 | 0 |
| 0 | 0 | 0 | 0 | 0 |
| 0 | 0 | 0 | 0 | 0 |
| 0 | 0 | 0 | 0 | 0 |
| 0 | 0 | 0 | 0 | 0 |
| 0 | 0 | 0 | 0 | 0 |
| 0 | 0 | 0 | 0 | 0 |
| 0 | 0 | 0 | 0 | 0 |
| 0 | 0 | 0 | 0 | 0 |
| 0 | 0 | 0 | 0 | 0 |
| 0 | 0.001620881 | 0.002254674 | 0.000896521 | 0 |
| 0 | 0 | 0 | 0 | 0 |
| 0 | 0 | 0 | 0 | 0.427559751 |
| 0 | 0 | 0 | 0 | 0 |
| 0 | 0 | 0 | 0 | 0 |
| 0 | 0 | 0 | 0 | 0 |
| 0 | 0 | 0 | 0 | 0 |
| 0 | 0 | 0 | 0 | 0 |
| 0.045111406 | 0.016836993 | 0.043306721 | 0.004358326 | 0.006069722 |
| 0 | 0 | 0 | 0 | 0 |
| 0 | 0 | 0 | 0 | 0 |
| 0 | 0 | 0 | 0 | 0.001186781 |
| 0 | 0 | 0 | 0 | 0 |
| 0 | 0 | 0 | 0 | 0 |
| 0.002754304 | 0 | 0.007142177 | 0.001171471 | 0.004418578 |
| 0 | 0 | 0 | 0 | 0 |
| 0 | 0 | 0 | 0 | 0 |
| 0.001813408 | 0 | 0.003581863 | 0.001880008 | 0 |
| 0.024287762 | 0 | 0.002892312 | 0 | 0 |
| 0 | 0 | 0 | 0 | 0 |
| 0 | 0 | 0 | 0 | 0 |
| 0 | 0 | 0.019674122 | 0 | 0.005564155 |
| 0.038583527 | 0.004382018 | 0.038650774 | 0.105795621 | 0 |
| 0 | 0 | 0 | 0 | 0 |
| 0 | 0 | 0 | 0 | 0 |
| 0 | 0 | 0 | 0 | 0 |
| 0 | 0.040774509 | 0 | 0 | 0 |
| 0 | 0 | 0 | 0 | 0 |
| 0 | 0.001349486 | 0 | 0 | 0 |
| 0 | 0.055798045 | 0 | 0 | 0 |
| 0 | 0.017763004 | 0 | 0 | 0 |
| 0 | 0 | 0 | 0 | 0 |
| 0 | 0 | 0 | 0 | 0 |
| 0 | 0.264097629 | 0 | 0.005192214 | 0 |
| 0 | 0 | 0.001889933 | 0 | 0 |
| 0 | 0.005152528 | 0.011905937 | 0 | 0 |

| | | | | |
|---|---|---|---|---|
| 0 | 0 | 0 | 0 | 0 |
| 0 | 0 | 0 | 0 | 0 |
| 0 | 0 | 0 | 0 | 0 |
| 0 | 0 | 0 | 0 | 0 |
| 0 | 0 | 0 | 0 | 0 |
| 0 | 0 | 0 | 0 | 0 |
| 0 | 0.003778409 | 0 | 0 | 0 |
| 0 | 0.004264552 | 0 | 0 | 0 |
| 0 | 0 | 0 | 0 | 0 |
| 0 | 0 | 0 | 0 | 0 |
| 0 | 0 | 0 | 0 | 0 |
| 0 | 0 | 0 | 0 | 0 |
| 0 | 0 | 0 | 0 | 0 |
| 0 | 0 | 0 | 0 | 0 |
| 0 | 0 | 0 | 0 | 0 |
| 0 | 0 | 0 | 0 | 0 |
| 0 | 0 | 0 | 0 | 0 |
| 0 | 0 | 0 | 0.059833588 | 0.094166465 |
| 0.011322405 | 0.047637158 | 0 | 0.013595802 | 0.058138283 |
| 0 | 0.078063691 | 0 | 0 | 0.005329904 |
| 0.096414357 | 0.000936929 | 0.011996137 | 0 | 0 |
| 0 | 0 | 0 | 0 | 0 |
| 0 | 0 | 0 | 0 | 0 |
| 0 | 0 | 0 | 0 | 0 |
| 0.014579731 | 0.004653518 | 0 | 0 | 0.00157274 |
| 0 | 0 | 0 | 0 | 0 |
| 0 | 0 | 0 | 0 | 0 |
| 0 | 0 | 0 | 0 | 0 |
| 0.020630583 | 0.204504339 | 0 | 0 | 0 |
| 0 | 0 | 0 | 0 | 0.002097702 |
| 1.818295733 | 0.063208717 | 0.893640478 | 0.080420273 | 0.024253542 |
| 0 | 0 | 0 | 0 | 0 |
| 0 | 0 | 0 | 0 | 0 |
| 0 | 0 | 0 | 0 | 0 |
| 0 | 0 | 0 | 0 | 0 |
| 0 | 0 | 0 | 0 | 0 |
| 0 | 0 | 0 | 0 | 0 |
| 0 | 0.012526588 | 0 | 0 | 0 |
| 0 | 0 | 0 | 0 | 0 |
| 0 | 0 | 0 | 0.007364889 | 0.000581422 |
| 0 | 0 | 0 | 0 | 0 |
| 0 | 0.013342613 | 0.112690871 | 0.00926371 | 0 |
| 0 | 0 | 0 | 0 | 0 |
| 0 | 0 | 0 | 0 | 0 |
| 0 | 0 | 0 | 0 | 0 |
| 0 | 0 | 0 | 0 | 0 |
| 0 | 0 | 0 | 0 | 0 |
| 0 | 0 | 0 | 0 | 0 |

| | | | | |
|---|---|---|---|---|
| 0 | 0 | 0 | 0 | 0 |
| 0 | 0 | 0 | 0 | 0 |
| 0.003572419 | 0.030862348 | 0.027347005 | 0.001097369 | 0.008195381 |
| 0 | 0 | 0 | 0 | 0 |
| 0 | 0 | 1.286562026 | 0 | 0 |
| 0 | 0 | 0 | 0 | 0 |
| 0 | 0 | 0 | 0 | 0 |
| 0 | 0 | 0 | 0 | 0 |
| 0 | 0 | 0 | 0 | 0 |
| 0 | 0 | 0 | 0.000602187 | 0 |
| 0 | 0 | 0 | 0 | 0 |
| 0 | 0 | 0 | 0 | 0 |
| 0 | 0 | 0 | 0 | 0 |
| 0.001316056 | 0 | 0 | 0 | 0 |
| 0 | 0.003820639 | 0.001144286 | 0 | 0.002038821 |
| 0 | 0 | 0 | 0 | 0.067667646 |
| 0.001485197 | 0 | 0 | 0.00364976 | 0 |
| 0 | 0 | 0 | 0 | 0 |
| 0 | 0 | 0 | 0 | 0 |
| 0 | 0 | 0 | 0 | 0 |
| 0 | 0 | 0 | 0 | 0 |
| 0 | 0.006825382 | 0.012240487 | 0 | 0 |
| 0 | 0 | 0 | 0 | 0 |
| 0.041914487 | 0 | 0 | 0 | 0 |
| 0 | 0 | 0 | 0 | 0 |
| 0 | 0 | 0 | 0 | 0 |
| 0 | 0 | 0 | 0 | 0 |
| 0 | 0 | 0 | 0 | 0 |
| 0 | 0 | 0 | 0 | 0 |
| 0 | 0 | 0 | 0 | 0 |
| 0 | 0 | 0 | 0 | 0 |
| 0 | 0 | 0 | 0 | 0 |
| 0 | 0.032531558 | 0 | 0 | 0 |
| 0 | 0 | 0 | 0 | 0 |
| 0 | 0 | 0 | 0 | 0 |
| 0 | 0.009501599 | 0 | 0.001233877 | 0 |
| 0 | 0 | 0 | 0 | 0 |
| 0.012633462 | 0.782438948 | 0.003548978 | 0.012159602 | 0.0073187 |
| 0 | 0 | 0 | 0 | 0 |
| 0 | 0 | 0 | 0 | 0 |
| 0 | 0 | 0 | 0 | 0 |
| 0 | 0 | 0 | 0 | 0 |
| 0 | 0 | 0 | 0 | 0 |
| 0 | 0 | 0 | 0 | 0 |
| 0 | 0 | 0 | 0 | 0 |
| 0 | 0 | 0 | 0 | 0 |
| 0 | 0 | 0 | 0 | 0 |
| 0 | 0 | 0 | 0 | 0 |

| | | | | |
|---|---|---|---|---|
| 0 | 1.584488736 | 0 | 0 | 0 |
| 0 | 0.039798804 | 0.234214968 | 0 | 0 |
| 0 | 0 | 0 | 0.004313666 | 0 |
| 0 | 0 | 0 | 0 | 0 |
| 0.003767597 | 0 | 0.002728659 | 0.002169982 | 0.006138589 |
| 0 | 0 | 0 | 0 | 0 |
| 0 | 0.010283926 | 0 | 0 | 0 |
| 0.004620977 | 0.008248964 | 0.009040476 | 0.001216683 | 0 |
| 0 | 0 | 0 | 0 | 0 |
| 0 | 0.056997472 | 0.036556072 | 0.03574075 | 0.001064272 |
| 0 | 0 | 0 | 0 | 0 |
| 0.003396698 | 0.455158844 | 0 | 0.007223479 | 0 |
| 0.07907921 | 0.001284155 | 0.008977803 | 0.017533649 | 0 |
| 0 | 0.014766554 | 0.044392916 | 0 | 0 |
| 0 | 0.001320724 | 0 | 0 | 0 |
| 0.04849598 | 0 | 0 | 0 | 0 |
| 0 | 0.051067469 | 0 | 0 | 0 |
| 0 | 0 | 0 | 0 | 0 |
| 0.510609712 | 0 | 0 | 0 | 0 |
| 0 | 0 | 0 | 0.006148235 | 0 |
| 0 | 0 | 0 | 0 | 0 |
| 0 | 0 | 0 | 0 | 0 |
| 0 | 0 | 0 | 0 | 0 |
| 0 | 0.064178009 | 0 | 0 | 0.055470352 |
| 0 | 0.036329139 | 0 | 0 | 0 |
| 0.00092338 | 0.005769178 | 0 | 0.0080838 | 0.002166441 |
| 0 | 0.00514304 | 0 | 25.43602765 | 0.015450513 |
| 0 | 0 | 0 | 0 | 0 |
| 0 | 0 | 0 | 0 | 0 |
| 0 | 0.002857957 | 0 | 0 | 0 |
| 0.010156075 | 0 | 0 | 0 | 0 |
| 0 | 0 | 0 | 0 | 0 |
| 0 | 0 | 0 | 0 | 0 |
| 0 | 0 | 0 | 0 | 0 |
| 0 | 0 | 0 | 0 | 0 |
| 0 | 0 | 0 | 0 | 0 |
| 0 | 0 | 0 | 0 | 0 |
| 0 | 0 | 0 | 0 | 0 |
| 0 | 0 | 0 | 0 | 0 |
| 0 | 0 | 0.001278829 | 0 | 0 |
| 0 | 0 | 0 | 0 | 0 |
| 0 | 0 | 0 | 0 | 0 |
| 0 | 0 | 0 | 0 | 0 |
| 0.006695165 | 0 | 0 | 0 | 0 |
| 0 | 0 | 0 | 0 | 0 |
| 0 | 0.003282968 | 0 | 0 | 0 |
| 0.001722443 | 0 | 0.002419334 | 0.002116388 | 0 |
| 0 | 0 | 0 | 0 | 0 |

| | | | | |
|---|---|---|---|---|
| 0 | 0 | 0 | 0 | 0 |
| 0.03304499 | 0 | 0 | 0.000886954 | 0 |
| 0 | 0 | 0 | 0 | 0 |
| 0 | 0.137269629 | 0 | 0 | 0.002577374 |
| 0.021848172 | 0 | 0 | 0 | 0 |
| 0.052682572 | 0.001674291 | 0.049067106 | 0 | 0 |
| 0 | 0 | 0 | 0.034753396 | 0 |
| 0 | 0 | 0 | 0 | 0 |
| 0 | 0 | 0 | 0 | 0 |
| 0 | 0.001638076 | 0 | 0 | 0 |
| 0.0159969 | 0.000970948 | 0.015470566 | 0 | 0.00355495 |
| 0.003440957 | 0 | 0.099165845 | 0 | 0 |
| 0 | 0 | 0 | 0 | 0 |
| 0.002229797 | 0.008255723 | 0.00626392 | 0 | 0 |
| 0.034845038 | 0 | 0.005017553 | 0 | 0 |
| 0 | 0 | 0 | 0 | 0 |
| 0 | 0 | 0 | 0 | 0 |
| 0 | 0 | 0.00126996 | 0 | 0 |
| 0 | 0 | 0 | 0 | 0 |
| 0.003268377 | 0.008250695 | 0 | 0 | 0 |
| 0.037639395 | 0 | 0 | 0 | 0 |
| 0 | 0.161270607 | 0.034229863 | 0.062726766 | 0.214103155 |
| 0 | 0.00239381 | 0 | 0 | 0 |
| 0 | 0 | 0 | 0 | 0 |
| 0 | 0 | 0 | 0 | 0 |
| 0 | 0 | 0 | 0 | 0 |
| 0 | 0 | 0 | 0 | 0 |
| 0 | 0 | 0 | 0 | 0 |
| 0 | 0 | 0 | 0 | 0 |
| 5.0664064 | 0.234291919 | 0.674847191 | 2.606012516 | 0.05680284 |
| 0 | 0 | 0 | 0 | 0.001226885 |
| 0 | 0.106593041 | 0 | 0 | 0 |
| 0 | 0 | 0 | 0 | 0 |
| 0.012379222 | 0 | 0 | 0 | 0 |
| 0 | 0 | 0 | 0 | 0 |
| 0 | 0 | 0 | 0 | 0 |
| 0 | 0 | 0 | 0.051864555 | 0.25080458 |
| 0.008080913 | 0.366744211 | 0 | 0 | 0 |
| 0 | 0 | 0.202760008 | 0 | 0 |
| 0 | 0 | 0.013897163 | 0 | 0 |
| 0.011406177 | 0.003582082 | 0.030468607 | 0 | 0 |
| 0 | 1.229053792 | 5.108913445 | 0.468791888 | 2.505253944 |
| 0.00277599 | 0.282858535 | 0 | 0 | 0 |
| 0 | 0 | 0 | 0 | 0 |
| 0 | 0 | 0 | 0 | 0 |
| 0 | 0 | 0 | 0 | 0 |
| 0 | 0.000791506 | 0.000675612 | 0.000788017 | 0.001560437 |
| 0 | 0 | 0 | 0 | 0 |

| | | | | |
|---|---|---|---|---|
| 0.001846368 | 0.005615365 | 0 | 0.016042642 | 0.008526408 |
| 0 | 0 | 0 | 0 | 0 |
| 5.551692244 | 0.001330672 | 0.002271667 | 0.015566491 | 0.776524373 |
| 0 | 0 | 0 | 0 | 0 |
| 0 | 0.033868852 | 0.341330369 | 0 | 0.427449821 |
| 0 | 0.006192777 | 0.099377235 | 0 | 0 |
| 0 | 0 | 0 | 0 | 0 |
| 0 | 0.001368192 | 0 | 0 | 0 |
| 0 | 0 | 0 | 0 | 0 |
| 0 | 0 | 0 | 0 | 0 |
| 0 | 0 | 0 | 0 | 0 |
| 0 | 0 | 0 | 0 | 0 |
| 0 | 0 | 0 | 0 | 0 |
| 0.013124279 | 0 | 0.01659086 | 0 | 0.0037324 |
| 0 | 0 | 0 | 0 | 0 |
| 0 | 0.011737374 | 0.064434459 | 0.008248691 | 0.068253258 |
| 0.09321881 | 0.047829613 | 0.02220953 | 0.052571181 | 0.043752931 |
| 0.011802859 | 0 | 0 | 0.003239881 | 0 |
| 0 | 0 | 0 | 0 | 0 |
| 0 | 0 | 0 | 0 | 0 |
| 0.001562956 | 0 | 0.003018565 | 0 | 0 |
| 0.002157784 | 0 | 0.002020541 | 0.009058599 | 0 |
| 0 | 0 | 0 | 0 | 0 |
| 0 | 0 | 0.039418735 | 0 | 0 |
| 0 | 0.005542973 | 0.000610499 | 0 | 0 |
| 0 | 0 | 0 | 0 | 0 |
| 0.001836794 | 0 | 0 | 0 | 0 |
| 0 | 0 | 0 | 0 | 0 |
| 0 | 0 | 0 | 0 | 0 |
| 0 | 0.003325167 | 0 | 0 | 0 |
| 0 | 0.013483815 | 0.040151856 | 0.001103375 | 0 |
| 0.008627798 | 0.004406074 | 0 | 0 | 0 |
| 0 | 0.004981774 | 0.01432366 | 0.030019958 | 0.029538246 |
| 0.01258456 | 0.490361879 | 0 | 0.002244601 | 0 |
| 0 | 0 | 0 | 0 | 0 |
| 0 | 0 | 0 | 0 | 0 |
| 0.001280478 | 0.00924478 | 0.001079131 | 0 | 0 |
| 0.032645419 | 0.002409949 | 0.142202331 | 0.00701342 | 0 |
| 0 | 0.01735193 | 0.253906954 | 0 | 0.019082559 |
| 0 | 0.004698597 | 0 | 0 | 0 |
| 0 | 0 | 0 | 0 | 0.025249789 |
| 0 | 0.003698872 | 0.035411041 | 0 | 0 |
| 0.017671467 | 0 | 0.010412301 | 0 | 0 |
| 0 | 0 | 0.153169131 | 0 | 0 |
| 0 | 0 | 0.152124459 | 0 | 0 |
| 0 | 0.001414561 | 0 | 0 | 0.002048898 |
| 0 | 0 | 0 | 0 | 0 |
| 0 | 0 | 0 | 0 | 0 |

| | | | | |
|---|---|---|---|---|
| 0.361384205 | 0 | 0 | 0 | 0 |
| 0.519338325 | 0.142216448 | 0.211880628 | 0.192390958 | 0.068582408 |
| 0 | 0 | 0 | 0 | 0 |
| 0 | 0 | 0 | 0 | 0 |
| 0 | 0 | 0 | 0 | 0 |
| 0 | 0 | 0 | 0 | 0 |
| 0 | 0 | 0 | 0 | 0 |
| 0 | 0 | 0 | 0 | 0 |
| 0 | 0.001156194 | 0 | 0 | 0 |
| 0 | 0 | 0 | 0 | 0 |
| 0.00121679 | 0.000563139 | 0.009827302 | 0 | 0 |
| 0.013291248 | 0.164966188 | 0.143516489 | 0.003062086 | 0 |
| 0 | 0.117895167 | 0 | 0 | 0 |
| 0 | 0 | 0.014374114 | 0 | 0 |
| 0 | 0.00818433 | 0.002451217 | 0 | 0 |
| 0 | 0 | 0 | 0 | 0 |
| 0.048205513 | 0.005700313 | 0.004921593 | 0.011154686 | 0.000885758 |
| 0.052377572 | 0 | 0.011035386 | 0 | 0 |
| 0 | 0 | 0 | 0 | 0 |
| 0 | 0 | 0 | 0 | 0 |
| 0.020366609 | 0.014189388 | 0.002076302 | 0.006861615 | 0.019410593 |
| 0.000817515 | 0 | 0.001148277 | 0 | 0 |
| 0 | 0 | 0 | 0 | 0 |
| 0 | 0 | 0 | 0 | 0 |
| 0.105602736 | 0 | 0.002325716 | 0 | 0 |
| 0 | 0 | 0.004654288 | 0 | 0 |
| 0 | 0.000600241 | 0 | 0 | 0.002704832 |
| 0 | 0.036410385 | 0.105190905 | 0.040754349 | 0.022815011 |
| 0 | 0.001470285 | 0 | 0 | 0 |
| 0 | 0 | 0.002957689 | 0 | 0 |
| 0.000999008 | 0 | 0.0007893 | 0 | 0 |
| 0 | 0 | 0 | 0 | 0 |
| 0 | 0 | 0 | 0 | 0 |
| 0 | 0 | 0 | 0 | 0 |
| 0 | 0.000563239 | 0.175854478 | 0 | 0.000634522 |
| 0.260389549 | 0.236644619 | 0.175954397 | 0.051186194 | 0.083387745 |
| 0 | 0.000675498 | 0 | 0 | 0 |
| 0 | 0 | 0.001575795 | 0 | 0 |
| 0.002871357 | 0.144316814 | 0 | 0.00238145 | 0 |
| 0 | 0 | 0 | 0 | 0 |
| 0 | 0 | 0.160578396 | 0 | 0 |
| 0 | 0 | 0 | 0 | 0 |
| 0.003488213 | 0.051982682 | 0.018373214 | 0.000964353 | 0 |
| 0.003704584 | 0 | 0 | 0 | 0 |
| 0 | 0 | 0 | 0 | 0 |
| 0 | 0.374256823 | 0 | 0 | 0.268684689 |
| 0 | 0 | 0 | 0 | 0 |
| 0.004899414 | 0 | 0 | 0 | 0 |

| | | | | |
|---|---|---|---|---|
| 0 | 0 | 0 | 0 | 0 |
| 0 | 0 | 0 | 0 | 0 |
| 0 | 0 | 0 | 0 | 0 |
| 0 | 0.367960823 | 5.101944986 | 0 | 0.994066796 |
| 0 | 0 | 0 | 0 | 0 |
| 0 | 0 | 0 | 0 | 0 |
| 0 | 0.00204078 | 0 | 0 | 0 |
| 0 | 0 | 0 | 0 | 0 |
| 0 | 0.477561345 | 0.615382792 | 2.43024671 | 0.478985961 |
| 0 | 0.003528971 | 0.650646996 | 0 | 0.086895189 |
| 0 | 0 | 0 | 0 | 0 |
| 0 | 0.003062485 | 0 | 0.001255466 | 0.001623563 |
| 0 | 0 | 0 | 0 | 0 |
| 0 | 0 | 0 | 0 | 0 |
| 0 | 0.040583379 | 0.392853022 | 0.012728165 | 0.02964238 |
| 0 | 0 | 0 | 0 | 0 |
| 0 | 0 | 0.049875956 | 0 | 0.058981863 |
| 0 | 2.28017749 | 0 | 0.000579805 | 5.28884755 |
| 0 | 0 | 0 | 0 | 0 |
| 0 | 0 | 0 | 0 | 0 |
| 0 | 0 | 0 | 0 | 0 |
| 0 | 0 | 0 | 0 | 0 |
| 0 | 0.004976524 | 0 | 0 | 0 |
| 0.276453515 | 0 | 0.242174578 | 0.017392781 | 0 |
| 0 | 0 | 0 | 0.007388459 | 0 |
| 0 | 0.344694363 | 0 | 0 | 0.102024259 |
| 0 | 0.057395263 | 0 | 0 | 0 |
| 0 | 1.974816337 | 2.540609096 | 1.356787653 | 1.909907433 |
| 0 | 0 | 0 | 0 | 0 |
| 0 | 0 | 0 | 0 | 0 |
| 0 | 0.00945717 | 0 | 0.001046166 | 0.042221676 |
| 0 | 0 | 0.196997933 | 0 | 0.12120086 |
| 0.000432002 | 0.003465511 | 0 | 0.001194313 | 0 |
| 0 | 0 | 0 | 0 | 0 |
| 0 | 0.064840865 | 0 | 0 | 0 |
| 0 | 0.345394743 | 0.1059634 | 0.025428237 | 0.063401492 |
| 0 | 0.45605172 | 0.720057031 | 0 | 0.453059181 |
| 0 | 0 | 0 | 0 | 0 |
| 0 | 0.005063368 | 0 | 0 | 0 |
| 0 | 0 | 0.004402548 | 0 | 0.002759995 |
| 0.204630216 | 0.125293073 | 0.113487283 | 0.001204395 | 0 |
| 0 | 0.021163424 | 0 | 0 | 0 |
| 0.185229183 | 0.020109695 | 0.013920925 | 5.010427455 | 0.071934747 |
| 0 | 0 | 0.003084091 | 0 | 0 |
| 0 | 0 | 0 | 0 | 0 |
| 0 | 0 | 0 | 0 | 0 |
| 0 | 0 | 0 | 0 | 0 |
| 0.024464234 | 0.006232413 | 0.002127942 | 0 | 0.002574435 |

| | | | | |
|---|---|---|---|---|
| 0 | 0 | 0 | 0 | 0 |
| 0 | 0.036149327 | 0 | 0.011117334 | 0.015778032 |
| 0.210204957 | 0 | 0.004533051 | 0.065429555 | 0.001794827 |
| 0 | 0.0036596 | 0 | 0 | 0 |
| 0 | 0 | 0 | 0 | 0 |
| 0 | 0.085030013 | 0 | 0 | 0.014492425 |
| 0 | 0 | 0.138072219 | 0 | 0 |
| 0.005668135 | 0 | 0 | 0 | 0 |
| 0 | 0.003558056 | 0 | 0 | 0 |
| 0 | 0 | 0 | 0 | 0 |
| 0.053021885 | 0.026200893 | 0.01891527 | 0.078256268 | 0.001101376 |
| 0 | 0.863038992 | 0.684644365 | 1.211630367 | 1.306172263 |
| 0 | 0 | 0 | 0 | 0 |
| 0.031005497 | 0.043892768 | 0.024977289 | 0.389092091 | 0.113159453 |
| 0.212715354 | 0.030598124 | 0.030849414 | 0.003145664 | 0 |
| 0 | 5.045993884 | 0 | 0.000797485 | 7.344788338 |
| 0 | 0 | 0 | 0 | 0 |
| 0 | 0 | 0 | 0 | 0 |
| 0 | 0.16306168 | 0 | 0 | 0.171281333 |
| 0.029755416 | 0.00144958 | 0.102286023 | 0 | 0.016511838 |
| 0.055063291 | 0.134547235 | 0.041965893 | 0 | 0 |
| 0.434680795 | 2.213424161 | 0.01415009 | 0.0379943 | 0 |
| 0.209372292 | 0.462824044 | 2.084772266 | 1.192799575 | 0.603706768 |
| 0 | 0 | 0 | 0 | 0 |
| 0 | 0.0673 | 0 | 0 | 0 |
| 0 | 0.212679495 | 0 | 0.001955145 | 0.389592669 |
| 1.766102543 | 0.016268886 | 0.007584745 | 0.014636012 | 0 |
| 0 | 0 | 0 | 0 | 0 |
| 0 | 0.031030042 | 0 | 0 | 0 |
| 0 | 0 | 0 | 0 | 0 |
| 0 | 0.005422733 | 0 | 0 | 0.002647247 |
| 0.069534824 | 0.093294532 | 0 | 0.003850886 | 0.207937864 |
| 1.984673439 | 0.00060244 | 0.009827511 | 0 | 0 |
| 0 | 0.043949971 | 0.09690224 | 0.006860741 | 0.0010351 |
| 0 | 0.015507105 | 0.001357593 | 0.000527821 | 0 |
| 0.042101366 | 0.106711169 | 0.139479268 | 0.218645912 | 0.022771388 |
| 0 | 0.001407256 | 0.01741746 | 0 | 0 |
| 0 | 0 | 0 | 0 | 0 |
| 0 | 0.190293755 | 0 | 0.612705892 | 2.91509914 |
| 0 | 0 | 0 | 0 | 0 |
| 0 | 0.058367062 | 0 | 0 | 0.023844852 |
| 0 | 0 | 0 | 0 | 0 |
| 0 | 0 | 0 | 0 | 0 |
| 0 | 0 | 0 | 0 | 0 |
| 0.030960899 | 0.016405573 | 0.273924005 | 0 | 0.046321623 |
| 0 | 0.25456735 | 0 | 0 | 0 |
| 0 | 0 | 0 | 0 | 0 |
| 0 | 0.223697684 | 0.157943634 | 0.099758937 | 0 |

| | | | | |
|---|---|---|---|---|
| 0 | 0 | 0 | 0 | 0 |
| 0 | 0 | 0 | 0 | 0 |
| 0 | 0 | 0 | 0 | 0 |
| 0 | 0.00141594 | 0 | 0 | 0.003389674 |
| 0 | 0 | 0 | 0 | 0 |
| 0 | 0 | 0 | 0 | 1.760846575 |
| 0 | 0.454785303 | 0.001764272 | 0 | 0.001528078 |
| 0 | 0.043973575 | 0.044485819 | 0 | 0.049538863 |
| 0 | 0 | 0 | 0 | 0 |
| 0 | 0 | 0 | 0 | 0 |
| 0.051963294 | 0.013207223 | 0.023332021 | 0.003728826 | 0 |
| 0 | 0 | 0 | 0 | 0 |
| 0 | 0.0094928 | 0 | 0 | 0.134358124 |
| 0 | 0 | 0 | 0 | 0 |
| 0 | 0 | 0 | 0 | 0 |
| 0 | 0 | 0 | 0 | 0 |
| 0 | 0 | 0 | 0 | 0 |
| 0 | 0.002133302 | 0.000933816 | 0.030551492 | 0.002218424 |
| 0 | 0.060782269 | 0.219517804 | 0 | 0 |
| 0 | 0 | 0 | 0 | 0 |
| 0 | 0 | 0 | 0 | 0 |
| 0 | 0 | 0 | 0 | 0 |
| 0 | 0.001135175 | 0 | 0 | 0.024297998 |
| 0 | 0.030638121 | 0 | 0 | 0 |
| 0 | 0 | 0 | 0 | 0 |
| 0 | 0 | 0 | 0 | 0 |
| 0 | 0 | 0 | 0 | 0 |
| 0 | 0 | 0 | 0 | 0 |
| 0 | 0 | 0 | 0 | 0 |
| 0 | 1.090471704 | 0.909201479 | 0 | 0.282946045 |
| 0 | 0.003881402 | 0 | 0 | 0 |
| 0 | 0 | 0 | 0 | 0 |
| 0 | 0.017660044 | 0 | 0 | 0.062003162 |
| 0 | 0 | 0 | 0 | 0 |
| 0 | 0 | 0 | 0.002964981 | 0 |
| 0 | 0 | 0 | 0 | 0 |
| 0 | 0.029926498 | 0.080329003 | 0.050172997 | 0.020355617 |
| 0.006309731 | 0.005339771 | 0.013483831 | 0.006313032 | 0 |
| 0 | 0 | 0 | 0 | 0 |
| 0 | 0 | 0 | 0 | 0 |
| 0 | 0 | 0.235026684 | 0 | 0 |
| 0 | 0 | 0 | 0 | 0 |
| 0 | 0 | 0 | 0 | 0 |
| 0 | 0 | 0.019918239 | 0 | 0.015152253 |
| 0 | 0.107622767 | 0 | 0.107903029 | 0.074496437 |
| 0 | 0 | 0 | 0 | 0 |
| 0 | 0 | 0 | 0 | 0.00142895 |
| 0 | 0 | 0 | 0 | 0 |

| | | | | |
|---|---|---|---|---|
| 0 | 0 | 0 | 0 | 0 |
| 0 | 0 | 0 | 0 | 0 |
| 0 | 0 | 0 | 0 | 0.051773016 |
| 0 | 0.041994101 | 0 | 0 | 1.222550869 |
| 0 | 0 | 0 | 0 | 0 |
| 0 | 0.187712923 | 0 | 0 | 0.556159203 |
| 0 | 0 | 0 | 0 | 0 |
| 0 | 0 | 0 | 0 | 0 |
| 0.141472463 | 0.005753465 | 0.048193631 | 0.01626783 | 0 |
| 0 | 0 | 0 | 0 | 0 |
| 0 | 0 | 0 | 0 | 0 |
| 0 | 0 | 0 | 0 | 0 |
| 0 | 0 | 0 | 0 | 0 |
| 0 | 0 | 0 | 0 | 0 |
| 0 | 0.257998546 | 0 | 0 | 0.51627779 |
| 0 | 0 | 0 | 0 | 0 |
| 0 | 0 | 0 | 0 | 0 |
| 0 | 0 | 0 | 0 | 0 |
| 0 | 0 | 0 | 0 | 0 |
| 0 | 0 | 0 | 0 | 0 |
| 0 | 0.001583766 | 0 | 0 | 0.101303302 |
| 0 | 0 | 0 | 0 | 0 |
| 0 | 0 | 0 | 0 | 0 |
| 0.19090298 | 0 | 0 | 0 | 0 |
| 1.683265742 | 0.343978939 | 0 | 10.26149527 | 0.793928768 |
| 0 | 1.647091218 | 22.14173021 | 0.197582398 | 1.100051586 |
| 0 | 0 | 0 | 0 | 0.170100505 |
| 0 | 0 | 0 | 0 | 0 |
| 0 | 0.040697549 | 0 | 0.004475847 | 0.010395818 |
| 0 | 0 | 0 | 0 | 0 |
| 0 | 0 | 0 | 0 | 0 |
| 0 | 0.014640476 | 0 | 0 | 0 |
| 0 | 0 | 0 | 0 | 0 |
| 0 | 0 | 0 | 0 | 0 |
| 0.006432264 | 0.004286731 | 0.002439375 | 0.218608196 | 0.049902342 |
| 0.002182479 | 0 | 0.091198554 | 0.007542112 | 0 |
| 0 | 0 | 0 | 0 | 0 |
| 0 | 1.489544623 | 1.598769523 | 0 | 0.298521848 |
| 0 | 0 | 0 | 0 | 1.486935944 |
| 0 | 0 | 0 | 0 | 0 |
| 0 | 0 | 0 | 0 | 0 |
| 0 | 0.007254348 | 0 | 0 | 0 |
| 0 | 0.085346924 | 0.144522469 | 0.259034946 | 0.29647921 |
| 0 | 0 | 0 | 0 | 0.962576709 |
| 0 | 0 | 0 | 0 | 0 |
| 0 | 0.002551763 | 0 | 0.001746605 | 0 |
| 0 | 0 | 0 | 0 | 0 |
| 1.785334032 | 0.056405541 | 0.438668763 | 0.018213067 | 0.001717411 |

| | | | | |
|---|---|---|---|---|
| 0 | 0.003599873 | 0 | 0 | 0 |
| 0 | 0 | 0 | 0 | 0 |
| 0 | 0 | 0 | 0 | 0 |
| 0 | 0 | 0 | 0 | 0 |
| 0 | 0 | 0 | 0 | 0 |
| 0 | 0.012352471 | 0 | 0 | 0 |
| 0 | 0 | 0 | 0 | 0.003721894 |
| 0 | 0 | 0 | 0 | 0 |
| 0 | 0 | 0 | 0 | 0 |
| 0 | 0.001893361 | 0 | 0 | 0 |
| 0 | 0 | 0 | 0 | 0 |
| 0 | 0.004524756 | 0 | 2.021448986 | 0.093321779 |
| 0 | 0.001827244 | 0.065767222 | 0.072312862 | 0.00102925 |
| 0 | 0 | 0 | 0 | 0 |
| 0.14212107 | 3.014724434 | 0 | 0.075801574 | 0.000983072 |
| 0 | 0 | 0 | 0 | 0 |
| 0 | 0 | 0.001381754 | 0 | 0 |
| 0.002832436 | 0 | 0 | 0 | 0 |
| 0 | 0 | 0.002393846 | 0 | 0.000646244 |
| 0 | 0.242346307 | 0 | 0 | 0 |
| 0 | 0 | 0 | 0 | 0.031923513 |
| 0 | 0 | 0 | 0 | 0 |
| 0 | 0.296558767 | 1.407829091 | 0 | 0.002094616 |
| 0 | 0 | 0 | 0 | 0 |
| 0 | 0 | 0 | 0 | 0 |
| 0 | 0 | 0 | 0 | 0 |
| 0 | 0 | 0 | 0 | 0 |
| 0 | 0 | 0 | 0 | 0 |
| 0 | 0.001179031 | 0 | 0 | 0 |
| 0 | 0 | 0 | 0 | 0 |
| 0 | 0 | 0 | 0 | 0 |
| 0.056580536 | 0.018967058 | 0.048005807 | 0.023956639 | 0 |
| 0 | 0 | 0 | 0 | 0 |
| 0 | 0.011597022 | 0 | 0 | 0 |
| 0.00376055 | 0 | 0.082566732 | 0.011673184 | 0 |
| 0 | 0.020782798 | 0.044886955 | 0.219608938 | 0.01090836 |
| 0 | 0 | 0 | 0 | 0 |
| 0 | 0.001226278 | 0 | 0 | 0.213141882 |
| 0 | 0 | 0 | 0 | 0 |
| 0 | 0 | 0 | 0 | 0 |
| 0 | 0.180905792 | 0 | 0 | 0.27062126 |
| 0 | 0 | 0 | 0 | 0 |
| 0 | 0 | 0 | 0 | 0 |
| 0 | 0.003877702 | 0 | 0 | 0 |
| 0.023786907 | 0.035552834 | 0.178899234 | 0.122359276 | 0 |
| 0 | 0 | 0 | 0 | 0 |
| 0 | 0.220174416 | 0 | 0 | 0.558174743 |
| 0 | 0 | 0.01146176 | 0 | 0 |

| | | | | |
|---|---|---|---|---|
| 0 | 0 | 0 | 0 | 0 |
| 0 | 0 | 0 | 0 | 0 |
| 0.004033052 | 0 | 0 | 0 | 0 |
| 0 | 0 | 0 | 0 | 0 |
| 0 | 0.002650096 | 0 | 0 | 0 |
| 0 | 0 | 0 | 0 | 0 |
| 0 | 0 | 0 | 0 | 0.180080923 |
| 0 | 0.121277726 | 0.025278161 | 0 | 0.000595756 |
| 0 | 0 | 0 | 0 | 0 |
| 0 | 0 | 0 | 0 | 0 |
| 0 | 0 | 0 | 0 | 0 |
| 0 | 0 | 0 | 0 | 0 |
| 0 | 0 | 0 | 0 | 0 |
| 0 | 0.139857529 | 0 | 0 | 0.421112296 |
| 0 | 0 | 0 | 0 | 0 |
| 0 | 0 | 0 | 0 | 0 |
| 0 | 0.580810428 | 0 | 0 | 0 |
| 0 | 0 | 0 | 0 | 0 |
| 0 | 6.04276136 | 0 | 0 | 2.221918455 |
| 0 | 0 | 0 | 0 | 0 |
| 0 | 0 | 0 | 0 | 0 |
| 0 | 0 | 0 | 0 | 0 |
| 0 | 0 | 0 | 0 | 0 |
| 0 | 0.000921312 | 0 | 0 | 0 |
| 0 | 0 | 0 | 0.003337296 | 0 |
| 0.005989963 | 0.002856202 | 0.035948455 | 0.021912561 | 0 |
| 0 | 0 | 0 | 0 | 0 |
| 0.055431833 | 0.308344186 | 0.102564574 | 0 | 0 |
| 0 | 0 | 0 | 0 | 0.346272354 |
| 0 | 0.32355498 | 0.42101938 | 0 | 0.349158316 |
| 0 | 0 | 0 | 0 | 0 |
| 0 | 0 | 0 | 0 | 0 |
| 0 | 0.072236418 | 0 | 0 | 0 |
| 0 | 0 | 0 | 0 | 0 |
| 0.00209237 | 0.00088033 | 0.007414122 | 0.001402321 | 0 |
| 0 | 0 | 0 | 0 | 0 |
| 0 | 0 | 0 | 0 | 0.604963261 |
| 0 | 0 | 0.001591622 | 0.001392321 | 0 |
| 0 | 0 | 0 | 0 | 0 |
| 0 | 0.003507507 | 0 | 0 | 0 |
| 4.29784741 | 1.805741698 | 6.494298124 | 3.098300242 | 0.730074753 |
| 0.404309246 | 0.236181437 | 0.52328478 | 0 | 0.019934845 |
| 0 | 0 | 0 | 0 | 0.057237907 |
| 0 | 0.020371394 | 0 | 0 | 0 |
| 0 | 0 | 0 | 0 | 0 |
| 0 | 0.00118034 | 0 | 0 | 0 |
| 0 | 0.002493835 | 0 | 0 | 0 |
| 0 | 0.003646861 | 0.008481479 | 0.000789302 | 0 |

| | | | | |
|---|---|---|---|---|
| 0 | 0 | 0 | 0 | 0 |
| 0 | 0.000815456 | 0.026171704 | 0 | 0 |
| 0 | 0 | 0.00867151 | 0 | 0 |
| 0 | 0 | 0 | 0 | 0 |
| 0 | 1.177380832 | 0.563661076 | 1.184456201 | 1.136117436 |
| 0 | 0 | 0 | 0 | 0 |
| 0 | 0 | 0 | 0 | 0 |
| 0 | 0 | 0 | 0 | 0 |
| 4.474106321 | 0 | 0.198871175 | 0 | 0 |
| 0 | 0 | 0 | 0 | 0 |
| 0 | 0 | 0 | 0 | 0 |
| 0 | 0 | 0 | 0 | 0 |
| 0 | 0.025672287 | 0.196949333 | 0 | 0.03695508 |
| 0.24289474 | 0.003896477 | 0.025498951 | 0.143146357 | 0.007462345 |
| 0 | 0 | 0 | 1.855071321 | 0.190574945 |
| 0 | 0.274498597 | 0.590973744 | 0 | 0.099057367 |
| 0.877907131 | 0.063221188 | 0.001151238 | 0 | 0 |
| 0 | 0 | 0 | 0 | 0 |
| 0 | 0 | 0 | 0 | 0.035932752 |
| 0 | 0 | 0 | 0 | 0 |
| 0 | 0 | 0 | 0 | 0 |
| 0 | 0 | 0 | 0 | 0 |
| 0 | 0 | 0 | 0 | 0 |
| 0 | 0 | 0 | 0 | 0 |
| 0 | 0 | 0 | 0 | 0 |
| 0 | 0 | 0 | 0 | 0 |
| 0 | 0.071831938 | 0.142315717 | 0 | 0.001651489 |
| 0.001429684 | 0.002481254 | 0.00602438 | 0.001317504 | 0 |
| 0 | 0 | 0 | 0 | 0 |
| 0 | 0 | 0 | 0 | 0 |
| 0 | 0 | 0 | 0 | 0 |
| 0 | 0 | 0 | 0 | 0 |
| 0 | 0.01765228 | 0 | 0 | 0 |
| 0 | 0 | 0 | 0 | 0.006446442 |
| 0 | 0.007178056 | 0 | 0.387744421 | 0.03766003 |
| 0 | 0.004592474 | 0.068165108 | 0 | 0 |
| 0 | 0.230986938 | 0.102839944 | 0 | 0 |
| 0 | 0 | 0 | 0 | 0 |
| 0 | 0 | 0 | 0 | 0 |
| 0 | 0.482245688 | 0 | 0 | 0 |
| 0 | 0.480200135 | 0 | 1.402985923 | 0.064156528 |
| 0 | 0.891555484 | 0 | 0 | 0.009603874 |
| 0 | 0 | 0 | 0 | 0.011915184 |
| 0 | 0.000809824 | 0 | 0 | 0 |
| 0 | 0 | 0 | 0 | 0 |
| 0 | 0 | 0 | 0 | 0 |
| 0 | 0 | 0 | 0 | 0 |
| 0 | 0 | 0 | 0 | 0 |

| | | | | |
|---|---|---|---|---|
| 0 | 0.723015358 | 0 | 0 | 0.048152989 |
| 0 | 0 | 0 | 0 | 0 |
| 0 | 0 | 0.017846621 | 0 | 0.001094328 |
| 0 | 0.00074954 | 0 | 0 | 0 |
| 0 | 0.006454467 | 0.07305051 | 0 | 0.127248508 |
| 0 | 0 | 0 | 0 | 0 |
| 0.711883067 | 0.227883758 | 0.022585953 | 0 | 0 |
| 0 | 0 | 0 | 0 | 0 |
| 0 | 0 | 0 | 0 | 0 |
| 0 | 0.007763716 | 0 | 0 | 0 |
| 0 | 0.028960284 | 0 | 0 | 0.00987955 |
| 0 | 0.084274833 | 0 | 0 | 0 |
| 0 | 0 | 0 | 0 | 0 |
| 0 | 0 | 0 | 0 | 0 |
| 0 | 0 | 0 | 0 | 0 |
| 0 | 0.000855494 | 0 | 0 | 0 |
| 0 | 1.724363163 | 0 | 0 | 0 |
| 0 | 0 | 0 | 0 | 0 |
| 0 | 0 | 0 | 0 | 0 |
| 0 | 0 | 0 | 0 | 0 |
| 0.014011726 | 0.022096095 | 0.184051837 | 0.007651727 | 0.007034858 |
| 0 | 0 | 0 | 0 | 0 |
| 0 | 0.012715782 | 0 | 0 | 0.025267866 |
| 0 | 0.00361196 | 0.012332369 | 0.000469049 | 0.000884585 |
| 0 | 0 | 0 | 0 | 0.000634522 |
| 0 | 0 | 0.001054306 | 0 | 0 |
| 0 | 0 | 0 | 0 | 0 |
| 0 | 0 | 0 | 0 | 0 |
| 0 | 0 | 0 | 0 | 0 |
| 0.028813684 | 0.649355107 | 0.149967627 | 0.002025214 | 0 |
| 0 | 0 | 0 | 0 | 0.110223772 |
| 0 | 0 | 0 | 0 | 0 |
| 0 | 0 | 0 | 0 | 0 |
| 0 | 0 | 0 | 0 | 0 |
| 0 | 0.017410392 | 0 | 0 | 0.037153154 |
| 0 | 0.17169023 | 0 | 0 | 0 |
| 0.009765406 | 0 | 0.010510963 | 0.001565071 | 0 |
| 0 | 0 | 0 | 0 | 0 |
| 0 | 0 | 0 | 0 | 0 |
| 0 | 0 | 0 | 0 | 0 |
| 0 | 0 | 0 | 0 | 0 |
| 0 | 0.039431897 | 0 | 0 | 0 |
| 0 | 0 | 0 | 0 | 0 |
| 0 | 0 | 0 | 0 | 0.006337183 |
| 0 | 0 | 0 | 0 | 0 |
| 0 | 0 | 0 | 0 | 0 |
| 0 | 0 | 0 | 0 | 0 |
| 0 | 0 | 0 | 0 | 0.00548597 |

| | | | | |
|---|---|---|---|---|
| 0.38374661 | 0.000506947 | 0.012885427 | 0 | 0 |
| 0 | 0 | 0 | 0.094297579 | 0.049469337 |
| 0 | 0 | 0 | 0 | 0.016741351 |
| 0 | 0 | 0 | 0 | 0 |
| 0 | 0 | 0 | 0 | 0 |
| 0 | 0 | 0 | 0 | 0 |
| 0 | 0 | 0 | 0 | 0 |
| 0 | 0.755101675 | 1.066529951 | 0 | 0.365716531 |
| 1.376631548 | 10.25855327 | 0.002716873 | 11.36602769 | 14.3131208 |
| 0.077204211 | 0 | 0.001471093 | 0.505561921 | 0 |
| 0 | 0 | 0 | 0 | 0.006102822 |
| 0 | 0 | 0 | 0 | 0 |
| 0 | 0 | 0 | 0 | 0 |
| 0 | 1.19193146 | 0 | 0 | 1.197885504 |
| 0 | 0 | 0 | 0 | 0 |
| 0 | 0 | 0 | 0 | 0 |
| 0 | 0.076220707 | 0 | 0 | 0 |
| 0 | 0 | 0 | 0 | 0.308277442 |
| 0 | 0 | 0 | 0 | 0 |
| 0 | 0.025111671 | 0.019517856 | 0 | 0 |
| 0 | 0 | 0 | 0 | 0 |
| 0 | 0 | 0 | 0 | 0 |
| 0 | 0.007178445 | 0 | 0 | 0 |
| 0 | 0 | 0 | 0 | 0 |
| 0 | 0.035403837 | 0 | 0 | 0.00794692 |
| 0 | 0 | 0 | 0 | 0 |
| 0 | 0 | 0 | 0 | 0 |
| 0 | 0 | 0 | 0 | 0 |
| 0 | 0 | 0 | 0 | 0 |
| 0 | 0 | 0 | 0 | 0 |
| 0 | 0 | 0.07448597 | 0 | 0 |
| 0 | 0 | 0 | 0 | 0 |
| 0 | 0 | 0 | 0 | 0 |
| 0 | 0 | 0 | 0 | 0 |
| 0 | 0 | 0.006286415 | 0 | 0 |
| 0 | 0 | 0 | 0 | 0.006072546 |
| 0 | 0 | 0 | 0 | 0 |
| 0 | 0 | 0 | 0 | 0 |
| 0 | 0 | 0 | 0 | 0.001810908 |
| 0 | 0 | 0 | 0 | 0 |
| 0 | 0 | 0 | 0 | 0 |
| 0 | 0.117825992 | 0 | 0 | 0.184696045 |
| 0 | 0 | 0 | 0 | 0 |
| 0 | 0 | 0 | 0 | 0 |
| 0 | 0 | 0.002432236 | 0 | 0 |
| 0 | 0 | 0 | 0 | 0 |
| 0 | 0 | 0 | 0 | 0 |
| 0 | 0 | 0 | 0 | 0 |

| | | | | |
|---|---|---|---|---|
| 0 | 0 | 0 | 0 | 0 |
| 0.005437435 | 0.001067599 | 0 | 0 | 0.001889979 |
| 0 | 0 | 0 | 0 | 0 |
| 0 | 0 | 0 | 0 | 0 |
| 0 | 0.153743249 | 2.089276068 | 1.476945889 | 0.371625376 |
| 0 | 0 | 0 | 0 | 0 |
| 0 | 0.002034245 | 0.024414669 | 0 | 0 |
| 0.032048135 | 0.00945815 | 0.033516327 | 0.004922244 | 0 |
| 0 | 0.001931648 | 0 | 0 | 0.001208954 |
| 0 | 0 | 0 | 0 | 0.043271403 |
| 0 | 0 | 0 | 0 | 0 |
| 0 | 0 | 0 | 0 | 0.184346642 |
| 0 | 0 | 0 | 0 | 0 |
| 0 | 0 | 0 | 0 | 0 |
| 0 | 0 | 0 | 0 | 0 |
| 0.042498318 | 0.044557112 | 0 | 0 | 0 |
| 0 | 0 | 0 | 0 | 0 |
| 0 | 0 | 0 | 0 | 0 |
| 0 | 0 | 0 | 0 | 0 |
| 0 | 0 | 0 | 0 | 0 |
| 0 | 0 | 0 | 0 | 0 |
| 0 | 0 | 0 | 0 | 0 |
| 0 | 0 | 0 | 0 | 0 |
| 0.002612719 | 0.017114614 | 0.011009433 | 0.004815423 | 0 |
| 0 | 0.209705148 | 0 | 0.148634903 | 0.354647781 |
| 0 | 0 | 0 | 0 | 0 |
| 0 | 0.034198963 | 0.001549135 | 0.417048645 | 0 |
| 0 | 0 | 0 | 0 | 0 |
| 0 | 0 | 0 | 0 | 0 |
| 0 | 0 | 0 | 0 | 0 |
| 0 | 0 | 0 | 0 | 0 |
| 0 | 0 | 0 | 0 | 0 |
| 0 | 0 | 0 | 0 | 0 |
| 0 | 0 | 0 | 0 | 0 |
| 0 | 0 | 0 | 0 | 0 |
| 0 | 0 | 0 | 0 | 0 |
| 0 | 0 | 0 | 0 | 0 |
| 0 | 0.133661437 | 0 | 0 | 0.011008284 |
| 0 | 0 | 0 | 0 | 0.007659446 |
| 0 | 0 | 0 | 0 | 0 |
| 0 | 0 | 0 | 0 | 0 |
| 0 | 0 | 0 | 0 | 0.017806809 |
| 0 | 0 | 0 | 0 | 0 |
| 0 | 0 | 0 | 0 | 0 |
| 0 | 0 | 0 | 0 | 0 |
| 0 | 0 | 0 | 0 | 0 |
| 0 | 0 | 0 | 0 | 0 |
| 0 | 0 | 0 | 0 | 0 |

| | | | | |
|---|---|---|---|---|
| 0 | 0.208647636 | 0 | 0 | 0.025596376 |
| 0 | 0 | 0 | 0 | 0 |
| 0 | 0 | 0 | 0 | 0 |
| 0 | 0 | 0 | 0 | 0 |
| 0 | 0 | 0 | 0 | 0 |
| 0.047665123 | 0.014273955 | 0 | 0 | 0.053671161 |
| 0 | 0 | 0 | 0 | 0 |
| 0 | 0 | 0 | 0 | 0 |
| 0 | 0 | 0 | 0 | 0 |
| 0 | 0.005182033 | 0 | 0 | 0 |
| 0 | 0 | 0 | 0 | 0 |
| 0 | 0 | 0.00097586 | 0 | 0 |
| 0 | 0 | 0 | 0 | 0.004030072 |
| 0 | 0 | 0 | 0 | 0 |
| 0 | 0 | 0 | 0 | 0 |
| 0 | 0 | 0 | 0 | 0 |
| 0 | 0 | 0 | 0 | 0 |
| 0 | 0 | 0 | 0 | 0 |
| 0 | 0 | 0 | 0 | 0 |
| 0 | 0 | 0.14103128 | 0 | 0 |
| 0 | 0 | 0.18975133 | 0.010328323 | 0 |
| 0 | 0 | 0 | 0 | 0 |
| 0.00101104 | 0.046089768 | 0.003461497 | 0.00652196 | 0.001054272 |
| 0 | 0.016273937 | 0 | 0 | 0 |
| 0 | 0 | 0 | 0 | 0 |
| 0 | 0 | 0 | 0 | 0 |
| 0 | 0 | 0 | 0 | 0 |
| 0 | 0 | 0 | 0 | 0 |
| 0 | 0.006076007 | 0 | 0 | 0 |
| 0 | 0 | 0 | 0 | 0 |
| 0 | 0 | 0 | 0 | 0 |
| 0 | 0 | 0 | 0 | 0 |
| 1.176442501 | 0.085887553 | 0.005735276 | 0.5060739 | 0 |
| 0 | 0 | 0 | 0 | 0.026299831 |
| 0 | 0.001763862 | 0.060892974 | 0.594533661 | 0.003091039 |
| 0 | 0.002285651 | 0 | 0 | 0 |
| 0 | 0 | 0 | 0 | 0 |
| 0 | 0 | 0 | 0 | 0 |
| 0 | 0.005490796 | 0.017358618 | 0 | 0 |
| 0 | 0 | 0 | 0 | 0 |
| 0 | 0 | 0 | 0 | 0.232495214 |
| 0 | 0 | 0.355284257 | 0 | 0.016973288 |
| 0 | 0.055405553 | 0.064139866 | 0.961891278 | 0.052773738 |
| 0 | 0 | 0 | 0 | 0 |
| 0 | 0 | 0 | 0 | 0 |
| 0 | 0.025683348 | 0.011821096 | 0 | 0.258064216 |
| 0 | 0 | 0 | 0 | 0 |
| 0 | 0 | 0 | 0 | 0 |

| | | | | |
|---|---|---|---|---|
| 0 | 0 | 0 | 0 | 0 |
| 0 | 0 | 0 | 0 | 0 |
| 0 | 0 | 0 | 0 | 0 |
| 0 | 0 | 0 | 0 | 0 |
| 0 | 0 | 0 | 0 | 0 |
| 0 | 0 | 0 | 0 | 0 |
| 0 | 0 | 0 | 0 | 0.35393487 |
| 2.024165285 | 0 | 0.010592166 | 0 | 0 |
| 0 | 0.010085193 | 0.176398864 | 0 | 0.007973034 |
| 0 | 0.005057299 | 0.156511786 | 0 | 0.01709205 |
| 0 | 0 | 0 | 0 | 0 |
| 0 | 0 | 0 | 0 | 0 |
| 0 | 0 | 0 | 0 | 0 |
| 0 | 0 | 0 | 0 | 0.136453043 |
| 0 | 0.001100343 | 0 | 0 | 0 |
| 0.150312872 | 0 | 0.019167109 | 0 | 0 |
| 0 | 0 | 0 | 0 | 0 |
| 0 | 0 | 0 | 0 | 0 |
| 0 | 0 | 0 | 0 | 0 |
| 0 | 0.588515846 | 0.812030346 | 0 | 0.18790668 |
| 0 | 0.067959389 | 0 | 0.002785995 | 0 |
| 0 | 0 | 0 | 0 | 0 |
| 0 | 0 | 0 | 0 | 0 |
| 0 | 0.015040233 | 0 | 0 | 0 |
| 0 | 0 | 0 | 0 | 0 |
| 0 | 0 | 0 | 0 | 0 |
| 0 | 0 | 0 | 0 | 0 |
| 0 | 0.111081328 | 0 | 0 | 0.105593313 |
| 0 | 0 | 0 | 0 | 0 |
| 0 | 0.15352168 | 0 | 0 | 0.007820798 |
| 0 | 0.015865745 | 0.180745866 | 0 | 0.782237699 |
| 0 | 0 | 0 | 0 | 0 |
| 0 | 0 | 0 | 0 | 0 |
| 0 | 0 | 0 | 0 | 0 |
| 0 | 0 | 0 | 0 | 0 |
| 0 | 0 | 0 | 0 | 0 |
| 0 | 0 | 0 | 0 | 0 |
| 0 | 0 | 0 | 0 | 0 |
| 0 | 0 | 0 | 0 | 0.00256742 |
| 0 | 0 | 0 | 0 | 0 |
| 0 | 0 | 0 | 0 | 0 |
| 0 | 0.932592355 | 0 | 0 | 0 |
| 0 | 0.001682372 | 0 | 0 | 0.001326705 |
| 0 | 0 | 0 | 0 | 0 |
| 0 | 0 | 0 | 0 | 0 |
| 0 | 0 | 0 | 0 | 0 |
| 25.19536426 | 0 | 0.169233106 | 0 | 0 |
| 0 | 0.00063687 | 0 | 0 | 0 |

| | | | | |
|---|---|---|---|---|
| 0 | 0 | 0 | 0 | 0 |
| 0 | 0 | 0 | 0 | 0 |
| 0 | 0 | 0 | 0 | 0 |
| 0 | 0 | 0 | 0 | 0 |
| 0 | 0.019612368 | 0.112937264 | 0 | 0.000494653 |
| 0 | 0 | 0 | 0 | 0 |
| 0 | 0 | 0 | 0 | 0 |
| 0 | 0 | 0 | 0 | 0 |
| 0 | 0 | 0 | 0 | 0 |
| 0 | 0 | 0 | 0 | 0 |
| 0 | 0.030104063 | 0 | 0 | 0.014026172 |
| 0.00241484 | 0.009753643 | 0.026132826 | 0.032975774 | 0.000686754 |
| 0 | 0 | 0.006983288 | 0 | 0 |
| 0 | 0 | 0 | 0 | 0 |
| 0 | 0 | 0 | 0 | 0 |
| 0 | 0 | 0 | 0 | 0 |
| 0 | 0 | 0 | 0 | 0 |
| 0 | 0 | 0 | 0 | 0 |
| 0.002659681 | 0 | 0.038700279 | 0.002757362 | 0 |
| 0.024960273 | 0.032947706 | 0.004344275 | 0 | 0 |
| 0 | 0 | 0 | 0 | 0 |
| 0 | 0 | 0 | 0 | 0.013890707 |
| 0 | 0 | 0 | 0 | 0 |
| 0 | 0 | 0 | 0 | 0 |
| 0 | 0 | 0 | 0 | 0 |
| 0 | 0 | 0 | 0 | 0 |
| 0 | 0.0035418 | 0.05643316 | 0.002724784 | 0 |
| 0 | 0 | 0 | 0 | 0 |
| 0 | 0 | 0 | 0 | 0 |
| 0 | 0 | 0 | 0 | 0 |
| 0.005520408 | 0 | 0 | 0.001090124 | 0 |
| 0 | 0.00177401 | 0 | 0 | 0 |
| 0 | 0 | 0 | 0 | 0 |
| 0 | 0 | 0 | 0 | 0 |
| 0 | 0.001852125 | 0 | 0 | 0 |
| 0 | 0 | 0 | 0 | 0 |
| 0 | 0 | 0 | 0 | 0 |
| 0.004848098 | 0.011923839 | 0.039690302 | 0.043783359 | 0 |
| 0 | 0.540675535 | 0 | 0 | 0.023453779 |
| 0 | 0 | 0 | 0 | 0 |
| 0.003871971 | 0 | 0 | 0 | 0.003172349 |
| 0 | 0.000986175 | 0 | 0 | 0 |
| 0 | 0 | 0 | 0 | 0 |
| 0.101058405 | 0.000570372 | 0.002596574 | 0 | 0 |
| 0 | 0 | 0 | 0 | 0 |
| 0 | 0 | 0.001160677 | 0 | 0 |
| 0 | 0 | 0 | 0 | 0 |
| 0 | 0 | 0 | 0 | 0 |

| | | | | |
|---|---|---|---|---|
| 0 | 0 | 0 | 0 | 0 |
| 0 | 0.407225826 | 0.736255845 | 0 | 0 |
| 0 | 0 | 0 | 0 | 0 |
| 0 | 0 | 0.023357153 | 0 | 0 |
| 0.002196188 | 0 | 0 | 0.001103926 | 0 |
| 0 | 0 | 0 | 0 | 0 |
| 0 | 0 | 0 | 0 | 0 |
| 0 | 0.002960798 | 0 | 0 | 0 |
| 0 | 0.007769649 | 0 | 0 | 0 |
| 0 | 0 | 0 | 0 | 0 |
| 0 | 0.018440143 | 0 | 0 | 0 |
| 0 | 0 | 0 | 0 | 0 |
| 0 | 0.01822007 | 0.092911722 | 0.045419741 | 0.011456372 |
| 0 | 0 | 0 | 0 | 0 |
| 0 | 0 | 0 | 0 | 0 |
| 0 | 0 | 0 | 0 | 0 |
| 0 | 0 | 0.218683779 | 0 | 0 |
| 0 | 0 | 0 | 0 | 0 |
| 0 | 0 | 0 | 0 | 0 |
| 0 | 0 | 0 | 0 | 0 |
| 0 | 0 | 0 | 0 | 0 |
| 0 | 0 | 0 | 0 | 0 |
| 0.033354033 | 0 | 0 | 0 | 0 |
| 0 | 0.102051111 | 0.005744801 | 0 | 0.006922725 |
| 0 | 0.002712538 | 0 | 0 | 0 |
| 0 | 0 | 0 | 0 | 0 |
| 0 | 0 | 0 | 0 | 0 |
| 0 | 0 | 0 | 0 | 0 |
| 0 | 0 | 0 | 0 | 0 |
| 0 | 0 | 0 | 0 | 0 |
| 0 | 0 | 0.006844214 | 0 | 0 |
| 0 | 0 | 0 | 0 | 0 |
| 0 | 0 | 0 | 0 | 0 |
| 0 | 0 | 0 | 0 | 0 |
| 0 | 0.002433946 | 0 | 0 | 0.189588719 |
| 0 | 0 | 0.003207304 | 0 | 0 |
| 0.350840659 | 0 | 0 | 0 | 0 |
| 0 | 0.007779499 | 0 | 0 | 0 |
| 0.000568644 | 0.02403642 | 0 | 0.001222726 | 0 |
| 0 | 0 | 0 | 0 | 0 |
| 0 | 0 | 0 | 0 | 0 |
| 0 | 0 | 0 | 0 | 0 |
| 0 | 0 | 0 | 0 | 0 |
| 0 | 0 | 0 | 0 | 0 |
| 0 | 0 | 0 | 0 | 0.015104438 |
| 0 | 0.304563211 | 0.388964804 | 0.589690249 | 0.373541528 |
| 0 | 0 | 0 | 0 | 0 |
| 0 | 0 | 0 | 0 | 0 |

| | | | | |
|---|---|---|---|---|
| 0 | 0 | 0 | 0 | 0.001592855 |
| 0 | 0 | 0 | 0 | 0 |
| 0 | 0.006175076 | 0 | 0 | 0.039242318 |
| 0 | 0 | 0 | 0 | 0.038333073 |
| 0 | 0 | 0.013118332 | 0 | 0 |
| 0 | 0 | 0 | 0 | 0 |
| 0 | 0 | 0 | 0 | 0 |
| 0 | 0 | 0 | 0 | 0 |
| 0 | 0 | 0 | 0 | 0.002377461 |
| 0 | 0.025229241 | 0 | 0 | 0 |
| 0 | 0 | 0.003077864 | 0 | 0 |
| 0.001377513 | 0 | 0.001934847 | 0 | 0 |
| 0 | 0 | 0 | 0 | 0 |
| 0 | 0 | 0 | 0 | 0 |
| 0 | 0 | 0 | 0 | 0 |
| 0 | 0 | 0 | 0 | 0 |
| 0 | 0.079924781 | 0 | 0 | 0 |
| 0 | 0 | 0 | 0 | 0 |
| 0 | 0 | 0 | 0 | 0 |
| 0 | 0 | 0 | 0 | 0 |
| 0 | 0 | 0 | 0 | 0 |
| 0 | 0 | 0 | 0 | 0 |
| 0 | 0 | 0 | 0 | 0 |
| 0 | 0 | 0 | 0 | 0 |
| 0 | 0 | 0 | 0 | 0 |
| 0 | 0 | 0 | 0 | 0 |
| 0 | 0 | 0 | 0 | 0 |
| 0 | 0.011489305 | 0 | 0 | 0.002353343 |
| 0 | 0 | 0 | 0 | 0 |
| 0 | 0 | 0 | 0 | 0 |
| 0.003503958 | 0 | 0.000553685 | 0.000968706 | 0.001096136 |
| 0 | 0 | 0 | 0 | 0 |
| 0 | 0.008849667 | 0.026358248 | 0.014059573 | 0.025454501 |
| 0.00353389 | 0 | 0.001532902 | 0.004022861 | 0 |
| 0 | 0 | 0 | 0 | 0 |
| 0 | 0 | 0 | 0 | 0 |
| 0 | 0 | 0 | 0 | 0 |
| 0 | 0 | 0 | 0 | 0 |
| 0 | 0 | 0 | 0 | 0 |
| 0 | 0 | 0 | 0 | 0 |
| 0 | 0 | 0 | 0 | 0 |
| 0.077986938 | 0.085972347 | 0.108247157 | 0.014342688 | 0 |
| 0 | 0.014153859 | 0.01030949 | 0.088118733 | 0.03678019 |
| 0 | 0 | 0 | 0 | 0 |
| 0 | 0 | 0 | 0 | 0 |
| 0 | 0.007363192 | 2.293039695 | 0.125165625 | 0.03840313 |
| 0 | 0 | 0.005290732 | 0 | 0 |
| 0 | 0 | 0 | 0 | 0 |

| | | | | |
|---|---|---|---|---|
| 0 | 0 | 0 | 0 | 0 |
| 0 | 0.003756438 | 0.001125058 | 0 | 0.180410537 |
| 0.005738557 | 0.00230943 | 0.00893647 | 0.010346633 | 0 |
| 0 | 0 | 0 | 0 | 0 |
| 0.01633569 | 0.065645274 | 0.940395941 | 0 | 0 |
| 0.280983413 | 0 | 0 | 0 | 0 |
| 0 | 0 | 0 | 0 | 0 |
| 0 | 0 | 0 | 0 | 0 |
| 0 | 0 | 0 | 0 | 0 |
| 0 | 0 | 0 | 0 | 0 |
| 0 | 0 | 0 | 0 | 0 |
| 0 | 0 | 0 | 0 | 0 |
| 0 | 0 | 0 | 0 | 0 |
| 0 | 0 | 0 | 0 | 0 |
| 0 | 0 | 0 | 0 | 0 |
| 0 | 0 | 0 | 0 | 0 |
| 0 | 0 | 0 | 0 | 0.860889368 |
| 0 | 0 | 0 | 0 | 0 |
| 0 | 0 | 0 | 0 | 0 |
| 0 | 0 | 0 | 0 | 0 |
| 0 | 0 | 0 | 0 | 0 |
| 0 | 0 | 0 | 0 | 0 |
| 0 | 0 | 0 | 0 | 0 |
| 0 | 0 | 0 | 0 | 0 |
| 0 | 0 | 0.001516304 | 0.001326435 | 0.007719031 |
| 0 | 0.057823017 | 0 | 0 | 0 |
| 0 | 0 | 0 | 0 | 0 |
| 0 | 0.238105367 | 0.003593457 | 0 | 0.086832742 |
| 0 | 0 | 0 | 0 | 0 |
| 0 | 0 | 0 | 0 | 0 |
| 0 | 0 | 0 | 0 | 0 |
| 0 | 0 | 0.08752092 | 0.001671652 | 0 |
| 0 | 0 | 0 | 0 | 0 |
| 0 | 0 | 0 | 0 | 0 |
| 0 | 0 | 0 | 0 | 0 |
| 0 | 0 | 0 | 0 | 0 |
| 0.000787982 | 0 | 0 | 0 | 0 |
| 0 | 0 | 0 | 0 | 0 |
| 0 | 0.607733944 | 0 | 0 | 0 |
| 0 | 0 | 0 | 0 | 0 |
| 0 | 0 | 0 | 0 | 0 |
| 0 | 0.001869409 | 0 | 0.008892258 | 0.025740013 |
| 0.314099682 | 0.001152948 | 0.007435662 | 0 | 0 |
| 0 | 0.001119152 | 0 | 0 | 0 |
| 0 | 0 | 0 | 0 | 0 |
| 0 | 0 | 0 | 0 | 0 |
| 0 | 0.001175856 | 0.001003686 | 0.00204868 | 0 |
| 0 | 0 | 0 | 0 | 0 |

| | | | | |
|---|---|---|---|---|
| 0 | 0 | 0 | 0 | 0 |
| 0.034463466 | 0.015022609 | 0.037149703 | 0.085122244 | 0 |
| 0 | 0 | 0 | 0 | 0.026012674 |
| 0 | 0 | 0 | 0 | 0 |
| 0 | 0 | 0 | 0 | 0 |
| 0 | 0 | 0 | 0 | 0 |
| 0 | 0 | 0 | 0 | 0 |
| 0 | 0 | 0 | 0 | 0 |
| 0 | 0 | 0 | 0 | 0 |
| 0 | 0 | 0 | 0 | 0 |
| 0 | 0 | 0 | 0 | 0 |
| 0 | 0 | 0 | 0 | 0 |
| 0 | 0.000784209 | 0.039493656 | 0 | 0.051682303 |
| 0 | 0 | 0 | 0 | 0 |
| 0.012741326 | 0 | 0.030792326 | 0.035915399 | 0 |
| 0 | 0 | 0 | 0 | 0 |
| 0 | 0 | 0 | 0 | 0 |
| 0 | 0 | 0 | 0 | 0 |
| 0 | 0 | 0 | 0 | 0 |
| 0 | 0 | 0 | 0 | 0 |
| 0 | 0 | 0 | 0 | 0 |
| 0.031405146 | 0.004087834 | 0.00180926 | 0 | 0 |
| 0 | 0 | 0.068520377 | 0.006949605 | 0 |
| 0.019711546 | 0 | 0 | 0 | 0 |
| 0 | 0 | 0 | 0 | 0 |
| 0 | 0 | 0 | 0 | 0 |
| 0 | 0 | 0 | 0 | 0 |
| 0 | 0 | 0 | 0 | 0 |
| 0 | 0.000918916 | 0 | 0 | 0 |
| 0 | 0 | 0 | 0 | 0 |
| 0 | 0 | 0 | 0 | 0 |
| 0 | 0 | 0 | 0 | 0 |
| 0 | 0 | 0 | 0 | 0 |
| 0 | 0 | 0 | 0 | 0 |
| 0 | 3.976280875 | 16.04362247 | 0.001026436 | 0.00116146 |
| 0 | 0 | 0 | 0 | 0 |
| 0 | 0 | 0 | 0 | 0 |
| 0 | 0 | 0 | 0 | 0 |
| 0.016660977 | 0.125187358 | 0.005678406 | 0.000903157 | 0 |
| 0 | 0 | 0 | 0 | 0 |
| 0.064368521 | 0.209039747 | 0.253877793 | 0.025913586 | 0.06574639 |
| 0 | 0 | 0 | 0 | 0 |
| 0 | 0 | 0 | 0 | 0 |
| 0 | 0 | 0 | 0 | 0 |
| 0 | 0 | 0 | 0 | 0 |
| 12.86462867 | 0.050288558 | 0.269475144 | 0.070000987 | 0 |
| 0 | 0 | 0 | 0 | 0 |
| 0 | 0 | 0 | 0 | 0 |

| | | | | |
|---|---|---|---|---|
| 0 | 0 | 0 | 0 | 0 |
| 0 | 0 | 0 | 0 | 0 |
| 0 | 0 | 0 | 0 | 0 |
| 0 | 0.013572568 | 0 | 0 | 0 |
| 0 | 0 | 0 | 0.001582166 | 0 |
| 0 | 0 | 0 | 0 | 0 |
| 0 | 0 | 0 | 0 | 0 |
| 0.001359329 | 0 | 0 | 0 | 0 |
| 0 | 0.001261742 | 0.076466744 | 0 | 0 |
| 0.004919887 | 0.047360722 | 0 | 0 | 0 |
| 0 | 0.011046335 | 0 | 0 | 0.022183419 |
| 0 | 0 | 0 | 0 | 0 |
| 0 | 0 | 0.009878279 | 0 | 0 |
| 0 | 0 | 0 | 0 | 0 |
| 0 | 0 | 0 | 0 | 0 |
| 0 | 0 | 0 | 0 | 0 |
| 0 | 0 | 0 | 0 | 0 |
| 0 | 0 | 0 | 0 | 0 |
| 0 | 0 | 0 | 0 | 0 |
| 0.250388741 | 1.224779836 | 0.376493588 | 1.163372968 | 0.205305633 |
| 0 | 0 | 0 | 0 | 0 |
| 0 | 0 | 0 | 0 | 0 |
| 0 | 0 | 0 | 0 | 0 |
| 0 | 0 | 0 | 0 | 0 |
| 0 | 0 | 0 | 0 | 0 |
| 0 | 0 | 0 | 0 | 0.014942807 |
| 0 | 0 | 0 | 0 | 0 |
| 0 | 0 | 0 | 0 | 0 |
| 0 | 0 | 0 | 0 | 0 |
| 0 | 0 | 0 | 0 | 0 |
| 0 | 0 | 0 | 0 | 0 |
| 0 | 0 | 0.005683009 | 0 | 0 |
| 0 | 0 | 0 | 0 | 0 |
| 0 | 0 | 0 | 0 | 0 |
| 0 | 0 | 0 | 0 | 0 |
| 0 | 0 | 0 | 0 | 0 |
| 0 | 0 | 0 | 0 | 0 |
| 0 | 0 | 0 | 0 | 0 |
| 0 | 0 | 0 | 0 | 0 |
| 0 | 0 | 0 | 0 | 0 |
| 0 | 0 | 0 | 0 | 0 |
| 0 | 0 | 0 | 0 | 0 |
| 0 | 0.005104422 | 0.057156692 | 0 | 0.000741992 |
| 0 | 0 | 0 | 0 | 0 |
| 0 | 0.001488414 | 0.019904169 | 0.006853578 | 0.004820765 |
| 0 | 0 | 0 | 0 | 0.003395083 |
| 0 | 0 | 0 | 0 | 0 |
| 0.000804844 | 0 | 0 | 0 | 0 |

| | | | | |
|---|---|---|---|---|
| 0 | 0 | 0 | 0 | 0 |
| 0 | 0 | 0 | 0 | 0 |
| 0 | 0 | 0 | 0 | 0 |
| 0 | 0 | 0 | 0 | 0 |
| 0 | 0 | 0 | 0 | 0 |
| 0.002100799 | 0.009583748 | 0.044577707 | 0 | 0.01423908 |
| 0 | 0 | 0 | 0 | 0 |
| 0 | 0 | 0 | 0 | 0 |
| 0 | 0 | 0 | 0 | 0 |
| 0 | 0 | 0 | 0 | 0 |
| 0 | 0 | 0 | 0 | 0 |
| 0 | 0 | 0 | 0 | 0 |
| 0 | 0 | 0 | 0 | 0 |
| 0 | 0 | 0 | 0 | 0 |
| 0 | 0 | 0 | 0 | 0 |
| 0 | 0 | 0 | 0 | 0 |
| 0 | 0.031842443 | 0 | 0 | 0 |
| 0.00110882 | 0 | 0.000778721 | 0 | 0 |
| 0 | 0 | 0 | 0 | 0 |
| 0 | 0 | 0 | 0 | 0 |
| 0 | 0 | 0 | 0.000481012 | 0 |
| 0 | 0 | 0 | 0 | 0 |
| 0 | 0 | 0 | 0 | 0 |
| 0 | 0 | 0 | 0 | 0 |
| 0 | 0 | 0 | 0 | 0 |
| 0 | 0 | 0 | 0 | 0 |
| 0 | 0 | 0 | 0 | 0 |
| 0 | 0.013008024 | 0 | 0 | 0 |
| 0 | 0 | 0 | 0 | 0 |
| 0 | 0 | 0 | 0 | 0 |
| 0 | 0 | 0 | 0 | 0 |
| 0 | 0 | 0 | 0 | 0 |
| 0 | 0 | 0 | 0 | 0 |
| 0 | 0 | 0 | 0 | 0 |
| 0 | 0 | 0 | 0 | 0 |
| 0 | 0 | 0 | 0 | 0 |
| 0 | 0 | 0 | 0 | 0 |
| 0 | 0 | 0 | 0 | 0 |
| 0 | 3.077202475 | 0 | 0 | 0 |
| 0 | 0 | 0 | 0 | 0 |
| 0 | 0.006647907 | 0 | 0 | 0 |
| 0 | 0 | 0 | 0 | 0 |
| 0 | 0 | 0 | 0 | 0 |
| 0 | 0 | 0 | 0 | 0 |
| 0 | 0 | 0 | 0 | 0 |
| 0 | 0 | 0 | 0 | 0 |
| 0 | 0 | 0 | 0 | 0 |
| 0 | 0 | 0 | 0 | 0 |

| | | | | |
|---|---|---|---|---|
| 0 | 0 | 0 | 0 | 0 |
| 0 | 0.011662802 | 0.279982226 | 0 | 0.005080353 |
| 0 | 0 | 0 | 0 | 0 |
| 0 | 0 | 0 | 0 | 0 |
| 0 | 0 | 0 | 0 | 0 |
| 0 | 0 | 0 | 0 | 0 |
| 0 | 0 | 0 | 0 | 0 |
| 0 | 0 | 0 | 0 | 0 |
| 0 | 0 | 0 | 0 | 0 |
| 0 | 0.001689098 | 0 | 0 | 0 |
| 0.002419824 | 0.002647063 | 0 | 0.009933419 | 0.028215012 |
| 0 | 0 | 0 | 0 | 0 |
| 0 | 0 | 0 | 0 | 0.011995989 |
| 0 | 0 | 0 | 0 | 0 |
| 0 | 0 | 0 | 0 | 0 |
| 0 | 0 | 0 | 0 | 0 |
| 0 | 0 | 0 | 0 | 0 |
| 0 | 0 | 0 | 0 | 0 |
| 0 | 0 | 0 | 0 | 0 |
| 0 | 0 | 0 | 0 | 0 |
| 0 | 0 | 0.014817811 | 0.003909278 | 0 |
| 0 | 0 | 0.00769115 | 0 | 0 |
| 0 | 0 | 0 | 0 | 0 |
| 0.003514186 | 0.451280365 | 2.448194155 | 0.904378165 | 0.190744305 |
| 0 | 0 | 0 | 0 | 0 |
| 0 | 0 | 0 | 0 | 0 |
| 0 | 0 | 0 | 0 | 0 |
| 0 | 0 | 0 | 0 | 0 |
| 0 | 0 | 0 | 0 | 0 |
| 0 | 0 | 0 | 0 | 0 |
| 0 | 0 | 0 | 0 | 0 |
| 0 | 0 | 0 | 0 | 0 |
| 0 | 0.001304334 | 0 | 0 | 0.00146941 |
| 0 | 0 | 0 | 0 | 0 |

| PHL018-0118_V3 | PHL018-0219_V2 | PHL018-0001_V1 | PHL018-0106_V3 | PHL018-0019_V2 |
|---|---|---|---|---|
| EYCF | REF | EYCF | CTRL | CTRL |
| 1.360023045 | 0.293163097 | 0.305774229 | 0.092729267 | 0.041823509 |
| 5.978815587 | 0.142874875 | 0.126958335 | 0.793893365 | 2.309412593 |
| 2.181941635 | 0 | 0 | 4.310357735 | 8.147012639 |
| 0 | 0 | 0 | 0 | 0 |
| 0 | 0 | 0 | 0 | 0 |
| 0 | 0 | 0 | 0 | 0 |
| 0 | 0 | 0 | 0 | 0 |
| 0 | 0 | 0 | 0 | 0 |
| 0 | 0 | 0 | 0 | 0 |
| 0 | 0 | 0 | 0 | 0 |
| 0 | 0 | 0 | 0 | 0 |
| 0 | 0 | 0 | 0 | 0 |
| 0 | 0 | 0 | 0 | 0 |
| 2.098520044 | 0.354780809 | 0.091973513 | 1.55352822 | 4.075844444 |
| 0 | 0.006596386 | 0 | 0 | 0 |
| 0 | 0 | 0 | 0 | 0 |
| 0 | 0 | 0 | 0 | 0 |
| 0 | 0 | 0 | 0 | 0 |
| 0 | 0 | 0 | 0 | 0 |
| 0 | 0 | 0 | 0 | 0 |
| 0 | 0 | 0 | 0 | 0 |
| 0 | 0 | 0 | 0 | 0 |
| 0 | 0 | 0 | 0 | 0 |
| 0 | 0 | 0 | 0 | 0 |
| 0.001531119 | 0 | 0.175542563 | 0 | 0 |
| 0 | 0 | 0 | 0 | 0 |
| 0 | 0 | 0 | 0 | 0 |
| 0 | 0 | 0 | 0 | 0 |
| 0 | 0 | 0 | 0 | 0 |
| 0 | 0 | 0 | 0 | 0 |
| 0 | 0 | 0 | 0 | 0 |
| 0 | 0 | 0 | 0 | 0 |
| 0 | 0 | 0 | 0 | 0 |
| 0 | 0 | 0 | 0 | 0 |
| 0 | 0 | 0 | 0 | 0 |
| 0 | 0.466355002 | 0.173389857 | 1.946636612 | 0 |
| 0 | 0 | 0 | 0 | 0 |
| 0 | 0 | 0 | 0 | 0 |
| 0 | 0 | 0 | 0 | 0 |
| 0 | 0 | 0 | 0 | 0 |
| 0 | 0 | 0 | 0 | 0 |
| 0.002969827 | 0.233864388 | 0.070015518 | 0.001599639 | 0.111780209 |
| 1.029448285 | 0.113542482 | 0.928732981 | 0.400984405 | 0.011286828 |
| 0.007866563 | 0.018044109 | 0.004651072 | 0.114475677 | 0.001279524 |
| 0.005671515 | 0.002674782 | 0 | 0.001350784 | 0 |
| 0 | 0 | 0 | 0 | 0 |

| | | | | |
|---|---|---|---|---|
| 0.00507528 | 0.354693673 | 0.001029291 | 0 | 0 |
| 0.002167202 | 0 | 0 | 0.013248169 | 0 |
| 0.029467133 | 0.008783916 | 0.014312075 | 0.0033858 | 0.003740429 |
| 3.763225273 | 0.201059733 | 0.199257772 | 0.673305834 | 0.921451414 |
| 0.004152029 | 0.060777821 | 0.021793741 | 0.00522516 | 0 |
| 0 | 0 | 0.069788527 | 0 | 0 |
| 0 | 0.344622303 | 0 | 0.0090363 | 7.487161943 |
| 0.068920889 | 0 | 0 | 0 | 0 |
| 0 | 0 | 0.0181002 | 0 | 0 |
| 0 | 0 | 0 | 0 | 0 |
| 0 | 0 | 0 | 0 | 0 |
| 0.264477495 | 0.336851467 | 0.043088801 | 0.002550859 | 0 |
| 0 | 0 | 0.000584457 | 0 | 0 |
| 0.090018244 | 0.015810335 | 0 | 0 | 0.038355744 |
| 0 | 0 | 0 | 0 | 0 |
| 0 | 0 | 0 | 0 | 0 |
| 0.37351063 | 0.011863825 | 0.024010833 | 0 | 0.000649156 |
| 0 | 0 | 0 | 0 | 0 |
| 0 | 0 | 0 | 0 | 0 |
| 0 | 0 | 0 | 0 | 0 |
| 0.209970595 | 0.083884996 | 0 | 0.236413153 | 0.594776655 |
| 0.005318247 | 0.005283679 | 0.000958725 | 0.090255128 | 0.017069452 |
| 0 | 0 | 0.02889929 | 0 | 0 |
| 0 | 0 | 0 | 0 | 0 |
| 0 | 0 | 0 | 0 | 0 |
| 0 | 0 | 0 | 0 | 0 |
| 0 | 0 | 0 | 0 | 0 |
| 0 | 0 | 0 | 0 | 0 |
| 0 | 0 | 0.027442649 | 0 | 0.281273577 |
| 0 | 0 | 0 | 0 | 0 |
| 0 | 0 | 0 | 0.631074676 | 1.989342908 |
| 0 | 0 | 0 | 0 | 0 |
| 0 | 0 | 0 | 0 | 0 |
| 0 | 0 | 0 | 0 | 0 |
| 0 | 0 | 0 | 0 | 0 |
| 0 | 0 | 0.00083587 | 0 | 0 |
| 0 | 0 | 0 | 0 | 0 |
| 0.011563606 | 0 | 0 | 0 | 0 |
| 0 | 0 | 0 | 0 | 0 |
| 0 | 0 | 0 | 0 | 0 |
| 0 | 0 | 0 | 0 | 0 |
| 0 | 0 | 0 | 0 | 0 |
| 0 | 0 | 0 | 0 | 0 |
| 0 | 0 | 0 | 0 | 0 |
| 0 | 0 | 0.053550064 | 0 | 0 |
| 0 | 0 | 0 | 0 | 0 |
| 0 | 0 | 0 | 0 | 0 |
| 0.089120193 | 0 | 0 | 0 | 0 |

| | | | | |
|---|---|---|---|---|
| 0 | 0 | 0 | 0 | 0 |
| 0 | 0 | 0 | 0 | 0 |
| 0 | 0 | 0 | 0 | 0 |
| 0 | 0 | 0 | 0 | 0 |
| 0 | 0 | 4.649805911 | 0 | 0.000713009 |
| 0 | 0 | 0 | 0 | 0 |
| 0 | 0 | 0 | 0 | 0 |
| 0 | 0 | 0 | 0 | 0 |
| 0 | 0 | 0 | 0 | 0 |
| 0 | 0 | 0 | 0 | 0 |
| 0 | 0 | 0 | 0 | 0 |
| 0 | 0.044654514 | 0 | 0.000823434 | 0 |
| 0 | 0 | 0 | 0 | 0 |
| 0 | 0 | 0 | 0 | 0 |
| 0 | 0 | 0 | 0 | 0 |
| 0 | 0 | 0 | 0 | 0 |
| 0 | 0 | 0 | 0 | 0 |
| 0 | 0 | 0 | 0 | 0 |
| 0 | 0 | 2.098166952 | 0 | 0 |
| 0 | 0.020856537 | 0 | 0 | 0 |
| 0 | 0 | 0 | 0 | 0 |
| 0 | 0 | 0 | 0 | 0 |
| 0 | 0 | 0 | 0 | 0 |
| 0 | 0 | 0 | 0 | 0 |
| 0 | 0 | 0 | 0 | 0 |
| 0 | 0 | 0 | 0 | 0 |
| 0 | 0 | 0 | 0 | 0 |
| 0 | 0 | 0.00155935 | 0 | 0 |
| 0 | 0 | 0 | 0 | 0 |
| 0 | 0 | 0.000536544 | 0 | 0 |
| 0 | 0.151082771 | 0.073211254 | 0.036590178 | 0.005602811 |
| 0.133363401 | 0.004648064 | 0.000674714 | 0 | 0.000881675 |
| 0 | 0 | 0 | 0 | 0 |
| 0 | 0 | 0.000427902 | 0 | 0 |
| 0 | 0 | 0 | 0 | 0 |
| 0 | 0 | 0 | 0 | 0 |
| 0 | 0 | 0 | 0 | 0 |
| 0 | 0 | 0 | 0 | 0 |
| 0.097257334 | 0 | 0.025738715 | 0.154736108 | 0 |
| 0 | 0 | 0 | 0 | 0 |
| 0 | 0 | 0 | 0 | 0 |
| 0 | 0 | 0.104407679 | 0 | 0 |
| 0 | 0 | 0 | 0 | 0 |
| 0 | 0 | 0 | 0 | 0 |
| 0 | 0 | 0 | 0 | 0 |
| 0 | 0 | 0 | 0 | 0 |
| 0 | 0 | 0 | 0 | 0 |
| 0 | 0 | 0 | 0 | 0 |

| | | | | |
|---|---|---|---|---|
| 0.003670065 | 0 | 0 | 0 | 0 |
| 0 | 0 | 0 | 0 | 0 |
| 0 | 0 | 0 | 0 | 0 |
| 0 | 0 | 0 | 0 | 0 |
| 0.003145333 | 0.241523144 | 0.007340781 | 0 | 0 |
| 0 | 0 | 0 | 0 | 0 |
| 0 | 0 | 0 | 0 | 0 |
| 0 | 0 | 0 | 0 | 0 |
| 0 | 0 | 0 | 0 | 0 |
| 0 | 0 | 0 | 0 | 0 |
| 0 | 0 | 0 | 0 | 0 |
| 0 | 0 | 0 | 0 | 0 |
| 0 | 0 | 0 | 0 | 0 |
| 0 | 0 | 0.000959438 | 0 | 0 |
| 0 | 0 | 0 | 0 | 0 |
| 0 | 0 | 0 | 0 | 0 |
| 0 | 0 | 0 | 0 | 0 |
| 0 | 0 | 0 | 0 | 0 |
| 0 | 0 | 0 | 0 | 0 |
| 0 | 0 | 0 | 0 | 0 |
| 0 | 0 | 0.001051997 | 0 | 0 |
| 0 | 0 | 0 | 0 | 0 |
| 0 | 0 | 0 | 0 | 0 |
| 0 | 0 | 0 | 0 | 0 |
| 0 | 0 | 0 | 0 | 0 |
| 0 | 0 | 0 | 0 | 0 |
| 0 | 0 | 0.056434872 | 0 | 0 |
| 0 | 0 | 0 | 0 | 0 |
| 0 | 0 | 0 | 0 | 0 |
| 0 | 0 | 0 | 0 | 0 |
| 0 | 0 | 0.001326541 | 0.02165967 | 0 |
| 0 | 0 | 0 | 0 | 0 |
| 0 | 0 | 0 | 0 | 0 |
| 0 | 0 | 0 | 0 | 0 |
| 0 | 0 | 0 | 0 | 0 |
| 0 | 0 | 0 | 0 | 0 |
| 0 | 0 | 0 | 0 | 0 |
| 0 | 0 | 0 | 0 | 0 |
| 0 | 0 | 0.002731294 | 0 | 0 |
| 0 | 0 | 0 | 0 | 0 |
| 0 | 0 | 0 | 0 | 0 |
| 0 | 0 | 0 | 0 | 0 |
| 0 | 0 | 0 | 0 | 0 |
| 0 | 0 | 0 | 0 | 0 |
| 0 | 0 | 0 | 0 | 0 |
| 0 | 0 | 0 | 0 | 0 |
| 0 | 0 | 0 | 0 | 0 |
| 0 | 0 | 0 | 0 | 0 |

| | | | | |
|---|---|---|---|---|
| 0 | 0 | 4.037619366 | 0 | 0 |
| 0 | 0 | 0 | 0 | 0 |
| 0 | 0 | 0 | 0 | 0 |
| 0 | 0 | 0 | 0 | 0 |
| 0 | 0 | 0 | 0 | 0 |
| 0 | 0 | 0 | 0 | 0 |
| 0 | 0 | 0 | 0 | 0 |
| 0 | 0 | 0 | 0 | 0 |
| 0 | 0 | 0 | 0 | 0 |
| 0 | 0 | 0 | 0 | 0 |
| 0 | 0 | 0 | 0 | 0 |
| 0 | 0 | 0.021284519 | 0 | 0 |
| 0.000569965 | 0.008723948 | 0 | 0 | 0 |
| 0 | 0 | 0 | 0 | 0 |
| 0 | 0 | 0.001751413 | 0 | 0 |
| 0 | 0 | 0 | 0 | 0 |
| 0 | 0 | 0 | 0 | 0 |
| 0 | 0 | 0 | 0 | 0 |
| 0 | 0 | 0 | 0 | 0 |
| 0 | 0 | 0 | 0 | 0 |
| 0.038720351 | 0.021429078 | 0.014160427 | 0.197926632 | 0.037007963 |
| 0 | 0 | 0 | 0 | 0 |
| 0 | 0 | 0 | 0 | 0 |
| 0.002222617 | 0 | 0 | 0.004817181 | 0.002214287 |
| 0 | 0 | 0 | 0 | 0 |
| 0 | 0 | 0 | 0 | 0 |
| 0.00682701 | 0 | 0.00229826 | 0.035387423 | 0.00459317 |
| 0 | 0 | 0 | 0 | 0 |
| 0 | 0 | 0 | 0 | 0 |
| 0 | 0 | 0 | 0 | 0 |
| 0 | 0 | 0 | 0.003919472 | 0 |
| 0 | 0 | 0 | 0 | 0 |
| 0 | 0 | 0 | 0 | 0 |
| 0 | 0.114542197 | 0.013903105 | 0 | 0 |
| 0.024268991 | 0.325056871 | 0 | 0.006473762 | 0.010658074 |
| 0 | 0 | 0 | 0 | 0 |
| 0 | 0 | 0 | 0 | 0 |
| 0 | 0 | 0 | 0 | 0 |
| 0 | 0 | 0 | 0 | 0 |
| 0 | 0 | 0 | 0 | 0 |
| 0 | 0 | 0 | 0 | 0 |
| 0.047551506 | 0 | 0 | 0 | 0 |
| 0 | 0 | 0.000784953 | 0 | 0 |
| 0 | 0 | 0 | 0 | 0 |
| 0 | 0 | 0 | 0 | 0 |
| 0 | 0 | 0 | 0 | 0 |
| 0 | 0 | 0 | 0 | 0 |
| 0 | 0.005635393 | 0 | 0.00049429 | 0 |

| | | | | |
|---|---|---|---|---|
| 0 | 0 | 0 | 0 | 0 |
| 0 | 0 | 0 | 0 | 0 |
| 0 | 0 | 0 | 0 | 0 |
| 0 | 0 | 0 | 0 | 0 |
| 0 | 0 | 0 | 0 | 0 |
| 0 | 0 | 0 | 0 | 0 |
| 0 | 0 | 0.001685407 | 0 | 0 |
| 0 | 0.031338004 | 0.007272265 | 0 | 0 |
| 0 | 0 | 0.006363126 | 0 | 0 |
| 0 | 0 | 0 | 0 | 0 |
| 0 | 0 | 0 | 0 | 0 |
| 0 | 0 | 0 | 0 | 0 |
| 0 | 0 | 0 | 0 | 0 |
| 0 | 0 | 0 | 0 | 0 |
| 0 | 0 | 0 | 0 | 0 |
| 0 | 0 | 0 | 0 | 0 |
| 0 | 0 | 0 | 0 | 0 |
| 0 | 0 | 0.006564617 | 0 | 0 |
| 0.002680173 | 0 | 0.011055998 | 0.00150807 | 0 |
| 0 | 0.215695226 | 0.062219349 | 0 | 0 |
| 0 | 0 | 0.006620373 | 0.062781951 | 0.006963079 |
| 0 | 0 | 0 | 0 | 0 |
| 0 | 0 | 0 | 0 | 0 |
| 0 | 0 | 0.010132274 | 0 | 0 |
| 0.011290871 | 0.005745911 | 0 | 0.002946369 | 0.001676804 |
| 0 | 0 | 0 | 0 | 0 |
| 0 | 0 | 0 | 0 | 0 |
| 0 | 0 | 0 | 0 | 0 |
| 0 | 0 | 0.00719437 | 0 | 0.025003093 |
| 0 | 0 | 0.013569858 | 0 | 0 |
| 0.310143504 | 0.118599558 | 0.035975392 | 0.321872579 | 0.470259596 |
| 0 | 0 | 0 | 0 | 0 |
| 0 | 0 | 0 | 0 | 0 |
| 0 | 0 | 0.003686662 | 0 | 0 |
| 0 | 0 | 0 | 0 | 0 |
| 0 | 0 | 0 | 0 | 0 |
| 0 | 0 | 0 | 0 | 0 |
| 0 | 0 | 0 | 0 | 0 |
| 0 | 0 | 0 | 0 | 0 |
| 0 | 0 | 0 | 0 | 0 |
| 0 | 0 | 0 | 0 | 0 |
| 0.081303657 | 0.238116594 | 0.007503623 | 0.010745084 | 0.008381929 |
| 0 | 0 | 0 | 0 | 0 |
| 0 | 0 | 0 | 0 | 0 |
| 0 | 0 | 0 | 0 | 0 |
| 0 | 0 | 0 | 0 | 0 |
| 0 | 0 | 0 | 0 | 0 |
| 0 | 0 | 0 | 0 | 0 |

| | | | | |
|---|---|---|---|---|
| 0 | 0 | 0 | 0 | 0 |
| 0 | 0 | 0 | 0 | 0 |
| 0 | 0.029313193 | 0.00440708 | 0.12212786 | 0 |
| 0 | 0 | 0 | 0 | 0 |
| 0 | 0 | 0.139795202 | 2.290709265 | 0.00999547 |
| 0 | 0 | 0 | 0 | 0 |
| 0 | 0 | 0 | 0 | 0 |
| 0 | 0 | 0 | 0 | 0 |
| 0 | 0 | 0 | 0 | 0 |
| 0 | 0 | 0 | 0 | 0 |
| 0 | 0 | 0 | 0 | 0 |
| 0 | 0 | 0 | 0 | 0 |
| 0 | 0 | 0 | 0 | 0 |
| 0.007453331 | 0 | 0.023681286 | 0.002570921 | 0.002414169 |
| 0.01187924 | 0 | 0.005267663 | 0.001060979 | 0.011412052 |
| 0 | 0 | 0.036361407 | 0 | 0 |
| 0.00314213 | 0 | 0.000789741 | 0.004110228 | 0.001651176 |
| 0 | 0 | 0.000612615 | 0 | 0 |
| 0 | 0 | 0 | 0 | 0 |
| 0 | 0 | 0 | 0 | 0 |
| 0 | 0 | 0 | 0 | 0 |
| 0 | 0.004125404 | 0 | 0.001527797 | 0.00074527 |
| 0 | 0 | 0 | 0 | 0 |
| 0 | 0 | 0 | 0 | 0 |
| 0 | 0 | 0 | 0 | 0 |
| 0 | 0 | 0 | 0 | 0 |
| 0 | 0 | 1.963237012 | 0 | 0 |
| 0 | 0 | 0 | 0 | 0 |
| 0 | 0 | 0 | 0 | 0 |
| 0 | 0 | 0 | 0 | 0 |
| 0 | 0 | 0 | 0 | 0 |
| 0 | 0 | 0 | 0 | 0 |
| 0 | 0 | 0.001475412 | 0 | 0 |
| 0 | 0 | 0 | 0 | 0.075617895 |
| 0.003230382 | 0 | 0 | 0 | 0 |
| 0 | 0 | 0 | 0 | 0 |
| 0 | 0 | 0 | 0 | 0 |
| 0.008223926 | 0.035294733 | 0.004299351 | 0 | 0 |
| 0 | 0 | 0 | 0 | 0 |
| 0 | 0 | 0 | 0 | 0 |
| 0 | 0 | 0.0147373 | 0 | 0 |
| 0 | 0 | 0 | 0 | 0 |
| 0 | 0 | 0 | 0 | 0 |
| 0 | 0 | 0 | 0 | 0 |
| 0 | 0 | 0 | 0 | 0 |
| 0 | 0 | 0 | 0 | 0 |
| 0 | 0 | 0 | 0 | 0 |
| 0.091643388 | 0 | 0 | 0.006310764 | 0 |

| | | | | |
|---|---|---|---|---|
| 0 | 0.325981458 | 0.000858275 | 0 | 0 |
| 0 | 0 | 0 | 0 | 0 |
| 0.005484843 | 0 | 0 | 0 | 0 |
| 0 | 0 | 0 | 0 | 0 |
| 0.004138711 | 0 | 0.002704574 | 0 | 0.008246401 |
| 0 | 0 | 0 | 0 | 0 |
| 0 | 0 | 0 | 0 | 0 |
| 0.093465617 | 0.015669803 | 0 | 0.01063908 | 0.018714807 |
| 0 | 0 | 0 | 0 | 0 |
| 0.061788582 | 0 | 0.008466299 | 0.070447793 | 0.129638514 |
| 0 | 0 | 0 | 0.003304873 | 0 |
| 0.00153078 | 0.095788545 | 0.002000675 | 0.002041681 | 0 |
| 0.022061873 | 0.004530298 | 0.001264653 | 0.003678133 | 0.005618741 |
| 0 | 0 | 0 | 0 | 0 |
| 0.005772052 | 0 | 0 | 0.001194592 | 0 |
| 0 | 0 | 0 | 0 | 0 |
| 0 | 0 | 0 | 0 | 0 |
| 0 | 0 | 0 | 0 | 0 |
| 0 | 0 | 0 | 3.777284316 | 0.040306585 |
| 0 | 0 | 0 | 0 | 0.034233879 |
| 0 | 0 | 0 | 0 | 0 |
| 0 | 0 | 0 | 0 | 0 |
| 0 | 0 | 0 | 0.00139829 | 0 |
| 0.013025891 | 0 | 0.075348618 | 0 | 0 |
| 0 | 0 | 0 | 0 | 0 |
| 0.003831928 | 0.008726755 | 0 | 0.008568174 | 0 |
| 0 | 0 | 0 | 0 | 0 |
| 0 | 0 | 0 | 0.063531151 | 0.081002115 |
| 0 | 0 | 0 | 0 | 0 |
| 0 | 0 | 0 | 0 | 0 |
| 0 | 0 | 0 | 0.029517952 | 0.005163601 |
| 0 | 0 | 0 | 0 | 0 |
| 0 | 0 | 0 | 0 | 0 |
| 0 | 0 | 0 | 0 | 0.04153571 |
| 0 | 0 | 0 | 0 | 0 |
| 0 | 0 | 0 | 0.014633408 | 0 |
| 0 | 0 | 0 | 0.592487057 | 0 |
| 0 | 0 | 0 | 0.193958985 | 0 |
| 0 | 0 | 0 | 0 | 0 |
| 0 | 0 | 0 | 0 | 0 |
| 0 | 0 | 0 | 0 | 0 |
| 0 | 0 | 0 | 0 | 0 |
| 0 | 0 | 0 | 0 | 0 |
| 0 | 0 | 0 | 0 | 0.004807188 |
| 0 | 0 | 0 | 0 | 0 |
| 0 | 0 | 0.007651696 | 0 | 0 |
| 0.004484993 | 0.018676238 | 0 | 0.0022432 | 0.000718101 |
| 0 | 0 | 0 | 0 | 0 |

| | | | | |
|---|---|---|---|---|
| 0 | 0 | 0 | 0 | 0 |
| 0 | 0.016359137 | 0 | 0 | 1.020013399 |
| 0 | 0 | 0 | 0 | 0 |
| 0 | 0 | 0.007294356 | 0 | 0 |
| 0 | 0 | 0 | 0 | 0.001419536 |
| 0.00728573 | 0 | 0 | 0 | 0.006787099 |
| 0 | 0 | 0 | 0 | 0 |
| 0 | 0 | 0 | 0 | 0.003657613 |
| 0 | 0 | 0 | 0 | 0 |
| 0 | 0 | 0 | 0 | 0 |
| 0.026887065 | 0.007608235 | 0.010374795 | 0.002817624 | 0.016181113 |
| 0 | 0 | 0 | 0 | 0 |
| 0 | 0 | 0 | 0 | 0 |
| 0.001935356 | 0 | 0.003003711 | 0.003871927 | 0.004338232 |
| 0.405598895 | 0 | 0 | 0 | 0 |
| 0 | 0 | 0 | 0 | 0.009269437 |
| 0 | 0 | 0 | 0 | 0 |
| 0 | 0 | 0 | 0 | 0 |
| 0 | 0 | 0 | 0.041725839 | 0.006376383 |
| 0 | 0 | 0 | 0 | 0 |
| 0 | 0.010560878 | 0 | 0 | 0 |
| 1.319989304 | 0 | 0.081325778 | 0.252534691 | 0 |
| 0.006887101 | 0 | 0 | 0 | 0 |
| 0 | 0 | 0 | 0 | 0 |
| 0 | 0 | 0 | 0 | 0 |
| 0 | 0 | 0 | 0 | 0 |
| 0 | 0 | 0 | 0.011807419 | 0.17516024 |
| 0 | 0 | 0 | 0 | 0 |
| 0 | 0 | 0 | 0 | 0 |
| 0.59998881 | 0.098973831 | 0.063956635 | 0.836416984 | 0.100228989 |
| 0 | 0.074771529 | 0.000643509 | 0 | 0 |
| 0 | 0 | 0 | 0 | 0 |
| 0 | 0 | 0 | 0 | 0 |
| 0 | 0 | 0 | 0.001395165 | 0 |
| 0 | 0 | 0 | 0 | 0 |
| 0 | 0 | 0 | 0 | 0 |
| 0 | 0.158706998 | 0.515643622 | 0 | 0 |
| 0 | 0.072597595 | 0 | 0 | 0 |
| 0 | 0 | 0 | 0 | 0 |
| 3.187137354 | 0 | 0 | 0 | 0 |
| 0.009745323 | 0 | 0 | 0 | 0.032607985 |
| 0.017446974 | 0.317251046 | 0.10831222 | 3.154786739 | 0 |
| 0 | 0.047183332 | 0 | 0 | 0 |
| 0 | 0 | 0 | 0 | 0 |
| 0 | 0 | 0 | 0 | 0 |
| 0 | 0 | 0 | 0 | 0 |
| 0 | 0 | 0.00054564 | 0 | 0.007843096 |
| 0 | 0 | 0 | 0 | 0 |

| | | | | |
|---|---|---|---|---|
| 0.001802878 | 0 | 0 | 0 | 0 |
| 0 | 0 | 0 | 0 | 0 |
| 0.790661397 | 0.025909504 | 0.002751974 | 0.001755236 | 0.092599872 |
| 0 | 0 | 0 | 0 | 0 |
| 0 | 0 | 0.000789551 | 0 | 0 |
| 0 | 0 | 0 | 0 | 0 |
| 0 | 0 | 0 | 0 | 0 |
| 0 | 0.016633794 | 0.00094319 | 0 | 0 |
| 0 | 0 | 0 | 0 | 0.038837597 |
| 0 | 0 | 0 | 0 | 0 |
| 0 | 0 | 0 | 0 | 0 |
| 0 | 0 | 0 | 0.000651728 | 0 |
| 0 | 0 | 0 | 0.000950435 | 0 |
| 0 | 0 | 0.010353889 | 0 | 0 |
| 0 | 0 | 0 | 0 | 0 |
| 0 | 0.381334309 | 0.020942411 | 0.03879682 | 0.025655872 |
| 0.049648968 | 0.207700359 | 0.003561005 | 0.001615107 | 0.001378757 |
| 0 | 0 | 0.003365042 | 0.182738469 | 0 |
| 0 | 0 | 0 | 0 | 0.001958209 |
| 0 | 0 | 0 | 0 | 0 |
| 0.028233629 | 0 | 0 | 0 | 0 |
| 0 | 0.05965076 | 0 | 0 | 0.001199465 |
| 0 | 0 | 0 | 0 | 0 |
| 0 | 0.077227396 | 0 | 0.25450547 | 0.359756673 |
| 0 | 0 | 0.000493052 | 0 | 0 |
| 0 | 0 | 0 | 0.002925476 | 0 |
| 0 | 0 | 0 | 0 | 0.005688614 |
| 0 | 0 | 0 | 0 | 0.007892307 |
| 0 | 0 | 0 | 0 | 0 |
| 0 | 0 | 0 | 0 | 0 |
| 0.001558828 | 0.034035004 | 0 | 0.05262695 | 0.079368674 |
| 0.028178756 | 0 | 0.017844799 | 0.017726221 | 0.002232622 |
| 0 | 0.016934544 | 0.003434283 | 3.196813628 | 0.011337375 |
| 0 | 0.01820164 | 0.067608094 | 0 | 0.061266855 |
| 0.001414614 | 0 | 0.005731434 | 0.002547104 | 0 |
| 0 | 0 | 0 | 0 | 0 |
| 0.002500635 | 0 | 0 | 0.010505961 | 0 |
| 0.022489541 | 0.011620975 | 0.00242812 | 0 | 0.013025665 |
| 0 | 0.106472098 | 0.035410991 | 0 | 0 |
| 0.000639565 | 0 | 0 | 0.000479824 | 0.003959545 |
| 0 | 0 | 0 | 0 | 0 |
| 0 | 0 | 0 | 0 | 0 |
| 0.041237072 | 0 | 0 | 0.004680852 | 0.001748193 |
| 0.056789385 | 0 | 0 | 0.169658076 | 0.079780768 |
| 0 | 0 | 0 | 0 | 0 |
| 0 | 0 | 0.02173202 | 0 | 0 |
| 0 | 0 | 0.002272169 | 0 | 0 |
| 0 | 0 | 0 | 0 | 0 |

| | | | | |
|---|---|---|---|---|
| 0 | 0 | 0 | 0 | 0 |
| 0.831674642 | 0.191940037 | 0.220632781 | 0.041850013 | 0.273290957 |
| 0 | 0 | 0 | 0 | 0 |
| 0 | 0 | 0 | 0 | 0 |
| 0 | 0 | 0 | 0 | 0.0033486 |
| 0 | 0 | 0 | 0 | 0 |
| 0 | 0 | 0 | 0.001726223 | 0 |
| 0.001637459 | 0 | 0 | 0 | 0 |
| 0.001219689 | 0.011420845 | 0.001115863 | 0 | 0 |
| 0 | 0 | 0 | 0.01239708 | 0.002976448 |
| 0.001188129 | 0.013371791 | 0 | 0 | 0.001183676 |
| 0.018287438 | 0.016199698 | 0.005782507 | 0.014162459 | 0.026446795 |
| 0 | 1.014035784 | 0.049939027 | 0 | 0 |
| 0 | 0 | 0 | 0 | 0 |
| 0 | 0.002045661 | 0 | 0.070682909 | 0 |
| 0 | 0 | 0 | 0 | 0 |
| 0.049558352 | 0.010641668 | 0.003387604 | 0.005185555 | 0.005489126 |
| 0.019111652 | 0 | 0 | 0.053852531 | 5.656152379 |
| 0 | 0 | 0 | 0 | 0 |
| 0.000524371 | 0 | 0 | 0.482047929 | 0 |
| 0.0102642 | 0.043319386 | 0.073921928 | 0.010267421 | 0.003469445 |
| 0 | 0 | 0 | 0 | 0 |
| 0 | 0 | 0 | 0 | 0 |
| 0 | 0 | 0 | 0 | 0 |
| 0 | 0.303754605 | 0 | 0 | 0.031907831 |
| 0 | 0 | 0 | 0 | 0 |
| 0.11207727 | 0 | 0.002068941 | 0.640159903 | 0.004866417 |
| 0 | 0.052590324 | 0.034308564 | 0.005911074 | 0.100727197 |
| 0.019720219 | 0 | 0.002895913 | 0 | 0 |
| 0 | 0 | 0 | 0 | 0 |
| 0 | 0 | 0 | 0 | 0 |
| 0 | 0 | 0 | 0.000627619 | 0 |
| 0 | 0 | 0 | 0 | 0 |
| 0 | 0 | 0 | 0 | 0 |
| 0.967308895 | 0 | 0 | 0 | 0 |
| 0.107703106 | 0.011082457 | 0.050775628 | 1.89732378 | 0.021678873 |
| 0 | 0 | 0 | 0 | 0 |
| 0 | 0 | 0 | 0 | 0.000831509 |
| 0.001401862 | 0.066138227 | 0.032796094 | 0.002804604 | 0.002873023 |
| 0 | 0 | 0 | 0 | 0 |
| 0 | 0 | 0 | 0 | 0 |
| 0 | 0 | 0 | 0 | 0 |
| 0.040021092 | 0.003680003 | 0 | 0.002384983 | 0.049590446 |
| 0.000493272 | 0 | 0 | 0 | 0.001684879 |
| 0 | 0 | 0 | 0 | 0 |
| 0.002038605 | 0 | 0.124893006 | 0 | 0 |
| 0 | 0 | 0 | 0 | 0 |
| 0 | 0 | 0 | 0.009039303 | 0 |

| | | | | |
|---|---|---|---|---|
| 0 | 0 | 0 | 0.000951964 | 0 |
| 0 | 0 | 0 | 0 | 0 |
| 0 | 0 | 0 | 0 | 0 |
| 0 | 0.52861152 | 2.133240439 | 0 | 0.014058198 |
| 0 | 0 | 0 | 0 | 0 |
| 0 | 0 | 0 | 0 | 0 |
| 0 | 0 | 0 | 0 | 0 |
| 0 | 0.003674558 | 0 | 0.004082494 | 0 |
| 0 | 5.407786078 | 3.891945434 | 0 | 0 |
| 0 | 0.09305324 | 0.048539445 | 0.015605079 | 0 |
| 0 | 0 | 0 | 0 | 0 |
| 0.015773256 | 0.183936371 | 0.001241872 | 0 | 0 |
| 0 | 0 | 0 | 0 | 0 |
| 0 | 0 | 0 | 3.536168224 | 0 |
| 0.081703664 | 0.535129531 | 0.053904285 | 0 | 0.017274922 |
| 0 | 0 | 0 | 0 | 0 |
| 0 | 0 | 0 | 0 | 0 |
| 0 | 3.861461477 | 3.571868785 | 0 | 0 |
| 0 | 0 | 0 | 0 | 0 |
| 0 | 0 | 0.010092135 | 0 | 0 |
| 0 | 0 | 0 | 0 | 0 |
| 0 | 0 | 0.041161654 | 0 | 0 |
| 0 | 0 | 0.000571777 | 0 | 0 |
| 0.107704835 | 0.093572882 | 0 | 0.611224069 | 0.385432939 |
| 0 | 0.00240295 | 0.001627794 | 0 | 0.000759679 |
| 0 | 0 | 0.245650313 | 0 | 0.050733789 |
| 0 | 0 | 0.223149735 | 0 | 0 |
| 0.53624792 | 1.069276478 | 2.153798934 | 0 | 1.107989282 |
| 0 | 0.006113109 | 0 | 0 | 0 |
| 0 | 0 | 0 | 0 | 0 |
| 0 | 0 | 0.042376643 | 0 | 0 |
| 0 | 0 | 0.015470158 | 0 | 0 |
| 0 | 0.108748009 | 0 | 0 | 0 |
| 0 | 0 | 0 | 0 | 0 |
| 0 | 0 | 0.050607196 | 0 | 0 |
| 0.088819297 | 0.341456166 | 0.091106217 | 0 | 0.015650661 |
| 0.047543196 | 2.668177611 | 1.638969391 | 0 | 0 |
| 0 | 0 | 0 | 0 | 0 |
| 0.007517572 | 0.015496341 | 0 | 0.00138525 | 0 |
| 0.00571305 | 0 | 0.00044445 | 0 | 0 |
| 0.002005395 | 0.011654176 | 0.043484443 | 0.007112269 | 0.087491566 |
| 0.00158338 | 0 | 0.344041205 | 0 | 0 |
| 3.357944745 | 0.526340592 | 0.005716704 | 6.336242701 | 19.5068513 |
| 0 | 0.054622289 | 0.014840906 | 0 | 0 |
| 0 | 0 | 0 | 0 | 0 |
| 0 | 0 | 0 | 0 | 0 |
| 0 | 0 | 0 | 0 | 0 |
| 0.125795505 | 0.004932977 | 0.002005003 | 0 | 0.004865743 |

| | | | | |
|---|---|---|---|---|
| 0 | 0 | 0.015776533 | 0 | 0 |
| 0 | 0.15028211 | 0.026746854 | 0 | 0 |
| 1.04160491 | 0.009079326 | 0 | 0.049805934 | 0.095978253 |
| 0 | 0.011450037 | 0 | 0 | 0 |
| 0 | 0 | 0 | 0 | 0 |
| 0.001985969 | 0.056771725 | 0.000540748 | 0.000827747 | 0 |
| 0 | 0 | 0 | 0 | 0 |
| 0 | 0 | 0 | 0.002013223 | 0 |
| 0 | 0 | 0 | 0 | 0 |
| 0 | 0 | 0 | 0 | 0 |
| 0.044553651 | 0.03120001 | 0.014018341 | 0.041266325 | 0.011272807 |
| 0.027027944 | 0.591537183 | 0.471074758 | 0.096849658 | 1.548414236 |
| 0 | 0 | 0 | 0 | 0 |
| 0.066338272 | 0.031467587 | 0.001891146 | 0.006680445 | 0.001520759 |
| 0.040874379 | 0.012003965 | 0.000458552 | 0.220229361 | 0.015879019 |
| 0 | 0 | 0 | 0 | 0 |
| 0 | 0 | 0 | 0 | 0 |
| 0 | 0 | 0 | 0 | 0 |
| 0 | 0 | 0.035580333 | 0 | 0 |
| 0.000679639 | 0 | 0.011103292 | 0 | 0.04758986 |
| 0.001901702 | 0.021636179 | 0.523188746 | 0.146822907 | 0.056394377 |
| 0 | 0 | 0 | 2.35065341 | 2.270176077 |
| 1.67569508 | 0.086154208 | 0.267196671 | 1.893466419 | 6.330549355 |
| 0 | 0 | 0 | 0 | 0 |
| 0 | 0 | 0.151806542 | 0 | 0 |
| 0 | 0.896427324 | 1.19417226 | 0 | 0 |
| 0 | 0 | 0.002297104 | 0 | 0 |
| 0 | 0 | 0 | 0.005591452 | 0 |
| 0 | 0 | 0 | 0 | 0 |
| 0 | 0 | 0.002871884 | 0 | 0 |
| 0 | 0 | 0.017320632 | 0 | 0 |
| 0.263427232 | 0.077443264 | 0.092151968 | 0.137305958 | 0.12822347 |
| 0 | 0 | 0.276592389 | 0 | 0.135854337 |
| 0.051856116 | 0.084360441 | 0.035470553 | 0 | 0 |
| 0 | 0.087616974 | 0 | 0.013986188 | 0.002387897 |
| 0.018825937 | 0.165523003 | 0.013808832 | 0.018639682 | 0.017060291 |
| 0.003896914 | 0.022388356 | 0.002425299 | 0 | 0.001584616 |
| 0 | 0 | 0 | 0 | 0 |
| 0.532320309 | 11.458992 | 0.051268487 | 0.020462985 | 0 |
| 0 | 0 | 0 | 0 | 0 |
| 0 | 0 | 0.086442078 | 0 | 0 |
| 0 | 0 | 0 | 0 | 0 |
| 0 | 0.166658038 | 0 | 0 | 0 |
| 0 | 0 | 0.002069391 | 0 | 0 |
| 0.260254966 | 0.431959068 | 0.00973476 | 0.006793296 | 0.023757945 |
| 0 | 0 | 0.079225417 | 0 | 0.37955057 |
| 0 | 0 | 0 | 0 | 0 |
| 0.685485482 | 0.316919097 | 0.001899567 | 0.041765879 | 0.019406593 |

| | | | | |
|---|---|---|---|---|
| 0 | 0 | 0 | 0 | 0 |
| 0 | 0 | 0.00097197 | 0 | 0 |
| 0 | 0 | 0 | 0 | 0 |
| 0 | 0.110783302 | 0.001098119 | 0 | 0 |
| 0 | 0 | 0 | 0 | 0 |
| 0 | 2.937223531 | 0.037608682 | 0 | 0 |
| 0 | 0 | 0.080148717 | 0 | 0 |
| 1.353684249 | 0 | 0.021668923 | 0 | 0.014964734 |
| 0 | 0 | 0 | 0 | 0 |
| 0 | 0 | 0 | 0 | 0 |
| 0.014764315 | 0.014042102 | 0 | 0.00124808 | 0 |
| 0 | 0 | 0 | 0 | 0 |
| 0.003459422 | 0.593436637 | 0.059015392 | 0 | 0 |
| 0 | 0 | 0 | 0 | 0 |
| 0 | 0 | 0 | 0 | 0 |
| 0 | 0 | 0 | 0 | 0 |
| 0 | 0.110563193 | 0.0039416 | 0 | 0 |
| 0 | 0.104895837 | 0 | 0.002770662 | 0.004878247 |
| 0 | 0 | 0.069399253 | 0 | 0 |
| 0 | 0.045963992 | 0 | 0 | 0 |
| 0 | 0 | 0.013165921 | 0 | 0 |
| 0 | 0 | 0.016488935 | 0 | 0 |
| 0.005189231 | 0 | 0.004043199 | 0 | 0 |
| 0 | 0 | 0.002417462 | 0 | 0 |
| 0 | 0 | 0 | 0 | 0 |
| 0 | 0 | 0 | 0 | 0 |
| 0 | 0 | 0 | 0 | 0 |
| 0 | 0 | 0 | 0 | 0 |
| 0 | 0 | 0 | 0 | 0 |
| 0.019238838 | 0 | 1.367094174 | 0.089321322 | 0.010178609 |
| 0 | 0 | 0.01252237 | 0 | 0.004195761 |
| 0 | 0 | 0 | 0 | 0 |
| 0 | 0 | 0.007801004 | 0 | 0 |
| 0 | 0 | 0 | 0 | 0 |
| 0 | 0.029030548 | 0 | 0 | 0 |
| 0 | 0 | 0 | 0 | 0 |
| 0 | 0.041966092 | 0.023744436 | 0.348719283 | 0 |
| 0 | 0.328395005 | 0 | 0.008099999 | 0.016685393 |
| 0 | 0 | 0.002503162 | 0 | 0 |
| 0.001736279 | 0 | 0 | 0 | 0 |
| 0.000990968 | 0.022454384 | 0.006044078 | 0 | 0 |
| 0 | 0 | 0 | 0 | 0 |
| 0 | 0 | 0 | 0 | 0 |
| 0 | 0 | 0.01362877 | 0 | 0 |
| 0 | 0.049490594 | 0.059431844 | 0 | 0 |
| 0 | 0 | 0.021886051 | 0 | 0 |
| 0 | 0 | 0.001202313 | 0 | 0 |
| 0 | 0 | 0 | 0 | 0 |

| | | | | |
|---|---|---|---|---|
| 0 | 0 | 0 | 0 | 0 |
| 0 | 0 | 0 | 0 | 0 |
| 0 | 2.424954312 | 0.103562814 | 0 | 0 |
| 0.0012257 | 0 | 0.455753555 | 0 | 0 |
| 0 | 0 | 0 | 0 | 0 |
| 0 | 0 | 0.569157936 | 0 | 0 |
| 0 | 0 | 0 | 0 | 0 |
| 0 | 0 | 0 | 0 | 0 |
| 0.421218304 | 0.003060213 | 0 | 0.015056905 | 0.109462217 |
| 0.001368551 | 0.002112323 | 0 | 0 | 0 |
| 0 | 0 | 0.001474721 | 0 | 0 |
| 0 | 0 | 0 | 0 | 0 |
| 0.000424895 | 0 | 0 | 0 | 0 |
| 0 | 0 | 0 | 0 | 0 |
| 1.828055233 | 0.14153101 | 0.315010868 | 0 | 3.327212562 |
| 0 | 0 | 0 | 0.297419154 | 0.352162362 |
| 0 | 0 | 0 | 0 | 0 |
| 0 | 0 | 0 | 0 | 0 |
| 0 | 0 | 0 | 0 | 0 |
| 0 | 0 | 0 | 0 | 0 |
| 0 | 0 | 0.186576428 | 0 | 0 |
| 0 | 0 | 0 | 0 | 0 |
| 0 | 0 | 0 | 0 | 0 |
| 0 | 0 | 0 | 0 | 0 |
| 0.042539348 | 0.036758391 | 0.039925657 | 0.017420902 | 0 |
| 0 | 6.658634237 | 2.734093679 | 1.689099004 | 0 |
| 0 | 0 | 2.48716339 | 0 | 0 |
| 0 | 0 | 0 | 0 | 0 |
| 0 | 0.780285754 | 0 | 0 | 0 |
| 0 | 0 | 0 | 0 | 0 |
| 0 | 0 | 0 | 0 | 0 |
| 0 | 0 | 0.092636528 | 0 | 0 |
| 0 | 0 | 0 | 0 | 0 |
| 0 | 0.006726823 | 0.000697477 | 0 | 0.000520812 |
| 0.024620568 | 0.060168547 | 0.001477571 | 0.004272255 | 0 |
| 0.003729376 | 0.021585723 | 0.000928412 | 0.001954096 | 0.001061543 |
| 0 | 0 | 0 | 0.945222695 | 0.203916308 |
| 0.090839671 | 0.46437006 | 0.578682234 | 3.231537066 | 0 |
| 0 | 0.707796602 | 2.182979402 | 0 | 0 |
| 0 | 0 | 0 | 0 | 0 |
| 0 | 0 | 0 | 0 | 0 |
| 0 | 0 | 0 | 0 | 0 |
| 0.853502095 | 0.167806716 | 1.46581415 | 0.106084006 | 1.941961979 |
| 0 | 0 | 0 | 0 | 0 |
| 0 | 0 | 0 | 0 | 0 |
| 0 | 0.011663939 | 0 | 0 | 0 |
| 0 | 0 | 0 | 0 | 0 |
| 0.007772935 | 0.006274459 | 0.637911943 | 0.157116185 | 0 |

| | | | | |
|---|---|---|---|---|
| 0.003597695 | 0.013856657 | 0 | 0.102166611 | 0 |
| 0 | 0 | 0.370637117 | 0 | 0 |
| 0 | 0 | 0.000457381 | 0 | 0 |
| 0 | 0 | 0 | 0 | 0 |
| 0 | 0 | 0 | 0 | 0.000849642 |
| 0 | 0.508290327 | 0.001878401 | 0 | 0 |
| 0 | 0.060333928 | 0 | 0 | 0 |
| 0 | 0 | 0.003019707 | 0 | 0 |
| 0 | 0 | 0 | 0 | 0 |
| 0 | 0 | 0 | 0 | 0 |
| 0 | 0.004249935 | 0.006467729 | 0 | 0 |
| 0 | 0.516377106 | 0.234901872 | 0.690868035 | 0.624100883 |
| 0 | 0.034710493 | 0.007085511 | 0.338880417 | 0.02962852 |
| 0 | 0 | 0 | 0 | 0 |
| 0.156954452 | 0.52506343 | 0.196711949 | 0.057782887 | 0.307557391 |
| 0 | 0 | 0 | 0 | 0 |
| 0 | 0 | 0 | 0 | 0 |
| 0 | 0 | 0 | 0 | 0 |
| 0 | 0 | 0.006195424 | 0.001008894 | 0.003789524 |
| 0 | 0.005034593 | 0.004238773 | 0 | 0 |
| 0 | 0 | 0.105422306 | 0 | 0 |
| 0 | 0 | 0 | 0 | 0 |
| 2.463922987 | 1.053756519 | 2.066303217 | 0.000981013 | 0.000669963 |
| 0 | 0 | 0 | 0 | 0 |
| 0 | 0 | 0.015672235 | 0 | 0 |
| 0 | 0 | 0 | 0 | 0 |
| 0 | 0 | 0 | 0 | 0 |
| 0 | 0 | 0 | 0 | 0 |
| 0 | 0 | 0.000677323 | 0 | 0 |
| 0 | 0 | 0.077360786 | 0 | 0 |
| 0 | 0 | 0 | 0 | 0 |
| 0.066894661 | 0.014788428 | 0.022637849 | 0.004182228 | 0.123682352 |
| 0 | 0 | 0 | 0 | 0 |
| 0.012681476 | 0.036671563 | 0.071171722 | 0 | 0 |
| 0.004251758 | 0.017574505 | 0.002146982 | 0.000966612 | 0.001485289 |
| 0.055806865 | 0.874149891 | 0.373153872 | 0.012211582 | 0 |
| 0 | 0 | 0.000560532 | 0 | 0 |
| 0.022176361 | 0 | 0 | 0 | 0 |
| 0 | 0.018391359 | 0 | 0 | 0 |
| 0 | 0 | 0 | 0 | 0 |
| 0 | 0 | 0.195082734 | 0 | 0 |
| 0 | 0 | 0.028953963 | 0 | 0 |
| 0 | 0 | 0 | 0 | 0 |
| 0 | 0 | 0 | 0 | 0 |
| 0.043787897 | 0.007542144 | 0.001119702 | 0.013140491 | 0.001625731 |
| 0 | 0 | 0 | 0 | 0 |
| 0 | 0.026536939 | 0.118703602 | 0 | 0 |
| 0.00216081 | 0.033722089 | 0 | 0 | 0.000768826 |

| | | | | |
|---|---|---|---|---|
| 0 | 0 | 0 | 0 | 0 |
| 0 | 0 | 0 | 0 | 0 |
| 0.001575222 | 0 | 0 | 0.010636088 | 0 |
| 0 | 0 | 0 | 0 | 0 |
| 0 | 0 | 0 | 0 | 0 |
| 0 | 0 | 0.019261041 | 0 | 0 |
| 0 | 0.060691574 | 0 | 0 | 0 |
| 0 | 0.019923871 | 0.03025826 | 0 | 0 |
| 0 | 0 | 0 | 0 | 0 |
| 0 | 0.066561037 | 0.000478318 | 0 | 0 |
| 0 | 0 | 0 | 0 | 0 |
| 0.001803094 | 0 | 0 | 0 | 0 |
| 0 | 0 | 0 | 0 | 0 |
| 0 | 0.112598132 | 0.223033802 | 0 | 0 |
| 0 | 0 | 0 | 0 | 0 |
| 0 | 0 | 0 | 0 | 0 |
| 0 | 0 | 0 | 0 | 0 |
| 0 | 0 | 0.019225511 | 0 | 0 |
| 0 | 0.174278361 | 2.083947273 | 0 | 0 |
| 0 | 0 | 0.000456434 | 0 | 0 |
| 0 | 0 | 0 | 0 | 0 |
| 0 | 0 | 0 | 0 | 0 |
| 0 | 0 | 0 | 0 | 0 |
| 0 | 0 | 0 | 0 | 0 |
| 0.001040045 | 0 | 0 | 0 | 0 |
| 0.005494397 | 0.010532177 | 0 | 0.00336859 | 0.009232317 |
| 0 | 0 | 0 | 0 | 0 |
| 0 | 0.005466844 | 0.519334025 | 0.184642057 | 0.000888846 |
| 0 | 0 | 0.025076744 | 0 | 0 |
| 0.068942851 | 0.867718468 | 0.390282816 | 0.466438945 | 0 |
| 0 | 0 | 0.005125976 | 0 | 0 |
| 0 | 0 | 0 | 0 | 0 |
| 0 | 0.002803793 | 0.00083794 | 0 | 0 |
| 0 | 0 | 0 | 0 | 0 |
| 0.003157496 | 0.012207668 | 0.000849622 | 0 | 0 |
| 0 | 0 | 0 | 0 | 0 |
| 0 | 0 | 0 | 0 | 0 |
| 0.095682734 | 0.007210697 | 0 | 0 | 0 |
| 0 | 0 | 0.888177763 | 0 | 0 |
| 0 | 0 | 0 | 0 | 0 |
| 5.267826125 | 1.054081666 | 0.617971534 | 16.39906144 | 0.787751942 |
| 0 | 0.139735055 | 0.178745025 | 0.040123711 | 0.201033572 |
| 0 | 0 | 0.370605212 | 0 | 0 |
| 0 | 0 | 0.112679059 | 0 | 0 |
| 0 | 0 | 0 | 0 | 0 |
| 0 | 0 | 0.329363728 | 0 | 0 |
| 0 | 0 | 0 | 0 | 0 |
| 0.011708674 | 0.009638413 | 0.001420976 | 0.001003916 | 0.001285508 |

| | | | | |
|---|---|---|---|---|
| 0 | 0 | 0 | 0 | 0.061415493 |
| 0 | 0 | 0.004834494 | 0.00103261 | 0.001175334 |
| 0 | 0 | 0 | 0 | 0 |
| 0 | 0 | 0 | 0 | 0 |
| 32.97986865 | 1.897540209 | 1.444968766 | 8.088550627 | 1.297659097 |
| 0 | 0 | 0 | 0 | 0 |
| 0 | 0 | 0 | 0 | 0 |
| 0 | 0 | 0.020145654 | 0 | 0 |
| 0.064594552 | 0.110284245 | 0 | 0.953068603 | 5.567711448 |
| 0 | 0 | 0.001499347 | 0 | 0 |
| 0 | 0 | 0 | 0 | 0 |
| 0 | 0 | 0 | 0 | 0 |
| 0 | 0.008127883 | 0.039218952 | 0 | 0 |
| 0.017263931 | 0.002775668 | 0.001477362 | 0.059003606 | 0.001755024 |
| 0.6367221 | 1.834101915 | 0 | 0 | 0 |
| 0 | 0.07956742 | 0.184491891 | 1.187504624 | 0.073448145 |
| 0 | 0 | 0 | 0 | 0 |
| 0 | 0 | 0 | 0 | 0 |
| 0 | 0 | 0 | 0 | 0 |
| 0 | 0 | 0.033086541 | 0 | 0 |
| 0 | 0 | 0.002516465 | 0 | 0 |
| 0 | 0 | 0 | 0 | 0 |
| 0 | 0 | 0 | 0 | 0 |
| 0 | 0 | 0 | 0 | 0 |
| 0 | 0 | 0 | 0 | 0 |
| 0 | 0 | 0 | 0 | 0 |
| 0 | 0.087441037 | 0.033147237 | 0 | 0 |
| 0.00610754 | 0.002985161 | 0 | 0 | 0.001490119 |
| 0 | 0 | 0 | 0 | 0 |
| 0 | 0 | 0 | 0 | 0 |
| 0 | 0 | 0 | 0 | 0 |
| 0 | 0 | 0 | 0 | 0 |
| 0 | 0 | 0.568252696 | 0 | 0 |
| 0 | 0 | 0.014583566 | 0 | 0 |
| 0 | 0.781314026 | 0.332386537 | 0 | 0 |
| 0 | 0 | 0.040365364 | 0 | 0 |
| 0.001856942 | 0 | 0 | 0 | 0 |
| 0 | 0 | 0 | 0 | 0 |
| 0 | 0 | 0 | 0 | 0 |
| 0 | 0 | 0.385441999 | 0 | 0 |
| 0 | 0.137563416 | 0.165759975 | 2.121886342 | 0 |
| 0 | 0 | 0.086870443 | 0 | 0 |
| 0 | 0.025887024 | 0.016264966 | 0 | 0 |
| 0 | 0 | 0 | 0 | 0 |
| 0 | 0 | 0 | 0 | 0 |
| 0 | 0 | 0 | 0 | 0 |
| 0 | 0 | 0 | 0 | 0 |
| 0 | 0 | 0 | 0 | 0 |

| | | | | |
|---|---|---|---|---|
| 0 | 0 | 0.587072326 | 0 | 0 |
| 0 | 0 | 0.006672832 | 0 | 0 |
| 0 | 0.011862382 | 0 | 0 | 0 |
| 0 | 0.010856722 | 0 | 0 | 0 |
| 0 | 0.010728342 | 0.025708292 | 0.001891962 | 0 |
| 0 | 0 | 0 | 0 | 0 |
| 0.001951574 | 0 | 0.004869399 | 0 | 0.000909005 |
| 0 | 0 | 0 | 0 | 0 |
| 0 | 0 | 0 | 0 | 0 |
| 0 | 0 | 0.017150945 | 0 | 0.025272889 |
| 0 | 0 | 0.03247719 | 0 | 0 |
| 0 | 0.018579288 | 0 | 0 | 0 |
| 0 | 0 | 0 | 0 | 0 |
| 0 | 0 | 0 | 0 | 0 |
| 0 | 0 | 0 | 0 | 0 |
| 0 | 0 | 0 | 0 | 0 |
| 0 | 0 | 0 | 0 | 0 |
| 0 | 0 | 0 | 0 | 0.001165217 |
| 0 | 0 | 0 | 0 | 0 |
| 0 | 0 | 0 | 0 | 0 |
| 0.000506729 | 0.076039738 | 0.166452294 | 0.001351702 | 0.00100966 |
| 0 | 0 | 0.000321268 | 0 | 0 |
| 0 | 0.205317592 | 0.003895943 | 0 | 0 |
| 0.027334888 | 0.160644162 | 0.006387326 | 0 | 0.002122009 |
| 0 | 0 | 0 | 0 | 0 |
| 0 | 0 | 0.002298999 | 0.00097755 | 0 |
| 0 | 0 | 0 | 0 | 0 |
| 0 | 0 | 0 | 0 | 0 |
| 0 | 0 | 0.04222194 | 0 | 0 |
| 0.005960805 | 0.189128379 | 0.006855692 | 0.751058569 | 0 |
| 0 | 0 | 0 | 0 | 0 |
| 0 | 0 | 0 | 0 | 0 |
| 0 | 0 | 0.12985531 | 0 | 0 |
| 0 | 0 | 0 | 0 | 0 |
| 0 | 0 | 0 | 0 | 0 |
| 0 | 0 | 0 | 0 | 0 |
| 0 | 0 | 0.347051096 | 0 | 0 |
| 0 | 0 | 0 | 0 | 0 |
| 0 | 0 | 0 | 0 | 0 |
| 0 | 0 | 0 | 0 | 0 |
| 0 | 0 | 0 | 0 | 0 |
| 0 | 0 | 0 | 0 | 0 |
| 0 | 0 | 0 | 0 | 0 |
| 0 | 0 | 0 | 0 | 0 |
| 0 | 0 | 0.022051022 | 0 | 0.000837643 |
| 0 | 0 | 0 | 0 | 0 |
| 0 | 0 | 0.000852485 | 0 | 0 |
| 0 | 0 | 0 | 0 | 0 |

| | | | | |
|---|---|---|---|---|
| 0.07041368 | 0 | 0.006640007 | 0.004101324 | 0.001826685 |
| 0 | 0.303065629 | 0.217092999 | 0 | 0 |
| 0 | 0 | 0.012597893 | 0 | 0 |
| 0 | 0 | 0 | 0 | 0 |
| 0 | 0 | 0 | 0 | 0 |
| 0 | 0.158573891 | 0 | 0 | 0 |
| 0 | 0 | 0 | 0 | 0 |
| 0.142218412 | 0.390570151 | 0.152024678 | 0.970391374 | 0 |
| 1.280968985 | 0.121681864 | 1.154153439 | 0.521449111 | 0.016486728 |
| 0.107916511 | 0.029990944 | 0 | 0.019095914 | 0.049736118 |
| 0 | 0 | 0.70506705 | 0 | 0 |
| 0 | 0 | 0 | 0 | 0 |
| 0 | 0 | 0.065152531 | 0 | 0 |
| 0 | 0 | 0.024095425 | 0 | 0.009552063 |
| 0 | 0 | 0 | 0 | 0 |
| 0 | 0 | 0 | 0 | 0 |
| 0 | 0 | 0 | 0.06637571 | 0 |
| 0 | 0 | 0 | 0 | 0 |
| 0 | 0 | 0.514045743 | 0 | 0 |
| 0.001453759 | 0 | 0.020477903 | 0 | 0 |
| 0 | 0 | 0 | 0 | 0 |
| 0 | 0 | 0 | 0 | 0 |
| 0 | 0 | 0 | 0 | 0 |
| 0 | 0 | 0 | 0 | 0 |
| 0 | 0 | 0.012111408 | 0 | 0 |
| 0 | 0 | 0.014602387 | 0 | 0 |
| 0 | 0 | 0 | 0 | 0 |
| 0 | 0 | 0.128098515 | 0 | 0 |
| 0 | 0 | 0 | 0 | 0 |
| 0 | 0 | 0 | 0 | 0 |
| 0 | 0 | 0.003829594 | 0 | 0 |
| 0 | 0 | 0 | 0 | 0 |
| 0 | 0 | 0 | 0 | 0 |
| 0 | 0 | 0.000833649 | 0 | 0 |
| 0.039277753 | 0 | 0 | 0 | 0 |
| 0 | 0 | 0 | 0 | 0 |
| 0 | 0 | 0.001104485 | 0 | 0 |
| 0 | 0 | 0 | 0 | 0 |
| 0 | 0.012595928 | 0 | 0 | 0 |
| 0 | 0 | 0 | 0 | 0 |
| 0 | 0 | 0 | 0 | 0 |
| 2.968798461 | 2.301379404 | 0.628208875 | 1.572652938 | 0 |
| 0 | 0 | 0.001939927 | 0 | 0 |
| 0 | 0 | 0 | 0 | 0 |
| 0 | 0.00202982 | 0 | 0 | 0 |
| 0 | 0 | 0 | 0 | 0 |
| 0 | 0 | 0 | 0 | 0 |
| 0 | 0 | 0 | 0 | 0 |

| | | | | |
|---|---|---|---|---|
| 0 | 0 | 0 | 0 | 0 |
| 0.002091566 | 0.008691512 | 0.040478338 | 0 | 0.000686943 |
| 0 | 0 | 0 | 0 | 0 |
| 0 | 0 | 0 | 0 | 0 |
| 2.011470723 | 7.279629386 | 0.577997943 | 0.177988435 | 0.068575633 |
| 0 | 0 | 0 | 0 | 0 |
| 0 | 0 | 0 | 0 | 0 |
| 0.018367786 | 0 | 0 | 0.000680502 | 0.033693304 |
| 0 | 0.032820524 | 0.002367323 | 0 | 0 |
| 0 | 0 | 0.014858621 | 0 | 0 |
| 0 | 0 | 0 | 0 | 0 |
| 0 | 0.615133202 | 0 | 0 | 0 |
| 0 | 0 | 0.055339231 | 0 | 0 |
| 0 | 0 | 0 | 0 | 0 |
| 0 | 0 | 0 | 0 | 0.000954209 |
| 0 | 0 | 0.003855604 | 0 | 0.002015307 |
| 0 | 0 | 0 | 0 | 0 |
| 0 | 0 | 0 | 0 | 0 |
| 0 | 0 | 0 | 0 | 0 |
| 0 | 0 | 0 | 0 | 0 |
| 0 | 0 | 0 | 0 | 0 |
| 0 | 0 | 0 | 0 | 0 |
| 0 | 0 | 0 | 0 | 0 |
| 0.009615981 | 0.012545032 | 0 | 0 | 0 |
| 0 | 0.04653271 | 0 | 0 | 0 |
| 0 | 0 | 0 | 0 | 0 |
| 0 | 0 | 1.24951305 | 0 | 0.018852233 |
| 0 | 0 | 0 | 0 | 0 |
| 0 | 0 | 0 | 0 | 0 |
| 0 | 0 | 0 | 0 | 0 |
| 0 | 0 | 0 | 0 | 0 |
| 0 | 0 | 0 | 0 | 0 |
| 0 | 0 | 0.012236257 | 0 | 0 |
| 0 | 0 | 0 | 0 | 0 |
| 0 | 0 | 0 | 0 | 0 |
| 0 | 0 | 0 | 0 | 0 |
| 0 | 0 | 0 | 0 | 0 |
| 0 | 1.699601786 | 0.010946386 | 0 | 0 |
| 0 | 0 | 0 | 0 | 0 |
| 0 | 0 | 0 | 0 | 0 |
| 0 | 0 | 0 | 0 | 0 |
| 1.015449612 | 0.578663666 | 0 | 0 | 0 |
| 0 | 0 | 0 | 0 | 0 |
| 0 | 0.669692137 | 0 | 0 | 0 |
| 0 | 0 | 0 | 0 | 0 |
| 0 | 0 | 0 | 0 | 0 |
| 0 | 0 | 0 | 0 | 0 |
| 0 | 0 | 0 | 0 | 0 |

| | | | | |
|---|---|---|---|---|
| 0.248800713 | 0.055014682 | 0.105339497 | 0 | 0.058808061 |
| 0 | 0 | 0.011525869 | 0 | 0 |
| 0 | 0 | 0 | 0 | 0 |
| 0 | 0 | 0 | 0 | 0 |
| 0 | 0 | 0 | 0 | 0 |
| 0.36451699 | 0.116030771 | 0 | 0.049686892 | 0 |
| 0 | 0 | 0 | 0 | 0 |
| 0 | 0 | 0 | 0 | 0 |
| 0 | 0 | 0 | 0 | 0 |
| 0 | 0 | 0.001004719 | 0 | 0 |
| 0 | 0 | 0 | 0 | 0 |
| 0 | 0 | 0.000591095 | 0 | 0 |
| 0 | 0 | 0 | 0 | 0 |
| 0 | 0 | 0 | 0 | 0 |
| 0 | 0 | 0 | 0 | 0 |
| 0 | 0 | 0 | 0 | 0 |
| 0 | 0 | 0 | 0 | 0 |
| 0 | 0 | 0 | 0 | 0 |
| 0 | 0 | 0 | 0 | 0 |
| 0 | 0 | 0 | 0 | 0 |
| 0 | 0 | 0 | 0.003323253 | 0.00250318 |
| 0 | 0 | 0 | 0 | 0 |
| 0.010612679 | 0.013110657 | 0.008064181 | 0.018763178 | 0.004636623 |
| 0 | 0.520048586 | 0.255226651 | 0 | 0 |
| 0 | 0 | 0.022454157 | 0 | 0 |
| 0 | 0.820391787 | 0 | 0 | 0 |
| 0 | 0 | 0.00070259 | 0 | 0 |
| 0 | 0 | 0 | 0 | 0 |
| 0 | 0 | 0 | 0 | 0 |
| 0 | 0 | 0 | 0 | 0 |
| 0 | 0 | 0 | 0 | 0 |
| 0 | 0 | 0 | 0 | 0 |
| 0 | 0 | 0 | 0.015953198 | 0.056250948 |
| 0 | 0 | 0 | 0 | 0 |
| 0 | 0.03704328 | 0.006349976 | 0.350133505 | 0.023657444 |
| 0 | 0 | 0 | 0.044898972 | 0.001900591 |
| 0 | 0 | 0 | 0 | 0 |
| 0 | 0 | 0.05733811 | 0 | 0 |
| 0.003003432 | 0.01854287 | 0 | 0.001287589 | 0.007144585 |
| 0 | 0 | 0 | 0 | 0 |
| 0 | 0 | 0 | 0 | 0 |
| 0 | 0.242604794 | 0.00514465 | 0 | 0 |
| 0 | 0.681859104 | 0.3614581 | 0 | 0 |
| 0 | 0 | 0 | 0 | 0 |
| 0 | 0 | 0 | 0 | 0 |
| 0.393856176 | 0.007354121 | 0.043872727 | 0 | 0 |
| 0 | 0 | 0 | 0 | 0 |
| 0 | 0 | 0 | 0 | 0 |

| | | | | |
|---|---|---|---|---|
| 0 | 0 | 0 | 0 | 0 |
| 0 | 0 | 0 | 0 | 0 |
| 0 | 0 | 0 | 0 | 0 |
| 0 | 0 | 0 | 0 | 0 |
| 0 | 0 | 0.004640544 | 0 | 0 |
| 0 | 0 | 0 | 0 | 0 |
| 0 | 0 | 0 | 0 | 0 |
| 0 | 0 | 0 | 0 | 0 |
| 0 | 0.042181034 | 0.008903981 | 0 | 0 |
| 0 | 0.422887891 | 0.061681528 | 0.001026289 | 0 |
| 0 | 0 | 0 | 0 | 0 |
| 0 | 0 | 0 | 0 | 0 |
| 0 | 0 | 0 | 0 | 0 |
| 0 | 0 | 0.068967356 | 0 | 0 |
| 0 | 0 | 0 | 0 | 0 |
| 0 | 0 | 0 | 0.065028239 | 0.010688681 |
| 0 | 0 | 0 | 0 | 0 |
| 0 | 0 | 0 | 0 | 0 |
| 0 | 0 | 0 | 0 | 0 |
| 0 | 0.672809173 | 0.408576663 | 0 | 0 |
| 0 | 0.02505986 | 0.004960495 | 0.001687388 | 0 |
| 0 | 0 | 0 | 0 | 0 |
| 0 | 0 | 0 | 0 | 0 |
| 0 | 0 | 0 | 0 | 0 |
| 0 | 0 | 0 | 0 | 0 |
| 0 | 0 | 0.164213515 | 0 | 0 |
| 0 | 0 | 0 | 0 | 0 |
| 0 | 0 | 0.037312956 | 0 | 0 |
| 0 | 0 | 0 | 0 | 0 |
| 0 | 0.033910595 | 0.013673533 | 1.35484269 | 0 |
| 0.313081157 | 0.018762823 | 0.149155552 | 0 | 0 |
| 0 | 0 | 0 | 0 | 0 |
| 0 | 0 | 0.081372324 | 0 | 0 |
| 0 | 0 | 0 | 0 | 0 |
| 0 | 0 | 0 | 0 | 0 |
| 0 | 0 | 0 | 0 | 0 |
| 0 | 0 | 0.000540737 | 0 | 0 |
| 0 | 0 | 0.005769921 | 0 | 0 |
| 0 | 0 | 0.006163416 | 0 | 0 |
| 0 | 0 | 0 | 0 | 0 |
| 0 | 0 | 0 | 0 | 0 |
| 0 | 0 | 0.013687473 | 0 | 0 |
| 0 | 0 | 0.042099871 | 0 | 0 |
| 0 | 0 | 0 | 0 | 0 |
| 0 | 0 | 0 | 0 | 0 |
| 0 | 0 | 0 | 0 | 0 |
| 0 | 0 | 0 | 0.230096967 | 0 |
| 0 | 0 | 0 | 0 | 0 |

| | | | | |
|---|---|---|---|---|
| 0 | 0 | 0 | 0 | 0 |
| 0 | 0 | 0 | 0 | 0 |
| 0 | 0 | 0 | 0 | 0 |
| 0.002037463 | 0 | 0 | 0 | 0 |
| 0 | 0.234517079 | 0.032892345 | 0.098382801 | 0.862666593 |
| 0 | 0 | 0 | 0 | 0 |
| 0 | 0 | 0 | 0 | 0 |
| 0 | 0 | 0 | 0 | 0 |
| 0 | 0 | 0 | 0 | 0 |
| 0 | 0 | 0 | 0 | 0 |
| 0 | 0.021709478 | 0.019856093 | 0 | 0 |
| 0.019078045 | 0 | 0.039922935 | 0.003859692 | 0.000915243 |
| 0.000924694 | 0.027973901 | 0.002296229 | 0 | 0 |
| 0 | 0 | 0 | 0 | 0 |
| 0 | 0 | 0.000320253 | 0 | 0.004045379 |
| 0 | 0 | 0 | 0 | 0 |
| 0.001318947 | 0.197426191 | 0 | 0.003793161 | 0 |
| 0 | 0 | 0.000880089 | 0 | 0 |
| 0.052589951 | 0.012860455 | 0 | 0.105375271 | 0.216778929 |
| 0.005510271 | 0 | 0.041825417 | 0 | 0.001447812 |
| 0 | 0 | 0 | 0 | 0 |
| 0 | 0 | 0 | 0 | 0 |
| 0 | 0 | 0 | 0 | 0 |
| 0 | 0 | 0 | 0 | 0 |
| 0 | 0 | 0 | 0 | 0 |
| 0 | 0 | 0 | 0 | 0 |
| 0.019021186 | 0.028899962 | 0.003107501 | 0 | 0.003190545 |
| 0 | 0 | 0.007296405 | 0 | 0 |
| 0 | 0 | 0 | 0 | 0 |
| 0 | 0 | 0.000317826 | 0 | 0 |
| 0.034844946 | 0.00259997 | 0 | 0 | 0 |
| 0 | 0 | 0.004212377 | 0 | 0 |
| 0 | 0 | 0 | 0 | 0 |
| 0 | 0 | 0 | 0 | 0 |
| 0 | 0 | 0.012884058 | 0 | 0 |
| 0 | 0 | 0 | 0 | 0 |
| 0 | 0 | 0 | 0 | 0 |
| 0.195307366 | 0.064663893 | 0.003888999 | 0.005682468 | 0.006698877 |
| 0 | 0 | 0.345190702 | 0 | 0 |
| 0 | 0 | 0 | 0 | 0 |
| 0.021874454 | 0.003768771 | 0 | 0.003646886 | 0 |
| 0 | 0 | 0.002039518 | 0 | 0 |
| 0 | 0 | 0 | 0 | 0 |
| 0 | 0 | 0 | 0.002206909 | 0.004795524 |
| 0 | 0 | 0 | 0 | 0 |
| 0 | 0 | 0.000585868 | 0 | 0 |
| 0 | 0 | 0.075171845 | 0 | 0 |
| 0 | 0 | 0.002030996 | 0 | 0 |

| | | | | |
|---|---|---|---|---|
| 0 | 0 | 0 | 0 | 0 |
| 0 | 0 | 0.01545914 | 0.004286953 | 0 |
| 0 | 0 | 0.009666081 | 0 | 0 |
| 0.001681545 | 0.002649492 | 0 | 0.001471814 | 0 |
| 0.00740813 | 0 | 0 | 0.001365084 | 0 |
| 0.001006411 | 0 | 0.001644179 | 0 | 0.001289108 |
| 0 | 0 | 0.0026659 | 0 | 0 |
| 0 | 0 | 0 | 0.007564278 | 0 |
| 0 | 0 | 0.004686638 | 0 | 0 |
| 0 | 0 | 0.027300623 | 0 | 0 |
| 0 | 0.033621639 | 0.030418168 | 0 | 0 |
| 0 | 0 | 0 | 0 | 0 |
| 0.007598866 | 0.656133419 | 0.00438152 | 0.016841986 | 0.009669739 |
| 0 | 0 | 0.019654821 | 0 | 0 |
| 0 | 0 | 0.000601833 | 0 | 0 |
| 0 | 0 | 0 | 0 | 0 |
| 0 | 0 | 0 | 0 | 0 |
| 0 | 0 | 0 | 0 | 0 |
| 0 | 0 | 0 | 0 | 0 |
| 0 | 0 | 0.003397236 | 0 | 0 |
| 0 | 0 | 0 | 0 | 0 |
| 0 | 0 | 0 | 0 | 0 |
| 0.060428122 | 0.010775418 | 0 | 0 | 0.000837685 |
| 0 | 0.011464672 | 0.340558931 | 0.075035072 | 0 |
| 0 | 0 | 0 | 0 | 0 |
| 0 | 0 | 0.001899424 | 0 | 0 |
| 0 | 0 | 0 | 0 | 0 |
| 0 | 0.003907718 | 0 | 0 | 0 |
| 0 | 0 | 0 | 0 | 0 |
| 0 | 0 | 0 | 0 | 0 |
| 0.001093784 | 0.018711173 | 0.006004053 | 0.016849557 | 0 |
| 0 | 0.005330855 | 0 | 0 | 0.000866736 |
| 0 | 0 | 0 | 0 | 0 |
| 0 | 0 | 0 | 0 | 0 |
| 0 | 0 | 0.037872324 | 0 | 0 |
| 0 | 0 | 0 | 0.004803837 | 0 |
| 0 | 0 | 0 | 0 | 0 |
| 0 | 0 | 0 | 0 | 0 |
| 0 | 0 | 0.001693282 | 0 | 0 |
| 0 | 0 | 0 | 0 | 0 |
| 0 | 0 | 0 | 0 | 0 |
| 0 | 0 | 0 | 0 | 0 |
| 0 | 0 | 0 | 0 | 0 |
| 0 | 0 | 0 | 0 | 0 |
| 0 | 0 | 0.003080924 | 0 | 0 |
| 0.37926093 | 0.381204362 | 0.171647198 | 0.106695516 | 0.022388091 |
| 0 | 0 | 0 | 0 | 0 |
| 0 | 0 | 0 | 0 | 0 |

| | | | | |
|---|---|---|---|---|
| 0 | 0 | 0 | 0 | 0 |
| 0 | 0 | 0 | 0 | 0 |
| 0 | 0 | 0 | 0 | 0 |
| 0 | 0 | 0.000963984 | 0 | 0 |
| 0 | 0 | 0 | 0 | 0 |
| 0 | 0 | 0 | 0 | 0 |
| 0 | 0 | 0 | 0 | 0 |
| 0 | 0 | 0 | 0 | 0 |
| 0 | 0.040705575 | 0.021262846 | 0 | 0 |
| 0 | 0 | 0 | 0 | 0 |
| 0 | 0.000540764 | 0 | 0 | 0 |
| 0 | 0.006257057 | 0 | 0 | 0 |
| 0 | 0 | 0 | 0 | 0 |
| 0 | 0 | 0 | 0 | 0 |
| 0 | 0 | 0 | 0 | 0 |
| 0 | 0 | 0 | 0 | 0 |
| 0.009049409 | 0 | 0.000721174 | 0 | 0 |
| 0 | 0 | 0 | 0 | 0 |
| 0 | 0 | 0 | 0 | 0 |
| 0 | 0 | 0 | 0 | 0.001190288 |
| 0 | 0 | 0 | 0 | 0 |
| 0 | 0 | 0 | 0 | 0 |
| 0 | 0 | 0 | 0 | 0 |
| 0 | 0 | 0 | 0 | 0 |
| 0 | 0 | 0 | 0 | 0 |
| 0 | 0 | 0 | 0 | 0 |
| 0 | 0 | 0 | 0 | 0 |
| 0 | 0 | 0.006192293 | 0 | 0 |
| 0 | 0 | 0 | 0 | 0 |
| 0 | 0 | 0 | 0 | 0 |
| 0.007184993 | 0.223953236 | 0.001006128 | 0.003080249 | 0.003359908 |
| 0 | 0 | 0 | 0 | 0 |
| 0 | 0.013055277 | 0.00817752 | 0.197501312 | 0.013230142 |
| 0.033694595 | 0 | 0.000795861 | 0 | 0.014386423 |
| 0 | 0 | 0 | 0 | 0 |
| 0 | 0 | 0 | 0 | 0 |
| 0 | 0 | 0 | 0 | 0 |
| 0 | 0 | 0 | 0 | 0 |
| 0 | 0 | 0 | 0 | 0 |
| 0 | 0 | 0 | 0 | 0 |
| 0 | 0 | 0 | 0 | 0 |
| 1.118228606 | 0 | 0.001281442 | 0.728065616 | 0.038792826 |
| 0 | 0.055745287 | 0.068821048 | 0.031066528 | 0.03230041 |
| 0 | 0 | 0 | 0 | 0 |
| 0 | 0 | 0 | 0 | 0 |
| 0.045166898 | 0.380244172 | 0.078019371 | 0 | 0 |
| 0 | 0 | 0 | 0 | 0 |
| 0 | 0 | 0 | 0 | 0 |

| | | | | |
|---|---|---|---|---|
| 0 | 0 | 0 | 0 | 0 |
| 0.04796989 | 0 | 0.000954053 | 0.008971098 | 0 |
| 0.020220928 | 0.003728961 | 0.012099581 | 0.00121851 | 0.007905496 |
| 0 | 0 | 0 | 0 | 0 |
| 0.018090671 | 0.028022547 | 0.032160736 | 0 | 0 |
| 0 | 0 | 0 | 0.003598738 | 0.065322731 |
| 0 | 0 | 0 | 0 | 0 |
| 0 | 0 | 0 | 0 | 0 |
| 0 | 0 | 0 | 0 | 0 |
| 0 | 0 | 0 | 0 | 0 |
| 0 | 0 | 0 | 0 | 0 |
| 0 | 0 | 0 | 0 | 0 |
| 0 | 0 | 0 | 0 | 0 |
| 0 | 0 | 0 | 0 | 0 |
| 0 | 0 | 0 | 0 | 0 |
| 0 | 0 | 0 | 0 | 0 |
| 0 | 0 | 0.207421307 | 0 | 0 |
| 0 | 0 | 0 | 0 | 0 |
| 0 | 0 | 0 | 0 | 0 |
| 0 | 0 | 0 | 0 | 0 |
| 0 | 0 | 0 | 0 | 0 |
| 0 | 0 | 0.00064031 | 0 | 0 |
| 0 | 0 | 0 | 0 | 0 |
| 0 | 0 | 0 | 0 | 0 |
| 0 | 0.016269818 | 0.010758999 | 0 | 0 |
| 0 | 0 | 0 | 0.004444992 | 0 |
| 0 | 0 | 0 | 0 | 0 |
| 0 | 0.124543199 | 0.224959799 | 0 | 0 |
| 0 | 0 | 0 | 0 | 0 |
| 0 | 0 | 0 | 0 | 0 |
| 0 | 0 | 0.002347592 | 0 | 0 |
| 0 | 0 | 0 | 0 | 0 |
| 0 | 0 | 0 | 0 | 0 |
| 0 | 0 | 0 | 0 | 0 |
| 0 | 0 | 0 | 0 | 0 |
| 0 | 0 | 0 | 0 | 0 |
| 0 | 0 | 0 | 0 | 0 |
| 0 | 0 | 0 | 0 | 0 |
| 2.269187808 | 0.670681219 | 0 | 0 | 0 |
| 0 | 0 | 0 | 0 | 0 |
| 0 | 0 | 0 | 0 | 0 |
| 0.005477976 | 0.168284848 | 0 | 0 | 0 |
| 0.054123757 | 0 | 0.007020795 | 0.003244389 | 0.001038605 |
| 0 | 0 | 0.055034339 | 0 | 0 |
| 0 | 0 | 0 | 0 | 0 |
| 0 | 0 | 0.002133593 | 0 | 0 |
| 0.004341508 | 0.073711013 | 0 | 0.008995945 | 0 |
| 0 | 0 | 0 | 0 | 0 |

| | | | | |
|---|---|---|---|---|
| 0 | 0 | 0 | 0 | 0 |
| 0.018782346 | 0.245735959 | 0.000767121 | 0.020158215 | 0.009523049 |
| 0 | 0.033836903 | 0.031077587 | 0 | 0 |
| 0 | 0 | 0 | 0 | 0 |
| 0 | 0 | 0 | 0 | 0 |
| 0 | 0 | 0 | 0 | 0 |
| 0 | 0 | 0 | 0 | 0 |
| 0 | 0 | 0 | 0 | 0 |
| 0 | 0 | 0 | 0 | 0 |
| 0 | 0 | 0 | 0 | 0 |
| 0 | 0 | 0 | 0 | 0 |
| 0 | 0 | 0 | 0 | 0 |
| 0.058529719 | 7.580287274 | 0.147316107 | 0.029791251 | 0.00317896 |
| 0 | 0 | 0 | 0 | 0 |
| 0.046593565 | 0.101912263 | 0 | 0.007076635 | 0 |
| 0 | 0 | 0 | 0 | 0 |
| 0 | 0 | 0 | 0 | 0 |
| 0 | 0 | 0.004700353 | 0 | 0 |
| 0 | 0 | 0 | 0 | 0 |
| 0 | 0 | 0 | 0 | 0 |
| 0 | 0 | 0 | 0 | 0 |
| 0.002156162 | 0 | 0.045714665 | 0 | 0 |
| 0.005338688 | 0.022044696 | 0 | 0 | 0 |
| 0 | 0 | 0 | 0 | 0 |
| 0 | 0 | 0 | 0 | 0 |
| 0 | 0 | 0 | 0 | 0 |
| 0 | 0 | 0 | 0 | 0 |
| 0 | 0 | 0.016921967 | 0 | 0 |
| 0 | 0 | 0 | 0 | 0 |
| 0 | 0 | 0 | 0 | 0 |
| 0 | 0 | 0 | 0 | 0 |
| 0 | 0 | 0 | 0 | 0 |
| 0 | 0 | 0 | 0 | 0 |
| 0 | 0 | 0 | 0 | 0 |
| 0 | 0 | 25.50069066 | 1.56206246 | 11.34874264 |
| 0 | 0 | 0.000585008 | 0 | 0 |
| 0 | 0 | 0 | 0 | 0 |
| 0 | 0 | 0 | 0 | 0 |
| 0.003349405 | 0.004092693 | 0.018917283 | 0 | 0.001634377 |
| 0 | 0 | 0 | 0 | 0 |
| 0.030031848 | 0 | 0.042334525 | 0.035620364 | 0.009708507 |
| 0 | 0 | 0 | 0 | 0 |
| 0 | 0 | 0 | 0 | 0 |
| 0 | 0 | 0 | 0 | 0 |
| 0 | 0 | 0 | 0 | 0 |
| 0.131397611 | 0.023882145 | 0 | 0.791826886 | 3.331737736 |
| 0 | 0 | 0 | 0 | 0 |
| 0 | 0 | 0 | 0 | 0 |

| | | | | |
|---|---|---|---|---|
| 0 | 0 | 0.000428849 | 0 | 0 |
| 0 | 0 | 0.002352716 | 0 | 0 |
| 0 | 0 | 0 | 0 | 0 |
| 0 | 0 | 0.005126856 | 0 | 0 |
| 0 | 0 | 0 | 0 | 0 |
| 0 | 0 | 0.010082481 | 0 | 0 |
| 0 | 0 | 0 | 0 | 0 |
| 0 | 0 | 0 | 0 | 0.000906745 |
| 0 | 0.039697275 | 0.003914129 | 0 | 0 |
| 0.003603002 | 0.613547903 | 0.003924163 | 0 | 0.005538084 |
| 0.022039305 | 0 | 0.013243487 | 0 | 0 |
| 0 | 0 | 0 | 0 | 0 |
| 0.022773214 | 0.026000015 | 0 | 0.001878793 | 0 |
| 0 | 0 | 0 | 0 | 0 |
| 0 | 0 | 0 | 0 | 0 |
| 0 | 0 | 0 | 0 | 0 |
| 0 | 0 | 0 | 0 | 0 |
| 0 | 0 | 0 | 0 | 0 |
| 0 | 0 | 0 | 0 | 0 |
| 0.236132348 | 1.618612391 | 0.018298504 | 0.155101928 | 0.08993529 |
| 0 | 0 | 0.001661742 | 0 | 0 |
| 0 | 0 | 0 | 0 | 0 |
| 0 | 0 | 0 | 0 | 0 |
| 0 | 0 | 0 | 0 | 0 |
| 0 | 0 | 0 | 0 | 0 |
| 0 | 0 | 0 | 0 | 0 |
| 0 | 0 | 0 | 0 | 0 |
| 0 | 0 | 0 | 0 | 0 |
| 0 | 0 | 0 | 0 | 0 |
| 0 | 0 | 0 | 0.011396401 | 0 |
| 0 | 0 | 0 | 0 | 0 |
| 0 | 0 | 0 | 0 | 0 |
| 0 | 0 | 0 | 0 | 0 |
| 0 | 0 | 0 | 0 | 0 |
| 0 | 0 | 0 | 0 | 0 |
| 0 | 0 | 0 | 0 | 0 |
| 0 | 0 | 0.01371908 | 0 | 0 |
| 0 | 0 | 0 | 0 | 0 |
| 0 | 0 | 0 | 0 | 0 |
| 0 | 0 | 0 | 0 | 0 |
| 0 | 0 | 0 | 0 | 0 |
| 0 | 0 | 0 | 0 | 0 |
| 0 | 0.037690866 | 0.016459056 | 0 | 0 |
| 0 | 0 | 0 | 0 | 0 |
| 0.024729917 | 0.05990564 | 0.004745561 | 0.001374315 | 0.002514004 |
| 0 | 0 | 0.016758776 | 0 | 0 |
| 0 | 0 | 0.021256047 | 0 | 0 |
| 0 | 0 | 0 | 0 | 0 |

| | | | | |
|---|---|---|---|---|
| 0 | 0 | 0 | 0 | 0 |
| 0 | 0 | 0 | 0 | 0 |
| 0 | 0 | 0 | 0 | 0 |
| 0 | 0 | 0 | 0 | 0 |
| 0 | 0 | 0 | 0 | 0 |
| 0.145789964 | 0.152634885 | 0.003255496 | 0 | 0.004879684 |
| 0 | 0 | 0.003326972 | 0 | 0 |
| 0 | 0 | 0 | 0 | 0 |
| 0 | 0 | 0 | 0 | 0 |
| 0 | 0 | 0 | 0 | 0 |
| 0 | 0 | 0 | 0 | 0 |
| 0 | 0 | 0 | 0 | 0 |
| 0 | 0 | 0 | 0 | 0 |
| 0 | 0.014505526 | 0 | 0 | 0 |
| 0 | 0 | 0 | 0 | 0 |
| 0 | 0 | 0.081152347 | 0 | 0 |
| 0 | 0 | 0.176654937 | 0 | 0 |
| 0 | 0 | 0 | 0 | 0 |
| 0 | 0 | 0 | 0 | 0 |
| 0 | 0 | 0 | 0 | 0 |
| 0 | 0 | 0 | 0 | 0 |
| 0 | 0 | 0 | 0 | 0 |
| 0 | 0 | 0 | 0 | 0 |
| 0 | 0 | 0 | 0 | 0 |
| 0 | 0 | 0 | 0 | 0 |
| 0 | 0.005984141 | 0.000748844 | 0 | 0 |
| 0 | 0 | 0 | 0 | 0 |
| 0 | 0 | 0.182261052 | 0 | 0 |
| 0 | 0 | 0 | 0 | 0 |
| 0 | 0 | 0 | 0 | 0 |
| 0 | 0 | 0 | 0 | 0 |
| 0 | 0 | 0 | 0 | 0 |
| 0 | 0 | 0 | 0 | 0 |
| 0 | 0 | 0 | 0 | 0 |
| 0 | 0 | 0 | 0 | 0 |
| 0 | 0 | 0 | 0 | 0 |
| 0 | 0 | 0 | 0 | 0 |
| 0 | 0 | 0 | 0 | 0 |
| 0 | 0 | 0 | 0 | 0 |
| 0 | 0 | 0 | 0 | 0 |
| 0 | 0 | 0 | 0 | 0 |
| 0 | 0 | 0 | 0 | 0 |
| 0 | 0 | 0 | 0 | 0 |
| 0 | 0 | 0 | 0 | 0 |
| 0 | 0.027838032 | 0 | 0.001220862 | 0 |
| 0 | 0 | 0 | 0 | 0 |
| 0 | 0 | 0 | 0 | 0 |
| 0 | 0 | 0 | 0 | 0 |

| | | | | |
|---|---|---|---|---|
| 0 | 0 | 0 | 0 | 0 |
| 0 | 0.060851743 | 0.015436759 | 0.032819061 | 0 |
| 0 | 0 | 0.0035436 | 0 | 0 |
| 0 | 0 | 0 | 0 | 0 |
| 0 | 0 | 0 | 0 | 0 |
| 0 | 0 | 0 | 0 | 0 |
| 0 | 0 | 0 | 0 | 0 |
| 0 | 0 | 0 | 0 | 0 |
| 0 | 0 | 0 | 0 | 0 |
| 0 | 0 | 0 | 0 | 0 |
| 0.020406228 | 0 | 0 | 0 | 0.001283984 |
| 0 | 0 | 0 | 0 | 0 |
| 0.006486726 | 0 | 0.016335503 | 1.179371948 | 0 |
| 0 | 0 | 0 | 0 | 0 |
| 0 | 0 | 0 | 0 | 0 |
| 0 | 0 | 0 | 0 | 0 |
| 0 | 0 | 0 | 0 | 0 |
| 0 | 0 | 0 | 0 | 0 |
| 0 | 0 | 0 | 0 | 0 |
| 0 | 0.009816744 | 0.000584017 | 0 | 0 |
| 0.053194808 | 0 | 0 | 0.00087232 | 0.0005585 |
| 0 | 0 | 0.002266375 | 0 | 0 |
| 0 | 0 | 0 | 0 | 0 |
| 2.441720693 | 1.014333009 | 0.068215154 | 4.836015569 | 0.678415585 |
| 0 | 0 | 0 | 0 | 0 |
| 0 | 0 | 0 | 0 | 0 |
| 0 | 0 | 0 | 0 | 0 |
| 0 | 0 | 0.143828042 | 0 | 0 |
| 0 | 0 | 0 | 0 | 0 |
| 0 | 0 | 0 | 0 | 0 |
| 0 | 0 | 0 | 0 | 0 |
| 0 | 0 | 0 | 0 | 0 |
| 0 | 0.097250649 | 0 | 0 | 0 |
| 0 | 0 | 0 | 0 | 0 |

| | | | | |
|---|---|---|---|---|
| PHL018-0020_V2 | PHL018-0245_V2 | PHL018-0117_V3 | PHL018-0210_V3 | PHL018-0008_V2 |
| EYCF | REF | EYCF | EYCF | CTRL |
| 0.164440145 | 0.064096191 | 0.063778557 | 0.10640613 | 0.024308066 |
| 6.189769176 | 8.411278129 | 0.590901928 | 1.455673993 | 0.497492286 |
| 0 | 0.065879707 | 3.773897704 | 0 | 1.753384221 |
| 0 | 0 | 0 | 0 | 0 |
| 0 | 0 | 0 | 0 | 0 |
| 0 | 0 | 0 | 0 | 0 |
| 0 | 0 | 0 | 0 | 0 |
| 0 | 0 | 0 | 0 | 0 |
| 0 | 0 | 0 | 0 | 0 |
| 0 | 0 | 0 | 0 | 0 |
| 0 | 0 | 0 | 0 | 0 |
| 0 | 0 | 0 | 0 | 0 |
| 0 | 0 | 0 | 0 | 0 |
| 0.05080631 | 3.010953374 | 1.850096066 | 0.224170343 | 0.556987116 |
| 0 | 0 | 0 | 0 | 0 |
| 0 | 0 | 0 | 0 | 0 |
| 0 | 0 | 0 | 0 | 0 |
| 0 | 0 | 0 | 0 | 0 |
| 0 | 0 | 0 | 0 | 0 |
| 0 | 0 | 0 | 0 | 0 |
| 0 | 0 | 0 | 0 | 0 |
| 0 | 0 | 0.007601999 | 0 | 0 |
| 0 | 0 | 0 | 0 | 0 |
| 0 | 0 | 0 | 0 | 0 |
| 0 | 0 | 0 | 0 | 0 |
| 0 | 0 | 0 | 0 | 0 |
| 0 | 0 | 0 | 0 | 0 |
| 0 | 0 | 0 | 0 | 0 |
| 0 | 0 | 0 | 0 | 0 |
| 0 | 0 | 0 | 0 | 0 |
| 0 | 0 | 0 | 0 | 0 |
| 0 | 0 | 0 | 0 | 0 |
| 0 | 0 | 0 | 0 | 0 |
| 0 | 0 | 0 | 0 | 0 |
| 0 | 0 | 0 | 0 | 0 |
| 0 | 5.780378691 | 6.177490193 | 1.078980567 | 0.751174427 |
| 0 | 0 | 0 | 0 | 0 |
| 0 | 0 | 0 | 0 | 0 |
| 0 | 0 | 0 | 0 | 0 |
| 0 | 0 | 0 | 0 | 0 |
| 0 | 0 | 0 | 0 | 0 |
| 0.008702847 | 0.013712425 | 0.003076061 | 0.001410258 | 0.061132029 |
| 0.12381817 | 0.0267925 | 0.384151567 | 0.139111698 | 0.012217844 |
| 0.039160924 | 0.001349098 | 0.039511146 | 0.01595606 | 0.150003995 |
| 0.011245604 | 0.003890608 | 0.004363834 | 0 | 0 |
| 0 | 0 | 0 | 0 | 0 |

| | | | | |
|---|---|---|---|---|
| 0 | 0 | 0 | 0.03781298 | 0 |
| 0.019200148 | 0.001445385 | 0.034585408 | 0.011000171 | 0.011966945 |
| 0 | 0 | 0 | 0.006882956 | 0.020169962 |
| 0.300683354 | 0.393767417 | 0.773015846 | 0.609995089 | 1.028248357 |
| 0.001898558 | 0 | 0.001803458 | 0.044978856 | 0 |
| 0 | 0 | 0 | 0 | 0 |
| 0.012576338 | 0.152443164 | 0 | 0.003505257 | 0.003206659 |
| 0 | 0 | 0 | 0 | 0 |
| 0 | 0 | 0 | 0 | 0 |
| 0 | 0 | 0 | 0 | 0 |
| 0 | 0 | 0 | 0 | 0 |
| 0 | 0.002623976 | 1.010477358 | 0.122338123 | 0.117677071 |
| 0 | 0 | 0 | 0 | 0.004701158 |
| 0.025546496 | 0.000665701 | 0.118637947 | 0.286636848 | 0 |
| 0 | 0 | 0 | 0 | 0 |
| 0 | 0 | 0 | 0 | 0 |
| 0.512975766 | 10.2853723 | 0.001137343 | 0 | 0 |
| 0 | 0 | 0 | 0 | 0 |
| 0 | 0 | 0 | 0 | 0 |
| 0 | 0 | 0 | 0 | 0 |
| 0.101749551 | 0.122164284 | 0.104148367 | 0.00361728 | 0.028933752 |
| 0 | 0.004182931 | 0.025516331 | 0.008603892 | 0.047847167 |
| 0 | 0 | 0.011632459 | 0.126977211 | 0 |
| 0 | 0 | 0 | 0 | 0 |
| 0 | 0 | 0.001999924 | 0 | 0 |
| 0 | 0 | 0 | 0 | 0 |
| 0 | 0 | 0 | 0 | 0 |
| 0 | 0 | 0 | 0 | 0 |
| 0.027227999 | 0.010702356 | 0 | 0.063144651 | 0 |
| 0 | 0 | 0 | 0 | 0 |
| 0.00073639 | 0 | 0 | 0 | 0.225404013 |
| 0 | 0 | 0 | 0 | 0 |
| 0 | 0 | 0 | 0 | 0 |
| 0 | 0 | 0 | 0 | 0 |
| 0 | 0 | 0 | 0 | 0 |
| 0 | 0 | 0 | 0 | 0 |
| 0 | 0 | 0 | 0 | 0 |
| 0 | 3.094068289 | 0 | 0 | 0 |
| 0 | 0 | 0 | 0 | 0 |
| 0 | 0 | 0 | 0 | 0 |
| 0 | 0 | 0 | 0 | 0 |
| 0 | 0 | 0 | 0 | 0 |
| 0 | 0 | 0 | 0 | 0 |
| 0 | 0 | 0 | 0 | 0 |
| 0 | 0 | 0 | 0 | 0 |
| 0 | 0.076675455 | 0 | 0 | 0 |
| 0 | 0 | 0 | 0 | 0 |
| 0 | 0.006672811 | 0 | 0 | 0 |

| | | | | |
|---|---|---|---|---|
| 0 | 0 | 0 | 0 | 0 |
| 0 | 0 | 0 | 0 | 0 |
| 0 | 0 | 0 | 0 | 0 |
| 0 | 0 | 0 | 0 | 0 |
| 0.081001224 | 0 | 0 | 0 | 0 |
| 0 | 0 | 0 | 0 | 0 |
| 0 | 0 | 0 | 0 | 0 |
| 0 | 0 | 0 | 0 | 0 |
| 0 | 0.001063982 | 0 | 0.017070373 | 0 |
| 0 | 0 | 0 | 0 | 0 |
| 0 | 0 | 0 | 0 | 0 |
| 0.004193392 | 0 | 0 | 0 | 0 |
| 0 | 0 | 0 | 0 | 0 |
| 0 | 0 | 0 | 0 | 0 |
| 0 | 0 | 0 | 0 | 0 |
| 0 | 0 | 0 | 0 | 0 |
| 0 | 0 | 0 | 0 | 0 |
| 0 | 0 | 0 | 0 | 0 |
| 0 | 0 | 0 | 0 | 0 |
| 0 | 0 | 0 | 0 | 0 |
| 0 | 0 | 0 | 0 | 0 |
| 0 | 0 | 0 | 0 | 0 |
| 0 | 0 | 0 | 0 | 0 |
| 0 | 0 | 0.000465768 | 0 | 0 |
| 0 | 0 | 0 | 0 | 0 |
| 0 | 0 | 0 | 0 | 0 |
| 0 | 0 | 0 | 0 | 0 |
| 0 | 0 | 0 | 0 | 0 |
| 0 | 0 | 0 | 0 | 0 |
| 0 | 0 | 0 | 0 | 0 |
| 0 | 0 | 0.008687432 | 0.101664173 | 0.011287011 |
| 0 | 0 | 0 | 0.004036727 | 0 |
| 0 | 0 | 0 | 0 | 0 |
| 0 | 0 | 0 | 0 | 0 |
| 0 | 0 | 0 | 0 | 0 |
| 0 | 0 | 0 | 0 | 0 |
| 0 | 0 | 0 | 0 | 0 |
| 0 | 0 | 0 | 0 | 0 |
| 0 | 0.013471836 | 0 | 0 | 0.208820142 |
| 0 | 0 | 0 | 0 | 0 |
| 0 | 0 | 0 | 0 | 0 |
| 0 | 0 | 0 | 0 | 0 |
| 0 | 0.000716327 | 0.005177825 | 0.000982277 | 0 |
| 0 | 0 | 0 | 0 | 0 |
| 0 | 0 | 0 | 0 | 0 |
| 0 | 0 | 0 | 0 | 0 |
| 0 | 0 | 0 | 0 | 0 |
| 0 | 0 | 0 | 0 | 0 |

| | | | | |
|---|---|---|---|---|
| 0 | 0 | 0 | 0 | 0 |
| 0 | 0 | 0 | 0 | 0 |
| 0 | 0 | 0 | 0 | 0 |
| 0 | 0 | 0 | 0 | 0 |
| 0.000796166 | 0 | 0 | 0.159957344 | 0 |
| 0 | 0 | 0 | 0 | 0 |
| 0 | 0 | 0 | 0 | 0 |
| 0 | 0 | 0 | 0 | 0 |
| 0 | 0 | 0 | 0 | 0 |
| 0 | 0 | 0 | 0 | 0 |
| 0 | 0 | 0 | 0 | 0 |
| 0 | 0 | 0 | 0 | 0 |
| 0 | 0 | 0 | 0 | 0 |
| 0 | 0 | 0 | 0 | 0 |
| 0 | 0 | 0 | 0 | 0 |
| 0 | 0 | 0 | 8.321777618 | 0 |
| 0 | 0 | 0 | 0 | 0 |
| 0 | 0 | 0 | 0 | 0 |
| 0 | 0 | 0 | 0 | 0 |
| 0 | 0 | 0 | 0 | 0 |
| 0 | 0 | 0 | 0 | 0 |
| 0 | 0 | 0 | 0 | 0 |
| 0 | 0 | 0 | 0 | 0 |
| 0 | 0 | 0 | 0 | 0 |
| 0 | 0 | 0 | 0 | 0 |
| 0 | 0 | 0 | 0 | 0 |
| 0 | 0 | 0 | 0 | 0 |
| 0 | 0 | 0 | 0 | 0 |
| 0 | 0 | 0 | 0 | 0 |
| 0 | 0 | 0 | 0 | 0 |
| 0 | 0 | 0 | 0 | 0.002802275 |
| 0 | 3.033486803 | 0.001247595 | 0 | 0 |
| 0 | 0 | 0 | 0 | 0 |
| 0 | 0 | 0 | 0 | 0 |
| 0 | 0 | 0 | 0 | 0 |
| 0 | 0 | 0 | 0 | 0 |
| 0 | 0 | 0 | 0 | 0 |
| 0 | 0 | 1.234530614 | 0 | 0 |
| 0 | 0 | 0 | 0 | 0 |
| 0 | 0 | 0 | 0 | 0 |
| 0 | 0 | 0 | 0 | 0 |
| 0 | 0 | 0 | 0 | 0 |
| 0 | 0 | 0 | 0 | 0 |
| 0 | 0 | 0 | 0 | 0 |
| 0 | 0 | 0 | 0 | 0 |
| 0 | 0 | 0 | 0 | 0 |
| 0 | 0 | 0 | 0 | 0 |
| 0 | 0 | 0 | 0.076035876 | 0 |

| | | | | |
|---|---|---|---|---|
| 0 | 0 | 0 | 0 | 0 |
| 0 | 0 | 0 | 0 | 0 |
| 0 | 0 | 0 | 0 | 0 |
| 0 | 0 | 0 | 0 | 0 |
| 0 | 0 | 0 | 0 | 0 |
| 0 | 0 | 0 | 0 | 0 |
| 0 | 0 | 0 | 0 | 0 |
| 0 | 0 | 0 | 0 | 0 |
| 0 | 0 | 0 | 0 | 0 |
| 0 | 0 | 0 | 0 | 0 |
| 0 | 0 | 0 | 0 | 0 |
| 0 | 0 | 0 | 0 | 0 |
| 0 | 0 | 0.001364373 | 0.001641972 | 0 |
| 0 | 0 | 0 | 0 | 0 |
| 0.012237468 | 0 | 0 | 0 | 0 |
| 0 | 0 | 0 | 0 | 0 |
| 0 | 0 | 0 | 0 | 0 |
| 0 | 0 | 0 | 0 | 0 |
| 0 | 0 | 0 | 0 | 0 |
| 0 | 0 | 0 | 0 | 0 |
| 0.011015038 | 0.154375322 | 0.026786013 | 0.01096462 | 0.080121272 |
| 0 | 0 | 0 | 0 | 0 |
| 0 | 0 | 0 | 0 | 0 |
| 0.005907327 | 0 | 0 | 0 | 0.008850675 |
| 0 | 0 | 0 | 0 | 0 |
| 0 | 0 | 0 | 0 | 0 |
| 0.004215505 | 0.007947363 | 0.005014139 | 0.004086743 | 0.00474516 |
| 0 | 0 | 0 | 0 | 0 |
| 0 | 0 | 0 | 0 | 0 |
| 0 | 0 | 0 | 0 | 0 |
| 0.001096211 | 0 | 0 | 0 | 0.407381485 |
| 0 | 0 | 0 | 0 | 0 |
| 0 | 0 | 0 | 0 | 0 |
| 0 | 0 | 0 | 0 | 0 |
| 0.008190824 | 0.003329662 | 0.017220927 | 0.001141467 | 0 |
| 0 | 0 | 0 | 0 | 0 |
| 0 | 0 | 0 | 0 | 0 |
| 0 | 0 | 0 | 0 | 0 |
| 0 | 0 | 0 | 0 | 0 |
| 0 | 0 | 0 | 0 | 0 |
| 0 | 0 | 0 | 0 | 0 |
| 0 | 0 | 0 | 0 | 0 |
| 0 | 0 | 0 | 0 | 0 |
| 0 | 0 | 0 | 0 | 0 |
| 0 | 0 | 0 | 0 | 0 |
| 0 | 0 | 0 | 0 | 0 |
| 0 | 0 | 0.00067394 | 0 | 0 |
| 0 | 0 | 0.003104989 | 0.00223696 | 0.001731572 |

| | | | | |
|---|---|---|---|---|
| 0 | 0 | 0 | 0 | 0 |
| 0 | 0 | 0 | 0 | 0 |
| 0 | 0 | 0.00081065 | 0 | 0.00373992 |
| 0 | 0 | 0 | 0 | 0 |
| 0 | 0 | 0 | 0 | 0 |
| 0 | 0 | 0 | 0 | 0 |
| 0 | 0 | 0 | 0 | 0 |
| 0.016571826 | 0.002273969 | 0 | 0 | 0 |
| 0 | 0 | 0 | 0.02043744 | 0 |
| 0 | 0 | 0 | 0 | 0 |
| 0 | 0 | 0 | 0 | 0 |
| 0 | 0 | 0 | 0 | 0 |
| 0 | 0 | 0 | 0 | 0 |
| 0 | 0 | 0 | 0 | 0 |
| 0 | 0 | 0 | 0 | 0 |
| 0 | 0 | 0 | 0 | 0 |
| 0 | 0 | 0 | 0 | 0 |
| 0 | 0 | 0 | 0 | 0 |
| 0.000890429 | 0 | 0 | 0 | 0 |
| 0 | 0 | 0 | 0 | 0 |
| 0 | 0.001186537 | 0 | 0 | 0.018183547 |
| 0 | 0 | 0 | 0 | 0 |
| 0 | 0 | 0 | 0 | 0 |
| 0 | 0 | 0 | 0 | 0 |
| 0.002174577 | 0.050682099 | 0.012338821 | 0.004242662 | 0 |
| 0 | 0 | 0 | 0 | 0 |
| 0 | 0 | 0 | 0 | 0 |
| 0 | 0 | 0 | 0 | 0 |
| 0.002282753 | 0 | 0 | 0 | 0 |
| 0.000745824 | 0 | 0 | 0 | 0 |
| 0.071790865 | 0.004144186 | 0.448189731 | 0.110365943 | 0.434102436 |
| 0 | 0 | 0 | 0 | 0 |
| 0 | 0 | 0 | 0 | 0 |
| 0 | 0 | 0 | 0 | 0 |
| 0 | 0 | 0 | 0 | 0 |
| 0 | 0 | 0 | 0 | 0 |
| 0 | 0 | 0 | 0 | 0 |
| 0 | 0 | 0 | 0 | 0 |
| 0 | 0 | 0 | 0 | 0 |
| 0 | 0 | 0 | 0 | 0 |
| 0 | 0 | 0 | 0 | 0 |
| 0.00262525 | 0.008003954 | 0.045053741 | 0 | 0.016814819 |
| 0 | 0 | 0 | 0 | 0 |
| 0 | 0 | 0 | 0 | 0 |
| 0 | 0 | 0 | 0 | 0 |
| 0 | 0 | 0 | 0 | 0 |
| 0 | 0 | 0 | 0 | 0 |
| 0 | 0 | 0 | 0 | 0 |

| | | | | |
|---|---|---|---|---|
| 0 | 0 | 0 | 0 | 0 |
| 0 | 0 | 0 | 0 | 0 |
| 0.029255849 | 0.000837523 | 0.038202017 | 0.01263318 | 0.141128903 |
| 0 | 0 | 0 | 0 | 0 |
| 0.075273121 | 0 | 0 | 0 | 0.532913347 |
| 0 | 0 | 0 | 0 | 0 |
| 0 | 0 | 0 | 0 | 0 |
| 0 | 0 | 0 | 0 | 0 |
| 0 | 0 | 0 | 0 | 0 |
| 0.002449601 | 0 | 0.0007637 | 0 | 0 |
| 0 | 0 | 0 | 0 | 0 |
| 0 | 0 | 0 | 0 | 0 |
| 0 | 0 | 0 | 0 | 0 |
| 0.00318776 | 0.005862226 | 0.001845689 | 0 | 0.001228133 |
| 0.003758684 | 0.010440974 | 0.000952109 | 0.004190457 | 0.001216396 |
| 0 | 0 | 0 | 0 | 0 |
| 0.007922964 | 0.012186713 | 0.001735748 | 0.003879403 | 0.002540952 |
| 0.40596667 | 0 | 0 | 0 | 0 |
| 0 | 0 | 0 | 0 | 0 |
| 0 | 0 | 0 | 0 | 0 |
| 0 | 0 | 0 | 0 | 0 |
| 0 | 0 | 0.004113074 | 0.001346921 | 0.001042616 |
| 0 | 0 | 0 | 0 | 0 |
| 0 | 0 | 0 | 0 | 0 |
| 0 | 0 | 0 | 0 | 0 |
| 0 | 0 | 0 | 0 | 0 |
| 24.39700164 | 0 | 0 | 0 | 0 |
| 0 | 0 | 0 | 0 | 0 |
| 0 | 0 | 0 | 0 | 0 |
| 0 | 0.026900932 | 0 | 0 | 0 |
| 0 | 0.003613001 | 0 | 0 | 0 |
| 0 | 0 | 0 | 0 | 0 |
| 0 | 0 | 0 | 0 | 0 |
| 0 | 0.008154168 | 0 | 0 | 0 |
| 0 | 0 | 0 | 0 | 0 |
| 0 | 0.017787814 | 0.001564815 | 0 | 0.001249486 |
| 0 | 0 | 0 | 0 | 0 |
| 0 | 0.007569066 | 0.005413731 | 0.006769066 | 0 |
| 0 | 0 | 0 | 0 | 0 |
| 0 | 0 | 0 | 0 | 0 |
| 0 | 0 | 0 | 0 | 0 |
| 0 | 0 | 0 | 0 | 0 |
| 0 | 0 | 0 | 0 | 0 |
| 0.023530654 | 0 | 0 | 0 | 0.001627149 |
| 0 | 0 | 0 | 0 | 0 |
| 0 | 0.003035823 | 0 | 0 | 0 |
| 0 | 0 | 0 | 0 | 0 |
| 0 | 0 | 0 | 0.002049759 | 0.004442661 |

| | | | | |
|---|---|---|---|---|
| 0 | 0 | 3.560342072 | 0 | 0 |
| 0 | 0 | 0 | 0 | 0 |
| 0 | 0 | 0 | 0 | 0 |
| 0 | 0 | 0 | 0 | 0 |
| 0 | 0.017665627 | 0.002476794 | 0.009368009 | 0 |
| 0 | 0.007651436 | 0 | 0 | 0 |
| 0 | 0 | 0 | 0 | 0 |
| 0.010850322 | 0.123037505 | 0.037610858 | 0.015120922 | 0.02402545 |
| 0 | 0 | 0 | 0 | 0 |
| 0.002648762 | 0.074176228 | 0.014611923 | 0.015175318 | 0.014921627 |
| 0 | 0 | 0 | 0 | 0 |
| 0.013335792 | 0.020520737 | 0.0020612 | 0 | 0.022432233 |
| 0.003600493 | 0.046697122 | 0.015982377 | 0.001433605 | 0.031072047 |
| 0 | 0 | 0 | 0.012700781 | 0 |
| 0 | 0 | 0.002858697 | 0 | 0 |
| 0.001029512 | 0.001743784 | 0 | 0 | 0 |
| 0 | 0 | 0 | 0 | 0 |
| 0 | 0 | 0 | 0 | 0 |
| 0.042094489 | 0 | 0 | 0 | 5.363952777 |
| 0 | 0 | 0 | 0 | 0 |
| 0 | 0 | 0 | 0 | 0 |
| 0 | 0 | 0 | 0 | 0 |
| 0 | 0 | 0 | 0 | 0.003816945 |
| 0 | 0 | 0 | 0 | 0 |
| 0 | 0 | 0 | 0 | 0 |
| 0.059310287 | 0 | 0.002225754 | 0 | 0 |
| 0 | 0.001447379 | 0 | 0 | 0 |
| 0 | 0 | 0 | 0 | 0.194858428 |
| 0 | 0 | 0 | 0 | 0 |
| 0 | 0 | 0.001040918 | 0 | 0 |
| 0 | 0 | 0 | 0 | 0.004623205 |
| 0 | 0 | 0 | 0 | 0 |
| 0 | 0 | 0 | 0 | 0 |
| 0.031340184 | 0 | 0 | 0 | 0 |
| 0 | 0 | 0 | 0 | 0 |
| 0 | 0 | 0 | 0 | 0.001310701 |
| 0 | 0.046626122 | 0.003669992 | 0 | 0.407820171 |
| 0 | 0 | 0 | 0 | 0 |
| 0 | 0 | 0 | 0 | 0 |
| 0 | 0 | 0 | 0 | 0 |
| 0 | 0 | 0 | 0 | 0 |
| 0 | 0 | 0 | 0 | 0 |
| 0 | 0 | 0 | 0 | 0 |
| 0.229581032 | 0 | 0.038145098 | 0 | 0 |
| 0 | 0 | 0 | 0 | 0 |
| 0 | 0 | 0 | 0 | 0 |
| 0.008195225 | 0 | 0.00654231 | 0.004499103 | 0.002411056 |
| 0 | 0 | 0 | 0 | 0 |

| | | | | |
|---|---|---|---|---|
| 0 | 0 | 0 | 0 | 0 |
| 0.0022203 | 0.017299393 | 0 | 0 | 0 |
| 0 | 0 | 0 | 0 | 0 |
| 0 | 0 | 0 | 0 | 0 |
| 0 | 0 | 0.005968975 | 0 | 0 |
| 0.007237124 | 0 | 0.03626822 | 0.01744536 | 0 |
| 0 | 0 | 0 | 0 | 0 |
| 0 | 0.044221168 | 0 | 0 | 0 |
| 0 | 0 | 0 | 0 | 0 |
| 0 | 0 | 0 | 0 | 0 |
| 0 | 0.053488432 | 0.003447953 | 0.023711344 | 0 |
| 0 | 0 | 0 | 0 | 0 |
| 0 | 0 | 0 | 0 | 0.002840362 |
| 0 | 0.006679673 | 0 | 0.001493419 | 0.006011284 |
| 0.0010843 | 0.021157447 | 0.020654663 | 0 | 0 |
| 0 | 0 | 0 | 0 | 0 |
| 0 | 0 | 0 | 0 | 0 |
| 0.045677789 | 0 | 0 | 0 | 0 |
| 0 | 0 | 0 | 0 | 0.00836673 |
| 0 | 0 | 0 | 0 | 0 |
| 0 | 0 | 0 | 0 | 0 |
| 0 | 0.204234919 | 0 | 0.077669719 | 0.21826928 |
| 0 | 0 | 0 | 0 | 0 |
| 0 | 0 | 0 | 0 | 0 |
| 0 | 0 | 0 | 0 | 0 |
| 0 | 0 | 0 | 0 | 0 |
| 0 | 0 | 0 | 0 | 0.009776736 |
| 0 | 0 | 0 | 0 | 0 |
| 0 | 0 | 0 | 0 | 0 |
| 0.354055927 | 0.872676278 | 0.490843643 | 0.233789421 | 0.859439058 |
| 0.003780492 | 0.002758382 | 0.000589313 | 0.001823699 | 0 |
| 0 | 0 | 0 | 0 | 0 |
| 0 | 0 | 0 | 0 | 0 |
| 0 | 0 | 0.037560091 | 0 | 0.003998837 |
| 0 | 0 | 0 | 0 | 0 |
| 0 | 0 | 0 | 0 | 0 |
| 0 | 0 | 0 | 0 | 0 |
| 0 | 0 | 0 | 0 | 0 |
| 0 | 0 | 0 | 0 | 0 |
| 0 | 0.115807312 | 0.080428488 | 0.008128646 | 0 |
| 0 | 0.000955029 | 0.028922191 | 0.001718854 | 0 |
| 1.154577474 | 4.887535378 | 0 | 1.439851648 | 3.866763261 |
| 0 | 0 | 0 | 0 | 0 |
| 0 | 0 | 0 | 0 | 0 |
| 0 | 0 | 0 | 0 | 0 |
| 0 | 0 | 0 | 0 | 0 |
| 0.000554803 | 0.002505929 | 0.005621466 | 0 | 0.004189427 |
| 0 | 0.034010719 | 0 | 0 | 0 |

| | | | | |
|---|---|---|---|---|
| 0.148087456 | 0.000927569 | 0.001618387 | 0 | 0 |
| 0.00131219 | 0 | 0 | 0 | 0.004089268 |
| 0.029536437 | 0.457947197 | 0 | 0 | 0.002347742 |
| 0 | 0 | 0 | 0 | 0 |
| 0 | 0 | 0 | 0 | 0 |
| 0 | 0.12129893 | 0 | 0 | 0.003537972 |
| 0 | 0 | 0 | 0 | 0 |
| 0.003068894 | 0 | 0 | 0 | 0 |
| 0 | 0 | 0 | 0 | 0 |
| 0 | 0 | 0 | 0 | 0 |
| 0 | 0 | 0 | 0 | 0 |
| 0 | 0 | 0 | 0 | 0 |
| 0 | 0 | 0 | 0 | 0 |
| 0 | 0.002237728 | 0.002300751 | 0 | 0 |
| 0 | 0 | 0 | 0 | 0 |
| 0.035812811 | 0.005683425 | 0.018633787 | 0.109708658 | 0.009745213 |
| 0.364583918 | 0.049911297 | 0.039857844 | 0 | 0 |
| 0 | 0 | 0 | 0 | 0.055774728 |
| 0 | 0 | 0 | 0 | 0 |
| 0 | 0.066886241 | 0 | 0 | 0 |
| 0.054984302 | 0 | 0.013014713 | 0.006594833 | 0 |
| 0.012444288 | 0 | 0 | 0 | 0 |
| 0 | 0 | 0 | 0 | 0 |
| 0 | 0 | 0 | 0 | 0.950477847 |
| 0 | 0 | 0 | 0 | 0 |
| 0 | 0.007340945 | 0 | 0 | 0.001270459 |
| 0 | 0 | 0.00344999 | 0 | 0 |
| 0 | 0 | 0 | 0 | 0 |
| 0 | 0 | 0 | 0 | 0 |
| 0 | 0 | 0 | 0.005142877 | 0.001676196 |
| 0 | 0.00163743 | 0.044078289 | 0.001202871 | 0.017318656 |
| 0.031656315 | 0.005579902 | 0.002868526 | 0.005379998 | 0.008329029 |
| 0.035042082 | 0.085669243 | 0.005462449 | 0.01092797 | 2.979172607 |
| 0 | 0 | 0 | 0 | 0 |
| 0.007018286 | 0 | 0 | 0.001047924 | 0 |
| 0 | 0 | 0 | 0 | 0 |
| 0.055163874 | 0.002701776 | 0.001122371 | 0.001234955 | 0.002628851 |
| 0.118853513 | 0.001643377 | 0 | 0.045633666 | 0 |
| 0.003217664 | 0 | 0 | 0.968388542 | 0 |
| 0.002691433 | 0 | 0.026265875 | 0 | 0 |
| 0 | 0 | 0 | 0 | 0 |
| 0 | 0 | 0 | 0 | 0 |
| 0 | 0 | 0.012601607 | 0 | 0.00391309 |
| 0 | 0.0106294 | 0 | 0 | 0.055440977 |
| 1.343973896 | 0 | 3.891404017 | 0 | 0.089078032 |
| 0 | 0 | 0 | 0 | 0 |
| 0 | 0 | 0 | 0 | 0 |
| 0 | 0 | 0 | 0 | 0 |

| | | | | |
|---|---|---|---|---|
| 0 | 0 | 0 | 0 | 0 |
| 0.053826941 | 1.187154283 | 0.428182316 | 0.244120585 | 0.027504673 |
| 0 | 0 | 0 | 0 | 0 |
| 0 | 0 | 0 | 0 | 0 |
| 0 | 0 | 0 | 0 | 0 |
| 0 | 0 | 0 | 0 | 0 |
| 0 | 0 | 0 | 0 | 0.409953756 |
| 0 | 0 | 0 | 0 | 0.000978078 |
| 0 | 0.06530409 | 0 | 0 | 0 |
| 0 | 0 | 0.0031289 | 0.001912643 | 0.013694871 |
| 0.004385888 | 0 | 0.001955335 | 0.000733457 | 0 |
| 1.037686667 | 0.053113995 | 0.016151283 | 0.003277514 | 0.006201645 |
| 0 | 0 | 0 | 0.004856309 | 0 |
| 0 | 0 | 0 | 0 | 0 |
| 0.049920005 | 0.001091024 | 0 | 0 | 0.016285537 |
| 0 | 0 | 0 | 0 | 0 |
| 0.002388182 | 0.146121317 | 0.028479109 | 0.012544572 | 0 |
| 0.004054095 | 0.004847163 | 0.147637637 | 0.046195054 | 0.09055357 |
| 0 | 0 | 0 | 0 | 0 |
| 0.00309708 | 0 | 0.020083675 | 0 | 0.321566637 |
| 0.002841714 | 0.051598426 | 0.026681773 | 0.015576745 | 0.002861113 |
| 0 | 0.00127773 | 0 | 0 | 0 |
| 0 | 0 | 0 | 0 | 0 |
| 0 | 0 | 0 | 0 | 0 |
| 0 | 0 | 0 | 0 | 0 |
| 0 | 0 | 0 | 0 | 0 |
| 0 | 0 | 0 | 0.023974616 | 0.117601977 |
| 0.016309739 | 0.035469633 | 0.031011046 | 0.015994567 | 0.006299091 |
| 0 | 0 | 0 | 0 | 0 |
| 0 | 0 | 0 | 0 | 0 |
| 0 | 0 | 0 | 0 | 0 |
| 0 | 0 | 0 | 0 | 0 |
| 0 | 0 | 0 | 0 | 0 |
| 0 | 0 | 0 | 0 | 0 |
| 0 | 0 | 0.125163642 | 0 | 0 |
| 0.349982075 | 0.077115768 | 0.323193078 | 0.189014044 | 1.918582768 |
| 0 | 0 | 0 | 0 | 0 |
| 0 | 0 | 0 | 0 | 0 |
| 0.016145589 | 0.014809642 | 0.007298765 | 0.01800029 | 0.008038586 |
| 0 | 0 | 0 | 0 | 0 |
| 1.165583261 | 0 | 3.682646079 | 0 | 0.044389439 |
| 0 | 0 | 0 | 0 | 0 |
| 0.010410589 | 0.001226673 | 0.141715322 | 0.032590691 | 0.002278621 |
| 0.00043701 | 0 | 0.000442794 | 0 | 0 |
| 0 | 0 | 0 | 0 | 0 |
| 0 | 0 | 0 | 0.429558801 | 0 |
| 0 | 0 | 0 | 0 | 0 |
| 0 | 0 | 0 | 0 | 0 |

| | | | | |
|---|---|---|---|---|
| 0 | 0 | 0 | 0 | 0.00864034 |
| 0 | 0 | 0 | 0 | 0 |
| 0 | 0 | 0 | 0 | 0 |
| 0 | 0 | 0.88349029 | 0.274276024 | 0 |
| 0 | 0 | 0 | 0 | 0 |
| 0 | 0 | 0 | 0 | 0 |
| 0 | 0 | 0 | 0 | 0 |
| 0 | 0 | 0.044918634 | 0.005257914 | 0.002543758 |
| 0 | 0.066598798 | 0.29533664 | 0.867675122 | 0 |
| 0 | 0 | 0 | 0.02692111 | 0.025413317 |
| 0 | 0 | 0 | 0 | 0 |
| 0.001852 | 0.011406956 | 0 | 0.005865757 | 0 |
| 0 | 0 | 0 | 0 | 0 |
| 0 | 0 | 0 | 0 | 0 |
| 0 | 0 | 0 | 0.061376749 | 0 |
| 0 | 0 | 0 | 0 | 0 |
| 0 | 0 | 0 | 0.009092055 | 0 |
| 0 | 0 | 0 | 0 | 0 |
| 0 | 0 | 0 | 0 | 0 |
| 0 | 0 | 0 | 0 | 0 |
| 0 | 0 | 0 | 0 | 0 |
| 0 | 0 | 0 | 0 | 0 |
| 0 | 0 | 0 | 0 | 0 |
| 0.028505616 | 0.062200848 | 0.277438869 | 0.792825448 | 0.525352463 |
| 0 | 0 | 0 | 0 | 0 |
| 0 | 0 | 0 | 0 | 0 |
| 0 | 0 | 0 | 0.005227561 | 0 |
| 0 | 0 | 0 | 0 | 0 |
| 0 | 0 | 0 | 0 | 0 |
| 0 | 0 | 0 | 0 | 0 |
| 0 | 0 | 0.033334793 | 0.004105812 | 0.017480101 |
| 0 | 0 | 0 | 0.050934408 | 0 |
| 0 | 0 | 0 | 0 | 0 |
| 0 | 0 | 0 | 0 | 0 |
| 0 | 0 | 0 | 0 | 0 |
| 0 | 0 | 0 | 0 | 0 |
| 0 | 0.003400205 | 0 | 11.60662487 | 0 |
| 0 | 0 | 0 | 0 | 0 |
| 0 | 0 | 0 | 0 | 0 |
| 0 | 0 | 0 | 0 | 0 |
| 0 | 0.183403598 | 0.075280267 | 0.148556636 | 0 |
| 0 | 0 | 0 | 0.118271814 | 0 |
| 0.475479425 | 1.235812903 | 0.042012816 | 0.076145093 | 2.582366057 |
| 0 | 0 | 0 | 0 | 0 |
| 0 | 0 | 0 | 0 | 0 |
| 0 | 0 | 0 | 0 | 0 |
| 0 | 0 | 0 | 0 | 0 |
| 0.083682695 | 0 | 0.035411289 | 0.002164632 | 0 |

| | | | | |
|---|---|---|---|---|
| 0 | 0 | 0 | 0 | 0 |
| 0 | 0 | 0 | 0 | 0 |
| 0.073518858 | 0.02244621 | 0.124844809 | 0.001556283 | 0.04798632 |
| 0 | 0 | 0 | 0.028318167 | 0 |
| 0 | 0 | 0 | 0 | 0 |
| 0.002639179 | 0.002781479 | 0.016341801 | 0 | 0.002530663 |
| 0 | 0 | 0 | 0 | 0 |
| 0.021842222 | 0 | 0 | 0 | 0 |
| 0 | 0 | 0 | 0 | 0 |
| 0 | 0 | 0 | 0 | 0 |
| 0.044954153 | 0.001485721 | 0.128685721 | 0.013497279 | 0.097776451 |
| 1.879696907 | 2.186813616 | 1.551613383 | 0.410265891 | 0.089522812 |
| 0 | 0 | 0 | 0 | 0 |
| 0.00216943 | 0.02057762 | 0.04056579 | 0.000824537 | 0.001914756 |
| 0.313477448 | 0.192274875 | 0.30581619 | 0.073423612 | 0.054991065 |
| 0 | 0 | 0 | 0 | 0 |
| 0 | 0 | 0 | 0 | 0 |
| 0 | 0 | 0 | 0 | 0 |
| 0 | 0 | 0 | 0 | 0 |
| 0 | 0 | 0.00899883 | 0.01153776 | 0 |
| 0.242917176 | 0.121412182 | 0.120117655 | 0.353895639 | 0.001652239 |
| 0.024046622 | 0.356237998 | 0.315926289 | 0 | 0.843489083 |
| 0.10126659 | 2.576992354 | 0.434760556 | 0.372164965 | 0.62891373 |
| 0 | 0 | 0 | 0 | 0 |
| 0 | 0 | 0 | 0 | 0 |
| 0 | 0.007709632 | 0.006508778 | 0 | 0 |
| 0 | 0.174009319 | 0.123248618 | 0 | 0 |
| 0 | 0 | 0 | 0.003116703 | 0.001033953 |
| 0 | 0 | 0 | 0 | 0 |
| 0 | 0 | 0 | 0 | 0 |
| 0 | 0 | 0 | 0 | 0 |
| 0 | 0.051335163 | 0.081607174 | 0.327857782 | 0.106069152 |
| 0.125182034 | 0.057983226 | 0.049632657 | 0 | 0 |
| 0 | 0 | 0 | 0.004786819 | 0 |
| 0.000619353 | 0 | 0.050580622 | 0.000863125 | 0 |
| 0.012083536 | 0.002306147 | 0.12398679 | 0.213933041 | 0.019460744 |
| 0 | 0 | 0 | 0.002749309 | 0 |
| 0 | 0 | 0 | 0 | 0 |
| 0.013343547 | 0 | 0 | 0.044407071 | 0.015468453 |
| 0 | 0 | 0 | 0 | 0 |
| 0 | 0 | 0 | 0 | 0 |
| 0 | 0 | 0 | 0 | 0 |
| 0 | 0 | 0 | 0 | 0 |
| 0 | 0 | 0 | 0 | 0 |
| 0.027171636 | 0 | 0.076104363 | 0.195822214 | 0.012980676 |
| 0 | 0 | 0 | 0.042647026 | 0 |
| 0 | 0 | 0 | 0 | 0 |
| 0 | 0.279169185 | 0.074011446 | 1.432951285 | 0.060107448 |

| | | | | |
|---|---|---|---|---|
| 0 | 0 | 0 | 0 | 0 |
| 0 | 0 | 0 | 0 | 0 |
| 0 | 0 | 0 | 0 | 0 |
| 0.000992497 | 0 | 0 | 0.002074705 | 0 |
| 0 | 0 | 0 | 0 | 0 |
| 0 | 0 | 0 | 0 | 0 |
| 0 | 0 | 0 | 0.744011891 | 0 |
| 0 | 0.001237525 | 0 | 0.009333397 | 0 |
| 0 | 0 | 0 | 0 | 0 |
| 0 | 0 | 0 | 0 | 0 |
| 0 | 0 | 0.010640103 | 0.005391568 | 0.001391157 |
| 0 | 0 | 0 | 0 | 0 |
| 0 | 0 | 0 | 0 | 0 |
| 0 | 0 | 0 | 0 | 0 |
| 0 | 0 | 0 | 0 | 0 |
| 0 | 0 | 0 | 0 | 0 |
| 0 | 0 | 0 | 0.264653846 | 0 |
| 0 | 0.004987639 | 0.001243178 | 0.006198207 | 0.008106743 |
| 0 | 0 | 0 | 0 | 0 |
| 0 | 0 | 0 | 0.001927502 | 0 |
| 0 | 0 | 0 | 0 | 0 |
| 0 | 0 | 0 | 0 | 0 |
| 0 | 0 | 0.029920037 | 0.001232084 | 0 |
| 0 | 0 | 0 | 0 | 0 |
| 0 | 0 | 0 | 0 | 0 |
| 0 | 0 | 0 | 0 | 0 |
| 0 | 0 | 0 | 0 | 0 |
| 0 | 0 | 0 | 0 | 0 |
| 0 | 0 | 0 | 0 | 0 |
| 0 | 0.069538818 | 0 | 0.053444395 | 0.18186765 |
| 0 | 0 | 0 | 0 | 0 |
| 0 | 0 | 0 | 0 | 0 |
| 0 | 0 | 0 | 0 | 0 |
| 0 | 0 | 0 | 0 | 0 |
| 0 | 0 | 0.00890578 | 0.005512001 | 0 |
| 0 | 0 | 0 | 0 | 0 |
| 0 | 0.032357383 | 0 | 0.034197318 | 0.517470051 |
| 0.0020274 | 0 | 0.010745232 | 0 | 0.044582027 |
| 0 | 0 | 0 | 0 | 0 |
| 0 | 0 | 0 | 0 | 0 |
| 0 | 0 | 0 | 0.128466432 | 0 |
| 0 | 0 | 0 | 0 | 0 |
| 0 | 0 | 0 | 0.004655144 | 0 |
| 3.830161639 | 0 | 0 | 0.145841783 | 0 |
| 0 | 0 | 0 | 0.779340073 | 0 |
| 0 | 0 | 0 | 0 | 0 |
| 0 | 0 | 0 | 0.002065056 | 0 |
| 0 | 0 | 0 | 0 | 0 |

| | | | | |
|---|---|---|---|---|
| 0 | 0 | 0 | 0 | 0 |
| 0 | 0 | 0 | 0 | 0 |
| 0 | 0 | 0 | 0 | 0 |
| 0 | 0.001261229 | 0 | 0.202349753 | 0 |
| 0 | 0 | 0 | 0 | 0 |
| 0 | 0 | 0 | 0 | 0 |
| 0 | 0 | 0 | 0 | 0 |
| 0 | 0 | 0 | 0 | 0 |
| 0.067106952 | 0.003205952 | 0.013729803 | 0.007793309 | 0.027146676 |
| 0.000519624 | 0 | 0 | 0.001448289 | 0 |
| 0 | 0 | 0 | 0.004072181 | 0 |
| 0 | 0 | 0 | 0 | 0 |
| 0 | 0 | 0 | 0 | 0 |
| 0 | 0 | 0 | 0 | 0 |
| 0 | 0.018431051 | 2.012003113 | 0.045665434 | 0 |
| 0 | 0 | 0.981936949 | 0 | 0.445993245 |
| 0 | 0 | 0 | 0 | 0 |
| 0 | 0 | 0 | 0 | 0 |
| 0 | 0 | 0 | 0 | 0 |
| 0 | 0 | 0 | 0 | 0 |
| 0 | 0 | 0 | 0.00366714 | 0 |
| 0 | 0 | 0 | 0 | 0 |
| 0 | 0 | 0 | 0 | 0 |
| 0 | 0 | 0.162418685 | 0 | 0 |
| 0.02453474 | 0.001713685 | 0.02896003 | 0.034543906 | 0.001364261 |
| 2.592147284 | 0.006783923 | 0 | 21.17568895 | 0.075609349 |
| 0 | 0 | 0 | 0 | 0 |
| 0 | 0 | 0 | 0 | 0 |
| 0 | 0 | 0 | 0 | 0 |
| 0 | 0 | 0 | 0.078259487 | 0 |
| 0 | 0 | 0 | 0 | 0 |
| 0 | 0 | 0 | 0 | 0 |
| 0 | 0 | 0 | 0 | 0 |
| 0 | 0.002745654 | 0 | 0.00225902 | 0 |
| 0.01691573 | 0.007539951 | 0.015786507 | 0 | 0 |
| 0.004720015 | 0 | 0.03124561 | 0.002411851 | 0.001188059 |
| 0.52784585 | 0 | 0 | 0 | 0.848597618 |
| 1.652566128 | 0.001444392 | 2.802374539 | 2.078694114 | 0.811622001 |
| 0 | 0 | 0 | 0 | 0 |
| 0 | 0 | 0 | 0 | 0 |
| 0 | 0 | 0 | 0 | 0 |
| 0 | 0 | 0.012112223 | 0 | 0 |
| 0.081914797 | 0.036416546 | 0.293001951 | 2.889849731 | 0.118515777 |
| 0 | 0 | 0 | 0 | 0 |
| 0 | 0 | 0 | 0 | 0 |
| 0.105679318 | 0 | 0 | 0 | 0 |
| 0 | 0 | 0 | 0 | 0 |
| 0.013297814 | 0.016731964 | 0.082767818 | 0.024819277 | 0.286642241 |

| | | | | |
|---|---|---|---|---|
| 0 | 0 | 0 | 0 | 0 |
| 0 | 0 | 0 | 0 | 0 |
| 0 | 0 | 0 | 0 | 0 |
| 0 | 0 | 0 | 0 | 0 |
| 0.000705194 | 0 | 0.002679482 | 0 | 0.002091989 |
| 0 | 0 | 0 | 0 | 0 |
| 0 | 0 | 0.01678044 | 0 | 0 |
| 0 | 0 | 0 | 0 | 0 |
| 0 | 0 | 0 | 0 | 0 |
| 0 | 0 | 0 | 0 | 0 |
| 0 | 0 | 0 | 0 | 0 |
| 0 | 0.068321638 | 0.618492769 | 3.787994065 | 6.772170831 |
| 0.006830933 | 2.60734784 | 0.005407306 | 0.522086938 | 0.963473166 |
| 0 | 0 | 0 | 0 | 0 |
| 0.061982346 | 2.299459032 | 0.036566091 | 0.000852415 | 0.531384937 |
| 0 | 0 | 0 | 0 | 0 |
| 0 | 0 | 0 | 0 | 0 |
| 0 | 0 | 0 | 0 | 0 |
| 0 | 0 | 0 | 0 | 0.029880933 |
| 0 | 0 | 0 | 0.012979175 | 0 |
| 0 | 0.00060145 | 0 | 0.100000966 | 0 |
| 0 | 0 | 0 | 0 | 0 |
| 5.120812803 | 0.054863155 | 1.784641332 | 0.624782493 | 0.171144191 |
| 0 | 0 | 0 | 0 | 0 |
| 0 | 0 | 0 | 0 | 0 |
| 0 | 0 | 0 | 0 | 0 |
| 0 | 0 | 0 | 0 | 0 |
| 0 | 0 | 0 | 0 | 0 |
| 0 | 0 | 0 | 0.001023748 | 0 |
| 0 | 0 | 0 | 0 | 0 |
| 0 | 0 | 0 | 0 | 0 |
| 0.054766999 | 0.164197081 | 0.088465387 | 0.034289977 | 0.009133063 |
| 0 | 0 | 0 | 0 | 0 |
| 0 | 0 | 0.023035377 | 0.031314149 | 0 |
| 0.00496534 | 0 | 0.032962143 | 0.003101916 | 0.00221641 |
| 0 | 0.033796721 | 0 | 0.033527943 | 0.006904952 |
| 0 | 0 | 0 | 0.002329863 | 0 |
| 0 | 0 | 0 | 0 | 0 |
| 0 | 0 | 0 | 0.000822379 | 0 |
| 0 | 0 | 0 | 0 | 0 |
| 0 | 0 | 0 | 0.052246984 | 0 |
| 0 | 0 | 0 | 0 | 0 |
| 0 | 0 | 0 | 0 | 0 |
| 0 | 0 | 0 | 0 | 0 |
| 0.026818133 | 0.001142754 | 0.050586423 | 0.01786408 | 0.013828112 |
| 0 | 0 | 0 | 0 | 0 |
| 0 | 0 | 1.8014195 | 0 | 0 |
| 0 | 0 | 0 | 0.000666956 | 0 |

| | | | | |
|---|---|---|---|---|
| 0 | 0 | 0 | 0 | 0 |
| 0 | 0 | 0 | 0 | 0 |
| 0 | 0 | 0.019619633 | 0 | 0.010538116 |
| 0 | 0 | 0 | 0 | 0 |
| 0 | 0 | 0 | 0 | 0 |
| 0 | 0 | 0 | 0 | 0 |
| 0 | 0 | 0 | 0 | 0 |
| 0.011532251 | 0.006250653 | 0 | 0.027091539 | 0 |
| 0 | 0 | 0 | 0 | 0 |
| 0 | 0 | 0 | 0 | 0 |
| 0 | 0 | 0 | 0 | 0 |
| 0 | 0 | 0 | 0 | 0 |
| 0 | 0 | 0 | 0 | 0 |
| 0 | 0 | 0 | 0 | 0 |
| 0 | 0 | 0 | 0 | 0 |
| 0 | 0 | 0 | 0 | 0 |
| 0 | 0 | 0 | 0 | 0 |
| 0 | 0 | 0 | 0 | 0 |
| 0 | 0.005304147 | 0 | 0 | 0 |
| 0 | 0 | 0 | 0 | 0 |
| 0 | 0 | 0 | 0 | 0 |
| 0 | 0 | 0 | 0 | 0 |
| 0 | 0 | 0 | 0 | 0 |
| 0 | 0 | 0 | 0 | 0 |
| 0.001842839 | 0.001248558 | 0 | 0 | 0 |
| 0.003140461 | 0 | 0.011932613 | 0.009190696 | 0.0040653 |
| 0 | 0 | 0 | 0 | 0 |
| 0.000461083 | 0.025199658 | 0.087519549 | 0.343770491 | 0 |
| 0 | 0 | 0 | 0 | 0 |
| 0.258764737 | 0.067064699 | 0 | 0.865150973 | 0.210628465 |
| 0 | 0 | 0 | 0 | 0 |
| 0 | 0 | 0 | 0 | 0 |
| 0 | 0 | 0 | 0 | 0 |
| 0 | 0 | 0 | 0 | 0 |
| 0 | 0 | 0.001834014 | 0.003669059 | 0 |
| 0 | 0 | 0 | 0 | 0 |
| 0 | 0 | 0 | 0 | 0 |
| 0 | 0 | 0 | 0 | 0 |
| 0 | 0 | 0 | 0 | 0 |
| 0 | 0 | 0 | 0 | 0 |
| 13.33000961 | 26.68559673 | 9.908168178 | 3.574485133 | 16.46631465 |
| 0.074352496 | 0 | 0.968990173 | 0.726081199 | 2.587125018 |
| 0 | 0 | 0 | 0.493847156 | 0 |
| 0 | 0 | 0 | 0 | 0 |
| 0 | 0 | 0 | 0 | 0 |
| 0 | 0 | 0 | 0 | 0 |
| 0 | 0 | 0 | 0 | 0 |
| 0.000889132 | 0 | 0.032582575 | 0.006401948 | 0.000479572 |

| | | | | |
|---|---|---|---|---|
| 0 | 0 | 0 | 0 | 0 |
| 0 | 0 | 0.012509785 | 0 | 0.000657706 |
| 0 | 0 | 0 | 0 | 0 |
| 0 | 0 | 0 | 0 | 0 |
| 0.967214042 | 0.385496061 | 2.954525913 | 2.799413905 | 6.320103032 |
| 0 | 0 | 0 | 0 | 0 |
| 0 | 0 | 0 | 0 | 0 |
| 0 | 0 | 0 | 0.002026956 | 0 |
| 0.148698905 | 0.17818794 | 0.267098802 | 0.052813895 | 5.901597422 |
| 0 | 0 | 0 | 0 | 0 |
| 0 | 0 | 0 | 0 | 0 |
| 0 | 0 | 0 | 0 | 0 |
| 0 | 0 | 0 | 0.149639627 | 0 |
| 0.013656105 | 0.066122874 | 0 | 0.009134898 | 0.05401522 |
| 0 | 0 | 0 | 0 | 0 |
| 0.347837364 | 0 | 0 | 0.365639756 | 1.697225354 |
| 0 | 0.164398136 | 0.210576763 | 0.14162834 | 0 |
| 0 | 0 | 0 | 0 | 0 |
| 0 | 0 | 0 | 0 | 0 |
| 0 | 0 | 0 | 0 | 0 |
| 0 | 0 | 0 | 0 | 0 |
| 0 | 0 | 0 | 0 | 0 |
| 0 | 0 | 0 | 0 | 0 |
| 0 | 0 | 0 | 0 | 0 |
| 0 | 0 | 0 | 0 | 0 |
| 0 | 0 | 0 | 0 | 0 |
| 0.081382564 | 0.002970409 | 0 | 0.594373841 | 0 |
| 0 | 0.001047429 | 0 | 0.001077232 | 0 |
| 0 | 0 | 0 | 0 | 0 |
| 0 | 0 | 0 | 0 | 0 |
| 0 | 0 | 0 | 0 | 0 |
| 0 | 0 | 0 | 0 | 0 |
| 0 | 0 | 0 | 0.148629377 | 0 |
| 0 | 0.000878389 | 0 | 0.015583322 | 0 |
| 0 | 0 | 0 | 0.12768085 | 0 |
| 0 | 0 | 0 | 0.10866283 | 0.006173433 |
| 0 | 0 | 0 | 0 | 0 |
| 0 | 0 | 0 | 0.003294769 | 0 |
| 0 | 0 | 0 | 0 | 0 |
| 0 | 0 | 0 | 0 | 0 |
| 0 | 5.976243706 | 6.43617075 | 1.210270859 | 0.832167933 |
| 0 | 0 | 0 | 0 | 0 |
| 0 | 0 | 0 | 0 | 0 |
| 0 | 0 | 0 | 0 | 0 |
| 0 | 0 | 0 | 0 | 0 |
| 0 | 0 | 0 | 0 | 0 |
| 0 | 0 | 0 | 0 | 0 |
| 0 | 0 | 0 | 0 | 0 |

| | | | | |
|---|---|---|---|---|
| 0 | 0 | 0 | 0 | 0 |
| 0 | 0 | 0 | 0 | 0 |
| 0.01893527 | 0 | 0 | 0.01409772 | 0 |
| 0 | 0 | 0 | 0 | 0.001889192 |
| 0 | 0 | 0 | 0 | 0.00144607 |
| 0 | 0 | 0 | 0 | 0 |
| 0.00047154 | 0.103297686 | 0.189997894 | 0.629973129 | 0 |
| 0 | 0 | 0 | 0 | 0 |
| 0 | 0 | 0 | 0 | 0 |
| 0 | 0 | 0 | 0 | 0 |
| 0 | 0 | 0 | 0 | 0 |
| 0 | 0 | 0 | 0 | 0 |
| 0 | 0 | 0 | 0 | 0 |
| 0 | 0 | 0 | 0 | 0 |
| 0 | 0 | 0 | 0 | 0 |
| 0 | 0 | 0 | 0 | 0 |
| 0 | 0 | 0 | 0.001877518 | 0 |
| 0 | 0 | 0 | 0 | 0 |
| 0 | 0 | 0 | 0 | 0 |
| 0 | 0 | 0 | 0 | 0 |
| 0 | 0.064279204 | 0 | 0.014389464 | 0.001937131 |
| 0 | 0 | 0 | 0 | 0 |
| 0 | 0.000670975 | 0.00735864 | 0.018631794 | 0 |
| 0.007925602 | 0.000596638 | 0.003122976 | 0.009204213 | 0 |
| 0 | 0 | 0 | 0 | 0 |
| 0 | 0 | 0 | 0 | 0.009152753 |
| 0 | 0 | 0 | 0 | 0 |
| 0 | 0 | 0 | 0 | 0 |
| 0 | 0 | 0 | 0 | 0 |
| 3.723897626 | 0.001144937 | 0.003424529 | 0 | 0.543243066 |
| 0 | 0 | 0 | 0.001154977 | 0 |
| 0 | 0 | 0 | 0 | 0 |
| 0 | 0 | 0 | 0 | 0 |
| 0 | 0 | 0 | 0 | 0 |
| 0 | 0 | 0 | 0 | 0 |
| 0 | 0 | 0 | 0 | 0 |
| 0.026261635 | 0.043050994 | 0.056381833 | 0.001535581 | 0 |
| 0 | 0 | 0 | 0 | 0 |
| 0 | 0 | 0 | 0 | 0 |
| 0 | 0 | 0 | 0 | 0 |
| 0 | 0 | 0 | 0 | 0 |
| 0 | 0 | 0 | 0 | 0 |
| 0 | 0 | 0 | 0 | 0 |
| 0 | 0 | 0 | 0 | 0 |
| 0 | 0 | 0 | 0 | 0 |
| 0 | 0 | 0 | 0 | 0 |
| 0 | 0 | 0 | 0 | 0 |
| 0 | 0 | 0 | 0 | 0 |

| | | | | |
|---|---|---|---|---|
| 0.000789651 | 0.003638023 | 0.0566473 | 0.001980812 | 0.005281347 |
| 0 | 0 | 0 | 0 | 0 |
| 0 | 0 | 0 | 0.066435058 | 0.001417242 |
| 0 | 0 | 0 | 0 | 0 |
| 0 | 0 | 0 | 0 | 0 |
| 0 | 0 | 0 | 0 | 0 |
| 0 | 0 | 0 | 0 | 0 |
| 6.250727294 | 0.151174925 | 2.615242879 | 1.297141951 | 2.016038339 |
| 0.153447083 | 0.022169877 | 0.531238085 | 0.16962315 | 0.009894357 |
| 0.124428125 | 0.013095518 | 0.198642635 | 0 | 0.081727026 |
| 0 | 0 | 0 | 0 | 0 |
| 0 | 0 | 0 | 0 | 0 |
| 0 | 0 | 0 | 0.16787496 | 0 |
| 0 | 0 | 0 | 0 | 0 |
| 0 | 0 | 0 | 0 | 0 |
| 0 | 0 | 0 | 0 | 0 |
| 0 | 0 | 0 | 0 | 0.217851174 |
| 0 | 0 | 0 | 0 | 0 |
| 0 | 0 | 0 | 0 | 0 |
| 0 | 0 | 0 | 0 | 0 |
| 0 | 0 | 0 | 0 | 0 |
| 0 | 0 | 0 | 0 | 0 |
| 0 | 0 | 0 | 0 | 0 |
| 0 | 0 | 0 | 0 | 0 |
| 0 | 0 | 0 | 0.029469766 | 0 |
| 0 | 0 | 0 | 0 | 0 |
| 0 | 0 | 0 | 0 | 0 |
| 0 | 0 | 0 | 0 | 0 |
| 0 | 0 | 0 | 0 | 0 |
| 0 | 0 | 0 | 0 | 0 |
| 0 | 0 | 0 | 0 | 0 |
| 0 | 0 | 0 | 0 | 0 |
| 0 | 0 | 0 | 0 | 0 |
| 0 | 0 | 0 | 0 | 0 |
| 0 | 0 | 0.005811825 | 0 | 0 |
| 0 | 0 | 0 | 0 | 0 |
| 0 | 0 | 0 | 0 | 0 |
| 0.055804495 | 0.088433663 | 0.008050212 | 0 | 0 |
| 0 | 0 | 0 | 0.002617045 | 0 |
| 0.049664088 | 0.001068202 | 0 | 0 | 0 |
| 0 | 0 | 0 | 0 | 0 |
| 0.548483318 | 0.007934685 | 0.390272903 | 0.571911398 | 1.176677072 |
| 0 | 0 | 0 | 0 | 0 |
| 0 | 0 | 0 | 0 | 0 |
| 0 | 0 | 0.002335101 | 0 | 0 |
| 0 | 0 | 0 | 0 | 0 |
| 0 | 0 | 0 | 0 | 0 |
| 0 | 0 | 0 | 0 | 0 |

| | | | | |
|---|---|---|---|---|
| 0 | 0 | 0 | 0 | 0 |
| 0.004133628 | 0.174217293 | 0 | 0 | 0.003536544 |
| 0 | 0 | 0 | 0 | 0 |
| 0 | 0 | 0 | 0 | 0 |
| 0 | 0.428290805 | 0 | 1.285603472 | 0.094807061 |
| 0 | 0 | 0 | 0 | 0 |
| 0 | 0 | 0 | 0 | 0 |
| 0.025313226 | 0.003538927 | 0.002442694 | 0.003359648 | 0.002817333 |
| 0.000601769 | 0 | 0 | 0 | 0 |
| 0 | 0 | 0 | 0 | 0 |
| 0 | 0 | 0 | 0 | 0 |
| 0 | 0 | 0 | 0 | 0 |
| 0 | 0 | 0 | 0 | 0 |
| 0 | 0 | 0 | 0 | 0 |
| 0 | 0 | 0 | 0 | 0 |
| 0 | 0.000944395 | 0 | 0 | 0 |
| 0 | 0 | 0 | 0 | 0 |
| 0 | 0 | 0 | 0 | 0 |
| 0 | 0 | 0 | 0 | 0 |
| 0 | 0 | 0 | 0 | 0 |
| 0 | 0 | 0 | 0 | 0 |
| 0 | 0 | 0 | 0 | 0 |
| 0 | 0 | 0 | 0 | 0 |
| 0 | 0 | 0.002466275 | 0.001211458 | 0 |
| 0 | 0.11444581 | 0.240745378 | 0.118348808 | 0 |
| 0 | 0 | 0 | 0 | 0 |
| 0.045637537 | 0.018099682 | 0.032546366 | 0 | 0 |
| 0 | 0 | 0 | 0.001866727 | 0 |
| 0 | 0 | 0 | 0 | 0 |
| 0 | 0 | 0 | 0 | 0 |
| 0 | 0 | 0 | 0 | 0 |
| 0 | 0 | 0 | 0 | 0 |
| 0 | 0 | 0 | 0 | 0 |
| 0 | 0 | 0 | 0 | 0 |
| 0 | 0 | 0 | 0 | 0 |
| 0 | 0 | 0 | 0 | 0 |
| 0 | 0 | 0 | 0 | 0 |
| 0 | 0 | 0 | 0 | 0 |
| 0 | 0 | 0 | 0 | 0 |
| 0 | 0 | 0 | 0 | 0 |
| 0 | 0 | 0 | 0 | 0 |
| 0 | 0 | 0 | 0 | 0 |
| 0 | 0 | 0 | 0 | 0 |
| 0 | 0 | 0 | 0 | 0 |
| 0 | 0 | 0 | 0 | 0 |
| 0 | 0 | 0 | 0 | 0 |
| 0 | 0 | 0 | 0 | 0 |
| 0 | 0 | 0 | 0 | 0 |

| | | | | |
|---|---|---|---|---|
| 0.001644947 | 0.011550094 | 0.552745189 | 0.480358934 | 0 |
| 0 | 0 | 0 | 0 | 0 |
| 0 | 0 | 0.028066646 | 0 | 0 |
| 0 | 0 | 0 | 0 | 0 |
| 0 | 0 | 0 | 0 | 0 |
| 0 | 0 | 0 | 0 | 0.067468665 |
| 0 | 0 | 0 | 0 | 0 |
| 0.000673111 | 0 | 0 | 0 | 0 |
| 0 | 0 | 0 | 0 | 0 |
| 0 | 0 | 0 | 0 | 0 |
| 0 | 0 | 0 | 0 | 0 |
| 0 | 0 | 0 | 0 | 0 |
| 0 | 0 | 0 | 0 | 0 |
| 0 | 0 | 0 | 0 | 0 |
| 0 | 0 | 0 | 0 | 0 |
| 0 | 0 | 0 | 0 | 0 |
| 0 | 0 | 0 | 0 | 0 |
| 0.380744131 | 0 | 0 | 0 | 0 |
| 0 | 0 | 0 | 0 | 0 |
| 0 | 0 | 0 | 0 | 0 |
| 0.003982092 | 0 | 0.114377829 | 0.002654066 | 0.00448242 |
| 0 | 0 | 0 | 0 | 0 |
| 0 | 0 | 0.020825739 | 0.004570762 | 0.009199076 |
| 0 | 0 | 0 | 1.663040823 | 0 |
| 0 | 0 | 0 | 0 | 0 |
| 0 | 0 | 0 | 0 | 0 |
| 0 | 0 | 0 | 0 | 0 |
| 0 | 0 | 0 | 0 | 0 |
| 0 | 0 | 0 | 0 | 0 |
| 0 | 0 | 0 | 0 | 0 |
| 0 | 0 | 0 | 0 | 0 |
| 0 | 0 | 0 | 0 | 0 |
| 0.014129171 | 0.041065835 | 0.307279429 | 0 | 0.013916372 |
| 0 | 0 | 0 | 0 | 0 |
| 0.413040056 | 2.338762771 | 0.142719333 | 1.999699951 | 0.92689159 |
| 0 | 0 | 0.003662887 | 0 | 0.066294795 |
| 0 | 0 | 0 | 0 | 0 |
| 0 | 0 | 0 | 0 | 0 |
| 0 | 0 | 0.025805401 | 0.003178424 | 0.006355863 |
| 0 | 0 | 0 | 0 | 0 |
| 0 | 0 | 0 | 0 | 0 |
| 1.34717515 | 0 | 0 | 0.29911691 | 0 |
| 0 | 0.024994833 | 0.023643999 | 0.073711186 | 0 |
| 0 | 0 | 0 | 0 | 0 |
| 0 | 0 | 0 | 0 | 0 |
| 0 | 0 | 0.519509067 | 0.017955382 | 0 |
| 0 | 0 | 0 | 0 | 0 |
| 0 | 0 | 0 | 0 | 0 |

| | | | | |
|---|---|---|---|---|
| 0 | 0 | 0 | 0 | 0 |
| 0 | 0 | 0 | 0 | 0 |
| 0 | 0 | 0 | 0 | 0 |
| 0 | 0 | 0 | 0 | 0 |
| 0 | 0 | 0 | 0 | 0 |
| 0 | 0 | 0 | 0 | 0 |
| 0 | 0 | 0 | 0 | 0 |
| 0.025923836 | 0 | 0.202877843 | 0 | 0 |
| 0 | 0 | 0 | 0.009448269 | 0 |
| 0 | 0 | 0 | 0.001773382 | 0 |
| 0 | 0 | 0 | 0 | 0 |
| 0 | 0 | 0 | 0 | 0 |
| 0 | 0 | 0 | 0 | 0 |
| 0 | 0 | 0 | 0.430562704 | 0.011806733 |
| 0 | 0 | 0 | 0 | 0 |
| 0 | 0 | 0.002355967 | 0 | 0.089526353 |
| 0 | 0 | 0 | 0 | 0 |
| 0 | 0 | 0 | 0 | 0 |
| 0 | 0 | 0 | 0 | 0 |
| 0.086105025 | 0.013798152 | 0 | 2.761560808 | 0 |
| 0 | 0.006834537 | 0.005678401 | 0 | 0 |
| 0 | 0 | 0 | 0 | 0 |
| 0 | 0 | 0 | 0 | 0 |
| 0 | 0 | 0 | 0 | 0 |
| 0 | 0 | 0 | 0 | 0 |
| 0 | 0 | 0 | 0 | 0 |
| 0 | 0 | 0 | 0 | 0 |
| 0 | 0 | 0 | 0 | 0 |
| 0 | 0 | 0 | 0 | 0 |
| 9.872910127 | 0.008289293 | 0 | 0.003616728 | 2.295283216 |
| 0 | 0 | 0.512447045 | 0.09562278 | 0 |
| 0 | 0 | 0 | 0 | 0 |
| 0 | 0 | 0 | 0 | 0 |
| 0 | 0 | 0 | 0 | 0 |
| 0 | 0 | 0 | 0 | 0 |
| 0 | 0 | 0 | 0 | 0 |
| 0 | 0 | 0 | 0 | 0 |
| 0 | 0 | 0 | 0 | 0 |
| 0 | 0 | 0 | 0 | 0 |
| 0 | 0 | 0 | 0 | 0 |
| 0 | 0 | 0 | 0 | 0 |
| 0 | 0 | 0 | 0.006206426 | 0 |
| 0 | 0 | 0 | 0 | 0 |
| 0 | 0 | 0 | 0 | 0 |
| 0 | 0 | 0 | 0 | 0 |
| 0 | 0 | 0 | 0 | 0 |
| 0 | 0.049621055 | 0.326147725 | 0 | 0.213514915 |
| 0 | 0 | 0 | 0 | 0 |

| | | | | |
|---|---|---|---|---|
| 0 | 0 | 0 | 0 | 0 |
| 0 | 0 | 0 | 0 | 0 |
| 0 | 0 | 0 | 0 | 0 |
| 0 | 0 | 0 | 0 | 0 |
| 0.035017781 | 0.000741413 | 1.396660214 | 0.024209623 | 0.001918271 |
| 0 | 0 | 0 | 0 | 0 |
| 0 | 0 | 0 | 0 | 0 |
| 0 | 0 | 0 | 0 | 0 |
| 0 | 0 | 0 | 0 | 0 |
| 0 | 0 | 0 | 0 | 0 |
| 0 | 0 | 0 | 0.059297346 | 0 |
| 0.008735884 | 0 | 0.015201518 | 0.003175893 | 0.004097292 |
| 0 | 0 | 0 | 0.001826664 | 0 |
| 0 | 0 | 0 | 0 | 0 |
| 0.019248436 | 0 | 0 | 0.055262433 | 0 |
| 0.19272547 | 0.140480832 | 0 | 0 | 0 |
| 0 | 0 | 0 | 0.020355325 | 0 |
| 0 | 0 | 0 | 0 | 0 |
| 0.002588424 | 0.089439005 | 0.215351646 | 0 | 0.09416066 |
| 0 | 0.122122969 | 0.016551405 | 0.399819305 | 0 |
| 0 | 0 | 0 | 0 | 0 |
| 0 | 0 | 0 | 0 | 0 |
| 0 | 0 | 0 | 0 | 0 |
| 0 | 0 | 0 | 0 | 0 |
| 0 | 0 | 0 | 0.001293632 | 0 |
| 0 | 0 | 0 | 0 | 0 |
| 0 | 0 | 0.007317737 | 0.003564588 | 0 |
| 0 | 0 | 0 | 0 | 0 |
| 0 | 0 | 0 | 0 | 0 |
| 0 | 0 | 0 | 0 | 0 |
| 0 | 0 | 0 | 0 | 0.000919929 |
| 0 | 0 | 0 | 0 | 0 |
| 0 | 0 | 0 | 0 | 0 |
| 0 | 0 | 0 | 0 | 0 |
| 0 | 0 | 0 | 0 | 0 |
| 0 | 0 | 0 | 0 | 0 |
| 0 | 0 | 0 | 0 | 0 |
| 0.002636206 | 0.003409787 | 0.021854453 | 0.020038882 | 0 |
| 0 | 0 | 0 | 0.244842936 | 0 |
| 0 | 0 | 0 | 0 | 0 |
| 0 | 0 | 0.00048484 | 0 | 0.003613294 |
| 0 | 0 | 0 | 0 | 0 |
| 0 | 0 | 0 | 0 | 0 |
| 0 | 0.002889304 | 0.006121394 | 0.00247626 | 0 |
| 0 | 0 | 0 | 0 | 0 |
| 0 | 0 | 0 | 0 | 0 |
| 0 | 0 | 0 | 0 | 0 |
| 0 | 0 | 0 | 0 | 0 |

| | | | | |
|---|---|---|---|---|
| 0 | 0 | 0 | 0 | 0 |
| 0 | 0 | 0 | 0 | 0.223956704 |
| 0 | 0 | 0 | 0 | 0 |
| 0 | 0 | 0.004151044 | 0.001038053 | 0.001205294 |
| 0.000518145 | 0.00912738 | 0.002450017 | 0 | 0 |
| 0 | 0 | 0.002484414 | 0 | 0 |
| 0 | 0 | 0 | 0.001007351 | 0 |
| 0 | 0 | 0 | 0 | 0 |
| 0 | 0 | 0 | 0.003794825 | 0 |
| 0 | 0 | 0 | 0 | 0 |
| 0.005847272 | 0 | 0 | 0.017798501 | 0 |
| 0 | 0 | 0 | 0 | 0 |
| 0.137811119 | 0.002861904 | 0.10151644 | 0.09455457 | 0.036168905 |
| 0 | 0 | 0 | 0 | 0 |
| 0 | 0 | 0 | 0 | 0 |
| 0 | 0 | 0 | 0 | 0 |
| 0 | 0 | 0 | 0 | 0 |
| 0 | 0 | 0 | 0 | 0 |
| 0 | 0 | 0 | 0 | 0 |
| 0 | 0 | 0 | 0 | 0 |
| 0 | 0 | 0 | 0 | 0 |
| 0 | 0 | 0 | 0 | 0 |
| 0.018250839 | 0.007065878 | 0.04350121 | 0 | 0 |
| 0.048201002 | 0.055030664 | 0.064218112 | 0.009432738 | 0.019692284 |
| 0 | 0 | 0 | 0 | 0 |
| 0 | 0 | 0 | 0 | 0 |
| 0 | 0 | 0 | 0 | 0 |
| 0 | 0 | 0 | 0 | 0 |
| 0 | 0 | 0 | 0 | 0 |
| 0 | 0 | 0 | 0 | 0 |
| 0.007364619 | 0.001575685 | 0 | 0.028899219 | 0.008780795 |
| 0 | 0 | 0 | 0 | 0 |
| 0 | 0 | 0 | 0 | 0 |
| 0 | 0 | 0 | 0 | 0 |
| 0 | 0 | 0 | 0 | 0 |
| 0 | 0 | 0 | 0 | 0.008304995 |
| 0 | 0 | 0 | 0 | 0 |
| 0 | 0 | 0 | 0 | 0 |
| 0 | 0.028218097 | 0.001993723 | 0 | 0.002476387 |
| 0 | 0 | 0 | 0 | 0 |
| 0 | 0 | 0 | 0 | 0 |
| 0 | 0 | 0 | 0 | 0 |
| 0 | 0 | 0 | 0 | 0 |
| 0 | 0 | 0 | 0 | 0 |
| 0 | 0 | 0 | 0 | 0 |
| 0.142826559 | 0.018571575 | 0.307560358 | 1.78192808 | 0.192202318 |
| 0 | 0 | 0 | 0 | 0 |
| 0 | 0 | 0 | 0 | 0 |

| | | | | |
|---|---|---|---|---|
| 0 | 0 | 0 | 0 | 0 |
| 0 | 0 | 0 | 0 | 0 |
| 0 | 0 | 0 | 0 | 0 |
| 0 | 0 | 0 | 0 | 0 |
| 0 | 0 | 0 | 0 | 0 |
| 0 | 0 | 0 | 0 | 0 |
| 0 | 0 | 0 | 0 | 0 |
| 0 | 0 | 0 | 0 | 0 |
| 0 | 0 | 0 | 0 | 0 |
| 0 | 0 | 0 | 0 | 0 |
| 0.000665131 | 0 | 0.000808722 | 0.002780762 | 0 |
| 0 | 0 | 0.00221361 | 0 | 0 |
| 0 | 0 | 0 | 0 | 0 |
| 0 | 0 | 0 | 0 | 0 |
| 0 | 0 | 0 | 0 | 0 |
| 0 | 0 | 0 | 0 | 0 |
| 0 | 0 | 0 | 0 | 0 |
| 0 | 0 | 0 | 0 | 0 |
| 0 | 0 | 0 | 0 | 0 |
| 0.001411324 | 0 | 0.002502509 | 0.001720959 | 0 |
| 0 | 0 | 0 | 0 | 0 |
| 0 | 0 | 0 | 0 | 0 |
| 0 | 0 | 0 | 0 | 0 |
| 0 | 0 | 0 | 0 | 0 |
| 0 | 0 | 0 | 0 | 0 |
| 0 | 0 | 0 | 0 | 0 |
| 0 | 0 | 0 | 0 | 0 |
| 0 | 0 | 0 | 0 | 0 |
| 0 | 0 | 0 | 0 | 0 |
| 0 | 0 | 0 | 0 | 0 |
| 0.007426399 | 0.006366419 | 0.008753221 | 0.016896931 | 0.004250827 |
| 0 | 0 | 0 | 0 | 0 |
| 0.021996953 | 0.036721853 | 0.024427776 | 0.006376194 | 0.194578235 |
| 0 | 0.019006549 | 0.004008583 | 0.001002427 | 0 |
| 0 | 0 | 0 | 0 | 0 |
| 0 | 0 | 0 | 0 | 0 |
| 0 | 0 | 0 | 0 | 0 |
| 0 | 0 | 0 | 0 | 0 |
| 0 | 0 | 0 | 0 | 0 |
| 0 | 0 | 0 | 0 | 0 |
| 0 | 0 | 0 | 0 | 0 |
| 0.00694913 | 0.053163047 | 0.493904723 | 0.039342275 | 0.319374631 |
| 0 | 0.003345927 | 0.034490952 | 0.264475197 | 0.269793293 |
| 0 | 0 | 0 | 0 | 0 |
| 0 | 0 | 0 | 0 | 0 |
| 0 | 0.006216553 | 0 | 0.05008187 | 0 |
| 0 | 0 | 0 | 0 | 0 |
| 0 | 0 | 0 | 0 | 0 |

| | | | | |
|---|---|---|---|---|
| 0 | 0 | 0 | 0 | 0 |
| 0 | 0.010015149 | 0 | 0 | 0.00896967 |
| 0.019857107 | 0.024274908 | 0.006779547 | 0.003308693 | 0.000698501 |
| 0 | 0 | 0 | 0 | 0 |
| 0 | 0.007706234 | 0 | 0.057880141 | 0 |
| 0.003187272 | 0.004773492 | 0 | 0 | 0.003257286 |
| 0 | 0 | 0 | 0 | 0 |
| 0 | 0 | 0 | 0 | 0 |
| 0 | 0 | 0 | 0 | 0 |
| 0 | 0 | 0 | 0 | 0 |
| 0 | 0 | 0 | 0 | 0 |
| 0 | 0 | 0 | 0 | 0 |
| 0 | 0 | 0 | 0 | 0 |
| 0 | 0 | 0 | 0 | 0 |
| 0 | 0 | 0 | 0 | 0 |
| 0 | 0 | 0 | 0 | 0 |
| 0 | 0 | 0 | 0 | 0 |
| 0 | 0 | 0 | 0 | 0 |
| 0 | 0 | 0 | 0 | 0 |
| 0 | 0 | 0 | 0 | 0 |
| 0 | 0 | 0 | 0 | 0 |
| 0 | 0 | 0 | 0 | 0 |
| 0 | 0 | 0 | 0.001700567 | 0 |
| 0 | 0 | 0 | 0 | 0 |
| 0 | 0 | 0 | 0 | 0 |
| 0.675334792 | 0 | 0.00344494 | 0 | 0.004246761 |
| 0 | 0 | 0 | 0 | 0 |
| 0.001215074 | 0 | 0 | 0.498560902 | 0 |
| 0 | 0 | 0 | 0 | 0 |
| 0 | 0 | 0 | 0 | 0 |
| 0 | 0 | 0 | 0 | 0 |
| 0 | 0 | 0 | 0 | 0 |
| 0 | 0 | 0 | 0 | 0 |
| 0 | 0 | 0 | 0 | 0 |
| 0 | 0 | 0 | 0 | 0 |
| 0 | 0 | 0 | 0 | 0 |
| 0 | 0 | 0 | 0 | 0 |
| 0 | 0 | 0 | 0.036490079 | 0 |
| 0 | 0.303805711 | 0 | 0 | 0 |
| 0 | 0 | 0 | 0 | 0 |
| 0 | 0 | 0 | 0 | 0 |
| 0 | 0 | 0 | 0 | 0 |
| 0.001257129 | 0.001216755 | 0.045673722 | 0.000750825 | 0.005037013 |
| 0 | 0 | 0 | 0 | 0 |
| 0 | 0 | 0 | 0 | 0 |
| 0 | 0 | 0 | 0 | 0 |
| 0 | 0 | 0.004453983 | 0 | 0 |
| 0 | 0 | 0 | 0 | 0 |

| | | | | |
|---|---|---|---|---|
| 0 | 0 | 0 | 0 | 0 |
| 0.011093376 | 0.004932356 | 0.020724145 | 0.001207784 | 0.011405951 |
| 0 | 0 | 0 | 0 | 0 |
| 0 | 0 | 0 | 0 | 0 |
| 0 | 0 | 0 | 0 | 0 |
| 0 | 0 | 0 | 0 | 0 |
| 0 | 0 | 0 | 0 | 0 |
| 0 | 0 | 0 | 0 | 0 |
| 0 | 0 | 0 | 0 | 0 |
| 0 | 0 | 0 | 0 | 0 |
| 0 | 0 | 0 | 0 | 0 |
| 0 | 0 | 0 | 0 | 0 |
| 0 | 0 | 0.01373845 | 0.430004396 | 0.073923862 |
| 0 | 0 | 0 | 0 | 0 |
| 0.003457942 | 0.00117141 | 0.005693534 | 0.00421659 | 0 |
| 0 | 0 | 0 | 0 | 0 |
| 0 | 0 | 0 | 0 | 0 |
| 0 | 0 | 0 | 0 | 0 |
| 0 | 0 | 0 | 0 | 0 |
| 0 | 0 | 0 | 0 | 0 |
| 0 | 0 | 0 | 0 | 0 |
| 0 | 0.006039692 | 0.025376807 | 0.198769177 | 0 |
| 0 | 0 | 0.004131356 | 0 | 0 |
| 0 | 0 | 0.004873185 | 0 | 0 |
| 0 | 0 | 0 | 0 | 0 |
| 0 | 0 | 0 | 0 | 0 |
| 0 | 0 | 0 | 0 | 0 |
| 0 | 0 | 0 | 0 | 0 |
| 0 | 0 | 0 | 0 | 0 |
| 0 | 0 | 0 | 0 | 0 |
| 0 | 0 | 0 | 0 | 0 |
| 0 | 0 | 0 | 0 | 0 |
| 0 | 0 | 0 | 0 | 0 |
| 0 | 0 | 0 | 0 | 0 |
| 0.001927096 | 0 | 20.58962443 | 0 | 0.19707411 |
| 0 | 0 | 0 | 0 | 0 |
| 0 | 0 | 0 | 0 | 0 |
| 0 | 0 | 0 | 0 | 0 |
| 0.000635867 | 0.053420644 | 0.002791895 | 0.066460404 | 0 |
| 0 | 0 | 0 | 0 | 0 |
| 0.02698656 | 0.006953033 | 0.061042074 | 0.223265899 | 0.024601314 |
| 0 | 0 | 0 | 0 | 0 |
| 0 | 0 | 0 | 0 | 0 |
| 0 | 0 | 0 | 0 | 0 |
| 0 | 0 | 0 | 0 | 0 |
| 0.483920067 | 3.478847622 | 0.058204106 | 0.00363878 | 0.514514103 |
| 0 | 0 | 0 | 0 | 0 |
| 0 | 0 | 0 | 0 | 0 |

| | | | | |
|---|---|---|---|---|
| 0 | 0 | 0 | 0 | 0 |
| 0 | 0 | 0 | 0 | 0 |
| 0 | 0 | 0 | 0 | 0 |
| 0 | 0 | 0 | 0 | 0 |
| 0 | 0 | 0 | 0 | 0 |
| 0 | 0 | 0 | 0 | 0 |
| 0 | 0 | 0 | 0 | 0 |
| 0 | 0.015296792 | 0 | 0 | 0 |
| 0 | 0 | 0.031364201 | 0 | 0 |
| 0.208972863 | 0.020473312 | 0.00064686 | 0 | 0.00091824 |
| 0 | 0 | 0 | 0.019704254 | 0 |
| 0 | 0 | 0 | 0 | 0 |
| 0 | 0 | 0.00442576 | 0 | 0 |
| 0 | 0 | 0 | 0 | 0 |
| 0 | 0 | 0 | 0 | 0 |
| 0 | 0 | 0 | 0 | 0 |
| 0 | 0 | 0 | 0 | 0 |
| 0 | 0 | 0 | 0 | 0 |
| 0 | 0 | 0 | 0 | 0 |
| 0.094972871 | 0.178864062 | 1.527486853 | 0.379774374 | 0.04753101 |
| 0 | 0 | 0 | 0 | 0 |
| 0 | 0 | 0 | 0 | 0 |
| 0 | 0 | 0 | 0 | 0 |
| 0 | 0 | 0 | 0 | 0 |
| 0 | 0 | 0 | 0 | 0 |
| 0 | 0 | 0 | 0 | 0 |
| 0 | 0 | 0 | 0 | 0 |
| 0 | 0 | 0 | 0 | 0 |
| 0 | 0 | 0 | 0 | 0 |
| 0 | 0 | 0 | 0.119423561 | 0.025832696 |
| 0 | 0 | 0 | 0 | 0 |
| 0 | 0 | 0 | 0 | 0 |
| 0 | 0 | 0 | 0 | 0 |
| 0 | 0 | 0 | 0 | 0 |
| 0 | 0 | 0 | 0 | 0 |
| 0 | 0 | 0 | 0.009685236 | 0 |
| 0 | 0 | 0 | 0 | 0 |
| 0.206515384 | 0 | 0 | 0 | 0 |
| 0 | 0 | 0 | 0 | 0 |
| 0 | 0 | 0 | 0 | 0 |
| 0 | 0 | 0 | 0 | 0 |
| 0 | 0 | 0 | 0 | 0 |
| 0 | 0 | 0 | 0.025520604 | 0 |
| 0 | 0 | 0 | 0 | 0 |
| 0.002608246 | 0 | 0.001938031 | 0.00945056 | 0 |
| 0 | 0 | 0 | 0.026952634 | 0 |
| 0 | 0 | 0 | 0 | 0 |
| 0 | 0.003302057 | 0 | 0 | 0 |

| | | | | |
|---|---|---|---|---|
| 0 | 0 | 0 | 0 | 0 |
| 0 | 0 | 0 | 0 | 0 |
| 0 | 0 | 0 | 0 | 0 |
| 0 | 0 | 0 | 0 | 0 |
| 0 | 0 | 0 | 0 | 0 |
| 0 | 0 | 0.014731215 | 0.001085417 | 0 |
| 0 | 0 | 0 | 0 | 0 |
| 0 | 0 | 0 | 0 | 0 |
| 0 | 0 | 0 | 0 | 0 |
| 0 | 0 | 0 | 0 | 0 |
| 0 | 0 | 0 | 0 | 0 |
| 0 | 0 | 0 | 0 | 0 |
| 0 | 0 | 0 | 0 | 0 |
| 0 | 0 | 0 | 0 | 0 |
| 0 | 0 | 0 | 0 | 0 |
| 0 | 0 | 0 | 0 | 0 |
| 0 | 0 | 0 | 0 | 0 |
| 0 | 0 | 0 | 0.000668374 | 0 |
| 0 | 0 | 0 | 0 | 0 |
| 0 | 0 | 0 | 0 | 0 |
| 0 | 0 | 0 | 0 | 0 |
| 0 | 0 | 0 | 0 | 0 |
| 0 | 0 | 0 | 0 | 0 |
| 0 | 0 | 0 | 0.000734183 | 0 |
| 0 | 0 | 0 | 0 | 0 |
| 0 | 0 | 0 | 0 | 0 |
| 0 | 0 | 0 | 0 | 0 |
| 0 | 0 | 0 | 0 | 0 |
| 0 | 0 | 0 | 0 | 0 |
| 0 | 0 | 0 | 0 | 0 |
| 0 | 0 | 0 | 0 | 0 |
| 0 | 0 | 0 | 0 | 0 |
| 0 | 0 | 0 | 0 | 0 |
| 0 | 0 | 0 | 0 | 0 |
| 0 | 0 | 0 | 0 | 0 |
| 0 | 0 | 0 | 0 | 0 |
| 0 | 0 | 0 | 0 | 0 |
| 0 | 0 | 0 | 0 | 0 |
| 0.626860996 | 0.131845118 | 0 | 0 | 1.524809866 |
| 0 | 0 | 0 | 0 | 0 |
| 0 | 0 | 0 | 0 | 0 |
| 0 | 0 | 0 | 0 | 0 |
| 0 | 0 | 0 | 0 | 0 |
| 0 | 0 | 0 | 0 | 0 |
| 0 | 0 | 0 | 0 | 0.000972013 |
| 0 | 0 | 0 | 0 | 0 |
| 0 | 0 | 0 | 0 | 0 |
| 0 | 0 | 0 | 0 | 0 |

| | | | | |
|---|---|---|---|---|
| 0 | 0 | 0 | 0 | 0 |
| 0.129201285 | 0 | 0.138127056 | 0 | 0.238928482 |
| 0 | 0 | 0 | 0 | 0 |
| 0 | 0 | 0 | 0 | 0 |
| 0 | 0 | 0 | 0 | 0 |
| 0 | 0 | 0 | 0 | 0 |
| 0 | 0 | 0 | 0 | 0 |
| 0 | 0 | 0 | 0 | 0 |
| 0 | 0 | 0 | 0 | 0 |
| 0 | 0 | 0 | 0 | 0 |
| 0 | 0.001805068 | 0.005206177 | 0 | 0 |
| 0 | 0 | 0 | 0 | 0 |
| 0.290146411 | 0 | 0 | 0.33519674 | 0.917508961 |
| 0 | 0 | 0 | 0 | 0 |
| 0 | 0 | 0 | 0 | 0 |
| 0 | 0.003020603 | 0 | 0 | 0 |
| 0 | 0 | 0 | 0 | 0 |
| 0 | 0 | 0 | 0 | 0 |
| 0 | 0 | 0 | 0 | 0 |
| 0 | 0 | 0 | 0 | 0 |
| 0.001545164 | 0.074590033 | 0.033465054 | 0 | 0.027919476 |
| 0 | 0 | 0 | 0 | 0 |
| 0 | 0 | 0 | 0 | 0 |
| 0.749287713 | 0.741484225 | 0 | 1.519365382 | 10.60147495 |
| 0 | 0 | 0 | 0 | 0 |
| 0 | 0 | 0 | 0 | 0 |
| 0 | 0 | 0 | 0 | 0 |
| 0 | 0 | 0 | 0 | 0 |
| 0 | 0 | 0 | 0 | 0 |
| 0 | 0 | 0 | 0 | 0 |
| 0 | 0 | 0 | 0 | 0 |
| 0 | 0 | 0 | 0 | 0 |
| 0.006095118 | 0 | 0.004323057 | 0 | 0 |
| 0 | 0 | 0 | 0 | 0 |

| PHL018-0012_V2 | PHL018-0232_V3 | PHL018-0262_V3 | PHL018-0145_V2 | PHL018-0022_V2 |
|---|---|---|---|---|
| EYCF | REF | REF | REF | CTRL |
| 0.268198523 | 1.155251979 | 0.001840296 | 0.013992851 | 0.217970388 |
| 0.196776975 | 3.207121701 | 2.898170746 | 1.839243077 | 3.643308367 |
| 0.003049074 | 3.690483348 | 8.77670851 | 2.517906386 | 0 |
| 0 | 0 | 0 | 0 | 0 |
| 0 | 0 | 0 | 0 | 0 |
| 0 | 0 | 0 | 0 | 0 |
| 0 | 0 | 0 | 0 | 0 |
| 0 | 0 | 0 | 0 | 0 |
| 0 | 0 | 0 | 0 | 0 |
| 0 | 0 | 0 | 0 | 0 |
| 0 | 0 | 0 | 0 | 0 |
| 0 | 0 | 0 | 0 | 0 |
| 0 | 0 | 0 | 0 | 0 |
| 1.466460337 | 0 | 0.913924994 | 2.31773903 | 0.292241796 |
| 0 | 0 | 0 | 0 | 0 |
| 0 | 0 | 0 | 0 | 0 |
| 0 | 0 | 0 | 0 | 0 |
| 0 | 0 | 0 | 0 | 0 |
| 0 | 0 | 0 | 0 | 0 |
| 0 | 0 | 0 | 0 | 0 |
| 0 | 0 | 0 | 0 | 0 |
| 0 | 0 | 0 | 0.000497414 | 0 |
| 0 | 0 | 0 | 0 | 0 |
| 0 | 0 | 0 | 0 | 0 |
| 0 | 0 | 0.222479645 | 0.382112287 | 0 |
| 0 | 0 | 0 | 0 | 0 |
| 0 | 0 | 0 | 0 | 0 |
| 0 | 0 | 0.00121851 | 0 | 0 |
| 0 | 0 | 0 | 0 | 0 |
| 0 | 0 | 0.017925057 | 0 | 0 |
| 0 | 0 | 0 | 0 | 0 |
| 0 | 0 | 0 | 0 | 0 |
| 0 | 0 | 0 | 0 | 0 |
| 0 | 0 | 0 | 0 | 0 |
| 0 | 0 | 0 | 0 | 0 |
| 0 | 2.233900355 | 0.730863222 | 0.059972761 | 0.008782593 |
| 0 | 0 | 0 | 0 | 0 |
| 0 | 0 | 0 | 0 | 0 |
| 0 | 0 | 0 | 0 | 0 |
| 0 | 0 | 0 | 0 | 0 |
| 0 | 0 | 0 | 0 | 0 |
| 0.001086244 | 0.002642765 | 0 | 0.036318596 | 0.006347241 |
| 0.171371487 | 0.031043653 | 0.010196327 | 0.004242333 | 1.093111002 |
| 0.008905853 | 0.00150005 | 0.012298236 | 0.061460356 | 0.079546074 |
| 0 | 0 | 0 | 0 | 0.005895785 |
| 0 | 0 | 0 | 0 | 0 |

| | | | | |
|---|---|---|---|---|
| 0.026490121 | 0 | 0 | 0.002741179 | 0.050708892 |
| 0 | 0.048427584 | 0.003425753 | 0 | 0.140976961 |
| 0.000788923 | 0 | 0.001960986 | 0 | 0.02094215 |
| 0.026695781 | 0.550273644 | 0.350819773 | 0.34023458 | 0.803025145 |
| 0 | 0.046482636 | 0.000586293 | 0.027586698 | 0.005581966 |
| 0 | 0 | 0 | 0 | 0 |
| 0 | 0.026871978 | 0 | 0.004648999 | 0.000768328 |
| 0 | 0 | 0 | 0 | 0 |
| 0 | 0 | 0 | 0 | 0 |
| 0 | 0 | 0 | 0 | 0 |
| 0 | 0 | 0.000778552 | 0 | 0 |
| 1.614387494 | 0.356106262 | 0.096676218 | 0.024678201 | 2.047094464 |
| 0 | 0 | 0.002202204 | 0 | 0 |
| 0.002636711 | 0.152478551 | 0.008900411 | 0.004777061 | 0.143982746 |
| 0 | 0 | 0 | 0 | 0 |
| 0 | 0 | 0 | 0 | 0 |
| 1.298664184 | 0.012627604 | 0.000831922 | 0 | 0.002263014 |
| 0 | 0 | 0 | 0 | 0 |
| 0 | 0 | 0 | 0 | 0 |
| 0 | 0 | 0 | 0 | 0 |
| 0 | 0.113151014 | 0.036123725 | 0.023862717 | 0.18102398 |
| 0 | 0.075068167 | 0.004816567 | 0.013356687 | 0.030571661 |
| 0.008313304 | 0 | 0 | 0.013531301 | 0 |
| 0 | 0 | 0 | 0 | 0 |
| 0 | 0 | 0 | 0 | 0 |
| 0 | 0 | 0 | 0 | 0 |
| 0 | 0 | 0 | 0 | 0 |
| 0 | 0 | 0 | 0 | 0 |
| 0 | 0 | 0 | 0 | 0 |
| 0 | 0 | 0 | 0 | 0 |
| 0 | 0 | 0 | 0 | 0 |
| 0 | 0 | 0 | 0 | 0 |
| 0 | 0 | 0 | 0 | 0 |
| 0 | 0 | 0 | 0 | 0 |
| 0 | 0 | 0 | 0 | 0 |
| 0 | 0 | 0 | 0 | 0 |
| 0 | 0 | 0 | 0 | 0 |
| 67.79458429 | 0.076978065 | 0 | 0 | 0 |
| 0 | 0 | 0 | 0 | 0 |
| 0 | 0 | 0 | 0 | 0 |
| 0 | 0 | 0 | 0 | 0 |
| 0 | 0 | 0 | 0 | 0 |
| 0 | 0 | 0 | 0 | 0 |
| 0 | 0 | 0 | 0 | 0 |
| 0 | 0 | 0.034364011 | 0 | 0 |
| 0 | 0 | 0 | 0 | 0 |
| 0 | 0 | 0 | 0.000887818 | 0 |
| 0 | 0 | 0 | 0 | 0 |

| | | | | |
|---|---|---|---|---|
| 0 | 0 | 0 | 0 | 0 |
| 0 | 0 | 0 | 0 | 0 |
| 0 | 0 | 0 | 0 | 0 |
| 0 | 0 | 0 | 0 | 0 |
| 0.005128171 | 0 | 0.009137515 | 0 | 0.005799754 |
| 0 | 0 | 0 | 0 | 0 |
| 0 | 0 | 0 | 0 | 0 |
| 0 | 0 | 0 | 0 | 0 |
| 0 | 0 | 0 | 0 | 0 |
| 0 | 0 | 0 | 0 | 0 |
| 0 | 0 | 0 | 0 | 0 |
| 0 | 0 | 0 | 0 | 0 |
| 0 | 0 | 0 | 0 | 0 |
| 0 | 0 | 0 | 0 | 0 |
| 0 | 0 | 0 | 0 | 0 |
| 0 | 0.0266672 | 0 | 0 | 0 |
| 0 | 0 | 0 | 0 | 0 |
| 0 | 0 | 0 | 0 | 0 |
| 0.004644491 | 0 | 0 | 0.511190293 | 0.002907762 |
| 0 | 0 | 0 | 0 | 0 |
| 0 | 0 | 0 | 0 | 0 |
| 0 | 0 | 0 | 0 | 0 |
| 0 | 0 | 0 | 0 | 0 |
| 0 | 0 | 0 | 0 | 0 |
| 0 | 0 | 0 | 0 | 0 |
| 0 | 0 | 0 | 0 | 0 |
| 0 | 0.006009897 | 0 | 0 | 0.001546524 |
| 0 | 0 | 0 | 0 | 0 |
| 0 | 0 | 0 | 0 | 0 |
| 0 | 0 | 0 | 0 | 0 |
| 0.003119774 | 0.109182355 | 0.055646892 | 0.020553637 | 0 |
| 0.054957697 | 0.014853683 | 0 | 0 | 0.008196254 |
| 0 | 0 | 0 | 0 | 0 |
| 0 | 0 | 0 | 0 | 0 |
| 0 | 0 | 0 | 0 | 0 |
| 0 | 0 | 0 | 0 | 0 |
| 0 | 0 | 0 | 0 | 0 |
| 0 | 0 | 0 | 0 | 0 |
| 0 | 0 | 0.085243085 | 0 | 0 |
| 0 | 0 | 0 | 0 | 0 |
| 0 | 0 | 0 | 0 | 0 |
| 0 | 0 | 0 | 0 | 0 |
| 0 | 0 | 0 | 0 | 0.002368393 |
| 0 | 0 | 0 | 0 | 0 |
| 0 | 0 | 0 | 0 | 0 |
| 0 | 0 | 0 | 0 | 0 |
| 0 | 0 | 0 | 0 | 0 |
| 0 | 0 | 0 | 0 | 0 |

| | | | | |
|---|---|---|---|---|
| 0 | 0 | 0 | 0 | 0 |
| 0 | 0 | 0 | 0 | 0 |
| 0 | 0 | 0 | 0 | 0 |
| 0 | 0 | 0 | 0 | 0 |
| 0.006172188 | 0 | 0 | 0.00715521 | 0.017537533 |
| 0 | 0 | 0 | 0 | 0 |
| 0 | 0 | 0 | 0 | 0 |
| 0 | 0 | 0 | 0 | 0 |
| 0 | 0 | 0 | 0 | 0 |
| 0 | 0 | 0 | 0 | 0 |
| 0 | 0 | 0 | 0 | 0 |
| 0 | 0 | 0 | 0 | 0 |
| 0 | 0 | 0 | 0 | 0 |
| 0 | 0 | 0 | 0 | 0 |
| 0 | 0 | 0 | 0 | 0 |
| 0.003216225 | 0 | 0 | 0 | 0 |
| 0 | 0 | 0 | 0 | 0 |
| 0 | 0 | 0 | 0 | 0 |
| 0 | 0 | 0 | 0 | 0 |
| 0 | 0 | 0 | 0 | 0 |
| 0 | 0 | 0 | 0 | 0 |
| 0 | 0 | 0 | 0 | 0 |
| 0 | 0 | 0 | 0 | 0 |
| 0 | 0 | 0 | 0 | 0 |
| 0 | 0 | 0.086338928 | 0 | 0 |
| 0 | 0 | 0 | 0 | 0 |
| 0 | 0 | 0 | 0.017274247 | 0 |
| 0 | 0 | 0 | 0 | 0 |
| 0 | 0 | 0 | 0 | 0 |
| 0 | 0 | 0 | 0 | 0 |
| 0 | 0 | 0 | 0 | 0 |
| 0 | 0 | 0.347427202 | 0 | 0 |
| 0 | 0 | 0 | 0 | 0 |
| 0 | 0 | 0 | 0 | 0 |
| 0 | 0 | 0 | 0 | 0 |
| 0 | 0 | 0 | 0 | 0 |
| 0 | 0 | 0 | 0 | 0 |
| 0 | 0 | 0 | 0 | 0 |
| 0 | 0 | 0 | 0 | 0 |
| 0 | 0 | 0 | 0 | 0 |
| 0 | 0 | 0 | 0 | 0 |
| 0 | 0 | 0 | 0 | 0 |
| 0 | 0 | 0 | 0 | 0 |
| 0 | 0 | 0 | 0 | 0 |
| 0 | 0 | 0 | 0 | 0 |
| 0 | 0 | 0 | 0 | 0 |
| 0 | 0 | 0 | 0 | 0 |
| 0 | 0 | 0 | 0 | 0.084298893 |

| | | | | |
|---|---|---|---|---|
| 0 | 0 | 0 | 0 | 0 |
| 0 | 0 | 0 | 0 | 0 |
| 0 | 0 | 0 | 0 | 0 |
| 0 | 0 | 0 | 0 | 0 |
| 0 | 0 | 0 | 0 | 0 |
| 0 | 0 | 0 | 0 | 0 |
| 0 | 0 | 0 | 0 | 0 |
| 0 | 0 | 0 | 0 | 0 |
| 0 | 0.020383238 | 0 | 0 | 0 |
| 0 | 0 | 0 | 0 | 0 |
| 0 | 0 | 0 | 0 | 0 |
| 0 | 0 | 0 | 0 | 0 |
| 0 | 0.002535979 | 0.000623741 | 0 | 0 |
| 0 | 0 | 0 | 0 | 0 |
| 0 | 0.002568426 | 0 | 0 | 0 |
| 0 | 0 | 0 | 0 | 0 |
| 0 | 0 | 0 | 0 | 0 |
| 0 | 0 | 0 | 0 | 0 |
| 0 | 0 | 0 | 0 | 0 |
| 0 | 0 | 0 | 0 | 0 |
| 0.010979079 | 0.013592793 | 0.110793289 | 0.074001361 | 0.050406674 |
| 0 | 0 | 0 | 0 | 0 |
| 0 | 0.000832105 | 0 | 0 | 0 |
| 0 | 0 | 0 | 0 | 0 |
| 0 | 0 | 0 | 0 | 0 |
| 0 | 0 | 0 | 0 | 0 |
| 0 | 0.000552288 | 0.001584785 | 0 | 0.001847559 |
| 0 | 0 | 0 | 0 | 0 |
| 0 | 0 | 0 | 0 | 0 |
| 0 | 0 | 0 | 0 | 0 |
| 0 | 0.031564118 | 0 | 0 | 0 |
| 0 | 0 | 0 | 0 | 0 |
| 0 | 0.001245573 | 0 | 0 | 0 |
| 0.001032266 | 0 | 0 | 0.060696758 | 0.019818877 |
| 0.00128218 | 0.076512574 | 0.003541166 | 0.002172146 | 0.066970647 |
| 0 | 0 | 0 | 0 | 0 |
| 0 | 0 | 0 | 0 | 0 |
| 0 | 0 | 0 | 0 | 0 |
| 0 | 0 | 0 | 0 | 0 |
| 0 | 0 | 0 | 0 | 0 |
| 0 | 0 | 0 | 0 | 0 |
| 0 | 0 | 0 | 0 | 0 |
| 0 | 0 | 0 | 0 | 0 |
| 0 | 0 | 0 | 0 | 0 |
| 0 | 0 | 0 | 0 | 0 |
| 0 | 0 | 0 | 0.003791552 | 0 |
| 0 | 0.000668087 | 0 | 0 | 0 |
| 0 | 0.005862895 | 0 | 0 | 0 |

| | | | | |
|---|---|---|---|---|
| 0 | 0 | 0 | 0 | 0 |
| 0 | 0 | 0 | 0 | 0 |
| 0 | 0 | 0 | 0 | 0.0020909 |
| 0 | 0 | 0 | 0.001639684 | 0 |
| 0 | 0 | 0 | 0 | 0 |
| 0 | 0 | 0 | 0 | 0 |
| 0 | 0 | 0 | 0.000550865 | 0 |
| 0 | 0 | 0 | 0 | 0 |
| 0 | 0 | 0.001065596 | 0 | 0 |
| 0 | 0 | 0 | 0 | 0 |
| 0 | 0 | 0 | 0 | 0 |
| 0 | 0 | 0 | 0 | 0 |
| 0 | 0 | 0 | 0 | 0 |
| 0 | 0 | 0 | 0 | 0 |
| 0 | 0 | 0 | 0 | 0 |
| 0 | 0 | 0 | 0 | 0 |
| 0 | 0 | 0 | 0 | 0 |
| 0 | 0 | 0 | 0 | 0 |
| 0.000530996 | 0 | 0 | 0 | 0.00349061 |
| 0 | 0 | 0 | 0.541350394 | 0 |
| 0 | 0.030343898 | 0.002974496 | 0.004644312 | 0 |
| 0 | 0 | 0 | 0 | 0 |
| 0 | 0 | 0 | 0 | 0 |
| 0 | 0 | 0 | 0 | 0 |
| 0 | 0 | 0.002148896 | 0 | 0.003409863 |
| 0 | 0 | 0 | 0 | 0 |
| 0 | 0 | 0 | 0 | 0 |
| 0 | 0 | 0 | 0 | 0 |
| 0 | 0 | 0 | 0 | 0.001162172 |
| 0 | 0 | 0 | 0 | 0 |
| 0.001295866 | 2.451393937 | 0.057064612 | 0.06090013 | 0.131250138 |
| 0 | 0 | 0 | 0 | 0 |
| 0 | 0.009247681 | 0.085294782 | 0 | 0 |
| 0 | 0 | 0 | 0 | 0 |
| 0 | 0 | 0 | 0 | 0 |
| 0 | 0 | 0 | 0 | 0 |
| 0 | 0 | 0 | 0 | 0 |
| 0 | 0 | 0 | 0 | 0 |
| 0 | 0 | 0 | 0 | 0 |
| 0 | 0 | 0 | 0 | 0 |
| 0 | 0 | 0 | 0 | 0 |
| 0 | 0.117177062 | 0.001621404 | 0.009717748 | 0.00128642 |
| 0 | 0 | 0 | 0 | 0 |
| 0 | 0 | 0 | 0 | 0 |
| 0 | 0 | 0 | 0 | 0 |
| 0 | 0 | 0 | 0 | 0 |
| 0 | 0 | 0 | 0 | 0 |
| 0 | 0 | 0 | 0 | 0 |

| | | | | |
|---|---|---|---|---|
| 0 | 0 | 0 | 0 | 0 |
| 0 | 0 | 0 | 0 | 0 |
| 0.008293163 | 0 | 0.00407188 | 0.060282669 | 0.080765606 |
| 0 | 0 | 0 | 0 | 0 |
| 0.002878815 | 0 | 0 | 0 | 0 |
| 0 | 0 | 0 | 0 | 0 |
| 0 | 0 | 0 | 0 | 0 |
| 0 | 0 | 0 | 0 | 0 |
| 0 | 0 | 0 | 0 | 0 |
| 0 | 0 | 0 | 0 | 0.001477352 |
| 0 | 0 | 0 | 0 | 0 |
| 0 | 0 | 0 | 0 | 0 |
| 0 | 0 | 0 | 0 | 0 |
| 0 | 0 | 0 | 0.032607142 | 0.004335514 |
| 0.003698383 | 0.000566304 | 0.004875012 | 0.001495163 | 0.001473458 |
| 0.011518856 | 0 | 0 | 0 | 0 |
| 0 | 0 | 0 | 0.001892893 | 0.003837426 |
| 0 | 0 | 0 | 0 | 0 |
| 0 | 0 | 0 | 0 | 0 |
| 0 | 0 | 0 | 0 | 0 |
| 0 | 0 | 0 | 0 | 0 |
| 0 | 0.006018945 | 0.00119387 | 0 | 0 |
| 0 | 0 | 0 | 0 | 0 |
| 0 | 0 | 0 | 0 | 0 |
| 0 | 0 | 0 | 0 | 0 |
| 0 | 0 | 0 | 0 | 0 |
| 0.009113788 | 0 | 0 | 0 | 0 |
| 0 | 0 | 0 | 0 | 0 |
| 0 | 0 | 0 | 0 | 0 |
| 0 | 0.001685121 | 0 | 0 | 0 |
| 0 | 0 | 0 | 0 | 0 |
| 0 | 0 | 0 | 0 | 0 |
| 0 | 0 | 0 | 0 | 0 |
| 0 | 0.142154473 | 0 | 0 | 0 |
| 0 | 0 | 0 | 0 | 0 |
| 0 | 0 | 0 | 0.021843005 | 0 |
| 0 | 0 | 0 | 0 | 0 |
| 0.001086217 | 0 | 0 | 0 | 0.461070023 |
| 0 | 0 | 0 | 0 | 0 |
| 0 | 0 | 0 | 0 | 0 |
| 0 | 0 | 0 | 0 | 0.001177795 |
| 0 | 0 | 0 | 0 | 0 |
| 0 | 0 | 0 | 0 | 0 |
| 0 | 0 | 0 | 0 | 0 |
| 0 | 0 | 0 | 0 | 0 |
| 0 | 0 | 0 | 0 | 0 |
| 0 | 0 | 0 | 0 | 0 |
| 0 | 0 | 0 | 0 | 0 |

| | | | | |
|---|---|---|---|---|
| 0 | 0.997524841 | 0 | 0.088286441 | 0 |
| 0 | 0 | 0 | 0 | 0 |
| 0 | 0 | 0.00350136 | 0 | 0 |
| 0 | 0 | 0 | 0 | 0 |
| 0 | 0.003887529 | 0.000754866 | 0.003781439 | 0.001368936 |
| 0 | 0 | 0 | 0 | 0 |
| 0 | 0 | 0 | 0 | 0 |
| 0.002298714 | 0.002868014 | 0 | 0.005048114 | 0.005756594 |
| 0 | 0 | 0 | 0 | 0 |
| 0.006844743 | 0.18416656 | 0.133055298 | 0.012747841 | 0.003955631 |
| 0 | 0 | 0 | 0 | 0 |
| 0 | 0 | 0 | 0.035964946 | 0.006075921 |
| 0 | 0.001377701 | 0.00360033 | 0.017884007 | 0.0011522 |
| 0 | 0.018046712 | 0 | 0 | 0 |
| 0 | 0.001948284 | 0 | 0 | 0 |
| 0 | 0 | 0 | 0 | 0 |
| 0 | 0 | 0 | 0 | 0.005031215 |
| 0 | 0 | 0 | 0 | 0 |
| 0 | 0.111544538 | 0.193961421 | 0 | 0 |
| 0 | 0 | 0.035646158 | 0 | 0 |
| 0 | 0 | 0 | 0.001790936 | 0.053610012 |
| 0 | 0 | 0 | 0 | 0 |
| 0 | 0 | 0 | 0 | 0 |
| 0 | 0 | 0 | 0 | 0 |
| 0 | 0 | 0 | 0 | 0 |
| 0.000893155 | 0 | 0 | 0.00088264 | 0.005368079 |
| 2.243189806 | 0 | 0.001319415 | 0 | 0 |
| 0 | 0.002425616 | 0 | 0 | 0 |
| 0 | 0 | 0 | 0 | 0 |
| 0 | 0 | 0.002030376 | 0.001271376 | 0.008054488 |
| 0 | 0 | 0 | 0.003599059 | 0 |
| 0 | 0 | 0 | 0 | 0 |
| 0 | 0 | 0 | 0 | 0 |
| 0 | 0 | 0 | 0 | 0 |
| 0 | 0 | 0 | 0 | 0 |
| 0 | 1.597321364 | 0.004752681 | 0 | 0 |
| 0 | 0.181223684 | 0 | 0 | 0 |
| 0 | 0 | 0 | 0 | 0 |
| 0 | 0 | 0 | 0 | 0 |
| 0 | 0 | 0 | 0 | 0 |
| 0 | 0 | 0 | 0 | 0 |
| 0 | 0.001648993 | 0 | 0 | 0 |
| 0 | 0.008833558 | 0 | 0 | 0 |
| 0 | 0.006463895 | 0 | 0 | 0 |
| 0 | 0 | 0 | 0 | 0 |
| 0 | 0 | 0 | 0 | 0 |
| 0 | 0.091545142 | 0 | 0 | 0.006397479 |
| 0 | 0 | 0 | 0 | 0 |

| | | | | |
|---|---|---|---|---|
| 0 | 0 | 0 | 0 | 0 |
| 0 | 0.004739067 | 0 | 0 | 0 |
| 0 | 0 | 0 | 0 | 0 |
| 0 | 0 | 0 | 0.099939491 | 0 |
| 0 | 0 | 0 | 0 | 0 |
| 0 | 0.035167349 | 0 | 0 | 0 |
| 0 | 0 | 0 | 0 | 0 |
| 0 | 0 | 0 | 0 | 0 |
| 0 | 0 | 0 | 3.192428278 | 0 |
| 0 | 0 | 0 | 0 | 0 |
| 0 | 0.008658948 | 0.001681362 | 0 | 0 |
| 0 | 0 | 0 | 0 | 0 |
| 0 | 0 | 0 | 0 | 0 |
| 0.001725448 | 0 | 0.011913495 | 0.001705135 | 0.142832507 |
| 0 | 0.020257444 | 0.019822653 | 0 | 0 |
| 0 | 0 | 0 | 0 | 0 |
| 0 | 0 | 0 | 0 | 0 |
| 0 | 0.369766836 | 0 | 0 | 0 |
| 0 | 0 | 0 | 0.001209902 | 0 |
| 0 | 0 | 0 | 0 | 0 |
| 0 | 0 | 0 | 0 | 0 |
| 0.073046854 | 0 | 0.573501299 | 0.551576319 | 0.138146567 |
| 0 | 0 | 0 | 0.010247911 | 0 |
| 0 | 0 | 0 | 0 | 0 |
| 0 | 0.07679703 | 0 | 0 | 0 |
| 0 | 0 | 0 | 0 | 0 |
| 0 | 0.002567594 | 0.001148212 | 0 | 0 |
| 0 | 0 | 0 | 0 | 0 |
| 0 | 0 | 0 | 0 | 0 |
| 0.275114831 | 0.05711482 | 14.2918525 | 0.289649234 | 0.66292273 |
| 0 | 0 | 0 | 0.000899732 | 0.014820077 |
| 0 | 0 | 0 | 0 | 0 |
| 0 | 0.001124508 | 0 | 0 | 0 |
| 0 | 0.007239917 | 0 | 0 | 0 |
| 0 | 0 | 0 | 0 | 0 |
| 0 | 0 | 0 | 0 | 0 |
| 0 | 0 | 2.711119501 | 0 | 0 |
| 0 | 0 | 0 | 0 | 0.005138272 |
| 0 | 0.00505402 | 0.154818628 | 0 | 0 |
| 0 | 0 | 0.008458028 | 0 | 0 |
| 0.002206565 | 0.032210615 | 0 | 0.051243804 | 0.000986755 |
| 0.001491081 | 0 | 1.699845414 | 0.322970191 | 3.91974962 |
| 0 | 0 | 0 | 0 | 0.006275986 |
| 0 | 0 | 0 | 0 | 0 |
| 0 | 0.059094695 | 0 | 0 | 0 |
| 0 | 0 | 0 | 0 | 0 |
| 0 | 0.001671792 | 0 | 0.000980864 | 0.002485609 |
| 0 | 0 | 0 | 0 | 0 |

| | | | | |
|---|---|---|---|---|
| 0 | 0 | 0 | 0 | 0.088937902 |
| 0 | 0 | 0 | 0 | 0 |
| 0.035876294 | 0.019986501 | 0 | 0.018688891 | 0.005571711 |
| 0 | 0 | 0 | 0 | 0 |
| 0.000683923 | 0 | 0 | 0 | 0 |
| 0 | 0 | 0 | 0 | 0 |
| 0 | 0 | 0 | 0 | 0 |
| 0 | 0 | 0 | 0 | 0 |
| 0 | 0.143283236 | 0 | 0 | 0 |
| 0 | 0 | 0 | 0 | 0 |
| 0 | 0 | 0 | 0 | 0 |
| 0 | 0 | 0 | 0 | 0 |
| 0 | 0 | 0 | 0 | 0 |
| 0 | 0 | 0 | 0 | 0 |
| 0 | 0 | 0 | 0 | 0 |
| 0.002308813 | 0.25763499 | 0.002191941 | 0.136184892 | 0.109313769 |
| 0.004798272 | 0 | 0.011705954 | 0.001896713 | 0.287586689 |
| 0 | 0.002618461 | 0 | 0 | 0 |
| 0 | 0.015211954 | 0 | 0 | 0 |
| 0 | 0 | 0.414143692 | 0 | 0 |
| 0 | 0 | 0 | 0.004979996 | 0 |
| 0 | 0.011666192 | 0.002305749 | 0 | 0.010918192 |
| 0 | 0.006108882 | 0 | 0 | 0 |
| 0 | 0 | 0 | 0 | 0 |
| 0 | 0.119510701 | 0 | 0 | 0 |
| 0 | 0 | 0.005528115 | 0.010202556 | 0 |
| 0 | 0.002964022 | 0 | 0.003009866 | 0 |
| 0 | 0 | 0 | 0 | 0 |
| 0 | 0 | 0 | 0 | 0 |
| 0 | 0 | 0 | 0 | 0 |
| 0.004323692 | 0.05999454 | 0.010448647 | 0.023195143 | 0.000580054 |
| 0.001611524 | 0 | 0.586546249 | 0.019565629 | 0.0141249 |
| 0.002849578 | 0.025105931 | 0.060841592 | 0.008448091 | 0.008233968 |
| 0 | 0 | 0 | 0.010865201 | 0.007473384 |
| 0 | 0 | 0.005573092 | 0.004874565 | 0 |
| 0 | 0.007947041 | 0 | 0 | 0 |
| 0 | 0 | 0 | 0.001370861 | 0.018362051 |
| 0 | 0.286444194 | 0.004494256 | 0.006891925 | 0 |
| 0.007022858 | 0 | 0 | 0.12059982 | 0.222145422 |
| 0 | 0.023903501 | 0 | 0 | 0.020466977 |
| 0 | 0 | 0 | 0 | 0 |
| 0 | 0 | 0 | 0 | 0 |
| 0 | 0.019779247 | 0 | 0 | 0 |
| 0.010162301 | 0 | 0.190618778 | 0 | 0 |
| 0 | 2.88888391 | 0 | 0 | 0.117685431 |
| 0 | 0 | 0 | 0 | 0.000846138 |
| 0 | 0 | 0 | 0 | 0 |
| 0 | 0 | 0 | 0 | 0 |

| | | | | |
|---|---|---|---|---|
| 0 | 0 | 0 | 0 | 0 |
| 0.009714178 | 0.061891021 | 0.039951774 | 0.048625144 | 0.029350932 |
| 0 | 0 | 0 | 0 | 0 |
| 0 | 0 | 0 | 0 | 0 |
| 0 | 0 | 0 | 0 | 0 |
| 0 | 0 | 0 | 0 | 0 |
| 0 | 0 | 0 | 0 | 0 |
| 0 | 0 | 0 | 0.000641189 | 0 |
| 0 | 0.0032561 | 0 | 0 | 0.002178516 |
| 0 | 0.001378544 | 0 | 0 | 0.001152905 |
| 0 | 0.005286413 | 0.000650114 | 0 | 0.003143921 |
| 0.003506235 | 0.00288723 | 0.090703347 | 0.347188597 | 0.105659271 |
| 0.00534364 | 0 | 0 | 0.626059883 | 0.14720094 |
| 0 | 0 | 0 | 0 | 0 |
| 0.001800555 | 0 | 0.002983698 | 0.004982199 | 0.006538152 |
| 0 | 0 | 0 | 0 | 0 |
| 0.002136244 | 0.002214255 | 0.007942236 | 0.005358931 | 0.005761243 |
| 0 | 0.076411372 | 0.09279108 | 0.000632421 | 0.010417025 |
| 0 | 0 | 0 | 0 | 0 |
| 0 | 0.000777705 | 0 | 0 | 0.004336071 |
| 0.073885444 | 0.011797854 | 0.018018989 | 0.448309081 | 0.25653664 |
| 0 | 0 | 0 | 0 | 0 |
| 0 | 0 | 0 | 0 | 0 |
| 0 | 0 | 0 | 0 | 0 |
| 0 | 0 | 0 | 0 | 0 |
| 0 | 0 | 0 | 0 | 0 |
| 0.009366979 | 0.192331503 | 0.036264232 | 0.239351893 | 0.001466089 |
| 0.011347127 | 0.032561931 | 0.092275529 | 0.032572658 | 0.023229148 |
| 0 | 0 | 0 | 0 | 0 |
| 0 | 0 | 0 | 0 | 0 |
| 0 | 0 | 0 | 0 | 0 |
| 0 | 0 | 0 | 0 | 0 |
| 0 | 0 | 0 | 0 | 0 |
| 0 | 0 | 0 | 0 | 0 |
| 0 | 0 | 0 | 0 | 0 |
| 0.153329847 | 2.140356288 | 0.248357921 | 0.158754519 | 0.403445062 |
| 0 | 0 | 0 | 0 | 0 |
| 0 | 0 | 0 | 0 | 0 |
| 0 | 0.002744451 | 0.001227301 | 0.012954858 | 0.001669267 |
| 0 | 0 | 0 | 0 | 0 |
| 0 | 2.24913741 | 0 | 0 | 0.118364534 |
| 0 | 0 | 0 | 0 | 0 |
| 0 | 0.045009581 | 0.001490963 | 0.000933608 | 0 |
| 0 | 0.002048426 | 0 | 0 | 0.036383876 |
| 0 | 0 | 0 | 0 | 0 |
| 0.410889376 | 0 | 0 | 0.394743233 | 0 |
| 0 | 0 | 0 | 0 | 0 |
| 0 | 0.004020649 | 0 | 0.000624434 | 0 |

| | | | | |
|---|---|---|---|---|
| 0 | 0 | 0 | 0 | 0 |
| 0 | 0.000749174 | 0 | 0 | 0 |
| 0.043289284 | 0 | 0.001366374 | 0.059524819 | 0 |
| 0.017133419 | 0.001412038 | 0 | 0.161394915 | 0.098803414 |
| 0 | 0 | 0 | 0 | 0 |
| 0 | 0 | 0 | 0 | 0 |
| 0 | 0 | 0 | 0 | 0 |
| 0 | 0 | 0 | 0.000555862 | 0.493541579 |
| 1.138823501 | 0 | 1.022003553 | 0.09309972 | 4.023037277 |
| 0.004074118 | 0 | 0 | 0.124116594 | 0 |
| 0 | 0 | 0 | 0 | 0 |
| 0 | 0 | 0.003743446 | 0.000744147 | 0.020554597 |
| 0 | 0 | 0 | 0 | 0 |
| 0.003962201 | 0 | 0 | 1.54981381 | 0 |
| 0.165786117 | 0.21448905 | 0.017504906 | 0.113971712 | 0.121999793 |
| 0 | 0 | 0 | 0 | 0 |
| 0.007943343 | 0 | 0 | 0.61244671 | 0 |
| 0 | 0 | 0.009412436 | 0.000641508 | 1.448247862 |
| 0 | 0 | 0 | 0 | 0 |
| 0 | 0 | 0 | 0.022993194 | 0 |
| 0 | 0 | 0 | 0 | 0 |
| 0 | 0 | 0 | 0.014370421 | 0 |
| 0 | 0 | 0 | 0 | 0 |
| 0.009936016 | 0.555569664 | 0.102988688 | 0.70070092 | 0.047666362 |
| 0.00493508 | 0 | 0 | 0 | 0 |
| 0.231490953 | 0 | 0 | 0.544556993 | 0.037847072 |
| 0.003187648 | 0 | 0 | 0 | 0 |
| 0.231780849 | 0 | 6.46353094 | 1.705143115 | 2.493106165 |
| 0 | 0 | 0 | 0 | 0 |
| 0 | 0 | 0 | 0 | 0 |
| 0.000585645 | 0 | 0 | 0.037184685 | 0.023282448 |
| 0 | 0 | 0 | 1.305423811 | 0 |
| 0.000557148 | 0 | 0.009386375 | 0.042395354 | 0 |
| 0 | 0 | 0 | 0 | 0 |
| 0 | 0 | 0 | 0 | 0 |
| 0.006951857 | 0 | 0 | 0 | 0 |
| 0.022125246 | 0 | 0 | 0.374552986 | 8.214367445 |
| 0 | 0 | 0 | 0 | 0 |
| 0.000627108 | 0.10316184 | 0 | 0.002788764 | 0 |
| 0 | 0 | 0.004465765 | 0.005006819 | 0 |
| 0.003611901 | 0.050616179 | 0.042894497 | 0.110365184 | 0.009768779 |
| 0 | 0 | 0 | 0 | 0 |
| 3.558886078 | 0 | 13.13594993 | 2.950405684 | 0.289064962 |
| 0 | 0 | 0 | 0 | 0 |
| 0 | 0 | 0 | 0 | 0 |
| 0 | 0 | 0 | 0 | 0 |
| 0 | 0 | 0 | 0 | 0 |
| 0.001389414 | 0.008970951 | 0.002638164 | 0 | 0 |

| | | | | |
|---|---|---|---|---|
| 0 | 0 | 0 | 0 | 0 |
| 0 | 0 | 0.011798714 | 0 | 0.107181935 |
| 0 | 1.59336925 | 0.008046747 | 0.000822644 | 0.002501595 |
| 0 | 0 | 0 | 0.017073379 | 0 |
| 0 | 0 | 0 | 0 | 0 |
| 0.002098455 | 0 | 0.001086671 | 0.012831331 | 0.026932352 |
| 0 | 0 | 0 | 0 | 0 |
| 0 | 0 | 0 | 0 | 0 |
| 0 | 0 | 0 | 0 | 0 |
| 0 | 0 | 0 | 0 | 0 |
| 0.058356103 | 3.497199149 | 0.001805824 | 0.104676656 | 0 |
| 0.256882677 | 1.817857925 | 1.647454125 | 0.442041314 | 0.848491186 |
| 0 | 0 | 0 | 0 | 0 |
| 0.003528307 | 0 | 0.001218074 | 0 | 0.079080633 |
| 0.014041229 | 0.295822978 | 0.001343845 | 0.140957402 | 0.075374138 |
| 0.041183432 | 0 | 0 | 0.051783158 | 0 |
| 0 | 0 | 0 | 0 | 0 |
| 0 | 0 | 0 | 0 | 0 |
| 0.004680505 | 0 | 0 | 0 | 0 |
| 0.00040395 | 0.014817384 | 0.001859404 | 0.026346838 | 0 |
| 0.008220337 | 0.035230013 | 0.017405781 | 0.005821885 | 0.001715494 |
| 0.001732337 | 0.002917848 | 9.916516763 | 0.016691439 | 0 |
| 0.390803502 | 2.226123205 | 1.428866244 | 0.253205689 | 0.281936596 |
| 0 | 0 | 0 | 0 | 0 |
| 0 | 0 | 0 | 0.005307923 | 0 |
| 1.903103091 | 0.004608747 | 0.022217608 | 8.594881041 | 6.236920654 |
| 0 | 0 | 0 | 0 | 0 |
| 0 | 0 | 0 | 0 | 0 |
| 0.025211876 | 0.301586316 | 0.073371476 | 0 | 0 |
| 0 | 0 | 0 | 0 | 0 |
| 0 | 0 | 0.001043376 | 0.017920153 | 0 |
| 0.016814696 | 0.161796743 | 0 | 0.130888767 | 0.137338471 |
| 0.021992309 | 0.145908105 | 0 | 0 | 0.050870452 |
| 0.019587315 | 0.351624277 | 0.013082251 | 0.087168477 | 0.013946121 |
| 0.000332409 | 0.027248003 | 0.000459029 | 0 | 0 |
| 0.005023807 | 0.142910536 | 0.036579316 | 0.022566649 | 0.07148271 |
| 0.006176456 | 0.01023813 | 0.001015377 | 0.003778506 | 0.004235148 |
| 0 | 0 | 0 | 0 | 0 |
| 0.088539333 | 0 | 0.024846689 | 0.433963841 | 0.110416952 |
| 0 | 0 | 0 | 0 | 0 |
| 0 | 0.004666313 | 0.001110687 | 0 | 0 |
| 0 | 0 | 0 | 0 | 0 |
| 0 | 0 | 0 | 0 | 0 |
| 0 | 0 | 0 | 0 | 0 |
| 0.00833321 | 0.030021387 | 0.008390859 | 0.002745035 | 0.155862257 |
| 0 | 0 | 0 | 0 | 0 |
| 0 | 0 | 0 | 0 | 0 |
| 2.843705362 | 0.472898359 | 0.062175968 | 0.083402414 | 0.286345358 |

| | | | | |
|---|---|---|---|---|
| 0 | 0 | 0 | 0 | 0 |
| 0 | 0 | 0 | 0 | 0 |
| 0 | 0 | 0 | 0 | 0 |
| 0.000591863 | 0 | 0.001838956 | 0.003509373 | 0 |
| 0 | 0 | 0 | 0 | 0 |
| 0 | 0 | 0 | 0 | 0 |
| 0.050542266 | 0 | 0 | 0.003201746 | 0 |
| 0.001089243 | 0.171234687 | 0.003384345 | 0.025834067 | 0 |
| 0 | 0 | 0 | 0 | 0 |
| 0 | 0 | 0 | 0 | 0 |
| 0 | 0.023130894 | 0.000682703 | 0.000651419 | 0 |
| 0 | 0 | 0 | 0 | 0 |
| 0 | 0 | 0 | 0 | 0 |
| 0 | 0 | 0 | 0.005697711 | 0 |
| 0 | 0 | 0 | 0 | 0 |
| 0 | 0 | 0 | 0 | 0 |
| 0 | 0 | 0 | 0 | 0 |
| 0.007682522 | 0.027882603 | 0 | 0.000813437 | 0.023018197 |
| 0 | 0 | 0 | 0 | 0 |
| 0 | 0 | 0 | 0 | 0 |
| 0 | 0 | 0 | 0 | 0 |
| 0 | 0 | 0 | 0 | 0 |
| 0 | 0 | 0 | 0.005783312 | 0.821103991 |
| 0 | 0 | 0 | 0 | 0 |
| 0 | 0 | 0 | 0 | 0 |
| 0 | 0 | 0 | 0 | 0 |
| 0 | 0 | 0 | 0 | 0 |
| 0 | 0 | 0 | 0 | 0 |
| 0 | 0 | 0 | 0 | 0 |
| 0.001325774 | 0 | 0 | 0.640834888 | 0.105827957 |
| 0 | 0 | 0 | 0.000513065 | 0.001462678 |
| 0 | 0 | 0 | 0 | 0 |
| 0 | 0 | 0 | 0.022380778 | 0 |
| 0 | 0 | 0 | 0 | 0 |
| 0 | 0.000735702 | 0.001447606 | 0 | 0.008039703 |
| 0 | 0 | 0 | 0 | 0 |
| 0.00165217 | 0 | 0 | 0.06752462 | 0 |
| 0 | 0.113568097 | 0.008668805 | 0 | 0.011703208 |
| 0 | 0 | 0 | 0 | 0 |
| 0 | 0.012515036 | 0.000950048 | 0 | 0 |
| 0.081411773 | 0 | 0.089468326 | 0 | 0 |
| 0 | 0 | 0 | 0 | 0 |
| 0 | 0 | 0 | 0.000466235 | 0 |
| 0.421049001 | 0 | 0 | 0 | 0 |
| 0.017265895 | 0 | 0.069122107 | 0 | 0.157284702 |
| 0 | 0 | 0 | 0 | 0 |
| 0.0005302 | 0.004762867 | 0 | 0.006811452 | 0 |
| 0 | 0 | 0 | 0 | 0 |

| | | | | |
|---|---|---|---|---|
| 0 | 0 | 0 | 0 | 0 |
| 0 | 0 | 0 | 0 | 0 |
| 0 | 0 | 0 | 0 | 0 |
| 0.00721569 | 0 | 0 | 0.096265001 | 0.000521251 |
| 0 | 0 | 0 | 0 | 0 |
| 0 | 0 | 0 | 0 | 0 |
| 0 | 0 | 0 | 0 | 0 |
| 0 | 0 | 0 | 0 | 0 |
| 0 | 2.072476659 | 0.002391146 | 0.007415111 | 0.002168151 |
| 0 | 0 | 0 | 0 | 0.000970001 |
| 0 | 0 | 0 | 0 | 0 |
| 0 | 0 | 0 | 0 | 0 |
| 0 | 0 | 0.000464983 | 0 | 0 |
| 0 | 0.002700986 | 0 | 0 | 0 |
| 0.158447013 | 0.721147471 | 0.007064289 | 4.293971215 | 0.054879406 |
| 0 | 0 | 0.045253989 | 0 | 0 |
| 0 | 0 | 0 | 0 | 0 |
| 0 | 0 | 0 | 0 | 0 |
| 0 | 0 | 0 | 0 | 0 |
| 0 | 0 | 0 | 0 | 0 |
| 0 | 0.000991164 | 0 | 0.055972244 | 0 |
| 0 | 0 | 0 | 0.002586783 | 0 |
| 0 | 0 | 0 | 0 | 0 |
| 0 | 0.083091081 | 0 | 0 | 0 |
| 0.005430056 | 0.016513737 | 0 | 0 | 1.55672349 |
| 0.050540786 | 0 | 0.010895889 | 0.631592773 | 5.228258262 |
| 0.25144758 | 0 | 0 | 0 | 0 |
| 0 | 0.002050169 | 0 | 0 | 0 |
| 0 | 0 | 1.451288279 | 0.007819218 | 0.834693797 |
| 0 | 0 | 0 | 0 | 0 |
| 0 | 0 | 0 | 0 | 0 |
| 0 | 0.071335891 | 0.004829044 | 0 | 0 |
| 0 | 0 | 0 | 0 | 0 |
| 0.00773333 | 0.078017732 | 0 | 0.0033435 | 0 |
| 0.004180996 | 0 | 0.002474402 | 0 | 0.027173007 |
| 0.001407361 | 0.009956001 | 0 | 0.001390792 | 0.000881102 |
| 0 | 3.470599182 | 0 | 0 | 0 |
| 0.127450117 | 0.693973267 | 0 | 0.74282032 | 0.429405903 |
| 0 | 0 | 0.107751886 | 10.06039277 | 0 |
| 0 | 0 | 0 | 0 | 0 |
| 0 | 0 | 0 | 0.281118858 | 0 |
| 0 | 0.00250893 | 0 | 0 | 0 |
| 0.034040349 | 1.0182194 | 0 | 0.044472686 | 0.072964913 |
| 0 | 0 | 0 | 0 | 0 |
| 0 | 0 | 0 | 0.279744918 | 0 |
| 0 | 0.042369417 | 0.000736469 | 0.001844641 | 0 |
| 0 | 0 | 0 | 0 | 0 |
| 1.451824417 | 0.001192572 | 0.01818589 | 0.084593577 | 0 |

| | | | | |
|---|---|---|---|---|
| 0 | 0.072922768 | 0.010936482 | 0.000939179 | 0 |
| 0 | 0 | 0 | 0 | 0 |
| 0 | 0 | 0 | 0 | 0 |
| 0 | 0 | 0 | 0 | 0 |
| 0.002050102 | 0.054186677 | 0 | 0.00218181 | 0 |
| 0 | 0 | 0 | 0 | 0 |
| 0 | 0 | 0 | 0 | 0 |
| 0 | 0 | 0 | 0 | 0 |
| 0 | 0 | 0 | 0 | 0 |
| 0 | 0 | 0 | 0 | 0 |
| 0 | 0 | 0 | 0.000953982 | 0 |
| 2.159921825 | 0 | 1.382443599 | 2.652660752 | 0.727591032 |
| 0.270190367 | 0.190613699 | 0 | 0.001320895 | 0.135325784 |
| 0 | 0 | 0 | 0 | 0 |
| 0.001459042 | 0.004300669 | 0 | 0.052267598 | 0.014158595 |
| 0 | 0 | 0 | 0 | 0 |
| 0 | 0.012394337 | 0 | 0 | 0 |
| 0 | 0 | 0 | 0 | 0 |
| 0 | 0 | 0.000882989 | 0 | 0.001200964 |
| 0 | 0 | 0 | 0.243482931 | 0 |
| 0 | 0.245504322 | 0 | 0.135496068 | 0 |
| 0 | 0 | 0 | 0 | 0 |
| 0.07616456 | 0 | 0.024040427 | 0.261287081 | 0.071818102 |
| 0 | 0 | 0 | 0 | 0 |
| 0 | 0 | 0 | 0 | 0 |
| 0 | 0 | 0 | 0 | 0 |
| 0 | 0 | 0 | 0 | 0 |
| 0 | 0 | 0 | 0 | 0 |
| 0 | 0 | 0 | 0 | 0 |
| 0 | 0 | 0 | 0 | 0 |
| 0 | 0 | 0 | 0 | 0 |
| 0.000946655 | 0.008769702 | 0.032681294 | 0.054025737 | 0.030522484 |
| 0 | 0 | 0 | 0 | 0 |
| 0.079098003 | 0.102493336 | 0.003755213 | 0.005725221 | 0 |
| 0.000765781 | 0.011006602 | 0 | 0.001362177 | 0.001150632 |
| 0.014610324 | 0.01928796 | 0.003271724 | 0.055802149 | 1.65289474 |
| 0 | 0 | 0 | 0 | 0 |
| 0 | 0 | 0 | 0 | 0 |
| 0 | 0 | 0 | 0.010432938 | 0.000881271 |
| 0 | 0 | 0 | 0 | 0 |
| 0.016572176 | 0 | 0 | 0.02862764 | 0 |
| 0 | 0 | 0 | 0.034699209 | 0 |
| 0 | 0 | 0 | 0 | 0 |
| 0 | 0 | 0 | 0.017003711 | 0 |
| 0.011768182 | 0.226508816 | 0 | 0.001789175 | 0.010768131 |
| 0 | 0 | 0 | 0 | 0 |
| 0.001142479 | 0 | 0.137958241 | 0.406324811 | 0.043869775 |
| 0 | 0.008652796 | 0.003744076 | 0 | 0 |

| | | | | |
|---|---|---|---|---|
| 0 | 0 | 0 | 0 | 0 |
| 0 | 0.016715958 | 0 | 0 | 0 |
| 0 | 0.002453056 | 0.002154804 | 0 | 0 |
| 0 | 0 | 0 | 0 | 0 |
| 0 | 0 | 0 | 0 | 0 |
| 0 | 0 | 0 | 0 | 0 |
| 0 | 0 | 0.000948785 | 0 | 0 |
| 0.006778869 | 0 | 0 | 0.047184696 | 0 |
| 0 | 0 | 0 | 0 | 0 |
| 0 | 0 | 0 | 0.001003151 | 0 |
| 0 | 0 | 0 | 0 | 0 |
| 0 | 0 | 0 | 0 | 0 |
| 0 | 0 | 0 | 0 | 0 |
| 0 | 0 | 0.438985945 | 0 | 0 |
| 0 | 1.121473163 | 0 | 0 | 0 |
| 0 | 0 | 0 | 0 | 0 |
| 0 | 0 | 0 | 9.734803837 | 0 |
| 0 | 0 | 0 | 0 | 0 |
| 0.107877915 | 0 | 0.022794535 | 2.236458662 | 0 |
| 0 | 0 | 0 | 0 | 0 |
| 0 | 0 | 0 | 0 | 0 |
| 0 | 0 | 0 | 0 | 0 |
| 0 | 0 | 0 | 0 | 0 |
| 0 | 0 | 0 | 0 | 0 |
| 0 | 0 | 0 | 0 | 0.000825622 |
| 0 | 0.06403417 | 0 | 0 | 0.01354217 |
| 0 | 0 | 0 | 0 | 0 |
| 0.018147433 | 0.002624395 | 0.043855144 | 0.00203793 | 0.019107988 |
| 0 | 0.012417443 | 0 | 0 | 0 |
| 0.023159462 | 0 | 0.090112979 | 0.669913305 | 0.628039056 |
| 0 | 0 | 0 | 0 | 0 |
| 0 | 0 | 0 | 0 | 0 |
| 0 | 0 | 0 | 0.01678465 | 0 |
| 0 | 0 | 0 | 0 | 0 |
| 0 | 0.004627852 | 0 | 0.000727293 | 0 |
| 0.159912846 | 0 | 0 | 0 | 0 |
| 0 | 0 | 0 | 0 | 0 |
| 0 | 0.002813173 | 0 | 0 | 0 |
| 0 | 0 | 0 | 0 | 0 |
| 0 | 0 | 0 | 0 | 0 |
| 2.191068789 | 1.811787737 | 9.761940508 | 4.280634654 | 7.849943619 |
| 0.316894725 | 0.164054008 | 0 | 0.410906877 | 0.059090548 |
| 0.170618614 | 0 | 0.007728166 | 0 | 0 |
| 0 | 0 | 0 | 0 | 0 |
| 0 | 0 | 0 | 0 | 0 |
| 0 | 0 | 0 | 0.001733597 | 0 |
| 0 | 0 | 0 | 0.002434904 | 0 |
| 0.001590668 | 0.020094207 | 0 | 0.001047961 | 0.004315412 |

| | | | | |
|---|---|---|---|---|
| 0 | 0 | 0 | 0 | 0 |
| 0 | 0.009798424 | 0.003765604 | 0.011318096 | 0.000512164 |
| 0 | 0.016271947 | 0 | 0.001888414 | 0 |
| 0 | 0 | 0 | 0.088221797 | 0 |
| 0.692713919 | 11.00727435 | 0 | 0.286550891 | 0.668355294 |
| 0 | 0 | 0 | 0 | 0 |
| 0 | 0 | 0 | 0 | 0 |
| 0 | 0 | 0 | 0 | 0 |
| 0 | 0.574044335 | 0.095693323 | 0.010628707 | 0.009205365 |
| 0 | 0 | 0 | 0 | 0 |
| 0 | 0 | 0 | 0 | 0 |
| 0 | 0 | 0 | 0 | 0 |
| 0.001324818 | 0 | 0 | 0.11390226 | 0.008294248 |
| 0.00553768 | 0.002194669 | 0.660796916 | 0.01416408 | 0.031610497 |
| 0 | 0 | 0 | 0.012890254 | 2.138496479 |
| 0.002405742 | 0 | 0 | 0.428331808 | 0.070956661 |
| 0 | 0 | 0.001946281 | 0.017688811 | 0 |
| 0 | 0 | 0 | 0 | 0 |
| 0 | 0 | 0 | 0 | 0 |
| 0 | 0 | 0 | 0 | 0 |
| 0 | 0 | 0 | 0 | 0 |
| 0 | 0 | 0 | 0 | 0 |
| 0 | 0 | 0 | 0 | 0 |
| 0 | 0 | 0 | 0 | 0 |
| 0 | 0 | 0 | 0 | 0 |
| 0 | 0 | 0 | 0 | 0 |
| 0.014706507 | 0 | 0.002369319 | 0.00169556 | 0 |
| 0 | 0.027485206 | 0 | 0 | 0 |
| 0 | 0 | 0 | 0 | 0 |
| 0 | 0 | 0 | 0.001479361 | 0 |
| 0 | 0 | 0 | 0 | 0 |
| 0 | 0.000648548 | 0 | 0 | 0 |
| 0 | 0 | 0 | 0 | 0 |
| 0 | 0 | 0 | 0 | 0 |
| 0.029147043 | 0 | 0.191777604 | 0.087089433 | 0 |
| 0 | 0 | 0.006185397 | 0.007113962 | 0.004607027 |
| 0 | 0.164142389 | 0 | 0 | 0 |
| 0 | 0 | 0 | 0 | 0 |
| 0 | 0 | 0 | 0 | 0 |
| 0 | 0 | 0 | 0 | 0 |
| 0.007053255 | 2.340158027 | 0.828437162 | 0.074929844 | 0.015945978 |
| 0 | 0 | 0 | 0 | 0 |
| 0 | 0 | 0 | 0 | 0 |
| 0 | 0 | 0.009582715 | 0 | 0 |
| 0 | 0 | 0 | 0 | 0 |
| 0 | 0 | 0 | 0 | 0 |
| 0 | 0 | 0 | 0.883190157 | 0 |
| 0 | 0 | 0 | 0 | 0 |

| | | | | |
|---|---|---|---|---|
| 0.054962996 | 0 | 0 | 0.063593022 | 0.203429622 |
| 0 | 0 | 0 | 0 | 0 |
| 0 | 0 | 0 | 0 | 0 |
| 0 | 0 | 0 | 0 | 0 |
| 0 | 0 | 0 | 0.01347916 | 0 |
| 0 | 0 | 0 | 0 | 0 |
| 0 | 0.002052405 | 0.008542811 | 0.077252477 | 0.004929349 |
| 0 | 0.001220875 | 0 | 0 | 0 |
| 0 | 0 | 0 | 0 | 0 |
| 0 | 0.196779152 | 0 | 0 | 0 |
| 0.000681995 | 0 | 0 | 0.005054745 | 0 |
| 0.001106605 | 0.003037468 | 0 | 0 | 0 |
| 0 | 0 | 0 | 0 | 0 |
| 0 | 0 | 0 | 0 | 0 |
| 0 | 0 | 0 | 0 | 0 |
| 0 | 0 | 0 | 0 | 0 |
| 0 | 0 | 0 | 0.027153344 | 0.005130519 |
| 0 | 0.1586127 | 0 | 0 | 0 |
| 0 | 0 | 0 | 0 | 0 |
| 0 | 0 | 0 | 0 | 0 |
| 0.036141568 | 0.01142341 | 0.117931873 | 0.00820147 | 0.001676077 |
| 0 | 0 | 0 | 0 | 0 |
| 0.002509957 | 0 | 0 | 0.136276512 | 0.010352759 |
| 0 | 0.005307174 | 0.001450371 | 0.003503018 | 0.002301445 |
| 0 | 0 | 0 | 0 | 0 |
| 0 | 0 | 0 | 0 | 0.000581826 |
| 0 | 0 | 0 | 0 | 0 |
| 0 | 0 | 0 | 0 | 0 |
| 0 | 0 | 0 | 0 | 0 |
| 0.228821884 | 0 | 0.850103455 | 4.371123128 | 0.902133727 |
| 0 | 0 | 0 | 0.038828775 | 0 |
| 0 | 0 | 0 | 0 | 0 |
| 0 | 0 | 0 | 0 | 0 |
| 0 | 3.301144657 | 0 | 0.554639728 | 0 |
| 0 | 0 | 0 | 0 | 0 |
| 0 | 0 | 0 | 0 | 0 |
| 0.028583692 | 0.003504787 | 0 | 0.00048702 | 0.049160657 |
| 0 | 0 | 0 | 0 | 0 |
| 0 | 0 | 0 | 0 | 0 |
| 0 | 0 | 0 | 0 | 0 |
| 0 | 0 | 0 | 0 | 0 |
| 0 | 0 | 0 | 0 | 0 |
| 0 | 0 | 0 | 0 | 0 |
| 0 | 0 | 0.000865874 | 0 | 0 |
| 0 | 0 | 0 | 0 | 0 |
| 0 | 0 | 0 | 0 | 0 |
| 0 | 0 | 0 | 0 | 0 |
| 0 | 0 | 0 | 0 | 0 |

| | | | | |
|---|---|---|---|---|
| 0 | 0.084232932 | 0.001170488 | 0 | 0 |
| 0 | 0 | 0 | 0 | 0 |
| 0.00386135 | 0 | 0 | 0 | 0 |
| 0 | 0 | 0 | 0 | 0 |
| 0 | 0 | 0 | 0 | 0 |
| 0 | 0 | 0 | 0 | 7.320497648 |
| 0 | 0 | 0 | 0 | 0 |
| 0.807648802 | 0.07883292 | 0.125991504 | 0.148647924 | 0.943356774 |
| 0.183825551 | 0.048304991 | 0.012554589 | 0.005259196 | 1.410198534 |
| 0 | 0.3993816 | 0.014708996 | 0 | 0.148642736 |
| 0 | 0.014645852 | 0 | 0 | 0 |
| 0 | 0 | 0 | 0 | 0 |
| 0 | 0.046687209 | 0 | 0.027215892 | 0 |
| 0 | 0 | 0.005440614 | 0.011355968 | 0 |
| 0 | 0 | 0 | 0 | 0 |
| 0 | 0 | 0 | 0 | 0 |
| 0 | 0 | 0 | 0 | 0 |
| 0 | 0 | 0 | 0 | 0 |
| 0 | 0 | 0 | 0 | 0 |
| 0 | 0 | 0 | 0.011764629 | 0 |
| 0 | 0 | 0 | 0 | 0 |
| 0 | 0 | 0 | 0 | 0 |
| 0 | 0 | 0 | 0 | 0 |
| 0 | 0 | 0 | 0 | 0 |
| 0 | 0 | 0 | 0.025620533 | 0 |
| 0 | 0 | 0 | 0 | 0 |
| 0 | 0 | 0 | 0 | 0 |
| 0 | 0 | 0 | 0 | 0 |
| 0 | 0 | 0 | 0 | 0 |
| 0 | 0 | 0 | 0 | 0 |
| 0.108992725 | 0 | 0 | 0 | 0 |
| 0 | 0 | 0 | 0 | 0 |
| 0 | 0 | 0 | 0 | 0 |
| 0 | 0 | 0 | 0 | 0 |
| 0 | 0 | 0 | 0 | 0 |
| 0.001201689 | 0 | 0 | 0 | 0 |
| 0 | 0 | 0 | 0 | 0 |
| 0 | 0 | 0 | 0 | 0 |
| 0 | 0 | 0 | 0 | 0 |
| 0 | 0.108003684 | 0 | 0 | 0.56808985 |
| 0 | 0 | 0 | 0 | 0 |
| 0.046559595 | 1.859097817 | 0.061481992 | 0.268160506 | 0 |
| 0 | 0 | 0 | 0 | 0 |
| 0 | 0 | 0 | 0 | 0 |
| 0 | 0 | 0 | 0 | 0 |
| 0 | 0 | 0.006071344 | 0 | 0 |
| 0 | 0 | 0 | 0 | 0 |
| 0 | 0 | 0 | 0 | 0 |

| | | | | |
|---|---|---|---|---|
| 0 | 0 | 0 | 0 | 0 |
| 0 | 0 | 0 | 0.027342185 | 0.000478948 |
| 0 | 0 | 0 | 0 | 0 |
| 0 | 0 | 0 | 0 | 0 |
| 0.315708395 | 1.079535473 | 0.062773311 | 0.12595934 | 1.94574228 |
| 0.001629158 | 0 | 0 | 0 | 0 |
| 0 | 0 | 0 | 0 | 0 |
| 0.001797051 | 0.207843505 | 0 | 0.009767418 | 0.00427528 |
| 0 | 0 | 0 | 0.003723645 | 0 |
| 0.001617099 | 0 | 0 | 0 | 0 |
| 0 | 0 | 0 | 0 | 0 |
| 0 | 0 | 0 | 0 | 0 |
| 0 | 0 | 0 | 0 | 0 |
| 0 | 0 | 0 | 0 | 0 |
| 0 | 0.000852317 | 0 | 0 | 0 |
| 0.039431784 | 0 | 0.01312874 | 0.007547077 | 0 |
| 0 | 0 | 0 | 0 | 0 |
| 0 | 0 | 0 | 0 | 0 |
| 0 | 0 | 0 | 0 | 0 |
| 0 | 0 | 0 | 0 | 0 |
| 0 | 0 | 0 | 0 | 0 |
| 0 | 0 | 0 | 0 | 0 |
| 0 | 0 | 0 | 0 | 0 |
| 0 | 0.004540444 | 0 | 0 | 0 |
| 0.023799493 | 0 | 0.246106116 | 0 | 0.028240837 |
| 0 | 0 | 0 | 0 | 0 |
| 0 | 0.543371545 | 0 | 0 | 0.007658484 |
| 0 | 0 | 0 | 0 | 0 |
| 0 | 0 | 0 | 0 | 0 |
| 0 | 0 | 0 | 0 | 0 |
| 0 | 0 | 0 | 0 | 0 |
| 0 | 0 | 0 | 0 | 0 |
| 0 | 0 | 0 | 0 | 0 |
| 0 | 0 | 0 | 0 | 0 |
| 0 | 0 | 0 | 0 | 0 |
| 0 | 0 | 0 | 0 | 0 |
| 0 | 0 | 0 | 0 | 0 |
| 0.004084531 | 0.08288455 | 0 | 0.482066742 | 0 |
| 0.03375919 | 0 | 0 | 0 | 0 |
| 0 | 0 | 0 | 0 | 0 |
| 0 | 0 | 0 | 0 | 0 |
| 0 | 0 | 0 | 0 | 0.187031045 |
| 0 | 0 | 0 | 0 | 0 |
| 0 | 0 | 0 | 0 | 0.046415489 |
| 0 | 0 | 0 | 0 | 0 |
| 0 | 0 | 0 | 0 | 0 |
| 0 | 0 | 0 | 0 | 0 |
| 0 | 0 | 0 | 0 | 0.000901317 |

| | | | | |
|---|---|---|---|---|
| 0.147676615 | 0.047464473 | 0 | 0.013219025 | 0.875814233 |
| 0 | 0 | 0 | 0 | 0 |
| 0 | 0 | 0 | 0 | 0 |
| 0 | 0 | 0 | 0 | 0 |
| 0 | 0 | 0 | 0.016774349 | 0 |
| 0 | 0 | 0 | 0.007351203 | 0 |
| 0 | 0 | 0 | 0 | 0 |
| 0 | 0.026536791 | 0 | 0 | 0 |
| 0 | 0 | 0 | 0 | 0 |
| 0 | 0 | 0 | 0 | 0 |
| 0 | 0 | 0 | 0 | 0 |
| 0 | 0.011268829 | 0 | 0 | 0 |
| 0 | 0 | 0 | 0.02475181 | 0 |
| 0 | 0 | 0 | 0 | 0 |
| 0 | 0 | 0 | 0 | 0.001051915 |
| 0 | 0 | 0 | 0 | 0 |
| 0 | 0 | 0 | 0 | 0 |
| 0 | 0.001915358 | 0 | 0 | 0 |
| 0 | 0 | 0 | 0 | 0 |
| 0 | 0 | 0 | 0 | 0 |
| 0 | 0.065735125 | 0.002138622 | 0 | 0 |
| 0 | 0 | 0 | 0 | 0 |
| 0.014669197 | 0.224897154 | 0 | 0.007344892 | 0.002938847 |
| 0 | 0 | 0 | 0.025297559 | 0 |
| 0 | 0 | 0 | 0 | 0 |
| 0 | 0 | 0 | 0 | 2.10207025 |
| 0 | 0 | 0 | 0 | 0 |
| 0 | 0 | 0 | 0 | 0 |
| 0 | 0 | 0.082770265 | 0 | 0 |
| 0 | 0 | 0 | 0 | 0 |
| 0 | 0 | 0 | 0 | 0 |
| 0 | 0 | 0 | 0 | 0 |
| 0.004396046 | 0 | 3.136714116 | 0 | 0.049081228 |
| 0 | 0 | 0 | 0.057006193 | 0 |
| 0.265426327 | 1.164218191 | 0 | 0.01149593 | 0.995406506 |
| 0 | 0 | 0 | 0 | 0 |
| 0 | 0 | 0 | 0 | 0 |
| 0 | 0 | 0 | 0 | 0.081592352 |
| 0.001360092 | 0.093925251 | 0 | 0 | 0.027461152 |
| 0 | 0 | 0 | 0 | 0 |
| 0 | 0 | 0 | 0 | 0 |
| 0 | 0 | 0 | 0 | 0 |
| 0.112318838 | 0 | 0.12600394 | 0.010608517 | 0.517249878 |
| 0 | 0 | 0 | 0 | 0 |
| 0 | 0 | 0 | 0 | 0 |
| 0 | 0 | 0 | 0.20953293 | 0.013047105 |
| 0 | 0 | 0 | 0 | 0 |
| 0 | 0 | 0 | 0 | 0 |

| | | | | |
|---|---|---|---|---|
| 0 | 0 | 0 | 0 | 0 |
| 0 | 0 | 0 | 0 | 0 |
| 0 | 0 | 0 | 0 | 0 |
| 0 | 0 | 0 | 0 | 0 |
| 0 | 0 | 0 | 0 | 0 |
| 0 | 0 | 0 | 0 | 0 |
| 0 | 0 | 0 | 0 | 0 |
| 0 | 1.088491833 | 0.000842687 | 0 | 0 |
| 0 | 0.368895716 | 0 | 0.047945377 | 0 |
| 0 | 0 | 0 | 0.001446277 | 0 |
| 0 | 0 | 0 | 0 | 0 |
| 0 | 0 | 0 | 0 | 0 |
| 0 | 0 | 0 | 0 | 0 |
| 0.022657511 | 0 | 0 | 0 | 0 |
| 0 | 0.001180499 | 0 | 0 | 0 |
| 0 | 0.084617117 | 0 | 0 | 0 |
| 0 | 0 | 0 | 0 | 0 |
| 0 | 0 | 0 | 0 | 0 |
| 0 | 0 | 0 | 0 | 0 |
| 0.024914774 | 0 | 0 | 0.295192718 | 0.466439908 |
| 0 | 0.001688724 | 0.002769019 | 0 | 0 |
| 0 | 0 | 0 | 0 | 0 |
| 0 | 0 | 0 | 0 | 0 |
| 0 | 0 | 0 | 0 | 0 |
| 0 | 0.008012645 | 0 | 0 | 0 |
| 0 | 0 | 0 | 0 | 0 |
| 0 | 0 | 0 | 0 | 0 |
| 0 | 0 | 0.003483532 | 0 | 0 |
| 0 | 0 | 0 | 0 | 0 |
| 0.003482209 | 0 | 0 | 0 | 0 |
| 0.003901111 | 0 | 0 | 1.196649078 | 0 |
| 0 | 0 | 0 | 0 | 0 |
| 0 | 0 | 0 | 0 | 0 |
| 0 | 0 | 0 | 0 | 0 |
| 0 | 0 | 0 | 0 | 0 |
| 0 | 0 | 0 | 0 | 0 |
| 0 | 0 | 0 | 0 | 0 |
| 0 | 0 | 0 | 0 | 0 |
| 0 | 0 | 0.02853596 | 0 | 0 |
| 0 | 0 | 0 | 0 | 0 |
| 0 | 0 | 0 | 0.007050788 | 0 |
| 0.092621598 | 0 | 0 | 1.056873396 | 0 |
| 0 | 0 | 0 | 0 | 0 |
| 0 | 0.001343274 | 0 | 0 | 0 |
| 0 | 1.01651186 | 0 | 0 | 0 |
| 0 | 0 | 0 | 0 | 0 |
| 0 | 4.072150047 | 0.017415102 | 0 | 0 |
| 0 | 0 | 0 | 0 | 0 |

| | | | | |
|---|---|---|---|---|
| 0 | 0 | 0 | 0 | 0 |
| 0 | 0 | 0 | 0 | 0 |
| 0 | 0 | 0 | 0 | 0 |
| 0 | 0 | 0 | 0 | 0 |
| 0.002569516 | 0.957643187 | 0.05609671 | 0.035307879 | 0.122107134 |
| 0 | 0 | 0 | 0 | 0 |
| 0 | 0 | 0.00137768 | 0 | 0 |
| 0 | 0 | 0 | 0 | 0 |
| 0 | 0 | 0 | 0 | 0 |
| 0 | 0 | 0 | 0 | 0 |
| 0 | 0 | 0 | 0 | 0 |
| 0.002887894 | 0.05970568 | 0 | 0.001175133 | 0.002127076 |
| 0 | 0.002468576 | 0 | 0.004779551 | 0 |
| 0 | 0 | 0 | 0 | 0 |
| 0 | 0 | 0 | 0.000511734 | 0 |
| 0 | 0 | 0 | 0 | 0 |
| 0.005095542 | 0.040932557 | 0.001804235 | 0.06313808 | 0 |
| 0 | 0.000998709 | 0 | 0 | 0 |
| 0 | 0.056764622 | 0.04724928 | 0 | 0 |
| 0.003023156 | 0 | 0.010900672 | 0.006307082 | 0 |
| 0 | 0 | 0 | 0 | 0 |
| 0 | 0 | 0 | 0 | 0 |
| 0 | 0 | 0 | 0 | 0 |
| 0 | 0 | 0 | 0 | 0 |
| 0 | 0 | 0 | 0 | 0 |
| 0 | 0 | 0 | 0 | 0 |
| 0 | 0.229413408 | 0.007620075 | 0.001995061 | 0 |
| 0 | 0 | 0 | 0 | 0 |
| 0 | 0 | 0 | 0 | 0 |
| 0 | 0 | 0 | 0 | 0 |
| 0.000610253 | 0.006338557 | 0.000842708 | 0 | 0 |
| 0 | 0.010177051 | 0 | 0 | 0 |
| 0 | 0 | 0 | 0 | 0 |
| 0 | 0 | 0 | 0 | 0 |
| 0 | 0 | 0 | 0 | 0 |
| 0 | 0 | 0 | 0 | 0 |
| 0 | 0 | 0 | 0 | 0 |
| 0.021115734 | 0.061503492 | 0.004588481 | 0.003495511 | 0.001207906 |
| 0 | 0 | 0 | 0 | 2.865530998 |
| 0 | 0.00782775 | 0 | 0 | 0 |
| 0 | 0 | 0 | 0.001268962 | 0 |
| 0 | 0 | 0 | 0 | 0 |
| 0 | 0 | 0 | 0 | 0 |
| 0 | 0 | 0.0037313 | 0 | 0 |
| 0 | 0 | 0 | 0 | 0 |
| 0 | 0 | 0 | 0 | 0 |
| 0 | 0 | 0 | 0 | 0 |
| 0 | 0 | 0 | 0 | 0 |

| | | | | |
|---|---|---|---|---|
| 0 | 0 | 0 | 0 | 0 |
| 0.000543402 | 0 | 0 | 0.001745264 | 0 |
| 0 | 0 | 0 | 0.006878054 | 0 |
| 0 | 0.013093148 | 0 | 0.00115229 | 0 |
| 0 | 0 | 0 | 0 | 0.01160688 |
| 0 | 0 | 0 | 0 | 0 |
| 0.000484942 | 0 | 0 | 0.002076676 | 0.003036062 |
| 0 | 0 | 0.000899496 | 0 | 0 |
| 0 | 0 | 0 | 0 | 0 |
| 0 | 0 | 0 | 0 | 0 |
| 0 | 0.014837585 | 0.025469777 | 0 | 0 |
| 0 | 0 | 0 | 0 | 0 |
| 0.006258107 | 0.01100485 | 0 | 0.019720167 | 0.455522168 |
| 0 | 0 | 0 | 0 | 0 |
| 0 | 0 | 0 | 0 | 0 |
| 0 | 0 | 0 | 0 | 0 |
| 0 | 0 | 0 | 0 | 0 |
| 0 | 0 | 0 | 0 | 0 |
| 0 | 0 | 0 | 0 | 0 |
| 0 | 0 | 0 | 0 | 0 |
| 0 | 0 | 0 | 0 | 0 |
| 0 | 0 | 0 | 0 | 0 |
| 0 | 0.118371055 | 0.001717648 | 0 | 0 |
| 0.003119037 | 0.018335827 | 0.05421914 | 0.017224711 | 0.006432507 |
| 0 | 0 | 0 | 0 | 0 |
| 0 | 0 | 0 | 0.00334753 | 0.000678638 |
| 0.000445645 | 0 | 0 | 0 | 0.001488021 |
| 0 | 0 | 0 | 0 | 0 |
| 0 | 0 | 0 | 0 | 0 |
| 0 | 0 | 0 | 0 | 0 |
| 0.000866802 | 0 | 0.002393962 | 0.020387021 | 0.049274996 |
| 0 | 0 | 0 | 0 | 0 |
| 0 | 0 | 0 | 0 | 0 |
| 0 | 0 | 0 | 0 | 0 |
| 0 | 0 | 0 | 0.260689207 | 0 |
| 0 | 0.00101749 | 0 | 0 | 0 |
| 0 | 0.02295678 | 0 | 0 | 0 |
| 0 | 0 | 0 | 0 | 0 |
| 0 | 0 | 0 | 0.034497667 | 0.000918283 |
| 0 | 0 | 0 | 0 | 0 |
| 0 | 0 | 0 | 0 | 0 |
| 0 | 0 | 0 | 0 | 0 |
| 0 | 0 | 0 | 0 | 0 |
| 0 | 0 | 0 | 0 | 0 |
| 0 | 8.35268679 | 0 | 0 | 0 |
| 0.005936288 | 0 | 0.145060386 | 0.277123673 | 0.162233878 |
| 0 | 0 | 0 | 0 | 0 |
| 0 | 0 | 0 | 0 | 0 |

| | | | | |
|---|---|---|---|---|
| 0 | 0 | 0 | 0 | 0 |
| 0 | 0 | 0 | 0 | 0 |
| 0.032033166 | 0 | 0.44253393 | 0 | 0 |
| 0 | 0 | 0 | 0.525260127 | 0 |
| 0 | 0 | 0 | 0 | 0 |
| 0 | 0 | 0 | 0.00305269 | 0 |
| 0 | 0 | 0 | 0 | 0 |
| 0 | 0 | 0 | 0 | 0 |
| 0 | 0 | 0 | 0 | 0 |
| 0 | 0 | 0 | 0 | 0 |
| 0 | 0 | 0 | 0.001763877 | 0.001638942 |
| 0 | 0.001595916 | 0 | 0 | 0 |
| 0 | 0 | 0 | 0 | 0 |
| 0 | 0 | 0 | 0 | 0 |
| 0 | 0 | 0 | 0 | 0 |
| 0 | 0 | 0 | 0 | 0 |
| 0 | 0 | 0 | 0 | 0 |
| 0 | 0 | 0 | 0 | 0 |
| 0 | 0 | 0 | 0 | 0 |
| 0 | 0 | 0 | 0 | 0.001778334 |
| 0 | 0 | 0 | 0 | 0 |
| 0 | 0 | 0 | 0 | 0 |
| 0 | 0 | 0 | 0 | 0 |
| 0 | 0 | 0 | 0 | 0 |
| 0 | 0 | 0 | 0 | 0 |
| 0 | 0 | 0 | 0 | 0 |
| 0 | 0 | 0 | 0 | 0 |
| 0.007160143 | 0 | 0.728544111 | 0.032963094 | 0.008746787 |
| 0 | 0 | 0 | 0 | 0 |
| 0 | 0.001910677 | 0 | 0 | 0 |
| 0.001491277 | 0.01019951 | 0.014040873 | 0 | 0.053472047 |
| 0 | 0 | 0 | 0 | 0 |
| 0.005666806 | 0.011842669 | 0.024128267 | 0.011977974 | 0.012023098 |
| 0 | 0.001083754 | 0.001110652 | 0.001589635 | 0 |
| 0 | 0 | 0 | 0 | 0 |
| 0 | 0 | 0 | 0 | 0 |
| 0 | 0 | 0 | 0 | 0 |
| 0 | 0 | 0 | 0 | 0 |
| 0 | 0 | 0 | 0 | 0 |
| 0 | 0 | 0 | 0 | 0 |
| 0 | 0 | 0 | 0 | 0 |
| 0 | 0.013232835 | 0 | 0 | 0 |
| 0.09261013 | 0 | 0.049019649 | 0.120051581 | 0.096007987 |
| 0 | 0 | 0 | 0 | 0 |
| 0 | 0 | 0 | 0 | 0 |
| 0.012539258 | 0 | 0.011648718 | 0.009688005 | 0.069654577 |
| 0 | 0.021316188 | 0 | 0 | 0 |
| 0 | 0 | 0 | 0 | 0 |

| | | | | |
|---|---|---|---|---|
| 0 | 0 | 0 | 0 | 0 |
| 0 | 0 | 0.015292254 | 0.018783824 | 0.002483484 |
| 0 | 0.001083978 | 0.007998344 | 0.039303886 | 0.003867962 |
| 0 | 0 | 0 | 0 | 0 |
| 0.098813954 | 0.982693474 | 0.037253372 | 0 | 0 |
| 0 | 0.270434994 | 0 | 0 | 0 |
| 0 | 0 | 0 | 0 | 0 |
| 0 | 0 | 0 | 0 | 0 |
| 0 | 0 | 0 | 0 | 0 |
| 0 | 0 | 0 | 0 | 0 |
| 0 | 0 | 0 | 0 | 0 |
| 0 | 0 | 0 | 0 | 0 |
| 0 | 0 | 0 | 0 | 0 |
| 0 | 0 | 0 | 0 | 0 |
| 0 | 0 | 0 | 0 | 0 |
| 0 | 0 | 0 | 0 | 0 |
| 0 | 0 | 0 | 0 | 0 |
| 0 | 0 | 0 | 0 | 0 |
| 0 | 0 | 0 | 0 | 0 |
| 0 | 0 | 0 | 0 | 0 |
| 0 | 0.002587652 | 0 | 0 | 0 |
| 0 | 0 | 0 | 0 | 0 |
| 0 | 0 | 0 | 0.001294432 | 0 |
| 0 | 0 | 0 | 0 | 0 |
| 0.001272925 | 0 | 0 | 0 | 0 |
| 0 | 0 | 0 | 0.005536374 | 0.004208919 |
| 0 | 0 | 0 | 0 | 0 |
| 0.009937289 | 0 | 0 | 0.007211782 | 0 |
| 0 | 0 | 0 | 0 | 0 |
| 0 | 0 | 0 | 0 | 0 |
| 0 | 0 | 0 | 0 | 0 |
| 0 | 0.001891433 | 0 | 0 | 0 |
| 0 | 0 | 0 | 0 | 0 |
| 0 | 0 | 0 | 0 | 0 |
| 0 | 0 | 0 | 0 | 0 |
| 0 | 0 | 0 | 0 | 0 |
| 0 | 0.000547749 | 0 | 0 | 0 |
| 0 | 0.536741737 | 0 | 0 | 0 |
| 0 | 0 | 3.014899251 | 1.69123287 | 1.888483593 |
| 0 | 0 | 0 | 0 | 0 |
| 0 | 0 | 0 | 0 | 0 |
| 0.00138918 | 0.112897961 | 0 | 0.004804889 | 0.00108715 |
| 0 | 0.062594129 | 0 | 0 | 0 |
| 0 | 0 | 0 | 0 | 0 |
| 0 | 0 | 0 | 0 | 0 |
| 0 | 0 | 0 | 0.001643757 | 0 |
| 0.001474526 | 0.00496721 | 0 | 0.003157193 | 0 |
| 0 | 0 | 0 | 0 | 0 |

| | | | | |
|---|---|---|---|---|
| 0 | 0 | 0 | 0 | 0 |
| 0.000465145 | 0.118738191 | 0.00342574 | 0.012411061 | 0.054941975 |
| 0 | 0 | 0 | 0 | 0 |
| 0 | 0 | 0 | 0 | 0 |
| 0 | 0 | 0 | 0 | 0 |
| 0 | 0 | 0 | 0 | 0 |
| 0 | 0 | 0 | 0 | 0 |
| 0 | 0 | 0 | 0 | 0 |
| 0 | 0 | 0 | 0 | 0 |
| 0 | 0 | 0 | 0 | 0 |
| 0 | 0 | 0 | 0 | 0 |
| 0 | 0 | 0 | 0 | 0 |
| 0.009506191 | 0.518446917 | 0 | 0.028506766 | 0.604386175 |
| 0 | 0.002615586 | 0 | 0 | 0 |
| 0 | 0.036252377 | 0.005072265 | 0 | 0 |
| 0 | 0 | 0 | 0 | 0 |
| 0 | 0 | 0 | 0 | 0 |
| 0 | 0 | 0 | 0 | 0 |
| 0 | 0.000960587 | 0.004429926 | 0 | 0 |
| 0 | 0 | 0 | 0 | 0 |
| 0 | 0 | 0 | 0 | 0 |
| 0 | 0 | 0.002359594 | 0.000750489 | 0 |
| 0.000729449 | 0.008190945 | 0.029614862 | 0.065021713 | 0 |
| 0 | 0 | 0 | 0 | 0.140058488 |
| 0 | 0 | 0 | 0 | 0 |
| 0 | 0 | 0 | 0 | 0 |
| 0 | 0 | 0 | 0 | 0 |
| 0 | 0 | 0 | 0 | 0 |
| 0 | 0 | 0 | 0 | 0 |
| 0 | 0 | 0 | 0 | 0 |
| 0 | 0 | 0 | 0 | 0 |
| 0 | 0 | 0 | 0 | 0 |
| 0 | 0 | 0 | 0 | 0 |
| 0 | 0 | 0 | 0 | 0 |
| 0 | 0 | 0 | 4.592820495 | 0 |
| 0 | 0 | 0 | 0 | 0 |
| 0 | 0.037692447 | 0 | 0 | 0 |
| 0 | 0 | 0 | 0 | 0 |
| 0.002275147 | 0 | 0.037701441 | 0.005433541 | 0.005934975 |
| 0 | 0 | 0 | 0 | 0 |
| 0.078538845 | 0.020616039 | 0.152354231 | 0.492564678 | 0.041081902 |
| 0 | 0 | 0 | 0 | 0 |
| 0 | 0 | 0 | 0 | 0 |
| 0 | 0 | 0 | 0 | 0 |
| 0 | 0 | 0 | 0 | 0 |
| 0.030557784 | 0.410009547 | 0.227652752 | 0.003077506 | 0.454762397 |
| 0 | 0 | 0 | 0 | 0 |
| 0 | 0 | 0 | 0 | 0 |

| | | | | |
|---|---|---|---|---|
| 0 | 0 | 0 | 0 | 0 |
| 0 | 0 | 0 | 0 | 0 |
| 0 | 0 | 0 | 0 | 0 |
| 0 | 0 | 0 | 0 | 0 |
| 0 | 0.001823333 | 0 | 0 | 0 |
| 0 | 0 | 0 | 0 | 0 |
| 0 | 0 | 0 | 0 | 0 |
| 0 | 0 | 0 | 0 | 0 |
| 0 | 0 | 0 | 0 | 0 |
| 0 | 0 | 0 | 0.003197919 | 0.007865501 |
| 0 | 0 | 0 | 0.056144905 | 0 |
| 0 | 0 | 0 | 0 | 0 |
| 0 | 0.00543192 | 0.004881598 | 0 | 0 |
| 0 | 0 | 0 | 0 | 0 |
| 0 | 0 | 0 | 0 | 0 |
| 0 | 0 | 0 | 0 | 0 |
| 0 | 0 | 0 | 0 | 0 |
| 0 | 0 | 0 | 0 | 0 |
| 0 | 0 | 0 | 0 | 0 |
| 0.002152826 | 0.457073841 | 0.094445854 | 0.0103101 | 0.596977449 |
| 0 | 0 | 0 | 0.000585729 | 0 |
| 0 | 0 | 0 | 0 | 0 |
| 0 | 0 | 0 | 0 | 0 |
| 0 | 0 | 0 | 0 | 0 |
| 0 | 0 | 0 | 0 | 0 |
| 0 | 0 | 0 | 0 | 0 |
| 0 | 0 | 0 | 0 | 0 |
| 0 | 0 | 0 | 0 | 0 |
| 0 | 0 | 0 | 0 | 0 |
| 0 | 0 | 0.001466791 | 0 | 0.027265009 |
| 0 | 0 | 0 | 0 | 0 |
| 0 | 0 | 0 | 0 | 0 |
| 0 | 0 | 0 | 0 | 0 |
| 0 | 0 | 0 | 0 | 0 |
| 0 | 0 | 0 | 0 | 0 |
| 0.002358048 | 0 | 0 | 0 | 0 |
| 0 | 0 | 0 | 0 | 0 |
| 0 | 0 | 0 | 0 | 0 |
| 0 | 0 | 0 | 0 | 0 |
| 0.019474931 | 0 | 0 | 0 | 0 |
| 0 | 0 | 0 | 0 | 0 |
| 0 | 0 | 0 | 0 | 0 |
| 0 | 0 | 0 | 0.38905813 | 0 |
| 0 | 0 | 0 | 0 | 0 |
| 0.003421869 | 0.019735949 | 0 | 0.000461125 | 0.013048667 |
| 0.002183509 | 0 | 0 | 0.052285229 | 0 |
| 0 | 0 | 0 | 0 | 0 |
| 0 | 0 | 0 | 0 | 0 |

| | | | | |
|---|---|---|---|---|
| 0 | 0 | 0 | 0 | 0 |
| 0 | 0 | 0 | 0 | 0 |
| 0 | 0 | 0 | 0 | 0 |
| 0 | 0 | 0 | 0 | 0 |
| 0 | 0 | 0 | 0 | 0 |
| 0.01463065 | 0.022687218 | 0.000641387 | 0.011015927 | 0.024426039 |
| 0 | 0 | 0 | 0 | 0 |
| 0 | 0.000776557 | 0 | 0 | 0 |
| 0 | 0 | 0 | 0 | 0 |
| 0 | 0.217892683 | 0 | 0 | 0 |
| 0 | 0 | 0 | 0 | 0 |
| 0 | 0 | 0 | 0 | 0 |
| 0 | 0 | 0 | 0 | 0 |
| 0 | 0 | 0 | 0 | 0 |
| 0 | 0 | 0 | 0 | 0 |
| 0 | 0 | 0 | 0 | 0 |
| 0 | 0 | 0 | 0.094927597 | 0 |
| 0 | 0 | 0 | 0 | 0 |
| 0 | 0 | 0 | 0 | 0 |
| 0 | 0 | 0 | 0 | 0 |
| 0 | 0 | 0 | 0 | 0 |
| 0 | 0 | 0 | 0 | 0 |
| 0 | 0 | 0 | 0 | 0 |
| 0 | 0 | 0 | 0 | 0 |
| 0 | 0 | 0 | 0 | 0 |
| 0 | 0 | 0 | 0 | 0 |
| 0 | 0 | 0 | 0 | 0 |
| 0 | 0.002136954 | 0 | 0.488168559 | 0 |
| 0 | 0 | 0 | 0 | 0 |
| 0 | 0.103714303 | 0 | 0 | 0 |
| 0 | 0 | 0 | 0 | 0 |
| 0 | 0 | 0 | 0 | 0 |
| 0 | 0 | 0 | 0 | 0 |
| 0 | 0 | 0 | 0 | 0 |
| 0 | 0 | 0 | 0 | 0 |
| 0 | 0 | 0 | 0 | 0 |
| 0 | 0 | 0 | 0 | 0 |
| 0 | 0 | 0 | 0 | 0 |
| 0.192264695 | 1.520269237 | 1.018321265 | 0.401869514 | 0 |
| 0 | 0 | 0 | 0 | 0 |
| 0 | 0 | 0 | 0 | 0 |
| 0 | 0 | 0 | 0 | 0 |
| 0 | 0 | 0 | 0 | 0 |
| 0 | 0 | 0 | 0 | 0 |
| 0 | 0 | 0 | 0 | 0 |
| 0 | 0 | 0 | 0 | 0 |
| 0 | 0 | 0 | 0 | 0 |
| 0 | 0 | 0 | 0 | 0 |

| | | | | |
|---|---|---|---|---|
| 0 | 0 | 0 | 0 | 0 |
| 0.000520006 | 0.455889241 | 0 | 0.583772346 | 0 |
| 0 | 0 | 0 | 0 | 0 |
| 0 | 0 | 0 | 0 | 0 |
| 0 | 0 | 0 | 0 | 0 |
| 0 | 0 | 0 | 0 | 0 |
| 0 | 0 | 0 | 0 | 0 |
| 0 | 0 | 0 | 0 | 0 |
| 0 | 0 | 0 | 0 | 0 |
| 0 | 0 | 0 | 0 | 0 |
| 0 | 0 | 0 | 0 | 0.016625432 |
| 0 | 0 | 0 | 0 | 0 |
| 0.06895931 | 0.334514987 | 0 | 0 | 0.311474562 |
| 0 | 0 | 0 | 0 | 0 |
| 0 | 0 | 0 | 0 | 0 |
| 0 | 0 | 0 | 0 | 0 |
| 0 | 0 | 0 | 0 | 0 |
| 0 | 0 | 0 | 0 | 0 |
| 0 | 0 | 0 | 0 | 0 |
| 0 | 0 | 0 | 0 | 0 |
| 0 | 0 | 0 | 0 | 0.389395694 |
| 0 | 0 | 0 | 0 | 0 |
| 0 | 0 | 0 | 0 | 0 |
| 0.015027859 | 0.656666864 | 2.317330354 | 1.733175144 | 6.408853007 |
| 0 | 0 | 0 | 0 | 0 |
| 0 | 0 | 0 | 0 | 0 |
| 0 | 0 | 0 | 0 | 0 |
| 0 | 0 | 0 | 0 | 0 |
| 0 | 0 | 0 | 0 | 0 |
| 0 | 0 | 0 | 0 | 0 |
| 0 | 0 | 0 | 0 | 0 |
| 0 | 0 | 0 | 0 | 0 |
| 0 | 0 | 0 | 0 | 0 |
| 0 | 0 | 0 | 0 | 0 |

| PHL018-0015_V2 | PHL018-0026_V2 | PHL018-0157_V2 | PHL018-0247_V2 | PHL018-0261_V3 |
|---|---|---|---|---|
| CTRL | EYCF | REF | REF | REF |
| 0.040809999 | 0.109361482 | 0.217650375 | 0.137372403 | 0.539810938 |
| 8.191622527 | 9.450228697 | 1.26491574 | 0.343161952 | 1.111645246 |
| 13.36969236 | 0 | 20.53120153 | 0 | 0 |
| 0 | 0 | 0 | 0 | 0 |
| 0 | 0 | 0 | 0 | 0 |
| 0 | 0 | 0 | 0 | 0 |
| 0 | 0 | 0 | 0 | 0 |
| 0 | 0 | 0 | 0 | 0 |
| 0 | 0 | 0 | 0 | 0 |
| 0 | 0 | 0 | 0 | 0 |
| 0 | 0 | 0 | 0 | 0 |
| 0 | 0 | 0 | 0 | 0 |
| 0 | 0 | 0 | 0 | 0 |
| 7.157047121 | 9.539207183 | 3.817241244 | 1.322089808 | 3.891025089 |
| 0 | 0 | 0 | 0 | 0 |
| 0 | 0 | 0 | 0 | 0 |
| 0 | 0 | 0 | 0 | 0 |
| 0 | 0 | 0 | 0 | 0 |
| 0 | 0 | 0 | 0 | 0 |
| 0 | 0 | 0 | 0 | 0 |
| 0 | 0 | 0 | 0 | 0 |
| 0.001666349 | 0 | 0 | 0 | 0 |
| 0 | 0 | 0 | 0 | 0 |
| 0 | 0 | 0 | 0 | 0 |
| 0 | 0 | 0 | 0 | 0 |
| 0 | 0 | 0 | 0 | 0 |
| 0 | 0 | 0 | 0 | 0 |
| 0 | 0 | 0 | 0 | 0 |
| 0 | 0 | 0 | 0 | 0 |
| 0 | 0 | 0 | 0 | 0 |
| 0 | 0 | 0 | 0 | 0 |
| 0 | 0 | 0 | 0 | 0 |
| 0 | 0 | 0 | 0 | 0 |
| 0 | 0 | 0 | 0 | 0 |
| 0 | 0 | 0 | 0 | 0 |
| 0 | 0 | 0 | 1.79228641 | 0 |
| 0 | 0 | 0 | 0 | 0 |
| 0 | 0 | 0 | 0 | 0 |
| 0 | 0 | 0 | 0 | 0 |
| 0 | 0 | 0 | 0 | 0 |
| 0 | 0 | 0 | 0 | 0 |
| 0.033363824 | 0.029147282 | 0 | 0.05457482 | 0.001235944 |
| 0.008716637 | 0.414956052 | 0.046136823 | 0.02371367 | 0.006056768 |
| 0.002129368 | 0.01206458 | 0 | 0.006676546 | 0.049483805 |
| 0 | 0 | 0 | 0 | 0.007549204 |
| 0 | 0 | 0 | 0 | 0 |

| | | | | |
|---|---|---|---|---|
| 0 | 0 | 0 | 0 | 0 |
| 0.176260092 | 0.01252793 | 0.011797229 | 0.002861223 | 0.054282099 |
| 0.001741197 | 0.058698397 | 0.038267147 | 0.001867707 | 0 |
| 0.838559141 | 1.670294092 | 0 | 0.126894378 | 0.502824609 |
| 0 | 0 | 0 | 0 | 0 |
| 0 | 0 | 0 | 0 | 0 |
| 6.505295075 | 0.007992809 | 0 | 0 | 0 |
| 0 | 0 | 0 | 0 | 0 |
| 0 | 0 | 0 | 0 | 0 |
| 0 | 0 | 0 | 0 | 0 |
| 0 | 0.056694481 | 0 | 0 | 0 |
| 0 | 0.048961527 | 0 | 0 | 0 |
| 0 | 0 | 0 | 0 | 0 |
| 0.016488205 | 0.029307897 | 0.082546689 | 0.103227225 | 0 |
| 0 | 0.014764849 | 0 | 0.000634394 | 0.00251542 |
| 0 | 0 | 0 | 0 | 0 |
| 0 | 0 | 0 | 6.211860405 | 0.004569779 |
| 0 | 0 | 0 | 0 | 0 |
| 0 | 0 | 0 | 0 | 0 |
| 0 | 0 | 0 | 0 | 0 |
| 0.079940616 | 0.168154281 | 0.50053985 | 0.508385677 | 0.26981649 |
| 0.022132029 | 0.003180326 | 0.005356141 | 0.094042811 | 0 |
| 0 | 0 | 0 | 0 | 0 |
| 0 | 0 | 0 | 0 | 0 |
| 0 | 0 | 0 | 0 | 0 |
| 0 | 0 | 0 | 0 | 0 |
| 0 | 0 | 0 | 0 | 0.003356574 |
| 0 | 0 | 0 | 0 | 0 |
| 0 | 0.004068557 | 0 | 0.000586868 | 0 |
| 0 | 0 | 0 | 0 | 0 |
| 2.235579301 | 0.002316639 | 1.226349408 | 0 | 0 |
| 0 | 0 | 0 | 0 | 0 |
| 0 | 0 | 0 | 0 | 0 |
| 0 | 0 | 0 | 0 | 0 |
| 0 | 0.00183132 | 0 | 0 | 0.018662892 |
| 0 | 0 | 0 | 0 | 0 |
| 0 | 0 | 0 | 0 | 0 |
| 0.000583422 | 0.00385648 | 0 | 0.13420184 | 0 |
| 0 | 0 | 0 | 0 | 0 |
| 0 | 0 | 0 | 0 | 0 |
| 0 | 0 | 0 | 0 | 0 |
| 0 | 0 | 0 | 0 | 0 |
| 0 | 0 | 0 | 0 | 0 |
| 0 | 0 | 0 | 0 | 0 |
| 0 | 0 | 0 | 0 | 0 |
| 0 | 0 | 0 | 0 | 0 |
| 0.001387965 | 0 | 0 | 0 | 0.001022206 |
| 0.00178236 | 0 | 0 | 0 | 1.524987849 |

| | | | | |
|---|---|---|---|---|
| 0 | 0 | 0 | 0 | 0 |
| 0 | 0 | 0.091449826 | 0 | 0 |
| 0 | 0 | 0 | 0 | 0 |
| 0 | 0 | 0 | 0 | 0 |
| 0.025374572 | 0 | 0 | 0 | 0.085264887 |
| 0 | 0 | 0 | 0 | 0 |
| 0 | 0 | 0 | 0 | 0 |
| 0 | 0 | 0 | 0 | 0 |
| 0 | 0 | 0 | 0.019399336 | 0 |
| 0 | 0 | 0 | 0 | 0 |
| 0 | 0 | 0 | 0 | 0 |
| 0.132279149 | 0.001784447 | 0 | 0 | 0 |
| 0 | 0 | 0 | 0 | 0 |
| 0 | 0 | 0 | 0 | 0 |
| 0 | 0 | 0 | 0 | 0 |
| 0 | 0 | 0 | 0 | 0 |
| 0 | 0 | 0 | 0 | 0 |
| 0 | 0 | 0 | 0 | 0 |
| 0 | 0 | 0 | 0.002748857 | 0.034349677 |
| 0 | 0 | 0 | 0 | 0 |
| 0 | 0 | 0 | 0 | 0 |
| 0 | 0 | 0 | 0 | 0 |
| 0 | 0 | 0 | 0 | 0 |
| 0 | 0 | 0.14469894 | 0 | 0 |
| 0 | 0 | 0 | 0 | 0 |
| 0 | 0 | 0 | 0 | 0 |
| 0 | 0 | 0 | 0 | 0 |
| 0 | 0 | 0 | 0 | 0 |
| 0 | 0 | 0 | 0 | 0 |
| 0 | 0 | 0 | 0 | 0 |
| 0.024673086 | 0 | 0 | 0.015181908 | 0 |
| 0 | 0 | 0.00551231 | 0.002044701 | 0 |
| 0 | 0 | 0 | 0 | 0 |
| 0 | 0 | 0 | 0 | 0 |
| 0 | 0 | 0 | 0 | 0 |
| 0 | 0 | 0 | 0 | 0 |
| 0 | 0 | 0 | 0 | 0 |
| 0 | 0 | 0 | 0 | 0 |
| 0 | 0 | 0 | 0 | 0 |
| 0 | 0 | 0 | 0 | 0 |
| 0 | 0 | 0 | 0 | 0 |
| 0 | 0 | 0 | 0 | 0 |
| 0 | 0.00418847 | 0.002436108 | 0.001741416 | 0 |
| 0 | 0 | 0 | 0 | 0 |
| 0 | 0 | 0 | 0.000530455 | 0 |
| 0 | 0 | 0 | 0 | 0 |
| 0 | 0.001808135 | 0 | 0 | 0 |
| 0 | 0 | 0 | 0 | 0 |

| | | | | |
|---|---|---|---|---|
| 0 | 0 | 0 | 0 | 0.003135107 |
| 0 | 0 | 0 | 0 | 0 |
| 0 | 0 | 0 | 0 | 0 |
| 0 | 0 | 0 | 0 | 0 |
| 0 | 0 | 0 | 0 | 0 |
| 0 | 0 | 0 | 0 | 0 |
| 0 | 0 | 0 | 0 | 0 |
| 0 | 0 | 0 | 0 | 0 |
| 0 | 0 | 0 | 0 | 0 |
| 0 | 0 | 0 | 0 | 0 |
| 0 | 0 | 0 | 0 | 0 |
| 0 | 0 | 0 | 0 | 0 |
| 0 | 0 | 0 | 0 | 0 |
| 0 | 0 | 0 | 0 | 0 |
| 0 | 0 | 0 | 0 | 0 |
| 0 | 0 | 0 | 0 | 0 |
| 0 | 0 | 0 | 0 | 0 |
| 0 | 0 | 0 | 0 | 0 |
| 0 | 0 | 0 | 0 | 0 |
| 0 | 0 | 0 | 0 | 0 |
| 0.005044751 | 0 | 0 | 0 | 0 |
| 0 | 0 | 0 | 0 | 0 |
| 0 | 0.002171722 | 0 | 0 | 0 |
| 0 | 0 | 0 | 0 | 0 |
| 0 | 0 | 0 | 0 | 0 |
| 0 | 0 | 0 | 0 | 0 |
| 0 | 0 | 0 | 0 | 0 |
| 0 | 0 | 0 | 0 | 0 |
| 0 | 0 | 0 | 0 | 0 |
| 0 | 0 | 0 | 0 | 0 |
| 0 | 0 | 0 | 0 | 0.264391458 |
| 0.002951754 | 0.011559273 | 0 | 0.00409746 | 0.001611246 |
| 0 | 0 | 0 | 0 | 0 |
| 0 | 0 | 0 | 0 | 0 |
| 0 | 0 | 0 | 0 | 0 |
| 0 | 0 | 0 | 0 | 0 |
| 0 | 0 | 0 | 0 | 0 |
| 0 | 0 | 0 | 0 | 0 |
| 0 | 0 | 0 | 0 | 0 |
| 0 | 0 | 0 | 0 | 0 |
| 0 | 0 | 0 | 0 | 0 |
| 0 | 0 | 0 | 0 | 0 |
| 0 | 0 | 0 | 0 | 0 |
| 0 | 0 | 0 | 0 | 0 |
| 0 | 0 | 0 | 0 | 0 |
| 0 | 0 | 0 | 0 | 0 |
| 0 | 0 | 0 | 0 | 0 |
| 0 | 0 | 0 | 0 | 0 |

| | | | | |
|---|---|---|---|---|
| 0 | 0 | 0 | 0 | 0 |
| 0 | 0 | 0 | 0 | 0 |
| 0 | 0 | 0 | 0 | 0 |
| 0 | 0 | 0 | 0 | 0 |
| 0 | 0 | 0 | 0 | 0 |
| 0 | 0 | 0 | 0 | 0 |
| 0 | 0 | 0 | 0 | 0 |
| 0 | 0 | 0 | 0 | 0 |
| 0 | 0 | 0 | 0 | 0 |
| 0 | 0 | 0 | 0 | 0 |
| 0 | 0 | 0 | 0 | 0 |
| 0 | 0 | 0 | 0 | 0 |
| 0.000830747 | 0.007766298 | 0.001252982 | 0.000594071 | 0 |
| 0 | 0 | 0 | 0 | 0 |
| 0 | 0 | 0.002913551 | 0 | 0 |
| 0 | 0 | 0 | 0 | 0 |
| 0 | 0 | 0 | 0 | 0 |
| 0 | 0 | 0 | 0 | 0 |
| 0 | 0 | 0 | 0 | 0 |
| 0 | 0 | 0 | 0 | 0 |
| 0.066170439 | 0.065124982 | 0.027778617 | 0.050428943 | 0.03267177 |
| 0 | 0 | 0 | 0 | 0 |
| 0 | 0 | 0 | 0 | 0.003372641 |
| 0 | 0 | 0.001396025 | 0 | 0.003006184 |
| 0 | 0 | 0 | 0 | 0 |
| 0 | 0 | 0 | 0 | 0 |
| 0.004523017 | 0.021013749 | 0.000584733 | 0.007762633 | 0.000559626 |
| 0 | 0 | 0 | 0 | 0 |
| 0 | 0 | 0 | 0 | 0 |
| 0 | 0.004660963 | 0 | 0 | 0 |
| 0.083861445 | 0.079956764 | 0.005828795 | 0 | 0.001673555 |
| 0 | 0 | 0 | 0 | 0 |
| 0 | 0 | 0 | 0 | 0 |
| 0 | 0 | 0 | 0.004616067 | 0.031712226 |
| 0.01334068 | 0.001145237 | 0.033099829 | 0.030065399 | 0.098161919 |
| 0 | 0 | 0 | 0 | 0 |
| 0 | 0 | 0 | 0 | 0 |
| 0 | 0 | 0 | 0 | 0 |
| 0 | 0 | 0 | 0 | 0 |
| 0.008010624 | 0 | 0 | 0 | 0 |
| 0 | 0 | 0 | 0 | 0 |
| 0 | 0 | 0 | 0 | 0 |
| 0 | 0 | 0 | 0.033653427 | 0 |
| 0 | 0 | 0 | 0 | 0 |
| 0 | 0 | 0 | 0 | 0 |
| 0 | 0 | 0 | 0 | 0 |
| 0 | 0 | 0 | 0.002191056 | 0 |
| 0 | 0.006120953 | 0 | 0 | 0.019307577 |

| | | | | |
|---|---|---|---|---|
| 0 | 0 | 0 | 0 | 0 |
| 0 | 0 | 0 | 0 | 0 |
| 0 | 0 | 0.00368688 | 0 | 0 |
| 0 | 0 | 0 | 0 | 0 |
| 0 | 0 | 0 | 0 | 0 |
| 0 | 0 | 0 | 0 | 0 |
| 0 | 0 | 0 | 0.003079207 | 0 |
| 0.000621199 | 0 | 0 | 0.000592297 | 0 |
| 0 | 0 | 0 | 0 | 0 |
| 0 | 0 | 0 | 0 | 0 |
| 0 | 0 | 0 | 0 | 0 |
| 0 | 0 | 0 | 0 | 0 |
| 0 | 0 | 0 | 0 | 0 |
| 0 | 0 | 0 | 0 | 0 |
| 0 | 0.017371286 | 0 | 0 | 0.001404999 |
| 0 | 0 | 0 | 0 | 0 |
| 0 | 0 | 0 | 0 | 0 |
| 0 | 0 | 0 | 0 | 0 |
| 0 | 0 | 0 | 0 | 0 |
| 0 | 0 | 0 | 0 | 0 |
| 0.010804545 | 0 | 0 | 0 | 0.156186168 |
| 0 | 0 | 0 | 0 | 0 |
| 0 | 0 | 0 | 0 | 0 |
| 0 | 0 | 0 | 0 | 0 |
| 0.001287931 | 0 | 0 | 0.001535009 | 0.00398384 |
| 0 | 0 | 0 | 0 | 0 |
| 0 | 0 | 0 | 0 | 0 |
| 0 | 0 | 0 | 0 | 0 |
| 0 | 0 | 0 | 0.007617382 | 0 |
| 0 | 0 | 0 | 0 | 0 |
| 0.526725474 | 0.783215946 | 2.085295012 | 0.360415586 | 0 |
| 0 | 0 | 0 | 0 | 0 |
| 0 | 0 | 0 | 0 | 0 |
| 0 | 0 | 0 | 0 | 0 |
| 0 | 0 | 0 | 0 | 0 |
| 0 | 0 | 0 | 0 | 0 |
| 0 | 0 | 0 | 0 | 0 |
| 0 | 0 | 0 | 0 | 0 |
| 0 | 0 | 0 | 0 | 0 |
| 0 | 0 | 0 | 0 | 0.000654567 |
| 0 | 0 | 0 | 0 | 0 |
| 0.021541147 | 0.032576694 | 0 | 0.017488949 | 0.057279522 |
| 0.000951254 | 0.023483324 | 0 | 0 | 0 |
| 0 | 0 | 0 | 0 | 0 |
| 0 | 0 | 0 | 0 | 0 |
| 0 | 0 | 0 | 0 | 0 |
| 0 | 0 | 0 | 0 | 0 |
| 0 | 0 | 0 | 0 | 0 |

| | | | | |
|---|---|---|---|---|
| 0 | 0 | 0 | 0 | 0 |
| 0 | 0 | 0 | 0 | 0 |
| 0 | 0.010658449 | 0.000876393 | 0.003490371 | 0.043405948 |
| 0 | 0 | 0 | 0 | 0 |
| 0.161313415 | 0 | 0 | 0 | 27.11085793 |
| 0 | 0 | 0 | 0 | 0 |
| 0 | 0 | 0 | 0 | 0 |
| 0 | 0 | 0 | 0 | 0 |
| 0 | 0 | 0 | 0 | 0 |
| 0 | 0 | 0.001001928 | 0 | 0 |
| 0 | 0 | 0 | 0 | 0 |
| 0 | 0 | 0 | 0 | 0 |
| 0 | 0 | 0 | 0 | 0 |
| 0.005619065 | 0.005412208 | 0 | 0.002893118 | 0 |
| 0.013171359 | 0.007357525 | 0 | 0.010082203 | 0.005929554 |
| 0 | 0 | 0 | 0 | 0.004914868 |
| 0.001479621 | 0.001197606 | 0 | 0.000906929 | 0.001743534 |
| 0.010001968 | 0 | 0 | 0 | 0 |
| 0 | 0 | 0 | 0 | 0 |
| 0 | 0 | 0 | 0 | 0 |
| 0 | 0 | 0 | 0 | 0 |
| 0 | 0.005675755 | 0 | 0.000818698 | 0.012394572 |
| 0 | 0 | 0 | 0 | 0 |
| 0 | 0 | 0.00318674 | 0 | 0.031109052 |
| 0 | 0 | 0 | 0.005032149 | 0 |
| 0 | 0 | 0 | 0 | 0 |
| 0 | 0 | 0 | 0 | 0 |
| 0 | 0 | 0 | 0 | 0 |
| 0 | 0 | 0 | 0 | 0 |
| 0 | 0.003909539 | 0 | 0.003552763 | 0 |
| 0 | 0 | 0 | 0 | 0 |
| 0 | 0 | 0 | 0 | 0 |
| 0 | 0 | 0 | 0.081046199 | 0 |
| 0.01254024 | 0 | 0 | 0 | 0 |
| 0 | 0 | 0 | 0 | 0 |
| 0 | 0.018354356 | 0 | 0.033794812 | 0 |
| 0 | 0 | 0 | 0 | 0 |
| 0 | 0 | 0.001033089 | 0.001028609 | 0.114692668 |
| 0 | 0 | 0 | 0 | 0 |
| 0 | 0 | 0 | 0 | 0 |
| 0 | 0 | 0 | 0 | 0 |
| 0 | 0 | 0 | 0 | 0 |
| 0 | 0 | 0 | 0 | 0 |
| 0 | 0 | 0 | 0 | 0 |
| 0 | 0 | 0 | 0 | 0 |
| 0 | 0 | 0 | 0.001317892 | 0 |
| 0 | 0 | 0 | 0.001300804 | 0 |
| 0 | 0.004935673 | 0.006569481 | 0.010590183 | 0.020359198 |

| | | | | |
|---|---|---|---|---|
| 0 | 0 | 0 | 0 | 0 |
| 0 | 0 | 0 | 0.072171169 | 0 |
| 0 | 0 | 0 | 0 | 0 |
| 0 | 0 | 0 | 0 | 0 |
| 0.001407545 | 0.125034599 | 0 | 0.010353002 | 0.007878361 |
| 0 | 0 | 0 | 0.00304805 | 0.002704502 |
| 0 | 0 | 0 | 0 | 0 |
| 0.030440323 | 0.111785383 | 0 | 0.000806223 | 0.013368153 |
| 0 | 0 | 0 | 0 | 0 |
| 0.106909112 | 0.011521962 | 0.082626422 | 0.028461453 | 0 |
| 0.001050648 | 0 | 0 | 0 | 0 |
| 0.004908581 | 0.114404552 | 0.001201853 | 0 | 0 |
| 0.004738787 | 0.013664241 | 0.029902061 | 0 | 0.004013517 |
| 0 | 0 | 0.021876024 | 0 | 0.041343369 |
| 0 | 0 | 0.005438135 | 0 | 0 |
| 0.006859645 | 0 | 0 | 0 | 0 |
| 0 | 0.003140341 | 0 | 0 | 0 |
| 0 | 0 | 0.014960017 | 0 | 0 |
| 1.188058523 | 0.126231342 | 0.003534089 | 0.013488598 | 0 |
| 0.058724588 | 0 | 0 | 0 | 0 |
| 0 | 0 | 0 | 0 | 0 |
| 0 | 0 | 0 | 0 | 0 |
| 0 | 0.002473637 | 0 | 0 | 0 |
| 0 | 0 | 0 | 0 | 0 |
| 0 | 0 | 0 | 0 | 0 |
| 0 | 0.001675301 | 0 | 0.001127716 | 0 |
| 0 | 0 | 0 | 0 | 0 |
| 0.092013908 | 0.044232242 | 0.728573933 | 0 | 1.080713874 |
| 0.001614614 | 0 | 0.696657683 | 0.001010291 | 0 |
| 0 | 0.014076668 | 0.003714491 | 0 | 0 |
| 0 | 0 | 0.081125965 | 0.007563126 | 0 |
| 0 | 0 | 0 | 0 | 0 |
| 0 | 0 | 0 | 0 | 0 |
| 0.16035302 | 0 | 0.034161741 | 0.018988628 | 0 |
| 0 | 0 | 0 | 0 | 0 |
| 0 | 0 | 0 | 0 | 0 |
| 0.003798829 | 0 | 0.603536112 | 0.017098349 | 0.054375204 |
| 0 | 0 | 0.205343399 | 0.00201793 | 0 |
| 0 | 0 | 0 | 0 | 0 |
| 0 | 0 | 0 | 0 | 0 |
| 0 | 0 | 0 | 0 | 0 |
| 0 | 0 | 0 | 0 | 0 |
| 0 | 0 | 0.013833914 | 0 | 0 |
| 0.002223343 | 0 | 0.021386348 | 0 | 0 |
| 0 | 0 | 0 | 0 | 0 |
| 0 | 0 | 0 | 0 | 0 |
| 0.007109047 | 0.002430597 | 0 | 0.008765022 | 0 |
| 0 | 0 | 0.009348791 | 0 | 0 |

| | | | | |
|---|---|---|---|---|
| 0 | 0 | 0 | 0 | 0 |
| 0.06246302 | 0.009313241 | 0 | 0.000979553 | 0 |
| 0 | 0 | 0.057364218 | 0 | 0 |
| 0 | 0 | 0 | 0 | 0 |
| 0.004845896 | 0 | 0 | 0 | 0 |
| 0.01332232 | 0.049774127 | 0.069058474 | 0.033551091 | 0 |
| 0 | 0.002736895 | 0 | 0 | 0 |
| 0.001498328 | 0 | 0 | 0 | 0 |
| 0 | 0 | 0 | 0 | 0 |
| 0 | 0 | 0 | 0 | 0 |
| 0.012316531 | 0 | 0.004825072 | 0.021618676 | 0.001385368 |
| 0 | 0 | 0 | 0 | 0 |
| 0 | 0 | 0.013767059 | 0 | 0 |
| 0.001904079 | 0.00599341 | 0 | 0.002571941 | 0.001526963 |
| 0 | 0.692234557 | 0.631663858 | 0 | 0 |
| 0 | 0 | 0.117378682 | 0.00046285 | 0 |
| 0 | 0.010465669 | 0 | 0 | 0 |
| 0 | 0.004374417 | 0 | 0.01869824 | 0 |
| 0.000900711 | 0 | 0 | 0 | 0 |
| 0 | 0.060376777 | 0 | 0 | 0 |
| 0.493287944 | 0 | 0 | 0 | 0 |
| 0 | 0 | 0 | 0 | 0 |
| 0 | 0 | 0 | 0 | 0 |
| 0 | 0 | 0 | 0 | 0 |
| 0 | 0 | 0 | 0 | 0 |
| 0 | 0 | 0 | 0 | 0 |
| 0.118824938 | 0 | 0.032374066 | 0.050195971 | 0 |
| 0 | 0 | 0 | 0 | 0 |
| 0 | 0 | 0 | 0 | 0.010812415 |
| 0.168079095 | 0.546871452 | 1.183344209 | 0.151684118 | 0.803197108 |
| 0 | 0.014637788 | 0 | 0.000513193 | 0 |
| 0.009874043 | 0.001427153 | 0 | 0 | 0 |
| 0 | 0 | 0 | 0 | 0.001595229 |
| 0 | 0 | 0 | 0 | 0 |
| 0 | 0.073340607 | 0 | 0 | 0 |
| 0 | 0 | 0 | 0 | 0 |
| 0 | 0 | 0 | 0 | 0 |
| 0.007547415 | 0 | 0 | 0 | 0 |
| 0 | 0 | 0 | 0 | 0 |
| 0 | 2.562244855 | 2.659170715 | 0 | 0 |
| 0.028524343 | 0.062822247 | 0.075513563 | 0 | 0 |
| 0 | 2.691530792 | 10.67174456 | 0.538485946 | 0 |
| 0 | 0 | 0 | 0 | 0 |
| 0 | 0 | 0 | 0 | 0 |
| 0 | 0 | 0.001902355 | 0.002946387 | 0 |
| 0 | 0 | 0 | 0 | 0 |
| 0.000730204 | 0.00181002 | 0.014946711 | 0.000913801 | 0.002635118 |
| 0 | 0 | 0 | 0 | 0 |

| | | | | |
|---|---|---|---|---|
| 0 | 0 | 0 | 0 | 0.002786823 |
| 0 | 0 | 0.017520536 | 0 | 0 |
| 0.077032623 | 0 | 0.001983811 | 14.17146663 | 0.005379451 |
| 0 | 0.03030201 | 0 | 0.000874181 | 0 |
| 0 | 0 | 0 | 0 | 0 |
| 0.142066906 | 0 | 0 | 0.000544733 | 0.009817755 |
| 0 | 0 | 0 | 0.000457799 | 0 |
| 0 | 0.002503033 | 0 | 0 | 0 |
| 0.05202691 | 0 | 0.05381619 | 0 | 0 |
| 0 | 0 | 0 | 0 | 0 |
| 0 | 0 | 0 | 0 | 0 |
| 0 | 0 | 0 | 0 | 0 |
| 0 | 0.004707815 | 0 | 0 | 0.04769152 |
| 0 | 0.008658751 | 0 | 0.026082855 | 0.001890876 |
| 0 | 0 | 0 | 0 | 0.020217673 |
| 0.014809311 | 0.299086567 | 0.085262174 | 0.006376899 | 0 |
| 0 | 0.003750071 | 0.001521195 | 0.072574522 | 0.273704959 |
| 0 | 0 | 0 | 0 | 0.007959754 |
| 0.00273468 | 0 | 0 | 0 | 0 |
| 0 | 0 | 0 | 0 | 0 |
| 0 | 0.297747858 | 0 | 0.001431619 | 0 |
| 0 | 0.004929872 | 0.002205634 | 0 | 0 |
| 0 | 0 | 0 | 0 | 0 |
| 0.471110393 | 0 | 0 | 0.157161109 | 0 |
| 0 | 0 | 0 | 0 | 0 |
| 0 | 0 | 0 | 0 | 0.00527412 |
| 0.005153592 | 0.004125979 | 0.007724676 | 0.052556404 | 0 |
| 0.005955626 | 0 | 0 | 0 | 0 |
| 0 | 0 | 0 | 0 | 0 |
| 0 | 0 | 0 | 0 | 0 |
| 0.068332007 | 0.100650827 | 0.033411757 | 0 | 0.003162572 |
| 0.001270257 | 0.005757622 | 0.007116119 | 0.00163506 | 0.005238896 |
| 0.027575613 | 0.033234837 | 0.099547832 | 0.111743543 | 0 |
| 0.043331735 | 0.020131745 | 0 | 0.017104672 | 0 |
| 0.005938144 | 0.004205545 | 0 | 0 | 0.008163522 |
| 0 | 0 | 0 | 0 | 0 |
| 0 | 0 | 0 | 0 | 0 |
| 0.01744434 | 0.007751858 | 0.024504967 | 0.013086025 | 0 |
| 0 | 0 | 0 | 0.177344016 | 0 |
| 0.006711779 | 0.007526299 | 0 | 0.000299977 | 0 |
| 0 | 0 | 0 | 0 | 0 |
| 0 | 0.023030147 | 0 | 0.044354561 | 0 |
| 0.001023058 | 0.003622779 | 0 | 0 | 0 |
| 0.078233627 | 0 | 0 | 0 | 0 |
| 1.12365772 | 3.816745586 | 0.282864717 | 0 | 0 |
| 0 | 0 | 0 | 0 | 0 |
| 0 | 0 | 0 | 0 | 0 |
| 0 | 0 | 0 | 0 | 0 |

| | | | | |
|---|---|---|---|---|
| 0 | 0.031247772 | 0.007375818 | 0 | 0 |
| 0.348861556 | 0.656879755 | 0.14561544 | 0.088156955 | 0.011293206 |
| 0.007272965 | 0 | 0 | 0 | 0 |
| 0.002117072 | 0 | 0 | 0 | 0 |
| 0.002286236 | 0 | 0.004433449 | 0.050518355 | 0 |
| 0 | 0 | 0 | 0 | 0.009099543 |
| 0.006467783 | 0.018933346 | 0 | 0 | 0 |
| 0 | 0 | 0 | 0 | 0 |
| 0.002559952 | 0.001510853 | 0 | 0 | 0 |
| 0.00541907 | 0.015351692 | 0 | 0.024220013 | 0 |
| 0.008312378 | 0.003679398 | 0.003544751 | 0.009907033 | 0 |
| 0.007222555 | 0.025575983 | 0.003056846 | 0.002582441 | 0.007180996 |
| 0 | 0 | 0 | 0 | 0 |
| 0 | 0 | 0 | 0 | 0 |
| 0 | 0.00168866 | 0 | 0.001705062 | 0 |
| 0 | 0 | 0 | 0 | 0 |
| 0.011243002 | 0.00102743 | 0.011721682 | 0.008947656 | 0.005235241 |
| 3.002561796 | 0.064686657 | 0.534601555 | 0.027270688 | 0.004126183 |
| 0 | 0 | 0 | 0 | 0 |
| 0.011005793 | 0.124496791 | 0 | 0 | 0 |
| 0.009724311 | 0.009535852 | 0.002619069 | 0.011901867 | 0.002892243 |
| 0 | 0 | 0 | 0 | 0 |
| 0 | 0 | 0 | 0 | 0 |
| 0 | 0 | 0 | 0 | 0 |
| 0.027021624 | 0 | 0 | 0 | 0 |
| 0 | 0 | 0 | 0 | 0.00424363 |
| 0.010521302 | 0 | 0 | 0.114309449 | 0 |
| 0.224002242 | 0.101352027 | 0.070662746 | 0.219881332 | 0 |
| 0 | 0 | 0 | 0 | 0 |
| 0 | 0.003735547 | 0 | 0 | 0 |
| 0.008530775 | 0 | 0 | 0 | 0 |
| 0 | 0 | 0 | 0 | 0 |
| 0 | 0 | 0 | 0 | 0 |
| 0 | 0 | 0.005676188 | 0 | 0 |
| 0 | 0.187682626 | 0 | 0.050658701 | 0 |
| 0.132873665 | 0.212985888 | 0.290283569 | 0.207116436 | 0.421064466 |
| 0 | 0 | 0 | 0 | 0 |
| 0 | 0 | 0 | 0 | 0 |
| 0.011033657 | 0.04827511 | 0.001584916 | 0.011396984 | 0 |
| 0 | 0 | 0 | 0 | 0 |
| 1.101941701 | 2.094785463 | 0.245320667 | 0 | 0 |
| 0 | 0 | 0 | 0 | 0 |
| 0.023680443 | 0.035018881 | 0.002888107 | 0 | 0.005528194 |
| 0.001581718 | 0.002240426 | 0 | 0.004935684 | 0.000741301 |
| 0 | 0 | 0 | 0 | 0 |
| 0 | 0 | 0 | 0.03718443 | 0 |
| 0 | 0 | 0 | 0 | 0 |
| 0 | 0.003292251 | 0.049830195 | 0.004155287 | 0.022527206 |

| | | | | |
|---|---|---|---|---|
| 0.01622895 | 0 | 0.014345772 | 0 | 0.013515262 |
| 0 | 0 | 0 | 0 | 0 |
| 0 | 0 | 0 | 0 | 0 |
| 0.069210746 | 0 | 0 | 0.024684442 | 0.111065794 |
| 0 | 0 | 0 | 0 | 0 |
| 0 | 0 | 0 | 0 | 0 |
| 0 | 0 | 0 | 0 | 0 |
| 0 | 0 | 0 | 0 | 0 |
| 0 | 8.972069116 | 0.139648394 | 0 | 0 |
| 0 | 0.000658782 | 0 | 0 | 0 |
| 0 | 0 | 0 | 0 | 0 |
| 0 | 0 | 0.003222816 | 0.003446533 | 0 |
| 0 | 0 | 0 | 0 | 0 |
| 0 | 0 | 0 | 0 | 0 |
| 0.015085792 | 0.000582771 | 0 | 0.035207848 | 0 |
| 0 | 0 | 0 | 0 | 0 |
| 0 | 0 | 0 | 0 | 0 |
| 0 | 0.002536705 | 0 | 0 | 0 |
| 0 | 0 | 0 | 0 | 0 |
| 0 | 0 | 0 | 0 | 0 |
| 0 | 0 | 0 | 0 | 0 |
| 0 | 0 | 0 | 0 | 0 |
| 0 | 0 | 0 | 0 | 0 |
| 0.414015199 | 1.445684364 | 0.641150828 | 0.309762118 | 0.001498298 |
| 0.000933599 | 0.024905153 | 0 | 0 | 0 |
| 0.621242223 | 0 | 0 | 0 | 0 |
| 0 | 0 | 0 | 0 | 0 |
| 0.557333137 | 0.002580907 | 0 | 0 | 0 |
| 0 | 0 | 0 | 0 | 0 |
| 0 | 0 | 0 | 0 | 0 |
| 0 | 0 | 0 | 0 | 0.049643417 |
| 0 | 0 | 0 | 0 | 0 |
| 0.002336348 | 0.065141381 | 0 | 0 | 0.022948301 |
| 0 | 0 | 0 | 0 | 0 |
| 0 | 0 | 0 | 0 | 0 |
| 0.005616681 | 0 | 0 | 0.033699537 | 0 |
| 0 | 0.001002429 | 0 | 0 | 0 |
| 0.015562813 | 0 | 0 | 0 | 0 |
| 0 | 0 | 0 | 0 | 0 |
| 0 | 0 | 0 | 0 | 0 |
| 0.054366743 | 0.228539323 | 0.006183432 | 0.491047385 | 0.072823366 |
| 0 | 0 | 0 | 0 | 0 |
| 9.101478709 | 4.999915905 | 10.33606548 | 5.269229884 | 0.111222031 |
| 0 | 0 | 0 | 0.039727555 | 0 |
| 0 | 0 | 0 | 0 | 0 |
| 0 | 0 | 0 | 0 | 0 |
| 0 | 0 | 0 | 0 | 0 |
| 0.009007882 | 0.167227251 | 0 | 0.001233494 | 0.006718802 |

| | | | | |
|---|---|---|---|---|
| 0 | 0 | 0 | 0 | 0 |
| 0 | 0.312611747 | 0 | 0 | 0 |
| 0.082492499 | 2.648175134 | 0.559554053 | 0.025356834 | 0.05891374 |
| 0 | 0 | 0 | 0 | 0 |
| 0 | 0 | 0 | 0 | 0 |
| 0 | 0 | 0 | 0 | 0 |
| 0 | 0 | 0 | 0.019081346 | 0 |
| 0 | 0 | 0 | 0 | 0 |
| 0 | 0 | 0 | 0 | 0 |
| 0 | 0 | 0 | 0 | 0 |
| 0.016054293 | 0.279269058 | 0.188309863 | 0.046953952 | 0.628087744 |
| 3.783795466 | 1.630203333 | 3.415537974 | 3.07697287 | 0 |
| 0 | 0 | 0 | 0 | 0 |
| 0.002141471 | 0.00716959 | 0.001887601 | 0.006125503 | 0.021879334 |
| 0.004909269 | 0.390278551 | 1.757565988 | 0.018321158 | 0.454612859 |
| 0 | 0 | 0 | 0 | 0 |
| 0 | 0 | 0 | 0 | 0.028397196 |
| 0 | 0 | 0 | 0 | 0 |
| 0 | 0 | 0 | 0 | 0 |
| 0.015750542 | 0.015785284 | 0.022571316 | 0 | 0 |
| 0.297692036 | 0.297886017 | 0.070853103 | 0.245423956 | 0.014341594 |
| 1.345186453 | 0 | 1.596977353 | 0.080770001 | 0.352494273 |
| 5.683238131 | 1.375046084 | 2.752898241 | 1.395142222 | 0 |
| 0 | 0 | 0 | 0 | 0 |
| 0 | 0 | 0 | 0 | 0 |
| 0 | 0 | 0 | 0.003238893 | 0 |
| 0 | 0 | 0 | 0 | 0 |
| 0 | 0 | 0 | 0.001578686 | 0 |
| 0 | 0 | 0 | 0 | 0 |
| 0 | 0 | 0 | 0 | 0 |
| 0.00083379 | 0 | 0 | 0 | 0 |
| 0.235654552 | 0.397478608 | 0 | 0 | 0.213689638 |
| 0.070028436 | 0 | 0 | 0.018944723 | 0.345548717 |
| 0 | 0 | 0 | 0.015659139 | 0 |
| 0.005746886 | 0.035851437 | 0 | 0.003235237 | 0 |
| 0.011591809 | 0.117076231 | 0 | 0.011172611 | 0 |
| 0.001947396 | 0 | 0 | 0.001044444 | 0 |
| 0 | 0 | 0 | 0 | 0 |
| 0.018086344 | 0.014479974 | 0 | 0 | 0 |
| 0 | 0 | 0 | 0 | 0 |
| 0 | 0 | 0 | 0 | 0 |
| 0 | 0 | 0 | 0 | 0 |
| 0 | 0 | 0 | 0 | 0 |
| 0 | 0 | 0 | 0 | 0 |
| 0.00536429 | 0.119401112 | 0.001238379 | 0 | 0.969892913 |
| 0 | 0 | 0 | 0 | 0 |
| 0 | 0 | 0 | 0 | 0 |
| 0.009706199 | 0.353856431 | 0.026390881 | 0 | 0.029785028 |

| | | | | |
|---|---|---|---|---|
| 0 | 0 | 0 | 0 | 0 |
| 0 | 0 | 0 | 0 | 0 |
| 0 | 0 | 0 | 0 | 0 |
| 0 | 0 | 0 | 0.005020913 | 0 |
| 0 | 0 | 0 | 0 | 0 |
| 0 | 0.001021344 | 0 | 0 | 0 |
| 0 | 0 | 0 | 0 | 0 |
| 0.0190819 | 0 | 0 | 0 | 0 |
| 0 | 0 | 0 | 0 | 0 |
| 0 | 0 | 0 | 0 | 0 |
| 0.000727422 | 0.010045988 | 0.006073463 | 0 | 0.153004718 |
| 0 | 0 | 0 | 0 | 0 |
| 0 | 0 | 0 | 0 | 0 |
| 0 | 0 | 0 | 0.417142887 | 0 |
| 0 | 0 | 0 | 0 | 0 |
| 0 | 0 | 0 | 0 | 0 |
| 0 | 0.003831575 | 0 | 0.241219181 | 0 |
| 0.004541718 | 0.00964968 | 0.00375124 | 0.002489983 | 0 |
| 0 | 0 | 0 | 0.068009261 | 0 |
| 0 | 0 | 0 | 0 | 0 |
| 0 | 0 | 0 | 0 | 0 |
| 0 | 0 | 0 | 0 | 0 |
| 0 | 0 | 0 | 0.018972058 | 0.023035589 |
| 0 | 0 | 0 | 0 | 0 |
| 0 | 0 | 0 | 0 | 0 |
| 0 | 0 | 0 | 0 | 0 |
| 0 | 0 | 0 | 0 | 0 |
| 0 | 0 | 0 | 0 | 0 |
| 0 | 0 | 0 | 0 | 0 |
| 0.060349903 | 0 | 0 | 0.03387131 | 0 |
| 0.005013103 | 0 | 0.006017987 | 0 | 0 |
| 0 | 0 | 0 | 0 | 0 |
| 0 | 0 | 0 | 0 | 0 |
| 0 | 0 | 0 | 0 | 0 |
| 0 | 0 | 0 | 0 | 0 |
| 0 | 0 | 0 | 0 | 0 |
| 0 | 0 | 0 | 0 | 0 |
| 0.006465644 | 0.029219232 | 0.0179116 | 0.020255825 | 0 |
| 0 | 0 | 0 | 0 | 0 |
| 0.001822101 | 0.019356875 | 0 | 0.002605986 | 0 |
| 0 | 0 | 0 | 0 | 0 |
| 0 | 0 | 0 | 0 | 0 |
| 0 | 0 | 0 | 0 | 0 |
| 0 | 0 | 0 | 0 | 0 |
| 0 | 0.249083618 | 0 | 0 | 0 |
| 0 | 0 | 0 | 0 | 0 |
| 0 | 0 | 0 | 0 | 0 |
| 0 | 0 | 0 | 0 | 0 |

| | | | | |
|---|---|---|---|---|
| 0 | 0 | 0 | 0 | 0 |
| 0 | 0 | 0 | 0 | 0 |
| 0 | 0 | 0 | 0 | 0 |
| 0.010259648 | 0 | 0 | 0 | 0 |
| 0 | 0 | 0 | 0 | 0 |
| 0 | 0 | 0 | 0 | 0 |
| 0.01684333 | 0 | 0 | 0 | 0 |
| 0 | 0 | 0 | 0 | 0 |
| 0.154140229 | 0.337722987 | 0.17406528 | 0.038108574 | 0.001313466 |
| 0 | 0 | 0 | 0 | 0 |
| 0 | 0 | 0 | 0 | 0 |
| 0 | 0 | 0 | 0 | 0 |
| 0 | 0 | 0 | 0 | 0 |
| 0 | 0 | 0 | 0 | 0 |
| 2.430209912 | 0.058567046 | 0 | 0.521530885 | 0 |
| 0.02732731 | 0 | 2.029101869 | 0 | 0.052472153 |
| 0 | 0 | 0 | 0 | 0.005475649 |
| 0 | 0 | 0 | 0 | 0 |
| 0 | 0 | 0 | 0 | 0 |
| 0 | 0 | 0 | 0 | 0 |
| 0 | 0 | 0 | 0.040284446 | 0 |
| 0 | 0 | 0 | 0 | 0 |
| 0 | 0 | 0 | 0 | 0 |
| 0 | 0 | 0.093611713 | 0 | 0 |
| 0 | 0.013438824 | 0.04330621 | 0.001785442 | 0 |
| 2.860197211 | 0.003333329 | 0 | 0 | 15.99807289 |
| 0 | 0 | 0 | 0.005976708 | 0.204138659 |
| 0 | 0 | 0 | 0 | 0 |
| 0 | 0 | 0.017167247 | 0.0120196 | 0 |
| 0 | 0 | 0 | 0 | 0 |
| 0 | 0 | 0 | 0 | 0 |
| 0 | 0 | 0 | 0 | 0 |
| 0 | 0 | 0 | 0 | 0 |
| 0.006267113 | 0.01492101 | 0 | 0 | 0.000687429 |
| 0.003075896 | 0.003423243 | 0 | 0.002513814 | 0.003171464 |
| 0.002018979 | 0.001539884 | 0.007027259 | 0 | 0.049160355 |
| 0.056095064 | 0.525316987 | 0 | 0 | 0 |
| 0 | 0.230017801 | 12.35118975 | 0.536110961 | 0 |
| 0 | 0 | 0 | 0 | 0 |
| 0 | 0 | 0.013582877 | 0 | 0 |
| 0 | 0 | 0 | 0 | 0 |
| 0 | 0 | 0 | 0 | 0 |
| 2.399031737 | 0 | 0.225438912 | 0.140383211 | 6.479990044 |
| 0 | 0 | 0 | 0 | 0 |
| 0 | 0 | 0 | 0 | 0 |
| 0 | 0.017297758 | 0 | 0.002946035 | 0 |
| 0 | 0 | 0 | 0 | 0 |
| 0.038441695 | 0.004316219 | 0.017171796 | 0.203491822 | 0 |

| | | | | |
|---|---|---|---|---|
| 0.000699171 | 0 | 0 | 0 | 0 |
| 0 | 0 | 0 | 0 | 0 |
| 0 | 0 | 0 | 0 | 0 |
| 0 | 0 | 0 | 0 | 0 |
| 0 | 0.006655497 | 0.003562189 | 0.000995577 | 0.011663173 |
| 0 | 0 | 0 | 0 | 0 |
| 0 | 0 | 0 | 0 | 0 |
| 0 | 0 | 0 | 0.00288983 | 0 |
| 0 | 0 | 0 | 0 | 0 |
| 0 | 0 | 0 | 0 | 0 |
| 0 | 0 | 0 | 0 | 0 |
| 0.176284456 | 0.816181271 | 0 | 1.08208813 | 0 |
| 0.92883401 | 0 | 0 | 0.063889866 | 0.977898148 |
| 0 | 0 | 0 | 0 | 0 |
| 0.05595072 | 0.381148119 | 0.001300949 | 0.285831261 | 0.001037574 |
| 0 | 0 | 0 | 0 | 0 |
| 0 | 0 | 0 | 0 | 0 |
| 0 | 0 | 0 | 0 | 0 |
| 0.000529215 | 0.037480362 | 0 | 0 | 0 |
| 0 | 0 | 0 | 0 | 0 |
| 0 | 0 | 0 | 0 | 0 |
| 0 | 0 | 0 | 0 | 0 |
| 0.030703861 | 0.004616311 | 0 | 0.089665822 | 0.018393373 |
| 0 | 0 | 0 | 0 | 0 |
| 0 | 0 | 0 | 0 | 0 |
| 0 | 0 | 0 | 0 | 0 |
| 0 | 0 | 0 | 0 | 0 |
| 0 | 0 | 0 | 0 | 0 |
| 0 | 0 | 0 | 0 | 0 |
| 0 | 0 | 0 | 0 | 0 |
| 0 | 0 | 0 | 0 | 0 |
| 0.047793204 | 0.055119188 | 0.018288435 | 0.013073224 | 0.041738308 |
| 0 | 0 | 0 | 0 | 0 |
| 0.006393204 | 0.090371783 | 0 | 0 | 0 |
| 0.00169012 | 0.002633368 | 0.004734116 | 0.000846029 | 0.043391523 |
| 0.002178769 | 0.030552581 | 0 | 0.004206729 | 0 |
| 0 | 0 | 0 | 0 | 0 |
| 0 | 0 | 0 | 0 | 0 |
| 0 | 0 | 0 | 0 | 0 |
| 0 | 0 | 0 | 0 | 0 |
| 0 | 0 | 0 | 0 | 0 |
| 0 | 0 | 0 | 0 | 0 |
| 0 | 0 | 0 | 0 | 0 |
| 0 | 0 | 0 | 0.021843063 | 0 |
| 0.004328837 | 0.038676148 | 0.035873685 | 0.014882555 | 0.838248314 |
| 0 | 0 | 0 | 0 | 0 |
| 0 | 0 | 0 | 0 | 0 |
| 0.001732206 | 0.02765859 | 0.002205456 | 0 | 0 |

| | | | | |
|---|---|---|---|---|
| 0 | 0 | 0 | 0.17005245 | 0 |
| 0 | 0 | 0 | 0 | 0 |
| 0 | 0 | 0.016696066 | 0 | 0 |
| 0 | 0 | 0 | 0 | 0 |
| 0 | 0 | 0 | 0 | 0 |
| 0 | 0 | 0 | 0 | 0 |
| 0 | 2.216101701 | 0 | 0 | 0 |
| 0 | 0 | 0 | 0.048377722 | 0.031858416 |
| 0 | 0 | 0 | 0 | 0 |
| 0 | 0 | 0.000862011 | 0 | 0 |
| 0 | 0 | 0 | 0 | 0 |
| 0 | 0 | 0 | 0 | 0 |
| 0 | 0 | 0 | 0 | 0 |
| 0 | 0 | 0 | 0 | 0 |
| 0 | 0 | 0 | 0 | 0 |
| 0 | 0 | 0 | 0 | 0 |
| 0 | 0 | 0 | 0 | 0 |
| 0 | 0 | 0 | 0 | 0 |
| 0 | 0.042220937 | 0 | 0 | 0 |
| 0 | 0 | 0 | 0 | 0 |
| 0 | 0 | 0 | 0.001006939 | 0.007420568 |
| 0 | 0 | 0 | 0 | 0 |
| 0 | 0 | 0 | 0 | 0 |
| 0 | 0 | 0 | 0 | 0 |
| 0 | 0 | 0 | 0 | 0 |
| 0.018909923 | 0.039957913 | 0.027385577 | 0.000997567 | 0.001438337 |
| 0 | 0 | 0 | 0 | 0 |
| 0.04187294 | 0.009240475 | 0 | 0.041660569 | 0.005005673 |
| 0 | 0 | 0 | 0 | 0 |
| 0 | 0.001200012 | 0 | 0.567868651 | 0 |
| 0 | 0 | 0 | 0 | 0 |
| 0 | 0 | 0 | 0 | 0 |
| 0 | 0 | 0 | 0 | 0 |
| 0 | 0 | 0 | 0 | 0 |
| 0 | 0.002990959 | 0.002099882 | 0.004646171 | 0.011890826 |
| 0 | 0 | 0 | 0 | 0 |
| 0 | 0 | 0 | 0 | 0 |
| 0 | 0.01984104 | 0.003375564 | 0 | 0 |
| 0 | 0 | 0 | 0 | 0 |
| 0 | 0 | 0 | 0 | 0 |
| 2.562349269 | 14.80937358 | 5.420906798 | 1.77345993 | 4.254335647 |
| 0.117781575 | 0.199319097 | 0.382267154 | 0.134974095 | 0.09055912 |
| 0 | 0.106093869 | 0 | 0.075190908 | 0 |
| 0 | 0 | 0 | 0 | 0 |
| 0 | 0 | 0 | 0 | 0 |
| 0 | 0 | 0 | 0 | 0 |
| 0 | 0 | 0 | 0 | 0 |
| 0.001609066 | 0.01616134 | 0.000787951 | 0 | 0.055050622 |

| | | | | |
|---|---|---|---|---|
| 0.059942869 | 0 | 0 | 0 | 0 |
| 0.006319311 | 0.035164615 | 0 | 0.020980925 | 0 |
| 0 | 0 | 0 | 0 | 0 |
| 0 | 0 | 0 | 0 | 0 |
| 1.579518205 | 0.001163649 | 0.138510629 | 2.187467413 | 4.149553712 |
| 0 | 0 | 0 | 0 | 0 |
| 0 | 0 | 0 | 0 | 0 |
| 0 | 0 | 0 | 0 | 0 |
| 3.083033112 | 0.79227179 | 1.158724395 | 0.203483518 | 0.049879676 |
| 0 | 0 | 0 | 0 | 0 |
| 0 | 0 | 0 | 0 | 0 |
| 0 | 0 | 0 | 0 | 0 |
| 0 | 0 | 0 | 0 | 0 |
| 0.008447533 | 0.047352887 | 0.013554334 | 0.002184996 | 0.025759351 |
| 0 | 0.002950994 | 0 | 0 | 0 |
| 0.267692587 | 0 | 0.024787483 | 0.005800855 | 0 |
| 0.001399793 | 0.007270038 | 0 | 0.016683306 | 0 |
| 0 | 0 | 0 | 0 | 0 |
| 0 | 0 | 0 | 0 | 0 |
| 0 | 0 | 0 | 0 | 0 |
| 0 | 0 | 0 | 0 | 0 |
| 0 | 0 | 0 | 0 | 0 |
| 0 | 0 | 0 | 0 | 0 |
| 0 | 0 | 0 | 0 | 0 |
| 0 | 0 | 0 | 0 | 0 |
| 0 | 0 | 0 | 0 | 0 |
| 0 | 0 | 0 | 0.010154774 | 0 |
| 0.001068238 | 0.003458529 | 0.004274559 | 0 | 0.028165082 |
| 0 | 0 | 0 | 0 | 0 |
| 0 | 0 | 0 | 0 | 0 |
| 0 | 0 | 0 | 0 | 0 |
| 0 | 0 | 0 | 0.005287054 | 0 |
| 0 | 0 | 0 | 0 | 0 |
| 0 | 0 | 0 | 0 | 0 |
| 0 | 0 | 0 | 0 | 0 |
| 0 | 0 | 0 | 0.00201983 | 0 |
| 0.001804378 | 0 | 0 | 0.04064504 | 0 |
| 0 | 0 | 0 | 0.008702022 | 0.003093772 |
| 0 | 0 | 0 | 0 | 0 |
| 0 | 0 | 0 | 0 | 0 |
| 0 | 0 | 0 | 1.918256056 | 0 |
| 0 | 0.039091126 | 0 | 0 | 0 |
| 0 | 0 | 0 | 0 | 0 |
| 0 | 0.892885566 | 0 | 0 | 0 |
| 0 | 0 | 0 | 0 | 0 |
| 0 | 0 | 0 | 0 | 0 |
| 0 | 0 | 0 | 0 | 0 |
| 0 | 0 | 0 | 0 | 0 |

| | | | | |
|---|---|---|---|---|
| 0 | 0 | 0 | 0.015891618 | 0 |
| 0 | 0 | 0 | 0 | 0 |
| 0 | 0 | 0 | 0 | 0.002111996 |
| 0 | 0.005142158 | 0 | 0 | 0 |
| 0 | 0.005154327 | 0 | 0 | 0 |
| 0 | 0 | 0 | 0 | 0 |
| 0 | 0.018680323 | 0 | 0.12792719 | 0.0006399 |
| 0 | 0 | 0 | 0 | 0 |
| 0 | 0 | 0 | 0 | 0.00033875 |
| 0.009929712 | 0 | 0.730766177 | 0 | 0 |
| 0 | 0 | 0 | 0 | 0 |
| 0 | 0 | 0 | 0 | 0 |
| 0 | 0 | 0 | 0 | 0 |
| 0 | 0 | 0 | 0 | 0 |
| 0 | 0 | 0 | 0 | 0 |
| 0 | 0 | 0 | 0 | 0 |
| 0 | 0 | 0 | 0 | 0 |
| 0.005369927 | 0.00971908 | 0 | 0 | 0 |
| 0 | 0 | 0 | 0 | 0 |
| 0 | 0 | 0 | 0 | 0 |
| 0.000738578 | 0.009415434 | 0 | 0.655658465 | 0 |
| 0 | 0 | 0 | 0 | 0 |
| 0 | 0 | 0 | 0 | 0 |
| 0.003477092 | 0.004309489 | 0.001248657 | 0.000725225 | 0 |
| 0 | 0 | 0 | 0 | 0 |
| 0 | 0.017189532 | 0 | 0 | 0 |
| 0 | 0 | 0 | 0 | 0 |
| 0 | 0 | 0 | 0 | 0 |
| 0 | 0 | 0 | 0 | 0 |
| 0 | 0.006497715 | 0 | 0.087825657 | 0.003869877 |
| 0 | 0 | 0 | 0 | 0 |
| 0 | 0 | 0 | 0 | 0 |
| 0 | 0 | 0 | 0 | 0 |
| 0 | 0 | 0 | 0 | 0 |
| 0 | 0 | 0 | 0 | 0 |
| 0 | 0 | 0 | 0 | 0 |
| 0 | 0 | 0 | 0.000518539 | 0 |
| 0 | 0 | 0 | 0 | 0 |
| 0 | 0 | 0 | 0 | 0 |
| 0 | 0 | 0 | 0 | 0 |
| 0 | 0 | 0 | 0 | 0 |
| 0 | 0 | 0 | 0 | 0 |
| 0 | 0 | 0 | 0 | 0 |
| 0 | 0 | 0 | 0 | 0 |
| 0.003602944 | 0 | 0 | 0 | 0 |
| 0 | 0 | 0 | 0 | 0 |
| 0 | 0 | 0 | 0 | 0 |
| 0 | 0 | 0 | 0 | 0 |

| | | | | |
|---|---|---|---|---|
| 0.000779474 | 0.022523363 | 0.056263209 | 0.000557405 | 0 |
| 0 | 0 | 0 | 0 | 0 |
| 0 | 0 | 0 | 0 | 0 |
| 0 | 0 | 0 | 0 | 0 |
| 0 | 0 | 0 | 0 | 0 |
| 0 | 0 | 0 | 0 | 0 |
| 0 | 0 | 0 | 0 | 0 |
| 1.834707485 | 0.950449814 | 0 | 1.531782309 | 0 |
| 0.009543311 | 0.578485226 | 0.060896811 | 0.03499726 | 0.003532243 |
| 0.032367067 | 0.169148931 | 0.002936418 | 0.020100335 | 0.181266495 |
| 0 | 0 | 0 | 0 | 0 |
| 0 | 0 | 0 | 0 | 0 |
| 0 | 0 | 0 | 0 | 0 |
| 0 | 0 | 0 | 0 | 0 |
| 0 | 0 | 0 | 0 | 0 |
| 0 | 0 | 0 | 0 | 0 |
| 0 | 0 | 0.00294194 | 0.020341547 | 0 |
| 0 | 0 | 0 | 0 | 0 |
| 0 | 0 | 0 | 0 | 0 |
| 0 | 0 | 0 | 0 | 0 |
| 0 | 0 | 0 | 0 | 0 |
| 0 | 0 | 0 | 0 | 0 |
| 0 | 0 | 0 | 0 | 0 |
| 0 | 0 | 0 | 0 | 0 |
| 0 | 0 | 0 | 0.047942515 | 0 |
| 0 | 0 | 0 | 0 | 0 |
| 0 | 0 | 0 | 0 | 0 |
| 0 | 0 | 0 | 0 | 0 |
| 0 | 0 | 0 | 0 | 0 |
| 0 | 0 | 0 | 0 | 0 |
| 0 | 0 | 0 | 0 | 0 |
| 0 | 0 | 0 | 0 | 0 |
| 0 | 0 | 0 | 0 | 0 |
| 0 | 0 | 0 | 0.002304738 | 0 |
| 0 | 0.01309919 | 0 | 0.002429344 | 0 |
| 0 | 0 | 0 | 0.010431276 | 0 |
| 0 | 0 | 0 | 0 | 0 |
| 0 | 0 | 0 | 0 | 0 |
| 0 | 0 | 0 | 0 | 0 |
| 0.880253522 | 0 | 0 | 0.002893619 | 0.221284362 |
| 0 | 0 | 0 | 0 | 0 |
| 0 | 0 | 0 | 0 | 0 |
| 0 | 0 | 0 | 0 | 0 |
| 0 | 0 | 0 | 0 | 0 |
| 0 | 0.00408155 | 0 | 0 | 0 |
| 0 | 0 | 0 | 0 | 0 |
| 0 | 0 | 0 | 0 | 0 |
| 0 | 0 | 0 | 0 | 0 |

| | | | | |
|---|---|---|---|---|
| 0 | 0 | 0 | 0 | 0 |
| 0.013507378 | 0 | 0 | 0.006439459 | 0.000725367 |
| 0 | 0 | 0 | 0 | 0 |
| 0 | 0 | 0 | 0 | 0 |
| 0.080065437 | 1.368149801 | 0.01229212 | 0.25715588 | 0 |
| 0 | 0 | 0 | 0.115063878 | 0 |
| 0 | 0 | 0 | 0.002928069 | 0 |
| 0.088247704 | 0.112358229 | 0.021578085 | 0.060553727 | 0 |
| 0 | 0 | 0 | 0.008495642 | 0 |
| 0 | 0 | 0 | 0 | 0 |
| 0 | 0 | 0 | 0 | 0 |
| 0 | 0 | 0 | 0 | 0 |
| 0 | 0 | 0 | 0 | 0 |
| 0 | 0 | 0 | 0 | 0 |
| 0.045650134 | 0.064661168 | 0.040607494 | 0 | 0 |
| 0.006363715 | 0 | 0.146380836 | 0 | 0 |
| 0 | 0 | 0 | 0 | 0 |
| 0 | 0 | 0 | 0 | 0 |
| 0 | 0 | 0 | 0 | 0 |
| 0 | 0 | 0 | 0 | 0 |
| 0 | 0 | 0 | 0 | 0 |
| 0 | 0 | 0 | 0 | 0 |
| 0 | 0 | 0 | 0 | 0 |
| 0 | 0.00486184 | 0.001479133 | 0.000613633 | 0.111657166 |
| 0 | 0 | 0 | 0 | 0 |
| 0 | 0 | 0 | 0 | 0 |
| 0 | 0.002223352 | 0.082353837 | 0.003255175 | 0.018773828 |
| 0 | 0.003745788 | 0 | 0.001536507 | 0 |
| 0 | 0 | 0 | 0 | 0 |
| 0 | 0 | 0 | 0 | 0 |
| 0 | 0 | 0 | 0 | 0 |
| 0 | 0 | 0 | 0 | 0 |
| 0 | 0 | 0 | 0 | 0 |
| 0 | 0 | 0 | 0 | 0 |
| 0 | 0 | 0 | 0 | 0 |
| 0 | 0 | 0 | 0 | 0 |
| 0 | 0 | 0 | 0 | 0 |
| 0 | 0.001824142 | 0 | 0 | 0 |
| 0 | 0 | 0 | 0 | 0 |
| 0 | 0 | 0 | 0 | 0 |
| 0 | 0 | 0 | 0 | 0 |
| 0 | 0 | 0 | 0 | 0 |
| 0 | 0 | 0 | 0 | 0 |
| 0 | 0.001632182 | 0 | 0 | 0 |
| 0 | 0 | 0 | 0 | 0 |
| 0 | 0 | 0 | 0 | 0 |
| 0 | 0 | 0 | 0 | 0 |
| 0 | 0 | 0 | 0 | 0 |

| | | | | |
|---|---|---|---|---|
| 0.383499591 | 0 | 0 | 0.036312303 | 0 |
| 0 | 0 | 0 | 0 | 0 |
| 0 | 0 | 0 | 0 | 0 |
| 0 | 0.001040151 | 0 | 0 | 0 |
| 0 | 0 | 0 | 0 | 0 |
| 0 | 0 | 0 | 0 | 2.022177439 |
| 0 | 0 | 0 | 0 | 0 |
| 0 | 0 | 0 | 0 | 0 |
| 0 | 0 | 0 | 0 | 0 |
| 0 | 0 | 0 | 0 | 0 |
| 0 | 0 | 0 | 0 | 0 |
| 0 | 0 | 0 | 0 | 0 |
| 0 | 0 | 0 | 0 | 0 |
| 0 | 0 | 0 | 0 | 0 |
| 0 | 0 | 0 | 0 | 0 |
| 0 | 0 | 0 | 0 | 0 |
| 0 | 0 | 0 | 0 | 0 |
| 0.247630026 | 0 | 0 | 0 | 0 |
| 0 | 0 | 0 | 0 | 0 |
| 0 | 0 | 0 | 0 | 0 |
| 0.048707376 | 0.181556842 | 0.033325492 | 0.001466563 | 0 |
| 0 | 0 | 0 | 0 | 0 |
| 0.013381964 | 0.064202059 | 0.018602285 | 0.004167365 | 0.288639817 |
| 0 | 0 | 0 | 0 | 0 |
| 0 | 0 | 0 | 0 | 0 |
| 0 | 0 | 0 | 0 | 0 |
| 0 | 0 | 0 | 0 | 0 |
| 0 | 0 | 0 | 0 | 0 |
| 0 | 0 | 0 | 0 | 0 |
| 0 | 0 | 0 | 0 | 0 |
| 0 | 0 | 0 | 0 | 0 |
| 0 | 0 | 0 | 0 | 0 |
| 0.039011384 | 0.018610108 | 0.140020988 | 0.003035449 | 0.064190809 |
| 0 | 0 | 0 | 0 | 0 |
| 0.813263977 | 0 | 0 | 0.42719063 | 0.860716338 |
| 0 | 0 | 0.046132655 | 0.009859274 | 0 |
| 0 | 0 | 0 | 0 | 0 |
| 0 | 0 | 0 | 0 | 0 |
| 0.017072724 | 0.004251899 | 0.013744168 | 0 | 0.002514739 |
| 0 | 0 | 0 | 0 | 0 |
| 0 | 0 | 0 | 0 | 0 |
| 0 | 0 | 0 | 0 | 0.045003385 |
| 0 | 0.968868539 | 0.016780068 | 0 | 0 |
| 0 | 0 | 0 | 0 | 0 |
| 0 | 0 | 0 | 0 | 0 |
| 0 | 0 | 0 | 0.02255022 | 0 |
| 0 | 0 | 0 | 0 | 0 |
| 0 | 0 | 0 | 0 | 0 |

| | | | | |
|---|---|---|---|---|
| 0 | 0 | 0 | 0 | 0 |
| 0 | 0 | 0 | 0 | 0 |
| 0 | 0 | 0 | 0 | 0 |
| 0 | 0 | 0 | 0 | 0 |
| 0 | 0 | 0 | 0 | 0 |
| 0 | 0 | 0 | 0 | 0 |
| 0 | 0 | 0 | 0 | 0 |
| 0 | 3.445095769 | 0 | 0 | 0 |
| 0 | 0 | 0 | 0 | 0 |
| 0 | 0.001779241 | 0 | 0.003208077 | 0 |
| 0 | 0 | 0 | 0 | 0 |
| 0 | 0 | 0 | 0 | 0 |
| 0 | 0 | 0 | 0 | 0 |
| 0 | 0 | 0 | 0 | 0 |
| 0 | 0 | 0 | 0 | 0 |
| 0.013418225 | 0 | 0.006276876 | 0.017549546 | 0.001092247 |
| 0 | 0 | 0 | 0 | 0 |
| 0 | 0 | 0 | 0 | 0 |
| 0 | 0 | 0 | 0 | 0 |
| 0 | 2.501634257 | 0 | 0.197880297 | 0 |
| 0 | 0 | 0 | 0.003032898 | 0 |
| 0 | 0 | 0 | 0.000609959 | 0 |
| 0 | 0 | 0 | 0 | 0 |
| 0 | 0 | 0 | 0 | 0 |
| 0 | 0 | 0 | 0 | 0 |
| 0 | 0 | 0 | 0 | 0 |
| 0 | 0 | 0 | 0 | 0 |
| 0 | 0 | 0 | 0 | 0 |
| 0 | 0 | 0 | 0 | 0 |
| 0 | 0 | 0 | 0.013347146 | 0 |
| 0 | 0 | 0 | 0 | 0 |
| 0 | 0 | 0 | 0 | 0 |
| 0 | 0 | 0 | 0 | 0 |
| 0 | 0 | 0 | 0 | 0 |
| 0 | 0 | 0 | 0 | 0 |
| 0 | 0 | 0 | 0 | 0 |
| 0 | 0 | 0 | 0 | 0 |
| 0 | 0 | 0 | 0 | 0 |
| 0 | 0 | 0 | 0 | 0 |
| 0 | 0 | 0 | 0 | 0 |
| 0 | 0 | 0 | 0 | 0 |
| 0 | 0 | 0 | 0 | 0 |
| 0 | 0 | 0 | 0 | 0 |
| 0 | 0 | 0 | 0 | 0 |
| 0 | 0 | 0 | 0 | 0 |
| 0 | 0 | 0 | 0 | 0 |
| 0 | 0.129795772 | 0.206592525 | 0.013570942 | 0.13529315 |
| 0 | 0 | 0 | 0 | 0 |

| | | | | |
|---|---|---|---|---|
| 0 | 0 | 0.001068808 | 0 | 0 |
| 0 | 0 | 0 | 0 | 0 |
| 0 | 0 | 0 | 0 | 0 |
| 0 | 0 | 0 | 0 | 0 |
| 0.029030437 | 0 | 0 | 0.056968755 | 0 |
| 0 | 0 | 0 | 0 | 0 |
| 0 | 0 | 0 | 0 | 0 |
| 0 | 0 | 0 | 0 | 0 |
| 0 | 0 | 0 | 0 | 0 |
| 0 | 0 | 0 | 0 | 0 |
| 0 | 0 | 0 | 0 | 0 |
| 0.00187463 | 0.005841705 | 0.022013551 | 0.002815168 | 0.145352136 |
| 0 | 0.002290873 | 0.001742399 | 0 | 0 |
| 0 | 0 | 0 | 0 | 0 |
| 0.017714624 | 0 | 0 | 0 | 0 |
| 0 | 0.035929055 | 0 | 0.551572264 | 0 |
| 0 | 0 | 0 | 0.004021083 | 0 |
| 0 | 0 | 0 | 0 | 0 |
| 0.089285313 | 0.232830261 | 1.742423469 | 0.162919853 | 0 |
| 0 | 0.007088208 | 0.000998365 | 0.265738937 | 0 |
| 0 | 0 | 0 | 0 | 0 |
| 0 | 0 | 0 | 0 | 0 |
| 0 | 0 | 0 | 0 | 0 |
| 0 | 0 | 0 | 0 | 0 |
| 0 | 0 | 0 | 0 | 0 |
| 0 | 0 | 0 | 0 | 0 |
| 0.004010098 | 0.065636782 | 0.006080271 | 0.002549013 | 0 |
| 0 | 0 | 0 | 0.020813484 | 0 |
| 0 | 0 | 0 | 0 | 0 |
| 0 | 0 | 0 | 0 | 0 |
| 0.001851934 | 0.004769406 | 0.000725506 | 0 | 0 |
| 0 | 0 | 0 | 0 | 0 |
| 0 | 0 | 0 | 0 | 0 |
| 0 | 0 | 0 | 0 | 0 |
| 0 | 0 | 0 | 0 | 0 |
| 0 | 0 | 0 | 0 | 0 |
| 0 | 0 | 0 | 0 | 0 |
| 0.010645488 | 0.097844718 | 0.024976251 | 0.006935959 | 0.003170914 |
| 0 | 0 | 0 | 0 | 0 |
| 0 | 0 | 0 | 0.015832476 | 0 |
| 0 | 0 | 0.002417105 | 0 | 0.098742195 |
| 0 | 0 | 0 | 0 | 0 |
| 0 | 0 | 0 | 0 | 0 |
| 0.001578591 | 0.00372666 | 0.029289158 | 0.002634001 | 0 |
| 0 | 0 | 0 | 0 | 0 |
| 0 | 0 | 0 | 0 | 0 |
| 0 | 0 | 0 | 0 | 0 |
| 0 | 0 | 0 | 0 | 0 |

| | | | | |
|---|---|---|---|---|
| 0 | 0 | 0 | 0 | 0 |
| 0 | 0 | 0 | 0 | 0 |
| 0 | 0 | 0 | 0 | 0 |
| 0 | 0.004686668 | 0.003168534 | 0.001314498 | 0 |
| 0.000852446 | 0 | 0.001469382 | 0.001341093 | 0 |
| 0 | 0 | 0 | 0 | 0.002722427 |
| 0 | 0.001263349 | 0 | 0 | 0 |
| 0 | 0 | 0 | 0 | 0 |
| 0 | 0 | 0 | 0 | 0 |
| 0 | 0 | 0 | 0 | 0 |
| 0 | 0 | 0 | 0 | 0 |
| 0 | 0 | 0 | 0 | 0 |
| 0.016678623 | 0.026946636 | 0.006878511 | 0.008013427 | 0.041782911 |
| 0 | 0 | 0 | 0 | 0 |
| 0.000966485 | 0 | 0 | 0 | 0 |
| 0 | 0 | 0 | 0 | 0 |
| 0 | 0 | 0 | 0 | 0 |
| 0 | 0 | 0 | 0 | 0 |
| 0 | 0 | 0 | 0 | 0 |
| 0 | 0 | 0 | 0 | 0 |
| 0 | 0 | 0 | 0 | 0 |
| 0 | 0 | 0 | 0 | 0 |
| 0.001372619 | 0 | 0.009611949 | 0 | 0 |
| 0 | 0.006309266 | 0.320423664 | 0.009266255 | 0 |
| 0 | 0 | 0 | 0 | 0 |
| 0 | 0 | 0 | 0.000641552 | 0 |
| 0 | 0 | 0 | 0 | 0 |
| 0 | 0 | 0 | 0 | 0 |
| 0 | 0 | 0 | 0 | 0 |
| 0 | 0 | 0 | 0 | 0 |
| 0 | 0 | 0 | 0.014638161 | 0 |
| 0.001183517 | 0 | 0 | 0 | 0 |
| 0 | 0 | 0 | 0 | 0 |
| 0 | 0 | 0 | 0 | 0 |
| 0 | 0 | 0 | 0 | 0 |
| 0 | 0 | 0 | 0.0034323 | 0 |
| 0 | 0 | 0 | 0.001889492 | 0 |
| 0.325064531 | 0 | 0 | 0 | 0 |
| 0.000647439 | 0.030263177 | 0.000871881 | 0.12674266 | 0 |
| 0 | 0 | 0 | 0 | 0 |
| 0 | 0 | 0 | 0 | 0 |
| 0 | 0 | 0 | 0 | 0 |
| 0 | 0 | 0 | 0 | 0 |
| 0 | 0 | 0 | 0 | 0 |
| 0 | 0 | 0 | 0 | 0 |
| 0.083879442 | 0.197480591 | 0.08376855 | 0.033402862 | 0.046399725 |
| 0 | 0 | 0 | 0 | 0 |
| 0 | 0 | 0 | 0 | 0 |

| | | | | |
|---|---|---|---|---|
| 0 | 0 | 0 | 0 | 0 |
| 0 | 0 | 0 | 0 | 0 |
| 0 | 0 | 0 | 0 | 0 |
| 0 | 0 | 0 | 0 | 0 |
| 0 | 0 | 0 | 0 | 0 |
| 0 | 0 | 0 | 0 | 0 |
| 0 | 0.001398273 | 0 | 0 | 0 |
| 0 | 0 | 0 | 0 | 0 |
| 0 | 0 | 0 | 0.000428386 | 0 |
| 0 | 0 | 0 | 0 | 0 |
| 0 | 0.055240974 | 0.002263453 | 0 | 0.020850295 |
| 0 | 0.006109256 | 0.000844836 | 0 | 0 |
| 0 | 0 | 0 | 0 | 0 |
| 0 | 0 | 0 | 0 | 0 |
| 0 | 0 | 0 | 0 | 0 |
| 0 | 0 | 0 | 0 | 0 |
| 0 | 0 | 0 | 0.322579142 | 0 |
| 0 | 0 | 0 | 0 | 0 |
| 0 | 0 | 0 | 0 | 0 |
| 0.001218993 | 0.003699951 | 0 | 0.001245297 | 0 |
| 0 | 0 | 0 | 0 | 0 |
| 0 | 0 | 0 | 0 | 0 |
| 0 | 0 | 0 | 0 | 0 |
| 0 | 0 | 0 | 0 | 0 |
| 0 | 0 | 0 | 0 | 0 |
| 0 | 0 | 0 | 0 | 0 |
| 0 | 0 | 0 | 0 | 0 |
| 0 | 0 | 0 | 0 | 0 |
| 0 | 0 | 0 | 0 | 0 |
| 0 | 0 | 0 | 0 | 0 |
| 0.005685023 | 0 | 0.005963463 | 0.003102532 | 0.00601591 |
| 0 | 0 | 0 | 0 | 0 |
| 0.051591175 | 0.05068596 | 0.070926136 | 0.031054788 | 0 |
| 0 | 0.001760043 | 0.000956185 | 0.002792647 | 0.002562359 |
| 0 | 0 | 0 | 0 | 0 |
| 0 | 0 | 0 | 0 | 0 |
| 0 | 0 | 0 | 0 | 0 |
| 0 | 0 | 0 | 0 | 0 |
| 0 | 0 | 0 | 0 | 0 |
| 0 | 0 | 0 | 0 | 0 |
| 0 | 0 | 0 | 0 | 0 |
| 0.079027967 | 0 | 0.090527625 | 0.025139717 | 0 |
| 0.007138178 | 0.041429936 | 0 | 0.044571384 | 0 |
| 0 | 0 | 0 | 0 | 0 |
| 0 | 0 | 0 | 0 | 0 |
| 0 | 0.551651343 | 0 | 0.005037556 | 0 |
| 0 | 0 | 0 | 0 | 0 |
| 0 | 0 | 0 | 0 | 0 |

| | | | | |
|---|---|---|---|---|
| 0 | 0 | 0 | 0 | 0 |
| 0 | 0 | 0 | 0 | 0 |
| 0.01129199 | 0.006639246 | 0.006197357 | 0.001675934 | 0.002636113 |
| 0 | 0 | 0 | 0 | 0 |
| 0 | 0 | 0 | 0.011313472 | 0 |
| 0.001159124 | 0 | 0.087056252 | 0 | 0.000682936 |
| 0 | 0 | 0 | 0 | 0 |
| 0 | 0 | 0 | 0 | 0 |
| 0 | 0 | 0 | 0 | 0 |
| 0 | 0 | 0 | 0 | 0 |
| 0 | 0 | 0 | 0 | 0 |
| 0 | 0 | 0 | 0 | 0 |
| 0 | 0 | 0 | 0 | 0 |
| 0 | 0 | 0 | 0 | 0 |
| 0 | 0 | 0 | 0 | 0 |
| 0 | 0 | 0 | 0 | 0 |
| 0 | 0 | 0 | 0 | 0 |
| 0 | 0 | 0 | 0 | 0 |
| 0 | 0 | 0 | 0 | 0 |
| 0 | 0 | 0 | 0 | 0 |
| 0 | 0 | 0 | 0 | 0 |
| 0 | 0 | 0.010154746 | 0 | 0 |
| 0 | 0 | 0 | 0 | 0 |
| 0 | 0 | 0 | 0 | 0 |
| 0 | 0.088044183 | 0 | 0 | 0 |
| 0 | 0 | 0 | 0 | 0 |
| 0 | 0 | 0 | 0 | 0 |
| 0.000685379 | 0 | 0 | 0.011272713 | 0 |
| 0 | 0 | 0 | 0 | 0 |
| 0 | 0 | 0 | 0 | 0 |
| 0 | 0 | 0 | 0 | 0 |
| 0 | 0 | 0.036713412 | 0 | 0 |
| 0 | 0 | 0 | 0 | 0 |
| 0 | 0 | 0 | 0 | 0 |
| 0 | 0 | 0 | 0 | 0 |
| 0 | 0 | 0 | 0 | 0 |
| 0 | 0.003621772 | 0 | 0 | 0.006660324 |
| 0 | 0 | 0 | 0 | 0 |
| 0 | 0 | 0 | 0 | 0 |
| 0 | 0 | 0.000842076 | 0 | 0 |
| 0 | 0 | 0 | 0 | 0 |
| 0 | 0 | 0 | 0 | 0 |
| 0.001240927 | 0.015066107 | 0.042207409 | 0.000633852 | 0 |
| 0 | 0 | 0 | 0 | 0 |
| 0 | 0 | 0 | 0 | 0 |
| 0 | 0 | 0 | 0 | 0 |
| 0 | 0.021127512 | 0.017822234 | 0.007757357 | 0 |
| 0 | 0 | 0 | 0.071766374 | 0 |

| | | | | |
|---|---|---|---|---|
| 0 | 0 | 0 | 0 | 0 |
| 0.018649909 | 0.019388388 | 0.026912324 | 0.0274074 | 0.074624027 |
| 0 | 0 | 0 | 0 | 0 |
| 0 | 0 | 0 | 0 | 0 |
| 0 | 0 | 0 | 0 | 0 |
| 0 | 0 | 0 | 0 | 0 |
| 0 | 0 | 0 | 0 | 0 |
| 0 | 0 | 0 | 0 | 0 |
| 0 | 0 | 0 | 0 | 0 |
| 0 | 0 | 0 | 0 | 0 |
| 0 | 0 | 0 | 0 | 0 |
| 0 | 0 | 0 | 0 | 0 |
| 0.02387457 | 0.003074287 | 0 | 0 | 0 |
| 0 | 0 | 0 | 0 | 0 |
| 0 | 0.00362616 | 0 | 0.04973377 | 0.01319786 |
| 0 | 0 | 0 | 0 | 0 |
| 0 | 0 | 0 | 0 | 0 |
| 0 | 0 | 0 | 0 | 0 |
| 0 | 0 | 0 | 0 | 0 |
| 0 | 0 | 0 | 0 | 0 |
| 0 | 0 | 0 | 0 | 0 |
| 0 | 0.007715879 | 0 | 0.115963453 | 0 |
| 0 | 0 | 0.000867214 | 0.025097721 | 0 |
| 0 | 0 | 0 | 0.010030643 | 0.00111251 |
| 0 | 0 | 0 | 0 | 0 |
| 0 | 0 | 0 | 0 | 0 |
| 0 | 0 | 0 | 0 | 0 |
| 0 | 0 | 0 | 0 | 0 |
| 0 | 0 | 0 | 0 | 0 |
| 0 | 0 | 0 | 0 | 0 |
| 0 | 0 | 0 | 0 | 0 |
| 0 | 0 | 0 | 0 | 0 |
| 0 | 0 | 0 | 0 | 0 |
| 0 | 0 | 0 | 0 | 0 |
| 0.030055704 | 0 | 0 | 40.33387656 | 0.001961363 |
| 0 | 0 | 0 | 0 | 0 |
| 0 | 0 | 0 | 0 | 0 |
| 0 | 0 | 0 | 0 | 0 |
| 0 | 0.001185421 | 0.001352417 | 0.00299234 | 11.61805582 |
| 0 | 0 | 0 | 0 | 0 |
| 0.029265115 | 0 | 0.044866969 | 0.02481801 | 6.946489922 |
| 0 | 0 | 0 | 0 | 0 |
| 0 | 0 | 0 | 0 | 0 |
| 0 | 0 | 0 | 0 | 0 |
| 0 | 0 | 0 | 0 | 0 |
| 2.154729873 | 0.410410768 | 1.205797841 | 0.240374972 | 0.087919236 |
| 0 | 0 | 0 | 0 | 0 |
| 0 | 0 | 0 | 0 | 0 |

| | | | | |
|---|---|---|---|---|
| 0 | 0 | 0 | 0 | 0 |
| 0 | 0 | 0 | 0 | 0 |
| 0 | 0 | 0 | 0 | 0 |
| 0 | 0 | 0 | 0 | 0 |
| 0 | 0.002768856 | 0 | 0 | 0 |
| 0 | 0 | 0 | 0 | 0 |
| 0 | 0 | 0 | 0 | 0 |
| 0 | 0 | 0 | 0.005478451 | 0.005026684 |
| 0 | 0.133550969 | 0 | 0 | 0 |
| 0.025417348 | 0.015769656 | 0 | 0.002253231 | 0 |
| 0 | 0 | 0 | 0.055923528 | 0 |
| 0 | 0 | 0 | 0 | 0 |
| 0.000821267 | 0 | 0 | 0 | 0 |
| 0 | 0 | 0 | 0 | 0 |
| 0 | 0 | 0 | 0 | 0 |
| 0 | 0 | 0 | 0 | 0 |
| 0 | 0 | 0 | 0 | 0 |
| 0 | 0 | 0 | 0 | 0 |
| 0 | 0 | 0 | 0 | 0 |
| 0.087169999 | 0.117000734 | 0.322879618 | 0.086119931 | 0.588448872 |
| 0 | 0 | 0 | 0 | 0 |
| 0 | 0 | 0 | 0 | 0 |
| 0 | 0 | 0 | 0 | 0 |
| 0 | 0 | 0 | 0 | 0 |
| 0 | 0 | 0 | 0 | 0 |
| 0 | 0 | 0 | 0 | 0 |
| 0 | 0 | 0 | 0 | 0 |
| 0 | 0 | 0 | 0 | 0 |
| 0 | 0 | 0 | 0 | 0 |
| 0 | 0 | 0 | 0 | 0 |
| 0 | 0 | 0 | 0 | 0 |
| 0 | 0 | 0 | 0 | 0 |
| 0 | 0 | 0 | 0 | 0 |
| 0 | 0 | 0 | 0 | 0 |
| 0 | 0 | 0 | 0 | 0 |
| 0.00520434 | 0 | 0 | 0 | 0.005122128 |
| 0 | 0 | 0 | 0 | 0 |
| 0 | 0 | 0 | 0 | 0 |
| 0 | 0 | 0 | 0 | 0 |
| 0 | 0 | 0 | 0 | 0 |
| 0 | 0 | 0 | 0 | 0 |
| 0 | 0 | 0 | 0 | 0 |
| 0 | 0 | 0 | 0 | 0 |
| 0 | 0 | 0 | 0 | 0 |
| 0.000686568 | 0.01239925 | 0 | 0.002454841 | 0.026192294 |
| 0 | 0 | 0 | 0 | 0 |
| 0 | 0 | 0 | 0 | 0 |
| 0 | 0 | 0 | 0 | 0 |

| | | | | |
|---|---|---|---|---|
| 0 | 0 | 0 | 0 | 0 |
| 0 | 0 | 0 | 0 | 0 |
| 0 | 0 | 0 | 0 | 0 |
| 0 | 0 | 0 | 0 | 0 |
| 0 | 0 | 0 | 0 | 0 |
| 0.003587849 | 0.010890026 | 0 | 0.0037569 | 0.022460177 |
| 0 | 0 | 0 | 0 | 0 |
| 0 | 0 | 0 | 0 | 0 |
| 0 | 0 | 0 | 0 | 0 |
| 0 | 0 | 0 | 0 | 0 |
| 0 | 0 | 0 | 0 | 0 |
| 0 | 0 | 0 | 0 | 0 |
| 0 | 0 | 0 | 0 | 0 |
| 0 | 0 | 0 | 0 | 0 |
| 0 | 0 | 0 | 0 | 0 |
| 0 | 0 | 0 | 0 | 0 |
| 0 | 0 | 0 | 0 | 0 |
| 0 | 0 | 0 | 0 | 0 |
| 0 | 0 | 0 | 0 | 0 |
| 0 | 0 | 0 | 0 | 0 |
| 0 | 0 | 0 | 0 | 0.00061276 |
| 0 | 0 | 0 | 0 | 0 |
| 0 | 0 | 0 | 0 | 0 |
| 0 | 0 | 0 | 0 | 0 |
| 0 | 0 | 0 | 0 | 0 |
| 0 | 0 | 0 | 0 | 0 |
| 0 | 0 | 0 | 0 | 0 |
| 0 | 0 | 0 | 0 | 0 |
| 0 | 0 | 0 | 0 | 0 |
| 0 | 0 | 0.078112872 | 0 | 0 |
| 0 | 0 | 0 | 0 | 0 |
| 0.00506954 | 0 | 0 | 0 | 0 |
| 0 | 0 | 0 | 0 | 0 |
| 0 | 0 | 0 | 0 | 0 |
| 0 | 0 | 0 | 0 | 0 |
| 0 | 0 | 0 | 0 | 0 |
| 0 | 0 | 0 | 0 | 0 |
| 0 | 0 | 0 | 0 | 0 |
| 0 | 0.029207271 | 0 | 0.982514992 | 0 |
| 0 | 0 | 0 | 0 | 0 |
| 0 | 0 | 0 | 0 | 0 |
| 0 | 0 | 0 | 0 | 0 |
| 0 | 0 | 0 | 0 | 0 |
| 0 | 0 | 0 | 0 | 0 |
| 0 | 0 | 0 | 0 | 0 |
| 0 | 0 | 0 | 0 | 0 |
| 0 | 0 | 0 | 0 | 0 |
| 0 | 0 | 0 | 0 | 0 |

| | | | | |
|---|---|---|---|---|
| 0 | 0 | 0 | 0 | 0 |
| 0 | 0 | 0 | 0 | 0 |
| 0 | 0.003234176 | 0 | 0 | 0 |
| 0 | 0 | 0 | 0 | 0 |
| 0 | 0 | 0 | 0 | 0 |
| 0 | 0 | 0 | 0 | 0 |
| 0 | 0 | 0 | 0 | 0 |
| 0 | 0 | 0 | 0 | 0 |
| 0 | 0 | 0 | 0 | 0 |
| 0 | 0 | 0 | 0 | 0 |
| 0.000939249 | 0.00239472 | 0 | 0.002149316 | 0.002130549 |
| 0 | 0 | 0 | 0 | 0 |
| 0 | 1.498277631 | 0 | 0.050856449 | 0 |
| 0 | 0 | 0 | 0 | 0 |
| 0 | 0 | 0 | 0 | 0 |
| 0 | 0 | 0 | 0 | 0 |
| 0 | 0 | 0 | 0 | 0 |
| 0 | 0 | 0 | 0 | 0 |
| 0 | 0 | 0 | 0 | 0 |
| 0 | 0 | 0 | 0 | 0 |
| 0.004003781 | 0.041048389 | 0 | 0 | 0.081384082 |
| 0 | 0 | 0 | 0 | 0 |
| 0 | 0 | 0 | 0 | 0 |
| 3.167951621 | 0.771811946 | 0 | 1.732437422 | 0 |
| 0 | 0.001816188 | 0 | 0 | 0.015297987 |
| 0 | 0 | 0 | 0 | 0 |
| 0 | 0 | 0 | 0 | 0 |
| 0 | 0 | 0 | 0 | 0 |
| 0 | 0 | 0 | 0 | 0 |
| 0 | 0 | 0 | 0 | 0 |
| 0 | 0 | 0 | 0 | 0 |
| 0 | 0 | 0 | 0 | 0 |
| 0 | 0 | 0 | 0.004302473 | 0.000827133 |
| 0 | 0 | 0 | 0 | 0 |

| PHL018-0193_V3 | PHL018-0256_V3 | PHL018-0213_V3 | PHL018-0162_V2 | PHL018-0016_V2 |
|---|---|---|---|---|
| REF | REF | REF | REF | CTRL |
| 2.247925923 | 0 | 0.015877016 | 4.6842074 | 0.106963111 |
| 0.006342883 | 7.470096412 | 5.143770694 | 0.179269253 | 7.182786403 |
| 9.927404504 | 0 | 7.811789037 | 0 | 5.182985577 |
| 0 | 0 | 0 | 0 | 0 |
| 0 | 0 | 0 | 0 | 0 |
| 0 | 0 | 0 | 0 | 0 |
| 0 | 0 | 0 | 0 | 0 |
| 0 | 0 | 0 | 0 | 0 |
| 0 | 0 | 0 | 0 | 0 |
| 0 | 0 | 0 | 0 | 0 |
| 0 | 0 | 0 | 0 | 0 |
| 0 | 0 | 0 | 0 | 0 |
| 0 | 0 | 0 | 0 | 0 |
| 0.295847309 | 0 | 11.2731239 | 1.784372178 | 1.713597654 |
| 0 | 0 | 0 | 0 | 0 |
| 0 | 0 | 0 | 0 | 0 |
| 0 | 0 | 0 | 0 | 0 |
| 0 | 0 | 0 | 0 | 0 |
| 0.073757727 | 0 | 0 | 0 | 0 |
| 0 | 0 | 0 | 0 | 0 |
| 0 | 0 | 0 | 0 | 0 |
| 0 | 0 | 0 | 0 | 0 |
| 0 | 0 | 0 | 0 | 0 |
| 0 | 0 | 0 | 0 | 0 |
| 0 | 0 | 0 | 0 | 0 |
| 0 | 0 | 0 | 0 | 0 |
| 0 | 0 | 0 | 0 | 0 |
| 0 | 0 | 0 | 0 | 0 |
| 0 | 0 | 0 | 0 | 0 |
| 0 | 0 | 0 | 0 | 0 |
| 0 | 0 | 0 | 0 | 0 |
| 0 | 0 | 0 | 0 | 0 |
| 0 | 0 | 0 | 0 | 0 |
| 0 | 0 | 0 | 0 | 0 |
| 0 | 0 | 0 | 0 | 0 |
| 0 | 0 | 0 | 0 | 3.02948349 |
| 0 | 0 | 0 | 0 | 0 |
| 0 | 0 | 0 | 0 | 0 |
| 0 | 0 | 0 | 0 | 0 |
| 0 | 0 | 0 | 0 | 0 |
| 0.00155058 | 0 | 0 | 0 | 0 |
| 0.002735749 | 0 | 0.067725825 | 0.017962554 | 0.005337848 |
| 0.333867468 | 0.557316038 | 0.763685274 | 1.949411505 | 0.017438823 |
| 0 | 0.008469158 | 0.014854292 | 0.003514413 | 0.150741705 |
| 0 | 0.001436697 | 0.002804848 | 0.003378356 | 0.008647612 |
| 0 | 0 | 0 | 0 | 0 |

| | | | | |
|---|---|---|---|---|
| 0 | 0 | 0.08625206 | 0.008913219 | 0 |
| 0.017302016 | 0.044834313 | 0.054336509 | 0 | 0 |
| 2.59076346 | 0 | 0 | 0 | 0.007581298 |
| 0 | 0 | 0 | 0.385351741 | 0.639976051 |
| 0.007485038 | 0 | 0.000632274 | 0.003829538 | 0.000927264 |
| 0 | 0 | 0 | 0 | 0 |
| 2.172079606 | 0.002402757 | 0.00389891 | 0 | 0.163051165 |
| 0 | 0 | 0 | 0 | 0 |
| 0 | 0 | 0 | 0 | 0 |
| 0 | 0 | 0 | 0 | 0 |
| 0 | 0 | 0 | 0 | 0 |
| 0 | 0 | 0.108686626 | 0 | 0.081809583 |
| 0 | 0.000892084 | 0 | 0 | 0.001547975 |
| 0.068066786 | 0.130533404 | 0.01361235 | 0.025269153 | 0.063664797 |
| 0 | 0.006070942 | 0 | 0 | 0 |
| 0 | 0 | 0 | 0 | 0 |
| 0 | 0 | 0.002392447 | 0.007924488 | 0 |
| 0 | 0 | 0 | 0 | 0 |
| 0 | 0 | 0 | 0 | 0 |
| 0 | 0 | 0 | 0 | 0 |
| 0.197259456 | 0.906049776 | 0.057351836 | 0.071728131 | 0.015888854 |
| 0.011163687 | 0.031461962 | 0.001038864 | 0 | 0.006506825 |
| 0 | 0 | 0 | 0.106595936 | 0 |
| 0 | 0 | 0 | 0 | 0 |
| 0 | 0 | 0 | 0 | 0 |
| 0 | 0 | 0 | 0 | 0 |
| 0 | 0 | 0 | 0 | 0 |
| 0 | 0 | 0 | 0 | 0 |
| 0 | 0 | 0.202673647 | 0 | 0 |
| 0 | 0 | 0 | 0 | 0 |
| 0 | 0.480582425 | 0.09495657 | 0 | 0 |
| 0 | 0 | 0 | 0 | 0 |
| 0 | 0 | 0 | 0 | 0 |
| 0 | 0 | 0 | 0.085750199 | 0 |
| 0 | 0 | 0 | 0 | 0 |
| 0 | 0 | 0 | 0 | 0 |
| 0 | 0 | 0 | 0 | 0 |
| 0 | 0.060331716 | 0.001049777 | 0 | 0 |
| 0 | 0 | 0 | 0 | 0 |
| 0 | 0 | 0 | 0 | 0 |
| 0 | 0 | 0 | 0 | 0 |
| 0 | 0 | 0 | 0 | 0 |
| 0 | 0 | 0 | 0 | 0 |
| 0 | 0 | 0 | 0 | 0 |
| 0 | 0 | 0 | 0 | 0 |
| 0 | 0 | 0 | 0 | 0 |
| 0 | 0 | 0 | 0 | 0 |
| 0 | 0 | 0.317500632 | 0 | 0.002886146 |

| | | | | |
|---|---|---|---|---|
| 0 | 0 | 0 | 0 | 0 |
| 0 | 0 | 0 | 0 | 0 |
| 0 | 0 | 0 | 0 | 0 |
| 0 | 0 | 0 | 0 | 0 |
| 0 | 0 | 0.002167913 | 0 | 0.002167745 |
| 0 | 0 | 0.000851266 | 0 | 0 |
| 0 | 0 | 0 | 0 | 0 |
| 0 | 0 | 0 | 0 | 0 |
| 0 | 0 | 0 | 0.013304071 | 0 |
| 0 | 0 | 0 | 0 | 0 |
| 0 | 0 | 0 | 0 | 0 |
| 0 | 0.001094758 | 0.384905245 | 0 | 0 |
| 0 | 0 | 0 | 0 | 0 |
| 0 | 0 | 0 | 0 | 0 |
| 0 | 0 | 0 | 0 | 0 |
| 0.018095943 | 0 | 0 | 0 | 0 |
| 0 | 0 | 0 | 0 | 0 |
| 0 | 0 | 0 | 0 | 0 |
| 0 | 0 | 0.469468411 | 0 | 0 |
| 0 | 0 | 0 | 0 | 0 |
| 0 | 0 | 0 | 0 | 0 |
| 0 | 0 | 0 | 0 | 0 |
| 0 | 0 | 0 | 0 | 0 |
| 0 | 0 | 0 | 0 | 0 |
| 0 | 0 | 0 | 0 | 0 |
| 0 | 0 | 0 | 0 | 0 |
| 0 | 0 | 0 | 0 | 0 |
| 0 | 0 | 0 | 0 | 0 |
| 0 | 0 | 0 | 0 | 0 |
| 0 | 0 | 0 | 0 | 0 |
| 0 | 0 | 0 | 0 | 0.007239495 |
| 0.001889013 | 0 | 0.003086921 | 0.007380298 | 0.002680539 |
| 0 | 0 | 0 | 0 | 0 |
| 0 | 0 | 0 | 0 | 0 |
| 0 | 0 | 0 | 0 | 0 |
| 0 | 0 | 0 | 0 | 0 |
| 0 | 0 | 0 | 0 | 0 |
| 0 | 0 | 0 | 0 | 0 |
| 0 | 0 | 0 | 0 | 0.047986088 |
| 0 | 0 | 0 | 0 | 0 |
| 0 | 0 | 0 | 0 | 0 |
| 0 | 0 | 0 | 0 | 0 |
| 0 | 0 | 0 | 0.000710871 | 0 |
| 0 | 0 | 0 | 0 | 0 |
| 0 | 0 | 0 | 0 | 0 |
| 0 | 0 | 0 | 0 | 0 |
| 0 | 0 | 0 | 0 | 0 |
| 0 | 0 | 0 | 0 | 0 |

| | | | | |
|---|---|---|---|---|
| 0.047034579 | 0 | 0 | 0 | 0 |
| 0 | 0 | 0 | 0 | 0 |
| 0 | 0 | 0 | 0 | 0 |
| 0 | 0 | 0 | 0 | 0 |
| 0 | 0 | 0 | 0.006825196 | 0.048735994 |
| 0 | 0 | 0 | 0 | 0 |
| 0 | 0 | 0 | 0 | 0 |
| 0 | 0 | 0 | 0 | 0 |
| 0 | 0 | 0 | 0 | 0 |
| 0 | 0 | 0 | 0 | 0 |
| 0 | 0 | 0 | 0 | 0 |
| 0 | 0 | 0 | 0 | 0 |
| 0 | 0 | 0 | 0 | 0 |
| 0 | 0 | 0 | 0 | 0 |
| 0 | 0 | 0 | 0 | 0 |
| 0 | 0 | 0 | 0 | 0 |
| 0 | 0 | 0 | 0 | 0 |
| 0 | 0 | 0 | 0 | 0 |
| 0 | 0 | 0 | 0 | 0 |
| 0 | 0 | 0 | 0 | 0 |
| 0 | 0 | 0 | 0 | 0 |
| 0 | 0 | 0 | 0 | 0 |
| 0 | 0 | 0 | 0 | 0.00455163 |
| 0 | 0 | 0 | 0 | 0 |
| 0 | 0 | 0 | 0 | 0 |
| 0 | 0 | 0 | 0 | 0 |
| 0 | 0 | 0 | 0.133783338 | 0 |
| 0 | 0 | 0 | 0 | 0 |
| 0 | 0 | 0 | 0 | 0 |
| 0 | 0 | 0 | 0 | 0 |
| 0 | 0 | 0 | 0 | 0 |
| 0 | 0 | 0.047238599 | 0.000709606 | 0.000687281 |
| 0 | 0 | 0 | 0 | 0 |
| 0 | 0 | 0 | 0 | 0 |
| 0 | 0 | 0 | 0 | 0 |
| 0 | 0 | 0 | 0 | 0 |
| 0 | 0 | 0 | 0 | 0 |
| 0 | 0 | 0 | 0 | 0 |
| 0 | 0 | 0 | 0 | 0 |
| 0 | 0 | 0 | 0 | 0 |
| 0 | 0 | 0 | 0 | 0 |
| 0 | 0 | 0 | 0 | 0 |
| 0 | 0 | 0 | 0 | 0 |
| 0 | 0 | 0 | 0 | 0 |
| 0 | 0 | 0 | 0 | 0 |
| 0 | 0 | 0 | 0 | 0 |
| 0 | 0 | 0 | 0 | 0 |
| 0 | 0 | 0 | 0 | 0 |

| | | | | |
|---|---|---|---|---|
| 0 | 0 | 0 | 0 | 0 |
| 0 | 0 | 0 | 0 | 0 |
| 0 | 0 | 0 | 0 | 0 |
| 0 | 0 | 0 | 0 | 0 |
| 0 | 0 | 0 | 0 | 0 |
| 0 | 0 | 0 | 0 | 0 |
| 0 | 0 | 0 | 0 | 0 |
| 0 | 0 | 0 | 0 | 0 |
| 0 | 0 | 0 | 0 | 0 |
| 0 | 0 | 0 | 0 | 0 |
| 0 | 0 | 0 | 0 | 0 |
| 0 | 0 | 0 | 0 | 0 |
| 0.001137589 | 0 | 0 | 0 | 0 |
| 0 | 0 | 0 | 0 | 0 |
| 0 | 0 | 0 | 0 | 0 |
| 0 | 0 | 0 | 0 | 0 |
| 0 | 0 | 0 | 0 | 0 |
| 0 | 0 | 0 | 0 | 0 |
| 0 | 0 | 0 | 0 | 0 |
| 0 | 0 | 0 | 0 | 0 |
| 0.037577333 | 0.032597737 | 0.112355547 | 0.010791688 | 0.038119675 |
| 0 | 0 | 0 | 0 | 0 |
| 0 | 0 | 0 | 0 | 0 |
| 0 | 0 | 0 | 0 | 0 |
| 0 | 0 | 0 | 0 | 0 |
| 0 | 0 | 0 | 0 | 0 |
| 0 | 0.018984153 | 0.052737224 | 0 | 0.008951613 |
| 0 | 0 | 0 | 0 | 0 |
| 0 | 0 | 0 | 0 | 0 |
| 0.017670583 | 0.143059081 | 0 | 0 | 0 |
| 0.080467744 | 0.12615985 | 0.095404919 | 0 | 0 |
| 0 | 0 | 0 | 0 | 0 |
| 0 | 0 | 0 | 0 | 0 |
| 0 | 0 | 0.014142885 | 0.023665163 | 0 |
| 0.298241622 | 0 | 0.009383564 | 0.034695199 | 0.003000326 |
| 0 | 0 | 0 | 0 | 0 |
| 0 | 0 | 0 | 0 | 0 |
| 0 | 0 | 0 | 0 | 0 |
| 0 | 0 | 0.004018799 | 0 | 0 |
| 0 | 0 | 0 | 0 | 0 |
| 0 | 0 | 0 | 0 | 0 |
| 0 | 0 | 0 | 0 | 0 |
| 0 | 0 | 0 | 0 | 0 |
| 0 | 0 | 0 | 0 | 0 |
| 0 | 0 | 0 | 0 | 0 |
| 0 | 0 | 0 | 0 | 0 |
| 0.001498452 | 0.000998461 | 0 | 0 | 0 |
| 0.007396811 | 0.001314321 | 0.000777556 | 0 | 0 |

| | | | | |
|---|---|---|---|---|
| 0 | 0 | 0 | 0 | 0 |
| 0 | 0 | 0 | 0 | 0 |
| 0 | 0 | 0 | 0 | 0 |
| 0 | 0 | 0 | 0 | 0.000724752 |
| 0 | 0 | 0 | 0 | 0 |
| 0 | 0 | 0 | 0 | 0 |
| 0 | 0 | 0 | 0 | 0 |
| 0 | 0 | 0.001117751 | 0.002961855 | 0.001434336 |
| 0 | 0 | 0 | 0 | 0 |
| 0 | 0 | 0 | 0 | 0 |
| 0 | 0 | 0.001346847 | 0 | 0 |
| 0 | 0 | 0 | 0 | 0 |
| 0 | 0 | 0 | 0 | 0 |
| 0 | 0 | 0 | 0 | 0 |
| 0 | 0 | 0 | 0 | 0 |
| 0 | 0 | 0 | 0 | 0 |
| 0 | 0 | 0 | 0 | 0 |
| 0 | 0 | 0 | 0 | 0 |
| 0 | 0.005123848 | 0.003795692 | 0.011225399 | 0 |
| 0 | 0 | 0 | 0.000857586 | 0 |
| 0.018000933 | 0.155435978 | 0.004432568 | 0 | 0 |
| 0 | 0 | 0 | 0 | 0 |
| 0 | 0 | 0 | 0 | 0 |
| 0 | 0 | 0 | 0 | 0 |
| 0.001469698 | 0 | 0.02271081 | 0.011404353 | 0 |
| 0 | 0 | 0 | 0.001064099 | 0 |
| 0 | 0 | 0 | 0 | 0 |
| 0 | 0 | 0 | 0 | 0 |
| 0 | 0 | 0.131587624 | 0 | 0 |
| 0 | 0 | 0 | 0 | 0 |
| 3.763397866 | 0.232208505 | 0.08851478 | 0.02987065 | 0.119497129 |
| 0 | 0 | 0 | 0 | 0 |
| 0 | 0 | 0 | 0 | 0 |
| 0 | 0 | 0 | 0 | 0.003865064 |
| 0 | 0 | 0 | 0 | 0 |
| 0 | 0 | 0 | 0 | 0 |
| 0 | 0 | 0 | 0 | 0 |
| 0 | 0 | 0 | 0 | 0 |
| 0 | 0 | 0 | 0 | 0 |
| 0 | 0 | 0 | 0 | 0 |
| 0 | 0 | 0 | 0 | 0 |
| 0 | 0 | 0 | 0 | 0.001923276 |
| 0 | 0 | 0 | 0 | 0 |
| 0 | 0 | 0 | 0 | 0 |
| 0 | 0 | 0 | 0 | 0 |
| 0 | 0 | 0 | 0 | 0 |
| 0 | 0 | 0 | 0 | 0 |
| 0 | 0 | 0 | 0 | 0 |

| | | | | |
|---|---|---|---|---|
| 0 | 0 | 0 | 0 | 0 |
| 0 | 0 | 0 | 0 | 0 |
| 0 | 0.005257674 | 0 | 0.000831145 | 0.144094342 |
| 0 | 0 | 0 | 0 | 0 |
| 0 | 0 | 0 | 0 | 0 |
| 0 | 0 | 0 | 0 | 0 |
| 0 | 0 | 0 | 0 | 0 |
| 0 | 0 | 0 | 0 | 0 |
| 0 | 0 | 0 | 0 | 0 |
| 0 | 0 | 0 | 0 | 0.001472487 |
| 0 | 0 | 0 | 0 | 0 |
| 0 | 0 | 0 | 0 | 0 |
| 0 | 0 | 0 | 0 | 0 |
| 0.016414956 | 0 | 0.00145593 | 0 | 0.002224166 |
| 0.001905243 | 0 | 0.008812319 | 0.005875705 | 0.002570061 |
| 0 | 0 | 0 | 0 | 0 |
| 0 | 0.003857337 | 0.009812647 | 0.002159626 | 0.002091681 |
| 0.005613227 | 0 | 0 | 0 | 0.010059825 |
| 0 | 0 | 0 | 0 | 0 |
| 0 | 0 | 0 | 0 | 0 |
| 0 | 0 | 0 | 0 | 0 |
| 0.010451528 | 0.001160692 | 0 | 0 | 0 |
| 0 | 0 | 0 | 0 | 0 |
| 0.007426033 | 0 | 0 | 0 | 0 |
| 0 | 0 | 0 | 0 | 0 |
| 0 | 0 | 0 | 0 | 0 |
| 0 | 0 | 0 | 0 | 0 |
| 0 | 0 | 0 | 0 | 0 |
| 0 | 0 | 0 | 0 | 0 |
| 0.046299585 | 0 | 0 | 0 | 0 |
| 0 | 0 | 0 | 0 | 0 |
| 0 | 0 | 0 | 0 | 0 |
| 0 | 0 | 0 | 0 | 0 |
| 0 | 0 | 0 | 0 | 0 |
| 0 | 0 | 0 | 0 | 0 |
| 0 | 0 | 0.004526023 | 0 | 0 |
| 0 | 0 | 0 | 0 | 0 |
| 0.001094272 | 0 | 0.094468659 | 0.032331777 | 0.000711695 |
| 0 | 0 | 0 | 0 | 0 |
| 0 | 0 | 0 | 0 | 0 |
| 0 | 0 | 0 | 0 | 0 |
| 0 | 0 | 0 | 0 | 0 |
| 0 | 0 | 0 | 0 | 0 |
| 0 | 0.005887124 | 0 | 0 | 0 |
| 0 | 0 | 0 | 0 | 0 |
| 0 | 0 | 0 | 0 | 0 |
| 0 | 0 | 0 | 0 | 0 |
| 0.02418924 | 0.082135436 | 0 | 0 | 0.001724082 |

| | | | | |
|---|---|---|---|---|
| 1.036319048 | 0 | 0.00454675 | 6.065141575 | 0 |
| 0 | 0 | 0 | 0 | 0 |
| 0.000684198 | 0 | 0.002157694 | 0 | 0.016415185 |
| 0 | 0 | 0 | 0 | 0 |
| 0 | 0 | 0.044719477 | 0.008628584 | 0.001591832 |
| 0 | 0 | 0 | 0 | 0 |
| 0 | 0 | 0 | 0 | 0 |
| 0.017689839 | 0.003429015 | 0.030429219 | 0.00201581 | 0.012272167 |
| 0 | 0 | 0 | 0 | 0 |
| 0.443566459 | 0 | 0.1260835 | 0.165031844 | 0.050884236 |
| 0.024877756 | 0.001997199 | 0 | 0 | 0 |
| 0 | 0 | 0.031795999 | 0.18487584 | 0.032014324 |
| 0.000579383 | 0.209759052 | 0.150191819 | 0.005014564 | 0.008206318 |
| 0.083887045 | 0 | 0 | 0 | 0.001780469 |
| 0.002582158 | 0 | 0 | 0 | 0 |
| 0 | 0 | 0 | 0 | 0.001257045 |
| 0 | 0 | 0.017752084 | 0 | 0 |
| 0 | 0.003016754 | 0 | 0 | 0 |
| 0.02183644 | 0.781971371 | 0.084407191 | 0 | 0 |
| 0 | 0 | 0 | 0 | 0 |
| 0 | 0 | 0 | 0 | 0 |
| 0 | 0 | 0 | 0 | 0 |
| 0 | 0 | 0.002639537 | 0 | 0 |
| 0 | 0 | 0 | 0 | 0 |
| 0 | 0 | 0 | 0 | 0 |
| 0.002474391 | 0.001199096 | 0.001915349 | 0 | 0.002730934 |
| 0 | 0 | 0 | 0 | 0 |
| 0 | 2.359215776 | 0 | 0 | 0 |
| 0.292955461 | 0 | 0 | 0 | 0.000798881 |
| 0 | 0 | 0 | 0 | 0.019467811 |
| 0.011586137 | 0 | 0 | 0 | 0 |
| 0 | 0 | 0 | 0 | 0 |
| 0 | 0 | 0 | 0 | 0 |
| 0 | 0.700592134 | 0 | 0 | 0 |
| 0 | 0 | 0 | 1.109480564 | 0 |
| 0 | 0 | 0 | 0 | 0 |
| 0 | 0 | 0 | 0 | 0 |
| 0 | 0 | 0 | 0 | 0 |
| 0 | 0 | 0 | 0 | 0 |
| 0 | 0 | 0 | 0 | 0 |
| 0 | 0 | 0 | 0 | 0 |
| 0 | 0 | 0 | 0 | 0 |
| 0 | 0 | 0 | 0 | 0 |
| 0.09604838 | 0 | 0.00342905 | 0 | 0 |
| 0 | 0 | 0 | 0 | 0.001557835 |
| 0 | 0 | 0 | 0 | 0 |
| 0.002517628 | 0 | 0 | 0.001001843 | 0.000970323 |
| 0.004570365 | 0 | 0 | 0 | 0 |

| | | | | |
|---|---|---|---|---|
| 0 | 0 | 0 | 0 | 0 |
| 0 | 0 | 0 | 0 | 0 |
| 0 | 0 | 0 | 0 | 0 |
| 0 | 0 | 0.07149277 | 0 | 0 |
| 0.285061424 | 0 | 0 | 0 | 0 |
| 0.18088794 | 0.068119696 | 0 | 0.001183609 | 0 |
| 0 | 0 | 0 | 0 | 0 |
| 0 | 0 | 0 | 0 | 0 |
| 0 | 0 | 0 | 0 | 0 |
| 0 | 0 | 0 | 0 | 0 |
| 0 | 0 | 0.131761195 | 0.005948742 | 0.001772796 |
| 0 | 0 | 0 | 0 | 0.002076892 |
| 0 | 0 | 0 | 0 | 0 |
| 0 | 0 | 0.003197686 | 0 | 0 |
| 0.362750494 | 0 | 0 | 0.022964724 | 0 |
| 0 | 0 | 0 | 0 | 0 |
| 0 | 0 | 0 | 0 | 0 |
| 0 | 0 | 0 | 0 | 0 |
| 0 | 0 | 0 | 0 | 0.005125029 |
| 0 | 0 | 0 | 0 | 0 |
| 0.821989008 | 0 | 0 | 0 | 0 |
| 0 | 0 | 0.079279042 | 0 | 1.394634951 |
| 0 | 0 | 0 | 0 | 0 |
| 0 | 0 | 0 | 0 | 0 |
| 0.145932363 | 0 | 0 | 0 | 0 |
| 0 | 0 | 0 | 0 | 0 |
| 0 | 0 | 0 | 0 | 0 |
| 0 | 0 | 0 | 0 | 0 |
| 0 | 0 | 0 | 0 | 0 |
| 0.008846879 | 2.677711275 | 0.131574663 | 0.341553888 | 0.582136506 |
| 0 | 0.010476993 | 0 | 0.011291672 | 0 |
| 0 | 0 | 0.003263289 | 0 | 0 |
| 0 | 0 | 0 | 0 | 0 |
| 0.01438259 | 0 | 0 | 0 | 0 |
| 0.017442125 | 0 | 0 | 0 | 0 |
| 0 | 0 | 0 | 0 | 0 |
| 0 | 0 | 0 | 0 | 0.210869532 |
| 0 | 0 | 0 | 0 | 0.036062846 |
| 0 | 0 | 0 | 0.694306169 | 0.006478874 |
| 2.146771655 | 0 | 0 | 0 | 0 |
| 0.059542729 | 0 | 0.00675989 | 0.023279259 | 0.006884539 |
| 0 | 0 | 8.359100102 | 2.627049386 | 3.495459033 |
| 0 | 0 | 0.00106633 | 0 | 0.035381645 |
| 0.000957767 | 0 | 0 | 0 | 0 |
| 0 | 0 | 0 | 0 | 0 |
| 0.0501615 | 0 | 0 | 0 | 0 |
| 0.002708089 | 0.001110447 | 0 | 0.001492107 | 0 |
| 0.909014604 | 0 | 0 | 0 | 0 |

| | | | | |
|---|---|---|---|---|
| 0.004112403 | 0 | 0 | 0.000920505 | 0 |
| 0 | 0 | 0 | 0.001176351 | 0 |
| 0 | 0.013534833 | 0.003313342 | 0 | 0.001518497 |
| 0 | 0 | 0 | 0 | 0 |
| 0 | 0 | 0 | 0 | 0.000896219 |
| 0 | 0 | 0 | 0 | 0 |
| 0 | 0 | 0 | 0 | 0 |
| 0 | 0 | 0 | 0 | 0 |
| 0.068619148 | 0 | 0 | 0 | 0 |
| 0 | 0 | 0 | 0.004515966 | 0 |
| 0 | 0 | 0 | 0 | 0 |
| 0 | 0 | 0 | 0 | 0 |
| 0 | 0 | 0 | 0 | 0 |
| 0 | 0 | 0.00087995 | 0 | 0 |
| 0 | 0 | 0 | 0 | 0 |
| 0 | 0.058603183 | 0 | 0.005857071 | 0.080364669 |
| 0 | 0.006173458 | 0.105756002 | 0.07303446 | 0.03720227 |
| 0.031077672 | 0.271322839 | 0.00092605 | 0 | 0 |
| 0 | 0 | 0 | 0 | 0 |
| 0 | 0 | 0 | 0 | 0 |
| 0 | 0 | 0.064600061 | 0.024090581 | 0.005502987 |
| 0.003504388 | 0.007783577 | 0 | 0.00146423 | 0 |
| 0 | 0 | 0 | 0 | 0 |
| 0 | 0 | 0 | 0 | 0.14363681 |
| 0.000941189 | 0 | 0 | 0 | 0 |
| 0 | 0.001060753 | 0.012299873 | 0 | 0 |
| 0.005113849 | 0 | 0 | 0.00160253 | 0 |
| 0.003883516 | 0 | 0 | 0 | 0 |
| 0 | 0 | 0 | 0 | 0.002621109 |
| 0 | 0 | 0 | 0 | 0 |
| 0.404851929 | 0 | 0 | 0.004352571 | 0.013658654 |
| 0 | 0.005022489 | 0.001919928 | 0.003114792 | 0.078185314 |
| 0.056034054 | 0.058488623 | 0.017496873 | 0.003707129 | 0.083538918 |
| 0 | 0 | 0.012724487 | 0 | 0 |
| 0 | 0 | 0.001602714 | 0 | 0 |
| 0 | 0 | 0 | 0 | 0 |
| 0 | 0 | 0.001180476 | 0 | 0.005842904 |
| 0.003903193 | 0.213006185 | 0.036927479 | 0.015726169 | 0.004400184 |
| 0 | 0 | 0 | 0.131363195 | 0.434619894 |
| 0 | 0 | 0 | 0 | 0.001383692 |
| 0 | 0 | 0 | 0 | 0 |
| 0 | 0 | 0 | 0 | 0 |
| 0.00612908 | 0 | 0.001380625 | 0 | 0 |
| 0 | 0 | 0 | 0.031372545 | 0.171876396 |
| 0 | 0 | 0 | 0 | 0 |
| 0 | 0 | 0 | 0 | 0 |
| 0 | 0 | 0 | 0 | 0 |
| 0 | 0 | 0 | 0 | 0 |

| | | | | |
|---|---|---|---|---|
| 0 | 0 | 0 | 0 | 0 |
| 0.139879071 | 0.032249772 | 1.65195226 | 0.554770415 | 0.520712905 |
| 0.001092027 | 0 | 0 | 0 | 0 |
| 0.000966343 | 0 | 0 | 0.000432605 | 0 |
| 0 | 0 | 0 | 0 | 0 |
| 0 | 0 | 0 | 0 | 0 |
| 0.003198256 | 0.023278044 | 0.097291697 | 0 | 0 |
| 0 | 0 | 0 | 0 | 0 |
| 0.017527453 | 0 | 0 | 0 | 0.039265095 |
| 0.015459678 | 0 | 0 | 0 | 0 |
| 0.010275975 | 0.010797466 | 0.001121762 | 0.000884668 | 0.001199569 |
| 0.035028009 | 0.450719953 | 0.035645777 | 0.011859637 | 0.040075187 |
| 0 | 0 | 0 | 0 | 0 |
| 0 | 0 | 0 | 0 | 0 |
| 0 | 0 | 0.002788676 | 0 | 0.002359466 |
| 0 | 0 | 0 | 0 | 0 |
| 0.001034657 | 0.000827305 | 0.043657689 | 0.033905311 | 0.008613406 |
| 0.083005267 | 0.012529493 | 0.001778995 | 0 | 0.006056587 |
| 0 | 0 | 0 | 0 | 0 |
| 0.122101936 | 0.053232671 | 0.039276347 | 0 | 0.003176517 |
| 0.001493789 | 0.004265802 | 0.076921152 | 0.003057043 | 0.026462729 |
| 0 | 0 | 0.005275686 | 0 | 0 |
| 0 | 0 | 0 | 0 | 0 |
| 0 | 0 | 0 | 0 | 0 |
| 0.002509844 | 0 | 0 | 0 | 0.006218506 |
| 0.004696616 | 0 | 0 | 0 | 0 |
| 0.541963591 | 0 | 0 | 0 | 0 |
| 0.287142738 | 0.09853715 | 0.077249602 | 0.009747556 | 0.012194478 |
| 0 | 0.004714843 | 0.017363479 | 0 | 0 |
| 0 | 0.001822989 | 0 | 0 | 0 |
| 0.001216842 | 0 | 0 | 0 | 0.015406162 |
| 0 | 0 | 0 | 0 | 0 |
| 0 | 0 | 0 | 0 | 0 |
| 0 | 0 | 0 | 0 | 0 |
| 0 | 0 | 0 | 0 | 0 |
| 0.021112526 | 2.383865129 | 0.222984918 | 0.10620045 | 0.420424736 |
| 0.001422262 | 0.00071077 | 0 | 0 | 0 |
| 0.003644038 | 0 | 0 | 0 | 0 |
| 0.006435325 | 0 | 0.008470758 | 0.058620417 | 0.013102188 |
| 0 | 0 | 0 | 0 | 0 |
| 0 | 0 | 0 | 0 | 0 |
| 0 | 0 | 0 | 0 | 0 |
| 0.17301197 | 0 | 0.013023959 | 0.027085618 | 0 |
| 0.00262538 | 0.001749365 | 0 | 0.009255569 | 0.005976252 |
| 0 | 0 | 0 | 0.001351075 | 0 |
| 0 | 0 | 0 | 0 | 0 |
| 0 | 0 | 0 | 0 | 0 |
| 0.004508956 | 0 | 0 | 0 | 0 |

| | | | | |
|---|---|---|---|---|
| 0 | 0 | 0 | 0 | 0 |
| 0 | 0 | 0 | 0 | 0 |
| 0 | 0 | 0 | 0 | 0 |
| 0 | 0 | 0 | 6.522260148 | 0.238078506 |
| 0 | 0 | 0 | 0 | 0 |
| 0 | 0 | 0 | 0.001690498 | 0 |
| 0 | 0 | 0 | 0 | 0 |
| 0 | 0 | 0 | 0 | 0.00230339 |
| 0 | 0 | 0 | 0 | 0.159783558 |
| 0 | 0 | 0.028118571 | 0.017106788 | 0 |
| 0 | 0 | 0 | 0 | 0 |
| 0 | 0 | 0 | 0.001188609 | 0.001480132 |
| 0 | 0 | 0 | 0 | 0 |
| 0 | 0 | 0 | 0 | 1.493349995 |
| 0.787345651 | 0 | 0.000888366 | 0 | 0.080748691 |
| 0 | 0 | 0 | 0 | 0 |
| 0 | 0 | 0.013326151 | 0.004021046 | 0.061073461 |
| 0 | 0 | 0 | 0 | 0 |
| 0 | 0 | 0 | 0 | 0 |
| 0 | 0 | 0 | 0 | 0 |
| 0 | 0 | 0 | 0 | 0 |
| 0 | 0 | 0 | 0 | 0 |
| 0 | 0 | 0 | 0 | 0 |
| 1.530549435 | 1.745986929 | 0.082720012 | 0 | 0.549700818 |
| 0 | 0 | 0 | 0.004292389 | 0 |
| 0.002744911 | 0 | 0 | 0 | 0 |
| 0 | 0 | 0 | 0 | 0 |
| 0 | 0 | 0 | 0 | 2.091117106 |
| 0 | 0 | 0 | 0 | 0 |
| 0 | 0 | 0 | 0 | 0 |
| 0 | 0 | 0 | 0.068671527 | 0 |
| 0 | 0 | 0 | 0 | 0 |
| 0 | 0.004861958 | 0 | 0.000879444 | 0.001946915 |
| 0 | 0 | 0 | 0 | 0 |
| 0 | 0 | 0 | 0 | 0 |
| 0 | 0 | 0 | 0 | 0 |
| 0 | 0 | 0 | 3.064355393 | 0.037116847 |
| 0 | 0 | 0 | 0 | 0 |
| 0 | 0 | 0 | 0 | 0 |
| 0 | 0 | 0 | 0 | 0 |
| 0.143910043 | 0.03491366 | 0.05284241 | 0.06369154 | 0.075098105 |
| 0 | 0 | 0 | 0 | 0 |
| 5.47311903 | 0 | 0.675206609 | 0.039668495 | 3.756689951 |
| 0.000792443 | 0 | 0 | 0 | 0 |
| 0 | 0 | 0 | 0 | 0 |
| 0 | 0 | 0 | 0.002001476 | 0 |
| 0 | 0 | 0 | 0 | 0 |
| 0.013341061 | 0 | 0.001034571 | 0.058940987 | 0.049121057 |

| | | | | |
|---|---|---|---|---|
| 0 | 0 | 0 | 0 | 0 |
| 0 | 0 | 0 | 0 | 0.020733903 |
| 0.307772531 | 1.126808943 | 0.013686188 | 0.004317397 | 0.025998437 |
| 0 | 0 | 0 | 0 | 0 |
| 0 | 0 | 0 | 0 | 0 |
| 0 | 0 | 0 | 0.039629965 | 0.001432207 |
| 0 | 0 | 0 | 0 | 0 |
| 0 | 0 | 0 | 0.0170835 | 0 |
| 0 | 0 | 0 | 0 | 0 |
| 0 | 0 | 0 | 0.01240693 | 0.001979204 |
| 0.049402405 | 0.208756388 | 0.000973725 | 0.860869055 | 0.016779233 |
| 0.910966612 | 2.234006824 | 2.269196645 | 0.276563018 | 0.96923757 |
| 0 | 0 | 0 | 0 | 0 |
| 0.001999391 | 0.002664501 | 0.018705737 | 0.020288304 | 0.008091181 |
| 1.157188226 | 0.466839826 | 0.146580242 | 0.192325772 | 0.011234188 |
| 0 | 0 | 0 | 0 | 0 |
| 0 | 0 | 0 | 0 | 0 |
| 0 | 0 | 0 | 0 | 0 |
| 0 | 0 | 0 | 0 | 0 |
| 0.139039814 | 0 | 0 | 0.003339938 | 0.002499664 |
| 0.112315106 | 0.011495965 | 0.014853479 | 0.13238183 | 0.093207882 |
| 3.45255531 | 0 | 3.232009028 | 0 | 0.381971329 |
| 7.86718855 | 0 | 3.074458383 | 6.056967528 | 1.150276114 |
| 0 | 0 | 0 | 0 | 0 |
| 0 | 0 | 0 | 0 | 0 |
| 0 | 0 | 0 | 0 | 0.004661262 |
| 0 | 0 | 0.015617981 | 0 | 0.232624813 |
| 0 | 0 | 0 | 0 | 0 |
| 0 | 0 | 0 | 0 | 0 |
| 0 | 0 | 0 | 0 | 0 |
| 0 | 0 | 0 | 0 | 0.002145227 |
| 0.416806735 | 0 | 0 | 0 | 0.076272083 |
| 1.491472777 | 0.001690394 | 0.035001445 | 0.007381987 | 0.00054998 |
| 0 | 0 | 0.001372696 | 0.184469039 | 0 |
| 0.18292502 | 0.061548387 | 0.009108548 | 0 | 0.000604989 |
| 0 | 0.027591788 | 0.018862567 | 0.017336027 | 0.063837586 |
| 0.520558638 | 0 | 0 | 0 | 0.006744745 |
| 0 | 0 | 0 | 0 | 0 |
| 0 | 0 | 0 | 0 | 0.044549366 |
| 0 | 0 | 0 | 0 | 0 |
| 0 | 0 | 0 | 0 | 0.008490386 |
| 0 | 0 | 0 | 0 | 0 |
| 0 | 0 | 0 | 0 | 0 |
| 0 | 0 | 0 | 0 | 0 |
| 0.501077039 | 0.218800389 | 0.009307478 | 0.074577057 | 0.002085403 |
| 0 | 0 | 0 | 0 | 0 |
| 0 | 0 | 0 | 0 | 0 |
| 0.024525538 | 0 | 4.265898564 | 0.01912542 | 0.21336572 |

| | | | | |
|---|---|---|---|---|
| 0 | 0 | 0 | 0 | 0 |
| 0 | 0 | 0 | 0 | 0 |
| 0 | 0 | 0 | 0 | 0 |
| 0 | 0 | 0 | 0.004671203 | 0.00145422 |
| 0 | 0 | 0 | 0 | 0 |
| 0.203566334 | 0 | 0 | 0 | 0 |
| 0 | 0 | 0 | 0 | 0 |
| 0 | 0 | 0.006164059 | 0 | 0.055012668 |
| 0 | 0 | 0 | 0 | 0 |
| 0 | 0 | 0 | 0 | 0 |
| 0.087158862 | 0.002765544 | 0.002355988 | 0 | 0 |
| 0 | 0 | 0 | 0 | 0 |
| 0 | 0 | 0.001547139 | 0 | 0 |
| 0 | 0 | 0 | 0 | 0 |
| 0 | 0 | 0 | 0 | 0 |
| 0 | 0 | 0 | 0 | 0 |
| 0 | 0 | 0 | 0.01824088 | 0 |
| 0 | 0 | 0 | 0.004176272 | 0.001348294 |
| 0 | 0 | 0 | 0.046712389 | 0 |
| 0 | 0.007682125 | 0 | 0 | 0 |
| 0 | 0 | 0 | 0 | 0 |
| 0 | 0 | 0 | 0 | 0 |
| 0 | 0 | 0 | 0.004279949 | 0.001036324 |
| 0 | 0 | 0 | 0 | 0 |
| 0 | 0 | 0 | 0 | 0 |
| 0 | 0 | 0 | 0 | 0 |
| 0 | 0 | 0 | 0 | 0 |
| 0 | 0 | 0 | 0 | 0 |
| 0 | 0 | 0 | 0 | 0 |
| 0 | 0 | 0 | 0.178066243 | 2.265739922 |
| 0 | 0 | 0 | 0 | 0 |
| 0 | 0 | 0 | 0 | 0 |
| 0 | 0 | 0 | 0 | 0 |
| 0 | 0 | 0 | 0 | 0 |
| 0 | 0 | 0.002966165 | 0 | 0 |
| 0 | 0 | 0 | 0 | 0 |
| 0 | 0 | 0 | 0 | 0.029769051 |
| 0.231709049 | 0.04096892 | 0.00265918 | 0.000786427 | 0.012186966 |
| 0 | 0 | 0 | 0.000684515 | 0 |
| 0 | 0 | 0 | 0 | 0.003305651 |
| 0 | 0 | 0 | 0.175759071 | 0 |
| 0 | 0 | 0 | 0 | 0 |
| 0 | 0 | 0 | 0 | 0 |
| 0 | 0 | 0 | 0 | 0.019675667 |
| 0 | 0 | 0 | 0 | 0 |
| 0 | 0 | 0 | 0 | 0 |
| 0 | 0 | 0 | 0.001046133 | 0 |
| 0 | 0 | 0 | 0 | 0 |

| | | | | |
|---|---|---|---|---|
| 0 | 0 | 0 | 0 | 0 |
| 0 | 0 | 0 | 0 | 0 |
| 0 | 0 | 0 | 0 | 0 |
| 0 | 0 | 0 | 0 | 0 |
| 0 | 0 | 0 | 0 | 0 |
| 0 | 0 | 0 | 0 | 0 |
| 0 | 0 | 0 | 0 | 0 |
| 0 | 0 | 0 | 0 | 0 |
| 1.571176739 | 0.328685921 | 0.131111476 | 0.002386152 | 0.010504913 |
| 0 | 0 | 0 | 0 | 0 |
| 0 | 0 | 0 | 0 | 0 |
| 0 | 0 | 0 | 0 | 0 |
| 0 | 0 | 0 | 0 | 0 |
| 0 | 0 | 0 | 0.001356009 | 0 |
| 1.105758627 | 0 | 0 | 0.094934899 | 1.731765364 |
| 2.65605971 | 1.507521874 | 0.97394336 | 0.005015656 | 0 |
| 0.005025488 | 0 | 0 | 0.071860384 | 0 |
| 0 | 0 | 0 | 0 | 0 |
| 0 | 0 | 0 | 0 | 0 |
| 0 | 0 | 0 | 0 | 0 |
| 0 | 0 | 0 | 0 | 0 |
| 0 | 0 | 0 | 0 | 0 |
| 0 | 0 | 0 | 0 | 0 |
| 0.060073687 | 0 | 0 | 0 | 0 |
| 0.017949502 | 0.035311249 | 0.056336229 | 0.12423135 | 0.005435531 |
| 0 | 0 | 1.995413677 | 11.98967298 | 8.396715282 |
| 0 | 0 | 0 | 0 | 0 |
| 0 | 0 | 0 | 0 | 0 |
| 0 | 0 | 0 | 0 | 0 |
| 0 | 0 | 0 | 0.993818954 | 0 |
| 0 | 0 | 0 | 0 | 0 |
| 0 | 0 | 0 | 0 | 0 |
| 0 | 0 | 0 | 0 | 0.000598219 |
| 0 | 0 | 0 | 0 | 0.002770971 |
| 0.004512859 | 0.004677627 | 0.045304598 | 0.004713993 | 0.066963388 |
| 0.302699775 | 0.002834156 | 0.003856397 | 0.001904127 | 0 |
| 0 | 0 | 0 | 0 | 0.441697451 |
| 0 | 0 | 0.65961788 | 0.204975076 | 1.955067741 |
| 0 | 0 | 0 | 0 | 0 |
| 0 | 0 | 0 | 0 | 0 |
| 0 | 0 | 0 | 0 | 0 |
| 0.002210716 | 0 | 0 | 0.032029543 | 0 |
| 0 | 0 | 0.33360344 | 0.291643423 | 0.090305747 |
| 0 | 0 | 0 | 0 | 0 |
| 0 | 0 | 0 | 0 | 0 |
| 0.000671593 | 0 | 0 | 0 | 0.002183959 |
| 0 | 0 | 0 | 0 | 0 |
| 0.005082156 | 0.153634905 | 1.03855938 | 0.003592328 | 0.350018508 |

| | | | | |
|---|---|---|---|---|
| 0 | 0 | 0 | 0 | 0 |
| 0 | 0 | 0 | 0 | 0 |
| 0 | 0 | 0 | 0 | 0 |
| 0 | 0 | 0 | 0 | 0 |
| 0.001191522 | 0.002117187 | 0 | 0.013335294 | 0 |
| 0 | 0 | 0 | 0 | 0 |
| 0 | 0 | 0 | 0 | 0 |
| 0 | 0 | 0 | 0 | 0 |
| 0 | 0 | 0 | 0 | 0 |
| 0 | 0 | 0 | 0 | 0 |
| 0 | 0 | 0 | 0.000952358 | 0 |
| 0 | 5.200501525 | 0 | 3.214930955 | 8.128030138 |
| 0 | 0 | 1.335137314 | 0.733706164 | 0.816132054 |
| 0 | 0 | 0 | 0 | 0 |
| 0.011712962 | 0.003060658 | 6.559563096 | 2.057536 | 0.370240024 |
| 0 | 0 | 0 | 0 | 0 |
| 0.00383438 | 0 | 0 | 0.009870141 | 0.000554181 |
| 0 | 0 | 0 | 0 | 0 |
| 0.074682806 | 0 | 0 | 0 | 0 |
| 0 | 0 | 0 | 0 | 0 |
| 0 | 0 | 0 | 0 | 0.32893389 |
| 0 | 0 | 0 | 0 | 0 |
| 0 | 0.538919905 | 0.132221925 | 0.396247543 | 0.043623065 |
| 0 | 0 | 0 | 0 | 0 |
| 0 | 0 | 0 | 0 | 0 |
| 0 | 0 | 0 | 0 | 0 |
| 0 | 0 | 0 | 0 | 0 |
| 0 | 0 | 0 | 0 | 0 |
| 0 | 0 | 0 | 0 | 0 |
| 0 | 0 | 0 | 0 | 0 |
| 0 | 0 | 0 | 0 | 0 |
| 0.002682213 | 0.01112057 | 0.142951369 | 0.033354246 | 0.056856593 |
| 0 | 0 | 4.689482228 | 0 | 0 |
| 0 | 0.015873271 | 0.00098602 | 0.052922344 | 0.024531223 |
| 0.287754394 | 0.003341295 | 0.005656457 | 0.000863403 | 0 |
| 0 | 0 | 0 | 0 | 0.008192905 |
| 0 | 0 | 0 | 0 | 0 |
| 0 | 0 | 0 | 0 | 0 |
| 0 | 0 | 0 | 0 | 0.001729288 |
| 0 | 0 | 0 | 0 | 0 |
| 0 | 0 | 0 | 0.128881055 | 0 |
| 0 | 0 | 0 | 0 | 0 |
| 0 | 0 | 0 | 0 | 0 |
| 0 | 0 | 0 | 0.002530402 | 0 |
| 0.118934511 | 0 | 0 | 0.003402154 | 0.031138865 |
| 0 | 0 | 0 | 0 | 0 |
| 0 | 0 | 0 | 0 | 0 |
| 0.001078186 | 0.003352655 | 0 | 0 | 0.001246637 |

| | | | | |
|---|---|---|---|---|
| 0 | 0 | 0 | 0.011882339 | 0 |
| 0 | 0 | 0 | 0.007931011 | 0 |
| 0.015326882 | 0.109983301 | 0 | 0 | 0 |
| 0 | 0 | 0 | 0 | 0 |
| 0 | 0 | 0 | 0 | 0 |
| 0 | 0 | 0 | 0 | 0 |
| 0 | 0 | 0 | 0 | 0 |
| 0 | 0 | 0 | 0.97648199 | 0 |
| 0 | 0 | 0 | 0 | 0 |
| 0 | 0 | 0 | 0 | 0 |
| 0 | 0 | 0 | 0.075267564 | 0 |
| 0.281904734 | 0.002397972 | 0.000567459 | 0 | 0 |
| 0 | 0 | 0 | 0 | 0 |
| 0 | 0 | 0 | 0 | 0 |
| 0 | 0 | 0 | 0 | 0 |
| 0 | 0 | 0 | 0 | 0 |
| 0 | 0 | 0 | 0.439275945 | 0 |
| 0 | 0 | 0 | 0 | 0 |
| 0 | 0 | 7.990278571 | 12.66722018 | 1.977171253 |
| 0 | 0 | 0 | 0 | 0 |
| 0 | 0 | 0 | 0.002397776 | 0 |
| 0 | 0 | 0 | 0 | 0 |
| 0 | 0 | 0 | 0 | 0 |
| 0 | 0 | 0 | 0 | 0 |
| 0 | 0 | 0 | 0.002230289 | 0 |
| 0.02546995 | 0.033471272 | 0.003179427 | 0 | 0 |
| 0 | 0 | 0 | 0 | 0 |
| 0.009348756 | 0 | 0.033085755 | 0.042316825 | 0.297557646 |
| 0 | 0 | 0 | 0 | 0 |
| 0 | 0 | 1.201072956 | 0.201393043 | 0.206793933 |
| 0 | 0 | 0 | 0 | 0 |
| 0 | 0 | 0 | 0 | 0 |
| 0 | 0 | 0 | 0 | 0 |
| 0 | 0 | 0 | 0 | 0 |
| 0.138644566 | 0.026677386 | 0.001052161 | 0 | 0.002411015 |
| 0 | 0 | 0 | 0 | 0 |
| 0 | 0 | 0 | 0 | 0 |
| 0.017877383 | 0 | 0.000596947 | 0.002259731 | 0 |
| 0 | 0 | 0 | 0 | 0 |
| 0 | 0 | 0 | 0 | 0 |
| 0.571463867 | 38.12687144 | 4.845455466 | 2.917233095 | 11.36375898 |
| 0.076824429 | 3.99879279 | 0.127740323 | 0.108483554 | 0.730286937 |
| 0 | 0 | 0 | 0 | 0 |
| 0 | 0 | 0 | 0 | 0 |
| 0 | 0 | 0 | 0 | 0 |
| 0 | 0 | 0 | 0 | 0 |
| 0 | 0 | 0 | 0 | 0 |
| 0.014021557 | 0.009342961 | 0.002526775 | 0.009415598 | 0.001158015 |

| | | | | |
|---|---|---|---|---|
| 0 | 0 | 0 | 0 | 0 |
| 0.003262194 | 0.011669293 | 0 | 0.00215216 | 0.00208445 |
| 0 | 0 | 0 | 0.002693144 | 0 |
| 0 | 0 | 0 | 0 | 0 |
| 0 | 0 | 3.102308993 | 2.791001793 | 0.159067178 |
| 0 | 0 | 0 | 0 | 0 |
| 0 | 0 | 0 | 0 | 0 |
| 0 | 0 | 0 | 0 | 0 |
| 0.592329555 | 0.725628126 | 0.011516044 | 0.000767495 | 0.01501565 |
| 0 | 0 | 0 | 0 | 0 |
| 0 | 0 | 0 | 0 | 0 |
| 0 | 0 | 0 | 0 | 0 |
| 0 | 0 | 0 | 0.034654008 | 0 |
| 0.000615303 | 0.029792879 | 0.004657025 | 0.003489086 | 0.031303132 |
| 0 | 0 | 0 | 0 | 0 |
| 0 | 0 | 0.064328938 | 1.056340162 | 0.338165143 |
| 0 | 0 | 0.00218288 | 0.463377396 | 0 |
| 0 | 0 | 0 | 0 | 0 |
| 0 | 0 | 0 | 0 | 0 |
| 0 | 0 | 0 | 0 | 0 |
| 0 | 0 | 0 | 0 | 0 |
| 0 | 0 | 0 | 0 | 0 |
| 0 | 0 | 0 | 0 | 0 |
| 0 | 0 | 0 | 0 | 0 |
| 0 | 0 | 0 | 0 | 0 |
| 0 | 0 | 0 | 0 | 0 |
| 0 | 0 | 0.744640854 | 0.036893402 | 0.048446571 |
| 0.145235114 | 0.011139484 | 0 | 0 | 0 |
| 0 | 0 | 0 | 0 | 0 |
| 0 | 0 | 0 | 0 | 0 |
| 0 | 0 | 0 | 0 | 0 |
| 0 | 0 | 0 | 0 | 0 |
| 0 | 0 | 0 | 0.115461792 | 0 |
| 0 | 0 | 0 | 0 | 0 |
| 0 | 0 | 0 | 0 | 0 |
| 0 | 0.228190842 | 0.009338706 | 0 | 0 |
| 0.001029516 | 0 | 0.002922025 | 0.005161923 | 0 |
| 0 | 0 | 0 | 0 | 0 |
| 0 | 0 | 0 | 0 | 0 |
| 0 | 0 | 0 | 0 | 0 |
| 0 | 0 | 0 | 0 | 3.115761027 |
| 0 | 0 | 0 | 0 | 0 |
| 0 | 0 | 0 | 0 | 0 |
| 0 | 0 | 0 | 0 | 0 |
| 0 | 0 | 0 | 0 | 0 |
| 0 | 0 | 0 | 0 | 0 |
| 0 | 0 | 0 | 0 | 0 |
| 0 | 0 | 0 | 0 | 0 |

| | | | | |
|---|---|---|---|---|
| 0 | 0 | 0 | 0 | 0 |
| 0 | 0 | 0 | 0 | 0 |
| 0 | 0 | 0 | 0 | 0 |
| 0.007890786 | 0 | 0 | 0 | 0 |
| 0 | 0 | 0.002678578 | 0.019772414 | 0 |
| 0 | 0 | 0 | 0 | 0 |
| 0 | 0.009437955 | 0.024455814 | 0.369198142 | 0.014278744 |
| 0 | 0 | 0 | 0 | 0 |
| 0 | 0 | 0 | 0 | 0.002167417 |
| 0 | 0 | 0 | 0 | 0 |
| 0 | 0 | 0 | 0 | 0 |
| 0 | 0 | 0 | 0.085119211 | 0 |
| 0 | 0 | 0 | 0 | 0 |
| 0 | 0 | 0 | 0 | 0 |
| 0 | 0 | 0 | 0 | 0 |
| 0 | 0 | 0 | 0 | 0 |
| 0 | 0 | 0.001148603 | 0 | 0 |
| 0 | 0 | 0 | 0 | 0 |
| 0 | 0 | 0 | 0 | 0 |
| 0 | 0 | 0 | 0 | 0 |
| 0.022418867 | 0 | 0.028067537 | 0 | 0.298340647 |
| 0 | 0 | 0 | 0 | 0 |
| 0 | 0 | 0 | 0.015314907 | 0.002902125 |
| 0.004629123 | 0 | 0.003910302 | 0 | 0.001433666 |
| 0 | 0 | 0 | 0 | 0 |
| 0.017749303 | 0 | 0 | 0 | 0 |
| 0 | 0 | 0 | 0 | 0 |
| 0 | 0 | 0 | 0 | 0 |
| 0 | 0 | 0 | 0 | 0.023164297 |
| 0.016418039 | 1.319118443 | 0.002026023 | 0 | 2.950429711 |
| 0 | 0 | 0 | 0 | 0 |
| 0 | 0 | 0 | 0 | 0 |
| 0 | 0 | 0 | 0 | 0 |
| 0.001235906 | 0 | 0 | 0 | 0 |
| 0 | 0 | 0 | 0 | 0 |
| 0 | 0 | 0 | 0 | 0 |
| 1.271808502 | 0.002481125 | 0.05734362 | 0.004630397 | 0 |
| 0 | 0 | 0 | 0 | 0 |
| 0 | 0 | 0 | 0 | 0 |
| 0 | 0 | 0 | 0 | 0 |
| 0 | 0 | 0 | 0 | 0 |
| 0 | 0 | 0 | 0.004088427 | 0 |
| 0 | 0 | 0 | 0 | 0 |
| 0 | 0 | 0 | 0 | 0 |
| 0 | 0 | 0 | 0.076778273 | 0.006960896 |
| 0 | 0 | 0 | 0 | 0 |
| 0 | 0 | 0 | 0 | 0 |
| 0 | 0 | 0 | 0 | 0 |

| | | | | |
|---|---|---|---|---|
| 0.50291323 | 0 | 0.00100983 | 0 | 0 |
| 0 | 0 | 0 | 0 | 0 |
| 0 | 0 | 0 | 0 | 0 |
| 0 | 0 | 0 | 0 | 0 |
| 0 | 0 | 0 | 0 | 0 |
| 0 | 0 | 0 | 0 | 0 |
| 0 | 0 | 0 | 0 | 0 |
| 0 | 5.910537041 | 3.124598182 | 1.751874742 | 1.21047703 |
| 0.492570855 | 0.841373745 | 0.870472725 | 2.359611365 | 0.025183198 |
| 0.722372393 | 0.114764462 | 0.013609644 | 0 | 0 |
| 0 | 0 | 0 | 0 | 0 |
| 0 | 0 | 0 | 0 | 0 |
| 0 | 0 | 0 | 0.005275648 | 0 |
| 0 | 0 | 0 | 0.58413809 | 0 |
| 0 | 0 | 0 | 0 | 0 |
| 0 | 0 | 0 | 0 | 0 |
| 0 | 0 | 0 | 0 | 0.012835782 |
| 0 | 0 | 0 | 0 | 0 |
| 0 | 0 | 0 | 0 | 0 |
| 0.004674717 | 0 | 0 | 0.003608176 | 0 |
| 0 | 0 | 0 | 0 | 0 |
| 0 | 0 | 0 | 0 | 0 |
| 0 | 0 | 0 | 0 | 0 |
| 0 | 0 | 0 | 0 | 0 |
| 0 | 0 | 0 | 0.038639843 | 0 |
| 0 | 0 | 0 | 0 | 0 |
| 0 | 0 | 0 | 0 | 0 |
| 0 | 0 | 0 | 0 | 0 |
| 0 | 0 | 0 | 0 | 0 |
| 0 | 0 | 0 | 0 | 0 |
| 0 | 0 | 0 | 0 | 0 |
| 0 | 0 | 0 | 0 | 0 |
| 0 | 0 | 0 | 0 | 0 |
| 0 | 0 | 0 | 0 | 0 |
| 0 | 0 | 0 | 0 | 0 |
| 0 | 0 | 0 | 0 | 0 |
| 0 | 0 | 0 | 0 | 0 |
| 0 | 0 | 0 | 0 | 0.07945572 |
| 0 | 0 | 0 | 0 | 0.001284053 |
| 0 | 0.061772165 | 0.248839162 | 0 | 0 |
| 0 | 0 | 0 | 0 | 0 |
| 0 | 0 | 0 | 0.121066709 | 1.225963746 |
| 0 | 0 | 0 | 0 | 0 |
| 0 | 0 | 0 | 0 | 0 |
| 0 | 0 | 0 | 0 | 0 |
| 0 | 0 | 0 | 0.039798341 | 0 |
| 0 | 0 | 0 | 0 | 0 |
| 0 | 0 | 0 | 0 | 0 |

| | | | | |
|---|---|---|---|---|
| 0 | 0 | 0 | 0 | 0 |
| 0 | 0 | 0.003797564 | 0 | 0.001392332 |
| 0 | 0 | 0 | 0 | 0 |
| 0 | 0 | 0 | 0 | 0 |
| 0 | 0 | 0 | 0 | 0.130427318 |
| 0 | 0 | 0 | 0.000734738 | 0 |
| 0 | 0 | 0 | 0 | 0 |
| 2.04866812 | 0.051569564 | 0.117752842 | 0.003444438 | 0.010008216 |
| 0 | 0 | 0 | 0 | 0.004310634 |
| 0 | 0 | 0 | 0 | 0 |
| 0 | 0 | 0 | 0 | 0 |
| 0 | 0 | 0 | 0 | 0 |
| 0 | 0 | 0 | 0 | 0 |
| 0 | 0 | 0 | 0 | 0 |
| 0 | 0.324817694 | 0.001507161 | 0.043788366 | 0 |
| 0 | 0 | 0 | 0.165006352 | 0.024508379 |
| 0 | 0 | 0 | 0 | 0 |
| 0 | 0 | 0 | 0 | 0 |
| 0 | 0 | 0 | 0 | 0 |
| 0 | 0 | 0 | 0 | 0 |
| 0 | 0 | 0 | 0 | 0 |
| 0 | 0 | 0 | 0 | 0 |
| 0 | 0 | 0 | 0 | 0 |
| 0.041126725 | 0.004436822 | 0 | 0 | 0 |
| 0 | 0 | 0 | 0 | 0.267598707 |
| 0 | 0 | 0 | 0 | 0 |
| 0.097440912 | 0.365217966 | 0.096254356 | 0.455300585 | 0.003106561 |
| 0.000943034 | 0 | 0 | 0 | 0 |
| 0.261978947 | 0.001774024 | 0 | 0 | 0 |
| 0 | 0 | 0 | 0 | 0 |
| 0 | 0 | 0 | 0.001408561 | 0 |
| 0 | 0 | 0 | 0 | 0 |
| 0 | 0 | 0 | 0 | 0 |
| 0 | 0 | 0 | 0 | 0 |
| 0 | 0 | 0 | 0 | 0 |
| 0 | 0 | 0 | 0 | 0 |
| 0 | 0 | 0 | 0 | 0 |
| 0 | 0 | 0 | 0 | 0 |
| 0 | 0 | 0 | 0 | 0.120752788 |
| 0 | 0 | 0 | 0 | 0 |
| 0 | 0 | 0 | 0 | 0 |
| 0 | 0 | 0 | 0 | 0 |
| 0 | 0 | 0 | 0 | 0 |
| 0 | 0 | 0 | 0 | 0 |
| 0 | 0 | 0 | 0 | 0 |
| 0 | 0 | 0 | 0 | 0 |
| 0 | 0 | 0 | 0 | 0 |
| 0 | 0 | 0 | 0 | 0 |

| | | | | |
|---|---|---|---|---|
| 0 | 0 | 0 | 0.182790843 | 1.091746838 |
| 0 | 0 | 0 | 0 | 0 |
| 0 | 0 | 0 | 0 | 0 |
| 0 | 0 | 0 | 0 | 0 |
| 0 | 0 | 0 | 0 | 0 |
| 16.97631831 | 0 | 0 | 0 | 0 |
| 0 | 0 | 0 | 0 | 0 |
| 0 | 0 | 0 | 0 | 0 |
| 0 | 0 | 0 | 0 | 0 |
| 0 | 0 | 0 | 0 | 0 |
| 0 | 0 | 0 | 0 | 0 |
| 0.004332832 | 0 | 0 | 0.008728603 | 0.000782777 |
| 0 | 0 | 0 | 0 | 0 |
| 0 | 0 | 0 | 0 | 0 |
| 0 | 0 | 0 | 0 | 0 |
| 0 | 0 | 0 | 0 | 0 |
| 0 | 0 | 0 | 0 | 0 |
| 0 | 0 | 0 | 0 | 0 |
| 0 | 0 | 0 | 0 | 0 |
| 0 | 0 | 0 | 0 | 0 |
| 0.021842562 | 0.071991891 | 0.002029588 | 0 | 0 |
| 0 | 0 | 0 | 0 | 0 |
| 0.011822385 | 0.005579959 | 0.002796242 | 0.091517117 | 0.020077014 |
| 0 | 0 | 0 | 0 | 0 |
| 0 | 0 | 0 | 0 | 0 |
| 0 | 0 | 0 | 0 | 0 |
| 0 | 0 | 0 | 0 | 0 |
| 0 | 0 | 0 | 0 | 0 |
| 0 | 0 | 0 | 0 | 0 |
| 0 | 0 | 0 | 0 | 0 |
| 0 | 0 | 0 | 0 | 0 |
| 0 | 0 | 0 | 0 | 0 |
| 0.01522347 | 0.055637367 | 0.032951584 | 0.001652152 | 0.009200999 |
| 0 | 0 | 0 | 0 | 0 |
| 0 | 0 | 1.172559846 | 0.695324862 | 0.770959687 |
| 0.004812441 | 0 | 0 | 0 | 0 |
| 0 | 0 | 0 | 0 | 0 |
| 0 | 0 | 0 | 0 | 0 |
| 0.034040368 | 0.116976666 | 0.002430569 | 0.003450325 | 0 |
| 0 | 0 | 0 | 0 | 0 |
| 0 | 0 | 0 | 0 | 0 |
| 0 | 0 | 0 | 0 | 0 |
| 0 | 0 | 0 | 0 | 0.01606431 |
| 0 | 0 | 0 | 0 | 0 |
| 0 | 0 | 0 | 0 | 0 |
| 0 | 0 | 0 | 0 | 0.00310326 |
| 0 | 0 | 0 | 0 | 0 |
| 0 | 0 | 0 | 0 | 0 |

| | | | | |
|---|---|---|---|---|
| 0 | 0 | 0 | 0 | 0 |
| 0 | 0 | 0 | 0 | 0 |
| 0 | 0 | 0 | 0 | 0 |
| 0 | 0 | 0 | 0 | 0 |
| 0 | 0 | 0 | 0 | 0 |
| 0 | 0 | 0 | 0 | 0 |
| 0 | 0 | 0 | 0 | 0 |
| 0 | 0 | 0 | 0 | 0 |
| 0 | 0 | 0 | 0.076215181 | 0.00597646 |
| 0 | 0 | 0 | 0.009533768 | 0 |
| 0 | 0 | 0 | 0 | 0 |
| 0 | 0 | 0 | 0 | 0 |
| 0 | 0 | 0 | 0 | 0 |
| 0 | 0 | 0 | 0 | 0 |
| 0 | 0 | 0 | 0 | 0 |
| 0.321752617 | 0.025775474 | 0 | 0 | 0 |
| 0 | 0 | 0 | 0 | 0 |
| 0 | 0 | 0 | 0 | 0 |
| 0 | 0 | 0 | 0 | 0 |
| 0 | 0 | 0 | 0 | 2.483164959 |
| 0 | 0 | 0 | 0 | 0.002007226 |
| 0 | 0 | 0 | 0 | 0 |
| 0 | 0 | 0 | 0 | 0 |
| 0 | 0 | 0 | 0 | 0 |
| 0.001633778 | 0 | 0 | 0.004875983 | 0 |
| 0 | 0 | 0 | 0 | 0 |
| 0 | 0 | 0 | 0 | 0 |
| 0 | 0 | 0 | 0 | 0 |
| 0 | 0 | 0 | 0 | 0 |
| 0 | 0 | 0 | 0 | 0.019556246 |
| 0 | 0 | 0.008365834 | 0.454797462 | 0.3878064 |
| 0 | 0 | 0 | 0 | 0 |
| 0 | 0 | 0 | 0 | 0 |
| 0 | 0 | 0 | 0 | 0 |
| 0 | 0 | 0 | 0 | 0 |
| 0 | 0 | 0 | 0 | 0 |
| 0 | 0 | 0 | 0 | 0 |
| 0 | 0 | 0 | 0 | 0 |
| 0 | 0 | 0 | 0 | 0 |
| 0 | 0 | 0 | 0 | 0 |
| 0 | 0 | 0 | 0 | 0 |
| 0 | 0 | 0 | 0 | 0 |
| 0 | 0 | 0 | 0.003012948 | 0 |
| 0 | 0 | 0 | 0 | 0 |
| 0 | 0 | 0 | 0 | 0 |
| 0 | 0 | 0 | 0 | 0 |
| 0.139628943 | 0 | 0.670650007 | 0 | 0 |
| 0 | 0 | 0 | 0 | 0 |

| | | | | |
|---|---|---|---|---|
| 0 | 0 | 0 | 0 | 0 |
| 0 | 0 | 0 | 0 | 0 |
| 0 | 0 | 0 | 0 | 0 |
| 0 | 0 | 0 | 0 | 0 |
| 0.229735678 | 0 | 0.040525205 | 0.018899997 | 0.049838739 |
| 0 | 0 | 0 | 0 | 0 |
| 0 | 0 | 0 | 0 | 0 |
| 0 | 0 | 0 | 0 | 0 |
| 0 | 0 | 0 | 0 | 0 |
| 0 | 0 | 0 | 0 | 0 |
| 0 | 0 | 0 | 0 | 0 |
| 0.0397891 | 0 | 0.004654881 | 0.018387081 | 0.007049238 |
| 0.043371357 | 0 | 0 | 0.096667702 | 0 |
| 0 | 0 | 0 | 0 | 0 |
| 0 | 0 | 0 | 0 | 0 |
| 0 | 0 | 0 | 0.021894673 | 0 |
| 0 | 0 | 0 | 0 | 0.012127488 |
| 0 | 0 | 0 | 0 | 0 |
| 1.866511294 | 0.052455327 | 0.061452429 | 0.009281868 | 0.004494925 |
| 0.00253798 | 0.004791529 | 0.005402569 | 0.149218784 | 0.003301314 |
| 0 | 0 | 0 | 0 | 0 |
| 0 | 0 | 0 | 0 | 0 |
| 0 | 0 | 0 | 0 | 0 |
| 0 | 0 | 0 | 0 | 0 |
| 0 | 0 | 0 | 0 | 0 |
| 0 | 0 | 0 | 0 | 0 |
| 0.327611197 | 0.020327672 | 0 | 0.006221586 | 0.005878876 |
| 0 | 0 | 0 | 0 | 0 |
| 0 | 0 | 0 | 0 | 0 |
| 0 | 0 | 0 | 0 | 0 |
| 0.012295565 | 0.019970153 | 0 | 0 | 0 |
| 0 | 0 | 0 | 0.002229517 | 0 |
| 0 | 0 | 0 | 0 | 0 |
| 0 | 0 | 0 | 0 | 0 |
| 0 | 0 | 0 | 0 | 0 |
| 0 | 0 | 0 | 0 | 0 |
| 0 | 0 | 0 | 0 | 0 |
| 0.311121242 | 0 | 0.006512662 | 0.116862611 | 0.008310449 |
| 0 | 0 | 0 | 0.262808358 | 0 |
| 0 | 0 | 0 | 0 | 0 |
| 1.244014136 | 0 | 0 | 0.010375722 | 0 |
| 0 | 0 | 0 | 0 | 0 |
| 0 | 0 | 0 | 0 | 0 |
| 0.03402606 | 3.138678007 | 0 | 0 | 0 |
| 0 | 0 | 0 | 0 | 0 |
| 0 | 0 | 0 | 0 | 0 |
| 0 | 0 | 0 | 0 | 0 |
| 0 | 0 | 0 | 0 | 0 |

| | | | | |
|---|---|---|---|---|
| 0 | 0 | 0 | 0 | 0 |
| 0 | 0 | 0 | 1.68608167 | 0 |
| 0 | 0 | 0 | 0 | 0 |
| 0.004824516 | 0.003913562 | 0 | 0 | 0 |
| 0 | 0 | 0.002944977 | 0 | 0 |
| 0.015818423 | 0 | 0 | 0.002248087 | 0 |
| 0 | 0 | 0 | 0.004556358 | 0 |
| 0.184886262 | 0 | 0 | 0 | 0 |
| 0 | 0 | 0 | 0 | 0 |
| 0 | 0 | 0 | 0 | 0 |
| 0 | 0 | 0 | 0 | 0.080263944 |
| 0 | 0 | 0 | 0 | 0 |
| 0 | 0.025165635 | 0 | 0.110231728 | 0.015473001 |
| 0 | 0 | 0 | 0 | 0 |
| 0 | 0 | 0 | 0 | 0 |
| 0 | 0 | 0 | 0 | 0 |
| 0 | 0 | 0 | 0 | 0 |
| 0 | 0 | 0 | 0 | 0 |
| 0 | 0 | 0 | 0 | 0 |
| 0 | 0 | 0 | 0 | 0 |
| 0 | 0 | 0 | 0 | 0 |
| 0 | 0 | 0 | 0 | 0 |
| 0.281157791 | 0.013307121 | 0 | 0 | 0.005942527 |
| 0 | 0 | 0 | 0.032477334 | 0.004207431 |
| 0 | 0 | 0 | 0 | 0 |
| 0 | 0 | 0 | 0 | 0 |
| 0 | 0 | 0 | 0 | 0 |
| 0 | 0 | 0 | 0 | 0 |
| 0 | 0 | 0 | 0 | 0 |
| 0 | 0 | 0 | 0 | 0 |
| 0 | 0 | 0.039448619 | 0.003909212 | 0.00643658 |
| 0 | 0 | 0 | 0 | 0 |
| 0 | 0 | 0 | 0 | 0 |
| 0 | 0 | 0 | 0 | 0 |
| 0 | 0 | 0 | 0 | 0 |
| 0.03309086 | 0 | 0 | 0 | 0 |
| 0.215700322 | 0 | 0 | 0 | 0 |
| 0 | 0 | 0 | 0 | 0 |
| 0 | 0 | 0.00646556 | 0.001488359 | 0 |
| 0 | 0 | 0 | 0 | 0 |
| 0 | 0 | 0 | 0 | 0 |
| 0.007744343 | 0 | 0 | 0 | 0 |
| 0 | 0 | 0 | 0 | 0 |
| 0 | 0 | 0 | 0 | 0 |
| 0 | 0 | 0 | 1.424990551 | 0 |
| 0 | 0 | 0.354232182 | 0.497031781 | 0.314963956 |
| 0 | 0 | 0 | 0 | 0 |
| 0 | 0 | 0 | 0 | 0 |

| | | | | |
|---|---|---|---|---|
| 0 | 0 | 0 | 0 | 0 |
| 0 | 0 | 0 | 0 | 0 |
| 0 | 0 | 0 | 0 | 0.51198737 |
| 0 | 0 | 0 | 0 | 0 |
| 0 | 0 | 0 | 0 | 0 |
| 0 | 0 | 0 | 0 | 0 |
| 0 | 0 | 0 | 0.00890925 | 0 |
| 0 | 0 | 0 | 0 | 0 |
| 0 | 0 | 0.006144048 | 0 | 0 |
| 0 | 0 | 0 | 0 | 0 |
| 0.004495317 | 0.002196595 | 0 | 0.000939134 | 0 |
| 0 | 0.001192554 | 0 | 0 | 0 |
| 0 | 0 | 0 | 0 | 0 |
| 0 | 0 | 0 | 0 | 0 |
| 0 | 0 | 0 | 0 | 0 |
| 0 | 0 | 0 | 0 | 0 |
| 0 | 0 | 0.003750989 | 0 | 0 |
| 0 | 0 | 0 | 0 | 0 |
| 0 | 0 | 0 | 0 | 0 |
| 0 | 0 | 0 | 0.009074013 | 0 |
| 0 | 0 | 0 | 0 | 0 |
| 0 | 0 | 0 | 0 | 0 |
| 0 | 0 | 0 | 0 | 0 |
| 0 | 0 | 0 | 0 | 0 |
| 0 | 0 | 0 | 0 | 0 |
| 0 | 0 | 0 | 0 | 0 |
| 0 | 0 | 0 | 0 | 0 |
| 0 | 0 | 0 | 0 | 0 |
| 0 | 0 | 0 | 0 | 0 |
| 0 | 0 | 0 | 0 | 0 |
| 0 | 0.001592578 | 0.004522431 | 0.008101223 | 0.000888266 |
| 0 | 0 | 0 | 0 | 0 |
| 0.01090228 | 0.065512564 | 0.025879946 | 0.028396513 | 0.036957324 |
| 0 | 0 | 0.007282369 | 0.00471545 | 0.00070263 |
| 0 | 0 | 0 | 0 | 0 |
| 0 | 0 | 0 | 0 | 0 |
| 0 | 0 | 0 | 0 | 0 |
| 0 | 0 | 0 | 0 | 0 |
| 0 | 0 | 0 | 0 | 0 |
| 0 | 0 | 0 | 0 | 0 |
| 0 | 0 | 0 | 0 | 0 |
| 1.43621467 | 0.759985415 | 0.120650113 | 0.083079525 | 0.076223263 |
| 0 | 0.265027853 | 0 | 0.169876351 | 0.347325855 |
| 0 | 0 | 0 | 0 | 0 |
| 0 | 0 | 0 | 0 | 0 |
| 0 | 0 | 0 | 0 | 0.042572724 |
| 0.009787889 | 0 | 0 | 0.016515895 | 0.001305819 |
| 0 | 0 | 0 | 0 | 0 |

| | | | | |
|---|---|---|---|---|
| 0 | 0 | 0 | 0 | 0 |
| 0 | 0 | 0.003544462 | 0 | 0.026171198 |
| 0.005591884 | 0.004212034 | 0.021851614 | 0.006312751 | 0.00822247 |
| 0 | 0 | 0 | 0 | 0 |
| 0 | 0 | 0 | 1.087689583 | 0 |
| 0.932701896 | 0.079070707 | 0.027351979 | 0 | 0 |
| 0 | 0 | 0 | 0 | 0 |
| 0 | 0 | 0 | 0 | 0 |
| 0 | 0 | 0 | 0 | 0 |
| 0 | 0 | 0 | 0 | 0 |
| 0 | 0 | 0 | 0 | 0 |
| 0 | 0 | 0 | 0 | 0 |
| 0 | 0 | 0 | 0 | 0 |
| 0.097885182 | 0 | 0 | 0 | 0.034378789 |
| 0 | 0 | 0 | 0 | 0 |
| 0 | 0.001027023 | 0 | 0 | 0 |
| 0 | 0 | 0 | 0 | 0 |
| 0 | 0 | 0 | 0 | 0 |
| 0 | 0 | 0 | 0 | 0 |
| 0 | 0 | 0 | 0 | 0 |
| 0 | 0 | 0 | 0 | 0 |
| 0 | 0 | 0 | 0 | 0 |
| 0 | 0 | 0 | 0 | 0 |
| 0 | 0 | 0 | 0 | 0 |
| 0 | 0 | 0 | 0 | 0 |
| 0.005240741 | 0 | 0 | 0 | 0.007690941 |
| 0 | 0 | 0 | 0 | 0 |
| 0 | 0 | 0 | 0.05549532 | 0 |
| 0 | 0 | 0 | 0 | 0 |
| 0 | 0 | 0 | 0 | 0 |
| 0 | 0 | 0 | 0 | 0 |
| 0.001414098 | 0 | 0 | 0 | 0 |
| 0 | 0 | 0 | 0 | 0 |
| 0 | 0 | 0 | 0 | 0 |
| 0 | 0 | 0 | 0 | 0 |
| 0 | 0 | 0 | 0 | 0 |
| 0 | 0 | 0 | 0 | 0 |
| 0.345507211 | 4.057883274 | 0.007403178 | 0 | 0 |
| 0 | 0 | 0 | 0.219400992 | 1.668881958 |
| 0 | 0 | 0 | 0 | 0 |
| 0 | 0 | 0 | 0 | 0 |
| 0 | 0 | 0 | 0 | 0.000758499 |
| 0.397508605 | 0 | 0.001339713 | 0 | 0 |
| 0 | 0 | 0 | 0.559264951 | 0 |
| 0 | 0 | 0 | 0 | 0 |
| 0 | 0 | 0 | 0 | 0 |
| 0.320302411 | 0.018146413 | 0 | 0.002770832 | 0.002415293 |
| 0 | 0 | 0 | 0 | 0 |

| | | | | |
|---|---|---|---|---|
| 0 | 0 | 0 | 0 | 0 |
| 0.07790383 | 0.052820221 | 0.007019386 | 0.026047291 | 0.013883762 |
| 0 | 0 | 0 | 0 | 0 |
| 0 | 0 | 0 | 0 | 0 |
| 0 | 0 | 0 | 0 | 0 |
| 0 | 0 | 0 | 0 | 0 |
| 0 | 0 | 0 | 0 | 0 |
| 0 | 0 | 0 | 0 | 0 |
| 0 | 0 | 0 | 0 | 0 |
| 0 | 0 | 0 | 0 | 0 |
| 0 | 0 | 0 | 0 | 0 |
| 0 | 0 | 0 | 0 | 0 |
| 0 | 0 | 0 | 0.054144619 | 0 |
| 0 | 0 | 0 | 0 | 0 |
| 0.009007445 | 0.008759552 | 0.00207287 | 0.009154604 | 0.010133244 |
| 0 | 0 | 0 | 0 | 0 |
| 0 | 0 | 0 | 0 | 0 |
| 0 | 0 | 0 | 0 | 0 |
| 0 | 0 | 0 | 0.001446766 | 0 |
| 0 | 0 | 0 | 0 | 0 |
| 0 | 0 | 0 | 0 | 0 |
| 0 | 0.038233651 | 0.002261913 | 0.077061799 | 0.001865934 |
| 0 | 0 | 0 | 0 | 0.000955878 |
| 0 | 0 | 0 | 0.474916836 | 0 |
| 0 | 0 | 0 | 0 | 0 |
| 0 | 0 | 0 | 0 | 0 |
| 0.089442544 | 0 | 0 | 0 | 0 |
| 0 | 0 | 0 | 0 | 0 |
| 0 | 0 | 0 | 0 | 0 |
| 0 | 0 | 0 | 0 | 0 |
| 0 | 0 | 0 | 0 | 0 |
| 0 | 0 | 0 | 0 | 0 |
| 0 | 0 | 0 | 0 | 0 |
| 0 | 0 | 0 | 0 | 0 |
| 0.001953656 | 0 | 0 | 0 | 0 |
| 0 | 0 | 0 | 0 | 0 |
| 0 | 0 | 0 | 0 | 0 |
| 0 | 0 | 0 | 0 | 0 |
| 0.00238752 | 0 | 0.023491455 | 0.051944983 | 0.004140801 |
| 0 | 0 | 0 | 0 | 0 |
| 0.018838399 | 0.728904258 | 0.028354303 | 0.019933568 | 0.36032489 |
| 0 | 0 | 0 | 0 | 0 |
| 0 | 0 | 0 | 0 | 0 |
| 0 | 0 | 0 | 0 | 0 |
| 0 | 0 | 0 | 0 | 0 |
| 0.442158039 | 2.752857823 | 0.393275107 | 0.174899952 | 0.006588857 |
| 0 | 0 | 0 | 0 | 0 |
| 0 | 0 | 0 | 0 | 0 |

| | | | | |
|---|---|---|---|---|
| 0 | 0 | 0 | 0 | 0 |
| 0 | 0 | 0 | 0 | 0 |
| 0 | 0 | 0 | 0 | 0 |
| 0 | 0 | 0 | 0 | 0 |
| 0 | 0 | 0 | 0.001497911 | 0 |
| 0 | 0 | 0 | 0 | 0 |
| 0 | 0 | 0 | 0 | 0 |
| 0 | 0 | 0 | 0 | 0 |
| 0 | 0 | 0 | 0 | 0 |
| 0 | 0 | 0.013153404 | 0.084560423 | 0.052174906 |
| 0 | 0 | 0 | 0 | 0.004384533 |
| 0 | 0 | 0 | 0 | 0 |
| 0.00140576 | 0 | 0.006649834 | 0 | 0 |
| 0 | 0 | 0 | 0 | 0 |
| 0 | 0 | 0 | 0 | 0 |
| 0 | 0 | 0 | 0 | 0 |
| 0 | 0 | 0 | 0 | 0 |
| 0 | 0 | 0 | 0 | 0 |
| 0 | 0 | 0 | 0 | 0 |
| 0.121160338 | 0.206486386 | 0.319221755 | 0.847303915 | 0.514848226 |
| 0 | 0 | 0 | 0 | 0 |
| 0 | 0 | 0 | 0 | 0 |
| 0 | 0 | 0 | 0 | 0 |
| 0 | 0 | 0 | 0 | 0 |
| 0 | 0 | 0 | 0 | 0 |
| 0 | 0 | 0 | 0 | 0 |
| 0 | 0 | 0 | 0 | 0 |
| 0 | 0 | 0 | 0 | 0 |
| 0 | 0 | 0 | 0 | 0 |
| 0.04191085 | 0 | 0 | 0 | 0.007346147 |
| 0 | 0 | 0 | 0 | 0 |
| 0 | 0 | 0 | 0 | 0 |
| 0 | 0 | 0 | 0 | 0 |
| 0 | 0 | 0 | 0 | 0 |
| 0 | 0 | 0 | 0 | 0 |
| 0 | 0 | 0.007406386 | 0.005559003 | 0.002106826 |
| 0 | 0 | 0 | 0 | 0 |
| 0 | 0 | 0 | 0 | 0 |
| 0 | 0 | 0 | 0 | 0 |
| 0 | 0 | 0 | 0 | 0 |
| 0 | 0 | 0 | 0 | 0 |
| 0 | 0 | 0 | 0 | 0 |
| 0 | 0 | 0 | 0 | 0.003457369 |
| 0 | 0 | 0 | 0 | 0 |
| 0.026833665 | 0 | 0 | 0.040159221 | 0.003566861 |
| 0 | 0 | 0 | 0 | 0 |
| 0 | 0 | 0 | 0 | 0 |
| 0.001960683 | 0 | 0 | 0 | 0 |

| | | | | |
|---|---|---|---|---|
| 0 | 0 | 0 | 0 | 0 |
| 0 | 0 | 0 | 0 | 0 |
| 0 | 0 | 0 | 0 | 0 |
| 0 | 0 | 0 | 0 | 0 |
| 0 | 0 | 0 | 0 | 0 |
| 0.085685921 | 0 | 0.045789876 | 0.012568211 | 0.0109048 |
| 0 | 0 | 0 | 0 | 0 |
| 0 | 0 | 0 | 0.000467837 | 0 |
| 0 | 0 | 0 | 0 | 0 |
| 0 | 0 | 0 | 0 | 0 |
| 0 | 0 | 0 | 0 | 0 |
| 0 | 0 | 0 | 0 | 0 |
| 0 | 0 | 0 | 0 | 0 |
| 0 | 0 | 0 | 0 | 0 |
| 0 | 0 | 0 | 0 | 0 |
| 0 | 0 | 0 | 0 | 0 |
| 0 | 0 | 0 | 0 | 0 |
| 0.008643832 | 0.023278504 | 0 | 0 | 0 |
| 0 | 0 | 0 | 0 | 0 |
| 0 | 0 | 0 | 0 | 0 |
| 0 | 0 | 0 | 0 | 0 |
| 0 | 0 | 0 | 0 | 0 |
| 0 | 0 | 0 | 0 | 0 |
| 0 | 0 | 0 | 0 | 0 |
| 0 | 0 | 0 | 0 | 0 |
| 0 | 0 | 0 | 0 | 0 |
| 0 | 0 | 0 | 0 | 0 |
| 0 | 0 | 0 | 0 | 0 |
| 0 | 0 | 0 | 0 | 0 |
| 0 | 0 | 0 | 0 | 0 |
| 0 | 0 | 0 | 0 | 0 |
| 0 | 0 | 0 | 0 | 0 |
| 0 | 0 | 0 | 0 | 0 |
| 0 | 0 | 0 | 0 | 0 |
| 0 | 0 | 0 | 0 | 0 |
| 0 | 0 | 0 | 0 | 0 |
| 0 | 0 | 0 | 0 | 0 |
| 0 | 0 | 0 | 0 | 0 |
| 0 | 0 | 0 | 0 | 0.735647134 |
| 0 | 0 | 0 | 0 | 0 |
| 0 | 0 | 0 | 0 | 0 |
| 0 | 0 | 0 | 0 | 0 |
| 0 | 0 | 0 | 0 | 0 |
| 0 | 0 | 0 | 0 | 0 |
| 0 | 0 | 0 | 0 | 0 |
| 0 | 0 | 0 | 0 | 0 |
| 0 | 0 | 0 | 0 | 0 |
| 0 | 0 | 0 | 0 | 0 |

| | | | | |
|---|---|---|---|---|
| 0 | 0 | 0 | 0 | 0 |
| 0.00163707 | 0 | 0.202428856 | 0.159033029 | 0.005678513 |
| 0 | 0 | 0 | 0 | 0 |
| 0 | 0 | 0 | 0 | 0 |
| 0 | 0 | 0 | 0 | 0 |
| 0 | 0 | 0 | 0 | 0 |
| 0 | 0 | 0 | 0 | 0 |
| 0 | 0 | 0 | 0 | 0 |
| 0 | 0 | 0 | 0 | 0 |
| 0 | 0 | 0 | 0 | 0 |
| 0 | 0 | 0.003650468 | 0.001919273 | 0.003160114 |
| 0 | 0 | 0 | 0 | 0 |
| 0 | 0 | 0 | 0.223496768 | 0.700876564 |
| 0 | 0 | 0 | 0 | 0 |
| 0 | 0 | 0 | 0 | 0 |
| 0 | 0 | 0 | 0 | 0 |
| 0 | 0 | 0 | 0 | 0 |
| 0 | 0 | 0 | 0 | 0 |
| 0 | 0 | 0 | 0 | 0 |
| 0 | 0 | 0 | 0 | 0 |
| 2.385587554 | 0 | 0 | 0 | 0 |
| 0 | 0 | 0 | 0 | 0 |
| 0 | 0 | 0 | 0 | 0 |
| 0 | 0 | 2.007443629 | 0.630625499 | 0.612504478 |
| 0 | 0 | 0 | 0 | 0 |
| 0 | 0 | 0 | 0 | 0 |
| 0 | 0 | 0 | 0 | 0 |
| 0 | 0 | 0 | 0 | 0.000866754 |
| 0 | 0 | 0 | 0 | 0 |
| 0 | 0 | 0 | 0 | 0 |
| 0 | 0 | 0 | 0 | 0 |
| 0 | 0 | 0 | 0 | 0 |
| 0 | 0 | 0 | 0 | 0 |
| 0 | 0 | 0 | 0 | 0 |

| PHL018-0038_V2 | PHL018-0089_V2 | PHL018-0254_V3 | PHL018-0264_V3 | PHL018-0220_V3 |
|---|---|---|---|---|
| CTRL | CTRL | REF | REF | REF |
| 0.748653362 | 0.007298851 | 0.020810557 | 0.166114693 | 0.33016286 |
| 0.762268635 | 0.050721065 | 7.066690126 | 1.124891415 | 1.095644789 |
| 0.598339973 | 0 | 5.269633535 | 2.583592292 | 0 |
| 0 | 0 | 0 | 0 | 0 |
| 0 | 0 | 0 | 0 | 0 |
| 0 | 0 | 0 | 0 | 0 |
| 0 | 0 | 0 | 0 | 0 |
| 0 | 0 | 0 | 0 | 0 |
| 0 | 0 | 0 | 0 | 0 |
| 0 | 0 | 0 | 0 | 0 |
| 0 | 0 | 0 | 0 | 0 |
| 0 | 0 | 0 | 0 | 0 |
| 0 | 0 | 0 | 0 | 0 |
| 0.04994762 | 0.31547312 | 2.745332929 | 2.285600411 | 0.496482719 |
| 0 | 0 | 0 | 0 | 0 |
| 0 | 0 | 0 | 0 | 0 |
| 0 | 0 | 0 | 0 | 0 |
| 0 | 0 | 0 | 0 | 0 |
| 0 | 0 | 0 | 0 | 0 |
| 0 | 0 | 0 | 0 | 0 |
| 0.045601313 | 0 | 0 | 0 | 0 |
| 0 | 0 | 0 | 0 | 0 |
| 0 | 0 | 0 | 0 | 0 |
| 0 | 0 | 0 | 0 | 0 |
| 0 | 0 | 0 | 0.014320012 | 0 |
| 0 | 0 | 0 | 0 | 0 |
| 0 | 0 | 0 | 0 | 0 |
| 0 | 0 | 0 | 0 | 0 |
| 0 | 0 | 0 | 0 | 0 |
| 0 | 0 | 0 | 0 | 0 |
| 0 | 0 | 0 | 0 | 0 |
| 0 | 0 | 0 | 0 | 0 |
| 0 | 0 | 0 | 0 | 0 |
| 0 | 0 | 0 | 0 | 0 |
| 0 | 0 | 0 | 0 | 0 |
| 0 | 0.467490671 | 7.878729241 | 4.233237107 | 0.030646367 |
| 0 | 0 | 0 | 0 | 0 |
| 0 | 0 | 0 | 0 | 0 |
| 0 | 0 | 0 | 0 | 0 |
| 0 | 0 | 0 | 0 | 0 |
| 0 | 0 | 0 | 0 | 0 |
| 0 | 0.079949441 | 0.00088693 | 0 | 0.000841308 |
| 0.010329248 | 0.393410815 | 0.217671417 | 0.53211263 | 0.242782448 |
| 0.006658642 | 0.09207611 | 0.01127117 | 0.078675311 | 0.076219349 |
| 0 | 0.019248783 | 0.009436779 | 0 | 0.000745947 |
| 0 | 0 | 0 | 0 | 0 |

| | | | | |
|---|---|---|---|---|
| 0.060803379 | 0.011993674 | 0 | 0 | 0 |
| 0 | 0 | 0.027812846 | 0.002755019 | 0.011971757 |
| 0 | 0.003456676 | 0.007729968 | 0.003742343 | 0.076073163 |
| 0.258631979 | 0.189599535 | 1.230477756 | 0.579276548 | 0.631059951 |
| 0.012012098 | 0 | 0 | 0.000875645 | 0 |
| 0 | 0 | 0 | 0.150758773 | 0 |
| 0.000625612 | 4.142744373 | 0 | 0 | 0.00166338 |
| 0 | 0 | 0 | 0 | 0 |
| 0.010408985 | 0 | 0 | 0.015090303 | 0 |
| 0 | 0 | 0 | 0 | 0 |
| 0 | 0.005417255 | 0 | 0 | 0 |
| 0.086388795 | 0.0013315 | 0 | 0.623402252 | 1.009042185 |
| 0 | 0 | 0 | 0 | 0.001852716 |
| 0.059440418 | 0 | 0 | 0.289425416 | 0.112148687 |
| 0 | 0 | 0 | 0 | 0 |
| 0 | 0 | 0 | 0 | 0 |
| 0 | 0 | 0 | 0.074549987 | 0.001633092 |
| 0 | 0 | 0 | 0 | 0 |
| 0 | 0 | 0 | 0 | 0 |
| 0 | 0 | 0 | 0 | 0 |
| 0.000946888 | 0 | 0.291762979 | 0.01707943 | 0.149796691 |
| 0.029930854 | 0 | 0.049305101 | 0.002397895 | 0.027014549 |
| 0.991672382 | 0.172162822 | 0 | 0 | 0 |
| 0 | 0 | 0 | 0 | 0 |
| 0 | 0 | 0 | 0 | 0 |
| 0 | 0 | 0 | 0 | 0 |
| 0 | 0 | 0 | 0 | 0 |
| 0 | 0 | 0 | 0 | 0 |
| 0 | 0.030655205 | 0 | 0 | 0 |
| 0 | 0 | 0 | 0 | 0 |
| 0 | 0 | 0 | 0 | 0.25062318 |
| 0 | 0 | 0 | 0 | 0 |
| 0 | 0 | 0 | 0 | 0 |
| 0 | 0 | 0 | 0 | 0 |
| 0 | 0 | 0 | 0 | 0 |
| 0 | 0 | 0 | 0 | 0 |
| 0 | 0 | 0 | 0 | 0 |
| 0 | 0 | 0 | 0 | 0 |
| 0 | 0 | 0 | 0 | 0 |
| 0 | 0 | 0 | 0 | 0 |
| 0 | 0 | 0 | 0 | 0 |
| 0 | 0 | 0 | 0 | 0 |
| 0 | 0 | 0 | 0 | 0 |
| 0 | 0 | 0 | 0 | 0 |
| 0 | 0 | 0 | 0 | 0 |
| 0 | 0 | 0 | 0 | 0 |
| 0 | 0 | 0 | 0 | 0 |
| 0 | 0 | 0 | 1.235702734 | 0 |

| | | | | |
|---|---|---|---|---|
| 0 | 0 | 0 | 0 | 0 |
| 0 | 0 | 0 | 0 | 0 |
| 0 | 0 | 0 | 0 | 0 |
| 0 | 0 | 0 | 0 | 0 |
| 0 | 0.026703567 | 0 | 0 | 0 |
| 0 | 0 | 0 | 0 | 0 |
| 0 | 0 | 0 | 0 | 0 |
| 0 | 0 | 0 | 0 | 0 |
| 0.001861865 | 0.008746419 | 0 | 0 | 0 |
| 0 | 0 | 0 | 0 | 0 |
| 0 | 0 | 0 | 0 | 0 |
| 0 | 0.003008717 | 0 | 0 | 0 |
| 0 | 0 | 0 | 0 | 0 |
| 0 | 0 | 0 | 0 | 0 |
| 0 | 0 | 0 | 0 | 0 |
| 0 | 0 | 0 | 0 | 0 |
| 0 | 0 | 0 | 0 | 0 |
| 0 | 0 | 0 | 0 | 0 |
| 0.088638755 | 0.108276155 | 0 | 0.025444161 | 0.102014569 |
| 0.000963347 | 0 | 0 | 0 | 0.00062727 |
| 0 | 0 | 0 | 0 | 0 |
| 0 | 0 | 0 | 0 | 0 |
| 0 | 0 | 0 | 0 | 0 |
| 0 | 0 | 0 | 0 | 0 |
| 0 | 0 | 0 | 0 | 0 |
| 0 | 0 | 0 | 0 | 0 |
| 0 | 0 | 0 | 0 | 0 |
| 0 | 0 | 0 | 0 | 0 |
| 0 | 0 | 0 | 0 | 0 |
| 0 | 0 | 0 | 0 | 0 |
| 0.27195558 | 0 | 0 | 0.172520832 | 0.141202448 |
| 0 | 0.000471721 | 0.001837254 | 0 | 0.001267454 |
| 0 | 0 | 0 | 0 | 0 |
| 0 | 0 | 0 | 0 | 0 |
| 0 | 0 | 0 | 0 | 0 |
| 0 | 0 | 0 | 0 | 0 |
| 0 | 0 | 0 | 0 | 0 |
| 0 | 0 | 0 | 0 | 0 |
| 0 | 0 | 0 | 0.0773229 | 0 |
| 0 | 0 | 0 | 0 | 0 |
| 0 | 0 | 0 | 0 | 0 |
| 0 | 0 | 0 | 0 | 0 |
| 0 | 0 | 0.000579157 | 0 | 0 |
| 0 | 0 | 0 | 0 | 0 |
| 0 | 0 | 0 | 0 | 0.000585701 |
| 0 | 0 | 0 | 0 | 0 |
| 0 | 0 | 0 | 0 | 0 |
| 0 | 0 | 0 | 0 | 0 |

| | | | | |
|---|---|---|---|---|
| 0 | 0 | 0 | 0 | 0 |
| 0 | 0 | 0 | 0 | 0 |
| 0 | 0 | 0 | 0 | 0 |
| 0 | 0 | 0 | 0 | 0 |
| 0.067769223 | 0.005200676 | 0 | 0.037454869 | 0.19870942 |
| 0 | 0 | 0 | 0 | 0 |
| 0 | 0 | 0 | 0 | 0 |
| 0 | 0 | 0 | 0 | 0 |
| 0 | 0 | 0 | 0 | 0 |
| 0 | 0 | 0 | 0 | 0 |
| 0 | 0 | 0 | 0 | 0 |
| 0 | 0 | 0 | 0 | 0 |
| 0 | 0 | 0 | 0 | 0 |
| 0 | 0 | 0 | 0 | 0 |
| 0 | 0 | 0 | 0 | 0 |
| 13.79371429 | 0.106801998 | 0 | 0 | 0 |
| 0 | 0 | 0 | 0 | 0 |
| 0 | 0 | 0 | 0 | 0 |
| 0 | 0 | 0 | 0 | 0 |
| 0 | 0 | 0 | 0 | 0 |
| 0.003143365 | 0 | 0 | 0 | 0 |
| 0 | 0 | 0 | 0 | 0 |
| 0 | 0 | 0 | 0 | 0 |
| 0 | 0 | 0 | 0 | 0 |
| 0 | 0 | 0 | 0.21337717 | 0 |
| 0 | 0 | 0 | 0 | 0 |
| 0 | 0 | 0 | 0 | 0 |
| 0 | 0 | 0 | 0 | 0 |
| 0 | 0 | 0 | 0 | 0 |
| 0 | 0 | 0 | 0 | 0 |
| 0 | 0 | 0 | 0 | 0.131448391 |
| 0 | 0 | 0 | 0 | 0 |
| 0 | 0 | 0 | 0 | 0 |
| 0 | 0 | 0 | 0 | 0 |
| 0 | 0 | 0 | 0 | 0 |
| 0 | 0 | 0 | 0 | 0 |
| 0 | 0 | 0 | 0 | 0 |
| 0.044108065 | 0.001524798 | 0 | 0 | 0 |
| 0 | 0 | 0 | 0 | 0 |
| 0 | 0 | 0 | 0 | 0 |
| 0 | 0 | 0 | 0 | 0 |
| 0 | 0 | 0 | 0 | 0 |
| 0 | 0 | 0 | 0 | 0 |
| 0 | 0 | 0 | 0 | 0 |
| 0 | 0 | 0 | 0 | 0 |
| 0 | 0 | 0 | 0 | 0 |
| 0 | 0 | 0 | 0 | 0 |
| 0 | 0 | 0 | 0 | 0 |

| | | | | |
|---|---|---|---|---|
| 0 | 0 | 0 | 0 | 0 |
| 0 | 0 | 0 | 0 | 0 |
| 0 | 0 | 0 | 0 | 0 |
| 0 | 0 | 0 | 0 | 0 |
| 0 | 0 | 0 | 0 | 0 |
| 0 | 0 | 0 | 0 | 0 |
| 0 | 0 | 0 | 0 | 0 |
| 0 | 0 | 0 | 0 | 0 |
| 0 | 0 | 0 | 0 | 0 |
| 0 | 0 | 0 | 0 | 0 |
| 0 | 0 | 0 | 0 | 0 |
| 0.011678898 | 0 | 0 | 0 | 0 |
| 0.002149078 | 0 | 0.008113742 | 0 | 0.00244885 |
| 0 | 0 | 0 | 0 | 0 |
| 0 | 0.001695442 | 0 | 0 | 0 |
| 0 | 0 | 0 | 0 | 0 |
| 0 | 0 | 0 | 0 | 0 |
| 0 | 0 | 0 | 0 | 0 |
| 0 | 0 | 0 | 0 | 0 |
| 0 | 0 | 0 | 0 | 0 |
| 0.019517263 | 0.009413253 | 0.018618646 | 0.095509874 | 0.073260185 |
| 0 | 0 | 0 | 0 | 0 |
| 0 | 0 | 0 | 0.002165149 | 0 |
| 0 | 0 | 0 | 0 | 0.00170526 |
| 0 | 0 | 0 | 0 | 0 |
| 0 | 0 | 0 | 0 | 0 |
| 0.000835764 | 0.003780729 | 0.011043858 | 0.001014396 | 0.010285314 |
| 0 | 0 | 0 | 0 | 0 |
| 0 | 0 | 0 | 0 | 0 |
| 0 | 0 | 0 | 0 | 0 |
| 0 | 0 | 0 | 0 | 0.002088515 |
| 0 | 0 | 0 | 0 | 0 |
| 0 | 0 | 0 | 0 | 0 |
| 0.519382858 | 0.177543165 | 0 | 0.171560799 | 0.033778931 |
| 0.094682036 | 0.029145428 | 0.139762938 | 0.006799931 | 0.167047233 |
| 0 | 0 | 0 | 0 | 0 |
| 0 | 0 | 0 | 0 | 0 |
| 0 | 0 | 0 | 0 | 0 |
| 0 | 0 | 0 | 0 | 0 |
| 0 | 0 | 0 | 0 | 0 |
| 0 | 0 | 0 | 0 | 0 |
| 0 | 0 | 0 | 0 | 0 |
| 0 | 0 | 0 | 0 | 0 |
| 0 | 0 | 0 | 0 | 0 |
| 0 | 0 | 0 | 0 | 0 |
| 0 | 0 | 0 | 0.241886109 | 0 |
| 0.002830802 | 0 | 0 | 0 | 0 |
| 0 | 0 | 0.013429058 | 0.026382769 | 0 |

| | | | | |
|---|---|---|---|---|
| 0 | 0 | 0 | 0 | 0 |
| 0 | 0 | 0 | 0 | 0 |
| 0 | 0 | 0 | 0 | 0 |
| 0 | 0 | 0 | 0 | 0 |
| 0 | 0 | 0 | 0 | 0 |
| 0 | 0 | 0 | 0 | 0 |
| 0 | 0 | 0 | 0 | 0 |
| 0 | 0 | 0 | 0 | 0 |
| 0 | 0 | 0 | 0.009681619 | 0 |
| 0 | 0 | 0 | 0 | 0 |
| 0 | 0 | 0 | 0 | 0 |
| 0 | 0 | 0 | 0 | 0 |
| 0 | 0 | 0 | 0 | 0 |
| 0 | 0 | 0 | 0 | 0 |
| 0 | 0 | 0 | 0 | 0 |
| 0 | 0 | 0 | 0 | 0 |
| 0 | 0 | 0 | 0 | 0 |
| 0 | 0 | 0 | 0 | 0 |
| 0 | 0.001530635 | 0.000975519 | 0 | 0 |
| 1.410929953 | 0.001052423 | 0 | 0 | 0 |
| 0 | 0 | 0.02110518 | 0 | 0 |
| 0 | 0 | 0 | 0 | 0 |
| 0 | 0 | 0 | 0 | 0 |
| 0 | 0 | 0 | 0 | 0 |
| 0 | 0.003139978 | 0.009529523 | 0 | 0 |
| 0 | 0 | 0 | 0 | 0 |
| 0 | 0 | 0 | 0 | 0 |
| 0 | 0 | 0 | 0 | 0 |
| 0 | 0.004708827 | 0.001591478 | 0 | 0 |
| 0 | 0 | 0 | 0 | 0 |
| 0.269231398 | 0 | 0.067364964 | 0.401045215 | 1.541625336 |
| 0 | 0 | 0 | 0 | 0 |
| 0 | 0 | 0 | 0 | 0.005407893 |
| 0 | 0.000859166 | 0.0045631 | 0 | 0 |
| 0 | 0 | 0 | 0 | 0 |
| 0 | 0 | 0 | 0 | 0 |
| 0 | 0 | 0 | 0 | 0 |
| 0 | 0 | 0 | 0 | 0 |
| 0 | 0 | 0 | 0 | 0 |
| 0 | 0 | 0.00880736 | 0 | 0 |
| 0 | 0 | 0 | 0 | 0 |
| 0.017956596 | 0.023522814 | 0.001258303 | 0.000908106 | 0.002216645 |
| 0 | 0 | 0 | 0 | 0 |
| 0 | 0 | 0 | 0 | 0 |
| 0 | 0 | 0 | 0 | 0 |
| 0 | 0 | 0 | 0 | 0 |
| 0 | 0 | 0 | 0 | 0 |
| 0 | 0 | 0 | 0 | 0 |

| | | | | |
|---|---|---|---|---|
| 0 | 0 | 0 | 0 | 0 |
| 0 | 0 | 0 | 0 | 0 |
| 0.005636865 | 0.108372301 | 0.00609431 | 0.083240157 | 0.076435413 |
| 0 | 0 | 0 | 0 | 0 |
| 0.037014828 | 0 | 0 | 0 | 10.24906221 |
| 0 | 0 | 0 | 0 | 0 |
| 0 | 0 | 0 | 0 | 0 |
| 0 | 0 | 0 | 0 | 0 |
| 0 | 0 | 0 | 0 | 0 |
| 0 | 0 | 0 | 0 | 0 |
| 0 | 0 | 0 | 0 | 0 |
| 0 | 0 | 0 | 0 | 0 |
| 0 | 0 | 0 | 0 | 0 |
| 0 | 0 | 0 | 0.001680281 | 0.001892996 |
| 0.001885344 | 0.001938338 | 0.001441253 | 0.001733565 | 0.001953025 |
| 0.222219525 | 0.103139775 | 0 | 0.000928005 | 0.004181943 |
| 0 | 0 | 0.003518954 | 0 | 0.000890119 |
| 0 | 0 | 0 | 0 | 0 |
| 0 | 0 | 0 | 0 | 0 |
| 0 | 0 | 0 | 0 | 0 |
| 0 | 0 | 0 | 0 | 0 |
| 0.000705161 | 0 | 0.010165156 | 0.03138221 | 0 |
| 0 | 0 | 0 | 0 | 0 |
| 0 | 0 | 0.013131926 | 0 | 0 |
| 0 | 0 | 0 | 0 | 0.002576805 |
| 0 | 0 | 0 | 0 | 0 |
| 0 | 0.00472881 | 0 | 18.48585893 | 0 |
| 0 | 0 | 0 | 0 | 0 |
| 0 | 0 | 0 | 0 | 0 |
| 0 | 0 | 0 | 0 | 0.002324608 |
| 0 | 0 | 0 | 0 | 0 |
| 0 | 0 | 0 | 0 | 0 |
| 0 | 0 | 0 | 0 | 0 |
| 0 | 0 | 0 | 0 | 0 |
| 0 | 0 | 0 | 0 | 0 |
| 0 | 0 | 0 | 0 | 0 |
| 0 | 0 | 0 | 0 | 0 |
| 0.001107453 | 0.228695525 | 0.040709048 | 0 | 0.012619297 |
| 0 | 0 | 0 | 0 | 0 |
| 0 | 0 | 0 | 0 | 0 |
| 0 | 0 | 0 | 0 | 0 |
| 0 | 0 | 0 | 0 | 0 |
| 0 | 0 | 0 | 0 | 0 |
| 0 | 0 | 0 | 0 | 0 |
| 0 | 0 | 0 | 0 | 0 |
| 0 | 0 | 0 | 0 | 0 |
| 0 | 0 | 0 | 0 | 0 |
| 0.001073122 | 0 | 0 | 0 | 0 |

| | | | | |
|---|---|---|---|---|
| 0.325429206 | 0 | 0 | 0 | 6.970664036 |
| 0 | 0 | 0 | 0 | 0.16627087 |
| 0 | 0 | 0 | 0.000747055 | 0.000631221 |
| 0 | 0 | 0 | 0 | 0 |
| 0.003343969 | 0.002731271 | 0.007364581 | 0.002818535 | 0 |
| 0 | 0.017814343 | 0 | 0 | 0 |
| 0 | 0 | 0 | 0 | 0 |
| 0 | 0.017964397 | 0.006882055 | 0.042405222 | 0.030266434 |
| 0 | 0 | 0 | 0 | 0 |
| 0 | 0 | 0 | 0.023075642 | 0.011316294 |
| 0 | 0 | 0.081042374 | 0 | 0 |
| 0 | 0.002098133 | 0 | 0 | 0 |
| 0.004847261 | 0.001945177 | 0.004320231 | 0.009647326 | 0.01924281 |
| 0 | 0 | 0 | 0 | 0.003247207 |
| 0.002251425 | 0 | 0 | 0 | 0 |
| 0 | 0 | 0 | 0 | 0 |
| 0 | 0.001158251 | 0 | 0 | 0 |
| 0 | 0 | 0 | 0 | 0 |
| 0 | 0 | 0 | 0 | 0.031369688 |
| 0 | 0 | 0 | 0 | 0 |
| 0 | 0 | 0 | 0 | 0 |
| 0 | 0 | 0 | 0 | 0 |
| 0 | 0 | 0 | 0 | 0 |
| 0 | 0 | 0 | 0 | 0 |
| 0 | 0 | 0 | 0 | 0 |
| 0 | 0.002677569 | 0.002406593 | 0.001105247 | 0.001452692 |
| 0 | 0 | 0 | 0 | 0 |
| 0 | 0 | 0 | 0 | 0.00150575 |
| 0 | 0 | 0 | 0 | 0 |
| 0 | 0 | 0.013055701 | 0 | 0.001494638 |
| 0 | 0 | 0 | 0 | 0 |
| 0 | 0 | 0 | 0 | 0 |
| 0 | 0 | 0 | 0 | 0 |
| 0 | 0 | 0 | 0 | 0 |
| 0 | 0 | 0 | 0 | 0 |
| 0 | 0 | 0 | 0 | 0 |
| 0 | 0 | 0.001240054 | 0 | 0.348410672 |
| 0 | 0 | 0 | 0 | 0 |
| 0 | 0 | 0 | 0 | 0 |
| 0 | 0 | 0 | 0 | 0 |
| 0 | 0 | 0 | 0 | 0 |
| 0 | 0 | 0 | 0 | 0 |
| 0 | 0 | 0 | 0 | 0 |
| 0.005575549 | 0 | 0 | 0 | 0 |
| 0 | 0 | 0 | 0 | 0 |
| 0 | 0 | 0 | 0 | 0 |
| 0.007926967 | 0.002305224 | 0.000816215 | 0.010308462 | 0 |
| 0 | 0 | 0 | 0 | 0 |

| | | | | |
|---|---|---|---|---|
| 0 | 0 | 0 | 0 | 0 |
| 0 | 0 | 0 | 0 | 0 |
| 0 | 0 | 0 | 0 | 0 |
| 0 | 0 | 0 | 0 | 0 |
| 0 | 0 | 0 | 0 | 0 |
| 0.027649549 | 0 | 0 | 0.006675756 | 0 |
| 0 | 0 | 0 | 0 | 0 |
| 0 | 0 | 0 | 0 | 0 |
| 0 | 0 | 0 | 0 | 0 |
| 0 | 0 | 0 | 0 | 0 |
| 0.001862061 | 0 | 0.014166759 | 0.005440854 | 0.015559844 |
| 0 | 0 | 0 | 0 | 0 |
| 0 | 0 | 0 | 0 | 0 |
| 0.000586394 | 0 | 0.001408846 | 0 | 0.001559108 |
| 0 | 0 | 0 | 0 | 0.00473261 |
| 0 | 0 | 0 | 0 | 0 |
| 0.152501549 | 0 | 0 | 0 | 0 |
| 0 | 0 | 0 | 0 | 0.00108376 |
| 0 | 0 | 0 | 0 | 0 |
| 0 | 0 | 0 | 0 | 0 |
| 0 | 0 | 0 | 0 | 0 |
| 0 | 0.21743591 | 0.171762727 | 0 | 0 |
| 0.018548655 | 0 | 0 | 0 | 0 |
| 0 | 0 | 0 | 0 | 0 |
| 0 | 0 | 0 | 0 | 0 |
| 0 | 0 | 0 | 0 | 0 |
| 0 | 0 | 0.039716628 | 0 | 0.003139473 |
| 0 | 0 | 0 | 0 | 0 |
| 0 | 0 | 0 | 0 | 0 |
| 0.257859127 | 0.056379467 | 2.090198944 | 0.577550119 | 0.758728016 |
| 0.002519537 | 0.012447349 | 0.025965638 | 0.010461743 | 0.001813253 |
| 0 | 0 | 0 | 0 | 0 |
| 0 | 0 | 0 | 0 | 0 |
| 0 | 0 | 0 | 0 | 0 |
| 0 | 0 | 0 | 0 | 0 |
| 0 | 0 | 0 | 0 | 0 |
| 0 | 0.012553825 | 0 | 0 | 0 |
| 0 | 0 | 0 | 0 | 0 |
| 0 | 0 | 0 | 0 | 0 |
| 0 | 0 | 0 | 0 | 0 |
| 0.009480896 | 0 | 0 | 0.016903362 | 0.003662161 |
| 0.394431073 | 0.178334149 | 2.820686859 | 0.968148595 | 4.704733895 |
| 0 | 0 | 0 | 0 | 0 |
| 0 | 0 | 0 | 0 | 0 |
| 0 | 0 | 0 | 0 | 0 |
| 0 | 0 | 0 | 0 | 0 |
| 0 | 0 | 0.00060782 | 0.000852947 | 0.007495215 |
| 0 | 0.002499472 | 0 | 0 | 0.004317275 |

| | | | | |
|---|---|---|---|---|
| 0.000624291 | 0.023534086 | 0.001499896 | 0.000841915 | 0 |
| 0.007978067 | 0 | 0 | 0 | 0 |
| 0 | 0 | 0 | 0.143396627 | 0.001615498 |
| 0 | 0 | 0 | 0 | 0 |
| 0.001394587 | 0 | 0 | 0.042034341 | 0 |
| 0 | 0 | 0 | 0 | 0.156782083 |
| 0 | 0 | 0 | 0 | 0 |
| 0 | 0.003165232 | 0 | 0 | 0 |
| 0.002960694 | 0 | 0 | 0 | 0.009128772 |
| 0 | 0 | 0 | 0 | 0 |
| 0 | 0 | 0 | 0 | 0 |
| 0 | 0 | 0 | 0 | 0 |
| 0 | 0 | 0 | 0 | 0 |
| 0.00602432 | 0 | 0.010629193 | 0 | 0 |
| 0 | 0 | 0 | 0 | 0 |
| 0.015889171 | 0.012811498 | 0.011487755 | 0.00565462 | 0.107627315 |
| 0.007990398 | 0.137022807 | 0.02468239 | 0.002803907 | 0.014121991 |
| 0 | 0 | 0.001190006 | 0 | 0 |
| 0 | 0 | 0 | 0 | 0 |
| 0 | 0 | 0 | 0 | 0 |
| 0 | 0 | 0 | 0 | 0.006557057 |
| 0.002648128 | 0.001925241 | 0.003408368 | 0 | 0.001508756 |
| 0 | 0 | 0 | 0 | 0 |
| 0.210609441 | 0 | 0 | 0.659815545 | 0 |
| 0 | 0 | 0 | 0.077228334 | 0 |
| 0 | 0 | 0 | 0.003259106 | 0 |
| 0.004968428 | 0 | 0 | 0 | 0 |
| 0 | 0 | 0 | 0.042005424 | 0.142715657 |
| 0 | 0 | 0 | 0 | 0 |
| 0 | 0 | 0 | 0 | 0 |
| 0.015113903 | 0 | 0 | 0.025159665 | 0 |
| 0 | 0.003716268 | 0 | 0.019704606 | 0.002942049 |
| 0.004246191 | 0.001010968 | 0 | 0.031419777 | 0.007894372 |
| 0 | 0.004346449 | 0 | 0 | 0 |
| 0 | 0.008014169 | 0 | 0.012022944 | 0 |
| 0 | 0 | 0 | 0 | 0 |
| 0 | 0 | 0.001213562 | 0 | 0 |
| 0.004898258 | 0 | 0 | 0.041712175 | 0.084982979 |
| 0 | 0.024027191 | 0 | 0.211327054 | 0.225373381 |
| 0 | 0 | 0 | 0 | 0 |
| 0 | 0 | 0 | 0 | 0 |
| 0 | 0 | 0 | 0 | 0 |
| 0 | 0 | 0 | 0.001195029 | 0 |
| 0.001603485 | 0 | 0 | 0.00299416 | 0.019958127 |
| 0 | 0 | 0 | 0 | 0.007701197 |
| 0 | 0 | 0.000827644 | 0 | 0 |
| 0 | 0 | 0 | 0 | 0 |
| 0 | 0 | 0 | 0 | 0 |

| | | | | |
|---|---|---|---|---|
| 0 | 0 | 0 | 0 | 0 |
| 0.123370555 | 0.423192915 | 0.117165418 | 0.086454934 | 1.018648245 |
| 0 | 0 | 0 | 0 | 0 |
| 0.00169517 | 0 | 0 | 0 | 0 |
| 0 | 0 | 0 | 0 | 0 |
| 0 | 0 | 0 | 0 | 0 |
| 0 | 0 | 0 | 0 | 0 |
| 0 | 0 | 0 | 0 | 0 |
| 0 | 0 | 0 | 0 | 0 |
| 0 | 0 | 0 | 0 | 0 |
| 0.01215971 | 0 | 0 | 0 | 0.00237008 |
| 0.00214487 | 0.008355103 | 0.021185292 | 0.020006857 | 0.013849643 |
| 0.241986897 | 0 | 0 | 0 | 0 |
| 0 | 0 | 0 | 0 | 0 |
| 0 | 0 | 0.001102629 | 0.00111406 | 0 |
| 0 | 0 | 0 | 0 | 0 |
| 0.000670156 | 0.009284189 | 0.037032099 | 0 | 0.00515454 |
| 0 | 0 | 0 | 0 | 0.00892174 |
| 0 | 0 | 0 | 0 | 0 |
| 0 | 0 | 0 | 0.00314253 | 0.022368621 |
| 0.003109951 | 0.027941444 | 0.043378219 | 0.009960894 | 0.491006832 |
| 0 | 0.027717657 | 0 | 0 | 0 |
| 0 | 0 | 0 | 0 | 0 |
| 0 | 0 | 0 | 0 | 0 |
| 0 | 0.054497728 | 0 | 0 | 0 |
| 0 | 0 | 0 | 0 | 0 |
| 0.018929677 | 0 | 0 | 0.011211816 | 0 |
| 0.013405311 | 0.009553084 | 0.026912758 | 0.039747534 | 0.007323726 |
| 0 | 0 | 0 | 0 | 0 |
| 0 | 0 | 0 | 0.007841445 | 0.001514422 |
| 0 | 0 | 0 | 0 | 0 |
| 0 | 0 | 0 | 0 | 0 |
| 0 | 0 | 0 | 0.000901929 | 0 |
| 0 | 0 | 0 | 0 | 0 |
| 0 | 0 | 0.687430168 | 0 | 0 |
| 0.381028862 | 0.023900558 | 0.795974122 | 0.112850694 | 0.249315949 |
| 0 | 0 | 0 | 0 | 0 |
| 0 | 0 | 0 | 0 | 0 |
| 0 | 0.007941929 | 0 | 0 | 0.049045161 |
| 0 | 0 | 0 | 0 | 0 |
| 0 | 0 | 0 | 0 | 0.00874689 |
| 0 | 0 | 0 | 0 | 0 |
| 0.117235157 | 0 | 0.034712182 | 0.015726754 | 0.001097555 |
| 0 | 0 | 0.000638361 | 0 | 0.000605525 |
| 0 | 0 | 0 | 0 | 0 |
| 0.357154694 | 0.114561093 | 0 | 0.947810145 | 0.180025652 |
| 0 | 0 | 0 | 0 | 0 |
| 0 | 0 | 0 | 0 | 0 |

| | | | | |
|---|---|---|---|---|
| 0 | 0 | 0 | 0 | 0.003066616 |
| 0 | 0.240168949 | 0 | 0 | 0 |
| 0 | 0.009840153 | 0 | 0.001658085 | 0 |
| 2.325538666 | 0.578885741 | 0 | 0.060623388 | 0.340576797 |
| 0 | 0 | 0 | 0 | 0 |
| 0 | 0 | 0 | 0 | 0 |
| 0.00136671 | 0 | 0 | 0 | 0 |
| 0.031828166 | 0 | 0.07388568 | 0 | 0.038554994 |
| 0 | 14.286903 | 0.712676558 | 2.599418169 | 3.623045192 |
| 0.059012154 | 0.004049636 | 0 | 0.032741236 | 0.005712448 |
| 0 | 0 | 0 | 0 | 0 |
| 0.001535467 | 0.015454757 | 0.021580955 | 0.001863649 | 0.012772424 |
| 0 | 0 | 0 | 0 | 0 |
| 0 | 0 | 0 | 0 | 0 |
| 0.237069021 | 0.001146364 | 0.031355417 | 0.697560263 | 1.147398372 |
| 0 | 0 | 0 | 0 | 0 |
| 0.225770206 | 0.088972256 | 0 | 0.017218509 | 0.005461651 |
| 0 | 0 | 0 | 0 | 0 |
| 0 | 0 | 0 | 0 | 0 |
| 0.00095985 | 0 | 0 | 0 | 0 |
| 0 | 0 | 0 | 0 | 0 |
| 0 | 0 | 0 | 0 | 0 |
| 0.002969201 | 0 | 0 | 0.003289202 | 0 |
| 0.267414687 | 0 | 0.00073309 | 0.217392529 | 0 |
| 0.000575036 | 0 | 0 | 0 | 0 |
| 0.139417264 | 0.020106658 | 0 | 3.411583815 | 0.010467154 |
| 0 | 0 | 0 | 0.002249113 | 0 |
| 0 | 4.441159632 | 0.697433053 | 0.082368248 | 0 |
| 0 | 0 | 0 | 0 | 0 |
| 0 | 0 | 0 | 0 | 0 |
| 0.126583983 | 0.18400987 | 0 | 0 | 0.012076783 |
| 0 | 0 | 0 | 0.103844753 | 0 |
| 0.019313384 | 0 | 0.007915563 | 0.024475493 | 0 |
| 0 | 0 | 0 | 0 | 0 |
| 0 | 0 | 0 | 0 | 0 |
| 0 | 0 | 0 | 0 | 0 |
| 0.291943741 | 1.088841655 | 0 | 0.697375475 | 0.460443323 |
| 0 | 0 | 0 | 0 | 0 |
| 0 | 0 | 0.019777564 | 0.009538622 | 0 |
| 0 | 0 | 0 | 0 | 0 |
| 0.149804712 | 0.044311645 | 0.017694931 | 0 | 0.224076228 |
| 0 | 0 | 0 | 0 | 0 |
| 0.335739792 | 0.003996786 | 5.167984208 | 4.859781865 | 0.483449788 |
| 0 | 0 | 0 | 0.078261386 | 0 |
| 0 | 0 | 0 | 0 | 0 |
| 0 | 0 | 0 | 0 | 0 |
| 0 | 0 | 0 | 0 | 0 |
| 0.005489239 | 0 | 0 | 0.167994729 | 0.008877958 |

| | | | | |
|---|---|---|---|---|
| 0 | 0 | 0 | 0 | 0 |
| 0 | 0.555704525 | 0 | 0 | 0 |
| 0.002376412 | 0 | 0.111742557 | 0.002060238 | 0.14970788 |
| 0.001100909 | 0 | 0 | 0.002598188 | 0 |
| 0 | 0 | 0 | 0 | 0 |
| 0.005883572 | 0.047635497 | 0 | 0.017041246 | 0.020569901 |
| 0 | 0 | 0 | 0 | 0 |
| 0 | 0 | 0 | 0 | 0 |
| 0 | 0 | 0 | 0 | 0 |
| 0 | 0 | 0 | 0 | 0 |
| 0.331649871 | 0.000942385 | 0.050451216 | 0.515642391 | 0.033423307 |
| 0.346264808 | 0.293994259 | 0.143744651 | 0.749926429 | 0.337979277 |
| 0 | 0 | 0 | 0 | 0 |
| 0.015468334 | 0.46276256 | 0.027008437 | 0.002001152 | 0.007788229 |
| 0.085894444 | 0 | 0.44968168 | 0.147519762 | 0.672370822 |
| 0 | 5.752527053 | 0 | 0 | 0 |
| 0 | 0 | 0 | 0 | 0 |
| 0 | 0 | 0 | 0 | 0 |
| 0 | 0.207943937 | 0 | 0 | 0 |
| 0.093489393 | 0 | 0 | 0.026382212 | 0.633705489 |
| 0.024724092 | 0.159077323 | 0.006879792 | 0 | 1.991675258 |
| 0 | 0.038949996 | 0.289436214 | 0 | 0.005869997 |
| 0.05171854 | 0.099372482 | 0.007290585 | 0.533513436 | 0.437785688 |
| 0 | 0 | 0 | 0 | 0 |
| 0 | 0 | 0 | 0 | 0 |
| 0 | 0.397334966 | 0 | 0.007110568 | 11.28749546 |
| 0.006849818 | 0 | 0.001003482 | 0 | 0 |
| 0 | 0 | 0 | 0.003094465 | 0 |
| 0.229006197 | 0 | 0 | 0 | 0 |
| 0 | 0 | 0 | 0 | 0 |
| 0.011709167 | 0 | 0 | 0.005142434 | 0 |
| 0.066991246 | 0.001491372 | 0.064950507 | 0.372135351 | 0 |
| 0 | 0 | 0.643366025 | 0 | 0.20225393 |
| 0 | 0 | 0 | 0.236444995 | 0 |
| 0 | 0 | 0 | 0.001942458 | 0.029993437 |
| 0.019091183 | 0.118836025 | 0.314361711 | 0.890600415 | 0.14944949 |
| 0.008996011 | 0 | 0.000720449 | 0.150891291 | 0.04988748 |
| 0 | 0 | 0 | 0 | 0 |
| 0 | 3.774602676 | 0 | 0 | 0.038702601 |
| 0 | 0 | 0 | 0 | 0 |
| 0.045102006 | 0 | 0 | 0 | 0 |
| 0 | 0 | 0 | 0 | 0 |
| 0 | 0 | 0 | 0 | 0 |
| 0 | 0 | 0 | 0 | 0 |
| 0.034161513 | 0.002001758 | 0.007654673 | 0.157724108 | 0.346507747 |
| 0 | 0.002475521 | 0 | 0 | 0 |
| 0 | 0 | 0 | 0 | 0 |
| 0.097148677 | 1.163145004 | 7.014478873 | 0.0207319 | 1.014788843 |

| | | | | |
|---|---|---|---|---|
| 0 | 0 | 0 | 0 | 0 |
| 0 | 0 | 0 | 0 | 0 |
| 0 | 0 | 0 | 0 | 0 |
| 0.004676618 | 0.002900353 | 0 | 0.001373267 | 0 |
| 0 | 0 | 0 | 0 | 0 |
| 0 | 0 | 0 | 0.015632431 | 0 |
| 0 | 0 | 0 | 0 | 0 |
| 0 | 0.007849558 | 0.082878945 | 0.057706892 | 0.132239061 |
| 0 | 0 | 0 | 0 | 0 |
| 0 | 0 | 0 | 0 | 0 |
| 0.002688265 | 0 | 0.030073417 | 0.019543052 | 0.008806838 |
| 0 | 0 | 0 | 0 | 0 |
| 0 | 0 | 0 | 0.018450628 | 0 |
| 0.600447692 | 0 | 0.088010351 | 0 | 0.169907224 |
| 0 | 0 | 0 | 0 | 0 |
| 0 | 0 | 0 | 0 | 0 |
| 0.039862273 | 0 | 0 | 0 | 0 |
| 0.020700756 | 0.006169086 | 0 | 0.000565883 | 0.000796901 |
| 0.146591588 | 0 | 0 | 0 | 0.066916406 |
| 0 | 0 | 0 | 0 | 0.00186855 |
| 0 | 0 | 0 | 0 | 0 |
| 0 | 0 | 0 | 0 | 0 |
| 0.007095443 | 0.001458977 | 0 | 0.015005728 | 0 |
| 0 | 0 | 0 | 0 | 0 |
| 0 | 0 | 0 | 0 | 0 |
| 0 | 0 | 0 | 0 | 0 |
| 0 | 0 | 0 | 0 | 0 |
| 0 | 0 | 0 | 0 | 0 |
| 0 | 0 | 0 | 0 | 0 |
| 0 | 0.065158986 | 0.002638614 | 0.082371626 | 0.102040855 |
| 0 | 0 | 0.020824749 | 0 | 0 |
| 0 | 0 | 0 | 0 | 0 |
| 0 | 0 | 0 | 0 | 0 |
| 0 | 0 | 0 | 0 | 0 |
| 0.002805571 | 0.053485605 | 0 | 0 | 0.008981812 |
| 0 | 0 | 0 | 0 | 0 |
| 0.06557405 | 0.016056439 | 0 | 0.025556253 | 0.095423369 |
| 0.000853374 | 0 | 0 | 0.000575427 | 0.117985729 |
| 0 | 0 | 0 | 0 | 0 |
| 0 | 0 | 0 | 0 | 0 |
| 0 | 0 | 0 | 0 | 0 |
| 0 | 0 | 0 | 0 | 0 |
| 0 | 0 | 0 | 0 | 0 |
| 0.119474831 | 0.234618346 | 0 | 0 | 0.007239826 |
| 0 | 0.058445756 | 0 | 0.092618282 | 0 |
| 0 | 0 | 0 | 0 | 0 |
| 0.001891979 | 0 | 0 | 0 | 0 |
| 0.187167786 | 0 | 0 | 0 | 0 |

| | | | | |
|---|---|---|---|---|
| 0 | 0 | 0 | 0 | 0 |
| 0 | 0 | 0 | 0 | 0 |
| 0 | 0 | 0 | 0 | 0 |
| 0.47069115 | 0.005759933 | 0 | 0.01116144 | 0.046589824 |
| 0 | 0 | 0 | 0 | 0 |
| 0 | 0 | 0 | 0 | 0 |
| 0 | 0 | 0 | 0 | 0 |
| 0 | 0 | 0 | 0 | 0 |
| 0.008827088 | 0 | 0.072812809 | 0.002380831 | 0.013858194 |
| 0 | 0.003215619 | 0 | 0 | 0 |
| 0 | 0 | 0 | 0 | 0 |
| 0 | 0 | 0 | 0 | 0 |
| 0 | 0 | 0 | 0 | 0 |
| 0.001498692 | 0 | 0 | 0 | 0 |
| 0.359881018 | 0.011637581 | 0 | 0.022954952 | 0.480274261 |
| 0 | 0 | 0 | 0 | 0 |
| 0 | 0 | 0 | 0 | 0 |
| 0 | 0 | 0 | 0 | 0 |
| 0 | 0 | 0 | 0 | 0 |
| 0 | 0 | 0 | 0 | 0 |
| 0.030298092 | 0 | 0 | 0 | 0 |
| 0 | 0 | 0 | 0 | 0 |
| 0 | 0 | 0 | 0 | 0 |
| 0 | 0 | 0 | 0 | 0 |
| 0 | 0.431488268 | 0.020228747 | 0.018665264 | 0.073861494 |
| 7.48125371 | 2.952354883 | 0 | 0.478523866 | 0.731603504 |
| 6.466742878 | 0.408691527 | 0 | 0.032108473 | 0.199441586 |
| 0 | 0 | 0 | 0 | 0 |
| 0 | 2.116581353 | 0 | 0 | 0 |
| 0.194726339 | 0 | 0 | 0 | 0.08208133 |
| 0 | 0 | 0 | 0 | 0 |
| 0 | 0 | 0 | 0 | 0 |
| 0 | 0 | 0 | 0 | 0 |
| 0.002587108 | 0 | 0.006067691 | 0.089591438 | 0 |
| 0.001217925 | 0.101237055 | 0.061448895 | 0 | 0.016653737 |
| 0.00172185 | 0 | 0.081702777 | 0.00624058 | 0.007848115 |
| 0 | 0 | 0 | 0 | 0 |
| 0.448477483 | 0.123866245 | 0 | 0 | 0.646178643 |
| 0 | 0 | 0 | 0 | 0 |
| 0 | 0 | 0 | 0 | 0 |
| 0 | 0 | 0 | 0 | 0 |
| 0.001139012 | 0 | 0 | 0 | 0.033930585 |
| 0.115686169 | 8.756635717 | 0.720767508 | 0.239176837 | 0.335143412 |
| 0 | 0 | 0 | 0 | 0 |
| 0 | 0 | 0 | 0 | 0 |
| 0.029362266 | 0 | 0.01371701 | 0 | 0 |
| 0 | 0 | 0 | 0 | 0 |
| 0.357978644 | 1.424182684 | 0.00910535 | 0 | 4.288387838 |

| | | | | |
|---|---|---|---|---|
| 0 | 0 | 0.011057788 | 0.00767696 | 0 |
| 0 | 0 | 0 | 0 | 0 |
| 0 | 0 | 0 | 0 | 0 |
| 0 | 0 | 0 | 0 | 0 |
| 0.006431331 | 0 | 0.000965729 | 0.008944291 | 0.000732843 |
| 0 | 0.00945553 | 0 | 0 | 0 |
| 0 | 0 | 0 | 0 | 0 |
| 0 | 0 | 0 | 0 | 0 |
| 0 | 0 | 0 | 0 | 0 |
| 0 | 0 | 0 | 0 | 0 |
| 0 | 0 | 0 | 0 | 0 |
| 1.985138179 | 0.37895288 | 0 | 1.456796449 | 0.778015478 |
| 0.367751173 | 0.048877254 | 0 | 0.107905864 | 0.090730941 |
| 0 | 0 | 0 | 0 | 0 |
| 0 | 0.097153883 | 0.023900919 | 0 | 0.001483182 |
| 0 | 0 | 0 | 0 | 0 |
| 0.009572062 | 0 | 0 | 0 | 0 |
| 0 | 0 | 0 | 0 | 0 |
| 0.009289909 | 0 | 0 | 0 | 0 |
| 0 | 0 | 0 | 0.031805137 | 0 |
| 0.081769526 | 0.013276079 | 0 | 0 | 0.076416166 |
| 0.025363901 | 0 | 0 | 0 | 0 |
| 0.030110582 | 0.832497586 | 0 | 0.028371428 | 0.323242589 |
| 0 | 0 | 0 | 0 | 0 |
| 0 | 0 | 0 | 0 | 0 |
| 0 | 0 | 0 | 0 | 0 |
| 0 | 0 | 0 | 0 | 0 |
| 0 | 0 | 0 | 0 | 0 |
| 0 | 0 | 0 | 0 | 0.001526825 |
| 0 | 0 | 0 | 0 | 0 |
| 0 | 0 | 0 | 0 | 0 |
| 0.017855514 | 0.055667604 | 0.024637913 | 0.001220262 | 0.045916324 |
| 0 | 0 | 0 | 0 | 0 |
| 0.059307988 | 0 | 0 | 0.173376433 | 0 |
| 0.001873805 | 0 | 0.071843282 | 0.005369877 | 0.006227605 |
| 0.028181548 | 1.463792449 | 0.438892122 | 0.286873337 | 0 |
| 0 | 0.004702697 | 0 | 0 | 0 |
| 0 | 0 | 0 | 0 | 0 |
| 0 | 0.001825919 | 0.001508515 | 0 | 0 |
| 0 | 0 | 0 | 0 | 0 |
| 0.008780686 | 0.077786953 | 0 | 0 | 0.02213336 |
| 0 | 0 | 0 | 0 | 0 |
| 0 | 0 | 0 | 0 | 0 |
| 0.034576853 | 0 | 0 | 0 | 0 |
| 0.049223608 | 0.001913585 | 0.024946038 | 0.318947868 | 0.088867184 |
| 0 | 0 | 0 | 0 | 0 |
| 0.098750398 | 0 | 0 | 0 | 0 |
| 0.001018428 | 0 | 0 | 0 | 0 |

| | | | | |
|---|---|---|---|---|
| 0.004806919 | 0 | 0 | 0 | 0 |
| 0 | 0 | 0 | 0 | 0 |
| 0.008909168 | 0 | 0 | 0 | 0 |
| 0 | 0 | 0 | 0 | 0 |
| 0 | 0 | 0 | 0 | 0 |
| 0 | 0 | 0 | 0 | 0 |
| 0 | 0 | 0 | 0 | 0 |
| 0.031101291 | 0.025317806 | 0 | 0.003951153 | 0.382302198 |
| 0 | 0 | 0 | 0 | 0 |
| 0 | 0 | 0 | 0 | 0 |
| 0 | 0 | 0 | 0 | 0 |
| 0 | 0 | 0.000777817 | 0 | 0 |
| 0 | 0 | 0 | 0 | 0 |
| 0 | 0 | 0 | 0 | 0 |
| 0 | 0 | 0 | 0 | 0 |
| 0 | 0 | 0 | 0 | 0 |
| 0 | 0 | 0 | 0 | 0 |
| 0 | 0 | 0 | 0 | 0 |
| 2.903179965 | 0.008459045 | 0 | 0.186899816 | 0 |
| 0 | 0 | 0 | 0 | 0 |
| 0 | 0.019943822 | 0 | 0 | 0.006753217 |
| 0 | 0 | 0 | 0 | 0 |
| 0 | 0 | 0 | 0 | 0 |
| 0 | 0 | 0 | 0 | 0 |
| 0 | 0 | 0.02665003 | 0 | 0 |
| 0 | 0 | 0.03509377 | 0 | 0.016807485 |
| 0 | 0 | 0 | 0 | 0 |
| 0.175911278 | 1.552694863 | 0 | 0 | 0.118352698 |
| 0 | 0 | 0 | 0 | 0 |
| 1.539100445 | 0.846251462 | 0 | 1.944641298 | 0.647078898 |
| 0 | 0 | 0 | 0 | 0 |
| 0 | 0 | 0 | 0 | 0 |
| 0.032117229 | 0.001171678 | 0 | 0 | 0 |
| 0 | 0 | 0 | 0 | 0 |
| 0.001350622 | 0 | 0.011357325 | 0.027169808 | 0.008721101 |
| 0 | 0 | 0 | 0 | 0 |
| 0 | 0 | 0 | 0 | 0 |
| 0.009365643 | 0 | 0.013296356 | 0.000688933 | 0.00756745 |
| 0 | 0 | 0 | 0 | 0 |
| 0 | 0 | 0 | 0 | 0 |
| 3.75910263 | 0.526749066 | 20.14962973 | 1.954163833 | 7.704720801 |
| 1.045680225 | 0.004792911 | 0 | 1.481388427 | 0.231376785 |
| 0 | 0 | 0 | 0 | 0 |
| 0.19222852 | 0 | 0 | 0 | 0 |
| 0 | 0 | 0 | 0 | 0 |
| 0 | 0 | 0 | 0 | 0 |
| 0 | 0 | 0 | 0 | 0 |
| 0.010271183 | 0 | 0 | 0.005604451 | 0.017555844 |

| | | | | |
|---|---|---|---|---|
| 0 | 0 | 0 | 0 | 0 |
| 0.011537829 | 0 | 0 | 0.00281202 | 0 |
| 0 | 0 | 0 | 0 | 0 |
| 0 | 0 | 0 | 0 | 0 |
| 0.20404659 | 6.848558828 | 13.32142526 | 1.35319005 | 0.818033026 |
| 0 | 0 | 0 | 0 | 0 |
| 0 | 0 | 0 | 0 | 0 |
| 0 | 0 | 0 | 0 | 0 |
| 0 | 0 | 0 | 0 | 0.015025843 |
| 0 | 0 | 0 | 0 | 0 |
| 0 | 0 | 0 | 0 | 0 |
| 0 | 0 | 0 | 0 | 0 |
| 0.03768506 | 0.001222044 | 0 | 0.002732357 | 0.080496444 |
| 0.00763863 | 0 | 0.209654735 | 0.006046478 | 0.023463311 |
| 0 | 10.54396328 | 3.224552886 | 0 | 0 |
| 1.231429583 | 0.025119132 | 0 | 0.019433311 | 0.32747027 |
| 0.034199819 | 0.013813302 | 0.00189882 | 0 | 0.019812634 |
| 0 | 0 | 0 | 0 | 0 |
| 0 | 0 | 0 | 0 | 0 |
| 0 | 0 | 0 | 0 | 0 |
| 0 | 0 | 0 | 0 | 0 |
| 0 | 0 | 0 | 0 | 0 |
| 0 | 0 | 0 | 0 | 0 |
| 0 | 0 | 0 | 0 | 0 |
| 0 | 0 | 0 | 0 | 0 |
| 0 | 0 | 0 | 0 | 0 |
| 0.012994821 | 0.047197082 | 0 | 0.002780366 | 0.005837553 |
| 0 | 0 | 0.017614615 | 0.015829268 | 0 |
| 0 | 0 | 0 | 0 | 0 |
| 0 | 0 | 0 | 0 | 0 |
| 0 | 0 | 0 | 0 | 0 |
| 0 | 0 | 0 | 0 | 0 |
| 0.45973383 | 0 | 0 | 0 | 0 |
| 0 | 0 | 0 | 0 | 0.001684137 |
| 0 | 0.00672148 | 0.01742917 | 0 | 0 |
| 0.006850152 | 0 | 0 | 0 | 0.013566999 |
| 0 | 0 | 0 | 0 | 0 |
| 0.014939121 | 0.101543842 | 0 | 0 | 0.085816772 |
| 0 | 0 | 0 | 0 | 0 |
| 0 | 0 | 0 | 0 | 0 |
| 0 | 0.479146336 | 0.951444197 | 4.453575987 | 0.022761757 |
| 0 | 0.558640152 | 0 | 0 | 0 |
| 0 | 0 | 0 | 0 | 0 |
| 0 | 0 | 0.009535606 | 0.576846108 | 0 |
| 0 | 0 | 0 | 0 | 0 |
| 0 | 0 | 0 | 0 | 0 |
| 0 | 0 | 0 | 0 | 0 |
| 0 | 0 | 0 | 0 | 0 |

| | | | | |
|---|---|---|---|---|
| 0.929945334 | 2.8582875 | 0 | 0.029042084 | 0.231141443 |
| 0 | 0 | 0 | 0 | 0 |
| 0 | 0 | 0 | 0 | 0 |
| 0 | 0 | 0 | 0 | 0.002547934 |
| 0.040191027 | 0.000518473 | 0 | 0.008037465 | 0.001044803 |
| 0 | 0 | 0 | 0 | 0 |
| 0.165852837 | 0.061765539 | 0.015498026 | 0 | 0.143414829 |
| 0 | 0 | 0 | 0 | 0 |
| 0 | 0 | 0 | 0 | 0 |
| 0 | 0 | 0 | 0 | 0 |
| 0.010603757 | 0 | 0 | 0.013714088 | 0 |
| 0.046759254 | 0 | 0 | 0.000380384 | 0 |
| 0 | 0 | 0 | 0 | 0 |
| 0 | 0 | 0 | 0 | 0 |
| 0 | 0 | 0 | 0 | 0 |
| 0 | 0 | 0 | 0 | 0 |
| 0 | 0.004446539 | 0 | 0 | 0 |
| 0 | 0 | 0 | 0 | 0 |
| 0 | 0 | 0 | 1.507497239 | 0 |
| 0 | 0 | 0 | 0 | 0 |
| 0.564732008 | 0.901819291 | 0 | 0 | 2.311986303 |
| 0 | 0 | 0 | 0 | 0 |
| 0.027848242 | 0.001021431 | 0 | 0.019793007 | 0 |
| 0.008968202 | 0.001967912 | 0.022189831 | 0.001489243 | 0.014794909 |
| 0 | 0 | 0 | 0 | 0 |
| 0.003158347 | 0 | 0 | 0 | 0 |
| 0 | 0 | 0 | 0 | 0 |
| 0 | 0 | 0 | 0 | 0 |
| 0 | 0 | 0 | 0 | 0 |
| 0.000770588 | 0 | 0.065261275 | 1.523741284 | 0.003073266 |
| 0 | 0 | 0 | 0 | 0 |
| 0 | 0 | 0 | 0 | 0 |
| 0 | 0 | 0 | 0 | 0 |
| 0 | 0 | 0 | 0 | 0 |
| 0 | 0 | 0 | 0 | 0 |
| 0 | 0 | 0 | 0 | 0 |
| 0 | 0 | 0.016699375 | 0 | 0.18760365 |
| 0 | 0 | 0 | 0 | 0 |
| 0 | 0 | 0 | 0 | 0 |
| 0 | 0 | 0 | 0 | 0 |
| 0 | 0 | 0 | 0 | 0 |
| 0 | 0 | 0 | 0 | 0 |
| 0 | 0 | 0 | 0 | 0 |
| 0 | 0 | 0 | 0 | 0 |
| 0 | 0 | 0 | 0 | 0 |
| 0 | 0 | 0 | 0 | 0 |
| 0 | 0 | 0 | 0 | 0 |
| 0 | 0 | 0 | 0 | 0 |

| | | | | |
|---|---|---|---|---|
| 0 | 0 | 0 | 0.001602479 | 0 |
| 0 | 0 | 0 | 0 | 0 |
| 0 | 0 | 0 | 0 | 0 |
| 0 | 0 | 0 | 0 | 0 |
| 0 | 0 | 0 | 0 | 0 |
| 0 | 0 | 0 | 0 | 0 |
| 0 | 0 | 0 | 0 | 0 |
| 1.821384741 | 0.118579596 | 0 | 0.549258854 | 2.472808738 |
| 0.015147286 | 0.471979838 | 0.288150755 | 0.77906653 | 0.326654981 |
| 0.000786947 | 0 | 0 | 0.000955144 | 0.430962127 |
| 0 | 0 | 0 | 0.001307652 | 0 |
| 0 | 0 | 0 | 0 | 0 |
| 0.018946198 | 0 | 0 | 0 | 0 |
| 0 | 0 | 0 | 0 | 0 |
| 0 | 0 | 0 | 0 | 0 |
| 0 | 0 | 0 | 0 | 0 |
| 0 | 0 | 0 | 0 | 0 |
| 0 | 0 | 0 | 0 | 0 |
| 0 | 0 | 0 | 0 | 0 |
| 0.054162079 | 0 | 0 | 0 | 0 |
| 0 | 0 | 0 | 0 | 0 |
| 0 | 0 | 0 | 0 | 0 |
| 0 | 0 | 0 | 0 | 0 |
| 0 | 0 | 0 | 0 | 0 |
| 0.006806681 | 0 | 0 | 0.004933953 | 0.019131802 |
| 0 | 0 | 0 | 0 | 0 |
| 0 | 0 | 0 | 0 | 0 |
| 0 | 0 | 0 | 0 | 0 |
| 0 | 0 | 0 | 0 | 0 |
| 0 | 0 | 0 | 0.104376586 | 0 |
| 0 | 0 | 0 | 0.045496944 | 0 |
| 0 | 0 | 0 | 0 | 0 |
| 0 | 0 | 0 | 0 | 0 |
| 0.001374312 | 0 | 0 | 0 | 0 |
| 0 | 0 | 0.035400019 | 0 | 0 |
| 0 | 0.432994618 | 0 | 0 | 0 |
| 0 | 0 | 0 | 0 | 0 |
| 0.007590876 | 0 | 0 | 0 | 0 |
| 0 | 0.00271161 | 0 | 0.000866124 | 0 |
| 0 | 0 | 0 | 0 | 0.006389966 |
| 0 | 0 | 0 | 0 | 0 |
| 0 | 0.004697401 | 0 | 0.33504181 | 0 |
| 0 | 0 | 0 | 0 | 0 |
| 0 | 0 | 0 | 0 | 0 |
| 0.008827468 | 0 | 0 | 0 | 0 |
| 0 | 0 | 0 | 0 | 0 |
| 0 | 0 | 0 | 0 | 0 |
| 0 | 0 | 0 | 0 | 0 |

| | | | | |
|---|---|---|---|---|
| 0 | 0 | 0 | 0 | 0 |
| 0 | 0 | 0 | 0.001314825 | 0.000444382 |
| 0 | 0 | 0 | 0 | 0 |
| 0 | 0 | 0 | 0 | 0 |
| 1.352407471 | 3.060983594 | 0.942327873 | 9.636234692 | 0 |
| 0 | 0 | 0 | 0 | 0 |
| 0 | 0 | 0 | 0 | 0.012393246 |
| 0.011909193 | 0 | 0.132498154 | 0.007041983 | 0.011900189 |
| 0.004756355 | 0.003723961 | 0 | 0 | 0 |
| 0 | 0 | 0 | 0 | 0 |
| 0 | 0 | 0 | 0 | 0 |
| 0 | 0 | 0 | 0 | 0 |
| 0 | 0 | 0 | 0 | 0 |
| 0 | 0 | 0 | 0 | 0 |
| 0.000386938 | 0 | 0 | 0 | 0 |
| 0 | 0 | 0 | 0 | 0 |
| 0 | 0 | 0 | 0 | 0 |
| 0 | 0 | 0 | 0 | 0 |
| 0 | 0 | 0 | 0 | 0 |
| 0 | 0 | 0 | 0 | 0 |
| 0 | 0 | 0 | 0 | 0 |
| 0 | 0 | 0 | 0 | 0 |
| 0 | 0 | 0 | 0 | 0 |
| 0.000792802 | 0 | 0.007619019 | 0.012188513 | 0 |
| 0 | 0 | 0.198957532 | 0.784503813 | 0.395290585 |
| 0 | 0 | 0 | 0 | 0 |
| 0.3618286 | 0.018204847 | 0.023518545 | 0.005427208 | 0.002809256 |
| 0 | 0 | 0 | 0 | 0 |
| 0 | 0 | 0.001035775 | 0 | 0 |
| 0 | 0 | 0 | 0 | 0 |
| 0 | 0 | 0 | 0 | 0 |
| 0 | 0 | 0 | 0 | 0 |
| 0 | 0 | 0 | 0 | 0 |
| 0 | 0 | 0 | 0 | 0 |
| 0 | 0 | 0 | 0 | 0 |
| 0 | 0 | 0 | 0 | 0 |
| 0 | 0 | 0 | 0 | 0 |
| 0.051013714 | 0 | 0 | 0.002707745 | 0 |
| 4.628854848 | 0.045876294 | 0 | 0 | 0.04722442 |
| 0.001658721 | 0 | 0 | 0 | 0 |
| 0 | 0 | 0 | 0 | 0 |
| 0 | 1.465961881 | 1.532378923 | 0 | 0 |
| 0 | 0 | 0 | 0 | 0 |
| 0 | 0 | 0 | 0 | 0 |
| 0 | 0 | 0 | 0 | 0 |
| 0 | 0 | 0 | 0 | 0 |
| 0 | 0 | 0 | 0 | 0 |
| 0 | 0 | 0 | 0 | 0 |

| | | | | |
|---|---|---|---|---|
| 2.611271813 | 0.024677607 | 0 | 0.76143494 | 0.119039231 |
| 0 | 0 | 0 | 0 | 0 |
| 0 | 0 | 0 | 0 | 0 |
| 0 | 0 | 0 | 0 | 0 |
| 0 | 0 | 0 | 0 | 0 |
| 0 | 0 | 0 | 0 | 0 |
| 0 | 0 | 0 | 0 | 0 |
| 0 | 0 | 0 | 0 | 0 |
| 0 | 0 | 0 | 0 | 0 |
| 0 | 0 | 0 | 0 | 0 |
| 0 | 0 | 0 | 0 | 0 |
| 0.006139031 | 0 | 0 | 0 | 0 |
| 0 | 0 | 0 | 0 | 0 |
| 0 | 0 | 0 | 0 | 0 |
| 0 | 0 | 0 | 0 | 0 |
| 0 | 0 | 0 | 0 | 0 |
| 0 | 0 | 0 | 0 | 0 |
| 0.002125543 | 0 | 0 | 0 | 0 |
| 0 | 0 | 0 | 0 | 0 |
| 0 | 0 | 0 | 0 | 0 |
| 0 | 0 | 0 | 0 | 0 |
| 0 | 0 | 0 | 0 | 0 |
| 0.145771002 | 0.001578642 | 0.004072368 | 0.007462732 | 0.008407473 |
| 0.023361702 | 0 | 0 | 0.00166142 | 0 |
| 0 | 0 | 0 | 0 | 0 |
| 0 | 0 | 0 | 0 | 0 |
| 0 | 0 | 0 | 0 | 0 |
| 0 | 0 | 0 | 0 | 0 |
| 0 | 0 | 0 | 0 | 0 |
| 0 | 0 | 0 | 0 | 0 |
| 0 | 0 | 0 | 0 | 0 |
| 0.0308785 | 0 | 0 | 0 | 0 |
| 0 | 0.00168959 | 0.009197882 | 0.19417581 | 0.160663262 |
| 0.2023825 | 0 | 0 | 0 | 0 |
| 0.338436434 | 0.044773159 | 0 | 0.55823889 | 0.436086615 |
| 0.002997174 | 0 | 0.000540067 | 0.000606296 | 0.013660984 |
| 0 | 0 | 0 | 0 | 0 |
| 0 | 0 | 0 | 0 | 0 |
| 0.006413406 | 0 | 0 | 0 | 0.067154626 |
| 0 | 0 | 0 | 0 | 0 |
| 0 | 0 | 0 | 0 | 0 |
| 0 | 0 | 0 | 0 | 0 |
| 0.014795706 | 1.438787602 | 0.097877583 | 0.433986359 | 0.646898207 |
| 0 | 0 | 0 | 0 | 0 |
| 0 | 0 | 0 | 0 | 0 |
| 0.094968638 | 0.003094631 | 0 | 0.001628061 | 0.006236158 |
| 0 | 0 | 0 | 0 | 0 |
| 0 | 0 | 0 | 0 | 0 |

| | | | | |
|---|---|---|---|---|
| 0 | 0 | 0 | 0 | 0 |
| 0 | 0 | 0 | 0 | 0 |
| 0 | 0 | 0 | 0 | 0 |
| 0 | 0 | 0 | 0 | 0 |
| 0 | 0 | 0 | 0 | 0 |
| 0 | 0 | 0 | 0 | 0 |
| 0.011763461 | 0 | 0 | 0 | 0 |
| 0 | 0 | 0 | 0 | 0.236081495 |
| 0 | 0 | 0 | 0 | 0 |
| 0 | 0.166121639 | 0 | 0.001006131 | 0 |
| 0 | 0 | 0 | 0 | 0 |
| 0 | 0 | 0 | 0 | 0 |
| 0 | 0 | 0 | 0 | 0 |
| 0 | 0 | 0 | 0 | 0 |
| 0 | 0 | 0 | 0 | 0 |
| 0 | 0 | 0 | 0 | 0 |
| 0 | 0 | 0 | 0 | 0 |
| 0 | 0 | 0 | 0 | 0 |
| 0 | 0 | 0 | 0 | 0 |
| 0.490010215 | 0.012664798 | 0 | 1.169842007 | 3.174316925 |
| 0 | 0.00115603 | 0 | 0.006203416 | 0 |
| 0 | 0 | 0 | 0 | 0 |
| 0 | 0 | 0 | 0 | 0 |
| 0 | 0 | 0 | 0.402171673 | 0 |
| 0.005291063 | 0 | 0 | 0 | 0 |
| 0 | 0 | 0 | 0 | 0 |
| 0 | 0 | 0 | 0 | 0 |
| 0 | 0 | 0 | 0 | 0 |
| 0 | 0 | 0 | 0 | 0 |
| 2.287568541 | 0.333252431 | 0 | 0 | 1.104039542 |
| 0.781024736 | 0.072119476 | 0 | 0.380030097 | 0.352452425 |
| 0 | 0 | 0 | 0 | 0 |
| 0 | 0 | 0 | 0 | 0 |
| 0 | 0 | 0 | 0 | 0 |
| 0 | 0 | 0 | 0 | 0 |
| 0 | 0 | 0 | 0 | 0 |
| 0 | 0 | 0 | 0 | 0 |
| 0 | 0 | 0 | 0 | 0 |
| 0 | 0 | 0 | 0 | 0 |
| 0 | 0 | 0 | 0 | 0 |
| 0 | 0 | 0 | 0 | 0 |
| 0 | 0 | 0 | 0.012324282 | 0 |
| 0.005018867 | 0 | 0 | 0.006961795 | 0 |
| 0 | 0 | 0 | 0 | 0 |
| 0 | 0 | 0 | 0 | 0 |
| 0 | 0 | 0 | 0 | 0 |
| 0 | 0 | 0.026679192 | 0 | 0.006088869 |
| 0 | 0 | 0 | 0 | 0.004175228 |

| | | | | |
|---|---|---|---|---|
| 0 | 0 | 0 | 0 | 0 |
| 0 | 0 | 0 | 0 | 0 |
| 0 | 0 | 0 | 0 | 0 |
| 0 | 0 | 0 | 0 | 0 |
| 0.297029853 | 0.00620762 | 0 | 0.026749702 | 0.128646887 |
| 0 | 0 | 0 | 0 | 0 |
| 0 | 0 | 0 | 0 | 0 |
| 0 | 0 | 0 | 0 | 0 |
| 0 | 0 | 0 | 0 | 0 |
| 0 | 0 | 0 | 0 | 0 |
| 0.177079128 | 0 | 0 | 0 | 0 |
| 0.031695103 | 0 | 0.00686593 | 0.074100958 | 0.016775289 |
| 0 | 0 | 0.004667053 | 0.001057951 | 0 |
| 0 | 0 | 0 | 0 | 0 |
| 0 | 0 | 0 | 0.010546426 | 0 |
| 0.400419812 | 0.121093127 | 0 | 0.846922938 | 1.364217299 |
| 0.026242225 | 0 | 0 | 0.044596914 | 0 |
| 0 | 0 | 0 | 0 | 0 |
| 0.004065525 | 0 | 0.071682097 | 0.001459117 | 0.001195515 |
| 0.011130384 | 0.061969573 | 0 | 0 | 0.033951272 |
| 0 | 0 | 0 | 0 | 0 |
| 0 | 0 | 0 | 0 | 0 |
| 0 | 0 | 0 | 0 | 0 |
| 0 | 0 | 0 | 0 | 0 |
| 0 | 0 | 0 | 0 | 0 |
| 0 | 0 | 0 | 0 | 0 |
| 0.004939913 | 0 | 0.016978462 | 0.000555161 | 0.01938869 |
| 0 | 0 | 0 | 0 | 0 |
| 0 | 0 | 0 | 0 | 0 |
| 0 | 0 | 0 | 0 | 0 |
| 0 | 0 | 0.014948333 | 0 | 0.001063456 |
| 0 | 0 | 0 | 0 | 0 |
| 0 | 0 | 0 | 0 | 0 |
| 0 | 0 | 0 | 0 | 0 |
| 0 | 0 | 0 | 0 | 0 |
| 0 | 0 | 0 | 0 | 0 |
| 0 | 0 | 0 | 0 | 0 |
| 0.014534506 | 0 | 0.044634626 | 0.043218309 | 0.071104116 |
| 0.864769094 | 0.050839129 | 0 | 0 | 0 |
| 0 | 0 | 0 | 0 | 0 |
| 0.006982314 | 0 | 0 | 0 | 0 |
| 0 | 0 | 0 | 0 | 0 |
| 0 | 0 | 0 | 0 | 0 |
| 0 | 0 | 0.00214136 | 0 | 0 |
| 0 | 0 | 0 | 0 | 0 |
| 0 | 0.255757196 | 0 | 0 | 0 |
| 0 | 0 | 0 | 0 | 0 |
| 0 | 0 | 0 | 0 | 0 |

| | | | | |
|---|---|---|---|---|
| 0 | 0 | 0 | 0 | 0 |
| 0.221333162 | 0.053257447 | 0 | 0.000980641 | 0.070706237 |
| 0 | 0 | 0 | 0 | 0 |
| 0.001698305 | 0 | 0 | 0.018723399 | 0.00154816 |
| 0 | 0.000712548 | 0.002649077 | 0 | 0.010230741 |
| 0 | 0 | 0 | 0 | 0 |
| 0.005768268 | 0 | 0 | 0 | 0 |
| 0.040377945 | 0 | 0 | 0 | 0 |
| 0 | 0 | 0 | 0 | 0 |
| 0 | 0 | 0 | 0 | 0 |
| 0.0162787 | 0 | 0 | 0 | 0 |
| 0 | 0 | 0 | 0 | 0 |
| 0.089205989 | 0.170047386 | 0 | 1.211915081 | 0.084501908 |
| 0 | 0 | 0 | 0 | 0 |
| 0 | 0 | 0 | 0 | 0 |
| 0 | 0 | 0 | 0 | 0 |
| 0 | 0 | 0 | 0 | 0 |
| 0 | 0 | 0 | 0 | 0 |
| 0 | 0 | 0 | 0 | 0 |
| 0 | 0 | 0 | 0 | 0 |
| 0 | 0 | 0 | 0 | 0 |
| 0 | 0 | 0 | 0 | 0 |
| 0 | 0 | 0 | 0 | 0.005599604 |
| 0.008604747 | 0 | 0 | 0.102357554 | 0.019396869 |
| 0 | 0 | 0 | 0 | 0 |
| 0 | 0 | 0 | 0 | 0 |
| 0.127371696 | 0 | 0 | 0 | 0.011735345 |
| 0 | 0 | 0 | 0 | 0 |
| 0 | 0 | 0 | 0 | 0 |
| 0 | 0 | 0 | 0 | 0 |
| 0.054969162 | 0.035780258 | 0 | 0.046659664 | 0.019334812 |
| 0.000683408 | 0 | 0 | 0 | 0 |
| 0 | 0 | 0 | 0 | 0 |
| 0 | 0 | 0 | 0 | 0 |
| 0.000889098 | 0 | 0 | 0 | 0 |
| 0 | 0 | 0 | 0 | 0 |
| 0 | 0 | 0 | 0 | 0 |
| 0 | 0 | 0 | 0 | 0 |
| 0 | 0 | 0 | 0 | 0.001022412 |
| 0 | 0 | 0 | 0 | 0 |
| 0 | 0 | 0 | 0 | 0 |
| 0 | 0 | 0 | 0 | 0 |
| 0 | 0 | 0 | 0 | 0 |
| 0 | 0 | 0 | 0 | 0 |
| 0 | 0 | 0 | 0 | 0 |
| 0.397349587 | 0.044490648 | 0.368585897 | 0.052699123 | 0.061319574 |
| 0 | 0 | 0 | 0 | 0 |
| 0 | 0 | 0 | 0 | 0 |

| | | | | |
|---|---|---|---|---|
| 0 | 0 | 0 | 0 | 0 |
| 0 | 0 | 0 | 0 | 0 |
| 0 | 0 | 0 | 0 | 0 |
| 0.013110701 | 0 | 0 | 0.016073644 | 0 |
| 0 | 0 | 0 | 0 | 0 |
| 0.004118511 | 0 | 0 | 0 | 0 |
| 0 | 0 | 0 | 0 | 0 |
| 0 | 0 | 0 | 0 | 0 |
| 0.083988527 | 0 | 0 | 0.003918607 | 0 |
| 0 | 0.003667199 | 0 | 0 | 0 |
| 0.000727914 | 0 | 0 | 0.001472491 | 0.003594283 |
| 0 | 0 | 0 | 0.032610097 | 0.000619186 |
| 0 | 0 | 0 | 0 | 0 |
| 0 | 0 | 0 | 0 | 0 |
| 0 | 0 | 0 | 0 | 0 |
| 0 | 0 | 0 | 0 | 0 |
| 0 | 0 | 0 | 0 | 0 |
| 0 | 0 | 0 | 0 | 0 |
| 0 | 0 | 0 | 0 | 0 |
| 0.001126229 | 0 | 0.001932739 | 0 | 0 |
| 0 | 0 | 0 | 0 | 0 |
| 0 | 0 | 0 | 0 | 0 |
| 0 | 0 | 0 | 0 | 0 |
| 0 | 0 | 0 | 0 | 0 |
| 0 | 0 | 0 | 0 | 0 |
| 0.002248317 | 0 | 0 | 0 | 0 |
| 0 | 0 | 0 | 0 | 0 |
| 0 | 0 | 0 | 0 | 0.143238982 |
| 0 | 0 | 0 | 0 | 0 |
| 0 | 0 | 0 | 0 | 0 |
| 0.002764421 | 0.021415417 | 0.00149438 | 0.00782897 | 0.004095035 |
| 0 | 0 | 0 | 0 | 0 |
| 0.005777594 | 0.003448497 | 0 | 0.010875813 | 0.005486253 |
| 0.000656008 | 0.000927366 | 0.016352035 | 0.000497637 | 0.001681907 |
| 0 | 0 | 0 | 0 | 0 |
| 0 | 0 | 0 | 0 | 0 |
| 0 | 0 | 0 | 0 | 0 |
| 0 | 0 | 0 | 0 | 0 |
| 0 | 0 | 0 | 0 | 0 |
| 0 | 0 | 0 | 0 | 0 |
| 0 | 0 | 0 | 0 | 0 |
| 0.130052138 | 0 | 0.37066734 | 0.152774077 | 0.586151946 |
| 0.092007898 | 0.05038956 | 0 | 0.080533516 | 0.048205304 |
| 0 | 0 | 0 | 0 | 0 |
| 0 | 0 | 0 | 0 | 0 |
| 0.062062474 | 0.176914978 | 0.354063713 | 0.197487443 | 0.026751565 |
| 0.016001654 | 0 | 0 | 0 | 0 |
| 0 | 0 | 0 | 0 | 0 |

| | | | | |
|---|---|---|---|---|
| 0 | 0 | 0 | 0 | 0 |
| 0 | 0.004764418 | 0.003846237 | 0 | 0 |
| 0.002755803 | 0.006678411 | 0.002128174 | 0.011547569 | 0.00919632 |
| 0 | 0 | 0 | 0 | 0 |
| 0 | 0 | 0.006041757 | 0.347823271 | 0 |
| 1.439158925 | 0 | 0 | 0 | 0.00767044 |
| 0 | 0 | 0 | 0 | 0 |
| 0 | 0 | 0 | 0 | 0 |
| 0 | 0 | 0 | 0 | 0 |
| 0 | 0 | 0 | 0 | 0 |
| 0.002359219 | 0 | 0 | 0 | 0 |
| 0 | 0 | 0 | 0 | 0 |
| 0 | 0 | 0 | 0 | 0 |
| 0 | 0 | 0 | 0 | 0 |
| 0 | 0 | 0 | 0 | 0 |
| 0 | 0 | 0 | 0 | 0 |
| 1.839305338 | 0 | 0 | 0.200371897 | 0 |
| 0 | 0 | 0 | 0 | 0 |
| 0 | 0 | 0 | 0 | 0 |
| 0 | 0 | 0 | 0 | 0 |
| 0 | 0 | 0 | 0 | 0 |
| 0 | 0 | 0 | 0 | 0 |
| 0.04629599 | 0 | 0 | 0 | 0 |
| 0 | 0 | 0 | 0 | 0 |
| 0 | 0.01541106 | 0.003507828 | 0.005086576 | 0.004991088 |
| 0 | 0.004244913 | 0.00098022 | 0.000495191 | 0.001115759 |
| 0 | 0 | 0 | 0 | 0 |
| 0.144533129 | 0.000716124 | 0 | 0.004323175 | 0.022007284 |
| 0 | 0 | 0 | 0 | 0 |
| 0 | 0 | 0 | 0 | 0 |
| 0 | 0 | 0 | 0.001381559 | 0 |
| 0 | 0 | 0 | 0 | 0.00195691 |
| 0 | 0 | 0 | 0 | 0 |
| 0 | 0 | 0 | 0 | 0 |
| 0 | 0 | 0 | 0 | 0 |
| 0 | 0 | 0 | 0 | 0 |
| 0 | 0 | 0 | 0 | 0 |
| 0 | 0 | 0 | 0 | 0.352074347 |
| 0 | 0 | 0 | 2.312415346 | 0 |
| 0 | 0 | 0 | 0 | 0 |
| 0 | 0 | 0 | 0 | 0 |
| 0 | 0.066473278 | 0 | 0 | 0 |
| 0.000818925 | 0 | 0.001180509 | 0.000662638 | 0 |
| 0 | 0 | 0 | 0 | 0 |
| 0 | 0 | 0 | 0 | 0 |
| 0.003203288 | 0 | 0 | 0.001334097 | 0 |
| 0.005512295 | 0 | 0 | 0.002280839 | 0.063097483 |
| 0 | 0 | 0 | 0 | 0 |

| | | | | |
|---|---|---|---|---|
| 0 | 0 | 0 | 0 | 0 |
| 0.077142842 | 0 | 0.097227724 | 0.00559611 | 0.104655473 |
| 0 | 0.001271862 | 0 | 0 | 0 |
| 0 | 0 | 0 | 0 | 0 |
| 0 | 0 | 0 | 0 | 0 |
| 0 | 0 | 0 | 0 | 0 |
| 0 | 0 | 0 | 0 | 0 |
| 0 | 0 | 0 | 0 | 0 |
| 0 | 0 | 0 | 0 | 0 |
| 0 | 0 | 0 | 0 | 0 |
| 0 | 0 | 0 | 0 | 0 |
| 0 | 0 | 0 | 0 | 0 |
| 1.035711458 | 0.422184316 | 0 | 0.118818732 | 0 |
| 0 | 0 | 0 | 0 | 0 |
| 0.001182609 | 0.001783247 | 0 | 0.001395501 | 0.051656813 |
| 0 | 0 | 0 | 0 | 0 |
| 0 | 0 | 0 | 0 | 0 |
| 0 | 0 | 0 | 0 | 0 |
| 0.003706769 | 0 | 0 | 0 | 0 |
| 0 | 0 | 0 | 0 | 0 |
| 0 | 0 | 0 | 0 | 0 |
| 0.005613506 | 0.015104869 | 0 | 0 | 0.003529118 |
| 0 | 0 | 0 | 0.046637779 | 0 |
| 0 | 0 | 0 | 0 | 0 |
| 0 | 0 | 0 | 0 | 0 |
| 0 | 0 | 0 | 0 | 0 |
| 0 | 0 | 0 | 0 | 0 |
| 0 | 0 | 0 | 0 | 0 |
| 0 | 0 | 0 | 0 | 0 |
| 0 | 0 | 0 | 0 | 0 |
| 0 | 0 | 0 | 0 | 0 |
| 0 | 0 | 0 | 0 | 0 |
| 0 | 0 | 0 | 0 | 0 |
| 0 | 0 | 0 | 0 | 0 |
| 8.340568987 | 0.003577665 | 0 | 0.016709584 | 1.983626384 |
| 0 | 0 | 0 | 0 | 0 |
| 0 | 0 | 0 | 0 | 0 |
| 0 | 0 | 0 | 0 | 0 |
| 0.032474774 | 0.311955432 | 0 | 0 | 0.010793019 |
| 0 | 0 | 0 | 0 | 0 |
| 0.02565151 | 0.033126049 | 0.14220546 | 0.083445045 | 0.082356385 |
| 0 | 0 | 0 | 0 | 0 |
| 0 | 0 | 0 | 0 | 0 |
| 0 | 0 | 0 | 0 | 0 |
| 0 | 0 | 0 | 0 | 0 |
| 0.004762578 | 0.016831541 | 0.489400251 | 0.005017799 | 0.84772762 |
| 0 | 0 | 0 | 0 | 0 |
| 0 | 0 | 0 | 0 | 0 |

| | | | | |
|---|---|---|---|---|
| 0 | 0 | 0 | 0 | 0 |
| 0 | 0 | 0 | 0 | 0 |
| 0 | 0 | 0 | 0 | 0 |
| 0 | 0 | 0 | 0 | 0.035393271 |
| 0 | 0 | 0 | 0 | 0 |
| 0 | 0 | 0 | 0 | 0 |
| 0 | 0 | 0 | 0 | 0 |
| 0 | 0.015281714 | 0 | 0 | 0 |
| 0.241705104 | 0 | 0 | 0 | 0 |
| 0 | 0.002194836 | 0 | 0 | 0.001548027 |
| 0 | 0 | 0 | 0 | 0 |
| 0 | 0 | 0 | 0 | 0 |
| 0.001896923 | 0 | 0 | 0.008633854 | 0 |
| 0 | 0 | 0 | 0 | 0 |
| 0 | 0 | 0 | 0 | 0 |
| 0 | 0 | 0 | 0 | 0 |
| 0 | 0.116215455 | 0 | 0 | 0 |
| 0 | 0 | 0 | 0 | 0 |
| 0 | 0 | 0 | 0 | 0 |
| 0.414838834 | 0.825259872 | 2.75170845 | 0.074285814 | 0.343225144 |
| 0.001692023 | 0 | 0 | 0 | 0 |
| 0 | 0 | 0 | 0 | 0 |
| 0 | 0 | 0 | 0 | 0 |
| 0 | 0 | 0 | 0 | 0 |
| 0 | 0 | 0 | 0 | 0 |
| 0 | 0 | 0 | 0 | 0 |
| 0 | 0 | 0 | 0 | 0 |
| 0 | 0 | 0 | 0 | 0 |
| 0 | 0.000706069 | 0 | 0 | 0 |
| 0 | 0 | 0 | 0 | 0.570731058 |
| 0 | 0 | 0 | 0 | 0 |
| 0 | 0.019431482 | 0 | 0 | 0 |
| 0 | 0 | 0 | 0 | 0 |
| 0 | 0 | 0 | 0 | 0 |
| 0 | 0 | 0 | 0 | 0 |
| 0.001748472 | 0 | 0 | 0 | 0 |
| 0 | 0 | 0 | 0 | 0 |
| 0 | 0 | 0 | 0 | 0 |
| 0 | 0 | 0 | 0 | 0 |
| 0 | 0 | 0 | 0.010131927 | 0 |
| 0 | 0 | 0 | 0 | 0 |
| 0 | 0 | 0 | 0 | 0 |
| 0.000842077 | 0 | 0 | 0 | 0 |
| 0 | 0 | 0 | 0 | 0 |
| 0.015540873 | 0 | 0 | 0.053892829 | 0.016443744 |
| 0 | 0.002517651 | 0 | 0 | 0 |
| 0 | 0 | 0 | 0 | 0 |
| 0 | 0 | 0 | 0 | 0 |

| | | | | |
|---|---|---|---|---|
| 0 | 0 | 0 | 0 | 0 |
| 0 | 0 | 0 | 0 | 0 |
| 0 | 0 | 0 | 0 | 0 |
| 0 | 0 | 0 | 0 | 0 |
| 0 | 0.001064535 | 0 | 0 | 0 |
| 0.013140889 | 0.000803313 | 0.046077734 | 0.109323849 | 0.060030473 |
| 0 | 0 | 0 | 0 | 0 |
| 0 | 0 | 0 | 0 | 0 |
| 0 | 0 | 0 | 0 | 0 |
| 0 | 0 | 0 | 0 | 0 |
| 0 | 0 | 0 | 0 | 0 |
| 0 | 0 | 0 | 0 | 0 |
| 0 | 0 | 0 | 0 | 0 |
| 0 | 0 | 0 | 0 | 0 |
| 0 | 0 | 0 | 0 | 0 |
| 0 | 0 | 0 | 0 | 0 |
| 0.055286713 | 0 | 0 | 0 | 0 |
| 0 | 0 | 0 | 0 | 0 |
| 0 | 0 | 0 | 0 | 0 |
| 0 | 0 | 0 | 0 | 0 |
| 0 | 0 | 0 | 0 | 0 |
| 0 | 0 | 0 | 0 | 0 |
| 0 | 0 | 0 | 0 | 0 |
| 0 | 0 | 0 | 0 | 0 |
| 0 | 0 | 0 | 0 | 0 |
| 0 | 0 | 0 | 0 | 0 |
| 0 | 0 | 0 | 0 | 0 |
| 0.10616114 | 0 | 0 | 0 | 0 |
| 0 | 0 | 0 | 0 | 0 |
| 0 | 0 | 0 | 0 | 0 |
| 0 | 0 | 0 | 0 | 0 |
| 0.016560419 | 0 | 0 | 0 | 0 |
| 0 | 0 | 0 | 0 | 0 |
| 0 | 0 | 0 | 0 | 0 |
| 0 | 0 | 0 | 0 | 0 |
| 0 | 0 | 0 | 0 | 0 |
| 0 | 0 | 0 | 0 | 0 |
| 0 | 0 | 0 | 0 | 0 |
| 3.221437182 | 0 | 0 | 0.020114052 | 0.212322043 |
| 0 | 0 | 0 | 0 | 0 |
| 0 | 0 | 0 | 0 | 0 |
| 0 | 0 | 0 | 0 | 0 |
| 0 | 0 | 0 | 0 | 0 |
| 0 | 0 | 0 | 0 | 0 |
| 0 | 0 | 0 | 0 | 0 |
| 0 | 0 | 0 | 0 | 0 |
| 0 | 0 | 0 | 0 | 0 |
| 0 | 0 | 0 | 0 | 0 |

| | | | | |
|---|---|---|---|---|
| 0 | 0 | 0 | 0 | 0 |
| 0.429307095 | 0.002098538 | 0 | 0.016355331 | 0.010421163 |
| 0 | 0 | 0 | 0 | 0 |
| 0 | 0 | 0 | 0 | 0 |
| 0 | 0 | 0 | 0 | 0 |
| 0 | 0 | 0 | 0 | 0 |
| 0 | 0 | 0 | 0 | 0 |
| 0 | 0 | 0 | 0 | 0 |
| 0 | 0 | 0 | 0 | 0 |
| 0 | 0 | 0 | 0 | 0 |
| 0.001388437 | 0.102456273 | 0.001459415 | 0 | 0.000791054 |
| 0 | 0 | 0 | 0 | 0 |
| 0.283019605 | 0.032816819 | 0 | 0.403658575 | 0.937340653 |
| 0 | 0 | 0 | 0.000530609 | 0 |
| 0 | 0 | 0 | 0 | 0 |
| 0 | 0 | 0 | 0.030501097 | 0 |
| 0 | 0 | 0 | 0 | 0 |
| 0 | 0 | 0 | 0 | 0 |
| 0 | 0 | 0 | 0 | 0 |
| 0 | 0 | 0 | 0 | 0 |
| 0 | 0.00159367 | 0.001692821 | 0 | 0.005218672 |
| 0.009184906 | 0 | 0 | 0 | 0 |
| 0 | 0 | 0 | 0 | 0 |
| 0.930389514 | 1.424160749 | 3.759476678 | 2.842891321 | 1.861446656 |
| 0 | 0 | 0 | 0 | 0 |
| 0 | 0.063912029 | 0 | 0 | 0 |
| 0 | 0 | 0 | 0 | 0 |
| 0 | 0 | 0 | 0 | 0 |
| 0 | 0 | 0 | 0 | 0 |
| 0 | 0 | 0 | 0 | 0 |
| 0 | 0 | 0 | 0 | 0 |
| 0 | 0 | 0 | 0 | 0 |
| 0 | 0.005448262 | 0 | 0 | 0 |
| 0 | 0 | 0 | 0 | 0 |

| PHL018-0159_V3 | PHL018-0009_V2 | PHL018-0011_V2 | PHL018-0208_V3 | PHL018-0192_V3 |
|---|---|---|---|---|
| REF | EYCF | CTRL | REF | CTRL |
| 0.015513362 | 0.016404472 | 0.030087449 | 0.568556366 | 0.009026585 |
| 2.031784889 | 0.636735623 | 2.834656172 | 0.358130516 | 0.137157257 |
| 20.9982686 | 0.154333768 | 3.78658742 | 0.902008766 | 0 |
| 0 | 0 | 0 | 0 | 0 |
| 0 | 0 | 0 | 0 | 0 |
| 0 | 0 | 0 | 0 | 0 |
| 0 | 0 | 0 | 0 | 0 |
| 0 | 0 | 0 | 0 | 0 |
| 0 | 0 | 0 | 0 | 0 |
| 0 | 0 | 0 | 0 | 0 |
| 0 | 0 | 0 | 0 | 0 |
| 0 | 0 | 0 | 0 | 0 |
| 0 | 0 | 0 | 0 | 0 |
| 18.46769034 | 0.768873082 | 3.752851747 | 0.096709042 | 0.014798371 |
| 0 | 0 | 0 | 0 | 0 |
| 0 | 0 | 0 | 0 | 0 |
| 0 | 0 | 0 | 0.000939522 | 0 |
| 0 | 0 | 0 | 0 | 0 |
| 0 | 0 | 0 | 0 | 0 |
| 0 | 0 | 0 | 0 | 0 |
| 0 | 0 | 0 | 0 | 0 |
| 0 | 0 | 0 | 0 | 0 |
| 0 | 0 | 0 | 0 | 0 |
| 0 | 0 | 0 | 0 | 0 |
| 0 | 0 | 0 | 0 | 0 |
| 0 | 0 | 0 | 0 | 0 |
| 0 | 0 | 0 | 0 | 0 |
| 0 | 0 | 0 | 0 | 0 |
| 0 | 0 | 0 | 0 | 0 |
| 0 | 0 | 0 | 0 | 0 |
| 0 | 0 | 0 | 0 | 0 |
| 0 | 0 | 0 | 0 | 0 |
| 0 | 0 | 0 | 0 | 0 |
| 0 | 0 | 0 | 0 | 0 |
| 0 | 0 | 0 | 0 | 0 |
| 0 | 0 | 0 | 1.053769125 | 0.915250625 |
| 0 | 0 | 0 | 0 | 0 |
| 0 | 0 | 0 | 0 | 0 |
| 0 | 0 | 0 | 0 | 0 |
| 0 | 0 | 0 | 0 | 0 |
| 0 | 0 | 0 | 0 | 0 |
| 0 | 0.007207153 | 0.183176478 | 0 | 0 |
| 0.043661607 | 0.016190283 | 2.649770951 | 0.050546564 | 7.610515632 |
| 0.003106274 | 0.010262948 | 0.479176718 | 0.010757648 | 0.01645156 |
| 0 | 0 | 0 | 0 | 0.004217242 |
| 0 | 0 | 0 | 0 | 0 |

| | | | | |
|---|---|---|---|---|
| 0 | 0.004940073 | 0 | 0 | 0.017991058 |
| 0.01757169 | 0 | 0.004350172 | 0.000638332 | 0.053425532 |
| 0 | 0 | 0 | 0 | 0 |
| 0 | 0.832183635 | 0 | 0.181251953 | 0.022534929 |
| 0.000777447 | 0.001334358 | 0 | 0.017279046 | 0 |
| 0 | 0 | 0 | 0 | 0 |
| 0 | 0.002142786 | 0.099351004 | 0.003420943 | 0.00688506 |
| 0 | 0 | 0 | 0 | 0.011151895 |
| 0 | 0 | 0 | 0 | 0 |
| 0 | 0 | 0 | 0 | 0 |
| 0 | 0 | 0 | 0.001885911 | 0 |
| 0 | 0.187817533 | 0 | 0.232250495 | 0.078335953 |
| 0 | 0 | 0.001864303 | 0 | 0 |
| 0.066981821 | 0.255970579 | 0.058308731 | 0.0545527 | 0.102706282 |
| 0 | 0 | 0 | 0 | 0 |
| 0 | 0 | 0 | 0 | 0 |
| 0 | 0.045441396 | 0.426085752 | 0.294665698 | 1.099026982 |
| 0 | 0 | 0 | 0 | 0 |
| 0 | 0 | 0 | 0 | 0 |
| 0 | 0 | 0 | 0 | 0 |
| 0.081144984 | 0.848094275 | 0.082514402 | 0.010182871 | 0.807358518 |
| 0.003649688 | 0.013093671 | 0.103467206 | 0.016204616 | 0.013721637 |
| 0 | 0 | 0 | 0 | 0 |
| 0 | 0 | 0 | 0 | 0 |
| 0 | 0 | 0 | 0 | 0 |
| 0 | 0 | 0 | 0 | 0 |
| 0 | 0 | 0 | 0 | 0 |
| 0 | 0 | 0 | 0 | 0 |
| 0 | 0 | 0.002608167 | 0 | 0 |
| 0 | 0 | 0 | 0 | 0 |
| 0.023606807 | 0 | 0 | 0 | 0.087475126 |
| 0 | 0 | 0 | 0 | 0 |
| 0 | 0 | 0 | 0 | 0 |
| 0 | 0 | 0 | 0 | 0 |
| 0 | 0 | 0 | 0 | 0 |
| 0 | 0 | 0 | 0 | 0 |
| 0 | 0 | 0 | 0 | 0 |
| 0 | 0 | 0.009270803 | 0.003340466 | 37.07109765 |
| 0 | 0 | 0 | 0 | 0 |
| 0 | 0 | 0 | 0 | 0 |
| 0 | 0 | 0 | 0 | 0 |
| 0 | 0 | 0 | 0 | 0 |
| 0 | 0 | 0 | 0 | 0 |
| 0 | 0 | 0 | 0 | 0 |
| 0 | 0 | 0 | 0 | 0 |
| 0 | 0 | 0 | 0 | 0 |
| 0.14251214 | 0 | 0.001575378 | 0 | 0 |
| 0 | 0 | 0 | 0 | 0 |

| | | | | |
|---|---|---|---|---|
| 0 | 0 | 0 | 0 | 0 |
| 0 | 0 | 0 | 0.016507167 | 0 |
| 0 | 0 | 0 | 0 | 0 |
| 0 | 0 | 0 | 0 | 0 |
| 0 | 0 | 0.259912089 | 0 | 0 |
| 0 | 0 | 0 | 0 | 0 |
| 0 | 0 | 0 | 0 | 0 |
| 0 | 0 | 0 | 0.000669526 | 0 |
| 0 | 0 | 0 | 0.006891734 | 0 |
| 0 | 0 | 0 | 0 | 0 |
| 0 | 0 | 0 | 0 | 0 |
| 0 | 0 | 0.002859819 | 0 | 0 |
| 0 | 0 | 0 | 0 | 0 |
| 0 | 0 | 0 | 0 | 0 |
| 0 | 0 | 0 | 0 | 0 |
| 0 | 0 | 0 | 0 | 0 |
| 0 | 0 | 0 | 0 | 0 |
| 0 | 0 | 0 | 0 | 0 |
| 0.010327197 | 0 | 0.013489059 | 0.585998297 | 0 |
| 0 | 0 | 0 | 0 | 0 |
| 0 | 0 | 0 | 0.041147014 | 0 |
| 0 | 0 | 0 | 0 | 0 |
| 0 | 0 | 0 | 0 | 0 |
| 0 | 0 | 0 | 0 | 0 |
| 0 | 0 | 0 | 0 | 0 |
| 0 | 0 | 0 | 0 | 0 |
| 0 | 0 | 0 | 0.001606691 | 0 |
| 0 | 0 | 0 | 0.006536418 | 0 |
| 0 | 0 | 0 | 0 | 0 |
| 0 | 0 | 0 | 0.001088257 | 0 |
| 0 | 0.017975864 | 0 | 0.237993324 | 0.037139856 |
| 0 | 0 | 0 | 0 | 0 |
| 0 | 0 | 0 | 0 | 0 |
| 0 | 0 | 0 | 0 | 0 |
| 0 | 0 | 0 | 0 | 0 |
| 0 | 0 | 0 | 0 | 0 |
| 0 | 0 | 0 | 0 | 0 |
| 0 | 0 | 0 | 0 | 0 |
| 0 | 0 | 0 | 0 | 0 |
| 0 | 0 | 0 | 0 | 0 |
| 0 | 0 | 0 | 0 | 0 |
| 0 | 0 | 0 | 0 | 0 |
| 0 | 0 | 0.002764009 | 0.008260369 | 0 |
| 0 | 0 | 0 | 0 | 0 |
| 0 | 0 | 0 | 0 | 0 |
| 0 | 0 | 0 | 0 | 0 |
| 0 | 0 | 0 | 0 | 0 |
| 0 | 0 | 0 | 0 | 0 |

| | | | | |
|---|---|---|---|---|
| 0 | 0 | 0 | 0 | 0 |
| 0 | 0 | 0 | 0 | 0 |
| 0 | 0 | 0 | 0 | 0 |
| 0 | 0 | 0 | 0 | 0 |
| 0 | 0 | 0.017483675 | 0.211094136 | 0 |
| 0 | 0 | 0 | 0 | 0 |
| 0 | 0 | 0 | 0 | 0 |
| 0 | 0 | 0 | 0 | 0 |
| 0 | 0 | 0 | 0 | 0 |
| 0 | 0 | 0 | 0 | 0 |
| 0 | 0 | 0 | 0 | 0 |
| 0 | 0 | 0 | 0 | 0 |
| 0 | 0 | 0 | 0 | 0 |
| 0 | 0 | 0 | 0 | 0 |
| 0 | 0 | 0 | 0 | 0 |
| 0 | 0 | 0 | 0 | 0 |
| 0 | 0 | 0 | 0 | 0 |
| 0 | 0 | 0 | 0 | 0 |
| 0 | 0 | 0 | 0 | 0 |
| 0 | 0 | 0 | 0 | 0 |
| 0 | 0 | 0 | 0 | 0 |
| 0 | 0 | 0 | 0 | 0 |
| 0 | 0 | 0 | 0 | 0 |
| 0 | 0 | 0 | 0 | 0 |
| 0 | 0 | 0 | 0 | 0.007351901 |
| 0 | 0 | 0 | 0 | 0 |
| 0 | 0 | 0 | 0 | 0 |
| 0 | 0 | 0 | 0 | 0 |
| 0 | 0 | 0 | 0 | 0 |
| 0 | 0 | 0 | 0 | 0 |
| 0 | 0 | 0 | 0 | 0 |
| 0.003622064 | 0 | 0.007173631 | 0.000701759 | 0 |
| 0 | 0 | 0 | 0 | 0 |
| 0 | 0 | 0 | 0 | 0 |
| 0 | 0 | 0 | 0 | 0 |
| 0 | 0 | 0 | 0 | 0 |
| 0 | 0 | 0 | 0 | 0 |
| 0 | 0 | 0 | 0 | 0 |
| 0 | 0 | 0 | 0 | 0 |
| 0 | 0 | 0 | 0 | 0 |
| 0 | 0 | 0 | 0 | 0 |
| 0 | 0 | 0 | 0 | 0 |
| 0 | 0 | 0 | 0 | 0 |
| 0 | 0 | 0 | 0 | 0 |
| 0 | 0 | 0 | 0 | 0 |
| 0 | 0 | 0 | 0 | 0 |
| 0 | 0 | 0 | 0 | 0 |
| 0 | 0 | 0 | 0 | 0.191954107 |

| | | | | |
|---|---|---|---|---|
| 0 | 0 | 0 | 0 | 0 |
| 0 | 0 | 0 | 0 | 0 |
| 0 | 0 | 0 | 0 | 0 |
| 0 | 0 | 0 | 0 | 0 |
| 0 | 0 | 0 | 0 | 0 |
| 0 | 0 | 0 | 0 | 0 |
| 0 | 0 | 0 | 0 | 0 |
| 0 | 0 | 0 | 0 | 0 |
| 0 | 0 | 0 | 0 | 0 |
| 0 | 0 | 0 | 0 | 0 |
| 0 | 0 | 0 | 0 | 0 |
| 0 | 0 | 0 | 0 | 0 |
| 0 | 0 | 0 | 0.003525449 | 0.000618066 |
| 0 | 0 | 0 | 0 | 0 |
| 0 | 0 | 0 | 0 | 0 |
| 0 | 0 | 0 | 0 | 0 |
| 0 | 0 | 0 | 0 | 0 |
| 0 | 0 | 0 | 0 | 0 |
| 0 | 0 | 0 | 0 | 0 |
| 0 | 0 | 0 | 0 | 0 |
| 0.053316951 | 0.012386668 | 0.192523414 | 0.046456096 | 0.021725921 |
| 0 | 0 | 0 | 0 | 0 |
| 0 | 0 | 0 | 0.002616502 | 0 |
| 0.010443984 | 0 | 0 | 0.002291287 | 0 |
| 0 | 0 | 0 | 0 | 0 |
| 0 | 0 | 0 | 0 | 0 |
| 0.007548196 | 0.003091587 | 0.00172494 | 0.000731214 | 0.00524951 |
| 0 | 0 | 0 | 0 | 0 |
| 0 | 0 | 0 | 0 | 0 |
| 0.001284771 | 0 | 0.009842598 | 0 | 0 |
| 0.080714898 | 0 | 0.002006051 | 0 | 0 |
| 0 | 0 | 0 | 0 | 0 |
| 0 | 0 | 0 | 0 | 0 |
| 0 | 0 | 0 | 0.001918316 | 0 |
| 0.014949648 | 0.007675644 | 0 | 0.011641407 | 0 |
| 0 | 0 | 0 | 0 | 0 |
| 0 | 0 | 0 | 0 | 0 |
| 0 | 0 | 0 | 0 | 0 |
| 0 | 0 | 0 | 0 | 0 |
| 0 | 0 | 0 | 0 | 0 |
| 0 | 0 | 0 | 0 | 0 |
| 0 | 0 | 0 | 0 | 0 |
| 0 | 0 | 0 | 0 | 0 |
| 0 | 0 | 0 | 0 | 0 |
| 0 | 0 | 0 | 0 | 0 |
| 0 | 0 | 0 | 0 | 0 |
| 0 | 0 | 0.001738843 | 0 | 0 |
| 0.000546335 | 0 | 0.001144461 | 0.000727718 | 0 |

| | | | | |
|---|---|---|---|---|
| 0 | 0 | 0 | 0 | 0 |
| 0 | 0 | 0 | 0 | 0 |
| 0 | 0 | 0.016314235 | 0 | 0 |
| 0 | 0 | 0 | 0 | 0 |
| 0 | 0 | 0 | 0 | 0 |
| 0 | 0 | 0 | 0 | 0 |
| 0 | 0 | 0 | 0 | 0 |
| 0 | 0 | 0 | 0 | 0 |
| 0 | 0 | 0 | 0 | 0 |
| 0 | 0 | 0 | 0 | 0 |
| 0 | 0 | 0 | 0 | 0 |
| 0 | 0 | 0 | 0 | 0 |
| 0 | 0 | 0 | 0 | 0 |
| 0 | 0 | 0 | 0 | 0 |
| 0 | 0 | 0 | 0.000688422 | 0 |
| 0 | 0 | 0 | 0 | 0 |
| 0 | 0 | 0 | 0 | 0 |
| 0 | 0 | 0 | 0 | 0 |
| 0 | 0 | 0.015130816 | 0 | 0.032478553 |
| 0 | 0 | 0 | 0 | 0 |
| 0 | 0.26545539 | 0.142844766 | 0.010261974 | 0.033761497 |
| 0 | 0 | 0 | 0 | 0 |
| 0 | 0 | 0 | 0 | 0 |
| 0 | 0 | 0 | 0 | 0 |
| 0.002035371 | 0.003260484 | 0.044342394 | 0.00260267 | 0.002235812 |
| 0 | 0 | 0 | 0 | 0 |
| 0 | 0 | 0 | 0 | 0 |
| 0 | 0 | 0 | 0 | 0 |
| 0 | 0 | 0.010741904 | 0 | 0 |
| 0 | 0 | 0 | 0 | 0 |
| 0.349987326 | 0.835663362 | 0.069938304 | 0.009793258 | 0.485266032 |
| 0 | 0 | 0 | 0 | 0 |
| 0 | 0 | 0 | 0 | 0 |
| 0 | 0 | 0.066311766 | 0 | 0 |
| 0 | 0 | 0 | 0 | 0 |
| 0 | 0 | 0 | 0 | 0 |
| 0 | 0 | 0 | 0 | 0 |
| 0 | 0 | 0 | 0 | 0 |
| 0 | 0 | 0 | 0 | 0 |
| 0 | 0 | 0 | 0 | 0 |
| 0 | 0 | 0 | 0 | 0 |
| 0 | 0.005842808 | 0.00488996 | 0.001636492 | 0.011808893 |
| 0 | 0 | 0 | 0 | 0 |
| 0 | 0 | 0 | 0 | 0 |
| 0 | 0 | 0 | 0 | 0 |
| 0 | 0 | 0 | 0 | 0 |
| 0 | 0 | 0 | 0 | 0 |
| 0 | 0 | 0 | 0 | 0 |

| | | | | |
|---|---|---|---|---|
| 0 | 0 | 0 | 0 | 0 |
| 0 | 0 | 0 | 0 | 0 |
| 0.001928382 | 0.013900938 | 0.541948995 | 0.006370142 | 0.015433234 |
| 0 | 0 | 0 | 0 | 0 |
| 0 | 0 | 0 | 0 | 0 |
| 0 | 0 | 0 | 0 | 0 |
| 0 | 0 | 0 | 0 | 0 |
| 0 | 0 | 0 | 0 | 0 |
| 0 | 0 | 0 | 0 | 0 |
| 0 | 0 | 0 | 0 | 0 |
| 0 | 0 | 0 | 0 | 0 |
| 0 | 0 | 0 | 0 | 0 |
| 0 | 0 | 0 | 0 | 0 |
| 0 | 0 | 0.039287218 | 0.005904648 | 0 |
| 0.008795177 | 0.002465591 | 0.028004666 | 0.001499543 | 0.005244714 |
| 0 | 0 | 0 | 0.019466183 | 0 |
| 0.003006395 | 0.001605326 | 0.028885866 | 0.004271486 | 0.267027524 |
| 0.010416762 | 0 | 0 | 0 | 0 |
| 0 | 0 | 0 | 0 | 0 |
| 0 | 0 | 0 | 0 | 0 |
| 0 | 0 | 0 | 0 | 0 |
| 0 | 0 | 0.000909617 | 0.000963983 | 0 |
| 0 | 0 | 0 | 0 | 0 |
| 0 | 0 | 0.050294075 | 0 | 0 |
| 0 | 0 | 0 | 0 | 0 |
| 0 | 0 | 0 | 0 | 0 |
| 3.251593685 | 0 | 0 | 0.017444744 | 0 |
| 0 | 0 | 0 | 0 | 0 |
| 0 | 0 | 0 | 0 | 0 |
| 0 | 0 | 0 | 0 | 0 |
| 0 | 0 | 0 | 0.00236389 | 0 |
| 0 | 0 | 0 | 0 | 0 |
| 0 | 0 | 0 | 0 | 0 |
| 0 | 0 | 0 | 0 | 0 |
| 0 | 0 | 0 | 0.001631113 | 0 |
| 0 | 0 | 0.011506597 | 0 | 0 |
| 0 | 0 | 0 | 0 | 0 |
| 0 | 0 | 0 | 0.001937832 | 0 |
| 0 | 0 | 0 | 0 | 0 |
| 0 | 0 | 0 | 0 | 0 |
| 0 | 0 | 0 | 0 | 0 |
| 0 | 0 | 0 | 0 | 0 |
| 0 | 0 | 0.003243888 | 0 | 0 |
| 0.040095582 | 0 | 0 | 0 | 0 |
| 0 | 0 | 0 | 0 | 0 |
| 0 | 0 | 0 | 0 | 0 |
| 0 | 0 | 0 | 0 | 0 |
| 0.002478041 | 0 | 0 | 0 | 0 |

| | | | | |
|---|---|---|---|---|
| 0.05953732 | 0 | 0 | 9.402713341 | 0 |
| 0 | 0 | 0 | 0 | 0 |
| 0 | 0 | 0.001587922 | 0 | 0 |
| 0 | 0 | 0 | 0 | 0 |
| 0.025167521 | 0 | 0.000958562 | 0.009142675 | 0 |
| 0 | 0 | 0 | 0 | 0 |
| 0 | 0 | 0 | 0 | 0 |
| 0.005612381 | 0 | 0.111073822 | 0.028763615 | 0.080104111 |
| 0 | 0 | 0 | 0 | 0 |
| 0.029199478 | 0.244355114 | 0.037854306 | 0.006164286 | 0.003674361 |
| 0.000747173 | 0.004987102 | 0 | 0.002123166 | 0 |
| 0 | 0 | 0.005672673 | 0 | 0 |
| 0.015245269 | 0 | 0.054055483 | 0.023598524 | 0.001133229 |
| 0 | 0.004392244 | 0 | 0 | 0 |
| 0.00231065 | 0.002974385 | 0 | 0 | 0 |
| 0 | 0 | 0.003229702 | 0 | 0 |
| 0 | 0 | 0.001467907 | 0 | 0 |
| 0 | 0 | 0 | 0 | 0 |
| 0.028253804 | 0.029066112 | 0 | 0 | 0.033354766 |
| 0.104509433 | 0.566231615 | 0 | 0 | 0 |
| 0 | 0 | 0 | 0 | 0 |
| 0.015044967 | 0 | 0 | 0 | 0 |
| 0 | 0 | 0 | 0 | 0 |
| 0 | 0 | 0 | 0 | 0 |
| 0 | 0 | 0 | 0 | 0 |
| 0.002429885 | 0.009356875 | 0 | 0 | 0.00073329 |
| 0 | 0 | 0 | 0.001598031 | 14.22103079 |
| 0 | 0 | 0 | 0 | 0.003901708 |
| 0 | 0 | 0 | 0 | 0 |
| 0 | 0 | 0 | 0 | 0 |
| 0.078422527 | 0.031727778 | 0 | 0 | 0 |
| 0 | 0 | 0 | 0 | 0 |
| 0.15364517 | 0 | 0 | 0 | 0 |
| 0.081890568 | 0 | 0 | 0.004167766 | 0.003452433 |
| 0 | 0 | 0 | 0.009873837 | 0 |
| 0 | 0 | 0 | 0 | 0 |
| 0 | 0.003818512 | 0 | 0 | 0.001995023 |
| 0 | 0 | 0.295132419 | 0 | 0 |
| 0 | 0 | 0.004182564 | 0 | 0 |
| 0 | 0 | 0 | 0 | 0 |
| 0 | 0 | 0 | 0 | 0 |
| 0 | 0 | 0 | 0.03087703 | 0 |
| 0 | 0 | 0 | 0 | 0 |
| 0.001204677 | 0 | 0 | 0.001444168 | 0 |
| 0 | 0 | 0 | 0 | 0 |
| 0 | 0 | 0 | 0 | 0 |
| 0.001627098 | 0 | 0 | 0 | 0.005289046 |
| 0 | 0 | 0 | 0 | 0 |

| | | | | |
|---|---|---|---|---|
| 0 | 0 | 0 | 0.042607548 | 0 |
| 0 | 0 | 0.06617078 | 0 | 0 |
| 0 | 0 | 0 | 0 | 0 |
| 0 | 0 | 0 | 0 | 0 |
| 0 | 0.008280707 | 0 | 0 | 0 |
| 0.038629185 | 0.133439056 | 0 | 0.111784602 | 0.033757057 |
| 0 | 0 | 0 | 0 | 0.001118093 |
| 0 | 0 | 0 | 0 | 0 |
| 0 | 0 | 0 | 0 | 0 |
| 0 | 0 | 0 | 0 | 0 |
| 0.018048743 | 0 | 0 | 0.001810141 | 0.002998915 |
| 0 | 0 | 0.326837218 | 0.004453349 | 0 |
| 0 | 0 | 0.011013505 | 0 | 0 |
| 0.001003032 | 0.040169211 | 0.09345915 | 0.002992727 | 0.000787007 |
| 0 | 0 | 0 | 0 | 0 |
| 0 | 0.004915659 | 0 | 0 | 0 |
| 0 | 0 | 0 | 0 | 0 |
| 0 | 0 | 0 | 0 | 0 |
| 0 | 0 | 0.08050864 | 0 | 0 |
| 0 | 0 | 0 | 0 | 0 |
| 0 | 0 | 0 | 0 | 0 |
| 0 | 0 | 0 | 0 | 0.371401032 |
| 0 | 0 | 0.016270149 | 0.030225235 | 0 |
| 0 | 0 | 0 | 0 | 0 |
| 0 | 0 | 0 | 0 | 0 |
| 0 | 0 | 0 | 0 | 0 |
| 0 | 0.016115002 | 0 | 0 | 0 |
| 0 | 0 | 0 | 0 | 0 |
| 0 | 0 | 0 | 0 | 0 |
| 0.715710364 | 1.524394126 | 1.126977162 | 0.518571924 | 0.036222632 |
| 0 | 0 | 0 | 0.000725117 | 0 |
| 0 | 0 | 0 | 0 | 0 |
| 0 | 0 | 0 | 0 | 0 |
| 0 | 0 | 0.003230311 | 0.00369725 | 0 |
| 0.021558721 | 0 | 0 | 0 | 0.015494093 |
| 0 | 0 | 0 | 0 | 0 |
| 0 | 0 | 0 | 0 | 0 |
| 0 | 0 | 0 | 0 | 0 |
| 0 | 0 | 0 | 0 | 0 |
| 0 | 0 | 0 | 0 | 0 |
| 0.002143964 | 0.006274458 | 0.083743087 | 0.027415299 | 0 |
| 0 | 0 | 0 | 0 | 0.345779117 |
| 0 | 0 | 0 | 0 | 0 |
| 0 | 0 | 0 | 0 | 0 |
| 0.001860387 | 0.012665743 | 0.004053021 | 0 | 0 |
| 0 | 0 | 0 | 0 | 0 |
| 0.015578612 | 0.00138642 | 0.019725471 | 0 | 0.00095071 |
| 0 | 0 | 0 | 0.000690589 | 0 |

| | | | | |
|---|---|---|---|---|
| 0 | 0 | 0.001073732 | 0 | 0.003519049 |
| 0 | 0 | 0 | 0 | 0 |
| 0 | 0.102556794 | 0.954390984 | 0.019846208 | 0.297060018 |
| 0 | 0 | 0 | 0 | 0 |
| 0 | 0 | 0 | 0.027452956 | 0 |
| 0 | 0 | 0 | 0 | 0 |
| 0 | 0 | 0 | 0 | 0 |
| 0 | 0 | 0 | 0 | 0 |
| 0.133697372 | 0 | 0 | 0 | 0 |
| 0 | 0 | 0 | 0 | 0 |
| 0 | 0 | 0 | 0 | 0 |
| 0.007383577 | 0 | 0 | 0 | 0 |
| 0 | 0 | 0 | 0 | 0 |
| 0.026276876 | 0 | 0 | 0 | 0 |
| 0 | 0 | 0 | 0 | 0 |
| 0.005133731 | 0.040816566 | 0.058957947 | 0.130648526 | 0.00047389 |
| 0.001004154 | 0.009048182 | 0.0016828 | 0.000713351 | 0.035586229 |
| 0 | 0.011400343 | 0.011926463 | 0.000866694 | 0.004148092 |
| 0.007951983 | 0 | 0.001448505 | 0 | 0 |
| 0 | 0 | 0 | 0 | 3.937329367 |
| 0 | 0 | 0.007776292 | 0 | 0.035746625 |
| 0.001553022 | 0 | 0 | 0 | 0 |
| 0 | 0 | 0 | 0.001166539 | 0 |
| 0 | 0 | 0 | 0.268323576 | 0 |
| 0 | 0.021924001 | 0 | 0 | 0 |
| 0 | 0 | 0.471437876 | 0.036413929 | 0.00467056 |
| 0.005099126 | 0 | 0.003916602 | 0.005660031 | 0 |
| 0 | 0 | 0 | 0.158893555 | 0 |
| 0 | 0 | 0 | 0 | 0 |
| 0 | 0 | 0 | 0 | 0 |
| 0 | 0.039795661 | 0.025453189 | 0.075069389 | 0 |
| 0.002649822 | 0 | 0.180856556 | 0 | 0.015876945 |
| 0.016284746 | 8.078616052 | 1.399892879 | 0.020285934 | 0.001619678 |
| 0 | 0 | 0.007711851 | 0 | 0 |
| 0 | 0 | 0.007470121 | 0 | 0.002577113 |
| 0 | 0 | 0 | 0 | 0 |
| 0 | 0.045673612 | 0.013205049 | 0 | 0.004067502 |
| 0.058217176 | 0.029873974 | 0.058700616 | 0.002246437 | 0.008397775 |
| 0.020720389 | 0.033047492 | 0 | 0 | 0.166358785 |
| 0 | 0.003185873 | 0 | 0 | 0.00353705 |
| 0 | 0 | 0.002069732 | 0 | 0 |
| 0 | 0 | 0 | 0 | 0 |
| 0.003880276 | 0 | 0 | 0 | 0.007040573 |
| 0 | 0.582754486 | 0.013577205 | 0.028192833 | 0 |
| 0 | 0.223217333 | 0 | 0 | 0 |
| 0 | 0 | 0.156712487 | 0 | 0 |
| 0 | 0 | 0 | 0 | 0 |
| 0 | 0 | 0 | 0 | 0 |

| | | | | |
|---|---|---|---|---|
| 0 | 0 | 0 | 0 | 0 |
| 0.063860376 | 0.003539746 | 0.639155664 | 0.708045766 | 0.013870344 |
| 0 | 0 | 0 | 0 | 0 |
| 0 | 0 | 0 | 0 | 0.002730132 |
| 0.002601385 | 0 | 0 | 0 | 0 |
| 0 | 0 | 0 | 0 | 0 |
| 0.003270819 | 0 | 0 | 0 | 0 |
| 0 | 0 | 0 | 0 | 0.002397125 |
| 0 | 0 | 0 | 0 | 0 |
| 0.003211494 | 0 | 0 | 0 | 0 |
| 0.00131364 | 0.006247244 | 0.006329165 | 0.001574792 | 0.002190276 |
| 0.021279155 | 0.34724008 | 1.774413908 | 0.008340256 | 0.032240965 |
| 0 | 0 | 0 | 0 | 0 |
| 0.023297261 | 0 | 0 | 0 | 0 |
| 0 | 0.026408166 | 0.013260907 | 0 | 0.007243549 |
| 0 | 0 | 0 | 0 | 0 |
| 0.007909531 | 0.004820248 | 0.061665022 | 0.010443884 | 0.002428449 |
| 0.063614874 | 0.041119717 | 0.054239294 | 0 | 0.02031587 |
| 0 | 0 | 0 | 0.002524702 | 0 |
| 0.020581617 | 0.054853132 | 0.006072442 | 0.02440296 | 0 |
| 0.00106392 | 0 | 0.07042687 | 0.013982467 | 0.176834498 |
| 0 | 0 | 0 | 0.000940483 | 0 |
| 0 | 0 | 0 | 0 | 0 |
| 0 | 0 | 0 | 0 | 0 |
| 0 | 0 | 0.005492123 | 0 | 0 |
| 0 | 0 | 0 | 0.001732745 | 0 |
| 0 | 0.147195569 | 0.004106362 | 0.028535314 | 0 |
| 0.040331516 | 2.99828667 | 0.579245032 | 0.005422366 | 0.002957498 |
| 0 | 0 | 0 | 0 | 0 |
| 0 | 0 | 0 | 0.000726738 | 0 |
| 0 | 0 | 0 | 0 | 0 |
| 0 | 0 | 0 | 0 | 0 |
| 0 | 0 | 0 | 0 | 0 |
| 0 | 0 | 0 | 0 | 0 |
| 0 | 0 | 0 | 0 | 0 |
| 1.276501183 | 0.725556025 | 0.71894992 | 0.239622461 | 0.058563041 |
| 0 | 0 | 0 | 0 | 0 |
| 0 | 0 | 0 | 0 | 0 |
| 0 | 0 | 0.003506582 | 0.012634956 | 0 |
| 0 | 0 | 0 | 0 | 0 |
| 0 | 0.215007158 | 0 | 0 | 0 |
| 0 | 0 | 0 | 0 | 0 |
| 0 | 0.012724948 | 0.014436339 | 0.000752418 | 0 |
| 0.00504476 | 0 | 0 | 0.004794541 | 0.002781479 |
| 0.071829348 | 0.018651312 | 0 | 0 | 0 |
| 0 | 0 | 0 | 0 | 0.006521409 |
| 0 | 0 | 0 | 0 | 0 |
| 0.022479874 | 0.003971776 | 0.001477359 | 0 | 0 |

| | | | | |
|---|---|---|---|---|
| 0 | 0 | 0 | 0 | 0 |
| 0 | 0 | 0 | 0 | 0 |
| 0 | 0 | 0 | 0 | 0 |
| 0.00213819 | 0.014491192 | 0.132580683 | 1.848703858 | 0 |
| 0 | 0 | 0 | 0 | 0 |
| 0 | 0 | 0 | 0 | 0 |
| 0 | 0 | 0 | 0 | 0 |
| 0 | 0 | 0 | 0 | 0 |
| 0 | 0 | 0.180082555 | 1.991861601 | 0.024388535 |
| 0 | 0.004120952 | 0 | 0.051692698 | 0 |
| 0 | 0 | 0 | 0 | 0 |
| 0 | 0 | 0 | 0.001511311 | 0 |
| 0 | 0 | 0 | 0 | 0 |
| 0 | 0 | 0 | 0 | 0 |
| 0.001170362 | 0.003385085 | 0 | 0 | 0.017957838 |
| 0 | 0 | 0 | 0 | 0 |
| 0 | 0 | 0 | 0 | 0.025820106 |
| 0 | 0 | 0 | 0 | 0.007594677 |
| 0 | 0 | 0 | 0 | 0 |
| 0 | 0 | 0 | 0 | 0 |
| 0 | 0 | 0 | 0 | 0 |
| 0 | 0 | 0 | 0 | 0 |
| 0 | 0 | 0 | 0 | 0 |
| 0.345347548 | 0.245806957 | 0.295146169 | 0.145625903 | 0.053564831 |
| 0 | 0 | 0 | 0.43644079 | 0 |
| 0 | 0 | 0 | 8.006596608 | 0 |
| 0 | 0 | 0 | 0 | 0 |
| 0 | 0 | 0 | 0 | 1.249031089 |
| 0 | 0 | 0 | 0 | 0 |
| 0 | 0.005688768 | 0 | 0 | 0 |
| 0 | 0 | 0 | 0 | 0 |
| 0 | 0 | 0 | 0 | 0 |
| 0 | 0 | 0 | 0.19431987 | 0.005214644 |
| 0 | 0 | 0 | 0 | 0 |
| 0 | 0 | 0 | 0 | 0 |
| 0 | 0 | 0 | 0 | 0.004280664 |
| 0 | 0 | 0.541761802 | 0.563296589 | 0 |
| 0 | 0 | 0 | 0 | 0 |
| 0 | 0 | 0 | 0 | 0 |
| 0 | 0 | 0.006426934 | 0 | 0 |
| 0.038700914 | 0 | 0 | 0.192505628 | 0.008540384 |
| 0 | 0 | 0 | 0 | 0 |
| 0.434522994 | 7.42767145 | 0.211831697 | 0.003865006 | 0.008964584 |
| 0 | 0 | 0 | 0.051502097 | 0.001343295 |
| 0 | 0 | 0 | 0 | 0 |
| 0 | 0 | 0 | 0 | 0 |
| 0 | 0 | 0 | 0 | 0 |
| 0.003089414 | 0 | 0.026800448 | 0.96709687 | 0.003422174 |

| | | | | |
|---|---|---|---|---|
| 0 | 0 | 0 | 0 | 0 |
| 0 | 0 | 0.0066589 | 0 | 0.005196162 |
| 0.369671369 | 0.082324845 | 0.770445716 | 0.276413598 | 0 |
| 0 | 0 | 0 | 0.009029916 | 0 |
| 0 | 12.52892901 | 0 | 0 | 0 |
| 0 | 0 | 0 | 0.050013303 | 0 |
| 0 | 0 | 0 | 0 | 0 |
| 0 | 0 | 0 | 0.013189628 | 0 |
| 0 | 0 | 0 | 0 | 0 |
| 0 | 0 | 0 | 0 | 0 |
| 0.022748624 | 0.002054959 | 0.010319014 | 0.682938274 | 0.004831362 |
| 5.372511085 | 18.87989004 | 4.133854272 | 0.670675158 | 0.141405341 |
| 0 | 0 | 0 | 0 | 0 |
| 0.001476766 | 0.003696326 | 0 | 0.000983527 | 0.025636499 |
| 0.552198709 | 0.676945633 | 0.508772759 | 0.065259772 | 0.006391765 |
| 0 | 0 | 0 | 0.003173336 | 0 |
| 0 | 0 | 0 | 0 | 0 |
| 0 | 0 | 0 | 0 | 0 |
| 0 | 0 | 0 | 0 | 0 |
| 0 | 0 | 0 | 0.107197447 | 0 |
| 0.002007021 | 0 | 0.088410502 | 0.299720944 | 0 |
| 0.892435969 | 0.719278226 | 1.838842925 | 0 | 0 |
| 15.34742288 | 0.278184398 | 0.06217556 | 0.387558443 | 0.010411819 |
| 0 | 0 | 0 | 0 | 0 |
| 0 | 0 | 0 | 0 | 0 |
| 0 | 0 | 0 | 0 | 3.915202867 |
| 0 | 0 | 0.027297806 | 1.314427438 | 0 |
| 0 | 0 | 0 | 0 | 0.001290497 |
| 0 | 0 | 0 | 0 | 0 |
| 0 | 0 | 0 | 0 | 0 |
| 0 | 0 | 0 | 0 | 0 |
| 0 | 0 | 0.356773032 | 0.218493228 | 0.168634022 |
| 0.001580984 | 0 | 0.005593334 | 0 | 0 |
| 0 | 0 | 0 | 0.379560928 | 0.005255361 |
| 0.015304212 | 0.007661247 | 0.012629397 | 0.033110529 | 0.000454853 |
| 0 | 0.004465612 | 0.006940808 | 0.241548104 | 0.00299971 |
| 0 | 0 | 0 | 0 | 0.00072442 |
| 0 | 0 | 0 | 0 | 0 |
| 0 | 0 | 0 | 0 | 0.172295601 |
| 0 | 0 | 0 | 0 | 0 |
| 0 | 0 | 0 | 0 | 0 |
| 0 | 0 | 0 | 0 | 0 |
| 0 | 0 | 0 | 0 | 0 |
| 0 | 0 | 0 | 0 | 0 |
| 0.009082942 | 0.001818758 | 0.005784187 | 0.093690629 | 0.004988709 |
| 0 | 0 | 0 | 0 | 0 |
| 0 | 0 | 0 | 0 | 0 |
| 0 | 0.020184061 | 0.068304197 | 0.033624744 | 0.026306149 |

| | | | | |
|---|---|---|---|---|
| 0 | 0 | 0 | 0 | 0 |
| 0 | 0 | 0 | 0 | 0 |
| 0 | 0 | 0 | 0 | 0 |
| 0 | 0 | 0 | 0.000659935 | 0 |
| 0 | 0 | 0 | 0 | 0 |
| 0 | 0 | 0 | 0 | 0 |
| 0 | 0 | 0 | 0 | 0 |
| 0 | 0.005420287 | 0.046079711 | 0 | 0.001788561 |
| 0 | 0 | 0 | 0 | 0 |
| 0 | 0 | 0 | 0 | 0 |
| 0 | 0.00656041 | 0.014448778 | 0.01249488 | 0 |
| 0 | 0 | 0 | 0 | 0 |
| 0 | 0.001511619 | 0 | 0 | 0.002724679 |
| 0 | 0 | 0 | 0 | 0 |
| 0 | 0 | 0 | 0 | 0 |
| 0 | 0 | 0 | 0 | 0 |
| 0 | 0 | 0 | 0 | 0 |
| 0 | 0 | 0 | 0 | 0 |
| 0 | 0 | 0 | 0.000965766 | 0 |
| 0 | 0 | 0 | 0.074769002 | 0 |
| 0 | 0 | 0 | 0 | 0.000500327 |
| 0 | 0 | 0 | 0.002306483 | 0 |
| 0.000993013 | 0 | 0 | 0 | 0.000909003 |
| 0 | 0 | 0 | 0 | 0 |
| 0 | 0 | 0 | 0 | 0 |
| 0 | 0 | 0 | 0 | 0 |
| 0 | 0 | 0 | 0 | 0 |
| 0 | 0 | 0 | 0 | 0 |
| 0 | 0 | 0 | 0 | 0 |
| 0 | 0 | 0 | 0.017739054 | 0.002449069 |
| 0 | 0.002719502 | 0 | 0.032273049 | 0.101021271 |
| 0 | 0 | 0 | 0 | 0 |
| 0 | 0 | 0 | 0 | 0 |
| 0 | 0 | 0 | 0 | 0 |
| 0 | 0 | 0 | 0 | 0 |
| 0 | 0 | 0 | 0 | 0 |
| 0 | 0 | 0 | 0 | 0.002325343 |
| 0.000583882 | 0 | 0.080970169 | 0.001711007 | 0.003092375 |
| 0 | 0 | 0 | 0 | 0 |
| 0 | 0 | 0 | 0.006597149 | 0.000903748 |
| 0 | 0 | 0.06724842 | 0.360327665 | 0 |
| 0 | 0 | 0 | 4.410279454 | 0 |
| 0 | 0 | 0 | 0 | 0 |
| 0 | 0.018450107 | 0 | 0 | 0 |
| 0 | 0 | 0 | 0 | 0.029651488 |
| 0 | 0 | 0 | 0 | 0 |
| 0 | 0 | 0 | 0 | 0 |
| 0 | 0 | 0 | 0 | 0 |

| | | | | |
|---|---|---|---|---|
| 0 | 0 | 0 | 0 | 0 |
| 0 | 0 | 0 | 0.507910468 | 0 |
| 0 | 0 | 0.051802147 | 0 | 0 |
| 0 | 0 | 0 | 0 | 0 |
| 0 | 0 | 0 | 0 | 0 |
| 0 | 0 | 0 | 0 | 0 |
| 0 | 0 | 0 | 0 | 0 |
| 0 | 0 | 0 | 0 | 0 |
| 0.303586907 | 0.137462602 | 0.016194038 | 0.120777034 | 0.003475106 |
| 0 | 0 | 0.002445185 | 0 | 0 |
| 0 | 0 | 0 | 0 | 0 |
| 0 | 0 | 0 | 0 | 0 |
| 0 | 0 | 0 | 0 | 0 |
| 0 | 0 | 0 | 0 | 0 |
| 5.560617782 | 0 | 0 | 2.838288305 | 0 |
| 0.21458774 | 0 | 0.183837607 | 0.24569471 | 0 |
| 0 | 0 | 0 | 0 | 0.007131178 |
| 0 | 0 | 0 | 0 | 0 |
| 0 | 0 | 0 | 0 | 0 |
| 0 | 0 | 0 | 0 | 0 |
| 0 | 0 | 0 | 0.001804378 | 0 |
| 0 | 0 | 0 | 0 | 0 |
| 0 | 0 | 0 | 0 | 0 |
| 0 | 0 | 0 | 0.001662246 | 0 |
| 0 | 0 | 0.116246087 | 0.019677385 | 0.380984919 |
| 0.980703996 | 1.112122036 | 0 | 0.013315619 | 0.077386456 |
| 0 | 0 | 0 | 0.642930327 | 0 |
| 0 | 0 | 0 | 0 | 0 |
| 0 | 0 | 0.030524383 | 0 | 0.010069023 |
| 0 | 0 | 0 | 0 | 0 |
| 0 | 0 | 0 | 0 | 0 |
| 0 | 0 | 0 | 0 | 0 |
| 0 | 0 | 0 | 0 | 0 |
| 0 | 0 | 0 | 0.004850302 | 0 |
| 0.007499778 | 0.005422977 | 0 | 0 | 0.004249941 |
| 0.002650717 | 0 | 0.004195387 | 0.040172153 | 0.003350831 |
| 0 | 0 | 0 | 0.347198606 | 0 |
| 0 | 0 | 0 | 0.011163133 | 0.149911497 |
| 0 | 0 | 0 | 0 | 0 |
| 0 | 0 | 0 | 0 | 0 |
| 0 | 0 | 0 | 0 | 0 |
| 0 | 0 | 0 | 0.040039052 | 0 |
| 0 | 0 | 0 | 0.095853496 | 1.142058237 |
| 0 | 0 | 0 | 0 | 0 |
| 0 | 0 | 0 | 0 | 0 |
| 0 | 0.001676148 | 0.013092808 | 0.00683856 | 0 |
| 0.110628249 | 0 | 0 | 0 | 0 |
| 0.054231401 | 0 | 0.083061177 | 0.003789442 | 0 |

| | | | | |
|---|---|---|---|---|
| 0 | 0 | 0 | 0 | 0.000390131 |
| 0 | 0 | 0 | 0 | 0 |
| 0 | 0 | 0 | 0 | 0 |
| 0 | 0 | 0 | 0 | 0 |
| 0.001320102 | 0.003965038 | 0 | 0.011605284 | 0 |
| 0 | 0 | 0 | 0 | 0.001371493 |
| 0 | 0 | 0 | 0 | 0 |
| 0 | 0 | 0 | 0 | 0 |
| 0 | 0 | 0 | 0 | 0 |
| 0 | 0 | 0 | 0.001941389 | 0 |
| 0 | 0 | 0 | 0 | 0 |
| 0 | 0 | 1.503172958 | 0 | 0.003225087 |
| 0 | 0.198839982 | 0 | 0 | 0.074935975 |
| 0 | 0 | 0 | 0 | 0 |
| 0.004580095 | 0.001528522 | 0.515537736 | 0 | 0.00119789 |
| 0 | 0 | 0 | 0 | 0 |
| 0 | 0 | 0 | 0 | 0 |
| 0 | 0 | 0 | 0 | 0 |
| 0 | 0 | 0 | 0 | 0.000918703 |
| 0 | 0 | 0 | 0.002964612 | 0 |
| 0 | 0 | 0 | 0 | 0 |
| 0 | 0 | 0 | 0.025780731 | 0 |
| 0.04570341 | 0.419475526 | 0.237133959 | 0.202778736 | 0.003700871 |
| 0 | 0 | 0 | 0 | 0 |
| 0 | 0 | 0 | 0 | 0 |
| 0 | 0 | 0 | 0 | 0 |
| 0 | 0 | 0 | 0 | 0 |
| 0 | 0 | 0 | 0 | 0 |
| 0 | 0 | 0 | 0 | 0 |
| 0 | 0 | 0 | 0 | 0 |
| 0 | 0 | 0 | 0 | 0 |
| 0.022535063 | 0.00446281 | 0.196710678 | 0.02770773 | 0.010881005 |
| 0 | 0 | 0 | 0.123162071 | 0 |
| 0 | 0.003220451 | 0.007878447 | 0.082131005 | 0.007183236 |
| 0.002564128 | 0 | 0.002954233 | 0.033129568 | 0.003772286 |
| 0 | 0 | 0.004500794 | 0 | 0 |
| 0 | 0 | 0 | 0 | 0 |
| 0 | 0 | 0 | 0 | 0 |
| 0 | 0 | 0 | 0 | 0.00043338 |
| 0 | 0 | 0 | 0 | 0 |
| 0 | 0 | 0 | 0.136767642 | 0 |
| 0 | 0 | 0 | 0 | 0 |
| 0 | 0 | 0 | 0 | 0 |
| 0 | 0 | 0 | 0.045321593 | 0 |
| 0.002841664 | 0.005057882 | 0.003439345 | 0.66550412 | 0.007679899 |
| 0 | 0 | 0 | 0 | 0 |
| 0 | 0 | 0 | 0 | 0 |
| 0 | 0.001195963 | 0 | 0 | 0 |

| | | | | |
|---|---|---|---|---|
| 0 | 0 | 0 | 0.299339636 | 0 |
| 0 | 0 | 0 | 0 | 0 |
| 0.009143529 | 0 | 0 | 0.009047393 | 0 |
| 0 | 0 | 0 | 0.005444149 | 0 |
| 0.002814975 | 0 | 0.023309728 | 0 | 0 |
| 0 | 0 | 0 | 0.1498301 | 0 |
| 0 | 0 | 0 | 0 | 0.07460112 |
| 0 | 0 | 0 | 0.016924518 | 0 |
| 0 | 0 | 0 | 0 | 0 |
| 0 | 0 | 0 | 0 | 0 |
| 0 | 0 | 0 | 0 | 0 |
| 0.004485529 | 0 | 0 | 0 | 0 |
| 0 | 0 | 0 | 0 | 0 |
| 0 | 0 | 0 | 0 | 0.26367304 |
| 0 | 0 | 0 | 0 | 0 |
| 0 | 0 | 0 | 0 | 0 |
| 0 | 0 | 0 | 0 | 0 |
| 0 | 0 | 0 | 0 | 0 |
| 0 | 0.53241114 | 0 | 2.850778085 | 0 |
| 0 | 0 | 0 | 0 | 0 |
| 0 | 0 | 0 | 0 | 0 |
| 0 | 0 | 0 | 0 | 0 |
| 0 | 0 | 0 | 0 | 0 |
| 0 | 0 | 0 | 0 | 0 |
| 0 | 0 | 0 | 0 | 0 |
| 0.008671301 | 0.001177174 | 0.0017241 | 0.020203017 | 0.005881201 |
| 0 | 0 | 0 | 0 | 0 |
| 0 | 0 | 0.005544811 | 0.069134926 | 0 |
| 0 | 0 | 0 | 0 | 0 |
| 0 | 0.331629755 | 0.247481669 | 0.863575375 | 3.101516705 |
| 0 | 0 | 0 | 0 | 0 |
| 0 | 0 | 0 | 0 | 0 |
| 0 | 0 | 0 | 0 | 0 |
| 0 | 0 | 0 | 0 | 0 |
| 0 | 0 | 0.002581072 | 0.013621981 | 0.000966766 |
| 0 | 0 | 0 | 0 | 0 |
| 0 | 0 | 0 | 0 | 0 |
| 0 | 0 | 0.002928761 | 0.003165883 | 0 |
| 0 | 0 | 0 | 0 | 0 |
| 0 | 0 | 0 | 0 | 0 |
| 12.0913473 | 23.34718975 | 34.21784939 | 17.44918705 | 0.634580707 |
| 1.42460563 | 0.005635554 | 0.835678997 | 0.164265648 | 0.147452063 |
| 0 | 0 | 0 | 0 | 0 |
| 0 | 0 | 0 | 0 | 0 |
| 0 | 0 | 0 | 0 | 0 |
| 0 | 0 | 0 | 0 | 0 |
| 0 | 0 | 0 | 0 | 0 |
| 0 | 0 | 0 | 0.009163673 | 0.000652978 |

| | | | | |
|---|---|---|---|---|
| 0 | 0 | 0 | 0 | 0 |
| 0 | 0 | 0.053316279 | 0.065979156 | 0.005373133 |
| 0 | 0 | 0 | 0.015802613 | 0 |
| 0 | 0 | 0 | 0 | 0 |
| 0 | 0 | 0 | 0.065990946 | 1.664819331 |
| 0 | 0 | 0 | 0 | 0 |
| 0 | 0 | 0 | 0 | 0 |
| 0 | 0 | 0 | 0 | 0 |
| 0.373235787 | 0.123229367 | 0.234436751 | 0 | 0.365505034 |
| 0 | 0 | 0 | 0 | 0 |
| 0 | 0 | 0 | 0 | 0 |
| 0 | 0 | 0 | 0 | 0 |
| 0 | 0 | 0 | 0.075927506 | 0 |
| 0 | 0.078148081 | 0.048552935 | 0.019613377 | 0.003476733 |
| 0 | 0 | 0 | 0 | 0 |
| 0 | 0.166620136 | 0.925276211 | 1.565646461 | 0.001207031 |
| 0 | 0 | 0 | 0.043530898 | 0 |
| 0 | 0 | 0 | 0 | 0 |
| 0 | 0 | 0 | 0 | 0 |
| 0 | 0 | 0 | 0 | 0 |
| 0 | 0 | 0 | 0 | 0 |
| 0 | 0 | 0 | 0 | 0 |
| 0 | 0 | 0 | 0 | 0 |
| 0 | 0 | 0 | 0 | 0 |
| 0 | 0 | 0 | 0 | 0 |
| 0 | 0 | 0 | 0 | 0 |
| 0.017183772 | 0 | 0 | 0.00259634 | 0 |
| 0.000434104 | 0 | 0.003394943 | 0 | 0.00034061 |
| 0 | 0 | 0 | 0 | 0 |
| 0 | 0 | 0 | 0 | 0 |
| 0 | 0 | 0 | 0 | 0 |
| 0 | 0 | 0 | 0.000386397 | 0 |
| 0 | 0 | 0 | 0 | 0 |
| 0 | 0 | 0 | 0 | 0 |
| 0 | 0 | 0 | 0 | 0.046170914 |
| 0 | 0.014077497 | 0.008415523 | 0.010345463 | 0.026793006 |
| 0 | 0.001370371 | 0.001720336 | 0 | 0 |
| 0 | 0 | 0 | 0 | 0 |
| 0 | 0 | 0 | 0 | 0 |
| 0 | 0 | 0 | 0 | 0 |
| 0 | 0 | 0 | 1.127240259 | 0.946152545 |
| 0 | 0 | 0 | 0 | 0.09617313 |
| 0 | 0 | 0 | 0 | 0 |
| 0 | 0 | 0 | 0 | 0 |
| 0 | 0 | 0 | 0 | 0 |
| 0 | 0 | 0 | 0 | 0 |
| 0 | 0 | 0 | 0 | 0 |
| 0 | 0 | 0 | 0 | 0 |

| | | | | |
|---|---|---|---|---|
| 0.015684995 | 0 | 0 | 0.144685258 | 0 |
| 0 | 0 | 0 | 0 | 0 |
| 0 | 0 | 0 | 0 | 0 |
| 0 | 0 | 0 | 0 | 0 |
| 0 | 0.048991775 | 0 | 0.006350793 | 0 |
| 0 | 0 | 0 | 0.005678741 | 0 |
| 0 | 0 | 0 | 0.086223022 | 0 |
| 0 | 0 | 0 | 0 | 0 |
| 0 | 0.001039657 | 0 | 0 | 0 |
| 0 | 0.122391937 | 0.023538715 | 0 | 0 |
| 0 | 0 | 0 | 0 | 0 |
| 0 | 0 | 0 | 0.014121049 | 0 |
| 0 | 0 | 0 | 0 | 0 |
| 0 | 0 | 0 | 0 | 0 |
| 0 | 0 | 0 | 0 | 0 |
| 0 | 0 | 0 | 0 | 0 |
| 0 | 0 | 0 | 0 | 0 |
| 0 | 0 | 0 | 0 | 0 |
| 0 | 0 | 0 | 0 | 0 |
| 0 | 0 | 0 | 0 | 0 |
| 0 | 0 | 0.001877807 | 0.000746265 | 0.000659392 |
| 0 | 0 | 0 | 0 | 0 |
| 0 | 0.001546754 | 0.026666882 | 0.040003992 | 0 |
| 0 | 0.002750779 | 0.006446107 | 0.001024707 | 0.001293457 |
| 0 | 0 | 0 | 0 | 0 |
| 0 | 0 | 0 | 0 | 0.000635829 |
| 0 | 0 | 0 | 0 | 0 |
| 0 | 0 | 0 | 0 | 0 |
| 0 | 0 | 0 | 0 | 0 |
| 0.206414087 | 0.18884509 | 4.34019453 | 0.006741912 | 0.313987938 |
| 0 | 0 | 0 | 0 | 0 |
| 0 | 0 | 0 | 0 | 0 |
| 0 | 0 | 0 | 0 | 0 |
| 0.212873428 | 0.0185073 | 0 | 0 | 0 |
| 0 | 0 | 0 | 0 | 0 |
| 0 | 0 | 0 | 0 | 0 |
| 0.001031349 | 0 | 0.012098625 | 0 | 0 |
| 0 | 0 | 0 | 0 | 0 |
| 0 | 0 | 0 | 0 | 0 |
| 0 | 0 | 0 | 0 | 0 |
| 0 | 0 | 0 | 0 | 0 |
| 0 | 0 | 0 | 0 | 0 |
| 0 | 0 | 0 | 0 | 0 |
| 0 | 0 | 0 | 0 | 0 |
| 0 | 0 | 0 | 0 | 0 |
| 0 | 0 | 0 | 0 | 0 |
| 0 | 0 | 0 | 0 | 0 |
| 0 | 0 | 0 | 0 | 0 |

| | | | | |
|---|---|---|---|---|
| 0.005469349 | 0.007991828 | 0.01808377 | 0.075293252 | 0.006843047 |
| 0 | 0 | 0 | 0 | 0 |
| 0 | 0 | 0 | 0 | 0 |
| 0 | 0 | 0 | 0 | 0 |
| 0 | 0 | 0 | 0 | 0 |
| 0 | 0 | 0 | 0 | 0 |
| 0 | 0 | 0 | 0 | 0 |
| 1.964765855 | 0.336437853 | 0 | 0.178393296 | 0.002262695 |
| 0.062182984 | 0.014867407 | 3.78261854 | 0.056619564 | 10.04264858 |
| 0.02713697 | 0.058867005 | 0 | 0 | 0.033332773 |
| 0 | 0 | 0.008576787 | 0 | 0 |
| 0 | 0 | 0 | 0 | 0 |
| 0 | 0 | 0 | 0 | 0 |
| 0 | 0 | 0 | 0 | 0 |
| 0 | 0 | 0 | 0 | 0 |
| 0 | 0 | 0 | 0 | 0 |
| 0.080269329 | 0 | 0 | 0 | 0 |
| 0 | 0 | 0 | 0 | 0 |
| 0 | 0 | 0 | 0 | 0 |
| 0 | 0 | 0 | 0.127173299 | 0 |
| 0 | 0 | 0 | 0 | 0 |
| 0 | 0 | 0 | 0 | 0 |
| 0 | 0 | 0 | 0.780663044 | 0 |
| 0 | 0 | 0 | 0 | 0 |
| 0 | 0.007460353 | 0 | 0.102851279 | 0 |
| 0 | 0 | 0 | 0 | 0 |
| 0 | 0 | 0 | 0 | 0 |
| 0 | 0 | 0 | 0 | 0 |
| 0 | 0 | 0 | 0 | 0 |
| 0 | 0 | 0 | 0 | 0 |
| 0 | 0 | 0 | 0 | 0 |
| 0 | 0 | 0 | 0 | 0 |
| 0 | 0 | 0 | 0.006692793 | 0 |
| 0 | 0 | 0 | 0 | 0 |
| 0 | 0 | 0 | 0 | 0 |
| 0 | 0 | 0 | 0 | 0 |
| 0 | 0 | 0 | 0 | 0 |
| 0 | 0 | 0.036496673 | 0 | 0 |
| 0 | 0 | 0 | 0 | 0.000827483 |
| 0 | 0 | 0 | 0 | 0.200082014 |
| 0 | 0 | 0 | 0 | 0 |
| 0 | 0 | 0 | 0.008111645 | 1.434786976 |
| 0 | 0 | 0 | 0 | 0 |
| 0 | 0 | 0 | 0 | 0 |
| 0 | 0 | 0 | 0 | 0 |
| 0.00118298 | 0 | 0 | 0.011625383 | 0 |
| 0 | 0 | 0 | 0 | 0 |
| 0 | 0 | 0 | 0 | 0 |

| | | | | |
|---|---|---|---|---|
| 0 | 0 | 0 | 0 | 0 |
| 0 | 0.000801442 | 0.048069856 | 0 | 0 |
| 0 | 0 | 0 | 0 | 0 |
| 0 | 0 | 0 | 0 | 0 |
| 0 | 0.02876022 | 0.097217757 | 0 | 0.00673247 |
| 0 | 0 | 0 | 0 | 0 |
| 0 | 0 | 0 | 0 | 0 |
| 0.022940641 | 0.257543332 | 0.036554086 | 0.006612321 | 0.00973763 |
| 0 | 0 | 0 | 0 | 0 |
| 0 | 0 | 0 | 0 | 0 |
| 0 | 0 | 0 | 0 | 0 |
| 0 | 0 | 0 | 0 | 0 |
| 0 | 0 | 0 | 0 | 0 |
| 0 | 0 | 0 | 0 | 0 |
| 0 | 0 | 0 | 0 | 0 |
| 0 | 0.084251864 | 0 | 0.037536982 | 0 |
| 0 | 0 | 0 | 0 | 0 |
| 0 | 0 | 0 | 0 | 0 |
| 0 | 0 | 0 | 0 | 0 |
| 0 | 0 | 0 | 0 | 0 |
| 0 | 0 | 0 | 0 | 0 |
| 0 | 0 | 0 | 0.043492091 | 0 |
| 0 | 0 | 0 | 0 | 0 |
| 0.000650926 | 0.000977555 | 0.013362871 | 0.006242639 | 0 |
| 0 | 0 | 0 | 0 | 0 |
| 0 | 0 | 0 | 0 | 0 |
| 0 | 0 | 0.0329242 | 0.705294465 | 0 |
| 0 | 0 | 0 | 0 | 0 |
| 0.002654723 | 0 | 0 | 0 | 0 |
| 0 | 0 | 0 | 0 | 0 |
| 0 | 0 | 0 | 0 | 0 |
| 0 | 0 | 0.05034462 | 0 | 0 |
| 0 | 0 | 0 | 0 | 0 |
| 0 | 0 | 0 | 0 | 0 |
| 0 | 0 | 0 | 0 | 0 |
| 0 | 0 | 0 | 0 | 0 |
| 0 | 0 | 0 | 0 | 0 |
| 0 | 0 | 0 | 0 | 0 |
| 0 | 0 | 0 | 0 | 0 |
| 0 | 0 | 0 | 0 | 0 |
| 0 | 0 | 0 | 0 | 0 |
| 0 | 0 | 0 | 0 | 0.051635505 |
| 0 | 0 | 0 | 0.17105855 | 0 |
| 0 | 0 | 0 | 0 | 0 |
| 0 | 0 | 0 | 0 | 0 |
| 0 | 0 | 0 | 0 | 0 |
| 0 | 0 | 0 | 0 | 0 |
| 0 | 0 | 0 | 0 | 0 |

| | | | | |
|---|---|---|---|---|
| 0.15200501 | 0 | 0 | 0 | 0.000658936 |
| 0 | 0 | 0 | 0 | 0 |
| 0 | 0 | 0 | 0 | 0 |
| 0 | 0 | 0 | 0.010684491 | 0 |
| 0 | 0 | 0 | 0 | 0 |
| 0 | 0 | 0 | 0 | 0 |
| 0 | 0 | 0 | 0 | 0 |
| 0 | 0 | 0 | 0 | 0 |
| 0 | 0 | 0 | 0 | 0 |
| 0 | 0 | 0 | 0 | 0 |
| 0 | 0 | 0 | 0 | 0 |
| 0.00060005 | 0 | 0 | 0.005275158 | 0 |
| 0 | 0 | 0 | 0 | 0 |
| 0 | 0 | 0 | 0 | 0 |
| 0 | 0 | 0 | 0 | 0 |
| 0 | 0 | 0 | 0 | 0 |
| 0 | 0 | 0 | 0 | 0 |
| 0 | 0 | 0 | 0 | 0 |
| 0 | 0 | 0 | 0 | 0 |
| 0 | 0 | 0 | 0 | 0 |
| 0.002430768 | 0.111624394 | 0.008147149 | 0.018131584 | 0.178518372 |
| 0 | 0 | 0 | 0 | 0 |
| 0 | 0 | 0 | 0.084834864 | 0.00979544 |
| 0 | 0 | 0 | 0.392018885 | 0 |
| 0 | 0 | 0 | 0 | 0 |
| 0 | 0 | 0 | 0 | 0 |
| 0 | 0 | 0 | 0 | 0 |
| 0 | 0 | 0 | 0 | 0 |
| 0 | 0 | 0 | 0 | 0 |
| 0 | 0 | 0 | 0 | 0 |
| 0 | 0 | 0 | 0 | 0 |
| 0 | 0 | 0 | 0 | 0 |
| 0.097364374 | 0.004989169 | 0.022162445 | 0.021853168 | 0.483933878 |
| 0 | 0 | 0 | 0 | 0 |
| 0 | 0.07003144 | 0 | 0 | 0.093354981 |
| 0 | 0 | 0.299500167 | 0 | 0 |
| 0 | 0 | 0 | 0 | 0 |
| 0 | 0 | 0 | 0 | 0 |
| 0.00106737 | 0.006411871 | 0.015502414 | 0.027486917 | 0 |
| 0 | 0 | 0 | 0 | 0 |
| 0 | 0 | 0 | 0 | 0 |
| 0 | 0 | 0 | 0 | 0 |
| 0 | 0 | 0.023355899 | 0.190182754 | 0.004896373 |
| 0 | 0 | 0 | 0 | 0 |
| 0 | 0 | 0 | 0 | 0 |
| 0 | 0 | 0 | 0 | 0 |
| 0 | 0 | 0 | 0 | 0 |
| 0 | 0 | 0 | 0 | 0 |

| | | | | |
|---|---|---|---|---|
| 0 | 0 | 0 | 0 | 0 |
| 0 | 0 | 0 | 0 | 0 |
| 0 | 0 | 0 | 0 | 0 |
| 0 | 0 | 0 | 0 | 0 |
| 0 | 0 | 0 | 0 | 0 |
| 0 | 0 | 0 | 0 | 0 |
| 0 | 0 | 0 | 0 | 0 |
| 0 | 0 | 0 | 0.021603417 | 0.014905105 |
| 0 | 0 | 0 | 0.420732708 | 0 |
| 0 | 0.036456063 | 0 | 0 | 0 |
| 0 | 0 | 0 | 0 | 0 |
| 0 | 0 | 0 | 0 | 0 |
| 0 | 0 | 0 | 0 | 0 |
| 0 | 0 | 0 | 0 | 0 |
| 0 | 0 | 0 | 0.005714537 | 0 |
| 0.05223221 | 0.013073651 | 0.001683322 | 0.001962324 | 0 |
| 0 | 0 | 0 | 0 | 0 |
| 0 | 0 | 0 | 0 | 0 |
| 0 | 0 | 0 | 0 | 0 |
| 0 | 2.272490394 | 0 | 0 | 0 |
| 0 | 0 | 0 | 0 | 0.01440508 |
| 0 | 0 | 0 | 0 | 0 |
| 0 | 0 | 0 | 0 | 0 |
| 0 | 0 | 0 | 0 | 0 |
| 0 | 0 | 0 | 0 | 0 |
| 0 | 0 | 0 | 0 | 0 |
| 0 | 0 | 0 | 0 | 0 |
| 0 | 0 | 0 | 0 | 0 |
| 0 | 0 | 0 | 0 | 0 |
| 0 | 0 | 0 | 0.544133529 | 0 |
| 0 | 0.819283883 | 1.357211842 | 0.22212914 | 0.022847038 |
| 0 | 0 | 0 | 0 | 0 |
| 0 | 0 | 0 | 0 | 0 |
| 0 | 0 | 0 | 0 | 0 |
| 0 | 0 | 0 | 0 | 0 |
| 0 | 0 | 0 | 0 | 0 |
| 0 | 0 | 0 | 0 | 0 |
| 0 | 0 | 0 | 0 | 0 |
| 0 | 0 | 0 | 0 | 0 |
| 0 | 0 | 0 | 0 | 0 |
| 0 | 0 | 0 | 0 | 0 |
| 0 | 0 | 0 | 0 | 0 |
| 0 | 0 | 0 | 0 | 0 |
| 0 | 0 | 0 | 0 | 0 |
| 0 | 0 | 0 | 0 | 0 |
| 0 | 0 | 0 | 0 | 0 |
| 0.281230229 | 0.015785551 | 0.006031217 | 0 | 0.033482385 |
| 0 | 0 | 0 | 0 | 0 |

| | | | | |
|---|---|---|---|---|
| 0 | 0 | 0 | 0 | 0 |
| 0 | 0 | 0 | 0 | 0 |
| 0 | 0 | 0 | 0 | 0 |
| 0 | 0 | 0 | 0 | 0 |
| 0 | 0.013843939 | 0 | 0.010368724 | 0 |
| 0 | 0 | 0 | 0 | 0 |
| 0 | 0 | 0 | 0 | 0 |
| 0 | 0 | 0 | 0 | 0 |
| 0 | 0 | 0 | 0 | 0 |
| 0 | 0 | 0 | 0 | 0 |
| 0 | 0 | 0 | 0 | 0 |
| 0.001422027 | 0 | 0.002383088 | 0.151341787 | 0.001394702 |
| 0 | 0 | 0 | 0 | 0 |
| 0 | 0 | 0 | 0 | 0 |
| 0 | 0 | 0 | 0 | 0 |
| 0 | 2.573013001 | 1.560384682 | 0.234495609 | 0 |
| 0 | 0 | 0 | 0.01180023 | 0 |
| 0 | 0 | 0 | 0 | 0 |
| 0.030957047 | 0.007815885 | 0.01195473 | 0.007027843 | 0.00623085 |
| 0 | 0 | 0 | 0.111238277 | 0 |
| 0 | 0 | 0 | 0 | 0 |
| 0 | 0 | 0 | 0 | 0 |
| 0 | 0 | 0 | 0 | 0 |
| 0 | 0 | 0 | 0 | 0 |
| 0 | 0 | 0 | 0 | 0 |
| 0 | 0 | 0 | 0 | 0 |
| 0.004084054 | 0.002255963 | 0 | 0.004502037 | 0 |
| 0 | 0 | 0 | 0.004743295 | 0 |
| 0 | 0 | 0 | 0 | 0 |
| 0 | 0 | 0 | 0 | 0 |
| 0 | 0 | 0 | 0.003572307 | 0 |
| 0 | 0 | 0.004334402 | 0.074965284 | 0 |
| 0 | 0 | 0 | 0 | 0 |
| 0 | 0 | 0 | 0 | 0 |
| 0 | 0 | 0 | 0 | 0 |
| 0 | 0 | 0 | 0 | 0 |
| 0 | 0 | 0 | 0 | 0 |
| 0.000448627 | 0 | 0.005074828 | 0.097284588 | 0.009064132 |
| 0 | 0 | 0 | 0 | 0 |
| 0 | 0 | 0 | 0.031280794 | 0 |
| 0 | 0 | 0 | 0.009187157 | 0 |
| 0 | 0 | 0 | 0 | 0 |
| 0 | 0 | 0 | 0 | 0 |
| 1.682102022 | 0.441592198 | 0.838098436 | 0 | 0 |
| 0 | 0 | 0 | 0 | 0 |
| 0 | 0 | 0 | 0 | 0 |
| 0 | 0 | 0 | 0 | 0 |
| 0 | 0 | 0 | 0 | 0 |

| | | | | |
|---|---|---|---|---|
| 0 | 0 | 0 | 0 | 0 |
| 0 | 0.296024918 | 0 | 0.085583327 | 0 |
| 0 | 0 | 0 | 0 | 0 |
| 0 | 0 | 0.002920954 | 0.016158687 | 0.002051386 |
| 0 | 0 | 0 | 0 | 0 |
| 0 | 0 | 0 | 0.002890193 | 0 |
| 0 | 0 | 0 | 0.005587404 | 0 |
| 0 | 0 | 0.003655103 | 0 | 0 |
| 0 | 0 | 0 | 0 | 0 |
| 0 | 0 | 0 | 0 | 0 |
| 0 | 0 | 0 | 0.232818947 | 0 |
| 0 | 0 | 0 | 0 | 0 |
| 0 | 0 | 0 | 0.066882196 | 0.00232663 |
| 0 | 0 | 0 | 0 | 0 |
| 0 | 0 | 0 | 0 | 0 |
| 0 | 0 | 0 | 0 | 0 |
| 0 | 0 | 0 | 0 | 0 |
| 0 | 0 | 0 | 0 | 0 |
| 0 | 0 | 0 | 0 | 0 |
| 0 | 0 | 0 | 0 | 0 |
| 0 | 0 | 0 | 0 | 0 |
| 0 | 0 | 0 | 0 | 0 |
| 0.004067264 | 0.008144245 | 0.025901096 | 0.005720981 | 0.003063636 |
| 0 | 0 | 0 | 0 | 0 |
| 0 | 0 | 0 | 0 | 0 |
| 0 | 0 | 0 | 0 | 0 |
| 0 | 0 | 0 | 0.043561587 | 0 |
| 0 | 0 | 0 | 0 | 0 |
| 0 | 0 | 0 | 0 | 0 |
| 0 | 0 | 0 | 0 | 0 |
| 0 | 0.011986652 | 0.003951946 | 0.02454898 | 0.055366705 |
| 0 | 0.000842668 | 0.005641964 | 0 | 0 |
| 0 | 0 | 0 | 0 | 0 |
| 0 | 0 | 0 | 0 | 0 |
| 0 | 0 | 0 | 0.21488835 | 0 |
| 0.319147028 | 0 | 0 | 0 | 0 |
| 0.022163193 | 0 | 0 | 0.03265321 | 0 |
| 0 | 0 | 0 | 0 | 0 |
| 0 | 0 | 0.02109053 | 0 | 0 |
| 0 | 0 | 0 | 0 | 0 |
| 0 | 0 | 0 | 0 | 0 |
| 0 | 0 | 0 | 0 | 0 |
| 0 | 0 | 0 | 0 | 0 |
| 0 | 0 | 0 | 0 | 0 |
| 0 | 0 | 0 | 0 | 0 |
| 0 | 0.098962835 | 0.15501201 | 0.006762927 | 0.027653282 |
| 0.01623227 | 0 | 0 | 0 | 0 |
| 0 | 0 | 0 | 0 | 0 |

| | | | | |
|---|---|---|---|---|
| 0 | 0 | 0 | 0 | 0 |
| 0 | 0 | 0 | 0 | 0 |
| 0 | 0 | 0 | 0 | 0 |
| 0 | 0 | 0 | 0 | 0 |
| 0 | 0 | 0 | 0 | 0 |
| 0 | 0 | 0 | 0 | 0 |
| 0 | 0 | 0 | 0.018120171 | 0 |
| 0 | 0 | 0 | 0 | 0 |
| 0.001136051 | 0 | 0 | 0.001361899 | 0 |
| 0 | 0 | 0 | 0 | 0 |
| 0 | 0 | 0 | 0 | 0 |
| 0 | 0 | 0 | 0.00237707 | 0 |
| 0 | 0 | 0 | 0 | 0 |
| 0 | 0 | 0 | 0 | 0 |
| 0 | 0 | 0 | 0 | 0 |
| 0 | 0 | 0 | 0 | 0 |
| 0 | 0 | 0 | 0 | 0 |
| 0 | 0 | 0 | 0 | 0 |
| 0 | 0 | 0 | 0 | 0 |
| 0 | 0 | 0 | 0 | 0.000388678 |
| 0 | 0 | 0 | 0 | 0 |
| 0 | 0 | 0 | 0 | 0 |
| 0 | 0 | 0 | 0 | 0 |
| 0 | 0 | 0 | 0 | 0 |
| 0 | 0 | 0 | 0 | 0 |
| 0 | 0 | 0 | 0 | 0 |
| 0 | 0 | 0 | 0 | 0 |
| 0 | 0 | 0 | 0 | 0.004444763 |
| 0 | 0 | 0 | 0 | 0 |
| 0 | 0 | 0 | 0 | 0 |
| 0 | 0.004544849 | 0 | 0.00256977 | 0.004229589 |
| 0 | 0 | 0 | 0 | 0 |
| 0.113320825 | 0.338059885 | 0.138842202 | 0.008073715 | 0.00180931 |
| 0 | 0 | 0.010154553 | 0.005918809 | 0 |
| 0 | 0 | 0 | 0 | 0 |
| 0 | 0 | 0 | 0 | 0 |
| 0 | 0 | 0 | 0 | 0 |
| 0 | 0 | 0 | 0 | 0 |
| 0 | 0 | 0 | 0 | 0 |
| 0 | 0 | 0 | 0 | 0 |
| 0 | 0 | 0 | 0 | 0 |
| 0.01463463 | 0.017365467 | 0.127168118 | 0.15248116 | 0 |
| 0 | 0 | 0.071100254 | 0.006508813 | 0 |
| 0 | 0 | 0 | 0 | 0 |
| 0 | 0 | 0 | 0 | 0 |
| 0 | 0 | 0.006396564 | 0 | 0.005053845 |
| 0 | 0 | 0 | 0 | 0 |
| 0 | 0 | 0 | 0 | 0 |

| | | | | |
|---|---|---|---|---|
| 0 | 0 | 0 | 0 | 0 |
| 0 | 0 | 0 | 0 | 0.006513579 |
| 0.003636381 | 0 | 0.038595263 | 0.008395679 | 0 |
| 0 | 0 | 0 | 0 | 1.249175956 |
| 0 | 0.120799673 | 0.275726216 | 2.515720227 | 0 |
| 0.008478677 | 0 | 0 | 0.052536152 | 0 |
| 0 | 0 | 0 | 0 | 0 |
| 0 | 0 | 0 | 0 | 0 |
| 0 | 0 | 0 | 0 | 0 |
| 0 | 0 | 0 | 0 | 0 |
| 0 | 0 | 0 | 0 | 0 |
| 0 | 0 | 0 | 0 | 0 |
| 0 | 0 | 0 | 0 | 0 |
| 0 | 0 | 0 | 0.089747843 | 0 |
| 0 | 0.026755493 | 0 | 0 | 0 |
| 0 | 0 | 0 | 0.002047123 | 0 |
| 0 | 0 | 0 | 0 | 0 |
| 0 | 0 | 0 | 0 | 0 |
| 0 | 0 | 0 | 0 | 0 |
| 0 | 0 | 0 | 0 | 0 |
| 0 | 0 | 0 | 0.003022927 | 0 |
| 0 | 0 | 0 | 0.007446004 | 0 |
| 0 | 0 | 0 | 0 | 0 |
| 0 | 0 | 0 | 0 | 0 |
| 0 | 0 | 0 | 0.002306407 | 0.002090171 |
| 0 | 0 | 0.214162173 | 0 | 0.002496912 |
| 0 | 0 | 0 | 0 | 0 |
| 0 | 0 | 0 | 0.180399508 | 0 |
| 0 | 0 | 0 | 0 | 0 |
| 0 | 0 | 0 | 0 | 0 |
| 0 | 0 | 0 | 0 | 0 |
| 0 | 0 | 0 | 0.00156513 | 0 |
| 0 | 0 | 0 | 0 | 0 |
| 0 | 0 | 0 | 0 | 0 |
| 0 | 0 | 0 | 0 | 0 |
| 0 | 0 | 0 | 0 | 0 |
| 0 | 0 | 0 | 0 | 0 |
| 0 | 0 | 0 | 0.006218032 | 0 |
| 0 | 0 | 0 | 0 | 0.799535056 |
| 0 | 0 | 0 | 0 | 0 |
| 0 | 0 | 0 | 0 | 0 |
| 0 | 0 | 0 | 0 | 0.001425666 |
| 0.003529961 | 0.003029295 | 0.011549593 | 0.054810822 | 0.005143738 |
| 0 | 0 | 0 | 0 | 0 |
| 0 | 0 | 0 | 0 | 0 |
| 0 | 0 | 0 | 0 | 0 |
| 0 | 0 | 0 | 0.010960762 | 0 |
| 0 | 0 | 0 | 0 | 0 |

| | | | | |
|---|---|---|---|---|
| 0 | 0 | 0 | 0 | 0 |
| 0.024497936 | 0.009745908 | 0.026100931 | 0.007088114 | 0 |
| 0 | 0 | 0 | 0 | 0 |
| 0 | 0 | 0 | 0 | 0 |
| 0 | 0 | 0 | 0 | 0 |
| 0 | 0 | 0 | 0 | 0 |
| 0 | 0 | 0 | 0 | 0 |
| 0 | 0 | 0 | 0 | 0 |
| 0 | 0 | 0 | 0 | 0 |
| 0 | 0 | 0 | 0 | 0 |
| 0 | 0 | 0 | 0 | 0 |
| 0 | 0 | 0 | 0 | 0 |
| 0 | 0 | 0 | 0.00511701 | 0 |
| 0 | 0 | 0 | 0 | 0 |
| 0.001011432 | 0 | 0 | 0 | 0.006031339 |
| 0 | 0 | 0 | 0 | 0 |
| 0 | 0 | 0 | 0 | 0 |
| 0 | 0 | 0 | 0 | 0 |
| 0 | 0 | 0 | 0.013353822 | 0 |
| 0 | 0 | 0 | 0 | 0 |
| 0 | 0 | 0 | 0 | 0 |
| 0 | 0 | 0 | 0.078326531 | 0 |
| 0 | 0 | 0.039141203 | 0.035299172 | 0 |
| 0 | 0 | 0.106302148 | 0 | 0 |
| 0 | 0 | 0 | 0 | 0 |
| 0 | 0 | 0 | 0 | 0 |
| 0 | 0 | 0 | 0 | 0 |
| 0 | 0 | 0 | 0 | 0 |
| 0 | 0 | 0 | 0 | 0 |
| 0 | 0 | 0 | 0 | 0 |
| 0 | 0 | 0 | 0 | 0 |
| 0 | 0 | 0 | 0 | 0 |
| 0 | 0 | 0 | 0 | 0 |
| 0 | 0 | 0 | 0 | 0 |
| 0 | 0 | 8.743649338 | 14.59456788 | 0.000566103 |
| 0 | 0 | 0 | 0 | 0 |
| 0 | 0 | 0 | 0 | 0 |
| 0 | 0 | 0 | 0 | 0 |
| 0 | 0 | 0.002659719 | 0.052638949 | 0 |
| 0 | 0 | 0 | 0 | 0 |
| 0 | 0.041317537 | 0.115364221 | 0.312756374 | 0.005443224 |
| 0 | 0 | 0 | 0 | 0 |
| 0 | 0 | 0 | 0 | 0 |
| 0 | 0 | 0 | 0 | 0 |
| 0 | 0 | 0 | 0 | 0 |
| 0.575749324 | 0.211244724 | 2.115394731 | 0.004991485 | 0.011185858 |
| 0 | 0 | 0 | 0 | 0 |
| 0 | 0 | 0 | 0 | 0 |

| | | | | |
|---|---|---|---|---|
| 0 | 0 | 0 | 0 | 0 |
| 0 | 0 | 0 | 0 | 0 |
| 0 | 0 | 0 | 0 | 0 |
| 0 | 0 | 0 | 0 | 0 |
| 0 | 0 | 0 | 0.040819327 | 0 |
| 0 | 0 | 0 | 0 | 0 |
| 0 | 0 | 0 | 0 | 0 |
| 0 | 0 | 0 | 0 | 0 |
| 0 | 0 | 0 | 0 | 0 |
| 0 | 0 | 0.002670357 | 0 | 0.000625131 |
| 0 | 0 | 0 | 0 | 0 |
| 0 | 0 | 0 | 0 | 0 |
| 0 | 0.001559319 | 0 | 0.004978884 | 0.000458259 |
| 0 | 0 | 0 | 0 | 0 |
| 0 | 0 | 0 | 0 | 0 |
| 0 | 0 | 0 | 0 | 0 |
| 0 | 0 | 0 | 0 | 0 |
| 0 | 0 | 0 | 0 | 0 |
| 0 | 0 | 0 | 0 | 0 |
| 0.223185947 | 0.087437958 | 0.095829041 | 0.036191062 | 0.000951728 |
| 0 | 0 | 0 | 0 | 0 |
| 0 | 0 | 0 | 0 | 0 |
| 0 | 0 | 0 | 0 | 0 |
| 0 | 0 | 0 | 0 | 0 |
| 0 | 0 | 0 | 0 | 0 |
| 0 | 0 | 0 | 0 | 0 |
| 0 | 0 | 0 | 0 | 0 |
| 0 | 0 | 0 | 0 | 0 |
| 0 | 0 | 0 | 0 | 0 |
| 0.014263496 | 0 | 0.004656491 | 0.068100236 | 0 |
| 0 | 0 | 0 | 0 | 0 |
| 0 | 0 | 0 | 0 | 0 |
| 0 | 0 | 0 | 0 | 0 |
| 0 | 0 | 0 | 0 | 0 |
| 0 | 0 | 0 | 0 | 0 |
| 0 | 0 | 0 | 0 | 0 |
| 0 | 0 | 0 | 0 | 0 |
| 0 | 0 | 0 | 0 | 0 |
| 0 | 0 | 0 | 0 | 0 |
| 0 | 0 | 0 | 0 | 0 |
| 0 | 0 | 0 | 0 | 0 |
| 0 | 0 | 0 | 0 | 0 |
| 0 | 0 | 0 | 0.069836551 | 0 |
| 0 | 0 | 0 | 0 | 0 |
| 0 | 0 | 0 | 0.022545698 | 0 |
| 0 | 0 | 0 | 0 | 0 |
| 0 | 0 | 0 | 0 | 0 |
| 0 | 0 | 0.078904904 | 0 | 0 |

| | | | | |
|---|---|---|---|---|
| 0 | 0 | 0 | 0 | 0 |
| 0 | 0 | 0 | 0 | 0 |
| 0 | 0 | 0 | 0 | 0 |
| 0 | 0 | 0 | 0 | 0 |
| 0 | 0 | 0 | 0 | 0 |
| 0 | 0.000729875 | 0 | 0.056190447 | 0.004480639 |
| 0 | 0 | 0 | 0 | 0 |
| 0 | 0 | 0 | 0 | 0 |
| 0 | 0 | 0 | 0 | 0 |
| 0 | 0 | 0 | 0 | 0 |
| 0 | 0 | 0 | 0 | 0 |
| 0 | 0 | 0 | 0 | 0 |
| 0 | 0 | 0 | 0 | 0 |
| 0 | 0 | 0 | 0 | 0 |
| 0 | 0 | 0 | 0 | 0 |
| 0 | 0 | 0 | 0 | 0 |
| 0 | 0 | 0 | 0 | 0 |
| 0 | 0 | 0 | 0 | 0 |
| 0 | 0 | 0 | 0 | 0 |
| 0 | 0 | 0 | 0 | 0 |
| 0 | 0 | 0 | 0 | 0 |
| 0 | 0 | 0 | 0 | 0 |
| 0 | 0 | 0 | 0 | 0 |
| 0 | 0 | 0 | 0 | 0 |
| 0 | 0 | 0 | 0 | 0 |
| 0 | 0 | 0 | 0 | 0 |
| 0 | 0 | 0 | 0 | 0 |
| 0 | 0 | 0 | 0.108219624 | 0 |
| 0 | 0 | 0 | 0 | 0 |
| 0 | 0 | 0 | 0.001170299 | 0 |
| 0 | 0 | 0 | 0 | 0 |
| 0 | 0 | 0 | 0 | 0 |
| 0 | 0 | 0 | 0 | 0 |
| 0 | 0 | 0 | 0 | 0 |
| 0 | 0 | 0 | 0 | 0 |
| 0 | 0 | 0 | 0 | 0 |
| 0 | 0 | 0 | 0 | 0 |
| 0 | 0 | 0 | 0 | 0 |
| 0 | 0.377083715 | 0 | 0.984711048 | 0.016955911 |
| 0 | 0 | 0 | 0 | 0 |
| 0 | 0 | 0 | 0 | 0 |
| 0 | 0 | 0 | 0 | 0 |
| 0 | 0 | 0 | 0 | 0 |
| 0 | 0 | 0 | 0 | 0 |
| 0 | 0 | 0 | 0 | 0 |
| 0 | 0 | 0 | 0 | 0 |
| 0 | 0 | 0 | 0 | 0 |
| 0 | 0 | 0 | 0 | 0 |

| | | | | |
|---|---|---|---|---|
| 0 | 0 | 0 | 0 | 0 |
| 0 | 0.001906691 | 0 | 0.00173944 | 0 |
| 0 | 0 | 0 | 0 | 0 |
| 0 | 0 | 0 | 0 | 0 |
| 0 | 0 | 0 | 0 | 0 |
| 0 | 0 | 0 | 0 | 0 |
| 0 | 0 | 0 | 0 | 0 |
| 0 | 0 | 0 | 0 | 0 |
| 0 | 0 | 0 | 0.019826859 | 0 |
| 0 | 0 | 0 | 0 | 0 |
| 0 | 0 | 0 | 0 | 0.002375886 |
| 0 | 0 | 0 | 0 | 0 |
| 0 | 0 | 0 | 0.175030703 | 0 |
| 0 | 0 | 0 | 0 | 0 |
| 0 | 0 | 0 | 0 | 0 |
| 0 | 0.003829755 | 0 | 0 | 0 |
| 0 | 0 | 0 | 0 | 0 |
| 0 | 0 | 0 | 0 | 0 |
| 0 | 0 | 0 | 0 | 0 |
| 0 | 0 | 0 | 0 | 0 |
| 0 | 0 | 0.004241443 | 0 | 0.023404584 |
| 0 | 0 | 0 | 0 | 0 |
| 0 | 0 | 0 | 0 | 0 |
| 0 | 1.888319204 | 0 | 1.382789489 | 0.229957554 |
| 0 | 0 | 0 | 0 | 0 |
| 0 | 0 | 0 | 0 | 0 |
| 0 | 0 | 0 | 0 | 0 |
| 0 | 0 | 0 | 0 | 0 |
| 0 | 0 | 0 | 0 | 0 |
| 0 | 0 | 0 | 0 | 0 |
| 0 | 0 | 0 | 0 | 0 |
| 0 | 0 | 0 | 0 | 0 |
| 0 | 0 | 0 | 0 | 0 |
| 0 | 0 | 0 | 0 | 0 |

| PHL018-0182_V2 | PHL018-0195_V2 | PHL018-0010_V2 | PHL018-0027_V2 | PHL018-0039_V2 |
|---|---|---|---|---|
| EYCF | CTRL | CTRL | EYCF | EYCF |
| 0.0043532 | 1.252492745 | 0.04201478 | 0.773244186 | 0.001219357 |
| 0.185797007 | 6.673819582 | 1.272135722 | 0 | 0.484957931 |
| 0 | 7.94403757 | 0 | 0 | 6.128706455 |
| 0 | 0 | 0 | 0 | 0 |
| 0 | 0 | 0 | 0 | 0 |
| 0 | 0 | 0 | 0 | 0 |
| 0 | 0 | 0 | 0 | 0 |
| 0 | 0 | 0 | 0 | 0 |
| 0 | 0 | 0 | 0 | 0 |
| 0 | 0 | 0 | 0 | 0 |
| 0 | 0 | 0 | 0 | 0 |
| 0 | 0 | 0 | 0 | 0 |
| 0 | 0 | 0 | 0 | 0 |
| 0.303486791 | 2.424225952 | 0.187667818 | 0 | 3.284431104 |
| 0 | 0.000820316 | 0 | 0 | 0 |
| 0 | 0 | 0 | 0 | 0 |
| 0 | 0 | 0 | 0 | 0 |
| 0 | 0 | 0 | 0 | 0 |
| 0 | 0 | 0 | 0 | 0 |
| 0 | 0 | 0 | 0 | 0 |
| 0 | 0 | 0 | 0 | 0 |
| 0 | 0 | 0.000868672 | 0 | 0 |
| 0 | 0 | 0 | 0 | 0 |
| 0 | 0 | 0 | 0 | 0 |
| 0 | 0 | 0 | 0 | 0 |
| 0 | 0 | 0 | 0 | 0 |
| 0 | 0 | 0 | 0 | 0 |
| 0 | 0 | 0 | 0 | 0 |
| 0 | 0 | 0 | 0 | 0 |
| 0 | 0 | 0 | 0 | 0 |
| 0 | 0 | 0 | 0 | 0 |
| 0 | 0 | 0 | 0 | 0 |
| 0 | 0 | 0 | 0 | 0 |
| 0 | 0 | 0 | 0 | 0 |
| 0 | 0 | 0 | 0 | 0 |
| 1.064059955 | 2.594012324 | 3.132121303 | 0 | 0 |
| 0 | 0 | 0 | 0 | 0 |
| 0 | 0 | 0 | 0 | 0 |
| 0 | 0 | 0 | 0 | 0 |
| 0 | 0 | 0 | 0 | 0 |
| 0 | 0 | 0 | 0 | 0 |
| 0 | 0.000818519 | 0.009507182 | 0.02559269 | 0.014577037 |
| 9.334813462 | 0.00155271 | 0.006985346 | 2.930088128 | 0.139518346 |
| 0.002685354 | 0.025836726 | 0.030849477 | 0.224815988 | 0.187527684 |
| 0.013767433 | 0 | 0 | 0.102869559 | 0 |
| 0 | 0 | 0 | 0 | 0 |

| | | | | |
|---|---|---|---|---|
| 0.005157711 | 0 | 0 | 0 | 0.005482071 |
| 0 | 0.007549287 | 0.000564576 | 0.017727395 | 0 |
| 0.003806104 | 0.062241864 | 0.000816921 | 0.015613577 | 0.000923964 |
| 0.558173873 | 0.421021529 | 0.65631676 | 0 | 0.186417658 |
| 0 | 0 | 0.003532783 | 0 | 0.000621552 |
| 0 | 0 | 0 | 0 | 0 |
| 0 | 0.003005458 | 0 | 0 | 0.006288173 |
| 0 | 0 | 0 | 0 | 0 |
| 0 | 0 | 0 | 0 | 0 |
| 0 | 0 | 0 | 0 | 0 |
| 0 | 0 | 0 | 0.002846438 | 5.253776645 |
| 0.285812737 | 0.085983222 | 0.053062074 | 0 | 0.001267921 |
| 0 | 0 | 0.000393175 | 0 | 0 |
| 0 | 0.009768651 | 0.021343888 | 0 | 0.001930025 |
| 0 | 0 | 0 | 0 | 0 |
| 0 | 0 | 0 | 0 | 0 |
| 0.339891901 | 0.031777118 | 0 | 6.206817636 | 0.157575649 |
| 0 | 0 | 0 | 0 | 0 |
| 0 | 0 | 0 | 0 | 0 |
| 0 | 0 | 0 | 0 | 0 |
| 0.003111525 | 0.601151812 | 0.033544658 | 0 | 0.076139167 |
| 0.002044993 | 0.16706 | 0.018166153 | 0 | 0 |
| 0.371742929 | 0 | 0 | 0 | 0 |
| 0 | 0 | 0 | 0 | 0 |
| 0 | 0 | 0 | 0 | 0 |
| 0 | 0 | 0 | 0 | 0 |
| 0 | 0 | 0 | 0.442434719 | 0 |
| 0 | 0 | 0 | 0 | 0 |
| 0 | 0 | 0 | 0.007321567 | 0.030484293 |
| 0 | 0 | 0 | 0 | 0 |
| 0 | 0.290367714 | 0 | 0 | 0.233606727 |
| 0 | 0 | 0 | 0 | 0 |
| 0 | 0 | 0 | 0 | 0 |
| 0 | 0 | 0 | 0 | 0.003540889 |
| 0 | 0 | 0 | 0 | 0 |
| 0 | 0 | 0 | 0 | 0 |
| 0 | 0 | 0 | 0 | 0 |
| 17.88356252 | 0 | 0 | 0.013701901 | 0 |
| 0 | 0 | 0 | 0 | 0 |
| 0 | 0 | 0 | 0 | 0 |
| 0 | 0 | 0 | 0 | 0 |
| 0 | 0 | 0 | 0 | 0 |
| 0 | 0 | 0 | 0 | 0 |
| 0 | 0 | 0 | 0 | 0 |
| 0 | 0 | 0 | 0 | 0 |
| 0 | 0 | 0 | 0 | 0 |
| 0 | 0.005077264 | 0 | 0 | 0 |
| 0 | 0.493740787 | 2.925404994 | 0.006375286 | 0 |

| | | | | |
|---|---|---|---|---|
| 0 | 0 | 0 | 0 | 0 |
| 0 | 0.000690039 | 0 | 0 | 0 |
| 0 | 0 | 0 | 0 | 0 |
| 0 | 0 | 0 | 0 | 0 |
| 0 | 1.697584178 | 0.016273077 | 0 | 0.000968704 |
| 0 | 0 | 0 | 0 | 0.00392264 |
| 0 | 0 | 0 | 0 | 0 |
| 0 | 0 | 0 | 0 | 0 |
| 0 | 0.004128212 | 0.046027465 | 0 | 0 |
| 0 | 0 | 0 | 0 | 0 |
| 0 | 0 | 0 | 0 | 0 |
| 0 | 0 | 0 | 0.001482092 | 0 |
| 0 | 0 | 0 | 0 | 0 |
| 0 | 0 | 0 | 0 | 0 |
| 0 | 0 | 0 | 0 | 0 |
| 0 | 0 | 0 | 0 | 0 |
| 0 | 0 | 0 | 0 | 0 |
| 0 | 0 | 0 | 0 | 0 |
| 0.006710435 | 0 | 0.056895932 | 0 | 0 |
| 0 | 0 | 0 | 0 | 0 |
| 0 | 0 | 0 | 0 | 0 |
| 0 | 0 | 0 | 0 | 0 |
| 0 | 0 | 0 | 0 | 0 |
| 0 | 0 | 0 | 0 | 0.003411147 |
| 0 | 0 | 0 | 0 | 0 |
| 0 | 0 | 0 | 0 | 0 |
| 0 | 0 | 0 | 0.015797196 | 0 |
| 0 | 0 | 0 | 0 | 0 |
| 0 | 0 | 0 | 0 | 0 |
| 0 | 0 | 0 | 0 | 0 |
| 0.018421957 | 0.012195019 | 0.089607643 | 0 | 0 |
| 0 | 0.001387262 | 0.002420761 | 0 | 0 |
| 0 | 0 | 0 | 0 | 0 |
| 0 | 0 | 0 | 0 | 0 |
| 0 | 0 | 0 | 0 | 0 |
| 0 | 0 | 0 | 0 | 0 |
| 0 | 0 | 0 | 0 | 0 |
| 0 | 0 | 0 | 0 | 0 |
| 0.329847319 | 0 | 0 | 0 | 0 |
| 0 | 0 | 0 | 0 | 0 |
| 0 | 0 | 0 | 0 | 0 |
| 0 | 0 | 0 | 0 | 0 |
| 0 | 0 | 0.00058292 | 0 | 0 |
| 0 | 0 | 0 | 0 | 0 |
| 0 | 0 | 0.001367243 | 0 | 0 |
| 0 | 0 | 0 | 0 | 0 |
| 0 | 0 | 0 | 0 | 0 |
| 0 | 0 | 0 | 0 | 0 |

| | | | | |
|---|---|---|---|---|
| 0 | 0.002595329 | 0 | 0 | 0 |
| 0 | 0 | 0 | 0 | 0 |
| 0 | 0 | 0 | 0 | 0 |
| 0 | 0 | 0 | 0 | 0 |
| 0.097706716 | 0 | 0 | 0 | 0 |
| 0 | 0 | 0 | 0 | 0 |
| 0 | 0 | 0 | 0 | 0 |
| 0 | 0 | 0 | 0 | 0 |
| 0 | 0 | 0 | 0 | 0 |
| 0 | 0 | 0 | 0 | 0 |
| 0 | 0 | 0 | 0 | 0 |
| 0 | 0 | 0 | 0 | 0 |
| 0 | 0 | 0 | 0 | 0 |
| 0 | 0 | 0 | 0 | 0 |
| 0 | 0 | 0 | 0 | 0 |
| 0 | 1.377047517 | 0 | 0 | 0 |
| 0 | 0 | 0 | 0 | 0 |
| 0 | 0 | 0 | 0 | 0 |
| 0 | 0 | 0 | 0 | 0 |
| 0 | 0 | 0 | 0 | 0 |
| 0 | 0 | 0 | 0 | 0 |
| 0 | 0 | 0 | 0 | 0 |
| 0 | 0 | 0 | 0 | 0 |
| 0 | 0 | 0 | 0 | 0 |
| 0 | 0 | 0 | 0 | 0 |
| 0 | 0 | 0 | 0 | 0 |
| 0 | 0 | 0 | 0 | 0 |
| 0 | 0 | 0 | 0 | 0 |
| 0 | 0 | 0 | 0 | 0 |
| 0 | 0 | 0 | 0 | 0 |
| 0 | 0 | 0.006478078 | 0 | 0.001308378 |
| 0.307273845 | 0 | 0 | 0.005242925 | 0.129914457 |
| 0 | 0 | 0 | 0 | 0 |
| 0 | 0 | 0 | 0 | 0 |
| 0 | 0 | 0 | 0 | 0 |
| 0 | 0 | 0 | 0 | 0 |
| 0 | 0 | 0 | 0 | 0 |
| 0 | 0 | 0 | 0 | 0 |
| 0 | 0 | 0 | 0 | 0 |
| 0 | 0 | 0 | 0 | 0 |
| 0 | 0 | 0 | 0 | 0 |
| 0 | 0 | 0 | 0 | 0 |
| 0 | 0 | 0 | 0 | 0 |
| 0 | 0 | 0 | 0 | 0 |
| 0 | 0 | 0 | 0 | 0 |
| 0 | 0 | 0 | 0 | 0 |
| 0 | 0 | 0 | 0 | 0 |
| 0 | 0 | 0 | 0 | 0 |

| | | | | |
|---|---|---|---|---|
| 0 | 0 | 0 | 0 | 0 |
| 0 | 0 | 0 | 0 | 0 |
| 0 | 0 | 0 | 0 | 0 |
| 0 | 0 | 0 | 0 | 0 |
| 0 | 0 | 0 | 0 | 0 |
| 0 | 0 | 0 | 0 | 0 |
| 0 | 0 | 0 | 0 | 0 |
| 0 | 0 | 0 | 0 | 0 |
| 0 | 0 | 0 | 0 | 0 |
| 0 | 0 | 0 | 0 | 0 |
| 0 | 0 | 0 | 0 | 0 |
| 0 | 0 | 0 | 0 | 0 |
| 0 | 0.001021079 | 0.000445444 | 0.00212841 | 0 |
| 0 | 0 | 0 | 0 | 0 |
| 0 | 0 | 0 | 0 | 0 |
| 0 | 0 | 0 | 0 | 0 |
| 0 | 0 | 0 | 0 | 0 |
| 0 | 0 | 0 | 0 | 0 |
| 0 | 0 | 0 | 0 | 0 |
| 0 | 0 | 0 | 0 | 0 |
| 0.014996224 | 0.10118611 | 0.05642711 | 0.304156246 | 0.237385454 |
| 0 | 0 | 0 | 0 | 0 |
| 0 | 0 | 0 | 0 | 0 |
| 0 | 0 | 0 | 0 | 0 |
| 0 | 0 | 0 | 0 | 0 |
| 0 | 0 | 0 | 0 | 0 |
| 0.054542849 | 0.012971662 | 0.00363784 | 0.326455252 | 0.008640481 |
| 0 | 0 | 0 | 0 | 0 |
| 0 | 0 | 0 | 0 | 0 |
| 0 | 0.041238031 | 0 | 0 | 0 |
| 0 | 7.738191698 | 0 | 0.030693816 | 0.023446751 |
| 0 | 0 | 0 | 0 | 0 |
| 0 | 0 | 0 | 0 | 0 |
| 0.00415008 | 0 | 0.00165425 | 0 | 0 |
| 0.00681173 | 0.0184261 | 0.011922047 | 0.000924768 | 0.013139455 |
| 0 | 0 | 0 | 0 | 0 |
| 0 | 0 | 0 | 0 | 0 |
| 0 | 0 | 0 | 0 | 0 |
| 0 | 0 | 0 | 0 | 0 |
| 0 | 0.002382081 | 0 | 0.001182233 | 0 |
| 0 | 0 | 0 | 0 | 0 |
| 0 | 0.000827062 | 0 | 0 | 0 |
| 0 | 0 | 0 | 0 | 0 |
| 0 | 0 | 0 | 0 | 0 |
| 0 | 0 | 0 | 0 | 0 |
| 0 | 0 | 0 | 0 | 0 |
| 0 | 0 | 0.001466864 | 0 | 0 |
| 0 | 0.000590154 | 0 | 0.004613101 | 0 |

| | | | | |
|---|---|---|---|---|
| 0 | 0 | 0 | 0 | 0 |
| 0 | 0 | 0 | 0 | 0 |
| 0 | 0 | 0 | 0 | 0 |
| 0 | 0 | 0.000368164 | 0 | 0 |
| 0 | 0 | 0 | 0 | 0 |
| 0 | 0 | 0 | 0 | 0 |
| 0 | 0.002730252 | 0 | 0 | 0 |
| 0 | 0.001484626 | 0 | 0 | 0 |
| 0 | 0 | 0 | 0 | 0 |
| 0 | 0 | 0 | 0 | 0 |
| 0 | 0 | 0 | 0 | 0 |
| 0 | 0 | 0 | 0 | 0 |
| 0 | 0 | 0 | 0 | 0 |
| 0 | 0 | 0 | 0.008901311 | 0 |
| 0 | 0 | 0 | 0.003324951 | 4.405755679 |
| 0 | 0 | 0 | 0 | 0.001770205 |
| 0 | 0 | 0 | 0 | 0 |
| 0 | 0 | 0 | 0 | 0 |
| 0.018546027 | 0 | 0 | 0.016621289 | 0 |
| 0 | 0 | 0.383793399 | 0 | 0 |
| 0 | 0.049799886 | 0.004200186 | 0.005338617 | 0.005446748 |
| 0 | 0 | 0 | 0 | 0 |
| 0 | 0 | 0 | 0 | 0 |
| 0 | 0 | 0 | 0 | 0 |
| 0 | 0.006156142 | 0.004891638 | 0.01257042 | 0 |
| 0 | 0 | 0 | 0.005717887 | 0 |
| 0 | 0 | 0 | 0 | 0 |
| 0 | 0 | 0 | 0 | 0 |
| 0 | 0 | 0 | 0.006935265 | 0.005978644 |
| 0 | 0 | 0 | 0 | 0 |
| 0.007670052 | 3.790678407 | 0.061128929 | 0 | 0.038574392 |
| 0 | 0 | 0 | 0 | 0 |
| 0 | 0 | 0 | 0 | 0 |
| 0 | 0.019160674 | 0.00220452 | 0.002069099 | 0.003817994 |
| 0 | 0 | 0 | 0 | 0 |
| 0 | 0 | 0 | 0 | 0 |
| 0 | 0 | 0 | 0 | 0 |
| 0 | 0 | 0 | 0 | 0 |
| 0 | 0 | 0 | 0 | 0 |
| 0 | 0 | 0 | 0 | 0 |
| 0 | 0 | 0 | 0 | 0 |
| 0.00250777 | 0.020902445 | 0 | 0 | 0 |
| 0 | 0 | 0 | 0 | 0 |
| 0 | 0 | 0 | 0 | 0 |
| 0 | 0 | 0 | 0 | 0 |
| 0 | 0 | 0 | 0 | 0 |
| 0 | 0 | 0 | 0 | 0 |
| 0 | 0 | 0 | 0 | 0 |

| | | | | |
|---|---|---|---|---|
| 0 | 0 | 0 | 0 | 0 |
| 0 | 0 | 0 | 0 | 0 |
| 0.000740922 | 0.020205608 | 0.027261825 | 0.261826418 | 0.191016676 |
| 0 | 0 | 0 | 0 | 0 |
| 0 | 0.002892369 | 0.804746075 | 0 | 0.107937517 |
| 0 | 0 | 0 | 0 | 0 |
| 0 | 0 | 0 | 0 | 0 |
| 0 | 0 | 0 | 0 | 0 |
| 0 | 0 | 0 | 0 | 0 |
| 0 | 0 | 0.000623336 | 0 | 0.000740264 |
| 0 | 0 | 0 | 0 | 0 |
| 0 | 0 | 0 | 0 | 0 |
| 0 | 0 | 0 | 0 | 0 |
| 0.002251844 | 0.005755372 | 0.000451939 | 0.001645291 | 0.017890502 |
| 0.000844819 | 0.002090135 | 0.002486774 | 0.014428452 | 0.007137029 |
| 0.058068304 | 0 | 0 | 0 | 0 |
| 0 | 0 | 0.001275055 | 0.007736436 | 0 |
| 0 | 0 | 0 | 0 | 0 |
| 0 | 0 | 0 | 0 | 0 |
| 0 | 0 | 0 | 0 | 0 |
| 0 | 0 | 0 | 0 | 0 |
| 0 | 0 | 0 | 0.001222164 | 0 |
| 0 | 0 | 0 | 0 | 0 |
| 0 | 0 | 0 | 0.052779063 | 0 |
| 0 | 0 | 0.000820257 | 0 | 0 |
| 0 | 0 | 0 | 0 | 0 |
| 0.986092574 | 0 | 0 | 0 | 0 |
| 0 | 0 | 0 | 0 | 0 |
| 0 | 0 | 0 | 0 | 0 |
| 0 | 0 | 0 | 0 | 0 |
| 0 | 0 | 0 | 0 | 0 |
| 0 | 0 | 0 | 0 | 0 |
| 0 | 0 | 0 | 0 | 0 |
| 0 | 0 | 0 | 0 | 0 |
| 0 | 0 | 0 | 0 | 0 |
| 0 | 0 | 0 | 0.003836012 | 0 |
| 0 | 0 | 0 | 0 | 0 |
| 0.001528443 | 0 | 0.001928168 | 0.050891602 | 0 |
| 0 | 0 | 0 | 0 | 0 |
| 0 | 0 | 0 | 0 | 0 |
| 0 | 0 | 0 | 0 | 0 |
| 0 | 0 | 0 | 0 | 0 |
| 0 | 0 | 0 | 0 | 0 |
| 0 | 0 | 0 | 0 | 0 |
| 0 | 0 | 0 | 0 | 0 |
| 0 | 0 | 0 | 0 | 0 |
| 0 | 0 | 0.001219204 | 0 | 0 |
| 0 | 0 | 0 | 0 | 0 |

| | | | | |
|---|---|---|---|---|
| 0 | 0 | 0 | 0 | 0 |
| 0 | 0 | 0 | 0 | 0 |
| 0 | 0 | 0.000401865 | 0.003840364 | 0 |
| 0 | 0 | 0 | 0 | 0 |
| 0.002380834 | 0 | 0.007277665 | 0.214531614 | 0 |
| 0 | 0 | 0 | 0 | 0 |
| 0 | 0 | 0 | 0 | 0 |
| 0.027211532 | 0.00692861 | 0.002455856 | 0.007737048 | 0.001869574 |
| 0 | 0 | 0 | 0 | 0 |
| 0 | 0 | 0.003894314 | 0.97371615 | 0.118318455 |
| 0 | 0 | 0 | 0 | 0 |
| 0 | 0.008684128 | 0 | 0.003470649 | 0 |
| 0 | 0.003987001 | 0.076794821 | 0.004026346 | 0.006062059 |
| 0 | 0.002632707 | 0 | 0 | 0.002386923 |
| 0 | 0.000534853 | 0 | 0 | 0 |
| 0 | 0.002081791 | 0 | 0.003022107 | 0 |
| 0 | 0 | 0 | 0.00118337 | 0.000980395 |
| 0 | 0 | 0 | 0 | 0 |
| 0 | 1.219678458 | 0 | 0 | 0.014868912 |
| 0 | 0 | 0 | 0 | 0 |
| 0 | 0 | 0 | 0 | 0 |
| 0 | 0 | 0 | 0 | 0 |
| 0 | 0 | 0 | 0 | 0 |
| 0.000512361 | 0 | 0 | 0 | 0 |
| 0 | 0 | 0 | 0 | 0 |
| 0.04991217 | 0.003432403 | 0 | 0.053209674 | 0 |
| 3.356821183 | 0 | 0 | 0 | 0.110852057 |
| 0 | 0 | 0 | 0 | 0.00126495 |
| 0 | 0.031628574 | 0 | 0 | 0.030523256 |
| 0 | 0 | 0 | 0 | 0 |
| 0 | 0 | 0.000708869 | 0 | 0 |
| 0 | 0 | 0 | 0 | 0 |
| 0 | 0 | 0 | 0 | 0 |
| 0 | 0.542899247 | 0 | 0 | 0.009234167 |
| 0 | 0 | 0 | 0 | 0 |
| 0 | 0 | 0 | 0 | 0 |
| 0 | 0.095214591 | 0 | 0.002249169 | 0.028755422 |
| 0 | 0 | 0 | 0 | 0 |
| 0 | 0 | 0 | 0 | 0 |
| 0 | 0.561250349 | 0 | 0 | 0 |
| 0 | 0 | 0 | 0 | 0 |
| 0 | 0.725123281 | 0 | 0 | 0 |
| 0 | 0 | 0 | 0 | 0 |
| 0 | 0.001626627 | 0.104100174 | 0 | 0 |
| 0 | 0 | 0 | 0 | 0 |
| 0 | 0 | 0 | 0 | 0 |
| 0.001562907 | 0.008034757 | 0.000821519 | 0.001121533 | 0 |
| 0 | 0 | 0 | 0 | 0 |

| | | | | |
|---|---|---|---|---|
| 0 | 0 | 0 | 0 | 0 |
| 0 | 0.012907892 | 0 | 0 | 0 |
| 0 | 0 | 0 | 0 | 0 |
| 0 | 0.004864128 | 0 | 0.00482816 | 0 |
| 0 | 0.01762024 | 0 | 0 | 0 |
| 0 | 0 | 0.084633564 | 0 | 0.003329824 |
| 0 | 0 | 0.005611937 | 0 | 0 |
| 0 | 0.001278895 | 0 | 0 | 0 |
| 0 | 0 | 0 | 0 | 0 |
| 0 | 0 | 0 | 0 | 0 |
| 0.005099028 | 0 | 0.00645399 | 0.112288384 | 0.016636461 |
| 0 | 0 | 0 | 0 | 0 |
| 0 | 0.071656063 | 0 | 0 | 0 |
| 0 | 0 | 0.004254004 | 0.084596517 | 0.009261976 |
| 0 | 0.729690285 | 0.000717381 | 0 | 0.038053787 |
| 0 | 0 | 0 | 0 | 0 |
| 0 | 0 | 0 | 0 | 0 |
| 0 | 0.042176136 | 0 | 0 | 0 |
| 0 | 0.065040506 | 0 | 0 | 0 |
| 0 | 0 | 0.00340113 | 0 | 0.001009782 |
| 0 | 0.00443615 | 0 | 0.000834318 | 0.000907218 |
| 0.449226122 | 0 | 0 | 0 | 0 |
| 0 | 0 | 0 | 0 | 0 |
| 0 | 0 | 0 | 0 | 0 |
| 0 | 0 | 0 | 0 | 0 |
| 0 | 0 | 0 | 0 | 0 |
| 0 | 2.106383777 | 0 | 0 | 0.171330011 |
| 0 | 0 | 0 | 0 | 0 |
| 0 | 0 | 0 | 0 | 0 |
| 0.1881093 | 0.351300615 | 0.473480656 | 0 | 1.41356173 |
| 0.000653632 | 0.000882068 | 0.001346802 | 0 | 0.006473924 |
| 0 | 0 | 0 | 0 | 0 |
| 0 | 0 | 0 | 0 | 0 |
| 0 | 0 | 0 | 0 | 0 |
| 0 | 0 | 0 | 0 | 0.002027734 |
| 0 | 0 | 0 | 0 | 0 |
| 0.146596251 | 0 | 0 | 0 | 0 |
| 0 | 0 | 0 | 0 | 0 |
| 0 | 0 | 0 | 0 | 0 |
| 0 | 3.040222543 | 0 | 0 | 0.206233872 |
| 0 | 0.060392202 | 0.152013969 | 0 | 0.056530795 |
| 0.420581167 | 1.884849418 | 4.316373389 | 0 | 3.913719821 |
| 0 | 0 | 0 | 0 | 0 |
| 0 | 0 | 0 | 0 | 0 |
| 0 | 0 | 0 | 0 | 0 |
| 0 | 0 | 0 | 0 | 0 |
| 0 | 0.000747917 | 0 | 0.001169258 | 0.003148288 |
| 0 | 0 | 0 | 0 | 0 |

| | | | | |
|---|---|---|---|---|
| 0.00882125 | 0.004152617 | 0 | 0.035654528 | 0 |
| 0 | 0.035083843 | 0 | 0 | 0 |
| 0.012019629 | 0.078586934 | 0.002674105 | 0.33417694 | 0.241029324 |
| 0 | 0 | 0 | 0 | 0 |
| 0 | 0 | 0 | 0 | 0 |
| 0 | 0 | 0 | 0 | 0.032590499 |
| 0 | 0 | 0 | 0 | 0 |
| 0 | 0 | 0 | 0 | 0 |
| 0 | 0 | 0.001642466 | 0 | 0 |
| 0 | 0 | 0 | 0 | 0.034595838 |
| 0 | 0 | 0 | 0 | 0 |
| 0 | 0.021398469 | 0 | 0.004865945 | 0 |
| 0 | 0 | 0 | 0 | 0 |
| 0 | 0 | 0.036328565 | 0.07141013 | 0.17003188 |
| 0 | 0 | 0 | 0 | 0 |
| 0.026976554 | 0 | 0.075031103 | 0 | 0.005281279 |
| 0.128605148 | 0.006327383 | 0.000354897 | 2.755369056 | 0.007024497 |
| 0 | 0.015814335 | 0 | 0.014323874 | 0 |
| 0 | 0.022781615 | 0.000366582 | 0 | 0 |
| 0 | 0.069435793 | 0.027924726 | 0 | 0 |
| 0 | 0.111868087 | 0.104512317 | 0 | 0 |
| 0.001305284 | 0 | 0 | 0.00337199 | 0 |
| 0 | 0 | 0 | 0 | 0 |
| 0 | 0 | 1.158308162 | 0 | 0 |
| 0 | 0 | 0 | 0 | 0.005470893 |
| 0 | 0 | 0.015272124 | 0 | 0.014188755 |
| 0 | 0.00780316 | 0.011413791 | 0 | 0.007134124 |
| 0 | 0 | 0.009789251 | 0 | 0.002633622 |
| 0 | 0 | 0 | 0 | 0 |
| 0 | 0 | 0 | 0 | 0 |
| 0 | 0.039794589 | 0.003883227 | 0 | 0 |
| 0.001619725 | 0.002862357 | 0.010727447 | 0 | 0.016851514 |
| 0.003745337 | 0.109014024 | 0.009403346 | 0 | 0.061933389 |
| 0 | 0 | 0.002555776 | 0.002854744 | 0.018395147 |
| 0.001577463 | 0.020527397 | 0.016915069 | 0 | 0 |
| 0 | 0 | 0 | 0 | 0 |
| 0 | 0 | 0.00161231 | 0.001400711 | 0 |
| 0 | 0.102650567 | 0.00263643 | 0 | 0.001134414 |
| 0.335804231 | 0 | 0 | 0 | 0 |
| 0.000636778 | 0.006731373 | 0 | 0 | 0 |
| 0 | 0 | 0 | 0 | 0.000863966 |
| 0 | 0 | 0 | 0 | 0 |
| 0 | 0 | 0.001371401 | 0 | 0 |
| 0 | 0 | 0.007337427 | 0 | 0.0053133 |
| 0 | 0 | 0 | 0 | 0 |
| 0 | 0.011457082 | 0 | 0.187132221 | 0.010634793 |
| 0 | 0 | 0 | 0 | 0 |
| 0 | 0 | 0 | 0 | 0 |

| | | | | |
|---|---|---|---|---|
| 0 | 0.025452694 | 0 | 0 | 0.022538257 |
| 0.269773004 | 0.017663323 | 0.615405882 | 0.847825042 | 0.099548425 |
| 0 | 0.00058811 | 0 | 0 | 0 |
| 0 | 0 | 0 | 0 | 0 |
| 0 | 0.002107526 | 0 | 0 | 0 |
| 0 | 0 | 0 | 0 | 0 |
| 0 | 7.068311401 | 0 | 0 | 0.02345283 |
| 0 | 0 | 0.000479895 | 0 | 0 |
| 0 | 0 | 0 | 0 | 0 |
| 0 | 0 | 0.000908027 | 0 | 0.002246581 |
| 0.001734995 | 0 | 0.013464063 | 0 | 0.000689211 |
| 0.006108404 | 0.19287016 | 0.020055103 | 0.016049824 | 0.27991961 |
| 0 | 0 | 0 | 0 | 0 |
| 0 | 0 | 0 | 0 | 0 |
| 0.001085831 | 0.167290233 | 0 | 0.013999376 | 0.007117056 |
| 0 | 0 | 0 | 0 | 0 |
| 0 | 0.002043114 | 0.035733209 | 0.019247624 | 0.007457607 |
| 0 | 1.215451291 | 0.011517772 | 1.013228404 | 0.191121755 |
| 0 | 0 | 0 | 0 | 0.007394611 |
| 0 | 0.141535905 | 0.003483388 | 0 | 0.00993647 |
| 0.30266664 | 0.008236322 | 0.033841794 | 0.020191383 | 0.001240431 |
| 0 | 0 | 0 | 0 | 0 |
| 0 | 0 | 0 | 0 | 0 |
| 0 | 0 | 0 | 0 | 0 |
| 0 | 0 | 0 | 0 | 0 |
| 0 | 0 | 0 | 0 | 0 |
| 0 | 0.144065166 | 0 | 0 | 0.064891487 |
| 0.009594579 | 0.196659507 | 0.058083001 | 0 | 0.314305453 |
| 0.003882734 | 0 | 0 | 0 | 0 |
| 0 | 0 | 0 | 0 | 0 |
| 0.010294992 | 0 | 0 | 0.002732028 | 0 |
| 0 | 0 | 0 | 0 | 0 |
| 0 | 0 | 0 | 0 | 0 |
| 0 | 0 | 0 | 0 | 0 |
| 0.399277574 | 0 | 0 | 0 | 0 |
| 0.162382534 | 0.152979825 | 0.301219015 | 0 | 0.024098099 |
| 0 | 0.000797872 | 0 | 0 | 0 |
| 0 | 0 | 0 | 0 | 0 |
| 0 | 0.004269385 | 0.016433917 | 0.000673065 | 0.005204442 |
| 0 | 0 | 0 | 0 | 0 |
| 0 | 0 | 0 | 0 | 0 |
| 0 | 0 | 0 | 0 | 0 |
| 0 | 0.018915758 | 0.001996442 | 0.012946276 | 0.00138305 |
| 0 | 0 | 0.001252893 | 0.002236732 | 0.008774887 |
| 0 | 0.277468545 | 0 | 0 | 0.008520819 |
| 0.290385434 | 0 | 0.00119492 | 0 | 0 |
| 0 | 0 | 0 | 0 | 0 |
| 0 | 0.002618746 | 0 | 0.002977474 | 0 |

| | | | | |
|---|---|---|---|---|
| 0 | 0.010442423 | 0 | 0 | 0 |
| 0 | 0 | 0 | 0 | 0 |
| 0 | 0 | 0 | 0 | 0 |
| 0.067934552 | 0 | 0.000581306 | 0 | 0 |
| 0 | 0 | 0 | 0 | 0 |
| 0 | 0 | 0 | 0 | 0 |
| 0 | 0 | 0 | 0 | 0 |
| 0 | 0 | 0.016225263 | 0 | 0 |
| 0.408098061 | 0 | 0 | 0 | 0.108346636 |
| 0.049844242 | 0 | 0.698683492 | 0 | 0 |
| 0 | 0 | 0 | 0 | 0 |
| 0 | 0 | 0.004455629 | 0 | 0 |
| 0 | 0 | 0 | 0 | 0 |
| 0 | 0 | 0 | 0 | 0 |
| 0.034100418 | 0.114342901 | 0.026289091 | 0 | 0.002183252 |
| 0 | 0 | 0 | 0 | 0 |
| 0.393322903 | 0 | 0 | 0 | 0 |
| 0 | 0 | 0 | 0 | 0 |
| 0 | 0 | 0.059980908 | 0 | 0 |
| 0 | 0 | 0 | 0 | 0 |
| 0 | 0 | 0 | 0 | 0 |
| 0 | 0 | 0 | 0 | 0 |
| 0 | 0 | 0 | 0 | 0 |
| 0.017446444 | 1.179623803 | 0.134761567 | 0 | 0 |
| 0 | 0.00143437 | 0 | 0 | 0 |
| 0 | 0 | 0 | 0 | 0 |
| 0.047046327 | 0 | 0 | 0 | 0 |
| 1.293003343 | 0 | 0 | 0 | 0 |
| 0 | 0 | 0 | 0 | 0 |
| 0 | 0 | 0 | 0 | 0 |
| 0.029284109 | 0 | 0.025664912 | 0 | 0 |
| 0 | 0 | 0 | 0 | 0 |
| 0 | 0.05025348 | 0 | 0 | 0.000489387 |
| 0 | 0 | 0 | 0 | 0 |
| 0 | 0 | 0 | 0 | 0 |
| 0.015721286 | 0 | 0 | 0 | 0 |
| 0.95381049 | 0 | 0.093918559 | 0 | 0 |
| 0 | 0 | 0 | 0 | 0 |
| 0 | 0 | 0 | 0 | 0.002754191 |
| 0 | 0 | 0 | 0 | 0 |
| 0 | 0.053725962 | 0.070634114 | 0 | 0.004441661 |
| 0 | 0 | 0.338753406 | 0 | 0.001102187 |
| 0.165488897 | 0.066801679 | 0.047558883 | 0 | 21.52868827 |
| 0 | 0.04779813 | 0 | 0 | 0 |
| 0 | 0 | 0 | 0 | 0 |
| 0 | 0 | 0 | 0 | 0 |
| 0 | 0 | 0 | 0 | 0 |
| 0 | 0.007459639 | 0.034169655 | 0.006137923 | 0 |

| | | | | |
|---|---|---|---|---|
| 0 | 0 | 0 | 0 | 0 |
| 0.081105169 | 0 | 0 | 0 | 0 |
| 0 | 0.169363569 | 0.011205819 | 0.216359125 | 0.014624013 |
| 0 | 0 | 0 | 0 | 0 |
| 0.001533434 | 0 | 0 | 0 | 0 |
| 0.012786633 | 0 | 0.002619152 | 0 | 0.001920039 |
| 0 | 0 | 0 | 0 | 0 |
| 0 | 0 | 0.000294918 | 0 | 0 |
| 0 | 0 | 0 | 0 | 0 |
| 0 | 0 | 0.054196331 | 0 | 0 |
| 0.00706466 | 0.161481202 | 0.036512555 | 0.475854702 | 0.002153728 |
| 0.317613606 | 9.309225561 | 1.417996009 | 0 | 4.835176773 |
| 0 | 0 | 0 | 0 | 0 |
| 0.182277999 | 0 | 0.000521932 | 0.162280538 | 0.003357454 |
| 0 | 1.125095161 | 0.089252563 | 1.67834532 | 0.30732014 |
| 0 | 0 | 0 | 0 | 0 |
| 0 | 0 | 0 | 0 | 0 |
| 0 | 0 | 0.01027708 | 0 | 0 |
| 0.0072035 | 0 | 0 | 0 | 0 |
| 0 | 0.022829178 | 0.004382041 | 0 | 0 |
| 0 | 0.075260507 | 0.785845934 | 0.001521748 | 0.284209481 |
| 0.072983363 | 0.357997714 | 0 | 22.46754529 | 0.564277498 |
| 0 | 0.599398884 | 0.051684756 | 0.022997246 | 3.092111811 |
| 0 | 0 | 0 | 0 | 0 |
| 0 | 0 | 0 | 0 | 0 |
| 6.696246218 | 0.00575252 | 0.033392878 | 0 | 0 |
| 0 | 0.144098202 | 0 | 0.004632935 | 0 |
| 0 | 0 | 0 | 0 | 0 |
| 0 | 0 | 0.002003458 | 0 | 0 |
| 0 | 0 | 0 | 0 | 0 |
| 0 | 0 | 0 | 0 | 0 |
| 0.116221882 | 0 | 0.178227913 | 0 | 0 |
| 0 | 0.110816607 | 0.00049668 | 1.401559855 | 0 |
| 0.003474177 | 0 | 0.00132565 | 0 | 0 |
| 0.001670504 | 0.155047494 | 0.001557121 | 0 | 0.002757597 |
| 0.434399345 | 0.004818031 | 0.006418168 | 0 | 0.004457377 |
| 0.002364911 | 0.00565147 | 0.015880351 | 0 | 0 |
| 0 | 0 | 0 | 0 | 0 |
| 1.245859328 | 0 | 0.017919916 | 0 | 0 |
| 0 | 0 | 0 | 0 | 0 |
| 0.020210135 | 0 | 0 | 0 | 0 |
| 0 | 0 | 0 | 0 | 0 |
| 0 | 0 | 0 | 0 | 0 |
| 0 | 0 | 0 | 0 | 0 |
| 0.005932732 | 0.026883397 | 0.013867944 | 0.657025673 | 0.001016626 |
| 0.171552929 | 0 | 0 | 0 | 0 |
| 0 | 0 | 0 | 0 | 0 |
| 0 | 0.063673916 | 0.007434902 | 0 | 0.178124334 |

| | | | | |
|---|---|---|---|---|
| 0 | 0 | 0 | 0 | 0 |
| 0 | 0 | 0 | 0 | 0 |
| 0 | 0 | 0 | 0 | 0 |
| 0 | 0 | 0.002736017 | 0 | 0.004332333 |
| 0 | 0 | 0 | 0 | 0 |
| 0.282003334 | 0 | 0 | 0 | 0 |
| 0.452799384 | 0 | 0 | 0 | 0 |
| 0.001915877 | 0 | 0.013091679 | 0 | 0 |
| 0 | 0 | 0 | 0 | 0 |
| 0 | 0 | 0 | 0 | 0 |
| 0 | 0.004842942 | 0.001096991 | 0.482395605 | 0.001206268 |
| 0 | 0 | 0 | 0 | 0 |
| 0.072367518 | 0.024137517 | 0 | 0 | 0.000633709 |
| 0.056677269 | 0 | 0.396175853 | 0 | 0 |
| 0 | 0 | 0 | 0 | 0 |
| 0 | 0 | 0 | 0 | 0 |
| 0.061717048 | 0 | 0 | 0 | 0 |
| 0.011444548 | 0 | 0.002435254 | 0 | 0 |
| 0 | 0 | 0.117022343 | 0 | 0 |
| 0 | 0 | 0 | 0 | 0 |
| 0 | 0 | 0 | 0 | 0 |
| 0 | 0 | 0 | 0 | 0 |
| 0.019394685 | 0 | 0.063055669 | 0 | 0 |
| 0 | 0 | 0 | 0 | 0 |
| 0 | 0 | 0 | 0 | 0 |
| 0 | 0 | 0 | 0 | 0 |
| 0 | 0 | 0 | 0 | 0 |
| 0 | 0 | 0 | 0 | 0 |
| 0 | 0 | 0 | 0 | 0 |
| 0.049136733 | 0 | 0 | 0 | 0.016740134 |
| 0 | 0 | 0.001920006 | 0 | 0 |
| 0 | 0 | 0 | 0 | 0 |
| 0 | 0 | 0 | 0 | 0 |
| 0 | 0 | 0 | 0 | 0 |
| 0 | 0 | 0 | 0 | 0 |
| 0 | 0 | 0 | 0 | 0 |
| 0.046377604 | 0 | 0.104481207 | 0 | 0 |
| 0.000981482 | 0.0312203 | 0.000619082 | 0 | 0.003471832 |
| 0 | 0 | 0 | 0 | 0 |
| 0 | 0 | 0.003969081 | 0 | 0.00120862 |
| 0 | 0 | 0.021394705 | 0 | 0 |
| 0 | 0 | 0 | 0 | 0 |
| 0 | 0 | 0 | 0 | 0 |
| 0 | 0 | 0.003406809 | 0 | 0 |
| 0.037095672 | 0 | 0 | 0 | 0 |
| 0 | 0 | 0 | 0 | 0 |
| 0 | 0 | 0.012352854 | 0 | 0 |
| 0 | 0 | 0 | 0 | 0 |

| | | | | |
|---|---|---|---|---|
| 0 | 0 | 0 | 0 | 0 |
| 0 | 0 | 0 | 0 | 0 |
| 0 | 0 | 0 | 0 | 0 |
| 0.08814473 | 0 | 0.019603117 | 0 | 0 |
| 0 | 0 | 0 | 0 | 0 |
| 0 | 0 | 0 | 0 | 0.128041587 |
| 0 | 0 | 0 | 0 | 0 |
| 0 | 0 | 0 | 0 | 0 |
| 0.001160252 | 0.312931687 | 0.003842176 | 0.673537485 | 0.031546023 |
| 0 | 0 | 0 | 0 | 0 |
| 0 | 0 | 0 | 0 | 0 |
| 0.002829383 | 0 | 0 | 0 | 0 |
| 0 | 0 | 0 | 0 | 0 |
| 0 | 0 | 0 | 0 | 0 |
| 0.400079069 | 0 | 0.062686948 | 0 | 0 |
| 0 | 1.159498886 | 0 | 0 | 0 |
| 0 | 0 | 0 | 0 | 0 |
| 0 | 0 | 0 | 0 | 0 |
| 0 | 0 | 0 | 0 | 0 |
| 0 | 0 | 0 | 0 | 0 |
| 0.002957263 | 0 | 0.00119692 | 0 | 0 |
| 0 | 0 | 0 | 0 | 0 |
| 0 | 0 | 0 | 0 | 0 |
| 0 | 0 | 0 | 0 | 0 |
| 10.71458709 | 0 | 0 | 0.116056227 | 0.005630829 |
| 0.976522124 | 0 | 0.023028397 | 0 | 0 |
| 0.138035804 | 0 | 0.002240715 | 0 | 0 |
| 0 | 0 | 0 | 0 | 0 |
| 0.045330046 | 0 | 0 | 0 | 0 |
| 0 | 0 | 0 | 0 | 0 |
| 0 | 0 | 0 | 0 | 0 |
| 0 | 0 | 0.001185475 | 0 | 0 |
| 0 | 0 | 0 | 0 | 0 |
| 0 | 0 | 0.000983097 | 0 | 0 |
| 0.067836734 | 0 | 0 | 2.408401999 | 0.001748808 |
| 0.003394856 | 0.005885721 | 0 | 0.002557936 | 0 |
| 0 | 0.610673273 | 0 | 0 | 0 |
| 0.205724711 | 0 | 1.087591093 | 0 | 0 |
| 0 | 0 | 0 | 0 | 0 |
| 0 | 0 | 0 | 0.001221506 | 0 |
| 0 | 0 | 0 | 0 | 0 |
| 0 | 0 | 0 | 0 | 0 |
| 0.821899517 | 0 | 0.110809096 | 0 | 0 |
| 0 | 0 | 0 | 0 | 0 |
| 0 | 0 | 0 | 0 | 0 |
| 0 | 0.003466151 | 0.000493077 | 0 | 0.001756709 |
| 0 | 0 | 0 | 0 | 0 |
| 0.001280948 | 0 | 0.317197026 | 0.001286881 | 3.133864709 |

| | | | | |
|---|---|---|---|---|
| 0 | 0 | 0 | 0 | 0.00115942 |
| 0 | 0 | 0 | 0 | 0 |
| 0 | 0 | 0 | 0 | 0 |
| 0 | 0 | 0 | 0 | 0 |
| 0 | 0.002317224 | 0 | 0.009394979 | 0 |
| 0 | 0 | 0 | 0 | 0 |
| 0 | 0 | 0 | 0 | 0 |
| 0 | 0 | 0.985725261 | 0 | 0 |
| 0 | 0 | 0 | 0 | 0 |
| 0 | 0 | 0 | 0 | 0 |
| 0.001334107 | 0 | 0 | 0 | 0 |
| 0.704384759 | 0 | 0.009469531 | 0 | 0 |
| 0.009979792 | 0 | 0.376946057 | 0 | 0.003354478 |
| 0 | 0 | 0 | 0 | 0 |
| 0 | 0.210678953 | 0.005935368 | 0.207361358 | 0.320931661 |
| 0 | 0 | 0 | 0 | 0 |
| 0 | 0 | 0 | 0 | 0 |
| 0 | 0 | 0 | 0 | 0.002139123 |
| 0.004820076 | 0.00379437 | 0.001064113 | 0 | 0 |
| 0 | 0 | 0 | 0 | 0 |
| 0 | 0 | 0 | 0 | 0 |
| 0 | 0 | 0 | 0 | 0 |
| 0.629290316 | 0 | 0.017589986 | 0 | 0.05506841 |
| 0 | 0.000924862 | 0 | 0 | 0 |
| 0 | 0 | 0 | 0 | 0 |
| 0 | 0 | 0 | 0 | 0 |
| 0 | 0 | 0 | 0 | 0 |
| 0 | 0 | 0 | 0 | 0 |
| 0 | 0 | 0 | 0 | 0 |
| 0 | 0 | 0 | 0 | 0 |
| 0 | 0 | 0 | 0 | 0 |
| 0.013796365 | 0.002942504 | 0.092423644 | 0.051617547 | 0.018709235 |
| 0.054533124 | 0 | 5.505046302 | 0 | 0 |
| 0.001069514 | 0.028865935 | 0.019588656 | 0 | 0.002423247 |
| 0.001231485 | 0.00276979 | 0 | 0.002319729 | 0 |
| 0.052983923 | 0.00200846 | 0.002190466 | 0 | 0 |
| 0 | 0 | 0 | 0 | 0 |
| 0 | 0 | 0 | 0 | 0 |
| 0 | 0 | 0.001041132 | 0 | 0 |
| 0 | 0 | 0 | 0 | 0 |
| 0.011934987 | 0 | 0 | 0 | 0 |
| 0 | 0 | 0 | 0 | 0 |
| 0 | 0 | 0 | 0 | 0 |
| 0 | 0 | 0 | 0 | 0 |
| 0.001819704 | 0.070429937 | 0.011047581 | 0.034429829 | 0 |
| 0 | 0 | 0 | 0 | 0 |
| 0 | 0 | 0.148347396 | 0 | 0 |
| 0 | 0.005161384 | 0 | 0 | 0 |

| | | | | |
|---|---|---|---|---|
| 0.017003712 | 0 | 0 | 0 | 0 |
| 0 | 0 | 0 | 0 | 0 |
| 0 | 0 | 0 | 0 | 0.001827514 |
| 0 | 0 | 0 | 0 | 0 |
| 0 | 0.012520763 | 0 | 0 | 0 |
| 0 | 0 | 0 | 0 | 0 |
| 0.776887035 | 0 | 0 | 0 | 0 |
| 0.000740584 | 0.346461669 | 0.004577896 | 0 | 0.031497969 |
| 0 | 0 | 0 | 0 | 0 |
| 0 | 0 | 0 | 0 | 0.001273776 |
| 0 | 0 | 0 | 0 | 0 |
| 0 | 0 | 0 | 0.001282526 | 0 |
| 0 | 0 | 0 | 0 | 0 |
| 0.104654893 | 0 | 0 | 0 | 0 |
| 0 | 0 | 0 | 0 | 0 |
| 0 | 0 | 0 | 0 | 0 |
| 0 | 0 | 32.72347193 | 0 | 0 |
| 0 | 0 | 0 | 0 | 0 |
| 0 | 3.173677706 | 0 | 0 | 0 |
| 0 | 0 | 0 | 0 | 0 |
| 0 | 0 | 0.003041049 | 0 | 0 |
| 0 | 0 | 0 | 0 | 0 |
| 0 | 0 | 0 | 0 | 0 |
| 0 | 0 | 0 | 0 | 0 |
| 0.00082841 | 0 | 0 | 0.003661888 | 0 |
| 0.001129384 | 0.008096727 | 0 | 0 | 0.006374393 |
| 0 | 0 | 0 | 0 | 0 |
| 0.001796343 | 0 | 1.550803605 | 0 | 0.006843062 |
| 0 | 0 | 0 | 0 | 0 |
| 0.149052082 | 0 | 0.384894112 | 0 | 0.027161153 |
| 0 | 0 | 0 | 0 | 0 |
| 0 | 0 | 0 | 0 | 0 |
| 0 | 0 | 0.009556016 | 0 | 0 |
| 0 | 0 | 0 | 0 | 0 |
| 0.001331465 | 0.109305072 | 0 | 0.333516234 | 0 |
| 0 | 0 | 0 | 0 | 0 |
| 0 | 0 | 0 | 0 | 0 |
| 0 | 0 | 0.001482398 | 0 | 0 |
| 0 | 0 | 0 | 0 | 0 |
| 0 | 0 | 0 | 0 | 0 |
| 1.768819109 | 3.880272143 | 9.356592138 | 0 | 19.41795519 |
| 0.006881336 | 0.395496018 | 0.570412334 | 0 | 1.275249596 |
| 0.174659988 | 0 | 0 | 0 | 0 |
| 0 | 0 | 0 | 0 | 0 |
| 0 | 0 | 0 | 0 | 0 |
| 0 | 0 | 0 | 0 | 0 |
| 0 | 0 | 0 | 0 | 0 |
| 0.000666151 | 0.002097581 | 0 | 0.0381465 | 0 |

| | | | | |
|---|---|---|---|---|
| 0 | 0 | 0 | 0 | 0 |
| 0 | 0 | 0.000705915 | 0 | 0 |
| 0 | 0 | 0.000706688 | 0 | 0 |
| 0 | 0 | 0 | 0 | 0 |
| 2.806781622 | 0 | 1.359672975 | 0 | 0.542928342 |
| 0 | 0 | 0 | 0 | 0 |
| 0 | 0 | 0 | 0 | 0 |
| 0 | 0 | 0 | 0 | 0 |
| 0 | 2.204827473 | 0.018930916 | 1.077765513 | 0.780294679 |
| 0 | 0 | 0 | 0 | 0 |
| 0 | 0 | 0 | 0.015709732 | 0 |
| 0 | 0 | 0 | 0 | 0 |
| 0.030492746 | 0 | 0.039489222 | 0 | 0 |
| 0.008348796 | 0.009941116 | 0.029032416 | 0 | 0.013829524 |
| 0.598239285 | 0 | 0 | 0 | 0 |
| 0.09860011 | 0.367394505 | 0.997238178 | 0 | 0 |
| 0 | 0 | 0.001980656 | 0 | 0 |
| 0 | 0 | 0 | 0 | 0 |
| 0 | 0 | 0 | 0 | 0 |
| 0 | 0 | 0 | 0 | 0 |
| 0 | 0 | 0 | 0 | 0 |
| 0 | 0 | 0 | 0 | 0 |
| 0 | 0 | 0 | 0 | 0 |
| 0 | 0 | 0 | 0 | 0 |
| 0 | 0 | 0 | 0 | 0 |
| 0 | 0 | 0 | 0 | 0 |
| 0.157175077 | 0 | 0.027465539 | 0 | 0.003408884 |
| 0 | 0.001406766 | 0 | 0.011170907 | 0.000607349 |
| 0 | 0 | 0 | 0 | 0 |
| 0 | 0 | 0 | 0 | 0 |
| 0 | 0 | 0 | 0 | 0 |
| 0 | 0 | 0 | 0 | 0 |
| 0 | 0 | 0 | 0 | 0 |
| 0.008887779 | 0 | 0 | 0 | 0 |
| 0.04514936 | 0 | 0 | 0 | 0 |
| 0.000482357 | 0 | 0 | 0 | 0 |
| 0 | 0 | 0 | 0.007264549 | 0.005984317 |
| 0 | 0 | 0 | 0 | 0 |
| 0 | 0 | 0 | 0 | 0 |
| 0 | 0 | 0 | 0 | 0 |
| 0.154976753 | 2.664731695 | 3.414416214 | 0 | 0 |
| 0 | 0 | 0 | 0 | 0 |
| 0.039137236 | 0 | 0 | 0 | 0 |
| 0 | 0 | 0 | 0 | 0 |
| 0 | 0 | 0 | 0 | 0 |
| 0 | 0 | 0 | 0 | 0 |
| 0 | 0 | 0 | 0 | 0 |
| 0 | 0 | 0 | 0 | 0 |

| | | | | |
|---|---|---|---|---|
| 0.313346496 | 0 | 0.010191131 | 0 | 0.00642824 |
| 0 | 0 | 0 | 0 | 0 |
| 0 | 0 | 0 | 0 | 0 |
| 0 | 0 | 0 | 0 | 0 |
| 0.020187112 | 0 | 0.103877496 | 0 | 0 |
| 0.048812419 | 0 | 0.059060357 | 0 | 0 |
| 0 | 0.018593416 | 0.471497705 | 0 | 0.003910792 |
| 0 | 0 | 0 | 0 | 0 |
| 0 | 0 | 0.000440408 | 0 | 0 |
| 0 | 0 | 0 | 0 | 0.049884544 |
| 0 | 0 | 0 | 0 | 0 |
| 0 | 0 | 0.047108256 | 0 | 0 |
| 0 | 0 | 0 | 0 | 0 |
| 0 | 0 | 0 | 0 | 0 |
| 0 | 0 | 0 | 0 | 0 |
| 0 | 0 | 0 | 0 | 0 |
| 0 | 0.014983624 | 0 | 39.28876478 | 0 |
| 0 | 0 | 0 | 0 | 0 |
| 0 | 0 | 0 | 0 | 0 |
| 0 | 0 | 0 | 0 | 0 |
| 0 | 0 | 0.618785969 | 0 | 0.001567702 |
| 0 | 0 | 0 | 0 | 0 |
| 0.002522731 | 0 | 0.001310436 | 0 | 0.002593755 |
| 0.004486478 | 0.005638946 | 0.001165253 | 0 | 0.002498593 |
| 0 | 0 | 0 | 0 | 0 |
| 0.001401098 | 0.001400566 | 0 | 0 | 0 |
| 0 | 0 | 0 | 0 | 0 |
| 0.021037657 | 0 | 0 | 0 | 0 |
| 0 | 0 | 0 | 0 | 0 |
| 0.005507521 | 0.900209238 | 0.005170974 | 0.261960376 | 4.106701932 |
| 0 | 0 | 0.00036555 | 0 | 0 |
| 0 | 0 | 0 | 0 | 0 |
| 0 | 0 | 0 | 0 | 0 |
| 0 | 0 | 0 | 0 | 0.607356799 |
| 0 | 0 | 0 | 0 | 0 |
| 0 | 0 | 0 | 0 | 0 |
| 0 | 0.079284707 | 0.001093525 | 0 | 0 |
| 0 | 0 | 0 | 0 | 0 |
| 0 | 0 | 0 | 0 | 0 |
| 0 | 0 | 0 | 0 | 0 |
| 0 | 0 | 0 | 0 | 0 |
| 0 | 0 | 0 | 0 | 0.04246855 |
| 0 | 0 | 0 | 0 | 0 |
| 0 | 0 | 0 | 0 | 0 |
| 0 | 0 | 0 | 0 | 0 |
| 0 | 0 | 0 | 0 | 0 |
| 0 | 0 | 0 | 0 | 0 |
| 0.060432225 | 0 | 0 | 0 | 0 |

| | | | | |
|---|---|---|---|---|
| 0 | 0.010059618 | 0.002612196 | 0 | 0 |
| 0 | 0 | 0 | 0 | 0 |
| 0 | 0 | 0 | 0 | 0 |
| 0 | 0 | 0 | 0 | 0 |
| 0 | 0 | 0 | 0 | 0 |
| 0 | 0 | 0 | 0 | 0 |
| 0 | 0 | 0 | 0.001425575 | 0 |
| 0.407337558 | 0 | 0.193864678 | 0 | 0 |
| 12.16035781 | 0.001754453 | 0.009348532 | 3.729354873 | 0.12790216 |
| 0 | 0.742086456 | 0.008791735 | 0 | 0.003163916 |
| 0 | 0 | 0 | 0 | 0 |
| 0 | 0 | 0 | 0 | 0 |
| 0 | 0 | 0 | 0 | 0 |
| 0 | 0 | 0 | 0 | 0 |
| 0 | 0 | 0 | 0 | 0 |
| 0 | 0 | 0 | 0 | 0 |
| 0 | 0.019346029 | 0 | 0 | 0.015698383 |
| 0.219027519 | 0 | 0 | 0 | 0 |
| 0 | 0 | 0 | 0 | 0 |
| 0.000900619 | 0 | 0 | 0 | 0 |
| 0 | 0 | 0 | 0 | 0 |
| 0 | 0 | 0 | 0 | 0 |
| 0 | 0 | 0 | 0 | 0 |
| 0 | 0 | 0 | 0 | 0 |
| 0.005591782 | 0 | 0 | 0 | 0 |
| 0 | 0 | 0 | 0 | 0 |
| 0 | 0 | 0 | 0 | 0 |
| 0 | 0 | 0 | 0 | 0 |
| 0 | 0 | 0 | 0 | 0 |
| 0 | 0 | 0 | 0 | 0 |
| 0 | 0 | 0 | 0 | 0 |
| 0 | 0 | 0 | 0 | 0 |
| 0 | 0 | 0 | 0 | 0 |
| 0 | 0 | 0 | 0 | 0 |
| 0.015642634 | 0 | 0 | 0 | 0 |
| 0 | 0 | 0 | 0 | 0 |
| 0 | 0 | 0 | 0 | 0 |
| 0 | 0 | 0 | 0 | 0 |
| 0 | 0 | 0.00298186 | 0 | 0 |
| 0 | 0 | 0 | 0.017658288 | 0 |
| 0 | 0 | 0 | 0 | 0 |
| 0.076629366 | 0 | 0 | 0 | 0.49140223 |
| 0 | 0 | 0 | 0 | 0 |
| 0 | 0 | 0 | 0 | 0 |
| 0 | 0 | 0 | 0 | 0 |
| 0 | 0 | 0 | 0 | 0 |
| 0 | 0 | 0 | 0 | 0 |
| 0 | 0 | 0 | 0 | 0 |

| | | | | |
|---|---|---|---|---|
| 0 | 0 | 0 | 0 | 0 |
| 0.024092358 | 2.409026407 | 0.001980401 | 0.001287444 | 0.381156013 |
| 0 | 0 | 0 | 0 | 0 |
| 0 | 0 | 0 | 0 | 0 |
| 0.103203584 | 0.105582351 | 0.090186365 | 0 | 0.040712004 |
| 0.009333464 | 0 | 0 | 0 | 0 |
| 0 | 0 | 0 | 0 | 0.001810663 |
| 0 | 0.158027392 | 0.010899118 | 0.029033002 | 0.005524715 |
| 0.001262394 | 0 | 0 | 0 | 0 |
| 0.154174131 | 0 | 0 | 0 | 0 |
| 0 | 0 | 0 | 0 | 0 |
| 0 | 0 | 0 | 0 | 0 |
| 0 | 0 | 0 | 0 | 0 |
| 0 | 0 | 0 | 0 | 0 |
| 0 | 0 | 0.056702347 | 0 | 0 |
| 0 | 0 | 0 | 0 | 0.001140844 |
| 0 | 0 | 0 | 0 | 0 |
| 0 | 0 | 0 | 0 | 0 |
| 0 | 0 | 0 | 0 | 0 |
| 0 | 0 | 0 | 0 | 0 |
| 0 | 0 | 0 | 0 | 0 |
| 0 | 0 | 0 | 0 | 0 |
| 0 | 0 | 0 | 0 | 0 |
| 0 | 0 | 0.000345084 | 0.008951021 | 0 |
| 0.038801471 | 0 | 0 | 0 | 0 |
| 0 | 0 | 0 | 0 | 0 |
| 0 | 0 | 0 | 0 | 0 |
| 0 | 0.00084645 | 0 | 0 | 0 |
| 0 | 0.004301476 | 0 | 0 | 0 |
| 0 | 0 | 0 | 0 | 0 |
| 0 | 0 | 0 | 0 | 0 |
| 0 | 0 | 0 | 0 | 0 |
| 0 | 0 | 0 | 0 | 0 |
| 0 | 0 | 0 | 0 | 0 |
| 0 | 0 | 0 | 0 | 0 |
| 0 | 0 | 0 | 0 | 0 |
| 0 | 0 | 0 | 0 | 0 |
| 0 | 0 | 0 | 0 | 0 |
| 0 | 0 | 0 | 0 | 0 |
| 0 | 0 | 0.001758701 | 0 | 0 |
| 0 | 0 | 0 | 0 | 0 |
| 0.044551481 | 0 | 0 | 0 | 0 |
| 0 | 0 | 0 | 0 | 0 |
| 0 | 0 | 0 | 0 | 0 |
| 0 | 0 | 0 | 0 | 0 |
| 0 | 0 | 0 | 0 | 0 |
| 0 | 0 | 0 | 0 | 0 |
| 0 | 0 | 0.004791669 | 0 | 0 |

| | | | | |
|---|---|---|---|---|
| 0.730578719 | 0 | 0.031165236 | 0 | 0.003133235 |
| 0 | 0 | 0 | 0 | 0 |
| 0 | 0 | 0 | 0 | 0 |
| 0 | 0 | 0 | 0 | 0 |
| 0 | 0 | 0 | 0 | 0 |
| 0.003115266 | 0.073745403 | 0 | 0 | 0 |
| 0 | 0 | 0 | 0 | 0 |
| 0 | 0 | 0 | 0 | 0 |
| 0 | 0 | 0 | 0 | 0 |
| 0 | 0 | 0 | 0 | 0 |
| 0 | 0 | 0 | 0 | 0 |
| 0 | 0 | 0 | 0 | 0 |
| 0 | 0 | 0 | 0 | 0 |
| 0 | 0 | 0 | 0 | 0 |
| 0 | 0 | 0 | 0 | 0 |
| 0 | 0 | 0 | 0 | 0 |
| 0 | 0 | 0 | 0 | 0 |
| 0 | 0 | 0.00042054 | 0 | 0 |
| 0 | 0 | 0 | 0 | 0 |
| 0 | 0 | 0 | 0 | 0 |
| 0 | 0.064067876 | 0 | 0 | 0.021085312 |
| 0 | 0 | 0 | 0 | 0 |
| 0 | 0 | 0 | 0.137456717 | 0 |
| 0 | 0 | 0.006223015 | 0 | 0 |
| 0 | 0 | 0 | 0 | 0 |
| 0 | 0 | 0 | 0 | 0 |
| 0 | 0 | 0 | 0 | 0 |
| 0 | 0 | 0 | 0 | 0 |
| 0 | 0 | 0 | 0 | 0.013385293 |
| 0 | 0 | 0 | 0 | 0 |
| 0 | 0 | 0 | 0 | 0 |
| 0.076891993 | 0 | 0.221390331 | 0 | 0 |
| 0.001288706 | 0.009109533 | 0 | 0.014981235 | 0 |
| 0 | 0 | 0.170481494 | 0 | 0 |
| 0.020255307 | 0 | 2.509604709 | 0 | 0.131683584 |
| 0.001772798 | 0 | 0.033701731 | 0.001335757 | 0.040238819 |
| 0 | 0 | 0 | 0 | 0 |
| 0 | 0 | 0 | 0 | 0 |
| 0 | 0.076481057 | 0 | 0 | 0.001742235 |
| 0 | 0 | 0 | 0 | 0 |
| 0 | 0 | 0 | 0 | 0 |
| 0.01230475 | 0 | 0 | 0 | 0 |
| 0.053947664 | 0 | 0 | 0 | 0.018625779 |
| 0 | 0 | 0 | 0 | 0 |
| 0 | 0 | 0 | 0 | 0 |
| 0.010314249 | 0.046931856 | 0.030243809 | 0 | 0.003235772 |
| 0 | 0 | 0 | 0 | 0 |
| 0 | 0 | 0 | 0 | 0 |

| | | | | |
|---|---|---|---|---|
| 0 | 0 | 0 | 0 | 0 |
| 0 | 0 | 0 | 0 | 0 |
| 0 | 0 | 0 | 0 | 0 |
| 0 | 0 | 0 | 0 | 0 |
| 0 | 0 | 0 | 0 | 0 |
| 0 | 0 | 0 | 0 | 0 |
| 0 | 0 | 0.070475957 | 0 | 0 |
| 0 | 0.332631778 | 0 | 0 | 0.195200435 |
| 0 | 0 | 0.034899887 | 0 | 0 |
| 0.007681653 | 0 | 0 | 0 | 0 |
| 0 | 0 | 0 | 0 | 0 |
| 0 | 0 | 0 | 0 | 0 |
| 0 | 0 | 0 | 0 | 0.001845163 |
| 0.003233179 | 0 | 0 | 0 | 0 |
| 0 | 0 | 0.000388789 | 0 | 0 |
| 0 | 0.023509003 | 0 | 2.070439478 | 0 |
| 0 | 0 | 0 | 0 | 0 |
| 0 | 0 | 0 | 0 | 0 |
| 0 | 0 | 0 | 0 | 0 |
| 0.283400648 | 0 | 4.636679473 | 0 | 0 |
| 0 | 0.003966332 | 0.002348271 | 0 | 0 |
| 0 | 0 | 0 | 0 | 0 |
| 0 | 0 | 0 | 0 | 0 |
| 0 | 0 | 0 | 0 | 0 |
| 0 | 0 | 0 | 0 | 0 |
| 0 | 0 | 0 | 0 | 0 |
| 0 | 0 | 0 | 0 | 0 |
| 0 | 0 | 0 | 0 | 0 |
| 0 | 0 | 0 | 0 | 0 |
| 1.253975491 | 0 | 1.585442489 | 0 | 0 |
| 0.055939113 | 0 | 0.623822784 | 0 | 6.077927331 |
| 0 | 0 | 0 | 0 | 0 |
| 0 | 0 | 0 | 0 | 0 |
| 0 | 0 | 0 | 0 | 0 |
| 0 | 0 | 0 | 0 | 0 |
| 0 | 0 | 0 | 0 | 0 |
| 0 | 0 | 0 | 0 | 0 |
| 0 | 0 | 0 | 0 | 0 |
| 0.003387963 | 0 | 0 | 0 | 0 |
| 0 | 0 | 0 | 0 | 0 |
| 0 | 0 | 0 | 0 | 0 |
| 0 | 0 | 0 | 0 | 0 |
| 0 | 0 | 0 | 0 | 0 |
| 0 | 0 | 0 | 0 | 0 |
| 0 | 0 | 0 | 0 | 0 |
| 0 | 0 | 0 | 0 | 0 |
| 0 | 0.186233777 | 0 | 0.089139135 | 0 |
| 0 | 0 | 0 | 0 | 0 |

| | | | | |
|---|---|---|---|---|
| 0 | 0 | 0 | 0 | 0 |
| 0 | 0 | 0 | 0 | 0 |
| 0 | 0 | 0 | 0 | 0 |
| 0 | 0 | 0 | 0 | 0 |
| 0.004058361 | 0.000829804 | 0.019276529 | 0 | 0.021853558 |
| 0 | 0 | 0 | 0 | 0 |
| 0 | 0 | 0 | 0 | 0 |
| 0 | 0 | 0 | 0 | 0 |
| 0 | 0 | 0 | 0 | 0 |
| 0 | 0 | 0 | 0 | 0 |
| 0 | 0 | 0 | 0 | 0 |
| 0 | 0.01728094 | 0.009046531 | 0.013894056 | 0 |
| 0 | 0 | 0 | 0 | 0 |
| 0 | 0 | 0 | 0 | 0 |
| 0.002602327 | 0.006145666 | 0 | 0 | 0.00170569 |
| 0.034388755 | 0 | 3.955504282 | 0 | 0.008509322 |
| 0 | 0 | 0.000676458 | 0 | 0 |
| 0 | 0 | 0 | 0 | 0 |
| 0 | 0.306775875 | 0.01227307 | 0 | 0 |
| 0.000506424 | 0.016705646 | 0.14347866 | 0 | 0.014015006 |
| 0 | 0 | 0 | 0 | 0 |
| 0 | 0 | 0 | 0 | 0 |
| 0 | 0 | 0 | 0 | 0 |
| 0 | 0 | 0 | 0 | 0 |
| 0 | 0 | 0 | 0 | 0 |
| 0 | 0 | 0 | 0 | 0 |
| 0 | 0.003498875 | 0.002090473 | 0 | 0.005319884 |
| 0 | 0 | 0 | 0 | 0 |
| 0 | 0 | 0 | 0 | 0 |
| 0 | 0 | 0 | 0 | 0 |
| 0 | 0.00310395 | 0 | 0 | 0 |
| 0 | 0 | 0 | 0 | 0.001929926 |
| 0 | 0 | 0 | 0 | 0 |
| 0 | 0 | 0 | 0 | 0 |
| 0 | 0 | 0 | 0 | 0 |
| 0 | 0 | 0 | 0 | 0 |
| 0 | 0 | 0 | 0 | 0 |
| 0.000646392 | 0.008238371 | 0.017124224 | 0 | 0.001255337 |
| 0.056755823 | 0 | 0.026222947 | 0 | 0 |
| 0 | 0 | 0 | 0 | 0 |
| 0 | 0.002177087 | 0 | 0.000648296 | 0 |
| 0 | 0 | 0 | 0 | 0 |
| 0 | 0 | 0 | 0 | 0 |
| 0 | 0.007365806 | 0 | 0 | 0 |
| 0 | 0 | 0 | 0 | 0 |
| 0 | 0 | 0 | 0 | 0 |
| 0 | 0 | 0 | 0 | 0 |
| 0 | 0 | 0 | 0 | 0 |

| | | | | |
|---|---|---|---|---|
| 0 | 0 | 0 | 0 | 0 |
| 0.01406381 | 0 | 0.061192124 | 0 | 0 |
| 0 | 0 | 0 | 0 | 0 |
| 0.000669687 | 0.001129668 | 0 | 0 | 0 |
| 0.003260901 | 0 | 0 | 0.019032026 | 0 |
| 0 | 0 | 0 | 0 | 0 |
| 0 | 0 | 0 | 0 | 0 |
| 0 | 0 | 0 | 0 | 0 |
| 0 | 0 | 0.008707698 | 0 | 0 |
| 0 | 0 | 0 | 0 | 0 |
| 0 | 0 | 0 | 0 | 0 |
| 0 | 0 | 0 | 0 | 0 |
| 0.137548317 | 0.002268864 | 0.002008687 | 0 | 0 |
| 0 | 0 | 0 | 0 | 0 |
| 0 | 0 | 0 | 0 | 0 |
| 0 | 0 | 0 | 0 | 0 |
| 0 | 0 | 0 | 0 | 0 |
| 0 | 0 | 0 | 0 | 0 |
| 0 | 0 | 0 | 0 | 0 |
| 0 | 0 | 0 | 0 | 0 |
| 0 | 0 | 0 | 0 | 0 |
| 0 | 0 | 0 | 0 | 0 |
| 0.009688874 | 0.027239621 | 0 | 0 | 0 |
| 0.003688131 | 0 | 0.003121164 | 0 | 0 |
| 0 | 0 | 0 | 0 | 0 |
| 0 | 0 | 0 | 0 | 0 |
| 0 | 0 | 0.08844664 | 0 | 0 |
| 0 | 0 | 0 | 0 | 0 |
| 0 | 0 | 0 | 0 | 0 |
| 0 | 0 | 0 | 0 | 0 |
| 0.004007584 | 0 | 0.010642541 | 0 | 0 |
| 0 | 0 | 0 | 0 | 0 |
| 0 | 0 | 0 | 0 | 0 |
| 0 | 0 | 0 | 0 | 0 |
| 0 | 0 | 0 | 0 | 0 |
| 0 | 0.754219095 | 0 | 0.025618928 | 0 |
| 0 | 0 | 0 | 0.385717609 | 0 |
| 0 | 0.170509102 | 0.020883909 | 0 | 0 |
| 0 | 0.003813083 | 0 | 0.004591233 | 0 |
| 0 | 0 | 0 | 0 | 0 |
| 0 | 0 | 0 | 0 | 0 |
| 0 | 0.004835601 | 0 | 0 | 0 |
| 0 | 0 | 0 | 0 | 0 |
| 0 | 0 | 0 | 0 | 0 |
| 0 | 0 | 0 | 0 | 0 |
| 0.381856044 | 0 | 0.046770329 | 0 | 0 |
| 0 | 0 | 0 | 0 | 0 |
| 0 | 0 | 0 | 0 | 0 |

| | | | | |
|---|---|---|---|---|
| 0 | 0 | 0 | 0 | 0 |
| 0 | 0 | 0 | 0 | 0 |
| 0.012372919 | 0 | 0 | 0 | 0 |
| 0 | 0 | 0 | 0 | 0 |
| 0 | 0 | 0 | 0 | 0 |
| 0 | 0 | 0 | 0 | 0 |
| 0 | 0 | 0 | 0 | 0 |
| 0 | 0 | 0 | 0 | 0 |
| 0 | 0 | 0 | 0 | 0 |
| 0 | 0 | 0 | 0 | 0 |
| 0 | 0 | 0 | 0.007689745 | 0 |
| 0 | 0 | 0.000788405 | 0 | 0 |
| 0 | 0 | 0 | 0 | 0 |
| 0 | 0 | 0 | 0 | 0 |
| 0 | 0 | 0 | 0 | 0 |
| 0 | 0 | 0 | 0 | 0 |
| 0 | 0.002570163 | 0 | 0.001766186 | 0 |
| 0 | 0 | 0 | 0 | 0 |
| 0 | 0 | 0 | 0 | 0 |
| 0 | 0 | 0 | 0 | 0 |
| 0 | 0 | 0 | 0 | 0 |
| 0 | 0 | 0 | 0 | 0 |
| 0 | 0 | 0 | 0 | 0 |
| 0 | 0 | 0 | 0 | 0 |
| 0 | 0 | 0 | 0 | 0 |
| 0 | 0 | 0 | 0 | 0 |
| 0 | 0 | 0 | 0 | 0 |
| 0.035280816 | 0 | 0 | 0 | 0 |
| 0 | 0 | 0 | 0 | 0 |
| 0 | 0 | 0 | 0 | 0 |
| 0.002997733 | 0.024364368 | 0.003409272 | 0 | 0.006152584 |
| 0 | 0 | 0 | 0 | 0 |
| 0.001582124 | 0.1094217 | 0.010245561 | 0 | 0.044245327 |
| 0 | 0 | 0.012730389 | 0.001136974 | 0.004474295 |
| 0 | 0 | 0 | 0 | 0 |
| 0 | 0 | 0 | 0 | 0 |
| 0 | 0 | 0 | 0 | 0 |
| 0 | 0 | 0 | 0 | 0 |
| 0 | 0 | 0 | 0 | 0 |
| 0 | 0 | 0 | 0 | 0 |
| 0 | 0 | 0 | 0 | 0 |
| 0 | 0.09807083 | 0.0005747 | 0 | 0.001516676 |
| 0.122575765 | 0 | 0.001400293 | 0 | 0 |
| 0 | 0 | 0 | 0 | 0 |
| 0 | 0 | 0 | 0 | 0 |
| 0.048808276 | 0 | 0 | 0 | 0.004338932 |
| 0 | 0 | 0 | 0 | 0 |
| 0 | 0 | 0 | 0 | 0 |

| | | | | |
|---|---|---|---|---|
| 0 | 0 | 0 | 0 | 0 |
| 0.011960976 | 0 | 0 | 0 | 0 |
| 0.004075074 | 0.01287526 | 0.017850021 | 0.008967698 | 0 |
| 0 | 0 | 0 | 0 | 0 |
| 0.000619762 | 0.312241574 | 0.001368229 | 0 | 0.017602938 |
| 0 | 1.007290462 | 0 | 0 | 0.075567957 |
| 0 | 0 | 0 | 0 | 0 |
| 0 | 0 | 0 | 0 | 0 |
| 0 | 0 | 0 | 0 | 0 |
| 0 | 0 | 0 | 0 | 0 |
| 0 | 0 | 0 | 0 | 0 |
| 0 | 0 | 0 | 0 | 0 |
| 0 | 0 | 0 | 0 | 0 |
| 0 | 0 | 0 | 0 | 0 |
| 0 | 0 | 0 | 0 | 0 |
| 0 | 0 | 0 | 0 | 0 |
| 0 | 0 | 0 | 0 | 0 |
| 0 | 0 | 0 | 0 | 0 |
| 0 | 0 | 0 | 0 | 0 |
| 0 | 0 | 0 | 0 | 0 |
| 0 | 0 | 0 | 0 | 0 |
| 0 | 0 | 0 | 0 | 0 |
| 0 | 0 | 0 | 0 | 0 |
| 0 | 0 | 0 | 0 | 0 |
| 0.027027399 | 0 | 0 | 0 | 0 |
| 0 | 0.000904613 | 0.000473563 | 0.007273187 | 0.007967275 |
| 0 | 0 | 0 | 0 | 0 |
| 0.004681811 | 0 | 0.034338106 | 0 | 0 |
| 0 | 0 | 0 | 0 | 0 |
| 0 | 0 | 0 | 0 | 0 |
| 0.00168319 | 0 | 0 | 0 | 0 |
| 0 | 0 | 0 | 0 | 0 |
| 0 | 0 | 0 | 0 | 0 |
| 0 | 0 | 0 | 0 | 0 |
| 0 | 0 | 0 | 0 | 0 |
| 0 | 0 | 0 | 0 | 0 |
| 0 | 0 | 0 | 0 | 0 |
| 0 | 0 | 0.038889856 | 0 | 0 |
| 0 | 0 | 0.620859376 | 0 | 0 |
| 0 | 0 | 0 | 0 | 0 |
| 0 | 0 | 0 | 0 | 0 |
| 0.011868129 | 0 | 0 | 0 | 0 |
| 0 | 0.006173573 | 0.000950544 | 0 | 0 |
| 0 | 0 | 0 | 0 | 0 |
| 0 | 0 | 0 | 0 | 0 |
| 0 | 0 | 0 | 0 | 0 |
| 0 | 0.062777206 | 0 | 0 | 0 |
| 0 | 0 | 0 | 0 | 0 |

| | | | | |
|---|---|---|---|---|
| 0 | 0 | 0 | 0 | 0 |
| 0.004363449 | 0.028215352 | 0.007683501 | 0.001565593 | 0.012257198 |
| 0.0859223 | 0 | 0 | 0 | 0 |
| 0 | 0 | 0 | 0 | 0 |
| 0 | 0 | 0 | 0 | 0 |
| 0 | 0 | 0 | 0 | 0 |
| 0 | 0 | 0 | 0 | 0 |
| 0 | 0 | 0 | 0 | 0 |
| 0 | 0 | 0 | 0 | 0 |
| 0 | 0 | 0 | 0 | 0 |
| 0 | 0 | 0 | 0 | 0 |
| 0 | 0 | 0 | 0 | 0 |
| 0.125362575 | 0 | 0.014304667 | 0 | 0 |
| 0 | 0 | 0 | 0 | 0 |
| 0 | 0.021851113 | 0 | 0 | 0.006226358 |
| 0 | 0 | 0 | 0 | 0 |
| 0 | 0 | 0 | 0 | 0 |
| 0 | 0 | 0 | 0 | 0 |
| 0 | 0 | 0 | 0 | 0 |
| 0 | 0 | 0 | 0 | 0 |
| 0 | 0 | 0 | 0 | 0 |
| 0 | 0.008798402 | 0.225663109 | 0 | 0.003335332 |
| 0 | 0.016984579 | 0 | 0 | 0 |
| 0 | 0 | 0 | 0 | 0 |
| 0 | 0 | 0 | 0 | 0 |
| 0 | 0 | 0 | 0 | 0 |
| 0 | 0 | 0 | 0 | 0 |
| 0 | 0 | 0 | 0 | 0 |
| 0 | 0 | 0 | 0 | 0 |
| 0 | 0 | 0 | 0 | 0 |
| 0 | 0 | 0 | 0 | 0 |
| 0 | 0 | 0 | 0 | 0 |
| 0 | 0 | 0 | 0 | 0 |
| 0 | 0 | 0 | 0 | 0 |
| 0 | 0 | 0 | 0 | 0 |
| 0 | 0 | 0 | 0 | 0 |
| 0 | 0 | 0 | 0 | 0 |
| 0 | 0 | 0 | 0 | 0 |
| 0 | 0.001714394 | 0.011919672 | 1.463391428 | 0 |
| 0 | 0 | 0 | 0 | 0 |
| 0.048866732 | 0.072055357 | 0.044007611 | 0 | 0.003484182 |
| 0 | 0 | 0 | 0 | 0 |
| 0 | 0 | 0 | 0 | 0 |
| 5.77627032 | 0 | 0 | 0 | 0 |
| 0 | 0 | 0 | 0 | 0 |
| 0.003912523 | 0.785165534 | 0.002159388 | 0.410752435 | 0.009972879 |
| 0 | 0 | 0 | 0 | 0 |
| 0 | 0 | 0 | 0 | 0 |

| | | | | |
|---|---|---|---|---|
| 0 | 0 | 0 | 0 | 0 |
| 0 | 0 | 0 | 0 | 0 |
| 0 | 0 | 0 | 0 | 0 |
| 0 | 0 | 0 | 0 | 0 |
| 0 | 0 | 0 | 0 | 0 |
| 0 | 0 | 0 | 0 | 0 |
| 0 | 0 | 0 | 0 | 0 |
| 0 | 0 | 0 | 0 | 0 |
| 0 | 0 | 0 | 0 | 0 |
| 0 | 0.043891902 | 0 | 0.003075342 | 0 |
| 0.01251018 | 0 | 0 | 0 | 0.01645832 |
| 0 | 0 | 0 | 0 | 0 |
| 0 | 0.010935457 | 0 | 0 | 0.002996154 |
| 0 | 0 | 0 | 0 | 0 |
| 0 | 0 | 0 | 0 | 0 |
| 0 | 0 | 0 | 0 | 0 |
| 0 | 0 | 0 | 0 | 0 |
| 0 | 0 | 0 | 0 | 0 |
| 0 | 0 | 0 | 0 | 0 |
| 0.11185092 | 0.124661475 | 0.063569695 | 0.386937275 | 0.124126879 |
| 0 | 0 | 0 | 0 | 0 |
| 0 | 0 | 0 | 0 | 0 |
| 0 | 0 | 0 | 0 | 0 |
| 0 | 0 | 0 | 0 | 0 |
| 0 | 0 | 0 | 0 | 0 |
| 0 | 0 | 0 | 0 | 0 |
| 0 | 0 | 0 | 0 | 0 |
| 0 | 0 | 0 | 0 | 0 |
| 0 | 0 | 0 | 0.01442764 | 0 |
| 0 | 0 | 0.002095014 | 0 | 0.021251716 |
| 0 | 0 | 0 | 0 | 0 |
| 0 | 0 | 0 | 0 | 0 |
| 0 | 0 | 0 | 0 | 0 |
| 0 | 0 | 0 | 0 | 0 |
| 0 | 0 | 0 | 0 | 0 |
| 0 | 0 | 0 | 0 | 0 |
| 0 | 0 | 0 | 0 | 0 |
| 0 | 0 | 0 | 0 | 0 |
| 0 | 0 | 0 | 0 | 0 |
| 0 | 0 | 0 | 0 | 0 |
| 0 | 0 | 0 | 0 | 0 |
| 0 | 0 | 0 | 0 | 0 |
| 0 | 0 | 0 | 0 | 0 |
| 0 | 0 | 0 | 0 | 0 |
| 0 | 0.0091419 | 0.001150424 | 0.02450061 | 0 |
| 0 | 0 | 0 | 0 | 0 |
| 0 | 0 | 0 | 0 | 0 |
| 0 | 0 | 0 | 0.018551634 | 0 |

| | | | | |
|---|---|---|---|---|
| 0 | 0 | 0 | 0 | 0 |
| 0 | 0 | 0 | 0 | 0 |
| 0 | 0 | 0 | 0 | 0 |
| 0 | 0 | 0 | 0 | 0 |
| 0 | 0 | 0 | 0 | 0 |
| 0.002567559 | 0.013518321 | 0.000429418 | 0 | 0 |
| 0 | 0 | 0 | 0 | 0 |
| 0 | 0 | 0 | 0 | 0 |
| 0 | 0 | 0 | 0 | 0 |
| 0 | 0 | 0 | 0 | 0 |
| 0 | 0 | 0 | 0 | 0 |
| 0 | 0 | 0 | 0 | 0 |
| 0 | 0 | 0 | 0 | 0 |
| 0 | 0 | 0 | 0 | 0 |
| 0 | 0 | 0 | 0 | 0 |
| 0 | 0 | 0 | 0 | 0 |
| 0 | 0 | 0 | 0 | 0 |
| 0 | 0 | 0 | 0 | 0 |
| 0 | 0 | 0 | 0 | 0 |
| 0 | 0 | 0 | 0 | 0 |
| 0 | 0 | 0 | 0 | 0 |
| 0 | 0 | 0 | 0 | 0 |
| 0 | 0 | 0 | 0 | 0 |
| 0 | 0 | 0 | 0 | 0 |
| 0 | 0 | 0 | 0 | 0 |
| 0 | 0 | 0 | 0 | 0 |
| 0 | 0 | 0 | 0 | 0 |
| 0 | 0 | 0 | 0 | 0 |
| 0 | 0 | 0 | 0 | 0 |
| 0 | 0 | 0 | 0 | 0 |
| 0 | 0 | 0 | 0 | 0 |
| 0 | 0 | 0 | 0 | 0 |
| 0 | 0 | 0 | 0 | 0 |
| 0 | 0 | 0 | 0 | 0 |
| 0 | 0 | 0 | 0 | 0 |
| 0 | 0 | 0 | 0 | 0 |
| 0 | 0 | 0 | 0 | 0 |
| 0 | 0 | 0 | 0 | 0 |
| 0.609712845 | 0 | 0.132628258 | 0 | 0 |
| 0 | 0 | 0 | 0 | 0 |
| 0 | 0 | 0 | 0 | 0 |
| 0 | 0 | 0 | 0 | 0 |
| 0 | 0 | 0 | 0 | 0 |
| 0 | 0 | 0 | 0 | 0 |
| 0 | 0 | 0 | 0 | 0 |
| 0 | 0 | 0 | 0 | 0 |
| 0 | 0 | 0 | 0 | 0 |
| 0 | 0 | 0 | 0 | 0 |

| | | | | |
|---|---|---|---|---|
| 0 | 0 | 0 | 0 | 0 |
| 0 | 0 | 0 | 0 | 0.003996668 |
| 0 | 0 | 0.000588604 | 0 | 0 |
| 0 | 0 | 0 | 0 | 0 |
| 0 | 0 | 0 | 0 | 0 |
| 0 | 0 | 0 | 0 | 0.000782946 |
| 0 | 0 | 0 | 0 | 0 |
| 0 | 0 | 0 | 0 | 0 |
| 0 | 0 | 0 | 0 | 0 |
| 0 | 0 | 0 | 0 | 0 |
| 0.014714002 | 0 | 0 | 0.113444445 | 0 |
| 0 | 0 | 0 | 0 | 0 |
| 0 | 0 | 0.268191936 | 0 | 0 |
| 0 | 0 | 0 | 0 | 0 |
| 0 | 0 | 0 | 0 | 0 |
| 0 | 0 | 0 | 0 | 0 |
| 0 | 0 | 0 | 0 | 0 |
| 0 | 0 | 0 | 0 | 0 |
| 0 | 0 | 0 | 0 | 0 |
| 0 | 0 | 0 | 0 | 0 |
| 0 | 0.008006529 | 0 | 0 | 0 |
| 0 | 0 | 0 | 0 | 0 |
| 0 | 0 | 0 | 0 | 0 |
| 1.219423954 | 1.121880887 | 0.203036269 | 0 | 1.100431064 |
| 0 | 0 | 0 | 0 | 0 |
| 0 | 0 | 0 | 0 | 0 |
| 0 | 0 | 0 | 0 | 0 |
| 0 | 0 | 0 | 0 | 0 |
| 0 | 0 | 0 | 0.109409281 | 0 |
| 0 | 0 | 0 | 0 | 0 |
| 0 | 0 | 0 | 0 | 0 |
| 0 | 0 | 0 | 0 | 0 |
| 0.023380776 | 0 | 0.001344192 | 0 | 0 |
| 0 | 0 | 0 | 0 | 0 |

| PHL018-0005_V2 | PHL018-0168_V3 | PHL018-0188_V3 | PHL018-0122_V3 | PHL018-0001_V2 |
|---|---|---|---|---|
| EYCF | EYCF | CTRL | CTRL | EYCF |
| 0.126407991 | 0.055656837 | 0.020561983 | 5.00521765 | 0.025952001 |
| 0.097908242 | 7.801588175 | 0.34381629 | 0.219718037 | 0.365107352 |
| 0.007198526 | 3.887857737 | 0 | 0 | 0 |
| 0 | 0 | 0 | 0 | 0 |
| 0 | 0 | 0 | 0 | 0 |
| 0 | 0 | 0.028841871 | 0 | 0 |
| 0 | 0 | 0 | 0 | 0 |
| 0 | 0 | 0 | 0 | 0 |
| 0 | 0 | 0 | 0 | 0 |
| 0 | 0 | 0 | 0 | 0 |
| 0 | 0 | 0 | 0 | 0 |
| 0 | 0 | 0 | 0 | 0 |
| 0 | 0 | 0 | 0 | 0 |
| 0.133602617 | 12.73210627 | 0.037951939 | 7.260317132 | 2.172040499 |
| 0 | 0 | 0 | 0 | 0 |
| 0 | 0 | 0 | 0 | 0 |
| 0 | 0 | 0 | 0 | 0 |
| 0 | 0 | 0 | 0 | 0 |
| 0 | 0 | 0 | 0 | 0 |
| 0 | 0 | 0 | 0 | 0 |
| 0 | 0 | 0 | 0 | 0 |
| 0 | 0 | 0 | 0 | 0 |
| 0 | 0 | 0 | 0 | 0 |
| 0 | 0 | 0 | 0 | 0 |
| 0.052748665 | 0 | 0 | 0 | 0 |
| 0 | 0 | 0 | 0 | 0 |
| 0 | 0 | 0 | 0 | 0 |
| 0 | 0 | 0 | 0 | 0 |
| 0 | 0 | 0 | 0 | 0 |
| 0 | 0 | 0.002801645 | 0 | 0 |
| 0 | 0 | 0 | 0 | 0 |
| 0 | 0 | 0 | 0 | 0 |
| 0 | 0 | 0 | 0 | 0 |
| 0 | 0 | 0 | 0 | 0 |
| 0 | 0 | 0 | 0 | 0 |
| 0.207853939 | 7.739604633 | 2.013953589 | 7.52114874 | 0.889326824 |
| 0 | 0 | 0 | 0 | 0 |
| 0 | 0 | 0 | 0 | 0 |
| 0 | 0 | 0 | 0 | 0 |
| 0 | 0 | 0 | 0 | 0 |
| 0 | 0 | 0 | 0 | 0 |
| 0.046244155 | 0.002119882 | 0.021785562 | 0.310652342 | 0 |
| 21.95411712 | 1.347291412 | 0.392869523 | 0.267739216 | 0.003343939 |
| 0.003514023 | 0.043914479 | 0.013185279 | 0.007885335 | 0.06262774 |
| 0.003200194 | 0 | 0 | 0.032594275 | 0.060434737 |
| 0 | 0 | 0 | 0 | 0 |

| | | | | |
|---|---|---|---|---|
| 0.003283454 | 0 | 0.001575832 | 0.289314676 | 0 |
| 0 | 0.089082185 | 0 | 0.009292931 | 0 |
| 0 | 0.000615856 | 0 | 0 | 0 |
| 0.138051784 | 0.493983728 | 0.256715639 | 0.497147945 | 0.544681609 |
| 0 | 0 | 0 | 0.022648991 | 0 |
| 0.003226706 | 0 | 0.033496399 | 0 | 0 |
| 0 | 5.912043547 | 0.607089387 | 0 | 0 |
| 0 | 0 | 0.000886123 | 0 | 0 |
| 0 | 0 | 0 | 0 | 0 |
| 0 | 0 | 0 | 0 | 0 |
| 0 | 0 | 0 | 0 | 0 |
| 0.079138904 | 0.094765633 | 0.153796497 | 0.005112287 | 0 |
| 0 | 0 | 0 | 0.003451556 | 0 |
| 0.003467931 | 0.015094136 | 0.009910555 | 0.016212303 | 0 |
| 0 | 0.222310843 | 0 | 0 | 0 |
| 0 | 0 | 0 | 0 | 0 |
| 0 | 0 | 0.902919964 | 0.004800676 | 0 |
| 0 | 0 | 0 | 0 | 0 |
| 0 | 0 | 0 | 0 | 0 |
| 0 | 0 | 0 | 0 | 0 |
| 0 | 0.372404922 | 0.018146839 | 0.087164623 | 0 |
| 0.001629777 | 0.005105236 | 0.012684763 | 0.003474301 | 0 |
| 0.004671518 | 0 | 0.05463674 | 0 | 0 |
| 0 | 0 | 0 | 0 | 0 |
| 0 | 0 | 0 | 0 | 0 |
| 0 | 0 | 0 | 0 | 0 |
| 0 | 0.01720379 | 0 | 0 | 0 |
| 0 | 0 | 0 | 0 | 0 |
| 0.007413179 | 0 | 0.002304494 | 0.00938312 | 0 |
| 0 | 0 | 0 | 0 | 0 |
| 0 | 0.048078079 | 0.00136078 | 0 | 0 |
| 0 | 0 | 0 | 0 | 0 |
| 0 | 0 | 0 | 0 | 0 |
| 0 | 0 | 0 | 0 | 0 |
| 0 | 0 | 0 | 0 | 0 |
| 0 | 0.002505763 | 0 | 0 | 0 |
| 0 | 0 | 0 | 0 | 0 |
| 0 | 0.000962988 | 40.83170867 | 0 | 0 |
| 0 | 0 | 0 | 0 | 0 |
| 0 | 0 | 0 | 0 | 0 |
| 0 | 0 | 0 | 0 | 0 |
| 0 | 0 | 0 | 0 | 0 |
| 0 | 0 | 0 | 0 | 0 |
| 0 | 0 | 0 | 0 | 0 |
| 0.076606155 | 0 | 0 | 0 | 0 |
| 0 | 0 | 0 | 0 | 0 |
| 0 | 0 | 0.00092797 | 0 | 0 |
| 0 | 0.102814946 | 0.001668318 | 0.027821334 | 0 |

| | | | | |
|---|---|---|---|---|
| 0 | 0 | 0 | 0 | 0 |
| 0 | 0 | 0 | 0 | 0 |
| 0 | 0 | 0 | 0 | 0 |
| 0 | 0 | 0 | 0 | 0 |
| 0.002267359 | 0.034220862 | 0.02187144 | 0.002929378 | 0 |
| 0 | 0 | 0 | 0 | 0 |
| 0 | 0 | 0 | 0 | 0 |
| 0.004863267 | 0 | 0 | 0 | 0 |
| 0 | 0 | 0 | 0.006716368 | 0 |
| 0 | 0 | 0 | 0 | 0 |
| 0 | 0 | 0 | 0 | 0 |
| 0.008738082 | 0 | 0 | 0.001443995 | 0 |
| 0 | 0 | 0 | 0 | 0 |
| 0 | 0 | 0 | 0 | 0 |
| 0 | 0 | 0 | 0 | 0 |
| 0 | 0 | 0.001953059 | 0 | 0 |
| 0 | 0 | 0 | 0 | 0 |
| 0 | 0 | 0 | 0 | 0 |
| 0.04647806 | 0 | 0.171356647 | 0 | 0 |
| 0 | 0 | 0 | 0 | 0 |
| 0 | 0 | 0 | 0 | 0 |
| 0 | 0 | 0 | 0 | 0 |
| 0 | 0 | 0 | 0 | 0 |
| 0 | 0 | 0 | 0 | 0 |
| 0 | 0 | 0 | 0 | 0 |
| 0 | 0 | 0 | 0 | 0 |
| 0 | 0 | 0 | 0 | 0 |
| 0 | 0 | 0 | 0 | 0 |
| 0 | 0 | 0 | 0 | 0 |
| 0 | 0 | 0.000504067 | 0 | 0 |
| 0.046323855 | 0 | 0.044913266 | 0 | 0 |
| 0.007816423 | 0.001277465 | 0.000845165 | 0 | 0 |
| 0 | 0 | 0 | 0 | 0 |
| 0 | 0 | 0 | 0 | 0 |
| 0 | 0 | 0 | 0 | 0 |
| 0 | 0 | 0 | 0 | 0 |
| 0 | 0 | 0 | 0 | 0 |
| 0 | 0 | 0 | 0 | 0 |
| 0 | 0 | 0 | 0 | 0 |
| 0 | 0 | 0 | 0 | 0 |
| 0 | 0 | 0 | 0 | 0 |
| 0 | 0 | 0 | 0 | 0 |
| 0.001767629 | 0.005414007 | 0.000325626 | 0 | 0 |
| 0 | 0 | 0 | 0 | 0 |
| 0 | 0 | 0 | 0 | 0 |
| 0 | 0 | 0 | 0 | 0 |
| 0 | 0 | 0 | 0 | 0 |
| 0 | 0 | 0 | 0 | 0 |

| | | | | |
|---|---|---|---|---|
| 0 | 0 | 0 | 0 | 0 |
| 0 | 0 | 0 | 0 | 0 |
| 0 | 0 | 0 | 0 | 0 |
| 0 | 0 | 0 | 0 | 0 |
| 0.050285345 | 0 | 0.027524455 | 0 | 0.005778647 |
| 0 | 0 | 0 | 0 | 0 |
| 0 | 0 | 0 | 0 | 0 |
| 0 | 0 | 0 | 0 | 0 |
| 0 | 0 | 0 | 0 | 0 |
| 0 | 0 | 0 | 0 | 0 |
| 0 | 0 | 0 | 0 | 0 |
| 0 | 0 | 0 | 0 | 0 |
| 0 | 0 | 0 | 0 | 0 |
| 0 | 0 | 0 | 0 | 0 |
| 0 | 0 | 0 | 0 | 0 |
| 0.003005623 | 0 | 1.986949696 | 0 | 0 |
| 0 | 0 | 0 | 0 | 0 |
| 0 | 0 | 0 | 0 | 0 |
| 0 | 0 | 0 | 0 | 0 |
| 0 | 0 | 0 | 0 | 0 |
| 0 | 0 | 0 | 0 | 0 |
| 0 | 0 | 0 | 0 | 0 |
| 0 | 0 | 0 | 0 | 0 |
| 0 | 0 | 0 | 0 | 0 |
| 0 | 0 | 0 | 0 | 0 |
| 0 | 0 | 0 | 0 | 0 |
| 0.012182348 | 0 | 0.001300606 | 0 | 0 |
| 0 | 0 | 0 | 0 | 0 |
| 0 | 0 | 0 | 0 | 0 |
| 0 | 0 | 0 | 0 | 0 |
| 0 | 0 | 0 | 0 | 0 |
| 2.880027232 | 0.01179135 | 0.148708484 | 0 | 0 |
| 0 | 0 | 0 | 0 | 0 |
| 0 | 0 | 0 | 0 | 0 |
| 0 | 0 | 0 | 0 | 0 |
| 0 | 0 | 0 | 0 | 0 |
| 0 | 0 | 0 | 0 | 0 |
| 0 | 0 | 0 | 0 | 0 |
| 0 | 0 | 0 | 0 | 0 |
| 0 | 0 | 0 | 0 | 0 |
| 0 | 0 | 0 | 0 | 0 |
| 0 | 0 | 0 | 0 | 0 |
| 0 | 0 | 0 | 0 | 0 |
| 0 | 0 | 0 | 0 | 0 |
| 0 | 0 | 0 | 0 | 0 |
| 0 | 0 | 0 | 0 | 0 |
| 0 | 0 | 0 | 0 | 0 |
| 0 | 0 | 0 | 0 | 0 |

| | | | | |
|---|---|---|---|---|
| 0 | 0 | 0.779079341 | 0 | 0 |
| 0 | 0 | 0 | 0 | 0 |
| 0 | 0 | 0 | 0 | 0 |
| 0 | 0 | 0 | 0 | 0 |
| 0 | 0 | 0 | 0 | 0 |
| 0 | 0 | 0 | 0 | 0 |
| 0 | 0 | 0 | 0 | 0 |
| 0 | 0 | 0 | 0 | 0 |
| 0 | 0 | 0 | 0 | 0 |
| 0 | 0 | 0 | 0 | 0 |
| 0 | 0 | 0 | 0 | 0 |
| 0.001332161 | 0 | 0.015539418 | 0 | 0 |
| 0.000750416 | 0 | 0 | 0.002799493 | 0 |
| 0 | 0 | 0 | 0 | 0 |
| 0 | 0 | 0 | 0 | 0 |
| 0 | 0 | 0 | 0 | 0 |
| 0 | 0 | 0 | 0 | 0 |
| 0 | 0 | 0 | 0 | 0 |
| 0 | 0 | 0 | 0 | 0 |
| 0 | 0 | 0 | 0 | 0 |
| 0.003156971 | 0.043292412 | 0.009595814 | 0.027916691 | 0.007309546 |
| 0 | 0 | 0 | 0 | 0 |
| 0 | 0 | 0 | 0 | 0 |
| 0 | 0.003666621 | 0 | 0.000779772 | 0 |
| 0 | 0 | 0 | 0 | 0 |
| 0 | 0 | 0 | 0 | 0 |
| 0 | 0.003199553 | 0.002286154 | 0.002467736 | 0.004256907 |
| 0 | 0 | 0 | 0 | 0 |
| 0 | 0 | 0 | 0 | 0 |
| 0 | 0 | 0 | 0 | 0 |
| 0 | 0 | 0 | 0.001591709 | 0 |
| 0 | 0 | 0 | 0 | 0 |
| 0 | 0 | 0 | 0 | 0 |
| 0.002358088 | 0 | 0.029499536 | 0.000761649 | 0 |
| 0 | 0.013154763 | 0.005959751 | 0.173045365 | 0 |
| 0 | 0 | 0 | 0 | 0 |
| 0 | 0 | 0 | 0 | 0 |
| 0 | 0 | 0 | 0 | 0 |
| 0 | 0 | 0 | 0.009903289 | 0 |
| 0 | 0 | 0 | 0 | 0 |
| 0 | 0 | 0 | 0 | 0 |
| 0 | 0 | 0 | 0 | 0 |
| 0 | 0 | 0 | 0 | 0 |
| 0 | 0 | 0 | 0 | 0 |
| 0 | 0 | 0 | 0 | 0 |
| 0.531304104 | 0 | 0 | 0 | 0 |
| 0 | 0 | 0.000512129 | 0 | 0 |
| 0 | 0 | 0 | 0.000809014 | 0.00169462 |

| | | | | |
|---|---|---|---|---|
| 0 | 0 | 0 | 0 | 0 |
| 0 | 0 | 0 | 0 | 0 |
| 0 | 0.001303547 | 0 | 0.002112171 | 0 |
| 0 | 0 | 0 | 0 | 0 |
| 0 | 0 | 0 | 0 | 0 |
| 0 | 0 | 0 | 0 | 0 |
| 0 | 0 | 0 | 0 | 0 |
| 0 | 0 | 0 | 0.004984166 | 0 |
| 0 | 0 | 0 | 0 | 0 |
| 0 | 0 | 0 | 0 | 0 |
| 0 | 0 | 0 | 0 | 0 |
| 0 | 0 | 0 | 0 | 0 |
| 0 | 0 | 0 | 0 | 0 |
| 0 | 0 | 0 | 0 | 0 |
| 0 | 0 | 0 | 0 | 0 |
| 0 | 0 | 0 | 0 | 0 |
| 0 | 0 | 0 | 0 | 0 |
| 0.00110217 | 0.00193341 | 0 | 0 | 0 |
| 0.268292756 | 0.00704668 | 0.001851108 | 0.001175374 | 0 |
| 0.057812951 | 0 | 0.095300672 | 0 | 0 |
| 0 | 0.001528613 | 0.000809059 | 0.055654848 | 0 |
| 0 | 0 | 0 | 0 | 0 |
| 0 | 0 | 0 | 0 | 0 |
| 0 | 0 | 0 | 0 | 0 |
| 0 | 0 | 0 | 0 | 0 |
| 0 | 0 | 0 | 0 | 0.00857689 |
| 0 | 0 | 0 | 0 | 0 |
| 0 | 0 | 0 | 0 | 0 |
| 0.008095514 | 0 | 0 | 0.079056824 | 0 |
| 0 | 0 | 0 | 0 | 0 |
| 0.001614682 | 0.00595714 | 0.141958109 | 0.196965779 | 0 |
| 0 | 0 | 0 | 0 | 0 |
| 0 | 0 | 0 | 0 | 0 |
| 0 | 0.025012068 | 0 | 0 | 0 |
| 0 | 0 | 0 | 0 | 0 |
| 0 | 0 | 0 | 0 | 0 |
| 0 | 0 | 0 | 0 | 0 |
| 0 | 0 | 0 | 0 | 0 |
| 0 | 0 | 0 | 0 | 0 |
| 0 | 0 | 0 | 0 | 0.019605193 |
| 0 | 0 | 0 | 0 | 0 |
| 0.001828777 | 0.002749727 | 0.003790013 | 0.003768571 | 0.058115639 |
| 0 | 0 | 0 | 0 | 0 |
| 0 | 0 | 0 | 0 | 0.007742476 |
| 0 | 0 | 0 | 0 | 0 |
| 0 | 0 | 0 | 0 | 0 |
| 0 | 0 | 0 | 0 | 0 |
| 0 | 0 | 0 | 0 | 0 |

| | | | | |
|---|---|---|---|---|
| 0 | 0 | 0 | 0 | 0 |
| 0 | 0 | 0 | 0 | 0 |
| 0.003903762 | 0.048626013 | 0.014276954 | 0.007179678 | 0.050842365 |
| 0 | 0 | 0 | 0 | 0 |
| 0 | 0 | 0.00198239 | 0 | 0 |
| 0 | 0 | 0 | 0 | 0 |
| 0 | 0 | 0 | 0 | 0 |
| 0 | 0 | 0 | 0 | 0 |
| 0 | 0 | 0 | 0 | 0 |
| 0 | 0 | 0 | 0.006118765 | 0 |
| 0 | 0 | 0 | 0 | 0 |
| 0 | 0 | 0 | 0 | 0 |
| 0 | 0 | 0 | 0 | 0 |
| 0 | 0 | 0.001157098 | 0 | 0 |
| 0.001885204 | 0 | 0.003299205 | 0.001041914 | 0.001636852 |
| 0.001569836 | 0 | 0.004740631 | 0 | 0 |
| 0 | 0 | 0 | 0 | 0.004351775 |
| 0 | 0 | 0 | 0 | 0 |
| 0 | 0 | 0 | 0 | 0 |
| 0 | 0 | 0 | 0 | 0 |
| 0 | 0 | 0 | 0 | 0 |
| 0 | 0 | 0 | 0.002755731 | 0 |
| 0 | 0 | 0 | 0 | 0 |
| 0 | 0.001255483 | 0 | 0 | 0 |
| 0 | 0 | 0 | 0 | 0 |
| 0 | 0 | 0 | 0 | 0 |
| 0.002649735 | 0 | 0 | 0 | 0 |
| 0 | 0 | 0 | 0 | 0 |
| 0 | 0 | 0 | 0 | 0 |
| 0 | 0 | 0 | 0 | 0 |
| 0 | 0 | 0 | 0 | 0 |
| 0 | 0 | 0 | 0 | 0 |
| 0 | 0 | 0 | 0 | 0 |
| 0 | 0.002956962 | 0 | 0 | 0 |
| 0 | 0 | 0 | 0 | 0 |
| 0 | 0 | 0 | 0.007338909 | 0 |
| 0 | 0 | 0 | 0 | 0 |
| 0.005684503 | 0 | 0 | 0.030775922 | 0.12973687 |
| 0 | 0 | 0 | 0 | 0 |
| 0 | 0 | 0 | 0 | 0 |
| 0 | 0.000979044 | 0.000647731 | 0 | 0 |
| 0 | 0 | 0 | 0 | 0 |
| 0 | 0 | 0 | 0 | 0 |
| 0 | 0.000842611 | 0 | 0 | 0 |
| 0 | 0 | 0 | 0 | 0 |
| 0 | 0 | 0 | 0 | 0 |
| 0 | 0 | 0 | 0 | 0 |
| 0 | 0.001437881 | 0 | 0 | 0 |

| | | | | |
|---|---|---|---|---|
| 0.031341859 | 0 | 0.247720382 | 0 | 0 |
| 0 | 0 | 0 | 0 | 0 |
| 0 | 0 | 0 | 0 | 0.004114704 |
| 0 | 0 | 0 | 0 | 0 |
| 0.002043386 | 0 | 0 | 0.001129337 | 0.003548391 |
| 0 | 0 | 0 | 0.001315348 | 0 |
| 0 | 0 | 0 | 0 | 0 |
| 0 | 0 | 0.005804057 | 0.010176525 | 0 |
| 0 | 0 | 0 | 0 | 0 |
| 0.001968166 | 0 | 0.002175406 | 0.066431379 | 0 |
| 0 | 0 | 0 | 0 | 0 |
| 0.009825202 | 0.000631327 | 0.000730945 | 0.075187223 | 0 |
| 0.001146579 | 0.029571085 | 0.002296998 | 0.0059748 | 0.007466489 |
| 0 | 0.05345664 | 0.001684126 | 0 | 0 |
| 0 | 0 | 0.000651699 | 0 | 0 |
| 0 | 0.043851942 | 0 | 0 | 0 |
| 0 | 0 | 0 | 0 | 0 |
| 0 | 0 | 0 | 0 | 0 |
| 0 | 0.026106162 | 0 | 0.119538037 | 0 |
| 0 | 0 | 0 | 0 | 0 |
| 0 | 0 | 0 | 0 | 0 |
| 0 | 0 | 0 | 0 | 0 |
| 0 | 0 | 0 | 0.001541305 | 0 |
| 0 | 0 | 0 | 0 | 0 |
| 0 | 0 | 0 | 0 | 0 |
| 0.000890315 | 0.000976111 | 0.006550159 | 0.00173978 | 0.053338877 |
| 2.263200453 | 0 | 4.488869949 | 0 | 0 |
| 0 | 0.048901853 | 0 | 0.020656248 | 0 |
| 0 | 0 | 0 | 0 | 0 |
| 0 | 0 | 0 | 0 | 0 |
| 0 | 0 | 0 | 0 | 0 |
| 0 | 0 | 0 | 0 | 0 |
| 0 | 0 | 0 | 0 | 0 |
| 0 | 0 | 0 | 0 | 0 |
| 0 | 0 | 0 | 0 | 0 |
| 0 | 0 | 0 | 0 | 0 |
| 0 | 0.136260261 | 0.000418325 | 0.015777747 | 0 |
| 0 | 0 | 0 | 0 | 0 |
| 0 | 0.000868912 | 0 | 0 | 0 |
| 0 | 0 | 0 | 0 | 0 |
| 0 | 0 | 0 | 0 | 0 |
| 0 | 0 | 0 | 0 | 0 |
| 0 | 0 | 0 | 0 | 0 |
| 0 | 0.056283016 | 0 | 0 | 0 |
| 0 | 0 | 0 | 0 | 0 |
| 0.000977085 | 0 | 0 | 0 | 0 |
| 0 | 0.005549125 | 0.002638729 | 0 | 0 |
| 0 | 0 | 0 | 0 | 0 |

| | | | | |
|---|---|---|---|---|
| 0 | 0 | 0 | 0 | 0 |
| 0 | 0.00096899 | 0.004872201 | 0 | 0 |
| 0 | 0 | 0 | 0 | 0 |
| 0.002681077 | 0 | 0.000740846 | 0 | 0 |
| 0 | 0.028109239 | 0 | 0 | 0 |
| 0 | 0 | 0.000361447 | 0 | 0 |
| 0 | 0 | 0 | 0 | 0 |
| 0 | 0 | 0 | 0 | 0 |
| 0 | 0 | 5.9259461 | 0 | 0 |
| 0 | 0 | 0 | 0 | 0 |
| 0 | 0 | 0 | 0 | 0 |
| 0 | 0 | 0 | 0 | 0 |
| 0 | 0 | 0 | 0 | 0 |
| 0 | 0 | 0 | 0.001697477 | 0 |
| 0 | 0 | 0 | 0 | 0 |
| 0 | 0 | 0 | 0 | 0 |
| 0 | 0 | 0 | 0 | 0 |
| 0 | 0 | 0 | 0 | 0 |
| 0 | 0 | 0 | 0 | 0 |
| 0 | 0 | 0 | 0 | 0 |
| 0 | 0 | 0 | 0 | 0 |
| 0.177174895 | 0 | 0.062689706 | 0 | 0 |
| 0.005667225 | 0 | 0.012026805 | 0 | 0 |
| 0 | 0 | 0 | 0 | 0 |
| 0 | 0 | 0 | 0 | 0 |
| 0 | 0 | 0 | 0 | 0 |
| 0 | 0.002271784 | 0 | 0.002024566 | 0 |
| 0 | 0 | 0 | 0 | 0 |
| 0 | 0 | 0 | 0 | 0 |
| 0.070730937 | 3.896434879 | 0.084758009 | 0.052251073 | 0.137723283 |
| 0.010534125 | 0 | 0.001343461 | 0.002303207 | 0 |
| 0 | 0 | 0 | 0.005012128 | 0 |
| 0 | 0 | 0 | 0 | 0 |
| 0 | 0 | 0 | 0 | 0 |
| 0 | 0 | 0 | 0 | 0 |
| 0 | 0 | 0 | 0 | 0 |
| 0 | 0 | 0.74171264 | 0 | 0 |
| 0 | 0 | 0 | 0.008131854 | 0 |
| 0 | 0 | 0 | 0 | 0 |
| 0 | 0 | 0 | 0 | 0 |
| 0 | 0.01980883 | 0 | 0.003488778 | 0 |
| 0.364490855 | 0.254488283 | 0.833300205 | 0 | 0.256053406 |
| 0 | 0 | 0 | 0.003645404 | 0 |
| 0 | 0 | 0 | 0 | 0 |
| 0 | 0 | 0.003581122 | 0 | 0 |
| 0 | 0 | 0 | 0 | 0 |
| 0.002061235 | 0.002195301 | 0 | 0.004101129 | 0 |
| 0 | 0 | 0 | 0 | 0 |

| | | | | |
|---|---|---|---|---|
| 0.001017287 | 0.000796657 | 0 | 0.015903189 | 0.02517327 |
| 0 | 0 | 0 | 0 | 0 |
| 0 | 0.063827736 | 0.035621002 | 0.001231211 | 0 |
| 0 | 0 | 0 | 0 | 0 |
| 0 | 0 | 0.002755105 | 0 | 0 |
| 0 | 0 | 0 | 0 | 0 |
| 0 | 0 | 0 | 0.000898887 | 0 |
| 0 | 0 | 0 | 0 | 0 |
| 0.003192669 | 0.095475759 | 0.000529327 | 0 | 0 |
| 0 | 0 | 0.261575433 | 0 | 0 |
| 0 | 0 | 0 | 0 | 0 |
| 0 | 0 | 0 | 0 | 0 |
| 0 | 0 | 0 | 0 | 0 |
| 0 | 0 | 0 | 0 | 0.002397223 |
| 0 | 0 | 0 | 0 | 0 |
| 0.003416246 | 0 | 0.280886924 | 0 | 0.001405042 |
| 0.027103744 | 0.002621967 | 0.004460607 | 1.535811108 | 0.076828873 |
| 0 | 0 | 0 | 0.004301408 | 0 |
| 0 | 0 | 0.005716676 | 0.000731386 | 0 |
| 0.001594691 | 0 | 0 | 0 | 0 |
| 0.001255825 | 0 | 0.000832835 | 0 | 0 |
| 0 | 0.000434478 | 0.003928463 | 0.000821328 | 0 |
| 0 | 0 | 0 | 0 | 0 |
| 0.018259468 | 0 | 0 | 0 | 0.004011257 |
| 0 | 0 | 0 | 0 | 0 |
| 0 | 0 | 0.004135008 | 0 | 0 |
| 0 | 0.000634021 | 0.00083893 | 0 | 0 |
| 0 | 0 | 0 | 0 | 0 |
| 0 | 0 | 0 | 0 | 0 |
| 0 | 0 | 0 | 0 | 0 |
| 0.001154449 | 0 | 0.005263532 | 0 | 0 |
| 0.007171423 | 0.004133431 | 0 | 0.00142686 | 0.014944029 |
| 0.002185013 | 0.022757948 | 0.023547085 | 0.002911207 | 0.040551898 |
| 0.011351093 | 0.002206987 | 0.001135658 | 0.397456684 | 0 |
| 0 | 0.035005045 | 0.007410934 | 0 | 0 |
| 0 | 0 | 0 | 0 | 0 |
| 0 | 0.000464092 | 0 | 0 | 0 |
| 0.002124177 | 0.020929633 | 0.02225116 | 0 | 0 |
| 0.037728237 | 0 | 0.031691867 | 0 | 0 |
| 0 | 0 | 0 | 0 | 0 |
| 0 | 0 | 0 | 0 | 0 |
| 0 | 0 | 0 | 0 | 0 |
| 0.001443956 | 0.012483908 | 0 | 0 | 0 |
| 0 | 0 | 0.233992082 | 0 | 0 |
| 0 | 0 | 0 | 0 | 0 |
| 0 | 0 | 0 | 0 | 0 |
| 0 | 0 | 0 | 0 | 0 |
| 0 | 0 | 0 | 0 | 0 |

| | | | | |
|---|---|---|---|---|
| 0 | 0 | 0 | 0 | 0 |
| 0.273394951 | 0.005934648 | 0.023776137 | 0.134250918 | 0.529132721 |
| 0 | 0 | 0.001074887 | 0 | 0 |
| 0.000956179 | 0.000798722 | 0 | 0 | 0 |
| 0 | 0 | 0 | 0 | 0 |
| 0 | 0 | 0 | 0 | 0 |
| 0 | 0 | 0 | 0 | 0 |
| 0 | 0 | 0 | 0 | 0 |
| 0 | 0 | 0 | 0 | 0 |
| 0 | 0 | 0 | 0 | 0 |
| 0 | 0.002082544 | 0.004619685 | 0 | 0 |
| 0.034077074 | 0.011130734 | 0.003138771 | 0.002069638 | 0.012138606 |
| 0.125213196 | 0 | 0.002606455 | 0 | 0 |
| 0 | 0 | 0 | 0 | 0 |
| 0 | 0.021083456 | 0 | 0 | 0 |
| 0 | 0 | 0 | 0 | 0 |
| 0.001433284 | 0 | 0.002291437 | 0 | 0.032533922 |
| 0 | 0.001165657 | 0 | 0.002266492 | 0 |
| 0 | 0 | 0 | 0 | 0 |
| 0 | 0 | 0.00264694 | 0 | 0 |
| 0.008446156 | 0.026060508 | 0.009277156 | 0.033309643 | 0.165553362 |
| 0 | 0.009711974 | 0 | 0 | 0 |
| 0 | 0 | 0 | 0 | 0 |
| 0 | 0 | 0 | 0 | 0 |
| 0 | 0.012224661 | 0.007058424 | 0 | 0 |
| 0 | 0 | 0 | 0 | 0 |
| 0.022509302 | 0.04243646 | 0 | 0 | 0 |
| 0.009425868 | 0.283311787 | 0.058231271 | 0.007654753 | 0.042089809 |
| 0 | 0.001370738 | 0 | 0 | 0 |
| 0 | 0.000678393 | 0 | 0.001731267 | 0 |
| 0 | 0 | 0 | 0 | 0.018190411 |
| 0 | 0 | 0 | 0 | 0 |
| 0 | 0 | 0 | 0 | 0 |
| 0 | 0 | 0 | 0 | 0 |
| 0 | 0 | 0.01353727 | 0 | 0.328067358 |
| 0.028487 | 0.248191709 | 0.184826144 | 0.025640526 | 0.062330179 |
| 0 | 0 | 0 | 0 | 0 |
| 0 | 0 | 0 | 0 | 0 |
| 0.002768539 | 0 | 0 | 0.035804687 | 0 |
| 0 | 0 | 0 | 0 | 0 |
| 0 | 0 | 0 | 0 | 0 |
| 0 | 0 | 0 | 0 | 0 |
| 0 | 0.0067427 | 0 | 0.00095597 | 0 |
| 0 | 0.003865282 | 0 | 0.000922971 | 0 |
| 0 | 0 | 0 | 0 | 0 |
| 0.034389113 | 0 | 0.613469891 | 0 | 0 |
| 0 | 0 | 0 | 0 | 0 |
| 0 | 0 | 0 | 0 | 0 |

| | | | | |
|---|---|---|---|---|
| 0 | 0 | 0.000876379 | 0.003004899 | 0 |
| 0 | 0 | 0.00902779 | 0 | 0 |
| 0 | 0 | 0 | 0 | 0 |
| 0.24599932 | 0 | 0.101800911 | 0 | 0 |
| 0 | 0 | 0 | 0 | 0 |
| 0 | 0 | 0.001055944 | 0 | 0 |
| 0 | 0 | 0 | 0 | 0 |
| 0 | 0 | 0.004575379 | 0 | 0 |
| 3.365504148 | 0.193329517 | 0.456831763 | 0 | 9.886468781 |
| 0.037110616 | 0 | 0.05100673 | 0 | 0.00820743 |
| 0 | 0 | 0 | 0 | 0 |
| 0.001125926 | 0 | 0 | 0 | 0.04105917 |
| 0 | 0 | 0 | 0 | 0 |
| 0.001410572 | 0 | 0 | 0 | 0 |
| 0.278270481 | 0 | 0.099678244 | 0.021677199 | 0 |
| 0 | 0 | 0 | 0 | 0 |
| 0.072717087 | 0 | 0.020093476 | 0 | 0 |
| 4.925337147 | 0 | 0.045679204 | 0 | 0 |
| 0 | 0 | 0 | 0 | 0 |
| 0 | 0 | 0 | 0 | 0 |
| 0 | 0 | 0 | 0 | 0 |
| 0 | 0 | 0.003394016 | 0 | 0 |
| 0 | 0 | 0.000859468 | 0 | 0 |
| 0.154383638 | 0 | 0.134347784 | 0.002119868 | 0 |
| 0.005622156 | 0 | 0 | 0 | 0 |
| 0.028790029 | 0.000680634 | 0.206314417 | 0 | 0 |
| 0.009616153 | 0 | 0 | 0 | 0 |
| 0.402217788 | 0 | 0.592558262 | 0 | 25.14728882 |
| 0 | 0 | 0 | 0 | 0 |
| 0 | 0 | 0 | 0 | 0 |
| 0.004925663 | 0.000685756 | 0.032968356 | 0 | 0 |
| 0.00687539 | 0 | 0 | 0 | 0 |
| 0.045818561 | 0.00304448 | 0 | 0 | 0 |
| 0 | 0 | 0 | 0 | 0 |
| 0.012127476 | 0 | 0 | 0 | 0 |
| 0 | 0 | 0.004103263 | 0 | 0 |
| 0.116067837 | 0 | 0.286994898 | 0 | 0.166516822 |
| 0 | 0 | 0 | 0 | 0 |
| 0 | 0 | 0 | 0 | 0 |
| 0.006178625 | 0 | 0 | 0 | 0 |
| 0 | 0.004060155 | 0.002984643 | 0.227186961 | 0 |
| 0.012664421 | 0 | 0 | 0 | 0 |
| 0.005182968 | 5.164516735 | 0.002058756 | 0.306912892 | 3.647956212 |
| 0.020073165 | 0 | 0 | 0 | 0 |
| 0 | 0 | 0 | 0 | 0 |
| 0 | 0 | 0 | 0 | 0 |
| 0 | 0 | 0 | 0 | 0 |
| 0.035274112 | 0 | 0.001166062 | 0.043057062 | 0.010146432 |

| | | | | |
|---|---|---|---|---|
| 0 | 0 | 0 | 0 | 0 |
| 0 | 0 | 0.018467949 | 0 | 0.259850319 |
| 0.075718955 | 0.534809484 | 0.000429924 | 1.067844622 | 0 |
| 0.002466669 | 0 | 0.013291198 | 0 | 0 |
| 0.001140613 | 0 | 0 | 0 | 0 |
| 0.016178627 | 0 | 0.000338677 | 0 | 0.008513514 |
| 0 | 0 | 0 | 0 | 0 |
| 0 | 0 | 0 | 0 | 0 |
| 0 | 0 | 0 | 0 | 0 |
| 0 | 0 | 0 | 0 | 0 |
| 0.186773018 | 0.016588445 | 0.000759796 | 0.084233658 | 0.012202393 |
| 0.54017302 | 1.88630921 | 0.691685179 | 0.410339105 | 1.021351137 |
| 0 | 0 | 0 | 0 | 0 |
| 0.03846814 | 0 | 0.00501113 | 0.684312181 | 0.447756883 |
| 0.003118049 | 1.091648483 | 0.003518167 | 0.188822494 | 0 |
| 0.074678775 | 0 | 1.126337208 | 0 | 0 |
| 0 | 0 | 0 | 0 | 0 |
| 0 | 0 | 0 | 0 | 0 |
| 0 | 0 | 0 | 0 | 0 |
| 0 | 0 | 0.003824774 | 0 | 0 |
| 0 | 0.001283405 | 0 | 0.124945388 | 0 |
| 0 | 0.745123479 | 0.000447341 | 0.330156744 | 0.033266615 |
| 0.244442905 | 1.982969565 | 0.356217542 | 1.45397766 | 0.031920097 |
| 0 | 0 | 0 | 0 | 0 |
| 0.075459008 | 0 | 0.057768276 | 0 | 0 |
| 0 | 0.011918762 | 0.001356627 | 0.010175285 | 2.776136225 |
| 0 | 0 | 0 | 0 | 0 |
| 0 | 0 | 0 | 0 | 0 |
| 0 | 0 | 0 | 0 | 0 |
| 0 | 0 | 0 | 0 | 0 |
| 0.009414569 | 0 | 0.00093653 | 0 | 0 |
| 0.033361282 | 0 | 0 | 0.274624604 | 0.003358418 |
| 0.013387708 | 0 | 0 | 0.116536024 | 0 |
| 0.370879783 | 0 | 0 | 0 | 0 |
| 0 | 0.003805823 | 0.005951433 | 0.007554174 | 0 |
| 0.018969058 | 0.011527497 | 0.009120394 | 0.002965421 | 0.733744068 |
| 0.009711641 | 0 | 0.003113942 | 0 | 0 |
| 0 | 0 | 0 | 0 | 0 |
| 0.164262454 | 0 | 0.503917207 | 0 | 0 |
| 0 | 0 | 0 | 0 | 0 |
| 0.017371339 | 0 | 0.003738553 | 0 | 0 |
| 0 | 0 | 0 | 0 | 0 |
| 0.051638702 | 0 | 0.184008667 | 0 | 0 |
| 0 | 0 | 0 | 0 | 0 |
| 0.098209542 | 0.021141702 | 0.00385546 | 0 | 0.002253877 |
| 0 | 0 | 0.203122815 | 0 | 0 |
| 0 | 0 | 0 | 0 | 0 |
| 0 | 0.09105761 | 0 | 19.34916346 | 33.80341474 |

| | | | | |
|---|---|---|---|---|
| 0 | 0 | 0 | 0 | 0 |
| 0 | 0 | 0.023567605 | 0 | 0 |
| 0 | 0 | 0 | 0 | 0 |
| 0.000737477 | 0 | 0.006189893 | 0 | 0 |
| 0 | 0 | 0 | 0 | 0 |
| 0.2282928 | 0 | 0 | 0 | 0 |
| 0.046828994 | 0 | 0.075715585 | 0 | 0 |
| 0.057342712 | 0 | 0.001406376 | 0 | 0 |
| 0 | 0 | 0 | 0 | 0 |
| 0 | 0 | 0 | 0 | 0 |
| 0.001026692 | 0.000514574 | 0 | 0.012402436 | 0.002139452 |
| 0 | 0 | 0 | 0 | 0 |
| 0 | 0 | 0.019971459 | 0 | 0.001404943 |
| 0.004730971 | 0 | 0.051129208 | 0 | 0 |
| 0 | 0 | 0 | 0 | 0 |
| 0 | 0 | 0 | 0 | 0 |
| 0.011453829 | 0 | 0 | 0 | 0 |
| 0.001538456 | 0 | 0.000566816 | 0.254839006 | 0.004897874 |
| 0.011691793 | 0 | 0.037948568 | 0 | 0 |
| 0 | 0 | 0.005495126 | 0 | 0 |
| 0.014882899 | 0 | 0.007994201 | 0 | 0 |
| 0 | 0 | 0 | 0 | 0 |
| 0 | 0 | 0.061673942 | 0 | 0 |
| 0 | 0 | 0 | 0 | 0 |
| 0 | 0 | 0 | 0 | 0 |
| 0 | 0 | 0 | 0 | 0 |
| 0 | 0 | 0 | 0 | 0 |
| 0 | 0 | 0 | 0 | 0 |
| 0 | 0 | 0 | 0 | 0 |
| 0.172009038 | 0.127297208 | 0.570362697 | 0 | 0.019901251 |
| 0 | 0 | 0 | 0 | 0 |
| 0 | 0 | 0 | 0 | 0 |
| 0 | 0 | 0 | 0 | 0 |
| 0 | 0 | 0 | 0 | 0 |
| 0 | 0 | 0.000406051 | 0.001276235 | 0 |
| 0 | 0 | 0 | 0 | 0 |
| 0.005440706 | 0 | 0.014201013 | 0 | 0.03102494 |
| 0 | 0.008494117 | 0 | 0.007658006 | 0 |
| 0 | 0 | 0 | 0 | 0 |
| 0 | 0 | 0 | 0 | 0 |
| 0.188367312 | 0.053028388 | 0.00033799 | 0 | 0 |
| 0 | 0 | 0 | 0 | 0 |
| 0 | 0 | 0 | 0 | 0 |
| 0 | 0 | 0.034423365 | 0.017752436 | 0 |
| 0.019145251 | 0 | 0.15003705 | 0.007293073 | 0 |
| 0 | 0 | 0 | 0 | 0 |
| 0.003963853 | 0 | 0 | 0 | 0 |
| 0 | 0 | 0 | 0 | 0 |

| | | | | |
|---|---|---|---|---|
| 0 | 0 | 0 | 0 | 0 |
| 0 | 0 | 0 | 0 | 0 |
| 0 | 0 | 0 | 0 | 0 |
| 0.241544794 | 0 | 0.247604872 | 0 | 0.009457861 |
| 0 | 0 | 0 | 0 | 0 |
| 0 | 0 | 0 | 0 | 0.015965266 |
| 0 | 0 | 0 | 0 | 0 |
| 0 | 0 | 0 | 0 | 0 |
| 0 | 0.130214581 | 0.008446014 | 0.15927679 | 0 |
| 0 | 0 | 0 | 0 | 0 |
| 0 | 0 | 0 | 0 | 0 |
| 0 | 0 | 0 | 0 | 0 |
| 0 | 0 | 0 | 0 | 0 |
| 0 | 0 | 0 | 0 | 0 |
| 0.194672503 | 5.610544658 | 0.081260276 | 0 | 0 |
| 0 | 0.150713632 | 0 | 0 | 0 |
| 0 | 0 | 0 | 0 | 0 |
| 0 | 0 | 0 | 0 | 0 |
| 0 | 0 | 0 | 0 | 0 |
| 0 | 0 | 0 | 0 | 0 |
| 0.007332329 | 0 | 0.013296271 | 0 | 0 |
| 0 | 0 | 0 | 0 | 0 |
| 0 | 0 | 0 | 0 | 0 |
| 0.002533033 | 0 | 0 | 0 | 0 |
| 8.409274065 | 1.951462905 | 0.01367148 | 0.129611032 | 0 |
| 0.076792396 | 0 | 6.945080985 | 0 | 0.808417293 |
| 0.058090266 | 0 | 0.067538976 | 0 | 0 |
| 0 | 0 | 0 | 0 | 0 |
| 0 | 0 | 0.011237659 | 0 | 0.629175449 |
| 0 | 0 | 1.449929125 | 0 | 0 |
| 0 | 0 | 0 | 0 | 0 |
| 0 | 0 | 0 | 0 | 0 |
| 0 | 0 | 0 | 0 | 0 |
| 0.002860664 | 0.000565955 | 0 | 0.037659437 | 0 |
| 0.009923097 | 0 | 0.000719772 | 0.008990323 | 0.195149102 |
| 0.001928967 | 0.008679142 | 0.00159906 | 0.00149531 | 0 |
| 0 | 0 | 0 | 0 | 0 |
| 0.070294437 | 1.254308097 | 0.132466478 | 0 | 0.087682697 |
| 0 | 0 | 0 | 0 | 0 |
| 0 | 0 | 0 | 0 | 0 |
| 0.003242576 | 0 | 0 | 0 | 0 |
| 0 | 0 | 0.000741824 | 0 | 0 |
| 0.011981673 | 0 | 0.064561066 | 0 | 0.401356983 |
| 0 | 0 | 0.211809894 | 0 | 0 |
| 0 | 0 | 0 | 0 | 0 |
| 0 | 0 | 0.000550876 | 0 | 0 |
| 0 | 0 | 0 | 0 | 0 |
| 0 | 0 | 0.013602974 | 1.596528658 | 0.277830379 |

| | | | | |
|---|---|---|---|---|
| 0 | 0 | 0 | 0 | 0 |
| 0 | 0 | 0 | 0 | 0 |
| 0.002591736 | 0 | 0.001145854 | 0 | 0 |
| 0 | 0 | 0 | 0 | 0 |
| 0.03792418 | 0 | 0 | 0.001396294 | 0 |
| 0 | 0 | 0.012627407 | 0 | 0.649630255 |
| 0 | 0 | 0.080269598 | 0 | 0 |
| 0 | 0 | 0.000260866 | 0 | 0 |
| 0 | 0 | 0 | 0 | 0 |
| 0 | 0 | 0 | 0 | 0 |
| 0 | 0 | 0 | 0 | 0 |
| 1.242871177 | 1.690184529 | 0.035240171 | 0.001674621 | 0.036359542 |
| 0 | 0 | 0.016370355 | 1.698951085 | 0 |
| 0 | 0 | 0 | 0 | 0 |
| 0.069084099 | 0.024914953 | 0.267976459 | 3.403865531 | 0 |
| 0 | 0 | 0 | 0 | 0 |
| 0 | 0 | 0 | 0 | 0 |
| 0 | 0 | 0 | 0 | 0 |
| 0.01314615 | 0 | 0 | 0 | 0 |
| 0.157033853 | 0 | 0.009337567 | 0 | 0 |
| 0.015830992 | 0 | 0 | 0 | 0 |
| 0 | 0 | 0 | 0 | 0 |
| 0.155524615 | 0 | 0.042948424 | 0 | 0.059530287 |
| 0 | 0 | 0 | 0 | 0 |
| 0 | 0 | 0 | 0 | 0 |
| 0 | 0 | 0 | 0 | 0 |
| 0 | 0 | 0 | 0 | 0 |
| 0 | 0 | 0 | 0 | 0 |
| 0 | 0 | 0 | 0 | 0 |
| 0 | 0 | 0 | 0 | 0 |
| 0 | 0 | 0 | 0 | 0 |
| 0.00943645 | 0 | 0.00281123 | 0.011105866 | 0.038406176 |
| 0 | 0 | 0 | 0 | 0 |
| 0.015910722 | 0.003691847 | 0 | 0 | 0 |
| 0.002290036 | 0.008727733 | 0.001977475 | 0.001491666 | 0.001491265 |
| 0.011316515 | 0.020498591 | 0.002549205 | 0.047024589 | 0.019224209 |
| 0.002202193 | 0 | 0.005687311 | 0 | 0 |
| 0 | 0 | 0.001437103 | 0 | 0 |
| 0 | 0 | 0 | 0 | 0.019987836 |
| 0 | 0 | 0 | 0 | 0 |
| 0.037233857 | 0 | 0.074199286 | 0 | 0.124091645 |
| 0.040809902 | 0 | 0 | 0 | 0 |
| 0 | 0 | 0 | 0 | 0 |
| 0 | 0 | 0 | 0 | 0 |
| 0.05978175 | 0.007184373 | 0.003428499 | 0.129043611 | 0 |
| 0 | 0 | 0 | 0 | 0 |
| 0.027680276 | 0.37913584 | 0 | 0.011127188 | 0 |
| 0 | 0 | 0.001916168 | 0 | 0 |

| | | | | |
|---|---|---|---|---|
| 0 | 0 | 0.300791156 | 0 | 0 |
| 0 | 0 | 0 | 0 | 0 |
| 0.00563852 | 0 | 0 | 0 | 0 |
| 0 | 0 | 0 | 0 | 0 |
| 0 | 0 | 0 | 0 | 0 |
| 0 | 0 | 0 | 0 | 0 |
| 0 | 0 | 0.152848964 | 0 | 0 |
| 0.006977659 | 0 | 0.008828643 | 0 | 0 |
| 0 | 0 | 0 | 0 | 0 |
| 0.00108415 | 0 | 0 | 0 | 0 |
| 0 | 0 | 0 | 0 | 0 |
| 0 | 0.000495757 | 0 | 0 | 0 |
| 0 | 0 | 0 | 0 | 0 |
| 0 | 0 | 0 | 0 | 0.00325736 |
| 0 | 0 | 0 | 0 | 0 |
| 0 | 0 | 0 | 0 | 0 |
| 0.001634551 | 0 | 0 | 0 | 0 |
| 0.003408205 | 0 | 0 | 0 | 0 |
| 1.128326519 | 8.35266088 | 2.049643373 | 0.030264028 | 0 |
| 0 | 0 | 0 | 0 | 0 |
| 0.001943245 | 0 | 0 | 0 | 0 |
| 0 | 0 | 0 | 0 | 0 |
| 0 | 0 | 0 | 0 | 0 |
| 0 | 0 | 0 | 0 | 0 |
| 0 | 0 | 0 | 0.001751445 | 0.002675099 |
| 0 | 0.020832657 | 0 | 0.293123753 | 0.013676255 |
| 0 | 0 | 0 | 0 | 0 |
| 0.005995647 | 0 | 0 | 0.35945897 | 0 |
| 0 | 0 | 0 | 0 | 0 |
| 0.12164693 | 0 | 0.258845159 | 0.020732209 | 0 |
| 0 | 0 | 0 | 0 | 0 |
| 0 | 0 | 0 | 0 | 0 |
| 0.002170589 | 0 | 0.00232417 | 0 | 0 |
| 0 | 0 | 0 | 0 | 0 |
| 0 | 0.000459607 | 0.013075175 | 0.002606496 | 0 |
| 0 | 0 | 0 | 0 | 0 |
| 0 | 0 | 0 | 0 | 0 |
| 0.009573061 | 0.001303799 | 0 | 0.014788048 | 0 |
| 0.037472695 | 0 | 0 | 0 | 0 |
| 0 | 0 | 0 | 0 | 0 |
| 0.560728563 | 6.003248261 | 1.334716703 | 1.207964517 | 1.03230003 |
| 0.066419138 | 0.786796178 | 0.046914319 | 0.248215373 | 0.012697818 |
| 0.331140476 | 0 | 0.075667065 | 0 | 0.261745955 |
| 0 | 0 | 0 | 0 | 0 |
| 0 | 0 | 0 | 0 | 0 |
| 0 | 0 | 0 | 0 | 0 |
| 0 | 0 | 0 | 0 | 0 |
| 0.012387571 | 0 | 0 | 0 | 0 |

| | | | | |
|---|---|---|---|---|
| 0 | 0 | 0 | 0 | 0 |
| 0.013421194 | 0 | 0.003802487 | 0 | 0 |
| 0 | 0 | 0 | 0 | 0 |
| 0 | 0 | 0 | 0 | 0 |
| 0.132184074 | 0 | 0.777931869 | 0 | 0.596806573 |
| 0 | 0 | 0 | 0 | 0 |
| 0 | 0 | 0 | 0 | 0 |
| 0.002161511 | 0 | 0.000821256 | 0 | 0 |
| 0 | 0.079920585 | 0.004851569 | 0.217820789 | 0 |
| 0 | 0 | 0.000512215 | 0 | 0 |
| 0 | 0 | 0 | 0 | 0 |
| 0 | 0 | 0 | 0 | 0 |
| 0.02360582 | 0 | 0.05692673 | 0 | 0 |
| 0.001420606 | 0.188426238 | 0.0063088 | 0.002163141 | 0.003700378 |
| 0 | 0 | 0.011789065 | 0 | 3.403544481 |
| 0.064615256 | 0 | 0.076204791 | 0 | 0.01474873 |
| 0 | 0 | 0 | 0.017345929 | 0 |
| 0 | 0 | 0 | 0 | 0 |
| 0 | 0 | 0 | 0 | 0 |
| 0 | 0 | 0 | 0 | 0 |
| 0 | 0 | 0 | 0 | 0 |
| 0 | 0 | 0 | 0 | 0 |
| 0 | 0 | 0 | 0 | 0 |
| 0 | 0 | 0 | 0 | 0 |
| 0.007703062 | 0 | 0 | 0 | 0 |
| 0 | 0 | 0 | 0 | 0 |
| 0.042910278 | 0 | 0.010190389 | 0.007704265 | 0 |
| 0 | 0.004318083 | 0 | 0.071628733 | 0 |
| 0 | 0 | 0 | 0 | 0 |
| 0 | 0 | 0 | 0 | 0 |
| 0 | 0 | 0 | 0 | 0 |
| 0 | 0 | 0.000656239 | 0 | 0 |
| 0.007243755 | 0 | 0 | 0 | 0 |
| 0.0057801 | 0 | 0 | 0 | 0 |
| 0 | 0 | 0.065437487 | 0 | 1.953779367 |
| 0 | 0 | 0.091872094 | 0.001274763 | 0 |
| 0 | 0.000382922 | 0.488184363 | 0.083099958 | 0 |
| 0.00075289 | 0 | 0.005695136 | 0 | 0 |
| 0 | 0 | 0 | 0 | 0 |
| 0 | 0 | 0 | 0 | 0 |
| 0.228501828 | 8.301779283 | 0.15269082 | 7.838547677 | 0.200307292 |
| 0 | 0 | 0 | 0 | 0.002102836 |
| 0.001483115 | 0 | 0 | 0 | 0 |
| 0.011810062 | 0 | 0 | 0 | 0 |
| 0 | 0 | 0 | 0 | 0 |
| 0 | 0 | 0 | 0 | 0 |
| 0.011680983 | 0 | 0 | 0 | 0 |
| 0 | 0 | 0 | 0 | 0 |

| | | | | |
|---|---|---|---|---|
| 0.014149848 | 0 | 0.00203317 | 0 | 0 |
| 0 | 0 | 0 | 0 | 0 |
| 0 | 0 | 0 | 0 | 0 |
| 0 | 0 | 0 | 0 | 0 |
| 0.020543957 | 0 | 0.088712717 | 0 | 0 |
| 0 | 0 | 0 | 0 | 0 |
| 0 | 0 | 0.001089203 | 0.984446515 | 0 |
| 0 | 0 | 0 | 0 | 0 |
| 0 | 0 | 0 | 0 | 0 |
| 0 | 0 | 0.014474845 | 0 | 0 |
| 0.008497829 | 0 | 0.005459458 | 0 | 0 |
| 0 | 0 | 0.004953147 | 0 | 0 |
| 0 | 0 | 0 | 0 | 0 |
| 0 | 0 | 0 | 0 | 0 |
| 0.001032828 | 0 | 0 | 0 | 0 |
| 0 | 0 | 0 | 0 | 0 |
| 0 | 0 | 0 | 2.233505631 | 0 |
| 0 | 0 | 0 | 0 | 0 |
| 0 | 0 | 0 | 0 | 0 |
| 0 | 0 | 0 | 0 | 0 |
| 0.024684917 | 0 | 0.236362688 | 0.035555688 | 0 |
| 0 | 0 | 0 | 0 | 0 |
| 0.006990813 | 0 | 0.018986867 | 0.003791054 | 0 |
| 0.000817935 | 0.003279564 | 0.000474631 | 0.008950704 | 0 |
| 0 | 0 | 0.000486369 | 0 | 0 |
| 0.005210887 | 0 | 0 | 0 | 0 |
| 0 | 0 | 0 | 0 | 0 |
| 0 | 0 | 0 | 0 | 0 |
| 0.016041107 | 0 | 0 | 0 | 0 |
| 2.977373526 | 0.427018044 | 0.002732424 | 0 | 0 |
| 0.022323615 | 0 | 0 | 0 | 0 |
| 0 | 0 | 0 | 0 | 0 |
| 0 | 0 | 0 | 0 | 0 |
| 0 | 0 | 0 | 0 | 0 |
| 0.028075959 | 0 | 0 | 0 | 0 |
| 0 | 0 | 0 | 0 | 0 |
| 0.011462619 | 0 | 0 | 0.0043635 | 0 |
| 0 | 0 | 0 | 0 | 0 |
| 0 | 0 | 0 | 0 | 0 |
| 0 | 0 | 0 | 0 | 0 |
| 0 | 0 | 0 | 0 | 0 |
| 0 | 0 | 0 | 0 | 0 |
| 0 | 0 | 0 | 0 | 0 |
| 0 | 0 | 0 | 0 | 0 |
| 0.019178604 | 0 | 0 | 0 | 0 |
| 0 | 0 | 0 | 0 | 0 |
| 0 | 0 | 0 | 0 | 0 |
| 0 | 0 | 0 | 0 | 0.222590104 |

| | | | | |
|---|---|---|---|---|
| 0.002288329 | 0.025915559 | 0.00029184 | 0 | 0 |
| 0 | 0 | 0.018267477 | 0 | 0 |
| 0.002719452 | 0 | 0 | 0 | 0 |
| 0 | 0 | 0 | 0 | 0 |
| 0 | 0 | 0 | 0 | 0 |
| 0 | 0 | 0 | 0 | 0 |
| 0.00094724 | 0 | 0 | 0 | 0 |
| 0.046449729 | 0 | 0.264744205 | 0.005650939 | 0.028247088 |
| 31.15538654 | 1.635962536 | 0.547299393 | 0.297249976 | 0 |
| 0 | 0.038200598 | 0.002630978 | 0.026379622 | 0 |
| 0.387335427 | 0 | 0 | 0 | 0 |
| 0 | 0 | 0 | 0 | 0 |
| 0.006663238 | 0 | 0 | 0 | 0 |
| 0 | 0.001537782 | 0.656894421 | 0.363228766 | 0 |
| 0 | 0 | 0 | 0 | 0 |
| 0 | 0 | 0 | 0 | 0 |
| 0 | 0 | 0 | 0 | 0 |
| 0 | 0 | 0 | 0 | 0 |
| 0 | 0 | 0 | 0 | 0 |
| 0.18757398 | 0 | 0.000198334 | 0 | 0 |
| 0 | 0 | 0 | 0 | 0 |
| 0 | 0 | 0 | 0 | 0 |
| 0 | 0 | 0 | 0 | 0 |
| 0 | 0 | 0 | 0 | 0 |
| 0.003466109 | 0 | 0.012470154 | 0 | 0 |
| 0 | 0 | 0 | 0 | 0 |
| 0 | 0 | 0 | 0 | 0 |
| 0.058253684 | 0 | 0 | 0 | 0 |
| 0 | 0 | 0 | 0 | 0 |
| 0 | 0 | 0 | 0 | 0 |
| 0.010488474 | 0.002379146 | 0 | 0 | 0 |
| 0 | 0 | 0 | 0 | 0 |
| 0 | 0 | 0 | 0 | 0 |
| 0 | 0 | 0.000464111 | 0 | 0 |
| 0 | 0 | 0 | 0 | 0.01665268 |
| 0.01815518 | 0 | 0 | 0 | 0 |
| 0 | 0 | 0 | 0 | 0 |
| 0 | 0 | 0 | 0 | 0 |
| 0.000837232 | 0 | 0 | 0 | 0.04906824 |
| 0 | 0.200148985 | 0 | 0 | 0 |
| 0 | 0 | 0 | 0 | 0 |
| 0.040972681 | 0.150721214 | 0.117225044 | 0 | 0 |
| 0 | 0 | 0 | 0 | 0 |
| 0 | 0 | 0 | 0 | 0 |
| 0 | 0 | 0 | 0 | 0 |
| 0 | 0 | 0 | 0 | 0 |
| 0 | 0 | 0 | 0 | 0 |
| 0 | 0 | 0 | 0 | 0 |

| | | | | |
|---|---|---|---|---|
| 0 | 0 | 0 | 0 | 0 |
| 0 | 0 | 0.092189477 | 0.011176261 | 0 |
| 0 | 0 | 0 | 0 | 0 |
| 0 | 0 | 0 | 0 | 0 |
| 0.514215657 | 0.247981632 | 0.137087816 | 1.528821009 | 0.76199558 |
| 0 | 0 | 0 | 0 | 0 |
| 0 | 0 | 0 | 0 | 0 |
| 0.000671751 | 0.026232891 | 0.010116339 | 0.021798443 | 0 |
| 0 | 0 | 0.003614044 | 0 | 0 |
| 0 | 0 | 0 | 0 | 0 |
| 0 | 0 | 0 | 0 | 0 |
| 0 | 0 | 0 | 0 | 0 |
| 0 | 0 | 0 | 0 | 0 |
| 0 | 0 | 0 | 0 | 0 |
| 0 | 0 | 0 | 0 | 0 |
| 0 | 0 | 0.000724444 | 0 | 0 |
| 0 | 0 | 0 | 0 | 0 |
| 0 | 0 | 0 | 0 | 0 |
| 0 | 0 | 0 | 0 | 0 |
| 0 | 0 | 0 | 0 | 0 |
| 0 | 0 | 0 | 0 | 0 |
| 0 | 0 | 0 | 0 | 0 |
| 0 | 0 | 0 | 0 | 0 |
| 0.000968909 | 0 | 0.000321279 | 0 | 0 |
| 0 | 0 | 0.088541557 | 0 | 0.025598423 |
| 0 | 0 | 0 | 0 | 0 |
| 0.065045379 | 0.003331111 | 0.001248847 | 10.07190569 | 0 |
| 0 | 0 | 0 | 0 | 0 |
| 0 | 0 | 0 | 0 | 0 |
| 0 | 0 | 0 | 0 | 0 |
| 0 | 0 | 0 | 0 | 0 |
| 0 | 0 | 0 | 0 | 0 |
| 0 | 0 | 0 | 0 | 0 |
| 0 | 0 | 0 | 0 | 0 |
| 0 | 0 | 0 | 0 | 0 |
| 0 | 0 | 0 | 0 | 0 |
| 0 | 0 | 0 | 0 | 0 |
| 0.008361202 | 0 | 0.065243706 | 0 | 0 |
| 0.003433861 | 0 | 0 | 0 | 0 |
| 0 | 0 | 0 | 0 | 0 |
| 0 | 0 | 0 | 0 | 0 |
| 0 | 0 | 0.002937156 | 0 | 0 |
| 0 | 0 | 0 | 0 | 0 |
| 0 | 0 | 0 | 0 | 0 |
| 0 | 0 | 0 | 0 | 0 |
| 0 | 0 | 0 | 0 | 0 |
| 0 | 0 | 0 | 0 | 0 |
| 0 | 0 | 0 | 0 | 0 |

| | | | | |
|---|---|---|---|---|
| 0.058002756 | 0 | 0.041243316 | 0.107185452 | 0 |
| 0 | 0 | 0.003380957 | 0 | 0 |
| 0 | 0 | 0 | 0 | 0 |
| 0 | 0 | 0 | 0 | 0 |
| 0 | 0 | 0 | 0 | 0 |
| 0 | 0 | 0.079477871 | 0 | 0 |
| 0 | 0 | 0 | 0 | 0 |
| 0 | 0 | 0 | 0 | 0 |
| 0 | 0 | 0 | 0 | 0 |
| 0.044364891 | 0 | 0 | 0 | 0 |
| 0 | 0 | 0 | 0 | 0 |
| 0 | 0 | 0 | 0 | 0 |
| 0.001242141 | 0 | 0 | 0 | 0 |
| 0 | 0 | 0 | 0 | 0 |
| 0 | 0 | 0 | 0 | 0 |
| 0 | 0 | 0 | 0 | 0 |
| 0 | 0 | 0 | 0 | 0 |
| 0 | 0 | 0.018793442 | 0 | 0 |
| 0.000565241 | 0 | 0 | 0 | 0 |
| 0 | 0 | 0 | 0 | 0 |
| 0 | 0.099255406 | 0.005598883 | 0.325256556 | 0 |
| 0 | 0 | 0 | 0 | 0 |
| 0.101870418 | 0.000763413 | 0.000404056 | 0 | 0.002539242 |
| 0.015390816 | 0 | 0.83023061 | 0 | 0 |
| 0.01619863 | 0 | 0 | 0 | 0 |
| 0 | 0 | 0 | 0 | 0 |
| 0 | 0 | 0 | 0 | 0 |
| 0 | 0 | 0 | 0 | 0 |
| 0 | 0 | 0 | 0.001499161 | 0 |
| 0 | 0 | 0 | 0 | 0 |
| 0 | 0 | 0 | 0 | 0 |
| 0 | 0 | 0 | 0 | 0 |
| 0 | 0.798293911 | 0.000662195 | 0.092928767 | 0 |
| 0 | 0 | 0 | 0 | 0 |
| 0 | 0 | 0.01226969 | 1.997595406 | 0 |
| 0 | 0 | 0 | 0 | 0 |
| 0 | 0 | 0 | 0 | 0 |
| 0 | 0 | 0.020601546 | 0 | 0 |
| 0 | 0.002388884 | 0.014487681 | 0.2200748 | 0 |
| 0 | 0 | 0 | 0 | 0 |
| 0 | 0 | 0 | 0 | 0 |
| 0.005573719 | 0 | 0.00188466 | 0 | 0 |
| 0.278417927 | 0.061763439 | 0.061396262 | 0 | 1.307847208 |
| 0 | 0 | 0 | 0 | 0 |
| 0 | 0 | 0 | 0 | 0 |
| 0.003540941 | 0 | 0.017612048 | 0 | 0 |
| 0 | 0 | 0 | 0 | 0 |
| 0 | 0 | 0 | 0 | 0 |

| | | | | |
|---|---|---|---|---|
| 0 | 0 | 0 | 0 | 0 |
| 0 | 0 | 0 | 0 | 0 |
| 0 | 0 | 0 | 0 | 0 |
| 0 | 0 | 0 | 0 | 0 |
| 0.003506069 | 0 | 0 | 0 | 0 |
| 0 | 0 | 0 | 0 | 0 |
| 0 | 0 | 0 | 0 | 0 |
| 0 | 0 | 0 | 0 | 0 |
| 0.003870456 | 0 | 0.033918426 | 0 | 0 |
| 0 | 0 | 0.008062318 | 0.012958036 | 0.008444446 |
| 0 | 0 | 0 | 0 | 0 |
| 0 | 0 | 0.013335725 | 0 | 0 |
| 0 | 0 | 0 | 0 | 0 |
| 0.018124133 | 0 | 0 | 0 | 0 |
| 0 | 0 | 0.001018039 | 0 | 0 |
| 0 | 0.018109674 | 0 | 0 | 0 |
| 0 | 0 | 0 | 0 | 0 |
| 0 | 0 | 0 | 0 | 0 |
| 0 | 0 | 0 | 0 | 0 |
| 0.117167687 | 0 | 0.209477061 | 0.000830208 | 0 |
| 0 | 0.013957426 | 0 | 0.017014523 | 0.003796422 |
| 0 | 0 | 0 | 0 | 0 |
| 0 | 0 | 0 | 0 | 0 |
| 0.020977955 | 0 | 0 | 0 | 0 |
| 0 | 0 | 0 | 0 | 0 |
| 0 | 0 | 0 | 0 | 0 |
| 0 | 0 | 0 | 0 | 0 |
| 0 | 0 | 0 | 0 | 0 |
| 0 | 0 | 0 | 0 | 0 |
| 0.17686261 | 0 | 0.299394314 | 0 | 0 |
| 0.041317496 | 0 | 0.160308329 | 0 | 0 |
| 0 | 0 | 0 | 0 | 0 |
| 0 | 0 | 0 | 0 | 0 |
| 0 | 0 | 0 | 0 | 0 |
| 0 | 0 | 0 | 0 | 0 |
| 0 | 0 | 0 | 0 | 0 |
| 0 | 0 | 0 | 0 | 0 |
| 0 | 0 | 0 | 0 | 0 |
| 0 | 0 | 0.00938565 | 0 | 0 |
| 0 | 0 | 0 | 0 | 0 |
| 0 | 0 | 0 | 0 | 0 |
| 0 | 0 | 0.311958423 | 0 | 0 |
| 0 | 0 | 0 | 0 | 0 |
| 0 | 0 | 0.000706081 | 0 | 0 |
| 0 | 0 | 0 | 0 | 0 |
| 0 | 0 | 0 | 0 | 0 |
| 0 | 0.058556852 | 0 | 0.191129629 | 0 |
| 0 | 0 | 0.000824927 | 0 | 0 |

| | | | | |
|---|---|---|---|---|
| 0 | 0 | 0 | 0 | 0 |
| 0 | 0 | 0 | 0 | 0 |
| 0 | 0 | 0 | 0 | 0 |
| 0 | 0 | 0.001743613 | 0 | 0 |
| 0.004878752 | 0 | 0.219658683 | 0 | 0 |
| 0.063043849 | 0 | 0 | 0 | 0 |
| 0 | 0 | 0 | 0 | 0 |
| 0 | 0 | 0 | 0 | 0 |
| 0 | 0 | 0 | 0 | 0 |
| 0 | 0 | 0 | 0 | 0 |
| 0.007162182 | 0 | 0 | 0 | 0 |
| 0.015875243 | 0.003447863 | 0.000614139 | 0.342784614 | 0 |
| 0 | 0 | 0 | 0 | 0 |
| 0 | 0 | 0 | 0 | 0 |
| 0 | 0 | 0 | 0 | 0 |
| 0.000658413 | 0 | 0.025516444 | 0 | 0 |
| 0.014977534 | 0 | 0.016059769 | 0 | 0 |
| 0 | 0 | 0 | 0 | 0 |
| 0 | 0.485296675 | 0.001129358 | 0 | 0 |
| 0 | 0 | 0.001040893 | 0.1656601 | 0 |
| 0 | 0 | 0 | 0 | 0 |
| 0 | 0 | 0 | 0 | 0 |
| 0 | 0 | 0 | 0 | 0 |
| 0 | 0 | 0 | 0 | 0 |
| 0 | 0 | 0 | 0 | 0 |
| 0 | 0 | 0 | 0 | 0 |
| 0 | 0.000945573 | 0.000347548 | 0.002025824 | 0 |
| 0 | 0 | 0.013533736 | 0 | 0 |
| 0 | 0 | 0 | 0 | 0 |
| 0 | 0 | 0 | 0 | 0 |
| 0.001900975 | 0.001071856 | 0 | 0 | 0 |
| 0.00082131 | 0 | 0.000425527 | 0 | 0 |
| 0 | 0 | 0 | 0 | 0 |
| 0 | 0 | 0 | 0 | 0 |
| 0.000526178 | 0 | 0 | 0 | 0 |
| 0 | 0 | 0 | 0 | 0 |
| 0 | 0 | 0 | 0 | 0 |
| 0.02631073 | 0.01656721 | 0 | 0.316694677 | 0.003478877 |
| 0.1559831 | 0 | 0.009055388 | 0 | 0 |
| 0 | 0 | 0 | 0 | 0 |
| 0.008533308 | 0.000501194 | 0.002597438 | 0.187878921 | 0 |
| 0 | 0 | 0 | 0 | 0 |
| 0 | 0 | 0 | 0 | 0 |
| 0 | 0.01215946 | 0 | 0 | 0 |
| 0 | 0 | 0 | 0 | 0 |
| 0 | 0 | 0.00550405 | 0 | 0 |
| 0 | 0 | 0 | 0 | 0 |
| 0 | 0 | 0 | 0 | 0 |

| | | | | |
|---|---|---|---|---|
| 0 | 0 | 0 | 0 | 0 |
| 0.023190404 | 0 | 0 | 0.109939012 | 0 |
| 0 | 0 | 0 | 0 | 0 |
| 0 | 0.001820453 | 0.000344115 | 0.013716227 | 0 |
| 0 | 0 | 0 | 0.114494233 | 0.006518625 |
| 0 | 0 | 0 | 0 | 0 |
| 0 | 0 | 0 | 0 | 0 |
| 0 | 0 | 0 | 0 | 0 |
| 0 | 0 | 0 | 0 | 0 |
| 0.009593689 | 0 | 0.017396967 | 0 | 0 |
| 0 | 0 | 0 | 0 | 0 |
| 0 | 0 | 0 | 0 | 0 |
| 0.089453519 | 0.013734019 | 0.113610001 | 0.045791602 | 0.060168042 |
| 0 | 0 | 0 | 0 | 0 |
| 0 | 0 | 0 | 0 | 0 |
| 0 | 0 | 0 | 0 | 0 |
| 0 | 0 | 0 | 0 | 0 |
| 0 | 0 | 0 | 0 | 0 |
| 0 | 0 | 0 | 0 | 0 |
| 0 | 0 | 0 | 0 | 0 |
| 0 | 0 | 0 | 0 | 0 |
| 0 | 0 | 0 | 0 | 0 |
| 0 | 0 | 0.003693782 | 0 | 0 |
| 0.03109118 | 0.028358353 | 0.074099437 | 0 | 0 |
| 0 | 0 | 0 | 0 | 0 |
| 0.011142904 | 0 | 0 | 0 | 0 |
| 0.000555285 | 0 | 0 | 0 | 0 |
| 0 | 0 | 0 | 0 | 0 |
| 0 | 0 | 0 | 0 | 0 |
| 0 | 0 | 0 | 0 | 0 |
| 0.003240172 | 0 | 0.007073164 | 0 | 0 |
| 0 | 0 | 0 | 0 | 0 |
| 0 | 0 | 0 | 0 | 0 |
| 0 | 0 | 0 | 0 | 0 |
| 0.002535388 | 0 | 0.006530486 | 0 | 0 |
| 0 | 0 | 0 | 0 | 0 |
| 0 | 0 | 0 | 0 | 0 |
| 0 | 0 | 0 | 0 | 0 |
| 0.00274141 | 0 | 0 | 0.012077681 | 0 |
| 0 | 0 | 0 | 0 | 0 |
| 0 | 0 | 0 | 0 | 0 |
| 0 | 0 | 0 | 0 | 0 |
| 0.021599764 | 0 | 0 | 0 | 0 |
| 0 | 0 | 0 | 0 | 0 |
| 0 | 0 | 0 | 0 | 0 |
| 0.068018059 | 0.079987456 | 0.174553767 | 0.521948723 | 0.086282648 |
| 0 | 0 | 0 | 0 | 0 |
| 0 | 0 | 0 | 0 | 0 |

| | | | | |
|---|---|---|---|---|
| 0.008837038 | 0 | 0 | 0 | 0 |
| 0 | 0 | 0 | 0 | 0 |
| 0 | 0 | 0.074584029 | 0 | 0 |
| 0.000971091 | 0 | 0 | 0 | 0 |
| 0 | 0 | 0 | 0 | 0 |
| 0 | 0 | 0.000245442 | 0 | 0 |
| 0 | 0 | 0 | 0 | 0 |
| 0 | 0 | 0 | 0 | 0 |
| 0 | 0 | 0 | 0 | 0 |
| 0 | 0 | 0 | 0 | 0 |
| 0.00207575 | 0 | 0 | 0 | 0 |
| 0 | 0 | 0 | 0.002674081 | 0 |
| 0 | 0 | 0 | 0 | 0 |
| 0 | 0 | 0 | 0 | 0 |
| 0 | 0 | 0 | 0 | 0 |
| 0 | 0 | 0 | 0 | 0 |
| 0 | 0 | 0 | 0.000464612 | 0 |
| 0 | 0 | 0 | 0 | 0 |
| 0 | 0 | 0 | 0 | 0 |
| 0.002162915 | 0 | 0 | 0 | 0 |
| 0 | 0 | 0 | 0 | 0 |
| 0 | 0 | 0 | 0 | 0 |
| 0 | 0 | 0 | 0 | 0 |
| 0 | 0 | 0 | 0 | 0 |
| 0 | 0 | 0 | 0 | 0 |
| 0 | 0 | 0 | 0 | 0 |
| 0 | 0 | 0 | 0 | 0 |
| 0 | 0 | 0.01109378 | 0 | 0.014170232 |
| 0 | 0 | 0 | 0 | 0 |
| 0 | 0 | 0 | 0 | 0 |
| 0.000675697 | 0 | 0.000910218 | 0 | 0.013200364 |
| 0 | 0 | 0 | 0 | 0 |
| 0.018633147 | 0.042393494 | 0.018779518 | 0.0073868 | 0.026566743 |
| 0 | 0 | 0 | 0 | 0 |
| 0 | 0 | 0.007824654 | 0 | 0 |
| 0 | 0 | 0 | 0 | 0 |
| 0 | 0 | 0 | 0 | 0 |
| 0 | 0 | 0 | 0 | 0 |
| 0 | 0 | 0 | 0 | 0 |
| 0 | 0 | 0 | 0 | 0 |
| 0 | 0 | 0 | 0 | 0 |
| 0 | 0.037606227 | 0.016252323 | 0.032334511 | 0 |
| 0.060154559 | 0.066998262 | 0.020533242 | 0 | 0.002560292 |
| 0 | 0 | 0 | 0 | 0 |
| 0 | 0 | 0 | 0 | 0 |
| 0.025850995 | 0.012636061 | 0.022842701 | 0.064797578 | 0.125053239 |
| 0 | 0.00058342 | 0 | 0 | 0 |
| 0 | 0 | 0 | 0 | 0 |

| | | | | |
|---|---|---|---|---|
| 0 | 0 | 0 | 0 | 0 |
| 0.00226542 | 0 | 0.090654739 | 0 | 0 |
| 0.020688866 | 0.025621507 | 0.000398849 | 0.002906063 | 0.006892921 |
| 0 | 0 | 0 | 0 | 0 |
| 0.201686245 | 0 | 0 | 0 | 0 |
| 0 | 0.015813461 | 0 | 0.101239153 | 0 |
| 0 | 0 | 0 | 0 | 0 |
| 0 | 0 | 0 | 0 | 0 |
| 0 | 0 | 0 | 0 | 0 |
| 0 | 0 | 0 | 0 | 0 |
| 0 | 0 | 0 | 0 | 0 |
| 0 | 0 | 0 | 0 | 0 |
| 0 | 0 | 0 | 0 | 0 |
| 0 | 0 | 0 | 0 | 0 |
| 0 | 0 | 0 | 0 | 0 |
| 0 | 0 | 0 | 0 | 0 |
| 0.01863942 | 0 | 0 | 0 | 0 |
| 0 | 0 | 0 | 0 | 0 |
| 0 | 0 | 0 | 0 | 0 |
| 0 | 0 | 0 | 0 | 0 |
| 0 | 0 | 0 | 0 | 0 |
| 0 | 0 | 0 | 0 | 0 |
| 0 | 0 | 0 | 0 | 0 |
| 0 | 0 | 0 | 0 | 0 |
| 0.004560027 | 0 | 0.003451427 | 0 | 0.015492916 |
| 0 | 0.01062096 | 0 | 0.001133797 | 0 |
| 0 | 0 | 0 | 0 | 0 |
| 0.001160823 | 0 | 0.079789843 | 0 | 0 |
| 0 | 0 | 0 | 0 | 0 |
| 0 | 0 | 0 | 0 | 0 |
| 0 | 0 | 0 | 0 | 0 |
| 0 | 0 | 0 | 0 | 0 |
| 0 | 0 | 0 | 0 | 0 |
| 0 | 0 | 0 | 0 | 0 |
| 0 | 0 | 0 | 0 | 0 |
| 0 | 0 | 0 | 0 | 0 |
| 0 | 0 | 0 | 0 | 0 |
| 0 | 0.002736353 | 0 | 0 | 0 |
| 0.009959517 | 0 | 0.00225168 | 0 | 0 |
| 0 | 0 | 0 | 0 | 0 |
| 0 | 0 | 0 | 0 | 0 |
| 0 | 0 | 0.005201555 | 0 | 0 |
| 0.002802334 | 0.022572619 | 0 | 0 | 0.002085568 |
| 0 | 0 | 0 | 0 | 0 |
| 0 | 0 | 0 | 0 | 0 |
| 0 | 0 | 0.001527197 | 0 | 0 |
| 0.005205671 | 0.001342899 | 0.002792289 | 0.000870374 | 0 |
| 0 | 0 | 0 | 0 | 0 |

| | | | | |
|---|---|---|---|---|
| 0 | 0 | 0 | 0 | 0 |
| 0 | 0.010530055 | 0.001841754 | 0.09046845 | 0 |
| 0.001145368 | 0 | 0 | 0 | 0 |
| 0 | 0 | 0 | 0 | 0 |
| 0 | 0 | 0 | 0 | 0 |
| 0 | 0 | 0 | 0 | 0 |
| 0 | 0 | 0 | 0 | 0 |
| 0 | 0 | 0 | 0 | 0 |
| 0 | 0 | 0 | 0 | 0 |
| 0 | 0 | 0 | 0 | 0 |
| 0 | 0 | 0 | 0 | 0 |
| 0 | 0 | 0 | 0 | 0 |
| 0.056774073 | 0 | 0.004740271 | 0 | 0.162779255 |
| 0 | 0 | 0 | 0 | 0 |
| 0.007708286 | 0.001961863 | 0.025659654 | 0 | 0 |
| 0 | 0 | 0 | 0 | 0 |
| 0 | 0 | 0 | 0 | 0 |
| 0 | 0 | 0 | 0 | 0 |
| 0 | 0 | 0 | 0 | 0 |
| 0 | 0 | 0 | 0 | 0 |
| 0 | 0 | 0 | 0 | 0 |
| 0 | 0 | 0 | 0.007396475 | 0 |
| 0.005089907 | 0 | 0.004822161 | 0 | 0 |
| 0 | 0 | 0 | 0 | 0 |
| 0 | 0 | 0 | 0 | 0 |
| 0 | 0 | 0 | 0 | 0 |
| 0 | 0 | 0 | 0 | 0 |
| 0 | 0 | 0 | 0 | 0 |
| 0 | 0 | 0 | 0 | 0 |
| 0 | 0 | 0 | 0 | 0 |
| 0 | 0 | 0 | 0 | 0 |
| 0 | 0 | 0 | 0 | 0 |
| 0 | 0 | 0 | 0 | 0 |
| 0 | 0 | 0 | 0 | 0 |
| 5.028226767 | 0.003633245 | 7.489690399 | 0 | 0.029652526 |
| 0 | 0 | 0 | 0 | 0 |
| 0 | 0 | 0 | 0 | 0 |
| 0 | 0 | 0 | 0 | 0 |
| 0 | 0 | 0.003525066 | 0.008057754 | 0 |
| 0 | 0 | 0 | 0 | 0 |
| 0.029231315 | 0.070997092 | 0.016769206 | 0.006472255 | 0.266494821 |
| 0 | 0 | 0 | 0 | 0 |
| 0 | 0 | 0 | 0 | 0 |
| 0 | 0 | 0 | 0 | 0 |
| 0 | 0 | 0 | 0 | 0 |
| 0.033710387 | 0.973314933 | 0.044531008 | 3.592612539 | 0 |
| 0 | 0 | 0 | 0 | 0 |
| 0 | 0 | 0 | 0 | 0 |

| | | | | |
|---|---|---|---|---|
| 0 | 0 | 0.003760311 | 0 | 0 |
| 0 | 0 | 0 | 0 | 0 |
| 0 | 0 | 0 | 0 | 0 |
| 0 | 0 | 0 | 0 | 0 |
| 0 | 0 | 0 | 0 | 0 |
| 0 | 0 | 0 | 0 | 0 |
| 0 | 0 | 0 | 0 | 0 |
| 0 | 0 | 0 | 0.001676402 | 0 |
| 0 | 0 | 0 | 0 | 0 |
| 0.005218085 | 0 | 0.005112128 | 0.084102115 | 0 |
| 0.023763888 | 0 | 0 | 0 | 0 |
| 0 | 0 | 0 | 0 | 0 |
| 0.001390974 | 0 | 0.004612315 | 0 | 0 |
| 0 | 0 | 0 | 0 | 0 |
| 0 | 0 | 0 | 0 | 0 |
| 0 | 0 | 0 | 0 | 0 |
| 0 | 0 | 0 | 0 | 0 |
| 0 | 0 | 0 | 0 | 0 |
| 0 | 0 | 0 | 0 | 0 |
| 0.0891407 | 0.061663502 | 0.013684286 | 7.503278158 | 0.251004524 |
| 0 | 0 | 0 | 0 | 0 |
| 0 | 0 | 0 | 0 | 0 |
| 0 | 0 | 0 | 0 | 0 |
| 0 | 0 | 0 | 0 | 0 |
| 0 | 0 | 0 | 0 | 0 |
| 0 | 0 | 0.002393341 | 0 | 0 |
| 0 | 0 | 0 | 0 | 0 |
| 0 | 0 | 0 | 0 | 0 |
| 0 | 0 | 0 | 0 | 0 |
| 0 | 0.009811885 | 0.007040073 | 0 | 0 |
| 0 | 0 | 0 | 0 | 0 |
| 0.002311783 | 0 | 0 | 0 | 0 |
| 0 | 0 | 0 | 0 | 0 |
| 0 | 0 | 0 | 0 | 0 |
| 0 | 0 | 0 | 0 | 0 |
| 0 | 0 | 0 | 0.012147432 | 0 |
| 0 | 0 | 0 | 0 | 0 |
| 0 | 0 | 0 | 0 | 0 |
| 0 | 0 | 0 | 0 | 0 |
| 0 | 0 | 0 | 0 | 0 |
| 0 | 0 | 0 | 0 | 0 |
| 0 | 0 | 0 | 0 | 0 |
| 0.005488686 | 0 | 0 | 0 | 0 |
| 0 | 0 | 0 | 0 | 0 |
| 0.007752243 | 0.002549789 | 0.000562309 | 0.045997234 | 0 |
| 0.01381282 | 0 | 0.003122848 | 0 | 0 |
| 0 | 0 | 0 | 0 | 0 |
| 0 | 0 | 0 | 0 | 0 |

| | | | | |
|---|---|---|---|---|
| 0 | 0 | 0 | 0 | 0 |
| 0 | 0 | 0 | 0 | 0 |
| 0 | 0 | 0.008538705 | 0 | 0 |
| 0 | 0 | 0 | 0 | 0 |
| 0 | 0 | 0 | 0 | 0 |
| 0.020400411 | 0.008883076 | 0.007496174 | 0.108876371 | 0.005276187 |
| 0 | 0 | 0 | 0 | 0 |
| 0 | 0 | 0 | 0 | 0 |
| 0 | 0 | 0 | 0 | 0 |
| 0 | 0 | 0 | 0 | 0 |
| 0 | 0 | 0 | 0 | 0 |
| 0 | 0 | 0 | 0 | 0 |
| 0 | 0 | 0 | 0 | 0 |
| 0 | 0 | 0 | 0 | 0 |
| 0 | 0 | 0 | 0 | 0 |
| 0 | 0 | 0 | 0 | 0 |
| 0.019831544 | 0 | 0.004291805 | 0 | 0 |
| 0 | 0 | 0 | 0 | 0 |
| 0 | 0 | 0 | 0 | 0 |
| 0 | 0 | 0 | 0 | 0 |
| 0 | 0.00557031 | 0 | 0 | 0 |
| 0 | 0 | 0 | 0 | 0 |
| 0 | 0 | 0 | 0 | 0 |
| 0 | 0 | 0 | 0 | 0 |
| 0 | 0 | 0 | 0 | 0 |
| 0.000678929 | 0 | 0 | 0 | 0 |
| 0 | 0 | 0 | 0 | 0 |
| 0.006266955 | 0 | 0.026256468 | 0 | 0 |
| 0 | 0 | 0 | 0 | 0 |
| 0.001569369 | 0 | 0.008781499 | 0 | 0 |
| 0 | 0 | 0 | 0 | 0 |
| 0 | 0 | 0 | 0 | 0 |
| 0 | 0 | 0 | 0 | 0 |
| 0 | 0 | 0 | 0 | 0 |
| 0 | 0 | 0 | 0 | 0 |
| 0 | 0 | 0 | 0 | 0 |
| 0 | 0 | 0 | 0 | 0 |
| 0 | 0 | 0 | 0 | 0 |
| 0 | 0 | 0.141453785 | 0 | 0 |
| 0 | 0 | 0 | 0 | 0 |
| 0 | 0 | 0 | 0 | 0 |
| 0 | 0 | 0 | 0 | 0 |
| 0 | 0 | 0 | 0 | 0 |
| 0 | 0 | 0 | 0 | 0 |
| 0 | 0 | 0 | 0 | 0 |
| 0 | 0 | 0 | 0 | 0 |
| 0 | 0 | 0 | 0 | 0 |
| 0 | 0 | 0 | 0 | 0 |

| | | | | |
|---|---|---|---|---|
| 0 | 0 | 0 | 0 | 0 |
| 0.00631742 | 0 | 0 | 0 | 0 |
| 0 | 0 | 0 | 0 | 0 |
| 0 | 0 | 0 | 0 | 0 |
| 0 | 0 | 0 | 0 | 0 |
| 0 | 0 | 0 | 0 | 0 |
| 0 | 0 | 0 | 0 | 0 |
| 0 | 0 | 0 | 0 | 0 |
| 0 | 0 | 0 | 0 | 0 |
| 0 | 0 | 0 | 0 | 0 |
| 0.002757387 | 0 | 0.000615407 | 0.057424501 | 0.055249297 |
| 0 | 0 | 0 | 0 | 0 |
| 0.004218137 | 0 | 0.060933543 | 0 | 0 |
| 0 | 0 | 0 | 0 | 0 |
| 0 | 0 | 0 | 0 | 0 |
| 0 | 0 | 0 | 0 | 0 |
| 0 | 0 | 0 | 0 | 0 |
| 0 | 0 | 0 | 0 | 0 |
| 0 | 0 | 0 | 0 | 0 |
| 0 | 0 | 0.000731555 | 0 | 0 |
| 0 | 0.00134869 | 0 | 0.01453236 | 0.049345746 |
| 0 | 0 | 0 | 0 | 0 |
| 0 | 0 | 0 | 0 | 0 |
| 0.137673972 | 0.465783341 | 0.792538201 | 0.348865988 | 1.174313325 |
| 0 | 0 | 0 | 0 | 0 |
| 0 | 0 | 0 | 0 | 0 |
| 0 | 0 | 0 | 0 | 0 |
| 0 | 0 | 0 | 0 | 0 |
| 0 | 0 | 0 | 0 | 0 |
| 0 | 0 | 0 | 0 | 0 |
| 0 | 0 | 0.001138513 | 0 | 0 |
| 0 | 0 | 0 | 0 | 0 |
| 0 | 0 | 0 | 0 | 0 |
| 0 | 0 | 0 | 0 | 0 |

| PHL018-0191_V3 | PHL018-0003_V2 | PHL018-0224_V3 | PHL018-0004_V2 | PHL018-0185_V3 |
|---|---|---|---|---|
| EYCF | CTRL | EYCF | CTRL | REF |
| 0.096379623 | 0.553912769 | 0.227974478 | 0.003340916 | 1.47707327 |
| 3.743511156 | 6.368494457 | 0.89189163 | 0.370823823 | 2.916937616 |
| 16.94742282 | 0 | 22.05226256 | 0 | 1.045652599 |
| 0 | 0 | 0 | 0 | 0 |
| 0 | 0 | 0 | 0 | 0 |
| 0 | 0 | 0 | 0 | 0 |
| 0 | 0 | 0 | 0 | 0 |
| 0 | 0 | 0 | 0 | 0 |
| 0 | 0 | 0 | 0 | 0 |
| 0 | 0 | 0 | 0 | 0 |
| 0 | 0 | 0 | 0 | 0 |
| 0 | 0 | 0 | 0 | 0 |
| 0 | 0 | 0 | 0 | 0 |
| 0.433816491 | 0 | 0.295382237 | 1.558938567 | 0.363699718 |
| 0 | 0 | 0 | 0 | 0 |
| 0 | 0 | 0 | 0 | 0 |
| 0 | 0 | 0 | 0 | 0 |
| 0 | 0 | 0 | 0 | 0 |
| 0 | 0 | 0.020352436 | 0 | 0 |
| 0 | 0 | 0 | 0 | 0 |
| 0 | 0 | 0 | 0 | 0 |
| 0 | 0 | 0 | 0 | 0 |
| 0 | 0 | 0 | 0 | 0 |
| 0 | 0 | 0 | 0 | 0 |
| 0 | 0 | 0 | 0 | 0 |
| 0 | 0 | 0 | 0 | 0 |
| 0 | 0 | 0 | 0 | 0 |
| 0 | 0 | 0 | 0 | 0 |
| 0 | 0 | 0 | 0 | 0 |
| 0 | 0 | 0 | 0 | 0 |
| 0 | 0 | 0 | 0 | 0 |
| 0 | 0 | 0 | 0 | 0 |
| 0 | 0 | 0 | 0 | 0 |
| 0 | 0 | 0 | 0 | 0 |
| 0 | 0 | 0 | 0 | 0 |
| 1.2818542 | 0 | 0 | 0.781722423 | 0 |
| 0 | 0 | 0 | 0 | 0 |
| 0 | 0 | 0 | 0 | 0 |
| 0 | 0 | 0 | 0 | 0 |
| 0 | 0 | 0 | 0 | 0 |
| 0 | 0 | 0 | 0 | 0 |
| 0.042902174 | 0.007137101 | 0.001498983 | 0 | 0.001554202 |
| 0.045999867 | 4.795637084 | 0.988534281 | 0.008179122 | 0.110560645 |
| 0.006823138 | 0.11140424 | 0 | 0.003721083 | 0.158735477 |
| 0 | 0.002596154 | 0 | 0 | 0 |
| 0 | 0 | 0 | 0 | 0 |

| | | | | |
|---|---|---|---|---|
| 0.005166308 | 0 | 0 | 0 | 0 |
| 0 | 0.145251755 | 0.056881146 | 0 | 0.010063455 |
| 0.013630853 | 0 | 0.019969668 | 0 | 0.021866354 |
| 1.700983501 | 0.949798584 | 0.780816012 | 0.213814034 | 0.577634386 |
| 0 | 0 | 0 | 0 | 0.007419853 |
| 0.075472773 | 0 | 0 | 0 | 0 |
| 0.21722585 | 0 | 0.058306136 | 0 | 0.001505079 |
| 0 | 0 | 0 | 0 | 0 |
| 0.015913375 | 0 | 0 | 0 | 0 |
| 0 | 0 | 0 | 0 | 0 |
| 0 | 0.001139806 | 0 | 0 | 0 |
| 0.819020634 | 0 | 0 | 0 | 0.022128597 |
| 0 | 0 | 0 | 0 | 0.001738485 |
| 0.092761706 | 0 | 0.015767174 | 0 | 0.168869487 |
| 0 | 0 | 0 | 0 | 0 |
| 0 | 0 | 0 | 0 | 0 |
| 0.024669341 | 0 | 0.024940529 | 0 | 0.00911232 |
| 0 | 0 | 0 | 0 | 0 |
| 0 | 0 | 0 | 0 | 0 |
| 0 | 0 | 0 | 0 | 0 |
| 0.037171751 | 2.063001991 | 0.049616964 | 0.000582069 | 0.109343822 |
| 0.08949771 | 0.003305389 | 0.005844675 | 0 | 0.044558678 |
| 0.073122626 | 0 | 0 | 0 | 0 |
| 0 | 0 | 0 | 0 | 0 |
| 0 | 0 | 0 | 0 | 0 |
| 0 | 0 | 0 | 0 | 0 |
| 0 | 0 | 0.001642058 | 0 | 0 |
| 0 | 0 | 0 | 0 | 0 |
| 0.008104427 | 0 | 0 | 0 | 0.001368082 |
| 0 | 0 | 0 | 0 | 0 |
| 0 | 0 | 0.576561092 | 0 | 0.061799636 |
| 0 | 0 | 0 | 0 | 0 |
| 0 | 0 | 0 | 0 | 0 |
| 0 | 0 | 0 | 0 | 0.002636712 |
| 0 | 0 | 0 | 0 | 0 |
| 0 | 0 | 0 | 0 | 0 |
| 0 | 0 | 0 | 0 | 0 |
| 0 | 0 | 0 | 0 | 0 |
| 0 | 0 | 0 | 0 | 0 |
| 0 | 0 | 0 | 0 | 0 |
| 0 | 0 | 0 | 0 | 0 |
| 0 | 0 | 0 | 0 | 0 |
| 0 | 0 | 0 | 0 | 0 |
| 0 | 0 | 0 | 0 | 0 |
| 0 | 0 | 0 | 0 | 0 |
| 0 | 0 | 0 | 0 | 0 |
| 0 | 0 | 0 | 0 | 0.000661077 |
| 0.496446288 | 0 | 0 | 0 | 0 |

| | | | | |
|---|---|---|---|---|
| 0 | 0 | 0 | 0 | 0 |
| 0 | 0 | 0.002166331 | 0 | 0 |
| 0 | 0 | 0 | 0 | 0 |
| 0 | 0 | 0 | 0 | 0 |
| 0 | 0.278166365 | 0 | 0 | 0.004666184 |
| 0 | 0 | 0 | 0 | 0 |
| 0 | 0 | 0 | 0 | 0 |
| 0 | 0 | 0 | 0 | 0 |
| 0.000668004 | 0 | 0 | 0 | 0 |
| 0 | 0 | 0 | 0 | 0 |
| 0 | 0 | 0 | 0 | 0 |
| 0 | 0 | 0.00115743 | 0 | 0.001200067 |
| 0 | 0 | 0 | 0 | 0 |
| 0 | 0 | 0 | 0 | 0 |
| 0 | 0 | 0 | 0 | 0 |
| 0 | 0 | 0.000536763 | 0 | 0 |
| 0 | 0 | 0 | 0 | 0 |
| 0 | 0 | 0 | 0 | 0 |
| 0.121934291 | 0 | 0 | 0 | 0.021360067 |
| 0 | 0 | 0 | 0 | 0 |
| 0 | 0 | 0 | 0 | 0 |
| 0 | 0 | 0 | 0 | 0 |
| 0 | 0 | 0 | 0 | 0 |
| 0 | 0 | 0 | 0 | 0 |
| 0 | 0 | 0 | 0 | 0 |
| 0 | 0 | 0 | 0 | 0 |
| 0 | 0 | 0 | 0 | 0 |
| 0.001648629 | 0 | 0 | 0 | 0 |
| 0 | 0 | 0 | 0 | 0 |
| 0 | 0 | 0 | 0 | 0 |
| 0.597546599 | 0 | 0.005535244 | 0 | 0 |
| 0.003999052 | 0.007650634 | 0.001451738 | 0.007350333 | 0.070912463 |
| 0 | 0 | 0 | 0 | 0 |
| 0 | 0 | 0 | 0 | 0 |
| 0 | 0 | 0 | 0 | 0 |
| 0 | 0 | 0 | 0 | 0 |
| 0 | 0 | 0 | 0 | 0 |
| 0 | 0 | 0 | 0 | 0 |
| 0 | 0 | 0 | 0 | 0 |
| 0 | 0 | 0 | 0 | 0 |
| 0 | 0 | 0 | 0 | 0 |
| 0 | 0 | 0 | 0 | 0 |
| 0.000749558 | 0 | 0 | 0.005334586 | 0 |
| 0 | 0 | 0 | 0 | 0 |
| 0.001198699 | 0 | 0 | 0 | 0 |
| 0 | 0 | 0 | 0 | 0 |
| 0 | 0 | 0 | 0 | 0 |
| 0 | 0 | 0 | 0 | 0 |

| | | | | |
|---|---|---|---|---|
| 0 | 0 | 0.001810635 | 0 | 0 |
| 0 | 0 | 0 | 0 | 0 |
| 0 | 0 | 0 | 0 | 0 |
| 0 | 0 | 0 | 0 | 0 |
| 0.136312609 | 0 | 0 | 0.000446346 | 0.002620243 |
| 0 | 0 | 0 | 0 | 0 |
| 0 | 0 | 0 | 0 | 0 |
| 0 | 0 | 0 | 0 | 0 |
| 0 | 0 | 0 | 0 | 0 |
| 0 | 0 | 0 | 0 | 0 |
| 0 | 0 | 0 | 0 | 0 |
| 0 | 0 | 0 | 0 | 0 |
| 0 | 0 | 0 | 0 | 0 |
| 0 | 0 | 0 | 0 | 0 |
| 0 | 0 | 0 | 0 | 0 |
| 0 | 0 | 0 | 0 | 0 |
| 0 | 0 | 0 | 0 | 0 |
| 0 | 0 | 0 | 0 | 0 |
| 0 | 0 | 0 | 0 | 0 |
| 0 | 0 | 0 | 0 | 0 |
| 0 | 0 | 0 | 0 | 0.017471338 |
| 0 | 0 | 0 | 0 | 0 |
| 0 | 0 | 0 | 0 | 0 |
| 0 | 0 | 0 | 0 | 0 |
| 0 | 0 | 0 | 0 | 0 |
| 0 | 0 | 0 | 0 | 0 |
| 0 | 0 | 0 | 0 | 0 |
| 0 | 0 | 0 | 0 | 0 |
| 0 | 0 | 0 | 0 | 0 |
| 0 | 0 | 0 | 0 | 0 |
| 0.060632923 | 0 | 0 | 0 | 0.001315277 |
| 0 | 0 | 0 | 0 | 0.001157797 |
| 0 | 0 | 0 | 0 | 0 |
| 0 | 0 | 0 | 0 | 0 |
| 0 | 0 | 0 | 0 | 0 |
| 0 | 0 | 0 | 0 | 0 |
| 0 | 0 | 0 | 0 | 0 |
| 0 | 0 | 0 | 0 | 0 |
| 0 | 0 | 0 | 0 | 0 |
| 0 | 0 | 0 | 0 | 0 |
| 0 | 0 | 0 | 0 | 0 |
| 0 | 0 | 0 | 0 | 0 |
| 0 | 0 | 0 | 0 | 0 |
| 0 | 0 | 0 | 0 | 0 |
| 0 | 0 | 0 | 0 | 0 |
| 0 | 0 | 0 | 0 | 0 |
| 0 | 0 | 0 | 0 | 0 |
| 0 | 0 | 0 | 0 | 0.001182903 |

| | | | | |
|---|---|---|---|---|
| 0 | 0 | 0 | 0 | 0 |
| 0 | 0 | 0 | 0 | 0 |
| 0 | 0 | 0 | 0 | 0 |
| 0 | 0 | 0 | 0 | 0 |
| 0 | 0 | 0 | 0 | 0 |
| 0 | 0 | 0 | 0 | 0 |
| 0 | 0 | 0 | 0 | 0 |
| 0 | 0 | 0 | 0 | 0 |
| 0 | 0 | 0 | 0 | 0 |
| 0 | 0 | 0 | 0 | 0 |
| 0 | 0 | 0 | 0 | 0 |
| 0.028446684 | 0 | 0 | 0 | 0 |
| 0.000357988 | 0 | 0 | 0 | 0.003692995 |
| 0 | 0 | 0 | 0 | 0 |
| 0 | 0 | 0 | 0 | 0 |
| 0 | 0 | 0 | 0 | 0 |
| 0 | 0 | 0 | 0 | 0 |
| 0 | 0 | 0 | 0 | 0 |
| 0 | 0 | 0 | 0 | 0 |
| 0 | 0 | 0 | 0 | 0 |
| 0.02766654 | 0.07000312 | 0.103558097 | 0.006704555 | 0.042120583 |
| 0 | 0 | 0 | 0 | 0 |
| 0 | 0 | 0.103283151 | 0 | 0.001666148 |
| 0 | 0 | 0.020834135 | 0 | 0.001800135 |
| 0 | 0 | 0 | 0 | 0 |
| 0 | 0 | 0 | 0 | 0 |
| 0.010784873 | 0.003728796 | 0.001357455 | 0 | 0 |
| 0 | 0 | 0 | 0 | 0 |
| 0 | 0 | 0 | 0 | 0 |
| 0 | 0.007446846 | 0 | 0 | 0 |
| 0 | 0 | 0 | 0 | 0.102418974 |
| 0 | 0 | 0 | 0 | 0 |
| 0 | 0 | 0 | 0 | 0 |
| 0.017726203 | 0.022434232 | 0 | 0 | 0 |
| 0.011468586 | 0 | 0.003249859 | 0.001607178 | 0.047847999 |
| 0 | 0 | 0 | 0 | 0 |
| 0 | 0 | 0 | 0 | 0 |
| 0 | 0 | 0 | 0 | 0 |
| 0 | 0 | 0 | 0 | 0 |
| 0 | 0 | 0 | 0 | 0 |
| 0 | 0 | 0 | 0 | 0 |
| 0 | 0 | 0 | 0 | 0 |
| 0 | 0 | 0 | 0 | 0 |
| 0 | 0 | 0 | 0 | 0 |
| 0 | 0 | 0 | 0 | 0 |
| 0.00898427 | 0 | 0 | 0 | 0 |
| 0 | 0 | 0 | 0 | 0 |
| 0.000724174 | 0 | 0 | 0 | 0.001760918 |

| | | | | |
|---|---|---|---|---|
| 0 | 0 | 0 | 0 | 0 |
| 0 | 0 | 0 | 0 | 0 |
| 0 | 0 | 0.001975172 | 0 | 0 |
| 0 | 0 | 0 | 0.001247871 | 0 |
| 0 | 0 | 0 | 0 | 0 |
| 0 | 0 | 0 | 0 | 0 |
| 0 | 0 | 0 | 0 | 0.025750077 |
| 0.000743581 | 0 | 0 | 0 | 0.001380737 |
| 0 | 0 | 0 | 0 | 0.040378227 |
| 0 | 0 | 0 | 0 | 0 |
| 0 | 0 | 0 | 0 | 0 |
| 0 | 0 | 0 | 0 | 0 |
| 0 | 0 | 0 | 0 | 0 |
| 0 | 0 | 0 | 0 | 0 |
| 0 | 0 | 0 | 0 | 0 |
| 0 | 0 | 0 | 0 | 0 |
| 0 | 0 | 0 | 0 | 0 |
| 0 | 0 | 0.009416436 | 0 | 0 |
| 0 | 0.026971705 | 0.005024626 | 0 | 0.000814019 |
| 0 | 0 | 0 | 0 | 0 |
| 0 | 0 | 0.017371487 | 0 | 0 |
| 0 | 0 | 0 | 0 | 0 |
| 0 | 0 | 0 | 0 | 0 |
| 0 | 0 | 0 | 0 | 0 |
| 0.002466664 | 0.005898755 | 0 | 0 | 0.002385566 |
| 0 | 0 | 0 | 0 | 0 |
| 0 | 0 | 0 | 0 | 0 |
| 0 | 0 | 0 | 0 | 0 |
| 0.003678083 | 0 | 0 | 0 | 0 |
| 0 | 0 | 0 | 0 | 0 |
| 0.207692556 | 1.772088766 | 0.569686824 | 0 | 1.033545135 |
| 0 | 0 | 0 | 0 | 0 |
| 0 | 0 | 0.025200282 | 0 | 0 |
| 0 | 0.003389459 | 0 | 0 | 0.00593996 |
| 0 | 0 | 0 | 0 | 0 |
| 0 | 0 | 0 | 0 | 0 |
| 0 | 0 | 0 | 0 | 0 |
| 0 | 0 | 0 | 0 | 0 |
| 0 | 0 | 0 | 0 | 0 |
| 0 | 0.001308417 | 0 | 0 | 0 |
| 0 | 0 | 0 | 0 | 0 |
| 0.008026293 | 0.024924379 | 0 | 0 | 0.005759931 |
| 0 | 0 | 0 | 0 | 0 |
| 0 | 0 | 0 | 0 | 0 |
| 0 | 0 | 0 | 0 | 0 |
| 0 | 0 | 0 | 0 | 0 |
| 0 | 0 | 0 | 0 | 0 |
| 0 | 0 | 0 | 0 | 0 |

| | | | | |
|---|---|---|---|---|
| 0 | 0 | 0 | 0 | 0 |
| 0 | 0 | 0 | 0 | 0 |
| 0.003505507 | 0.115685864 | 0 | 0.00924023 | 0.158211843 |
| 0 | 0 | 0 | 0 | 0 |
| 6.61977417 | 0 | 0 | 0 | 0.067865729 |
| 0 | 0 | 0 | 0 | 0 |
| 0 | 0 | 0 | 0 | 0 |
| 0 | 0 | 0 | 0 | 0 |
| 0 | 0 | 0 | 0 | 0 |
| 0 | 0 | 0.000797476 | 0 | 0 |
| 0 | 0 | 0 | 0 | 0 |
| 0 | 0 | 0 | 0 | 0 |
| 0 | 0 | 0 | 0 | 0 |
| 0.001452832 | 0.023779566 | 0.002168232 | 0 | 0.001498737 |
| 0.000932651 | 0.024215029 | 0.002584966 | 0 | 0.006803562 |
| 0 | 0 | 0 | 0 | 0 |
| 0.001062672 | 0.004356451 | 0 | 0 | 0.003288748 |
| 0 | 0 | 0 | 0 | 0 |
| 0 | 0 | 0 | 0 | 0 |
| 0 | 0 | 0 | 0 | 0 |
| 0 | 0 | 0 | 0 | 0 |
| 0.001370414 | 0 | 0 | 0 | 0.0014844 |
| 0 | 0 | 0 | 0 | 0 |
| 0 | 0.041456041 | 0 | 0 | 0 |
| 0 | 0 | 0 | 0 | 0 |
| 0 | 0 | 0 | 0 | 0 |
| 0 | 0 | 0 | 0 | 0 |
| 0 | 0 | 0 | 0 | 0 |
| 0 | 0 | 0 | 0 | 0 |
| 0 | 0 | 0.018638197 | 0 | 0 |
| 0 | 0 | 0 | 0 | 0 |
| 0 | 0 | 0 | 0 | 0 |
| 0 | 0 | 0 | 0 | 0 |
| 0 | 0 | 0 | 0 | 0 |
| 0 | 0 | 0 | 0 | 0 |
| 0 | 0 | 0 | 0 | 0.011435961 |
| 0 | 0 | 0 | 0 | 0 |
| 0 | 0.008234908 | 0 | 0.001633852 | 0.002664279 |
| 0 | 0 | 0 | 0 | 0 |
| 0 | 0 | 0 | 0 | 0 |
| 0 | 0 | 0 | 0 | 0.002307185 |
| 0 | 0 | 0 | 0 | 0 |
| 0 | 0 | 0 | 0 | 0 |
| 0 | 0.005319102 | 0.002553496 | 0 | 0.000992835 |
| 0 | 0 | 0 | 0 | 0 |
| 0 | 0 | 0 | 0 | 0 |
| 0 | 0 | 0 | 0 | 0 |
| 0.003545368 | 0.020348059 | 0.002178718 | 0 | 0.006776932 |

| | | | | |
|---|---|---|---|---|
| 0 | 7.992899869 | 0 | 0 | 0 |
| 0 | 0 | 0 | 0 | 0.028296716 |
| 0.001722482 | 0 | 0 | 0 | 0 |
| 0 | 0 | 0 | 0 | 0 |
| 0.003465965 | 0 | 0.001939751 | 0 | 0 |
| 0 | 0 | 0 | 0 | 0 |
| 0 | 0 | 0 | 0 | 0 |
| 0 | 0.010069038 | 0.017824476 | 0 | 0.004385344 |
| 0 | 0 | 0 | 0 | 0 |
| 0.0317146 | 0 | 5.57670283 | 0.020459289 | 0 |
| 0 | 2.176947215 | 0 | 0 | 0.03707245 |
| 0.005448299 | 0 | 0 | 0 | 0.003223493 |
| 0.001337059 | 0.016277823 | 0 | 0 | 0.014106542 |
| 0 | 0 | 0 | 0 | 0 |
| 0 | 0.016263046 | 0 | 0 | 0.001740991 |
| 0 | 0 | 0 | 0 | 0 |
| 0 | 0 | 0 | 0 | 0 |
| 0 | 0 | 0.00401635 | 0 | 0 |
| 0 | 1.727806408 | 0.029887359 | 0.001242053 | 0.002126651 |
| 0 | 0 | 0 | 0 | 0 |
| 0 | 0 | 0 | 0 | 0 |
| 0 | 0 | 0 | 0 | 0 |
| 0 | 0 | 0 | 0 | 0 |
| 0 | 0 | 0 | 0 | 0 |
| 0 | 0 | 0 | 0 | 0 |
| 0 | 0 | 0 | 0.007612914 | 0.000876295 |
| 0 | 0 | 0 | 0 | 0 |
| 0 | 3.391933679 | 0 | 0 | 0.077356918 |
| 0 | 0 | 0.099512102 | 0 | 0 |
| 0.030880683 | 0 | 0 | 0 | 0 |
| 0 | 0 | 0.001813807 | 0 | 0 |
| 0 | 0 | 0 | 0 | 0 |
| 0 | 0 | 0.102813549 | 0 | 0 |
| 0 | 0 | 0 | 0 | 0 |
| 0 | 0 | 0 | 0 | 0 |
| 0 | 0 | 0 | 0 | 0 |
| 0 | 0 | 0 | 0 | 0 |
| 0 | 0 | 0 | 0 | 0 |
| 0 | 0 | 0 | 0 | 0 |
| 0 | 0 | 0 | 0 | 0 |
| 0 | 0 | 0 | 0 | 0 |
| 0 | 0 | 0 | 0 | 0 |
| 0 | 0 | 0 | 0 | 0 |
| 0 | 0 | 0.002212897 | 0 | 0.525950519 |
| 0 | 0 | 0 | 0 | 0.001093474 |
| 0 | 0 | 0 | 0 | 0 |
| 0.004577575 | 0.003031413 | 0.000788268 | 0 | 0 |
| 0 | 0 | 0.022657166 | 0 | 0 |

| | | | | |
|---|---|---|---|---|
| 0 | 0 | 0 | 0 | 0 |
| 0.019282483 | 0 | 0.018206191 | 0 | 0 |
| 0 | 0 | 0 | 0 | 0 |
| 0 | 0 | 0 | 0 | 0 |
| 0 | 0 | 0.008050912 | 0 | 0 |
| 0.064342437 | 0 | 0.008278088 | 0 | 0.286887954 |
| 0.000462577 | 0 | 0 | 0 | 0 |
| 0 | 0 | 0 | 0 | 0 |
| 0 | 0 | 0 | 0 | 0 |
| 0 | 0 | 0 | 0 | 0 |
| 0.003377488 | 0.000923073 | 0.001440178 | 0.001271826 | 0 |
| 0 | 0 | 0 | 0 | 0 |
| 0 | 0 | 0 | 0 | 0 |
| 0.00136752 | 0.001744146 | 0.00226768 | 0 | 0 |
| 0.002152398 | 0 | 0.382949473 | 0 | 0.003330612 |
| 0 | 0 | 0 | 0 | 0 |
| 0 | 0 | 0 | 0 | 0 |
| 0 | 0 | 0 | 0 | 0 |
| 0 | 0 | 0 | 0 | 0 |
| 0.069974392 | 0 | 0 | 0 | 0 |
| 0.000654865 | 0 | 0.003176332 | 0 | 0 |
| 0 | 0 | 0 | 0.449932123 | 0 |
| 0 | 0 | 0 | 0 | 0 |
| 0 | 0 | 0 | 0 | 0 |
| 0 | 0 | 0 | 0 | 0 |
| 0 | 0 | 0 | 0 | 0 |
| 0 | 0.019541534 | 0.000983506 | 0 | 0 |
| 0 | 0 | 0 | 0 | 0 |
| 0 | 0 | 0 | 0 | 0 |
| 0.322724214 | 0.0217307 | 0.007645071 | 0.405787668 | 0.371520949 |
| 0.007009695 | 0.018537811 | 0.003230725 | 0.003912773 | 0.00239267 |
| 0 | 0 | 0 | 0 | 0 |
| 0 | 0.001214747 | 0 | 0 | 0 |
| 0 | 0 | 0 | 0 | 0.011973903 |
| 0 | 0 | 0.000614376 | 0 | 0.003185042 |
| 0 | 0 | 0 | 0 | 0 |
| 0 | 0 | 0 | 1.163893591 | 0 |
| 0.002787735 | 0 | 0 | 0 | 0 |
| 0 | 0 | 0 | 0 | 0.000752714 |
| 0.008809799 | 0 | 2.436118805 | 0 | 0 |
| 0.06920386 | 0 | 0.007829975 | 0 | 0.021649105 |
| 2.747508094 | 0 | 0 | 4.968560611 | 2.481001008 |
| 0.003688743 | 0 | 0 | 0 | 0 |
| 0.004068896 | 0 | 1.765296636 | 0 | 0 |
| 0 | 0 | 0 | 0 | 0 |
| 0 | 0 | 0.185679998 | 0 | 0 |
| 0.061883474 | 0.001254131 | 0.007239773 | 0.001658845 | 0.01095539 |
| 0 | 0.003286851 | 0 | 0 | 0 |

| | | | | |
|---|---|---|---|---|
| 0 | 0 | 0 | 0 | 0 |
| 0 | 0 | 0 | 0 | 0 |
| 0.004408387 | 0 | 0.051317421 | 0 | 0.030014684 |
| 0 | 0 | 0 | 0 | 0 |
| 0.714963001 | 0 | 0 | 0 | 0 |
| 0.005470951 | 0 | 0 | 0 | 0.000634929 |
| 0 | 0.001319428 | 0 | 0 | 0 |
| 0 | 0 | 0 | 0 | 0 |
| 0 | 0 | 0.030913605 | 0 | 0 |
| 0 | 0 | 0 | 0 | 0 |
| 0 | 0 | 0 | 0 | 0 |
| 0 | 0 | 0 | 0 | 0 |
| 0 | 0 | 0 | 0 | 0 |
| 0.019756728 | 0 | 0 | 0 | 0 |
| 0 | 0 | 0 | 0 | 0 |
| 0.05867026 | 0.343073435 | 0 | 0 | 0.054330088 |
| 0.004056444 | 0.394569428 | 0.016837438 | 0.005814022 | 0.025500003 |
| 0 | 0 | 0.029880764 | 0 | 0 |
| 0 | 0 | 0 | 0 | 0.019653377 |
| 0.02011765 | 0 | 0 | 0.24330716 | 0 |
| 0.059110662 | 0 | 0 | 0 | 0.028676324 |
| 0 | 0.008720394 | 0.001097222 | 0 | 0 |
| 0 | 0 | 0 | 0 | 0 |
| 0.322401918 | 0 | 0 | 0 | 0.594805782 |
| 0 | 0 | 0 | 0 | 0 |
| 0 | 0 | 0 | 0 | 0 |
| 0.027357718 | 0 | 0.000720514 | 0 | 0 |
| 0 | 0 | 0 | 0 | 0 |
| 0 | 0 | 0 | 0 | 0 |
| 0 | 0 | 0.031442282 | 0 | 0.001491528 |
| 0.081998019 | 0 | 0.033242253 | 0 | 0.021020978 |
| 0 | 0.014311815 | 0 | 0.002020002 | 0.013269963 |
| 0.012682122 | 0.002326524 | 0.145452922 | 0.100561875 | 0.006451812 |
| 0.003485562 | 0 | 0 | 0 | 0 |
| 0 | 0.137004137 | 0 | 0 | 0 |
| 0 | 0 | 0 | 0 | 0 |
| 0 | 0 | 0.001640813 | 0 | 0 |
| 0.111406468 | 0 | 0.020164447 | 0.007187669 | 0.024328452 |
| 0.391264693 | 0 | 0 | 0 | 0.832058201 |
| 0.001104683 | 0.131221875 | 0 | 0 | 0 |
| 0 | 0 | 0 | 0 | 0 |
| 0 | 0 | 0 | 0 | 0 |
| 0.018920225 | 0 | 0 | 0 | 0 |
| 0.087158247 | 0 | 0.004436054 | 0 | 0 |
| 0 | 3.385411607 | 0 | 0 | 0 |
| 0 | 0.007684645 | 0 | 0 | 0.00248625 |
| 0 | 0 | 0 | 0 | 0 |
| 0 | 0 | 0 | 0 | 0 |

| | | | | |
|---|---|---|---|---|
| 0.001274809 | 0 | 0 | 0 | 0 |
| 0.148792769 | 0.487023216 | 0.003497024 | 0.02514704 | 0.285405962 |
| 0 | 0 | 0.000820592 | 0 | 0 |
| 0 | 0 | 0 | 0 | 0 |
| 0.008045739 | 0 | 0 | 0 | 0 |
| 0 | 0 | 0 | 0 | 0 |
| 0 | 0 | 0 | 0 | 0.002156388 |
| 0 | 0.000983787 | 0 | 0 | 0.008355098 |
| 0 | 0 | 0.012576291 | 0 | 0 |
| 0.001216252 | 0.114479843 | 0 | 0 | 0 |
| 0.002860623 | 0 | 0.001113716 | 0 | 0.000769829 |
| 0.002408379 | 0.019493267 | 0.029860414 | 0.003222979 | 0.042140558 |
| 0.67221282 | 0 | 0 | 0 | 0 |
| 0 | 0 | 0 | 0 | 0 |
| 0 | 0 | 0 | 0 | 0 |
| 0 | 0 | 0 | 0 | 0 |
| 0.023833643 | 0.021178668 | 0 | 0.001853809 | 0.008867341 |
| 0.006086432 | 0.095416316 | 0.198575419 | 0.003743445 | 0.007063588 |
| 0 | 0.001056378 | 0 | 0 | 0 |
| 0 | 0 | 0.014582034 | 0 | 0 |
| 0.010073155 | 0.009507063 | 0 | 0.014444296 | 0.002909603 |
| 0 | 0 | 0 | 0 | 0 |
| 0 | 0 | 0 | 0 | 0 |
| 0 | 0 | 0 | 0 | 0 |
| 0.015345133 | 0 | 0 | 0 | 0 |
| 0 | 0 | 0.001470508 | 0 | 0.003049357 |
| 0.171014567 | 0 | 0.031655843 | 0 | 0 |
| 0.055815389 | 0.216293151 | 0.24563883 | 0.03499388 | 0.071126994 |
| 0 | 0 | 0 | 0 | 0 |
| 0 | 0.003953033 | 0 | 0 | 0 |
| 0 | 0 | 0 | 0 | 0.002844198 |
| 0 | 0.004146515 | 0 | 0 | 0 |
| 0 | 0 | 0 | 0 | 0 |
| 0 | 0 | 0 | 0 | 0 |
| 0 | 0 | 0 | 0.001377181 | 0 |
| 0.191827591 | 0.013403831 | 0.017307021 | 0.008406137 | 0.349919222 |
| 0 | 0 | 0 | 0 | 0 |
| 0 | 0 | 0 | 0 | 0 |
| 0.007924434 | 0 | 0 | 0 | 0.003542423 |
| 0 | 0 | 0 | 0 | 0 |
| 0 | 2.813365973 | 0 | 0 | 0 |
| 0 | 0 | 0 | 0 | 0 |
| 0 | 0 | 0.064173837 | 0.002030046 | 0 |
| 0 | 0.017979062 | 0.000462378 | 0 | 0.001598036 |
| 0 | 0 | 0 | 0 | 0.133524968 |
| 0.31220982 | 0 | 0 | 0.090283673 | 5.708846525 |
| 0 | 0 | 0 | 0 | 0 |
| 0 | 0 | 0 | 0 | 0 |

| | | | | |
|---|---|---|---|---|
| 0 | 0 | 0 | 0 | 0 |
| 0 | 0 | 0 | 0 | 0 |
| 0 | 0 | 0 | 0 | 0 |
| 0.105021197 | 0.106059551 | 0 | 0 | 0.049157632 |
| 0 | 0 | 0 | 0 | 0 |
| 0 | 0 | 0 | 0 | 0 |
| 0 | 0 | 0 | 0 | 0.057838769 |
| 0 | 0 | 0 | 0 | 0.211433429 |
| 0.145161661 | 0 | 0 | 0.150642301 | 0 |
| 0.036743571 | 0 | 0 | 0 | 0 |
| 0 | 0 | 0 | 0 | 0 |
| 0.005848705 | 0 | 0 | 0.000755104 | 0 |
| 0 | 0 | 0 | 0 | 0 |
| 0 | 0 | 0 | 0 | 0 |
| 0 | 0 | 0.492888006 | 0.013864282 | 0 |
| 0 | 0 | 0 | 0 | 0 |
| 0.013490511 | 0 | 0 | 0 | 0.00954293 |
| 0 | 0 | 0 | 0 | 0 |
| 0 | 0 | 0 | 0 | 0 |
| 0 | 0 | 0 | 0 | 0 |
| 0 | 0 | 0 | 0 | 0 |
| 0 | 0 | 0 | 0 | 0 |
| 0 | 0 | 0 | 0 | 0 |
| 0 | 0 | 0.350312997 | 0 | 0.465253517 |
| 0.062469761 | 0 | 0 | 0 | 0 |
| 0 | 0 | 0 | 0.002185821 | 0 |
| 0.776136919 | 0 | 0 | 0 | 0 |
| 0 | 0 | 0 | 0.066229704 | 0.75405241 |
| 0 | 0 | 0 | 0 | 0 |
| 0 | 0 | 0 | 0 | 0 |
| 0 | 0 | 0 | 0 | 0 |
| 0.006013193 | 0 | 0 | 0 | 0 |
| 0.070121812 | 0 | 0 | 0 | 0.030611318 |
| 0 | 0 | 0 | 0 | 0 |
| 0 | 0 | 0 | 0 | 0 |
| 0 | 0 | 0 | 0 | 0 |
| 0.442709691 | 0 | 0 | 0.010249301 | 0 |
| 0 | 0 | 0 | 0 | 0 |
| 0 | 0 | 0 | 0 | 0.039608007 |
| 0.032807291 | 0 | 0 | 0 | 0.002511852 |
| 0.079352557 | 0.691580365 | 0.088521336 | 0.000724387 | 0.011517081 |
| 0.019293329 | 0 | 0 | 0 | 0.023391088 |
| 2.628319475 | 0 | 5.18640287 | 13.13915287 | 0.014028709 |
| 0 | 0 | 0 | 0 | 0 |
| 0 | 0 | 0 | 0 | 0 |
| 0 | 0 | 0 | 0 | 0 |
| 0 | 0 | 0 | 0 | 0 |
| 0.015416707 | 0.06267451 | 0 | 0 | 0.010223891 |

| | | | | |
|---|---|---|---|---|
| 0 | 0 | 0 | 0 | 0 |
| 0 | 0 | 0 | 0 | 0 |
| 0.00870885 | 0.98956271 | 0.233753488 | 0 | 0.456143425 |
| 0 | 0 | 0 | 0 | 0 |
| 0 | 0 | 0 | 0 | 0 |
| 0.008627594 | 0 | 0 | 0 | 0.008364044 |
| 0 | 0 | 0 | 0 | 0 |
| 0 | 0 | 0 | 0 | 0 |
| 0 | 0 | 0 | 0 | 0 |
| 1.312829686 | 0 | 0 | 0 | 0 |
| 0.041845855 | 0.127147455 | 0.013147687 | 0.023972812 | 0.480127766 |
| 1.288426007 | 2.736062558 | 0.533947471 | 2.091591899 | 2.023391808 |
| 0 | 0 | 0 | 0 | 0 |
| 0.001258377 | 0.015514459 | 0 | 0.006191694 | 0.006707047 |
| 0.212984241 | 0.131745569 | 0.466679279 | 0 | 0.085760042 |
| 1.190044788 | 0 | 0 | 0 | 0.001026575 |
| 0.000358017 | 0 | 0 | 0 | 0 |
| 0.003303738 | 0 | 0 | 0 | 0.01495789 |
| 0 | 0 | 0 | 0 | 0 |
| 0.076196778 | 0 | 0.043798738 | 0 | 0 |
| 0.050926414 | 0.002700561 | 0.004537523 | 0 | 0.018482653 |
| 0 | 2.411497114 | 37.87568703 | 66.89947531 | 0 |
| 1.3468345 | 2.55937859 | 7.459128768 | 1.329114873 | 0.19234786 |
| 0 | 0 | 0 | 0 | 0 |
| 0.013515851 | 0 | 0 | 0 | 0 |
| 0.003773408 | 0 | 0 | 0.031897063 | 0 |
| 0.208835531 | 0 | 0 | 0 | 0 |
| 0 | 0 | 0 | 0.001612029 | 0 |
| 0 | 0 | 0 | 0 | 0 |
| 0 | 0 | 0 | 0 | 0 |
| 0 | 0 | 0 | 0 | 0 |
| 0.308255575 | 0.155913968 | 0.101280541 | 0.006323031 | 0.504911812 |
| 0.016897992 | 0.782229746 | 0.563552375 | 0 | 1.269098683 |
| 0 | 0 | 0 | 0 | 0 |
| 0.005444715 | 0.470247799 | 0.003931835 | 0 | 0.044163983 |
| 0.027509693 | 0.137343543 | 0 | 0.034157583 | 0.02837892 |
| 0 | 0 | 0.126458144 | 0 | 0 |
| 0 | 0 | 0 | 0 | 0 |
| 0 | 0 | 0 | 0.027466291 | 0.003277211 |
| 0 | 0 | 0 | 0 | 0 |
| 0.001381174 | 0 | 0 | 0 | 0 |
| 0 | 0 | 0 | 0 | 0 |
| 0 | 0 | 0 | 0 | 0 |
| 0 | 0 | 0 | 0 | 0 |
| 0.037150676 | 0 | 0.114790323 | 0.017409093 | 0.012349015 |
| 0 | 0 | 0 | 0 | 0 |
| 0 | 0 | 0 | 0 | 0 |
| 0.063403324 | 0 | 0.012385376 | 0.003675022 | 0.177728117 |

| | | | | |
|---|---|---|---|---|
| 0 | 0 | 0 | 0 | 0 |
| 0 | 0 | 0 | 0 | 0 |
| 0 | 0 | 0 | 0 | 0 |
| 0.008443565 | 0 | 0 | 0 | 0.002903453 |
| 0 | 0 | 0 | 0 | 0 |
| 0 | 0 | 0 | 0 | 0 |
| 0.0512277 | 0 | 0 | 0 | 0 |
| 0.048344232 | 0.247941966 | 0 | 0 | 0.003172646 |
| 0 | 0 | 0 | 0 | 0 |
| 0 | 0 | 0 | 0 | 0 |
| 0.032129979 | 1.220619435 | 0.00506803 | 0 | 0.028900992 |
| 0 | 0 | 0 | 0 | 0 |
| 0 | 0 | 0 | 0 | 0 |
| 0 | 0 | 0 | 0 | 0 |
| 0 | 0 | 0 | 0 | 0 |
| 0 | 0 | 0 | 0 | 0 |
| 0.109087328 | 0 | 0 | 0 | 0 |
| 0.001848403 | 0 | 0 | 0 | 0.006056916 |
| 0.163952888 | 0 | 0 | 0 | 0.000708165 |
| 0.003431962 | 0 | 0 | 0 | 0 |
| 0 | 0 | 0 | 0 | 0 |
| 0 | 0 | 0 | 0 | 0 |
| 0.013538601 | 0 | 0 | 0 | 7.559303917 |
| 0 | 0 | 0 | 0 | 0 |
| 0 | 0 | 0 | 0 | 0 |
| 0 | 0 | 0 | 0 | 0 |
| 0 | 0 | 0 | 0 | 0 |
| 0 | 0 | 0 | 0 | 0 |
| 0 | 0 | 0 | 0 | 0 |
| 0.000656722 | 0 | 0 | 0.007062745 | 0 |
| 0 | 0 | 0 | 0 | 0 |
| 0 | 0 | 0 | 0 | 0 |
| 0 | 0 | 0 | 0 | 0 |
| 0 | 0 | 0 | 0 | 0 |
| 0.002388652 | 0.00496713 | 0 | 0 | 0 |
| 0 | 0 | 0 | 0 | 0 |
| 0 | 0 | 0 | 0.008431731 | 0.096245781 |
| 0 | 0.566873882 | 0.007755318 | 0.00104917 | 0.040376127 |
| 0 | 0 | 0 | 0 | 0 |
| 0 | 0 | 0 | 0 | 0.008943701 |
| 0.130914586 | 0 | 0 | 0 | 0 |
| 0 | 0 | 0 | 0 | 0 |
| 0 | 0 | 0 | 0 | 0 |
| 0.003221104 | 0 | 0 | 0.053830194 | 0.118294584 |
| 0 | 0.135701073 | 0 | 0.03693096 | 0 |
| 0 | 0 | 0 | 0 | 0 |
| 0 | 0 | 0 | 0 | 0.013167333 |
| 0 | 0 | 0 | 0 | 0 |

| | | | | |
|---|---|---|---|---|
| 0 | 0 | 0 | 0 | 0 |
| 0 | 0 | 0 | 0 | 0 |
| 0 | 0 | 0 | 0 | 0 |
| 0.104369262 | 0 | 0 | 0 | 0 |
| 0 | 0 | 0 | 0 | 0 |
| 1.394273256 | 0 | 0 | 0 | 0 |
| 0 | 0 | 0 | 0 | 0 |
| 0 | 0 | 0 | 0 | 0 |
| 0.009759063 | 0.677085721 | 0.33362041 | 0.006993738 | 0.063471943 |
| 0.001719141 | 0 | 0 | 0 | 0 |
| 0 | 0 | 0 | 0 | 0 |
| 0 | 0 | 0 | 0 | 0 |
| 0 | 0 | 0 | 0 | 0.017619432 |
| 0 | 0 | 0 | 0 | 0 |
| 1.71569405 | 0 | 1.228833365 | 0 | 0.193302524 |
| 0 | 0 | 0.131897788 | 0 | 0 |
| 0.016000769 | 0 | 0 | 0 | 0.104364256 |
| 0 | 0 | 0 | 0 | 0 |
| 0 | 0 | 0 | 0 | 0 |
| 0 | 0 | 0 | 0 | 0 |
| 0 | 0 | 0 | 0 | 0 |
| 0 | 0 | 0 | 0 | 0 |
| 0 | 0 | 0 | 0 | 0 |
| 0 | 0 | 0 | 0 | 0.019446522 |
| 0.003048413 | 0.128646089 | 0.093131091 | 0.005388424 | 0.205332662 |
| 10.09307456 | 0 | 0 | 0 | 16.4530556 |
| 0.021342668 | 0.001275966 | 0 | 0 | 0 |
| 0 | 0 | 0 | 0 | 0 |
| 0.057693417 | 0 | 0 | 0 | 0 |
| 0 | 0 | 0 | 0 | 0 |
| 0 | 0 | 0 | 0 | 0 |
| 0 | 0 | 0 | 0 | 0 |
| 0.01233059 | 0 | 0 | 0 | 0 |
| 0 | 0 | 0.003858981 | 0 | 0.016152743 |
| 0.000946768 | 0.037734861 | 0.000941983 | 0.011479775 | 0.005127587 |
| 0.002230834 | 0.045665841 | 0.019976024 | 0 | 0.083710594 |
| 0 | 0 | 0 | 0 | 0 |
| 0.372811616 | 0 | 0 | 0.044022348 | 0.74625022 |
| 0 | 0 | 0 | 0 | 0 |
| 0 | 0 | 0 | 0 | 0 |
| 0 | 0 | 0 | 0 | 0 |
| 0 | 0.024440787 | 0 | 0 | 0 |
| 0.851896251 | 0 | 0 | 0.004339186 | 0.320297647 |
| 0 | 0 | 0 | 0 | 0 |
| 0 | 0 | 0 | 0 | 0 |
| 0.000422687 | 0 | 0 | 0 | 0 |
| 0 | 0 | 0 | 0 | 0 |
| 0.046295644 | 0.116266612 | 0 | 0.001597504 | 0.048713622 |

| | | | | |
|---|---|---|---|---|
| 0 | 0 | 0 | 0 | 0.026030253 |
| 0 | 0 | 0 | 0 | 0 |
| 0 | 0 | 0 | 0 | 0 |
| 0 | 0 | 0 | 0 | 0 |
| 0.000624934 | 0.247960875 | 0 | 0.017592892 | 0.624114634 |
| 0 | 0 | 0 | 0 | 0 |
| 0 | 0 | 0 | 0 | 0 |
| 0 | 0 | 0 | 0 | 0 |
| 0 | 0 | 0 | 0 | 0 |
| 0 | 0 | 0 | 0 | 0 |
| 0 | 0 | 0 | 0 | 0 |
| 1.813453059 | 0 | 0 | 0.022613371 | 1.274686124 |
| 0.348075225 | 0 | 0 | 0.02106256 | 0.025525435 |
| 0 | 0 | 0 | 0 | 0 |
| 0.08470465 | 0.011337933 | 0.037104672 | 0 | 0.151649406 |
| 0 | 0 | 0 | 0 | 0 |
| 0 | 0 | 0 | 0 | 0 |
| 0.057324801 | 0 | 0 | 0 | 0 |
| 0.002660597 | 0 | 0.009454102 | 0 | 0 |
| 0 | 0 | 0 | 0 | 0 |
| 0.029264845 | 0 | 0 | 0.008460488 | 0.005843135 |
| 0 | 0 | 0 | 0 | 0 |
| 0.005174138 | 0 | 0 | 0.001169023 | 0.147928487 |
| 0 | 0 | 0 | 0 | 0 |
| 0 | 0 | 0 | 0 | 0 |
| 0 | 0 | 0 | 0 | 0 |
| 0 | 0 | 0 | 0 | 0 |
| 0 | 0 | 0 | 0 | 0 |
| 0 | 0 | 0 | 0.001235512 | 0 |
| 0 | 0 | 0 | 0 | 0 |
| 0 | 0 | 0 | 0 | 0 |
| 0.017256457 | 0.112317785 | 0 | 0.001483259 | 0.055436852 |
| 0 | 0 | 0 | 0 | 0.009320065 |
| 0.845891572 | 0 | 0 | 0 | 0.201552151 |
| 0.001213853 | 0.048999207 | 0.015217244 | 0 | 0.078137754 |
| 0.001095363 | 0 | 0 | 0.006187044 | 0.027361494 |
| 0 | 0 | 0 | 0 | 0 |
| 0 | 0 | 0 | 0 | 0 |
| 0 | 0 | 0 | 0 | 0 |
| 0 | 0 | 0 | 0 | 0 |
| 0 | 0 | 0 | 0 | 0 |
| 0 | 0 | 0 | 0 | 0 |
| 0 | 0 | 0 | 0 | 0 |
| 0.037748795 | 0 | 0 | 0 | 0.004281531 |
| 0.001434921 | 0.19124641 | 0.010707508 | 0.000756467 | 0.510134595 |
| 0 | 0 | 0 | 0 | 0 |
| 1.716291507 | 0 | 0 | 0 | 0 |
| 0.00248816 | 0 | 0 | 0 | 0 |

| | | | | |
|---|---|---|---|---|
| 0 | 0 | 0 | 0 | 0 |
| 0 | 0 | 0 | 0 | 0.00521624 |
| 0.013480275 | 0 | 0.01291995 | 0 | 0.076547963 |
| 0 | 0 | 0 | 0 | 0 |
| 0 | 0 | 0 | 0 | 0.005822302 |
| 0 | 0 | 0 | 0 | 0 |
| 0 | 0 | 0 | 0 | 0 |
| 0.095306254 | 0 | 0 | 0 | 0.083136254 |
| 0 | 0 | 0 | 0 | 0 |
| 0.001494124 | 0 | 0 | 0 | 0 |
| 0 | 0 | 0 | 0 | 0 |
| 0 | 0.088108498 | 0.004694905 | 0 | 0 |
| 0 | 0 | 0 | 0 | 0 |
| 0 | 0 | 0 | 0 | 0 |
| 0 | 0 | 0 | 0 | 0 |
| 0 | 0 | 0 | 0 | 0 |
| 0 | 0 | 0 | 0 | 0 |
| 0 | 0 | 0 | 0 | 0 |
| 0 | 0 | 0 | 0 | 0 |
| 0 | 0 | 0 | 0 | 0 |
| 0 | 0 | 0 | 0 | 0 |
| 0 | 0 | 0 | 0 | 0 |
| 0 | 0 | 0 | 0 | 0 |
| 0 | 0 | 0 | 0 | 0 |
| 0 | 0 | 0 | 0 | 0 |
| 0 | 0.269194471 | 0.015783118 | 0 | 0.00396194 |
| 0 | 0 | 0 | 0 | 0 |
| 0.093706901 | 0.05461536 | 0 | 0 | 0.029506151 |
| 0 | 0 | 0 | 0 | 0 |
| 0.75357937 | 0 | 0 | 0.039935975 | 0.995040765 |
| 0 | 0 | 0 | 0 | 0 |
| 0 | 0 | 0 | 0 | 0 |
| 0 | 0 | 0 | 0 | 0 |
| 0 | 0 | 0 | 0 | 0 |
| 0.001633211 | 0.104448105 | 0 | 0 | 0.00270774 |
| 0 | 0 | 0 | 0.004264818 | 0 |
| 0 | 0 | 0 | 0 | 0 |
| 0 | 0 | 0 | 0 | 0.005735318 |
| 0 | 0 | 0 | 0 | 0 |
| 0 | 0 | 0 | 0 | 0 |
| 3.204195039 | 0.219396157 | 0.58557283 | 1.492807291 | 11.46393738 |
| 0.013371833 | 2.7040719 | 0 | 0 | 1.911416381 |
| 0 | 0 | 0 | 0.069969571 | 0 |
| 0 | 0 | 0 | 0 | 0 |
| 0 | 0 | 0 | 0 | 0 |
| 0 | 0 | 0 | 0 | 0 |
| 0 | 0 | 0 | 0 | 0 |
| 0.000525291 | 0 | 0.004233351 | 0.017778578 | 0 |

| | | | | |
|---|---|---|---|---|
| 0 | 0 | 0 | 0 | 0 |
| 0.018694562 | 0 | 0 | 0 | 0.005350823 |
| 0.006689088 | 0 | 0 | 0 | 0 |
| 0 | 0 | 0 | 0 | 0 |
| 7.729014296 | 0 | 0 | 0.084294769 | 1.62742248 |
| 0 | 0 | 0 | 0 | 0 |
| 0 | 0 | 0 | 0 | 0 |
| 0 | 0 | 0 | 0 | 0 |
| 0.006366202 | 6.996054577 | 0.242517958 | 0.023379337 | 0.201695745 |
| 0 | 0 | 0 | 0 | 0 |
| 0 | 0 | 0.046097633 | 0 | 0 |
| 0 | 0 | 0 | 0 | 0 |
| 0.138809642 | 0 | 0 | 0 | 0 |
| 0.024655519 | 0.001728698 | 0 | 0.005103922 | 0.016978569 |
| 0 | 0 | 0 | 0.957770206 | 0 |
| 0.608129421 | 0.666721945 | 0 | 0 | 0.209643013 |
| 0.150912554 | 0 | 0 | 0 | 0 |
| 0 | 0 | 0 | 0 | 0 |
| 0 | 0 | 0 | 0 | 0 |
| 0 | 0 | 0 | 0 | 0 |
| 0 | 0 | 0 | 0 | 0 |
| 0 | 0 | 0 | 0 | 0 |
| 0 | 0 | 0 | 0 | 0 |
| 0 | 0 | 0 | 0 | 0 |
| 0 | 0 | 0 | 0 | 0 |
| 0 | 0 | 0 | 0 | 0 |
| 0.016220962 | 0 | 0 | 0 | 0.022394831 |
| 0 | 0.05367833 | 0.000654288 | 0 | 0.014585405 |
| 0 | 0 | 0 | 0 | 0 |
| 0 | 0 | 0 | 0 | 0 |
| 0 | 0 | 0 | 0 | 0 |
| 0 | 0 | 0 | 0 | 0 |
| 0 | 0 | 0 | 0 | 0 |
| 0.032514492 | 0 | 0 | 0 | 0 |
| 0 | 0 | 0 | 0.005157888 | 0 |
| 0 | 0.007034164 | 0.025355338 | 0 | 0 |
| 0.001036731 | 0 | 0 | 0 | 0 |
| 0.002035286 | 0 | 0 | 0 | 0 |
| 0 | 0 | 0 | 0 | 0 |
| 0 | 0 | 0 | 0 | 0 |
| 1.322219646 | 0 | 0 | 0.101917849 | 0 |
| 0 | 0 | 0 | 0 | 0 |
| 0 | 0 | 0 | 0 | 0 |
| 0 | 0 | 0 | 0 | 0 |
| 0 | 0 | 0 | 0 | 0 |
| 0 | 0 | 0 | 0 | 0 |
| 0 | 0 | 0 | 0 | 0 |
| 0 | 0 | 0 | 0 | 0 |

| | | | | |
|---|---|---|---|---|
| 0.000840028 | 0 | 0 | 0 | 0.064621383 |
| 0 | 0 | 0 | 0 | 0 |
| 0 | 0 | 0 | 0 | 0 |
| 0 | 0.01567681 | 0 | 0 | 0.000768484 |
| 0.001187946 | 0 | 0 | 0 | 0 |
| 0 | 0 | 0 | 0 | 0 |
| 0.518050692 | 0 | 0 | 0 | 0.009656108 |
| 0 | 0 | 0 | 0 | 0 |
| 0 | 0 | 0 | 0 | 0.003164411 |
| 0 | 0 | 0 | 0 | 0 |
| 0.009594259 | 0 | 0 | 0 | 0.003136503 |
| 0.027578354 | 0 | 0 | 0 | 0 |
| 0 | 0 | 0 | 0 | 0 |
| 0 | 0 | 0 | 0 | 0 |
| 0 | 0 | 0 | 0 | 0.040154273 |
| 0 | 0 | 0 | 0 | 0 |
| 0 | 0 | 0 | 0 | 0 |
| 0 | 0 | 0 | 0 | 0 |
| 0 | 0 | 0 | 0 | 0 |
| 0 | 0 | 0 | 0 | 0 |
| 0.000848721 | 0 | 0 | 0.046980357 | 0.079619272 |
| 0 | 0 | 0 | 0 | 0 |
| 0.018254663 | 0 | 0 | 0 | 0.007242868 |
| 0.002393214 | 0 | 0.005124579 | 0 | 0.005635379 |
| 0 | 0 | 0 | 0 | 0 |
| 0.000736553 | 0 | 0.002564906 | 0 | 0 |
| 0 | 0 | 0 | 0 | 0 |
| 0 | 0 | 0 | 0 | 0 |
| 0 | 0 | 0 | 0 | 0 |
| 0.001048295 | 0.010027524 | 0.089176141 | 0.647303716 | 0.068592773 |
| 0 | 0 | 0 | 0 | 0 |
| 0 | 0 | 0 | 0 | 0 |
| 0 | 0 | 0 | 0 | 0 |
| 0 | 0 | 0 | 0 | 0 |
| 0 | 0 | 0 | 0 | 0 |
| 0 | 0 | 0 | 0 | 0 |
| 0.020440927 | 0.684973883 | 0.610322502 | 0 | 0.003424926 |
| 0 | 0 | 0 | 0 | 0 |
| 0 | 0 | 0 | 0 | 0 |
| 0 | 0 | 0 | 0 | 0 |
| 0 | 0 | 0 | 0 | 0 |
| 0 | 0 | 0 | 0 | 0 |
| 0 | 0 | 0 | 0 | 0 |
| 0 | 0 | 0 | 0 | 0 |
| 0 | 0 | 0 | 0 | 0 |
| 0 | 0 | 0 | 0 | 0 |
| 0 | 0 | 0 | 0 | 0 |
| 0 | 0 | 0 | 0 | 0 |

| | | | | |
|---|---|---|---|---|
| 0.071097454 | 0 | 0.002005174 | 0 | 0 |
| 0 | 0 | 0 | 0 | 0 |
| 0.016100099 | 0 | 0 | 0 | 0 |
| 0 | 0 | 0 | 0 | 0 |
| 0 | 0 | 0 | 0 | 0 |
| 0 | 0 | 0 | 0 | 0 |
| 0 | 0 | 0 | 0 | 0 |
| 1.357933867 | 0.024852725 | 0 | 0.117091198 | 1.574878399 |
| 0.055008213 | 6.254701245 | 1.424732962 | 0.019734437 | 0.152290428 |
| 0 | 0.624957019 | 0.078881064 | 0 | 0.398521711 |
| 0 | 0 | 0 | 0 | 0 |
| 0 | 0 | 0 | 0 | 0 |
| 0 | 0 | 0 | 0 | 0 |
| 0 | 0 | 0 | 0 | 0 |
| 0 | 0 | 0 | 0 | 0.011751457 |
| 0 | 0 | 0 | 0 | 0 |
| 0.056386121 | 0 | 0 | 0 | 0 |
| 0 | 0 | 0 | 0 | 0 |
| 0 | 0 | 0 | 0 | 0 |
| 0.052248957 | 0 | 0.003936718 | 0 | 0.011303274 |
| 0 | 0 | 0 | 0 | 0 |
| 0 | 0 | 0 | 0 | 0 |
| 0 | 0 | 0 | 0 | 0 |
| 0 | 0 | 0 | 0 | 0 |
| 0 | 0 | 0 | 0 | 0 |
| 0 | 0 | 0 | 0 | 0 |
| 0 | 0 | 0 | 0 | 0 |
| 0 | 0 | 0 | 0 | 0 |
| 0 | 0 | 0 | 0 | 0 |
| 0 | 0 | 0 | 0 | 0 |
| 0.032666888 | 0 | 0 | 0 | 0 |
| 0 | 0 | 0 | 0 | 0 |
| 0 | 0 | 0 | 0 | 0 |
| 0 | 0 | 0 | 0 | 0 |
| 0 | 0 | 0 | 0 | 0 |
| 0 | 0 | 0 | 0 | 0 |
| 0 | 0 | 0 | 0 | 0 |
| 0 | 0.058497541 | 0.006204611 | 0 | 0 |
| 0 | 0 | 0 | 0 | 0 |
| 0 | 0.411426797 | 0.203515487 | 0 | 0.000518883 |
| 0 | 0 | 0 | 0 | 0 |
| 0.118891207 | 0 | 0 | 0.075504809 | 0.237325329 |
| 0 | 0 | 0 | 0 | 0 |
| 0 | 0 | 0 | 0 | 0 |
| 0 | 0 | 0 | 0 | 0 |
| 0 | 0 | 0 | 0 | 0 |
| 0 | 0 | 0 | 0 | 0 |
| 0 | 0 | 0 | 0 | 0 |

| | | | | |
|---|---|---|---|---|
| 0 | 0 | 0 | 0 | 0 |
| 0.002728427 | 0.037891783 | 0 | 0 | 0.004691062 |
| 0 | 0 | 0 | 0 | 0 |
| 0 | 0 | 0 | 0 | 0 |
| 0.084912944 | 0 | 0 | 0.0288252 | 1.211622492 |
| 0 | 0 | 0 | 0 | 0 |
| 0 | 0 | 0 | 0 | 0.003223291 |
| 0.019512491 | 1.482011596 | 0.036985536 | 0 | 0.006832116 |
| 0.00071104 | 0 | 0 | 0 | 0 |
| 0 | 0 | 0 | 0 | 0 |
| 0 | 0 | 0 | 0 | 0 |
| 0 | 0 | 0 | 0 | 0 |
| 0 | 0 | 0 | 0 | 0 |
| 0 | 0 | 0 | 0 | 0 |
| 0.001203163 | 0 | 0 | 0 | 0.442635464 |
| 0.002470517 | 0 | 0 | 0 | 0 |
| 0 | 0 | 0 | 0 | 0 |
| 0 | 0 | 0 | 0 | 0 |
| 0 | 0 | 0 | 0 | 0 |
| 0 | 0 | 0 | 0 | 0 |
| 0 | 0 | 0 | 0 | 0 |
| 0 | 0 | 0 | 0 | 0 |
| 0 | 0 | 0 | 0 | 0 |
| 0.005669904 | 0.131109174 | 0 | 0 | 0.002860952 |
| 1.279360927 | 0 | 0 | 0.018731738 | 0 |
| 0 | 0 | 0 | 0 | 0 |
| 0 | 0.22645671 | 0 | 0 | 0.233057642 |
| 0.000830938 | 0 | 0 | 0 | 0 |
| 0 | 0.051291421 | 0.006001864 | 0 | 0 |
| 0 | 0 | 0 | 0 | 0 |
| 0 | 0 | 0 | 0 | 0 |
| 0 | 0 | 0 | 0 | 0 |
| 0 | 0 | 0 | 0 | 0 |
| 0 | 0 | 0 | 0 | 0 |
| 0 | 0 | 0 | 0 | 0 |
| 0 | 0 | 0 | 0 | 0 |
| 0 | 0 | 0 | 0 | 0 |
| 0.002427925 | 0 | 0 | 0 | 0 |
| 0 | 0 | 0 | 0 | 0 |
| 0 | 0 | 0 | 0 | 0 |
| 0 | 0 | 0 | 0 | 0 |
| 0 | 0 | 0 | 0 | 0 |
| 0 | 0 | 0 | 0 | 0 |
| 0 | 0 | 0 | 0 | 0 |
| 0 | 0 | 0 | 0 | 0 |
| 0 | 0 | 0 | 0 | 0 |
| 0 | 0 | 0 | 0 | 0 |
| 0 | 0 | 0 | 0 | 0.039019577 |

| | | | | |
|---|---|---|---|---|
| 0.072515461 | 0 | 0 | 0.009724913 | 4.353063314 |
| 0 | 0 | 0 | 0 | 0 |
| 0 | 0.208339885 | 0 | 0 | 0.004387513 |
| 0.001054808 | 0 | 0 | 0 | 0 |
| 0 | 0 | 0 | 0 | 0 |
| 0 | 0 | 0.333254967 | 0 | 0 |
| 0 | 0 | 0 | 0 | 0 |
| 0 | 0 | 0 | 0 | 0 |
| 0 | 0 | 0 | 0 | 0 |
| 0 | 0 | 0 | 0 | 0 |
| 0 | 0 | 0 | 0 | 0 |
| 0 | 0 | 0 | 0 | 0 |
| 0 | 0 | 0 | 0 | 0 |
| 0 | 0 | 0 | 0 | 0 |
| 0 | 0 | 0 | 0 | 0 |
| 0 | 0 | 0 | 0 | 0 |
| 0 | 0 | 0 | 0 | 0 |
| 0 | 0 | 0 | 0 | 0 |
| 0 | 0 | 0 | 0 | 0.015438435 |
| 0 | 0 | 0 | 0 | 0 |
| 0 | 0.174707658 | 0.002930955 | 0 | 0.019183214 |
| 0 | 0 | 0 | 0 | 0 |
| 0.000620065 | 0 | 0.002313489 | 0.022555266 | 0.004077812 |
| 0.026956982 | 0 | 0 | 0 | 0 |
| 0 | 0 | 0 | 0 | 0 |
| 0 | 0 | 0 | 0 | 0 |
| 0 | 0 | 0 | 0 | 0 |
| 0 | 0 | 0 | 0 | 0 |
| 0 | 0 | 0 | 0 | 0 |
| 0 | 0 | 0 | 0 | 0 |
| 0 | 0 | 0 | 0 | 0 |
| 0.018704232 | 0 | 0 | 0 | 0.20959446 |
| 0.001742064 | 0.002221842 | 0.016465954 | 0 | 0.027181258 |
| 0 | 0 | 0 | 0 | 0 |
| 0.319574745 | 0 | 0 | 0.025055546 | 0.145278222 |
| 0 | 0 | 0.001738584 | 0.057114983 | 0.035511806 |
| 0 | 0 | 0 | 0 | 0 |
| 0 | 0 | 0 | 0 | 0 |
| 0 | 0.557322099 | 0.002413137 | 0 | 0.007297593 |
| 0 | 0 | 0 | 0 | 0 |
| 0 | 0 | 0 | 0 | 0 |
| 0 | 0 | 0 | 0 | 0 |
| 0.013864962 | 0 | 0 | 0.022924878 | 0 |
| 0 | 0 | 0 | 0 | 0 |
| 0 | 0 | 0 | 0 | 0 |
| 0.013013218 | 0 | 0 | 0 | 0.247447801 |
| 0 | 0 | 0 | 0 | 0 |
| 0 | 0 | 0 | 0 | 0 |

| | | | | |
|---|---|---|---|---|
| 0 | 0 | 0 | 0 | 0 |
| 0 | 0 | 0 | 0 | 0 |
| 0 | 0 | 0 | 0 | 0 |
| 0 | 0 | 0 | 0 | 0 |
| 0 | 0 | 0 | 0 | 0 |
| 0 | 0 | 0 | 0 | 0 |
| 0 | 0 | 0 | 0 | 0 |
| 0 | 0 | 0 | 0 | 0.164647426 |
| 0.16729076 | 0 | 0 | 0 | 0 |
| 0 | 0 | 0 | 0.001426807 | 0 |
| 0 | 0 | 0 | 0 | 0 |
| 0 | 0 | 0 | 0 | 0 |
| 0 | 0 | 0 | 0 | 0 |
| 0.085242343 | 0 | 0 | 0 | 0 |
| 0.010102768 | 0 | 0 | 0 | 0 |
| 0.001268032 | 0.845421735 | 0.041255062 | 0 | 0 |
| 0 | 0 | 0 | 0 | 0 |
| 0 | 0 | 0 | 0 | 0 |
| 0 | 0 | 0 | 0 | 0 |
| 1.066944744 | 0 | 0 | 0.001511129 | 0.002464158 |
| 0.001589243 | 0 | 0 | 0 | 0 |
| 0 | 0 | 0 | 0 | 0 |
| 0 | 0 | 0 | 0 | 0 |
| 0 | 0 | 0 | 0 | 0 |
| 0 | 0 | 0 | 0 | 0 |
| 0 | 0 | 0 | 0 | 0 |
| 0 | 0 | 0 | 0 | 0 |
| 0 | 0 | 0 | 0 | 0 |
| 0 | 0 | 0 | 0 | 0 |
| 0.000919953 | 0 | 0 | 0 | 0 |
| 0.475322346 | 0 | 0 | 0.121270472 | 0.801854523 |
| 0 | 0 | 0 | 0 | 0 |
| 0 | 0 | 0 | 0 | 0 |
| 0 | 0 | 0 | 0 | 0 |
| 0 | 0 | 0 | 0 | 0 |
| 0 | 0 | 0 | 0 | 0 |
| 0 | 0 | 0 | 0 | 0 |
| 0 | 0 | 0 | 0 | 0 |
| 0 | 0 | 0 | 0 | 0 |
| 0 | 0 | 0 | 0 | 0 |
| 0 | 0 | 0 | 0 | 0 |
| 0 | 0 | 0 | 0 | 0 |
| 0.007691479 | 0 | 0 | 0 | 0 |
| 0 | 0 | 0 | 0 | 0.002515025 |
| 0 | 0 | 0 | 0 | 0 |
| 0 | 0 | 0 | 0 | 0 |
| 0 | 9.265924677 | 0.308994714 | 0.001231783 | 0.007431953 |
| 0 | 0 | 0 | 0 | 0 |

| | | | | |
|---|---|---|---|---|
| 0 | 0 | 0 | 0 | 0 |
| 0 | 0 | 0 | 0 | 0 |
| 0 | 0 | 0 | 0 | 0 |
| 0 | 0 | 0 | 0 | 0 |
| 0.04625751 | 0.018181526 | 0.021564562 | 0 | 0.189375856 |
| 0 | 0 | 0 | 0 | 0 |
| 0 | 0 | 0 | 0 | 0 |
| 0 | 0 | 0 | 0 | 0 |
| 0 | 0 | 0 | 0 | 0 |
| 0 | 0 | 0 | 0 | 0 |
| 0 | 0 | 0 | 0 | 0 |
| 0.001077096 | 0.041469663 | 0.003616823 | 0.001490546 | 0.644801851 |
| 0.000580789 | 0 | 0 | 0 | 0.001977162 |
| 0 | 0 | 0 | 0 | 0 |
| 0.003488494 | 0 | 0 | 0 | 0 |
| 0.00764305 | 0 | 0 | 0 | 0 |
| 0.004866934 | 0 | 0 | 0 | 0 |
| 0 | 0 | 0 | 0 | 0 |
| 0 | 0 | 0.240090867 | 0 | 0 |
| 0.125791641 | 0 | 0 | 0 | 0.007827173 |
| 0 | 0.065579386 | 0 | 0 | 0 |
| 0 | 0 | 0 | 0 | 0 |
| 0 | 0 | 0 | 0 | 0 |
| 0 | 0 | 0 | 0 | 0 |
| 0 | 0 | 0 | 0 | 0 |
| 0 | 0 | 0 | 0 | 0 |
| 0.015040389 | 0 | 0.000477586 | 0 | 0.022943313 |
| 0 | 0 | 0 | 0 | 0 |
| 0 | 0 | 0 | 0 | 0 |
| 0 | 0 | 0 | 0 | 0 |
| 0 | 0.002775896 | 0.002345932 | 0 | 0.013471485 |
| 0 | 0 | 0 | 0 | 0 |
| 0 | 0 | 0 | 0 | 0 |
| 0 | 0 | 0 | 0 | 0 |
| 0 | 0 | 0 | 0 | 0 |
| 0 | 0 | 0 | 0 | 0 |
| 0 | 0 | 0 | 0 | 0 |
| 0.011638376 | 0 | 0.011790854 | 0 | 0.121989137 |
| 0.077656361 | 0 | 0 | 0 | 0.158270108 |
| 0 | 0 | 0 | 0 | 0 |
| 0 | 0 | 0.02442814 | 0 | 0.002493432 |
| 0 | 0 | 0 | 0 | 0 |
| 0 | 0 | 0 | 0 | 0 |
| 0 | 0 | 0.002444048 | 0 | 0.023781379 |
| 0 | 0 | 0 | 0 | 0 |
| 0 | 0 | 0 | 0 | 0 |
| 0 | 0 | 0 | 0 | 0 |
| 0 | 0 | 0 | 0 | 0 |

| | | | | |
|---|---|---|---|---|
| 0 | 0 | 0 | 0 | 0 |
| 0.038976376 | 0 | 0 | 0 | 0 |
| 0 | 0 | 0 | 0 | 0 |
| 0.000924138 | 0.024751723 | 0.002758402 | 0 | 0.001430008 |
| 0 | 0.00210828 | 0 | 0 | 0 |
| 0.017699204 | 0 | 0.019810959 | 0 | 0.001711729 |
| 0.007558794 | 0 | 0 | 0 | 0 |
| 0 | 0 | 0.00354415 | 0 | 0 |
| 0.116581144 | 0 | 0 | 0 | 0 |
| 0 | 0 | 0 | 0 | 0 |
| 0.091090142 | 0 | 0 | 0 | 0 |
| 0 | 0 | 0 | 0 | 0 |
| 0.017500102 | 0 | 0 | 0 | 0.080369883 |
| 0 | 0 | 0 | 0 | 0 |
| 0 | 0 | 0 | 0 | 0 |
| 0 | 0 | 0 | 0 | 0 |
| 0 | 0 | 0 | 0 | 0 |
| 0 | 0 | 0 | 0 | 0 |
| 0 | 0 | 0 | 0 | 0 |
| 0 | 0 | 0 | 0 | 0 |
| 0 | 0 | 0 | 0 | 0 |
| 0 | 0 | 0 | 0 | 0 |
| 0.000862595 | 0.009901442 | 0.009563186 | 0 | 0.028983693 |
| 0.002472022 | 0 | 0 | 0 | 0.002025104 |
| 0 | 0 | 0 | 0 | 0 |
| 0 | 0 | 0 | 0 | 0 |
| 0.007770412 | 0 | 0 | 0 | 0 |
| 0 | 0 | 0 | 0 | 0 |
| 0 | 0 | 0 | 0 | 0 |
| 0 | 0 | 0 | 0 | 0 |
| 0.015113832 | 0 | 0 | 0.001955724 | 0.076964746 |
| 0.01285638 | 0 | 0 | 0 | 0 |
| 0 | 0 | 0 | 0 | 0 |
| 0 | 0 | 0 | 0 | 0 |
| 0 | 0 | 0 | 0 | 0 |
| 0 | 0.009067915 | 0 | 0 | 0 |
| 0 | 0 | 0.014596416 | 0 | 0.001807059 |
| 0 | 0 | 0 | 0 | 0 |
| 0.001394984 | 0.000889587 | 0 | 0 | 0.028421456 |
| 0 | 0 | 0 | 0 | 0 |
| 0 | 0 | 0 | 0 | 0 |
| 0 | 0 | 0 | 0 | 0 |
| 0 | 0 | 0 | 0 | 0 |
| 0 | 0 | 0 | 0 | 0 |
| 0 | 0 | 0 | 0 | 2.973188954 |
| 0.237902333 | 1.331941423 | 0.002137015 | 0.016176039 | 0.798298751 |
| 0 | 0 | 0.18022146 | 0 | 0 |
| 0 | 0 | 0 | 0 | 0 |

| | | | | |
|---|---|---|---|---|
| 0 | 0 | 0 | 0 | 0 |
| 0 | 0 | 0 | 0 | 0 |
| 0 | 0 | 0 | 0 | 0 |
| 0.000926523 | 0 | 0 | 0 | 0 |
| 0 | 0 | 0 | 0 | 0 |
| 0 | 0 | 0 | 0 | 0 |
| 0 | 0 | 0 | 0 | 0 |
| 0 | 0 | 0 | 0 | 0 |
| 0 | 0.00679061 | 0 | 0 | 0 |
| 0 | 0 | 0 | 0 | 0 |
| 0.000377235 | 0 | 0.001266733 | 0 | 0.004523923 |
| 0 | 0.003501843 | 0 | 0 | 0 |
| 0 | 0 | 0 | 0 | 0 |
| 0 | 0 | 0 | 0 | 0 |
| 0 | 0 | 0 | 0 | 0 |
| 0 | 0 | 0 | 0 | 0 |
| 0 | 0 | 0 | 0 | 0 |
| 0 | 0 | 0 | 0 | 0 |
| 0 | 0 | 0 | 0 | 0 |
| 0.014758219 | 0 | 0 | 0.00098902 | 0 |
| 0 | 0 | 0 | 0 | 0 |
| 0 | 0 | 0 | 0 | 0 |
| 0 | 0 | 0 | 0 | 0 |
| 0 | 0 | 0 | 0 | 0 |
| 0 | 0 | 0 | 0 | 0 |
| 0 | 0 | 0 | 0 | 0 |
| 0 | 0 | 0 | 0 | 0 |
| 0 | 0 | 0 | 0.154545938 | 0 |
| 0 | 0 | 0 | 0 | 0 |
| 0 | 0 | 0.001006331 | 0 | 0 |
| 0.005372389 | 0 | 0 | 0 | 0.003824077 |
| 0 | 0 | 0 | 0 | 0 |
| 0.017840366 | 0.06635183 | 0.027928621 | 0.018744529 | 0.146524713 |
| 0.001402378 | 0.000975603 | 0 | 0 | 0 |
| 0 | 0 | 0 | 0 | 0 |
| 0 | 0 | 0 | 0 | 0 |
| 0 | 0 | 0 | 0 | 0 |
| 0 | 0 | 0 | 0 | 0 |
| 0 | 0 | 0 | 0 | 0 |
| 0 | 0 | 0 | 0 | 0 |
| 0 | 0 | 0 | 0 | 0 |
| 0.03202278 | 1.06307295 | 0.009956537 | 0 | 0.319546297 |
| 0.080151246 | 0 | 0 | 0.003361897 | 0.104483422 |
| 0 | 0 | 0 | 0 | 0 |
| 0 | 0 | 0 | 0 | 0 |
| 0.003252471 | 0 | 0 | 0 | 0.027960335 |
| 0 | 0 | 0 | 0 | 0 |
| 0 | 0 | 0 | 0 | 0 |

| | | | | |
|---|---|---|---|---|
| 0 | 0 | 0 | 0 | 0 |
| 0 | 0 | 0 | 0.016159915 | 0 |
| 0.00183622 | 0.010538681 | 0 | 0 | 0.013496452 |
| 0 | 0 | 0 | 0 | 0 |
| 0.016616173 | 0 | 0 | 0 | 0.005671719 |
| 0.195635057 | 0 | 0.023073562 | 0 | 0 |
| 0 | 0 | 0 | 0 | 0 |
| 0 | 0 | 0 | 0 | 0 |
| 0 | 0 | 0 | 0 | 0 |
| 0 | 0 | 0 | 0 | 0 |
| 0 | 0 | 0 | 0 | 0 |
| 0 | 0 | 0 | 0 | 0 |
| 0 | 0 | 0 | 0 | 0 |
| 0 | 0 | 0.05470536 | 0 | 0 |
| 0 | 0 | 0 | 0 | 0 |
| 0 | 0 | 0 | 0 | 0 |
| 0 | 0 | 0 | 0 | 0 |
| 0 | 0 | 0 | 0 | 0 |
| 0 | 0 | 0 | 0 | 0 |
| 0 | 0 | 0 | 0 | 0 |
| 0 | 0 | 0 | 0 | 0 |
| 0 | 0 | 0 | 0 | 0 |
| 0 | 0 | 0 | 0 | 0 |
| 0 | 0 | 0 | 0 | 0 |
| 0.000504434 | 0 | 0 | 0 | 0 |
| 0 | 0.006310244 | 0.000946658 | 0 | 0.005692877 |
| 0 | 0 | 0 | 0 | 0 |
| 1.441042168 | 0 | 0 | 0 | 0 |
| 0 | 0 | 0 | 0 | 0 |
| 0 | 0 | 0 | 0 | 0 |
| 0 | 0 | 0 | 0 | 0 |
| 0 | 0 | 0 | 0 | 0 |
| 0 | 0 | 0 | 0 | 0 |
| 0 | 0 | 0 | 0 | 0 |
| 0 | 0 | 0 | 0 | 0 |
| 0 | 0 | 0 | 0 | 0 |
| 0 | 0 | 0 | 0 | 0 |
| 0 | 0 | 0 | 0 | 0.01191978 |
| 2.66206241 | 0.007271685 | 0 | 0.020038102 | 4.092172839 |
| 0 | 0 | 0 | 0 | 0 |
| 0 | 0 | 0 | 0 | 0 |
| 0 | 0 | 0 | 0 | 0.333072358 |
| 0.055256933 | 0 | 0 | 0 | 0 |
| 0 | 0 | 0 | 0 | 0 |
| 0 | 0 | 0 | 0 | 0 |
| 0 | 0 | 0 | 0 | 0 |
| 0.039344524 | 0 | 0.002325484 | 0 | 0 |
| 0 | 0 | 0 | 0 | 0 |

| | | | | |
|---|---|---|---|---|
| 0 | 0 | 0 | 0 | 0 |
| 0.009339262 | 0.034323552 | 0.005135084 | 0.001166094 | 0.031945497 |
| 0 | 0 | 0 | 0 | 0 |
| 0 | 0 | 0 | 0 | 0 |
| 0 | 0 | 0 | 0 | 0 |
| 0 | 0 | 0 | 0 | 0 |
| 0 | 0 | 0 | 0 | 0 |
| 0 | 0 | 0 | 0 | 0 |
| 0 | 0 | 0 | 0 | 0 |
| 0 | 0 | 0 | 0 | 0 |
| 0 | 0 | 0 | 0 | 0 |
| 0 | 0 | 0 | 0 | 0 |
| 0.032864793 | 3.495348613 | 0 | 0 | 0.162011857 |
| 0 | 0 | 0 | 0 | 0 |
| 0.003217601 | 0 | 0.010747348 | 0 | 0.009009443 |
| 0 | 0 | 0 | 0 | 0 |
| 0 | 0 | 0 | 0 | 0 |
| 0 | 0 | 0 | 0 | 0 |
| 0 | 0 | 0 | 0 | 0 |
| 0 | 0 | 0 | 0 | 0 |
| 0 | 0 | 0 | 0 | 0 |
| 0.066208235 | 0 | 0 | 0 | 0 |
| 0.064519524 | 0 | 0 | 0 | 0 |
| 0 | 0.700942538 | 0 | 0 | 0 |
| 0 | 0 | 0 | 0 | 0 |
| 0 | 0 | 0 | 0 | 0 |
| 0 | 0 | 0 | 0 | 0 |
| 0 | 0 | 0 | 0 | 0 |
| 0 | 0 | 0 | 0 | 0 |
| 0 | 0 | 0 | 0 | 0 |
| 0 | 0 | 0 | 0 | 0 |
| 0 | 0 | 0 | 0 | 0 |
| 0 | 0 | 0 | 0 | 0 |
| 0 | 0 | 0 | 0 | 0 |
| 0 | 0 | 0 | 0 | 0.356855495 |
| 0 | 0 | 0 | 0 | 0 |
| 0 | 0 | 0 | 0 | 0 |
| 0 | 0 | 0 | 0 | 0 |
| 0 | 0 | 0.001569815 | 0 | 0 |
| 0 | 0 | 0 | 0 | 0 |
| 0.068040229 | 0.012372459 | 0.01307008 | 0 | 0.123631877 |
| 0 | 0 | 0 | 0 | 0 |
| 0 | 0 | 0 | 0 | 0 |
| 0 | 0 | 0 | 0 | 0 |
| 0 | 0 | 0 | 0 | 0 |
| 0.002468164 | 0.271212659 | 0.844680142 | 0.015126169 | 0.64783631 |
| 0 | 0 | 0 | 0 | 0 |
| 0 | 0 | 0 | 0 | 0 |

| | | | | |
|---|---|---|---|---|
| 0 | 0 | 0 | 0 | 0 |
| 0 | 0 | 0 | 0 | 0 |
| 0 | 0 | 0 | 0 | 0 |
| 0 | 0 | 0 | 0 | 0 |
| 0.005264763 | 0 | 0 | 0 | 0 |
| 0 | 0 | 0 | 0 | 0 |
| 0 | 0 | 0 | 0 | 0 |
| 0 | 0.00223286 | 0 | 0 | 0 |
| 0 | 0 | 0 | 0 | 0 |
| 0.001056067 | 0.001443125 | 0 | 0 | 0.007236969 |
| 0 | 0 | 0 | 0 | 0 |
| 0 | 0 | 0 | 0 | 0 |
| 0.001474596 | 0 | 0 | 0 | 0.001140892 |
| 0 | 0 | 0 | 0 | 0 |
| 0 | 0 | 0 | 0 | 0 |
| 0 | 0 | 0 | 0 | 0 |
| 0 | 0 | 0 | 0 | 0 |
| 0 | 0 | 0 | 0 | 0 |
| 0 | 0 | 0 | 0 | 0 |
| 0.087674617 | 4.104647459 | 0.010969225 | 0.114998025 | 0.712050356 |
| 0 | 0 | 0 | 0 | 0 |
| 0 | 0 | 0 | 0 | 0 |
| 0 | 0 | 0 | 0 | 0 |
| 0 | 0 | 0 | 0 | 0 |
| 0 | 0 | 0 | 0 | 0 |
| 0 | 0 | 0 | 0 | 0 |
| 0 | 0 | 0 | 0 | 0 |
| 0 | 0 | 0 | 0 | 0 |
| 0 | 0 | 0 | 0 | 0 |
| 0.038584634 | 0 | 0.011307483 | 0 | 0.020842716 |
| 0 | 0 | 0 | 0 | 0 |
| 0 | 0 | 0 | 0 | 0 |
| 0 | 0 | 0 | 0 | 0 |
| 0 | 0 | 0 | 0 | 0 |
| 0 | 0 | 0 | 0 | 0 |
| 0 | 0 | 0 | 0 | 0 |
| 0 | 0 | 0 | 0 | 0 |
| 0 | 0 | 0 | 0 | 0 |
| 0 | 0 | 0 | 0 | 0 |
| 0 | 0 | 0 | 0 | 0 |
| 0 | 0 | 0 | 0 | 0 |
| 0 | 0 | 0 | 0 | 0 |
| 0.009273474 | 0 | 0 | 0 | 0.003038767 |
| 0 | 0 | 0 | 0 | 0 |
| 0.00345168 | 0 | 0.002575675 | 0 | 0.022508983 |
| 0 | 0 | 0 | 0 | 0 |
| 0 | 0 | 0 | 0 | 0 |
| 0 | 0 | 0.042358319 | 0 | 0 |

| | | | | |
|---|---|---|---|---|
| 0 | 0 | 0 | 0 | 0 |
| 0 | 0 | 0 | 0 | 0 |
| 0 | 0 | 0 | 0 | 0 |
| 0 | 0 | 0 | 0 | 0 |
| 0 | 0 | 0 | 0 | 0 |
| 0.005705801 | 0 | 0.009476861 | 0 | 0.022500045 |
| 0 | 0 | 0 | 0 | 0 |
| 0 | 0 | 0 | 0 | 0 |
| 0 | 0 | 0 | 0 | 0 |
| 0 | 0 | 0 | 0 | 0 |
| 0 | 0 | 0 | 0 | 0 |
| 0 | 0 | 0 | 0 | 0 |
| 0 | 0 | 0 | 0 | 0 |
| 0 | 0 | 0 | 0 | 0 |
| 0 | 0 | 0 | 0 | 0 |
| 0 | 0 | 0 | 0 | 0 |
| 0 | 0 | 0 | 0 | 0 |
| 0 | 0 | 0 | 0 | 0 |
| 0 | 0 | 0 | 0 | 0 |
| 0 | 0 | 0 | 0 | 0 |
| 0 | 0 | 0.000955505 | 0 | 0 |
| 0 | 0 | 0 | 0 | 0 |
| 0 | 0 | 0 | 0 | 0 |
| 0 | 0 | 0 | 0 | 0.077059108 |
| 0 | 0 | 0 | 0 | 0 |
| 0 | 0 | 0 | 0 | 0.005345901 |
| 0 | 0 | 0 | 0 | 0 |
| 0 | 0 | 0 | 0 | 0 |
| 0 | 0 | 0 | 0 | 0 |
| 0 | 0 | 0 | 0 | 0 |
| 0 | 0 | 0 | 0 | 0 |
| 0 | 0 | 0 | 0 | 0 |
| 0 | 0 | 0 | 0 | 0 |
| 0 | 0 | 0 | 0 | 0 |
| 0 | 0 | 0 | 0 | 0 |
| 0 | 0 | 0 | 0 | 0 |
| 0 | 0 | 0 | 0 | 0 |
| 0 | 0 | 0 | 0 | 0 |
| 1.102392918 | 0 | 0 | 0 | 0 |
| 0 | 0 | 0 | 0 | 0 |
| 0 | 0 | 0 | 0 | 0 |
| 0 | 0 | 0 | 0 | 0 |
| 0 | 0 | 0 | 0 | 0 |
| 0 | 0 | 0 | 0 | 0 |
| 0 | 0 | 0 | 0 | 0 |
| 0 | 0 | 0 | 0 | 0 |
| 0 | 0 | 0 | 0 | 0 |
| 0 | 0 | 0 | 0 | 0 |

| | | | | |
|---|---|---|---|---|
| 0 | 0 | 0 | 0 | 0 |
| 0.020606787 | 0 | 0 | 0 | 0.43429894 |
| 0 | 0 | 0 | 0 | 0 |
| 0 | 0 | 0 | 0 | 0 |
| 0 | 0 | 0 | 0 | 0 |
| 0 | 0 | 0 | 0 | 0 |
| 0 | 0 | 0 | 0 | 0 |
| 0 | 0 | 0 | 0 | 0 |
| 0 | 0 | 0 | 0 | 0 |
| 0 | 0 | 0 | 0 | 0 |
| 0 | 0.002839178 | 0 | 0.000640124 | 0 |
| 0 | 0 | 0 | 0 | 0 |
| 0.020669283 | 0 | 0 | 0.460639408 | 0.751613633 |
| 0 | 0 | 0 | 0 | 0 |
| 0 | 0 | 0 | 0 | 0 |
| 0 | 0 | 0 | 0 | 0 |
| 0 | 0 | 0 | 0 | 0 |
| 0 | 0 | 0 | 0 | 0 |
| 0 | 0 | 0 | 0 | 0 |
| 0 | 0 | 0 | 0 | 0 |
| 0 | 0.155082176 | 0.042301976 | 0 | 0.001906969 |
| 0 | 0 | 0 | 0 | 0 |
| 0 | 0 | 0 | 0 | 0 |
| 0.641804281 | 0 | 0 | 0.492812285 | 1.717695819 |
| 0 | 0 | 0 | 0 | 0 |
| 0 | 0 | 0 | 0 | 0 |
| 0 | 0 | 0 | 0 | 0 |
| 0 | 0 | 0 | 0 | 0 |
| 0 | 0 | 0 | 0 | 0 |
| 0 | 0 | 0 | 0 | 0 |
| 0 | 0 | 0 | 0 | 0 |
| 0 | 0 | 0 | 0 | 0 |
| 0.002160565 | 0 | 0 | 0 | 0 |
| 0 | 0 | 0 | 0 | 0 |

| PHL018-0047_V2 | PHL018-0114_V3 | PHL018-0002_V2 | PHL018-0034_V2 | PHL018-0041_V2 |
|---|---|---|---|---|
| EYCF | EYCF | CTRL | CTRL | EYCF |
| 0.235920591 | 0.021990751 | 0.028317287 | 0.018596776 | 0.003304245 |
| 0.748539633 | 0.104625562 | 0 | 0.610390563 | 0.018755062 |
| 6.349143326 | 10.29990276 | 0 | 0 | 3.851769593 |
| 0 | 0 | 0 | 0 | 0 |
| 0 | 0 | 0 | 0 | 0 |
| 0 | 0 | 0 | 0 | 0 |
| 0 | 0 | 0 | 0 | 0 |
| 0 | 0 | 0 | 0 | 0 |
| 0 | 0 | 0 | 0 | 0 |
| 0 | 0 | 0 | 0 | 0 |
| 0 | 0 | 0 | 0 | 0 |
| 0 | 0 | 0 | 0 | 0 |
| 0 | 0 | 0 | 0 | 0 |
| 3.745354458 | 2.670061549 | 35.07404729 | 0.100385876 | 10.19356626 |
| 0 | 0 | 0 | 0 | 0 |
| 0 | 0 | 0 | 0 | 0 |
| 0 | 0 | 0 | 0 | 0 |
| 0 | 0 | 0 | 0 | 0 |
| 0 | 0 | 0 | 0 | 0 |
| 0 | 0 | 0 | 0 | 0 |
| 0 | 0 | 0 | 0 | 0 |
| 0 | 0.00238159 | 0 | 0 | 0 |
| 0 | 0 | 0 | 0 | 0 |
| 0 | 0 | 0 | 0 | 0 |
| 0 | 0 | 0 | 0 | 0 |
| 0 | 0 | 0 | 0 | 0 |
| 0 | 0 | 0 | 0 | 0 |
| 0 | 0 | 0 | 0 | 0 |
| 0 | 0 | 0 | 0 | 0 |
| 0 | 0 | 0 | 0 | 0 |
| 0 | 0 | 0 | 0 | 0 |
| 0 | 0 | 0 | 0 | 0 |
| 0 | 0 | 0 | 0 | 0 |
| 0 | 0 | 0 | 0 | 0 |
| 0 | 0 | 0 | 0 | 0 |
| 0 | 1.020738787 | 0 | 0 | 0 |
| 0 | 0 | 0 | 0 | 0 |
| 0 | 0 | 0 | 0 | 0 |
| 0 | 0 | 0 | 0 | 0 |
| 0 | 0 | 0 | 0 | 0 |
| 0 | 0 | 0 | 0 | 0 |
| 0.116298804 | 0 | 0 | 0 | 0 |
| 0.031303622 | 0.318086435 | 0.117810552 | 13.83218453 | 0 |
| 0.337592546 | 0.001474854 | 0.005455489 | 0.069703499 | 0 |
| 0 | 0 | 0 | 0.007012153 | 0 |
| 0 | 0 | 0 | 0 | 0 |

| | | | | |
|---|---|---|---|---|
| 0 | 0 | 0 | 0 | 0 |
| 0.001656581 | 0.005417539 | 0.028347536 | 0.097023914 | 0.025898634 |
| 0.002348092 | 0.004031474 | 0 | 0 | 0 |
| 2.572195772 | 0.244442307 | 0.497311605 | 0.112846025 | 0.001177589 |
| 0.001447935 | 0 | 0 | 0 | 0 |
| 0 | 0 | 0 | 0 | 0 |
| 0.092330561 | 0.003774373 | 0.001566185 | 0 | 0 |
| 0 | 0 | 0 | 0 | 0 |
| 0 | 0 | 0 | 0 | 0 |
| 0 | 0 | 0 | 0 | 0 |
| 0.004661204 | 0 | 0 | 0 | 0 |
| 0.07088838 | 0.161664344 | 0 | 0 | 0 |
| 0 | 0 | 0 | 0 | 0 |
| 0.017439361 | 0.009148911 | 0.140879529 | 0.021685491 | 0.007493319 |
| 0.002990867 | 0.015405185 | 0 | 0 | 0 |
| 0 | 0 | 0 | 0 | 0 |
| 0 | 0 | 0.001230133 | 25.56651999 | 0 |
| 0 | 0 | 0 | 0 | 0 |
| 0 | 0 | 0 | 0 | 0 |
| 0 | 0 | 0 | 0 | 0 |
| 0.51310557 | 0.314635821 | 0.048357836 | 0.022351405 | 0.015997856 |
| 0.03586587 | 0.02145453 | 0.000890261 | 0 | 0.009861897 |
| 0 | 0 | 0 | 0 | 0 |
| 0 | 0 | 0 | 0 | 0 |
| 0 | 0 | 0 | 0 | 0 |
| 0 | 0 | 0 | 0 | 0 |
| 0 | 0 | 0 | 0 | 0 |
| 0 | 0 | 0 | 0 | 0 |
| 0.000691701 | 0 | 0.001138901 | 0 | 0 |
| 0 | 0 | 0 | 0 | 0 |
| 0 | 0 | 0 | 0 | 0.435982014 |
| 0 | 0 | 0 | 0 | 0 |
| 0 | 0 | 0 | 0 | 0 |
| 0 | 0 | 0 | 0 | 0 |
| 0 | 0 | 0 | 0 | 0 |
| 0 | 0.038849352 | 0 | 0 | 0 |
| 0 | 0 | 0 | 0 | 0 |
| 0 | 0.002251374 | 0 | 0 | 0 |
| 0 | 0 | 0 | 0 | 0 |
| 0 | 0 | 0 | 0 | 0 |
| 0 | 0 | 0 | 0 | 0 |
| 0 | 0 | 0 | 0 | 0 |
| 0 | 0 | 0 | 0 | 0 |
| 0 | 0 | 0 | 0 | 0 |
| 0 | 0 | 0 | 0 | 0 |
| 0 | 0 | 0 | 0 | 0 |
| 0 | 0 | 0 | 0 | 0 |
| 0 | 0 | 0.450175431 | 0 | 0 |

| | | | | |
|---|---|---|---|---|
| 0 | 0 | 0 | 0 | 0 |
| 0 | 0 | 0 | 0 | 0 |
| 0 | 0 | 0 | 0 | 0 |
| 0 | 0 | 0 | 0 | 0 |
| 0 | 0 | 0 | 0 | 0 |
| 0.086616958 | 0 | 0.004650547 | 0 | 0 |
| 0 | 0 | 0 | 0 | 0 |
| 0 | 0 | 0 | 0 | 0 |
| 0.004877793 | 0 | 0.00129076 | 0 | 0 |
| 0 | 0 | 0 | 0 | 0 |
| 0 | 0 | 0 | 0 | 0 |
| 0.003640514 | 0 | 0 | 0 | 0 |
| 0 | 0 | 0 | 0 | 0 |
| 0 | 0 | 0 | 0 | 0 |
| 0 | 0 | 0 | 0 | 0 |
| 0 | 0 | 0 | 0 | 0 |
| 0 | 0 | 0 | 0 | 0 |
| 0 | 0 | 0 | 0 | 0 |
| 0.545201006 | 0 | 0 | 0 | 0 |
| 0 | 0 | 0 | 0 | 0 |
| 0 | 0 | 0 | 0 | 0 |
| 0 | 0 | 0 | 0 | 0 |
| 0 | 0 | 0 | 0 | 0 |
| 0 | 0 | 0 | 0 | 0 |
| 0 | 0 | 0 | 0 | 0 |
| 0 | 0 | 0 | 0 | 0 |
| 0 | 0 | 0 | 0 | 0 |
| 0 | 0 | 0 | 0 | 0 |
| 0 | 0 | 0 | 0 | 0 |
| 0 | 0.001616312 | 0 | 0 | 0 |
| 0.176520556 | 0.137281331 | 0 | 0 | 0 |
| 0.106037689 | 0 | 0 | 0.000827395 | 0 |
| 0 | 0 | 0 | 0 | 0 |
| 0 | 0 | 0 | 0 | 0 |
| 0 | 0 | 0 | 0 | 0 |
| 0 | 0 | 0 | 0 | 0 |
| 0 | 0 | 0 | 0 | 0 |
| 0 | 0 | 0 | 0 | 0 |
| 0.105962923 | 0.856428922 | 0 | 0 | 0 |
| 0 | 0 | 0 | 0 | 0 |
| 0 | 0 | 0 | 0 | 0 |
| 0 | 0 | 0 | 0 | 0 |
| 0.000586425 | 0 | 0.001206952 | 0 | 0.000822773 |
| 0 | 0 | 0 | 0 | 0 |
| 0 | 0 | 0 | 0 | 0 |
| 0 | 0 | 0 | 0 | 0 |
| 0 | 0 | 0 | 0 | 0 |
| 0 | 0 | 0 | 0 | 0 |

| | | | | |
|---|---|---|---|---|
| 0 | 0 | 0 | 0 | 0 |
| 0 | 0 | 0 | 0 | 0 |
| 0 | 0 | 0 | 0 | 0 |
| 0 | 0 | 0 | 0 | 0 |
| 0.107896909 | 0 | 0 | 0 | 0 |
| 0 | 0 | 0 | 0 | 0 |
| 0 | 0 | 0 | 0 | 0 |
| 0 | 0 | 0 | 0 | 0 |
| 0 | 0 | 0 | 0 | 0 |
| 0 | 0 | 0 | 0 | 0 |
| 0 | 0 | 0 | 0 | 0 |
| 0 | 0 | 0 | 0 | 0 |
| 0 | 0 | 0 | 0 | 0 |
| 0 | 0.002642519 | 0 | 0 | 0 |
| 0 | 0 | 0 | 0 | 0 |
| 0 | 0 | 0 | 0 | 0 |
| 0 | 0 | 0 | 0 | 0 |
| 0 | 0 | 0 | 0 | 0 |
| 0 | 0 | 0 | 0 | 0 |
| 0 | 0 | 0 | 0 | 0 |
| 0 | 0 | 0 | 0 | 0 |
| 0 | 0 | 0 | 0 | 0 |
| 0 | 0 | 0 | 0 | 0 |
| 0 | 0 | 0 | 0 | 0 |
| 0.056516688 | 0 | 0 | 0 | 0.016860562 |
| 0 | 0 | 0 | 0 | 0 |
| 0 | 0 | 0 | 0 | 0 |
| 0 | 0 | 0 | 0 | 0 |
| 0 | 0 | 0 | 0 | 0 |
| 0 | 0 | 0 | 0 | 0 |
| 0 | 0 | 0 | 0 | 0 |
| 0.085660741 | 0 | 0 | 0 | 0 |
| 0 | 0 | 0 | 0 | 0 |
| 0 | 0 | 0 | 0 | 0 |
| 0 | 0 | 0 | 0 | 0 |
| 0 | 0 | 0 | 0 | 0 |
| 0 | 0 | 0 | 0 | 0 |
| 0 | 0 | 0 | 0 | 0 |
| 0 | 0 | 0 | 0 | 0 |
| 0 | 0 | 0 | 0 | 0 |
| 0 | 0 | 0 | 0 | 0 |
| 0 | 0 | 0 | 0 | 0 |
| 0 | 0 | 0 | 0 | 0 |
| 0 | 0 | 0 | 0 | 0 |
| 0 | 0 | 0 | 0 | 0 |
| 0 | 0 | 0 | 0 | 0 |
| 0 | 0 | 0 | 0 | 0 |
| 0 | 0 | 0.0454951 | 0 | 0 |

| | | | | |
|---|---|---|---|---|
| 0 | 0 | 0 | 0 | 0 |
| 0 | 0 | 0 | 0 | 0 |
| 0 | 0 | 0 | 0 | 0 |
| 0 | 0 | 0 | 0 | 0 |
| 0 | 0 | 0 | 0 | 0 |
| 0 | 0 | 0 | 0 | 0 |
| 0 | 0 | 0 | 0 | 0 |
| 0 | 0 | 0 | 0 | 0 |
| 0 | 0 | 0 | 0 | 0 |
| 0 | 0 | 0 | 0 | 0 |
| 0 | 0 | 0 | 0 | 0 |
| 0 | 0 | 0 | 0 | 0 |
| 0 | 0.001282311 | 0.00161403 | 0.001278889 | 0 |
| 0 | 0 | 0 | 0 | 0 |
| 0.006281033 | 0 | 0 | 0 | 0 |
| 0 | 0 | 0 | 0 | 0 |
| 0 | 0 | 0 | 0 | 0 |
| 0 | 0 | 0 | 0 | 0 |
| 0 | 0 | 0 | 0 | 0 |
| 0 | 0 | 0 | 0 | 0 |
| 0.057953362 | 0.123077492 | 0.177190847 | 0.008198452 | 0.027919844 |
| 0 | 0 | 0 | 0 | 0 |
| 0 | 0 | 0 | 0 | 0 |
| 0 | 0 | 0 | 0 | 0 |
| 0 | 0 | 0 | 0 | 0 |
| 0 | 0 | 0 | 0 | 0 |
| 0.014028894 | 0.002559908 | 0.030631134 | 0 | 0.000513469 |
| 0 | 0 | 0 | 0 | 0 |
| 0 | 0 | 0 | 0 | 0 |
| 0 | 0 | 0 | 0 | 0 |
| 0 | 0.016006707 | 0.027780971 | 0.056518481 | 0 |
| 0 | 0 | 0 | 0 | 0 |
| 0 | 0 | 0 | 0 | 0 |
| 0.036270929 | 0.036143387 | 0 | 0 | 0 |
| 0.010449079 | 0.007800076 | 0.006171231 | 0.002000381 | 0 |
| 0 | 0 | 0 | 0 | 0 |
| 0 | 0 | 0 | 0 | 0 |
| 0 | 0 | 0 | 0 | 0 |
| 0 | 0 | 0 | 0 | 0 |
| 0 | 0 | 0 | 0 | 0 |
| 0 | 0 | 0 | 0 | 0 |
| 0 | 0 | 0 | 0 | 0 |
| 0 | 0 | 0 | 0 | 0 |
| 0 | 0 | 0 | 0 | 0 |
| 0 | 0 | 0 | 0 | 0 |
| 0 | 0 | 0 | 0 | 0 |
| 0 | 0 | 0 | 0 | 0 |
| 0 | 0.00574383 | 0.004997484 | 0 | 0 |

| | | | | |
|---|---|---|---|---|
| 0 | 0 | 0 | 0 | 0 |
| 0 | 0 | 0 | 0 | 0 |
| 0.00266254 | 0 | 0 | 0 | 0 |
| 0 | 0.001413127 | 0 | 0 | 0 |
| 0 | 0 | 0 | 0 | 0 |
| 0 | 0 | 0 | 0 | 0 |
| 0 | 0 | 0 | 0 | 0 |
| 0 | 0 | 0 | 0 | 0 |
| 0 | 0 | 0 | 0 | 0 |
| 0 | 0 | 0 | 0 | 0 |
| 0 | 0 | 0 | 0 | 0 |
| 0 | 0 | 0 | 0 | 0 |
| 0 | 0 | 0 | 0 | 0 |
| 0 | 0 | 0 | 0 | 0.017005581 |
| 0.003573147 | 0 | 0 | 0 | 0 |
| 0.006426665 | 0 | 0 | 0 | 0 |
| 0 | 0 | 0 | 0 | 0 |
| 0 | 0 | 0 | 0.085653265 | 0 |
| 0 | 0.001319034 | 0 | 0.065560931 | 0 |
| 0 | 0 | 0 | 0 | 0 |
| 0.015056162 | 0.014731801 | 0.008096835 | 0.007128431 | 0.169062365 |
| 0 | 0 | 0 | 0 | 0 |
| 0 | 0 | 0 | 0 | 0 |
| 0 | 0 | 0 | 0 | 0 |
| 0.005789468 | 0 | 0.086983955 | 0 | 0 |
| 0 | 0 | 0 | 0 | 0 |
| 0 | 0 | 0 | 0 | 0 |
| 0 | 0 | 0 | 0 | 0 |
| 0 | 0 | 0 | 0 | 0 |
| 0 | 0 | 0 | 0 | 0 |
| 0.897626798 | 0.532212759 | 0.275188592 | 0.001223024 | 0.055917622 |
| 0 | 0 | 0 | 0 | 0 |
| 0.045293459 | 0 | 0 | 0 | 0.020746609 |
| 0 | 0.179809421 | 0 | 0 | 0 |
| 0 | 0 | 0 | 0 | 0 |
| 0 | 0 | 0 | 0 | 0 |
| 0 | 0 | 0 | 0 | 0 |
| 0 | 0 | 0 | 0 | 0 |
| 0 | 0 | 0 | 0 | 0 |
| 0 | 0 | 0 | 0.01518313 | 0 |
| 0 | 0 | 0 | 0 | 0 |
| 0 | 0.009375042 | 0 | 0 | 0.000459667 |
| 0 | 0 | 0 | 0 | 0 |
| 0 | 0 | 0 | 0 | 0 |
| 0 | 0 | 0 | 0 | 0 |
| 0 | 0 | 0 | 0 | 0 |
| 0 | 0 | 0 | 0 | 0 |
| 0 | 0 | 0 | 0 | 0 |

| | | | | |
|---|---|---|---|---|
| 0 | 0 | 0 | 0 | 0 |
| 0 | 0 | 0 | 0 | 0 |
| 0.310139297 | 0.001046392 | 0.005080171 | 0.049645515 | 0 |
| 0 | 0 | 0 | 0 | 0 |
| 0 | 0 | 0 | 0 | 0 |
| 0 | 0 | 0 | 0 | 0 |
| 0 | 0 | 0 | 0 | 0 |
| 0 | 0 | 0 | 0 | 0 |
| 0 | 0 | 0 | 0 | 0 |
| 0 | 0 | 0 | 0.00184076 | 0 |
| 0 | 0 | 0 | 0 | 0 |
| 0 | 0 | 0 | 0 | 0 |
| 0 | 0 | 0 | 0 | 0 |
| 0.010608634 | 0.011275406 | 0.017779272 | 0 | 0 |
| 0.003752588 | 0.014317494 | 0.023427648 | 0 | 0 |
| 0 | 0 | 0 | 0 | 0 |
| 0 | 0 | 0.031093765 | 0.00859984 | 0.001399766 |
| 0 | 0.036487596 | 0 | 0 | 0 |
| 0 | 0 | 0 | 0 | 0 |
| 0 | 0 | 0 | 0 | 0 |
| 0 | 0 | 0 | 0 | 0 |
| 0 | 0.004908812 | 0.006355199 | 0 | 0 |
| 0 | 0 | 0 | 0 | 0 |
| 0 | 0 | 0 | 0 | 0.001679016 |
| 0 | 0 | 0 | 0 | 0 |
| 0 | 0 | 0 | 0 | 0 |
| 0 | 0 | 0 | 0 | 0 |
| 0 | 0 | 0 | 0 | 0 |
| 0 | 0 | 0 | 0 | 0 |
| 0.001550887 | 0 | 0 | 0 | 0 |
| 0 | 0 | 0 | 0 | 0 |
| 0 | 0 | 0 | 0 | 0 |
| 0 | 0 | 0 | 0 | 0 |
| 0.000741717 | 0 | 0 | 0 | 0.011863444 |
| 0 | 0 | 0 | 0 | 0 |
| 0.011992312 | 0 | 0 | 0 | 0 |
| 0 | 0 | 0 | 0 | 0 |
| 0.004579991 | 0 | 0.002328856 | 0.230397228 | 0 |
| 0 | 0 | 0 | 0 | 0 |
| 0 | 0 | 0 | 0 | 0 |
| 0 | 0.00222533 | 0 | 0 | 0 |
| 0 | 0 | 0 | 0 | 0 |
| 0 | 0 | 0 | 0 | 0 |
| 0 | 0 | 0 | 0.000982343 | 0 |
| 0 | 0 | 0 | 0 | 0 |
| 0 | 0 | 0 | 0 | 0 |
| 0 | 0 | 0 | 0 | 0 |
| 0 | 0 | 0.041103509 | 0 | 0 |

| | | | | |
|---|---|---|---|---|
| 0 | 0 | 0 | 0.007367453 | 0 |
| 0 | 0 | 0 | 0 | 0 |
| 0.001796804 | 0 | 0 | 0.004175557 | 0 |
| 0 | 0 | 0 | 0 | 0 |
| 0.017173702 | 0 | 0.008650495 | 0.000663318 | 0.009321059 |
| 0 | 0 | 0 | 0 | 0 |
| 0 | 0 | 0 | 0 | 0 |
| 0.113711944 | 0.009063773 | 0.02190426 | 0 | 0.006399436 |
| 0 | 0 | 0 | 0 | 0 |
| 0.007835456 | 0.123691088 | 0 | 0.006069553 | 2.412225856 |
| 0 | 0 | 0 | 0 | 0 |
| 0.010029502 | 0 | 0 | 0 | 0 |
| 0.034615187 | 0.005660125 | 0.037109175 | 0.005955186 | 0 |
| 0.00751022 | 0.005290012 | 0 | 0 | 0 |
| 0 | 0 | 0 | 0 | 0 |
| 0 | 0 | 0 | 0 | 0 |
| 0 | 0 | 0 | 0 | 0 |
| 0 | 0.003402265 | 0 | 0 | 0 |
| 0 | 0 | 0.24165879 | 0 | 0.344216064 |
| 0 | 0 | 0 | 0.0118064 | 0 |
| 0 | 0 | 0 | 0 | 0 |
| 0 | 0 | 0.395783226 | 0 | 0 |
| 0.004710119 | 0 | 0 | 0 | 0 |
| 0 | 0 | 0 | 0 | 0 |
| 0 | 0 | 0 | 0 | 0 |
| 0 | 0 | 0 | 0 | 0 |
| 0.007405654 | 0 | 0 | 0 | 0 |
| 0 | 0 | 0.044328805 | 0.001572728 | 0 |
| 0 | 0.314647971 | 0 | 0 | 0.000763737 |
| 0.002507155 | 0 | 0 | 0 | 0 |
| 0 | 0.093597384 | 0.056360915 | 0 | 0 |
| 0 | 0 | 0 | 0 | 0 |
| 0 | 0.002577186 | 0 | 0 | 0 |
| 0 | 0 | 0 | 0 | 0 |
| 0 | 0 | 0 | 0 | 0 |
| 0 | 0 | 0.006935151 | 0 | 0 |
| 0 | 0.003161819 | 0 | 0.008845849 | 0 |
| 0 | 0 | 0 | 0 | 0 |
| 0 | 0 | 0 | 0 | 0 |
| 0 | 0.013067415 | 0 | 0.002279745 | 0.046287855 |
| 0 | 0.012552781 | 0 | 0 | 0 |
| 0 | 0.270987902 | 0 | 0 | 0 |
| 0 | 0 | 0 | 0 | 0 |
| 0 | 0 | 0 | 0 | 0 |
| 0 | 0 | 0 | 0 | 0 |
| 0 | 0 | 0 | 0 | 0 |
| 0.006060692 | 0.002207267 | 0 | 0 | 0 |
| 0 | 0.004865586 | 0 | 0 | 0 |

| | | | | |
|---|---|---|---|---|
| 0 | 0.017881886 | 0 | 0 | 0 |
| 0 | 0 | 0 | 0 | 0.032656049 |
| 0 | 0 | 0 | 0 | 0 |
| 0 | 0 | 0 | 0 | 0 |
| 0 | 0 | 0.018493616 | 0 | 0 |
| 0 | 0.237179777 | 0.030545742 | 0 | 0 |
| 0.043428289 | 0 | 0 | 0 | 0 |
| 0 | 0 | 0 | 0 | 0 |
| 0 | 0 | 0 | 0 | 0 |
| 0 | 0 | 0 | 0 | 0 |
| 0.022649278 | 0.002304407 | 0.091988277 | 0 | 0.016524384 |
| 0 | 0 | 0 | 0 | 0 |
| 0 | 0 | 0 | 0 | 0 |
| 0.004755089 | 0 | 0.029360101 | 0.006513835 | 0.003402486 |
| 0 | 0.256353393 | 0 | 0 | 0 |
| 0 | 0 | 0 | 0 | 0 |
| 0 | 0 | 0 | 0 | 0 |
| 0 | 0 | 0.108002615 | 0 | 0 |
| 0.043778177 | 0 | 0 | 0 | 0 |
| 0.049898023 | 0.048301919 | 0 | 0 | 0 |
| 0 | 0 | 0 | 0 | 0 |
| 0.642717947 | 0 | 0 | 0 | 0.011766926 |
| 0.001353706 | 0 | 0 | 0 | 0 |
| 0 | 0 | 0 | 0 | 0 |
| 0 | 0 | 0 | 0 | 0 |
| 0 | 0 | 0 | 0 | 0 |
| 0 | 0.124518477 | 0 | 0.006810482 | 0 |
| 0 | 0 | 0 | 0 | 0 |
| 0 | 0 | 0 | 0 | 0 |
| 0.734982719 | 0.877168758 | 0.98853138 | 0.39078264 | 0.143752518 |
| 0.004032437 | 0 | 0 | 0 | 0 |
| 0.001132283 | 0 | 0 | 0 | 0 |
| 0 | 0 | 0 | 0 | 0 |
| 0 | 0 | 0 | 0 | 0 |
| 0 | 0 | 0 | 0 | 0.001626751 |
| 0 | 0 | 0 | 0 | 0 |
| 0 | 0 | 0 | 0 | 0 |
| 0.004604367 | 0 | 0 | 0 | 0 |
| 0.305471778 | 0 | 0 | 0 | 2.534997684 |
| 0 | 0 | 0 | 0 | 0 |
| 0.076229407 | 0.013423525 | 0.092445468 | 0 | 0.000658168 |
| 3.695763499 | 0 | 0 | 0 | 0 |
| 0 | 0 | 0 | 0 | 0 |
| 0 | 0 | 0 | 0 | 0.046052907 |
| 0.001763913 | 0.003280861 | 0.001089121 | 0 | 0 |
| 0 | 0.054718969 | 0 | 0 | 0 |
| 0.002666941 | 0.005400765 | 0.021787 | 0.001806603 | 0.001726986 |
| 0 | 0 | 0 | 0 | 0 |

| | | | | |
|---|---|---|---|---|
| 0 | 0 | 0 | 0.003962746 | 0 |
| 0 | 0 | 0 | 0 | 0 |
| 0 | 0 | 0.00468499 | 1.526383927 | 0 |
| 0 | 0 | 0 | 0 | 0 |
| 0 | 0.188336832 | 0 | 0 | 0 |
| 0 | 0.093820205 | 0 | 0 | 0 |
| 0 | 0 | 0 | 0 | 0 |
| 0.00283696 | 0 | 0 | 0 | 0 |
| 0.117782259 | 0 | 0.264998521 | 0.001450953 | 0 |
| 0 | 0.660654307 | 0 | 0 | 0 |
| 0 | 0 | 0 | 0 | 0 |
| 0 | 0 | 0 | 0 | 0 |
| 0 | 0 | 0 | 0 | 0 |
| 0.050607037 | 0.00917369 | 0.001979455 | 0 | 0 |
| 0 | 0 | 0 | 0 | 0 |
| 0.128845877 | 0.325681082 | 0.062318366 | 0 | 0 |
| 0.00614886 | 0.001589236 | 0.005143765 | 0.000970405 | 0 |
| 0 | 0.002482533 | 0 | 0 | 0.482014922 |
| 0.001639047 | 0 | 0 | 0.001603768 | 0 |
| 0 | 0 | 0 | 0 | 0 |
| 0 | 0 | 0 | 0 | 0.00273567 |
| 0 | 0 | 0 | 0 | 0 |
| 0.01790473 | 0 | 0 | 0 | 0 |
| 0 | 0.48018148 | 0 | 0 | 0.006819823 |
| 0.02409908 | 0 | 0 | 0 | 0.001716598 |
| 0 | 0 | 0 | 0 | 0.006818768 |
| 0.008057834 | 0 | 0.008706734 | 0 | 0 |
| 0.254967171 | 0 | 0 | 0 | 0 |
| 0 | 0 | 0 | 0 | 0 |
| 0 | 0 | 0 | 0 | 0 |
| 0.090387409 | 0.288894137 | 0.007803848 | 0 | 0.001289659 |
| 0.00256951 | 0 | 0.043012688 | 0.012850374 | 0.001682382 |
| 0.045124723 | 0.064718311 | 0 | 0 | 0.075973717 |
| 0.018309651 | 0 | 0 | 0 | 0 |
| 0 | 0 | 0 | 0.012787145 | 0 |
| 0 | 0 | 0 | 0 | 0 |
| 0 | 0 | 0.006069696 | 0.001683279 | 0 |
| 0.101286396 | 0.005499686 | 0.006408158 | 0 | 0 |
| 0 | 0 | 0 | 0 | 0 |
| 0 | 0.002518071 | 0 | 0 | 0 |
| 0 | 0 | 0 | 0 | 0 |
| 0 | 0 | 0 | 0 | 0 |
| 0 | 0 | 0 | 0.003937355 | 0 |
| 0 | 0 | 0 | 0 | 0.02917513 |
| 0.22695002 | 0 | 0 | 0 | 0.023726889 |
| 0 | 0.56090218 | 0 | 0 | 0 |
| 0 | 0 | 0 | 0 | 0 |
| 0 | 0 | 0 | 0 | 0 |

| | | | | |
|---|---|---|---|---|
| 0 | 0 | 0.002463267 | 0 | 0 |
| 0.723861514 | 0 | 1.072733482 | 1.684857989 | 0.051367946 |
| 0 | 0 | 0 | 0 | 0 |
| 0.00095166 | 0 | 0 | 0 | 0 |
| 0 | 0.003528953 | 0 | 0 | 0 |
| 0 | 0 | 0 | 0 | 0 |
| 0 | 0.012756618 | 0 | 0.001185333 | 0 |
| 0 | 0 | 0 | 0.002558773 | 0 |
| 0.003356331 | 0 | 0 | 0 | 0 |
| 0 | 0 | 0 | 0 | 0 |
| 0.005449143 | 0 | 0 | 0 | 0 |
| 0.026089242 | 0.283025361 | 0.045462319 | 0.044815281 | 0.002684296 |
| 0 | 0 | 0 | 0 | 0 |
| 0 | 0 | 0 | 0 | 0 |
| 0.001116466 | 0.020191101 | 0 | 0 | 0 |
| 0 | 0 | 0 | 0 | 0 |
| 0.011615865 | 0 | 0.151496334 | 0.055433412 | 0.008234491 |
| 0.026718926 | 0.078425043 | 0.00653366 | 0.007765492 | 4.310543893 |
| 0 | 0 | 0 | 0 | 0 |
| 0 | 0.160836881 | 0 | 0 | 0.003325995 |
| 0.370715051 | 0.005532569 | 0.064101399 | 0.091095193 | 0.00159222 |
| 0 | 0 | 0 | 0 | 0 |
| 0 | 0 | 0 | 0 | 0 |
| 0 | 0 | 0 | 0 | 0 |
| 0 | 0 | 0 | 0 | 0 |
| 0 | 0 | 0 | 0 | 0 |
| 0.124875508 | 0 | 0 | 0 | 0 |
| 0.064552405 | 0.093062207 | 0 | 0.055950587 | 0.14403306 |
| 0 | 0 | 0 | 0 | 0 |
| 0 | 0 | 0 | 0 | 0 |
| 0 | 0 | 0.032852357 | 0.010787544 | 0 |
| 0 | 0 | 0 | 0 | 0 |
| 0 | 0 | 0 | 0 | 0 |
| 0 | 0 | 0 | 0 | 0 |
| 0.455668174 | 0 | 0 | 0 | 0 |
| 0.45817414 | 0.661163218 | 2.29866093 | 2.825995126 | 8.256903314 |
| 0.007178339 | 0.009619195 | 0 | 0 | 0 |
| 0 | 0.002190733 | 0 | 0 | 0 |
| 0.013501723 | 0.002523135 | 0.007145643 | 0 | 0 |
| 0 | 0 | 0 | 0 | 0 |
| 0.071237333 | 0 | 0 | 0 | 0.023580026 |
| 0 | 0 | 0 | 0 | 0 |
| 0 | 0.012069152 | 0.004547057 | 0.005076807 | 0.001690748 |
| 0.005494163 | 0.041986132 | 0 | 0.005692136 | 0 |
| 0 | 1.207446074 | 0 | 0 | 0 |
| 0 | 0 | 0 | 0 | 0 |
| 0 | 0 | 0 | 0 | 0 |
| 0.003134431 | 0.028103652 | 0.029675242 | 0 | 0 |

| | | | | |
|---|---|---|---|---|
| 0 | 0 | 0 | 0 | 0 |
| 0 | 0 | 0 | 0 | 0 |
| 0.000460154 | 0 | 0 | 0 | 0 |
| 1.942903501 | 0 | 0 | 0 | 0 |
| 0 | 0 | 0 | 0 | 0 |
| 0 | 0 | 0 | 0 | 0 |
| 0 | 0 | 0 | 0 | 0 |
| 0.055106936 | 0 | 0 | 0 | 0 |
| 0 | 0.117342194 | 0 | 0 | 0 |
| 0 | 0 | 0 | 0 | 0 |
| 0 | 0 | 0 | 0 | 0 |
| 0.004855952 | 0 | 0 | 0 | 0 |
| 0 | 0 | 0 | 0 | 0 |
| 0 | 1.644953606 | 0 | 0 | 0 |
| 0.06087776 | 0.009349638 | 0 | 0.007690983 | 0 |
| 0 | 0 | 0 | 0 | 0 |
| 0 | 0 | 0 | 0 | 0 |
| 0.111698581 | 0 | 0 | 0 | 0 |
| 0 | 0 | 0 | 0 | 0 |
| 0 | 0 | 0 | 0 | 0 |
| 0 | 0 | 0 | 0 | 0 |
| 0 | 0 | 0 | 0 | 0 |
| 0 | 0 | 0 | 0 | 0 |
| 0.047951914 | 0 | 0.110364356 | 0.002614725 | 0.093731059 |
| 0.018710233 | 0 | 0.004102777 | 0.015912135 | 0 |
| 0 | 5.583429393 | 0 | 0 | 0 |
| 0 | 0 | 0 | 0 | 0 |
| 0 | 0.358303347 | 0 | 0 | 0 |
| 0 | 0 | 0 | 0 | 0 |
| 0 | 0 | 0 | 0 | 0 |
| 0 | 0 | 0 | 0 | 0 |
| 1.728686272 | 0 | 0 | 0 | 0 |
| 0.047536637 | 0.008066731 | 0 | 0 | 0 |
| 0 | 0 | 0 | 0 | 0 |
| 0 | 0 | 0 | 0 | 0 |
| 0.147792795 | 0 | 0 | 0 | 0 |
| 0 | 0 | 0 | 0 | 0 |
| 0 | 0 | 0 | 0 | 0 |
| 0 | 0 | 0 | 0.00114144 | 0 |
| 0.001069465 | 0 | 0 | 0 | 0 |
| 0.049092555 | 0.261066323 | 0.016151648 | 0.01227194 | 0 |
| 0 | 0 | 0 | 0 | 0 |
| 1.918736338 | 0.028046069 | 1.202366546 | 0.041010444 | 0.306870009 |
| 0.074762515 | 0 | 0 | 0 | 0 |
| 0 | 0 | 0 | 0 | 0 |
| 0 | 0 | 0 | 0 | 0 |
| 0 | 0 | 0 | 0 | 0 |
| 0.001723062 | 0 | 0.044683706 | 0 | 0 |

| | | | | |
|---|---|---|---|---|
| 0 | 0 | 0 | 0 | 0 |
| 0.000784879 | 0.030544931 | 0 | 0 | 0 |
| 0.11397073 | 1.00320416 | 2.016275299 | 0.783047518 | 0.164771244 |
| 0 | 0 | 0 | 0 | 0 |
| 0 | 0 | 0 | 0 | 0 |
| 0.000650592 | 0 | 0 | 0 | 0 |
| 0 | 0 | 0 | 0 | 0 |
| 0 | 0 | 0 | 0 | 0 |
| 0 | 0 | 0 | 0 | 0 |
| 0 | 1.164702728 | 0 | 0 | 0 |
| 0.019663419 | 0.029235817 | 0.007509957 | 0.007735743 | 0 |
| 3.562118937 | 7.133860534 | 0 | 0.902203718 | 1.51717286 |
| 0 | 0 | 0 | 0 | 0 |
| 0.008313602 | 0 | 0.002971852 | 0.086056127 | 0 |
| 0.33308887 | 0.004933446 | 2.750920516 | 0.066465965 | 0.00682134 |
| 0 | 0 | 0 | 0 | 0 |
| 0 | 0 | 0 | 0 | 0 |
| 0 | 0 | 0 | 0 | 0 |
| 0 | 0 | 0 | 0 | 0 |
| 0.00651239 | 0.01720187 | 0 | 0 | 0.006044537 |
| 0.305830039 | 0.148384846 | 0.393403142 | 0.001662485 | 0 |
| 0 | 0.278686981 | 0.328966931 | 0 | 9.041485853 |
| 1.331020584 | 3.302689954 | 0 | 0.416432251 | 39.02005592 |
| 0 | 0 | 0 | 0 | 0 |
| 0 | 0 | 0 | 0 | 0 |
| 6.857175458 | 0.00209736 | 0 | 0 | 0 |
| 0.00162572 | 0 | 0 | 0 | 0 |
| 0 | 0 | 0 | 0 | 0 |
| 0.248191208 | 0 | 0 | 0 | 0 |
| 0 | 0 | 0 | 0 | 0 |
| 0 | 0 | 0 | 0 | 0 |
| 0.320166163 | 0 | 0 | 0.006591956 | 0 |
| 0.068912302 | 0 | 0.317001092 | 0.103893658 | 0 |
| 0.043342639 | 0 | 0 | 0 | 0 |
| 0 | 0.004089325 | 0.002715 | 0.000537813 | 0 |
| 0.003587087 | 0.002809241 | 0 | 0.336763523 | 0 |
| 0.002735587 | 0 | 0 | 0 | 0 |
| 0 | 0 | 0 | 0 | 0 |
| 0.016569536 | 0 | 0 | 0 | 0 |
| 0 | 0 | 0 | 0 | 0 |
| 0 | 0 | 0 | 0 | 0 |
| 0 | 0 | 0 | 0 | 0 |
| 0 | 0 | 0 | 0 | 0 |
| 0 | 0 | 0 | 0 | 0 |
| 0.006889541 | 0.000739297 | 0.00132935 | 0 | 0 |
| 0 | 0 | 0 | 0 | 0 |
| 0 | 0 | 0 | 0 | 0 |
| 0.009089795 | 0.228299333 | 0.006414236 | 13.5879659 | 0.003060786 |

| | | | | |
|---|---|---|---|---|
| 0 | 0 | 0 | 0 | 0 |
| 0 | 0 | 0 | 0 | 0 |
| 0 | 0 | 0.002519655 | 0 | 0 |
| 0.012661345 | 0 | 0 | 0 | 0 |
| 0 | 0 | 0 | 0 | 0 |
| 0.0351679 | 0 | 0 | 0 | 0 |
| 0 | 0 | 0 | 0 | 0 |
| 0.108233589 | 0 | 0.001251077 | 0.003139119 | 0 |
| 0 | 0 | 0 | 0 | 0 |
| 0 | 0 | 0 | 0 | 0 |
| 0.000817472 | 0.010058623 | 0.015394708 | 0.014397772 | 0 |
| 0 | 0 | 0 | 0 | 0 |
| 0.005905023 | 0 | 0 | 0 | 0 |
| 0 | 0 | 0 | 0 | 0 |
| 0 | 0 | 0 | 0 | 0 |
| 0 | 0 | 0 | 0 | 0 |
| 0 | 0 | 0 | 0 | 0 |
| 0 | 0 | 0 | 0.001331757 | 0 |
| 0 | 0 | 0 | 0 | 0 |
| 0 | 0 | 0 | 0 | 0 |
| 0.011940735 | 0 | 0 | 0 | 0 |
| 0 | 0 | 0 | 0 | 0 |
| 0 | 0 | 0 | 0 | 0 |
| 0 | 0 | 0 | 0 | 0 |
| 0 | 0 | 0 | 0 | 0 |
| 0 | 0 | 0 | 0 | 0 |
| 0 | 0 | 0.066419595 | 0 | 0 |
| 0 | 0 | 0 | 0 | 0 |
| 0 | 0 | 0 | 0 | 0 |
| 0.100087014 | 0 | 0 | 0 | 0.001903094 |
| 0 | 0 | 0 | 0 | 0 |
| 0 | 0 | 0 | 0 | 0 |
| 0 | 0 | 0 | 0 | 0 |
| 0 | 0 | 0 | 0 | 0 |
| 0 | 0 | 0 | 0 | 0 |
| 0 | 0 | 0 | 0 | 0 |
| 0 | 0 | 0 | 0 | 0 |
| 0.002595015 | 0 | 0.002777288 | 0.000677109 | 0.009029406 |
| 0 | 0 | 0 | 0 | 0 |
| 0 | 0 | 0 | 0 | 0 |
| 0.015095552 | 0.028983314 | 0 | 0.012229449 | 0 |
| 0 | 0 | 0 | 0 | 0 |
| 0 | 0 | 0 | 0 | 0 |
| 0 | 0.010384175 | 0 | 0 | 0 |
| 0 | 0 | 0 | 0 | 0.065485606 |
| 0 | 0 | 0 | 0 | 0 |
| 0.001479419 | 0 | 0 | 0 | 0 |
| 0 | 0 | 0 | 0 | 0 |

| | | | | |
|---|---|---|---|---|
| 0 | 0 | 0 | 0 | 0 |
| 0 | 0 | 0 | 0 | 0 |
| 0 | 0 | 0 | 0 | 0 |
| 0 | 0.103015565 | 0 | 0 | 0 |
| 0 | 0 | 0 | 0 | 0 |
| 0 | 0 | 0 | 0 | 0 |
| 0 | 0 | 0 | 0 | 0 |
| 0 | 0 | 0 | 0 | 0 |
| 0.081600274 | 0.001911705 | 5.59053442 | 0.004669234 | 0.365356673 |
| 0 | 0 | 0 | 0 | 0 |
| 0 | 0 | 0 | 0 | 0 |
| 0 | 0 | 0 | 0 | 0 |
| 0 | 0 | 0 | 0 | 0 |
| 0 | 0 | 0 | 0 | 0 |
| 0.352697976 | 2.909319778 | 0 | 0 | 0 |
| 0.000735574 | 0 | 5.854572901 | 0 | 0.002012467 |
| 0 | 0 | 0 | 0 | 0 |
| 0 | 0 | 0 | 0 | 0 |
| 0 | 0 | 0 | 0 | 0 |
| 0 | 0 | 0 | 0 | 0 |
| 0 | 0 | 0 | 0 | 0 |
| 0 | 0 | 0 | 0 | 0 |
| 0 | 0 | 0 | 0 | 0 |
| 0 | 0 | 0 | 0 | 0 |
| 0.008136925 | 0.015415618 | 0.003118414 | 0.541950428 | 0 |
| 1.45559752 | 0 | 0 | 0 | 0 |
| 0 | 0 | 0 | 0 | 0 |
| 0 | 0 | 0 | 0 | 0 |
| 0.02256855 | 0 | 0 | 0 | 0 |
| 0 | 0 | 0 | 0 | 0 |
| 0 | 0 | 0 | 0 | 0 |
| 0.036320178 | 0 | 0 | 0 | 0 |
| 0 | 0 | 0 | 0 | 0 |
| 0.003146843 | 0 | 0.001295337 | 0.000879746 | 0 |
| 0.001728334 | 0 | 0.006098017 | 0.047593327 | 0 |
| 0.005585019 | 0.006392666 | 0 | 0.004781705 | 0 |
| 0 | 0 | 0 | 0 | 0 |
| 6.305663563 | 3.671922688 | 0 | 0 | 0.31222923 |
| 0 | 0 | 0 | 0 | 0 |
| 0 | 0 | 0 | 0 | 0 |
| 0 | 0 | 0 | 0 | 0 |
| 0 | 0 | 0 | 0 | 0 |
| 0.645383372 | 0 | 0 | 0 | 0 |
| 0 | 0 | 0 | 0 | 0 |
| 0 | 0 | 0 | 0 | 0 |
| 0.000661388 | 0 | 0 | 0 | 0 |
| 0 | 0 | 0 | 0 | 0 |
| 1.6329258 | 0.007236248 | 0 | 0.009536667 | 0 |

| | | | | |
|---|---|---|---|---|
| 0.004714339 | 0 | 0 | 0.012108776 | 0 |
| 0 | 0 | 0 | 0 | 0 |
| 0 | 0 | 0 | 0 | 0 |
| 0 | 0 | 0 | 0 | 0 |
| 0 | 0 | 0 | 0.001339521 | 0 |
| 0 | 0 | 0 | 0 | 0 |
| 0 | 0 | 0 | 0 | 0 |
| 0 | 0 | 0 | 0 | 0 |
| 0 | 0 | 0 | 0 | 0 |
| 0 | 0 | 0 | 0 | 0 |
| 0 | 0 | 0 | 0 | 0 |
| 0.395131751 | 0 | 0 | 0 | 0.331718184 |
| 0 | 0 | 0 | 0 | 0 |
| 0 | 0 | 0 | 0 | 0 |
| 0.359620255 | 0 | 0.697699435 | 0.000885232 | 0.003046392 |
| 0 | 0 | 0 | 0 | 0 |
| 0.000786691 | 0 | 0 | 0 | 0 |
| 0 | 0 | 0 | 0 | 0 |
| 0.000991213 | 0 | 0 | 0 | 0 |
| 0 | 0 | 0 | 0 | 0 |
| 0 | 0 | 0 | 0 | 0 |
| 0 | 0 | 0 | 0 | 0 |
| 0.121634131 | 0.069942755 | 0 | 0 | 0 |
| 0 | 0 | 0 | 0 | 0 |
| 0 | 0 | 0 | 0 | 0 |
| 0 | 0 | 0 | 0 | 0 |
| 0 | 0 | 0 | 0 | 0 |
| 0 | 0 | 0 | 0 | 0 |
| 0.001833548 | 0 | 0 | 0 | 0 |
| 0 | 0 | 0 | 0 | 0 |
| 0 | 0 | 0 | 0 | 0 |
| 0.083059178 | 0.006718751 | 0.186654016 | 0.099650794 | 0.01012987 |
| 0 | 0 | 0 | 0 | 0 |
| 0.147065082 | 0 | 0.00778585 | 0 | 0 |
| 0.003608753 | 0.004782816 | 0 | 0.004274463 | 0 |
| 0.006855757 | 0.012611515 | 0 | 0 | 0.001648945 |
| 0.001180192 | 0 | 0 | 0 | 0 |
| 0 | 0 | 0 | 0 | 0 |
| 0.00065462 | 0 | 0 | 0 | 0 |
| 0 | 0 | 0 | 0 | 0 |
| 0 | 0 | 0 | 0 | 0 |
| 0 | 0 | 0 | 0 | 0 |
| 0 | 0 | 0 | 0 | 0 |
| 0 | 0 | 0 | 0 | 0 |
| 0.118998401 | 0.007709814 | 0.003234752 | 0 | 0 |
| 0 | 0 | 0 | 0 | 0 |
| 0 | 0 | 0 | 0 | 0.002650475 |
| 0.003539346 | 0.001012792 | 0 | 0 | 0 |

| | | | | |
|---|---|---|---|---|
| 0 | 0 | 0 | 0 | 0 |
| 0 | 0 | 0 | 0 | 0 |
| 0.012771843 | 0 | 0 | 0 | 0 |
| 0 | 0 | 0 | 0 | 0 |
| 0 | 0.06020072 | 0 | 0 | 0 |
| 0 | 0.003620475 | 0 | 0 | 0 |
| 0 | 0 | 0 | 0 | 0 |
| 0 | 0.078234372 | 0 | 0 | 0 |
| 0 | 0 | 0 | 0 | 0 |
| 0 | 0 | 0 | 0 | 0 |
| 0 | 0.83592524 | 0 | 0 | 0 |
| 0 | 0.006986389 | 0.005349157 | 0 | 0 |
| 0 | 0 | 0 | 0 | 0 |
| 0 | 0 | 0 | 0 | 0.03143229 |
| 0 | 0 | 0 | 0 | 0 |
| 0 | 0 | 0 | 0 | 0 |
| 0 | 0 | 0 | 0 | 0 |
| 0 | 0 | 0 | 0 | 0 |
| 0 | 0.037158188 | 0 | 0 | 0 |
| 0 | 0 | 0 | 0 | 0 |
| 0 | 0 | 0 | 0 | 0 |
| 0 | 0 | 0 | 0 | 0 |
| 0 | 0 | 0 | 0 | 0 |
| 0 | 0 | 0 | 0 | 0 |
| 0 | 0 | 0 | 0 | 0 |
| 0.001393497 | 0.003389396 | 0.01613267 | 0.006647078 | 0.005278827 |
| 0 | 0 | 0 | 0 | 0 |
| 0.071266723 | 0.043323233 | 0 | 0.001167774 | 0 |
| 0 | 0 | 0 | 0 | 0 |
| 0.586794492 | 0.243922865 | 0 | 0.101697557 | 0.287729011 |
| 0 | 0 | 0 | 0 | 0 |
| 0 | 0 | 0 | 0 | 0 |
| 0 | 0 | 0 | 0 | 0 |
| 0 | 0 | 0 | 0 | 0 |
| 0 | 0.022355936 | 0.006762416 | 0.015538928 | 0 |
| 0 | 0 | 0 | 0 | 0 |
| 0 | 0 | 0 | 0 | 0 |
| 0 | 0 | 0.001278894 | 0.001216009 | 0 |
| 0 | 0 | 0 | 0 | 0 |
| 0 | 0 | 0 | 0 | 0 |
| 14.04130429 | 25.6765757 | 1.178806718 | 14.2943452 | 6.274465075 |
| 0.4833565 | 0.103605058 | 0 | 0 | 0 |
| 0 | 0 | 0 | 0 | 0 |
| 0 | 0 | 0 | 0 | 0 |
| 0 | 0 | 0 | 0 | 0 |
| 0 | 0 | 0 | 0 | 0 |
| 0 | 0 | 0 | 0 | 0 |
| 0 | 0 | 0 | 0 | 0 |

| | | | | |
|---|---|---|---|---|
| 0 | 0 | 0 | 0 | 0 |
| 0.194617502 | 0.00193537 | 0 | 0 | 0 |
| 0.01145398 | 0 | 0 | 0 | 0 |
| 0 | 0 | 0 | 0 | 0 |
| 1.89260784 | 0 | 0 | 0 | 0 |
| 0 | 0 | 0 | 0 | 0 |
| 0 | 0 | 0 | 0 | 0 |
| 0 | 0 | 0 | 0 | 0 |
| 0.09150927 | 0.114404969 | 0.128015658 | 0.385582 | 4.052257975 |
| 0 | 0 | 0 | 0 | 0 |
| 0 | 0 | 0 | 0 | 0.01897605 |
| 0 | 0 | 0 | 0 | 0 |
| 0 | 0 | 0 | 0 | 0 |
| 0.054737815 | 0.024968894 | 0 | 0.036760483 | 0.006121241 |
| 0.343955063 | 0 | 0 | 0 | 0 |
| 0.391826065 | 5.255693493 | 0 | 0 | 0 |
| 0.002272222 | 0 | 0 | 0 | 0 |
| 0 | 0 | 0 | 0 | 0 |
| 0 | 0 | 0 | 0 | 0 |
| 0 | 0 | 0 | 0 | 0 |
| 0 | 0 | 0 | 0 | 0 |
| 0 | 0 | 0 | 0 | 0 |
| 0 | 0 | 0 | 0 | 0 |
| 0 | 0 | 0 | 0 | 0 |
| 0 | 0 | 0 | 0 | 0 |
| 0 | 0 | 0 | 0 | 0 |
| 0.002279758 | 0.009567921 | 0 | 0 | 0 |
| 0.000685987 | 0.00314075 | 0.0116479 | 0.005034164 | 0 |
| 0 | 0 | 0 | 0 | 0 |
| 0 | 0 | 0 | 0 | 0 |
| 0 | 0 | 0 | 0 | 0 |
| 0.000859515 | 0 | 0 | 0 | 0 |
| 0 | 0 | 0 | 0 | 0 |
| 0 | 0 | 0 | 0 | 0 |
| 0 | 0.052081798 | 0 | 0 | 0 |
| 0.002975793 | 0 | 0 | 0 | 0 |
| 0.006894336 | 0 | 0 | 0 | 0 |
| 0 | 0 | 0 | 0 | 0 |
| 0 | 0 | 0 | 0 | 0 |
| 0 | 0 | 0 | 0 | 0 |
| 0.043006055 | 1.101308266 | 0 | 0 | 0 |
| 0 | 0 | 0 | 0 | 0 |
| 0 | 0 | 0 | 0 | 0 |
| 0 | 0 | 0 | 0 | 0 |
| 0 | 0 | 0 | 0 | 0 |
| 0 | 0 | 0 | 0 | 0 |
| 0 | 0 | 0 | 0 | 0 |
| 0 | 0 | 0 | 0 | 0 |

| | | | | |
|---|---|---|---|---|
| 0 | 0 | 0 | 0 | 0 |
| 0 | 0 | 0 | 0 | 0 |
| 0 | 0 | 0 | 0 | 0 |
| 0 | 0 | 0 | 0 | 0 |
| 0 | 0 | 0 | 0 | 0 |
| 0 | 0 | 0 | 0 | 0 |
| 0.021969524 | 0.004590239 | 0.001507221 | 0 | 0 |
| 0 | 0 | 0 | 0 | 0 |
| 0 | 0 | 0 | 0 | 0 |
| 0 | 0.038946037 | 0 | 0 | 0 |
| 0 | 0 | 0 | 0 | 0 |
| 0 | 0 | 0 | 0 | 0 |
| 0 | 0 | 0 | 0 | 0 |
| 0 | 0 | 0 | 0 | 0 |
| 0 | 0 | 0 | 0 | 0 |
| 0 | 0 | 0 | 0 | 0 |
| 0 | 0 | 0 | 0 | 0 |
| 0 | 0 | 0 | 0 | 0 |
| 0 | 0 | 0 | 0 | 0 |
| 0 | 0 | 0 | 0 | 0 |
| 0.013446136 | 0.544180353 | 0.000819976 | 0 | 0 |
| 0 | 0 | 0 | 0 | 0 |
| 0.015929636 | 0 | 0 | 0 | 0 |
| 0.005535668 | 0.003540806 | 0 | 0 | 0 |
| 0 | 0 | 0 | 0 | 0 |
| 0 | 0 | 0 | 0 | 0 |
| 0 | 0 | 0 | 0 | 0 |
| 0 | 0 | 0 | 0 | 0 |
| 0 | 0 | 0 | 0 | 0 |
| 0.00164029 | 0.366915557 | 0.003472425 | 0.007107788 | 0 |
| 0 | 0 | 0 | 0 | 0 |
| 0 | 0 | 0 | 0 | 0 |
| 0 | 0 | 0 | 0 | 0 |
| 0 | 0 | 1.127050423 | 0 | 1.854966726 |
| 0 | 0 | 0 | 0 | 0 |
| 0 | 0 | 0 | 0 | 0 |
| 0.020983385 | 0 | 0.004779922 | 0 | 0 |
| 0 | 0 | 0 | 0 | 0 |
| 0 | 0 | 0 | 0 | 0 |
| 0 | 0 | 0 | 0 | 0 |
| 0 | 0 | 0 | 0 | 0 |
| 0 | 0 | 0 | 0 | 0 |
| 0 | 0 | 0 | 0 | 0 |
| 0 | 0 | 0 | 0 | 0 |
| 0.016967075 | 0 | 0 | 0 | 0 |
| 2.657579571 | 0 | 0 | 0 | 0 |
| 0 | 0 | 0 | 0 | 0 |
| 0 | 0 | 0 | 0 | 0 |

| | | | | |
|---|---|---|---|---|
| 0.002978288 | 0 | 0.005192277 | 0.00479983 | 0 |
| 0 | 0 | 0 | 0 | 0 |
| 0 | 0 | 0 | 0 | 0 |
| 0 | 0 | 0 | 0 | 0 |
| 0 | 0 | 0 | 0 | 0 |
| 0 | 0 | 0 | 0 | 0 |
| 0 | 0 | 0 | 0 | 0 |
| 0.359048487 | 0.261283246 | 0 | 0 | 0 |
| 0.035749003 | 0.52344398 | 0.141267686 | 17.51775111 | 0 |
| 0 | 0.006354645 | 0 | 0.001311246 | 0.000966954 |
| 0 | 0 | 0 | 0 | 0 |
| 0 | 0 | 0 | 0 | 0 |
| 0 | 0 | 0 | 0 | 0 |
| 0 | 0 | 0 | 0 | 0.008226189 |
| 0 | 0 | 0 | 0 | 0 |
| 0 | 0 | 0 | 0 | 0 |
| 0 | 0 | 0 | 0 | 0.002798676 |
| 0 | 0 | 0 | 0 | 0 |
| 0 | 0 | 0 | 0 | 0 |
| 0.030003339 | 0 | 0 | 0 | 0 |
| 0 | 0 | 0 | 0 | 0 |
| 0 | 0 | 0 | 0 | 0 |
| 0 | 0 | 0 | 0 | 0 |
| 0 | 0 | 0 | 0 | 0 |
| 0 | 0 | 0 | 0 | 0 |
| 0 | 0 | 0 | 0 | 0 |
| 0 | 0 | 0 | 0 | 0 |
| 0 | 0 | 0 | 0 | 0 |
| 0 | 0 | 0 | 0 | 0 |
| 0 | 0 | 0 | 0 | 0 |
| 0.345383659 | 0 | 0 | 0 | 0 |
| 0 | 0 | 0 | 0 | 0 |
| 0 | 0 | 0 | 0 | 0 |
| 0 | 0 | 0 | 0 | 0 |
| 0.019724952 | 0 | 0 | 0 | 0 |
| 0 | 0 | 0 | 0 | 0 |
| 0 | 0 | 0 | 0 | 0 |
| 0 | 0 | 0 | 0 | 0 |
| 0.003333098 | 0 | 0 | 0 | 0 |
| 0 | 0 | 0 | 0.449738392 | 0 |
| 0 | 0 | 0 | 0 | 0 |
| 0.352576216 | 0 | 0 | 0 | 0 |
| 0 | 0 | 0 | 0 | 0 |
| 0 | 0 | 0 | 0 | 0 |
| 0 | 0 | 0 | 0 | 0 |
| 0 | 0 | 0 | 0 | 0 |
| 0 | 0 | 0.001456917 | 0 | 0 |
| 0 | 0 | 0 | 0 | 0 |

| | | | | |
|---|---|---|---|---|
| 0 | 0 | 0 | 0 | 0 |
| 0.006482907 | 0 | 0.009177254 | 0 | 0 |
| 0 | 0 | 0 | 0 | 0 |
| 0 | 0 | 0 | 0 | 0 |
| 0.322325252 | 0.011386931 | 0 | 0 | 0.030893259 |
| 0 | 0 | 0 | 0 | 0 |
| 0 | 0 | 0 | 0 | 0 |
| 0.009805782 | 0.001785603 | 0.411983913 | 0.001526433 | 0.031330321 |
| 0.007565548 | 0 | 0 | 0 | 0 |
| 0 | 0 | 0 | 0 | 0 |
| 0 | 0 | 0 | 0 | 0 |
| 0.101890626 | 0 | 0 | 0 | 0 |
| 0 | 0 | 0 | 0 | 0 |
| 0 | 0 | 0 | 0 | 0 |
| 0 | 0 | 0 | 0 | 0 |
| 0 | 0 | 0 | 0.000756494 | 0 |
| 0 | 0 | 0 | 0 | 0 |
| 0 | 0 | 0 | 0 | 0 |
| 0 | 0 | 0 | 0 | 0 |
| 0 | 0 | 0 | 0 | 0 |
| 0 | 0 | 0 | 0 | 0 |
| 0 | 0 | 0 | 0 | 0 |
| 0 | 0 | 0 | 0 | 0 |
| 0.000771463 | 0.005739658 | 0.002381684 | 0 | 0 |
| 0 | 0 | 0 | 0 | 0.151964822 |
| 0 | 0 | 0 | 0 | 0 |
| 0 | 0.016152519 | 0.04530907 | 0.054714477 | 0 |
| 0 | 0 | 0 | 0 | 0 |
| 0 | 0.007502727 | 0.001618902 | 0 | 0 |
| 0 | 0 | 0 | 0 | 0 |
| 0 | 0 | 0 | 0 | 0 |
| 0 | 0 | 0 | 0 | 0 |
| 0 | 0 | 0 | 0 | 0 |
| 0 | 0 | 0 | 0 | 0 |
| 0 | 0 | 0 | 0 | 0 |
| 0 | 0 | 0 | 0 | 0 |
| 0 | 0 | 0 | 0 | 0 |
| 0 | 0 | 0 | 0 | 0 |
| 0 | 0 | 0 | 0 | 0 |
| 0 | 0 | 0 | 0 | 0 |
| 0 | 0 | 0 | 0 | 0 |
| 0 | 0 | 0 | 0 | 0 |
| 0 | 0 | 0 | 0 | 0 |
| 0 | 0 | 0 | 0 | 0 |
| 0 | 0 | 0 | 0 | 0 |
| 0 | 0 | 0 | 0 | 0 |
| 0 | 0 | 0 | 0 | 0 |
| 0 | 0 | 0 | 0 | 0 |

| | | | | |
|---|---|---|---|---|
| 0.038485743 | 0 | 0 | 0 | 0 |
| 0 | 0 | 0 | 0 | 0 |
| 0 | 0 | 0 | 0 | 0 |
| 0 | 0 | 0 | 0.001453461 | 0 |
| 0 | 0 | 0 | 0 | 0 |
| 0.023062738 | 0 | 0 | 0 | 0.002103254 |
| 0 | 0 | 0 | 0 | 0 |
| 0 | 0 | 0 | 0 | 0 |
| 0 | 0 | 0 | 0 | 0 |
| 0 | 0 | 0 | 0 | 0 |
| 0 | 0 | 0 | 0 | 0 |
| 0 | 0 | 0 | 0 | 0 |
| 0 | 0 | 0 | 0 | 0 |
| 0 | 0 | 0 | 0 | 0 |
| 0 | 0 | 0 | 0 | 0 |
| 0 | 0 | 0 | 0 | 0 |
| 0 | 0 | 0 | 0 | 0 |
| 0 | 0 | 0 | 0 | 0 |
| 0 | 0 | 0 | 0 | 0 |
| 0 | 0 | 0 | 0 | 0 |
| 0.001344417 | 0 | 0.027037702 | 0.001127554 | 0 |
| 0 | 0 | 0 | 0 | 0 |
| 0.005578817 | 0 | 0.003893916 | 0 | 0 |
| 0 | 0 | 0 | 0 | 0 |
| 0 | 0 | 0 | 0 | 0 |
| 0 | 0 | 0 | 0 | 0 |
| 0 | 0 | 0 | 0 | 0 |
| 0 | 0 | 0 | 0 | 0 |
| 0 | 0.013699517 | 0 | 0 | 0 |
| 0 | 0 | 0 | 0 | 0 |
| 0 | 0 | 0 | 0 | 0 |
| 0 | 0 | 0 | 0 | 0 |
| 0.001817231 | 0.002340023 | 0.032258706 | 0.007556996 | 0.013959243 |
| 0 | 0 | 0 | 0 | 0 |
| 0 | 0 | 0 | 0 | 0 |
| 0 | 0 | 0 | 0 | 0 |
| 0 | 0 | 0 | 0 | 0 |
| 0 | 0 | 0 | 0 | 0 |
| 0.002319211 | 0.001447959 | 0.009633369 | 0.001031496 | 0.012424086 |
| 0 | 0 | 0 | 0 | 0 |
| 0 | 0 | 0 | 0 | 0 |
| 0 | 0 | 0 | 0 | 0 |
| 0.002958385 | 0.008747671 | 0 | 0 | 0 |
| 0 | 0 | 0 | 0 | 0 |
| 0 | 0 | 0 | 0 | 0 |
| 0.050317826 | 0 | 0 | 0 | 0 |
| 0 | 0 | 0 | 0 | 0 |
| 0 | 0 | 0 | 0 | 0 |

| | | | | |
|---|---|---|---|---|
| 0 | 0 | 0 | 0 | 0 |
| 0 | 0 | 0 | 0 | 0 |
| 0 | 0 | 0 | 0 | 0 |
| 0 | 0 | 0 | 0 | 0 |
| 0 | 0 | 0 | 0 | 0 |
| 0 | 0 | 0 | 0 | 0 |
| 0 | 0 | 0 | 0 | 0 |
| 0 | 0 | 0 | 0 | 0 |
| 0.263414576 | 0 | 0 | 0 | 0 |
| 0.00846975 | 0 | 0 | 0 | 0 |
| 0 | 0 | 0 | 0 | 0 |
| 0 | 0 | 0 | 0 | 0 |
| 0 | 0 | 0 | 0 | 0 |
| 0 | 0 | 0 | 0 | 0 |
| 0.022978682 | 0.045887933 | 0 | 0 | 0 |
| 0.001587295 | 0 | 0.032342268 | 0 | 0 |
| 0 | 0 | 0 | 0 | 0 |
| 0 | 0 | 0 | 0 | 0 |
| 0 | 0 | 0 | 0 | 0 |
| 0 | 2.352586709 | 0 | 0 | 0 |
| 0.01429867 | 0.002134747 | 0 | 0 | 0 |
| 0 | 0 | 0 | 0 | 0 |
| 0 | 0 | 0 | 0 | 0 |
| 0 | 0 | 0 | 0 | 0 |
| 0 | 0 | 0 | 0 | 0 |
| 0 | 4.336250199 | 0 | 0 | 0 |
| 0 | 0 | 0 | 0 | 0 |
| 0 | 0 | 0 | 0 | 0 |
| 0 | 0 | 0 | 0 | 0 |
| 0 | 0 | 0 | 0 | 0 |
| 0.405416658 | 0.064604346 | 0 | 0.644266859 | 0 |
| 0 | 0 | 0 | 0 | 0 |
| 0 | 0 | 0 | 0 | 0 |
| 0 | 0 | 0 | 0 | 0 |
| 0 | 0 | 0 | 0 | 0 |
| 0 | 0 | 0 | 0 | 0 |
| 0 | 0 | 0 | 0 | 0 |
| 0 | 0 | 0 | 0 | 0 |
| 0 | 0 | 0 | 0 | 0 |
| 0 | 0 | 0 | 0 | 0 |
| 0 | 0 | 0 | 0 | 0 |
| 0 | 0 | 0 | 0 | 0 |
| 0 | 0 | 0 | 0 | 0 |
| 0 | 0 | 0 | 0 | 0 |
| 0 | 0 | 0 | 0 | 0 |
| 0 | 0.060815289 | 0 | 0 | 0 |
| 0.0095463 | 0.004184727 | 5.877519032 | 0.105531437 | 0.233906432 |
| 0 | 0 | 0 | 0 | 0 |

| | | | | |
|---|---|---|---|---|
| 0 | 0 | 0 | 0 | 0 |
| 0 | 0 | 0 | 0 | 0 |
| 0 | 0 | 0 | 0 | 0 |
| 0 | 0 | 0 | 0 | 0 |
| 0.344905953 | 0.012678898 | 0 | 0 | 0 |
| 0 | 0 | 0 | 0 | 0 |
| 0 | 0 | 0 | 0 | 0 |
| 0 | 0 | 0 | 0 | 0 |
| 0 | 0 | 0 | 0 | 0 |
| 0 | 0 | 0 | 0 | 0 |
| 0 | 0 | 0 | 0 | 0 |
| 0.015800211 | 0.003134746 | 0.002081232 | 0 | 0 |
| 0.016176167 | 0 | 0 | 0 | 0.000765023 |
| 0 | 0 | 0 | 0 | 0 |
| 0.01123814 | 0 | 0.023394153 | 0 | 0 |
| 0 | 0 | 0 | 0 | 0 |
| 0.029975519 | 0 | 0 | 0 | 0 |
| 0 | 0 | 0 | 0 | 0 |
| 0.190307523 | 0.316425484 | 0 | 0.012955187 | 0 |
| 0.001874562 | 0.001192023 | 0 | 0.002038015 | 0 |
| 0 | 0 | 0 | 0 | 0 |
| 0 | 0 | 0 | 0 | 0 |
| 0 | 0 | 0 | 0 | 0 |
| 0 | 0 | 0 | 0 | 0 |
| 0 | 0 | 0 | 0 | 0 |
| 0 | 0 | 0 | 0 | 0 |
| 0.003004345 | 0.004202981 | 0 | 0 | 0 |
| 0 | 0 | 0 | 0 | 0 |
| 0 | 0 | 0 | 0 | 0 |
| 0 | 0 | 0 | 0 | 0 |
| 0 | 0.005197417 | 0 | 0.004257914 | 0 |
| 0 | 0.022455265 | 0 | 0 | 0 |
| 0 | 0 | 0 | 0 | 0 |
| 0 | 0 | 0 | 0 | 0 |
| 0 | 0 | 0 | 0 | 0 |
| 0 | 0 | 0 | 0 | 0 |
| 0 | 0 | 0 | 0 | 0 |
| 0.00345607 | 0.007759554 | 0.010669682 | 0.006763362 | 0.001566593 |
| 0 | 0 | 0 | 0 | 0 |
| 0 | 0.004344234 | 0 | 0 | 0 |
| 0.001725136 | 0 | 0 | 0 | 0 |
| 0 | 0 | 0 | 0 | 0 |
| 0 | 0 | 0 | 0 | 0 |
| 0.002168233 | 0.016244378 | 0.097851654 | 0.001157216 | 0 |
| 0 | 0 | 0 | 0 | 0 |
| 0 | 0 | 0 | 0 | 0 |
| 0 | 0 | 0 | 0 | 0 |
| 0 | 0 | 0 | 0 | 0 |

| | | | | |
|---|---|---|---|---|
| 0 | 0 | 0 | 0 | 0 |
| 0 | 0 | 0 | 0 | 0 |
| 0 | 0 | 0 | 0 | 0 |
| 0.001652595 | 0 | 0.001785679 | 0 | 0.003651863 |
| 0.000957972 | 0 | 0.00709795 | 0 | 0 |
| 0.001483626 | 0.00650966 | 0 | 0 | 0 |
| 0 | 0.005965799 | 0 | 0 | 0 |
| 0 | 0 | 0 | 0 | 0 |
| 0 | 0 | 0 | 0 | 0 |
| 0 | 0 | 0 | 0 | 0 |
| 0 | 0 | 0 | 0 | 0 |
| 0 | 0 | 0 | 0 | 0 |
| 0.001903618 | 0.010894495 | 0 | 0.090266583 | 0 |
| 0 | 0 | 0 | 0 | 0 |
| 0 | 0 | 0 | 0 | 0 |
| 0 | 0 | 0 | 0 | 0 |
| 0 | 0 | 0 | 0 | 0 |
| 0 | 0 | 0 | 0 | 0 |
| 0 | 0 | 0 | 0 | 0 |
| 0 | 0 | 0 | 0 | 0 |
| 0 | 0 | 0 | 0 | 0 |
| 0 | 0 | 0 | 0 | 0 |
| 0 | 0.00375191 | 0.046669282 | 0.000566001 | 0.001947808 |
| 0.131512987 | 0.007552595 | 0 | 0 | 0.002490021 |
| 0 | 0 | 0 | 0 | 0 |
| 0 | 0 | 0 | 0 | 0 |
| 0 | 0 | 0 | 0 | 0 |
| 0 | 0 | 0 | 0 | 0 |
| 0 | 0 | 0 | 0 | 0 |
| 0 | 0 | 0 | 0 | 0 |
| 0.015909297 | 0.019194255 | 0 | 0.003576171 | 0.008325224 |
| 0 | 0.00323502 | 0 | 0 | 0 |
| 0 | 0 | 0 | 0 | 0 |
| 0 | 0 | 0 | 0 | 0 |
| 0 | 0.229827219 | 0 | 0 | 0 |
| 0 | 0 | 0 | 0 | 0 |
| 0 | 0.044706615 | 0 | 0 | 0 |
| 0 | 0 | 0.079679496 | 0 | 0 |
| 0.022373316 | 0.001041005 | 0.001796984 | 0 | 0 |
| 0 | 0 | 0 | 0 | 0 |
| 0 | 0 | 0 | 0 | 0 |
| 0 | 0 | 0 | 0 | 0 |
| 0 | 0 | 0 | 0 | 0 |
| 0 | 0 | 0 | 0 | 0 |
| 0 | 0 | 0 | 0 | 0 |
| 0.146595908 | 0.008609573 | 0 | 0 | 0 |
| 0 | 0.190332663 | 0 | 0 | 0 |
| 0 | 0 | 0 | 0 | 0 |

| | | | | |
|---|---|---|---|---|
| 0 | 0 | 0 | 0 | 0 |
| 0 | 0 | 0 | 0 | 0 |
| 0 | 0 | 0 | 0 | 0 |
| 0 | 0 | 0 | 0 | 0 |
| 0 | 0.000927337 | 0 | 0 | 0 |
| 0.001768085 | 0 | 0 | 0 | 0 |
| 0 | 0 | 0 | 0 | 0 |
| 0 | 0 | 0 | 0 | 0 |
| 0 | 0.021190533 | 0 | 0 | 0 |
| 0 | 0 | 0 | 0 | 0 |
| 0 | 0 | 0 | 0 | 0 |
| 0 | 0 | 0 | 0 | 0.001483744 |
| 0 | 0 | 0 | 0 | 0 |
| 0 | 0 | 0 | 0 | 0 |
| 0 | 0 | 0 | 0 | 0 |
| 0 | 0 | 0 | 0 | 0 |
| 0 | 0 | 0 | 0 | 0 |
| 0 | 0 | 0 | 0 | 0 |
| 0 | 0 | 0 | 0 | 0 |
| 0.00293549 | 0 | 0 | 0 | 0 |
| 0 | 0 | 0 | 0 | 0 |
| 0 | 0 | 0 | 0 | 0 |
| 0 | 0 | 0 | 0 | 0 |
| 0 | 0 | 0 | 0 | 0 |
| 0 | 0 | 0 | 0 | 0 |
| 0 | 0 | 0 | 0 | 0 |
| 0 | 0 | 0 | 0 | 0 |
| 0.022956884 | 0 | 0 | 0 | 0 |
| 0 | 0 | 0 | 0 | 0 |
| 0 | 0 | 0 | 0 | 0 |
| 0.001008756 | 0.002501702 | 0 | 0 | 0 |
| 0 | 0 | 0 | 0 | 0 |
| 0.037675794 | 0.168250925 | 0 | 0.012415624 | 0.061789088 |
| 0.001994848 | 0 | 0.018722008 | 0.014053759 | 0.000839651 |
| 0 | 0 | 0 | 0 | 0 |
| 0 | 0 | 0 | 0 | 0 |
| 0 | 0 | 0 | 0 | 0 |
| 0 | 0 | 0 | 0 | 0 |
| 0 | 0 | 0 | 0 | 0 |
| 0 | 0 | 0 | 0 | 0 |
| 0 | 0 | 0 | 0 | 0 |
| 0.11771879 | 0.090808424 | 0.002776507 | 0.002199986 | 0 |
| 0.026413345 | 0 | 0 | 0 | 0.015317624 |
| 0 | 0 | 0 | 0 | 0 |
| 0 | 0 | 0 | 0 | 0 |
| 0 | 0.003559823 | 0 | 0 | 0 |
| 0.001482949 | 0 | 0 | 0 | 0 |
| 0 | 0 | 0 | 0 | 0 |

| | | | | |
|---|---|---|---|---|
| 0 | 0 | 0 | 0 | 0 |
| 0.019677498 | 0 | 0 | 0 | 0 |
| 0.010534973 | 0.003562718 | 0.015966268 | 0.031627482 | 0 |
| 0 | 0 | 0 | 0 | 0 |
| 0.006308752 | 0 | 0.015345172 | 0 | 0 |
| 0 | 0.030671713 | 0.007123752 | 0.000910413 | 0 |
| 0 | 0 | 0 | 0 | 0 |
| 0 | 0 | 0 | 0 | 0 |
| 0 | 0 | 0 | 0 | 0 |
| 0 | 0 | 0 | 0 | 0 |
| 0 | 0 | 0 | 0 | 0 |
| 0 | 0 | 0 | 0 | 0 |
| 0 | 0 | 0 | 0 | 0 |
| 0 | 0 | 0 | 0 | 0 |
| 0 | 0 | 0 | 0 | 0 |
| 0 | 0 | 0 | 0 | 0 |
| 0 | 0 | 0 | 0 | 0 |
| 0 | 0 | 0 | 0 | 0 |
| 0 | 0.010598746 | 0 | 0 | 0 |
| 0 | 0 | 0 | 0 | 0 |
| 0 | 0 | 0 | 0 | 0 |
| 0 | 0.000661337 | 0 | 0 | 0 |
| 0 | 0 | 0 | 0 | 0 |
| 0 | 0 | 0 | 0 | 0 |
| 0 | 0 | 0 | 0 | 0 |
| 0.0093297 | 0.021357728 | 0 | 0 | 0 |
| 0 | 0 | 0 | 0 | 0 |
| 0 | 0 | 0 | 0 | 0 |
| 0 | 0 | 0 | 0 | 0 |
| 0 | 0 | 0 | 0 | 0 |
| 0 | 0 | 0 | 0 | 0 |
| 0 | 0 | 0 | 0 | 0 |
| 0 | 0 | 0 | 0 | 0 |
| 0 | 0 | 0 | 0 | 0 |
| 0 | 0 | 0 | 0 | 0 |
| 0 | 0 | 0 | 0 | 0 |
| 0 | 0.002769679 | 0 | 0 | 0 |
| 0 | 0.001809347 | 0 | 0 | 0 |
| 12.63512287 | 0.60999572 | 0 | 0 | 0.011227127 |
| 0 | 0 | 0 | 0 | 0 |
| 0 | 0 | 0 | 0 | 0 |
| 0 | 0 | 0 | 0 | 0 |
| 0.003984414 | 0 | 0.006888448 | 0.004678383 | 0 |
| 0 | 0 | 0 | 0 | 0 |
| 0 | 0 | 0 | 0 | 0 |
| 0 | 0 | 0 | 0 | 0 |
| 0.003047687 | 0 | 0.003387207 | 0 | 0 |
| 0 | 0 | 0 | 0 | 0 |

| | | | | |
|---|---|---|---|---|
| 0 | 0 | 0 | 0 | 0 |
| 0.013075108 | 0.004181647 | 0.005936156 | 0.001316996 | 0.001618658 |
| 0 | 0 | 0 | 0 | 0 |
| 0 | 0 | 0 | 0 | 0 |
| 0 | 0 | 0 | 0 | 0 |
| 0 | 0 | 0 | 0 | 0 |
| 0 | 0 | 0 | 0 | 0 |
| 0 | 0 | 0 | 0 | 0 |
| 0 | 0 | 0 | 0 | 0 |
| 0 | 0 | 0 | 0 | 0 |
| 0 | 0 | 0 | 0 | 0 |
| 0 | 0 | 0 | 0 | 0 |
| 0.227242502 | 0 | 0 | 0.052107369 | 0 |
| 0 | 0 | 0 | 0 | 0 |
| 0.041475958 | 0 | 0.003848776 | 0 | 0 |
| 0 | 0 | 0 | 0 | 0 |
| 0 | 0 | 0 | 0 | 0 |
| 0 | 0 | 0 | 0 | 0 |
| 0 | 0 | 0 | 0 | 0 |
| 0 | 0.001370661 | 0 | 0 | 0 |
| 0 | 0 | 0 | 0 | 0 |
| 0.001883591 | 0.001616983 | 0 | 0.001612668 | 0 |
| 0 | 0 | 0 | 0 | 0 |
| 0 | 0 | 0.001796851 | 0 | 0 |
| 0 | 0 | 0 | 0 | 0 |
| 0 | 0 | 0 | 0 | 0 |
| 0 | 0 | 0 | 0 | 0 |
| 0 | 0 | 0 | 0 | 0 |
| 0 | 0 | 0 | 0 | 0 |
| 0 | 0 | 0 | 0 | 0 |
| 0 | 0 | 0 | 0 | 0 |
| 0 | 0 | 0 | 0 | 0 |
| 0 | 0 | 0 | 0 | 0 |
| 0 | 0 | 0 | 0 | 0 |
| 0.025011619 | 0.000734065 | 0 | 0 | 0 |
| 0 | 0 | 0 | 0 | 0 |
| 0.00103552 | 0 | 0 | 0 | 0 |
| 0 | 0 | 0 | 0 | 0 |
| 0.00611323 | 0 | 0 | 0.003450951 | 0 |
| 0 | 0 | 0 | 0 | 0 |
| 0.363453649 | 0.585651049 | 25.78406101 | 0.073023918 | 0.051112025 |
| 0 | 0 | 0 | 0 | 0 |
| 0 | 0 | 0 | 0 | 0 |
| 0 | 0 | 0 | 0 | 0 |
| 0 | 0 | 0 | 0 | 0 |
| 0.156169322 | 0.162452406 | 1.589913043 | 0.485348137 | 0.253788953 |
| 0 | 0 | 0 | 0 | 0 |
| 0 | 0 | 0 | 0 | 0 |

| | | | | |
|---|---|---|---|---|
| 0 | 0 | 0 | 0 | 0 |
| 0 | 0 | 0 | 0 | 0 |
| 0 | 0 | 0 | 0 | 0 |
| 0 | 0 | 0 | 0 | 0 |
| 0.003112093 | 0 | 0 | 0.00179124 | 0 |
| 0 | 0 | 0 | 0 | 0 |
| 0 | 0 | 0 | 0 | 0 |
| 0 | 0 | 0 | 0 | 0 |
| 0 | 0 | 0 | 0 | 0 |
| 0.003304905 | 0 | 0.002332115 | 0 | 0 |
| 0 | 0.064117457 | 0 | 0 | 0 |
| 0 | 0 | 0 | 0 | 0 |
| 0.005999067 | 0 | 0 | 0 | 0 |
| 0 | 0 | 0 | 0 | 0 |
| 0 | 0 | 0 | 0 | 0 |
| 0 | 0 | 0 | 0 | 0 |
| 0 | 0 | 0 | 0 | 0 |
| 0 | 0 | 0 | 0 | 0 |
| 0 | 0 | 0 | 0 | 0 |
| 0.135748881 | 0.120119378 | 0.022317543 | 0.313278802 | 0.020227373 |
| 0 | 0 | 0 | 0 | 0 |
| 0 | 0 | 0 | 0 | 0 |
| 0 | 0 | 0 | 0 | 0 |
| 0 | 0 | 0 | 0 | 0 |
| 0 | 0 | 0 | 0 | 0 |
| 0 | 0 | 0 | 0 | 0 |
| 0 | 0 | 0 | 0 | 0 |
| 0 | 0 | 0 | 0 | 0 |
| 0 | 0 | 0 | 0 | 0 |
| 0 | 0 | 0 | 0 | 0 |
| 0 | 0 | 0 | 0 | 0 |
| 0 | 0 | 0 | 0 | 0 |
| 0 | 0 | 0 | 0 | 0 |
| 0 | 0 | 0 | 0 | 0 |
| 0 | 0 | 0 | 0 | 0 |
| 0 | 0 | 0 | 0 | 0 |
| 0 | 0 | 0 | 0 | 0 |
| 0 | 0 | 0 | 0 | 0 |
| 0 | 0 | 0 | 0 | 0 |
| 0 | 0 | 0 | 0 | 0 |
| 0 | 0 | 0 | 0 | 0 |
| 0 | 0 | 0 | 0 | 0 |
| 0 | 0 | 0 | 0 | 0 |
| 0 | 0 | 0 | 0 | 0 |
| 0 | 0 | 0 | 0 | 0 |
| 0.005015142 | 0 | 0 | 0 | 0 |
| 0 | 0 | 0 | 0 | 0 |
| 0 | 0 | 0 | 0 | 0 |
| 0 | 0 | 0 | 0 | 0 |

| | | | | |
|---|---|---|---|---|
| 0 | 0 | 0 | 0 | 0 |
| 0 | 0 | 0 | 0 | 0 |
| 0 | 0 | 0 | 0 | 0 |
| 0 | 0 | 0 | 0 | 0 |
| 0 | 0 | 0 | 0 | 0 |
| 0.016847993 | 0 | 0 | 0 | 0.000727331 |
| 0 | 0 | 0 | 0 | 0 |
| 0 | 0 | 0 | 0 | 0 |
| 0 | 0 | 0 | 0 | 0 |
| 0 | 0 | 0 | 0 | 0 |
| 0 | 0.444538229 | 0 | 0 | 0 |
| 0 | 0 | 0 | 0 | 0 |
| 0 | 0 | 0 | 0 | 0 |
| 0 | 0 | 0 | 0 | 0 |
| 0 | 0 | 0 | 0 | 0 |
| 0 | 0 | 0 | 0 | 0 |
| 0 | 0 | 0 | 0 | 0 |
| 0 | 0.00507473 | 0 | 0 | 0 |
| 0 | 0 | 0 | 0 | 0 |
| 0 | 0 | 0 | 0 | 0 |
| 0 | 0 | 0 | 0 | 0 |
| 0 | 0 | 0 | 0 | 0 |
| 0 | 0 | 0 | 0 | 0 |
| 0 | 0 | 0 | 0 | 0 |
| 0 | 0 | 0 | 0 | 0 |
| 0 | 0 | 0 | 0 | 0 |
| 0 | 0 | 0 | 0 | 0 |
| 0 | 0 | 0 | 0 | 0 |
| 0 | 0 | 0 | 0 | 0 |
| 0 | 0.02711534 | 0 | 0 | 0 |
| 0 | 0 | 0 | 0 | 0 |
| 0 | 0.001630242 | 0 | 0 | 0 |
| 0 | 0 | 0 | 0 | 0 |
| 0 | 0 | 0 | 0 | 0 |
| 0 | 0 | 0 | 0 | 0 |
| 0 | 0 | 0 | 0 | 0 |
| 0 | 0 | 0 | 0 | 0 |
| 0 | 0 | 0 | 0 | 0 |
| 0 | 0.513022887 | 0 | 0 | 0 |
| 0 | 0 | 0 | 0 | 0 |
| 0 | 0 | 0 | 0 | 0 |
| 0 | 0 | 0 | 0 | 0 |
| 0 | 0 | 0 | 0 | 0 |
| 0 | 0 | 0 | 0 | 0 |
| 0 | 0 | 0 | 0 | 0 |
| 0 | 0 | 0 | 0 | 0 |
| 0 | 0 | 0 | 0 | 0 |
| 0 | 0 | 0 | 0 | 0 |

| | | | | |
|---|---|---|---|---|
| 0 | 0 | 0 | 0 | 0 |
| 0 | 0 | 0 | 0 | 0 |
| 0 | 0 | 0 | 0 | 0 |
| 0 | 0 | 0 | 0 | 0 |
| 0 | 0 | 0 | 0 | 0 |
| 0 | 0 | 0 | 0 | 0 |
| 0 | 0 | 0 | 0 | 0 |
| 0 | 0 | 0 | 0 | 0 |
| 0 | 0 | 0 | 0 | 0 |
| 0 | 0 | 0 | 0 | 0 |
| 0 | 0 | 0.001564145 | 0.04027927 | 0 |
| 0 | 0 | 0 | 0 | 0 |
| 0.430704315 | 0 | 0 | 0 | 0 |
| 0 | 0 | 0 | 0 | 0 |
| 0 | 0 | 0 | 0 | 0 |
| 0 | 0 | 0 | 0 | 0 |
| 0 | 0 | 0 | 0 | 0 |
| 0 | 0 | 0 | 0 | 0 |
| 0 | 0 | 0 | 0 | 0 |
| 0 | 0 | 0 | 0 | 0 |
| 0 | 0 | 0 | 0.013836658 | 0 |
| 0.015148626 | 0 | 0 | 0 | 0 |
| 0 | 0 | 0 | 0 | 0 |
| 2.77134902 | 0 | 0 | 0.235150447 | 1.341751895 |
| 0 | 0 | 0 | 0 | 0 |
| 0 | 0 | 0 | 0 | 0 |
| 0 | 0 | 0 | 0 | 0 |
| 0 | 0.509817416 | 0 | 0 | 0 |
| 0 | 0 | 0 | 0 | 0 |
| 0 | 0 | 0 | 0 | 0 |
| 0 | 0 | 0 | 0 | 0 |
| 0 | 0 | 0 | 0 | 0 |
| 0 | 0 | 0 | 0 | 0 |
| 0 | 0 | 0 | 0 | 0 |

| PHL018-0152_V3 | PHL018-0013_V2 | PHL018-0007_V2 | PHL018-0135_V3 | PHL018-0112_V3 |
|---|---|---|---|---|
| REF | EYCF | EYCF | EYCF | CTRL |
| 0.011853654 | 0.625099234 | 0.006536909 | 0.12940613 | 0.105836707 |
| 1.453459216 | 0.808400014 | 0.05719406 | 0.008385935 | 0.910846681 |
| 5.917471218 | 0 | 20.57846246 | 0 | 0.013731682 |
| 0 | 0 | 0 | 0 | 0 |
| 0 | 0 | 0 | 0 | 0 |
| 0 | 0 | 0 | 0.129698184 | 0 |
| 0 | 0 | 0 | 0 | 0 |
| 0 | 0 | 0 | 0 | 0 |
| 0 | 0 | 0 | 0 | 0 |
| 0 | 0 | 0 | 0 | 0 |
| 0 | 0 | 0 | 0 | 0 |
| 0 | 0 | 0 | 0 | 0 |
| 0 | 0 | 0 | 0 | 0 |
| 3.680926897 | 0.456207955 | 1.405735187 | 0 | 1.012064458 |
| 0 | 0 | 0 | 0.000925501 | 0 |
| 0 | 0 | 0 | 0 | 0 |
| 0 | 0 | 0 | 0 | 0 |
| 0 | 0 | 0 | 0 | 0 |
| 0 | 0 | 0 | 0 | 0 |
| 0 | 0 | 0 | 0 | 0 |
| 0 | 0 | 0 | 0.000872126 | 0 |
| 0 | 0 | 0 | 0 | 0 |
| 0 | 0 | 0 | 0 | 0 |
| 0 | 0 | 0 | 0 | 0 |
| 0 | 0 | 0 | 1.585164091 | 0 |
| 0 | 0 | 0 | 0 | 0 |
| 0 | 0 | 0 | 0 | 0 |
| 0 | 0 | 0 | 0 | 0 |
| 0 | 0 | 0 | 0 | 0 |
| 0 | 0 | 0 | 0 | 0 |
| 0 | 0 | 0 | 0.003241831 | 0 |
| 0 | 0 | 0 | 0 | 0 |
| 0 | 0 | 0 | 0 | 0 |
| 0 | 0 | 0 | 0 | 0 |
| 0 | 0 | 0 | 0 | 0 |
| 0.017461969 | 0.03063224 | 0 | 0.704062081 | 7.085546874 |
| 0 | 0 | 0 | 0 | 0 |
| 0 | 0 | 0 | 0 | 0 |
| 0 | 0 | 0 | 0 | 0 |
| 0 | 0 | 0 | 0 | 0 |
| 0 | 0 | 0 | 0 | 0 |
| 0.012882436 | 0.052983152 | 0 | 0.057139925 | 0.162076883 |
| 2.150514859 | 0.035064145 | 0.1141209 | 0.265690708 | 0.001285658 |
| 0.088126662 | 0.130318801 | 0 | 0 | 0.003492741 |
| 0.01570558 | 0.004069625 | 0 | 0 | 0.005494129 |
| 0 | 0 | 0 | 0 | 0 |

| | | | | |
|---|---|---|---|---|
| 0 | 0.00424239 | 0 | 0.001179092 | 0.035899244 |
| 0 | 0.003155249 | 0 | 0 | 0.10940309 |
| 0 | 0.137748503 | 0.000526949 | 0 | 0.019070519 |
| 0.306847171 | 0.816276477 | 0.179147554 | 0.232282491 | 0.356183624 |
| 0 | 0.034134622 | 0 | 0 | 0.013055828 |
| 0 | 0 | 0 | 0.004002828 | 0 |
| 0 | 0.001690959 | 0.02886062 | 0.015649999 | 0 |
| 0 | 0 | 0 | 0.000759719 | 0 |
| 0 | 0 | 0 | 0.001084942 | 0 |
| 0 | 0 | 0 | 0 | 0 |
| 0 | 0.000776832 | 0 | 0.031954045 | 0 |
| 0 | 0.377934873 | 0 | 0.062739478 | 0.714843519 |
| 0 | 0 | 0 | 0 | 0 |
| 0.001791532 | 0.171721472 | 0 | 0 | 0.187700867 |
| 0 | 0 | 0 | 0 | 0 |
| 0 | 0 | 0 | 0 | 0 |
| 0.001339637 | 0.015273554 | 0 | 0 | 0.001933107 |
| 0 | 0 | 0 | 0 | 0 |
| 0 | 0 | 0 | 0 | 0 |
| 0 | 0 | 0 | 0.100350697 | 0 |
| 0.00757239 | 0.093159723 | 0.01395746 | 0 | 0.608436478 |
| 0.001131094 | 0.056469662 | 0 | 0 | 0.017565352 |
| 2.924429534 | 0 | 0 | 0 | 0.158135781 |
| 0 | 0 | 0 | 0 | 0 |
| 0 | 0 | 0 | 0 | 0 |
| 0 | 0 | 0 | 0 | 0 |
| 0 | 0 | 0.199650075 | 0 | 0 |
| 0 | 0 | 0 | 0 | 0 |
| 0.001653711 | 0.016446362 | 0 | 0.010669118 | 0.002585178 |
| 0 | 0 | 0 | 0 | 0 |
| 0 | 0 | 0 | 0 | 0 |
| 0 | 0 | 0 | 0 | 0 |
| 0 | 0 | 0 | 0 | 0 |
| 0 | 0 | 0 | 0 | 0 |
| 0 | 0 | 0 | 0 | 0 |
| 0 | 0 | 0 | 0 | 0 |
| 0 | 0 | 0 | 0 | 0 |
| 0 | 0 | 0 | 0 | 0 |
| 0 | 0 | 0 | 0.001827521 | 0 |
| 0 | 0 | 0 | 0 | 0 |
| 0 | 0 | 0 | 0 | 0 |
| 0 | 0 | 0 | 0 | 0 |
| 0 | 0 | 0 | 0 | 0 |
| 0 | 0 | 0 | 0 | 0 |
| 0 | 0 | 0 | 3.301382206 | 0 |
| 0 | 0 | 0 | 0 | 0 |
| 0 | 0 | 0 | 0 | 0 |
| 0 | 0 | 0 | 0 | 0 |

| | | | | |
|---|---|---|---|---|
| 0 | 0 | 0 | 0 | 0 |
| 0 | 0 | 0 | 0 | 0 |
| 0 | 0 | 0 | 0 | 0 |
| 0 | 0 | 0 | 0 | 0 |
| 0 | 0.0020514 | 0 | 0.013360459 | 0 |
| 0 | 0 | 0 | 0 | 0 |
| 0 | 0 | 0 | 0 | 0 |
| 0 | 0 | 0 | 0 | 0 |
| 0 | 0 | 0 | 0.004627696 | 0.003005007 |
| 0 | 0 | 0 | 0 | 0 |
| 0 | 0 | 0 | 0.051495652 | 0 |
| 0.005983795 | 0 | 0 | 0 | 0 |
| 0 | 0 | 0 | 0 | 0 |
| 0 | 0 | 0 | 0 | 0 |
| 0 | 0 | 0 | 0 | 0 |
| 0 | 0 | 0 | 0 | 0 |
| 0 | 0 | 0 | 0 | 0 |
| 0 | 0 | 0 | 0 | 0 |
| 0 | 0.021478277 | 0 | 0.807288608 | 0 |
| 0 | 0 | 0 | 0 | 0 |
| 0 | 0 | 0 | 0 | 0 |
| 0 | 0 | 0 | 0 | 0 |
| 0 | 0 | 0 | 0.007837393 | 0 |
| 0 | 0 | 0 | 0 | 0 |
| 0 | 0 | 0 | 0 | 0 |
| 0 | 0 | 0 | 0 | 0 |
| 0 | 0 | 0 | 0 | 0 |
| 0 | 0 | 0 | 0.001406706 | 0 |
| 0 | 0 | 0 | 0.008472627 | 0 |
| 0 | 0 | 0 | 0 | 0 |
| 0 | 0.177426008 | 0 | 0.071280149 | 0 |
| 0 | 0.001352892 | 0 | 0.000724604 | 0.000583449 |
| 0 | 0 | 0 | 0 | 0 |
| 0 | 0 | 0 | 0 | 0 |
| 0 | 0 | 0 | 0 | 0 |
| 0 | 0 | 0 | 0 | 0 |
| 0 | 0 | 0 | 0 | 0 |
| 0 | 0 | 0 | 0 | 0 |
| 0 | 0 | 0 | 0.200874524 | 0 |
| 0 | 0 | 0 | 0 | 0 |
| 0 | 0 | 0 | 0 | 0 |
| 0 | 0 | 0 | 0 | 0 |
| 0 | 0 | 0 | 0 | 0 |
| 0 | 0 | 0 | 0 | 0 |
| 0 | 0 | 0 | 0 | 0 |
| 0 | 0 | 0 | 0 | 0 |
| 0 | 0 | 0 | 0 | 0 |
| 0 | 0 | 0 | 0 | 0 |

| | | | | |
|---|---|---|---|---|
| 0 | 0 | 0 | 0 | 0 |
| 0 | 0 | 0 | 0 | 0 |
| 0 | 0 | 0 | 0 | 0 |
| 0 | 0 | 0 | 0 | 0 |
| 0 | 0.737531951 | 0 | 0.001892056 | 0 |
| 0 | 0 | 0 | 0 | 0 |
| 0 | 0 | 0 | 0 | 0 |
| 0 | 0 | 0 | 0 | 0 |
| 0 | 0 | 0 | 0 | 0 |
| 0 | 0 | 0 | 0 | 0 |
| 0 | 0 | 0 | 0 | 0 |
| 0 | 0 | 0 | 0 | 0 |
| 0 | 0 | 0 | 0 | 0 |
| 0 | 0 | 0 | 0.003462075 | 0 |
| 0 | 0 | 0 | 0 | 0 |
| 0 | 0 | 0 | 0.457650592 | 0 |
| 0 | 0 | 0 | 0 | 0 |
| 0 | 0 | 0 | 0 | 0 |
| 0 | 0 | 0 | 0 | 0 |
| 0 | 0 | 0 | 0.005183303 | 0 |
| 0 | 0 | 0 | 0 | 0 |
| 0 | 0 | 0 | 0 | 0 |
| 0 | 0.051400916 | 0 | 0.004798548 | 0 |
| 0 | 0 | 0 | 0 | 0 |
| 0 | 0 | 0 | 0 | 0 |
| 0 | 0 | 0 | 0 | 0 |
| 0 | 0 | 0 | 0.438264758 | 0 |
| 0 | 0 | 0 | 0.000611009 | 0 |
| 0 | 0 | 0 | 0 | 0 |
| 0 | 0 | 0 | 0.233795637 | 0 |
| 0 | 0 | 0 | 0 | 0 |
| 0 | 0 | 0 | 0.002842523 | 0 |
| 0 | 0 | 0 | 0 | 0 |
| 0 | 0 | 0 | 0 | 0 |
| 0 | 0 | 0 | 0 | 0 |
| 0 | 0 | 0 | 0 | 0 |
| 0 | 0 | 0 | 0 | 0 |
| 0 | 0 | 0 | 0 | 0 |
| 0 | 0 | 0 | 0 | 0 |
| 0 | 0 | 0 | 0 | 0 |
| 0 | 0 | 0 | 0 | 0 |
| 0 | 0 | 0 | 0 | 0 |
| 0 | 0 | 0 | 0 | 0 |
| 0 | 0 | 0 | 0 | 0 |
| 0 | 0 | 0 | 0 | 0 |
| 0 | 0 | 0 | 0 | 0 |
| 0 | 0 | 0 | 0 | 0 |
| 0.008793695 | 0.007655001 | 0 | 0 | 0 |

| | | | | |
|---|---|---|---|---|
| 0 | 0 | 0 | 9.212887149 | 0 |
| 0 | 0 | 0 | 0 | 0 |
| 0 | 0 | 0 | 0 | 0 |
| 0 | 0 | 0 | 0 | 0 |
| 0 | 0 | 0 | 0 | 0 |
| 0 | 0 | 0 | 0 | 0 |
| 0 | 0 | 0 | 0 | 0 |
| 0 | 0 | 0 | 0 | 0 |
| 0 | 0 | 0 | 0 | 0 |
| 0 | 0 | 0 | 0 | 0 |
| 0 | 0 | 0 | 0 | 0 |
| 0 | 0 | 0 | 0.104039181 | 0 |
| 0 | 0.00286287 | 0 | 0 | 0.001127283 |
| 0 | 0 | 0 | 0 | 0 |
| 0 | 0 | 0 | 0 | 0 |
| 0 | 0 | 0 | 0 | 0 |
| 0 | 0 | 0 | 0 | 0 |
| 0 | 0 | 0 | 0 | 0 |
| 0 | 0 | 0 | 0 | 0 |
| 0 | 0 | 0 | 0 | 0 |
| 0.028169919 | 0.119857942 | 0.007051259 | 0.005684102 | 0.017464199 |
| 0 | 0 | 0 | 0 | 0 |
| 0 | 0 | 0 | 0 | 0 |
| 0 | 0 | 0 | 0 | 0 |
| 0 | 0 | 0 | 0 | 0 |
| 0 | 0 | 0 | 0 | 0 |
| 0.030988161 | 0.001219848 | 0 | 0 | 0.004559296 |
| 0 | 0 | 0 | 0 | 0 |
| 0 | 0 | 0 | 0 | 0 |
| 0 | 0 | 0 | 0 | 0 |
| 0 | 0 | 0 | 0 | 0.069221473 |
| 0 | 0 | 0 | 0 | 0 |
| 0 | 0 | 0 | 0 | 0 |
| 0 | 0.213206494 | 0 | 0.00603339 | 0 |
| 0 | 0.370849764 | 0.009787573 | 0.003114425 | 0.006073402 |
| 0 | 0 | 0 | 0 | 0 |
| 0 | 0 | 0 | 0 | 0 |
| 0 | 0 | 0 | 0 | 0 |
| 0 | 0 | 0 | 0.011697497 | 0 |
| 0 | 0 | 0 | 0 | 0 |
| 0 | 0 | 0 | 0 | 0 |
| 0 | 0 | 0 | 0.001555187 | 0 |
| 0 | 0 | 0 | 0.007081147 | 0 |
| 0 | 0 | 0 | 0 | 0 |
| 0 | 0 | 0 | 0 | 0 |
| 0 | 0 | 0 | 0.005189226 | 0 |
| 0 | 0.002951233 | 0 | 0 | 0.001484876 |
| 0.012626247 | 0.004748147 | 0 | 0 | 0 |

| | | | | |
|---|---|---|---|---|
| 0 | 0 | 0 | 0 | 0 |
| 0 | 0 | 0 | 0 | 0 |
| 0 | 0 | 0.001195033 | 0 | 0 |
| 0 | 0.000411512 | 0 | 0 | 0 |
| 0 | 0 | 0 | 0 | 0 |
| 0 | 0 | 0 | 0 | 0 |
| 0 | 0 | 0 | 0 | 0 |
| 0 | 0 | 0 | 0 | 0 |
| 0 | 0 | 0 | 0 | 0 |
| 0 | 0 | 0 | 0 | 0 |
| 0 | 0 | 0 | 0 | 0 |
| 0 | 0 | 0 | 0.001197213 | 0 |
| 0 | 0 | 0 | 0 | 0 |
| 0 | 0 | 0 | 0 | 0 |
| 0 | 0 | 0 | 0.019902241 | 0 |
| 0 | 0 | 0 | 0 | 0 |
| 0 | 0 | 0 | 0 | 0 |
| 0 | 0 | 0 | 0 | 0 |
| 0.007379776 | 0 | 0 | 0.001974997 | 0 |
| 0 | 0 | 0 | 0.001778272 | 0 |
| 0 | 0 | 0.677348839 | 0 | 0.025133558 |
| 0 | 0 | 0 | 0 | 0 |
| 0 | 0 | 0 | 0 | 0 |
| 0 | 0 | 0 | 0 | 0 |
| 0.025087531 | 0.008040562 | 0 | 0 | 0.002080547 |
| 0 | 0.000624196 | 0 | 0 | 0 |
| 0 | 0 | 0 | 0 | 0 |
| 0 | 0 | 0 | 0 | 0 |
| 0 | 0.019029599 | 0 | 0.002564484 | 0 |
| 0 | 0 | 0 | 0 | 0 |
| 0.0027215 | 0.402220459 | 0.022480367 | 0.1202672 | 0.191737211 |
| 0 | 0 | 0 | 0 | 0 |
| 0.71342096 | 0 | 0.091780104 | 0 | 0 |
| 0 | 0 | 0 | 0 | 0 |
| 0 | 0 | 0 | 0 | 0 |
| 0 | 0 | 0 | 0 | 0 |
| 0 | 0 | 0 | 0 | 0 |
| 0 | 0 | 0 | 0 | 0 |
| 0 | 0 | 0 | 0 | 0 |
| 0.001918871 | 0 | 0 | 0 | 0 |
| 0 | 0 | 0 | 0 | 0 |
| 0.00342685 | 0.072680713 | 0 | 0 | 0.005965365 |
| 0 | 0 | 0 | 0 | 0 |
| 0 | 0 | 0 | 0 | 0 |
| 0 | 0 | 0 | 0 | 0 |
| 0 | 0 | 0 | 0 | 0 |
| 0 | 0 | 0 | 0 | 0 |
| 0 | 0 | 0 | 0 | 0 |

| | | | | |
|---|---|---|---|---|
| 0 | 0 | 0 | 0 | 0 |
| 0 | 0 | 0 | 0 | 0 |
| 0.080732001 | 0.13011395 | 0 | 0 | 0.004927959 |
| 0 | 0 | 0 | 0 | 0 |
| 0 | 0 | 0 | 0 | 0 |
| 0 | 0 | 0 | 0 | 0 |
| 0 | 0 | 0 | 0 | 0 |
| 0 | 0 | 0 | 0 | 0 |
| 0 | 0 | 0 | 0 | 0 |
| 0 | 0 | 0.001407276 | 0 | 0 |
| 0 | 0 | 0 | 0 | 0 |
| 0 | 0 | 0 | 0 | 0 |
| 0 | 0 | 0 | 0 | 0 |
| 0 | 0.005556652 | 0 | 0.007142691 | 0.000653554 |
| 0.003551277 | 0.021819615 | 0 | 0.00125053 | 0.00161827 |
| 0 | 0 | 0 | 0.013388576 | 0 |
| 0.007879757 | 0.004592251 | 0 | 0 | 0.004302372 |
| 0 | 0 | 0 | 0 | 0 |
| 0 | 0 | 0 | 0 | 0 |
| 0 | 0 | 0 | 0 | 0 |
| 0 | 0 | 0 | 0 | 0 |
| 0.008266659 | 0.00500318 | 0 | 0 | 0 |
| 0 | 0 | 0 | 0 | 0 |
| 0 | 0 | 0 | 0 | 0 |
| 0 | 0 | 0 | 0 | 0 |
| 0 | 0 | 0 | 0 | 0 |
| 0 | 0 | 0 | 0 | 0 |
| 0 | 0 | 0 | 0 | 0 |
| 0 | 0 | 0 | 0 | 0 |
| 0 | 0 | 0 | 0 | 0 |
| 0 | 0 | 0 | 0 | 0 |
| 0 | 0 | 0 | 0 | 0 |
| 0 | 0 | 0 | 0.017873223 | 0 |
| 0 | 0 | 0.001997438 | 0 | 0 |
| 0 | 0 | 0 | 0 | 0 |
| 0 | 0.002569665 | 0 | 0 | 0 |
| 0 | 0 | 0 | 0 | 0 |
| 0.255548852 | 0.063039372 | 0 | 0 | 0.003485436 |
| 0 | 0 | 0 | 0 | 0 |
| 0 | 0 | 0 | 0 | 0 |
| 0 | 0.001814488 | 0 | 0 | 0 |
| 0 | 0 | 0 | 0 | 0 |
| 0 | 0 | 0 | 0 | 0 |
| 0 | 0 | 0 | 0 | 0 |
| 0 | 0 | 0 | 0 | 0 |
| 0 | 0 | 0 | 0 | 0 |
| 0 | 0 | 0 | 0 | 0 |
| 0 | 0 | 0.002636367 | 0.006711178 | 0.001125797 |

| | | | | |
|---|---|---|---|---|
| 0 | 1.097108877 | 0 | 0.135495329 | 0 |
| 0 | 0 | 0 | 0 | 0 |
| 0.005034117 | 0.002994538 | 0 | 0 | 0.000774856 |
| 0 | 0 | 0 | 0 | 0 |
| 0.005672584 | 0.002259595 | 0.000456401 | 0.000968184 | 0.000584685 |
| 0 | 0 | 0.001488406 | 0 | 0.001588969 |
| 0 | 0 | 0 | 0 | 0 |
| 0.001419888 | 0.063768708 | 0 | 0.000814274 | 0.01857679 |
| 0 | 0 | 0 | 0 | 0 |
| 0.027513981 | 0.002394059 | 0.644014309 | 0.005595266 | 0.02534228 |
| 0 | 0 | 0 | 0 | 0 |
| 0.003821561 | 0.064018751 | 0 | 0.001718886 | 0 |
| 0.007673268 | 0.004818013 | 0.000896331 | 0.001792775 | 0 |
| 0 | 0 | 0 | 0 | 0 |
| 0 | 0.000521602 | 0 | 0 | 0 |
| 0 | 0 | 0 | 0 | 0 |
| 0 | 0 | 0 | 0.017435884 | 0 |
| 0 | 0 | 0 | 0 | 0 |
| 0 | 0 | 0 | 0 | 1.040772614 |
| 0 | 0 | 0 | 0 | 0 |
| 0 | 0 | 0 | 0 | 0 |
| 0 | 0 | 0.002770306 | 0 | 0 |
| 0 | 0 | 0 | 0 | 0 |
| 0 | 0 | 0 | 0.479290095 | 0 |
| 0 | 0 | 0 | 0 | 0 |
| 0.280435772 | 0.0013291 | 0 | 0 | 0.000573189 |
| 0 | 0 | 0 | 0.008376741 | 0 |
| 0 | 0 | 0 | 0 | 0.668191162 |
| 0 | 0 | 0 | 0 | 0 |
| 0 | 0.048621398 | 0 | 0 | 0 |
| 0 | 0 | 0 | 0 | 0.003075313 |
| 0 | 0 | 0 | 0 | 0 |
| 0 | 0 | 0 | 0 | 0 |
| 0 | 0 | 0 | 0 | 0.0009995 |
| 0 | 0 | 0 | 0 | 0 |
| 0 | 0 | 0 | 0 | 0 |
| 0 | 0 | 0 | 0 | 0.183234579 |
| 0 | 0 | 0 | 0 | 0 |
| 0 | 0 | 0 | 0 | 0 |
| 0 | 0.001553192 | 0 | 0 | 0 |
| 0 | 0 | 0 | 0 | 0 |
| 0 | 0 | 0 | 0 | 0 |
| 0 | 0 | 0 | 0 | 0 |
| 0 | 0 | 0 | 0 | 0 |
| 0 | 0 | 0 | 0 | 0.000572197 |
| 0 | 0 | 0 | 0.82359903 | 0 |
| 0 | 0.019650444 | 0.002225644 | 0 | 0.001425611 |
| 0 | 0 | 0 | 0 | 0 |

| | | | | |
|---|---|---|---|---|
| 0 | 0 | 0 | 0 | 0 |
| 0 | 0 | 0.003109141 | 0 | 0 |
| 0 | 0 | 0 | 0 | 0 |
| 0 | 0 | 1.763561666 | 0.10619969 | 0 |
| 0 | 0 | 0 | 0 | 0 |
| 0 | 0.012005605 | 0 | 0 | 0 |
| 0 | 0 | 0 | 0 | 0.004161792 |
| 0 | 0 | 0 | 0 | 0 |
| 0 | 0 | 0 | 2.828144743 | 0 |
| 0 | 0 | 0 | 0 | 0 |
| 0.072425155 | 0.004361885 | 0.001524857 | 0 | 0.019317548 |
| 0 | 0 | 0 | 0 | 0 |
| 0 | 0 | 0 | 0 | 0 |
| 0.006821049 | 0.001267966 | 0 | 0 | 0.001025295 |
| 0 | 0 | 0 | 0 | 0 |
| 0 | 0 | 0 | 0 | 0 |
| 0 | 0.015277411 | 0 | 0 | 0 |
| 0 | 0 | 0 | 0 | 0.034722656 |
| 0 | 0 | 0 | 0 | 0 |
| 0 | 0 | 0 | 0 | 0 |
| 0 | 0 | 0 | 0 | 0 |
| 0.410660552 | 0.141206962 | 0 | 0.064957259 | 0.177590508 |
| 0 | 0 | 0.005164482 | 0 | 0.005448552 |
| 0 | 0 | 0 | 0 | 0 |
| 0 | 0 | 0 | 0 | 0 |
| 0 | 0 | 0 | 0 | 0 |
| 0 | 0 | 0 | 0 | 0.002668059 |
| 0 | 0 | 0 | 0 | 0 |
| 0 | 0 | 0 | 0 | 0 |
| 0.821104543 | 0.435370678 | 2.983733828 | 0.019411167 | 0.693423048 |
| 0.000433832 | 0.004623652 | 0 | 0.000552873 | 0 |
| 0 | 0 | 0 | 0.001552433 | 0 |
| 0 | 0 | 0 | 0 | 0 |
| 0 | 0 | 0 | 0 | 0.003546651 |
| 0.002310009 | 0 | 0 | 0 | 0.039629971 |
| 0 | 0 | 0 | 0 | 0 |
| 0 | 0.043816509 | 0 | 0.726087566 | 0 |
| 0.000651794 | 0.066396808 | 0 | 0.001038303 | 0 |
| 0 | 0 | 0 | 0 | 0 |
| 0 | 0 | 0 | 0 | 0 |
| 0.000876194 | 0.014072483 | 0.004079383 | 0 | 0.001854389 |
| 1.404040797 | 1.424111354 | 0 | 0.082685113 | 3.625244665 |
| 0 | 0.064375525 | 0 | 0 | 0 |
| 0 | 0 | 0 | 0 | 0 |
| 0 | 0 | 0 | 0 | 0.001901679 |
| 0 | 0 | 0 | 0 | 0 |
| 0.000735703 | 0 | 0.002900442 | 0 | 0 |
| 0 | 0 | 0 | 0 | 0 |

| | | | | |
|---|---|---|---|---|
| 0.013616013 | 0.003374778 | 0 | 0 | 0.003056359 |
| 0 | 0 | 0 | 0 | 0 |
| 0 | 0.030119478 | 0 | 0 | 0 |
| 0 | 0 | 0 | 0 | 0 |
| 0 | 0 | 0 | 0.022094667 | 0 |
| 0 | 0 | 0 | 0 | 0.124223975 |
| 0 | 0 | 0 | 0 | 0 |
| 0.002034772 | 0.001512977 | 0 | 0 | 0 |
| 0 | 0 | 0 | 0 | 0 |
| 0 | 0 | 0 | 0 | 0 |
| 0 | 0 | 0 | 0 | 0 |
| 0 | 0 | 0 | 0 | 0 |
| 0 | 0 | 0 | 0 | 0.004563749 |
| 0.47096062 | 0.003511038 | 0.003237522 | 0 | 0.03653764 |
| 0 | 0 | 0 | 0 | 0 |
| 0 | 0.011333151 | 0 | 0.011194343 | 0.100477576 |
| 0.235447134 | 0.106310942 | 0 | 0 | 0.112737506 |
| 0 | 0 | 0.053216207 | 0 | 0.001662782 |
| 0 | 0 | 0 | 0 | 0.026682623 |
| 0 | 0 | 0 | 0 | 0 |
| 0.003361749 | 0 | 0.022383499 | 0 | 0 |
| 0 | 0.030215283 | 0.003717556 | 0 | 0.002579674 |
| 0 | 0 | 0 | 0 | 0.018128165 |
| 0 | 0 | 0 | 0 | 0 |
| 0.016786147 | 0 | 0 | 0.003177056 | 0 |
| 0 | 0 | 0.01073075 | 0 | 0 |
| 0 | 0.001510775 | 0.000678115 | 0 | 0 |
| 0 | 0 | 0 | 0 | 0 |
| 0 | 0 | 0 | 0 | 0 |
| 0 | 0 | 0 | 0 | 0.007804928 |
| 0.001545187 | 0.282001032 | 0.005414896 | 0 | 0.005615585 |
| 0.01151842 | 0.008564646 | 0 | 0 | 0 |
| 0.349240979 | 0.009060778 | 0.01738621 | 0.009084663 | 0.649436001 |
| 0 | 0.02441176 | 0 | 0.009347123 | 0 |
| 0 | 0 | 0.008236698 | 0 | 0 |
| 0 | 0 | 0 | 0 | 0 |
| 0.002203335 | 0 | 0 | 0 | 0 |
| 0.02205581 | 0 | 0.00504638 | 0 | 0.080395617 |
| 0 | 1.035645487 | 0 | 0.002936312 | 0 |
| 0 | 0.012256245 | 0 | 0 | 0 |
| 0 | 0 | 0 | 0 | 0 |
| 0 | 0 | 0 | 0 | 0 |
| 0 | 0.046560944 | 0 | 0 | 0.011403402 |
| 0 | 0 | 0.047473877 | 0 | 0.044901577 |
| 0 | 0.002152943 | 0 | 0 | 0.497833796 |
| 0 | 0 | 0 | 0 | 0 |
| 0 | 0 | 0 | 0.002013145 | 0 |
| 0 | 0 | 0 | 0 | 0 |

| | | | | |
|---|---|---|---|---|
| 0 | 0 | 0 | 0 | 0.010076703 |
| 1.043666537 | 0.48534581 | 0.020805316 | 0.064632011 | 0.215710333 |
| 0 | 0 | 0 | 0 | 0 |
| 0 | 0 | 0 | 0 | 0 |
| 0 | 0 | 0 | 0 | 0 |
| 0 | 0 | 0 | 0 | 0 |
| 0 | 0 | 0 | 0 | 0.021314278 |
| 0 | 0.002413792 | 0 | 0 | 0 |
| 0 | 0.006073085 | 0 | 0.00082174 | 0 |
| 0 | 0.020045046 | 0 | 0 | 0.008535214 |
| 0.001221352 | 0.015179067 | 0 | 0 | 0.003692691 |
| 0.02702868 | 0.006956816 | 0 | 0 | 0.006000402 |
| 0 | 0.452813477 | 0 | 0.174144892 | 0 |
| 0 | 0 | 0 | 0 | 0 |
| 0.001201156 | 0.000744277 | 0 | 0 | 0 |
| 0 | 0 | 0 | 0 | 0 |
| 0.04823399 | 0.027034531 | 0 | 0.000523885 | 0.00703051 |
| 0.01043573 | 0.046204892 | 0 | 0 | 0.013233531 |
| 0 | 0 | 0 | 0 | 0 |
| 0 | 0.003206432 | 0 | 0 | 0.005037372 |
| 0.376828879 | 0.40474195 | 0 | 0.028273464 | 0.035163613 |
| 0 | 0 | 0 | 0 | 0 |
| 0 | 0 | 0 | 0 | 0 |
| 0 | 0 | 0 | 0.387877615 | 0 |
| 0 | 0 | 0 | 0 | 0 |
| 0 | 0 | 0 | 0 | 0 |
| 0.005021311 | 0.038027915 | 0.003770678 | 0 | 0.033098067 |
| 0.019825417 | 0.030078418 | 0.336549946 | 0.059526823 | 0.003275018 |
| 0 | 0 | 0 | 0 | 0 |
| 0 | 0 | 0 | 0 | 0 |
| 0.004297513 | 0 | 0 | 0 | 0 |
| 0 | 0 | 0 | 0 | 0 |
| 0 | 0 | 0 | 0 | 0 |
| 0 | 0 | 0 | 0 | 0 |
| 0 | 0 | 0 | 0.007839401 | 0.046502594 |
| 0.089136638 | 0.458153357 | 0.058791534 | 0.027211029 | 0.662524262 |
| 0 | 0 | 0 | 0 | 0 |
| 0 | 0 | 0 | 0 | 0 |
| 0.002964469 | 0.028839107 | 0 | 0.01794504 | 0.004040101 |
| 0 | 0 | 0 | 0 | 0 |
| 0 | 0.001152655 | 0 | 0 | 0.501071664 |
| 0 | 0 | 0 | 0 | 0 |
| 0 | 0 | 0 | 0 | 0.008949951 |
| 0 | 0 | 0 | 0.005815803 | 0.001811824 |
| 0 | 0 | 0 | 0 | 0 |
| 0 | 0 | 0 | 0.032906007 | 0 |
| 0 | 0 | 0 | 0 | 0 |
| 0 | 0 | 0 | 0 | 0 |

| | | | | |
|---|---|---|---|---|
| 0 | 0 | 0 | 0 | 0 |
| 0 | 0 | 0 | 0 | 0 |
| 0 | 0 | 0 | 0.024315939 | 0 |
| 0.005243009 | 0.485492674 | 0 | 0.043597844 | 0 |
| 0 | 0 | 0 | 0.012097045 | 0 |
| 0 | 0 | 0 | 0 | 0 |
| 0 | 0 | 0 | 0 | 0 |
| 0 | 0.821335118 | 0 | 0 | 0 |
| 1.118601349 | 2.027236969 | 0.00524378 | 0.015152521 | 0.125820449 |
| 0 | 0 | 0 | 0.06359927 | 0 |
| 0 | 0 | 0 | 0 | 0 |
| 0.017749226 | 0.01531423 | 0 | 0.009922713 | 0.03559946 |
| 0 | 0 | 0 | 0 | 0 |
| 0 | 0 | 0 | 0 | 0 |
| 0.015613942 | 0.232506534 | 0.007885877 | 0 | 0.014488993 |
| 0 | 0 | 0 | 0 | 0 |
| 0 | 0 | 0 | 0 | 0 |
| 0 | 0.091233141 | 0 | 0.042253091 | 0 |
| 0 | 0 | 0 | 0 | 0 |
| 0 | 0 | 0 | 0.009147099 | 0 |
| 0 | 0 | 0 | 0 | 0 |
| 0 | 0 | 0 | 0.034026857 | 0 |
| 0 | 0 | 0 | 0.003389584 | 0 |
| 0 | 0 | 0 | 0 | 0.405766133 |
| 0 | 0 | 0 | 0 | 0 |
| 0 | 0.292773695 | 0 | 0.149022734 | 0 |
| 0 | 0 | 0 | 1.787878851 | 0 |
| 0 | 0.86765491 | 0 | 0.652516851 | 1.441772855 |
| 0 | 0 | 0 | 0 | 0 |
| 0 | 0 | 0 | 0 | 0 |
| 0 | 0.000968328 | 0 | 0.006068003 | 0 |
| 0 | 0 | 0 | 0.086344144 | 0 |
| 0 | 0.32223956 | 0 | 0 | 0.001311031 |
| 0 | 0 | 0 | 0 | 0 |
| 0 | 0 | 0 | 0.776737982 | 0 |
| 0 | 0 | 0.000414588 | 0 | 0.044614028 |
| 0.977749138 | 3.726934257 | 0 | 0.272095219 | 0 |
| 0 | 0 | 0 | 0 | 0 |
| 0 | 0 | 0 | 0 | 0 |
| 0 | 0 | 0 | 0 | 0.000691796 |
| 0 | 0.001672177 | 0.001930012 | 0.003377734 | 0.041414269 |
| 0 | 0 | 0 | 0.176016951 | 0 |
| 22.79297271 | 0.405351472 | 15.75479629 | 0 | 0.397081233 |
| 0 | 0 | 0 | 0.002006215 | 0 |
| 0 | 0 | 0 | 0 | 0 |
| 0 | 0 | 0 | 0 | 0 |
| 0 | 0 | 0 | 0 | 0 |
| 0 | 0.003876714 | 0 | 0 | 0 |

| | | | | |
|---|---|---|---|---|
| 0 | 0 | 0 | 0.053370668 | 0 |
| 0 | 0.021714008 | 0 | 0.001681437 | 0 |
| 0.009995869 | 0.217883432 | 0.098554247 | 0 | 0.46958532 |
| 0 | 0 | 0 | 1.423463946 | 0 |
| 0 | 0 | 0 | 0 | 0 |
| 0.06970267 | 0.029817464 | 0 | 0.000836254 | 0.002805622 |
| 0 | 0 | 0 | 0 | 0 |
| 0 | 0 | 0 | 0 | 0 |
| 0 | 0 | 0 | 0 | 0 |
| 0 | 0 | 0 | 0 | 0 |
| 0.003634878 | 0 | 0.005322636 | 0.00069484 | 0.021679992 |
| 1.309150605 | 1.090230509 | 6.440285056 | 0.1459256 | 0.038130659 |
| 0 | 0 | 0 | 0 | 0 |
| 0.141224709 | 0.044628893 | 0 | 0 | 0.146237071 |
| 0.027204341 | 0.126080716 | 0.027393017 | 0 | 0.021561111 |
| 0 | 0 | 0 | 0 | 0 |
| 0 | 0 | 0 | 0 | 0 |
| 0 | 0 | 0 | 0 | 0 |
| 0 | 0 | 0 | 0.016787016 | 0 |
| 0 | 0.021818299 | 0 | 0.027044273 | 0.253188968 |
| 0.000913969 | 0.003964284 | 0 | 0.794946696 | 0.120163237 |
| 0.252155895 | 0 | 23.81588963 | 0 | 0 |
| 2.00220115 | 0.062905281 | 14.84312804 | 0.211777195 | 0.595444484 |
| 0 | 0 | 0 | 0 | 0 |
| 0 | 0 | 0 | 0.217042108 | 0 |
| 6.609038272 | 9.479085723 | 0 | 0.461637148 | 0.012291966 |
| 0 | 0 | 0 | 0 | 0 |
| 0 | 0 | 0 | 0 | 0 |
| 0 | 0 | 0 | 0 | 0 |
| 0 | 0 | 0 | 0 | 0 |
| 0 | 0 | 0 | 0.01220462 | 0 |
| 0.311347469 | 0.437989685 | 0 | 0.039311413 | 0.107771141 |
| 0 | 0.686593545 | 0.365413556 | 0.019802931 | 0.501701381 |
| 0 | 0.13864782 | 0 | 0.001768617 | 0.006846567 |
| 0.022791093 | 0 | 0 | 0 | 0.043257744 |
| 1.264391089 | 0.029702248 | 0.006228152 | 0.021193237 | 0.156969561 |
| 0 | 0.008388753 | 0 | 0 | 0.000471879 |
| 0 | 0 | 0 | 0 | 0 |
| 0.274526877 | 2.275593722 | 0 | 0.710502137 | 0 |
| 0 | 0 | 0 | 0 | 0 |
| 0 | 0 | 0 | 0.021510829 | 0 |
| 0 | 0 | 0 | 0 | 0 |
| 0 | 0 | 0 | 1.17912209 | 0 |
| 0 | 0 | 0 | 0.078080479 | 0 |
| 0 | 0.264948264 | 0 | 0.00332086 | 0.003713817 |
| 0 | 0 | 0 | 0.002994557 | 0 |
| 0 | 0 | 0 | 0 | 0 |
| 2.561013061 | 0.24411485 | 0.001923329 | 0 | 1.879308753 |

| | | | | |
|---|---|---|---|---|
| 0 | 0 | 0 | 0 | 0 |
| 0 | 0 | 0 | 0 | 0 |
| 0 | 0 | 0 | 0 | 0 |
| 0 | 0 | 0 | 0 | 0 |
| 0 | 0 | 0 | 0 | 0 |
| 0 | 1.957536974 | 0 | 0.004860629 | 0 |
| 0 | 0 | 0 | 0.00051645 | 0 |
| 0.001665211 | 0 | 0.002841966 | 0 | 0.004223302 |
| 0 | 0 | 0 | 0 | 0 |
| 0 | 0 | 0 | 0 | 0 |
| 0 | 0.003405977 | 0 | 0 | 0.00334901 |
| 0 | 0 | 0 | 0 | 0 |
| 0 | 0 | 0 | 0.015180308 | 0 |
| 0 | 0 | 0 | 0 | 0 |
| 0 | 0 | 0 | 0 | 0 |
| 0 | 0 | 0 | 0 | 0 |
| 0 | 0 | 0 | 0 | 0 |
| 0.000610123 | 0.002948814 | 0 | 0 | 0.007336775 |
| 0 | 0 | 0 | 0.15448185 | 0 |
| 0 | 0 | 0 | 0 | 0 |
| 0.001084093 | 0 | 0 | 0.002072343 | 0 |
| 0 | 0 | 0 | 0 | 0 |
| 0 | 1.112771522 | 0 | 0.000840418 | 0 |
| 0 | 0 | 0 | 0 | 0 |
| 0 | 0 | 0 | 0 | 0 |
| 0 | 0 | 0 | 0 | 0 |
| 0 | 0 | 0 | 0 | 0 |
| 0 | 0 | 0 | 0 | 0 |
| 0 | 0 | 0 | 0 | 0 |
| 0 | 0.143170685 | 0.017018792 | 0.116761598 | 0.087564052 |
| 0 | 0.101830983 | 0 | 0 | 0 |
| 0 | 0 | 0 | 0 | 0 |
| 0 | 0 | 0 | 0.225439499 | 0 |
| 0 | 0 | 0 | 0 | 0 |
| 0 | 0.000541653 | 0 | 0 | 0 |
| 0 | 0 | 0 | 0 | 0 |
| 0.14341308 | 0 | 0 | 0.025834556 | 0 |
| 0.002481654 | 0.00472848 | 0.007454242 | 0 | 0.006266832 |
| 0 | 0 | 0 | 0.011613234 | 0 |
| 0 | 0.003412618 | 0 | 0 | 0 |
| 0 | 0 | 0 | 0.000417278 | 0 |
| 0 | 0 | 0 | 3.684817103 | 0 |
| 0 | 0 | 0 | 0.842463111 | 0 |
| 0 | 0.001343975 | 0 | 0.00129569 | 0 |
| 0 | 0.06272727 | 0.266358253 | 0.068511578 | 0 |
| 0 | 0 | 0 | 0.026190542 | 0 |
| 0 | 0 | 0 | 0.002535473 | 0 |
| 0 | 0 | 0 | 0.364823478 | 0 |

| | | | | |
|---|---|---|---|---|
| 0 | 0 | 0 | 0 | 0 |
| 0 | 0 | 0 | 0 | 0 |
| 0 | 0 | 0 | 0.429041883 | 0 |
| 0 | 1.624755732 | 0 | 0.210281486 | 0 |
| 0 | 0 | 0 | 0 | 0 |
| 0.154336394 | 2.348928852 | 0 | 1.063070498 | 0 |
| 0 | 0 | 0 | 0 | 0 |
| 0 | 0 | 0 | 0 | 0 |
| 0.001925221 | 0.033561164 | 0.401427044 | 0 | 0.225747147 |
| 0 | 0 | 0 | 0 | 0 |
| 0 | 0 | 0 | 0 | 0 |
| 0 | 0 | 0 | 0 | 0 |
| 0 | 0 | 0 | 0 | 0 |
| 0 | 0 | 0 | 0 | 0 |
| 0 | 0.522250512 | 0 | 0.133998903 | 0 |
| 0 | 0 | 0.020799394 | 0 | 0 |
| 0 | 0 | 0 | 0 | 0 |
| 0 | 0 | 0 | 0 | 0 |
| 0 | 0 | 0 | 0 | 0 |
| 0 | 0 | 0 | 0 | 0 |
| 0 | 0 | 0 | 0.288442042 | 0 |
| 0 | 0 | 0 | 0 | 0 |
| 0 | 0 | 0 | 0 | 0 |
| 0 | 0 | 0 | 0 | 0 |
| 0.067668593 | 0.008604174 | 0.005667068 | 0.016349669 | 0 |
| 0.558138378 | 3.261547073 | 0 | 4.760632395 | 20.65797697 |
| 0 | 0 | 0 | 0.228578134 | 0 |
| 0 | 0.00104498 | 0 | 0 | 0 |
| 0 | 0.03156183 | 0 | 0 | 0 |
| 0 | 0 | 0 | 0 | 0 |
| 0 | 0 | 0 | 0 | 0 |
| 0 | 0 | 0 | 0.069988724 | 0 |
| 0 | 0 | 0 | 0 | 0.000468752 |
| 0 | 0 | 0 | 0 | 0 |
| 0.777864671 | 0.023043414 | 0 | 0 | 0.005536728 |
| 0 | 0.007446354 | 0 | 0 | 0.006021224 |
| 0 | 0 | 0 | 0 | 0 |
| 1.684192335 | 4.502996521 | 0 | 0.157056453 | 7.323036083 |
| 0 | 0 | 0 | 4.264293004 | 0 |
| 0 | 0 | 0 | 0 | 0 |
| 0 | 0 | 0 | 0 | 0 |
| 0 | 0.004485995 | 0 | 0 | 0 |
| 0.085530796 | 0.139119201 | 0 | 0.194973723 | 0 |
| 0 | 0 | 0 | 3.423826155 | 0 |
| 0 | 0 | 0 | 0.001227853 | 0 |
| 0 | 0.012235124 | 0 | 0.00085013 | 0 |
| 0 | 0 | 0 | 0 | 0 |
| 0.119972324 | 2.4774181 | 0.000709381 | 0.406533424 | 0.006134211 |

| | | | | |
|---|---|---|---|---|
| 0 | 0 | 0 | 0 | 0 |
| 0 | 0 | 0 | 0 | 0 |
| 0 | 0 | 0 | 0.008841601 | 0 |
| 0 | 0 | 0 | 0 | 0 |
| 0 | 0 | 0 | 0 | 0.036096518 |
| 0 | 0 | 0 | 0.002743517 | 0 |
| 0 | 0 | 0 | 0.002668276 | 0 |
| 0 | 0 | 0 | 0 | 0 |
| 0 | 0 | 0 | 0 | 0 |
| 0 | 0 | 0 | 0.025735858 | 0 |
| 0 | 0.00089783 | 0 | 0.292627966 | 0 |
| 3.847939979 | 1.192798729 | 0 | 0.021181471 | 0 |
| 0.063053856 | 0.051462313 | 0.005420464 | 0 | 0 |
| 0 | 0 | 0 | 0 | 0 |
| 0.02189996 | 0.325980597 | 0.005481815 | 0.07752553 | 0.037649088 |
| 0 | 0 | 0 | 0 | 0 |
| 0 | 0.001887973 | 0 | 0 | 0 |
| 0 | 0 | 0 | 0 | 0 |
| 0 | 0.01665163 | 0 | 0 | 0 |
| 0 | 0 | 0 | 0.087402107 | 0 |
| 0 | 0 | 0 | 0.008438561 | 0 |
| 0 | 0 | 0 | 0 | 0 |
| 0 | 0.578137487 | 0 | 0.364557603 | 2.200060071 |
| 0 | 0 | 0 | 0 | 0 |
| 0 | 0 | 0 | 0.006363524 | 0 |
| 0 | 0 | 0 | 0 | 0 |
| 0 | 0 | 0 | 0.159044195 | 0 |
| 0 | 0 | 0 | 0 | 0 |
| 0 | 0 | 0 | 0.000872886 | 0 |
| 0 | 0 | 0 | 0 | 0 |
| 0 | 0 | 0 | 0 | 0 |
| 0.109989194 | 0.043044029 | 0 | 0.001509003 | 0.039235846 |
| 0 | 0 | 0 | 0 | 0 |
| 0 | 0 | 0 | 0.002261616 | 0.016692935 |
| 0 | 0.007850265 | 0 | 0 | 0.006224971 |
| 0.009658787 | 0.132701841 | 0.006099215 | 0.009022015 | 0.020695388 |
| 0 | 0 | 0 | 0.005778995 | 0 |
| 0 | 0 | 0 | 0.008871139 | 0 |
| 0 | 0 | 0 | 0 | 0 |
| 0 | 0 | 0 | 0.004401383 | 0 |
| 0 | 0 | 0 | 0.427797212 | 0 |
| 0 | 0 | 0 | 1.056859224 | 0 |
| 0 | 0 | 0 | 0 | 0 |
| 0 | 0 | 0 | 0.000794998 | 0 |
| 0.000670991 | 0.027066543 | 0.001889514 | 0.001603326 | 0.009359704 |
| 0 | 0 | 0 | 0.032452621 | 0 |
| 0.088917647 | 0.062862367 | 0.000953877 | 0.082019322 | 0 |
| 0 | 0 | 0 | 0 | 0.001068418 |

| | | | | |
|---|---|---|---|---|
| 0 | 0 | 0 | 0 | 0 |
| 0 | 0 | 0 | 0 | 0 |
| 0 | 0 | 0 | 0 | 0 |
| 0 | 0 | 0 | 0 | 0 |
| 0 | 0 | 0 | 0 | 0 |
| 0 | 0 | 0 | 0.007132837 | 0 |
| 0 | 0 | 0 | 0.028658091 | 0 |
| 0 | 0.003045774 | 0 | 0.007203842 | 0 |
| 0 | 0 | 0 | 0 | 0 |
| 0 | 0 | 0 | 0.000462317 | 0 |
| 0 | 0 | 0 | 0 | 0 |
| 0 | 0 | 0 | 0 | 0 |
| 0 | 0 | 0 | 0.105716592 | 0 |
| 0 | 0 | 0 | 0.128117092 | 0 |
| 0 | 0 | 0 | 0 | 0 |
| 0 | 0 | 0 | 0 | 0 |
| 0 | 0 | 0 | 0 | 0 |
| 0 | 0 | 0 | 0.023530737 | 0 |
| 14.69809584 | 0 | 0 | 1.587785043 | 0 |
| 0 | 0 | 0 | 0 | 0 |
| 0 | 0 | 0 | 0.001958657 | 0 |
| 0 | 0 | 0 | 0 | 0 |
| 0 | 0 | 0 | 0 | 0 |
| 0 | 0 | 0 | 0.00941279 | 0 |
| 0.002565905 | 0 | 0 | 0 | 0 |
| 0.079332715 | 0.005341496 | 0 | 0 | 0.043868122 |
| 0 | 0 | 0 | 0 | 0 |
| 0.001375706 | 0.002273158 | 0 | 0.631879623 | 0.120432732 |
| 0 | 0 | 0 | 0.072365649 | 0 |
| 0.779173921 | 2.291747974 | 0 | 0.64886939 | 1.999080492 |
| 0 | 0 | 0 | 0.032315818 | 0 |
| 0 | 0 | 0 | 0 | 0 |
| 0 | 0 | 0 | 0.000617073 | 0 |
| 0 | 0 | 0 | 0 | 0 |
| 0.003109409 | 0.001460232 | 0 | 0 | 0.001574352 |
| 0 | 0 | 0 | 0 | 0 |
| 0 | 0.191508385 | 0 | 0.082016541 | 0 |
| 0 | 0.002347326 | 0 | 0 | 0 |
| 0 | 0 | 0 | 0 | 0 |
| 0 | 0 | 0 | 0.002389015 | 0 |
| 2.598342844 | 7.325222125 | 2.282969172 | 0.235647449 | 7.186246064 |
| 0.058042976 | 0.351837653 | 0 | 0.039626601 | 3.109124925 |
| 0 | 0 | 0 | 0.110557836 | 0.102987957 |
| 0 | 0 | 0 | 0.07339019 | 0 |
| 0 | 0 | 0 | 0 | 0 |
| 0 | 0 | 0 | 0.419101089 | 0 |
| 0 | 0 | 0 | 0.064653762 | 0 |
| 0 | 0.008657322 | 0.000442693 | 0.006198089 | 0.000708904 |

| | | | | |
|---|---|---|---|---|
| 0 | 0 | 0 | 0 | 0 |
| 0 | 0.026488873 | 0 | 0 | 0.002479166 |
| 0 | 0 | 0 | 0 | 0 |
| 0 | 0 | 0 | 0.605657046 | 0 |
| 1.114514381 | 1.429905504 | 0 | 0.648062327 | 0 |
| 0 | 0 | 0 | 0 | 0 |
| 0 | 0 | 0 | 0 | 0 |
| 0 | 0 | 0 | 0.064521686 | 0 |
| 0.023462271 | 0.012605871 | 0.001364024 | 0 | 0.397683556 |
| 0 | 0 | 0 | 0 | 0 |
| 0 | 0 | 0.025465079 | 0 | 0 |
| 0 | 0 | 0 | 0 | 0 |
| 0 | 0.153883099 | 0 | 0.04181381 | 0.128523628 |
| 0.034950068 | 0.024910299 | 0.006119353 | 0 | 0.021950218 |
| 0 | 0.401972151 | 0 | 0.008754661 | 0 |
| 0.019763781 | 0.133475583 | 0 | 0.069453229 | 0.61027016 |
| 0 | 0.000349557 | 0 | 0.004043979 | 0.00075375 |
| 0 | 0 | 0 | 0 | 0 |
| 0 | 0 | 0 | 0.007282069 | 0 |
| 0 | 0 | 0 | 0.370706098 | 0 |
| 0 | 0 | 0 | 0 | 0 |
| 0 | 0 | 0 | 0 | 0 |
| 0 | 0 | 0 | 0 | 0 |
| 0 | 0 | 0 | 0 | 0 |
| 0 | 0 | 0 | 0.163973636 | 0 |
| 0 | 0 | 0 | 0 | 0 |
| 0.041695721 | 0 | 0 | 0.014586573 | 0 |
| 0.000768774 | 0 | 0 | 0 | 0 |
| 0 | 0 | 0 | 0 | 0 |
| 0 | 0 | 0 | 0.048737298 | 0 |
| 0 | 0 | 0 | 0.015101936 | 0 |
| 0 | 0 | 0 | 0 | 0 |
| 0 | 0.06531404 | 0 | 0.002534541 | 0 |
| 0 | 0 | 0 | 0.02310771 | 0 |
| 0 | 0 | 0 | 0.000911009 | 0.069503097 |
| 0 | 0.014018656 | 0 | 0.001020001 | 0 |
| 0 | 0.000405531 | 0.080272304 | 0.002780169 | 0 |
| 0 | 0 | 0 | 0.005297434 | 0 |
| 0 | 0 | 0 | 0 | 0 |
| 0 | 0 | 0 | 0 | 0 |
| 0.032239671 | 0.039359688 | 0 | 0.066834253 | 7.46575148 |
| 0 | 0 | 0 | 0 | 0 |
| 0 | 0 | 0 | 0 | 0 |
| 0 | 0 | 0 | 0.398039174 | 0 |
| 0 | 0 | 0 | 0 | 0 |
| 0 | 0 | 0 | 0.000519467 | 0 |
| 0 | 1.066508833 | 0 | 0 | 0 |
| 0 | 0 | 0 | 0 | 0 |

| | | | | |
|---|---|---|---|---|
| 0 | 0.176248205 | 0 | 0.006114407 | 0 |
| 0 | 0 | 0 | 0 | 0 |
| 0 | 0 | 0 | 0.000493139 | 0 |
| 0 | 0 | 0 | 0 | 0 |
| 0 | 0 | 0 | 0.072633418 | 0 |
| 0 | 0 | 0 | 0 | 0 |
| 0 | 0.007439078 | 0 | 0.010608309 | 0.01112838 |
| 0 | 0 | 0 | 0 | 0 |
| 0 | 0 | 0 | 0 | 0 |
| 0 | 0 | 0 | 0.019594794 | 0 |
| 0 | 0 | 0 | 0.007066312 | 0 |
| 0 | 0 | 0 | 0.001175979 | 0 |
| 0 | 0 | 0 | 0 | 0 |
| 0 | 0 | 0 | 0 | 0 |
| 0 | 0 | 0 | 0.008955441 | 0 |
| 0 | 0 | 0 | 0 | 0 |
| 0 | 0 | 2.795168896 | 0 | 0 |
| 0 | 0 | 0 | 0 | 0 |
| 0 | 0 | 0 | 0 | 0 |
| 0 | 0 | 0 | 0 | 0 |
| 0.055215235 | 0.01283688 | 0 | 0.180372137 | 0.119836207 |
| 0 | 0 | 0 | 0 | 0 |
| 0 | 0.025876856 | 0 | 0.27473278 | 0 |
| 0 | 0.003690278 | 0.001315368 | 0 | 0.004774415 |
| 0 | 0 | 0 | 0.000667183 | 0 |
| 0 | 0.011780595 | 0 | 0 | 0 |
| 0 | 0 | 0 | 0.959747252 | 0 |
| 0 | 0 | 0 | 0.00151659 | 0 |
| 0 | 0 | 0 | 0 | 0 |
| 0.006302552 | 0.007185707 | 0 | 0 | 0.002627339 |
| 0 | 0 | 0 | 0.219539687 | 0 |
| 0 | 0 | 0 | 0 | 0 |
| 0 | 0 | 0 | 0.123675464 | 0 |
| 0 | 0 | 0 | 0 | 0 |
| 0 | 0 | 0 | 0.116006829 | 0 |
| 0 | 0 | 0 | 0 | 0 |
| 0 | 0.869582638 | 0 | 0.027233324 | 0.466325716 |
| 0 | 0 | 0 | 0 | 0 |
| 0 | 0 | 0 | 0.014960545 | 0 |
| 0 | 0 | 0 | 0 | 0 |
| 0 | 0 | 0 | 0 | 0 |
| 0 | 0 | 0 | 0 | 0 |
| 0 | 0 | 0 | 0 | 0 |
| 0 | 0 | 0 | 0.020989642 | 0 |
| 0 | 0 | 0 | 0.03023522 | 0 |
| 0 | 0 | 0 | 0.941251449 | 0 |
| 0 | 0 | 0 | 0 | 0 |
| 0 | 0 | 0 | 0.000998368 | 0 |

| | | | | |
|---|---|---|---|---|
| 0 | 0.039708699 | 0 | 0 | 0.007101755 |
| 0 | 0 | 0 | 0.000996651 | 0 |
| 0 | 0 | 0 | 0.011239806 | 0 |
| 0 | 0 | 0 | 0 | 0 |
| 0 | 0 | 0 | 0 | 0 |
| 0 | 0.420143877 | 0 | 0 | 0 |
| 0 | 0 | 0 | 0 | 0 |
| 0.024674965 | 1.679680704 | 0.000484426 | 0.059191796 | 1.532384489 |
| 2.465679109 | 0.048029815 | 0.195875947 | 0.327310968 | 0.00118588 |
| 0 | 0.016718458 | 0.000386662 | 0 | 0.039132209 |
| 0 | 0 | 0 | 5.513639298 | 0 |
| 0 | 0 | 0 | 0 | 0 |
| 0 | 0 | 0 | 0.012502252 | 0 |
| 2.698014948 | 0 | 0.002467096 | 0.146714262 | 0 |
| 0 | 0 | 0 | 0 | 0 |
| 0 | 0 | 0 | 0 | 0 |
| 0.008941856 | 0 | 0 | 0 | 0 |
| 0 | 0 | 0 | 0.458462792 | 0 |
| 0 | 0 | 0 | 0 | 0 |
| 0 | 0.09429193 | 0 | 0 | 0 |
| 0 | 0 | 0 | 0.214425101 | 0 |
| 0 | 0 | 0 | 0.024111676 | 0 |
| 0 | 0 | 0 | 0 | 0 |
| 0 | 0 | 0 | 0 | 0 |
| 0 | 0 | 0 | 0.087501231 | 0 |
| 0 | 0 | 0 | 0.006729804 | 0 |
| 0 | 0 | 0 | 0 | 0 |
| 0 | 0 | 0 | 0.071064873 | 0 |
| 0 | 0 | 0 | 0 | 0 |
| 0 | 0 | 0 | 0 | 0 |
| 0 | 0 | 0 | 0 | 0 |
| 0 | 0 | 0 | 0 | 0 |
| 0 | 0 | 0 | 0 | 0 |
| 0 | 0 | 0 | 0.000716232 | 0 |
| 0 | 0 | 0 | 0.000545245 | 0.002926868 |
| 0 | 0 | 0 | 0.007821772 | 0 |
| 0 | 0 | 0 | 0 | 0 |
| 0 | 0 | 0 | 0 | 0 |
| 0.001307371 | 0 | 0 | 0 | 0 |
| 0 | 0 | 0 | 0 | 0.320397522 |
| 0 | 0 | 0 | 0 | 0 |
| 0 | 2.398997299 | 0.011054643 | 0.173174605 | 0 |
| 0 | 0 | 0 | 0 | 0 |
| 0 | 0 | 0 | 0 | 0 |
| 0 | 0 | 0 | 0 | 0 |
| 0 | 0 | 0 | 0 | 0 |
| 0 | 0 | 0 | 0 | 0 |
| 0 | 0 | 0 | 0.000559209 | 0 |

| | | | | |
|---|---|---|---|---|
| 0 | 0 | 0 | 0 | 0 |
| 0.000708808 | 0.030146787 | 0.01117764 | 0.009891145 | 0 |
| 0 | 0 | 0 | 0 | 0 |
| 0 | 0 | 0 | 0 | 0 |
| 0.133588554 | 0.499898336 | 0.126065159 | 0.052664666 | 0.42100379 |
| 0 | 0 | 0 | 0 | 0 |
| 0 | 0 | 0 | 0 | 0 |
| 0 | 0.036250033 | 0.033008598 | 0 | 0.022680973 |
| 0 | 0 | 0 | 0 | 0 |
| 0 | 0 | 0 | 0.045662387 | 0 |
| 0 | 0 | 0 | 0 | 0 |
| 0 | 0 | 0 | 0.004993311 | 0 |
| 0 | 0 | 0 | 0.001671499 | 0 |
| 0 | 0 | 0 | 0 | 0 |
| 0 | 0 | 0 | 0 | 0 |
| 0.004158904 | 0 | 0 | 0 | 0 |
| 0 | 0 | 0 | 0 | 0 |
| 0 | 0 | 0 | 0 | 0 |
| 0 | 0 | 0 | 0 | 0 |
| 0 | 0 | 0 | 0 | 0 |
| 0 | 0 | 0 | 0.015322172 | 0 |
| 0 | 0 | 0 | 0 | 0 |
| 0 | 0 | 0 | 0 | 0 |
| 0 | 0.001671428 | 0 | 0 | 0.00149709 |
| 0.25353519 | 0.041982046 | 0 | 0.114841077 | 0 |
| 0 | 0 | 0 | 0 | 0 |
| 0 | 0.159691464 | 0.015913751 | 0.132867725 | 0.045185539 |
| 0 | 0 | 0 | 0 | 0 |
| 0 | 0 | 0 | 0 | 0 |
| 0 | 0 | 0 | 0 | 0 |
| 0 | 0 | 0 | 0 | 0 |
| 0 | 0 | 0 | 0 | 0 |
| 0 | 0 | 0 | 0 | 0 |
| 0 | 0 | 0 | 0 | 0 |
| 0 | 0 | 0 | 0 | 0 |
| 0 | 0 | 0 | 0 | 0 |
| 0 | 0 | 0 | 0 | 0 |
| 0 | 0 | 0 | 1.080032726 | 0 |
| 0 | 0 | 0 | 0 | 0 |
| 0 | 0 | 0 | 0 | 0 |
| 0 | 0 | 0 | 0 | 0 |
| 0 | 0.233698822 | 0 | 0.001422029 | 0.032203525 |
| 0 | 0 | 0 | 0 | 0 |
| 0 | 0.087477121 | 0 | 0 | 0 |
| 0 | 0 | 0 | 0 | 0 |
| 0 | 0 | 0 | 0.03135454 | 0 |
| 0 | 0 | 0 | 0 | 0 |
| 0 | 0.243869386 | 0 | 0.000611961 | 0 |

| | | | | |
|---|---|---|---|---|
| 0.028109055 | 0.289293499 | 0 | 0.00312732 | 0.817241142 |
| 0 | 0 | 0 | 0.018050098 | 0 |
| 0 | 0 | 0 | 0 | 0 |
| 0 | 0 | 0 | 0 | 0 |
| 0 | 0 | 0 | 0 | 0 |
| 0 | 0 | 0 | 0 | 0 |
| 0 | 0 | 0 | 0 | 0 |
| 0 | 0 | 0 | 0 | 0 |
| 0 | 0 | 0 | 0 | 0 |
| 0 | 0 | 0 | 0 | 0 |
| 0 | 0 | 0 | 0 | 0 |
| 0 | 0.002014879 | 0 | 0 | 0 |
| 0 | 0 | 0 | 0.003840246 | 0 |
| 0 | 0 | 0 | 0 | 0 |
| 0 | 0.025465926 | 0 | 0.00148794 | 0 |
| 0 | 0 | 0 | 0.151926617 | 0 |
| 0 | 0 | 0 | 0 | 0 |
| 0 | 0 | 0 | 0 | 0.076626619 |
| 0 | 0 | 0 | 0 | 0 |
| 0 | 0 | 0 | 0 | 0 |
| 0 | 0.008962374 | 0 | 0 | 0.100382673 |
| 0 | 0 | 0 | 0 | 0 |
| 0.01217797 | 0.009701851 | 0.00440913 | 0.027644178 | 0.009623261 |
| 0 | 0.001775928 | 0 | 0.117375633 | 0 |
| 0 | 0 | 0 | 0.026510331 | 0 |
| 0 | 0.197667944 | 0 | 0 | 0 |
| 0 | 0 | 0 | 0.187125946 | 0 |
| 0 | 0 | 0 | 0 | 0 |
| 0.008659711 | 0 | 0 | 0 | 0 |
| 0 | 0 | 0 | 0.015838412 | 0 |
| 0 | 0 | 0 | 0 | 0 |
| 0 | 0 | 0 | 0 | 0 |
| 0.044600204 | 0.005602875 | 0.625640016 | 0 | 0.749967148 |
| 0 | 0 | 0 | 0 | 0 |
| 0.066126782 | 0.164981234 | 0.058001461 | 0.000522345 | 0 |
| 0.00571983 | 0 | 0 | 0 | 0.110050168 |
| 0 | 0 | 0 | 0 | 0 |
| 0 | 0.141849658 | 0 | 0.004935899 | 0 |
| 0.009451269 | 0.00449766 | 0 | 0 | 0.173296824 |
| 0 | 0 | 0 | 0 | 0 |
| 0 | 0 | 0 | 0.007110405 | 0 |
| 0 | 0 | 0 | 0.006880252 | 0 |
| 0.072939012 | 0.247835647 | 0 | 0.001901313 | 0.016159842 |
| 0 | 0 | 0 | 0 | 0 |
| 0 | 0 | 0 | 0 | 0 |
| 0 | 0.089145308 | 0 | 0.101168146 | 0 |
| 0 | 0 | 0 | 0 | 0 |
| 0 | 0 | 0 | 0 | 0 |

| | | | | |
|---|---|---|---|---|
| 0 | 0 | 0 | 0 | 0 |
| 0 | 0 | 0 | 0.071657636 | 0 |
| 0 | 0 | 0 | 0 | 0 |
| 0 | 0 | 0 | 0 | 0 |
| 0 | 0 | 0 | 0.061387104 | 0 |
| 0 | 0 | 0 | 0 | 0 |
| 0 | 0 | 0 | 0 | 0 |
| 0 | 0 | 0 | 0 | 0 |
| 0 | 0 | 0 | 0.013832662 | 0 |
| 0 | 0 | 0 | 0.012269232 | 0 |
| 0 | 0 | 0 | 0 | 0 |
| 0 | 0 | 0 | 0 | 0 |
| 0 | 0 | 0 | 0 | 0 |
| 0 | 0 | 0 | 0.000743146 | 0 |
| 0 | 0 | 0 | 0 | 0 |
| 0 | 0 | 0.001870124 | 0 | 0.016770416 |
| 0 | 0 | 0 | 0 | 0 |
| 0 | 0 | 0 | 0 | 0 |
| 0 | 0 | 0 | 0 | 0 |
| 0 | 1.169851355 | 0 | 0.029181314 | 0 |
| 0 | 0.036332171 | 0 | 0.007635768 | 0.008042836 |
| 0 | 0 | 0 | 0 | 0 |
| 0 | 0 | 0 | 0 | 0 |
| 0 | 0 | 0 | 0 | 0 |
| 0 | 0.001787645 | 0 | 0 | 0 |
| 0 | 0 | 0 | 0.801140912 | 0 |
| 0 | 0 | 0 | 0 | 0 |
| 0 | 0 | 0 | 0.058474288 | 0 |
| 0 | 0 | 0 | 0.043086932 | 0 |
| 0 | 0 | 0 | 0.035066755 | 0 |
| 0.181592748 | 0 | 0 | 0.093001319 | 0.549254444 |
| 0 | 0 | 0 | 0.039321191 | 0 |
| 0 | 0 | 0 | 0.294467684 | 0 |
| 0 | 0 | 0 | 0 | 0 |
| 0 | 0 | 0 | 0 | 0 |
| 0 | 0 | 0 | 0 | 0 |
| 0 | 0 | 0 | 0 | 0 |
| 0 | 0 | 0 | 0.003054017 | 0 |
| 0 | 0 | 0 | 0.001868935 | 0 |
| 0 | 0 | 0 | 0 | 0 |
| 0 | 0.003202025 | 0 | 0.001949675 | 0 |
| 0 | 0 | 0 | 2.909627263 | 0 |
| 0 | 0 | 0 | 0.011060318 | 0 |
| 0 | 0 | 0 | 0 | 0 |
| 0 | 0 | 0 | 0 | 0 |
| 0 | 0 | 0 | 0 | 0 |
| 0 | 0 | 0 | 0 | 0.12980923 |
| 0 | 0 | 0 | 0.004384968 | 0 |

| | | | | |
|---|---|---|---|---|
| 0 | 0 | 0 | 0 | 0 |
| 0 | 0 | 0 | 0 | 0 |
| 0 | 0 | 0 | 0 | 0 |
| 0 | 0.000728107 | 0 | 0 | 0 |
| 0 | 0.621500529 | 0 | 0.027175984 | 0.07237299 |
| 0 | 0 | 0 | 0.024240677 | 0 |
| 0 | 0 | 0 | 0.024536079 | 0 |
| 0 | 0 | 0 | 0 | 0 |
| 0 | 0 | 0 | 0 | 0 |
| 0 | 0 | 0 | 0 | 0 |
| 0 | 0 | 0 | 0.001981094 | 0 |
| 0 | 0.011796956 | 0.001843826 | 0.005776827 | 0.005632659 |
| 0 | 0.020113272 | 0 | 0 | 0.002194642 |
| 0 | 0 | 0 | 0 | 0 |
| 0.000719679 | 0 | 0 | 0.002476319 | 0 |
| 0 | 0 | 0 | 0 | 0 |
| 0 | 0.127025627 | 0 | 0 | 0.001816722 |
| 0 | 0 | 0 | 0 | 0 |
| 0.000715086 | 0.005529781 | 0.002255328 | 0 | 0.15588185 |
| 0 | 0 | 0 | 0.048904143 | 0.138144572 |
| 0 | 0 | 0 | 0 | 0 |
| 0 | 0 | 0 | 0 | 0 |
| 0 | 0 | 0 | 0.005731979 | 0 |
| 0 | 0 | 0 | 0 | 0 |
| 0 | 0 | 0 | 0 | 0 |
| 0 | 0 | 0 | 0 | 0 |
| 0.000598563 | 0.001891539 | 0 | 0 | 0.001439552 |
| 0 | 0 | 0 | 0.008137291 | 0 |
| 0 | 0 | 0 | 0 | 0 |
| 0 | 0 | 0 | 0 | 0 |
| 0 | 0.007819873 | 0 | 0 | 0.001468627 |
| 0 | 0 | 0 | 0 | 0 |
| 0 | 0 | 0 | 0 | 0 |
| 0 | 0 | 0 | 0 | 0 |
| 0 | 0 | 0 | 0.017950341 | 0 |
| 0 | 0 | 0 | 0.001413735 | 0 |
| 0 | 0 | 0 | 0 | 0 |
| 0.004171092 | 0.028622051 | 0.001968823 | 0.003644995 | 0.003324737 |
| 0 | 0.055636572 | 0 | 0.002466102 | 0 |
| 0 | 0 | 0 | 0 | 0 |
| 0 | 0 | 0 | 0 | 0 |
| 0 | 0 | 0 | 0 | 0 |
| 0 | 0 | 0 | 0 | 0 |
| 0 | 0 | 0 | 0 | 0.003060102 |
| 0 | 0 | 0 | 0 | 0 |
| 0 | 0 | 0 | 0.000755025 | 0 |
| 0 | 0 | 0 | 0 | 0 |
| 0 | 0 | 0 | 3.76268251 | 0 |

| | | | | |
|---|---|---|---|---|
| 0 | 0 | 0 | 0 | 0 |
| 0 | 0 | 0 | 0.00678525 | 0.097063703 |
| 0 | 0 | 0 | 0.060338414 | 0 |
| 0 | 0.00068855 | 0 | 0 | 0.002138 |
| 0.061323021 | 0.000510895 | 0 | 0 | 0 |
| 0 | 0.000989039 | 0 | 0 | 0 |
| 0 | 0.031137437 | 0 | 0.024049368 | 0 |
| 0 | 0 | 0 | 0 | 0 |
| 0 | 0 | 0 | 0 | 0 |
| 0 | 0 | 0 | 0.039274178 | 0 |
| 0 | 0 | 0 | 0.048269628 | 0 |
| 0 | 0 | 0 | 0 | 0 |
| 0 | 0.041194368 | 0 | 0.036765604 | 0 |
| 0 | 0 | 0 | 0.00318836 | 0 |
| 0 | 0 | 0 | 0 | 0 |
| 0 | 0 | 0 | 0 | 0 |
| 0 | 0 | 0 | 0 | 0 |
| 0 | 0 | 0 | 0 | 0 |
| 0 | 0 | 0 | 0 | 0 |
| 0 | 0 | 0 | 0.112989128 | 0 |
| 0 | 0 | 0 | 0 | 0 |
| 0 | 0 | 0 | 0 | 0 |
| 0 | 0.150005102 | 0 | 0 | 0.024778929 |
| 0 | 0.084031015 | 0 | 0.002924622 | 0.010400801 |
| 0 | 0 | 0 | 0 | 0 |
| 0 | 0.00179228 | 0 | 0.003311788 | 0 |
| 0 | 0.011421141 | 0 | 0.003157223 | 0 |
| 0 | 0 | 0 | 0 | 0 |
| 0 | 0 | 0 | 0.188675955 | 0 |
| 0 | 0 | 0 | 0 | 0 |
| 0.021202375 | 0.142603897 | 0 | 0.016949029 | 0.066753172 |
| 0 | 0 | 0 | 0 | 0 |
| 0 | 0 | 0 | 0 | 0 |
| 0 | 0 | 0 | 0 | 0 |
| 0 | 0 | 0 | 0.025021402 | 0 |
| 0 | 0 | 0 | 0 | 0.001938388 |
| 0 | 0 | 0 | 0 | 0 |
| 0 | 0 | 0 | 0 | 0 |
| 0 | 0.003152736 | 0 | 0 | 0 |
| 0 | 0 | 0 | 0 | 0 |
| 0 | 0 | 0 | 0 | 0 |
| 0 | 0 | 0 | 0 | 0 |
| 0 | 0 | 0 | 0 | 0 |
| 0 | 0 | 0 | 0 | 0 |
| 0 | 0 | 0 | 0.183002946 | 0 |
| 0.109477143 | 0.428245474 | 0 | 0.030582217 | 0.210911304 |
| 0 | 0 | 0 | 0 | 0 |
| 0 | 0 | 0 | 0.063177967 | 0 |

| | | | | |
|---|---|---|---|---|
| 0 | 0 | 0 | 0 | 0 |
| 0 | 0 | 0 | 0 | 0 |
| 0 | 0.083386884 | 0 | 0.398930914 | 0 |
| 0 | 0.013047178 | 0 | 0 | 0 |
| 0 | 0 | 0 | 0 | 0 |
| 0 | 0 | 0 | 0 | 0 |
| 0 | 0 | 0 | 0 | 0 |
| 0 | 0 | 0 | 0 | 0 |
| 0 | 0 | 0 | 0.014854799 | 0 |
| 0 | 0 | 0 | 0 | 0 |
| 0 | 0.002065846 | 0 | 0.029463421 | 0 |
| 0 | 0 | 0 | 0 | 0.001140123 |
| 0 | 0 | 0 | 0 | 0 |
| 0 | 0 | 0 | 0 | 0 |
| 0 | 0 | 0 | 0 | 0 |
| 0 | 0 | 0 | 0 | 0 |
| 0 | 0.002747494 | 0 | 0.053347459 | 0 |
| 0 | 0 | 0 | 0 | 0 |
| 0 | 0 | 0 | 0 | 0 |
| 0 | 0.000521841 | 0 | 0 | 0 |
| 0 | 0 | 0 | 0 | 0 |
| 0 | 0 | 0 | 0 | 0 |
| 0 | 0 | 0 | 0 | 0 |
| 0 | 0 | 0 | 0 | 0 |
| 0 | 0 | 0 | 0 | 0 |
| 0 | 0 | 0 | 0 | 0 |
| 0 | 0 | 0 | 0 | 0 |
| 0.005436729 | 0.01472639 | 0 | 0.005567575 | 0 |
| 0 | 0 | 0 | 0 | 0 |
| 0 | 0 | 0 | 0 | 0 |
| 0 | 0.006388497 | 0 | 0.002737324 | 0.021605842 |
| 0 | 0 | 0 | 0.001640498 | 0 |
| 0.072456607 | 0.008979316 | 0.096729241 | 0.014218761 | 0.038555889 |
| 0.004471183 | 0 | 0 | 0.004957305 | 0 |
| 0 | 0 | 0 | 0 | 0 |
| 0 | 0 | 0 | 0 | 0 |
| 0 | 0 | 0 | 0 | 0 |
| 0 | 0 | 0 | 0 | 0 |
| 0 | 0 | 0 | 0 | 0 |
| 0 | 0 | 0 | 0 | 0 |
| 0 | 0 | 0 | 0 | 0 |
| 0.001583824 | 0.017022682 | 0.004216783 | 0 | 0 |
| 0.202075962 | 0.072519276 | 0 | 0.01240678 | 0 |
| 0 | 0 | 0 | 0 | 0 |
| 0 | 0 | 0 | 0 | 0 |
| 0 | 0.023371635 | 0.000761401 | 0.001453675 | 0.011704955 |
| 0 | 0.003707203 | 0 | 0 | 0 |
| 0 | 0 | 0 | 0 | 0 |

| | | | | |
|---|---|---|---|---|
| 0 | 0 | 0 | 0 | 0 |
| 0.006248131 | 0.002732862 | 0 | 0.087120036 | 0.007071455 |
| 0.021680768 | 0.010534495 | 0 | 0.003282755 | 0.002478062 |
| 0 | 0 | 0 | 0 | 0 |
| 0 | 0 | 0 | 0 | 0.003297688 |
| 0 | 0 | 0 | 0 | 0 |
| 0 | 0 | 0 | 0 | 0 |
| 0 | 0 | 0 | 0 | 0 |
| 0 | 0 | 0 | 0 | 0 |
| 0 | 0 | 0 | 0 | 0 |
| 0 | 0 | 0 | 0.564712423 | 0 |
| 0 | 0 | 0 | 0 | 0 |
| 0 | 0 | 0 | 0 | 0 |
| 0 | 0 | 0.042814791 | 0 | 0 |
| 0 | 0 | 0 | 0 | 0 |
| 0 | 0 | 0 | 0 | 0 |
| 0 | 0 | 0 | 0 | 0 |
| 0 | 0 | 0 | 0 | 0 |
| 0 | 0 | 0 | 0 | 0 |
| 0 | 0 | 0 | 0 | 0 |
| 0 | 0 | 0 | 0 | 0 |
| 0 | 0 | 0 | 0 | 0 |
| 0 | 0 | 0 | 0.004871892 | 0 |
| 0 | 0 | 0 | 0 | 0.044339222 |
| 0 | 0.001052352 | 0 | 0.001690907 | 0 |
| 0.008720441 | 0.005690197 | 0 | 0 | 0 |
| 0 | 0 | 0 | 0 | 0 |
| 0 | 0.192418954 | 0 | 0.054946387 | 0 |
| 0 | 0 | 0 | 0 | 0 |
| 0 | 0 | 0 | 0 | 0 |
| 0 | 0 | 0 | 0 | 0 |
| 0 | 0 | 0 | 0 | 0 |
| 0 | 0 | 0 | 0 | 0 |
| 0 | 0 | 0 | 0 | 0 |
| 0 | 0 | 0 | 0 | 0 |
| 0 | 0 | 0 | 0 | 0 |
| 0 | 0 | 0 | 0 | 0 |
| 0 | 0 | 0 | 0 | 0.077882656 |
| 0 | 5.902142497 | 0 | 1.181918499 | 0 |
| 0 | 0 | 0 | 0 | 0 |
| 0 | 0 | 0 | 0 | 0 |
| 0.003668293 | 0.028998682 | 0 | 0.007381346 | 0 |
| 0 | 0.027491208 | 0 | 0 | 0.006529323 |
| 0 | 0 | 0 | 0.531269958 | 0 |
| 0 | 0 | 0 | 0 | 0 |
| 0 | 0.001069534 | 0 | 0 | 0 |
| 0 | 0.030069144 | 0.000820739 | 0 | 0 |
| 0 | 0 | 0 | 0 | 0 |

| | | | | |
|---|---|---|---|---|
| 0 | 0 | 0 | 0 | 0 |
| 0.00275822 | 0.288921138 | 0.008284975 | 0.002965832 | 0.035489532 |
| 0 | 0 | 0 | 0.001172213 | 0 |
| 0 | 0 | 0 | 0 | 0 |
| 0 | 0 | 0 | 0 | 0 |
| 0 | 0 | 0 | 0 | 0 |
| 0 | 0 | 0 | 0 | 0 |
| 0 | 0 | 0 | 0 | 0 |
| 0 | 0 | 0 | 0 | 0 |
| 0 | 0 | 0 | 0 | 0 |
| 0 | 0 | 0 | 0 | 0 |
| 0 | 0 | 0 | 0 | 0 |
| 0 | 0.098778932 | 0 | 0 | 0.014199822 |
| 0 | 0 | 0 | 0 | 0 |
| 0 | 0.027489562 | 0 | 0 | 0.033704513 |
| 0 | 0 | 0 | 0 | 0 |
| 0 | 0 | 0 | 0 | 0 |
| 0 | 0 | 0 | 0 | 0 |
| 0 | 0.01343723 | 0 | 0 | 0 |
| 0 | 0 | 0 | 0 | 0 |
| 0 | 0 | 0 | 0 | 0 |
| 0 | 0.001883506 | 0 | 0.00665807 | 0.021728544 |
| 0 | 0 | 0 | 0 | 0.043692058 |
| 0 | 0 | 0 | 0 | 0 |
| 0 | 0 | 0 | 0 | 0 |
| 0 | 0 | 0 | 0 | 0 |
| 0 | 0 | 0 | 0 | 0 |
| 0 | 0 | 0 | 0.015330262 | 0 |
| 0 | 0 | 0 | 0 | 0 |
| 0 | 0 | 0 | 0 | 0 |
| 0 | 0 | 0 | 0 | 0 |
| 0 | 0 | 0 | 0 | 0 |
| 0 | 0 | 0 | 0 | 0 |
| 0 | 0 | 0 | 0 | 0 |
| 1.092071161 | 0 | 0 | 2.703834069 | 0 |
| 0 | 0 | 0 | 0.283623633 | 0 |
| 0 | 0 | 0 | 0 | 0 |
| 0 | 0 | 0 | 0 | 0 |
| 0.030565954 | 0.001567424 | 0.001741265 | 0.021357037 | 0.001216743 |
| 0 | 0 | 0 | 0 | 0 |
| 0.042905224 | 0.0404756 | 0.006954769 | 0.006322918 | 0.057457799 |
| 0 | 0 | 0 | 0.016995596 | 0 |
| 0 | 0 | 0 | 0 | 0 |
| 0 | 0 | 0 | 0 | 0 |
| 0 | 0 | 0 | 0 | 0 |
| 0.006492101 | 0.376527189 | 1.176372543 | 0.003929912 | 7.166638908 |
| 0 | 0 | 0 | 0 | 0 |
| 0 | 0 | 0 | 0.00899194 | 0 |

| | | | | |
|---|---|---|---|---|
| 0 | 0 | 0 | 0.001473787 | 0 |
| 0 | 0 | 0 | 0 | 0 |
| 0 | 0 | 0 | 0 | 0 |
| 0 | 0 | 0 | 0.016352649 | 0 |
| 0 | 0 | 0 | 0 | 0 |
| 0 | 0 | 0 | 0.063993434 | 0 |
| 0 | 0 | 0 | 0 | 0 |
| 0 | 0 | 0.001580803 | 0 | 0.00202513 |
| 0 | 0 | 0 | 0.013871697 | 0.000705142 |
| 0.012910212 | 0.012746914 | 0 | 0.001011437 | 0.00386843 |
| 0 | 0 | 0 | 0.077442747 | 0 |
| 0 | 0 | 0 | 0 | 0 |
| 0 | 0 | 0 | 0.002372626 | 0.002786048 |
| 0 | 0 | 0 | 0 | 0 |
| 0 | 0 | 0 | 0 | 0 |
| 0 | 0 | 0 | 0 | 0 |
| 0 | 0 | 0 | 0.010375338 | 0 |
| 0 | 0 | 0 | 0 | 0 |
| 0 | 0 | 0 | 0 | 0 |
| 0.215423563 | 0.07283422 | 0.040649666 | 0.009679105 | 0.148255354 |
| 0 | 0 | 0 | 0 | 0 |
| 0 | 0 | 0 | 0 | 0 |
| 0 | 0 | 0 | 0 | 0 |
| 0 | 0 | 0 | 0 | 0 |
| 0 | 0 | 0 | 0 | 0 |
| 0 | 0 | 0 | 0.073342029 | 0 |
| 0 | 0 | 0 | 0 | 0 |
| 0 | 0 | 0 | 0 | 0 |
| 0 | 0 | 0 | 0 | 0 |
| 0 | 0.017123557 | 0 | 0 | 0.256382197 |
| 0 | 0 | 0 | 0.008236245 | 0 |
| 0 | 0 | 0 | 0 | 0 |
| 0 | 0 | 0 | 0 | 0 |
| 0 | 0 | 0 | 0 | 0 |
| 0 | 0 | 0 | 0 | 0 |
| 0 | 0 | 0 | 0 | 0 |
| 0 | 0 | 0 | 0.004541698 | 0 |
| 0 | 0 | 0 | 0 | 0 |
| 0 | 0 | 0 | 0 | 0 |
| 0 | 0 | 0 | 0 | 0 |
| 0 | 0 | 0 | 0 | 0 |
| 0 | 0 | 0 | 0 | 0 |
| 0 | 0 | 0 | 0.003218262 | 0 |
| 0 | 0 | 0 | 0 | 0 |
| 0 | 0.010287004 | 0 | 0 | 0 |
| 0 | 0 | 0 | 0.006068727 | 0 |
| 0 | 0 | 0 | 0.056228339 | 0 |
| 0 | 0 | 0 | 0 | 0 |

| | | | | |
|---|---|---|---|---|
| 0 | 0 | 0 | 0 | 0 |
| 0 | 0 | 0 | 0 | 0 |
| 0 | 0 | 0 | 0 | 0 |
| 0 | 0 | 0 | 0 | 0 |
| 0 | 0 | 0 | 0 | 0 |
| 0.020269102 | 0.041470101 | 0.000872529 | 0.004812432 | 0.000620987 |
| 0 | 0 | 0 | 0.003991872 | 0 |
| 0 | 0 | 0 | 0 | 0 |
| 0 | 0 | 0 | 0 | 0 |
| 0 | 0 | 0 | 0 | 0 |
| 0 | 0 | 0 | 0 | 0 |
| 0 | 0 | 0 | 0 | 0 |
| 0 | 0 | 0 | 0 | 0 |
| 0 | 0 | 0 | 0 | 0 |
| 0 | 0 | 0 | 0 | 0 |
| 0 | 0 | 0 | 0.424902553 | 0 |
| 0 | 0 | 0 | 0.678490915 | 0 |
| 0 | 0 | 0 | 0 | 0 |
| 0 | 0 | 0 | 0 | 0 |
| 0 | 0 | 0 | 0 | 0 |
| 0 | 0 | 0 | 0 | 0 |
| 0 | 0 | 0 | 0 | 0 |
| 0 | 0 | 0 | 0 | 0 |
| 0 | 0 | 0 | 0 | 0 |
| 0 | 0 | 0 | 0 | 0 |
| 0.001413566 | 0.001651686 | 0 | 0 | 0 |
| 0 | 0 | 0 | 0 | 0 |
| 0 | 0 | 0 | 4.733239845 | 0 |
| 0 | 0 | 0 | 0 | 0 |
| 0 | 0 | 0 | 0 | 0 |
| 0 | 0 | 0 | 0 | 0 |
| 0 | 0 | 0 | 0 | 0 |
| 0 | 0 | 0 | 0 | 0 |
| 0 | 0 | 0 | 0 | 0 |
| 0 | 0 | 0 | 0.131247145 | 0 |
| 0 | 0 | 0 | 0 | 0 |
| 0 | 0 | 0 | 0 | 0 |
| 0 | 0 | 0 | 0 | 0 |
| 0 | 0 | 0 | 0.622185592 | 0 |
| 0 | 0 | 0 | 0 | 0 |
| 0 | 0 | 0 | 0.004614118 | 0 |
| 0 | 0 | 0 | 0 | 0 |
| 0 | 0 | 0 | 0 | 0 |
| 0 | 0 | 0 | 0 | 0 |
| 0 | 0 | 0 | 0 | 0 |
| 0 | 0 | 0 | 0 | 0 |
| 0 | 0 | 0 | 0 | 0 |
| 0 | 0 | 0 | 0 | 0 |

| | | | | |
|---|---|---|---|---|
| 0 | 0 | 0 | 0 | 0 |
| 0 | 0 | 0 | 0.011742872 | 0 |
| 0 | 0 | 0 | 0 | 0 |
| 0 | 0 | 0 | 0 | 0 |
| 0 | 0 | 0 | 0 | 0 |
| 0 | 0 | 0 | 0 | 0 |
| 0 | 0 | 0 | 0 | 0 |
| 0 | 0 | 0 | 0 | 0 |
| 0 | 0 | 0 | 0 | 0 |
| 0 | 0 | 0 | 0.00225092 | 0 |
| 0.025929252 | 0.011680569 | 0 | 0 | 0.010196122 |
| 0 | 0 | 0 | 0.030630899 | 0 |
| 0 | 0.599849051 | 0 | 0.229307392 | 0 |
| 0 | 0 | 0 | 0 | 0 |
| 0 | 0 | 0 | 0 | 0 |
| 0 | 0 | 0 | 0 | 0 |
| 0 | 0 | 0 | 0 | 0 |
| 0 | 0 | 0 | 0 | 0 |
| 0 | 0 | 0 | 0 | 0 |
| 0 | 0 | 0 | 0 | 0 |
| 0 | 0.043849495 | 0 | 0 | 0.026240714 |
| 0 | 0 | 0 | 0 | 0 |
| 0 | 0 | 0 | 0.001102468 | 0 |
| 1.327614044 | 8.20067227 | 0.802890923 | 0.04243449 | 4.780784445 |
| 0 | 0 | 0 | 0 | 0 |
| 0 | 0 | 0 | 0 | 0 |
| 0 | 0 | 0 | 0 | 0 |
| 0 | 0.003280936 | 0 | 0 | 0 |
| 0 | 0 | 0 | 0 | 0 |
| 0 | 0 | 0 | 0 | 0 |
| 0 | 0 | 0 | 0.002162451 | 0 |
| 0 | 0 | 0 | 0 | 0 |
| 0 | 0 | 0 | 0 | 0 |
| 0 | 0 | 0 | 0.003264759 | 0 |

| PHL018-0006_V2 | PHL018-0028_V2 | PHL018-0070_V1 | PHL018-0274_V3 | PHL018-0066_V1 |
|---|---|---|---|---|
| EYCF | CTRL | EYCF | REF | CTRL |
| 0.013365686 | 0 | 0.063096014 | 0.163023455 | 0.205813926 |
| 2.834091213 | 0.003811683 | 0.101792833 | 9.071407162 | 3.631951748 |
| 16.81439848 | 2.930293177 | 0 | 0 | 8.125527763 |
| 0 | 0 | 0 | 0 | 0 |
| 0 | 0 | 0 | 0 | 0 |
| 0.033223892 | 0 | 0 | 0 | 0 |
| 0 | 0 | 0 | 0 | 0 |
| 0 | 0 | 0 | 0 | 0 |
| 0 | 0 | 0 | 0 | 0 |
| 0 | 0 | 0 | 0 | 0 |
| 0 | 0 | 0 | 0 | 0 |
| 0 | 0 | 0 | 0 | 0 |
| 0 | 0 | 0 | 0 | 0 |
| 1.645922484 | 0.119833016 | 3.115920042 | 18.69260993 | 5.194733337 |
| 0 | 0 | 0 | 0 | 0 |
| 0 | 0 | 0 | 0 | 0 |
| 0 | 0 | 0.000789983 | 0 | 0.005470985 |
| 0 | 0 | 0 | 0 | 0 |
| 0 | 0 | 0 | 0 | 0 |
| 0 | 0 | 0 | 0 | 0 |
| 0 | 0 | 0 | 0 | 0 |
| 0 | 0 | 0 | 0 | 0 |
| 0 | 0 | 0 | 0 | 0 |
| 0 | 0 | 0 | 0 | 0 |
| 0.002403828 | 0 | 0 | 0 | 0 |
| 0 | 0 | 0 | 0 | 0 |
| 0 | 0 | 0 | 0 | 0 |
| 0 | 0 | 0 | 0 | 0 |
| 0 | 0 | 0 | 0 | 0 |
| 0 | 0 | 0 | 0 | 0 |
| 0 | 0 | 0 | 0 | 0 |
| 0 | 0 | 0 | 0 | 0 |
| 0 | 0 | 0 | 0 | 0 |
| 0 | 0 | 0 | 0 | 0 |
| 0 | 0 | 0 | 0 | 0 |
| 3.595490226 | 0 | 0.394560264 | 0 | 0 |
| 0 | 0 | 0 | 0 | 0 |
| 0 | 0 | 0 | 0 | 0 |
| 0 | 0 | 0 | 0 | 0 |
| 0 | 0 | 0 | 0 | 0 |
| 0 | 0 | 0 | 0 | 0 |
| 0.005533599 | 0.485201388 | 0 | 0 | 0.163447347 |
| 0.092724408 | 14.26951385 | 0.02125064 | 0.016901156 | 0.020174854 |
| 0.001323252 | 0.031391644 | 0.00055664 | 0.013218037 | 0.033409843 |
| 0 | 0.001278659 | 0 | 0 | 0.003705741 |
| 0 | 0 | 0 | 0 | 0 |

| | | | | |
|---|---|---|---|---|
| 0.000883163 | 0 | 0.000866861 | 0.002860173 | 0.009605442 |
| 0 | 0.492966753 | 0.001908377 | 0.046151331 | 0 |
| 0 | 0 | 0.005128222 | 0.008578508 | 0.132694331 |
| 0.319236993 | 0 | 0.472209878 | 0.395806687 | 0.279936504 |
| 0.000540714 | 0 | 0.001857565 | 0 | 0.003413024 |
| 0 | 0 | 0 | 0 | 0 |
| 0.027612193 | 0.003177128 | 0.001789788 | 0 | 0.167713161 |
| 0 | 0 | 0 | 0 | 0 |
| 0 | 0 | 0 | 0 | 0 |
| 0 | 0 | 0 | 0 | 0 |
| 0 | 0 | 0 | 0 | 0 |
| 0 | 0 | 0.122881508 | 0 | 0.087832348 |
| 0 | 0 | 0 | 0.000498297 | 0 |
| 0.003358016 | 0 | 0.002014242 | 0.083348517 | 0.008876974 |
| 0 | 0 | 0 | 0 | 0 |
| 0 | 0 | 0 | 0 | 0 |
| 0.745764509 | 0.001919555 | 7.09079819 | 0 | 4.231471094 |
| 0 | 3.964898668 | 0 | 0 | 0 |
| 0 | 0 | 0 | 0 | 0 |
| 0 | 0 | 0 | 0 | 0 |
| 0.021093201 | 0.001479602 | 0.105260693 | 0.213872579 | 0.157932403 |
| 0.003701768 | 0 | 0.013988739 | 0.018255046 | 0.006111063 |
| 0.001245589 | 0 | 0.038511824 | 0 | 0 |
| 0 | 0 | 0 | 0 | 0 |
| 0 | 0 | 0 | 0 | 0 |
| 0 | 0 | 0 | 0 | 0 |
| 0 | 0.009330506 | 0 | 0.008458602 | 0 |
| 0 | 0 | 0 | 0 | 0 |
| 0 | 0.032433749 | 0 | 0 | 0.09473354 |
| 0 | 0 | 0 | 0 | 0 |
| 0 | 0 | 0.006793194 | 0 | 0 |
| 0 | 0 | 0 | 0 | 0 |
| 0 | 0 | 0 | 0 | 0 |
| 0 | 0 | 0 | 0 | 0 |
| 0 | 0 | 0 | 0 | 0 |
| 0 | 0 | 0 | 0 | 0 |
| 0 | 0 | 0 | 0 | 0 |
| 0.028952653 | 0.974093393 | 0 | 0 | 0.102000682 |
| 0 | 0 | 0 | 0 | 0 |
| 0 | 0 | 0 | 0 | 0 |
| 0 | 0 | 0 | 0 | 0 |
| 0 | 0 | 0 | 0 | 0 |
| 0 | 0 | 0 | 0 | 0 |
| 0 | 0 | 0 | 0 | 0 |
| 0 | 0 | 0 | 0 | 0 |
| 0 | 0 | 0 | 0 | 0 |
| 0 | 0 | 0 | 0 | 0 |
| 0 | 0 | 0.62259295 | 0.017480038 | 0 |

| | | | | |
|---|---|---|---|---|
| 0 | 0 | 0 | 0 | 0 |
| 0 | 0 | 0 | 0 | 0.017931784 |
| 0 | 0 | 0 | 0 | 0 |
| 0 | 0 | 0 | 0 | 0 |
| 0 | 0 | 0 | 0 | 0 |
| 0 | 0 | 0 | 0 | 0 |
| 0 | 0 | 0 | 0 | 0 |
| 0 | 0 | 0 | 0 | 0.003713101 |
| 0.013596587 | 0 | 0.117125309 | 0 | 0 |
| 0 | 0 | 0 | 0 | 0 |
| 0 | 0 | 0 | 0 | 0 |
| 0 | 0.002143533 | 0 | 0.000917258 | 0.001613577 |
| 0 | 0 | 0 | 0 | 0 |
| 0 | 0 | 0 | 0 | 0 |
| 0 | 0 | 0 | 0 | 0 |
| 0 | 0 | 0 | 0 | 0 |
| 0 | 0 | 0 | 0 | 0 |
| 0 | 0 | 0 | 0 | 0 |
| 0.056932341 | 0 | 0.435439851 | 0 | 0 |
| 0 | 0 | 0 | 0 | 0 |
| 0 | 0 | 0 | 0 | 0 |
| 0 | 0 | 0 | 0 | 0 |
| 0 | 0 | 0 | 0 | 0 |
| 0 | 0 | 0 | 0 | 0 |
| 0 | 0 | 0 | 0 | 0 |
| 0 | 0 | 0 | 0 | 0 |
| 0 | 0 | 0.015696882 | 0 | 0 |
| 0 | 0 | 0 | 0 | 0 |
| 0 | 0 | 0 | 0 | 0 |
| 0 | 0 | 0 | 0 | 0 |
| 0.014071845 | 0 | 0.208806422 | 0 | 0.03456441 |
| 0 | 0 | 0 | 0 | 0 |
| 0 | 0 | 0 | 0 | 0 |
| 0 | 0 | 0 | 0 | 0 |
| 0 | 0 | 0 | 0 | 0 |
| 0 | 0 | 0 | 0 | 0 |
| 0 | 0 | 0 | 0 | 0 |
| 0 | 0 | 0 | 0 | 0 |
| 0 | 0 | 0 | 0 | 0.855303175 |
| 0 | 0 | 0 | 0 | 0 |
| 0 | 0 | 0 | 0 | 0 |
| 0 | 0 | 0 | 0 | 0 |
| 0.002609669 | 0 | 0 | 0 | 0 |
| 0 | 0 | 0 | 0 | 0 |
| 0 | 0 | 0 | 0 | 0 |
| 0 | 0 | 0 | 0 | 0 |
| 0 | 0 | 0 | 0 | 0 |
| 0 | 0 | 0 | 0 | 0 |

| | | | | |
|---|---|---|---|---|
| 0 | 0 | 0 | 0 | 0 |
| 0 | 0 | 0 | 0 | 0 |
| 0 | 0 | 0 | 0 | 0 |
| 0 | 0 | 0 | 0 | 0 |
| 0.001814 | 0 | 0 | 0 | 0 |
| 0 | 0 | 0 | 0 | 0 |
| 0 | 0 | 0 | 0 | 0 |
| 0 | 0 | 0 | 0 | 0 |
| 0 | 0 | 0 | 0 | 0 |
| 0 | 0 | 0 | 0 | 0 |
| 0 | 0 | 0 | 0 | 0 |
| 0 | 0 | 0 | 0 | 0 |
| 0 | 0 | 0 | 0 | 0 |
| 0 | 0 | 0 | 0 | 0 |
| 0 | 0 | 0 | 0 | 0 |
| 0.316216096 | 0 | 0 | 0 | 0 |
| 0 | 0 | 0 | 0 | 0 |
| 0 | 0 | 0 | 0 | 0 |
| 0 | 0 | 0 | 0 | 0 |
| 0 | 0 | 0 | 0 | 0 |
| 0 | 0 | 0 | 0 | 0 |
| 0 | 0 | 0 | 0 | 0 |
| 0 | 0 | 0 | 0 | 0 |
| 0 | 0 | 0 | 0 | 0 |
| 0 | 0 | 0 | 0 | 0 |
| 0 | 0 | 0 | 0 | 0 |
| 0.004716508 | 0 | 0 | 0 | 0 |
| 0 | 0 | 0 | 0 | 0 |
| 0 | 0 | 0 | 0 | 0 |
| 0 | 0 | 0 | 0 | 0 |
| 0.002276424 | 0 | 0 | 0 | 0.004642276 |
| 0 | 0 | 0.000590063 | 0 | 0 |
| 0 | 0 | 0 | 0 | 0 |
| 0 | 0 | 0 | 0 | 0 |
| 0 | 0 | 0 | 0 | 0 |
| 0 | 0 | 0 | 0 | 0 |
| 0 | 0 | 0 | 0 | 0 |
| 0 | 0 | 0 | 0 | 0 |
| 0 | 0 | 0 | 0 | 0 |
| 0 | 0 | 0 | 0 | 0 |
| 0 | 0 | 0 | 0 | 0 |
| 0 | 0 | 0 | 0 | 0 |
| 0 | 0 | 0 | 0 | 0 |
| 0 | 0 | 0 | 0 | 0 |
| 0 | 0 | 0 | 0 | 0 |
| 0 | 0 | 0 | 0 | 0 |
| 0 | 0 | 0 | 0 | 0 |
| 0 | 0 | 0 | 0 | 0 |

| | | | | |
|---|---|---|---|---|
| 0 | 0 | 0 | 0 | 0 |
| 0 | 0 | 0 | 0 | 0 |
| 0 | 0 | 0 | 0 | 0.001850664 |
| 0 | 0 | 0 | 0 | 0 |
| 0 | 0 | 0 | 0 | 0 |
| 0 | 0 | 0 | 0 | 0 |
| 0 | 0 | 0 | 0 | 0 |
| 0 | 0 | 0 | 0 | 0 |
| 0 | 0 | 0 | 0 | 0 |
| 0 | 0 | 0 | 0 | 0 |
| 0 | 0 | 0 | 0 | 0 |
| 0 | 0 | 0 | 0 | 0 |
| 0 | 0 | 0 | 0 | 0 |
| 0 | 0 | 0 | 0 | 0 |
| 0 | 0 | 0 | 0 | 0 |
| 0 | 0 | 0 | 0 | 0 |
| 0 | 0 | 0 | 0 | 0 |
| 0 | 0 | 0 | 0 | 0 |
| 0 | 0 | 0 | 0 | 0 |
| 0 | 0 | 0 | 0 | 0 |
| 0.03692825 | 0.027246005 | 0.016187804 | 0.036416607 | 0.033075291 |
| 0 | 0 | 0 | 0 | 0 |
| 0 | 0 | 0 | 0 | 0 |
| 0 | 0 | 0 | 0 | 0 |
| 0 | 0 | 0 | 0 | 0 |
| 0 | 0 | 0 | 0 | 0 |
| 0.034451587 | 0.044859775 | 0.006763131 | 0.004917837 | 0.002027607 |
| 0 | 0 | 0 | 0 | 0 |
| 0 | 0 | 0 | 0 | 0 |
| 0 | 0 | 0 | 0 | 0 |
| 0 | 0.007810931 | 0.014300562 | 0 | 0 |
| 0 | 0 | 0 | 0 | 0 |
| 0 | 0 | 0 | 0 | 0 |
| 0 | 0 | 0 | 0 | 0.123621899 |
| 0.001632936 | 0 | 0.002976618 | 0.005322697 | 0.002038789 |
| 0 | 0 | 0.002041212 | 0 | 0 |
| 0 | 0 | 0 | 0 | 0 |
| 0 | 0 | 0 | 0 | 0 |
| 0 | 0 | 0 | 0 | 0 |
| 0 | 0 | 0 | 0 | 0 |
| 0 | 0 | 0 | 0 | 0 |
| 0 | 0 | 0 | 0 | 0 |
| 0 | 0 | 0 | 0 | 0 |
| 0 | 0 | 0 | 0 | 0 |
| 0 | 0 | 0 | 0 | 0 |
| 0 | 0 | 0 | 0 | 0.019842855 |
| 0 | 0 | 0.003222886 | 0.001487243 | 0 |
| 0 | 0 | 0 | 0 | 0.000484299 |

| | | | | |
|---|---|---|---|---|
| 0 | 0 | 0 | 0 | 0 |
| 0 | 0 | 0 | 0 | 0 |
| 0 | 0 | 0 | 0.001118082 | 0 |
| 0 | 0 | 0 | 0 | 0 |
| 0 | 0 | 0 | 0 | 0 |
| 0 | 0 | 0 | 0 | 0 |
| 0 | 0 | 0 | 0 | 0 |
| 0 | 0.001793631 | 0 | 0 | 0 |
| 0 | 0 | 0 | 0 | 0 |
| 0 | 0 | 0 | 0 | 0 |
| 0 | 0.021419592 | 0 | 0 | 0 |
| 0 | 0 | 0 | 0 | 0 |
| 0 | 0 | 0 | 0 | 0 |
| 0 | 0 | 0 | 0.038083726 | 0 |
| 0 | 0 | 0.001029064 | 0 | 0 |
| 0 | 0 | 0 | 0 | 0 |
| 0 | 0 | 0 | 0 | 0 |
| 0 | 0 | 0 | 0 | 0 |
| 0 | 0.066460239 | 0 | 0 | 0 |
| 0.001356178 | 0 | 0.104780137 | 0 | 0 |
| 0 | 0 | 0 | 0.223022525 | 0 |
| 0 | 0 | 0 | 0 | 0 |
| 0 | 0 | 0 | 0 | 0 |
| 0 | 0 | 0 | 0 | 0 |
| 0.03170942 | 0.012550713 | 0 | 0.010940281 | 0.003367945 |
| 0 | 0.001578764 | 0 | 0 | 0 |
| 0 | 0 | 0 | 0 | 0 |
| 0 | 0 | 0 | 0 | 0 |
| 0 | 0.107352348 | 0 | 0 | 0.011249275 |
| 0 | 0 | 0 | 0 | 0 |
| 0.151100217 | 0 | 1.055816729 | 0.897279587 | 0.018519523 |
| 0 | 0 | 0 | 0 | 0 |
| 0 | 0 | 0 | 0 | 0 |
| 0 | 0 | 0 | 0 | 0 |
| 0 | 0 | 0 | 0 | 0 |
| 0 | 0 | 0 | 0 | 0 |
| 0 | 0 | 0 | 0 | 0 |
| 0 | 0.108043611 | 0 | 0 | 0 |
| 0 | 0 | 0 | 0 | 0 |
| 0 | 0 | 0 | 0 | 0 |
| 0 | 0 | 0 | 0 | 0 |
| 0 | 0 | 0.020915475 | 0 | 0.008349696 |
| 0 | 0.046790617 | 0 | 0.000692605 | 0 |
| 0 | 0 | 0 | 0 | 0 |
| 0 | 0 | 0 | 0 | 0 |
| 0 | 0 | 0 | 0 | 0 |
| 0 | 0 | 0 | 0 | 0 |
| 0 | 0 | 0 | 0 | 0 |

| | | | | |
|---|---|---|---|---|
| 0 | 0 | 0 | 0 | 0 |
| 0 | 0 | 0 | 0 | 0 |
| 0 | 0.039856834 | 0 | 0.006910139 | 0.03122533 |
| 0 | 0.00355296 | 0 | 0 | 0 |
| 0.191872299 | 0 | 0 | 0 | 0.502800262 |
| 0 | 0 | 0 | 0 | 0 |
| 0 | 0 | 0 | 0 | 0 |
| 0 | 0 | 0 | 0 | 0 |
| 0 | 0 | 0 | 0 | 0 |
| 0 | 0 | 0 | 0 | 0 |
| 0 | 0 | 0 | 0 | 0 |
| 0 | 0 | 0 | 0 | 0 |
| 0 | 0 | 0 | 0 | 0 |
| 0.008560026 | 0.011194773 | 0 | 0 | 0 |
| 0.006851084 | 0.008838088 | 0.000840578 | 0.000472748 | 0 |
| 0 | 0 | 0.002474861 | 0 | 0 |
| 0.005610704 | 0.015105313 | 0 | 0.001795512 | 0.000947563 |
| 0.031412378 | 0 | 0 | 0 | 0 |
| 0 | 0 | 0 | 0 | 0 |
| 0 | 0 | 0 | 0 | 0 |
| 0 | 0 | 0 | 0 | 0 |
| 0 | 0 | 0 | 0 | 0 |
| 0 | 0 | 0 | 0 | 0 |
| 0 | 0.003843379 | 0 | 0.001507599 | 0 |
| 0 | 0 | 0.0048521 | 0 | 0 |
| 0 | 0 | 0 | 0 | 0 |
| 0.64012477 | 0 | 0 | 0 | 13.40600995 |
| 0 | 0 | 0 | 0 | 0 |
| 0 | 0 | 0 | 0 | 0 |
| 0 | 0 | 0.003126582 | 0 | 0 |
| 0 | 0 | 0 | 0 | 0 |
| 0 | 0 | 0 | 0 | 0 |
| 0 | 0 | 0 | 0 | 0 |
| 0 | 0 | 0 | 0 | 0.010109054 |
| 0 | 0 | 0 | 0 | 0 |
| 0 | 0.015681072 | 0 | 0 | 0 |
| 0 | 0 | 0 | 0 | 0 |
| 0 | 0.23465587 | 0 | 0 | 0.001880716 |
| 0 | 0 | 0 | 0 | 0 |
| 0 | 0 | 0 | 0 | 0 |
| 0 | 0 | 0 | 0 | 0 |
| 0 | 0 | 0 | 0 | 0 |
| 0 | 0 | 0 | 0 | 0 |
| 0 | 0 | 0 | 0 | 0 |
| 0 | 0 | 0 | 0 | 0 |
| 0 | 0 | 0.002087642 | 0 | 0 |
| 0 | 0 | 0 | 0 | 0 |
| 0 | 0 | 0 | 0 | 0 |

| | | | | |
|---|---|---|---|---|
| 0 | 0 | 0.523475156 | 0 | 0 |
| 0 | 0 | 0 | 0 | 0.090775155 |
| 0 | 0 | 0 | 0.001867468 | 0.001119928 |
| 0 | 0 | 0 | 0 | 0 |
| 0.072170826 | 0.130196344 | 0.020499931 | 0.026816361 | 0.010140804 |
| 0 | 0 | 0.007428221 | 0 | 0 |
| 0 | 0 | 0 | 0 | 0 |
| 0.002602271 | 0 | 0.00798199 | 0.020111329 | 0.028429137 |
| 0 | 0 | 0 | 0 | 0 |
| 0.133440316 | 0.010065846 | 0.040806963 | 0.02763891 | 0.013023345 |
| 0 | 0 | 0 | 0 | 0 |
| 0 | 1.7471294 | 0 | 0 | 0.009751996 |
| 0.053908261 | 0.015576199 | 0.007093435 | 0.001293861 | 0.135615978 |
| 0 | 0 | 0 | 0 | 0 |
| 0 | 0 | 0 | 0.000443569 | 0 |
| 0 | 0 | 0 | 0 | 0 |
| 0 | 0 | 0 | 0 | 0 |
| 0 | 0 | 0 | 0 | 0 |
| 0 | 0 | 0.053262586 | 0 | 0.010722889 |
| 0 | 0 | 0.003971466 | 0.203070073 | 0 |
| 0 | 0 | 0 | 0 | 0 |
| 0 | 0 | 0 | 0 | 0 |
| 0 | 0 | 0 | 0 | 0 |
| 0.004869141 | 0 | 0 | 0 | 0 |
| 0 | 0 | 0 | 0 | 0 |
| 0.000909991 | 0.001067194 | 0 | 0 | 0.001767363 |
| 0 | 0 | 0 | 0 | 0 |
| 0 | 0 | 0 | 0 | 0.014159167 |
| 0 | 0 | 0 | 0 | 0 |
| 0 | 0 | 0 | 0 | 0 |
| 0 | 0 | 0 | 0 | 0 |
| 0 | 0 | 0 | 0 | 0 |
| 0 | 0 | 0 | 0 | 0 |
| 0 | 0 | 0.004964567 | 0 | 0.049405867 |
| 0 | 0 | 0 | 0 | 0 |
| 0 | 0 | 0.002037929 | 0 | 0 |
| 0 | 0 | 0 | 0 | 0.070372469 |
| 0 | 0 | 0 | 0 | 0 |
| 0 | 0 | 0 | 0 | 0 |
| 0.00406758 | 0 | 0.008689554 | 0.001937219 | 0.187507777 |
| 0 | 0 | 0 | 0 | 0.178966571 |
| 0 | 0 | 0.116699958 | 0 | 0.538367406 |
| 0 | 0 | 0 | 0 | 0 |
| 0 | 0 | 0 | 0.062863766 | 0.042359528 |
| 0 | 0 | 0 | 0 | 0 |
| 0.001165125 | 0 | 0 | 0 | 0 |
| 0 | 0 | 0 | 0.006455221 | 0.011538739 |
| 0 | 0 | 0 | 0 | 0 |

| | | | | |
|---|---|---|---|---|
| 0 | 0 | 0.093960926 | 0 | 0 |
| 0.135638354 | 0 | 0 | 0 | 0.965308941 |
| 0 | 0 | 0 | 0 | 0 |
| 0 | 0 | 0 | 0 | 0 |
| 0 | 0 | 0 | 0.017288582 | 0 |
| 0.000668481 | 0 | 0.030619952 | 0 | 0 |
| 0 | 0 | 0 | 0.007477113 | 0 |
| 0.003170191 | 0 | 0 | 0 | 0.013993336 |
| 0.108660414 | 0 | 0 | 0 | 0 |
| 0 | 0 | 0 | 0 | 0 |
| 0.025068858 | 0.025459415 | 0.004312413 | 0.006657785 | 0.031371219 |
| 0 | 0 | 0 | 0 | 0 |
| 0 | 0 | 0 | 0 | 0 |
| 0.013428957 | 0.148210913 | 0.002636215 | 0.000898563 | 0.001422622 |
| 0 | 0 | 0.067654357 | 0.003273063 | 0.010076062 |
| 0 | 0 | 0 | 0 | 0 |
| 0 | 0 | 0 | 0 | 0 |
| 0 | 0 | 0.003265135 | 0 | 0 |
| 0 | 0 | 0 | 0 | 0 |
| 0 | 0 | 0 | 0 | 0 |
| 0 | 0 | 0 | 0 | 0.000766408 |
| 0.399172024 | 0 | 0.389907151 | 0 | 0.274773118 |
| 0 | 0 | 0 | 0 | 0 |
| 0 | 0 | 0 | 0 | 0 |
| 0 | 0 | 0 | 0.019143906 | 0 |
| 0 | 0 | 0 | 0 | 0 |
| 0 | 0 | 0.001039402 | 0 | 0.122114464 |
| 0 | 0 | 0 | 0 | 0 |
| 0 | 0 | 0 | 0 | 0 |
| 0.978117732 | 0.024847319 | 0.219553377 | 0.002370789 | 0.624189424 |
| 0.001656451 | 0 | 0 | 0 | 0 |
| 0 | 0 | 0 | 0 | 0 |
| 0 | 0 | 0 | 0 | 0 |
| 0 | 0 | 0 | 0 | 0 |
| 0 | 0 | 0 | 0 | 0.023125607 |
| 0 | 0 | 0 | 0 | 0 |
| 0 | 0 | 1.701531573 | 0 | 0.018546476 |
| 0 | 0 | 0 | 0 | 0 |
| 0 | 0 | 0.114061485 | 0 | 0 |
| 0 | 0 | 0.132996349 | 0 | 0 |
| 0.021076485 | 0.003201505 | 0 | 0.013296897 | 0.017153397 |
| 0.306944892 | 0 | 0.688637957 | 0 | 13.5466165 |
| 0 | 0 | 0 | 0 | 0 |
| 0 | 0 | 0 | 0 | 0 |
| 0 | 0 | 0 | 0 | 0.00197897 |
| 0 | 0 | 0 | 0 | 0 |
| 0 | 0.029648901 | 0 | 0.005427352 | 0 |
| 0 | 0 | 0 | 0 | 0 |

| | | | | |
|---|---|---|---|---|
| 0 | 0 | 0 | 0 | 0.002019415 |
| 0.000498287 | 0 | 0.006521185 | 0 | 0 |
| 1.727745809 | 0 | 0.400149224 | 0 | 9.625813767 |
| 0 | 0 | 0 | 0 | 0 |
| 0 | 0 | 0 | 0 | 0 |
| 0 | 0 | 0.004961667 | 0 | 0 |
| 0 | 0.287838167 | 0 | 0 | 0 |
| 0 | 0 | 0 | 0 | 0 |
| 0.001740388 | 0 | 0.000640598 | 0.004803704 | 0 |
| 0 | 0 | 0 | 0 | 0 |
| 0 | 0 | 0 | 0 | 0 |
| 0 | 0 | 0 | 0 | 0 |
| 0 | 0 | 0 | 0 | 0 |
| 0.042094235 | 0.030226403 | 0.003923983 | 0.013154734 | 0 |
| 0 | 0 | 0 | 0 | 0 |
| 0.001653985 | 0 | 0.003607676 | 0 | 0.095477566 |
| 0.004481326 | 0.021213051 | 0 | 0.008545896 | 0.044308844 |
| 0 | 0 | 0 | 0.01803351 | 0 |
| 0 | 0 | 0 | 0.008207808 | 0 |
| 0 | 0 | 0 | 0 | 0 |
| 0 | 0 | 0.028221433 | 0 | 0.013461851 |
| 0 | 0 | 0 | 0.006956347 | 0.001606124 |
| 0 | 0 | 0 | 0 | 0.428537224 |
| 0 | 0 | 0 | 0 | 0 |
| 0 | 0 | 0 | 0 | 0 |
| 0 | 0 | 0 | 0.824380465 | 0.004429797 |
| 0.00801643 | 0 | 0 | 0 | 0.00125559 |
| 0.064805284 | 0 | 0 | 0 | 0 |
| 0 | 0 | 0 | 0 | 0 |
| 0 | 0 | 0 | 0.001140033 | 0 |
| 0.006883114 | 0 | 0.078563334 | 0 | 0.019479403 |
| 0.012314253 | 0.007152016 | 0.004604552 | 0.001510622 | 0.002562476 |
| 0.004745766 | 0.03692945 | 0.019180683 | 0.038012641 | 0.030633346 |
| 0 | 0.568646158 | 0 | 0 | 0.191142311 |
| 0.002569917 | 0 | 0 | 0 | 0 |
| 0 | 0 | 0 | 0 | 0 |
| 0.0042905 | 0.010418566 | 0 | 0.000557287 | 0 |
| 0.009473985 | 0 | 0.00271224 | 0.033558484 | 0.026066873 |
| 0.023899665 | 0 | 0 | 0 | 0 |
| 0 | 0 | 0 | 0.011640147 | 0.004074416 |
| 0 | 0 | 0 | 0 | 0 |
| 0 | 0 | 0.007276221 | 0 | 0 |
| 0 | 0 | 0 | 0 | 0.006879347 |
| 0.026038721 | 0.008239562 | 0.85778365 | 0 | 0.025435509 |
| 0 | 0 | 0 | 0.000887688 | 0.019909904 |
| 0 | 0 | 0 | 0 | 0 |
| 0 | 0 | 0.000645841 | 0 | 0 |
| 0 | 0 | 0 | 0 | 0 |

| | | | | |
|---|---|---|---|---|
| 0 | 0 | 0 | 0.010985112 | 0 |
| 3.842998689 | 0.614165325 | 0.289854031 | 1.031149553 | 0.188043995 |
| 0 | 0 | 0 | 0 | 0 |
| 0 | 0 | 0 | 0 | 0.003954395 |
| 0 | 0 | 0 | 0 | 0 |
| 0 | 0 | 0 | 0 | 0 |
| 0 | 0 | 0.014653155 | 0 | 0 |
| 0 | 0 | 0 | 0 | 0.000802431 |
| 0 | 0 | 0 | 0 | 0 |
| 0 | 0 | 0 | 0.007480216 | 0 |
| 0 | 0 | 0.022363232 | 0.001471025 | 0.00407567 |
| 0.034830183 | 0.359455194 | 0.011980153 | 0.025198033 | 0.140785339 |
| 0 | 0 | 0 | 0 | 0 |
| 0 | 0 | 0 | 0 | 0 |
| 0.001605183 | 0.003442253 | 0 | 0 | 0 |
| 0 | 0 | 0 | 0 | 0 |
| 0.113848304 | 0.072648343 | 0.009243763 | 0.012631078 | 0.005283961 |
| 0.006791858 | 0.010195415 | 0.678648618 | 19.56728182 | 0.025590507 |
| 0 | 0 | 0.000870915 | 0 | 0 |
| 0.006174389 | 0 | 0.033765185 | 0 | 0.007537671 |
| 0.138340907 | 0.019438383 | 0.090242432 | 0.003971289 | 0.003772452 |
| 0 | 0 | 0 | 0 | 0 |
| 0 | 0 | 0 | 0 | 0 |
| 0 | 0 | 0 | 0 | 0 |
| 0 | 0 | 0 | 0 | 0 |
| 0 | 0 | 0 | 0 | 0 |
| 0.015550866 | 0 | 0.370095326 | 0 | 0.060405037 |
| 0.021103445 | 0.037446489 | 0.149500354 | 0.293943058 | 0.11404968 |
| 0 | 0 | 0 | 0 | 0 |
| 0 | 0.001297967 | 0 | 0 | 0 |
| 0 | 0 | 0 | 0 | 0.003107188 |
| 0 | 0 | 0 | 0 | 0 |
| 0 | 0 | 0.000819996 | 0 | 0 |
| 0 | 0 | 0 | 0 | 0 |
| 0 | 0 | 0 | 0 | 0 |
| 0.506240046 | 0.006556133 | 0.43402149 | 0 | 0.437693098 |
| 0 | 0 | 0 | 0.000661698 | 0 |
| 0 | 0 | 0 | 0 | 0 |
| 0.010752951 | 0.593359525 | 0.025275169 | 0 | 0.006320195 |
| 0 | 0 | 0 | 0 | 0 |
| 0 | 0 | 0 | 0 | 0.021726537 |
| 0 | 0 | 0 | 0 | 0 |
| 0.013063013 | 0.081113676 | 0.002361927 | 0.0182176 | 0.02754053 |
| 0 | 0.000622773 | 0.018406052 | 0.00061072 | 0 |
| 0 | 0 | 0.013909589 | 0 | 0.008468586 |
| 0.139567718 | 0 | 0.254965729 | 0 | 0 |
| 0 | 0 | 0 | 0 | 0 |
| 0 | 0 | 0 | 0 | 0 |

| | | | | |
|---|---|---|---|---|
| 0 | 0 | 0 | 0 | 0 |
| 0 | 0 | 0 | 0 | 0 |
| 0 | 0 | 0 | 0 | 0 |
| 0.040638134 | 0 | 0.045389804 | 0 | 0.130636811 |
| 0 | 0 | 0 | 0 | 0 |
| 0.001215154 | 0 | 0 | 0 | 0 |
| 0 | 0 | 0 | 0 | 0 |
| 0 | 0 | 0 | 0 | 0.021217223 |
| 0 | 12.76643022 | 0.110127246 | 0 | 0 |
| 0.025048619 | 0 | 0 | 0 | 0 |
| 0 | 0 | 0 | 0 | 0 |
| 0.002301618 | 0 | 0.002070872 | 0 | 0.005587683 |
| 0 | 0 | 0 | 0 | 0 |
| 0 | 0 | 0 | 0 | 0 |
| 0.002690676 | 0 | 0.025322625 | 0 | 0.074544024 |
| 0 | 0 | 0 | 0 | 0 |
| 0 | 0 | 0 | 0 | 0 |
| 0 | 0 | 0 | 0 | 0 |
| 0 | 0 | 0 | 0 | 0 |
| 0 | 0 | 0 | 0 | 0 |
| 0 | 0 | 0 | 0 | 0 |
| 0 | 0 | 0 | 0 | 0 |
| 0 | 0 | 0 | 0 | 0 |
| 0 | 0 | 0.419904617 | 0.186838993 | 0.143906105 |
| 0 | 0 | 0 | 0 | 0.001220683 |
| 0.531894132 | 0 | 2.592396614 | 0 | 0 |
| 0 | 0 | 0 | 0 | 0.000622468 |
| 0 | 0 | 0 | 0 | 0 |
| 0 | 0 | 0 | 0 | 0 |
| 0 | 0 | 0 | 0 | 0 |
| 0 | 0 | 0 | 0 | 0 |
| 0.003162302 | 0 | 0.332120516 | 0 | 0 |
| 0 | 0 | 0 | 0 | 0.001929335 |
| 0 | 0 | 0 | 0 | 0 |
| 0.001377277 | 0 | 0 | 0 | 0 |
| 0 | 0 | 0.137180964 | 0 | 0 |
| 0.006329813 | 0 | 0.623301554 | 0 | 0.14038511 |
| 0 | 0 | 0 | 0 | 0 |
| 0 | 0 | 0 | 0 | 0 |
| 0.002196582 | 0 | 0.002560293 | 0 | 0.000399951 |
| 0.024471899 | 0.030382605 | 0.355065225 | 0.109562881 | 0.01608115 |
| 0 | 0 | 0.151993903 | 0 | 0 |
| 5.932551393 | 0.052600607 | 0.052863894 | 0.399338789 | 7.184728218 |
| 0.002083736 | 0.040302358 | 0 | 0 | 0.045762026 |
| 0.01355452 | 0 | 0 | 0 | 0 |
| 0 | 0 | 0 | 0 | 0 |
| 0 | 0 | 0 | 0 | 0 |
| 0.002875446 | 0 | 0 | 0.004558468 | 0.015250317 |

| | | | | |
|---|---|---|---|---|
| 0 | 0 | 0 | 0 | 0 |
| 0 | 0 | 0.026305954 | 0 | 0 |
| 0 | 1.028868901 | 0.032466719 | 0.340220594 | 0.240083508 |
| 0 | 0 | 0 | 0 | 0 |
| 0 | 0 | 0 | 0 | 0 |
| 0.000668128 | 0 | 0 | 0 | 0 |
| 0 | 0 | 0 | 0 | 0 |
| 0 | 0 | 0 | 0 | 0 |
| 0 | 0 | 0 | 0 | 0 |
| 0 | 0 | 0 | 0 | 0.102789803 |
| 0.001040898 | 0 | 0.030854876 | 0.006742017 | 0.310923227 |
| 1.310492752 | 0.496079884 | 2.637159222 | 1.891183307 | 0.697627801 |
| 0 | 0 | 0 | 0 | 0 |
| 0.002471436 | 0.012647485 | 0 | 0.005457195 | 0.049308707 |
| 0.255664921 | 0.081560854 | 0.074553811 | 1.042018998 | 0.03438686 |
| 0 | 0 | 0.014858486 | 0 | 0 |
| 0 | 0 | 0 | 0 | 0 |
| 0 | 0 | 0 | 0.001013041 | 0 |
| 0 | 0 | 0.057046127 | 0 | 0 |
| 0.001543367 | 0 | 0 | 0 | 0.004329708 |
| 0.418589987 | 0.007039201 | 1.267870627 | 0.000642137 | 0.004574893 |
| 0.293062105 | 0.107634688 | 0 | 1.082715909 | 1.324214075 |
| 1.872520056 | 0.415002319 | 1.19737277 | 0.589046082 | 0.136674702 |
| 0 | 0 | 0 | 0 | 0 |
| 0 | 0 | 0 | 0 | 0 |
| 1.912296933 | 0 | 3.904427838 | 0 | 0 |
| 0 | 0 | 0 | 0 | 0.258997819 |
| 0 | 0 | 0 | 0 | 0.001413787 |
| 0 | 0 | 0 | 0 | 0 |
| 0 | 0 | 0 | 0 | 0.000957167 |
| 0 | 0 | 0 | 0 | 0.001495108 |
| 0.040028758 | 0 | 0.41779858 | 0 | 0.05502789 |
| 0 | 0.000601779 | 0 | 0.850893071 | 0 |
| 0 | 0 | 0 | 0 | 0.018366239 |
| 0.000705573 | 0 | 0.002908705 | 0.001246384 | 0.004248074 |
| 0.000775531 | 1.005367901 | 0.00095152 | 0.036104277 | 0.008472475 |
| 0 | 0 | 0.002022141 | 0 | 0.002728099 |
| 0 | 0 | 0 | 0 | 0 |
| 0 | 0 | 0.053001188 | 0 | 0 |
| 0 | 0 | 0 | 0 | 0 |
| 0 | 0 | 0 | 0 | 0 |
| 0 | 0 | 0 | 0 | 0 |
| 0.011201495 | 0 | 0 | 0 | 0 |
| 0 | 0 | 0 | 0 | 0 |
| 0 | 0.201007096 | 0.023004127 | 0.005207631 | 0.027482728 |
| 0 | 0 | 0.288679511 | 0 | 0 |
| 0 | 0 | 0 | 0 | 0 |
| 0.055208677 | 14.74810022 | 0.036388186 | 0.022967925 | 0.096606067 |

| | | | | |
|---|---|---|---|---|
| 0 | 0 | 0 | 0 | 0 |
| 0 | 0 | 0 | 0 | 0 |
| 0 | 0 | 0 | 0 | 0 |
| 0.000942219 | 0 | 0 | 0 | 0.0040259 |
| 0 | 0 | 0 | 0 | 0 |
| 0 | 0 | 0 | 0 | 0.066102752 |
| 0 | 0 | 0 | 0 | 0 |
| 0 | 0 | 0.016509553 | 0 | 0.029973231 |
| 0 | 0 | 0 | 0 | 0 |
| 0 | 0 | 0 | 0 | 0 |
| 0 | 0.279016465 | 0.001236014 | 0.002162672 | 0 |
| 0 | 0 | 0 | 0 | 0 |
| 0 | 0 | 0 | 0 | 0.002855204 |
| 0 | 0 | 0.232918771 | 0 | 0 |
| 0 | 0 | 0 | 0 | 0 |
| 0 | 0 | 0 | 0 | 0 |
| 0.100992044 | 0 | 0 | 0 | 0 |
| 0 | 0 | 0 | 0.000771589 | 0.002544989 |
| 0.007537808 | 0 | 0 | 0 | 0 |
| 0 | 0 | 0.006804936 | 0 | 0.002601179 |
| 0 | 0 | 0 | 0 | 0 |
| 0 | 0 | 0 | 0 | 0 |
| 0 | 0 | 0 | 0 | 0.003325415 |
| 0 | 0 | 0 | 0 | 0 |
| 0 | 0 | 0 | 0 | 0 |
| 0 | 0 | 0 | 0 | 0 |
| 0 | 0 | 0 | 0 | 0 |
| 0 | 0 | 0 | 0 | 0 |
| 0 | 0 | 0 | 0 | 0 |
| 0.779222379 | 0.40216657 | 1.87978034 | 0 | 0 |
| 0 | 0 | 0 | 0 | 0 |
| 0 | 0 | 0 | 0 | 0 |
| 0 | 0 | 0 | 0 | 0 |
| 0 | 0 | 0 | 0 | 0 |
| 0 | 0 | 0 | 0 | 0.022202024 |
| 0 | 0 | 0 | 0 | 0 |
| 0.079656804 | 0 | 0 | 0 | 0 |
| 0 | 0.025002716 | 0 | 0.004707616 | 0.003278021 |
| 0 | 0 | 0 | 0 | 0 |
| 0 | 0 | 0 | 0 | 0 |
| 0.004834093 | 0 | 0 | 0.074596679 | 0.009550533 |
| 0 | 0 | 0 | 0 | 0 |
| 0 | 0 | 0 | 0 | 0 |
| 0.008626643 | 0 | 0 | 0 | 0 |
| 0 | 0 | 0.108312534 | 0 | 0 |
| 0 | 0 | 0 | 0 | 0 |
| 0 | 0 | 0 | 0 | 0.00147537 |
| 0 | 0 | 0 | 0 | 0 |

| | | | | |
|---|---|---|---|---|
| 0 | 0 | 0 | 0 | 0 |
| 0 | 0 | 0 | 0 | 0 |
| 0 | 0 | 0 | 0 | 0 |
| 0.013784447 | 0 | 0.028621163 | 0 | 0 |
| 0 | 0 | 0 | 0 | 0 |
| 0.128196261 | 0 | 0 | 0 | 0 |
| 0 | 0 | 0 | 0 | 0 |
| 0 | 0 | 0 | 0 | 0 |
| 0.000980114 | 0.064598005 | 0.039923918 | 0.434282019 | 0.076380073 |
| 0 | 0 | 0 | 0 | 0 |
| 0 | 0 | 0 | 0 | 0 |
| 0 | 0 | 0 | 0 | 0 |
| 0 | 0 | 0.012627577 | 0 | 0 |
| 0 | 0 | 0 | 0 | 0 |
| 2.379245543 | 0 | 1.433654896 | 0 | 2.496497048 |
| 0 | 0 | 0 | 0 | 0.014121044 |
| 0 | 0 | 0 | 0 | 0 |
| 0 | 0 | 0 | 0 | 0 |
| 0 | 0 | 0 | 0 | 0 |
| 0 | 0 | 0 | 0 | 0 |
| 0 | 0 | 0 | 0 | 0 |
| 0 | 0 | 0 | 0 | 0 |
| 0 | 0 | 0 | 0 | 0 |
| 0 | 0 | 0 | 0 | 0 |
| 0.006339222 | 2.096212888 | 0.030262559 | 0.001484603 | 0.006715573 |
| 2.090428513 | 1.97245553 | 0.037587336 | 0 | 0 |
| 0 | 0 | 0.057015764 | 0 | 0.007493386 |
| 0 | 0 | 0 | 0 | 0 |
| 0 | 0 | 0 | 0 | 0 |
| 0.041435867 | 0 | 0 | 0 | 0 |
| 0 | 0 | 0 | 0 | 0 |
| 0.00092784 | 0 | 0 | 0 | 0 |
| 0 | 0 | 0 | 0 | 0.000677505 |
| 0 | 0.009240235 | 0.004229348 | 0 | 0 |
| 0.00101424 | 0.006660933 | 0 | 0.005412247 | 0.017728491 |
| 0 | 0 | 0.000527783 | 0.001846939 | 0.002610811 |
| 0 | 0 | 0 | 0 | 0.081187786 |
| 3.514073368 | 0.12930922 | 3.625810674 | 0 | 0 |
| 0 | 0 | 0 | 0 | 0 |
| 0 | 0 | 0 | 0 | 0 |
| 0 | 0 | 0.295011952 | 0.000457387 | 0 |
| 0 | 0 | 0.001995034 | 0 | 0 |
| 0.146957644 | 0 | 0.131022547 | 0 | 0 |
| 0 | 0 | 0 | 0 | 0 |
| 0 | 0 | 0 | 0 | 0 |
| 0 | 0 | 0 | 0 | 0.001813838 |
| 0 | 0 | 0 | 0 | 0 |
| 0.001082071 | 0 | 0.141790008 | 0.153514374 | 0.004203145 |

| | | | | |
|---|---|---|---|---|
| 0 | 0 | 0.011682107 | 0 | 0.002154824 |
| 0 | 0 | 0 | 0 | 0 |
| 0 | 0 | 0 | 0 | 0 |
| 0 | 0 | 0 | 0 | 0 |
| 0 | 0 | 0 | 0 | 0.007411302 |
| 0 | 0 | 0 | 0 | 0.055405452 |
| 0 | 0 | 0 | 0 | 0 |
| 0 | 0 | 0 | 0 | 0 |
| 0 | 0 | 0 | 0 | 0 |
| 0 | 0 | 0 | 0 | 0 |
| 0 | 0 | 0 | 0 | 0 |
| 0.099499993 | 0 | 0 | 0 | 0.450368688 |
| 0.185305616 | 0 | 0.248480861 | 0 | 0 |
| 0 | 0 | 0 | 0 | 0 |
| 0.052725879 | 5.472153862 | 0.038757477 | 0 | 0.102628491 |
| 0 | 0 | 0 | 0 | 0 |
| 0 | 0 | 0 | 0 | 0.001098353 |
| 0 | 0 | 0 | 0 | 0 |
| 0.000610758 | 0 | 0 | 0.003596319 | 0 |
| 0 | 0 | 0 | 0 | 0 |
| 0.034384327 | 0 | 0 | 0 | 0 |
| 0 | 0 | 0 | 0 | 0 |
| 0.009502071 | 0 | 0.170211837 | 0 | 0 |
| 0 | 0 | 0 | 0 | 0 |
| 0 | 0 | 0 | 0 | 0 |
| 0 | 0 | 0 | 0 | 0 |
| 0 | 0 | 0 | 0 | 0 |
| 0 | 0 | 0 | 0 | 0 |
| 0 | 0 | 0 | 0 | 0 |
| 0 | 0 | 0 | 0 | 0 |
| 0 | 0 | 0 | 0 | 0 |
| 0.157032617 | 0.139693247 | 0.045559764 | 0.030836554 | 0.014927287 |
| 0 | 0 | 0 | 0 | 0 |
| 0.001054041 | 0 | 0 | 0 | 0 |
| 0 | 0 | 0 | 0.003158465 | 0.001894145 |
| 0 | 0 | 0.001974453 | 0.001665673 | 0.001953426 |
| 0 | 0 | 0 | 0 | 0 |
| 0 | 0 | 0 | 0 | 0 |
| 0 | 0 | 0 | 0 | 0.00282892 |
| 0 | 0 | 0 | 0 | 0 |
| 0.010303569 | 0 | 0 | 0 | 0 |
| 0 | 0 | 0 | 0 | 0 |
| 0 | 0 | 0 | 0 | 0 |
| 0 | 0 | 0 | 0 | 0 |
| 0 | 0.010455848 | 0.007166831 | 0.021779835 | 0.205251444 |
| 0 | 0 | 0 | 0 | 0 |
| 0.646351273 | 0 | 0 | 0 | 0.643520387 |
| 0 | 0 | 0.001962209 | 0 | 0.000882414 |

| | | | | |
|---|---|---|---|---|
| 2.403573996 | 0 | 0 | 0 | 0 |
| 0 | 0 | 0 | 0 | 0 |
| 0.003378388 | 0 | 0.00429133 | 0 | 0.016210577 |
| 0 | 0 | 0 | 0 | 0 |
| 0 | 0 | 0 | 0 | 0.003914254 |
| 0 | 0 | 0 | 0 | 0 |
| 0 | 0 | 0 | 0 | 0 |
| 0.06587592 | 0 | 0.003131678 | 0 | 0 |
| 0 | 0 | 0 | 0 | 0 |
| 0 | 0 | 0 | 0 | 0 |
| 0 | 0 | 0 | 0 | 0 |
| 0 | 0 | 0 | 0 | 0 |
| 0 | 0 | 0 | 0 | 0 |
| 0 | 0 | 0 | 0 | 0 |
| 0 | 0 | 0 | 0 | 0 |
| 0 | 0 | 0 | 0 | 0 |
| 0 | 0 | 0 | 0 | 0 |
| 0 | 0 | 0 | 0 | 0 |
| 5.85889739 | 1.854191066 | 11.69408812 | 0 | 0 |
| 0 | 0 | 0 | 0 | 0 |
| 0 | 0 | 0 | 0 | 0 |
| 0 | 0 | 0 | 0 | 0 |
| 0 | 0 | 0 | 0 | 0 |
| 0 | 0 | 0 | 0 | 0 |
| 0.001259628 | 0.000590893 | 0 | 0 | 0 |
| 0 | 0.692120938 | 0 | 0.002106619 | 0.004690178 |
| 0 | 0 | 0 | 0 | 0 |
| 0.026613687 | 0 | 0.896755685 | 0 | 0.000510084 |
| 0 | 0 | 0 | 0 | 0 |
| 0.307497773 | 0.533876283 | 0.591967362 | 0 | 0 |
| 0 | 0 | 0 | 0 | 0 |
| 0 | 0 | 0 | 0 | 0 |
| 0 | 0 | 0.00544402 | 0 | 0 |
| 0 | 0 | 0 | 0 | 0 |
| 0 | 0.024446336 | 0.006807897 | 0.00386331 | 0.001092226 |
| 0 | 0 | 0 | 0 | 0 |
| 0 | 0 | 0 | 0 | 0 |
| 0 | 0.016364269 | 0 | 0.00767152 | 0 |
| 0 | 0 | 0 | 0 | 0 |
| 0 | 0 | 0 | 0 | 0 |
| 17.82256816 | 0.644808649 | 9.983551439 | 0 | 9.248082635 |
| 0.111484175 | 0 | 0.025828281 | 0 | 1.548464103 |
| 0 | 0 | 0 | 0 | 0 |
| 0 | 0 | 0 | 0 | 0 |
| 0 | 0 | 0 | 0 | 0 |
| 0 | 0 | 0.012198044 | 0 | 0 |
| 0 | 0 | 0 | 0 | 0 |
| 0 | 0.005701868 | 0 | 0.000869792 | 0 |

| | | | | |
|---|---|---|---|---|
| 0 | 0 | 0 | 0 | 0 |
| 0 | 0 | 0.00238613 | 0.009202142 | 0.001686227 |
| 0 | 0 | 0 | 0 | 0 |
| 0 | 0 | 0 | 0 | 0 |
| 2.215569943 | 0 | 0.592331375 | 0 | 0 |
| 0 | 0 | 0 | 0 | 0 |
| 0 | 0 | 0 | 0 | 0 |
| 0 | 0 | 0 | 0 | 0 |
| 0.008149193 | 0.180728704 | 0.65453766 | 20.94356946 | 0.07240087 |
| 0 | 0 | 0 | 0 | 0 |
| 0 | 0.024479576 | 0 | 0 | 0 |
| 0 | 0 | 0 | 0 | 0 |
| 0.065802307 | 0 | 0.231356402 | 0 | 0 |
| 0.038166905 | 0 | 0.012623198 | 0 | 0.032229063 |
| 0 | 0 | 0 | 0 | 0 |
| 0.454302761 | 0.03497335 | 0.474319303 | 0 | 0 |
| 0.004846432 | 0 | 0 | 0 | 0.003834773 |
| 0 | 0 | 0 | 0 | 0 |
| 0 | 0 | 0 | 0 | 0 |
| 0 | 0 | 0 | 0 | 0 |
| 0 | 0 | 0 | 0 | 0 |
| 0 | 0 | 0 | 0 | 0 |
| 0 | 0 | 0 | 0 | 0 |
| 0 | 0 | 0 | 0 | 0 |
| 0 | 0 | 0 | 0 | 0 |
| 0 | 0 | 0 | 0 | 0 |
| 0.002809448 | 0 | 0.00520878 | 0 | 0 |
| 0 | 0.113351539 | 0.002074421 | 0 | 0 |
| 0 | 0 | 0 | 0 | 0 |
| 0 | 0 | 0 | 0 | 0 |
| 0 | 0 | 0 | 0 | 0 |
| 0 | 0 | 0 | 0 | 0 |
| 0 | 0 | 0 | 0 | 0.025809476 |
| 0.00055386 | 0 | 0 | 0 | 0 |
| 0 | 0 | 0.042007849 | 0 | 0 |
| 0 | 0 | 0 | 0.001799462 | 0.001582745 |
| 0 | 0 | 0 | 0 | 0 |
| 0 | 0 | 0.01076337 | 0 | 0 |
| 0 | 0 | 0 | 0 | 0 |
| 0 | 0 | 0 | 0 | 0 |
| 3.676194463 | 0 | 0.494483962 | 0 | 0 |
| 0 | 0 | 0 | 0 | 0 |
| 0 | 0 | 0 | 0 | 0 |
| 0 | 0 | 0 | 0 | 0 |
| 0 | 0 | 0 | 0 | 0 |
| 0 | 0 | 0 | 0 | 0 |
| 0 | 0 | 0 | 0 | 0 |
| 0 | 0 | 0 | 0 | 0 |

| | | | | |
|---|---|---|---|---|
| 0.001349839 | 0 | 0.013438504 | 0 | 0.028276098 |
| 0 | 0 | 0 | 0 | 0 |
| 0 | 0 | 0 | 0 | 0 |
| 0 | 0 | 0.003720694 | 0 | 0 |
| 0.005726722 | 0 | 0 | 0 | 0 |
| 0 | 0 | 0 | 0 | 0 |
| 0.034558776 | 0 | 0.481219363 | 0 | 0.000521652 |
| 0 | 0 | 0 | 0 | 0 |
| 0 | 0 | 0 | 0 | 0 |
| 0.00544806 | 0 | 0 | 0 | 0 |
| 0 | 0 | 0 | 0 | 0 |
| 0.000782961 | 0 | 0.000768509 | 0 | 0 |
| 0 | 0 | 0 | 0 | 0 |
| 0 | 0 | 0 | 0 | 0 |
| 0 | 0 | 0 | 0 | 0 |
| 0 | 0 | 0 | 0 | 0 |
| 0 | 0 | 0 | 0.15544231 | 0 |
| 0 | 0 | 0 | 0 | 0.00133737 |
| 0 | 0 | 0 | 0 | 0 |
| 0 | 0 | 0 | 0 | 0 |
| 0.016706655 | 0 | 0.025434055 | 0 | 0.012416044 |
| 0 | 0 | 0 | 0 | 0 |
| 0.003948725 | 0 | 0.009966437 | 0 | 0 |
| 0 | 0 | 0 | 0.006399474 | 0.005682882 |
| 0 | 0 | 0 | 0 | 0 |
| 0 | 0.001110424 | 0 | 0.001451912 | 0 |
| 0 | 0 | 0 | 0 | 0 |
| 0 | 0 | 0 | 0 | 0 |
| 0 | 0 | 0 | 0 | 0 |
| 0.007219296 | 0.119207573 | 0 | 0.437137092 | 0.139510375 |
| 0 | 0 | 0 | 0 | 0 |
| 0 | 0 | 0 | 0 | 0 |
| 0 | 0 | 0 | 0 | 0 |
| 0 | 0 | 0 | 4.124208839 | 0 |
| 0 | 0 | 0.019419308 | 0 | 0 |
| 0 | 0 | 0 | 0 | 0 |
| 0 | 0.000588853 | 0 | 0.022482315 | 0 |
| 0 | 0 | 0 | 0 | 0 |
| 0 | 0 | 0 | 0 | 0 |
| 0 | 0 | 0 | 0 | 0 |
| 0 | 0 | 0 | 0 | 0 |
| 0 | 0 | 0 | 0 | 0 |
| 0 | 0 | 0 | 0 | 0 |
| 0 | 0 | 0 | 0 | 0 |
| 0 | 0 | 0 | 0 | 0 |
| 0 | 0 | 0 | 0 | 0 |
| 0 | 0 | 0 | 0 | 0 |
| 0 | 0 | 0 | 0 | 0 |

| | | | | |
|---|---|---|---|---|
| 0 | 0 | 0.000706378 | 0.048599704 | 0.009609259 |
| 0 | 0 | 0 | 0 | 0 |
| 0.04981813 | 0 | 0 | 0 | 0 |
| 0 | 0 | 0 | 0 | 0 |
| 0 | 0 | 0 | 0 | 0 |
| 0 | 0 | 0 | 0 | 0 |
| 0 | 0 | 0 | 0 | 0 |
| 0.319217913 | 0 | 0.877420428 | 0 | 0.149791981 |
| 0.123410744 | 17.09207918 | 0.036589297 | 0.019330958 | 0.029892112 |
| 0 | 0 | 0.035313969 | 0.00781411 | 0.053838695 |
| 0 | 0 | 0 | 0 | 0 |
| 0 | 0 | 0 | 0 | 0 |
| 0 | 0 | 0 | 0 | 0 |
| 0 | 0 | 0 | 0 | 0 |
| 0 | 0 | 0 | 0 | 0 |
| 0 | 0 | 0 | 0 | 0 |
| 0 | 0 | 0.065476247 | 0 | 0 |
| 0 | 0 | 0 | 0 | 0 |
| 0 | 0 | 0 | 0 | 0 |
| 0 | 0 | 0 | 0 | 0 |
| 0 | 0 | 0 | 0 | 0 |
| 0 | 0 | 0 | 0 | 0 |
| 0 | 0 | 0 | 0 | 0 |
| 0 | 0 | 0 | 0 | 0 |
| 0.088000931 | 0 | 0 | 0 | 0 |
| 0 | 0 | 0 | 0 | 0 |
| 0 | 0 | 0 | 0 | 0 |
| 0 | 0 | 0 | 0 | 0 |
| 0 | 0 | 0 | 0 | 0 |
| 0 | 0 | 0 | 0 | 0 |
| 0 | 0 | 0 | 0 | 0 |
| 0 | 0 | 0 | 0 | 0 |
| 0 | 0 | 0 | 0 | 0 |
| 0 | 0 | 0 | 0 | 0 |
| 0 | 0 | 0 | 0 | 0 |
| 0 | 0 | 0 | 0 | 0 |
| 0 | 0 | 0 | 0 | 0 |
| 0 | 0 | 0 | 0 | 0 |
| 0 | 0 | 0 | 0 | 0.003531722 |
| 0 | 1.811844963 | 0 | 0 | 0 |
| 0 | 0 | 0 | 0 | 0 |
| 0 | 0 | 0.432695572 | 0 | 0.115704154 |
| 0 | 0 | 0 | 0 | 0 |
| 0 | 0 | 0 | 0 | 0 |
| 0 | 0 | 0 | 0 | 0 |
| 0 | 0 | 0.000785142 | 0 | 0.007301736 |
| 0 | 0 | 0 | 0 | 0 |
| 0 | 0 | 0 | 0 | 0 |

| | | | | |
|---|---|---|---|---|
| 0 | 0 | 0 | 0 | 0 |
| 0 | 0 | 0.005419066 | 0.000836631 | 0 |
| 0 | 0 | 0 | 0 | 0 |
| 0 | 0 | 0 | 0 | 0 |
| 0.058801381 | 0 | 0.135710564 | 0.045452268 | 0.094140003 |
| 0.030292584 | 0 | 0 | 0 | 0 |
| 0 | 0 | 0 | 0 | 0.005544904 |
| 0 | 0.00300611 | 0.03459472 | 0.031163851 | 0.01289042 |
| 0 | 0 | 0.002915845 | 0 | 0 |
| 0 | 0 | 0 | 0 | 0 |
| 0 | 0 | 0 | 0 | 0 |
| 0 | 0 | 0 | 0 | 0 |
| 0 | 0 | 0 | 0 | 0 |
| 0 | 0 | 0 | 0 | 0 |
| 0.001288906 | 0 | 0 | 0 | 0.04192999 |
| 0.002580414 | 0 | 0 | 0 | 0.001542037 |
| 0 | 0 | 0 | 0 | 0 |
| 0 | 0 | 0 | 0 | 0 |
| 0 | 0 | 0 | 0 | 0 |
| 0 | 0 | 0 | 0 | 0 |
| 0 | 0 | 0 | 0 | 0 |
| 0 | 0 | 0 | 0 | 0 |
| 0 | 0 | 0 | 0 | 0 |
| 0 | 0.021741446 | 0 | 0.003352996 | 0 |
| 0.129699189 | 0.237581452 | 0 | 0 | 0 |
| 0 | 0 | 0 | 0 | 0.004031859 |
| 0 | 0.272271808 | 0 | 0.000800007 | 0.073180563 |
| 0 | 0 | 0 | 0 | 0.000926163 |
| 0 | 0 | 0 | 0 | 0 |
| 0 | 0 | 0 | 0 | 0 |
| 0 | 0 | 0 | 0 | 0 |
| 0 | 0 | 0 | 0 | 0 |
| 0 | 0 | 0 | 0 | 0 |
| 0 | 0 | 0 | 0 | 0 |
| 0 | 0 | 0 | 0 | 0 |
| 0 | 0 | 0 | 0 | 0 |
| 0 | 0 | 0 | 0 | 0 |
| 0 | 0 | 0 | 0 | 0.096700271 |
| 0 | 0 | 0 | 0 | 0 |
| 0 | 0 | 0 | 0 | 0 |
| 0 | 0 | 0 | 0 | 0 |
| 0 | 0 | 0 | 0 | 0 |
| 0 | 0 | 0 | 0 | 0 |
| 0 | 0 | 0 | 0 | 0 |
| 0 | 0 | 0 | 0 | 0 |
| 0 | 0 | 0 | 0 | 0 |
| 0 | 0 | 0 | 0 | 0 |
| 0 | 0 | 0 | 0 | 0 |

| | | | | |
|---|---|---|---|---|
| 0.003577521 | 0 | 0 | 0 | 0.013730916 |
| 0 | 0 | 0 | 0 | 0 |
| 0 | 0 | 0 | 0 | 0 |
| 0 | 0 | 0 | 0 | 0 |
| 0 | 0 | 0 | 0 | 0 |
| 0 | 0 | 0 | 0.015544017 | 0 |
| 0 | 0 | 0 | 0 | 0 |
| 0 | 0 | 0 | 0 | 0 |
| 0 | 0 | 0 | 0 | 0 |
| 0 | 0 | 0 | 0 | 0 |
| 0 | 0 | 0 | 0 | 0 |
| 0 | 0 | 0 | 0 | 0.002127658 |
| 0 | 0 | 0 | 0 | 0 |
| 0 | 0 | 0 | 0 | 0 |
| 0 | 0 | 0 | 0 | 0 |
| 0 | 0 | 0 | 0 | 0 |
| 0 | 0 | 0 | 0 | 0 |
| 0.001930984 | 0 | 0 | 0 | 0.037268712 |
| 0 | 0 | 0 | 0 | 0 |
| 0 | 0 | 0 | 0 | 0 |
| 0 | 0 | 0.001161574 | 0.000435519 | 0.00995976 |
| 0 | 0 | 0 | 0 | 0 |
| 0 | 0.002337015 | 0.004156452 | 0 | 0 |
| 0.030549971 | 0 | 0 | 0 | 0 |
| 0 | 0 | 0 | 0 | 0 |
| 0 | 0 | 0 | 0 | 0 |
| 0 | 0 | 0 | 0 | 0 |
| 0 | 0 | 0 | 0 | 0 |
| 0 | 0 | 0 | 0 | 0 |
| 0 | 0 | 0 | 0 | 0 |
| 0 | 0 | 0 | 0 | 0 |
| 0 | 0 | 0.053347653 | 0 | 0 |
| 0 | 0.071129673 | 0.0038925 | 3.667509787 | 0.068186155 |
| 0 | 0 | 0 | 0 | 0 |
| 0.235373883 | 0 | 0.42130059 | 0 | 0 |
| 0 | 0.002634396 | 0.059899017 | 0 | 0.001090695 |
| 0 | 0 | 0 | 0 | 0 |
| 0 | 0 | 0 | 0 | 0 |
| 0.000649561 | 0 | 0.000850094 | 0.0014343 | 0.001682082 |
| 0 | 0 | 0 | 0 | 0 |
| 0 | 0 | 0 | 0 | 0 |
| 0 | 0 | 0 | 0 | 0 |
| 0 | 1.076253865 | 0.013170699 | 0 | 0 |
| 0 | 0 | 0 | 0 | 0 |
| 0 | 0 | 0 | 0 | 0 |
| 0 | 0 | 0.002170906 | 0 | 0.326463434 |
| 0 | 0 | 0 | 0 | 0 |
| 0 | 0 | 0 | 0 | 0 |

| | | | | |
|---|---|---|---|---|
| 0 | 0 | 0 | 0 | 0 |
| 0 | 0 | 0 | 0 | 0 |
| 0 | 0 | 0 | 0 | 0 |
| 0 | 0 | 0 | 0 | 0 |
| 0 | 0 | 0 | 0 | 0 |
| 0 | 0 | 0 | 0 | 0 |
| 2.73163956 | 0 | 0 | 0 | 0 |
| 0 | 0 | 0.079143623 | 0.279436212 | 0.395843286 |
| 0.000565142 | 0 | 0 | 0 | 0 |
| 0 | 0 | 0 | 0 | 0.003619956 |
| 0 | 0 | 0 | 0 | 0 |
| 0 | 0 | 0 | 0 | 0 |
| 0 | 0 | 0 | 0 | 0 |
| 0.329703676 | 0 | 0 | 0 | 0 |
| 0 | 0 | 0.095113931 | 0 | 0 |
| 0 | 0 | 0 | 0.119829641 | 0 |
| 0 | 0 | 0 | 0 | 0 |
| 0 | 0 | 0 | 0 | 0 |
| 0 | 0 | 0 | 0 | 0 |
| 0.890331006 | 0 | 0 | 0 | 0 |
| 0.007874069 | 0 | 0.010861992 | 0 | 0 |
| 0 | 0 | 0 | 0 | 0 |
| 0 | 0 | 0 | 0 | 0 |
| 0 | 0 | 0 | 0 | 0 |
| 0 | 0 | 0 | 0 | 0.002406824 |
| 0 | 0 | 0 | 0 | 0 |
| 0 | 0 | 0 | 0 | 0 |
| 0 | 0 | 0 | 0 | 0 |
| 0 | 0 | 0 | 0 | 0 |
| 0.00886962 | 0 | 0 | 0 | 0 |
| 0.03030668 | 0 | 0.655577581 | 0 | 0 |
| 0 | 0 | 0 | 0 | 0 |
| 0 | 0 | 0 | 0 | 0 |
| 0 | 0 | 0 | 0 | 0 |
| 0 | 0 | 0 | 0 | 0 |
| 0 | 0 | 0 | 0 | 0 |
| 0 | 0 | 0 | 0 | 0 |
| 0 | 0 | 0 | 0 | 0 |
| 0 | 0 | 0 | 0 | 0 |
| 0 | 0 | 0 | 0 | 0 |
| 0 | 0 | 0 | 0 | 0 |
| 0.022196656 | 0 | 0 | 0 | 0 |
| 0 | 0 | 0.002637236 | 0 | 0 |
| 0 | 0 | 0.001068136 | 0 | 0 |
| 0 | 0 | 0 | 0 | 0 |
| 0 | 0 | 0 | 0 | 0 |
| 0 | 0.017417047 | 0.004708956 | 2.478399622 | 0.001215341 |
| 0 | 0 | 0 | 0 | 0 |

| | | | | |
|---|---|---|---|---|
| 0 | 0 | 0 | 0 | 0 |
| 0 | 0 | 0 | 0 | 0 |
| 0 | 0 | 0 | 0 | 0 |
| 0 | 0 | 0 | 0 | 0 |
| 0.00779152 | 0 | 0.018966295 | 0 | 0.156772735 |
| 0 | 0 | 0 | 0 | 0 |
| 0 | 0 | 0 | 0 | 0 |
| 0 | 0 | 0 | 0 | 0 |
| 0 | 0 | 0 | 0 | 0 |
| 0 | 0 | 0 | 0 | 0 |
| 0.020972149 | 0 | 0 | 0 | 0 |
| 0 | 0.0119757 | 0.001698835 | 0.004458713 | 0.045800228 |
| 0 | 0 | 0 | 0 | 0.005437714 |
| 0 | 0 | 0 | 0 | 0.025294133 |
| 0 | 0 | 0 | 0 | 0.003042002 |
| 0 | 0 | 0.032366581 | 0 | 0 |
| 0.003993532 | 0 | 0 | 0 | 0.00307014 |
| 0 | 0 | 0 | 0 | 0 |
| 0.001474378 | 0 | 0.269815581 | 0.325558568 | 0.017340098 |
| 0.060533355 | 0.010636053 | 0.129959359 | 0 | 0.000623144 |
| 0 | 0 | 0 | 0 | 0 |
| 0 | 0 | 0 | 0 | 0 |
| 0 | 0 | 0 | 0 | 0 |
| 0 | 0 | 0 | 0 | 0 |
| 0 | 0 | 0 | 0 | 0 |
| 0 | 0 | 0 | 0 | 0 |
| 0 | 0 | 0.005552018 | 0.006055757 | 0.00565934 |
| 0 | 0 | 0 | 0 | 0 |
| 0 | 0 | 0 | 0 | 0 |
| 0 | 0 | 0 | 0 | 0 |
| 0 | 0 | 0.000572137 | 0.004147313 | 0.000754725 |
| 0 | 0 | 0.01009358 | 0 | 0 |
| 0 | 0 | 0.001629787 | 0 | 0 |
| 0 | 0 | 0 | 0 | 0 |
| 0 | 0 | 0 | 0 | 0 |
| 0 | 0 | 0 | 0 | 0 |
| 0 | 0 | 0 | 0 | 0 |
| 0.005187328 | 0.256017264 | 0.009781188 | 0.071638844 | 0.004241981 |
| 0.001641919 | 0 | 0 | 0 | 0 |
| 0 | 0 | 0.019766323 | 0 | 0 |
| 0 | 0 | 0 | 0 | 0 |
| 0 | 0 | 0 | 0 | 0 |
| 0 | 0 | 0 | 0 | 0 |
| 0 | 0 | 0.025432129 | 0.011323395 | 0 |
| 0 | 0 | 0 | 0 | 0 |
| 0 | 0 | 0 | 0 | 0 |
| 0 | 0 | 0 | 0 | 0 |
| 0 | 0 | 0 | 0 | 0 |

| | | | | |
|---|---|---|---|---|
| 0 | 0 | 0 | 0 | 0 |
| 0.014014133 | 0 | 0 | 0 | 0 |
| 0 | 0 | 0 | 0 | 0 |
| 0.000848569 | 0.004776776 | 0.000832906 | 0.000780722 | 0.001030044 |
| 0 | 0 | 0 | 0.000434464 | 0 |
| 0 | 0 | 0 | 0.001682153 | 0 |
| 0 | 0 | 0.000606204 | 0 | 0 |
| 0.000995482 | 0 | 0 | 0 | 0 |
| 0 | 0 | 0 | 0 | 0 |
| 0 | 0 | 0 | 0 | 0 |
| 0.004908302 | 0 | 0 | 0 | 0 |
| 0 | 0 | 0 | 0 | 0 |
| 0.001052653 | 0 | 0.00634694 | 0 | 0.018545972 |
| 0 | 0 | 0 | 0 | 0 |
| 0 | 0 | 0 | 0 | 0 |
| 0 | 0 | 0 | 0 | 0 |
| 0 | 0 | 0 | 0 | 0 |
| 0 | 0 | 0 | 0 | 0 |
| 0 | 0 | 0 | 0 | 0 |
| 0 | 0 | 0 | 0 | 0 |
| 0 | 0 | 0 | 0 | 0 |
| 0 | 0 | 0 | 0 | 0 |
| 0 | 0.005759106 | 0 | 0.024631037 | 0.04576491 |
| 0.009346559 | 0 | 0.022705734 | 0 | 0 |
| 0 | 0 | 0 | 0 | 0 |
| 0 | 0 | 0 | 0 | 0.00083787 |
| 0 | 0 | 0 | 0 | 0 |
| 0 | 0 | 0 | 0 | 0 |
| 0 | 0 | 0 | 0 | 0 |
| 0 | 0 | 0 | 0 | 0 |
| 0.009272984 | 0.007457084 | 0.016253245 | 0 | 0 |
| 0 | 0 | 0 | 0 | 0.004642354 |
| 0 | 0 | 0 | 0 | 0 |
| 0 | 0 | 0 | 0 | 0 |
| 0 | 0 | 0.063771343 | 0 | 0 |
| 0 | 0 | 0 | 0.001528913 | 0 |
| 0 | 0 | 0 | 0.016919761 | 0 |
| 0.007070707 | 0 | 0 | 0.001784329 | 0 |
| 0 | 0.018401864 | 0.000733407 | 0 | 0 |
| 0 | 0 | 0 | 0 | 0 |
| 0 | 0 | 0 | 0 | 0 |
| 0 | 0 | 0 | 0 | 0 |
| 0 | 0 | 0 | 0 | 0 |
| 0 | 0 | 0 | 0 | 0 |
| 0 | 0 | 0 | 0 | 0 |
| 0.134933717 | 0 | 0.394425325 | 0.209761277 | 0.220568729 |
| 0 | 0 | 0 | 0 | 0 |
| 0 | 0 | 0 | 0 | 0 |

| | | | | |
|---|---|---|---|---|
| 0 | 0 | 0 | 0 | 0 |
| 0 | 0 | 0 | 0 | 0 |
| 0.194203551 | 0 | 0.276331098 | 0 | 0.230169928 |
| 0 | 0 | 0 | 0 | 0 |
| 0 | 0 | 0 | 0 | 0 |
| 0 | 0 | 0 | 0 | 0 |
| 0 | 0 | 0 | 0 | 0 |
| 0 | 0 | 0 | 0 | 0 |
| 0 | 0 | 0 | 0 | 0 |
| 0 | 0 | 0 | 0 | 0 |
| 0 | 0 | 0 | 0 | 0.001471631 |
| 0 | 0.007005054 | 0 | 0 | 0.000659145 |
| 0 | 0 | 0 | 0 | 0 |
| 0 | 0 | 0 | 0 | 0 |
| 0 | 0 | 0 | 0 | 0 |
| 0 | 0 | 0 | 0 | 0 |
| 0 | 0 | 0 | 0 | 0 |
| 0 | 0 | 0 | 0 | 0 |
| 0 | 0 | 0 | 0 | 0 |
| 0 | 0 | 0 | 0.00177509 | 0 |
| 0 | 0 | 0 | 0 | 0 |
| 0 | 0 | 0 | 0 | 0 |
| 0 | 0 | 0 | 0 | 0 |
| 0 | 0 | 0 | 0 | 0 |
| 0 | 0 | 0 | 0 | 0 |
| 0 | 0 | 0 | 0 | 0 |
| 0 | 0 | 0 | 0 | 0 |
| 0.005560298 | 0 | 0.035147344 | 0 | 0 |
| 0 | 0 | 0 | 0 | 0 |
| 0 | 0 | 0.001488933 | 0 | 0 |
| 0.001208603 | 0.064147133 | 0 | 0.00177915 | 0.007041961 |
| 0 | 0 | 0 | 0 | 0 |
| 0.035884681 | 0.007899538 | 0.076544548 | 0.04662749 | 0.007592403 |
| 0.03871887 | 0.056699496 | 0.010255094 | 0.000753928 | 0.002785143 |
| 0 | 0 | 0 | 0 | 0 |
| 0 | 0 | 0 | 0 | 0 |
| 0 | 0 | 0 | 0 | 0 |
| 0 | 0 | 0 | 0 | 0 |
| 0 | 0 | 0 | 0 | 0 |
| 0 | 0 | 0 | 0 | 0 |
| 0 | 0 | 0 | 0 | 0 |
| 0.030841421 | 0.789768398 | 0.031567194 | 0 | 0.001761749 |
| 0.006027842 | 0 | 0.007494329 | 0 | 0.023609377 |
| 0 | 0 | 0 | 0 | 0 |
| 0 | 0 | 0 | 0 | 0 |
| 0 | 0.055844384 | 0.007409882 | 0 | 0.003101563 |
| 0 | 0 | 0.000747404 | 0 | 0 |
| 0 | 0 | 0 | 0 | 0 |

| | | | | |
|---|---|---|---|---|
| 0 | 0 | 0 | 0 | 0 |
| 0.012419428 | 0 | 0.017975354 | 0 | 0.006745624 |
| 0.04499692 | 0.008304826 | 0.003861538 | 0.01031586 | 0.002626534 |
| 0 | 0 | 0 | 0 | 0 |
| 0.014135557 | 0 | 0.126413333 | 0 | 0 |
| 0.033443149 | 0 | 0.000562729 | 0.057529596 | 0.093345991 |
| 0 | 0 | 0 | 0 | 0 |
| 0 | 0 | 0 | 0 | 0 |
| 0 | 0 | 0 | 0 | 0 |
| 0 | 0 | 0 | 0 | 0 |
| 0 | 0 | 0 | 0 | 0 |
| 0 | 0 | 0 | 0 | 0 |
| 0 | 0 | 0 | 0 | 0 |
| 0 | 0 | 0 | 0 | 0 |
| 0 | 0 | 0 | 0 | 0 |
| 0 | 0 | 0 | 0 | 0 |
| 0 | 0 | 0.021951813 | 0 | 0 |
| 0 | 0 | 0 | 0 | 0 |
| 0 | 0 | 0 | 0 | 0 |
| 0 | 0 | 0 | 0 | 0 |
| 0 | 0 | 0 | 0 | 0 |
| 0 | 0 | 0.011453968 | 0 | 0 |
| 0 | 0 | 0 | 0 | 0 |
| 0 | 0 | 0 | 0 | 0 |
| 0 | 0.006464103 | 0 | 0 | 0 |
| 0 | 0.004207652 | 0 | 0 | 0.001979609 |
| 0 | 0 | 0 | 0 | 0 |
| 0 | 0 | 0.024456104 | 0 | 0 |
| 0 | 0 | 0 | 0 | 0 |
| 0 | 0 | 0 | 0 | 0 |
| 0 | 0 | 0 | 0 | 0 |
| 0 | 0 | 0 | 0 | 0 |
| 0 | 0 | 0 | 0 | 0 |
| 0 | 0 | 0 | 0 | 0 |
| 0 | 0 | 0 | 0 | 0 |
| 0 | 0 | 0 | 0 | 0 |
| 0 | 0 | 0 | 0 | 0 |
| 0 | 0 | 0 | 0 | 0 |
| 0 | 0 | 0 | 0.702666545 | 1.356803755 |
| 0 | 0 | 0 | 0 | 0 |
| 0 | 0 | 0 | 0 | 0 |
| 0 | 0 | 0 | 0 | 0 |
| 0 | 0 | 0 | 0.03297831 | 0.005562909 |
| 0 | 0 | 0 | 0 | 0 |
| 0 | 0 | 0 | 0 | 0 |
| 0 | 0 | 0 | 0 | 0.001371414 |
| 0 | 0 | 0 | 0.007602109 | 0.001823606 |
| 0 | 0 | 0 | 0 | 0 |

| | | | | |
|---|---|---|---|---|
| 0 | 0 | 0 | 0 | 0 |
| 0.003554347 | 0.004260999 | 0.007365115 | 0.018458225 | 0.005177375 |
| 0 | 0 | 0 | 0 | 0 |
| 0 | 0 | 0 | 0 | 0 |
| 0 | 0 | 0 | 0 | 0 |
| 0 | 0 | 0 | 0 | 0 |
| 0 | 0 | 0 | 0 | 0 |
| 0 | 0 | 0 | 0 | 0 |
| 0 | 0 | 0 | 0 | 0 |
| 0 | 0 | 0 | 0 | 0 |
| 0 | 0 | 0 | 0 | 0 |
| 0 | 0 | 0 | 0 | 0 |
| 0 | 0 | 0 | 0 | 0.042770048 |
| 0 | 0 | 0 | 0 | 0 |
| 0.001477245 | 0 | 0.007491548 | 0.019027839 | 0.00836812 |
| 0 | 0 | 0 | 0 | 0 |
| 0 | 0 | 0 | 0 | 0 |
| 0 | 0 | 0 | 0 | 0 |
| 0 | 0 | 0 | 0 | 0 |
| 0 | 0 | 0 | 0 | 0 |
| 0 | 0 | 0 | 0 | 0 |
| 0.002176157 | 0 | 0.43028289 | 0 | 0 |
| 0.005202395 | 0 | 0.037933003 | 0 | 0.005412839 |
| 0.001743335 | 0.003738516 | 0 | 0 | 0 |
| 0 | 0 | 0 | 0 | 0 |
| 0 | 0 | 0 | 0 | 0 |
| 0 | 0 | 0 | 0 | 0 |
| 0 | 0 | 0 | 0 | 0 |
| 0 | 0 | 0 | 0 | 0 |
| 0 | 0 | 0 | 0 | 0 |
| 0 | 0 | 0 | 0 | 0 |
| 0 | 0 | 0 | 0 | 0 |
| 0 | 0 | 0 | 0 | 0 |
| 0 | 0 | 0 | 0 | 0 |
| 0 | 0 | 21.18554812 | 0 | 2.297115076 |
| 0 | 0 | 0 | 0 | 0 |
| 0 | 0 | 0 | 0 | 0 |
| 0 | 0 | 0 | 0 | 0 |
| 0.002897541 | 0.008155436 | 0.036735729 | 0 | 0.001875845 |
| 0 | 0 | 0 | 0 | 0 |
| 0.160428282 | 0.012187378 | 0.129628699 | 0 | 0.050248048 |
| 0 | 0 | 0 | 0 | 0 |
| 0 | 0 | 0 | 0 | 0 |
| 0 | 0 | 0 | 0 | 0 |
| 0 | 0 | 0 | 0 | 0 |
| 0.005205491 | 3.048654992 | 0.433569476 | 5.253799097 | 0.121079036 |
| 0 | 0 | 0 | 0 | 0 |
| 0 | 0 | 0 | 0 | 0 |

| | | | | |
|---|---|---|---|---|
| 0 | 0 | 0 | 0 | 0 |
| 0 | 0 | 0.000990719 | 0 | 0 |
| 0 | 0 | 0 | 0 | 0.000746005 |
| 0 | 0 | 0 | 0 | 0 |
| 0 | 0 | 0 | 0 | 0 |
| 0 | 0 | 0 | 0 | 0 |
| 0 | 0 | 0 | 0 | 0 |
| 0 | 0.003267766 | 0.016304655 | 0 | 0 |
| 0 | 0 | 0.002307514 | 0 | 0 |
| 0 | 0.035026681 | 0 | 0 | 0.001177092 |
| 0 | 0 | 0 | 0 | 0 |
| 0 | 0 | 0 | 0 | 0 |
| 0.001421715 | 0 | 0.002093208 | 0 | 0.001610712 |
| 0 | 0 | 0 | 0 | 0 |
| 0 | 0 | 0 | 0 | 0 |
| 0 | 0 | 0 | 0 | 0 |
| 0 | 0 | 0 | 0 | 0 |
| 0 | 0 | 0 | 0 | 0 |
| 0 | 0 | 0 | 0 | 0 |
| 0.014552407 | 0 | 0.054236998 | 0.181467628 | 0.17285692 |
| 0 | 0 | 0 | 0 | 0 |
| 0 | 0 | 0 | 0 | 0 |
| 0 | 0 | 0 | 0 | 0 |
| 0 | 0 | 0 | 0 | 0 |
| 0 | 0 | 0 | 0 | 0 |
| 0 | 0 | 0 | 0 | 0 |
| 0 | 0 | 0 | 0 | 0 |
| 0 | 0 | 0 | 0 | 0 |
| 0 | 0 | 0 | 0 | 0 |
| 0 | 0 | 0 | 0.046133215 | 0.010071321 |
| 0 | 0 | 0 | 0 | 0 |
| 0 | 0 | 0 | 0 | 0 |
| 0 | 0 | 0 | 0 | 0 |
| 0 | 0 | 0 | 0 | 0 |
| 0 | 0 | 0 | 0 | 0 |
| 0 | 0 | 0 | 0 | 0 |
| 0 | 0 | 0 | 0 | 0 |
| 0 | 0 | 0 | 0 | 0 |
| 0 | 0 | 0 | 0 | 0 |
| 0 | 0 | 0 | 0 | 0 |
| 0 | 0 | 0 | 0 | 0 |
| 0 | 0 | 0 | 0 | 0 |
| 0.014375592 | 0 | 0.026327639 | 0 | 0 |
| 0 | 0 | 0 | 0 | 0 |
| 0 | 0 | 0.001361029 | 0.00189541 | 0.002693069 |
| 0.007272953 | 0 | 0 | 0 | 0 |
| 0 | 0 | 0 | 0 | 0 |
| 0 | 0.005395144 | 0 | 0 | 0 |

| | | | | |
|---|---|---|---|---|
| 0 | 0 | 0 | 0 | 0 |
| 0 | 0 | 0 | 0 | 0 |
| 0 | 0 | 0 | 0 | 0 |
| 0 | 0 | 0 | 0 | 0 |
| 0 | 0 | 0 | 0 | 0 |
| 0.001774576 | 0.086020362 | 0.00362879 | 0.001306151 | 0.002872115 |
| 0 | 0 | 0 | 0 | 0 |
| 0 | 0 | 0 | 0 | 0 |
| 0 | 0 | 0 | 0 | 0 |
| 0 | 0 | 0 | 0 | 0 |
| 0 | 0 | 0 | 0 | 0 |
| 0 | 0 | 0 | 0 | 0 |
| 0 | 0 | 0 | 0 | 0 |
| 0 | 0 | 0 | 0 | 0 |
| 0 | 0 | 0 | 0 | 0 |
| 0 | 0 | 0 | 0 | 0 |
| 0.001614411 | 0 | 0 | 0 | 0 |
| 0 | 0 | 0 | 0 | 0 |
| 0 | 0 | 0 | 0 | 0 |
| 0 | 0 | 0 | 0 | 0 |
| 0 | 0 | 0 | 0 | 0 |
| 0 | 0 | 0 | 0 | 0 |
| 0 | 0 | 0 | 0 | 0 |
| 0 | 0 | 0 | 0 | 0 |
| 0 | 0 | 0 | 0 | 0 |
| 0 | 0 | 0 | 0 | 0 |
| 0 | 0 | 0 | 0 | 0 |
| 0.003289288 | 0 | 0 | 0 | 0 |
| 0 | 0 | 0 | 0 | 0 |
| 0 | 0 | 0 | 0 | 0 |
| 0 | 0 | 0 | 0 | 0 |
| 0 | 0 | 0 | 0 | 0 |
| 0 | 0 | 0 | 0 | 0 |
| 0 | 0 | 0 | 0 | 0 |
| 0 | 0 | 0 | 0 | 0 |
| 0 | 0 | 0 | 0 | 0 |
| 0 | 0 | 0 | 0 | 0 |
| 0 | 0 | 0 | 0 | 0 |
| 0.226282519 | 0 | 0 | 0 | 0 |
| 0 | 0 | 0 | 0 | 0 |
| 0 | 0 | 0 | 0 | 0 |
| 0 | 0 | 0 | 0 | 0 |
| 0 | 0 | 0 | 0 | 0 |
| 0 | 0 | 0 | 0 | 0 |
| 0 | 0 | 0 | 0 | 0 |
| 0 | 0 | 0 | 0 | 0 |
| 0 | 0 | 0 | 0 | 0 |
| 0 | 0 | 0 | 0 | 0 |

| | | | | |
|---|---|---|---|---|
| 0 | 0 | 0 | 0 | 0 |
| 0.004801386 | 0.014290676 | 0 | 0 | 0.246472998 |
| 0 | 0 | 0 | 0 | 0 |
| 0 | 0 | 0 | 0 | 0 |
| 0 | 0 | 0 | 0 | 0 |
| 0 | 0 | 0 | 0 | 0 |
| 0 | 0 | 0 | 0 | 0 |
| 0 | 0 | 0 | 0 | 0 |
| 0 | 0 | 0 | 0 | 0 |
| 0 | 0 | 0 | 0 | 0 |
| 0 | 0 | 0 | 0.002712662 | 0.002736842 |
| 0 | 0 | 0 | 0 | 0 |
| 0.026985913 | 0 | 0 | 0 | 0 |
| 0 | 0 | 0 | 0 | 0 |
| 0 | 0 | 0 | 0 | 0 |
| 0 | 0 | 0 | 0 | 0 |
| 0 | 0 | 0 | 0 | 0 |
| 0 | 0 | 0 | 0 | 0 |
| 0 | 0 | 0 | 0 | 0 |
| 0 | 0 | 0 | 0 | 0 |
| 0.005720862 | 0 | 0.00388749 | 0 | 0.003846088 |
| 0 | 0 | 0 | 0 | 0 |
| 0 | 0 | 0 | 0 | 0 |
| 0.492508064 | 0 | 0.778619886 | 0.823615544 | 0.082484527 |
| 0 | 0 | 0 | 0 | 0 |
| 0 | 0 | 0 | 0 | 0 |
| 0 | 0 | 0 | 0 | 0 |
| 0 | 0 | 0 | 0 | 0 |
| 0 | 0 | 0 | 0 | 0 |
| 0 | 0 | 0 | 0 | 0 |
| 0 | 0 | 0 | 0 | 0 |
| 0 | 0 | 0 | 0 | 0 |
| 0 | 0 | 0 | 0 | 0 |
| 0 | 0 | 0 | 0 | 0 |

| PHL018-0131_V3 | PHL018-0014_V2 | PHL018-0065_V1 | PHL018-0119_V3 | PHL018-0021_V2 |
|---|---|---|---|---|
| EYCF | CTRL | EYCF | REF | EYCF |
| 0.007448864 | 0.110389972 | 0.000623164 | 0.072147712 | 0.126761291 |
| 0.057046366 | 0.936503511 | 0.60338055 | 0.286670948 | 4.71882019 |
| 0 | 0 | 0 | 5.878423913 | 1.258244629 |
| 0 | 0 | 0 | 0 | 0 |
| 0 | 0 | 0 | 0 | 0 |
| 0 | 0 | 0 | 0 | 0 |
| 0 | 0 | 0 | 0 | 0 |
| 0 | 0 | 0 | 0 | 0 |
| 0 | 0 | 0 | 0 | 0 |
| 0 | 0 | 0 | 0 | 0 |
| 0 | 0 | 0 | 0 | 0 |
| 0 | 0 | 0 | 0 | 0 |
| 0 | 0 | 0 | 0 | 0 |
| 0.052758188 | 0.270935129 | 12.99025474 | 4.597184448 | 7.81677689 |
| 0 | 0 | 0 | 0 | 0 |
| 0 | 0 | 0 | 0 | 0 |
| 0 | 0 | 0 | 0 | 0 |
| 0 | 0 | 0 | 0 | 0 |
| 0 | 0 | 0 | 0 | 0 |
| 0 | 0 | 0 | 0 | 0 |
| 0 | 0 | 0 | 0 | 0 |
| 0 | 0 | 0 | 0 | 0 |
| 0 | 0 | 0 | 0 | 0 |
| 0 | 0 | 0 | 0 | 0 |
| 0 | 0.239058208 | 0 | 0.197242601 | 0 |
| 0 | 0 | 0 | 0 | 0 |
| 0 | 0 | 0 | 0 | 0 |
| 0 | 0 | 0 | 0 | 0 |
| 0 | 0 | 0 | 0 | 0 |
| 0 | 0 | 0 | 0 | 0 |
| 0 | 0 | 0 | 0 | 0 |
| 0 | 0 | 0 | 0 | 0 |
| 0 | 0 | 0 | 0 | 0 |
| 0 | 0 | 0 | 0 | 0 |
| 0 | 0 | 0 | 0 | 0 |
| 0 | 1.702114617 | 0.269423032 | 0 | 1.723426544 |
| 0 | 0 | 0 | 0 | 0 |
| 0 | 0 | 0 | 0 | 0 |
| 0 | 0 | 0 | 0 | 0 |
| 0 | 0 | 0 | 0 | 0 |
| 0 | 0 | 0 | 0 | 0 |
| 0 | 0.04916834 | 0.015802469 | 0.006791368 | 0 |
| 20.76736065 | 0.316500087 | 13.31638139 | 0.147179205 | 0.037501446 |
| 0.000106621 | 0.032070378 | 0 | 0 | 0.096086167 |
| 0 | 0.003100104 | 0 | 0 | 0.003353694 |
| 0 | 0 | 0 | 0 | 0 |

| | | | | |
|---|---|---|---|---|
| 0.001818166 | 0.013225267 | 0 | 0.000472976 | 0 |
| 0 | 0 | 0 | 0 | 0 |
| 0.00011334 | 0 | 0 | 0 | 0.016225165 |
| 0.043304027 | 0.206198573 | 0.303815516 | 0.133781389 | 0.742474839 |
| 0.002211085 | 0.024855112 | 0.00092648 | 0.002533807 | 0 |
| 0 | 0.0022733 | 0.041097805 | 0 | 0 |
| 0.000404043 | 0.000740667 | 0.02397471 | 0.033713973 | 0 |
| 0.000975708 | 0.02640549 | 0 | 0 | 0 |
| 0 | 0.008831687 | 0 | 0 | 0 |
| 0 | 0 | 0 | 0 | 0 |
| 0 | 0 | 0 | 0.00234995 | 0 |
| 0.046374704 | 0.054187335 | 0.331371441 | 0.004102208 | 0 |
| 0.000127282 | 0 | 0 | 0 | 0 |
| 0.002683184 | 0.042081753 | 0 | 0 | 0.075028612 |
| 0 | 0 | 0 | 0.002284618 | 0 |
| 0 | 0 | 0 | 0 | 0 |
| 0 | 0.009210949 | 0.135720031 | 0.003424147 | 0 |
| 0 | 0 | 0 | 0 | 0 |
| 0 | 0 | 0 | 0 | 0 |
| 0 | 0 | 0 | 0 | 0 |
| 0.004503122 | 0.003363085 | 0.001930142 | 0.110676715 | 0.464323495 |
| 0.000260987 | 0.003683877 | 0.053822784 | 0.000826031 | 0.020068425 |
| 0 | 0.015347049 | 0 | 0 | 0 |
| 0 | 0 | 0 | 0 | 0 |
| 0 | 0 | 0 | 0 | 0 |
| 0 | 0 | 0 | 0 | 0 |
| 0 | 0 | 0 | 0 | 0 |
| 0 | 0 | 0 | 0 | 0 |
| 0 | 0.006956908 | 0 | 0 | 0 |
| 0 | 0 | 0 | 0 | 0 |
| 0 | 0 | 0 | 0.00056159 | 0 |
| 0 | 0 | 0 | 0 | 0 |
| 0 | 0 | 0 | 0 | 0 |
| 0 | 0 | 0 | 0 | 0 |
| 0 | 0 | 0 | 0 | 0 |
| 0 | 0 | 0 | 0 | 0 |
| 0 | 0 | 0 | 0 | 0 |
| 0 | 0 | 2.399671318 | 0 | 0.000690357 |
| 0 | 0 | 0 | 0 | 0 |
| 0 | 0 | 0 | 0 | 0 |
| 0 | 0 | 0 | 0 | 0 |
| 0 | 0 | 0 | 0 | 0 |
| 0 | 0 | 0 | 0 | 0 |
| 0 | 0 | 0 | 0 | 0 |
| 0.000238976 | 0 | 0 | 0 | 0 |
| 0 | 0 | 0 | 0 | 0 |
| 0 | 0 | 0 | 0.003472268 | 0 |
| 0 | 0.01240636 | 0 | 0.180932226 | 0 |

| | | | | |
|---|---|---|---|---|
| 0 | 0 | 0 | 0 | 0 |
| 0.000109633 | 0 | 0 | 0 | 0 |
| 0 | 0 | 0 | 0 | 0 |
| 0 | 0 | 0 | 0 | 0 |
| 0 | 0.000798707 | 0 | 0 | 0 |
| 0 | 0 | 0 | 0.000609179 | 0 |
| 0 | 0 | 0 | 0 | 0 |
| 0 | 0 | 0 | 0 | 0 |
| 0 | 0.002882517 | 0 | 0 | 0 |
| 0 | 0 | 0 | 0 | 0 |
| 0 | 0 | 0 | 0 | 0 |
| 0 | 0 | 0 | 0 | 0.000798594 |
| 0 | 0 | 0 | 0 | 0 |
| 0 | 0 | 0 | 0 | 0 |
| 0 | 0 | 0 | 0 | 0 |
| 0 | 0 | 0 | 0 | 0 |
| 0 | 0 | 0 | 0 | 0 |
| 0 | 0 | 0 | 0 | 0 |
| 0.002841023 | 0.137525923 | 0 | 0 | 0.002984988 |
| 0.001939216 | 0 | 0 | 0 | 0 |
| 0 | 0 | 0 | 0 | 0 |
| 0 | 0 | 0 | 0 | 0 |
| 0.000193144 | 0.00055638 | 0 | 0 | 0 |
| 0 | 0 | 0 | 0 | 0 |
| 0 | 0 | 0 | 0 | 0 |
| 0 | 0 | 0 | 0 | 0 |
| 0 | 0 | 0 | 0 | 0 |
| 0 | 0 | 0 | 0 | 0 |
| 0 | 0 | 0 | 0 | 0 |
| 0 | 0 | 0 | 0.001479304 | 0 |
| 0.005765926 | 0.053975559 | 0.145558955 | 0.077282418 | 0.014258212 |
| 0.003085704 | 0.002798332 | 0.000510147 | 0.012866744 | 0.002537536 |
| 0 | 0 | 0 | 0 | 0 |
| 0 | 0 | 0 | 0 | 0 |
| 0 | 0 | 0 | 0 | 0 |
| 0 | 0 | 0 | 0 | 0 |
| 0 | 0 | 0 | 0 | 0 |
| 0 | 0 | 0 | 0 | 0 |
| 0 | 0 | 0 | 0 | 0 |
| 0 | 0 | 0 | 0 | 0 |
| 0 | 0 | 0 | 0 | 0 |
| 0 | 0 | 0 | 0 | 0 |
| 0.000141531 | 0 | 0 | 0.003046056 | 0 |
| 0 | 0 | 0 | 0 | 0 |
| 0 | 0 | 0 | 0 | 0 |
| 0 | 0 | 0 | 0 | 0 |
| 0 | 0 | 0 | 0 | 0 |
| 0 | 0 | 0 | 0 | 0 |

| | | | | |
|---|---|---|---|---|
| 0 | 0 | 0 | 0 | 0 |
| 0 | 0.020147631 | 0 | 0 | 0 |
| 0 | 0 | 0 | 0 | 0 |
| 0 | 0 | 0 | 0 | 0 |
| 0.004625468 | 0.013181062 | 0.026937534 | 0 | 0.00453352 |
| 0 | 0 | 0 | 0 | 0 |
| 0 | 0 | 0 | 0 | 0 |
| 0 | 0 | 0 | 0 | 0 |
| 0 | 0 | 0 | 0 | 0 |
| 0 | 0 | 0 | 0 | 0 |
| 0 | 0 | 0 | 0 | 0 |
| 0 | 0 | 0 | 0 | 0 |
| 0 | 0 | 0 | 0 | 0 |
| 0 | 0 | 0 | 0 | 0 |
| 0 | 0 | 0 | 0 | 0 |
| 0 | 6.06994884 | 0.194107585 | 0 | 0 |
| 0 | 0 | 0 | 0 | 0 |
| 0 | 0 | 0 | 0 | 0 |
| 0 | 0 | 0 | 0 | 0 |
| 0 | 0 | 0 | 0 | 0 |
| 0 | 0 | 0 | 0 | 0 |
| 0 | 0 | 0 | 0 | 0 |
| 0 | 0.003144471 | 0 | 0 | 0 |
| 0 | 0 | 0 | 0 | 0 |
| 0.014960996 | 0.080486943 | 0 | 0 | 0 |
| 0 | 0 | 0 | 0 | 0 |
| 0 | 0.005185434 | 0 | 0 | 0 |
| 0 | 0 | 0 | 0 | 0 |
| 0 | 0 | 0 | 0 | 0 |
| 0 | 0 | 0 | 0 | 0 |
| 0 | 0.007119895 | 0 | 0 | 0 |
| 0 | 0.00208914 | 0 | 0.004113798 | 0 |
| 0 | 0 | 0 | 0 | 0 |
| 0 | 0 | 0 | 0 | 0 |
| 0 | 0 | 0 | 0 | 0 |
| 0 | 0 | 0 | 0 | 0 |
| 0 | 0 | 0 | 0 | 0 |
| 0 | 0 | 0 | 0 | 0 |
| 0 | 0 | 0 | 0 | 0 |
| 0 | 0 | 0 | 0 | 0 |
| 0 | 0 | 0 | 0 | 0 |
| 0 | 0 | 0 | 0 | 0 |
| 0 | 0 | 0 | 0 | 0 |
| 0 | 0 | 0 | 0 | 0 |
| 0 | 0 | 0 | 0 | 0 |
| 0 | 0 | 0 | 0 | 0 |
| 0 | 0 | 0 | 0 | 0 |
| 0 | 0 | 0 | 0 | 0 |

| | | | | |
|---|---|---|---|---|
| 0 | 0 | 0 | 0 | 0 |
| 0 | 0 | 0 | 0 | 0 |
| 0 | 0 | 0 | 0 | 0 |
| 0 | 0 | 0 | 0 | 0 |
| 0 | 0 | 0 | 0 | 0 |
| 0 | 0 | 0 | 0 | 0 |
| 0 | 0 | 0 | 0 | 0 |
| 0 | 0 | 0 | 0 | 0 |
| 0 | 0 | 0 | 0 | 0 |
| 0 | 0 | 0 | 0 | 0 |
| 0 | 0 | 0 | 0 | 0 |
| 0 | 0 | 0 | 0 | 0 |
| 0 | 0 | 0 | 0 | 0 |
| 0 | 0 | 0 | 0 | 0 |
| 0 | 0 | 0 | 0 | 0 |
| 0 | 0 | 0 | 0 | 0 |
| 0.000190947 | 0 | 0 | 0 | 0 |
| 0 | 0 | 0 | 0 | 0 |
| 0 | 0 | 0 | 0 | 0 |
| 0 | 0 | 0 | 0 | 0 |
| 0.000252773 | 0.023446454 | 0.024923574 | 0.024641045 | 0.030051277 |
| 0 | 0 | 0 | 0 | 0 |
| 0 | 0 | 0 | 0 | 0 |
| 0 | 0 | 0 | 0.007341615 | 0 |
| 0 | 0 | 0 | 0 | 0 |
| 0 | 0 | 0 | 0 | 0 |
| 0 | 0.004947341 | 0.001839917 | 0.004845577 | 0.004014026 |
| 0 | 0 | 0 | 0 | 0 |
| 0 | 0 | 0 | 0 | 0 |
| 0 | 0 | 0 | 0.008573885 | 0 |
| 0 | 0 | 0 | 0 | 0 |
| 0 | 0 | 0 | 0 | 0 |
| 0 | 0 | 0 | 0 | 0 |
| 0.000957773 | 0.080782426 | 0 | 0 | 0.156864738 |
| 0.000263149 | 0.068097071 | 0.003654446 | 0.004372576 | 0.157141241 |
| 0 | 0 | 0 | 0 | 0 |
| 0 | 0 | 0 | 0 | 0 |
| 0 | 0 | 0 | 0 | 0 |
| 0 | 0 | 0 | 0 | 0 |
| 0 | 0 | 0 | 0 | 0 |
| 0 | 0 | 0 | 0 | 0 |
| 0 | 0 | 0 | 0 | 0 |
| 0 | 0 | 0 | 0 | 0 |
| 0 | 0 | 0 | 0 | 0 |
| 0 | 0 | 0 | 0 | 0 |
| 0.000125273 | 0.009683273 | 0 | 0.22025142 | 0 |
| 0 | 0 | 0 | 0 | 0.001359584 |
| 0 | 0 | 0 | 0 | 0.000894842 |

| | | | | |
|---|---|---|---|---|
| 0 | 0 | 0 | 0 | 0 |
| 0 | 0 | 0 | 0 | 0 |
| 0 | 0 | 0 | 0.000813529 | 0 |
| 0 | 0 | 0 | 0 | 0 |
| 0 | 0 | 0 | 0 | 0 |
| 0 | 0 | 0 | 0 | 0 |
| 0 | 0 | 0 | 0 | 0 |
| 0 | 0 | 0 | 0 | 0 |
| 0 | 0 | 0 | 0 | 0 |
| 0 | 0 | 0 | 0 | 0 |
| 0 | 0 | 0 | 0 | 0 |
| 0 | 0 | 0 | 0 | 0 |
| 0 | 0 | 0 | 0 | 0 |
| 0 | 0 | 0 | 0 | 0 |
| 0 | 0 | 0 | 0.001169744 | 0 |
| 0 | 0 | 0 | 0 | 0 |
| 0 | 0 | 0 | 0 | 0 |
| 0.131462645 | 0 | 0 | 0 | 0 |
| 0.147683802 | 0.001762588 | 0.079620904 | 0.001358129 | 0 |
| 0 | 0.224753655 | 0.005311379 | 0.097704395 | 0 |
| 0 | 0 | 0 | 0.005787537 | 0 |
| 0 | 0 | 0 | 0 | 0 |
| 0 | 0 | 0 | 0 | 0 |
| 0 | 0 | 0 | 0 | 0 |
| 0 | 0.001408757 | 0 | 0 | 0.001523994 |
| 0 | 0 | 0 | 0 | 0 |
| 0 | 0 | 0 | 0 | 0 |
| 0 | 0 | 0 | 0 | 0 |
| 0 | 0.073717797 | 0 | 0 | 0 |
| 0 | 0 | 0 | 0 | 0 |
| 0.037699483 | 0.196912992 | 0.030163215 | 0.386111477 | 0.129871079 |
| 0 | 0 | 0 | 0 | 0 |
| 0 | 0 | 0 | 0.013839235 | 0.016306252 |
| 0 | 0 | 0 | 0.00232936 | 0 |
| 0 | 0 | 0 | 0 | 0 |
| 0 | 0 | 0 | 0 | 0 |
| 0 | 0 | 0 | 0 | 0 |
| 0 | 0 | 0 | 0 | 0 |
| 0 | 0 | 0 | 0 | 0 |
| 0 | 0 | 0 | 0 | 0 |
| 0 | 0 | 0 | 0 | 0 |
| 0.000316283 | 0.004960425 | 0 | 0.003837329 | 0.023285428 |
| 0 | 0 | 0 | 0 | 0 |
| 0 | 0 | 0 | 0 | 0 |
| 0 | 0 | 0 | 0 | 0 |
| 0 | 0 | 0 | 0 | 0 |
| 0 | 0 | 0 | 0 | 0 |
| 0 | 0 | 0 | 0 | 0 |

| | | | | |
|---|---|---|---|---|
| 0 | 0 | 0 | 0 | 0 |
| 0 | 0 | 0 | 0 | 0 |
| 0 | 0.035147268 | 0 | 0 | 0.101974431 |
| 0 | 0 | 0 | 0 | 0 |
| 0 | 1.027214683 | 0 | 0 | 0 |
| 0 | 0 | 0 | 0 | 0 |
| 0 | 0 | 0 | 0 | 0 |
| 0 | 0 | 0 | 0 | 0 |
| 0 | 0 | 0 | 0 | 0 |
| 0 | 0 | 0 | 0 | 0 |
| 0 | 0 | 0 | 0 | 0 |
| 0 | 0 | 0 | 0 | 0 |
| 0 | 0 | 0 | 0 | 0 |
| 0 | 0 | 0.001015902 | 0.001620713 | 0 |
| 0 | 0 | 0.004821339 | 0.007452824 | 0.000823179 |
| 0 | 0.01911784 | 0 | 0 | 0 |
| 0 | 0 | 0.001194235 | 0 | 0.001125526 |
| 0 | 0.001614142 | 0 | 0 | 0 |
| 0 | 0 | 0 | 0 | 0 |
| 0 | 0 | 0 | 0 | 0 |
| 0 | 0 | 0 | 0 | 0 |
| 0 | 0 | 0 | 0 | 0.001016029 |
| 0 | 0 | 0 | 0 | 0 |
| 0 | 0 | 0 | 0 | 0 |
| 0 | 0 | 0 | 0 | 0 |
| 0 | 0 | 0 | 0 | 0 |
| 0 | 0.04161983 | 0 | 0 | 0 |
| 0 | 0 | 0 | 0 | 0 |
| 0 | 0 | 0 | 0 | 0 |
| 0 | 0 | 0 | 0 | 0 |
| 0 | 0.001919268 | 0 | 0 | 0 |
| 0 | 0 | 0 | 0 | 0 |
| 0 | 0 | 0 | 0 | 0 |
| 0 | 0 | 0 | 0 | 0 |
| 0 | 0 | 0 | 0 | 0 |
| 0 | 0 | 0 | 0.001570373 | 0 |
| 0 | 0 | 0 | 0 | 0 |
| 0.000936306 | 0.000786673 | 0 | 0 | 0.113398895 |
| 0 | 0 | 0 | 0 | 0 |
| 0 | 0 | 0 | 0 | 0 |
| 0 | 0 | 0 | 0 | 0 |
| 0 | 0 | 0 | 0 | 0 |
| 0 | 0 | 0 | 0 | 0 |
| 0 | 0 | 0 | 0 | 0 |
| 0 | 0 | 0 | 0 | 0 |
| 0 | 0 | 0 | 0 | 0 |
| 0 | 0 | 0 | 0 | 0 |
| 0 | 0.002858577 | 0.004593664 | 0 | 0 |

| | | | | |
|---|---|---|---|---|
| 0.033114901 | 0.197455157 | 0 | 1.302476572 | 0 |
| 0 | 0 | 0 | 0.0019006 | 0 |
| 0 | 0 | 0 | 0 | 0.00124158 |
| 0 | 0 | 0 | 0 | 0 |
| 0.000327221 | 0 | 0.009770127 | 0 | 0.002676748 |
| 0 | 0 | 0 | 0 | 0 |
| 0 | 0 | 0 | 0 | 0 |
| 0 | 0.043161567 | 0.007325224 | 0.004645468 | 0.004002188 |
| 0 | 0 | 0 | 0 | 0 |
| 0.015553921 | 0.026692537 | 0.050225688 | 0.477322131 | 0.010312855 |
| 0 | 0 | 0 | 0 | 0 |
| 0 | 0.00219644 | 0.014622741 | 0.004596711 | 0 |
| 0.000110166 | 0.002961916 | 0.011856825 | 0.000697354 | 0.004205515 |
| 0 | 0 | 0 | 0 | 0 |
| 0 | 0 | 0.00078674 | 0 | 0 |
| 0 | 0 | 0 | 0 | 0 |
| 0 | 1.360447578 | 0 | 0 | 0 |
| 0 | 0 | 0 | 0 | 0 |
| 0 | 0 | 0 | 0.292018825 | 0.025958758 |
| 0 | 0 | 0 | 0 | 0 |
| 0 | 0 | 0 | 0 | 0 |
| 0 | 0 | 0 | 0 | 0 |
| 0 | 0 | 0.023877421 | 0 | 0 |
| 0 | 0 | 0 | 0 | 0 |
| 0 | 0 | 0 | 0 | 0 |
| 0 | 0.001293704 | 0 | 0 | 0 |
| 0 | 0 | 0.411039852 | 0 | 0 |
| 0 | 0 | 0 | 0 | 0 |
| 0 | 0 | 0 | 0 | 0 |
| 0 | 0.004880549 | 0 | 0.002298153 | 0.012959457 |
| 0 | 0 | 0 | 0 | 0 |
| 0 | 0 | 0 | 0 | 0 |
| 0 | 0 | 0 | 0 | 0 |
| 0 | 0 | 0 | 0.010888163 | 0 |
| 0 | 0 | 0 | 0 | 0 |
| 0 | 0 | 0 | 0 | 0 |
| 0 | 0 | 0 | 0.003222648 | 0 |
| 0 | 0 | 0 | 0 | 0 |
| 0 | 0 | 0 | 0 | 0 |
| 0 | 0 | 0 | 0 | 0.00165594 |
| 0 | 0 | 0 | 0 | 0 |
| 0 | 0 | 0 | 0.044595453 | 0.031229182 |
| 0 | 0 | 0 | 0 | 0 |
| 0 | 0 | 0 | 0 | 0 |
| 0 | 0 | 0 | 0 | 0 |
| 0 | 0 | 0 | 0 | 0 |
| 0.000757956 | 0.006971567 | 0 | 0 | 0.003045745 |
| 0 | 0 | 0 | 0 | 0 |

| | | | | |
|---|---|---|---|---|
| 0 | 0 | 0 | 0 | 0 |
| 0 | 0.004494931 | 0 | 0.019472477 | 0 |
| 0 | 0 | 0 | 0 | 0 |
| 0 | 0 | 0 | 0 | 0 |
| 0 | 0 | 0 | 0 | 0 |
| 0 | 0 | 0 | 0.035800349 | 0.000514048 |
| 0 | 0 | 0 | 0 | 0 |
| 0 | 0 | 0.010722764 | 0 | 0 |
| 0 | 0.077046838 | 0 | 0 | 0 |
| 0 | 0 | 0 | 0 | 0 |
| 0.000400863 | 0.000979782 | 0.001265212 | 0.006459036 | 0.002186102 |
| 0 | 0 | 0 | 0 | 0 |
| 0 | 0 | 0 | 0 | 0 |
| 0.000723001 | 0.001157061 | 0.000956247 | 0 | 0.000938782 |
| 0 | 0 | 0 | 0.169352291 | 0 |
| 0 | 0 | 0 | 0 | 0 |
| 0 | 0 | 0 | 0 | 0 |
| 0 | 0 | 0 | 0.001060429 | 0 |
| 0.000329796 | 0 | 0.002544445 | 0 | 0 |
| 0 | 0.084784027 | 0 | 0.025744198 | 0 |
| 0 | 0.912573811 | 0 | 0 | 0 |
| 0.028960412 | 0.193087762 | 0.41948264 | 0.003203185 | 0 |
| 0 | 0 | 0 | 0 | 0 |
| 0 | 0 | 0 | 0 | 0 |
| 0 | 0 | 0 | 0 | 0 |
| 0 | 0 | 0 | 0 | 0 |
| 0 | 0 | 0 | 0.00921566 | 0 |
| 0 | 0 | 0 | 0 | 0 |
| 0 | 0 | 0 | 0 | 0 |
| 0.164163106 | 0.196397813 | 0.796141569 | 6.891423464 | 0.645921188 |
| 7.00712E-05 | 0.001726942 | 0.011677275 | 0.001330663 | 0 |
| 0 | 0.121669483 | 0 | 0 | 0 |
| 0 | 0 | 0 | 0 | 0 |
| 0 | 0 | 0 | 0 | 0.011726681 |
| 0 | 0 | 0 | 0 | 0 |
| 0 | 0 | 0 | 0 | 0 |
| 0.228133705 | 0.358058307 | 0 | 1.254477493 | 0 |
| 0 | 0.002122836 | 0 | 0 | 0 |
| 0 | 0 | 0 | 0.46280232 | 0.013824823 |
| 0 | 0 | 0 | 0.026560876 | 0 |
| 0 | 0.002092706 | 0.006878716 | 0 | 0 |
| 0.583299649 | 1.761855493 | 6.810314581 | 1.14905202 | 8.431261228 |
| 0 | 0.002160728 | 0 | 0 | 0 |
| 0 | 0 | 0 | 0 | 0 |
| 0 | 0 | 0 | 0 | 0 |
| 0 | 0 | 0 | 0 | 0 |
| 0 | 0 | 0 | 0.000752191 | 0 |
| 0 | 0 | 0 | 0 | 0 |

| | | | | |
|---|---|---|---|---|
| 0 | 0 | 0 | 0 | 0.001399233 |
| 0 | 0 | 0 | 0 | 0 |
| 0 | 0.020477384 | 0.203566648 | 0.002529152 | 0 |
| 0 | 0 | 0 | 0 | 0 |
| 0.046622258 | 0 | 0 | 0 | 0 |
| 0 | 0 | 0 | 0.002550242 | 0.005577249 |
| 0 | 0 | 0 | 0 | 0 |
| 0 | 0 | 0 | 0 | 0 |
| 0 | 0.004742316 | 0 | 0 | 0 |
| 0 | 0 | 0 | 0 | 0 |
| 0 | 0 | 0 | 0 | 0 |
| 0 | 0 | 0 | 0 | 0 |
| 0 | 0 | 0 | 0 | 0 |
| 0 | 0 | 0.00990076 | 0.018891176 | 0.008137613 |
| 0 | 0 | 0 | 0 | 0 |
| 0.001528907 | 0.058349935 | 0.001259556 | 0.082345447 | 0.390891385 |
| 8.61678E-05 | 0.003282006 | 0.003789378 | 0 | 0.059522665 |
| 0 | 0 | 0 | 0 | 0.001903098 |
| 0 | 0 | 0 | 0 | 0 |
| 0 | 0 | 0 | 0.001939794 | 0 |
| 0 | 0.002919714 | 0.038205554 | 0.002291392 | 0.004343002 |
| 0.001399303 | 0.007166035 | 0.001156706 | 0 | 0.008902939 |
| 0 | 0 | 0 | 0 | 0 |
| 0 | 0 | 0.014090095 | 0.320520112 | 0.14870202 |
| 0 | 0 | 0 | 0 | 0 |
| 0 | 0 | 0 | 0.024190487 | 0 |
| 0 | 0 | 0 | 0 | 0 |
| 0 | 0 | 0 | 0 | 0 |
| 0 | 0 | 0 | 0 | 0 |
| 0 | 0 | 0 | 0 | 0 |
| 0.000138652 | 0.011556203 | 0 | 0.342117764 | 0.006049115 |
| 0 | 0.01556157 | 0.001722426 | 0 | 0.001578235 |
| 0.005274754 | 0.055298109 | 0.031159662 | 0.021429042 | 0.134813069 |
| 0 | 0.006825151 | 0 | 0.002499644 | 0 |
| 0 | 0 | 0 | 0.001274364 | 0 |
| 0 | 0 | 0 | 0 | 0 |
| 0 | 0 | 0 | 0 | 0 |
| 0.001725532 | 0.004489617 | 0.015073584 | 0.008280112 | 0.106851114 |
| 0 | 0.093524601 | 0.107526747 | 0 | 0 |
| 0 | 0 | 0 | 0 | 0.001365027 |
| 0.001324759 | 0 | 0 | 0 | 0 |
| 0 | 0 | 0 | 0 | 0 |
| 0.00029135 | 0 | 0 | 0 | 0.007036457 |
| 0 | 0 | 0 | 0 | 0.019075404 |
| 0 | 0 | 0 | 0.007266284 | 0 |
| 0 | 0 | 0 | 0.002688595 | 0 |
| 0 | 0 | 0 | 0 | 0 |
| 0 | 0 | 0 | 0 | 0 |

| | | | | |
|---|---|---|---|---|
| 0 | 0.018028324 | 0 | 0 | 0 |
| 0.00087224 | 0.069843428 | 0.046082594 | 0.080659323 | 0.227704 |
| 0 | 0 | 0 | 0 | 0 |
| 0 | 0.0029332 | 0 | 0 | 0 |
| 0 | 0 | 0 | 0 | 0 |
| 0 | 0 | 0 | 0 | 0 |
| 0 | 0 | 0 | 0 | 0 |
| 0 | 0 | 0 | 0.004056568 | 0 |
| 0 | 0.001633399 | 0 | 0 | 0 |
| 0 | 0 | 0 | 0 | 0 |
| 0.000197269 | 0 | 0 | 0.002675835 | 0.012141243 |
| 0.000251861 | 0.046836515 | 0.002040321 | 0 | 0.016940123 |
| 0 | 0.030458632 | 0.001295637 | 0 | 0 |
| 0 | 0 | 0 | 0 | 0 |
| 0 | 0 | 0 | 0 | 0 |
| 0 | 0 | 0 | 0 | 0 |
| 0 | 0.00852909 | 0.001434353 | 0 | 0.002413981 |
| 0 | 0 | 0 | 0.017701837 | 0.005431714 |
| 0 | 0 | 0 | 0 | 0 |
| 0 | 0.000668794 | 0 | 0.009762578 | 0 |
| 0.015976877 | 0.025568732 | 0.001901803 | 0.005008559 | 0.155505989 |
| 0 | 0 | 0 | 0 | 0 |
| 0 | 0 | 0 | 0 | 0 |
| 0 | 0.010358882 | 0 | 0 | 0 |
| 0 | 0 | 0.006745811 | 0 | 0 |
| 0 | 0 | 0 | 0 | 0 |
| 0 | 0.039774483 | 0 | 0.051148265 | 0 |
| 0.009606429 | 0.036306852 | 0.065131939 | 0.204539843 | 0.119241429 |
| 0.000788334 | 0 | 0 | 0 | 0 |
| 0 | 0 | 0 | 0 | 0 |
| 0 | 0 | 0 | 0 | 0 |
| 0 | 0 | 0 | 0 | 0 |
| 0 | 0 | 0 | 0 | 0 |
| 0 | 0 | 0 | 0 | 0 |
| 0.001493874 | 0.170319554 | 0.012525893 | 0 | 0.210280631 |
| 0.007937558 | 0.116271312 | 0.215522411 | 0.055096362 | 0.07851445 |
| 0 | 0 | 0.004577147 | 0 | 0 |
| 0 | 0 | 0 | 0 | 0 |
| 0.000558614 | 0.012336969 | 0.003324725 | 0.002778329 | 0 |
| 0 | 0 | 0 | 0 | 0 |
| 0 | 0 | 0 | 0 | 0 |
| 0 | 0 | 0 | 0 | 0 |
| 0 | 0 | 0 | 0.01718283 | 0 |
| 0 | 0 | 0 | 0 | 0 |
| 0 | 0 | 0 | 0 | 0 |
| 0.037280776 | 0.158117521 | 0.288076829 | 0 | 0 |
| 0 | 0 | 0 | 0 | 0 |
| 0 | 0 | 0 | 0 | 0 |

| | | | | |
|---|---|---|---|---|
| 0 | 0 | 0 | 0 | 0 |
| 0 | 0 | 0 | 0 | 0 |
| 0.000510858 | 0.001492929 | 0 | 0 | 0 |
| 0.001637215 | 0.248740785 | 0.104862928 | 0 | 0.012469174 |
| 0 | 0 | 0 | 0 | 0 |
| 0 | 0 | 0 | 0 | 0 |
| 0 | 0 | 0 | 0 | 0 |
| 0.000883849 | 0 | 0 | 0 | 0 |
| 0.220057339 | 3.35469981 | 0.069214493 | 0.262231687 | 2.191034262 |
| 0.004970064 | 0.022553502 | 0.004204337 | 0.008144657 | 0.04994331 |
| 0 | 0 | 0 | 0 | 0 |
| 0 | 0.002544993 | 0.002065737 | 0 | 0.00103244 |
| 0 | 0 | 0 | 0 | 0 |
| 0 | 0 | 0 | 0 | 0 |
| 0.000245496 | 0.226514523 | 0.059197908 | 0.042381924 | 0.122684018 |
| 0 | 0 | 0 | 0 | 0 |
| 0.010305539 | 0.018393394 | 0 | 0 | 0 |
| 0 | 0 | 0.041282526 | 0.022137765 | 0 |
| 0 | 0 | 0 | 0 | 0 |
| 0 | 0 | 0 | 0 | 0 |
| 0 | 0.442839767 | 0 | 0 | 0 |
| 0 | 0 | 0 | 0 | 0 |
| 0.000547891 | 0.005021803 | 0 | 0 | 0 |
| 0.008061682 | 0 | 0.089415266 | 0 | 0 |
| 0 | 0.003403948 | 0 | 0.008655398 | 0 |
| 0 | 0.080336054 | 0 | 0 | 0 |
| 0 | 0 | 0 | 0.006293581 | 0 |
| 0.893365302 | 2.247394169 | 5.547489424 | 10.01193571 | 8.646962026 |
| 0 | 0 | 0 | 0 | 0 |
| 0 | 0 | 0 | 0 | 0 |
| 0 | 0.01590536 | 0 | 0 | 0 |
| 0 | 0.099239293 | 0 | 0 | 0 |
| 0 | 0.002286528 | 0.010837922 | 0.019000241 | 0.023025992 |
| 0 | 0 | 0 | 0 | 0 |
| 0 | 0 | 0 | 0 | 0 |
| 0.011748532 | 0.324262571 | 0.046748792 | 0 | 0 |
| 0 | 0.069475271 | 0.019992159 | 0 | 2.192258642 |
| 0 | 0 | 0 | 0 | 0 |
| 0 | 0 | 0 | 0.000891086 | 0 |
| 0 | 0 | 0 | 0 | 0 |
| 0.000934025 | 0.144744043 | 0.00108093 | 0.024635091 | 0.003678784 |
| 0 | 0.110414954 | 0.036769872 | 0 | 0 |
| 0.097481559 | 0.666519828 | 1.470683839 | 29.16311289 | 2.25596998 |
| 0.000632845 | 0 | 0.004237343 | 0.002002969 | 0 |
| 0 | 0 | 0 | 0 | 0 |
| 0 | 0.141182273 | 0 | 0 | 0 |
| 0 | 0 | 0 | 0 | 0 |
| 0 | 0.128298017 | 0 | 0 | 0.007483927 |

| | | | | |
|---|---|---|---|---|
| 0 | 0 | 0 | 0 | 0 |
| 0.004176872 | 0.098930491 | 0 | 0 | 0.07406903 |
| 0.000149491 | 0.237535905 | 0.005812911 | 0.004731427 | 0.03929499 |
| 0 | 0.002389522 | 0 | 0 | 0 |
| 0 | 0 | 0 | 0 | 0 |
| 0.000895001 | 0.064748516 | 0.000490627 | 0 | 0.00565155 |
| 0 | 0 | 0 | 0.012788974 | 0 |
| 0 | 0 | 0 | 0 | 0 |
| 0 | 0 | 0 | 0 | 0 |
| 0 | 0 | 0 | 0 | 0 |
| 0.000821933 | 0.018546958 | 0.028332411 | 0.106844703 | 0.037620197 |
| 0.157626002 | 0.872485078 | 4.665235546 | 3.118416622 | 3.149374829 |
| 0 | 0 | 0 | 0.001085381 | 0 |
| 9.50425E-05 | 0.011924803 | 0 | 0 | 0.052176925 |
| 9.98628E-05 | 0.147838124 | 0.264191835 | 0.005847262 | 0.095986025 |
| 0 | 5.340255863 | 0 | 0 | 0 |
| 0 | 0 | 0 | 0 | 0 |
| 0 | 0 | 0 | 0.0008775 | 0 |
| 0.00220639 | 0.006002725 | 0 | 0 | 0 |
| 0.000314348 | 0.027792616 | 0 | 0.003979669 | 0 |
| 0.001057955 | 0.055400027 | 0 | 0.056222827 | 0.021065568 |
| 0.308320757 | 0 | 0 | 2.418559594 | 1.419732437 |
| 1.69935263 | 0.335515473 | 9.321902005 | 5.902909175 | 1.665006337 |
| 0 | 0.004255097 | 0 | 0 | 0 |
| 0 | 0 | 0 | 0 | 0 |
| 0.270472344 | 0.696357845 | 0 | 0 | 3.021587572 |
| 0 | 0 | 0 | 0 | 0.006579691 |
| 0 | 0 | 0 | 0 | 0 |
| 0 | 0.08614045 | 0 | 0 | 0 |
| 0 | 0 | 0 | 0.216280315 | 0 |
| 0.001112622 | 0.017510628 | 0.00301491 | 0 | 0.0113954 |
| 0.013611421 | 0.169707867 | 0.15967301 | 0.073222083 | 0.244857066 |
| 0 | 0 | 0 | 0 | 0 |
| 0.001954171 | 0.056866705 | 0.050126084 | 0.011496818 | 0.012412344 |
| 0 | 0 | 0 | 0.002645075 | 0 |
| 0.038465672 | 0.27655551 | 0.026196833 | 0.00449945 | 0.070520489 |
| 0 | 0.016685709 | 0.003667505 | 0.000835848 | 0.004896706 |
| 0 | 0 | 0 | 0 | 0 |
| 0.042037357 | 3.73879182 | 0.002665703 | 0.043539697 | 0.149867737 |
| 0 | 0 | 0 | 0 | 0 |
| 0.000361098 | 0.026374705 | 0 | 0 | 0 |
| 0 | 0 | 0 | 0 | 0 |
| 0 | 0 | 0 | 0 | 0 |
| 0 | 0 | 0 | 0 | 0 |
| 0.003335916 | 0.01571662 | 0.003030748 | 0.015195987 | 0.027373612 |
| 0 | 0.002591511 | 0 | 0 | 0 |
| 0 | 0 | 0 | 0 | 0 |
| 0.017224335 | 0.019464099 | 0 | 0.003570879 | 0.199761088 |

| | | | | |
|---|---|---|---|---|
| 0 | 0 | 0 | 0 | 0 |
| 0 | 0 | 0 | 0 | 0 |
| 0 | 0 | 0 | 0 | 0 |
| 0 | 0.00125022 | 0.002767601 | 0.00336402 | 0 |
| 0 | 0 | 0 | 0 | 0 |
| 0 | 0.101585123 | 0 | 0.004902284 | 0 |
| 0 | 1.651888727 | 0.507827303 | 0 | 0 |
| 0.000391215 | 0.008217346 | 0 | 0.023371082 | 0 |
| 0 | 0 | 0 | 0 | 0.001275063 |
| 0 | 0 | 0 | 0 | 0 |
| 0.00073985 | 0.001790246 | 0 | 0.002060645 | 0.002743643 |
| 0 | 0 | 0 | 0 | 0 |
| 0.008188148 | 0.043367514 | 0 | 0 | 0.046491059 |
| 0 | 0.827826545 | 0 | 0 | 0 |
| 0 | 0 | 0 | 0 | 0 |
| 0 | 0 | 0 | 0 | 0 |
| 0 | 0.121015993 | 0.033775799 | 0 | 0 |
| 0.001724543 | 0.003643051 | 0.000855335 | 0.000935695 | 0.002687082 |
| 0 | 0.057850335 | 0 | 0 | 0 |
| 0 | 0.011499001 | 0 | 0 | 0.002180973 |
| 0.001247584 | 0.104452743 | 0.005471267 | 0 | 0.0054907 |
| 0 | 0 | 0 | 0 | 0 |
| 0 | 0.647590966 | 0.003944562 | 0 | 0.005421523 |
| 0 | 0 | 0 | 0 | 0 |
| 0 | 0 | 0 | 0 | 0 |
| 0 | 0 | 0 | 0 | 0 |
| 0 | 0 | 0 | 0 | 0 |
| 0 | 0 | 0 | 0 | 0 |
| 0 | 0 | 0 | 0 | 0 |
| 0.000922574 | 0.097817187 | 0.026037587 | 0 | 0.057940586 |
| 0 | 0 | 0 | 0 | 0 |
| 0 | 0 | 0 | 0 | 0 |
| 0.001261298 | 0 | 0 | 0 | 0 |
| 0 | 0 | 0 | 0 | 0 |
| 0 | 0.005852235 | 0 | 0 | 0 |
| 0 | 0 | 0 | 0 | 0 |
| 0.003623958 | 0.054984578 | 0.011330999 | 0.110808723 | 0.13002129 |
| 0 | 0 | 0 | 0 | 0.000956341 |
| 0 | 0 | 0 | 0 | 0 |
| 0.000345936 | 0 | 0 | 0.005005243 | 0 |
| 0 | 0 | 0 | 0.057134066 | 0 |
| 0 | 0 | 0 | 0 | 0 |
| 0 | 0.054479849 | 0 | 0 | 0 |
| 0.001046117 | 0.450995975 | 0.013007476 | 0 | 0 |
| 0.009728567 | 0.026768091 | 0.345859257 | 0.09849564 | 0 |
| 0 | 0 | 0 | 0 | 0 |
| 0.001166369 | 0.03279897 | 0 | 0 | 0 |
| 0 | 0 | 0 | 0 | 0 |

| | | | | |
|---|---|---|---|---|
| 0 | 0 | 0 | 0 | 0 |
| 0 | 0 | 0 | 0 | 0 |
| 0 | 1.182559337 | 0 | 0 | 0 |
| 0.015549604 | 0.550220404 | 0 | 0.005363157 | 0.142148198 |
| 0 | 0 | 0 | 0 | 0 |
| 0 | 0.368704451 | 0 | 0.042278735 | 0.452012582 |
| 0 | 0 | 0 | 0 | 0 |
| 0 | 0 | 0 | 0 | 0 |
| 0 | 0 | 0.000959637 | 0.110009792 | 0.000753688 |
| 0 | 0 | 0 | 0 | 0 |
| 0.000222343 | 0.008468728 | 0 | 0 | 0 |
| 0 | 0 | 0 | 0 | 0 |
| 0 | 0 | 0 | 0 | 0 |
| 0 | 0 | 0 | 0 | 0 |
| 0.014251859 | 0.467955139 | 0.060686594 | 0.980733727 | 0 |
| 0 | 0 | 0 | 0.001854202 | 0 |
| 0 | 0 | 0 | 0 | 0 |
| 0 | 0 | 0 | 0 | 0 |
| 0 | 0 | 0 | 0 | 0 |
| 0 | 0 | 0 | 0 | 0 |
| 0 | 0.153424861 | 0 | 0 | 0 |
| 0 | 0.003385269 | 0 | 0 | 0 |
| 0 | 0 | 0 | 0 | 0 |
| 0 | 0 | 0 | 0 | 0 |
| 1.345116743 | 0.023213401 | 0.468611968 | 0.004886678 | 0.001551052 |
| 0.197416069 | 10.01467824 | 0.006647898 | 0.913664739 | 7.343487228 |
| 0 | 0.149317591 | 0 | 0 | 0 |
| 0 | 0 | 0 | 0 | 0 |
| 0 | 0.20963697 | 0 | 0 | 0.142386578 |
| 0 | 0.161360267 | 0 | 0.007582659 | 0 |
| 0 | 0 | 0 | 0 | 0 |
| 0 | 0.000703509 | 0 | 0.039752185 | 0 |
| 0 | 0 | 0 | 0 | 0 |
| 0 | 0.000875114 | 0 | 0 | 0.002011734 |
| 0 | 0.000961274 | 0 | 0 | 0.012673863 |
| 0 | 0 | 0.000701966 | 0.000479948 | 0.010612832 |
| 0.00083461 | 0.00070123 | 0 | 0.349181282 | 0 |
| 0.006706469 | 0.73351758 | 0.430800374 | 0 | 1.194325746 |
| 0 | 1.223339673 | 0 | 0 | 0 |
| 0 | 0 | 0 | 0 | 0 |
| 0 | 0 | 0 | 0 | 0 |
| 0.000143302 | 0 | 0 | 0.004898379 | 0 |
| 0.193578244 | 0.089605531 | 0.049401328 | 0.045472744 | 0.70403649 |
| 0.019904571 | 3.180403086 | 0 | 0 | 0 |
| 0 | 0.726149269 | 0 | 0 | 0 |
| 0 | 0.006115578 | 0.006151479 | 0.005607844 | 0 |
| 0 | 0 | 0 | 0 | 0 |
| 0.00575987 | 0.192036671 | 0 | 0.190269369 | 1.380438567 |

| | | | | |
|---|---|---|---|---|
| 0 | 0 | 0 | 0 | 0 |
| 0 | 0.038593467 | 0 | 0 | 0 |
| 0 | 0 | 0 | 0 | 0 |
| 0 | 0 | 0 | 0 | 0 |
| 0 | 0 | 0 | 0.001971932 | 0 |
| 0 | 0.176157803 | 0 | 0 | 0.05815186 |
| 0 | 0.029246772 | 0 | 0 | 0 |
| 0 | 0 | 0 | 0 | 0 |
| 0 | 0 | 0 | 0 | 0 |
| 0 | 0 | 0 | 0 | 0 |
| 0 | 0 | 0 | 0 | 0 |
| 0.082639552 | 1.046474705 | 0.56892536 | 1.595136641 | 0.665681581 |
| 0.064843304 | 0 | 0 | 0 | 0 |
| 0 | 0 | 0 | 0 | 0 |
| 0 | 3.476495663 | 0.047758177 | 0.041671672 | 0 |
| 0 | 0 | 0 | 0 | 0 |
| 0 | 0 | 0 | 0 | 0.001615264 |
| 0 | 0 | 0 | 0 | 0 |
| 0.000143535 | 0 | 0 | 0 | 0 |
| 0 | 0 | 0 | 0 | 0 |
| 0.004705823 | 0.175883161 | 0.001485263 | 0 | 0 |
| 0 | 0 | 0 | 0 | 0 |
| 0.004494102 | 0.617839063 | 0.033337726 | 0 | 0.589727806 |
| 0 | 0 | 0 | 0 | 0 |
| 0 | 0 | 0 | 0 | 0 |
| 0 | 0 | 0 | 0 | 0 |
| 0 | 0 | 0 | 0 | 0 |
| 0 | 0 | 0 | 0 | 0 |
| 0 | 0 | 0 | 0 | 0 |
| 0 | 0 | 0 | 0 | 0 |
| 0 | 0 | 0 | 0 | 0 |
| 0.000297502 | 0.015997302 | 0.005902182 | 0 | 0.063738198 |
| 0 | 0.21417943 | 0 | 0 | 0 |
| 0.002972539 | 0.011702537 | 0.000884589 | 0.003225654 | 0.004747428 |
| 0 | 0.000924339 | 0 | 0 | 0.007799609 |
| 0.004502897 | 0.836492429 | 0.003692921 | 0.002468811 | 0.004060528 |
| 0 | 0 | 0 | 0 | 0 |
| 0.011677254 | 0 | 0 | 0 | 0 |
| 0 | 0.000637157 | 0 | 0 | 0 |
| 0 | 0 | 0 | 0 | 0 |
| 0 | 0.091039618 | 0 | 0 | 0.126260612 |
| 0 | 0 | 0 | 0 | 0 |
| 0 | 0 | 0 | 0 | 0 |
| 0 | 0 | 0 | 0.003118155 | 0 |
| 0.000704447 | 0.00728453 | 0.003762664 | 0.01372056 | 0.023788978 |
| 0 | 0 | 0 | 0 | 0 |
| 0 | 0.134072731 | 0.239810903 | 0 | 0 |
| 0.000384393 | 0.001377972 | 0 | 0.002919869 | 0 |

| | | | | |
|---|---|---|---|---|
| 0 | 0.762252718 | 0 | 0 | 0 |
| 0 | 0 | 0 | 0 | 0 |
| 0 | 0 | 0.000778307 | 0.002660717 | 0 |
| 0 | 0 | 0 | 0.009900556 | 0 |
| 0 | 0 | 0 | 0 | 0 |
| 0 | 0 | 0 | 0.196090029 | 0 |
| 0.007341381 | 0 | 0 | 0 | 0 |
| 0.000423428 | 0.001956676 | 0.000918798 | 0 | 0.005772906 |
| 0 | 0 | 0 | 0 | 0 |
| 0.000312502 | 0 | 0 | 0 | 0 |
| 0 | 0 | 0 | 0 | 0 |
| 0 | 0 | 0 | 0 | 0 |
| 0 | 0 | 0 | 0 | 0 |
| 0.016108049 | 0 | 0 | 0.920534451 | 0.359735187 |
| 0 | 0 | 0 | 0 | 0 |
| 0 | 0 | 0 | 0 | 0 |
| 0.043346079 | 0 | 0 | 0 | 0 |
| 0 | 0 | 0 | 0 | 0 |
| 0.415841813 | 1.803682013 | 0 | 0 | 0.00342912 |
| 0 | 0 | 0 | 0 | 0 |
| 0 | 0.005989671 | 0 | 0 | 0 |
| 0 | 0 | 0 | 0 | 0 |
| 0 | 0 | 0 | 0 | 0 |
| 0 | 0 | 0 | 0 | 0 |
| 0 | 0 | 0 | 0 | 0 |
| 0.000100894 | 0 | 0 | 0 | 0.023522207 |
| 0 | 0 | 0 | 0 | 0 |
| 0.012764153 | 0.178558412 | 0 | 0.010627161 | 0.001750329 |
| 0.005199875 | 0 | 0 | 0 | 0 |
| 0.001470585 | 0.802192511 | 0.07897793 | 0.019199543 | 0 |
| 0 | 0 | 0 | 0 | 0 |
| 0 | 0 | 0 | 0 | 0 |
| 0 | 0.004205403 | 0 | 0.004455542 | 0 |
| 0 | 0 | 0 | 0 | 0 |
| 7.92983E-05 | 0 | 0 | 0 | 0.00144151 |
| 0 | 0 | 0 | 0 | 0 |
| 0.02290847 | 0 | 0 | 0 | 0 |
| 0.000179961 | 0.001612809 | 0.000833061 | 0 | 0.000981415 |
| 0 | 0 | 0 | 0 | 0 |
| 0 | 0 | 0 | 0 | 0 |
| 0.137005049 | 1.271656906 | 6.671539879 | 4.321586334 | 6.068751806 |
| 0.001514919 | 0.161887536 | 0.135986417 | 0.40748447 | 0.283408845 |
| 0 | 1.16985594 | 0.018967106 | 0 | 0.15388412 |
| 0 | 0.019747422 | 0 | 0 | 0 |
| 0 | 0 | 0 | 0 | 0 |
| 0 | 0 | 0 | 0 | 0 |
| 0 | 0 | 0 | 0 | 0 |
| 0.000166631 | 0 | 0 | 0.001205467 | 0.00103854 |

| | | | | |
|---|---|---|---|---|
| 0 | 0 | 0 | 0 | 0 |
| 0 | 0.031927568 | 0.003570321 | 0.005269669 | 0.006676402 |
| 0 | 0.00345986 | 0 | 0 | 0 |
| 0 | 0 | 0 | 0 | 0 |
| 0.815885533 | 1.082358455 | 0.720051115 | 0.435683665 | 1.713206551 |
| 0 | 0 | 0 | 0 | 0 |
| 0 | 0 | 0 | 0 | 0 |
| 0 | 0 | 0 | 0 | 0 |
| 0 | 0.002136322 | 0.001527882 | 0.021047599 | 0.028132877 |
| 0 | 0 | 0 | 0 | 0 |
| 0 | 0 | 0 | 0 | 0 |
| 0 | 0 | 0 | 0 | 0 |
| 0.002688407 | 0.049732866 | 0.029405393 | 0 | 0.174896591 |
| 0.006200761 | 0.003538736 | 0.018278565 | 0.312712802 | 0.005901818 |
| 0.798734627 | 1.532819136 | 0 | 0 | 0.759739449 |
| 0.011184658 | 0.064530254 | 0.018499085 | 0 | 0.23625932 |
| 0 | 0.05767195 | 0 | 0.00112151 | 0 |
| 0 | 0 | 0 | 0 | 0 |
| 0 | 0 | 0 | 0 | 0 |
| 0 | 0 | 0 | 0 | 0 |
| 0 | 0 | 0 | 0 | 0 |
| 0 | 0 | 0 | 0 | 0 |
| 0 | 0 | 0 | 0 | 0 |
| 0 | 0 | 0 | 0 | 0 |
| 0 | 0 | 0 | 0 | 0 |
| 0 | 0 | 0 | 0 | 0 |
| 0.000220084 | 0.059319766 | 0 | 0.009055423 | 0.020763928 |
| 0 | 0 | 0 | 0 | 0.000406297 |
| 0 | 0 | 0 | 0 | 0 |
| 0 | 0 | 0 | 0 | 0 |
| 0 | 0 | 0 | 0 | 0 |
| 0 | 0.000836587 | 0 | 0 | 0 |
| 0.034853273 | 0.001439425 | 0 | 0 | 0 |
| 0 | 0.032721037 | 0.002349918 | 0 | 0 |
| 0.062687874 | 0.173012958 | 0 | 0 | 0.300897595 |
| 0.001493846 | 0 | 0 | 0 | 0 |
| 37.60818882 | 0.415059492 | 0 | 0.002973934 | 0 |
| 0 | 0.000729342 | 0 | 0 | 0 |
| 0 | 0 | 0 | 0 | 0 |
| 0 | 0 | 0 | 0 | 0 |
| 0.002040682 | 0.255882822 | 0.272159157 | 0 | 0.581002631 |
| 0 | 0.001251276 | 0.007432669 | 0 | 0 |
| 0 | 0 | 0.000848126 | 0 | 0 |
| 0 | 0.0104192 | 0.012874131 | 0.006541592 | 0 |
| 0 | 0 | 0 | 0 | 0 |
| 0 | 0 | 0 | 0 | 0 |
| 0 | 0.169734728 | 0 | 0 | 0 |
| 0 | 0 | 0 | 0 | 0 |

| | | | | |
|---|---|---|---|---|
| 0.152921753 | 0.259707577 | 0 | 0 | 0 |
| 0 | 0 | 0 | 0 | 0 |
| 0 | 0.00280065 | 0 | 0 | 0 |
| 0 | 0 | 0 | 0 | 0 |
| 0.001157426 | 0.161925214 | 0.032334149 | 0.000851926 | 0.006018436 |
| 0.014307002 | 0 | 0 | 0 | 0 |
| 0.000277737 | 0.128979659 | 0 | 0.001118783 | 0 |
| 0 | 0 | 0 | 0 | 0 |
| 0 | 0.002395759 | 0 | 0 | 0 |
| 0.000264901 | 0 | 0 | 0 | 0 |
| 0.003612962 | 0.060505569 | 0 | 0 | 0 |
| 0 | 0.047195865 | 0 | 0 | 0 |
| 0 | 0 | 0 | 0 | 0 |
| 0 | 0 | 0 | 0 | 0 |
| 0 | 0 | 0 | 0 | 0 |
| 0 | 0 | 0 | 0 | 0 |
| 0 | 0 | 0 | 0 | 0 |
| 0 | 0 | 0 | 0.006760869 | 0 |
| 0.002274668 | 0 | 0 | 0 | 0 |
| 0 | 0 | 0 | 0 | 0 |
| 0.005552843 | 0.274351721 | 0 | 0.06816922 | 0.172517934 |
| 0 | 0 | 0 | 0.001181024 | 0 |
| 0.000106057 | 0.005881093 | 0.005339075 | 0.007216937 | 0.004771626 |
| 0.001626789 | 0.019016469 | 0.001145963 | 0 | 0.001285751 |
| 0 | 0.003220726 | 0 | 0 | 0 |
| 0.000111261 | 0 | 0 | 0 | 0 |
| 0 | 0 | 0 | 0 | 0 |
| 0 | 0 | 0 | 0 | 0 |
| 0 | 0 | 0.111094582 | 0 | 0 |
| 0.002782451 | 0.097616663 | 0 | 0 | 0 |
| 0.000244075 | 0 | 0 | 0 | 0 |
| 0 | 0 | 0 | 0 | 0 |
| 0 | 0 | 0 | 0 | 0 |
| 0.001863205 | 0 | 0.000699325 | 0 | 0 |
| 0 | 0.060308284 | 0.00069806 | 0 | 0 |
| 0 | 0 | 0 | 0 | 0 |
| 0 | 0 | 0 | 0 | 0 |
| 0 | 0 | 0 | 0 | 0 |
| 0.000271843 | 0.034351216 | 0 | 0 | 0 |
| 0 | 0 | 0 | 0 | 0 |
| 0 | 0 | 0 | 0 | 0 |
| 0 | 0 | 0 | 0 | 0 |
| 0 | 0 | 0 | 0 | 0 |
| 0 | 0 | 0 | 0 | 0 |
| 0 | 0 | 0 | 0 | 0 |
| 0 | 0 | 0 | 0 | 0 |
| 0 | 0.003188995 | 0 | 0 | 0 |
| 0.002643143 | 0.111320386 | 0 | 0 | 0.082805946 |

| | | | | |
|---|---|---|---|---|
| 0.000507386 | 0 | 0 | 0.004335901 | 0.005810759 |
| 0 | 0.244036092 | 0 | 0 | 0 |
| 0 | 0.004255566 | 0 | 0 | 0 |
| 0 | 0 | 0 | 0 | 0 |
| 0 | 0 | 0 | 0 | 0 |
| 0.00179263 | 0 | 0 | 0 | 0 |
| 0 | 0 | 0 | 0 | 0 |
| 0.015498886 | 0.510374137 | 0.079765038 | 0.050785663 | 1.395838639 |
| 29.2063515 | 0.448057315 | 18.72404677 | 0.174431342 | 0.05363934 |
| 0 | 0 | 0 | 0.007896711 | 0.156625163 |
| 0.000325314 | 0.108892839 | 0 | 0 | 0 |
| 0 | 0 | 0 | 0 | 0 |
| 0.00247284 | 0.042601932 | 0 | 0 | 0 |
| 0 | 0 | 0 | 0 | 0 |
| 0 | 0 | 0 | 0 | 0 |
| 0 | 0 | 0 | 0 | 0 |
| 0 | 0 | 0 | 0 | 0 |
| 0 | 0 | 0 | 0 | 0 |
| 0 | 0 | 0 | 0 | 0 |
| 0 | 0.001236105 | 0 | 0.000582057 | 0 |
| 0 | 0 | 0 | 0 | 0 |
| 0 | 0 | 0 | 0 | 0 |
| 0 | 0 | 0 | 0 | 0 |
| 0 | 0 | 0 | 0 | 0 |
| 0 | 0.012356346 | 0.029414536 | 0 | 0 |
| 0 | 0.001981785 | 0 | 0 | 0 |
| 0 | 0 | 0 | 0 | 0 |
| 0 | 0 | 0 | 0 | 0 |
| 0 | 0 | 0 | 0 | 0 |
| 0 | 0 | 0 | 0 | 0 |
| 0 | 0 | 0.000904853 | 0.005444257 | 0 |
| 0 | 0 | 0 | 0 | 0 |
| 0 | 0 | 0 | 0 | 0 |
| 0 | 0.003932061 | 0 | 0 | 0 |
| 0.000245705 | 0.003715892 | 0 | 0 | 0.010384619 |
| 0 | 0 | 0 | 0 | 0 |
| 0 | 0 | 0 | 0.001522585 | 0 |
| 0 | 0 | 0 | 0 | 0.057669329 |
| 0 | 0 | 0.003560889 | 0 | 0.000987064 |
| 0 | 0 | 0 | 0 | 0 |
| 0 | 0 | 0 | 0 | 0 |
| 0.025109773 | 0.493857104 | 0.025763106 | 0.085509987 | 0 |
| 0 | 0 | 0 | 0 | 0 |
| 0 | 0 | 0 | 0 | 0 |
| 0 | 0 | 0 | 0 | 0 |
| 0 | 0 | 0 | 0 | 0 |
| 0 | 0 | 0 | 0 | 0 |
| 0 | 0 | 0 | 0 | 0 |

| | | | | |
|---|---|---|---|---|
| 0 | 0 | 0 | 0 | 0 |
| 0 | 0 | 0 | 0.015943237 | 0 |
| 0 | 0 | 0 | 0 | 0 |
| 0 | 0 | 0 | 0 | 0 |
| 0.183146955 | 4.361837847 | 0.013115059 | 0.036273351 | 0.140539557 |
| 0 | 0 | 0 | 0 | 0 |
| 0 | 0 | 0 | 0 | 0 |
| 0.000322716 | 0.000650741 | 0.001568588 | 0.027169336 | 0.002991881 |
| 0.00016111 | 0.003248705 | 0.00581723 | 0 | 0.01194913 |
| 0 | 0 | 0 | 0 | 0 |
| 0 | 0 | 0 | 0 | 0 |
| 0.028809894 | 0 | 0 | 0 | 0 |
| 0 | 0 | 0 | 0 | 0 |
| 0 | 0 | 0 | 0 | 0 |
| 0 | 0 | 0 | 0 | 0 |
| 0 | 0.001693149 | 0 | 0.003366252 | 0 |
| 0 | 0 | 0 | 0 | 0 |
| 0 | 0 | 0 | 0 | 0 |
| 0 | 0 | 0 | 0 | 0 |
| 0 | 0 | 0 | 0 | 0 |
| 0 | 0 | 0 | 0 | 0 |
| 0 | 0 | 0 | 0 | 0 |
| 0 | 0 | 0 | 0 | 0 |
| 8.37852E-05 | 0.000938605 | 0 | 0.00194467 | 0 |
| 0.026289668 | 0.062298109 | 0 | 0.995814973 | 0.257199781 |
| 0 | 0 | 0 | 0 | 0 |
| 0 | 0.121386347 | 0 | 0 | 0 |
| 0 | 0 | 0 | 0.000510772 | 0 |
| 0 | 0 | 0 | 0 | 0 |
| 0 | 0 | 0 | 0 | 0 |
| 0 | 0 | 0 | 0 | 0 |
| 0 | 0 | 0 | 0 | 0 |
| 0 | 0 | 0 | 0 | 0 |
| 0 | 0 | 0 | 0 | 0 |
| 0 | 0 | 0 | 0 | 0 |
| 0 | 0 | 0 | 0 | 0 |
| 0 | 0 | 0 | 0 | 0 |
| 0.009771318 | 0.169917545 | 0.058572083 | 0.045602002 | 0 |
| 0 | 0 | 0 | 0 | 0 |
| 0 | 0.000755297 | 0 | 0 | 0 |
| 0 | 0 | 0 | 0 | 0 |
| 0.051485217 | 0.308706518 | 0 | 0 | 0 |
| 0 | 0 | 0 | 0 | 0 |
| 0 | 0 | 0 | 0 | 0 |
| 0 | 0 | 0 | 0 | 0 |
| 0 | 0 | 0 | 0 | 0 |
| 0 | 0 | 0 | 0 | 0 |
| 0 | 0 | 0 | 0 | 0 |

| | | | | |
|---|---|---|---|---|
| 0.000216195 | 0.165498208 | 0.001834788 | 0.017486689 | 0 |
| 0 | 0 | 0 | 0 | 0 |
| 0 | 0.013754558 | 0 | 0 | 0 |
| 0 | 0 | 0 | 0 | 0 |
| 0 | 0 | 0 | 0 | 0 |
| 0.000779253 | 0 | 0 | 0 | 0.011231228 |
| 0 | 0 | 0 | 0 | 0 |
| 0 | 0 | 0 | 0 | 0 |
| 0 | 0 | 0 | 0 | 0 |
| 0 | 0.032192146 | 0 | 0 | 0 |
| 0 | 0 | 0 | 0 | 0 |
| 0 | 0 | 0 | 0 | 0.000702016 |
| 0 | 0 | 0 | 0 | 0 |
| 0 | 0 | 0 | 0 | 0 |
| 0 | 0 | 0 | 0 | 0 |
| 0 | 0 | 0 | 0 | 0 |
| 0 | 0 | 0 | 0 | 0 |
| 0 | 0 | 0 | 0 | 0 |
| 0 | 0 | 0 | 0 | 0 |
| 0 | 0 | 0 | 0 | 0 |
| 0 | 0 | 0 | 0.005105424 | 0.060516638 |
| 0 | 0 | 0 | 0 | 0 |
| 0.00119422 | 0.005666085 | 0.006097274 | 0.002001035 | 0.006512671 |
| 0.002443203 | 0.082542183 | 0 | 0 | 0 |
| 0 | 0.015692001 | 0 | 0 | 0 |
| 0.080895049 | 0 | 0 | 0 | 0 |
| 0 | 0 | 0 | 0 | 0 |
| 0 | 0 | 0 | 0 | 0 |
| 0 | 0 | 0 | 0 | 0 |
| 0 | 0 | 0 | 0 | 0 |
| 0 | 0 | 0 | 0 | 0 |
| 0.006992817 | 0 | 0 | 0 | 0 |
| 0.01424292 | 0 | 0.001142014 | 0.912827198 | 0.00143508 |
| 0 | 0.000894595 | 0 | 0 | 0 |
| 0.080295777 | 0 | 0 | 0 | 0 |
| 0 | 0 | 0 | 0.001503775 | 0 |
| 0 | 0 | 0 | 0 | 0 |
| 0.053259905 | 0 | 0 | 0.015995016 | 0 |
| 0 | 0.010671082 | 0 | 0 | 0.002497496 |
| 0 | 0 | 0 | 0 | 0 |
| 0 | 0 | 0 | 0 | 0 |
| 0 | 0.063656024 | 0 | 0 | 0 |
| 0.025882269 | 0.376010457 | 0.003718314 | 0.023727967 | 0.231289397 |
| 0 | 0 | 0 | 0 | 0 |
| 0 | 0 | 0 | 0 | 0 |
| 0.001247476 | 0.066698233 | 0.032089118 | 0 | 0.564503283 |
| 0 | 0 | 0 | 0 | 0 |
| 0 | 0 | 0 | 0 | 0 |

| | | | | |
|---|---|---|---|---|
| 0 | 0 | 0 | 0 | 0 |
| 0 | 0 | 0 | 0 | 0 |
| 0 | 0 | 0 | 0 | 0 |
| 0 | 0 | 0 | 0 | 0 |
| 0 | 0 | 0 | 0 | 0 |
| 0 | 0 | 0 | 0 | 0 |
| 0 | 0 | 0 | 0 | 0 |
| 0 | 0 | 0 | 0 | 0 |
| 0.002603169 | 0.098198666 | 0 | 0.007398367 | 0.008112212 |
| 0 | 0.094215008 | 0 | 0 | 0.003025804 |
| 0 | 0 | 0 | 0 | 0 |
| 0 | 0 | 0 | 0 | 0 |
| 0 | 0 | 0 | 0 | 0 |
| 0 | 0.012323862 | 0 | 0 | 0 |
| 0 | 0 | 0 | 0 | 0 |
| 0 | 0 | 0 | 0 | 0 |
| 0 | 0 | 0 | 0 | 0 |
| 0 | 0 | 0 | 0 | 0 |
| 0 | 0 | 0 | 0 | 0 |
| 0.201123316 | 0.161847078 | 0.920168678 | 0 | 0 |
| 0.000780209 | 0 | 0 | 0 | 0.00736419 |
| 0 | 0 | 0 | 0 | 0 |
| 0 | 0 | 0 | 0 | 0 |
| 0 | 0 | 0 | 0.01065931 | 0 |
| 0 | 0 | 0 | 0 | 0.000847068 |
| 0 | 0 | 0 | 1.378884934 | 0 |
| 0 | 0 | 0 | 0 | 0 |
| 0 | 0.26376827 | 0 | 0 | 0 |
| 0 | 0 | 0 | 0 | 0 |
| 0 | 0.028221586 | 0.087670211 | 0 | 0 |
| 0.029096805 | 0.079265687 | 0.140260004 | 3.355373621 | 0 |
| 0 | 0 | 0 | 0 | 0 |
| 0 | 0.21933451 | 0 | 0.000749086 | 0 |
| 0 | 0 | 0 | 0 | 0 |
| 0 | 0 | 0 | 0 | 0 |
| 0 | 0 | 0 | 0 | 0 |
| 0 | 0 | 0 | 0 | 0 |
| 0 | 0 | 0 | 0 | 0 |
| 0.002131824 | 0.003538041 | 0 | 0.015327135 | 0 |
| 0 | 0 | 0 | 0 | 0 |
| 0 | 0 | 0 | 0 | 0 |
| 0.052373309 | 0.057101787 | 0 | 0.102269101 | 0 |
| 0 | 0.002546519 | 0 | 0 | 0 |
| 0 | 0 | 0 | 0 | 0 |
| 0 | 0 | 0 | 0 | 0 |
| 0 | 0 | 0 | 0 | 0 |
| 0 | 0 | 0 | 0.062929138 | 0.041383082 |
| 0 | 0 | 0 | 0 | 0 |

| | | | | |
|---|---|---|---|---|
| 0 | 0 | 0 | 0 | 0 |
| 0 | 0 | 0 | 0 | 0 |
| 0 | 0 | 0 | 0 | 0 |
| 0 | 0 | 0 | 0 | 0 |
| 0.001362335 | 0.042240089 | 0 | 0.105848268 | 0.066625724 |
| 0 | 0 | 0 | 0 | 0 |
| 0 | 0 | 0 | 0 | 0 |
| 0 | 0 | 0 | 0 | 0 |
| 0 | 0 | 0 | 0 | 0 |
| 0 | 0 | 0 | 0 | 0 |
| 0 | 0.003375329 | 0 | 0 | 0 |
| 0.000213546 | 0.006561591 | 0.001270967 | 0.001737969 | 0.006321951 |
| 0 | 0 | 0 | 0.007163939 | 0 |
| 0 | 0 | 0 | 0 | 0 |
| 0 | 0.005312915 | 0 | 0.010448467 | 0 |
| 0 | 0 | 0 | 0 | 0 |
| 0 | 0.013405143 | 0 | 0.001485227 | 0 |
| 0 | 0 | 0 | 0 | 0 |
| 0.002656466 | 0.000621059 | 0 | 0.014037333 | 0 |
| 0 | 0.058994005 | 0 | 0.003627512 | 0 |
| 0 | 0 | 0 | 0 | 0 |
| 0 | 0 | 0 | 0 | 0 |
| 0 | 0 | 0 | 0 | 0 |
| 0 | 0 | 0 | 0 | 0 |
| 0 | 0 | 0 | 0 | 0 |
| 0 | 0 | 0 | 0 | 0 |
| 0 | 0.002599291 | 0 | 0.003977851 | 0.003822445 |
| 0 | 0 | 0 | 0.134821841 | 0 |
| 0 | 0 | 0 | 0 | 0 |
| 0 | 0 | 0 | 0 | 0 |
| 0 | 0.003683041 | 0 | 0 | 0 |
| 0 | 0 | 0 | 0 | 0 |
| 0 | 0 | 0 | 0 | 0 |
| 0 | 0 | 0 | 0 | 0 |
| 0 | 0.004417582 | 0 | 0 | 0 |
| 0 | 0 | 0 | 0 | 0 |
| 0 | 0 | 0 | 0 | 0 |
| 0.001193416 | 0.006339614 | 0.001871193 | 0.003777189 | 0.009972377 |
| 0 | 0.006535934 | 0 | 0 | 0 |
| 0 | 0 | 0 | 0 | 0 |
| 0 | 0 | 0 | 0 | 0 |
| 0 | 0 | 0 | 0 | 0 |
| 0 | 0 | 0 | 0 | 0 |
| 0 | 0 | 0 | 0 | 0 |
| 0 | 0 | 0 | 0 | 0 |
| 0 | 0 | 0 | 0 | 0 |
| 0 | 0 | 0 | 0 | 0 |
| 0 | 0 | 0 | 0 | 0 |

| | | | | |
|---|---|---|---|---|
| 0 | 0 | 0 | 0 | 0 |
| 0.000219565 | 0.003935492 | 0 | 0 | 0 |
| 0 | 0 | 0 | 0 | 0 |
| 0.000239308 | 0.001407447 | 0 | 0 | 0 |
| 0 | 0 | 0 | 0 | 0 |
| 0 | 0 | 0 | 0.001359948 | 0 |
| 0.000638633 | 0.017170309 | 0 | 0 | 0 |
| 0 | 0 | 0 | 0 | 0 |
| 0.00041312 | 0.001960085 | 0 | 0 | 0 |
| 0 | 0.007899591 | 0 | 0 | 0 |
| 0 | 0.039043405 | 0 | 0 | 0 |
| 0 | 0 | 0 | 0 | 0 |
| 0.00850299 | 0.228611596 | 0.002208559 | 0.002952955 | 0.007863414 |
| 0 | 0.006035814 | 0 | 0 | 0 |
| 0 | 0 | 0 | 0 | 0 |
| 0 | 0 | 0 | 0 | 0 |
| 0 | 0 | 0 | 0 | 0 |
| 0 | 0 | 0 | 0 | 0 |
| 0 | 0 | 0 | 0 | 0 |
| 0 | 0 | 0 | 0 | 0 |
| 0 | 0 | 0 | 0 | 0 |
| 0 | 0 | 0 | 0 | 0 |
| 0 | 0 | 0 | 0.007069749 | 0.021165417 |
| 0 | 0.1284477 | 0.001830258 | 0 | 0.010421607 |
| 0 | 0 | 0 | 0 | 0 |
| 0.000194661 | 0.025186928 | 0 | 0 | 0 |
| 0 | 0.02241326 | 0 | 0 | 0 |
| 0 | 0 | 0 | 0 | 0 |
| 0 | 0 | 0 | 0 | 0 |
| 0 | 0 | 0 | 0 | 0 |
| 0 | 0.019460758 | 0.002161726 | 0.000788274 | 0 |
| 0 | 0 | 0 | 0 | 0 |
| 0 | 0 | 0 | 0 | 0 |
| 0 | 0 | 0 | 0 | 0 |
| 0.003105967 | 0.023332865 | 0 | 0 | 0 |
| 0 | 0 | 0 | 0 | 0 |
| 0 | 0 | 0 | 0.020099598 | 0 |
| 0 | 0 | 0 | 0 | 0 |
| 0 | 0 | 0.001097382 | 0.003501405 | 0 |
| 0 | 0 | 0 | 0 | 0 |
| 0 | 0 | 0 | 0 | 0 |
| 0 | 0 | 0 | 0 | 0 |
| 0 | 3.670471899 | 3.95927384 | 0.331359952 | 0 |
| 0 | 0 | 0 | 0 | 0 |
| 0 | 0 | 0 | 0 | 0 |
| 0.018099574 | 0.372601917 | 0.176850174 | 0.036234408 | 0.185531784 |
| 0 | 0 | 0 | 0 | 0 |
| 0 | 0 | 0 | 0 | 0 |

| | | | | |
|---|---|---|---|---|
| 0 | 0 | 0 | 0 | 0 |
| 0 | 0 | 0 | 0 | 0 |
| 0.01371578 | 0.029718317 | 0 | 0.222997392 | 0 |
| 0.000979696 | 0 | 0.001457724 | 0 | 0 |
| 0 | 0 | 0 | 0 | 0 |
| 0 | 0 | 0 | 0 | 0 |
| 0 | 0 | 0 | 0 | 0 |
| 0 | 0 | 0 | 0 | 0 |
| 0 | 0.007699229 | 0 | 0 | 0 |
| 0 | 0 | 0 | 0 | 0 |
| 0 | 0.00229809 | 0 | 0 | 0 |
| 0 | 0 | 0 | 0 | 0 |
| 0 | 0 | 0 | 0 | 0 |
| 0 | 0 | 0 | 0 | 0 |
| 0 | 0 | 0 | 0 | 0 |
| 0 | 0 | 0 | 0 | 0 |
| 0 | 0 | 0 | 0 | 0 |
| 0 | 0 | 0 | 0 | 0 |
| 0 | 0 | 0 | 0 | 0 |
| 0.000368403 | 0 | 0 | 0 | 0 |
| 0 | 0 | 0 | 0 | 0 |
| 0 | 0 | 0 | 0 | 0 |
| 0 | 0 | 0 | 0 | 0 |
| 0 | 0 | 0 | 0 | 0 |
| 0 | 0 | 0 | 0 | 0 |
| 0 | 0 | 0 | 0 | 0 |
| 0 | 0 | 0 | 0 | 0 |
| 0.001223102 | 0.007799492 | 0 | 0 | 0.020010541 |
| 0 | 0 | 0 | 0 | 0 |
| 0 | 0 | 0 | 0 | 0 |
| 0 | 0.005072873 | 0.006254865 | 0 | 0.008231754 |
| 0 | 0 | 0 | 0 | 0 |
| 0.005003447 | 0.017860372 | 0.165468033 | 0.05135525 | 0.065517894 |
| 0 | 0.000776654 | 0.001002909 | 0 | 0.00346576 |
| 0.002384163 | 0 | 0 | 0 | 0 |
| 0 | 0 | 0 | 0 | 0 |
| 0 | 0 | 0 | 0 | 0 |
| 0 | 0 | 0 | 0 | 0 |
| 0 | 0 | 0 | 0 | 0 |
| 0 | 0 | 0 | 0 | 0 |
| 0 | 0 | 0 | 0 | 0 |
| 0 | 0 | 0.000645927 | 0.44222123 | 0 |
| 0.004958138 | 0.058082728 | 0.027542229 | 0.069226883 | 0.048052492 |
| 0 | 0 | 0 | 0 | 0 |
| 0 | 0 | 0 | 0 | 0 |
| 0.006386956 | 0.291153205 | 0 | 0 | 0.021657883 |
| 0 | 0 | 0 | 0 | 0.004098831 |
| 0 | 0 | 0 | 0 | 0 |

| | | | | |
|---|---|---|---|---|
| 0 | 0 | 0 | 0 | 0 |
| 0 | 0.005586171 | 0.009068443 | 0 | 0 |
| 0 | 0.001398265 | 0.001203739 | 0.001755774 | 0.006239655 |
| 0 | 0 | 0 | 0 | 0 |
| 0.001993209 | 0.02363147 | 0.000768902 | 0.027687543 | 0 |
| 0 | 0 | 0 | 0 | 0 |
| 0 | 0 | 0 | 0 | 0 |
| 0 | 0 | 0 | 0 | 0 |
| 0 | 0 | 0 | 0 | 0 |
| 0 | 0 | 0 | 0 | 0 |
| 0 | 0 | 0 | 0 | 0 |
| 0 | 0 | 0 | 0 | 0 |
| 0 | 0 | 0 | 0 | 0 |
| 0 | 0 | 0 | 0 | 0 |
| 0 | 0 | 0 | 0 | 0 |
| 0 | 0 | 0 | 0 | 0 |
| 0.000525594 | 1.088882613 | 0 | 0 | 0 |
| 0 | 0 | 0 | 0 | 0 |
| 0 | 0 | 0 | 0 | 0 |
| 0 | 0 | 0 | 0 | 0 |
| 0 | 0 | 0 | 0 | 0 |
| 0 | 0 | 0 | 0 | 0 |
| 0 | 0 | 0 | 0 | 0 |
| 0 | 0 | 0 | 0 | 0 |
| 0.001085817 | 0.004033285 | 0 | 0 | 0.002493262 |
| 0 | 0 | 0 | 0 | 0.002978443 |
| 0 | 0 | 0 | 0 | 0 |
| 0.004935389 | 0.094271995 | 0 | 0 | 0.360388907 |
| 0 | 0 | 0 | 0 | 0 |
| 0 | 0 | 0 | 0 | 0 |
| 0.001112731 | 0 | 0 | 0 | 0 |
| 0.00010083 | 0 | 0 | 0 | 0.002474449 |
| 0 | 0 | 0 | 0 | 0 |
| 0 | 0 | 0 | 0 | 0 |
| 0 | 0 | 0 | 0 | 0 |
| 0 | 0 | 0 | 0 | 0 |
| 0 | 0 | 0 | 0 | 0 |
| 0 | 0 | 0 | 0 | 0 |
| 0.170342355 | 0.094024366 | 0.006342603 | 0.915881219 | 0 |
| 0 | 0 | 0 | 0 | 0 |
| 0 | 0 | 0 | 0 | 0 |
| 0.000623676 | 0.034374725 | 0 | 0 | 0.001360485 |
| 0.00043273 | 0 | 0 | 0.002191359 | 0.004719778 |
| 0 | 0 | 0 | 0 | 0 |
| 0 | 0 | 0 | 0 | 0 |
| 0 | 0.000892329 | 0 | 0 | 0.003981954 |
| 0 | 0.026400847 | 0.002451555 | 0.000838088 | 0.007461015 |
| 0 | 0 | 0 | 0 | 0 |

| | | | | |
|---|---|---|---|---|
| 0 | 0 | 0 | 0 | 0 |
| 0.001030218 | 0.047162242 | 0.001933383 | 0.002467529 | 0.130739075 |
| 0 | 0 | 0.002750934 | 0 | 0 |
| 0 | 0 | 0 | 0 | 0 |
| 0 | 0 | 0 | 0 | 0 |
| 0 | 0 | 0 | 0 | 0 |
| 0 | 0 | 0 | 0 | 0 |
| 0 | 0 | 0 | 0 | 0 |
| 0 | 0 | 0 | 0 | 0 |
| 0 | 0 | 0 | 0 | 0 |
| 0 | 0 | 0 | 0 | 0 |
| 0 | 0 | 0 | 0 | 0 |
| 0.241205755 | 0.132352276 | 0 | 0 | 0.339006141 |
| 0 | 0 | 0 | 0 | 0 |
| 0.00260377 | 0.060904358 | 0.001928509 | 0.010328709 | 0.007951801 |
| 0 | 0 | 0 | 0 | 0 |
| 0 | 0 | 0 | 0 | 0 |
| 0 | 0 | 0 | 0 | 0 |
| 0 | 0 | 0 | 0.047163572 | 0 |
| 0 | 0 | 0 | 0 | 0 |
| 0 | 0 | 0 | 0 | 0 |
| 0 | 0.009395894 | 0 | 0.001294929 | 0 |
| 0.000419185 | 0.050011538 | 0 | 0.00597028 | 0.042862893 |
| 0 | 0 | 0 | 0 | 0 |
| 0 | 0 | 0 | 0 | 0 |
| 0 | 0 | 0 | 0 | 0 |
| 0 | 0 | 0 | 0 | 0 |
| 0 | 0 | 0 | 0 | 0 |
| 0 | 0 | 0 | 0 | 0 |
| 0 | 0.00088749 | 0 | 0 | 0 |
| 0 | 0 | 0 | 0 | 0 |
| 0 | 0 | 0 | 0 | 0 |
| 0 | 0 | 0 | 0 | 0 |
| 0 | 0 | 0 | 0 | 0 |
| 0.093828081 | 1.141066434 | 0 | 0 | 12.44957198 |
| 0 | 0 | 0 | 0 | 0 |
| 0 | 0 | 0 | 0 | 0 |
| 0 | 0 | 0 | 0 | 0 |
| 0 | 0.048287798 | 0 | 0 | 0 |
| 0 | 0 | 0 | 0 | 0 |
| 0.233755247 | 0.051914638 | 0.111077087 | 0.000579737 | 0.124531512 |
| 0 | 0 | 0 | 0 | 0 |
| 0 | 0 | 0 | 0 | 0 |
| 0 | 0 | 0 | 0 | 0 |
| 0 | 0 | 0 | 0 | 0 |
| 0.006641038 | 0.070480704 | 0.001698912 | 0.001770024 | 0.012580598 |
| 0 | 0 | 0 | 0 | 0 |
| 0 | 0 | 0 | 0 | 0 |

| | | | | |
|---|---|---|---|---|
| 0 | 0 | 0 | 0 | 0 |
| 0 | 0 | 0 | 0 | 0 |
| 0 | 0 | 0 | 0.015428106 | 0 |
| 0 | 0 | 0 | 0 | 0 |
| 0 | 0 | 0 | 0 | 0 |
| 0 | 0 | 0 | 0 | 0 |
| 0 | 0 | 0 | 0 | 0 |
| 0 | 0 | 0 | 0 | 0 |
| 0 | 0 | 0 | 0.013939188 | 0 |
| 0 | 0.002527442 | 0.008545056 | 0.002813016 | 0 |
| 0.003893598 | 0.035621423 | 0.04856457 | 0.018484992 | 0 |
| 0 | 0 | 0 | 0 | 0 |
| 0.000200471 | 0.00269494 | 0 | 0 | 0.002733175 |
| 0 | 0 | 0 | 0 | 0 |
| 0 | 0 | 0 | 0 | 0 |
| 0 | 0 | 0 | 0 | 0 |
| 0 | 0 | 0 | 0 | 0 |
| 0 | 0 | 0 | 0 | 0 |
| 0 | 0 | 0 | 0 | 0 |
| 0.16124463 | 0.00339814 | 0.039234654 | 0.185048752 | 0.531143651 |
| 0 | 0 | 0 | 0 | 0 |
| 0 | 0 | 0 | 0 | 0 |
| 0 | 0 | 0 | 0 | 0 |
| 0 | 0 | 0 | 0 | 0 |
| 0 | 0 | 0 | 0 | 0 |
| 0 | 0.04614755 | 0 | 0 | 0 |
| 0 | 0 | 0 | 0 | 0 |
| 0 | 0 | 0 | 0 | 0 |
| 0 | 0 | 0 | 0 | 0 |
| 0 | 0 | 0 | 0.001006206 | 0 |
| 0 | 0 | 0 | 0 | 0 |
| 0 | 0 | 0 | 0 | 0 |
| 0 | 0 | 0 | 0 | 0 |
| 0 | 0 | 0 | 0 | 0 |
| 0 | 0 | 0 | 0 | 0 |
| 0.000346468 | 0 | 0 | 0 | 0 |
| 0 | 0 | 0 | 0 | 0 |
| 0 | 0 | 0 | 0 | 0 |
| 0 | 0 | 0 | 0 | 0 |
| 0 | 0 | 0 | 0 | 0 |
| 0 | 0 | 0 | 0 | 0 |
| 0 | 0 | 0 | 0 | 0 |
| 0 | 0.02708358 | 0 | 0 | 0 |
| 0 | 0 | 0 | 0 | 0 |
| 0 | 0.027598453 | 0 | 0.001768104 | 0.002589552 |
| 0 | 0 | 0 | 0 | 0 |
| 0 | 0 | 0 | 0 | 0 |
| 0 | 0 | 0 | 0 | 0 |

| | | | | |
|---|---|---|---|---|
| 0 | 0 | 0 | 0 | 0 |
| 0 | 0 | 0 | 0 | 0 |
| 0.00199839 | 0 | 0 | 0 | 0 |
| 0 | 0 | 0 | 0 | 0 |
| 0 | 0 | 0 | 0 | 0 |
| 0.000375341 | 0.042047589 | 0 | 0.000527983 | 0.011485491 |
| 0 | 0.054577803 | 0 | 0 | 0 |
| 0 | 0 | 0 | 0 | 0 |
| 0 | 0 | 0 | 0 | 0 |
| 0 | 0 | 0 | 0 | 0 |
| 0 | 0 | 0 | 0 | 0 |
| 0 | 0 | 0 | 0 | 0 |
| 0 | 0 | 0 | 0 | 0 |
| 0 | 0 | 0 | 0 | 0 |
| 0 | 0 | 0 | 0 | 0 |
| 0 | 0 | 0 | 0 | 0 |
| 0.003642286 | 0.014790995 | 0 | 0 | 0 |
| 0 | 0 | 0 | 0 | 0 |
| 0 | 0 | 0 | 0 | 0 |
| 0 | 0 | 0 | 0 | 0 |
| 0 | 0 | 0 | 0 | 0 |
| 0 | 0 | 0 | 0 | 0 |
| 0 | 0 | 0 | 0 | 0 |
| 0 | 0 | 0 | 0 | 0 |
| 0 | 0 | 0 | 0 | 0 |
| 0 | 0 | 0 | 0 | 0 |
| 0 | 0 | 0 | 0 | 0 |
| 0.001415847 | 0.010008863 | 0 | 0 | 0 |
| 0 | 0 | 0 | 0 | 0 |
| 0 | 0 | 0 | 0 | 0 |
| 0 | 0 | 0 | 0 | 0 |
| 0 | 0 | 0 | 0 | 0 |
| 0 | 0 | 0 | 0 | 0 |
| 0 | 0 | 0 | 0 | 0 |
| 0 | 0 | 0 | 0 | 0 |
| 0 | 0 | 0 | 0 | 0 |
| 0 | 0 | 0 | 0 | 0 |
| 0 | 0 | 0 | 0.012245216 | 0 |
| 0.260292314 | 0.018004029 | 0.285323546 | 0 | 0 |
| 0 | 0 | 0 | 0 | 0 |
| 0 | 0 | 0 | 0 | 0 |
| 0 | 0 | 0 | 0 | 0 |
| 0 | 0 | 0 | 0 | 0 |
| 0 | 0 | 0 | 0 | 0 |
| 0 | 0.01576084 | 0 | 0 | 0 |
| 0 | 0 | 0 | 0 | 0 |
| 0 | 0 | 0 | 0 | 0 |
| 0 | 0 | 0 | 0 | 0 |

| | | | | |
|---|---|---|---|---|
| 0 | 0 | 0 | 0 | 0 |
| 0.001237327 | 0.044721881 | 0 | 0.023497012 | 0 |
| 0 | 0 | 0 | 0 | 0 |
| 0 | 0 | 0 | 0 | 0 |
| 0 | 0 | 0 | 0 | 0 |
| 0 | 0 | 0 | 0 | 0 |
| 0 | 0 | 0 | 0 | 0 |
| 0 | 0 | 0 | 0 | 0 |
| 0 | 0 | 0 | 0 | 0 |
| 0 | 0 | 0 | 0 | 0 |
| 0 | 0.001232837 | 0 | 0 | 0.00383434 |
| 0 | 0 | 0 | 0 | 0 |
| 0 | 0.020582394 | 0 | 0.277357641 | 0 |
| 0 | 0 | 0 | 0 | 0 |
| 0 | 0 | 0 | 0 | 0 |
| 0 | 0 | 0 | 0 | 0 |
| 0 | 0 | 0 | 0 | 0 |
| 0 | 0 | 0 | 0 | 0 |
| 0 | 0 | 0 | 0 | 0 |
| 0 | 0.000683907 | 0 | 0 | 0 |
| 0 | 0.001876883 | 0 | 0 | 0 |
| 0 | 0.004054744 | 0 | 0 | 0 |
| 0 | 0 | 0 | 0 | 0 |
| 0.073070967 | 1.288764048 | 1.24748591 | 0.513140843 | 3.626937599 |
| 0 | 0 | 0 | 0 | 0 |
| 0 | 0 | 0 | 0 | 0 |
| 0 | 0 | 0 | 0 | 0 |
| 0 | 0 | 0 | 0 | 0 |
| 0 | 0 | 0 | 0 | 0 |
| 0 | 0 | 0 | 0 | 0 |
| 0 | 0.005321795 | 0 | 0 | 0 |
| 0 | 0 | 0 | 0 | 0 |
| 0 | 0 | 0 | 0 | 0.00160185 |
| 0 | 0 | 0 | 0 | 0 |

| PHL018-0173_V3 | PHL018-0057_V1 | PHL018-0167_V3 | PHL018-0150_V3 | PHL018-0048_V2 |
|---|---|---|---|---|
| CTRL | REF | REF | EYCF | CTRL |
| 0.010777332 | 0.806385066 | 0.011858751 | 0.008062876 | 0.015920155 |
| 0.070112119 | 1.767144758 | 0.038418427 | 4.368549792 | 0 |
| 0 | 0 | 21.62912553 | 3.719217998 | 8.12220071 |
| 0 | 0 | 0 | 0 | 0 |
| 0 | 0 | 0 | 0 | 0 |
| 0 | 0 | 0 | 0 | 0 |
| 0 | 0 | 0 | 0 | 0 |
| 0 | 0 | 0 | 0 | 0 |
| 0 | 0 | 0 | 0 | 0 |
| 0 | 0 | 0 | 0 | 0 |
| 0 | 0 | 0 | 0 | 0 |
| 0 | 0 | 0 | 0 | 0 |
| 0 | 0 | 0 | 0 | 0 |
| 2.230771679 | 0.023588075 | 2.118195686 | 14.5151646 | 0 |
| 0 | 0 | 0 | 0 | 0 |
| 0 | 0 | 0 | 0 | 0 |
| 0 | 0 | 0 | 0.001551555 | 0 |
| 0 | 0 | 0 | 0 | 0 |
| 0 | 0 | 0 | 0 | 0 |
| 0 | 0 | 0 | 0 | 0 |
| 0 | 0 | 0 | 0 | 0 |
| 0 | 0 | 0 | 0 | 0 |
| 0 | 0 | 0 | 0 | 0 |
| 0 | 0 | 0 | 0 | 0 |
| 0 | 0 | 0 | 0 | 0 |
| 0 | 0 | 0 | 0 | 0 |
| 0 | 0 | 0 | 0 | 0 |
| 0 | 0 | 0 | 0 | 0 |
| 0 | 0 | 0 | 0 | 0 |
| 0 | 0 | 0 | 0 | 0 |
| 0 | 0 | 0 | 0 | 0 |
| 0 | 0 | 0 | 0 | 0 |
| 0 | 0 | 0 | 0 | 0 |
| 0 | 0 | 0 | 0 | 0 |
| 0 | 0 | 0 | 0 | 0 |
| 1.017524946 | 0 | 0 | 0 | 0 |
| 0 | 0 | 0 | 0 | 0 |
| 0 | 0 | 0 | 0 | 0 |
| 0 | 0 | 0 | 0 | 0 |
| 0 | 0 | 0 | 0 | 0 |
| 0 | 0.000805699 | 0 | 0 | 0 |
| 0.102858801 | 3.004130976 | 0.006443988 | 0.008572163 | 0.000950652 |
| 0.014341165 | 0.058398295 | 0.001746298 | 0.125030437 | 0.016410613 |
| 0.002618989 | 0.017482133 | 0.082079357 | 0.001561802 | 0 |
| 0.019385503 | 0 | 0 | 0 | 0 |
| 0 | 0 | 0 | 0 | 0 |

| | | | | |
|---|---|---|---|---|
| 0.002773432 | 0.073916144 | 0 | 0 | 0 |
| 0 | 0.084284332 | 0.626357203 | 0.053812337 | 0.023047226 |
| 0.010950521 | 0.035309246 | 0.019857304 | 0 | 0 |
| 0.106001007 | 0.257405247 | 0.31641909 | 0 | 0 |
| 0 | 0 | 0 | 0 | 0 |
| 0 | 0 | 0 | 0 | 0 |
| 0.00649616 | 10.29084009 | 0.130266572 | 0.004519556 | 0 |
| 0 | 0 | 0 | 0 | 0 |
| 0 | 0 | 0 | 0 | 0 |
| 0 | 0 | 0 | 0 | 0 |
| 0 | 0 | 0.050886068 | 0 | 0 |
| 0.016470259 | 1.31355884 | 0 | 0 | 0 |
| 0 | 0 | 0 | 0.000559333 | 0 |
| 0 | 0.233560132 | 0.021228307 | 0.001541316 | 0.033690666 |
| 0.000709228 | 0 | 0 | 0 | 0 |
| 0 | 0 | 0 | 0 | 0 |
| 0.051259198 | 0 | 0.091645017 | 0.09106949 | 0 |
| 0 | 0 | 0 | 0 | 0 |
| 0 | 0 | 0 | 0 | 0 |
| 0 | 0 | 0 | 0 | 0 |
| 0 | 0.630562085 | 0.187521833 | 0.450171713 | 0.058359071 |
| 0 | 0.025247665 | 0.014410334 | 0.00122335 | 0.005494611 |
| 0 | 0 | 0.33983472 | 0 | 0 |
| 0 | 0 | 0 | 0 | 0 |
| 0 | 0 | 0.004284823 | 0 | 0 |
| 0 | 0 | 0 | 0 | 0 |
| 0 | 0 | 0 | 0 | 0 |
| 0 | 0 | 0 | 0 | 0 |
| 0.004592669 | 0.049543482 | 0.265180173 | 0.017410828 | 0 |
| 0 | 0 | 0 | 0 | 0 |
| 0 | 0 | 0 | 1.452034394 | 0.075230838 |
| 0 | 0 | 0 | 0 | 0 |
| 0 | 0 | 0 | 0 | 0 |
| 0 | 0 | 0 | 0 | 0 |
| 0 | 0 | 0 | 0 | 0 |
| 0 | 0 | 0 | 0 | 0 |
| 0 | 0 | 0 | 0 | 0 |
| 0.001865688 | 0 | 0 | 6.921361648 | 0 |
| 0 | 0 | 0 | 0 | 0 |
| 0 | 0 | 0 | 0 | 0 |
| 0 | 0 | 0 | 0 | 0 |
| 0 | 0 | 0 | 0 | 0 |
| 0 | 0 | 0 | 0 | 0 |
| 0 | 0 | 0 | 0 | 0 |
| 0 | 0 | 0 | 0 | 0 |
| 0 | 0 | 0 | 0 | 0 |
| 0.003381702 | 0 | 0 | 0 | 0 |
| 0 | 0 | 0 | 0 | 0 |

| | | | | |
|---|---|---|---|---|
| 0 | 0 | 0 | 0 | 0 |
| 0 | 0 | 0 | 0.012847326 | 0 |
| 0 | 0 | 0 | 0 | 0 |
| 0 | 0 | 0 | 0 | 0 |
| 0 | 0 | 0.001051456 | 0 | 0 |
| 0 | 0 | 0.001589554 | 0 | 0 |
| 0 | 0 | 0 | 0 | 0 |
| 0 | 0 | 0 | 0.002369305 | 0 |
| 0 | 0 | 0 | 0.001625887 | 0 |
| 0 | 0 | 0 | 0 | 0 |
| 0 | 0 | 0 | 0 | 0 |
| 0.00115104 | 0.538795506 | 0 | 0.018361424 | 0 |
| 0 | 0 | 0 | 0 | 0 |
| 0 | 0 | 0 | 0 | 0 |
| 0 | 0 | 0 | 0 | 0 |
| 0 | 0 | 0 | 0 | 0 |
| 0 | 0 | 0 | 0 | 0 |
| 0 | 0 | 0 | 0 | 0 |
| 0 | 0 | 0 | 0 | 0 |
| 0 | 0 | 0 | 0 | 0 |
| 0 | 0 | 0 | 0 | 0 |
| 0 | 0 | 0 | 0 | 0 |
| 0 | 0 | 0 | 0 | 0 |
| 0 | 0 | 0 | 0 | 0 |
| 0 | 0 | 0 | 0 | 0 |
| 0 | 0 | 0 | 0 | 0 |
| 0 | 0 | 0 | 0 | 0 |
| 0 | 0 | 0 | 0 | 0 |
| 0 | 0 | 0 | 0 | 0 |
| 0 | 0 | 0 | 0 | 0 |
| 0.001681987 | 0 | 0 | 0 | 0 |
| 0.001443724 | 0 | 0 | 0 | 0 |
| 0 | 0 | 0 | 0 | 0 |
| 0 | 0 | 0 | 0 | 0 |
| 0 | 0 | 0 | 0 | 0 |
| 0 | 0 | 0 | 0 | 0 |
| 0 | 0 | 0 | 0 | 0 |
| 0 | 0 | 0 | 0 | 0 |
| 0 | 0 | 0 | 0 | 0 |
| 0 | 0 | 0 | 0 | 0 |
| 0 | 0 | 0 | 0 | 0 |
| 0 | 0 | 0 | 0 | 0 |
| 0 | 0 | 0.000801498 | 0 | 0 |
| 0 | 0 | 0 | 0 | 0 |
| 0 | 0 | 0 | 0 | 0 |
| 0 | 0 | 0 | 0 | 0 |
| 0 | 0 | 0 | 0 | 0 |
| 0 | 0 | 0 | 0 | 0 |

| | | | | |
|---|---|---|---|---|
| 0 | 0 | 0 | 0 | 0 |
| 0 | 0 | 0 | 0 | 0 |
| 0 | 0 | 0 | 0 | 0 |
| 0 | 0 | 0 | 0 | 0 |
| 0.00069811 | 0.009261724 | 0.033497251 | 0 | 0 |
| 0 | 0 | 0 | 0 | 0 |
| 0 | 0 | 0 | 0 | 0 |
| 0 | 0 | 0 | 0 | 0 |
| 0 | 0 | 0 | 0 | 0 |
| 0 | 0 | 0 | 0 | 0 |
| 0 | 0 | 0 | 0 | 0 |
| 0 | 0 | 0 | 0 | 0 |
| 0 | 0 | 0 | 0 | 0 |
| 0 | 0 | 0 | 0 | 0 |
| 0 | 0 | 0 | 0 | 0 |
| 0 | 0 | 0 | 0.004060966 | 0 |
| 0 | 0 | 0 | 0 | 0 |
| 0 | 0 | 0 | 0 | 0 |
| 0 | 0 | 0 | 0 | 0 |
| 0 | 0 | 0 | 0 | 0 |
| 0 | 0.002504311 | 0 | 0 | 0 |
| 0 | 0 | 0 | 0 | 0 |
| 0 | 0 | 0 | 0 | 0 |
| 0 | 0 | 0 | 0 | 0 |
| 0 | 0.10358466 | 0 | 0 | 0 |
| 0 | 0 | 0 | 0 | 0 |
| 0 | 0 | 0 | 0 | 0 |
| 0 | 0 | 0 | 0 | 0 |
| 0 | 0 | 0 | 0 | 0 |
| 0 | 0 | 0 | 0 | 0 |
| 0 | 0 | 0 | 0 | 0 |
| 0 | 0 | 0 | 0.023178095 | 0 |
| 0 | 0 | 0 | 0 | 0 |
| 0 | 0 | 0 | 0 | 0 |
| 0 | 0 | 0 | 0 | 0 |
| 0 | 0 | 0 | 0 | 0 |
| 0 | 0 | 0 | 0 | 0 |
| 0 | 0 | 0 | 0 | 0 |
| 0 | 0 | 0 | 0 | 0 |
| 0 | 0 | 0 | 0 | 0 |
| 0 | 0 | 0 | 0 | 0 |
| 0 | 0 | 0 | 0 | 0 |
| 0 | 0 | 0 | 0 | 0 |
| 0 | 0 | 0 | 0 | 0 |
| 0 | 0 | 0 | 0 | 0 |
| 0 | 0 | 0 | 0 | 0 |
| 0 | 0 | 0 | 0 | 0 |
| 0 | 0 | 0 | 0 | 0 |

| | | | | |
|---|---|---|---|---|
| 0 | 0 | 0 | 0 | 0 |
| 0 | 0 | 0 | 0 | 0 |
| 0 | 0 | 0 | 0 | 0 |
| 0 | 0 | 0 | 0 | 0 |
| 0 | 0 | 0 | 0 | 0 |
| 0 | 0 | 0 | 0 | 0 |
| 0 | 0 | 0 | 0 | 0 |
| 0 | 0 | 0 | 0 | 0 |
| 0 | 0 | 0 | 0 | 0 |
| 0 | 0 | 0 | 0 | 0 |
| 0 | 0 | 0 | 0 | 0 |
| 0 | 0 | 0 | 0 | 0 |
| 0 | 0.005763271 | 0.001531179 | 0 | 0 |
| 0 | 0 | 0 | 0 | 0 |
| 0 | 0 | 0 | 0 | 0 |
| 0 | 0 | 0 | 0 | 0 |
| 0 | 0 | 0 | 0 | 0 |
| 0.002981938 | 0 | 0 | 0 | 0 |
| 0 | 0 | 0 | 0 | 0 |
| 0 | 0 | 0 | 0 | 0 |
| 0.009603231 | 0.041031431 | 0.336752895 | 0.078792445 | 0.083077422 |
| 0 | 0.001508496 | 0 | 0 | 0 |
| 0 | 0 | 0 | 0 | 0 |
| 0 | 0 | 0 | 0 | 0 |
| 0 | 0 | 0 | 0 | 0 |
| 0 | 0 | 0 | 0 | 0 |
| 0.053612735 | 0 | 0.00187572 | 0.005865238 | 0.002066149 |
| 0 | 0 | 0 | 0 | 0 |
| 0 | 0 | 0 | 0 | 0 |
| 0 | 0 | 0 | 0 | 0 |
| 0 | 0 | 0.175135532 | 0.565405102 | 0 |
| 0 | 0 | 0 | 0 | 0 |
| 0 | 0 | 0 | 0 | 0 |
| 0 | 0 | 0 | 0 | 0 |
| 0.000861845 | 0.005393381 | 0.031900091 | 0.000578194 | 0.013273193 |
| 0 | 0 | 0 | 0 | 0 |
| 0 | 0 | 0 | 0 | 0 |
| 0 | 0 | 0 | 0 | 0 |
| 0 | 0 | 0 | 0.013263382 | 0 |
| 0 | 0 | 0 | 0 | 0 |
| 0 | 0 | 0 | 0 | 0 |
| 0 | 0 | 0 | 0 | 0 |
| 0 | 0.061944252 | 0 | 0 | 0 |
| 0 | 0 | 0 | 0 | 0 |
| 0 | 0 | 0 | 0 | 0 |
| 0 | 0 | 0 | 0 | 0 |
| 0 | 0.004087718 | 0 | 0 | 0 |
| 0.000614174 | 0.01056954 | 0 | 0 | 0 |

| | | | | |
|---|---|---|---|---|
| 0 | 0 | 0 | 0 | 0 |
| 0 | 0 | 0 | 0 | 0 |
| 0 | 0 | 0 | 0 | 0 |
| 0 | 0 | 0 | 0 | 0 |
| 0 | 0 | 0 | 0 | 0 |
| 0 | 0 | 0 | 0 | 0 |
| 0 | 0.013404268 | 0 | 0.009188959 | 0 |
| 0.002427937 | 0.077903307 | 0 | 0 | 0 |
| 0 | 0.194268134 | 0 | 0 | 0 |
| 0 | 0 | 0 | 0 | 0 |
| 0 | 0.002377672 | 0 | 0 | 0 |
| 0 | 0 | 0 | 0 | 0 |
| 0 | 0 | 0 | 0 | 0 |
| 0 | 0 | 0 | 0 | 0 |
| 0 | 0 | 0.01543569 | 0 | 0 |
| 0 | 0 | 0 | 0 | 0 |
| 0 | 0 | 0 | 0 | 0 |
| 0 | 0 | 0 | 0 | 0 |
| 0 | 0 | 0 | 0.000838078 | 0 |
| 0 | 0 | 0 | 0 | 0 |
| 0 | 0.100037539 | 0.027879937 | 0.102196652 | 0.009665551 |
| 0 | 0 | 0 | 0 | 0 |
| 0 | 0 | 0 | 0 | 0 |
| 0 | 0 | 0 | 0 | 0 |
| 0.005949077 | 0.005154788 | 0.049454811 | 0.018011192 | 0.021245496 |
| 0 | 0 | 0 | 0 | 0 |
| 0 | 0 | 0 | 0 | 0 |
| 0 | 0 | 0 | 0 | 0 |
| 0.014848298 | 0.337517889 | 0.009675066 | 0 | 0.000779803 |
| 0 | 0 | 0 | 0 | 0 |
| 0.002032437 | 0.710773786 | 0.029285866 | 0.008635633 | 0.061997893 |
| 0 | 0 | 0 | 0 | 0 |
| 0 | 0 | 0 | 0 | 0 |
| 0 | 0 | 0 | 0 | 0 |
| 0 | 0 | 0 | 0 | 0 |
| 0 | 0 | 0 | 0 | 0 |
| 0 | 0 | 0 | 0 | 0 |
| 0 | 0 | 0 | 0 | 0 |
| 0 | 0 | 0 | 0 | 0 |
| 0.000676707 | 0 | 0 | 0 | 0 |
| 0 | 0 | 0 | 0 | 0 |
| 0.144503249 | 0.032411975 | 0.030971524 | 0 | 0 |
| 0 | 0 | 0 | 0 | 0 |
| 0 | 0 | 0 | 0 | 0 |
| 0 | 0 | 0 | 0 | 0 |
| 0 | 0 | 0 | 0 | 0 |
| 0 | 0 | 0 | 0 | 0 |
| 0 | 0 | 0 | 0 | 0 |

| | | | | |
|---|---|---|---|---|
| 0 | 0 | 0 | 0 | 0 |
| 0 | 0 | 0 | 0 | 0 |
| 0 | 0.018450018 | 0.064660223 | 0 | 0 |
| 0 | 0 | 0 | 0 | 0 |
| 0 | 0 | 0 | 0 | 0 |
| 0 | 0 | 0 | 0 | 0 |
| 0 | 0 | 0 | 0 | 0 |
| 0 | 0 | 0 | 0 | 0 |
| 0 | 0 | 0 | 0 | 0 |
| 0 | 0 | 0 | 0.001241466 | 0 |
| 0 | 0 | 0 | 0 | 0 |
| 0 | 0 | 0 | 0 | 0 |
| 0.00377409 | 0 | 0 | 0 | 0 |
| 0 | 0 | 0.044533805 | 0.006214992 | 0.000641708 |
| 0.001977458 | 0.001237483 | 0.027140214 | 0.018572915 | 0.012181861 |
| 0 | 0 | 0 | 0 | 0 |
| 0.002703763 | 0.016356013 | 0.011687751 | 0.00806178 | 0.004425551 |
| 0 | 0 | 0.002833245 | 0 | 0 |
| 0 | 0 | 0 | 0 | 0 |
| 0 | 0 | 0 | 0 | 0 |
| 0 | 0 | 0 | 0 | 0 |
| 0.000610182 | 0.006364143 | 0 | 0 | 0.000907957 |
| 0 | 0 | 0 | 0 | 0 |
| 0 | 0 | 0 | 0 | 0 |
| 0 | 0 | 0 | 0 | 0 |
| 0 | 0 | 0 | 0 | 0 |
| 0 | 0 | 0 | 0 | 0 |
| 0 | 0 | 0 | 0 | 0 |
| 0 | 0 | 0 | 0 | 0 |
| 0 | 0 | 0 | 0 | 0 |
| 0 | 0 | 0 | 0 | 0 |
| 0 | 0 | 0 | 0 | 0 |
| 0 | 0.042147149 | 0 | 0 | 0 |
| 0.010084036 | 0 | 0.012164927 | 0 | 0 |
| 0 | 0 | 0 | 0 | 0 |
| 0 | 0.011694554 | 0 | 0.032342031 | 0.012694602 |
| 0 | 0 | 0 | 0 | 0 |
| 0.05596399 | 0.015671939 | 0.006627933 | 0.000914341 | 0 |
| 0 | 0 | 0 | 0 | 0 |
| 0 | 0 | 0 | 0 | 0 |
| 0 | 0 | 0 | 0 | 0 |
| 0 | 0 | 0 | 0 | 0 |
| 0 | 0 | 0 | 0 | 0 |
| 0 | 0 | 0 | 0.003123324 | 0 |
| 0 | 0 | 0 | 0 | 0 |
| 0 | 0 | 0 | 0 | 0 |
| 0 | 0 | 0 | 0 | 0 |
| 0 | 0 | 0.012376638 | 0.001661244 | 0.044215643 |

| | | | | |
|---|---|---|---|---|
| 0 | 0 | 0 | 0 | 0 |
| 0 | 0 | 0 | 0 | 0 |
| 0 | 0 | 0.00207207 | 0 | 0 |
| 0 | 0 | 0 | 0 | 0 |
| 0.013717632 | 0 | 0.018299052 | 0.019747856 | 0.046309687 |
| 0 | 0 | 0 | 0 | 0 |
| 0 | 0 | 0 | 0 | 0 |
| 0.02403535 | 0 | 0.111042418 | 0.076861713 | 0.003487074 |
| 0 | 0 | 0 | 0 | 0 |
| 0.103740105 | 0.001162747 | 0.102071394 | 0.356226644 | 0.528624241 |
| 0 | 0.001752123 | 0 | 0 | 0 |
| 0.008561911 | 0.508414788 | 0 | 0.010849805 | 0.13207357 |
| 0.003608063 | 0.013999029 | 0.034118074 | 0.027433209 | 0.001288521 |
| 0 | 0.074756117 | 0 | 0 | 0 |
| 0 | 0 | 0 | 0 | 0 |
| 0 | 0.008473736 | 0 | 0 | 0 |
| 0 | 0 | 0 | 0 | 0 |
| 0 | 0 | 0 | 0 | 0 |
| 0 | 0 | 0 | 5.47923607 | 0.060357102 |
| 0 | 0 | 0 | 0 | 0 |
| 0 | 0 | 0.158097918 | 0 | 0 |
| 0 | 0 | 0 | 0 | 0 |
| 0 | 0.075254776 | 0 | 0 | 0 |
| 0 | 0 | 0 | 0 | 0 |
| 0 | 0 | 0 | 0 | 0 |
| 0.012607421 | 0 | 0 | 0.000751828 | 0 |
| 0 | 0 | 0.011538735 | 0 | 0 |
| 0 | 0 | 0 | 0.102736114 | 0 |
| 0 | 0 | 0 | 0 | 0 |
| 0 | 0 | 0.033409935 | 0 | 0 |
| 0 | 0 | 0 | 0 | 0 |
| 0 | 0 | 0 | 0 | 0 |
| 0 | 0 | 0 | 0 | 0 |
| 0 | 0 | 0 | 0 | 0 |
| 0 | 0 | 0 | 0 | 0 |
| 0 | 0 | 0 | 0.187739491 | 0 |
| 0 | 0 | 0 | 0 | 0.126747485 |
| 0 | 0 | 0.207452288 | 0 | 0 |
| 0 | 0 | 0 | 0 | 0 |
| 0 | 0 | 0 | 0 | 0.105756523 |
| 0 | 0 | 0 | 0 | 0 |
| 0 | 0 | 0 | 0 | 0 |
| 0 | 0 | 0 | 0 | 0 |
| 0 | 0.007415554 | 0.083056122 | 0 | 0 |
| 0 | 0 | 0 | 0 | 0 |
| 0 | 0 | 0 | 0 | 0 |
| 0 | 0.002943425 | 0.003388691 | 0 | 0.003499427 |
| 0 | 0 | 0 | 0 | 0 |

| | | | | |
|---|---|---|---|---|
| 0 | 0 | 0 | 0 | 0 |
| 0.003504325 | 0.082968081 | 0.026509694 | 0 | 0 |
| 0 | 0 | 0 | 0 | 0 |
| 0 | 0 | 0 | 0 | 0 |
| 0 | 0.00161626 | 0 | 0.002079242 | 0 |
| 0 | 0 | 0 | 0 | 0 |
| 0 | 0.021768311 | 0 | 0 | 0 |
| 0.039928393 | 0 | 0 | 0 | 0 |
| 0 | 0 | 0 | 0 | 0 |
| 0 | 0 | 0 | 0 | 0 |
| 0.035805682 | 0 | 0.001547798 | 0.504767891 | 0.04411521 |
| 0 | 0.007280166 | 0 | 0 | 0 |
| 0 | 0 | 0 | 0 | 0 |
| 0.003382741 | 0 | 0.01949709 | 0.008674201 | 0.004429528 |
| 0 | 0 | 0.028358354 | 0.646824771 | 0 |
| 0 | 0 | 0 | 0 | 0 |
| 0 | 0 | 0 | 0 | 0 |
| 0 | 0 | 0 | 0 | 0 |
| 0 | 0.00751039 | 0 | 0 | 0.03214467 |
| 0 | 0.013709172 | 0 | 0 | 0 |
| 0 | 0 | 0 | 0 | 0 |
| 1.183317418 | 0.443574504 | 0 | 0 | 0 |
| 0 | 0 | 0 | 0 | 0 |
| 0 | 0 | 0 | 0 | 0 |
| 0 | 0 | 0 | 0 | 0 |
| 0 | 0 | 0 | 0 | 0 |
| 0 | 0 | 0 | 0 | 0.001309849 |
| 0 | 0 | 0 | 0 | 0 |
| 0 | 0 | 0 | 0 | 0 |
| 0.413199131 | 1.198146795 | 1.681139066 | 0.71438096 | 0.547457753 |
| 0.093173784 | 0.001340407 | 0 | 0 | 0 |
| 0 | 0.044896496 | 0 | 0 | 0 |
| 0 | 0 | 0 | 0 | 0 |
| 0 | 0 | 0 | 0 | 0 |
| 0 | 0 | 0 | 0.006740529 | 0 |
| 0 | 0 | 0 | 0 | 0 |
| 0 | 0 | 0.002350297 | 0 | 0 |
| 0 | 0.449662057 | 0 | 0.001644897 | 0 |
| 0 | 0 | 0 | 0 | 0 |
| 0 | 0 | 0 | 2.812997198 | 0 |
| 0 | 0.012762376 | 0.310756986 | 0.040465057 | 0 |
| 0.034281909 | 0 | 0 | 0 | 6.68103364 |
| 0 | 0.389781825 | 0 | 0.001318478 | 0 |
| 0 | 0 | 0 | 0 | 0 |
| 0 | 0 | 0 | 0 | 0 |
| 0 | 0 | 0 | 0 | 0 |
| 0.001556714 | 0 | 0 | 0.00121843 | 0.002432228 |
| 0 | 0 | 0 | 0 | 0 |

| | | | | |
|---|---|---|---|---|
| 0.007682897 | 0.030650479 | 0 | 0 | 0 |
| 0 | 0 | 0 | 0 | 0 |
| 0.083748281 | 0.157227768 | 0.012727432 | 0.019020978 | 0 |
| 0 | 0 | 0 | 0 | 0 |
| 0 | 0 | 0 | 0 | 0 |
| 0 | 0 | 0 | 0.15452622 | 0 |
| 0 | 0 | 0 | 0 | 0 |
| 0 | 0 | 0 | 0 | 0 |
| 0 | 0 | 0 | 0.013839749 | 0 |
| 0 | 0 | 0 | 0 | 0 |
| 0 | 0 | 0 | 0 | 0 |
| 0 | 0 | 0 | 0 | 0 |
| 0 | 0.124527013 | 0 | 0 | 0 |
| 0.001737631 | 0 | 0.005633536 | 0.081407619 | 0.012604863 |
| 0 | 0 | 0 | 0 | 0 |
| 0 | 0.456121979 | 0.030450701 | 0 | 0 |
| 0.025398934 | 0.044269198 | 0.015655782 | 0.008919513 | 0 |
| 0 | 0.005721846 | 0.001482169 | 0.00429385 | 0.012652987 |
| 0 | 0 | 0 | 0 | 0.010930704 |
| 0 | 0 | 0 | 0 | 0 |
| 0 | 0.0320506 | 0 | 0 | 0 |
| 0 | 0 | 0.005896069 | 0.000780843 | 0 |
| 0 | 0 | 0 | 0 | 0 |
| 0.132917246 | 0 | 2.398160357 | 0 | 0 |
| 0 | 0.015185081 | 0 | 0 | 0 |
| 0 | 0 | 0.08838743 | 0 | 0 |
| 0 | 0.00089681 | 0.018842667 | 0.002563787 | 0 |
| 0 | 0 | 0 | 0 | 0 |
| 0 | 0 | 0 | 0 | 0 |
| 0 | 0 | 0 | 0 | 0 |
| 0 | 0.008184259 | 0.002355583 | 0.000974876 | 0 |
| 0 | 0.025081326 | 0.029558363 | 0.001695658 | 0 |
| 5.336060308 | 0.016135684 | 0.331871067 | 0.018220737 | 0.132367222 |
| 0 | 0.688632994 | 0 | 0.001322131 | 0.001319618 |
| 0 | 0 | 0.003420252 | 0 | 0.001883744 |
| 0 | 0 | 0 | 0 | 0 |
| 0 | 0.064186192 | 0.005290278 | 0.001876648 | 0.002913682 |
| 0.000911506 | 0.060692226 | 0.057921394 | 0 | 0 |
| 0.05268072 | 0 | 0 | 0 | 0 |
| 0 | 0.007461969 | 0 | 0 | 0 |
| 0 | 0 | 0 | 0 | 0 |
| 0 | 0 | 0 | 0 | 0 |
| 0 | 0.024568004 | 0.000883891 | 0 | 0 |
| 0 | 0 | 0.041985159 | 0 | 0 |
| 0 | 0 | 0.579037871 | 0.136509754 | 0.00124316 |
| 0 | 0 | 0 | 0 | 0 |
| 0 | 0 | 0 | 0 | 0 |
| 0 | 0 | 0 | 0 | 0 |

| | | | | |
|---|---|---|---|---|
| 0 | 0 | 0 | 0 | 0 |
| 0.634435066 | 0.009948975 | 0.576369063 | 1.625859062 | 0.019073133 |
| 0 | 0 | 0 | 0 | 0 |
| 0 | 0 | 0 | 0 | 0 |
| 0 | 0 | 0 | 0 | 0 |
| 0 | 0 | 0 | 0 | 0 |
| 0 | 0 | 0.004470376 | 0.012334001 | 0 |
| 0.004006881 | 0.004059739 | 0 | 0 | 0 |
| 0.002046577 | 0.005122951 | 0 | 0 | 0 |
| 0 | 0.007681584 | 0 | 0 | 0.006124218 |
| 0 | 0.018251904 | 0.006583173 | 0 | 0.014008616 |
| 0.009623562 | 0.837626567 | 0.110538369 | 0.035171215 | 0 |
| 0 | 0 | 0 | 0 | 0 |
| 0 | 0 | 0 | 0 | 0 |
| 0.003812386 | 0 | 0 | 0 | 0 |
| 0 | 0 | 0 | 0 | 0 |
| 0.007538591 | 0.001935431 | 0.171084942 | 0.062591834 | 0.076624179 |
| 0 | 0.005024913 | 0.017897515 | 0.013691778 | 0.040549206 |
| 0 | 0 | 0 | 0 | 0 |
| 0 | 0 | 0 | 0.033959655 | 0.050042564 |
| 0.090100855 | 0.011726014 | 0.014648789 | 0.015334601 | 0 |
| 0 | 0 | 0 | 0 | 0 |
| 0 | 0 | 0 | 0 | 0 |
| 0 | 0 | 0 | 0 | 0 |
| 0 | 0.125756754 | 0 | 0 | 0 |
| 0 | 0.013727342 | 0 | 0 | 0 |
| 0 | 0.03201364 | 0.122264335 | 0 | 0 |
| 0.026907116 | 0.106333444 | 0.351916493 | 0.00644356 | 0.674720402 |
| 0 | 0 | 0 | 0 | 0 |
| 0 | 0 | 0 | 0 | 0 |
| 0.010912009 | 0.003129812 | 0 | 0.001220108 | 0 |
| 0 | 0.001098044 | 0 | 0 | 0 |
| 0 | 0 | 0 | 0.00138043 | 0 |
| 0 | 0 | 0 | 0 | 0 |
| 0 | 0 | 0 | 0 | 0 |
| 0.043745826 | 1.333341908 | 0.041463208 | 20.12099625 | 0.340944516 |
| 0 | 0 | 0 | 0 | 0 |
| 0 | 0 | 0 | 0 | 0 |
| 0.001045447 | 0.235524744 | 0 | 0.012157068 | 0.06206988 |
| 0 | 0 | 0 | 0 | 0 |
| 0 | 0 | 0.298472067 | 0.066897159 | 0 |
| 0 | 0 | 0 | 0 | 0 |
| 0 | 0.219758093 | 0.001029393 | 0 | 0.010063367 |
| 0 | 0.001151025 | 0 | 0.005758433 | 0 |
| 0 | 0 | 0 | 1.177116248 | 0 |
| 0 | 0.011892463 | 0 | 0 | 0 |
| 0 | 0 | 0 | 0 | 0 |
| 0 | 0 | 0.00535499 | 0 | 0 |

| | | | | |
|---|---|---|---|---|
| 0 | 0 | 0 | 0 | 0 |
| 0 | 0 | 0 | 0 | 0 |
| 0 | 0.002549298 | 0 | 0 | 0 |
| 0 | 0 | 0 | 0.019351072 | 0 |
| 0 | 0 | 0 | 0 | 0 |
| 0 | 0 | 0 | 0 | 0 |
| 0 | 0 | 0 | 0 | 0 |
| 0 | 0 | 0.00858048 | 0 | 0 |
| 16.78181465 | 3.364250896 | 0.305981568 | 0 | 0 |
| 0.003800857 | 0 | 0 | 0 | 0 |
| 0 | 0 | 0 | 0 | 0 |
| 0.002302996 | 0.003547578 | 0.011104018 | 0 | 0 |
| 0 | 0 | 0 | 0 | 0 |
| 0 | 0 | 0 | 0 | 0 |
| 0.006724628 | 0 | 0.006635322 | 0 | 0 |
| 0 | 0 | 0 | 0 | 0 |
| 0.004576533 | 0.163485024 | 0 | 0 | 0 |
| 0 | 0 | 0 | 0 | 0 |
| 0 | 0 | 0 | 0 | 0 |
| 0 | 0 | 0 | 0 | 0 |
| 0 | 0 | 0 | 0 | 0 |
| 0 | 0 | 0 | 0 | 0 |
| 0 | 0 | 0 | 0 | 0 |
| 0 | 0 | 0.221572558 | 0.006927851 | 0.066003791 |
| 0 | 0.006227699 | 0 | 0 | 0 |
| 0 | 0 | 0 | 0 | 0 |
| 0 | 0 | 0 | 0 | 0 |
| 0.741455176 | 0 | 0 | 0 | 0 |
| 0 | 0 | 0 | 0 | 0 |
| 0 | 0 | 0 | 0 | 0 |
| 0 | 0 | 0 | 0 | 0 |
| 0 | 0.076565327 | 0 | 0 | 0 |
| 0 | 0.002296725 | 0.00184147 | 0 | 0 |
| 0 | 0 | 0 | 0 | 0 |
| 0 | 0 | 0 | 0 | 0 |
| 0 | 0.242894642 | 0 | 0 | 0 |
| 0 | 0 | 0 | 0 | 0 |
| 0 | 0 | 0 | 0 | 0 |
| 0 | 0 | 0 | 0 | 0 |
| 0 | 0 | 0 | 0 | 0 |
| 0 | 0.008082758 | 0.211209807 | 0 | 0.084361834 |
| 0 | 0.065396879 | 0 | 0 | 0 |
| 12.76832948 | 0.191118403 | 0.054860566 | 0.245463667 | 0 |
| 0 | 0 | 0 | 0 | 0 |
| 0 | 0 | 0 | 0 | 0 |
| 0 | 0 | 0 | 0 | 0 |
| 0 | 0 | 0 | 0 | 0 |
| 0 | 0.006648065 | 0.021636561 | 0 | 0.001276777 |

| | | | | |
|---|---|---|---|---|
| 0 | 0 | 0 | 0 | 0 |
| 0 | 0 | 0.004022759 | 0 | 0 |
| 0.000783363 | 0.023285672 | 0.534400436 | 0.442506843 | 0.985617567 |
| 0 | 0 | 0 | 0 | 0 |
| 0 | 0 | 0 | 0 | 0 |
| 0.019593027 | 0.083801083 | 0.005335193 | 0 | 0 |
| 0 | 0 | 0 | 0 | 0 |
| 0 | 0 | 0 | 0.001118805 | 0 |
| 0 | 0 | 0 | 0 | 0 |
| 0 | 0 | 0 | 0 | 0 |
| 0.061914449 | 0.026231597 | 0.004779322 | 0.002923941 | 0.000858348 |
| 0.431944151 | 0.849695249 | 5.626205595 | 0.300704035 | 7.206569336 |
| 0 | 0 | 0 | 0 | 0 |
| 0.200046758 | 0 | 0.002466888 | 0.001856257 | 0 |
| 0.012101343 | 0.267305011 | 0.683582024 | 0.282759527 | 0.528965663 |
| 0 | 0 | 0 | 0 | 0 |
| 0 | 0 | 0 | 0 | 0 |
| 0 | 0 | 0 | 0 | 0 |
| 0 | 0 | 0 | 0 | 0 |
| 0 | 0 | 0 | 0.009209192 | 0 |
| 0.004351312 | 1.290110812 | 0.029085416 | 0.288749325 | 0.000575538 |
| 3.743343108 | 0 | 0.912803603 | 0.226176504 | 21.31433107 |
| 7.673981284 | 0.049153152 | 1.646996783 | 2.901593592 | 14.75491551 |
| 0 | 0 | 0 | 0 | 0 |
| 0 | 0 | 0 | 0 | 0 |
| 0.001158706 | 0 | 0 | 0 | 0 |
| 0 | 0 | 0 | 0 | 0 |
| 0.094800872 | 0 | 0.016348506 | 0 | 0 |
| 0 | 0 | 0 | 0 | 0 |
| 0 | 0 | 0 | 0 | 0 |
| 0 | 0.001557335 | 0 | 0 | 0 |
| 0 | 0.105686326 | 0 | 0 | 0 |
| 0 | 0 | 0.001493888 | 0.019430941 | 0.036495999 |
| 0 | 0 | 0 | 0 | 0 |
| 0 | 0.008645849 | 0 | 0.001399054 | 0 |
| 4.885369338 | 0.431001208 | 0.072378745 | 0.000961107 | 0 |
| 0 | 0 | 0 | 0 | 0 |
| 0 | 0 | 0 | 0 | 0 |
| 0.704314199 | 0.148973271 | 0.061598362 | 0 | 0 |
| 0 | 0 | 0 | 0 | 0 |
| 0 | 0 | 0 | 0 | 0 |
| 0 | 0 | 0 | 0 | 0 |
| 0 | 0 | 0 | 0 | 0 |
| 0 | 0 | 0 | 0 | 0 |
| 0 | 0.180194023 | 0.001103473 | 0 | 0.001093951 |
| 0 | 0 | 0 | 0 | 0 |
| 0 | 0 | 0 | 0 | 0 |
| 21.95357175 | 0 | 0.150945536 | 0.441586874 | 0.01913425 |

| | | | | |
|---|---|---|---|---|
| 0 | 0 | 0 | 0 | 0 |
| 0 | 0 | 0 | 0 | 0 |
| 0 | 0 | 0 | 0 | 0 |
| 0 | 0 | 0.003385746 | 0 | 0 |
| 0 | 0 | 0 | 0 | 0 |
| 0 | 0 | 0 | 0 | 0 |
| 0 | 0 | 0 | 0 | 0 |
| 0.000960957 | 0.144326956 | 0.006577166 | 0.001432638 | 0.004575728 |
| 0 | 0 | 0 | 0 | 0 |
| 0 | 0 | 0 | 0 | 0 |
| 0 | 0.142659587 | 0 | 0.000866992 | 0.011076405 |
| 0 | 0 | 0 | 0 | 0 |
| 0.01833075 | 0 | 0 | 0 | 0 |
| 0.005624149 | 0 | 0 | 0 | 0 |
| 0 | 0 | 0 | 0 | 0 |
| 0 | 0 | 0 | 0 | 0 |
| 0 | 0 | 0 | 0 | 0 |
| 0.024367492 | 0.001615794 | 0.007847815 | 0 | 0 |
| 0 | 0 | 0 | 0 | 0 |
| 0 | 0 | 0 | 0 | 0 |
| 0 | 0 | 0 | 0 | 0 |
| 0 | 0 | 0 | 0 | 0 |
| 0 | 0.000698586 | 0 | 0 | 0 |
| 0 | 0 | 0 | 0 | 0 |
| 0 | 0 | 0 | 0 | 0 |
| 0 | 0 | 0 | 0 | 0 |
| 0 | 0 | 0 | 0 | 0 |
| 0 | 0 | 0 | 0 | 0 |
| 0 | 0 | 0 | 0 | 0 |
| 0.004873462 | 0 | 0 | 0 | 0 |
| 0 | 0 | 0 | 0 | 0 |
| 0 | 0 | 0 | 0 | 0 |
| 0 | 0 | 0 | 0 | 0 |
| 0 | 0 | 0 | 0 | 0 |
| 0.165082406 | 0.006173385 | 0 | 0 | 0 |
| 0 | 0 | 0 | 0 | 0 |
| 0.001665813 | 0 | 0 | 0 | 0 |
| 0.000492287 | 0.001232284 | 0.003014732 | 0.006458531 | 0.003955656 |
| 0 | 0 | 0 | 0 | 0 |
| 0 | 0 | 0.006646793 | 0 | 0 |
| 0 | 0.073610599 | 0 | 0.002203527 | 0 |
| 0 | 0 | 0 | 0 | 0 |
| 0 | 0 | 0 | 0 | 0 |
| 0 | 0 | 0 | 0 | 0 |
| 0 | 0 | 0 | 0 | 0 |
| 0 | 0 | 0 | 0 | 0 |
| 0 | 0.00195146 | 0 | 0 | 0 |
| 0 | 0 | 0 | 0 | 0 |

| | | | | |
|---|---|---|---|---|
| 0 | 0 | 0 | 0 | 0 |
| 0 | 0 | 0 | 0 | 0 |
| 0 | 0 | 0 | 0 | 0 |
| 0 | 0 | 0.044099631 | 0 | 0 |
| 0 | 0 | 0 | 0 | 0 |
| 0 | 0 | 0 | 0 | 0 |
| 0 | 0 | 0 | 0 | 0 |
| 0 | 0 | 0 | 0 | 0 |
| 0 | 0.00198278 | 0.993718796 | 0.090288568 | 5.730306973 |
| 0 | 0 | 0.001969573 | 0 | 0 |
| 0 | 0 | 0 | 0 | 0 |
| 0 | 0 | 0 | 0 | 0 |
| 0 | 0 | 0.000570728 | 0 | 0 |
| 0 | 0 | 0 | 0 | 0 |
| 0.11726078 | 0 | 2.420389851 | 0 | 0 |
| 0 | 0 | 0.000942514 | 0.007957362 | 0 |
| 0 | 0 | 0 | 0 | 0 |
| 0 | 0 | 0 | 0 | 0 |
| 0 | 0 | 0 | 0 | 0 |
| 0 | 0 | 0 | 0 | 0 |
| 0 | 0.065842903 | 0 | 0 | 0 |
| 0 | 0 | 0 | 0 | 0 |
| 0 | 0 | 0 | 0 | 0 |
| 0 | 0 | 0 | 0 | 0 |
| 0 | 0 | 0 | 0.006903872 | 0 |
| 1.36336226 | 0 | 0 | 0 | 0 |
| 0 | 0 | 0 | 0 | 0 |
| 0 | 0 | 0 | 0 | 0 |
| 0.002559513 | 0 | 0.006537931 | 0 | 0 |
| 0 | 0 | 0 | 0 | 0 |
| 0 | 0 | 0 | 0 | 0 |
| 0 | 0 | 0 | 0 | 0 |
| 0 | 0 | 0 | 0 | 0 |
| 0 | 0.020635961 | 0 | 0 | 0.000761401 |
| 0.075176762 | 0 | 0.002277841 | 0.001466426 | 0 |
| 0 | 0.027764535 | 0.001610158 | 0 | 0.00339945 |
| 0 | 0 | 0 | 0 | 0 |
| 0.019440941 | 0.875019303 | 1.295328109 | 0 | 0 |
| 0 | 0 | 0 | 0 | 0 |
| 0 | 0 | 0 | 0 | 0.003529044 |
| 0 | 0 | 0 | 0 | 0 |
| 0 | 0 | 0 | 0 | 0 |
| 0.433445324 | 0.081261119 | 1.04979183 | 0 | 0 |
| 0 | 0 | 0 | 0 | 0 |
| 0 | 0 | 0 | 0 | 0 |
| 0 | 0 | 0.003050844 | 0 | 0 |
| 0 | 0 | 0 | 0 | 0 |
| 0.001249291 | 0 | 0.145810817 | 1.697481249 | 0 |

| | | | | |
|---|---|---|---|---|
| 0 | 0.006529479 | 0 | 0 | 0 |
| 0 | 0 | 0 | 0 | 0 |
| 0 | 0 | 0 | 0 | 0 |
| 0 | 0 | 0 | 0 | 0 |
| 0.001669524 | 0 | 0 | 0 | 0 |
| 0 | 0 | 0.00193051 | 0 | 0 |
| 0 | 0 | 0 | 0 | 0 |
| 0 | 0 | 0 | 0 | 0 |
| 0 | 0 | 0 | 0 | 0 |
| 0 | 0 | 0 | 0 | 0 |
| 0 | 0 | 0 | 0 | 0 |
| 0.713303511 | 0.012423293 | 0.457190765 | 0 | 0.043087676 |
| 0.025830293 | 0.808785664 | 0 | 0 | 0 |
| 0 | 0 | 0 | 0 | 0 |
| 0.828958857 | 12.50014202 | 0.054019942 | 1.018999415 | 0.027006747 |
| 0 | 0 | 0 | 0 | 0 |
| 0 | 0.000933932 | 0 | 0 | 0 |
| 0 | 0 | 0 | 0 | 0 |
| 0 | 0 | 0 | 0 | 0 |
| 0 | 0 | 0 | 0 | 0 |
| 0.007316883 | 0 | 0 | 0 | 0 |
| 0 | 0 | 0 | 0 | 0 |
| 0 | 1.534618998 | 0.428584853 | 0.002289742 | 0 |
| 0 | 0 | 0 | 0 | 0 |
| 0 | 0 | 0 | 0 | 0 |
| 0 | 0 | 0 | 0 | 0 |
| 0 | 0 | 0 | 0 | 0 |
| 0 | 0 | 0 | 0 | 0 |
| 0 | 0 | 0 | 0 | 0 |
| 0 | 0 | 0 | 0 | 0 |
| 0 | 0 | 0 | 0 | 0 |
| 0.030901034 | 0.014982372 | 0.2313549 | 0.08740589 | 0.005965112 |
| 0 | 0 | 0 | 0 | 0 |
| 0 | 0.040906039 | 0.009168317 | 0 | 0 |
| 0 | 0.028185421 | 0.003309815 | 0 | 0.002090995 |
| 0.021598776 | 0.05057752 | 0.003513796 | 0 | 0 |
| 0 | 0 | 0.010196657 | 0 | 0 |
| 0 | 0 | 0 | 0 | 0 |
| 0 | 0 | 0 | 0 | 0 |
| 0 | 0 | 0 | 0 | 0 |
| 0 | 0 | 0 | 0 | 0 |
| 0 | 0 | 0 | 0 | 0 |
| 0 | 0 | 0 | 0 | 0 |
| 0 | 0 | 0 | 0 | 0 |
| 0.001419786 | 0.110395488 | 0.000575382 | 0 | 0 |
| 0 | 0 | 0 | 0 | 0 |
| 0 | 0 | 0 | 0 | 0 |
| 0 | 0 | 0 | 0 | 0 |

| | | | | |
|---|---|---|---|---|
| 0 | 0 | 0 | 0 | 0 |
| 0 | 0 | 0 | 0 | 0 |
| 0 | 0 | 0 | 0 | 0 |
| 0 | 0 | 0 | 0 | 0 |
| 0 | 0 | 0 | 0.029638888 | 0 |
| 0 | 0 | 0 | 0 | 0 |
| 0 | 0 | 0 | 0 | 0 |
| 0 | 0 | 0 | 0 | 0 |
| 0 | 0 | 0 | 0 | 0 |
| 0 | 0 | 0 | 0 | 0 |
| 0 | 0 | 0 | 0 | 0 |
| 0 | 0.001168731 | 0 | 0 | 0 |
| 0 | 0 | 0 | 0 | 0 |
| 0 | 0 | 0 | 0 | 0 |
| 0 | 0 | 0 | 0 | 0 |
| 0 | 0 | 0 | 0 | 0 |
| 0 | 0 | 0 | 0 | 0 |
| 0 | 0 | 0 | 0 | 0 |
| 0 | 0 | 0.001271745 | 0.824921298 | 0 |
| 0 | 0 | 0 | 0 | 0 |
| 0 | 0 | 0 | 0 | 0 |
| 0 | 0 | 0 | 0 | 0 |
| 0 | 0 | 0 | 0 | 0 |
| 0 | 0 | 0 | 0 | 0 |
| 0.017839252 | 0 | 0 | 0 | 0 |
| 0.056340316 | 0.020678879 | 0.004463826 | 0.017291566 | 0.008260574 |
| 0 | 0 | 0 | 0 | 0 |
| 0 | 0.130551284 | 0.012583289 | 0 | 0 |
| 0 | 0 | 0 | 0 | 0 |
| 0.030704224 | 1.233875515 | 0 | 0 | 0 |
| 0 | 0 | 0 | 0 | 0 |
| 0 | 0 | 0 | 0 | 0 |
| 0 | 0 | 0 | 0 | 0 |
| 0 | 0 | 0 | 0 | 0 |
| 0 | 0.02102008 | 0.001684013 | 0 | 0 |
| 0 | 0 | 0 | 0 | 0 |
| 0 | 0 | 0 | 0 | 0 |
| 0 | 0.002213038 | 0 | 0 | 0 |
| 0 | 0 | 0 | 0 | 0 |
| 0 | 0 | 0 | 0 | 0 |
| 1.090638621 | 17.01857995 | 17.06789749 | 18.6975924 | 1.912255646 |
| 0 | 1.692671942 | 0 | 0.042323107 | 0 |
| 0 | 0 | 0 | 0 | 0 |
| 0 | 0 | 0 | 0 | 0 |
| 0 | 0 | 0 | 0 | 0 |
| 0 | 0 | 0 | 0 | 0 |
| 0 | 0 | 0 | 0 | 0 |
| 0 | 0.041177578 | 0 | 0 | 0 |

| | | | | |
|---|---|---|---|---|
| 0 | 0 | 0 | 0 | 0 |
| 0 | 0.00501831 | 0.000866618 | 0 | 0 |
| 0 | 0 | 0 | 0 | 0 |
| 0 | 0 | 0 | 0 | 0 |
| 0.511409108 | 2.908831848 | 7.760022276 | 0 | 0 |
| 0 | 0 | 0 | 0 | 0 |
| 0 | 0 | 0 | 0 | 0 |
| 0 | 0 | 0 | 0 | 0 |
| 0 | 0.079573275 | 1.98093723 | 0.120760742 | 0.424218397 |
| 0 | 0 | 0 | 0 | 0 |
| 0 | 0 | 0 | 0 | 0.005823896 |
| 0 | 0 | 0 | 0 | 0 |
| 0.013090423 | 0 | 0 | 0 | 0 |
| 0.022990491 | 0.01750459 | 0.056316835 | 0.024849552 | 0.005644666 |
| 1.814563838 | 0 | 0 | 0 | 0 |
| 0.014149373 | 0.220971636 | 0.064562541 | 0 | 0 |
| 0 | 0 | 0.033145937 | 0 | 0 |
| 0 | 0 | 0 | 0 | 0 |
| 0 | 0 | 0 | 0 | 0 |
| 0 | 0 | 0 | 0 | 0 |
| 0 | 0 | 0 | 0 | 0 |
| 0 | 0 | 0 | 0 | 0 |
| 0 | 0 | 0 | 0 | 0 |
| 0 | 0 | 0 | 0 | 0 |
| 0 | 0 | 0 | 0 | 0 |
| 0 | 0 | 0 | 0 | 0 |
| 0.005766421 | 0 | 0 | 0 | 0.001544489 |
| 0 | 0 | 0.001230567 | 0 | 0.002323711 |
| 0 | 0 | 0 | 0 | 0 |
| 0 | 0 | 0 | 0 | 0 |
| 0 | 0 | 0 | 0 | 0 |
| 0 | 0 | 0 | 0 | 0 |
| 0.005611002 | 0 | 0 | 0 | 0 |
| 0 | 0.007896608 | 0 | 0 | 0 |
| 2.420971314 | 0 | 0 | 0 | 0 |
| 0 | 0 | 0 | 0 | 0.001680032 |
| 0 | 0 | 0 | 0.004645256 | 0 |
| 0 | 0 | 0 | 0 | 0 |
| 0 | 0 | 0 | 0 | 0 |
| 0 | 0 | 0 | 0 | 0 |
| 0.201196035 | 0 | 0 | 0 | 0 |
| 0 | 0 | 0.10196411 | 0 | 0 |
| 0 | 0 | 0 | 0 | 0 |
| 0 | 0.021928063 | 0.013124648 | 0 | 0 |
| 0 | 0 | 0 | 0 | 0 |
| 0 | 0 | 0 | 0 | 0 |
| 0 | 0 | 0 | 0 | 0 |
| 0 | 0 | 0 | 0 | 0 |

| | | | | |
|---|---|---|---|---|
| 0.003205932 | 0 | 0 | 0.007169319 | 0 |
| 0 | 0 | 0 | 0 | 0 |
| 0 | 0 | 0 | 0 | 0 |
| 0 | 0 | 0 | 0 | 0 |
| 0.001057874 | 0.004413416 | 0 | 0 | 0 |
| 0 | 0 | 0 | 0 | 0 |
| 0 | 0.00186296 | 0.010723894 | 0 | 0 |
| 0 | 0 | 0 | 0 | 0 |
| 0 | 0 | 0 | 0 | 0 |
| 0 | 0 | 0 | 0 | 0 |
| 0 | 0 | 0 | 0 | 0 |
| 0 | 0 | 0 | 0 | 0 |
| 0 | 0 | 0 | 0 | 0 |
| 0 | 0 | 0 | 0 | 0 |
| 0 | 0 | 0 | 0 | 0 |
| 0 | 0 | 0 | 0 | 0 |
| 0 | 0 | 0 | 0 | 14.64385691 |
| 0 | 0 | 0 | 0 | 0 |
| 0 | 0 | 0 | 0 | 0 |
| 0 | 0 | 0 | 0 | 0 |
| 0 | 0 | 0.155358283 | 0 | 0 |
| 0 | 0 | 0 | 0 | 0 |
| 0.000694697 | 0 | 0 | 0 | 0 |
| 0 | 0.001739581 | 0.015688092 | 0.000414424 | 0.001930302 |
| 0 | 0 | 0 | 0 | 0 |
| 0 | 0 | 0 | 0 | 0 |
| 0 | 0 | 0 | 0 | 0 |
| 0 | 0 | 0 | 0 | 0 |
| 0 | 0 | 0 | 0 | 0 |
| 0.014224973 | 4.69186677 | 0.243563258 | 0.011332609 | 0 |
| 0 | 0 | 0 | 0 | 0 |
| 0 | 0 | 0 | 0 | 0 |
| 0 | 0 | 0 | 0 | 0 |
| 0.612201885 | 0 | 0.128862729 | 0 | 0 |
| 0 | 0 | 0 | 0 | 0 |
| 0 | 0 | 0 | 0 | 0 |
| 0 | 0 | 0.002714769 | 0 | 0.002932874 |
| 0 | 0 | 0 | 0 | 0 |
| 0 | 0 | 0 | 0 | 0 |
| 0 | 0 | 0 | 0 | 0 |
| 0 | 0 | 0 | 0 | 0 |
| 0 | 0 | 0 | 0 | 0 |
| 0 | 0 | 0 | 0 | 0 |
| 0 | 0 | 0 | 0 | 0 |
| 0 | 0 | 0 | 0 | 0 |
| 0 | 0 | 0 | 0 | 0 |
| 0 | 0 | 0 | 0 | 0 |
| 0 | 0 | 0 | 0 | 0 |

| | | | | |
|---|---|---|---|---|
| 0 | 0.110439174 | 0.002693768 | 0.00089187 | 0 |
| 0.037823992 | 0 | 0 | 0 | 0 |
| 0 | 0 | 0 | 0 | 0 |
| 0 | 0 | 0 | 0 | 0 |
| 0 | 0 | 0 | 0 | 0 |
| 0 | 0 | 0 | 0 | 0 |
| 0 | 0 | 0 | 0 | 0 |
| 0.023887431 | 0 | 0.148436127 | 0 | 0.961384005 |
| 0.015911065 | 0.003264612 | 0.002818849 | 0.166357546 | 0.027479494 |
| 0 | 0.048181761 | 0.00883079 | 0.0021116 | 0.01280764 |
| 0 | 0 | 0 | 0 | 0 |
| 0 | 0 | 0 | 0 | 0 |
| 0 | 0 | 0 | 0 | 0 |
| 0 | 0 | 0 | 0 | 0 |
| 0 | 0.00099346 | 0 | 0 | 0 |
| 0 | 0 | 0 | 0 | 0 |
| 0 | 0 | 0 | 0 | 0.001082846 |
| 0 | 0 | 0 | 0 | 0 |
| 0 | 0 | 0 | 0 | 0 |
| 0 | 0 | 0 | 0 | 0 |
| 0 | 0 | 0 | 0 | 0 |
| 0 | 0 | 0 | 0 | 0 |
| 0 | 0 | 0 | 0 | 0 |
| 0 | 0 | 0 | 0 | 0 |
| 0 | 0 | 0 | 0 | 0 |
| 0 | 0 | 0 | 0 | 0 |
| 0 | 0 | 0 | 0 | 0 |
| 0 | 0 | 0 | 0 | 0 |
| 0 | 0 | 0 | 0.000837523 | 0 |
| 0 | 0 | 0 | 0 | 0 |
| 0 | 0 | 0 | 0 | 0 |
| 0 | 0 | 0 | 0 | 0 |
| 0 | 0 | 0 | 0 | 0 |
| 0 | 0 | 0 | 0 | 0 |
| 0 | 0 | 0 | 0 | 0 |
| 0 | 0 | 0 | 0 | 0 |
| 0 | 0 | 0 | 0 | 0 |
| 0 | 0 | 0 | 0 | 0 |
| 0 | 0 | 0 | 0 | 0 |
| 0 | 0.477970122 | 3.240156024 | 0 | 0.047395884 |
| 0 | 0 | 0 | 0 | 0 |
| 0.006674856 | 0 | 0.267823193 | 0 | 0 |
| 0 | 0 | 0 | 0 | 0 |
| 0 | 0 | 0 | 0 | 0 |
| 0 | 0.002630331 | 0 | 0 | 0 |
| 0 | 0 | 0 | 0 | 0 |
| 0 | 0 | 0 | 0 | 0 |
| 0 | 0 | 0 | 0 | 0 |

| | | | | |
|---|---|---|---|---|
| 0 | 0 | 0 | 0 | 0 |
| 0 | 0 | 0.048949028 | 0 | 0 |
| 0 | 0 | 0 | 0 | 0 |
| 0 | 0 | 0 | 0 | 0 |
| 0.569528432 | 1.86287505 | 0.149339052 | 0 | 0 |
| 0 | 0 | 0 | 0 | 0 |
| 0 | 0 | 0 | 0 | 0 |
| 0 | 0.006349678 | 0.068533351 | 0.011534314 | 0.776425833 |
| 0 | 0 | 0 | 0 | 0 |
| 0 | 0 | 0 | 0 | 0 |
| 0 | 0 | 0 | 0 | 0 |
| 0 | 0 | 0 | 0 | 0 |
| 0 | 0 | 0 | 0 | 0 |
| 0 | 0 | 0 | 0 | 0 |
| 0 | 0 | 0.021710277 | 0.018635492 | 0 |
| 0 | 0.062871907 | 0.000990641 | 0 | 0 |
| 0 | 0 | 0 | 0 | 0 |
| 0 | 0 | 0 | 0 | 0 |
| 0 | 0 | 0 | 0 | 0 |
| 0 | 0 | 0 | 0 | 0 |
| 0 | 0 | 0 | 0 | 0 |
| 0 | 0 | 0 | 0 | 0 |
| 0 | 0 | 0 | 0 | 0 |
| 0 | 0.039610682 | 0 | 0 | 0.000816641 |
| 0 | 0 | 0 | 0 | 0 |
| 0 | 0 | 0 | 0 | 0 |
| 0 | 0 | 0 | 0.000748334 | 0.026291279 |
| 0 | 0 | 0 | 0 | 0 |
| 0 | 0 | 0 | 0 | 0 |
| 0 | 0 | 0 | 0 | 0 |
| 0 | 0 | 0 | 0 | 0 |
| 0 | 0 | 0 | 0 | 0 |
| 0 | 0 | 0 | 0 | 0 |
| 0 | 0 | 0 | 0 | 0 |
| 0 | 0 | 0 | 0 | 0 |
| 0 | 0 | 0 | 0 | 0 |
| 0 | 0 | 0 | 0 | 0 |
| 0 | 0 | 0 | 0 | 0 |
| 0 | 0 | 0 | 0 | 0 |
| 0 | 0 | 0 | 0 | 0 |
| 0 | 0 | 0 | 0 | 0 |
| 0.539900491 | 0 | 0.093346162 | 0 | 0 |
| 0 | 0 | 0 | 0 | 0 |
| 0 | 0 | 0 | 0 | 0 |
| 0 | 0 | 0 | 0 | 0 |
| 0 | 0 | 0 | 0 | 0 |
| 0 | 0 | 0 | 0 | 0 |
| 0 | 0 | 0 | 0 | 0 |

| | | | | |
|---|---|---|---|---|
| 0.004405739 | 0.566188008 | 0 | 0 | 0 |
| 0 | 0 | 0 | 0 | 0 |
| 0 | 0 | 0 | 0 | 0 |
| 0 | 0 | 0 | 0 | 0 |
| 0 | 0 | 0 | 0 | 0 |
| 0 | 0.005247693 | 0.004334145 | 0 | 0 |
| 0 | 0 | 0 | 0 | 0 |
| 0 | 0 | 0 | 0 | 0 |
| 0 | 0 | 0 | 0 | 0 |
| 0 | 0 | 0 | 0 | 0 |
| 0 | 0 | 0 | 0 | 0 |
| 0 | 0.001055339 | 0 | 0 | 0 |
| 0 | 0 | 0 | 0 | 0 |
| 0 | 0 | 0 | 0 | 0 |
| 0 | 0 | 0 | 0 | 0 |
| 0 | 0 | 0 | 0 | 0 |
| 0 | 0 | 0 | 0 | 0 |
| 0 | 0 | 0 | 0 | 0 |
| 0 | 0 | 0.00057667 | 0 | 0 |
| 0 | 0 | 0 | 0 | 0 |
| 0 | 0 | 0.003740596 | 0.025258066 | 0.001789101 |
| 0 | 0 | 0 | 0 | 0 |
| 0.00138043 | 0.016125503 | 0.001740458 | 0 | 0 |
| 0 | 0 | 0 | 0 | 0 |
| 0 | 0 | 0 | 0 | 0 |
| 0 | 0 | 0 | 0 | 0 |
| 0 | 0 | 0 | 0 | 0 |
| 0 | 0 | 0 | 0 | 0 |
| 0 | 0 | 0 | 0 | 0 |
| 0 | 0 | 0 | 0 | 0 |
| 0 | 0 | 0 | 0 | 0 |
| 0 | 0 | 0 | 0 | 0 |
| 0.052357065 | 0.039911019 | 0.137845647 | 0.029487911 | 0.078677334 |
| 0 | 0 | 0 | 0 | 0 |
| 0.041436734 | 0.870313405 | 0 | 0 | 0 |
| 0 | 0.010170711 | 0.006539759 | 0.001546589 | 0 |
| 0 | 0 | 0 | 0 | 0 |
| 0 | 0 | 0 | 0 | 0 |
| 0 | 0 | 0.013183435 | 0.005545512 | 0 |
| 0 | 0 | 0 | 0 | 0 |
| 0 | 0 | 0 | 0 | 0 |
| 0 | 0 | 0 | 0 | 0 |
| 1.623563927 | 0.333355818 | 0.038917531 | 0 | 0 |
| 0 | 0 | 0 | 0 | 0 |
| 0 | 0 | 0 | 0 | 0 |
| 0 | 0.010924398 | 0 | 0 | 0 |
| 0 | 0 | 0 | 0 | 0 |
| 0 | 0 | 0 | 0 | 0 |

| | | | | |
|---|---|---|---|---|
| 0 | 0 | 0 | 0 | 0 |
| 0 | 0 | 0 | 0 | 0 |
| 0 | 0 | 0 | 0 | 0 |
| 0 | 0 | 0 | 0 | 0 |
| 0 | 0 | 0 | 0 | 0 |
| 0 | 0 | 0 | 0 | 0 |
| 0 | 0 | 0 | 0 | 0 |
| 0 | 0 | 1.399251491 | 1.978300935 | 0 |
| 0 | 0 | 0 | 0 | 0 |
| 0 | 0.004548685 | 0 | 0 | 0 |
| 0 | 0 | 0 | 0 | 0 |
| 0 | 0 | 0 | 0 | 0 |
| 0 | 0 | 0 | 0 | 0 |
| 0 | 0 | 0 | 0 | 0 |
| 0 | 0.018186318 | 0 | 0 | 0 |
| 0 | 0 | 0.002033849 | 0.001515104 | 0.289675013 |
| 0 | 0 | 0 | 0 | 0 |
| 0 | 0 | 0 | 0 | 0 |
| 0 | 0 | 0 | 0 | 0 |
| 0.03261613 | 0 | 0 | 0 | 0 |
| 0 | 0 | 0 | 0 | 0 |
| 0 | 0 | 0 | 0 | 0 |
| 0 | 0 | 0 | 0 | 0 |
| 0 | 0 | 0 | 0 | 0 |
| 0 | 0 | 0 | 0 | 0 |
| 0 | 0 | 0 | 0 | 0 |
| 0 | 0 | 0 | 0 | 0 |
| 0 | 0 | 0 | 0 | 0 |
| 0 | 0 | 0 | 0 | 0 |
| 0 | 0.007323808 | 0 | 0 | 0 |
| 0.035028427 | 0.292338677 | 0.349125362 | 0 | 0 |
| 0 | 0 | 0 | 0 | 0 |
| 0 | 0 | 0 | 0 | 0 |
| 0 | 0 | 0 | 0 | 0 |
| 0 | 0 | 0 | 0 | 0 |
| 0 | 0 | 0 | 0 | 0 |
| 0 | 0 | 0 | 0 | 0 |
| 0 | 0 | 0 | 0 | 0 |
| 0 | 0 | 0 | 0 | 0 |
| 0 | 0 | 0 | 0 | 0 |
| 0 | 0 | 0 | 0 | 0 |
| 0 | 0 | 0 | 0 | 0 |
| 0 | 0 | 0 | 0 | 0 |
| 0 | 0 | 0 | 0 | 0 |
| 0 | 0 | 0 | 0 | 0 |
| 0 | 0 | 0 | 0 | 0 |
| 0 | 0 | 0 | 0.01688868 | 2.341000512 |
| 0 | 0 | 0 | 0 | 0 |

| | | | | |
|---|---|---|---|---|
| 0 | 0 | 0 | 0 | 0 |
| 0 | 0 | 0 | 0 | 0 |
| 0 | 0 | 0 | 0 | 0 |
| 0 | 0 | 0.027243246 | 0.001081148 | 0 |
| 0 | 0 | 0.190074401 | 0 | 0 |
| 0 | 0 | 0 | 0 | 0 |
| 0 | 0 | 0 | 0 | 0 |
| 0.000575186 | 0 | 0 | 0 | 0 |
| 0 | 0 | 0 | 0 | 0 |
| 0 | 0 | 0 | 0 | 0 |
| 0 | 0 | 0 | 0 | 0 |
| 0 | 0.018507351 | 0 | 0 | 0.001427242 |
| 0 | 0 | 0 | 0 | 0 |
| 0 | 0 | 0 | 0 | 0 |
| 0 | 0 | 0 | 0 | 0 |
| 0.224852938 | 3.38894803 | 0 | 0 | 0 |
| 0 | 0 | 0 | 0 | 0 |
| 0 | 0 | 0 | 0 | 0 |
| 0 | 0.008332573 | 0.265226877 | 0 | 0 |
| 0 | 0.001483609 | 0.005337631 | 0 | 0 |
| 0 | 0 | 0 | 0 | 0 |
| 0 | 0 | 0 | 0 | 0 |
| 0 | 0 | 0 | 0 | 0 |
| 0 | 0 | 0 | 0 | 0 |
| 0 | 0 | 0 | 0 | 0 |
| 0 | 0 | 0 | 0 | 0 |
| 0 | 0 | 0 | 0 | 0 |
| 0 | 0 | 0 | 0 | 0 |
| 0 | 0 | 0 | 0 | 0 |
| 0 | 0 | 0 | 0 | 0 |
| 0 | 0.001347663 | 0 | 0 | 0.000640892 |
| 0 | 0 | 0 | 0 | 0 |
| 0 | 0 | 0 | 0 | 0 |
| 0 | 0 | 0 | 0 | 0 |
| 0 | 0 | 0 | 0 | 0 |
| 0 | 0 | 0 | 0 | 0 |
| 0 | 0 | 0 | 0 | 0 |
| 0.004917254 | 0.061701663 | 0.003815209 | 0.000676695 | 0.000675409 |
| 0 | 0 | 0 | 0 | 0 |
| 0 | 0 | 0 | 0 | 0 |
| 0 | 0.0007877 | 0 | 0 | 0 |
| 0 | 0 | 0 | 0 | 0 |
| 0 | 0 | 0 | 0 | 0 |
| 0 | 0.519723893 | 0.311767109 | 0.000668968 | 0 |
| 0 | 0 | 0 | 0 | 0 |
| 0 | 0 | 0 | 0.006130135 | 0 |
| 0 | 0 | 0 | 0 | 0 |
| 0 | 0 | 0 | 0 | 0 |

| | | | | |
|---|---|---|---|---|
| 0 | 0 | 0 | 0 | 0 |
| 0 | 3.56089176 | 0 | 0 | 0 |
| 0 | 0 | 0 | 0 | 0 |
| 0 | 0.008092845 | 0 | 0 | 0.002274185 |
| 0.010176997 | 0 | 0 | 0 | 0 |
| 0 | 0 | 0 | 0 | 0 |
| 0 | 0 | 0 | 0 | 0 |
| 0 | 0.009014465 | 0.001821692 | 0 | 0 |
| 0 | 0 | 0 | 0 | 0 |
| 0 | 0 | 0 | 0 | 0 |
| 0 | 0 | 0 | 0 | 0 |
| 0 | 0 | 0 | 0 | 0 |
| 0.050002037 | 0.049370283 | 0.073850362 | 0 | 0 |
| 0 | 0 | 0 | 0 | 0 |
| 0 | 0 | 0 | 0 | 0 |
| 0 | 0 | 0 | 0 | 0 |
| 0 | 0 | 0 | 0 | 0 |
| 0 | 0 | 0 | 0 | 0 |
| 0 | 0 | 0 | 0 | 0 |
| 0 | 0 | 0 | 0 | 0 |
| 0 | 0 | 0 | 0 | 0 |
| 0 | 0 | 0 | 0 | 0 |
| 0.001097353 | 0 | 0.009882511 | 0.002781169 | 0 |
| 0 | 0.004591987 | 0.034052173 | 0 | 0 |
| 0 | 0 | 0 | 0 | 0 |
| 0 | 0 | 0 | 0 | 0 |
| 0 | 0 | 0 | 0 | 0 |
| 0 | 0 | 0 | 0 | 0 |
| 0 | 0 | 0 | 0 | 0 |
| 0 | 0 | 0 | 0 | 0 |
| 0.001427467 | 0.036497762 | 0 | 0 | 0 |
| 0 | 0 | 0 | 0 | 0 |
| 0 | 0 | 0 | 0 | 0 |
| 0 | 0 | 0 | 0 | 0 |
| 0 | 0 | 0 | 0 | 0 |
| 0 | 0 | 0 | 0 | 0.002283904 |
| 0 | 0 | 0 | 0 | 0.002514594 |
| 0 | 0 | 0 | 0 | 0 |
| 0.000690135 | 0.01986662 | 0 | 0.066671677 | 0.026340711 |
| 0 | 0 | 0 | 0 | 0 |
| 0 | 0 | 0 | 0 | 0 |
| 0 | 0 | 0 | 0 | 0 |
| 0 | 0 | 0 | 0 | 0 |
| 0 | 0 | 0 | 0 | 0 |
| 0 | 0 | 0 | 0 | 0 |
| 0.036432289 | 0.358896527 | 0.006890121 | 0.009505103 | 0 |
| 0 | 0 | 0 | 0 | 0 |
| 0 | 0 | 0 | 0 | 0 |

| | | | | |
|---|---|---|---|---|
| 0 | 0 | 0 | 0 | 0 |
| 0 | 0 | 0 | 0 | 0 |
| 0 | 0 | 0 | 0 | 0 |
| 0 | 0 | 0 | 0 | 0 |
| 0 | 0 | 0 | 0 | 0 |
| 0 | 0 | 0 | 0 | 0 |
| 0 | 0 | 0 | 0 | 0 |
| 0 | 0 | 0 | 0 | 0 |
| 0 | 0 | 0 | 0 | 0 |
| 0 | 0 | 0 | 0.006693074 | 0 |
| 0 | 0.002627796 | 0 | 0 | 0 |
| 0 | 0.002353985 | 0 | 0 | 0 |
| 0 | 0 | 0 | 0 | 0 |
| 0 | 0 | 0 | 0 | 0 |
| 0 | 0 | 0 | 0 | 0 |
| 0 | 0 | 0 | 0 | 0 |
| 0 | 0.001802615 | 0 | 0 | 0 |
| 0 | 0 | 0 | 0 | 0 |
| 0 | 0 | 0 | 0 | 0 |
| 0.102651023 | 0 | 0.007823572 | 0 | 0 |
| 0 | 0 | 0 | 0 | 0 |
| 0 | 0 | 0 | 0 | 0 |
| 0 | 0 | 0 | 0 | 0 |
| 0 | 0 | 0 | 0 | 0 |
| 0 | 0 | 0 | 0 | 0 |
| 0 | 0 | 0 | 0 | 0 |
| 0 | 0 | 0 | 0 | 0 |
| 0.001848836 | 0 | 0 | 0 | 0 |
| 0 | 0 | 0 | 0 | 0 |
| 0 | 0 | 0 | 0 | 0 |
| 0.108919174 | 0.004590643 | 0.091684854 | 0 | 0.01295634 |
| 0 | 0 | 0 | 0 | 0 |
| 0.255339621 | 0.009269921 | 0.06903596 | 0.039916692 | 0.072573117 |
| 0.000756866 | 0 | 0.009610787 | 0.012355638 | 0.127544875 |
| 0 | 0 | 0 | 0 | 0 |
| 0 | 0 | 0 | 0 | 0 |
| 0 | 0 | 0 | 0 | 0 |
| 0 | 0 | 0 | 0 | 0 |
| 0 | 0 | 0 | 0 | 0 |
| 0 | 0 | 0 | 0 | 0 |
| 0 | 0 | 0 | 0 | 0 |
| 0 | 0 | 0.077044002 | 0.001090095 | 0.048145004 |
| 0.034147294 | 0 | 0.020858071 | 0 | 0.002651037 |
| 0 | 0 | 0 | 0 | 0 |
| 0 | 0 | 0 | 0 | 0 |
| 1.313816308 | 0.096667789 | 0.008114982 | 0 | 0 |
| 0 | 0.001100315 | 0 | 0 | 0 |
| 0 | 0 | 0 | 0 | 0 |

| | | | | |
|---|---|---|---|---|
| 0 | 0 | 0 | 0 | 0 |
| 0.030913245 | 0.016303632 | 0 | 0 | 0 |
| 0.010674006 | 0.001421221 | 0.024052376 | 0.012392045 | 0.000811049 |
| 0 | 0 | 0 | 0 | 0 |
| 0 | 0 | 0 | 0.006488165 | 0 |
| 0 | 0 | 0 | 0.000631554 | 0.003466945 |
| 0 | 0 | 0 | 0 | 0 |
| 0 | 0 | 0 | 0 | 0 |
| 0 | 0 | 0 | 0 | 0 |
| 0 | 0 | 0 | 0 | 0 |
| 0 | 0 | 0 | 0 | 0 |
| 0 | 0 | 0 | 0 | 0 |
| 0 | 0 | 0 | 0 | 0 |
| 0 | 0 | 0 | 0 | 0 |
| 0 | 0 | 0 | 0 | 0 |
| 0 | 0 | 0 | 0 | 0 |
| 0 | 0 | 0 | 0 | 0 |
| 0 | 0 | 0 | 0 | 0 |
| 0 | 0 | 0 | 0 | 0 |
| 0 | 0 | 0 | 0 | 0 |
| 0 | 0 | 0 | 0 | 0 |
| 0 | 0 | 0 | 0 | 0 |
| 0 | 0 | 0 | 0 | 0 |
| 0 | 0.038356145 | 0 | 0 | 0 |
| 0.020588446 | 0.009136031 | 0 | 0 | 0 |
| 0 | 0.397318072 | 0.002441756 | 0.000842117 | 0 |
| 0 | 0 | 0 | 0 | 0 |
| 0 | 0 | 0 | 0 | 0 |
| 0 | 0 | 0 | 0 | 0 |
| 0 | 0 | 0 | 0 | 0 |
| 0 | 0 | 0 | 0 | 0 |
| 0 | 0 | 0 | 0 | 0 |
| 0 | 0 | 0 | 0 | 0 |
| 0 | 0 | 0 | 0 | 0 |
| 0 | 0 | 0 | 0 | 0 |
| 0 | 0 | 0 | 0 | 0 |
| 0 | 0 | 0 | 0 | 0 |
| 0 | 0 | 0 | 0.001676513 | 0 |
| 0 | 1.758308792 | 0.268801847 | 0.074928417 | 0 |
| 0 | 0 | 0 | 0 | 0 |
| 0 | 0 | 0 | 0 | 0 |
| 0.008170463 | 0.003067821 | 0 | 0 | 0 |
| 0 | 0.088690402 | 0.001225284 | 0 | 0 |
| 0 | 0 | 0 | 0 | 0 |
| 0 | 0 | 0 | 0 | 0 |
| 0 | 0 | 0 | 0 | 0 |
| 0 | 0.001085432 | 0.026867047 | 0 | 0.001290466 |
| 0 | 0 | 0 | 0 | 0 |

| | | | | |
|---|---|---|---|---|
| 0 | 0 | 0 | 0 | 0 |
| 0.000729533 | 0.005934999 | 0.199662718 | 0.000489429 | 0.03272944 |
| 0 | 0 | 0 | 0 | 0 |
| 0 | 0 | 0 | 0 | 0 |
| 0 | 0 | 0 | 0 | 0 |
| 0 | 0 | 0 | 0 | 0 |
| 0 | 0 | 0 | 0 | 0 |
| 0 | 0 | 0 | 0 | 0 |
| 0 | 0 | 0 | 0 | 0 |
| 0 | 0 | 0 | 0 | 0 |
| 0 | 0 | 0 | 0 | 0 |
| 0 | 0 | 0 | 0 | 0 |
| 0 | 0.017373654 | 0 | 0 | 0 |
| 0 | 0 | 0 | 0 | 0 |
| 0 | 0 | 0.023100924 | 0 | 0 |
| 0 | 0 | 0 | 0 | 0 |
| 0 | 0 | 0 | 0 | 0 |
| 0 | 0 | 0 | 0 | 0 |
| 0 | 0 | 0 | 0 | 0 |
| 0 | 0 | 0 | 0 | 0 |
| 0 | 0 | 0 | 0 | 0 |
| 0 | 0.005590332 | 0 | 0 | 0 |
| 0 | 0 | 0 | 0 | 0 |
| 0 | 0 | 0 | 0 | 0 |
| 0 | 0 | 0 | 0 | 0 |
| 0 | 0 | 0 | 0 | 0 |
| 0 | 0 | 0 | 0 | 0 |
| 0 | 0 | 0 | 0 | 0 |
| 0 | 0 | 0 | 0 | 0 |
| 0 | 0 | 0 | 0 | 0 |
| 0 | 0 | 0 | 0 | 0 |
| 0 | 0 | 0 | 0 | 0 |
| 0 | 0 | 0 | 0 | 0 |
| 0 | 0 | 0 | 0 | 0 |
| 0.014396706 | 0 | 0 | 0 | 0 |
| 0 | 0 | 0 | 0 | 0 |
| 0 | 0 | 0 | 0 | 0 |
| 0 | 0 | 0 | 0 | 0 |
| 0 | 0.001953918 | 0.00096407 | 0.020946868 | 0 |
| 0 | 0 | 0 | 0 | 0 |
| 0.017797156 | 0.002753058 | 4.557202573 | 0.161700959 | 0 |
| 0 | 0 | 0 | 0 | 0 |
| 0 | 0 | 0 | 0 | 0 |
| 0 | 0 | 0 | 0 | 0 |
| 0 | 0 | 0 | 0 | 0 |
| 0.001831598 | 0.189983506 | 2.973794463 | 1.994519115 | 6.01040873 |
| 0 | 0 | 0 | 0 | 0 |
| 0 | 0 | 0 | 0 | 0 |

| | | | | |
|---|---|---|---|---|
| 0 | 0 | 0 | 0 | 0 |
| 0 | 0 | 0 | 0 | 0 |
| 0 | 0 | 0 | 0 | 0 |
| 0 | 0 | 0 | 0 | 0 |
| 0 | 0 | 0 | 0 | 0 |
| 0 | 0 | 0 | 0 | 0 |
| 0 | 0 | 0 | 0 | 0 |
| 0 | 0 | 0 | 0 | 0 |
| 0 | 0.110647719 | 0 | 0 | 0 |
| 0.009404357 | 0.915008462 | 0.00483963 | 0.014220725 | 0.002398935 |
| 0 | 0 | 0 | 0 | 0 |
| 0 | 0 | 0 | 0 | 0 |
| 0 | 0 | 0.00614943 | 0 | 0 |
| 0 | 0 | 0 | 0 | 0 |
| 0 | 0 | 0 | 0 | 0 |
| 0 | 0 | 0 | 0 | 0 |
| 0 | 0 | 0 | 0 | 0 |
| 0 | 0 | 0 | 0 | 0 |
| 0 | 0 | 0 | 0 | 0 |
| 1.043141229 | 0.039252886 | 0.174518109 | 0.003659203 | 0.092697529 |
| 0 | 0 | 0 | 0 | 0 |
| 0 | 0 | 0 | 0 | 0 |
| 0 | 0 | 0 | 0 | 0 |
| 0 | 0 | 0 | 0 | 0 |
| 0 | 0 | 0 | 0 | 0 |
| 0 | 0 | 0 | 0 | 0 |
| 0 | 0 | 0 | 0 | 0 |
| 0 | 0 | 0 | 0 | 0 |
| 0 | 0 | 0 | 0 | 0 |
| 0 | 0 | 0 | 0.029803787 | 0 |
| 0 | 0 | 0 | 0 | 0 |
| 0 | 0 | 0 | 0 | 0 |
| 0 | 0 | 0 | 0 | 0 |
| 0 | 0 | 0 | 0 | 0 |
| 0 | 0 | 0 | 0 | 0 |
| 0 | 0.006627647 | 0 | 0 | 0 |
| 0 | 0 | 0 | 0 | 0 |
| 0 | 0 | 0 | 0 | 0 |
| 0 | 0 | 0 | 0 | 0 |
| 0 | 0 | 0 | 0 | 0 |
| 0 | 0 | 0 | 0 | 0 |
| 0 | 0 | 0 | 0 | 0 |
| 0 | 0 | 0 | 0 | 0 |
| 0 | 0 | 0 | 0 | 0 |
| 0 | 0.020838296 | 0 | 0 | 0 |
| 0 | 0 | 0 | 0 | 0 |
| 0 | 0 | 0 | 0 | 0 |
| 0 | 0 | 0 | 0 | 0 |

| | | | | |
|---|---|---|---|---|
| 0 | 0 | 0 | 0 | 0 |
| 0 | 0 | 0 | 0 | 0 |
| 0 | 0 | 0 | 0 | 0 |
| 0 | 0 | 0 | 0 | 0 |
| 0 | 0 | 0 | 0 | 0 |
| 0.021990597 | 0.023078469 | 0.003395012 | 0.054735957 | 0.000609731 |
| 0 | 0 | 0 | 0 | 0 |
| 0 | 0 | 0 | 0 | 0 |
| 0 | 0 | 0 | 0 | 0 |
| 0 | 0 | 0 | 0 | 0 |
| 0 | 0 | 0 | 0 | 0 |
| 0 | 0 | 0 | 0 | 0 |
| 0 | 0 | 0 | 0 | 0 |
| 0.000698531 | 0 | 0 | 0 | 0 |
| 0 | 0 | 0 | 0 | 0 |
| 0 | 0 | 0 | 0 | 0 |
| 0 | 0 | 0 | 0 | 0 |
| 0 | 0 | 0 | 0 | 0 |
| 0 | 0 | 0 | 0 | 0 |
| 0 | 0 | 0 | 0 | 0 |
| 0 | 0 | 0 | 0 | 0 |
| 0 | 0 | 0 | 0 | 0 |
| 0 | 0 | 0 | 0 | 0 |
| 0 | 0 | 0 | 0 | 0 |
| 0 | 0 | 0 | 0 | 0 |
| 0.079262615 | 0.005882735 | 0 | 0 | 0 |
| 0 | 0 | 0 | 0 | 0 |
| 0 | 0 | 0 | 0 | 0 |
| 0 | 0 | 0 | 0 | 0 |
| 0 | 0 | 0 | 0 | 0 |
| 0 | 0 | 0 | 0 | 0 |
| 0 | 0 | 0 | 0 | 0 |
| 0 | 0 | 0 | 0 | 0 |
| 0 | 0 | 0 | 0 | 0 |
| 0 | 0 | 0 | 0 | 0 |
| 0 | 0 | 0 | 0 | 0 |
| 0 | 0 | 0 | 0 | 0 |
| 0 | 0 | 0 | 0 | 0 |
| 0 | 0 | 0 | 0 | 0 |
| 0 | 0 | 0 | 0 | 0 |
| 0 | 0 | 0 | 0 | 0 |
| 0 | 0 | 0 | 0 | 0 |
| 0 | 0 | 0 | 0 | 0 |
| 0 | 0 | 0 | 0 | 0 |
| 0 | 0 | 0 | 0 | 0 |
| 0 | 0 | 0 | 0 | 0 |
| 0 | 0 | 0 | 0 | 0 |
| 0 | 0 | 0 | 0 | 0 |

| | | | | |
|---|---|---|---|---|
| 0 | 0 | 0 | 0 | 0 |
| 0 | 0 | 0 | 0 | 0 |
| 0 | 0 | 0 | 0 | 0 |
| 0 | 0 | 0 | 0 | 0 |
| 0 | 0 | 0 | 0 | 0 |
| 0 | 0 | 0 | 0 | 0 |
| 0 | 0.002653495 | 0 | 0 | 0 |
| 0 | 0 | 0 | 0 | 0 |
| 0 | 0 | 0 | 0 | 0 |
| 0 | 0 | 0 | 0 | 0 |
| 0.025430181 | 0 | 0 | 0 | 0 |
| 0 | 0 | 0 | 0 | 0 |
| 0 | 1.17030655 | 0 | 0 | 0 |
| 0 | 0 | 0 | 0 | 0 |
| 0 | 0 | 0 | 0 | 0 |
| 0 | 0 | 0 | 0 | 0 |
| 0 | 0 | 0 | 0 | 0 |
| 0 | 0 | 0 | 0 | 0 |
| 0 | 0 | 0 | 0 | 0 |
| 0 | 0 | 0 | 0 | 0 |
| 0.001219375 | 0 | 0.031846107 | 0 | 0.00127011 |
| 0 | 0 | 0 | 0 | 0 |
| 0 | 0 | 0 | 0 | 0 |
| 0.34293903 | 0.777314248 | 3.95774107 | 0 | 2.733357909 |
| 0 | 0 | 0 | 0 | 0 |
| 0 | 0 | 0 | 0 | 0 |
| 0 | 0 | 0 | 0 | 0 |
| 0 | 0 | 0 | 0 | 0 |
| 0 | 0 | 0 | 0 | 0 |
| 0 | 0 | 0 | 0 | 0 |
| 0 | 0 | 0 | 0 | 0 |
| 0 | 0 | 0 | 0 | 0 |
| 0 | 0 | 0 | 0 | 0 |
| 0 | 0 | 0 | 0 | 0 |

| PHL018-0206_V3 | PHL018-0273_V3 | PHL018-0049_V1 | PHL018-0058_V1 | PHL018-0042_V2 |
|---|---|---|---|---|
| REF | REF | EYCF | EYCF | CTRL |
| 0.144654623 | 0.092595996 | 0.045204628 | 0.010776199 | 0.115981215 |
| 0.079252019 | 3.590876236 | 0.226389789 | 0.458663586 | 2.330664792 |
| 0.927048725 | 22.70009853 | 0 | 0.067261818 | 2.003974615 |
| 0 | 0 | 0 | 0 | 0 |
| 0 | 0 | 0 | 0 | 0 |
| 0 | 0 | 0 | 0 | 0 |
| 0 | 0.161726258 | 0 | 0 | 0 |
| 0 | 0 | 0 | 0 | 0 |
| 0 | 0 | 0 | 0 | 0 |
| 0 | 0 | 0 | 0 | 0 |
| 0 | 0 | 0 | 0 | 0 |
| 0 | 0 | 0 | 0 | 0 |
| 0 | 0 | 0 | 0 | 0 |
| 0.833803797 | 1.30035158 | 2.579240869 | 0.302920128 | 3.136195373 |
| 0 | 0.004695818 | 0 | 0 | 0 |
| 0 | 0 | 0 | 0 | 0 |
| 0 | 0 | 0 | 0 | 0 |
| 0 | 0.00331957 | 0 | 0 | 0 |
| 0 | 0 | 0 | 0 | 0 |
| 0 | 0 | 0 | 0 | 0 |
| 0 | 0 | 0 | 0 | 0 |
| 0 | 0 | 0 | 0 | 0 |
| 0 | 0 | 0 | 0 | 0 |
| 0 | 0 | 0 | 0 | 0 |
| 0.03624968 | 0 | 0.061355236 | 0.000545092 | 0 |
| 0 | 0 | 0 | 0 | 0 |
| 0 | 0 | 0 | 0 | 0 |
| 0 | 0 | 0 | 0 | 0 |
| 0 | 0 | 0 | 0 | 0 |
| 0 | 0 | 0.005932472 | 0 | 0 |
| 0 | 0 | 0 | 0 | 0 |
| 0 | 0 | 0 | 0 | 0 |
| 0 | 0 | 0 | 0 | 0 |
| 0 | 0 | 0 | 0 | 0 |
| 0 | 0 | 0 | 0 | 0 |
| 4.73102417 | 0 | 0.604565581 | 0.625536564 | 0.018795316 |
| 0 | 0 | 0 | 0 | 0 |
| 0 | 0 | 0 | 0 | 0 |
| 0 | 0 | 0 | 0 | 0 |
| 0 | 0 | 0 | 0 | 0 |
| 0 | 0 | 0 | 0 | 0 |
| 0.003064859 | 0.001064893 | 0 | 0.056117451 | 0 |
| 1.310535667 | 0.052522001 | 0.005258057 | 7.679921439 | 0.006280089 |
| 0.001625932 | 0.304050205 | 0.039024999 | 0.348070626 | 0.008645433 |
| 0 | 0 | 0.00271187 | 0.008364888 | 0 |
| 0 | 0 | 0 | 0 | 0 |

| | | | | |
|---|---|---|---|---|
| 0.058145752 | 0 | 0.001903764 | 0.03401855 | 0 |
| 0.002850506 | 0.003648032 | 0 | 0 | 0 |
| 0 | 0.017401868 | 0.000666414 | 0 | 0.01859422 |
| 0.485260128 | 0.672891067 | 0.471385492 | 0.469402523 | 0.664813364 |
| 0 | 0.01716919 | 0 | 0.000858287 | 0.004313229 |
| 0 | 0 | 0.019016064 | 0.031092801 | 0 |
| 0.001425799 | 0.005113211 | 0 | 0.00771839 | 0 |
| 0.01509271 | 0 | 0 | 0 | 0 |
| 0.042192292 | 0 | 0 | 0.005274364 | 0 |
| 0 | 0 | 0 | 0 | 0 |
| 0.000935738 | 0.140112457 | 0 | 0 | 0 |
| 0.081216524 | 0 | 0.489102197 | 0.009726901 | 0.063545971 |
| 0 | 0.000781698 | 0 | 0 | 0 |
| 0.006564293 | 0.00150785 | 0.013611063 | 0.01026567 | 0.004365227 |
| 0 | 0 | 0 | 0 | 0 |
| 0 | 0 | 0 | 0 | 0 |
| 0.001599813 | 2.601005144 | 0.550874183 | 0.021650994 | 0 |
| 0 | 0 | 0 | 0 | 0 |
| 0 | 0 | 0 | 0 | 0 |
| 0 | 0 | 0 | 0 | 0 |
| 0 | 0.150911371 | 0 | 0.001043043 | 0.012160756 |
| 0.013025275 | 0.030560819 | 0.010895237 | 0 | 0.003149728 |
| 0.007304682 | 0 | 0.004819265 | 0.262302038 | 0 |
| 0 | 0 | 0 | 0 | 0 |
| 0 | 0 | 0 | 0 | 0 |
| 0 | 0 | 0 | 0 | 0 |
| 0 | 0 | 0 | 0 | 0 |
| 0 | 0 | 0 | 0 | 0 |
| 0 | 0 | 0 | 0.133551522 | 0 |
| 0 | 0 | 0 | 0 | 0 |
| 0 | 0.026879683 | 0 | 0 | 0.008253288 |
| 0 | 0 | 0 | 0 | 0 |
| 0 | 0 | 0 | 0 | 0 |
| 0 | 0 | 0 | 0 | 0 |
| 0 | 0 | 0 | 0 | 0 |
| 0 | 0 | 0 | 0 | 0 |
| 0 | 0 | 0 | 0 | 0 |
| 0 | 0.122577146 | 0.014886333 | 0 | 0 |
| 0 | 0 | 0 | 0 | 0 |
| 0 | 0 | 0 | 0 | 0 |
| 0 | 0 | 0 | 0 | 0 |
| 0 | 0 | 0 | 0 | 0 |
| 0 | 0 | 0 | 0 | 0 |
| 0 | 0 | 0 | 0 | 0 |
| 0.260268596 | 0.498613792 | 0.504720051 | 0 | 0 |
| 0 | 0 | 0 | 0 | 0 |
| 0 | 0 | 0 | 0 | 0 |
| 0 | 0 | 0 | 0 | 0 |

| | | | | |
|---|---|---|---|---|
| 0 | 0 | 0 | 0 | 0 |
| 0 | 0 | 0 | 0 | 0 |
| 0 | 0 | 0 | 0 | 0 |
| 0 | 0 | 0 | 0 | 0 |
| 0.002635762 | 0 | 0 | 0.017166644 | 0 |
| 0 | 0 | 0 | 0 | 0 |
| 0 | 0 | 0 | 0 | 0 |
| 0 | 0 | 0 | 0 | 0 |
| 0 | 0.053501479 | 0 | 0 | 0 |
| 0 | 0 | 0 | 0 | 0 |
| 0 | 0 | 0 | 0 | 0 |
| 0 | 0 | 0 | 0.010989674 | 0 |
| 0 | 0 | 0 | 0 | 0 |
| 0 | 0 | 0 | 0 | 0 |
| 0 | 0 | 0 | 0 | 0 |
| 0 | 0 | 0 | 0 | 0 |
| 0 | 0 | 0 | 0 | 0 |
| 0 | 0 | 0 | 0 | 0 |
| 0.494888338 | 0.085159075 | 0.213480654 | 0.099563411 | 0.045414459 |
| 0 | 0 | 0.006128633 | 0 | 0 |
| 0 | 0 | 0 | 0 | 0 |
| 0 | 0 | 0 | 0 | 0 |
| 0 | 0 | 0 | 0 | 0 |
| 0 | 0 | 0 | 0 | 0 |
| 0 | 0 | 0 | 0 | 0 |
| 0 | 0 | 0 | 0 | 0 |
| 0 | 0 | 0 | 0 | 0 |
| 0 | 0.006673898 | 0 | 0 | 0 |
| 0 | 0 | 0 | 0 | 0 |
| 0 | 0 | 0 | 0 | 0 |
| 0.112749704 | 0.150819965 | 0.094432098 | 0.004379714 | 0.004049802 |
| 0 | 0.002005365 | 0.00791964 | 0.118910883 | 0 |
| 0 | 0 | 0 | 0 | 0 |
| 0 | 0 | 0 | 0 | 0 |
| 0 | 0 | 0 | 0 | 0 |
| 0 | 0 | 0 | 0 | 0 |
| 0 | 0 | 0 | 0 | 0 |
| 0 | 0 | 0 | 0 | 0 |
| 0 | 0 | 0 | 2.053028994 | 0 |
| 0 | 0 | 0 | 0 | 0 |
| 0 | 0 | 0 | 0 | 0 |
| 0 | 0 | 0 | 0 | 0 |
| 0.0004709 | 0.001390731 | 0 | 0 | 0 |
| 0 | 0 | 0 | 0 | 0 |
| 0 | 0 | 0.00124209 | 0 | 0 |
| 0 | 0 | 0 | 0 | 0 |
| 0 | 0 | 0 | 0 | 0 |
| 0 | 0 | 0 | 0 | 0 |

| | | | | |
|---|---|---|---|---|
| 0 | 0 | 0 | 0 | 0 |
| 0 | 0 | 0 | 0 | 0 |
| 0 | 0 | 0 | 0 | 0 |
| 0 | 0 | 0 | 0 | 0 |
| 0.000472804 | 0 | 0.080712524 | 0.005118935 | 0.001286233 |
| 0 | 0 | 0 | 0 | 0 |
| 0 | 0 | 0 | 0 | 0 |
| 0 | 0 | 0 | 0 | 0 |
| 0 | 0 | 0 | 0 | 0 |
| 0 | 0 | 0 | 0 | 0 |
| 0 | 0 | 0 | 0 | 0 |
| 0 | 0 | 0 | 0 | 0 |
| 0 | 0 | 0 | 0 | 0 |
| 0 | 0 | 0.001719962 | 0 | 0 |
| 0 | 0 | 0 | 0 | 0 |
| 0 | 0 | 0 | 0.788231224 | 0 |
| 0 | 0 | 0 | 0 | 0 |
| 0 | 0 | 0 | 0 | 0 |
| 0 | 0 | 0 | 0 | 0 |
| 0 | 0 | 0 | 0 | 0 |
| 0 | 0 | 0 | 0 | 0 |
| 0 | 0 | 0.006654515 | 0 | 0 |
| 0 | 0 | 0.006601632 | 0 | 0 |
| 0 | 0 | 0 | 0 | 0 |
| 0.017197752 | 0 | 0 | 0.051936598 | 0 |
| 0 | 0 | 0 | 0 | 0 |
| 0 | 0 | 0.03190875 | 0.017568588 | 0 |
| 0 | 0.000845494 | 0 | 0 | 0 |
| 0 | 0 | 0 | 0 | 0 |
| 0 | 0 | 0 | 0 | 0 |
| 0 | 0 | 0 | 0 | 0 |
| 0 | 0.009023657 | 0.005980945 | 0.008482071 | 0 |
| 0 | 0 | 0 | 0 | 0 |
| 0 | 0 | 0 | 0 | 0 |
| 0 | 0 | 0 | 0 | 0 |
| 0 | 0 | 0 | 0 | 0 |
| 0 | 0 | 0 | 0 | 0 |
| 0 | 0 | 0 | 0 | 0 |
| 0 | 0 | 0.010223104 | 0 | 0 |
| 0 | 0 | 0 | 0 | 0 |
| 0 | 0 | 0 | 0 | 0 |
| 0 | 0 | 0 | 0 | 0 |
| 0 | 0 | 0 | 0 | 0 |
| 0 | 0 | 0 | 0 | 0 |
| 0 | 0 | 0 | 0 | 0 |
| 0 | 0 | 0 | 0 | 0 |
| 0 | 0 | 0 | 0 | 0 |
| 0 | 0 | 0 | 0 | 0 |

| | | | | |
|---|---|---|---|---|
| 0 | 0 | 2.148670568 | 0.001084415 | 0.036239935 |
| 0 | 0 | 0 | 0 | 0 |
| 0 | 0 | 0 | 0 | 0 |
| 0 | 0 | 0 | 0 | 0 |
| 0 | 0 | 0 | 0 | 0 |
| 0 | 0 | 0 | 0 | 0 |
| 0 | 0 | 0 | 0 | 0 |
| 0 | 0 | 0 | 0 | 0 |
| 0 | 0 | 0 | 0 | 0 |
| 0 | 0 | 0 | 0 | 0 |
| 0 | 0 | 0 | 0 | 0 |
| 0 | 0 | 0.007579648 | 0 | 0 |
| 0.001949145 | 0 | 0.000476932 | 0.001623302 | 0 |
| 0 | 0 | 0 | 0 | 0 |
| 0 | 0.03147323 | 0 | 0 | 0 |
| 0 | 0 | 0 | 0 | 0 |
| 0 | 0 | 0 | 0 | 0 |
| 0 | 0 | 0 | 0 | 0 |
| 0 | 0 | 0 | 0 | 0 |
| 0 | 0 | 0 | 0 | 0 |
| 0.020745235 | 0.156481504 | 0.007431228 | 0.09598772 | 0.003431925 |
| 0 | 0 | 0 | 0 | 0 |
| 0.000737945 | 0 | 0 | 0 | 0 |
| 0.001754036 | 0 | 0 | 0.003560723 | 0 |
| 0 | 0 | 0 | 0 | 0 |
| 0 | 0 | 0 | 0 | 0 |
| 0.002612224 | 0 | 0.002250441 | 0.012373341 | 0.003553196 |
| 0 | 0 | 0 | 0 | 0 |
| 0 | 0.015285129 | 0 | 0 | 0 |
| 0 | 0 | 0 | 0 | 0 |
| 0 | 0 | 0 | 0 | 0 |
| 0 | 0 | 0 | 0 | 0 |
| 0 | 0 | 0 | 0 | 0 |
| 0.017818009 | 0.011637729 | 0.019619302 | 0.28403408 | 0 |
| 0.007296192 | 0.014275677 | 0 | 0 | 0.054088068 |
| 0 | 0 | 0 | 0 | 0 |
| 0 | 0 | 0 | 0 | 0 |
| 0 | 0 | 0 | 0 | 0 |
| 0 | 0 | 0.006373713 | 0 | 0 |
| 0 | 0 | 0 | 0 | 0 |
| 0 | 0 | 0 | 0 | 0 |
| 0 | 0 | 0 | 0.003698032 | 0 |
| 0 | 0 | 0 | 0 | 0 |
| 0 | 0 | 0 | 0 | 0 |
| 0 | 0 | 0 | 0 | 0 |
| 0.125389191 | 0.540089639 | 0 | 0 | 0 |
| 0 | 0 | 0 | 0 | 0 |
| 0.003899598 | 0 | 0 | 0 | 0 |

| | | | | |
|---|---|---|---|---|
| 0 | 0 | 0 | 0 | 0 |
| 0 | 0 | 0 | 0 | 0 |
| 0.002138023 | 0 | 0 | 0 | 0 |
| 0 | 0.000975963 | 0 | 0 | 0 |
| 0 | 0 | 0 | 0 | 0 |
| 0 | 0 | 0 | 0 | 0 |
| 0 | 0 | 0 | 0.013272006 | 0 |
| 0 | 0 | 0 | 0.049818046 | 0 |
| 0 | 0 | 0 | 0 | 0 |
| 0 | 0 | 0 | 0 | 0 |
| 0 | 0 | 0 | 0 | 0 |
| 0 | 0 | 0 | 0 | 0 |
| 0 | 0 | 0 | 0 | 0 |
| 0 | 0 | 0 | 0 | 0 |
| 0 | 0.125897426 | 0 | 0 | 0 |
| 0 | 0 | 0 | 0 | 0 |
| 0 | 0 | 0 | 0 | 0 |
| 0 | 0 | 0 | 0 | 0 |
| 0.002379519 | 0 | 0 | 0.051882777 | 0 |
| 0.027268196 | 0 | 0.058146879 | 1.312731536 | 0 |
| 0.002925027 | 0.010654302 | 0 | 0 | 0.005039657 |
| 0 | 0 | 0 | 0 | 0 |
| 0 | 0 | 0 | 0 | 0 |
| 0 | 0 | 0 | 0 | 0 |
| 0.013559432 | 0 | 0 | 0.013107564 | 0 |
| 0 | 0 | 0 | 0 | 0 |
| 0 | 0 | 0 | 0 | 0 |
| 0 | 0 | 0 | 0 | 0 |
| 0 | 0 | 0 | 0.032272546 | 0 |
| 0 | 0 | 0 | 0 | 0 |
| 0.255081499 | 0.280333872 | 0.079969191 | 0.026002573 | 0.034521129 |
| 0 | 0 | 0 | 0 | 0 |
| 0 | 0 | 0 | 0.025576187 | 0.03807411 |
| 0 | 0 | 0 | 0 | 0 |
| 0 | 0 | 0 | 0 | 0 |
| 0 | 0 | 0 | 0 | 0 |
| 0 | 0 | 0 | 0 | 0 |
| 0 | 0 | 0 | 0 | 0 |
| 0 | 0 | 0 | 0 | 0 |
| 0 | 0 | 0 | 0.000775312 | 0 |
| 0 | 0 | 0 | 0 | 0 |
| 0.055831946 | 0.020719306 | 0.001239776 | 0.001780207 | 0 |
| 0 | 0 | 0 | 0 | 0 |
| 0 | 0 | 0 | 0 | 0 |
| 0 | 0 | 0 | 0 | 0 |
| 0 | 0 | 0 | 0 | 0 |
| 0 | 0 | 0 | 0 | 0 |
| 0 | 0 | 0 | 0 | 0 |

| | | | | |
|---|---|---|---|---|
| 0 | 0 | 0 | 0 | 0 |
| 0 | 0 | 0 | 0 | 0 |
| 0.002018765 | 0.313011 | 0.003891858 | 0.329092085 | 0.007738619 |
| 0 | 0 | 0 | 0 | 0 |
| 0 | 0 | 0 | 0 | 0 |
| 0 | 0 | 0 | 0 | 0 |
| 0 | 0 | 0 | 0 | 0 |
| 0 | 0 | 0 | 0 | 0 |
| 0 | 0 | 0 | 0 | 0 |
| 0 | 0 | 0 | 0 | 0 |
| 0 | 0 | 0 | 0 | 0 |
| 0 | 0 | 0 | 0 | 0 |
| 0 | 0 | 0 | 0 | 0 |
| 0 | 0 | 0 | 0.001372476 | 0 |
| 0 | 0.033867225 | 0.000532512 | 0.002718718 | 0.003187948 |
| 0.026705516 | 0.005293334 | 0.000570125 | 0.087322414 | 0 |
| 0 | 0.001126676 | 0 | 0.004646604 | 0 |
| 0 | 0 | 0 | 0 | 0 |
| 0 | 0 | 0 | 0 | 0 |
| 0 | 0 | 0 | 0.001577835 | 0 |
| 0 | 0 | 0 | 0 | 0 |
| 0.0030994 | 0 | 0 | 0 | 0 |
| 0 | 0 | 0 | 0 | 0 |
| 0 | 0 | 0 | 0 | 0 |
| 0 | 0.007746338 | 0 | 0 | 0 |
| 0 | 0 | 0 | 0 | 0 |
| 0 | 0 | 0 | 0.253022995 | 0 |
| 0 | 0 | 0 | 0 | 0 |
| 0 | 0 | 0 | 0 | 0 |
| 0 | 0.050020703 | 0 | 0 | 0 |
| 0 | 0.003896974 | 0 | 0 | 0 |
| 0 | 0 | 0 | 0 | 0 |
| 0 | 0 | 0 | 0 | 0 |
| 0 | 0 | 0 | 0.001880788 | 0 |
| 0 | 0 | 0 | 0 | 0 |
| 0 | 0.004570756 | 0 | 0.097743704 | 0 |
| 0 | 0 | 0 | 0 | 0 |
| 0.001514363 | 0 | 0.012616196 | 0.116233372 | 0 |
| 0 | 0 | 0 | 0 | 0 |
| 0 | 0 | 0 | 0 | 0 |
| 0 | 0 | 0 | 0 | 0 |
| 0 | 0 | 0 | 0 | 0 |
| 0 | 0 | 0 | 0 | 0 |
| 0 | 0.00476183 | 0 | 0 | 0 |
| 0 | 0 | 0 | 0 | 0 |
| 0 | 0.007367469 | 0 | 0 | 0 |
| 0 | 0.003462829 | 0 | 0 | 0 |
| 0 | 0 | 0 | 0 | 0 |

| | | | | |
|---|---|---|---|---|
| 0 | 0.091833771 | 1.252318701 | 0 | 0 |
| 0 | 0 | 0 | 0 | 0 |
| 0 | 0 | 0.001721089 | 0.023431849 | 0 |
| 0 | 0 | 0 | 0 | 0 |
| 0.001088724 | 0 | 0.013660252 | 0.001473419 | 0 |
| 0 | 0.009612108 | 0 | 0 | 0 |
| 0 | 0 | 0 | 0 | 0 |
| 0.015769554 | 0.039436794 | 0.002427192 | 0.03132403 | 0.015222609 |
| 0 | 0 | 0 | 0 | 0 |
| 0.072880933 | 0.235760474 | 0 | 0.017030166 | 0.054559359 |
| 0 | 0 | 0 | 0 | 0 |
| 0.000604028 | 0 | 0.00106743 | 0.264311297 | 0 |
| 0.003665411 | 0.139374856 | 0.012954937 | 0.00992113 | 0.00186966 |
| 0 | 0 | 0 | 0 | 0.002524028 |
| 0 | 0 | 0 | 0 | 0 |
| 0 | 0 | 0 | 0 | 0 |
| 0 | 0 | 0 | 0.105371135 | 0 |
| 0 | 0 | 0 | 0 | 0 |
| 0 | 0.016028305 | 0 | 0 | 1.22241371 |
| 0 | 0.007299817 | 0 | 0 | 0 |
| 0 | 0 | 0 | 0 | 0 |
| 0.095455265 | 0 | 0 | 0 | 0 |
| 0 | 0 | 0 | 0.007464716 | 0 |
| 0.533276318 | 0 | 0.362785635 | 0.248011582 | 0 |
| 0 | 0 | 0 | 0 | 0 |
| 0 | 0 | 0 | 0.003611121 | 0 |
| 0 | 0.012996482 | 0.001513299 | 0.00128768 | 0 |
| 0 | 0 | 0 | 0 | 0 |
| 0.006738803 | 0.027970417 | 0 | 0 | 0 |
| 0 | 0 | 0 | 0 | 0 |
| 0 | 0 | 0 | 0 | 0 |
| 0 | 0 | 0 | 0 | 0 |
| 0 | 0 | 0 | 0 | 0 |
| 0 | 0.090350239 | 0 | 0 | 0 |
| 0 | 0.002804729 | 0 | 0 | 0 |
| 0 | 0.0037292 | 0 | 0 | 0.029443745 |
| 0 | 0.005955491 | 0 | 0 | 0.001920038 |
| 0 | 0 | 0 | 0 | 0 |
| 0 | 0 | 0 | 0 | 0 |
| 0 | 0 | 0 | 0 | 0 |
| 0 | 0.001477749 | 0 | 0 | 0 |
| 0 | 0.005552957 | 0 | 0 | 0 |
| 0 | 0.005302102 | 0 | 0 | 0 |
| 0 | 0 | 0 | 0 | 0 |
| 0 | 0 | 0.000564866 | 0 | 0 |
| 0.081473022 | 0 | 0.01379982 | 0 | 0 |
| 0.001548509 | 0.013066528 | 0.000703673 | 0 | 0.000902704 |
| 0 | 0 | 0.003832228 | 0 | 0.00109248 |

| | | | | |
|---|---|---|---|---|
| 0 | 0 | 0 | 0 | 0 |
| 0 | 0 | 0 | 0 | 0 |
| 0 | 0 | 0 | 0 | 0 |
| 0 | 0 | 0 | 0 | 0 |
| 0 | 0 | 0 | 0 | 0 |
| 0.084677741 | 0.097768999 | 0 | 0 | 0.001658975 |
| 0 | 0.024603638 | 0 | 0 | 0 |
| 0 | 0 | 0 | 0 | 0 |
| 0 | 0 | 0.006065824 | 0 | 0 |
| 0 | 0 | 0 | 0 | 0 |
| 0 | 0 | 0.007713729 | 0.035826779 | 0.018141809 |
| 0 | 0 | 0 | 0 | 0 |
| 0 | 0 | 0 | 0 | 0 |
| 0.006682097 | 0 | 0.001214591 | 0.073120588 | 0 |
| 0 | 0.043073447 | 0 | 0 | 0.01418896 |
| 0 | 0 | 0 | 0 | 0 |
| 0 | 0 | 0 | 0 | 0 |
| 0 | 0.007407608 | 0 | 0 | 0.004548947 |
| 0 | 0 | 0 | 0 | 0 |
| 0.119517605 | 0.025019445 | 0 | 0 | 0 |
| 0 | 0 | 0 | 0 | 0 |
| 0.13598703 | 0.286869269 | 0.253589136 | 0.189587699 | 0 |
| 0 | 0.03798936 | 0.050521792 | 0 | 0 |
| 0 | 0 | 0 | 0 | 0 |
| 0 | 0 | 0 | 0 | 0 |
| 0 | 0 | 0 | 0 | 0 |
| 0 | 0.104541985 | 0 | 0 | 0.030128155 |
| 0 | 0 | 0 | 0 | 0 |
| 0 | 0 | 0 | 0 | 0 |
| 0.254658414 | 0.054134298 | 0.226696043 | 0.550288923 | 0.990069528 |
| 0.020464483 | 0 | 0.000824003 | 0.001928168 | 0.001409426 |
| 0 | 0 | 0 | 0.01993401 | 0 |
| 0 | 0 | 0 | 0 | 0 |
| 0 | 0 | 0 | 0 | 0 |
| 0 | 0.002291411 | 0 | 0 | 0.003517839 |
| 0 | 0 | 0 | 0 | 0 |
| 0.74300366 | 0 | 0.431867296 | 0 | 0.374262318 |
| 0 | 0 | 0 | 0.049247315 | 0 |
| 0 | 0 | 0 | 0 | 0 |
| 0 | 0.239848428 | 0 | 0 | 0.055310122 |
| 0.002511273 | 0.080424445 | 0 | 0 | 0.015157971 |
| 0.031182129 | 0 | 1.484248065 | 0.916712107 | 1.32366328 |
| 0 | 0 | 0 | 0.032327483 | 0 |
| 0 | 0 | 0.063443532 | 0 | 0.897074485 |
| 0 | 0 | 0 | 0 | 0 |
| 0 | 0 | 0 | 0 | 0 |
| 0.001976821 | 0.011919741 | 0 | 0 | 0.009859333 |
| 0.060443722 | 0 | 0 | 0 | 0 |

| | | | | |
|---|---|---|---|---|
| 0 | 0.001200568 | 0.001293086 | 0.001925523 | 0 |
| 0 | 0 | 0 | 0 | 0 |
| 0.000830853 | 0.253968215 | 1.153769786 | 0.005622157 | 0 |
| 0 | 0 | 0 | 0 | 0 |
| 0 | 0.04928025 | 0 | 0.018065785 | 0 |
| 0 | 0.045171124 | 0 | 0 | 0 |
| 0 | 0 | 0 | 0 | 0 |
| 0 | 0 | 0 | 0.030830304 | 0 |
| 0 | 0 | 0.012625633 | 0 | 0.000694146 |
| 0 | 0.889382088 | 0 | 0 | 0 |
| 0 | 0 | 0 | 0 | 0 |
| 0 | 0 | 0 | 0 | 0 |
| 0 | 0 | 0 | 0 | 0 |
| 0.0009194 | 0 | 0.000584911 | 0 | 0.009254334 |
| 0 | 0 | 0 | 0 | 0 |
| 0.071562164 | 0.143232796 | 0.007770666 | 0.063204209 | 0.024237315 |
| 0.002866959 | 0.002822375 | 0.014186073 | 0.022848798 | 0.00844933 |
| 0 | 0.027146853 | 0 | 0 | 0 |
| 0 | 0 | 0 | 0 | 0 |
| 0 | 0.480223641 | 0 | 0 | 0 |
| 0 | 0.33998955 | 0 | 0.001901622 | 0 |
| 0 | 0 | 0 | 0 | 0.001759115 |
| 0 | 0 | 0 | 0 | 0 |
| 0 | 0 | 0 | 0 | 0 |
| 0 | 0.16859828 | 0.038669916 | 0 | 0.01822319 |
| 0 | 0 | 0 | 0 | 0 |
| 0 | 0.005075957 | 0 | 0 | 0.001375191 |
| 0 | 0 | 0.006105579 | 0 | 0.009050914 |
| 0 | 0 | 0 | 0 | 0 |
| 0 | 0 | 0 | 0 | 0 |
| 0.007842458 | 0.052226923 | 0.004239253 | 0 | 0.010667457 |
| 0 | 0.021327947 | 0.018231363 | 0.006825825 | 0 |
| 0.005457108 | 0.10862681 | 0.014349877 | 0.019792738 | 0.209324649 |
| 0 | 0 | 0 | 0.008043784 | 0 |
| 0 | 0 | 0.00213068 | 0.001813015 | 0 |
| 0 | 0 | 0 | 0 | 0 |
| 0.015590182 | 0.00203989 | 0 | 0.009080537 | 0.001879018 |
| 0 | 0.094349914 | 0.002781887 | 0.001462053 | 0.030019353 |
| 0.0369169 | 0 | 0.489779458 | 0.029635431 | 0.016686369 |
| 0 | 0.023850263 | 0 | 0 | 0 |
| 0 | 0 | 0 | 0 | 0 |
| 0 | 0 | 0 | 0 | 0 |
| 0.00577009 | 0.001022465 | 0 | 0 | 0.003139433 |
| 0 | 0.385159445 | 0 | 0 | 0.027202879 |
| 0 | 0 | 0 | 0 | 0 |
| 0 | 0 | 0 | 0 | 0 |
| 0 | 0 | 0 | 0 | 0 |
| 0 | 0.000926069 | 0 | 0 | 0 |

| | | | | |
|---|---|---|---|---|
| 0 | 0 | 0 | 0 | 0 |
| 0.007991294 | 0.06925051 | 0.396013912 | 1.102273974 | 0.062072891 |
| 0 | 0 | 0 | 0 | 0 |
| 0 | 0 | 0 | 0 | 0 |
| 0 | 0 | 0 | 0 | 0 |
| 0 | 0 | 0 | 0 | 0 |
| 0 | 0 | 0 | 0 | 0 |
| 0 | 0 | 0 | 0.005683767 | 0 |
| 0.001732588 | 0 | 0 | 0.010681819 | 0 |
| 0 | 0.003610618 | 0 | 0 | 0 |
| 0 | 0.001384593 | 0.000662796 | 0 | 0.001487966 |
| 0.101256821 | 0.386010721 | 0.010859796 | 0.203190439 | 0.370133377 |
| 0.053952664 | 0 | 0 | 0.074016794 | 0 |
| 0 | 0 | 0 | 0 | 0 |
| 0.030123186 | 0.015621697 | 0 | 0.028148686 | 0.000975574 |
| 0 | 0 | 0 | 0 | 0 |
| 0.016855049 | 0.005799474 | 0.005205327 | 0.025689717 | 0.003561406 |
| 0 | 0.036701529 | 0 | 0 | 0.164412444 |
| 0.000693797 | 0 | 0 | 0 | 0 |
| 0 | 0.113049491 | 0 | 0 | 0.011632992 |
| 0.024638291 | 0.003737951 | 0.032029098 | 0.167177308 | 0.121199368 |
| 0 | 0 | 0 | 0.005115377 | 0 |
| 0 | 0 | 0 | 0 | 0 |
| 0.000872058 | 0 | 0 | 0 | 0 |
| 0 | 0 | 0 | 0 | 0 |
| 0 | 0 | 0 | 0 | 0 |
| 0.018655843 | 0.364280985 | 0.138997037 | 0 | 0.007476865 |
| 0.009506191 | 0.348289998 | 0.035563642 | 0.032526999 | 0.056845396 |
| 0 | 0 | 0 | 0.002129838 | 0 |
| 0.001730822 | 0 | 0.002294015 | 0 | 0 |
| 0 | 0 | 0 | 0.00234413 | 0 |
| 0 | 0 | 0 | 0 | 0 |
| 0 | 0.00482306 | 0 | 0 | 0 |
| 0 | 0 | 0 | 0 | 0 |
| 0.00687727 | 0 | 0.073749202 | 0 | 0 |
| 0.35486123 | 0.154200954 | 0.137115826 | 0.352073537 | 0.41249457 |
| 0 | 0 | 0 | 0 | 0 |
| 0 | 0 | 0 | 0 | 0 |
| 0 | 0.002287135 | 0 | 0.105404801 | 0 |
| 0 | 0 | 0 | 0 | 0 |
| 0 | 0 | 0 | 0 | 0 |
| 0 | 0 | 0 | 0 | 0 |
| 0 | 0.010518538 | 0 | 0.002000765 | 0.025045182 |
| 0 | 0 | 0 | 0 | 0.00476553 |
| 0 | 0.067716471 | 0 | 0 | 0.01205784 |
| 0.540562895 | 0 | 0.540328061 | 1.088461545 | 0 |
| 0 | 0 | 0 | 0 | 0 |
| 0 | 0 | 0 | 0 | 0 |

| | | | | |
|---|---|---|---|---|
| 0 | 0 | 0 | 0 | 0 |
| 0 | 0 | 0 | 0 | 0 |
| 0 | 0 | 0 | 0.001000132 | 0 |
| 0.546922464 | 0.346488697 | 0.006306965 | 0.621119761 | 0.001277506 |
| 0 | 0 | 0 | 0 | 0 |
| 0 | 0 | 0 | 0 | 0 |
| 0 | 0 | 0 | 0 | 0 |
| 0 | 0 | 0.017520691 | 0 | 0 |
| 2.373460421 | 0 | 0.72804432 | 1.19667769 | 0.325348718 |
| 0.38528742 | 0 | 0.087668246 | 1.025434839 | 0.011417758 |
| 0 | 0 | 0 | 0 | 0 |
| 0.007498719 | 0 | 0.001590197 | 0 | 0 |
| 0 | 0 | 0 | 0 | 0 |
| 17.09306103 | 0 | 0.690502183 | 0 | 0 |
| 0.101358823 | 0 | 0.226098082 | 0.03366528 | 0.043094945 |
| 0 | 0 | 0 | 0 | 0 |
| 0.436999384 | 0 | 0.00753146 | 0.013326955 | 0 |
| 0 | 0 | 2.992426975 | 0.059053147 | 0.191908279 |
| 0 | 0 | 0 | 0 | 0 |
| 0 | 0 | 0.006532412 | 0.184880224 | 0 |
| 0 | 0 | 0 | 0 | 0 |
| 0.004594927 | 0 | 0 | 0 | 0 |
| 0.001657213 | 0 | 0.047736272 | 0.001308288 | 0 |
| 0 | 0.363868592 | 0.409542023 | 0.014842801 | 0.079617716 |
| 0 | 0.001036731 | 0 | 0.000570087 | 0 |
| 0.66162178 | 0 | 0.31547186 | 0.550911482 | 0.09251457 |
| 0.003174357 | 0.010608529 | 0.071922057 | 0 | 0 |
| 0.883384003 | 0 | 2.511861441 | 0.417793397 | 2.261448282 |
| 0 | 0 | 0 | 0 | 0 |
| 0 | 0 | 0 | 0 | 0 |
| 0 | 0.004306001 | 0.001855132 | 0.011247161 | 0.000793283 |
| 1.269585395 | 0.887682881 | 0.002184848 | 0 | 0 |
| 0 | 0.026381322 | 0 | 0.001651914 | 0 |
| 0 | 0 | 0 | 0 | 0 |
| 0 | 0 | 0.354839207 | 0.001457455 | 0 |
| 0.108046213 | 0 | 0.368986487 | 0.009536335 | 0.032790001 |
| 7.410109687 | 0 | 0.123242267 | 0.639363498 | 0.017808072 |
| 0 | 0.001612985 | 0 | 0 | 0 |
| 0.014987842 | 0 | 0 | 0 | 0.000849448 |
| 0 | 0 | 0.057821622 | 0.000435835 | 0.023946672 |
| 0.172648339 | 0.556418082 | 0 | 0.029401306 | 0.160669344 |
| 0 | 0 | 0.030870864 | 0.602536366 | 0.003059427 |
| 0.778744415 | 8.585896719 | 0.038979782 | 0.100439104 | 17.63864785 |
| 0.015791923 | 0 | 0.040664913 | 0.000508855 | 0.001022879 |
| 0 | 0 | 0 | 0 | 0 |
| 0 | 0 | 0 | 0 | 0 |
| 0 | 0 | 0 | 0 | 0 |
| 0 | 0 | 0.010819645 | 0.002106581 | 0 |

| | | | | |
|---|---|---|---|---|
| 0.217466285 | 0 | 0.137623358 | 0 | 0 |
| 0.014338389 | 0 | 0.435880037 | 0.073354358 | 0 |
| 0.008952908 | 0.080792093 | 0 | 0.010321354 | 0.020973069 |
| 0 | 0 | 0.0022803 | 0 | 0 |
| 0 | 0 | 0 | 0.03927242 | 0 |
| 0.022725545 | 0 | 0.018418347 | 0.156252095 | 0 |
| 0 | 0.026420893 | 0 | 0 | 0 |
| 0 | 0 | 0 | 0 | 0 |
| 0 | 0 | 0 | 0 | 0 |
| 0 | 0 | 0 | 0 | 0 |
| 0.025393817 | 0.011057216 | 0.02140223 | 0.008151052 | 0.018156242 |
| 0.717324213 | 0.764992728 | 2.021521918 | 0.582472278 | 1.111443839 |
| 0 | 0 | 0 | 0 | 0 |
| 0 | 0.002075374 | 0.034647238 | 0.054683725 | 0 |
| 0.008998755 | 0.155574467 | 0.002055091 | 0.001311522 | 0.075324765 |
| 1.115151696 | 0 | 0.406215716 | 12.68739691 | 0 |
| 0 | 0 | 0 | 0 | 0 |
| 0.001076201 | 0 | 0 | 0 | 0 |
| 0.074138857 | 0 | 0 | 0.034299349 | 0 |
| 0.024806392 | 0.42531502 | 0.010236661 | 0 | 0.002371068 |
| 0 | 0.044524656 | 0 | 0.001107856 | 0.019114795 |
| 0 | 0.233089871 | 0.00243888 | 0 | 22.4533757 |
| 1.961849123 | 5.043128757 | 0.0122974 | 0.232300195 | 5.609831236 |
| 0 | 0 | 0.002047221 | 0 | 0 |
| 0.021395041 | 0 | 0.017297643 | 0 | 0 |
| 0.442401978 | 0 | 0.810930117 | 3.350058162 | 2.536034642 |
| 0.067230917 | 0 | 0 | 0.073761001 | 0 |
| 0 | 0 | 0 | 0 | 0.003290471 |
| 0 | 0 | 0 | 0 | 0 |
| 0 | 0 | 0 | 0 | 0 |
| 0.006320309 | 0 | 0.014998604 | 0.000814625 | 0 |
| 0.091579988 | 0 | 0.047254266 | 0.247181369 | 0.084097082 |
| 0.084426004 | 0 | 0.001240843 | 0.000678757 | 0 |
| 0.025115106 | 0.018825797 | 0.040417917 | 0.006901402 | 0.033988623 |
| 0.75296706 | 0.002281128 | 0.000467984 | 0 | 0.001350791 |
| 0.297897544 | 0.006044393 | 0.048705829 | 0.110586383 | 0.011547849 |
| 0.001903789 | 0 | 0.010714166 | 0 | 0 |
| 0 | 0 | 0 | 0 | 0 |
| 0.360308996 | 0 | 0.45107453 | 0.019687363 | 0.058638137 |
| 0 | 0 | 0 | 0 | 0 |
| 0.025096688 | 0 | 0.019108486 | 0.007949133 | 0 |
| 0 | 0 | 0 | 0 | 0 |
| 0 | 0 | 0 | 0.007634511 | 0 |
| 0 | 0 | 0 | 0 | 0 |
| 0.007261155 | 0.006382356 | 0.06800867 | 0 | 0.012463528 |
| 0.002138019 | 0 | 0 | 0 | 0 |
| 0 | 0 | 0 | 0 | 0 |
| 0.152238675 | 0.636542182 | 0.009287212 | 0.012418334 | 0.084249492 |

| | | | | |
|---|---|---|---|---|
| 0 | 0 | 0 | 0 | 0 |
| 0 | 0 | 0 | 0 | 0 |
| 0 | 0 | 0 | 0 | 0 |
| 0.001178791 | 0 | 0 | 0.002193553 | 0 |
| 0 | 0 | 0 | 0 | 0 |
| 0.252857359 | 0 | 0.607968718 | 0.080311208 | 0 |
| 0.001935828 | 0.077658253 | 0.179943068 | 0.025980106 | 0 |
| 0.098436631 | 0 | 0.004025428 | 0 | 0 |
| 0 | 0 | 0 | 0 | 0 |
| 0 | 0 | 0 | 0 | 0 |
| 0.022646854 | 0.000969334 | 0.00226207 | 0.008217487 | 0 |
| 0 | 0 | 0 | 0 | 0 |
| 0.134061902 | 0 | 0 | 0.000972304 | 0 |
| 0.007562039 | 0 | 0.084932737 | 0.081493341 | 0 |
| 0 | 0 | 0 | 0 | 0 |
| 0 | 0 | 0 | 0 | 0 |
| 0.003905692 | 0 | 0.307099918 | 0.004955389 | 0 |
| 0.010656037 | 0 | 0.003186822 | 0.003143103 | 0 |
| 0 | 0.003608775 | 0.080328732 | 0 | 0 |
| 0.206737799 | 0 | 0 | 0 | 0.000837928 |
| 0 | 0 | 0 | 0 | 0 |
| 0 | 0 | 0 | 0 | 0 |
| 0.160185958 | 0 | 0.071813466 | 0.038156193 | 0 |
| 0 | 0 | 0 | 0 | 0 |
| 0 | 0 | 0 | 0 | 0 |
| 0 | 0 | 0 | 0 | 0 |
| 0 | 0 | 0 | 0 | 0 |
| 0 | 0 | 0 | 0 | 0 |
| 0 | 0 | 0 | 0 | 0 |
| 1.151254479 | 1.562826181 | 0.045670873 | 0.182248322 | 2.978372607 |
| 0.036190852 | 0 | 0 | 0 | 0 |
| 0 | 0 | 0 | 0 | 0 |
| 0 | 0 | 0 | 0 | 0 |
| 0 | 0 | 0 | 0 | 0 |
| 0.010439232 | 0 | 0 | 0 | 0 |
| 0 | 0 | 0 | 0 | 0 |
| 0.076858144 | 0 | 0.056447445 | 0.028628142 | 0.131079478 |
| 0 | 0.010462115 | 0 | 0 | 0.004346116 |
| 0 | 0 | 0 | 0 | 0 |
| 0 | 0 | 0.000726436 | 0 | 0 |
| 0 | 0 | 0.025014664 | 0 | 0 |
| 0 | 0 | 0 | 0 | 0 |
| 0 | 0 | 0.022085037 | 0 | 0 |
| 0.11305271 | 0 | 0.01459053 | 0.130177224 | 0 |
| 0.084392082 | 0 | 0.298217558 | 0.057212541 | 0 |
| 0 | 0 | 0 | 0 | 0 |
| 0.010295786 | 0 | 0.004758584 | 0.023044279 | 0 |
| 0 | 0 | 0.157432881 | 0 | 0 |

| | | | | |
|---|---|---|---|---|
| 0 | 0 | 0 | 0 | 0 |
| 0 | 0 | 0.182310034 | 0 | 0 |
| 0 | 0 | 1.756492489 | 0.004351138 | 0 |
| 0.048364635 | 0 | 0.230767355 | 0.021506326 | 0 |
| 0 | 0 | 0 | 0 | 0 |
| 1.234300472 | 0 | 0 | 0 | 0 |
| 0 | 0 | 0 | 0 | 0 |
| 0 | 0 | 0 | 0 | 0 |
| 0.00249072 | 0.061111076 | 0 | 0.002592926 | 0.003648534 |
| 0.000771449 | 0 | 0 | 0.001044036 | 0 |
| 0 | 0 | 0.022079238 | 0.001483218 | 0 |
| 0 | 0 | 0.000866948 | 0 | 0 |
| 0 | 0 | 0 | 0 | 0 |
| 0 | 0 | 0 | 0 | 0 |
| 0.749416237 | 4.742118196 | 0.147448406 | 0.533848357 | 1.119565173 |
| 0.007678669 | 0.003052778 | 0 | 0 | 0 |
| 0 | 0 | 0 | 0 | 0 |
| 0 | 0 | 0 | 0 | 0 |
| 0 | 0 | 0 | 0 | 0 |
| 0 | 0 | 0 | 0 | 0 |
| 0.366398967 | 0.026176435 | 0.179724735 | 0.008723721 | 0 |
| 0 | 0 | 0.19307822 | 0 | 0 |
| 0 | 0 | 0 | 0 | 0 |
| 0 | 0.029893998 | 0 | 0 | 0 |
| 1.362668415 | 0 | 0 | 0.274733563 | 0.004290572 |
| 1.150370684 | 0 | 2.715565125 | 4.228810672 | 0.092146397 |
| 1.197021729 | 0.22695309 | 0.006219911 | 1.642892674 | 0 |
| 0 | 0 | 0 | 0 | 0 |
| 0.038411965 | 0 | 0.126781109 | 0.028258317 | 0 |
| 0 | 0 | 0.00057496 | 0 | 0 |
| 0 | 0 | 0 | 0 | 0 |
| 0 | 0 | 0.019231447 | 0 | 0 |
| 0 | 0 | 0 | 0 | 0 |
| 0 | 0.003731434 | 0.002168976 | 0 | 0.001309395 |
| 0 | 0 | 0.014084926 | 0.025758798 | 0.006472406 |
| 0.019620895 | 0.002483457 | 0.002377632 | 0 | 0 |
| 0 | 0.416250955 | 0.000920119 | 0 | 0.059215313 |
| 1.586800069 | 1.059306146 | 0.036579826 | 2.452466762 | 0.672251663 |
| 0 | 0 | 0 | 0.956502765 | 0 |
| 0 | 0 | 0 | 0 | 0 |
| 0 | 0 | 0 | 0.108283978 | 0 |
| 0 | 0 | 0.003201802 | 0 | 0 |
| 0.060902229 | 0.033371264 | 0.019088327 | 0.773675641 | 0.149268922 |
| 0 | 0 | 10.3830636 | 0 | 0 |
| 0 | 0 | 1.29278975 | 0 | 0 |
| 0 | 0.001176382 | 0 | 0.000718755 | 0.004153833 |
| 0 | 0 | 0 | 0 | 0 |
| 0.018614166 | 0.007496469 | 0 | 0.138266368 | 0 |

| | | | | |
|---|---|---|---|---|
| 0.006940302 | 0 | 0 | 0 | 0.000858212 |
| 0.083915951 | 0 | 2.887030772 | 0.006702956 | 0 |
| 0 | 0 | 0 | 0 | 0 |
| 0 | 0 | 0 | 0 | 0 |
| 0.00062817 | 0 | 0 | 0 | 0 |
| 0 | 0 | 0.290074507 | 0 | 0 |
| 0 | 0 | 0.062568411 | 0 | 0 |
| 0 | 0 | 0 | 0 | 0 |
| 0 | 0 | 0 | 0 | 0 |
| 0.000630501 | 0.011638049 | 2.170208624 | 0 | 0 |
| 0 | 0.001419558 | 0.001528952 | 0 | 0 |
| 1.136887666 | 0.918672085 | 0.082033096 | 1.28135071 | 1.777185816 |
| 0.003993172 | 0.012074017 | 0.011492316 | 0.680149573 | 0 |
| 0 | 0 | 0 | 0 | 0 |
| 0.002542677 | 0.00938676 | 0.022531127 | 1.104107542 | 0 |
| 0 | 0 | 0 | 0 | 0 |
| 0.000631714 | 0 | 0.002411331 | 0 | 0 |
| 0.001576351 | 0 | 0 | 0 | 0 |
| 0.03613592 | 0.010108025 | 0 | 0 | 0 |
| 0 | 0 | 0.053902476 | 0 | 0 |
| 0.024381882 | 0 | 0.017188364 | 0.106125801 | 0 |
| 0 | 0 | 0.018233067 | 0 | 0 |
| 0.391153686 | 0.082515201 | 0.032004477 | 0.031213105 | 0.058321765 |
| 0 | 0.039506557 | 0 | 0 | 0 |
| 0 | 0 | 0 | 0 | 0 |
| 0 | 0 | 0 | 0 | 0 |
| 0 | 0 | 0 | 0 | 0 |
| 0 | 0 | 0 | 0 | 0 |
| 0 | 0 | 0.002428443 | 0 | 0 |
| 0.014813617 | 0 | 0 | 0 | 0 |
| 0 | 0 | 0 | 0 | 0 |
| 0.011783852 | 0.056726954 | 0.038983051 | 0.046567042 | 0.015066898 |
| 0 | 0 | 0 | 0 | 0 |
| 0.157300892 | 0.069719406 | 0.026216696 | 0.002230803 | 0.009609144 |
| 0.022115038 | 0 | 0.00258746 | 0 | 0 |
| 0.024969921 | 0.004355011 | 0.703800925 | 0.0303338 | 0.014174183 |
| 0 | 0 | 0 | 0 | 0 |
| 0.093193097 | 0 | 0.02164935 | 0 | 0 |
| 0 | 0 | 0 | 0.001185666 | 0 |
| 0 | 0 | 0 | 0 | 0 |
| 0.294196541 | 0 | 0.228168358 | 0.136292313 | 0 |
| 0.020568338 | 0 | 0.161385634 | 0 | 0 |
| 0 | 0 | 0 | 0 | 0 |
| 0.000620816 | 0 | 0.007109203 | 0 | 0 |
| 0.157757276 | 0.003993532 | 0.018798191 | 0.001626667 | 0.017371116 |
| 0 | 0 | 0 | 0 | 0 |
| 0 | 0 | 0.157761622 | 1.240159586 | 0.256891809 |
| 0.001563154 | 0.002518111 | 0.000452027 | 0 | 0 |

| | | | | |
|---|---|---|---|---|
| 0 | 0 | 0 | 0 | 0 |
| 0 | 0 | 0 | 0 | 0 |
| 0 | 0.021569405 | 0 | 0 | 0 |
| 0 | 0 | 0 | 0 | 0 |
| 0 | 0.001861659 | 0 | 0 | 0.001500487 |
| 0 | 0 | 0 | 0 | 0 |
| 0.004276278 | 0 | 0 | 0.065512008 | 0 |
| 0.342813211 | 0.011051964 | 0.009180592 | 0.029790959 | 0.001596921 |
| 0 | 0 | 0.005105409 | 0 | 0 |
| 0.002454967 | 0 | 0 | 0.004104162 | 0 |
| 0 | 0 | 0 | 0 | 0 |
| 0 | 0 | 0 | 0 | 0 |
| 0 | 0 | 0 | 0 | 0 |
| 0.086825411 | 0 | 0.214193362 | 0.350992561 | 0 |
| 0 | 0 | 0 | 0 | 0 |
| 0 | 0 | 0 | 0 | 0 |
| 0.008584538 | 0 | 0.131983267 | 0 | 0 |
| 0 | 0 | 0.116763366 | 0 | 0 |
| 0.047155454 | 0.39744879 | 3.355425818 | 7.868440057 | 0.197845601 |
| 0 | 0 | 0.001636478 | 0 | 0 |
| 0.000988306 | 0 | 0 | 0 | 0 |
| 0 | 0 | 0 | 0 | 0 |
| 0 | 0 | 0 | 0 | 0 |
| 0 | 0 | 0 | 0 | 0 |
| 0 | 0 | 0 | 0 | 0 |
| 0 | 0.004337465 | 0 | 0 | 0.003995399 |
| 0 | 0 | 0 | 0 | 0 |
| 0 | 0.221373328 | 0 | 0.007040747 | 0.007076506 |
| 0 | 0 | 0.669708455 | 0.061770916 | 0 |
| 0.716988394 | 0.209982926 | 0.548253228 | 0.11019061 | 0.023397943 |
| 0 | 0 | 0 | 0 | 0 |
| 0 | 0 | 0 | 0 | 0 |
| 0.000578249 | 0 | 0.002912343 | 0.027389968 | 0 |
| 0 | 0 | 0 | 0 | 0 |
| 0.000732888 | 0.002164474 | 0.000466254 | 0.002777182 | 0.00259191 |
| 0 | 0.003263013 | 0 | 0 | 0 |
| 0.231704326 | 0 | 1.267636381 | 0 | 0 |
| 0.019293435 | 0 | 0.002468957 | 0 | 0 |
| 0 | 0 | 1.69153605 | 0 | 0 |
| 0 | 0 | 0 | 0 | 0 |
| 5.164078417 | 4.357264779 | 2.110139943 | 5.625364614 | 9.431923417 |
| 0.001095741 | 2.208820515 | 0.266058584 | 0.028834466 | 0.487873003 |
| 0 | 0 | 0 | 0 | 0 |
| 0.796962623 | 0 | 0 | 0.046988438 | 0 |
| 0 | 0 | 0 | 0 | 0 |
| 0 | 0 | 0.863633503 | 0 | 0 |
| 0.004907276 | 0 | 0.009205744 | 0 | 0 |
| 0.001584035 | 0.000779701 | 0.004198931 | 0.006252592 | 0.001975081 |

| | | | | |
|---|---|---|---|---|
| 0 | 0 | 0 | 0 | 0 |
| 0.001765087 | 0.011829305 | 0.004750842 | 0 | 0.001477479 |
| 0.001744364 | 0.007259234 | 0.014292144 | 0 | 0 |
| 0 | 0 | 0 | 0 | 0 |
| 0.196457556 | 5.233544279 | 0.198218636 | 0.63308313 | 1.231084734 |
| 0 | 0 | 0 | 0 | 0 |
| 0 | 0 | 0 | 0 | 0 |
| 0 | 0 | 0.004884484 | 0.008572273 | 0 |
| 0.006914361 | 0.211212165 | 0 | 0 | 0.14734567 |
| 0 | 0.008993647 | 0 | 0 | 0 |
| 0 | 0 | 0 | 0 | 0 |
| 0 | 0 | 0 | 0 | 0 |
| 0.069394874 | 0 | 0.087848679 | 0.127481857 | 0.127590974 |
| 0.018003512 | 0.01532847 | 0.008082879 | 0.007243638 | 0.020738188 |
| 0 | 0 | 0.412823207 | 0 | 0 |
| 0.566319484 | 1.341175262 | 0.178378685 | 0.148421792 | 0.097398646 |
| 0.002526364 | 0.012435387 | 0 | 0.002469309 | 0 |
| 0 | 0 | 0 | 0 | 0 |
| 0 | 0 | 0 | 0 | 0 |
| 0 | 0 | 0 | 0 | 0 |
| 0 | 0 | 0.000743606 | 0 | 0 |
| 0 | 0 | 0 | 0 | 0 |
| 0 | 0 | 0 | 0 | 0 |
| 0 | 0 | 0 | 0 | 0 |
| 0.125765061 | 0 | 0 | 0 | 0 |
| 0 | 0 | 0 | 0 | 0 |
| 0.00683442 | 0 | 0.009964105 | 0.006441496 | 0.001328043 |
| 0 | 0 | 0 | 0 | 0 |
| 0 | 0 | 0 | 0 | 0 |
| 0 | 0 | 0 | 0 | 0 |
| 0 | 0 | 0 | 0 | 0 |
| 0 | 0 | 0 | 0 | 0 |
| 0 | 0 | 0 | 0 | 0 |
| 0.009960774 | 0 | 0.023878253 | 0 | 0.003534477 |
| 0.085027664 | 0 | 0.22412252 | 0.012246593 | 0 |
| 0 | 0.000940962 | 0 | 0 | 0 |
| 0.000651313 | 0.007213324 | 0 | 0.001322176 | 0 |
| 0 | 0 | 0 | 0.011807732 | 0 |
| 0 | 0 | 0 | 0 | 0 |
| 0 | 0 | 0 | 0 | 0 |
| 1.295704313 | 0 | 0.06226914 | 0.097697324 | 0.010615516 |
| 0 | 0 | 0.000820931 | 0.008731727 | 0 |
| 0.000677322 | 0 | 0.002513606 | 0 | 0 |
| 0 | 0 | 0.116342313 | 0 | 0 |
| 0 | 0 | 0 | 0 | 0 |
| 0 | 0 | 0 | 0 | 0 |
| 0 | 0 | 0 | 0 | 0 |
| 0 | 0 | 0 | 0 | 0 |

| | | | | |
|---|---|---|---|---|
| 1.250735525 | 0 | 0.051480044 | 0.69870101 | 0 |
| 0 | 0 | 0 | 0 | 0 |
| 0 | 0 | 0 | 0 | 0 |
| 0 | 0 | 0 | 0 | 0 |
| 0.156576105 | 0 | 0.085937709 | 0.068479127 | 0.070857223 |
| 0 | 0 | 0 | 0 | 0 |
| 0.003780332 | 0.369673462 | 0 | 0.26148922 | 0.032376046 |
| 0 | 0 | 0 | 0 | 0 |
| 0 | 0 | 0 | 0 | 0 |
| 0 | 0 | 0 | 0 | 0.005195066 |
| 0.007131089 | 0 | 0.00594093 | 0 | 0 |
| 0 | 0.055327543 | 0.035183466 | 0 | 0 |
| 0 | 0 | 0 | 0 | 0 |
| 0 | 0 | 0 | 0 | 0 |
| 0 | 0 | 0 | 0.002792771 | 0 |
| 0 | 0 | 0 | 0 | 0 |
| 0 | 0.053519071 | 0 | 0 | 0.779959216 |
| 0 | 0.0047705 | 0 | 0 | 0 |
| 0 | 0 | 0 | 0 | 0 |
| 0 | 0 | 0 | 0 | 0 |
| 0.008397869 | 0.046651056 | 0 | 0.061877277 | 0 |
| 0 | 0 | 0 | 0 | 0 |
| 0.004704925 | 0 | 0.112869137 | 0.003780636 | 0.005199777 |
| 0.00339923 | 0.006370972 | 0.00512912 | 0.000530807 | 0.007646881 |
| 0 | 0 | 0 | 0 | 0 |
| 0.012493711 | 0.003644274 | 0 | 0 | 0 |
| 0 | 0 | 3.204591565 | 0 | 0 |
| 0 | 0 | 0 | 0 | 0 |
| 0 | 0 | 0 | 0 | 0.068563828 |
| 0.697903816 | 1.686877516 | 0 | 0.608363863 | 0.113895733 |
| 0 | 0 | 0.322897196 | 0.003330379 | 0 |
| 0 | 0 | 0 | 0 | 0 |
| 0 | 0 | 0.042881446 | 0 | 0 |
| 0 | 0.977687522 | 0 | 0 | 0.110151558 |
| 0.405286103 | 0 | 0.217672719 | 0.006978749 | 0 |
| 0 | 0 | 5.206663928 | 0.005519999 | 0 |
| 0.077377652 | 0 | 0.001908012 | 0 | 0 |
| 0 | 0 | 0 | 0 | 0 |
| 0 | 0 | 0.20411159 | 0 | 0 |
| 0.038670366 | 0 | 0.775896841 | 0 | 0 |
| 0 | 0 | 0 | 0 | 0 |
| 0 | 0 | 0 | 0 | 0 |
| 0 | 0 | 0 | 0 | 0 |
| 0 | 0 | 0.055779766 | 0 | 0 |
| 0 | 0 | 0 | 0 | 0 |
| 0 | 0 | 0 | 0 | 0 |
| 0 | 0 | 0 | 0 | 0 |
| 0 | 0 | 0.107133899 | 0 | 0 |

| | | | | |
|---|---|---|---|---|
| 0.005205177 | 0.003739305 | 0 | 0 | 0.003061699 |
| 0 | 0 | 0.160282914 | 0 | 0 |
| 0 | 0 | 0 | 0.017874432 | 0 |
| 0 | 0 | 0 | 0 | 0 |
| 0 | 0 | 0 | 0 | 0 |
| 0 | 0 | 0.063617781 | 0 | 0 |
| 0 | 0 | 0 | 0 | 0 |
| 0.131880149 | 2.411369681 | 0.214882114 | 0.61363269 | 0.666065678 |
| 1.608667276 | 0.074345741 | 0.006321707 | 9.309004538 | 0.011413766 |
| 0 | 0.038136939 | 0 | 0 | 0.02153763 |
| 0.034816559 | 0 | 0.629683485 | 0 | 0 |
| 0 | 0 | 0 | 0 | 0 |
| 0.062305986 | 0 | 0.04121931 | 0 | 0 |
| 0 | 0.005552226 | 0 | 0.001106199 | 0.370235143 |
| 0 | 0 | 0 | 0 | 0 |
| 0 | 0 | 0.403999704 | 0 | 0 |
| 0 | 0.4800341 | 0 | 0 | 0 |
| 0 | 0 | 0 | 0 | 0 |
| 0 | 0 | 0 | 0 | 0 |
| 0.045763403 | 0 | 0 | 0 | 0.001387147 |
| 0 | 0 | 0 | 0 | 0 |
| 0 | 0 | 0 | 0 | 0 |
| 0 | 0 | 0.00052805 | 0 | 0 |
| 0 | 0 | 0 | 0 | 0 |
| 0.012299396 | 0 | 0.030546967 | 0.01634541 | 0.024265843 |
| 0 | 0 | 0 | 0 | 0 |
| 0 | 0 | 0 | 0 | 0 |
| 0 | 0 | 0 | 0 | 0 |
| 0 | 0 | 0 | 0 | 0 |
| 0 | 0 | 0 | 0 | 0 |
| 0 | 0 | 0 | 0 | 0 |
| 0 | 0 | 0 | 0 | 0 |
| 0 | 0 | 0 | 0 | 0 |
| 0.001118616 | 0 | 0.00533737 | 0 | 0 |
| 0 | 0 | 0.002528198 | 0 | 0 |
| 0 | 0 | 0 | 0 | 0 |
| 0 | 0 | 0 | 0 | 0 |
| 0 | 0 | 0 | 0 | 0 |
| 0 | 0 | 0.001064215 | 0.002037489 | 0 |
| 0 | 0.002073887 | 0 | 0 | 0 |
| 0 | 0 | 0 | 0 | 0 |
| 0 | 0 | 0.03364516 | 0.075886413 | 0.130076003 |
| 0 | 0 | 0 | 0 | 0 |
| 0 | 0 | 0 | 0 | 0 |
| 0 | 0 | 0 | 0 | 0 |
| 0 | 0 | 0 | 0 | 0 |
| 0 | 0 | 0 | 0 | 0 |
| 0 | 0 | 0 | 0 | 0 |

| | | | | |
|---|---|---|---|---|
| 0 | 0 | 0 | 0 | 0 |
| 0 | 0 | 0 | 0.018042631 | 0 |
| 0 | 0 | 0 | 0 | 0 |
| 0 | 0 | 0 | 0 | 0 |
| 0.243028017 | 0.238549695 | 3.508591023 | 0.040361816 | 0.370413631 |
| 0 | 0 | 0.088935077 | 0 | 0 |
| 0 | 0 | 0 | 0 | 0 |
| 0.00250538 | 0.039903148 | 0 | 0.010898498 | 0.001703932 |
| 0 | 0 | 0.001136744 | 0.001692716 | 0 |
| 0 | 0 | 0 | 0.024751395 | 0 |
| 0 | 0 | 0 | 0 | 0 |
| 0 | 0 | 0 | 0 | 0 |
| 0 | 0 | 0 | 0 | 0 |
| 0 | 0 | 0 | 0 | 0 |
| 0 | 0.021951445 | 0 | 0 | 0 |
| 0.30172236 | 0 | 0.011026051 | 0.012392866 | 0 |
| 0 | 0 | 0 | 0 | 0 |
| 0 | 0 | 0 | 0 | 0 |
| 0 | 0 | 0 | 0 | 0 |
| 0 | 0 | 0 | 0 | 0 |
| 0 | 0 | 0 | 0 | 0 |
| 0 | 0 | 0 | 0 | 0 |
| 0 | 0 | 0 | 0 | 0 |
| 0.007588701 | 0 | 0 | 0.001676756 | 0.000842636 |
| 0 | 0 | 0.078457201 | 0.833940175 | 0.091464615 |
| 0 | 0 | 0 | 0 | 0.000575989 |
| 0.004391073 | 0 | 0.061127599 | 0.003833959 | 0 |
| 0 | 0.001541724 | 0 | 0 | 0 |
| 0 | 0 | 0 | 0 | 0 |
| 0 | 0 | 0 | 0 | 0 |
| 0 | 0 | 0 | 0 | 0 |
| 0 | 0 | 0 | 0 | 0 |
| 0 | 0 | 0 | 0 | 0 |
| 0 | 0 | 0 | 0 | 0 |
| 0 | 0 | 0 | 0 | 0 |
| 0 | 0.000934967 | 0 | 0 | 0 |
| 0 | 0 | 0 | 0 | 0 |
| 0.004793839 | 0 | 0.152951118 | 0.035190952 | 0 |
| 0 | 0 | 0 | 0 | 0 |
| 0 | 0 | 0 | 0 | 0 |
| 0 | 0 | 0 | 0 | 0 |
| 0.233031473 | 0 | 0.326916793 | 0 | 0 |
| 0 | 0 | 0 | 0 | 0 |
| 0 | 0 | 0.114298961 | 0 | 0 |
| 0 | 0 | 0 | 0 | 0 |
| 0 | 0 | 0 | 0 | 0 |
| 0 | 0 | 0 | 0 | 0 |
| 0 | 0 | 0 | 0 | 0 |

| | | | | |
|---|---|---|---|---|
| 0.259088172 | 0 | 0.077541513 | 0.255630537 | 0.01594481 |
| 0 | 0 | 0.015132325 | 0 | 0 |
| 0 | 0 | 0 | 0 | 0.000717919 |
| 0 | 0 | 0 | 0 | 0 |
| 0.001727355 | 0 | 0 | 0 | 0 |
| 0 | 0.025523975 | 0 | 0 | 0.012805138 |
| 0 | 0 | 0 | 0 | 0 |
| 0 | 0 | 0 | 0 | 0 |
| 0 | 0 | 0 | 0 | 0 |
| 0 | 0 | 0 | 0 | 0 |
| 0 | 0 | 0 | 0 | 0 |
| 0 | 0 | 0.002573416 | 0 | 0 |
| 0 | 0 | 0.09012888 | 0 | 0 |
| 0 | 0 | 0 | 0 | 0 |
| 0 | 0 | 0 | 0 | 0 |
| 0 | 0 | 0 | 0 | 0 |
| 0 | 0 | 0 | 0 | 0 |
| 0 | 0 | 0 | 0 | 0 |
| 0 | 0 | 0 | 0 | 0 |
| 0 | 0 | 0.000365023 | 0 | 0 |
| 0 | 0.009337283 | 0 | 0 | 0.013635597 |
| 0 | 0 | 0 | 0 | 0 |
| 0.017140089 | 0.008628518 | 0.009499969 | 0.005535517 | 0.030202574 |
| 0.000713068 | 0 | 0.143087259 | 0.074307111 | 0 |
| 0 | 0 | 0.00537944 | 0 | 0 |
| 0 | 0 | 1.27117744 | 0.030910253 | 0 |
| 0 | 0 | 0 | 0 | 0 |
| 0 | 0 | 0 | 0 | 0 |
| 0 | 0 | 0 | 0 | 0.001376097 |
| 0 | 0 | 0 | 0 | 0 |
| 0 | 0 | 0 | 0 | 0 |
| 0 | 0.009832764 | 0.198312199 | 0.170954426 | 0 |
| 0.005472144 | 0.002962876 | 0.003674716 | 0 | 0 |
| 0 | 0 | 0 | 0 | 0 |
| 0.004079006 | 0.255149573 | 0.011591057 | 0.686504756 | 0 |
| 0 | 0 | 0 | 0 | 0 |
| 0 | 0 | 0 | 0 | 0 |
| 0 | 0 | 0.159101824 | 0 | 0 |
| 0.131717433 | 0.000750014 | 0.000897569 | 0 | 0 |
| 0 | 0 | 0 | 0 | 0 |
| 0 | 0 | 0 | 0 | 0 |
| 0 | 0 | 0.005220394 | 0.001110521 | 0 |
| 0.317734604 | 0 | 0.105372465 | 0.207098757 | 0.027466186 |
| 0 | 0 | 0 | 0 | 0 |
| 0 | 0 | 0 | 0 | 0 |
| 0.205170703 | 0 | 0.011669085 | 0.007659772 | 0.002138521 |
| 0 | 0 | 0 | 0 | 0 |
| 0 | 0 | 0 | 0 | 0 |

| | | | | |
|---|---|---|---|---|
| 0 | 0 | 0 | 0 | 0 |
| 0 | 0 | 0 | 0 | 0 |
| 0 | 0 | 0 | 0 | 0 |
| 0 | 0 | 0 | 0 | 0 |
| 0 | 0 | 0.006291679 | 0 | 0 |
| 0 | 0 | 0 | 0 | 0 |
| 0 | 0 | 0 | 0 | 0 |
| 0 | 0.003140776 | 0 | 0 | 0 |
| 0.015024555 | 0.195762065 | 0.342042058 | 0.005980414 | 0 |
| 0.040969242 | 0 | 0.034340083 | 0 | 0.020044143 |
| 0 | 0 | 0 | 0 | 0 |
| 0.006033393 | 0 | 0 | 0 | 0 |
| 0 | 0 | 0 | 0 | 0 |
| 0 | 0 | 0 | 0.098769377 | 0 |
| 0.019891415 | 0.055847601 | 0 | 0 | 0 |
| 0 | 0.008940296 | 0 | 0 | 0 |
| 0 | 0 | 0 | 0 | 0 |
| 0 | 0 | 0 | 0 | 0 |
| 0 | 0 | 0 | 0 | 0 |
| 0.17260907 | 0 | 0.521902826 | 0.626964052 | 0 |
| 0 | 0 | 0 | 0 | 0 |
| 0 | 0 | 0 | 0 | 0 |
| 0 | 0 | 0 | 0 | 0 |
| 0.006366722 | 0.016609444 | 0 | 0 | 0 |
| 0 | 0 | 0.001369915 | 0 | 0 |
| 0 | 0 | 0 | 0 | 0 |
| 0 | 0 | 0 | 0 | 0 |
| 0 | 0 | 0 | 0 | 0 |
| 0 | 0 | 0 | 0 | 0 |
| 0.18742034 | 0 | 0.041145531 | 0.096094346 | 0 |
| 0.260025603 | 0.673864971 | 0.073320869 | 1.411122414 | 0.032233851 |
| 0 | 0 | 0 | 0 | 0 |
| 0 | 0 | 0.520457824 | 0 | 0 |
| 0.220005177 | 0 | 0 | 0.011197169 | 0 |
| 0 | 0 | 0 | 0 | 0 |
| 0 | 0 | 0 | 0 | 0 |
| 0 | 0 | 0 | 0 | 0 |
| 0 | 0 | 0 | 0 | 0 |
| 0.00890269 | 0 | 0.005106688 | 0.011455866 | 0.014094785 |
| 0 | 0 | 0 | 0 | 0 |
| 0 | 0 | 0 | 0 | 0 |
| 0 | 0 | 0.184905487 | 0.026173257 | 0 |
| 0 | 0 | 0.00059403 | 0 | 0 |
| 0 | 0 | 0 | 0 | 0.001388907 |
| 0 | 0 | 0 | 0 | 0 |
| 0 | 0 | 0 | 0 | 0 |
| 0 | 0.348262083 | 0 | 0 | 0.201884707 |
| 0.001855714 | 0 | 0 | 0 | 0 |

| | | | | |
|---|---|---|---|---|
| 0 | 0 | 0 | 0 | 0 |
| 0 | 0 | 0 | 0 | 0 |
| 0 | 0 | 0 | 0 | 0 |
| 0 | 0 | 0 | 0 | 0 |
| 0.050688644 | 0.066393916 | 0.036821048 | 0.058540202 | 0.207671605 |
| 0 | 0 | 0 | 0 | 0 |
| 0 | 0 | 0.006320504 | 0 | 0 |
| 0 | 0 | 0 | 0 | 0 |
| 0 | 0 | 0 | 0 | 0 |
| 0 | 0 | 0 | 0 | 0 |
| 0.023205636 | 0 | 0.009021917 | 0.116827458 | 0.001052158 |
| 0.028758527 | 0 | 0.013452807 | 0.001602597 | 0.004141893 |
| 0.010654291 | 0.001077597 | 0.014237163 | 0 | 0.01985228 |
| 0 | 0 | 0 | 0 | 0 |
| 0 | 0.001713324 | 0 | 0 | 0.000876781 |
| 0 | 0.142003563 | 0.014367078 | 0 | 0 |
| 0.014312113 | 0.003842604 | 0.04304271 | 0 | 0.004247477 |
| 0 | 0.00287735 | 0 | 0 | 0 |
| 0 | 0.17799418 | 0.001629847 | 0 | 0.059937517 |
| 0 | 0.489757095 | 0 | 0.022182375 | 0.007735013 |
| 0 | 0 | 0 | 0 | 0 |
| 0.015250971 | 0 | 0 | 0.031555117 | 0 |
| 0 | 0 | 0 | 0 | 0 |
| 0 | 0 | 0 | 0 | 0 |
| 0 | 0 | 0 | 0 | 0 |
| 0 | 0 | 0 | 0 | 0 |
| 0 | 0.019989382 | 0 | 0.000906926 | 0.029351345 |
| 0 | 0 | 0 | 0 | 0 |
| 0 | 0 | 0 | 0 | 0 |
| 0 | 0 | 0 | 0 | 0 |
| 0.017623536 | 0 | 0 | 0 | 0 |
| 0 | 0.027139901 | 0 | 0 | 0 |
| 0 | 0 | 0 | 0 | 0 |
| 0 | 0 | 0 | 0 | 0 |
| 0 | 0 | 0.007431453 | 0 | 0 |
| 0 | 0 | 0 | 0 | 0 |
| 0 | 0 | 0 | 0 | 0 |
| 0.050808015 | 0.015289088 | 0.119967908 | 0 | 0.003048971 |
| 0.031454515 | 0 | 0.000408387 | 0.296417922 | 0 |
| 0 | 0.035003549 | 0 | 0 | 0 |
| 0 | 0 | 0 | 0 | 0.001449455 |
| 0 | 0 | 0.00101561 | 0 | 0 |
| 0 | 0 | 0 | 0 | 0 |
| 0 | 0.002804756 | 0 | 0.001071045 | 0 |
| 0 | 0 | 0 | 0 | 0 |
| 0 | 0 | 0 | 0 | 0 |
| 0 | 0 | 0 | 0 | 0 |
| 0 | 0 | 0.016904273 | 0 | 0 |

| | | | | |
|---|---|---|---|---|
| 0 | 0 | 0 | 0 | 0 |
| 0.051678478 | 0 | 0.004590186 | 0.010435896 | 0.034594966 |
| 0 | 0 | 0 | 0 | 0 |
| 0.014763266 | 0 | 0.00175884 | 0 | 0.000676897 |
| 0 | 0 | 0 | 0.001665702 | 0 |
| 0 | 0 | 0 | 0 | 0 |
| 0 | 0 | 0.000682728 | 0 | 0 |
| 0.009081243 | 0 | 0 | 0 | 0 |
| 0.009163495 | 0 | 0.005572509 | 0 | 0 |
| 0 | 0 | 0.02174713 | 0 | 0 |
| 0 | 0 | 0.034299226 | 0.043592799 | 0 |
| 0 | 0 | 0 | 0 | 0 |
| 0.323241552 | 0.009723554 | 0.031792638 | 0.288031203 | 0.004638303 |
| 0 | 0 | 0 | 0 | 0 |
| 0 | 0 | 0 | 0 | 0 |
| 0 | 0 | 0 | 0 | 0 |
| 0 | 0 | 0 | 0 | 0 |
| 0 | 0 | 0 | 0 | 0 |
| 0 | 0 | 0 | 0 | 0 |
| 0 | 0 | 0 | 0 | 0 |
| 0 | 0 | 0 | 0 | 0 |
| 0 | 0 | 0 | 0 | 0 |
| 0 | 0.006173203 | 0 | 0 | 0.016637881 |
| 0.001096246 | 0.025361172 | 0.058311919 | 0.081845236 | 0.006710106 |
| 0 | 0 | 0 | 0 | 0.011847928 |
| 0 | 0 | 0.004578223 | 0 | 0 |
| 0.003550298 | 0 | 0.003293873 | 0.001000995 | 0 |
| 0 | 0 | 0 | 0 | 0 |
| 0 | 0 | 0 | 0 | 0 |
| 0 | 0 | 0 | 0 | 0 |
| 0.018299584 | 0.005608447 | 0.013179587 | 0.010747358 | 0 |
| 0 | 0 | 0.006369909 | 0 | 0 |
| 0 | 0 | 0 | 0 | 0 |
| 0 | 0 | 0 | 0 | 0 |
| 0.009118335 | 0.038898329 | 0.258895206 | 0 | 0 |
| 0 | 0 | 0 | 0 | 0 |
| 0 | 0.011646285 | 0 | 0 | 0 |
| 0.006002657 | 2.123153103 | 0 | 0 | 0 |
| 0 | 0.005823575 | 0 | 0.182453226 | 0 |
| 0 | 0 | 0 | 0 | 0 |
| 0 | 0 | 0 | 0 | 0 |
| 0 | 0 | 0 | 0 | 0 |
| 0 | 0 | 3.894624668 | 0 | 0 |
| 0 | 0 | 0 | 0 | 0 |
| 0 | 0 | 0 | 0 | 0 |
| 0.22116853 | 0 | 0.087071748 | 0.365233223 | 0.304858036 |
| 0 | 0 | 0 | 0 | 0 |
| 0 | 0 | 0 | 0 | 0 |

| | | | | |
|---|---|---|---|---|
| 0 | 0 | 0 | 0 | 0 |
| 0 | 0 | 0 | 0 | 0 |
| 0.274705616 | 0 | 0.084838398 | 0.064043166 | 0.134474455 |
| 0 | 0 | 0 | 0 | 0 |
| 0 | 0.005764109 | 0 | 0 | 0 |
| 0 | 0 | 0 | 0 | 0 |
| 0 | 0 | 0 | 0 | 0 |
| 0 | 0 | 0 | 0 | 0 |
| 0 | 0 | 0 | 0 | 0 |
| 0 | 0 | 0 | 0 | 0 |
| 0.000829476 | 0 | 0 | 0 | 0 |
| 0.002653735 | 0 | 0.000750344 | 0 | 0 |
| 0 | 0 | 0 | 0 | 0 |
| 0 | 0 | 0 | 0 | 0 |
| 0 | 0 | 0 | 0 | 0 |
| 0 | 0 | 0 | 0 | 0 |
| 0 | 0 | 0 | 0.001885857 | 0 |
| 0 | 0 | 0 | 0 | 0 |
| 0 | 0 | 0 | 0 | 0 |
| 0.000628587 | 0 | 0 | 0 | 0 |
| 0 | 0 | 0 | 0 | 0 |
| 0 | 0 | 0 | 0 | 0 |
| 0 | 0 | 0 | 0 | 0 |
| 0 | 0 | 0 | 0 | 0 |
| 0 | 0 | 0 | 0 | 0 |
| 0 | 0 | 0 | 0.005696253 | 0 |
| 0 | 0 | 0 | 0 | 0 |
| 0.013738846 | 0 | 0.005347553 | 0.014827647 | 0.00875253 |
| 0 | 0 | 0 | 0 | 0 |
| 0 | 0 | 0 | 0 | 0 |
| 0.010260404 | 0.009369865 | 0.000572591 | 0.000730835 | 0 |
| 0 | 0 | 0 | 0 | 0 |
| 0.017556552 | 0.061572588 | 0.035568775 | 0.036488681 | 0.10405183 |
| 0 | 0.002838519 | 0.002208022 | 0.01105617 | 0 |
| 0 | 0 | 0 | 0.010103967 | 0 |
| 0 | 0 | 0 | 0 | 0 |
| 0 | 0 | 0 | 0 | 0 |
| 0 | 0 | 0 | 0 | 0 |
| 0 | 0 | 0 | 0 | 0 |
| 0 | 0 | 0 | 0 | 0 |
| 0 | 0 | 0 | 0 | 0 |
| 0.114517239 | 0.134826648 | 0 | 0 | 0.144366505 |
| 0.088296204 | 0.052896281 | 0.010661533 | 0.068890486 | 0.071591132 |
| 0 | 0 | 0 | 0 | 0 |
| 0 | 0 | 0 | 0 | 0 |
| 0.017254725 | 0.007710932 | 0.191138823 | 0.007988704 | 0.012043917 |
| 0.000744256 | 0 | 0.006628808 | 0 | 0 |
| 0 | 0 | 0 | 0 | 0 |

| | | | | |
|---|---|---|---|---|
| 0 | 0 | 0 | 0 | 0 |
| 0.002962697 | 0.938910108 | 0.00872607 | 0.026062082 | 0 |
| 0 | 0.011356419 | 0.012027706 | 0.011969147 | 0 |
| 0 | 0 | 0 | 0 | 0 |
| 1.699544053 | 0.191098698 | 0.329938707 | 0.003947369 | 0.209437871 |
| 0 | 0 | 0 | 0 | 0.000813023 |
| 0 | 0 | 0 | 0 | 0 |
| 0 | 0 | 0 | 0 | 0 |
| 0 | 0 | 0 | 0 | 0 |
| 0 | 0 | 0 | 0 | 0 |
| 0 | 0 | 0 | 0 | 0 |
| 0 | 0 | 0 | 0 | 0 |
| 0 | 0 | 0.062441125 | 0 | 0 |
| 0 | 0.086146921 | 0 | 0 | 0.001318888 |
| 0 | 0 | 0 | 0 | 0 |
| 0 | 0 | 0 | 0 | 0 |
| 0.029469621 | 0 | 0.227141742 | 0 | 0 |
| 0 | 0 | 0 | 0 | 0 |
| 0 | 0 | 0 | 0 | 0 |
| 0 | 0 | 0 | 0 | 0 |
| 0 | 0 | 0 | 0 | 0 |
| 0 | 0.000685119 | 0 | 0 | 0 |
| 0 | 0 | 0 | 0.002353722 | 0 |
| 0 | 0 | 0 | 0 | 0 |
| 0.035176378 | 0 | 0.001848097 | 0.003645487 | 0 |
| 0.047500937 | 0.007767557 | 0 | 0.013590486 | 0 |
| 0 | 0 | 0 | 0 | 0 |
| 0.006339531 | 0.004794873 | 0.07016992 | 0.003766642 | 0 |
| 0 | 0 | 0 | 0 | 0 |
| 0 | 0 | 0 | 0 | 0 |
| 0 | 0 | 0.00424384 | 0 | 0 |
| 0 | 0 | 0 | 0 | 0 |
| 0 | 0 | 0 | 0 | 0 |
| 0 | 0 | 0 | 0 | 0 |
| 0 | 0 | 0 | 0 | 0 |
| 0 | 0 | 0 | 0 | 0 |
| 0 | 0 | 0 | 0 | 0 |
| 0 | 0.001171508 | 0 | 0 | 0 |
| 2.092967729 | 0 | 3.408594478 | 0 | 0 |
| 0 | 0 | 0 | 0 | 0 |
| 0 | 0 | 0 | 0 | 0 |
| 0.004323087 | 0 | 0.003850412 | 0 | 0 |
| 0.003839386 | 0.00141738 | 0.000848114 | 0 | 0.001958402 |
| 0 | 0 | 0 | 0 | 0 |
| 0 | 0 | 0.102043093 | 0 | 0 |
| 0 | 0 | 0 | 0.002739838 | 0 |
| 0.007831345 | 0.008311871 | 0.019980717 | 0 | 0.003994628 |
| 0 | 0 | 0 | 0 | 0 |

| | | | | |
|---|---|---|---|---|
| 0 | 0 | 0 | 0 | 0 |
| 0.00756569 | 0.022800115 | 0.000654857 | 0 | 0.047884616 |
| 0.002563077 | 0 | 0.007958869 | 0 | 0 |
| 0 | 0 | 0 | 0 | 0 |
| 0 | 0 | 0 | 0 | 0 |
| 0 | 0 | 0 | 0 | 0 |
| 0 | 0 | 0 | 0 | 0 |
| 0 | 0 | 0 | 0 | 0 |
| 0 | 0 | 0 | 0 | 0 |
| 0 | 0 | 0 | 0 | 0 |
| 0 | 0 | 0 | 0 | 0 |
| 0 | 0 | 0 | 0 | 0 |
| 0.07524277 | 0 | 0.017825225 | 0.098736992 | 0 |
| 0 | 0 | 0 | 0 | 0 |
| 0 | 0.009097058 | 0 | 0 | 0.01309318 |
| 0 | 0 | 0 | 0 | 0 |
| 0 | 0 | 0 | 0 | 0 |
| 0 | 0 | 0 | 0 | 0 |
| 0.002981613 | 0.009749211 | 0 | 0 | 0.001738134 |
| 0 | 0 | 0 | 0 | 0 |
| 0 | 0 | 0 | 0 | 0 |
| 0 | 0.386955134 | 0 | 0.004093938 | 0.001285854 |
| 0.002615068 | 0.014373741 | 0.001078307 | 0 | 0.002173761 |
| 0 | 0.001150251 | 0 | 0 | 0 |
| 0 | 0 | 0 | 0 | 0 |
| 0 | 0 | 0 | 0 | 0 |
| 0 | 0 | 0 | 0 | 0 |
| 0 | 0 | 0 | 0 | 0 |
| 0 | 0 | 0 | 0 | 0 |
| 0 | 0 | 0.001682085 | 0 | 0 |
| 0 | 0 | 0 | 0 | 0 |
| 0 | 0 | 0 | 0 | 0 |
| 0 | 0 | 0 | 0 | 0 |
| 0 | 0 | 0 | 0 | 0 |
| 9.774386305 | 0 | 0 | 0 | 0 |
| 0 | 0 | 0 | 0 | 0 |
| 0 | 0 | 0 | 0 | 0 |
| 0 | 0 | 0 | 0 | 0 |
| 0.001321635 | 0.093956748 | 0 | 0.00459933 | 0.001027264 |
| 0 | 0 | 0 | 0 | 0 |
| 0.143895359 | 0.106993207 | 0.056542155 | 1.943276722 | 0.154050348 |
| 0 | 0 | 0 | 0 | 0 |
| 0 | 0 | 0 | 0 | 0 |
| 0 | 0 | 0 | 0 | 0 |
| 0 | 0 | 0 | 0 | 0 |
| 0.015505919 | 0.132231245 | 0 | 0 | 0.379118628 |
| 0 | 0 | 0 | 0 | 0 |
| 0 | 0 | 0 | 0 | 0 |

| | | | | |
|---|---|---|---|---|
| 0 | 0 | 0 | 0 | 0 |
| 0 | 0 | 0 | 0 | 0 |
| 0 | 0 | 0 | 0 | 0 |
| 0 | 0 | 0 | 0 | 0 |
| 0 | 0 | 0 | 0 | 0 |
| 0 | 0 | 0 | 0 | 0 |
| 0 | 0 | 0 | 0 | 0 |
| 0 | 0.012993026 | 0 | 0 | 0 |
| 0 | 0 | 0 | 0.001776975 | 0.0019646 |
| 0 | 0 | 0 | 0.322984245 | 0 |
| 0.051179077 | 0 | 0.069528085 | 0.041665992 | 0 |
| 0 | 0 | 0 | 0 | 0 |
| 0.019083748 | 0.00082079 | 0 | 0 | 0.002016161 |
| 0 | 0 | 0 | 0 | 0 |
| 0 | 0 | 0 | 0 | 0 |
| 0 | 0 | 0 | 0 | 0 |
| 0 | 0 | 0 | 0 | 0 |
| 0 | 0 | 0 | 0 | 0 |
| 0 | 0 | 0 | 0 | 0 |
| 0.221476116 | 0.152443811 | 0.001049146 | 0.201310276 | 0.373036941 |
| 0 | 0 | 0.001877501 | 0 | 0 |
| 0 | 0 | 0 | 0 | 0 |
| 0 | 0 | 0 | 0 | 0 |
| 0 | 0 | 0 | 0 | 0 |
| 0 | 0 | 0 | 0 | 0 |
| 0 | 0 | 0.159551152 | 0 | 0 |
| 0 | 0 | 0 | 0 | 0 |
| 0 | 0 | 0 | 0 | 0 |
| 0 | 0 | 0 | 0 | 0 |
| 0 | 0 | 0 | 0 | 0.003357159 |
| 0.005145367 | 0 | 0.002386868 | 0 | 0 |
| 0.003387248 | 0 | 0 | 0 | 0 |
| 0 | 0 | 0 | 0 | 0 |
| 0 | 0 | 0 | 0 | 0 |
| 0 | 0 | 0 | 0 | 0 |
| 0 | 0 | 0 | 0 | 0 |
| 0 | 0 | 0.00112816 | 0 | 0 |
| 0 | 0 | 0 | 0 | 0 |
| 0 | 0 | 0 | 0 | 0 |
| 0 | 0 | 0.008151263 | 0 | 0 |
| 0 | 0 | 0 | 0 | 0 |
| 0 | 0 | 0 | 0 | 0 |
| 0.019054248 | 0.277320623 | 0.048110399 | 0 | 0 |
| 0 | 0 | 0 | 0 | 0 |
| 0.002788037 | 0.001829787 | 0.016094815 | 0 | 0.002528226 |
| 0 | 0 | 0 | 0.085112508 | 0 |
| 0 | 0 | 0 | 0 | 0 |
| 0 | 0 | 0 | 0 | 0 |

| | | | | |
|---|---|---|---|---|
| 0 | 0 | 0 | 0 | 0 |
| 0 | 0 | 0 | 0 | 0 |
| 0 | 0 | 0 | 0.003856456 | 0 |
| 0 | 0 | 0 | 0 | 0 |
| 0 | 0 | 0 | 0 | 0 |
| 0.701308655 | 0.003073516 | 0.089870325 | 0.006103115 | 0.001730134 |
| 0 | 0 | 0 | 0 | 0 |
| 0 | 0 | 0 | 0 | 0 |
| 0 | 0 | 0 | 0 | 0 |
| 0 | 0 | 0 | 0 | 0 |
| 0 | 0 | 0 | 0 | 0 |
| 0 | 0 | 0 | 0 | 0 |
| 0 | 0 | 0 | 0 | 0 |
| 0 | 0 | 0 | 0 | 0 |
| 0 | 0 | 0 | 0 | 0 |
| 0 | 0 | 0 | 0 | 0 |
| 0.058629064 | 0 | 0.16076678 | 0.044038567 | 0 |
| 0 | 0 | 0 | 0 | 0 |
| 0 | 0 | 0 | 0 | 0 |
| 0 | 0 | 0 | 0 | 0 |
| 0 | 0 | 0 | 0.000907242 | 0 |
| 0 | 0 | 0 | 0 | 0 |
| 0 | 0 | 0 | 0 | 0 |
| 0 | 0 | 0 | 0 | 0 |
| 0 | 0 | 0 | 0 | 0 |
| 0 | 0 | 0 | 0.014441831 | 0 |
| 0 | 0 | 0 | 0 | 0 |
| 0.077971507 | 1.825224809 | 0.004880073 | 0.108453524 | 0.011415997 |
| 0 | 0 | 0 | 0 | 0 |
| 0 | 0.026546997 | 0 | 0 | 0 |
| 0 | 0 | 0 | 0 | 0 |
| 0 | 0 | 0 | 0 | 0 |
| 0 | 0 | 0 | 0 | 0 |
| 0 | 0 | 0 | 0 | 0 |
| 0 | 0 | 0 | 0 | 0 |
| 0 | 0 | 0 | 0 | 0 |
| 0 | 0 | 0 | 0 | 0 |
| 0 | 0 | 0 | 0 | 0 |
| 0 | 0 | 0.20595283 | 0.034460373 | 6.796308443 |
| 0 | 0 | 0 | 0 | 0 |
| 0 | 0 | 0.005868282 | 0 | 0 |
| 0 | 0 | 0 | 0 | 0 |
| 0 | 0 | 0 | 0 | 0 |
| 0 | 0 | 0 | 0 | 0 |
| 0 | 0 | 0 | 0 | 0 |
| 0 | 0 | 0 | 0 | 0 |
| 0 | 0 | 0 | 0 | 0 |
| 0 | 0 | 0 | 0 | 0 |

| | | | | |
|---|---|---|---|---|
| 0 | 0 | 0 | 0 | 0 |
| 0 | 0.008984961 | 0.011255934 | 0 | 0 |
| 0 | 0 | 0 | 0 | 0 |
| 0 | 0 | 0 | 0 | 0 |
| 0 | 0 | 0 | 0 | 0 |
| 0 | 0 | 0 | 0 | 0 |
| 0 | 0 | 0 | 0 | 0 |
| 0 | 0 | 0 | 0 | 0 |
| 0 | 0 | 0 | 0 | 0 |
| 0 | 0 | 0 | 0 | 0 |
| 0 | 0 | 0.002516373 | 0.004588297 | 0 |
| 0 | 0 | 0 | 0 | 0 |
| 0.019977242 | 0 | 0.153040833 | 0.134504598 | 0 |
| 0 | 0 | 0 | 0 | 0 |
| 0 | 0 | 0 | 0 | 0 |
| 0 | 0 | 0 | 0 | 0 |
| 0 | 0 | 0 | 0 | 0 |
| 0 | 0 | 0 | 0 | 0 |
| 0 | 0 | 0 | 0 | 0 |
| 0 | 0 | 0 | 0 | 0 |
| 0 | 0 | 0 | 0 | 0 |
| 0 | 0 | 0 | 0 | 0 |
| 0 | 0 | 0 | 0 | 0 |
| 0.994401705 | 1.154605223 | 0.914589058 | 1.039850701 | 0.003101857 |
| 0 | 0 | 0 | 0 | 0 |
| 0 | 0 | 0 | 0 | 0 |
| 0 | 0 | 0 | 0 | 0 |
| 0 | 0 | 0 | 0 | 0 |
| 0 | 0 | 0 | 0 | 0 |
| 0 | 0 | 0 | 0 | 0 |
| 0 | 0 | 0 | 0 | 0 |
| 0 | 0 | 0 | 0 | 0 |
| 0 | 0 | 0 | 0 | 0 |
| 0 | 0 | 0 | 0 | 0 |

| PHL018-0142_V3 EYCF | PHL018-0056_V1 REF | PHL018-0109_V3 REF | PHL018-0241_V3 REF | PHL018-0062_V1 EYCF |
|---|---|---|---|---|
| 0.001002336 | 0.063476617 | 0.106133906 | 0.332752161 | 0 |
| 0.025458345 | 0.580978442 | 0.214283151 | 0.167689956 | 2.252963848 |
| 0 | 0 | 0 | 0 | 0.010396873 |
| 0 | 0 | 0 | 0 | 0 |
| 0 | 0 | 0 | 0 | 0 |
| 0 | 0 | 0 | 0 | 0 |
| 0 | 0 | 0 | 0 | 0 |
| 0 | 0 | 0 | 0 | 0 |
| 0 | 0 | 0 | 0 | 0 |
| 0 | 0 | 0 | 0 | 0 |
| 0 | 0 | 0 | 0 | 0 |
| 0 | 0 | 0 | 0 | 0 |
| 0 | 0 | 0 | 0 | 0 |
| 1.234342639 | 0.153883994 | 0.279836551 | 0.108897802 | 14.33926154 |
| 0 | 0 | 0 | 0 | 0 |
| 0 | 0 | 0 | 0 | 0 |
| 0 | 0 | 0 | 0 | 0 |
| 0 | 0 | 0 | 0 | 0 |
| 0 | 0 | 0 | 0 | 0.010687674 |
| 0 | 0 | 0 | 0 | 0 |
| 0 | 0 | 0 | 0 | 0 |
| 0 | 0 | 0 | 0 | 0 |
| 0 | 0 | 0 | 0 | 0 |
| 0 | 0 | 0 | 0 | 0 |
| 0.00129795 | 0.094728997 | 1.357610509 | 0.110010479 | 0 |
| 0 | 0 | 0 | 0 | 0 |
| 0 | 0 | 0 | 0.001426349 | 0 |
| 0 | 0 | 0 | 0 | 0 |
| 0 | 0 | 0 | 0 | 0 |
| 0 | 0.002560394 | 0 | 0 | 0 |
| 0 | 0 | 0 | 0 | 0 |
| 0 | 0 | 0 | 0 | 0 |
| 0 | 0 | 0 | 0 | 0 |
| 0 | 0 | 0 | 0 | 0 |
| 0 | 0 | 0 | 0 | 0 |
| 1.761747334 | 5.344790975 | 0.957324571 | 1.110971565 | 2.695395376 |
| 0 | 0 | 0 | 0 | 0 |
| 0 | 0 | 0 | 0 | 0 |
| 0 | 0 | 0 | 0 | 0 |
| 0 | 0 | 0 | 0 | 0 |
| 0 | 0 | 0 | 0 | 0 |
| 0.654033338 | 0.090718282 | 0.006032321 | 0.032752835 | 0.306944529 |
| 9.656433423 | 1.489243643 | 1.308162058 | 3.76054688 | 0.013953273 |
| 0.001071738 | 0.008009908 | 0.013338548 | 0.012596114 | 0.003748767 |
| 0.01236296 | 0.011314013 | 0.008860259 | 0.004590574 | 0.001175099 |
| 0 | 0 | 0 | 0 | 0 |

| | | | | |
|---|---|---|---|---|
| 0 | 0 | 0 | 0.054838724 | 0 |
| 0 | 0.003152418 | 0.00289022 | 0 | 0.029755697 |
| 0 | 0.006429446 | 0.002389738 | 0 | 0.054230721 |
| 0.361680548 | 0.398903676 | 0.327760846 | 0.588949826 | 0.451962627 |
| 0 | 0.003857521 | 0.006644666 | 0.037613325 | 0.002331061 |
| 0 | 0 | 0 | 0 | 0 |
| 0.029537181 | 0 | 0 | 0.007248187 | 0.003818213 |
| 0 | 0 | 0 | 0 | 0 |
| 0 | 0 | 0 | 0 | 0 |
| 0 | 0 | 0 | 0 | 0 |
| 0.018544951 | 0 | 0.000569265 | 0.002278312 | 0 |
| 0.222174452 | 0.053255271 | 0.384631012 | 0.235569786 | 0.023775969 |
| 0 | 0 | 0 | 0.000495724 | 0 |
| 0 | 0.049042302 | 0.044667466 | 0.115839549 | 0.019784795 |
| 0 | 0 | 0 | 0 | 0 |
| 0 | 0 | 0 | 0 | 0 |
| 0.010633093 | 0.624105746 | 0.002838679 | 0.010299772 | 0 |
| 0 | 0 | 0 | 0 | 0 |
| 0 | 0 | 0 | 0 | 0 |
| 0 | 0 | 0 | 0 | 0 |
| 0 | 0.181208667 | 0.024693912 | 0.01775475 | 1.800842963 |
| 0 | 0.010403503 | 0.005576182 | 0.040929544 | 0.001436276 |
| 0.001765464 | 0 | 0 | 0.210230917 | 0 |
| 0 | 0 | 0 | 0 | 0 |
| 0 | 0 | 0 | 0 | 0 |
| 0 | 0 | 0 | 0 | 0 |
| 0 | 0 | 0 | 0 | 0 |
| 0 | 0 | 0 | 0 | 0 |
| 0.140507781 | 0 | 0 | 0 | 0.008370434 |
| 0 | 0 | 0 | 0 | 0 |
| 0 | 0 | 0 | 0 | 0.004882378 |
| 0 | 0 | 0 | 0 | 0 |
| 0 | 0 | 0 | 0 | 0 |
| 0 | 0 | 0.002713525 | 0 | 0 |
| 0.011480729 | 0 | 0 | 0 | 0 |
| 0.011385264 | 0 | 0 | 0 | 0 |
| 0 | 0 | 0 | 0 | 0 |
| 0 | 32.63860371 | 0 | 0 | 0.010836844 |
| 0 | 0 | 0 | 0 | 0 |
| 0 | 0 | 0 | 0 | 0 |
| 0 | 0 | 0 | 0 | 0 |
| 0 | 0 | 0 | 0 | 0 |
| 0 | 0 | 0 | 0 | 0 |
| 0 | 0 | 0 | 0 | 0 |
| 0 | 0.076355417 | 0.592553859 | 0 | 0 |
| 0 | 0 | 0 | 0 | 0 |
| 0.002378498 | 0 | 0 | 0.000502677 | 0 |
| 0.000883492 | 0 | 0.093401617 | 0 | 0 |

| | | | | |
|---|---|---|---|---|
| 0 | 0 | 0 | 0 | 0 |
| 0 | 0 | 0 | 0 | 0 |
| 0 | 0 | 0 | 0 | 0 |
| 0 | 0 | 0 | 0 | 0 |
| 2.860688333 | 0 | 0 | 0 | 0 |
| 0 | 0 | 0.000901823 | 0 | 0 |
| 0 | 0 | 0 | 0 | 0 |
| 0 | 0 | 0 | 0 | 0 |
| 0.079114288 | 0 | 0.002269397 | 0.001571985 | 0.000771263 |
| 0 | 0 | 0 | 0 | 0 |
| 0 | 0 | 0 | 0 | 0 |
| 0.042784887 | 0.010237714 | 0 | 0.000608348 | 0.102078001 |
| 0 | 0 | 0 | 0 | 0 |
| 0 | 0 | 0.030011146 | 0 | 0 |
| 0 | 0 | 0 | 0 | 0 |
| 0 | 0 | 0 | 0 | 0 |
| 0 | 0 | 0 | 0 | 0 |
| 0 | 0 | 0 | 0 | 0 |
| 0.015719676 | 0 | 0.828294109 | 0 | 0 |
| 0 | 0 | 0 | 0 | 0 |
| 0 | 0 | 0 | 0 | 0 |
| 0 | 0 | 0 | 0 | 0 |
| 0 | 0.001353694 | 0 | 0 | 0 |
| 0 | 0 | 0 | 0 | 0 |
| 0 | 0 | 0 | 0 | 0 |
| 0 | 0 | 0 | 0 | 0 |
| 0 | 0 | 0 | 0 | 0 |
| 0 | 0 | 0.002864072 | 0 | 0 |
| 0 | 0 | 0 | 0 | 0 |
| 0 | 0 | 0 | 0 | 0 |
| 0 | 0.007444276 | 0.058407178 | 0.055641077 | 0 |
| 0.028555231 | 0.017872183 | 0 | 0.333828721 | 0 |
| 0 | 0 | 0 | 0 | 0 |
| 0 | 0 | 0 | 0 | 0 |
| 0 | 0 | 0 | 0 | 0 |
| 0 | 0 | 0 | 0 | 0 |
| 0 | 0 | 0 | 0 | 0 |
| 0 | 0 | 0 | 0 | 0 |
| 0 | 0 | 0.053524975 | 0.03821443 | 0 |
| 0 | 0 | 0 | 0 | 0 |
| 0 | 0 | 0 | 0 | 0 |
| 0 | 0 | 0 | 0 | 0 |
| 0 | 0.001388734 | 0 | 0 | 0 |
| 0 | 0 | 0 | 0 | 0 |
| 0 | 0 | 0 | 0 | 0 |
| 0 | 0 | 0 | 0 | 0 |
| 0 | 0 | 0 | 0 | 0 |
| 0 | 0 | 0 | 0 | 0 |

| | | | | |
|---|---|---|---|---|
| 0 | 0 | 0 | 0 | 0 |
| 0 | 0 | 0 | 0 | 0 |
| 0 | 0 | 0 | 0 | 0 |
| 0 | 0 | 0 | 0 | 0 |
| 0.004856548 | 0.06588302 | 0.084372852 | 0 | 0 |
| 0 | 0 | 0 | 0 | 0 |
| 0 | 0 | 0 | 0 | 0 |
| 0 | 0 | 0 | 0 | 0 |
| 0 | 0 | 0 | 0 | 0 |
| 0 | 0 | 0 | 0 | 0 |
| 0 | 0 | 0 | 0 | 0 |
| 0 | 0 | 0 | 0 | 0 |
| 0 | 0 | 0 | 0 | 0 |
| 0 | 0 | 0 | 0 | 0 |
| 0 | 0 | 0 | 0 | 0 |
| 0.038317276 | 0 | 0.562456128 | 0 | 0 |
| 0 | 0 | 0 | 0 | 0 |
| 0 | 0 | 0 | 0 | 0 |
| 0 | 0 | 0 | 0 | 0 |
| 0 | 0 | 0 | 0 | 0 |
| 0 | 0 | 0 | 0 | 0 |
| 0 | 0 | 0 | 0 | 0 |
| 0 | 0 | 0.0168343 | 0 | 0 |
| 0 | 0 | 0 | 0 | 0 |
| 0 | 0 | 0 | 0 | 0 |
| 0 | 0 | 0 | 0 | 0 |
| 0 | 0.011419986 | 0 | 0 | 0 |
| 0 | 0 | 0 | 0 | 0 |
| 0 | 0 | 0 | 0 | 0 |
| 0 | 0 | 0 | 0 | 0 |
| 0 | 0 | 0 | 0 | 0 |
| 0.001893481 | 0 | 0.000953221 | 0.00234768 | 0.003628304 |
| 0 | 0 | 0 | 0 | 0 |
| 0 | 0 | 0 | 0 | 0 |
| 0 | 0 | 0 | 0 | 0 |
| 0 | 0 | 0 | 0 | 0 |
| 0 | 0 | 0 | 0 | 0 |
| 0 | 0 | 0 | 0 | 0 |
| 0 | 0 | 0.013892102 | 0 | 0 |
| 0 | 0 | 0 | 0 | 0 |
| 0 | 0 | 0 | 0 | 0 |
| 0 | 0 | 0 | 0 | 0 |
| 0 | 0 | 0 | 0 | 0 |
| 0 | 0 | 0 | 0 | 0 |
| 0 | 0 | 0 | 0 | 0 |
| 0 | 0 | 0 | 0 | 0 |
| 0 | 0 | 0 | 0 | 0 |
| 0 | 0 | 0 | 0 | 0 |

| | | | | |
|---|---|---|---|---|
| 0 | 0 | 0 | 0 | 0 |
| 0 | 0 | 0 | 0 | 0 |
| 0 | 0 | 0 | 0 | 0 |
| 0 | 0 | 0 | 0 | 0 |
| 0 | 0 | 0 | 0 | 0 |
| 0 | 0 | 0 | 0 | 0 |
| 0 | 0 | 0 | 0 | 0 |
| 0 | 0 | 0 | 0 | 0 |
| 0 | 0 | 0 | 0 | 0 |
| 0 | 0 | 0 | 0 | 0 |
| 0 | 0 | 0 | 0 | 0 |
| 0 | 0 | 0.002775867 | 0 | 0 |
| 0 | 0.002155589 | 0.005016765 | 0.001544469 | 0 |
| 0 | 0 | 0 | 0 | 0 |
| 0 | 0 | 0 | 0 | 0 |
| 0 | 0 | 0 | 0 | 0 |
| 0 | 0 | 0 | 0 | 0 |
| 0 | 0 | 0 | 0 | 0 |
| 0 | 0 | 0 | 0 | 0 |
| 0 | 0 | 0 | 0 | 0 |
| 0.037265499 | 0.031313373 | 0.011938366 | 0.061693596 | 0.023802818 |
| 0 | 0 | 0 | 0 | 0 |
| 0 | 0 | 0 | 0 | 0 |
| 0 | 0 | 0 | 0 | 0 |
| 0 | 0 | 0 | 0 | 0 |
| 0 | 0 | 0 | 0 | 0 |
| 0.004735101 | 0.008486139 | 0.001820925 | 0.005962668 | 0.002160343 |
| 0 | 0 | 0 | 0 | 0 |
| 0 | 0 | 0 | 0 | 0 |
| 0 | 0 | 0 | 0 | 0 |
| 0 | 0 | 0 | 0 | 0.029072183 |
| 0 | 0 | 0 | 0 | 0 |
| 0 | 0 | 0 | 0 | 0 |
| 0 | 0.054378795 | 0 | 0.033692305 | 0 |
| 0 | 0.026829325 | 0.022563427 | 0.002903827 | 0.005229506 |
| 0 | 0 | 0 | 0 | 0 |
| 0 | 0 | 0 | 0 | 0 |
| 0 | 0 | 0 | 0 | 0 |
| 0.159811797 | 0 | 0 | 0.000496691 | 0.025928792 |
| 0.002536162 | 0 | 0 | 0 | 0 |
| 0 | 0 | 0 | 0 | 0 |
| 0 | 0 | 0 | 0.000682365 | 0 |
| 0 | 0.000629009 | 0 | 0 | 0 |
| 0 | 0 | 0 | 0 | 0 |
| 0 | 0 | 0 | 0 | 0 |
| 0 | 0.038988882 | 0 | 0 | 0 |
| 0 | 0 | 0.01421757 | 0 | 0 |
| 0 | 0 | 0.005667297 | 0.010711908 | 0 |

| | | | | |
|---|---|---|---|---|
| 0 | 0 | 0 | 0 | 0 |
| 0 | 0 | 0 | 0 | 0 |
| 0.001435382 | 0 | 0 | 0 | 0.001047806 |
| 0 | 0 | 0 | 0 | 0 |
| 0 | 0 | 0 | 0 | 0 |
| 0 | 0 | 0 | 0 | 0 |
| 0.057242654 | 0 | 0 | 0 | 0 |
| 0.01806465 | 0 | 0 | 0 | 0.005769272 |
| 0 | 0 | 0 | 0.016431058 | 0 |
| 0 | 0 | 0 | 0 | 0 |
| 0 | 0 | 0 | 0 | 0 |
| 0 | 0 | 0 | 0 | 0 |
| 0 | 0 | 0 | 0 | 0 |
| 0 | 0 | 0 | 0 | 0 |
| 0.001733663 | 0 | 0 | 0.001343453 | 0 |
| 0 | 0 | 0 | 0 | 0 |
| 0 | 0 | 0 | 0 | 0 |
| 0 | 0 | 0 | 0 | 0 |
| 0.075083096 | 0.010233792 | 0.006567827 | 0.030577307 | 0 |
| 0 | 0 | 0.393524683 | 0 | 0 |
| 0 | 0.008410564 | 0.000593156 | 0 | 0.222766287 |
| 0 | 0 | 0 | 0 | 0 |
| 0 | 0 | 0 | 0 | 0 |
| 0 | 0 | 0 | 0 | 0 |
| 0 | 0.005569788 | 0.002356859 | 0.010521001 | 0.004271934 |
| 0 | 0 | 0 | 0 | 0 |
| 0 | 0 | 0 | 0 | 0 |
| 0 | 0 | 0 | 0 | 0 |
| 0.173350199 | 0 | 0.001312025 | 0.005193281 | 0.03852546 |
| 0 | 0 | 0 | 0 | 0 |
| 0.022352204 | 0.109731408 | 0.069928956 | 0.215239573 | 0.405705307 |
| 0 | 0 | 0 | 0 | 0 |
| 0 | 0 | 0 | 0 | 0.293712904 |
| 0 | 0 | 0 | 0 | 0 |
| 0 | 0 | 0 | 0 | 0 |
| 0 | 0 | 0 | 0 | 0 |
| 0 | 0 | 0 | 0 | 0 |
| 0 | 0 | 0 | 0 | 0 |
| 0 | 0 | 0 | 0 | 0 |
| 0 | 0 | 0 | 0.00053648 | 0 |
| 0 | 0 | 0 | 0 | 0 |
| 0.001059737 | 0.002747832 | 0.017931381 | 0.017519228 | 0 |
| 0 | 0.000678098 | 0.002735471 | 0.000689029 | 0 |
| 0 | 0 | 0 | 0 | 0 |
| 0 | 0 | 0 | 0 | 0 |
| 0 | 0 | 0 | 0 | 0 |
| 0 | 0 | 0 | 0 | 0 |
| 0 | 0 | 0 | 0 | 0 |

| | | | | |
|---|---|---|---|---|
| 0 | 0 | 0 | 0 | 0 |
| 0 | 0 | 0 | 0 | 0 |
| 0 | 0.006291824 | 0.007815401 | 0.011686583 | 0.005059226 |
| 0 | 0 | 0 | 0 | 0 |
| 0 | 0 | 0.001776348 | 0 | 0 |
| 0 | 0 | 0 | 0 | 0 |
| 0 | 0 | 0 | 0 | 0 |
| 0 | 0 | 0 | 0 | 0 |
| 0 | 0 | 0 | 0 | 0 |
| 0 | 0 | 0 | 0 | 0 |
| 0 | 0 | 0 | 0 | 0 |
| 0 | 0 | 0 | 0 | 0 |
| 0 | 0 | 0 | 0 | 0 |
| 0.002205954 | 0.004710504 | 0 | 0.00284907 | 0 |
| 0.002023029 | 0 | 0.000848698 | 0.004232762 | 0 |
| 0.00216592 | 0 | 0.029440059 | 0 | 0 |
| 0 | 0.002742331 | 0.001934032 | 0.005001477 | 0 |
| 0 | 0 | 0 | 0 | 0 |
| 0 | 0 | 0 | 0 | 0 |
| 0.002113353 | 0 | 0 | 0 | 0 |
| 0 | 0 | 0 | 0 | 0 |
| 0 | 0.001142558 | 0.007158103 | 0.012577239 | 0 |
| 0 | 0 | 0 | 0 | 0 |
| 0 | 0 | 0 | 0 | 0 |
| 0 | 0 | 0 | 0 | 0 |
| 0 | 0 | 0 | 0 | 0 |
| 0 | 0 | 0 | 0 | 0.001334363 |
| 0 | 0 | 0 | 0 | 0 |
| 0 | 0 | 0 | 0 | 0 |
| 0.00240793 | 0 | 0 | 0 | 0 |
| 0.00350803 | 0 | 0 | 0 | 0 |
| 0 | 0 | 0 | 0 | 0 |
| 0.00258382 | 0 | 0 | 0 | 0 |
| 0 | 0 | 0 | 0 | 0 |
| 0 | 0 | 0 | 0 | 0 |
| 0.360336036 | 0 | 0 | 0.001932403 | 0.014790287 |
| 0 | 0 | 0 | 0 | 0 |
| 0.496983016 | 0 | 0.172410341 | 0.005267333 | 0 |
| 0 | 0 | 0 | 0 | 0 |
| 0 | 0 | 0 | 0 | 0 |
| 0 | 0 | 0 | 0 | 0 |
| 0 | 0 | 0 | 0 | 0 |
| 0 | 0 | 0 | 0 | 0 |
| 0 | 0 | 0 | 0 | 0.002666869 |
| 0.008447906 | 0 | 0 | 0 | 0 |
| 0.009646757 | 0.001471378 | 0 | 0 | 0 |
| 0.001133533 | 0.004668107 | 0 | 0 | 0 |
| 0 | 0 | 0.006642245 | 0 | 0.009535223 |

| | | | | |
|---|---|---|---|---|
| 0 | 0 | 0.284277716 | 0 | 0.372212475 |
| 0 | 0 | 0 | 0 | 0.021113387 |
| 0 | 0 | 0.000914336 | 0.000675573 | 0.001193244 |
| 0 | 0 | 0 | 0 | 0 |
| 0.053284429 | 0 | 0.002943712 | 0.009515685 | 0.023410102 |
| 0.090818163 | 0 | 0 | 0 | 0 |
| 0 | 0 | 0 | 0 | 0 |
| 0.004610485 | 0.057945109 | 0.066535953 | 0.018340199 | 0.012620927 |
| 0 | 0.006827905 | 0 | 0 | 0 |
| 0.002534468 | 0.004348928 | 0.003721398 | 0.003437025 | 0.215365643 |
| 0 | 0.005242658 | 0 | 0 | 0 |
| 1.362545831 | 0.000890671 | 0.01143227 | 0.002451119 | 0.225125375 |
| 0.014395727 | 0.003040225 | 0.001393679 | 0 | 0.008083578 |
| 0 | 0 | 0.005174007 | 0.002172106 | 0 |
| 0 | 0 | 0 | 0 | 0 |
| 0.001108224 | 0 | 0 | 0 | 0 |
| 0.003626573 | 0 | 0 | 0 | 0 |
| 0 | 0 | 0 | 0 | 0.001754634 |
| 0 | 0.003000992 | 0 | 0 | 0.212991916 |
| 0 | 0 | 0 | 0 | 0.045134515 |
| 0 | 0 | 0 | 0 | 0 |
| 0 | 0 | 0 | 0 | 0 |
| 0 | 0 | 0 | 0 | 0 |
| 0 | 0.041123087 | 0.289397424 | 0 | 0 |
| 0 | 0 | 0 | 0 | 0 |
| 0 | 0.008655991 | 0.002344739 | 0 | 0 |
| 0 | 1.205095995 | 0.023394846 | 0 | 0.001836067 |
| 0 | 0.010229601 | 0 | 0 | 0.098676276 |
| 0 | 0 | 0.001108004 | 0 | 0.231759668 |
| 0 | 0 | 0 | 0 | 0.000807264 |
| 0 | 0 | 0 | 0 | 0 |
| 0 | 0 | 0 | 0.004761176 | 0 |
| 0 | 0 | 0 | 0 | 0 |
| 0 | 0 | 0 | 0 | 0.001539187 |
| 0 | 0 | 0 | 0 | 0 |
| 0 | 0 | 0 | 0 | 0 |
| 0 | 0 | 0 | 0 | 0.759577193 |
| 0 | 0 | 0 | 0 | 0 |
| 0 | 0 | 0.005940591 | 0 | 0 |
| 0 | 0 | 0 | 0 | 0.037959559 |
| 0 | 0 | 0 | 0 | 0 |
| 0 | 0 | 0 | 0 | 0 |
| 0 | 0 | 0 | 0 | 0 |
| 0 | 0 | 0 | 0 | 0 |
| 0 | 0 | 0 | 0 | 0 |
| 0 | 0 | 0 | 0 | 0 |
| 0 | 0.007094724 | 0.011663474 | 0.000828631 | 0 |
| 0 | 0 | 0 | 0 | 0.015277245 |

| | | | | |
|---|---|---|---|---|
| 0 | 0 | 0 | 0 | 0 |
| 0 | 0.001093634 | 0 | 0 | 0 |
| 0 | 0 | 0 | 0 | 0 |
| 0 | 0 | 0 | 0 | 0 |
| 0 | 0 | 0 | 0 | 0 |
| 0 | 0.002697633 | 0 | 0.005221179 | 0.007300752 |
| 0 | 0.035566918 | 0 | 0.058600917 | 0 |
| 0 | 0 | 0 | 0 | 0 |
| 0 | 0 | 0.114720843 | 0 | 0 |
| 0 | 0 | 0 | 0 | 0 |
| 0.018071212 | 0.007598515 | 0 | 0.043525259 | 0.03030525 |
| 0 | 0 | 0 | 0.003015141 | 0 |
| 0 | 0 | 0 | 0 | 0 |
| 0.000922853 | 0.002744795 | 0.001742193 | 0.001609062 | 0.00189469 |
| 0 | 0 | 0 | 0 | 0 |
| 0 | 0 | 0 | 0 | 0 |
| 0 | 0 | 0.006490048 | 0 | 0 |
| 0 | 0 | 0 | 0 | 0.033598858 |
| 0 | 0 | 0.001236198 | 0 | 0 |
| 0 | 0 | 0 | 0 | 0 |
| 0 | 0.001364956 | 0 | 0 | 0 |
| 0 | 0.071375633 | 0.265914594 | 0 | 0 |
| 0 | 0 | 0 | 0.101455553 | 0 |
| 0 | 0 | 0 | 0 | 0 |
| 0 | 0 | 0 | 0 | 0 |
| 0 | 0 | 0 | 0 | 0 |
| 0 | 0 | 0 | 0 | 0.265923678 |
| 0 | 0.001386739 | 0 | 0 | 0 |
| 0 | 0 | 0 | 0 | 0 |
| 0.426441384 | 0.147314479 | 0.105836245 | 0.247909609 | 0.103374273 |
| 0 | 0.002148606 | 0 | 0.002789698 | 0 |
| 0 | 0 | 0.001290649 | 0.001021734 | 0.024864163 |
| 0 | 0 | 0 | 0 | 0 |
| 0 | 0 | 0.001860034 | 0 | 0.000809139 |
| 0 | 0 | 0.004020802 | 0 | 0 |
| 0 | 0 | 0 | 0 | 0 |
| 0.013843974 | 0 | 0.246416911 | 0 | 0 |
| 0.000940633 | 0 | 0.017954891 | 0.004920189 | 0.035619805 |
| 0 | 0 | 0 | 0 | 0 |
| 0 | 0 | 0 | 0 | 0 |
| 0 | 0.022912359 | 0.008275349 | 0.070403641 | 0.033575831 |
| 0.128379637 | 0.947958575 | 0.14631217 | 1.413636343 | 0 |
| 0 | 0 | 0.018797534 | 0.003171741 | 0.009631761 |
| 0 | 0 | 0.016319025 | 0 | 0.138847797 |
| 0 | 0 | 0 | 0 | 0.002538035 |
| 0 | 0 | 0 | 0 | 0.005643992 |
| 0.000995368 | 0 | 0 | 0 | 0.002724761 |
| 0 | 0 | 0 | 0 | 0 |

| | | | | |
|---|---|---|---|---|
| 0.009579316 | 0.0094409 | 0.004327833 | 0.002284068 | 0 |
| 0 | 0 | 0.01316837 | 0.066405076 | 0 |
| 0.001673404 | 0.032159873 | 0.005616194 | 0.015042387 | 0 |
| 0 | 0 | 0 | 0 | 0 |
| 0.005925873 | 0.075320655 | 0.001104895 | 0.244146082 | 0 |
| 0 | 0 | 0 | 0 | 0 |
| 0 | 0 | 0 | 0 | 0 |
| 0.002202353 | 0 | 0 | 0 | 0 |
| 0 | 0.000940617 | 0.001034859 | 0.010832175 | 0 |
| 0 | 0 | 0 | 0 | 0 |
| 0 | 0 | 0 | 0 | 0 |
| 0 | 0 | 0 | 0 | 0 |
| 0 | 0 | 0 | 0 | 0 |
| 0.001555466 | 0 | 0 | 0.079554049 | 0 |
| 0 | 0 | 0 | 0 | 0 |
| 0 | 0.066110325 | 0.888922911 | 0.403029219 | 0.007922735 |
| 0.100857424 | 0.333971296 | 0.054908153 | 0.011335695 | 0.023885751 |
| 0 | 0.000642029 | 0 | 0 | 0.029020256 |
| 0 | 0 | 0 | 0.006316652 | 0 |
| 0 | 0 | 0 | 0 | 0 |
| 0 | 0.007547764 | 0.005291757 | 0 | 0.007968399 |
| 0 | 0 | 0 | 0.001557097 | 0 |
| 0 | 0 | 0.002535274 | 0 | 0 |
| 0.41410271 | 0 | 0 | 0.291754737 | 0.108181708 |
| 0.001079325 | 0 | 0 | 0 | 0 |
| 0 | 0 | 0.0068077 | 0 | 0.909253659 |
| 0 | 0.002907031 | 0 | 0.004544449 | 0.002229644 |
| 0 | 0 | 0 | 0 | 0 |
| 0 | 0 | 0 | 0 | 0 |
| 0 | 0 | 0 | 0 | 0 |
| 0 | 0.010373701 | 0.013564701 | 0.064656763 | 0 |
| 0.010250694 | 0.001267698 | 0.003486777 | 0.004508466 | 0 |
| 0.207862615 | 0.007483523 | 0.009074347 | 0.000613238 | 0.004091876 |
| 0.393528911 | 0 | 0.001903086 | 0.019334272 | 0.002759557 |
| 0.129781987 | 0 | 0 | 0.003554481 | 0.000984811 |
| 0 | 0 | 0 | 0 | 0 |
| 0 | 0.005019654 | 0 | 0.002032836 | 0.000652824 |
| 0.000746009 | 0.022188185 | 0.081370857 | 0 | 0.068412217 |
| 0.009005552 | 0.269432594 | 0.058123039 | 0.269803407 | 0.000379264 |
| 0 | 0.011163759 | 0 | 0 | 0 |
| 0 | 0 | 0 | 0 | 0 |
| 0 | 0 | 0 | 0 | 0 |
| 0 | 0.005104984 | 0.000702058 | 0.031555931 | 0 |
| 0 | 0 | 0.055457831 | 0 | 0.080106252 |
| 0 | 0 | 0 | 0 | 0.17079817 |
| 0 | 0 | 0 | 0 | 0 |
| 0 | 0 | 0 | 0 | 0 |
| 0 | 0 | 0 | 0 | 0.007088159 |

| | | | | |
|---|---|---|---|---|
| 0 | 0 | 0 | 0 | 0 |
| 0.627708646 | 0.213965231 | 0.359714194 | 0.424194294 | 0 |
| 0 | 0 | 0 | 0 | 0 |
| 0 | 0 | 0 | 0 | 0 |
| 0 | 0 | 0 | 0 | 0 |
| 0 | 0 | 0 | 0 | 0 |
| 0 | 0 | 0 | 0 | 0.026172625 |
| 0 | 0 | 0 | 0 | 0 |
| 0.002326379 | 0 | 0 | 0 | 0.006792878 |
| 0 | 0 | 0 | 0 | 0 |
| 0 | 0.009159757 | 0.044049401 | 0.004097603 | 0 |
| 0.010689226 | 0.057406589 | 0.025961912 | 0.013732866 | 0.037731441 |
| 0.202027389 | 0 | 0.811123099 | 0 | 0 |
| 0 | 0 | 0 | 0 | 0 |
| 0 | 0.002181254 | 0 | 0 | 0.000988582 |
| 0 | 0 | 0 | 0 | 0 |
| 0 | 0.003800441 | 0.007964215 | 0.049956959 | 0.005954671 |
| 0 | 0.010101922 | 0 | 0 | 0.467781243 |
| 0 | 0 | 0 | 0 | 0 |
| 0 | 0 | 0 | 0 | 0.007909434 |
| 0.156823697 | 0.021275728 | 0.017623977 | 0.020069984 | 0.007629439 |
| 0 | 0 | 0 | 0 | 0 |
| 0 | 0 | 0 | 0 | 0 |
| 0 | 0 | 0 | 0 | 0 |
| 0 | 0 | 0 | 0.001327611 | 0 |
| 0 | 0 | 0.005962562 | 0 | 0 |
| 0 | 0.015289624 | 0.034791491 | 0.019498108 | 0 |
| 0.01083748 | 0.012298275 | 0.067288578 | 0.05223681 | 0.191648522 |
| 0.006761969 | 0 | 0.002304878 | 0 | 0 |
| 0 | 0 | 0.001974303 | 0 | 0 |
| 0 | 0.002341236 | 0 | 0 | 0 |
| 0 | 0 | 0 | 0 | 0 |
| 0 | 0 | 0 | 0 | 0 |
| 0 | 0 | 0 | 0 | 0 |
| 0.067053175 | 0.371122478 | 0 | 0 | 0 |
| 0.136462902 | 0.06747796 | 0.046476129 | 0.161464839 | 0.102762211 |
| 0 | 0 | 0 | 0 | 0 |
| 0 | 0 | 0 | 0 | 0 |
| 0.646001845 | 0.004649269 | 0.006281686 | 0.012432144 | 0.072951029 |
| 0 | 0 | 0 | 0 | 0 |
| 0 | 0 | 0 | 0 | 0.051135678 |
| 0 | 0 | 0 | 0 | 0 |
| 0 | 0 | 0 | 0.006041179 | 0 |
| 0 | 0 | 0.000394702 | 0 | 0 |
| 0 | 0 | 0 | 0 | 0.025189137 |
| 0 | 0 | 0.414333097 | 0 | 0 |
| 0 | 0 | 0 | 0 | 0 |
| 0 | 0 | 0 | 0.001178502 | 0 |

| | | | | |
|---|---|---|---|---|
| 0 | 0 | 0 | 0 | 0 |
| 0.009152956 | 0 | 0 | 0 | 0 |
| 0 | 0 | 0 | 0 | 0 |
| 0.388157371 | 0.151125917 | 2.265624506 | 1.983729855 | 0 |
| 0 | 0 | 0 | 0 | 0 |
| 0 | 0 | 0 | 0 | 0 |
| 0 | 0 | 0 | 0.086255646 | 0 |
| 0 | 0 | 0.063609538 | 0.048257929 | 0 |
| 2.787921808 | 0.331634278 | 6.428263336 | 1.17107273 | 0 |
| 0 | 0 | 0.2132008 | 0.008253669 | 0 |
| 0 | 0 | 0 | 0 | 0 |
| 0.001087417 | 0.005307487 | 0.006386683 | 0.000842663 | 0 |
| 0 | 0 | 0 | 0 | 0 |
| 0 | 0.03553207 | 0.264424063 | 0 | 0 |
| 0 | 0.075003246 | 0.084450393 | 0.142782161 | 0 |
| 0 | 0 | 0 | 0 | 10.37056229 |
| 0 | 0 | 0.179559403 | 0.185642121 | 0 |
| 0 | 0.601419734 | 3.92745768 | 8.411073909 | 0 |
| 0 | 0 | 0 | 0 | 0 |
| 0 | 0 | 0 | 0 | 0 |
| 0 | 0 | 0 | 0 | 0 |
| 0 | 0 | 0 | 0 | 0 |
| 0 | 0 | 0 | 0 | 0 |
| 0.00115249 | 0 | 0.310038653 | 0.415063447 | 0.0763742 |
| 0 | 0 | 0.078588598 | 0.001446405 | 0 |
| 0 | 0 | 0.559619986 | 0.027103112 | 0 |
| 0 | 0 | 0.06521865 | 0 | 0 |
| 4.207500208 | 0 | 1.004266559 | 0 | 0.001359832 |
| 0 | 0 | 0 | 0 | 0 |
| 0 | 0 | 0 | 0 | 0 |
| 0 | 0 | 0.248801282 | 0.054750678 | 0 |
| 0 | 0 | 2.734307366 | 0 | 0 |
| 0 | 0 | 0.017101675 | 0.12053958 | 0 |
| 0 | 0 | 0 | 0 | 0 |
| 0 | 0 | 0.228350216 | 0 | 0 |
| 0.001493914 | 0.055643491 | 0 | 0 | 0 |
| 0.23152784 | 0.405757187 | 1.562987795 | 2.227599233 | 0.000704215 |
| 0 | 0 | 0 | 0 | 0 |
| 0 | 0.009150937 | 0.01693794 | 0 | 0 |
| 0 | 0 | 0 | 0.03478198 | 0 |
| 0.033034093 | 0.006205369 | 0.042596407 | 0.015763498 | 0.003490237 |
| 0 | 0 | 0.077665537 | 0.026211539 | 0 |
| 0.034831321 | 0.358843883 | 0.009099932 | 0.002262765 | 0.175852622 |
| 0 | 0 | 0.0921322 | 0 | 0 |
| 0 | 0 | 0 | 0 | 0 |
| 0 | 0 | 0 | 0 | 0 |
| 0 | 0 | 0 | 0 | 0 |
| 0 | 0 | 0.116966599 | 0.206501326 | 0 |

| | | | | |
|---|---|---|---|---|
| 0 | 0 | 0 | 0 | 0 |
| 0.049696646 | 0 | 1.07630277 | 0 | 0 |
| 0.031455574 | 0.027503214 | 0.000504313 | 0 | 0.742793657 |
| 0 | 0 | 0 | 0 | 0 |
| 0 | 0 | 0 | 0 | 0 |
| 0.002525298 | 0 | 0.013110197 | 0.060908793 | 0 |
| 0 | 0 | 0 | 0 | 0 |
| 0 | 0 | 0 | 0.002974712 | 0 |
| 0 | 0 | 0 | 0 | 0 |
| 0 | 0 | 0 | 0 | 0 |
| 0.005901362 | 0.027543344 | 0.006767 | 0.272708906 | 0.009692776 |
| 1.014618592 | 0.544969482 | 0.585854057 | 0.249132009 | 5.729866019 |
| 0 | 0 | 0 | 0 | 0 |
| 0.150308179 | 0.16184101 | 0.225331141 | 0.049025675 | 0.010460839 |
| 0.106068387 | 0.007349066 | 0.004912992 | 0.361578991 | 0.276006009 |
| 0 | 0 | 0.016227549 | 0 | 0 |
| 0 | 0 | 0 | 0 | 0 |
| 0 | 0 | 0 | 0 | 0 |
| 0 | 0 | 0.102641234 | 0 | 0 |
| 0 | 0.007711113 | 0 | 0.015068107 | 0.003252861 |
| 4.473682567 | 0.092945053 | 0.0397022 | 0.173248307 | 0.094027473 |
| 0 | 0 | 0 | 0 | 4.85661575 |
| 0.312388745 | 0.099475034 | 0.304439995 | 0.054304389 | 34.78217103 |
| 0 | 0 | 0 | 0 | 0 |
| 0 | 0 | 0.111312838 | 0 | 0 |
| 4.410708495 | 0.009401875 | 0 | 0 | 0.008292741 |
| 0 | 0 | 0.007941856 | 0.262989418 | 0 |
| 0 | 0 | 0 | 0 | 0 |
| 0 | 0 | 0.055225973 | 0 | 0 |
| 0 | 0 | 0 | 0 | 0 |
| 0 | 0.009153213 | 0.032652149 | 0 | 0 |
| 0.017277527 | 0.066665122 | 0.063732325 | 0.313368772 | 0 |
| 0.009091279 | 0 | 0 | 0.002661456 | 0 |
| 0 | 0.01297115 | 0 | 0 | 0 |
| 0 | 0.010616419 | 0.012082858 | 0 | 0.017033976 |
| 0.006778502 | 0.118199415 | 0.151100827 | 0.079218051 | 0.003677715 |
| 0 | 0.003401077 | 0.003266683 | 0.00548556 | 0 |
| 0 | 0 | 0 | 0 | 0 |
| 0 | 0 | 7.266466105 | 1.243656922 | 0.002151835 |
| 0 | 0 | 0 | 0 | 0 |
| 0.003065066 | 0.007233966 | 0 | 0 | 0 |
| 0 | 0 | 0 | 0 | 0 |
| 0 | 0 | 0 | 0 | 0 |
| 0 | 0 | 0 | 0 | 0 |
| 0.00104461 | 0.029635344 | 0.081511397 | 0.307282781 | 0 |
| 0 | 0 | 0 | 0 | 0 |
| 0 | 0 | 0 | 0 | 0 |
| 0.015625051 | 0.131230941 | 0.013955809 | 0.034371651 | 0 |

| | | | | |
|---|---|---|---|---|
| 0 | 0 | 0 | 0 | 0 |
| 0 | 0 | 0 | 0 | 0 |
| 0 | 0 | 0 | 0 | 0 |
| 0 | 0.000814777 | 0.001195214 | 0.003449629 | 0 |
| 0 | 0 | 0 | 0 | 0 |
| 0.103479974 | 0.2340158 | 0 | 0 | 0 |
| 0 | 0 | 0 | 0 | 0 |
| 0 | 0.006597732 | 0.01869683 | 0.205312659 | 0.009269639 |
| 0 | 0 | 0 | 0 | 0 |
| 0 | 0 | 0 | 0 | 0 |
| 0 | 0.003629783 | 0.004991821 | 0.01029649 | 0 |
| 0 | 0 | 0 | 0 | 0 |
| 0 | 0.005085054 | 0 | 0.052746711 | 0 |
| 0 | 0 | 0 | 0 | 0.305283152 |
| 0 | 0 | 0 | 0 | 0 |
| 0 | 0 | 0 | 0 | 0 |
| 0 | 0 | 0.036630992 | 0 | 0 |
| 0.00165093 | 0.014202043 | 0.022301597 | 0.003198355 | 0 |
| 0 | 0 | 0 | 0 | 0 |
| 0 | 0.000545017 | 0.062960392 | 0 | 0 |
| 0 | 0 | 0 | 0 | 0 |
| 0 | 0 | 0 | 0 | 0 |
| 0.007613618 | 0.551836607 | 0.002714947 | 1.214211771 | 0 |
| 0 | 0 | 0 | 0 | 0 |
| 0 | 0 | 0 | 0 | 0 |
| 0 | 0 | 0 | 0 | 0 |
| 0 | 0 | 0 | 0 | 0 |
| 0 | 0 | 0 | 0 | 0 |
| 0 | 0 | 0 | 0 | 0 |
| 0.634190309 | 0 | 1.017366413 | 0 | 0 |
| 0 | 0 | 0 | 0 | 0 |
| 0 | 0 | 0 | 0 | 0 |
| 0 | 0 | 0 | 0 | 0 |
| 0 | 0 | 0 | 0 | 0 |
| 0 | 0 | 0 | 0.003421544 | 0 |
| 0 | 0 | 0 | 0 | 0 |
| 0.075836979 | 0 | 0 | 0.085070155 | 0 |
| 0 | 0 | 0.001126844 | 0.003512482 | 0.010876123 |
| 0 | 0.00989561 | 0 | 0 | 0 |
| 0 | 0 | 0 | 0.003592833 | 0 |
| 0 | 0 | 0 | 0.163192105 | 0 |
| 0 | 0 | 0 | 0 | 0 |
| 0 | 0.00225153 | 0 | 0 | 0 |
| 0 | 0 | 0 | 0.001389681 | 0 |
| 0 | 0.167820433 | 0.070853385 | 0.112856575 | 0 |
| 0 | 0 | 0 | 0 | 0 |
| 0 | 0 | 0 | 0 | 0 |
| 0 | 0 | 0 | 0 | 0 |

| | | | | |
|---|---|---|---|---|
| 0 | 0 | 0 | 0 | 0 |
| 0 | 0 | 0 | 0 | 0 |
| 0 | 0.217426591 | 0 | 0.919494152 | 0 |
| 0 | 0.102695399 | 0.26929123 | 0 | 0 |
| 0 | 0 | 0 | 0 | 0 |
| 0 | 0.33653429 | 0 | 0 | 0 |
| 0 | 0 | 0 | 0 | 0 |
| 0 | 0 | 0 | 0 | 0 |
| 0 | 0.004025845 | 0.016706636 | 0.006279647 | 1.005421874 |
| 0 | 0 | 0.00688333 | 0 | 0 |
| 0 | 0 | 0 | 0 | 0 |
| 0 | 0 | 0 | 0 | 0 |
| 0 | 0 | 0 | 0.022713254 | 0 |
| 0 | 0 | 0 | 0 | 0 |
| 0.657550816 | 0 | 0.603362733 | 0.120425159 | 0.001062757 |
| 0 | 0 | 0.00074862 | 0 | 0.017911268 |
| 0 | 0 | 0 | 0 | 0.004640015 |
| 0 | 0 | 0 | 0 | 0 |
| 0 | 0 | 0 | 0 | 0 |
| 0 | 0 | 0 | 0 | 0 |
| 0 | 0.018307997 | 0.082589397 | 0 | 0 |
| 0 | 0 | 0 | 0 | 0 |
| 0 | 0 | 0 | 0 | 0 |
| 0 | 0.012537452 | 0 | 0 | 0 |
| 5.676638746 | 0.058804729 | 0.055055968 | 6.662463776 | 0.007453336 |
| 0.289478085 | 6.87727485 | 1.521678895 | 11.45995815 | 0 |
| 0.005063488 | 0 | 0.499618457 | 0 | 0 |
| 0 | 0 | 0 | 0 | 0 |
| 0 | 0.049233668 | 1.069747879 | 0.416156414 | 0 |
| 0 | 0 | 0 | 0 | 0 |
| 0 | 0 | 0 | 0 | 0 |
| 0 | 0.070766617 | 0.063997972 | 0 | 0 |
| 0 | 0 | 0 | 0 | 0 |
| 0 | 0.00705293 | 0.02440118 | 0.006760965 | 0 |
| 0.016531891 | 0.162036224 | 0.050055769 | 0.005755633 | 0.001530365 |
| 0.001016178 | 0.002634884 | 0.004689361 | 0.004331021 | 0.002781726 |
| 0 | 0 | 0.081094782 | 0 | 0 |
| 1.038050915 | 0.357203778 | 0.41686871 | 0.106552998 | 0 |
| 0 | 0 | 0.000897341 | 0 | 0 |
| 0 | 0 | 0 | 0 | 0 |
| 0 | 0 | 0 | 0 | 0 |
| 0 | 0 | 0.001611446 | 0 | 0.0017525 |
| 2.68907206 | 1.476040847 | 0.363707956 | 1.081994317 | 0.001689457 |
| 0 | 0 | 3.877178413 | 0 | 0 |
| 0 | 0 | 0 | 0 | 0 |
| 0 | 0.001762039 | 0.000807742 | 0.002238054 | 0 |
| 0 | 0 | 0 | 0.520587259 | 0 |
| 3.847791981 | 0.113221292 | 0.024664238 | 0 | 0 |

| | | | | |
|---|---|---|---|---|
| 0 | 0.004884336 | 0.014553788 | 0 | 0 |
| 0 | 0 | 0 | 0 | 0 |
| 0 | 0 | 0 | 0 | 0 |
| 0 | 0 | 0 | 0 | 0 |
| 0 | 0 | 0.013375368 | 0.001764755 | 0.013333963 |
| 0 | 0 | 0 | 0 | 0 |
| 0 | 0 | 0 | 0 | 0 |
| 0 | 0 | 0 | 0 | 0 |
| 0 | 0 | 0 | 0 | 0 |
| 0 | 0 | 0.00127857 | 0 | 0 |
| 0 | 0 | 0.001340233 | 0 | 0 |
| 0.109951502 | 1.502448733 | 0.83424714 | 5.268391784 | 0.000479277 |
| 0 | 0.357915203 | 0 | 0.79418045 | 0 |
| 0 | 0 | 0 | 0 | 0 |
| 6.685954245 | 0.272963394 | 0.033699476 | 0.097454819 | 1.236662223 |
| 0 | 0 | 0 | 0 | 0 |
| 0 | 0 | 0.00294637 | 0 | 0 |
| 0 | 0 | 0 | 0 | 0 |
| 0 | 0 | 0.003228139 | 0.001043511 | 0 |
| 0 | 0 | 0 | 0.003801856 | 0 |
| 0.203860501 | 0 | 0.07216028 | 0 | 0 |
| 0 | 0 | 0 | 0 | 0 |
| 0.169097909 | 0.808332446 | 0.647874415 | 1.126576178 | 0.000682737 |
| 0 | 0 | 0 | 0 | 0 |
| 0 | 0 | 0 | 0 | 0 |
| 0 | 0 | 0 | 0 | 0 |
| 0 | 0.013491847 | 0 | 0 | 0 |
| 0 | 0 | 0 | 0 | 0 |
| 0 | 0 | 0 | 0 | 0 |
| 0 | 0 | 0 | 0 | 0 |
| 0 | 0 | 0 | 0 | 0 |
| 0.076327264 | 0.009382998 | 0.132384153 | 0.045684956 | 0.010914846 |
| 0 | 0 | 0 | 0 | 0 |
| 0.00256109 | 0.000911475 | 0 | 0.039251866 | 0 |
| 0 | 0.0023614 | 0.003247491 | 0.002570856 | 0.002186322 |
| 0.053696937 | 0.002609255 | 0.616199599 | 0.010310674 | 0 |
| 0 | 0 | 0 | 0.000633908 | 0 |
| 0.001233697 | 0 | 0 | 0 | 0 |
| 0 | 0 | 0.003197904 | 0 | 0 |
| 0 | 0 | 0 | 0 | 0 |
| 0.308097209 | 0 | 0.004216189 | 0 | 0 |
| 0 | 0.022909316 | 0 | 0 | 0 |
| 0 | 0 | 0 | 0 | 0 |
| 0 | 0 | 0.0552671 | 0 | 0 |
| 0.002178758 | 0.003323165 | 0.04174069 | 0.033485952 | 0.015904586 |
| 0 | 0 | 0 | 0 | 0 |
| 0 | 0 | 0.238403625 | 0.169992978 | 0 |
| 0 | 0.000471468 | 0.008501001 | 0.00598835 | 0 |

| | | | | |
|---|---|---|---|---|
| 0 | 0 | 0 | 0 | 0 |
| 0 | 0 | 0 | 0 | 0 |
| 0 | 0 | 0 | 0.002764808 | 0.003084248 |
| 0 | 0 | 0 | 0 | 0 |
| 0 | 0 | 0 | 0 | 0.000695083 |
| 0 | 0 | 0 | 0 | 0 |
| 0.052296678 | 0.555964918 | 0.273158974 | 0 | 0.000943077 |
| 0.001418738 | 0.001298364 | 0.092998016 | 0.00192397 | 0 |
| 0 | 0 | 0 | 0 | 0 |
| 0 | 0 | 0 | 0 | 0 |
| 0 | 0 | 0 | 0 | 0 |
| 0 | 0.001049115 | 0.000961856 | 0 | 0 |
| 0 | 0 | 0 | 0 | 0 |
| 0.8149342 | 0 | 0 | 0 | 0 |
| 0 | 0 | 0 | 0 | 0 |
| 0 | 0 | 0 | 0 | 0 |
| 0 | 0 | 0 | 0 | 0 |
| 0 | 0 | 0 | 0 | 0 |
| 0.311209615 | 0 | 1.872260969 | 0.052399733 | 0 |
| 0 | 0 | 0 | 0 | 0 |
| 0 | 0 | 0 | 0 | 0 |
| 0 | 0 | 0 | 0 | 0 |
| 0 | 0 | 0 | 0 | 0 |
| 0 | 0 | 0 | 0 | 0 |
| 0.022614581 | 0.000544627 | 0.001997314 | 0.001383516 | 0 |
| 0.00219737 | 0.005878108 | 0 | 0.008775918 | 0.022518322 |
| 0 | 0 | 0 | 0 | 0 |
| 0.142613777 | 0.155798695 | 0.127570737 | 0.039487796 | 0 |
| 0.120120382 | 0 | 0 | 0 | 0 |
| 0.021095188 | 1.070671748 | 0.567422428 | 0.687056508 | 0 |
| 0 | 0 | 0 | 0 | 0 |
| 0 | 0 | 0 | 0 | 0 |
| 0 | 0 | 0.007768547 | 0 | 0 |
| 0 | 0 | 0 | 0 | 0 |
| 0 | 0.107311837 | 0.070445798 | 0.008235778 | 0.001616288 |
| 0 | 0 | 0.053024953 | 0 | 0 |
| 0 | 0 | 0 | 0 | 0 |
| 0 | 0.002207266 | 0.011804798 | 0.001246026 | 0 |
| 0 | 0 | 0 | 0 | 0 |
| 0 | 0 | 0 | 0 | 0 |
| 4.410888904 | 2.103510937 | 0.732951735 | 4.492207493 | 4.464040822 |
| 0.68855528 | 0.149993477 | 0.27170294 | 0.416828219 | 0.08605466 |
| 0 | 0.414366285 | 2.577450726 | 0 | 0 |
| 0 | 0 | 0 | 0 | 0 |
| 0 | 0 | 0 | 0 | 0 |
| 0 | 0 | 1.174907914 | 0 | 0 |
| 0 | 0 | 0.003061936 | 0 | 0 |
| 0 | 0 | 0.015659516 | 0.00840578 | 0.000727787 |

| | | | | |
|---|---|---|---|---|
| 0 | 0 | 0 | 0 | 0 |
| 0 | 0.004354542 | 0.02367882 | 0.017800667 | 0 |
| 0 | 0 | 0.045181412 | 0.008761574 | 0 |
| 0 | 0 | 0 | 0 | 0 |
| 1.999607114 | 1.697812992 | 2.005727362 | 2.255001078 | 0.002724913 |
| 0 | 0 | 0 | 0 | 0 |
| 0 | 0 | 0 | 0 | 0 |
| 0 | 0 | 0.041599546 | 0 | 0 |
| 0 | 0.13434198 | 0.0065983 | 0.002031363 | 0.486613226 |
| 0 | 0 | 0 | 0 | 0 |
| 0 | 0 | 0 | 0 | 0.003044701 |
| 0 | 0 | 0 | 0 | 0 |
| 0 | 0.071638253 | 0.041200389 | 0.086729002 | 0 |
| 0.009800131 | 0.006816159 | 0.001644535 | 0.009416977 | 0.014665576 |
| 0.294525963 | 0 | 2.602020601 | 0 | 0 |
| 0.025412456 | 0 | 0.100402339 | 0.163648584 | 0.023335123 |
| 0.01289035 | 0.034303435 | 0.000996163 | 0.014063482 | 0 |
| 0 | 0 | 0 | 0 | 0 |
| 0 | 0 | 0 | 0 | 0 |
| 0.001024243 | 0 | 0 | 0 | 0 |
| 0 | 0 | 0 | 0 | 0 |
| 0 | 0 | 0 | 0 | 0 |
| 0 | 0 | 0 | 0 | 0 |
| 0 | 0 | 0 | 0 | 0 |
| 0 | 0 | 0 | 0 | 0 |
| 0 | 0 | 0 | 0 | 0 |
| 0.047046997 | 0.017411058 | 0.162351534 | 0 | 0 |
| 0 | 0 | 0 | 0 | 0 |
| 0 | 0 | 0 | 0 | 0 |
| 0 | 0 | 0 | 0.002157468 | 0 |
| 0 | 0 | 0 | 0 | 0 |
| 0 | 0 | 0 | 0 | 0 |
| 0 | 0 | 0.028446477 | 0 | 0 |
| 0 | 0 | 0 | 0.039068562 | 0 |
| 1.192576828 | 0.185045627 | 1.794531423 | 2.636237131 | 0 |
| 0 | 0.023255375 | 0.158454846 | 0.001193447 | 0.002634935 |
| 0 | 0 | 0 | 0.000609923 | 0 |
| 0.099436288 | 0 | 0 | 0 | 0 |
| 0 | 0 | 0 | 0 | 0 |
| 0 | 0 | 0 | 0 | 0 |
| 1.842119794 | 5.742113629 | 1.031460038 | 1.189247596 | 2.870197829 |
| 0 | 0 | 0 | 0 | 0 |
| 0 | 0 | 0 | 0 | 0 |
| 0 | 0 | 0.207980345 | 0 | 0 |
| 0 | 0 | 0 | 0 | 0 |
| 0 | 0 | 0 | 0 | 0 |
| 0 | 0 | 0 | 0 | 0 |
| 0 | 0 | 0 | 0 | 0 |

| | | | | |
|---|---|---|---|---|
| 0.018403401 | 0 | 0.418439667 | 0.391123986 | 0 |
| 0 | 0 | 0 | 0 | 0 |
| 0 | 0 | 0 | 0 | 0 |
| 0 | 0 | 0 | 0 | 0 |
| 0 | 0 | 0.030873731 | 0.047244612 | 0 |
| 0 | 0 | 0 | 0 | 0 |
| 0.050082444 | 0.004800094 | 0.025837264 | 0.296450599 | 0 |
| 0 | 0 | 0 | 0 | 0 |
| 0 | 0 | 0 | 0 | 0 |
| 0 | 0 | 0 | 0 | 0 |
| 0 | 0.006947525 | 0 | 0 | 0 |
| 0 | 0 | 0 | 0 | 0 |
| 0 | 0 | 0 | 0 | 0 |
| 0 | 0 | 0 | 0 | 0 |
| 0 | 0 | 0 | 0 | 0 |
| 0 | 0 | 0 | 0 | 0 |
| 0 | 0 | 0 | 0 | 0 |
| 0 | 0 | 0 | 0 | 0 |
| 0 | 0 | 0 | 0 | 0 |
| 0 | 0 | 0 | 0 | 0 |
| 0.017558308 | 0.606624908 | 0.015002391 | 0 | 0.001028911 |
| 0 | 0 | 0 | 0 | 0 |
| 0 | 0.035934931 | 0.022659756 | 0.046537707 | 0 |
| 0 | 0.028917386 | 0.029826251 | 0.011630982 | 0.003027454 |
| 0 | 0 | 0.000950875 | 0 | 0 |
| 0 | 0 | 0 | 0 | 0 |
| 0 | 0 | 0 | 0 | 0 |
| 0 | 0 | 0 | 0 | 0 |
| 0.053343376 | 0 | 0 | 0 | 0 |
| 0.007731171 | 0.087607781 | 0 | 0.00070483 | 0.046269481 |
| 0 | 0 | 0 | 0 | 0 |
| 0 | 0 | 0 | 0 | 0 |
| 0 | 0 | 0 | 0 | 0 |
| 0 | 0 | 0 | 0 | 0 |
| 0 | 0 | 0 | 0 | 0 |
| 0 | 0 | 0 | 0 | 0 |
| 0.009489062 | 0 | 0 | 0.002297901 | 0 |
| 0 | 0 | 0 | 0 | 0 |
| 0 | 0 | 0.00794795 | 0 | 0 |
| 0 | 0 | 0 | 0 | 0 |
| 0 | 0 | 0 | 0 | 0 |
| 0 | 0 | 0 | 0 | 0 |
| 0 | 0 | 0 | 0 | 0 |
| 0 | 0 | 0 | 0.007601543 | 0 |
| 0 | 0 | 0 | 0 | 0 |
| 0 | 0 | 0 | 0 | 0 |
| 0 | 0 | 0 | 0 | 0 |
| 0 | 0.183994238 | 0 | 0 | 0 |

| | | | | |
|---|---|---|---|---|
| 0 | 0.005756485 | 0.002567528 | 0.097619727 | 0.001085883 |
| 0 | 0 | 0.633108557 | 0.333220703 | 0 |
| 0 | 0 | 0 | 0.020509427 | 0 |
| 0 | 0 | 0 | 0 | 0 |
| 0 | 0 | 0 | 0 | 0 |
| 0 | 0 | 0 | 0 | 0 |
| 0 | 0 | 0.000657932 | 0 | 0 |
| 0.039100678 | 0.245050648 | 0.466620032 | 0.546075872 | 0.142555084 |
| 11.81550499 | 2.006162382 | 1.753326718 | 5.2356283 | 0.014122633 |
| 0 | 0 | 0 | 0.127115429 | 0.021867176 |
| 0.004141997 | 0.187333725 | 0 | 0 | 0 |
| 0 | 0 | 0 | 0 | 0 |
| 0 | 0 | 0 | 0 | 0 |
| 0 | 0 | 0.022211017 | 0 | 0.293467573 |
| 0 | 0 | 0 | 0 | 0 |
| 0 | 0 | 0 | 0 | 0 |
| 0 | 0 | 0 | 0 | 0.008378366 |
| 0 | 0 | 0 | 0 | 0 |
| 0 | 0 | 0 | 0 | 0 |
| 0 | 0 | 0 | 0.034260315 | 0 |
| 0 | 0 | 0 | 0 | 0 |
| 0 | 0 | 0 | 0 | 0 |
| 0 | 0 | 0 | 0 | 0 |
| 0 | 0 | 0 | 0 | 0 |
| 0 | 0 | 0.023031574 | 0 | 0 |
| 0 | 0 | 0 | 0 | 0 |
| 0 | 0 | 0 | 0 | 0 |
| 0 | 0 | 0 | 0 | 0 |
| 0 | 0 | 0 | 0 | 0 |
| 0 | 0 | 0 | 0 | 0 |
| 0 | 0 | 0 | 0 | 0 |
| 0 | 0 | 0 | 0 | 0 |
| 0 | 0 | 0 | 0 | 0 |
| 0 | 0 | 0 | 0 | 0 |
| 0.002469778 | 0.019588639 | 0 | 0 | 0 |
| 0 | 0 | 0 | 0 | 0 |
| 0 | 0 | 0 | 0 | 0 |
| 0 | 0 | 0 | 0 | 0 |
| 0 | 0 | 0 | 0 | 0 |
| 0.000678872 | 0 | 0 | 0 | 0 |
| 0 | 0 | 0 | 0 | 0 |
| 0.117488317 | 0.128350895 | 0.011752858 | 0.97204351 | 0 |
| 0 | 0 | 0 | 0 | 0 |
| 0 | 0 | 0 | 0 | 0 |
| 0 | 0 | 0 | 0 | 0 |
| 0 | 0 | 0.008244363 | 0 | 0 |
| 0 | 0 | 0 | 0 | 0 |
| 0 | 0 | 0 | 0 | 0 |

| | | | | |
|---|---|---|---|---|
| 0 | 0 | 0 | 0 | 0 |
| 0 | 0.005897571 | 0 | 0.001189016 | 0 |
| 0 | 0 | 0 | 0 | 0 |
| 0 | 0 | 0 | 0 | 0 |
| 0.180202081 | 0.052222346 | 0.715302452 | 0.601127925 | 0 |
| 0 | 0 | 0 | 0 | 0 |
| 0 | 0 | 0 | 0 | 0.001358137 |
| 0 | 0.001583279 | 0.021410982 | 0.002513752 | 0.057620829 |
| 0 | 0.00869465 | 0.004710423 | 0 | 0 |
| 0 | 0 | 0 | 0 | 0 |
| 0 | 0 | 0 | 0 | 0 |
| 0 | 0 | 0.47497631 | 0 | 0 |
| 0 | 0 | 0 | 0 | 0 |
| 0 | 0 | 0 | 0 | 0 |
| 0 | 0 | 0.003321076 | 0 | 0 |
| 0 | 0.009955454 | 0.016523783 | 0.018022644 | 0 |
| 0 | 0 | 0 | 0 | 0 |
| 0 | 0 | 0 | 0 | 0 |
| 0 | 0 | 0 | 0 | 0 |
| 0 | 0 | 0 | 0 | 0 |
| 0 | 0 | 0 | 0 | 0 |
| 0 | 0 | 0 | 0 | 0 |
| 0 | 0 | 0 | 0 | 0 |
| 0 | 0 | 0.008165517 | 0.001450297 | 0 |
| 0.358006294 | 0 | 0.259305369 | 0 | 0 |
| 0 | 0 | 0 | 0 | 0 |
| 0 | 0 | 1.12541413 | 0 | 0.000937155 |
| 0 | 0.000494843 | 0.000604913 | 0 | 0 |
| 0 | 0.002794093 | 0.001707799 | 0 | 0 |
| 0 | 0 | 0 | 0 | 0 |
| 0 | 0 | 0 | 0 | 0 |
| 0 | 0 | 0 | 0 | 0 |
| 0 | 0 | 0 | 0 | 0 |
| 0 | 0 | 0 | 0 | 0 |
| 0 | 0 | 0 | 0 | 0 |
| 0 | 0 | 0 | 0 | 0 |
| 0 | 0 | 0 | 0 | 0 |
| 0.001053291 | 0.040002824 | 0.198401986 | 0 | 0 |
| 0 | 0 | 0 | 0 | 0 |
| 0 | 0 | 0 | 0 | 0 |
| 0 | 0 | 0 | 0 | 0 |
| 0 | 0.072027023 | 0.240838186 | 0 | 0 |
| 0 | 0 | 0 | 0 | 0 |
| 0 | 0 | 0 | 0 | 0 |
| 0 | 0 | 0 | 0 | 0 |
| 0 | 0 | 0 | 0 | 0 |
| 0 | 0 | 0 | 0 | 0 |
| 0 | 0.346113034 | 0 | 0 | 0 |

| | | | | |
|---|---|---|---|---|
| 0 | 0.921166751 | 0.032685287 | 0.153058739 | 0.049500423 |
| 0 | 0 | 0 | 0 | 0 |
| 0 | 0 | 0 | 0 | 0 |
| 0 | 0 | 0.001075043 | 0 | 0 |
| 0 | 0 | 0 | 0 | 0 |
| 0.016225294 | 0.154118689 | 0 | 0 | 0.022803501 |
| 0 | 0 | 0 | 0 | 0 |
| 0 | 0 | 0 | 0 | 0 |
| 0 | 0 | 0 | 0 | 0 |
| 0 | 0 | 0 | 0 | 0 |
| 0 | 0 | 0 | 0 | 0 |
| 0 | 0 | 0.00173707 | 0 | 0 |
| 0 | 0.001921274 | 0 | 0 | 0 |
| 0 | 0 | 0 | 0 | 0 |
| 0 | 0 | 0 | 0 | 0 |
| 0 | 0 | 0 | 0 | 0 |
| 0 | 0 | 0 | 0 | 0 |
| 0 | 0 | 0 | 0 | 0 |
| 0 | 0 | 0 | 0.007826177 | 0 |
| 0 | 0 | 0 | 0 | 0 |
| 0 | 0 | 0 | 0 | 0.013604829 |
| 0 | 0 | 0 | 0 | 0 |
| 0 | 0.00064621 | 0.174973822 | 0.011673362 | 0.010738693 |
| 0 | 0 | 0 | 0.531699392 | 0 |
| 0 | 0 | 0 | 0 | 0 |
| 0 | 0 | 0 | 0 | 0 |
| 0 | 0 | 0 | 0 | 0 |
| 0 | 0 | 0 | 0 | 0 |
| 0 | 0 | 0 | 0 | 0 |
| 0 | 0 | 0 | 0 | 0 |
| 0 | 0 | 0 | 0 | 0 |
| 0 | 0 | 0.126491255 | 0 | 0 |
| 0 | 0.020374149 | 0.00665806 | 0.000854067 | 0.007441989 |
| 0 | 0 | 0 | 0 | 0 |
| 0 | 0.403466937 | 0.000661734 | 0.916597686 | 0 |
| 0 | 0 | 0.004749168 | 0.000685353 | 0 |
| 0 | 0 | 0 | 0 | 0 |
| 0 | 0 | 0 | 0 | 0 |
| 0 | 0 | 0.007553101 | 0 | 0.004480494 |
| 0 | 0 | 0 | 0 | 0 |
| 0 | 0 | 0 | 0 | 0 |
| 0 | 0 | 0 | 0 | 0 |
| 0.302148505 | 0.046851672 | 0.661785705 | 1.123805097 | 0 |
| 0 | 0 | 0 | 0 | 0 |
| 0 | 0 | 0 | 0 | 0 |
| 0 | 0.034948033 | 0.592046326 | 0.117340719 | 0 |
| 0 | 0 | 0 | 0 | 0 |
| 0 | 0 | 0 | 0 | 0 |

| | | | | |
|---|---|---|---|---|
| 0 | 0 | 0 | 0 | 0 |
| 0 | 0 | 0 | 0 | 0 |
| 0 | 0 | 0 | 0 | 0.495862348 |
| 0 | 0 | 0.003266838 | 0 | 0 |
| 0 | 0.00236971 | 0 | 0 | 0 |
| 0 | 0 | 0 | 0 | 0 |
| 0 | 0 | 0 | 0 | 0 |
| 0 | 0 | 0 | 0 | 0 |
| 0 | 0 | 0.89372106 | 0.279464391 | 0 |
| 0 | 0.022385627 | 0.027419708 | 0.124347439 | 0 |
| 0 | 0 | 0 | 0 | 0 |
| 0 | 0 | 0 | 0 | 0 |
| 0 | 0 | 0 | 0 | 0 |
| 0 | 0 | 0 | 0.131731448 | 0 |
| 0 | 0 | 0 | 0 | 0 |
| 0 | 0 | 0 | 0 | 0.066233253 |
| 0 | 0 | 0 | 0 | 0 |
| 0 | 0 | 0 | 0 | 0 |
| 0 | 0 | 0 | 0 | 0 |
| 1.721261578 | 0 | 0.357060838 | 1.422158361 | 0 |
| 0.005831607 | 0.009569469 | 0 | 0.000779145 | 0.006055188 |
| 0 | 0 | 0 | 0 | 0 |
| 0 | 0 | 0 | 0 | 0 |
| 0 | 0.001408211 | 0 | 0 | 0 |
| 0 | 0 | 0.001746656 | 0 | 0 |
| 0 | 0 | 0 | 0 | 0 |
| 0 | 0 | 0 | 0 | 0 |
| 0 | 0 | 0.183550739 | 0 | 0 |
| 0 | 0 | 0 | 0 | 0 |
| 0.009578328 | 0.048393672 | 0 | 0 | 0 |
| 0.024568559 | 0 | 0.122370651 | 0.065119683 | 0 |
| 0 | 0 | 0 | 0 | 0 |
| 0 | 0 | 0.013639951 | 0 | 0 |
| 0 | 0 | 0 | 0 | 0 |
| 0 | 0 | 0 | 0 | 0 |
| 0 | 0 | 0 | 0 | 0 |
| 0 | 0 | 0 | 0 | 0 |
| 0 | 0 | 0 | 0 | 0 |
| 0.246209295 | 0 | 0.002663624 | 0 | 0 |
| 0 | 0 | 0 | 0 | 0 |
| 0 | 0.000898008 | 0 | 0 | 0 |
| 0 | 0 | 0 | 0 | 0 |
| 0 | 0 | 0.00511241 | 0.072662377 | 0 |
| 0 | 0 | 0.001380422 | 0 | 0 |
| 0 | 0 | 0 | 0 | 0 |
| 0 | 0 | 0 | 0 | 0 |
| 0 | 0.013347714 | 0 | 0 | 0.045501411 |
| 0 | 0 | 0.010886218 | 0 | 0 |

| | | | | |
|---|---|---|---|---|
| 0 | 0 | 0 | 0 | 0 |
| 0 | 0 | 0 | 0 | 0 |
| 0 | 0 | 0 | 0 | 0 |
| 0 | 0 | 0 | 0 | 0 |
| 0 | 0.787632634 | 0.397445295 | 0.063670334 | 0.028484213 |
| 0 | 0 | 0 | 0 | 0 |
| 0 | 0 | 0 | 0 | 0 |
| 0 | 0 | 0 | 0 | 0 |
| 0 | 0 | 0 | 0 | 0 |
| 0 | 0 | 0 | 0 | 0 |
| 0 | 0 | 0 | 0.050270971 | 0 |
| 0 | 0.001309597 | 0.050428262 | 0.004435691 | 0.002238463 |
| 0 | 0.003228149 | 0.14694672 | 0 | 0 |
| 0 | 0 | 0 | 0 | 0 |
| 0 | 0 | 0 | 0 | 0 |
| 0 | 0 | 0.012671553 | 1.541256414 | 0 |
| 0 | 0 | 0 | 0.052757542 | 0 |
| 0 | 0 | 0 | 0 | 0 |
| 0 | 0.203851653 | 0.05532866 | 0.051820421 | 0.045623137 |
| 3.132800675 | 0.09470333 | 0.014923307 | 0.014566056 | 0 |
| 0 | 0 | 0 | 0 | 0 |
| 0 | 0 | 0.064678861 | 0 | 0 |
| 0 | 0 | 0 | 0 | 0 |
| 0 | 0 | 0 | 0 | 0 |
| 0 | 0 | 0 | 0 | 0 |
| 0 | 0 | 0 | 0 | 0 |
| 0 | 0.002519789 | 0.003125577 | 0.004643877 | 0.004581484 |
| 0 | 0.004872377 | 0.02439737 | 0 | 0.007660914 |
| 0 | 0 | 0 | 0 | 0 |
| 0 | 0 | 0 | 0 | 0 |
| 0 | 0.001176129 | 0 | 0.004979534 | 0.001172692 |
| 0 | 0 | 0 | 0 | 0 |
| 0 | 0 | 0 | 0 | 0 |
| 0 | 0 | 0 | 0 | 0 |
| 0 | 0 | 0 | 0 | 0 |
| 0 | 0.002197655 | 0 | 0 | 0 |
| 0 | 0 | 0 | 0 | 0 |
| 0.000644945 | 0.011686442 | 0.041991942 | 0.037483672 | 0 |
| 0.029012566 | 0 | 0 | 0 | 0 |
| 0 | 0 | 0.026438033 | 0 | 0 |
| 0.000643863 | 0.007542197 | 0.020960747 | 0 | 0.001997542 |
| 0 | 0 | 0 | 0 | 0 |
| 0 | 0 | 0 | 0 | 0 |
| 0 | 0.003675957 | 0.001925837 | 0 | 0.000872671 |
| 0 | 0 | 0 | 0 | 0 |
| 0 | 0 | 0 | 0 | 0 |
| 0 | 0 | 0 | 0 | 0 |
| 0 | 0 | 0 | 0 | 0 |

| | | | | |
|---|---|---|---|---|
| 0 | 0 | 0 | 0 | 0 |
| 0 | 0 | 0 | 0 | 0 |
| 0 | 0.376596879 | 0 | 0 | 0 |
| 0 | 0.002017934 | 0.005382095 | 0.001864057 | 0 |
| 0 | 0.00425364 | 0.001091958 | 0.001296663 | 0 |
| 0 | 0 | 0 | 0.000743763 | 0.001313686 |
| 0 | 0.025635296 | 0.009303317 | 0 | 0 |
| 0 | 0 | 0.004866962 | 0.023447155 | 0 |
| 0 | 0.014903013 | 0 | 0 | 0 |
| 0 | 0.017382808 | 0 | 0 | 0 |
| 0 | 0 | 0 | 0 | 0 |
| 0 | 0 | 0 | 0 | 0 |
| 0.179608604 | 0.21274413 | 0.077614415 | 0.065186811 | 0 |
| 0 | 0.005323448 | 0 | 0 | 0 |
| 0 | 0 | 0 | 0 | 0 |
| 0.109604635 | 0 | 0 | 0 | 0 |
| 0 | 0 | 0 | 0 | 0 |
| 0 | 0 | 0 | 0 | 0 |
| 0 | 0 | 0 | 0 | 0 |
| 0 | 0 | 0 | 0 | 0 |
| 0 | 0 | 0 | 0 | 0 |
| 0 | 0 | 0 | 0 | 0 |
| 0 | 0 | 0.005808619 | 0 | 0.00256097 |
| 0.015455482 | 0.008284429 | 0.002593546 | 0.002395358 | 0 |
| 0 | 0 | 0 | 0 | 0 |
| 0 | 0.003282899 | 0 | 0 | 0 |
| 0 | 0 | 0 | 0 | 0 |
| 0 | 0 | 0 | 0 | 0 |
| 0 | 0 | 0 | 0 | 0 |
| 0 | 0 | 0 | 0 | 0 |
| 0 | 0.030547607 | 0.012953169 | 0.017945019 | 0 |
| 0 | 0 | 0.083518158 | 0 | 0 |
| 0 | 0 | 0 | 0 | 0 |
| 0 | 0 | 0 | 0 | 0 |
| 0.001574146 | 0.133974543 | 0.645268409 | 0 | 0 |
| 0 | 0 | 0 | 0 | 0.000796008 |
| 0 | 0 | 0 | 0 | 0 |
| 0 | 0 | 0 | 0 | 0.119142588 |
| 0.740811841 | 0.000646135 | 0.001036689 | 0.003419534 | 0.018361037 |
| 0 | 0 | 0 | 0 | 0 |
| 0 | 0 | 0 | 0 | 0 |
| 0 | 0 | 0 | 0 | 0 |
| 0 | 0 | 0 | 0 | 0 |
| 0 | 0 | 0 | 0 | 0 |
| 0 | 0 | 0 | 0 | 0 |
| 0.015374651 | 0.319350336 | 0.341912712 | 0.231062398 | 0 |
| 0 | 0 | 0 | 0 | 0.217401097 |
| 0 | 0 | 0 | 0 | 0 |

| | | | | |
|---|---|---|---|---|
| 0 | 0 | 0 | 0 | 0 |
| 0 | 0 | 0 | 0 | 0 |
| 0 | 0 | 0.089280218 | 0 | 0 |
| 0 | 0.00643727 | 0 | 0 | 0 |
| 0 | 0 | 0 | 0 | 0 |
| 0 | 0 | 0.149953594 | 0.007534118 | 0 |
| 0 | 0 | 0 | 0.001807467 | 0 |
| 0 | 0 | 0 | 0 | 0 |
| 0 | 0 | 0.009181036 | 0 | 0 |
| 0.001611548 | 0 | 0 | 0 | 0 |
| 0 | 0 | 0.001682064 | 0.003218023 | 0 |
| 0 | 0.000978269 | 0.001076283 | 0 | 0.001170492 |
| 0 | 0 | 0 | 0 | 0 |
| 0 | 0 | 0 | 0 | 0 |
| 0 | 0 | 0 | 0 | 0 |
| 0 | 0 | 0 | 0 | 0 |
| 0.017681419 | 0 | 0 | 0 | 0 |
| 0 | 0 | 0 | 0 | 0 |
| 0 | 0 | 0 | 0 | 0 |
| 0 | 0 | 0 | 0 | 0 |
| 0 | 0 | 0 | 0 | 0 |
| 0 | 0 | 0 | 0 | 0 |
| 0 | 0 | 0 | 0 | 0 |
| 0 | 0 | 0 | 0 | 0 |
| 0 | 0 | 0 | 0 | 0 |
| 0 | 0 | 0 | 0 | 0 |
| 0 | 0 | 0 | 0 | 0 |
| 0.008616574 | 0 | 0 | 0 | 0 |
| 0 | 0 | 0 | 0 | 0 |
| 0 | 0 | 0 | 0 | 0 |
| 0.003099789 | 0.004180524 | 0.028198568 | 0.000884982 | 0 |
| 0 | 0 | 0 | 0 | 0 |
| 0.006061761 | 0.009361314 | 0.016847521 | 0.01893636 | 0.072424623 |
| 0.003677962 | 0 | 0.014780065 | 0.007950372 | 0 |
| 0 | 0 | 0 | 0 | 0 |
| 0 | 0 | 0 | 0 | 0 |
| 0 | 0 | 0 | 0 | 0 |
| 0 | 0 | 0 | 0 | 0 |
| 0 | 0 | 0 | 0 | 0 |
| 0 | 0 | 0 | 0 | 0 |
| 0 | 0 | 0 | 0 | 0 |
| 0 | 0.054338134 | 0.007060909 | 0.130909985 | 0.215011019 |
| 0.007404538 | 0.06637292 | 0.04731198 | 0.227457779 | 0 |
| 0 | 0 | 0 | 0 | 0 |
| 0 | 0 | 0 | 0 | 0 |
| 0.478485257 | 0.01556711 | 0.277850182 | 0.228128617 | 0 |
| 0 | 0.00049385 | 0.004074975 | 0.000975744 | 0 |
| 0 | 0 | 0 | 0 | 0 |

| | | | | |
|---|---|---|---|---|
| 0 | 0 | 0 | 0 | 0 |
| 0 | 0.003640546 | 0.010680797 | 0 | 0 |
| 0.014172755 | 0.00297678 | 0.003508959 | 0.006841731 | 0 |
| 0 | 0 | 0 | 0 | 0 |
| 0 | 0.008318854 | 0.027394741 | 1.769944991 | 0 |
| 0 | 0 | 0.011060279 | 0.007976175 | 0 |
| 0 | 0 | 0 | 0 | 0 |
| 0 | 0 | 0 | 0 | 0 |
| 0.007759721 | 0 | 0 | 0 | 0 |
| 0 | 0 | 0 | 0 | 0 |
| 0 | 0 | 0 | 0 | 0 |
| 0 | 0 | 0 | 0 | 0 |
| 0 | 0 | 0 | 0 | 0 |
| 0 | 0.001965905 | 0 | 0 | 0 |
| 0 | 0 | 0 | 0 | 0 |
| 0 | 0 | 0 | 0 | 0 |
| 0.000978365 | 0 | 1.196521434 | 0 | 0 |
| 0 | 0 | 0 | 0 | 0 |
| 0 | 0 | 0 | 0 | 0 |
| 0 | 0 | 0 | 0 | 0 |
| 0 | 0 | 0 | 0 | 0 |
| 0 | 0 | 0 | 0 | 0 |
| 0 | 0 | 0 | 0 | 0 |
| 0 | 0 | 0 | 0 | 0 |
| 0.010148495 | 0.021729097 | 3.431049693 | 0.005341783 | 0 |
| 0 | 0.000528846 | 0 | 0 | 0 |
| 0 | 0 | 0 | 0 | 0 |
| 0.028028002 | 0 | 0.570981664 | 0.113955052 | 0 |
| 0 | 0 | 0 | 0 | 0 |
| 0 | 0 | 0 | 0 | 0 |
| 0 | 0 | 0.005072756 | 0 | 0 |
| 0 | 0 | 0 | 0 | 0 |
| 0 | 0 | 0 | 0 | 0 |
| 0 | 0 | 0 | 0 | 0 |
| 0 | 0 | 0 | 0 | 0 |
| 0 | 0 | 0 | 0 | 0 |
| 0 | 0 | 0.005417886 | 0.000909796 | 0 |
| 0 | 0 | 0 | 0 | 0.06596034 |
| 0 | 0 | 0.454741622 | 4.971196479 | 0 |
| 0 | 0 | 0 | 0 | 0 |
| 0 | 0 | 0 | 0 | 0 |
| 0 | 0 | 0.070659073 | 0.015383782 | 0 |
| 0 | 0.006722889 | 0.001784235 | 0.076402274 | 0.001058407 |
| 0 | 0 | 0 | 0 | 0 |
| 0 | 0 | 0 | 0 | 0 |
| 0 | 0 | 0 | 0.001206445 | 0 |
| 0 | 0.002165201 | 0.010421846 | 0.00275013 | 0.004857468 |
| 0 | 0 | 0 | 0 | 0 |

| | | | | |
|---|---|---|---|---|
| 0 | 0 | 0 | 0 | 0 |
| 0 | 0.016221757 | 0.017220831 | 0.015181942 | 0.004937436 |
| 0 | 0 | 0 | 0 | 0 |
| 0 | 0 | 0 | 0 | 0 |
| 0 | 0 | 0 | 0 | 0 |
| 0 | 0 | 0 | 0 | 0 |
| 0 | 0 | 0 | 0 | 0 |
| 0 | 0 | 0 | 0 | 0 |
| 0 | 0 | 0 | 0 | 0 |
| 0 | 0 | 0 | 0 | 0 |
| 0 | 0 | 0 | 0 | 0 |
| 0 | 0 | 0 | 0 | 0 |
| 0.103352921 | 0 | 0.183032381 | 0.017882791 | 0 |
| 0 | 0 | 0 | 0 | 0 |
| 0 | 0.008303341 | 0.044309947 | 0.024878895 | 0 |
| 0 | 0 | 0 | 0 | 0 |
| 0 | 0 | 0 | 0 | 0 |
| 0 | 0 | 0 | 0 | 0 |
| 0 | 0 | 0.006046307 | 0 | 0 |
| 0 | 0 | 0.003899939 | 0 | 0 |
| 0 | 0 | 0 | 0 | 0 |
| 1.411752683 | 0 | 0.014185789 | 0.043377496 | 0 |
| 0 | 0.013656871 | 0 | 0.01891084 | 0.002883593 |
| 0 | 0 | 0 | 0 | 0 |
| 0 | 0 | 0 | 0 | 0 |
| 0 | 0 | 0 | 0 | 0 |
| 0 | 0 | 0 | 0 | 0 |
| 0 | 0 | 0 | 0 | 0 |
| 0 | 0 | 0 | 0 | 0 |
| 0 | 0 | 0 | 0 | 0 |
| 0 | 0 | 0 | 0 | 0 |
| 0 | 0 | 0 | 0 | 0 |
| 0 | 0 | 0 | 0 | 0 |
| 0 | 0 | 0 | 0 | 0 |
| 0 | 14.36463701 | 7.318571762 | 0 | 0 |
| 0 | 0 | 0 | 0 | 0 |
| 0 | 0 | 0 | 0 | 0 |
| 0 | 0 | 0 | 0 | 0 |
| 0.468870541 | 0.007516884 | 0.001340048 | 0.003005716 | 0 |
| 0 | 0 | 0 | 0 | 0 |
| 0.255106741 | 0.030516682 | 0.073808312 | 0.14426296 | 0.303901808 |
| 0 | 0 | 0 | 0 | 0 |
| 0 | 0 | 0 | 0 | 0 |
| 0 | 0 | 0 | 0 | 0.004401039 |
| 0 | 0 | 0 | 0 | 0 |
| 0.015693144 | 0.070736459 | 0.201044465 | 0.001815069 | 0.706152652 |
| 0 | 0 | 0 | 0 | 0 |
| 0 | 0 | 0 | 0 | 0 |

| | | | | |
|---|---|---|---|---|
| 0 | 0 | 0 | 0 | 0 |
| 0 | 0 | 0.002880836 | 0 | 0 |
| 0 | 0 | 0 | 0 | 0 |
| 0 | 0 | 0 | 0 | 0 |
| 0 | 0 | 0.006558151 | 0 | 0 |
| 0 | 0 | 0 | 0 | 0 |
| 0 | 0 | 0 | 0 | 0 |
| 0.097721781 | 0.000926741 | 0 | 0 | 0 |
| 0 | 0 | 0 | 0 | 0 |
| 0.335592568 | 0.002096375 | 0.004420627 | 0.004082823 | 0.139001587 |
| 0 | 0 | 0 | 0 | 0 |
| 0 | 0 | 0 | 0 | 0 |
| 0 | 0.002458836 | 0.016343861 | 0.00381711 | 0.002451651 |
| 0 | 0 | 0 | 0 | 0 |
| 0 | 0 | 0 | 0 | 0 |
| 0 | 0 | 0 | 0 | 0 |
| 0 | 0 | 0 | 0 | 0 |
| 0 | 0 | 0 | 0 | 0 |
| 0 | 0 | 0 | 0 | 0 |
| 0.020526539 | 0.439896021 | 0.289773325 | 0.198908039 | 0.247673201 |
| 0 | 0 | 0 | 0 | 0 |
| 0 | 0 | 0 | 0 | 0 |
| 0 | 0 | 0 | 0 | 0 |
| 0 | 0 | 0 | 0 | 0 |
| 0 | 0 | 0 | 0 | 0 |
| 0 | 0 | 0.03743279 | 0 | 0 |
| 0 | 0 | 0 | 0 | 0 |
| 0 | 0 | 0 | 0 | 0 |
| 0 | 0 | 0 | 0 | 0 |
| 0 | 0 | 0.004111232 | 0 | 0 |
| 0 | 0 | 0 | 0 | 0 |
| 0 | 0 | 0 | 0 | 0 |
| 0 | 0 | 0 | 0 | 0 |
| 0 | 0 | 0 | 0.001607204 | 0 |
| 0 | 0 | 0 | 0 | 0 |
| 0 | 0.001888674 | 0 | 0.00819624 | 0 |
| 0 | 0 | 0 | 0 | 0 |
| 0 | 0 | 0 | 0 | 0 |
| 0 | 0 | 0 | 0 | 0 |
| 0 | 0 | 0 | 0 | 0 |
| 0 | 0 | 0 | 0 | 0 |
| 0 | 0 | 0 | 0 | 0 |
| 0 | 0 | 0 | 0 | 0 |
| 0 | 0 | 0 | 0 | 0 |
| 0 | 0.002226851 | 0.008480641 | 0.018130992 | 0 |
| 0 | 0 | 0.010005942 | 0 | 0 |
| 0 | 0 | 0 | 0 | 0 |
| 0 | 0 | 0 | 0 | 0 |

| | | | | |
|---|---|---|---|---|
| 0 | 0 | 0 | 0 | 0 |
| 0 | 0 | 0 | 0 | 0 |
| 0 | 0 | 0 | 0 | 0 |
| 0 | 0 | 0 | 0 | 0 |
| 0 | 0 | 0 | 0 | 0 |
| 0.00265497 | 0.005882448 | 0.130022615 | 0.093557309 | 0.000637528 |
| 0 | 0 | 0 | 0 | 0 |
| 0 | 0 | 0 | 0 | 0 |
| 0 | 0 | 0 | 0 | 0 |
| 0 | 0 | 0 | 0 | 0 |
| 0 | 0 | 0 | 0 | 0 |
| 0 | 0 | 0 | 0 | 0 |
| 0 | 0 | 0 | 0 | 0 |
| 0.004287773 | 0 | 0.037025325 | 0 | 0 |
| 0 | 0 | 0 | 0 | 0 |
| 0 | 0 | 0 | 0 | 0 |
| 0 | 0.02807612 | 0 | 0 | 0 |
| 0 | 0 | 0.014005941 | 0 | 0 |
| 0 | 0 | 0 | 0 | 0 |
| 0 | 0 | 0 | 0 | 0 |
| 0 | 0 | 0 | 0 | 0 |
| 0 | 0 | 0 | 0 | 0 |
| 0 | 0 | 0 | 0 | 0 |
| 0 | 0 | 0 | 0 | 0 |
| 0 | 0 | 0 | 0 | 0 |
| 0.003934249 | 0 | 0 | 0 | 0.001994402 |
| 0 | 0 | 0 | 0 | 0 |
| 0 | 0 | 0.317466593 | 0 | 0 |
| 0 | 0 | 0 | 0 | 0 |
| 0 | 0 | 0 | 0 | 0 |
| 0 | 0 | 0 | 0 | 0 |
| 0 | 0 | 0 | 0.003366054 | 0 |
| 0 | 0 | 0 | 0 | 0 |
| 0 | 0 | 0 | 0 | 0 |
| 0 | 0 | 0 | 0 | 0 |
| 0 | 0 | 0 | 0 | 0 |
| 0 | 0 | 0 | 0 | 0 |
| 0 | 0 | 0 | 0 | 0 |
| 0 | 0 | 0 | 0 | 0 |
| 0 | 0 | 0 | 0 | 0 |
| 0 | 0 | 0 | 0 | 0 |
| 0 | 0 | 0 | 0 | 0 |
| 0 | 0 | 0 | 0 | 0 |
| 0 | 0 | 0 | 0 | 0 |
| 0 | 0 | 0 | 0 | 0 |
| 0 | 0 | 0 | 0 | 0 |
| 0 | 0 | 0 | 0 | 0 |
| 0 | 0 | 0 | 0 | 0 |

| | | | | |
|---|---|---|---|---|
| 0 | 0 | 0 | 0 | 0 |
| 0 | 0 | 0.079676712 | 0 | 0 |
| 0 | 0 | 0 | 0 | 0 |
| 0 | 0 | 0 | 0 | 0 |
| 0 | 0 | 0 | 0 | 0 |
| 0 | 0 | 0 | 0 | 0 |
| 0 | 0 | 0 | 0 | 0 |
| 0 | 0 | 0 | 0 | 0 |
| 0 | 0 | 0 | 0 | 0 |
| 0 | 0 | 0 | 0 | 0 |
| 0.021919183 | 0.0228715 | 0.006187631 | 0.002539911 | 0.001682312 |
| 0 | 0 | 0 | 0 | 0 |
| 0.022632599 | 0 | 0.061652893 | 0.312769874 | 0 |
| 0 | 0 | 0 | 0 | 0 |
| 0 | 0 | 0 | 0 | 0 |
| 0 | 0 | 0 | 0 | 0 |
| 0 | 0 | 0 | 0 | 0 |
| 0 | 0 | 0 | 0 | 0 |
| 0 | 0 | 0 | 0 | 0 |
| 0 | 0 | 0 | 0.000924654 | 0 |
| 0.004989903 | 0.007040068 | 0.008896771 | 0.00370567 | 0 |
| 0 | 0 | 0.003854325 | 0 | 0 |
| 0 | 0 | 0 | 0 | 0 |
| 0.374955033 | 0.390585649 | 0.031503484 | 1.793416004 | 0.00204309 |
| 0 | 0 | 0 | 0 | 0 |
| 0 | 0 | 0 | 0 | 0 |
| 0 | 0 | 0 | 0 | 0 |
| 0.010029325 | 0 | 0 | 0 | 0 |
| 0 | 0 | 0 | 0 | 0 |
| 0 | 0 | 0 | 0 | 0 |
| 0 | 0 | 0 | 0 | 0.002606888 |
| 0 | 0 | 0 | 0 | 0 |
| 0 | 0.003602665 | 0.00183501 | 0 | 0 |
| 0 | 0 | 0 | 0 | 0 |

| PHL018-0141_V3 | PHL018-0077_V2 | PHL018-0186_V3 | PHL018-0183_V2 | PHL018-0050_V1 |
|---|---|---|---|---|
| EYCF | EYCF | REF | REF | EYCF |
| 0.000911267 | 0.41179855 | 0.023314625 | 0.058491479 | 0.058786867 |
| 1.059355937 | 3.506575191 | 2.776743922 | 0.171171678 | 0.408570896 |
| 0.090947217 | 5.906798934 | 0.744353578 | 0.109482662 | 0.005458535 |
| 0 | 0 | 0 | 0 | 0 |
| 0 | 0 | 0 | 0 | 0 |
| 0 | 0 | 0 | 0 | 0 |
| 0 | 0 | 0 | 0 | 0 |
| 0 | 0 | 0 | 0 | 0 |
| 0 | 0 | 0 | 0 | 0 |
| 0 | 0 | 0 | 0 | 0 |
| 0 | 0 | 0 | 0 | 0 |
| 0 | 0 | 0 | 0 | 0 |
| 0 | 0 | 0 | 0 | 0 |
| 3.602887659 | 2.950467411 | 10.06031051 | 0.077343504 | 4.834878121 |
| 0 | 0 | 0 | 0 | 0 |
| 0 | 0 | 0 | 0 | 0 |
| 0 | 0 | 0 | 0 | 0 |
| 0 | 0 | 0 | 0 | 0 |
| 0 | 0 | 0 | 0 | 0 |
| 0 | 0 | 0 | 0 | 0 |
| 0 | 0 | 0 | 0.003475619 | 0 |
| 0 | 0 | 0 | 0.001291926 | 0 |
| 0 | 0 | 0 | 0 | 0 |
| 0 | 0 | 0 | 0 | 0 |
| 0 | 0 | 0 | 0.412138183 | 0 |
| 0 | 0 | 0 | 0 | 0 |
| 0 | 0 | 0 | 0 | 0 |
| 0 | 0 | 0 | 0 | 0 |
| 0 | 0 | 0 | 0 | 0 |
| 0 | 0 | 0 | 0 | 0 |
| 0 | 0 | 0 | 0 | 0 |
| 0 | 0 | 0 | 0 | 0 |
| 0 | 0 | 0 | 0 | 0 |
| 0 | 0 | 0 | 0 | 0 |
| 0 | 0 | 0 | 0 | 0 |
| 7.855425916 | 0 | 0.872880904 | 0.080309619 | 0.285690715 |
| 0 | 0 | 0 | 0 | 0 |
| 0 | 0 | 0 | 0 | 0 |
| 0 | 0 | 0 | 0 | 0 |
| 0 | 0 | 0 | 0 | 0 |
| 0 | 0 | 0 | 0 | 0 |
| 0.003367212 | 0.060076518 | 0.001709934 | 0.040148192 | 0.005901393 |
| 0 | 0.839638442 | 0.010763195 | 25.585842 | 0.175222948 |
| 0.008606874 | 0.016304012 | 0.041038406 | 0.04343153 | 0.009676818 |
| 0 | 0.003711984 | 0.017642082 | 0.002636851 | 0.004549985 |
| 0 | 0 | 0 | 0 | 0 |

| | | | | |
|---|---|---|---|---|
| 0.000758693 | 0.006093696 | 0 | 0 | 0 |
| 0.002714151 | 0.080976332 | 0 | 0.000881645 | 0 |
| 0 | 0.017696323 | 0.00054192 | 0 | 0.00670867 |
| 0.539576528 | 0.386485005 | 0.455642484 | 0.139935622 | 0.332521197 |
| 0 | 0.003927207 | 0 | 0.000817306 | 0.003159053 |
| 0.009916215 | 0.033752676 | 0 | 0 | 0 |
| 0.001118897 | 0.045406987 | 0.00193187 | 0 | 0 |
| 0 | 0 | 0 | 0 | 0 |
| 0 | 0 | 0 | 0 | 0 |
| 0 | 0 | 0 | 0 | 0 |
| 0 | 0.002172929 | 0 | 0.061646119 | 0 |
| 0.04579878 | 0.051405306 | 0 | 0.074914826 | 0.049303543 |
| 0 | 0 | 0 | 0 | 0 |
| 0.011058201 | 0.099758979 | 0.025155353 | 0.01218179 | 0.067679658 |
| 0 | 0 | 0 | 0 | 0 |
| 0 | 0 | 0 | 0 | 0 |
| 0 | 1.117987376 | 0.00155184 | 0 | 0.679070564 |
| 0 | 0 | 0 | 0 | 0 |
| 0 | 0 | 0 | 0 | 0 |
| 0 | 0 | 0 | 0 | 0 |
| 0.145301838 | 0.214766424 | 0.006645365 | 0.005244311 | 0 |
| 0.007950132 | 0.107375825 | 0 | 0.024171859 | 0.003295562 |
| 0.00820364 | 0 | 0 | 0 | 0.027953702 |
| 0 | 0 | 0 | 0 | 0 |
| 0 | 0 | 0 | 0 | 0 |
| 0 | 0 | 0 | 0 | 0 |
| 0 | 0 | 0 | 0 | 0 |
| 0 | 0 | 0 | 0 | 0 |
| 0 | 0.001504778 | 0.001436748 | 0 | 0.004742974 |
| 0 | 0 | 0 | 0 | 0 |
| 0.025403623 | 0.038461767 | 0 | 0.000760816 | 0 |
| 0 | 0 | 0 | 0 | 0 |
| 0 | 0 | 0 | 0 | 0 |
| 0 | 0 | 0 | 0.000620809 | 0 |
| 0 | 0 | 0 | 0 | 0 |
| 0 | 0 | 0 | 0 | 0 |
| 0 | 0 | 0 | 0 | 0 |
| 0 | 0.028119277 | 0 | 0 | 0 |
| 0 | 0 | 0 | 0 | 0 |
| 0 | 0 | 0 | 0 | 0 |
| 0 | 0 | 0 | 0 | 0 |
| 0 | 0 | 0 | 0 | 0 |
| 0 | 0 | 0 | 0 | 0 |
| 0 | 0 | 0 | 0 | 0 |
| 0 | 0 | 0 | 0.001639503 | 0 |
| 0 | 0.02103476 | 0 | 0 | 0 |
| 0.000786326 | 0 | 0 | 0 | 0 |
| 2.117128925 | 0.006790882 | 0 | 0 | 0 |

| | | | | |
|---|---|---|---|---|
| 0 | 0 | 0 | 0 | 0 |
| 0 | 0 | 0 | 0 | 0 |
| 0 | 0 | 0 | 0 | 0 |
| 0 | 0 | 0 | 0 | 0 |
| 0.025881066 | 1.542212064 | 0.000994281 | 0 | 0.002344506 |
| 0 | 0 | 0 | 0.00137548 | 0 |
| 0 | 0 | 0 | 0 | 0 |
| 0 | 0 | 0 | 0 | 0 |
| 0 | 0.006334427 | 0 | 0 | 0 |
| 0 | 0 | 0 | 0 | 0 |
| 0 | 0 | 0 | 0 | 0 |
| 0 | 0.002451382 | 0 | 0.001506952 | 0 |
| 0 | 0 | 0 | 0 | 0 |
| 0 | 0 | 0 | 0 | 0 |
| 0 | 0 | 0 | 0 | 0 |
| 0 | 0 | 0 | 0 | 0.00085753 |
| 0 | 0 | 0 | 0 | 0 |
| 0 | 0 | 0 | 0 | 0 |
| 0.049226065 | 0.041282867 | 0 | 0.032633845 | 0.007405273 |
| 0 | 0 | 0 | 0 | 0 |
| 0 | 0 | 0 | 0 | 0 |
| 0 | 0 | 0 | 0 | 0 |
| 0 | 0 | 0 | 0 | 0 |
| 0 | 0 | 0 | 0.003588371 | 0 |
| 0 | 0 | 0 | 0 | 0 |
| 0 | 0 | 0 | 0 | 0 |
| 0 | 0 | 0 | 0 | 0 |
| 0 | 0 | 0 | 0.002329784 | 0 |
| 0 | 0 | 0 | 0 | 0 |
| 0 | 0 | 0 | 0 | 0 |
| 0 | 0.011122189 | 0 | 0.00140132 | 0.003684617 |
| 0 | 0.002522842 | 0.007376908 | 0.02583187 | 0.001932742 |
| 0 | 0 | 0 | 0 | 0 |
| 0 | 0 | 0 | 0 | 0 |
| 0 | 0 | 0 | 0 | 0 |
| 0 | 0 | 0 | 0 | 0 |
| 0 | 0 | 0 | 0 | 0 |
| 0 | 0 | 0 | 0 | 0 |
| 0.083943404 | 0 | 0 | 0 | 0.070928532 |
| 0 | 0 | 0 | 0 | 0 |
| 0 | 0 | 0 | 0 | 0 |
| 0 | 0 | 0 | 0 | 0 |
| 0 | 0.000729002 | 0.000406026 | 0.001456466 | 0 |
| 0 | 0 | 0 | 0 | 0 |
| 0 | 0 | 0 | 0 | 0 |
| 0 | 0 | 0 | 0 | 0 |
| 0 | 0 | 0 | 0 | 0 |
| 0 | 0 | 0 | 0 | 0 |

| | | | | |
|---|---|---|---|---|
| 0 | 0 | 0 | 0 | 0 |
| 0 | 0 | 0 | 0 | 0 |
| 0 | 0 | 0 | 0 | 0 |
| 0 | 0 | 0 | 0 | 0 |
| 0.01610285 | 0.016331623 | 0 | 0.019010624 | 0.089831264 |
| 0 | 0 | 0 | 0 | 0 |
| 0 | 0 | 0 | 0 | 0 |
| 0 | 0 | 0 | 0 | 0 |
| 0 | 0 | 0 | 0 | 0 |
| 0 | 0 | 0 | 0 | 0 |
| 0 | 0 | 0 | 0 | 0 |
| 0 | 0 | 0 | 0 | 0 |
| 0 | 0 | 0 | 0 | 0 |
| 0 | 0 | 0 | 0 | 0 |
| 0 | 0 | 0 | 0 | 0 |
| 0 | 6.451043727 | 0 | 0.643155056 | 0 |
| 0 | 0 | 0 | 0 | 0 |
| 0 | 0 | 0 | 0 | 0 |
| 0 | 0 | 0 | 0 | 0 |
| 0 | 0 | 0 | 0 | 0 |
| 0 | 0 | 0 | 0 | 0 |
| 0 | 0 | 0 | 0 | 0 |
| 0 | 0 | 0 | 0 | 0 |
| 0 | 0 | 0 | 0 | 0 |
| 0 | 0 | 0 | 0.00861902 | 0.00498581 |
| 0 | 0 | 0 | 0 | 0 |
| 0 | 0.008564006 | 0 | 0 | 0 |
| 0 | 0 | 0 | 0 | 0 |
| 0 | 0 | 0 | 0 | 0 |
| 0 | 0 | 0 | 0 | 0 |
| 0 | 0.003306736 | 0 | 0.002752708 | 0 |
| 0 | 0.080229348 | 0.000405303 | 0.003150059 | 0 |
| 0 | 0 | 0 | 0 | 0 |
| 0 | 0 | 0 | 0 | 0 |
| 0 | 0 | 0 | 0 | 0 |
| 0 | 0 | 0 | 0 | 0 |
| 0 | 0 | 0 | 0 | 0 |
| 0 | 0 | 0 | 0 | 0 |
| 0 | 0 | 0 | 0.003531423 | 0 |
| 0 | 0 | 0 | 0 | 0 |
| 0 | 0 | 0 | 0 | 0 |
| 0 | 0 | 0 | 0 | 0 |
| 0 | 0 | 0 | 0 | 0 |
| 0 | 0 | 0 | 0 | 0 |
| 0 | 0 | 0 | 0 | 0 |
| 0 | 0 | 0 | 0 | 0 |
| 0 | 0 | 0 | 0 | 0 |
| 0 | 0 | 0 | 0 | 0 |

| | | | | |
|---|---|---|---|---|
| 0 | 0 | 0 | 1.067540273 | 0 |
| 0 | 0 | 0 | 0 | 0 |
| 0 | 0 | 0 | 0 | 0 |
| 0 | 0 | 0 | 0 | 0 |
| 0 | 0 | 0 | 0 | 0 |
| 0 | 0 | 0 | 0 | 0 |
| 0 | 0 | 0 | 0 | 0 |
| 0 | 0 | 0 | 0 | 0 |
| 0 | 0 | 0 | 0 | 0 |
| 0 | 0 | 0 | 0 | 0 |
| 0 | 0 | 0 | 0 | 0 |
| 0 | 0 | 0 | 0 | 0 |
| 0 | 0 | 0 | 0 | 0 |
| 0 | 0 | 0 | 0 | 0 |
| 0 | 0 | 0 | 0 | 0 |
| 0 | 0 | 0 | 0 | 0 |
| 0 | 0 | 0 | 0 | 0 |
| 0 | 0 | 0 | 0 | 0 |
| 0 | 0 | 0 | 0 | 0 |
| 0 | 0 | 0 | 0 | 0 |
| 0.066219371 | 0.024086829 | 0.007976743 | 0.025578824 | 0.028726603 |
| 0 | 0 | 0 | 0 | 0 |
| 0 | 0 | 0 | 0 | 0 |
| 0 | 0 | 0 | 0 | 0 |
| 0 | 0 | 0 | 0 | 0 |
| 0 | 0 | 0 | 0 | 0 |
| 0 | 0.001137373 | 0.002815434 | 0.01211919 | 0.001858855 |
| 0 | 0 | 0 | 0 | 0 |
| 0 | 0 | 0 | 0 | 0 |
| 0 | 0 | 0 | 0 | 0 |
| 0 | 0.017195479 | 0 | 0 | 0 |
| 0 | 0 | 0 | 0 | 0 |
| 0 | 0 | 0 | 0 | 0 |
| 0 | 0 | 0 | 0.003974285 | 0 |
| 0 | 0.005718226 | 0.008964708 | 0 | 0.025700206 |
| 0 | 0 | 0 | 0 | 0 |
| 0 | 0 | 0 | 0 | 0 |
| 0 | 0 | 0 | 0 | 0 |
| 0 | 0 | 0 | 0 | 0 |
| 0 | 0 | 0 | 0 | 0 |
| 0 | 0 | 0 | 0 | 0 |
| 0 | 0 | 0 | 0 | 0 |
| 0 | 0 | 0 | 0.002345556 | 0 |
| 0 | 0 | 0 | 0 | 0 |
| 0 | 0 | 0 | 0 | 0 |
| 0 | 0.0278268 | 0 | 0 | 0 |
| 0 | 0 | 0 | 0 | 0 |
| 0 | 0 | 0.010647488 | 0.00361837 | 0 |

| | | | | |
|---|---|---|---|---|
| 0 | 0 | 0 | 0 | 0 |
| 0 | 0 | 0 | 0 | 0 |
| 0 | 0 | 0 | 0 | 0 |
| 0 | 0 | 0 | 0 | 0 |
| 0 | 0 | 0 | 0 | 0 |
| 0 | 0 | 0 | 0 | 0 |
| 0 | 0 | 0 | 0.007726056 | 0 |
| 0 | 0.01952611 | 0 | 0.002600737 | 0 |
| 0 | 0 | 0.002650304 | 0 | 0 |
| 0 | 0 | 0 | 0 | 0 |
| 0 | 0 | 0 | 0 | 0 |
| 0 | 0 | 0 | 0 | 0 |
| 0 | 0 | 0 | 0 | 0 |
| 0 | 0 | 0 | 0 | 0 |
| 0 | 0 | 0 | 0.044688932 | 0 |
| 0 | 0 | 0 | 0 | 0 |
| 0 | 0 | 0 | 0 | 0 |
| 0 | 0 | 0 | 0 | 0 |
| 0 | 0.003837231 | 0 | 0.142492299 | 0 |
| 0 | 0 | 0 | 0.074030879 | 0 |
| 0.008355323 | 0.01788657 | 0 | 0 | 0 |
| 0 | 0 | 0 | 0 | 0 |
| 0 | 0 | 0 | 0 | 0 |
| 0 | 0 | 0 | 0 | 0 |
| 0.011492042 | 0 | 0.00534464 | 0.004193855 | 0.011578649 |
| 0 | 0 | 0 | 0 | 0 |
| 0 | 0 | 0 | 0 | 0 |
| 0 | 0 | 0 | 0 | 0 |
| 0 | 0.014595037 | 0 | 0 | 0 |
| 0 | 0.001028502 | 0 | 0 | 0 |
| 0.009136734 | 0.150332211 | 0.048463423 | 0.088252702 | 0.024487779 |
| 0 | 0 | 0 | 0 | 0 |
| 0 | 0 | 0 | 0 | 0.09261459 |
| 0.004687595 | 0.001723112 | 0 | 0 | 0.006072328 |
| 0 | 0 | 0 | 0 | 0 |
| 0 | 0 | 0 | 0 | 0 |
| 0 | 0 | 0 | 0 | 0 |
| 0 | 0 | 0 | 0 | 0 |
| 0 | 0 | 0 | 0 | 0 |
| 0 | 0 | 0.015559785 | 0 | 0.003261318 |
| 0 | 0 | 0 | 0 | 0 |
| 0.002408633 | 0.006787982 | 0.004788814 | 0 | 0.160583841 |
| 0 | 0 | 0 | 0 | 0 |
| 0 | 0 | 0 | 0 | 0 |
| 0 | 0 | 0 | 0 | 0 |
| 0 | 0 | 0 | 0 | 0 |
| 0 | 0 | 0 | 0 | 0 |
| 0 | 0 | 0 | 0 | 0 |

| | | | | |
|---|---|---|---|---|
| 0 | 0 | 0 | 0 | 0 |
| 0 | 0 | 0 | 0 | 0 |
| 0.007258644 | 0.009588857 | 0.021837209 | 0.034909214 | 0.008880493 |
| 0 | 0 | 0 | 0 | 0 |
| 0 | 0.387965651 | 0 | 0.144825742 | 0 |
| 0 | 0 | 0 | 0 | 0 |
| 0 | 0 | 0 | 0 | 0 |
| 0 | 0 | 0 | 0 | 0 |
| 0 | 0 | 0 | 0 | 0 |
| 0 | 0 | 0 | 0 | 0 |
| 0 | 0 | 0 | 0 | 0 |
| 0 | 0 | 0 | 0 | 0 |
| 0 | 0 | 0 | 0 | 0 |
| 0.002451201 | 0.011539411 | 0.001748845 | 0.003136665 | 0.012123854 |
| 0.016553009 | 0.003109967 | 0.000721721 | 0.002588907 | 0.002382537 |
| 0 | 0 | 0 | 0 | 0 |
| 0.005867764 | 0.000885882 | 0.009703572 | 0.001769896 | 0.001900281 |
| 0 | 0.009620711 | 0 | 0 | 0.009055063 |
| 0 | 0 | 0 | 0 | 0 |
| 0 | 0 | 0 | 0 | 0 |
| 0 | 0 | 0 | 0 | 0 |
| 0 | 0 | 0.009650359 | 0.002396567 | 0 |
| 0 | 0 | 0 | 0 | 0 |
| 0 | 0 | 0 | 0 | 0 |
| 0 | 0 | 0 | 0 | 0 |
| 0 | 0 | 0 | 0 | 0 |
| 0 | 0.861280756 | 0 | 0.000935696 | 0 |
| 0 | 0 | 0 | 0 | 0 |
| 0 | 0.013026149 | 0 | 0 | 0 |
| 0 | 0 | 0 | 0 | 0 |
| 0 | 0 | 0 | 0 | 0 |
| 0 | 0 | 0 | 0 | 0 |
| 0 | 0 | 0 | 0 | 0 |
| 0 | 0 | 0 | 0 | 0 |
| 0 | 0 | 0 | 0 | 0 |
| 0 | 0 | 0 | 0.032443876 | 0 |
| 0 | 0 | 0 | 0 | 0 |
| 0 | 0.001255923 | 0.201642702 | 0.031783179 | 0.017857676 |
| 0 | 0 | 0 | 0 | 0 |
| 0 | 0 | 0 | 0 | 0 |
| 0 | 0.005075426 | 0 | 0 | 0 |
| 0 | 0 | 0 | 0 | 0 |
| 0 | 0 | 0 | 0 | 0 |
| 0 | 0 | 0 | 0 | 0 |
| 0 | 0 | 0 | 0 | 0 |
| 0 | 0 | 0 | 0 | 0 |
| 0 | 0 | 0 | 0 | 0 |
| 0 | 0.003650971 | 0.004066899 | 0 | 0 |

| | | | | |
|---|---|---|---|---|
| 0.034731728 | 0 | 0.121841055 | 0.760045404 | 0 |
| 0 | 0 | 0 | 0 | 0 |
| 0 | 0.001465839 | 0 | 0 | 0 |
| 0 | 0.009777388 | 0 | 0 | 0 |
| 0.000797419 | 0 | 0.00547595 | 0 | 0 |
| 0 | 0 | 0 | 0 | 0 |
| 0 | 0 | 0 | 0 | 0 |
| 0.013133653 | 0.025397294 | 0.00153515 | 0.108955565 | 0.005067817 |
| 0 | 0 | 0 | 0 | 0 |
| 0.010080849 | 0.045658503 | 0.05063399 | 0.035272018 | 0.748454137 |
| 0 | 0 | 0 | 0 | 0 |
| 0 | 0.013558909 | 0.000520813 | 0.080644947 | 0.001146201 |
| 0.008054018 | 0.038125111 | 0.009086267 | 0.00425134 | 0.001304151 |
| 0.00178383 | 0 | 0 | 0 | 0 |
| 0 | 0 | 0 | 0 | 0 |
| 0 | 0.002129567 | 0 | 0 | 0 |
| 0 | 0 | 0 | 0 | 0.00071184 |
| 0 | 0 | 0 | 0 | 0 |
| 0 | 0.042390968 | 0 | 0 | 0 |
| 0 | 0 | 0 | 0 | 0 |
| 0 | 0 | 0 | 0 | 0 |
| 0 | 0 | 0 | 0 | 0 |
| 0 | 0.000548933 | 0 | 0 | 0 |
| 0 | 0 | 0 | 0.073402318 | 0 |
| 0 | 0 | 0 | 0 | 0 |
| 0 | 0.001239238 | 0.004908143 | 0.001650576 | 0.007595015 |
| 0 | 0 | 0 | 4.017020713 | 0 |
| 0 | 0.010490084 | 0 | 0 | 0 |
| 0 | 0.001057334 | 0 | 0 | 0 |
| 0 | 0 | 0 | 0 | 0 |
| 0 | 0.000886513 | 0 | 0 | 0 |
| 0 | 0 | 0 | 0 | 0 |
| 0 | 0 | 0 | 0 | 0 |
| 0.001277965 | 0.010534523 | 0 | 0 | 0 |
| 0 | 0 | 0 | 0 | 0 |
| 0 | 0.003141634 | 0 | 0 | 0 |
| 0 | 0.119642426 | 0 | 0 | 0 |
| 0 | 0 | 0 | 0 | 0 |
| 0 | 0 | 0 | 0 | 0 |
| 0.001849928 | 0 | 0 | 0 | 0 |
| 0 | 0.090862362 | 0 | 0 | 0 |
| 0 | 0.012855956 | 0 | 0 | 0 |
| 0 | 0 | 0 | 0 | 0 |
| 0 | 0 | 0 | 0 | 0 |
| 0 | 0.00082473 | 0 | 0 | 0 |
| 0 | 0 | 0 | 0.046493624 | 0 |
| 0 | 0.002825333 | 0.003242572 | 0 | 0.038409673 |
| 0 | 0.001398806 | 0 | 0 | 0 |

| | | | | |
|---|---|---|---|---|
| 0 | 0 | 0 | 0 | 0 |
| 0.007061934 | 0.026121194 | 0.000639494 | 0 | 0 |
| 0 | 0 | 0 | 0 | 0 |
| 0 | 0 | 0.002052817 | 0 | 0 |
| 0 | 0 | 0 | 0 | 0 |
| 0 | 0.027714998 | 0.000901383 | 0.001616688 | 0 |
| 0 | 0 | 0 | 0 | 0 |
| 0 | 0 | 0 | 0 | 0 |
| 0 | 0 | 0 | 0 | 0 |
| 0 | 0 | 0 | 0 | 0 |
| 0 | 0 | 0.019340867 | 0.0021876 | 0.003451233 |
| 0 | 0 | 0 | 0 | 0 |
| 0 | 0 | 0 | 0 | 0 |
| 0 | 0.000886678 | 0.001646151 | 0 | 0.004619115 |
| 0.001910076 | 0 | 0 | 0 | 0 |
| 0 | 0 | 0 | 0 | 0 |
| 0 | 0 | 0 | 0 | 0 |
| 0 | 0 | 0 | 0 | 0 |
| 0.002902224 | 0 | 0 | 0 | 0 |
| 0 | 0 | 0 | 0 | 0 |
| 0.003389976 | 0.04108045 | 0 | 0 | 0 |
| 0.23145508 | 0.251623925 | 0.304670653 | 0 | 0.455578509 |
| 0.005971319 | 0 | 0 | 0 | 0 |
| 0 | 0 | 0 | 0 | 0 |
| 0 | 0 | 0 | 0 | 0 |
| 0 | 0 | 0 | 0 | 0 |
| 0 | 0.001922788 | 0 | 0 | 0 |
| 0 | 0 | 0 | 0 | 0 |
| 0 | 0 | 0 | 0 | 0 |
| 0.160797127 | 0.592981011 | 2.438007175 | 0.076600539 | 0.497444019 |
| 0.003272891 | 0.000601541 | 0 | 0 | 0.068388447 |
| 0 | 0 | 0 | 0 | 0.008799241 |
| 0 | 0 | 0 | 0 | 0 |
| 0 | 0.001064984 | 0 | 0 | 0 |
| 0 | 0.012211445 | 0 | 0 | 0 |
| 0 | 0 | 0 | 0 | 0 |
| 0 | 0 | 0 | 0 | 0 |
| 0 | 0 | 0 | 0.000902808 | 0.010800955 |
| 0 | 0.5547085 | 0 | 0.002205467 | 0 |
| 0 | 0 | 0 | 0 | 0 |
| 0.022761869 | 0.015854573 | 0.000947321 | 0.0075245 | 0 |
| 9.228383521 | 2.899211147 | 0.427037985 | 0.883031269 | 0.540992917 |
| 0 | 0.001448832 | 0 | 0.001378385 | 0.001775913 |
| 0 | 0 | 0 | 0 | 0 |
| 0 | 0.001027861 | 0 | 0 | 0 |
| 0 | 0 | 0 | 0 | 0 |
| 0.001809865 | 0.001338892 | 0.000994281 | 0 | 0 |
| 0 | 0 | 0 | 0 | 0 |

| | | | | |
|---|---|---|---|---|
| 0 | 0.002359954 | 0.021205701 | 0 | 0.002024909 |
| 0 | 0 | 0 | 0 | 0 |
| 0.001521364 | 2.633269688 | 0.010029458 | 0 | 0.026408463 |
| 0 | 0 | 0 | 0 | 0 |
| 0 | 0 | 0 | 0.001474559 | 0 |
| 0 | 0.308879122 | 0 | 0 | 0 |
| 0 | 0 | 0 | 0 | 0 |
| 0 | 0.002380527 | 0 | 0 | 0 |
| 0 | 0 | 0 | 0.009207276 | 0 |
| 0 | 0.009725399 | 0 | 0 | 0 |
| 0 | 0 | 0 | 0 | 0 |
| 0 | 0 | 0 | 0 | 0 |
| 0 | 0 | 0 | 0 | 0 |
| 0 | 0 | 0.011573976 | 0 | 0 |
| 0 | 0 | 0 | 0 | 0 |
| 0.001263002 | 0.040710387 | 0 | 0.012666916 | 0.053991225 |
| 0.000874941 | 0.109849148 | 0.131290065 | 0.046060908 | 0.210359476 |
| 0 | 0 | 0 | 0 | 0 |
| 0 | 0.016045679 | 0 | 0 | 0 |
| 0 | 0 | 0 | 0 | 0 |
| 0 | 0.181558385 | 0 | 0 | 0.001428409 |
| 0 | 0.001501574 | 0.004938263 | 0 | 0 |
| 0 | 0 | 0.010779808 | 0 | 0 |
| 0 | 0.052872477 | 0 | 0.050301654 | 0 |
| 0 | 0 | 0 | 0.010589439 | 0 |
| 0.039879302 | 0.053595144 | 0 | 0 | 0 |
| 0.001555043 | 0.007277922 | 0 | 0 | 0 |
| 0 | 0 | 0 | 0 | 0 |
| 0 | 0 | 0 | 0 | 0 |
| 0 | 0 | 0 | 0 | 0 |
| 0.003378874 | 0.027852739 | 0 | 0.012128138 | 0 |
| 0.013097465 | 0.01064744 | 0.005732538 | 0 | 0.00326279 |
| 0.022303112 | 0.002787896 | 2.120397586 | 0.005738698 | 0.04815256 |
| 0 | 0.003995326 | 0 | 0 | 0.005342502 |
| 0.009076186 | 0 | 0 | 0.001726449 | 0.005719781 |
| 0 | 0 | 0 | 0 | 0 |
| 0.002384936 | 0 | 0.00102094 | 0 | 0.001404298 |
| 0.001017344 | 0.017919169 | 0.01503696 | 0.002386704 | 0.005930421 |
| 0.260987114 | 0.664382531 | 0 | 0.010106159 | 0.17834242 |
| 0 | 0 | 0.010335875 | 0 | 0 |
| 0 | 0 | 0 | 0 | 0 |
| 0 | 0 | 0 | 0 | 0 |
| 0 | 0 | 0 | 0.002855449 | 0 |
| 0 | 0.02909529 | 0 | 0.00111787 | 0 |
| 0 | 0.26798519 | 0 | 0 | 0 |
| 0.007208422 | 0.004883339 | 0 | 0 | 0.000718294 |
| 0 | 0 | 0 | 0 | 0 |
| 0 | 0 | 0 | 0 | 0 |

| | | | | |
|---|---|---|---|---|
| 0.007793314 | 0.005047954 | 0 | 0 | 0 |
| 1.045471215 | 0.260775069 | 1.072539487 | 0.268955835 | 0.426793896 |
| 0 | 0 | 0 | 0 | 0 |
| 0 | 0 | 0 | 0 | 0 |
| 0 | 0 | 0 | 0 | 0 |
| 0 | 0 | 0 | 0 | 0 |
| 0 | 0.009713286 | 0 | 0 | 0 |
| 0 | 0 | 0 | 0 | 0.005977132 |
| 0 | 0 | 0.001991861 | 0 | 0.007123466 |
| 0 | 0 | 0 | 0 | 0 |
| 0 | 0.001451568 | 0 | 0 | 0 |
| 0.061888557 | 0.045134931 | 0.022679761 | 0.005039704 | 0.014576486 |
| 0.001786375 | 0 | 0 | 0 | 0.512883662 |
| 0 | 0 | 0 | 0 | 0 |
| 0.011228621 | 0 | 0 | 0.000831869 | 0.002041488 |
| 0 | 0 | 0 | 0 | 0 |
| 0.039013283 | 0.003040003 | 0.016085059 | 0.008098129 | 0.023289381 |
| 0.062547286 | 0.044889667 | 0 | 0 | 0.018442028 |
| 0 | 0 | 0 | 0 | 0 |
| 0 | 0.007207171 | 0 | 0 | 0 |
| 0.287004458 | 0.00313501 | 0.031720442 | 0.004175607 | 0.316787225 |
| 0.026992456 | 0.001950509 | 0 | 0 | 0 |
| 0 | 0 | 0 | 0 | 0 |
| 0 | 0 | 0 | 0 | 0 |
| 0 | 0.001975276 | 0 | 0 | 0 |
| 0 | 0 | 0 | 0 | 0 |
| 0.079789054 | 0.136540927 | 0.006750248 | 0.007727874 | 0 |
| 0.066802133 | 0.049356039 | 0.012990772 | 0.081549242 | 0.09087529 |
| 0 | 0.002030284 | 0 | 0 | 0.000995453 |
| 0 | 0 | 0 | 0 | 0 |
| 0 | 0.00111728 | 0.000497825 | 0.002678645 | 0 |
| 0 | 0 | 0 | 0 | 0 |
| 0 | 0 | 0 | 0 | 0 |
| 0 | 0 | 0 | 0 | 0 |
| 0.002060652 | 0 | 0 | 0 | 0.049828352 |
| 0.06436671 | 0.229449075 | 0.132963598 | 0.023002871 | 0.374133909 |
| 0 | 0 | 0.003272992 | 0 | 0 |
| 0 | 0 | 0 | 0 | 0 |
| 0 | 0.02928703 | 0.010302152 | 0.028742889 | 0.012281148 |
| 0 | 0 | 0 | 0 | 0 |
| 0 | 0.249873099 | 0 | 0 | 0 |
| 0 | 0 | 0 | 0 | 0 |
| 0 | 0.000468142 | 0 | 0.006027467 | 0 |
| 0.025803419 | 0.002862553 | 0 | 0 | 0.001662059 |
| 0 | 0 | 0 | 0 | 0 |
| 0 | 0 | 0 | 0.410311253 | 0.42645025 |
| 0 | 0 | 0 | 0 | 0 |
| 0 | 0 | 0 | 0 | 0 |

| | | | | |
|---|---|---|---|---|
| 0.058583639 | 0 | 0 | 0 | 0 |
| 0 | 0 | 0 | 0 | 0.070917804 |
| 0 | 0 | 0 | 0 | 0 |
| 0.044197321 | 0.032714226 | 0.001619605 | 0.273783576 | 0.117625619 |
| 0 | 0 | 0 | 0 | 0 |
| 0 | 0 | 0 | 0 | 0 |
| 0 | 0 | 0 | 0 | 0.00276341 |
| 0 | 0 | 0 | 0 | 0 |
| 0.010771394 | 0 | 0.199969182 | 0.211221071 | 6.04418679 |
| 0.09391221 | 0 | 0 | 0.217002007 | 0.007765164 |
| 0 | 0 | 0 | 0 | 0 |
| 0.002636313 | 0.004353298 | 0.002715572 | 0.004870553 | 0.006616738 |
| 0 | 0 | 0 | 0 | 0 |
| 0 | 1.651864571 | 0 | 0.115645184 | 0 |
| 0.00475889 | 0.015950045 | 0.033826879 | 0.036172738 | 0.666490437 |
| 0 | 0 | 0 | 0 | 0 |
| 0 | 0 | 0.604376255 | 0.017725269 | 0.008960295 |
| 0 | 0 | 0 | 0.271420417 | 0 |
| 0 | 0 | 0 | 0 | 0 |
| 0 | 0 | 0 | 0 | 0 |
| 0 | 0 | 0 | 0.002444821 | 0 |
| 0 | 0 | 0 | 0 | 0 |
| 0 | 0 | 0 | 0 | 0 |
| 0.107004374 | 0.585077025 | 0.003186443 | 0.097525259 | 0 |
| 0 | 0.016629261 | 0 | 0 | 0 |
| 3.041752713 | 0 | 0 | 0 | 0.011121477 |
| 0 | 0 | 0 | 0.81335243 | 0 |
| 0.892626742 | 1.908886456 | 9.073036074 | 0.448807123 | 1.070897701 |
| 0 | 0 | 0 | 0 | 0 |
| 0 | 0.000553992 | 0 | 0 | 0 |
| 0 | 0.013373587 | 0 | 0.00067643 | 0.019297792 |
| 0 | 0 | 0 | 0 | 0 |
| 0 | 0.002319102 | 0 | 0.001372834 | 0 |
| 0 | 0 | 0 | 0 | 0 |
| 0 | 0 | 0 | 0.08743549 | 0 |
| 0 | 0 | 0.028992925 | 0.203605568 | 0.312820361 |
| 0.105947631 | 0 | 0 | 0.626654115 | 3.918525612 |
| 0 | 0.016124977 | 0 | 0 | 0 |
| 0 | 0.002537799 | 0 | 0 | 0 |
| 0 | 0.003240614 | 0 | 0 | 0.000611108 |
| 0.052478233 | 0.781285582 | 0 | 0.009789844 | 0.012285633 |
| 0 | 0 | 0 | 0 | 0 |
| 0.58953085 | 0.098794793 | 10.70540696 | 0.020552209 | 14.32447519 |
| 0.001101576 | 0 | 0 | 0 | 0.00053512 |
| 0 | 0 | 0 | 0 | 0 |
| 0 | 0 | 0 | 0 | 0 |
| 0 | 0 | 0 | 0 | 0 |
| 0 | 0.004819473 | 0 | 0.074623086 | 0.086889424 |

| | | | | |
|---|---|---|---|---|
| 0 | 0 | 0 | 0 | 0 |
| 0 | 0 | 0 | 0.015838507 | 0.326950207 |
| 0.01165763 | 0.013860028 | 0 | 0.00153838 | 0.002359584 |
| 0 | 0 | 0 | 0 | 0.002040478 |
| 0 | 0 | 0 | 0 | 0 |
| 0 | 0 | 0 | 0.048273067 | 0.002974063 |
| 0 | 0 | 0 | 0 | 0 |
| 0 | 0.011218436 | 0 | 0 | 0 |
| 0 | 0 | 0 | 0 | 0 |
| 0 | 0 | 0 | 0 | 0 |
| 0 | 0.096957641 | 0.002105327 | 0.028697903 | 0.034055391 |
| 2.221577288 | 0.913781274 | 0.18970727 | 0.231579328 | 1.676470038 |
| 0 | 0 | 0 | 0 | 0 |
| 0 | 0.052830264 | 0.213582592 | 0.020376281 | 0.049505342 |
| 0.012624295 | 0.095802453 | 0.004177939 | 0.006208819 | 0.00472874 |
| 0 | 0 | 0 | 1.759871515 | 0 |
| 0 | 0.00600643 | 0 | 0 | 0 |
| 0 | 0.001606568 | 0 | 0 | 0 |
| 0 | 0 | 0 | 0 | 0.000829188 |
| 0.003240966 | 0.003736495 | 0 | 0.060135619 | 0 |
| 0.009593204 | 0.390007584 | 0 | 0.022154143 | 0.000970865 |
| 0 | 0.607407038 | 6.159659494 | 0 | 0.038669043 |
| 0.424451837 | 0.749610801 | 2.681137927 | 0.34795493 | 10.32254825 |
| 0 | 0 | 0 | 0 | 0 |
| 0 | 0 | 0 | 0 | 0 |
| 7.271285351 | 0 | 0.436430632 | 0 | 7.333762408 |
| 0 | 0 | 0 | 0 | 0 |
| 0 | 0 | 0.000613473 | 0 | 0 |
| 0 | 0 | 0 | 0 | 0 |
| 0 | 0 | 0 | 0 | 0 |
| 0 | 0 | 0 | 0 | 0 |
| 0.26207882 | 0 | 0 | 0.03067564 | 0.234642183 |
| 0.055101836 | 0.0463921 | 0 | 0.006204946 | 0 |
| 0 | 0.013766167 | 0 | 0.039234109 | 0.01451523 |
| 0 | 0.002946633 | 0 | 0.009043879 | 0 |
| 0.002831478 | 0.001056129 | 0.836845032 | 0.048765157 | 0.390524316 |
| 0 | 0.001360275 | 0.00063135 | 0.00067942 | 0.109837618 |
| 0 | 0 | 0 | 0 | 0 |
| 0.009940204 | 0 | 0.604056165 | 0.018930243 | 0.297423726 |
| 0 | 0 | 0 | 0 | 0 |
| 0 | 0 | 0 | 0.009496385 | 0 |
| 0 | 0 | 0 | 0 | 0 |
| 0.001467883 | 0 | 0 | 0 | 0 |
| 0 | 0 | 0 | 0 | 0 |
| 0 | 0.034726724 | 0.013266961 | 0.008555558 | 0.126715224 |
| 0 | 0 | 0 | 0.001033246 | 0.022250646 |
| 0 | 0 | 0 | 0 | 0 |
| 0.002291195 | 0.038257845 | 0.008990775 | 0.000967532 | 1.047712143 |

| | | | | |
|---|---|---|---|---|
| 0 | 0 | 0 | 0 | 0 |
| 0 | 0 | 0 | 0 | 0 |
| 0 | 0 | 0 | 0 | 0 |
| 0 | 0.001197584 | 0.002795082 | 0 | 0.00482325 |
| 0 | 0 | 0 | 0 | 0 |
| 0 | 0 | 0 | 0 | 0.082419115 |
| 0.006557633 | 0 | 0 | 0 | 0 |
| 0 | 0.026920152 | 0 | 0.015306818 | 0 |
| 0 | 0 | 0 | 0 | 0 |
| 0 | 0 | 0 | 0 | 0 |
| 0 | 0 | 0.002122487 | 0.004060605 | 0 |
| 0 | 0 | 0 | 0 | 0 |
| 0 | 0.00400402 | 0 | 0 | 0.024539749 |
| 0 | 0 | 0 | 0.013643557 | 0 |
| 0 | 0 | 0 | 0 | 0 |
| 0 | 0 | 0 | 0 | 0 |
| 0 | 0.232550822 | 0 | 0.013023839 | 0.009032677 |
| 0 | 0.003172433 | 0.020731886 | 0.002957816 | 0.008554979 |
| 0 | 0 | 0 | 0.010226478 | 0 |
| 0 | 0 | 0 | 0.02743673 | 0 |
| 0 | 0 | 0 | 0 | 0 |
| 0 | 0 | 0 | 0 | 0 |
| 0 | 0.106070048 | 0 | 0 | 0.013449926 |
| 0 | 0 | 0.001722323 | 0 | 0 |
| 0 | 0 | 0 | 0 | 0 |
| 0 | 0 | 0 | 0 | 0 |
| 0 | 0 | 0 | 0 | 0 |
| 0 | 0 | 0 | 0 | 0 |
| 0 | 0 | 0 | 0 | 0 |
| 0.005983272 | 0 | 0.002419016 | 0.377718713 | 0 |
| 0 | 0 | 0 | 0 | 0 |
| 0 | 0 | 0 | 0 | 0 |
| 0 | 0 | 0 | 0 | 0 |
| 0 | 0 | 0 | 0 | 0 |
| 0.000716817 | 0 | 0 | 0 | 0 |
| 0 | 0 | 0 | 0 | 0 |
| 0.375978902 | 0 | 0 | 0 | 0 |
| 0.001220997 | 0.003548532 | 0 | 0 | 0.003163369 |
| 0 | 0 | 0 | 0 | 0 |
| 0 | 0.00254551 | 0 | 0 | 0 |
| 0 | 0.01982507 | 0 | 0.02398673 | 0.000927507 |
| 0 | 0 | 0 | 0 | 0 |
| 0 | 0 | 0 | 0 | 0 |
| 0 | 0 | 0.002326441 | 0 | 0.00499201 |
| 0.213163574 | 0 | 0.354715783 | 0 | 0.070267968 |
| 0 | 0 | 0 | 0 | 0 |
| 0 | 0 | 0 | 0 | 0 |
| 0 | 0 | 0 | 0 | 0 |

| | | | | |
|---|---|---|---|---|
| 0 | 0 | 0 | 0 | 0 |
| 0 | 0 | 0 | 0.193540423 | 0 |
| 0 | 0 | 0.007069025 | 0.267496776 | 0 |
| 0.00804629 | 0 | 0 | 0.025002743 | 0.060375867 |
| 0 | 0 | 0 | 0 | 0 |
| 0 | 0 | 0 | 0 | 0.125317337 |
| 0 | 0 | 0 | 0 | 0 |
| 0 | 0 | 0 | 0 | 0 |
| 0.005051877 | 0.269615901 | 0.018502254 | 0.001185178 | 0.001908731 |
| 0.001017064 | 0 | 0 | 0 | 0.000878339 |
| 0 | 0 | 0 | 0 | 0 |
| 0 | 0 | 0 | 0.000903177 | 0 |
| 0 | 0 | 0 | 0 | 0 |
| 0 | 0 | 0 | 0 | 0 |
| 0 | 2.091158686 | 0 | 0.105823733 | 0.270217934 |
| 0 | 0.782337919 | 0 | 0 | 0 |
| 0 | 0.001764292 | 0 | 0 | 0.001663531 |
| 0 | 0 | 0 | 0 | 0 |
| 0 | 0 | 0 | 0 | 0 |
| 0 | 0 | 0 | 0 | 0 |
| 0 | 0 | 0 | 0.050296232 | 0 |
| 0 | 0 | 0 | 0 | 0 |
| 0 | 0 | 0 | 0 | 0 |
| 0 | 0 | 0 | 0 | 0 |
| 0 | 0.047611414 | 0.000971345 | 0.796519767 | 0.008337136 |
| 8.63071434 | 0.806037358 | 0.081665539 | 2.919595302 | 2.41696969 |
| 0 | 0 | 0 | 0.028252092 | 0 |
| 0 | 0.002273432 | 0 | 0 | 0 |
| 0.002813424 | 0 | 0.155579506 | 0 | 0 |
| 0 | 0 | 0 | 0.800845897 | 0 |
| 0 | 0 | 0 | 0 | 0 |
| 0.010202531 | 0.035884664 | 0 | 0.012341633 | 0.00165205 |
| 0 | 0 | 0 | 0 | 0 |
| 0 | 0.00307366 | 0 | 0 | 0 |
| 0 | 0.021178487 | 0.307540277 | 0.024528881 | 0.012697654 |
| 0 | 0.000650897 | 0.001570938 | 0 | 0.002592981 |
| 0 | 0.005205732 | 0 | 0.004920661 | 0 |
| 0 | 0 | 0.197307909 | 0.631656726 | 0.986484905 |
| 0 | 0 | 10.92891576 | 0 | 0 |
| 0 | 0 | 0 | 0 | 0 |
| 0 | 0 | 0 | 0 | 0 |
| 0 | 0 | 0 | 0.002949376 | 0 |
| 16.62511199 | 0.621956219 | 0.004128298 | 0.224796616 | 0.277199319 |
| 0 | 0 | 0 | 0 | 0.002457701 |
| 0 | 0 | 0 | 0 | 0 |
| 0 | 0.001079125 | 0.001488267 | 0 | 0 |
| 0 | 0 | 0 | 0.000600074 | 0 |
| 0.097139786 | 0.687915812 | 0 | 0.014393138 | 0.003311454 |

| | | | | |
|---|---|---|---|---|
| 0 | 0.001648274 | 0.001360037 | 0 | 0 |
| 0 | 0 | 0 | 0 | 0 |
| 0 | 0 | 0 | 0 | 0 |
| 0 | 0 | 0 | 0 | 0 |
| 0 | 0.001458709 | 0 | 0 | 0 |
| 0 | 0 | 0 | 0 | 0.16185273 |
| 0 | 0 | 0 | 0 | 0 |
| 0 | 0 | 0 | 0 | 0 |
| 0 | 0 | 0 | 0 | 0 |
| 0 | 0 | 0 | 0 | 0 |
| 0 | 0 | 0 | 0 | 0 |
| 0.00238762 | 1.360892135 | 0.099562829 | 0.940809517 | 0.017320416 |
| 1.15547982 | 0.334040369 | 0.455138122 | 0.009115959 | 0 |
| 0 | 0 | 0 | 0 | 0 |
| 0 | 0.593823034 | 0.043221237 | 0.035670464 | 0.021970866 |
| 0 | 0 | 0 | 0 | 0 |
| 0 | 0 | 0 | 0 | 0 |
| 0 | 0 | 0 | 0.00337539 | 0 |
| 0 | 0 | 0 | 0 | 0 |
| 0 | 0 | 0 | 0.009554154 | 0 |
| 0 | 0 | 0 | 0.068482042 | 0.051206287 |
| 0 | 0 | 0 | 0 | 0 |
| 0.083499454 | 0.04924408 | 0 | 0.088091079 | 0.592376915 |
| 0 | 0 | 0 | 0 | 0 |
| 0 | 0 | 0 | 0 | 0 |
| 0 | 0 | 0 | 0 | 0 |
| 0.015199541 | 0 | 0 | 0 | 0 |
| 0 | 0 | 0 | 0 | 0 |
| 0 | 0 | 0 | 0 | 0.00442369 |
| 0 | 0 | 0 | 0 | 0 |
| 0 | 0 | 0 | 0 | 0 |
| 0.135159863 | 0.013681968 | 0.103636434 | 0.026241684 | 0.050647658 |
| 0 | 0 | 0 | 0.059605816 | 0 |
| 0 | 0.004511282 | 0.000507598 | 0.042060907 | 0.002178378 |
| 0 | 0 | 0.001446563 | 0 | 0.002821813 |
| 0 | 0.005250523 | 0.025259499 | 0.002736515 | 0.091501106 |
| 0 | 0.000785963 | 0 | 0.001465584 | 0 |
| 0 | 0 | 0 | 0.113898024 | 0 |
| 0 | 0 | 0 | 0 | 0.005735574 |
| 0 | 0 | 0 | 0 | 0 |
| 0 | 0 | 0 | 0.018086127 | 0 |
| 0 | 0 | 0 | 0 | 0 |
| 0 | 0 | 0 | 0 | 0 |
| 0 | 0 | 0 | 0.00422433 | 0 |
| 0 | 0.040645952 | 0.001165917 | 0.023467315 | 0.079971794 |
| 0 | 0 | 0 | 0 | 0 |
| 0 | 0 | 0 | 0.225208801 | 0.064947628 |
| 0 | 0.001319959 | 0 | 0.000879045 | 0 |

| | | | | |
|---|---|---|---|---|
| 0 | 0 | 0 | 0 | 0 |
| 0 | 0 | 0 | 0 | 0 |
| 0.00324369 | 0 | 0 | 0.004325538 | 0.001548064 |
| 0 | 0 | 0 | 0 | 0 |
| 0.001558217 | 0 | 0 | 0 | 0 |
| 0 | 0.015831865 | 0 | 0 | 0 |
| 0 | 0 | 0 | 0 | 0 |
| 0.002095984 | 0.024025095 | 0 | 0.016113318 | 0 |
| 0 | 0 | 0 | 0 | 0 |
| 0 | 0 | 0 | 0 | 0.006576757 |
| 0 | 0 | 0 | 0 | 0 |
| 0 | 0 | 0 | 0 | 0 |
| 0 | 0 | 0 | 0 | 0 |
| 0 | 0 | 0 | 0 | 0 |
| 0 | 0 | 0 | 0 | 0 |
| 0 | 0 | 0 | 0 | 0 |
| 0 | 0 | 0 | 0 | 0.006905539 |
| 0 | 0 | 0 | 0.025073612 | 0 |
| 0.124045913 | 0 | 0.781540856 | 0.831699538 | 0 |
| 0 | 0 | 0 | 0 | 0 |
| 0 | 0 | 0.033720829 | 0 | 0 |
| 0 | 0 | 0 | 0 | 0 |
| 0 | 0 | 0 | 0 | 0 |
| 0 | 0 | 0 | 0 | 0 |
| 0 | 0 | 0.004812381 | 0 | 0 |
| 0 | 0.000649611 | 0.010010027 | 0 | 0 |
| 0 | 0 | 0 | 0 | 0 |
| 0 | 0.906707888 | 0.001534599 | 0.00063517 | 0.001948461 |
| 0 | 0 | 0 | 0 | 0 |
| 1.088697609 | 1.648388761 | 0.031860913 | 0.14975824 | 0.783582295 |
| 0 | 0 | 0 | 0 | 0 |
| 0 | 0 | 0 | 0 | 0 |
| 0 | 0 | 0 | 0.063491449 | 0 |
| 0 | 0 | 0 | 0 | 0 |
| 0 | 0.002382633 | 0.001011074 | 0.007707058 | 0 |
| 0 | 0 | 0 | 0 | 0 |
| 0 | 0 | 0 | 0 | 0 |
| 0 | 0.000965568 | 0 | 0 | 0 |
| 0 | 0 | 0 | 0 | 0 |
| 0 | 0.001808217 | 0 | 0 | 0 |
| 3.914713992 | 13.95312923 | 2.534971848 | 1.043869016 | 9.2433058 |
| 0.018592599 | 2.342618366 | 0 | 0.018828133 | 0.146415544 |
| 0 | 0 | 0.380110664 | 0 | 0.456980945 |
| 0 | 0 | 0 | 0 | 0 |
| 0 | 0 | 0 | 0 | 0 |
| 0 | 0 | 0 | 0.454035109 | 0 |
| 0 | 0 | 0 | 0 | 0 |
| 0 | 0.001839188 | 0.017983143 | 0.001020694 | 0.036446042 |

| | | | | |
|---|---|---|---|---|
| 0 | 0 | 0 | 0 | 0 |
| 0 | 0.004256452 | 0 | 0.002519683 | 0 |
| 0 | 0 | 0 | 0.001716661 | 0 |
| 0 | 0 | 0 | 0 | 0 |
| 11.5787116 | 0.384044247 | 0.00298301 | 0.290822624 | 1.813341722 |
| 0 | 0 | 0 | 0 | 0 |
| 0 | 0 | 0 | 0 | 0 |
| 0 | 0 | 0 | 0 | 0 |
| 0.159674894 | 0.25689954 | 0.000935184 | 0 | 0.017944443 |
| 0 | 0 | 0 | 0 | 0 |
| 0 | 0 | 0 | 0 | 0 |
| 0 | 0 | 0 | 0 | 0 |
| 0.075371117 | 0.114425688 | 0 | 0.038152691 | 0.042246361 |
| 0.011048057 | 0.017513723 | 0.116074642 | 0.002257453 | 0.028392557 |
| 0 | 0 | 0 | 0 | 0 |
| 0.619848159 | 2.628463801 | 0.078954226 | 0.610690846 | 0.028791923 |
| 0.000616799 | 0.124013632 | 0 | 0.003906951 | 0 |
| 0 | 0 | 0 | 0 | 0 |
| 0 | 0 | 0 | 0 | 0 |
| 0 | 0 | 0 | 0 | 0 |
| 0 | 0 | 0 | 0 | 0 |
| 0 | 0 | 0 | 0 | 0 |
| 0 | 0 | 0 | 0 | 0 |
| 0 | 0 | 0 | 0 | 0 |
| 0 | 0 | 0 | 0.017486801 | 0 |
| 0 | 0 | 0 | 0 | 0 |
| 0.003217991 | 0.024514384 | 0.163632288 | 0.035293696 | 0.069476628 |
| 0 | 0.000799472 | 0.003680941 | 0 | 0 |
| 0 | 0 | 0 | 0 | 0 |
| 0 | 0 | 0 | 0 | 0 |
| 0 | 0 | 0 | 0 | 0 |
| 0.0018957 | 0.002938342 | 0 | 0 | 0 |
| 0.025115302 | 0 | 0 | 0 | 0 |
| 0 | 0 | 0 | 0.026789658 | 0.006163542 |
| 0 | 0 | 0.120422937 | 0.084229363 | 0.875751375 |
| 0 | 0 | 0 | 0 | 0 |
| 0 | 0.015691664 | 0 | 0 | 0 |
| 0 | 0.009606251 | 0 | 0 | 0 |
| 0 | 0 | 0 | 0 | 0 |
| 0 | 0 | 0 | 0 | 0 |
| 1.321923528 | 0 | 0.915663206 | 0.090819863 | 0.075527736 |
| 0 | 0 | 0 | 0 | 0 |
| 0 | 0 | 0 | 0 | 0 |
| 0 | 0 | 0 | 0 | 0 |
| 0 | 0 | 0 | 0 | 0 |
| 0 | 0 | 0 | 0 | 0 |
| 0 | 0 | 0 | 0 | 0 |
| 0 | 0 | 0 | 0 | 0 |

| | | | | |
|---|---|---|---|---|
| 0.00099394 | 0 | 0 | 0.193772456 | 0.257940003 |
| 0 | 0 | 0 | 0 | 0 |
| 0 | 0 | 0 | 0 | 0 |
| 0 | 0 | 0 | 0 | 0 |
| 0.014102899 | 0 | 0 | 0.030469537 | 0.001274578 |
| 0 | 0 | 0 | 0 | 0 |
| 0.001230598 | 0.773810139 | 0 | 0.034644005 | 0.006177219 |
| 0 | 0 | 0 | 0 | 0 |
| 0 | 0 | 0 | 0 | 0 |
| 0 | 0.006651735 | 0 | 0.00590642 | 0 |
| 0 | 0 | 0 | 0.004988869 | 0.000966566 |
| 0 | 0 | 0 | 0.001514842 | 0 |
| 0 | 0 | 0 | 0 | 0 |
| 0 | 0 | 0 | 0 | 0 |
| 0 | 0 | 0 | 0 | 0 |
| 0 | 0 | 0 | 0 | 0 |
| 0 | 0 | 0 | 0 | 0 |
| 0 | 0 | 0 | 0.00104083 | 0 |
| 0 | 0 | 0 | 0 | 0 |
| 0 | 0 | 0 | 0 | 0 |
| 0.024457102 | 1.070242074 | 0.424571122 | 0.165739693 | 0 |
| 0 | 0 | 0 | 0 | 0 |
| 0.002907603 | 0.017412621 | 0.005324477 | 0.012733056 | 0.048337013 |
| 0.000718186 | 0.005161161 | 0.016909215 | 0 | 0.002418888 |
| 0 | 0 | 0 | 0 | 0 |
| 0 | 0 | 0 | 0 | 0 |
| 0 | 0 | 0 | 0 | 0 |
| 0 | 0 | 0 | 0 | 0 |
| 0 | 0 | 0.157518944 | 0 | 0 |
| 0.21148245 | 0.288604327 | 0.010870225 | 0 | 0.221467186 |
| 0 | 0 | 0 | 0 | 0 |
| 0 | 0 | 0 | 0 | 0 |
| 0 | 0 | 0 | 0 | 0 |
| 0 | 0 | 0 | 0.356486382 | 0 |
| 0 | 0 | 0 | 0 | 0 |
| 0 | 0 | 0 | 2.682700742 | 0 |
| 0.057692505 | 0.045205642 | 0 | 0.003541803 | 0 |
| 0 | 0 | 0 | 0 | 0 |
| 0 | 0 | 0 | 0 | 0 |
| 0 | 0 | 0 | 0 | 0 |
| 0 | 0 | 0 | 0 | 0 |
| 0 | 0 | 0 | 0 | 0 |
| 0 | 0 | 0 | 0 | 0 |
| 0 | 0 | 0 | 0 | 0 |
| 0 | 0 | 0 | 0.144377538 | 0 |
| 0 | 0 | 0 | 0 | 0.001526722 |
| 0 | 0 | 0 | 0 | 0 |
| 0 | 0 | 0 | 0 | 0 |

| | | | | |
|---|---|---|---|---|
| 0 | 0.002776796 | 0.000485198 | 0.000870233 | 0 |
| 0 | 0 | 0 | 0.010935564 | 0 |
| 0 | 0 | 0.027677246 | 0 | 0 |
| 0 | 0 | 0 | 0 | 0 |
| 0 | 0 | 0 | 0 | 0 |
| 0 | 0 | 0 | 0 | 0 |
| 0 | 0 | 0 | 0 | 0 |
| 2.99127903 | 4.395790169 | 0.014945653 | 0.072152785 | 0.424515788 |
| 0 | 1.082727165 | 0.016564599 | 35.92138319 | 0.230673587 |
| 0 | 0.023560646 | 0.203860609 | 0 | 0 |
| 0 | 0.001466182 | 0 | 0 | 0 |
| 0 | 0 | 0 | 0 | 0 |
| 0 | 0 | 0 | 0 | 0 |
| 0 | 0.002087895 | 0 | 0 | 0 |
| 0 | 0 | 0 | 0 | 0 |
| 0 | 0 | 0 | 0 | 0 |
| 0 | 0 | 0 | 0 | 0 |
| 0 | 0 | 0 | 0 | 0 |
| 0 | 0 | 0 | 0 | 0 |
| 0 | 0 | 0 | 0 | 0 |
| 0 | 0 | 0 | 0 | 0 |
| 0 | 0 | 0 | 0 | 0 |
| 0 | 0 | 0 | 0.150181956 | 0 |
| 0 | 0 | 0 | 0 | 0 |
| 0.096171824 | 0 | 0 | 0.022447812 | 0.010723458 |
| 0 | 0 | 0 | 0 | 0 |
| 0 | 0 | 0 | 0 | 0 |
| 0 | 0 | 0 | 0 | 0 |
| 0 | 0 | 0 | 0 | 0 |
| 0 | 0 | 0 | 0 | 0 |
| 0 | 0 | 0 | 0.006705113 | 0 |
| 0 | 0 | 0 | 0 | 0 |
| 0 | 0 | 0 | 0 | 0 |
| 0 | 0 | 0 | 0 | 0 |
| 0 | 0 | 0 | 0 | 0.004605405 |
| 0 | 0 | 0 | 0 | 0 |
| 0 | 0 | 0 | 0 | 0 |
| 0 | 0 | 0.031237719 | 0 | 0 |
| 0 | 0 | 0 | 0 | 0.012379773 |
| 0 | 0.227802392 | 0 | 0 | 0 |
| 0 | 0 | 0 | 0 | 0 |
| 0.347025185 | 0.299787501 | 0.210008687 | 0.114724567 | 0.048046925 |
| 0 | 0 | 0 | 0 | 0 |
| 0 | 0 | 0 | 0 | 0 |
| 0 | 0 | 0 | 0 | 0.002532099 |
| 0 | 0.000580973 | 0 | 0 | 0.000534098 |
| 0 | 0 | 0 | 0 | 0 |
| 0 | 0 | 0 | 0 | 0 |

| | | | | |
|---|---|---|---|---|
| 0 | 0 | 0 | 0 | 0 |
| 0 | 0.19503977 | 0 | 0 | 0.009035188 |
| 0 | 0 | 0 | 0 | 0 |
| 0 | 0 | 0 | 0 | 0 |
| 0 | 0.204880469 | 0.591752021 | 0.034591179 | 4.545665514 |
| 0 | 0 | 0 | 0 | 0 |
| 0 | 0 | 0 | 0 | 0 |
| 0.00078644 | 0.168510809 | 0.010646815 | 0.00138375 | 0 |
| 0 | 0 | 0.019103998 | 0 | 0.066626041 |
| 0 | 0 | 0 | 0 | 0.067118621 |
| 0 | 0 | 0 | 0 | 0 |
| 0 | 0 | 0 | 0 | 0 |
| 0 | 0 | 0 | 0 | 0 |
| 0 | 0 | 0 | 0 | 0 |
| 0 | 0.156363546 | 0 | 0 | 0 |
| 0.001364147 | 0 | 0 | 0.01440139 | 0 |
| 0 | 0 | 0 | 0 | 0 |
| 0 | 0 | 0 | 0 | 0 |
| 0 | 0 | 0 | 0 | 0 |
| 0 | 0 | 0 | 0 | 0 |
| 0 | 0 | 0 | 0 | 0 |
| 0 | 0 | 0 | 0 | 0 |
| 0 | 0 | 0 | 0 | 0 |
| 0 | 0 | 0 | 0.003113556 | 0 |
| 0.06010008 | 0 | 0 | 0 | 0.264938497 |
| 0 | 0 | 0 | 0 | 0 |
| 0 | 0.004604986 | 0 | 0 | 0 |
| 0 | 0.0006927 | 0 | 0 | 0.001273621 |
| 0 | 0 | 0 | 0 | 0 |
| 0 | 0 | 0 | 0 | 0 |
| 0 | 0 | 0 | 0 | 0 |
| 0 | 0 | 0 | 0 | 0 |
| 0 | 0 | 0 | 0 | 0 |
| 0 | 0 | 0 | 0 | 0 |
| 0 | 0 | 0 | 0 | 0 |
| 0 | 0 | 0 | 0 | 0 |
| 0 | 0 | 0 | 0 | 0 |
| 0 | 0.326708447 | 0 | 0.04987287 | 0 |
| 0 | 0 | 0 | 0 | 0.012205524 |
| 0 | 0 | 0 | 0 | 0 |
| 0 | 0 | 0 | 0 | 0 |
| 0 | 0 | 0 | 0 | 1.501313853 |
| 0 | 0 | 0 | 0 | 0 |
| 0 | 0 | 0.343693084 | 0 | 0 |
| 0 | 0 | 0 | 0 | 0 |
| 0 | 0 | 0 | 0 | 0 |
| 0 | 0 | 0 | 0 | 0 |
| 0 | 0 | 0 | 0 | 0 |

| | | | | |
|---|---|---|---|---|
| 0.030147768 | 0.001855968 | 0 | 0.043054266 | 0.045499263 |
| 0 | 0 | 0 | 0.006811906 | 0 |
| 0 | 0.001072423 | 0 | 0 | 0 |
| 0 | 0 | 0 | 0 | 0 |
| 0 | 0 | 0 | 0 | 0 |
| 0 | 0 | 0 | 0 | 0 |
| 0 | 0 | 0 | 0 | 0 |
| 0 | 0 | 0 | 0 | 0 |
| 0 | 0 | 0 | 0 | 0 |
| 0 | 0 | 0 | 0 | 0 |
| 0.280500243 | 0 | 0 | 1.067608685 | 0 |
| 0 | 0 | 0 | 0 | 0 |
| 0 | 0 | 0 | 0 | 0 |
| 0 | 0 | 0 | 0 | 0 |
| 0 | 0 | 0 | 0 | 0 |
| 0 | 0 | 0 | 0 | 0 |
| 0 | 0 | 0 | 0 | 0 |
| 0 | 0 | 0 | 0 | 0 |
| 0 | 0 | 0 | 0 | 0 |
| 0 | 0 | 0 | 0 | 0 |
| 0.006099785 | 0.005192917 | 0.054653618 | 0 | 0 |
| 0 | 0 | 0 | 0 | 0 |
| 0 | 0.003392212 | 0.094214613 | 0.01174727 | 0.40748617 |
| 0 | 0.182351075 | 0.092685854 | 0.60430142 | 0 |
| 0 | 0 | 0 | 0.011633638 | 0 |
| 0 | 0 | 0 | 0 | 0 |
| 0 | 0 | 0 | 0 | 0 |
| 0 | 0 | 0 | 0 | 0 |
| 0 | 0 | 0 | 0 | 0 |
| 0 | 0 | 0 | 0 | 0 |
| 0 | 0 | 0 | 0 | 0 |
| 0 | 0 | 0 | 0.042098715 | 0 |
| 0.044689047 | 0.007412515 | 0.390043419 | 0 | 0 |
| 0 | 0 | 0 | 0 | 0 |
| 1.036459545 | 0.314094343 | 0.540923261 | 0.008831293 | 0 |
| 0 | 0.03449975 | 0.000378621 | 0 | 0 |
| 0 | 0 | 0 | 0 | 0 |
| 0 | 0 | 0 | 0 | 0 |
| 0.001860046 | 0.002358882 | 0.000729892 | 0 | 0 |
| 0 | 0 | 0 | 0 | 0 |
| 0 | 0 | 0 | 0 | 0 |
| 0 | 0 | 0 | 0 | 0 |
| 0.004349902 | 0 | 0.036016121 | 0.346559518 | 4.085287443 |
| 0 | 0 | 0 | 0 | 0 |
| 0 | 0 | 0 | 0 | 0 |
| 0.017963797 | 0.002555607 | 0 | 0.00316075 | 0.003356304 |
| 0 | 0 | 0 | 0 | 0 |
| 0 | 0 | 0 | 0 | 0 |

| | | | | |
|---|---|---|---|---|
| 0 | 0 | 0 | 0 | 0 |
| 0 | 0 | 0 | 0 | 0 |
| 0 | 0 | 0 | 0 | 0 |
| 0 | 0 | 0 | 0 | 0 |
| 0 | 0 | 0 | 0 | 0 |
| 0 | 0 | 0 | 0 | 0 |
| 0 | 0 | 0 | 0 | 0 |
| 0.061756979 | 0.008996132 | 0 | 0 | 0 |
| 0.000809154 | 0 | 0 | 0.025740787 | 0.001886727 |
| 0 | 0 | 0 | 0 | 0.046323205 |
| 0 | 0 | 0 | 0 | 0 |
| 0 | 0 | 0 | 0 | 0 |
| 0 | 0 | 0 | 0 | 0 |
| 0 | 0 | 0.189262131 | 0 | 0 |
| 0 | 0 | 0 | 0 | 0 |
| 0 | 0 | 0 | 0 | 0 |
| 0 | 0 | 0 | 0 | 0 |
| 0 | 0 | 0 | 0 | 0 |
| 0 | 0 | 0 | 0 | 0 |
| 0.224223294 | 0 | 0.085226087 | 0.248575493 | 0.265590288 |
| 0.014991202 | 0 | 0.003443491 | 0 | 0.007341579 |
| 0 | 0 | 0 | 0 | 0 |
| 0 | 0 | 0 | 0 | 0 |
| 0 | 0 | 0 | 0 | 0 |
| 0 | 0 | 0 | 0 | 0 |
| 0 | 0.190041605 | 0 | 0 | 0 |
| 0 | 0 | 0 | 0 | 0 |
| 0 | 0 | 0 | 0 | 0 |
| 0 | 0 | 0 | 0 | 0 |
| 0.02902691 | 0 | 0 | 0 | 0.313268081 |
| 0.494102609 | 0 | 0.027243772 | 0.264543531 | 0 |
| 0 | 0 | 0 | 0 | 0 |
| 0 | 0 | 0 | 0 | 0 |
| 0 | 0 | 0 | 0 | 0 |
| 0 | 0 | 0 | 0 | 0 |
| 0 | 0 | 0 | 0 | 0 |
| 0 | 0 | 0 | 0 | 0 |
| 0 | 0 | 0 | 0 | 0 |
| 0 | 0 | 0 | 0 | 0 |
| 0 | 0 | 0 | 0 | 0 |
| 0 | 0 | 0 | 0 | 0 |
| 0 | 0 | 0 | 0 | 0 |
| 0 | 0 | 0 | 0 | 0 |
| 0 | 0 | 0 | 0 | 0 |
| 0 | 0 | 0 | 0 | 0 |
| 0 | 0 | 0 | 0 | 0 |
| 0 | 0.197703839 | 0 | 0 | 0 |
| 0 | 0 | 0 | 0 | 0 |

| | | | | |
|---|---|---|---|---|
| 0 | 0 | 0 | 0 | 0 |
| 0 | 0 | 0 | 0 | 0 |
| 0 | 0 | 0 | 0 | 0 |
| 0 | 0 | 0 | 0 | 0 |
| 0.073761358 | 0.17104285 | 0 | 0.071039618 | 0.149829118 |
| 0 | 0 | 0 | 0 | 0 |
| 0 | 0 | 0 | 0 | 0 |
| 0 | 0 | 0 | 0 | 0 |
| 0 | 0 | 0 | 0 | 0 |
| 0 | 0 | 0 | 0 | 0 |
| 0 | 0 | 0 | 0.029427221 | 0.014531528 |
| 0 | 0.00903518 | 0.005105177 | 0.006801943 | 0.038280709 |
| 0 | 0 | 0 | 0 | 0 |
| 0 | 0 | 0 | 0 | 0 |
| 0 | 0.007484172 | 0.000666943 | 0 | 0 |
| 0 | 0 | 0 | 0.140577967 | 0 |
| 0 | 0.00740233 | 0 | 0.01006058 | 0 |
| 0 | 0 | 0 | 0 | 0 |
| 0.002251705 | 0.428188053 | 0.001104477 | 0.00713142 | 0 |
| 0 | 0.076123858 | 0 | 0.081475292 | 0.001190169 |
| 0 | 0 | 0 | 0 | 0 |
| 0 | 0 | 0 | 0 | 0.028174944 |
| 0 | 0 | 0 | 0 | 0 |
| 0 | 0 | 0 | 0 | 0 |
| 0 | 0 | 0 | 0 | 0 |
| 0 | 0 | 0 | 0 | 0 |
| 0.001177995 | 0.003579172 | 0 | 0.002279964 | 0 |
| 0 | 0.001770718 | 0 | 0 | 0 |
| 0 | 0 | 0 | 0 | 0 |
| 0 | 0 | 0 | 0 | 0 |
| 0 | 0 | 0 | 0 | 0 |
| 0 | 0 | 0 | 0 | 0 |
| 0 | 0 | 0 | 0 | 0 |
| 0 | 0 | 0 | 0 | 0 |
| 0 | 0 | 0 | 0 | 0 |
| 0 | 0 | 0 | 0 | 0 |
| 0 | 0 | 0 | 0 | 0 |
| 0 | 0.007931705 | 0.005337996 | 0.010894596 | 0.004101608 |
| 0 | 0 | 0 | 0.006551983 | 0.193864118 |
| 0 | 0 | 0 | 0.004189492 | 0 |
| 0 | 0 | 0 | 0 | 0 |
| 0 | 0 | 0 | 0 | 0 |
| 0 | 0 | 0 | 0 | 0 |
| 0 | 0.005145749 | 0 | 0 | 0 |
| 0 | 0 | 0 | 0 | 0 |
| 0 | 0 | 0 | 0 | 0 |
| 0 | 0 | 0 | 0 | 0 |
| 0 | 0 | 0 | 0 | 0 |

| | | | | |
|---|---|---|---|---|
| 0 | 0 | 0 | 0 | 0 |
| 0 | 0 | 0.71889179 | 0.01861994 | 0.253569646 |
| 0 | 0 | 0 | 0 | 0 |
| 0 | 0 | 0 | 0 | 0 |
| 0 | 0 | 0.001591861 | 0.011420419 | 0 |
| 0 | 0.001613796 | 0 | 0 | 0 |
| 0 | 0 | 0 | 0 | 0.00366555 |
| 0 | 0 | 0 | 0.013040749 | 0 |
| 0.000444154 | 0.003442201 | 0 | 0 | 0 |
| 0 | 0 | 0 | 0 | 0 |
| 0 | 0 | 0 | 0 | 0.000975379 |
| 0 | 0 | 0 | 0 | 0 |
| 0.016018952 | 0.012150768 | 0.033761496 | 0.040732624 | 0.149106057 |
| 0 | 0 | 0 | 0 | 0 |
| 0 | 0 | 0 | 0 | 0 |
| 0 | 0 | 0 | 0 | 0 |
| 0 | 0 | 0 | 0 | 0 |
| 0 | 0 | 0 | 0 | 0 |
| 0 | 0 | 0 | 0 | 0 |
| 0 | 0 | 0 | 0 | 0 |
| 0 | 0 | 0 | 0 | 0 |
| 0 | 0 | 0 | 0 | 0 |
| 0 | 0.009168403 | 0.007209099 | 0 | 0.033053589 |
| 0.249510013 | 0 | 0.005986397 | 0.003108075 | 0.025742885 |
| 0 | 0 | 0 | 0 | 0 |
| 0 | 0 | 0 | 0 | 0 |
| 0 | 0 | 0 | 0 | 0 |
| 0 | 0 | 0 | 0 | 0 |
| 0 | 0 | 0 | 0 | 0 |
| 0 | 0 | 0 | 0 | 0 |
| 0.031864316 | 0.057327457 | 0.001786247 | 0.00293677 | 0.01769021 |
| 0 | 0 | 0 | 0 | 0 |
| 0 | 0 | 0 | 0 | 0 |
| 0 | 0 | 0 | 0 | 0 |
| 0 | 0 | 0 | 0.004700415 | 0 |
| 0 | 0 | 0 | 0 | 0 |
| 0 | 0 | 0 | 0 | 0 |
| 0 | 0 | 0 | 0 | 0 |
| 0 | 0.000508773 | 0 | 0.079510762 | 0.002702401 |
| 0 | 0 | 0 | 0 | 0 |
| 0 | 0 | 0 | 0 | 0 |
| 0 | 0 | 0 | 0 | 0 |
| 0 | 0 | 0 | 0 | 0 |
| 0 | 0 | 0 | 0 | 0 |
| 0 | 0 | 0 | 0 | 0 |
| 0.023719837 | 0 | 0.137289958 | 0.176480755 | 0.161681327 |
| 0 | 0 | 0 | 0 | 0 |
| 0 | 0 | 0 | 0 | 0 |

| | | | | |
|---|---|---|---|---|
| 0 | 0 | 0 | 0 | 0 |
| 0 | 0 | 0 | 0 | 0 |
| 0 | 0 | 0 | 0 | 0.096051671 |
| 0 | 0 | 0 | 0 | 0.004970456 |
| 0 | 0 | 0 | 0 | 0 |
| 0 | 0 | 0 | 0 | 0 |
| 0 | 0 | 0 | 0 | 0 |
| 0 | 0 | 0 | 0 | 0 |
| 0 | 0 | 0 | 0.000627005 | 0 |
| 0 | 0 | 0 | 0 | 0 |
| 0 | 0.000550335 | 0.001021717 | 0 | 0.004216099 |
| 0 | 0 | 0 | 0 | 0 |
| 0 | 0 | 0 | 0 | 0 |
| 0 | 0 | 0 | 0 | 0 |
| 0 | 0 | 0 | 0 | 0 |
| 0 | 0 | 0 | 0 | 0 |
| 0 | 0 | 0 | 0 | 0 |
| 0 | 0 | 0 | 0 | 0 |
| 0 | 0 | 0 | 0 | 0 |
| 0 | 0 | 0.000677486 | 0 | 0.001565556 |
| 0 | 0 | 0 | 0 | 0 |
| 0 | 0 | 0 | 0 | 0 |
| 0 | 0 | 0 | 0 | 0 |
| 0 | 0 | 0 | 0 | 0 |
| 0 | 0 | 0 | 0 | 0 |
| 0 | 0 | 0 | 0 | 0 |
| 0 | 0 | 0 | 0 | 0 |
| 0.0240743 | 0 | 0.001199605 | 0 | 0.026235791 |
| 0 | 0 | 0 | 0 | 0 |
| 0 | 0 | 0 | 0 | 0 |
| 0.001631562 | 0.00470255 | 0.048541361 | 0.002087819 | 0.000768557 |
| 0 | 0 | 0 | 0 | 0 |
| 0.054593434 | 0.01019226 | 0.123670835 | 0.003878672 | 0.038818217 |
| 0.00422374 | 0 | 0.006629675 | 0.000743172 | 0 |
| 0 | 0 | 0 | 0 | 0 |
| 0 | 0 | 0 | 0 | 0 |
| 0 | 0 | 0 | 0 | 0 |
| 0 | 0 | 0 | 0 | 0 |
| 0 | 0 | 0 | 0 | 0 |
| 0 | 0 | 0 | 0 | 0 |
| 0 | 0 | 0 | 0 | 0 |
| 0.0158685 | 0.176386147 | 0 | 0.009572846 | 0 |
| 0 | 0.065487958 | 0.030344413 | 0.047864538 | 0.042931107 |
| 0 | 0 | 0 | 0 | 0 |
| 0 | 0 | 0 | 0 | 0 |
| 0 | 0 | 0.017618218 | 0.009128717 | 0.206148546 |
| 0 | 0 | 0 | 0 | 0 |
| 0 | 0 | 0 | 0 | 0 |

| | | | | |
|---|---|---|---|---|
| 0 | 0 | 0 | 0 | 0 |
| 0.005244121 | 0.004013242 | 0.011779255 | 0 | 0.015226259 |
| 0.010139068 | 0.002455561 | 0.017572263 | 0.00267597 | 0.005746207 |
| 0 | 0 | 0 | 0 | 0 |
| 0.012817966 | 0.067731462 | 0 | 0.067660057 | 0.002840234 |
| 0 | 0.436348532 | 0 | 0.002079786 | 0 |
| 0 | 0 | 0 | 0 | 0 |
| 0 | 0 | 0 | 0 | 0 |
| 0 | 0 | 0 | 0 | 0 |
| 0 | 0 | 0 | 0 | 0 |
| 0 | 0 | 0 | 0 | 0 |
| 0 | 0 | 0 | 0 | 0 |
| 0 | 0 | 0 | 0 | 0 |
| 0.005503889 | 0.00375266 | 0.000975374 | 0 | 0 |
| 0 | 0 | 0 | 0 | 0 |
| 0 | 0 | 0 | 0 | 0 |
| 0 | 0 | 0 | 0.36033453 | 0 |
| 0 | 0 | 0 | 0 | 0 |
| 0 | 0 | 0 | 0 | 0 |
| 0 | 0 | 0 | 0 | 0 |
| 0 | 0 | 0 | 0 | 0 |
| 0 | 0 | 0 | 0 | 0 |
| 0 | 0 | 0 | 0.001681003 | 0 |
| 0 | 0 | 0 | 0 | 0 |
| 0 | 0 | 0.000819737 | 0 | 0.023903921 |
| 0.001751241 | 0.001295523 | 0.001099514 | 0.000986025 | 0 |
| 0 | 0 | 0 | 0 | 0 |
| 0 | 0 | 0 | 0.247249675 | 0.001100293 |
| 0 | 0 | 0 | 0 | 0 |
| 0 | 0 | 0 | 0 | 0 |
| 0 | 0 | 0 | 0 | 0 |
| 0 | 0 | 0.000361578 | 0 | 0 |
| 0 | 0 | 0 | 0 | 0 |
| 0 | 0 | 0 | 0 | 0 |
| 0 | 0 | 0 | 0 | 0 |
| 0 | 0 | 0 | 0 | 0 |
| 0 | 0 | 0 | 0 | 0 |
| 0 | 0.084559903 | 0 | 0 | 0 |
| 1.070221633 | 0 | 0 | 0 | 1.61828894 |
| 0 | 0 | 0 | 0 | 0 |
| 0 | 0 | 0 | 0 | 0 |
| 0 | 0 | 0.00656042 | 0 | 0.002461042 |
| 0 | 0.001671684 | 0.000413806 | 0.002226563 | 0 |
| 0 | 0 | 0 | 0 | 0 |
| 0 | 0 | 0 | 0 | 0 |
| 0 | 0 | 0 | 0.001423101 | 0 |
| 0 | 0.0034098 | 0.001477098 | 0 | 0 |
| 0 | 0 | 0 | 0 | 0 |

| | | | | |
|---|---|---|---|---|
| 0 | 0 | 0 | 0 | 0 |
| 0.002714141 | 0.008784354 | 0.011715481 | 0 | 0.014942611 |
| 0 | 0 | 0 | 0 | 0 |
| 0 | 0 | 0 | 0 | 0 |
| 0 | 0 | 0 | 0 | 0 |
| 0 | 0 | 0 | 0 | 0 |
| 0 | 0 | 0 | 0 | 0 |
| 0 | 0 | 0 | 0 | 0 |
| 0 | 0 | 0 | 0 | 0 |
| 0 | 0 | 0 | 0 | 0 |
| 0 | 0 | 0 | 0 | 0 |
| 0 | 0 | 0 | 0 | 0 |
| 0.002869087 | 0 | 0 | 0.01110602 | 0 |
| 0 | 0 | 0 | 0 | 0 |
| 0.002961109 | 0.012517446 | 0.004315834 | 0.007740726 | 0 |
| 0 | 0 | 0 | 0 | 0 |
| 0 | 0 | 0 | 0 | 0 |
| 0 | 0 | 0 | 0 | 0 |
| 0 | 0 | 0 | 0.005599046 | 0 |
| 0 | 0 | 0 | 0 | 0 |
| 0 | 0 | 0 | 0 | 0 |
| 0.001246304 | 0.151249277 | 0 | 0.025729859 | 0.007803258 |
| 0 | 0.008602841 | 0 | 0.00359478 | 0.014473501 |
| 0 | 0 | 0 | 0 | 0 |
| 0 | 0 | 0 | 0 | 0 |
| 0 | 0 | 0 | 0 | 0 |
| 0 | 0 | 0 | 0 | 0 |
| 0 | 0 | 0 | 0 | 0 |
| 0 | 0 | 0 | 0 | 0 |
| 0 | 0 | 0 | 0 | 0 |
| 0 | 0 | 0 | 0 | 0 |
| 0 | 0 | 0 | 0 | 0 |
| 0 | 0 | 0 | 0 | 0 |
| 0 | 0 | 0 | 0 | 0 |
| 0.006412257 | 0 | 0 | 0.332898239 | 0.018323078 |
| 0 | 0 | 0 | 0 | 0 |
| 0 | 0 | 0 | 0 | 0 |
| 0 | 0 | 0 | 0 | 0 |
| 0 | 0.078041351 | 0 | 0.015767004 | 0 |
| 0 | 0 | 0 | 0 | 0 |
| 0.026782451 | 0.164125331 | 0.018391765 | 0.027750809 | 0.062401174 |
| 0 | 0 | 0 | 0 | 0 |
| 0 | 0 | 0 | 0 | 0 |
| 0 | 0 | 0 | 0 | 0 |
| 0 | 0 | 0 | 0 | 0 |
| 0.148635626 | 0.139527722 | 0.082223818 | 0 | 0.000827549 |
| 0 | 0 | 0 | 0 | 0 |
| 0 | 0 | 0 | 0 | 0 |

| | | | | |
|---|---|---|---|---|
| 0 | 0 | 0 | 0.000800915 | 0 |
| 0 | 0 | 0 | 0 | 0 |
| 0 | 0 | 0 | 0 | 0 |
| 0 | 0 | 0 | 0 | 0 |
| 0 | 0.000512038 | 0 | 0 | 0 |
| 0 | 0 | 0 | 0 | 0 |
| 0 | 0 | 0 | 0 | 0 |
| 0 | 0 | 0 | 0 | 0 |
| 0 | 0.00853733 | 0 | 0 | 0.037934464 |
| 0 | 0.349289311 | 0.000817226 | 0.038695721 | 0.001618689 |
| 0 | 0 | 0 | 0 | 0 |
| 0 | 0 | 0 | 0 | 0 |
| 0.005088929 | 0.00752932 | 0 | 0 | 0 |
| 0 | 0 | 0 | 0 | 0 |
| 0 | 0 | 0 | 0 | 0 |
| 0 | 0 | 0 | 0 | 0 |
| 0 | 0 | 0 | 0 | 0 |
| 0 | 0 | 0 | 0 | 0 |
| 0 | 0 | 0 | 0 | 0 |
| 0.001811802 | 0.286446658 | 6.39267866 | 0.024227936 | 0.545915889 |
| 0 | 0 | 0 | 0 | 0 |
| 0 | 0 | 0 | 0 | 0 |
| 0 | 0 | 0 | 0 | 0 |
| 0 | 0 | 0 | 0 | 0 |
| 0 | 0 | 0 | 0 | 0 |
| 0 | 0 | 0 | 0 | 0 |
| 0 | 0 | 0 | 0 | 0 |
| 0 | 0 | 0 | 0 | 0 |
| 0 | 0 | 0 | 0 | 0 |
| 0.003292638 | 0.012076705 | 0 | 0 | 0 |
| 0 | 0 | 0 | 0 | 0 |
| 0 | 0 | 0 | 0 | 0 |
| 0 | 0 | 0 | 0 | 0 |
| 0 | 0 | 0 | 0 | 0 |
| 0 | 0 | 0 | 0 | 0 |
| 0.009381347 | 0.0012393 | 0 | 0 | 0 |
| 0 | 0 | 0 | 0 | 0 |
| 0 | 0 | 0 | 0 | 0 |
| 0 | 0 | 0 | 0 | 0 |
| 0 | 0 | 0 | 0 | 0 |
| 0 | 0 | 0 | 0 | 0 |
| 0 | 0 | 0 | 0 | 0 |
| 0.004668729 | 0 | 0 | 0.004027843 | 0 |
| 0 | 0 | 0 | 0 | 0 |
| 0 | 0.005215371 | 0.005074983 | 0 | 0.026011962 |
| 0 | 0 | 0 | 0 | 0.005475074 |
| 0 | 0 | 0 | 0 | 0 |
| 0 | 0 | 0 | 0 | 0 |

| | | | | |
|---|---|---|---|---|
| 0 | 0 | 0 | 0 | 0 |
| 0 | 0 | 0 | 0 | 0 |
| 0 | 0 | 0 | 0 | 0 |
| 0 | 0 | 0 | 0 | 0 |
| 0 | 0 | 0 | 0 | 0 |
| 0 | 0.00308793 | 0.000997019 | 0.032187913 | 0.012342547 |
| 0 | 0 | 0 | 0 | 0 |
| 0 | 0 | 0 | 0 | 0 |
| 0 | 0 | 0 | 0 | 0 |
| 0 | 0 | 0 | 0 | 0 |
| 0 | 0 | 0 | 0 | 0 |
| 0 | 0 | 0 | 0 | 0 |
| 0 | 0 | 0 | 0 | 0 |
| 0 | 0 | 0 | 0 | 0 |
| 0 | 0 | 0 | 0 | 0 |
| 0 | 0 | 0 | 0 | 0 |
| 0 | 0 | 0 | 0.059216297 | 0 |
| 0 | 0 | 0 | 0 | 0 |
| 0 | 0 | 0 | 0 | 0 |
| 0 | 0 | 0 | 0 | 0 |
| 0 | 0 | 0 | 0 | 0 |
| 0 | 0 | 0 | 0 | 0 |
| 0 | 0 | 0 | 0 | 0 |
| 0.01752977 | 0 | 0 | 0 | 0 |
| 0 | 0 | 0 | 0 | 0 |
| 0 | 0.026670228 | 0 | 0 | 0 |
| 0 | 0 | 0 | 0 | 0 |
| 0 | 0 | 0 | 0.146748404 | 0 |
| 0 | 0 | 0 | 0 | 0 |
| 0 | 0 | 0 | 0 | 0 |
| 0 | 0 | 0 | 0 | 0 |
| 0 | 0 | 0 | 0 | 0 |
| 0 | 0 | 0 | 0 | 0 |
| 0 | 0 | 0 | 0 | 0 |
| 0 | 0 | 0 | 0 | 0 |
| 0 | 0 | 0 | 0 | 0 |
| 0 | 0 | 0 | 0 | 0 |
| 0 | 0 | 0 | 0 | 0 |
| 0.003407222 | 0 | 16.02824661 | 0.081027757 | 1.437481622 |
| 0 | 0 | 0 | 0 | 0 |
| 0 | 0 | 0 | 0 | 0 |
| 0 | 0 | 0 | 0 | 0 |
| 0 | 0 | 0 | 0 | 0 |
| 0 | 0 | 0 | 0 | 0 |
| 0 | 0 | 0 | 0 | 0 |
| 0 | 0 | 0 | 0 | 0 |
| 0 | 0 | 0 | 0 | 0 |
| 0 | 0 | 0 | 0 | 0 |

| | | | | |
|---|---|---|---|---|
| 0 | 0 | 0 | 0 | 0 |
| 0.00696931 | 0.014129373 | 0 | 0 | 0 |
| 0 | 0 | 0 | 0 | 0 |
| 0 | 0 | 0 | 0 | 0 |
| 0 | 0 | 0 | 0 | 0 |
| 0 | 0 | 0 | 0 | 0 |
| 0 | 0 | 0 | 0 | 0 |
| 0 | 0 | 0 | 0 | 0 |
| 0 | 0 | 0 | 0 | 0 |
| 0 | 0 | 0 | 0 | 0 |
| 0 | 0.000984111 | 0.02850182 | 0.003670143 | 0.005548888 |
| 0 | 0 | 0 | 0 | 0 |
| 0.143896752 | 0.002319513 | 0 | 0.018343452 | 0 |
| 0 | 0 | 0 | 0 | 0 |
| 0 | 0 | 0 | 0 | 0 |
| 0 | 0 | 0 | 0 | 0 |
| 0 | 0 | 0 | 0 | 0 |
| 0 | 0 | 0 | 0 | 0 |
| 0 | 0 | 0 | 0 | 0 |
| 0 | 0 | 0 | 0 | 0 |
| 0.001323157 | 0 | 0 | 0 | 0 |
| 0 | 0 | 0 | 0 | 0 |
| 0 | 0 | 0 | 0 | 0 |
| 2.594317845 | 11.79300853 | 0.355399188 | 0.095879182 | 1.027395655 |
| 0 | 0 | 0 | 0 | 0 |
| 0 | 0 | 0 | 0 | 0 |
| 0 | 0 | 0 | 0 | 0 |
| 0 | 0 | 0 | 0.004278236 | 0 |
| 0 | 0 | 0 | 0 | 0 |
| 0 | 0 | 0 | 0 | 0 |
| 0 | 0 | 0 | 0.005484089 | 0 |
| 0 | 0 | 0 | 0 | 0 |
| 0 | 0 | 0 | 0 | 0.01493904 |
| 0 | 0 | 0 | 0 | 0 |

| PHL018-0160_V3 | PHL018-0269_V2 | PHL018-0061_V1 | PHL018-0229_V3 | PHL018-0262_V2 |
|---|---|---|---|---|
| REF | REF | REF | REF | REF |
| 0.067337909 | 0.001359582 | 0.111665646 | 0.252098941 | 1.197088093 |
| 0.012199623 | 0 | 1.570455545 | 0.084530844 | 0.305631508 |
| 0.000141854 | 5.697860748 | 2.986396612 | 0 | 7.101148725 |
| 0 | 0 | 0 | 0 | 0 |
| 0 | 0 | 0 | 0 | 0 |
| 0 | 0 | 0 | 0 | 0 |
| 0 | 0 | 0 | 0 | 0 |
| 0 | 0 | 0 | 0 | 0 |
| 0 | 0 | 0 | 0 | 0 |
| 0 | 0 | 0 | 0 | 0 |
| 0 | 0 | 0 | 0 | 0 |
| 0 | 0 | 0 | 0 | 0 |
| 0 | 0 | 0 | 0 | 0 |
| 0.065419905 | 3.005679496 | 9.559799818 | 0.866410235 | 4.527761067 |
| 0 | 0 | 0 | 0 | 0 |
| 0 | 0 | 0 | 0 | 0 |
| 0 | 0 | 0 | 0 | 0 |
| 0 | 0 | 0 | 0 | 0 |
| 0 | 0 | 0 | 0 | 0 |
| 0 | 0 | 0 | 0 | 0 |
| 0 | 0 | 0 | 0 | 0 |
| 0 | 0 | 0 | 0 | 0 |
| 0 | 0 | 0 | 0 | 0 |
| 0 | 0 | 0 | 0 | 0 |
| 0 | 0 | 0 | 1.159235324 | 0 |
| 0 | 0 | 0 | 0 | 0 |
| 0 | 0 | 0 | 0 | 0 |
| 0 | 0 | 0 | 0 | 0 |
| 0 | 0 | 0 | 0 | 0 |
| 0 | 0 | 0 | 0 | 0 |
| 0 | 0 | 0 | 0 | 0 |
| 0 | 0 | 0 | 0 | 0 |
| 0 | 0 | 0 | 0 | 0 |
| 0 | 0 | 0 | 0 | 0 |
| 0 | 0 | 0 | 0 | 0 |
| 0 | 0 | 0 | 0.032965273 | 0.018364455 |
| 0 | 0 | 0 | 0 | 0 |
| 0 | 0 | 0 | 0 | 0 |
| 0 | 0 | 0 | 0 | 0 |
| 0 | 0 | 0 | 0 | 0 |
| 0 | 0 | 0 | 0 | 0 |
| 0 | 0.006402849 | 0.001531857 | 0.010901957 | 0 |
| 0.012583182 | 0.028029345 | 0.015013796 | 3.776270435 | 1.405915104 |
| 0.009000701 | 0.026449609 | 0.009907913 | 0.001172229 | 0.013790621 |
| 0.000132432 | 0 | 0 | 0.002535408 | 0 |
| 0 | 0 | 0 | 0 | 0 |

| | | | | |
|---|---|---|---|---|
| 0 | 0 | 0 | 0.008899432 | 0.004454338 |
| 0.001518154 | 0 | 0.010943968 | 0 | 0 |
| 0 | 0 | 0 | 0.008099653 | 0.013392786 |
| 0.018512599 | 0 | 0.247800311 | 0.878579165 | 0.737631698 |
| 0 | 0 | 0.001596639 | 0 | 0 |
| 0 | 0 | 0 | 0.081340405 | 0.004860567 |
| 0.001265613 | 0.558400632 | 0.001922978 | 0.002019172 | 0.002346115 |
| 0 | 0 | 0 | 0 | 0 |
| 0 | 0 | 0 | 0 | 0 |
| 0 | 0 | 0 | 0 | 0 |
| 0 | 0.713480979 | 0 | 0 | 0 |
| 0.007413353 | 0 | 0 | 1.382801285 | 0.528669279 |
| 0 | 0 | 0 | 0 | 0 |
| 0.003392496 | 0 | 0.025477768 | 0.000723034 | 0.234893931 |
| 0 | 0 | 0 | 0 | 0 |
| 0 | 0 | 0 | 0 | 0 |
| 0 | 0.666238121 | 0.001510371 | 0.010506746 | 2.247653422 |
| 0 | 0 | 0 | 0 | 0 |
| 0 | 0 | 0 | 0 | 0 |
| 0 | 0 | 0 | 0 | 0 |
| 0.002887014 | 0 | 0.212321206 | 0 | 0.042611253 |
| 0.001002032 | 0 | 0.002049509 | 0.011047096 | 0.011168848 |
| 0 | 0 | 0 | 0.003218339 | 0.007665878 |
| 0 | 0 | 0 | 0 | 0 |
| 0 | 0 | 0 | 0 | 0 |
| 0 | 0 | 0 | 0 | 0 |
| 0.000935694 | 0 | 0 | 0 | 0 |
| 0 | 0 | 0 | 0 | 0 |
| 0 | 0 | 0.000699177 | 0 | 0 |
| 0.00016904 | 0 | 0 | 0 | 0 |
| 0 | 0 | 0 | 0 | 0.009633319 |
| 0 | 0 | 0 | 0 | 0 |
| 0 | 0 | 0 | 0 | 0 |
| 0 | 0 | 0 | 0 | 0 |
| 0 | 0 | 0 | 0 | 0 |
| 0 | 0 | 0 | 0 | 0 |
| 0 | 0 | 0 | 0 | 0 |
| 0 | 0 | 0 | 0 | 0.004042807 |
| 0 | 0 | 0 | 0 | 0 |
| 0 | 0 | 0 | 0 | 0 |
| 0 | 0 | 0 | 0 | 0 |
| 0 | 0 | 0 | 0 | 0 |
| 0 | 0 | 0 | 0 | 0 |
| 0 | 0 | 0 | 0 | 0 |
| 0 | 0 | 0 | 0 | 0 |
| 0 | 0 | 0 | 0 | 0 |
| 0 | 0.000977644 | 0 | 0 | 0 |
| 0.012719603 | 0 | 0 | 0 | 1.692622672 |

| | | | | |
|---|---|---|---|---|
| 0 | 0 | 0 | 0 | 0 |
| 0 | 0 | 0 | 0 | 0.003151143 |
| 0 | 0 | 0 | 0 | 0 |
| 0 | 0 | 0 | 0 | 0 |
| 0.000555664 | 0 | 0 | 0 | 0.238701969 |
| 0 | 0 | 0 | 0 | 0 |
| 0 | 0 | 0 | 0 | 0 |
| 0 | 0 | 0 | 0 | 0 |
| 0 | 0 | 0 | 0 | 0.000805639 |
| 0 | 0 | 0 | 0 | 0 |
| 0 | 0 | 0 | 0 | 0 |
| 0 | 0 | 0 | 0 | 0 |
| 0 | 0 | 0 | 0 | 0 |
| 0 | 0 | 0 | 0 | 0 |
| 0 | 0 | 0 | 0 | 0 |
| 0.000227309 | 0 | 0.000995489 | 0 | 0 |
| 0 | 0 | 0 | 0 | 0 |
| 0 | 0 | 0 | 0 | 0 |
| 0 | 0 | 0 | 0.181536147 | 0.098722777 |
| 0 | 0 | 0 | 0 | 0 |
| 0 | 0 | 0 | 0 | 0 |
| 0 | 0 | 0 | 0 | 0 |
| 0 | 0 | 0 | 0 | 0 |
| 0 | 0 | 0 | 0 | 0 |
| 0 | 0 | 0 | 0 | 0 |
| 0 | 0 | 0 | 0 | 0 |
| 0 | 0 | 0 | 0 | 0 |
| 0 | 0 | 0 | 0 | 0 |
| 0 | 0 | 0 | 0 | 0 |
| 0 | 0 | 0 | 0 | 0 |
| 0 | 0 | 0 | 0.196336806 | 0.025794824 |
| 9.64365E-05 | 0 | 0 | 0.000673119 | 0.006569727 |
| 0 | 0 | 0 | 0 | 0 |
| 0 | 0 | 0 | 0 | 0 |
| 0 | 0 | 0 | 0 | 0 |
| 0 | 0 | 0 | 0 | 0 |
| 0 | 0 | 0 | 0 | 0 |
| 0 | 0 | 0 | 0 | 0 |
| 0 | 0 | 0 | 0 | 0 |
| 0 | 0 | 0 | 0 | 0 |
| 0 | 0 | 0 | 0 | 0 |
| 0 | 0 | 0 | 0 | 0 |
| 0 | 0 | 0 | 0 | 0.003796782 |
| 0 | 0 | 0 | 0 | 0 |
| 0 | 0 | 0 | 0 | 0 |
| 0 | 0 | 0 | 0 | 0 |
| 0.000189898 | 0 | 0 | 0 | 0.26138881 |
| 0 | 0 | 0 | 0 | 0 |

| | | | | |
|---|---|---|---|---|
| 0 | 0 | 0 | 0 | 0 |
| 0 | 0 | 0 | 0 | 0 |
| 0 | 0 | 0 | 0 | 0 |
| 0 | 0 | 0 | 0 | 0 |
| 0 | 0 | 0 | 0.178691541 | 0.000408443 |
| 0 | 0 | 0 | 0 | 0 |
| 0 | 0 | 0 | 0 | 0 |
| 0 | 0 | 0 | 0 | 0 |
| 0 | 0 | 0 | 0 | 0 |
| 0 | 0 | 0 | 0 | 0 |
| 0 | 0 | 0 | 0 | 0 |
| 0 | 0 | 0 | 0 | 0 |
| 0 | 0 | 0 | 0 | 0 |
| 0 | 0 | 0 | 0 | 0 |
| 0 | 0 | 0 | 0 | 0 |
| 0 | 0 | 0 | 0 | 0 |
| 0 | 0 | 0 | 0 | 0 |
| 0 | 0 | 0 | 0 | 0 |
| 0 | 0 | 0 | 0 | 0 |
| 0 | 0 | 0 | 0 | 0 |
| 0 | 0 | 0 | 0 | 0 |
| 0 | 0 | 0 | 0 | 0 |
| 0 | 0 | 0 | 0 | 0 |
| 0 | 0 | 0 | 0 | 0 |
| 0 | 0 | 0 | 0.049408914 | 0 |
| 0 | 0 | 0 | 0 | 0 |
| 0 | 0 | 0 | 0.000438712 | 0 |
| 0 | 0 | 0 | 0 | 0 |
| 0 | 0 | 0 | 0 | 0 |
| 0 | 0 | 0 | 0 | 0 |
| 0.000363403 | 0 | 0 | 0 | 0 |
| 95.28449115 | 0 | 0 | 0 | 0 |
| 0 | 0 | 0 | 0 | 0 |
| 0 | 0 | 0 | 0 | 0 |
| 0 | 0 | 0 | 0 | 0 |
| 0 | 0 | 0 | 0 | 0 |
| 0 | 0 | 0 | 0 | 0 |
| 0 | 0 | 0 | 0 | 0 |
| 0 | 0 | 0 | 0 | 0 |
| 0 | 0 | 0 | 0 | 0 |
| 0 | 0 | 0 | 0 | 0 |
| 0 | 0 | 0 | 0 | 0 |
| 0 | 0 | 0 | 0 | 0 |
| 0 | 0 | 0 | 0 | 0 |
| 0 | 0 | 0 | 0 | 0 |
| 0 | 0 | 0 | 0 | 0 |
| 0 | 0 | 0 | 0 | 0 |
| 0 | 0 | 0 | 0 | 0 |

| | | | | |
|---|---|---|---|---|
| 0 | 0 | 0 | 0 | 0 |
| 0 | 0 | 0 | 0 | 0 |
| 0 | 0 | 0 | 0 | 0 |
| 0 | 0 | 0 | 0 | 0 |
| 0 | 0 | 0 | 0 | 0 |
| 0 | 0 | 0 | 0 | 0 |
| 0 | 0 | 0 | 0 | 0 |
| 0 | 0 | 0 | 0 | 0 |
| 0 | 0 | 0 | 0 | 0 |
| 0 | 0 | 0 | 0 | 0 |
| 0 | 0 | 0 | 0 | 0 |
| 0 | 0 | 0 | 0 | 0 |
| 0 | 0 | 0 | 0 | 0.001726991 |
| 0 | 0 | 0 | 0 | 0 |
| 0 | 0 | 0 | 0 | 0 |
| 0 | 0 | 0 | 0 | 0 |
| 0.000469949 | 0 | 0 | 0 | 0 |
| 0 | 0 | 0 | 0 | 0 |
| 0 | 0 | 0 | 0 | 0 |
| 0 | 0 | 0 | 0 | 0 |
| 0.00126911 | 0.050930346 | 0.069872259 | 0.008337213 | 0.064581159 |
| 0 | 0 | 0 | 0 | 0 |
| 0.000152842 | 0 | 0 | 0 | 0 |
| 0 | 0 | 0 | 0 | 0 |
| 0 | 0 | 0 | 0 | 0 |
| 0 | 0 | 0 | 0 | 0 |
| 5.79686E-05 | 0 | 0.005548903 | 0 | 0.002444685 |
| 0 | 0 | 0 | 0 | 0 |
| 0 | 0 | 0 | 0.001558661 | 0 |
| 0.000914791 | 0 | 0 | 0 | 0 |
| 0 | 0.002667664 | 0 | 0 | 0 |
| 0 | 0 | 0 | 0 | 0 |
| 0 | 0 | 0 | 0 | 0 |
| 0 | 0 | 0 | 0.006623743 | 0.125113538 |
| 0.006006933 | 0.007723894 | 0.015498619 | 0.001808212 | 0.003991895 |
| 0 | 0 | 0 | 0 | 0 |
| 0 | 0 | 0 | 0 | 0 |
| 0 | 0 | 0 | 0 | 0 |
| 0 | 0 | 0 | 0 | 0 |
| 0 | 0 | 0 | 0 | 0 |
| 0 | 0 | 0 | 0 | 0 |
| 0 | 0 | 0 | 0 | 0 |
| 0 | 0 | 0 | 0 | 0 |
| 0 | 0 | 0 | 0 | 0 |
| 0 | 0 | 0 | 0 | 0 |
| 0 | 0 | 0 | 0.270982138 | 0 |
| 8.76538E-05 | 0 | 0 | 0 | 0.004094678 |
| 0.003380717 | 0 | 0.001227193 | 0.000386574 | 0.002545287 |

| | | | | |
|---|---|---|---|---|
| 0 | 0 | 0 | 0 | 0 |
| 0 | 0 | 0 | 0 | 0 |
| 0 | 0 | 0.001569936 | 0 | 0 |
| 0 | 0 | 0 | 0 | 0 |
| 0 | 0 | 0 | 0 | 0 |
| 0 | 0 | 0 | 0 | 0 |
| 0 | 0 | 0.001921583 | 0 | 0 |
| 0 | 0 | 0 | 0 | 0 |
| 0 | 0 | 0.005682919 | 0 | 0 |
| 0 | 0 | 0 | 0 | 0 |
| 0 | 0 | 0 | 0 | 0 |
| 0 | 0 | 0 | 0 | 0 |
| 0 | 0 | 0 | 0 | 0 |
| 0 | 0 | 0 | 0 | 0 |
| 0 | 0.073626311 | 0 | 0 | 0 |
| 0 | 0 | 0 | 0 | 0 |
| 0 | 0 | 0 | 0 | 0 |
| 0 | 0 | 0 | 0 | 0 |
| 0 | 0 | 0 | 0.014415258 | 0.011572295 |
| 0 | 0 | 0 | 0.168946599 | 0 |
| 0.013328215 | 0 | 0.005891188 | 0 | 0.001123047 |
| 0 | 0 | 0 | 0 | 0 |
| 0 | 0 | 0 | 0 | 0 |
| 0 | 0 | 0 | 0 | 0 |
| 0 | 0.008996234 | 0.001828764 | 0.005760733 | 0 |
| 0 | 0 | 0 | 0 | 0 |
| 0 | 0 | 0 | 0 | 0 |
| 0 | 0 | 0 | 0 | 0 |
| 0.000410221 | 0 | 0 | 0 | 0 |
| 0 | 0 | 0 | 0 | 0 |
| 0.024118751 | 0 | 0.06524756 | 0.074623455 | 0.416191071 |
| 0 | 0 | 0 | 0 | 0 |
| 0 | 0 | 0.115467495 | 0 | 0.190366421 |
| 0.000181133 | 0.014950393 | 0 | 0 | 0.021794736 |
| 0 | 0 | 0 | 0 | 0 |
| 0 | 0 | 0 | 0 | 0 |
| 0 | 0 | 0 | 0 | 0 |
| 0 | 0 | 0 | 0 | 0 |
| 0 | 0 | 0 | 0 | 0 |
| 0 | 0 | 0 | 0 | 0 |
| 0 | 0 | 0 | 0 | 0 |
| 0.001894152 | 0 | 0 | 0.00507108 | 0.022727014 |
| 0 | 0 | 0 | 0 | 0 |
| 0 | 0 | 0 | 0 | 0 |
| 0 | 0 | 0 | 0 | 0 |
| 0 | 0 | 0 | 0 | 0 |
| 0 | 0 | 0 | 0 | 0 |
| 0 | 0 | 0 | 0 | 0 |

| | | | | |
|---|---|---|---|---|
| 0 | 0 | 0 | 0 | 0 |
| 0 | 0 | 0 | 0 | 0 |
| 0.009448516 | 0.027324856 | 0.007970124 | 0 | 0.016488337 |
| 0 | 0 | 0 | 0 | 0 |
| 0.020598171 | 0 | 0 | 0 | 0 |
| 0 | 0 | 0 | 0 | 0 |
| 0 | 0 | 0 | 0 | 0 |
| 0 | 0 | 0 | 0 | 0 |
| 0 | 0 | 0 | 0 | 0 |
| 0 | 0 | 0 | 0 | 0 |
| 0 | 0 | 0 | 0 | 0 |
| 0 | 0 | 0 | 0 | 0 |
| 0 | 0 | 0 | 0 | 0 |
| 0 | 0 | 0 | 0.004825592 | 0.009344918 |
| 0.00011888 | 0.002286719 | 0 | 0.004148848 | 0.003470853 |
| 0 | 0 | 0 | 0.002132107 | 0.06647522 |
| 0 | 0.004950472 | 0.001440651 | 0 | 0.001757656 |
| 0 | 0 | 0 | 0 | 0 |
| 0 | 0 | 0 | 0 | 0 |
| 0 | 0 | 0 | 0 | 0 |
| 0 | 0 | 0 | 0 | 0 |
| 0.002919316 | 0 | 0.000812811 | 0.001365552 | 0.002776658 |
| 0 | 0 | 0 | 0 | 0 |
| 0 | 0 | 0 | 0 | 0 |
| 0 | 0 | 0 | 0 | 0 |
| 0 | 0 | 0 | 0 | 0 |
| 0 | 0 | 0 | 0 | 0 |
| 0 | 0 | 0 | 0 | 0 |
| 0 | 0 | 0 | 0 | 0 |
| 0 | 0.022454769 | 0 | 0 | 0 |
| 0 | 0 | 0 | 0 | 0 |
| 0 | 0 | 0 | 0 | 0 |
| 0 | 0 | 0 | 0 | 0 |
| 0 | 0 | 0 | 0 | 0 |
| 0 | 0 | 0 | 0 | 0 |
| 0 | 0 | 0.001623474 | 0 | 0 |
| 0 | 0 | 0 | 0 | 0 |
| 0.000384065 | 0 | 0 | 0.024877277 | 0 |
| 0 | 0 | 0 | 0 | 0 |
| 0 | 0 | 0 | 0 | 0 |
| 0 | 0 | 0 | 0 | 0 |
| 0 | 0 | 0 | 0 | 0 |
| 0 | 0 | 0 | 0 | 0 |
| 0 | 0 | 0.001522206 | 0 | 0 |
| 0 | 0 | 0 | 0 | 0 |
| 0 | 0 | 0 | 0 | 0 |
| 0 | 0 | 0 | 0 | 0 |
| 0.00016282 | 0 | 0 | 0 | 0.000905475 |

| | | | | |
|---|---|---|---|---|
| 0.203376001 | 0 | 0 | 0 | 0 |
| 0 | 0 | 0 | 0 | 0 |
| 0 | 0 | 0 | 0 | 0 |
| 0 | 0 | 0 | 0 | 0 |
| 0 | 0 | 0.002398329 | 0 | 0.002508054 |
| 0 | 0 | 0 | 0 | 0 |
| 0 | 0 | 0 | 0 | 0 |
| 0 | 0.002084574 | 0.181776537 | 0.001512837 | 0.025780991 |
| 0 | 0 | 0 | 0 | 0 |
| 0.000232708 | 0.015040309 | 2.431980615 | 0.003378512 | 0.136790688 |
| 0 | 0 | 0 | 0 | 0 |
| 0 | 0.005500521 | 0.005132317 | 0 | 0 |
| 0.001179433 | 0.00208617 | 0 | 0.000756997 | 0.023220658 |
| 0.00615615 | 0 | 0 | 0 | 0 |
| 0 | 0 | 0 | 0 | 0.001447389 |
| 8.14034E-05 | 0 | 0 | 0 | 0 |
| 0.523063659 | 0 | 0 | 0 | 0 |
| 0 | 0 | 0.000938923 | 0 | 0 |
| 0 | 0 | 0.010713278 | 0 | 0 |
| 0 | 0 | 0.067952121 | 0 | 0.104086119 |
| 0 | 0 | 0 | 0 | 0 |
| 0 | 0 | 0 | 0 | 0 |
| 0 | 0 | 0 | 0 | 0 |
| 0 | 0 | 0 | 0.207332074 | 0 |
| 0 | 0 | 0 | 0 | 0 |
| 0 | 0.008990469 | 0.000671764 | 0 | 0 |
| 0 | 0.018845421 | 0 | 5.363143152 | 0 |
| 0 | 0 | 0 | 0 | 0 |
| 0 | 0 | 0 | 0 | 0 |
| 0 | 0 | 0 | 0.001088601 | 0 |
| 0 | 0 | 0.037844041 | 0 | 0 |
| 0.00097783 | 0 | 0 | 0 | 0 |
| 0 | 0 | 0 | 0 | 0 |
| 0 | 0 | 0.011201428 | 0 | 0 |
| 0 | 0 | 0 | 0 | 0 |
| 0 | 0 | 0 | 0 | 0 |
| 0 | 0.001377246 | 0.011803449 | 0 | 0 |
| 0 | 0 | 0 | 0 | 0 |
| 0 | 0 | 0 | 0.000627902 | 0 |
| 0 | 0 | 0.001059785 | 0 | 0 |
| 0 | 0 | 0 | 0 | 0 |
| 0 | 0 | 0 | 0 | 0 |
| 0 | 0 | 0 | 0 | 0 |
| 0.005342845 | 0 | 0 | 0 | 0 |
| 0 | 0 | 0 | 0 | 0 |
| 0 | 0 | 0 | 0.014063034 | 0 |
| 7.85449E-05 | 0 | 0 | 0.003289425 | 0 |
| 0 | 0 | 0 | 0 | 0 |

| | | | | |
|---|---|---|---|---|
| 0 | 0 | 0 | 0 | 0 |
| 0 | 0.33364579 | 0 | 0 | 0 |
| 0 | 0 | 0.164752202 | 0 | 0 |
| 0 | 0 | 0 | 0 | 0.004936135 |
| 0.000815151 | 0 | 0 | 0 | 0 |
| 0.001252739 | 0 | 0.162190023 | 0 | 0 |
| 0 | 0 | 0 | 0 | 0 |
| 0 | 0 | 0 | 0 | 0 |
| 0 | 0 | 0 | 0 | 0 |
| 0 | 0 | 0 | 0 | 0 |
| 0 | 0 | 0.006295875 | 0 | 0.003491471 |
| 0 | 0 | 0 | 0 | 0.006326453 |
| 0 | 0 | 0 | 0 | 0 |
| 0 | 0.015907902 | 0 | 0 | 0 |
| 0 | 0.003958017 | 0.041945962 | 0 | 0.003337556 |
| 0 | 0 | 0 | 0 | 0 |
| 0 | 0 | 0 | 0 | 0 |
| 0 | 0 | 0 | 0 | 0 |
| 0 | 0 | 0 | 0 | 0.026916259 |
| 0 | 0 | 0 | 0 | 0 |
| 0 | 0 | 0 | 0 | 0 |
| 0 | 0.131206485 | 0 | 0.341295578 | 0 |
| 0 | 0 | 0 | 0 | 0 |
| 0 | 0 | 0 | 0 | 0 |
| 0 | 0 | 0 | 0 | 0 |
| 0 | 0 | 0 | 0 | 0 |
| 0 | 0 | 0.037327392 | 0 | 0 |
| 0 | 0 | 0 | 0 | 0 |
| 0 | 0 | 0 | 0 | 0 |
| 0.388727996 | 0 | 0.391542639 | 0.256053887 | 1.186510698 |
| 0 | 0 | 0.002934737 | 0.003595131 | 0.0120842 |
| 0.049328406 | 0 | 0 | 0 | 0 |
| 0 | 0 | 0 | 0 | 0 |
| 0 | 0 | 0 | 0 | 0 |
| 0 | 0 | 0 | 0 | 0.001588748 |
| 0 | 0 | 0 | 0 | 0 |
| 0 | 0 | 0 | 0.21354358 | 0 |
| 0 | 0 | 0 | 0 | 0 |
| 0 | 0 | 0.109827481 | 0 | 0.159338114 |
| 0 | 0.019748846 | 0.13756164 | 0 | 0 |
| 0.000905587 | 0 | 0.014669775 | 0 | 0.071410212 |
| 0 | 0 | 0 | 2.255194719 | 0.149730158 |
| 0 | 0 | 0 | 0 | 0 |
| 0 | 0 | 0 | 0 | 0 |
| 0 | 0 | 0.000501463 | 0 | 0 |
| 0 | 0 | 0 | 0 | 0 |
| 0 | 0.008550827 | 0.002799451 | 0 | 0.002276966 |
| 0 | 0 | 0 | 0 | 0 |

| | | | | |
|---|---|---|---|---|
| 0.000108252 | 0.005552782 | 0 | 0.002619381 | 0 |
| 0.000115283 | 0 | 0.005885434 | 0 | 0 |
| 0.000221253 | 0.041991803 | 0.002091739 | 0.002745467 | 0.106865421 |
| 0 | 0 | 0 | 0 | 0 |
| 0 | 0 | 0 | 0.026196122 | 0 |
| 0 | 0 | 0 | 0 | 0 |
| 0 | 0 | 0 | 0 | 0 |
| 0 | 0 | 0 | 0 | 0 |
| 0.009437214 | 0 | 0 | 0 | 0 |
| 0 | 0 | 0 | 0 | 0.077413368 |
| 0 | 0 | 0 | 0 | 0 |
| 0 | 0 | 0 | 0 | 0 |
| 0 | 0 | 0 | 0 | 0 |
| 0 | 0 | 0 | 0 | 0 |
| 0 | 0 | 0 | 0 | 0 |
| 0.008699235 | 0 | 0.000949656 | 0.028917663 | 0.182896587 |
| 0.001074497 | 0.009137694 | 0.018645965 | 0.07594638 | 0.002384962 |
| 0.001580308 | 0 | 0 | 0 | 0.001337371 |
| 0 | 0 | 0.003106431 | 0 | 0 |
| 0 | 0 | 0 | 0 | 0 |
| 0 | 0 | 0 | 0 | 0 |
| 0 | 0 | 0 | 0 | 0.00085121 |
| 0 | 0 | 0 | 0 | 0 |
| 0 | 0 | 0 | 0 | 1.144211384 |
| 0 | 0 | 0 | 0 | 0 |
| 9.31224E-05 | 0 | 0 | 0 | 0 |
| 5.38454E-05 | 0 | 0.00057269 | 0 | 0.005356751 |
| 0 | 0 | 0 | 0 | 0 |
| 0 | 0 | 0 | 0 | 0 |
| 0 | 0 | 0 | 0 | 0 |
| 0 | 0 | 0.002032471 | 0.006249984 | 0.02993353 |
| 0.000488402 | 0 | 0.001731522 | 0 | 0.00369693 |
| 0.000445649 | 48.56350183 | 0.008243218 | 0.010603071 | 0.159907513 |
| 0 | 0 | 0.00443 | 0 | 0 |
| 0.000158553 | 0.00548975 | 0 | 0 | 0.007972465 |
| 0 | 0 | 0 | 0 | 0 |
| 0 | 0.021295583 | 0.004844084 | 0.001173785 | 0 |
| 0.012137602 | 0 | 0.006410966 | 0.004130793 | 0.012621302 |
| 0 | 0 | 0 | 0.126723528 | 0.422975777 |
| 0.001332871 | 0 | 0 | 0 | 0.014824771 |
| 0 | 0 | 0 | 0 | 0 |
| 0 | 0 | 0 | 0 | 0 |
| 0.017680557 | 0 | 0 | 0 | 0 |
| 0 | 0 | 0 | 0 | 0.062861607 |
| 0 | 0 | 0.030270525 | 0 | 0 |
| 0 | 0 | 0 | 0 | 0.05367655 |
| 0 | 0 | 0 | 0 | 0 |
| 0 | 0 | 0 | 0 | 0 |

| | | | | |
|---|---|---|---|---|
| 0 | 0 | 0 | 0 | 0 |
| 0.012152262 | 0 | 0.008338675 | 0.195754684 | 0.182154784 |
| 0 | 0 | 0 | 0 | 0 |
| 0.011203736 | 0 | 0 | 0 | 0 |
| 0 | 0 | 0 | 0 | 0 |
| 0 | 0 | 0 | 0 | 0 |
| 0 | 0 | 0 | 0 | 0 |
| 0 | 0 | 0.000914997 | 0 | 0 |
| 0.000290499 | 0 | 0 | 0.009160246 | 0 |
| 0.00037982 | 0 | 0.006348102 | 0 | 0 |
| 0.000124845 | 0 | 0 | 0 | 0 |
| 0.000599205 | 0.428293003 | 0.119769368 | 0.00207678 | 0.174276004 |
| 0 | 0 | 0 | 0.461722952 | 0 |
| 0 | 0 | 0 | 0 | 0 |
| 0 | 0.00759262 | 0.013373126 | 0 | 0 |
| 0 | 0 | 0 | 0 | 0 |
| 0.000261462 | 0.023693926 | 0.000772462 | 0.026928609 | 0.006408567 |
| 0 | 0 | 0.09967456 | 0 | 0 |
| 0 | 0 | 0 | 0 | 0 |
| 0 | 0 | 0.014715795 | 0 | 0.00746755 |
| 0.000269633 | 0 | 0.001911847 | 0.119445274 | 0.347158965 |
| 0 | 0 | 0 | 0 | 0.019107897 |
| 0 | 0 | 0 | 0 | 0 |
| 0 | 0 | 0 | 0 | 0 |
| 0 | 0.025174852 | 0 | 0 | 0 |
| 0 | 0 | 0 | 0 | 0 |
| 0.007984233 | 0 | 0 | 0.059774847 | 0.047773319 |
| 0.008183449 | 0.054145366 | 0.225383932 | 0.071655372 | 0.098752426 |
| 0 | 0 | 0 | 0 | 0 |
| 0.000556933 | 0 | 0 | 0 | 0.004734838 |
| 0 | 0 | 0 | 0 | 0 |
| 0 | 0 | 0 | 0 | 0 |
| 0 | 0 | 0 | 0 | 0 |
| 0 | 0 | 0 | 0 | 0 |
| 0 | 0 | 0 | 1.04974957 | 0 |
| 0.008945914 | 0 | 0.964082616 | 0.054166351 | 0.050349567 |
| 0 | 0 | 0 | 0 | 0 |
| 0 | 0 | 0 | 0 | 0 |
| 0 | 0.002720133 | 0.000835571 | 0 | 0 |
| 0 | 0 | 0 | 0 | 0 |
| 0 | 0 | 0.034081139 | 0 | 0 |
| 0 | 0 | 0 | 0 | 0 |
| 0 | 0.001835833 | 0.023219894 | 0 | 0.003560508 |
| 0.000264917 | 0 | 0.014333056 | 0 | 0 |
| 0 | 0 | 0.000965657 | 0 | 0 |
| 0 | 0 | 0 | 1.007945539 | 0 |
| 0 | 0 | 0.013643524 | 0 | 0.017521768 |
| 0 | 0 | 0.008910875 | 0 | 0 |

| | | | | |
|---|---|---|---|---|
| 0 | 0 | 0 | 0 | 0.000648797 |
| 0 | 0 | 0 | 0 | 0 |
| 0 | 0 | 0 | 0.000610493 | 0 |
| 0 | 0 | 0 | 0.236638683 | 0.528274546 |
| 0 | 0 | 0 | 0 | 0 |
| 0 | 0 | 0 | 0 | 0 |
| 0 | 0 | 0 | 0 | 0 |
| 0 | 0 | 0 | 0 | 0 |
| 0.979370038 | 0.036251164 | 0 | 1.209857405 | 1.971370215 |
| 0 | 0 | 0.005019018 | 0.034394231 | 0.028844507 |
| 0 | 0 | 0 | 0 | 0 |
| 0 | 0 | 0 | 0.000743362 | 0 |
| 0 | 0 | 0 | 0 | 0 |
| 0 | 0 | 0 | 0 | 0 |
| 0 | 0.002535755 | 0.047203396 | 0.237026997 | 0.432211241 |
| 0 | 0 | 0 | 0 | 0 |
| 0 | 0 | 0 | 0.018883723 | 0.017292776 |
| 0 | 0 | 0 | 0 | 0 |
| 0 | 0 | 0 | 0 | 0 |
| 0 | 0 | 0 | 0 | 0 |
| 0 | 0 | 0 | 0 | 0 |
| 0 | 0 | 0 | 0.002208502 | 0 |
| 0 | 0 | 0 | 0.018344876 | 0 |
| 0.012380779 | 0 | 0.020933733 | 0 | 0.273800271 |
| 0 | 0 | 0.00066282 | 0 | 0 |
| 0 | 0 | 0 | 0.29379646 | 0.588726346 |
| 0 | 0 | 0 | 0.011096601 | 0 |
| 0 | 0.820635471 | 0 | 8.898343768 | 0 |
| 0 | 0 | 0 | 0 | 0 |
| 0 | 0 | 0 | 0 | 0 |
| 0 | 0 | 0 | 0.005636859 | 0 |
| 0 | 0 | 0 | 0.005719503 | 0 |
| 0.000344744 | 0 | 0.000314283 | 0.001980029 | 0.010864112 |
| 0 | 0 | 0 | 0 | 0 |
| 0 | 0 | 0 | 0 | 0 |
| 0 | 0.000675455 | 0 | 0.659805915 | 0 |
| 0 | 0 | 0 | 1.891523033 | 0 |
| 0 | 0 | 0 | 0 | 0 |
| 0 | 0 | 0 | 0.012690968 | 0 |
| 0 | 0 | 0 | 0 | 0 |
| 0.000140481 | 0 | 0.001086639 | 0.032803639 | 0.039275228 |
| 0 | 0 | 0 | 0.073084279 | 1.022183329 |
| 0.029997229 | 19.52443409 | 0.276842942 | 0.015056414 | 9.05272805 |
| 0 | 0 | 0.000828279 | 0.022861006 | 0.048938807 |
| 0 | 0 | 0 | 0 | 0 |
| 0 | 0 | 0 | 0 | 0 |
| 0 | 0 | 0 | 0 | 0 |
| 0.000245635 | 0 | 0.001142982 | 0.003943374 | 0.01095668 |

| | | | | |
|---|---|---|---|---|
| 0 | 0 | 0 | 0 | 0 |
| 0 | 0 | 0 | 0.034519481 | 0 |
| 0.025886824 | 0.127954964 | 0.565369408 | 0.003451459 | 0.006874831 |
| 0 | 0 | 0 | 0.003908522 | 0 |
| 0 | 0 | 0 | 0 | 0 |
| 0 | 0 | 0 | 0.075612038 | 0.052652902 |
| 0 | 0 | 0 | 0 | 0 |
| 0 | 0 | 0 | 0.001705707 | 0 |
| 0 | 0 | 0 | 0 | 0 |
| 0 | 0 | 0 | 0 | 0 |
| 0.014521519 | 0 | 0.021976253 | 0.054380892 | 0.167621652 |
| 0.050796076 | 2.716603107 | 0.642600311 | 0.814936835 | 2.058188601 |
| 0 | 0 | 0 | 0 | 0 |
| 0.002510673 | 0 | 0.004146446 | 0.010623426 | 0.001821182 |
| 0.012878728 | 0.018721586 | 0.521240869 | 0.010293033 | 0.319403184 |
| 0 | 0 | 0 | 3.699664031 | 0 |
| 0 | 0 | 0 | 0 | 0 |
| 0 | 0 | 0 | 0 | 0 |
| 0 | 0 | 0 | 0.035953443 | 0 |
| 0 | 0 | 0 | 0.005981601 | 0.01837926 |
| 0.000145327 | 0.004659101 | 0.012107816 | 0.024209721 | 0.024515318 |
| 0 | 1.458242593 | 35.68148697 | 0 | 2.961685332 |
| 0.012628405 | 0.432670274 | 18.16689337 | 0.189576179 | 2.25942407 |
| 0 | 0 | 0 | 0 | 0 |
| 0 | 0 | 0 | 0 | 0 |
| 0 | 0 | 0 | 2.084866898 | 7.093720816 |
| 0 | 0 | 0 | 0 | 0.35317325 |
| 0 | 0 | 0 | 0 | 0.001311232 |
| 0 | 0 | 0 | 0.39112401 | 0 |
| 0 | 0 | 0 | 0 | 0 |
| 0 | 0 | 0 | 0.162603007 | 0 |
| 0 | 0 | 0 | 0.075079495 | 0 |
| 0.035081239 | 0 | 0 | 0.02949972 | 0.004428961 |
| 0 | 0 | 0 | 0.714684628 | 0 |
| 0.023026725 | 0 | 0.001770925 | 0 | 0.033934185 |
| 0.042927702 | 0.075377912 | 0.006011293 | 0.021490842 | 0.046798116 |
| 0 | 0 | 0.000967804 | 0.009000779 | 0.016868037 |
| 0 | 0 | 0 | 0 | 0 |
| 0.020723527 | 0.004604231 | 0 | 0 | 0.094609637 |
| 0 | 0 | 0 | 0 | 0 |
| 0 | 0 | 0 | 0.028848822 | 0 |
| 0 | 0 | 0 | 0 | 0 |
| 0 | 0 | 0 | 0 | 0 |
| 0 | 0 | 0 | 0 | 0 |
| 0.006813684 | 0 | 0 | 0.062040892 | 0.113904705 |
| 0 | 0 | 0 | 0.019340047 | 0 |
| 0 | 0 | 0 | 0 | 0 |
| 0.009959258 | 0.293981622 | 0.032880077 | 0.582580787 | 0.006245464 |

| | | | | |
|---|---|---|---|---|
| 0 | 0 | 0 | 0 | 0 |
| 0 | 0 | 0 | 0 | 0 |
| 0 | 0 | 0 | 0 | 0 |
| 0 | 0 | 0 | 0.001314626 | 0.001357769 |
| 0 | 0 | 0 | 0 | 0 |
| 0 | 0 | 0 | 0.553056263 | 0.101180256 |
| 0 | 0 | 0 | 0 | 0 |
| 0 | 0.001666867 | 0.029313645 | 0.017473363 | 0 |
| 0 | 0 | 0 | 0 | 0 |
| 0 | 0 | 0 | 0 | 0 |
| 0.029104951 | 0 | 0.001084529 | 0.009923655 | 0.010018296 |
| 0 | 0 | 0 | 0 | 0 |
| 0 | 0 | 0 | 0.042874914 | 0.024329337 |
| 0 | 0 | 0 | 0 | 0 |
| 0 | 0 | 0 | 0 | 0 |
| 0 | 0 | 0 | 0 | 0 |
| 0 | 0 | 0 | 0.021294808 | 0 |
| 0 | 0 | 0 | 0.000541719 | 0.011172457 |
| 0 | 0 | 0 | 0 | 0 |
| 0 | 0 | 0 | 0 | 0 |
| 0 | 0 | 0 | 0.025026287 | 0 |
| 0 | 0 | 0 | 0 | 0 |
| 0 | 0 | 0 | 0.037317662 | 0 |
| 0 | 0 | 0 | 0 | 0.011503995 |
| 0 | 0 | 0 | 0 | 0 |
| 0 | 0 | 0 | 0 | 0 |
| 0 | 0 | 0 | 0 | 0 |
| 0 | 0 | 0 | 0 | 0 |
| 0 | 0 | 0 | 0 | 0 |
| 0.017315097 | 0 | 1.125061928 | 0.016686992 | 0.650670271 |
| 0 | 0 | 0.000610134 | 0.012556826 | 0.00952818 |
| 0 | 0 | 0 | 0 | 0 |
| 0 | 0 | 0 | 0 | 0 |
| 0 | 0 | 0 | 0 | 0 |
| 0 | 0 | 0 | 0 | 0 |
| 0 | 0 | 0 | 0 | 0 |
| 0 | 0 | 0 | 0.220767121 | 0.137258517 |
| 0.001085151 | 0 | 0.002885371 | 0 | 0.070085481 |
| 0 | 0 | 0 | 0 | 0 |
| 0 | 0 | 0 | 0 | 0 |
| 0.01309698 | 0 | 0 | 0 | 0 |
| 0 | 0 | 0 | 0 | 0 |
| 0 | 0 | 0 | 0.014127554 | 0 |
| 0 | 0 | 0 | 0.004145832 | 0.012275897 |
| 0 | 0.052359145 | 0 | 0.177639396 | 0 |
| 0 | 0 | 0 | 0.11016965 | 0 |
| 0 | 0 | 0 | 0.057967111 | 0 |
| 0 | 0 | 0 | 0 | 0 |

| | | | | |
|---|---|---|---|---|
| 0 | 0 | 0 | 0 | 0 |
| 0 | 0 | 0 | 0 | 0 |
| 0 | 0 | 0 | 0 | 0 |
| 0 | 0 | 0 | 2.194088194 | 0 |
| 0 | 0 | 0 | 0 | 0 |
| 0 | 0 | 0 | 0.542091354 | 0 |
| 0 | 0 | 0 | 0 | 0 |
| 0 | 0 | 0 | 0 | 0 |
| 0.00146259 | 0.16644742 | 0.08971757 | 0.001329513 | 0.047888423 |
| 0 | 0 | 0 | 0 | 0 |
| 0 | 0 | 0 | 0.053371835 | 0 |
| 0 | 0 | 0 | 0 | 0 |
| 0 | 0 | 0 | 0 | 0 |
| 3.54373E-05 | 0 | 0 | 0 | 0 |
| 0.040661197 | 0 | 0 | 0.013785394 | 1.686121773 |
| 0 | 0 | 0 | 0 | 0.000850436 |
| 0 | 0 | 0 | 0 | 0 |
| 0 | 0 | 0 | 0 | 0 |
| 0 | 0 | 0 | 0 | 0 |
| 0 | 0 | 0 | 0 | 0 |
| 0 | 0 | 0 | 0.144648163 | 0 |
| 0 | 0 | 0 | 0 | 0 |
| 0 | 0 | 0 | 0 | 0 |
| 0 | 0 | 0 | 0 | 0 |
| 0.005279891 | 0.001846579 | 0 | 2.397469179 | 2.310748941 |
| 0 | 0 | 0.004811765 | 5.599603991 | 0.527127227 |
| 0 | 0 | 0 | 0.030072795 | 2.118924539 |
| 0 | 0 | 0 | 0 | 0 |
| 0 | 0 | 0 | 0.064245223 | 0 |
| 0 | 0 | 0 | 0 | 0 |
| 0 | 0 | 0 | 0 | 0 |
| 0 | 0 | 0 | 0 | 0 |
| 0 | 0 | 0 | 0 | 0 |
| 0.002841163 | 0 | 0.002272041 | 0.010974197 | 0.014691527 |
| 5.27971E-05 | 0.018145084 | 0.001684622 | 0.003930872 | 0 |
| 0.035529702 | 0 | 0.000661569 | 0 | 0.03664425 |
| 0 | 0 | 0.021710517 | 0 | 0 |
| 0.004958708 | 0 | 0.223061812 | 0.063065274 | 2.641974715 |
| 0 | 0 | 0.0026737 | 0 | 0 |
| 0 | 0 | 0 | 0 | 0 |
| 0 | 0 | 0 | 0.285639139 | 0 |
| 0.000691267 | 0 | 0 | 0 | 0 |
| 0 | 0 | 0 | 0.504821032 | 0.053825131 |
| 0 | 0 | 0 | 0 | 0 |
| 0 | 0 | 0 | 0 | 0 |
| 0.001390712 | 0 | 0 | 0.012898867 | 0 |
| 0 | 0 | 0 | 0 | 0 |
| 0.001520857 | 0.030627047 | 0 | 0.086391528 | 0 |

| | | | | |
|---|---|---|---|---|
| 0 | 0 | 0 | 0.053320363 | 0 |
| 0 | 0 | 0 | 4.694406733 | 0 |
| 0 | 0 | 0 | 0 | 0 |
| 0 | 0 | 0 | 0 | 0 |
| 0.000167279 | 0 | 0 | 0.000467038 | 0.001989754 |
| 0 | 0.007643233 | 0 | 0 | 0 |
| 0 | 0 | 0 | 0 | 0 |
| 0 | 0 | 0 | 0 | 0 |
| 0 | 0 | 0 | 0 | 0 |
| 0 | 0 | 0 | 0 | 0 |
| 0 | 0 | 0 | 0.001667706 | 0 |
| 0.012436065 | 0 | 0 | 1.384247854 | 6.304041685 |
| 0 | 0 | 0 | 0.011310011 | 0.159667115 |
| 0 | 0 | 0 | 0 | 0 |
| 0.00801239 | 1.033524378 | 0.018003902 | 0.010622535 | 0.001255173 |
| 0 | 0 | 0 | 0 | 0 |
| 0.000280371 | 0 | 0 | 0 | 0 |
| 0 | 0 | 0 | 0 | 0 |
| 0.000169565 | 0 | 0 | 0 | 0 |
| 0 | 0 | 0 | 0.001049897 | 0 |
| 0 | 0 | 0 | 0.041415966 | 0 |
| 0 | 0 | 0 | 0 | 0 |
| 0.074882663 | 0 | 0 | 0.385762931 | 0.558409943 |
| 0 | 0 | 0 | 0 | 0 |
| 0 | 0 | 0 | 0 | 0 |
| 0 | 0 | 0 | 0 | 0 |
| 0 | 0 | 0 | 0 | 0 |
| 0 | 0 | 0 | 0 | 0 |
| 0 | 0 | 0 | 0 | 0 |
| 0 | 0 | 0 | 0 | 0 |
| 0 | 0 | 0 | 0 | 0 |
| 0.003158907 | 0.01191125 | 0.004005014 | 0.009578868 | 0.048862872 |
| 0 | 0 | 0 | 0 | 0 |
| 0.005769078 | 0 | 0 | 0.150566194 | 0 |
| 0.032248093 | 0 | 0.001295915 | 0 | 0.038824079 |
| 0.000942432 | 0 | 0.011694138 | 0.138383667 | 0.13202957 |
| 0 | 0 | 0 | 0.000671048 | 0 |
| 0 | 0 | 0 | 0.188093254 | 0 |
| 0 | 0 | 0 | 0.002779182 | 0 |
| 0 | 0 | 0 | 0 | 0 |
| 0 | 0 | 0 | 0.143116015 | 0 |
| 0 | 0 | 0 | 0 | 0 |
| 0 | 0 | 0 | 0 | 0 |
| 0 | 0 | 0 | 0 | 0 |
| 0.033528025 | 0 | 0.010780227 | 0.04617159 | 0.268584547 |
| 0 | 0 | 0 | 0 | 0 |
| 0.013038862 | 0 | 0 | 0.012907473 | 0 |
| 0 | 0 | 0 | 0.001408712 | 0 |

| | | | | |
|---|---|---|---|---|
| 0 | 0 | 0 | 0 | 0 |
| 0 | 0 | 0 | 0 | 0 |
| 0 | 0 | 0 | 0 | 0.013065854 |
| 0 | 0 | 0 | 0 | 0 |
| 0 | 0 | 0 | 0 | 0 |
| 0 | 0 | 0 | 0 | 0 |
| 0 | 0 | 0 | 0.059008998 | 0 |
| 0 | 0 | 0 | 0.00160026 | 0.073867832 |
| 0 | 0 | 0 | 0 | 0 |
| 0 | 0 | 0 | 0.00071578 | 0 |
| 0 | 0 | 0 | 0 | 0 |
| 0.00248408 | 0 | 0 | 0 | 0 |
| 0 | 0 | 0 | 0 | 0 |
| 0 | 0 | 0 | 0 | 0 |
| 0 | 0 | 0 | 0 | 0 |
| 0 | 0 | 0 | 0 | 0 |
| 0 | 0 | 0 | 0 | 0 |
| 0 | 0 | 0 | 0 | 0 |
| 0.006529199 | 0 | 0 | 1.734377385 | 3.300985243 |
| 0 | 0 | 0 | 0 | 0 |
| 0 | 0 | 0 | 0 | 0 |
| 0 | 0 | 0 | 0 | 0 |
| 0 | 0 | 0 | 0 | 0 |
| 0 | 0 | 0 | 0 | 0 |
| 0 | 0 | 0 | 0.026036888 | 0 |
| 0.00588509 | 0 | 0.004093625 | 0 | 0.012083218 |
| 0 | 0 | 0 | 0 | 0 |
| 0 | 0.068445678 | 0 | 0.069759402 | 0.023811829 |
| 0 | 0 | 0 | 0 | 0 |
| 0.002759737 | 0 | 0 | 0.395107242 | 0.550811475 |
| 0 | 0 | 0 | 0.022556249 | 0 |
| 0 | 0 | 0 | 0 | 0 |
| 0 | 0 | 0 | 0.15405524 | 0 |
| 0 | 0 | 0 | 0 | 0 |
| 0.000364308 | 0 | 0 | 0.001162441 | 0.006584482 |
| 0 | 0 | 0 | 0 | 0 |
| 0 | 0 | 0 | 0 | 0 |
| 0.001417312 | 0 | 0 | 0.000989272 | 0.003639941 |
| 0 | 0 | 0 | 0 | 0 |
| 0 | 0 | 0 | 0 | 0 |
| 0.294957199 | 0 | 18.0013442 | 0.67602774 | 1.594466507 |
| 0.135931316 | 0 | 0 | 0.003379375 | 0.584076252 |
| 0 | 0 | 0 | 0.748948967 | 0 |
| 0 | 0 | 0 | 0 | 0 |
| 0 | 0 | 0 | 0 | 0 |
| 0 | 0 | 0 | 0 | 0 |
| 0 | 0 | 0 | 0 | 0 |
| 0.000210911 | 0.001442487 | 0.00087236 | 0.00418742 | 0.008362486 |

| | | | | |
|---|---|---|---|---|
| 0 | 0 | 0 | 0 | 0 |
| 0.000349513 | 0 | 0 | 0.020054967 | 0.00156391 |
| 0 | 0 | 0 | 0 | 0 |
| 0 | 0 | 0 | 0 | 0 |
| 0 | 0.001518981 | 0 | 2.173015091 | 0.518892604 |
| 0 | 0 | 0 | 0 | 0 |
| 0 | 0 | 0 | 0 | 0 |
| 0 | 0 | 0 | 0.015555154 | 0 |
| 0.000156447 | 0 | 0.390515542 | 0 | 0.000624642 |
| 0 | 0 | 0 | 0 | 0 |
| 0 | 0 | 0.07791437 | 0 | 0 |
| 0 | 0 | 0 | 0 | 0 |
| 0 | 0 | 0 | 0.020271523 | 0.1274949 |
| 0.01972411 | 0 | 0.072581647 | 0.002411785 | 0.060342873 |
| 0 | 0 | 0 | 0.477452819 | 0 |
| 0 | 0 | 0 | 0.074677781 | 0.259388968 |
| 0 | 0 | 0 | 0.005704537 | 0 |
| 0 | 0 | 0 | 0 | 0 |
| 0 | 0 | 0 | 0 | 0 |
| 0 | 0 | 0 | 0 | 0 |
| 0 | 0 | 0 | 0 | 0 |
| 0 | 0 | 0 | 0 | 0 |
| 0 | 0 | 0 | 0 | 0 |
| 0 | 0 | 0 | 0 | 0 |
| 0 | 0 | 0 | 0 | 0 |
| 0 | 0 | 0 | 0 | 0 |
| 0 | 0 | 0 | 0.006775084 | 0.000702866 |
| 0.001197956 | 0 | 0 | 0 | 0.013006941 |
| 0 | 0 | 0 | 0 | 0 |
| 0 | 0 | 0 | 0 | 0 |
| 0 | 0 | 0 | 0 | 0 |
| 0 | 0 | 0 | 0 | 0 |
| 0 | 0 | 0 | 0 | 0 |
| 0 | 0 | 0 | 0.026188349 | 0 |
| 0 | 0.013060424 | 0 | 0.022680297 | 0 |
| 0.000523205 | 0 | 0 | 0.007106457 | 0.026973301 |
| 0.000173442 | 0 | 0 | 0 | 0 |
| 0 | 0 | 0 | 0 | 0.010222831 |
| 0 | 0 | 0 | 0 | 0 |
| 0 | 0 | 0 | 0 | 0 |
| 0 | 0 | 0 | 0.043324492 | 0.017079503 |
| 0 | 0 | 0 | 0 | 0 |
| 0 | 0 | 0 | 0 | 0 |
| 0 | 0 | 0 | 0 | 0 |
| 0 | 0 | 0 | 0 | 0 |
| 0 | 0 | 0 | 0 | 0 |
| 0 | 0 | 0 | 0 | 0 |
| 0 | 0 | 0 | 0 | 0 |

| | | | | |
|---|---|---|---|---|
| 0 | 0 | 0 | 1.253030366 | 0 |
| 0 | 0 | 0 | 0 | 0 |
| 0 | 0 | 0 | 0 | 0 |
| 0 | 0 | 0 | 0 | 0.002874988 |
| 0 | 0 | 0 | 0.003107294 | 0.053812546 |
| 0 | 0 | 0 | 0 | 0 |
| 0 | 0 | 0 | 0.007772617 | 0.007579721 |
| 0 | 0 | 0 | 0.000939158 | 0 |
| 0 | 0 | 0 | 0 | 0 |
| 0 | 0 | 0 | 0 | 0 |
| 0 | 0 | 0 | 0.055711853 | 0 |
| 0 | 0 | 0 | 0.000606902 | 0 |
| 0 | 0 | 0.000628197 | 0 | 0 |
| 0 | 0 | 0 | 0 | 0 |
| 0 | 0 | 0 | 0 | 0 |
| 0 | 0 | 0 | 0 | 0 |
| 0 | 0 | 0 | 0 | 0.008293875 |
| 0 | 0 | 0 | 0 | 0 |
| 0 | 0 | 0 | 0 | 0 |
| 0 | 0 | 0 | 0 | 0 |
| 0 | 0.011835327 | 0 | 0.229134605 | 0 |
| 0 | 0 | 0 | 0 | 0 |
| 0 | 0 | 0 | 0.198514775 | 0.004741873 |
| 0.001241748 | 0 | 0 | 0.003499323 | 0 |
| 0 | 0 | 0 | 0 | 0 |
| 0.000109531 | 0 | 0 | 0 | 0 |
| 0 | 0 | 0 | 0 | 0 |
| 0 | 0 | 0 | 0 | 0 |
| 0 | 0 | 0 | 0 | 0 |
| 0.000116917 | 0.432831816 | 0.429012649 | 0 | 0.019289366 |
| 0 | 0 | 0 | 0 | 0 |
| 0 | 0 | 0 | 0 | 0 |
| 0 | 0 | 0 | 0 | 0 |
| 0 | 0 | 1.611846194 | 0.159586551 | 0 |
| 0 | 0 | 0 | 0 | 0 |
| 0 | 0 | 0 | 0 | 0 |
| 0.000174252 | 0 | 0 | 0.029514731 | 0.006971807 |
| 0 | 0 | 0 | 0 | 0 |
| 0 | 0 | 0 | 0 | 0 |
| 0 | 0 | 0 | 0 | 0 |
| 0 | 0 | 0 | 0 | 0 |
| 0 | 0 | 0 | 0 | 0 |
| 0 | 0 | 0 | 0 | 0 |
| 0 | 0 | 0 | 0 | 0 |
| 0 | 0 | 0 | 0.093162712 | 0 |
| 0 | 0 | 0 | 0 | 0 |
| 0 | 0 | 0 | 0 | 0 |
| 0 | 0 | 0 | 0 | 0 |

| | | | | |
|---|---|---|---|---|
| 0.06388618 | 0 | 0.006773579 | 0 | 0.002916725 |
| 0 | 0 | 0 | 0 | 0 |
| 0 | 0 | 0 | 0 | 0 |
| 0 | 0 | 0 | 0 | 0 |
| 0 | 0 | 0 | 0 | 0 |
| 0 | 0 | 0 | 0 | 0 |
| 0 | 0 | 0 | 0 | 0 |
| 0 | 0 | 0.151508477 | 0.199754188 | 0.648877124 |
| 0.015876848 | 0.041323356 | 0.016260934 | 4.833136996 | 1.956688118 |
| 0.037252654 | 0.070345308 | 0.01785136 | 0 | 0.491896148 |
| 0 | 0 | 0 | 3.157073472 | 0 |
| 0 | 0 | 0 | 0 | 0 |
| 0 | 0 | 0 | 0.046191812 | 0 |
| 0.001189902 | 0 | 0 | 0 | 0 |
| 0 | 0 | 0 | 0 | 0 |
| 0 | 0 | 0 | 0 | 0 |
| 0 | 0 | 0.001938746 | 0 | 0.025545803 |
| 0 | 0 | 0 | 0 | 0 |
| 0 | 0 | 0 | 0 | 0 |
| 0 | 0 | 0 | 0 | 0 |
| 0 | 0 | 0 | 0 | 0 |
| 0 | 0 | 0 | 0 | 0 |
| 0 | 0 | 0 | 0 | 0 |
| 0 | 0 | 0 | 0 | 0 |
| 0 | 0 | 0 | 0.008457939 | 0 |
| 0 | 0 | 0 | 0.040172562 | 0 |
| 0 | 0 | 0 | 0 | 0 |
| 0 | 0 | 0 | 0 | 0 |
| 0 | 0 | 0 | 0 | 0 |
| 0 | 0 | 0 | 0 | 0 |
| 0.001693399 | 0 | 0 | 0 | 0 |
| 0 | 0 | 0 | 0 | 0 |
| 0 | 0 | 0 | 0 | 0 |
| 0 | 0 | 0 | 0 | 0 |
| 0 | 0 | 0 | 0.220329492 | 0 |
| 0 | 0 | 0 | 0 | 0 |
| 0 | 0 | 0 | 0 | 0 |
| 0 | 0 | 0 | 0 | 0 |
| 0 | 0 | 0 | 0.005306488 | 0 |
| 0 | 0 | 0 | 0 | 0 |
| 0 | 0 | 0 | 0 | 0 |
| 0.019819367 | 0 | 0 | 0.010054387 | 0.044059308 |
| 0 | 0 | 0 | 0 | 0 |
| 0 | 0 | 0 | 0 | 0 |
| 0 | 0 | 0 | 0 | 0 |
| 0 | 0 | 0 | 0 | 0.00043226 |
| 0 | 0 | 0 | 0 | 0 |
| 0 | 0 | 0 | 0 | 0 |

| | | | | |
|---|---|---|---|---|
| 0 | 0 | 0 | 0 | 0 |
| 0 | 0 | 0 | 0.001636283 | 0.005557441 |
| 0 | 0 | 0 | 0 | 0 |
| 0 | 0 | 0 | 0 | 0 |
| 0.040875724 | 0.005334748 | 0.141284556 | 0.267139249 | 0.555560983 |
| 0 | 0 | 0 | 0 | 0 |
| 0 | 0 | 0 | 0 | 0 |
| 0.001032527 | 0.108778732 | 0.012333397 | 0.001064412 | 0.022880094 |
| 0 | 0 | 0 | 0.00867934 | 0 |
| 0 | 0 | 0 | 0 | 0 |
| 0 | 0 | 0 | 0 | 0 |
| 0 | 0 | 0 | 0 | 0 |
| 0 | 0 | 0 | 0 | 0 |
| 0 | 0 | 0 | 0 | 0 |
| 0 | 0 | 0 | 0 | 0 |
| 0.008665716 | 0 | 0 | 0 | 0.028961127 |
| 0 | 0 | 0 | 0 | 0 |
| 0 | 0 | 0 | 0 | 0 |
| 0 | 0 | 0 | 0 | 0 |
| 0 | 0 | 0 | 0 | 0 |
| 0 | 0 | 0 | 0 | 0 |
| 0 | 0 | 0 | 0 | 0 |
| 0 | 0 | 0 | 0 | 0 |
| 0.001718399 | 0 | 0 | 0 | 0.003567718 |
| 0 | 0 | 0 | 0.089060517 | 0 |
| 0 | 0 | 0 | 0 | 0 |
| 0.002866728 | 0 | 0 | 1.194533814 | 0.113555304 |
| 0 | 0 | 0 | 0 | 0 |
| 0.001476412 | 0 | 0 | 0 | 0 |
| 0 | 0 | 0 | 0 | 0 |
| 0 | 0 | 0 | 0 | 0 |
| 0 | 0 | 0 | 0 | 0 |
| 0 | 0 | 0 | 0 | 0 |
| 0 | 0 | 0 | 0 | 0 |
| 0 | 0 | 0 | 0 | 0 |
| 0 | 0 | 0 | 0 | 0 |
| 0 | 0 | 0 | 0 | 0 |
| 0 | 0 | 0 | 0.383633671 | 0 |
| 0 | 0 | 0 | 0 | 0 |
| 0 | 0 | 0 | 0 | 0 |
| 0 | 0 | 0 | 0 | 0 |
| 0 | 0 | 0 | 0 | 0 |
| 0 | 0 | 0 | 0 | 0 |
| 0 | 0 | 0 | 0 | 0 |
| 0 | 0 | 0 | 0 | 0 |
| 0 | 0 | 0 | 0 | 0 |
| 0 | 0 | 0 | 0 | 0 |
| 0 | 0 | 0 | 0.010090525 | 0 |

| | | | | |
|---|---|---|---|---|
| 0 | 0 | 0.003898557 | 0.16796861 | 0 |
| 0 | 0 | 0 | 0 | 0 |
| 0 | 0 | 0 | 0 | 0 |
| 0 | 0 | 0 | 0 | 0 |
| 0 | 0 | 0 | 0 | 0 |
| 0 | 0 | 0.066439259 | 0 | 0 |
| 0 | 0 | 0 | 0 | 0 |
| 0 | 0 | 0 | 0 | 0 |
| 0 | 0 | 0 | 0 | 0 |
| 0 | 0 | 0 | 0 | 0 |
| 0 | 0 | 0 | 0 | 0 |
| 0.000190091 | 0 | 0 | 0 | 0 |
| 0 | 0 | 0 | 0 | 0 |
| 0 | 0 | 0 | 0 | 0 |
| 0 | 0 | 0 | 0 | 0 |
| 0 | 0 | 0 | 0 | 0 |
| 0 | 0 | 0 | 0 | 0 |
| 0 | 0 | 0 | 0 | 0 |
| 0 | 0 | 0 | 0 | 0 |
| 0 | 0 | 0 | 0 | 0 |
| 0.025558714 | 0 | 0 | 0 | 0 |
| 0 | 0 | 0 | 0 | 0 |
| 0.000345782 | 0 | 0.006252066 | 0.021625266 | 0.050926691 |
| 0 | 0 | 0 | 0.002650791 | 0.055440113 |
| 0 | 0 | 0 | 0 | 0 |
| 0 | 0 | 0 | 0 | 0 |
| 0 | 0 | 0 | 0 | 0 |
| 0 | 0 | 0 | 0 | 0 |
| 0 | 0 | 0 | 0 | 0 |
| 0 | 0 | 0 | 0 | 0 |
| 0 | 0 | 0 | 0 | 0 |
| 0 | 0 | 0 | 0 | 0 |
| 0.08668765 | 0.151238831 | 0.006716068 | 0 | 0.221024905 |
| 0 | 0 | 0 | 0 | 0 |
| 0 | 0 | 0 | 0.012616008 | 1.091891901 |
| 0 | 0.043586648 | 0 | 0.010737505 | 0 |
| 0 | 0 | 0 | 0 | 0 |
| 0 | 0 | 0 | 0 | 0 |
| 0.007709454 | 0 | 0 | 0 | 0.052652213 |
| 0 | 0 | 0 | 0 | 0 |
| 0 | 0 | 0 | 0 | 0 |
| 0 | 0 | 0 | 0 | 0 |
| 0.199710782 | 0.011357353 | 0 | 0.09596467 | 0.19290747 |
| 0 | 0 | 0 | 0 | 0 |
| 0 | 0 | 0 | 0 | 0 |
| 0 | 0 | 0 | 0.002961225 | 0.057405473 |
| 0 | 0 | 0 | 0 | 0 |
| 0 | 0 | 0 | 0 | 0 |

| | | | | |
|---|---|---|---|---|
| 0 | 0 | 0 | 0 | 0 |
| 0 | 0 | 0 | 0 | 0 |
| 0 | 0 | 0 | 0 | 0 |
| 0 | 0 | 0 | 0 | 0 |
| 0 | 0 | 0 | 0.025163082 | 0 |
| 0 | 0 | 0 | 0 | 0 |
| 0 | 0 | 0 | 0 | 0 |
| 0 | 0 | 0.0767348 | 0 | 0 |
| 0 | 0 | 0 | 0.000438062 | 0 |
| 0 | 0 | 0 | 0.013965925 | 0 |
| 0 | 0 | 0 | 0 | 0 |
| 0 | 0 | 0 | 0 | 0 |
| 0 | 0 | 0 | 0 | 0 |
| 0 | 0 | 0 | 0 | 0 |
| 0 | 0 | 0 | 0 | 0 |
| 0.00022628 | 0 | 0.005715859 | 0 | 0 |
| 0 | 0 | 0 | 0 | 0 |
| 0 | 0 | 0 | 0 | 0 |
| 0 | 0 | 0 | 0 | 0 |
| 0.007873967 | 0 | 0 | 0.167672417 | 1.97833352 |
| 0 | 0 | 0 | 0.003299178 | 0 |
| 0 | 0 | 0 | 0 | 0 |
| 0 | 0 | 0 | 0 | 0 |
| 0 | 0 | 0 | 0.014937091 | 0 |
| 0.00019114 | 0 | 0 | 0 | 0 |
| 0 | 0 | 0 | 0 | 0 |
| 0 | 0 | 0 | 0 | 0 |
| 0 | 0 | 0 | 0 | 0 |
| 0 | 0 | 0 | 0 | 0 |
| 0 | 0 | 0 | 0.22999067 | 0 |
| 0.026725015 | 0 | 0 | 0.041720432 | 0.155420867 |
| 0 | 0 | 0 | 0 | 0 |
| 0 | 0 | 0 | 0 | 0 |
| 0 | 0 | 0 | 0 | 0 |
| 0 | 0 | 0 | 0 | 0 |
| 0 | 0 | 0 | 0 | 0 |
| 0 | 0 | 0 | 0 | 0 |
| 0 | 0 | 0 | 0 | 0 |
| 0 | 0 | 0 | 0 | 0 |
| 0 | 0 | 0 | 0 | 0 |
| 0 | 0 | 0 | 0.003219817 | 0 |
| 0 | 0 | 0 | 0 | 0 |
| 0 | 0 | 0 | 0.002082661 | 0 |
| 0 | 0 | 0 | 0.001012226 | 0 |
| 0 | 0 | 0 | 0 | 0 |
| 0 | 0 | 0 | 0 | 0 |
| 0.00095552 | 0 | 0.151671428 | 0 | 0 |
| 0 | 0 | 0 | 0 | 0 |

| | | | | |
|---|---|---|---|---|
| 0 | 0 | 0 | 0 | 0 |
| 0 | 0 | 0 | 0 | 0 |
| 0 | 0 | 0 | 0 | 0 |
| 0 | 0 | 0 | 0 | 0 |
| 0.000508344 | 0 | 0.002645815 | 0.008213709 | 0.090805223 |
| 0 | 0 | 0 | 0 | 0 |
| 0 | 0 | 0 | 0 | 0 |
| 0 | 0 | 0 | 0 | 0 |
| 0 | 0 | 0 | 0 | 0 |
| 0 | 0 | 0 | 0 | 0 |
| 0 | 0 | 0 | 0.030518861 | 0 |
| 0.006006484 | 0 | 0.003034489 | 0.047123449 | 0.051246318 |
| 0 | 0 | 0 | 0 | 0 |
| 0 | 0 | 0 | 0 | 0 |
| 0 | 0 | 0 | 0 | 0 |
| 0 | 0 | 0 | 0 | 0 |
| 0 | 0 | 0 | 0.053913855 | 0.047657077 |
| 0 | 0 | 0 | 0 | 0 |
| 0.010915545 | 0 | 0 | 0 | 0.010918237 |
| 0 | 0 | 0 | 0.00215541 | 0.001733825 |
| 0 | 0 | 0 | 0 | 0 |
| 0 | 0 | 0 | 0 | 0 |
| 0 | 0 | 0 | 0 | 0 |
| 0 | 0 | 0 | 0 | 0 |
| 0 | 0 | 0 | 0 | 0 |
| 0 | 0 | 0 | 0 | 0 |
| 0.008506327 | 0 | 0.001898012 | 0 | 0.01034326 |
| 0 | 0 | 0 | 0 | 0 |
| 0 | 0 | 0 | 0 | 0 |
| 0 | 0 | 0 | 0 | 0 |
| 0.003357972 | 0 | 0 | 0.001054256 | 0.011724628 |
| 0 | 0 | 0 | 0 | 0 |
| 0 | 0 | 0 | 0 | 0 |
| 0 | 0 | 0 | 0 | 0 |
| 0 | 0 | 0 | 0 | 0 |
| 0 | 0 | 0 | 0 | 0 |
| 0 | 0 | 0 | 0 | 0 |
| 0.01108546 | 0.00145802 | 0.001612351 | 0.064757449 | 0.01020452 |
| 0 | 0 | 0 | 0.033981395 | 0 |
| 0 | 0 | 0 | 0 | 0 |
| 0 | 0 | 0.003621703 | 0.02408485 | 0 |
| 0 | 0 | 0 | 0 | 0 |
| 0 | 0 | 0 | 0 | 0 |
| 0.000295047 | 0 | 0.020023883 | 0 | 0 |
| 0 | 0 | 0 | 0 | 0 |
| 0 | 0 | 0 | 0 | 0 |
| 0 | 0 | 0 | 0 | 0 |
| 0 | 0 | 0 | 0.016716099 | 0 |

| | | | | |
|---|---|---|---|---|
| 0 | 0 | 0 | 0 | 0 |
| 0 | 0 | 0 | 0.007778348 | 0.043007544 |
| 0 | 0 | 0 | 0.035619664 | 0 |
| 0.010601505 | 0 | 0.001409448 | 0 | 0.008024739 |
| 0 | 0.013659996 | 0.000435747 | 0 | 0 |
| 0 | 0 | 0.000562373 | 0.000590505 | 0.008233432 |
| 0 | 0 | 0 | 0 | 0 |
| 0.000253345 | 0 | 0 | 0 | 0 |
| 0 | 0 | 0 | 0 | 0 |
| 0 | 0 | 0 | 0.102610315 | 0 |
| 0 | 0 | 0 | 0.070249292 | 0 |
| 0 | 0 | 0 | 0 | 0 |
| 0.043879103 | 0.036781232 | 0 | 0.064460046 | 0 |
| 0 | 0 | 0 | 0 | 0 |
| 0 | 0 | 0 | 0 | 0 |
| 0 | 0 | 0 | 0 | 0 |
| 0 | 0 | 0 | 0 | 0 |
| 0 | 0 | 0 | 0 | 0 |
| 0 | 0 | 0 | 0 | 0 |
| 0 | 0 | 0 | 0 | 0 |
| 0 | 0 | 0 | 0 | 0 |
| 0 | 0 | 0 | 0 | 0 |
| 0 | 0 | 0.003508227 | 0 | 0.042980204 |
| 0 | 0 | 0.003967336 | 0 | 0 |
| 0 | 0 | 0 | 0 | 0 |
| 0 | 0 | 0 | 0 | 0 |
| 0 | 0 | 0 | 0 | 0 |
| 0 | 0 | 0 | 0 | 0 |
| 0 | 0 | 0 | 0 | 0 |
| 0 | 0 | 0 | 0 | 0 |
| 0 | 0 | 0 | 0.024643982 | 0.019089555 |
| 0 | 0 | 0 | 0 | 0.01291681 |
| 0 | 0 | 0 | 0 | 0 |
| 0 | 0 | 0 | 0 | 0 |
| 0 | 0 | 0 | 0.008034818 | 0 |
| 0 | 0 | 0 | 0 | 0 |
| 0.00039074 | 0 | 0 | 0 | 0 |
| 0 | 0 | 0 | 0 | 0 |
| 0 | 0 | 0.003447431 | 0 | 0 |
| 0 | 0 | 0 | 0 | 0 |
| 0 | 0 | 0 | 0 | 0 |
| 0 | 0 | 0 | 0 | 0 |
| 0 | 0 | 0 | 0 | 0 |
| 0 | 0 | 0 | 0 | 0 |
| 0 | 0 | 0 | 0 | 0 |
| 0.037530206 | 0 | 0 | 0.349064802 | 0.178664775 |
| 0 | 0 | 0 | 0 | 0 |
| 0 | 0 | 0 | 0 | 0 |

| | | | | |
|---|---|---|---|---|
| 0 | 0 | 0 | 0.002479625 | 0 |
| 0 | 0 | 0 | 0 | 0 |
| 0 | 0 | 0 | 0.148051485 | 0 |
| 0 | 0 | 0 | 0 | 0 |
| 0 | 0 | 0 | 0 | 0 |
| 0 | 0 | 0 | 0 | 0 |
| 0 | 0 | 0 | 0 | 0 |
| 0 | 0 | 0 | 0 | 0 |
| 0 | 0 | 0 | 0 | 0 |
| 0 | 0 | 0 | 0 | 0 |
| 5.25918E-05 | 0 | 0 | 0.000469871 | 0.002593274 |
| 0.001209204 | 0 | 0.001169173 | 0 | 0.001018886 |
| 0 | 0 | 0 | 0 | 0 |
| 0 | 0 | 0 | 0 | 0 |
| 0 | 0 | 0 | 0 | 0 |
| 0 | 0 | 0 | 0 | 0 |
| 0 | 0 | 0 | 0 | 0 |
| 0 | 0 | 0 | 0 | 0 |
| 0 | 0 | 0 | 0 | 0 |
| 0 | 0 | 0 | 0.00062313 | 0.003620134 |
| 0 | 0 | 0 | 0 | 0 |
| 0 | 0 | 0 | 0 | 0 |
| 0 | 0 | 0 | 0 | 0 |
| 0 | 0 | 0 | 0 | 0 |
| 0 | 0 | 0 | 0 | 0 |
| 0 | 0 | 0 | 0 | 0 |
| 0 | 0 | 0 | 0 | 0 |
| 0 | 0 | 0 | 0.014826364 | 0.008813838 |
| 0 | 0 | 0 | 0 | 0 |
| 0 | 0 | 0 | 0 | 0 |
| 0 | 0 | 0 | 0.001204499 | 0.002954566 |
| 0 | 0 | 0 | 0 | 0 |
| 0.001266208 | 2.896416646 | 0.029154235 | 0.025484615 | 0.032680521 |
| 7.10949E-05 | 0 | 0 | 0.017626335 | 0.00276762 |
| 0 | 0 | 0 | 0 | 0 |
| 0 | 0 | 0 | 0 | 0 |
| 0 | 0 | 0 | 0 | 0 |
| 0 | 0 | 0 | 0 | 0 |
| 0 | 0 | 0 | 0 | 0 |
| 0 | 0 | 0 | 0 | 0 |
| 0 | 0 | 0 | 0 | 0 |
| 0 | 0 | 0.070006759 | 0 | 0.08600539 |
| 0.000543892 | 0 | 0 | 0.090644558 | 0.2725335 |
| 0 | 0 | 0 | 0 | 0 |
| 0 | 0 | 0 | 0 | 0 |
| 0.027367607 | 0 | 0.003643826 | 0.021156087 | 0.021966605 |
| 0.000396384 | 0 | 0 | 0 | 0 |
| 0 | 0 | 0 | 0 | 0 |

| | | | | |
|---|---|---|---|---|
| 0 | 0 | 0 | 0 | 0 |
| 0 | 0 | 0 | 0.007342438 | 0 |
| 8.53315E-05 | 0.004989865 | 0 | 0.001905943 | 0.020816796 |
| 0 | 0 | 0 | 0 | 0 |
| 0 | 0 | 0.101741213 | 0.181551171 | 0.402799107 |
| 0.000623411 | 0 | 0.010157363 | 0.026367399 | 0.003270221 |
| 0 | 0 | 0 | 0 | 0 |
| 0 | 0 | 0 | 0 | 0 |
| 0 | 0 | 0 | 0 | 0 |
| 0 | 0 | 0 | 0 | 0 |
| 0 | 0 | 0 | 0 | 0 |
| 0 | 0 | 0 | 0 | 0 |
| 0 | 0 | 0 | 0 | 0 |
| 0 | 0 | 0 | 0 | 0 |
| 0 | 0 | 0 | 0 | 0 |
| 0 | 0 | 0 | 0 | 0 |
| 0 | 0 | 0 | 0 | 0 |
| 0 | 0 | 0 | 0 | 0 |
| 0 | 0 | 0 | 0 | 0 |
| 0 | 0 | 0 | 0 | 0 |
| 0 | 0 | 0 | 0 | 0 |
| 0 | 0 | 0 | 0 | 0 |
| 0 | 0 | 0 | 0 | 0 |
| 0 | 0 | 0 | 0 | 0 |
| 0.001167398 | 0 | 0 | 0.004555218 | 0.000912551 |
| 8.48945E-05 | 0.021337834 | 0.002708773 | 0 | 0.000734404 |
| 0 | 0 | 0 | 0 | 0 |
| 6.86256E-05 | 0 | 0 | 0.009196817 | 0.738400134 |
| 0 | 0 | 0 | 0 | 0 |
| 0 | 0 | 0 | 0 | 0 |
| 0 | 0 | 0 | 0 | 0 |
| 4.96317E-05 | 0 | 0 | 0 | 0.061504761 |
| 0 | 0 | 0 | 0 | 0 |
| 0 | 0 | 0 | 0 | 0 |
| 0 | 0 | 0 | 0 | 0 |
| 0 | 0 | 0 | 0 | 0 |
| 5.3899E-05 | 0 | 0 | 0 | 0 |
| 0 | 0 | 0.002696356 | 0 | 0.031800135 |
| 0 | 0 | 0 | 8.535187751 | 4.491605107 |
| 0 | 0 | 0 | 0 | 0 |
| 0 | 0 | 0 | 0 | 0 |
| 0.000168845 | 0 | 0 | 0 | 0.001394252 |
| 0.05371919 | 0 | 0.003473702 | 0 | 0.002211164 |
| 0 | 0 | 0 | 0 | 0 |
| 0 | 0 | 0 | 0 | 0 |
| 0 | 0 | 0 | 0 | 0 |
| 0.001781324 | 0 | 0.002310472 | 0.002911258 | 0.026215525 |
| 0 | 0 | 0 | 0 | 0 |

| | | | | |
|---|---|---|---|---|
| 0 | 0 | 0 | 0 | 0 |
| 0.003837541 | 0.004007231 | 0.012827716 | 0.001530613 | 0.00640242 |
| 0 | 0 | 0 | 0 | 0 |
| 0 | 0 | 0 | 0 | 0 |
| 0 | 0 | 0 | 0 | 0 |
| 0 | 0 | 0 | 0 | 0 |
| 0 | 0 | 0 | 0 | 0 |
| 0 | 0 | 0 | 0 | 0 |
| 0 | 0 | 0 | 0 | 0 |
| 0 | 0 | 0 | 0 | 0 |
| 0 | 0 | 0 | 0 | 0 |
| 0 | 0 | 0 | 0 | 0 |
| 0 | 0 | 0 | 0.150473433 | 0 |
| 0 | 0 | 0 | 0 | 0 |
| 0.002392418 | 0 | 0 | 0 | 0.020178878 |
| 0 | 0 | 0 | 0 | 0 |
| 0 | 0 | 0 | 0 | 0 |
| 0 | 0 | 0 | 0 | 0 |
| 0 | 0 | 0 | 0 | 0 |
| 0 | 0 | 0 | 0 | 0.018607469 |
| 0 | 0 | 0 | 0 | 0 |
| 0 | 0 | 0 | 0.002061667 | 0.001742176 |
| 0 | 0 | 0.0030175 | 0 | 0 |
| 0 | 0 | 0 | 0 | 0.000897216 |
| 0 | 0 | 0 | 0 | 0 |
| 0 | 0 | 0 | 0 | 0 |
| 0 | 0 | 0 | 0 | 0 |
| 0 | 0 | 0 | 0 | 0 |
| 0 | 0 | 0 | 0 | 0 |
| 0 | 0 | 0 | 0.034960809 | 0 |
| 0 | 0 | 0 | 0 | 0 |
| 0 | 0 | 0 | 0 | 0 |
| 0 | 0 | 0 | 0 | 0 |
| 0 | 0 | 0 | 0 | 0 |
| 0 | 0 | 0 | 0.938662199 | 0 |
| 0 | 0 | 0 | 0 | 0 |
| 0 | 0 | 0 | 0 | 0 |
| 0 | 0 | 0 | 0 | 0 |
| 0 | 0 | 0 | 0.089652426 | 0 |
| 0 | 0 | 0 | 0 | 0 |
| 0.001427559 | 0 | 0.007671554 | 0.140296608 | 0.027103905 |
| 0 | 0 | 0 | 0 | 0 |
| 0 | 0 | 0 | 0 | 0 |
| 0 | 0 | 0 | 0 | 0 |
| 0 | 0 | 0 | 0 | 0 |
| 0.211842945 | 8.315193069 | 0.183902578 | 0.003650683 | 0.091756863 |
| 0 | 0 | 0 | 0 | 0 |
| 0 | 0 | 0 | 0 | 0 |

| | | | | |
|---|---|---|---|---|
| 0 | 0 | 0.002444717 | 0 | 0 |
| 0 | 0 | 0 | 0 | 0 |
| 0 | 0 | 0 | 0 | 0 |
| 0 | 0 | 0 | 0 | 0 |
| 0 | 0 | 0 | 0 | 0 |
| 0 | 0 | 0 | 0 | 0 |
| 0 | 0 | 0 | 0 | 0 |
| 0 | 0 | 0 | 0 | 0 |
| 0 | 0 | 0.001859423 | 0 | 0 |
| 0 | 0 | 0.002505471 | 0 | 0 |
| 0 | 0 | 0 | 0.041816528 | 0 |
| 0 | 0 | 0 | 0 | 0 |
| 0.000296034 | 0 | 0 | 0 | 0 |
| 0 | 0 | 0 | 0 | 0 |
| 0 | 0 | 0 | 0 | 0 |
| 0 | 0 | 0 | 0 | 0 |
| 0 | 0 | 0 | 0 | 0 |
| 0 | 0 | 0 | 0 | 0 |
| 0 | 0 | 0 | 0 | 0 |
| 0.033609756 | 0.222559523 | 0.148218378 | 0.03661946 | 1.229928495 |
| 0 | 0 | 0 | 0 | 0 |
| 0 | 0 | 0 | 0 | 0 |
| 0 | 0 | 0 | 0 | 0 |
| 0 | 0 | 0 | 0 | 0 |
| 0 | 0 | 0 | 0 | 0 |
| 0 | 0 | 0 | 0 | 0 |
| 0 | 0 | 0 | 0 | 0 |
| 0 | 0 | 0 | 0 | 0 |
| 0 | 0 | 0 | 0 | 0 |
| 0 | 0 | 0 | 0 | 0 |
| 0 | 0 | 0 | 0 | 0 |
| 0 | 0 | 0 | 0 | 0 |
| 0 | 0 | 0 | 0 | 0 |
| 0 | 0 | 0 | 0 | 0 |
| 0 | 0 | 0 | 0 | 0 |
| 0 | 0 | 0 | 0 | 0 |
| 0 | 0 | 0 | 0 | 0 |
| 0 | 0 | 0 | 0 | 0 |
| 0 | 0 | 0 | 0 | 0 |
| 0 | 0 | 0 | 0 | 0 |
| 0 | 0 | 0 | 0 | 0 |
| 0 | 0 | 0 | 0 | 0 |
| 0 | 0 | 0 | 0 | 0 |
| 0 | 0 | 0 | 0 | 0 |
| 0.001072416 | 0 | 0 | 0.007677309 | 0.017662426 |
| 0 | 0 | 0 | 0 | 0 |
| 0 | 0 | 0 | 0 | 0 |
| 0 | 0 | 0 | 0 | 0 |

| | | | | |
|---|---|---|---|---|
| 0 | 0 | 0 | 0 | 0 |
| 0 | 0 | 0 | 0 | 0 |
| 0 | 0 | 0 | 0 | 0 |
| 0 | 0 | 0 | 0 | 0 |
| 0 | 0 | 0 | 0 | 0 |
| 0.001436974 | 0 | 0 | 0.1923461 | 0.439522931 |
| 0 | 0 | 0 | 0 | 0 |
| 0 | 0 | 0 | 0 | 0 |
| 0 | 0 | 0 | 0 | 0 |
| 0 | 0 | 0 | 0 | 0 |
| 0 | 0 | 0 | 0 | 0 |
| 0 | 0 | 0 | 0 | 0 |
| 0 | 0 | 0 | 0 | 0 |
| 0 | 0 | 0 | 0 | 0 |
| 0 | 0 | 0 | 0 | 0 |
| 0 | 0 | 0 | 0 | 0 |
| 0 | 0 | 0 | 0.369715832 | 0.004685154 |
| 0 | 0 | 0 | 0 | 0 |
| 0 | 0 | 0 | 0 | 0 |
| 0 | 0 | 0 | 0 | 0 |
| 0 | 0 | 0 | 0 | 0 |
| 0 | 0 | 0 | 0 | 0 |
| 0 | 0 | 0 | 0 | 0 |
| 0 | 0 | 0 | 0 | 0 |
| 0 | 0 | 0 | 0 | 0 |
| 0 | 0 | 0 | 0 | 0 |
| 0 | 0 | 0 | 0 | 0 |
| 0 | 0 | 0 | 0 | 0 |
| 0 | 0 | 0 | 0 | 0 |
| 0 | 0 | 0 | 0 | 0.048156129 |
| 0 | 0 | 0 | 0 | 0 |
| 0 | 0 | 0 | 0.020245841 | 0 |
| 0 | 0 | 0 | 0 | 0 |
| 0 | 0 | 0 | 0 | 0 |
| 0 | 0 | 0 | 0 | 0 |
| 0 | 0 | 0 | 0 | 0 |
| 0 | 0 | 0 | 0 | 0 |
| 0 | 0 | 0 | 0 | 0 |
| 0 | 0 | 0 | 0.19465498 | 0 |
| 0 | 0 | 0 | 0 | 0 |
| 0 | 0 | 0 | 0 | 0 |
| 0 | 0 | 0 | 0 | 0 |
| 0 | 0 | 0 | 0 | 0 |
| 0 | 0 | 0 | 0 | 0 |
| 0 | 0 | 0 | 0 | 0 |
| 0 | 0 | 0 | 0 | 0 |
| 0 | 0 | 0 | 0 | 0 |
| 0 | 0 | 0 | 0 | 0 |

| | | | | |
|---|---|---|---|---|
| 0 | 0 | 0 | 0 | 0 |
| 0 | 0 | 0 | 0.001026684 | 0.052786908 |
| 0 | 0 | 0 | 0 | 0 |
| 0 | 0 | 0 | 0 | 0 |
| 0 | 0 | 0 | 0 | 0 |
| 0 | 0 | 0 | 0.000703945 | 0 |
| 0 | 0 | 0 | 0 | 0 |
| 0 | 0 | 0 | 0 | 0 |
| 0 | 0 | 0 | 0 | 0 |
| 0 | 0 | 0 | 0 | 0 |
| 7.5236E-05 | 0 | 0 | 0.01529211 | 0 |
| 0 | 0 | 0 | 0 | 0 |
| 0 | 0 | 0 | 0.898597482 | 0.279720536 |
| 0 | 0 | 0 | 0 | 0 |
| 0 | 0 | 0 | 0 | 0 |
| 0 | 0 | 0 | 0 | 0 |
| 0 | 0 | 0 | 0 | 0 |
| 0 | 0 | 0 | 0 | 0 |
| 0 | 0 | 0 | 0 | 0 |
| 0 | 0 | 0 | 0 | 0 |
| 0.000274896 | 0.002820154 | 0.007796662 | 0 | 0.001189032 |
| 0 | 0 | 0 | 0 | 0 |
| 0 | 0 | 0 | 0.011606865 | 0 |
| 0 | 0 | 0 | 1.462051235 | 1.381289941 |
| 0 | 0 | 0 | 0 | 0 |
| 0 | 0 | 0 | 0 | 0 |
| 0 | 0 | 0 | 0 | 0 |
| 0 | 0 | 0 | 0 | 0 |
| 0 | 0 | 0 | 0 | 0 |
| 0 | 0 | 0 | 0 | 0 |
| 0 | 0 | 0 | 0 | 0.001556046 |
| 0 | 0 | 0 | 0 | 0 |
| 0 | 0 | 0 | 0 | 0 |
| 0 | 0 | 0 | 0 | 0 |

| PHL018-0063_V1 | PHL018-0128_V3 | PHL018-0275_V3 | PHL018-0153_V2 | PHL018-0059_V1 |
|---|---|---|---|---|
| EYCF | CTRL | REF | CTRL | CTRL |
| 0.113601062 | 0.020834349 | 0.0083753 | 0.030100394 | 0.467863678 |
| 0.125264727 | 1.706654137 | 1.86300031 | 51.61349583 | 3.871501401 |
| 11.7695801 | 4.95293323 | 4.863498313 | 0 | 0.590729678 |
| 0 | 0 | 0 | 0 | 0 |
| 0 | 0 | 0 | 0 | 0 |
| 0 | 0 | 0 | 0 | 0 |
| 0.050809672 | 0 | 0 | 0 | 0 |
| 0 | 0 | 0 | 0 | 0 |
| 0.059653754 | 0 | 0 | 0 | 0 |
| 0 | 0 | 0 | 0 | 0 |
| 0 | 0 | 0 | 0 | 0 |
| 0 | 0 | 0 | 0 | 0 |
| 0 | 0 | 0 | 0 | 0 |
| 0.22569513 | 0.45260767 | 5.813576834 | 1.480290655 | 0 |
| 0 | 0 | 0 | 0 | 0 |
| 0 | 0 | 0 | 0 | 0 |
| 0.001298866 | 0 | 0.002246578 | 0 | 0 |
| 0 | 0 | 0 | 0 | 0 |
| 0 | 0 | 0 | 0 | 0 |
| 0 | 0 | 0 | 0 | 0 |
| 0 | 0 | 0 | 0 | 0 |
| 0 | 0 | 0 | 0 | 0 |
| 0 | 0 | 0 | 0 | 0 |
| 0 | 0 | 0 | 0 | 0 |
| 0 | 0.003976845 | 0 | 0 | 0 |
| 0 | 0 | 0 | 0 | 0 |
| 0 | 0 | 0 | 0 | 0 |
| 0 | 0 | 0 | 0 | 0 |
| 0 | 0 | 0 | 0 | 0 |
| 0 | 0 | 0 | 0 | 0 |
| 0 | 0 | 0 | 0 | 0 |
| 0 | 0 | 0 | 0 | 0 |
| 0 | 0 | 0 | 0 | 0 |
| 0 | 0 | 0 | 0 | 0 |
| 0 | 0 | 0 | 0 | 0 |
| 1.546783977 | 0 | 0 | 0.914106909 | 0 |
| 0 | 0 | 0 | 0 | 0 |
| 0 | 0 | 0 | 0 | 0 |
| 0 | 0 | 0 | 0 | 0 |
| 0 | 0 | 0 | 0 | 0 |
| 0 | 0 | 0 | 0 | 0.001120827 |
| 0.620651487 | 0.001708873 | 0 | 0.002320058 | 0.025460563 |
| 0.261306333 | 0.007699015 | 0.006802011 | 0 | 0.349105952 |
| 0.002379548 | 0.00893178 | 0.000904566 | 0 | 0.041546331 |
| 0 | 0 | 0 | 0 | 0.001461143 |
| 0 | 0 | 0 | 0 | 0 |

| | | | | |
|---|---|---|---|---|
| 0 | 0.001636418 | 0 | 0.020439533 | 0 |
| 0.007530471 | 0 | 0.100595251 | 0.036927078 | 0.005471662 |
| 0.021014313 | 0.02718073 | 0.001682746 | 0.008290317 | 0 |
| 0.710964001 | 0.979254941 | 0 | 1.154981114 | 0.318808883 |
| 0.001047138 | 0.001628074 | 0 | 0 | 0.01521707 |
| 0 | 0 | 0 | 0 | 0.071358783 |
| 0.003279027 | 0 | 0.00155812 | 0 | 0 |
| 0 | 0 | 0 | 0 | 0 |
| 0 | 0 | 0 | 0 | 0 |
| 0 | 0 | 0 | 0 | 0 |
| 0.01181941 | 0.008647828 | 0 | 0 | 0 |
| 0 | 0.460362818 | 0.044424029 | 0 | 0.034490795 |
| 0.000874046 | 0 | 0 | 0 | 0 |
| 0.043715174 | 0.048394019 | 0.015399093 | 0.018584371 | 0.06560221 |
| 0.000991364 | 0 | 0 | 0 | 0 |
| 0 | 0 | 0 | 0 | 0 |
| 0.023030518 | 0 | 0 | 0 | 0.016177095 |
| 0 | 0 | 0 | 0 | 0 |
| 0 | 0 | 0 | 0 | 0 |
| 0 | 0 | 0 | 0 | 0 |
| 0.025578206 | 0.207472449 | 0.149514647 | 0.141829686 | 0.053047728 |
| 0.013979138 | 0.029150871 | 0.005978315 | 0 | 0.00833418 |
| 0 | 0 | 0.007202072 | 0 | 0 |
| 0 | 0 | 0 | 0 | 0 |
| 0.001781404 | 0 | 0 | 0 | 0 |
| 0 | 0 | 0 | 0 | 0 |
| 0 | 0 | 0 | 0.003335723 | 0 |
| 0 | 0 | 0 | 0 | 0 |
| 0.06717724 | 0 | 0 | 0 | 0 |
| 0 | 0 | 0 | 0 | 0 |
| 0.014215301 | 0 | 0.007777659 | 0 | 0 |
| 0 | 0 | 0 | 0 | 0 |
| 0 | 0 | 0 | 0 | 0 |
| 0 | 0 | 0 | 0 | 0 |
| 0 | 0 | 0 | 0 | 0 |
| 0 | 0 | 0 | 0 | 0 |
| 0 | 0 | 0 | 0 | 0 |
| 0.00391181 | 0 | 0 | 0 | 0.764692597 |
| 0 | 0 | 0 | 0 | 0 |
| 0 | 0 | 0 | 0 | 0 |
| 0 | 0 | 0 | 0 | 0 |
| 0 | 0 | 0 | 0 | 0 |
| 0 | 0 | 0 | 0 | 0 |
| 0 | 0 | 0 | 0 | 0 |
| 0 | 0.014445293 | 0 | 0 | 0 |
| 0 | 0 | 0 | 0 | 0 |
| 0 | 0 | 0 | 0 | 0 |
| 0 | 0 | 0 | 0 | 0 |

| | | | | |
|---|---|---|---|---|
| 0 | 0 | 0 | 0 | 0 |
| 0.003199608 | 0 | 0 | 0 | 0 |
| 0 | 0 | 0 | 0 | 0 |
| 0 | 0 | 0 | 0 | 0 |
| 0 | 0 | 0 | 0 | 0 |
| 0.005066541 | 0 | 0 | 0 | 0 |
| 0 | 0 | 0 | 0 | 0 |
| 0.008700669 | 0 | 0 | 0 | 0 |
| 0.072236872 | 0.002486171 | 0.017549548 | 0 | 0 |
| 0 | 0 | 0 | 0 | 0 |
| 0 | 0 | 0 | 0 | 0 |
| 0.002413398 | 0 | 0 | 0.000626996 | 0 |
| 0 | 0 | 0 | 0 | 0 |
| 0 | 0 | 0 | 0 | 0 |
| 0 | 0 | 0 | 0 | 0 |
| 0 | 0.000892385 | 0 | 0 | 0 |
| 0 | 0 | 0 | 0 | 0 |
| 0 | 0 | 0 | 0 | 0 |
| 0.260601113 | 0.520258849 | 0.023660715 | 0 | 0 |
| 0 | 0 | 0 | 0 | 0 |
| 0 | 0 | 0 | 0 | 0 |
| 0 | 0 | 0 | 0 | 0 |
| 0 | 0.000906436 | 0 | 0 | 0 |
| 0 | 0 | 0 | 0 | 0 |
| 0 | 0 | 0 | 0 | 0 |
| 0 | 0 | 0 | 0 | 0 |
| 0 | 0 | 0 | 0 | 0 |
| 0 | 0 | 0 | 0 | 0 |
| 0 | 0 | 0 | 0 | 0 |
| 0 | 0.000959653 | 0 | 0 | 0 |
| 0.271551427 | 0.054294865 | 0.002178515 | 0 | 0 |
| 0.111833489 | 0 | 0 | 0 | 0.269244167 |
| 0 | 0 | 0 | 0 | 0 |
| 0 | 0 | 0 | 0 | 0 |
| 0 | 0 | 0 | 0 | 0 |
| 0 | 0 | 0 | 0 | 0 |
| 0 | 0 | 0 | 0 | 0 |
| 0 | 0 | 0 | 0 | 0 |
| 0.14300032 | 0 | 0 | 0 | 0 |
| 0 | 0 | 0 | 0 | 0 |
| 0 | 0 | 0 | 0 | 0 |
| 0 | 0 | 0 | 0 | 0 |
| 0.026824259 | 0.001115878 | 0 | 0 | 0.000860868 |
| 0 | 0 | 0 | 0 | 0 |
| 0 | 0 | 0 | 0 | 0 |
| 0 | 0 | 0 | 0 | 0 |
| 0 | 0 | 0.001762391 | 0 | 0 |
| 0 | 0 | 0 | 0 | 0 |

| | | | | |
|---|---|---|---|---|
| 0 | 0 | 0 | 0 | 0 |
| 0 | 0 | 0 | 0 | 0 |
| 0 | 0 | 0 | 0 | 0 |
| 0 | 0 | 0 | 0 | 0 |
| 0 | 0.005601952 | 0 | 0 | 0 |
| 0 | 0 | 0 | 0 | 0.340823051 |
| 0 | 0 | 0 | 0 | 0 |
| 0 | 0 | 0 | 0 | 0 |
| 0 | 0 | 0 | 0 | 0 |
| 0 | 0 | 0 | 0 | 0 |
| 0 | 0 | 0 | 0 | 0 |
| 0 | 0 | 0 | 0 | 0 |
| 0 | 0 | 0 | 0 | 0 |
| 0 | 0 | 0 | 0 | 0 |
| 0 | 0 | 0 | 0 | 0 |
| 2.532807307 | 4.060446842 | 0 | 0 | 0 |
| 0 | 0 | 0 | 0 | 0 |
| 0 | 0 | 0 | 0 | 0 |
| 0 | 0 | 0 | 0 | 0 |
| 0 | 0 | 0 | 0 | 0 |
| 0 | 0 | 0 | 0 | 0 |
| 0 | 0 | 0 | 0 | 0 |
| 0 | 0 | 0 | 0 | 0 |
| 0 | 0 | 0 | 0 | 0 |
| 0 | 0 | 0 | 0 | 0 |
| 0 | 0 | 0 | 0 | 0 |
| 0 | 0 | 0 | 0 | 0 |
| 0 | 0 | 0 | 0 | 0 |
| 0 | 0 | 0 | 0 | 0 |
| 0 | 0 | 0 | 0 | 0 |
| 0 | 0.009490497 | 0 | 0 | 0 |
| 0.034925877 | 0.000742594 | 0 | 0.004234381 | 0.065524307 |
| 0 | 0 | 0 | 0 | 0 |
| 0 | 0 | 0 | 0 | 0 |
| 0 | 0 | 0 | 0 | 0 |
| 0 | 0 | 0 | 0 | 0 |
| 0 | 0 | 0 | 0 | 0 |
| 0 | 0 | 0 | 0 | 0 |
| 0 | 0 | 0 | 0 | 0 |
| 0 | 0 | 0 | 0 | 0 |
| 0 | 0 | 0 | 0 | 0 |
| 0 | 0 | 0 | 0 | 0 |
| 0 | 0 | 0 | 0 | 0 |
| 0 | 0 | 0 | 0 | 0 |
| 0 | 0 | 0 | 0 | 0 |
| 0 | 0 | 0 | 0 | 0 |
| 0 | 0 | 0 | 0 | 0 |
| 0 | 0 | 0 | 0 | 0 |

| | | | | |
|---|---|---|---|---|
| 0 | 0 | 0 | 0 | 0 |
| 0 | 0 | 0 | 0 | 0 |
| 0 | 0 | 0 | 0 | 0 |
| 0 | 0 | 0 | 0 | 0 |
| 0 | 0 | 0 | 0 | 0 |
| 0 | 0 | 0 | 0 | 0 |
| 0 | 0 | 0 | 0 | 0 |
| 0 | 0 | 0 | 0 | 0 |
| 0 | 0 | 0 | 0 | 0 |
| 0 | 0 | 0 | 0 | 0 |
| 0 | 0 | 0 | 0 | 0 |
| 0.007345533 | 0 | 0 | 0 | 0 |
| 0.001299692 | 0 | 0 | 0 | 0.001027874 |
| 0 | 0 | 0 | 0 | 0 |
| 0.000805911 | 0 | 0 | 0 | 0 |
| 0 | 0 | 0 | 0 | 0 |
| 0 | 0 | 0 | 0 | 0 |
| 0 | 0 | 0 | 0 | 0 |
| 0 | 0 | 0 | 0 | 0 |
| 0 | 0 | 0 | 0 | 0 |
| 0.062835726 | 0.063442201 | 0.063906278 | 0.108950899 | 0.013837523 |
| 0 | 0 | 0 | 0 | 0 |
| 0 | 0 | 0 | 0 | 0.002023594 |
| 0 | 0 | 0 | 0 | 0 |
| 0 | 0 | 0 | 0 | 0 |
| 0 | 0 | 0 | 0 | 0 |
| 0.00202177 | 0.001934411 | 0 | 0.002101008 | 0 |
| 0 | 0 | 0 | 0 | 0 |
| 0 | 0 | 0 | 0 | 0 |
| 0 | 0 | 0 | 0.041809755 | 0 |
| 0.001007681 | 0 | 0 | 0 | 0 |
| 0 | 0 | 0 | 0 | 0 |
| 0 | 0 | 0 | 0 | 0 |
| 0 | 0 | 0 | 0 | 0 |
| 0 | 0.011238228 | 0.002511594 | 0.021830137 | 0.068025913 |
| 0 | 0 | 0 | 0 | 0 |
| 0 | 0 | 0 | 0 | 0 |
| 0 | 0 | 0 | 0 | 0 |
| 0 | 0 | 0 | 0 | 0 |
| 0 | 0 | 0 | 0 | 0 |
| 0 | 0 | 0 | 0 | 0 |
| 0 | 0 | 0 | 0 | 0 |
| 0 | 0 | 0 | 0 | 0 |
| 0 | 0 | 0 | 0 | 0 |
| 0 | 0 | 0 | 0 | 0 |
| 0 | 0.075517431 | 0 | 0 | 0 |
| 0 | 0.002808003 | 0 | 0 | 0.002166293 |
| 0.018189402 | 0 | 0 | 0 | 0 |

| | | | | |
|---|---|---|---|---|
| 0 | 0 | 0 | 0 | 0 |
| 0 | 0 | 0 | 0 | 0 |
| 0 | 0.002814674 | 0 | 0 | 0 |
| 0 | 0 | 0 | 0 | 0 |
| 0 | 0 | 0 | 0 | 0 |
| 0 | 0 | 0 | 0 | 0 |
| 0 | 0 | 0 | 0 | 0 |
| 0 | 0.003763736 | 0 | 0 | 0 |
| 0 | 0 | 0 | 0 | 0 |
| 0 | 0 | 0 | 0 | 0 |
| 0 | 0 | 0 | 0 | 0 |
| 0 | 0.001139361 | 0 | 0 | 0 |
| 0 | 0 | 0 | 0 | 0 |
| 0 | 0 | 0 | 0 | 0 |
| 0.00964415 | 0.003885234 | 0 | 0 | 0 |
| 0 | 0 | 0 | 0 | 0 |
| 0 | 0 | 0 | 0 | 0 |
| 0 | 0 | 0 | 0 | 0 |
| 0.001473332 | 0 | 0 | 0 | 0.002718799 |
| 0 | 0 | 0 | 0 | 0 |
| 0.005795506 | 0.001617316 | 0 | 0.004015096 | 0 |
| 0 | 0 | 0 | 0 | 0 |
| 0 | 0 | 0 | 0 | 0 |
| 0 | 0 | 0 | 0 | 0 |
| 0.003358249 | 0.005508241 | 0.00237085 | 0 | 0.006374174 |
| 0 | 0 | 0 | 0 | 0 |
| 0 | 0 | 0 | 0 | 0 |
| 0 | 0 | 0 | 0 | 0 |
| 0.0439519 | 0 | 0 | 0 | 0 |
| 0 | 0 | 0 | 0 | 0 |
| 1.19160325 | 0.140666795 | 0.009871568 | 0.036534692 | 0.157865695 |
| 0 | 0 | 0 | 0 | 0 |
| 0 | 0 | 0 | 0 | 0.036178648 |
| 0 | 0 | 0 | 0 | 0 |
| 0 | 0 | 0 | 0 | 0 |
| 0 | 0 | 0 | 0 | 0 |
| 0 | 0 | 0 | 0 | 0 |
| 0 | 0 | 0 | 0 | 0 |
| 0 | 0 | 0 | 0 | 0 |
| 0 | 0 | 0 | 0 | 0 |
| 0 | 0 | 0 | 0 | 0 |
| 0.002714889 | 0.023897735 | 0 | 0.003949808 | 0.038475993 |
| 0 | 0 | 0 | 0 | 0 |
| 0 | 0 | 0 | 0 | 0 |
| 0 | 0 | 0 | 0 | 0 |
| 0 | 0 | 0 | 0 | 0 |
| 0 | 0 | 0 | 0 | 0 |
| 0 | 0 | 0 | 0 | 0 |

| | | | | |
|---|---|---|---|---|
| 0 | 0 | 0 | 0 | 0 |
| 0 | 0 | 0 | 0 | 0 |
| 0 | 0.009785067 | 0 | 0 | 0.040764044 |
| 0 | 0 | 0 | 0 | 0 |
| 0 | 0.698588076 | 0 | 0 | 0 |
| 0 | 0 | 0 | 0 | 0 |
| 0 | 0 | 0 | 0 | 0 |
| 0 | 0 | 0 | 0 | 0 |
| 0 | 0 | 0 | 0 | 0 |
| 0 | 0 | 0 | 0 | 0 |
| 0 | 0 | 0 | 0 | 0 |
| 0 | 0 | 0 | 0 | 0 |
| 0 | 0 | 0 | 0 | 0 |
| 0.004018719 | 0.000720951 | 0.003103108 | 0.001566083 | 0.001668579 |
| 0.023218487 | 0.005752157 | 0.008708112 | 0.001077164 | 0.002524855 |
| 0.001109753 | 0 | 0 | 0 | 0 |
| 0 | 0.001808018 | 0 | 0.002700129 | 0.001046126 |
| 0.010993869 | 0 | 0 | 0 | 0 |
| 0 | 0 | 0 | 0 | 0 |
| 0 | 0 | 0 | 0 | 0 |
| 0 | 0 | 0 | 0 | 0 |
| 0.019830309 | 0 | 0 | 0 | 0 |
| 0 | 0 | 0 | 0 | 0 |
| 0 | 0 | 0 | 0.006526669 | 0 |
| 0.002735204 | 0 | 0.001689619 | 0 | 0 |
| 0 | 0 | 0 | 0 | 0 |
| 0 | 0 | 0 | 0 | 0 |
| 0 | 0 | 0 | 0 | 0 |
| 0 | 0 | 0 | 0 | 0 |
| 0.001850627 | 0 | 0.002667442 | 0 | 0 |
| 0 | 0 | 0 | 0 | 0 |
| 0 | 0 | 0 | 0 | 0 |
| 0 | 0 | 0 | 0 | 0 |
| 0 | 0 | 0 | 0 | 0 |
| 0 | 0 | 0 | 0 | 0 |
| 0.054940438 | 0.002037461 | 0 | 0 | 0.018390553 |
| 0 | 0 | 0 | 0 | 0 |
| 0 | 0 | 0 | 0 | 0.037670791 |
| 0 | 0 | 0 | 0 | 0 |
| 0 | 0 | 0 | 0 | 0 |
| 0.002319935 | 0.001479795 | 0 | 0 | 0 |
| 0 | 0 | 0 | 0 | 0 |
| 0 | 0 | 0 | 0 | 0 |
| 0 | 0 | 0 | 0 | 0 |
| 0 | 0 | 0 | 0 | 0 |
| 0 | 0 | 0.005597651 | 0 | 0 |
| 0 | 0 | 0.00466402 | 0 | 0 |
| 0 | 0.004036152 | 0 | 0.007755878 | 0 |

| | | | | |
|---|---|---|---|---|
| 0 | 0.019444518 | 0.327194963 | 0 | 0 |
| 0 | 0 | 0 | 0 | 0 |
| 0 | 0 | 0 | 0 | 0 |
| 0 | 0 | 0 | 0 | 0 |
| 0.002471727 | 0.006449795 | 0.00222089 | 0 | 0.003483082 |
| 0 | 0 | 0 | 0.001142273 | 0 |
| 0 | 0 | 0 | 0 | 0 |
| 0.006929339 | 0 | 0.0023348 | 0 | 0.023016584 |
| 0 | 0 | 0 | 0 | 0 |
| 0.02597171 | 0.072684737 | 0.050136009 | 0.017205866 | 1.303365236 |
| 0.001174089 | 0 | 0 | 0 | 0 |
| 0.025182467 | 0.001908463 | 0 | 0.001295516 | 0 |
| 0.041229617 | 0.036371874 | 0.005374151 | 0.001375773 | 0.000837608 |
| 0.002010643 | 0.009618818 | 0.031228326 | 0 | 0 |
| 0 | 0 | 0 | 0 | 0 |
| 0 | 0 | 0 | 0 | 0 |
| 0 | 0 | 0 | 0 | 0 |
| 0 | 0 | 0 | 0 | 0 |
| 0 | 0 | 0 | 0 | 0 |
| 0 | 0 | 0 | 0 | 0 |
| 0 | 0 | 0 | 0 | 0 |
| 0 | 0 | 0 | 0 | 0 |
| 0 | 0 | 0 | 0 | 0 |
| 0 | 0 | 0 | 0 | 0 |
| 0 | 0 | 0 | 0 | 0 |
| 0 | 0 | 0 | 0.00206026 | 0.002438998 |
| 0 | 0 | 0 | 0 | 0 |
| 0.002930235 | 0.001784123 | 0 | 0.060624721 | 0.003834255 |
| 0 | 0 | 0 | 0 | 0 |
| 0 | 0.001734813 | 0 | 0 | 0 |
| 0 | 0 | 0.001460174 | 0 | 0 |
| 0 | 0 | 0 | 0 | 0 |
| 0 | 0 | 0 | 0 | 0 |
| 0 | 0 | 0 | 0 | 0.02392321 |
| 0 | 0 | 0 | 0 | 0 |
| 0 | 0 | 0 | 0 | 0 |
| 0 | 0 | 0 | 0.087971089 | 0 |
| 0 | 0 | 0 | 0 | 0 |
| 0 | 0 | 0 | 0 | 0 |
| 0 | 0 | 0 | 0 | 0 |
| 0 | 0 | 0 | 0 | 0.035948973 |
| 0 | 0 | 0 | 0 | 0 |
| 0 | 0 | 0 | 0 | 0 |
| 0 | 0 | 0 | 0 | 0 |
| 0 | 0 | 0 | 0 | 0 |
| 0 | 0 | 0 | 0 | 0 |
| 0 | 0.001048417 | 0 | 0.003416128 | 0.012738965 |
| 0 | 0 | 0 | 0 | 0 |

| | | | | |
|---|---|---|---|---|
| 0.063434907 | 0 | 0 | 0 | 0 |
| 0 | 0 | 0 | 0 | 0.009152154 |
| 0 | 0 | 0 | 0 | 0 |
| 0.524496629 | 0 | 0 | 0 | 0 |
| 0.002166096 | 0 | 0 | 0 | 0 |
| 0.017692462 | 0 | 0.123419735 | 0 | 0 |
| 0 | 0 | 0 | 0 | 0 |
| 0 | 0 | 0 | 0 | 0 |
| 0 | 0 | 0 | 0 | 0 |
| 0 | 0 | 0 | 0 | 0 |
| 0.007257171 | 0.00287321 | 0.008965955 | 0 | 0.001385374 |
| 0 | 0 | 0 | 0 | 0.008829198 |
| 0 | 0 | 0.002990294 | 0.11419218 | 0.008575559 |
| 0 | 0.002035849 | 0.001752535 | 0 | 0.001047066 |
| 0.094250953 | 0 | 0.128560627 | 0 | 0 |
| 0 | 0 | 0 | 0 | 0 |
| 0 | 0 | 0 | 0 | 0 |
| 0 | 0 | 0 | 0 | 0 |
| 0 | 0.003852174 | 0 | 0 | 0 |
| 0 | 0.008274075 | 0 | 0 | 0 |
| 0 | 0 | 0 | 0 | 0 |
| 0 | 0.370069858 | 0 | 0.211387274 | 0.640743261 |
| 0 | 0 | 0 | 0 | 0 |
| 0 | 0 | 0 | 0 | 0 |
| 0 | 0 | 0 | 0 | 0 |
| 0 | 0 | 0 | 0 | 0 |
| 0 | 0 | 0 | 0.221103386 | 0 |
| 0 | 0 | 0 | 0 | 0 |
| 0 | 0 | 0 | 0 | 0 |
| 0.537489348 | 0.774215326 | 0.219636556 | 0 | 1.362500016 |
| 0.000801964 | 0 | 0 | 0.007000519 | 0.016515657 |
| 0 | 0 | 0 | 0 | 0 |
| 0 | 0 | 0 | 0 | 0 |
| 0 | 0 | 0 | 0.001180373 | 0 |
| 0 | 0 | 0 | 0 | 0 |
| 0 | 0 | 0 | 0 | 0 |
| 1.035612842 | 0 | 0 | 0 | 0 |
| 0 | 0 | 0 | 0 | 0 |
| 0.177360721 | 0 | 0 | 0 | 0 |
| 0 | 0 | 0.727867618 | 0 | 0 |
| 0.001360547 | 0.045561535 | 0.033137805 | 0 | 0.001936804 |
| 2.467130658 | 5.82302769 | 10.03671858 | 0.306081398 | 0.636495259 |
| 0 | 0 | 0 | 0 | 0 |
| 0 | 0 | 0 | 0 | 0 |
| 0 | 0 | 0 | 0 | 0 |
| 0 | 0 | 0 | 0 | 0 |
| 0.007343956 | 0 | 0 | 0 | 0.019198809 |
| 0 | 0 | 0 | 0 | 0 |

| | | | | |
|---|---|---|---|---|
| 0 | 0 | 0 | 0.01595546 | 0 |
| 0 | 0 | 0 | 0 | 0.035079911 |
| 0.043213074 | 0.00196885 | 0 | 0 | 0 |
| 0 | 0 | 0 | 0 | 0 |
| 0 | 0 | 0 | 0 | 0 |
| 0.00148969 | 0.1582106 | 0.008939348 | 0 | 0.023091471 |
| 0 | 0 | 0 | 0 | 0 |
| 0 | 0.001889407 | 0 | 0 | 0 |
| 0 | 0 | 0 | 0.001751251 | 0.001399399 |
| 0.317352127 | 0 | 0 | 0 | 0 |
| 0 | 0 | 0 | 0 | 0 |
| 0 | 0 | 0.00104885 | 0 | 0 |
| 0 | 0 | 0 | 0 | 0 |
| 0.018671964 | 0.034423106 | 0 | 0 | 0 |
| 0 | 0 | 0 | 0 | 0 |
| 0.002313337 | 0.018388069 | 0.011652004 | 0 | 0.313468211 |
| 0 | 0.000566146 | 0.001218399 | 0.00737885 | 0.089318512 |
| 0 | 0 | 0 | 0.012700439 | 0 |
| 0 | 0 | 0 | 0 | 0 |
| 0.004997799 | 0 | 0 | 0 | 0 |
| 0 | 0 | 0 | 0 | 0.011284196 |
| 0 | 0 | 0 | 0.004279541 | 0 |
| 0 | 0 | 0 | 0 | 0.000857088 |
| 0.741356595 | 0 | 0 | 0 | 0.005062408 |
| 0.004792785 | 0.020459237 | 0 | 0 | 0 |
| 0.024423661 | 0.004474786 | 0 | 0 | 0 |
| 0 | 0.002635332 | 0.002474828 | 0 | 0 |
| 0 | 0.004396799 | 0 | 0 | 0 |
| 0 | 0 | 0 | 0 | 0 |
| 0 | 0 | 0 | 0 | 0 |
| 0.041131752 | 0.005648095 | 0.019526785 | 0 | 0.004427622 |
| 0.0017034 | 0 | 0.005261212 | 0.004425399 | 0 |
| 0.041357607 | 0.016293761 | 0.002337713 | 0.024719658 | 0.2409282 |
| 0.148754785 | 0 | 0 | 0.001610258 | 0 |
| 0 | 0.098410832 | 0 | 0 | 0 |
| 0 | 0 | 0 | 0 | 0 |
| 0 | 0 | 0.001207688 | 0.026665555 | 0 |
| 0.040325566 | 0.008228626 | 0 | 0.0059582 | 0.010157037 |
| 0 | 0.135975816 | 0 | 0.76395652 | 0 |
| 0.001406315 | 0.002242581 | 0 | 0 | 0 |
| 0 | 0 | 0 | 0 | 0.056606645 |
| 0 | 0 | 0 | 0 | 0 |
| 0 | 0 | 0 | 0 | 0.007594916 |
| 0.343435212 | 0.032260029 | 0 | 0 | 0.070203087 |
| 0 | 0.066481685 | 0.159065514 | 0 | 0 |
| 0 | 0 | 0 | 0 | 0 |
| 0 | 0 | 0 | 0 | 0 |
| 0 | 0 | 0 | 0 | 0 |

| | | | | |
|---|---|---|---|---|
| 0 | 0 | 0 | 0.069494396 | 0 |
| 0.265628785 | 0.154483987 | 0.064991346 | 0 | 0.117642137 |
| 0 | 0 | 0 | 0 | 0 |
| 0 | 0.006338028 | 0 | 0 | 0 |
| 0 | 0 | 0 | 0 | 0 |
| 0 | 0 | 0 | 0 | 0 |
| 0 | 0 | 0 | 0 | 0 |
| 0 | 0 | 0 | 0.001662959 | 0 |
| 0.001668754 | 0 | 0 | 0 | 0 |
| 0 | 0 | 0 | 0.003146547 | 0 |
| 0.002902812 | 0.007221196 | 0.002390875 | 0 | 0.018212702 |
| 0.007494623 | 0.004688589 | 0.004273531 | 0.570047523 | 0.003829883 |
| 0 | 0.105082288 | 0 | 0 | 0 |
| 0 | 0 | 0 | 0 | 0 |
| 0 | 0 | 0 | 0.004153377 | 0 |
| 0 | 0 | 0 | 0 | 0 |
| 0.011550721 | 0.002132769 | 0.01427047 | 0.000842345 | 0.01099406 |
| 0.008944047 | 0.008557578 | 0.097500219 | 0.154454087 | 0.011067933 |
| 0 | 0 | 0 | 0 | 0 |
| 0.064910667 | 0.008662139 | 0.005275962 | 0 | 0 |
| 0.012329662 | 0.004598792 | 0.001032734 | 0.002823178 | 0.001156903 |
| 0.000780116 | 0 | 0 | 0 | 0 |
| 0 | 0 | 0 | 0 | 0 |
| 0 | 0 | 0 | 0 | 0 |
| 0 | 0 | 0 | 0 | 0 |
| 0 | 0 | 0 | 0 | 0 |
| 0 | 0.028419576 | 0.075432621 | 0 | 0.069885568 |
| 0.586153672 | 0.225053036 | 0 | 0.160187043 | 0.058775696 |
| 0 | 0 | 0 | 0 | 0 |
| 0 | 0 | 0 | 0 | 0 |
| 0 | 0 | 0 | 0 | 0 |
| 0 | 0 | 0 | 0 | 0 |
| 0 | 0 | 0 | 0 | 0 |
| 0 | 0 | 0 | 0 | 0 |
| 0 | 0 | 0 | 0 | 0.091722598 |
| 0.188331599 | 0.237021476 | 0.778839067 | 0 | 0.853648161 |
| 0 | 0 | 0 | 0 | 0 |
| 0 | 0 | 0 | 0 | 0 |
| 0.024112004 | 0.009962103 | 0.011848077 | 0 | 0.013955206 |
| 0 | 0 | 0 | 0 | 0 |
| 0 | 0.016450715 | 0.153367856 | 0 | 0 |
| 0 | 0 | 0 | 0 | 0 |
| 0.001664318 | 0.028344784 | 0.0010281 | 0 | 0 |
| 0.002731675 | 0 | 0.003573402 | 0 | 0 |
| 0.04158772 | 0 | 0 | 0 | 0 |
| 0 | 0 | 0 | 0 | 0 |
| 0 | 0 | 0 | 0 | 0 |
| 0 | 0 | 0.001925376 | 0 | 0 |

| | | | | |
|---|---|---|---|---|
| 0 | 0 | 0.004308836 | 0.000966485 | 0 |
| 0 | 0 | 0 | 0 | 0 |
| 0 | 0 | 0 | 0 | 0 |
| 1.428991398 | 0.072516671 | 0 | 0 | 0 |
| 0 | 0 | 0 | 0 | 0 |
| 0 | 0 | 0 | 0 | 0 |
| 0 | 0 | 0 | 0 | 0 |
| 0 | 0 | 0 | 0 | 0.048768365 |
| 0.282172882 | 0.249351249 | 0 | 0.068739802 | 1.517847543 |
| 0.12681581 | 0.35323407 | 0 | 0 | 0.000959092 |
| 0 | 0 | 0 | 0 | 0 |
| 0.002042919 | 0.006397022 | 0 | 0 | 0.001850669 |
| 0 | 0 | 0 | 0 | 0 |
| 0 | 4.668305034 | 0 | 0 | 0 |
| 0.149271893 | 0.024194678 | 0.002840139 | 0.00318526 | 0 |
| 0 | 0 | 0 | 0 | 0 |
| 0.129941343 | 0 | 0 | 0 | 0.026118232 |
| 0 | 0 | 0 | 0 | 0 |
| 0 | 0 | 0 | 0 | 0 |
| 0 | 0 | 0 | 0 | 0 |
| 0 | 0 | 0 | 0 | 0 |
| 0 | 0 | 0 | 0 | 0 |
| 0 | 0 | 0 | 0 | 0 |
| 0.075288929 | 0.229525596 | 0.097940975 | 0.484165402 | 0.01830656 |
| 0 | 0.00083184 | 0 | 0 | 0.013861595 |
| 1.314273964 | 0 | 0 | 0 | 0 |
| 0 | 0.001979522 | 0 | 0 | 0 |
| 0 | 0 | 0 | 0.473713702 | 0 |
| 0 | 0 | 0 | 0 | 0 |
| 0 | 0 | 0 | 0 | 0 |
| 0 | 0.010537743 | 0 | 0 | 0 |
| 0 | 0 | 0 | 0 | 0 |
| 0 | 0.018143598 | 0 | 0 | 0.232932719 |
| 0 | 0 | 0 | 0 | 0 |
| 0 | 0.031261574 | 0 | 0 | 0 |
| 0 | 0 | 0 | 0 | 0 |
| 0.971167495 | 0.083424034 | 0 | 0 | 1.368618888 |
| 0 | 0 | 0 | 0 | 0 |
| 0.00174001 | 0.001294864 | 0 | 0 | 0 |
| 0 | 0 | 0 | 0 | 0 |
| 0.014787702 | 0.115576382 | 0.147477778 | 0.000555443 | 0 |
| 0 | 0.120219023 | 0 | 0 | 0 |
| 0.019022049 | 7.446095992 | 0.135196808 | 1.040795438 | 0 |
| 0 | 0 | 0 | 0 | 0 |
| 0 | 0 | 0 | 0 | 0 |
| 0 | 0 | 0.131321577 | 0 | 0 |
| 0 | 0 | 0 | 0 | 0 |
| 0.003855148 | 0 | 0 | 0 | 0.003082759 |

| | | | | |
|---|---|---|---|---|
| 0 | 0 | 0 | 0 | 0 |
| 0 | 0 | 0 | 0 | 0 |
| 0.001847797 | 0.005303864 | 0.009892489 | 0.055472908 | 0.038417248 |
| 0 | 0 | 0 | 0 | 0 |
| 0 | 0 | 0 | 0 | 0 |
| 0 | 0.0029402 | 0 | 0 | 0.00644669 |
| 0 | 0 | 0 | 0 | 0 |
| 0 | 0 | 0 | 0 | 0 |
| 0 | 0 | 0 | 0 | 0 |
| 0 | 0 | 0.183234778 | 0 | 0 |
| 0.030438396 | 0.017936784 | 0.005727985 | 0.004189581 | 0.640775346 |
| 1.753431607 | 6.515065384 | 0.356281227 | 5.764821741 | 1.442033596 |
| 0 | 0 | 0 | 0 | 0 |
| 0 | 0.00124891 | 0 | 0.000904313 | 0.015656847 |
| 0.168182004 | 0.640215053 | 0.431026652 | 0.023338752 | 0.016134563 |
| 0 | 0 | 0 | 0 | 0 |
| 0 | 0.003064668 | 0 | 0 | 0 |
| 0 | 0 | 0 | 0 | 0 |
| 0.279396493 | 0 | 0 | 0 | 0 |
| 1.207500066 | 0.002700841 | 0 | 0 | 0 |
| 0.562950415 | 0.399442905 | 0.537908944 | 0 | 0.008792063 |
| 0 | 1.484438186 | 0.070601209 | 1.244130721 | 0 |
| 3.693636941 | 0.968630737 | 0.922290362 | 0.257706329 | 13.17417976 |
| 0 | 0 | 0 | 0 | 0 |
| 0 | 0 | 0 | 0 | 0 |
| 0.002227016 | 0 | 0 | 0.001683125 | 0 |
| 0 | 0.141720943 | 0 | 0 | 0.75892152 |
| 0 | 0.002191806 | 0 | 0 | 0 |
| 0 | 0 | 0 | 0 | 0 |
| 0.235898834 | 0 | 0 | 0 | 0 |
| 0 | 0 | 0 | 0 | 0 |
| 0 | 0.389313697 | 0 | 0 | 0.08830193 |
| 0.067076639 | 0.004357785 | 0.27060832 | 0 | 0 |
| 0.008367315 | 0.025678873 | 0 | 0 | 0.235104146 |
| 0.013527377 | 0.000392208 | 0.002701019 | 0.001135963 | 0 |
| 0.011264114 | 0.008442294 | 0.004870733 | 0.049553613 | 0.09831838 |
| 0.005259141 | 0.000867568 | 0 | 0 | 0 |
| 0 | 0 | 0 | 0 | 0 |
| 0 | 0.015764663 | 0 | 0.015752573 | 0 |
| 0 | 0 | 0 | 0 | 0 |
| 0 | 0.005377689 | 0 | 0 | 0 |
| 0 | 0 | 0 | 0 | 0 |
| 0 | 0 | 0 | 0 | 0 |
| 0 | 0 | 0 | 0 | 0 |
| 0 | 0.002662919 | 0.007406023 | 0 | 0.037452665 |
| 0 | 0 | 0 | 0 | 0 |
| 0 | 0.005946533 | 0 | 0 | 0 |
| 0.010588329 | 0.018037786 | 0.056473672 | 0.00885632 | 0.066337534 |

| | | | | |
|---|---|---|---|---|
| 0 | 0 | 0 | 0 | 0 |
| 0 | 0 | 0 | 0 | 0 |
| 0 | 0 | 0 | 0 | 0 |
| 0.003466904 | 0.00122209 | 0 | 0 | 0.022627369 |
| 0 | 0 | 0 | 0 | 0 |
| 0 | 0 | 0 | 0 | 0 |
| 0 | 0.464844021 | 0 | 0 | 0 |
| 0.009906348 | 0.005622719 | 0.002074392 | 0 | 0 |
| 0 | 0 | 0 | 0 | 0 |
| 0 | 0 | 0 | 0 | 0 |
| 0.009551423 | 0.000583322 | 0.009289688 | 0 | 0.002025072 |
| 0 | 0 | 0 | 0 | 0 |
| 0.029181594 | 0 | 0 | 0.000924551 | 0 |
| 0 | 0.688077605 | 0 | 0 | 0.255751728 |
| 0 | 0 | 0 | 0 | 0 |
| 0 | 0 | 0 | 0 | 0 |
| 0 | 0.056109705 | 0 | 0 | 0 |
| 0 | 0.001456807 | 0 | 0 | 0.007679876 |
| 0 | 0 | 0 | 0 | 0 |
| 0.057518918 | 0.001605753 | 0 | 0 | 0.000675705 |
| 0.000450903 | 0 | 0 | 0 | 0 |
| 0 | 0 | 0 | 0 | 0 |
| 0 | 0.002985944 | 0 | 0 | 0.005398992 |
| 0 | 0 | 0 | 0 | 0 |
| 0 | 0 | 0 | 0 | 0 |
| 0 | 0 | 0 | 0 | 0 |
| 0 | 0 | 0 | 0 | 0 |
| 0 | 0 | 0 | 0 | 0 |
| 0 | 0 | 0 | 0 | 0 |
| 1.101526762 | 0.937000694 | 0 | 0.043961494 | 0.036203791 |
| 0 | 0.003522304 | 0 | 0.002162327 | 0 |
| 0 | 0 | 0 | 0 | 0 |
| 0 | 0 | 0 | 0 | 0 |
| 0 | 0 | 0 | 0 | 0 |
| 0 | 0.000927658 | 0 | 0 | 0 |
| 0 | 0 | 0 | 0 | 0 |
| 0 | 0 | 0 | 0 | 0.097001364 |
| 0 | 0 | 0.001275224 | 0.005541964 | 0.007999876 |
| 0 | 0 | 0 | 0 | 0 |
| 0 | 0 | 0 | 0 | 0 |
| 0 | 0.111191833 | 0 | 0 | 0 |
| 0 | 0 | 0 | 0 | 0 |
| 0 | 0 | 0 | 0 | 0 |
| 0 | 0.013906392 | 0 | 0 | 0.446707556 |
| 0 | 0 | 0 | 0.123826899 | 0 |
| 0 | 0 | 0 | 0 | 0 |
| 0.000653832 | 0 | 0 | 0 | 0 |
| 0 | 0 | 0 | 0 | 0 |

| | | | | |
|---|---|---|---|---|
| 0 | 0 | 0 | 0 | 0 |
| 0 | 0 | 0 | 0 | 0 |
| 0 | 0 | 0 | 0 | 0 |
| 0.003080164 | 0.083991527 | 0 | 0 | 0 |
| 0 | 0 | 0 | 0 | 0 |
| 0 | 0 | 0 | 0 | 0 |
| 0 | 0 | 0 | 0 | 0 |
| 0 | 0 | 0 | 0 | 0 |
| 0.151608617 | 0.024289782 | 0.084126752 | 0.304252393 | 0.017863219 |
| 0 | 0 | 0 | 0 | 0 |
| 0 | 0.000649269 | 0 | 0 | 0 |
| 0 | 0 | 0 | 0 | 0 |
| 0 | 0.002118909 | 0 | 0 | 0 |
| 0 | 0 | 0 | 0 | 0 |
| 0.001875508 | 0.872927062 | 0 | 0 | 0.200105874 |
| 0.094905508 | 0 | 0.285110105 | 0 | 0 |
| 0 | 0 | 0 | 0 | 0 |
| 0 | 0 | 0 | 0 | 0 |
| 0 | 0 | 0 | 0 | 0 |
| 0 | 0 | 0 | 0 | 0 |
| 0 | 0.017184385 | 0 | 0 | 0 |
| 0 | 0 | 0 | 0 | 0 |
| 0 | 0 | 0 | 0 | 0 |
| 0.037269576 | 0.090507139 | 0 | 0 | 0 |
| 0.006138221 | 0 | 0 | 0 | 0.01637282 |
| 0.252459449 | 2.954719603 | 3.852338256 | 0 | 4.645925108 |
| 0.002698155 | 0 | 0 | 0 | 0 |
| 0 | 0.001657109 | 0 | 0 | 0 |
| 0.00268328 | 0 | 0 | 0 | 0 |
| 0 | 0 | 0 | 0 | 0 |
| 0 | 0 | 0 | 0 | 0 |
| 0 | 0.003610319 | 0 | 0 | 0 |
| 0 | 0 | 0 | 0 | 0 |
| 0 | 0 | 0 | 0 | 0.023262704 |
| 0 | 0 | 0 | 0.002041141 | 0.014407576 |
| 0.00399172 | 0.000498162 | 0.000857672 | 0.002524969 | 0.000768635 |
| 0 | 0.06271886 | 0 | 0 | 0.459466727 |
| 0.271616315 | 0.466508772 | 0.055202236 | 0 | 1.025015167 |
| 5.927981128 | 0 | 0 | 0 | 0 |
| 0 | 0 | 0 | 0 | 0 |
| 0.214210824 | 0 | 0 | 0 | 0 |
| 0 | 0 | 0 | 0 | 0 |
| 0.173129213 | 0.080328114 | 0.184886862 | 0 | 0.316156243 |
| 0 | 0 | 0 | 0 | 0 |
| 0 | 0 | 0 | 0 | 0 |
| 0 | 0.000943891 | 0 | 0 | 0.592378388 |
| 0 | 0 | 0 | 0 | 0 |
| 0.422771653 | 1.932546037 | 0.099026847 | 0 | 0 |

| | | | | |
|---|---|---|---|---|
| 0.001171975 | 0.065224289 | 0.000723965 | 0 | 0 |
| 0 | 0 | 0 | 0 | 0 |
| 0 | 0 | 0 | 0 | 0 |
| 0 | 0 | 0 | 0 | 0 |
| 0.066704178 | 0 | 0 | 0 | 0.009043488 |
| 0 | 0 | 0 | 0 | 0 |
| 0 | 0 | 0 | 0 | 0 |
| 0 | 0 | 0 | 0 | 0 |
| 0 | 0 | 0 | 0 | 0 |
| 0.00136636 | 0 | 0 | 0 | 0 |
| 0 | 0 | 0 | 0 | 0 |
| 1.297698475 | 0.787753248 | 0.409181719 | 0 | 0.82081148 |
| 0.148347668 | 0.3100098 | 0.182986907 | 1.916499309 | 1.026178812 |
| 0 | 0 | 0 | 0 | 0 |
| 1.565583263 | 0.054227788 | 0 | 0.013789622 | 0.431549759 |
| 0 | 0 | 0 | 0 | 0 |
| 0 | 0 | 0 | 0 | 0.008141745 |
| 0.037689878 | 0 | 0 | 0 | 0 |
| 0 | 0 | 0 | 0 | 0.051073818 |
| 0 | 0 | 0 | 0 | 0 |
| 0 | 0.026546218 | 0 | 0 | 0 |
| 0 | 0 | 0 | 0 | 0 |
| 0.059805098 | 0.122144708 | 1.197346569 | 0 | 0.250858527 |
| 0 | 0 | 0 | 0 | 0 |
| 0 | 0 | 0 | 0 | 0 |
| 0 | 0 | 0 | 0 | 0 |
| 0 | 0 | 0 | 0 | 0 |
| 0 | 0 | 0 | 0 | 0 |
| 0 | 0 | 0 | 0 | 0 |
| 0 | 0 | 0 | 0 | 0 |
| 0 | 0 | 0 | 0 | 0 |
| 0.021013152 | 0.030157774 | 0.003966255 | 0.015467663 | 0.011956063 |
| 0 | 0 | 0 | 0 | 0 |
| 0.051760028 | 0.037805263 | 0.003782811 | 0 | 0 |
| 0.002644169 | 0 | 0.000933362 | 0.003729156 | 0.002300285 |
| 0.020695399 | 0.001397729 | 0 | 0 | 0.042862725 |
| 0 | 0 | 0 | 0 | 0 |
| 0 | 0 | 0 | 0 | 0 |
| 0 | 0 | 0 | 0 | 0 |
| 0 | 0 | 0 | 0 | 0 |
| 0 | 0.728404014 | 0 | 0 | 0 |
| 0 | 0 | 0 | 0 | 0 |
| 0 | 0 | 0 | 0 | 0 |
| 0.006919061 | 0.009268127 | 0 | 0 | 0 |
| 0.027350096 | 0.00498444 | 0 | 0 | 0.52653853 |
| 0 | 0 | 0 | 0 | 0 |
| 0 | 1.438022453 | 0 | 0 | 0.689658512 |
| 0.001231824 | 0 | 0 | 0 | 0.007793603 |

| | | | | |
|---|---|---|---|---|
| 0 | 0 | 0 | 0 | 0 |
| 0 | 0 | 0 | 0 | 0.05159617 |
| 0.016548766 | 0.008100963 | 0 | 0 | 0.001704453 |
| 0 | 0 | 0 | 0 | 0 |
| 0.001756344 | 0 | 0 | 0 | 0 |
| 0 | 0.083154962 | 0 | 0 | 0 |
| 0 | 0.061610933 | 0 | 0 | 0 |
| 0.00926822 | 0.033036692 | 0.672213347 | 0 | 0 |
| 0 | 0 | 0 | 0 | 0 |
| 0 | 0 | 0 | 0 | 0 |
| 0 | 0 | 0 | 0 | 0 |
| 0 | 0 | 0 | 0 | 0 |
| 0 | 0 | 0 | 0 | 0 |
| 0.651152026 | 0 | 0 | 0 | 0 |
| 0.010617413 | 0.420676506 | 0 | 0 | 0 |
| 0 | 0 | 0 | 0 | 0 |
| 0 | 1.395896212 | 0 | 0 | 0 |
| 0 | 0 | 0 | 0 | 0 |
| 23.28447125 | 3.824245368 | 17.21888387 | 0 | 1.978802779 |
| 0 | 0 | 0 | 0 | 0 |
| 0 | 0 | 0 | 0 | 0 |
| 0 | 0 | 0 | 0 | 0 |
| 0 | 0 | 0 | 0 | 0 |
| 0 | 0 | 0 | 0 | 0 |
| 0 | 0 | 0 | 0 | 0 |
| 0.000866051 | 0.001491533 | 0.00235394 | 0.007379947 | 0.039122944 |
| 0 | 0 | 0 | 0 | 0 |
| 0.185814062 | 2.693541796 | 0.057810444 | 0 | 0.038105866 |
| 0 | 0 | 0 | 0 | 0 |
| 0 | 1.037321552 | 1.292466699 | 0 | 0.759965145 |
| 0 | 0 | 0 | 0 | 0 |
| 0 | 0 | 0 | 0 | 0 |
| 0 | 0 | 0 | 0 | 0 |
| 0 | 0 | 0 | 0 | 0 |
| 0.010164763 | 0 | 0 | 0 | 0.003818481 |
| 0 | 0 | 0 | 0 | 0 |
| 0 | 0 | 0 | 0 | 0 |
| 0 | 0 | 0 | 0 | 0.012086391 |
| 0 | 0 | 0 | 0 | 0 |
| 0 | 0 | 0 | 0 | 0 |
| 8.795008262 | 7.086392794 | 25.34705039 | 0 | 25.01544871 |
| 0.01221214 | 2.335737732 | 0.341241177 | 0.025381518 | 0.197478259 |
| 0 | 0 | 0 | 0.057190976 | 0 |
| 0 | 0 | 0 | 0 | 0 |
| 0 | 0 | 0 | 0 | 0 |
| 0 | 0 | 0.00250564 | 0 | 0 |
| 0 | 0 | 0 | 0 | 0 |
| 0.001471186 | 0.003910045 | 0.003837141 | 0 | 0.127054434 |

| | | | | |
|---|---|---|---|---|
| 0 | 0 | 0 | 0 | 0 |
| 0.017150005 | 0.000804361 | 0 | 0 | 0.006143358 |
| 0 | 0.001315227 | 0 | 0 | 0 |
| 0 | 0 | 0 | 0 | 0 |
| 0.577909753 | 0.764773837 | 1.665776472 | 0 | 1.752723028 |
| 0 | 0 | 0 | 0 | 0 |
| 0 | 0 | 0 | 0 | 0 |
| 0 | 0 | 0 | 0 | 0 |
| 0.00570824 | 0.008513654 | 0.422516374 | 11.97348229 | 0.24146837 |
| 0 | 0 | 0 | 0 | 0 |
| 0 | 0 | 0 | 0 | 0 |
| 0 | 0 | 0 | 0 | 0 |
| 0.198826219 | 0.088161505 | 0 | 0 | 0.115225947 |
| 0.010444292 | 0.021331209 | 0.028536632 | 0 | 0.056040758 |
| 0 | 0 | 0 | 0.016935749 | 1.96493737 |
| 0.541590359 | 0.447067649 | 0 | 0 | 0.048723941 |
| 0 | 0.056374335 | 0.018037378 | 0 | 0 |
| 0 | 0 | 0 | 0 | 0 |
| 0 | 0 | 0 | 0 | 0 |
| 0 | 0 | 0 | 0 | 0 |
| 0 | 0 | 0 | 0 | 0 |
| 0 | 0 | 0 | 0 | 0 |
| 0 | 0 | 0 | 0 | 0 |
| 0 | 0 | 0 | 0 | 0 |
| 0 | 0 | 0 | 0 | 0 |
| 0 | 0 | 0 | 0 | 0 |
| 0.00695205 | 0.006507053 | 0.002240606 | 0 | 0 |
| 0 | 0 | 0 | 0 | 0 |
| 0 | 0 | 0 | 0 | 0 |
| 0 | 0 | 0 | 0 | 0 |
| 0 | 0 | 0 | 0 | 0 |
| 0 | 0 | 0 | 0 | 0.001892643 |
| 0 | 0 | 0 | 0 | 0 |
| 0 | 0.014538553 | 0 | 0 | 0 |
| 0 | 0 | 0 | 0 | 0.516891253 |
| 0 | 0 | 0.019741639 | 0 | 0 |
| 0.001613099 | 0.001157548 | 0 | 0 | 0.007367375 |
| 0 | 0 | 0 | 0 | 0 |
| 0 | 0 | 0 | 0 | 0 |
| 0 | 0 | 0 | 0 | 0 |
| 1.671453922 | 0 | 0 | 0.96966548 | 0 |
| 0 | 0 | 0 | 0 | 0 |
| 0 | 0 | 0 | 0 | 0 |
| 0 | 0 | 0 | 0 | 0.00161767 |
| 0 | 0 | 0 | 0 | 0 |
| 0 | 0 | 0 | 0 | 0 |
| 0 | 0 | 0 | 0 | 0 |
| 0 | 0 | 0 | 0 | 0 |

| | | | | |
|---|---|---|---|---|
| 0 | 0 | 0 | 0 | 0 |
| 0 | 0 | 0 | 0 | 0 |
| 0 | 0.019710973 | 0 | 0 | 0 |
| 0 | 0 | 0 | 0 | 0 |
| 0.017005102 | 0.081528446 | 0 | 0 | 0.018009509 |
| 0 | 0 | 0 | 0 | 0 |
| 0.009882856 | 0.537322929 | 0.058050518 | 0 | 0 |
| 0 | 0 | 0 | 0 | 0 |
| 0 | 0 | 0 | 0 | 0 |
| 0.195186686 | 0.01096498 | 0 | 0 | 0 |
| 0 | 0.006035122 | 0 | 0 | 0 |
| 0 | 0.001450753 | 0 | 0 | 0 |
| 0 | 0 | 0 | 0 | 0 |
| 0 | 0 | 0 | 0 | 0 |
| 0 | 0 | 0 | 0 | 0 |
| 0 | 0.001547066 | 0 | 0 | 0 |
| 0 | 0 | 0 | 0 | 0 |
| 0 | 0 | 0 | 0.001039337 | 0 |
| 0 | 0 | 0 | 0 | 0 |
| 0 | 0 | 0 | 0 | 0 |
| 0.017332423 | 0.416640601 | 0.051800374 | 0 | 0.013159223 |
| 0 | 0 | 0 | 0 | 0 |
| 0.007829113 | 0.001567846 | 0 | 0 | 0 |
| 0 | 0.00216867 | 0 | 0.000504738 | 0.013802798 |
| 0 | 0 | 0 | 0 | 0 |
| 0 | 0 | 0 | 0 | 0.015861232 |
| 0 | 0 | 0 | 0 | 0 |
| 0 | 0 | 0 | 0 | 0 |
| 0 | 0 | 0 | 0 | 0 |
| 0.006524364 | 0.000668834 | 0 | 0.810460202 | 0 |
| 0 | 0 | 0 | 0 | 0 |
| 0 | 0 | 0 | 0 | 0 |
| 0 | 0 | 0 | 0 | 0 |
| 0.155783036 | 0 | 0 | 0 | 0 |
| 0 | 0 | 0 | 0 | 0 |
| 0 | 0 | 0 | 0 | 0 |
| 0.0826525 | 0.002325915 | 0 | 0 | 0 |
| 0 | 0 | 0 | 0 | 0 |
| 0 | 0 | 0 | 0 | 0 |
| 0 | 0 | 0 | 0 | 0 |
| 0 | 0 | 0 | 0 | 0 |
| 0 | 0 | 0 | 0 | 0 |
| 0 | 0 | 0 | 0 | 0 |
| 0 | 0.004808885 | 0 | 0 | 0 |
| 0 | 0 | 0 | 0 | 0 |
| 0 | 0 | 0 | 0 | 0 |
| 0 | 0 | 0 | 0 | 0 |
| 0 | 0 | 0 | 0 | 0 |

| | | | | |
|---|---|---|---|---|
| 0.002787374 | 0 | 0.001291385 | 0 | 0.021024685 |
| 0 | 0 | 0 | 0 | 0 |
| 0 | 0.165798401 | 0 | 0 | 0 |
| 0 | 0 | 0 | 0 | 0 |
| 0 | 0 | 0 | 0 | 0 |
| 0 | 0 | 0 | 0 | 0 |
| 0 | 0 | 0 | 0 | 0 |
| 0.650211272 | 0.87951958 | 0.099447024 | 0 | 0.163986537 |
| 0.347555086 | 0.013604991 | 0.00810809 | 0 | 0.426596091 |
| 0 | 0.001457136 | 0.00235192 | 0.035015673 | 0.433566256 |
| 0 | 0.016458017 | 0 | 0 | 0 |
| 0 | 0.002710397 | 0 | 0 | 0 |
| 0 | 0.012303729 | 0 | 0 | 0 |
| 0 | 0 | 0 | 0 | 0 |
| 0 | 0 | 0 | 0 | 0 |
| 0 | 0 | 0 | 0 | 0 |
| 0.007629029 | 0 | 0.10902013 | 0 | 0 |
| 0 | 0 | 0 | 0 | 0 |
| 0 | 0 | 0 | 0 | 0 |
| 0.052882271 | 0.029905241 | 0 | 0 | 0 |
| 0 | 0 | 0 | 0 | 0 |
| 0 | 0 | 0 | 0 | 0 |
| 2.446292328 | 0 | 0 | 0 | 0 |
| 0 | 0 | 0 | 0 | 0 |
| 0 | 0.019430356 | 0 | 0 | 0 |
| 0 | 0 | 0 | 0 | 0 |
| 0 | 0 | 0 | 0 | 0 |
| 0 | 0 | 0 | 0 | 0 |
| 0 | 0 | 0 | 0 | 0 |
| 0 | 0 | 0 | 0 | 0 |
| 0.059597571 | 0.028939288 | 0 | 0 | 0 |
| 0 | 0 | 0 | 0 | 0 |
| 0 | 0 | 0 | 0 | 0 |
| 0 | 0 | 0 | 0 | 0 |
| 0 | 0 | 0 | 0 | 0.002101652 |
| 0 | 0 | 0 | 0 | 0 |
| 0 | 0 | 0 | 0 | 0 |
| 0 | 0 | 0 | 0 | 0 |
| 0 | 0 | 0 | 0 | 0 |
| 0 | 0 | 0.544474855 | 0 | 0 |
| 0 | 0 | 0 | 0 | 0 |
| 0 | 0.322745825 | 0 | 0.038596874 | 0.007151815 |
| 0 | 0 | 0 | 0 | 0 |
| 0 | 0 | 0 | 0 | 0 |
| 0 | 0 | 0 | 0 | 0 |
| 0.003717813 | 0 | 0 | 0 | 0 |
| 0 | 0 | 0 | 0 | 0 |
| 0 | 0 | 0 | 0 | 0 |

| | | | | |
|---|---|---|---|---|
| 0 | 0 | 0 | 0 | 0 |
| 0 | 0.005114924 | 0 | 0.004575066 | 0.000696356 |
| 0 | 0 | 0 | 0 | 0 |
| 0 | 0 | 0 | 0 | 0 |
| 0.215437052 | 0.091495639 | 0 | 0.003198292 | 1.185848973 |
| 0 | 0 | 0 | 0 | 0 |
| 0 | 0 | 0 | 0 | 0 |
| 0.230473181 | 0.034773445 | 0.02628376 | 0.07691824 | 0.002208296 |
| 0 | 0.001058536 | 0 | 0 | 0.002204902 |
| 0 | 0 | 0 | 0 | 0 |
| 0 | 0 | 0 | 0 | 0 |
| 0 | 0 | 0 | 0 | 0 |
| 0 | 0 | 0 | 0 | 0 |
| 0 | 0 | 0 | 0 | 0 |
| 0 | 0.011941112 | 0 | 0 | 0 |
| 0 | 0 | 0.003324374 | 0 | 0.05469071 |
| 0 | 0 | 0 | 0 | 0 |
| 0 | 0 | 0 | 0 | 0 |
| 0 | 0 | 0 | 0 | 0 |
| 0 | 0 | 0 | 0 | 0 |
| 0 | 0 | 0 | 0 | 0 |
| 0 | 0 | 0 | 0 | 0 |
| 0 | 0 | 0 | 0 | 0 |
| 0.000958921 | 0.000550492 | 0 | 0 | 0.00297282 |
| 0 | 0 | 0 | 0 | 0 |
| 0 | 0 | 0 | 0 | 0 |
| 0.09261564 | 0.002853094 | 0.004117496 | 0 | 0.006991666 |
| 0 | 0.00123703 | 0 | 0 | 0 |
| 0 | 0 | 0 | 0 | 0 |
| 0 | 0 | 0 | 0 | 0 |
| 0 | 0 | 0 | 0 | 0 |
| 0 | 0.000806547 | 0 | 0 | 0 |
| 0 | 0 | 0 | 0 | 0 |
| 0 | 0 | 0 | 0 | 0 |
| 0 | 0 | 0 | 0 | 0 |
| 0.007840667 | 0 | 0 | 0 | 0 |
| 0 | 0 | 0 | 0 | 0 |
| 0 | 0.010327114 | 0 | 0 | 0 |
| 0 | 2.773426779 | 0 | 0 | 0 |
| 0 | 0 | 0 | 0 | 0 |
| 0 | 0 | 0 | 0 | 0 |
| 0 | 0 | 0 | 0 | 0.002968995 |
| 0 | 0 | 0 | 0 | 0 |
| 0 | 0 | 0 | 0 | 0 |
| 0 | 0 | 0 | 0 | 0 |
| 0 | 0 | 0 | 0 | 0 |
| 0 | 0 | 0 | 0 | 0 |
| 0 | 0 | 0 | 0 | 0 |

| | | | | |
|---|---|---|---|---|
| 0 | 0.047979952 | 0 | 0 | 0.045294855 |
| 0 | 0 | 0 | 0 | 0 |
| 0 | 0 | 0 | 0 | 0 |
| 0 | 0 | 0 | 0 | 0 |
| 0 | 0 | 0 | 0 | 0 |
| 0 | 0 | 0 | 0 | 0 |
| 0 | 0 | 0 | 0 | 0 |
| 0 | 0 | 0 | 0 | 0 |
| 0 | 0 | 0 | 0 | 0 |
| 0 | 0 | 0 | 0 | 0 |
| 0 | 0 | 0 | 0 | 0 |
| 0 | 0 | 0 | 0 | 0.009787378 |
| 0 | 0 | 0 | 0 | 0 |
| 0 | 0 | 0 | 0 | 0 |
| 0 | 0 | 0 | 0 | 0 |
| 0 | 0 | 0 | 0 | 0 |
| 0 | 0 | 0 | 0 | 0 |
| 0 | 0 | 0 | 0 | 0 |
| 0 | 0.000535243 | 0.000863919 | 0 | 0 |
| 0 | 0 | 0 | 0 | 0 |
| 0.078302815 | 0 | 0.000707854 | 0.004167817 | 0.309361045 |
| 0 | 0 | 0 | 0 | 0 |
| 0.024119696 | 0.021462069 | 0 | 0 | 0.101213587 |
| 0 | 0.059985699 | 0 | 0 | 0 |
| 0 | 0 | 0 | 0 | 0 |
| 0 | 0 | 0 | 0 | 0 |
| 0 | 0 | 0 | 0 | 0 |
| 0 | 0 | 0 | 0 | 0 |
| 0.007725601 | 0 | 0 | 0 | 0 |
| 0 | 0 | 0 | 0 | 0 |
| 0 | 0 | 0 | 0 | 0 |
| 0 | 2.729301609 | 0 | 0 | 0 |
| 0.005195239 | 0.002161198 | 11.37891524 | 2.841204053 | 0 |
| 0 | 0 | 0 | 0 | 0 |
| 0.13092746 | 0.339273291 | 0.194206843 | 1.694855601 | 0.912285425 |
| 0.053652723 | 0.001387417 | 0.047922893 | 0 | 0 |
| 0 | 0 | 0 | 0 | 0 |
| 0 | 0 | 0 | 0 | 0 |
| 0.019917219 | 0.002607746 | 0.001554124 | 0.002178717 | 0.001392785 |
| 0 | 0 | 0 | 0 | 0 |
| 0 | 0 | 0 | 0 | 0 |
| 0 | 0.409732395 | 0 | 0 | 0 |
| 0.023534346 | 0.02720857 | 0 | 0.063179822 | 0.166386911 |
| 0 | 0 | 0 | 0 | 0 |
| 0 | 0 | 0 | 0 | 0 |
| 0.011097397 | 0.001490229 | 0 | 0 | 0.160163244 |
| 0 | 0 | 0 | 0 | 0 |
| 0 | 0 | 0 | 0 | 0 |

| | | | | |
|---|---|---|---|---|
| 0 | 0 | 0 | 0 | 0 |
| 0 | 0 | 0 | 0 | 0 |
| 0 | 0 | 0 | 0 | 0 |
| 0 | 0 | 0 | 0 | 0 |
| 0 | 0 | 0 | 0 | 0 |
| 0 | 0 | 0 | 0 | 0 |
| 0 | 0 | 0 | 0 | 0 |
| 0 | 0 | 0 | 0 | 0 |
| 0 | 0.0317637 | 0 | 0 | 0 |
| 0.003208476 | 0 | 0 | 0.004792945 | 0 |
| 0 | 0 | 0 | 0 | 0 |
| 0 | 0 | 0 | 0 | 0 |
| 0 | 0 | 0 | 0 | 0.001475068 |
| 0 | 1.158451514 | 0 | 0 | 0 |
| 0 | 0 | 0 | 0 | 0 |
| 0 | 0.000755096 | 0 | 0.321694694 | 0 |
| 0 | 0 | 0 | 0 | 0 |
| 0 | 0 | 0 | 0 | 0 |
| 0 | 0 | 0 | 0 | 0 |
| 0 | 3.433739584 | 0 | 0 | 0.00658417 |
| 0.001854584 | 0 | 0 | 0.001498987 | 0 |
| 0 | 0 | 0 | 0 | 0 |
| 0 | 0 | 0 | 0 | 0 |
| 0 | 0.003771761 | 0 | 0 | 0 |
| 0 | 0 | 0 | 0 | 0.002361933 |
| 0 | 0 | 0 | 0 | 0 |
| 0 | 0 | 0 | 0 | 0 |
| 0 | 0 | 0 | 0 | 0 |
| 0 | 0 | 0 | 0 | 0 |
| 0.009610813 | 0.012521578 | 0 | 0 | 0 |
| 1.731558443 | 0.167238839 | 0 | 0 | 0 |
| 0 | 0 | 0 | 0 | 0 |
| 0 | 0.139952528 | 0 | 0 | 0 |
| 0 | 0 | 0 | 0 | 0 |
| 0 | 0 | 0 | 0 | 0 |
| 0 | 0 | 0 | 0 | 0 |
| 0 | 0 | 0 | 0 | 0 |
| 0 | 0 | 0 | 0 | 0 |
| 0.016085066 | 0 | 0 | 0 | 0 |
| 0 | 0 | 0 | 0 | 0 |
| 0 | 0 | 0 | 0 | 0 |
| 0 | 0 | 0 | 0 | 0 |
| 0 | 0 | 0 | 0 | 0 |
| 0 | 0 | 0 | 0 | 0 |
| 0 | 0.38025568 | 0 | 0 | 0 |
| 0 | 0 | 0 | 0 | 0 |
| 0.003433552 | 0 | 0.057392056 | 0 | 0 |
| 0 | 0 | 0 | 0 | 0 |

| | | | | |
|---|---|---|---|---|
| 0 | 0 | 0 | 0 | 0 |
| 0 | 0 | 0 | 0 | 0 |
| 0 | 0 | 0 | 0 | 0 |
| 0 | 0 | 0 | 0 | 0 |
| 0.063373554 | 0.269682309 | 0.036724356 | 0 | 0.442389215 |
| 0 | 0 | 0 | 0 | 0 |
| 0 | 0 | 0 | 0 | 0 |
| 0 | 0 | 0 | 0 | 0 |
| 0 | 0 | 0 | 0 | 0 |
| 0 | 0 | 0 | 0 | 0 |
| 0 | 0.211893818 | 0 | 0 | 0 |
| 0.007541568 | 0.001603489 | 0 | 0 | 0.108782682 |
| 0.003072501 | 0.000518778 | 0 | 0 | 0 |
| 0 | 0 | 0 | 0 | 0 |
| 0.005427912 | 0 | 0 | 0 | 0 |
| 0 | 0.400267702 | 0 | 0 | 0.354969948 |
| 0.001127848 | 0.005087252 | 0.003184943 | 0 | 0 |
| 0 | 0 | 0 | 0 | 0 |
| 0.326292672 | 0.030202502 | 0.485039858 | 0 | 0.012821052 |
| 0.051572029 | 0.014862537 | 0.138945312 | 0 | 0 |
| 0 | 0 | 0 | 0 | 0 |
| 0 | 0 | 0 | 0 | 0 |
| 0 | 0 | 0 | 0 | 0 |
| 0 | 0 | 0 | 0 | 0 |
| 0 | 0 | 0 | 0 | 0 |
| 0 | 0 | 0 | 0 | 0 |
| 0.001991664 | 0.004605212 | 0 | 0 | 0 |
| 0 | 0.013451749 | 0 | 0 | 0 |
| 0 | 0 | 0 | 0 | 0 |
| 0 | 0 | 0 | 0 | 0 |
| 0.00846621 | 0 | 0 | 0 | 0.001666458 |
| 0.125990785 | 0 | 0 | 0 | 0 |
| 0 | 0 | 0 | 0 | 0 |
| 0 | 0 | 0 | 0 | 0 |
| 0 | 0 | 0 | 0 | 0 |
| 0 | 0 | 0 | 0 | 0 |
| 0 | 0 | 0 | 0 | 0 |
| 0.017712159 | 0.002529377 | 0.001469733 | 0 | 0.130105676 |
| 0 | 0.021752601 | 0 | 0 | 0 |
| 0.016144761 | 0 | 0 | 0 | 0 |
| 0 | 0 | 0 | 0 | 0 |
| 0 | 0 | 0 | 0 | 0 |
| 0 | 0 | 0 | 0 | 0 |
| 0.007840264 | 0 | 0.042862074 | 0.000814754 | 0 |
| 0 | 0 | 0 | 0 | 0 |
| 0 | 0 | 0 | 0 | 0 |
| 0 | 0 | 0 | 0 | 0 |
| 0 | 0 | 0 | 0 | 0 |

| | | | | |
|---|---|---|---|---|
| 0 | 0 | 0 | 0 | 0 |
| 0 | 0 | 0 | 0 | 0 |
| 0 | 0 | 0 | 0 | 0 |
| 0 | 0 | 0.00076135 | 0 | 0.000909749 |
| 0 | 0 | 0 | 0 | 0.001687555 |
| 0.059749859 | 0 | 0 | 0 | 0 |
| 0 | 0.011825076 | 0 | 0 | 0.001986395 |
| 0 | 0 | 0 | 0 | 0 |
| 0 | 0.018840503 | 0 | 0 | 0 |
| 0 | 0 | 0 | 0 | 0 |
| 0 | 0 | 0 | 0 | 0 |
| 0 | 0 | 0 | 0 | 0 |
| 0.007862988 | 0.004040254 | 0.005756693 | 0.004085577 | 0.016283323 |
| 0 | 0 | 0 | 0 | 0 |
| 0 | 0 | 0 | 0 | 0 |
| 0 | 0 | 0 | 0 | 0 |
| 0 | 0 | 0 | 0 | 0 |
| 0 | 0 | 0 | 0 | 0 |
| 0 | 0 | 0.000858434 | 0 | 0 |
| 0 | 0 | 0 | 0 | 0 |
| 0 | 0 | 0 | 0 | 0 |
| 0 | 0 | 0 | 0 | 0 |
| 0.007861176 | 0.000917256 | 0.004263884 | 0.009763275 | 0.058167713 |
| 0.006561397 | 0 | 0 | 0 | 0.01703472 |
| 0 | 0 | 0 | 0 | 0 |
| 0 | 0 | 0 | 0 | 0 |
| 0 | 0 | 0 | 0 | 0 |
| 0 | 0 | 0 | 0 | 0 |
| 0 | 0 | 0 | 0 | 0 |
| 0 | 0 | 0 | 0 | 0 |
| 0 | 0.051750449 | 0.041995534 | 0 | 0.025090565 |
| 0 | 0 | 0 | 0 | 0 |
| 0 | 0 | 0 | 0 | 0 |
| 0 | 0 | 0 | 0 | 0 |
| 0 | 0 | 0 | 0 | 0 |
| 0 | 0 | 0.064332901 | 0 | 0 |
| 0 | 0 | 0.004209168 | 0 | 0 |
| 0 | 0 | 0 | 0 | 0 |
| 0.118112519 | 0.005018773 | 0 | 0 | 0.026635581 |
| 0 | 0 | 0 | 0 | 0 |
| 0 | 0 | 0 | 0 | 0 |
| 0 | 0 | 0 | 0 | 0 |
| 0 | 0 | 0 | 0 | 0 |
| 0 | 0 | 0 | 0 | 0 |
| 0 | 0 | 0 | 0 | 9.771522175 |
| 0.012094766 | 0.077198465 | 0.021037632 | 0 | 0.09268243 |
| 0 | 0 | 0 | 0 | 0 |
| 0 | 0 | 0 | 0 | 0 |

| | | | | |
|---|---|---|---|---|
| 0 | 0 | 0 | 0 | 0 |
| 0 | 0 | 0 | 0 | 0 |
| 0.465054338 | 0 | 0 | 0 | 0 |
| 0 | 0.000919553 | 0 | 0 | 0 |
| 0 | 0 | 0 | 0 | 0 |
| 0.108566927 | 0 | 0 | 0 | 0 |
| 0 | 0.001055482 | 0 | 0 | 0 |
| 0 | 0 | 0 | 0 | 0 |
| 0.001673591 | 0 | 0 | 0 | 0 |
| 0 | 0 | 0 | 0 | 0 |
| 0 | 0.00940673 | 0 | 0 | 0.000649882 |
| 0 | 0 | 0 | 0 | 0 |
| 0 | 0 | 0 | 0 | 0 |
| 0 | 0 | 0 | 0 | 0 |
| 0 | 0 | 0 | 0 | 0 |
| 0 | 0 | 0 | 0 | 0 |
| 0 | 0 | 0 | 0 | 0 |
| 0 | 0 | 0 | 0 | 0 |
| 0 | 0 | 0 | 0 | 0 |
| 0.001362213 | 0 | 0 | 0 | 0 |
| 0 | 0 | 0 | 0 | 0 |
| 0 | 0 | 0 | 0 | 0 |
| 0 | 0 | 0 | 0 | 0 |
| 0 | 0 | 0 | 0 | 0 |
| 0 | 0 | 0 | 0 | 0 |
| 0 | 0 | 0 | 0 | 0 |
| 0 | 0 | 0 | 0 | 0 |
| 0 | 0 | 0 | 0 | 0 |
| 0 | 0 | 0 | 0 | 0 |
| 0 | 0 | 0 | 0 | 0 |
| 0.001003098 | 0.002879265 | 0 | 0 | 0.008514868 |
| 0 | 0 | 0 | 0 | 0 |
| 0.047170328 | 0.107908479 | 1.355962322 | 0.104896689 | 0.030519594 |
| 0.021026805 | 0 | 0.074012243 | 0 | 0 |
| 0 | 0 | 0 | 0 | 0 |
| 0 | 0 | 0 | 0 | 0 |
| 0 | 0 | 0 | 0 | 0 |
| 0 | 0 | 0 | 0 | 0 |
| 0 | 0 | 0 | 0 | 0 |
| 0 | 0 | 0 | 0 | 0 |
| 0 | 0 | 0 | 0 | 0 |
| 0.045673613 | 0.020169265 | 0.023084177 | 0 | 0.794090801 |
| 0.068094995 | 0.040953087 | 0.015864277 | 0 | 0.049114505 |
| 0 | 0 | 0 | 0 | 0 |
| 0 | 0 | 0 | 0 | 0 |
| 0.005060674 | 0.012374012 | 0 | 0 | 0.214789357 |
| 0 | 0 | 0 | 0 | 0.017551728 |
| 0 | 0 | 0 | 0 | 0 |

| | | | | |
|---|---|---|---|---|
| 0 | 0 | 0 | 0 | 0 |
| 0 | 0.010335913 | 0 | 0.004871692 | 0.0227932 |
| 0.007380741 | 0.003872612 | 0 | 0 | 0 |
| 0 | 0 | 0 | 0 | 0 |
| 0.119954449 | 0.095127374 | 0 | 0 | 0.023994938 |
| 0.001110266 | 0 | 0.003429226 | 0.003653634 | 0 |
| 0 | 0 | 0 | 0 | 0 |
| 0 | 0 | 0 | 0 | 0 |
| 0 | 0 | 0 | 0 | 0 |
| 0 | 0 | 0 | 0 | 0 |
| 0 | 0 | 0 | 0 | 0 |
| 0 | 0 | 0 | 0 | 0 |
| 0 | 0 | 0 | 0 | 0 |
| 0 | 0 | 0 | 0 | 0 |
| 0 | 0 | 0 | 0 | 0 |
| 0 | 0 | 0 | 0 | 0 |
| 0 | 0 | 0 | 0 | 0 |
| 0 | 0 | 0 | 0 | 0 |
| 0 | 0 | 0 | 0 | 0 |
| 0 | 0 | 0 | 0 | 0 |
| 0 | 0 | 0 | 0 | 0 |
| 0.003255748 | 0 | 0.006861658 | 0 | 0 |
| 0 | 0 | 0 | 0 | 0 |
| 0 | 0 | 0 | 0 | 0 |
| 0.00235462 | 0.003003837 | 0 | 0 | 0 |
| 0 | 0.000755447 | 0 | 0.037538269 | 0.001748417 |
| 0 | 0 | 0 | 0 | 0 |
| 0.001340333 | 0.39919835 | 0 | 0 | 0 |
| 0 | 0 | 0 | 0 | 0 |
| 0 | 0 | 0 | 0 | 0 |
| 0 | 0 | 0 | 0 | 0 |
| 0.001038601 | 0 | 0 | 0 | 0.001916573 |
| 0 | 0 | 0 | 0 | 0 |
| 0 | 0 | 0 | 0 | 0 |
| 0 | 0 | 0 | 0 | 0 |
| 0 | 0 | 0 | 0 | 0 |
| 0 | 0 | 0 | 0 | 0.179496588 |
| 0.029669408 | 0 | 0.145165133 | 0 | 0 |
| 0 | 0.480806607 | 0 | 3.789039306 | 0 |
| 0 | 0 | 0 | 0 | 0 |
| 0 | 0 | 0 | 0 | 0 |
| 0 | 0 | 0 | 0 | 0 |
| 0.002575344 | 0 | 0.000734247 | 0 | 0.017108594 |
| 0 | 0 | 0 | 0 | 0 |
| 0 | 0 | 0 | 0 | 0 |
| 0 | 0 | 0 | 0 | 0 |
| 0.005152011 | 0.002899644 | 0 | 0 | 0.00268439 |
| 0 | 0 | 0 | 0 | 0 |

| | | | | |
|---|---|---|---|---|
| 0 | 0 | 0 | 0 | 0 |
| 0 | 0.011159392 | 0.001889788 | 0.014703523 | 0.072401534 |
| 0 | 0 | 0 | 0 | 0 |
| 0 | 0 | 0 | 0 | 0 |
| 0 | 0 | 0 | 0 | 0 |
| 0 | 0 | 0 | 0 | 0 |
| 0 | 0 | 0 | 0 | 0 |
| 0 | 0 | 0 | 0 | 0 |
| 0 | 0 | 0 | 0 | 0 |
| 0 | 0 | 0 | 0 | 0 |
| 0 | 0 | 0 | 0 | 0 |
| 0 | 0 | 0 | 0 | 0 |
| 0.009499552 | 0.041964478 | 0 | 0 | 0.005818438 |
| 0 | 0 | 0 | 0 | 0 |
| 0.013350449 | 0 | 0 | 0 | 0.054111489 |
| 0 | 0 | 0 | 0 | 0 |
| 0 | 0 | 0 | 0 | 0 |
| 0 | 0 | 0 | 0 | 0 |
| 0 | 0 | 0 | 0 | 0 |
| 0 | 0 | 0 | 0 | 0 |
| 0 | 0 | 0 | 0 | 0 |
| 0.093885518 | 0.005152273 | 0.05524803 | 0 | 0 |
| 0.005757074 | 0.003614831 | 0 | 0 | 0 |
| 0 | 0 | 0 | 0 | 0 |
| 0 | 0 | 0 | 0 | 0 |
| 0 | 0 | 0 | 0 | 0 |
| 0 | 0 | 0 | 0 | 0 |
| 0 | 0 | 0 | 0 | 0 |
| 0 | 0 | 0 | 0 | 0 |
| 0 | 0 | 0 | 0 | 0 |
| 0 | 0 | 0 | 0 | 0 |
| 0 | 0 | 0 | 0 | 0 |
| 0 | 0 | 0 | 0 | 0 |
| 0 | 0 | 0 | 0 | 0 |
| 0.002338332 | 0 | 0 | 0.0320315 | 0 |
| 0 | 0 | 0 | 0 | 0 |
| 0 | 0 | 0 | 0 | 0 |
| 0 | 0 | 0 | 0 | 0 |
| 0.007247974 | 0.002684436 | 0.049394687 | 0 | 0 |
| 0 | 0 | 0 | 0 | 0 |
| 0.582792673 | 0.185937007 | 0.747989795 | 0 | 0.077757426 |
| 0 | 0 | 0 | 0 | 0 |
| 0 | 0 | 0 | 0 | 0 |
| 0 | 0 | 0 | 0 | 0 |
| 0 | 0 | 0 | 0 | 0 |
| 0.19609703 | 0.014467928 | 0.309287569 | 4.2789331 | 0.016742384 |
| 0 | 0 | 0 | 0 | 0 |
| 0 | 0 | 0 | 0 | 0 |

| | | | | |
|---|---|---|---|---|
| 0.039121519 | 0 | 0 | 0 | 0 |
| 0.001368285 | 0 | 0 | 0 | 0 |
| 0.017480652 | 0 | 0 | 0 | 0 |
| 0 | 0 | 0 | 0 | 0 |
| 0 | 0 | 0 | 0 | 0.001612423 |
| 0 | 0 | 0 | 0 | 0 |
| 0 | 0 | 0 | 0 | 0 |
| 0.001556572 | 0 | 0 | 0.001078385 | 0 |
| 0 | 0 | 0 | 0 | 0 |
| 0.011502297 | 0.111400466 | 0.001450066 | 0.000731822 | 0.00129953 |
| 0 | 0.023213295 | 0 | 0 | 0 |
| 0 | 0 | 0 | 0 | 0 |
| 0.002064954 | 0 | 0 | 0 | 0.002540362 |
| 0 | 0 | 0 | 0 | 0 |
| 0 | 0 | 0 | 0 | 0 |
| 0 | 0 | 0 | 0 | 0 |
| 0 | 0 | 0 | 0 | 0 |
| 0 | 0 | 0 | 0 | 0 |
| 0 | 0 | 0 | 0 | 0 |
| 0.125593577 | 0.121729875 | 0.005550654 | 0.050720792 | 0.524801701 |
| 0 | 0 | 0 | 0 | 0 |
| 0 | 0 | 0 | 0 | 0 |
| 0 | 0 | 0 | 0 | 0 |
| 0 | 0 | 0 | 0 | 0 |
| 0 | 0 | 0 | 0 | 0 |
| 0 | 0 | 0 | 0 | 0 |
| 0 | 0 | 0 | 0 | 0 |
| 0 | 0 | 0 | 0 | 0 |
| 0 | 0 | 0 | 0 | 0 |
| 0 | 0 | 0 | 0 | 0 |
| 0 | 0 | 0 | 0 | 0 |
| 0 | 0 | 0 | 0 | 0 |
| 0 | 0 | 0 | 0 | 0 |
| 0 | 0 | 0 | 0 | 0 |
| 0 | 0 | 0 | 0 | 0 |
| 0 | 0.003793977 | 0.002939398 | 0.018405909 | 0 |
| 0 | 0 | 0 | 0 | 0 |
| 0 | 0 | 0 | 0 | 0 |
| 0 | 0 | 0 | 0 | 0 |
| 0 | 0 | 0 | 0 | 0 |
| 0 | 0 | 0 | 0 | 0 |
| 0 | 0 | 0 | 0 | 0 |
| 0 | 0.001624184 | 0 | 0 | 0 |
| 0 | 0 | 0 | 0 | 0 |
| 0.001534466 | 0 | 0 | 0 | 0.008919581 |
| 0 | 0.025366774 | 0 | 0 | 0 |
| 0 | 0 | 0 | 0 | 0 |
| 0 | 0 | 0 | 0 | 0 |

| | | | | |
|---|---|---|---|---|
| 0 | 0 | 0 | 0 | 0 |
| 0 | 0 | 0 | 0 | 0 |
| 0 | 0 | 0 | 0 | 0 |
| 0 | 0 | 0 | 0 | 0 |
| 0 | 0 | 0 | 0 | 0 |
| 0.326621753 | 0.003151117 | 0 | 0.001190435 | 0 |
| 0 | 0 | 0 | 0 | 0 |
| 0 | 0 | 0 | 0 | 0 |
| 0 | 0 | 0 | 0 | 0 |
| 0 | 0 | 0 | 0 | 0 |
| 0 | 0 | 0 | 0 | 0 |
| 0 | 0 | 0 | 0 | 0 |
| 0 | 0 | 0 | 0 | 0 |
| 0 | 0 | 0 | 0 | 0 |
| 0 | 0 | 0 | 0 | 0 |
| 0 | 0 | 0 | 0 | 0 |
| 0 | 0.018114294 | 0 | 0 | 0 |
| 0 | 0 | 0 | 0 | 0 |
| 0 | 0 | 0 | 0 | 0 |
| 0 | 0 | 0 | 0 | 0 |
| 0 | 0 | 0 | 0 | 0.061031199 |
| 0 | 0 | 0 | 0 | 0 |
| 0 | 0 | 0 | 0 | 0 |
| 0 | 0 | 0 | 0 | 0.011152921 |
| 0 | 0 | 0 | 0 | 0 |
| 0 | 0 | 0 | 0 | 0 |
| 0 | 0 | 0 | 0 | 0 |
| 0.21674715 | 0.012831034 | 0 | 0 | 0 |
| 0 | 0 | 0 | 0 | 0 |
| 0.020968087 | 0 | 0 | 0 | 0 |
| 0 | 0 | 0 | 0 | 0 |
| 0 | 0 | 0 | 0 | 0 |
| 0 | 0 | 0 | 0 | 0 |
| 0 | 0 | 0 | 0 | 0 |
| 0 | 0 | 0 | 0 | 0 |
| 0 | 0 | 0 | 0 | 0 |
| 0 | 0 | 0 | 0 | 0 |
| 0 | 0 | 0 | 0 | 0 |
| 0 | 0 | 0 | 0 | 0.449386243 |
| 0 | 0 | 0 | 0 | 0 |
| 0 | 0 | 0 | 0 | 0 |
| 0 | 0 | 0 | 0 | 0 |
| 0 | 0 | 0 | 0 | 0 |
| 0 | 0 | 0 | 0 | 0 |
| 0 | 0 | 0 | 0 | 0 |
| 0 | 0 | 0 | 0 | 0 |
| 0 | 0 | 0 | 0 | 0 |
| 0 | 0 | 0 | 0 | 0 |

| | | | | |
|---|---|---|---|---|
| 0 | 0 | 0 | 0 | 0 |
| 0.001763469 | 0.006595693 | 0.739172864 | 0 | 0.140226487 |
| 0 | 0 | 0 | 0 | 0 |
| 0 | 0 | 0 | 0 | 0 |
| 0 | 0 | 0 | 0 | 0 |
| 0 | 0 | 0 | 0 | 0 |
| 0 | 0 | 0 | 0 | 0 |
| 0 | 0 | 0 | 0 | 0 |
| 0 | 0 | 0 | 0 | 0 |
| 0 | 0 | 0 | 0 | 0 |
| 0 | 0 | 0 | 0 | 0.002091822 |
| 0 | 0 | 0 | 0 | 0 |
| 0 | 0 | 0 | 0 | 0.324067413 |
| 0 | 0 | 0 | 0 | 0 |
| 0 | 0 | 0 | 0 | 0 |
| 0 | 0 | 0 | 0 | 0 |
| 0 | 0 | 0 | 0 | 0 |
| 0 | 0 | 0 | 0 | 0 |
| 0 | 0.069737559 | 0 | 0 | 0 |
| 0.001222743 | 0 | 0 | 0 | 0 |
| 0.001917508 | 0 | 0 | 0.013062988 | 0.012502543 |
| 0 | 0.002161916 | 0 | 0 | 0 |
| 0 | 0 | 0 | 0 | 0 |
| 0.258389964 | 2.228233884 | 0 | 2.205542666 | 2.799153122 |
| 0 | 0 | 0 | 0 | 0 |
| 0 | 0 | 0 | 0 | 0 |
| 0 | 0 | 0 | 0 | 0 |
| 0.156853517 | 0 | 0 | 0 | 0 |
| 0 | 0 | 0 | 0 | 0 |
| 0 | 0 | 0 | 0 | 0 |
| 0 | 0 | 0 | 0 | 0 |
| 0 | 0 | 0 | 0 | 0 |
| 0 | 0 | 0 | 0 | 0.000992565 |
| 0 | 0 | 0 | 0 | 0 |

| PHL018-0181_V2 | PHL018-0067_V1 | PHL018-0239_V3 | PHL018-0275_V2 | PHL018-0143_V2 |
|---|---|---|---|---|
| CTRL | EYCF | REF | REF | CTRL |
| 0.063698919 | 0.049494579 | 0.123735231 | 0.050497504 | 0.127115904 |
| 0.032100975 | 0.61774802 | 0.845715337 | 8.527753128 | 0 |
| 0 | 11.55173228 | 1.185705871 | 2.835410204 | 1.289629262 |
| 0 | 0 | 0 | 0 | 0 |
| 0 | 0 | 0 | 0 | 0 |
| 0 | 0 | 0 | 0 | 0 |
| 0 | 0 | 0 | 0 | 0 |
| 0 | 0 | 0 | 0 | 0 |
| 0 | 0 | 0 | 0 | 0 |
| 0 | 0 | 0 | 0 | 0 |
| 0 | 0 | 0 | 0 | 0 |
| 0 | 0 | 0 | 0 | 0 |
| 0 | 0 | 0 | 0 | 0 |
| 0.348154425 | 10.66594459 | 0.970269568 | 5.90145516 | 0.631361047 |
| 0 | 0 | 0 | 0 | 0 |
| 0 | 0 | 0 | 0 | 0 |
| 0 | 0 | 0 | 0 | 0 |
| 0 | 0.002923637 | 0 | 0 | 0 |
| 0 | 0 | 0 | 0 | 0 |
| 0 | 0 | 0 | 0 | 0 |
| 0 | 0 | 0 | 0 | 0 |
| 0 | 0 | 0 | 0 | 0 |
| 0 | 0 | 0 | 0 | 0 |
| 0 | 0 | 0 | 0 | 0 |
| 0.949235752 | 0 | 0 | 0 | 0 |
| 0 | 0 | 0 | 0 | 0 |
| 0 | 0 | 0 | 0 | 0 |
| 0 | 0 | 0 | 0 | 0 |
| 0 | 0 | 0 | 0 | 0 |
| 0 | 0 | 0 | 0 | 0 |
| 0 | 0 | 0 | 0 | 0 |
| 0 | 0 | 0 | 0 | 0 |
| 0 | 0 | 0 | 0 | 0 |
| 0 | 0 | 0 | 0 | 0 |
| 0 | 0 | 0 | 0 | 0 |
| 3.824155353 | 2.397061073 | 0.654793848 | 8.280801003 | 0.553743642 |
| 0 | 0 | 0 | 0 | 0 |
| 0 | 0 | 0 | 0 | 0 |
| 0 | 0 | 0 | 0 | 0 |
| 0 | 0 | 0 | 0 | 0 |
| 0 | 0 | 0 | 0 | 0 |
| 0.136806462 | 0 | 0.035683584 | 0.052484632 | 0.00116091 |
| 0.045511282 | 0.063434092 | 0.195238079 | 0.022721635 | 0 |
| 0.003453519 | 0 | 0.051062737 | 0.006070746 | 0.000317267 |
| 0.003680671 | 0 | 0 | 0 | 0 |
| 0 | 0 | 0 | 0 | 0 |

| | | | | |
|---|---|---|---|---|
| 0.001403009 | 0 | 0 | 0 | 0 |
| 0 | 0.046303881 | 0.00801017 | 0.116383726 | 0.006526293 |
| 0.002074999 | 0.016408159 | 0.007199016 | 0 | 0.159411936 |
| 0.894858644 | 0.269729613 | 0.299298962 | 0.715532929 | 0.024557018 |
| 0.078167883 | 0.022706325 | 0.012978678 | 0.00408579 | 0.008091907 |
| 0 | 0 | 0.414069053 | 0 | 0 |
| 0.006414256 | 0.001418008 | 0.000559931 | 0.002460445 | 0 |
| 0.004628452 | 0 | 0 | 0 | 0 |
| 0.009811447 | 0 | 0 | 0 | 0 |
| 0 | 0 | 0 | 0 | 0 |
| 0 | 0 | 0 | 0.003843144 | 0 |
| 0.059285217 | 0.153825655 | 0.116044616 | 0.050008287 | 0.037333367 |
| 0 | 0 | 0 | 0 | 0 |
| 0.077944522 | 0.089221822 | 0.050927638 | 0.032246312 | 0.040286293 |
| 0 | 0 | 0 | 0 | 0.005441393 |
| 0 | 0 | 0 | 0 | 0 |
| 0.005078597 | 1.157504966 | 0.001099471 | 0.529025957 | 0 |
| 0 | 0 | 0 | 0 | 0 |
| 0 | 0 | 0 | 0 | 0 |
| 0 | 0 | 0 | 0 | 0 |
| 0.039302666 | 0.031886566 | 0.11101957 | 0.184150833 | 0.012462262 |
| 0.004998592 | 0.000575738 | 0.008354839 | 0.016083692 | 0.012943418 |
| 0.000989383 | 0 | 0 | 0 | 0 |
| 0 | 0 | 0 | 0 | 0 |
| 0 | 0 | 0 | 0 | 0 |
| 0 | 0 | 0 | 0 | 0 |
| 0 | 0 | 0 | 0 | 0.022996885 |
| 0 | 0 | 0 | 0 | 0 |
| 0.012036979 | 0.001104803 | 0.000848274 | 0.014537175 | 0 |
| 0 | 0 | 0 | 0 | 0 |
| 0 | 0 | 0 | 0 | 0.464511712 |
| 0 | 0 | 0 | 0 | 0 |
| 0 | 0 | 0 | 0 | 0 |
| 0 | 0 | 0 | 0 | 0 |
| 0.04182042 | 0 | 0 | 0 | 0 |
| 0 | 0 | 0 | 0 | 0 |
| 0 | 0 | 0 | 0 | 0 |
| 0 | 0.025656752 | 0 | 0.019503168 | 0 |
| 0 | 0 | 0 | 0 | 0 |
| 0 | 0 | 0 | 0 | 0 |
| 0 | 0 | 0 | 0 | 0 |
| 0 | 0 | 0 | 0 | 0 |
| 0 | 0 | 0 | 0 | 0 |
| 0 | 0 | 0 | 0 | 0 |
| 0 | 0 | 0 | 0 | 0 |
| 0 | 0 | 0 | 0 | 0 |
| 0 | 0 | 0 | 0 | 0 |
| 0.014259377 | 0 | 0.001473841 | 0 | 0.042474229 |

| | | | | |
|---|---|---|---|---|
| 0 | 0 | 0 | 0 | 0 |
| 0 | 0 | 0 | 0 | 0.001957373 |
| 0 | 0 | 0 | 0 | 0 |
| 0 | 0 | 0 | 0 | 0 |
| 0 | 0.033422342 | 0.052682299 | 0.001193963 | 0 |
| 0 | 0 | 0 | 0 | 0 |
| 0 | 0 | 0 | 0 | 0 |
| 0 | 0 | 0 | 0 | 0 |
| 0.003126285 | 0 | 0.007562869 | 0.001013879 | 0 |
| 0 | 0 | 0 | 0 | 0 |
| 0 | 0 | 0 | 0 | 0 |
| 0.033490894 | 0 | 0 | 0 | 0 |
| 0 | 0 | 0 | 0 | 0 |
| 0 | 0 | 0 | 0 | 0 |
| 0 | 0 | 0 | 0 | 0 |
| 0 | 0 | 0 | 0 | 0 |
| 0 | 0 | 0 | 0 | 0 |
| 0 | 0 | 0 | 0 | 0 |
| 0.519317496 | 0 | 0.00821142 | 0.08642357 | 0 |
| 0 | 0 | 0.054072804 | 0 | 0 |
| 0 | 0 | 0 | 0 | 0 |
| 0 | 0 | 0 | 0 | 0 |
| 0 | 0 | 0 | 0 | 0 |
| 0 | 0 | 0 | 0 | 0 |
| 0 | 0 | 0 | 0 | 0 |
| 0 | 0 | 0 | 0 | 0 |
| 0 | 0 | 0 | 0 | 0 |
| 0 | 0.001872856 | 0 | 0 | 0.001751546 |
| 0 | 0 | 0 | 0 | 0 |
| 0 | 0 | 0 | 0 | 0 |
| 0 | 0.014933988 | 0.117194528 | 0.002345536 | 0.017504659 |
| 0.00717359 | 0.080091061 | 0 | 0.002624716 | 0 |
| 0 | 0 | 0 | 0 | 0 |
| 0 | 0 | 0 | 0 | 0 |
| 0 | 0 | 0 | 0 | 0 |
| 0 | 0 | 0 | 0 | 0 |
| 0 | 0 | 0 | 0 | 0 |
| 0 | 0 | 0 | 0 | 0 |
| 0 | 0.634675643 | 0 | 0 | 0 |
| 0 | 0 | 0 | 0 | 0 |
| 0 | 0 | 0 | 0 | 0 |
| 0 | 0 | 0 | 0 | 0 |
| 0 | 0 | 0.004171174 | 0.012703856 | 0 |
| 0 | 0 | 0 | 0 | 0 |
| 0.003399973 | 0 | 0 | 0 | 0 |
| 0 | 0 | 0 | 0 | 0 |
| 0 | 0 | 0 | 0 | 0 |
| 0 | 0 | 0 | 0 | 0 |

| | | | | |
|---|---|---|---|---|
| 0 | 0 | 0 | 0 | 0 |
| 0.226607454 | 0 | 0 | 0 | 0 |
| 0 | 0 | 0 | 0 | 0 |
| 0 | 0 | 0 | 0 | 0 |
| 0.010926648 | 0 | 0.046357288 | 0.004854601 | 0 |
| 0 | 0 | 0 | 0 | 0 |
| 0 | 0 | 0 | 0 | 0 |
| 0 | 0 | 0 | 0 | 0 |
| 0 | 0 | 0 | 0 | 0 |
| 0 | 0 | 0 | 0 | 0 |
| 0 | 0 | 0 | 0 | 0 |
| 0 | 0 | 0 | 0 | 0 |
| 0 | 0 | 0 | 0 | 0 |
| 0 | 0 | 0 | 0 | 0 |
| 0 | 0 | 0 | 0 | 0 |
| 0 | 0 | 0 | 0 | 0 |
| 0 | 0 | 0 | 0 | 0 |
| 0 | 0 | 0 | 0 | 0 |
| 0 | 0 | 0 | 0 | 0 |
| 0 | 0 | 0 | 0 | 0 |
| 0 | 0 | 0 | 0 | 0 |
| 0 | 0 | 0 | 0 | 0 |
| 0 | 0 | 0 | 0 | 0 |
| 0 | 0 | 0 | 0 | 0 |
| 0.044969407 | 0 | 0 | 0 | 0 |
| 0 | 0 | 0 | 0 | 0 |
| 0.018581972 | 0 | 0 | 0 | 0 |
| 0 | 0 | 0 | 0 | 0 |
| 0 | 0 | 0 | 0 | 0 |
| 0 | 0 | 0 | 0 | 0 |
| 0.011210741 | 0 | 0 | 0 | 0 |
| 0.218857041 | 0 | 0 | 0 | 0 |
| 0 | 0 | 0 | 0 | 0 |
| 0 | 0 | 0 | 0 | 0 |
| 0 | 0 | 0 | 0 | 0 |
| 0 | 0 | 0 | 0 | 0 |
| 0 | 0 | 0 | 0 | 0 |
| 0 | 0 | 0.001896291 | 0 | 0 |
| 0 | 0 | 0 | 0 | 0 |
| 0 | 0 | 0 | 0 | 0 |
| 0 | 0 | 0 | 0 | 0 |
| 0 | 0 | 0 | 0 | 0 |
| 0 | 0 | 0 | 0 | 0 |
| 0 | 0 | 0 | 0 | 0 |
| 0 | 0 | 0 | 0 | 0 |
| 0 | 0 | 0 | 0 | 0 |
| 0 | 0 | 0 | 0 | 0 |
| 0 | 0.000764208 | 0 | 0 | 0 |

| | | | | |
|---|---|---|---|---|
| 0 | 0 | 0 | 0 | 0 |
| 0 | 0 | 0 | 0 | 0 |
| 0 | 0.002037781 | 0 | 0 | 0 |
| 0 | 0 | 0 | 0 | 0 |
| 0 | 0 | 0 | 0 | 0 |
| 0 | 0 | 0 | 0 | 0 |
| 0 | 0 | 0 | 0 | 0 |
| 0 | 0 | 0 | 0 | 0 |
| 0 | 0 | 0 | 0 | 0.035621771 |
| 0 | 0 | 0 | 0 | 0 |
| 0 | 0 | 0 | 0 | 0 |
| 0 | 0 | 0.003205518 | 0 | 0 |
| 0 | 0 | 0.002198234 | 0 | 0 |
| 0 | 0 | 0 | 0 | 0 |
| 0 | 0 | 0 | 0 | 0 |
| 0 | 0 | 0 | 0 | 0 |
| 0.004087391 | 0 | 0 | 0 | 0 |
| 0 | 0 | 0 | 0 | 0 |
| 0.001572907 | 0 | 0 | 0 | 0 |
| 0.000494561 | 0 | 0 | 0 | 0 |
| 0.012530366 | 0.014498081 | 0.091965047 | 0.168304823 | 0.007621928 |
| 0 | 0 | 0 | 0 | 0 |
| 0 | 0 | 0 | 0 | 0 |
| 0 | 0.024422325 | 0 | 0.006709564 | 0 |
| 0 | 0 | 0 | 0 | 0 |
| 0 | 0 | 0 | 0 | 0 |
| 0.010448535 | 0.003085067 | 0.004039311 | 0.006113722 | 0 |
| 0 | 0 | 0 | 0 | 0 |
| 0 | 0 | 0 | 0 | 0 |
| 0 | 0 | 0 | 0 | 0 |
| 0 | 0 | 0 | 0.182043902 | 0 |
| 0 | 0 | 0 | 0 | 0 |
| 0 | 0 | 0 | 0 | 0 |
| 0.639772807 | 0 | 0.061070093 | 0 | 0 |
| 0.004632343 | 0.017959408 | 0.002172865 | 0.049576096 | 0.004306702 |
| 0 | 0 | 0 | 0 | 0 |
| 0 | 0 | 0 | 0 | 0 |
| 0 | 0 | 0 | 0 | 0 |
| 0 | 0 | 0.000810033 | 0 | 0 |
| 0 | 0 | 0 | 0 | 0 |
| 0.002119484 | 0 | 0 | 0 | 0 |
| 0 | 0 | 0 | 0 | 0 |
| 0.001348144 | 0 | 0 | 0 | 0 |
| 0 | 0 | 0 | 0 | 0 |
| 0 | 0 | 0 | 0 | 0 |
| 2.08757553 | 0.005699771 | 0.000954833 | 0 | 0 |
| 0.004814972 | 0.001178501 | 0 | 0 | 0.000989124 |
| 0.000660227 | 0 | 0.002263108 | 0 | 0 |

| | | | | |
|---|---|---|---|---|
| 0 | 0 | 0 | 0 | 0.002432529 |
| 0 | 0 | 0 | 0 | 0 |
| 0 | 0 | 0 | 0.003443536 | 0 |
| 0 | 0 | 0 | 0 | 0 |
| 0 | 0 | 0 | 0 | 0 |
| 0 | 0 | 0 | 0 | 0 |
| 0.013533468 | 0 | 0 | 0.002235145 | 0 |
| 0.007972337 | 0 | 0 | 0 | 0 |
| 0 | 0 | 0.012205227 | 0 | 0 |
| 0 | 0 | 0 | 0 | 0 |
| 0 | 0 | 0 | 0 | 0 |
| 0 | 0 | 0 | 0 | 0 |
| 0 | 0 | 0 | 0 | 0 |
| 0 | 0 | 0 | 0 | 0 |
| 0 | 0 | 0 | 0.00470377 | 0 |
| 0 | 0 | 0 | 0 | 0 |
| 0 | 0 | 0 | 0 | 0 |
| 0 | 0 | 0 | 0 | 0 |
| 0 | 0.000525893 | 0.001695891 | 0 | 0 |
| 0.233911684 | 0 | 0.233348356 | 0 | 0 |
| 0 | 0.079707844 | 0.00053606 | 0.033684357 | 0.089571181 |
| 0 | 0 | 0 | 0 | 0 |
| 0 | 0 | 0 | 0 | 0 |
| 0 | 0 | 0 | 0 | 0 |
| 0 | 0.007320616 | 0 | 0.011231513 | 0 |
| 0 | 0 | 0 | 0 | 0 |
| 0 | 0 | 0 | 0 | 0 |
| 0 | 0 | 0 | 0 | 0 |
| 0.013896043 | 0 | 0.004573534 | 0.026572689 | 0 |
| 0 | 0 | 0 | 0 | 0 |
| 0.274269658 | 1.012469414 | 0.41426679 | 0.031349825 | 0.314022618 |
| 0 | 0 | 0 | 0 | 0 |
| 0 | 0.094330816 | 0 | 0 | 0.019899428 |
| 0 | 0.002767413 | 0.00594954 | 0 | 0 |
| 0 | 0 | 0 | 0 | 0 |
| 0 | 0 | 0 | 0 | 0 |
| 0 | 0 | 0 | 0 | 0 |
| 0 | 0 | 0 | 0 | 0 |
| 0 | 0 | 0 | 0 | 0 |
| 0 | 0 | 0 | 0 | 0 |
| 0 | 0 | 0 | 0 | 0 |
| 0.003563322 | 0.067155553 | 0.00241071 | 0.002295178 | 0 |
| 0 | 0 | 0 | 0 | 0 |
| 0 | 0 | 0 | 0 | 0 |
| 0 | 0 | 0 | 0 | 0 |
| 0 | 0.009964109 | 0 | 0 | 0 |
| 0 | 0 | 0 | 0 | 0 |
| 0 | 0 | 0 | 0 | 0 |

| | | | | |
|---|---|---|---|---|
| 0 | 0 | 0 | 0 | 0 |
| 0 | 0 | 0 | 0 | 0 |
| 0.005965783 | 0 | 0.053141432 | 0.006650707 | 0 |
| 0 | 0 | 0 | 0 | 0 |
| 1.982571319 | 0 | 0 | 0 | 0 |
| 0 | 0 | 0 | 0 | 0 |
| 0 | 0 | 0 | 0 | 0 |
| 0 | 0 | 0 | 0 | 0 |
| 0 | 0 | 0 | 0 | 0 |
| 0 | 0 | 0 | 0.001216537 | 0 |
| 0 | 0 | 0 | 0 | 0 |
| 0 | 0 | 0 | 0 | 0 |
| 0 | 0 | 0 | 0 | 0 |
| 0 | 0.002420628 | 0.007248431 | 0.001960062 | 0 |
| 0 | 0.008990602 | 0.002301012 | 0.007279985 | 0 |
| 0.086301329 | 0 | 0 | 0 | 0 |
| 0 | 0.002276441 | 0 | 0.020506813 | 0.000545895 |
| 0 | 0 | 0 | 0.016086216 | 0 |
| 0 | 0 | 0 | 0 | 0 |
| 0 | 0 | 0 | 0 | 0 |
| 0.001263602 | 0 | 0 | 0 | 0 |
| 0 | 0 | 0.003628993 | 0 | 0 |
| 0 | 0 | 0 | 0 | 0 |
| 0 | 0 | 0 | 0 | 0.15832513 |
| 0 | 0 | 0 | 0 | 0 |
| 0 | 0 | 0 | 0 | 0 |
| 0.494166696 | 0 | 0 | 0 | 0 |
| 0 | 0 | 0 | 0 | 0 |
| 0 | 0 | 0 | 0 | 0 |
| 0.002530176 | 0 | 0 | 0.003008707 | 0 |
| 0 | 0 | 0 | 0 | 0 |
| 0 | 0 | 0 | 0 | 0 |
| 0.002533996 | 0 | 0 | 0 | 0 |
| 0.001008388 | 0 | 0 | 0 | 0.004687505 |
| 0 | 0 | 0 | 0.010998242 | 0 |
| 0.081403172 | 0 | 0.001890886 | 0.009139811 | 0 |
| 0 | 0 | 0 | 0 | 0 |
| 0.001977875 | 0 | 0 | 0 | 0 |
| 0 | 0 | 0 | 0 | 0 |
| 0 | 0 | 0 | 0 | 0 |
| 0 | 0 | 0.002288897 | 0 | 0 |
| 0 | 0 | 0 | 0 | 0 |
| 0 | 0 | 0 | 0 | 0 |
| 0 | 0.001870797 | 0 | 0 | 0.001538129 |
| 0.003873508 | 0 | 0 | 0 | 0 |
| 0 | 0 | 0 | 0 | 0 |
| 0 | 0 | 0 | 0 | 0 |
| 0.000798567 | 0.005472752 | 0 | 0 | 0.002249789 |

| | | | | |
|---|---|---|---|---|
| 0 | 0 | 0.515410039 | 0 | 0.107653482 |
| 0 | 0 | 0 | 0 | 0 |
| 0 | 0 | 0 | 0 | 0 |
| 0 | 0 | 0 | 0 | 0 |
| 0 | 0 | 0 | 0.001534327 | 0 |
| 0 | 0 | 0 | 0 | 0 |
| 0 | 0 | 0 | 0 | 0 |
| 0.047282806 | 0 | 0.018645363 | 0.206159704 | 0 |
| 0 | 0 | 0 | 0 | 0 |
| 0 | 0.086562095 | 0.054291357 | 0.082970548 | 1.496809753 |
| 0 | 0.00165013 | 0 | 0 | 0 |
| 0.143171951 | 0 | 0.003136444 | 0.129876407 | 0.002016778 |
| 0.008377792 | 0.020657171 | 0.004758208 | 0.015496878 | 0.000437085 |
| 0 | 0 | 0.001487807 | 0 | 0.000995729 |
| 0 | 0 | 0 | 0 | 0 |
| 0 | 0 | 0 | 0.004431121 | 0.001640343 |
| 0.423409596 | 0 | 0.006110959 | 0.005034888 | 0 |
| 0 | 0.004747636 | 0 | 0 | 0.001423116 |
| 0 | 0.143279393 | 0 | 0.703557928 | 0 |
| 0 | 0 | 0.055082562 | 0 | 0 |
| 0 | 0 | 0 | 0 | 0 |
| 0 | 0 | 0 | 0 | 0 |
| 0.000640355 | 0 | 0 | 0 | 0 |
| 0 | 0 | 0 | 0 | 0 |
| 0.052113393 | 0 | 0 | 0 | 0 |
| 0 | 0.001769142 | 0.002608047 | 0.004082718 | 0 |
| 0 | 0 | 0 | 0 | 0 |
| 0 | 0.163449014 | 0 | 0.005976539 | 0 |
| 0 | 0.013222831 | 0 | 0 | 0 |
| 0.000929606 | 0 | 0.000670838 | 0 | 0.005106975 |
| 0 | 0 | 0 | 0 | 0 |
| 0 | 0 | 0 | 0 | 0 |
| 0 | 0 | 0 | 0 | 0 |
| 0 | 0.014807765 | 0 | 0 | 0 |
| 0 | 0 | 0 | 0 | 0 |
| 0 | 0 | 0 | 0 | 0 |
| 0 | 0 | 0 | 5.717993658 | 0.005049701 |
| 0 | 0 | 0 | 0 | 0 |
| 0 | 0 | 0 | 0 | 0 |
| 0 | 0 | 0 | 0 | 0.034401709 |
| 0.002439786 | 0 | 0 | 0 | 0 |
| 0.010398908 | 0.044255513 | 0 | 0 | 0 |
| 0 | 0.000486948 | 0 | 0 | 0 |
| 0 | 0 | 0.03650721 | 0 | 0 |
| 0 | 0 | 0 | 0 | 0 |
| 0 | 0 | 0.00190815 | 0 | 0 |
| 0.003595506 | 0.007260226 | 0 | 0.001068881 | 0 |
| 0 | 0 | 0 | 0 | 0 |

| | | | | |
|---|---|---|---|---|
| 0 | 0 | 0 | 0 | 0 |
| 0.002762533 | 0.014506509 | 0 | 0.004778195 | 0 |
| 0 | 0 | 0 | 0 | 0 |
| 0 | 0 | 0 | 0 | 0 |
| 0 | 0 | 0 | 0 | 0 |
| 0.039646554 | 0 | 0.02346948 | 0.011575752 | 0.010471451 |
| 0 | 0 | 0.017752905 | 0 | 0 |
| 0 | 0 | 0 | 0 | 0 |
| 0 | 0 | 10.77396675 | 0 | 0 |
| 0.001847095 | 0 | 0 | 0 | 0 |
| 0.008006026 | 0 | 0 | 0 | 0 |
| 0 | 0 | 0 | 0 | 0 |
| 0 | 0 | 0 | 0 | 0 |
| 0.001551526 | 0.229937292 | 0 | 0.001844965 | 0 |
| 0 | 0.028625621 | 0.052395374 | 0 | 0 |
| 0 | 0.006583418 | 0 | 0 | 0.038307096 |
| 0 | 0 | 0 | 0 | 0.011830797 |
| 0 | 0.00277167 | 0 | 0 | 0 |
| 0 | 0 | 0 | 0.005891046 | 0 |
| 0 | 0 | 0 | 0.002074322 | 0 |
| 0.197470265 | 0 | 0 | 0.004224234 | 0 |
| 0.271896149 | 0 | 0 | 0.317061896 | 0 |
| 0 | 0 | 0 | 0 | 0 |
| 0 | 0 | 0 | 0 | 0 |
| 0 | 0 | 0 | 0 | 0 |
| 0 | 0 | 0 | 0 | 0 |
| 0 | 0 | 0 | 0.036007759 | 0 |
| 0 | 0 | 0 | 0 | 0 |
| 0 | 0 | 0 | 0 | 0 |
| 0.102055289 | 1.943476213 | 0.182322125 | 0.05205841 | 0.053258982 |
| 0 | 0 | 0.001305538 | 0.002346867 | 0 |
| 0.148888313 | 0 | 0.000666522 | 0 | 0 |
| 0 | 0 | 0 | 0 | 0 |
| 0 | 0 | 0 | 0 | 0 |
| 0 | 0.015261123 | 0 | 0 | 0.007278176 |
| 0 | 0 | 0 | 0 | 0 |
| 0.55028481 | 0.065105195 | 0 | 0 | 0 |
| 0.111885367 | 0 | 0 | 0 | 0 |
| 0 | 0 | 0.174552245 | 0 | 0 |
| 0 | 0 | 0.264963781 | 0 | 0 |
| 0 | 0.031843789 | 0 | 0.001895953 | 0.008646878 |
| 0.243260636 | 0 | 0.002226483 | 6.041697602 | 0.086729075 |
| 0.014124647 | 0 | 0 | 0 | 0 |
| 0 | 0.009039161 | 0 | 0 | 0.002709511 |
| 0 | 0.017080299 | 0 | 0 | 0.002786894 |
| 0 | 0.000636306 | 0 | 0 | 0 |
| 0.048362465 | 0.034569187 | 0.000603809 | 0.051738406 | 0 |
| 0 | 0 | 0 | 0 | 0 |

| | | | | |
|---|---|---|---|---|
| 0 | 0 | 0.000744998 | 0.004664976 | 0 |
| 0 | 0 | 0 | 0 | 0 |
| 0.017724262 | 2.652467224 | 0.000761338 | 1.208717162 | 0 |
| 0 | 0 | 0 | 0 | 0 |
| 0 | 0 | 0 | 0 | 0 |
| 0 | 0.046317612 | 0 | 0.074940581 | 0 |
| 0 | 0 | 0 | 0 | 0 |
| 0.002082746 | 0 | 0 | 0 | 0 |
| 0.006393628 | 0 | 0.086821944 | 0 | 0 |
| 0 | 0 | 0 | 0 | 0.033674678 |
| 0 | 0 | 0 | 0 | 0 |
| 0 | 0 | 0 | 0 | 0 |
| 0.005839708 | 0 | 0 | 0 | 0 |
| 0 | 0 | 0 | 0 | 0.005130904 |
| 0 | 0 | 0 | 0 | 0 |
| 0.052988933 | 0.005287817 | 0.258743715 | 0.081410515 | 0 |
| 0.000970789 | 0.014098065 | 0.021600501 | 0.002501185 | 0.006267672 |
| 0 | 0.003271755 | 0 | 0.002805085 | 0.204773647 |
| 0 | 0 | 0.004371884 | 0.015103746 | 0 |
| 0 | 0 | 0 | 0 | 0 |
| 0.001427381 | 0.001197814 | 0 | 0.023035345 | 0.020250291 |
| 0.002064451 | 0.009370834 | 0 | 0 | 0.000660925 |
| 0 | 0.005180782 | 0 | 0 | 0 |
| 0 | 0.295337648 | 0.134204398 | 0 | 0 |
| 0 | 0 | 0 | 0 | 0 |
| 0 | 0.321770635 | 0 | 0 | 0.001050834 |
| 0 | 0.002614257 | 0 | 0 | 0 |
| 0 | 0 | 0 | 0 | 0 |
| 0 | 0 | 0 | 0 | 0 |
| 0 | 0 | 0 | 0 | 0 |
| 0.03842745 | 0.022787089 | 0 | 0.000928764 | 0.019363742 |
| 0.016069318 | 0.00091202 | 0.012184426 | 0.026585737 | 0 |
| 0.056544082 | 0.02778773 | 0 | 0.063810398 | 0.006090904 |
| 0.027328839 | 0 | 0.02522517 | 0.013225737 | 0 |
| 0 | 0 | 0.004091899 | 0.001078836 | 0 |
| 0 | 0 | 0 | 0 | 0 |
| 0 | 0.011775948 | 0 | 0.030989958 | 0 |
| 0.054871782 | 0.028241969 | 0.137035631 | 0.012117792 | 0.00761904 |
| 0.057417027 | 0 | 0.245201623 | 0 | 0 |
| 0 | 0.008282532 | 0 | 0 | 0 |
| 0 | 0 | 0 | 0 | 0 |
| 0 | 0 | 0 | 0 | 0 |
| 0 | 0.00275451 | 0 | 0.002788021 | 0 |
| 0 | 0.192546 | 0.039521574 | 0.047006091 | 0.01456731 |
| 0 | 0 | 0 | 0 | 0 |
| 0 | 0.000501946 | 0 | 0.001016106 | 0 |
| 0 | 0 | 0 | 0 | 0 |
| 0 | 0 | 0 | 0 | 0 |

| | | | | |
|---|---|---|---|---|
| 0.007438322 | 0 | 0 | 0 | 0 |
| 0.194187759 | 0 | 0.195207328 | 0.049536086 | 0.001805549 |
| 0 | 0 | 0 | 0 | 0 |
| 0 | 0 | 0 | 0 | 0.000455629 |
| 0 | 0 | 0 | 0 | 0 |
| 0 | 0 | 0 | 0 | 0 |
| 0 | 0 | 0 | 0.110690632 | 0 |
| 0 | 0.000803239 | 0 | 0.001170737 | 0 |
| 0.038133958 | 0 | 0 | 0 | 0 |
| 0 | 0 | 0 | 0.017721553 | 0.003717496 |
| 0.003016229 | 0.006527639 | 0.00114559 | 0.002642821 | 0.004025162 |
| 0.003073993 | 0.134039916 | 0.004479272 | 0.555054618 | 0 |
| 0 | 0 | 0 | 0 | 0 |
| 0 | 0.025006118 | 0 | 0 | 0 |
| 0 | 0.028888646 | 0.004929082 | 0.030539665 | 0 |
| 0 | 0 | 0 | 0 | 0 |
| 0.004654524 | 0.000488241 | 0.001499498 | 0.017988201 | 0.000468325 |
| 0.000863174 | 0.019859217 | 0.007591567 | 0.033102208 | 0.535908456 |
| 0 | 0 | 0 | 0 | 0 |
| 0 | 0.000823119 | 0.002022387 | 0.002666027 | 0.000592157 |
| 0.040285617 | 0.015105002 | 0.006030808 | 0.042197003 | 0.002173326 |
| 0.007892648 | 0 | 0 | 0 | 0 |
| 0 | 0 | 0 | 0 | 0 |
| 0 | 0 | 0 | 0 | 0 |
| 0 | 0 | 0 | 0 | 0 |
| 0 | 0 | 0 | 0 | 0.001328667 |
| 0.003384163 | 0 | 0 | 0 | 0 |
| 0.040813433 | 0.091866771 | 0 | 0.107464714 | 0.016682787 |
| 0 | 0 | 0 | 0 | 0 |
| 0 | 0 | 0 | 0 | 0 |
| 0 | 0 | 0.001234474 | 0.000929917 | 0 |
| 0 | 0 | 0 | 0 | 0 |
| 0 | 0.008662183 | 0 | 0 | 0 |
| 0 | 0 | 0 | 0 | 0 |
| 0.082881634 | 0.190734161 | 0 | 0.028509779 | 0 |
| 0.056431236 | 0.037435728 | 0.139637124 | 0.046936159 | 2.033978745 |
| 0 | 0 | 0 | 0 | 0 |
| 0 | 0 | 0 | 0 | 0 |
| 0.053269913 | 0 | 0.001216504 | 0.043566267 | 0 |
| 0 | 0.00135742 | 0 | 0 | 0 |
| 0 | 0 | 0 | 0 | 0 |
| 0 | 0 | 0 | 0 | 0 |
| 0 | 0.000801986 | 0 | 0.165433187 | 0.00086543 |
| 0.000395443 | 0 | 0.000475611 | 0.163641239 | 0.002692349 |
| 0 | 0.002034507 | 0 | 0 | 0 |
| 0.580993121 | 0 | 0 | 0 | 0 |
| 0 | 0 | 0 | 0.006201776 | 0 |
| 0 | 0 | 0 | 0 | 0 |

| | | | | |
|---|---|---|---|---|
| 0.001144389 | 0 | 0 | 0.001360827 | 0 |
| 0 | 0 | 0 | 0 | 0 |
| 0 | 0 | 0 | 0 | 0 |
| 0.34378188 | 0 | 0.501736622 | 0.000567254 | 0 |
| 0 | 0 | 0 | 0 | 0 |
| 0 | 0 | 0 | 0 | 0 |
| 0 | 0 | 0 | 0 | 0 |
| 0.00526334 | 0 | 0.003207956 | 0 | 0 |
| 0.279208687 | 0.126253795 | 0 | 0 | 0 |
| 0.006537371 | 0 | 0.084481077 | 0 | 0 |
| 0 | 0 | 0 | 0 | 0 |
| 0.006551036 | 0 | 0 | 0.00471025 | 0.002038756 |
| 0 | 0 | 0 | 0 | 0 |
| 0 | 0 | 0 | 0 | 0 |
| 0.976964981 | 0.199640688 | 0.04014524 | 0.036327664 | 0.078806538 |
| 0 | 0 | 0 | 0 | 0 |
| 0.01213525 | 0 | 0.359165312 | 0 | 0 |
| 0 | 0 | 0 | 0 | 0 |
| 0 | 0 | 0 | 0 | 0 |
| 0 | 0 | 0 | 0 | 0 |
| 0 | 0 | 0 | 0 | 0 |
| 0 | 0 | 0 | 0 | 0 |
| 0 | 0 | 0 | 0 | 0 |
| 0.293425168 | 0.062125708 | 0.089684173 | 0.832484246 | 0.056549352 |
| 0 | 0 | 0 | 0 | 0 |
| 0 | 0 | 0 | 0 | 0 |
| 0.020196379 | 0 | 0.081292368 | 0 | 0 |
| 1.563411803 | 0.232210579 | 0 | 0.347091381 | 0 |
| 0 | 0 | 0 | 0 | 0 |
| 0 | 0 | 0 | 0 | 0 |
| 0.820972956 | 0 | 0 | 0 | 0 |
| 0.228438376 | 0 | 0 | 0 | 0 |
| 0 | 0 | 0 | 0.000938291 | 0 |
| 0 | 0 | 0 | 0 | 0 |
| 0 | 0 | 0 | 0 | 0 |
| 0.508852586 | 0.021392814 | 0 | 0 | 0 |
| 0.440664863 | 0 | 0 | 0 | 0 |
| 0 | 0 | 0 | 0 | 0 |
| 0.005550861 | 0 | 0 | 0 | 0 |
| 0 | 0 | 0 | 0.005964911 | 0 |
| 0.126275569 | 0.0045788 | 0.115752082 | 0.063608592 | 0 |
| 0 | 0 | 0 | 0 | 0 |
| 0.112384991 | 16.4982384 | 0.152934499 | 4.138887855 | 0.008273614 |
| 0 | 0 | 0 | 0 | 0 |
| 0 | 0 | 0 | 0 | 0 |
| 0 | 0 | 0 | 0 | 0 |
| 0 | 0 | 0 | 0 | 0 |
| 0.040233423 | 0 | 0 | 0.009819253 | 0.000494973 |

| | | | | |
|---|---|---|---|---|
| 0 | 0 | 0 | 0 | 0 |
| 0 | 0 | 0 | 0 | 0 |
| 0.004715772 | 0.012531541 | 0.020053834 | 0.302613531 | 0.112690818 |
| 0 | 0 | 0 | 0 | 0 |
| 0 | 0 | 0 | 0 | 0 |
| 0.001724777 | 0 | 0.002154223 | 0 | 0 |
| 0 | 0 | 0 | 0 | 0 |
| 0 | 0 | 0 | 0.00687496 | 0 |
| 0 | 0 | 0 | 0 | 0 |
| 0 | 0 | 0.935030169 | 0 | 0.158005978 |
| 0.038197928 | 0.011494294 | 0.002983229 | 0.012387894 | 0 |
| 0.827079911 | 4.135713823 | 0.300139215 | 2.145428045 | 0.648590183 |
| 0 | 0 | 0 | 0 | 0 |
| 0.016061595 | 0 | 0.004024527 | 0.000636644 | 0.00138264 |
| 0.055269568 | 0.006884298 | 0.09260727 | 0.035620682 | 0.361044894 |
| 1.595998437 | 0 | 0 | 0 | 0 |
| 0 | 0 | 0 | 0 | 0 |
| 0 | 0 | 0 | 0 | 0 |
| 0 | 0 | 0.868217514 | 0 | 0 |
| 0.300756738 | 0.008668137 | 0.006634967 | 0.003077507 | 0 |
| 0.294653372 | 0.000814133 | 0.030629714 | 0.08899611 | 0 |
| 0 | 17.24909691 | 0.055063765 | 0.001562905 | 0.956563971 |
| 0.035082946 | 1.217533684 | 0.434548028 | 2.197364063 | 27.95175265 |
| 0 | 0 | 0 | 0 | 0 |
| 0 | 0 | 0 | 0 | 0 |
| 0.166410766 | 0.0040649 | 0.000898863 | 0.017774006 | 0.001988533 |
| 0 | 0 | 0 | 0 | 0 |
| 0.007083061 | 0 | 0 | 0 | 0 |
| 0 | 0 | 0 | 0 | 0.039637929 |
| 0 | 0 | 0 | 0 | 0 |
| 0.016815359 | 0 | 0 | 0 | 0 |
| 0 | 0 | 0.172202483 | 0.20180155 | 0 |
| 0 | 0 | 0 | 0 | 0 |
| 0.226637242 | 0 | 0 | 0.00261801 | 0 |
| 0.015468225 | 0.004389525 | 0 | 0.004398498 | 0 |
| 0.010626198 | 0.05261994 | 0.142251698 | 0.034977702 | 0.002494749 |
| 0.059803395 | 0.008593061 | 0.002952237 | 0.005307013 | 0.088956488 |
| 0 | 0 | 0 | 0 | 0 |
| 0.67940848 | 0 | 0 | 0 | 0 |
| 0 | 0 | 0 | 0 | 0 |
| 0.026985848 | 0 | 0.025810365 | 0 | 0 |
| 0 | 0 | 0 | 0 | 0 |
| 0 | 0 | 0 | 0 | 0 |
| 0 | 0 | 0 | 0 | 0 |
| 0.505793622 | 0.125000397 | 0.063843195 | 0.001044191 | 0.087575832 |
| 0.018325246 | 0 | 0 | 0 | 0 |
| 0 | 0 | 0 | 0 | 0 |
| 0.359084177 | 0.164680883 | 0.037455133 | 0.114873675 | 0 |

| | | | | |
|---|---|---|---|---|
| 0 | 0 | 0 | 0 | 0 |
| 0 | 0 | 0 | 0 | 0 |
| 0 | 0 | 0 | 0 | 0 |
| 0.001496826 | 0 | 0 | 0.002135903 | 0 |
| 0 | 0 | 0 | 0 | 0 |
| 0 | 0 | 0 | 0 | 0 |
| 0.600434078 | 0 | 0.278052861 | 0 | 0 |
| 0.015701799 | 0 | 0.085355202 | 0.006878963 | 0 |
| 0 | 0 | 0 | 0 | 0 |
| 0 | 0 | 0 | 0 | 0 |
| 0.001333654 | 0.001795321 | 0.003909807 | 0 | 0.003405033 |
| 0 | 0 | 0 | 0 | 0 |
| 0.620934298 | 0 | 0 | 0 | 0 |
| 0.335154351 | 0 | 0 | 0 | 0 |
| 0 | 0 | 0 | 0 | 0 |
| 0 | 0 | 0 | 0 | 0 |
| 0.06906017 | 0 | 0.016776154 | 0 | 0 |
| 0.002498032 | 0.001358694 | 0.002503711 | 0.00264043 | 0 |
| 0.511115235 | 0 | 0 | 0 | 0.04566715 |
| 0.039299046 | 0 | 0 | 0 | 0 |
| 0 | 0 | 0 | 0 | 0 |
| 0 | 0 | 0 | 0 | 0 |
| 0.147042688 | 0 | 0.140288765 | 0 | 0 |
| 0 | 0 | 0 | 0 | 0 |
| 0 | 0 | 0 | 0 | 0 |
| 0 | 0 | 0 | 0.008620583 | 0 |
| 0 | 0 | 0 | 0 | 0 |
| 0 | 0 | 0 | 0 | 0 |
| 0 | 0 | 0 | 0 | 0 |
| 0.375020731 | 0.023306323 | 0 | 0.056017629 | 0 |
| 0 | 0.062216659 | 0 | 0 | 0 |
| 0 | 0 | 0 | 0 | 0 |
| 0 | 0 | 0 | 0 | 0 |
| 0 | 0 | 0 | 0 | 0 |
| 0 | 0 | 0.004543759 | 0 | 0 |
| 0 | 0 | 0 | 0 | 0 |
| 0.023279368 | 0.076087565 | 0 | 0 | 0 |
| 0.002116803 | 0.004697472 | 0.026223164 | 0.001174671 | 0.004373328 |
| 0 | 0 | 0 | 0 | 0 |
| 0 | 0 | 0.000502235 | 0 | 0 |
| 0 | 0 | 0 | 0 | 0 |
| 0 | 0 | 0 | 0 | 0 |
| 0 | 0 | 0 | 0 | 0 |
| 0.003289066 | 0 | 0 | 0 | 0 |
| 0.037175801 | 0.352353203 | 0.252952575 | 0 | 0 |
| 0 | 0 | 0 | 0 | 0 |
| 0.135428697 | 0 | 0 | 0 | 0 |
| 0 | 0 | 0 | 0 | 0 |

| | | | | |
|---|---|---|---|---|
| 0 | 0 | 0 | 0 | 0 |
| 0 | 0 | 0 | 0 | 0 |
| 0 | 0 | 0 | 0 | 0 |
| 0.653998891 | 0 | 0 | 0 | 0 |
| 0 | 0 | 0 | 0 | 0 |
| 0 | 0 | 0 | 0 | 0 |
| 0 | 0 | 0 | 0 | 0 |
| 0 | 0 | 0 | 0 | 0 |
| 0.002919425 | 0.491230207 | 0.024403355 | 0.018977937 | 0.278959144 |
| 0 | 0 | 0.01442083 | 0 | 0 |
| 0 | 0 | 0 | 0 | 0 |
| 0 | 0 | 0 | 0 | 0 |
| 0 | 0 | 0 | 0.00783097 | 0 |
| 0 | 0 | 0 | 0 | 0.001999448 |
| 0.349802482 | 0 | 0.115693041 | 0 | 0 |
| 0 | 0.010427052 | 0 | 0.003567518 | 0 |
| 0 | 0 | 0 | 0 | 0 |
| 0 | 0 | 0 | 0 | 0 |
| 0 | 0 | 0 | 0 | 0 |
| 0 | 0 | 0 | 0 | 0 |
| 0.259693101 | 0 | 0.122161703 | 0 | 0 |
| 0 | 0 | 0 | 0 | 0 |
| 0 | 0 | 0 | 0 | 0 |
| 0 | 0 | 0.155452094 | 0 | 0 |
| 0.018767944 | 0.001792623 | 0.05945988 | 0.001360824 | 0.000806015 |
| 0.280875841 | 0 | 13.42453851 | 1.507350661 | 0 |
| 1.61863942 | 0 | 0 | 0 | 0 |
| 0.005682993 | 0.000695478 | 0 | 0 | 0 |
| 0.112258492 | 0 | 0 | 0 | 0 |
| 0 | 0 | 0 | 0 | 0 |
| 0 | 0 | 0 | 0 | 0 |
| 0.321033466 | 0.027418403 | 0.003855841 | 0 | 0 |
| 0 | 0 | 0 | 0.000790777 | 0 |
| 0.025058217 | 0 | 0.009151728 | 0.004505958 | 0.031423845 |
| 0.00120843 | 0 | 0.008175447 | 0.002873959 | 0 |
| 0 | 0.001951369 | 0.003981123 | 0.005586737 | 0.0015041 |
| 0 | 0.375278303 | 0.001325293 | 0 | 0.253213807 |
| 0.98191672 | 0.081841784 | 0.779150173 | 5.417188279 | 0 |
| 0.003056693 | 0 | 0.002270699 | 0 | 0 |
| 0 | 0 | 0 | 0 | 0 |
| 0 | 0 | 0 | 0 | 0 |
| 0 | 0 | 0 | 0 | 0 |
| 0.149414147 | 0 | 2.380910269 | 0.756728604 | 0 |
| 0 | 0 | 0 | 0 | 0 |
| 0 | 0 | 0 | 0 | 0 |
| 0 | 0.001452532 | 0.000851655 | 0 | 0.002754895 |
| 0 | 0 | 0 | 0 | 0 |
| 0.892804483 | 0 | 0.314968704 | 0.133378228 | 0 |

| | | | | |
|---|---|---|---|---|
| 0.001602323 | 0 | 0 | 0.002286443 | 0 |
| 0 | 0 | 0 | 0 | 0 |
| 0 | 0 | 0.003289914 | 0 | 0 |
| 0 | 0 | 0 | 0 | 0 |
| 0.00047859 | 0.000468554 | 0 | 0 | 0 |
| 0.034411967 | 0 | 0 | 0 | 0 |
| 0 | 0 | 0.007790068 | 0 | 0 |
| 0 | 0 | 0 | 0 | 0 |
| 0 | 0 | 0 | 0 | 0 |
| 0 | 0 | 0 | 0 | 0 |
| 0 | 0 | 0 | 0 | 0 |
| 1.222889152 | 0.080546423 | 0.031464105 | 0.335322693 | 0.216648045 |
| 0 | 0 | 1.061654146 | 0 | 0.002176774 |
| 0 | 0 | 0 | 0 | 0 |
| 0.450355343 | 0.017159604 | 0.217349888 | 1.198746461 | 0.052757442 |
| 0 | 0 | 0 | 0 | 0 |
| 0 | 0 | 0 | 0 | 0.008135561 |
| 0 | 0 | 0 | 0 | 0 |
| 0 | 0 | 0 | 0 | 0 |
| 0.003227596 | 0 | 0 | 0 | 0 |
| 0.41329158 | 0 | 0 | 0.034228471 | 0 |
| 0 | 0 | 0 | 0 | 0 |
| 0.068557205 | 0 | 4.186232966 | 0 | 0 |
| 0 | 0 | 0 | 0 | 0 |
| 0 | 0 | 0 | 0 | 0 |
| 0 | 0 | 0 | 0 | 0 |
| 0 | 0 | 0 | 0 | 0 |
| 0 | 0 | 0 | 0 | 0 |
| 0 | 0 | 0 | 0 | 0 |
| 0 | 0 | 0 | 0 | 0 |
| 0 | 0 | 0 | 0 | 0 |
| 0.017955726 | 0.004453397 | 0.011661762 | 0.02818422 | 0 |
| 0.575145983 | 0 | 5.678989872 | 0 | 0 |
| 0.028226768 | 0 | 0.021901502 | 0.001991161 | 0 |
| 0 | 0.002881999 | 0.002096351 | 0.002947767 | 0.002073327 |
| 0.098665389 | 0 | 0.01639487 | 0 | 0 |
| 0 | 0 | 0 | 0 | 0 |
| 0 | 0 | 0 | 0 | 0 |
| 0.001957948 | 0 | 0 | 0.001058297 | 0 |
| 0 | 0 | 0 | 0 | 0 |
| 0.175432702 | 0 | 0.092639003 | 0.186636479 | 0 |
| 0 | 0 | 0 | 0 | 0 |
| 0 | 0 | 0 | 0 | 0 |
| 0 | 0 | 0.391385541 | 0 | 0.000799522 |
| 0.010531097 | 0.004930995 | 0.00936186 | 0.016152645 | 0.107281597 |
| 0 | 0 | 0 | 0 | 0 |
| 0 | 0.579955885 | 0 | 0 | 0 |
| 0.010826667 | 0.001695942 | 0.000520861 | 0 | 0.000610035 |

| | | | | |
|---|---|---|---|---|
| 0 | 0 | 0 | 0.044371048 | 0 |
| 0 | 0 | 0 | 0 | 0 |
| 0 | 0.008654347 | 0 | 0 | 0.001778854 |
| 0 | 0 | 0 | 0 | 0 |
| 0 | 0 | 0 | 0 | 0 |
| 0 | 0 | 0 | 0 | 0 |
| 0 | 0 | 0 | 0 | 0 |
| 0.001490765 | 0 | 0 | 0 | 0 |
| 0 | 0 | 0 | 0 | 0 |
| 0 | 0 | 0.054915874 | 0 | 0 |
| 0 | 0 | 0 | 0 | 0 |
| 0 | 0 | 0 | 0 | 0 |
| 0 | 0 | 0 | 0 | 0 |
| 0 | 0 | 0.255968768 | 0.0445182 | 0 |
| 0 | 0 | 0 | 0 | 0 |
| 0 | 0 | 0 | 0 | 0 |
| 0 | 0 | 0 | 0 | 0 |
| 0 | 0 | 0 | 0 | 0 |
| 0 | 0.036329282 | 0 | 0 | 0 |
| 0 | 0 | 0 | 0 | 0 |
| 0.001864495 | 0 | 0 | 0 | 0 |
| 0 | 0 | 0 | 0 | 0 |
| 0 | 0 | 0 | 0 | 0 |
| 0 | 0 | 0 | 0 | 0 |
| 0 | 0 | 0 | 0 | 0 |
| 0 | 0.01154598 | 0.001922545 | 0.002703371 | 0.000600452 |
| 0 | 0 | 0 | 0 | 0 |
| 0.304992851 | 0 | 0.081544202 | 0.133295404 | 0 |
| 0 | 0.004755536 | 0 | 0 | 0 |
| 0.427531595 | 0 | 0.973064978 | 1.436206667 | 0 |
| 0 | 0 | 0 | 0 | 0 |
| 0 | 0 | 0 | 0 | 0 |
| 0.002790185 | 0 | 0.004238948 | 0 | 0 |
| 0 | 0 | 0 | 0 | 0 |
| 0 | 0.004810634 | 0 | 0.013279527 | 0 |
| 0.000598587 | 0 | 0 | 0 | 0 |
| 0 | 0 | 0 | 0 | 0 |
| 0.004899751 | 0.00082707 | 0.000762035 | 0 | 0.012732976 |
| 0 | 0 | 0 | 0 | 0 |
| 0 | 0 | 0 | 0 | 0 |
| 1.66689374 | 1.488633995 | 5.193588826 | 0.791839196 | 1.699983262 |
| 0.475291617 | 0 | 3.186003609 | 0.421938763 | 0 |
| 0.184065996 | 0 | 0 | 0 | 0 |
| 0 | 0 | 0 | 0 | 0 |
| 0 | 0 | 0 | 0 | 0 |
| 0 | 0 | 0 | 0 | 0 |
| 0 | 0 | 0 | 0 | 0 |
| 0.015554854 | 0.002494352 | 0.033626482 | 0 | 0.002644455 |

| | | | | |
|---|---|---|---|---|
| 0 | 0 | 0 | 0 | 0 |
| 0.012413367 | 0 | 0.001990654 | 0.000656049 | 0 |
| 0 | 0 | 0 | 0 | 0 |
| 0 | 0 | 0 | 0 | 0 |
| 1.724703253 | 0 | 2.027437993 | 5.248059132 | 0 |
| 0 | 0 | 0 | 0 | 0 |
| 0 | 0 | 0 | 0 | 0 |
| 0 | 0 | 0.000527652 | 0 | 0 |
| 0.00137723 | 0.447382327 | 0.074539466 | 0.647220652 | 1.327944507 |
| 0 | 0 | 0 | 0 | 0.011188393 |
| 0 | 0.071550794 | 0 | 0 | 0 |
| 0 | 0 | 0 | 0 | 0 |
| 0.143132195 | 0 | 0 | 0 | 0 |
| 0.004448591 | 0.112108775 | 0.014713723 | 0.001763317 | 0 |
| 0.091324355 | 0 | 0 | 0 | 0 |
| 0.175493511 | 0 | 0.01487904 | 1.147603914 | 0 |
| 0.116912722 | 0.002372974 | 0.226997636 | 0.001356335 | 0 |
| 0 | 0 | 0 | 0 | 0 |
| 0 | 0 | 0 | 0 | 0 |
| 0 | 0 | 0 | 0 | 0 |
| 0 | 0 | 0 | 0 | 0 |
| 0 | 0 | 0 | 0 | 0 |
| 0 | 0 | 0 | 0 | 0 |
| 0 | 0 | 0 | 0 | 0 |
| 0 | 0 | 0 | 0 | 0 |
| 0 | 0 | 0 | 0 | 0 |
| 0.083435928 | 0 | 0.155446712 | 0 | 0 |
| 0.00041968 | 0.001780476 | 0 | 0 | 0.000591178 |
| 0 | 0 | 0 | 0 | 0 |
| 0 | 0 | 0 | 0 | 0 |
| 0 | 0 | 0 | 0 | 0 |
| 0.002804491 | 0 | 0 | 0 | 0.001481445 |
| 0 | 0.029378617 | 0 | 0 | 0 |
| 0.042380519 | 0 | 0 | 0 | 0 |
| 0.079436631 | 0.044489011 | 0 | 0 | 0 |
| 0 | 0 | 0 | 0 | 0 |
| 0.122566637 | 0 | 0 | 0.001376833 | 0 |
| 0 | 0 | 0 | 0.017989483 | 0 |
| 0 | 0 | 0 | 0 | 0 |
| 0 | 0 | 0 | 0 | 0 |
| 0.422952182 | 2.388447757 | 0.708158385 | 8.928331905 | 0.548046371 |
| 0 | 0 | 0 | 0.011534715 | 0 |
| 0 | 0 | 0 | 0 | 0 |
| 0 | 0 | 0.0054056 | 0 | 0 |
| 0 | 0 | 0 | 0 | 0 |
| 0 | 0 | 0 | 0 | 0 |
| 0.591305326 | 0 | 0 | 0.003967815 | 0 |
| 0 | 0 | 0 | 0 | 0 |

| | | | | |
|---|---|---|---|---|
| 0.109363333 | 0 | 0.006079319 | 0.004371333 | 0 |
| 0 | 0 | 0 | 0 | 0 |
| 0 | 0 | 0.014714831 | 0 | 0 |
| 0.002852488 | 0 | 0.000428846 | 0 | 0 |
| 0.074145242 | 0 | 0.003009025 | 0 | 0 |
| 0 | 0 | 0 | 0 | 0 |
| 0.176507188 | 0.001810215 | 0.431080716 | 0.000845648 | 0 |
| 0.001237356 | 0 | 0 | 0 | 0 |
| 0.001204695 | 0 | 0 | 0 | 0 |
| 0 | 0 | 0 | 0.000709778 | 0 |
| 0.006381646 | 0 | 0 | 0 | 0 |
| 0 | 0 | 0 | 0 | 0 |
| 0 | 0 | 0 | 0 | 0 |
| 0 | 0 | 0 | 0 | 0 |
| 0 | 0 | 0 | 0 | 0 |
| 0 | 0 | 0 | 0 | 0 |
| 0 | 0.223192296 | 0.001979442 | 0 | 51.67232942 |
| 0 | 0 | 0.001233449 | 0 | 0.006548948 |
| 0 | 0 | 0 | 0 | 0 |
| 0 | 0 | 0 | 0 | 0 |
| 0.222750603 | 0 | 0.235010225 | 0.212386491 | 0 |
| 0 | 0 | 0 | 0 | 0 |
| 0.010156312 | 0 | 0 | 0.005683374 | 0 |
| 0.012085734 | 0.004420859 | 0 | 0.003632357 | 0.001309573 |
| 0 | 0 | 0 | 0 | 0 |
| 0 | 0 | 0 | 0 | 0 |
| 0 | 0 | 0 | 0 | 0 |
| 0 | 0 | 0 | 0 | 0 |
| 0 | 0 | 0 | 0 | 0 |
| 0.21847885 | 0.262927259 | 0.07483101 | 1.614485291 | 0.069602345 |
| 0 | 0 | 0 | 0 | 0 |
| 0 | 0 | 0 | 0 | 0 |
| 0 | 0 | 0 | 0 | 0 |
| 0 | 0 | 5.877445599 | 0.540043585 | 0.693678516 |
| 0 | 0 | 0 | 0 | 0 |
| 0 | 0 | 0 | 0 | 0 |
| 0 | 0 | 0 | 0 | 0 |
| 0 | 0 | 0 | 0 | 0 |
| 0 | 0 | 0 | 0 | 0 |
| 0 | 0 | 0 | 0 | 0 |
| 0 | 0 | 0 | 0 | 0 |
| 0 | 0 | 0 | 0 | 0 |
| 0 | 0 | 0 | 0 | 0 |
| 0 | 0 | 0 | 0 | 0 |
| 0 | 0 | 0 | 0 | 0 |
| 0 | 0 | 0 | 0 | 0 |
| 0 | 0 | 0 | 0 | 0 |
| 0.009345325 | 0 | 0 | 0 | 0 |

| | | | | |
|---|---|---|---|---|
| 0.008002879 | 0.038195921 | 0.001160191 | 0.003738609 | 0.009662725 |
| 0.064514117 | 0 | 0 | 0 | 0 |
| 0.032438027 | 0 | 0 | 0 | 0 |
| 0 | 0 | 0 | 0 | 0 |
| 0 | 0 | 0 | 0 | 0 |
| 0 | 0 | 0 | 0 | 0 |
| 0 | 0 | 0 | 0 | 0 |
| 0.540717687 | 0.014653095 | 0.190997692 | 1.499014762 | 0 |
| 0.052490047 | 0.10541404 | 0.213472662 | 0.032809176 | 0.000789953 |
| 0.008120444 | 0.094331706 | 0 | 0.001299881 | 0.000549942 |
| 0.076966591 | 0 | 0 | 0 | 0 |
| 0 | 0 | 0 | 0 | 0 |
| 0 | 0 | 0 | 0 | 0 |
| 0 | 0 | 0 | 0 | 0.224218476 |
| 0 | 0 | 0.002832477 | 0 | 0 |
| 0 | 0 | 0 | 0 | 0 |
| 0 | 0.032064672 | 0 | 0 | 0.003526245 |
| 0 | 0 | 0 | 0 | 0 |
| 0 | 0 | 0 | 0 | 0 |
| 0 | 0 | 0.000817664 | 0 | 0 |
| 0 | 0 | 0 | 0 | 0 |
| 0 | 0 | 0 | 0 | 0 |
| 0 | 0 | 0 | 0 | 0 |
| 0 | 0 | 0 | 0 | 0 |
| 0 | 0 | 0.010559595 | 0 | 0 |
| 0 | 0 | 0 | 0 | 0 |
| 0 | 0 | 0 | 0 | 0 |
| 0 | 0 | 0 | 0 | 0 |
| 0 | 0 | 0 | 0 | 0 |
| 0 | 0 | 0 | 0 | 0 |
| 0 | 0 | 0 | 0.061801597 | 0 |
| 0 | 0 | 0 | 0 | 0 |
| 0 | 0 | 0 | 0 | 0 |
| 0.001249965 | 0 | 0.042230983 | 0 | 0 |
| 0.002768175 | 0.007960998 | 0 | 0.000617197 | 0 |
| 0 | 0 | 0 | 0 | 0 |
| 0 | 0 | 0 | 0 | 0 |
| 0 | 0 | 0 | 0.086711956 | 0 |
| 0 | 0 | 0 | 0.001818613 | 0 |
| 0 | 0 | 0 | 0.309102491 | 0 |
| 0 | 0 | 0 | 0 | 0 |
| 0 | 0.145109071 | 0.131972688 | 0.698871596 | 0.027782666 |
| 0 | 0 | 0 | 0 | 0 |
| 0 | 0 | 0 | 0 | 0 |
| 0 | 0 | 0 | 0 | 0 |
| 0 | 0 | 0 | 0 | 0 |
| 0 | 0 | 0 | 0 | 0 |
| 0 | 0 | 0 | 0 | 0 |

| | | | | |
|---|---|---|---|---|
| 0 | 0 | 0 | 0 | 0 |
| 0 | 0 | 0.01512511 | 0.003681009 | 0 |
| 0 | 0 | 0 | 0 | 0 |
| 0 | 0 | 0 | 0 | 0 |
| 0.132906069 | 0.010063483 | 0.405534969 | 0.053395638 | 0 |
| 0 | 0 | 0 | 0 | 0 |
| 0 | 0 | 0 | 0 | 0 |
| 0.005999071 | 0.038977169 | 0.055754212 | 0.018806952 | 0.029832946 |
| 0.000544532 | 0 | 0.003765811 | 0 | 0 |
| 0 | 0 | 0 | 0 | 0 |
| 0 | 0 | 0 | 0 | 0 |
| 0 | 0 | 0 | 0 | 0 |
| 0 | 0 | 0 | 0 | 0 |
| 0 | 0 | 0 | 0 | 0 |
| 0 | 0 | 0 | 0 | 0 |
| 0 | 0 | 0.102968156 | 0 | 0.01499131 |
| 0 | 0 | 0 | 0 | 0 |
| 0 | 0 | 0 | 0 | 0 |
| 0 | 0 | 0 | 0 | 0 |
| 0 | 0 | 0 | 0 | 0 |
| 0 | 0 | 0 | 0 | 0 |
| 0 | 0 | 0 | 0 | 0 |
| 0 | 0 | 0 | 0 | 0 |
| 0.000471973 | 0.000924152 | 0.001561051 | 0 | 0.006205174 |
| 0.032194142 | 0.523627166 | 0 | 0 | 0 |
| 0 | 0 | 0 | 0 | 0 |
| 0.005180073 | 0.005071449 | 0.01414779 | 0.095989865 | 0 |
| 0 | 0 | 0.000546685 | 0.00090084 | 0.002027553 |
| 0 | 0 | 0 | 0 | 0 |
| 0 | 0 | 0 | 0 | 0 |
| 0 | 0 | 0 | 0 | 0 |
| 0 | 0 | 0 | 0 | 0 |
| 0 | 0 | 0 | 0 | 0 |
| 0 | 0 | 0 | 0 | 0 |
| 0 | 0 | 0 | 0 | 0 |
| 0 | 0 | 0 | 0 | 0 |
| 0 | 0 | 0 | 0 | 0 |
| 0 | 0 | 0 | 0.006843647 | 0 |
| 0 | 0 | 1.526608916 | 0 | 0 |
| 0 | 0 | 0 | 0 | 0 |
| 0 | 0 | 0 | 0 | 0 |
| 0.030119052 | 0.070571121 | 0 | 0 | 0 |
| 0 | 0 | 0 | 0 | 0 |
| 0 | 0 | 0 | 0 | 0 |
| 0 | 0 | 0 | 0 | 0 |
| 0 | 0 | 0 | 0 | 0 |
| 0 | 0 | 0 | 0 | 0 |
| 0 | 0 | 0 | 0 | 0 |

| | | | | |
|---|---|---|---|---|
| 0 | 0 | 1.716804542 | 0.019148223 | 0 |
| 0 | 0 | 0 | 0 | 0 |
| 0.007104067 | 0 | 0 | 0 | 0 |
| 0 | 0 | 0 | 0 | 0 |
| 0 | 0 | 0 | 0 | 0 |
| 0 | 0.496524346 | 0 | 0 | 0.054988217 |
| 0 | 0 | 0 | 0 | 0 |
| 0 | 0 | 0 | 0 | 0 |
| 0 | 0 | 0 | 0 | 0 |
| 0 | 0 | 0 | 0 | 0 |
| 0 | 0 | 0 | 0 | 0 |
| 0 | 0 | 0 | 0 | 0.0117468 |
| 0 | 0 | 0 | 0 | 0 |
| 0 | 0 | 0 | 0 | 0 |
| 0 | 0 | 0 | 0 | 0 |
| 0 | 0 | 0 | 0 | 0 |
| 0 | 0 | 0 | 0 | 0 |
| 0 | 0 | 0 | 0 | 0 |
| 0 | 0 | 0 | 0.004774787 | 0 |
| 0 | 0 | 0 | 0 | 0 |
| 0 | 0.076230546 | 0 | 0 | 0.003641335 |
| 0 | 0 | 0 | 0 | 0 |
| 0.01780727 | 0.003293062 | 0.024094466 | 0.001646956 | 0.002229692 |
| 0.201734772 | 0 | 0 | 0 | 0 |
| 0 | 0 | 0 | 0 | 0 |
| 0 | 0 | 0 | 0 | 0 |
| 0 | 0 | 0 | 0 | 0 |
| 0 | 0 | 0 | 0 | 0 |
| 0 | 0 | 0 | 0 | 0.00168893 |
| 0 | 0 | 0 | 0 | 0 |
| 0 | 0 | 0 | 0 | 0 |
| 0 | 0 | 0 | 0 | 0 |
| 0.002038231 | 0.434472544 | 0.027411528 | 0.039220201 | 0.005089738 |
| 0 | 0 | 0 | 0 | 0 |
| 0 | 0 | 1.017408995 | 0 | 0.012374132 |
| 0 | 0 | 0.011266553 | 0.00406668 | 0 |
| 0 | 0 | 0 | 0 | 0 |
| 0 | 0 | 0 | 0 | 0 |
| 0.009975073 | 0.007408613 | 0.011480175 | 0.004703759 | 0 |
| 0 | 0 | 0 | 0 | 0 |
| 0 | 0 | 0 | 0 | 0 |
| 0 | 0 | 0.315483845 | 0 | 0 |
| 0.034388246 | 0.010237959 | 0 | 0 | 0 |
| 0 | 0 | 0 | 0 | 0 |
| 0 | 0 | 0 | 0 | 0 |
| 0.033858298 | 0 | 0.003457556 | 0 | 0 |
| 0 | 0 | 0 | 0 | 0 |
| 0 | 0 | 0 | 0 | 0 |

| | | | | |
|---|---|---|---|---|
| 0 | 0 | 0 | 0 | 0 |
| 0 | 0 | 0 | 0 | 0 |
| 0 | 0 | 0 | 0 | 0 |
| 0 | 0 | 0 | 0 | 0 |
| 0 | 0 | 0 | 0 | 0 |
| 0 | 0 | 0 | 0 | 0 |
| 0 | 0.117993428 | 0 | 0 | 0 |
| 0 | 0.626130212 | 0 | 0 | 0 |
| 0.003291915 | 0 | 0.000539902 | 0 | 0 |
| 0.000493495 | 0.002576784 | 0 | 0 | 0 |
| 0 | 0 | 0 | 0 | 0 |
| 0 | 0 | 0 | 0 | 0.014697483 |
| 0 | 0 | 0 | 0 | 0 |
| 0.216611809 | 0 | 0 | 0 | 0 |
| 0 | 0.000911047 | 0.000719493 | 0 | 0 |
| 0 | 0 | 0 | 0.000962293 | 0.007865519 |
| 0 | 0 | 0 | 0 | 0 |
| 0 | 0 | 0 | 0 | 0 |
| 0 | 0 | 0 | 0 | 0 |
| 0.935116663 | 0 | 0 | 0 | 0 |
| 0 | 0.004302853 | 0 | 0.013266615 | 0.000595288 |
| 0 | 0 | 0 | 0 | 0 |
| 0 | 0 | 0 | 0 | 0 |
| 0.107145878 | 0 | 0 | 0 | 0 |
| 0 | 0 | 0 | 0 | 0.007549167 |
| 0 | 0 | 0 | 0 | 0 |
| 0 | 0 | 0 | 0 | 0 |
| 0 | 0 | 0 | 0 | 0 |
| 0 | 0 | 0 | 0 | 0 |
| 0.008219424 | 0 | 0.192317828 | 0 | 0 |
| 0.125231558 | 0 | 0.0143579 | 1.041007845 | 0 |
| 0 | 0 | 0 | 0 | 0 |
| 0.004999605 | 0 | 0 | 0 | 0 |
| 0 | 0 | 0 | 0 | 0 |
| 0 | 0 | 0 | 0 | 0 |
| 0 | 0 | 0 | 0 | 0 |
| 0 | 0 | 0 | 0 | 0 |
| 0 | 0 | 0 | 0 | 0 |
| 0.006671575 | 0.005370487 | 0 | 0 | 0 |
| 0 | 0 | 0 | 0 | 0 |
| 0 | 0 | 0 | 0 | 0 |
| 0 | 0 | 0 | 0 | 0 |
| 0.01678884 | 0 | 0.016941089 | 0 | 0 |
| 0.003111788 | 0 | 0 | 0 | 0 |
| 0 | 0 | 0 | 0 | 0 |
| 0 | 0 | 0 | 0 | 0 |
| 0 | 0.063585771 | 0.040651387 | 0.17357324 | 0.001633714 |
| 0 | 0 | 0.003157981 | 0 | 0 |

| | | | | |
|---|---|---|---|---|
| 0 | 0 | 0 | 0 | 0 |
| 0 | 0 | 0 | 0 | 0 |
| 0 | 0 | 0 | 0 | 0 |
| 0 | 0 | 0 | 0 | 0 |
| 0.065230753 | 0.042898406 | 0.03584062 | 0.206062478 | 0.063466319 |
| 0 | 0 | 0 | 0 | 0 |
| 0 | 0 | 0 | 0 | 0 |
| 0 | 0 | 0 | 0 | 0 |
| 0 | 0 | 0 | 0 | 0 |
| 0 | 0 | 0 | 0 | 0 |
| 0 | 0 | 0 | 0 | 0 |
| 0.00704573 | 0.014805427 | 0.005270475 | 0.005517407 | 0.018397376 |
| 0 | 0 | 0.001069904 | 0 | 0.005952113 |
| 0 | 0 | 0 | 0 | 0 |
| 0.002619189 | 0.001666773 | 0 | 0 | 0.014665747 |
| 0 | 0 | 0.054004312 | 0 | 0 |
| 0.007005052 | 0.001552791 | 0.001311466 | 0.002514691 | 0.000651634 |
| 0.008817306 | 0 | 0 | 0 | 0.004055645 |
| 0.001171112 | 0.008025881 | 0.082164227 | 0.392250136 | 0.300149409 |
| 0.166333822 | 0.001330712 | 0.070345977 | 0.001818317 | 0 |
| 0 | 0 | 0 | 0 | 0 |
| 0 | 0 | 0 | 0 | 0 |
| 0 | 0 | 0 | 0 | 0 |
| 0 | 0 | 0 | 0 | 0 |
| 0 | 0 | 0 | 0 | 0 |
| 0 | 0 | 0 | 0 | 0 |
| 0.018107959 | 0.036122955 | 0.001228136 | 0.003238001 | 0.001821972 |
| 0 | 0 | 0 | 0 | 0 |
| 0 | 0 | 0 | 0 | 0 |
| 0 | 0 | 0 | 0 | 0 |
| 0 | 0.000604388 | 0 | 0 | 0.0008696 |
| 0 | 0 | 0 | 0 | 0 |
| 0 | 0 | 0 | 0 | 0 |
| 0 | 0 | 0 | 0 | 0 |
| 0 | 0 | 0 | 0 | 0 |
| 0 | 0 | 0 | 0 | 0 |
| 0 | 0 | 0 | 0 | 0 |
| 0.031770008 | 0.008386349 | 0.012715158 | 0.005931119 | 0.07109999 |
| 0.017737003 | 0 | 0 | 0 | 0 |
| 0 | 0 | 0 | 0 | 0 |
| 0 | 0.027236402 | 0 | 0 | 0.003812068 |
| 0 | 0 | 0 | 0 | 0 |
| 0 | 0 | 0 | 0 | 0 |
| 0 | 0 | 0.015664126 | 0.010515877 | 0.010758395 |
| 0 | 0 | 0 | 0 | 0 |
| 0 | 0 | 0 | 0 | 0 |
| 0 | 0 | 0 | 0 | 0 |
| 0 | 0 | 0 | 0 | 0 |

| | | | | |
|---|---|---|---|---|
| 0 | 0 | 0 | 0 | 0 |
| 0 | 0 | 1.988276474 | 0 | 0 |
| 0 | 0 | 0 | 0 | 0 |
| 0 | 0 | 0.001064005 | 0.001001881 | 0.000712096 |
| 0 | 0.000918059 | 0.000422935 | 0.000557537 | 0 |
| 0 | 0.0045419 | 0 | 0.003357923 | 0 |
| 0.009320802 | 0 | 0.001180041 | 0 | 0 |
| 0 | 0 | 0 | 0.004544633 | 0 |
| 0.045858673 | 0 | 0 | 0 | 0 |
| 0 | 0 | 0.008255345 | 0 | 0 |
| 0 | 0 | 0 | 0 | 0 |
| 0 | 0 | 0 | 0 | 0 |
| 0.004777898 | 0 | 0.051287618 | 0.000852231 | 0 |
| 0 | 0 | 0.000456297 | 0 | 0 |
| 0 | 0 | 0 | 0 | 0 |
| 0 | 0 | 0.065974093 | 0 | 0 |
| 0 | 0 | 0 | 0 | 0 |
| 0 | 0 | 0 | 0 | 0 |
| 0 | 0 | 0 | 0 | 0 |
| 0 | 0 | 0 | 0 | 0 |
| 0 | 0 | 0 | 0 | 0 |
| 0 | 0 | 0 | 0 | 0 |
| 0.016986754 | 0.085308546 | 0 | 0.013653331 | 0.011299442 |
| 0.02078736 | 0 | 0.01942084 | 0 | 0 |
| 0 | 0 | 0 | 0 | 0 |
| 0.014803392 | 0 | 0 | 0 | 0 |
| 0.002554624 | 0 | 0 | 0 | 0 |
| 0.003350543 | 0 | 0 | 0 | 0 |
| 0 | 0 | 0 | 0 | 0 |
| 0 | 0 | 0 | 0 | 0 |
| 0.011223796 | 0 | 0.028000241 | 0.04671291 | 0 |
| 0 | 0 | 0 | 0 | 0 |
| 0 | 0 | 0 | 0 | 0 |
| 0 | 0 | 0 | 0 | 0 |
| 2.501824782 | 0 | 0 | 0 | 0 |
| 0 | 0 | 0 | 0 | 0 |
| 0 | 0 | 0 | 0 | 0.040418453 |
| 0 | 0.01319653 | 0 | 0 | 0 |
| 0.137248463 | 0 | 0.00307838 | 0.018878985 | 0 |
| 0 | 0 | 0 | 0 | 0 |
| 0 | 0 | 0 | 0 | 0 |
| 0 | 0 | 0 | 0 | 0 |
| 0 | 0 | 0 | 0 | 0 |
| 0 | 0 | 0 | 0 | 0 |
| 0 | 0 | 0 | 0 | 0 |
| 0.545425362 | 0 | 1.032278931 | 0.26964714 | 0.002482566 |
| 0 | 0 | 0 | 0 | 0 |
| 0 | 0 | 0 | 0 | 0 |

| | | | | |
|---|---|---|---|---|
| 0 | 0 | 0 | 0 | 0 |
| 0 | 0 | 0 | 0 | 0 |
| 0 | 0 | 0 | 0 | 0 |
| 0 | 0 | 0 | 0 | 0 |
| 0 | 0.031542168 | 0 | 0 | 0.005352896 |
| 0 | 0 | 0 | 0 | 0 |
| 0 | 0 | 0 | 0 | 0 |
| 0 | 0 | 0 | 0 | 0 |
| 0.019494886 | 0 | 0.030340807 | 0 | 0 |
| 0.008466817 | 0 | 0 | 0 | 0 |
| 0.003490825 | 0.001649887 | 0.00195448 | 0 | 0.001017376 |
| 0 | 0 | 0 | 0 | 0.000886052 |
| 0 | 0 | 0 | 0 | 0 |
| 0 | 0 | 0 | 0 | 0 |
| 0 | 0 | 0 | 0 | 0 |
| 0 | 0 | 0 | 0 | 0 |
| 0.002477209 | 0 | 0 | 0 | 0 |
| 0 | 0 | 0 | 0 | 0 |
| 0 | 0 | 0 | 0 | 0 |
| 0 | 0.017035404 | 0 | 0 | 0 |
| 0 | 0 | 0 | 0 | 0 |
| 0 | 0 | 0 | 0 | 0 |
| 0 | 0 | 0 | 0 | 0 |
| 0 | 0 | 0 | 0 | 0 |
| 0 | 0 | 0 | 0 | 0 |
| 0 | 0 | 0 | 0 | 0 |
| 0 | 0 | 0 | 0 | 0 |
| 0.010953156 | 0 | 0 | 0 | 0 |
| 0 | 0 | 0 | 0 | 0 |
| 0 | 0 | 0 | 0 | 0 |
| 0.002880014 | 0.006042045 | 0.001608227 | 0 | 0 |
| 0 | 0 | 0 | 0 | 0 |
| 0.010828157 | 0.230729678 | 1.069971732 | 0.030486011 | 0.072003036 |
| 0.000650894 | 0 | 0 | 0 | 0 |
| 0 | 0 | 0 | 0 | 0 |
| 0 | 0 | 0 | 0 | 0 |
| 0 | 0 | 0 | 0 | 0 |
| 0 | 0 | 0 | 0 | 0 |
| 0 | 0 | 0 | 0 | 0 |
| 0 | 0 | 0 | 0 | 0 |
| 0 | 0 | 0 | 0 | 0 |
| 0 | 0.159165975 | 0.00685392 | 0.119483189 | 0.05729861 |
| 0.057296039 | 0.007187603 | 0.001439656 | 0.013854221 | 0.009667157 |
| 0 | 0 | 0 | 0 | 0 |
| 0 | 0 | 0 | 0 | 0 |
| 0.024216322 | 0.00338693 | 0 | 0 | 0 |
| 0 | 0 | 0 | 0 | 0.020873865 |
| 0 | 0 | 0 | 0 | 0 |

| | | | | |
|---|---|---|---|---|
| 0 | 0 | 0 | 0 | 0 |
| 0.00601924 | 0 | 0 | 0.00795295 | 0 |
| 0.003124937 | 0 | 0.015503603 | 0 | 0 |
| 0 | 0 | 0 | 0 | 0 |
| 0 | 0.011145229 | 0.001828698 | 0.029670106 | 0 |
| 0 | 0 | 0.009721779 | 0 | 0.004062678 |
| 0 | 0 | 0 | 0 | 0 |
| 0 | 0 | 0 | 0 | 0 |
| 0 | 0 | 0 | 0 | 0 |
| 0 | 0 | 0 | 0 | 0 |
| 0 | 0 | 0 | 0 | 0 |
| 0 | 0 | 0 | 0 | 0 |
| 0 | 0 | 0 | 0 | 0 |
| 0 | 0 | 0 | 0 | 0.565278677 |
| 0 | 0 | 0.00257438 | 0 | 0 |
| 0 | 0 | 0 | 0 | 0 |
| 0 | 0 | 0.524203524 | 0 | 0 |
| 0 | 0 | 0 | 0 | 0 |
| 0 | 0 | 0 | 0 | 0 |
| 0 | 0 | 0 | 0 | 0 |
| 0 | 0 | 0 | 0 | 0 |
| 0 | 0 | 0 | 0 | 0.000442604 |
| 0 | 0 | 0 | 0 | 0 |
| 0 | 0 | 0 | 0 | 0 |
| 0 | 0 | 0 | 0 | 0 |
| 0 | 0.014584592 | 0.001752751 | 0.09280823 | 0 |
| 0 | 0 | 0 | 0 | 0 |
| 0.000471215 | 0 | 1.029205826 | 0 | 0 |
| 0 | 0 | 0 | 0 | 0 |
| 0 | 0 | 0 | 0 | 0 |
| 0 | 0 | 0 | 0 | 0 |
| 0 | 0.152921802 | 0 | 0 | 0.003300386 |
| 0 | 0 | 0 | 0 | 0 |
| 0 | 0 | 0 | 0 | 0 |
| 0 | 0 | 0 | 0 | 0 |
| 0 | 0 | 0 | 0 | 0 |
| 0 | 0 | 0 | 0 | 0.000781997 |
| 0 | 0 | 0 | 0 | 0 |
| 0.265801258 | 7.988033606 | 0.217618539 | 2.893947607 | 1.48141705 |
| 0 | 0 | 0 | 0 | 0 |
| 0 | 0 | 0 | 0 | 0 |
| 0.007377813 | 0 | 0 | 0 | 0 |
| 0.004387718 | 0.027206156 | 0.002198847 | 0.002512161 | 0.00503614 |
| 0 | 0 | 0 | 0 | 0 |
| 0 | 0 | 0 | 0 | 0 |
| 0.005608793 | 0 | 0 | 0 | 0 |
| 0.013921911 | 0.013143188 | 0.001569776 | 0.002956242 | 0.004027252 |
| 0 | 0 | 0 | 0 | 0 |

| | | | | |
|---|---|---|---|---|
| 0 | 0 | 0.005716656 | 0 | 0 |
| 0.002195861 | 0.019809007 | 0.00452747 | 0.044762799 | 0.009058596 |
| 0 | 0 | 0 | 0 | 0 |
| 0 | 0 | 0 | 0 | 0 |
| 0 | 0 | 0 | 0 | 0 |
| 0 | 0 | 0 | 0 | 0 |
| 0 | 0 | 0 | 0 | 0 |
| 0 | 0 | 0 | 0 | 0 |
| 0 | 0 | 0 | 0 | 0 |
| 0 | 0 | 0 | 0 | 0 |
| 0 | 0 | 0 | 0 | 0 |
| 0 | 0 | 0 | 0 | 0 |
| 0.007792667 | 0 | 0 | 0.118689931 | 0 |
| 0 | 0 | 0 | 0 | 0 |
| 0.001564519 | 0.022592743 | 0.00123486 | 0.006744006 | 0.000964182 |
| 0 | 0 | 0 | 0 | 0 |
| 0 | 0 | 0 | 0 | 0 |
| 0 | 0 | 0 | 0 | 0 |
| 0.015576868 | 0 | 0 | 0 | 0.001066635 |
| 0 | 0 | 0 | 0 | 0 |
| 0 | 0 | 0 | 0 | 0 |
| 0.104288655 | 0 | 0.01905718 | 0 | 0 |
| 0.099765768 | 0.056494703 | 0 | 0 | 0.004573595 |
| 0 | 0 | 0 | 0 | 0 |
| 0 | 0 | 0 | 0 | 0 |
| 0 | 0 | 0 | 0 | 0 |
| 0 | 0.002153408 | 0 | 0 | 0 |
| 0 | 0 | 0 | 0 | 0 |
| 0 | 0 | 0 | 0 | 0 |
| 0 | 0 | 0 | 0 | 0 |
| 0 | 0 | 0 | 0 | 0 |
| 0 | 0 | 0 | 0 | 0 |
| 0 | 0 | 0 | 0 | 0 |
| 0 | 0 | 0 | 0 | 0 |
| 43.43088785 | 0 | 10.65038132 | 0 | 0 |
| 0 | 0 | 0 | 0 | 0 |
| 0 | 0 | 0 | 0 | 0 |
| 0 | 0 | 0 | 0 | 0 |
| 0.09321251 | 0.000938867 | 0.001730082 | 0.001368416 | 0 |
| 0 | 0 | 0 | 0 | 0 |
| 0.255547185 | 0.010101821 | 0.07601111 | 0.036400001 | 0.014776864 |
| 0 | 0 | 0 | 0 | 0 |
| 0 | 0 | 0 | 0 | 0 |
| 0 | 0 | 0 | 0 | 0 |
| 0 | 0 | 0 | 0 | 0 |
| 0.00472545 | 1.603226138 | 0.137468005 | 1.250733997 | 0.092497065 |
| 0 | 0 | 0 | 0 | 0 |
| 0 | 0 | 0 | 0 | 0 |

| | | | | |
|---|---|---|---|---|
| 0 | 0 | 0 | 0 | 0 |
| 0 | 0 | 0 | 0 | 0 |
| 0 | 0 | 0 | 0 | 0 |
| 0 | 0 | 0 | 0 | 0 |
| 0 | 0 | 0 | 0 | 0.000525877 |
| 0 | 0 | 0 | 0 | 0 |
| 0 | 0 | 0 | 0 | 0 |
| 0 | 0 | 0 | 0.001265319 | 0 |
| 0 | 0 | 0 | 0.00301356 | 0 |
| 0.152701994 | 0 | 0 | 0.029080666 | 0 |
| 0 | 0 | 0 | 0 | 0 |
| 0 | 0 | 0 | 0 | 0 |
| 0.001317495 | 0.001105601 | 0 | 0 | 0 |
| 0 | 0 | 0 | 0 | 0 |
| 0 | 0 | 0 | 0 | 0 |
| 0 | 0 | 0 | 0 | 0 |
| 0 | 0 | 0 | 0 | 0 |
| 0 | 0 | 0 | 0 | 0 |
| 0 | 0 | 0 | 0 | 0 |
| 0.072035035 | 1.183991212 | 0.044729674 | 0.079085148 | 0.101707444 |
| 0 | 0 | 0 | 0 | 0 |
| 0 | 0 | 0 | 0 | 0 |
| 0 | 0 | 0 | 0 | 0 |
| 0 | 0 | 0 | 0 | 0 |
| 0 | 0 | 0 | 0 | 0 |
| 0 | 0 | 0 | 0 | 0 |
| 0 | 0 | 0 | 0 | 0 |
| 0 | 0 | 0 | 0 | 0 |
| 0 | 0 | 0 | 0 | 0 |
| 0.009491513 | 0 | 0 | 0 | 0.000882933 |
| 0 | 0 | 0 | 0 | 0 |
| 0 | 0 | 0 | 0 | 0 |
| 0 | 0 | 0 | 0 | 0 |
| 0 | 0 | 0 | 0 | 0 |
| 0 | 0 | 0 | 0 | 0 |
| 0 | 0 | 0.009976278 | 0 | 0 |
| 0 | 0 | 0.128138477 | 0 | 0 |
| 0 | 0 | 0 | 0 | 0 |
| 0 | 0 | 0 | 0 | 0 |
| 0 | 0 | 0 | 0.139295068 | 0 |
| 0 | 0 | 0 | 0 | 0 |
| 0 | 0 | 0 | 0 | 0 |
| 0 | 0 | 0.046978783 | 0 | 0 |
| 0 | 0 | 0 | 0 | 0 |
| 0.011800806 | 0.003697071 | 0.004683743 | 0.001122615 | 0.004765286 |
| 0 | 0 | 0 | 0 | 0 |
| 0 | 0 | 0 | 0 | 0 |
| 0 | 0 | 0 | 0 | 0 |

| | | | | |
|---|---|---|---|---|
| 0 | 0 | 0 | 0 | 0 |
| 0 | 0 | 0 | 0 | 0 |
| 0 | 0 | 0 | 0 | 0 |
| 0 | 0 | 0 | 0 | 0 |
| 0 | 0 | 0 | 0 | 0 |
| 0.228466448 | 0.004255011 | 0.0383566 | 0.001396793 | 0.000413659 |
| 0 | 0 | 0 | 0 | 0 |
| 0 | 0 | 0 | 0 | 0 |
| 0 | 0 | 0 | 0 | 0 |
| 0 | 0 | 0 | 0 | 0 |
| 0 | 0 | 0 | 0 | 0 |
| 0 | 0 | 0 | 0 | 0 |
| 0 | 0 | 0 | 0 | 0 |
| 0 | 0 | 0 | 0 | 0 |
| 0 | 0 | 0 | 0 | 0 |
| 0 | 0 | 0 | 0 | 0 |
| 0.062264792 | 0 | 0.010025001 | 0 | 0 |
| 0.000433988 | 0 | 0 | 0 | 0 |
| 0 | 0 | 0 | 0 | 0 |
| 0 | 0 | 0 | 0 | 0 |
| 0.000953382 | 0 | 0 | 0 | 0 |
| 0 | 0 | 0 | 0 | 0 |
| 0 | 0 | 0 | 0 | 0 |
| 0 | 0 | 0 | 0 | 0 |
| 0 | 0 | 0 | 0 | 0 |
| 0.004409582 | 0 | 0 | 0 | 0 |
| 0 | 0 | 0 | 0 | 0 |
| 0 | 0 | 0 | 0 | 0 |
| 0 | 0 | 0 | 0 | 0 |
| 0 | 0 | 0 | 0 | 0.006132122 |
| 0 | 0 | 0 | 0 | 0 |
| 0 | 0 | 0 | 0 | 0.008884254 |
| 0 | 0 | 0 | 0 | 0 |
| 0 | 0 | 0 | 0 | 0 |
| 0 | 0 | 0 | 0 | 0 |
| 0 | 0 | 0 | 0 | 0 |
| 0 | 0 | 0 | 0 | 0 |
| 0 | 0 | 0 | 0 | 0 |
| 0.001326482 | 0 | 0.014059434 | 0 | 0 |
| 0 | 0 | 0 | 0 | 0 |
| 0 | 0 | 0 | 0 | 0 |
| 0 | 0 | 0 | 0 | 0 |
| 0 | 0 | 0 | 0 | 0 |
| 0 | 0 | 0 | 0 | 0 |
| 0.004239852 | 0 | 0 | 0.001939128 | 0 |
| 0 | 0 | 0 | 0 | 0 |
| 0 | 0 | 0 | 0 | 0 |
| 0 | 0 | 0 | 0 | 0 |

| | | | | |
|---|---|---|---|---|
| 0 | 0 | 0 | 0 | 0 |
| 0.035113122 | 0 | 0 | 0 | 0 |
| 0 | 0 | 0 | 0 | 0.001020606 |
| 0 | 0 | 0 | 0 | 0 |
| 0 | 0 | 0 | 0 | 0 |
| 0.00595119 | 0 | 0 | 0 | 0 |
| 0 | 0 | 0.000632562 | 0 | 0 |
| 0 | 0 | 0 | 0 | 0 |
| 0 | 0 | 0 | 0 | 0 |
| 0 | 0 | 0 | 0 | 0 |
| 0.001722017 | 0 | 0.001708672 | 0 | 0 |
| 0 | 0 | 0 | 0 | 0 |
| 0.005961257 | 0 | 0.213605777 | 0.193355756 | 0 |
| 0 | 0 | 0 | 0 | 0 |
| 0 | 0 | 0 | 0 | 0 |
| 0 | 0 | 0 | 0 | 0 |
| 0 | 0 | 0 | 0 | 0 |
| 0 | 0 | 0 | 0 | 0 |
| 0 | 0 | 0 | 0 | 0 |
| 0.008024316 | 0 | 0 | 0 | 0 |
| 0 | 0 | 0 | 0.002493952 | 0 |
| 0.002625867 | 0 | 0.00222932 | 0 | 0 |
| 0 | 0 | 0 | 0 | 0 |
| 0.581593307 | 1.331712487 | 1.991664196 | 3.554877725 | 0 |
| 0.03123207 | 0 | 0 | 0 | 0 |
| 0 | 0 | 0 | 0 | 0 |
| 0 | 0 | 0 | 0 | 0 |
| 0 | 0 | 0 | 0 | 0 |
| 0 | 0 | 0 | 0 | 0 |
| 0 | 0 | 0 | 0 | 0 |
| 0.003087737 | 0 | 0 | 0 | 0 |
| 0 | 0 | 0 | 0 | 0 |
| 0.00091923 | 0 | 0 | 0 | 0 |
| 0 | 0 | 0 | 0 | 0 |

| PHL018-0047_V1 | PHL018-0050_V3 | PHL018-0167_V2 | PHL018-0044_V1 | PHL018-0153_V3 |
|---|---|---|---|---|
| EYCF | EYCF | REF | CTRL | CTRL |
| 0.779768383 | 2.919508872 | 0.00953553 | 0.145068918 | 0.121223529 |
| 1.166649513 | 0.920981268 | 1.344428503 | 0.712679424 | 0 |
| 0 | 1.395055608 | 1.133870278 | 0 | 0 |
| 0 | 0 | 0 | 0 | 0 |
| 0 | 0 | 0 | 0 | 0 |
| 0 | 0 | 0 | 0 | 0 |
| 0 | 0 | 0 | 0 | 0 |
| 0 | 0 | 0 | 0 | 0 |
| 0 | 0 | 0 | 0 | 0 |
| 0 | 0 | 0 | 0 | 0 |
| 0 | 0 | 0 | 0 | 0 |
| 0 | 0 | 0 | 0 | 0 |
| 0 | 0 | 0 | 0 | 0 |
| 0.491393576 | 0.1120238 | 0.299495265 | 3.236074515 | 0 |
| 0 | 0 | 0 | 0 | 0 |
| 0 | 0 | 0 | 0 | 0 |
| 0 | 0 | 0 | 0.003505362 | 0 |
| 0 | 0 | 0 | 0 | 0 |
| 0 | 0 | 0 | 0 | 0 |
| 0 | 0 | 0 | 0 | 0 |
| 0 | 0 | 0 | 0 | 0 |
| 0.010085379 | 0 | 0 | 0.003722584 | 0 |
| 0 | 0 | 0 | 0 | 0 |
| 0 | 0 | 0 | 0 | 0 |
| 0 | 0.016354781 | 0 | 0 | 0 |
| 0 | 0 | 0 | 0 | 0 |
| 0 | 0 | 0 | 0 | 0 |
| 0 | 0 | 0 | 0 | 0 |
| 0 | 0 | 0 | 0 | 0 |
| 0 | 0 | 0 | 0 | 0 |
| 0 | 0.000188379 | 0 | 0 | 0 |
| 0 | 0 | 0 | 0 | 0 |
| 0 | 0 | 0 | 0 | 0 |
| 0 | 0 | 0 | 0 | 0 |
| 0 | 0 | 0 | 0 | 0 |
| 0.074690783 | 0.010943425 | 1.450376201 | 0 | 0 |
| 0 | 0 | 0 | 0 | 0 |
| 0 | 0 | 0 | 0 | 0 |
| 0 | 0 | 0 | 0 | 0 |
| 0 | 0 | 0 | 0 | 0 |
| 0 | 0 | 0 | 0 | 0 |
| 0.248213311 | 0.001614831 | 0.001759775 | 0.516661196 | 0 |
| 0.526197606 | 0.004382868 | 0.14632667 | 1.622774243 | 0.136270105 |
| 0.024488829 | 0.00044811 | 0.139310073 | 0.003293279 | 0 |
| 0 | 0.002173389 | 0 | 0 | 0 |
| 0 | 0 | 0 | 0 | 0 |

| | | | | |
|---|---|---|---|---|
| 0 | 0.047237808 | 0 | 0 | 0 |
| 0.004149047 | 0.014559874 | 0 | 0.007409479 | 0.641715847 |
| 0.02320687 | 0.408403842 | 0 | 0.068922254 | 0.00290074 |
| 1.217752933 | 0.160465641 | 0.273298852 | 0.375927216 | 0 |
| 0.049820411 | 0.065311582 | 0 | 0 | 0 |
| 0 | 0 | 0 | 0 | 0 |
| 0.012818139 | 0 | 0 | 0.003403614 | 0 |
| 0 | 0.450195836 | 0 | 0 | 0 |
| 0 | 0 | 0 | 0 | 0 |
| 0 | 0 | 0 | 0 | 0 |
| 0 | 0 | 0 | 0 | 0 |
| 2.674503172 | 1.600473997 | 0.060021911 | 0 | 0 |
| 0 | 0 | 0 | 0 | 0 |
| 0.283828994 | 0.025629354 | 0.061542928 | 0.083689608 | 0 |
| 0 | 0.016520027 | 0 | 0 | 0 |
| 0 | 0 | 0 | 0 | 0 |
| 0.052136764 | 0 | 0 | 0 | 0 |
| 0 | 0 | 0 | 0 | 0 |
| 0 | 0 | 0 | 0 | 0 |
| 0 | 0 | 0 | 0 | 0 |
| 0.392766313 | 0.020731262 | 0.031930841 | 0.156476918 | 1.332479 |
| 0.030854689 | 0.042638718 | 0.012870973 | 0.028214432 | 0.01536282 |
| 0 | 0 | 0 | 0 | 0 |
| 0 | 0 | 0 | 0 | 0 |
| 0 | 0 | 0 | 0 | 0 |
| 0 | 0 | 0 | 0 | 0 |
| 0 | 0 | 0 | 0 | 0 |
| 0 | 0 | 0 | 0 | 0 |
| 0.005706805 | 0 | 0 | 0.005414163 | 0 |
| 0 | 0 | 0 | 0 | 0 |
| 0.028011536 | 0.003525277 | 0 | 0.025804409 | 0 |
| 0 | 0 | 0 | 0 | 0 |
| 0 | 0 | 0 | 0 | 0 |
| 0 | 0 | 0 | 0 | 0 |
| 0 | 0 | 0 | 0 | 0 |
| 0 | 0 | 0 | 0 | 0 |
| 0 | 0 | 0 | 0 | 0 |
| 0.0020285 | 0 | 0 | 0 | 0 |
| 0 | 0 | 0 | 0 | 0 |
| 0 | 0 | 0 | 0 | 0 |
| 0 | 0 | 0 | 0 | 0 |
| 0 | 0 | 0 | 0 | 0 |
| 0 | 0 | 0 | 0 | 0 |
| 0 | 0 | 0 | 0 | 0 |
| 0 | 0 | 0 | 0 | 0 |
| 0 | 0 | 0 | 0 | 0 |
| 0 | 0 | 0 | 0 | 0 |
| 0 | 0.004916395 | 0 | 0 | 0 |

| | | | | |
|---|---|---|---|---|
| 0 | 0 | 0 | 0 | 0 |
| 0 | 0 | 0.004450649 | 0.111112944 | 0 |
| 0 | 0 | 0 | 0 | 0 |
| 0 | 0 | 0 | 0 | 0 |
| 0 | 0 | 0 | 0.003211538 | 0 |
| 0 | 0 | 0 | 0 | 0 |
| 0 | 0 | 0 | 0 | 0 |
| 0 | 0 | 0 | 0 | 0 |
| 0.002874552 | 0.004105945 | 0 | 0 | 0 |
| 0 | 0 | 0 | 0 | 0 |
| 0 | 0 | 0 | 0 | 0 |
| 0 | 0 | 0 | 0.030983162 | 0 |
| 0 | 0 | 0 | 0 | 0 |
| 0 | 0 | 0 | 0 | 0 |
| 0 | 0 | 0 | 0 | 0 |
| 0.023795672 | 0 | 0 | 0 | 0 |
| 0 | 0 | 0 | 0 | 0 |
| 0 | 0 | 0 | 0 | 0 |
| 0 | 0.399735663 | 0.322605651 | 0.002012669 | 0 |
| 0 | 0 | 0 | 0 | 0 |
| 0 | 0 | 0 | 0 | 0 |
| 0 | 0 | 0 | 0 | 0 |
| 0 | 0 | 0 | 0 | 0 |
| 0 | 0 | 0 | 0 | 0 |
| 0 | 0 | 0 | 0 | 0 |
| 0 | 0 | 0 | 0 | 0 |
| 0 | 0 | 0 | 0 | 0 |
| 0 | 0 | 0.001429667 | 0 | 0 |
| 0 | 0 | 0 | 0 | 0 |
| 0 | 0 | 0 | 0 | 0 |
| 0.055694302 | 0.958389192 | 0.014601038 | 0 | 0 |
| 0.003488249 | 0.028407475 | 0 | 0.000945535 | 0 |
| 0 | 0 | 0 | 0 | 0 |
| 0 | 0 | 0 | 0 | 0 |
| 0 | 0 | 0 | 0 | 0 |
| 0 | 0 | 0 | 0 | 0 |
| 0 | 0 | 0 | 0 | 0 |
| 0 | 0 | 0 | 0 | 0 |
| 0 | 0.080569212 | 0 | 0 | 0 |
| 0 | 0 | 0 | 0 | 0 |
| 0 | 0 | 0 | 0 | 0 |
| 0 | 0 | 0 | 0 | 0 |
| 0 | 0 | 0 | 0.00808738 | 0 |
| 0 | 0 | 0 | 0 | 0 |
| 0 | 0.00470439 | 0 | 0 | 0 |
| 0 | 0 | 0 | 0 | 0 |
| 0 | 0 | 0 | 0 | 0 |
| 0 | 0 | 0 | 0 | 0 |

| | | | | |
|---|---|---|---|---|
| 0 | 0 | 0 | 0 | 0 |
| 0 | 0 | 0 | 0 | 0 |
| 0 | 0 | 0 | 0 | 0 |
| 0 | 0 | 0 | 0 | 0 |
| 0 | 0.595252382 | 0.004358662 | 0 | 0 |
| 0 | 0 | 0 | 0 | 0 |
| 0 | 0 | 0 | 0 | 0 |
| 0 | 0 | 0 | 0 | 0 |
| 0 | 0 | 0 | 0 | 0 |
| 0 | 0 | 0 | 0 | 0 |
| 0 | 0 | 0 | 0 | 0 |
| 0 | 0 | 0 | 0 | 0 |
| 0 | 0 | 0 | 0 | 0 |
| 0 | 0 | 0 | 0 | 0 |
| 0 | 0 | 0 | 0 | 0 |
| 0 | 0 | 0 | 0 | 0 |
| 0 | 0 | 0 | 0 | 0 |
| 0 | 0 | 0 | 0 | 0 |
| 0 | 0 | 0 | 0 | 0 |
| 0 | 0 | 0 | 0 | 0 |
| 0 | 0 | 0 | 0 | 0 |
| 0 | 0 | 0 | 0 | 0 |
| 0 | 0 | 0 | 0 | 0 |
| 0 | 0 | 0 | 0 | 0 |
| 0 | 0 | 0 | 0 | 0 |
| 0 | 0 | 0 | 0 | 0 |
| 0 | 0 | 0 | 0 | 0 |
| 0 | 0 | 0 | 0 | 0 |
| 0 | 0 | 0 | 0 | 0 |
| 0 | 0 | 0 | 0 | 0 |
| 0 | 0 | 0 | 0 | 0 |
| 0.008049374 | 0 | 0.002549047 | 0 | 0 |
| 0 | 0 | 0 | 0 | 0 |
| 0 | 0 | 0 | 0 | 0 |
| 0 | 0 | 0 | 0 | 0 |
| 0 | 0 | 0 | 0 | 0 |
| 0 | 0 | 0 | 0 | 0 |
| 0 | 0 | 0 | 0 | 0 |
| 0 | 0 | 0 | 0 | 0 |
| 0 | 0 | 0 | 0 | 0 |
| 0 | 0 | 0 | 0 | 0 |
| 0 | 0 | 0 | 0 | 0 |
| 0 | 0 | 0 | 0 | 0 |
| 0 | 0 | 0 | 0 | 0 |
| 0 | 0 | 0 | 0 | 0 |
| 0 | 0 | 0 | 0 | 0 |
| 0 | 0 | 0 | 0 | 0 |
| 0 | 0 | 0 | 0 | 0.060584624 |

| | | | | |
|---|---|---|---|---|
| 0 | 0 | 0 | 0 | 0 |
| 0 | 0 | 0 | 0 | 0 |
| 0 | 0 | 0 | 0 | 0 |
| 0 | 0 | 0 | 0 | 0 |
| 0 | 0 | 0 | 0 | 0 |
| 0 | 0 | 0 | 0 | 0 |
| 0 | 0 | 0 | 0 | 0 |
| 0 | 0 | 0 | 0 | 0 |
| 0 | 0 | 0 | 0 | 0 |
| 0 | 0 | 0 | 0 | 0 |
| 0 | 0 | 0 | 0 | 0 |
| 0 | 0 | 0 | 0 | 0 |
| 0.0075099 | 0.009979299 | 0.000813063 | 0.001670282 | 0 |
| 0 | 0 | 0 | 0 | 0 |
| 0 | 0 | 0 | 0 | 0 |
| 0 | 0 | 0 | 0 | 0 |
| 0 | 0 | 0 | 0 | 0 |
| 0 | 0 | 0 | 0 | 0 |
| 0 | 0 | 0 | 0 | 0 |
| 0 | 0 | 0 | 0 | 0 |
| 0.035284293 | 0.004249437 | 0.027060124 | 0.041770465 | 0.050029033 |
| 0 | 0 | 0 | 0 | 0 |
| 0.000631832 | 0.001813464 | 0 | 0 | 0 |
| 0 | 0 | 0 | 0 | 0 |
| 0 | 0 | 0 | 0 | 0 |
| 0 | 0 | 0 | 0 | 0 |
| 0.015306715 | 0.000427461 | 0.005312083 | 0.002955512 | 0.003516381 |
| 0 | 0 | 0 | 0 | 0 |
| 0 | 0 | 0 | 0 | 0 |
| 0 | 0.000770246 | 0 | 0 | 0.042726965 |
| 0 | 0 | 0 | 0 | 1.31896982 |
| 0 | 0 | 0 | 0 | 0 |
| 0 | 0 | 0 | 0 | 0 |
| 0 | 0.151074099 | 0 | 0 | 0 |
| 0.06161135 | 0.27400259 | 0.002373945 | 0.033528112 | 0.061455091 |
| 0 | 0 | 0 | 0 | 0 |
| 0 | 0 | 0 | 0 | 0 |
| 0 | 0 | 0 | 0 | 0 |
| 0 | 0 | 0 | 0 | 0 |
| 0 | 0 | 0 | 0 | 0 |
| 0 | 0 | 0 | 0 | 0 |
| 0 | 0 | 0 | 0 | 0 |
| 0 | 0 | 0 | 0 | 0 |
| 0 | 0 | 0 | 0 | 0 |
| 0 | 0 | 0 | 0 | 0 |
| 0 | 0 | 0 | 0 | 0 |
| 0 | 0 | 0 | 0.006325354 | 0.0009115 |
| 0.027211649 | 0.006054347 | 0.000563912 | 0.002896126 | 0.001399826 |

| | | | | |
|---|---|---|---|---|
| 0 | 0 | 0 | 0 | 0 |
| 0 | 0 | 0 | 0 | 0 |
| 0 | 0 | 0 | 0 | 0 |
| 0.000636618 | 0 | 0 | 0 | 0 |
| 0 | 0 | 0 | 0 | 0 |
| 0 | 0 | 0 | 0 | 0 |
| 0 | 0 | 0 | 0.001803637 | 0 |
| 0 | 0 | 0 | 0.007025457 | 0 |
| 0 | 0 | 0 | 0 | 0 |
| 0 | 0 | 0 | 0 | 0 |
| 0 | 0 | 0 | 0 | 0 |
| 0 | 0 | 0 | 0 | 0 |
| 0 | 0 | 0 | 0 | 0 |
| 0 | 0 | 0 | 0 | 0 |
| 0 | 0 | 0 | 0 | 0 |
| 0 | 0 | 0 | 0 | 0 |
| 0 | 0 | 0 | 0 | 0 |
| 0 | 0 | 0 | 0 | 0 |
| 0.002037355 | 0 | 0.001003617 | 0.005706598 | 0 |
| 0 | 0 | 0 | 0 | 0 |
| 0.034811456 | 0 | 0.006133238 | 0.018193338 | 0.141898339 |
| 0 | 0 | 0 | 0 | 0 |
| 0 | 0 | 0 | 0 | 0 |
| 0 | 0 | 0 | 0 | 0 |
| 0.00398045 | 0 | 0.004201715 | 0.131092739 | 0.007152077 |
| 0 | 0.000207219 | 0 | 0 | 0 |
| 0 | 0 | 0 | 0 | 0 |
| 0 | 0 | 0 | 0 | 0 |
| 0.011870628 | 0 | 0 | 0.021880277 | 0 |
| 0 | 0 | 0 | 0 | 0 |
| 0.87581739 | 0.35419177 | 0.034834066 | 1.829729923 | 0.637938458 |
| 0 | 0 | 0 | 0 | 0 |
| 0 | 0 | 0 | 0 | 0 |
| 0 | 0 | 0 | 0 | 0 |
| 0 | 0 | 0 | 0 | 0 |
| 0 | 0 | 0 | 0 | 0 |
| 0 | 0 | 0 | 0 | 0 |
| 0 | 0 | 0 | 0 | 0 |
| 0 | 0 | 0 | 0 | 0 |
| 0 | 0.000157888 | 0 | 0 | 0 |
| 0 | 0 | 0 | 0 | 0 |
| 0.017269348 | 0.051277857 | 0.002219221 | 0.042740303 | 0.068804733 |
| 0 | 0 | 0 | 0.000512296 | 0 |
| 0 | 0 | 0 | 0 | 0 |
| 0 | 0 | 0 | 0 | 0 |
| 0 | 0 | 0 | 0 | 0 |
| 0 | 0 | 0 | 0 | 0 |
| 0 | 0 | 0 | 0 | 0 |

| | | | | |
|---|---|---|---|---|
| 0 | 0 | 0 | 0 | 0 |
| 0 | 0 | 0 | 0 | 0 |
| 0.023098715 | 0 | 0.139130822 | 0.004855629 | 0 |
| 0 | 0 | 0 | 0 | 0 |
| 0 | 0 | 0 | 0 | 0 |
| 0 | 0 | 0 | 0 | 0 |
| 0 | 0 | 0 | 0 | 0 |
| 0 | 0 | 0 | 0 | 0 |
| 0 | 0 | 0 | 0 | 0 |
| 0 | 0 | 0 | 0 | 0 |
| 0 | 0 | 0 | 0 | 0 |
| 0 | 0 | 0 | 0 | 0 |
| 0 | 0 | 0 | 0 | 0 |
| 0 | 0.001481338 | 0.003739627 | 0.019488314 | 0 |
| 0.004945054 | 0.00041524 | 0 | 0.034268154 | 0.00540843 |
| 0 | 0 | 0.018661179 | 0 | 0 |
| 0.004409566 | 0.000131425 | 0.003103123 | 0.003453002 | 0.003521384 |
| 0 | 0 | 0 | 0 | 0 |
| 0 | 0 | 0 | 0 | 0 |
| 0 | 0 | 0 | 0 | 0 |
| 0 | 0 | 0 | 0 | 0 |
| 0.027200628 | 0.00609804 | 0.000746996 | 0.002157976 | 0.001854303 |
| 0 | 0 | 0 | 0 | 0 |
| 0 | 0 | 0 | 0 | 0 |
| 0 | 0 | 0 | 0 | 0 |
| 0 | 0 | 0 | 0 | 0 |
| 0 | 0 | 0.098268191 | 0 | 0 |
| 0 | 0 | 0 | 0 | 0 |
| 0 | 0 | 0 | 0 | 0 |
| 0 | 0 | 0 | 0 | 0 |
| 0 | 0 | 0 | 0 | 0 |
| 0 | 0 | 0 | 0 | 0 |
| 0 | 0 | 0 | 0 | 0 |
| 0 | 0 | 0 | 0 | 0 |
| 0 | 0 | 0 | 0 | 0 |
| 0 | 0 | 0 | 0.01819883 | 0 |
| 0 | 0 | 0 | 0 | 0 |
| 0 | 0.016157835 | 0.018301199 | 0.005121293 | 0.00133128 |
| 0 | 0 | 0 | 0 | 0 |
| 0 | 0 | 0 | 0 | 0 |
| 0 | 0 | 0 | 0 | 0 |
| 0 | 0 | 0 | 0 | 0 |
| 0 | 0 | 0 | 0 | 0 |
| 0 | 0 | 0 | 0 | 0 |
| 0 | 0 | 0 | 0 | 0 |
| 0 | 0 | 0 | 0 | 0 |
| 0 | 0 | 0 | 0 | 0 |
| 0 | 0.034592621 | 0 | 0 | 0.011690723 |

| | | | | |
|---|---|---|---|---|
| 3.086995851 | 0 | 3.060466402 | 0 | 0 |
| 0 | 0 | 0 | 0 | 0 |
| 0 | 0 | 0 | 0.005525205 | 0 |
| 0 | 0 | 0 | 0 | 0 |
| 0.014448635 | 0.001650298 | 0.002558367 | 0.002021414 | 0.005024773 |
| 0 | 0 | 0 | 0 | 0 |
| 0 | 0 | 0 | 0 | 0 |
| 0 | 0.003960575 | 0.04303338 | 0.006375278 | 0.039520878 |
| 0 | 0 | 0 | 0 | 0 |
| 0.020875146 | 0.019435847 | 0.007676903 | 0.028109864 | 39.57992446 |
| 0 | 0 | 0 | 0 | 0.001458603 |
| 0.001292928 | 0.010237971 | 0 | 0.01542045 | 0 |
| 0.003922926 | 0.000463005 | 0.010932222 | 0.000850688 | 0.001174786 |
| 0.00953266 | 0 | 0 | 0.17118676 | 0 |
| 0.001412124 | 0.002012987 | 0 | 0.002187283 | 0 |
| 0 | 0 | 0 | 0 | 0 |
| 0 | 0 | 0 | 0 | 0 |
| 0 | 0 | 0 | 0 | 0 |
| 0.30161712 | 0.029302836 | 0.005350982 | 0.049123044 | 8.353103459 |
| 0 | 0.000463221 | 0 | 0 | 0 |
| 0 | 0 | 0 | 0 | 0 |
| 0 | 0 | 0 | 0 | 0 |
| 0 | 0 | 0 | 0.047620652 | 0 |
| 0 | 0.100435472 | 0 | 0 | 0 |
| 0 | 0 | 0 | 0 | 0 |
| 0.002284604 | 0.000759899 | 0 | 0 | 0 |
| 0 | 0 | 0 | 0 | 0 |
| 0.115758029 | 0.001926787 | 0 | 0.00419368 | 0.006287811 |
| 0 | 0 | 0 | 0 | 0 |
| 0 | 0.002522067 | 0 | 0.001784027 | 0 |
| 0.001225752 | 0 | 0 | 0 | 0.192639483 |
| 0 | 0 | 0 | 0 | 0 |
| 0 | 0 | 0 | 0 | 0 |
| 0 | 0 | 0.004289346 | 0 | 0 |
| 0 | 0 | 0.002942804 | 0 | 0 |
| 0 | 0 | 0 | 0 | 0 |
| 0.031336978 | 0.030427706 | 0 | 0.001404016 | 2.444593938 |
| 0 | 0 | 0 | 0 | 0 |
| 0 | 0 | 0 | 0 | 0 |
| 0 | 0.002603909 | 0 | 0 | 0 |
| 0.24702339 | 0 | 0 | 0 | 0 |
| 0 | 0.020670155 | 0.04984746 | 0 | 0 |
| 0 | 0 | 0 | 0 | 0 |
| 0 | 0 | 0.005906326 | 0 | 0 |
| 0 | 0 | 0 | 0 | 0 |
| 0 | 0 | 0 | 0 | 0 |
| 0.002272869 | 0.026581788 | 0 | 0.002156317 | 0 |
| 0 | 0 | 0 | 0 | 0 |

| | | | | |
|---|---|---|---|---|
| 0 | 0 | 0.004308514 | 0 | 0 |
| 0 | 0 | 0 | 0 | 0 |
| 0 | 0 | 0 | 0 | 0 |
| 0 | 0 | 0 | 0 | 0 |
| 0 | 0 | 0 | 0 | 0 |
| 0.021258142 | 0.028499404 | 0.065004673 | 0.016740984 | 0 |
| 0 | 0 | 0 | 0 | 0 |
| 0 | 0 | 0 | 0 | 0 |
| 0 | 1.643463245 | 0.056759977 | 0 | 0 |
| 0 | 0 | 0 | 0 | 0 |
| 0.018427015 | 0.000835413 | 0.00547925 | 0.012100277 | 0.079276708 |
| 0 | 0 | 0.003902832 | 0 | 0 |
| 0 | 0 | 0 | 0 | 0 |
| 0.003187548 | 0.00018416 | 0 | 0.00186098 | 0.001468562 |
| 0.003721415 | 0.006255586 | 0 | 0.044922454 | 0 |
| 0 | 0 | 0 | 0 | 0 |
| 0.008905735 | 0 | 0 | 0 | 0 |
| 0.003817845 | 0 | 0 | 0 | 0 |
| 0 | 0 | 0 | 0 | 0 |
| 0 | 0 | 0 | 0 | 0 |
| 0 | 0 | 0 | 0 | 0 |
| 0 | 0.537515739 | 0 | 0 | 0 |
| 0 | 0.021245682 | 0.023971876 | 0 | 0 |
| 0 | 0.000185486 | 0 | 0 | 0 |
| 0 | 0 | 0 | 0 | 0 |
| 0 | 0 | 0 | 0 | 0 |
| 0.002658574 | 0.00068461 | 0 | 0.089359484 | 0.02133695 |
| 0 | 0.001407024 | 0 | 0 | 0 |
| 0 | 0 | 0 | 0 | 0 |
| 1.005258626 | 0.072999403 | 0.435230101 | 2.005717215 | 0 |
| 0.394073707 | 0.002730783 | 0 | 0 | 0 |
| 0 | 0 | 0 | 0.005824087 | 0 |
| 0 | 0.000109939 | 0 | 0 | 0 |
| 0.021439866 | 0 | 0.002387516 | 0 | 0 |
| 0 | 0 | 0 | 0 | 0 |
| 0 | 0 | 0 | 0 | 0 |
| 0 | 0.001691408 | 0 | 0 | 0 |
| 0.00099968 | 0 | 0 | 0.006232455 | 0 |
| 0 | 0 | 0 | 0.167099398 | 0 |
| 0 | 0.049029338 | 0 | 0.120167124 | 0 |
| 0 | 0.00741857 | 0.065026521 | 0.019233379 | 0.17554397 |
| 0.147360614 | 0.126117555 | 0 | 0 | 0 |
| 0 | 0 | 0 | 0 | 0 |
| 0 | 0.003157212 | 0 | 0 | 3.928210553 |
| 0.002501293 | 0 | 0 | 0.005177515 | 0.01198487 |
| 0 | 0 | 0 | 0 | 0 |
| 0 | 0.000794523 | 0 | 0.008258241 | 0 |
| 0 | 0 | 0 | 0 | 0 |

| | | | | |
|---|---|---|---|---|
| 0 | 0.000756237 | 0 | 0 | 0 |
| 0.060547802 | 0 | 0 | 0 | 0 |
| 0.1269814 | 0.000114492 | 0.00090111 | 0 | 0 |
| 0.005667125 | 0 | 0 | 0 | 0.004525644 |
| 0 | 0.449045077 | 0.008568472 | 0 | 0 |
| 0.019864038 | 0.002320783 | 0 | 0 | 0 |
| 0 | 0 | 0 | 0 | 0 |
| 0 | 0 | 0 | 0.005075609 | 0 |
| 0 | 0 | 0 | 0 | 0.004972242 |
| 0.109112665 | 0 | 0 | 0 | 0 |
| 0 | 0 | 0 | 0.069855521 | 0 |
| 0 | 0 | 0 | 0 | 0 |
| 0 | 0 | 0 | 0 | 0.108433986 |
| 0.068722099 | 0.000304066 | 0 | 0.045065717 | 0 |
| 0 | 0 | 0 | 0 | 0 |
| 0.010519563 | 0.013108837 | 0.002337752 | 0.01760904 | 0 |
| 0.006750433 | 0.024626 | 0.004836822 | 0.002439723 | 0.001715238 |
| 0.005716206 | 0 | 0.000839507 | 0.001616819 | 0.020839478 |
| 0.036553784 | 0 | 0 | 0 | 0 |
| 0 | 0 | 0 | 0 | 0 |
| 0.057489845 | 0.048323962 | 0 | 0 | 0 |
| 0.003796437 | 0.002749561 | 0 | 0.001286342 | 0.005400309 |
| 0.01123965 | 0 | 0 | 0 | 0 |
| 0.653259526 | 0 | 0.493498708 | 0 | 0 |
| 0.087177969 | 0 | 0 | 0 | 0 |
| 0 | 0 | 0.027762237 | 0 | 0 |
| 0 | 0 | 0.007894781 | 0 | 0.016486853 |
| 0 | 0 | 0.186523164 | 0 | 0 |
| 0 | 0 | 0 | 0 | 0 |
| 0 | 0 | 0 | 0 | 0 |
| 0.015009434 | 0.000614514 | 0.048865549 | 0.031263225 | 0 |
| 0.016782919 | 0.000252735 | 0.004226939 | 0.001276976 | 0 |
| 0.042892631 | 0.039825624 | 0.046638214 | 0.018541774 | 0 |
| 0.002410562 | 0.000560394 | 0 | 0.137216944 | 0 |
| 0 | 0 | 0 | 0 | 0 |
| 0 | 0 | 0 | 0 | 0 |
| 0 | 0 | 0.000856125 | 0.003847255 | 0 |
| 0.250537287 | 0.065013581 | 0 | 0.063398284 | 0.006173158 |
| 0.286684367 | 0 | 0 | 0 | 0 |
| 0 | 0.005807606 | 0 | 0 | 0 |
| 0 | 0 | 0 | 0 | 0 |
| 0 | 0 | 0 | 0 | 0 |
| 0.058395244 | 0.026747956 | 0 | 0.398853335 | 0 |
| 0.042240537 | 0.404780387 | 0.007186442 | 0 | 0 |
| 0 | 0 | 0 | 0 | 2.369864052 |
| 0 | 0 | 0 | 0 | 0 |
| 0 | 0 | 0 | 0 | 0 |
| 0 | 0 | 0 | 0 | 0 |

| | | | | |
|---|---|---|---|---|
| 0.002938795 | 0 | 0 | 0 | 0 |
| 1.488444525 | 0.040281014 | 1.949169293 | 0.187125637 | 0.099322901 |
| 0 | 0 | 0 | 0 | 0 |
| 0 | 0 | 0 | 0 | 0 |
| 0 | 0 | 0 | 0 | 0 |
| 0 | 0 | 0 | 0 | 0 |
| 0.000749594 | 0 | 0 | 0 | 0.035018679 |
| 0 | 0.000957139 | 0 | 0 | 0 |
| 0.013103805 | 0.043134567 | 0 | 0 | 0 |
| 0.002355195 | 0.000758109 | 0 | 0 | 0.027741919 |
| 0.001404925 | 0.00133515 | 0 | 0.039823208 | 0.002644578 |
| 0.046337803 | 0.001539672 | 0.009593383 | 0.009724229 | 0.003939174 |
| 0 | 0 | 0 | 0 | 0 |
| 0 | 0 | 0 | 0 | 0 |
| 0 | 0 | 0 | 0 | 0 |
| 0 | 0 | 0 | 0.002566143 | 0 |
| 0.006304966 | 0.000744147 | 0.022362301 | 0.057879586 | 0.017370782 |
| 0.030283453 | 0.003015109 | 0 | 0.100279535 | 0.120918547 |
| 0 | 0.000262916 | 0 | 0 | 0 |
| 0 | 0 | 0.012117951 | 0.055896284 | 0 |
| 0.407459343 | 0.245034235 | 0.030016131 | 0.00540486 | 0.005711601 |
| 0 | 0 | 0 | 0.001462061 | 0 |
| 0 | 0 | 0 | 0 | 0 |
| 0 | 0 | 0 | 0 | 0 |
| 0 | 0 | 0 | 0 | 0 |
| 0.009142541 | 0 | 0.001007035 | 0 | 0 |
| 0.344636374 | 0.077743506 | 0.043718512 | 0.069585222 | 0 |
| 0.059662183 | 0.01188852 | 0.012644369 | 0.160847916 | 0 |
| 0 | 0 | 0 | 0 | 0 |
| 0.006668717 | 0.002891919 | 0 | 0.001205095 | 0 |
| 0.000988688 | 0 | 0 | 0 | 0 |
| 0 | 0 | 0 | 0 | 0 |
| 0 | 0 | 0 | 0 | 0 |
| 0 | 0 | 0 | 0 | 0 |
| 0.011241392 | 2.703469297 | 0 | 0.081619426 | 0 |
| 0.07368423 | 0.025558615 | 0.283465578 | 0.198673891 | 0 |
| 0 | 0 | 0.000762394 | 0.004502803 | 0 |
| 0 | 0 | 0 | 0 | 0 |
| 0.001705019 | 0.004604031 | 0.008399054 | 0.015405582 | 0.002723182 |
| 0 | 0 | 0 | 0 | 0 |
| 0 | 0 | 0 | 0 | 1.285452966 |
| 0 | 0 | 0 | 0 | 0 |
| 0.026236579 | 0.258246443 | 0.006413583 | 0.073924927 | 0 |
| 0 | 0 | 0.002251707 | 0 | 0.003194009 |
| 0 | 0 | 0 | 0 | 0 |
| 0 | 0.909454839 | 0 | 0 | 0 |
| 0 | 0 | 0 | 0.165227463 | 0 |
| 0 | 0 | 0 | 0 | 0.047111176 |

| | | | | |
|---|---|---|---|---|
| 0.001205693 | 0 | 0 | 0.002876005 | 0.033506843 |
| 0 | 0 | 0 | 0 | 0 |
| 0 | 0 | 0 | 0 | 0 |
| 0.002814489 | 0.737378559 | 0.358040848 | 2.57360162 | 0 |
| 0 | 0 | 0 | 0 | 0 |
| 0 | 0 | 0 | 0 | 0 |
| 0 | 0 | 0 | 0 | 0 |
| 0 | 0.017926811 | 0 | 0 | 0 |
| 2.545889706 | 3.202655942 | 0.106148938 | 1.615462386 | 0 |
| 0 | 0.048543231 | 0.138682041 | 0 | 0 |
| 0 | 0 | 0 | 0 | 0 |
| 0.005393167 | 0.014549291 | 0 | 0.006891757 | 0 |
| 0 | 0 | 0 | 0 | 0 |
| 0 | 0 | 6.057889973 | 0 | 0 |
| 0.103155184 | 2.576010157 | 0 | 0 | 0 |
| 0 | 0 | 0 | 0 | 0 |
| 0 | 0.164973561 | 0 | 0 | 0 |
| 0 | 0 | 0 | 0 | 0 |
| 0 | 0 | 0 | 0 | 0 |
| 0 | 0.001181174 | 0 | 0 | 0 |
| 0 | 0 | 0 | 0 | 0 |
| 0 | 0 | 0 | 0 | 0 |
| 0 | 0 | 0 | 0 | 0 |
| 0.14698041 | 0.271086668 | 0.145841711 | 0.335327972 | 0.286836059 |
| 0.02272222 | 0 | 0 | 0 | 0 |
| 0 | 0 | 1.855286247 | 0 | 0 |
| 0 | 0 | 0 | 0 | 0 |
| 0.553541588 | 0.000538132 | 0 | 0 | 0 |
| 0 | 0 | 0 | 0 | 0 |
| 0 | 0 | 0 | 0 | 0 |
| 0 | 0 | 0 | 0 | 0 |
| 0 | 1.831788343 | 0 | 0 | 0 |
| 0.146503472 | 0 | 0.002647658 | 0 | 0 |
| 0 | 0 | 0 | 0 | 0 |
| 0 | 0 | 0 | 0 | 0 |
| 0 | 0.737697298 | 0 | 0 | 0 |
| 0 | 0.027831521 | 0.507012377 | 0 | 0 |
| 0 | 0 | 0 | 0 | 0 |
| 0 | 0.05866851 | 0 | 0 | 0 |
| 0 | 0.039658975 | 0 | 0 | 0 |
| 0.131561816 | 0.04022678 | 0.206440137 | 0.009015557 | 0 |
| 0 | 0.004442533 | 0 | 0 | 0 |
| 3.471918384 | 0.538841641 | 0.011792871 | 6.849669406 | 0 |
| 0 | 0.000656297 | 0 | 0 | 0 |
| 0 | 0 | 0 | 0 | 0 |
| 0 | 0 | 0 | 0 | 0 |
| 0 | 0 | 0 | 0 | 0 |
| 0.008218594 | 0.098358886 | 0.009754018 | 0 | 0 |

| | | | | |
|---|---|---|---|---|
| 0 | 0 | 0 | 0 | 0 |
| 0 | 0 | 0 | 0 | 0 |
| 0.016821555 | 0.000502626 | 0.03830038 | 0.530079637 | 1.134776227 |
| 0 | 0 | 0.028572437 | 0 | 0 |
| 2.041014878 | 0 | 0 | 0 | 0 |
| 0.051328101 | 0.010330683 | 0.003824564 | 0 | 0 |
| 0 | 0 | 0 | 0 | 0 |
| 0 | 0 | 0 | 0 | 0 |
| 0 | 0 | 0 | 0 | 0 |
| 0 | 0 | 0 | 0 | 0 |
| 0.823071406 | 0.975763821 | 0.009003818 | 0.090896396 | 0.012020488 |
| 0.927643628 | 0.479523155 | 1.464060776 | 1.183604512 | 0 |
| 0 | 0 | 0 | 0 | 0 |
| 0.001805013 | 0.012104415 | 0.006859262 | 0 | 0.005135146 |
| 0.090501458 | 0.071826735 | 0.116815863 | 0.134947867 | 1.557337541 |
| 0 | 0 | 0 | 0 | 0 |
| 0 | 0 | 0 | 0 | 0 |
| 0 | 0 | 0 | 0 | 0 |
| 0 | 0 | 0 | 0 | 0 |
| 0 | 0.007021462 | 0.044500658 | 0 | 0 |
| 0.034869483 | 0.015755051 | 0.451311811 | 0.077324484 | 0.092563506 |
| 0.032495098 | 0 | 0 | 0.032029884 | 0 |
| 0.414419192 | 0.520516309 | 0.36226382 | 3.048801189 | 0.030927079 |
| 0 | 0 | 0 | 0 | 0 |
| 0 | 0 | 0 | 0 | 0 |
| 11.46921045 | 4.996159459 | 0.001950451 | 0 | 0 |
| 0 | 0 | 1.366287998 | 0 | 0 |
| 0 | 0 | 0 | 0 | 0 |
| 0 | 0 | 0 | 0 | 0 |
| 0 | 0 | 0 | 0 | 0 |
| 0 | 0.00398146 | 0 | 0 | 0 |
| 0 | 0.342012689 | 0.474687169 | 0 | 0 |
| 0.005582477 | 0 | 0 | 1.142118837 | 1.31117955 |
| 0.31158482 | 0.385036557 | 0.00069134 | 0 | 0.007845241 |
| 0.503076485 | 0.002067892 | 0.005026196 | 0.031805791 | 0.00373454 |
| 0.020638538 | 0.117532008 | 0.011345055 | 0.183874464 | 0 |
| 0.005078177 | 0.091114011 | 0 | 0 | 0 |
| 0 | 0 | 0 | 0 | 0 |
| 0.020277943 | 0.733444825 | 0 | 0 | 0.055403692 |
| 0 | 0 | 0 | 0 | 0 |
| 0 | 0.014937112 | 0 | 0 | 0 |
| 0 | 0 | 0 | 0 | 0 |
| 0 | 0 | 0 | 0 | 0 |
| 0 | 0 | 0 | 0 | 0 |
| 0.33905033 | 2.185820841 | 0.040500773 | 0.019500257 | 0.002925678 |
| 0 | 0 | 0 | 0 | 0 |
| 0 | 0 | 0 | 0 | 0 |
| 0.137400466 | 0.939698785 | 0 | 0.019166595 | 0.151112546 |

| | | | | |
|---|---|---|---|---|
| 0 | 0 | 0 | 0 | 0 |
| 0 | 0 | 0 | 0 | 0 |
| 0 | 0 | 0 | 0 | 0 |
| 0.008137369 | 0 | 0 | 0 | 0 |
| 0 | 0 | 0 | 0 | 0 |
| 0 | 0 | 0 | 0 | 0 |
| 0.297100439 | 0.000444598 | 0 | 0 | 0 |
| 0.007487853 | 1.52753727 | 0 | 0.216373283 | 0 |
| 0 | 0 | 0 | 0 | 0 |
| 0 | 0 | 0 | 0 | 0 |
| 0.054588011 | 0.006173671 | 0.00284775 | 0.052102978 | 0.009341315 |
| 0 | 0 | 0 | 0 | 0 |
| 0 | 0.451847031 | 0 | 0 | 0 |
| 0 | 0 | 0 | 0 | 0 |
| 0 | 0 | 0 | 0 | 0 |
| 0 | 0 | 0 | 0 | 0 |
| 0 | 0 | 0 | 0 | 0 |
| 0 | 0.010956609 | 0 | 0 | 0 |
| 0 | 0.46111093 | 0.010757927 | 0 | 0 |
| 0 | 0.153614632 | 0.012560558 | 0 | 0 |
| 0.036475782 | 0 | 0 | 0 | 0 |
| 0 | 0 | 0 | 0 | 0 |
| 0 | 0.208214032 | 0 | 0 | 0 |
| 0 | 0 | 0 | 0 | 0 |
| 0 | 0 | 0 | 0 | 0 |
| 0 | 0 | 0 | 0 | 0 |
| 0 | 0 | 0 | 0 | 0 |
| 0 | 0 | 0 | 0 | 0 |
| 0 | 0 | 0.001212646 | 0 | 0 |
| 0 | 0.023241696 | 0.003758697 | 0 | 0 |
| 0 | 0 | 0.000841095 | 0 | 0 |
| 0 | 0 | 0 | 0 | 0 |
| 0 | 0 | 0 | 0 | 0 |
| 0 | 0 | 0 | 0 | 0 |
| 0 | 0.000413579 | 0 | 0 | 0 |
| 0 | 0 | 0 | 0 | 0 |
| 0 | 0 | 0 | 0 | 0 |
| 0.227837047 | 0.020138751 | 0 | 0 | 0.013464269 |
| 0 | 0 | 0 | 0 | 0 |
| 0 | 0.046694921 | 0.004309676 | 0 | 0 |
| 0 | 0.108054296 | 0.026152127 | 0 | 0 |
| 0 | 0 | 0 | 0 | 0 |
| 0 | 0 | 0 | 0 | 0 |
| 0 | 0 | 0 | 0 | 0 |
| 0 | 0 | 0 | 0 | 0 |
| 0 | 0 | 0 | 0 | 0 |
| 0 | 0.008967345 | 0 | 0 | 0 |
| 0 | 0 | 0 | 0 | 0 |

| | | | | |
|---|---|---|---|---|
| 0 | 0 | 0 | 0 | 0 |
| 0 | 0 | 0 | 0 | 0 |
| 0 | 0 | 0 | 0 | 0 |
| 0 | 0.584493341 | 0 | 0 | 0 |
| 0 | 0 | 0 | 0 | 0 |
| 0 | 0 | 0 | 0 | 0 |
| 0 | 0 | 0 | 0 | 0 |
| 0 | 0 | 0 | 0 | 0 |
| 0.004183109 | 0.141038456 | 0.017039176 | 0.110720642 | 2.287582509 |
| 0 | 0.000212613 | 0 | 0 | 0 |
| 0 | 0 | 0 | 0 | 0 |
| 0 | 0 | 0 | 0 | 0 |
| 0 | 0.003126671 | 0 | 0 | 0 |
| 0 | 0 | 0 | 0 | 0 |
| 0 | 0.065513892 | 3.545766023 | 3.946320334 | 0 |
| 0 | 0.000569761 | 0.163676626 | 0.947532246 | 0 |
| 0 | 0 | 0 | 0.085289595 | 0 |
| 0 | 0 | 0 | 0 | 0 |
| 0 | 0 | 0 | 0 | 0 |
| 0 | 0 | 0 | 0 | 0 |
| 0 | 0 | 0 | 0 | 0 |
| 0 | 0 | 0 | 0 | 0 |
| 0 | 0 | 0 | 0 | 0 |
| 0 | 0 | 0 | 0 | 0 |
| 0.002893657 | 0.000217334 | 0.236296896 | 0.077808848 | 0.006239193 |
| 1.572341898 | 0.280376668 | 0.176425305 | 4.395553433 | 0 |
| 0 | 0.007737115 | 0.522059207 | 0 | 0 |
| 0 | 0.00406656 | 0 | 0 | 0 |
| 0 | 0 | 0 | 0 | 0 |
| 0 | 0 | 0 | 0 | 0 |
| 0 | 0 | 0 | 0 | 0 |
| 0 | 0.191131612 | 0 | 0 | 0 |
| 0 | 0 | 0 | 0 | 0 |
| 0.021635584 | 0 | 0 | 0 | 0 |
| 0.004838025 | 0.00278675 | 0.007956108 | 0 | 0 |
| 0.076137716 | 0.010795795 | 0.006384005 | 0.162567853 | 0.001293658 |
| 0 | 0 | 0 | 0 | 0.084106219 |
| 1.977370744 | 1.788509493 | 0 | 0.478861387 | 0 |
| 0 | 0 | 0 | 0 | 0 |
| 0 | 0 | 0 | 0 | 0 |
| 0 | 0 | 0 | 0 | 0 |
| 0.001837038 | 0 | 0.013444809 | 0 | 0 |
| 0.280647009 | 0.364092149 | 0.041542172 | 0.709392514 | 0 |
| 0 | 0 | 0 | 0 | 0 |
| 0 | 0 | 0 | 0 | 0 |
| 0.008867159 | 0.005690166 | 0 | 0 | 0 |
| 0 | 0 | 0 | 0 | 0 |
| 0.050800878 | 0.01900468 | 0.005505861 | 0 | 0 |

| | | | | |
|---|---|---|---|---|
| 0 | 0.034625298 | 0 | 0 | 0 |
| 0 | 0 | 0 | 0 | 0 |
| 0 | 0 | 0 | 0 | 0 |
| 0 | 0 | 0 | 0 | 0 |
| 0.022791066 | 0.013590305 | 0 | 0.002186837 | 0 |
| 0 | 0 | 0 | 0 | 0 |
| 0 | 0 | 0 | 0 | 0 |
| 0 | 0 | 0 | 0 | 0 |
| 0 | 0 | 0 | 0 | 0 |
| 0 | 0 | 0 | 0 | 0 |
| 0 | 0.002069858 | 0 | 0 | 0 |
| 3.321955516 | 0.691442508 | 0 | 3.053481018 | 0 |
| 0.032236018 | 0.017870437 | 0 | 0 | 0 |
| 0 | 0 | 0 | 0 | 0 |
| 0.348483654 | 0.462005202 | 0.002757683 | 0.800958601 | 0 |
| 0 | 0 | 0 | 0 | 0 |
| 0.000973577 | 0 | 0 | 0 | 0 |
| 0 | 0 | 0 | 0 | 0 |
| 0.002657818 | 0 | 0 | 0 | 0 |
| 0 | 0 | 0.015123855 | 0 | 0 |
| 0 | 0 | 0 | 0 | 0 |
| 0 | 0 | 0.21115597 | 0 | 0 |
| 2.078427728 | 0.025062946 | 0.110631993 | 0.688671237 | 0 |
| 0 | 0 | 0 | 0 | 0 |
| 0 | 0 | 0 | 0 | 0 |
| 0 | 0 | 0 | 0 | 0 |
| 0 | 0 | 0 | 0 | 0 |
| 0 | 0 | 0 | 0 | 0 |
| 0 | 0 | 0 | 0 | 0 |
| 0 | 0 | 0 | 0 | 0 |
| 0 | 0 | 0 | 0 | 0 |
| 0.115927043 | 0.007307215 | 0.042941746 | 0.031833758 | 0.027192931 |
| 0 | 0 | 0.496855273 | 0 | 0 |
| 0 | 0.169012743 | 0.006895593 | 1.340984726 | 0.00072454 |
| 0.068949489 | 0.010403439 | 0.004135355 | 0.155463936 | 0 |
| 0.017423333 | 0.362538456 | 0.001279434 | 0.002190294 | 0 |
| 0 | 0 | 0 | 0 | 0 |
| 0 | 0.472162957 | 0 | 0 | 0 |
| 0.002250363 | 0 | 0 | 0 | 0 |
| 0 | 0 | 0 | 0 | 0 |
| 0 | 0 | 0.127544829 | 0 | 0 |
| 0 | 0 | 0 | 0 | 0 |
| 0 | 0 | 0 | 0 | 0 |
| 0 | 0.094480387 | 0.198896358 | 0 | 0 |
| 0.029330022 | 0.739274156 | 0.018576252 | 0.154193649 | 0.001849131 |
| 0 | 0 | 0 | 0 | 0 |
| 0 | 0 | 0 | 0 | 0 |
| 0.003832632 | 0.094895267 | 0 | 0 | 0 |

| | | | | |
|---|---|---|---|---|
| 0 | 0 | 1.085215464 | 0 | 0 |
| 0 | 0 | 0 | 0 | 0 |
| 0 | 0.062868609 | 0.004044735 | 0 | 0 |
| 0 | 0 | 0.03990663 | 0 | 0 |
| 0 | 0 | 0 | 0 | 0 |
| 0 | 0 | 0.043060613 | 0 | 0 |
| 0 | 0 | 0 | 0 | 0 |
| 0 | 0.007704807 | 0.100271704 | 0.1307867 | 0 |
| 0 | 0 | 0 | 0 | 0 |
| 0 | 0.022387276 | 0 | 0 | 0 |
| 0 | 0 | 0 | 0 | 0 |
| 0.003045859 | 8.71489E-05 | 0 | 0.06252692 | 0 |
| 0 | 0 | 0 | 0 | 0 |
| 0 | 0 | 0 | 0 | 0 |
| 0 | 0 | 0 | 0 | 0 |
| 0 | 0 | 0 | 0 | 0 |
| 0 | 0 | 0 | 0 | 0 |
| 0 | 0 | 0 | 0 | 0 |
| 0.161640989 | 0.003546479 | 1.914976193 | 0 | 0 |
| 0 | 0 | 0 | 0 | 0 |
| 0.055667351 | 0.009533264 | 0 | 0 | 0 |
| 0 | 0 | 0 | 0 | 0 |
| 0 | 0 | 0 | 0 | 0 |
| 0 | 0 | 0 | 0 | 0 |
| 0 | 0.000113104 | 0 | 0 | 0 |
| 0.392511418 | 0.004240401 | 0.002427198 | 0.091154245 | 0.03485978 |
| 0 | 0 | 0 | 0 | 0 |
| 0.011604842 | 0.002697831 | 0.017224147 | 0 | 0 |
| 0 | 0 | 0 | 0 | 0 |
| 0 | 0.592564065 | 0.706999135 | 0 | 0 |
| 0 | 0 | 0 | 0 | 0 |
| 0 | 0 | 0 | 0 | 0 |
| 0 | 0 | 0 | 0 | 0 |
| 0 | 0 | 0 | 0 | 0 |
| 0.032567364 | 0.009533708 | 0 | 0.026738558 | 0 |
| 0 | 0 | 0 | 0 | 0 |
| 0 | 0 | 0 | 0 | 0 |
| 0.151662644 | 0.085512357 | 0 | 0.013201562 | 0 |
| 0 | 0 | 0 | 0 | 0 |
| 0 | 0 | 0 | 0 | 0 |
| 18.10515768 | 1.188889768 | 20.06849749 | 12.09899373 | 0 |
| 0.699880869 | 0.042781655 | 1.228800907 | 0.188355495 | 0 |
| 0.853948418 | 0 | 0 | 0 | 0.188403161 |
| 0 | 0 | 0 | 0 | 0 |
| 0 | 0 | 0 | 0 | 0 |
| 0 | 0 | 0 | 0 | 0 |
| 0 | 0 | 0 | 0 | 0 |
| 0.009832868 | 0.112505893 | 0.000715825 | 0.053122713 | 0 |

| | | | | |
|---|---|---|---|---|
| 0 | 0 | 0 | 0 | 0 |
| 0.017263409 | 0.004902031 | 0.008540903 | 0.004726738 | 0 |
| 0.005498915 | 0 | 0.002063854 | 0.001104113 | 0 |
| 0 | 0 | 0 | 0 | 0 |
| 1.056852907 | 1.675537211 | 0.031625453 | 2.854929721 | 0 |
| 0 | 0 | 0 | 0 | 0 |
| 0 | 0 | 0 | 0 | 0 |
| 0 | 0 | 0 | 0 | 0 |
| 0.630491335 | 0.045323637 | 0 | 0.697204364 | 10.8429081 |
| 0 | 0 | 0 | 0 | 0 |
| 0 | 0 | 0 | 0 | 0 |
| 0 | 0 | 0 | 0 | 0 |
| 0 | 0 | 0.228507101 | 0 | 0 |
| 0.054784627 | 0.002682054 | 0.023219952 | 0.010841126 | 0 |
| 0 | 2.700688678 | 0 | 0 | 0.011569287 |
| 0.178797865 | 0 | 2.050339311 | 0.148639831 | 0 |
| 0.021090072 | 0.002765742 | 0.002131104 | 0 | 0 |
| 0 | 0 | 0 | 0 | 0 |
| 0 | 0 | 0 | 0 | 0 |
| 0 | 0 | 0 | 0 | 0 |
| 0 | 0 | 0 | 0 | 0 |
| 0 | 0 | 0 | 0 | 0 |
| 0 | 0 | 0 | 0 | 0 |
| 0 | 0 | 0 | 0 | 0 |
| 0 | 0 | 0 | 0 | 0 |
| 0 | 0 | 0 | 0 | 0 |
| 0 | 0.006138447 | 0.043547441 | 0 | 0 |
| 0.000884323 | 0.001935639 | 0 | 0.000767063 | 0 |
| 0 | 0 | 0 | 0 | 0 |
| 0 | 0 | 0 | 0 | 0 |
| 0 | 0 | 0 | 0 | 0 |
| 0 | 0 | 0.000998071 | 0 | 0 |
| 0 | 0 | 0 | 0 | 0 |
| 0 | 0 | 0 | 0 | 0 |
| 0 | 0 | 0 | 0 | 0.001050675 |
| 0 | 0 | 0 | 0 | 0 |
| 0 | 0 | 0 | 0 | 0 |
| 0 | 0.004850627 | 0.001631485 | 0 | 0 |
| 0 | 0 | 0 | 0 | 0 |
| 0 | 0 | 0 | 0 | 0 |
| 0.065150009 | 0.017959777 | 1.495199672 | 0 | 0 |
| 0 | 0 | 0 | 0 | 0 |
| 0 | 0 | 0 | 0 | 0 |
| 0 | 0 | 0 | 0 | 0 |
| 0 | 0 | 0 | 0 | 0 |
| 0 | 0 | 0 | 0 | 0 |
| 0 | 0 | 0 | 0 | 0 |
| 0 | 0 | 0 | 0 | 0 |

| | | | | |
|---|---|---|---|---|
| 0 | 0 | 0.002289445 | 0 | 0 |
| 0 | 0 | 0 | 0 | 0 |
| 0 | 0 | 0 | 0 | 0 |
| 0.006210939 | 0.0007309 | 0 | 0.006516989 | 0 |
| 0 | 0.015611471 | 0.012950696 | 0 | 0 |
| 0 | 0 | 0.013751524 | 0 | 0 |
| 0.024275526 | 0.004977806 | 0.039936966 | 0.002924532 | 0 |
| 0.001390549 | 0.001529303 | 0 | 0 | 0 |
| 0 | 0 | 0 | 0 | 0 |
| 0.097536752 | 0 | 0 | 0.102890653 | 0 |
| 0 | 0.004960146 | 0 | 0 | 0 |
| 0 | 0 | 0.008897404 | 0 | 0 |
| 0 | 0 | 0 | 0 | 0 |
| 0 | 0 | 0 | 0 | 0 |
| 0 | 0 | 0 | 0 | 0 |
| 0 | 0 | 0 | 0 | 0 |
| 0 | 0 | 0 | 0.334564621 | 0 |
| 0 | 0 | 0 | 0 | 0 |
| 0 | 0 | 0 | 0 | 0 |
| 0 | 0 | 0 | 0 | 0 |
| 1.136408593 | 0.050661131 | 0.000578285 | 0 | 0 |
| 0 | 0 | 0 | 0 | 0 |
| 0 | 0.054643147 | 0.002232461 | 0 | 0 |
| 0.050876652 | 0.033978245 | 0 | 0 | 0.003620405 |
| 0 | 0 | 0 | 0 | 0 |
| 0.002179053 | 0 | 0 | 0 | 0 |
| 0 | 0 | 0 | 0 | 0 |
| 0 | 0 | 0 | 0 | 0 |
| 0 | 0 | 0 | 0 | 0 |
| 0.039390822 | 0 | 0.012040482 | 0.000786066 | 0.053553511 |
| 0 | 0 | 0 | 0 | 0 |
| 0 | 0 | 0 | 0 | 0 |
| 0 | 0 | 0 | 0 | 0 |
| 0.00286908 | 0.01394903 | 0 | 0 | 0 |
| 0 | 0 | 0 | 0 | 0 |
| 0 | 0 | 0 | 0 | 0 |
| 0.008403943 | 0 | 0 | 0.628271797 | 0.002265032 |
| 0 | 0 | 0 | 0 | 0 |
| 0 | 0 | 0 | 0 | 0 |
| 0 | 0 | 0 | 0 | 0 |
| 0 | 0 | 0 | 0 | 0 |
| 0 | 0 | 0 | 0 | 0 |
| 0 | 0 | 0 | 0 | 0 |
| 0 | 0.098693445 | 0 | 0 | 0 |
| 0 | 0 | 0 | 0 | 0 |
| 0 | 0 | 0 | 0 | 0 |
| 0 | 0 | 0 | 0 | 0 |
| 0 | 0 | 0 | 0 | 0 |

| | | | | |
|---|---|---|---|---|
| 0.062514188 | 0.092005457 | 0 | 0.369074116 | 0 |
| 0 | 0 | 0 | 0 | 0 |
| 0 | 0.000875452 | 0 | 0 | 0 |
| 0 | 0 | 0 | 0 | 0 |
| 0 | 0 | 0 | 0 | 0 |
| 0 | 0 | 0 | 0 | 0 |
| 0 | 0 | 0 | 0 | 0 |
| 0.252484942 | 0.452339594 | 0.239378501 | 0.88402883 | 0 |
| 0.713819803 | 0.005416375 | 0.194945948 | 2.168749799 | 0.171895564 |
| 2.132478431 | 0.073020587 | 0 | 1.114007618 | 0.016081938 |
| 0 | 0 | 0 | 0 | 0 |
| 0 | 0 | 0 | 0 | 0 |
| 0 | 0 | 0 | 0 | 0 |
| 0 | 0 | 0 | 0 | 0.02389218 |
| 0 | 0 | 0 | 0 | 0 |
| 0 | 0 | 0 | 0 | 0 |
| 0 | 0 | 0 | 0.086931881 | 0 |
| 0 | 0 | 0 | 0 | 0 |
| 0 | 0 | 0 | 0 | 0 |
| 0 | 0.865289668 | 0.011889825 | 0.110056185 | 0 |
| 0 | 0 | 0 | 0 | 0 |
| 0 | 0 | 0 | 0 | 0 |
| 0 | 0 | 4.201633112 | 0 | 0 |
| 0 | 0 | 0 | 0 | 0 |
| 0.113988758 | 0 | 0.035091084 | 0 | 0 |
| 0 | 0 | 0 | 0 | 0 |
| 0 | 0 | 0 | 0 | 0 |
| 0 | 0 | 0 | 0 | 0 |
| 0 | 0 | 0 | 0.00507735 | 0 |
| 0 | 0 | 0 | 0 | 0 |
| 0 | 0 | 0 | 0 | 0 |
| 0 | 0 | 0 | 0 | 0 |
| 0 | 0.000422977 | 0 | 0 | 0 |
| 0 | 0 | 0 | 0 | 0 |
| 0 | 0.102584549 | 0 | 0.002371636 | 0 |
| 0 | 0 | 0 | 0 | 0 |
| 0 | 0 | 0 | 0 | 0 |
| 0 | 0 | 0 | 0 | 0 |
| 0 | 0 | 0 | 0 | 0 |
| 0.001082232 | 0.043118917 | 0 | 0 | 2.413842911 |
| 0 | 0 | 0 | 0 | 0 |
| 0.682934906 | 0.933701923 | 0.078571977 | 0 | 0 |
| 0 | 0 | 0 | 0 | 0 |
| 0 | 0 | 0 | 0 | 0 |
| 0 | 0 | 0 | 0 | 0 |
| 0 | 0.680521768 | 0.000407014 | 0.045464689 | 0 |
| 0 | 0 | 0 | 0 | 0 |
| 0 | 0 | 0 | 0 | 0 |

| | | | | |
|---|---|---|---|---|
| 0 | 0 | 0 | 0 | 0 |
| 0 | 0.000489905 | 0 | 0.002475307 | 0 |
| 0 | 0 | 0 | 0 | 0 |
| 0 | 0 | 0 | 0 | 0 |
| 0.792503827 | 11.27986352 | 0.087602687 | 0.136949849 | 0.010104902 |
| 0 | 0.365436694 | 0 | 0 | 0 |
| 0 | 0 | 0 | 0 | 0 |
| 0.001608838 | 0.019678889 | 0.001552704 | 0.082982929 | 0.120586093 |
| 0.055075383 | 0.000541692 | 0 | 0 | 0 |
| 0 | 0 | 0 | 0 | 0 |
| 0 | 0 | 0 | 0 | 0 |
| 0 | 0 | 0 | 0 | 0 |
| 0 | 0 | 0 | 0 | 0 |
| 0 | 0 | 0 | 0 | 0 |
| 0 | 0 | 0 | 0 | 0 |
| 0 | 0 | 0.017338104 | 0 | 0 |
| 0 | 0 | 0 | 0 | 0 |
| 0 | 0 | 0 | 0 | 0 |
| 0 | 0 | 0 | 0 | 0 |
| 0 | 0 | 0 | 0 | 0 |
| 0 | 0 | 0 | 0 | 0 |
| 0 | 0 | 0 | 0 | 0 |
| 0 | 0 | 0 | 0 | 0 |
| 0.040377193 | 0.003201214 | 0.005542918 | 0.004744527 | 0.000953035 |
| 0 | 0 | 0 | 0 | 0 |
| 0 | 0 | 0 | 0 | 0 |
| 0.124614408 | 0.068278009 | 0.03994252 | 0.005128392 | 0.015907771 |
| 0 | 0 | 0 | 0.00062308 | 0 |
| 0 | 0 | 0 | 0.029318161 | 0 |
| 0 | 0 | 0 | 0 | 0 |
| 0 | 0 | 0 | 0 | 0 |
| 0 | 0 | 0 | 0 | 0 |
| 0 | 0 | 0 | 0 | 0 |
| 0 | 0 | 0 | 0 | 0 |
| 0 | 0 | 0 | 0 | 0 |
| 0 | 0 | 0 | 0 | 0 |
| 0 | 0 | 0 | 0 | 0 |
| 0 | 0.017275567 | 0 | 0 | 0 |
| 0 | 0 | 0 | 0 | 0 |
| 0 | 0 | 0 | 0 | 0 |
| 0 | 0 | 0 | 0 | 0 |
| 0 | 8.699560308 | 0 | 0 | 0 |
| 0 | 0 | 0 | 0 | 0 |
| 0 | 0 | 0 | 0 | 0 |
| 0 | 0 | 0 | 0 | 0 |
| 0 | 0 | 0 | 0 | 0 |
| 0 | 0 | 0 | 0 | 0 |
| 0 | 0 | 0 | 0 | 0 |

| | | | | |
|---|---|---|---|---|
| 0 | 0.375950961 | 0 | 0 | 0 |
| 0 | 0 | 0 | 0 | 0 |
| 0 | 0 | 0 | 0 | 0 |
| 0.001021285 | 0 | 0 | 0 | 0 |
| 0 | 0 | 0 | 0 | 0 |
| 0 | 0 | 0 | 0.478806233 | 0 |
| 0 | 0 | 0 | 0 | 0 |
| 0 | 0 | 0 | 0 | 0 |
| 0 | 0 | 0 | 0 | 0 |
| 0 | 0 | 0 | 0 | 0 |
| 0 | 0 | 0 | 0 | 0 |
| 0 | 0 | 0 | 0 | 0 |
| 0 | 0 | 0 | 0 | 0 |
| 0 | 0 | 0 | 0 | 0 |
| 0 | 0.000845412 | 0 | 0 | 0 |
| 0 | 0 | 0 | 0 | 0 |
| 0 | 0 | 0 | 0 | 0 |
| 0 | 0 | 0 | 0 | 0 |
| 0 | 0.003610542 | 0 | 0 | 0 |
| 0 | 0 | 0 | 0 | 0 |
| 0.361479495 | 0 | 0.012544879 | 0.046602662 | 0.024675327 |
| 0 | 0 | 0 | 0 | 0 |
| 0.019261488 | 0.08808887 | 0.004436115 | 0.125848278 | 0 |
| 0 | 0.002358275 | 1.717449351 | 0 | 0 |
| 0 | 0 | 0 | 0 | 0 |
| 0 | 0 | 0 | 0 | 0 |
| 0 | 0 | 0 | 0 | 0 |
| 0 | 0 | 0 | 0 | 0 |
| 0 | 0 | 0 | 0 | 0 |
| 0 | 0 | 0 | 0 | 0 |
| 0 | 0 | 0 | 0 | 0 |
| 0 | 0.029362503 | 0 | 0 | 0 |
| 0.001405582 | 0.013171026 | 0.01266104 | 0.009144031 | 0.208217706 |
| 0 | 0 | 0 | 0 | 0 |
| 0.012782413 | 0.144547237 | 0 | 0 | 0 |
| 0 | 0.00816894 | 0 | 0.003465026 | 0.003828115 |
| 0 | 0 | 0 | 0 | 0 |
| 0 | 0 | 0 | 0 | 0 |
| 0.141768571 | 0.043253918 | 0.007161127 | 0.141453536 | 0.129448652 |
| 0 | 0 | 0 | 0 | 0 |
| 0 | 0 | 0 | 0 | 0 |
| 0 | 0 | 0 | 0 | 0 |
| 0.249163407 | 2.942756284 | 0.010305851 | 0.19600086 | 0 |
| 0 | 0 | 0 | 0 | 0 |
| 0 | 0 | 0 | 0 | 0 |
| 0.076324741 | 0.08211001 | 0 | 0 | 0 |
| 0 | 0 | 0 | 0 | 0 |
| 0 | 0 | 0 | 0 | 0 |

| | | | | |
|---|---|---|---|---|
| 0 | 0 | 0 | 0 | 0 |
| 0 | 0 | 0 | 0 | 0 |
| 0 | 0 | 0 | 0 | 0 |
| 0 | 0 | 0 | 0 | 0 |
| 0 | 0 | 0 | 0 | 0 |
| 0 | 0.015006167 | 0 | 0 | 0 |
| 0 | 0 | 0 | 0 | 0 |
| 0.235505112 | 0 | 0.047618459 | 0 | 0 |
| 0.150017991 | 0 | 0.03418758 | 0.017845125 | 0 |
| 0 | 0 | 0 | 0.006313851 | 0.002242112 |
| 0 | 0 | 0 | 0 | 0 |
| 0 | 0 | 0 | 0 | 0 |
| 0 | 0 | 0 | 0 | 0 |
| 0 | 0 | 0 | 0 | 0 |
| 0 | 0.042847017 | 0 | 0 | 0 |
| 0 | 0 | 0.001209581 | 0.026401559 | 0.389725049 |
| 0 | 0 | 0 | 0 | 0 |
| 0 | 0 | 0 | 0 | 0 |
| 0 | 0 | 0 | 0 | 0 |
| 0 | 0 | 0 | 0 | 0 |
| 0 | 0.003966987 | 0 | 0 | 0 |
| 0 | 0 | 0 | 0 | 0 |
| 0 | 0 | 0 | 0 | 0 |
| 0 | 0 | 0 | 0 | 0 |
| 0 | 0 | 0 | 0 | 0 |
| 0 | 0 | 0 | 0 | 0 |
| 0 | 0 | 0 | 0 | 0 |
| 0 | 0 | 0 | 0 | 0 |
| 0 | 0 | 0 | 0 | 0 |
| 0 | 1.913154867 | 0.341168539 | 0 | 0 |
| 0.582753925 | 0.119647395 | 1.095047793 | 0.50165879 | 0 |
| 0 | 0 | 0 | 0 | 0 |
| 0 | 0 | 0 | 0 | 0 |
| 0 | 0 | 0 | 0 | 0 |
| 0 | 0 | 0 | 0 | 0 |
| 0 | 0 | 0 | 0 | 0 |
| 0 | 0 | 0 | 0 | 0 |
| 0 | 0 | 0 | 0 | 0 |
| 0 | 0 | 0 | 0 | 0 |
| 0 | 0 | 0 | 0 | 0 |
| 0 | 0.002321819 | 0 | 0 | 0 |
| 0 | 0 | 0 | 0 | 0 |
| 0 | 0 | 0 | 0 | 0 |
| 0 | 0.000633182 | 0 | 0 | 0 |
| 0 | 0 | 0 | 0 | 0 |
| 0 | 0 | 0 | 0 | 0 |
| 0.025555352 | 0.000157327 | 0.194403532 | 0.072223635 | 4.768434568 |
| 0 | 0 | 0 | 0 | 0 |

| | | | | |
|---|---|---|---|---|
| 0 | 0 | 0 | 0 | 0 |
| 0 | 0 | 0 | 0 | 0 |
| 0 | 0 | 0 | 0 | 0 |
| 0 | 8.05718E-05 | 0 | 0 | 0 |
| 0.237082054 | 0.186494563 | 0.10624941 | 0.288107077 | 0 |
| 0 | 0 | 0 | 0 | 0 |
| 0 | 0 | 0 | 0 | 0 |
| 0 | 0 | 0 | 0 | 0 |
| 0 | 0 | 0 | 0 | 0 |
| 0 | 0 | 0 | 0 | 0 |
| 0 | 0 | 0 | 0 | 0 |
| 0.008907796 | 0.280903286 | 0.005320701 | 0.03085943 | 0 |
| 0 | 0.013474993 | 0 | 0.128241902 | 0 |
| 0 | 0 | 0 | 0 | 0 |
| 0 | 0 | 0 | 0.001795194 | 0 |
| 0 | 0 | 0.04694035 | 0 | 0 |
| 0 | 0.016996969 | 0.009595622 | 0 | 0 |
| 0 | 0 | 0 | 0 | 0 |
| 0.040634719 | 0.051630881 | 0.004167792 | 0.135920658 | 0.102276637 |
| 0.014391861 | 0.006430277 | 0.00689303 | 0.001397409 | 0 |
| 0 | 0 | 0 | 0 | 0 |
| 0 | 0 | 0 | 0 | 0 |
| 0 | 0 | 0 | 0 | 0 |
| 0 | 0 | 0 | 0 | 0 |
| 0 | 0 | 0 | 0 | 0 |
| 0 | 0 | 0 | 0 | 0 |
| 0.038385481 | 0.059729035 | 0.000581442 | 0.011384715 | 0.007010513 |
| 0 | 0 | 0.019664487 | 0 | 0 |
| 0 | 0 | 0 | 0 | 0 |
| 0 | 0 | 0 | 0 | 0 |
| 0.037658333 | 0.006196962 | 0 | 0.076373232 | 0 |
| 0.028030188 | 0.000723615 | 0.005339248 | 0.029249236 | 0 |
| 0 | 0 | 0 | 0 | 0 |
| 0 | 0 | 0 | 0 | 0 |
| 0 | 0 | 0 | 0 | 0 |
| 0 | 0 | 0 | 0 | 0 |
| 0 | 0 | 0 | 0 | 0 |
| 0.277736796 | 0.83406268 | 0.0056725 | 0.106126144 | 0.00410528 |
| 0 | 0 | 0 | 0 | 0 |
| 0 | 0 | 0.002570873 | 0 | 0 |
| 0.002326557 | 0.001101309 | 0 | 0.013799962 | 0 |
| 0 | 0 | 0 | 0 | 0 |
| 0 | 0 | 0 | 0 | 0 |
| 0 | 0.001788526 | 0 | 0.001983676 | 0.00389607 |
| 0 | 0 | 0 | 0 | 0 |
| 0 | 0 | 0 | 0 | 0 |
| 0 | 0 | 0 | 0 | 0 |
| 0 | 0 | 0 | 0 | 0 |

| | | | | |
|---|---|---|---|---|
| 0 | 0 | 0 | 0 | 0 |
| 0 | 0.078148327 | 0.064851758 | 0 | 0 |
| 0 | 0 | 0 | 0 | 0 |
| 0.05901208 | 0.011132007 | 0.001079436 | 0.002078901 | 0 |
| 0 | 0.000254409 | 0.0008343 | 0 | 0 |
| 0 | 0.000273616 | 0 | 0 | 0 |
| 0 | 0.031787198 | 0.002444191 | 0 | 0 |
| 0 | 0 | 0 | 0 | 0 |
| 0 | 0 | 0 | 0 | 0 |
| 0 | 0 | 0 | 0 | 0 |
| 0 | 0 | 0 | 0 | 0 |
| 0 | 0 | 0 | 0 | 0 |
| 0.097706944 | 0.467936212 | 0.012625273 | 0 | 0 |
| 0 | 0 | 0 | 0 | 0 |
| 0 | 0 | 0 | 0 | 0 |
| 0 | 0 | 0 | 0 | 0 |
| 0 | 0 | 0 | 0 | 0 |
| 0 | 0 | 0 | 0 | 0 |
| 0 | 0 | 0 | 0 | 0 |
| 0 | 0 | 0 | 0 | 0 |
| 0 | 0 | 0 | 0 | 0 |
| 0 | 0 | 0 | 0 | 0 |
| 0.160672773 | 0.077663269 | 0.001847175 | 0.08818291 | 0.004525779 |
| 0.038952352 | 0.013444672 | 0 | 0 | 0 |
| 0 | 0 | 0 | 0 | 0 |
| 0 | 0.007270125 | 0 | 0 | 0 |
| 0.004179699 | 0 | 0.055030167 | 0 | 0 |
| 0 | 0.001975367 | 0 | 0 | 0 |
| 0 | 0 | 0 | 0 | 0 |
| 0 | 0 | 0 | 0 | 0 |
| 0 | 0.01298913 | 0.025838517 | 0 | 0 |
| 0.000857286 | 0 | 0 | 0 | 0 |
| 0 | 0 | 0 | 0 | 0 |
| 0 | 0 | 0 | 0 | 0 |
| 0 | 0.256827626 | 0.06137689 | 0 | 0 |
| 0 | 0 | 0 | 0 | 0.024154793 |
| 0 | 0 | 0.090709415 | 0 | 0 |
| 0 | 0 | 0 | 0 | 0.095991877 |
| 0 | 0 | 0 | 0.024000607 | 0 |
| 0 | 0 | 0 | 0 | 0 |
| 0 | 0 | 0 | 0 | 0 |
| 0 | 0 | 0 | 0 | 0 |
| 0 | 0 | 0 | 0 | 0 |
| 0 | 0 | 0 | 0 | 0 |
| 0 | 0 | 0 | 0 | 0 |
| 0.250213287 | 0.212012384 | 0.420783509 | 0.06478134 | 0 |
| 0 | 0 | 0 | 0 | 0.007059217 |
| 0 | 0 | 0 | 0 | 0 |

| | | | | |
|---|---|---|---|---|
| 0 | 0 | 0 | 0 | 0 |
| 0 | 0 | 0 | 0 | 0 |
| 0 | 0 | 0 | 0 | 0 |
| 0 | 0 | 0 | 0 | 0 |
| 0 | 0 | 0 | 0 | 0 |
| 0 | 0 | 0.001539828 | 0 | 0 |
| 0 | 0 | 0 | 0 | 0 |
| 0 | 0 | 0 | 0 | 0 |
| 0.00885211 | 0.007374883 | 0.000879452 | 0 | 0 |
| 0 | 0 | 0 | 0 | 0 |
| 0.002130602 | 0.013585668 | 0 | 0 | 0 |
| 0.008634131 | 0.002120985 | 0 | 0 | 0.001905206 |
| 0 | 0 | 0 | 0 | 0 |
| 0 | 0 | 0 | 0 | 0 |
| 0 | 0 | 0 | 0 | 0 |
| 0 | 0 | 0 | 0 | 0 |
| 0 | 0 | 0 | 0 | 0 |
| 0 | 0 | 0 | 0 | 0 |
| 0 | 0 | 0 | 0 | 0 |
| 0.001614597 | 0 | 0 | 0 | 0 |
| 0 | 0 | 0 | 0 | 0 |
| 0 | 0 | 0 | 0 | 0 |
| 0 | 0 | 0 | 0 | 0 |
| 0 | 0 | 0 | 0 | 0 |
| 0 | 0 | 0 | 0 | 0 |
| 0 | 0.000277394 | 0 | 0 | 0 |
| 0 | 0 | 0 | 0 | 0 |
| 0.046457433 | 0.007285323 | 0 | 0 | 0 |
| 0 | 0 | 0 | 0 | 0 |
| 0 | 0 | 0 | 0 | 0 |
| 0.028088892 | 0.026845363 | 0 | 0.004699913 | 0 |
| 0 | 0 | 0 | 0 | 0 |
| 0.011475319 | 0.009461225 | 0.015810925 | 0.024884272 | 0 |
| 0.004114572 | 0.000353182 | 0.001389854 | 0.002007555 | 0.002957227 |
| 0 | 0 | 0 | 0 | 0 |
| 0 | 0 | 0 | 0 | 0 |
| 0 | 0 | 0 | 0 | 0 |
| 0 | 0 | 0 | 0 | 0 |
| 0 | 0 | 0 | 0 | 0 |
| 0 | 0 | 0 | 0 | 0 |
| 0 | 0 | 0 | 0 | 0 |
| 0.001159377 | 1.315119319 | 0.033987351 | 1.193665051 | 0 |
| 0.13983235 | 0.041329838 | 0 | 0.134329991 | 0 |
| 0 | 0 | 0 | 0 | 0 |
| 0 | 0 | 0 | 0 | 0 |
| 0.034115287 | 0.281039859 | 0.00196987 | 0.007587603 | 0 |
| 0 | 0 | 0 | 0 | 0 |
| 0 | 0 | 0 | 0 | 0 |

| | | | | |
|---|---|---|---|---|
| 0 | 0 | 0 | 0 | 0 |
| 0 | 0.01975536 | 0 | 0 | 0 |
| 0.019507096 | 0.001006775 | 0.027733304 | 0.008835063 | 0.001478919 |
| 0 | 0 | 0 | 0 | 0 |
| 0 | 0.250635556 | 0.209116524 | 2.085956685 | 0 |
| 0 | 0 | 0.020906296 | 0 | 0 |
| 0 | 0 | 0 | 0 | 0 |
| 0 | 0 | 0 | 0 | 0 |
| 0 | 0 | 0 | 0 | 0 |
| 0 | 0 | 0 | 0 | 0 |
| 0 | 0 | 0 | 0 | 0 |
| 0 | 0 | 0 | 0 | 0 |
| 0 | 0 | 0 | 0 | 0 |
| 0 | 0 | 0.034527676 | 0 | 0 |
| 0 | 0 | 0 | 0 | 0 |
| 0 | 0 | 0.002148151 | 0 | 0 |
| 0 | 0 | 0 | 0 | 0 |
| 0 | 0 | 0 | 0 | 0 |
| 0 | 0 | 0 | 0 | 0 |
| 0 | 0 | 0 | 0 | 0 |
| 0 | 0 | 0 | 0 | 0 |
| 0 | 0 | 0 | 0 | 0 |
| 0 | 0 | 0 | 0 | 0 |
| 0 | 0 | 0 | 0 | 0 |
| 0 | 0.026376498 | 0 | 0 | 0 |
| 0.001023587 | 0 | 0.005877843 | 0 | 0 |
| 0.036310148 | 0 | 0 | 0 | 0 |
| 0.102865971 | 0.035945159 | 0.001341579 | 0 | 0 |
| 0 | 0 | 0 | 0 | 0 |
| 0 | 0 | 0 | 0 | 0 |
| 0 | 0 | 0 | 0 | 0 |
| 0.004488124 | 9.63115E-05 | 0 | 0.001167902 | 0 |
| 0 | 0 | 0 | 0 | 0 |
| 0 | 0 | 0 | 0 | 0 |
| 0 | 0 | 0 | 0 | 0 |
| 0 | 0 | 0 | 0 | 0 |
| 0.55446807 | 0 | 0 | 0 | 0 |
| 0 | 0.029648468 | 0.005678813 | 0 | 0.009031704 |
| 2.759199369 | 2.673253191 | 0 | 0 | 0 |
| 0 | 0 | 0 | 0 | 0 |
| 0 | 0 | 0 | 0 | 0 |
| 0 | 0.008626101 | 0 | 0 | 0.001596183 |
| 0.057322276 | 0.070256198 | 0 | 0.296724661 | 0.001230546 |
| 0 | 0 | 0 | 0 | 0 |
| 0 | 0 | 0 | 0 | 0 |
| 0 | 0.00180067 | 0 | 0 | 0 |
| 0.005971847 | 0.020672782 | 0 | 0.002385522 | 0.002635493 |
| 0 | 0 | 0 | 0 | 0 |

| | | | | |
|---|---|---|---|---|
| 0 | 0 | 0 | 0 | 0 |
| 0.056713597 | 0.237767255 | 0.001004747 | 0.019565581 | 0.05605856 |
| 0 | 0 | 0 | 0 | 0 |
| 0 | 0 | 0 | 0 | 0 |
| 0 | 0 | 0 | 0 | 0 |
| 0 | 0 | 0 | 0 | 0 |
| 0 | 0 | 0 | 0 | 0 |
| 0 | 0 | 0 | 0 | 0 |
| 0 | 0 | 0 | 0 | 0 |
| 0 | 0 | 0 | 0 | 0 |
| 0 | 0 | 0 | 0 | 0 |
| 0 | 0 | 0 | 0 | 0 |
| 0.314429881 | 0.170349635 | 0 | 0.035228524 | 0 |
| 0 | 0 | 0 | 0 | 0 |
| 0.16194831 | 0.076694914 | 0 | 0.091415469 | 0.00770047 |
| 0 | 0 | 0 | 0 | 0 |
| 0 | 0 | 0 | 0 | 0 |
| 0 | 0 | 0 | 0 | 0 |
| 0.010393828 | 0 | 0 | 0.003558798 | 0 |
| 0 | 0 | 0 | 0 | 0 |
| 0 | 0 | 0 | 0 | 0 |
| 0.011169644 | 0.001094244 | 0.01476381 | 0 | 0 |
| 0.025189114 | 0.044404198 | 0.001890797 | 0 | 0 |
| 0 | 0 | 0 | 0 | 0 |
| 0 | 0 | 0 | 0 | 0 |
| 0 | 0 | 0 | 0 | 0 |
| 0.000926944 | 0 | 0 | 0 | 0 |
| 0 | 0 | 0 | 0 | 0 |
| 0 | 0 | 0 | 0 | 0 |
| 0 | 0 | 0 | 0 | 0 |
| 0 | 0 | 0 | 0 | 0 |
| 0 | 0 | 0 | 0 | 0 |
| 0 | 0 | 0 | 0 | 0 |
| 0 | 0 | 0 | 0 | 0 |
| 0 | 0 | 21.72811634 | 17.88809487 | 0 |
| 0 | 0 | 0 | 0 | 0 |
| 0 | 0 | 0 | 0 | 0 |
| 0 | 0 | 0 | 0 | 0 |
| 0.018428747 | 0.000494332 | 0 | 0 | 0 |
| 0 | 0 | 0 | 0 | 0 |
| 0.007827067 | 0.00898133 | 0.453317084 | 0.366805932 | 0 |
| 0 | 0 | 0 | 0 | 0 |
| 0 | 0 | 0 | 0 | 0 |
| 0 | 0 | 0 | 0 | 0 |
| 0 | 0 | 0 | 0 | 0 |
| 0.075674546 | 0.062419215 | 0.000630641 | 0.158432593 | 9.297093466 |
| 0 | 0 | 0 | 0 | 0 |
| 0 | 0 | 0 | 0 | 0 |

| | | | | |
|---|---|---|---|---|
| 0 | 0 | 0 | 0 | 0 |
| 0 | 0 | 0 | 0 | 0 |
| 0 | 0 | 0 | 0 | 0 |
| 0 | 0 | 0 | 0 | 0 |
| 0.00226553 | 0 | 0 | 0 | 0 |
| 0 | 0 | 0 | 0 | 0 |
| 0 | 0 | 0 | 0 | 0 |
| 0 | 0 | 0 | 0 | 0 |
| 0 | 0 | 0 | 0 | 0 |
| 0.001704174 | 0.004466787 | 0 | 0.06572723 | 0 |
| 0 | 0.005675077 | 0 | 0 | 0 |
| 0 | 0 | 0 | 0 | 0 |
| 0.064247896 | 0.012714751 | 0 | 0.013932181 | 0 |
| 0 | 0 | 0 | 0 | 0 |
| 0 | 0 | 0 | 0 | 0 |
| 0 | 0 | 0 | 0 | 0 |
| 0 | 0 | 0 | 0 | 0 |
| 0 | 0 | 0 | 0 | 0 |
| 0 | 0 | 0 | 0 | 0 |
| 2.117121776 | 0.545103388 | 0.003398268 | 1.345697357 | 0 |
| 0 | 0 | 0 | 0 | 0 |
| 0 | 0 | 0 | 0 | 0 |
| 0 | 0 | 0 | 0 | 0 |
| 0 | 0 | 0 | 0 | 0 |
| 0 | 0 | 0 | 0 | 0 |
| 0 | 0 | 0 | 0 | 0 |
| 0 | 0 | 0 | 0 | 0 |
| 0 | 0 | 0 | 0 | 0 |
| 0 | 0 | 0 | 0 | 0 |
| 0.024679159 | 0.001263254 | 0.05372725 | 0.65889554 | 0 |
| 0 | 0 | 0 | 0 | 0 |
| 0 | 0 | 0 | 0 | 0 |
| 0 | 0 | 0 | 0 | 0 |
| 0 | 0 | 0 | 0 | 0 |
| 0 | 0 | 0 | 0 | 0 |
| 0 | 0 | 0 | 0 | 0 |
| 0 | 0 | 0 | 0 | 0 |
| 0 | 0 | 0 | 0 | 0 |
| 0 | 0 | 0 | 0 | 0 |
| 0 | 0 | 0 | 0 | 0 |
| 0 | 0 | 0 | 0 | 0 |
| 0 | 0 | 0 | 0 | 0 |
| 0 | 0.021005541 | 0.277571247 | 0 | 0 |
| 0 | 0 | 0 | 0 | 0 |
| 0.02088738 | 0.123454325 | 0 | 0 | 0.001429732 |
| 0 | 0 | 0 | 0 | 0 |
| 0 | 0 | 0 | 0 | 0 |
| 0 | 0 | 0 | 0 | 0.018367162 |

| | | | | |
|---|---|---|---|---|
| 0 | 0 | 0 | 0 | 0 |
| 0 | 0 | 0 | 0 | 0 |
| 0 | 0 | 0 | 0 | 0 |
| 0 | 0 | 0 | 0 | 0 |
| 0 | 0 | 0 | 0 | 0 |
| 0.007425353 | 0.077440246 | 0.005768852 | 0 | 0.00302416 |
| 0 | 0 | 0 | 0 | 0 |
| 0 | 0 | 0 | 0 | 0 |
| 0 | 0 | 0 | 0 | 0 |
| 0 | 0 | 0 | 0 | 0 |
| 0 | 0 | 0 | 0 | 0 |
| 0 | 0 | 0 | 0 | 0 |
| 0 | 0 | 0 | 0 | 0 |
| 0 | 0 | 0 | 0 | 0 |
| 0 | 0 | 0 | 0 | 0 |
| 0 | 0 | 0 | 0 | 0 |
| 0 | 0 | 0 | 0 | 0 |
| 0 | 0 | 0 | 0 | 0 |
| 0 | 0 | 0 | 0 | 0 |
| 0 | 0 | 0 | 0 | 0 |
| 0.000516576 | 0 | 0 | 0 | 0 |
| 0 | 0 | 0 | 0 | 0 |
| 0 | 0 | 0 | 0 | 0 |
| 0 | 0 | 0 | 0.007406174 | 0 |
| 0 | 0 | 0 | 0 | 0 |
| 0 | 0 | 0 | 0.004029773 | 0 |
| 0 | 0 | 0 | 0 | 0 |
| 0 | 0 | 0.054027441 | 0 | 0 |
| 0 | 0 | 0 | 0 | 0 |
| 0 | 0 | 0 | 0 | 0 |
| 0 | 0 | 0 | 0 | 0 |
| 0 | 0 | 0 | 0 | 0 |
| 0 | 0 | 0 | 0 | 0 |
| 0 | 0 | 0 | 0 | 0 |
| 0 | 0 | 0 | 0 | 0 |
| 0 | 0 | 0 | 0 | 0 |
| 0 | 0 | 0 | 0 | 0 |
| 0 | 0 | 0 | 0 | 0 |
| 2.971100555 | 1.557339978 | 2.91953254 | 0 | 0 |
| 0 | 0 | 0 | 0 | 0 |
| 0 | 0 | 0 | 0 | 0 |
| 0 | 0 | 0 | 0 | 0 |
| 0 | 0 | 0 | 0 | 0 |
| 0 | 0 | 0 | 0 | 0 |
| 0 | 0 | 0 | 0 | 0 |
| 0 | 0 | 0 | 0 | 0 |
| 0 | 0 | 0 | 0 | 0 |
| 0 | 0 | 0 | 0 | 0 |

| | | | | |
|---|---|---|---|---|
| 0 | 0 | 0 | 0 | 0 |
| 0.092609952 | 0.004388549 | 0.528068456 | 0 | 0 |
| 0 | 0 | 0 | 0 | 0 |
| 0 | 0 | 0 | 0 | 0 |
| 0 | 0 | 0 | 0 | 0 |
| 0 | 0.006164786 | 0 | 0 | 0 |
| 0 | 0 | 0 | 0 | 0 |
| 0 | 0 | 0 | 0 | 0 |
| 0 | 0 | 0 | 0 | 0 |
| 0 | 0 | 0 | 0 | 0 |
| 0.00261254 | 0.009390551 | 0.001654659 | 0 | 0 |
| 0 | 0 | 0 | 0 | 0 |
| 0.264426455 | 0.20732642 | 0.229394016 | 0 | 0 |
| 0 | 0 | 0 | 0 | 0 |
| 0 | 0 | 0 | 0 | 0 |
| 0 | 0 | 0 | 0 | 0 |
| 0 | 0 | 0 | 0 | 0 |
| 0 | 0 | 0 | 0 | 0 |
| 0 | 0 | 0 | 0 | 0 |
| 0 | 0.003265554 | 0 | 0 | 0 |
| 0.001325785 | 0.00116172 | 0 | 0.028270544 | 0.091051937 |
| 0 | 0 | 0 | 0.028584592 | 0 |
| 0 | 0 | 0 | 0 | 0 |
| 3.511201304 | 1.504967149 | 2.209067022 | 0.825629809 | 0 |
| 0 | 0 | 0 | 0 | 0 |
| 0 | 0 | 0 | 0 | 0 |
| 0 | 0 | 0 | 0 | 0 |
| 0 | 0 | 0 | 0 | 0 |
| 0 | 0 | 0 | 0 | 0 |
| 0 | 0 | 0 | 0 | 0 |
| 0.006289412 | 0 | 0 | 0 | 0 |
| 0 | 0 | 0 | 0 | 0 |
| 0 | 0.001546227 | 0 | 0 | 0 |
| 0 | 0 | 0 | 0 | 0 |

| PHL018-0272_V2 | PHL018-0038_V1 | PHL018-0148_V3 | PHL018-0226_V2 | PHL018-0045_V1 |
|---|---|---|---|---|
| REF | CTRL | CTRL | REF | EYCF |
| 0 | 0.055115968 | 0.483529587 | 2.144040524 | 0.030442763 |
| 1.479989408 | 0.049409193 | 7.234570538 | 0.08738417 | 1.612110339 |
| 0 | 0.000652284 | 0.185247558 | 3.787771596 | 7.994122799 |
| 0 | 0 | 0 | 0 | 0 |
| 0 | 0 | 0 | 0 | 0 |
| 0 | 0 | 0 | 0 | 0 |
| 0 | 0 | 0 | 0 | 0 |
| 0 | 0 | 0 | 0 | 0 |
| 0 | 0 | 0 | 0 | 0 |
| 0 | 0 | 0 | 0 | 0 |
| 0 | 0 | 0 | 0 | 0 |
| 0 | 0 | 0 | 0 | 0 |
| 0 | 0 | 0 | 0 | 0 |
| 0 | 0.199001072 | 6.688028253 | 0.418044486 | 0 |
| 0 | 0 | 0 | 0.001900499 | 0 |
| 0 | 0 | 0 | 0 | 0 |
| 0.0353102 | 0 | 0.001568823 | 0 | 0 |
| 0 | 0 | 0 | 0 | 0 |
| 0 | 0 | 0 | 0 | 0 |
| 0 | 0 | 0 | 0 | 0 |
| 0 | 0 | 0 | 0 | 0 |
| 0 | 0 | 0 | 0.075377491 | 0 |
| 0 | 0 | 0 | 0 | 0 |
| 0 | 0 | 0 | 0 | 0 |
| 0 | 0.757769272 | 0 | 0 | 0 |
| 0 | 0 | 0 | 0 | 0 |
| 0 | 0 | 0 | 0 | 0 |
| 0 | 0 | 0 | 0 | 0 |
| 0 | 0 | 0 | 0 | 0 |
| 0 | 0 | 0 | 0 | 0 |
| 0 | 0 | 0 | 0 | 0 |
| 0 | 0 | 0 | 0 | 0 |
| 0 | 0 | 0 | 0 | 0 |
| 0 | 0 | 0 | 0 | 0 |
| 0 | 0 | 0 | 0 | 0 |
| 0 | 0.088008593 | 0 | 0.067755261 | 0 |
| 0 | 0 | 0 | 0 | 0 |
| 0 | 0 | 0 | 0 | 0 |
| 0 | 0 | 0 | 0 | 0 |
| 0 | 0 | 0 | 0 | 0 |
| 0 | 0 | 0 | 0 | 0 |
| 0 | 0.006769968 | 0.740547194 | 0 | 0 |
| 0.057380925 | 0.652173851 | 6.055922204 | 1.253575644 | 1.353042061 |
| 0.000963112 | 0 | 0.039242716 | 0.001821984 | 0.053762263 |
| 0 | 0 | 0.005578825 | 0 | 0.028894335 |
| 0 | 0 | 0 | 0 | 0 |

| | | | | |
|---|---|---|---|---|
| 0 | 0.00259317 | 0 | 0.015094933 | 0 |
| 0.006810209 | 0 | 0 | 0.00281091 | 0.026390967 |
| 0 | 0.017059601 | 0 | 0 | 0 |
| 0 | 0.473872919 | 0.84286341 | 0.121024701 | 0.593035062 |
| 0.006198435 | 0 | 0 | 0 | 0 |
| 0 | 0 | 0 | 0 | 0 |
| 0 | 0 | 0 | 0 | 0 |
| 0 | 0 | 0 | 0 | 0 |
| 0 | 0 | 0 | 0 | 0 |
| 0 | 0 | 0 | 0 | 0 |
| 0 | 0 | 0 | 0 | 0 |
| 0 | 1.049295513 | 0 | 0 | 0 |
| 0 | 0 | 0 | 0 | 0.00145864 |
| 0.272312149 | 0 | 0.00354552 | 0.024693707 | 0.176776548 |
| 0 | 0 | 0 | 0 | 0.017206022 |
| 0 | 0 | 0 | 0 | 0 |
| 0 | 0.741959866 | 0 | 0.033688126 | 0.007273564 |
| 0 | 0 | 0 | 0 | 0.688198448 |
| 0 | 0 | 0 | 0 | 0 |
| 0 | 0 | 0 | 0 | 0 |
| 0.070975006 | 0.000377496 | 0.335746033 | 0.005066727 | 0.025229248 |
| 0.014930821 | 0.012286973 | 0 | 0.010822546 | 0.002631978 |
| 0 | 0.002252708 | 0 | 0 | 0 |
| 0 | 0 | 0 | 0 | 0 |
| 0 | 0 | 0 | 0 | 0 |
| 0 | 0 | 0 | 0 | 0 |
| 0.01576364 | 0 | 0.002907804 | 0.000994038 | 0.13284943 |
| 0 | 0 | 0 | 0 | 0 |
| 0 | 0 | 0.033231156 | 0 | 0.002142676 |
| 0 | 0 | 0 | 0 | 0 |
| 0.77093877 | 0 | 0.090448153 | 0 | 0 |
| 0 | 0 | 0 | 0 | 0.000698165 |
| 0 | 0 | 0 | 0 | 0 |
| 0 | 0 | 0 | 0.024301373 | 0.128533478 |
| 0 | 0 | 0.004051086 | 0 | 0 |
| 0 | 0 | 0 | 0 | 0 |
| 0 | 0 | 0 | 0 | 0 |
| 0.002096391 | 42.54537324 | 0 | 1.289129007 | 0 |
| 0 | 0 | 0 | 0 | 0 |
| 0 | 0 | 0 | 0 | 0 |
| 0 | 0 | 0 | 0 | 0 |
| 0 | 0 | 0 | 0 | 0 |
| 0 | 0 | 0 | 0 | 0 |
| 0 | 0 | 0 | 0 | 0 |
| 0 | 0.014171597 | 0 | 0 | 0 |
| 0 | 0 | 0 | 0 | 0 |
| 0 | 0 | 0 | 0 | 0.136077289 |
| 0.085587309 | 0 | 0 | 0.023911214 | 0.240048718 |

| | | | | |
|---|---|---|---|---|
| 0 | 0 | 0 | 0 | 0 |
| 0 | 0 | 0 | 0 | 0 |
| 0 | 0 | 0 | 0 | 0 |
| 0 | 0 | 0 | 0 | 0 |
| 0 | 0 | 0 | 0 | 0 |
| 0 | 0 | 0 | 0 | 0 |
| 0 | 0 | 0 | 0 | 0 |
| 0 | 0 | 0 | 0 | 0 |
| 0 | 0 | 0.009677079 | 0 | 0 |
| 0 | 0 | 0 | 0 | 0 |
| 0 | 0 | 0 | 0 | 0 |
| 0 | 0 | 0.030971879 | 0 | 0 |
| 0 | 0 | 0 | 0 | 0 |
| 0 | 0 | 0 | 0 | 0 |
| 0 | 0 | 0 | 0 | 0 |
| 0 | 0 | 0 | 0 | 0 |
| 0 | 0 | 0 | 0 | 0 |
| 0 | 0 | 0 | 0 | 0 |
| 0 | 0.02395172 | 0 | 0.006413733 | 0.041100506 |
| 0 | 0 | 0 | 0 | 0 |
| 0 | 0 | 0 | 0 | 0 |
| 0 | 0 | 0 | 0 | 0 |
| 0 | 0 | 0 | 0 | 0 |
| 0 | 0 | 0 | 0 | 0 |
| 0 | 0 | 0 | 0 | 0 |
| 0 | 0 | 0 | 0 | 0 |
| 0 | 0 | 0 | 0 | 0 |
| 0.003067543 | 0 | 0 | 0 | 0 |
| 0 | 0 | 0 | 0 | 0 |
| 0 | 0 | 0 | 0 | 0 |
| 0 | 1.592282295 | 0 | 0.021487816 | 0 |
| 0 | 0 | 0 | 0.001785552 | 0.001347117 |
| 0 | 0 | 0 | 0 | 0 |
| 0 | 0 | 0 | 0 | 0 |
| 0 | 0 | 0 | 0 | 0 |
| 0 | 0 | 0 | 0 | 0 |
| 0 | 0 | 0 | 0 | 0 |
| 0 | 0 | 0 | 0 | 0 |
| 0 | 0 | 0 | 1.465507477 | 0 |
| 0 | 0 | 0 | 0 | 0 |
| 0 | 0 | 0 | 0 | 0 |
| 0 | 0 | 0 | 0 | 0 |
| 0 | 0 | 0 | 0.548716927 | 0 |
| 0 | 0 | 0 | 0 | 0 |
| 0 | 0 | 0 | 0 | 0 |
| 0 | 0 | 0 | 0 | 0 |
| 0 | 0 | 0 | 0 | 0 |
| 0 | 0 | 0 | 0 | 0 |

| | | | | |
|---|---|---|---|---|
| 0 | 0 | 0 | 0 | 0 |
| 0 | 0 | 0 | 0 | 0 |
| 0 | 0 | 0 | 0 | 0 |
| 0 | 0 | 0 | 0 | 0 |
| 0 | 0.170886154 | 0 | 0 | 0 |
| 0 | 0 | 0 | 0 | 0 |
| 0 | 0 | 0 | 0 | 0 |
| 0 | 0 | 0 | 0 | 0 |
| 0 | 0 | 0 | 0 | 0 |
| 0 | 0 | 0 | 0 | 0 |
| 0 | 0 | 0 | 0 | 0 |
| 0 | 0 | 0 | 0 | 0 |
| 0 | 0 | 0 | 0 | 0 |
| 0 | 0.003972586 | 0 | 0 | 0 |
| 0 | 0 | 0 | 0 | 0 |
| 0 | 0 | 0 | 0 | 0 |
| 0 | 0 | 0 | 0 | 0 |
| 0 | 0 | 0 | 0 | 0 |
| 0 | 0 | 0 | 0 | 0 |
| 0 | 0 | 0 | 0 | 0 |
| 0 | 0 | 0 | 0 | 0 |
| 0 | 0 | 0 | 0 | 0 |
| 0 | 0 | 0 | 0 | 0 |
| 0 | 0 | 0 | 0 | 0 |
| 0 | 0.129341595 | 0 | 0 | 0 |
| 0 | 0 | 0 | 0 | 0 |
| 0 | 0.027817933 | 0 | 0 | 0 |
| 0 | 0 | 0 | 0 | 0 |
| 0 | 0 | 0 | 0 | 0 |
| 0 | 0 | 0 | 0.009872921 | 0 |
| 0 | 0 | 0 | 0 | 0 |
| 0 | 0 | 0 | 0.012618366 | 0 |
| 0 | 0 | 0 | 0 | 0 |
| 0 | 0 | 0 | 0 | 0 |
| 0 | 0 | 0 | 0 | 0 |
| 0 | 0 | 0 | 0 | 0 |
| 0 | 0 | 0 | 0 | 0 |
| 0 | 0 | 0 | 0 | 0 |
| 0 | 0 | 0 | 0 | 0 |
| 0 | 0 | 0 | 0 | 0 |
| 0 | 0 | 0 | 0 | 0 |
| 0 | 0 | 0 | 0 | 0 |
| 0 | 0 | 0 | 0 | 0 |
| 0 | 0 | 0 | 0 | 0 |
| 0 | 0 | 0 | 0 | 0 |
| 0 | 0 | 0 | 0 | 0 |
| 0 | 0.00668642 | 0 | 0 | 0 |
| 0 | 0 | 0 | 0 | 0 |

| | | | | |
|---|---|---|---|---|
| 0 | 0 | 0 | 0 | 0 |
| 0 | 0 | 0 | 0 | 0 |
| 0 | 0 | 0 | 0 | 0 |
| 0 | 0 | 0 | 0 | 0 |
| 0 | 0 | 0 | 0 | 0 |
| 0 | 0 | 0 | 0 | 0 |
| 0 | 0 | 0 | 0 | 0 |
| 0 | 0 | 0 | 0 | 0 |
| 0 | 0 | 0 | 0 | 0 |
| 0 | 0 | 0 | 0 | 0 |
| 0 | 0 | 0 | 0 | 0 |
| 0 | 0 | 0 | 0 | 0 |
| 0 | 0 | 0 | 0.001848144 | 0.004214005 |
| 0 | 0 | 0 | 0 | 0 |
| 0 | 0 | 0 | 0 | 0.001075947 |
| 0 | 0 | 0 | 0 | 0 |
| 0 | 0 | 0 | 0 | 0 |
| 0 | 0 | 0 | 0 | 0 |
| 0 | 0 | 0 | 0 | 0 |
| 0 | 0 | 0 | 0 | 0 |
| 0.008524333 | 0.006865546 | 0.085434906 | 0.017277924 | 0.030589743 |
| 0 | 0 | 0 | 0 | 0 |
| 0 | 0 | 0 | 0.00697374 | 0 |
| 0 | 0 | 0 | 0 | 0.002899904 |
| 0 | 0 | 0 | 0 | 0 |
| 0 | 0 | 0 | 0 | 0 |
| 0.000531897 | 0 | 0.035103415 | 0.00080498 | 0.029421321 |
| 0 | 0 | 0 | 0 | 0 |
| 0 | 0 | 0 | 0 | 0 |
| 0.014416398 | 0 | 0 | 0.001291855 | 0.032054992 |
| 0 | 0 | 0 | 0.000401214 | 0.01237698 |
| 0 | 0 | 0 | 0 | 0 |
| 0 | 0 | 0 | 0 | 0 |
| 0 | 0.003076917 | 0 | 0 | 0.007931022 |
| 0.054077255 | 0.001786836 | 0.004092405 | 0.036423879 | 0.014776705 |
| 0 | 0 | 0 | 0 | 0 |
| 0 | 0 | 0 | 0 | 0 |
| 0 | 0 | 0 | 0 | 0 |
| 0 | 0 | 0 | 0 | 0 |
| 0 | 0 | 0 | 0 | 0 |
| 0 | 0 | 0 | 0 | 0 |
| 0 | 0 | 0 | 0 | 0 |
| 0 | 0 | 0 | 0 | 0 |
| 0 | 0 | 0 | 0 | 0 |
| 0 | 0 | 0 | 0.199808589 | 0 |
| 0 | 0.203584421 | 0 | 0 | 0 |
| 0.001286841 | 0 | 0 | 0.000486881 | 0 |
| 0.000988126 | 0 | 0 | 0.019013507 | 0.076505703 |

| | | | | |
|---|---|---|---|---|
| 0 | 0 | 0 | 0 | 0 |
| 0 | 0 | 0 | 0 | 0 |
| 0 | 0 | 0.001776605 | 0.000585645 | 0 |
| 0 | 0 | 0 | 0.001086231 | 0 |
| 0 | 0 | 0 | 0 | 0 |
| 0 | 0 | 0 | 0 | 0 |
| 0 | 0 | 0.00172975 | 0 | 0 |
| 0 | 0 | 0.003842959 | 0 | 0 |
| 0 | 0 | 0 | 0 | 0 |
| 0 | 0 | 0 | 0 | 0 |
| 0 | 0 | 0 | 0 | 0 |
| 0 | 0 | 0 | 0 | 0 |
| 0 | 0 | 0 | 0 | 0 |
| 0 | 0 | 0 | 0 | 0 |
| 0 | 0 | 0 | 0 | 0 |
| 0 | 0 | 0 | 0 | 0 |
| 0 | 0 | 0 | 0 | 0 |
| 0 | 0 | 0 | 0 | 0 |
| 0 | 0.004316621 | 0.038556926 | 0.005323 | 0.006993779 |
| 0 | 0.014963613 | 0 | 0.004108105 | 0 |
| 0.007411779 | 0.004298032 | 0.173542858 | 0.030286156 | 1.122245739 |
| 0 | 0 | 0 | 0 | 0 |
| 0 | 0 | 0 | 0.002135423 | 0 |
| 0 | 0 | 0 | 0 | 0 |
| 0 | 0 | 0.0296974 | 0 | 0.001921498 |
| 0 | 0 | 0 | 0 | 0 |
| 0 | 0 | 0 | 0 | 0 |
| 0 | 0 | 0 | 0 | 0 |
| 0 | 0 | 0.097464221 | 0 | 0 |
| 0 | 0 | 0 | 0 | 0 |
| 3.91270096 | 0.075028842 | 0.386034938 | 0.887123263 | 0.329268749 |
| 0 | 0 | 0 | 0 | 0 |
| 0.032333566 | 0 | 0 | 0 | 0 |
| 0 | 0 | 0 | 0 | 0 |
| 0 | 0 | 0 | 0 | 0 |
| 0 | 0 | 0 | 0 | 0 |
| 0 | 0 | 0 | 0 | 0 |
| 0 | 0 | 0 | 0 | 0 |
| 0 | 0 | 0 | 0 | 0 |
| 0 | 0 | 0.001071098 | 0 | 0 |
| 0 | 0 | 0 | 0 | 0 |
| 0 | 0 | 0.004645468 | 0.011169816 | 0.002174739 |
| 0 | 0 | 0.001146388 | 0.004232465 | 0 |
| 0 | 0 | 0 | 0 | 0 |
| 0 | 0 | 0 | 0 | 0 |
| 0.048960533 | 0 | 0 | 0 | 0 |
| 0 | 0 | 0 | 0 | 0 |
| 0 | 0 | 0 | 0 | 0 |

| | | | | |
|---|---|---|---|---|
| 0 | 0 | 0 | 0 | 0 |
| 0 | 0 | 0 | 0 | 0 |
| 0 | 0 | 0.008235034 | 0.001809747 | 0.061896838 |
| 0 | 0 | 0 | 0 | 0 |
| 0 | 0 | 0.000893324 | 0 | 0 |
| 0 | 0 | 0 | 0 | 0 |
| 0 | 0 | 0 | 0 | 0 |
| 0 | 0 | 0 | 0 | 0 |
| 0 | 0 | 0 | 0 | 0 |
| 0 | 0 | 0.001569104 | 0 | 0 |
| 0 | 0 | 0 | 0 | 0 |
| 0 | 0 | 0 | 0 | 0 |
| 0 | 0 | 0 | 0 | 0 |
| 0 | 0 | 0.00568825 | 0 | 0 |
| 0 | 0.002596542 | 0.012206754 | 0 | 0.008026343 |
| 0 | 0.001316814 | 0 | 0 | 0 |
| 0.002278573 | 0 | 0.016048271 | 0 | 0.009460653 |
| 0 | 0 | 0 | 0 | 0.003914021 |
| 0 | 0 | 0 | 0 | 0 |
| 0 | 0 | 0 | 0 | 0 |
| 0 | 0 | 0 | 0 | 0 |
| 0.002430884 | 0 | 0.001609668 | 0.01980965 | 0.085972055 |
| 0 | 0 | 0 | 0 | 0 |
| 0 | 0 | 0.002495352 | 0.004167715 | 0 |
| 0 | 0 | 0 | 0 | 0 |
| 0 | 0 | 0 | 0 | 0 |
| 0 | 0 | 0 | 0 | 0 |
| 0 | 0 | 0 | 0 | 0 |
| 0 | 0 | 0 | 0 | 0 |
| 0 | 0 | 0 | 0 | 0 |
| 0 | 0 | 0.002467028 | 0 | 0 |
| 0 | 0 | 0 | 0 | 0 |
| 0 | 0 | 0 | 0 | 0 |
| 0 | 0 | 0 | 0 | 0 |
| 0 | 0.04756822 | 0 | 0 | 0 |
| 0 | 0 | 0.076518958 | 0 | 0 |
| 0 | 0 | 0 | 0 | 0 |
| 0 | 0.001942637 | 0.265740829 | 0 | 0.001788328 |
| 0 | 0 | 0 | 0 | 0 |
| 0 | 0 | 0 | 0 | 0 |
| 0 | 0 | 0 | 0 | 0 |
| 0 | 0 | 0 | 0 | 0 |
| 0 | 0 | 0 | 0 | 0 |
| 0.0248052 | 0 | 0.010550875 | 0 | 0.004442765 |
| 0 | 0 | 0 | 0 | 0 |
| 0 | 0 | 0 | 0 | 0 |
| 0 | 0 | 0 | 0 | 0 |
| 0 | 0 | 0.005389143 | 0.010766624 | 0 |

| | | | | |
|---|---|---|---|---|
| 0 | 0 | 0 | 0 | 0 |
| 0 | 0 | 0 | 0 | 0 |
| 0 | 0 | 0.011464809 | 0 | 0 |
| 0 | 0 | 0 | 0 | 0 |
| 0.001379368 | 0 | 0.023408663 | 0 | 0.022799497 |
| 0 | 0 | 0.010510566 | 0 | 0 |
| 0 | 0 | 0 | 0 | 0 |
| 0 | 0.001868691 | 0.01711951 | 0 | 0.031538007 |
| 0 | 0 | 0 | 0 | 0 |
| 1.946197457 | 0 | 0 | 0 | 0.345006636 |
| 0.002831445 | 0 | 0 | 0 | 0.006269962 |
| 0 | 0 | 0.183703064 | 0.001323638 | 0 |
| 0 | 0.001496097 | 0.022557947 | 0 | 0.014139845 |
| 0.00075567 | 0 | 0 | 0.001906071 | 0 |
| 0.023880852 | 0 | 0 | 0.018070815 | 0 |
| 0 | 0 | 0 | 0 | 0 |
| 0 | 0 | 0 | 0 | 0 |
| 0.003672064 | 0 | 0 | 0.001634516 | 0 |
| 0.238159449 | 0 | 0 | 0 | 0.001631382 |
| 0 | 0 | 0.074679239 | 0 | 0 |
| 0 | 0 | 0 | 0 | 0 |
| 0 | 0 | 0 | 0 | 0 |
| 0 | 0 | 0 | 0 | 0 |
| 0 | 0.099767648 | 0 | 0 | 0 |
| 0 | 0 | 0 | 0 | 0 |
| 0 | 0 | 0 | 0.003508303 | 0.021468929 |
| 0 | 4.051586314 | 0 | 0.016028653 | 0 |
| 0 | 0 | 0.341860111 | 0 | 0 |
| 0 | 0 | 0 | 0 | 0.41733952 |
| 0 | 0.001195258 | 0 | 0 | 0.003328453 |
| 0.071307982 | 0.003895572 | 0.002141294 | 0 | 0.008836235 |
| 0 | 0 | 0 | 0 | 0 |
| 0.059639848 | 0 | 0 | 0 | 0 |
| 0 | 0 | 0 | 0 | 0 |
| 0 | 0 | 0 | 0 | 0 |
| 0.241731908 | 0 | 0 | 0 | 0 |
| 0 | 0 | 0.010933574 | 0 | 0.006334348 |
| 0 | 0 | 0 | 0 | 0 |
| 0 | 0 | 0 | 0 | 0 |
| 0 | 0 | 0 | 0 | 0 |
| 0 | 0 | 0 | 0 | 0 |
| 0 | 0 | 0 | 0 | 0 |
| 0 | 0 | 0 | 0 | 0 |
| 0 | 0 | 0.01420096 | 0 | 0 |
| 0 | 0 | 0 | 0 | 0 |
| 0 | 0.061196865 | 0 | 0 | 0 |
| 0 | 0.001264098 | 0.004963157 | 0 | 0.002925842 |
| 0 | 0 | 0 | 0 | 0 |

| | | | | |
|---|---|---|---|---|
| 0 | 0 | 0 | 0 | 0 |
| 0 | 0 | 0 | 0 | 0 |
| 0 | 0 | 0 | 0 | 0 |
| 0 | 0 | 0 | 0 | 0.123201194 |
| 0 | 0 | 0 | 0 | 0.004337833 |
| 0 | 0 | 0.021174222 | 0.012599675 | 0 |
| 0 | 0 | 0 | 0 | 0 |
| 0 | 0 | 0 | 0 | 0 |
| 0 | 0 | 0 | 0 | 0 |
| 0 | 0 | 0 | 0 | 0 |
| 0.019092505 | 0.000989798 | 0.109191013 | 0 | 0.010022924 |
| 0 | 0 | 0 | 0.074706435 | 0 |
| 0 | 0 | 0 | 0 | 0 |
| 0.004768571 | 0.000623407 | 0.018918282 | 0.010197688 | 0.003787662 |
| 0 | 0 | 0.114851409 | 0 | 0 |
| 0 | 0 | 0 | 0 | 0 |
| 0 | 0 | 0 | 0.003506641 | 0 |
| 0 | 0 | 0 | 0 | 0 |
| 0.013460873 | 0 | 0 | 0 | 0.01511769 |
| 0 | 0 | 0 | 0.112890212 | 0 |
| 0 | 0 | 0 | 0 | 0 |
| 0 | 0.280522178 | 0 | 0.00212854 | 0 |
| 0 | 0 | 0 | 0.021289661 | 0 |
| 0 | 0 | 0 | 0 | 0 |
| 0 | 0 | 0 | 0 | 0 |
| 0.005993349 | 0 | 0 | 0 | 0 |
| 0 | 0 | 0.00503135 | 0 | 0.018480728 |
| 0 | 0 | 0 | 0 | 0 |
| 0 | 0 | 0 | 0 | 0 |
| 0.103022493 | 0.128158288 | 0.227074033 | 0.203288382 | 1.81362694 |
| 0 | 0.001057331 | 0 | 0.003831685 | 0 |
| 0 | 0 | 0 | 0 | 0 |
| 0 | 0 | 0 | 0 | 0 |
| 0 | 0 | 0.0020579 | 0 | 0 |
| 0 | 0 | 0 | 0 | 0 |
| 0 | 0 | 0 | 0 | 0 |
| 0 | 0.057280451 | 0 | 0.035134603 | 0 |
| 0 | 0 | 0.003274433 | 0 | 0 |
| 0 | 0 | 0 | 0 | 0 |
| 0 | 0 | 0.81177916 | 0 | 0 |
| 0.016874268 | 0.000768763 | 0.05135387 | 0.002579572 | 0.000778468 |
| 0.971903261 | 1.315725573 | 0 | 0.011869194 | 0 |
| 0 | 0 | 0.004332744 | 0 | 0 |
| 0 | 0 | 0 | 0 | 0 |
| 0 | 0 | 0 | 0 | 0 |
| 0 | 0 | 0 | 0 | 0 |
| 0.002146758 | 0 | 0.022175824 | 0.000812234 | 0.00326823 |
| 0 | 0 | 0 | 0 | 0 |

| | | | | |
|---|---|---|---|---|
| 0 | 0 | 0.003040138 | 0.002505401 | 0.006384709 |
| 0 | 0 | 0 | 0 | 0 |
| 0 | 0 | 0 | 0 | 0 |
| 0 | 0 | 0 | 0 | 0 |
| 0 | 0 | 0 | 0 | 0 |
| 0 | 0 | 0 | 0 | 0 |
| 0 | 0 | 0 | 0 | 0 |
| 0 | 0 | 0 | 0 | 0 |
| 0.006650272 | 0 | 0 | 0.055774763 | 0.01546782 |
| 0 | 0 | 0 | 0 | 0.076164763 |
| 0 | 0 | 0 | 0 | 0 |
| 0 | 0 | 0 | 0 | 0 |
| 0 | 0 | 0 | 0 | 0 |
| 0.218058531 | 0 | 0.448992418 | 0 | 0.077521141 |
| 0 | 0 | 0 | 0 | 0 |
| 0 | 0.001055757 | 0 | 0.001062772 | 0.000950297 |
| 0.030269325 | 0.001040174 | 0.047944186 | 0.004581004 | 1.02986912 |
| 0 | 0 | 0.008321474 | 0.057088602 | 0.292737023 |
| 0.016972932 | 0 | 0 | 0 | 0.010335855 |
| 0 | 0 | 0 | 0.039099882 | 0 |
| 0 | 0 | 0 | 0 | 0 |
| 0 | 0 | 0.00379963 | 0 | 0.004276227 |
| 0 | 0 | 0 | 0 | 0 |
| 0 | 0 | 0 | 0.011703002 | 0 |
| 0 | 0 | 0 | 0 | 0 |
| 0.028937494 | 0 | 0.45505153 | 0 | 0 |
| 0.001537086 | 0 | 0.005670702 | 0.001827767 | 0.001002885 |
| 0 | 0 | 0 | 0.000630921 | 0 |
| 0 | 0 | 0 | 0 | 0 |
| 0 | 0 | 0.004205233 | 0 | 0.006292939 |
| 0.067799077 | 0.041550593 | 0.006325077 | 0.054084081 | 0 |
| 0 | 0 | 0.021860254 | 0 | 0.001137078 |
| 0 | 0.010691558 | 0.629723038 | 0.010224476 | 0 |
| 0 | 0 | 0.036261945 | 0 | 0 |
| 0 | 0 | 0 | 0 | 0 |
| 0 | 0 | 0 | 0 | 0 |
| 0.000857235 | 0 | 0.006272405 | 0 | 0 |
| 0.127040302 | 0.001637824 | 0.02827768 | 0.000760942 | 0.022198371 |
| 0.005104685 | 0.018156757 | 0 | 0 | 0.244514331 |
| 0 | 0 | 0.030433327 | 0.000622148 | 0 |
| 0 | 0.000599693 | 0 | 0 | 0 |
| 0 | 0 | 0 | 0 | 0 |
| 0.144120618 | 0 | 0 | 0.038122578 | 0.087000681 |
| 0.074312544 | 0 | 0 | 0.050688763 | 0.032266982 |
| 0 | 0 | 0 | 0 | 0 |
| 0 | 0 | 0 | 0 | 0 |
| 0 | 0 | 0 | 0 | 0 |
| 0.09534596 | 0 | 0 | 0 | 0 |

| | | | | |
|---|---|---|---|---|
| 0 | 0 | 0.001711282 | 0 | 0 |
| 0 | 0.107810331 | 7.113858559 | 0.000691253 | 0.054585645 |
| 0 | 0 | 0 | 0 | 0 |
| 0 | 0 | 0 | 0 | 0 |
| 0 | 0 | 0 | 0 | 0 |
| 0.002402409 | 0 | 0 | 0 | 0 |
| 0 | 0 | 0 | 0 | 0.008041215 |
| 0 | 0 | 0 | 0 | 0 |
| 0 | 0.001414368 | 0 | 0 | 0 |
| 0 | 0 | 0 | 0 | 0 |
| 0.00712773 | 0 | 0 | 0.000770515 | 0 |
| 0.037159348 | 0 | 0.030899716 | 0.002677976 | 0.138542426 |
| 0 | 0.044043324 | 0 | 0 | 0 |
| 0 | 0 | 0 | 0 | 0 |
| 0 | 0 | 0.002011426 | 0.000736725 | 0.00266796 |
| 0.010467988 | 0 | 0 | 0 | 0 |
| 0.001954795 | 0.001469443 | 0.216614322 | 0 | 0.016773734 |
| 3.569010465 | 0 | 0.017078246 | 0.183991595 | 0.003512036 |
| 0 | 0 | 0.00227003 | 0 | 0 |
| 0.054676365 | 0 | 0 | 0 | 0 |
| 0 | 0.114754553 | 0.074461338 | 0 | 0.003068991 |
| 0 | 0 | 0 | 0 | 0 |
| 0 | 0 | 0 | 0 | 0 |
| 0 | 0 | 0 | 0 | 0 |
| 0 | 0 | 0 | 0 | 0 |
| 0 | 0 | 0 | 0 | 0 |
| 0 | 0.014005586 | 0 | 0.023406508 | 0.076557354 |
| 0 | 0.030162344 | 0.105633661 | 0.017834449 | 0 |
| 0 | 0 | 0 | 0 | 0 |
| 0 | 0 | 0 | 0.001244526 | 0.046857473 |
| 0 | 0 | 0.019070747 | 0 | 0 |
| 0.006452538 | 0 | 0 | 0 | 0 |
| 0 | 0 | 0 | 0 | 0 |
| 0 | 0 | 0 | 0 | 0 |
| 0 | 0.094930005 | 0 | 0.001798188 | 0 |
| 0.008241377 | 0.03147132 | 0.208858891 | 0.041159647 | 4.012934599 |
| 0.000458029 | 0 | 0 | 0 | 0 |
| 0 | 0 | 0 | 0 | 0 |
| 0 | 0.001083746 | 0.090597128 | 0 | 0 |
| 0 | 0 | 0 | 0 | 0 |
| 0 | 0 | 0 | 0 | 0 |
| 0 | 0 | 0 | 0 | 0 |
| 0.017660216 | 0.003071992 | 0.092973193 | 0 | 0 |
| 0 | 0 | 0.005094706 | 0 | 0.0160896 |
| 0 | 0 | 0 | 0 | 0 |
| 0 | 0.363135986 | 0 | 0 | 0 |
| 0 | 0 | 0 | 0 | 0 |
| 0.021411011 | 0.001198557 | 0 | 0 | 0.005201518 |

| | | | | |
|---|---|---|---|---|
| 0 | 0 | 0.004212505 | 0 | 0 |
| 0 | 0 | 0 | 0 | 0 |
| 0 | 0.001709282 | 0 | 0 | 0 |
| 0 | 0.002938989 | 2.287433716 | 0 | 0 |
| 0 | 0 | 0 | 0 | 0 |
| 0 | 0 | 0 | 0 | 0 |
| 0 | 0 | 0 | 0 | 0 |
| 0 | 0.000342944 | 0 | 0 | 0 |
| 0 | 0.384443272 | 0 | 0.01694679 | 0 |
| 0 | 0.02706677 | 0 | 0 | 0.577312191 |
| 0 | 0 | 0 | 0 | 0 |
| 0 | 0.002571009 | 0 | 0 | 0 |
| 0 | 0 | 0 | 0 | 0 |
| 0 | 0 | 0 | 0 | 0 |
| 0 | 0.122042695 | 0 | 0.003966303 | 0 |
| 0 | 0 | 0 | 0 | 0 |
| 0 | 0.022271346 | 0 | 0 | 0 |
| 0 | 0 | 0 | 0 | 0 |
| 0 | 0 | 0 | 0 | 0 |
| 0 | 0 | 0 | 0 | 0 |
| 0 | 0 | 0 | 0 | 0 |
| 0 | 0 | 0 | 0 | 0 |
| 0 | 0.002480201 | 0 | 0 | 0 |
| 0.192118478 | 0 | 0.008246741 | 0.122845912 | 0 |
| 0 | 0 | 0 | 0 | 0 |
| 0 | 0.095569495 | 0 | 0.001664851 | 4.095930574 |
| 0 | 0.004637119 | 0 | 0 | 0 |
| 0.000669607 | 1.037240148 | 0 | 0 | 0 |
| 0 | 0 | 0 | 0 | 0 |
| 0.01606273 | 0 | 0 | 0 | 0 |
| 0 | 0.000714131 | 0 | 0 | 0 |
| 0 | 0 | 0 | 0 | 0 |
| 0 | 0 | 0 | 0.073222626 | 0 |
| 0 | 0 | 0 | 0 | 0 |
| 0 | 0 | 0 | 0.004756732 | 0 |
| 0 | 0.386916656 | 0 | 0 | 0 |
| 0 | 0.018507591 | 0 | 0 | 0 |
| 0 | 0 | 0 | 0.004872761 | 0 |
| 0.320778103 | 0.011342925 | 0 | 0.015523759 | 0 |
| 0 | 0 | 0 | 0.03696156 | 0 |
| 0.023436251 | 0.003875808 | 0.009952771 | 0.027902095 | 0.036392999 |
| 0 | 0.139136313 | 0 | 0 | 0 |
| 0 | 0.012539582 | 0.134239489 | 0.238984325 | 0 |
| 0 | 0 | 0 | 0 | 0 |
| 0 | 0 | 0 | 0 | 0 |
| 0 | 0 | 0 | 0 | 0 |
| 0 | 0 | 0 | 0 | 0 |
| 0 | 0.001411869 | 0.003556975 | 0.975972946 | 0 |

| | | | | |
|---|---|---|---|---|
| 0 | 0 | 0 | 0 | 0 |
| 0 | 0.032542182 | 0 | 0 | 0 |
| 1.77724464 | 0 | 0.793234753 | 0.07466113 | 0.003563354 |
| 0 | 0.0004389 | 0 | 0 | 0 |
| 0 | 0 | 0 | 0 | 0 |
| 0 | 0.009169185 | 0 | 0.000536635 | 0 |
| 0 | 0 | 0 | 0 | 0 |
| 0 | 0 | 0 | 0 | 0 |
| 0 | 0 | 0 | 0 | 0 |
| 1.634394825 | 0.003788709 | 0 | 0 | 1.260515365 |
| 0 | 0.023786121 | 0.063912204 | 2.804412658 | 0.003767703 |
| 0 | 0.328446874 | 7.659837858 | 0.581302835 | 0 |
| 0 | 0 | 0 | 0 | 0 |
| 0 | 0 | 0.008211507 | 0.002454225 | 0.164973932 |
| 1.097056902 | 0.034695621 | 0.209659822 | 0.041524615 | 0.917135881 |
| 0 | 0.299543565 | 0 | 0 | 0 |
| 0 | 0 | 0 | 0 | 0 |
| 0 | 0 | 0 | 0 | 0 |
| 0 | 0 | 0 | 0 | 0 |
| 0.005970235 | 0.008756873 | 0 | 0.00198339 | 0.174466421 |
| 0.005333864 | 0.031519358 | 1.344807839 | 0.002690783 | 0.000676691 |
| 32.50168009 | 0 | 0.214486756 | 2.003458642 | 2.312568356 |
| 30.6406165 | 0.136756312 | 0.189855924 | 0.151431605 | 6.679978086 |
| 0 | 0 | 0 | 0 | 0 |
| 0 | 0 | 0 | 0 | 0 |
| 0 | 2.220534813 | 0 | 0 | 0 |
| 0 | 0 | 0.004881496 | 39.79778351 | 0 |
| 0 | 0 | 0 | 0 | 0 |
| 0 | 0.725979702 | 0 | 0 | 0 |
| 0 | 0 | 0 | 0 | 0 |
| 0 | 0.035747792 | 0 | 0 | 0 |
| 0 | 0.058987643 | 0 | 0.088804833 | 0 |
| 0.003449484 | 0.073559847 | 1.420001152 | 0 | 0.007186269 |
| 0 | 0.283493495 | 0 | 0.095544852 | 0 |
| 0.010065421 | 0 | 0.050543365 | 0.114611367 | 0.277284633 |
| 0 | 0.005692846 | 0 | 0.634610079 | 0 |
| 0 | 0.004184182 | 0 | 0.003971302 | 0 |
| 0 | 0 | 0 | 0 | 0 |
| 0 | 0 | 0 | 0.137182421 | 0 |
| 0 | 0 | 0 | 0 | 0 |
| 0 | 0.005666685 | 0 | 0 | 0 |
| 0 | 0 | 0 | 0 | 0 |
| 0 | 0.016481443 | 0 | 0 | 0 |
| 0 | 0 | 0 | 0 | 0 |
| 0 | 0.077057741 | 0.03329314 | 0.155423759 | 0 |
| 0 | 0.003708837 | 0 | 0 | 0 |
| 0 | 0 | 0 | 0 | 0 |
| 0 | 0.047327562 | 0.710024258 | 0.335722329 | 0 |

| | | | | |
|---|---|---|---|---|
| 0 | 0 | 0 | 0 | 0 |
| 0 | 0 | 0 | 0 | 0 |
| 0 | 0 | 0 | 0 | 0.004186294 |
| 0 | 0 | 0 | 0 | 0 |
| 0 | 0 | 0 | 0 | 0 |
| 0 | 0.0456813 | 0 | 0 | 0 |
| 0 | 0 | 0 | 0 | 0 |
| 0.308327682 | 0.000553423 | 0.888524176 | 0.095821338 | 0 |
| 0 | 0 | 0 | 0 | 0 |
| 0 | 0 | 0 | 0 | 0 |
| 0.029227278 | 0.005760557 | 0.003375068 | 0.073631746 | 0.075425613 |
| 0 | 0 | 0 | 0 | 0 |
| 0 | 0.001407577 | 0 | 0.180658675 | 0 |
| 0 | 0 | 0 | 0 | 0 |
| 0 | 0 | 0 | 0 | 0 |
| 0 | 0 | 0 | 0 | 0 |
| 0 | 0.006476324 | 0 | 0 | 0 |
| 0 | 0 | 0 | 0.003592483 | 0 |
| 0.025758823 | 0 | 0 | 0 | 0 |
| 0 | 0 | 0 | 0 | 0 |
| 0 | 0.045279691 | 0 | 0 | 0 |
| 0 | 0 | 0 | 0 | 0 |
| 0 | 0 | 0 | 0 | 0 |
| 0 | 0 | 0 | 0.035859822 | 0 |
| 0 | 0 | 0 | 0 | 0 |
| 0 | 0 | 0 | 0 | 0 |
| 0 | 0 | 0 | 0 | 0 |
| 0 | 0 | 0 | 0 | 0 |
| 0 | 0 | 0 | 0 | 0 |
| 0 | 0.073018238 | 0 | 0 | 0 |
| 0 | 0.013505726 | 0 | 0.000743503 | 0 |
| 0 | 0 | 0 | 0 | 0 |
| 0 | 0 | 0 | 0 | 0 |
| 0 | 0 | 0 | 0 | 0 |
| 0 | 0 | 0 | 0 | 0 |
| 0 | 0 | 0 | 0 | 0 |
| 0 | 0.039813232 | 0.01779747 | 0 | 0 |
| 0.010862079 | 0.00068043 | 0.052465911 | 0.000570793 | 0.002985749 |
| 0 | 0 | 0 | 0 | 0 |
| 0.001636824 | 0 | 0 | 0 | 0 |
| 0 | 0 | 0 | 0.170410358 | 0 |
| 0 | 0 | 0 | 0 | 0 |
| 0 | 0 | 0 | 0 | 0 |
| 0 | 0.002422852 | 0 | 0 | 0 |
| 0 | 0.054856074 | 0 | 0.00998119 | 0 |
| 0 | 0.03339172 | 0 | 0 | 0 |
| 0 | 0 | 0 | 0 | 0 |
| 0 | 0 | 0 | 0 | 0 |

| | | | | |
|---|---|---|---|---|
| 0 | 0 | 0 | 0 | 0 |
| 0 | 0 | 0 | 0 | 0 |
| 0 | 0 | 0 | 0 | 0 |
| 0 | 0.71348952 | 0 | 0.008403017 | 0 |
| 0 | 0 | 0 | 0 | 0 |
| 0 | 0.036964509 | 0 | 0 | 0 |
| 0 | 0 | 0 | 0.007806455 | 0 |
| 0 | 0 | 0 | 0.000648887 | 0 |
| 0.857640555 | 0.042854789 | 0.5122462 | 0.006927522 | 0.127653258 |
| 0 | 0.000503807 | 0 | 0 | 0 |
| 0 | 0 | 0 | 0 | 0 |
| 0 | 0 | 0 | 0 | 0 |
| 0 | 0 | 0 | 0.010654564 | 0 |
| 0 | 0 | 0 | 0 | 0 |
| 0 | 0.027312298 | 0 | 0.794378342 | 0 |
| 0.172548275 | 0.000482181 | 0.72665727 | 0.108362184 | 1.382285495 |
| 0 | 0 | 0 | 0.048890818 | 0 |
| 0 | 0 | 0 | 0 | 0 |
| 0 | 0 | 0 | 0 | 0 |
| 0 | 0 | 0 | 0 | 0 |
| 0 | 0 | 0 | 0 | 0 |
| 0 | 0 | 0 | 0 | 0 |
| 0 | 0 | 0 | 0 | 0 |
| 0 | 0 | 0 | 0 | 0 |
| 0.006116939 | 0.944097549 | 0.205148561 | 0.864276913 | 0.632873688 |
| 0 | 0.293496635 | 0 | 0 | 1.756526586 |
| 0 | 0.010398315 | 0 | 0 | 0 |
| 0 | 0 | 0 | 0 | 0 |
| 0 | 0.019778995 | 0 | 0 | 0 |
| 0 | 0 | 0 | 0 | 0 |
| 0 | 0 | 0 | 0 | 0 |
| 0 | 0 | 0 | 0 | 0 |
| 0 | 0 | 0 | 0 | 0 |
| 0 | 0.005893731 | 0 | 0.006328423 | 0 |
| 0 | 0 | 0.00667236 | 0.012545276 | 0.039006596 |
| 0.001826366 | 0 | 0.08568398 | 0.044455115 | 0.032438746 |
| 0 | 0 | 0 | 0 | 0 |
| 0 | 0.165617574 | 0 | 0 | 0 |
| 0 | 0 | 0 | 1.327256712 | 0 |
| 0 | 0 | 0 | 0 | 0 |
| 0 | 0.053681169 | 0 | 0 | 0 |
| 0 | 0 | 0 | 0 | 0 |
| 0.44466146 | 1.190381418 | 0 | 0.18806918 | 0 |
| 0 | 0 | 0 | 0 | 0 |
| 0 | 0 | 0 | 0 | 0 |
| 0.008507073 | 0.000758713 | 0 | 0.005455394 | 0 |
| 0 | 0 | 0 | 0 | 0 |
| 0 | 0.03332617 | 3.731727659 | 0.004345548 | 0 |

| | | | | |
|---|---|---|---|---|
| 0.190136013 | 0.354228478 | 0 | 0.009462202 | 0 |
| 0 | 0 | 0 | 0 | 0 |
| 0 | 0 | 0 | 0 | 0 |
| 0 | 0 | 0 | 0 | 0 |
| 0 | 0.000512796 | 0.001761691 | 0.042973961 | 0 |
| 0 | 0 | 0 | 0 | 0 |
| 0 | 0 | 0 | 0 | 0 |
| 0 | 0 | 0 | 0 | 0 |
| 0 | 0 | 0 | 0 | 0 |
| 0 | 0 | 0 | 0 | 0 |
| 0 | 0 | 0 | 0 | 0 |
| 0 | 0.010762731 | 5.236628864 | 0.004405136 | 0 |
| 0 | 0.057899495 | 0 | 0 | 0 |
| 0 | 0 | 0 | 0 | 0 |
| 0.001380629 | 0.023870207 | 4.205185505 | 0.001790969 | 0.016814981 |
| 0 | 0 | 0 | 0 | 0 |
| 0 | 0 | 0 | 0.003945279 | 0 |
| 0 | 0.077704548 | 0 | 0.008071203 | 0 |
| 0 | 0 | 0 | 0.651457756 | 0 |
| 0 | 0 | 0 | 0 | 0 |
| 0 | 0.006885614 | 0 | 0 | 0 |
| 0 | 0 | 0 | 0 | 0 |
| 0 | 0.12964529 | 0 | 0.002416793 | 0.791785873 |
| 0 | 0 | 0 | 0 | 0 |
| 0 | 0 | 0 | 0 | 0 |
| 0 | 0 | 0 | 0 | 0 |
| 0 | 0 | 0 | 0 | 0 |
| 0 | 0 | 0 | 0.253797387 | 0 |
| 0 | 0 | 0 | 0 | 0 |
| 0 | 0 | 0 | 0 | 0 |
| 0 | 0 | 0 | 0 | 0 |
| 0.00255937 | 0.002308691 | 0.142765542 | 0 | 0.028054015 |
| 0 | 0 | 0 | 0 | 0 |
| 0 | 0.208281567 | 0 | 0.0910454 | 0 |
| 0.002318801 | 0.00124505 | 0.076136349 | 0.044117003 | 0.026728267 |
| 0 | 0.009148618 | 0.034186726 | 0.037645496 | 0 |
| 0 | 0 | 0 | 0 | 0 |
| 0 | 0.143104916 | 0 | 0 | 0 |
| 0 | 0.000429114 | 0 | 0.000719942 | 0 |
| 0 | 0 | 0 | 0 | 0 |
| 0 | 0.196033505 | 0 | 0 | 0 |
| 0 | 0.000719597 | 0 | 0 | 0 |
| 0 | 0 | 0 | 0 | 0 |
| 0 | 0 | 0 | 0 | 0 |
| 0 | 0.03397403 | 0.048596759 | 2.149525141 | 0.001241982 |
| 0 | 0 | 0 | 0 | 0 |
| 0 | 0.091224275 | 0 | 0 | 0 |
| 0.011111106 | 0.001508064 | 0 | 0 | 0 |

| | | | | |
|---|---|---|---|---|
| 0 | 0 | 0 | 0 | 0 |
| 0 | 0 | 0 | 0 | 0 |
| 0.005737469 | 0 | 0 | 0 | 0 |
| 0 | 0 | 0 | 0 | 0 |
| 0 | 0 | 0 | 0 | 0 |
| 0 | 0 | 0 | 0 | 0 |
| 0 | 0.412759984 | 0 | 0 | 0 |
| 0 | 0.001557382 | 0 | 0 | 0 |
| 0 | 0 | 0 | 0 | 0 |
| 0 | 0 | 0 | 0 | 0 |
| 0 | 0 | 0 | 0 | 0 |
| 0.016998113 | 0 | 0 | 0.030922202 | 0.004182299 |
| 0 | 0 | 0 | 0 | 0 |
| 0 | 0 | 0 | 0 | 0 |
| 0 | 0 | 0 | 0 | 0 |
| 0 | 0 | 0 | 0 | 0 |
| 0 | 0 | 0 | 0 | 0 |
| 0 | 0.172909301 | 0 | 0 | 0 |
| 0 | 0.376406111 | 0 | 0.000955114 | 3.313889846 |
| 0 | 0 | 0 | 0.001811849 | 0 |
| 0 | 0 | 0 | 0 | 0 |
| 0 | 0 | 0 | 0 | 0 |
| 0 | 0 | 0 | 0 | 0 |
| 0 | 0 | 0 | 0 | 0 |
| 0 | 0.001608068 | 0.008593601 | 0 | 0 |
| 0 | 0 | 0.129187414 | 0.005862002 | 0.008324917 |
| 0 | 0 | 0 | 0 | 0 |
| 0 | 0.201730238 | 0.001791784 | 0.001687569 | 0 |
| 0 | 0 | 0 | 0 | 0 |
| 0 | 0.100479979 | 0 | 0.000628247 | 0 |
| 0 | 0.007921549 | 0 | 0 | 0 |
| 0 | 0 | 0 | 0 | 0 |
| 0 | 0.013453244 | 0 | 0 | 0 |
| 0 | 0 | 0 | 0 | 0 |
| 0.015917817 | 0.015914267 | 0.006029087 | 0.040230731 | 0.066156984 |
| 0 | 1.583182744 | 0 | 0 | 0 |
| 0 | 0 | 0 | 0 | 0 |
| 0 | 0.001357747 | 0.003109658 | 0.976556691 | 0 |
| 0 | 7.839748418 | 0 | 0 | 0 |
| 0 | 0 | 0 | 0 | 0 |
| 0 | 0.56138212 | 6.752030729 | 1.384916987 | 38.32226376 |
| 0.083295513 | 0.007454076 | 0.048712478 | 0.150672491 | 0.657395571 |
| 0 | 0.338527916 | 0 | 0 | 0 |
| 0 | 0.066170593 | 0 | 0 | 0 |
| 0 | 0 | 0 | 0 | 0 |
| 0 | 0.00053478 | 0 | 0 | 0 |
| 0 | 0 | 0 | 0 | 0 |
| 0 | 0.007435322 | 0.000987199 | 0.011606758 | 0 |

| | | | | |
|---|---|---|---|---|
| 0 | 0 | 0 | 0 | 0 |
| 0.008699422 | 0.015074009 | 0 | 0.000892599 | 0 |
| 0 | 0.008414437 | 0 | 0 | 0 |
| 0 | 0 | 0 | 0 | 0 |
| 1.413584818 | 2.216600652 | 0 | 0.113515998 | 0 |
| 0 | 0 | 0 | 0 | 0 |
| 0 | 0 | 0 | 0 | 0 |
| 0 | 0 | 0 | 0 | 0 |
| 3.353026059 | 0 | 0.593395724 | 0.230842251 | 0.194332861 |
| 0 | 0 | 0 | 0 | 0 |
| 0.041979506 | 0 | 0 | 0.015757044 | 0 |
| 0 | 0 | 0 | 0 | 0 |
| 0 | 0.049110425 | 0 | 0 | 0 |
| 0 | 0.00225087 | 0.02153048 | 0.011995557 | 0.150700892 |
| 0 | 0.15445119 | 0 | 0 | 0 |
| 0 | 0.131031721 | 0 | 0 | 0 |
| 0 | 0.000916602 | 0 | 0 | 0 |
| 0 | 0 | 0 | 0 | 0 |
| 0 | 0 | 0 | 0 | 0 |
| 0 | 0 | 0 | 0 | 0 |
| 0 | 0 | 0 | 0 | 0 |
| 0 | 0 | 0 | 0 | 0 |
| 0 | 0 | 0 | 0 | 0 |
| 0 | 0 | 0 | 0 | 0 |
| 0 | 0 | 0 | 0 | 0 |
| 0 | 0 | 0 | 0 | 0 |
| 0 | 0.00139479 | 0 | 0 | 0.037327645 |
| 0.002093707 | 0 | 0.013131169 | 0.00950594 | 0.042120072 |
| 0 | 0 | 0 | 0 | 0 |
| 0 | 0 | 0 | 0 | 0 |
| 0 | 0 | 0 | 0 | 0 |
| 0 | 0 | 0 | 0 | 0 |
| 0 | 0 | 0 | 0 | 0 |
| 0 | 0.003653195 | 0 | 0 | 0 |
| 0 | 0.003066343 | 0 | 0 | 0 |
| 0 | 0 | 0 | 0.014530956 | 0.014485567 |
| 0 | 0 | 0 | 0 | 0 |
| 0 | 0 | 0 | 0 | 0.001243498 |
| 0 | 0 | 0 | 0 | 0 |
| 0 | 0 | 0 | 0 | 0 |
| 0 | 0.097474506 | 0 | 0.004809914 | 0 |
| 0 | 0 | 0 | 0 | 0 |
| 0 | 0 | 0 | 0 | 0 |
| 0 | 0 | 0 | 0 | 0 |
| 0 | 0 | 0 | 0.085602276 | 0 |
| 0 | 0 | 0 | 0 | 0 |
| 0 | 0 | 0 | 0 | 0 |
| 0 | 0 | 0 | 0 | 0 |

| | | | | |
|---|---|---|---|---|
| 0 | 0.024617633 | 0 | 0 | 0 |
| 0 | 0 | 0 | 0 | 0 |
| 0 | 0 | 0 | 0 | 0 |
| 0 | 0 | 0 | 0 | 0 |
| 0 | 0.003411734 | 0 | 0 | 0.079213697 |
| 0 | 0 | 0 | 0 | 0 |
| 0 | 0 | 0.329835833 | 0 | 0 |
| 0 | 0 | 0 | 0 | 0 |
| 0 | 0 | 0 | 0 | 0 |
| 0 | 0 | 0 | 0 | 0.205944476 |
| 0 | 0.025780226 | 0 | 0 | 0 |
| 0 | 0 | 0 | 0 | 0 |
| 0 | 0 | 0 | 0 | 0 |
| 0 | 0 | 0 | 0 | 0 |
| 0 | 0 | 0 | 0 | 0 |
| 0 | 0 | 0 | 0 | 0 |
| 0 | 0 | 0 | 9.232713045 | 0.45515122 |
| 0 | 0 | 0 | 0 | 0 |
| 0 | 0 | 0 | 0 | 0 |
| 0 | 0 | 0 | 0 | 0 |
| 0 | 0.009358241 | 0 | 0.216998478 | 0 |
| 0 | 0 | 0 | 0 | 0 |
| 0 | 0.027725659 | 0 | 0 | 0 |
| 0.002129679 | 0.000640362 | 0 | 0.00257847 | 0 |
| 0 | 0 | 0 | 0 | 0 |
| 0 | 0 | 0 | 0.225362611 | 0 |
| 0 | 0 | 0 | 0 | 0 |
| 0 | 0 | 0 | 0 | 0 |
| 0 | 0 | 0 | 0 | 0 |
| 0.011238728 | 0 | 0.01512754 | 0.057056978 | 3.320558932 |
| 0 | 0.069908754 | 0 | 0 | 0 |
| 0 | 0 | 0 | 0 | 0 |
| 0 | 0 | 0 | 0 | 0 |
| 0 | 0 | 0 | 0.049336241 | 0 |
| 0 | 0 | 0 | 0 | 0 |
| 0 | 0 | 0 | 0 | 0 |
| 0.004085996 | 0.093880832 | 0.00183513 | 0 | 0 |
| 0 | 0 | 0 | 0 | 0 |
| 0 | 0 | 0 | 0 | 0 |
| 0 | 0 | 0 | 0 | 0 |
| 0 | 0 | 0 | 0 | 0 |
| 0 | 0 | 0 | 0 | 0 |
| 0 | 0 | 0 | 0 | 0 |
| 0 | 0 | 0 | 0 | 0 |
| 0 | 0.00594026 | 0 | 0 | 0 |
| 0 | 0 | 0 | 0 | 0 |
| 0 | 0 | 0 | 0 | 0 |
| 0 | 0 | 0 | 0 | 0 |

| | | | | |
|---|---|---|---|---|
| 0.793966594 | 0 | 0.007364664 | 0.306816258 | 0.073031243 |
| 0 | 0 | 0 | 0 | 0 |
| 0 | 0 | 0 | 0 | 0 |
| 0 | 0 | 0 | 0 | 0 |
| 0 | 0 | 0 | 0 | 0 |
| 0 | 0 | 0 | 0 | 0 |
| 0 | 0 | 0 | 0 | 0 |
| 0.200002101 | 0.031955465 | 0.295722165 | 0.007834229 | 2.722203952 |
| 0.083930621 | 0.873909541 | 7.557727615 | 1.745098498 | 1.615821324 |
| 0 | 0 | 0.021556306 | 0.029307827 | 0.037868971 |
| 0 | 0.045062695 | 0 | 0 | 0 |
| 0 | 0 | 0 | 0 | 0 |
| 0 | 0.005434019 | 0 | 0 | 0 |
| 0 | 0 | 0 | 0 | 0 |
| 0 | 0 | 0.002261461 | 0 | 0 |
| 0 | 0 | 0 | 0 | 0 |
| 0.002899117 | 0 | 0 | 0 | 0 |
| 0 | 0 | 0 | 0 | 0 |
| 0 | 0 | 0 | 0 | 0 |
| 0 | 0 | 0 | 0 | 0 |
| 0 | 0 | 0 | 0 | 0 |
| 0 | 0 | 0 | 0 | 0 |
| 0 | 0 | 0 | 0 | 0 |
| 0 | 0 | 0 | 0 | 0 |
| 0 | 0.024060776 | 0 | 0 | 0 |
| 0 | 0 | 0 | 0 | 0 |
| 0 | 0 | 0 | 0 | 0 |
| 0 | 0 | 0 | 0 | 0 |
| 0.003270994 | 0 | 0 | 0 | 0 |
| 0 | 0 | 0 | 0 | 0 |
| 0 | 0 | 0 | 0 | 0 |
| 0 | 0 | 0 | 0 | 0 |
| 0 | 0 | 0 | 0 | 0 |
| 0 | 0 | 0 | 0 | 0 |
| 0 | 0.02794553 | 0 | 0 | 0 |
| 0 | 0 | 0 | 0 | 0 |
| 0 | 0 | 0 | 0 | 0 |
| 0 | 0 | 0 | 0 | 0 |
| 0 | 0.001229006 | 0 | 0.001787027 | 0 |
| 0 | 0 | 0 | 0 | 0 |
| 0 | 0 | 0 | 0 | 0 |
| 0 | 0.038086197 | 0 | 0.412087839 | 0 |
| 0 | 0 | 0 | 0 | 0 |
| 0 | 0 | 0 | 0 | 0 |
| 0 | 0 | 0 | 0 | 0 |
| 0 | 0 | 0 | 0 | 0 |
| 0 | 0 | 0 | 0 | 0 |
| 0 | 0 | 0 | 0 | 0 |

| | | | | |
|---|---|---|---|---|
| 0 | 0 | 0 | 0 | 0 |
| 0.088798093 | 0.0008292 | 0 | 0.000939049 | 0 |
| 0 | 0 | 0 | 0 | 0 |
| 0 | 0 | 0 | 0 | 0 |
| 0 | 0.237809213 | 1.066079367 | 0.507225692 | 0.027881861 |
| 0 | 0 | 0 | 0 | 0 |
| 0 | 0 | 0 | 0 | 0 |
| 0.053054716 | 0.058434969 | 0.041488566 | 0.010882308 | 0.036687031 |
| 0 | 0 | 0 | 0 | 0 |
| 0 | 0 | 0 | 0 | 0 |
| 0 | 0 | 0 | 0 | 0 |
| 0 | 0 | 0 | 0 | 0 |
| 0 | 0 | 0 | 0 | 0 |
| 0 | 0 | 0 | 0 | 0 |
| 0 | 0 | 0 | 0.003312761 | 0.007914534 |
| 0.283837023 | 0.000760203 | 0 | 0.020789426 | 0 |
| 0 | 0 | 0 | 0 | 0 |
| 0 | 0 | 0 | 0 | 0 |
| 0 | 0 | 0 | 0 | 0 |
| 0 | 0 | 0 | 0 | 0 |
| 0 | 0 | 0 | 0 | 0 |
| 0 | 0 | 0 | 0 | 0 |
| 0 | 0 | 0 | 0 | 0 |
| 0.018668528 | 0.000632133 | 0.005501563 | 0.034616562 | 0.028421017 |
| 0 | 0.023396727 | 0 | 0 | 0 |
| 0 | 0 | 0 | 0 | 0 |
| 0.403173261 | 0.860067136 | 0 | 0.914437679 | 0 |
| 0 | 0 | 0 | 0 | 0 |
| 0.00320097 | 0 | 0 | 0.007093578 | 0.002436583 |
| 0 | 0 | 0 | 0 | 0 |
| 0 | 0 | 0 | 0 | 0 |
| 0 | 0 | 0 | 0 | 0 |
| 0 | 0 | 0 | 0 | 0 |
| 0 | 0 | 0 | 0 | 0 |
| 0 | 0 | 0 | 0 | 0 |
| 0 | 0 | 0 | 0 | 0 |
| 0 | 0 | 0 | 0 | 0 |
| 0 | 0.13590057 | 0 | 0 | 0 |
| 0 | 0 | 0 | 0 | 0 |
| 0 | 0 | 0 | 0 | 0 |
| 0 | 0 | 0 | 0 | 0 |
| 0 | 0 | 0 | 0 | 0 |
| 0 | 0 | 0 | 0 | 0 |
| 0 | 0 | 0 | 0 | 0 |
| 0 | 0 | 0 | 0 | 0 |
| 0 | 0 | 0 | 0 | 0 |
| 0 | 0 | 0 | 0 | 0 |
| 0 | 0.004447267 | 0 | 0 | 0 |

| | | | | |
|---|---|---|---|---|
| 0 | 0.009460507 | 0 | 0 | 0 |
| 0 | 0 | 0 | 0 | 0 |
| 0 | 0 | 0 | 0 | 0 |
| 0 | 0 | 0 | 0 | 0 |
| 0 | 0 | 0 | 0 | 0 |
| 0 | 0 | 0.672876506 | 0.009638356 | 0 |
| 0 | 0 | 0 | 0 | 0 |
| 0 | 0 | 0 | 0 | 0 |
| 0 | 0 | 0 | 0 | 0 |
| 0 | 0 | 0 | 0 | 0 |
| 0 | 0 | 0 | 0 | 0 |
| 0 | 0 | 0 | 0.004575467 | 0 |
| 0 | 0 | 0 | 0 | 0 |
| 0 | 0 | 0 | 0 | 0 |
| 0 | 0 | 0 | 0 | 0 |
| 0 | 0 | 0 | 0 | 0 |
| 0 | 0 | 0 | 0 | 0 |
| 0 | 0 | 0 | 0 | 0 |
| 0 | 0 | 0 | 0.005382748 | 0 |
| 0 | 0 | 0 | 0 | 0 |
| 0 | 0 | 0.17416069 | 0.006843677 | 0 |
| 0 | 0 | 0 | 0 | 0 |
| 0 | 0.060261066 | 0 | 0.139249393 | 0 |
| 0 | 0.002037353 | 0 | 0 | 0 |
| 0 | 0 | 0 | 0 | 0 |
| 0 | 0 | 0 | 0 | 0 |
| 0 | 0 | 0 | 0 | 0 |
| 0 | 0 | 0 | 0 | 0 |
| 0 | 0 | 0 | 0 | 0 |
| 0 | 0 | 0 | 0 | 0 |
| 0 | 0 | 0 | 0 | 0 |
| 0 | 0 | 0 | 0 | 0 |
| 0.01683724 | 0.001637933 | 0.120043884 | 0.028779362 | 0.008443831 |
| 0 | 0 | 0 | 0 | 0 |
| 0 | 0.072057431 | 0 | 0.000402234 | 0 |
| 0.00127164 | 0.017684286 | 0.001915657 | 0.004931581 | 0 |
| 0 | 0 | 0 | 0 | 0 |
| 0 | 0 | 0 | 0 | 0 |
| 0 | 0 | 0.365600475 | 0.019060219 | 0 |
| 0 | 0 | 0 | 0 | 0 |
| 0 | 0 | 0 | 0 | 0 |
| 0 | 0 | 0 | 0 | 0 |
| 0 | 0.052845064 | 0 | 0.000976078 | 0 |
| 0 | 0 | 0 | 0 | 0 |
| 0 | 0 | 0 | 0 | 0 |
| 0 | 0.006545491 | 0 | 0 | 0 |
| 0 | 0 | 0 | 0 | 0 |
| 0 | 0 | 0 | 0 | 0 |

| | | | | |
|---|---|---|---|---|
| 0 | 0 | 0 | 0 | 0 |
| 0 | 0 | 0 | 0 | 0 |
| 0.379414786 | 0 | 0 | 0 | 0 |
| 0 | 0 | 0 | 0 | 0 |
| 0 | 0 | 0 | 0 | 0 |
| 0 | 0 | 0 | 0 | 0 |
| 0 | 0 | 0 | 0 | 0 |
| 0 | 0 | 0 | 0 | 0 |
| 0 | 0.2206504 | 0 | 0 | 0 |
| 0 | 0 | 0 | 0 | 0 |
| 0 | 0 | 0 | 0 | 0 |
| 0 | 0 | 0 | 0 | 0 |
| 0 | 0 | 0 | 0 | 0 |
| 0 | 0.000454789 | 0 | 0.000343358 | 0 |
| 0 | 0 | 0 | 0 | 0 |
| 0.039102801 | 0 | 0 | 0.003404088 | 0.027131024 |
| 0 | 0 | 0 | 0 | 0 |
| 0 | 0 | 0 | 0 | 0 |
| 0 | 0 | 0 | 0 | 0 |
| 0 | 0.097458466 | 0 | 0 | 0 |
| 0 | 0.005161946 | 0 | 0 | 0 |
| 0 | 0 | 0 | 0 | 0 |
| 0 | 0 | 0 | 0 | 0 |
| 0 | 0.010221482 | 0 | 0 | 0 |
| 0 | 0 | 0 | 0.011147887 | 0 |
| 0 | 0 | 0 | 0 | 0 |
| 0 | 0 | 0 | 0 | 0 |
| 0 | 0 | 0 | 0 | 0 |
| 0 | 0 | 0 | 0 | 0 |
| 0 | 0.005526777 | 0 | 0 | 0 |
| 0 | 0.037315933 | 0 | 0 | 0 |
| 0 | 0 | 0 | 0 | 0 |
| 0 | 0 | 0 | 0 | 0 |
| 0 | 0 | 0 | 0 | 0 |
| 0 | 0 | 0 | 0 | 0 |
| 0 | 0 | 0 | 0 | 0 |
| 0 | 0 | 0 | 0 | 0 |
| 0 | 0 | 0 | 0 | 0 |
| 0 | 0 | 0 | 0 | 0 |
| 0 | 0 | 0 | 0 | 0 |
| 0 | 0 | 0 | 0 | 0 |
| 0 | 0 | 0 | 0 | 0 |
| 0 | 0.006974471 | 0 | 0 | 0 |
| 0.006098777 | 0.010558297 | 0 | 0 | 0 |
| 0 | 0 | 0 | 0 | 0 |
| 0 | 0 | 0 | 0 | 0 |
| 1.198220774 | 0 | 0.022870502 | 0.006567394 | 0.376160151 |
| 0 | 0 | 0 | 0 | 0 |

| | | | | |
|---|---|---|---|---|
| 0 | 0 | 0 | 0 | 0 |
| 0 | 0 | 0 | 0 | 0 |
| 0 | 0 | 0 | 0 | 0 |
| 0 | 0 | 0 | 0 | 0 |
| 0 | 0.007857993 | 0 | 0.001702197 | 0.070506741 |
| 0 | 0 | 0 | 0 | 0 |
| 0 | 0 | 0 | 0 | 0 |
| 0 | 0 | 0 | 0 | 0 |
| 0 | 0 | 0 | 0 | 0 |
| 0 | 0 | 0 | 0 | 0 |
| 0 | 0.016291427 | 0 | 0 | 0 |
| 0 | 0.035490984 | 0.003795422 | 0.386322423 | 0 |
| 0 | 0.001072288 | 0 | 0 | 0 |
| 0 | 0 | 0 | 0 | 0 |
| 0 | 0 | 0 | 0 | 0 |
| 0 | 0 | 0 | 0 | 0 |
| 0.001836835 | 0 | 0 | 0 | 0 |
| 0 | 0 | 0 | 0 | 0 |
| 0 | 0 | 0 | 0.293262391 | 0 |
| 0 | 0 | 2.188933618 | 0.000549765 | 0 |
| 0 | 0 | 0 | 0 | 0 |
| 0 | 0 | 0 | 0 | 0 |
| 0 | 0 | 0 | 0 | 0 |
| 0 | 0 | 0 | 0 | 0 |
| 0 | 0 | 0 | 0 | 0 |
| 0 | 0 | 0 | 0 | 0 |
| 0.032457421 | 0 | 0 | 0.002092621 | 0.004210099 |
| 0 | 0 | 0 | 0 | 0 |
| 0 | 0 | 0 | 0 | 0 |
| 0 | 0 | 0 | 0 | 0 |
| 0 | 0 | 0.001420255 | 0.017478605 | 0.013563572 |
| 0.011405159 | 0 | 0 | 0.030610835 | 0 |
| 0 | 0 | 0 | 0 | 0 |
| 0 | 0 | 0 | 0 | 0 |
| 0 | 0 | 0 | 0 | 0 |
| 0 | 0 | 0 | 0 | 0 |
| 0 | 0 | 0 | 0 | 0 |
| 0 | 0.029190214 | 0.042507541 | 1.731825277 | 0.006176445 |
| 0 | 0.002934554 | 0 | 0.001371526 | 0 |
| 0 | 0 | 0 | 0 | 0 |
| 0 | 0.015310026 | 0.05897241 | 0.018651739 | 0 |
| 0 | 0 | 0 | 0 | 0 |
| 0 | 0 | 0 | 0 | 0 |
| 0 | 0 | 0 | 0.145807062 | 0 |
| 0 | 0 | 0 | 0 | 0 |
| 0 | 0 | 0 | 0 | 0 |
| 0 | 0 | 0 | 0 | 0 |
| 0 | 0 | 0 | 0 | 0 |

| | | | | |
|---|---|---|---|---|
| 0 | 0 | 0 | 0 | 0 |
| 0 | 0.001214804 | 0 | 0 | 0 |
| 0 | 0 | 0 | 0 | 0 |
| 0.001981531 | 0 | 0.001240548 | 0.023718398 | 0.058962537 |
| 0 | 0 | 0.014094707 | 0.030721952 | 0.283099622 |
| 0.000646884 | 0 | 0.023759115 | 0 | 0.00131309 |
| 0 | 0 | 0 | 0 | 0 |
| 0 | 0 | 0 | 0.002985032 | 0 |
| 0 | 0 | 0 | 0 | 0 |
| 0 | 0 | 0 | 0 | 0 |
| 0 | 0.03524651 | 0 | 0 | 0 |
| 0 | 0 | 0 | 0 | 0 |
| 0 | 0.001055875 | 0 | 0.072566477 | 0 |
| 0 | 0 | 0 | 0 | 0 |
| 0 | 0 | 0 | 0 | 0 |
| 0 | 0 | 0 | 0 | 0 |
| 0 | 0 | 0 | 0 | 0 |
| 0 | 0 | 0 | 0 | 0 |
| 0 | 0.00160314 | 0 | 0 | 0 |
| 0 | 0 | 0 | 0 | 0 |
| 0 | 0 | 0 | 0 | 0 |
| 0 | 0 | 0 | 0 | 0 |
| 0.027575398 | 0 | 0 | 0.012850714 | 0.00895933 |
| 0 | 0 | 0 | 0 | 0 |
| 0 | 0 | 0 | 0 | 0 |
| 0 | 0 | 0 | 0 | 0 |
| 0 | 0 | 0 | 0 | 0 |
| 0 | 0 | 0 | 0 | 0 |
| 0 | 0 | 0 | 0 | 0 |
| 0 | 0 | 0 | 0 | 0 |
| 0 | 0.003523243 | 0 | 0 | 0 |
| 0 | 0 | 0 | 0.00164559 | 0 |
| 0 | 0 | 0 | 0 | 0 |
| 0 | 0 | 0 | 0 | 0 |
| 0 | 0.383167909 | 0 | 0 | 0 |
| 0 | 0 | 0 | 0 | 0 |
| 0 | 0 | 0.015774417 | 0 | 0 |
| 0 | 0 | 0 | 0 | 0 |
| 0 | 0.002027017 | 0.139274821 | 0 | 0.003139279 |
| 0 | 0 | 0 | 0 | 0 |
| 0 | 0 | 0 | 0 | 0 |
| 0 | 0 | 0 | 0 | 0 |
| 0 | 0.605411753 | 0 | 0 | 0 |
| 0 | 0 | 0 | 0 | 0 |
| 0 | 0 | 0 | 0 | 0 |
| 0 | 0.022135642 | 0 | 0.037489916 | 0 |
| 0.805592196 | 0 | 0 | 0 | 0.362114624 |
| 0 | 0 | 0 | 0 | 0 |

| | | | | |
|---|---|---|---|---|
| 0 | 0 | 0 | 0 | 0 |
| 0 | 0 | 0 | 0 | 0 |
| 0 | 0.120949293 | 0 | 0 | 0 |
| 0 | 0 | 0 | 0 | 0 |
| 0 | 0 | 0 | 0 | 0 |
| 0 | 0 | 0 | 0.001944516 | 0 |
| 0 | 0 | 0 | 0 | 0 |
| 0 | 0 | 0 | 0 | 0 |
| 0 | 0 | 0 | 0 | 0 |
| 0 | 0 | 0 | 0 | 0 |
| 0 | 0.000773861 | 0 | 0.001558006 | 0 |
| 0 | 0 | 0 | 0.004216072 | 0.008774712 |
| 0 | 0 | 0 | 0 | 0 |
| 0 | 0 | 0 | 0 | 0 |
| 0 | 0 | 0 | 0 | 0 |
| 0 | 0 | 0 | 0 | 0 |
| 0 | 0 | 0 | 0 | 0 |
| 0 | 0 | 0 | 0 | 0 |
| 0 | 0 | 0 | 0 | 0 |
| 0 | 0 | 0 | 0 | 0 |
| 0 | 0 | 0 | 0 | 0 |
| 0 | 0 | 0 | 0 | 0 |
| 0 | 0 | 0 | 0 | 0 |
| 0 | 0 | 0 | 0 | 0 |
| 0 | 0 | 0 | 0 | 0 |
| 0 | 0 | 0 | 0 | 0 |
| 0 | 0 | 0 | 0 | 0 |
| 0 | 0.014622701 | 0 | 0 | 0 |
| 0 | 0 | 0 | 0 | 0 |
| 0 | 0 | 0 | 0 | 0 |
| 0 | 0.000661256 | 0.002019305 | 0.003106366 | 0 |
| 0 | 0 | 0 | 0 | 0 |
| 0 | 0.008829528 | 0.090267833 | 0.056678393 | 0 |
| 0 | 0.008368992 | 0.034441729 | 0 | 0.008209807 |
| 0 | 0 | 0 | 0 | 0 |
| 0 | 0 | 0 | 0 | 0 |
| 0 | 0 | 0 | 0 | 0 |
| 0 | 0 | 0 | 0 | 0 |
| 0 | 0 | 0 | 0 | 0 |
| 0 | 0 | 0 | 0 | 0 |
| 0 | 0 | 0 | 0 | 0 |
| 0.27183111 | 0 | 0.028210116 | 0.002861313 | 0.33260422 |
| 0 | 0.00782353 | 0.232937823 | 0.002711244 | 0 |
| 0 | 0 | 0 | 0 | 0 |
| 0 | 0 | 0 | 0 | 0 |
| 0 | 0.01390026 | 0 | 0.004477642 | 0 |
| 0 | 0 | 0 | 0.008562368 | 0 |
| 0 | 0 | 0 | 0 | 0 |

| | | | | |
|---|---|---|---|---|
| 0 | 0 | 0 | 0 | 0 |
| 0 | 0.005643275 | 0 | 0.013796171 | 0 |
| 0 | 0.002511216 | 0.086271864 | 0 | 0.001907187 |
| 0 | 0 | 0 | 0 | 0 |
| 0 | 0.121182316 | 0 | 0.396743557 | 0 |
| 0 | 0.005611246 | 0 | 0.063607403 | 0.041256841 |
| 0 | 0 | 0 | 0 | 0 |
| 0 | 0 | 0 | 0 | 0 |
| 0 | 0 | 0 | 0 | 0 |
| 0 | 0 | 0 | 0 | 0 |
| 0 | 0 | 0 | 0 | 0 |
| 0 | 0 | 0 | 0 | 0 |
| 0 | 0 | 0 | 0 | 0 |
| 2.349521391 | 0 | 0 | 0 | 0 |
| 0 | 0 | 0 | 0 | 0 |
| 0 | 0 | 0 | 0 | 0 |
| 0 | 0 | 0 | 0 | 0 |
| 0 | 0 | 0 | 0 | 0 |
| 0 | 0 | 0 | 0 | 0 |
| 0 | 0 | 0 | 0 | 0 |
| 0 | 0 | 0 | 0 | 0 |
| 0.319270142 | 0 | 0.024288509 | 0 | 0.004090964 |
| 0 | 0 | 0 | 0 | 0 |
| 0 | 0 | 0 | 0 | 0 |
| 0 | 0.011641471 | 0 | 0 | 0 |
| 0.009693702 | 0 | 0 | 0.009038132 | 0.001844715 |
| 0 | 0 | 0 | 0 | 0 |
| 0 | 0.107416259 | 0 | 0 | 0.02914227 |
| 0 | 0 | 0 | 0 | 0 |
| 0 | 0 | 0 | 0 | 0 |
| 0 | 0 | 0 | 0 | 0 |
| 0.006375602 | 0 | 0 | 0 | 0 |
| 0 | 0 | 0 | 0 | 0 |
| 0 | 0 | 0 | 0 | 0 |
| 0 | 0 | 0 | 0 | 0 |
| 0 | 0 | 0 | 0 | 0 |
| 0 | 0 | 0 | 0 | 0 |
| 0 | 0 | 0 | 0.000521548 | 0 |
| 0 | 4.99347405 | 0 | 0 | 0 |
| 0 | 0 | 0 | 0 | 0 |
| 0 | 0 | 0 | 0 | 0 |
| 0 | 0 | 0 | 0.000568405 | 0 |
| 0.618292856 | 0 | 0.010169288 | 0.226637327 | 0.067707338 |
| 0 | 0 | 0 | 0 | 0 |
| 0 | 0 | 0 | 0 | 0 |
| 0 | 0 | 0 | 0 | 0 |
| 0.031094815 | 0.002397369 | 0 | 0.006435467 | 0.001618422 |
| 0 | 0 | 0 | 0 | 0 |

| | | | | |
|---|---|---|---|---|
| 0 | 0 | 0 | 0 | 0 |
| 0.039403612 | 0.001386476 | 0.016454641 | 0.024234244 | 0.013273977 |
| 0 | 0 | 0 | 0 | 0 |
| 0 | 0 | 0 | 0 | 0 |
| 0 | 0 | 0 | 0 | 0 |
| 0 | 0 | 0 | 0 | 0 |
| 0 | 0 | 0 | 0 | 0 |
| 0 | 0 | 0 | 0 | 0 |
| 0 | 0 | 0 | 0 | 0 |
| 0 | 0 | 0 | 0 | 0 |
| 0 | 0 | 0 | 0 | 0 |
| 0 | 0 | 0 | 0 | 0 |
| 0 | 0.00359745 | 0.008544428 | 0.008583956 | 0 |
| 0 | 0 | 0 | 0 | 0 |
| 0 | 0 | 0 | 0 | 0 |
| 0 | 0 | 0 | 0 | 0 |
| 0 | 0 | 0 | 0 | 0 |
| 0 | 0 | 0 | 0 | 0 |
| 0 | 0 | 0 | 0 | 0 |
| 0 | 0 | 0 | 0 | 0 |
| 0 | 0 | 0 | 0 | 0 |
| 0 | 0 | 0.03782315 | 0 | 0 |
| 0 | 0 | 0 | 0 | 0.00552438 |
| 0 | 0 | 0.001820452 | 0.000960159 | 0 |
| 0 | 0 | 0 | 0 | 0 |
| 0 | 0 | 0 | 0 | 0 |
| 0 | 0 | 0 | 0 | 0 |
| 0 | 0 | 0 | 0 | 0 |
| 0 | 0 | 0 | 0 | 0 |
| 0 | 0.059671179 | 0 | 0 | 0 |
| 0 | 0 | 0 | 0 | 0 |
| 0 | 0 | 0 | 0 | 0 |
| 0 | 0 | 0 | 0 | 0 |
| 0 | 0 | 0 | 0 | 0 |
| 0 | 6.048824447 | 0 | 8.410225987 | 0 |
| 0 | 0 | 0 | 0 | 0 |
| 0 | 0 | 0 | 0 | 0 |
| 0 | 0 | 0 | 0 | 0 |
| 0 | 0.000770639 | 0.260513941 | 0 | 0 |
| 0 | 0 | 0 | 0 | 0 |
| 0 | 0.057351285 | 0.041779467 | 0.011685594 | 0.294434607 |
| 0 | 0 | 0 | 0 | 0 |
| 0 | 0 | 0 | 0 | 0 |
| 0 | 0 | 0 | 0 | 0 |
| 0 | 0 | 0 | 0 | 0 |
| 0.651664713 | 0.004272071 | 8.827666611 | 0.0060525 | 0.75272407 |
| 0 | 0 | 0 | 0 | 0 |
| 0 | 0 | 0 | 0 | 0 |

| | | | | |
|---|---|---|---|---|
| 0.16010201 | 0 | 0 | 0 | 0 |
| 0 | 0 | 0 | 0 | 0 |
| 0 | 0 | 0 | 0 | 0 |
| 0 | 0 | 0 | 0 | 0 |
| 0 | 0 | 0 | 0 | 0 |
| 0 | 0 | 0 | 0 | 0 |
| 0 | 0 | 0 | 0 | 0 |
| 0 | 0 | 0.009008768 | 0 | 0 |
| 0 | 0.000321559 | 0 | 0 | 0 |
| 0 | 0 | 0.047491184 | 0.002959677 | 0 |
| 0 | 0.015830383 | 0 | 0 | 0 |
| 0 | 0 | 0 | 0 | 0 |
| 0 | 0.000756247 | 0.002424854 | 0.000761272 | 0 |
| 0 | 0 | 0 | 0 | 0 |
| 0 | 0 | 0 | 0 | 0 |
| 0 | 0 | 0 | 0 | 0 |
| 0 | 0 | 0 | 0 | 0 |
| 0 | 0 | 0 | 0 | 0 |
| 0 | 0 | 0 | 0 | 0 |
| 0.604780344 | 0.002827075 | 1.274316769 | 0.639912401 | 0 |
| 0 | 0 | 0 | 0 | 0 |
| 0 | 0 | 0 | 0 | 0 |
| 0 | 0 | 0 | 0 | 0 |
| 0 | 0 | 0 | 0 | 0 |
| 0 | 0 | 0 | 0 | 0 |
| 0 | 0 | 0 | 0 | 0 |
| 0 | 0 | 0 | 0 | 0 |
| 0 | 0 | 0 | 0 | 0 |
| 0 | 0 | 0 | 0 | 0 |
| 0 | 0 | 0 | 0.003187143 | 0 |
| 0 | 0 | 0 | 0 | 0 |
| 0 | 0 | 0 | 0 | 0 |
| 0 | 0 | 0 | 0 | 0 |
| 0 | 0 | 0 | 0 | 0 |
| 0 | 0 | 0 | 0 | 0 |
| 0 | 0 | 0 | 0 | 0 |
| 0 | 0 | 0 | 0 | 0 |
| 0 | 0 | 0 | 0 | 0 |
| 0 | 0 | 0 | 0 | 0 |
| 0 | 0 | 0 | 0 | 0 |
| 0 | 0 | 0 | 0 | 0 |
| 0 | 0 | 0 | 0 | 0 |
| 0 | 0.006938026 | 0 | 0.057900698 | 0 |
| 0 | 0 | 0 | 0 | 0 |
| 0 | 0.004299046 | 0.00984614 | 0.027238513 | 0 |
| 0 | 0 | 0 | 0 | 0 |
| 0 | 0 | 0 | 0 | 0 |
| 0.001010278 | 0 | 0 | 0 | 0 |

| | | | | |
|---|---|---|---|---|
| 0 | 0 | 0 | 0 | 0 |
| 0 | 0 | 0 | 0 | 0 |
| 0 | 0 | 0 | 0 | 0 |
| 0 | 0 | 0 | 0 | 0 |
| 0 | 0 | 0 | 0 | 0 |
| 0.003516027 | 0.018973247 | 0.070694765 | 0.220354944 | 0 |
| 0 | 0 | 0 | 0 | 0 |
| 0 | 0 | 0 | 0 | 0 |
| 0 | 0 | 0 | 0 | 0 |
| 0 | 0 | 0 | 0 | 0 |
| 0 | 0 | 0 | 0 | 0 |
| 0 | 0 | 0 | 0 | 0 |
| 0 | 0 | 0 | 0 | 0 |
| 0 | 0 | 0 | 0 | 0 |
| 0 | 0 | 0 | 0 | 0 |
| 0 | 0 | 0 | 0 | 0 |
| 0 | 0.068127247 | 0 | 0 | 0 |
| 0 | 0 | 0.001065008 | 0 | 0.004708763 |
| 0 | 0 | 0 | 0 | 0 |
| 0 | 0 | 0 | 0 | 0 |
| 0 | 0 | 0 | 0 | 0 |
| 0 | 0 | 0 | 0 | 0 |
| 0 | 0 | 0 | 0 | 0 |
| 0 | 0 | 0 | 0 | 0 |
| 0 | 0 | 0 | 0 | 0 |
| 0 | 0 | 0 | 0 | 0 |
| 0 | 0 | 0 | 0 | 0 |
| 0 | 0.000994542 | 0 | 0.008454164 | 0 |
| 0 | 0 | 0 | 0 | 0 |
| 0 | 0 | 0 | 0 | 0 |
| 0 | 0 | 0 | 0 | 0 |
| 0 | 0.003601171 | 0 | 0 | 0 |
| 0 | 0 | 0 | 0 | 0 |
| 0 | 0 | 0 | 0 | 0 |
| 0 | 0 | 0 | 0 | 0 |
| 0 | 0 | 0 | 0 | 0 |
| 0 | 0 | 0 | 0 | 0 |
| 0 | 0 | 0 | 0 | 0 |
| 0 | 0.327385329 | 0 | 0 | 0 |
| 0 | 0 | 0 | 0 | 0 |
| 0 | 0 | 0 | 0 | 0 |
| 0 | 0 | 0 | 0 | 0 |
| 0 | 0 | 0 | 0 | 0 |
| 0 | 0 | 0 | 0 | 0 |
| 0 | 0 | 0 | 0 | 0 |
| 0 | 0 | 0 | 0 | 0 |
| 0 | 0 | 0 | 0 | 0 |
| 0 | 0 | 0 | 0 | 0 |

| | | | | |
|---|---|---|---|---|
| 0 | 0 | 0 | 0 | 0 |
| 0 | 0.000634092 | 0 | 0 | 0.04088014 |
| 0 | 0 | 0 | 0 | 0 |
| 0 | 0 | 0 | 0 | 0 |
| 0 | 0 | 0 | 0 | 0 |
| 0 | 0 | 0 | 0.003938881 | 0 |
| 0 | 0 | 0 | 0 | 0 |
| 0 | 0 | 0 | 0 | 0 |
| 0 | 0 | 0 | 0 | 0 |
| 0 | 0 | 0 | 0 | 0 |
| 0 | 0 | 0 | 0 | 0.001681549 |
| 0 | 0 | 0 | 0 | 0 |
| 0 | 0.222605235 | 0 | 0 | 0 |
| 0 | 0 | 0 | 0 | 0 |
| 0 | 0 | 0 | 0 | 0 |
| 0 | 0 | 0 | 0 | 0 |
| 0 | 0 | 0 | 0 | 0 |
| 0 | 0 | 0 | 0 | 0 |
| 0 | 0 | 0 | 0 | 0 |
| 0 | 0 | 0 | 0.002434213 | 0 |
| 0 | 0 | 0.044069177 | 0.000848297 | 0 |
| 0 | 0 | 0 | 0.002623991 | 0 |
| 0 | 0 | 0 | 0 | 0 |
| 0 | 1.081341831 | 1.052837325 | 0.105526513 | 0 |
| 0 | 0 | 0.001545235 | 0 | 0 |
| 0 | 0 | 0 | 0 | 0 |
| 0 | 0 | 0 | 0 | 0 |
| 0 | 0 | 0 | 0 | 0 |
| 0 | 0 | 0 | 0 | 0 |
| 0 | 0 | 0 | 0 | 0 |
| 0 | 0 | 0 | 0 | 0 |
| 0 | 0 | 0 | 0 | 0 |
| 0 | 0 | 0 | 0 | 0 |
| 0 | 0 | 0 | 0 | 0 |

| PHL018-0053_V1 | PHL018-0268_V3 | PHL018-0260_V2 | PHL018-0060_V1 | PHL018-0159_V2 |
|---|---|---|---|---|
| CTRL | REF | REF | CTRL | REF |
| 0.271388417 | 0.003959862 | 0.009649965 | 0.30969785 | 0.012949593 |
| 0.290619562 | 0.393948316 | 0.935810203 | 2.554432099 | 0.165658171 |
| 0 | 0 | 0 | 0 | 3.333562295 |
| 0 | 0 | 0 | 0 | 0 |
| 0 | 0 | 0 | 0 | 0 |
| 0 | 0 | 0 | 0 | 0 |
| 0 | 0 | 0 | 0 | 0 |
| 0 | 0 | 0 | 0 | 0 |
| 0 | 0 | 0 | 0 | 0 |
| 0 | 0 | 0 | 0 | 0 |
| 0 | 0 | 0 | 0 | 0 |
| 0 | 0 | 0 | 0 | 0 |
| 0 | 0 | 0 | 0 | 0 |
| 0.010111931 | 0.835386444 | 0.606677497 | 0.34602686 | 1.558565054 |
| 0 | 0 | 0 | 0 | 0 |
| 0 | 0 | 0 | 0 | 0 |
| 0 | 0 | 0 | 0 | 0 |
| 0 | 0 | 0 | 0 | 0 |
| 0 | 0 | 0 | 0 | 0 |
| 0 | 0 | 0 | 0 | 0 |
| 0 | 0 | 0 | 0 | 0 |
| 0 | 0 | 0 | 0 | 0 |
| 0 | 0 | 0 | 0 | 0 |
| 0 | 0 | 0 | 0 | 0 |
| 0 | 0 | 0.047472471 | 0 | 0 |
| 0 | 0 | 0 | 0 | 0 |
| 0 | 0 | 0 | 0 | 0 |
| 0 | 0 | 0 | 0 | 0 |
| 0 | 0 | 0 | 0 | 0 |
| 0 | 0 | 0 | 0 | 0 |
| 0 | 0 | 0 | 0 | 0 |
| 0 | 0 | 0 | 0 | 0 |
| 0 | 0 | 0 | 0 | 0 |
| 0 | 0.000888139 | 0 | 0 | 0 |
| 0 | 0 | 0 | 0 | 0 |
| 0 | 0.501904731 | 5.073273576 | 2.539996948 | 0 |
| 0 | 0 | 0 | 0 | 0 |
| 0 | 0 | 0 | 0 | 0 |
| 0 | 0 | 0 | 0 | 0 |
| 0 | 0 | 0 | 0 | 0 |
| 0 | 0 | 0 | 0 | 0 |
| 0.100539406 | 0.066578288 | 0.028583546 | 0.006291152 | 0.001391884 |
| 1.171046033 | 11.48105746 | 0.030036809 | 0.006238312 | 0.409403989 |
| 0.002627245 | 0.005188776 | 0.060357655 | 0.049235097 | 0.037785464 |
| 0.001578459 | 0.005985479 | 0.041084691 | 0.005747096 | 0.01554073 |
| 0 | 0 | 0 | 0 | 0 |

| | | | | |
|---|---|---|---|---|
| 0.091341285 | 0.003684723 | 0 | 0.01161068 | 0 |
| 0.06178386 | 0.000800511 | 0 | 0.008138426 | 0.004564442 |
| 0 | 0 | 0 | 0.008722962 | 0.001617447 |
| 0 | 0.74044084 | 0.324865593 | 0.793652346 | 0.320338271 |
| 0 | 0 | 0 | 0 | 0 |
| 0 | 0.007571264 | 0 | 0 | 0 |
| 0.001810178 | 0 | 0 | 0 | 0.00104836 |
| 0 | 0 | 0.003410518 | 0 | 0 |
| 0 | 0.004996528 | 0 | 0 | 0 |
| 0 | 0 | 0 | 0 | 0 |
| 0.007623013 | 0 | 0 | 0 | 0 |
| 0 | 0.144720617 | 0.237576818 | 0.028272456 | 0 |
| 0.000653405 | 0 | 0 | 0 | 0 |
| 0 | 0 | 0.029782982 | 0.184639326 | 0 |
| 0.00088933 | 0 | 0 | 0 | 0.001030106 |
| 0 | 0 | 0 | 0 | 0 |
| 0.014513935 | 0.02807984 | 0.017791545 | 0.040283662 | 0.901983919 |
| 0 | 0 | 0 | 0 | 0 |
| 0 | 0 | 0 | 0 | 0 |
| 0 | 0 | 0 | 0 | 0 |
| 0.003538876 | 0 | 0.004571271 | 0.115890569 | 0.002644556 |
| 0 | 0.002235385 | 0.005612586 | 0.014921196 | 0 |
| 0 | 0.003191039 | 0.017589445 | 0 | 0 |
| 0 | 0 | 0 | 0 | 0 |
| 0 | 0 | 0 | 0 | 0 |
| 0 | 0 | 0 | 0 | 0 |
| 0 | 0 | 0 | 0 | 0.009042535 |
| 0 | 0 | 0 | 0 | 0 |
| 0.003153702 | 0.000779919 | 0.053428546 | 0 | 0.001429402 |
| 0 | 0 | 0 | 0 | 0 |
| 0 | 0 | 0 | 0.001820793 | 0 |
| 0 | 0 | 0 | 0 | 0 |
| 0 | 0 | 0 | 0 | 0 |
| 0 | 0 | 0 | 0 | 0 |
| 0 | 0 | 0 | 0 | 0 |
| 0 | 0 | 0 | 0 | 0 |
| 0 | 0 | 0 | 0 | 0 |
| 1.01493673 | 0.921124602 | 0.001801549 | 0.004175427 | 0.001806525 |
| 0 | 0 | 0 | 0 | 0 |
| 0 | 0 | 0 | 0 | 0 |
| 0 | 0 | 0 | 0 | 0 |
| 0 | 0 | 0 | 0 | 0 |
| 0 | 0 | 0 | 0 | 0 |
| 0 | 0 | 0 | 0 | 0 |
| 0 | 0 | 0 | 0 | 0 |
| 0 | 0 | 0 | 0.115869863 | 0 |
| 0 | 0 | 0 | 0 | 0 |
| 0 | 0 | 0 | 0 | 0.002884906 |

| | | | | |
|---|---|---|---|---|
| 0 | 0 | 0 | 0 | 0 |
| 0 | 0 | 0 | 0 | 0 |
| 0 | 0 | 0 | 0 | 0 |
| 0 | 0 | 0 | 0 | 0 |
| 0 | 0 | 0 | 0 | 0 |
| 0 | 0 | 0 | 0 | 0 |
| 0 | 0 | 0 | 0 | 0 |
| 0 | 0 | 0 | 0 | 0 |
| 0.012846443 | 0 | 0 | 0 | 0 |
| 0 | 0 | 0 | 0 | 0 |
| 0 | 0 | 0 | 0 | 0 |
| 0.001082502 | 0 | 0 | 0 | 0 |
| 0 | 0 | 0 | 0 | 0 |
| 0 | 0 | 0 | 0 | 0 |
| 0 | 0 | 0 | 0 | 0 |
| 0 | 0 | 0 | 0 | 0 |
| 0 | 0 | 0 | 0 | 0 |
| 0 | 0 | 0 | 0 | 0 |
| 0.213227178 | 0.211879657 | 0.109961086 | 0 | 0 |
| 0 | 0 | 0 | 0 | 0 |
| 0 | 0 | 0 | 0 | 0 |
| 0 | 0 | 0 | 0 | 0 |
| 0 | 0 | 0 | 0 | 0 |
| 0 | 0 | 0 | 0 | 0 |
| 0 | 0 | 0 | 0 | 0 |
| 0 | 0 | 0 | 0 | 0 |
| 0.001518168 | 0 | 0 | 0 | 0 |
| 0 | 0 | 0 | 0 | 0 |
| 0 | 0 | 0 | 0 | 0 |
| 0 | 0 | 0 | 0 | 0 |
| 0 | 0.009815362 | 0.014617469 | 0 | 0 |
| 0.001106321 | 0.002478948 | 0.002788181 | 0 | 0.005824755 |
| 0 | 0 | 0 | 0 | 0 |
| 0 | 0 | 0 | 0 | 0 |
| 0 | 0 | 0 | 0 | 0 |
| 0 | 0 | 0 | 0 | 0 |
| 0 | 0 | 0 | 0 | 0 |
| 0 | 0 | 0 | 0 | 0 |
| 0 | 0.04340195 | 0.279473527 | 0 | 0 |
| 0 | 0 | 0 | 0 | 0 |
| 0 | 0 | 0 | 0 | 0 |
| 0 | 0 | 0 | 0 | 0 |
| 0 | 0 | 0 | 0 | 0.00067325 |
| 0 | 0 | 0 | 0 | 0 |
| 0 | 0 | 0 | 0 | 0 |
| 0 | 0 | 0 | 0 | 0 |
| 0 | 0 | 0 | 0 | 0 |
| 0 | 0 | 0 | 0 | 0 |

| | | | | |
|---|---|---|---|---|
| 0 | 0 | 0 | 0 | 0 |
| 0 | 0 | 0 | 0 | 0 |
| 0 | 0 | 0 | 0 | 0 |
| 0 | 0 | 0 | 0 | 0 |
| 0 | 0 | 0.031952819 | 0.010124201 | 0 |
| 0 | 0 | 0 | 0 | 0 |
| 0 | 0 | 0 | 0 | 0 |
| 0 | 0 | 0 | 0 | 0 |
| 0 | 0 | 0 | 0 | 0 |
| 0 | 0 | 0 | 0 | 0 |
| 0 | 0 | 0 | 0 | 0 |
| 0 | 0 | 0 | 0 | 0 |
| 0 | 0 | 0 | 0 | 0 |
| 0 | 0 | 0 | 0 | 0 |
| 0 | 0 | 0 | 0 | 0 |
| 0 | 0 | 7.689952084 | 0 | 0 |
| 0 | 0 | 0 | 0 | 0 |
| 0 | 0 | 0 | 0 | 0 |
| 0 | 0 | 0 | 0 | 0 |
| 0 | 0 | 0 | 0 | 0 |
| 0 | 0 | 0 | 0 | 0 |
| 0 | 0 | 0 | 0 | 0 |
| 0 | 0 | 0 | 0 | 0 |
| 0 | 0 | 0 | 0 | 0 |
| 0 | 0.095475316 | 0.008173351 | 0 | 0 |
| 0 | 0 | 0 | 0 | 0 |
| 0 | 0.000828554 | 0 | 0 | 0 |
| 0 | 0 | 0 | 0 | 0 |
| 0 | 0 | 0 | 0 | 0 |
| 0 | 0 | 0 | 0 | 0 |
| 0 | 0 | 0 | 0 | 0 |
| 0.005337907 | 0 | 0 | 0 | 0 |
| 0 | 0 | 0 | 0 | 0 |
| 0 | 0 | 0 | 0 | 0 |
| 0 | 0 | 0 | 0 | 0 |
| 0 | 0 | 0 | 0 | 0 |
| 0 | 0 | 0 | 0 | 0 |
| 0 | 0.006868277 | 0 | 0 | 0 |
| 0 | 0 | 0 | 0 | 0 |
| 0 | 0 | 0 | 0 | 0 |
| 0 | 0 | 0 | 0 | 0 |
| 0 | 0 | 0 | 0 | 0 |
| 0 | 0 | 0 | 0 | 0 |
| 0 | 0 | 0 | 0 | 0 |
| 0 | 0 | 0 | 0 | 0 |
| 0 | 0 | 0 | 0 | 0 |
| 0 | 0 | 0 | 0 | 0 |
| 0 | 0 | 0 | 0 | 0 |

| | | | | |
|---|---|---|---|---|
| 0 | 1.673348264 | 0 | 0 | 0 |
| 0 | 0 | 0 | 0 | 0 |
| 0 | 0 | 0 | 0 | 0 |
| 0 | 0 | 0 | 0 | 0 |
| 0 | 0 | 0 | 0 | 0 |
| 0 | 0 | 0 | 0 | 0 |
| 0 | 0 | 0 | 0 | 0 |
| 0 | 0 | 0 | 0 | 0 |
| 0 | 0 | 0 | 0 | 0 |
| 0 | 0 | 0 | 0 | 0 |
| 0 | 0 | 0 | 0 | 0 |
| 0 | 0 | 0 | 0 | 0 |
| 0 | 0 | 0.000855087 | 0.000713457 | 0 |
| 0 | 0 | 0 | 0 | 0 |
| 0 | 0 | 0 | 0 | 0 |
| 0 | 0 | 0 | 0 | 0 |
| 0 | 0 | 0 | 0 | 0 |
| 0 | 0 | 0 | 0 | 0 |
| 0 | 0 | 0 | 0 | 0 |
| 0 | 0 | 0 | 0 | 0 |
| 0.034464698 | 0.014958413 | 0.008633564 | 0.05158109 | 0.018998207 |
| 0 | 0 | 0 | 0 | 0 |
| 0 | 0 | 0 | 0 | 0 |
| 0 | 0 | 0 | 0 | 0 |
| 0 | 0 | 0 | 0 | 0 |
| 0 | 0 | 0 | 0 | 0 |
| 0.007133821 | 0.013067141 | 0.025139921 | 0.00595613 | 0.008683224 |
| 0 | 0 | 0 | 0 | 0 |
| 0 | 0 | 0 | 0 | 0 |
| 0 | 0 | 0 | 0 | 0 |
| 0.001084761 | 0 | 0 | 0 | 0 |
| 0 | 0 | 0 | 0 | 0 |
| 0 | 0 | 0 | 0 | 0 |
| 0 | 0.002886838 | 1.079491526 | 0 | 0 |
| 0 | 0 | 0.012899346 | 0.253735806 | 0.000782357 |
| 0 | 0 | 0 | 0 | 0 |
| 0 | 0 | 0 | 0 | 0 |
| 0 | 0 | 0 | 0 | 0 |
| 0 | 0 | 0 | 0 | 0.582232728 |
| 0 | 0 | 0 | 0 | 0 |
| 0 | 0 | 0 | 0 | 0 |
| 0 | 0 | 0 | 0 | 0 |
| 0 | 0 | 0 | 0 | 0 |
| 0 | 0 | 0 | 0 | 0 |
| 0 | 0 | 0 | 0 | 0 |
| 0 | 0 | 0.054078579 | 0 | 0 |
| 0 | 0 | 0 | 0 | 0 |
| 0 | 0.000912607 | 0 | 0 | 0 |

| | | | | |
|---|---|---|---|---|
| 0 | 0 | 0 | 0 | 0 |
| 0 | 0 | 0 | 0 | 0 |
| 0 | 0 | 0.002438664 | 0 | 0 |
| 0 | 0 | 0 | 0 | 0 |
| 0 | 0 | 0 | 0 | 0 |
| 0 | 0 | 0 | 0 | 0 |
| 0.000411105 | 0 | 0 | 0 | 0 |
| 0.000691929 | 0 | 0 | 0 | 0 |
| 0 | 0 | 0 | 0 | 0 |
| 0 | 0 | 0 | 0 | 0 |
| 0 | 0 | 0 | 0 | 0 |
| 0 | 0 | 0 | 0 | 0 |
| 0 | 0 | 0 | 0 | 0 |
| 0 | 0 | 0 | 0 | 0 |
| 0.000455344 | 0 | 0 | 0 | 0 |
| 0 | 0 | 0 | 0 | 0 |
| 0 | 0 | 0 | 0 | 0 |
| 0 | 0.06406961 | 0 | 0 | 0 |
| 0.004699349 | 0.028957202 | 0 | 0 | 0.001814411 |
| 0 | 0.08232091 | 0.386362903 | 0 | 0 |
| 0 | 0 | 0 | 0.002474427 | 0 |
| 0 | 0 | 0 | 0 | 0 |
| 0 | 0.000912212 | 0 | 0 | 0 |
| 0.401353728 | 0 | 0 | 0 | 0 |
| 0.012624259 | 0 | 0.003535112 | 0.004301475 | 0.007643639 |
| 0.001809719 | 0 | 0 | 0 | 0 |
| 0 | 0 | 0 | 0 | 0 |
| 0 | 0 | 0 | 0 | 0 |
| 0.026833099 | 0.002336107 | 0 | 0 | 0 |
| 0 | 0 | 0 | 0 | 0 |
| 0 | 0.020133455 | 0.043012876 | 0.478059134 | 0 |
| 0 | 0 | 0 | 0 | 0 |
| 0 | 0 | 0 | 0 | 0 |
| 0 | 0 | 0 | 0 | 0 |
| 0 | 0 | 0 | 0 | 0 |
| 0.07358304 | 0 | 0 | 0 | 0 |
| 0 | 0 | 0 | 0 | 0 |
| 0 | 0 | 0 | 0 | 0 |
| 0 | 0 | 0 | 0 | 0 |
| 0 | 0 | 0.001633608 | 0.003861916 | 0 |
| 0 | 0 | 0 | 0 | 0 |
| 0.788332578 | 0 | 0.002834054 | 0.005332037 | 0 |
| 0 | 0 | 0 | 0 | 0 |
| 0 | 0 | 0 | 0 | 0 |
| 0 | 0 | 0 | 0 | 0 |
| 0 | 0 | 0 | 0 | 0 |
| 0 | 0 | 0 | 0 | 0 |
| 0 | 0 | 0 | 0 | 0 |

| | | | | |
|---|---|---|---|---|
| 0 | 0 | 0 | 0 | 0 |
| 0 | 0 | 0 | 0 | 0 |
| 0.00040775 | 0.005669321 | 0.060287219 | 0.053270899 | 0.038413312 |
| 0 | 0 | 0 | 0 | 0 |
| 0 | 0 | 0 | 0 | 0 |
| 0 | 0 | 0 | 0 | 0 |
| 0 | 0 | 0 | 0 | 0 |
| 0 | 0 | 0 | 0 | 0 |
| 0 | 0 | 0 | 0 | 0 |
| 0.000372925 | 0 | 0.000957261 | 0 | 0 |
| 0 | 0 | 0 | 0 | 0 |
| 0 | 0 | 0 | 0 | 0 |
| 0 | 0 | 0 | 0 | 0 |
| 0.004356162 | 0 | 0 | 0 | 0 |
| 0.004339326 | 0 | 0.003055162 | 0.005576217 | 0.000718031 |
| 0 | 0.005536722 | 0.079116196 | 0 | 0 |
| 0.01115993 | 0 | 0.004568921 | 0.003328076 | 0.007035937 |
| 0.006794059 | 0 | 0 | 0 | 0 |
| 0 | 0 | 0 | 0 | 0 |
| 0 | 0 | 0 | 0 | 0 |
| 0 | 0 | 0 | 0 | 0 |
| 0 | 0.00145068 | 0 | 0 | 0 |
| 0 | 0 | 0 | 0 | 0 |
| 0 | 0 | 0 | 0 | 0 |
| 0.006747641 | 0 | 0 | 0 | 0 |
| 0 | 0 | 0 | 0 | 0 |
| 0 | 0 | 0 | 0 | 0 |
| 0 | 0 | 0 | 0 | 0 |
| 0 | 0 | 0 | 0 | 0 |
| 0.001844616 | 0 | 0 | 0 | 0 |
| 0.243523429 | 0 | 0 | 0 | 0 |
| 0 | 0 | 0 | 0 | 0 |
| 0 | 0 | 0 | 0 | 0 |
| 0.001029227 | 0 | 0 | 0 | 0 |
| 0 | 0 | 0 | 0 | 0 |
| 0 | 0 | 0 | 0 | 0 |
| 0 | 0 | 0 | 0 | 0 |
| 0.196106952 | 0.012150862 | 0.030104496 | 0 | 0.076273297 |
| 0 | 0 | 0 | 0 | 0 |
| 0 | 0 | 0 | 0 | 0 |
| 0 | 0 | 0 | 0 | 0 |
| 0 | 0 | 0 | 0 | 0 |
| 0 | 0 | 0 | 0 | 0 |
| 0 | 0 | 0 | 0 | 0 |
| 0 | 0 | 0 | 0 | 0 |
| 0.002463328 | 0 | 0 | 0 | 0 |
| 0.001736706 | 0 | 0 | 0 | 0 |
| 0 | 0 | 0 | 0 | 0.001573486 |

| | | | | |
|---|---|---|---|---|
| 0 | 0 | 0.164331612 | 0 | 0 |
| 0 | 0 | 0 | 0 | 0 |
| 0.008949142 | 0 | 0 | 0 | 0 |
| 0 | 0 | 0 | 0 | 0 |
| 0.001075069 | 0.007134109 | 0.008692715 | 0.005756285 | 0.000466967 |
| 0.003067744 | 0 | 0 | 0 | 0 |
| 0 | 0 | 0 | 0 | 0 |
| 0.002448786 | 0.014642893 | 0.038585052 | 0.04639503 | 0.006981948 |
| 0 | 0 | 0 | 0 | 0 |
| 0.002135707 | 0.003681158 | 0.00149513 | 0 | 0.039130668 |
| 0 | 0 | 0 | 0 | 0 |
| 0.08469613 | 0 | 0.000918618 | 0.006387212 | 0 |
| 0.002488363 | 0.002144503 | 0.008535858 | 0.004118188 | 0.053452796 |
| 0 | 0 | 0 | 0 | 0 |
| 0 | 0 | 0 | 0 | 0 |
| 0 | 0 | 0 | 0 | 0 |
| 0 | 0 | 0 | 0 | 0 |
| 0 | 0 | 0 | 0 | 0 |
| 0 | 0 | 0 | 0 | 0 |
| 0 | 0 | 0 | 0 | 0 |
| 0 | 0 | 0 | 0.01145024 | 0 |
| 0 | 0 | 0 | 0 | 0 |
| 0 | 0 | 0 | 0 | 0 |
| 0 | 0.032789091 | 0.242321209 | 0 | 0 |
| 0 | 0 | 0 | 0 | 0 |
| 0.001053931 | 0.004995599 | 0.104493484 | 0.002539397 | 0.016022506 |
| 0.002113978 | 0.179361444 | 0 | 0 | 0 |
| 0.012107703 | 0 | 0 | 0.168213641 | 0 |
| 0 | 0 | 0 | 0 | 0 |
| 0 | 0 | 0 | 0 | 0 |
| 0 | 0 | 0 | 0 | 0.006058493 |
| 0 | 0 | 0 | 0 | 0 |
| 0 | 0 | 0 | 0 | 0 |
| 0 | 0 | 0 | 0.116975292 | 0 |
| 0 | 0 | 0 | 0 | 0 |
| 0 | 0 | 0 | 0 | 0 |
| 0 | 0 | 0 | 0 | 0 |
| 0 | 0 | 0 | 0 | 0 |
| 0 | 0 | 0 | 0 | 0 |
| 0 | 0 | 0 | 0 | 0 |
| 0 | 0 | 0 | 0 | 0 |
| 0 | 0 | 0 | 0 | 0 |
| 0 | 0 | 0 | 0 | 0 |
| 0 | 0 | 0 | 0 | 0 |
| 0 | 0 | 0 | 0 | 0 |
| 0 | 0 | 0.000593799 | 0 | 0 |
| 0 | 0 | 0.002018577 | 0.008070294 | 0 |
| 0 | 0 | 0 | 0 | 0 |

| | | | | |
|---|---|---|---|---|
| 0 | 0 | 0 | 0 | 0 |
| 0 | 0 | 0 | 0.001176407 | 0 |
| 0 | 0 | 0 | 0 | 0.001207242 |
| 0 | 0 | 0 | 0 | 0.005310037 |
| 0 | 0 | 0 | 0 | 0 |
| 0 | 0.00146791 | 0.001192406 | 0 | 0 |
| 0 | 0 | 0 | 0 | 0 |
| 0 | 0 | 0 | 0.001072321 | 0 |
| 0 | 0 | 0 | 0 | 0 |
| 0 | 0 | 0 | 0 | 0 |
| 0.021251816 | 0.012485212 | 0.003918471 | 0.004166939 | 0.021668899 |
| 0 | 0 | 0 | 0 | 0 |
| 0 | 0 | 0 | 0 | 0 |
| 0.001413919 | 0.001072309 | 0.001959869 | 0.002422594 | 0.002947923 |
| 0 | 0 | 0 | 0 | 0.077220552 |
| 0 | 0 | 0 | 0 | 0 |
| 0 | 0 | 0 | 0.004653356 | 0 |
| 0 | 0 | 0 | 0 | 0 |
| 0 | 0 | 0 | 0 | 0 |
| 0 | 0 | 0 | 0 | 0 |
| 0 | 0 | 0 | 0 | 0 |
| 0.087922371 | 0.492521965 | 0.125564491 | 0 | 0 |
| 0 | 0 | 0 | 0 | 0 |
| 0 | 0 | 0 | 0 | 0 |
| 0 | 0 | 0 | 0 | 0 |
| 0 | 0 | 0 | 0 | 0 |
| 0 | 0 | 0 | 0 | 0 |
| 0 | 0 | 0 | 0 | 0 |
| 0 | 0 | 0 | 0 | 0 |
| 0.160412186 | 0.108059917 | 0.243865138 | 1.395548935 | 0.508202012 |
| 0.005947236 | 0 | 0 | 0.000821769 | 0 |
| 0.000808039 | 0.013532945 | 0 | 0 | 0 |
| 0 | 0 | 0 | 0 | 0 |
| 0 | 0 | 0 | 0 | 0 |
| 0 | 0 | 0 | 0 | 0 |
| 0 | 0 | 0 | 0 | 0 |
| 0 | 0.893922771 | 0.038734051 | 0.063274048 | 0 |
| 0.003458784 | 0.019673436 | 0 | 0 | 0 |
| 0 | 0 | 0 | 0.030699422 | 0 |
| 0 | 0 | 0 | 0 | 0.313351886 |
| 0 | 0 | 0 | 0.030372445 | 0 |
| 0 | 2.588596732 | 1.413898062 | 0 | 4.801239795 |
| 0.003432509 | 0.005506765 | 0 | 0 | 0 |
| 0 | 0 | 0 | 0 | 0 |
| 0 | 0 | 0 | 0 | 0 |
| 0 | 0 | 0 | 0 | 0 |
| 0 | 0.001156569 | 0.001127398 | 0 | 0.001695767 |
| 0 | 0 | 0 | 0 | 0 |

| | | | | |
|---|---|---|---|---|
| 0 | 0.00599345 | 0.034311816 | 0.004513529 | 0.024933152 |
| 0 | 0 | 0 | 0 | 0 |
| 0.000820433 | 0.003499943 | 0 | 0.106746921 | 0.0567806 |
| 0 | 0 | 0 | 0 | 0 |
| 0 | 0.16028082 | 0 | 0 | 0 |
| 0 | 0 | 0 | 0 | 0 |
| 0 | 0 | 0 | 0 | 0 |
| 0 | 0 | 0 | 0 | 0 |
| 0 | 0 | 0 | 0 | 0 |
| 0 | 0 | 0 | 0 | 0 |
| 0 | 0 | 0 | 0 | 0 |
| 0 | 0 | 0 | 0 | 0 |
| 0 | 0 | 0 | 0 | 0 |
| 0.065638935 | 0.002323767 | 0 | 0 | 0 |
| 0 | 0 | 0 | 0 | 0 |
| 0 | 0.007263934 | 0.689059224 | 0.395001348 | 0 |
| 0.114891443 | 0.072909121 | 0.168410213 | 0.089180431 | 0.527120703 |
| 0 | 0 | 0 | 0 | 0 |
| 0 | 0 | 0 | 0 | 0 |
| 0 | 0 | 0 | 0 | 0 |
| 0 | 0 | 0 | 0 | 0 |
| 0 | 0 | 0 | 0.008791331 | 0 |
| 0 | 0 | 0 | 0.233668458 | 0 |
| 0 | 0 | 0 | 1.246306507 | 0 |
| 0 | 0 | 0 | 0 | 0 |
| 0 | 0 | 0 | 0.006656062 | 0 |
| 0 | 0 | 0 | 0.002245062 | 0 |
| 0 | 0 | 0 | 0 | 0 |
| 0 | 0 | 0 | 0 | 0 |
| 0 | 0 | 0 | 0.00164658 | 0 |
| 0 | 0 | 0 | 0.004390363 | 0.000527644 |
| 0.009168482 | 0 | 0.061189867 | 0.009818239 | 0.006686544 |
| 0.023279026 | 0.011953703 | 0.036848854 | 2.428478183 | 0.100553159 |
| 0.009266976 | 0.012738303 | 0 | 0.003473299 | 0 |
| 0.001322853 | 0 | 0 | 0 | 0.926438714 |
| 0 | 0 | 0 | 0 | 0 |
| 0 | 0 | 0 | 0 | 0 |
| 0 | 0 | 0.04013573 | 0.068787386 | 0 |
| 0.212609879 | 0.018674227 | 0.149339668 | 0.014366214 | 0 |
| 0 | 0 | 0 | 0 | 0 |
| 0 | 0 | 0 | 0 | 0 |
| 0 | 0 | 0 | 0.016376956 | 0 |
| 0 | 0 | 0 | 0 | 0 |
| 0 | 0.184729397 | 0 | 0 | 0 |
| 0 | 0 | 0 | 0 | 0 |
| 0 | 0 | 0 | 0 | 0 |
| 0 | 0 | 0 | 0 | 0 |
| 0 | 0 | 0 | 0 | 0 |

| | | | | |
|---|---|---|---|---|
| 0 | 0 | 0 | 0 | 0 |
| 0.933689521 | 0.419607182 | 2.339917441 | 0.098735003 | 0.467051178 |
| 0 | 0 | 0 | 0 | 0 |
| 0 | 0 | 0 | 0 | 0 |
| 0 | 0 | 0 | 0 | 0 |
| 0 | 0 | 0 | 0 | 0 |
| 0 | 0 | 0 | 0 | 0 |
| 0 | 0 | 0.003500642 | 0 | 0 |
| 0 | 0.005135961 | 0 | 0 | 0 |
| 0 | 0 | 0 | 0 | 0 |
| 0 | 0 | 0.000891243 | 0.00371812 | 0 |
| 0.022410841 | 0.001634257 | 0.03690537 | 0.032860255 | 0.01337854 |
| 0 | 0.757164344 | 0.177536691 | 0 | 0 |
| 0 | 0 | 0 | 0 | 0 |
| 0 | 0 | 0.000613554 | 0.001422026 | 0 |
| 0 | 0 | 0 | 0 | 0 |
| 0.022056873 | 0 | 0.01530546 | 0.012458907 | 0.01473944 |
| 0 | 0 | 0 | 0.016847273 | 0 |
| 0 | 0 | 0 | 0 | 0 |
| 0 | 0 | 0 | 0.003938306 | 0 |
| 0.029745032 | 0.014928411 | 0.319333026 | 0.011777586 | 0.005645744 |
| 0 | 0 | 0 | 0 | 0 |
| 0 | 0 | 0 | 0 | 0 |
| 0 | 0 | 0 | 0 | 0 |
| 0 | 0 | 0 | 0 | 0 |
| 0 | 0 | 0 | 0 | 0 |
| 0 | 0 | 0 | 0.122327402 | 0 |
| 0.008899902 | 0.019140777 | 0.02045834 | 0.02361319 | 0.064929493 |
| 0.001424518 | 0 | 0 | 0 | 0 |
| 0 | 0 | 0 | 0 | 0 |
| 0.007126569 | 0 | 0.005268436 | 0 | 0.004787708 |
| 0 | 0 | 0 | 0 | 0 |
| 0 | 0 | 0 | 0.001036129 | 0 |
| 0 | 0 | 0 | 0 | 0 |
| 0 | 0 | 0.037795401 | 0 | 0 |
| 0.081382752 | 0.068625452 | 0.092101224 | 0.577310053 | 0.144783049 |
| 0 | 0 | 0 | 0 | 0 |
| 0 | 0 | 0 | 0 | 0 |
| 0.053911928 | 0 | 0 | 0.004211496 | 0.001328636 |
| 0 | 0 | 0 | 0 | 0 |
| 0 | 0 | 0 | 0 | 0 |
| 0 | 0 | 0 | 0 | 0 |
| 0 | 0 | 0 | 0.040503673 | 0.000807034 |
| 0 | 0 | 0 | 0 | 0 |
| 0 | 0 | 0 | 0 | 0 |
| 0 | 0.271272012 | 0.579721246 | 0 | 0 |
| 0 | 0 | 0 | 0 | 0 |
| 0 | 0 | 0 | 0 | 0.003778434 |

| | | | | |
|---|---|---|---|---|
| 0 | 0 | 0 | 0 | 0 |
| 0 | 0 | 0.003182863 | 0 | 0 |
| 0 | 0.000691788 | 0 | 0 | 0 |
| 0.345460889 | 0.032309891 | 0.007320259 | 0.043697986 | 0 |
| 0 | 0 | 0 | 0 | 0 |
| 0 | 0 | 0 | 0 | 0 |
| 0 | 0 | 0 | 0 | 0 |
| 0 | 0 | 0 | 0 | 0 |
| 15.36874071 | 1.370253155 | 0.722063681 | 0.989632377 | 0.591620212 |
| 0 | 0.133188242 | 0.104041885 | 0 | 0 |
| 0 | 0 | 0 | 0 | 0 |
| 0 | 0.002527049 | 0.002908076 | 0 | 0 |
| 0 | 0 | 0 | 0 | 0 |
| 0 | 0 | 0 | 0 | 0 |
| 0 | 0.0076462 | 0.008046226 | 0.130736779 | 0 |
| 0 | 0 | 0 | 0 | 0 |
| 0.154108022 | 0.141219905 | 0.067208648 | 0.06631904 | 0 |
| 0 | 0.13207148 | 0.025622723 | 2.684394197 | 0 |
| 0 | 0 | 0 | 0 | 0 |
| 0 | 0 | 0 | 0 | 0 |
| 0 | 0 | 0 | 0 | 0 |
| 0 | 0.00117309 | 0 | 0 | 0 |
| 0 | 0 | 0 | 0 | 0 |
| 0 | 0 | 0.140326256 | 0.066370109 | 0 |
| 0 | 0 | 0 | 0 | 0 |
| 0 | 0.140612835 | 0.395154964 | 0 | 0 |
| 0 | 0.113543632 | 0.00171508 | 0 | 0 |
| 2.526496148 | 2.239835386 | 6.098128745 | 4.501526313 | 0 |
| 0 | 0 | 0 | 0 | 0 |
| 0 | 0 | 0 | 0 | 0 |
| 0.436991014 | 0.00982689 | 0.035422472 | 0 | 0 |
| 0 | 0.002314688 | 0 | 0 | 0 |
| 0 | 0 | 0 | 0.011088493 | 0 |
| 0 | 0 | 0 | 0 | 0 |
| 0 | 0 | 0 | 0 | 0 |
| 0 | 0.04964545 | 0.031162344 | 0 | 0 |
| 0.716077421 | 0.580194922 | 0.841166638 | 0.003545425 | 0 |
| 0 | 0 | 0 | 0 | 0 |
| 0 | 0 | 0 | 0.212224041 | 0 |
| 0 | 0 | 0 | 0 | 0 |
| 0.001917933 | 0.023434433 | 0 | 0 | 0.002221532 |
| 0 | 0 | 0 | 0 | 0 |
| 0.077715777 | 0.02544664 | 0.378568839 | 1.531693398 | 8.856535908 |
| 0 | 0 | 0.000714783 | 0.028030412 | 0 |
| 0 | 0 | 0 | 0 | 0 |
| 0 | 0 | 0 | 0 | 0 |
| 0 | 0 | 0 | 0 | 0 |
| 0.001665138 | 0 | 0 | 0.019203085 | 0 |

| | | | | |
|---|---|---|---|---|
| 0 | 0 | 0 | 0 | 0 |
| 0 | 0.017699632 | 0.236512756 | 0 | 0 |
| 0 | 0 | 0.007375215 | 0.003681665 | 0.164267335 |
| 0 | 0.019628482 | 0 | 0 | 0 |
| 0 | 0 | 0 | 0.005783662 | 0 |
| 0.134933266 | 0.042180158 | 0 | 0.063391698 | 0 |
| 0 | 0 | 0 | 0 | 0 |
| 0 | 0 | 0 | 0 | 0 |
| 0 | 0 | 0 | 0 | 0 |
| 0 | 0 | 0 | 0 | 0 |
| 0 | 0.008914219 | 0.000557012 | 0.120577471 | 0.002373839 |
| 0.788753478 | 0.628957713 | 0.859851443 | 0.501229257 | 3.019672291 |
| 0 | 0 | 0 | 0 | 0 |
| 0.008456971 | 0.138141784 | 0.608363608 | 0.052665891 | 0.195913324 |
| 0 | 0.001943947 | 0 | 0.144051844 | 0.049285148 |
| 0 | 0.545436219 | 2.465956886 | 0 | 0 |
| 0 | 0 | 0 | 0 | 0 |
| 0 | 0 | 0 | 0 | 0 |
| 0 | 0.007567373 | 0.015284656 | 0 | 0 |
| 0 | 0.008660939 | 0 | 0 | 0 |
| 2.697628948 | 0 | 0.001400574 | 0 | 0.016268122 |
| 0 | 0 | 0 | 0.006840749 | 50.46275929 |
| 0.184956826 | 0.179861625 | 0.018520261 | 0.228520165 | 1.462277847 |
| 0 | 0 | 0 | 0 | 0 |
| 0 | 0 | 0 | 0 | 0 |
| 0 | 0.025022521 | 5.65123387 | 0.006742307 | 0 |
| 0 | 0 | 0 | 0 | 0 |
| 0 | 0 | 0 | 0 | 0 |
| 0 | 0 | 0 | 0 | 0 |
| 0 | 0 | 0 | 0 | 0 |
| 0 | 0.006972978 | 0.002574661 | 0 | 0 |
| 0 | 0.155540945 | 0.140892967 | 0.086420594 | 0 |
| 0.745846708 | 0 | 0 | 0 | 0 |
| 0 | 0.004117294 | 0.002472051 | 0 | 0 |
| 0 | 0 | 0 | 0.041304261 | 0 |
| 0.06321098 | 0.106223403 | 0.020058668 | 0.149328211 | 0 |
| 0 | 0 | 0 | 0.005342556 | 0 |
| 0 | 0 | 0 | 0 | 0 |
| 0 | 0.113342262 | 0.02812676 | 0.08807556 | 0 |
| 0 | 0 | 0 | 0 | 0 |
| 0 | 0.0037489 | 0.018728517 | 0.005293519 | 0 |
| 0 | 0 | 0 | 0 | 0 |
| 0 | 0.243887912 | 0 | 0 | 0 |
| 0 | 0 | 0 | 0 | 0 |
| 0 | 0.032043922 | 0.013803658 | 0.037842639 | 0 |
| 0 | 0.00656712 | 0.097089168 | 0 | 0 |
| 0 | 0 | 0 | 0 | 0 |
| 7.470797433 | 0.048024326 | 0 | 0.074096219 | 7.642086174 |

| | | | | |
|---|---|---|---|---|
| 0 | 0 | 0 | 0 | 0 |
| 0 | 0 | 0 | 0 | 0 |
| 0 | 0 | 0 | 0 | 0 |
| 0 | 0 | 0.001512617 | 0.003505775 | 0 |
| 0 | 0 | 0 | 0 | 0 |
| 0 | 0.00475107 | 0 | 0 | 0 |
| 0 | 0 | 0.912839424 | 0 | 0 |
| 0 | 0.006853902 | 0 | 0.003441011 | 0 |
| 0 | 0 | 0 | 0 | 0 |
| 0 | 0 | 0 | 0 | 0 |
| 0 | 0.002304327 | 0 | 0.004164805 | 0.000703878 |
| 0 | 0 | 0 | 0 | 0 |
| 0 | 0.255632252 | 0.014914368 | 0 | 0 |
| 0 | 0.001573079 | 0 | 0.996438065 | 0 |
| 0 | 0 | 0 | 0 | 0 |
| 0 | 0 | 0 | 0 | 0 |
| 0 | 0.003599006 | 0.055129357 | 0 | 0 |
| 0.037233059 | 0.002493789 | 0 | 0.102496296 | 0.002929814 |
| 0 | 0 | 0.06214487 | 0 | 0 |
| 0 | 0 | 0.009134421 | 0 | 0 |
| 0 | 0.003749368 | 0.003184107 | 0 | 0 |
| 0 | 0 | 0 | 0 | 0 |
| 0 | 0.001548164 | 0.000898283 | 0.697283233 | 0 |
| 0 | 0 | 0 | 0 | 0 |
| 0 | 0 | 0 | 0 | 0 |
| 0 | 0 | 0 | 0 | 0 |
| 0 | 0 | 0 | 0 | 0 |
| 0 | 0 | 0 | 0 | 0 |
| 0 | 0 | 0 | 0 | 0 |
| 0.073210175 | 0.110071217 | 0.034070865 | 0.112035208 | 0 |
| 0 | 0 | 0 | 0 | 0 |
| 0 | 0 | 0 | 0 | 0 |
| 0 | 0.004810311 | 0.022793678 | 0 | 0 |
| 0 | 0 | 0 | 0 | 0 |
| 0.208936357 | 0 | 0.001934914 | 0 | 0 |
| 0 | 0 | 0 | 0 | 0 |
| 0 | 0.020132016 | 0.086727365 | 0 | 0 |
| 0 | 0 | 0 | 0.016966855 | 0.000476676 |
| 0 | 0 | 0 | 0 | 0 |
| 0 | 0 | 0 | 0 | 0 |
| 0 | 0 | 0 | 0 | 0 |
| 0 | 0 | 0 | 0 | 0 |
| 0 | 0 | 0 | 0 | 0 |
| 0 | 0.695971423 | 0 | 0.075964591 | 0 |
| 0 | 0.053158713 | 0.099317716 | 0 | 0 |
| 0 | 0 | 0 | 0 | 0 |
| 0 | 0.002409481 | 0.025143231 | 0 | 0 |
| 0 | 0 | 0 | 0 | 0 |

| | | | | |
|---|---|---|---|---|
| 0 | 0 | 0 | 0 | 0 |
| 0 | 0 | 0 | 0 | 0 |
| 0 | 0.02302395 | 0 | 0 | 0 |
| 0 | 0.353333744 | 0.115847571 | 0.023671704 | 0 |
| 0 | 0 | 0 | 0 | 0 |
| 0 | 0.531973592 | 0 | 0 | 0 |
| 0 | 0 | 0 | 0 | 0 |
| 0 | 0 | 0 | 0 | 0 |
| 0 | 0 | 0.000874141 | 0.041330095 | 0.182597381 |
| 0.002628116 | 0 | 0 | 0 | 0 |
| 0 | 0.006540347 | 0.00281267 | 0 | 0 |
| 0 | 0 | 0 | 0 | 0 |
| 0 | 0 | 0 | 0 | 0 |
| 0 | 0 | 0 | 0 | 0 |
| 0.228918489 | 2.183279773 | 0.884478203 | 1.15200171 | 0 |
| 0 | 0 | 0 | 0 | 0.000760034 |
| 0 | 0 | 0 | 0 | 0 |
| 0 | 0 | 0 | 0 | 0 |
| 0 | 0 | 0 | 0 | 0 |
| 0 | 0 | 0 | 0 | 0 |
| 0 | 0.028182299 | 0.061827544 | 0 | 0 |
| 0 | 0 | 0 | 0 | 0 |
| 0 | 0 | 0 | 0 | 0 |
| 0 | 0 | 0 | 0 | 0 |
| 0.056065725 | 10.584436 | 0.004882831 | 0 | 0.081755628 |
| 0.03381405 | 1.905737529 | 14.3515801 | 0.35267246 | 0 |
| 0 | 0.036477846 | 0.97975115 | 0 | 0 |
| 0 | 0 | 0 | 0 | 0 |
| 0 | 0 | 0.230300828 | 0.264018594 | 0 |
| 0 | 0 | 0 | 0 | 0 |
| 0 | 0 | 0 | 0 | 0 |
| 0 | 0 | 0.060077846 | 0 | 0 |
| 0 | 0 | 0 | 0.001730593 | 0 |
| 0 | 0.000506884 | 0 | 0.125395843 | 0 |
| 0.014242864 | 0.104676448 | 0.568752799 | 0.003144791 | 0.723506111 |
| 0 | 0 | 0.000959139 | 0 | 0 |
| 0 | 0 | 0 | 0.004129308 | 0 |
| 1.220178597 | 0.101772858 | 2.211014317 | 0.236705737 | 2.190106401 |
| 0 | 0.075804247 | 0.415290901 | 0 | 0 |
| 0 | 0 | 0 | 0 | 0.004248421 |
| 0 | 0.36411231 | 0 | 0 | 0 |
| 0 | 0 | 0 | 0 | 0 |
| 10.43088759 | 0.445867578 | 0.055267054 | 0.372731868 | 1.815214393 |
| 0 | 1.014584228 | 0 | 0 | 0 |
| 0 | 0 | 0 | 0 | 0 |
| 0 | 0 | 0 | 0.045910798 | 0 |
| 0 | 0 | 0 | 0 | 0 |
| 13.23739451 | 0.019602892 | 0.02581573 | 0 | 0.019596727 |

| | | | | |
|---|---|---|---|---|
| 0 | 0 | 0 | 0.141608072 | 0 |
| 0 | 0 | 0 | 0 | 0 |
| 0 | 0 | 0 | 0 | 0 |
| 0 | 0 | 0 | 0 | 0 |
| 0 | 0 | 0 | 0.001743658 | 0 |
| 0 | 0 | 0.003277407 | 0 | 0 |
| 0 | 0 | 0 | 0 | 0 |
| 0 | 0 | 0 | 0 | 0 |
| 0 | 0 | 0 | 0 | 0 |
| 0 | 0 | 0 | 0 | 0 |
| 0 | 0 | 0 | 0 | 0 |
| 0 | 0.423758547 | 0.20111488 | 0.74387934 | 0 |
| 0.446836436 | 0.002403015 | 0 | 0.007238622 | 0 |
| 0 | 0 | 0 | 0 | 0 |
| 0.094826995 | 0.043353816 | 2.594453506 | 0.33647426 | 0.016826188 |
| 0 | 0 | 0 | 0 | 0 |
| 0 | 0 | 0 | 0.006813578 | 0 |
| 0 | 0 | 0 | 0 | 0 |
| 0 | 0.002682315 | 0 | 0 | 0 |
| 0 | 0 | 0 | 0 | 0 |
| 0 | 0.025353117 | 0.100869006 | 0 | 0 |
| 0 | 0 | 0 | 0 | 0 |
| 0 | 0.128235862 | 1.134547363 | 0.531061234 | 0 |
| 0 | 0 | 0 | 0 | 0 |
| 0 | 0 | 0 | 0 | 0 |
| 0 | 0 | 0 | 0 | 0 |
| 0 | 0 | 0 | 0 | 0 |
| 0 | 0 | 0 | 0 | 0 |
| 0 | 0 | 0.005597923 | 0 | 0 |
| 0 | 0 | 0 | 0 | 0 |
| 0 | 0 | 0 | 0 | 0 |
| 0.056202148 | 0.006287618 | 0.127150522 | 0.017943387 | 0.041040458 |
| 0 | 0 | 0.348892871 | 0 | 0 |
| 0 | 0.005125093 | 0.002820225 | 0.005976138 | 0 |
| 0 | 0 | 0 | 0 | 0 |
| 0 | 0.0196014 | 0.465789475 | 0.342110161 | 0.007758364 |
| 0 | 0 | 0 | 0 | 0 |
| 0 | 0.018225867 | 0.011977195 | 0 | 0 |
| 0 | 0 | 0 | 0 | 0 |
| 0 | 0 | 0 | 0 | 0 |
| 0 | 0.101632272 | 0.085669987 | 0 | 0 |
| 0 | 0 | 0 | 0 | 0 |
| 0 | 0 | 0 | 0 | 0 |
| 0 | 0 | 0 | 0 | 0 |
| 0 | 0.010548371 | 0 | 0.030503942 | 0.003350984 |
| 0 | 0 | 0 | 0 | 0 |
| 0 | 0.238919344 | 0.067624415 | 0.405833 | 0 |
| 0 | 0 | 0 | 0.002704807 | 0 |

| | | | | |
|---|---|---|---|---|
| 0 | 0 | 0 | 0 | 0 |
| 0 | 0.037514177 | 0.037162592 | 0 | 0 |
| 0 | 0 | 0 | 0 | 0 |
| 0 | 0 | 0 | 0 | 0 |
| 0 | 0 | 0 | 0 | 0 |
| 0 | 0 | 0 | 0 | 0 |
| 0.004687135 | 0.038669373 | 0.881088669 | 0 | 0 |
| 0 | 0.000618189 | 0.002678209 | 0.001396632 | 0 |
| 0 | 0 | 0 | 0 | 0 |
| 0 | 0 | 0 | 0 | 0.000495514 |
| 0 | 0 | 0 | 0 | 0 |
| 0 | 0 | 0 | 0 | 0 |
| 0 | 0 | 0 | 0 | 0 |
| 0 | 0.589233386 | 0.22614068 | 0 | 0 |
| 0 | 0 | 0 | 0 | 0 |
| 0 | 0.006300445 | 0 | 0 | 0 |
| 0 | 0.012054029 | 0 | 0 | 0 |
| 0 | 0 | 0 | 0 | 0 |
| 0 | 0.993437204 | 0 | 0 | 0 |
| 0 | 0 | 0 | 0 | 0 |
| 0.002614046 | 0 | 0 | 0 | 0 |
| 0 | 0 | 0 | 0 | 0 |
| 0 | 0 | 0 | 0 | 0 |
| 0 | 0 | 0 | 0 | 0 |
| 0.004376615 | 0 | 0.001310671 | 0 | 0.007604116 |
| 0.010876802 | 0 | 0 | 0.135556159 | 0.053993757 |
| 0 | 0 | 0 | 0 | 0 |
| 0.091760324 | 0 | 0.000780794 | 0 | 0.025015264 |
| 0 | 0.207733247 | 0 | 0 | 0 |
| 1.95639332 | 0.098225185 | 0.408977135 | 0.203117418 | 0 |
| 0 | 0 | 0 | 0 | 0 |
| 0 | 0 | 0 | 0 | 0 |
| 0 | 0.001420916 | 0.012201871 | 0 | 0 |
| 0 | 0 | 0 | 0 | 0 |
| 0 | 0.005145447 | 0 | 0.003719921 | 0 |
| 0 | 0 | 0 | 0 | 0 |
| 0 | 0.115569822 | 0 | 0 | 0 |
| 0 | 0.001401258 | 0 | 0.001055255 | 0 |
| 0 | 0 | 0 | 0 | 0 |
| 0 | 0 | 0 | 0 | 0 |
| 0.440838913 | 0.974528494 | 2.965101446 | 21.21020196 | 2.310558784 |
| 0 | 0.011624909 | 0 | 0 | 0 |
| 0 | 0 | 0 | 0 | 0 |
| 0 | 0 | 0 | 0 | 0 |
| 0 | 0 | 0 | 0 | 0 |
| 0 | 0 | 0 | 0 | 0 |
| 0 | 0 | 0 | 0 | 0 |
| 0 | 0.002594937 | 0.001204518 | 0.10866677 | 0 |

| | | | | |
|---|---|---|---|---|
| 0 | 0 | 0 | 0.113653482 | 0 |
| 0 | 0.007244719 | 0.002477892 | 0 | 0 |
| 0 | 0 | 0 | 0 | 0 |
| 0 | 0 | 0 | 0 | 0 |
| 7.191394857 | 1.303814767 | 0.405322038 | 4.333567173 | 1.223988315 |
| 0 | 0 | 0 | 0 | 0 |
| 0 | 0 | 0 | 0 | 0 |
| 0 | 0 | 0 | 0 | 0 |
| 0 | 0 | 0.001855679 | 0.013762831 | 0 |
| 0 | 0 | 0 | 0 | 0 |
| 0 | 0 | 0 | 0 | 0 |
| 0 | 0 | 0 | 0 | 0 |
| 0 | 0.0513141 | 0.033707721 | 0.09186542 | 0 |
| 0.001201196 | 0.004554907 | 0.005365028 | 0.083353753 | 0.00528709 |
| 0 | 0.04631792 | 0.155177671 | 0 | 0 |
| 0 | 0.027846044 | 0.071806342 | 0.051510725 | 0.015184918 |
| 0 | 0.000985391 | 0 | 0 | 0 |
| 0 | 0 | 0 | 0 | 0 |
| 0 | 0 | 0 | 0 | 0 |
| 0 | 0 | 0 | 0 | 0 |
| 0 | 0 | 0 | 0 | 0 |
| 0 | 0 | 0 | 0 | 0 |
| 0 | 0 | 0 | 0 | 0 |
| 0 | 0 | 0 | 0 | 0 |
| 0 | 0 | 0 | 0 | 0 |
| 0 | 0 | 0 | 0 | 0 |
| 0 | 0.165027071 | 0.120829521 | 0 | 0 |
| 0 | 0 | 0 | 0.004368658 | 0.001771994 |
| 0 | 0 | 0 | 0 | 0 |
| 0 | 0 | 0 | 0 | 0 |
| 0 | 0 | 0 | 0 | 0 |
| 0 | 0 | 0 | 0 | 0 |
| 0 | 0 | 0 | 0 | 0 |
| 0 | 0.023310798 | 0.041823218 | 0 | 0 |
| 0 | 0.045071712 | 0.005647661 | 0 | 0 |
| 0 | 0 | 0 | 0 | 0 |
| 0.001326484 | 0 | 0 | 0 | 0 |
| 0 | 0 | 0.080814741 | 0 | 0 |
| 0 | 0 | 0 | 0 | 0 |
| 0 | 0 | 0 | 0 | 0 |
| 0 | 0.051504547 | 0.555242497 | 2.666082604 | 0 |
| 0 | 0 | 0.087574626 | 0 | 0 |
| 0 | 0 | 0 | 0 | 0 |
| 0 | 0 | 0 | 0 | 0 |
| 0 | 0 | 0 | 0 | 0 |
| 0 | 0 | 0 | 0 | 0 |
| 0 | 0 | 0 | 0 | 0 |
| 0 | 0 | 0 | 0 | 0 |

| | | | | |
|---|---|---|---|---|
| 0.321380591 | 0.08659398 | 0.419813692 | 0.002391635 | 0 |
| 0 | 0 | 0 | 0 | 0 |
| 0 | 0 | 0 | 0 | 0 |
| 0 | 0 | 0 | 0 | 0 |
| 0 | 0.052396865 | 0.035412212 | 0 | 0 |
| 0 | 0 | 0.007024547 | 0 | 0 |
| 0.041787943 | 0 | 0 | 0 | 0 |
| 0 | 0 | 0 | 0 | 0 |
| 0 | 0 | 0 | 0 | 0 |
| 0 | 0 | 0 | 0 | 0 |
| 0 | 0.000715226 | 0.006197211 | 0 | 0 |
| 0 | 0 | 0.012010845 | 0 | 0 |
| 0 | 0 | 0 | 0 | 0 |
| 0 | 0 | 0 | 0 | 0 |
| 0 | 0 | 0.00141227 | 0 | 0 |
| 0 | 0 | 0 | 0 | 0 |
| 0 | 0 | 0.029840992 | 0 | 0 |
| 0 | 0 | 0 | 0 | 0 |
| 0 | 0 | 0 | 0 | 0 |
| 0 | 0 | 0 | 0 | 0 |
| 0.000888486 | 0.064761955 | 0 | 0.002325772 | 0.027443448 |
| 0 | 0 | 0 | 0 | 0 |
| 0 | 0.002683869 | 0.018782805 | 0.022155022 | 0 |
| 0 | 0 | 0 | 0.025299557 | 0 |
| 0 | 0 | 0 | 0 | 0 |
| 0 | 0.00129949 | 0 | 0 | 0 |
| 0 | 0 | 0 | 0 | 0 |
| 0 | 0 | 0 | 0 | 0 |
| 0 | 0 | 0 | 0.198725957 | 0 |
| 0.005156087 | 0 | 0.01523831 | 0.754504274 | 0 |
| 0 | 0 | 0.001122755 | 0 | 0 |
| 0 | 0 | 0 | 0 | 0 |
| 0 | 0.00898425 | 0 | 0 | 0 |
| 0 | 0 | 0.000637021 | 0 | 0 |
| 0 | 0 | 0 | 0 | 0 |
| 0 | 0 | 0 | 0 | 0 |
| 0.000726918 | 0 | 0 | 0 | 0 |
| 0 | 0 | 0 | 0 | 0 |
| 0 | 0 | 0 | 0 | 0 |
| 0 | 0 | 0.002261952 | 0 | 0 |
| 0 | 0 | 0 | 0 | 0 |
| 0 | 0 | 0 | 0 | 0 |
| 0 | 0 | 0 | 0 | 0 |
| 0 | 0.008548556 | 0.013176021 | 0 | 0 |
| 0 | 0 | 0 | 0 | 0 |
| 0 | 4.755039346 | 0 | 0 | 0 |
| 0 | 0 | 0 | 0 | 0 |
| 0 | 0.002189441 | 0.051043398 | 0 | 0 |

| | | | | |
|---|---|---|---|---|
| 0 | 0 | 0 | 0.051545608 | 0 |
| 0 | 0 | 0.049915674 | 0 | 0 |
| 0 | 0 | 0.007437055 | 0 | 0 |
| 0 | 0 | 0 | 0 | 0 |
| 0 | 0 | 0 | 0 | 0 |
| 0 | 0 | 0 | 0 | 0 |
| 0 | 0 | 0 | 0 | 0 |
| 0 | 0.378473599 | 0.187989685 | 0.827565332 | 0 |
| 1.322524954 | 14.688683 | 0.008311709 | 0.005604057 | 0.544405269 |
| 0 | 0 | 0.001052066 | 0.032186355 | 0 |
| 0.00227121 | 0.023050759 | 0.284982394 | 0 | 0 |
| 0 | 0 | 0 | 0 | 0 |
| 0 | 0.023741215 | 0 | 0 | 0 |
| 0 | 0 | 0 | 0 | 0 |
| 0 | 0 | 0 | 0 | 0 |
| 0 | 0 | 0 | 0 | 0 |
| 0 | 0 | 0 | 0 | 0 |
| 0 | 0.225867807 | 0 | 0 | 0 |
| 0 | 0 | 0 | 0 | 0 |
| 0 | 0 | 0 | 0.003841699 | 0 |
| 0 | 0 | 0 | 0 | 0 |
| 0 | 0 | 0 | 0 | 0 |
| 0 | 0 | 0 | 0 | 0 |
| 0 | 0 | 0 | 0 | 0 |
| 0 | 0.026605548 | 0.019463514 | 0 | 0 |
| 0 | 0.030009131 | 0 | 0 | 0 |
| 0 | 0 | 0 | 0 | 0 |
| 0 | 0 | 0 | 0 | 0 |
| 0 | 0 | 0 | 0 | 0 |
| 0 | 0 | 0 | 0 | 0 |
| 0 | 0 | 0 | 0 | 0 |
| 0 | 0 | 0 | 0 | 0 |
| 0 | 0 | 0 | 0 | 0 |
| 0 | 0 | 0 | 0 | 0 |
| 0 | 0 | 0.001942624 | 0 | 0 |
| 0 | 0 | 0.333218709 | 0 | 0 |
| 0 | 0 | 0 | 0 | 0 |
| 0 | 0 | 0 | 0.028064095 | 0 |
| 0 | 0 | 0 | 0 | 0 |
| 0.361023081 | 0 | 0 | 0.029404948 | 0 |
| 0 | 0 | 0 | 0 | 0 |
| 0 | 0.337728669 | 0.052224146 | 0.449300807 | 0 |
| 0 | 0 | 0 | 0 | 0 |
| 0 | 0 | 0 | 0 | 0 |
| 0 | 0 | 0 | 0 | 0 |
| 0 | 0 | 0 | 0 | 0 |
| 0 | 0 | 0 | 0 | 0 |
| 0 | 0 | 0 | 0 | 0 |

| | | | | |
|---|---|---|---|---|
| 0 | 0 | 0 | 0 | 0 |
| 0.001880667 | 0 | 0 | 0 | 0 |
| 0 | 0 | 0 | 0 | 0 |
| 0 | 0 | 0 | 0 | 0 |
| 0 | 0.199415164 | 0.428660511 | 1.408453385 | 0.266952318 |
| 0 | 0 | 0 | 0 | 0 |
| 0 | 0 | 0 | 0 | 0 |
| 0 | 0 | 0.00061236 | 0.023559692 | 0.017960994 |
| 0 | 0 | 0.005910379 | 0.019839079 | 0 |
| 0 | 0.009690821 | 0 | 0 | 0 |
| 0 | 0 | 0 | 0 | 0 |
| 0 | 0 | 0.030345318 | 0 | 0 |
| 0 | 0 | 0 | 0 | 0 |
| 0 | 0 | 0 | 0 | 0 |
| 0 | 0 | 0 | 0 | 0 |
| 0 | 0.014383695 | 0 | 0.025110612 | 0 |
| 0 | 0 | 0 | 0 | 0 |
| 0 | 0 | 0 | 0 | 0 |
| 0 | 0 | 0 | 0 | 0 |
| 0 | 0 | 0 | 0 | 0 |
| 0 | 0 | 0 | 0 | 0 |
| 0 | 0 | 0 | 0 | 0 |
| 0 | 0 | 0 | 0 | 0 |
| 0 | 0 | 0 | 0 | 0 |
| 0 | 0.251139238 | 0.029076092 | 0.151680568 | 0 |
| 0 | 0 | 0 | 0 | 0 |
| 0 | 0 | 0.000646262 | 0.011458428 | 0.054921995 |
| 0 | 0 | 0 | 0 | 0 |
| 0 | 0 | 0 | 0 | 0 |
| 0 | 0 | 0 | 0 | 0 |
| 0 | 0 | 0 | 0 | 0 |
| 0 | 0 | 0 | 0 | 0 |
| 0 | 0 | 0 | 0 | 0 |
| 0 | 0 | 0 | 0 | 0 |
| 0 | 0 | 0 | 0 | 0 |
| 0 | 0 | 0 | 0 | 0 |
| 0 | 0 | 0 | 0 | 0 |
| 0 | 0.009790976 | 0.087486916 | 0 | 0 |
| 0 | 0 | 0.352937275 | 0 | 0 |
| 0 | 0 | 0 | 0 | 0 |
| 0 | 0 | 0 | 0 | 0 |
| 5.358663001 | 0.053794763 | 0.138694567 | 0 | 0 |
| 0 | 0 | 0 | 0 | 0 |
| 0 | 0 | 0 | 0 | 0 |
| 0 | 0 | 0 | 0 | 0 |
| 0 | 0 | 0 | 0 | 0 |
| 0 | 0 | 0 | 0 | 0 |
| 0 | 0 | 0 | 0 | 0 |

| | | | | |
|---|---|---|---|---|
| 0 | 0.027308393 | 0.020511691 | 0.060428235 | 0 |
| 0 | 0.003216217 | 0 | 0 | 0 |
| 0 | 0 | 0 | 0 | 0 |
| 0 | 0 | 0 | 0 | 0 |
| 0 | 0 | 0 | 0 | 0 |
| 0 | 0.003467217 | 0 | 0 | 0 |
| 0 | 0 | 0 | 0 | 0 |
| 0 | 0 | 0 | 0 | 0 |
| 0 | 0 | 0 | 0 | 0 |
| 0 | 0 | 0 | 0 | 0 |
| 0 | 0 | 0 | 0 | 0 |
| 0 | 0.0006014 | 0 | 0 | 0 |
| 0 | 0.088863811 | 0 | 0 | 0 |
| 0 | 0 | 0 | 0 | 0 |
| 0 | 0 | 0 | 0 | 0 |
| 0 | 0 | 0 | 0 | 0 |
| 0 | 0 | 0 | 0 | 0 |
| 0 | 0 | 0 | 0 | 0 |
| 0 | 0 | 0 | 0 | 0 |
| 0 | 0 | 0 | 0 | 0 |
| 0 | 0 | 0 | 0.211843859 | 0 |
| 0 | 0 | 0 | 0 | 0 |
| 0.003894699 | 0.008751766 | 0 | 0.154779707 | 0 |
| 0 | 0.003754721 | 0 | 0 | 0 |
| 0 | 0.007860867 | 0 | 0 | 0 |
| 0 | 0 | 0.174698707 | 0.413421198 | 0 |
| 0 | 0 | 0 | 0 | 0 |
| 0 | 0 | 0 | 0 | 0 |
| 0 | 0 | 0 | 0 | 0 |
| 0 | 0 | 0 | 0 | 0 |
| 0 | 0 | 0 | 0 | 0 |
| 0 | 0 | 0 | 0 | 0 |
| 0.067408797 | 0 | 0.001872486 | 0.023724443 | 0.023314249 |
| 0 | 0 | 0 | 0 | 0 |
| 0.42679052 | 0.005040252 | 0 | 0.042442802 | 0 |
| 0 | 0 | 0.003172137 | 0 | 0.01469073 |
| 0 | 0 | 0 | 0 | 0 |
| 0 | 0 | 0 | 0 | 0 |
| 0 | 0 | 0 | 0.102850955 | 0 |
| 0 | 0 | 0 | 0 | 0 |
| 0 | 0 | 0 | 0 | 0 |
| 0 | 0 | 0 | 0 | 0 |
| 1.693220902 | 0.281866026 | 0.087611462 | 0.124973648 | 0.057908557 |
| 0 | 0 | 0 | 0 | 0 |
| 0 | 0 | 0 | 0 | 0 |
| 0 | 0.001986839 | 0.041065777 | 0.023191758 | 0 |
| 0 | 0 | 0 | 0 | 0 |
| 0 | 0 | 0 | 0 | 0 |

| | | | | |
|---|---|---|---|---|
| 0 | 0 | 0 | 0 | 0 |
| 0 | 0 | 0 | 0 | 0 |
| 0 | 0 | 0 | 0 | 0 |
| 0 | 0 | 0 | 0 | 0 |
| 0 | 0 | 0.000940026 | 0 | 0 |
| 0 | 0 | 0 | 0 | 0 |
| 0 | 0 | 0 | 0 | 0 |
| 0 | 0 | 0 | 0.004578507 | 0 |
| 0 | 0.008893756 | 0 | 0 | 0 |
| 0 | 0 | 0.021056296 | 0.038270979 | 0 |
| 0 | 0 | 0 | 0 | 0 |
| 0 | 0 | 0 | 0 | 0 |
| 0 | 0 | 0 | 0 | 0 |
| 0 | 0 | 0.045434763 | 0 | 0 |
| 0 | 0 | 0 | 0 | 0 |
| 0 | 0 | 0 | 0 | 0 |
| 0 | 0 | 0 | 0 | 0 |
| 0 | 0 | 0 | 0 | 0 |
| 0 | 0 | 0 | 0 | 0 |
| 0.297454028 | 0.24948754 | 1.595890712 | 3.950204239 | 0 |
| 0 | 0 | 0.033025669 | 0.004223072 | 0 |
| 0 | 0 | 0 | 0 | 0 |
| 0 | 0 | 0 | 0 | 0 |
| 0 | 0 | 0.001936531 | 0 | 0 |
| 0 | 0 | 0 | 0 | 0 |
| 0 | 0 | 0 | 0 | 0 |
| 0 | 0 | 0 | 0 | 0 |
| 0 | 0 | 0 | 0 | 0 |
| 0 | 0 | 0 | 0 | 0 |
| 0.006970685 | 0.009042755 | 0.332998935 | 2.161616269 | 0 |
| 0 | 0.388390815 | 0.191572031 | 0.346508093 | 0 |
| 0 | 0 | 0 | 0 | 0 |
| 0 | 0 | 0 | 0 | 0 |
| 0 | 0 | 0 | 0 | 0 |
| 0 | 0 | 0 | 0 | 0 |
| 0 | 0 | 0 | 0 | 0 |
| 0 | 0 | 0 | 0 | 0 |
| 0 | 0 | 0 | 0 | 0 |
| 0 | 0 | 0.001165277 | 0 | 0 |
| 0 | 0 | 0 | 0 | 0 |
| 0 | 0 | 0 | 0 | 0 |
| 0 | 0 | 0 | 0 | 0 |
| 0 | 0.00609665 | 0 | 0 | 0 |
| 0 | 0 | 0 | 0.004049023 | 0 |
| 0 | 0 | 0 | 0 | 0 |
| 0 | 0 | 0 | 0 | 0 |
| 0.229985274 | 0 | 0 | 0 | 0 |
| 0 | 0 | 0.001663089 | 0 | 0 |

| | | | | |
|---|---|---|---|---|
| 0 | 0 | 0 | 0 | 0 |
| 0 | 0 | 0 | 0 | 0 |
| 0 | 0 | 0 | 0 | 0 |
| 0 | 0 | 0.001500559 | 0 | 0 |
| 0 | 0.003079677 | 0.454747544 | 0.0575943 | 0 |
| 0 | 0 | 0 | 0 | 0 |
| 0 | 0 | 0 | 0 | 0 |
| 0 | 0 | 0 | 0 | 0 |
| 0 | 0 | 0 | 0 | 0 |
| 0 | 0 | 0 | 0 | 0 |
| 0 | 0.022591801 | 0.010058119 | 0 | 0 |
| 0 | 0.00237538 | 0 | 0.012074694 | 0.002902329 |
| 0 | 0 | 0 | 0.004166967 | 0 |
| 0 | 0 | 0 | 0 | 0 |
| 0 | 0 | 0 | 0 | 0 |
| 0 | 0 | 0 | 0.035472525 | 0 |
| 0 | 0 | 0 | 0 | 0 |
| 0 | 0 | 0 | 0 | 0 |
| 0 | 0 | 0 | 0.170060305 | 0.011427831 |
| 0.042238412 | 0.271250613 | 0 | 0 | 0 |
| 0 | 0 | 0 | 0 | 0 |
| 0 | 0.003912308 | 0.006188788 | 0 | 0 |
| 0 | 0 | 0 | 0 | 0 |
| 0 | 0 | 0 | 0 | 0 |
| 0 | 0 | 0 | 0 | 0 |
| 0 | 0 | 0 | 0 | 0 |
| 0 | 0 | 0 | 0.000850353 | 0 |
| 0 | 0 | 0 | 0 | 0 |
| 0 | 0 | 0 | 0 | 0 |
| 0 | 0 | 0 | 0 | 0 |
| 0 | 0.004479914 | 0 | 0.0024098 | 0 |
| 0 | 0 | 0 | 0 | 0 |
| 0 | 0 | 0 | 0 | 0 |
| 0 | 0 | 0 | 0 | 0 |
| 0 | 0 | 0 | 0 | 0 |
| 0 | 0 | 0 | 0 | 0 |
| 0 | 0 | 0 | 0 | 0 |
| 0 | 0.020533413 | 0 | 0.027935414 | 0.000457819 |
| 0 | 0.002523831 | 0.484859397 | 0 | 0 |
| 0 | 0 | 0 | 0 | 0 |
| 0 | 0 | 0 | 0 | 0 |
| 0 | 0 | 0 | 0 | 0 |
| 0 | 0 | 0 | 0 | 0 |
| 0 | 0 | 0 | 0.004267997 | 0 |
| 0 | 0 | 0 | 0 | 0 |
| 0 | 0 | 0 | 0 | 0 |
| 0 | 0 | 0 | 0 | 0 |
| 0 | 0 | 0 | 0 | 0 |

| | | | | |
|---|---|---|---|---|
| 0 | 0 | 0 | 0 | 0 |
| 0 | 0 | 0.008641201 | 0.031972641 | 0 |
| 0 | 0.034574796 | 0 | 0 | 0 |
| 0 | 0.001863364 | 0.000946024 | 0.001052442 | 0 |
| 0.001936991 | 0.00129618 | 0.033166558 | 0 | 0.044608175 |
| 0.004999745 | 0 | 0 | 0 | 0 |
| 0 | 0 | 0 | 0 | 0 |
| 0 | 0 | 0 | 0.003292404 | 0 |
| 0 | 0 | 0 | 0 | 0 |
| 0 | 0 | 0 | 0 | 0 |
| 0 | 0.015012336 | 0.032519318 | 0 | 0 |
| 0 | 0 | 0 | 0 | 0 |
| 1.989841156 | 0.088002488 | 0.254961312 | 0.030027848 | 0 |
| 0 | 0 | 0 | 0 | 0 |
| 0 | 0 | 0 | 0 | 0 |
| 0 | 0 | 0.017369051 | 0 | 0 |
| 0 | 0 | 0 | 0 | 0 |
| 0 | 0 | 0 | 0 | 0 |
| 0 | 0 | 0 | 0 | 0 |
| 0 | 0 | 0 | 0 | 0 |
| 0 | 0 | 0 | 0 | 0 |
| 0 | 0 | 0 | 0 | 0 |
| 0 | 0 | 0 | 0 | 0 |
| 0.010419032 | 0.005131002 | 0.012920767 | 0.005216453 | 0.05485596 |
| 0 | 0 | 0 | 0 | 0 |
| 0 | 0 | 0 | 0 | 0 |
| 0 | 0.014124653 | 0 | 0 | 0 |
| 0 | 0 | 0 | 0 | 0 |
| 0 | 0 | 0 | 0 | 0 |
| 0 | 0 | 0 | 0 | 0 |
| 0.133224114 | 0 | 0.010239492 | 0.004928943 | 0 |
| 0 | 0 | 0 | 0 | 0 |
| 0 | 0 | 0 | 0 | 0 |
| 0 | 0 | 0 | 0 | 0 |
| 0 | 0 | 0 | 0 | 0 |
| 0 | 0 | 0 | 0 | 0 |
| 0 | 0 | 0 | 0 | 0 |
| 0 | 0 | 0 | 0 | 0 |
| 0.006057719 | 0 | 0 | 0 | 0.000751781 |
| 0 | 0 | 0 | 0 | 0 |
| 0 | 0 | 0 | 0 | 0 |
| 0 | 0 | 0 | 0 | 0 |
| 0 | 0 | 0 | 8.909810167 | 0 |
| 0 | 0 | 0 | 0 | 0 |
| 0 | 11.52930204 | 12.5269056 | 0 | 0 |
| 0.200438244 | 0.256961317 | 0.169156229 | 0.484730344 | 0.503248082 |
| 0 | 0 | 0 | 0 | 0 |
| 0 | 0 | 0 | 0 | 0 |

| | | | | |
|---|---|---|---|---|
| 0 | 0 | 0.002013933 | 0 | 0 |
| 0 | 0 | 0 | 0 | 0 |
| 0 | 0.217481835 | 0.049916903 | 0 | 0 |
| 0 | 0 | 0 | 0 | 0 |
| 0 | 0 | 0 | 0 | 0 |
| 0 | 0 | 0 | 0 | 0 |
| 0 | 0 | 0 | 0 | 0 |
| 0 | 0 | 0 | 0 | 0 |
| 0 | 0 | 0 | 0 | 0 |
| 0 | 0.00093627 | 0 | 0 | 0 |
| 0 | 0 | 0 | 0.033267853 | 0 |
| 0 | 0 | 0 | 0 | 0 |
| 0 | 0 | 0 | 0 | 0 |
| 0 | 0 | 0 | 0 | 0 |
| 0 | 0 | 0 | 0 | 0 |
| 0 | 0 | 0 | 0 | 0 |
| 0.002580001 | 0 | 0 | 0 | 0 |
| 0 | 0 | 0 | 0 | 0 |
| 0 | 0 | 0 | 0 | 0 |
| 0 | 0 | 0 | 0.00324037 | 0 |
| 0 | 0 | 0 | 0 | 0 |
| 0 | 0 | 0 | 0 | 0 |
| 0 | 0 | 0 | 0 | 0 |
| 0 | 0 | 0 | 0 | 0 |
| 0 | 0 | 0 | 0 | 0 |
| 0 | 0 | 0 | 0 | 0 |
| 0 | 0 | 0 | 0 | 0 |
| 0 | 0.000976784 | 0.029357874 | 0 | 0 |
| 0 | 0 | 0 | 0 | 0 |
| 0 | 0 | 0 | 0 | 0 |
| 0.35214376 | 0.00132698 | 0.003079786 | 0.001498976 | 0.041112901 |
| 0 | 0 | 0 | 0 | 0 |
| 0.010448248 | 0.015407584 | 0.011621823 | 0.14445854 | 0.068713327 |
| 0.006050286 | 0 | 0.004933209 | 0.001016323 | 0.00068706 |
| 0 | 0 | 0 | 0 | 0 |
| 0 | 0 | 0 | 0 | 0 |
| 0 | 0 | 0 | 0 | 0 |
| 0 | 0 | 0 | 0 | 0 |
| 0 | 0 | 0 | 0 | 0 |
| 0 | 0 | 0 | 0 | 0 |
| 0 | 0 | 0 | 0 | 0 |
| 0 | 0 | 0 | 0 | 0 |
| 0.009657475 | 0.031105711 | 0.040499894 | 0.029655044 | 0 |
| 0 | 0 | 0 | 0 | 0 |
| 0 | 0 | 0 | 0 | 0 |
| 0.096849349 | 0.019765211 | 0.109540458 | 0.019446172 | 0.005258442 |
| 0 | 0 | 0 | 0 | 0 |
| 0 | 0 | 0 | 0 | 0 |

| | | | | |
|---|---|---|---|---|
| 0 | 0 | 0 | 0 | 0 |
| 0 | 0.01040022 | 0.121654859 | 0 | 0 |
| 0.031040813 | 0.004319486 | 0.025657978 | 0.011893436 | 0.011544995 |
| 0 | 0 | 0 | 0 | 0 |
| 0 | 0.021124525 | 0.004027292 | 1.187772502 | 0 |
| 0 | 0 | 0 | 0.006636486 | 0 |
| 0 | 0 | 0 | 0 | 0 |
| 0 | 0 | 0 | 0 | 0 |
| 0 | 0 | 0.00696689 | 0 | 0 |
| 0 | 0 | 0 | 0 | 0 |
| 0 | 0 | 0 | 0 | 0 |
| 0 | 0 | 0 | 0 | 0 |
| 0 | 0 | 0 | 0 | 0 |
| 0 | 0 | 0 | 0 | 0 |
| 0 | 0 | 0 | 0 | 0 |
| 0 | 0 | 0 | 0 | 0 |
| 0 | 0 | 0 | 0 | 0 |
| 0 | 0 | 0 | 0 | 0 |
| 0 | 0 | 0 | 0 | 0 |
| 0 | 0 | 0 | 0 | 0 |
| 0 | 0 | 0 | 0 | 0 |
| 0 | 0 | 0 | 0 | 0 |
| 0 | 0 | 0 | 0 | 0 |
| 0 | 0 | 0 | 0 | 0 |
| 0.014785923 | 0.002669898 | 0.032351237 | 0.001005319 | 0 |
| 0 | 0 | 0.001999946 | 0.002275487 | 0 |
| 0 | 0 | 0 | 0 | 0 |
| 0 | 0.059706451 | 0.058729624 | 0.002943069 | 0 |
| 0 | 0 | 0 | 0 | 0 |
| 0 | 0 | 0 | 0 | 0 |
| 0 | 0 | 0.002282626 | 0 | 0 |
| 0 | 0 | 0 | 0 | 0 |
| 0 | 0 | 0 | 0 | 0 |
| 0 | 0 | 0 | 0 | 0 |
| 0 | 0 | 0 | 0 | 0 |
| 0 | 0 | 0 | 0 | 0 |
| 0 | 0 | 0 | 0 | 0 |
| 0 | 0 | 0 | 0 | 0 |
| 0 | 0.055476892 | 0.005117238 | 4.464776009 | 0 |
| 0 | 0 | 0 | 0 | 0 |
| 0 | 0 | 0 | 0 | 0 |
| 0 | 0.001456869 | 0 | 0 | 0 |
| 0 | 0 | 0 | 0.039837866 | 0 |
| 0 | 0 | 0 | 0 | 0 |
| 0 | 0 | 0 | 0 | 0 |
| 0 | 0 | 0.007137439 | 0 | 0 |
| 0 | 0 | 0 | 0 | 0 |
| 0 | 0 | 0 | 0 | 0 |

| | | | | |
|---|---|---|---|---|
| 0 | 0 | 0 | 0 | 0 |
| 0 | 0.000650413 | 0.008981779 | 0.207679773 | 0.000397349 |
| 0 | 0 | 0 | 0 | 0 |
| 0 | 0 | 0 | 0 | 0 |
| 0 | 0 | 0 | 0 | 0 |
| 0 | 0 | 0 | 0 | 0 |
| 0 | 0 | 0 | 0 | 0 |
| 0 | 0 | 0 | 0 | 0 |
| 0 | 0 | 0 | 0 | 0 |
| 0 | 0 | 0 | 0 | 0 |
| 0 | 0 | 0 | 0 | 0 |
| 0 | 0 | 0 | 0 | 0 |
| 0.000725262 | 0.006187895 | 0.019919909 | 0.097600287 | 0 |
| 0 | 0 | 0 | 0 | 0 |
| 0 | 0 | 0.00746595 | 0.022291288 | 0 |
| 0 | 0 | 0 | 0 | 0 |
| 0 | 0 | 0 | 0 | 0 |
| 0 | 0 | 0 | 0 | 0 |
| 0 | 0 | 0 | 0 | 0 |
| 0 | 0 | 0 | 0 | 0 |
| 0 | 0 | 0 | 0 | 0 |
| 0.004900728 | 0 | 0 | 0 | 0.000486556 |
| 0 | 0 | 0 | 0.001382635 | 0 |
| 0 | 0 | 0 | 0 | 0 |
| 0 | 0 | 0 | 0 | 0 |
| 0 | 0 | 0 | 0 | 0 |
| 0 | 0 | 0 | 0 | 0 |
| 0 | 0 | 0 | 0 | 0 |
| 0 | 0 | 0 | 0 | 0 |
| 0 | 0 | 0 | 0 | 0 |
| 0 | 0 | 0 | 0 | 0 |
| 0 | 0 | 0 | 0 | 0 |
| 0 | 0 | 0 | 0 | 0 |
| 0 | 0 | 0 | 0 | 0 |
| 0 | 8.150975168 | 0.044446505 | 0 | 0 |
| 0 | 0 | 0 | 0 | 0 |
| 0 | 0 | 0 | 0 | 0 |
| 0 | 0 | 0 | 0 | 0 |
| 2.41300688 | 0.12274669 | 0 | 0 | 0.007450239 |
| 0 | 0 | 0 | 0 | 0 |
| 0.997851986 | 0.045402161 | 0.166639664 | 0.031416631 | 0.021056915 |
| 0 | 0 | 0 | 0 | 0 |
| 0 | 0 | 0 | 0 | 0 |
| 0 | 0 | 0 | 0 | 0 |
| 0 | 0 | 0 | 0 | 0 |
| 2.539306512 | 0 | 0.100369812 | 0.061487017 | 0.322558245 |
| 0 | 0 | 0 | 0 | 0 |
| 0 | 0 | 0 | 0 | 0 |

| | | | | |
|---|---|---|---|---|
| 0 | 0 | 0 | 0 | 0 |
| 0 | 0 | 0 | 0 | 0 |
| 0 | 0 | 0 | 0 | 0 |
| 0 | 0 | 0.045550244 | 0 | 0 |
| 0 | 0 | 0 | 0 | 0 |
| 0 | 0 | 0 | 0 | 0 |
| 0 | 0 | 0 | 0 | 0 |
| 0.00387879 | 0 | 0 | 0 | 0 |
| 0 | 0 | 0 | 0 | 0 |
| 0.017407986 | 0.010913052 | 0.00237837 | 0 | 0 |
| 0 | 0.023299461 | 0 | 0 | 0 |
| 0 | 0 | 0 | 0 | 0 |
| 0 | 0 | 0 | 0.001469408 | 0 |
| 0 | 0 | 0 | 0 | 0 |
| 0 | 0 | 0 | 0 | 0 |
| 0 | 0 | 0 | 0 | 0 |
| 0 | 0 | 0 | 0 | 0 |
| 0 | 0 | 0 | 0 | 0 |
| 0 | 0 | 0 | 0 | 0 |
| 0.743297368 | 0.006946842 | 0.040065467 | 1.75203156 | 0.853176923 |
| 0 | 0 | 0 | 0 | 0 |
| 0 | 0 | 0 | 0 | 0 |
| 0 | 0 | 0 | 0 | 0 |
| 0 | 0 | 0 | 0 | 0 |
| 0 | 0 | 0 | 0 | 0 |
| 0 | 0.028002441 | 0 | 0 | 0 |
| 0 | 0 | 0 | 0 | 0 |
| 0 | 0 | 0 | 0 | 0 |
| 0 | 0 | 0 | 0 | 0 |
| 0 | 0 | 0 | 0 | 0 |
| 0 | 0 | 0 | 0 | 0 |
| 0 | 0 | 0 | 0 | 0 |
| 0 | 0 | 0 | 0 | 0 |
| 0 | 0 | 0 | 0 | 0 |
| 0 | 0 | 0 | 0 | 0 |
| 0 | 0 | 0 | 0 | 0 |
| 0 | 0 | 0 | 0 | 0 |
| 0 | 0 | 0 | 0 | 0 |
| 0 | 0 | 0 | 0 | 0 |
| 0 | 0 | 0 | 0 | 0 |
| 0 | 0 | 0 | 0 | 0 |
| 0 | 0 | 0 | 0 | 0 |
| 0 | 0.010009115 | 0 | 0 | 0 |
| 0 | 0 | 0 | 0 | 0 |
| 0 | 0 | 0 | 0.006142029 | 0 |
| 0 | 0 | 0 | 0 | 0 |
| 0 | 0 | 0 | 0 | 0 |
| 0 | 0 | 0 | 0 | 0 |

| | | | | |
|---|---|---|---|---|
| 0 | 0 | 0 | 0 | 0 |
| 0 | 0 | 0 | 0 | 0 |
| 0 | 0 | 0.000902853 | 0 | 0 |
| 0 | 0 | 0 | 0 | 0 |
| 0 | 0 | 0 | 0 | 0 |
| 0 | 0.000811827 | 0 | 0.007703243 | 0 |
| 0 | 0 | 0 | 0 | 0 |
| 0 | 0 | 0 | 0 | 0 |
| 0 | 0 | 0 | 0 | 0 |
| 0 | 0 | 0 | 0 | 0 |
| 0 | 0 | 0 | 0 | 0 |
| 0 | 0 | 0 | 0 | 0 |
| 0 | 0 | 0 | 0 | 0 |
| 0 | 0 | 0 | 0 | 0 |
| 0 | 0 | 0 | 0 | 0 |
| 0 | 0 | 0 | 0 | 0 |
| 0 | 0.137469831 | 0.005119486 | 0 | 0 |
| 0 | 0 | 0 | 0 | 0 |
| 0 | 0 | 0 | 0 | 0 |
| 0 | 0 | 0 | 0 | 0 |
| 0 | 0 | 0 | 0 | 0 |
| 0 | 0 | 0 | 0 | 0 |
| 0 | 0 | 0 | 0 | 0 |
| 0 | 0 | 0 | 0 | 0 |
| 0 | 0 | 0 | 0 | 0 |
| 0 | 0 | 0 | 0 | 0 |
| 0 | 0 | 0 | 0 | 0 |
| 0 | 0.123739891 | 0 | 0 | 0 |
| 0 | 0 | 0 | 0 | 0 |
| 0 | 0 | 0 | 0 | 0 |
| 0 | 0 | 0 | 0 | 0 |
| 0 | 0 | 0 | 0 | 0 |
| 0 | 0 | 0 | 0 | 0 |
| 0 | 0 | 0 | 0 | 0 |
| 0 | 0 | 0 | 0 | 0 |
| 0 | 0 | 0 | 0 | 0 |
| 0 | 0 | 0 | 0 | 0 |
| 0 | 0 | 0 | 0 | 0 |
| 0 | 0 | 1.766449631 | 4.641566713 | 0 |
| 0 | 0 | 0 | 0 | 0 |
| 0 | 0 | 0 | 0 | 0 |
| 0 | 0 | 0 | 0 | 0 |
| 0 | 0 | 0 | 0 | 0 |
| 0 | 0 | 0 | 0 | 0 |
| 0 | 0 | 0 | 0 | 0.070918412 |
| 0 | 0 | 0 | 0 | 0 |
| 0 | 0 | 0 | 0 | 0 |
| 0 | 0 | 0 | 0 | 0 |

| | | | | |
|---|---|---|---|---|
| 0 | 0 | 0 | 0 | 0 |
| 0 | 0 | 0 | 0 | 0 |
| 0 | 0 | 0 | 0 | 0 |
| 0 | 0 | 0 | 0 | 0 |
| 0 | 0 | 0 | 0 | 0 |
| 0 | 0 | 0 | 0 | 0 |
| 0 | 0 | 0 | 0 | 0 |
| 0 | 0 | 0 | 0 | 0 |
| 0 | 0 | 0 | 0 | 0 |
| 0 | 0.001480893 | 0 | 0 | 0 |
| 0.001129888 | 0.009045069 | 0.066900413 | 0.002688802 | 0.017595322 |
| 0 | 0 | 0 | 0 | 0 |
| 0.249758275 | 0.002805116 | 0.014953559 | 0.168337302 | 0 |
| 0 | 0 | 0 | 0 | 0 |
| 0 | 0 | 0 | 0 | 0 |
| 0 | 0 | 0 | 0 | 0 |
| 0 | 0 | 0 | 0 | 0 |
| 0 | 0 | 0 | 0 | 0 |
| 0 | 0 | 0 | 0 | 0 |
| 0 | 0 | 0 | 0 | 0 |
| 0.883648392 | 0 | 0 | 0.000818692 | 0 |
| 0 | 0 | 0 | 0 | 0 |
| 0 | 0 | 0 | 0 | 0 |
| 4.399683616 | 0.228946738 | 1.697363884 | 1.861217588 | 0 |
| 0 | 0 | 0 | 0 | 0 |
| 0 | 0 | 0 | 0 | 0 |
| 0 | 0 | 0 | 0 | 0 |
| 0 | 0 | 0 | 0 | 0 |
| 0 | 0 | 0 | 0 | 0 |
| 0 | 0 | 0 | 0 | 0 |
| 0 | 0 | 0 | 0 | 0 |
| 0 | 0 | 0 | 0 | 0 |
| 0 | 0 | 0.000825713 | 0 | 0 |
| 0 | 0 | 0 | 0 | 0 |

| PHL018-0266_V2 | PHL018-0048_V1 | PHL018-0218_V3 | PHL018-0240_V2 | PHL018-0068_V1 |
|---|---|---|---|---|
| REF | CTRL | REF | REF | REF |
| 0.013986094 | 3.298012414 | 0.010518152 | 0.049063664 | 0.209011029 |
| 1.370041156 | 9.44801132 | 1.033836235 | 0.100564463 | 0.398059945 |
| 2.392483575 | 4.424828623 | 0 | 1.560554375 | 6.275223176 |
| 0 | 0 | 0 | 0 | 0 |
| 0 | 0 | 0 | 0 | 0 |
| 0 | 0 | 0 | 0 | 0 |
| 0 | 0 | 0 | 0 | 0 |
| 0 | 0 | 0 | 0 | 0 |
| 0 | 0 | 0 | 0 | 0 |
| 0 | 0 | 0 | 0 | 0 |
| 0 | 0 | 0 | 0 | 0 |
| 0 | 0 | 0 | 0 | 0 |
| 0 | 0 | 0 | 0 | 0 |
| 5.045546239 | 11.34012963 | 0.538727102 | 0.041561192 | 0.177679993 |
| 0 | 0 | 0 | 0 | 0 |
| 0 | 0 | 0 | 0 | 0 |
| 0 | 0 | 0 | 0 | 0 |
| 0 | 0 | 0 | 0 | 0 |
| 0 | 0 | 0 | 0 | 0 |
| 0 | 0 | 0 | 0 | 0 |
| 0 | 0 | 0 | 0 | 0.043167397 |
| 0 | 0 | 0 | 0 | 0 |
| 0 | 0 | 0 | 0 | 0 |
| 0 | 0 | 0 | 0 | 0 |
| 0 | 0 | 0 | 0 | 0 |
| 0 | 0 | 0 | 0 | 0 |
| 0 | 0 | 0 | 0 | 0 |
| 0 | 0 | 0 | 0 | 0 |
| 0 | 0 | 0 | 0 | 0 |
| 0 | 0 | 0 | 0.010309543 | 0 |
| 0 | 0 | 0 | 0 | 0 |
| 0 | 0 | 0 | 0 | 0 |
| 0 | 0 | 0 | 0 | 0 |
| 0 | 0 | 0 | 0 | 0 |
| 0 | 0 | 0 | 0 | 0 |
| 3.434357355 | 0 | 8.472255946 | 0.301350304 | 0 |
| 0 | 0 | 0 | 0 | 0 |
| 0 | 0 | 0 | 0 | 0 |
| 0 | 0 | 0 | 0 | 0 |
| 0 | 0 | 0 | 0 | 0 |
| 0 | 0 | 0 | 0 | 0 |
| 0.003848089 | 0 | 0.001096126 | 0.00672125 | 0 |
| 0.019344287 | 0.003417386 | 0.028070873 | 0.055037703 | 0.006388602 |
| 0.010884593 | 0.029736919 | 0.011297778 | 0.227771026 | 0.031610562 |
| 0 | 0.00596324 | 0 | 0.309624483 | 0 |
| 0 | 0 | 0 | 0 | 0 |

| | | | | |
|---|---|---|---|---|
| 0.013413151 | 0 | 0 | 0.035528218 | 0.055692248 |
| 0.000608423 | 0.06405613 | 0.003136065 | 0 | 0 |
| 0 | 0 | 0 | 0.001562094 | 0 |
| 0.254943415 | 0.957978245 | 0.656934 | 0.476161101 | 2.167629131 |
| 0.001128044 | 0 | 0 | 0 | 0 |
| 0 | 0 | 0.018733281 | 0 | 0 |
| 0 | 0 | 0.924372968 | 0.003543687 | 0.003673692 |
| 0 | 0 | 0.007172823 | 0 | 0 |
| 0 | 0 | 0 | 0 | 0.019210159 |
| 0 | 0 | 0 | 0 | 0 |
| 0 | 0 | 0.001137845 | 0 | 0 |
| 0.38413492 | 0 | 0.022098866 | 0.133975196 | 0.105371397 |
| 0 | 0.010344157 | 0 | 0 | 0 |
| 0.02568697 | 0 | 0.016597684 | 0.001268939 | 0.04078023 |
| 0 | 0 | 0 | 0 | 0 |
| 0 | 0 | 0 | 0 | 0 |
| 0.450313841 | 0 | 0.249248178 | 0 | 0 |
| 0 | 0 | 0 | 0 | 0 |
| 0 | 0 | 0 | 0 | 0 |
| 0 | 0 | 0 | 0 | 0 |
| 0.144928327 | 0.005339574 | 0.287714028 | 0.007853711 | 0.025815571 |
| 0.019924508 | 0.003277953 | 0.031928525 | 0 | 0.015848123 |
| 0 | 0 | 0.015508945 | 0.67194022 | 0.68780798 |
| 0 | 0 | 0 | 0 | 0 |
| 0 | 0 | 0 | 0 | 0 |
| 0 | 0 | 0 | 0 | 0 |
| 0 | 0.006216339 | 0 | 0 | 0 |
| 0 | 0 | 0 | 0 | 0 |
| 0 | 0 | 0 | 0.002300808 | 0 |
| 0 | 0 | 0 | 0 | 0 |
| 0 | 0 | 0 | 0 | 0 |
| 0 | 0 | 0 | 0 | 0 |
| 0 | 0 | 0 | 0 | 0 |
| 0 | 0 | 0 | 0 | 0 |
| 0 | 0 | 0 | 0 | 0 |
| 0 | 0 | 0 | 0 | 0 |
| 0 | 0 | 0 | 0 | 0 |
| 0.007117064 | 0 | 0.004572838 | 0 | 0 |
| 0 | 0 | 0 | 0 | 0 |
| 0 | 0 | 0 | 0 | 0 |
| 0 | 0 | 0 | 0 | 0 |
| 0 | 0 | 0 | 0 | 0 |
| 0 | 0 | 0 | 0 | 0 |
| 0 | 0 | 0 | 0 | 0 |
| 0 | 0 | 0 | 0 | 0 |
| 0 | 0 | 0 | 0 | 0 |
| 0 | 0 | 0.0228455 | 0 | 0 |
| 0 | 0.143770793 | 0 | 0.008903652 | 0.031081599 |

| | | | | |
|---|---|---|---|---|
| 0 | 0 | 0 | 0 | 0 |
| 0 | 0 | 0 | 0 | 0 |
| 0 | 0 | 0 | 0 | 0 |
| 0 | 0 | 0 | 0 | 0 |
| 0.357770132 | 0 | 0.019173073 | 0 | 0.003183405 |
| 0 | 0 | 0 | 0 | 0 |
| 0 | 0 | 0 | 0 | 0 |
| 0 | 0.859615434 | 0 | 0 | 0 |
| 0 | 0.062995216 | 0.002673037 | 0.000869198 | 0 |
| 0 | 0 | 0 | 0 | 0 |
| 0 | 0 | 0 | 0 | 0 |
| 0 | 0 | 0 | 0.000605473 | 0 |
| 0 | 0 | 0 | 0 | 0 |
| 0 | 0 | 0 | 0 | 0 |
| 0 | 0 | 0 | 0 | 0 |
| 0 | 0 | 0 | 0 | 0 |
| 0 | 0 | 0 | 0 | 0 |
| 0 | 0 | 0 | 0 | 0 |
| 0 | 0.123628304 | 0.004644896 | 8.177654554 | 0.026254702 |
| 0 | 0 | 0 | 0 | 0 |
| 0 | 0 | 0 | 0 | 0 |
| 0 | 0 | 0 | 0 | 0 |
| 0 | 0 | 0 | 0 | 0 |
| 0 | 0 | 0 | 0 | 0 |
| 0 | 0 | 0 | 0 | 0 |
| 0 | 0 | 0 | 0 | 0 |
| 0 | 0 | 0 | 0 | 0 |
| 0 | 0 | 0 | 0 | 0 |
| 0 | 0 | 0 | 0 | 0 |
| 0 | 0 | 0 | 0 | 0 |
| 0 | 0 | 0 | 0.028795026 | 0.023347469 |
| 0.000724657 | 0.00398053 | 0 | 0.050122467 | 0 |
| 0 | 0 | 0 | 0 | 0 |
| 0 | 0 | 0 | 0 | 0 |
| 0 | 0 | 0 | 0 | 0 |
| 0 | 0 | 0 | 0 | 0 |
| 0 | 0 | 0 | 0 | 0 |
| 0 | 0 | 0 | 0 | 0 |
| 0 | 0 | 0.038762147 | 0.419505234 | 0 |
| 0 | 0 | 0 | 0 | 0 |
| 0 | 0 | 0 | 0 | 0 |
| 0 | 0 | 0 | 0 | 0 |
| 0 | 0 | 0.002726704 | 0.004486444 | 0 |
| 0 | 0 | 0 | 0 | 0 |
| 0 | 0 | 0 | 0 | 0 |
| 0 | 0 | 0 | 0 | 0 |
| 0 | 0 | 0 | 0 | 0 |
| 0 | 0 | 0 | 0 | 0 |

| | | | | |
|---|---|---|---|---|
| 0 | 0.014419883 | 0 | 0 | 0 |
| 0 | 0 | 0 | 0 | 0 |
| 0 | 0 | 0 | 0 | 0 |
| 0 | 0 | 0 | 0 | 0 |
| 0 | 0 | 0.008726514 | 0.001615775 | 0.038526198 |
| 0 | 0 | 0 | 0 | 0 |
| 0 | 0 | 0 | 0 | 0 |
| 0 | 0 | 0 | 0 | 0 |
| 0 | 0 | 0 | 0 | 0 |
| 0 | 0 | 0 | 0 | 0 |
| 0 | 0 | 0 | 0 | 0 |
| 0 | 0 | 0 | 0 | 0 |
| 0 | 0 | 0 | 0 | 0 |
| 0 | 0 | 0 | 0 | 0 |
| 0 | 0 | 0 | 0 | 0 |
| 0 | 0 | 0 | 0 | 5.041551641 |
| 0 | 0 | 0 | 0 | 0 |
| 0 | 0 | 0 | 0 | 0 |
| 0 | 0 | 0 | 0 | 0 |
| 0 | 0 | 0 | 0 | 0 |
| 0 | 0.000564213 | 0 | 0 | 0 |
| 0 | 0 | 0 | 0 | 0 |
| 0 | 0 | 0 | 0 | 0 |
| 0 | 0 | 0 | 0 | 0 |
| 0 | 0 | 0 | 0.073217828 | 0 |
| 0 | 0 | 0 | 0 | 0 |
| 0 | 0 | 0 | 0 | 0 |
| 0 | 0 | 0 | 0 | 0 |
| 0 | 0 | 0 | 0 | 0 |
| 0 | 0 | 0 | 0 | 0 |
| 0 | 0.004031592 | 0 | 0 | 0 |
| 0 | 0 | 0.014153619 | 0 | 0 |
| 0 | 0 | 0 | 0 | 0 |
| 0 | 0 | 0 | 0 | 0 |
| 0 | 0 | 0 | 0 | 0 |
| 0 | 0 | 0 | 0 | 0 |
| 0 | 0 | 0 | 0 | 0 |
| 0 | 0 | 0 | 0 | 0.851435655 |
| 0 | 0 | 0 | 0 | 0 |
| 0 | 0 | 0 | 0 | 0 |
| 0 | 0 | 0 | 0 | 0 |
| 0 | 0 | 0 | 0 | 0 |
| 0 | 0 | 0 | 0 | 0 |
| 0 | 0 | 0 | 0 | 0 |
| 0 | 0 | 0 | 0 | 0 |
| 0 | 0 | 0 | 0 | 0 |
| 0 | 0 | 0 | 0 | 0 |
| 0 | 0 | 0 | 0.002148527 | 0 |

| | | | | |
|---|---|---|---|---|
| 0 | 0 | 0 | 0 | 0 |
| 0 | 0 | 0 | 0 | 0 |
| 0 | 0 | 0 | 0 | 0 |
| 0 | 0 | 0 | 0 | 0 |
| 0 | 0 | 0 | 0 | 0 |
| 0 | 0 | 0 | 0 | 0 |
| 0 | 0 | 0 | 0 | 0 |
| 0 | 0 | 0 | 0 | 0 |
| 0 | 0 | 0 | 0 | 0 |
| 0 | 0 | 0 | 0 | 0 |
| 0 | 0 | 0 | 0 | 0 |
| 0 | 0 | 0 | 0 | 0.005094542 |
| 0 | 0 | 0 | 0 | 0.004056343 |
| 0 | 0 | 0 | 0 | 0 |
| 0 | 0 | 0 | 0 | 0 |
| 0 | 0 | 0 | 0 | 0 |
| 0 | 0 | 0 | 0 | 0 |
| 0 | 0 | 0 | 0 | 0 |
| 0 | 0 | 0 | 0 | 0 |
| 0 | 0 | 0 | 0 | 0 |
| 0.038594894 | 0.042953039 | 0.070369625 | 0.023689873 | 0.050201414 |
| 0 | 0 | 0 | 0 | 0 |
| 0 | 0 | 0 | 0 | 0 |
| 0 | 0 | 0 | 0 | 0 |
| 0 | 0 | 0 | 0 | 0 |
| 0 | 0 | 0 | 0 | 0 |
| 0.002439338 | 0.007830732 | 0.001134439 | 0.003246217 | 0.001472323 |
| 0 | 0 | 0 | 0 | 0 |
| 0 | 0 | 0 | 0 | 0 |
| 0 | 0.084012529 | 0 | 0 | 0 |
| 0 | 0.004770278 | 0.000424066 | 0.000808982 | 0 |
| 0 | 0.482725537 | 0 | 0 | 0 |
| 0 | 0 | 0 | 0 | 0 |
| 0 | 0 | 0 | 0 | 0.002427913 |
| 0.016157266 | 0.020413853 | 0.001425869 | 0.001133375 | 0.005639778 |
| 0 | 0 | 0 | 0 | 0 |
| 0 | 0 | 0 | 0 | 0 |
| 0 | 0 | 0 | 0 | 0 |
| 0 | 0 | 0 | 0 | 0 |
| 0 | 0 | 0 | 0 | 0 |
| 0 | 0 | 0 | 0 | 0 |
| 0 | 0 | 0 | 0 | 0 |
| 0 | 0 | 0 | 0 | 0 |
| 0 | 0 | 0 | 0 | 0 |
| 0 | 0 | 0 | 0 | 0 |
| 0 | 0 | 0 | 0 | 0 |
| 0 | 0 | 0 | 0 | 0 |
| 0 | 0.27986548 | 0 | 0 | 0.001004765 |

| | | | | |
|---|---|---|---|---|
| 0 | 0 | 0 | 0 | 0 |
| 0 | 0 | 0 | 0 | 0 |
| 0 | 0.002848536 | 0 | 0 | 0 |
| 0 | 0 | 0 | 0 | 0 |
| 0 | 0 | 0 | 0 | 0 |
| 0 | 0 | 0 | 0 | 0 |
| 0 | 0 | 0 | 0 | 0 |
| 0 | 0 | 0 | 0.000928836 | 0 |
| 0 | 0 | 0.001446117 | 0 | 0 |
| 0 | 0 | 0 | 0 | 0 |
| 0 | 0 | 0 | 0 | 0 |
| 0 | 0 | 0 | 0 | 0 |
| 0 | 0 | 0 | 0 | 0 |
| 0 | 0 | 0 | 0 | 0 |
| 0 | 0 | 0.001780081 | 0 | 0 |
| 0 | 0 | 0 | 0 | 0 |
| 0 | 0 | 0 | 0 | 0 |
| 0 | 0 | 0 | 0 | 0 |
| 0 | 0 | 0 | 0 | 0 |
| 0 | 0 | 0 | 0 | 0.153398446 |
| 0.067219414 | 0.003117664 | 0.013549692 | 0 | 0 |
| 0 | 0 | 0 | 0 | 0 |
| 0 | 0 | 0 | 0 | 0 |
| 0 | 0 | 0 | 0 | 0 |
| 0.00330763 | 0.004129266 | 0.015815105 | 0.004332945 | 0.001996402 |
| 0 | 0 | 0 | 0 | 0 |
| 0 | 0 | 0 | 0 | 0 |
| 0 | 0 | 0 | 0 | 0 |
| 0 | 0 | 0 | 0 | 0 |
| 0 | 0 | 0 | 0 | 0 |
| 0.067483114 | 0.788209483 | 0.04358849 | 0.003563692 | 0.106214883 |
| 0 | 0 | 0 | 0 | 0 |
| 0 | 0 | 0 | 0 | 0 |
| 0 | 0 | 0.004350376 | 0.008068587 | 0 |
| 0 | 0 | 0 | 0 | 0 |
| 0 | 0 | 0 | 0 | 0 |
| 0 | 0 | 0 | 0 | 0 |
| 0 | 0.002939163 | 0 | 0 | 0 |
| 0 | 0 | 0 | 0 | 0 |
| 0 | 0 | 0 | 0.001601835 | 0 |
| 0 | 0 | 0 | 0 | 0 |
| 0.007331128 | 0 | 0.002158088 | 0.009263109 | 0.012050745 |
| 0 | 0 | 0 | 0 | 0 |
| 0 | 0 | 0 | 0 | 0 |
| 0 | 0 | 0 | 0 | 0 |
| 0 | 0 | 0 | 0 | 0 |
| 0 | 0 | 0 | 0 | 0 |
| 0 | 0 | 0 | 0 | 0 |

| | | | | |
|---|---|---|---|---|
| 0 | 0 | 0 | 0 | 0 |
| 0 | 0 | 0 | 0 | 0 |
| 0.01096818 | 0.021272656 | 0.013867975 | 0.245398834 | 0.035701339 |
| 0 | 0 | 0 | 0 | 0 |
| 0 | 0.471922532 | 0 | 0 | 0 |
| 0 | 0 | 0 | 0 | 0 |
| 0 | 0 | 0 | 0 | 0 |
| 0 | 0 | 0 | 0 | 0 |
| 0 | 0 | 0 | 0 | 0 |
| 0 | 0.024822929 | 0 | 0 | 0 |
| 0 | 0 | 0 | 0 | 0 |
| 0 | 0 | 0 | 0 | 0 |
| 0 | 0 | 0 | 0 | 0 |
| 0.003896299 | 0.002702313 | 0.004051864 | 0 | 0.001829106 |
| 0.001965263 | 0.008252502 | 0.110652159 | 0.002912525 | 0.020057276 |
| 0 | 0 | 0 | 0 | 0.024013893 |
| 0 | 0.002033078 | 0.008118045 | 0.008533443 | 0.000737209 |
| 0 | 0 | 0.019663012 | 0 | 0.002096839 |
| 0 | 0 | 0 | 0 | 0 |
| 0 | 0 | 0 | 0 | 0 |
| 0 | 0 | 0 | 0 | 0 |
| 0 | 0.295940387 | 0 | 0.000641939 | 0.001109148 |
| 0 | 0 | 0 | 0 | 0 |
| 0 | 0 | 0 | 0 | 0 |
| 0 | 0 | 0 | 0 | 0 |
| 0 | 0 | 0 | 0 | 0 |
| 0 | 0 | 0 | 0 | 0 |
| 0 | 0 | 0 | 0 | 0 |
| 0 | 0 | 0 | 0 | 0 |
| 0 | 0.001659227 | 0.001514346 | 0 | 0 |
| 0 | 0 | 0 | 0 | 0 |
| 0 | 0 | 0 | 0 | 0 |
| 0 | 0 | 0 | 0 | 0 |
| 0 | 0 | 0 | 0 | 0 |
| 0 | 0 | 0 | 0 | 0 |
| 0 | 0 | 0.002091028 | 0.001709575 | 0 |
| 0 | 0 | 0 | 0 | 0 |
| 0 | 0.015564493 | 0.001127415 | 0.457571063 | 0.002229648 |
| 0 | 0 | 0 | 0 | 0 |
| 0 | 0 | 0 | 0 | 0 |
| 0.001332897 | 0 | 0 | 0 | 0 |
| 0 | 0 | 0 | 0 | 0 |
| 0 | 0 | 0 | 0 | 0 |
| 0 | 0 | 0 | 0 | 0 |
| 0 | 0 | 0 | 0 | 0 |
| 0 | 0 | 0 | 0 | 0 |
| 0 | 0 | 0 | 0 | 0 |
| 0 | 0.006284169 | 0 | 0 | 0.002700663 |

| | | | | |
|---|---|---|---|---|
| 0 | 0 | 0 | 0 | 0 |
| 0 | 0 | 0 | 0 | 0 |
| 0 | 0.005286387 | 0 | 0.004706665 | 0.001161746 |
| 0 | 0 | 0 | 0 | 0 |
| 0.002517464 | 0.00556037 | 0 | 0 | 0.014493478 |
| 0 | 0 | 0 | 0 | 0 |
| 0 | 0 | 0 | 0 | 0 |
| 0.029044574 | 0.066758008 | 0.018336084 | 0.002844706 | 0.030801374 |
| 0 | 0 | 0 | 0 | 0 |
| 0.050081309 | 0.172196954 | 0.009108144 | 0.002280522 | 0.032760866 |
| 0 | 0.093476143 | 0 | 0 | 0 |
| 0.004727284 | 0.001073013 | 0.002448298 | 0.032276941 | 0 |
| 0.016136156 | 0.005290469 | 0.012203937 | 0.115583505 | 0.03738346 |
| 0.03935918 | 0 | 0.000805854 | 0.000864737 | 0 |
| 0 | 0 | 0 | 0 | 0 |
| 0.000734032 | 0 | 0 | 0 | 0 |
| 0 | 0 | 0 | 0 | 0 |
| 0 | 0 | 0 | 0 | 0 |
| 0 | 0.101558957 | 0 | 0.001839369 | 0 |
| 0 | 0 | 0 | 0 | 0 |
| 0 | 0 | 0 | 0 | 0 |
| 0 | 0 | 0 | 0 | 0 |
| 0 | 0 | 0 | 0 | 0 |
| 0 | 0 | 0 | 0 | 0 |
| 0 | 0 | 0 | 0 | 0 |
| 0 | 0 | 0 | 0.043548747 | 0 |
| 0 | 0 | 0.000619814 | 0 | 0 |
| 0.016529298 | 0.24790993 | 0 | 0 | 0 |
| 0 | 0.090510565 | 0 | 0 | 0 |
| 0 | 0 | 0 | 0 | 0 |
| 0 | 0.052134746 | 0 | 0 | 0 |
| 0 | 0.023708613 | 0 | 0 | 0 |
| 0 | 0 | 0 | 0 | 0 |
| 0.014648546 | 0.029445708 | 0 | 0 | 0 |
| 0 | 0 | 0 | 0 | 0 |
| 0.005390321 | 0.031483559 | 0 | 0 | 0 |
| 0 | 0.004298653 | 0 | 0 | 0 |
| 0 | 0 | 0 | 0 | 0 |
| 0 | 0 | 0 | 0 | 0 |
| 0.000998334 | 0 | 0 | 0 | 0 |
| 0 | 0 | 0 | 0 | 0 |
| 0 | 0 | 0 | 0 | 0 |
| 0 | 0 | 0 | 0 | 0 |
| 0 | 0.000763748 | 0 | 0 | 0.004984929 |
| 0 | 0 | 0 | 0 | 0 |
| 0 | 0 | 0 | 0 | 0 |
| 0.001652598 | 0 | 0 | 0 | 0 |
| 0 | 0 | 0 | 0 | 0 |

| | | | | |
|---|---|---|---|---|
| 0 | 0 | 0 | 0 | 0 |
| 0 | 0 | 0 | 0 | 0 |
| 0 | 0.225805373 | 0 | 0 | 0 |
| 0 | 0 | 0 | 0 | 0 |
| 0 | 0 | 0 | 0 | 0 |
| 0 | 0 | 0 | 0 | 0.008305184 |
| 0 | 0 | 0 | 0 | 0 |
| 0 | 0 | 0 | 0 | 0 |
| 0 | 0 | 0 | 0 | 0 |
| 0 | 0 | 0 | 0 | 0 |
| 0.003234988 | 0.037693407 | 0 | 0.004018048 | 0.023691025 |
| 0.005457458 | 0 | 0 | 0 | 0 |
| 0 | 0 | 0 | 0 | 0 |
| 0.007538761 | 0.004324173 | 0 | 0.001423519 | 0.004673184 |
| 0.002267745 | 0 | 0.0040268 | 0 | 0 |
| 0 | 0 | 0 | 0 | 0 |
| 0 | 0 | 0 | 0 | 0.017179568 |
| 0 | 0 | 0 | 0 | 0 |
| 0 | 0 | 0 | 0 | 0 |
| 0 | 0 | 0 | 0 | 0 |
| 0 | 0 | 0.217617351 | 0 | 0 |
| 0 | 0 | 0.418458017 | 0.307939935 | 0 |
| 0 | 0 | 0 | 0 | 0.01120327 |
| 0 | 0 | 0 | 0 | 0 |
| 0 | 0 | 0 | 0 | 0 |
| 0 | 0 | 0 | 0 | 0 |
| 0 | 0.008549706 | 0 | 0 | 0 |
| 0 | 0 | 0 | 0 | 0 |
| 0 | 0 | 0 | 0 | 0 |
| 22.62821209 | 0.667016222 | 0.144622001 | 0.091809527 | 0.358358359 |
| 0 | 0 | 0.000787485 | 0 | 0.001501761 |
| 0 | 0 | 0 | 0 | 0 |
| 0 | 0 | 0 | 0 | 0 |
| 0.002153566 | 0 | 0 | 0 | 0 |
| 0.001840049 | 0 | 0 | 0 | 0 |
| 0 | 0 | 0 | 0 | 0 |
| 0 | 0 | 0 | 0 | 0 |
| 0.001453731 | 0 | 0.001014109 | 0 | 0 |
| 0 | 0 | 0 | 0.014937568 | 0 |
| 0 | 0 | 0 | 0 | 0 |
| 0 | 0.0708897 | 0.046759397 | 0.000975239 | 0.011930003 |
| 0 | 0 | 7.576844763 | 0.62654401 | 0.365545145 |
| 0.002092689 | 0 | 0.000774158 | 0 | 0 |
| 0.003839489 | 0 | 0 | 0 | 0 |
| 0.001133726 | 0 | 0 | 0 | 0 |
| 0 | 0 | 0 | 0 | 0 |
| 0 | 0.006145449 | 0.001287744 | 0.004299051 | 0 |
| 0 | 0 | 0 | 0 | 0 |

| | | | | |
|---|---|---|---|---|
| 0 | 0.001624811 | 0 | 0.226317145 | 0 |
| 0 | 0 | 0 | 0 | 0.002007792 |
| 1.020890198 | 0 | 0.582610209 | 0.004818351 | 0 |
| 0 | 0 | 0 | 0 | 0 |
| 0 | 0 | 0 | 0 | 0 |
| 0 | 0 | 0 | 0 | 0 |
| 0 | 0 | 0 | 0 | 0 |
| 0 | 0 | 0 | 0 | 0 |
| 0.000907705 | 0 | 0 | 0 | 0.004602089 |
| 0.001915547 | 0 | 0 | 0 | 0 |
| 0 | 0 | 0 | 0 | 0 |
| 0 | 0 | 0 | 0 | 0 |
| 0 | 0.132868229 | 0 | 0 | 0 |
| 0.002354884 | 0.002204889 | 0 | 0 | 0.053300559 |
| 0 | 0 | 0 | 0 | 0 |
| 0.02039667 | 0 | 0.014103709 | 0.040893736 | 0.018882905 |
| 0.028726928 | 0.014854445 | 0.000968384 | 0.74838113 | 0.001231161 |
| 0.00640216 | 0 | 0.001176549 | 0 | 0 |
| 0.001404627 | 0 | 0 | 0 | 0 |
| 0 | 0 | 0 | 0.002903646 | 0 |
| 0.008354793 | 0 | 0 | 0 | 0 |
| 0.003549061 | 0.002215335 | 0.000481404 | 0 | 0 |
| 0 | 0 | 0 | 0 | 0 |
| 0 | 0 | 0 | 0.034740249 | 0.452109536 |
| 0.013821207 | 0 | 0 | 0 | 0 |
| 0.007837238 | 0 | 0 | 0 | 0 |
| 0 | 0.00700434 | 0.00386374 | 0 | 0.00442841 |
| 0 | 0 | 0 | 0 | 0 |
| 0 | 0 | 0 | 0 | 0 |
| 0 | 0 | 0.000601099 | 0 | 0 |
| 0.003610424 | 0 | 0.02110279 | 0 | 0.027140454 |
| 0.001468009 | 0.003054452 | 0.005177237 | 0.053276136 | 0 |
| 0.028653731 | 0.014832083 | 0.011944387 | 0.012478074 | 0.016310417 |
| 0 | 0 | 0 | 0.002721219 | 0 |
| 0.003097695 | 0 | 0.000969644 | 0.314923283 | 0 |
| 0 | 0 | 0 | 0 | 0 |
| 0.001684875 | 0.001051704 | 0.003770922 | 0 | 0 |
| 0.014164738 | 0.052638799 | 0 | 0.003260414 | 0.025051957 |
| 0 | 0 | 0.018422215 | 0.085484582 | 0 |
| 0 | 0 | 0 | 0 | 0 |
| 0 | 0 | 0 | 0.001278077 | 0 |
| 0 | 0 | 0 | 0 | 0 |
| 0.006157949 | 0 | 0 | 0 | 0 |
| 0 | 0 | 0 | 0.00209601 | 0.00124166 |
| 0 | 0.415562245 | 0 | 0 | 0 |
| 0.0560175 | 0 | 0.001022856 | 0 | 0.001300413 |
| 0 | 0 | 0 | 0 | 0 |
| 0 | 0 | 0 | 0 | 0 |

| | | | | |
|---|---|---|---|---|
| 0 | 0 | 0 | 0 | 0 |
| 0.002917631 | 1.040602475 | 0.954195728 | 4.539160479 | 0.083173778 |
| 0 | 0 | 0 | 0 | 0 |
| 0 | 0.035974264 | 0 | 0 | 0 |
| 0 | 0 | 0 | 0 | 0 |
| 0 | 0 | 0 | 0 | 0 |
| 0 | 0 | 0 | 0.000967083 | 0 |
| 0 | 0 | 0 | 0 | 0 |
| 0 | 0.003590792 | 0 | 0 | 0 |
| 0 | 0 | 0.001327327 | 0 | 0 |
| 0.001334227 | 0.015615537 | 0.001085868 | 0 | 0.001409285 |
| 0.017886322 | 0.017677437 | 0.077030155 | 0.033555229 | 0.008996442 |
| 0 | 0 | 0.010784501 | 0 | 0.187573194 |
| 0 | 0 | 0 | 0 | 0 |
| 0.005166648 | 0 | 0.000622949 | 0 | 0 |
| 0 | 0.016808851 | 0 | 0 | 0 |
| 0.001397128 | 0.017877882 | 0.073340399 | 0.050229279 | 0.012438251 |
| 0 | 0 | 0.044281846 | 0 | 0 |
| 0 | 0 | 0 | 0 | 0 |
| 0.002502612 | 0 | 0 | 0.001885606 | 0 |
| 0.013867621 | 0.023832687 | 0.324955717 | 0.11513232 | 0.003478502 |
| 0 | 0 | 0 | 0 | 0 |
| 0 | 0 | 0 | 0 | 0 |
| 0 | 0 | 0 | 0 | 0 |
| 0 | 0 | 0.910715853 | 0 | 0 |
| 0 | 0.001237089 | 0 | 0 | 0 |
| 0 | 0 | 0.184751735 | 0.002276967 | 0.05257478 |
| 0.096283567 | 0.040624592 | 0.027418526 | 0.018723074 | 0.057536645 |
| 0 | 0 | 0 | 0 | 0 |
| 0 | 0.072351152 | 0 | 0 | 0 |
| 0 | 0 | 0.003176034 | 0 | 0 |
| 0 | 0.183230221 | 0 | 0 | 0 |
| 0 | 0 | 0 | 0 | 0 |
| 0 | 0 | 0 | 0 | 0 |
| 0 | 0 | 0.015612123 | 2.960745012 | 0 |
| 1.118649757 | 0.341346856 | 0.058268935 | 0.065261542 | 0.247940697 |
| 0 | 0.015172273 | 0 | 0 | 0 |
| 0 | 0 | 0 | 0 | 0 |
| 0.003069772 | 0.013855318 | 0.014413559 | 0.012098431 | 0.004275776 |
| 0 | 0 | 0 | 0 | 0 |
| 0 | 0.413625917 | 0 | 0 | 0 |
| 0 | 0 | 0 | 0 | 0 |
| 0 | 0.001074374 | 0 | 0.003841404 | 0.025971719 |
| 0 | 0 | 0.00586061 | 0 | 0 |
| 0 | 0 | 0 | 0 | 0 |
| 0 | 0 | 0 | 1.174411795 | 0.004490695 |
| 0 | 0 | 0 | 0 | 0 |
| 0 | 0.019002553 | 0 | 0 | 0 |

| | | | | |
|---|---|---|---|---|
| 0 | 0.015759702 | 0 | 0 | 0 |
| 0 | 0 | 0 | 0 | 0 |
| 0 | 0 | 0 | 0.001071428 | 0 |
| 0 | 0.00417102 | 0.008429374 | 8.05284624 | 2.347511173 |
| 0 | 0 | 0 | 0 | 0 |
| 0 | 0 | 0 | 0 | 0 |
| 0 | 0 | 0 | 0 | 0.001563416 |
| 0 | 0 | 0 | 0.003306405 | 0 |
| 0.05123371 | 0 | 0.05153376 | 1.250929706 | 0 |
| 0 | 0.290400771 | 0.060634835 | 0.027643002 | 1.127540501 |
| 0 | 0 | 0 | 0 | 0 |
| 0.002720944 | 0 | 0.004168385 | 0 | 0.001738898 |
| 0 | 0 | 0 | 0 | 0 |
| 0 | 0 | 0 | 0 | 0 |
| 0.048208546 | 0 | 0.007416987 | 0.010304327 | 0.017697632 |
| 0 | 0 | 0 | 0 | 0 |
| 0 | 0 | 0 | 0.031696904 | 0.358129613 |
| 0 | 0 | 0 | 0 | 0 |
| 0 | 0 | 0 | 0 | 0 |
| 0 | 0 | 0 | 0 | 0 |
| 0 | 0 | 0 | 0 | 0 |
| 0 | 0 | 0 | 0 | 0 |
| 0 | 0 | 0 | 0 | 0 |
| 0.026717285 | 0 | 0.12042038 | 0.001777739 | 0.663157479 |
| 0.009590583 | 0 | 0.001097627 | 0 | 0 |
| 0 | 0 | 1.912137859 | 0.484471628 | 0.016592357 |
| 0 | 0 | 0 | 0 | 0 |
| 0 | 0 | 0.86688714 | 2.336302017 | 0 |
| 0 | 0 | 0 | 0 | 0 |
| 0 | 0 | 0 | 0 | 0 |
| 0 | 0 | 0.003545833 | 0.69503284 | 0 |
| 0 | 0 | 0 | 0 | 0 |
| 0.006039622 | 0.036664621 | 0.00212036 | 0.001103172 | 0.006718905 |
| 0 | 0 | 0 | 0 | 0 |
| 0 | 0 | 0 | 0 | 0 |
| 0 | 0 | 0 | 0 | 0 |
| 0 | 0 | 0.73206017 | 0.945220847 | 0.753846373 |
| 0 | 0 | 0 | 0 | 0 |
| 0 | 0 | 0 | 0 | 0 |
| 0 | 0 | 0 | 0 | 0 |
| 0.003224438 | 0.120187173 | 0.43524661 | 0.01269424 | 0.022427509 |
| 0 | 0 | 0 | 0 | 0 |
| 20.38014335 | 0.429548439 | 0.189334592 | 0 | 1.198030733 |
| 0 | 0 | 0.004898665 | 0 | 0 |
| 0 | 0 | 0 | 0 | 0 |
| 0 | 0 | 0 | 0 | 0 |
| 0 | 0 | 0 | 0 | 0 |
| 0 | 5.227043707 | 0 | 0 | 0.002228135 |

| | | | | |
|---|---|---|---|---|
| 0 | 0 | 0 | 0 | 0 |
| 0 | 0 | 0 | 2.828222809 | 0 |
| 0.046181109 | 0.558414683 | 0.02318446 | 0 | 0.008543678 |
| 0 | 0 | 0 | 0 | 0.004386771 |
| 0.268995911 | 0 | 0 | 0 | 0 |
| 0 | 0.308345916 | 0 | 0 | 0.065452944 |
| 0 | 0 | 0 | 0 | 0 |
| 0 | 0 | 0.001839369 | 0 | 0 |
| 0 | 0 | 0 | 0.053462917 | 0 |
| 0 | 0 | 0 | 0 | 0 |
| 0.0220628 | 1.026910978 | 0.008483116 | 0.006473223 | 0.024955419 |
| 1.938957205 | 1.501297455 | 1.194562715 | 0.453517049 | 1.441519712 |
| 0 | 0 | 0 | 0 | 0 |
| 0.006187129 | 0.037215879 | 0.001983654 | 6.408715688 | 0 |
| 0.378826735 | 0.856216598 | 0.062648052 | 0.003096769 | 0.386137078 |
| 0 | 0 | 0 | 0 | 0 |
| 0 | 0 | 0 | 0 | 0 |
| 0 | 0 | 0 | 0 | 0 |
| 0 | 0 | 0 | 0 | 0 |
| 0 | 0 | 0 | 0.001999587 | 0.01226492 |
| 0.003640133 | 0.713101665 | 0.538353107 | 0.06561481 | 0.045699515 |
| 0.987736195 | 0 | 0 | 0 | 0.00259286 |
| 2.761282188 | 3.906645481 | 0.295489137 | 0.558893993 | 0.811488826 |
| 0 | 0 | 0 | 0 | 0 |
| 0 | 0 | 0 | 0 | 0 |
| 0.003664056 | 0 | 8.559367238 | 1.016655359 | 0 |
| 0.526631844 | 0 | 0 | 0 | 0 |
| 0 | 0 | 0.002842687 | 0.014677286 | 0 |
| 0 | 0 | 0 | 0 | 0.012296385 |
| 0 | 0 | 0 | 0 | 0 |
| 0 | 0 | 0 | 0 | 0.011050411 |
| 0 | 0 | 0.322646532 | 0 | 0.263675269 |
| 0.043355544 | 0.321410902 | 0.003775365 | 0 | 0.04996747 |
| 0 | 0 | 0.002768277 | 0.001109006 | 0 |
| 0 | 0 | 0.003066816 | 0.013437863 | 0 |
| 0.014884893 | 0 | 0.002633511 | 0.029578164 | 0.135934009 |
| 0.001094024 | 0 | 0 | 0 | 0 |
| 0 | 0 | 0 | 0 | 0 |
| 0.016661012 | 0 | 0.003235827 | 0 | 0 |
| 0 | 0 | 0 | 0 | 0 |
| 0 | 0 | 0 | 0 | 0.006191211 |
| 0 | 0 | 0 | 0 | 0 |
| 0 | 0 | 0 | 0 | 0 |
| 0 | 0 | 0 | 0 | 0 |
| 0 | 0.017505698 | 0.00150161 | 0.008808612 | 0.00445452 |
| 0 | 0 | 0 | 0 | 0 |
| 0 | 0 | 0 | 0 | 0 |
| 0.020253358 | 1.608200065 | 0.023185308 | 2.495459388 | 0.151797668 |

| | | | | |
|---|---|---|---|---|
| 0 | 0 | 0 | 0 | 0 |
| 0 | 0 | 0 | 0 | 0 |
| 0 | 0 | 0 | 0 | 0 |
| 0 | 0 | 0 | 0 | 0.001138971 |
| 0 | 0 | 0 | 0 | 0 |
| 0 | 0.140788844 | 0 | 0 | 0 |
| 0 | 0 | 0.061510707 | 0.003808958 | 0 |
| 0.0085374 | 0 | 0 | 0 | 0 |
| 0 | 0 | 0 | 0 | 0 |
| 0 | 0 | 0 | 0 | 0 |
| 0 | 0.317252922 | 0 | 0 | 0.006976816 |
| 0 | 0 | 0 | 0 | 0 |
| 0 | 0 | 0.001497634 | 0 | 0 |
| 0 | 0 | 0 | 0 | 0.393473355 |
| 0 | 0 | 0 | 0 | 0 |
| 0 | 0 | 0 | 0 | 0 |
| 0 | 0 | 0 | 0.020931208 | 0.219349548 |
| 0 | 0 | 0 | 0.041424583 | 0.001232008 |
| 0 | 0 | 0 | 0 | 0.009815597 |
| 0 | 0.007058393 | 0.000642066 | 0.011023706 | 0.001111068 |
| 0 | 0 | 0 | 0 | 0 |
| 0 | 0 | 0 | 0 | 0 |
| 0 | 0 | 0 | 0.00097868 | 0 |
| 0 | 0 | 0 | 0.702685743 | 0 |
| 0 | 0 | 0 | 0 | 0 |
| 0 | 0 | 0 | 0 | 0 |
| 0 | 0 | 0 | 0 | 0 |
| 0 | 0 | 0 | 0 | 0 |
| 0 | 0 | 0 | 0 | 0 |
| 0.031173936 | 0 | 0.008313686 | 0.557828823 | 0.793452648 |
| 0 | 0 | 0 | 0 | 0 |
| 0 | 0 | 0 | 0 | 0 |
| 0 | 0 | 0 | 0 | 0 |
| 0 | 0 | 0 | 0 | 0 |
| 0 | 0 | 0.000340017 | 0.001297286 | 0.000672439 |
| 0 | 0 | 0 | 0 | 0 |
| 0 | 0 | 0.31060959 | 0 | 0.117638013 |
| 0.000593032 | 0.00444207 | 0.004585103 | 0.002934816 | 0 |
| 0 | 0 | 0 | 0 | 0 |
| 0 | 0 | 0 | 0 | 0 |
| 0 | 0 | 0 | 0 | 0 |
| 0 | 0 | 0 | 0 | 0 |
| 0 | 0 | 0 | 0 | 0 |
| 0 | 0 | 0 | 0 | 0.17101987 |
| 0.315984447 | 0 | 0.24373878 | 0 | 0 |
| 0 | 0 | 0 | 0 | 0 |
| 0 | 0 | 0 | 0 | 0.000510153 |
| 0 | 0 | 0 | 0 | 0 |

| | | | | |
|---|---|---|---|---|
| 0 | 0 | 0 | 0 | 0 |
| 0 | 0 | 0 | 0 | 0.00508381 |
| 0 | 0 | 0 | 0 | 0 |
| 0 | 0 | 0.005160943 | 0 | 0.037206659 |
| 0 | 0 | 0 | 0 | 0 |
| 0 | 0 | 0 | 0 | 0 |
| 0 | 0 | 0 | 0 | 0 |
| 0 | 0 | 0 | 0 | 0 |
| 0.07320119 | 0.781365712 | 0.054083617 | 0 | 0.024189238 |
| 0 | 0 | 0.002680203 | 0 | 0 |
| 0 | 0 | 0 | 0 | 0 |
| 0 | 0 | 0 | 0 | 0 |
| 0 | 0 | 0 | 0 | 0 |
| 0 | 0 | 0 | 0 | 0 |
| 0 | 0 | 0 | 0.050474726 | 1.804819189 |
| 0 | 0.855030825 | 0 | 0 | 0.000760952 |
| 0 | 0 | 0 | 0 | 0 |
| 0 | 0 | 0 | 0 | 0 |
| 0 | 0 | 0 | 0 | 0 |
| 0 | 0 | 0 | 0 | 0 |
| 0 | 0 | 0 | 0 | 0.004718403 |
| 0 | 0 | 0 | 0 | 0 |
| 0 | 0 | 0 | 0 | 0 |
| 0 | 0 | 0 | 0 | 0 |
| 0 | 0.005103131 | 0.002152642 | 0.002519928 | 0 |
| 0 | 0 | 3.739548546 | 3.119874402 | 5.575130606 |
| 0 | 0 | 0.004803932 | 0 | 0.553407846 |
| 0.001044827 | 0 | 0 | 0 | 0 |
| 0.005255638 | 0 | 0.009410124 | 5.934273286 | 0 |
| 0 | 0 | 0 | 0 | 0.091863699 |
| 0 | 0 | 0 | 0 | 0 |
| 0 | 0 | 0 | 0 | 0 |
| 0.007102845 | 0 | 0 | 0 | 0 |
| 0 | 0 | 0.001985758 | 0 | 0.003410429 |
| 0.003808657 | 0.005283056 | 0 | 1.142976736 | 0 |
| 0.001046989 | 0.941089067 | 0 | 0.000522491 | 0.004152725 |
| 0 | 0 | 0 | 0 | 0 |
| 0.268873358 | 0 | 2.379024121 | 0.781531062 | 2.375153263 |
| 0 | 0 | 0 | 6.158026733 | 0 |
| 0 | 0 | 0 | 0 | 0 |
| 0 | 0 | 0 | 0 | 0.137716883 |
| 0 | 0 | 0 | 0 | 0 |
| 0 | 0 | 17.77184761 | 0.09258121 | 2.243015338 |
| 0 | 0 | 0 | 0 | 0 |
| 0 | 0 | 0 | 0 | 0 |
| 0.001700385 | 0 | 0 | 0 | 0 |
| 0 | 0 | 0 | 0 | 0 |
| 0.045148613 | 0.00281819 | 0.029946721 | 0.014983144 | 1.608055505 |

| | | | | |
|---|---|---|---|---|
| 0 | 0 | 0.000685009 | 0 | 0 |
| 0 | 0 | 0 | 0 | 0 |
| 0 | 0 | 0 | 0 | 0 |
| 0 | 0 | 0 | 0 | 0 |
| 0 | 0.016320051 | 0 | 0 | 0 |
| 0.042608328 | 0 | 0 | 0 | 0 |
| 0 | 0 | 0 | 0 | 0.000644249 |
| 0 | 0 | 0 | 0 | 0 |
| 0 | 0 | 0 | 0 | 0 |
| 0 | 0 | 0 | 0 | 0 |
| 0 | 0 | 0 | 0 | 0 |
| 0.011441926 | 0 | 0.256962887 | 0.628177038 | 3.102126041 |
| 0.342753508 | 0 | 0.035839277 | 0.009215747 | 0.508803114 |
| 0 | 0 | 0 | 0 | 0 |
| 0.134710493 | 0.009195093 | 0.002681716 | 0.102467502 | 0 |
| 0 | 0 | 0 | 0 | 0 |
| 0 | 0 | 0 | 0 | 0 |
| 0 | 0.140875506 | 0 | 0 | 0 |
| 0 | 0 | 0 | 0 | 0.004101642 |
| 0 | 0 | 0 | 0.009432307 | 0.008186519 |
| 0 | 0 | 0 | 0 | 0.159092463 |
| 0 | 0 | 0 | 0 | 0.062893114 |
| 0 | 0.399673521 | 0.170206701 | 1.809892745 | 0.046264161 |
| 0 | 0 | 0 | 0 | 0 |
| 0 | 0 | 0 | 0 | 0 |
| 0 | 0 | 0 | 0 | 0 |
| 0 | 0 | 0.180463192 | 0 | 0 |
| 0 | 0 | 0 | 0 | 0 |
| 0 | 0 | 0 | 0 | 0 |
| 0 | 0 | 0 | 0 | 0 |
| 0 | 0 | 0 | 0 | 0 |
| 0.003521266 | 0.070335522 | 0.124048258 | 0.110414425 | 0.037952624 |
| 0 | 0 | 0 | 0 | 0 |
| 0.010429328 | 0 | 0 | 0.001170528 | 0.01789869 |
| 0.000976617 | 0.922742667 | 0 | 0.000758136 | 0.002554333 |
| 0.002517956 | 0 | 0 | 0.086493585 | 0.003546141 |
| 0 | 0 | 0 | 0 | 0 |
| 0 | 0 | 0 | 0 | 0 |
| 0.000560995 | 0 | 0 | 0 | 0 |
| 0 | 0 | 0 | 0 | 0 |
| 0 | 0 | 0 | 0.023738084 | 0.111158771 |
| 0 | 0 | 0 | 0 | 0 |
| 0 | 0 | 0 | 0 | 0 |
| 0 | 0 | 0 | 0 | 0.009757477 |
| 0.01330858 | 0 | 0.001696462 | 0.00056013 | 0.001548476 |
| 0 | 0 | 0 | 0 | 0 |
| 0.014300471 | 0 | 0 | 0 | 2.104922083 |
| 0 | 0 | 0 | 0 | 0 |

| | | | | |
|---|---|---|---|---|
| 0 | 0 | 0 | 0 | 0 |
| 0 | 0 | 0 | 0 | 0 |
| 0 | 0 | 0 | 0 | 0.004203969 |
| 0 | 0 | 0 | 0 | 0 |
| 0 | 0 | 0 | 0.001175017 | 0 |
| 0 | 0 | 0 | 0 | 0 |
| 0 | 0 | 0 | 0.150921632 | 0 |
| 0 | 0.04281917 | 0.000637317 | 0.048874991 | 0.044239997 |
| 0 | 0 | 0 | 0 | 0 |
| 0 | 0 | 0 | 0 | 0 |
| 0 | 0 | 0 | 0 | 0 |
| 0 | 0 | 0 | 0 | 0 |
| 0 | 0 | 0 | 0 | 0 |
| 0.116527113 | 0 | 0 | 0 | 0 |
| 0 | 0 | 0 | 0 | 0 |
| 0 | 0 | 0 | 0 | 0 |
| 0 | 0 | 0 | 0 | 0 |
| 0 | 0 | 0 | 0 | 0 |
| 0 | 0 | 0 | 7.76762184 | 5.184073245 |
| 0 | 0 | 0 | 0 | 0 |
| 0 | 0 | 0 | 0 | 0 |
| 0 | 0 | 0 | 0 | 0 |
| 0 | 0 | 0 | 0 | 0 |
| 0 | 0 | 0 | 0 | 0 |
| 0 | 0 | 0 | 0.020195678 | 0 |
| 0.005075333 | 0.139766417 | 0 | 0.002085838 | 0 |
| 0 | 0 | 0 | 0 | 0 |
| 0.016510436 | 0.014045162 | 0.060050792 | 0.098167921 | 0.313950455 |
| 0 | 0 | 0 | 0 | 0 |
| 0 | 0 | 1.672800405 | 0.318747048 | 1.661724278 |
| 0 | 0 | 0 | 0.076096243 | 0 |
| 0 | 0 | 0 | 0 | 0 |
| 0 | 0 | 0 | 0 | 0.005400917 |
| 0 | 0 | 0 | 0 | 0 |
| 0.001251444 | 0.018357142 | 0 | 0 | 0.003587864 |
| 0 | 0 | 0 | 0 | 0 |
| 0 | 0 | 0 | 0 | 0 |
| 0 | 0.015733282 | 0 | 0.003720414 | 0.003214085 |
| 0 | 0 | 0 | 0 | 0 |
| 0 | 0 | 0 | 0 | 0 |
| 16.20299878 | 17.01722198 | 3.215289502 | 0.771441032 | 4.642397028 |
| 0 | 3.28602849 | 1.2139701 | 0.040396504 | 1.781012613 |
| 0 | 0 | 0 | 0 | 0 |
| 0 | 0 | 0 | 0 | 0.098233533 |
| 0 | 0 | 0 | 0 | 0 |
| 0 | 0 | 0 | 0 | 0 |
| 0 | 0 | 0 | 0 | 0 |
| 0 | 0.262398741 | 0 | 0 | 0.001700584 |

| | | | | |
|---|---|---|---|---|
| 0 | 0 | 0 | 0 | 0 |
| 0.001159221 | 0 | 0 | 0 | 0.002273948 |
| 0 | 0 | 0 | 0 | 0 |
| 0 | 0 | 0 | 0 | 0 |
| 0 | 0 | 13.36848992 | 0.683433702 | 2.70922881 |
| 0 | 0 | 0 | 0 | 0 |
| 0 | 0 | 0 | 0 | 0 |
| 0 | 0 | 0 | 0 | 0 |
| 0.002025645 | 0.065027014 | 0.796500869 | 0 | 0.001921273 |
| 0 | 0 | 0 | 0 | 0 |
| 0 | 0.021868262 | 0 | 0 | 0 |
| 0 | 0 | 0 | 0 | 0 |
| 0 | 0 | 0.179675338 | 0.099681006 | 0.184580574 |
| 1.106950791 | 0.045595935 | 0.009861275 | 0.002418704 | 0.010447654 |
| 0 | 0 | 0 | 0 | 0 |
| 0 | 0 | 0.444486982 | 0.294076879 | 2.237623262 |
| 0 | 0 | 0.011093425 | 0 | 0.015731034 |
| 0 | 0 | 0 | 0 | 0 |
| 0 | 0 | 0 | 0 | 0 |
| 0 | 0 | 0 | 0 | 0 |
| 0 | 0 | 0 | 0 | 0 |
| 0 | 0 | 0 | 0 | 0 |
| 0 | 0 | 0 | 0 | 0 |
| 0 | 0 | 0 | 0 | 0 |
| 0 | 0 | 0 | 0 | 0 |
| 0 | 0 | 0 | 0 | 0 |
| 0 | 0 | 0.004982098 | 0.158488254 | 0.029244289 |
| 0 | 0.264023348 | 0.001315726 | 0 | 0 |
| 0 | 0 | 0 | 0 | 0 |
| 0 | 0 | 0 | 0 | 0 |
| 0 | 0 | 0 | 0 | 0 |
| 0 | 0 | 0 | 0 | 0 |
| 0 | 0 | 0.00193397 | 0 | 0.422442843 |
| 0 | 0 | 0 | 0 | 0 |
| 0 | 0 | 0 | 1.000626244 | 0 |
| 0 | 0.006367343 | 0 | 0 | 0 |
| 0 | 0 | 0.012728373 | 0 | 0 |
| 0 | 0 | 0.000914595 | 0.007477512 | 0.004263509 |
| 0 | 0 | 0 | 0 | 0 |
| 0 | 0 | 0 | 0 | 0 |
| 3.584119696 | 0 | 1.489573928 | 0.330715066 | 0 |
| 0 | 0 | 0 | 0 | 0 |
| 0 | 0 | 0 | 0 | 0 |
| 0 | 0 | 0 | 0 | 0 |
| 0 | 0 | 0 | 0 | 0 |
| 0 | 0 | 0 | 0 | 0 |
| 0 | 0 | 0 | 0 | 0 |
| 0 | 0 | 0 | 0 | 0 |

| | | | | |
|---|---|---|---|---|
| 0 | 0 | 0.001440597 | 0.169576185 | 0.646467848 |
| 0 | 0 | 0 | 0 | 0 |
| 0 | 0 | 0 | 0 | 0 |
| 0 | 0 | 0 | 0 | 0 |
| 0 | 0.042358348 | 0.007519323 | 0.000556466 | 0.302093026 |
| 0 | 0 | 0 | 0 | 0 |
| 0 | 1.916154893 | 0.008755873 | 0.007481726 | 0.040765159 |
| 0 | 0 | 0 | 0 | 0 |
| 0 | 0 | 0 | 0 | 0 |
| 0 | 0 | 0 | 0 | 0 |
| 0 | 0 | 0 | 0 | 0 |
| 0 | 0 | 0 | 0 | 0.000473229 |
| 0 | 0 | 0 | 0 | 0 |
| 0 | 0 | 0 | 0 | 0 |
| 0 | 0 | 0 | 0 | 0 |
| 0 | 0 | 0 | 0 | 0 |
| 0 | 0 | 0 | 0 | 0 |
| 0.000718276 | 0 | 0 | 0 | 0 |
| 0 | 0 | 0 | 0 | 0 |
| 0 | 0 | 0 | 0 | 0 |
| 0.003841013 | 0.004084753 | 0 | 0.006129111 | 0.269785906 |
| 0 | 0 | 0 | 0 | 0 |
| 0 | 0 | 0.002298666 | 0.005298682 | 0.023487614 |
| 0.000837167 | 0.007838431 | 0.00124911 | 0.008773404 | 0.001010585 |
| 0 | 0 | 0 | 0 | 0 |
| 0 | 0 | 0 | 0 | 0.001788395 |
| 0 | 0 | 0 | 0 | 0 |
| 0 | 0 | 0 | 0 | 0 |
| 0 | 0 | 0 | 0 | 0 |
| 0.204427835 | 0 | 0.080735855 | 0.093533227 | 0 |
| 0 | 0 | 0 | 0 | 0 |
| 0 | 0 | 0 | 0 | 0 |
| 0 | 0 | 0 | 0 | 0 |
| 0 | 0 | 0 | 0 | 0 |
| 0 | 0 | 0 | 0 | 0 |
| 0 | 0 | 0 | 0 | 0 |
| 0.039805471 | 0 | 0 | 0 | 0.04215013 |
| 0 | 0 | 0 | 0 | 0 |
| 0 | 0 | 0 | 0 | 0 |
| 0 | 0 | 0 | 0 | 0 |
| 0 | 0 | 0 | 0 | 0 |
| 0 | 0 | 0 | 0 | 0 |
| 0 | 0 | 0 | 0 | 0 |
| 0 | 0 | 0 | 0 | 0 |
| 0 | 0 | 0 | 0 | 0 |
| 0 | 0 | 0 | 0 | 0 |
| 0 | 0 | 0 | 0 | 0 |
| 0 | 0 | 0 | 0.262169969 | 0 |

| | | | | |
|---|---|---|---|---|
| 0.007056432 | 0 | 0 | 0 | 0 |
| 0 | 0 | 0 | 0 | 0 |
| 0 | 0 | 0 | 0 | 0 |
| 0 | 0 | 0 | 0 | 0 |
| 0 | 0 | 0 | 0 | 0 |
| 0 | 0 | 0 | 0 | 0 |
| 0 | 0.021469216 | 0 | 0 | 0 |
| 0 | 0 | 3.523828547 | 0.314851253 | 6.689379293 |
| 0.017203351 | 0.004707237 | 0.035098497 | 0.078756381 | 0.005689593 |
| 0.083014892 | 0 | 0 | 0 | 0 |
| 0 | 0 | 0 | 0 | 0 |
| 0 | 0 | 0 | 0 | 0 |
| 0 | 0 | 0 | 0 | 0.046651686 |
| 0.003140369 | 0.434952849 | 0 | 1.445225923 | 0 |
| 0 | 0.003043994 | 0.000466924 | 0 | 0 |
| 0 | 0 | 0 | 0 | 0 |
| 0 | 0.05745599 | 0 | 0 | 0 |
| 0 | 0 | 0 | 0 | 0 |
| 0 | 0 | 0 | 0 | 0 |
| 0.002720867 | 0 | 0 | 0 | 0.020528083 |
| 0 | 0 | 0 | 0 | 0 |
| 0 | 0 | 0 | 0 | 0 |
| 0 | 0 | 0 | 0 | 0 |
| 0 | 0 | 0 | 0 | 0 |
| 0 | 0 | 0.066887716 | 0.011979736 | 0.068091625 |
| 0 | 0 | 0 | 0 | 0 |
| 0 | 0 | 0 | 0 | 0 |
| 0 | 0 | 0 | 0 | 0 |
| 0 | 0 | 0 | 0 | 0 |
| 0 | 0 | 0 | 0 | 0 |
| 0 | 0 | 0 | 0 | 0 |
| 0 | 0 | 0 | 0 | 0 |
| 0 | 0 | 0 | 0 | 0 |
| 0 | 0 | 0 | 0 | 0 |
| 0 | 0 | 0.003846123 | 0.115560368 | 0 |
| 0 | 0 | 0 | 0 | 0 |
| 0 | 0 | 0 | 0 | 0 |
| 0 | 0.092271395 | 0 | 0 | 0.004992961 |
| 0 | 0 | 0 | 0 | 0 |
| 0 | 0.024325029 | 0 | 0 | 0 |
| 0 | 0 | 0 | 0 | 0 |
| 0.060306297 | 0 | 0.093630693 | 0 | 0 |
| 0 | 0 | 0 | 0 | 0 |
| 0 | 0 | 0 | 0 | 0 |
| 0 | 0 | 0 | 0 | 0 |
| 0 | 0 | 0 | 0 | 0 |
| 0 | 0 | 0 | 0 | 0 |
| 0 | 0 | 0 | 0 | 0 |

| | | | | |
|---|---|---|---|---|
| 0 | 0 | 0 | 0 | 0 |
| 0.023577883 | 0.010149922 | 0.003198165 | 0.000788931 | 0.002126473 |
| 0 | 0 | 0 | 0 | 0 |
| 0 | 0 | 0 | 0 | 0 |
| 0.05816273 | 0 | 0.014031208 | 2.671335034 | 0.240821792 |
| 0 | 0 | 0 | 0 | 0 |
| 0 | 0 | 0 | 0 | 0 |
| 0.003628714 | 0.062229392 | 0.004196722 | 0.000667166 | 0.045187282 |
| 0 | 0 | 0 | 0 | 0 |
| 0 | 0 | 0 | 0 | 0 |
| 0 | 0 | 0 | 0 | 0 |
| 0 | 0 | 0 | 0 | 0 |
| 0 | 0 | 0 | 0 | 0 |
| 0 | 0 | 0 | 0 | 0 |
| 0.000537787 | 0 | 0 | 0 | 0.001947566 |
| 0.020539338 | 0 | 0.012941459 | 0 | 0 |
| 0 | 0 | 0 | 0 | 0 |
| 0 | 0 | 0 | 0 | 0 |
| 0 | 0 | 0 | 0 | 0 |
| 0 | 0 | 0 | 0 | 0 |
| 0 | 0 | 0 | 0 | 0 |
| 0 | 0 | 0 | 0 | 0 |
| 0 | 0 | 0 | 0 | 0 |
| 0 | 0.169197775 | 0.000538062 | 0 | 0.004588954 |
| 0 | 0 | 0.060558605 | 0 | 0 |
| 0 | 0 | 0 | 0 | 0 |
| 0.003476885 | 1.016824464 | 0.002583622 | 0 | 0.116971896 |
| 0 | 0 | 0 | 0 | 0 |
| 0 | 0 | 0 | 0 | 0 |
| 0 | 0 | 0 | 0 | 0 |
| 0 | 0 | 0 | 0 | 0 |
| 0 | 0 | 0 | 0 | 0 |
| 0 | 0 | 0 | 0 | 0 |
| 0 | 0 | 0 | 0 | 0 |
| 0 | 0 | 0 | 0 | 0 |
| 0 | 0 | 0 | 0 | 0 |
| 0 | 0 | 0 | 0 | 0 |
| 0.003100652 | 0 | 0 | 0 | 0 |
| 0 | 0 | 0 | 0 | 2.895386395 |
| 0 | 0 | 0 | 0 | 0.001284432 |
| 0 | 0 | 0 | 0 | 0 |
| 0 | 0 | 0 | 0.242807481 | 0 |
| 0 | 0 | 0 | 0 | 0 |
| 0 | 0 | 0 | 0 | 0 |
| 0 | 0 | 0 | 0 | 0 |
| 0 | 0 | 0 | 0 | 0 |
| 0 | 0 | 0 | 0 | 0 |
| 0 | 0 | 0 | 0 | 0.001846957 |

| | | | | |
|---|---|---|---|---|
| 0 | 0 | 0.038759088 | 0 | 1.32860434 |
| 0 | 0 | 0 | 0 | 0 |
| 0 | 0 | 0 | 0 | 0 |
| 0 | 0 | 0 | 0 | 0 |
| 0 | 0 | 0 | 0 | 0 |
| 0 | 0 | 0 | 0 | 0 |
| 0 | 0 | 0 | 0 | 0 |
| 0 | 0 | 0 | 0 | 0 |
| 0 | 0 | 0 | 0 | 0 |
| 0 | 0 | 0 | 0 | 0 |
| 0 | 0 | 0.090590006 | 0 | 0 |
| 0 | 0 | 0 | 0 | 0 |
| 0 | 0 | 0 | 0 | 0 |
| 0 | 0 | 0 | 0 | 0 |
| 0 | 0 | 0 | 0 | 0 |
| 0 | 0 | 0 | 0 | 0 |
| 0 | 0 | 0 | 0 | 0 |
| 0 | 0 | 0 | 0 | 0 |
| 0 | 0 | 0 | 0 | 0 |
| 0 | 0 | 0 | 0 | 0 |
| 0.000822954 | 0 | 0.001607438 | 0.02989812 | 0 |
| 0 | 0 | 0 | 0 | 0 |
| 0.002702261 | 1.492652444 | 0 | 0 | 0.016310162 |
| 0 | 0 | 0 | 0.912937276 | 0.241143165 |
| 0 | 0 | 0 | 0 | 0 |
| 0 | 0 | 0 | 0 | 0 |
| 0 | 0 | 0 | 0 | 0 |
| 0 | 0 | 0 | 0 | 0 |
| 0 | 0 | 0 | 0.002723953 | 0 |
| 0 | 0 | 0 | 0 | 0 |
| 0 | 0 | 0 | 0 | 0 |
| 0 | 0 | 0 | 0 | 0.296206598 |
| 3.174610761 | 0.016768522 | 0.04451879 | 0 | 0.005874769 |
| 0 | 0 | 0 | 0 | 0.040251489 |
| 0.335021661 | 0 | 0.018281203 | 0.010137982 | 0.45150556 |
| 0 | 0.056944384 | 0 | 0 | 0.000565711 |
| 0 | 0 | 0 | 0 | 0 |
| 0 | 0 | 0 | 0 | 0 |
| 0 | 0.586470846 | 0.000882299 | 0.007153355 | 0.016140237 |
| 0 | 0 | 0 | 0 | 0 |
| 0 | 0 | 0 | 0 | 0 |
| 0 | 0 | 0 | 0 | 0 |
| 0.05809994 | 0 | 0.014787294 | 0.175652719 | 0.004335637 |
| 0 | 0 | 0 | 0 | 0 |
| 0 | 0 | 0 | 0 | 0 |
| 0 | 0 | 0.035913799 | 0.076001405 | 0.013165359 |
| 0 | 0 | 0 | 0 | 0 |
| 0 | 0 | 0 | 0 | 0 |

| | | | | |
|---|---|---|---|---|
| 0 | 0 | 0 | 0 | 0 |
| 0 | 0 | 0 | 0 | 0 |
| 0 | 0 | 0 | 0 | 0 |
| 0 | 0 | 0 | 0 | 0 |
| 0 | 0 | 0 | 0 | 0 |
| 0 | 0 | 0 | 0 | 0 |
| 0 | 0 | 0 | 0 | 0 |
| 0.123871453 | 0 | 0.268132112 | 0 | 0 |
| 0 | 0 | 0 | 0 | 0 |
| 0 | 0 | 0 | 0.038234744 | 0 |
| 0 | 0 | 0 | 0 | 0 |
| 0 | 0 | 0 | 0 | 0 |
| 0 | 0 | 0 | 0 | 0 |
| 0 | 0 | 0 | 0 | 0 |
| 0 | 0 | 0 | 0 | 0 |
| 0.020064079 | 0.227768076 | 0 | 0 | 0 |
| 0 | 0 | 0 | 0 | 0 |
| 0 | 0 | 0 | 0 | 0 |
| 0 | 0 | 0 | 0 | 0 |
| 0 | 0 | 0.356614269 | 0 | 1.98518523 |
| 0.003906959 | 0 | 0.020957111 | 0 | 0 |
| 0 | 0 | 0 | 0 | 0 |
| 0 | 0 | 0 | 0 | 0 |
| 0 | 0 | 0 | 0 | 0 |
| 0 | 0 | 0 | 0 | 0 |
| 0 | 0 | 0 | 0 | 0 |
| 0 | 0 | 0 | 0 | 0 |
| 0 | 0 | 0 | 0 | 0 |
| 0 | 0 | 0 | 0 | 0 |
| 0 | 0 | 0.116173384 | 0 | 0.137638414 |
| 0 | 3.522522304 | 0.193455737 | 0.438677582 | 3.65318254 |
| 0 | 0 | 0 | 0 | 0 |
| 0 | 0 | 0 | 0 | 0 |
| 0 | 0 | 0 | 0 | 0 |
| 0 | 0 | 0 | 0 | 0 |
| 0 | 0 | 0 | 0 | 0 |
| 0 | 0 | 0 | 0 | 0 |
| 0 | 0 | 0 | 0 | 0 |
| 0 | 0 | 0 | 0 | 0 |
| 0 | 0 | 0 | 0 | 0 |
| 0 | 0 | 0 | 0 | 0 |
| 0 | 0 | 0 | 0 | 0 |
| 0 | 0 | 0 | 0.003655112 | 0.003067451 |
| 0 | 0 | 0 | 0 | 0 |
| 0 | 0 | 0 | 0 | 0 |
| 0 | 0 | 0 | 0 | 0 |
| 0.114533552 | 0.988069962 | 0.004674854 | 0.001621476 | 0 |
| 0 | 0 | 0 | 0 | 0 |

| | | | | |
|---|---|---|---|---|
| 0 | 0 | 0 | 0 | 0 |
| 0 | 0 | 0 | 0 | 0 |
| 0 | 0 | 0 | 0 | 0 |
| 0.001559999 | 0 | 0 | 0 | 0 |
| 0.052535421 | 0 | 0.210500383 | 0.685783212 | 0.232480708 |
| 0 | 0 | 0 | 0 | 0 |
| 0 | 0 | 0 | 0 | 0 |
| 0 | 0 | 0 | 0 | 0 |
| 0 | 0 | 0 | 0 | 0 |
| 0 | 0 | 0 | 0 | 0 |
| 0 | 0 | 0 | 0 | 0.1243659 |
| 0.002347008 | 0.081364383 | 0 | 0 | 0.003487002 |
| 0 | 0 | 0 | 0 | 0 |
| 0 | 0 | 0 | 0 | 0 |
| 0.006191252 | 0.393845561 | 0 | 0 | 0 |
| 0 | 0 | 0 | 0 | 0.83772681 |
| 0 | 0 | 0 | 0 | 0.002681918 |
| 0 | 0 | 0 | 0 | 0 |
| 0.005468209 | 0 | 0.004672848 | 0 | 0.005610803 |
| 0 | 0.371911151 | 0.483311366 | 0.005611829 | 0.004955823 |
| 0 | 0 | 0 | 0 | 0 |
| 0 | 0 | 0 | 0 | 0 |
| 0 | 0 | 0 | 0 | 0 |
| 0 | 0 | 0 | 0 | 0 |
| 0 | 0 | 0 | 0 | 0 |
| 0 | 0 | 0 | 0 | 0 |
| 0 | 0 | 0.00116411 | 0 | 0.004489328 |
| 0 | 0 | 0 | 0 | 0 |
| 0 | 0 | 0 | 0 | 0 |
| 0 | 0 | 0 | 0 | 0 |
| 0.000486418 | 0.044329068 | 0.001187622 | 0 | 0 |
| 0 | 0 | 0 | 0 | 0 |
| 0 | 0 | 0 | 0 | 0 |
| 0 | 0 | 0 | 0 | 0 |
| 0 | 0 | 0 | 0 | 0.003611722 |
| 0 | 0 | 0 | 0 | 0 |
| 0 | 0 | 0 | 0 | 0 |
| 0.009113155 | 0.76054706 | 0.001854198 | 0.01233603 | 0.007150611 |
| 0 | 0 | 0 | 0 | 0.843035889 |
| 0 | 0 | 0 | 0 | 0 |
| 0 | 0.129195347 | 0 | 0 | 0.00260834 |
| 0 | 0 | 0 | 0 | 0 |
| 0 | 0 | 0 | 0 | 0 |
| 0 | 0 | 0 | 0 | 0 |
| 0 | 0 | 0 | 0 | 0 |
| 0 | 0.001319706 | 0 | 0.253220905 | 0 |
| 0 | 0 | 0 | 0 | 0 |
| 0 | 0 | 0 | 0 | 0 |

| | | | | |
|---|---|---|---|---|
| 0 | 0 | 0 | 0 | 0 |
| 0 | 0 | 0 | 0 | 0.644681489 |
| 0 | 0 | 0 | 0 | 0 |
| 0.000531089 | 0.081771806 | 0 | 0 | 0.000641104 |
| 0 | 0.003074677 | 0 | 0.009559519 | 0 |
| 0 | 0.017459932 | 0 | 0.002220746 | 0 |
| 0 | 0 | 0 | 0 | 0.019701174 |
| 0 | 0.017630184 | 0 | 0 | 0.004846852 |
| 0 | 0 | 0 | 0 | 0 |
| 0 | 0 | 0 | 0 | 0 |
| 0 | 0 | 0 | 0 | 0.054388065 |
| 0 | 0 | 0 | 0 | 0 |
| 0.006399943 | 0 | 0.011132167 | 0.288009129 | 0.149211808 |
| 0 | 0 | 0 | 0 | 0 |
| 0 | 0 | 0 | 0 | 0 |
| 0 | 0 | 0 | 0 | 0 |
| 0.098125425 | 0 | 0 | 0 | 0 |
| 0 | 0 | 0 | 0 | 0 |
| 0 | 0 | 0 | 0 | 0 |
| 0 | 0 | 0 | 0 | 0 |
| 0 | 0 | 0 | 0 | 0 |
| 0 | 0 | 0 | 0 | 0 |
| 0.001652402 | 0 | 0.000672408 | 0 | 0 |
| 0 | 0 | 0.015393217 | 0.001135257 | 0.020713528 |
| 0 | 0 | 0 | 0 | 0 |
| 0 | 0 | 0 | 0 | 0 |
| 0 | 0 | 0.007709134 | 0 | 0.126351855 |
| 0 | 0 | 0 | 0 | 0 |
| 0 | 0 | 0 | 0 | 0 |
| 0 | 0 | 0 | 0 | 0 |
| 0 | 0 | 0.045433768 | 0.019522882 | 0.065387873 |
| 0.003419413 | 0 | 0 | 0 | 0 |
| 0 | 0 | 0 | 0 | 0 |
| 0 | 0 | 0 | 0 | 0 |
| 0 | 0 | 0 | 0 | 0 |
| 0 | 0.009858251 | 0 | 0 | 0 |
| 0 | 0 | 0 | 0 | 0 |
| 0 | 0 | 0 | 0 | 0 |
| 0.000623526 | 0 | 0.004694008 | 0.005808426 | 0 |
| 0 | 0 | 0 | 0 | 0 |
| 0 | 0 | 0 | 0 | 0 |
| 0 | 0 | 0 | 0 | 0 |
| 0 | 0 | 0 | 0 | 0 |
| 0 | 0 | 0 | 0 | 0 |
| 0 | 0 | 0 | 0 | 0 |
| 0.063500474 | 0.345177444 | 0.084273334 | 0.158583828 | 0.693698894 |
| 0 | 0 | 0 | 0 | 0 |
| 0 | 0 | 0 | 0 | 0 |

| | | | | |
|---|---|---|---|---|
| 0 | 0 | 0 | 0 | 0 |
| 0 | 0 | 0 | 0 | 0 |
| 0 | 0 | 0 | 0 | 0 |
| 0 | 0 | 0 | 0 | 0.001499769 |
| 0 | 0 | 0 | 0 | 0 |
| 0 | 0 | 0 | 0 | 0.000914542 |
| 0 | 0 | 0 | 0 | 0 |
| 0 | 0 | 0 | 0 | 0 |
| 0 | 0 | 0 | 0.094050937 | 0.001044657 |
| 0 | 0 | 0 | 0 | 0 |
| 0 | 1.443140924 | 0 | 0 | 0 |
| 0 | 0.016499615 | 0 | 0 | 0 |
| 0 | 0 | 0 | 0 | 0 |
| 0 | 0 | 0 | 0 | 0 |
| 0 | 0 | 0 | 0 | 0 |
| 0 | 0 | 0 | 0 | 0 |
| 0 | 0 | 0 | 0 | 0 |
| 0 | 0 | 0 | 0 | 0 |
| 0 | 0 | 0 | 0 | 0 |
| 0.00067084 | 0 | 0 | 0.003710447 | 0.002429412 |
| 0 | 0 | 0 | 0 | 0 |
| 0 | 0 | 0 | 0 | 0 |
| 0 | 0 | 0 | 0 | 0 |
| 0 | 0 | 0 | 0 | 0 |
| 0 | 0 | 0 | 0 | 0 |
| 0 | 0 | 0 | 0 | 0 |
| 0 | 0 | 0 | 0 | 0 |
| 0 | 0 | 0.024923434 | 0.003674001 | 0 |
| 0 | 0 | 0 | 0 | 0 |
| 0 | 0 | 0 | 0 | 0 |
| 0.007924413 | 0 | 0.001055344 | 0.03573532 | 0.004174233 |
| 0 | 0 | 0 | 0 | 0 |
| 0.057046511 | 0.023182144 | 0.030089449 | 0.010324386 | 0.02504133 |
| 0.001880495 | 0.009603907 | 0.003060902 | 0.001194385 | 0.002270038 |
| 0 | 0 | 0 | 0 | 0 |
| 0 | 0 | 0 | 0 | 0 |
| 0 | 0 | 0 | 0 | 0 |
| 0 | 0 | 0 | 0 | 0 |
| 0 | 0 | 0 | 0 | 0 |
| 0 | 0 | 0 | 0 | 0 |
| 0 | 0 | 0 | 0 | 0 |
| 0.04349091 | 0 | 0.397525201 | 0.008333519 | 0.227610864 |
| 0 | 0 | 0.017330464 | 0.046467559 | 0.1518034 |
| 0 | 0 | 0 | 0 | 0 |
| 0 | 0 | 0 | 0 | 0 |
| 0.002180673 | 0 | 0 | 0.452831554 | 0.007312216 |
| 0 | 0 | 0 | 0 | 0 |
| 0 | 0 | 0 | 0 | 0 |

| | | | | |
|---|---|---|---|---|
| 0 | 0 | 0 | 0 | 0 |
| 0 | 0 | 0.011865706 | 0.008795294 | 0 |
| 0 | 0.01972409 | 0.018703187 | 0.051130236 | 0.001733841 |
| 0 | 0 | 0 | 0 | 0 |
| 0.02228117 | 0 | 0.058401022 | 0 | 0 |
| 0 | 0 | 0 | 0 | 0.089708725 |
| 0 | 0 | 0 | 0 | 0 |
| 0 | 0 | 0 | 0 | 0 |
| 0 | 0 | 0 | 0 | 0 |
| 0 | 0 | 0 | 0 | 0 |
| 0 | 0 | 0 | 0 | 0 |
| 0 | 0 | 0 | 0 | 0 |
| 0 | 0 | 0 | 0 | 0 |
| 0 | 0 | 0 | 0 | 0 |
| 0 | 0 | 0 | 0 | 0 |
| 0 | 0 | 0 | 0 | 0 |
| 0 | 0 | 0 | 0 | 0.080311624 |
| 0 | 0 | 0 | 0 | 0 |
| 0 | 0 | 0 | 0 | 0 |
| 0 | 0 | 0 | 0 | 0 |
| 0 | 0 | 0 | 0 | 0 |
| 0 | 0 | 0 | 0 | 0 |
| 0 | 0 | 0 | 0 | 0.03024788 |
| 0 | 0 | 0 | 0 | 0 |
| 0 | 0 | 0 | 0.805751181 | 0 |
| 0.005103413 | 0.000637113 | 0.00069224 | 0.012281313 | 0.002053527 |
| 0 | 0 | 0 | 0 | 0 |
| 0 | 0 | 0.003491784 | 0.20886708 | 0.003386388 |
| 0 | 0 | 0 | 0 | 0 |
| 0 | 0 | 0 | 0 | 0 |
| 0 | 0 | 0 | 0.013678073 | 0 |
| 0 | 0 | 0 | 0 | 0 |
| 0 | 0 | 0 | 0 | 0 |
| 0 | 0 | 0 | 0 | 0 |
| 0 | 0 | 0 | 0 | 0 |
| 0 | 0 | 0 | 0 | 0 |
| 0 | 0 | 0 | 0 | 0 |
| 0 | 0.040588328 | 0.03541803 | 0 | 0 |
| 2.290028961 | 0 | 1.374402725 | 0.517777322 | 0 |
| 0 | 0 | 0 | 0 | 0 |
| 0 | 0 | 0 | 0 | 0 |
| 0 | 0 | 0 | 0 | 0 |
| 0.006146198 | 0.003197063 | 0 | 0 | 0.000824375 |
| 0 | 0 | 0 | 0 | 0 |
| 0 | 0 | 0 | 0 | 0 |
| 0 | 0 | 0 | 0 | 0 |
| 0.00444006 | 0.045648221 | 0 | 0 | 0.003783399 |
| 0 | 0 | 0 | 0 | 0.019048556 |

| | | | | |
|---|---|---|---|---|
| 0 | 0 | 0 | 0 | 0 |
| 0.011040297 | 0.014605627 | 0.002548043 | 0 | 0.005768528 |
| 0 | 0 | 0 | 0 | 0 |
| 0.008553948 | 0 | 0 | 0 | 0 |
| 0 | 0 | 0 | 0 | 0 |
| 0 | 0 | 0 | 0 | 0 |
| 0 | 0 | 0 | 0 | 0 |
| 0 | 0 | 0 | 0 | 0 |
| 0 | 0 | 0 | 0 | 0 |
| 0 | 0 | 0 | 0 | 0 |
| 0 | 0 | 0 | 0 | 0 |
| 0 | 0 | 0 | 0 | 0 |
| 0 | 0 | 0.012049903 | 0.028598954 | 0.456707545 |
| 0 | 0 | 0 | 0 | 0 |
| 0.034516695 | 0 | 0.005685205 | 0.001674679 | 0.00074405 |
| 0 | 0 | 0 | 0 | 0 |
| 0 | 0 | 0 | 0 | 0 |
| 0 | 0 | 0 | 0 | 0 |
| 0 | 0 | 0 | 0 | 0.004389931 |
| 0 | 0 | 0 | 0 | 0 |
| 0 | 0 | 0 | 0 | 0 |
| 0 | 0.131490001 | 0.050578307 | 0.001409715 | 0 |
| 0.000930283 | 0 | 0 | 0 | 0 |
| 0 | 0.996556113 | 0 | 0 | 0 |
| 0 | 0 | 0 | 0 | 0 |
| 0 | 0 | 0 | 0 | 0 |
| 0 | 0 | 0 | 0 | 0 |
| 0 | 0 | 0 | 0 | 0 |
| 0 | 0 | 0 | 0 | 0 |
| 0 | 0 | 0 | 0 | 0 |
| 0 | 0 | 0 | 0 | 0 |
| 0 | 0 | 0 | 0 | 0 |
| 0 | 0 | 0 | 0 | 0 |
| 0 | 0 | 0 | 0 | 0 |
| 0 | 0 | 0 | 0.053330939 | 0.004865296 |
| 0 | 0 | 0 | 0 | 0 |
| 0 | 0 | 0 | 0 | 0 |
| 0 | 0 | 0 | 0 | 0 |
| 0 | 0.062132649 | 0.000983933 | 0 | 0 |
| 0 | 0 | 0 | 0 | 0 |
| 0.288526886 | 0.087737828 | 0.016468219 | 0.048807103 | 0.206098194 |
| 0 | 0 | 0 | 0 | 0 |
| 0 | 0 | 0 | 0 | 0 |
| 0 | 0 | 0 | 0 | 0 |
| 0 | 0 | 0 | 0 | 0 |
| 0.801347085 | 1.963627292 | 0.244946529 | 0.013488472 | 0.05568355 |
| 0 | 0 | 0 | 0 | 0 |
| 0 | 0 | 0 | 0 | 0 |

| | | | | |
|---|---|---|---|---|
| 0 | 0 | 0 | 0 | 0 |
| 0 | 0 | 0 | 0 | 0 |
| 0 | 0 | 0 | 0 | 0 |
| 0 | 0 | 0 | 0 | 0 |
| 0.000941292 | 0 | 0 | 0 | 0 |
| 0 | 0 | 0 | 0 | 0 |
| 0 | 0 | 0 | 0 | 0 |
| 0 | 0 | 0 | 0 | 0 |
| 0 | 0 | 0 | 0.000979019 | 0.009134422 |
| 0.007889786 | 0 | 0.000823224 | 0.023321124 | 0 |
| 0 | 0 | 0 | 0 | 0.046359581 |
| 0 | 0 | 0 | 0 | 0 |
| 0 | 0.004443328 | 0 | 0 | 0 |
| 0 | 0 | 0 | 0 | 0 |
| 0 | 0 | 0 | 0 | 0 |
| 0 | 0 | 0 | 0 | 0 |
| 0 | 0 | 0 | 0 | 0 |
| 0 | 0 | 0 | 0 | 0 |
| 0 | 0 | 0 | 0 | 0 |
| 0.523942024 | 1.063861495 | 0.000716179 | 0.086073084 | 0.206080613 |
| 0 | 0 | 0 | 0 | 0 |
| 0 | 0 | 0 | 0 | 0 |
| 0 | 0 | 0 | 0 | 0 |
| 0 | 0 | 0 | 0 | 0 |
| 0 | 0 | 0 | 0 | 0 |
| 0 | 0 | 0 | 0 | 0 |
| 0 | 0 | 0 | 0 | 0 |
| 0 | 0 | 0 | 0 | 0 |
| 0 | 0 | 0 | 0 | 0 |
| 0 | 0 | 0.021896362 | 0 | 0 |
| 0 | 0 | 0 | 0 | 0 |
| 0 | 0 | 0 | 0 | 0 |
| 0 | 0 | 0 | 0 | 0 |
| 0 | 0 | 0 | 0 | 0 |
| 0 | 0 | 0 | 0 | 0 |
| 0 | 0 | 0 | 0 | 0.004400274 |
| 0 | 0 | 0 | 0 | 0 |
| 0 | 0 | 0 | 0 | 0 |
| 0 | 0 | 0 | 0 | 0 |
| 0 | 0 | 0 | 0 | 0 |
| 0 | 0 | 0 | 0 | 0 |
| 0 | 0 | 0 | 0 | 0 |
| 0 | 0 | 0.003690961 | 0.002384911 | 0 |
| 0 | 0 | 0 | 0 | 0 |
| 0 | 0 | 0 | 0.016746051 | 0.003392262 |
| 0 | 0 | 0 | 0 | 0 |
| 0 | 0 | 0 | 0 | 0 |
| 0 | 0 | 0 | 0 | 0 |

| | | | | |
|---|---|---|---|---|
| 0 | 0 | 0 | 0 | 0 |
| 0 | 0 | 0 | 0 | 0 |
| 0 | 0 | 0 | 0 | 0 |
| 0 | 0 | 0 | 0 | 0 |
| 0 | 0 | 0 | 0 | 0 |
| 0.016783047 | 0 | 0.000803469 | 0 | 0.313875521 |
| 0 | 0 | 0 | 0 | 0 |
| 0 | 0 | 0 | 0 | 0 |
| 0 | 0 | 0 | 0 | 0 |
| 0 | 0 | 0 | 0 | 0 |
| 0 | 0 | 0 | 0 | 0 |
| 0 | 0 | 0 | 0 | 0 |
| 0 | 0 | 0 | 0 | 0 |
| 0 | 0 | 0 | 0 | 0 |
| 0 | 0 | 0 | 0 | 0 |
| 0 | 0 | 0 | 0 | 0 |
| 0 | 0 | 0 | 0 | 0.022406441 |
| 0 | 0 | 0 | 0 | 0 |
| 0 | 0 | 0 | 0 | 0 |
| 0 | 0 | 0 | 0 | 0 |
| 0 | 0 | 0 | 0 | 0 |
| 0 | 0 | 0 | 0 | 0 |
| 0 | 0 | 0 | 0 | 0 |
| 0 | 0 | 0 | 0 | 0 |
| 0 | 0 | 0 | 0 | 0 |
| 0.036095815 | 0 | 0 | 0 | 0 |
| 0 | 0 | 0 | 0 | 0 |
| 0 | 0 | 0 | 0 | 0.097659835 |
| 0 | 0 | 0 | 0 | 0 |
| 0 | 0 | 0 | 0 | 0 |
| 0 | 0 | 0 | 0 | 0 |
| 0 | 0 | 0 | 0 | 0 |
| 0 | 0 | 0 | 0 | 0 |
| 0 | 0 | 0 | 0 | 0 |
| 0 | 0 | 0 | 0 | 0 |
| 0 | 0 | 0 | 0 | 0 |
| 0 | 0 | 0 | 0 | 0 |
| 0 | 0 | 0 | 0 | 0 |
| 0 | 0 | 0.045083201 | 0.759433732 | 0.300492478 |
| 0 | 0 | 0 | 0 | 0 |
| 0 | 0 | 0 | 0 | 0 |
| 0 | 0 | 0 | 0 | 0 |
| 0 | 0 | 0 | 0 | 0 |
| 0 | 0 | 0 | 0 | 0 |
| 0 | 0 | 0 | 0 | 0 |
| 0 | 0 | 0 | 0 | 0 |
| 0 | 0 | 0 | 0 | 0 |
| 0 | 0 | 0 | 0 | 0 |

| | | | | |
|---|---|---|---|---|
| 0 | 0 | 0 | 0 | 0 |
| 0.001657939 | 0 | 0.018328297 | 0 | 0.362750096 |
| 0 | 0 | 0 | 0 | 0 |
| 0 | 0 | 0 | 0 | 0 |
| 0 | 0 | 0 | 0 | 0 |
| 0 | 0 | 0 | 0 | 0 |
| 0 | 0 | 0 | 0 | 0 |
| 0 | 0 | 0 | 0 | 0 |
| 0 | 0 | 0 | 0 | 0 |
| 0 | 0 | 0 | 0 | 0 |
| 0 | 0.002032661 | 0 | 1.286916245 | 0 |
| 0 | 0 | 0 | 0 | 0 |
| 0 | 0 | 0.124279908 | 0 | 0.242727677 |
| 0 | 0 | 0 | 0 | 0 |
| 0 | 0 | 0 | 0 | 0 |
| 0 | 0 | 0 | 0 | 0 |
| 0 | 0 | 0 | 0 | 0 |
| 0 | 0 | 0 | 0 | 0 |
| 0 | 0 | 0 | 0 | 0 |
| 0 | 0 | 0 | 0 | 0 |
| 0.01248578 | 0 | 0.001046049 | 0.006414188 | 0 |
| 0 | 0 | 0 | 0 | 0 |
| 0 | 0 | 0 | 0 | 0 |
| 1.739485432 | 0 | 3.536129234 | 0.402587011 | 2.865308762 |
| 0 | 0 | 0 | 0 | 0 |
| 0 | 0 | 0 | 0 | 0 |
| 0 | 0 | 0 | 0 | 0 |
| 0 | 0 | 0 | 0 | 0 |
| 0 | 0 | 0 | 0 | 0 |
| 0 | 0 | 0 | 0 | 0 |
| 0 | 0 | 0 | 0 | 0 |
| 0 | 0 | 0 | 0 | 0 |
| 0 | 0 | 0 | 0 | 0 |
| 0 | 0 | 0 | 0 | 0 |

| PHL018-0278_V2 | PHL018-0052_V1 | PHL018-0145_V3 | PHL018-0228_V2 | PHL018-0249_V2 |
|---|---|---|---|---|
| REF | EYCF | REF | REF | REF |
| 0.524220228 | 0.056426763 | 0.072821119 | 0.108518057 | 0.010545488 |
| 0.826101557 | 1.637395111 | 0.32418168 | 1.928496229 | 4.356745957 |
| 12.88073217 | 13.2342648 | 0.612047368 | 12.00548721 | 20.53861587 |
| 0 | 0 | 0 | 0 | 0 |
| 0 | 0 | 0 | 0 | 0 |
| 0 | 0 | 0 | 0 | 0 |
| 0 | 0 | 0 | 0 | 0 |
| 0 | 0 | 0 | 0 | 0 |
| 0 | 0 | 0 | 0 | 0 |
| 0 | 0 | 0 | 0 | 0 |
| 0 | 0 | 0 | 0 | 0 |
| 0 | 0 | 0 | 0 | 0 |
| 0 | 0 | 0 | 0 | 0 |
| 16.01455863 | 15.88338992 | 1.685966398 | 8.982860458 | 7.903768459 |
| 0 | 0 | 0 | 0 | 0 |
| 0 | 0 | 0 | 0 | 0 |
| 0 | 0 | 0 | 0 | 0.027234113 |
| 0 | 0 | 0 | 0 | 0 |
| 0 | 0 | 0 | 0 | 0 |
| 0 | 0 | 0 | 0 | 0 |
| 0 | 0 | 0 | 0 | 0 |
| 0 | 0 | 0 | 0 | 0 |
| 0 | 0 | 0 | 0 | 0 |
| 0 | 0 | 0 | 0 | 0 |
| 0.101775693 | 0.001062041 | 0 | 0 | 0 |
| 0 | 0 | 0 | 0 | 0 |
| 0 | 0 | 0 | 0 | 0 |
| 0 | 0 | 0 | 0 | 0 |
| 0 | 0 | 0 | 0 | 0 |
| 0 | 0 | 0 | 0 | 0 |
| 0 | 0 | 0 | 0 | 0 |
| 0 | 0 | 0 | 0 | 0 |
| 0 | 0 | 0 | 0 | 0 |
| 0 | 0 | 0 | 0 | 0 |
| 0 | 0 | 0 | 0 | 0 |
| 0 | 0 | 1.656813707 | 0 | 0 |
| 0 | 0 | 0 | 0 | 0 |
| 0 | 0 | 0 | 0 | 0 |
| 0 | 0 | 0 | 0 | 0 |
| 0 | 0 | 0 | 0 | 0 |
| 0 | 0 | 0 | 0 | 0 |
| 0.012736763 | 0.00876058 | 0.122344964 | 0.001998922 | 0.088296073 |
| 0.040077128 | 0.051401737 | 0.299885659 | 0.160614285 | 0.00122483 |
| 0.001491795 | 0.036247007 | 0.0064403 | 0.014749805 | 0 |
| 0 | 0 | 0.005339713 | 0 | 0 |
| 0 | 0 | 0 | 0 | 0 |

| | | | | |
|---|---|---|---|---|
| 0 | 0.006555231 | 0 | 0 | 0 |
| 0 | 0.018039016 | 0 | 0 | 0 |
| 0.00422881 | 0 | 0 | 0.003733167 | 0.026307774 |
| 0.760405084 | 2.033415183 | 0.298278155 | 1.997458734 | 2.005099338 |
| 0.01789808 | 0.003822305 | 0 | 0.000837103 | 0 |
| 0 | 0 | 0 | 0 | 0 |
| 0.00639551 | 0 | 0.00221869 | 9.139930195 | 0.005471091 |
| 0 | 0 | 0 | 0 | 0 |
| 0 | 0 | 0 | 0 | 0 |
| 0 | 0 | 0 | 0 | 0 |
| 0 | 0 | 0 | 0 | 0 |
| 0.669606095 | 0.803114052 | 0.32255193 | 0.320123383 | 0 |
| 0 | 0 | 0.001922071 | 0.001676951 | 0 |
| 0.115651638 | 0.295074216 | 0.00143006 | 0.009011066 | 0.009403735 |
| 0 | 0 | 0 | 0 | 0 |
| 0 | 0 | 0 | 0 | 0 |
| 0 | 0 | 0.017208495 | 0.003167491 | 0.018442043 |
| 0 | 0 | 0 | 0 | 0 |
| 0 | 0 | 0 | 0 | 0 |
| 0 | 0 | 0 | 0 | 0 |
| 0.04407791 | 0.086921501 | 0.001007422 | 0.02929822 | 0.263740624 |
| 0.093156513 | 0.013574545 | 0 | 0.014212556 | 0.001295797 |
| 0 | 0 | 0 | 0 | 0 |
| 0 | 0 | 0 | 0 | 0 |
| 0 | 0 | 0 | 0 | 0 |
| 0 | 0 | 0 | 0 | 0 |
| 0 | 0 | 0 | 0 | 0 |
| 0 | 0 | 0 | 0 | 0 |
| 0 | 0 | 0.14359178 | 0 | 0.003149628 |
| 0 | 0 | 0 | 0 | 0 |
| 0 | 0.001853188 | 0 | 0 | 0.007341403 |
| 0 | 0 | 0 | 0 | 0 |
| 0 | 0 | 0 | 0 | 0 |
| 0 | 0.005292559 | 0.000728788 | 0 | 0.000479234 |
| 0 | 0 | 0.00181553 | 0 | 0 |
| 0 | 0 | 0 | 0 | 0 |
| 0 | 0 | 0 | 0 | 0 |
| 0 | 0.004363041 | 0 | 0.001945799 | 0 |
| 0 | 0 | 0 | 0 | 0 |
| 0 | 0 | 0 | 0 | 0 |
| 0 | 0 | 0 | 0 | 0 |
| 0 | 0 | 0 | 0 | 0 |
| 0 | 0 | 0 | 0 | 0 |
| 0 | 0 | 0 | 0 | 0 |
| 0 | 0 | 0 | 0 | 0 |
| 0 | 0 | 0 | 0 | 0 |
| 0 | 0 | 0 | 0.006801895 | 0 |
| 0 | 0 | 0 | 0.012969673 | 0 |

| | | | | |
|---|---|---|---|---|
| 0 | 0 | 0 | 0 | 0 |
| 0 | 0 | 0 | 0 | 0.516429113 |
| 0 | 0 | 0 | 0 | 0 |
| 0 | 0 | 0 | 0 | 0 |
| 0 | 0 | 0 | 0.77208879 | 0 |
| 0 | 0 | 0 | 0 | 0 |
| 0 | 0 | 0 | 0 | 0 |
| 0.000670549 | 0 | 0 | 0 | 0 |
| 0 | 0.024818979 | 0.0025904 | 0 | 0.014528873 |
| 0 | 0 | 0 | 0 | 0 |
| 0 | 0 | 0 | 0 | 0 |
| 0.001456979 | 0 | 0.003007403 | 0.006173815 | 0 |
| 0 | 0 | 0 | 0 | 0 |
| 0 | 0 | 0 | 0 | 0 |
| 0 | 0 | 0 | 0 | 0 |
| 0 | 0 | 0 | 0 | 0 |
| 0 | 0 | 0 | 0 | 0 |
| 0 | 0 | 0 | 0 | 0 |
| 0.006159054 | 0 | 1.20455917 | 0.282961963 | 0 |
| 0 | 0 | 0 | 0.001703285 | 0 |
| 0 | 0 | 0 | 0 | 0 |
| 0 | 0 | 0 | 0 | 0 |
| 0 | 0 | 0 | 0 | 0 |
| 0 | 0 | 0 | 0 | 0 |
| 0 | 0 | 0 | 0 | 0 |
| 0 | 0 | 0 | 0 | 0 |
| 0 | 0 | 0 | 0 | 0 |
| 0 | 0 | 0 | 0 | 0 |
| 0 | 0 | 0 | 0 | 0 |
| 0 | 0 | 0 | 0 | 0 |
| 0.095516875 | 0.218111071 | 0 | 0.070532914 | 0 |
| 0.000609152 | 0 | 0.000591705 | 0.007528578 | 0 |
| 0 | 0 | 0 | 0 | 0 |
| 0 | 0 | 0 | 0 | 0 |
| 0 | 0 | 0 | 0 | 0 |
| 0 | 0 | 0 | 0 | 0 |
| 0 | 0 | 0 | 0 | 0 |
| 0 | 0 | 0 | 0 | 0 |
| 0 | 0 | 0 | 0 | 0 |
| 0 | 0 | 0 | 0 | 0 |
| 0 | 0 | 0 | 0 | 0 |
| 0 | 0 | 0 | 0 | 0 |
| 0 | 0 | 0.002051752 | 0.001988993 | 0 |
| 0 | 0 | 0 | 0 | 0 |
| 0 | 0 | 0 | 0 | 0 |
| 0 | 0 | 0 | 0 | 0 |
| 0 | 0 | 0.011293032 | 0.012511543 | 0 |
| 0 | 0 | 0 | 0 | 0 |

| | | | | |
|---|---|---|---|---|
| 0 | 0.011714013 | 0 | 0 | 0 |
| 0 | 0 | 0 | 0 | 0 |
| 0 | 0 | 0 | 0 | 0 |
| 0 | 0 | 0 | 0 | 0 |
| 0.079397146 | 0 | 0.000386259 | 0.041750541 | 0 |
| 0 | 0 | 0 | 0 | 0 |
| 0 | 0 | 0 | 0 | 0 |
| 0 | 0 | 0 | 0 | 0 |
| 0 | 0 | 0 | 0 | 0 |
| 0 | 0 | 0 | 0 | 0 |
| 0 | 0 | 0 | 0 | 0 |
| 0 | 0 | 0 | 0 | 0 |
| 0 | 0 | 0 | 0 | 0 |
| 0 | 0 | 0 | 0 | 0 |
| 0 | 0 | 0 | 0 | 0 |
| 0.01077482 | 0 | 0 | 5.16181402 | 0 |
| 0 | 0 | 0 | 0 | 0 |
| 0 | 0 | 0 | 0 | 0 |
| 0 | 0 | 0 | 0 | 0 |
| 0 | 0 | 0 | 0 | 0 |
| 0 | 0 | 0 | 0 | 0 |
| 0 | 0 | 0 | 0 | 0 |
| 0 | 0.001172651 | 0 | 0 | 0 |
| 0 | 0 | 0 | 0 | 0 |
| 0 | 0 | 0 | 0 | 0 |
| 0 | 0 | 0 | 0 | 0 |
| 0.000992553 | 0.001500345 | 0 | 0 | 0 |
| 0 | 0 | 0 | 0 | 0 |
| 0 | 0 | 0 | 0 | 0 |
| 0 | 0 | 0 | 0 | 0 |
| 0 | 0 | 0 | 0 | 0 |
| 0 | 0 | 0.045058195 | 0.016876346 | 0 |
| 0 | 0 | 0 | 0 | 0 |
| 0 | 0 | 0 | 0 | 0 |
| 0 | 0 | 0 | 0 | 0 |
| 0 | 0 | 0 | 0 | 0 |
| 0 | 0 | 0 | 0 | 0 |
| 0 | 0 | 0 | 0 | 0 |
| 0 | 0 | 0 | 0 | 0 |
| 0 | 0 | 0 | 0 | 0 |
| 0 | 0 | 0 | 0 | 0 |
| 0 | 0 | 0 | 0 | 0 |
| 0 | 0 | 0 | 0 | 0 |
| 0 | 0 | 0 | 0 | 0 |
| 0 | 0 | 0 | 0 | 0 |
| 0 | 0 | 0 | 0 | 0 |
| 0 | 0 | 0 | 0 | 0 |
| 0 | 0 | 0 | 0 | 0 |

| | | | | |
|---|---|---|---|---|
| 0 | 0 | 0 | 0 | 0 |
| 0 | 0 | 0 | 0 | 0 |
| 0 | 0 | 0 | 0 | 0 |
| 0 | 0 | 0 | 0 | 0 |
| 0 | 0 | 0 | 0 | 0 |
| 0 | 0 | 0 | 0 | 0 |
| 0 | 0 | 0 | 0 | 0 |
| 0 | 0 | 0 | 0 | 0 |
| 0 | 0 | 0 | 0 | 0 |
| 0 | 0 | 0 | 0 | 0 |
| 0 | 0 | 0 | 0 | 0 |
| 0 | 0 | 0 | 0 | 0 |
| 0.001345079 | 0.001355482 | 0.001306554 | 0.001662398 | 0 |
| 0 | 0 | 0 | 0 | 0 |
| 0 | 0 | 0 | 0 | 0 |
| 0 | 0 | 0 | 0 | 0 |
| 0 | 0 | 0 | 0 | 0 |
| 0 | 0 | 0 | 0 | 0 |
| 0 | 0 | 0 | 0 | 0 |
| 0 | 0 | 0 | 0 | 0 |
| 0.045583912 | 0.106445847 | 0.0293153 | 0.065718021 | 0.018825234 |
| 0 | 0 | 0 | 0 | 0 |
| 0 | 0 | 0 | 0 | 0 |
| 0.003933919 | 0.001982172 | 0 | 0.022226165 | 0.006020153 |
| 0 | 0 | 0 | 0 | 0 |
| 0 | 0 | 0 | 0 | 0 |
| 0.003478572 | 0.001106992 | 0 | 0.008275162 | 0.00175414 |
| 0 | 0 | 0 | 0 | 0 |
| 0 | 0 | 0 | 0 | 0 |
| 0 | 0 | 0 | 0 | 0 |
| 0 | 0 | 0 | 0 | 0.241012517 |
| 0 | 0 | 0 | 0 | 0 |
| 0 | 0 | 0 | 0 | 0 |
| 0 | 0 | 0.00317257 | 0.010583433 | 0 |
| 0.078136974 | 0.007008386 | 0 | 0.015601493 | 0.003103004 |
| 0 | 0 | 0 | 0 | 0 |
| 0 | 0 | 0 | 0 | 0 |
| 0 | 0 | 0 | 0 | 0 |
| 0 | 0 | 0 | 0 | 0 |
| 0 | 0 | 0 | 0 | 0 |
| 0 | 0 | 0 | 0 | 0 |
| 0 | 0 | 0 | 0 | 0 |
| 0 | 0 | 0 | 0 | 0 |
| 0 | 0 | 0 | 0 | 0 |
| 0 | 0 | 0 | 0 | 0 |
| 0 | 0 | 0 | 0 | 0 |
| 0.00088588 | 0.053563892 | 0 | 0 | 0 |
| 0.001749189 | 0.001175145 | 0.002973407 | 0.003706015 | 0 |

| | | | | |
|---|---|---|---|---|
| 0 | 0 | 0 | 0 | 0 |
| 0 | 0 | 0 | 0 | 0 |
| 0 | 0 | 0 | 0 | 0.001063487 |
| 0 | 0 | 0 | 0 | 0 |
| 0 | 0 | 0 | 0 | 0 |
| 0 | 0 | 0 | 0 | 0 |
| 0.000622487 | 0 | 0 | 0 | 0 |
| 0.002095409 | 0 | 0 | 0 | 0 |
| 0 | 0 | 0 | 0 | 0 |
| 0 | 0 | 0 | 0 | 0 |
| 0 | 0 | 0 | 0 | 0 |
| 0 | 0 | 0 | 0 | 0 |
| 0 | 0 | 0 | 0 | 0 |
| 0 | 0 | 0 | 0 | 0 |
| 0 | 0 | 0 | 0 | 0 |
| 0 | 0 | 0.000749509 | 0 | 0 |
| 0 | 0 | 0 | 0 | 0 |
| 0 | 0 | 0 | 0 | 0 |
| 0.000592971 | 0.000597557 | 0.002591944 | 0 | 0 |
| 0 | 0 | 0 | 0.020155766 | 0 |
| 0.144761685 | 0 | 0 | 0.016678945 | 0.005587032 |
| 0 | 0 | 0 | 0 | 0 |
| 0 | 0 | 0 | 0 | 0 |
| 0 | 0 | 0 | 0 | 0 |
| 0.003041084 | 0 | 0.012237929 | 0 | 0 |
| 0 | 0 | 0 | 0 | 0 |
| 0 | 0 | 0 | 0 | 0 |
| 0 | 0 | 0 | 0 | 0 |
| 0 | 0 | 0.035640578 | 0 | 0.003641247 |
| 0 | 0 | 0 | 0 | 0 |
| 0.39618741 | 0.499388409 | 0.015930876 | 0.268445801 | 0.460753832 |
| 0 | 0 | 0 | 0 | 0 |
| 0.019993018 | 0 | 0 | 0 | 0.001436668 |
| 0 | 0 | 0 | 0 | 0 |
| 0 | 0 | 0 | 0 | 0 |
| 0 | 0 | 0 | 0 | 0 |
| 0 | 0 | 0 | 0 | 0 |
| 0 | 0 | 0 | 0 | 0 |
| 0 | 0 | 0 | 0 | 0 |
| 0 | 0 | 0 | 0 | 0 |
| 0 | 0 | 0 | 0 | 0 |
| 0.072443367 | 0 | 0 | 0.004861551 | 0 |
| 0 | 0 | 0 | 0 | 0 |
| 0 | 0 | 0 | 0 | 0 |
| 0 | 0 | 0 | 0 | 0 |
| 0 | 0 | 0 | 0 | 0 |
| 0 | 0 | 0 | 0 | 0 |
| 0 | 0 | 0 | 0 | 0 |

| | | | | |
|---|---|---|---|---|
| 0 | 0 | 0 | 0 | 0 |
| 0 | 0 | 0 | 0 | 0 |
| 0 | 0.0437601 | 0 | 0.009011371 | 0 |
| 0 | 0 | 0 | 0 | 0 |
| 0 | 0 | 0 | 0 | 0 |
| 0 | 0 | 0 | 0 | 0 |
| 0 | 0 | 0 | 0 | 0 |
| 0 | 0 | 0 | 0 | 0 |
| 0 | 0 | 0 | 0 | 0 |
| 0 | 0 | 0.00091417 | 0 | 0 |
| 0 | 0 | 0 | 0 | 0 |
| 0 | 0 | 0 | 0 | 0 |
| 0 | 0.002166375 | 0 | 0 | 0 |
| 0.002047037 | 0.00389653 | 0.00110467 | 0 | 0 |
| 0.000563187 | 0.005864608 | 0.003282339 | 0.003712262 | 0 |
| 0 | 0 | 0 | 0 | 0 |
| 0.000855602 | 0 | 0.007064314 | 0 | 0 |
| 0 | 0 | 0 | 0.002578022 | 0 |
| 0 | 0 | 0 | 0 | 0 |
| 0 | 0 | 0 | 0 | 0 |
| 0 | 0 | 0 | 0 | 0 |
| 0.000772364 | 0.000778337 | 0.002438288 | 0.003545558 | 0 |
| 0 | 0 | 0 | 0 | 0 |
| 0 | 0 | 0 | 0 | 0 |
| 0 | 0.001872028 | 0.015337865 | 0 | 0.003213617 |
| 0 | 0 | 0 | 0 | 0 |
| 0 | 0 | 0 | 0 | 0 |
| 0 | 0 | 0 | 0 | 0 |
| 0 | 0 | 0 | 0 | 0 |
| 0 | 0.007881106 | 0.00434093 | 0 | 0.002453079 |
| 0 | 0 | 0 | 0 | 0 |
| 0 | 0 | 0 | 0 | 0 |
| 0 | 0 | 0.001164504 | 0 | 0 |
| 0 | 0 | 0 | 0 | 0.002310836 |
| 0 | 0 | 0 | 0 | 0 |
| 0 | 0 | 0.002997005 | 0 | 0 |
| 0 | 0 | 0 | 0 | 0 |
| 0 | 0 | 0.916444619 | 0.004454626 | 0 |
| 0 | 0 | 0 | 0 | 0 |
| 0 | 0 | 0 | 0 | 0 |
| 0 | 0 | 0 | 0 | 0 |
| 0 | 0 | 0 | 0 | 0 |
| 0 | 0 | 0 | 0 | 0 |
| 0 | 0 | 0 | 0 | 0 |
| 0 | 0 | 0 | 0 | 0 |
| 0 | 0 | 0.001811544 | 0 | 0 |
| 0 | 0 | 0 | 0 | 0 |
| 0 | 0.003257327 | 0 | 0 | 0.002346164 |

| | | | | |
|---|---|---|---|---|
| 0 | 0.925329683 | 0 | 1.367290564 | 0 |
| 0 | 0 | 0 | 0 | 0 |
| 0 | 0.000815248 | 0 | 0 | 0 |
| 0 | 0 | 0 | 0 | 0 |
| 0.009970558 | 0.005536471 | 0.026880776 | 0.003736336 | 0.001949576 |
| 0 | 0.003343599 | 0.002578326 | 0 | 0 |
| 0 | 0 | 0 | 0 | 0 |
| 0 | 0.025293749 | 0.0036017 | 0.133349639 | 0.00250503 |
| 0 | 0 | 0.001921472 | 0 | 0 |
| 1.299122433 | 0.223083225 | 0.519443478 | 0.04152458 | 0.923882413 |
| 0 | 0 | 0 | 0 | 0 |
| 0.000677351 | 0.003640477 | 0.090358531 | 0.005421513 | 0.000721087 |
| 0.002397708 | 0 | 0.099483038 | 0.015723921 | 0.001230683 |
| 0 | 0 | 0 | 0 | 0 |
| 0 | 0 | 0 | 0 | 0 |
| 0 | 0 | 0.051195339 | 0 | 0.001026367 |
| 0 | 0 | 0 | 0 | 0 |
| 0 | 0 | 0 | 0 | 0.001958984 |
| 0 | 0 | 0.001612271 | 0 | 0.415462294 |
| 0 | 0 | 0 | 0 | 0 |
| 0 | 0 | 0 | 0 | 0 |
| 0 | 0 | 0 | 0 | 0 |
| 0 | 0 | 0 | 0 | 0.021165106 |
| 0 | 0 | 0 | 0 | 0 |
| 0 | 0 | 0 | 0 | 0 |
| 0 | 0.002010227 | 0 | 0.001408797 | 0 |
| 0 | 0 | 0 | 0 | 0 |
| 0 | 0 | 0 | 0 | 0 |
| 0 | 0 | 0 | 0 | 0.045985681 |
| 0 | 0 | 0 | 0.00173937 | 0 |
| 0 | 0 | 0 | 0 | 0.004443547 |
| 0.048462361 | 0 | 0 | 0 | 0 |
| 0 | 0 | 0 | 0 | 0 |
| 0 | 0.262900731 | 0 | 0 | 0.038743104 |
| 0 | 0 | 0 | 0 | 0 |
| 0 | 0 | 0 | 0 | 0.007267886 |
| 0 | 0 | 0 | 0 | 0.057956286 |
| 0 | 0 | 0 | 0 | 0 |
| 0 | 0 | 0 | 0 | 0 |
| 0 | 0.223687024 | 0 | 0 | 0.044558076 |
| 0 | 0 | 0 | 0 | 0 |
| 0 | 0.002360859 | 0.000910256 | 0 | 0.247468285 |
| 0.000915098 | 0 | 0 | 0 | 0 |
| 0.002089205 | 0 | 0 | 0 | 0 |
| 0 | 0 | 0 | 0 | 0 |
| 0 | 0 | 0 | 0 | 0 |
| 0.004217174 | 0.004749764 | 0 | 0.00245273 | 0 |
| 0 | 0 | 0 | 0 | 0 |

| | | | | |
|---|---|---|---|---|
| 0 | 0 | 0 | 0 | 0 |
| 0 | 0 | 0 | 0 | 0.01571597 |
| 0 | 0 | 0 | 0 | 0 |
| 0 | 0 | 0 | 0 | 0 |
| 0 | 0.001482519 | 0 | 0 | 0 |
| 0.014263063 | 0.071079278 | 0 | 0 | 0 |
| 0.003041594 | 0 | 0 | 0 | 0 |
| 0 | 0 | 0 | 0 | 0.031273897 |
| 0.009741659 | 0 | 0 | 0 | 0 |
| 0 | 0 | 0 | 0 | 0 |
| 0.006798391 | 0.010048088 | 0.019370779 | 0.019845213 | 0.003618682 |
| 0 | 0.047942861 | 0 | 0 | 0 |
| 0 | 0 | 0 | 0 | 0 |
| 0.00085637 | 0.00258898 | 0.005199017 | 0.001814396 | 0.002222187 |
| 0 | 0 | 0 | 0 | 1.084100567 |
| 0 | 0 | 0 | 0 | 0 |
| 0 | 0 | 0 | 0 | 0 |
| 0 | 0.000629874 | 0 | 0 | 0 |
| 0 | 0 | 0 | 0.017702169 | 0 |
| 0 | 0.017206234 | 0.000748202 | 0.004170569 | 0 |
| 0.001384056 | 0 | 0 | 0.003258229 | 0 |
| 0 | 0 | 0 | 1.316980497 | 0 |
| 0 | 0 | 0 | 0 | 0.036822002 |
| 0 | 0 | 0 | 0 | 0 |
| 0 | 0 | 0 | 0 | 0 |
| 0 | 0 | 0 | 0 | 0 |
| 0 | 0 | 0 | 0 | 0.234642269 |
| 0 | 0 | 0 | 0 | 0 |
| 0 | 0 | 0 | 0 | 0 |
| 0.047699868 | 0.457602068 | 0.166740245 | 0.484919987 | 0.922806914 |
| 0.001161959 | 0.003659203 | 0.002257356 | 0.006359772 | 0 |
| 0 | 0 | 0.006734673 | 0 | 0 |
| 0 | 0 | 0 | 0 | 0 |
| 0 | 0 | 0.001398771 | 0 | 0 |
| 0 | 0.026498279 | 0 | 0.003277127 | 0 |
| 0 | 0 | 0 | 0 | 0 |
| 0 | 0 | 0 | 0 | 0 |
| 0 | 0 | 0.008266725 | 0 | 0 |
| 0 | 0 | 0 | 0 | 0 |
| 0 | 0 | 0 | 0 | 4.886061079 |
| 0 | 0.021461605 | 0.00324836 | 0.026103542 | 0.006042755 |
| 0 | 0.055221115 | 0.000735191 | 0.32862741 | 8.777169383 |
| 0 | 0 | 0 | 0 | 0 |
| 0 | 0 | 0.004950101 | 0 | 0.00356023 |
| 0 | 0.001200486 | 0 | 0 | 0 |
| 0 | 0 | 0 | 0 | 0 |
| 0.005172508 | 0.0033509 | 0.003768269 | 0.001565574 | 0.002064936 |
| 0 | 0 | 0 | 0 | 0 |

| | | | | |
|---|---|---|---|---|
| 0 | 0 | 0.005313614 | 0 | 0 |
| 0.002621516 | 0 | 0 | 0 | 0 |
| 0 | 0 | 0.041329507 | 0.03772574 | 0.044138307 |
| 0 | 0 | 0 | 0 | 0 |
| 0 | 0 | 0.006825218 | 0 | 0 |
| 0 | 0.598343842 | 0.001871944 | 0.052807325 | 0 |
| 0 | 0 | 0 | 0 | 0 |
| 0.001788234 | 0 | 0 | 0.001804162 | 0 |
| 0.019838621 | 0.037677325 | 0.000926462 | 0 | 0 |
| 0 | 0 | 0 | 0 | 0 |
| 0 | 0 | 0 | 0 | 0 |
| 0.001153164 | 0 | 0 | 0 | 0 |
| 0 | 0 | 0 | 0 | 0 |
| 0.028455798 | 0 | 0.000801182 | 0.006116319 | 0.003457375 |
| 0 | 0 | 0 | 0 | 0 |
| 0.003222857 | 0.320231346 | 0.005791517 | 0.040059259 | 0 |
| 0 | 0.001799914 | 0.032443451 | 0.003027376 | 0 |
| 0.012803251 | 0 | 0 | 0 | 0 |
| 0 | 0.003718351 | 0 | 0.007817627 | 0 |
| 0 | 0 | 0 | 0 | 0 |
| 0 | 0 | 0 | 0.027343377 | 0.096153221 |
| 0 | 0 | 0 | 0 | 0 |
| 0 | 0 | 0 | 0 | 0 |
| 0 | 0 | 0.057263287 | 0.029241122 | 0.148762307 |
| 0 | 0 | 0 | 0.215504091 | 0 |
| 0.476927304 | 0 | 0.001828386 | 0.042206693 | 0 |
| 0 | 0.002970507 | 0 | 0.001921639 | 0 |
| 0 | 0.082424826 | 0 | 0 | 0.001489417 |
| 0 | 0 | 0 | 0 | 0 |
| 0 | 0 | 0 | 0 | 0 |
| 0.049662831 | 0.004691898 | 0.002010017 | 0.06698088 | 0.018311594 |
| 0 | 0.00362706 | 0.017230961 | 0 | 0 |
| 6.711922821 | 0.018253959 | 0.019497228 | 0.331917315 | 0 |
| 0.001052389 | 0.00707019 | 0.102565491 | 0.001486468 | 0.070301485 |
| 0 | 0 | 0 | 0.017329017 | 0 |
| 0 | 0 | 0 | 0 | 0 |
| 0 | 0 | 0.003224422 | 0.003125793 | 0 |
| 0.06316935 | 0.109003243 | 0 | 0.098758408 | 0.052232507 |
| 0.125866165 | 0 | 0.072682325 | 0 | 0 |
| 0 | 0 | 0 | 0 | 0 |
| 0 | 0 | 0 | 0 | 0 |
| 0 | 0 | 0 | 0 | 0 |
| 0 | 0 | 0 | 0.005118076 | 0 |
| 0.024453082 | 0.243154516 | 0.000918613 | 0 | 0 |
| 0 | 0.039163993 | 0 | 0 | 0.328238635 |
| 0 | 0 | 0 | 0 | 0 |
| 0 | 0 | 0 | 0 | 0 |
| 0 | 0 | 0 | 0 | 0 |

| | | | | |
|---|---|---|---|---|
| 0 | 0 | 0 | 0 | 0 |
| 0.019337679 | 0.272345977 | 0.270990929 | 0.005662635 | 0.01054421 |
| 0 | 0 | 0 | 0 | 0 |
| 0 | 0 | 0 | 0 | 0 |
| 0 | 0 | 0 | 0 | 0 |
| 0 | 0 | 0 | 0 | 0 |
| 0 | 0 | 0 | 0 | 0.226275772 |
| 0 | 0 | 0 | 0 | 0 |
| 0.000863515 | 0 | 0 | 0.003354144 | 0 |
| 0.001713697 | 0 | 0 | 0 | 0 |
| 0.001577196 | 0.083708085 | 0 | 0.002722796 | 0 |
| 0.001044124 | 0.007102342 | 0.004310428 | 0.01106095 | 0 |
| 0 | 0.078676267 | 0 | 0 | 0 |
| 0 | 0 | 0 | 0 | 0 |
| 0 | 0 | 0 | 0.011644252 | 0 |
| 0 | 0 | 0 | 0 | 0 |
| 0.002202069 | 0.00258895 | 0.08270795 | 0.005183928 | 0.00073258 |
| 0.004287885 | 0.126990782 | 0.000694179 | 0.011776545 | 0.33722125 |
| 0 | 0 | 0 | 0 | 0 |
| 0 | 0 | 0.001352291 | 0.003495805 | 0.053354036 |
| 0.002649371 | 0.026507901 | 0.009558672 | 0.067091511 | 0.002719711 |
| 0 | 0 | 0 | 0 | 0 |
| 0 | 0 | 0 | 0 | 0 |
| 0 | 0 | 0 | 0 | 0 |
| 0 | 0 | 0 | 0.126199511 | 0 |
| 0 | 0 | 0 | 0.004412137 | 0 |
| 0 | 0.161881839 | 0 | 0.30842353 | 0.294846529 |
| 0.09352777 | 0.206136281 | 0.103157562 | 0.261085234 | 0 |
| 0 | 0 | 0 | 0 | 0 |
| 0 | 0 | 0 | 0 | 0 |
| 0 | 0.000869948 | 0 | 0.036275534 | 0 |
| 0 | 0 | 0 | 0 | 0.000554161 |
| 0 | 0 | 0 | 0 | 0 |
| 0 | 0 | 0 | 0 | 0 |
| 0 | 0.06853271 | 0 | 0.061892711 | 0 |
| 0.025871082 | 0.334122567 | 0.106802898 | 0.140398678 | 8.50300991 |
| 0 | 0.008738195 | 0 | 0 | 0 |
| 0 | 0 | 0 | 0 | 0 |
| 0.00297747 | 0.002500415 | 0.053505528 | 0.009112104 | 0 |
| 0 | 0 | 0 | 0 | 0 |
| 0 | 0.011296966 | 0 | 0 | 0.319486221 |
| 0 | 0 | 0 | 0 | 0 |
| 0.001959274 | 0.002581943 | 0.002195951 | 0 | 0.015282384 |
| 0 | 0 | 0.002968209 | 0.001849768 | 0 |
| 0 | 0.156236004 | 0 | 0 | 0.017247589 |
| 0.343382977 | 0 | 0.00540336 | 0 | 0 |
| 0 | 0 | 0 | 0 | 0 |
| 0 | 0 | 0 | 0 | 0 |

| | | | | |
|---|---|---|---|---|
| 0 | 0.018247329 | 0 | 0 | 0 |
| 0 | 0 | 0 | 0 | 0 |
| 0 | 0 | 0 | 0 | 0 |
| 0.001228734 | 0.133022012 | 1.199508792 | 0.004958713 | 0 |
| 0 | 0 | 0 | 0 | 0 |
| 0 | 0 | 0 | 0 | 0 |
| 0 | 0 | 0 | 0 | 0 |
| 0 | 0.007279406 | 0 | 0 | 0 |
| 0 | 0.220059055 | 0.11894685 | 0.352619118 | 0 |
| 0 | 0 | 0.00502888 | 0 | 0 |
| 0 | 0 | 0 | 0 | 0 |
| 0.000840899 | 0.006779223 | 0.007678059 | 0.009977058 | 0 |
| 0 | 0 | 0 | 0 | 0 |
| 0.001053488 | 0.002547925 | 0 | 0 | 0 |
| 0.197209889 | 0.102374442 | 0.007279606 | 0.021170813 | 0.003102617 |
| 0 | 0 | 0 | 0 | 0 |
| 0 | 0 | 0 | 0 | 0 |
| 0 | 0 | 0 | 0 | 0 |
| 0 | 0.01016411 | 0 | 0 | 0 |
| 0 | 0 | 0 | 0 | 0 |
| 0 | 0 | 0 | 0 | 0 |
| 0 | 0 | 0 | 0 | 0 |
| 0 | 0 | 0 | 0 | 0 |
| 0.277526092 | 0.877411667 | 0.010648044 | 0.216769178 | 0.459499619 |
| 0 | 0 | 0.001529495 | 0.000889626 | 0 |
| 0.120735077 | 0.039902142 | 0.105139493 | 0 | 0 |
| 0 | 0 | 0 | 0 | 0 |
| 0 | 0 | 0 | 1.237419954 | 0 |
| 0 | 0 | 0 | 0.006030265 | 0 |
| 0 | 0 | 0 | 0 | 0 |
| 0 | 0 | 0.0009529 | 0 | 0 |
| 0 | 0.038964803 | 0 | 0 | 0 |
| 0.006968368 | 0.00551749 | 0 | 0.075225658 | 0 |
| 0 | 0 | 0 | 0 | 0 |
| 0 | 0 | 0 | 0 | 0 |
| 0 | 0.070409533 | 0 | 0.066183685 | 0 |
| 0 | 0 | 0 | 0 | 0 |
| 0.039371983 | 0 | 0 | 0 | 0 |
| 0 | 0.020112938 | 0 | 0.011622561 | 0 |
| 0 | 0 | 0 | 0 | 0 |
| 0.297024246 | 0.022762083 | 0.609631267 | 0.011394291 | 0.023959988 |
| 0 | 0 | 0 | 0 | 0 |
| 0.323027239 | 0.747986972 | 0.034968361 | 1.969681315 | 0.226698398 |
| 0.000562188 | 0 | 0 | 0.00833777 | 0 |
| 0 | 0 | 0 | 0 | 0 |
| 0 | 0 | 0 | 0 | 0 |
| 0 | 0 | 0 | 0 | 0 |
| 0.004460791 | 0.024626372 | 0.00150714 | 0.153409137 | 0 |

| | | | | |
|---|---|---|---|---|
| 0 | 0 | 0 | 0 | 0 |
| 0 | 0 | 0 | 0 | 0 |
| 0.134234606 | 0.0256681 | 0.024922173 | 0.258667995 | 0.113411262 |
| 0 | 0 | 0 | 0.00596118 | 0 |
| 0 | 0 | 0 | 0 | 0 |
| 0.000585844 | 0.004132624 | 0.009674097 | 0.003103076 | 0 |
| 0 | 0 | 0 | 0 | 0 |
| 0 | 0 | 0 | 0 | 0 |
| 0 | 0 | 0 | 0 | 0 |
| 0 | 0 | 0 | 0 | 0 |
| 0.012777868 | 0.030168245 | 0.003191629 | 0.007992842 | 0.010202202 |
| 0.505412665 | 4.83368356 | 1.059850722 | 9.282292884 | 0 |
| 0 | 0.007790071 | 0 | 0 | 0 |
| 0 | 0.001588234 | 0.002104994 | 0.00918273 | 0 |
| 0.252588512 | 0.161298587 | 0.004524044 | 0.520797424 | 0.104493469 |
| 0 | 0.259235216 | 0 | 0 | 0 |
| 0 | 0 | 0 | 0 | 0 |
| 0 | 0.001042436 | 0 | 0 | 0 |
| 0 | 0 | 0 | 0 | 0 |
| 0.009773771 | 0.006061145 | 0.001314531 | 0.009345029 | 0.005162414 |
| 0.071755223 | 0.037928148 | 0.536942967 | 0.638150769 | 0.015513862 |
| 2.869386438 | 0.063033442 | 0.003758237 | 0.336548023 | 3.289310362 |
| 40.2733991 | 5.624376691 | 12.38144777 | 1.259383848 | 8.060193372 |
| 0 | 0 | 0 | 0 | 0 |
| 0 | 0 | 0 | 0 | 0 |
| 0 | 0 | 0 | 1.598533237 | 0 |
| 0 | 9.033856809 | 0 | 0.042647334 | 0 |
| 0 | 0 | 0 | 0.002479301 | 0 |
| 0.032808108 | 0 | 0 | 0.022841575 | 0 |
| 0 | 0.080563627 | 0 | 0 | 0 |
| 0 | 0.005271735 | 0 | 0 | 0 |
| 0.159173047 | 0.879456806 | 0 | 0.772409698 | 0 |
| 0.752896905 | 0.236344214 | 0.009105239 | 0.101154298 | 0.034726844 |
| 0.020586771 | 0 | 0 | 0.023020236 | 0 |
| 0.002227237 | 0.020823621 | 0.001923063 | 0.005767492 | 0.007804706 |
| 0.008500243 | 0.007538065 | 0.107503308 | 0.033377614 | 0 |
| 0.008046896 | 0.004633788 | 0 | 0.005103101 | 0 |
| 0 | 0 | 0 | 0 | 0 |
| 0.013726861 | 0 | 0.047730788 | 0 | 0 |
| 0 | 0 | 0 | 0 | 0 |
| 0 | 0.001810267 | 0 | 0 | 0 |
| 0 | 0 | 0 | 0 | 0 |
| 0 | 0 | 0 | 0 | 0 |
| 0 | 0 | 0 | 0 | 0 |
| 0.010469047 | 0.032040781 | 0.002824777 | 0.007667444 | 0.003869804 |
| 0 | 0 | 0.008587764 | 0 | 0 |
| 0 | 0 | 0 | 0 | 0 |
| 0 | 0.076592863 | 0.004770434 | 0.717130318 | 0 |

| | | | | |
|---|---|---|---|---|
| 0 | 0 | 0 | 0 | 0 |
| 0 | 0 | 0 | 0 | 0 |
| 0 | 0 | 0 | 0 | 0 |
| 0.002478535 | 0.01165595 | 0 | 0.014703594 | 0 |
| 0 | 0 | 0 | 0 | 0 |
| 0 | 0 | 0 | 0 | 0 |
| 0 | 0 | 0.008258854 | 0 | 0 |
| 0.258175342 | 0 | 0.023335316 | 0.18125863 | 0 |
| 0 | 0 | 0 | 0 | 0 |
| 0 | 0 | 0 | 0 | 0 |
| 0.020795252 | 0 | 0 | 0.002079481 | 0 |
| 0 | 0 | 0 | 0 | 0 |
| 0 | 0.0029228 | 0.001408649 | 0.020711009 | 0 |
| 0 | 0.023104725 | 0 | 0 | 0 |
| 0 | 0 | 0 | 0 | 0 |
| 0 | 0 | 0 | 0 | 0 |
| 0.222548932 | 0 | 0 | 0 | 0 |
| 0.001225598 | 0 | 0 | 0 | 0 |
| 0 | 0 | 0 | 0.035748854 | 0 |
| 0 | 0.003898421 | 0 | 0 | 0 |
| 0 | 0 | 0 | 0.001153473 | 0 |
| 0 | 0 | 0 | 0 | 0 |
| 0 | 0 | 0 | 0 | 0 |
| 0 | 0 | 0.010042315 | 0 | 0 |
| 0 | 0 | 0 | 0 | 0 |
| 0 | 0 | 0 | 0 | 0 |
| 0 | 0 | 0 | 0 | 0 |
| 0 | 0 | 0 | 0 | 0 |
| 0 | 0 | 0 | 0 | 0 |
| 0 | 0.01827653 | 0 | 0 | 0 |
| 0 | 0 | 0 | 0 | 0 |
| 0 | 0 | 0 | 0 | 0 |
| 0 | 0 | 0 | 0 | 0 |
| 0 | 0 | 0 | 0 | 0 |
| 0 | 0 | 0 | 0.006820695 | 0 |
| 0 | 0 | 0 | 0 | 0 |
| 0 | 0.275172295 | 0 | 0.275120343 | 0 |
| 0.024458023 | 0.004238687 | 0 | 0.013202377 | 0.00062191 |
| 0 | 0 | 0 | 0 | 0 |
| 0.000921938 | 0.002787204 | 0 | 0 | 0.000858784 |
| 0 | 0.067460637 | 0.006814906 | 0.011767742 | 0 |
| 0 | 0 | 0 | 0 | 0 |
| 0 | 0 | 0 | 0 | 0 |
| 0 | 0.004421176 | 0 | 0.641053591 | 0 |
| 0 | 0 | 0 | 0 | 0 |
| 0 | 0 | 0 | 0 | 0 |
| 0 | 0 | 0 | 0 | 0 |
| 0 | 0 | 0 | 0 | 0 |

| | | | | |
|---|---|---|---|---|
| 0 | 0 | 0 | 0 | 0 |
| 0 | 0 | 0 | 0 | 0 |
| 0 | 0 | 0 | 0 | 0 |
| 0.083367946 | 0 | 0.00195677 | 0 | 0 |
| 0 | 0 | 0 | 0 | 0 |
| 0 | 0 | 0 | 0 | 0 |
| 0 | 0 | 0 | 0 | 0 |
| 0.034755389 | 0.129946409 | 0 | 0 | 0 |
| 0.014609908 | 0.099163053 | 0 | 0.003034714 | 0.215458546 |
| 0 | 0 | 0 | 0 | 0 |
| 0 | 0 | 0 | 0 | 0 |
| 0 | 0 | 0 | 0 | 0 |
| 0 | 0 | 0 | 0 | 0 |
| 0 | 0 | 0 | 0 | 0 |
| 0 | 1.839098761 | 0.70383651 | 2.184314687 | 0 |
| 0 | 0.000834364 | 0.138812923 | 0 | 0.088979545 |
| 0 | 0 | 0 | 0 | 0 |
| 0 | 0 | 0 | 0 | 0 |
| 0 | 0 | 0 | 0 | 0 |
| 0 | 0 | 0 | 0 | 0 |
| 0.005914254 | 0 | 0 | 0 | 0 |
| 0 | 0 | 0 | 0 | 0 |
| 0 | 0 | 0 | 0 | 0 |
| 0 | 0 | 0 | 0 | 0 |
| 0 | 0.004328427 | 0.009571489 | 0.008208095 | 0 |
| 0 | 1.192239121 | 2.701137691 | 3.488475926 | 0 |
| 0.006014356 | 0 | 0 | 0 | 0 |
| 0 | 0 | 0 | 0 | 0 |
| 0 | 0 | 0 | 0 | 0 |
| 0 | 0 | 0.001328993 | 0 | 0 |
| 0 | 0 | 0 | 0 | 0 |
| 0 | 0.007050814 | 0 | 0.033784809 | 0 |
| 0 | 0 | 0 | 0 | 0 |
| 0.027527007 | 0 | 0 | 0 | 0 |
| 0 | 0.002016467 | 0 | 0.025123026 | 0 |
| 0.008486752 | 0 | 0 | 0.0033298 | 0 |
| 0 | 1.086393894 | 0 | 0.446943084 | 0.102171157 |
| 0.263710219 | 6.663951431 | 0.198409742 | 2.373512034 | 2.57609246 |
| 0 | 0 | 8.403725356 | 0 | 0 |
| 0 | 0 | 0 | 0 | 0 |
| 0 | 0 | 0 | 0 | 0 |
| 0 | 0.003053224 | 0 | 0.017117957 | 0 |
| 0.135731329 | 0.342818347 | 2.051088551 | 0.071893651 | 4.84517239 |
| 0 | 0 | 0 | 0 | 0 |
| 0 | 0 | 0 | 0 | 0 |
| 0.004168957 | 0 | 0 | 0.000841218 | 0 |
| 0 | 0 | 0 | 0 | 0 |
| 0.101759543 | 0.679579976 | 14.44450199 | 0.132675953 | 0.00113633 |

| | | | | |
|---|---|---|---|---|
| 0 | 0.018894455 | 0 | 0.005746318 | 0 |
| 0 | 0 | 0 | 0 | 0 |
| 0 | 0 | 0 | 0 | 0 |
| 0 | 0 | 0 | 0 | 0 |
| 0 | 0.000887341 | 0 | 0 | 0 |
| 0 | 0 | 0 | 0 | 0 |
| 0 | 0 | 0 | 0 | 0 |
| 0 | 0 | 0 | 0 | 0 |
| 0 | 0 | 0 | 0 | 0 |
| 0 | 0.004096908 | 0 | 0 | 0 |
| 0 | 0 | 0 | 0 | 0 |
| 2.140043389 | 0 | 5.237589489 | 0.224663603 | 0.362437218 |
| 0.006397009 | 0.339084983 | 0.096935102 | 0.002108302 | 0 |
| 0 | 0 | 0 | 0 | 0 |
| 0.075356685 | 0.058493927 | 1.075817947 | 0.009780885 | 0.100657524 |
| 0 | 0 | 0 | 0 | 0 |
| 0 | 0 | 0.000963352 | 0 | 0 |
| 0 | 0 | 0 | 0 | 0 |
| 0.002677697 | 0 | 0 | 0 | 0 |
| 0 | 0 | 0 | 0.003914231 | 0 |
| 0.103159178 | 0 | 0 | 0 | 0 |
| 0 | 0 | 0 | 0 | 0 |
| 0.022565372 | 0.08326298 | 0.065757193 | 0.085321318 | 0 |
| 0 | 0 | 0 | 0 | 0 |
| 0 | 0 | 0 | 0 | 0 |
| 0 | 0 | 0 | 0 | 0 |
| 0 | 0 | 0 | 0 | 0 |
| 0 | 0 | 0 | 0 | 0 |
| 0 | 0 | 0 | 0 | 0 |
| 0 | 0 | 0 | 0 | 0 |
| 0 | 0 | 0 | 0 | 0 |
| 0.002114288 | 0.03142694 | 0.006417912 | 0.028370501 | 0 |
| 0 | 0 | 0 | 0 | 0 |
| 0.068260934 | 0.029404907 | 0.001026009 | 0.015665351 | 0 |
| 0.005815078 | 0 | 0.000498399 | 0.005556274 | 0 |
| 0.000881922 | 0.000666557 | 0.015848262 | 0.073184218 | 0 |
| 0 | 0 | 0 | 0 | 0 |
| 0 | 0 | 0 | 0 | 0 |
| 0 | 0 | 0 | 0 | 0 |
| 0 | 0 | 0 | 0 | 0 |
| 0 | 0 | 0 | 0 | 0 |
| 0 | 0 | 0 | 0 | 0 |
| 0 | 0 | 0 | 0 | 0 |
| 0 | 0 | 0 | 0 | 0 |
| 0.028305207 | 0.037862368 | 0.001800235 | 0.009043152 | 0.001614261 |
| 0 | 0 | 0 | 0 | 0 |
| 0 | 0 | 0 | 0 | 0 |
| 0 | 0.003693514 | 0 | 0.003826433 | 0 |

| | | | | |
|---|---|---|---|---|
| 0 | 0 | 0 | 0 | 0 |
| 0 | 0 | 0 | 0 | 0 |
| 0.012546286 | 0 | 0 | 0 | 0.000834775 |
| 0 | 0 | 0 | 0 | 0 |
| 0 | 0.06678204 | 0 | 0 | 0 |
| 0 | 0 | 0 | 0 | 0 |
| 0 | 0 | 0 | 0 | 0 |
| 0 | 0 | 0.004635741 | 0.029288112 | 0 |
| 0 | 0 | 0 | 0 | 0 |
| 0 | 0 | 0 | 0 | 0 |
| 0 | 0 | 0 | 0 | 0 |
| 0.005673574 | 0 | 0 | 0 | 0 |
| 0 | 0 | 0 | 0 | 0 |
| 0 | 0 | 0 | 0 | 0 |
| 0 | 0 | 0 | 0 | 0 |
| 0 | 0 | 0 | 0 | 0 |
| 0 | 0 | 0 | 0 | 0 |
| 0 | 0 | 0 | 0 | 0 |
| 0 | 0 | 16.08958469 | 0 | 0.005351907 |
| 0 | 0 | 0 | 0 | 0 |
| 0.009974496 | 0 | 0 | 0 | 0 |
| 0 | 0 | 0 | 0 | 0 |
| 0 | 0 | 0 | 0 | 0 |
| 0 | 0 | 0 | 0 | 0 |
| 0 | 0 | 0.290924361 | 0 | 0 |
| 0 | 0.006954848 | 0.000914155 | 0.001107741 | 0.007639316 |
| 0 | 0 | 0 | 0 | 0 |
| 0.015045604 | 0.037750198 | 2.301512963 | 0.009107769 | 0 |
| 0 | 0 | 0 | 0 | 0 |
| 0.060869784 | 0 | 0 | 0.093646018 | 0 |
| 0 | 0 | 0 | 0 | 0 |
| 0 | 0 | 0 | 0 | 0 |
| 0 | 0 | 0 | 0 | 0 |
| 0 | 0 | 0 | 0 | 0 |
| 0.004109271 | 0.001987705 | 0 | 0 | 0 |
| 0 | 0 | 0 | 0.012133398 | 0 |
| 0 | 0 | 0 | 0 | 0 |
| 0 | 0 | 0.001087024 | 0 | 0 |
| 0 | 0 | 0 | 0 | 0 |
| 0 | 0 | 0 | 0.006629396 | 0 |
| 0.757108645 | 9.343834387 | 1.171248892 | 7.598193462 | 7.979758474 |
| 0.03822789 | 0.106765238 | 0.053179737 | 0.058816788 | 0 |
| 0 | 0 | 0 | 0 | 0 |
| 0 | 0.01358171 | 0 | 0 | 0 |
| 0 | 0 | 0 | 0 | 0 |
| 0 | 0 | 0 | 0 | 0 |
| 0 | 0 | 0 | 0 | 0 |
| 0.008585557 | 0.002088403 | 0 | 0 | 0 |

| | | | | |
|---|---|---|---|---|
| 0 | 0 | 0 | 0 | 0 |
| 0.002436125 | 0.002761837 | 0 | 0.011828277 | 0 |
| 0 | 0 | 0 | 0 | 0 |
| 0 | 0 | 0 | 0 | 0 |
| 0.718141087 | 1.337781737 | 2.000927835 | 0.637028306 | 3.180879813 |
| 0 | 0 | 0 | 0 | 0 |
| 0 | 0 | 0 | 0 | 0 |
| 0 | 0 | 0 | 0 | 0 |
| 0.032687169 | 0.776923599 | 0.000738394 | 0.022547963 | 0.353359345 |
| 0 | 0 | 0 | 0 | 0 |
| 0 | 0 | 0 | 0 | 0.00315894 |
| 0 | 0 | 0 | 0 | 0 |
| 0 | 0.05352281 | 0.017532262 | 0.068750153 | 0 |
| 0.003273887 | 0.033541938 | 0.000706693 | 0.02491966 | 0.010891508 |
| 0 | 0 | 0 | 0 | 0 |
| 0.055969337 | 0.260352539 | 0.083071688 | 1.006844455 | 0 |
| 0 | 0.01221285 | 0.443667249 | 0 | 0 |
| 0 | 0 | 0 | 0 | 0 |
| 0 | 0 | 0 | 0 | 0 |
| 0 | 0 | 0 | 0 | 0 |
| 0 | 0 | 0 | 0 | 0 |
| 0 | 0 | 0 | 0 | 0 |
| 0 | 0 | 0 | 0 | 0 |
| 0 | 0 | 0 | 0 | 0 |
| 0 | 0 | 0 | 0 | 0 |
| 0 | 0 | 0 | 0 | 0 |
| 0.002326588 | 0 | 0.180397256 | 0.006765779 | 0 |
| 0.001081004 | 0 | 0.001050042 | 0 | 0 |
| 0 | 0 | 0 | 0 | 0 |
| 0 | 0 | 0 | 0 | 0 |
| 0 | 0 | 0 | 0 | 0 |
| 0 | 0.000649967 | 0 | 0 | 0 |
| 0 | 0 | 0 | 0 | 0 |
| 0 | 0 | 0.004874636 | 0 | 0 |
| 0 | 0 | 0 | 0 | 0 |
| 0.005180629 | 0 | 0 | 0 | 0 |
| 0 | 0.005704192 | 0 | 0.037396741 | 0 |
| 0 | 0 | 0 | 0 | 0 |
| 0 | 0 | 0 | 0 | 0 |
| 0 | 0 | 0 | 0 | 0 |
| 0 | 0 | 0.151742456 | 0.007416855 | 0 |
| 0 | 0 | 0 | 0 | 0 |
| 0 | 0 | 0 | 0 | 0 |
| 0 | 0 | 0 | 0.053126593 | 0 |
| 0 | 0 | 0 | 0 | 0 |
| 0 | 0 | 0 | 0 | 0 |
| 0 | 0 | 0 | 0 | 0 |
| 0 | 0 | 0 | 0 | 0 |

| | | | | |
|---|---|---|---|---|
| 0 | 0 | 0 | 0 | 0.006615119 |
| 0 | 0 | 0 | 0 | 0 |
| 0 | 0.090082251 | 0 | 0 | 0 |
| 0 | 0.002644362 | 0 | 0 | 0 |
| 0 | 0 | 0.025201023 | 0 | 0 |
| 0 | 0 | 0 | 0 | 0 |
| 0.002826153 | 0.03306856 | 1.012066461 | 0 | 0 |
| 0 | 0 | 0 | 0 | 0 |
| 0 | 0.003238713 | 0 | 0 | 0 |
| 0 | 0.056108486 | 0 | 0.011400955 | 0.006050055 |
| 0 | 0 | 0 | 0 | 0 |
| 0 | 0.000553475 | 0 | 0 | 0 |
| 0 | 0 | 0 | 0 | 0 |
| 0 | 0 | 0 | 0 | 0 |
| 0 | 0 | 0 | 0 | 0 |
| 0 | 0 | 0 | 0 | 0 |
| 0 | 0 | 0 | 0 | 0 |
| 0 | 0.002281719 | 0 | 0 | 0 |
| 0 | 0 | 0 | 0 | 0 |
| 0 | 0 | 0 | 0 | 0 |
| 0.000448442 | 2.072462455 | 4.228789336 | 0.034837632 | 0 |
| 0 | 0 | 0 | 0 | 0 |
| 0 | 0.004319951 | 0 | 0.013274363 | 0 |
| 0.000733051 | 0.002954881 | 0 | 0.008904546 | 0.000936461 |
| 0 | 0 | 0 | 0 | 0 |
| 0.001383736 | 0 | 0 | 0 | 0 |
| 0 | 0 | 0 | 0 | 0 |
| 0 | 0 | 0 | 0 | 0 |
| 0 | 0 | 0 | 0 | 0 |
| 0.031861971 | 0 | 0.005329035 | 0.984426824 | 0 |
| 0 | 0 | 0 | 0 | 0 |
| 0 | 0 | 0 | 0 | 0 |
| 0 | 0 | 0 | 0 | 0 |
| 0.34022845 | 0 | 0 | 0 | 0.15038761 |
| 0 | 0 | 0 | 0 | 0 |
| 0 | 0 | 0 | 0 | 0 |
| 0.913934525 | 0.251232994 | 0.014433641 | 0.124892848 | 0.037056757 |
| 0 | 0 | 0 | 0 | 0 |
| 0 | 0 | 0 | 0 | 0 |
| 0 | 0 | 0 | 0 | 0 |
| 0 | 0 | 0 | 0 | 0 |
| 0 | 0 | 0 | 0 | 0 |
| 0 | 0 | 0 | 0 | 0 |
| 0 | 0.004233766 | 0 | 0 | 0 |
| 0 | 0 | 0 | 0 | 0 |
| 0 | 0 | 0 | 0 | 0 |
| 0 | 0 | 0 | 0 | 0 |
| 0 | 0 | 0 | 0 | 0 |

| | | | | |
|---|---|---|---|---|
| 0.006783584 | 0.000635911 | 0 | 0.006462014 | 0.000629791 |
| 0 | 0 | 0 | 0 | 0 |
| 0 | 0 | 0 | 0 | 0.021255801 |
| 0 | 0 | 0 | 0 | 0 |
| 0 | 0 | 0 | 0 | 0 |
| 0 | 0 | 0 | 0 | 0 |
| 0 | 0 | 0 | 0 | 0 |
| 0.572928816 | 0.160296098 | 0.09298922 | 0.403135164 | 0.246762696 |
| 0.049277277 | 0.065628916 | 0.381483999 | 0.232591823 | 0.001581682 |
| 0.00137911 | 0.112398125 | 0.000669805 | 0.012174695 | 0 |
| 0 | 0 | 0 | 0 | 0 |
| 0 | 0 | 0 | 0 | 0 |
| 0 | 0 | 0 | 0 | 0 |
| 0 | 0 | 3.953513876 | 0 | 0 |
| 0 | 0 | 0 | 0 | 0 |
| 0 | 0 | 0 | 0 | 0 |
| 0 | 0.002088584 | 0 | 0 | 0.042105086 |
| 0 | 0 | 0 | 0 | 0 |
| 0 | 0 | 0 | 0 | 0 |
| 0.013151299 | 0.004321633 | 0 | 0 | 0 |
| 0 | 0 | 0 | 0 | 0 |
| 0 | 0 | 0 | 0 | 0 |
| 0 | 0 | 0 | 0 | 0 |
| 0 | 0 | 0 | 0 | 0 |
| 0 | 0 | 0 | 0 | 0 |
| 0 | 0 | 0 | 0 | 0 |
| 0 | 0 | 0 | 0 | 0 |
| 0 | 0 | 0 | 0 | 0 |
| 0 | 0 | 0 | 0 | 0 |
| 0 | 0 | 0 | 0 | 0 |
| 0 | 0 | 0 | 0 | 0 |
| 0 | 0 | 0 | 0 | 0 |
| 0 | 0 | 0 | 0 | 0 |
| 0 | 0 | 0 | 0 | 0 |
| 0 | 0.101814016 | 0 | 0.001618589 | 0 |
| 0 | 0 | 0 | 0 | 0 |
| 0 | 0 | 0 | 0 | 0 |
| 0 | 0 | 0 | 0 | 0 |
| 0 | 0 | 0 | 0 | 0 |
| 0 | 0.040788387 | 0.002141732 | 0 | 0 |
| 0 | 0 | 0 | 0 | 0 |
| 0.588504708 | 1.338060376 | 0 | 0.483783747 | 0 |
| 0 | 0 | 0 | 0 | 0 |
| 0 | 0.000964003 | 0 | 0 | 0 |
| 0 | 0.004639768 | 0 | 0 | 0 |
| 0 | 0.000848182 | 0 | 0 | 0.000560012 |
| 0 | 0 | 0 | 0 | 0 |
| 0 | 0 | 0 | 0 | 0 |

| | | | | |
|---|---|---|---|---|
| 0 | 0 | 0 | 0 | 0 |
| 0 | 0.007030738 | 0.004840682 | 0.021921218 | 0.044222586 |
| 0 | 0 | 0 | 0 | 0 |
| 0 | 0 | 0 | 0 | 0 |
| 0.027670987 | 0.050754696 | 0.43334244 | 1.265604569 | 0 |
| 0 | 0 | 0 | 0 | 0 |
| 0 | 0 | 0 | 0 | 0 |
| 0.02026861 | 0.008291485 | 0.003118905 | 0.001984174 | 0.030763755 |
| 0 | 0.006461456 | 0 | 0 | 0 |
| 0 | 0 | 0 | 0 | 0 |
| 0 | 0 | 0 | 0 | 0 |
| 0 | 0 | 0 | 0 | 0 |
| 0 | 0 | 0 | 0 | 0 |
| 0 | 0 | 0 | 0 | 0 |
| 0 | 0.458411187 | 0.001646699 | 0 | 0 |
| 0.031328579 | 0.012979135 | 0.003888441 | 0.022371173 | 0 |
| 0 | 0 | 0 | 0 | 0 |
| 0 | 0 | 0 | 0 | 0 |
| 0 | 0 | 0 | 0 | 0 |
| 0 | 0 | 0 | 0 | 0 |
| 0 | 0 | 0 | 0 | 0 |
| 0 | 0 | 0 | 0 | 0 |
| 0 | 0 | 0 | 0 | 0 |
| 0.003994442 | 0.000700058 | 0 | 0.000981222 | 0 |
| 0 | 0 | 0 | 0 | 0 |
| 0 | 0 | 0 | 0 | 0 |
| 0.004765266 | 0.078114492 | 0.0084861 | 0.061250234 | 0.008624032 |
| 0 | 0.000505648 | 0 | 0 | 0.000534166 |
| 0.008263477 | 0 | 0 | 0 | 0.000565525 |
| 0 | 0 | 0 | 0 | 0 |
| 0 | 0 | 0 | 0 | 0 |
| 0 | 0 | 0 | 0 | 0 |
| 0 | 0 | 0 | 0 | 0 |
| 0 | 0 | 0 | 0 | 0 |
| 0 | 0 | 0 | 0 | 0 |
| 0 | 0 | 0 | 0 | 0 |
| 0 | 0 | 0 | 0 | 0 |
| 0.003746746 | 0.1321503 | 0 | 0.042567419 | 0 |
| 0 | 0 | 0 | 0 | 0 |
| 0 | 0 | 0 | 0 | 0 |
| 0 | 0 | 0 | 0 | 0 |
| 0 | 0 | 0 | 0 | 0 |
| 0 | 0 | 0 | 0 | 0 |
| 0 | 0 | 0 | 0 | 0 |
| 0 | 0 | 0 | 0 | 0 |
| 0 | 0 | 0 | 0 | 0 |
| 0 | 0 | 0 | 0 | 0 |
| 0 | 0 | 0 | 0 | 0 |

| | | | | |
|---|---|---|---|---|
| 0.054373379 | 0.001655859 | 0 | 0.019622171 | 0.017412989 |
| 0 | 0 | 0 | 0 | 0 |
| 0 | 0 | 0 | 0 | 0 |
| 0 | 0 | 0 | 0 | 0 |
| 0 | 0 | 0 | 0 | 0 |
| 0.70522919 | 0 | 0 | 0.306290064 | 0 |
| 0 | 0 | 0 | 0 | 0 |
| 0 | 0 | 0 | 0 | 0 |
| 0 | 0 | 0 | 0 | 0 |
| 0 | 0 | 0 | 0 | 0 |
| 0 | 0 | 0 | 0 | 0 |
| 0 | 0.000645342 | 0.000622048 | 0 | 0 |
| 0 | 0.001542535 | 0 | 0 | 0 |
| 0 | 0 | 0 | 0 | 0 |
| 0 | 0 | 0 | 0 | 0 |
| 0 | 0 | 0 | 0 | 0 |
| 0 | 0 | 0 | 0 | 0 |
| 0 | 0.126690217 | 0 | 0 | 0 |
| 0 | 0 | 0 | 0 | 0 |
| 0 | 0 | 0.002124961 | 0 | 0 |
| 0 | 0 | 0 | 0.006839845 | 0.000828506 |
| 0 | 0 | 0 | 0 | 0 |
| 0.125371623 | 0.006823307 | 0 | 0.002159556 | 0 |
| 0 | 0 | 0.04738026 | 0.437097399 | 0 |
| 0 | 0 | 0 | 0 | 0 |
| 0 | 0 | 0 | 0 | 0 |
| 0 | 0 | 0 | 0 | 0 |
| 0 | 0 | 0 | 0 | 0 |
| 0 | 0 | 0.007713916 | 0.016024205 | 0 |
| 0 | 0 | 0 | 0 | 0 |
| 0 | 0 | 0 | 0 | 0 |
| 0 | 0 | 0 | 0 | 0 |
| 0 | 0.007420644 | 0.013113441 | 0 | 0.005389428 |
| 0 | 0 | 0 | 0 | 0 |
| 0.005488991 | 0.309023234 | 0.119787273 | 0.005168689 | 0 |
| 0 | 0.066991 | 0.045599611 | 0 | 0 |
| 0 | 0 | 0 | 0 | 0 |
| 0 | 0 | 0 | 0 | 0 |
| 0 | 0.000765291 | 0.002213 | 0 | 0.001667435 |
| 0 | 0 | 0 | 0 | 0 |
| 0 | 0 | 0 | 0 | 0 |
| 0 | 0 | 0 | 0 | 0 |
| 0 | 0.020581647 | 0.015957236 | 0.042017519 | 0 |
| 0 | 0 | 0 | 0 | 0 |
| 0 | 0 | 0 | 0 | 0 |
| 0 | 0 | 0 | 0 | 0 |
| 0 | 0 | 0 | 0 | 0 |
| 0 | 0 | 0 | 0 | 0 |

| | | | | |
|---|---|---|---|---|
| 0 | 0 | 0 | 0 | 0 |
| 0 | 0 | 0 | 0 | 0 |
| 0 | 0 | 0 | 0 | 0 |
| 0 | 0 | 0 | 0 | 0 |
| 0 | 0 | 0 | 0 | 0 |
| 0 | 0 | 0 | 0 | 0 |
| 0 | 0 | 0 | 0.300590478 | 0 |
| 0 | 0 | 0 | 0 | 0 |
| 0 | 0 | 0 | 0 | 0 |
| 0 | 0 | 0 | 0 | 0 |
| 0 | 0 | 0 | 0 | 0 |
| 0 | 0 | 0 | 0 | 0 |
| 0 | 0 | 0 | 0 | 0 |
| 0 | 0 | 0 | 0 | 0.144654844 |
| 0 | 0 | 0 | 0 | 0 |
| 0.004466637 | 0 | 0 | 0 | 0.010984162 |
| 0 | 0 | 0 | 0 | 0 |
| 0 | 0 | 0 | 0 | 0 |
| 0 | 0 | 0 | 0 | 0 |
| 0 | 0.497144684 | 0 | 0.009085188 | 0 |
| 0 | 0 | 0 | 0 | 0 |
| 0 | 0 | 0 | 0 | 0 |
| 0 | 0 | 0 | 0 | 0 |
| 0 | 0 | 0 | 0 | 0 |
| 0 | 0 | 0 | 0 | 0 |
| 0 | 0 | 0 | 0 | 0 |
| 0 | 0 | 0 | 0 | 0 |
| 0 | 0 | 0 | 0 | 0 |
| 0 | 0 | 0 | 0 | 0 |
| 0.042774978 | 0 | 0 | 0 | 0 |
| 0.151313291 | 0 | 0.526135624 | 0.923819046 | 0 |
| 0 | 0 | 0 | 0 | 0 |
| 0 | 0 | 0 | 0 | 0 |
| 0 | 0 | 0 | 0 | 0 |
| 0 | 0 | 0 | 0 | 0 |
| 0 | 0 | 0 | 0 | 0 |
| 0 | 0 | 0 | 0 | 0 |
| 0 | 0 | 0 | 0 | 0 |
| 0 | 0 | 0 | 0 | 0 |
| 0 | 0 | 0 | 0 | 0 |
| 0 | 0 | 0 | 0 | 0 |
| 0 | 0 | 0 | 0 | 0 |
| 0 | 0 | 0 | 0 | 0 |
| 0 | 0 | 0 | 0 | 0 |
| 0 | 0 | 0 | 0 | 0 |
| 0 | 0 | 0 | 0 | 0 |
| 0 | 0.000552939 | 0 | 0.002841726 | 0.165745471 |
| 0 | 0 | 0 | 0.00398908 | 0 |

| | | | | |
|---|---|---|---|---|
| 0 | 0 | 0 | 0 | 0 |
| 0 | 0 | 0 | 0 | 0 |
| 0 | 0 | 0 | 0 | 0 |
| 0 | 0 | 0 | 0.009492725 | 0 |
| 0 | 0.062238424 | 0.006238091 | 0.25224877 | 0 |
| 0 | 0 | 0 | 0 | 0 |
| 0 | 0 | 0 | 0 | 0 |
| 0 | 0 | 0 | 0 | 0 |
| 0 | 0 | 0 | 0 | 0 |
| 0 | 0 | 0 | 0 | 0 |
| 0 | 0 | 0 | 0 | 0 |
| 0.012141005 | 0.01108788 | 0 | 0.005091051 | 0 |
| 0.042225845 | 0 | 0 | 0.008091068 | 0.001568106 |
| 0 | 0 | 0 | 0 | 0 |
| 0 | 0.020541606 | 0.003651088 | 0 | 0 |
| 0 | 0 | 0 | 0.235248766 | 0 |
| 0.001313139 | 0 | 0 | 0.001648683 | 0.003145338 |
| 0 | 0 | 0 | 0 | 0 |
| 0.052717477 | 0.001302798 | 0.005720736 | 0.007304159 | 0.051725015 |
| 0 | 0.004158138 | 0.172710306 | 0 | 0.130731478 |
| 0 | 0 | 0 | 0 | 0 |
| 0 | 0 | 0 | 0 | 0 |
| 0 | 0 | 0 | 0 | 0 |
| 0 | 0 | 0 | 0 | 0 |
| 0 | 0 | 0 | 0 | 0 |
| 0 | 0 | 0 | 0 | 0 |
| 0.060419403 | 0.026353936 | 0 | 0.023564191 | 0.00276003 |
| 0 | 0 | 0 | 0 | 0 |
| 0 | 0 | 0 | 0 | 0 |
| 0 | 0 | 0 | 0 | 0 |
| 0.000851848 | 0 | 0 | 0.002647061 | 0.000408083 |
| 0 | 0 | 0 | 0 | 0 |
| 0 | 0 | 0 | 0 | 0 |
| 0 | 0 | 0 | 0 | 0 |
| 0 | 0 | 0 | 0 | 0 |
| 0 | 0 | 0 | 0 | 0 |
| 0 | 0 | 0 | 0 | 0 |
| 0.011610581 | 0.019299589 | 0.003255516 | 0.006086448 | 0 |
| 0.008206325 | 0 | 0 | 0 | 0 |
| 0 | 0.000956692 | 0 | 0 | 0 |
| 0.043018525 | 0.001324621 | 0 | 0.010970983 | 0 |
| 0 | 0 | 0 | 0 | 0 |
| 0 | 0 | 0 | 0 | 0 |
| 0 | 0 | 0 | 0 | 0 |
| 0 | 0 | 0 | 0 | 0 |
| 0 | 0 | 0 | 0 | 0 |
| 0 | 0 | 0 | 0 | 0 |
| 0 | 0 | 0 | 0 | 0 |

| | | | | |
|---|---|---|---|---|
| 0 | 0 | 0 | 0 | 0 |
| 0 | 0 | 1.051716096 | 0 | 0 |
| 0 | 0 | 0 | 0 | 0 |
| 0.000744062 | 0 | 0 | 0.001576449 | 0 |
| 0 | 0 | 0 | 0.004142704 | 0 |
| 0.002671944 | 0.001570688 | 0 | 0.005975564 | 0 |
| 0.003971309 | 0 | 0 | 0 | 0 |
| 0 | 0 | 0 | 0 | 0.000813088 |
| 0.026596339 | 0.047969546 | 0 | 0 | 0 |
| 0 | 0 | 0 | 0 | 0 |
| 0 | 0 | 0 | 0 | 0 |
| 0 | 0 | 0 | 0 | 0 |
| 0.001318589 | 0.012490594 | 0.126032925 | 0.007263621 | 0 |
| 0 | 0 | 0 | 0 | 0 |
| 0 | 0 | 0 | 0 | 0 |
| 0 | 0 | 0 | 0 | 0 |
| 0 | 0 | 0 | 0 | 0 |
| 0 | 0 | 0 | 0 | 0 |
| 0 | 0 | 0 | 0 | 0 |
| 0 | 0 | 0 | 0 | 0 |
| 0 | 0 | 0 | 0 | 0 |
| 0 | 0 | 0 | 0 | 0 |
| 0.021182587 | 0.004374265 | 0 | 0.002207196 | 0.007624606 |
| 0.001024434 | 0.008052381 | 0.000796074 | 0 | 0.023065798 |
| 0 | 0 | 0 | 0 | 0 |
| 0 | 0 | 0 | 0 | 0 |
| 0 | 0 | 0 | 0 | 0 |
| 0 | 0 | 0 | 0 | 0 |
| 0 | 0 | 0 | 0 | 0 |
| 0 | 0 | 0 | 0 | 0 |
| 0.003097507 | 0 | 0 | 0.003007905 | 0 |
| 0 | 0 | 0 | 0 | 0 |
| 0 | 0 | 0 | 0 | 0 |
| 0 | 0 | 0 | 0 | 0 |
| 0 | 0 | 0.006936879 | 0 | 0 |
| 0.071237097 | 0 | 0 | 0 | 0 |
| 0 | 0 | 0 | 0 | 0 |
| 0 | 0 | 0.034929504 | 0 | 0 |
| 0 | 0 | 0.007636934 | 0.001156771 | 0 |
| 0 | 0 | 0 | 0 | 0 |
| 0 | 0 | 0 | 0 | 0 |
| 0 | 0 | 0 | 0 | 0 |
| 0 | 0 | 0 | 0 | 0 |
| 0 | 0 | 0 | 0 | 0 |
| 0 | 0 | 0 | 0 | 0 |
| 0.059374044 | 0.102094089 | 0.017218053 | 0.044985325 | 0 |
| 0 | 0 | 0 | 0 | 0 |
| 0 | 0 | 0 | 0 | 0 |

| | | | | |
|---|---|---|---|---|
| 0 | 0 | 0 | 0 | 0 |
| 0 | 0 | 0 | 0 | 0 |
| 0 | 0 | 0 | 0.004384552 | 0 |
| 0 | 0 | 0 | 0 | 0 |
| 0 | 0 | 0 | 0 | 0 |
| 0 | 0 | 0 | 0 | 0 |
| 0 | 0 | 0 | 0 | 0 |
| 0 | 0 | 0 | 0 | 0 |
| 0 | 0.002290878 | 0 | 0 | 0 |
| 0 | 0 | 0 | 0 | 0 |
| 0.003454903 | 0 | 0 | 0 | 0 |
| 0 | 0.001599408 | 0 | 0 | 0 |
| 0 | 0 | 0 | 0 | 0 |
| 0 | 0 | 0 | 0 | 0 |
| 0 | 0 | 0 | 0 | 0 |
| 0 | 0 | 0 | 0 | 0 |
| 0 | 0 | 0 | 0 | 0 |
| 0 | 0 | 0 | 0 | 0 |
| 0 | 0 | 0 | 0 | 0 |
| 0 | 0 | 0.004621741 | 0.003235824 | 0 |
| 0 | 0 | 0 | 0 | 0 |
| 0 | 0 | 0 | 0 | 0 |
| 0 | 0 | 0 | 0 | 0 |
| 0 | 0 | 0 | 0 | 0 |
| 0 | 0 | 0 | 0 | 0 |
| 0 | 0 | 0 | 0 | 0 |
| 0 | 0 | 0 | 0 | 0 |
| 0 | 0 | 0.011555519 | 0.008263814 | 0 |
| 0 | 0 | 0 | 0 | 0 |
| 0 | 0 | 0 | 0 | 0 |
| 0.002876481 | 0.004576937 | 0.024852728 | 0.010050435 | 0 |
| 0 | 0 | 0 | 0 | 0 |
| 0.372836839 | 0.144902977 | 0.008708189 | 0.30961627 | 0 |
| 0.000718527 | 0.004163482 | 0.096142204 | 0 | 0 |
| 0 | 0 | 0 | 0 | 0 |
| 0 | 0 | 0 | 0 | 0 |
| 0 | 0 | 0 | 0 | 0 |
| 0 | 0 | 0 | 0 | 0 |
| 0 | 0 | 0 | 0 | 0 |
| 0 | 0 | 0 | 0 | 0 |
| 0 | 0 | 0 | 0 | 0 |
| 0.000578462 | 0.593428614 | 0.016013978 | 0.589100355 | 0.013393941 |
| 0.10377147 | 0 | 0.210326538 | 0.012940325 | 0.018708968 |
| 0 | 0 | 0 | 0 | 0 |
| 0 | 0 | 0 | 0 | 0 |
| 0 | 0.004105039 | 0.008655636 | 0.032724421 | 0 |
| 0 | 0 | 0.001621394 | 0 | 0 |
| 0 | 0 | 0 | 0 | 0 |

| | | | | |
|---|---|---|---|---|
| 0 | 0 | 0 | 0 | 0 |
| 0 | 0 | 0 | 0.050576944 | 0 |
| 0 | 0.002389969 | 0.010052511 | 0 | 0 |
| 0 | 0 | 0 | 0 | 0 |
| 0.198142598 | 0.204358957 | 0 | 0.067596856 | 0 |
| 0.002681089 | 0 | 0 | 0 | 0.01551975 |
| 0 | 0 | 0 | 0 | 0 |
| 0 | 0 | 0 | 0 | 0 |
| 0 | 0 | 0 | 0 | 0 |
| 0 | 0 | 0 | 0 | 0 |
| 0 | 0 | 0 | 0 | 0 |
| 0 | 0 | 0 | 0 | 0 |
| 0 | 0 | 0 | 0 | 0 |
| 0 | 0 | 0 | 0 | 0 |
| 0 | 0 | 0 | 0 | 0 |
| 0 | 0 | 0 | 0 | 0 |
| 0 | 0 | 0.002116513 | 0 | 0 |
| 0 | 0 | 0 | 0 | 0 |
| 0 | 0 | 0 | 0 | 0 |
| 0 | 0 | 0 | 0 | 0 |
| 0 | 0 | 0 | 0 | 0 |
| 0 | 0 | 0 | 0 | 0 |
| 0 | 0 | 0 | 0 | 0 |
| 0 | 0 | 0 | 0 | 0 |
| 0 | 0 | 0 | 0 | 0 |
| 0 | 0.000900656 | 0 | 0 | 0 |
| 0 | 0 | 0 | 0 | 0 |
| 0 | 0 | 0 | 0.001714381 | 0 |
| 0 | 0 | 0 | 0 | 0 |
| 0 | 0 | 0 | 0 | 0 |
| 0 | 0 | 0 | 0 | 0 |
| 0 | 0 | 0 | 0 | 0.001376706 |
| 0 | 0 | 0 | 0 | 0 |
| 0 | 0 | 0 | 0 | 0 |
| 0 | 0 | 0 | 0 | 0 |
| 0 | 0 | 0 | 0 | 0 |
| 0 | 0 | 0 | 0 | 0 |
| 0 | 0 | 0.000518499 | 0 | 0 |
| 0 | 0 | 0.000788208 | 0 | 0 |
| 0 | 0 | 0 | 0 | 0 |
| 0 | 0 | 0 | 0 | 0 |
| 0 | 0 | 0 | 0 | 0 |
| 0.004664239 | 0 | 0 | 0.004814379 | 0.001002631 |
| 0 | 0 | 0 | 0 | 0 |
| 0 | 0 | 0 | 0 | 0 |
| 0 | 0 | 0 | 0 | 0 |
| 0.024973807 | 0.002212479 | 0 | 0 | 0.001095592 |
| 0 | 0 | 0 | 0 | 0 |

| | | | | |
|---|---|---|---|---|
| 0 | 0.008862911 | 0 | 0 | 0 |
| 0.049519408 | 0.039607837 | 0 | 0.014429145 | 0.002488384 |
| 0 | 0 | 0 | 0 | 0 |
| 0 | 0 | 0 | 0 | 0 |
| 0 | 0 | 0 | 0 | 0 |
| 0 | 0 | 0 | 0 | 0 |
| 0 | 0 | 0 | 0 | 0 |
| 0 | 0 | 0 | 0 | 0 |
| 0 | 0 | 0 | 0 | 0 |
| 0 | 0 | 0 | 0 | 0 |
| 0 | 0 | 0 | 0 | 0 |
| 0 | 0 | 0 | 0 | 0 |
| 0.003843617 | 0 | 0.006578124 | 0 | 0 |
| 0 | 0 | 0 | 0 | 0 |
| 0.005613022 | 0.007396873 | 0 | 0.00182959 | 0 |
| 0 | 0 | 0 | 0 | 0 |
| 0 | 0 | 0 | 0 | 0 |
| 0 | 0 | 0 | 0 | 0 |
| 0.006209455 | 0 | 0.001855879 | 0 | 0 |
| 0 | 0 | 0 | 0 | 0 |
| 0 | 0 | 0 | 0 | 0 |
| 0 | 0.004059469 | 0.306444767 | 0 | 0 |
| 0.046105614 | 0 | 0.000633005 | 0 | 0 |
| 0 | 0 | 0 | 0 | 0 |
| 0 | 0 | 0 | 0 | 0 |
| 0 | 0 | 0 | 0 | 0 |
| 0 | 0 | 0 | 0 | 0 |
| 0 | 0 | 0 | 0 | 0 |
| 0 | 0 | 0 | 0 | 0 |
| 0 | 0 | 0 | 0 | 0 |
| 0 | 0 | 0 | 0 | 0 |
| 0 | 0 | 0 | 0 | 0 |
| 0 | 0 | 0 | 0 | 0 |
| 0 | 0 | 0 | 0 | 0 |
| 0 | 0 | 0 | 0 | 0 |
| 0 | 0 | 0 | 0 | 0 |
| 0 | 0 | 0 | 0 | 0 |
| 0 | 0 | 0 | 0 | 0 |
| 0.000635173 | 0.001493531 | 0.06066972 | 0.00149527 | 0.001690464 |
| 0 | 0 | 0 | 0 | 0 |
| 0.033221777 | 0.152088998 | 1.382088872 | 0.024132732 | 0.101401986 |
| 0 | 0 | 0 | 0 | 0 |
| 0 | 0 | 0 | 0 | 0 |
| 0 | 0 | 0 | 0 | 0 |
| 0 | 0 | 0 | 0 | 0 |
| 0.008911449 | 0.030226609 | 0.03948077 | 0.001228015 | 0.068895447 |
| 0 | 0 | 0 | 0 | 0 |
| 0 | 0 | 0 | 0 | 0 |

| | | | | |
|---|---|---|---|---|
| 0 | 0 | 0 | 0 | 0 |
| 0 | 0 | 0 | 0 | 0 |
| 0 | 0.062111531 | 0 | 0 | 0 |
| 0 | 0 | 0 | 0 | 0 |
| 0 | 0 | 0 | 0 | 0 |
| 0 | 0 | 0 | 0 | 0 |
| 0 | 0 | 0 | 0 | 0 |
| 0 | 0.007339069 | 0.00319478 | 0 | 0 |
| 0 | 0.085466923 | 0 | 0.007903009 | 0 |
| 0.036349643 | 0 | 0.009498202 | 0.002702251 | 0 |
| 0 | 0.203102366 | 0 | 0 | 0 |
| 0 | 0 | 0 | 0 | 0 |
| 0 | 0.001675017 | 0 | 0 | 0 |
| 0 | 0 | 0 | 0 | 0 |
| 0 | 0 | 0 | 0 | 0 |
| 0 | 0 | 0 | 0 | 0 |
| 0 | 0 | 0 | 0 | 0 |
| 0 | 0 | 0 | 0 | 0 |
| 0 | 0 | 0 | 0 | 0 |
| 0.22099159 | 0.12206608 | 0.770268496 | 0.557679771 | 0.078846901 |
| 0 | 0 | 0 | 0 | 0 |
| 0 | 0 | 0 | 0 | 0 |
| 0 | 0 | 0 | 0 | 0 |
| 0 | 0 | 0 | 0 | 0 |
| 0 | 0 | 0 | 0 | 0 |
| 0 | 0 | 0 | 0 | 0 |
| 0 | 0 | 0 | 0 | 0 |
| 0 | 0 | 0 | 0 | 0 |
| 0 | 0 | 0 | 0 | 0 |
| 0 | 0 | 0 | 0.008935549 | 0.00615591 |
| 0 | 0 | 0 | 0 | 0 |
| 0 | 0 | 0 | 0 | 0 |
| 0 | 0 | 0 | 0 | 0 |
| 0 | 0 | 0 | 0 | 0 |
| 0 | 0 | 0 | 0 | 0 |
| 0 | 0.002894872 | 0 | 0 | 0 |
| 0 | 0 | 0 | 0 | 0 |
| 0 | 0 | 0 | 0 | 0 |
| 0 | 0 | 0 | 0 | 0 |
| 0 | 0 | 0 | 0 | 0 |
| 0 | 0 | 0 | 0 | 0 |
| 0 | 0 | 0 | 0 | 0 |
| 0 | 0 | 0 | 0 | 0 |
| 0 | 0 | 0 | 0 | 0 |
| 0.00590557 | 0.000875183 | 0 | 0.005397402 | 0 |
| 0 | 0 | 0 | 0 | 0 |
| 0 | 0 | 0 | 0 | 0 |
| 0 | 0 | 0 | 0 | 0 |

| | | | | |
|---|---|---|---|---|
| 0 | 0 | 0 | 0 | 0 |
| 0 | 0 | 0 | 0 | 0 |
| 0 | 0 | 0 | 0 | 0 |
| 0 | 0 | 0 | 0 | 0 |
| 0 | 0 | 0 | 0 | 0 |
| 0.001815363 | 0.004442834 | 0.002015277 | 0.001465226 | 0.001656498 |
| 0 | 0 | 0 | 0 | 0 |
| 0 | 0 | 0 | 0 | 0 |
| 0 | 0 | 0 | 0 | 0 |
| 0 | 0 | 0 | 0 | 0 |
| 0 | 0 | 0 | 0 | 0 |
| 0 | 0 | 0 | 0 | 0 |
| 0 | 0 | 0 | 0 | 0 |
| 0 | 0 | 0 | 0 | 0 |
| 0 | 0 | 0 | 0 | 0 |
| 0 | 0 | 0 | 0 | 0 |
| 0 | 0.00681566 | 0 | 0 | 0 |
| 0 | 0.001770219 | 0 | 0 | 0 |
| 0 | 0 | 0 | 0 | 0 |
| 0 | 0 | 0 | 0 | 0 |
| 0 | 0 | 0 | 0 | 0 |
| 0 | 0 | 0 | 0 | 0 |
| 0 | 0 | 0 | 0 | 0 |
| 0 | 0 | 0 | 0 | 0 |
| 0 | 0 | 0 | 0 | 0 |
| 0.000608471 | 0 | 0 | 0.002578341 | 0 |
| 0 | 0 | 0 | 0 | 0 |
| 0 | 0.561871731 | 0 | 0 | 0 |
| 0 | 0 | 0 | 0 | 0 |
| 0 | 0 | 0 | 0 | 0 |
| 0 | 0 | 0 | 0 | 0 |
| 0 | 0 | 0 | 0 | 0 |
| 0 | 0 | 0 | 0 | 0 |
| 0 | 0 | 0 | 0 | 0 |
| 0 | 0 | 0 | 0 | 0 |
| 0 | 0 | 0 | 0 | 0 |
| 0 | 0 | 0 | 0 | 0 |
| 0 | 0 | 0 | 0 | 0 |
| 0.186150739 | 0 | 0 | 0.059980622 | 0 |
| 0 | 0 | 0 | 0 | 0 |
| 0 | 0 | 0 | 0 | 0 |
| 0 | 0 | 0 | 0 | 0 |
| 0 | 0 | 0 | 0 | 0 |
| 0 | 0 | 0 | 0 | 0 |
| 0 | 0 | 0 | 0 | 0 |
| 0 | 0 | 0 | 0 | 0 |
| 0 | 0 | 0 | 0 | 0 |
| 0 | 0 | 0 | 0 | 0 |

| | | | | |
|---|---|---|---|---|
| 0 | 0 | 0 | 0 | 0 |
| 0 | 0.017701984 | 0 | 0.006766808 | 0 |
| 0 | 0 | 0 | 0 | 0 |
| 0 | 0 | 0 | 0 | 0 |
| 0 | 0 | 0 | 0 | 0 |
| 0 | 0 | 0 | 0 | 0 |
| 0 | 0 | 0 | 0 | 0 |
| 0 | 0 | 0 | 0 | 0 |
| 0 | 0 | 0 | 0 | 0 |
| 0 | 0 | 0 | 0 | 0 |
| 0 | 0 | 0 | 0 | 0 |
| 0 | 0 | 0 | 0 | 0 |
| 0.003500358 | 0.578921755 | 0 | 0 | 0 |
| 0 | 0 | 0 | 0 | 0 |
| 0 | 0 | 0 | 0 | 0 |
| 0 | 0 | 0 | 0 | 0 |
| 0 | 0 | 0 | 0 | 0 |
| 0 | 0 | 0 | 0 | 0 |
| 0 | 0 | 0 | 0 | 0 |
| 0 | 0 | 0 | 0 | 0 |
| 0.021222811 | 0 | 0 | 0.001362572 | 0 |
| 0 | 0 | 0 | 0 | 0 |
| 0 | 0 | 0 | 0 | 0 |
| 0.134574317 | 1.135011389 | 0.368406554 | 4.379565166 | 1.40396817 |
| 0 | 0 | 0.00126037 | 0 | 0 |
| 0 | 0 | 0 | 0 | 0 |
| 0 | 0 | 0 | 0 | 0 |
| 0 | 0 | 0 | 0 | 0 |
| 0 | 0 | 0 | 0 | 0 |
| 0 | 0 | 0 | 0 | 0 |
| 0 | 0 | 0 | 0 | 0.003175083 |
| 0 | 0 | 0 | 0 | 0 |
| 0 | 0 | 0 | 0 | 0 |
| 0 | 0 | 0 | 0 | 0 |

| PHL018-0064_V1 | PHL018-0270_V3 | PHL018-0268_V2 | PHL018-0064_V3 | PHL018-0046_V1 |
|---|---|---|---|---|
| CTRL | REF | REF | CTRL | CTRL |
| 0.079809765 | 0.145212017 | 0.032759339 | 0.122222576 | 0.461777121 |
| 0.053800796 | 2.167789164 | 3.879370866 | 1.080146427 | 0 |
| 0 | 8.88912531 | 1.053489405 | 0.094973927 | 1.051812847 |
| 0 | 0 | 0 | 0 | 0 |
| 0 | 0 | 0 | 0 | 0 |
| 0 | 0 | 0 | 0 | 0 |
| 0 | 0 | 0 | 0 | 0 |
| 0 | 0 | 0 | 0 | 0 |
| 0 | 0 | 0 | 0 | 0 |
| 0 | 0 | 0 | 0 | 0 |
| 0 | 0 | 0 | 0 | 0 |
| 0 | 0 | 0 | 0 | 0 |
| 0 | 0 | 0 | 0 | 0 |
| 2.339575658 | 3.28575782 | 0 | 1.460352675 | 0 |
| 0 | 0 | 0 | 0 | 0 |
| 0 | 0 | 0 | 0 | 0 |
| 0 | 0 | 0 | 0 | 0 |
| 0 | 0 | 0 | 0 | 0 |
| 0 | 0 | 0 | 0 | 0 |
| 0 | 0.025482994 | 0 | 0 | 0 |
| 0 | 0 | 0 | 0 | 0 |
| 0 | 0 | 0 | 0 | 0 |
| 0 | 0 | 0 | 0 | 0 |
| 0 | 0 | 0 | 0 | 0 |
| 0.00566231 | 0 | 0 | 3.341158949 | 0 |
| 0 | 0 | 0 | 0 | 0 |
| 0 | 0 | 0 | 0 | 0 |
| 0 | 0 | 0 | 0 | 0 |
| 0 | 0 | 0 | 0 | 0 |
| 0 | 0 | 0 | 0 | 0 |
| 0 | 0 | 0 | 0 | 0 |
| 0 | 0 | 0 | 0 | 0 |
| 0 | 0 | 0 | 0 | 0 |
| 0 | 0 | 0 | 0 | 0 |
| 0 | 0 | 0 | 0 | 0 |
| 1.991176315 | 0 | 0 | 0.259613616 | 0 |
| 0 | 0 | 0 | 0 | 0 |
| 0 | 0 | 0 | 0 | 0 |
| 0 | 0 | 0 | 0 | 0 |
| 0 | 0 | 0 | 0 | 0 |
| 0 | 0 | 0 | 0 | 0 |
| 0.118558626 | 0.004311885 | 0.221487105 | 0.006942351 | 0.143589635 |
| 1.785066069 | 0.085476213 | 0.032411038 | 0.14969298 | 0.108916001 |
| 0.003232415 | 0.00353521 | 0.003080315 | 0.008917255 | 0.048983171 |
| 0.00133169 | 0.002548762 | 0 | 0.001915029 | 0.003351945 |
| 0 | 0 | 0 | 0 | 0 |

| | | | | |
|---|---|---|---|---|
| 0.003451798 | 0 | 0 | 0 | 0 |
| 0 | 0.007726545 | 0.00961761 | 0.000975696 | 0.018358808 |
| 0 | 0.018414153 | 0 | 0 | 0 |
| 2.244144881 | 0.545371627 | 0 | 0.516015547 | 0 |
| 0.005943805 | 0 | 0.005349448 | 0.028084493 | 0 |
| 0 | 0 | 0 | 0 | 0 |
| 0.302573006 | 0.006820149 | 0.022638332 | 0 | 0.002288102 |
| 0 | 0 | 0 | 0 | 0 |
| 0 | 0 | 0 | 0 | 0 |
| 0 | 0 | 0 | 0 | 0 |
| 0.022509426 | 0 | 0.023400276 | 0 | 0 |
| 0.10171468 | 0 | 0 | 0.116979273 | 0 |
| 0 | 0 | 0 | 0 | 0 |
| 0.022330116 | 0.265500728 | 0 | 0.123391301 | 0 |
| 0.001563117 | 0 | 0.002383303 | 0.00308274 | 0 |
| 0 | 0 | 0 | 0 | 0 |
| 0.005310284 | 0 | 0.020043194 | 0.002310179 | 0.003594302 |
| 0 | 0 | 0 | 0 | 0 |
| 0 | 0 | 0 | 0 | 0 |
| 0 | 0 | 0 | 0 | 0 |
| 0.444713424 | 0.067833984 | 0.063786113 | 0.023149064 | 0 |
| 0.141290798 | 0.094841973 | 0.005170269 | 0.018948213 | 0 |
| 0.133118252 | 0 | 0 | 0.003542105 | 0 |
| 0 | 0 | 0 | 0 | 0 |
| 0 | 0.745068213 | 0 | 0 | 0 |
| 0 | 0 | 0 | 0 | 0 |
| 0 | 0 | 0.167198482 | 0 | 0.20363492 |
| 0 | 0 | 0 | 0 | 0 |
| 0.234543527 | 0 | 0.003307134 | 0 | 0.014350845 |
| 0 | 0 | 0 | 0 | 0 |
| 0 | 0 | 0.063955125 | 0.003367908 | 0 |
| 0 | 0 | 0 | 0 | 0 |
| 0 | 0 | 0 | 0 | 0 |
| 0 | 0.04512525 | 0 | 0 | 0 |
| 0 | 0 | 0 | 0 | 0 |
| 0 | 0 | 0 | 0 | 0 |
| 0 | 0 | 0 | 0 | 0 |
| 0 | 0 | 0 | 0 | 0 |
| 0 | 0 | 0 | 0 | 0 |
| 0 | 0 | 0 | 0 | 0 |
| 0 | 0 | 0 | 0 | 0 |
| 0 | 0 | 0 | 0 | 0 |
| 0 | 0 | 0 | 0 | 0 |
| 0 | 0 | 0 | 0 | 0 |
| 0 | 0 | 0 | 0 | 0 |
| 0 | 0 | 0 | 0 | 0 |
| 0 | 0 | 0.000532684 | 0 | 0.000603002 |
| 0 | 0 | 0 | 0 | 0 |

| | | | | |
|---|---|---|---|---|
| 0 | 0 | 0 | 0 | 0 |
| 0 | 0 | 0 | 0 | 0 |
| 0 | 0 | 0 | 0 | 0 |
| 0 | 0 | 0 | 0 | 0 |
| 0 | 0 | 0.132742016 | 0.15562798 | 0.007217155 |
| 0 | 0.013188931 | 0 | 0 | 0 |
| 0 | 0 | 0 | 0 | 0 |
| 0 | 0 | 0 | 0 | 0 |
| 0 | 0 | 0.001527005 | 0 | 0 |
| 0 | 0 | 0 | 0 | 0 |
| 0 | 0 | 0 | 0 | 0 |
| 0.035769665 | 0 | 0.002256319 | 0 | 0.029920263 |
| 0 | 0 | 0 | 0 | 0 |
| 0 | 0 | 0 | 0 | 0 |
| 0 | 0 | 0 | 0 | 0 |
| 0 | 0 | 0 | 0 | 0 |
| 0 | 0 | 0 | 0 | 0 |
| 0 | 0 | 0 | 0 | 0 |
| 0 | 0 | 0 | 0 | 0 |
| 0 | 0 | 0 | 0 | 0 |
| 0 | 0 | 0 | 0 | 0 |
| 0 | 0 | 0 | 0 | 0 |
| 0 | 0 | 0 | 0 | 0 |
| 0 | 0 | 0 | 0 | 0 |
| 0 | 0 | 0 | 0 | 0 |
| 0 | 0 | 0 | 0 | 0 |
| 0 | 0 | 0 | 0 | 0 |
| 0 | 0 | 0 | 0.007251514 | 0 |
| 0 | 0 | 0 | 0 | 0 |
| 0 | 0 | 0 | 0 | 0 |
| 0.022242286 | 0.116719722 | 0 | 0.466571352 | 0 |
| 0.076578259 | 0 | 0 | 0.014991012 | 0 |
| 0 | 0 | 0 | 0 | 0 |
| 0 | 0 | 0 | 0 | 0 |
| 0 | 0 | 0 | 0 | 0 |
| 0 | 0 | 0 | 0 | 0 |
| 0 | 0 | 0 | 0 | 0 |
| 0 | 0 | 0 | 0 | 0 |
| 0.298099409 | 0 | 0 | 2.101886881 | 0 |
| 0 | 0 | 0 | 0 | 0 |
| 0 | 0 | 0 | 0 | 0 |
| 0 | 0 | 0 | 0 | 0 |
| 0.002451866 | 0 | 0 | 0.001410357 | 0 |
| 0 | 0 | 0 | 0 | 0 |
| 0 | 0 | 0 | 0 | 0 |
| 0 | 0 | 0 | 0 | 0 |
| 0 | 0 | 0 | 0 | 0 |
| 0 | 0 | 0 | 0 | 0 |

| | | | | |
|---|---|---|---|---|
| 0 | 0 | 0 | 0 | 0 |
| 0 | 0 | 0 | 0 | 0 |
| 0 | 0 | 0 | 0 | 0 |
| 0 | 0 | 0 | 0 | 0 |
| 0 | 0 | 0 | 0 | 0 |
| 0 | 0 | 0 | 0 | 0 |
| 0 | 0 | 0 | 0 | 0 |
| 0 | 0 | 0 | 0 | 0 |
| 0 | 0 | 0 | 0 | 0 |
| 0 | 0 | 0 | 0 | 0 |
| 0 | 0 | 0 | 0 | 0 |
| 0 | 0 | 0 | 0 | 0 |
| 0 | 0 | 0 | 0 | 0 |
| 0 | 0 | 0 | 0 | 0 |
| 0 | 0 | 0 | 0 | 0 |
| 0 | 0 | 0 | 0 | 0 |
| 0 | 0 | 0 | 0 | 0 |
| 0 | 0 | 0 | 0 | 0 |
| 0 | 0 | 0 | 0 | 0 |
| 0 | 0 | 0 | 0 | 0 |
| 0 | 0 | 0 | 0 | 0 |
| 0 | 0 | 0 | 0 | 0 |
| 0 | 0 | 0 | 0 | 0 |
| 0 | 0 | 0 | 0 | 0 |
| 0 | 0 | 0 | 0 | 0 |
| 0 | 0 | 0 | 0 | 0 |
| 0 | 0 | 0 | 0.052829146 | 0 |
| 0 | 0 | 0 | 0 | 0 |
| 0 | 0 | 0 | 0 | 0 |
| 0 | 0 | 0 | 0 | 0 |
| 0 | 0 | 0 | 0 | 0 |
| 0.000979 | 0 | 0 | 0 | 0 |
| 0 | 0 | 0 | 0 | 0 |
| 0 | 0 | 0 | 0 | 0 |
| 0 | 0 | 0 | 0 | 0 |
| 0 | 0 | 0 | 0 | 0 |
| 0 | 0 | 0 | 0 | 0 |
| 0 | 0 | 0 | 0 | 0 |
| 0 | 0 | 0 | 0 | 0 |
| 0 | 0 | 0 | 0 | 0 |
| 0 | 0 | 0 | 0 | 0 |
| 0 | 0 | 0 | 0 | 0 |
| 0 | 0 | 0 | 0 | 0 |
| 0 | 0 | 0 | 0 | 0 |
| 0 | 0 | 0 | 0 | 0 |
| 0 | 0 | 0 | 0 | 0 |
| 0 | 0 | 0 | 0 | 0 |
| 0 | 0 | 0 | 0 | 0 |

| | | | | |
|---|---|---|---|---|
| 0 | 0 | 0 | 0 | 0 |
| 0 | 0 | 0 | 0 | 0 |
| 0 | 0 | 0 | 0 | 0 |
| 0 | 0 | 0 | 0 | 0 |
| 0 | 0 | 0 | 0 | 0 |
| 0 | 0 | 0 | 0 | 0 |
| 0 | 0 | 0 | 0 | 0 |
| 0 | 0 | 0 | 0 | 0 |
| 0 | 0 | 0 | 0 | 0 |
| 0 | 0 | 0 | 0 | 0 |
| 0 | 0 | 0 | 0 | 0 |
| 0 | 0 | 0 | 0 | 0 |
| 0 | 0 | 0 | 0.001539623 | 0.026611735 |
| 0 | 0 | 0 | 0 | 0 |
| 0 | 0.048640843 | 0 | 0.006891648 | 0 |
| 0 | 0 | 0 | 0 | 0 |
| 0 | 0 | 0 | 0 | 0 |
| 0 | 0 | 0 | 0 | 0.001012799 |
| 0 | 0 | 0 | 0 | 0 |
| 0 | 0 | 0 | 0 | 0 |
| 0.079698015 | 0.023467148 | 0.108496934 | 0.03346519 | 0.045978497 |
| 0 | 0 | 0 | 0 | 0 |
| 0 | 0 | 0 | 0 | 0 |
| 0 | 0 | 0 | 0 | 0 |
| 0 | 0 | 0 | 0 | 0 |
| 0 | 0 | 0 | 0 | 0 |
| 0.01071098 | 0 | 0 | 0.002933879 | 0.001834027 |
| 0 | 0 | 0 | 0 | 0 |
| 0 | 0 | 0 | 0 | 0 |
| 0 | 0.064335962 | 0.024960665 | 0 | 0 |
| 0 | 0.015958832 | 0 | 0 | 0 |
| 0 | 0 | 0 | 0 | 0 |
| 0 | 0 | 0 | 0 | 0 |
| 0.003211416 | 0 | 0 | 0 | 0 |
| 0 | 0.161414904 | 0.019368067 | 0.078902339 | 0.110443538 |
| 0 | 0 | 0 | 0 | 0 |
| 0 | 0 | 0 | 0 | 0 |
| 0 | 0 | 0 | 0 | 0 |
| 0 | 0 | 0 | 0 | 0 |
| 0 | 0 | 0 | 0 | 0 |
| 0 | 0 | 0 | 0 | 0 |
| 0.000853654 | 0 | 0 | 0 | 0 |
| 0 | 0 | 0 | 0 | 0 |
| 0 | 0 | 0 | 0 | 0 |
| 0 | 0 | 0 | 0 | 0 |
| 0 | 0 | 0 | 0.203970662 | 0 |
| 0 | 0.008502296 | 0 | 0.00228152 | 0 |
| 0.001421301 | 0 | 0.001805896 | 0.011345697 | 0.010075415 |

| | | | | |
|---|---|---|---|---|
| 0 | 0 | 0 | 0 | 0 |
| 0 | 0 | 0 | 0 | 0 |
| 0 | 0 | 0 | 0.002439403 | 0 |
| 0 | 0 | 0 | 0 | 0 |
| 0 | 0 | 0 | 0 | 0 |
| 0 | 0 | 0 | 0 | 0 |
| 0 | 0 | 0 | 0 | 0 |
| 0.001751268 | 0 | 0.001112575 | 0 | 0.002099071 |
| 0.00420115 | 0 | 0 | 0 | 0 |
| 0 | 0 | 0 | 0 | 0 |
| 0.001256078 | 0 | 0 | 0 | 0 |
| 0 | 0 | 0 | 0 | 0 |
| 0 | 0 | 0 | 0 | 0 |
| 0 | 0 | 0 | 0 | 0 |
| 0.019848154 | 0 | 0.023185156 | 0 | 0 |
| 0 | 0 | 0 | 0 | 0 |
| 0 | 0 | 0 | 0 | 0 |
| 0 | 0 | 0 | 0 | 0 |
| 0.009292203 | 0 | 0 | 0 | 0 |
| 0 | 0 | 0 | 0 | 0 |
| 0 | 0.128722187 | 0.249435251 | 0 | 0.110622977 |
| 0 | 0 | 0 | 0 | 0 |
| 0 | 0 | 0 | 0 | 0 |
| 0 | 0 | 0 | 0 | 0 |
| 0.011497848 | 0 | 0 | 0.001989103 | 0 |
| 0 | 0 | 0 | 0 | 0 |
| 0 | 0 | 0 | 0 | 0 |
| 0 | 0 | 0 | 0 | 0 |
| 0.083737981 | 0.002210602 | 0.007154278 | 0 | 0.023257782 |
| 0 | 0 | 0 | 0 | 0 |
| 0.154988185 | 1.305886983 | 0.166761906 | 0.755877513 | 0.566165785 |
| 0 | 0 | 0 | 0 | 0 |
| 0 | 0 | 0.077139021 | 0.03592897 | 0.047065703 |
| 0 | 0 | 0.005890856 | 0 | 0.002292293 |
| 0 | 0 | 0 | 0 | 0 |
| 0 | 0 | 0 | 0 | 0 |
| 0 | 0 | 0 | 0 | 0 |
| 0 | 0 | 0 | 0 | 0 |
| 0 | 0 | 0 | 0 | 0 |
| 0 | 0.001027626 | 0 | 0.002206038 | 0 |
| 0 | 0 | 0 | 0 | 0 |
| 0.001826413 | 0 | 0.005076375 | 0.007128969 | 0 |
| 0 | 0 | 0 | 0 | 0 |
| 0 | 0 | 0 | 0 | 0 |
| 0 | 0 | 0 | 0 | 0 |
| 0 | 0 | 0 | 0 | 0 |
| 0 | 0 | 0 | 0 | 0 |
| 0 | 0 | 0 | 0 | 0 |

| | | | | |
|---|---|---|---|---|
| 0 | 0 | 0 | 0 | 0 |
| 0 | 0 | 0 | 0 | 0 |
| 0.001433351 | 0 | 0.001821207 | 0.002591251 | 0.064528661 |
| 0 | 0 | 0 | 0 | 0 |
| 0 | 0 | 0 | 0 | 0 |
| 0 | 0 | 0 | 0 | 0 |
| 0 | 0 | 0 | 0 | 0 |
| 0 | 0 | 0 | 0 | 0 |
| 0 | 0 | 0 | 0 | 0 |
| 0.001310932 | 0 | 0 | 0 | 0 |
| 0 | 0 | 0 | 0 | 0 |
| 0 | 0 | 0 | 0 | 0 |
| 0 | 0 | 0 | 0 | 0 |
| 0.006970091 | 0 | 0.005031906 | 0 | 0.005468306 |
| 0.01516673 | 0.00280271 | 0.005482197 | 0.007091076 | 0.004325313 |
| 0 | 0 | 0 | 0 | 0 |
| 0.005065158 | 0 | 0.004921463 | 0.000979351 | 0.011142259 |
| 0 | 0 | 0 | 0 | 0 |
| 0 | 0 | 0 | 0 | 0 |
| 0 | 0 | 0 | 0 | 0 |
| 0 | 0 | 0 | 0 | 0 |
| 0.00188275 | 0 | 0.001366977 | 0.009945837 | 0.006383139 |
| 0 | 0 | 0 | 0 | 0 |
| 0 | 0 | 0 | 0 | 0 |
| 0 | 0 | 0 | 0 | 0 |
| 0 | 0 | 0 | 0 | 0 |
| 0 | 0 | 0 | 2.388151217 | 0 |
| 0 | 0 | 0 | 0 | 0 |
| 0 | 0 | 0 | 0 | 0 |
| 0 | 0 | 0 | 0 | 0 |
| 0 | 0 | 0 | 0 | 0 |
| 0 | 0 | 0 | 0 | 0 |
| 0 | 0 | 0 | 0 | 0 |
| 0 | 0.004748308 | 0.002167166 | 0 | 0.002007202 |
| 0 | 0 | 0 | 0 | 0 |
| 0.009669922 | 0 | 0 | 0 | 0 |
| 0 | 0 | 0 | 0 | 0 |
| 0.001689629 | 0 | 0 | 0.011385166 | 0 |
| 0 | 0 | 0 | 0 | 0 |
| 0 | 0 | 0 | 0 | 0 |
| 0.002926334 | 0 | 0 | 0 | 0.001753753 |
| 0 | 0 | 0 | 0 | 0 |
| 0 | 0 | 0 | 0 | 0 |
| 0 | 0 | 0 | 0 | 0 |
| 0 | 0 | 0 | 0 | 0 |
| 0 | 0 | 0 | 0 | 0 |
| 0 | 0 | 0 | 0 | 0 |
| 0.003274503 | 0.008774039 | 0.009361267 | 0 | 0.008830848 |

| | | | | |
|---|---|---|---|---|
| 0 | 0.300782359 | 0 | 0 | 0 |
| 0 | 0 | 0 | 0 | 0 |
| 0.001690315 | 0 | 0 | 0 | 0 |
| 0 | 0 | 0 | 0 | 0 |
| 0.052152332 | 0.003471859 | 0 | 0.005124044 | 0 |
| 0 | 0 | 0 | 0 | 0 |
| 0 | 0 | 0 | 0 | 0 |
| 0.044497461 | 0 | 0.026502281 | 0.006529523 | 0.007714482 |
| 0 | 0 | 0 | 0 | 0 |
| 0.020065717 | 0.173054879 | 1.121016087 | 0.056533011 | 1.045472845 |
| 0 | 0 | 0 | 0 | 0 |
| 0.09277842 | 0 | 0.018781469 | 0.006719431 | 0.036640869 |
| 0.00262418 | 0 | 0.002728036 | 0.00588107 | 0.001544078 |
| 0.013043297 | 0 | 0 | 0 | 0 |
| 0 | 0 | 0 | 0 | 0 |
| 0 | 0 | 0.001137565 | 0 | 0 |
| 0 | 0 | 0 | 0 | 0 |
| 0 | 0 | 0 | 0.009446503 | 0 |
| 0 | 0.296772709 | 2.441664134 | 0 | 0 |
| 0 | 0.853631153 | 0 | 0 | 0 |
| 0 | 0 | 0 | 0 | 0 |
| 0 | 0 | 0 | 0 | 0 |
| 0.029293627 | 0 | 0 | 0 | 0 |
| 0 | 0 | 0 | 0 | 0 |
| 0 | 0 | 0 | 0 | 0 |
| 0 | 0 | 0 | 0 | 0.01099057 |
| 0 | 0 | 0 | 0 | 0 |
| 0 | 0.506769265 | 0 | 0 | 0 |
| 0 | 0 | 0.000542211 | 0.00490935 | 0 |
| 0 | 0.007003742 | 0 | 0 | 0 |
| 0 | 0 | 0 | 0 | 0 |
| 0.027796251 | 0 | 0 | 0 | 0 |
| 0 | 0 | 0.002467024 | 0 | 0 |
| 0 | 0.160802377 | 0.004848133 | 0 | 0.022213817 |
| 0 | 0 | 0 | 0 | 0 |
| 0 | 1.463405168 | 0 | 0 | 0 |
| 0 | 1.087082695 | 0.134874284 | 0 | 0.003624418 |
| 0 | 0 | 0 | 0 | 0 |
| 0 | 0 | 0 | 0 | 0 |
| 0 | 0 | 0 | 0 | 0 |
| 0 | 0 | 0 | 0 | 0 |
| 0 | 0.722505494 | 0.062194889 | 0 | 0 |
| 0 | 0.006635832 | 0 | 0 | 0 |
| 0.000895427 | 0 | 0 | 0 | 0 |
| 0 | 0 | 0 | 0 | 0 |
| 0 | 0 | 0 | 0.006515201 | 0 |
| 0 | 0.011639758 | 0.001975715 | 0.005962915 | 0 |
| 0 | 0 | 0 | 0 | 0 |

| | | | | |
|---|---|---|---|---|
| 0 | 0.017002209 | 0 | 0 | 0 |
| 0.085729998 | 0 | 0.006378667 | 0 | 0.026105588 |
| 0 | 0 | 0 | 0 | 0 |
| 0 | 0 | 0.004961475 | 0 | 0 |
| 0 | 0.014642324 | 0 | 0 | 0 |
| 0.025855092 | 0.303368173 | 0.051178898 | 0.003131012 | 0.252094945 |
| 0.017249664 | 0.008803909 | 0 | 0 | 0 |
| 0 | 0.002155874 | 0 | 0.002314044 | 0 |
| 0 | 0 | 0 | 0 | 0 |
| 0 | 0 | 0 | 0 | 0 |
| 0.020202141 | 0.003383231 | 0.001002684 | 0.001037556 | 0 |
| 0 | 0.003170858 | 0 | 0 | 0.192759855 |
| 0 | 0 | 0 | 0 | 0 |
| 0.003876837 | 0 | 0.001136743 | 0 | 0.004074869 |
| 0 | 0 | 0 | 0 | 0 |
| 0 | 0 | 0 | 0 | 0 |
| 0 | 0 | 0 | 0 | 0 |
| 0 | 0.008442846 | 0 | 0 | 0 |
| 0 | 0 | 0 | 0 | 0 |
| 0 | 0.057772276 | 0 | 0 | 0 |
| 0 | 0 | 0 | 0 | 0 |
| 0 | 0.082084225 | 0 | 0.451614891 | 0 |
| 0.007074871 | 0 | 0 | 0.037207693 | 0 |
| 0 | 0 | 0 | 0 | 0 |
| 0 | 0 | 0 | 0 | 0 |
| 0 | 0 | 0 | 0 | 0 |
| 0 | 0.295074691 | 0.003286747 | 0 | 0 |
| 0 | 0 | 0 | 0 | 0 |
| 0 | 0 | 0 | 0 | 0.008858718 |
| 0.69328101 | 4.814960297 | 0 | 1.857799588 | 0 |
| 0.001618539 | 0 | 0 | 0.010307636 | 0 |
| 0 | 0 | 0 | 0 | 0 |
| 0 | 0 | 0.000791708 | 0 | 0 |
| 0 | 0.01798877 | 0 | 0 | 0 |
| 0 | 0.063504179 | 0 | 0 | 0 |
| 0 | 0 | 0 | 0 | 0 |
| 0 | 0 | 0 | 0.044114169 | 0 |
| 0.00395153 | 0 | 0.012165759 | 0 | 0.040440772 |
| 0 | 0 | 0 | 0.003922615 | 0 |
| 0 | 0 | 0 | 0 | 0 |
| 0.319698868 | 0.027778666 | 0.075385116 | 0.000805857 | 0.008639436 |
| 3.389700074 | 0 | 0 | 0.387079943 | 0 |
| 0.001949154 | 0 | 0.01149842 | 0 | 0.036445611 |
| 0 | 0 | 0 | 0 | 0 |
| 0 | 0.005928391 | 0.010717642 | 0 | 0.012331337 |
| 0 | 0 | 0 | 0 | 0 |
| 0 | 0 | 0 | 0 | 0 |
| 0 | 0 | 0 | 0 | 0 |

| | | | | |
|---|---|---|---|---|
| 0 | 0 | 0 | 0 | 0.013927956 |
| 0.004057351 | 0 | 0 | 0.025005643 | 0 |
| 0 | 0 | 0.039301167 | 0.009598219 | 0.007466717 |
| 0 | 0 | 0 | 0 | 0 |
| 0.002127707 | 0 | 0 | 0.026051469 | 0 |
| 0 | 0.023017067 | 0 | 0 | 0 |
| 0 | 0 | 0 | 0 | 0 |
| 0 | 0 | 0 | 0 | 0.003070901 |
| 0.03773106 | 0.004882116 | 0 | 0 | 0 |
| 0 | 0 | 0 | 0 | 0 |
| 0 | 0 | 0 | 0 | 0 |
| 0 | 0 | 0 | 0 | 0 |
| 0 | 0 | 0 | 0 | 0 |
| 0.058881392 | 0 | 0 | 0.012509337 | 0 |
| 0 | 0 | 0 | 0 | 0 |
| 0.009202958 | 0.504528629 | 0 | 0 | 0 |
| 0.016917935 | 0.233896098 | 0.004741726 | 0.010426603 | 0.129181833 |
| 0 | 0.016661705 | 0.013826429 | 0 | 0.007825806 |
| 0.003340799 | 0.012984855 | 0.011265043 | 0 | 0.052302115 |
| 0 | 0 | 0 | 0 | 0 |
| 0 | 0 | 0.017728574 | 0.018808932 | 0 |
| 0 | 0.010162978 | 0.006966864 | 0 | 0 |
| 0 | 0 | 0 | 0 | 0 |
| 0.666267906 | 0 | 0 | 0 | 0 |
| 0 | 0.203645523 | 0 | 0 | 0 |
| 0 | 0 | 0.959653421 | 0 | 0.467390064 |
| 0.027478509 | 0.003385229 | 0.005618349 | 0 | 0.001362859 |
| 0 | 0.003978592 | 0 | 0 | 0 |
| 0 | 0 | 0 | 0 | 0 |
| 0 | 0 | 0 | 0 | 0 |
| 0.060530192 | 0.142698496 | 0.07813007 | 0.03691028 | 0.634307694 |
| 0.009668902 | 0.002193257 | 0.005687615 | 0.008533862 | 0.002832904 |
| 0.004773535 | 0 | 6.436934995 | 0.093862273 | 4.731568343 |
| 0.006840941 | 0.008229939 | 0.004346028 | 0 | 0.016516249 |
| 0.02580851 | 0 | 0.007976465 | 0.001719557 | 0.006772059 |
| 0 | 0 | 0 | 0 | 0 |
| 0.002157802 | 0 | 0.006462555 | 0 | 0.028154194 |
| 0 | 0.143955028 | 0.071522008 | 0.008122022 | 0.094312828 |
| 0.132701871 | 0 | 0 | 0 | 0 |
| 0 | 0.012675163 | 0 | 0 | 0 |
| 0 | 0 | 0 | 0 | 0 |
| 0 | 0 | 0 | 0 | 0 |
| 0.011897233 | 0 | 0 | 0 | 0 |
| 0 | 0.184697809 | 0.125292646 | 0.365104992 | 0.027608545 |
| 0 | 1.2280904 | 0.003275375 | 0 | 0 |
| 0 | 0 | 0.034224389 | 0 | 0.005858583 |
| 0 | 0 | 0 | 0 | 0 |
| 0 | 0 | 0 | 0 | 0 |

| | | | | |
|---|---|---|---|---|
| 0 | 0.022438304 | 0 | 0 | 0 |
| 0.869186278 | 0.069912362 | 0.001460824 | 0.072342255 | 0 |
| 0 | 0 | 0 | 0 | 0 |
| 0 | 0.036858499 | 0 | 0 | 0 |
| 0 | 0 | 0 | 0 | 0 |
| 0 | 0 | 0 | 0 | 0.008946414 |
| 0 | 0 | 0 | 0 | 0 |
| 0.001261577 | 0 | 0 | 0 | 0.028125571 |
| 0 | 0.010589765 | 0.000955188 | 0 | 0 |
| 0 | 0 | 0 | 0 | 0 |
| 0.001708731 | 0.008222702 | 0 | 0.013640142 | 0 |
| 0.026542839 | 0 | 0.037883065 | 0.022707657 | 0.036608198 |
| 0 | 0 | 0 | 0.649836814 | 0 |
| 0 | 0 | 0 | 0 | 0 |
| 0 | 0 | 0 | 0 | 0.01510665 |
| 0 | 0 | 0 | 0 | 0 |
| 0.055212325 | 0.007436217 | 0.009581004 | 0.004410987 | 0.006985406 |
| 0 | 0.042677468 | 0.137866163 | 0 | 0 |
| 0 | 0 | 0 | 0 | 0 |
| 0.001077334 | 0.011546882 | 0.009855752 | 0.000708231 | 0 |
| 0.047975399 | 0.001210834 | 0.00167465 | 0.001299669 | 0.003033144 |
| 0 | 0 | 0 | 0 | 0 |
| 0 | 0 | 0.234789833 | 0 | 0.372067448 |
| 0 | 0 | 0 | 0 | 0 |
| 0.003100293 | 0 | 0.028699982 | 0 | 0.005096247 |
| 0 | 0 | 0 | 0 | 0 |
| 0.050216179 | 0.429458563 | 0 | 0.097709497 | 0 |
| 0.298940377 | 0.018446109 | 0.010679432 | 0.122480236 | 0.00805946 |
| 0 | 0 | 0 | 0 | 0 |
| 0 | 0 | 0 | 0 | 0.002430399 |
| 0 | 0 | 0 | 0 | 0.001080976 |
| 0 | 0 | 0 | 0 | 0 |
| 0 | 0.020831112 | 0 | 0.000838478 | 0 |
| 0 | 0 | 0 | 0 | 0 |
| 0 | 0 | 0 | 0.07904662 | 0 |
| 0.788543718 | 0.061711856 | 0.002409907 | 0.041302241 | 0 |
| 0 | 0 | 0 | 0 | 0 |
| 0 | 0.007811109 | 0 | 0 | 0 |
| 0.0504605 | 0 | 0.005928423 | 0.003976133 | 0.020630159 |
| 0 | 0 | 0 | 0 | 0 |
| 0 | 0.625623514 | 0.004960155 | 0 | 0 |
| 0 | 0 | 0 | 0 | 0 |
| 0.021413348 | 0.002250083 | 0.002800789 | 0 | 0.025968008 |
| 0 | 0 | 0.000772603 | 0 | 0 |
| 0 | 0 | 0 | 0 | 0 |
| 0 | 0 | 0 | 0.622956882 | 0 |
| 0 | 0 | 0 | 0 | 0 |
| 0 | 0.035617022 | 0 | 0 | 0 |

| | | | | |
|---|---|---|---|---|
| 0 | 0.003143415 | 0.001490576 | 0 | 0.014975167 |
| 0 | 0 | 0 | 0 | 0 |
| 0 | 0 | 0 | 0 | 0 |
| 0.169688268 | 0.507654442 | 0 | 0.002411059 | 0 |
| 0 | 0 | 0 | 0 | 0 |
| 0 | 0 | 0 | 0 | 0 |
| 0.00151649 | 0 | 0 | 0 | 0 |
| 0 | 0 | 0 | 0 | 0 |
| 0.114330298 | 0 | 0.294392878 | 3.761244017 | 0.398576483 |
| 0.000655589 | 0 | 0 | 0 | 0 |
| 0 | 0 | 0 | 0 | 0 |
| 0 | 0 | 0 | 0.010010232 | 0 |
| 0 | 0 | 0 | 0 | 0 |
| 0 | 0 | 0 | 0 | 0 |
| 0.025324358 | 0.016279685 | 0.045195136 | 0.868937804 | 0.367860176 |
| 0 | 0 | 0 | 0 | 0 |
| 0.039337491 | 0 | 0 | 0.004351466 | 0 |
| 0.440005023 | 0 | 0 | 0.272369954 | 0 |
| 0 | 0.728189685 | 0 | 0 | 0 |
| 0 | 0 | 0 | 0 | 0 |
| 0 | 0 | 0 | 0 | 0 |
| 0 | 0 | 0 | 0.263978719 | 0 |
| 0 | 0 | 0 | 0 | 0 |
| 0.30110573 | 2.5649246 | 0.305333 | 0.080028544 | 0.0448621 |
| 0 | 0 | 0 | 0.025773373 | 0 |
| 0 | 0 | 0 | 0 | 0 |
| 0 | 0 | 0 | 0.004503448 | 0 |
| 0 | 0 | 0 | 0.60704858 | 0 |
| 0 | 0 | 0 | 0 | 0 |
| 0 | 0 | 0 | 0 | 0 |
| 0.002277451 | 0 | 0 | 0.017404686 | 0 |
| 0 | 0 | 0 | 0 | 0 |
| 0.021493011 | 0.021496946 | 0 | 0 | 0 |
| 0 | 0 | 0 | 0 | 0 |
| 0 | 0 | 0 | 0 | 0 |
| 0 | 0 | 0 | 0 | 0 |
| 0.470354741 | 0 | 0 | 0 | 0 |
| 0 | 0.036947806 | 0.000619404 | 0 | 0.001402339 |
| 0 | 0.070945868 | 0 | 0 | 0 |
| 0.001173762 | 0 | 0.004580652 | 0.001515685 | 0.026529592 |
| 0.025844521 | 0.010323051 | 0.2541367 | 0.04229026 | 0 |
| 0.001951859 | 0 | 0 | 0.100565767 | 0 |
| 0.042596354 | 22.57973396 | 0 | 11.27078034 | 0 |
| 0 | 0 | 0 | 0.027187852 | 0 |
| 0 | 0.028407976 | 0 | 0 | 0 |
| 0 | 0 | 0 | 0 | 0 |
| 0 | 0 | 0 | 0 | 0 |
| 0.00486283 | 0.033919275 | 0 | 0 | 0.003108582 |

| | | | | |
|---|---|---|---|---|
| 0 | 0 | 0 | 0 | 0 |
| 0 | 0 | 0 | 0.0962697 | 0 |
| 0.005179524 | 0.341015746 | 0.868372279 | 0.00212811 | 0.644409178 |
| 0 | 0 | 0 | 0.002070346 | 0 |
| 0 | 0 | 0 | 0 | 0 |
| 0.001224069 | 0 | 0 | 0.003185242 | 0 |
| 0 | 0 | 0 | 0 | 0 |
| 0.002894448 | 0.005698051 | 0 | 0 | 0 |
| 0 | 0 | 0 | 0 | 0 |
| 0 | 0.194561349 | 0 | 0 | 0 |
| 0.007119523 | 0.040878818 | 0.005976836 | 0.197450807 | 0.20498604 |
| 1.413496326 | 0.779890856 | 0.407531392 | 4.443577394 | 0.301808179 |
| 0 | 0 | 0 | 0 | 0 |
| 0.000823251 | 0 | 0 | 0.039011401 | 0.017958845 |
| 0.194626101 | 0.003973342 | 0.221462577 | 0.014038474 | 0.034836329 |
| 0 | 0 | 0 | 0 | 0 |
| 0 | 0.002151756 | 0 | 0 | 0 |
| 0 | 0.004780181 | 0 | 0 | 0 |
| 0 | 0 | 0 | 0 | 0 |
| 0.008378019 | 0 | 0.004790277 | 0.010326825 | 0 |
| 0.178376922 | 0.009136659 | 0.097887727 | 0.017687596 | 0.001226109 |
| 0 | 0.603075538 | 51.82472715 | 1.502050721 | 38.46164698 |
| 0.825699402 | 4.809878123 | 15.88501343 | 2.180759299 | 15.05371485 |
| 0 | 0 | 0 | 0 | 0 |
| 0 | 0 | 0 | 0.088050985 | 0 |
| 0 | 0 | 0 | 0 | 0 |
| 0.105526526 | 0.390671324 | 0 | 0 | 0.017964419 |
| 0 | 0 | 0 | 0.020274361 | 0 |
| 0 | 0.006203271 | 0 | 0.001065341 | 0 |
| 0 | 0 | 0 | 0 | 0 |
| 0 | 0 | 0 | 0.004828951 | 0 |
| 0.369195616 | 0 | 0 | 0.036868847 | 0 |
| 0.021413526 | 0 | 0.00464525 | 0 | 0 |
| 0 | 0.003658943 | 0 | 0.157586421 | 0 |
| 0.00086178 | 0.183411855 | 0.009416763 | 0 | 0 |
| 0.005683359 | 0.01196531 | 0.057920369 | 0.59409361 | 1.183939223 |
| 0.00114376 | 0 | 0.001453254 | 0.041354402 | 0.01562839 |
| 0 | 0 | 0 | 0 | 0 |
| 0.039904067 | 0 | 0.020949727 | 1.695327448 | 0.087487293 |
| 0 | 0 | 0 | 0 | 0 |
| 0 | 0 | 0 | 0 | 0 |
| 0 | 0 | 0 | 0 | 0 |
| 0 | 0 | 0 | 0 | 0 |
| 0 | 0 | 0 | 0 | 0 |
| 0.010531992 | 0.008269744 | 0.001544064 | 0.295586342 | 0.031267819 |
| 0 | 0 | 0 | 0 | 0 |
| 0 | 0 | 0 | 0 | 0 |
| 0.029317986 | 0.052371748 | 0.020488188 | 0.512886595 | 12.34792404 |

| | | | | |
|---|---|---|---|---|
| 0 | 0 | 0 | 0 | 0 |
| 0 | 0 | 0 | 0 | 0 |
| 0 | 0 | 0 | 0 | 0 |
| 0 | 0 | 0 | 0.000945672 | 0 |
| 0 | 0 | 0 | 0 | 0 |
| 0 | 0 | 0 | 0.049945653 | 0 |
| 0 | 0 | 0 | 0 | 0 |
| 0.008471681 | 0 | 0.01318598 | 0.036896034 | 0.308280395 |
| 0 | 0 | 0 | 0 | 0 |
| 0 | 0 | 0 | 0 | 0 |
| 0.001538047 | 0.007064915 | 0.000488558 | 0.014955842 | 0.254772249 |
| 0 | 0 | 0 | 0 | 0 |
| 0.001571127 | 0 | 0 | 0.346925512 | 0 |
| 0 | 0 | 0 | 0 | 0 |
| 0 | 0 | 0 | 0 | 0 |
| 0 | 0 | 0 | 0 | 0 |
| 0 | 0 | 0 | 0.005796602 | 0 |
| 0 | 0 | 0.002033568 | 0.000876789 | 0.016114087 |
| 0 | 0 | 0 | 0 | 0 |
| 0 | 0 | 0 | 0 | 0 |
| 0 | 0 | 0 | 0 | 0 |
| 0 | 0 | 0 | 0 | 0 |
| 0.024357245 | 0 | 0 | 0 | 0 |
| 0 | 0 | 0 | 0 | 0 |
| 0 | 0 | 0 | 0 | 0 |
| 0 | 0 | 0 | 0 | 0 |
| 0 | 0 | 0 | 0 | 0 |
| 0 | 0 | 0 | 0 | 0 |
| 0 | 0 | 0 | 0 | 0 |
| 0 | 0 | 0 | 0.020759393 | 0 |
| 0 | 0 | 0 | 0.006304463 | 0 |
| 0 | 0 | 0 | 0 | 0 |
| 0 | 0 | 0 | 0.262369519 | 0 |
| 0 | 0 | 0 | 0 | 0 |
| 0 | 0 | 0 | 0 | 0 |
| 0 | 0 | 0 | 0 | 0 |
| 0.182652117 | 0 | 0 | 0.020967693 | 0 |
| 0.001952975 | 0.066118571 | 0.006479311 | 0 | 0.012172355 |
| 0 | 0 | 0 | 0.065031948 | 0 |
| 0 | 0.003932605 | 0.001767678 | 0.000527641 | 0.000615699 |
| 0.026467626 | 0.154309616 | 0 | 0.147896578 | 0 |
| 0 | 0 | 0 | 0 | 0 |
| 0 | 0 | 0 | 0 | 0 |
| 0.00168584 | 0 | 0 | 0 | 0 |
| 0.298388135 | 0 | 0 | 0.161980083 | 0 |
| 0 | 0 | 0 | 0 | 0 |
| 0 | 0 | 0 | 0.001186005 | 0 |
| 0 | 0 | 0 | 0 | 0 |

| | | | | |
|---|---|---|---|---|
| 0 | 0 | 0 | 0 | 0 |
| 0 | 0 | 0 | 0 | 0 |
| 0 | 0 | 0 | 0.465714355 | 0 |
| 0 | 0.002313468 | 0 | 0 | 0 |
| 0 | 0 | 0 | 0 | 0 |
| 0 | 0 | 0 | 0 | 0 |
| 0 | 0 | 0 | 0 | 0 |
| 0 | 0 | 0 | 0 | 0 |
| 0.011671742 | 0.007331743 | 0.109894426 | 0 | 0.225988416 |
| 0 | 0 | 0 | 0 | 0 |
| 0 | 0 | 0 | 0.003692747 | 0 |
| 0 | 0 | 0 | 0 | 0 |
| 0.00104755 | 0 | 0 | 0.007460382 | 0 |
| 0 | 0 | 0 | 0.001195688 | 0 |
| 0.096565188 | 0 | 0 | 0 | 0 |
| 0 | 3.722803253 | 0.305970355 | 0.005307187 | 0.836872081 |
| 0 | 0 | 0 | 0 | 0 |
| 0 | 0 | 0 | 0 | 0 |
| 0 | 0 | 0 | 0 | 0 |
| 0 | 0 | 0 | 0 | 0 |
| 0 | 0 | 0 | 0.013163225 | 0 |
| 0 | 0 | 0 | 0 | 0 |
| 0 | 0 | 0 | 0 | 0 |
| 0 | 0 | 0 | 0 | 0 |
| 0.074963093 | 0.078675027 | 0.00223586 | 0.006651659 | 0.030625209 |
| 1.700052467 | 2.451161507 | 0 | 0 | 0 |
| 0 | 0 | 0 | 0 | 0 |
| 0 | 0 | 0 | 0 | 0 |
| 0.067385353 | 0 | 0 | 0.151960282 | 0 |
| 0 | 0 | 0 | 0 | 0 |
| 0 | 0 | 0 | 0 | 0 |
| 0 | 0 | 0 | 0 | 0 |
| 0 | 0 | 0 | 0 | 0 |
| 0.005262831 | 0 | 0.002029958 | 0.000617811 | 0.039470262 |
| 0.000929087 | 0.006164457 | 0.000786995 | 0.004580808 | 0.039421612 |
| 0 | 0.021452425 | 0.00111262 | 0 | 0.052741303 |
| 0 | 0.973393763 | 0 | 0 | 0 |
| 0.481042576 | 0.531402929 | 0 | 0.37924444 | 0 |
| 0 | 0 | 0 | 0.182893381 | 0 |
| 0 | 0 | 0 | 0 | 0 |
| 0 | 0 | 0 | 0 | 0 |
| 0 | 0.001520451 | 0 | 0 | 0 |
| 4.482232005 | 0 | 0 | 0.021386959 | 0 |
| 0 | 0 | 0 | 2.773891544 | 0 |
| 0 | 0 | 0 | 0 | 0 |
| 0 | 0.040964513 | 0 | 0 | 0 |
| 0.031113706 | 0 | 0 | 0 | 0 |
| 0 | 0 | 0.016792568 | 0.030680544 | 0 |

| | | | | |
|---|---|---|---|---|
| 0 | 0.051306245 | 0 | 0 | 0 |
| 0 | 0 | 0 | 0 | 0 |
| 0 | 0 | 0 | 0 | 0 |
| 0 | 0 | 0 | 0 | 0 |
| 0 | 0 | 0.007947927 | 0.002620502 | 0.283320702 |
| 0 | 0 | 0 | 0.003882315 | 0 |
| 0 | 0 | 0 | 0 | 0 |
| 0 | 0 | 0 | 0 | 0 |
| 0 | 0 | 0 | 0 | 0 |
| 0 | 0 | 0 | 0 | 0 |
| 0 | 0 | 0 | 0.000616967 | 0 |
| 0.681645159 | 0 | 0 | 0.228740319 | 0 |
| 1.01135901 | 0 | 0 | 0.391007392 | 0 |
| 0 | 0 | 0 | 0 | 0 |
| 3.204341756 | 0 | 0.291072958 | 0.324275359 | 1.234949107 |
| 0 | 0 | 0 | 0 | 0 |
| 0 | 0 | 0 | 0.003729941 | 0 |
| 0 | 0 | 0.018049579 | 0 | 0.157192084 |
| 0 | 0 | 0 | 0 | 0 |
| 0 | 0 | 0 | 0 | 0 |
| 0 | 0 | 0 | 0.029942804 | 0 |
| 0 | 0 | 0 | 0 | 0 |
| 0.000967145 | 0 | 0 | 0.043114726 | 0 |
| 0 | 0 | 0 | 0 | 0 |
| 0 | 0 | 0 | 0 | 0 |
| 0 | 0 | 0 | 0 | 0 |
| 0 | 0 | 0 | 0 | 0 |
| 0 | 0 | 0 | 0 | 0 |
| 0 | 0 | 0 | 0 | 0 |
| 0 | 0 | 0 | 0 | 0 |
| 0 | 0 | 0 | 0 | 0 |
| 0.133264765 | 0.004509339 | 0.025726233 | 0.016638098 | 0.003176975 |
| 0 | 0 | 0 | 0 | 0 |
| 0.008460033 | 0.034918216 | 0.016941777 | 0.051988451 | 0.144696267 |
| 0.000952948 | 0.017144435 | 0.000605405 | 0 | 0.052941158 |
| 0.001228465 | 0.010345269 | 0 | 0.151421802 | 0.001766927 |
| 0 | 0 | 0 | 0 | 0 |
| 0 | 0 | 0 | 0.283982892 | 0 |
| 0.001642195 | 0 | 0 | 0.030362772 | 0 |
| 0 | 0 | 0 | 0 | 0 |
| 0 | 0 | 0 | 0 | 0 |
| 0 | 0 | 0 | 0 | 0 |
| 0 | 0 | 0 | 0 | 0 |
| 0 | 0 | 0 | 0 | 0 |
| 0.005397808 | 0.028747212 | 0.004472883 | 0.005399852 | 0.131140368 |
| 0 | 0 | 0 | 0 | 0 |
| 0.688742553 | 0 | 0 | 0.987197097 | 0 |
| 0 | 0.007817044 | 0 | 0.018605141 | 0 |

| | | | | |
|---|---|---|---|---|
| 0 | 0 | 0 | 0 | 0 |
| 0 | 0 | 0 | 0.051021099 | 0 |
| 0 | 0.891582689 | 0.003238257 | 0 | 0 |
| 0 | 0 | 0 | 0 | 0 |
| 0 | 0 | 0 | 0 | 0 |
| 0 | 0 | 0 | 0 | 0 |
| 0 | 0 | 0 | 1.562376173 | 0 |
| 0 | 0 | 0 | 0 | 0 |
| 0 | 0 | 0 | 0 | 0 |
| 0 | 0 | 0 | 0 | 0 |
| 0 | 0 | 0 | 0 | 0 |
| 0 | 0 | 0 | 0 | 0 |
| 0 | 0 | 0 | 0 | 0 |
| 0 | 0 | 0 | 0 | 0 |
| 0 | 0 | 0 | 0 | 0 |
| 0 | 0 | 0 | 0 | 0 |
| 0 | 0 | 0 | 0 | 0 |
| 0 | 0 | 0 | 0 | 0 |
| 0.300598292 | 0.197764607 | 0 | 0.027919561 | 0 |
| 0 | 0 | 0 | 0 | 0 |
| 0 | 0 | 0 | 0 | 0 |
| 0 | 0 | 0 | 0 | 0 |
| 0 | 0 | 0 | 0 | 0 |
| 0 | 0 | 0 | 0 | 0 |
| 0 | 0.018452685 | 0 | 0 | 0 |
| 0.00393273 | 0.003345321 | 0.006662537 | 0.001974916 | 0.200963863 |
| 0 | 0 | 0 | 0 | 0 |
| 0.058168114 | 0.000982321 | 0.001630325 | 0.037255137 | 0 |
| 0 | 0 | 0 | 0 | 0 |
| 0.585952621 | 0 | 0 | 0.040070522 | 0 |
| 0 | 0 | 0 | 0 | 0 |
| 0 | 0 | 0 | 0 | 0 |
| 0 | 0 | 0 | 0 | 0 |
| 0 | 0 | 0 | 0 | 0 |
| 0 | 0.010867637 | 0.001454568 | 0.006585057 | 0.024698714 |
| 0 | 0 | 0 | 0 | 0 |
| 0 | 0 | 0 | 0 | 0 |
| 0 | 0.003182345 | 0 | 0 | 0.001868387 |
| 0 | 0 | 0 | 0 | 0 |
| 0 | 0 | 0 | 0 | 0 |
| 24.0351353 | 3.359198095 | 0 | 1.510658786 | 0 |
| 0.247608648 | 0.837160691 | 0.387584074 | 0 | 0 |
| 0 | 0 | 0 | 2.658898569 | 0 |
| 0 | 0 | 0 | 0 | 0 |
| 0 | 0 | 0 | 0 | 0 |
| 0 | 0 | 0 | 0 | 0 |
| 0 | 0 | 0 | 0 | 0 |
| 0 | 0 | 0 | 0.010674505 | 0.000593142 |

| | | | | |
|---|---|---|---|---|
| 0 | 0 | 0 | 0 | 0 |
| 0.000636258 | 0.01104097 | 0.002425277 | 0.009062535 | 0.001677763 |
| 0.001238523 | 0 | 0 | 0 | 0 |
| 0 | 0 | 0 | 0 | 0 |
| 3.150173473 | 0 | 0 | 0.351768432 | 0 |
| 0 | 0 | 0 | 0 | 0 |
| 0 | 0 | 0 | 0 | 0 |
| 0 | 0 | 0 | 0 | 0 |
| 0.012071083 | 0.518160505 | 1.470376464 | 0.001044135 | 0.00319828 |
| 0 | 0 | 0 | 0 | 0 |
| 0 | 0 | 0 | 0 | 0 |
| 0 | 0 | 0 | 0 | 0 |
| 0.194125254 | 0 | 0 | 0 | 0 |
| 0.046363352 | 0.246715826 | 0 | 0.099930762 | 0 |
| 0 | 0 | 0 | 1.09427529 | 0 |
| 0.302284466 | 0 | 0 | 0.016571331 | 0 |
| 0 | 0 | 0.000835682 | 0.003242797 | 0 |
| 0 | 0 | 0 | 0 | 0 |
| 0 | 0 | 0 | 0 | 0 |
| 0 | 0 | 0 | 0 | 0 |
| 0 | 0 | 0 | 0 | 0 |
| 0 | 0 | 0 | 0 | 0 |
| 0 | 0 | 0 | 0 | 0 |
| 0 | 0 | 0 | 0 | 0 |
| 0 | 0 | 0 | 0 | 0 |
| 0 | 0 | 0 | 0 | 0 |
| 0 | 0 | 0 | 0.026474272 | 0 |
| 0 | 0 | 0.000819954 | 0.003005002 | 0.00448627 |
| 0 | 0.001975662 | 0 | 0 | 0 |
| 0 | 0 | 0 | 0 | 0 |
| 0 | 0 | 0 | 0 | 0 |
| 0 | 0 | 0 | 0 | 0 |
| 0 | 0 | 0 | 0.174151664 | 0 |
| 0.001578453 | 0 | 0 | 0 | 0 |
| 0.140816749 | 0 | 0 | 0.342103502 | 0 |
| 0.126409805 | 0 | 0.003635981 | 0 | 0 |
| 0 | 0 | 0 | 0 | 0 |
| 0 | 0 | 0 | 0 | 0 |
| 0 | 0 | 0 | 0 | 0 |
| 0 | 0 | 0 | 0 | 0 |
| 2.118710761 | 0 | 0 | 0.271973104 | 0 |
| 0 | 0 | 0 | 0.300788451 | 0 |
| 0 | 0 | 0 | 0 | 0 |
| 0 | 0 | 0 | 1.84347408 | 0 |
| 0 | 0 | 0 | 0 | 0 |
| 0 | 0 | 0 | 0 | 0 |
| 0 | 0 | 0 | 0 | 0 |
| 0 | 0 | 0 | 0 | 0 |

| | | | | |
|---|---|---|---|---|
| 0.000942103 | 0 | 0 | 0 | 0 |
| 0 | 0 | 0 | 0 | 0 |
| 0 | 0 | 0 | 0 | 0 |
| 0 | 0 | 0 | 0 | 0.000700974 |
| 0.009792392 | 0 | 0 | 0 | 0 |
| 0 | 0 | 0 | 0 | 0 |
| 0.006561104 | 0.001674333 | 0 | 0.019050034 | 0 |
| 0 | 0.036253608 | 0 | 0 | 0 |
| 0 | 0.010163556 | 0 | 0 | 0 |
| 0 | 0.650311 | 0 | 0.078815028 | 0 |
| 0 | 0 | 0 | 0.012422389 | 0 |
| 0 | 0 | 0 | 0 | 0 |
| 0 | 0 | 0 | 0 | 0 |
| 0 | 0 | 0 | 0 | 0 |
| 0 | 0 | 0 | 0 | 0 |
| 0 | 0 | 0 | 0 | 0 |
| 0 | 0 | 3.413896611 | 0 | 3.907155296 |
| 0 | 0.004507147 | 0 | 0 | 0 |
| 0 | 0 | 0 | 0 | 0 |
| 0 | 0 | 0 | 0 | 0 |
| 0 | 0.000956435 | 0.175006679 | 0.003422018 | 0 |
| 0 | 0 | 0 | 0 | 0 |
| 0.004822936 | 0.012659333 | 0 | 0.005850434 | 0 |
| 0.001429535 | 0.01125681 | 0.000518959 | 0.00167815 | 0.007196454 |
| 0 | 0 | 0 | 0 | 0 |
| 0 | 0 | 0 | 0 | 0 |
| 0 | 0 | 0 | 0 | 0 |
| 0 | 0 | 0 | 0 | 0 |
| 0 | 0 | 0 | 0 | 0 |
| 0.001763516 | 0 | 0.500799328 | 0.583524881 | 0.259357362 |
| 0 | 0 | 0 | 0 | 0 |
| 0 | 0 | 0 | 0 | 0 |
| 0 | 0 | 0 | 0 | 0 |
| 0 | 0 | 0 | 0 | 0 |
| 0 | 0 | 0 | 0 | 0 |
| 0 | 0 | 0 | 0 | 0 |
| 0.029897147 | 0 | 0.001948058 | 0 | 0 |
| 0 | 0 | 0 | 0 | 0 |
| 0 | 0 | 0 | 0 | 0 |
| 0 | 0 | 0 | 0 | 0 |
| 0 | 0 | 0 | 0 | 0 |
| 0 | 0 | 0 | 0 | 0 |
| 0 | 0 | 0 | 0 | 0 |
| 0.002438385 | 0 | 0 | 0 | 0 |
| 0 | 0 | 0 | 0 | 0 |
| 0 | 0 | 0 | 0 | 0 |
| 0 | 0 | 0 | 0 | 0 |
| 0 | 0 | 0 | 0 | 0 |

| | | | | |
|---|---|---|---|---|
| 0.033181822 | 0.00942101 | 0 | 0.075119179 | 0 |
| 0.021882763 | 0 | 0 | 0.561378833 | 0 |
| 0 | 0 | 0 | 0 | 0 |
| 0 | 0 | 0 | 0 | 0 |
| 0 | 0 | 0 | 0 | 0 |
| 0 | 0 | 0 | 0 | 0 |
| 0 | 0 | 0 | 0 | 0 |
| 0.711413509 | 0 | 0 | 0.03819439 | 0 |
| 2.217875324 | 0.091894932 | 0.039662627 | 0.184519118 | 0.109641311 |
| 0.031456671 | 0.245603232 | 0.008848022 | 0 | 0 |
| 0 | 0 | 0 | 0.005788849 | 0 |
| 0 | 0 | 0 | 0 | 0 |
| 0 | 0 | 0 | 0 | 0 |
| 0 | 1.389952444 | 0 | 0.013009784 | 0 |
| 0 | 0 | 0 | 0 | 0 |
| 0 | 0 | 0 | 0 | 0 |
| 0 | 0.245082771 | 0 | 0 | 0 |
| 0 | 0 | 0 | 0 | 0 |
| 0 | 0 | 0 | 0 | 0 |
| 0.003185909 | 0 | 0 | 0 | 0 |
| 0 | 0 | 0 | 0 | 0 |
| 0 | 0 | 0 | 0 | 0 |
| 0 | 0 | 0 | 0 | 0 |
| 0 | 0 | 0 | 0 | 0 |
| 0 | 0 | 0 | 0.001422278 | 0 |
| 0 | 0 | 0 | 0 | 0 |
| 0 | 0 | 0 | 0 | 0 |
| 0 | 0 | 0 | 0 | 0 |
| 0 | 0.237350625 | 0 | 0 | 0 |
| 0 | 0 | 0 | 0 | 0 |
| 0 | 0.02212221 | 0 | 0 | 0 |
| 0 | 0 | 0 | 0 | 0 |
| 0 | 0 | 0 | 0 | 0 |
| 0 | 0 | 0 | 0 | 0 |
| 0 | 0 | 0 | 0.003935005 | 0 |
| 0 | 0 | 0 | 0 | 0 |
| 0 | 0 | 0 | 0 | 0 |
| 0 | 0 | 0 | 0 | 0 |
| 0 | 0 | 0 | 0.060765116 | 0 |
| 0 | 0 | 0 | 0 | 0 |
| 0 | 0 | 0 | 0 | 0 |
| 0.048886369 | 0 | 0 | 0.751548653 | 0 |
| 0 | 0 | 0 | 0 | 0 |
| 0 | 0 | 0 | 0 | 0 |
| 0 | 0 | 0 | 0 | 0 |
| 0.002540199 | 0 | 0 | 0.030347319 | 0 |
| 0 | 0 | 0 | 0 | 0 |
| 0 | 0 | 0.001430052 | 0 | 0.007652643 |

| | | | | |
|---|---|---|---|---|
| 0 | 0 | 0 | 0 | 0 |
| 0.011899886 | 0 | 0.031877833 | 0.015319818 | 0.00741687 |
| 0 | 0 | 0 | 0 | 0 |
| 0 | 0 | 0 | 0 | 0 |
| 0.016637713 | 0.109965967 | 0.019025798 | 1.483397672 | 0.068400975 |
| 0 | 0 | 0 | 0 | 0 |
| 0 | 0 | 0 | 0 | 0 |
| 0.019287867 | 0.013268229 | 0.055939993 | 0 | 0.465987577 |
| 0 | 0 | 0 | 0 | 0 |
| 0 | 0 | 0 | 0 | 0 |
| 0 | 0 | 0 | 0 | 0 |
| 0 | 0 | 0 | 0.25139708 | 0 |
| 0 | 0 | 0 | 0 | 0 |
| 0 | 0 | 0 | 0 | 0 |
| 0 | 0.028925002 | 0 | 0 | 0 |
| 0 | 0.012621059 | 0.002926382 | 0 | 0.014994257 |
| 0 | 0 | 0 | 0 | 0 |
| 0 | 0 | 0 | 0 | 0 |
| 0 | 0 | 0 | 0 | 0 |
| 0 | 0 | 0 | 0 | 0 |
| 0 | 0 | 0 | 0 | 0 |
| 0 | 0 | 0 | 0 | 0 |
| 0 | 0 | 0 | 0 | 0 |
| 0 | 0.001481621 | 0 | 0.009343142 | 0.009046696 |
| 0 | 0 | 0 | 0 | 0 |
| 0 | 0 | 0 | 0 | 0 |
| 0.003982639 | 0.00135511 | 0 | 0.005272671 | 0 |
| 0 | 0 | 0 | 0 | 0 |
| 0 | 0 | 0 | 0 | 0 |
| 0 | 0 | 0 | 0 | 0 |
| 0 | 0 | 0 | 0 | 0 |
| 0 | 0 | 0 | 0 | 0 |
| 0 | 0 | 0 | 0 | 0 |
| 0 | 0 | 0 | 0 | 0 |
| 0 | 0 | 0 | 0 | 0 |
| 0 | 0 | 0 | 0 | 0 |
| 0 | 0 | 0 | 0 | 0 |
| 0 | 0 | 0 | 0.427560156 | 0 |
| 0 | 0 | 0 | 0 | 0 |
| 0 | 0 | 0 | 0 | 0 |
| 0 | 0 | 0 | 0 | 0 |
| 0 | 0 | 0 | 0 | 0 |
| 0 | 0 | 0 | 0 | 0 |
| 0 | 0 | 0 | 0 | 0 |
| 0 | 0 | 0 | 0 | 0 |
| 0 | 0 | 0 | 0 | 0 |
| 0 | 0 | 0 | 0 | 0 |
| 0 | 0 | 0 | 0.054470053 | 0 |

| | | | | |
|---|---|---|---|---|
| 0.020599309 | 0 | 0 | 0 | 0 |
| 0 | 0 | 0 | 0 | 0 |
| 0 | 0 | 0 | 0 | 0 |
| 0 | 0 | 0 | 0.001020709 | 0 |
| 0 | 0 | 0 | 0 | 0 |
| 0 | 0 | 0.000459445 | 0.251578279 | 0.231962138 |
| 0 | 0 | 0 | 0 | 0 |
| 0 | 0 | 0 | 0 | 0 |
| 0 | 0 | 0 | 0 | 0 |
| 0 | 0 | 0 | 0 | 0 |
| 0 | 0 | 0 | 0 | 0 |
| 0 | 0 | 0 | 0.002199035 | 0 |
| 0 | 0 | 0 | 0 | 0 |
| 0 | 0 | 0 | 0 | 0 |
| 0 | 0 | 0 | 0.008411732 | 0 |
| 0 | 0 | 0 | 0 | 0 |
| 0 | 0 | 0 | 0 | 0 |
| 0 | 3.804380895 | 0 | 0 | 0 |
| 0 | 0 | 0 | 0.001304664 | 0 |
| 0 | 0 | 0 | 0 | 0 |
| 0 | 0.01586084 | 0.008264414 | 0 | 0 |
| 0 | 0 | 0 | 0 | 0 |
| 0.00365091 | 0 | 0.002126128 | 0.080502614 | 0.004375983 |
| 0.094675922 | 0 | 0 | 0 | 0 |
| 0 | 0 | 0 | 0 | 0 |
| 0 | 0 | 0 | 0 | 0 |
| 0 | 0 | 0 | 0 | 0 |
| 0 | 0 | 0 | 0 | 0 |
| 0 | 0.009275185 | 0 | 0.396062681 | 0 |
| 0 | 0 | 0 | 0 | 0 |
| 0 | 0 | 0 | 0 | 0 |
| 0 | 0 | 0 | 0 | 0 |
| 0 | 0.732910917 | 0.076386207 | 0.379527276 | 0.497508184 |
| 0 | 0.151217648 | 0 | 0 | 0 |
| 1.133113076 | 0 | 0 | 0.348909677 | 0 |
| 0.004115481 | 0.064764348 | 0.083375172 | 0 | 0.595884508 |
| 0 | 0 | 0 | 0 | 0 |
| 0 | 0 | 0 | 0 | 0 |
| 0 | 0.027129645 | 0 | 0.00282508 | 0 |
| 0 | 0 | 0 | 0 | 0 |
| 0 | 0 | 0 | 0 | 0 |
| 0 | 0 | 0 | 0 | 0 |
| 0.095242257 | 0 | 0.028289039 | 0.339230157 | 0.040474002 |
| 0 | 0 | 0 | 0 | 0 |
| 0 | 0 | 0 | 0 | 0 |
| 0.001473484 | 0 | 0 | 0 | 0 |
| 0 | 0 | 0 | 0 | 0 |
| 0 | 0 | 0 | 0 | 0 |

| | | | | |
|---|---|---|---|---|
| 0 | 0 | 0 | 0 | 0 |
| 0 | 0 | 0 | 0 | 0 |
| 0 | 0 | 0 | 0 | 0 |
| 0 | 0 | 0 | 0 | 0 |
| 0 | 0 | 0 | 0 | 0 |
| 0 | 0 | 0 | 0 | 0 |
| 0 | 0 | 0 | 0 | 0 |
| 0.078311905 | 0.053776399 | 0 | 0 | 0 |
| 0.089503607 | 0 | 0 | 0 | 0 |
| 0.001770618 | 0.021301282 | 0 | 0.001039277 | 0 |
| 0 | 0 | 0 | 0 | 0 |
| 0 | 0 | 0 | 0 | 0 |
| 0 | 0 | 0 | 0 | 0 |
| 0 | 0 | 0 | 0 | 0 |
| 0 | 0 | 0 | 0 | 0 |
| 0 | 0.044964665 | 0.369495071 | 0 | 0.177545906 |
| 0 | 0 | 0 | 0 | 0 |
| 0 | 0 | 0 | 0 | 0 |
| 0 | 0 | 0 | 0 | 0 |
| 1.25038124 | 0 | 0 | 0 | 0 |
| 0 | 0 | 0 | 0 | 0 |
| 0 | 0 | 0 | 0 | 0 |
| 0 | 0 | 0 | 0 | 0 |
| 0 | 0 | 0 | 0.013620353 | 0 |
| 0 | 0 | 0 | 0.002487566 | 0 |
| 0 | 0 | 0 | 0 | 0 |
| 0 | 0 | 0 | 0 | 0 |
| 0 | 0 | 0 | 0 | 0 |
| 0 | 0 | 0 | 0 | 0 |
| 0 | 0 | 0 | 0 | 0 |
| 0.12176946 | 0 | 0 | 0.287555856 | 0 |
| 0 | 0 | 0 | 0 | 0 |
| 0 | 0 | 0 | 0 | 0 |
| 0 | 0 | 0 | 0.017619501 | 0 |
| 0 | 0 | 0 | 0 | 0 |
| 0 | 0 | 0 | 0 | 0 |
| 0 | 0 | 0 | 0 | 0 |
| 0 | 0 | 0 | 0 | 0 |
| 0 | 0 | 0 | 0 | 0 |
| 0 | 0 | 0 | 0 | 0 |
| 0 | 0 | 0 | 0 | 0 |
| 0 | 0 | 0 | 0 | 0 |
| 0.007000897 | 0 | 0 | 0 | 0 |
| 0 | 0 | 0 | 0 | 0 |
| 0 | 0 | 0 | 0 | 0 |
| 0 | 0 | 0 | 0 | 0 |
| 0 | 0.02223483 | 0.419683664 | 0.000628056 | 0 |
| 0 | 0 | 0 | 0.002211822 | 0 |

| | | | | |
|---|---|---|---|---|
| 0 | 0 | 0 | 0 | 0 |
| 0 | 0 | 0 | 0 | 0 |
| 0 | 0 | 0 | 0 | 0 |
| 0 | 0 | 0 | 0.001501014 | 0 |
| 0.017510343 | 0 | 0 | 0.017360572 | 0 |
| 0 | 0 | 0 | 0 | 0 |
| 0 | 0 | 0 | 0 | 0 |
| 0 | 0 | 0 | 0 | 0 |
| 0 | 0 | 0 | 0 | 0 |
| 0 | 0 | 0 | 0 | 0 |
| 0.143541718 | 0 | 0 | 0 | 0 |
| 0.001321226 | 0.004450574 | 0.000671497 | 0.004994238 | 0.029075314 |
| 0 | 0 | 0 | 0.001873378 | 0.002131377 |
| 0 | 0 | 0 | 0 | 0 |
| 0 | 0.054266372 | 0 | 0.010425016 | 0 |
| 3.366512275 | 0 | 0 | 0 | 0 |
| 0.005690607 | 0.004045387 | 0.001032921 | 0 | 0.002630867 |
| 0 | 0 | 0 | 0.001818985 | 0 |
| 0.032614347 | 1.357803716 | 0.111607233 | 0.002630724 | 0 |
| 0 | 0.00160007 | 0.025559973 | 0.013310331 | 0 |
| 0 | 0 | 0 | 0 | 0 |
| 0 | 0 | 0 | 0 | 0 |
| 0 | 0 | 0 | 0 | 0 |
| 0 | 0 | 0 | 0 | 0 |
| 0 | 0 | 0 | 0 | 0 |
| 0 | 0 | 0 | 0 | 0 |
| 0.001256129 | 0.09135744 | 0 | 0.040772325 | 0 |
| 0 | 0 | 0 | 0 | 0 |
| 0 | 0 | 0 | 0 | 0 |
| 0 | 0 | 0 | 0 | 0 |
| 0 | 0.091567478 | 0 | 0.002730152 | 0.001536006 |
| 0 | 0 | 0 | 0 | 0 |
| 0 | 0 | 0 | 0 | 0 |
| 0 | 0 | 0 | 0 | 0 |
| 0 | 0 | 0 | 0 | 0 |
| 0 | 0 | 0 | 0 | 0 |
| 0 | 0 | 0 | 0 | 0 |
| 0.001667304 | 0.054376408 | 0.004342855 | 0.032060098 | 0.058394173 |
| 0 | 0 | 0 | 0 | 0 |
| 0 | 0 | 0 | 0.004346635 | 0 |
| 0 | 0 | 0 | 0.0091642 | 0.008499036 |
| 0 | 0 | 0 | 0.008606267 | 0 |
| 0 | 0 | 0 | 0 | 0 |
| 0 | 0.028392013 | 0.752840444 | 0 | 0 |
| 0 | 0 | 0 | 0 | 0 |
| 0 | 0 | 0 | 0 | 0 |
| 0 | 0 | 0 | 0 | 0 |
| 0 | 0 | 0 | 0 | 0 |

| | | | | |
|---|---|---|---|---|
| 0 | 0 | 0 | 0 | 0 |
| 0 | 0 | 0 | 0 | 0 |
| 0 | 0 | 0 | 0 | 0 |
| 0 | 0.001190198 | 0.001152277 | 0.007239275 | 0.027019402 |
| 0 | 0 | 0 | 0 | 0.001901108 |
| 0 | 0 | 0 | 0 | 0 |
| 0 | 0 | 0 | 0.002479471 | 0 |
| 0.001215872 | 0 | 0.0012874 | 0 | 0.135824607 |
| 0 | 0 | 0 | 0 | 0 |
| 0 | 0 | 0 | 0 | 0 |
| 0 | 0 | 0 | 0.28273395 | 0 |
| 0 | 0 | 0 | 0 | 0 |
| 0.009183578 | 0 | 0 | 0.006942789 | 0 |
| 0 | 0 | 0 | 0 | 0 |
| 0 | 0.011820357 | 0 | 0 | 0 |
| 0 | 0 | 0 | 0 | 0 |
| 0 | 0 | 0 | 0 | 0 |
| 0 | 0 | 0 | 0 | 0 |
| 0 | 0 | 0 | 0 | 0 |
| 0 | 0 | 0 | 0 | 0 |
| 0 | 0 | 0 | 0 | 0 |
| 0 | 0 | 0 | 0 | 0 |
| 0 | 0.019256227 | 0.018591474 | 0 | 0.399519973 |
| 0 | 0 | 0 | 0.000703561 | 0 |
| 0 | 0 | 0 | 0 | 0 |
| 0 | 0 | 0 | 0.007101018 | 0 |
| 0 | 0 | 0 | 0 | 0 |
| 0 | 0 | 0 | 0.002442487 | 0 |
| 0 | 0 | 0 | 0 | 0 |
| 0 | 0 | 0 | 0 | 0 |
| 0.010516941 | 0 | 0 | 0 | 0 |
| 0 | 0 | 0 | 0 | 0 |
| 0 | 0 | 0 | 0 | 0 |
| 0 | 0 | 0 | 0 | 0 |
| 0.001130406 | 0 | 0 | 0 | 0 |
| 0 | 0 | 0 | 0 | 0 |
| 0 | 0 | 0 | 0 | 0 |
| 0 | 0 | 0 | 0 | 0 |
| 0.02646603 | 0 | 0 | 0.001124906 | 0 |
| 0 | 0 | 0 | 0 | 0 |
| 0 | 0 | 0 | 0 | 0 |
| 0 | 0 | 0 | 0 | 0 |
| 0 | 0 | 0 | 0 | 0 |
| 0 | 0 | 0 | 0 | 0 |
| 0 | 0 | 0 | 20.30577449 | 0 |
| 0.139312927 | 0 | 0 | 0.050146399 | 0 |
| 0 | 0 | 0 | 0 | 0 |
| 0 | 0 | 0 | 0 | 0 |

| | | | | |
|---|---|---|---|---|
| 0 | 0 | 0 | 0 | 0 |
| 0 | 0 | 0 | 0 | 0 |
| 0 | 0 | 0 | 0.219618148 | 0 |
| 0 | 0 | 0 | 0 | 0 |
| 0 | 0 | 0 | 0 | 0 |
| 0 | 0 | 0 | 0 | 0 |
| 0 | 0 | 0 | 0 | 0 |
| 0 | 0 | 0 | 0 | 0 |
| 0 | 0 | 0 | 0 | 0 |
| 0 | 0 | 0 | 0 | 0 |
| 0 | 0 | 0 | 0 | 0 |
| 0 | 0.00465441 | 0.001580065 | 0 | 0.005564673 |
| 0 | 0 | 0 | 0 | 0 |
| 0 | 0 | 0 | 0 | 0 |
| 0 | 0 | 0 | 0 | 0 |
| 0 | 0 | 0 | 0 | 0 |
| 0.00136041 | 0 | 0 | 0 | 0 |
| 0 | 0 | 0 | 0 | 0 |
| 0 | 0 | 0 | 0 | 0 |
| 0 | 0 | 0.005302132 | 0.018355681 | 0.013063285 |
| 0 | 0 | 0 | 0 | 0 |
| 0 | 0 | 0 | 0 | 0 |
| 0 | 0 | 0 | 0 | 0 |
| 0 | 0 | 0 | 0 | 0 |
| 0 | 0 | 0 | 0 | 0 |
| 0 | 0 | 0 | 0 | 0 |
| 0 | 0 | 0 | 0 | 0 |
| 0 | 0 | 0 | 0 | 0 |
| 0 | 0 | 0 | 0 | 0 |
| 0 | 0 | 0 | 0 | 0 |
| 0.000632647 | 0.010978309 | 0 | 0.003292518 | 0 |
| 0 | 0 | 0 | 0 | 0 |
| 0.091820985 | 0.031867336 | 0.453446421 | 0.085714541 | 0.304284766 |
| 0.055548004 | 0.061681868 | 0 | 0.001028063 | 0 |
| 0 | 0 | 0 | 0 | 0 |
| 0 | 0 | 0 | 0 | 0 |
| 0 | 0 | 0 | 0 | 0 |
| 0 | 0 | 0 | 0 | 0 |
| 0 | 0 | 0 | 0 | 0 |
| 0 | 0 | 0 | 0 | 0 |
| 0 | 0 | 0 | 0 | 0 |
| 0.064662489 | 0 | 0.065651022 | 0 | 0.710288975 |
| 0.032148928 | 0 | 0 | 0.028233028 | 0 |
| 0 | 0 | 0 | 0 | 0 |
| 0 | 0 | 0 | 0 | 0 |
| 0.013121566 | 0 | 0 | 0.085094408 | 0.002550408 |
| 0 | 0 | 0 | 0.0112727 | 0.000668848 |
| 0 | 0 | 0 | 0 | 0 |

| | | | | |
|---|---|---|---|---|
| 0 | 0 | 0 | 0 | 0 |
| 0 | 0.003936592 | 0 | 0.038873734 | 0 |
| 0.027629571 | 0 | 0.004769832 | 0.003208218 | 0.002159793 |
| 0 | 0 | 0 | 0 | 0 |
| 0.421794488 | 0.389920198 | 0 | 0.019507629 | 0 |
| 0 | 0 | 0 | 0 | 0.001678609 |
| 0 | 0 | 0 | 0 | 0 |
| 0 | 0 | 0 | 0 | 0 |
| 0 | 0 | 0 | 0 | 0 |
| 0 | 0 | 0 | 0 | 0 |
| 0 | 0 | 0 | 0 | 0 |
| 0 | 0 | 0 | 0 | 0 |
| 0 | 0 | 0 | 0 | 0 |
| 0 | 1.551425722 | 0 | 0 | 0 |
| 0 | 0 | 0 | 0 | 0 |
| 0 | 0 | 0 | 0 | 0 |
| 0 | 0 | 0 | 0.017042791 | 0 |
| 0 | 0 | 0 | 0 | 0 |
| 0 | 0 | 0 | 0 | 0 |
| 0 | 0 | 0 | 0 | 0 |
| 0 | 0 | 0 | 0 | 0 |
| 0 | 0 | 0 | 0 | 0 |
| 0 | 0 | 0 | 0 | 0 |
| 0 | 0 | 0 | 0 | 0 |
| 0 | 0 | 0 | 0.066914116 | 0 |
| 0 | 0 | 0 | 0 | 0.003581222 |
| 0 | 0 | 0 | 0 | 0 |
| 0 | 0 | 0 | 0 | 0 |
| 0 | 0 | 0 | 0 | 0 |
| 0 | 0 | 0 | 0 | 0 |
| 0 | 0 | 0 | 0.00663589 | 0 |
| 0 | 0.001002959 | 0 | 0 | 0 |
| 0 | 0 | 0 | 0 | 0 |
| 0 | 0 | 0 | 0 | 0 |
| 0 | 0 | 0 | 0 | 0 |
| 0 | 0 | 0 | 0 | 0 |
| 0 | 0 | 0 | 0 | 0 |
| 0 | 0 | 0.00062982 | 0 | 0 |
| 5.096748014 | 1.930023424 | 0 | 0.254866054 | 0 |
| 0 | 0 | 0 | 0 | 0 |
| 0 | 0 | 0 | 0 | 0 |
| 0 | 0 | 0 | 0.018422804 | 0.002913805 |
| 0.031235612 | 0.008800027 | 0 | 0.053799163 | 0 |
| 0 | 0 | 0 | 0.000996607 | 0 |
| 0 | 0 | 0 | 0 | 0 |
| 0 | 0 | 0 | 0 | 0 |
| 0 | 0.022242092 | 0.008257176 | 0.021046744 | 0 |
| 0 | 0 | 0 | 0 | 0 |

| | | | | |
|---|---|---|---|---|
| 0 | 0 | 0 | 0 | 0 |
| 0.005305968 | 0.085673415 | 0.011185143 | 0.076500358 | 0.129391741 |
| 0 | 0 | 0 | 0 | 0 |
| 0 | 0 | 0 | 0 | 0 |
| 0 | 0 | 0 | 0 | 0 |
| 0 | 0 | 0 | 0 | 0 |
| 0 | 0 | 0 | 0 | 0 |
| 0 | 0 | 0 | 0 | 0 |
| 0 | 0 | 0 | 0 | 0 |
| 0 | 0 | 0 | 0 | 0 |
| 0 | 0 | 0 | 0 | 0 |
| 0 | 0 | 0 | 0 | 0 |
| 0 | 0 | 0 | 0.635209162 | 0 |
| 0 | 0 | 0 | 0 | 0 |
| 0.017441502 | 0.007367023 | 0 | 0.011614165 | 0 |
| 0 | 0 | 0 | 0 | 0 |
| 0 | 0 | 0 | 0 | 0 |
| 0 | 0 | 0 | 0.001863882 | 0 |
| 0.056886446 | 0 | 0 | 0 | 0 |
| 0 | 0 | 0 | 0 | 0 |
| 0 | 0 | 0 | 0 | 0 |
| 0 | 0 | 0.009193378 | 0.052661867 | 0 |
| 0.006354157 | 0.081307746 | 0 | 0.002983693 | 0 |
| 0 | 0 | 0 | 0 | 0 |
| 0 | 0 | 0 | 0 | 0 |
| 0 | 0 | 0 | 0 | 0 |
| 0 | 0 | 0 | 0 | 0 |
| 0 | 0 | 0 | 0 | 0 |
| 0 | 0 | 0 | 0 | 0 |
| 0 | 0 | 0 | 0 | 0 |
| 0 | 0 | 0 | 0 | 0 |
| 0 | 0 | 0 | 0 | 0 |
| 0 | 0 | 0 | 0 | 0 |
| 0 | 0 | 0 | 0 | 0 |
| 0 | 1.537140328 | 0 | 0.133086096 | 0 |
| 0 | 0 | 0 | 0 | 0 |
| 0 | 0.028240258 | 0 | 0 | 0 |
| 0 | 0 | 0 | 0 | 0 |
| 0 | 0.005418766 | 0.001873612 | 0.004604588 | 0 |
| 0 | 0 | 0 | 0 | 0 |
| 0.480741068 | 0.004453754 | 0 | 0.007388061 | 0 |
| 0 | 0 | 0 | 0 | 0 |
| 0 | 0 | 0 | 0 | 0 |
| 0 | 0 | 0 | 0 | 0 |
| 0 | 0 | 0 | 0 | 0 |
| 0.570095139 | 0.512242314 | 1.333121265 | 0.009453983 | 5.401066814 |
| 0 | 0 | 0 | 0 | 0 |
| 0 | 0 | 0 | 0 | 0 |

| | | | | |
|---|---|---|---|---|
| 0 | 0 | 0 | 0 | 0 |
| 0 | 0 | 0 | 0 | 0 |
| 0 | 0 | 0 | 0 | 0 |
| 0 | 0 | 0 | 0 | 0 |
| 0 | 0 | 0 | 0.00245294 | 0 |
| 0 | 0 | 0 | 0 | 0 |
| 0 | 0 | 0 | 0 | 0 |
| 0 | 0 | 0 | 0 | 0 |
| 0 | 0 | 0 | 0 | 0 |
| 0.03553185 | 0 | 0.00921742 | 0.009246011 | 0.047273237 |
| 0 | 0 | 0 | 0 | 0 |
| 0 | 0 | 0 | 0 | 0 |
| 0 | 0.047858143 | 0.002022485 | 0 | 0 |
| 0 | 0 | 0 | 0 | 0 |
| 0 | 0 | 0 | 0 | 0 |
| 0 | 0 | 0 | 0 | 0 |
| 0 | 0 | 0 | 0 | 0 |
| 0 | 0 | 0 | 0 | 0 |
| 0 | 0 | 0 | 0 | 0 |
| 0.01030387 | 0.366414153 | 0.089025826 | 1.39389601 | 1.232676057 |
| 0 | 0 | 0 | 0 | 0 |
| 0 | 0 | 0 | 0 | 0 |
| 0 | 0 | 0 | 0 | 0 |
| 0 | 0 | 0 | 0 | 0 |
| 0 | 0 | 0 | 0 | 0 |
| 0 | 0 | 0 | 0.052042007 | 0 |
| 0 | 0 | 0 | 0 | 0 |
| 0 | 0 | 0 | 0 | 0 |
| 0 | 0 | 0 | 0 | 0 |
| 0 | 0 | 0 | 0 | 0 |
| 0 | 0 | 0 | 0 | 0 |
| 0 | 0 | 0 | 0 | 0 |
| 0 | 0 | 0 | 0 | 0 |
| 0 | 0 | 0 | 0 | 0 |
| 0 | 0 | 0 | 0 | 0 |
| 0.009336686 | 0 | 0 | 0.001918081 | 0 |
| 0 | 0 | 0 | 0 | 0 |
| 0 | 0 | 0 | 0 | 0 |
| 0 | 0 | 0 | 0 | 0 |
| 0 | 0 | 0 | 0 | 0 |
| 0 | 0 | 0 | 0 | 0 |
| 0 | 0 | 0 | 0 | 0 |
| 0 | 0 | 0 | 0 | 0 |
| 0 | 0 | 0 | 0 | 0 |
| 0 | 0.00222271 | 0 | 0.019285078 | 0.004697913 |
| 0 | 0 | 0 | 0 | 0 |
| 0 | 0 | 0 | 0 | 0 |
| 0 | 0 | 0.006770063 | 0 | 0.0132374 |

| | | | | |
|---|---|---|---|---|
| 0 | 0 | 0 | 0 | 0 |
| 0 | 0 | 0 | 0 | 0 |
| 0 | 0 | 0 | 0 | 0 |
| 0 | 0 | 0 | 0 | 0 |
| 0 | 0 | 0 | 0 | 0 |
| 0.001264346 | 0.060012783 | 0.000573739 | 0.070649494 | 0.015587444 |
| 0 | 0 | 0 | 0 | 0 |
| 0 | 0 | 0 | 0 | 0 |
| 0 | 0 | 0 | 0 | 0 |
| 0 | 0 | 0 | 0 | 0 |
| 0 | 0 | 0 | 0 | 0 |
| 0 | 0 | 0 | 0 | 0 |
| 0 | 0 | 0 | 0 | 0 |
| 0 | 0 | 0 | 0.572521746 | 0 |
| 0 | 0 | 0 | 0 | 0 |
| 0 | 0 | 0 | 0 | 0 |
| 0 | 0 | 0 | 0.355031834 | 0 |
| 0 | 0 | 0 | 0 | 0 |
| 0 | 0 | 0 | 0 | 0 |
| 0 | 0 | 0 | 0 | 0 |
| 0 | 0 | 0 | 0 | 0 |
| 0 | 0 | 0 | 0 | 0 |
| 0 | 0 | 0 | 0 | 0 |
| 0 | 0 | 0 | 0 | 0 |
| 0 | 0 | 0 | 0 | 0 |
| 0.001695128 | 0 | 0.001256394 | 0 | 0 |
| 0 | 0 | 0 | 0 | 0 |
| 0 | 0 | 0 | 0.296425922 | 0 |
| 0 | 0 | 0 | 0 | 0 |
| 0 | 0 | 0 | 0 | 0 |
| 0 | 0 | 0 | 0 | 0 |
| 0 | 0 | 0 | 0 | 0 |
| 0 | 0 | 0 | 0 | 0 |
| 0 | 0 | 0 | 0 | 0 |
| 0 | 0 | 0 | 0 | 0 |
| 0 | 0 | 0 | 0 | 0 |
| 0 | 0 | 0 | 0 | 0 |
| 0 | 0 | 0 | 0 | 0 |
| 0 | 0 | 0 | 0 | 0 |
| 0 | 0 | 0 | 0 | 0 |
| 0 | 0 | 0 | 0 | 0 |
| 0 | 0 | 0 | 0 | 0 |
| 0 | 0 | 0 | 0 | 0 |
| 0 | 0 | 0 | 0 | 0 |
| 0 | 0 | 0 | 0 | 0 |
| 0 | 0 | 0 | 0 | 0 |
| 0 | 0 | 0 | 0 | 0 |
| 0 | 0 | 0 | 0 | 0 |

| | | | | |
|---|---|---|---|---|
| 0 | 0 | 0 | 0 | 0 |
| 0 | 0 | 0 | 0 | 0 |
| 0 | 0 | 0 | 0 | 0 |
| 0 | 0 | 0 | 0 | 0 |
| 0 | 0 | 0 | 0 | 0 |
| 0 | 0 | 0 | 0.005241034 | 0 |
| 0 | 0 | 0 | 0 | 0 |
| 0 | 0 | 0 | 0 | 0 |
| 0 | 0 | 0 | 0 | 0 |
| 0 | 0 | 0 | 0 | 0 |
| 0 | 0 | 0 | 0.001523122 | 0 |
| 0 | 0 | 0 | 0 | 0 |
| 0.244179587 | 0 | 0 | 0.001282122 | 0 |
| 0 | 0 | 0 | 0 | 0 |
| 0 | 0 | 0 | 0 | 0 |
| 0 | 0 | 0 | 0 | 0 |
| 0 | 0 | 0 | 0 | 0 |
| 0 | 0 | 0 | 0 | 0 |
| 0 | 0 | 0 | 0 | 0 |
| 0 | 0 | 0 | 0.003259056 | 0 |
| 0.005106183 | 0 | 0.007341553 | 0 | 0.086779048 |
| 0 | 0 | 0 | 0 | 0 |
| 0 | 0 | 0 | 0 | 0 |
| 14.37028602 | 0 | 0 | 1.17559456 | 0 |
| 0 | 0 | 0 | 0 | 0 |
| 0 | 0 | 0 | 0 | 0 |
| 0 | 0 | 0 | 0 | 0 |
| 0 | 0 | 0 | 0 | 0 |
| 0 | 0 | 0 | 0 | 0 |
| 0 | 0 | 0 | 0 | 0 |
| 0 | 1.278674078 | 0 | 0 | 0 |
| 0 | 0 | 0 | 0 | 0 |
| 0 | 0 | 0 | 0 | 0 |
| 0 | 0 | 0 | 0 | 0 |

| PHL018-0244_V2 | PHL018-0212_V2 | PHL018-0155_V3 | PHL018-0043_V1 | PHL018-0041_V1 |
|---|---|---|---|---|
| REF | REF | CTRL | EYCF | EYCF |
| 0.001326442 | 0.007211329 | 0.034988237 | 0.023542351 | 0.017033307 |
| 0.212302374 | 0.907242497 | 1.926325515 | 1.554661213 | 0.232963322 |
| 0.096092589 | 0 | 0 | 3.369929208 | 9.660719641 |
| 0 | 0 | 0 | 0 | 0 |
| 0 | 0 | 0 | 0 | 0 |
| 0 | 0 | 0 | 0 | 0 |
| 0 | 0 | 0 | 0 | 0 |
| 0 | 0 | 0 | 0 | 0 |
| 0 | 0 | 0 | 0 | 0 |
| 0 | 0 | 0 | 0 | 0 |
| 0 | 0 | 0 | 0 | 0 |
| 0 | 0 | 0 | 0 | 0 |
| 0 | 0 | 0 | 0 | 0 |
| 5.20756661 | 8.381955126 | 0 | 1.336024391 | 3.43695974 |
| 0 | 0 | 0 | 0 | 0 |
| 0 | 0 | 0 | 0 | 0 |
| 0 | 0 | 0.042664309 | 0 | 0 |
| 0 | 0 | 0 | 0 | 0.001445229 |
| 0 | 0 | 0 | 0 | 0 |
| 0 | 0 | 0 | 0 | 0 |
| 0 | 0 | 0 | 0 | 0 |
| 0 | 0 | 0 | 0 | 0 |
| 0 | 0 | 0 | 0 | 0 |
| 0 | 0 | 0 | 0 | 0 |
| 0 | 0 | 0 | 0.006941967 | 0 |
| 0 | 0 | 0 | 0 | 0 |
| 0 | 0 | 0 | 0 | 0 |
| 0 | 0 | 0 | 0 | 0 |
| 0 | 0 | 0 | 0 | 0 |
| 0 | 0 | 0 | 0 | 0 |
| 0 | 0 | 0 | 0 | 0 |
| 0 | 0 | 0 | 0 | 0 |
| 0 | 0 | 0 | 0 | 0 |
| 0 | 0 | 0 | 0 | 0 |
| 0 | 0 | 0 | 0 | 0 |
| 0 | 0.011001395 | 0 | 0 | 0 |
| 0 | 0 | 0 | 0 | 0 |
| 0 | 0 | 0 | 0 | 0 |
| 0 | 0 | 0 | 0 | 0 |
| 0 | 0 | 0 | 0 | 0 |
| 0 | 0 | 0 | 0 | 0 |
| 0 | 0.118428835 | 0.002263385 | 0 | 0.010116823 |
| 0.48170189 | 0.223169924 | 0.040145041 | 0.000564363 | 0.001224981 |
| 0.008036943 | 0.001427896 | 0.016330115 | 0.001138287 | 0 |
| 0.001192955 | 0 | 0 | 0 | 0 |
| 0 | 0 | 0 | 0 | 0 |

| | | | | |
|---|---|---|---|---|
| 0 | 0.012274694 | 0 | 0.006533541 | 0 |
| 0 | 0.035981047 | 0 | 0 | 0 |
| 0 | 0.000607151 | 0.156035513 | 0.051166535 | 0.006566029 |
| 0.969135863 | 0.138533673 | 0.593860602 | 0.879588552 | 1.256056303 |
| 0 | 0 | 0 | 0.018954155 | 0.005426574 |
| 0 | 0 | 0 | 0 | 0 |
| 0 | 0.001574117 | 0 | 0 | 0 |
| 0 | 0 | 0 | 0 | 0 |
| 0 | 0 | 0 | 0 | 0 |
| 0 | 0 | 0 | 0 | 0 |
| 0 | 0 | 0 | 0 | 0.000670332 |
| 0.072411719 | 0.000833171 | 0.099255731 | 0.031469751 | 0.253490185 |
| 0 | 0 | 0 | 0 | 0 |
| 0 | 0.062848837 | 0.400517185 | 0.087140396 | 0.007489082 |
| 0 | 0 | 0 | 0 | 0 |
| 0 | 0 | 0 | 0 | 0 |
| 0 | 5.820375624 | 0.001004237 | 0 | 0 |
| 0 | 0 | 0 | 0 | 0 |
| 0 | 0 | 0 | 0 | 0 |
| 0 | 0 | 0 | 0 | 0 |
| 0.02945738 | 0.080855703 | 0.149395705 | 0.003391549 | 0.022820767 |
| 0 | 0.017475973 | 0.019259597 | 0.058114101 | 0.057022075 |
| 0 | 0 | 0 | 0 | 0 |
| 0 | 0 | 0 | 0 | 0 |
| 0 | 0 | 0 | 0 | 0 |
| 0 | 0 | 0 | 0 | 0 |
| 0 | 0 | 0 | 0 | 0 |
| 0 | 0 | 0 | 0 | 0 |
| 0 | 0.00232511 | 0 | 0 | 0 |
| 0 | 0 | 0 | 0 | 0 |
| 0 | 0 | 3.140077224 | 0 | 0 |
| 0 | 0 | 0 | 0 | 0 |
| 0 | 0 | 0 | 0 | 0 |
| 0 | 0 | 0 | 0 | 0 |
| 0 | 0 | 0 | 0 | 0 |
| 0 | 0 | 0 | 0 | 0 |
| 0 | 0 | 0 | 0 | 0 |
| 0 | 0.124605753 | 0 | 0 | 0 |
| 0 | 0 | 0 | 0 | 0 |
| 0 | 0 | 0 | 0 | 0 |
| 0 | 0 | 0 | 0 | 0 |
| 0 | 0 | 0 | 0 | 0 |
| 0 | 0 | 0 | 0 | 0 |
| 0 | 0 | 0 | 0 | 0 |
| 0 | 0 | 0 | 0.017932039 | 0 |
| 0 | 0.014113722 | 0 | 0 | 0 |
| 0 | 0 | 0 | 0 | 0 |
| 0 | 0 | 0.00055071 | 0 | 0 |

| | | | | |
|---|---|---|---|---|
| 0 | 0 | 0 | 0 | 0 |
| 0 | 0 | 1.421541939 | 0 | 0 |
| 0 | 0 | 0 | 0 | 0 |
| 0 | 0 | 0 | 0 | 0 |
| 0 | 0 | 0 | 0 | 0 |
| 0 | 0 | 0 | 0 | 0 |
| 0 | 0 | 0 | 0 | 0 |
| 0 | 0 | 0 | 0 | 0 |
| 0 | 0 | 0 | 0 | 0.001002115 |
| 0 | 0 | 0 | 0 | 0 |
| 0 | 0 | 0 | 0 | 0 |
| 0 | 0.009099578 | 0 | 0 | 0.000775624 |
| 0 | 0 | 0 | 0 | 0 |
| 0 | 0 | 0 | 0 | 0 |
| 0 | 0 | 0 | 0 | 0 |
| 0 | 0 | 0 | 0.004308656 | 0 |
| 0 | 0 | 0 | 0 | 0 |
| 0 | 0 | 0 | 0 | 0 |
| 0.101008345 | 0 | 0 | 0.060104854 | 0 |
| 0 | 0 | 0 | 0 | 0 |
| 0 | 0 | 0 | 0 | 0 |
| 0 | 0 | 0 | 0 | 0 |
| 0 | 0 | 0 | 0 | 0 |
| 0 | 0 | 0 | 0 | 0 |
| 0 | 0 | 0 | 0 | 0 |
| 0 | 0 | 0 | 0 | 0 |
| 0 | 0 | 0 | 0 | 0 |
| 0 | 0 | 0 | 0.001035853 | 0 |
| 0 | 0 | 0 | 0 | 0 |
| 0 | 0 | 0 | 0 | 0 |
| 0 | 0 | 0 | 0.019795038 | 0.01576457 |
| 0 | 0 | 0 | 0.001643409 | 0 |
| 0 | 0 | 0 | 0 | 0 |
| 0 | 0 | 0 | 0 | 0 |
| 0 | 0 | 0 | 0 | 0 |
| 0 | 0 | 0 | 0 | 0 |
| 0 | 0 | 0 | 0 | 0 |
| 0 | 0 | 0 | 0 | 0 |
| 0.000833101 | 0 | 0 | 0 | 0 |
| 0 | 0 | 0 | 0 | 0 |
| 0 | 0 | 0 | 0 | 0 |
| 0 | 0 | 0 | 0 | 0 |
| 0 | 0.001364698 | 0.001182375 | 0 | 0 |
| 0 | 0 | 0 | 0 | 0 |
| 0 | 0 | 0 | 0 | 0 |
| 0 | 0 | 0 | 0 | 0 |
| 0 | 0 | 0 | 0 | 0 |
| 0 | 0 | 0 | 0 | 0 |

| | | | | |
|---|---|---|---|---|
| 0 | 0 | 0 | 0 | 0 |
| 0 | 0 | 0 | 0 | 0 |
| 0 | 0 | 0 | 0 | 0 |
| 0 | 0 | 0 | 0 | 0 |
| 0.022777734 | 0 | 0 | 0.009622704 | 0 |
| 0 | 0 | 0 | 0 | 0 |
| 0 | 0 | 0 | 0 | 0 |
| 0 | 0 | 0 | 0 | 0 |
| 0 | 0 | 0 | 0 | 0 |
| 0 | 0 | 0 | 0 | 0 |
| 0 | 0 | 0 | 0 | 0 |
| 0 | 0 | 0 | 0 | 0 |
| 0 | 0 | 0 | 0 | 0 |
| 0 | 0 | 0 | 0 | 0 |
| 0 | 0 | 0 | 0 | 0 |
| 0 | 0 | 0 | 0.19625977 | 0 |
| 0 | 0 | 0 | 0 | 0 |
| 0 | 0 | 0 | 0 | 0 |
| 0 | 0 | 0 | 0 | 0 |
| 0 | 0 | 0 | 0 | 0 |
| 0 | 0 | 0 | 0.000815297 | 0 |
| 0 | 0 | 0 | 0 | 0 |
| 0 | 0 | 0 | 0 | 0 |
| 0 | 0 | 0 | 0 | 0 |
| 0 | 0.040149062 | 0 | 0 | 0 |
| 0 | 0 | 0 | 0 | 0 |
| 0.00129751 | 0 | 0 | 0 | 0 |
| 0 | 0 | 0 | 0 | 0 |
| 0 | 0 | 0 | 0 | 0 |
| 0 | 0 | 0 | 0 | 0 |
| 0 | 0 | 0 | 0.000587467 | 0 |
| 0 | 0 | 0.002950677 | 0 | 0 |
| 0 | 0 | 0 | 0 | 0 |
| 0 | 0 | 0 | 0 | 0 |
| 0 | 0 | 0 | 0 | 0 |
| 0 | 0 | 0 | 0 | 0 |
| 0 | 0 | 0 | 0 | 0 |
| 0.001053003 | 0 | 0 | 0 | 0 |
| 0 | 0 | 0 | 0 | 0 |
| 0 | 0 | 0 | 0 | 0 |
| 0 | 0 | 0 | 0 | 0 |
| 0 | 0 | 0 | 0 | 0 |
| 0 | 0 | 0 | 0 | 0 |
| 0 | 0 | 0 | 0 | 0 |
| 0 | 0 | 0 | 0 | 0 |
| 0 | 0 | 0 | 0 | 0 |
| 0 | 0 | 0 | 0 | 0 |
| 0 | 0.012062335 | 0.003376417 | 0 | 0 |

| | | | | |
|---|---|---|---|---|
| 0.017299087 | 0 | 0 | 0 | 0 |
| 0 | 0 | 0 | 0 | 0 |
| 0 | 0 | 0 | 0 | 0 |
| 0 | 0 | 0 | 0 | 0 |
| 0 | 0 | 0 | 0 | 0 |
| 0 | 0 | 0 | 0 | 0 |
| 0 | 0 | 0 | 0 | 0 |
| 0 | 0 | 0 | 0 | 0 |
| 0 | 0 | 0 | 0 | 0 |
| 0 | 0 | 0 | 0 | 0 |
| 0 | 0 | 0 | 0 | 0 |
| 0 | 0 | 0 | 0 | 0 |
| 0 | 0.000434519 | 0 | 0 | 0.000716054 |
| 0 | 0 | 0 | 0 | 0 |
| 0 | 0 | 0 | 0 | 0 |
| 0 | 0 | 0 | 0 | 0 |
| 0 | 0 | 0 | 0 | 0 |
| 0 | 0 | 0 | 0 | 0 |
| 0 | 0 | 0 | 0 | 0 |
| 0 | 0 | 0 | 0 | 0 |
| 0.014122108 | 0.053350664 | 0.080597035 | 0.046570174 | 0.04920267 |
| 0 | 0 | 0 | 0 | 0 |
| 0 | 0 | 0 | 0 | 0 |
| 0 | 0 | 0 | 0 | 0 |
| 0 | 0 | 0 | 0 | 0 |
| 0 | 0 | 0 | 0 | 0 |
| 0.00861602 | 0.002838899 | 0.001639749 | 0.001257282 | 0 |
| 0 | 0 | 0 | 0 | 0 |
| 0 | 0 | 0 | 0 | 0 |
| 0 | 0.000506216 | 0.013815447 | 0 | 0.003753917 |
| 0 | 1.071431568 | 0.110332211 | 0 | 0.001554484 |
| 0 | 0 | 0 | 0 | 0 |
| 0 | 0 | 0 | 0 | 0 |
| 0 | 0 | 0 | 0 | 0 |
| 0.001458507 | 0.013920356 | 0.126407342 | 0.001806022 | 0.003266722 |
| 0 | 0 | 0 | 0 | 0 |
| 0 | 0 | 0 | 0 | 0 |
| 0 | 0 | 0 | 0 | 0 |
| 0 | 0.003586624 | 0 | 0 | 0 |
| 0 | 0 | 0 | 0 | 0 |
| 0 | 0 | 0 | 0 | 0 |
| 0 | 0 | 0 | 0 | 0 |
| 0 | 0 | 0 | 0 | 0 |
| 0 | 0 | 0 | 0 | 0 |
| 0 | 0 | 0 | 0 | 0 |
| 0 | 0 | 0 | 0.546428341 | 0.015551378 |
| 0 | 0 | 0.002479448 | 0 | 0 |
| 0 | 0 | 0.001142335 | 0 | 0 |

| | | | | |
|---|---|---|---|---|
| 0 | 0 | 0 | 0 | 0 |
| 0 | 0 | 0 | 0 | 0 |
| 0 | 0 | 0 | 0 | 0 |
| 0 | 0 | 0.001659494 | 0 | 0.001578205 |
| 0 | 0 | 0 | 0 | 0 |
| 0 | 0 | 0 | 0 | 0 |
| 0 | 0 | 0 | 0 | 0 |
| 0 | 0 | 0 | 0.001644547 | 0 |
| 0 | 0 | 0 | 0 | 0 |
| 0 | 0 | 0 | 0 | 0 |
| 0 | 0 | 0 | 0 | 0 |
| 0 | 0 | 0 | 0 | 0 |
| 0 | 0 | 0 | 0 | 0 |
| 0 | 0 | 0 | 0 | 0 |
| 0 | 0 | 0 | 0 | 0.001957553 |
| 0 | 0 | 0 | 0 | 0 |
| 0 | 0 | 0 | 0 | 0 |
| 0.002817341 | 0 | 0 | 0 | 0 |
| 0.002325475 | 0.000638519 | 0 | 0 | 0 |
| 0.027981322 | 0 | 0 | 0 | 0 |
| 0.005613239 | 0.084957593 | 0.027663943 | 0.002788857 | 0.597886032 |
| 0 | 0 | 0 | 0 | 0 |
| 0 | 0 | 0 | 0 | 0 |
| 0 | 0 | 0 | 0 | 0 |
| 0.00805414 | 0.004116737 | 0.001945496 | 0 | 0 |
| 0 | 0 | 0 | 0 | 0 |
| 0 | 0 | 0 | 0 | 0 |
| 0 | 0 | 0 | 0 | 0 |
| 0 | 0 | 0 | 0 | 0 |
| 0 | 0 | 0 | 0 | 0 |
| 0.001605105 | 0.248214979 | 2.10181289 | 0.10742229 | 0.818990539 |
| 0 | 0 | 0 | 0 | 0 |
| 0.011404697 | 0 | 0 | 0.047073548 | 0.002980123 |
| 0 | 0 | 0 | 0 | 0 |
| 0 | 0 | 0 | 0 | 0 |
| 0 | 0 | 0 | 0 | 0 |
| 0 | 0 | 0 | 0 | 0 |
| 0 | 0 | 0 | 0 | 0 |
| 0 | 0 | 0 | 0 | 0 |
| 0 | 0 | 0 | 0 | 0 |
| 0 | 0 | 0 | 0 | 0 |
| 0.000467468 | 0.005506441 | 0 | 0.027495384 | 0 |
| 0 | 0 | 0 | 0 | 0 |
| 0 | 0 | 0 | 0 | 0 |
| 0 | 0 | 0 | 0 | 0 |
| 0 | 0 | 0 | 0 | 0 |
| 0 | 0 | 0 | 0 | 0 |
| 0 | 0 | 0 | 0 | 0 |

| | | | | |
|---|---|---|---|---|
| 0 | 0 | 0 | 0 | 0 |
| 0 | 0 | 0 | 0 | 0 |
| 0.007777522 | 0.00106373 | 0.012441815 | 0.000807602 | 0 |
| 0 | 0 | 0 | 0 | 0 |
| 0 | 0 | 0 | 0.060098677 | 0 |
| 0 | 0 | 0 | 0 | 0 |
| 0 | 0 | 0 | 0 | 0 |
| 0 | 0 | 0 | 0 | 0 |
| 0 | 0 | 0 | 0 | 0 |
| 0 | 0 | 0 | 0 | 0 |
| 0 | 0 | 0 | 0 | 0 |
| 0 | 0 | 0 | 0 | 0 |
| 0 | 0 | 0 | 0 | 0 |
| 0 | 0.003330904 | 0.002037104 | 0 | 0 |
| 0.000401576 | 0.001778911 | 0.003572896 | 0.001657529 | 0.001199252 |
| 0 | 0 | 0 | 0 | 0 |
| 0 | 0.003869554 | 0.001915762 | 0 | 0 |
| 0 | 0 | 0 | 0 | 0 |
| 0 | 0 | 0 | 0 | 0 |
| 0 | 0 | 0 | 0 | 0 |
| 0 | 0 | 0 | 0 | 0 |
| 0 | 0.000499015 | 0.001080867 | 0 | 0 |
| 0 | 0 | 0 | 0 | 0 |
| 0 | 0 | 0.004021415 | 0 | 0 |
| 0 | 0 | 0 | 0 | 0 |
| 0 | 0 | 0 | 0 | 0 |
| 0 | 0 | 0 | 0 | 0 |
| 0 | 0 | 0 | 0 | 0 |
| 0 | 0 | 0 | 0 | 0 |
| 0 | 0 | 0 | 0 | 0 |
| 0 | 0 | 0 | 0 | 0 |
| 0 | 0 | 0 | 0 | 0 |
| 0 | 0 | 0 | 0 | 0 |
| 0 | 0 | 0 | 0 | 0 |
| 0 | 0 | 0 | 0 | 0 |
| 0 | 0.013289478 | 0 | 0 | 0 |
| 0 | 0 | 0 | 0 | 0 |
| 0 | 0 | 0.003530792 | 0 | 0.000774886 |
| 0 | 0 | 0 | 0 | 0 |
| 0 | 0 | 0 | 0 | 0 |
| 0 | 0 | 0 | 0 | 0 |
| 0 | 0 | 0 | 0 | 0 |
| 0 | 0 | 0 | 0 | 0 |
| 0 | 0 | 0.002361582 | 0 | 0 |
| 0 | 0 | 0 | 0 | 0 |
| 0 | 0 | 0 | 0 | 0 |
| 0 | 0 | 0 | 0 | 0 |
| 0 | 0 | 0.019738498 | 0 | 0.008447231 |

| | | | | |
|---|---|---|---|---|
| 0 | 0 | 0 | 1.67221818 | 0.044138118 |
| 0 | 0.00225177 | 0 | 0 | 0 |
| 0 | 0 | 0 | 0 | 0 |
| 0 | 0 | 0 | 0 | 0 |
| 0.003046899 | 0.003330481 | 0.008656592 | 0.003193969 | 0.00173317 |
| 0 | 0 | 0 | 0 | 0 |
| 0 | 0 | 0 | 0 | 0 |
| 0.044539383 | 0.026781888 | 0.008302292 | 0 | 0 |
| 0 | 0 | 0 | 0 | 0 |
| 0.067289378 | 0.040014112 | 0.879103413 | 0.152569977 | 0.780639057 |
| 0 | 0 | 0 | 0 | 0 |
| 0 | 0.113588741 | 0 | 0 | 0.001682745 |
| 0.000854834 | 0.138536414 | 0.001917381 | 0 | 0 |
| 0.002040142 | 0.211969566 | 0.000873602 | 0.000765528 | 0 |
| 0 | 0 | 0 | 0 | 0 |
| 0 | 0 | 0 | 0 | 0 |
| 0 | 0 | 0 | 0 | 0 |
| 0 | 0 | 0 | 0.007221108 | 0 |
| 0 | 0.05600221 | 1.846766962 | 0 | 0 |
| 0 | 0 | 0 | 0 | 0 |
| 0 | 0 | 0 | 0 | 0 |
| 0 | 0 | 0 | 0 | 0.002849211 |
| 0 | 0 | 0 | 0 | 0 |
| 0.000304432 | 0 | 0 | 0 | 0.010303621 |
| 0 | 0 | 0 | 0 | 0 |
| 0.000568951 | 0 | 0 | 0 | 0 |
| 0 | 0 | 0 | 0 | 0 |
| 0 | 0.325422159 | 0.018094232 | 0 | 0 |
| 0 | 0 | 0 | 0 | 0.212016833 |
| 0.000878068 | 0.003005664 | 0.00160842 | 0.001208092 | 0.004151861 |
| 0.054335834 | 0 | 0 | 0 | 0 |
| 0 | 0 | 0 | 0 | 0 |
| 0 | 0 | 0 | 0 | 0 |
| 0 | 0 | 0 | 0 | 0.004166438 |
| 0 | 0 | 0 | 0 | 0 |
| 0 | 0 | 0 | 0 | 0 |
| 0 | 0 | 0 | 0 | 0.003129904 |
| 0 | 0 | 0 | 0 | 0 |
| 0 | 0 | 0 | 0 | 0 |
| 0 | 0 | 0 | 0 | 0 |
| 0 | 0 | 0 | 0 | 0 |
| 0 | 0 | 0 | 0 | 0 |
| 0 | 0 | 0.001229388 | 0 | 0 |
| 0 | 0 | 0.057034407 | 0 | 0.000513318 |
| 0 | 0 | 0 | 0 | 0 |
| 0 | 0 | 0 | 0 | 0 |
| 0 | 0.000641097 | 0.001388617 | 0.00121683 | 0 |
| 0 | 0 | 0 | 0 | 0 |

| | | | | |
|---|---|---|---|---|
| 0 | 0 | 0 | 0 | 0 |
| 0 | 0 | 0 | 0 | 0 |
| 0 | 0 | 0 | 0 | 0 |
| 0 | 0 | 0 | 0 | 0 |
| 0 | 0 | 0 | 0.001924335 | 0.001827381 |
| 0 | 0.019187784 | 0.139119198 | 0 | 0.003744469 |
| 0 | 0 | 0 | 0 | 0.000925254 |
| 0 | 0 | 0 | 0 | 0.003228671 |
| 0 | 0 | 0 | 0 | 0.563374306 |
| 0 | 0 | 0 | 0 | 0 |
| 0 | 0.015617262 | 0.007611078 | 0.018896926 | 0.000723826 |
| 0 | 0 | 0 | 0 | 0.004522596 |
| 0 | 0.227329987 | 0 | 0 | 0 |
| 0.0032058 | 0.003504171 | 0.001438113 | 0.004620742 | 0.000683834 |
| 0.012511388 | 0 | 0.078090989 | 0 | 0 |
| 0 | 0 | 0 | 0 | 0 |
| 0 | 0 | 0 | 0 | 0 |
| 0 | 0 | 0 | 0 | 0.013309618 |
| 0 | 0 | 0 | 0.005141618 | 0 |
| 0 | 0 | 0 | 0.027833508 | 0.012028148 |
| 0 | 0 | 0 | 0 | 0 |
| 0.377121019 | 0 | 0.592926975 | 0.221555668 | 0 |
| 0 | 0 | 0 | 0 | 0.135192864 |
| 0 | 0 | 0 | 0 | 0 |
| 0 | 0 | 0 | 0 | 0 |
| 0 | 0 | 0 | 0 | 0 |
| 0 | 0 | 0.027287661 | 0 | 0.012110466 |
| 0 | 0 | 0 | 0 | 0 |
| 0 | 0 | 0 | 0 | 0 |
| 0.634816682 | 13.92310607 | 0.223796361 | 0.343577815 | 0.423663053 |
| 0 | 0 | 0 | 0 | 0 |
| 0 | 0 | 0 | 0 | 0 |
| 0 | 0.005202233 | 0 | 0 | 0 |
| 0 | 0 | 0 | 0 | 0.00065708 |
| 0 | 0 | 0.068833526 | 0 | 0 |
| 0 | 0 | 0 | 0 | 0 |
| 3.944616051 | 0.297616533 | 0 | 0 | 0 |
| 0 | 0 | 0 | 0 | 0 |
| 0.020200157 | 0 | 0 | 0 | 0 |
| 0 | 0 | 0.52269181 | 0 | 0 |
| 0.000878505 | 0 | 0.051429403 | 0.0428223 | 0.137735555 |
| 0.124126351 | 0 | 0 | 1.008638983 | 0.003706861 |
| 0 | 0 | 0 | 0 | 0 |
| 0 | 0.00384125 | 2.958171026 | 0 | 0 |
| 0 | 0.000855185 | 0 | 0 | 0 |
| 0 | 0 | 0 | 0 | 0 |
| 0 | 0.001432239 | 0 | 0 | 0.016914895 |
| 0 | 0 | 0 | 0 | 0 |

| | | | | |
|---|---|---|---|---|
| 0.001300183 | 0 | 0 | 0 | 0 |
| 0 | 0 | 0 | 0 | 0 |
| 0 | 13.23578257 | 0.002781571 | 0 | 0 |
| 0 | 0 | 0 | 0 | 0 |
| 0 | 0 | 0 | 0 | 0 |
| 0 | 0 | 0.003667771 | 0.123834569 | 0 |
| 0 | 0 | 0 | 0 | 0 |
| 0 | 0.003301015 | 0 | 0 | 0 |
| 0 | 0.033522648 | 0 | 0 | 0 |
| 0 | 0 | 0 | 0 | 0 |
| 0 | 0 | 0 | 0 | 0 |
| 0 | 0 | 0 | 0 | 0 |
| 0 | 0 | 0 | 0 | 0 |
| 0 | 0.006217127 | 0.001154255 | 0.027309439 | 0.002195429 |
| 0 | 0 | 0 | 0 | 0 |
| 0.006204692 | 0 | 0 | 0.012963201 | 0.015098072 |
| 0.006622541 | 0.000923182 | 0.005798877 | 0 | 0 |
| 0 | 0.003177951 | 0.004615958 | 0 | 0.057299085 |
| 0 | 0.044500259 | 0 | 0 | 0 |
| 0 | 0 | 0 | 0 | 0 |
| 0 | 0 | 0.602551401 | 0.007949949 | 0.140203269 |
| 0 | 0.00374795 | 0.001623613 | 0 | 0 |
| 0.091303558 | 0 | 0.266305118 | 0 | 0 |
| 0.000421757 | 0.013417842 | 0 | 0 | 0 |
| 0.000595135 | 0 | 0 | 0.122495202 | 0 |
| 0.019796839 | 0.002432258 | 0 | 0.011772154 | 0.005260724 |
| 0.001131761 | 0 | 0.005330902 | 0.002446929 | 0.015933569 |
| 0 | 0 | 0 | 0.003660143 | 0 |
| 0 | 0 | 0 | 0 | 0 |
| 0 | 0 | 0.001954902 | 0 | 0 |
| 0.001475488 | 0 | 0 | 0.006936017 | 0.061137963 |
| 0 | 0.006422571 | 0 | 0 | 0 |
| 0.011868718 | 0.070429724 | 0.016990697 | 0.004562684 | 0.005994253 |
| 0 | 0.005137291 | 0 | 0 | 0 |
| 0 | 0 | 0 | 0 | 0 |
| 0 | 0 | 0 | 0 | 0 |
| 0.016095209 | 0 | 0.001486525 | 0 | 0 |
| 0.014931874 | 0.066195205 | 0.005231393 | 0.022751271 | 0.006449252 |
| 0.174362154 | 0 | 0 | 0.003405473 | 0 |
| 0 | 0 | 0 | 0.000832902 | 0.003766373 |
| 0 | 0 | 0 | 0 | 0 |
| 0 | 0 | 0 | 0 | 0 |
| 0 | 0 | 0 | 0 | 0.003031237 |
| 0 | 0.005004417 | 0.15775362 | 0.0075547 | 0.462017379 |
| 0 | 0.029607187 | 2.514654225 | 0.39708757 | 0 |
| 0 | 0 | 0 | 0 | 0 |
| 0 | 0 | 0 | 0 | 0 |
| 0 | 0 | 0 | 0 | 0 |

| | | | | |
|---|---|---|---|---|
| 0 | 0 | 0.029302027 | 0 | 0 |
| 0.002690456 | 0.134892742 | 0.187979496 | 0.015500747 | 0.00426842 |
| 0 | 0 | 0 | 0 | 0 |
| 0 | 0 | 0 | 0.024660597 | 0 |
| 0.000989799 | 0 | 0 | 0 | 0 |
| 0 | 0 | 0 | 0 | 0 |
| 0 | 1.008495715 | 0.089639502 | 0 | 0.00185828 |
| 0 | 0 | 0 | 0 | 0 |
| 0 | 0 | 0 | 0.001694288 | 0 |
| 0 | 0.207267442 | 0.001534843 | 0 | 0 |
| 0 | 0.002264459 | 0.001569543 | 0.001719216 | 0.000559748 |
| 0.035736221 | 0.006521087 | 0.00905928 | 0.000768246 | 0 |
| 0.003545795 | 0 | 0 | 0.093016573 | 0 |
| 0 | 0 | 0 | 0 | 0 |
| 0 | 0 | 0 | 0 | 0 |
| 0 | 0 | 0 | 0 | 0.001523231 |
| 0.004187117 | 0.009642943 | 0.033487086 | 0 | 0.002149167 |
| 0.000679432 | 0.240712529 | 6.18139683 | 0.002570696 | 0.061124519 |
| 0 | 0 | 0 | 0 | 0 |
| 0 | 0 | 0.07896831 | 0.002124532 | 0.009716901 |
| 0.040750883 | 0.305176114 | 0.005932158 | 0 | 0 |
| 0 | 0 | 0 | 0 | 0 |
| 0 | 0 | 0 | 0 | 0 |
| 0 | 0 | 0 | 0 | 0 |
| 0 | 0 | 0 | 0 | 0 |
| 0 | 0 | 0 | 0 | 0 |
| 0.020244768 | 0.018826505 | 0 | 0.014843153 | 0.115944492 |
| 0.028970394 | 0.597773607 | 0.611135438 | 0.220605714 | 0.243487033 |
| 0.000978737 | 0 | 0 | 0 | 0 |
| 0 | 0 | 0.002172948 | 0 | 0 |
| 0 | 0.003346495 | 0 | 0 | 0 |
| 0 | 0 | 0 | 0 | 0 |
| 0 | 0 | 0 | 0 | 0 |
| 0 | 0 | 0 | 0 | 0 |
| 0 | 0 | 0 | 0 | 0 |
| 0.179374556 | 0.8207935 | 4.177256768 | 0.118888082 | 0.006523809 |
| 0 | 0 | 0 | 0.001392012 | 0 |
| 0 | 0 | 0 | 0 | 0 |
| 0 | 0.050443845 | 0 | 0.001460511 | 0.004490998 |
| 0 | 0 | 0 | 0 | 0 |
| 0 | 0.005633299 | 2.39746622 | 0.08675992 | 0 |
| 0 | 0 | 0 | 0 | 0 |
| 0.001289581 | 0.003635297 | 0.000506189 | 0.015820601 | 0 |
| 0 | 0 | 0.081615642 | 0 | 0 |
| 0 | 0 | 0.205887264 | 0 | 0 |
| 0.466001921 | 0 | 0 | 0 | 0 |
| 0 | 0 | 0 | 0 | 0 |
| 0.01794526 | 0 | 0.005266481 | 0 | 0.001252127 |

| | | | | |
|---|---|---|---|---|
| 0 | 0 | 0.005657261 | 0 | 0 |
| 0.000425002 | 0 | 0 | 0 | 0 |
| 0 | 0 | 0 | 0 | 0 |
| 0.003754884 | 0 | 0 | 0.002755289 | 0 |
| 0 | 0 | 0 | 0 | 0 |
| 0 | 0 | 0 | 0 | 0 |
| 0 | 0 | 0 | 0 | 0.000869353 |
| 0.008285894 | 0 | 0 | 0.004775746 | 0 |
| 0.50591335 | 0.007184947 | 0 | 0.033101543 | 0.070637724 |
| 0.000671188 | 0 | 0 | 0.001539094 | 0 |
| 0 | 0 | 0 | 0 | 0 |
| 0.00395734 | 0 | 0 | 0.010229468 | 0.001432491 |
| 0 | 0 | 0 | 0 | 0 |
| 0 | 0 | 0 | 2.323755418 | 0 |
| 0.016229079 | 0.004184395 | 0 | 0.067394916 | 0.014776186 |
| 0 | 0 | 0 | 0 | 0 |
| 0.0014199 | 0 | 0 | 0 | 0 |
| 0 | 0 | 0 | 0 | 0 |
| 0 | 0 | 0 | 0 | 0 |
| 0 | 0 | 0 | 0 | 0 |
| 0 | 0 | 0 | 0 | 0 |
| 0 | 0 | 0 | 0 | 0 |
| 0 | 0 | 0 | 0 | 0 |
| 0 | 0.047100964 | 0.047144064 | 0.293248363 | 0.255059922 |
| 0 | 0 | 0.007580135 | 0.002317112 | 0 |
| 0.048135918 | 0 | 0 | 0 | 0 |
| 0 | 0 | 0 | 0.00955762 | 0 |
| 4.336232659 | 0 | 0 | 0 | 5.550371082 |
| 0 | 0 | 0 | 0 | 0 |
| 0 | 0 | 0 | 0 | 0 |
| 0 | 0 | 0 | 0 | 0 |
| 0.000988578 | 0 | 0 | 0 | 0 |
| 0.000976003 | 0 | 0 | 0.023194445 | 0.004107074 |
| 0 | 0 | 0 | 0 | 0 |
| 0 | 0 | 0 | 0.061311389 | 0 |
| 0.002570063 | 0 | 0 | 0 | 0.016973862 |
| 0.001914952 | 0 | 0 | 0.027752092 | 0 |
| 0 | 0 | 0 | 0 | 0 |
| 0 | 0 | 0 | 0.00085878 | 0 |
| 0 | 0.000414801 | 0 | 0 | 0.005895697 |
| 0 | 0.00055594 | 0.046781902 | 0.013137222 | 0.063214059 |
| 0.001998303 | 0 | 0 | 0.425190079 | 0 |
| 38.56676442 | 0.110875213 | 0 | 15.46483276 | 27.08351716 |
| 0 | 0 | 0 | 0 | 0.00299281 |
| 0 | 0 | 0 | 0 | 0 |
| 0 | 0 | 0 | 0 | 0 |
| 0 | 0 | 0 | 0 | 0 |
| 0 | 0 | 0 | 0 | 0 |

| | | | | |
|---|---|---|---|---|
| 0 | 0 | 0 | 0 | 0 |
| 0.004661579 | 0 | 0 | 0 | 0 |
| 0.005435338 | 0.603807717 | 0.095330697 | 0.00127677 | 0.094422049 |
| 0 | 0 | 0 | 0 | 0 |
| 0 | 0 | 0 | 0 | 0 |
| 0.005639378 | 0 | 0.000983815 | 0.00201158 | 0 |
| 0 | 0 | 0 | 0 | 0 |
| 0 | 0 | 0 | 0 | 0 |
| 0 | 0 | 0 | 0 | 0 |
| 0 | 0 | 0 | 0 | 0 |
| 0 | 0.001100749 | 0.008378846 | 0.021489629 | 0.006996657 |
| 0.763983721 | 5.008440176 | 2.570721553 | 4.453315442 | 3.384097091 |
| 0 | 0 | 0 | 0 | 0 |
| 0.03427551 | 0.001866815 | 0.003087783 | 0.000773083 | 0 |
| 0.005424223 | 0.033300856 | 0.082036673 | 0.073563181 | 0.172565624 |
| 0 | 0 | 0 | 0 | 0 |
| 0 | 0 | 0 | 0.003340453 | 0 |
| 0 | 0 | 0 | 0 | 0 |
| 0 | 0 | 0 | 0 | 0 |
| 0 | 0 | 0 | 0.013128834 | 0.002561513 |
| 0 | 0.12520864 | 0.08735562 | 0.012908223 | 0.191239025 |
| 0.609924513 | 1.120303025 | 10.38420137 | 2.532997875 | 10.98210986 |
| 0.604849448 | 1.420147029 | 8.290748733 | 21.88090267 | 17.49063572 |
| 0 | 0 | 0 | 0 | 0 |
| 0 | 0 | 0 | 0 | 0 |
| 1.089469614 | 5.626248599 | 0 | 0 | 0 |
| 0 | 0.102779472 | 0 | 0.433903096 | 0.27821768 |
| 0 | 0 | 0 | 0.00156547 | 0.012062672 |
| 0 | 0 | 0 | 0 | 0 |
| 0 | 0 | 0 | 0.073624985 | 0 |
| 0 | 0 | 0 | 0.011754162 | 0 |
| 0.153664011 | 0 | 0 | 0.706383305 | 0.175161803 |
| 0 | 0 | 0.282505472 | 0 | 0.034531517 |
| 0 | 0.001108404 | 0 | 0.024965087 | 0.003196486 |
| 0 | 0.01748106 | 0.021056079 | 0.004491413 | 0.002766565 |
| 0.768296687 | 0.042470692 | 0.017890776 | 0.003168854 | 0.043079041 |
| 0 | 0.002970856 | 0 | 0.00306107 | 0.001223934 |
| 0 | 0 | 0 | 0 | 0 |
| 0.106389509 | 0.003598914 | 0 | 0.023029869 | 0 |
| 0 | 0 | 0 | 0 | 0 |
| 0 | 0 | 0 | 0.005286952 | 0 |
| 0 | 0 | 0 | 0 | 0 |
| 0 | 0 | 0 | 0 | 0 |
| 0 | 0 | 0 | 0 | 0 |
| 0.025021156 | 0 | 0 | 0.003613718 | 0 |
| 0 | 0 | 0 | 0 | 0 |
| 0 | 0 | 0 | 0 | 0 |
| 0.489420998 | 0.085016173 | 0 | 0.00802998 | 0.006473813 |

| | | | | |
|---|---|---|---|---|
| 0 | 0 | 0 | 0 | 0 |
| 0 | 0 | 0 | 0 | 0 |
| 0 | 0 | 0 | 0 | 0 |
| 0.002945501 | 0 | 0 | 0.000648412 | 0 |
| 0 | 0 | 0.064746911 | 0 | 0 |
| 0 | 0 | 0 | 0 | 0 |
| 0 | 0 | 0 | 0.107193057 | 0 |
| 0 | 0.003143535 | 0 | 0.007010714 | 0.004370877 |
| 0 | 0 | 0 | 0 | 0 |
| 0 | 0 | 0 | 0 | 0 |
| 0.001180983 | 0.003487695 | 0.004944666 | 0 | 0.000587807 |
| 0 | 0 | 0 | 0 | 0 |
| 0 | 0.003053749 | 0 | 0 | 0.001544013 |
| 0 | 0 | 0 | 0.29795172 | 0 |
| 0 | 0 | 0 | 0 | 0 |
| 0 | 0 | 0 | 0 | 0 |
| 0 | 0 | 0 | 0.097751556 | 0 |
| 0.01004989 | 0 | 0 | 0.00225443 | 0.001304896 |
| 0 | 0 | 0.104857484 | 0 | 0 |
| 0 | 0 | 0.001082737 | 0 | 0 |
| 0 | 0 | 0 | 0.000534102 | 0 |
| 0 | 0 | 0 | 0 | 0 |
| 0 | 0 | 0 | 0.009530385 | 0 |
| 0 | 0 | 0 | 0 | 0 |
| 0 | 0 | 0 | 0 | 0 |
| 0 | 0 | 0 | 0 | 0 |
| 0 | 0 | 0 | 0 | 0 |
| 0 | 0 | 0 | 0 | 0 |
| 0 | 0 | 0 | 0 | 0 |
| 0.164024358 | 0 | 0 | 0.836244118 | 0 |
| 0 | 0 | 0 | 0.004550248 | 0 |
| 0 | 0 | 0 | 0 | 0 |
| 0 | 0 | 0 | 0 | 0 |
| 0 | 0 | 0 | 0 | 0 |
| 0 | 0 | 0 | 0 | 0 |
| 0 | 0 | 0 | 0 | 0 |
| 0.695746312 | 0 | 0 | 0.060635822 | 0 |
| 0 | 0.003220794 | 0.045868807 | 0.000611321 | 0.0044783 |
| 0 | 0 | 0 | 0 | 0 |
| 0 | 0 | 0 | 0 | 0 |
| 0 | 0.00831023 | 0 | 0.107544374 | 0 |
| 0 | 0 | 0 | 0 | 0 |
| 0.001502679 | 0 | 0 | 0 | 0 |
| 0 | 0 | 0 | 0 | 0 |
| 0 | 0 | 0 | 0 | 0.532048317 |
| 0 | 0 | 0 | 0 | 0 |
| 0.00042218 | 0 | 0 | 0 | 0 |
| 0 | 0 | 0 | 0 | 0 |

| | | | | |
|---|---|---|---|---|
| 0 | 0 | 0 | 0 | 0 |
| 0 | 0 | 0 | 0 | 0 |
| 0 | 0 | 0 | 0 | 0 |
| 0.018894193 | 0 | 0 | 0.114015899 | 0 |
| 0 | 0 | 0 | 0 | 0 |
| 0.895330072 | 0 | 0 | 0 | 0 |
| 0 | 0 | 0 | 0 | 0.442505379 |
| 0 | 0 | 0 | 0 | 0 |
| 0.00091919 | 0.243940539 | 2.930439754 | 0.004847901 | 0.341756591 |
| 0 | 0 | 0 | 0 | 0 |
| 0 | 0 | 0 | 0 | 0 |
| 0 | 0 | 0 | 0 | 0 |
| 0 | 0 | 0 | 0.001844461 | 0 |
| 0 | 0 | 0 | 0 | 0 |
| 0.007486279 | 0 | 1.278953466 | 0.386402123 | 0 |
| 0 | 0.30526947 | 0.223391493 | 0 | 0.270982471 |
| 0 | 0 | 0 | 0.001285909 | 0 |
| 0 | 0 | 0 | 0 | 0 |
| 0 | 0 | 0 | 0 | 0 |
| 0 | 0 | 0 | 0 | 0 |
| 0 | 0 | 0 | 0.110240439 | 0 |
| 0 | 0 | 0 | 0 | 0 |
| 0 | 0 | 0 | 0.000795587 | 0 |
| 0 | 0 | 0 | 0.007423735 | 0 |
| 0.025762429 | 0.015018082 | 0.029983424 | 0 | 0 |
| 0.003905523 | 0 | 0.064519544 | 0.01051322 | 0 |
| 0 | 0 | 0 | 0 | 0 |
| 0 | 0 | 0 | 0 | 0.002504775 |
| 0.002047612 | 0 | 0 | 0 | 0.006350097 |
| 0 | 0 | 0 | 0 | 0 |
| 0 | 0 | 0 | 0 | 0 |
| 0 | 0 | 0 | 0.004788875 | 0.001732418 |
| 0 | 0 | 0 | 0 | 0 |
| 0 | 0 | 0 | 0.000794269 | 0.000718334 |
| 0.008085155 | 0.000574585 | 0.001244551 | 0 | 0 |
| 0 | 0 | 0.003518987 | 0 | 0.004015935 |
| 0 | 0.005728351 | 0 | 0.220847544 | 0.335518389 |
| 0.066306307 | 0 | 0 | 0.324171741 | 0 |
| 0 | 0 | 0 | 0 | 0 |
| 0 | 0 | 0 | 0 | 0 |
| 0.175355646 | 0 | 0 | 0 | 0 |
| 0 | 0 | 0 | 0.006993767 | 0 |
| 0.073811744 | 0 | 0 | 0.0459311 | 0 |
| 0.005868473 | 0 | 0 | 0 | 0 |
| 0 | 0 | 0 | 0 | 0 |
| 0 | 0 | 0 | 0.01519113 | 0 |
| 0 | 0 | 0 | 0 | 0 |
| 0 | 0 | 0.04248913 | 0.006050323 | 0.005556077 |

| | | | | |
|---|---|---|---|---|
| 0 | 0 | 0 | 0.040085005 | 0 |
| 0 | 0 | 0 | 0 | 0 |
| 0 | 0 | 0 | 0 | 0 |
| 0 | 0 | 0 | 0 | 0 |
| 0 | 0 | 0 | 0 | 0 |
| 0 | 0 | 0 | 0 | 0 |
| 0 | 0 | 0 | 0 | 0 |
| 0 | 0 | 0 | 0 | 0 |
| 0 | 0 | 0 | 0 | 0 |
| 0 | 0 | 0 | 0 | 0 |
| 0.001441259 | 0 | 0 | 0 | 0 |
| 0.294599917 | 0 | 0 | 0.637077595 | 0 |
| 0 | 0 | 0 | 0.210864544 | 0.008173084 |
| 0 | 0 | 0 | 0 | 0 |
| 0 | 0.414585204 | 0.005700331 | 0.200005283 | 0.021901264 |
| 0 | 0 | 0 | 0 | 0 |
| 0 | 0 | 0 | 0 | 0 |
| 0 | 0.000468551 | 0 | 0 | 0 |
| 0 | 0 | 0 | 0 | 0 |
| 0 | 0 | 0 | 0.000970949 | 0 |
| 0.000316147 | 0 | 0 | 0.020298681 | 0 |
| 0 | 0 | 0 | 0 | 0 |
| 0.606100507 | 0 | 0.934725764 | 0.115455908 | 0 |
| 0 | 0 | 0 | 0 | 0 |
| 0 | 0 | 0 | 0 | 0 |
| 0 | 0 | 0 | 0 | 0 |
| 0 | 0 | 0 | 0 | 0 |
| 0 | 0 | 0 | 0 | 0 |
| 0 | 0 | 0 | 0 | 0 |
| 0 | 0 | 0 | 0 | 0 |
| 0 | 0 | 0 | 0 | 0 |
| 0.014887327 | 0.012294136 | 0.04734065 | 0.002852031 | 0 |
| 0 | 0 | 0 | 0 | 0 |
| 0.004236523 | 0 | 0 | 0.031216632 | 0.0080128 |
| 0 | 0 | 0.003063632 | 0.000503368 | 0.002549367 |
| 0.001100484 | 0.000949665 | 0 | 0 | 0.003286444 |
| 0 | 0 | 0 | 0 | 0 |
| 0 | 0 | 0 | 0 | 0 |
| 0.001401059 | 0 | 0 | 0 | 0.00502088 |
| 0 | 0 | 0 | 0 | 0 |
| 0 | 0 | 0 | 0.198996142 | 0 |
| 0 | 0 | 0 | 0 | 0 |
| 0 | 0 | 0 | 0 | 0 |
| 0 | 0 | 0 | 0.009561709 | 0 |
| 0.006847741 | 0 | 0.023200757 | 0.015041314 | 0.012736296 |
| 0 | 0 | 0 | 0 | 0 |
| 0 | 0 | 0 | 0.314160419 | 2.263195785 |
| 0 | 0 | 0.02408462 | 0.000625336 | 0 |

| | | | | |
|---|---|---|---|---|
| 0 | 0 | 0 | 0 | 0 |
| 0 | 0 | 0 | 0 | 0 |
| 0 | 0 | 0 | 0.007692766 | 0.003339512 |
| 0 | 0 | 0 | 0 | 0 |
| 0 | 0 | 8.428939326 | 0 | 0 |
| 0 | 0 | 0 | 0.092063873 | 0.029208309 |
| 0.000957406 | 0 | 0 | 0.087314872 | 0 |
| 0 | 0 | 0.043111935 | 0.003874719 | 0 |
| 0 | 0 | 0 | 0 | 0 |
| 0 | 0 | 0 | 0.002065305 | 0 |
| 0 | 0 | 0 | 0 | 0 |
| 0 | 0 | 0 | 0 | 0 |
| 0 | 0 | 0 | 0 | 0 |
| 0 | 0.098357305 | 0 | 0 | 0 |
| 0 | 0 | 0 | 0.010611435 | 0 |
| 0 | 0 | 0 | 0 | 0 |
| 0 | 0 | 0 | 16.01187897 | 0 |
| 0 | 0 | 0 | 0 | 0 |
| 0.01301104 | 0.519049848 | 0 | 0 | 0 |
| 0 | 0 | 0 | 0 | 0.00039487 |
| 0 | 0 | 0 | 0 | 0 |
| 0 | 0 | 0 | 0 | 0 |
| 0 | 0 | 0 | 0 | 0 |
| 0 | 0 | 0 | 0 | 0 |
| 0 | 0 | 0 | 0 | 0 |
| 0 | 0.018781657 | 0.04829051 | 0.002000414 | 0.003173003 |
| 0 | 0 | 0 | 0 | 0 |
| 0 | 0 | 0.056548286 | 0.062505607 | 0.012259494 |
| 0.000695166 | 0 | 0 | 0 | 0 |
| 0.028221971 | 0 | 0 | 0.208865043 | 0.034533911 |
| 0 | 0 | 0 | 0 | 0 |
| 0 | 0 | 0 | 0 | 0 |
| 0 | 0 | 0 | 0 | 0 |
| 0 | 0 | 0 | 0 | 0 |
| 0 | 0.002973542 | 0.00055206 | 0 | 0 |
| 0 | 0 | 0 | 0 | 0 |
| 0 | 0 | 0 | 0 | 0 |
| 0 | 0 | 0.001670469 | 0 | 0 |
| 0 | 0 | 0 | 0 | 0 |
| 0 | 0 | 0 | 0 | 0 |
| 2.920628116 | 8.588030434 | 3.639621063 | 4.430773985 | 0.262747129 |
| 0 | 0 | 3.078223806 | 0.223009754 | 0 |
| 0 | 0 | 0 | 0 | 0 |
| 0 | 0 | 0 | 0 | 0 |
| 0 | 0 | 0 | 0 | 0 |
| 0 | 0 | 0 | 0 | 0 |
| 0 | 0 | 0 | 0 | 0 |
| 0.000844395 | 0 | 0 | 0.001452205 | 0 |

| | | | | |
|---|---|---|---|---|
| 0 | 0 | 0 | 0 | 0 |
| 0 | 0 | 0 | 0.001941826 | 0 |
| 0 | 0 | 0 | 0 | 0 |
| 0 | 0 | 0 | 0 | 0 |
| 1.388394921 | 0 | 0 | 0.491251672 | 0 |
| 0 | 0 | 0 | 0 | 0 |
| 0 | 0 | 0 | 0 | 0 |
| 0 | 0 | 0 | 0 | 0 |
| 0.003035375 | 3.904712899 | 7.841121834 | 0.00149151 | 0.130629158 |
| 0 | 0 | 0 | 0.004165064 | 0 |
| 0 | 0.000592745 | 0.007510738 | 0 | 0 |
| 0 | 0 | 0 | 0 | 0 |
| 0.013713786 | 0 | 0 | 0.071408685 | 0.066787041 |
| 0.027234908 | 0.645532984 | 0.004479751 | 0.011062932 | 0.000968253 |
| 0 | 0 | 0 | 0 | 0 |
| 0.012770725 | 0 | 0 | 0.027966595 | 0.001589083 |
| 0 | 0 | 0 | 0.001544073 | 0.010054476 |
| 0 | 0 | 0 | 0 | 0 |
| 0 | 0 | 0 | 0 | 0 |
| 0 | 0 | 0 | 0 | 0 |
| 0 | 0 | 0 | 0 | 0 |
| 0 | 0 | 0 | 0 | 0 |
| 0 | 0 | 0 | 0 | 0 |
| 0 | 0 | 0 | 0 | 0 |
| 0 | 0 | 0 | 0 | 0 |
| 0 | 0 | 0 | 0 | 0 |
| 0 | 0 | 0.041062497 | 0.005504803 | 0.001019991 |
| 0 | 0 | 0.001210228 | 0 | 0.000986525 |
| 0 | 0 | 0 | 0 | 0 |
| 0 | 0 | 0 | 0 | 0 |
| 0 | 0 | 0 | 0 | 0 |
| 0 | 0 | 0 | 0.000632751 | 0 |
| 0 | 0 | 0 | 0 | 0 |
| 0.009926927 | 0 | 0 | 0.011116967 | 0 |
| 0.029488794 | 0 | 0 | 0 | 0 |
| 0 | 0.025853766 | 0 | 0 | 0 |
| 0.00143217 | 0.000943774 | 0 | 0 | 0 |
| 0 | 0 | 0 | 0 | 0 |
| 0 | 0 | 0 | 0 | 0 |
| 0 | 0 | 0 | 0 | 0 |
| 0.004992232 | 0.018093835 | 0 | 0 | 0 |
| 0.013103813 | 0 | 0 | 0 | 0 |
| 0 | 0 | 0 | 0 | 0 |
| 0.116711328 | 0 | 0 | 0 | 0 |
| 0 | 0 | 0 | 0 | 0 |
| 0 | 0 | 0 | 0 | 0 |
| 0 | 0 | 0 | 0 | 0 |
| 0 | 0 | 0 | 0 | 0 |

| | | | | |
|---|---|---|---|---|
| 0 | 0 | 0 | 0 | 0 |
| 0 | 0 | 0 | 0 | 0.000515428 |
| 0 | 0 | 0 | 0.002033535 | 0 |
| 0 | 0 | 0 | 0 | 0 |
| 0 | 0 | 0 | 0.002627328 | 0 |
| 0 | 0 | 0 | 0 | 0 |
| 0 | 0.014607534 | 0.00140622 | 0 | 0.039785777 |
| 0 | 0 | 0 | 0 | 0 |
| 0 | 0 | 0 | 0 | 0.000589965 |
| 0 | 0 | 0 | 0.006464178 | 0.027710913 |
| 0 | 0 | 0 | 0.007097975 | 0 |
| 0 | 0 | 0 | 0.028287786 | 0 |
| 0 | 0 | 0 | 0 | 0 |
| 0 | 0 | 0 | 0 | 0 |
| 0 | 0 | 0 | 0 | 0 |
| 0 | 0 | 0 | 0.001436465 | 0 |
| 0 | 0.032229147 | 0 | 0 | 2.96629301 |
| 0 | 0 | 0 | 0.003850221 | 0 |
| 0 | 0 | 0 | 0 | 0 |
| 0 | 0 | 0 | 0 | 0 |
| 0 | 0 | 1.418457916 | 0.064671275 | 0.053952672 |
| 0 | 0 | 0 | 0 | 0 |
| 0.003056669 | 0 | 0 | 0.005661285 | 0 |
| 0.000627236 | 0.001136676 | 0 | 0.001438307 | 0.005307265 |
| 0 | 0 | 0 | 0 | 0 |
| 0 | 0 | 0 | 0.001187815 | 0 |
| 0 | 0 | 0 | 0 | 0 |
| 0 | 0 | 0 | 0 | 0 |
| 0 | 0 | 0 | 0 | 0 |
| 0.280451008 | 0.007634402 | 0 | 0 | 0 |
| 0 | 0 | 0 | 0 | 0 |
| 0 | 0 | 0 | 0 | 0 |
| 0 | 0 | 0 | 0 | 0 |
| 0 | 0 | 0 | 0 | 0 |
| 0 | 0 | 0 | 0 | 0 |
| 0 | 0 | 0 | 0 | 0 |
| 0 | 0 | 0.003080652 | 0.000719878 | 0.041211798 |
| 0 | 0 | 0 | 0 | 0 |
| 0 | 0 | 0 | 0 | 0 |
| 0 | 0 | 0 | 0 | 0 |
| 0 | 0 | 0 | 0 | 0 |
| 0 | 0 | 0 | 0 | 0 |
| 0 | 0 | 0 | 0 | 0 |
| 0 | 0 | 0 | 0.002060812 | 0 |
| 0 | 0 | 0 | 0 | 0 |
| 0 | 0 | 0 | 0 | 0 |
| 0 | 0 | 0 | 0 | 0 |
| 0 | 0 | 0 | 0 | 0 |

| | | | | |
|---|---|---|---|---|
| 0 | 0.002038506 | 0.005475105 | 0 | 0.002855403 |
| 0.004053959 | 0 | 0 | 0 | 0 |
| 0 | 0 | 0 | 0.243700712 | 0 |
| 0 | 0 | 0 | 0 | 0 |
| 0 | 0 | 0 | 0 | 0 |
| 0 | 0 | 0 | 0 | 0 |
| 0 | 0 | 0 | 0 | 0 |
| 0.095171787 | 0 | 1.326357385 | 0.316867785 | 0.051738592 |
| 0.568898208 | 0.255126058 | 0.051841222 | 0.001700509 | 0 |
| 0.008112753 | 0.002970085 | 0.024124534 | 0.0338241 | 0.005322733 |
| 0 | 0 | 0 | 0.010518376 | 0 |
| 0 | 0 | 0 | 0.000444111 | 0 |
| 0 | 0 | 0 | 0.020944222 | 0 |
| 0.086795085 | 0.00094753 | 0 | 0 | 0.015614547 |
| 0 | 0 | 0 | 0 | 0 |
| 0 | 0 | 0 | 0 | 0 |
| 0 | 0 | 0 | 0 | 0.004413304 |
| 0 | 0 | 0 | 0 | 0 |
| 0 | 0 | 0 | 0 | 0 |
| 0 | 0 | 0 | 0.012341013 | 0 |
| 0 | 0 | 0 | 0 | 0 |
| 0 | 0 | 0 | 0 | 0 |
| 0 | 0 | 0 | 0 | 0 |
| 0 | 0 | 0 | 0 | 0 |
| 0.004695794 | 0 | 0 | 0.021210667 | 0 |
| 0 | 0 | 0 | 0 | 0 |
| 0 | 0 | 0 | 0 | 0 |
| 0 | 0 | 0 | 0 | 0 |
| 0 | 0 | 0 | 0 | 0 |
| 0 | 0 | 0 | 0 | 0 |
| 0 | 0 | 0 | 0.004928887 | 0 |
| 0 | 0 | 0 | 0 | 0 |
| 0 | 0 | 0 | 0 | 0 |
| 0 | 0 | 0 | 0 | 0 |
| 0 | 0 | 0 | 0 | 0 |
| 0.143768664 | 0 | 0 | 0 | 0 |
| 0 | 0 | 0 | 0 | 0 |
| 0 | 0 | 0 | 0 | 0 |
| 0 | 0 | 0 | 0 | 0 |
| 0 | 0 | 0 | 0 | 0 |
| 0 | 0 | 0 | 0 | 0 |
| 0.015640501 | 0 | 0 | 0.466723743 | 0.091859434 |
| 0 | 0 | 0 | 0 | 0 |
| 0 | 0 | 0 | 0 | 0 |
| 0 | 0 | 0 | 0 | 0 |
| 0 | 0 | 0 | 0 | 0 |
| 0 | 0 | 0 | 0 | 0 |
| 0 | 0 | 0 | 0.005188473 | 0 |

| | | | | |
|---|---|---|---|---|
| 0 | 0 | 0 | 0 | 0 |
| 0 | 0.000981249 | 0.316735593 | 0.001396839 | 0 |
| 0 | 0 | 0 | 0 | 0 |
| 0 | 0 | 0 | 0 | 0 |
| 0.016039975 | 0.001543414 | 0 | 0.043941987 | 0.058498762 |
| 0 | 0 | 0 | 0 | 0 |
| 0 | 0 | 0 | 0 | 0 |
| 0.001574022 | 0.04788642 | 0.374747399 | 0.011024983 | 0.043160026 |
| 0 | 0 | 0 | 0.003931442 | 0 |
| 0 | 0 | 0 | 0 | 0 |
| 0 | 0 | 0 | 0 | 0 |
| 0.00406214 | 0 | 0 | 0 | 0 |
| 0 | 0 | 0 | 0 | 0 |
| 0 | 0 | 0 | 0 | 0 |
| 0 | 0 | 0.574274756 | 0.002356078 | 0 |
| 0 | 0.001049533 | 0 | 0 | 0 |
| 0 | 0 | 0 | 0 | 0 |
| 0 | 0 | 0 | 0 | 0 |
| 0 | 0 | 0 | 0 | 0 |
| 0 | 0 | 0 | 0 | 0 |
| 0 | 0 | 0 | 0 | 0 |
| 0 | 0 | 0 | 0 | 0 |
| 0 | 0 | 0 | 0 | 0 |
| 0 | 0.000448828 | 0.000972161 | 0.000681515 | 0 |
| 0.145435648 | 0.17543933 | 0 | 0 | 0 |
| 0 | 0 | 0 | 0 | 0 |
| 0.00090609 | 0.001094677 | 0.073443943 | 0.003116616 | 0.004566226 |
| 0 | 0 | 0 | 0.002625356 | 0.000712309 |
| 0 | 0 | 0 | 0 | 0 |
| 0 | 0 | 0 | 0 | 0 |
| 0 | 0 | 0 | 0 | 0 |
| 0 | 0.001052152 | 0 | 0.000898663 | 0 |
| 0 | 0 | 0 | 0 | 0 |
| 0 | 0 | 0 | 0 | 0 |
| 0 | 0 | 0 | 0 | 0 |
| 0 | 0 | 0 | 0 | 0 |
| 0 | 0 | 0 | 0 | 0 |
| 0.000464624 | 0 | 0 | 0.414556488 | 0 |
| 0.002194389 | 0 | 0 | 3.29201208 | 0 |
| 0 | 0 | 0 | 0 | 0 |
| 0 | 0 | 0 | 0 | 0 |
| 0.09243335 | 0 | 0 | 0 | 0 |
| 0 | 0 | 0 | 0 | 0 |
| 0 | 0 | 0 | 0 | 0 |
| 0 | 0 | 0 | 0 | 0 |
| 0 | 0 | 0 | 0 | 0 |
| 0 | 0 | 0 | 0 | 0 |
| 0 | 0 | 0 | 0 | 0 |

| | | | | |
|---|---|---|---|---|
| 0.002769323 | 0 | 0 | 0.109909068 | 0 |
| 0 | 0 | 0 | 0 | 0 |
| 0 | 0 | 0 | 0 | 0 |
| 0 | 0 | 0 | 0 | 0 |
| 0 | 0 | 0 | 0 | 0 |
| 0 | 0 | 0 | 0 | 0 |
| 0 | 0 | 0 | 0 | 0 |
| 0 | 0 | 0 | 0 | 0 |
| 0 | 0 | 0 | 0 | 0 |
| 0 | 0 | 0 | 0 | 0 |
| 0 | 0 | 0 | 0 | 0 |
| 0 | 0 | 0 | 0 | 0 |
| 0 | 0 | 0 | 0 | 0 |
| 0 | 0 | 0 | 0 | 0 |
| 0 | 0 | 0 | 0 | 0 |
| 0 | 0 | 0 | 0 | 0 |
| 0 | 0 | 0 | 0 | 0 |
| 0 | 0.109575955 | 1.801506225 | 0 | 0.233001686 |
| 0 | 0 | 0 | 0.000745466 | 0 |
| 0 | 0 | 0 | 0 | 0 |
| 0 | 0.181462274 | 0.128176094 | 0 | 0.001657217 |
| 0 | 0 | 0 | 0 | 0 |
| 0.000778705 | 0 | 0 | 0.025927465 | 0.009301973 |
| 0 | 0 | 0 | 0.008236917 | 0 |
| 0 | 0 | 0 | 0.000769856 | 0 |
| 0.003461569 | 0 | 0 | 0 | 0 |
| 0 | 0 | 0 | 0 | 0 |
| 0 | 0 | 0 | 0 | 0 |
| 3.10522543 | 0 | 0 | 0 | 0 |
| 0 | 0 | 0 | 0 | 0 |
| 0 | 0 | 0 | 0 | 0 |
| 0 | 0 | 0 | 0.002800914 | 0 |
| 0.105304232 | 11.84780844 | 0.117705387 | 0 | 0.084717001 |
| 0 | 0 | 0 | 0 | 0 |
| 0 | 0 | 0 | 0.249322134 | 0.059415376 |
| 0.032536871 | 0 | 0.037854961 | 0.000966171 | 0.004718534 |
| 0 | 0 | 0 | 0 | 0 |
| 0 | 0 | 0 | 0 | 0 |
| 0 | 0.099274868 | 0.038258956 | 0 | 0 |
| 0 | 0 | 0 | 0 | 0 |
| 0 | 0 | 0 | 0 | 0 |
| 0 | 0 | 0 | 0.048232073 | 0 |
| 0.051920119 | 0.000764956 | 0 | 0.00522691 | 0.010163485 |
| 0 | 0 | 0 | 0 | 0 |
| 0 | 0 | 0 | 0 | 0 |
| 0 | 0 | 0 | 0.001037768 | 0 |
| 0 | 0 | 0 | 0 | 0 |
| 0 | 0 | 0 | 0 | 0 |

| | | | | |
|---|---|---|---|---|
| 0 | 0 | 0 | 0 | 0 |
| 0 | 0 | 0 | 0 | 0 |
| 0 | 0 | 0.07146087 | 0 | 0 |
| 0 | 0 | 0 | 0 | 0 |
| 0 | 0 | 0 | 0 | 0 |
| 0 | 0 | 0 | 0 | 0 |
| 0 | 0 | 0 | 0 | 0 |
| 0 | 0 | 0 | 0 | 0 |
| 0 | 0 | 0 | 0.028358534 | 0 |
| 0 | 0.000625726 | 0 | 0 | 0 |
| 0 | 0 | 0 | 0 | 0 |
| 0 | 0 | 0 | 0 | 0 |
| 0 | 0 | 0 | 0 | 0 |
| 0 | 0 | 0 | 1.642561252 | 0 |
| 0 | 0 | 0 | 0.002447458 | 0.005781074 |
| 0 | 0.008157302 | 0.191222309 | 0 | 0.006848113 |
| 0 | 0 | 0 | 0 | 0 |
| 0 | 0 | 0 | 0 | 0 |
| 0 | 0 | 0 | 0 | 0 |
| 0.072315499 | 0 | 0 | 1.186167984 | 0 |
| 0 | 0 | 0 | 0 | 0 |
| 0 | 0 | 0 | 0 | 0 |
| 0 | 0 | 0 | 0 | 0 |
| 0 | 0 | 0 | 0.027549958 | 0 |
| 0 | 0 | 0 | 0 | 0 |
| 0 | 0 | 0 | 0 | 0 |
| 0 | 0 | 0 | 0 | 0 |
| 0 | 0 | 0 | 0 | 0 |
| 0 | 0 | 0 | 0 | 0 |
| 0 | 0 | 0 | 0.002361612 | 0 |
| 0.023297391 | 0 | 0 | 0.055987263 | 0 |
| 0 | 0 | 0 | 0 | 0 |
| 0 | 0 | 0 | 0.022921235 | 0 |
| 0 | 0 | 0 | 0 | 0 |
| 0 | 0 | 0 | 0 | 0 |
| 0 | 0 | 0 | 0 | 0 |
| 0 | 0 | 0 | 0 | 0 |
| 0 | 0 | 0 | 0 | 0 |
| 0.051347162 | 0 | 0 | 0 | 0 |
| 0 | 0 | 0 | 0 | 0 |
| 0 | 0 | 0 | 0 | 0 |
| 0.000925196 | 0 | 0 | 0 | 0 |
| 0 | 0 | 0 | 0 | 0 |
| 0 | 0 | 0 | 0 | 0 |
| 0 | 0 | 0 | 0.008761061 | 0 |
| 0 | 0 | 0 | 0 | 0 |
| 0 | 0.170792806 | 0.036652475 | 0 | 0.169806578 |
| 0 | 0 | 0 | 0 | 0 |

| | | | | |
|---|---|---|---|---|
| 0 | 0 | 0 | 0 | 0 |
| 0 | 0 | 0 | 0 | 0 |
| 0 | 0 | 0 | 0 | 0 |
| 0 | 0 | 0 | 0 | 0 |
| 0.178685385 | 0 | 0.642026958 | 0.095844604 | 0.030841671 |
| 0 | 0 | 0 | 0 | 0 |
| 0 | 0 | 0 | 0 | 0 |
| 0 | 0 | 0 | 0 | 0 |
| 0 | 0 | 0 | 0 | 0 |
| 0 | 0 | 0 | 0.000456902 | 0 |
| 0 | 0 | 0 | 0.010041498 | 0 |
| 0.001352664 | 0 | 0.007220937 | 0.00334992 | 0.003029658 |
| 0 | 0 | 0.013375854 | 0.001766195 | 0 |
| 0 | 0 | 0 | 0 | 0 |
| 0 | 0.041346384 | 0.003560673 | 0 | 0 |
| 0 | 0 | 0 | 0.093926094 | 0.00078533 |
| 0 | 0 | 0 | 0 | 0.000466033 |
| 0 | 0 | 0 | 0 | 0 |
| 0 | 0.007053317 | 0.171429579 | 0.051718027 | 0.076469158 |
| 0 | 0.003716107 | 0 | 0 | 0.025760118 |
| 0 | 0 | 0 | 0 | 0 |
| 0 | 0 | 0 | 0 | 0 |
| 0 | 0 | 0 | 0 | 0 |
| 0 | 0 | 0 | 0 | 0 |
| 0 | 0 | 0 | 0 | 0 |
| 0 | 0 | 0 | 0 | 0 |
| 0 | 0.003884201 | 0.031297044 | 0.003391295 | 0.006400857 |
| 0 | 0 | 0 | 0.002609865 | 0 |
| 0 | 0 | 0 | 0 | 0 |
| 0 | 0 | 0 | 0 | 0 |
| 0.001214807 | 0.000587059 | 0.001525886 | 0 | 0 |
| 0 | 0 | 0 | 0 | 0 |
| 0 | 0 | 0 | 0 | 0 |
| 0 | 0 | 0 | 0 | 0 |
| 0 | 0 | 0 | 0 | 0 |
| 0 | 0 | 0 | 0 | 0 |
| 0 | 0 | 0 | 0 | 0 |
| 0 | 0.001237353 | 0.081341264 | 0.002231121 | 0.015165507 |
| 0 | 0 | 0 | 0.001977376 | 0 |
| 0 | 0 | 0 | 0 | 0.002223703 |
| 0 | 0 | 0 | 0 | 0 |
| 0 | 0 | 0 | 0 | 0 |
| 0 | 0 | 0 | 0 | 0 |
| 0 | 0.279506845 | 0.001390991 | 0.00139304 | 0.001133875 |
| 0 | 0 | 0 | 0 | 0 |
| 0 | 0 | 0 | 0 | 0 |
| 0 | 0 | 0 | 0 | 0 |
| 0 | 0 | 0 | 0 | 0 |

| | | | | |
|---|---|---|---|---|
| 0 | 0 | 0 | 0 | 0 |
| 0 | 0 | 0 | 0 | 0.000807609 |
| 0 | 0 | 0 | 0 | 0 |
| 0 | 0.001121702 | 0.002082521 | 0.001094934 | 0.002970766 |
| 0 | 0 | 0 | 0 | 0 |
| 0 | 0 | 0 | 0 | 0 |
| 0.000514857 | 0 | 0 | 0.000885459 | 0 |
| 0.020539261 | 0 | 0 | 0 | 0 |
| 0 | 0 | 0 | 0.002965012 | 0 |
| 0 | 0 | 0 | 0 | 0 |
| 0 | 0 | 0 | 0 | 0 |
| 0 | 0 | 0 | 0 | 0 |
| 0.149775431 | 0 | 0 | 0 | 0.000561561 |
| 0 | 0 | 0 | 0 | 0 |
| 0 | 0 | 0 | 0 | 0 |
| 0 | 0 | 0 | 0 | 0 |
| 0 | 0 | 0 | 0 | 0 |
| 0 | 0 | 0 | 0 | 0 |
| 0 | 0 | 0 | 0 | 0 |
| 0 | 0 | 0 | 0 | 0 |
| 0 | 0 | 0 | 0 | 0 |
| 0 | 0 | 0 | 0 | 0 |
| 0.004333139 | 0.003141004 | 0.00971917 | 0.005450753 | 0 |
| 0 | 0 | 0 | 0.041808492 | 0 |
| 0 | 0 | 0 | 0 | 0 |
| 0 | 0 | 0 | 0 | 0 |
| 0 | 0 | 0 | 0 | 0 |
| 0 | 0 | 0 | 0 | 0 |
| 0 | 0 | 0 | 0 | 0 |
| 0 | 0 | 0 | 0 | 0 |
| 0.002070613 | 0 | 0 | 0.00778688 | 0.00164896 |
| 0 | 0 | 0 | 0 | 0 |
| 0 | 0 | 0 | 0 | 0 |
| 0 | 0 | 0 | 0 | 0 |
| 0 | 0 | 0 | 0 | 0 |
| 0 | 0 | 0.001585994 | 0 | 0 |
| 0 | 0.025337128 | 0.042369092 | 0 | 0 |
| 0 | 0 | 0 | 0 | 0 |
| 0 | 0.02045956 | 0 | 0 | 0.000697567 |
| 0 | 0 | 0 | 0 | 0 |
| 0 | 0 | 0 | 0 | 0 |
| 0 | 0 | 0 | 0 | 0 |
| 0 | 0 | 0 | 0 | 0 |
| 0 | 0 | 0 | 0 | 0 |
| 0 | 0 | 0 | 0 | 0 |
| 0.261210341 | 0 | 0 | 0.009331053 | 0 |
| 0 | 0 | 0 | 0.00286056 | 0 |
| 0 | 0 | 0 | 0 | 0 |

| | | | | |
|---|---|---|---|---|
| 0 | 0 | 0 | 0 | 0 |
| 0 | 0 | 0 | 0 | 0 |
| 0.230826835 | 0 | 0 | 0 | 0 |
| 0 | 0 | 0 | 0.00128072 | 0 |
| 0 | 0 | 0 | 0 | 0 |
| 0 | 0 | 0 | 0 | 0 |
| 0 | 0 | 0 | 0.004900123 | 0 |
| 0 | 0 | 0 | 0 | 0 |
| 0 | 0 | 0.011876895 | 0 | 0 |
| 0 | 0 | 0 | 0 | 0 |
| 0 | 0 | 0 | 0.000912532 | 0 |
| 0 | 0 | 0 | 0 | 0 |
| 0 | 0 | 0 | 0 | 0 |
| 0 | 0 | 0 | 0 | 0 |
| 0 | 0 | 0 | 0 | 0 |
| 0 | 0 | 0 | 0 | 0 |
| 0 | 0 | 0 | 0 | 0 |
| 0 | 0 | 0 | 0 | 0 |
| 0 | 0 | 0 | 0 | 0 |
| 0.000879581 | 0 | 0.039063175 | 0.001210174 | 0.000938124 |
| 0 | 0 | 0 | 0 | 0 |
| 0 | 0 | 0 | 0 | 0 |
| 0 | 0 | 0 | 0 | 0 |
| 0 | 0 | 0 | 0 | 0 |
| 0 | 0 | 0 | 0 | 0 |
| 0 | 0 | 0 | 0 | 0 |
| 0 | 0 | 0 | 0 | 0 |
| 0.004588909 | 0.006048012 | 0 | 0 | 0 |
| 0 | 0 | 0 | 0 | 0 |
| 0 | 0 | 0 | 0 | 0 |
| 0.015868672 | 0.000521672 | 0 | 0.010693692 | 0.003062592 |
| 0 | 0 | 0 | 0 | 0 |
| 0.021809354 | 0.040734317 | 0.065336555 | 0.038456552 | 0.063820511 |
| 0.00179319 | 0 | 0.001005525 | 0.000528678 | 0 |
| 0 | 0 | 0 | 0 | 0 |
| 0 | 0 | 0 | 0 | 0 |
| 0 | 0 | 0 | 0 | 0 |
| 0 | 0 | 0 | 0 | 0 |
| 0 | 0 | 0 | 0 | 0 |
| 0 | 0 | 0 | 0 | 0 |
| 0 | 0 | 0 | 0 | 0 |
| 0 | 0.398652828 | 0.264549477 | 0.006100573 | 0.030948433 |
| 0.025753244 | 0 | 0 | 0.032667192 | 0 |
| 0 | 0 | 0 | 0 | 0 |
| 0 | 0 | 0 | 0 | 0 |
| 0.004901518 | 0 | 0 | 0.00274746 | 0 |
| 0 | 0 | 0 | 0 | 0 |
| 0 | 0 | 0 | 0 | 0 |

| | | | | |
|---|---|---|---|---|
| 0 | 0 | 0 | 0 | 0 |
| 0.007896567 | 0 | 0.037608245 | 0.007243009 | 0 |
| 0 | 0.003157423 | 0.002655134 | 0 | 0 |
| 0 | 0 | 0 | 0 | 0 |
| 0 | 0.014978003 | 0 | 0.065189427 | 0.066899541 |
| 0.000358449 | 0 | 0 | 0.008219559 | 0.195358841 |
| 0 | 0 | 0 | 0 | 0 |
| 0 | 0 | 0 | 0 | 0 |
| 0 | 0 | 0 | 0 | 0 |
| 0 | 0 | 0 | 0 | 0 |
| 0 | 0 | 0 | 0 | 0 |
| 0 | 0 | 0 | 0 | 0 |
| 0.00027973 | 0 | 0 | 0 | 0 |
| 0 | 0 | 0 | 0 | 0.267807137 |
| 0 | 0 | 0 | 0 | 0 |
| 0 | 0 | 0 | 0 | 0 |
| 0 | 0 | 0 | 0 | 0 |
| 0 | 0 | 0 | 0 | 0 |
| 0 | 0 | 0 | 0 | 0 |
| 0 | 0 | 0 | 0 | 0 |
| 0 | 0 | 0 | 0 | 0 |
| 0 | 0 | 0.151832301 | 0 | 0 |
| 0 | 0 | 0 | 0 | 0 |
| 0 | 0 | 0 | 0 | 0 |
| 0 | 0 | 0 | 0.004183634 | 0 |
| 0.000382366 | 0 | 0 | 0 | 0 |
| 0 | 0 | 0 | 0 | 0 |
| 0 | 0 | 0.00058236 | 0.271998188 | 0 |
| 0 | 0 | 0 | 0 | 0 |
| 0 | 0 | 0 | 0 | 0 |
| 0 | 0 | 0 | 0 | 0 |
| 0 | 0 | 0 | 0 | 0 |
| 0 | 0 | 0 | 0 | 0 |
| 0 | 0 | 0 | 0 | 0 |
| 0 | 0 | 0 | 0 | 0 |
| 0 | 0 | 0 | 0 | 0 |
| 0 | 0 | 0 | 0 | 0 |
| 0 | 0 | 0.22270464 | 0 | 0 |
| 0 | 0 | 0 | 0.824882421 | 0 |
| 0 | 0 | 0 | 0 | 0 |
| 0 | 0 | 0 | 0 | 0 |
| 0.000967886 | 0 | 0 | 0 | 0 |
| 0 | 0.001081771 | 0.006627678 | 0 | 0.001528007 |
| 0 | 0 | 0 | 0 | 0 |
| 0 | 0 | 0 | 0 | 0 |
| 0 | 0 | 0 | 0 | 0 |
| 0.000587057 | 0.002600555 | 0.005223147 | 0.000807703 | 0.020745765 |
| 0 | 0 | 0 | 0 | 0 |

| | | | | |
|---|---|---|---|---|
| 0 | 0 | 0 | 0 | 0.001702533 |
| 0.001851894 | 0.007905244 | 0.106807316 | 0.004416417 | 0.004793034 |
| 0 | 0 | 0 | 0 | 0 |
| 0 | 0 | 0 | 0 | 0 |
| 0 | 0 | 0 | 0 | 0 |
| 0 | 0 | 0 | 0 | 0 |
| 0 | 0 | 0 | 0 | 0 |
| 0 | 0 | 0 | 0 | 0 |
| 0 | 0 | 0 | 0 | 0 |
| 0 | 0 | 0 | 0 | 0 |
| 0 | 0 | 0 | 0 | 0 |
| 0 | 0 | 0 | 0 | 0 |
| 0 | 0 | 0 | 0.002513832 | 0 |
| 0 | 0 | 0 | 0 | 0 |
| 0.001847226 | 0.005765209 | 0.014259864 | 0 | 0.002528388 |
| 0 | 0 | 0 | 0 | 0 |
| 0 | 0 | 0 | 0 | 0 |
| 0 | 0 | 0 | 0 | 0 |
| 0 | 0 | 0 | 0 | 0 |
| 0 | 0 | 0 | 0 | 0 |
| 0 | 0 | 0 | 0 | 0 |
| 0 | 0 | 0.009969171 | 0 | 0.001354406 |
| 0 | 0 | 0 | 0.005434175 | 0.001387667 |
| 0 | 0 | 0 | 0 | 0 |
| 0 | 0 | 0 | 0 | 0 |
| 0 | 0 | 0 | 0 | 0 |
| 0 | 0 | 0 | 0 | 0 |
| 0 | 0 | 0 | 0 | 0 |
| 0 | 0 | 0 | 0 | 0 |
| 0 | 0 | 0 | 0 | 0 |
| 0 | 0 | 0 | 0 | 0 |
| 0 | 0 | 0 | 0 | 0 |
| 0 | 0 | 0 | 0 | 0 |
| 0 | 0 | 0 | 0 | 0 |
| 22.7521013 | 0 | 4.252458607 | 0 | 0 |
| 0 | 0 | 0 | 0 | 0 |
| 0 | 0 | 0 | 0 | 0 |
| 0 | 0 | 0 | 0 | 0 |
| 0 | 0 | 0 | 0 | 0.039899869 |
| 0 | 0 | 0 | 0 | 0 |
| 0.041556465 | 0.139823749 | 0 | 0.112488747 | 0 |
| 0 | 0 | 0 | 0 | 0 |
| 0 | 0 | 0 | 0 | 0 |
| 0 | 0 | 0 | 0 | 0 |
| 0 | 0 | 0 | 0 | 0 |
| 0.010073798 | 6.763044457 | 0.394445948 | 0.003198476 | 0.190917695 |
| 0 | 0 | 0 | 0 | 0 |
| 0 | 0 | 0 | 0 | 0 |

| | | | | |
|---|---|---|---|---|
| 0 | 0 | 0 | 0 | 0 |
| 0 | 0 | 0 | 0 | 0 |
| 0 | 0 | 0 | 0.003138963 | 0 |
| 0 | 0 | 0 | 0 | 0 |
| 0 | 0 | 0.000738205 | 0.001293761 | 0 |
| 0 | 0 | 0 | 0 | 0 |
| 0 | 0 | 0 | 0 | 0 |
| 0 | 0.001214268 | 0 | 0 | 0 |
| 0 | 0 | 0 | 0 | 0 |
| 0 | 0.012452111 | 0.007615423 | 0.018977227 | 0 |
| 0.008472288 | 0 | 0 | 0.030571135 | 0 |
| 0 | 0 | 0 | 0 | 0 |
| 0 | 0 | 0.007443435 | 0 | 0 |
| 0 | 0 | 0 | 0 | 0 |
| 0 | 0 | 0 | 0 | 0 |
| 0 | 0 | 0 | 0 | 0 |
| 0 | 0 | 0 | 0 | 0 |
| 0 | 0 | 0 | 0 | 0 |
| 0 | 0 | 0 | 0 | 0 |
| 0.249880553 | 0.073122417 | 0 | 0.167635942 | 0.125225479 |
| 0 | 0 | 0 | 0 | 0 |
| 0 | 0 | 0 | 0 | 0 |
| 0 | 0 | 0 | 0 | 0 |
| 0 | 0 | 0 | 0 | 0 |
| 0 | 0 | 0 | 0 | 0 |
| 0 | 0 | 0 | 0 | 0 |
| 0 | 0 | 0 | 0 | 0 |
| 0 | 0 | 0 | 0 | 0 |
| 0 | 0 | 0 | 0 | 0 |
| 0 | 0 | 0.049798313 | 0 | 0 |
| 0 | 0 | 0 | 0 | 0 |
| 0 | 0 | 0 | 0 | 0 |
| 0 | 0 | 0 | 0 | 0 |
| 0 | 0 | 0 | 0 | 0 |
| 0 | 0 | 0 | 0 | 0 |
| 0 | 0 | 0 | 0.00422729 | 0 |
| 0 | 0 | 0 | 0 | 0 |
| 0 | 0 | 0 | 0 | 0 |
| 0 | 0 | 0 | 0 | 0 |
| 0 | 0 | 0 | 0 | 0 |
| 0 | 0 | 0 | 0 | 0 |
| 0 | 0 | 0 | 0 | 0 |
| 0 | 0 | 0 | 0.001206455 | 0 |
| 0 | 0 | 0 | 0 | 0 |
| 0 | 0 | 0 | 0.000681601 | 0 |
| 0 | 0 | 0 | 0.004600249 | 0 |
| 0 | 0 | 0 | 0 | 0 |
| 0 | 0.057366662 | 0 | 0 | 0 |

| | | | | |
|---|---|---|---|---|
| 0 | 0 | 0 | 0 | 0 |
| 0 | 0 | 0 | 0 | 0 |
| 0 | 0 | 0 | 0 | 0 |
| 0 | 0 | 0 | 0 | 0 |
| 0 | 0 | 0 | 0 | 0 |
| 0.081918968 | 0 | 0 | 0.002162577 | 0 |
| 0 | 0 | 0 | 0 | 0 |
| 0 | 0 | 0 | 0 | 0 |
| 0 | 0 | 0 | 0 | 0 |
| 0 | 0 | 0 | 0 | 0 |
| 0 | 0 | 0 | 0 | 0 |
| 0 | 0 | 0 | 0 | 0 |
| 0 | 0 | 0 | 0 | 0 |
| 0 | 0 | 0 | 0 | 0 |
| 0 | 0 | 0 | 0 | 0 |
| 0 | 0 | 0 | 0 | 0 |
| 0.000336458 | 0 | 0 | 0.009875542 | 0 |
| 0 | 0 | 0 | 0 | 0 |
| 0 | 0 | 0 | 0 | 0 |
| 0 | 0 | 0 | 0 | 0 |
| 0 | 0 | 0 | 0 | 0 |
| 0 | 0 | 0 | 0 | 0 |
| 0 | 0 | 0 | 0 | 0 |
| 0 | 0 | 0 | 0 | 0 |
| 0 | 0 | 0 | 0.001126975 | 0 |
| 0 | 0.000873612 | 0 | 0 | 0 |
| 0 | 0 | 0 | 0 | 0 |
| 0 | 0 | 0 | 0.140879844 | 0 |
| 0 | 0 | 0 | 0 | 0 |
| 0 | 0 | 0 | 0 | 0 |
| 0 | 0 | 0 | 0 | 0 |
| 0 | 0 | 0.005538262 | 0 | 0 |
| 0 | 0 | 0 | 0 | 0 |
| 0 | 0 | 0 | 0 | 0 |
| 0 | 0 | 0 | 0 | 0 |
| 0 | 0 | 0 | 0 | 0 |
| 0 | 0 | 0 | 0 | 0 |
| 0 | 0 | 0 | 0.062183646 | 0 |
| 0 | 0 | 0 | 0 | 0 |
| 0 | 0 | 0 | 0 | 0 |
| 0 | 0 | 0 | 0 | 0 |
| 0 | 0 | 0 | 0 | 0 |
| 0 | 0 | 0 | 0 | 0 |
| 0 | 0 | 0 | 0 | 0 |
| 0 | 0 | 0 | 0 | 0 |
| 0 | 0 | 0 | 0 | 0 |
| 0 | 0 | 0 | 0 | 0 |
| 0 | 0 | 0 | 0 | 0 |

| | | | | |
|---|---|---|---|---|
| 0 | 0 | 0 | 0 | 0 |
| 0 | 0.001000485 | 0 | 0.007548383 | 0 |
| 0 | 0 | 0 | 0 | 0 |
| 0 | 0 | 0 | 0 | 0 |
| 0 | 0 | 0 | 0 | 0 |
| 0 | 0 | 0 | 0 | 0 |
| 0 | 0 | 0 | 0 | 0 |
| 0 | 0 | 0 | 0 | 0 |
| 0 | 0 | 0 | 0.001099715 | 0 |
| 0 | 0 | 0 | 0 | 0 |
| 0.001355456 | 0 | 0.000851275 | 0 | 0 |
| 0 | 0 | 0 | 0 | 0 |
| 0 | 0 | 0 | 0 | 0 |
| 0 | 0 | 0 | 0 | 0 |
| 0 | 0 | 0 | 0 | 0 |
| 0 | 0 | 0 | 0 | 0 |
| 0 | 0 | 0 | 0 | 0 |
| 0 | 0 | 0 | 0 | 0 |
| 0 | 0 | 0 | 0.097738777 | 0 |
| 0 | 0 | 0 | 0 | 0 |
| 0 | 0.005152309 | 0 | 0 | 0 |
| 0 | 0 | 0 | 0 | 0 |
| 0 | 0 | 0 | 0 | 0 |
| 1.390353793 | 0 | 0 | 0.336172512 | 3.896506354 |
| 0 | 0 | 0 | 0 | 0 |
| 0 | 0 | 0 | 0 | 0 |
| 0 | 0 | 0 | 0 | 0 |
| 0 | 0 | 0 | 0 | 0 |
| 0 | 0 | 0 | 0 | 0 |
| 0 | 0 | 0 | 0 | 0 |
| 0 | 0 | 0 | 0 | 0 |
| 0 | 0 | 0 | 0 | 0 |
| 0 | 0 | 0 | 0 | 0 |
| 0 | 0 | 0 | 0 | 0 |

| PHL018-0182_V1 | PHL018-0072_V1 | PHL018-0258_V3 | PHL018-0211_V1 | PHL018-0054_V1 |
|---|---|---|---|---|
| EYCF | CTRL | REF | REF | CTRL |
| 0.013258359 | 0.003642277 | 0.831361868 | 0.501716153 | 0.203665125 |
| 11.93551222 | 1.808249571 | 6.685575081 | 0.37949373 | 9.067374705 |
| 12.41617853 | 7.772963035 | 0 | 6.316156791 | 2.039515435 |
| 0 | 0 | 0 | 0 | 0 |
| 0 | 0 | 0 | 0 | 0 |
| 0 | 0 | 0 | 0 | 0 |
| 0 | 0 | 0 | 0 | 0 |
| 0 | 0 | 0 | 0 | 0 |
| 0 | 0 | 0 | 0 | 0 |
| 0 | 0 | 0 | 0 | 0 |
| 0 | 0 | 0 | 0 | 0 |
| 0 | 0 | 0 | 0 | 0 |
| 0 | 0 | 0 | 0 | 0 |
| 8.405089074 | 10.03393206 | 0.481151798 | 0.162369851 | 8.399779035 |
| 0 | 0 | 0 | 0 | 0 |
| 0 | 0 | 0 | 0 | 0 |
| 0.012799768 | 0 | 0.001364971 | 0 | 0 |
| 0 | 0 | 0 | 0 | 0 |
| 0 | 0 | 0 | 0 | 0 |
| 0 | 0 | 0 | 0 | 0 |
| 0 | 0 | 0 | 0 | 0 |
| 0 | 0 | 0 | 0 | 0 |
| 0 | 0 | 0 | 0 | 0 |
| 0 | 0 | 0 | 0 | 0 |
| 0 | 0 | 0 | 0 | 0.007775089 |
| 0 | 0 | 0 | 0 | 0 |
| 0 | 0 | 0 | 0 | 0 |
| 0 | 0 | 0 | 0 | 0 |
| 0 | 0 | 0 | 0 | 0 |
| 0 | 0 | 0 | 0 | 0 |
| 0 | 0 | 0 | 0 | 0 |
| 0 | 0 | 0 | 0 | 0 |
| 0 | 0 | 0 | 0 | 0 |
| 0 | 0 | 0 | 0 | 0 |
| 0 | 0 | 0 | 0 | 0 |
| 0 | 1.838187031 | 0 | 0 | 2.312485252 |
| 0 | 0 | 0 | 0 | 0 |
| 0 | 0 | 0 | 0 | 0 |
| 0 | 0 | 0 | 0 | 0 |
| 0 | 0 | 0 | 0 | 0 |
| 0 | 0 | 0 | 0 | 0 |
| 0.036255044 | 0 | 0.037538966 | 0.3376193 | 0.043067584 |
| 0 | 0.150715802 | 0.001335197 | 1.332647717 | 0.093581528 |
| 0 | 0.75826197 | 0.007694322 | 0.024272467 | 0.080892152 |
| 0 | 0.243674594 | 0 | 0 | 0 |
| 0 | 0 | 0 | 0 | 0 |

| | | | | |
|---|---|---|---|---|
| 0 | 0.17393243 | 0 | 0 | 0 |
| 0.005062715 | 0 | 0.121179062 | 0.000854953 | 0 |
| 0.006616634 | 0.002464218 | 0 | 0.039409991 | 0.013876656 |
| 1.310829165 | 0.113724388 | 0.759005432 | 0.637036466 | 0.75206506 |
| 0 | 0 | 0.015956266 | 0.102002608 | 0.002142425 |
| 0 | 0 | 0 | 0 | 0 |
| 3.262149641 | 0.072512881 | 0.003976053 | 0.005727322 | 0.048952317 |
| 0 | 0 | 0 | 0 | 0 |
| 0 | 0 | 0 | 0 | 0 |
| 0 | 0 | 0 | 0 | 0 |
| 0 | 0 | 0.000730645 | 0 | 0 |
| 0 | 0.130527987 | 0.052191635 | 1.48354528 | 0.142974199 |
| 0 | 0 | 0 | 0 | 0 |
| 0.001629708 | 0 | 0.077072989 | 0 | 0.017317861 |
| 0 | 0 | 0 | 0 | 0 |
| 0 | 0 | 0 | 0 | 0 |
| 1.257314788 | 0.994186163 | 0.003383167 | 0.021592454 | 0.022003811 |
| 0 | 0 | 0 | 0 | 0 |
| 0 | 0 | 0 | 0 | 0 |
| 0 | 0 | 0 | 0 | 0 |
| 1.288527742 | 0 | 0.15666632 | 0 | 0.070297422 |
| 0.00572128 | 0 | 0.015443971 | 0.030113938 | 0.000628595 |
| 0 | 2.323772322 | 0 | 0 | 0.099880966 |
| 0 | 0 | 0 | 0 | 0 |
| 0 | 0 | 0 | 0 | 0 |
| 0 | 0 | 0.001745764 | 0 | 0 |
| 0 | 0 | 0 | 0 | 0 |
| 0 | 0 | 0 | 0 | 0 |
| 0.004455148 | 0 | 0.00265037 | 0.013327372 | 0.001072206 |
| 0 | 0 | 0 | 0 | 0 |
| 0.147977906 | 0 | 1.400404692 | 0 | 0 |
| 0 | 0 | 0 | 0 | 0 |
| 0 | 0 | 0 | 0 | 0 |
| 0.00034499 | 0 | 0 | 0 | 0 |
| 0 | 0 | 0 | 0 | 0 |
| 0 | 0 | 0 | 0 | 0 |
| 0 | 0 | 0 | 0 | 0 |
| 0.02714734 | 0.019265973 | 0 | 0.002368626 | 0.000762237 |
| 0 | 0 | 0 | 0 | 0 |
| 0 | 0 | 0 | 0 | 0 |
| 0 | 0 | 0 | 0 | 0 |
| 0 | 0 | 0 | 0 | 0 |
| 0 | 0 | 0 | 0 | 0 |
| 0 | 0 | 0 | 0 | 0 |
| 0 | 0 | 0 | 0 | 0 |
| 0 | 0 | 0 | 0 | 0 |
| 0 | 0 | 0.001164269 | 0 | 0 |
| 0.001884895 | 0.021785003 | 0.003615424 | 0 | 0 |

| | | | | |
|---|---|---|---|---|
| 0 | 0 | 0 | 0 | 0 |
| 0.300940007 | 0 | 0.002175704 | 0 | 0 |
| 0 | 0 | 0 | 0 | 0 |
| 0 | 0 | 0 | 0 | 0 |
| 0 | 0.183689732 | 0 | 0.002593974 | 0.321977044 |
| 0 | 0 | 0 | 0 | 0 |
| 0 | 0 | 0 | 0 | 0 |
| 0 | 0 | 0 | 0 | 0 |
| 0 | 0 | 0.043327054 | 0.013059032 | 0.002835401 |
| 0 | 0 | 0 | 0 | 0 |
| 0 | 0 | 0 | 0 | 0 |
| 0 | 0 | 0.000845409 | 0.002374658 | 0.000940527 |
| 0 | 0 | 0 | 0 | 0 |
| 0 | 0 | 0 | 0 | 0 |
| 0 | 0 | 0 | 0 | 0 |
| 0 | 0 | 0 | 0 | 0 |
| 0 | 0 | 0 | 0 | 0 |
| 0 | 0 | 0 | 0 | 0 |
| 0 | 0 | 0.079939816 | 0.430144875 | 0.362222577 |
| 0 | 0 | 0 | 0 | 0 |
| 0 | 0 | 0 | 0 | 0 |
| 0 | 0 | 0 | 0 | 0 |
| 0 | 0 | 0 | 0 | 0 |
| 0 | 0 | 0 | 0 | 0 |
| 0 | 0 | 0 | 0 | 0 |
| 0 | 0 | 0 | 0 | 0 |
| 0 | 0 | 0 | 0 | 0 |
| 0 | 0 | 0.003185868 | 0 | 0 |
| 0 | 0 | 0 | 0 | 0 |
| 0 | 0 | 0 | 0 | 0 |
| 0 | 0 | 0 | 0.007279788 | 0 |
| 0 | 0 | 0 | 0.020620255 | 0.009437458 |
| 0 | 0 | 0 | 0 | 0 |
| 0 | 0 | 0 | 0 | 0 |
| 0 | 0 | 0 | 0 | 0 |
| 0 | 0 | 0 | 0 | 0 |
| 0 | 0 | 0 | 0 | 0 |
| 0 | 0 | 0 | 0 | 0 |
| 0 | 0 | 0 | 0.002539035 | 0 |
| 0 | 0 | 0 | 0 | 0 |
| 0 | 0 | 0 | 0 | 0 |
| 0 | 0 | 0 | 0 | 0 |
| 0 | 0 | 0 | 0.006179121 | 0.002272543 |
| 0 | 0 | 0 | 0 | 0 |
| 0 | 0 | 0 | 0 | 0 |
| 0 | 0 | 0 | 0 | 0 |
| 0 | 0 | 0 | 0 | 0 |
| 0 | 0 | 0 | 0 | 0 |

| | | | | |
|---|---|---|---|---|
| 0 | 0 | 0.000991893 | 0.0054865 | 0 |
| 0 | 0 | 0 | 0 | 0 |
| 0 | 0 | 0 | 0 | 0 |
| 0 | 0 | 0 | 0 | 0 |
| 0 | 0 | 0 | 0.035762251 | 0.061777923 |
| 0 | 0 | 0 | 0 | 0 |
| 0 | 0 | 0 | 0 | 0 |
| 0 | 0 | 0 | 0 | 0 |
| 0 | 0 | 0 | 0 | 0 |
| 0 | 0 | 0 | 0 | 0 |
| 0 | 0 | 0 | 0 | 0 |
| 0 | 0 | 0 | 0 | 0 |
| 0 | 0 | 0 | 0 | 0 |
| 0 | 0 | 0 | 0 | 0 |
| 0 | 0 | 0 | 0 | 0 |
| 0 | 0 | 0 | 9.997541014 | 0 |
| 0 | 0 | 0 | 0 | 0 |
| 0 | 0 | 0 | 0 | 0 |
| 0 | 0 | 0 | 0 | 0 |
| 0 | 0 | 0 | 0 | 0 |
| 0 | 0 | 0 | 0 | 0 |
| 0 | 0 | 0 | 0 | 0 |
| 0 | 0 | 0 | 0 | 0 |
| 0 | 0 | 0 | 0 | 0 |
| 0 | 0.008990411 | 0 | 0 | 0 |
| 0 | 0 | 0 | 0 | 0 |
| 0 | 0 | 0 | 0.001825113 | 0 |
| 0 | 0 | 0 | 0 | 0 |
| 0 | 0 | 0 | 0 | 0 |
| 0 | 0 | 0 | 0 | 0 |
| 0 | 0 | 0 | 0 | 0 |
| 0 | 0 | 0.005301607 | 0 | 0 |
| 0 | 0 | 0 | 0 | 0 |
| 0 | 0 | 0 | 0 | 0 |
| 0 | 0 | 0 | 0 | 0 |
| 0 | 0 | 0 | 0 | 0 |
| 0 | 0 | 0 | 0 | 0 |
| 0 | 0.233675656 | 0 | 0 | 0 |
| 0 | 0 | 0 | 0 | 0 |
| 0 | 0 | 0 | 0 | 0 |
| 0 | 0 | 0 | 0 | 0 |
| 0 | 0 | 0 | 0 | 0 |
| 0 | 0 | 0 | 0 | 0 |
| 0 | 0 | 0 | 0 | 0 |
| 0 | 0 | 0 | 0 | 0 |
| 0 | 0 | 0 | 0 | 0 |
| 0 | 0 | 0 | 0 | 0 |
| 0 | 0 | 0 | 0 | 0.005006204 |

| | | | | |
|---|---|---|---|---|
| 0 | 0 | 0 | 0 | 0 |
| 0 | 0 | 0 | 0 | 0 |
| 0 | 0 | 0 | 0 | 0 |
| 0 | 0 | 0 | 0 | 0 |
| 0 | 0 | 0 | 0 | 0 |
| 0 | 0 | 0 | 0 | 0 |
| 0 | 0 | 0 | 0 | 0 |
| 0 | 0 | 0 | 0 | 0 |
| 0 | 0 | 0 | 0 | 0 |
| 0 | 0 | 0 | 0 | 0 |
| 0 | 0 | 0 | 0 | 0 |
| 0 | 0 | 0 | 0 | 0 |
| 0 | 0 | 0 | 0.001180457 | 0 |
| 0 | 0 | 0 | 0 | 0 |
| 0 | 0 | 0 | 0 | 0 |
| 0 | 0 | 0 | 0 | 0 |
| 0 | 0 | 0 | 0 | 0 |
| 0 | 0 | 0 | 0 | 0 |
| 0 | 0 | 0 | 0 | 0 |
| 0 | 0 | 0 | 0 | 0 |
| 0.010359674 | 0.039569291 | 0.096314419 | 0.060854877 | 0.10146859 |
| 0 | 0 | 0 | 0 | 0 |
| 0 | 0 | 0.002080737 | 0 | 0 |
| 0.002763572 | 0 | 0 | 0 | 0 |
| 0 | 0 | 0 | 0 | 0 |
| 0 | 0 | 0 | 0 | 0 |
| 0.009119999 | 0.039175178 | 0.006798939 | 0.009181463 | 0.001654602 |
| 0 | 0 | 0 | 0 | 0 |
| 0 | 0 | 0 | 0 | 0 |
| 0 | 0 | 0.004546301 | 0 | 0 |
| 0.692321482 | 0 | 0.184260232 | 0 | 0 |
| 0 | 0 | 0 | 0 | 0 |
| 0 | 0 | 0 | 0 | 0 |
| 0 | 0.048633278 | 0.068671623 | 0.028712008 | 0 |
| 0.002037843 | 0 | 0.003323266 | 0.037749008 | 0 |
| 0 | 0 | 0 | 0 | 0 |
| 0 | 0 | 0 | 0 | 0 |
| 0 | 0 | 0 | 0 | 0 |
| 0 | 0 | 0 | 0.112351665 | 0 |
| 0 | 0 | 0 | 0 | 0 |
| 0 | 0 | 0 | 0 | 0 |
| 0 | 0 | 0 | 0 | 0 |
| 0.000814657 | 0 | 0 | 0 | 0 |
| 0 | 0 | 0 | 0 | 0 |
| 0 | 0 | 0 | 0 | 0 |
| 0 | 0 | 0 | 0 | 0 |
| 0 | 0 | 0.002056122 | 0 | 0 |
| 0 | 0 | 0.006597265 | 0.002046812 | 0 |

| | | | | |
|---|---|---|---|---|
| 0 | 0 | 0 | 0 | 0 |
| 0 | 0 | 0 | 0 | 0 |
| 0 | 0 | 0 | 0 | 0.002407536 |
| 0 | 0 | 0 | 0 | 0 |
| 0 | 0 | 0 | 0 | 0 |
| 0 | 0 | 0 | 0 | 0 |
| 0 | 0 | 0 | 0.049947628 | 0 |
| 0 | 0 | 0 | 0.004623662 | 0 |
| 0 | 0 | 0 | 0 | 0 |
| 0 | 0 | 0 | 0 | 0 |
| 0 | 0 | 0 | 0 | 0 |
| 0 | 0 | 0 | 0 | 0 |
| 0 | 0 | 0 | 0 | 0 |
| 0 | 0 | 0 | 0 | 0 |
| 0 | 0 | 0.002133682 | 0 | 0 |
| 0 | 0 | 0 | 0 | 0 |
| 0 | 0 | 0 | 0 | 0 |
| 0 | 0 | 0 | 0.00693516 | 0 |
| 0 | 0 | 0 | 0.00446056 | 0.000956955 |
| 0 | 0 | 0 | 0.161015304 | 0 |
| 0.107755801 | 0 | 0.008374394 | 0 | 0 |
| 0 | 0 | 0 | 0 | 0 |
| 0 | 0 | 0 | 0 | 0 |
| 0 | 0 | 0 | 0 | 0 |
| 0.005992899 | 0.02491073 | 0.024704172 | 0.013943599 | 0.011217819 |
| 0 | 0.00294793 | 0 | 0 | 0 |
| 0 | 0 | 0 | 0 | 0 |
| 0 | 0 | 0 | 0 | 0 |
| 0.00301842 | 0 | 0 | 0.024534006 | 0 |
| 0 | 0 | 0 | 0 | 0 |
| 0.016019072 | 0 | 0.769151252 | 0.194169472 | 0.025948844 |
| 0 | 0 | 0 | 0 | 0 |
| 0 | 0 | 0 | 0 | 0 |
| 0 | 0.000872796 | 0 | 0 | 0 |
| 0 | 0 | 0 | 0 | 0 |
| 0 | 0 | 0 | 0 | 0 |
| 0 | 0 | 0 | 0 | 0 |
| 0 | 0 | 0 | 0 | 0 |
| 0 | 0 | 0 | 0 | 0 |
| 0 | 0.04402438 | 0 | 0 | 0 |
| 0 | 0 | 0 | 0 | 0 |
| 0 | 0 | 0.005325718 | 0.059179781 | 0 |
| 0 | 0 | 0 | 0 | 0 |
| 0 | 0 | 0 | 0 | 0 |
| 0 | 0 | 0 | 0 | 0 |
| 0 | 0 | 0 | 0 | 0 |
| 0 | 0 | 0 | 0 | 0 |
| 0 | 0 | 0 | 0 | 0 |

| | | | | |
|---|---|---|---|---|
| 0 | 0 | 0 | 0 | 0 |
| 0 | 0 | 0 | 0 | 0 |
| 0 | 0.745455849 | 0.010986304 | 0.030962481 | 0.067488805 |
| 0 | 0 | 0 | 0 | 0 |
| 0 | 0 | 0 | 0 | 0 |
| 0 | 0 | 0 | 0 | 0 |
| 0 | 0 | 0 | 0 | 0 |
| 0 | 0 | 0 | 0 | 0 |
| 0 | 0 | 0 | 0 | 0 |
| 0 | 0 | 0 | 0.000566361 | 0.003402151 |
| 0 | 0 | 0 | 0 | 0 |
| 0 | 0 | 0 | 0 | 0 |
| 0 | 0 | 0 | 0 | 0 |
| 0.00087154 | 0.014632322 | 0 | 0.006615699 | 0.002642858 |
| 0.0027335 | 0 | 0.002178589 | 0.00320092 | 0.002181333 |
| 0 | 0 | 0 | 0.264081182 | 0 |
| 0.001147476 | 0.005384635 | 0.004219921 | 0.006650706 | 0.003037752 |
| 0 | 0 | 0.027150493 | 0 | 0.001071107 |
| 0 | 0 | 0 | 0 | 0 |
| 0 | 0 | 0 | 0 | 0 |
| 0 | 0 | 0 | 0 | 0 |
| 0 | 0 | 0.006722443 | 0.001355672 | 0 |
| 0 | 0 | 0 | 0 | 0 |
| 0 | 0 | 0.005558037 | 0 | 0 |
| 0.00403365 | 0 | 0.006108125 | 0 | 0 |
| 0 | 0 | 0 | 0 | 0 |
| 0 | 0.032262724 | 0 | 0 | 0 |
| 0 | 0 | 0 | 0 | 0 |
| 0 | 0 | 0 | 0 | 0 |
| 0 | 0 | 0.057047908 | 0 | 0 |
| 0 | 0 | 0 | 0 | 0.002292446 |
| 0 | 0 | 0 | 0 | 0 |
| 0.001837493 | 0 | 0 | 0.003206453 | 0 |
| 0.003071119 | 0 | 0 | 0 | 0 |
| 0 | 0 | 0 | 0 | 0 |
| 0 | 0 | 0 | 0.831282893 | 0 |
| 0 | 0 | 0 | 0 | 0 |
| 0 | 0.310782394 | 0.002533816 | 0.169352846 | 0.009709536 |
| 0 | 0 | 0 | 0 | 0 |
| 0 | 0 | 0 | 0 | 0 |
| 0 | 0.003917439 | 0 | 0 | 0 |
| 0 | 0 | 0 | 0 | 0 |
| 0 | 0 | 0 | 0 | 0 |
| 0 | 0 | 0 | 0 | 0 |
| 0 | 0 | 0 | 0 | 0 |
| 0.142637603 | 0 | 0 | 0 | 0 |
| 0 | 0 | 0 | 0 | 0 |
| 0.006530627 | 0 | 0 | 0 | 0 |

| | | | | |
|---|---|---|---|---|
| 0 | 0.049578748 | 0 | 0 | 2.084865446 |
| 0 | 0 | 0 | 0 | 0.081476193 |
| 0.002789911 | 0.099280294 | 0.001642955 | 0.000811406 | 0 |
| 0 | 0 | 0 | 0 | 0 |
| 0 | 0 | 0.002833664 | 0.007959439 | 0.008406619 |
| 0.001017097 | 0 | 0.002310273 | 0 | 0 |
| 0 | 0 | 0 | 0 | 0 |
| 0.004153099 | 0.014759056 | 0.001324001 | 0.005149341 | 0.027986352 |
| 0 | 0 | 0 | 0 | 0 |
| 0.645257615 | 0.039881362 | 0.029681724 | 0.046883249 | 0.00341599 |
| 0 | 0 | 0 | 0 | 0 |
| 0.00173032 | 0 | 0 | 0.147650362 | 0.001166005 |
| 0.052894124 | 0.029700393 | 0.087053501 | 0.00755815 | 0.031840425 |
| 0 | 0 | 0.011470431 | 0.022696877 | 0 |
| 0 | 0 | 0.004905893 | 0 | 0 |
| 0.004137618 | 0.019416144 | 0 | 0 | 0.00298736 |
| 0 | 0 | 0 | 0 | 0 |
| 0 | 0 | 0 | 0 | 0 |
| 0.421885317 | 0 | 2.342906435 | 0 | 0 |
| 0 | 0 | 0.469624806 | 0 | 0 |
| 0 | 0 | 0 | 0 | 0 |
| 0 | 0 | 0 | 0 | 0 |
| 0.030201782 | 0 | 0 | 0 | 0.002281611 |
| 0 | 0 | 0 | 0.082361361 | 0 |
| 0 | 0 | 0 | 0 | 0 |
| 0.001528739 | 0.318593921 | 0 | 0 | 0 |
| 0 | 0 | 0 | 0 | 0 |
| 0 | 0 | 1.118651031 | 0 | 0 |
| 0.054312714 | 0 | 0 | 0 | 0 |
| 0 | 0 | 0 | 0 | 0 |
| 0.004265091 | 0 | 0.029560494 | 0 | 0 |
| 0 | 0 | 0 | 0 | 0 |
| 0 | 0 | 0 | 0 | 0 |
| 0.133153998 | 0 | 0.265515162 | 0 | 0 |
| 0 | 0 | 0 | 0 | 0 |
| 0.02867912 | 0 | 0 | 0 | 0.027944212 |
| 0.271038371 | 0 | 0 | 0 | 0 |
| 0 | 0 | 0 | 0 | 0 |
| 0 | 0 | 0 | 0 | 0 |
| 0.090521493 | 0 | 0 | 0 | 0 |
| 0 | 0 | 0 | 0 | 0 |
| 0.405937135 | 0 | 0 | 0 | 0 |
| 0 | 0 | 0 | 0 | 0 |
| 0 | 0 | 0 | 0 | 0.001867361 |
| 0 | 0 | 0 | 0 | 0 |
| 0 | 0 | 0 | 0 | 0 |
| 0 | 0 | 0.004318249 | 0.01169405 | 0 |
| 0 | 0 | 0 | 0 | 0 |

| | | | | |
|---|---|---|---|---|
| 0 | 0 | 0 | 0 | 0 |
| 0.082647735 | 0.003877211 | 0 | 0 | 0.006442699 |
| 0 | 0 | 0 | 0 | 0 |
| 0 | 0 | 0 | 0 | 0 |
| 0.002818568 | 0 | 0 | 0 | 0 |
| 0.009583055 | 0 | 0 | 0.022144348 | 0 |
| 0 | 0 | 0 | 0.068422257 | 0 |
| 0.012588158 | 0 | 0 | 0 | 0 |
| 0 | 0 | 0 | 0 | 0 |
| 0 | 0 | 0 | 0 | 0 |
| 0.0059047 | 0.010141613 | 0.008678459 | 0.017046713 | 0.007899447 |
| 0 | 0 | 0 | 0 | 0 |
| 0 | 0 | 0 | 0 | 0 |
| 0.004265884 | 0.000898246 | 0.002484537 | 0.014172292 | 0.001934852 |
| 0.467323368 | 0 | 0.4786475 | 0 | 0 |
| 0 | 0 | 0 | 0 | 0 |
| 0 | 0 | 0 | 0 | 0 |
| 0 | 0 | 0 | 0 | 0 |
| 0 | 0 | 0 | 0 | 0 |
| 0 | 0 | 0 | 0.15361134 | 0.009447377 |
| 0 | 0.010323443 | 0 | 0 | 0 |
| 0 | 0.951922035 | 0 | 0.502063817 | 0 |
| 0.009030422 | 0 | 0 | 0 | 0 |
| 0 | 0 | 0 | 0 | 0 |
| 0 | 0 | 0 | 0 | 0 |
| 0 | 0 | 0 | 0 | 0 |
| 0.234825357 | 0 | 0 | 0 | 0 |
| 0 | 0 | 0 | 0 | 0 |
| 0 | 0 | 0 | 0 | 0 |
| 0.803977717 | 0.127275191 | 0.363452724 | 0.452927698 | 0.790458166 |
| 0.002337513 | 0 | 0 | 0.003204925 | 0.004500491 |
| 0 | 0 | 0 | 0 | 0 |
| 0 | 0 | 0 | 0 | 0 |
| 0 | 0 | 0.004297197 | 0 | 0 |
| 0.014520883 | 0 | 0 | 0 | 0 |
| 0 | 0 | 0 | 0 | 0 |
| 0 | 0 | 0 | 1.23534985 | 0 |
| 0.00284297 | 0 | 0 | 0 | 0 |
| 0 | 0 | 0 | 0 | 0.207948022 |
| 2.297061211 | 0 | 2.54855002 | 0 | 0 |
| 0.007418698 | 0 | 0.001429792 | 0 | 0.02931357 |
| 0 | 0.139722662 | 0 | 1.778334211 | 1.945734872 |
| 0.003217132 | 0 | 0 | 0 | 0 |
| 0 | 0 | 0 | 0 | 0 |
| 0.000760789 | 0 | 0.012211803 | 0 | 0 |
| 0 | 0 | 0 | 0 | 0 |
| 0.000849432 | 0 | 0.00321572 | 0.001482271 | 0.000715505 |
| 0 | 0 | 0 | 0 | 0 |

| | | | | |
|---|---|---|---|---|
| 0 | 0.06024669 | 0 | 0 | 0 |
| 0 | 0 | 0 | 0.078734172 | 0 |
| 2.934418418 | 2.24164157 | 0.009370813 | 0.002180483 | 0.011227064 |
| 0 | 0 | 0 | 0 | 0 |
| 0 | 0 | 0 | 0 | 0 |
| 0.001033821 | 0 | 0 | 0 | 0 |
| 0 | 0 | 0 | 0 | 0 |
| 0 | 0 | 0 | 0 | 0 |
| 0 | 0 | 0.085228071 | 0 | 0 |
| 0 | 0 | 0 | 0 | 0 |
| 0 | 0 | 0 | 0 | 0.008626369 |
| 0 | 0 | 0 | 0 | 0 |
| 0 | 0 | 0 | 0 | 0 |
| 0.000790125 | 0 | 0 | 0.118506125 | 0 |
| 0 | 0 | 0 | 0 | 0 |
| 0 | 0 | 0.014960457 | 0.202029869 | 0.086339037 |
| 0.0041064 | 0.399916591 | 0 | 0.003045435 | 0.007609706 |
| 0.014801043 | 0 | 0.024679601 | 0 | 0 |
| 0 | 0 | 0 | 0 | 0 |
| 0 | 0 | 0 | 0 | 0 |
| 0.232885442 | 0 | 0 | 0.002031943 | 0.016855846 |
| 0 | 0 | 0 | 0.018493919 | 0.001337404 |
| 0 | 0 | 0 | 0 | 0 |
| 0 | 0 | 0.35968465 | 0 | 0.16647589 |
| 0 | 0 | 0 | 0 | 0 |
| 0 | 0 | 0 | 0 | 0 |
| 0 | 0 | 0.04868071 | 0 | 0 |
| 0 | 0 | 0 | 0 | 0 |
| 0 | 0 | 0 | 0 | 0 |
| 0 | 0 | 0.017787439 | 0 | 0 |
| 0.017840491 | 0 | 0 | 0.034072163 | 0.007346846 |
| 0.001773199 | 0.019415421 | 0.002386793 | 0.013666309 | 0.022902242 |
| 0 | 0.017116765 | 0.003124755 | 0.394784455 | 0 |
| 0.046508828 | 0 | 0.055772436 | 0.08725723 | 0.005887713 |
| 0 | 0.021710136 | 0.001743387 | 0 | 0 |
| 0 | 0 | 0 | 0 | 0 |
| 0 | 0.005570905 | 0.004879535 | 0.000887842 | 0 |
| 0.012865282 | 0.004598745 | 0.01586662 | 0.039056251 | 0.01362986 |
| 0 | 0.015463113 | 0 | 0.000773699 | 0 |
| 0 | 0 | 0 | 0 | 0.00310565 |
| 0 | 0 | 0 | 0 | 0 |
| 0 | 0 | 0 | 0 | 0 |
| 0 | 0 | 0.002402887 | 0 | 0 |
| 0 | 0 | 0.004389923 | 0.271683899 | 0.021628417 |
| 0.08246169 | 0 | 0 | 0 | 0 |
| 0 | 0 | 0 | 0 | 0 |
| 0 | 0 | 0 | 0 | 0 |
| 0 | 0 | 0 | 0 | 0 |

| | | | | |
|---|---|---|---|---|
| 0 | 0 | 0.004168975 | 0 | 0 |
| 0.482181297 | 2.750608061 | 0.539959731 | 0.838023386 | 0.335216849 |
| 0 | 0 | 0 | 0 | 0 |
| 0 | 0 | 0 | 0 | 0 |
| 0 | 0 | 0.040810102 | 0 | 0 |
| 0 | 0 | 0 | 0 | 0 |
| 0.66176411 | 0 | 0.165835869 | 0 | 0 |
| 0.001249361 | 0 | 0 | 0 | 0 |
| 0 | 0 | 0 | 0.003897416 | 0.010033682 |
| 0 | 0 | 0 | 0 | 0 |
| 0 | 0 | 0.039860531 | 0.01107333 | 0 |
| 0.006201371 | 0.165372145 | 0.013934571 | 0.032726269 | 0.020557479 |
| 0 | 0 | 0 | 0 | 0 |
| 0 | 0 | 0 | 0 | 0 |
| 0.000770464 | 0.01518499 | 0 | 0 | 0 |
| 0 | 0 | 0 | 0 | 0 |
| 0.026720058 | 0.049274606 | 0.116913833 | 0.03809909 | 0.006396782 |
| 1.34564693 | 0 | 0.384816514 | 0 | 0.001230208 |
| 0 | 0 | 0 | 0 | 0 |
| 0.075157324 | 0 | 0.026926695 | 0 | 0.004992707 |
| 0.02001372 | 0.039169549 | 0.010541434 | 0.013666008 | 0.015880935 |
| 0 | 0.000987977 | 0.006558561 | 0 | 0.000912059 |
| 0 | 0 | 0 | 0 | 0 |
| 0 | 0 | 0 | 0 | 0 |
| 0.015351357 | 0.012450947 | 0 | 0 | 0 |
| 0 | 0 | 0.002013915 | 0 | 0 |
| 0.092331009 | 0 | 0 | 0.04196588 | 0.026045075 |
| 0 | 0.04500697 | 0.001167082 | 0.041355758 | 0.062322792 |
| 0 | 0 | 0 | 0 | 0 |
| 0 | 0 | 0.002533994 | 0 | 0 |
| 0.002150132 | 0.007243878 | 0 | 0 | 0 |
| 0 | 0 | 0 | 0 | 0 |
| 0 | 0 | 0 | 0 | 0 |
| 0 | 0 | 0.008485644 | 0 | 0 |
| 0 | 0.010540494 | 0 | 0 | 0 |
| 7.507505327 | 0.09045379 | 0.252432722 | 0.267677307 | 0.7669101 |
| 0 | 0 | 0.008782097 | 0 | 0 |
| 0 | 0 | 0 | 0 | 0 |
| 0.002852275 | 0 | 0.004607124 | 0.138865658 | 0.009930609 |
| 0 | 0 | 0 | 0 | 0 |
| 0.08424075 | 0 | 0 | 0 | 0 |
| 0 | 0 | 0 | 0 | 0 |
| 0.013975626 | 0 | 0.001224317 | 0.03113964 | 0.026073822 |
| 0.002230282 | 0 | 0.002195244 | 0 | 0 |
| 0.037065424 | 0 | 0 | 0 | 0 |
| 0 | 0 | 0 | 0.10540333 | 0 |
| 0 | 0 | 0 | 0 | 0.011011937 |
| 0.000721015 | 0 | 0.010645326 | 0 | 0 |

| | | | | |
|---|---|---|---|---|
| 0 | 0 | 0 | 0 | 0 |
| 0 | 0 | 0 | 0 | 0 |
| 0 | 0 | 0 | 0 | 0 |
| 0 | 0.750950762 | 0 | 5.286651933 | 0.104927382 |
| 0 | 0 | 0 | 0 | 0 |
| 0 | 0 | 0 | 0 | 0 |
| 0 | 0 | 0 | 0 | 0 |
| 0 | 0 | 0 | 0 | 0.212471919 |
| 0 | 2.474197391 | 0 | 0.697182238 | 1.264035481 |
| 0 | 0 | 0 | 0.31281583 | 0 |
| 0 | 0 | 0 | 0 | 0 |
| 0 | 0 | 0 | 0.004891776 | 0.007382458 |
| 0 | 0 | 0 | 0 | 0 |
| 0 | 0 | 0 | 0 | 0 |
| 0.00404159 | 0.002911374 | 0 | 1.031030353 | 0.071312508 |
| 0 | 0 | 0 | 0 | 0 |
| 0 | 0.076712179 | 0 | 0.102587032 | 0.383536 |
| 0 | 0.016727992 | 0 | 0 | 0 |
| 0 | 0 | 0 | 0 | 0 |
| 0 | 0 | 0 | 0 | 0 |
| 0 | 0 | 0 | 0 | 0 |
| 0 | 0 | 0 | 0 | 0 |
| 0 | 0 | 0 | 0 | 0 |
| 0.342284309 | 0 | 0.238758525 | 0.069454554 | 0.271489616 |
| 0 | 0 | 0.03800805 | 0.132186899 | 0 |
| 0 | 0 | 0 | 0 | 0 |
| 0 | 0 | 0 | 0 | 0 |
| 0 | 0 | 0 | 2.554909053 | 2.249921375 |
| 0 | 0 | 0 | 0 | 0 |
| 0 | 0 | 0 | 0 | 0 |
| 0 | 0 | 0 | 0 | 0.021108848 |
| 0 | 0 | 0 | 0.051914262 | 0 |
| 0 | 0 | 0 | 0 | 0.002409807 |
| 0 | 0 | 0 | 0 | 0 |
| 0 | 0 | 0 | 0 | 0 |
| 0 | 0 | 0 | 0 | 0 |
| 0 | 0.126949629 | 0 | 2.317055838 | 3.618236267 |
| 0 | 0 | 0 | 0 | 0 |
| 0 | 0 | 0 | 0 | 0.03277486 |
| 0 | 0 | 0 | 0 | 0 |
| 0.053414099 | 0 | 0.094927025 | 0.042720521 | 0.007915343 |
| 0 | 0 | 0 | 0.081936786 | 0 |
| 0.218780632 | 14.83971968 | 5.842971133 | 1.257141444 | 0.142181504 |
| 0 | 0 | 0 | 0 | 0 |
| 0 | 0 | 0 | 0 | 0 |
| 0 | 0 | 0 | 0 | 0 |
| 0 | 0 | 0 | 0 | 0 |
| 0 | 0 | 0.006977334 | 0.047075386 | 0.000751196 |

| | | | | |
|---|---|---|---|---|
| 0 | 0 | 0 | 0 | 0 |
| 0 | 0 | 0 | 0 | 0 |
| 0.161005156 | 0.010660641 | 0.99077028 | 0.008204922 | 0.069850306 |
| 0 | 0 | 0 | 0 | 0 |
| 0 | 0 | 0 | 0 | 0 |
| 0 | 0.049978592 | 0 | 0 | 0.024581784 |
| 0 | 0.000786737 | 0 | 0 | 0 |
| 0 | 0 | 0 | 0 | 0 |
| 0 | 0 | 0 | 0 | 0 |
| 0 | 0 | 0 | 0 | 0 |
| 0.00223828 | 0.001276446 | 0.327501757 | 0.700303478 | 0.011547938 |
| 0 | 0.948327946 | 0.774181017 | 2.323129529 | 0.116755691 |
| 0 | 0 | 0 | 0 | 0 |
| 0 | 0.994346482 | 0 | 0.000592781 | 0.0043239 |
| 0.195239941 | 0 | 0.001621483 | 0.030519443 | 0.070152333 |
| 0 | 0.002091664 | 0 | 0 | 0 |
| 0 | 0 | 0 | 0 | 0 |
| 0 | 0 | 0 | 0 | 0 |
| 0 | 0 | 0 | 0 | 0 |
| 0.001037111 | 0 | 0.012912916 | 0 | 0 |
| 0.147735637 | 0.00534925 | 0.286923309 | 0.217442796 | 0.300835173 |
| 0.010995563 | 0.878609741 | 0 | 0 | 0.007284664 |
| 1.027943849 | 4.396075556 | 0.739564692 | 1.715509927 | 0.288437221 |
| 0 | 0 | 0 | 0 | 0 |
| 0 | 0 | 0 | 0 | 0 |
| 0 | 0.002820402 | 0 | 0 | 0.003787165 |
| 0 | 0 | 0 | 0 | 0 |
| 0 | 0 | 0 | 0 | 0 |
| 0 | 0 | 0 | 0 | 0 |
| 0 | 0 | 0 | 0 | 0 |
| 0 | 0 | 0 | 0 | 0 |
| 0 | 0 | 0 | 0.954991774 | 0.094546167 |
| 0.061779488 | 0 | 0 | 0.041367472 | 0 |
| 0 | 0 | 0 | 0.284199884 | 0 |
| 0.000853435 | 0 | 0.050688847 | 0 | 0.016773792 |
| 0 | 0.268665521 | 0 | 0.065306227 | 0.03226463 |
| 0 | 0 | 0 | 0.04381355 | 0.002120217 |
| 0 | 0 | 0 | 0 | 0 |
| 0 | 0.074563983 | 0 | 1.032787325 | 0 |
| 0 | 0 | 0 | 0 | 0 |
| 0 | 0 | 0 | 0 | 0 |
| 0 | 0 | 0 | 0 | 0 |
| 0 | 0 | 0 | 0 | 0 |
| 0 | 0 | 0 | 0 | 0 |
| 0.005200145 | 0 | 0.189888921 | 0.159643425 | 0.176461729 |
| 0 | 0 | 0 | 0 | 0 |
| 0 | 0 | 0 | 0 | 0 |
| 0 | 6.603399575 | 0 | 0.003049277 | 0.406398577 |

| | | | | |
|---|---|---|---|---|
| 0 | 0 | 0 | 0 | 0 |
| 0 | 0 | 0 | 0 | 0 |
| 0 | 0 | 0 | 0 | 0 |
| 0 | 0 | 0 | 0.028339645 | 0.002986619 |
| 0 | 0 | 0 | 0 | 0 |
| 0 | 0 | 0 | 0.529715681 | 0 |
| 0 | 0 | 0 | 0 | 0 |
| 0 | 0 | 0.13180791 | 1.425575666 | 0.001570417 |
| 0 | 0 | 0 | 0 | 0 |
| 0 | 0 | 0 | 0 | 0 |
| 0 | 0 | 0.020502215 | 0.001845781 | 0 |
| 0 | 0 | 0 | 0 | 0 |
| 0 | 0.00090139 | 0 | 0 | 0 |
| 0 | 0 | 0 | 0 | 2.107128671 |
| 0 | 0 | 0 | 0 | 0 |
| 0 | 0 | 0 | 0 | 0 |
| 0 | 0.188161454 | 0 | 0 | 0 |
| 0 | 0.007284658 | 0 | 0 | 0.000791163 |
| 0 | 0 | 0 | 0 | 0 |
| 0.000741169 | 0 | 0 | 0 | 0.005351237 |
| 0 | 0 | 0 | 0 | 0.004217317 |
| 0 | 0 | 0 | 0 | 0 |
| 0 | 0 | 0 | 0.042340194 | 0.028732871 |
| 0 | 0 | 0 | 0 | 0 |
| 0 | 0 | 0 | 0 | 0 |
| 0 | 0 | 0 | 0 | 0 |
| 0 | 0 | 0 | 0 | 0 |
| 0 | 0 | 0 | 0 | 0 |
| 0 | 0 | 0 | 0 | 0 |
| 0.003403824 | 0.029764036 | 0 | 0.47443548 | 0.013618984 |
| 0 | 0 | 0.013624687 | 0 | 0 |
| 0 | 0 | 0 | 0 | 0 |
| 0 | 0 | 0 | 0 | 0 |
| 0 | 0 | 0 | 0 | 0 |
| 0 | 0.010437053 | 0 | 0 | 0.011713191 |
| 0 | 0 | 0 | 0 | 0 |
| 0 | 0.001843068 | 0 | 0.055118787 | 0 |
| 0.001910186 | 0 | 0.02729873 | 0 | 0 |
| 0 | 0 | 0 | 0 | 0 |
| 0.000706538 | 0 | 0 | 0 | 0 |
| 0 | 0 | 0 | 0 | 0 |
| 0 | 0 | 0 | 0 | 0 |
| 0 | 0 | 0 | 0.00722184 | 0 |
| 0 | 0.007828306 | 0 | 0 | 0 |
| 0 | 0.066299742 | 0 | 0 | 0 |
| 0 | 0 | 0 | 0 | 0 |
| 0 | 0 | 0 | 0 | 0.004395393 |
| 0 | 0 | 0 | 0 | 0 |

| | | | | |
|---|---|---|---|---|
| 0 | 0 | 0 | 0 | 0 |
| 0 | 0 | 0 | 0 | 0 |
| 0 | 0 | 0 | 0 | 0 |
| 0 | 0 | 0 | 0.990035465 | 0.08322584 |
| 0 | 0 | 0 | 0 | 0 |
| 0 | 0 | 0 | 0 | 0 |
| 0 | 0 | 0 | 0 | 0 |
| 0 | 0 | 0 | 0 | 0 |
| 0.296542069 | 0 | 0.109208505 | 0.008188732 | 0.013591978 |
| 0 | 0 | 0 | 0 | 0 |
| 0 | 0 | 0 | 0 | 0 |
| 0 | 0 | 0 | 0 | 0 |
| 0 | 0 | 0 | 0 | 0 |
| 0 | 0 | 0 | 0 | 0 |
| 0 | 0.122000704 | 7.998879567 | 1.637942751 | 0.016345703 |
| 0.388324737 | 0 | 0.395676018 | 0 | 0 |
| 0 | 0 | 0.024185983 | 0.002497876 | 0 |
| 0 | 0 | 0 | 0 | 0 |
| 0 | 0 | 0 | 0 | 0 |
| 0 | 0 | 0 | 0 | 0 |
| 0 | 0 | 0 | 0.11534268 | 0 |
| 0 | 0 | 0 | 0 | 0 |
| 0 | 0 | 0 | 0 | 0 |
| 0 | 0 | 0.392717914 | 0 | 0 |
| 0 | 0.009187152 | 0 | 0.049162231 | 0.002283397 |
| 0.748262233 | 2.000625161 | 0 | 3.765037406 | 3.782947175 |
| 0 | 0.00131426 | 0 | 0.008294426 | 0 |
| 0 | 0 | 0 | 0.002044973 | 0 |
| 0 | 0.328393308 | 0 | 0.006646678 | 0 |
| 0 | 0 | 0 | 1.054203593 | 0 |
| 0 | 0 | 0 | 0 | 0 |
| 0 | 0 | 0 | 0 | 0 |
| 0 | 0 | 0 | 0 | 0 |
| 0 | 0 | 0.15346124 | 0.014210607 | 0.004529499 |
| 0.003066969 | 3.087000684 | 0.003354208 | 0 | 0.001435228 |
| 0 | 0 | 0 | 0.004413674 | 0 |
| 0 | 0 | 0 | 0 | 0 |
| 4.883796954 | 0 | 0 | 0.969171265 | 0.455712523 |
| 0 | 0.017765912 | 0 | 0 | 0 |
| 0 | 0 | 0 | 0 | 0 |
| 0 | 0 | 0 | 0 | 0 |
| 0 | 0 | 0 | 0 | 0.013345655 |
| 6.42401794 | 0.155884362 | 0 | 3.90293881 | 1.576864153 |
| 0 | 0.002708276 | 0 | 0 | 0 |
| 0 | 0 | 0 | 0 | 0 |
| 0 | 0 | 0 | 0.001642694 | 0.00115337 |
| 0 | 0 | 0 | 0 | 0.216473309 |
| 0.006362385 | 0.113453039 | 0.017617392 | 0.206743626 | 0.563792995 |

| | | | | |
|---|---|---|---|---|
| 0 | 0 | 0 | 0 | 0.014843763 |
| 0 | 0 | 0 | 0 | 0 |
| 0 | 0 | 0 | 0 | 0 |
| 0 | 0 | 0 | 0 | 0 |
| 0 | 0 | 0.00408741 | 0.012364227 | 0.000909458 |
| 0 | 0.061902285 | 0 | 0.002211219 | 0 |
| 0 | 0 | 0 | 0 | 0 |
| 0 | 0 | 0 | 0 | 0 |
| 0 | 0 | 0 | 0 | 0 |
| 0 | 0 | 0 | 0 | 0 |
| 0 | 0 | 0 | 0 | 0 |
| 0.101351296 | 2.490992619 | 0 | 1.370770078 | 2.027941992 |
| 0 | 0.053380287 | 0 | 0.222211097 | 0 |
| 0 | 0 | 0 | 0 | 0 |
| 0.078197314 | 0 | 0.400857217 | 0.842224544 | 0.042860348 |
| 0 | 0 | 0 | 0 | 0 |
| 0 | 0 | 0.009207447 | 0.002415743 | 0 |
| 0 | 0 | 0 | 0 | 0 |
| 0 | 0 | 0.005179097 | 0 | 0 |
| 0 | 0 | 0 | 0 | 0 |
| 0 | 0.021496015 | 0 | 0.074709784 | 0 |
| 0 | 0.019883469 | 0 | 0 | 0 |
| 0 | 0.004369621 | 0 | 0.292484541 | 0.118326247 |
| 0 | 0 | 0 | 0 | 0 |
| 0 | 0 | 0 | 0 | 0 |
| 0 | 0 | 0 | 0 | 0 |
| 0 | 0.975575907 | 0 | 0 | 0 |
| 0 | 0 | 0 | 0 | 0 |
| 0 | 0 | 0 | 0 | 0 |
| 0 | 0 | 0 | 0 | 0 |
| 0 | 0 | 0 | 0 | 0 |
| 0.023697042 | 0.186284595 | 0.100291883 | 0.121669414 | 0.107140067 |
| 0 | 0 | 0 | 0 | 0 |
| 0 | 0 | 0 | 0.097995898 | 0.032047013 |
| 0 | 0 | 0 | 0.005660895 | 0 |
| 0 | 0.023434526 | 0 | 0.005749608 | 0.003700512 |
| 0 | 0 | 0.003699906 | 0 | 0 |
| 0 | 0 | 0 | 0 | 0 |
| 0 | 0.003847168 | 0 | 0 | 0 |
| 0 | 0 | 0 | 0 | 0 |
| 0 | 0 | 0 | 0.250362888 | 0.343719896 |
| 0 | 0 | 0 | 0 | 0 |
| 0 | 0 | 0 | 0 | 0 |
| 0 | 0 | 0 | 0.002234419 | 0 |
| 0 | 0 | 0.472582354 | 0.173379965 | 0.014138992 |
| 0 | 0 | 0 | 0 | 0 |
| 0 | 0 | 0 | 0 | 0 |
| 0 | 0 | 0.001109586 | 0.003356447 | 0.002057378 |

| | | | | |
|---|---|---|---|---|
| 0 | 0 | 0 | 0 | 0 |
| 0 | 0 | 0 | 0 | 0.014151175 |
| 0.00080125 | 0 | 0 | 0.013457609 | 0 |
| 0 | 0 | 0 | 0 | 0 |
| 0.001083444 | 0 | 0 | 0 | 0 |
| 0 | 0 | 0 | 0 | 0 |
| 0 | 0 | 0 | 0.509250365 | 0 |
| 0 | 0 | 0 | 0.005446909 | 0.063102439 |
| 0 | 0 | 0 | 0 | 0 |
| 0 | 0 | 0 | 0 | 0 |
| 0 | 0 | 0 | 0 | 0 |
| 0 | 0 | 0 | 0 | 0 |
| 0 | 0 | 0 | 0 | 0 |
| 0 | 0.013714791 | 0 | 0 | 0 |
| 0 | 0 | 0 | 0 | 0 |
| 0 | 0 | 0 | 0 | 0 |
| 0 | 0 | 0 | 0 | 0 |
| 0 | 0 | 0 | 0 | 0 |
| 0.004566196 | 0 | 0 | 1.849217347 | 0.027164206 |
| 0 | 0 | 0 | 0 | 0 |
| 0 | 0 | 0 | 0 | 0 |
| 0 | 0 | 0 | 0 | 0 |
| 0 | 0 | 0 | 0 | 0 |
| 0 | 0 | 0 | 0 | 0 |
| 0 | 0.0028319 | 0 | 0 | 0 |
| 0.0069719 | 0.000877448 | 0.001820258 | 0 | 0.000810023 |
| 0 | 0 | 0 | 0 | 0 |
| 0.018825112 | 0.041439359 | 0.094072193 | 0.143667882 | 0.201384151 |
| 0 | 0 | 0 | 0 | 0 |
| 0 | 0.470803433 | 0.200310122 | 0.679878551 | 0.584604877 |
| 0 | 0 | 0 | 0 | 0 |
| 0 | 0 | 0 | 0 | 0 |
| 0 | 0 | 0 | 0.003577084 | 0 |
| 0 | 0 | 0 | 0 | 0 |
| 0 | 0 | 0.005341049 | 0.012364612 | 0 |
| 0 | 0 | 0 | 0 | 0 |
| 0 | 0 | 0 | 0 | 0 |
| 0 | 0 | 0.0041125 | 0.130013312 | 0.002408 |
| 0 | 0 | 0 | 0 | 0 |
| 0 | 0 | 0 | 0 | 0 |
| 14.41514224 | 2.66960353 | 31.10694827 | 5.536596368 | 14.6105372 |
| 0 | 0.067592378 | 0.081913717 | 0.145425748 | 2.901702744 |
| 0 | 0 | 0 | 0 | 0.486186311 |
| 0 | 0 | 0 | 0 | 0 |
| 0 | 0 | 0 | 0 | 0 |
| 0 | 0 | 0 | 0 | 0 |
| 0 | 0 | 0 | 0 | 0 |
| 0 | 0 | 0.005668892 | 0.007126498 | 0.011275622 |

| | | | | |
|---|---|---|---|---|
| 0 | 0 | 0 | 0 | 0 |
| 0 | 0 | 0.050534671 | 0 | 0.001179448 |
| 0 | 0 | 0 | 0 | 0 |
| 0 | 0 | 0 | 0 | 0 |
| 4.11976031 | 1.492077774 | 0 | 4.012499323 | 4.994857684 |
| 0 | 0 | 0 | 0 | 0 |
| 0 | 0 | 0 | 0 | 0 |
| 0 | 0 | 0 | 0 | 0 |
| 1.363896487 | 0 | 3.007496473 | 0.004117154 | 0.012562233 |
| 0 | 0 | 0 | 0 | 0 |
| 0.000527317 | 0 | 0 | 0 | 0 |
| 0 | 0 | 0 | 0 | 0 |
| 0 | 0.015931677 | 0 | 0.129986187 | 0.093401853 |
| 0.035404541 | 0.007631056 | 0.116090771 | 0.016418301 | 0.04179836 |
| 0 | 0 | 0 | 0 | 0 |
| 0.022189865 | 0.013776376 | 0.370487427 | 0.321549372 | 0.35378515 |
| 0 | 0 | 0 | 0.001105027 | 0.00182882 |
| 0 | 0 | 0 | 0 | 0 |
| 0 | 0 | 0 | 0 | 0 |
| 0 | 0 | 0 | 0 | 0 |
| 0 | 0 | 0 | 0 | 0 |
| 0 | 0 | 0 | 0 | 0 |
| 0 | 0 | 0 | 0 | 0 |
| 0 | 0 | 0 | 0 | 0 |
| 0 | 0 | 0 | 0 | 0 |
| 0 | 0 | 0 | 0 | 0 |
| 0 | 0.012632423 | 0 | 0.0016472 | 0 |
| 0 | 0 | 0.00734779 | 0 | 0 |
| 0 | 0 | 0 | 0 | 0 |
| 0 | 0 | 0 | 0 | 0 |
| 0 | 0 | 0 | 0 | 0 |
| 0 | 0 | 0 | 0 | 0 |
| 0 | 0 | 0 | 0 | 0 |
| 0 | 0 | 0 | 0 | 0.066253412 |
| 0 | 0 | 0 | 0.207206704 | 0 |
| 0 | 0 | 0.001451197 | 0 | 0 |
| 0 | 0 | 0 | 0 | 0 |
| 0 | 0.010380344 | 0 | 0 | 0 |
| 0 | 0 | 0 | 0 | 0 |
| 0 | 0 | 0 | 0 | 0 |
| 0 | 1.905986759 | 0 | 0 | 2.398480816 |
| 0 | 3.135351588 | 0 | 0.158632882 | 0 |
| 0 | 0 | 0 | 0 | 0 |
| 0 | 0 | 0 | 0 | 0 |
| 0 | 0 | 0 | 0 | 0 |
| 0 | 0 | 0 | 0 | 0 |
| 0 | 0 | 0 | 0 | 0 |
| 0 | 0 | 0 | 0 | 0 |

| | | | | |
|---|---|---|---|---|
| 0.000518323 | 0.001891769 | 0 | 0.323917127 | 0.208476665 |
| 0 | 0 | 0 | 0 | 0 |
| 0 | 0 | 0.017289639 | 0 | 0 |
| 0 | 0 | 0.000812059 | 0 | 0 |
| 0 | 0.001638616 | 0 | 0.032568906 | 0 |
| 0 | 0 | 0 | 0 | 0 |
| 0 | 0 | 0.21245428 | 0.069604914 | 0.024527684 |
| 0 | 0 | 0 | 0.000913335 | 0 |
| 0 | 0 | 0 | 0 | 0 |
| 0.001893613 | 0 | 0 | 0 | 0 |
| 0 | 0 | 0 | 0 | 0 |
| 0 | 0 | 0 | 0 | 0.027299902 |
| 0 | 0 | 0 | 0 | 0 |
| 0 | 0 | 0 | 0 | 0 |
| 0 | 0 | 0 | 0 | 0 |
| 0 | 0 | 0 | 0 | 0 |
| 0 | 0 | 0 | 0 | 0 |
| 0 | 0 | 0 | 0 | 0 |
| 0 | 0 | 0 | 0 | 0 |
| 0 | 0 | 0 | 0 | 0 |
| 0.241141331 | 0.007734984 | 0.001214304 | 0.001799125 | 0.002508863 |
| 0 | 0 | 0 | 0 | 0 |
| 0 | 0 | 0 | 0.065651499 | 0.005534534 |
| 0.000337069 | 0 | 0.001361125 | 0.013381353 | 0.001514266 |
| 0 | 0 | 0 | 0 | 0 |
| 0 | 0 | 0.005018191 | 0 | 0 |
| 0 | 0 | 0 | 0 | 0 |
| 0 | 0 | 0 | 0 | 0 |
| 0 | 0 | 0 | 0 | 0 |
| 0 | 5.911424882 | 0 | 0.001269818 | 0.016072955 |
| 0 | 0 | 0 | 0 | 0 |
| 0 | 0 | 0 | 0 | 0 |
| 0 | 0 | 0 | 0 | 0 |
| 0 | 0 | 0 | 0.157666384 | 0 |
| 0 | 0 | 0 | 0 | 0 |
| 0 | 0 | 0 | 0 | 0 |
| 0.003092916 | 0 | 0 | 0.039374915 | 0 |
| 0 | 0 | 0 | 0 | 0 |
| 0 | 0 | 0 | 0 | 0 |
| 0 | 0 | 0 | 0 | 0 |
| 0 | 0 | 0 | 0 | 0 |
| 0 | 0 | 0 | 0 | 0 |
| 0 | 0 | 0 | 0 | 0 |
| 0 | 0 | 0 | 0 | 0 |
| 0 | 0 | 0 | 0 | 0 |
| 0 | 0 | 0 | 0 | 0 |
| 0 | 0 | 0 | 0 | 0 |
| 0 | 0 | 0 | 0 | 0 |

| | | | | |
|---|---|---|---|---|
| 0.000362699 | 0 | 0.13858953 | 0 | 0.009980099 |
| 0 | 0 | 0 | 0 | 0 |
| 0.030890582 | 0 | 0 | 0 | 0 |
| 0 | 0 | 0 | 0 | 0 |
| 0 | 0 | 0 | 0 | 0 |
| 0 | 0 | 0 | 0 | 0 |
| 0 | 0 | 0 | 0 | 0 |
| 0 | 0.420451347 | 0 | 0.396414862 | 0.651645027 |
| 0 | 0.183890933 | 0.000862102 | 1.741774627 | 0.138110088 |
| 0 | 0 | 0.051814641 | 0 | 0.02604011 |
| 0 | 0 | 0 | 0 | 0 |
| 0 | 0 | 0 | 0 | 0 |
| 0 | 0 | 0 | 0 | 0 |
| 0 | 0 | 0 | 0 | 0.007810406 |
| 0 | 0 | 0 | 0 | 0 |
| 0 | 0 | 0 | 0 | 0 |
| 0.079416384 | 0 | 0.043293467 | 0 | 0 |
| 0 | 0 | 0 | 0 | 0 |
| 0 | 0 | 0 | 0 | 0 |
| 0 | 0 | 0 | 0 | 0.005721245 |
| 0 | 0 | 0 | 0 | 0 |
| 0 | 0 | 0 | 0 | 0 |
| 0 | 0 | 0 | 0 | 0 |
| 0 | 0 | 0 | 0 | 0 |
| 0 | 0 | 0 | 0.034646315 | 0.013315083 |
| 0 | 0 | 0 | 0 | 0 |
| 0 | 0 | 0 | 0 | 0 |
| 0 | 0 | 0 | 0 | 0 |
| 0 | 0 | 0 | 0 | 0 |
| 0 | 0 | 0 | 0 | 0 |
| 0 | 0 | 0 | 0 | 0 |
| 0 | 0 | 0 | 0 | 0 |
| 0 | 0 | 0 | 0 | 0 |
| 0 | 0 | 0 | 0 | 0 |
| 0 | 0.000801308 | 0 | 0 | 0 |
| 0 | 0 | 0 | 0 | 0 |
| 0 | 0 | 0 | 0 | 0 |
| 0 | 0 | 0 | 0 | 0.100512031 |
| 0 | 0 | 0 | 0 | 0 |
| 0 | 0 | 0 | 0 | 0 |
| 0 | 0 | 0 | 0 | 0 |
| 0 | 0.011588943 | 0 | 0.331309104 | 0.868460092 |
| 0 | 0 | 0 | 0 | 0 |
| 0 | 0 | 0 | 0 | 0 |
| 0 | 0 | 0 | 0 | 0 |
| 0 | 0 | 0 | 0 | 0 |
| 0 | 0 | 0 | 0 | 0 |
| 0 | 0 | 0 | 0 | 0 |

| | | | | |
|---|---|---|---|---|
| 0 | 0 | 0 | 0 | 0 |
| 0.04495627 | 0 | 0 | 0.016708573 | 0.008272172 |
| 0 | 0 | 0 | 0 | 0 |
| 0 | 0 | 0 | 0 | 0 |
| 0 | 1.136740673 | 0 | 0.278733882 | 0.343114558 |
| 0 | 0.003562097 | 0 | 0 | 0 |
| 0 | 0 | 0 | 0 | 0.062634546 |
| 0.044138597 | 0 | 0.078250236 | 0.010869031 | 0.011399959 |
| 0 | 0 | 0 | 0 | 0.00439776 |
| 0 | 0 | 0 | 0.179829229 | 0 |
| 0 | 0 | 0 | 0 | 0 |
| 0 | 0 | 0 | 0 | 0 |
| 0 | 0 | 0 | 0 | 0 |
| 0 | 0 | 0 | 0 | 0 |
| 0 | 0 | 0.081480496 | 0 | 0 |
| 0 | 0 | 0 | 0 | 0.036851916 |
| 0 | 0 | 0 | 0 | 0 |
| 0 | 0 | 0 | 0 | 0 |
| 0 | 0 | 0 | 0 | 0 |
| 0 | 0 | 0 | 0 | 0 |
| 0 | 0 | 0 | 0 | 0 |
| 0 | 0 | 0 | 0 | 0 |
| 0 | 0 | 0 | 0 | 0 |
| 0 | 0 | 0.006046351 | 0.002264468 | 0 |
| 0 | 0 | 0 | 0.11592514 | 0 |
| 0 | 0 | 0 | 0 | 0 |
| 0.008034224 | 0 | 0.013087843 | 0.00398291 | 0.001435529 |
| 0.000640892 | 0 | 0 | 0 | 0 |
| 0 | 0 | 0 | 0 | 0 |
| 0 | 0 | 0 | 0 | 0 |
| 0 | 0 | 0 | 0 | 0 |
| 0 | 0 | 0.206940289 | 0 | 0 |
| 0 | 0 | 0 | 0 | 0 |
| 0 | 0 | 0 | 0 | 0 |
| 0 | 0 | 0 | 0 | 0 |
| 0 | 0 | 0 | 0 | 0 |
| 0 | 0 | 0 | 0 | 0 |
| 0 | 0 | 0 | 0 | 0.003365073 |
| 0 | 0 | 0 | 0 | 0 |
| 0 | 0 | 0 | 0 | 0 |
| 0 | 0 | 0 | 0 | 0 |
| 0 | 0 | 0 | 0 | 0 |
| 0 | 0 | 0 | 0 | 0 |
| 0 | 0 | 0 | 0 | 0 |
| 0 | 0 | 0 | 0 | 0 |
| 0 | 0 | 0 | 0 | 0 |
| 0 | 0 | 0 | 0 | 0 |
| 0 | 0 | 0 | 0.001612478 | 0 |

| | | | | |
|---|---|---|---|---|
| 0.001831635 | 0.01838399 | 0 | 0.015431802 | 0 |
| 0 | 0 | 0 | 0 | 0 |
| 0 | 0 | 0 | 0 | 0 |
| 0 | 0 | 0.001379805 | 0.0070061 | 0 |
| 0 | 0 | 0 | 0 | 0 |
| 0 | 0.009923359 | 0.125523916 | 1.780920457 | 0 |
| 0 | 0 | 0 | 0 | 0 |
| 0 | 0 | 0 | 0 | 0 |
| 0 | 0 | 0 | 0 | 0 |
| 0 | 0 | 0 | 0 | 0 |
| 0 | 0 | 0 | 0 | 0 |
| 0 | 0 | 0.005759564 | 0 | 0 |
| 0 | 0 | 0 | 0 | 0 |
| 0 | 0 | 0 | 0 | 0 |
| 0 | 0 | 0 | 0 | 0 |
| 0 | 0 | 0 | 0 | 0 |
| 0 | 0 | 0 | 0 | 0 |
| 0 | 0 | 0 | 0 | 0 |
| 0 | 0 | 0 | 0 | 0 |
| 0 | 0 | 0 | 0 | 0 |
| 0.005434092 | 0 | 0 | 0.010234239 | 0.059170188 |
| 0 | 0 | 0 | 0 | 0 |
| 0 | 0 | 0.005829872 | 0.003286042 | 0.029045082 |
| 0 | 0 | 0 | 0.01744383 | 0 |
| 0 | 0 | 0 | 0 | 0 |
| 0 | 0 | 0 | 0 | 0 |
| 0 | 0 | 0 | 0 | 0 |
| 0 | 0 | 0 | 0 | 0 |
| 0 | 0 | 0 | 0 | 0 |
| 0 | 0 | 0 | 0 | 0 |
| 0 | 0 | 0 | 0 | 0 |
| 0 | 0.08047158 | 0 | 0 | 0 |
| 0.043421721 | 0 | 0.338260965 | 0.00697536 | 0 |
| 0 | 0 | 0 | 0.119397701 | 0 |
| 0 | 0.057062172 | 0 | 0.234896057 | 0 |
| 0 | 0 | 0.02476297 | 0.001481674 | 0 |
| 0 | 0 | 0 | 0 | 0 |
| 0 | 0 | 0 | 0 | 0 |
| 0.002909944 | 0 | 0.04428539 | 0.004189265 | 0 |
| 0 | 0 | 0 | 0 | 0 |
| 0 | 0 | 0 | 0 | 0 |
| 0 | 0 | 0.33348089 | 0 | 0 |
| 0 | 0.27197107 | 0 | 0.08505587 | 0.14043991 |
| 0 | 0 | 0 | 0 | 0 |
| 0 | 0 | 0 | 0 | 0 |
| 0 | 0 | 0 | 0.131561622 | 1.159949919 |
| 0 | 0 | 0 | 0 | 0 |
| 0 | 0 | 0 | 0 | 0 |

| | | | | |
|---|---|---|---|---|
| 0 | 0 | 0 | 0 | 0 |
| 0 | 0 | 0 | 0 | 0 |
| 0 | 0 | 0 | 0 | 0 |
| 0 | 0 | 0 | 0 | 0 |
| 0 | 0 | 0 | 0 | 0 |
| 0 | 0 | 0 | 0 | 0 |
| 0 | 0 | 0 | 0 | 0 |
| 0 | 0 | 0 | 0 | 0.028153949 |
| 0 | 0 | 0 | 0.026673425 | 0 |
| 0 | 0 | 0 | 0 | 0 |
| 0 | 0 | 0 | 0 | 0 |
| 0 | 0 | 0 | 0 | 0 |
| 0 | 0 | 0 | 0 | 0 |
| 0.195939586 | 0 | 0 | 0 | 0 |
| 0 | 0 | 0 | 0 | 0 |
| 0.001369225 | 0 | 0.049139735 | 0 | 0.004382706 |
| 0 | 0 | 0 | 0 | 0 |
| 0 | 0 | 0 | 0 | 0 |
| 0 | 0 | 0 | 0 | 0 |
| 0 | 0.313379278 | 0 | 0.889083262 | 0 |
| 0 | 0.002348736 | 0 | 0 | 0.006504762 |
| 0 | 0 | 0 | 0 | 0 |
| 0 | 0 | 0 | 0 | 0 |
| 0 | 0 | 0 | 0 | 0 |
| 0 | 0 | 0.003082491 | 0 | 0 |
| 0 | 0 | 0 | 0 | 0 |
| 0 | 0 | 0 | 0 | 0 |
| 0 | 0 | 0 | 0 | 0 |
| 0 | 0 | 0 | 0 | 0 |
| 0 | 0 | 0.021919053 | 0.053860491 | 0 |
| 0 | 0.202071288 | 0 | 0.302656076 | 1.051768717 |
| 0 | 0 | 0 | 0 | 0 |
| 0 | 0 | 0 | 0 | 0 |
| 0 | 0 | 0 | 0 | 0 |
| 0 | 0 | 0 | 0 | 0 |
| 0 | 0 | 0 | 0 | 0 |
| 0 | 0 | 0 | 0 | 0 |
| 0 | 0 | 0 | 0 | 0 |
| 0 | 0 | 0 | 0.012475457 | 0 |
| 0 | 0 | 0 | 0 | 0 |
| 0 | 0 | 0 | 0 | 0 |
| 0 | 0 | 0 | 0 | 0 |
| 0 | 0 | 0 | 0 | 0 |
| 0 | 0 | 0 | 0 | 0 |
| 0 | 0 | 0 | 0 | 0 |
| 0 | 0 | 0 | 0 | 0 |
| 0.180113991 | 0.000767368 | 6.540792014 | 0.011923895 | 0 |
| 0 | 0 | 0 | 0 | 0 |

| | | | | |
|---|---|---|---|---|
| 0 | 0 | 0 | 0 | 0 |
| 0 | 0 | 0 | 0 | 0 |
| 0 | 0 | 0 | 0 | 0 |
| 0 | 0 | 0 | 0 | 0 |
| 0 | 0.00764375 | 0.175852943 | 0.127316446 | 0.022933254 |
| 0 | 0 | 0 | 0 | 0 |
| 0 | 0 | 0 | 0 | 0 |
| 0 | 0 | 0 | 0 | 0 |
| 0 | 0 | 0 | 0 | 0 |
| 0 | 0 | 0 | 0 | 0 |
| 0 | 0 | 0 | 0.042630433 | 0.078168652 |
| 0.000726907 | 0 | 0.081675607 | 0.038434438 | 0.004163625 |
| 0.000376282 | 0 | 0 | 0.002462319 | 0 |
| 0 | 0 | 0 | 0 | 0 |
| 0 | 0 | 0.223133549 | 0.008264923 | 0.021650875 |
| 0 | 0.002682068 | 9.07039886 | 0 | 0.058090098 |
| 0 | 0 | 0 | 0.023707051 | 0 |
| 0 | 0 | 0 | 0 | 0 |
| 0 | 0 | 0.066512803 | 0 | 0.050628986 |
| 0.045132989 | 0 | 0.058332285 | 0.012979979 | 0.011138745 |
| 0 | 0 | 0 | 0 | 0 |
| 0 | 0 | 0 | 0 | 0 |
| 0 | 0 | 0 | 0 | 0 |
| 0 | 0 | 0 | 0 | 0 |
| 0 | 0 | 0 | 0 | 0 |
| 0 | 0 | 0 | 0 | 0 |
| 0.001497365 | 0 | 0 | 0.001959694 | 0 |
| 0 | 0 | 0 | 0 | 0 |
| 0 | 0 | 0 | 0 | 0 |
| 0 | 0 | 0 | 0 | 0 |
| 0 | 0 | 0.008897097 | 0.003075807 | 0 |
| 0 | 0 | 0 | 0 | 0 |
| 0 | 0 | 0 | 0 | 0 |
| 0 | 0 | 0 | 0 | 0 |
| 0 | 0 | 0 | 0 | 0 |
| 0 | 0 | 0 | 0 | 0 |
| 0 | 0 | 0 | 0 | 0 |
| 0 | 0 | 0.05570195 | 0.306978141 | 0.029980025 |
| 0 | 0 | 0 | 0 | 0.109108855 |
| 0 | 0 | 0 | 0 | 0 |
| 0 | 0 | 0.008320474 | 0.048420456 | 0 |
| 0 | 0 | 0 | 0 | 0 |
| 0 | 0 | 0 | 0 | 0 |
| 0 | 0 | 0 | 0 | 0.002062418 |
| 0 | 0 | 0 | 0 | 0 |
| 0 | 0 | 0 | 0 | 0 |
| 0 | 0 | 0 | 0 | 0 |
| 0 | 0 | 0 | 0 | 0 |

| | | | | |
|---|---|---|---|---|
| 0 | 0 | 0 | 0 | 0 |
| 0 | 0 | 0 | 0 | 0.020173839 |
| 0 | 0 | 0 | 0 | 0 |
| 0 | 0 | 0.008850688 | 0.000932854 | 0.000960633 |
| 0.001718831 | 0.071419977 | 0 | 0 | 0 |
| 0 | 0.001245599 | 0 | 0.001116633 | 0 |
| 0 | 0 | 0 | 0 | 0 |
| 0 | 0 | 0 | 0 | 0 |
| 0 | 0 | 0 | 0 | 0 |
| 0 | 0 | 0 | 0 | 0 |
| 0 | 0 | 0 | 0 | 0 |
| 0 | 0 | 0 | 0 | 0 |
| 0 | 0.002212905 | 0 | 0.010976958 | 0.028259568 |
| 0 | 0 | 0 | 0 | 0 |
| 0 | 0 | 0 | 0 | 0 |
| 0 | 0 | 0 | 0 | 0 |
| 0 | 0 | 0 | 0 | 0 |
| 0 | 0 | 0 | 0 | 0 |
| 0 | 0 | 0 | 0 | 0 |
| 0 | 0 | 0 | 0 | 0 |
| 0 | 0 | 0 | 0 | 0 |
| 0 | 0 | 0 | 0 | 0 |
| 0.012773928 | 0 | 0.014507597 | 0 | 0.003586635 |
| 0.042237691 | 0 | 0 | 0.117545056 | 0.007142095 |
| 0 | 0 | 0 | 0 | 0 |
| 0 | 0 | 0 | 0 | 0 |
| 0 | 0 | 0 | 0.017969202 | 0 |
| 0 | 0 | 0 | 0 | 0 |
| 0 | 0 | 0 | 0 | 0 |
| 0 | 0 | 0 | 0 | 0 |
| 0 | 0.037633905 | 0 | 0.022135619 | 0.052238011 |
| 0 | 0 | 0 | 0 | 0 |
| 0 | 0 | 0 | 0 | 0 |
| 0 | 0 | 0 | 0 | 0 |
| 0 | 0 | 0 | 0 | 0 |
| 0 | 0 | 0.001409154 | 0 | 0 |
| 0 | 0 | 0.283803006 | 0 | 0 |
| 0 | 0 | 0 | 0 | 0 |
| 0 | 0 | 0 | 1.70640811 | 0 |
| 0 | 0 | 0 | 0 | 0 |
| 0 | 0 | 0 | 0 | 0 |
| 0 | 0 | 0 | 0 | 0 |
| 0 | 0 | 0 | 0 | 0 |
| 0 | 0 | 0 | 0 | 0 |
| 0 | 0 | 0 | 0 | 6.087997495 |
| 0 | 0.244272291 | 0.548049333 | 0.232857082 | 0.587197828 |
| 0 | 0 | 0 | 0 | 0 |
| 0 | 0 | 0 | 0 | 0 |

| | | | | |
|---|---|---|---|---|
| 0 | 0 | 0 | 0 | 0 |
| 0 | 0 | 0 | 0 | 0 |
| 0 | 0.122589604 | 0 | 0 | 0 |
| 0 | 0 | 0 | 0 | 0 |
| 0 | 0 | 0 | 0 | 0 |
| 0 | 0 | 0 | 0 | 0 |
| 0 | 0 | 0.046178529 | 0.010520429 | 0 |
| 0 | 0 | 0 | 0 | 0 |
| 0 | 0 | 0 | 0.000608022 | 0 |
| 0 | 0 | 0 | 0 | 0 |
| 0 | 0 | 0.002158909 | 0.011195316 | 0.001544021 |
| 0 | 0 | 0.002762793 | 0 | 0 |
| 0 | 0 | 0 | 0 | 0 |
| 0 | 0 | 0 | 0 | 0 |
| 0 | 0 | 0 | 0 | 0 |
| 0 | 0 | 0 | 0 | 0 |
| 0 | 0 | 0 | 0 | 0 |
| 0 | 0 | 0 | 0 | 0 |
| 0 | 0 | 0 | 0 | 0 |
| 0 | 0 | 0 | 0 | 0.002502667 |
| 0 | 0 | 0 | 0 | 0 |
| 0 | 0 | 0 | 0 | 0 |
| 0 | 0 | 0 | 0 | 0 |
| 0 | 0 | 0 | 0 | 0 |
| 0 | 0 | 0 | 0 | 0 |
| 0 | 0 | 0 | 0 | 0 |
| 0 | 0 | 0 | 0 | 0 |
| 0 | 0.007364039 | 0 | 0 | 0 |
| 0 | 0 | 0 | 0 | 0 |
| 0 | 0 | 0 | 0 | 0 |
| 0 | 0.001270373 | 0 | 0.001670302 | 0.001172755 |
| 0 | 0 | 0 | 0 | 0 |
| 0 | 0.00983355 | 0.08343442 | 0.032617027 | 0.001361687 |
| 0.007846787 | 0.003768308 | 0.005211548 | 0.021079646 | 0.003478742 |
| 0 | 0 | 0 | 0 | 0 |
| 0 | 0 | 0 | 0 | 0 |
| 0 | 0 | 0 | 0 | 0 |
| 0 | 0 | 0 | 0 | 0 |
| 0 | 0 | 0 | 0 | 0 |
| 0 | 0 | 0 | 0 | 0 |
| 0 | 0 | 0 | 0 | 0 |
| 0.033248355 | 0 | 0.030040813 | 0.41614042 | 0.001493664 |
| 0.004860704 | 0.106443363 | 0 | 0.079165358 | 0.104632965 |
| 0 | 0 | 0 | 0 | 0 |
| 0 | 0 | 0 | 0 | 0 |
| 0 | 0.03240149 | 0 | 0.038814098 | 0.012490593 |
| 0 | 0 | 0.012010045 | 0 | 0 |
| 0 | 0 | 0 | 0 | 0 |

| | | | | |
|---|---|---|---|---|
| 0 | 0 | 0 | 0 | 0 |
| 0 | 0.03071785 | 0 | 0.012218281 | 0 |
| 0.006443954 | 0.034977112 | 0.010258443 | 0.014272243 | 0.005567136 |
| 0 | 0 | 0 | 0 | 0 |
| 0 | 0 | 0 | 0 | 0.021114177 |
| 0.007448233 | 0 | 0.013028962 | 0 | 0.03245118 |
| 0 | 0 | 0 | 0 | 0 |
| 0 | 0 | 0 | 0 | 0 |
| 0 | 0 | 0 | 0 | 0 |
| 0 | 0 | 0 | 0 | 0 |
| 0 | 0 | 0 | 0 | 0 |
| 0 | 0 | 0 | 0 | 0 |
| 0 | 0 | 0 | 0 | 0 |
| 0 | 0 | 0 | 0 | 0 |
| 0 | 0 | 0 | 0 | 0 |
| 0 | 0 | 0 | 0 | 0 |
| 0 | 0 | 0 | 0 | 0 |
| 0 | 0 | 0 | 0 | 0 |
| 0 | 0 | 0 | 0 | 0 |
| 0 | 0 | 0 | 0 | 0 |
| 0 | 0 | 0 | 0 | 0 |
| 0 | 0 | 0.008251731 | 0 | 0 |
| 0 | 0 | 0 | 0 | 0 |
| 0 | 0 | 0 | 0 | 0 |
| 0 | 0.001988004 | 0 | 0.019603891 | 0.002294051 |
| 0 | 0.030748232 | 0 | 0.000896411 | 0.00300009 |
| 0 | 0 | 0 | 0 | 0 |
| 0 | 0 | 0 | 0.02643433 | 0 |
| 0 | 0 | 0 | 0 | 0 |
| 0 | 0 | 0 | 0 | 0 |
| 0 | 0 | 0 | 0 | 0 |
| 0.000840902 | 0 | 0 | 0 | 0 |
| 0 | 0 | 0 | 0 | 0 |
| 0 | 0 | 0 | 0 | 0 |
| 0 | 0 | 0 | 0 | 0 |
| 0 | 0 | 0 | 0 | 0 |
| 0 | 0 | 0 | 0 | 0 |
| 0 | 0 | 0.124098272 | 0 | 0 |
| 0 | 0.030186651 | 0 | 0 | 0 |
| 0 | 0 | 0 | 0 | 0 |
| 0 | 0 | 0 | 0 | 0 |
| 0 | 0.000813588 | 0 | 0.016142971 | 0 |
| 0.000687403 | 0 | 0.097847364 | 0.000899647 | 0.00717988 |
| 0 | 0 | 0 | 0 | 0 |
| 0 | 0 | 0 | 0 | 0 |
| 0 | 0 | 0.002634615 | 0 | 0 |
| 0 | 0 | 0 | 0.00082577 | 0 |
| 0 | 0 | 0 | 0 | 0 |

| | | | | |
|---|---|---|---|---|
| 0 | 0 | 0 | 0 | 0 |
| 0.002786524 | 0 | 0.026523274 | 0.032301065 | 0.003129576 |
| 0 | 0 | 0 | 0 | 0 |
| 0 | 0 | 0 | 0 | 0 |
| 0 | 0 | 0 | 0 | 0 |
| 0 | 0 | 0 | 0 | 0 |
| 0 | 0 | 0 | 0 | 0 |
| 0 | 0 | 0 | 0 | 0 |
| 0 | 0 | 0 | 0 | 0 |
| 0 | 0 | 0 | 0 | 0 |
| 0 | 0 | 0 | 0 | 0 |
| 0 | 0 | 0 | 0 | 0 |
| 0 | 0.009215003 | 0 | 0.055807026 | 0 |
| 0 | 0 | 0 | 0 | 0 |
| 0.000827232 | 0 | 0 | 0 | 0.003065943 |
| 0 | 0 | 0 | 0 | 0 |
| 0 | 0 | 0 | 0 | 0 |
| 0 | 0 | 0 | 0 | 0 |
| 0 | 0 | 0 | 0 | 0 |
| 0 | 0 | 0 | 0 | 0 |
| 0 | 0 | 0 | 0 | 0 |
| 0 | 0 | 0.084393243 | 0.005316236 | 0.009032994 |
| 0 | 0 | 0 | 0 | 0 |
| 0 | 0.001832434 | 0 | 0 | 0 |
| 0 | 0 | 0 | 0 | 0 |
| 0 | 0 | 0 | 0 | 0 |
| 0 | 0 | 0 | 0 | 0 |
| 0 | 0 | 0 | 0 | 0 |
| 0 | 0 | 0 | 0 | 0 |
| 0 | 0 | 0 | 0 | 0 |
| 0 | 0 | 0 | 0 | 0 |
| 0 | 0 | 0 | 0 | 0 |
| 0 | 0 | 0 | 0 | 0 |
| 0 | 0 | 0 | 0 | 0 |
| 0 | 0.037959482 | 0 | 0 | 0 |
| 0 | 0 | 0 | 0 | 0 |
| 0 | 0 | 0 | 0 | 0 |
| 0 | 0 | 0 | 0 | 0 |
| 0 | 0.000888309 | 0.000982821 | 0.021447976 | 0.321185811 |
| 0 | 0 | 0 | 0 | 0 |
| 0.350764853 | 0.019115668 | 0.257318762 | 0.270756509 | 0.300485358 |
| 0 | 0 | 0 | 0 | 0 |
| 0 | 0 | 0 | 0 | 0 |
| 0 | 0 | 0 | 0 | 0 |
| 0 | 0 | 0 | 0 | 0 |
| 0.249023151 | 0.00364769 | 1.194845268 | 0.25658747 | 0.010102178 |
| 0 | 0 | 0 | 0 | 0 |
| 0 | 0 | 0 | 0 | 0 |

| | | | | |
|---|---|---|---|---|
| 0 | 0 | 0 | 0 | 0 |
| 0 | 0 | 0 | 0 | 0 |
| 0 | 0 | 0 | 0 | 0 |
| 0 | 0 | 0 | 0 | 0 |
| 0 | 0 | 0.005739074 | 0.011242947 | 0 |
| 0 | 0 | 0 | 0 | 0 |
| 0 | 0 | 0 | 0 | 0 |
| 0.005761259 | 0 | 0 | 0 | 0 |
| 0 | 0 | 0 | 0 | 0 |
| 0 | 0 | 0.000986752 | 0.101699055 | 0.003293315 |
| 0 | 0.002194723 | 0 | 0 | 0.021129067 |
| 0 | 0 | 0 | 0 | 0 |
| 0 | 0 | 0 | 0.009377572 | 0.001341224 |
| 0 | 0 | 0 | 0 | 0 |
| 0 | 0 | 0 | 0 | 0 |
| 0 | 0 | 0 | 0 | 0 |
| 0 | 0 | 0 | 0 | 0 |
| 0 | 0 | 0 | 0 | 0 |
| 0 | 0 | 0 | 0 | 0 |
| 0.028202978 | 0.727528051 | 0.125547336 | 0.634349267 | 0.311338954 |
| 0 | 0 | 0 | 0 | 0 |
| 0 | 0 | 0 | 0 | 0 |
| 0 | 0 | 0 | 0 | 0 |
| 0 | 0 | 0 | 0 | 0 |
| 0 | 0 | 0 | 0 | 0 |
| 0 | 0 | 0 | 0 | 0 |
| 0 | 0 | 0 | 0 | 0 |
| 0 | 0 | 0 | 0 | 0 |
| 0 | 0 | 0 | 0.005029734 | 0 |
| 0.004393637 | 0 | 0.914707301 | 0 | 0 |
| 0 | 0 | 0 | 0 | 0 |
| 0 | 0.011831765 | 0 | 0 | 0 |
| 0 | 0 | 0 | 0 | 0 |
| 0 | 0 | 0 | 0 | 0 |
| 0 | 0 | 0 | 0 | 0 |
| 0 | 0 | 0 | 0 | 0 |
| 0 | 0 | 0 | 0 | 0 |
| 0 | 0 | 0 | 0 | 0 |
| 0 | 0 | 0 | 0 | 0 |
| 0 | 0 | 0 | 0 | 0 |
| 0 | 0 | 0 | 0 | 0 |
| 0 | 0 | 0 | 0 | 0 |
| 0 | 0 | 0 | 0 | 0 |
| 0 | 0 | 0 | 0 | 0 |
| 0 | 0 | 0.013303645 | 0.009581646 | 0.006278978 |
| 0 | 0 | 0 | 0 | 0 |
| 0 | 0 | 0 | 0 | 0 |
| 0 | 0 | 0 | 0 | 0 |

| | | | | |
|---|---|---|---|---|
| 0 | 0 | 0 | 0 | 0 |
| 0 | 0 | 0 | 0 | 0 |
| 0 | 0 | 0 | 0 | 0 |
| 0 | 0 | 0 | 0 | 0 |
| 0 | 0 | 0 | 0 | 0 |
| 0 | 0.000544037 | 0.000752401 | 0.14709315 | 0 |
| 0 | 0 | 0 | 0 | 0 |
| 0 | 0 | 0.000716986 | 0 | 0 |
| 0 | 0 | 0 | 0 | 0 |
| 0 | 0 | 0 | 0 | 0 |
| 0 | 0 | 0 | 0 | 0 |
| 0 | 0 | 0 | 0 | 0 |
| 0 | 0 | 0 | 0 | 0 |
| 0 | 0 | 0 | 0 | 0 |
| 0 | 0 | 0 | 0 | 0 |
| 0 | 0 | 0 | 0 | 0 |
| 0 | 0 | 0 | 0 | 0 |
| 0 | 0 | 0 | 0 | 0 |
| 0 | 0 | 0 | 0 | 0 |
| 0 | 0 | 0 | 0 | 0 |
| 0 | 0 | 0 | 0 | 0 |
| 0 | 0 | 0 | 0 | 0 |
| 0 | 0 | 0 | 0 | 0 |
| 0 | 0 | 0 | 0.00738953 | 0 |
| 0 | 0 | 0 | 0 | 0 |
| 0 | 0 | 0 | 0.001424004 | 0 |
| 0 | 0 | 0 | 0 | 0 |
| 0 | 0 | 0 | 0.055724474 | 0 |
| 0 | 0 | 0 | 0 | 0 |
| 0 | 0 | 0 | 0 | 0 |
| 0 | 0 | 0 | 0 | 0 |
| 0 | 0 | 0 | 0 | 0 |
| 0 | 0 | 0 | 0 | 0 |
| 0 | 0 | 0 | 0 | 0 |
| 0 | 0 | 0 | 0 | 0 |
| 0 | 0 | 0 | 0 | 0 |
| 0 | 0 | 0 | 0 | 0 |
| 0 | 0 | 0 | 0 | 0 |
| 0 | 0.11263378 | 0 | 0 | 0 |
| 0 | 0 | 0 | 0 | 0 |
| 0 | 0 | 0 | 0 | 0 |
| 0 | 0 | 0 | 0 | 0 |
| 0 | 0 | 0 | 0 | 0 |
| 0 | 0 | 0 | 0 | 0 |
| 0 | 0.030966144 | 0 | 0 | 0 |
| 0 | 0 | 0 | 0 | 0 |
| 0 | 0 | 0 | 0 | 0 |
| 0 | 0 | 0 | 0 | 0 |

| | | | | |
|---|---|---|---|---|
| 0 | 0 | 0 | 0 | 0 |
| 0 | 0.001015156 | 0 | 0 | 0 |
| 0 | 0 | 0 | 0 | 0 |
| 0 | 0 | 0 | 0 | 0 |
| 0 | 0 | 0 | 0 | 0 |
| 0 | 0 | 0 | 0 | 0 |
| 0 | 0 | 0 | 0 | 0 |
| 0 | 0 | 0 | 0 | 0 |
| 0 | 0 | 0 | 0 | 0 |
| 0 | 0 | 0 | 0 | 0 |
| 0 | 0.025521919 | 0 | 0 | 0.000736274 |
| 0 | 0 | 0 | 0 | 0 |
| 0 | 0.022127029 | 0 | 0.220899999 | 0.528564382 |
| 0 | 0 | 0 | 0 | 0 |
| 0 | 0 | 0 | 0 | 0 |
| 0 | 0 | 0 | 0 | 0 |
| 0 | 0 | 0 | 0 | 0 |
| 0 | 0 | 0 | 0 | 0 |
| 0 | 0 | 0 | 0 | 0 |
| 0 | 0 | 0 | 0 | 0 |
| 0.002513583 | 0 | 0 | 0.011610635 | 0 |
| 0 | 0 | 0 | 0 | 0 |
| 0 | 0 | 0 | 0 | 0 |
| 1.823316872 | 1.210824213 | 0.849492142 | 1.519362108 | 6.171012843 |
| 0 | 0 | 0 | 0 | 0 |
| 0 | 0 | 0 | 0 | 0 |
| 0 | 0 | 0 | 0 | 0 |
| 0 | 0 | 0 | 0 | 0 |
| 0 | 0 | 0 | 0 | 0 |
| 0 | 0 | 0 | 0 | 0 |
| 0.002176835 | 0 | 0 | 0 | 0 |
| 0 | 0 | 0 | 0 | 0 |
| 0 | 0 | 0 | 0 | 0 |
| 0 | 0 | 0 | 0 | 0 |

| PHL018-0247_V3 | PHL018-0210_V1 | PHL018-0266_V3 | PHL018-0227_V3 | PHL018-0216_V1 |
|---|---|---|---|---|
| REF | EYCF | REF | REF | CTRL |
| 0.135334961 | 4.616653511 | 0.155957274 | 0.142270761 | 0.054212819 |
| 11.57867098 | 7.575780919 | 0.35059937 | 0.444248254 | 1.186270098 |
| 0.743162954 | 0 | 0 | 10.95700944 | 29.09128524 |
| 0 | 0 | 0 | 0 | 0 |
| 0 | 0 | 0 | 0 | 0 |
| 0 | 0 | 0 | 0 | 0 |
| 0 | 0 | 0 | 0 | 0 |
| 0 | 0 | 0 | 0 | 0 |
| 0 | 0 | 0 | 0 | 0 |
| 0 | 0 | 0 | 0 | 0 |
| 0 | 0 | 0 | 0 | 0 |
| 0 | 0 | 0 | 0 | 0 |
| 0 | 0 | 0 | 0 | 0 |
| 9.942881102 | 0.095130488 | 0.047833748 | 7.350612078 | 6.164396493 |
| 0 | 0 | 0 | 0 | 0 |
| 0 | 0 | 0 | 0 | 0 |
| 0 | 0 | 0 | 0 | 0.001922102 |
| 0 | 0 | 0 | 0 | 0.000528149 |
| 0 | 0 | 0 | 0 | 0 |
| 0 | 0 | 0 | 0 | 0 |
| 0 | 0 | 0 | 0 | 0 |
| 0 | 0 | 0 | 0 | 0 |
| 0 | 0 | 0 | 0 | 0 |
| 0 | 0 | 0 | 0 | 0 |
| 0 | 0 | 0 | 0 | 0 |
| 0 | 0 | 0 | 0 | 0 |
| 0 | 0 | 0 | 0 | 0 |
| 0 | 0 | 0 | 0 | 0 |
| 0 | 0 | 0 | 0 | 0 |
| 0 | 0 | 0 | 0 | 0 |
| 0 | 0 | 0 | 0 | 0 |
| 0 | 0 | 0 | 0 | 0 |
| 0 | 0 | 0.000407608 | 0 | 0 |
| 0 | 0 | 0 | 0 | 0 |
| 0 | 0 | 0 | 0 | 0 |
| 0 | 0.812836998 | 0.176141234 | 0.029363172 | 0 |
| 0 | 0 | 0 | 0 | 0 |
| 0 | 0 | 0 | 0 | 0 |
| 0 | 0 | 0 | 0 | 0 |
| 0 | 0 | 0 | 0 | 0 |
| 0 | 0 | 0 | 0 | 0 |
| 0.005326867 | 0.02337363 | 0 | 0.258025744 | 0 |
| 0.018525719 | 0.008751159 | 0.288057887 | 1.228528735 | 0.169529095 |
| 0.001941053 | 0.044756793 | 0.084870493 | 0.266939367 | 0.007855272 |
| 0 | 0.007089885 | 0.026857692 | 0.004657081 | 0 |
| 0 | 0 | 0 | 0 | 0 |

| | | | | |
|---|---|---|---|---|
| 0 | 0 | 0 | 0.149835703 | 0 |
| 0.087966568 | 0.080233724 | 0 | 0.122908701 | 0.007777463 |
| 0 | 0.313741832 | 0.090523956 | 0 | 0.000863823 |
| 0.760081294 | 0.640863946 | 0.100236747 | 0.130630331 | 0.177714556 |
| 0 | 0.012711964 | 0 | 0 | 0 |
| 0 | 0 | 0 | 0 | 0 |
| 0 | 0.003127186 | 0 | 0.443949166 | 0 |
| 0 | 0 | 0 | 0 | 0 |
| 0 | 0 | 0 | 0 | 0 |
| 0 | 0 | 0 | 0 | 0 |
| 0 | 0 | 0 | 0.002044628 | 0 |
| 0 | 0 | 0 | 0 | 0.483640052 |
| 0 | 0 | 0 | 0 | 0.000754507 |
| 0.054354945 | 0.2164939 | 0 | 0.183753192 | 0.033325709 |
| 0.019711556 | 0 | 0 | 0.001749251 | 0 |
| 0 | 0 | 0 | 0 | 0 |
| 0 | 0.005287642 | 0.000604101 | 1.278756981 | 0.000488621 |
| 0 | 0 | 0 | 0 | 0 |
| 0 | 0 | 0 | 0 | 0 |
| 0 | 0 | 0 | 0 | 0 |
| 0.345883747 | 0.37601995 | 0.001430193 | 0.119230477 | 0.048867891 |
| 0.001187824 | 0.004320494 | 0 | 0.01027751 | 0.022808486 |
| 0 | 0 | 0.502448569 | 0 | 0 |
| 0 | 0 | 0 | 0 | 0 |
| 0 | 0 | 0 | 0 | 0 |
| 0 | 0 | 0 | 0 | 0 |
| 0.045385723 | 0 | 0 | 0.188795507 | 0 |
| 0 | 0 | 0 | 0 | 0 |
| 0 | 0 | 0 | 0.007686469 | 0 |
| 0 | 0 | 0 | 0 | 0.001163264 |
| 0.326793229 | 0 | 0 | 0 | 0 |
| 0 | 0 | 0 | 0 | 0 |
| 0 | 0 | 0 | 0 | 0 |
| 0 | 0 | 0 | 0.000877155 | 0 |
| 0 | 0 | 0 | 0 | 0 |
| 0 | 0 | 0.000580831 | 0.001470441 | 0.043779364 |
| 0 | 0 | 0 | 0 | 0 |
| 0 | 0 | 0.157452845 | 0.029910147 | 0 |
| 0 | 0 | 0 | 0 | 0 |
| 0 | 0 | 0 | 0 | 0 |
| 0 | 0 | 0 | 0 | 0 |
| 0 | 0 | 0 | 0 | 0 |
| 0 | 0 | 0 | 0 | 0 |
| 0 | 0 | 0 | 0 | 0 |
| 0 | 0 | 0 | 0 | 0 |
| 0 | 0 | 0 | 0 | 0 |
| 0 | 0 | 0 | 0.004691632 | 0 |
| 0.330983951 | 0 | 0 | 0 | 0.046534493 |

| | | | | |
|---|---|---|---|---|
| 0 | 0 | 0 | 0 | 0 |
| 0 | 0 | 0 | 0 | 0 |
| 0 | 0 | 0 | 0 | 0 |
| 0 | 0 | 0 | 0 | 0 |
| 0 | 0.003512749 | 0.014573782 | 0.002339697 | 0 |
| 0 | 0 | 0 | 0.001360412 | 0 |
| 0 | 0 | 0 | 0 | 0 |
| 0 | 0 | 0 | 0 | 0 |
| 0 | 0.000357951 | 0.00060369 | 0 | 0 |
| 0 | 0 | 0 | 0 | 0 |
| 0 | 0 | 0 | 0 | 0 |
| 0 | 0 | 0 | 0.001064603 | 0 |
| 0 | 0 | 0 | 0 | 0 |
| 0 | 0 | 0 | 0 | 0 |
| 0 | 0 | 0 | 0 | 0 |
| 0 | 0 | 0 | 0 | 0 |
| 0 | 0 | 0 | 0 | 0 |
| 0 | 0 | 0 | 0 | 0 |
| 0 | 0 | 0 | 0 | 0.014744237 |
| 0 | 0 | 0 | 0 | 0 |
| 0 | 0 | 0 | 0 | 0 |
| 0 | 0 | 0 | 0 | 0 |
| 0 | 0 | 0 | 0 | 0 |
| 0 | 0 | 0 | 0 | 0 |
| 0 | 0 | 0 | 0 | 0 |
| 0 | 0 | 0 | 0 | 0 |
| 0 | 0 | 0.000265396 | 0 | 0 |
| 0 | 0 | 0 | 0 | 0.000690188 |
| 0 | 0 | 0 | 0 | 0 |
| 0 | 0 | 0 | 0 | 0 |
| 0 | 0 | 0 | 0 | 0.386826377 |
| 0 | 0 | 0 | 0 | 0 |
| 0 | 0 | 0 | 0 | 0 |
| 0 | 0 | 0 | 0 | 0 |
| 0 | 0 | 0 | 0 | 0 |
| 0 | 0 | 0 | 0 | 0 |
| 0 | 0 | 0 | 0 | 0 |
| 0 | 0 | 0 | 0 | 0 |
| 0 | 0 | 0 | 0 | 0.736679463 |
| 0 | 0 | 0 | 0 | 0 |
| 0 | 0 | 0 | 0 | 0 |
| 0 | 0 | 0 | 0 | 0 |
| 0 | 0 | 0 | 0 | 0 |
| 0 | 0 | 0 | 0 | 0 |
| 0 | 0 | 0 | 0 | 0 |
| 0 | 0 | 0 | 0 | 0 |
| 0 | 0 | 0 | 0.000898946 | 0 |
| 0 | 0 | 0 | 0 | 0 |

| | | | | |
|---|---|---|---|---|
| 0 | 0 | 0 | 0 | 0 |
| 0 | 0 | 0 | 0 | 0 |
| 0 | 0 | 0 | 0 | 0 |
| 0 | 0 | 0 | 0 | 0 |
| 0 | 0 | 0 | 0 | 0 |
| 0 | 0 | 0 | 0 | 0 |
| 0 | 0 | 0 | 0 | 0 |
| 0 | 0 | 0 | 0 | 0 |
| 0 | 0 | 0 | 0 | 0 |
| 0 | 0 | 0 | 0 | 0 |
| 0 | 0 | 0 | 0 | 0 |
| 0 | 0 | 0 | 0 | 0 |
| 0 | 0 | 0 | 0 | 0 |
| 0 | 0 | 0 | 0 | 0.001557084 |
| 0 | 0 | 0 | 0 | 0 |
| 0 | 0 | 0 | 0 | 0 |
| 0 | 0 | 0 | 0 | 0 |
| 0 | 0 | 0 | 0 | 0 |
| 0 | 0 | 0 | 0 | 0 |
| 0 | 0 | 0 | 0 | 0 |
| 0 | 0 | 0 | 0 | 0 |
| 0 | 0 | 0 | 0 | 0 |
| 0 | 0 | 0 | 0 | 0 |
| 0 | 0 | 0 | 0 | 0 |
| 0 | 0 | 0 | 0.148571658 | 0 |
| 0 | 0 | 0 | 0 | 0 |
| 0 | 0 | 0 | 0 | 0 |
| 0 | 0 | 0 | 0 | 0 |
| 0 | 0 | 0 | 0 | 0 |
| 0 | 0 | 0 | 0 | 0 |
| 0 | 0 | 0 | 0 | 0 |
| 0 | 0 | 0 | 0.000855921 | 0.000574271 |
| 0 | 0 | 0 | 0 | 0 |
| 0 | 0 | 0 | 0 | 0 |
| 0 | 0 | 0 | 0 | 0 |
| 0 | 0 | 0 | 0 | 0 |
| 0 | 0 | 0 | 0 | 0 |
| 0 | 0 | 0 | 0 | 0 |
| 0 | 0 | 0 | 0 | 0 |
| 0 | 0 | 0 | 0 | 0 |
| 0 | 0 | 0 | 0 | 0 |
| 0 | 0 | 0 | 0 | 0 |
| 0 | 0 | 0 | 0 | 0 |
| 0 | 0 | 0 | 0 | 0 |
| 0 | 0 | 0 | 0 | 0 |
| 0 | 0 | 0 | 0 | 0 |
| 0 | 0 | 0 | 0 | 0 |
| 0 | 0 | 0 | 0 | 0 |

| | | | | |
|---|---|---|---|---|
| 0 | 0 | 0 | 0 | 0 |
| 0 | 0 | 0 | 0 | 0 |
| 0 | 0 | 0 | 0 | 0 |
| 0 | 0 | 0 | 0 | 0 |
| 0 | 0 | 0 | 0 | 0 |
| 0 | 0 | 0 | 0 | 0 |
| 0 | 0 | 0 | 0 | 0 |
| 0 | 0 | 0 | 0 | 0 |
| 0 | 0 | 0 | 0 | 0 |
| 0 | 0 | 0 | 0 | 0 |
| 0 | 0 | 0 | 0 | 0 |
| 0 | 0 | 0 | 0 | 0 |
| 0 | 0.000511543 | 0 | 0 | 0.003113953 |
| 0 | 0 | 0 | 0 | 0 |
| 0 | 0 | 0 | 0 | 0 |
| 0 | 0 | 0 | 0 | 0 |
| 0 | 0 | 0 | 0 | 0 |
| 0 | 0 | 0 | 0 | 0 |
| 0 | 0 | 0 | 0 | 0 |
| 0 | 0 | 0 | 0 | 0 |
| 0.049238883 | 0.064800286 | 0.001451778 | 0.116692186 | 0.137510086 |
| 0 | 0 | 0 | 0 | 0 |
| 0 | 0.001678472 | 0 | 0 | 0 |
| 0 | 0 | 0 | 0 | 0 |
| 0 | 0 | 0 | 0 | 0 |
| 0 | 0 | 0 | 0 | 0 |
| 0.005627915 | 0.004595422 | 0.00151482 | 0.004815976 | 0.00428836 |
| 0 | 0 | 0 | 0 | 0 |
| 0 | 0 | 0 | 0 | 0 |
| 0 | 0.031883222 | 0 | 0 | 0 |
| 0.004274343 | 0 | 0 | 0 | 0.011134277 |
| 0 | 0 | 0 | 0 | 0 |
| 0 | 0 | 0 | 0 | 0 |
| 0 | 0 | 0 | 0 | 0.00617462 |
| 0.024552058 | 0.202100048 | 0.009013096 | 0.005978437 | 0.038551807 |
| 0 | 0 | 0 | 0 | 0 |
| 0 | 0 | 0 | 0 | 0 |
| 0 | 0 | 0 | 0 | 0 |
| 0 | 0 | 0 | 0 | 0 |
| 0 | 0 | 0 | 0 | 0 |
| 0 | 0 | 0 | 0 | 0 |
| 0 | 0 | 0 | 0 | 0 |
| 0 | 0 | 0 | 0 | 0 |
| 0 | 0 | 0 | 0 | 0 |
| 0 | 0 | 0 | 0 | 0 |
| 0 | 0 | 0 | 0 | 0 |
| 0 | 0 | 0 | 0 | 0 |
| 0 | 0.071290314 | 0 | 0 | 0.006113947 |

| | | | | |
|---|---|---|---|---|
| 0 | 0 | 0 | 0 | 0 |
| 0 | 0 | 0 | 0 | 0 |
| 0.005069334 | 0 | 0 | 0 | 0 |
| 0.001084892 | 0 | 0 | 0 | 0.000706511 |
| 0 | 0 | 0 | 0 | 0 |
| 0 | 0 | 0 | 0 | 0 |
| 0 | 0 | 0 | 0.015970166 | 0 |
| 0 | 0 | 0 | 0.028988815 | 0 |
| 0 | 0.245898491 | 0 | 0 | 0 |
| 0 | 0 | 0 | 0 | 0 |
| 0 | 0 | 0 | 0 | 0 |
| 0 | 0 | 0 | 0 | 0 |
| 0 | 0 | 0 | 0 | 0 |
| 0 | 0 | 0 | 0 | 0 |
| 0 | 0 | 0 | 0.001119539 | 0 |
| 0 | 0 | 0 | 0 | 0 |
| 0 | 0 | 0 | 0 | 0 |
| 0 | 0 | 0 | 0 | 0 |
| 0 | 0 | 0.002281965 | 0.007510177 | 0 |
| 0 | 0 | 0 | 0 | 0 |
| 0.12835738 | 0.096468978 | 0 | 1.276345268 | 0.874313376 |
| 0 | 0 | 0 | 0 | 0 |
| 0 | 0 | 0 | 0 | 0 |
| 0 | 0 | 0 | 0 | 0 |
| 0 | 0.004295738 | 0.004179706 | 0.007618617 | 0.004496343 |
| 0 | 0 | 0.001419575 | 0 | 0 |
| 0 | 0 | 0 | 0 | 0 |
| 0 | 0 | 0 | 0 | 0 |
| 0 | 0.00252276 | 0 | 0.017570512 | 0 |
| 0 | 0 | 0 | 0 | 0 |
| 0.529099197 | 0.073257287 | 0 | 2.152698078 | 0.4493169 |
| 0 | 0 | 0 | 0 | 0 |
| 0 | 0 | 0 | 0 | 0 |
| 0 | 0 | 0.000400281 | 0.003242742 | 0.125555105 |
| 0 | 0 | 0 | 0 | 0 |
| 0 | 0 | 0 | 0 | 0 |
| 0 | 0 | 0 | 0 | 0 |
| 0 | 0 | 0 | 0 | 0 |
| 0 | 0 | 0 | 0 | 0 |
| 0 | 0 | 0.025742771 | 0 | 0 |
| 0 | 0 | 0 | 0 | 0 |
| 0.001919319 | 0.045626955 | 0.12797752 | 0.028742337 | 0.005535329 |
| 0 | 0 | 0 | 0 | 0 |
| 0 | 0 | 0 | 0 | 0 |
| 0 | 0 | 0 | 0 | 0 |
| 0 | 0 | 0 | 0 | 0 |
| 0 | 0 | 0 | 0 | 0 |
| 0 | 0 | 0 | 0 | 0 |

| | | | | |
|---|---|---|---|---|
| 0 | 0 | 0 | 0 | 0 |
| 0 | 0 | 0 | 0 | 0 |
| 0 | 0.041795093 | 0.083120418 | 0.273287055 | 0.007623148 |
| 0 | 0 | 0 | 0 | 0 |
| 0 | 0 | 0 | 0 | 0 |
| 0 | 0 | 0 | 0 | 0 |
| 0 | 0 | 0 | 0 | 0 |
| 0 | 0 | 0 | 0 | 0 |
| 0 | 0 | 0 | 0 | 0 |
| 0 | 0 | 0 | 0 | 0 |
| 0 | 0 | 0 | 0 | 0 |
| 0 | 0 | 0 | 0 | 0 |
| 0 | 0 | 0 | 0 | 0 |
| 0.001664693 | 0.003114008 | 0 | 0.001329559 | 0.00136286 |
| 0.009068307 | 0.002855785 | 0 | 0.002011858 | 0.037836292 |
| 0 | 0 | 0.059325624 | 0 | 0 |
| 0.002191747 | 0.002765824 | 0.001152431 | 0.002083938 | 0.001631229 |
| 0 | 0 | 0 | 0 | 0.015636384 |
| 0 | 0 | 0 | 0 | 0 |
| 0 | 0 | 0 | 0 | 0 |
| 0 | 0 | 0 | 0 | 0 |
| 0 | 0.077105509 | 0 | 0 | 0.00904557 |
| 0 | 0 | 0 | 0 | 0 |
| 0.0179648 | 0 | 0 | 0 | 0 |
| 0 | 0 | 0 | 0 | 0 |
| 0 | 0 | 0 | 0 | 0 |
| 0 | 0 | 0 | 0 | 0 |
| 0 | 0 | 0 | 0 | 0 |
| 0 | 0 | 0 | 0 | 0 |
| 0.0024531 | 0 | 0 | 0 | 0 |
| 0 | 0 | 0 | 0 | 0 |
| 0 | 0 | 0 | 0 | 0 |
| 0 | 0 | 0 | 0 | 0 |
| 0 | 0 | 0 | 0 | 0 |
| 0 | 0 | 0 | 0 | 0 |
| 0 | 0 | 0 | 0.005636148 | 0 |
| 0 | 0 | 0 | 0 | 0 |
| 0 | 0.001660716 | 0.572254343 | 0.06546991 | 0.000792895 |
| 0 | 0 | 0 | 0 | 0 |
| 0 | 0 | 0 | 0 | 0 |
| 0 | 0.000798961 | 0 | 0 | 0 |
| 0 | 0 | 0 | 0 | 0 |
| 0 | 0 | 0 | 0 | 0 |
| 0.019408794 | 0 | 0 | 0 | 0 |
| 0 | 0 | 0 | 0 | 0 |
| 0 | 0 | 0 | 0 | 0 |
| 0 | 0 | 0 | 0 | 0 |
| 0.022366963 | 0 | 0 | 0.010878767 | 0.003521445 |

| | | | | |
|---|---|---|---|---|
| 0 | 0 | 0 | 0 | 0 |
| 0 | 0 | 0 | 0 | 0 |
| 0 | 0 | 0.001673391 | 0 | 0 |
| 0 | 0 | 0 | 0 | 0 |
| 0.145948718 | 0 | 0.000900527 | 0.021410181 | 0 |
| 0 | 0 | 0 | 0 | 0 |
| 0 | 0 | 0 | 0 | 0 |
| 0.006680139 | 0.020826676 | 0.001207404 | 0.095312971 | 0.007613006 |
| 0 | 0 | 0 | 0 | 0 |
| 0 | 0.052100998 | 0 | 0.051555119 | 0.10104873 |
| 0 | 0 | 0 | 0 | 0 |
| 0 | 0.001202139 | 0 | 0.049493499 | 0 |
| 0.00927579 | 0.028919138 | 0.002438627 | 0.012847904 | 0.013713904 |
| 0 | 0 | 0 | 0 | 0.010343098 |
| 0 | 0 | 0 | 0 | 0 |
| 0 | 0 | 0 | 0 | 0 |
| 0 | 0 | 0 | 0.001275042 | 0 |
| 0 | 0 | 0 | 0 | 0.001215005 |
| 0 | 0 | 0 | 0 | 0 |
| 0.650463466 | 0 | 0 | 0 | 0 |
| 0 | 0 | 0 | 0 | 0 |
| 0 | 0 | 0 | 0.002206071 | 0.001681977 |
| 0 | 0 | 0 | 0 | 0 |
| 0 | 0 | 0 | 0 | 0 |
| 0 | 0 | 0 | 0 | 0 |
| 0 | 0.037476602 | 0.004145417 | 0.001360412 | 0 |
| 0 | 0 | 0 | 0.003378415 | 0 |
| 3.810376341 | 0 | 0 | 0 | 0 |
| 0 | 0 | 0 | 0 | 1.963066744 |
| 0 | 0 | 0 | 0 | 0 |
| 0.003759958 | 0 | 0 | 0 | 0.021221093 |
| 0 | 0 | 0 | 0 | 0 |
| 0 | 0 | 0 | 0 | 0.001932747 |
| 0 | 0 | 0 | 0 | 0.086686445 |
| 0 | 0 | 0 | 0 | 0 |
| 0 | 0 | 0 | 0 | 0 |
| 2.126737774 | 0.000687993 | 0 | 0 | 0.005789396 |
| 0 | 0 | 0 | 0 | 0 |
| 0 | 0 | 0 | 0 | 0 |
| 0 | 0 | 0 | 0 | 0.093141301 |
| 0 | 0 | 0 | 0 | 0 |
| 0 | 0 | 0 | 0 | 0.047472547 |
| 0 | 0 | 0 | 0 | 0 |
| 0.405795845 | 0.156096299 | 0 | 0.581190956 | 0 |
| 0 | 0 | 0 | 0 | 0 |
| 0 | 0 | 0 | 0 | 0 |
| 0.001089368 | 0.04075589 | 0 | 0.002658511 | 0.002162061 |
| 0.006921529 | 0 | 0 | 0 | 0 |

| | | | | |
|---|---|---|---|---|
| 0 | 0 | 0 | 0 | 0 |
| 0 | 0 | 0 | 0 | 0 |
| 0 | 0 | 0 | 0 | 0 |
| 0 | 0 | 0 | 0 | 0 |
| 0 | 0 | 0 | 0 | 0 |
| 0.053196581 | 0 | 0 | 0 | 1.631576822 |
| 0 | 0.010245402 | 0 | 0 | 0 |
| 0 | 0 | 0 | 0 | 0 |
| 0 | 0 | 0 | 0 | 0 |
| 0 | 0 | 0 | 0 | 0 |
| 0.03881071 | 0 | 0.000130813 | 0.001766233 | 0 |
| 0 | 0 | 0 | 0 | 0 |
| 0.000962503 | 0 | 0 | 0 | 0 |
| 0.008461479 | 0 | 0.000411952 | 0.113050938 | 0 |
| 0.30979931 | 0.031635181 | 0 | 0 | 3.762770729 |
| 0 | 0 | 0 | 0 | 0.00796818 |
| 0 | 0 | 0 | 0 | 0 |
| 0.03537736 | 0 | 0 | 0 | 0 |
| 0 | 0 | 0 | 0.071928803 | 0 |
| 0.001315428 | 0 | 0 | 0 | 0.019930061 |
| 0 | 0 | 0 | 0 | 0 |
| 0 | 0 | 0.13609413 | 0.212005338 | 0 |
| 0 | 0 | 0 | 0 | 0 |
| 0 | 0 | 0 | 0 | 0 |
| 0 | 0 | 0 | 0 | 0 |
| 0 | 0 | 0 | 0 | 0 |
| 0.331640576 | 0.002295326 | 0 | 0 | 0.203960845 |
| 0 | 0 | 0 | 0 | 0 |
| 0 | 0 | 0 | 0 | 0 |
| 0.849359073 | 0.222460826 | 0.033633656 | 0.142473049 | 0.366157959 |
| 0 | 0.090037266 | 0.003465514 | 0 | 0 |
| 0 | 0.000775518 | 0 | 0 | 0 |
| 0 | 0.001360975 | 0 | 0 | 0 |
| 0.006323668 | 0 | 0 | 0.001603364 | 0 |
| 0 | 0 | 0 | 0 | 0 |
| 0 | 0 | 0 | 0 | 0.003515683 |
| 0 | 0 | 0 | 1.671771344 | 0.015245347 |
| 0 | 0.000829896 | 0 | 0 | 0 |
| 0 | 1.032557717 | 0 | 0 | 0 |
| 2.157421029 | 0.113105183 | 0 | 0 | 0.105150358 |
| 0.085020909 | 0 | 0 | 0 | 0.001534004 |
| 0 | 0 | 0.095478367 | 0 | 0 |
| 0 | 0 | 0 | 0 | 0 |
| 0 | 0 | 0 | 0 | 0 |
| 0.005231346 | 0 | 0 | 0 | 0 |
| 0 | 0 | 0 | 0 | 0.235795254 |
| 0.004596983 | 0 | 0 | 0 | 0.003320722 |
| 0 | 0 | 0 | 0 | 0 |

| | | | | |
|---|---|---|---|---|
| 0 | 0.034846512 | 0.022235785 | 0.002442672 | 0.003848897 |
| 0 | 0 | 0 | 0 | 0 |
| 0 | 0.011338756 | 0 | 0.073828353 | 0.005582767 |
| 0 | 0 | 0 | 0 | 0 |
| 0 | 0 | 0 | 0 | 0 |
| 0 | 0 | 0 | 0 | 0 |
| 0 | 0 | 0 | 0 | 0 |
| 0 | 0 | 0 | 0.001493313 | 0 |
| 0 | 0 | 0 | 0 | 0 |
| 0 | 0 | 0 | 0 | 0.004994742 |
| 0 | 0 | 0 | 0 | 0 |
| 0 | 0 | 0 | 0 | 0 |
| 0 | 0 | 0 | 0 | 0 |
| 0.052217805 | 0 | 0.000634829 | 0.001205358 | 0 |
| 0 | 0 | 0 | 0 | 0 |
| 0 | 0 | 0.017270764 | 0 | 0 |
| 0.002614489 | 0.067247348 | 0.090593411 | 0.047157245 | 0 |
| 0.004764752 | 0.003796297 | 0 | 0.135730203 | 0.095747777 |
| 0 | 0.026661964 | 0 | 0 | 0 |
| 0 | 0 | 0 | 0 | 0 |
| 0 | 0.067800634 | 0 | 0 | 0 |
| 0.001819603 | 0.084937066 | 0 | 0.006459035 | 0 |
| 0 | 0 | 0 | 0 | 0 |
| 0 | 0.258539479 | 0 | 0 | 0.30587288 |
| 0 | 0 | 0 | 0 | 0 |
| 0.33373599 | 0.003579254 | 0 | 0.006647673 | 0.008843475 |
| 0.006638229 | 0 | 0 | 0.000662729 | 0.000864599 |
| 0 | 0 | 0 | 0 | 0 |
| 0 | 0 | 0 | 0 | 0 |
| 0 | 0 | 0 | 0 | 0 |
| 0.017164404 | 0 | 0 | 0 | 0 |
| 0 | 0.023074215 | 0.00247341 | 0 | 0.000700207 |
| 0.67183342 | 0 | 0.001648515 | 0.113039716 | 0.053735336 |
| 0 | 0.001600923 | 0 | 0.1165421 | 0 |
| 0 | 0.00285663 | 0.001204437 | 0.005122609 | 0.038052071 |
| 0 | 0 | 0 | 0 | 0 |
| 0 | 0 | 0.000255492 | 0.003665246 | 0.029051831 |
| 0.131227282 | 0.084379241 | 0 | 0.401294968 | 0.00056564 |
| 0 | 0 | 0 | 0.002254613 | 0 |
| 0.014913133 | 0 | 0 | 0 | 0.002235265 |
| 0 | 0 | 0 | 0 | 0 |
| 0 | 0 | 0 | 0 | 0 |
| 0 | 0 | 0 | 0 | 0.022275733 |
| 0 | 0.000924829 | 0 | 0 | 0 |
| 0 | 4.793755177 | 0 | 0 | 0.125002564 |
| 0 | 0 | 0 | 0 | 0.259894341 |
| 0 | 0 | 0 | 0 | 0 |
| 0 | 0 | 0 | 0 | 0.000957703 |

| | | | | |
|---|---|---|---|---|
| 0.08113198 | 0 | 0 | 0 | 0 |
| 0.560605486 | 0.263318024 | 0.646123101 | 0.017231575 | 0 |
| 0 | 0 | 0 | 0 | 0 |
| 0 | 0.011297984 | 0 | 0 | 0 |
| 0 | 0 | 0 | 0.001126999 | 0 |
| 0 | 0 | 0 | 0 | 0 |
| 0 | 0 | 0 | 0 | 0 |
| 0 | 0 | 0 | 0 | 0 |
| 0 | 0 | 0 | 0 | 0 |
| 0 | 0 | 0 | 0 | 0 |
| 0.000769565 | 0.001466223 | 0 | 0 | 0.003341078 |
| 0.002674674 | 0.036532034 | 0.016826046 | 0.037383722 | 2.589981039 |
| 0 | 0 | 0 | 0 | 0 |
| 0 | 0 | 0 | 0 | 0 |
| 0 | 0.025540482 | 0 | 0.007052195 | 0.060130711 |
| 0 | 0 | 0 | 0 | 0 |
| 0.001611691 | 0.006420541 | 0.009639571 | 0.008402738 | 0.000699718 |
| 0.1931796 | 0.001087141 | 0 | 0.006730416 | 0.067995063 |
| 0 | 0 | 0 | 0 | 0.000529755 |
| 0 | 0.02764903 | 0 | 0 | 0.001348166 |
| 0.00249309 | 0.007916649 | 0.012416933 | 0.000921845 | 0.003401763 |
| 0 | 0 | 0 | 0 | 0 |
| 0 | 0 | 0 | 0 | 0 |
| 0 | 0 | 0 | 0 | 0 |
| 0 | 0 | 0 | 0 | 0 |
| 0 | 0 | 0 | 0 | 0 |
| 0 | 0 | 0 | 0.069334918 | 0 |
| 0.133667033 | 0 | 0.011804097 | 0.059766893 | 0.089841481 |
| 0 | 0 | 0 | 0 | 0 |
| 0 | 0 | 0 | 0.003072811 | 0 |
| 0 | 0 | 0 | 0 | 0 |
| 0 | 0 | 0 | 0 | 0 |
| 0 | 0 | 0 | 0 | 0 |
| 0 | 0 | 0 | 0 | 0 |
| 0 | 0 | 0 | 0 | 0 |
| 0.903897337 | 0.239500691 | 0.025610794 | 0.003979747 | 0.246063581 |
| 0 | 0 | 0 | 0 | 0.026880227 |
| 0 | 0 | 0 | 0 | 0 |
| 0.001816005 | 0.001887253 | 0.004105914 | 0.024415226 | 0 |
| 0 | 0 | 0 | 0 | 0 |
| 0 | 4.885314529 | 0 | 0 | 0.115344744 |
| 0 | 0 | 0 | 0 | 0 |
| 0.062213095 | 0.044707514 | 0.011715977 | 0.036267869 | 0.009194864 |
| 0.000638995 | 0 | 0 | 0 | 0.008718942 |
| 0 | 0 | 0 | 0 | 1.977908822 |
| 0 | 0 | 0.083719471 | 0 | 0 |
| 0 | 0 | 0 | 0 | 0 |
| 0.00309866 | 0 | 0 | 0.011686755 | 0.008327974 |

| | | | | |
|---|---|---|---|---|
| 0 | 0 | 0 | 0 | 0 |
| 0 | 0 | 0 | 0 | 0 |
| 0 | 0 | 0 | 0.0014802 | 0 |
| 0.025694528 | 0.918970619 | 0.000202655 | 0 | 0.193050778 |
| 0 | 0 | 0 | 0 | 0 |
| 0 | 0 | 0 | 0 | 0 |
| 0 | 0 | 0 | 0 | 0 |
| 0 | 0.97452549 | 0.064450809 | 0 | 0 |
| 0 | 0 | 16.20991619 | 0 | 0.065361203 |
| 0 | 0 | 0 | 0 | 0 |
| 0 | 0 | 0 | 0 | 0 |
| 0 | 0 | 0 | 0 | 0 |
| 0 | 0 | 0 | 0 | 0 |
| 0 | 0 | 0 | 0 | 0 |
| 0 | 0.22019779 | 0.003524957 | 0.047729021 | 0.01330524 |
| 0 | 0 | 0 | 0 | 0 |
| 0 | 0 | 0 | 0 | 0 |
| 0 | 0 | 0 | 0 | 0 |
| 0 | 0 | 0 | 0 | 0 |
| 0 | 0 | 0 | 0 | 0 |
| 0 | 0 | 0 | 0 | 0 |
| 0 | 0 | 0 | 0 | 0 |
| 0 | 0 | 0 | 0 | 0 |
| 0.272002001 | 0.088462613 | 0 | 0.009377394 | 0 |
| 0 | 0 | 0 | 0 | 0 |
| 0 | 0 | 0 | 0 | 1.23145413 |
| 0 | 0 | 0 | 0 | 0 |
| 0 | 0 | 0 | 0.125972615 | 1.094039621 |
| 0 | 0 | 0 | 0 | 0 |
| 0 | 0 | 0 | 0 | 0 |
| 0 | 0 | 0.304943933 | 0 | 0 |
| 0 | 0 | 0 | 0 | 0 |
| 0 | 0 | 0 | 0 | 0.011929744 |
| 0 | 0 | 0 | 0 | 0 |
| 0 | 0 | 0 | 0 | 0 |
| 0 | 0 | 0 | 0 | 0 |
| 0 | 0 | 0.044682408 | 0 | 0 |
| 0 | 0.00186336 | 0 | 0 | 0 |
| 0 | 0 | 0 | 0 | 0 |
| 0 | 0 | 0 | 0 | 0 |
| 0.008738174 | 0.017302994 | 0.001396998 | 0.786554149 | 0.019949839 |
| 0 | 0 | 0 | 0 | 0 |
| 0.209366925 | 0.964835528 | 0.001266214 | 7.679220054 | 0.084553696 |
| 0 | 0.00277945 | 0 | 0 | 0 |
| 0 | 0 | 0 | 0 | 0 |
| 0 | 0 | 0 | 0 | 0 |
| 0 | 0 | 0 | 0 | 0 |
| 0 | 0.037617355 | 0.00070906 | 0.008125044 | 0 |

| | | | | |
|---|---|---|---|---|
| 0 | 0 | 0 | 0 | 0 |
| 0 | 0 | 0 | 0.066619303 | 0.030895581 |
| 1.112548648 | 0.038888754 | 0 | 0.311550859 | 0.016305292 |
| 0 | 0 | 0 | 0 | 0 |
| 0 | 0 | 0 | 0 | 0 |
| 0 | 0.088451777 | 0.090914189 | 0 | 0 |
| 0 | 0 | 0 | 0 | 0 |
| 0 | 0.09550795 | 0 | 0 | 0 |
| 0 | 0 | 0 | 0 | 0 |
| 0 | 0 | 0 | 0 | 0 |
| 0.005611261 | 0.104826562 | 0.005900855 | 0.042148479 | 0.018892497 |
| 8.162737198 | 0.081993776 | 0.213761745 | 0.859827121 | 6.074929809 |
| 0 | 0 | 0 | 0 | 0 |
| 0.000865129 | 0.194798713 | 0.554016625 | 0.026294957 | 0 |
| 0.361557229 | 0.043454815 | 0 | 0.171216526 | 0.001353074 |
| 0 | 0 | 0.361886995 | 0 | 0 |
| 0 | 0 | 0 | 0 | 0 |
| 0 | 0 | 0 | 0 | 0 |
| 0 | 0 | 0 | 0 | 0 |
| 0 | 0 | 0 | 0 | 0.001474337 |
| 0.056884667 | 0.001629194 | 0.021726128 | 0.085857131 | 0 |
| 0.401637099 | 0.069647545 | 0 | 0.246898921 | 0.102041066 |
| 0.038498694 | 0.569580659 | 0.009342828 | 2.19855227 | 2.409688525 |
| 0 | 0 | 0 | 0 | 0 |
| 0 | 0 | 0 | 0 | 0 |
| 0 | 0.000976131 | 0.003210206 | 0 | 0 |
| 0.188306332 | 0.224366416 | 0 | 0 | 0 |
| 0 | 0 | 0 | 0 | 0 |
| 0 | 0 | 0 | 0 | 0 |
| 0 | 0 | 0 | 0 | 0 |
| 0 | 0 | 0 | 0 | 0 |
| 0 | 0 | 0.004446857 | 0.006895364 | 0 |
| 2.298578311 | 8.071904007 | 0.001262569 | 0.016620972 | 0.00204243 |
| 0 | 0 | 0 | 0.000557133 | 0 |
| 0.002354605 | 0.040845826 | 0 | 0 | 0 |
| 0 | 1.427560217 | 0.089172812 | 0.011428352 | 0.030096785 |
| 0 | 0.007869314 | 0.000157997 | 0.003199893 | 0 |
| 0 | 0 | 0 | 0 | 0 |
| 0 | 0 | 0.523588409 | 0.233551357 | 0 |
| 0 | 0 | 0 | 0 | 0 |
| 0 | 0 | 0 | 0 | 0 |
| 0 | 0 | 0 | 0 | 0 |
| 0 | 0 | 0 | 0 | 0 |
| 0 | 0 | 0 | 0 | 0 |
| 0.001418941 | 0.5380855 | 0.031745943 | 0.019265788 | 0.000844848 |
| 0 | 0 | 0 | 0 | 0 |
| 0 | 0 | 0 | 0 | 0 |
| 0.171847664 | 1.480303757 | 15.75064989 | 0.315104409 | 0.058344509 |

| | | | | |
|---|---|---|---|---|
| 0 | 0 | 0 | 0 | 0 |
| 0 | 0 | 0 | 0 | 0 |
| 0 | 0 | 0 | 0 | 0 |
| 0 | 0 | 0 | 0 | 0 |
| 0 | 0 | 0 | 0 | 0 |
| 0 | 0 | 0 | 0 | 0 |
| 0 | 0 | 0 | 0 | 0 |
| 0.011128276 | 0.161792666 | 0 | 0 | 0 |
| 0 | 0 | 0 | 0 | 0 |
| 0 | 0 | 0 | 0 | 0 |
| 0 | 0.04801135 | 0 | 0.460957666 | 0.05914434 |
| 0 | 0 | 0 | 0 | 0 |
| 0 | 0 | 0 | 0.002197768 | 0 |
| 0 | 0 | 0.025774866 | 0 | 0 |
| 0 | 0 | 0 | 0 | 0 |
| 0 | 0 | 0 | 0 | 0 |
| 0 | 0 | 0 | 0 | 0 |
| 0 | 0.238062231 | 0.010700646 | 0 | 0 |
| 0 | 0 | 0 | 0 | 0 |
| 0 | 0 | 0 | 0 | 0 |
| 0 | 0 | 0 | 0 | 0 |
| 0 | 0 | 0 | 0 | 0 |
| 0 | 0 | 0.000713715 | 0 | 0 |
| 0 | 0 | 0 | 0 | 0.026843834 |
| 0 | 0 | 0 | 0 | 0 |
| 0 | 0 | 0 | 0 | 0 |
| 0 | 0 | 0 | 0 | 0 |
| 0 | 0 | 0 | 0 | 0 |
| 0 | 0 | 0 | 0 | 0 |
| 0 | 0 | 0.022896973 | 0.121522004 | 0 |
| 0.003819012 | 0 | 0 | 0 | 0.030949897 |
| 0 | 0 | 0 | 0 | 0 |
| 0 | 0 | 0 | 0 | 0 |
| 0 | 0 | 0 | 0 | 0 |
| 0 | 0.001335614 | 0.000534976 | 0 | 0 |
| 0 | 0 | 0 | 0 | 0 |
| 0.145452528 | 0 | 0.046582581 | 0 | 0 |
| 0.026224086 | 0.003910206 | 0 | 0 | 0.000424294 |
| 0 | 0 | 0 | 0 | 0 |
| 0 | 0.005493034 | 0 | 0 | 0 |
| 0 | 0 | 0 | 0 | 0.044188322 |
| 0 | 0 | 0 | 0 | 0 |
| 0 | 0 | 0 | 0 | 0 |
| 0 | 0 | 0 | 0 | 0 |
| 0 | 0 | 0.030735332 | 0.32295921 | 0 |
| 0 | 0 | 0 | 0 | 0 |
| 0 | 0 | 0 | 0 | 0 |
| 0 | 0 | 0 | 0 | 0 |

| | | | | |
|---|---|---|---|---|
| 0 | 0 | 0 | 0 | 0 |
| 0 | 0 | 0 | 0 | 0 |
| 0 | 0 | 0 | 0 | 0 |
| 0 | 0 | 0 | 0 | 0.067612152 |
| 0 | 0 | 0 | 0 | 0 |
| 0 | 0 | 0 | 0 | 0 |
| 0 | 0 | 0 | 0 | 0 |
| 0 | 0 | 0 | 0 | 0 |
| 0.120767648 | 0.061935808 | 0 | 1.352629082 | 0.107788793 |
| 0 | 0 | 0.000366211 | 0 | 0 |
| 0 | 0 | 0 | 0 | 0 |
| 0 | 0 | 0 | 0 | 0 |
| 0 | 0 | 0 | 0 | 0 |
| 0 | 0 | 0 | 0 | 0 |
| 0 | 0 | 0.009209768 | 0 | 0 |
| 1.255354329 | 0 | 0 | 1.696537151 | 0 |
| 0 | 0 | 0 | 0 | 0 |
| 0 | 0 | 0 | 0 | 0 |
| 0 | 0 | 0 | 0 | 0 |
| 0 | 0 | 0 | 0 | 0 |
| 0 | 0 | 0 | 0 | 0 |
| 0 | 0 | 0 | 0 | 0 |
| 0 | 0 | 0 | 0 | 0 |
| 0 | 0 | 0 | 0 | 0 |
| 0.007396839 | 0 | 0.0351981 | 0.359386226 | 0.005092283 |
| 0 | 0 | 0 | 0.001160056 | 2.73543339 |
| 0 | 0 | 0.461822755 | 0 | 0 |
| 0 | 0 | 0 | 0 | 0 |
| 0 | 0 | 0 | 0.076184275 | 0 |
| 0 | 0 | 0 | 0 | 0 |
| 0 | 0 | 0 | 0 | 0 |
| 0 | 0 | 0 | 0 | 0 |
| 0 | 0 | 0 | 0 | 0 |
| 0 | 0.083749656 | 0 | 0.003549489 | 0.000367514 |
| 0 | 0.152536356 | 0.209668302 | 0.005848436 | 0 |
| 0.00414096 | 0.250435445 | 0 | 0.024804813 | 0.020717556 |
| 0 | 0 | 0 | 0 | 0 |
| 0 | 0 | 0.063820873 | 0.125348565 | 0 |
| 0 | 0 | 0 | 0.446704724 | 0 |
| 0 | 0 | 0 | 0 | 0 |
| 0 | 0 | 0 | 0.004954695 | 0 |
| 0 | 0 | 0 | 0 | 0 |
| 0 | 1.860263746 | 6.518619829 | 0.005021662 | 0.000818911 |
| 0 | 0 | 0 | 0 | 0 |
| 0 | 0 | 0 | 0 | 0 |
| 0 | 0 | 0 | 0 | 0.024574656 |
| 0 | 0 | 0 | 0 | 0 |
| 0.029860504 | 0 | 0.010041205 | 0.789652559 | 0.030751778 |

| | | | | |
|---|---|---|---|---|
| 0 | 0 | 0 | 0 | 0 |
| 0 | 0 | 0 | 0 | 0 |
| 0 | 0 | 0 | 0 | 0 |
| 0 | 0 | 0 | 0 | 0 |
| 0 | 0.001339487 | 0 | 0.000857862 | 0.000383716 |
| 0 | 0 | 0 | 0 | 0 |
| 0 | 0 | 0 | 0 | 0 |
| 0 | 0 | 0 | 0 | 0 |
| 0 | 0 | 0 | 0 | 0 |
| 0 | 0 | 0 | 0 | 0 |
| 0 | 0 | 0 | 0 | 0 |
| 4.703401985 | 0 | 0.059897907 | 0 | 0 |
| 0 | 5.835375814 | 0.278921601 | 0 | 0 |
| 0 | 0 | 0 | 0 | 0 |
| 0.009838182 | 0.063668692 | 0 | 0.512924947 | 0 |
| 0 | 0 | 0 | 0 | 0 |
| 0 | 0 | 0 | 0 | 0 |
| 0 | 0 | 0 | 0 | 0 |
| 0 | 0 | 0 | 0.00139134 | 0.001166882 |
| 0 | 0 | 0 | 0 | 0 |
| 0 | 0 | 0.011033911 | 0 | 0 |
| 0 | 0 | 0 | 0 | 0 |
| 0.037858747 | 2.235219302 | 0.083332609 | 0 | 0.109492184 |
| 0 | 0 | 0 | 0 | 0 |
| 0 | 0 | 0 | 0 | 0 |
| 0 | 0 | 0 | 0 | 0 |
| 0 | 0 | 0 | 0 | 0 |
| 0 | 0 | 0 | 0 | 0 |
| 0 | 0 | 0 | 0 | 0 |
| 0 | 0 | 0 | 0 | 0 |
| 0 | 0 | 0 | 0 | 0 |
| 0.017795611 | 0.053069159 | 0.08756946 | 0.026778163 | 0.046787842 |
| 0 | 0 | 0 | 0 | 0 |
| 0.007730765 | 0.075620701 | 0 | 0.004502181 | 0 |
| 0.005758181 | 0.237802748 | 0 | 0.015163213 | 0.019005628 |
| 0.006454777 | 0 | 0.707257147 | 0.150363233 | 0.003362822 |
| 0 | 0 | 0 | 0 | 0 |
| 0 | 0 | 0 | 0 | 0 |
| 0 | 0 | 0.003478356 | 0 | 0 |
| 0 | 0 | 0 | 0 | 0 |
| 0 | 0 | 0 | 0 | 0 |
| 0 | 0 | 0 | 0 | 0 |
| 0 | 0 | 0 | 0 | 0 |
| 0 | 0 | 0 | 0 | 0 |
| 0.038720195 | 0.346519583 | 0.030765824 | 0.033321635 | 0.02872611 |
| 0 | 0 | 0 | 0 | 0 |
| 0 | 0 | 0 | 0.101371805 | 0 |
| 0 | 0 | 0 | 0 | 0.001302065 |

| | | | | |
|---|---|---|---|---|
| 0 | 0 | 0 | 0 | 0 |
| 0 | 0 | 0 | 0 | 0 |
| 0.00102029 | 0 | 0 | 0 | 0.006169805 |
| 0 | 0 | 0 | 0 | 0 |
| 0 | 0 | 0 | 0 | 0.082400859 |
| 0 | 0 | 0 | 0 | 0 |
| 0 | 0 | 0.002805058 | 0 | 0 |
| 0 | 0 | 0 | 0 | 0.007081808 |
| 0 | 0 | 0 | 0 | 0 |
| 0 | 0 | 0 | 0 | 0 |
| 0 | 0 | 0 | 0 | 1.947430313 |
| 0 | 0.001078849 | 0 | 0.002763758 | 0 |
| 0 | 0 | 0 | 0 | 0 |
| 0 | 0 | 0 | 0.794356024 | 0 |
| 0 | 0 | 0 | 0 | 0 |
| 0 | 0 | 0 | 0 | 0 |
| 0 | 0 | 0 | 0 | 0 |
| 0 | 0 | 0 | 0 | 0 |
| 0 | 0 | 0 | 0.735043751 | 0.935140538 |
| 0 | 0 | 0 | 0 | 0 |
| 0 | 0 | 0 | 0 | 0 |
| 0 | 0 | 0 | 0 | 0 |
| 0 | 0 | 0 | 0 | 0 |
| 0 | 0 | 0 | 0 | 0 |
| 0.001077839 | 0.000980111 | 0.001239728 | 0 | 0 |
| 0.01561273 | 0.024934429 | 0 | 0.058069209 | 0.005724523 |
| 0 | 0 | 0 | 0 | 0 |
| 0 | 0 | 0.063247812 | 0.061774155 | 0 |
| 0 | 0 | 0 | 0 | 0 |
| 0 | 0 | 0.23248901 | 0.025892758 | 0 |
| 0 | 0 | 0 | 0 | 0 |
| 0 | 0 | 0 | 0 | 0 |
| 0 | 0 | 0 | 0 | 0 |
| 0 | 0 | 0 | 0 | 0 |
| 0 | 0.025754671 | 0 | 0.004003483 | 0.00411868 |
| 0 | 0 | 0 | 0 | 0 |
| 0 | 0 | 0 | 0 | 0 |
| 0 | 0.015463189 | 0.004809042 | 0.00145369 | 0.000914379 |
| 0 | 0 | 0 | 0 | 0 |
| 0 | 0 | 0 | 0 | 0 |
| 25.01032659 | 5.678796543 | 0.020858158 | 0.123926399 | 10.85738086 |
| 1.808838054 | 0.789950268 | 0.088242456 | 0 | 0 |
| 0 | 0 | 10.52915649 | 0 | 0 |
| 0 | 0 | 0 | 0 | 0 |
| 0 | 0 | 0 | 0 | 0 |
| 0 | 0 | 0 | 0 | 0 |
| 0 | 0 | 0 | 0 | 0 |
| 0 | 0.023981929 | 0.008744281 | 0 | 0.000403168 |

| | | | | |
|---|---|---|---|---|
| 0 | 0 | 0 | 0 | 0 |
| 0.003120243 | 0 | 0 | 0 | 0.011030791 |
| 0 | 0 | 0 | 0 | 0 |
| 0 | 0 | 0 | 0 | 0 |
| 0 | 1.118065012 | 4.780795451 | 0.044411728 | 0 |
| 0 | 0 | 0 | 0 | 0 |
| 0 | 0 | 0 | 0 | 0 |
| 0 | 0 | 0 | 0 | 0 |
| 5.303047188 | 0.07354616 | 0 | 0.004739822 | 0.103934084 |
| 0 | 0 | 0 | 0 | 0 |
| 0 | 0 | 0 | 0 | 0 |
| 0 | 0 | 0 | 0 | 0 |
| 0 | 0 | 0 | 0 | 0 |
| 0.089988857 | 0.023933232 | 0 | 0.001594803 | 0.013771506 |
| 0 | 0 | 16.36902202 | 0 | 0 |
| 0 | 0 | 0 | 0 | 0 |
| 0.001151944 | 0.006464549 | 0.000999401 | 0.003680145 | 0 |
| 0 | 0 | 0 | 0 | 0 |
| 0 | 0 | 0 | 0 | 0 |
| 0 | 0 | 0 | 0 | 0 |
| 0 | 0 | 0 | 0 | 0 |
| 0 | 0 | 0 | 0 | 0 |
| 0 | 0 | 0 | 0 | 0 |
| 0 | 0 | 0 | 0 | 0 |
| 0 | 0 | 0 | 0 | 0 |
| 0 | 0 | 0 | 0 | 0 |
| 0 | 0 | 0.054195249 | 0 | 0 |
| 0.00158237 | 0.000822226 | 0 | 0.049649654 | 0.005383745 |
| 0 | 0 | 0 | 0 | 0 |
| 0 | 0 | 0 | 0 | 0 |
| 0 | 0 | 0 | 0 | 0 |
| 0 | 0 | 0 | 0 | 0 |
| 0 | 0 | 0 | 0 | 0 |
| 0 | 0 | 0 | 0 | 0 |
| 0 | 0 | 0.002689404 | 0 | 0.067478833 |
| 0.048638289 | 0.001087023 | 0 | 0.043684911 | 0 |
| 0 | 0 | 0 | 0.016544085 | 0.001193557 |
| 0 | 0 | 0 | 0 | 0 |
| 0 | 0 | 0 | 0 | 0 |
| 0 | 0 | 0 | 0 | 0 |
| 0 | 0.868521786 | 0.192056915 | 0.001278951 | 0 |
| 0 | 0 | 0 | 0 | 0 |
| 0 | 0 | 0 | 0 | 0 |
| 0 | 0.000718809 | 0 | 0 | 0 |
| 0 | 0 | 0 | 0 | 0 |
| 0 | 0 | 0 | 0 | 0 |
| 0 | 0 | 0 | 0 | 0 |
| 0 | 0 | 0 | 0 | 0 |

| | | | | |
|---|---|---|---|---|
| 0 | 0 | 0.044898354 | 0 | 0 |
| 0 | 0 | 0 | 0 | 0 |
| 0 | 0 | 0 | 0 | 0 |
| 0 | 0 | 0 | 0 | 0 |
| 0.005635281 | 0 | 0 | 0.00211994 | 0 |
| 0 | 0 | 0 | 0 | 0 |
| 0.00781417 | 0.00214961 | 0.009879073 | 0.061951435 | 0 |
| 0 | 0 | 0 | 0 | 0 |
| 0 | 0 | 0 | 0 | 0 |
| 0 | 0 | 0 | 0 | 0.031674963 |
| 0 | 0 | 0 | 0 | 0 |
| 0 | 0 | 0 | 0 | 0 |
| 0 | 0 | 0 | 0 | 0 |
| 0 | 0 | 0 | 0 | 0 |
| 0 | 0 | 0 | 0 | 0 |
| 0 | 0 | 0 | 0 | 0 |
| 0 | 0.005732262 | 0 | 0 | 0 |
| 0 | 0 | 0 | 0 | 0 |
| 0 | 0 | 0 | 0 | 0 |
| 0 | 0 | 0 | 0 | 0 |
| 0.000656429 | 0 | 0.218395758 | 0.232575325 | 0 |
| 0 | 0 | 0 | 0 | 0 |
| 0 | 0 | 0 | 0 | 0 |
| 0 | 0.02386393 | 0 | 0 | 0.001197928 |
| 0 | 0 | 0 | 0 | 0 |
| 0 | 0 | 0 | 0 | 0 |
| 0 | 0 | 0 | 0 | 0 |
| 0 | 0 | 0 | 0 | 0 |
| 0 | 0 | 0 | 0 | 0 |
| 0.001235482 | 1.097781321 | 0 | 0.602743966 | 0.714926672 |
| 0 | 0 | 0 | 0 | 0 |
| 0 | 0 | 0 | 0 | 0 |
| 0 | 0 | 0 | 0 | 0 |
| 0 | 0 | 0 | 0.020287837 | 0 |
| 0 | 0 | 0 | 0 | 0 |
| 0 | 0 | 0 | 0 | 0 |
| 0 | 1.166352238 | 0.001270747 | 0.000714899 | 0 |
| 0 | 0 | 0 | 0 | 0 |
| 0 | 0 | 0 | 0 | 0 |
| 0 | 0 | 0 | 0 | 0 |
| 0 | 0 | 0 | 0 | 0 |
| 0 | 0 | 0 | 0 | 0 |
| 0 | 0 | 0 | 0 | 0 |
| 0 | 0 | 0 | 0 | 0 |
| 0 | 0 | 0 | 0 | 0 |
| 0 | 0 | 0 | 0 | 0 |
| 0 | 0 | 0 | 0 | 0 |
| 0 | 0 | 0 | 0 | 0 |

| | | | | |
|---|---|---|---|---|
| 0.007389611 | 0 | 0 | 0.001075875 | 0.021397622 |
| 0 | 0 | 0 | 0 | 0 |
| 0 | 0 | 0 | 0 | 0 |
| 0 | 0 | 0 | 0 | 0 |
| 0 | 0 | 0 | 0 | 0 |
| 0 | 0 | 0 | 0 | 0 |
| 0 | 0.000645713 | 0 | 0 | 0 |
| 1.13124286 | 7.628630287 | 0.098304316 | 0 | 0.62982163 |
| 0.024285623 | 0.007157169 | 0.377319506 | 1.527110794 | 0.247786821 |
| 0.02851466 | 0.111715277 | 0 | 0 | 0.044134906 |
| 0 | 0 | 0 | 0 | 0 |
| 0 | 0 | 0 | 0 | 0 |
| 0 | 0 | 0 | 0 | 0 |
| 0 | 0 | 0 | 0 | 0 |
| 0 | 0.076115747 | 0 | 0 | 0 |
| 0 | 0 | 0 | 0 | 0 |
| 0 | 0 | 0 | 0 | 0.00150529 |
| 0 | 0 | 0 | 0 | 0 |
| 0 | 0 | 0 | 0 | 0 |
| 0 | 0 | 0 | 0 | 0 |
| 0 | 0 | 0 | 0 | 0 |
| 0 | 0 | 0 | 0 | 0 |
| 0 | 0 | 0 | 0 | 0 |
| 0 | 0 | 0 | 0 | 0 |
| 0 | 0 | 0 | 0 | 0 |
| 0 | 0 | 0 | 0 | 0 |
| 0 | 0 | 0 | 0 | 0 |
| 0 | 0 | 0 | 0 | 0 |
| 0 | 0 | 0 | 0 | 0.001743072 |
| 0 | 0 | 0 | 0 | 0 |
| 0 | 0 | 0 | 0 | 0 |
| 0 | 0 | 0 | 0 | 0 |
| 0 | 0 | 0 | 0 | 0 |
| 0 | 0 | 0 | 0 | 0 |
| 0 | 0 | 0 | 0 | 0 |
| 0 | 0 | 0 | 0 | 0 |
| 0 | 0 | 0 | 0 | 0 |
| 0 | 0 | 0 | 0.004970611 | 0 |
| 0 | 0 | 0.008662788 | 0 | 0 |
| 0.004610716 | 0 | 0.000181825 | 0 | 0.002230521 |
| 0 | 0 | 0 | 0 | 0 |
| 1.464847882 | 0 | 0 | 0.002532764 | 0 |
| 0 | 0 | 0 | 0 | 0 |
| 0 | 0 | 0 | 0 | 0 |
| 0 | 0 | 0 | 0 | 0 |
| 0 | 0 | 0 | 0 | 0 |
| 0 | 0 | 0 | 0 | 0 |
| 0 | 0 | 0 | 0 | 0 |

| | | | | |
|---|---|---|---|---|
| 0 | 0 | 0 | 0 | 0 |
| 0 | 0 | 0 | 0.00124846 | 0 |
| 0 | 0 | 0 | 0 | 0 |
| 0 | 0 | 0 | 0 | 0 |
| 0.132295944 | 0 | 8.457660985 | 0.511244286 | 0.002168779 |
| 0 | 0 | 0 | 0 | 0 |
| 0 | 0 | 0 | 0 | 0 |
| 0.022031531 | 0.038770405 | 0 | 0.096583457 | 0.20310645 |
| 0 | 0 | 0.001773526 | 0 | 0 |
| 0 | 0 | 0 | 0 | 0 |
| 0 | 0 | 0 | 0 | 0 |
| 0 | 0 | 0 | 0 | 0 |
| 0 | 0 | 0 | 0 | 0 |
| 0 | 0 | 0 | 0 | 0 |
| 0 | 0 | 0 | 0 | 0 |
| 0 | 0 | 0 | 0.002882639 | 0 |
| 0 | 0 | 0 | 0 | 0 |
| 0 | 0 | 0 | 0 | 0 |
| 0 | 0 | 0 | 0 | 0 |
| 0 | 0 | 0 | 0 | 0 |
| 0 | 0 | 0 | 0 | 0 |
| 0 | 0 | 0 | 0 | 0 |
| 0 | 0 | 0 | 0 | 0 |
| 0.001016879 | 0.028004526 | 0 | 0.092552652 | 0.039733154 |
| 0 | 0 | 0 | 2.083336662 | 0 |
| 0 | 0 | 0 | 0 | 0 |
| 0 | 1.061986147 | 0.017238137 | 0.008047157 | 0.151591712 |
| 0 | 0 | 0 | 0 | 0 |
| 0 | 0 | 0 | 0 | 0 |
| 0 | 0 | 0 | 0 | 0 |
| 0 | 0 | 0 | 0 | 0 |
| 0 | 0 | 0 | 0 | 0 |
| 0 | 0 | 0 | 0 | 0 |
| 0 | 0 | 0 | 0 | 0 |
| 0 | 0 | 0 | 0 | 0 |
| 0 | 0 | 0 | 0 | 0 |
| 0 | 0 | 0 | 0 | 0 |
| 0 | 0 | 0 | 0 | 0 |
| 0 | 0 | 0 | 0 | 0 |
| 0 | 0 | 0 | 0 | 0 |
| 0 | 0 | 0 | 0 | 0 |
| 0 | 0 | 3.082579347 | 0 | 0 |
| 0 | 0 | 0 | 0 | 0 |
| 0 | 0 | 0 | 0 | 0.015562081 |
| 0 | 0 | 0 | 0 | 0 |
| 0 | 0 | 0 | 0 | 0 |
| 0 | 0 | 0 | 0 | 0 |
| 0 | 0 | 0 | 0 | 0 |

| | | | | |
|---|---|---|---|---|
| 0 | 2.240670059 | 0.027075789 | 0 | 0 |
| 0 | 0 | 0 | 0 | 0 |
| 0 | 0 | 0 | 0 | 0 |
| 0 | 0 | 0 | 0 | 0 |
| 0 | 0 | 0 | 0 | 0 |
| 0.023306192 | 0 | 0 | 0 | 0 |
| 0 | 0 | 0 | 0 | 0 |
| 0 | 0 | 0 | 0 | 0 |
| 0 | 0 | 0 | 0 | 0 |
| 0 | 0 | 0 | 0 | 0 |
| 0 | 0 | 0 | 0 | 0 |
| 0 | 0 | 0 | 0 | 0 |
| 0 | 0 | 0 | 0 | 0 |
| 0 | 0 | 0 | 0 | 0 |
| 0 | 0 | 0 | 0 | 0 |
| 0 | 0 | 0 | 0 | 0 |
| 0 | 0 | 0 | 0 | 0 |
| 0 | 0 | 0 | 0 | 0 |
| 0 | 0 | 0 | 0 | 0 |
| 0 | 0 | 0 | 0 | 0 |
| 0.153666276 | 0.006051052 | 0 | 0 | 0.039001814 |
| 0 | 0 | 0 | 0 | 0 |
| 0 | 0.050503907 | 0.088131482 | 0 | 0.007138616 |
| 0 | 0 | 0 | 0 | 0 |
| 0 | 0 | 0 | 0 | 0 |
| 0 | 0 | 0 | 0 | 0 |
| 0 | 0 | 0 | 0 | 0 |
| 0 | 0 | 0 | 0 | 0 |
| 0 | 0 | 0 | 0 | 0.027808993 |
| 0 | 0 | 0 | 0 | 0 |
| 0 | 0 | 0 | 0 | 0 |
| 0 | 0 | 0 | 0 | 0 |
| 0.068566114 | 0.254064186 | 0.001692415 | 12.67787445 | 0.006462442 |
| 0 | 0 | 0 | 0 | 0 |
| 0 | 5.240460952 | 0.254046418 | 0 | 0 |
| 0.001201341 | 0.001623017 | 0.000631671 | 0.000959489 | 0 |
| 0 | 0 | 0 | 0 | 0 |
| 0 | 0 | 0 | 0 | 0 |
| 0.0269571 | 0.052563714 | 0 | 0 | 0.00206836 |
| 0 | 0 | 0 | 0 | 0 |
| 0 | 0 | 0 | 0 | 0 |
| 0 | 0 | 0 | 0 | 0 |
| 0 | 0 | 1.728849221 | 0.005623323 | 0.00713467 |
| 0 | 0 | 0 | 0 | 0 |
| 0 | 0 | 0 | 0 | 0 |
| 0 | 0 | 0.00033924 | 0 | 0.00086433 |
| 0 | 0 | 0 | 0 | 0 |
| 0 | 0 | 0 | 0 | 0 |

| | | | | |
|---|---|---|---|---|
| 0 | 0 | 0 | 0 | 0 |
| 0 | 0 | 0 | 0 | 0 |
| 0 | 0 | 0 | 0 | 0 |
| 0 | 0 | 0 | 0 | 0 |
| 0 | 0 | 0 | 0 | 0 |
| 0 | 0 | 0 | 0 | 0 |
| 0 | 0 | 0 | 0 | 0 |
| 0 | 0.027730591 | 0 | 0 | 0 |
| 0 | 0 | 0 | 0 | 0 |
| 0 | 0 | 0.014954897 | 0.010614971 | 0.001582668 |
| 0 | 0 | 0 | 0 | 0 |
| 0 | 0 | 0 | 0 | 0 |
| 0 | 0 | 0 | 0 | 0 |
| 0 | 0 | 0 | 0 | 0 |
| 0 | 0 | 0 | 0 | 0 |
| 0.0026153 | 0 | 0 | 0.258487968 | 0.000389293 |
| 0 | 0 | 0 | 0 | 0 |
| 0 | 0 | 0 | 0 | 0 |
| 0 | 0 | 0 | 0 | 0 |
| 0 | 0 | 0 | 0 | 0 |
| 0 | 0.001561264 | 0 | 0 | 0 |
| 0 | 0 | 0 | 0 | 0 |
| 0 | 0 | 0 | 0 | 0 |
| 0 | 0 | 0 | 0 | 0 |
| 0 | 0 | 0 | 0 | 0 |
| 0 | 0 | 0 | 0 | 2.086521867 |
| 0 | 0 | 0 | 0 | 0 |
| 0 | 0 | 0 | 0 | 0 |
| 0 | 0 | 0 | 0 | 0 |
| 0 | 0 | 0 | 0 | 0 |
| 0 | 0 | 0.033194879 | 0.01216693 | 0.649027898 |
| 0 | 0 | 0 | 0 | 0 |
| 0 | 0 | 0 | 0 | 0 |
| 0 | 0 | 0 | 0 | 0 |
| 0 | 0 | 0 | 0 | 0 |
| 0 | 0 | 0 | 0 | 0 |
| 0 | 0 | 0 | 0 | 0 |
| 0 | 0 | 0 | 0 | 0 |
| 0 | 0 | 0 | 0.011958669 | 0.003031117 |
| 0 | 0 | 0 | 0 | 0 |
| 0 | 0 | 0 | 0 | 0 |
| 0 | 0 | 0 | 0 | 0 |
| 0 | 0 | 0 | 0 | 0 |
| 0 | 0 | 0 | 0 | 0 |
| 0 | 0 | 0 | 0 | 0 |
| 0 | 0 | 0 | 0 | 0 |
| 0.1595649 | 0.436960576 | 0 | 0 | 0.007272916 |
| 0 | 0 | 0 | 0 | 0.00048581 |

| | | | | |
|---|---|---|---|---|
| 0 | 0 | 0 | 0 | 0 |
| 0 | 0 | 0 | 0 | 0 |
| 0 | 0 | 0 | 0 | 0 |
| 0 | 0 | 0 | 0 | 0 |
| 0 | 0 | 0 | 0 | 0.03059085 |
| 0 | 0 | 0 | 0 | 0 |
| 0 | 0 | 0 | 0 | 0 |
| 0 | 0 | 0.000350233 | 0 | 0 |
| 0 | 0 | 0 | 0 | 0 |
| 0 | 0 | 0 | 0 | 0 |
| 0 | 0 | 0 | 0 | 0 |
| 0.005553739 | 0.094943361 | 0.011096692 | 0.005174947 | 0.011780262 |
| 0 | 0.014315962 | 0.00847142 | 0 | 0 |
| 0 | 0 | 0 | 0 | 0.00102819 |
| 0 | 0 | 0 | 0 | 0 |
| 0 | 0.207694359 | 0 | 0 | 0 |
| 0 | 0 | 0 | 0 | 0 |
| 0 | 0 | 0 | 0 | 0 |
| 0 | 0.023599693 | 0 | 0.000559786 | 0 |
| 0.013727177 | 0.001854547 | 0.013822114 | 0.004020002 | 0 |
| 0 | 0 | 0 | 0 | 0 |
| 0 | 0 | 0.00914459 | 0 | 0 |
| 0 | 0 | 0 | 0 | 0 |
| 0 | 0 | 0 | 0 | 0 |
| 0 | 0 | 0 | 0 | 0 |
| 0 | 0 | 0 | 0 | 0 |
| 0.003300065 | 0 | 0 | 0.008639241 | 0.006140264 |
| 0 | 0 | 0 | 0 | 0 |
| 0 | 0 | 0 | 0 | 0 |
| 0 | 0 | 0 | 0 | 0 |
| 0 | 0.050538305 | 0 | 0.004481558 | 0.017075966 |
| 0 | 0 | 0 | 0 | 0.003508376 |
| 0 | 0 | 0 | 0 | 0 |
| 0 | 0 | 0 | 0 | 0 |
| 0 | 0 | 0 | 0 | 0 |
| 0 | 0 | 0 | 0 | 0 |
| 0 | 0 | 0 | 0 | 0 |
| 0 | 0.377190897 | 0.061149378 | 0.019474751 | 0.017734811 |
| 0 | 0 | 0.089368304 | 0 | 0 |
| 0 | 0 | 0 | 0 | 0 |
| 0 | 0.020450295 | 0.000896732 | 0 | 0 |
| 0 | 0 | 0 | 0 | 0 |
| 0 | 0 | 0 | 0 | 0 |
| 0 | 0 | 0 | 0 | 0 |
| 0 | 0 | 0 | 0 | 0 |
| 0 | 0 | 0 | 0 | 0 |
| 0 | 0 | 0 | 0 | 0 |
| 0 | 0 | 0 | 0 | 0 |

| | | | | |
|---|---|---|---|---|
| 0 | 0 | 0 | 0 | 0 |
| 0 | 0 | 0 | 0 | 0 |
| 0 | 0 | 0 | 0 | 0 |
| 0 | 0 | 0 | 0.009605025 | 0.003850423 |
| 0 | 0.002230842 | 0.000863127 | 0.015127653 | 0 |
| 0 | 0 | 0 | 0 | 0.010188279 |
| 0 | 0 | 0 | 0 | 0.032153934 |
| 0.000638861 | 0 | 0 | 0 | 0 |
| 0 | 0 | 0 | 0 | 0 |
| 0 | 0 | 0 | 0 | 0 |
| 0 | 0 | 0 | 0 | 0 |
| 0 | 0 | 0 | 0 | 0 |
| 0 | 0.631248237 | 0.127494295 | 0.000770786 | 0 |
| 0 | 0 | 0 | 0 | 0 |
| 0 | 0 | 0 | 0 | 0 |
| 0 | 0 | 0 | 0 | 0 |
| 0 | 0 | 0 | 0 | 0 |
| 0 | 0 | 0 | 0 | 0 |
| 0 | 0 | 0 | 0 | 0 |
| 0 | 0 | 0 | 0 | 0 |
| 0 | 0 | 0 | 0 | 0 |
| 0 | 0 | 0 | 0 | 0 |
| 0.00482896 | 0 | 0 | 0 | 0.032629746 |
| 0 | 0 | 0.000473086 | 0 | 0 |
| 0 | 0 | 0 | 0 | 0 |
| 0 | 0 | 0 | 0 | 0 |
| 0 | 0 | 0 | 0 | 0 |
| 0 | 0 | 0 | 0 | 0 |
| 0 | 0 | 0 | 0 | 0 |
| 0 | 0 | 0 | 0 | 0 |
| 0 | 0 | 0.010542025 | 0.001320272 | 0 |
| 0 | 0 | 0 | 0 | 0.00546379 |
| 0 | 0 | 0 | 0 | 0 |
| 0 | 0 | 0 | 0 | 0 |
| 0 | 0 | 0 | 0 | 0 |
| 0 | 0 | 0 | 0 | 0 |
| 0.018666541 | 0 | 0 | 0 | 0.005377842 |
| 0 | 0 | 0 | 0 | 0 |
| 0 | 0.000498336 | 0 | 0.019628025 | 0 |
| 0 | 0 | 0 | 0 | 0 |
| 0 | 0 | 0 | 0 | 0 |
| 0 | 0 | 0 | 0 | 0 |
| 0 | 0 | 0 | 2.768915952 | 0 |
| 0 | 0 | 0 | 0 | 0 |
| 0 | 0 | 0 | 0 | 0 |
| 0 | 0 | 0.209275821 | 0.000982812 | 0.031321874 |
| 0 | 0 | 0 | 0 | 0.046528549 |
| 0 | 0 | 0 | 0 | 0 |

| | | | | |
|---|---|---|---|---|
| 0 | 0 | 0 | 0 | 0 |
| 0 | 0 | 0 | 0 | 0 |
| 0 | 0 | 0 | 0 | 0 |
| 0 | 0 | 0 | 0 | 0 |
| 0 | 0 | 0 | 0 | 0 |
| 0 | 0 | 0 | 0 | 0 |
| 0 | 0 | 0 | 0 | 0 |
| 0 | 0 | 0 | 0 | 0 |
| 0 | 0 | 0 | 0 | 0 |
| 0 | 0 | 0 | 0 | 0 |
| 0 | 0.02395383 | 0.002761415 | 0 | 0 |
| 0 | 0 | 0 | 0.008504508 | 0.000540342 |
| 0 | 0 | 0 | 0 | 0 |
| 0 | 0 | 0 | 0 | 0 |
| 0 | 0 | 0 | 0 | 0 |
| 0 | 0 | 0 | 0 | 0 |
| 0 | 0 | 0 | 0 | 0 |
| 0 | 0 | 0 | 0 | 0 |
| 0 | 0 | 0 | 0 | 0 |
| 0 | 0.001474413 | 0 | 0.001716863 | 0 |
| 0 | 0 | 0 | 0 | 0 |
| 0 | 0 | 0 | 0 | 0 |
| 0 | 0 | 0 | 0 | 0 |
| 0 | 0 | 0 | 0 | 0 |
| 0 | 0 | 0 | 0 | 0 |
| 0 | 0 | 0 | 0 | 0 |
| 0 | 0 | 0 | 0 | 0 |
| 0 | 0 | 0 | 0.002279999 | 0 |
| 0 | 0 | 0 | 0 | 0 |
| 0 | 0 | 0 | 0 | 0 |
| 0.005097022 | 0.041915366 | 0.001631327 | 0 | 0.012370137 |
| 0 | 0 | 0 | 0 | 0 |
| 0.104983039 | 0.35725527 | 0.004329457 | 0.039731901 | 0.239766164 |
| 0.005258886 | 0.000956412 | 0.001693653 | 0.000875037 | 0 |
| 0 | 0 | 0 | 0 | 0 |
| 0 | 0 | 0 | 0 | 0 |
| 0 | 0 | 0 | 0 | 0 |
| 0 | 0 | 0 | 0 | 0 |
| 0 | 0 | 0 | 0 | 0 |
| 0 | 0 | 0 | 0 | 0 |
| 0 | 0 | 0 | 0 | 0 |
| 0.295304383 | 0 | 0 | 0.000845355 | 0.403802386 |
| 0.194968879 | 0 | 0.016170608 | 0 | 0 |
| 0 | 0 | 0 | 0 | 0 |
| 0 | 0 | 0 | 0 | 0 |
| 0 | 0 | 0.286241493 | 0.022571791 | 0.010956048 |
| 0 | 0 | 0 | 0 | 0 |
| 0 | 0 | 0 | 0 | 0 |

| | | | | |
|---|---|---|---|---|
| 0 | 0 | 0 | 0 | 0 |
| 0 | 0 | 0.003089842 | 0.006473623 | 0 |
| 0.004102778 | 0.022794634 | 0.004770863 | 0.000630156 | 0 |
| 0 | 0 | 0 | 0 | 0 |
| 0.012095521 | 0 | 0 | 0.00570236 | 0 |
| 0 | 0.011215998 | 0 | 0 | 0.029482829 |
| 0 | 0 | 0 | 0 | 0 |
| 0 | 0 | 0 | 0 | 0 |
| 0 | 0 | 0 | 0 | 0 |
| 0 | 0 | 0 | 0 | 0 |
| 0 | 0 | 0 | 0 | 0 |
| 0 | 0 | 0 | 0 | 0 |
| 0 | 0 | 0 | 0 | 0 |
| 0.003183223 | 0 | 0 | 0 | 0.023395304 |
| 0 | 0 | 0 | 0 | 0 |
| 0 | 0 | 0 | 0 | 0 |
| 0 | 0 | 0 | 0 | 0 |
| 0 | 0 | 0 | 0 | 0 |
| 0 | 0 | 0 | 0 | 0 |
| 0 | 0 | 0 | 0 | 0 |
| 0 | 0 | 0 | 0 | 0 |
| 0.002284761 | 0 | 0 | 0 | 0.021161228 |
| 0 | 0 | 0 | 0 | 0 |
| 0 | 0 | 0 | 0 | 0 |
| 0 | 0 | 0.02030892 | 0 | 0 |
| 0 | 0.001903421 | 0.003370654 | 0.011667851 | 0.215573764 |
| 0 | 0 | 0 | 0 | 0 |
| 0 | 0 | 0 | 0 | 0.082077745 |
| 0 | 0 | 0 | 0 | 0 |
| 0 | 0 | 0 | 0 | 0 |
| 0 | 0 | 0 | 0 | 0 |
| 0 | 0.001073049 | 0 | 0 | 0 |
| 0 | 0 | 0 | 0 | 0 |
| 0 | 0 | 0 | 0 | 0 |
| 0 | 0 | 0 | 0 | 0 |
| 0 | 0 | 0 | 0 | 0 |
| 0 | 0 | 0 | 0 | 0 |
| 0 | 0.033563187 | 0 | 0 | 0.002229209 |
| 0 | 0 | 0 | 0.001423006 | 0 |
| 0 | 0 | 0 | 0 | 0 |
| 0 | 0 | 0 | 0 | 0 |
| 0 | 0 | 0.008880412 | 0.003211686 | 0 |
| 0.00814048 | 0 | 0 | 0.00157298 | 0.017198698 |
| 0 | 0 | 0 | 0 | 0 |
| 0 | 0 | 0 | 0 | 0 |
| 0 | 0 | 0 | 0 | 0 |
| 0 | 0.004174816 | 0 | 0 | 0.021676381 |
| 0 | 0 | 0 | 0 | 0 |

| | | | | |
|---|---|---|---|---|
| 0 | 0 | 0 | 0 | 0 |
| 0.016220723 | 0.160142655 | 0.002032285 | 0.021423299 | 0.021126757 |
| 0 | 0 | 0 | 0 | 0 |
| 0 | 0 | 0 | 0 | 0 |
| 0 | 0 | 0 | 0 | 0 |
| 0 | 0 | 0 | 0 | 0 |
| 0 | 0 | 0 | 0 | 0 |
| 0 | 0 | 0 | 0 | 0 |
| 0 | 0 | 0 | 0 | 0 |
| 0 | 0 | 0 | 0 | 0 |
| 0 | 0 | 0 | 0 | 0 |
| 0 | 0 | 0 | 0 | 0 |
| 0 | 0.002784292 | 0.04106432 | 0 | 0 |
| 0 | 0 | 0 | 0 | 0 |
| 0 | 0 | 0 | 0 | 0 |
| 0 | 0 | 0 | 0 | 0 |
| 0 | 0 | 0 | 0 | 0 |
| 0 | 0 | 0 | 0 | 0 |
| 0 | 0 | 0 | 0 | 0 |
| 0 | 0 | 0 | 0 | 0.002819437 |
| 0 | 0 | 0 | 0 | 0 |
| 0.008069078 | 0 | 0.000815915 | 0 | 0 |
| 0 | 0 | 0 | 0 | 0 |
| 0 | 0 | 0.000168078 | 0 | 0.000951635 |
| 0 | 0 | 0 | 0 | 0 |
| 0 | 0 | 0 | 0 | 0 |
| 0 | 0 | 0 | 0 | 0 |
| 0 | 0 | 0 | 0 | 0 |
| 0 | 0 | 0 | 0 | 0 |
| 0 | 0 | 0 | 0 | 0 |
| 0 | 0 | 0 | 0 | 0 |
| 0 | 0 | 0 | 0 | 0 |
| 0 | 0 | 0 | 0 | 0 |
| 0 | 0 | 0 | 0 | 0 |
| 0 | 0 | 0 | 0.008251918 | 7.008230293 |
| 0 | 0 | 0 | 0 | 0 |
| 0 | 0 | 0 | 0 | 0 |
| 0 | 0 | 0 | 0 | 0 |
| 0.009917478 | 0.001932483 | 0.011692238 | 0.000825094 | 0 |
| 0 | 0 | 0 | 0 | 0 |
| 0.167564863 | 0.036387264 | 0.418687994 | 0.004808741 | 0.23587696 |
| 0 | 0 | 0 | 0 | 0 |
| 0 | 0 | 0 | 0 | 0 |
| 0 | 0 | 0 | 0 | 0 |
| 0 | 0 | 0 | 0 | 0 |
| 2.524916231 | 10.31389322 | 0.021873157 | 23.46203616 | 0.145864839 |
| 0 | 0 | 0 | 0 | 0 |
| 0 | 0 | 0 | 0 | 0 |

| | | | | |
|---|---|---|---|---|
| 0 | 0 | 0 | 0 | 0.001687235 |
| 0 | 0 | 0 | 0 | 0.000578523 |
| 0 | 0 | 0 | 0 | 0 |
| 0 | 0 | 0 | 0 | 0 |
| 0 | 0 | 0 | 0 | 0 |
| 0 | 0 | 0 | 0 | 0 |
| 0 | 0 | 0 | 0 | 0 |
| 0 | 0 | 0 | 0 | 0 |
| 0 | 0 | 0 | 0 | 0 |
| 0 | 0.002101897 | 0 | 0.039555841 | 0 |
| 0 | 0 | 0 | 0 | 0 |
| 0 | 0 | 0 | 0 | 0 |
| 0 | 0.003634756 | 0 | 0.055261066 | 0 |
| 0 | 0 | 0 | 0 | 0 |
| 0 | 0 | 0 | 0 | 0 |
| 0 | 0 | 0 | 0 | 0 |
| 0 | 0 | 0 | 0 | 0 |
| 0 | 0 | 0 | 0 | 0 |
| 0 | 0 | 0 | 0 | 0 |
| 0.229283118 | 1.48523328 | 1.391791747 | 1.433427812 | 0.020751547 |
| 0 | 0 | 0 | 0 | 0 |
| 0 | 0 | 0 | 0 | 0 |
| 0 | 0 | 0 | 0 | 0 |
| 0 | 0 | 0 | 0 | 0 |
| 0 | 0 | 0 | 0 | 0 |
| 0 | 0 | 0 | 0 | 0 |
| 0 | 0 | 0 | 0 | 0 |
| 0 | 0 | 0 | 0 | 0 |
| 0 | 0 | 0 | 0 | 0 |
| 0 | 0 | 0 | 0 | 0 |
| 0 | 0 | 0 | 0 | 0 |
| 0 | 0 | 0 | 0 | 0 |
| 0 | 0 | 0 | 0 | 0 |
| 0 | 0.010779506 | 0 | 0 | 0 |
| 0 | 0 | 0 | 0 | 0 |
| 0 | 0.035505926 | 0 | 0 | 0 |
| 0 | 0 | 0 | 0 | 0 |
| 0 | 0 | 0 | 0 | 0 |
| 0 | 0 | 0 | 0 | 0 |
| 0 | 0 | 0 | 0 | 0 |
| 0 | 0 | 0 | 0 | 0 |
| 0 | 0 | 0 | 0 | 0 |
| 0 | 0 | 0 | 0 | 0 |
| 0 | 0 | 0 | 0 | 0 |
| 0 | 0.049278308 | 0.040607335 | 0.002199882 | 0 |
| 0 | 0 | 0 | 0 | 0 |
| 0 | 0 | 0 | 0 | 0 |
| 0 | 0 | 0 | 0.028797377 | 0 |

| | | | | |
|---|---|---|---|---|
| 0 | 0 | 0 | 0 | 0 |
| 0 | 0 | 0 | 0 | 0 |
| 0 | 0 | 0 | 0 | 0 |
| 0 | 0 | 0 | 0 | 0 |
| 0 | 0 | 0 | 0 | 0 |
| 0.002277705 | 0 | 0.003942192 | 0.00833782 | 0 |
| 0 | 0 | 0 | 0 | 0 |
| 0 | 0 | 0 | 0 | 0 |
| 0 | 0 | 0 | 0 | 0 |
| 0 | 0 | 0 | 0 | 0 |
| 0 | 0 | 0 | 0 | 0.428001067 |
| 0 | 0 | 0 | 0 | 0 |
| 0 | 0 | 0 | 0 | 0 |
| 0 | 0 | 0 | 0 | 0 |
| 0 | 0 | 0 | 0 | 0 |
| 0 | 0 | 0 | 0 | 0 |
| 0 | 0 | 0 | 0 | 0 |
| 0 | 0.001214654 | 0 | 0 | 0.02600969 |
| 0 | 0 | 0 | 0 | 0 |
| 0 | 0 | 0 | 0 | 0 |
| 0 | 0 | 0 | 0 | 0 |
| 0 | 0 | 0 | 0 | 0 |
| 0 | 0 | 0 | 0 | 0 |
| 0 | 0 | 0 | 0 | 0 |
| 0 | 0 | 0 | 0 | 0 |
| 0 | 0 | 0 | 0.000988011 | 0 |
| 0 | 0 | 0 | 0 | 0 |
| 0 | 0 | 0 | 0 | 0 |
| 0 | 0 | 0 | 0 | 0 |
| 0 | 0 | 0 | 0 | 0.463012321 |
| 0 | 0 | 0 | 0 | 0 |
| 0 | 0 | 0 | 0 | 0 |
| 0 | 0 | 0 | 0 | 0 |
| 0 | 0 | 0 | 0 | 0 |
| 0 | 0 | 0 | 0 | 0 |
| 0 | 0 | 0 | 0 | 0 |
| 0 | 0 | 0 | 0 | 0 |
| 0 | 0 | 0 | 0 | 0 |
| 0 | 0 | 0 | 0 | 1.055383004 |
| 0 | 0 | 0 | 0 | 0 |
| 0 | 0 | 0 | 0 | 0 |
| 0 | 0 | 0 | 0 | 0 |
| 0 | 0 | 0 | 0 | 0 |
| 0 | 0 | 0 | 0 | 0 |
| 0 | 0 | 0 | 0 | 0 |
| 0 | 0 | 0 | 0 | 0 |
| 0 | 0 | 0 | 0 | 0 |
| 0 | 0 | 0 | 0 | 0 |

| | | | | |
|---|---|---|---|---|
| 0 | 0 | 0 | 0 | 0 |
| 0 | 0 | 0 | 0 | 0 |
| 0 | 0 | 0 | 0 | 0 |
| 0 | 0 | 0 | 0 | 0 |
| 0 | 0 | 0 | 0 | 0 |
| 0 | 0 | 0 | 0 | 0 |
| 0 | 0.01165833 | 0 | 0 | 0 |
| 0 | 0 | 0 | 0 | 0 |
| 0 | 0 | 0 | 0 | 0 |
| 0 | 0 | 0 | 0 | 0 |
| 0 | 0.005638954 | 0.077873712 | 0.002222411 | 0 |
| 0 | 0 | 0 | 0 | 0 |
| 0 | 0 | 0.067945839 | 0 | 0 |
| 0 | 0 | 0 | 0 | 0 |
| 0 | 0 | 0 | 0 | 0 |
| 0 | 0 | 0 | 0 | 0 |
| 0 | 0 | 0 | 0 | 0 |
| 0 | 0 | 0 | 0 | 0 |
| 0 | 0 | 0 | 0 | 0 |
| 0 | 0 | 0 | 0 | 0 |
| 0.025982377 | 0.335467527 | 1.746167455 | 0.059773757 | 0 |
| 0 | 0 | 0 | 0 | 0 |
| 0 | 0 | 0 | 0 | 0 |
| 1.569598993 | 0.703505692 | 1.331752111 | 0 | 0 |
| 0 | 0 | 0 | 0 | 0 |
| 0 | 0 | 0 | 0 | 0 |
| 0 | 0 | 0 | 0 | 0 |
| 0 | 0 | 0 | 0 | 0.114911668 |
| 0 | 0 | 0 | 0 | 0 |
| 0 | 0 | 0 | 0 | 0 |
| 0 | 0 | 0 | 0 | 0.005776486 |
| 0 | 0 | 0 | 0 | 0 |
| 0 | 0 | 0 | 0 | 0 |
| 0 | 0 | 0 | 0 | 0 |

| PHL018-0035_V1 | PHL018-0215_V1 | PHL018-0276_V3 | PHL018-0176_V3 | PHL018-0223_V1 |
|---|---|---|---|---|
| CTRL | CTRL | REF | REF | REF |
| 0.315355669 | 0.410157007 | 0.100501155 | 0.098082156 | 0.008944367 |
| 6.718749615 | 5.466497212 | 4.138746095 | 1.227715548 | 1.422018369 |
| 0 | 3.195073077 | 0.004184373 | 3.801636341 | 0.283222287 |
| 0 | 0 | 0 | 0 | 0 |
| 0 | 0 | 0 | 0 | 0 |
| 0 | 0.010048458 | 0 | 0 | 0 |
| 0 | 0 | 0 | 0 | 0 |
| 0 | 0 | 0 | 0 | 0 |
| 0 | 0 | 0 | 0 | 0 |
| 0 | 0 | 0 | 0 | 0 |
| 0 | 0 | 0 | 0 | 0 |
| 0 | 0 | 0 | 0 | 0 |
| 0 | 0 | 0 | 0 | 0 |
| 0.114907098 | 0.228513814 | 4.399238553 | 0.537802833 | 3.668526957 |
| 0 | 0 | 0 | 0 | 0 |
| 0 | 0 | 0 | 0 | 0 |
| 0 | 0 | 0 | 0 | 0 |
| 0 | 0 | 0 | 0 | 0 |
| 0 | 0 | 0 | 0 | 0 |
| 0 | 0 | 0 | 0 | 0 |
| 0 | 0 | 0 | 0 | 0 |
| 0 | 0 | 0 | 0 | 0 |
| 0 | 0 | 0 | 0 | 0 |
| 0 | 0 | 0 | 0 | 0 |
| 0 | 0 | 0 | 0 | 0 |
| 0 | 0 | 0 | 0 | 0 |
| 0 | 0 | 0 | 0 | 0 |
| 0 | 0 | 0 | 0 | 0 |
| 0 | 0 | 0 | 0 | 0 |
| 0 | 0 | 0 | 0 | 0 |
| 0 | 0 | 0 | 0 | 0 |
| 0 | 0 | 0 | 0 | 0 |
| 0 | 0 | 0 | 0 | 0 |
| 0 | 0 | 0 | 0 | 0 |
| 0 | 0 | 0 | 0 | 0 |
| 2.368888631 | 0.878171716 | 9.916735029 | 2.445212511 | 0.150750542 |
| 0 | 0 | 0 | 0 | 0 |
| 0 | 0 | 0 | 0 | 0 |
| 0 | 0 | 0 | 0 | 0 |
| 0 | 0 | 0 | 0 | 0 |
| 0 | 0 | 0 | 0 | 0 |
| 0.004906091 | 0 | 0.006511099 | 0.073701639 | 0.006128236 |
| 0.646818065 | 0.053182611 | 0.045545772 | 0.036203839 | 0.265774195 |
| 0.004597011 | 0.117926142 | 0.118376617 | 0.019613273 | 0.175160712 |
| 0 | 0.001773572 | 0.007889876 | 0 | 0 |
| 0 | 0 | 0 | 0 | 0 |

| | | | | |
|---|---|---|---|---|
| 0 | 0.427393573 | 0 | 0 | 0 |
| 0.084219287 | 0 | 0.017329452 | 0.011866217 | 0 |
| 0 | 0 | 0 | 0 | 0 |
| 0.332849892 | 0.691699864 | 0.852829422 | 0.888697093 | 1.095151278 |
| 0 | 0 | 0.002863034 | 0.006084517 | 0 |
| 0 | 0 | 0 | 0 | 0 |
| 7.575774438 | 0.012076474 | 0 | 0.046105179 | 0.026500807 |
| 0 | 0 | 0 | 0 | 0 |
| 0 | 0 | 0 | 0 | 0 |
| 0.017800286 | 0 | 0 | 0 | 0 |
| 0 | 0 | 0 | 0 | 0.020181246 |
| 1.551470459 | 0.029006999 | 0.051914496 | 0.048666148 | 0 |
| 0 | 0 | 0.000477955 | 0 | 0 |
| 0.069562941 | 0.01244212 | 0.017517001 | 0.08425558 | 0 |
| 0 | 0 | 0 | 0 | 0 |
| 0 | 0 | 0 | 0 | 0 |
| 0 | 0.375042233 | 0.00866669 | 0.277837502 | 0.699914628 |
| 0 | 0 | 0 | 0 | 0 |
| 0 | 0 | 0 | 0 | 0 |
| 0 | 0 | 0 | 0 | 0 |
| 0.187162202 | 0.001603352 | 0.026164619 | 20.46627738 | 0.004697569 |
| 0.04125929 | 0.00361294 | 0.072522048 | 0.00465972 | 0 |
| 0.105595943 | 0.011312764 | 0 | 0 | 0 |
| 0 | 0 | 0 | 0 | 0 |
| 0 | 0 | 0.002597669 | 0 | 0 |
| 0 | 0 | 0 | 0 | 0 |
| 0 | 0 | 0 | 0 | 0 |
| 0 | 0 | 0 | 0 | 0 |
| 0 | 0 | 0.000668661 | 0.013186742 | 0.156684577 |
| 0 | 0 | 0 | 0 | 0 |
| 0 | 0 | 0 | 0 | 0.020951688 |
| 0 | 0 | 0.001143844 | 0 | 0 |
| 0 | 0 | 0 | 0 | 0 |
| 0 | 0 | 0 | 0 | 0 |
| 0 | 0 | 0 | 0 | 0 |
| 0 | 0 | 0.00182281 | 0 | 0 |
| 0 | 0 | 0 | 0 | 0 |
| 0 | 0.01241303 | 0 | 0.004623055 | 0.014604883 |
| 0 | 0 | 0 | 0 | 0 |
| 0 | 0 | 0 | 0 | 0 |
| 0 | 0 | 0 | 0 | 0 |
| 0 | 0 | 0 | 0 | 0 |
| 0 | 0 | 0 | 0 | 0 |
| 0 | 0 | 0 | 0 | 0 |
| 0 | 0 | 0 | 0 | 0 |
| 0 | 0 | 0 | 0 | 0 |
| 0.002782393 | 0 | 0.001777084 | 0.000698303 | 0 |
| 0 | 0 | 0 | 0.016548721 | 0 |

| | | | | |
|---|---|---|---|---|
| 0 | 0 | 0 | 0 | 0 |
| 0 | 0 | 0 | 0 | 0 |
| 0 | 0 | 0 | 0 | 0 |
| 0 | 0 | 0 | 0 | 0 |
| 0 | 0 | 0 | 0 | 0 |
| 0 | 0 | 0 | 0 | 0 |
| 0 | 0 | 0 | 0 | 0 |
| 0 | 0 | 0 | 0 | 0 |
| 0 | 0.001600587 | 0.000757819 | 0 | 0.004263151 |
| 0 | 0 | 0 | 0 | 0 |
| 0 | 0 | 0 | 0 | 0 |
| 0.002315014 | 0 | 0.002346167 | 0.055617945 | 0.000951814 |
| 0 | 0 | 0 | 0 | 0 |
| 0 | 0 | 0 | 0 | 0 |
| 0 | 0 | 0 | 0 | 0 |
| 0 | 0 | 0 | 0 | 0 |
| 0 | 0 | 0 | 0 | 0 |
| 0 | 0 | 0 | 0 | 0 |
| 0.015920147 | 0.019694723 | 0.324941746 | 0.003102399 | 0.054381907 |
| 0.000522561 | 0 | 0 | 0 | 0 |
| 0 | 0 | 0 | 0 | 0 |
| 0 | 0 | 0 | 0 | 0 |
| 0 | 0 | 0.001519617 | 0 | 0 |
| 0 | 0 | 0 | 0 | 0 |
| 0 | 0 | 0 | 0 | 0 |
| 0 | 0 | 0 | 0 | 0 |
| 0 | 0 | 0 | 0 | 0 |
| 0 | 0 | 0 | 0 | 0 |
| 0 | 0 | 0 | 0 | 0 |
| 0 | 0 | 0 | 0 | 0 |
| 0.277954981 | 0.005789897 | 0.029803697 | 0.01528214 | 0.004400188 |
| 0 | 0.000423775 | 0.000858301 | 0.005299933 | 0 |
| 0 | 0 | 0 | 0 | 0 |
| 0 | 0 | 0 | 0 | 0 |
| 0 | 0 | 0 | 0 | 0 |
| 0 | 0 | 0 | 0 | 0 |
| 0 | 0 | 0 | 0 | 0 |
| 0 | 0 | 0 | 0 | 0 |
| 0 | 0 | 0 | 0 | 0 |
| 0 | 0 | 0 | 0 | 0 |
| 0 | 0 | 0 | 0 | 0 |
| 0 | 0 | 0 | 0 | 0 |
| 0 | 0.0029389 | 0 | 0.000459441 | 0 |
| 0 | 0 | 0 | 0 | 0 |
| 0 | 0 | 0 | 0 | 0 |
| 0 | 0 | 0 | 0 | 0 |
| 0 | 0 | 0 | 0 | 0 |
| 0 | 0 | 0 | 0 | 0 |

| | | | | |
|---|---|---|---|---|
| 0 | 0 | 0 | 0 | 0 |
| 0 | 0 | 0 | 0 | 0 |
| 0 | 0 | 0 | 0 | 0 |
| 0 | 0 | 0 | 0 | 0 |
| 0.060655613 | 0.266554191 | 0.002347883 | 0.029177171 | 0 |
| 0 | 0 | 0 | 0 | 0 |
| 0 | 0 | 0 | 0 | 0 |
| 0 | 0 | 0 | 0 | 0 |
| 0 | 0 | 0 | 0 | 0 |
| 0 | 0 | 0 | 0 | 0 |
| 0 | 0 | 0 | 0 | 0 |
| 0 | 0 | 0 | 0 | 0 |
| 0 | 0 | 0 | 0 | 0 |
| 0 | 0 | 0 | 0 | 0 |
| 0 | 0 | 0 | 0 | 0 |
| 0 | 0 | 0 | 0 | 0 |
| 0 | 0 | 0 | 0 | 0 |
| 0 | 0 | 0 | 0 | 0 |
| 0 | 0 | 0 | 0 | 0 |
| 0 | 0 | 0 | 0 | 0 |
| 0 | 0 | 0 | 0 | 0 |
| 0 | 0 | 0 | 0 | 0 |
| 0 | 0 | 0 | 0 | 0 |
| 0 | 0 | 0 | 0 | 0 |
| 0.025587291 | 0 | 0 | 0 | 0 |
| 0 | 0 | 0 | 0 | 0 |
| 0 | 0 | 0 | 0 | 0 |
| 0 | 0 | 0 | 0 | 0 |
| 0 | 0 | 0 | 0 | 0 |
| 0 | 0 | 0 | 0 | 0 |
| 0.02099009 | 0.005369345 | 0 | 0 | 0.009388726 |
| 0.001015215 | 0 | 0 | 0 | 0.029936218 |
| 0 | 0 | 0 | 0 | 0 |
| 0 | 0 | 0 | 0 | 0 |
| 0 | 0 | 0 | 0 | 0 |
| 0 | 0 | 0 | 0 | 0 |
| 0 | 0 | 0 | 0 | 0 |
| 0 | 0 | 0 | 0 | 0 |
| 0 | 0 | 0 | 0 | 0 |
| 0 | 0 | 0 | 0 | 0 |
| 0 | 0 | 0 | 0 | 0 |
| 0 | 0 | 0 | 0 | 0 |
| 0 | 0 | 0 | 0 | 0 |
| 0 | 0 | 0 | 0 | 0 |
| 0 | 0 | 0 | 0 | 0 |
| 0 | 0 | 0 | 0 | 0 |
| 0 | 0 | 0 | 0 | 0 |
| 0 | 0 | 0 | 0 | 0 |

| | | | | |
|---|---|---|---|---|
| 0 | 0 | 0 | 0 | 0 |
| 0 | 0 | 0 | 0 | 0 |
| 0 | 0 | 0 | 0 | 0 |
| 0 | 0 | 0 | 0 | 0 |
| 0 | 0 | 0 | 0 | 0 |
| 0 | 0 | 0 | 0 | 0 |
| 0 | 0 | 0 | 0 | 0 |
| 0 | 0 | 0 | 0 | 0 |
| 0 | 0 | 0 | 0 | 0 |
| 0 | 0 | 0 | 0 | 0 |
| 0 | 0 | 0 | 0 | 0 |
| 0 | 0 | 0 | 0 | 0 |
| 0.001165754 | 0.001091702 | 0 | 0.000877715 | 0 |
| 0 | 0 | 0 | 0 | 0 |
| 0 | 0 | 0 | 0 | 0 |
| 0 | 0 | 0 | 0 | 0 |
| 0 | 0 | 0 | 0 | 0 |
| 0 | 0 | 0 | 0 | 0 |
| 0 | 0 | 0 | 0 | 0 |
| 0 | 0 | 0 | 0 | 0 |
| 0.02833583 | 0.018662582 | 0.04935759 | 0.027898982 | 0.082477653 |
| 0.002908886 | 0 | 0 | 0 | 0 |
| 0.000478133 | 0 | 0 | 0 | 0 |
| 0 | 0 | 0 | 0.001711354 | 0 |
| 0 | 0 | 0 | 0 | 0 |
| 0 | 0 | 0 | 0 | 0 |
| 0.005923838 | 0.006623082 | 0.051593045 | 0 | 0.024494937 |
| 0 | 0 | 0 | 0 | 0 |
| 0 | 0 | 0 | 0 | 0 |
| 0 | 0 | 0 | 0 | 0.001842663 |
| 0.072547884 | 0 | 0 | 0 | 0 |
| 0 | 0 | 0 | 0 | 0 |
| 0 | 0 | 0 | 0 | 0 |
| 0.00111007 | 0.004455239 | 0.064814558 | 0 | 0 |
| 0.158367393 | 0.048950947 | 0.006093554 | 0.007296619 | 0.000855209 |
| 0 | 0 | 0 | 0 | 0 |
| 0 | 0 | 0 | 0 | 0 |
| 0 | 0 | 0 | 0 | 0 |
| 0 | 0 | 0 | 0 | 0 |
| 0 | 0 | 0 | 0 | 0 |
| 0 | 0 | 0 | 0 | 0 |
| 0 | 0 | 0 | 0 | 0 |
| 0 | 0 | 0 | 0 | 0 |
| 0 | 0 | 0 | 0 | 0 |
| 0 | 0 | 0 | 0 | 0 |
| 0 | 0 | 0 | 0.021349858 | 0 |
| 0 | 0 | 0.000534949 | 0.003853804 | 0 |
| 0.007243054 | 0 | 0 | 0 | 0 |

| | | | | |
|---|---|---|---|---|
| 0 | 0 | 0 | 0 | 0 |
| 0 | 0 | 0 | 0 | 0 |
| 0 | 0 | 0 | 0 | 0.00278449 |
| 0.001070566 | 0 | 0.000447551 | 0.002901768 | 0 |
| 0 | 0 | 0 | 0 | 0 |
| 0 | 0 | 0 | 0 | 0 |
| 0 | 0 | 0 | 0 | 0 |
| 0.004116381 | 0 | 0 | 0.002187735 | 0 |
| 0 | 0 | 0 | 0 | 0 |
| 0 | 0 | 0 | 0 | 0 |
| 0.001042034 | 0 | 0 | 0 | 0 |
| 0 | 0 | 0.000578821 | 0 | 0 |
| 0 | 0 | 0 | 0 | 0 |
| 0 | 0 | 0 | 0 | 0 |
| 0 | 0 | 0 | 0 | 0.021620085 |
| 0 | 0 | 0 | 0 | 0 |
| 0 | 0 | 0 | 0 | 0 |
| 0 | 0 | 0 | 0 | 0 |
| 0.003940027 | 0 | 0 | 0 | 0 |
| 0 | 0.00252121 | 0.016960486 | 0 | 0 |
| 0.002274238 | 0 | 0 | 0.005517448 | 0 |
| 0 | 0 | 0 | 0 | 0 |
| 0 | 0 | 0 | 0 | 0 |
| 0 | 0 | 0 | 0 | 0 |
| 0.00301217 | 0.012895214 | 0.006296202 | 0 | 0.012714739 |
| 0 | 0 | 0 | 0 | 0 |
| 0 | 0 | 0 | 0 | 0 |
| 0 | 0 | 0 | 0 | 0 |
| 0 | 0 | 0 | 0.061502239 | 0.005200226 |
| 0 | 0 | 0 | 0 | 0 |
| 0.37421154 | 0.139897828 | 0.23950106 | 0.858400402 | 0 |
| 0 | 0 | 0 | 0 | 0 |
| 0 | 0 | 0 | 0 | 0 |
| 0 | 0 | 0 | 0 | 0 |
| 0 | 0 | 0 | 0 | 0 |
| 0 | 0 | 0 | 0 | 0 |
| 0 | 0 | 0 | 0 | 0 |
| 0 | 0 | 0 | 0 | 0 |
| 0 | 0 | 0 | 0 | 0 |
| 0 | 0 | 0 | 0 | 0 |
| 0 | 0 | 0 | 0 | 0 |
| 0.142237476 | 0.019003541 | 0.023621373 | 0.002139007 | 0 |
| 0.001271292 | 0 | 0 | 0.005862706 | 0 |
| 0 | 0 | 0 | 0 | 0 |
| 0 | 0 | 0 | 0 | 0 |
| 0 | 0 | 0 | 0 | 0 |
| 0 | 0 | 0 | 0 | 0 |
| 0 | 0 | 0 | 0 | 0 |

| | | | | |
|---|---|---|---|---|
| 0 | 0 | 0 | 0 | 0 |
| 0 | 0 | 0 | 0 | 0 |
| 0.002853836 | 0.112819882 | 0.125767235 | 0.02614252 | 0.189515566 |
| 0 | 0 | 0 | 0 | 0 |
| 2.807475059 | 0.379207943 | 0 | 0 | 0.713311932 |
| 0 | 0 | 0 | 0 | 0 |
| 0 | 0 | 0 | 0 | 0 |
| 0 | 0 | 0 | 0 | 0 |
| 0 | 0 | 0 | 0 | 0 |
| 0 | 0 | 0 | 0 | 0 |
| 0 | 0 | 0 | 0 | 0 |
| 0 | 0 | 0 | 0 | 0 |
| 0 | 0 | 0 | 0 | 0 |
| 0.004205338 | 0.002109752 | 0 | 0.001583135 | 0.007132199 |
| 0.002386282 | 0.037960864 | 0.005894837 | 0.000653335 | 0.104783203 |
| 0 | 0 | 0 | 0 | 0 |
| 0.002224599 | 0.002777716 | 0.006372195 | 0.002791563 | 0.024370086 |
| 0 | 0 | 0 | 0 | 0 |
| 0 | 0 | 0 | 0 | 0 |
| 0 | 0 | 0 | 0 | 0 |
| 0 | 0 | 0 | 0 | 0 |
| 0.008478976 | 0 | 0.000777334 | 0 | 0 |
| 0 | 0 | 0 | 0 | 0 |
| 0 | 0 | 0 | 0 | 0 |
| 0.00143111 | 0 | 0 | 0 | 0.015210099 |
| 0 | 0 | 0 | 0 | 0 |
| 0 | 0 | 0 | 0 | 0 |
| 0 | 0 | 0 | 0 | 0 |
| 0 | 0 | 0 | 0 | 0 |
| 0 | 0 | 0 | 0 | 0.005838916 |
| 0 | 0 | 0 | 0 | 0 |
| 0 | 0 | 0 | 0 | 0 |
| 0 | 0 | 0 | 0 | 0 |
| 0 | 0 | 0 | 0 | 0 |
| 0 | 0 | 0 | 0 | 0 |
| 0 | 0 | 0 | 0 | 0.018946752 |
| 0 | 0 | 0 | 0 | 0 |
| 0 | 0.006075761 | 0.011915008 | 0.00379932 | 0.00177503 |
| 0 | 0 | 0 | 0 | 0 |
| 0 | 0 | 0 | 0 | 0 |
| 0 | 0 | 0.000845741 | 0 | 0 |
| 0 | 0 | 0 | 0 | 0.018168594 |
| 0 | 0 | 0 | 0 | 0 |
| 0 | 0 | 0 | 0 | 0 |
| 0 | 0 | 0 | 0 | 0 |
| 0 | 0 | 0 | 0 | 0.044055169 |
| 0 | 0 | 0 | 0 | 0.003848136 |
| 0 | 0 | 0 | 0 | 0 |

| | | | | |
|---|---|---|---|---|
| 0 | 0 | 0 | 0 | 0 |
| 0 | 0 | 0 | 0 | 0 |
| 0 | 0.002813994 | 0 | 0 | 0.015432121 |
| 0 | 0 | 0 | 0 | 0 |
| 0.001175685 | 0.029443955 | 0.003768139 | 0.00123927 | 0.055936863 |
| 0.004217536 | 0 | 0 | 0 | 0.014565876 |
| 0 | 0 | 0 | 0 | 0 |
| 0.007580717 | 0.011376307 | 0.049603712 | 0 | 0.042334263 |
| 0 | 0 | 0 | 0 | 0.007581322 |
| 0.032953045 | 0.017452362 | 0.017752646 | 0 | 0.126295212 |
| 0.00337868 | 0 | 0 | 0.002081343 | 0 |
| 0.006000922 | 0 | 0.000727157 | 0.067576464 | 0.006607991 |
| 0.005937277 | 0.021287335 | 0.011583062 | 0.021904345 | 0.054957339 |
| 0 | 0 | 0.005026195 | 0 | 0 |
| 0 | 0 | 0 | 0 | 0 |
| 0 | 0 | 0 | 0.001342125 | 0 |
| 0 | 0 | 0 | 0 | 0 |
| 0 | 0 | 0 | 0 | 0 |
| 0.015344271 | 0 | 0 | 0.02045836 | 0.019373187 |
| 0 | 0 | 0 | 0 | 0.014833489 |
| 0 | 0 | 0 | 0 | 0 |
| 0 | 0 | 0 | 0 | 0 |
| 0 | 0 | 0.001992037 | 0.005996569 | 0 |
| 0 | 0 | 0 | 0 | 0 |
| 0 | 0 | 0 | 0 | 0 |
| 0.001613597 | 0.001110193 | 0.008191155 | 0.001388461 | 0 |
| 0 | 0 | 0 | 0 | 0.005297477 |
| 0 | 0 | 0 | 0.002308674 | 0 |
| 0 | 0 | 0 | 0 | 0 |
| 0.006640752 | 0 | 0 | 0 | 0 |
| 0 | 0 | 0 | 0 | 0 |
| 0.019490837 | 0 | 0 | 0 | 0 |
| 0 | 0 | 0 | 0 | 0 |
| 0 | 0 | 0.007561783 | 0 | 0 |
| 0 | 0 | 0 | 0 | 0 |
| 0 | 0 | 0 | 0 | 0 |
| 0 | 0 | 0 | 0 | 0 |
| 0 | 0 | 0 | 0 | 0 |
| 0.001641586 | 0 | 0 | 0 | 0 |
| 0 | 0 | 0.001689215 | 0.791485814 | 0.010964738 |
| 0 | 0 | 0 | 0 | 0 |
| 0.001624326 | 0 | 0.006224635 | 0 | 0 |
| 0 | 0 | 0 | 0 | 0 |
| 0 | 0 | 0.004140601 | 0.023490461 | 0 |
| 0 | 0 | 0 | 0 | 0 |
| 0 | 0 | 0 | 0 | 0 |
| 0.022932971 | 0.001840818 | 0.01038608 | 0 | 0 |
| 0 | 0 | 0.054568766 | 0 | 0 |

| | | | | |
|---|---|---|---|---|
| 0 | 0 | 0 | 0 | 0 |
| 0.048695479 | 0.005914572 | 0 | 0.002894497 | 0.001738672 |
| 0 | 0 | 0 | 0 | 0 |
| 0 | 0 | 0 | 0 | 0 |
| 0 | 0 | 0 | 0 | 0 |
| 0 | 0 | 0.008180296 | 0 | 0.0016338 |
| 0.012552811 | 0 | 0 | 0 | 0 |
| 0 | 0 | 0 | 0.003517867 | 0 |
| 0 | 0 | 0 | 0 | 0 |
| 0 | 0 | 0 | 0 | 0 |
| 0 | 0.001891802 | 0.005656168 | 0 | 0.052584392 |
| 1.421531099 | 0 | 0 | 0 | 0 |
| 0 | 0 | 0 | 0 | 0 |
| 0.032161979 | 0.003375971 | 0.003275149 | 0.118468635 | 0.008951187 |
| 0 | 0 | 0 | 0.018470375 | 0 |
| 0 | 0 | 0 | 0 | 0 |
| 0 | 0 | 0 | 0.002472025 | 0 |
| 0 | 0 | 0.003187249 | 0 | 0.009364272 |
| 0.013692593 | 0 | 0 | 0.022006561 | 0 |
| 0 | 0 | 0 | 0 | 0 |
| 0.058910463 | 0.004707315 | 0 | 0.074058239 | 0 |
| 0.141254735 | 0.192416754 | 0.183001745 | 0 | 0.425305101 |
| 0 | 0 | 0.009160302 | 0 | 0 |
| 0 | 0 | 0 | 0 | 0 |
| 0 | 0 | 0 | 0 | 0 |
| 0.002298757 | 0 | 0 | 0 | 0 |
| 0 | 0 | 0 | 0 | 0 |
| 0 | 0 | 0 | 0 | 0 |
| 0 | 0 | 0 | 0 | 0 |
| 0.54431716 | 0.229084914 | 0.814094596 | 0.055972157 | 0.374920171 |
| 0.002349776 | 0.002829229 | 0 | 0 | 0 |
| 0 | 0 | 0 | 0 | 0 |
| 0 | 0 | 0 | 0 | 0 |
| 0.001426323 | 0 | 0 | 0 | 0 |
| 0.020108198 | 0 | 0.005759839 | 0 | 0 |
| 0 | 0 | 0 | 0 | 0 |
| 0 | 0 | 0.89711045 | 0 | 0.552463419 |
| 0.001260832 | 0 | 0 | 0 | 0 |
| 0 | 0 | 0 | 0 | 0.01695489 |
| 0 | 0 | 0 | 0 | 0 |
| 0.003254233 | 0 | 0.017997428 | 0.018682518 | 0 |
| 0.483139073 | 0.247262507 | 5.368618491 | 10.29231824 | 6.783180022 |
| 0.000924003 | 0 | 0 | 0 | 0 |
| 0 | 0 | 0 | 0 | 0 |
| 0.003441511 | 0 | 0 | 0 | 0 |
| 0 | 0 | 0 | 0 | 0 |
| 0.00106736 | 0 | 0.002974739 | 0.003696711 | 0.00559964 |
| 0 | 0 | 0 | 0 | 0 |

| | | | | |
|---|---|---|---|---|
| 0 | 0.002325627 | 0.002936263 | 0.000793239 | 0 |
| 0 | 0.000810549 | 0 | 0 | 0 |
| 0.093310654 | 0.855011993 | 0.003250718 | 0.577713395 | 1.488720531 |
| 0 | 0 | 0 | 0 | 0 |
| 0 | 0 | 0.405853109 | 0 | 0 |
| 0 | 0 | 0.003568756 | 0 | 0.03807098 |
| 0 | 0 | 0 | 0 | 0 |
| 0 | 0 | 0 | 0 | 0 |
| 0 | 0 | 0 | 0 | 0.001196323 |
| 0 | 0 | 0 | 0 | 0 |
| 0 | 0 | 0 | 0 | 0 |
| 0 | 0 | 0 | 0 | 0 |
| 0 | 0 | 0 | 0 | 0 |
| 0 | 0.010902214 | 0 | 0 | 0.003448503 |
| 0 | 0 | 0 | 0 | 0 |
| 0.054001623 | 0.001943133 | 0.044761632 | 0.004206074 | 0 |
| 0.011764637 | 0.032140736 | 0.025885358 | 0.002175597 | 0 |
| 0 | 0 | 0 | 0 | 0 |
| 0 | 0 | 0 | 0 | 0 |
| 0 | 0 | 0 | 0 | 0 |
| 0.072573233 | 0 | 0.004349722 | 0.074813933 | 0.129641817 |
| 0 | 0.007686924 | 0 | 0 | 0 |
| 0 | 0 | 0 | 0 | 0 |
| 0 | 0 | 0 | 0 | 0 |
| 0 | 0 | 0 | 0 | 0 |
| 0 | 0 | 0 | 0 | 0 |
| 0 | 0 | 0.000912824 | 0.001775532 | 0 |
| 0 | 0.013044023 | 0 | 0 | 0 |
| 0 | 0 | 0 | 0 | 0 |
| 0 | 0 | 0 | 0 | 0 |
| 0.019727494 | 0.003358972 | 0.005275921 | 0 | 0.003604856 |
| 0 | 0 | 0.060235097 | 0.001342075 | 0.351485661 |
| 0.009617366 | 0 | 0.013204746 | 0.186351181 | 0.002302719 |
| 0 | 0 | 0 | 0 | 0 |
| 0.002314656 | 0.000928981 | 0.028021361 | 0 | 0 |
| 0 | 0 | 0 | 0 | 0 |
| 0.021225449 | 0.002052718 | 0 | 0.009819646 | 0.00693939 |
| 0.006199733 | 0.012682031 | 0.086393482 | 0.117148771 | 0 |
| 0 | 3.518363083 | 1.309771576 | 0.282220237 | 0 |
| 0.016351339 | 0 | 0 | 0 | 0 |
| 0 | 0 | 0 | 0 | 0 |
| 0 | 0 | 0 | 0 | 0.004034056 |
| 0 | 0 | 0 | 0.0608006 | 0 |
| 0 | 0.010025203 | 0.079188507 | 0.001763157 | 0.101108353 |
| 0 | 0 | 0 | 0.084877921 | 0 |
| 0 | 0 | 0.00227845 | 0 | 0 |
| 0 | 0 | 0 | 0 | 0 |
| 0 | 0 | 0 | 0 | 0 |

| | | | | |
|---|---|---|---|---|
| 0 | 0 | 0 | 0 | 0 |
| 2.510123072 | 0.555831714 | 3.375016068 | 0.020723183 | 17.32594135 |
| 0 | 0 | 0 | 0 | 0 |
| 0 | 0 | 0 | 0.007455889 | 0 |
| 0 | 0 | 0 | 0 | 0 |
| 0 | 0 | 0 | 0 | 0 |
| 0 | 0 | 0 | 0 | 0 |
| 0 | 0 | 0 | 0 | 0.000757339 |
| 0.012972118 | 0 | 0 | 0 | 0 |
| 0 | 0 | 0 | 0 | 0 |
| 0.009315342 | 0.00097531 | 0 | 0.019821255 | 0 |
| 0.692868353 | 0.022517652 | 0.062840397 | 0.189182771 | 0.589884062 |
| 0.913294767 | 0 | 0 | 0 | 0 |
| 0 | 0 | 0 | 0 | 0 |
| 0.041590999 | 0 | 0.002428374 | 0.004723424 | 0 |
| 0 | 0 | 0 | 0 | 0 |
| 0.002544654 | 0.0905544 | 0.024526388 | 0.000638637 | 0.161119471 |
| 0.00137638 | 0.00419829 | 0.003644182 | 0.164978872 | 0.12275426 |
| 0 | 0 | 0 | 0 | 0 |
| 0.008758744 | 0 | 0.004857226 | 0 | 0.013904822 |
| 0.016182445 | 0.427777932 | 0.117627191 | 0.008067804 | 0.313122719 |
| 0 | 0 | 0 | 0 | 0 |
| 0 | 0 | 0 | 0 | 0 |
| 0 | 0 | 0 | 0 | 0 |
| 0.233315531 | 0 | 0 | 0.021024727 | 0 |
| 0.002406447 | 0 | 0 | 0 | 0 |
| 0.090009348 | 0.013167871 | 0.232207831 | 0.047454922 | 0.408445847 |
| 0.195935417 | 0.013432854 | 0.187368565 | 0.022924692 | 0.052769179 |
| 0 | 0 | 0 | 0.002559107 | 0 |
| 0 | 0 | 0 | 0 | 0 |
| 0.003242118 | 0 | 0.000521296 | 0 | 0.002481415 |
| 0.005453901 | 0 | 0 | 0 | 0 |
| 0 | 0 | 0 | 0 | 0 |
| 0 | 0 | 0 | 0 | 0 |
| 0 | 0 | 0 | 0 | 0 |
| 0.113683276 | 0.143983924 | 0.131832296 | 0.122911831 | 0.435513276 |
| 0 | 0 | 0 | 0 | 0 |
| 0 | 0 | 0 | 0 | 0 |
| 0.029245838 | 0.001611062 | 0.001997754 | 0.03151836 | 0.026453633 |
| 0 | 0 | 0 | 0 | 0 |
| 0 | 0 | 0 | 0.080737321 | 0 |
| 0 | 0 | 0 | 0 | 0 |
| 0.185132589 | 0.090169518 | 0.0249974 | 0.069760261 | 0 |
| 0.072472074 | 0 | 0 | 0 | 0.097188643 |
| 0.039321688 | 0 | 0 | 0.008316278 | 0.00079927 |
| 0 | 0.385869088 | 0 | 0 | 0.020900337 |
| 0 | 0 | 0 | 0 | 0 |
| 0 | 0 | 0 | 0.002660348 | 0 |

| | | | | |
|---|---|---|---|---|
| 0 | 0 | 0.015257144 | 0 | 0 |
| 0 | 0 | 0 | 0 | 0 |
| 0.000638429 | 0 | 0 | 0 | 0 |
| 0.00324547 | 0.463385968 | 0.002543947 | 0.000916338 | 0.00348604 |
| 0 | 0 | 0 | 0 | 0 |
| 0 | 0 | 0 | 0 | 0 |
| 0 | 0.001262308 | 0 | 0 | 0 |
| 0.000907319 | 0.005389421 | 0 | 0.004918575 | 0 |
| 0.187942907 | 0 | 0.645742958 | 0 | 0.028135085 |
| 0.011421316 | 0.016007323 | 0 | 0 | 0 |
| 0 | 0 | 0 | 0 | 0 |
| 0.006024673 | 0 | 0.002437375 | 0.045068112 | 0 |
| 0 | 0 | 0 | 0 | 0 |
| 0.046747509 | 0 | 0 | 0 | 0 |
| 0 | 0.305478406 | 0.044919655 | 0.2763917 | 0 |
| 0 | 0 | 0 | 0 | 0 |
| 0 | 0 | 0 | 0 | 0 |
| 0 | 0 | 0 | 0 | 0 |
| 0 | 0 | 0 | 0 | 0 |
| 0 | 0 | 0 | 0 | 0 |
| 0 | 0 | 0 | 0 | 0 |
| 0 | 0 | 0 | 0 | 0 |
| 0 | 0 | 0 | 0 | 0 |
| 0.382339719 | 0 | 0.162420729 | 1.338391523 | 0.025990127 |
| 0.009461708 | 0 | 0.00063389 | 0 | 0 |
| 0 | 0.178974249 | 0 | 0 | 2.51683577 |
| 0 | 0 | 0 | 0 | 0 |
| 0 | 0 | 0 | 0 | 0 |
| 0 | 0 | 0 | 0 | 0 |
| 0 | 0 | 0 | 0 | 0 |
| 0.000944679 | 0.0068246 | 0 | 0 | 0.004101538 |
| 0.085624042 | 0 | 0.329301097 | 0 | 0.003542375 |
| 0.0240136 | 0 | 0.003506605 | 0.000541324 | 0 |
| 0 | 0 | 0 | 0 | 0 |
| 0 | 0 | 0 | 0 | 0 |
| 0.477705734 | 0 | 0 | 0 | 0.00304294 |
| 0 | 0.382632583 | 0 | 0 | 0.002807124 |
| 0 | 0 | 0 | 0.047501132 | 0 |
| 0 | 0 | 0 | 0.008682489 | 0 |
| 0 | 0.005806321 | 0 | 0.012463532 | 0 |
| 0.44931705 | 0.030229965 | 1.746264628 | 0.025547835 | 0.084150683 |
| 0.835046672 | 0 | 0 | 0 | 0.0477477 |
| 1.06035309 | 0.864065843 | 4.05944144 | 1.135651199 | 0.053003321 |
| 0 | 0.015253007 | 0 | 0 | 0 |
| 0 | 0.002090832 | 0 | 0 | 0 |
| 0 | 0 | 0 | 0 | 0 |
| 0 | 0 | 0 | 0 | 0 |
| 0.004930653 | 0.199689475 | 0 | 0.029530219 | 0 |

| | | | | |
|---|---|---|---|---|
| 0 | 0 | 0 | 0 | 0 |
| 0 | 0 | 0 | 0 | 0.005171219 |
| 0.05091824 | 0.002069458 | 0.014520273 | 0.084115345 | 0.030704595 |
| 0.003019082 | 0 | 0 | 0 | 0 |
| 0 | 0 | 0 | 0 | 0.001246579 |
| 0.001353972 | 0.011819237 | 0 | 0.006243997 | 0 |
| 0 | 0 | 0 | 0 | 0 |
| 0 | 0 | 0.003585107 | 0 | 0 |
| 0 | 0 | 0 | 0 | 0 |
| 0 | 0 | 0 | 0 | 0 |
| 0.109688585 | 0.023196854 | 0.017195783 | 0.056857527 | 0 |
| 0.851167431 | 0.848715819 | 0.711240667 | 1.78288536 | 0.911006697 |
| 0 | 0 | 0 | 0 | 0 |
| 0.001365928 | 0.05134914 | 0.037592636 | 0.001542644 | 0 |
| 0.140829436 | 0.041329115 | 0.029874434 | 0.606345615 | 0.153185275 |
| 0.00064811 | 0.05809285 | 0 | 0 | 0 |
| 0 | 0 | 0 | 0.002633359 | 0 |
| 0 | 0 | 0 | 0 | 0 |
| 0 | 0 | 0 | 0 | 0.000682911 |
| 0 | 0.01771335 | 0 | 0.002354864 | 0 |
| 0.794355376 | 0.025402817 | 0.076128084 | 0.003993434 | 2.465149099 |
| 0 | 0.059813828 | 0 | 0.001542875 | 0.05830268 |
| 0.560085829 | 0.328135611 | 1.417618652 | 0.033444139 | 4.911753858 |
| 0 | 0 | 0 | 0 | 0 |
| 0 | 0 | 0 | 0 | 0 |
| 0.002118576 | 9.166409794 | 0.00560926 | 0.003509243 | 2.748559893 |
| 0.125395285 | 0 | 0 | 0 | 0 |
| 0.00073759 | 0 | 0 | 0 | 0 |
| 0 | 0 | 0 | 0 | 0 |
| 0 | 0 | 0 | 0 | 0 |
| 0.002145045 | 0 | 0.005842385 | 0 | 0 |
| 0.267725688 | 0.189312053 | 0.15141701 | 0.284140469 | 0.255299832 |
| 0.120452091 | 0 | 0 | 0.328507938 | 0 |
| 0 | 0 | 0.008863234 | 0.003482799 | 0 |
| 0.010294948 | 0 | 0.022316068 | 0.017763251 | 0 |
| 0.041451621 | 0.038003757 | 0.010129052 | 0.031209612 | 0 |
| 0 | 0.011576948 | 0.00158668 | 0.014716825 | 0 |
| 0 | 0 | 0 | 0 | 0 |
| 0.063449611 | 0 | 1.546821189 | 0.005538813 | 0.008941483 |
| 0 | 0 | 0 | 0 | 0 |
| 0 | 0.002360547 | 0 | 0 | 0 |
| 0 | 0 | 0 | 0 | 0 |
| 0 | 0 | 0 | 0 | 0 |
| 0 | 0 | 0 | 0 | 0 |
| 0.087372695 | 0.017803146 | 0.001404855 | 0.008771247 | 0 |
| 0 | 0 | 0 | 0 | 0.014253088 |
| 0 | 0 | 0 | 0 | 0 |
| 0 | 0.032538574 | 0.01374541 | 0.054937317 | 0.004888865 |

| | | | | |
|---|---|---|---|---|
| 0 | 0 | 0 | 0 | 0 |
| 0 | 0 | 0 | 0 | 0 |
| 0 | 0 | 0 | 0 | 0 |
| 0.004200725 | 0.000919609 | 0.000532157 | 0 | 0 |
| 0 | 0 | 0 | 0 | 0 |
| 0 | 0 | 0 | 0 | 0 |
| 0 | 0 | 0 | 0 | 0 |
| 1.347101378 | 0.049785165 | 0.00954877 | 0.194463905 | 0.000635706 |
| 0 | 0 | 0 | 0 | 0 |
| 0 | 0 | 0 | 0 | 0 |
| 0.015736756 | 0.001365601 | 0.004296942 | 0.007365262 | 0 |
| 0 | 0 | 0 | 0 | 0 |
| 0 | 0 | 0 | 0 | 0 |
| 0 | 0 | 0 | 0 | 0.065328721 |
| 0 | 0 | 0 | 0 | 0 |
| 0 | 0 | 0 | 0 | 0 |
| 0 | 0.650649364 | 0 | 0 | 0 |
| 0.001239236 | 0.00426312 | 0 | 0.006531287 | 0 |
| 0.031296108 | 0.016747481 | 0 | 0 | 0 |
| 0.017721709 | 0 | 0.004227117 | 0 | 0 |
| 0 | 0 | 0 | 0.001776286 | 0 |
| 0 | 0 | 0 | 0 | 0 |
| 0 | 0 | 0.007252811 | 0 | 0 |
| 0 | 0 | 0 | 0 | 0 |
| 0 | 0 | 0 | 0 | 0 |
| 0 | 0 | 0 | 0 | 0 |
| 0 | 0 | 0 | 0 | 0 |
| 0 | 0 | 0 | 0 | 0 |
| 0 | 0 | 0 | 0 | 0 |
| 0.100084174 | 0.216807016 | 1.254463615 | 0 | 2.357423513 |
| 0 | 0 | 0 | 0.003404914 | 0 |
| 0 | 0 | 0 | 0 | 0 |
| 0 | 0 | 0 | 0 | 0 |
| 0 | 0 | 0 | 0 | 0 |
| 0 | 0 | 0 | 0.000636576 | 0 |
| 0 | 0 | 0 | 0 | 0 |
| 0 | 0.243015316 | 0.106213304 | 0 | 0 |
| 0.034923774 | 0 | 0.001505146 | 0.009352234 | 0 |
| 0 | 0 | 0 | 0 | 0 |
| 0 | 0 | 0 | 0 | 0 |
| 0.03800313 | 0.029013562 | 0 | 0 | 0 |
| 0 | 0 | 0 | 0 | 0 |
| 0 | 0 | 0 | 0 | 0 |
| 1.625303552 | 0 | 0.025335705 | 0 | 0 |
| 0 | 0 | 0 | 0 | 0.052694916 |
| 0 | 0 | 0 | 0 | 0 |
| 0.002394681 | 0 | 0.006912348 | 0.005924126 | 0 |
| 0 | 0 | 0 | 0 | 0 |

| | | | | |
|---|---|---|---|---|
| 0 | 0 | 0 | 0 | 0 |
| 0 | 0 | 0 | 0 | 0 |
| 0 | 0 | 0 | 0 | 0 |
| 0 | 0.556257599 | 0 | 0 | 0 |
| 0 | 0 | 0 | 0 | 0 |
| 0 | 0.441548956 | 0 | 0 | 0 |
| 0 | 0 | 0 | 0 | 0 |
| 0 | 0 | 0 | 0 | 0 |
| 0.070510637 | 0.002391487 | 0.016952759 | 0.153284062 | 0.005165179 |
| 0 | 0 | 0 | 0 | 0 |
| 0 | 0 | 0 | 0 | 0 |
| 0 | 0 | 0 | 0 | 0 |
| 0 | 0 | 0 | 0 | 0.000393035 |
| 0 | 0 | 0 | 0 | 0 |
| 0.898871528 | 0.792267895 | 2.632333327 | 0.190620106 | 5.373767249 |
| 0 | 0 | 0 | 0 | 0 |
| 0 | 0 | 0 | 0 | 0 |
| 0 | 0 | 0 | 0 | 0 |
| 0 | 0 | 0 | 0 | 0 |
| 0 | 0 | 0 | 0 | 0 |
| 0.051637374 | 0 | 0.023676978 | 0 | 0 |
| 0 | 0 | 0 | 0 | 0 |
| 0 | 0 | 0 | 0 | 0 |
| 0 | 0 | 0 | 0 | 0 |
| 0.034452053 | 0 | 0.013222831 | 0.001318959 | 0.060476943 |
| 2.996550939 | 7.126485052 | 1.510907534 | 0.181296146 | 0.014935032 |
| 0.00738441 | 0.023884045 | 0 | 0 | 0.003143244 |
| 0 | 0 | 0 | 0 | 0 |
| 0 | 0 | 0.057624825 | 0.005765735 | 0 |
| 0 | 0 | 0 | 0 | 0 |
| 0 | 0 | 0 | 0 | 0 |
| 0 | 0.003320441 | 0.022926552 | 0.009202014 | 0 |
| 0 | 0 | 0 | 0 | 0 |
| 0.010447235 | 0.000625816 | 0.001086437 | 0.005048262 | 0 |
| 0.005395353 | 0.017117055 | 0.113492664 | 0 | 0.000929569 |
| 0.042678911 | 0.001312014 | 0.001645003 | 0.001914346 | 0 |
| 0 | 0 | 1.014510734 | 0 | 0 |
| 2.047026468 | 6.296827813 | 5.467174206 | 0.233764532 | 1.208671301 |
| 0 | 0 | 0 | 0 | 0 |
| 0 | 0 | 0 | 0 | 0 |
| 0 | 0 | 0 | 0 | 0.336979148 |
| 0 | 0 | 0 | 0 | 0 |
| 0.012657005 | 0.503403733 | 2.025727668 | 0 | 0.190809913 |
| 0 | 0 | 0 | 0 | 0 |
| 0 | 0 | 0 | 0 | 0 |
| 0.00103233 | 0 | 0.010789161 | 0 | 0 |
| 0 | 0 | 0 | 0 | 0 |
| 0.034537122 | 0.54059395 | 0.440023708 | 0.011350765 | 9.495144898 |

| | | | | |
|---|---|---|---|---|
| 0.043741549 | 0 | 0.008829784 | 0.006617526 | 0.001294195 |
| 0 | 0 | 0 | 0 | 0 |
| 0 | 0 | 0 | 0 | 0 |
| 0 | 0 | 0 | 0 | 0 |
| 0.004070073 | 0 | 0 | 0.00122577 | 0 |
| 0 | 0 | 0 | 0 | 0 |
| 0 | 0.047985571 | 0 | 0 | 0 |
| 0 | 0 | 0 | 0 | 0 |
| 0 | 0 | 0 | 0 | 0 |
| 0 | 0 | 0 | 0 | 0 |
| 0 | 0 | 0 | 0 | 0 |
| 0.070779901 | 0.138796544 | 0 | 10.03432481 | 0 |
| 0.287064421 | 1.161817223 | 2.526854552 | 0.361400915 | 0.156017288 |
| 0 | 0 | 0 | 0 | 0 |
| 0.08072572 | 0.12392875 | 0.02115366 | 2.52847227 | 0.055775098 |
| 0 | 0 | 0 | 0 | 0 |
| 0.00130977 | 0 | 0 | 0.004560933 | 0 |
| 0 | 0 | 0 | 0 | 0 |
| 0.061885544 | 0.003808432 | 0 | 0 | 0 |
| 0.001601161 | 0 | 0.003506208 | 0 | 0 |
| 0 | 0.048666372 | 0.065328177 | 0.754034134 | 0 |
| 0 | 0 | 0 | 0 | 0 |
| 0.564444148 | 0.010465516 | 0.686907734 | 1.16454007 | 0.236589636 |
| 0 | 0 | 0 | 0 | 0 |
| 0 | 0 | 0 | 0 | 0 |
| 0 | 0 | 0 | 0 | 0 |
| 0 | 0 | 0 | 0 | 0 |
| 0 | 0 | 0 | 0 | 0 |
| 0 | 0 | 0 | 0 | 0 |
| 0 | 0 | 0 | 0 | 0 |
| 0 | 0 | 0 | 0 | 0 |
| 0.014353897 | 0.084084612 | 0.288755209 | 0.010577345 | 2.62625392 |
| 0 | 0 | 0 | 0 | 0 |
| 0.051417307 | 0.012614414 | 0.004358542 | 0.010338768 | 0 |
| 0.033203484 | 0.003172888 | 0.000688529 | 0.002232094 | 0 |
| 0 | 0.031903856 | 0.00763334 | 0 | 0 |
| 0 | 0 | 0 | 0 | 0 |
| 0 | 0 | 0 | 0 | 0 |
| 0 | 0 | 0 | 0.001196695 | 0 |
| 0 | 0 | 0 | 0 | 0 |
| 0.128649824 | 0.23302554 | 0 | 0 | 0 |
| 0 | 0 | 0.081711764 | 0 | 0 |
| 0 | 0 | 0 | 0 | 0 |
| 0.025260826 | 0 | 0 | 0.011750774 | 0 |
| 0.05801911 | 0.005157262 | 0.058195592 | 0.003811316 | 0 |
| 0 | 0 | 0 | 0 | 0 |
| 0 | 0.223008761 | 1.048683996 | 0 | 0 |
| 0 | 0 | 0.001667958 | 0.00138647 | 0 |

| | | | | |
|---|---|---|---|---|
| 0 | 4.494435918 | 0 | 0 | 0 |
| 0 | 0 | 0 | 0 | 0 |
| 0 | 0.0014547 | 0.005331409 | 0 | 0 |
| 0 | 0 | 0 | 0 | 0 |
| 0.016336924 | 0 | 0 | 0.002767577 | 0 |
| 0 | 0 | 0 | 0 | 0 |
| 0 | 0 | 0 | 0 | 0 |
| 0 | 0.13326131 | 0.012190043 | 0.003006807 | 0.001032077 |
| 0 | 0 | 0 | 0 | 0 |
| 0 | 0 | 0 | 0 | 0 |
| 0 | 0 | 0 | 0 | 0 |
| 0 | 0 | 0 | 0 | 0 |
| 0 | 0 | 0 | 0 | 0 |
| 0 | 0 | 0 | 0 | 0 |
| 0 | 0 | 0 | 0 | 0 |
| 0 | 0 | 0 | 0 | 0 |
| 1.15958451 | 0 | 0 | 0 | 0 |
| 0 | 0 | 0 | 0 | 0 |
| 0 | 0.518649592 | 0 | 0 | 0.016737399 |
| 0 | 0 | 0 | 0 | 0 |
| 0 | 0 | 0 | 0 | 0 |
| 0 | 0 | 0 | 0 | 0 |
| 0 | 0 | 0 | 0 | 0 |
| 0 | 0 | 0 | 0 | 0 |
| 0 | 0.000683001 | 0 | 0 | 0 |
| 0.068151386 | 0 | 0.005430419 | 0.005322287 | 0 |
| 0 | 0 | 0 | 0 | 0 |
| 2.242124088 | 0.041013252 | 0.111497681 | 0.001736487 | 0.087939318 |
| 0 | 0 | 0 | 0 | 0 |
| 11.78302829 | 1.812986964 | 2.849090481 | 0.397447157 | 0.012246867 |
| 0 | 0 | 0 | 0 | 0 |
| 0 | 0 | 0 | 0 | 0 |
| 0 | 0 | 0.000522093 | 0 | 0 |
| 0 | 0 | 0 | 0 | 0 |
| 0.061541419 | 0 | 0.000794057 | 0.007722592 | 0 |
| 0 | 0 | 0 | 0 | 0 |
| 0 | 0 | 0 | 0 | 0 |
| 0.001293176 | 0.001730042 | 0 | 0.002271858 | 0 |
| 0 | 0 | 0 | 0 | 0 |
| 0 | 0 | 0 | 0 | 0 |
| 7.567744113 | 3.017577171 | 4.686982399 | 2.227868827 | 15.06940758 |
| 0.228133114 | 0.956257668 | 0.85137219 | 1.516902909 | 0.12472477 |
| 0 | 0 | 0 | 0 | 0 |
| 0 | 0 | 0 | 0 | 0 |
| 0 | 0 | 0 | 0 | 0 |
| 0 | 0 | 0 | 0 | 0 |
| 0 | 0 | 0 | 0 | 0 |
| 0.010776525 | 0.002196892 | 0 | 0.001030327 | 0 |

| | | | | |
|---|---|---|---|---|
| 0 | 0 | 0 | 0 | 0 |
| 0.008973204 | 0 | 0.003677703 | 0 | 0 |
| 0 | 0 | 0.00257721 | 0 | 0 |
| 0 | 0 | 0 | 0 | 0 |
| 0.541181583 | 7.12267707 | 1.323981291 | 0 | 0.634629015 |
| 0 | 0 | 0 | 0 | 0 |
| 0 | 0 | 0 | 0 | 0 |
| 0 | 0 | 0 | 0 | 0 |
| 0.013176433 | 0.007051102 | 0.103925538 | 0.220373027 | 0.205155929 |
| 0 | 0 | 0 | 0 | 0 |
| 0 | 0 | 0 | 0 | 0 |
| 0 | 0 | 0 | 0 | 0 |
| 0.313883887 | 0.103882561 | 0.27182537 | 0 | 0.043298776 |
| 0.006305365 | 0.008435472 | 0.021527039 | 0 | 0.015399095 |
| 0 | 0 | 0.39088079 | 0 | 0 |
| 0.149705019 | 0.250579556 | 0.83301571 | 0 | 0.524488037 |
| 0.111672582 | 0.001459918 | 0.006462884 | 0.002327952 | 0 |
| 0 | 0 | 0 | 0 | 0 |
| 0 | 0 | 0 | 0 | 0 |
| 0 | 0 | 0 | 0 | 0 |
| 0 | 0 | 0 | 0 | 0 |
| 0 | 0 | 0 | 0 | 0 |
| 0 | 0 | 0 | 0 | 0 |
| 0 | 0 | 0 | 0 | 0 |
| 0 | 0 | 0 | 0 | 0 |
| 0 | 0 | 0 | 0 | 0 |
| 0.011861226 | 0.093813031 | 0.048373869 | 0 | 0.000572202 |
| 0 | 0 | 0 | 0.000806167 | 0 |
| 0 | 0 | 0 | 0 | 0 |
| 0 | 0 | 0 | 0 | 0 |
| 0 | 0 | 0 | 0 | 0 |
| 0 | 0 | 0.001350189 | 0 | 0 |
| 0 | 0.483289302 | 0 | 0 | 0 |
| 0.01197233 | 0.065468986 | 0.021897069 | 0.012535324 | 0 |
| 0 | 0 | 0 | 0 | 0.023414922 |
| 0.174643579 | 0.104062038 | 0.003452006 | 0 | 0 |
| 0.017513075 | 0 | 0 | 0 | 0.135507682 |
| 0.00292399 | 0 | 0.004889508 | 0 | 0.002292183 |
| 0 | 0 | 0 | 0 | 0 |
| 0 | 0 | 0 | 0 | 0 |
| 2.530819653 | 0.950998495 | 0.996884439 | 2.598974763 | 0.210624066 |
| 0 | 0 | 0 | 0 | 0 |
| 0 | 0 | 0 | 0 | 0 |
| 0 | 0 | 0 | 0 | 0 |
| 0 | 0 | 0 | 0 | 0 |
| 0 | 0 | 0 | 0 | 0 |
| 0 | 0 | 0 | 0 | 0 |
| 0 | 0 | 0 | 0 | 0 |

| | | | | |
|---|---|---|---|---|
| 0 | 0.013958676 | 0 | 0 | 0.003357984 |
| 0 | 0 | 0 | 0 | 0 |
| 0.132640378 | 0 | 0 | 0 | 0 |
| 0 | 0 | 0 | 0 | 0 |
| 0.008123702 | 0.028878063 | 0 | 0 | 0 |
| 0 | 0 | 0 | 0 | 0 |
| 0.344907785 | 0.077333172 | 0.772522551 | 0.003278529 | 1.525919643 |
| 0 | 0 | 0 | 0 | 0 |
| 0 | 0 | 0 | 0 | 0 |
| 0 | 0.001222374 | 0 | 0 | 0 |
| 0 | 0 | 0 | 0 | 0 |
| 0.11439982 | 0 | 0 | 0 | 0 |
| 0 | 0 | 0 | 0 | 0 |
| 0 | 0 | 0 | 0 | 0 |
| 0 | 0 | 0 | 0 | 0 |
| 0 | 0 | 0 | 0 | 0 |
| 0 | 0 | 0 | 0 | 0 |
| 0 | 0 | 0 | 0 | 0 |
| 0 | 0 | 0 | 0 | 0 |
| 0 | 0 | 0 | 0 | 0 |
| 0.102087069 | 0.075427997 | 0.027440758 | 0.00858369 | 0.001093711 |
| 0 | 0 | 0 | 0 | 0 |
| 0.004572657 | 0.01896398 | 0.017788535 | 0.002438671 | 0 |
| 0.003388378 | 0.004215742 | 0.004957802 | 0.002806283 | 0.001685683 |
| 0 | 0 | 0 | 0 | 0 |
| 0.017988861 | 0.001123077 | 0 | 0 | 0 |
| 0 | 0 | 0 | 0 | 0 |
| 0 | 0 | 0 | 0 | 0 |
| 0 | 0 | 0 | 0 | 0 |
| 0.681514732 | 0 | 0.028881541 | 0.29135147 | 0.014997663 |
| 0 | 0 | 0 | 0 | 0 |
| 0 | 0 | 0 | 0 | 0 |
| 0 | 0 | 0 | 0 | 0 |
| 0 | 0 | 0 | 0 | 0 |
| 0 | 0 | 0 | 0 | 0.001633591 |
| 0 | 0 | 0 | 0 | 0 |
| 0.11214109 | 0 | 0 | 0.003990212 | 0.004410199 |
| 0 | 0 | 0 | 0 | 0 |
| 0 | 0 | 0 | 0 | 0 |
| 0 | 0 | 0 | 0 | 0 |
| 0 | 0 | 0 | 0 | 0 |
| 0 | 0 | 0 | 0 | 0 |
| 0 | 0 | 0 | 0 | 0 |
| 0 | 0 | 0 | 0 | 0 |
| 0 | 0 | 0 | 0 | 0 |
| 0 | 0 | 0 | 0 | 0 |
| 0 | 0 | 0 | 0 | 0 |
| 0 | 0 | 0 | 0 | 0 |

| | | | | |
|---|---|---|---|---|
| 0 | 0 | 0.00152422 | 0.061491255 | 0 |
| 0 | 0 | 0 | 0 | 0 |
| 0 | 0.481879316 | 0 | 0 | 0 |
| 0 | 0 | 0 | 0 | 0 |
| 0 | 0 | 0 | 0 | 0 |
| 0 | 0 | 0 | 0 | 0 |
| 0 | 0 | 0 | 0 | 0 |
| 0.581199127 | 2.027319949 | 1.010748379 | 1.828736096 | 1.312497108 |
| 0.879047978 | 0.074419509 | 0.063799805 | 0.045674529 | 0.361066185 |
| 0.056176643 | 0 | 0.002081977 | 0.057744982 | 0 |
| 0 | 0 | 0 | 0.000880035 | 0 |
| 0 | 0 | 0 | 0 | 0 |
| 0 | 0 | 0 | 0 | 0 |
| 0 | 0 | 0 | 0 | 0 |
| 0 | 0 | 0 | 0 | 0 |
| 0 | 0 | 0 | 0 | 0 |
| 0 | 0 | 0 | 0 | 0.390661898 |
| 0 | 0 | 0 | 0 | 0 |
| 0 | 0 | 0 | 0 | 0 |
| 0 | 0 | 0.002301901 | 0 | 0 |
| 0 | 0 | 0 | 0 | 0 |
| 0 | 0 | 0 | 0 | 0 |
| 0 | 0 | 0 | 0 | 0 |
| 0 | 0 | 0 | 0 | 0 |
| 0.131956567 | 0.01129515 | 0.076734181 | 0.012216331 | 0 |
| 0 | 0 | 0 | 0 | 0 |
| 0 | 0 | 0 | 0 | 0 |
| 0 | 0 | 0 | 0 | 0 |
| 0 | 0 | 0 | 0 | 0 |
| 0 | 0 | 0 | 0 | 0 |
| 0 | 0 | 0 | 0 | 0 |
| 0 | 0 | 0 | 0 | 0 |
| 0 | 0 | 0 | 0 | 0 |
| 0 | 0 | 0 | 0 | 0 |
| 0 | 0 | 0 | 0 | 0 |
| 0 | 0 | 0 | 0 | 0 |
| 0 | 0 | 0 | 0 | 0 |
| 0 | 0 | 0 | 0 | 0 |
| 0 | 0 | 0 | 0.002774563 | 0 |
| 0.285911403 | 0 | 0.044761847 | 0 | 0 |
| 0 | 0 | 0 | 0 | 0 |
| 0 | 0 | 0.328098898 | 0.571544211 | 0.008491619 |
| 0 | 0 | 0 | 0 | 0 |
| 0 | 0 | 0 | 0 | 0 |
| 0.000709393 | 0 | 0 | 0 | 0 |
| 0.40817391 | 0.000780713 | 0.014344032 | 0 | 0.000586504 |
| 0 | 0 | 0 | 0 | 0 |
| 0 | 0 | 0 | 0 | 0 |

| | | | | |
|---|---|---|---|---|
| 0 | 0 | 0 | 0 | 0 |
| 0.009872065 | 0 | 0.000458558 | 0 | 0.071733997 |
| 0 | 0 | 0 | 0 | 0 |
| 0 | 0 | 0 | 0 | 0 |
| 0 | 0.256668644 | 0.548325784 | 0 | 0.097173023 |
| 0 | 0 | 0 | 0 | 0 |
| 0 | 0 | 0 | 0 | 0 |
| 0.015073485 | 0.002792171 | 0.011633512 | 0.011174476 | 0.000839038 |
| 0 | 0 | 0 | 0 | 0 |
| 0 | 0 | 0 | 0 | 0 |
| 0 | 0 | 0 | 0 | 0 |
| 0 | 0 | 0 | 0 | 0 |
| 0 | 0 | 0 | 0 | 0 |
| 0 | 0 | 0 | 0 | 0 |
| 0 | 0 | 0 | 0 | 0 |
| 0 | 0.046656719 | 0 | 0.000605721 | 0 |
| 0 | 0 | 0 | 0 | 0 |
| 0 | 0 | 0 | 0 | 0 |
| 0 | 0 | 0 | 0 | 0 |
| 0 | 0 | 0 | 0 | 0 |
| 0 | 0 | 0 | 0 | 0 |
| 0 | 0 | 0 | 0 | 0 |
| 0 | 0 | 0 | 0 | 0 |
| 0.002207592 | 0 | 0.003775442 | 0.007404044 | 0 |
| 0 | 0.030672098 | 0 | 0 | 0 |
| 0 | 0 | 0 | 0 | 0 |
| 0 | 0.217617891 | 0 | 0 | 0 |
| 0 | 0 | 0.001346656 | 0 | 0 |
| 0 | 0 | 0 | 0 | 0 |
| 0 | 0 | 0 | 0 | 0 |
| 0 | 0 | 0 | 0 | 0 |
| 0 | 0 | 0 | 0 | 0 |
| 0 | 0 | 0 | 0 | 0 |
| 0 | 0 | 0 | 0 | 0 |
| 0 | 0 | 0 | 0 | 0 |
| 0 | 0 | 0 | 0 | 0 |
| 0 | 0 | 0 | 0 | 0 |
| 0 | 0 | 0 | 0.003968524 | 0 |
| 0 | 0 | 0 | 0 | 0 |
| 0 | 0 | 0 | 0 | 0 |
| 0 | 0 | 0 | 0 | 0 |
| 0 | 0 | 0 | 0 | 0 |
| 0 | 0 | 0 | 0 | 0 |
| 0 | 0 | 0 | 0 | 0 |
| 0 | 0 | 0 | 0 | 0 |
| 0 | 0 | 0 | 0 | 0 |
| 0 | 0 | 0 | 0 | 0 |
| 0 | 0 | 0 | 0 | 0 |

| | | | | |
|---|---|---|---|---|
| 0.160015633 | 0.028404488 | 0.016958202 | 0 | 0 |
| 0 | 0 | 0 | 0 | 0 |
| 0 | 0 | 0 | 0 | 0 |
| 0 | 0 | 0 | 0 | 0 |
| 0 | 0 | 0 | 0 | 0 |
| 2.382037717 | 0 | 0 | 0 | 0 |
| 0 | 0 | 0 | 0 | 0 |
| 0 | 0 | 0 | 0 | 0 |
| 0 | 0 | 0 | 0 | 0 |
| 0 | 0 | 0 | 0 | 0 |
| 0 | 0 | 0 | 0 | 0 |
| 0 | 0 | 0 | 0 | 0 |
| 0 | 0 | 0 | 0 | 0 |
| 0 | 0 | 0 | 0 | 0 |
| 0 | 0 | 0 | 0 | 0 |
| 0 | 0 | 0 | 0 | 0 |
| 0 | 0 | 0 | 0 | 0 |
| 0.014918953 | 0 | 0 | 11.90890045 | 0 |
| 0 | 0.000469891 | 0 | 0.000661127 | 0 |
| 0 | 0 | 0 | 0 | 0 |
| 0 | 0.002406293 | 0.065306684 | 0 | 0 |
| 0 | 0 | 0 | 0 | 0 |
| 0.1532053 | 0.019449424 | 0.017937556 | 0.001900343 | 0.003196203 |
| 0.06329602 | 0.029668545 | 0.205539235 | 0 | 0 |
| 0 | 0 | 0 | 0 | 0 |
| 0 | 0 | 0 | 0 | 0 |
| 0 | 0 | 0 | 0 | 0 |
| 0 | 0 | 0 | 0 | 0 |
| 0 | 0 | 0 | 0 | 0 |
| 0 | 0 | 0 | 0 | 0 |
| 0 | 0 | 0 | 0 | 0 |
| 0 | 0 | 0 | 0 | 0 |
| 0.063819696 | 0 | 0.014657469 | 0.017796632 | 0 |
| 0 | 0 | 0 | 0 | 0 |
| 0.352244323 | 1.288983054 | 2.748603504 | 2.373841682 | 0.561878014 |
| 0.004172881 | 0.003958559 | 0 | 0.001285295 | 0.001715673 |
| 0 | 0 | 0 | 0 | 0 |
| 0 | 0 | 0 | 0 | 0 |
| 0.004387815 | 0 | 0.001834331 | 0.010736892 | 0 |
| 0 | 0 | 0 | 0 | 0 |
| 0 | 0 | 0 | 0 | 0 |
| 2.941202703 | 0 | 0 | 0 | 0 |
| 0.019240016 | 0 | 0.031637011 | 0 | 0.000696124 |
| 0 | 0 | 0 | 0 | 0 |
| 0 | 0 | 0 | 0 | 0 |
| 0.007741821 | 0.130500702 | 0.068562187 | 0 | 0 |
| 0 | 0 | 0 | 0 | 0 |
| 0 | 0 | 0 | 0 | 0 |

| | | | | |
|---|---|---|---|---|
| 0 | 0 | 0 | 0 | 0 |
| 0 | 0 | 0 | 0 | 0 |
| 0 | 0 | 0 | 0 | 0 |
| 0 | 0 | 0 | 0 | 0 |
| 0 | 0 | 0 | 0 | 0 |
| 0 | 0 | 0 | 0 | 0 |
| 0 | 0.192564413 | 0 | 0 | 0 |
| 0 | 0 | 0.01042487 | 0.83762806 | 0 |
| 0 | 0 | 0.072216092 | 0 | 0 |
| 0.000839367 | 0.001010634 | 0 | 0 | 0 |
| 0 | 0 | 0 | 0 | 0 |
| 0 | 0 | 0 | 0 | 0 |
| 0 | 0 | 0 | 0 | 0 |
| 0 | 3.083478173 | 0 | 0 | 0 |
| 0.008987785 | 0 | 0 | 0 | 0 |
| 0 | 0 | 0 | 0.015233908 | 0 |
| 0 | 0 | 0 | 0 | 0 |
| 0 | 0 | 0 | 0 | 0 |
| 0 | 0 | 0 | 0 | 0 |
| 0.029558347 | 0.818072764 | 1.88412557 | 0 | 0 |
| 0.002803243 | 0.010039123 | 0 | 0.003084735 | 0.006241494 |
| 0 | 0 | 0 | 0 | 0 |
| 0 | 0 | 0 | 0 | 0 |
| 0 | 0 | 0 | 0.00144942 | 0 |
| 0 | 0 | 0 | 0 | 0 |
| 0 | 0 | 0 | 0 | 0 |
| 0 | 0 | 0 | 0 | 0 |
| 0 | 0 | 0 | 0 | 0 |
| 0 | 0 | 0 | 0 | 0 |
| 0 | 0.831669484 | 0 | 0 | 0 |
| 0.162100453 | 0.389812761 | 1.900986069 | 0.62590721 | 0.256060993 |
| 0 | 0 | 0 | 0 | 0 |
| 0 | 0 | 0 | 0 | 0 |
| 0 | 0 | 0 | 0 | 0 |
| 0 | 0 | 0 | 0 | 0 |
| 0 | 0 | 0 | 0 | 0 |
| 0 | 0 | 0 | 0 | 0 |
| 0 | 0 | 0 | 0 | 0 |
| 0 | 0 | 0.007379248 | 0 | 0 |
| 0 | 0 | 0 | 0 | 0 |
| 0 | 0 | 0 | 0 | 0 |
| 0 | 0 | 0 | 0 | 0 |
| 0 | 0 | 0 | 0 | 0 |
| 0 | 0 | 0 | 0 | 0 |
| 0 | 0 | 0 | 0 | 0 |
| 0 | 0 | 0 | 0 | 0 |
| 0 | 0 | 0.012958917 | 0.004773931 | 0.001911742 |
| 0 | 0 | 0 | 0 | 0 |

| | | | | |
|---|---|---|---|---|
| 0 | 0 | 0 | 0 | 0 |
| 0 | 0 | 0 | 0 | 0 |
| 0 | 0 | 0 | 0 | 0 |
| 0 | 0 | 0 | 0 | 0 |
| 0.1334224 | 0.36032909 | 0.045656029 | 0.119120529 | 0 |
| 0 | 0 | 0 | 0 | 0 |
| 0 | 0 | 0 | 0 | 0 |
| 0 | 0 | 0 | 0 | 0 |
| 0 | 0 | 0 | 0 | 0 |
| 0 | 0 | 0 | 0 | 0 |
| 0.065153878 | 0.016735576 | 0 | 0 | 0 |
| 0.014468261 | 0.009326071 | 0.009011611 | 0.002970926 | 0 |
| 0 | 0 | 0 | 0 | 0 |
| 0 | 0 | 0 | 0 | 0 |
| 0 | 0 | 0.003957529 | 0 | 0 |
| 0.005114147 | 0.213328325 | 0 | 0 | 0.033614737 |
| 0 | 0.000812021 | 0.004111609 | 0.004062212 | 0 |
| 0 | 0 | 0 | 0 | 0 |
| 0.002489875 | 0 | 0.004626199 | 0.018621704 | 0.049397326 |
| 0.620077538 | 0.045406609 | 0.372089014 | 0 | 0.041560929 |
| 0 | 0 | 0 | 0 | 0 |
| 0 | 0 | 0 | 0 | 0 |
| 0 | 0 | 0 | 0 | 0 |
| 0 | 0 | 0 | 0 | 0 |
| 0 | 0 | 0 | 0 | 0 |
| 0 | 0 | 0 | 0 | 0 |
| 0.047588135 | 0 | 0.003872363 | 0.001830725 | 0.001728073 |
| 0 | 0 | 0 | 0 | 0 |
| 0 | 0 | 0 | 0 | 0 |
| 0 | 0 | 0 | 0 | 0 |
| 0.005709364 | 0 | 0 | 0.001334072 | 0 |
| 0 | 0 | 0 | 0 | 0 |
| 0 | 0 | 0 | 0 | 0 |
| 0 | 0 | 0 | 0 | 0 |
| 0 | 0 | 0 | 0 | 0 |
| 0 | 0 | 0 | 0 | 0 |
| 0 | 0 | 0 | 0 | 0 |
| 0.198625229 | 0.034307484 | 0.003951317 | 0.013850915 | 0.001126014 |
| 0 | 0.025506766 | 0 | 0 | 0 |
| 0 | 0.000704473 | 0 | 0 | 0 |
| 0.09086082 | 0.007648042 | 0 | 0.003950765 | 0 |
| 0 | 0 | 0 | 0 | 0 |
| 0 | 0 | 0 | 0 | 0 |
| 0.135781929 | 0 | 0 | 0 | 0.004452629 |
| 0 | 0 | 0 | 0 | 0 |
| 0 | 0 | 0 | 0 | 0 |
| 0 | 0 | 0 | 0 | 0 |
| 0 | 0 | 0 | 0 | 0 |

| | | | | |
|---|---|---|---|---|
| 0 | 0 | 0 | 0 | 0 |
| 1.331812769 | 10.26865762 | 1.750717815 | 0 | 0 |
| 0 | 0 | 0 | 0 | 0 |
| 0.010532785 | 0.00224306 | 0.001048391 | 0.00161843 | 0.001166594 |
| 0 | 0.001120217 | 0.011529478 | 0 | 0 |
| 0 | 0 | 0.004661173 | 0.000968636 | 0 |
| 0 | 0 | 0 | 0 | 0 |
| 0 | 0 | 0 | 0 | 0 |
| 0 | 0 | 0 | 0 | 0 |
| 0 | 0 | 0 | 0.001683171 | 0 |
| 0.013321537 | 0 | 0 | 0 | 0 |
| 0 | 0 | 0 | 0 | 0 |
| 0.015542015 | 0.247552931 | 0.026116817 | 0.045545398 | 0 |
| 0 | 0 | 0 | 0 | 0 |
| 0 | 0 | 0 | 0 | 0 |
| 0 | 0 | 0 | 0 | 0 |
| 0 | 0 | 0 | 0 | 0 |
| 0 | 0 | 0 | 0 | 0 |
| 0 | 0 | 0 | 0 | 0 |
| 0 | 0 | 0 | 0 | 0 |
| 0 | 0 | 0 | 0 | 0 |
| 0 | 0 | 0 | 0 | 0 |
| 0 | 0.015622088 | 0.002236737 | 0.019638446 | 0 |
| 0.017757133 | 0.097494334 | 0.045530107 | 0.005882641 | 0.067031227 |
| 0 | 0 | 0 | 0 | 0 |
| 0 | 0 | 0 | 0 | 0 |
| 0 | 0 | 0 | 0 | 0 |
| 0 | 0 | 0 | 0 | 0 |
| 0 | 0 | 0 | 0 | 0 |
| 0 | 0 | 0 | 0 | 0 |
| 0.220356715 | 0.042917892 | 0.064686873 | 0.009600896 | 0 |
| 0 | 0 | 0.001084831 | 0 | 0 |
| 0 | 0 | 0 | 0 | 0 |
| 0 | 0 | 0 | 0 | 0 |
| 0 | 0.002709896 | 1.523593052 | 0 | 0 |
| 0 | 0 | 0 | 0 | 0 |
| 0 | 0 | 0 | 0 | 0 |
| 0 | 0.233977995 | 0 | 0 | 0 |
| 0.005110458 | 0 | 0 | 0 | 0.03954843 |
| 0 | 0 | 0 | 0 | 0 |
| 0 | 0 | 0 | 0 | 0 |
| 0 | 0 | 0 | 0 | 0 |
| 0 | 0 | 0 | 0 | 0 |
| 0 | 0 | 0 | 0 | 0 |
| 0 | 0 | 0 | 0 | 0 |
| 0.013156012 | 1.945627934 | 0.829507769 | 0.042003823 | 0.002862016 |
| 0 | 0 | 0 | 0 | 0 |
| 0 | 0 | 0 | 0 | 0 |

| | | | | |
|---|---|---|---|---|
| 0 | 0 | 0 | 0 | 0 |
| 0 | 0 | 0 | 0 | 0 |
| 0 | 0.158638394 | 0 | 0 | 0.146626333 |
| 0 | 0 | 0 | 0 | 0 |
| 0 | 0 | 0 | 0 | 0 |
| 0 | 0 | 0 | 0 | 0 |
| 0 | 0 | 0 | 0 | 0 |
| 0 | 0 | 0 | 0 | 0 |
| 0 | 0.008153443 | 0 | 0 | 0 |
| 0 | 0 | 0 | 0 | 0 |
| 0.001535537 | 0 | 0 | 0 | 0 |
| 0.001146281 | 0 | 0 | 0.001380886 | 0 |
| 0 | 0 | 0 | 0 | 0 |
| 0 | 0 | 0 | 0 | 0 |
| 0 | 0 | 0 | 0 | 0 |
| 0 | 0 | 0 | 0 | 0 |
| 0 | 0 | 0 | 0 | 0 |
| 0 | 0 | 0 | 0 | 0 |
| 0 | 0 | 0 | 0 | 0 |
| 0 | 0 | 0 | 0 | 0 |
| 0 | 0 | 0 | 0 | 0 |
| 0 | 0 | 0 | 0 | 0 |
| 0 | 0 | 0 | 0 | 0 |
| 0 | 0 | 0 | 0 | 0 |
| 0 | 0 | 0 | 0 | 0 |
| 0 | 0 | 0 | 0 | 0 |
| 0 | 0 | 0 | 0 | 0 |
| 0 | 0.024782713 | 0.021197743 | 0 | 0.021811367 |
| 0 | 0 | 0 | 0 | 0 |
| 0 | 0 | 0 | 0 | 0 |
| 0.010496798 | 0.011796014 | 0.000609472 | 0.004610208 | 0 |
| 0 | 0 | 0 | 0 | 0 |
| 0.009344019 | 0.008315656 | 0.003962889 | 0.038541027 | 0.018373722 |
| 0 | 0.001999458 | 0.001157041 | 0 | 0.088998007 |
| 0 | 0 | 0 | 0 | 0 |
| 0 | 0 | 0 | 0 | 0 |
| 0 | 0 | 0 | 0 | 0 |
| 0 | 0 | 0 | 0 | 0 |
| 0 | 0 | 0 | 0 | 0 |
| 0 | 0 | 0 | 0 | 0 |
| 0 | 0 | 0 | 0 | 0 |
| 0.081384474 | 0.00737778 | 0.011410815 | 0.003271394 | 0.001511589 |
| 0.004682613 | 0.014707991 | 0.004539295 | 0.425220039 | 0 |
| 0 | 0 | 0 | 0.000944376 | 0 |
| 0 | 0 | 0 | 0 | 0 |
| 0 | 0 | 0.018243878 | 0 | 0.001596694 |
| 0 | 0 | 0 | 0 | 0 |
| 0 | 0 | 0 | 0 | 0 |

| | | | | |
|---|---|---|---|---|
| 0 | 0 | 0 | 0 | 0 |
| 0.006825263 | 0.006677047 | 0.005349953 | 0 | 0.014855299 |
| 0.013204665 | 0.007399522 | 0.039926144 | 0.001875851 | 0.265020883 |
| 0 | 0 | 0 | 0 | 0 |
| 0 | 0.095648358 | 0.001524653 | 0.007339107 | 0.013675245 |
| 0 | 0.579449076 | 0.001888839 | 0.001020549 | 0.00052545 |
| 0 | 0 | 0 | 0 | 0 |
| 0 | 0 | 0 | 0 | 0 |
| 0 | 0 | 0 | 0 | 0 |
| 0 | 0 | 0 | 0 | 0 |
| 0 | 0 | 0 | 0 | 0 |
| 0 | 0 | 0 | 0 | 0 |
| 0 | 0 | 0 | 0 | 0 |
| 0 | 0 | 0 | 0 | 0 |
| 0 | 0 | 0 | 0 | 0 |
| 0 | 0 | 0 | 0 | 0 |
| 0 | 0 | 0 | 0 | 0 |
| 0 | 0 | 0 | 0 | 0 |
| 0 | 0 | 0 | 0 | 0 |
| 0 | 0 | 0 | 0 | 0 |
| 0 | 0 | 0 | 0 | 0 |
| 0 | 0 | 0 | 0 | 0.001087646 |
| 0 | 0 | 0 | 0 | 0 |
| 0.090271883 | 0 | 0 | 0 | 0 |
| 0 | 0 | 0 | 0 | 0 |
| 0.05101966 | 0.002984445 | 0.003741898 | 0.023483586 | 0.009528664 |
| 0 | 0 | 0 | 0 | 0 |
| 0 | 0.002090845 | 0.020522171 | 0.0125214 | 0 |
| 0 | 0 | 0 | 0 | 0 |
| 0 | 0 | 0 | 0 | 0 |
| 0 | 0 | 0 | 0 | 0 |
| 0 | 0 | 0 | 0 | 0 |
| 0 | 0 | 0 | 0 | 0 |
| 0 | 0 | 0 | 0 | 0 |
| 0 | 0 | 0 | 0 | 0 |
| 0 | 0 | 0 | 0 | 0 |
| 0 | 0 | 0 | 0.019905774 | 0 |
| 0.006785145 | 0 | 0.002865189 | 0.047835524 | 0 |
| 1.81986704 | 0 | 1.937142529 | 4.011360279 | 0 |
| 0 | 0 | 0 | 0 | 0 |
| 0 | 0 | 0 | 0 | 0 |
| 0 | 0 | 0.021077563 | 0 | 0 |
| 0 | 0 | 0.001299947 | 0.045263702 | 0 |
| 0 | 0 | 0 | 0 | 0 |
| 0 | 0 | 0 | 0 | 0 |
| 0 | 0 | 0.000498514 | 0 | 0 |
| 0.013953841 | 0 | 0.004640218 | 0.001671422 | 0 |
| 0 | 0 | 0 | 0 | 0 |

| | | | | |
|---|---|---|---|---|
| 0 | 0 | 0 | 0 | 0 |
| 0.126851734 | 0.051072215 | 0.007667395 | 0.005875145 | 0.001085875 |
| 0 | 0 | 0 | 0 | 0 |
| 0 | 0 | 0 | 0 | 0 |
| 0 | 0 | 0 | 0 | 0 |
| 0 | 0 | 0 | 0 | 0 |
| 0 | 0 | 0 | 0 | 0 |
| 0 | 0 | 0 | 0 | 0 |
| 0 | 0 | 0 | 0 | 0 |
| 0 | 0 | 0 | 0 | 0 |
| 0 | 0 | 0 | 0 | 0 |
| 0 | 0 | 0 | 0 | 0 |
| 0.035955677 | 0 | 0.512538064 | 0.142046406 | 0 |
| 0 | 0 | 0 | 0 | 0 |
| 0.004989432 | 0.002202745 | 0.010081545 | 0 | 0 |
| 0 | 0 | 0 | 0 | 0 |
| 0 | 0 | 0 | 0 | 0 |
| 0 | 0 | 0 | 0 | 0 |
| 0 | 0 | 0 | 0 | 0 |
| 0 | 0 | 0 | 0 | 0 |
| 0 | 0 | 0 | 0 | 0 |
| 0.222215726 | 0.023402642 | 0.099520846 | 0 | 0.475131174 |
| 0 | 0.008613713 | 0 | 0.029766739 | 0 |
| 0 | 0.001823038 | 0 | 0 | 0 |
| 0 | 0 | 0 | 0 | 0 |
| 0 | 0 | 0 | 0 | 0 |
| 0 | 0 | 0 | 0 | 0 |
| 0 | 0 | 0 | 0 | 0 |
| 0 | 0 | 0 | 0 | 0 |
| 0 | 0 | 0 | 0 | 0 |
| 0 | 0 | 0 | 0 | 0 |
| 0 | 0 | 0 | 0 | 0 |
| 0 | 0 | 0 | 0 | 0 |
| 0 | 0 | 0 | 0 | 0 |
| 0 | 0 | 0.052541738 | 0 | 0 |
| 0 | 0 | 0 | 0 | 0 |
| 0 | 0 | 0 | 0 | 0 |
| 0 | 0 | 0 | 0 | 0 |
| 0.002935955 | 0.013943666 | 0.049606569 | 0.000736843 | 0.002655649 |
| 0 | 0 | 0 | 0 | 0 |
| 0.021717896 | 0.023948146 | 0.112802032 | 0.002147201 | 0.301611146 |
| 0 | 0 | 0 | 0 | 0 |
| 0 | 0 | 0 | 0 | 0 |
| 0 | 0 | 0 | 0 | 0 |
| 0 | 0 | 0 | 0 | 0 |
| 0.202691606 | 0.053426722 | 0.272301369 | 1.380864006 | 0.001590309 |
| 0 | 0 | 0 | 0 | 0 |
| 0 | 0 | 0 | 0 | 0 |

| | | | | |
|---|---|---|---|---|
| 0 | 0 | 0 | 0 | 0 |
| 0 | 0 | 0 | 0 | 0 |
| 0 | 0 | 0 | 0 | 0 |
| 0 | 0 | 0 | 0.004530583 | 0 |
| 0 | 0 | 0 | 0 | 0 |
| 0 | 0 | 0 | 0 | 0 |
| 0 | 0 | 0 | 0 | 0 |
| 0.001357379 | 0 | 0 | 0 | 0.013505637 |
| 0.039644541 | 0 | 0 | 0 | 0.001385132 |
| 0.053304211 | 0 | 0.003936476 | 0.072499449 | 0 |
| 0 | 0 | 0.027617007 | 0 | 0 |
| 0 | 0 | 0 | 0 | 0 |
| 0.000960376 | 0.001541777 | 0.002843859 | 0.002892331 | 0 |
| 0 | 0 | 0 | 0 | 0 |
| 0 | 0 | 0 | 0 | 0 |
| 0 | 0 | 0 | 0 | 0 |
| 0 | 0 | 0 | 0 | 0 |
| 0 | 0 | 0 | 0 | 0 |
| 0 | 0 | 0 | 0 | 0 |
| 0.09808854 | 0.032591895 | 0.628193026 | 0.27787199 | 0.034600299 |
| 0 | 0 | 0 | 0 | 0 |
| 0 | 0 | 0 | 0 | 0 |
| 0 | 0 | 0 | 0 | 0 |
| 0 | 0 | 0 | 0 | 0 |
| 0 | 0 | 0 | 0 | 0 |
| 0 | 0 | 0 | 0 | 0 |
| 0 | 0 | 0 | 0 | 0 |
| 0 | 0 | 0 | 0 | 0 |
| 0 | 0 | 0 | 0 | 0 |
| 0.001599145 | 0 | 0.00716276 | 0.117994244 | 0 |
| 0 | 0 | 0 | 0 | 0 |
| 0 | 0 | 0 | 0 | 0 |
| 0 | 0 | 0 | 0 | 0 |
| 0 | 0 | 0 | 0 | 0 |
| 0 | 0 | 0 | 0 | 0 |
| 0 | 0.007771743 | 0.015226673 | 0 | 0 |
| 0 | 0 | 0 | 0 | 0 |
| 0 | 0 | 0 | 0 | 0 |
| 0 | 0 | 0 | 0 | 0 |
| 0 | 0 | 0 | 0 | 0 |
| 0 | 0 | 0 | 0 | 0 |
| 0 | 0 | 0 | 0 | 0 |
| 0.002486908 | 0 | 0.012871912 | 0 | 0.003213534 |
| 0 | 0 | 0 | 0 | 0 |
| 0.006824323 | 0.003705603 | 0.003356371 | 0.001208975 | 0 |
| 0.068708582 | 0.025923188 | 0 | 0 | 0 |
| 0 | 0 | 0 | 0 | 0 |
| 0 | 0 | 0 | 0 | 0 |

| | | | | |
|---|---|---|---|---|
| 0 | 0 | 0 | 0 | 0 |
| 0 | 0 | 0 | 0 | 0 |
| 0 | 0 | 0 | 0 | 0 |
| 0 | 0 | 0 | 0 | 0 |
| 0 | 0 | 0 | 0 | 0 |
| 0.017831179 | 0.01491438 | 0.035392463 | 0.039035167 | 0 |
| 0 | 0 | 0 | 0 | 0 |
| 0 | 0 | 0 | 0 | 0 |
| 0 | 0 | 0 | 0 | 0 |
| 0 | 0 | 0 | 0 | 0 |
| 0 | 0 | 0 | 0 | 0 |
| 0 | 0 | 0 | 0 | 0 |
| 0 | 0 | 0 | 0 | 0 |
| 0 | 0 | 0 | 0 | 0 |
| 0 | 0 | 0 | 0 | 0 |
| 0 | 0 | 0 | 0 | 0 |
| 0 | 0 | 0 | 0.021348277 | 0 |
| 0 | 0 | 0 | 0 | 0 |
| 0 | 0 | 0 | 0 | 0 |
| 0 | 0 | 0 | 0 | 0 |
| 0.001042439 | 0 | 0 | 0 | 0.000785761 |
| 0 | 0 | 0 | 0 | 0 |
| 0 | 0 | 0 | 0 | 0 |
| 0.009121858 | 0 | 0 | 0 | 0 |
| 0 | 0 | 0 | 0 | 0 |
| 0 | 0 | 0 | 0.000705868 | 0 |
| 0 | 0 | 0 | 0 | 0 |
| 0 | 0.313150672 | 0 | 0 | 0.110908941 |
| 0 | 0 | 0 | 0 | 0 |
| 0 | 0 | 0 | 0 | 0 |
| 0 | 0 | 0 | 0 | 0 |
| 0 | 0 | 0 | 0 | 0 |
| 0 | 0 | 0 | 0 | 0 |
| 0 | 0 | 0 | 0 | 0 |
| 0 | 0 | 0 | 0 | 0 |
| 0 | 0 | 0 | 0 | 0 |
| 0 | 0 | 0 | 0 | 0 |
| 0 | 0 | 0.078973228 | 0 | 0 |
| 0.006980001 | 0 | 0 | 0 | 0 |
| 0 | 0 | 0 | 0 | 0 |
| 0 | 0 | 0 | 0 | 0 |
| 0 | 0 | 0 | 0 | 0 |
| 0 | 0 | 0 | 0 | 0 |
| 0 | 0 | 0 | 0 | 0 |
| 0 | 0 | 0 | 0 | 0 |
| 0 | 0 | 0 | 0 | 0 |
| 0 | 0 | 0 | 0 | 0 |
| 0 | 0 | 0 | 0 | 0 |

| | | | | |
|---|---|---|---|---|
| 0 | 0 | 0 | 0 | 0 |
| 0 | 0.028682598 | 0.330089073 | 0 | 0 |
| 0 | 0 | 0 | 0 | 0 |
| 0 | 0 | 0 | 0 | 0 |
| 0 | 0 | 0 | 0 | 0 |
| 0 | 0 | 0 | 0 | 0 |
| 0 | 0 | 0 | 0 | 0 |
| 0 | 0 | 0 | 0 | 0 |
| 0 | 0 | 0 | 0 | 0 |
| 0 | 0 | 0 | 0 | 0 |
| 0 | 0.002821227 | 0.003214141 | 0.000661568 | 0 |
| 0 | 0 | 0 | 0 | 0 |
| 0.728895105 | 0.068962847 | 0 | 0 | 0 |
| 0 | 0 | 0 | 0 | 0 |
| 0 | 0 | 0 | 0 | 0 |
| 0 | 0 | 0 | 0 | 0 |
| 0 | 0 | 0 | 0 | 0 |
| 0 | 0 | 0 | 0 | 0 |
| 0 | 0 | 0 | 0 | 0 |
| 0 | 0 | 0 | 0 | 0 |
| 0 | 0.004474025 | 0 | 0.02232577 | 0 |
| 0 | 0 | 0 | 0 | 0 |
| 0 | 0 | 0 | 0 | 0 |
| 5.716214106 | 1.361983904 | 3.163319441 | 0 | 0.176748096 |
| 0 | 0 | 0 | 0 | 0 |
| 0 | 0 | 0 | 0 | 0 |
| 0 | 0 | 0 | 0 | 0 |
| 0 | 0 | 0 | 0 | 0 |
| 0 | 0 | 0 | 0 | 0 |
| 0 | 0 | 0 | 0 | 0 |
| 0 | 0 | 0 | 0 | 0 |
| 0 | 0 | 0 | 0 | 0 |
| 0 | 0 | 0 | 0 | 0 |
| 0 | 0 | 0 | 0 | 0 |

| | | | | |
|---|---|---|---|---|
| PHL018-0206_V1 | PHL018-0073_V1 | PHL018-0255_V3 | PHL018-0201_V3 | PHL018-0187_V3 |
| REF | CTRL | REF | CTRL | CTRL |
| 0.017034092 | 0.532446699 | 0.080106348 | 0.048723917 | 7.342077357 |
| 0.970875199 | 0.191630366 | 0.386288339 | 0.493576288 | 2.027209303 |
| 0.000159577 | 1.207166633 | 0.310857248 | 1.060542841 | 0.986073588 |
| 0 | 0 | 0 | 0 | 0 |
| 0 | 0 | 0 | 0 | 0 |
| 0 | 0 | 0 | 0 | 0 |
| 0 | 0 | 0 | 0 | 0 |
| 0 | 0 | 0 | 0 | 0 |
| 0 | 0 | 0 | 0 | 0 |
| 0 | 0 | 0 | 0 | 0 |
| 0 | 0 | 0 | 0 | 0 |
| 0 | 0 | 0 | 0 | 0 |
| 0 | 0 | 0 | 0 | 0 |
| 1.399553994 | 0.299131587 | 0.123225061 | 0.21117957 | 1.216470129 |
| 0 | 0 | 0 | 0 | 0 |
| 0 | 0 | 0 | 0 | 0 |
| 0 | 0 | 0 | 0 | 0 |
| 0 | 0 | 0 | 0 | 0 |
| 0 | 0 | 0 | 0 | 0 |
| 0 | 0 | 0 | 0 | 0 |
| 0 | 0 | 0 | 0 | 0 |
| 0 | 0 | 0 | 0 | 0 |
| 0 | 0 | 0 | 0 | 0 |
| 0 | 0 | 0 | 0 | 0 |
| 0 | 0 | 0.215140063 | 0 | 0 |
| 0 | 0 | 0 | 0 | 0 |
| 0 | 0 | 0 | 0 | 0 |
| 0 | 0 | 0 | 0 | 0 |
| 0 | 0 | 0 | 0 | 0 |
| 0 | 0 | 0 | 0 | 0 |
| 0 | 0 | 0 | 0 | 0 |
| 0 | 0 | 0 | 0 | 0 |
| 0 | 0 | 0 | 0 | 0 |
| 0 | 0 | 0 | 0 | 0 |
| 0 | 0 | 0 | 0 | 0 |
| 2.642305957 | 0.011576046 | 1.42855596 | 0.34507767 | 0 |
| 0 | 0 | 0 | 0 | 0 |
| 0 | 0 | 0 | 0 | 0 |
| 0 | 0 | 0 | 0 | 0 |
| 0 | 0 | 0 | 0 | 0 |
| 0 | 0 | 0 | 0 | 0 |
| 0.002910391 | 0.021664728 | 0.001299385 | 0.008052712 | 0.037712894 |
| 0.762914309 | 10.20211366 | 0.709754811 | 0.374488507 | 0.041896613 |
| 0.029962282 | 0.003055894 | 0.001893928 | 0.004201414 | 0.003416562 |
| 0.000673946 | 0.019277928 | 0.002048182 | 0 | 0.006650696 |
| 0 | 0 | 0 | 0 | 0 |

| | | | | |
|---|---|---|---|---|
| 0 | 0 | 0.002765093 | 0 | 0.158502039 |
| 0.000537648 | 0 | 0 | 0 | 0 |
| 0 | 0 | 0.012582989 | 0.005742228 | 0.433463115 |
| 0.754999856 | 0.226102353 | 0.648374666 | 0.366416087 | 0.151023574 |
| 0.00369411 | 0 | 0.005586635 | 0.038628063 | 0.014414655 |
| 0 | 0 | 0.022528915 | 0 | 0 |
| 0.000169492 | 0.003421466 | 0.00163115 | 0 | 0.007768261 |
| 0 | 0 | 0 | 0 | 0 |
| 0 | 0 | 0 | 0 | 0 |
| 0 | 0 | 0 | 0 | 0 |
| 0 | 0 | 0 | 0 | 0.040013418 |
| 1.205977655 | 0.231060149 | 0.078968494 | 0.120240578 | 0.590588882 |
| 0 | 0 | 0 | 0 | 0 |
| 0.026000571 | 0 | 0.024414931 | 0.032973004 | 0.056308194 |
| 0 | 0 | 0 | 0 | 0 |
| 0 | 0 | 0 | 0 | 0 |
| 0.541449783 | 1.327903946 | 0.000800725 | 0 | 0.000924462 |
| 0 | 0 | 0 | 0 | 0 |
| 0 | 0 | 0 | 0 | 0 |
| 0 | 0 | 0.004996063 | 0 | 0 |
| 0.042609995 | 0 | 0 | 0.017525474 | 0.005700639 |
| 0.010067894 | 0 | 0.000927189 | 0.006660241 | 0.016224294 |
| 0.029932747 | 0.001887731 | 0.030028566 | 0.001977416 | 0.035644467 |
| 0 | 0 | 0 | 0 | 0 |
| 0 | 0 | 0 | 0 | 0 |
| 0 | 0 | 0 | 0 | 0 |
| 0 | 0 | 0 | 0 | 0 |
| 0 | 0 | 0 | 0 | 0 |
| 0 | 0 | 0 | 0 | 0.024179163 |
| 0 | 0 | 0 | 0 | 0 |
| 0 | 0 | 0 | 0 | 0 |
| 0 | 0 | 0 | 0 | 0 |
| 0 | 0 | 0 | 0 | 0 |
| 0.000534471 | 0 | 0 | 0 | 0 |
| 0 | 0 | 0 | 0 | 0 |
| 0.000503985 | 0 | 0 | 0 | 0 |
| 0 | 0 | 0 | 0 | 0 |
| 27.83341407 | 0.028345395 | 0 | 0 | 0 |
| 0 | 0 | 0 | 0 | 0 |
| 0 | 0 | 0 | 0 | 0 |
| 0 | 0 | 0 | 0 | 0 |
| 0 | 0 | 0 | 0 | 0 |
| 0 | 0 | 0.000867622 | 0 | 0 |
| 0 | 0 | 0 | 0 | 0 |
| 0 | 0 | 0.002122482 | 0.006457277 | 0 |
| 0 | 0 | 0 | 0 | 0 |
| 0 | 0 | 0 | 0 | 0 |
| 0 | 0 | 0 | 0 | 0 |

| | | | | |
|---|---|---|---|---|
| 0 | 0 | 0 | 0 | 0 |
| 0 | 0 | 0 | 0 | 0 |
| 0 | 0 | 0 | 0 | 0 |
| 0 | 0 | 0 | 0 | 0 |
| 0.050527695 | 0.002128604 | 0 | 0.025641925 | 0 |
| 0 | 0 | 0 | 0 | 0 |
| 0 | 0 | 0 | 0 | 0 |
| 0 | 0 | 0 | 0 | 0 |
| 0 | 0 | 0 | 0 | 0 |
| 0 | 0 | 0 | 0 | 0 |
| 0 | 0 | 0 | 0 | 0 |
| 0 | 0.010282793 | 0 | 0 | 0.002440053 |
| 0 | 0 | 0 | 0 | 0 |
| 0 | 0 | 0 | 0 | 0 |
| 0 | 0 | 0 | 0 | 0 |
| 0 | 0 | 0 | 0 | 0 |
| 0 | 0 | 0 | 0 | 0 |
| 0 | 0 | 0 | 0 | 0 |
| 0.10997594 | 0.538800509 | 0.394000527 | 0.00097816 | 0.742163321 |
| 0 | 0 | 0 | 0 | 0 |
| 0 | 0 | 0.008630665 | 0 | 0 |
| 0 | 0 | 0 | 0 | 0 |
| 0 | 0 | 0.00073518 | 0 | 0.001945141 |
| 0 | 0 | 0 | 0 | 0 |
| 0 | 0 | 0 | 0 | 0 |
| 0 | 0 | 0 | 0 | 0 |
| 0 | 0.002119038 | 0 | 0 | 0 |
| 0 | 0 | 0 | 0 | 0 |
| 0 | 0 | 0 | 0 | 0 |
| 0 | 0 | 0 | 0 | 0 |
| 0.009285227 | 0 | 0.065930772 | 0.058871237 | 0.224258338 |
| 0.000813634 | 0 | 0.005981434 | 0.002573377 | 0 |
| 0 | 0 | 0 | 0 | 0 |
| 0 | 0 | 0 | 0 | 0 |
| 0 | 0 | 0 | 0 | 0 |
| 0 | 0 | 0 | 0 | 0 |
| 0 | 0 | 0 | 0 | 0 |
| 0 | 0 | 0 | 0 | 0 |
| 0 | 0 | 0 | 0 | 0 |
| 0 | 0 | 0 | 0 | 0 |
| 0 | 0 | 0 | 0 | 0 |
| 0 | 0 | 0 | 0 | 0 |
| 0 | 0 | 0 | 0 | 0 |
| 0 | 0 | 0 | 0 | 0 |
| 0 | 0 | 0 | 0 | 0.003094502 |
| 0 | 0 | 0 | 0 | 0 |
| 0 | 0 | 0 | 0 | 0 |
| 0 | 0 | 0 | 0 | 0 |

| | | | | |
|---|---|---|---|---|
| 0 | 0 | 0 | 0 | 0 |
| 0 | 0 | 0 | 0 | 0 |
| 0 | 0 | 0 | 0 | 0 |
| 0 | 0 | 0 | 0 | 0 |
| 0.03261547 | 0 | 0.05840366 | 0.011679133 | 0.048495247 |
| 0 | 0 | 0 | 0 | 0 |
| 0 | 0 | 0 | 0 | 0 |
| 0 | 0 | 0 | 0 | 0 |
| 0 | 0 | 0 | 0 | 0 |
| 0 | 0 | 0 | 0 | 0 |
| 0 | 0 | 0 | 0 | 0 |
| 0 | 0 | 0 | 0 | 0 |
| 0 | 0 | 0 | 0 | 0 |
| 0 | 0 | 0 | 0 | 0.00535632 |
| 0 | 0 | 0 | 0 | 0 |
| 0 | 0 | 0 | 0 | 0 |
| 0 | 0 | 0 | 0 | 0 |
| 0 | 0 | 0 | 0 | 0 |
| 0 | 0 | 0 | 0 | 0 |
| 0 | 0 | 0 | 0 | 0 |
| 0 | 0 | 0 | 0 | 0 |
| 0 | 0 | 0 | 0 | 0 |
| 0 | 0 | 0 | 0 | 0.01459108 |
| 0 | 0 | 0 | 0 | 0 |
| 0.006731311 | 0 | 0.030732562 | 0 | 0 |
| 0 | 0 | 0 | 0 | 0 |
| 0 | 0 | 0.03201465 | 0 | 0 |
| 0 | 0 | 0 | 0 | 0 |
| 0 | 0 | 0 | 0 | 0 |
| 0 | 0.01369787 | 0 | 0 | 0 |
| 0 | 0 | 0 | 0 | 0 |
| 0 | 0 | 0.000878345 | 0.000848321 | 0.001267597 |
| 0 | 0 | 0 | 0 | 0 |
| 0 | 0 | 0 | 0 | 0 |
| 0 | 0 | 0 | 0 | 0 |
| 0 | 0 | 0 | 0 | 0 |
| 0 | 0 | 0 | 0 | 0 |
| 0 | 0 | 0 | 0 | 0 |
| 0 | 0.001311396 | 0 | 0 | 0 |
| 0 | 0 | 0 | 0 | 0 |
| 0 | 0 | 0 | 0 | 0 |
| 0 | 0 | 0 | 0 | 0 |
| 0 | 0 | 0 | 0 | 0 |
| 0 | 0 | 0 | 0 | 0 |
| 0 | 0 | 0 | 0 | 0 |
| 0 | 0 | 0 | 0 | 0 |
| 0 | 0 | 0 | 0 | 0 |
| 0.005051335 | 0 | 0 | 0 | 0 |

| | | | | |
|---|---|---|---|---|
| 0 | 0 | 0.791195712 | 0 | 0 |
| 0 | 0 | 0 | 0 | 0 |
| 0 | 0 | 0 | 0 | 0 |
| 0 | 0 | 0 | 0 | 0 |
| 0 | 0 | 0 | 0 | 0 |
| 0 | 0 | 0 | 0 | 0 |
| 0 | 0 | 0 | 0 | 0 |
| 0 | 0 | 0 | 0 | 0 |
| 0 | 0 | 0 | 0 | 0 |
| 0 | 0 | 0 | 0 | 0 |
| 0 | 0 | 0 | 0 | 0 |
| 0.000278438 | 0 | 0.010596692 | 0 | 0 |
| 0.000848394 | 0 | 0.00072042 | 0 | 0.001906089 |
| 0 | 0 | 0 | 0 | 0 |
| 0 | 0 | 0.007556962 | 0 | 0 |
| 0 | 0 | 0 | 0 | 0 |
| 0 | 0 | 0 | 0 | 0 |
| 0 | 0 | 0 | 0 | 0 |
| 0 | 0 | 0 | 0 | 0 |
| 0 | 0 | 0 | 0 | 0 |
| 0.016609426 | 0.005795826 | 0.009653591 | 0.012521813 | 0.008423832 |
| 0 | 0 | 0 | 0 | 0 |
| 0 | 0 | 0 | 0 | 0.001847847 |
| 0 | 0 | 0 | 0 | 0.00101358 |
| 0 | 0 | 0 | 0 | 0 |
| 0 | 0 | 0 | 0 | 0 |
| 0.000815135 | 0 | 0.001830426 | 0.004198659 | 0.001698176 |
| 0 | 0 | 0 | 0 | 0 |
| 0 | 0 | 0 | 0.004078983 | 0 |
| 0 | 0 | 0 | 0 | 0 |
| 0 | 0 | 0 | 0 | 0 |
| 0 | 0 | 0 | 0 | 0 |
| 0 | 0 | 0 | 0 | 0 |
| 0.176039637 | 0 | 0.416681695 | 0.001391373 | 0.309877394 |
| 0.006587399 | 0.022155055 | 0.002629315 | 0.012097593 | 0.035837268 |
| 0 | 0 | 0 | 0 | 0 |
| 0 | 0 | 0 | 0 | 0 |
| 0 | 0 | 0 | 0 | 0 |
| 0 | 0 | 0 | 0 | 0.005721203 |
| 0 | 0 | 0 | 0 | 0.001347493 |
| 0 | 0 | 0 | 0 | 0 |
| 0 | 0 | 0 | 0 | 0 |
| 0 | 0 | 0 | 0 | 0 |
| 0 | 0 | 0 | 0 | 0 |
| 0 | 0 | 0 | 0 | 0 |
| 0 | 0 | 0.20374845 | 0.019745531 | 0 |
| 0 | 0 | 0 | 0 | 0 |
| 0.001622474 | 0 | 0.001561434 | 0.001055641 | 0 |

| | | | | |
|---|---|---|---|---|
| 0 | 0 | 0 | 0 | 0 |
| 0 | 0 | 0 | 0 | 0 |
| 0 | 0 | 0 | 0 | 0 |
| 0 | 0 | 0 | 0 | 0 |
| 0 | 0 | 0 | 0 | 0 |
| 0 | 0 | 0 | 0 | 0 |
| 0 | 0 | 0 | 0 | 0.151617077 |
| 0 | 0 | 0 | 0 | 0.004535038 |
| 0 | 0 | 0 | 0 | 0.213439278 |
| 0 | 0 | 0 | 0 | 0 |
| 0 | 0 | 0 | 0 | 0 |
| 0 | 0 | 0 | 0 | 0 |
| 0 | 0 | 0 | 0 | 0 |
| 0 | 0 | 0 | 0 | 0 |
| 0 | 0 | 0 | 0 | 0.026054416 |
| 0 | 0 | 0 | 0 | 0 |
| 0 | 0 | 0 | 0 | 0 |
| 0 | 0.05508852 | 0 | 0 | 0 |
| 0.003410094 | 0.049536282 | 0.002011426 | 0.002147161 | 0 |
| 0 | 0 | 0.513894105 | 0.411731632 | 0.626607091 |
| 0.002855876 | 0 | 0 | 0 | 0 |
| 0 | 0 | 0 | 0 | 0 |
| 0 | 0 | 0 | 0 | 0 |
| 0 | 0 | 0 | 0 | 0 |
| 0.002901376 | 0.00851003 | 0.006515182 | 0.003146236 | 0.002238686 |
| 0 | 0 | 0 | 0 | 0 |
| 0 | 0 | 0 | 0 | 0 |
| 0 | 0 | 0 | 0 | 0 |
| 0 | 0 | 0 | 0 | 0 |
| 0.000626543 | 0 | 0 | 0 | 0 |
| 0.03899178 | 0.005187639 | 0.086003994 | 0.121102813 | 0.443609762 |
| 0 | 0 | 0 | 0 | 0 |
| 0 | 0 | 0 | 0 | 0 |
| 0.006359859 | 0 | 0 | 0 | 0 |
| 0 | 0 | 0 | 0 | 0 |
| 0 | 0 | 0 | 0 | 0 |
| 0 | 0 | 0 | 0 | 0 |
| 0 | 0 | 0 | 0 | 0 |
| 0 | 0 | 0 | 0 | 0 |
| 0 | 0.002220742 | 0.000573472 | 0 | 0 |
| 0 | 0 | 0 | 0 | 0 |
| 0.009145875 | 0.096127114 | 0.003160217 | 0.002967408 | 0.013175397 |
| 0 | 0 | 0 | 0 | 0 |
| 0 | 0 | 0 | 0 | 0 |
| 0 | 0 | 0 | 0 | 0 |
| 0 | 0 | 0 | 0 | 0 |
| 0 | 0 | 0 | 0 | 0 |
| 0 | 0 | 0 | 0 | 0 |

| | | | | |
|---|---|---|---|---|
| 0 | 0 | 0 | 0 | 0 |
| 0 | 0 | 0 | 0 | 0 |
| 0.027549744 | 0.001185692 | 0.001175755 | 0 | 0 |
| 0 | 0 | 0 | 0 | 0 |
| 0 | 0 | 0 | 0 | 0.001656604 |
| 0 | 0 | 0 | 0 | 0 |
| 0 | 0 | 0 | 0 | 0 |
| 0 | 0 | 0 | 0 | 0 |
| 0 | 0 | 0 | 0 | 0 |
| 0 | 0 | 0 | 0 | 0.001357902 |
| 0 | 0 | 0 | 0 | 0 |
| 0 | 0 | 0 | 0 | 0 |
| 0 | 0 | 0 | 0 | 0 |
| 0.000573843 | 0.003669129 | 0.001624277 | 0.000823596 | 0 |
| 0.024573163 | 0.00194682 | 0.004155947 | 0.001586131 | 0.000773899 |
| 0 | 0 | 0.136355325 | 0 | 0 |
| 0.001555495 | 0.005175861 | 0.000763763 | 0.002581792 | 0 |
| 0 | 0 | 0 | 0 | 0 |
| 0 | 0 | 0 | 0 | 0 |
| 0 | 0 | 0 | 0 | 0.002694839 |
| 0 | 0 | 0 | 0 | 0 |
| 0.001490138 | 0 | 0.001103136 | 0 | 0 |
| 0 | 0 | 0 | 0 | 0 |
| 0 | 0 | 0 | 0 | 0 |
| 0 | 0 | 0 | 0 | 0 |
| 0 | 0 | 0 | 0 | 0 |
| 0 | 0 | 5.650183801 | 0 | 0 |
| 0 | 0 | 0 | 0 | 0 |
| 0 | 0 | 0 | 0 | 0 |
| 0 | 0 | 0 | 0 | 0 |
| 0 | 0.001704989 | 0 | 0 | 0 |
| 0 | 0 | 0 | 0 | 0 |
| 0 | 0 | 0 | 0 | 0 |
| 0 | 0 | 0 | 0 | 0 |
| 0 | 0 | 0 | 0 | 0 |
| 0 | 0 | 0 | 0 | 0.01987379 |
| 0 | 0 | 0 | 0 | 0 |
| 0 | 0.644055066 | 0 | 0.023718294 | 0.1757668 |
| 0 | 0 | 0 | 0 | 0 |
| 0 | 0 | 0 | 0 | 0 |
| 0.005352275 | 0 | 0 | 0 | 0 |
| 0 | 0 | 0 | 0 | 0 |
| 0 | 0 | 0 | 0 | 0 |
| 0 | 0 | 0 | 0 | 0 |
| 0 | 0 | 0 | 0 | 0 |
| 0 | 0 | 0 | 0 | 0 |
| 0 | 0 | 0 | 0 | 0 |
| 0 | 0 | 0 | 0 | 0 |

| | | | | |
|---|---|---|---|---|
| 1.639307973 | 0 | 0.133360177 | 0 | 0.00099824 |
| 0 | 0 | 0 | 0 | 0 |
| 0.000300154 | 0 | 0 | 0 | 0 |
| 0 | 0 | 0 | 0 | 0 |
| 0 | 0 | 0.001162493 | 0.004420848 | 0.005871839 |
| 0 | 0 | 0 | 0 | 0 |
| 0 | 0 | 0 | 0 | 0 |
| 0.008889248 | 0 | 0.001425803 | 0.001721331 | 0.001763716 |
| 0 | 0 | 0 | 0 | 0 |
| 0.002748702 | 0.017276192 | 0.004828706 | 0.006032324 | 0.012422319 |
| 0 | 0 | 0 | 0 | 0 |
| 0 | 0.02731703 | 0 | 0.008720715 | 0.056079254 |
| 0.017232928 | 0.024273098 | 0.001834579 | 0.003720922 | 0.007589786 |
| 0 | 0 | 0 | 0 | 0 |
| 0.000209129 | 0 | 0 | 0 | 0 |
| 0 | 0 | 0 | 0 | 0 |
| 0 | 0.001723435 | 0 | 0 | 0 |
| 0 | 0 | 0 | 0 | 0 |
| 0 | 0 | 0 | 0 | 0 |
| 0 | 0 | 0 | 0 | 0 |
| 0 | 0 | 0 | 0 | 0 |
| 0 | 0.009724989 | 0 | 0 | 0 |
| 0 | 0 | 0 | 0 | 0 |
| 0 | 0 | 0.17135138 | 0 | 0 |
| 0 | 0 | 0 | 0 | 0 |
| 0.001480234 | 0.006895603 | 0.000427363 | 0 | 0 |
| 3.07796366 | 0.003073606 | 0 | 0 | 0 |
| 0 | 0 | 0 | 0 | 0 |
| 0 | 0 | 0 | 0 | 0 |
| 0 | 0 | 0 | 0 | 0 |
| 0 | 0 | 0 | 0 | 0 |
| 0 | 0 | 0 | 0 | 0 |
| 0 | 0 | 0 | 0 | 0 |
| 0 | 0 | 0 | 0 | 0 |
| 0 | 0 | 0 | 0 | 0 |
| 0 | 0 | 0 | 0 | 0 |
| 0.007450304 | 0 | 0 | 0 | 0 |
| 0 | 0 | 0 | 0 | 0 |
| 0 | 0 | 0 | 0 | 0 |
| 0.00090296 | 0 | 0 | 0 | 0 |
| 0 | 0 | 0 | 0 | 0 |
| 0 | 0 | 0 | 0 | 0 |
| 0 | 0 | 0 | 0 | 0 |
| 0 | 0 | 0 | 0 | 0 |
| 0.000443302 | 0 | 0 | 0 | 0 |
| 0 | 0 | 0.083380264 | 0 | 0 |
| 0.00051542 | 0 | 0 | 0.001497105 | 0.001278308 |
| 0 | 0 | 0 | 0 | 0 |

| | | | | |
|---|---|---|---|---|
| 0 | 0 | 0 | 0 | 0 |
| 0.000576018 | 0 | 0 | 0 | 0 |
| 0 | 0 | 0 | 0 | 0 |
| 0 | 0.125822222 | 0 | 0.000966748 | 0 |
| 0 | 0 | 0 | 0 | 0 |
| 0 | 0 | 0.003209184 | 0.001179152 | 0 |
| 0 | 0 | 0 | 0 | 0 |
| 0 | 0 | 0 | 0 | 0 |
| 0 | 0 | 0 | 0 | 0 |
| 0 | 0 | 0 | 0 | 0 |
| 0 | 0 | 0.00161831 | 0 | 0.006772915 |
| 0 | 0 | 0 | 0 | 0 |
| 0 | 0 | 0 | 0 | 0 |
| 0 | 0.004440439 | 0.001376008 | 0.003100935 | 0 |
| 0 | 0.001497642 | 0 | 0 | 0 |
| 0 | 0 | 0 | 0 | 0 |
| 0 | 0 | 0 | 0 | 0 |
| 0 | 0 | 0 | 0 | 0 |
| 0.023232845 | 0 | 0 | 0 | 0 |
| 0 | 0 | 0.002842018 | 0 | 0.004763033 |
| 0 | 0 | 0 | 0 | 0 |
| 0.04533507 | 0.220897518 | 0.090404993 | 0.014900256 | 0.264930016 |
| 0 | 0 | 0.001160681 | 0 | 0 |
| 0 | 0 | 0 | 0 | 0 |
| 0 | 0 | 0 | 0 | 0 |
| 0 | 0 | 0 | 0 | 0 |
| 0 | 0 | 0 | 0 | 0 |
| 0 | 0 | 0 | 0 | 0 |
| 0 | 0 | 0 | 0 | 0 |
| 0.708189722 | 0.387632461 | 0.377391845 | 0.717024307 | 0.089587231 |
| 0.000711337 | 0 | 0.002281919 | 0 | 0.006586369 |
| 0 | 0 | 0 | 0 | 0.003993047 |
| 0 | 0 | 0 | 0 | 0 |
| 0 | 0 | 0 | 0 | 0 |
| 0 | 0 | 0 | 0 | 0.004981563 |
| 0 | 0 | 0 | 0 | 0 |
| 0.628450795 | 0.661364888 | 0.027850168 | 0.757025051 | 0 |
| 0 | 0 | 0 | 0 | 0.002923651 |
| 0 | 0.009477022 | 0 | 0 | 0 |
| 0 | 0 | 0 | 0 | 0 |
| 0.005145863 | 0 | 0 | 0.008285243 | 0.038092811 |
| 0 | 0 | 2.141145986 | 2.691123821 | 1.426214072 |
| 0 | 0 | 0 | 0 | 0 |
| 0 | 0 | 0 | 0 | 0 |
| 0 | 0 | 0 | 0 | 0 |
| 0 | 0 | 0 | 0 | 0 |
| 0.000301576 | 0 | 0.00092346 | 0 | 0 |
| 0 | 0 | 0 | 0 | 0 |

| | | | | |
|---|---|---|---|---|
| 0.003179714 | 0.029940278 | 0.000813853 | 0 | 0.003288668 |
| 0 | 0 | 0 | 0 | 0 |
| 0.001613204 | 2.823865112 | 0.001330725 | 0.002624024 | 0.001280303 |
| 0 | 0 | 0 | 0 | 0 |
| 0 | 0 | 0 | 0 | 0 |
| 0 | 0 | 0 | 0 | 0 |
| 0 | 0 | 0 | 0 | 0 |
| 0 | 0 | 0 | 0 | 0.000789842 |
| 0.018373161 | 0 | 0.010897954 | 0 | 0 |
| 0 | 0 | 0 | 0 | 0 |
| 0 | 0 | 0 | 0 | 0 |
| 0 | 0 | 0 | 0 | 0 |
| 0 | 0 | 0 | 0 | 0 |
| 0 | 0 | 0.00854076 | 0 | 0 |
| 0 | 0 | 0 | 0 | 0 |
| 0.02623486 | 0 | 0.184583242 | 0.050764633 | 0.788523316 |
| 0.060330819 | 0.082116528 | 0.003571419 | 0.004958421 | 0.02190512 |
| 0 | 0 | 0 | 0 | 0 |
| 0 | 0 | 0 | 0 | 0 |
| 0 | 0 | 0 | 0 | 0 |
| 0 | 0 | 0 | 0 | 0.003247845 |
| 0.001168393 | 0 | 0 | 0.002250591 | 0.007686696 |
| 0.060166155 | 0 | 0 | 0 | 0 |
| 0 | 0 | 0 | 0 | 0 |
| 0 | 0 | 0.012397677 | 0 | 0 |
| 0.006215549 | 0 | 0 | 0 | 0 |
| 0 | 0 | 0 | 0 | 0.007711664 |
| 0 | 0 | 0.077163005 | 0 | 0 |
| 0 | 0 | 0 | 0 | 0 |
| 0 | 0 | 0 | 0 | 0 |
| 0.011900165 | 0 | 0.008989565 | 0.004370871 | 0 |
| 0.181537506 | 0 | 0.000550781 | 0 | 0.004239295 |
| 0.092200264 | 0.012974454 | 0.002796896 | 0.052295274 | 0.008325038 |
| 0 | 0 | 0 | 0.002117069 | 0.00361533 |
| 0.00787763 | 0.00375054 | 0.001251623 | 0 | 0 |
| 0 | 0 | 0 | 0 | 0 |
| 0.000919602 | 0.005354909 | 0 | 0 | 0 |
| 0.008989647 | 0.003190695 | 0.002348248 | 0 | 0 |
| 0 | 0 | 0.112011593 | 0 | 0.364642675 |
| 0 | 0 | 0 | 0 | 0 |
| 0 | 0 | 0 | 0 | 0 |
| 0 | 0 | 0 | 0 | 0 |
| 0.000652994 | 0 | 0 | 0 | 0.005068108 |
| 0 | 0.001712377 | 0 | 0 | 0.004037809 |
| 0 | 0 | 0 | 0 | 0 |
| 0.009155485 | 0 | 0 | 0 | 0 |
| 0 | 0 | 0 | 0 | 0 |
| 0 | 0 | 0 | 0 | 0 |

| | | | | |
|---|---|---|---|---|
| 0.003313125 | 0 | 0 | 0 | 0 |
| 0.156409053 | 0.001358665 | 0.059280245 | 0.385975603 | 0.424353204 |
| 0 | 0 | 0 | 0 | 0 |
| 0 | 0 | 0 | 0 | 0 |
| 0 | 0 | 0 | 0 | 0 |
| 0 | 0 | 0 | 0 | 0 |
| 0 | 0 | 0 | 0 | 0 |
| 0 | 0 | 0 | 0 | 0 |
| 0 | 0 | 0 | 0 | 0 |
| 0 | 0 | 0 | 0 | 0 |
| 0.002340705 | 0 | 0.000876028 | 0 | 0.001264253 |
| 0.043969233 | 0.089029813 | 0.002050508 | 0.005041057 | 0.012912965 |
| 0 | 0 | 0.116005849 | 0 | 0.004954134 |
| 0 | 0 | 0 | 0 | 0 |
| 0.000298407 | 0.001621804 | 0 | 0 | 0 |
| 0 | 0 | 0 | 0 | 0 |
| 0.009205079 | 0.011629585 | 0.006814436 | 0.014840011 | 0.008699621 |
| 0.012479765 | 0 | 0 | 0.001437641 | 0.002455067 |
| 0 | 0 | 0.000972309 | 0 | 0 |
| 0 | 0 | 0 | 0 | 0.002550708 |
| 0.016092753 | 0.012211091 | 0.01202767 | 0.058017374 | 0.009946668 |
| 0 | 0 | 0 | 0 | 0 |
| 0 | 0 | 0 | 0 | 0 |
| 0 | 0 | 0 | 0 | 0 |
| 0 | 0 | 0.001135322 | 0 | 0.001310765 |
| 0 | 0 | 0 | 0 | 0 |
| 0.07326761 | 0 | 0.005602475 | 0.008792818 | 0 |
| 0.018447484 | 0.045425055 | 0.01666182 | 0.048068713 | 0.008291639 |
| 0 | 0.001807566 | 0 | 0 | 0 |
| 0.001296232 | 0 | 0 | 0.001171356 | 0 |
| 0 | 0 | 0.000924732 | 0 | 0.002891504 |
| 0 | 0 | 0 | 0 | 0 |
| 0 | 0 | 0 | 0 | 0 |
| 0 | 0 | 0 | 0 | 0 |
| 0.211273172 | 0 | 0.36211423 | 0 | 0 |
| 0.045821503 | 0.013728428 | 0.237018609 | 0.1483262 | 0.072972141 |
| 0 | 0 | 0 | 0 | 0 |
| 0 | 0 | 0 | 0 | 0 |
| 0.000994239 | 0.015438734 | 0.00460698 | 0.004492283 | 0.031197425 |
| 0 | 0 | 0 | 0 | 0 |
| 0 | 0 | 0 | 0 | 0 |
| 0 | 0 | 0 | 0 | 0 |
| 0 | 0 | 0.007211133 | 0.094776194 | 0 |
| 0 | 0.000503001 | 0 | 0 | 0.001499645 |
| 0.001106471 | 0 | 0 | 0 | 0 |
| 0 | 0 | 0.405299858 | 0 | 0 |
| 0 | 0 | 0 | 0 | 0 |
| 0 | 0 | 0 | 0 | 0 |

| | | | | |
|---|---|---|---|---|
| 0 | 0 | 0 | 0 | 0 |
| 0 | 0 | 0 | 0 | 0 |
| 0.000204982 | 0 | 0 | 0 | 0 |
| 0.421110981 | 0 | 0.437859961 | 0.398739361 | 2.245222297 |
| 0 | 0 | 0 | 0 | 0 |
| 0 | 0 | 0.007174241 | 0 | 0 |
| 0 | 0 | 0.000971838 | 0 | 0 |
| 0.020322155 | 0 | 0.078387396 | 0.001137243 | 0.036413926 |
| 0.385156403 | 34.43764737 | 1.713765704 | 1.766085087 | 0 |
| 0.036880268 | 0 | 0.347869445 | 0.00624887 | 0.021180778 |
| 0 | 0 | 0 | 0 | 0 |
| 0.001397729 | 0 | 0.004083474 | 0 | 0.004159854 |
| 0 | 0 | 0 | 0 | 0 |
| 0 | 0 | 0 | 0 | 0 |
| 0.015241488 | 0 | 0.114767786 | 0.1090499 | 0.806832653 |
| 0 | 0 | 0 | 0 | 0 |
| 0 | 0.004171875 | 0.065156372 | 0.03583464 | 0.288624101 |
| 0 | 0 | 0.769278067 | 0 | 0 |
| 0 | 0 | 0 | 0 | 0 |
| 0 | 0 | 0.919171134 | 0 | 0.119959095 |
| 0 | 0 | 0 | 0 | 0.067236136 |
| 0 | 0 | 0 | 0 | 0 |
| 0 | 0 | 0.00962166 | 0 | 0 |
| 0.076720546 | 0.000924229 | 0 | 0 | 0.241656686 |
| 0 | 0 | 0 | 0 | 0.110024882 |
| 0.202230065 | 0 | 0.494261058 | 0.001175226 | 0 |
| 0 | 0 | 0.00709107 | 0.001130678 | 0 |
| 2.204502323 | 3.110713247 | 1.428356292 | 3.325824496 | 0 |
| 0 | 0 | 0 | 0 | 0 |
| 0 | 0 | 0 | 0 | 0 |
| 0.100262343 | 0.142426254 | 0.125516781 | 0 | 0 |
| 0 | 0 | 0 | 0 | 1.239928657 |
| 0.000221608 | 0.006882339 | 0 | 0 | 0.029880766 |
| 0 | 0 | 0 | 0 | 0 |
| 0 | 0 | 0.150385002 | 0 | 0 |
| 0.044583589 | 0 | 0.249478111 | 0 | 0 |
| 1.525058173 | 0 | 1.052399763 | 0 | 3.500718413 |
| 0 | 0 | 0 | 0 | 0.002164107 |
| 0 | 0 | 0 | 0.011622436 | 0.00739778 |
| 0.000357306 | 0 | 0 | 0.016273361 | 0.005210647 |
| 0.00665647 | 0.105965558 | 0.014402071 | 0.105157813 | 0.056700286 |
| 0.022397143 | 0 | 0.318514602 | 0.115517093 | 0 |
| 0.09188785 | 0.125334573 | 0.066620315 | 1.092596618 | 0.107513829 |
| 0 | 0 | 0.00080295 | 0 | 0.005938791 |
| 0 | 0 | 0 | 0 | 0 |
| 0 | 0 | 0.123162807 | 0 | 0 |
| 0 | 0 | 0 | 0 | 0 |
| 0.000115134 | 0.004246092 | 0 | 0.00280915 | 0.054168408 |

| | | | | |
|---|---|---|---|---|
| 0 | 0 | 0 | 0 | 0 |
| 0.158201384 | 0.568437977 | 0.205169728 | 0.040307641 | 0 |
| 0.000386293 | 0 | 0.007435191 | 0 | 0.015137256 |
| 0 | 0 | 0.005597256 | 0 | 0.691166575 |
| 0 | 0 | 0 | 0 | 0 |
| 0.001152017 | 0 | 0.041836837 | 0.00618728 | 0.168905916 |
| 0 | 0 | 0 | 0 | 0 |
| 0 | 0 | 0 | 0 | 0 |
| 0.042301757 | 0 | 0 | 0 | 0 |
| 0 | 0 | 0 | 0 | 0 |
| 0.000406362 | 0.164061513 | 0.014730437 | 0.011457065 | 0.161225572 |
| 1.617593967 | 0.960483297 | 0.407377044 | 0.824785118 | 0.291423453 |
| 0 | 0 | 0 | 0 | 0 |
| 0.007514029 | 0.548891806 | 0.016038375 | 0.003804594 | 0.049946672 |
| 0.005806142 | 0.001609979 | 0.012084519 | 0.003747708 | 0.220639398 |
| 0 | 0 | 1.887799861 | 0 | 0 |
| 0 | 0 | 0 | 0 | 0 |
| 0 | 0 | 0 | 0 | 0.000710731 |
| 0 | 0 | 0.182897577 | 0 | 0 |
| 0.009037845 | 0 | 0.01342257 | 0.017060288 | 0.031768407 |
| 0.029041047 | 0.155665777 | 0.142692349 | 0.004616686 | 0.046199902 |
| 0 | 0.165509491 | 0 | 0.111495601 | 0 |
| 0.173029724 | 0.075929473 | 0.082351878 | 0.170236732 | 0.201951103 |
| 0 | 0 | 0 | 0 | 0 |
| 0.005585793 | 0 | 0.012528896 | 0 | 0.020862993 |
| 0.004298969 | 0.225615658 | 0.045038249 | 1.296741804 | 0 |
| 0 | 0 | 0 | 0.031142793 | 0 |
| 0 | 0 | 0 | 0 | 0 |
| 0 | 0 | 0.005732684 | 0 | 0.13776689 |
| 0 | 0 | 0 | 0 | 0 |
| 0.000951679 | 0 | 0.006989576 | 0 | 0.009739278 |
| 0 | 0 | 0.094071833 | 0.215484522 | 0.088572246 |
| 0.001836313 | 0 | 0.035344559 | 0.003846807 | 0 |
| 0.002121714 | 0 | 0.02419641 | 0.044519997 | 0.273461681 |
| 0.030501738 | 0 | 0 | 0 | 0 |
| 0.035599949 | 0.259946519 | 0.044677203 | 0.099520542 | 0.081822163 |
| 0 | 0 | 0.003752832 | 0.002576827 | 0.027926018 |
| 0 | 0 | 0 | 0 | 0 |
| 1.976963376 | 0.788555637 | 0.751828535 | 0.251887896 | 0 |
| 0 | 0 | 0 | 0 | 0 |
| 0 | 0 | 0.052062055 | 0 | 0 |
| 0 | 0 | 0 | 0 | 0 |
| 0 | 0 | 0 | 0 | 0 |
| 0 | 0 | 0.025794789 | 0 | 0 |
| 0.004344616 | 0.005137987 | 0.023259453 | 0.006786113 | 0.341066968 |
| 0 | 0 | 0.000749072 | 0 | 0 |
| 0 | 0 | 0 | 0 | 0 |
| 0.119184676 | 0.227702374 | 0 | 0.045445893 | 0 |

| | | | | |
|---|---|---|---|---|
| 0 | 0 | 0 | 0 | 0 |
| 0 | 0 | 0 | 0 | 0 |
| 0 | 0 | 0 | 0 | 0 |
| 0 | 0 | 0 | 0.002792168 | 0.017710462 |
| 0 | 0 | 0.00041027 | 0 | 0 |
| 0 | 0 | 0.179565071 | 0 | 0.601004863 |
| 0.064326548 | 0 | 1.088013943 | 0 | 0 |
| 0.001286213 | 0 | 0.255491134 | 0 | 0 |
| 0 | 0 | 0 | 0 | 0 |
| 0 | 0 | 0 | 0 | 0 |
| 0.004042114 | 0.002968695 | 0.002496986 | 0 | 0.012327978 |
| 0 | 0 | 0 | 0 | 0 |
| 0 | 0 | 0 | 0.038897381 | 0 |
| 0 | 0 | 0.173375778 | 0 | 0.057616232 |
| 0 | 0 | 0 | 0 | 0 |
| 0 | 0 | 0 | 0 | 0 |
| 0.209280958 | 0 | 0.052780751 | 0.216414294 | 0 |
| 0 | 0 | 0.007548906 | 0 | 0.054155799 |
| 0.079692781 | 0 | 0 | 0 | 0.148197513 |
| 0.227864801 | 0 | 0 | 0 | 0.00469885 |
| 0 | 0 | 0.001846747 | 0.006078388 | 0 |
| 0 | 0 | 0 | 0 | 0 |
| 0.071536061 | 0 | 0.130676361 | 0.05948046 | 0 |
| 0 | 0 | 0 | 0 | 0 |
| 0 | 0 | 0 | 0 | 0 |
| 0 | 0 | 0 | 0 | 0 |
| 0 | 0 | 0 | 0 | 0 |
| 0 | 0 | 0 | 0 | 0 |
| 0 | 0 | 0 | 0 | 0 |
| 0.001538052 | 0 | 0.412073569 | 0 | 0.179665824 |
| 0 | 0 | 0 | 0 | 0.006273931 |
| 0 | 0 | 0 | 0 | 0 |
| 0 | 0 | 0 | 0.001932027 | 0 |
| 0 | 0 | 0 | 0 | 0 |
| 0 | 0 | 0.066879394 | 0 | 0.036043436 |
| 0 | 0 | 0 | 0 | 0 |
| 0.022374646 | 0 | 0.065502092 | 0.024178111 | 0 |
| 0.00085543 | 0.00287206 | 0 | 0 | 0 |
| 0 | 0 | 0 | 0 | 0 |
| 0 | 0 | 0.001316766 | 0 | 0 |
| 0 | 0.002526284 | 0.004905845 | 0.014995733 | 1.069372564 |
| 0 | 0 | 0 | 0 | 0 |
| 0 | 0 | 0 | 0 | 0 |
| 1.918118911 | 0 | 0.115491082 | 0.115221346 | 0.746362987 |
| 0.076603957 | 0 | 0.036633609 | 0.30873285 | 0 |
| 0 | 0 | 0.034571334 | 0 | 0 |
| 0.001713474 | 0 | 0.009090696 | 0 | 0 |
| 0 | 0 | 0 | 0 | 0 |

| | | | | |
|---|---|---|---|---|
| 0 | 0 | 0 | 0 | 0 |
| 0 | 0 | 0 | 0 | 1.458966239 |
| 0 | 0 | 0.095521701 | 0 | 0 |
| 0.071268799 | 0 | 0.074364032 | 0.062657246 | 0.131913617 |
| 0 | 0 | 0 | 0 | 0 |
| 0 | 0 | 0.440788781 | 0.181442081 | 0 |
| 0 | 0 | 0 | 0 | 0 |
| 0 | 0 | 0 | 0 | 0 |
| 0.018398203 | 0 | 0.000767159 | 0.001037309 | 0.017492763 |
| 0 | 0 | 0.00061779 | 0 | 0 |
| 0 | 0 | 0.000987377 | 0.004821104 | 0 |
| 0 | 0 | 0.000935395 | 0 | 0 |
| 0.001878769 | 0 | 0.00116362 | 0 | 0.033456731 |
| 0 | 0 | 0 | 0 | 0 |
| 4.703691412 | 0.423419093 | 0.491208937 | 0.203877225 | 0.150690007 |
| 0 | 0.000572417 | 0 | 0 | 0 |
| 0 | 0.002416543 | 0 | 0 | 0 |
| 0 | 0 | 0 | 0 | 0 |
| 0 | 0 | 0 | 0 | 0.411414345 |
| 0 | 0 | 0.12091107 | 0 | 0 |
| 0.000463247 | 0 | 0.068632561 | 0.045409075 | 0.002878321 |
| 0 | 0 | 0 | 0 | 0 |
| 0 | 0 | 0 | 0 | 0 |
| 0 | 0 | 0 | 0 | 0 |
| 0.024260552 | 0.596234463 | 0.035725423 | 0.047878958 | 0.003124707 |
| 4.449794392 | 0.048294798 | 12.07302364 | 47.87640422 | 11.41963962 |
| 0 | 0 | 0.654320603 | 0 | 0.009297649 |
| 0.001280187 | 0 | 0 | 0 | 0 |
| 0.110413452 | 2.298923687 | 0.376703816 | 0.735881051 | 0 |
| 0 | 0 | 0 | 0 | 0.019270744 |
| 0 | 0 | 0 | 0 | 0 |
| 0 | 0 | 0.057867022 | 0.012176608 | 0 |
| 0 | 0 | 0 | 0 | 0 |
| 0.000300387 | 0 | 0.003565393 | 0 | 0.198447467 |
| 0.021909472 | 0.023569066 | 0.00349303 | 0.00161014 | 0.008478075 |
| 0.001819296 | 0 | 0.002244681 | 0 | 0.014901442 |
| 0 | 0 | 0 | 0 | 0.177848483 |
| 0.490266516 | 0 | 1.160227971 | 1.202514251 | 1.025124286 |
| 0 | 0 | 0.394905606 | 0.618348102 | 0 |
| 0 | 0 | 0 | 0 | 0 |
| 0 | 0 | 0 | 0.86967615 | 0 |
| 0.006162839 | 0 | 0.001018194 | 0 | 0 |
| 0.293997312 | 0 | 1.179260403 | 0.049511498 | 0.590086831 |
| 0.109593955 | 0 | 0.017171279 | 0 | 0 |
| 0 | 0 | 0.056053158 | 0 | 0.386074655 |
| 0.001458392 | 0 | 0.000850621 | 0 | 0.001227587 |
| 0 | 0 | 0 | 0.196601995 | 0.209873862 |
| 0.120605263 | 0.365154012 | 0.107733757 | 0.00143152 | 0.206814496 |

| | | | | |
|---|---|---|---|---|
| 0 | 0 | 0.009978989 | 0.006618415 | 0.002916724 |
| 0 | 0 | 0.067088074 | 0 | 0 |
| 0 | 0 | 0 | 0 | 0 |
| 0 | 0 | 0 | 0 | 0 |
| 0 | 0 | 0.00050305 | 0.014454166 | 0.003266929 |
| 0.103945358 | 0 | 0 | 0 | 0 |
| 0 | 0 | 0.098916766 | 0 | 0 |
| 0 | 0 | 0 | 0 | 0 |
| 0 | 0 | 0 | 0 | 0 |
| 0 | 0 | 0.184420667 | 0 | 0 |
| 0 | 0 | 0 | 0 | 0 |
| 0.052202319 | 1.033758326 | 0.066249682 | 1.469191134 | 2.424220863 |
| 0.001139251 | 0 | 0.212577822 | 0.028311186 | 0.22789102 |
| 0 | 0 | 0 | 0 | 0 |
| 0.006075432 | 0.054914582 | 0 | 0.077927187 | 0.135385619 |
| 0 | 0 | 0 | 0 | 0 |
| 0 | 0 | 0 | 0 | 0.001022111 |
| 0 | 0 | 0 | 0 | 0 |
| 0 | 0 | 0.001019852 | 0.001508266 | 0.191519808 |
| 0.001645085 | 0 | 0.003675271 | 0 | 0.01897208 |
| 0.013050599 | 0 | 0.142588544 | 0.171964641 | 0.025589069 |
| 0 | 0 | 0 | 0 | 0 |
| 1.447708225 | 0.019600944 | 2.205467905 | 0.007332918 | 0.848665554 |
| 0 | 0 | 0 | 0 | 0 |
| 0 | 0 | 0 | 0 | 0 |
| 0 | 0 | 0 | 0 | 0 |
| 0 | 0 | 0 | 0 | 0 |
| 0 | 0 | 0 | 0 | 0 |
| 0.000735102 | 0 | 0 | 0 | 0.012856553 |
| 0 | 0 | 0 | 0 | 0 |
| 0 | 0 | 0 | 0 | 0 |
| 0.03498647 | 0.054814884 | 0.009059251 | 0.035727443 | 0.042490475 |
| 0 | 0 | 0 | 0 | 0 |
| 0.020574629 | 0.006998279 | 0.014897576 | 0.021832925 | 0.437885411 |
| 0.00170064 | 0 | 0.002687052 | 0 | 0.010752247 |
| 0.038643935 | 0.020578191 | 0.148634365 | 0.110440452 | 0.025222384 |
| 0 | 0 | 0 | 0.000549742 | 0 |
| 0 | 0 | 0.018336495 | 0.184175204 | 0 |
| 0 | 0 | 0 | 0 | 0 |
| 0 | 0 | 0 | 0 | 0 |
| 0 | 0 | 0.455274142 | 0 | 0 |
| 0.009242319 | 0 | 0.046826105 | 0 | 0 |
| 0 | 0 | 0 | 0 | 0 |
| 0.005537911 | 0 | 0 | 0 | 0.00674436 |
| 0.00172531 | 0.079596262 | 0.018529112 | 0.01098147 | 0.519009999 |
| 0 | 0 | 0 | 0 | 0 |
| 0 | 0 | 0.00951596 | 0 | 0.342158675 |
| 0.000260148 | 0 | 0.0003414 | 0 | 0 |

| | | | | |
|---|---|---|---|---|
| 0 | 0 | 0 | 0 | 0 |
| 0 | 0 | 0.012523949 | 0 | 0 |
| 0 | 0 | 0 | 0.006520097 | 0 |
| 0 | 0 | 0 | 0 | 0 |
| 0 | 0 | 0 | 0 | 0 |
| 0 | 0 | 0 | 0 | 0 |
| 0.410154665 | 0 | 0.365875757 | 0 | 0 |
| 0.16246318 | 0 | 0.030673194 | 0 | 0.127880925 |
| 0 | 0 | 0 | 0 | 0 |
| 0 | 0 | 0 | 0.001368237 | 0.18225062 |
| 0 | 0 | 0 | 0 | 0 |
| 0 | 0 | 0 | 0 | 0 |
| 0 | 0 | 0 | 0 | 0 |
| 0 | 0 | 0.175979334 | 0 | 0 |
| 0 | 0 | 0 | 0 | 0 |
| 0 | 0 | 0 | 0 | 0 |
| 5.047091163 | 0 | 0 | 0 | 0 |
| 0 | 0 | 0.014334345 | 0 | 0 |
| 0.83507608 | 0.002145469 | 0.096401853 | 2.004680055 | 0.019189484 |
| 0 | 0 | 0 | 0 | 0 |
| 0.002255707 | 0.003283785 | 0.001017583 | 0 | 0 |
| 0 | 0 | 0 | 0 | 0 |
| 0 | 0 | 0 | 0 | 0 |
| 0 | 0 | 0 | 0 | 0 |
| 0 | 0 | 0.002629177 | 0 | 0 |
| 0.001536364 | 0.002891751 | 0 | 0 | 0.012285574 |
| 0 | 0 | 0 | 0 | 0 |
| 0.002665818 | 0.052894011 | 0.146805038 | 0.00463269 | 0.037499429 |
| 0 | 0 | 0.08911688 | 0 | 0 |
| 0.225247254 | 0.042807245 | 0.505096311 | 0.151079641 | 0.933186887 |
| 0 | 0 | 0 | 0 | 0 |
| 0 | 0 | 0 | 0 | 0 |
| 0 | 0 | 0.457862686 | 0.023284588 | 0.015704805 |
| 0 | 0 | 0 | 0 | 0 |
| 0.000243942 | 0.000757594 | 0 | 0 | 0.008978285 |
| 0 | 0 | 0.031026481 | 0 | 0 |
| 0 | 0 | 0 | 0 | 0 |
| 0.000581285 | 0.003008765 | 0 | 0 | 0.021528774 |
| 0 | 0 | 0 | 0 | 0 |
| 0 | 0 | 0 | 0 | 0 |
| 10.73184771 | 0.26054558 | 2.371101005 | 3.937259945 | 1.084538044 |
| 0.036998499 | 0 | 0.067956641 | 0.098379882 | 1.819760887 |
| 0.07895615 | 0 | 0 | 0.349312603 | 0 |
| 0 | 0 | 0.130681507 | 0 | 0 |
| 0 | 0 | 0.042773112 | 0 | 0 |
| 0 | 0.119494753 | 0 | 0 | 0 |
| 0 | 0 | 0 | 0 | 0 |
| 0.000285591 | 0.023367529 | 0.01057103 | 0 | 0.045004426 |

| | | | | |
|---|---|---|---|---|
| 0 | 0 | 0 | 0 | 0 |
| 0.001400981 | 0 | 0.006523906 | 0.001286433 | 0.005178289 |
| 0 | 0 | 0.000888948 | 0 | 0.004215238 |
| 0 | 0 | 0 | 0 | 0 |
| 0.932045262 | 0.028737755 | 2.652687027 | 0.385319257 | 2.011591706 |
| 0 | 0 | 0 | 0 | 0 |
| 0 | 0 | 0 | 0 | 0 |
| 0 | 0 | 0.022365071 | 0 | 0 |
| 0.021525253 | 0 | 0 | 0.001651547 | 0.002506984 |
| 0 | 0 | 0 | 0 | 0 |
| 0 | 0 | 0 | 0 | 0 |
| 0 | 0 | 0 | 0 | 0 |
| 0.12796146 | 0 | 0.072265587 | 0.083749287 | 0.037355748 |
| 0.008880719 | 0.019281055 | 0.008702476 | 0.02107522 | 0.005623481 |
| 0 | 0 | 0.942901682 | 0.052281917 | 0 |
| 0.096088073 | 0.010662581 | 0.187696028 | 0.247883147 | 0.528062652 |
| 0 | 0.008161025 | 0.01584807 | 0.050532613 | 0.014598751 |
| 0 | 0 | 0 | 0 | 0 |
| 0 | 0 | 0 | 0 | 0 |
| 0 | 0 | 0.056821009 | 0 | 0 |
| 0 | 0 | 0.000936034 | 0 | 0 |
| 0 | 0 | 0 | 0 | 0 |
| 0 | 0 | 0 | 0 | 0 |
| 0 | 0 | 0 | 0 | 0 |
| 0 | 0 | 0.025052961 | 0 | 0 |
| 0 | 0 | 0 | 0 | 0 |
| 0.136652501 | 0.054878416 | 0.079067119 | 0.001817075 | 0.087307985 |
| 0.00176474 | 0.002313259 | 0 | 0 | 0 |
| 0 | 0 | 0 | 0 | 0 |
| 0 | 0 | 0.002279098 | 0 | 0 |
| 0 | 0 | 0 | 0 | 0 |
| 0 | 0 | 0 | 0 | 0 |
| 0 | 0 | 1.675903531 | 1.300960538 | 2.346036516 |
| 0 | 0 | 0.009248425 | 0 | 0.040041004 |
| 0.154634873 | 0 | 0.674900649 | 0.137734836 | 0 |
| 0.001087 | 0.005969449 | 0.019136064 | 0 | 0 |
| 0 | 0.002314361 | 0 | 0 | 0 |
| 0 | 0 | 0.182264745 | 0 | 0 |
| 0 | 0 | 0 | 0 | 0 |
| 0 | 0 | 0 | 0 | 0 |
| 2.749156498 | 0.011848926 | 0.142182977 | 0.411704076 | 0 |
| 0.000622784 | 0 | 0 | 0 | 0 |
| 0 | 0 | 0.006373337 | 0 | 0 |
| 0.085497954 | 0 | 0 | 0 | 0 |
| 0 | 0 | 0 | 0 | 0 |
| 0 | 0 | 0 | 0 | 0 |
| 0 | 0 | 1.901960715 | 0 | 0 |
| 0 | 0 | 0 | 0 | 0 |

| | | | | |
|---|---|---|---|---|
| 0.007001181 | 0.03955067 | 0.519461408 | 0 | 1.035281271 |
| 0 | 0 | 0 | 0 | 0 |
| 0 | 0 | 0.013877536 | 0 | 0 |
| 0 | 0 | 0 | 0 | 0 |
| 0.019028159 | 0 | 0.090246576 | 0.036163433 | 0.001725042 |
| 0 | 0 | 0 | 0 | 0.001256032 |
| 0 | 0.02641361 | 0.04810135 | 0.174728493 | 0.036408133 |
| 0 | 0 | 0 | 0 | 0.001876975 |
| 0 | 0 | 0.000395709 | 0 | 0 |
| 0 | 0 | 0.009410936 | 0 | 0 |
| 0.000565138 | 0 | 0.007342334 | 0 | 0.005494325 |
| 0.009414459 | 0 | 0.009217158 | 0.013092696 | 0.054339585 |
| 0 | 0 | 0 | 0 | 0 |
| 0 | 0 | 0 | 0 | 0 |
| 0 | 0 | 0 | 0 | 0.038635941 |
| 0 | 0 | 0 | 0 | 0 |
| 0 | 0.006202771 | 0 | 0 | 0 |
| 0 | 0 | 0 | 0 | 0 |
| 0 | 0 | 0 | 0 | 0 |
| 0 | 0 | 0 | 0 | 0 |
| 0.006921515 | 0.001722413 | 0.123508174 | 0 | 0.102601213 |
| 0 | 0 | 0.003210571 | 0.009277465 | 0 |
| 0.005676866 | 0 | 0.003532301 | 0.006965261 | 0.098725317 |
| 0.001000882 | 0.005574807 | 0.007538301 | 0.002477433 | 0.051373106 |
| 0 | 0.003635336 | 0 | 0 | 0 |
| 0 | 0 | 0.000617604 | 0.001252634 | 0.062926107 |
| 0 | 0 | 0 | 0 | 0 |
| 0 | 0 | 0 | 0 | 0 |
| 0.005770391 | 0.021271465 | 0 | 0 | 0 |
| 0.170230033 | 0.617805998 | 0 | 0.001782803 | 0.00239211 |
| 0 | 0 | 0 | 0 | 0 |
| 0 | 0 | 0 | 0 | 0 |
| 0 | 0 | 0 | 0 | 0 |
| 0 | 0 | 0.04170581 | 0 | 4.642894347 |
| 0 | 0 | 1.263307742 | 0 | 0 |
| 0 | 0 | 0.597318136 | 0 | 0 |
| 0.004573848 | 0 | 0.039170524 | 0.007749775 | 0 |
| 0 | 0 | 0 | 0 | 0 |
| 0.000376315 | 0 | 2.098342397 | 0 | 0 |
| 0.15178368 | 0 | 0 | 0 | 0 |
| 0 | 0 | 0 | 0 | 0 |
| 0 | 0 | 0 | 0 | 0 |
| 0 | 0 | 0 | 0 | 0 |
| 0 | 0 | 0.014501223 | 0 | 0.034278053 |
| 0 | 0 | 0.127582044 | 0 | 0 |
| 0 | 0 | 0 | 0 | 0 |
| 0 | 0 | 0 | 0 | 0 |
| 0 | 0 | 0 | 0 | 0 |

| | | | | |
|---|---|---|---|---|
| 0.005244448 | 0 | 0.000901276 | 0.010091978 | 0.007641548 |
| 0.023070495 | 0 | 0.246172557 | 2.500427862 | 0 |
| 0 | 0.000864103 | 0 | 0 | 0 |
| 0 | 0 | 0 | 0 | 0 |
| 0 | 0 | 0 | 0 | 0 |
| 0 | 0 | 0 | 0.590817058 | 0 |
| 0 | 0 | 0 | 0 | 0 |
| 0.208447089 | 0.033782823 | 0.199540722 | 0.321423106 | 1.562384095 |
| 0.946159671 | 12.60039867 | 0.877990286 | 0.48927382 | 0.047978231 |
| 0.211606008 | 0 | 0 | 0.001248446 | 0 |
| 0.004113079 | 0 | 0.033515855 | 0.004395089 | 0 |
| 0 | 0 | 0.001939689 | 0 | 0 |
| 0 | 0 | 0.033690293 | 0 | 0 |
| 0 | 0.394795973 | 0 | 0 | 0 |
| 0 | 0 | 0 | 0 | 0 |
| 0 | 0 | 0 | 0 | 0 |
| 0 | 0 | 0 | 0 | 0 |
| 0 | 0 | 0 | 0 | 0 |
| 0 | 0 | 0 | 0 | 0 |
| 0 | 0 | 0.057064953 | 0.038303942 | 0 |
| 0 | 0 | 0 | 0 | 0 |
| 0 | 0 | 0 | 0 | 0 |
| 0 | 0 | 0 | 0 | 0 |
| 0 | 0 | 0 | 0 | 0 |
| 0.10371083 | 0 | 0.013310239 | 0 | 0 |
| 0 | 0 | 0.043020816 | 0 | 0 |
| 0 | 0 | 0 | 0 | 0 |
| 0 | 0 | 0 | 0 | 0 |
| 0 | 0 | 0 | 0 | 0 |
| 0 | 0 | 0 | 0 | 0 |
| 0 | 0 | 0 | 0 | 0 |
| 0 | 0 | 0 | 0 | 0 |
| 0 | 0 | 0.001034987 | 0 | 0 |
| 0 | 0 | 0.033055292 | 0 | 0 |
| 0.006944375 | 0 | 0.015957643 | 0 | 0 |
| 0 | 0 | 0 | 0 | 0 |
| 0 | 0 | 0.002492357 | 0.014643562 | 0 |
| 0.01065657 | 0 | 0 | 0 | 0 |
| 0 | 0 | 0.000669804 | 0.001584926 | 0 |
| 0 | 0 | 0 | 0 | 0 |
| 0 | 0 | 0 | 0 | 0 |
| 0.599681789 | 0 | 0.330923683 | 0.867221242 | 0.596469397 |
| 0 | 0 | 0 | 0 | 0 |
| 0 | 0 | 0 | 0 | 0 |
| 0 | 0 | 0 | 0 | 0 |
| 0 | 0 | 0 | 0 | 0 |
| 0 | 0 | 0 | 0 | 0 |
| 0 | 0 | 0 | 0 | 0 |

| | | | | |
|---|---|---|---|---|
| 0 | 0 | 0 | 0 | 0 |
| 0.021722668 | 0 | 0 | 0.007389876 | 0 |
| 0 | 0 | 0 | 0 | 0 |
| 0 | 0 | 0 | 0 | 0 |
| 0.066267436 | 1.115031746 | 0.558274233 | 0.778270324 | 1.303000906 |
| 0 | 0 | 0.00545672 | 0 | 0 |
| 0 | 0 | 0 | 0 | 0 |
| 0.006760666 | 0 | 0.00143311 | 0.000726664 | 0.048603014 |
| 0.000594738 | 0 | 0.000858544 | 0 | 0.00660811 |
| 0 | 0 | 0 | 0 | 0 |
| 0 | 0 | 0 | 0 | 0 |
| 0 | 0 | 0 | 0 | 0 |
| 0 | 0 | 0 | 0 | 0 |
| 0 | 0 | 0 | 0 | 0 |
| 0.000251591 | 0 | 0 | 0 | 0 |
| 0.003409598 | 0 | 0.110496058 | 0 | 0.05811737 |
| 0 | 0 | 0 | 0 | 0 |
| 0 | 0 | 0 | 0 | 0 |
| 0 | 0 | 0 | 0 | 0 |
| 0 | 0 | 0 | 0 | 0 |
| 0 | 0 | 0 | 0 | 0 |
| 0 | 0 | 0.001237186 | 0 | 0 |
| 0 | 0 | 0 | 0 | 0 |
| 0.004046588 | 0.00460265 | 0 | 0 | 0.005548592 |
| 0.018345627 | 0 | 0.014345039 | 0.295983933 | 0 |
| 0 | 0 | 0 | 0 | 0 |
| 0.000165016 | 0.168751879 | 0.000907471 | 0 | 1.329602049 |
| 0 | 0 | 0 | 0 | 0 |
| 0 | 0 | 0 | 0 | 0 |
| 0 | 0 | 0 | 0 | 0 |
| 0 | 0 | 0 | 0 | 0 |
| 0 | 0 | 0 | 0 | 0 |
| 0 | 0 | 0 | 0 | 0 |
| 0 | 0 | 0 | 0 | 0 |
| 0 | 0 | 0 | 0 | 0 |
| 0 | 0 | 0 | 0 | 0 |
| 0 | 0 | 0 | 0 | 0 |
| 0.024998157 | 0 | 0.009306647 | 0.001179743 | 0.343666354 |
| 0 | 0 | 0 | 0 | 0 |
| 0 | 0 | 0.000399209 | 0 | 0 |
| 0 | 0 | 0 | 0 | 0 |
| 0.161005781 | 0 | 0.485281067 | 0 | 0 |
| 0 | 0 | 0 | 0 | 0 |
| 0 | 0 | 0 | 0 | 0 |
| 0 | 0 | 0 | 0 | 0 |
| 0 | 0 | 0 | 0 | 0 |
| 0 | 0 | 0 | 0 | 0 |
| 0 | 0 | 0.021354485 | 0 | 0 |

| | | | | |
|---|---|---|---|---|
| 0.050279278 | 0 | 0.135050276 | 0.421900915 | 1.05901717 |
| 0 | 0 | 0.005361719 | 0 | 0 |
| 0 | 0 | 0 | 0 | 0 |
| 0 | 0 | 0 | 0 | 0.002603908 |
| 0 | 0 | 0 | 0 | 0 |
| 0.000333893 | 0.003390029 | 0.108511062 | 0.014622121 | 0 |
| 0 | 0 | 0 | 0 | 0 |
| 0 | 0 | 0 | 0 | 0 |
| 0 | 0 | 0.059555616 | 0 | 0 |
| 0 | 0 | 0 | 0 | 0 |
| 0 | 0 | 0 | 0 | 0 |
| 0 | 0 | 0 | 0 | 0 |
| 0 | 0 | 0 | 0.001847561 | 0 |
| 0 | 0 | 0 | 0 | 0 |
| 0 | 0 | 0 | 0 | 0 |
| 0 | 0 | 0 | 0 | 0 |
| 0 | 0 | 0 | 0 | 0 |
| 0 | 0 | 0 | 0 | 0 |
| 0.001691581 | 0 | 0.000452206 | 0 | 0.016967796 |
| 0 | 0 | 0 | 0 | 0.000928353 |
| 0.094530468 | 0 | 0 | 0 | 0.00124767 |
| 0 | 0 | 0 | 0 | 0 |
| 0.000324152 | 0.391102085 | 0.136481014 | 0.00395447 | 0.015532065 |
| 0.098319948 | 0 | 0.034405032 | 0 | 0.02801946 |
| 0 | 0 | 0.020594739 | 0 | 0 |
| 0 | 0 | 0 | 0 | 0 |
| 0 | 0 | 0 | 0 | 0 |
| 0 | 0 | 0 | 0 | 0 |
| 0 | 5.891863925 | 0 | 0 | 0 |
| 0 | 0 | 0 | 0 | 0 |
| 0 | 0 | 0 | 0 | 0 |
| 0 | 0 | 1.100518576 | 0 | 0 |
| 0 | 0.077307002 | 0.010663332 | 0.062709973 | 0.004005342 |
| 0 | 0 | 0.007289525 | 0 | 0 |
| 0.01105164 | 0 | 0.228396542 | 0.032463719 | 0.763316274 |
| 0 | 0 | 0 | 0.002179305 | 0.001691642 |
| 0 | 0 | 0 | 0 | 0 |
| 0 | 0 | 0 | 0 | 0 |
| 0.001014341 | 0 | 0 | 0 | 0 |
| 0 | 0 | 0 | 0 | 0 |
| 0 | 0 | 0 | 0 | 0 |
| 0 | 0 | 0.196388255 | 0 | 0 |
| 0.053241322 | 2.884641022 | 0.178668167 | 0.181939033 | 0 |
| 0 | 0 | 0 | 0 | 0 |
| 0 | 0 | 0 | 0 | 0 |
| 0.378923356 | 0 | 0.104877813 | 0.002127999 | 0.018169984 |
| 0 | 0 | 0 | 0 | 0 |
| 0 | 0 | 0 | 0 | 0 |

| | | | | |
|---|---|---|---|---|
| 0 | 0 | 0 | 0 | 0 |
| 0 | 0 | 0 | 0 | 0 |
| 0 | 0 | 0 | 0 | 0 |
| 0 | 0 | 0 | 0 | 0 |
| 0 | 0 | 0 | 0 | 0 |
| 0 | 0 | 0 | 0 | 0 |
| 0 | 0 | 0 | 0 | 0 |
| 0 | 0 | 0 | 0 | 0 |
| 0.000563829 | 0 | 0.011560093 | 0 | 0.073541911 |
| 0.048267076 | 0.027201288 | 0.047203323 | 0.00284936 | 0 |
| 0 | 0 | 0.123777101 | 0 | 0 |
| 0 | 0 | 0 | 0 | 0 |
| 0 | 0 | 0 | 0 | 0 |
| 0 | 0.010078114 | 0 | 0 | 0 |
| 0 | 0 | 0 | 0 | 0 |
| 0.002969749 | 0 | 0 | 0 | 0 |
| 0 | 0 | 0 | 0 | 0 |
| 0 | 0 | 0 | 0 | 0 |
| 0 | 0 | 0 | 0 | 0 |
| 0.041846457 | 0 | 0.0954994 | 0.001986043 | 0.09342254 |
| 0.005067943 | 0 | 0 | 0.007432642 | 0 |
| 0 | 0 | 0 | 0 | 0 |
| 0 | 0 | 0 | 0 | 0 |
| 0 | 0 | 0.013256322 | 0.000861752 | 0 |
| 0 | 0 | 0 | 0 | 0 |
| 0 | 0 | 0 | 0 | 0 |
| 0 | 0 | 0 | 0 | 0 |
| 0 | 0.005274726 | 0 | 0 | 0 |
| 0 | 0 | 0 | 0 | 0 |
| 0 | 0 | 0.540454683 | 0.069957104 | 0.1045806 |
| 1.268802263 | 0.032461071 | 0.245150997 | 1.264255828 | 0.171359269 |
| 0 | 0 | 0 | 0 | 0 |
| 0 | 0 | 0.001445162 | 0 | 0 |
| 0 | 0 | 0 | 0 | 0 |
| 0 | 0 | 0 | 0 | 0 |
| 0 | 0 | 0 | 0 | 0 |
| 0 | 0 | 0 | 0 | 0 |
| 0 | 0 | 0 | 0 | 0 |
| 0.005975091 | 0.005091713 | 0 | 0 | 0 |
| 0 | 0 | 0 | 0 | 0 |
| 0 | 0 | 0 | 0 | 0.014780476 |
| 0 | 0 | 1.89146809 | 0 | 0 |
| 0.005174701 | 0 | 0 | 0 | 0.025575318 |
| 0 | 0 | 0 | 0 | 0.002950218 |
| 0 | 0 | 0 | 0 | 0 |
| 0 | 0 | 0 | 0 | 0 |
| 0 | 0 | 0 | 0 | 0 |
| 0 | 0 | 0.003502925 | 0 | 0 |

| | | | | |
|---|---|---|---|---|
| 0 | 0 | 0 | 0 | 0 |
| 0 | 0 | 0 | 0 | 0 |
| 0 | 0 | 0 | 0 | 0 |
| 0 | 0 | 0 | 0 | 0 |
| 0.116986535 | 0 | 0.085478174 | 0.059867427 | 0.428517937 |
| 0 | 0 | 0.002031975 | 0 | 0 |
| 0 | 0 | 0 | 0 | 0 |
| 0 | 0 | 0 | 0 | 0 |
| 0 | 0 | 0 | 0 | 0 |
| 0 | 0 | 0 | 0 | 0 |
| 0 | 0 | 0.021804627 | 0 | 0.076702446 |
| 0.001717377 | 0.038690086 | 0.010160446 | 0.001602811 | 0.136465492 |
| 0 | 0 | 0.001402545 | 0.001185276 | 0.035759156 |
| 0 | 0 | 0 | 0 | 0 |
| 0.008667859 | 0 | 0 | 0 | 0 |
| 0 | 0 | 0.564355445 | 0 | 0.81582623 |
| 0.015395673 | 0 | 0 | 0 | 0.066764216 |
| 0 | 0 | 0 | 0 | 0 |
| 0.036986997 | 0.00811029 | 0 | 0.010229402 | 0.000740204 |
| 0 | 0.004322273 | 0.020894493 | 0.002037429 | 0.004445811 |
| 0 | 0 | 0 | 0 | 0 |
| 0 | 0 | 0.046739172 | 0 | 0 |
| 0 | 0 | 0 | 0 | 0 |
| 0 | 0 | 0 | 0 | 0 |
| 0 | 0 | 0 | 0 | 0 |
| 0 | 0 | 0 | 0 | 0 |
| 0.002565129 | 0.006407725 | 0 | 0 | 0 |
| 0 | 0 | 0 | 0 | 0 |
| 0 | 0 | 0 | 0 | 0 |
| 0 | 0 | 0 | 0 | 0 |
| 0.000758533 | 0 | 0.008394947 | 0 | 0.001229088 |
| 0 | 0 | 0 | 0 | 0 |
| 0 | 0 | 0 | 0 | 0 |
| 0 | 0 | 0 | 0 | 0 |
| 0 | 0 | 0 | 0 | 0 |
| 0 | 0 | 0 | 0 | 0 |
| 0 | 0 | 0 | 0 | 0 |
| 0.003126482 | 0.063582107 | 0.014104023 | 0.000722374 | 0.365516205 |
| 0.010448255 | 0 | 0.008122277 | 0.18437368 | 0 |
| 0 | 0 | 0 | 0 | 0 |
| 0 | 0.002891506 | 0.006229493 | 0.001730788 | 0 |
| 0 | 0 | 0.004821493 | 0 | 0 |
| 0 | 0 | 0 | 0 | 0 |
| 0.00068489 | 0 | 0 | 0 | 0 |
| 0 | 0 | 0 | 0 | 0 |
| 0 | 0 | 0 | 0 | 0 |
| 0 | 0 | 0 | 0 | 0 |
| 0 | 0 | 0 | 0 | 0 |

| | | | | |
|---|---|---|---|---|
| 0 | 0 | 0 | 0 | 0 |
| 0 | 0 | 0.083528468 | 0 | 0.208257325 |
| 0 | 0 | 0 | 0 | 0 |
| 0.002180909 | 0 | 0.001195553 | 0 | 0.002683924 |
| 0.000332859 | 0 | 0 | 0 | 0 |
| 0 | 0 | 0 | 0 | 0.002294769 |
| 0 | 0 | 0.006703338 | 0.004139751 | 0.059160362 |
| 0.007945351 | 0 | 0.051426415 | 0.010711338 | 0 |
| 0 | 0 | 0.006582879 | 0 | 0 |
| 0 | 0 | 0.014736327 | 0 | 0 |
| 0 | 0 | 0.044455672 | 0 | 0 |
| 0 | 0 | 0 | 0 | 0 |
| 0.18663033 | 0 | 0.117328704 | 0.05904634 | 0.387163674 |
| 0 | 0 | 0 | 0 | 0 |
| 0 | 0 | 0 | 0 | 0 |
| 0 | 0 | 0.347013331 | 0 | 0 |
| 0 | 0 | 0 | 0 | 0 |
| 0 | 0 | 0 | 0 | 0 |
| 0 | 0 | 0 | 0 | 0 |
| 0 | 0 | 0 | 0 | 0 |
| 0 | 0 | 0 | 0 | 0 |
| 0 | 0 | 0 | 0 | 0 |
| 0.022804654 | 0 | 0 | 0.009640214 | 0.061377056 |
| 0.006020604 | 0 | 0.011705249 | 0.019783973 | 0.015836816 |
| 0 | 0 | 0 | 0 | 0 |
| 0 | 0 | 0.017829155 | 0 | 0 |
| 0 | 0 | 0.018361961 | 0 | 0 |
| 0 | 0 | 0 | 0 | 0 |
| 0 | 0 | 0 | 0 | 0 |
| 0 | 0 | 0 | 0 | 0 |
| 0.016089457 | 0 | 0.010092343 | 0.005140729 | 0.026137038 |
| 0 | 0 | 0 | 0 | 0 |
| 0 | 0 | 0 | 0 | 0 |
| 0 | 0 | 0 | 0 | 0 |
| 0.116005734 | 0 | 0.131785987 | 0 | 0 |
| 0 | 0 | 0 | 0 | 0 |
| 0 | 0 | 0 | 0 | 0 |
| 0 | 0 | 0 | 0 | 0 |
| 0 | 0.001509872 | 0 | 0.004349413 | 0.055875192 |
| 0 | 0 | 0 | 0 | 0 |
| 0 | 0 | 0 | 0 | 0 |
| 0 | 0 | 0 | 0 | 0 |
| 0 | 0 | 6.602754136 | 0 | 0 |
| 0 | 0 | 0 | 0 | 0 |
| 0 | 0 | 2.058889312 | 0 | 0 |
| 0.284090448 | 0 | 0.514880865 | 0.24369534 | 0.353929053 |
| 0 | 0 | 0 | 0 | 0 |
| 0 | 0 | 0 | 0 | 0 |

| | | | | |
|---|---|---|---|---|
| 0 | 0 | 0.004006238 | 0 | 0 |
| 0 | 0 | 0 | 0 | 0 |
| 0.057432466 | 0 | 0.028079233 | 0.072258921 | 0 |
| 0 | 0 | 0.001243032 | 0.002626185 | 0.666882473 |
| 0 | 0 | 0 | 0 | 0 |
| 0 | 0 | 0 | 0 | 0 |
| 0 | 0 | 0 | 0 | 0 |
| 0 | 0 | 0 | 0 | 0 |
| 0 | 0 | 0 | 0 | 0 |
| 0 | 0 | 0 | 0 | 0 |
| 0 | 0.002756048 | 0.000569365 | 0 | 0.003834541 |
| 0.00047109 | 0 | 0 | 0 | 0.003680345 |
| 0 | 0 | 0 | 0 | 0 |
| 0 | 0 | 0 | 0 | 0 |
| 0 | 0 | 0 | 0 | 0 |
| 0 | 0 | 0 | 0 | 0 |
| 0 | 0 | 0 | 0 | 0 |
| 0 | 0 | 0 | 0 | 0 |
| 0 | 0 | 0 | 0 | 0 |
| 0.000915359 | 0.042641592 | 0 | 0 | 0.000726466 |
| 0 | 0 | 0 | 0 | 0 |
| 0 | 0 | 0 | 0 | 0 |
| 0 | 0 | 0 | 0 | 0 |
| 0 | 0 | 0 | 0 | 0 |
| 0 | 0 | 0 | 0 | 0 |
| 0 | 0 | 0 | 0 | 0 |
| 0 | 0 | 0 | 0 | 0 |
| 0 | 0 | 0.007520569 | 0.004472428 | 0 |
| 0 | 0 | 0 | 0 | 0 |
| 0 | 0 | 0 | 0 | 0 |
| 0.003707258 | 0.609686784 | 0.003135326 | 0.011694926 | 0.002808491 |
| 0 | 0 | 0 | 0 | 0 |
| 0.844491547 | 0.015798815 | 0.011297894 | 0.014215516 | 0.002717454 |
| 0.000986383 | 0.004967592 | 0.003591847 | 0.022548932 | 0.006849793 |
| 0 | 0 | 0 | 0 | 0 |
| 0 | 0 | 0 | 0 | 0 |
| 0 | 0 | 0 | 0 | 0 |
| 0 | 0 | 0 | 0 | 0 |
| 0 | 0 | 0 | 0 | 0 |
| 0 | 0 | 0 | 0 | 0 |
| 0 | 0 | 0 | 0 | 0 |
| 0.002275002 | 0.006665406 | 0.131571286 | 0 | 0.185109714 |
| 0.011923083 | 0.059887537 | 0.016859485 | 0.070388278 | 0.115299959 |
| 0 | 0 | 0.000484459 | 0 | 0 |
| 0 | 0 | 0 | 0 | 0 |
| 0.033741463 | 0.15812187 | 0.067089509 | 0.097106471 | 0 |
| 0 | 0 | 0 | 0 | 0 |
| 0 | 0 | 0 | 0 | 0 |

| | | | | |
|---|---|---|---|---|
| 0 | 0 | 0 | 0 | 0 |
| 0.001753124 | 0.005954996 | 0.002372581 | 0 | 0.006467603 |
| 0.016542661 | 0 | 0 | 0.006505841 | 0.006443837 |
| 0 | 0 | 0 | 0 | 0 |
| 0 | 0.005950803 | 0.024095472 | 0.023687381 | 2.706674088 |
| 0 | 0 | 0 | 0 | 2.64312251 |
| 0 | 0 | 0 | 0 | 0 |
| 0 | 0 | 0 | 0 | 0 |
| 0 | 0 | 0 | 0 | 0 |
| 0 | 0 | 0 | 0 | 0 |
| 0 | 0 | 0 | 0 | 0 |
| 0 | 0 | 0 | 0 | 0 |
| 0 | 0 | 0 | 0 | 0 |
| 0 | 0 | 0 | 0 | 0 |
| 0 | 0 | 0 | 0 | 0 |
| 0 | 0 | 0 | 0 | 0 |
| 0 | 0 | 0.451248939 | 0 | 0 |
| 0 | 0 | 0 | 0 | 0 |
| 0 | 0 | 0 | 0 | 0 |
| 0 | 0 | 0 | 0 | 0 |
| 0 | 0 | 0 | 0 | 0 |
| 0 | 0 | 0 | 0 | 0 |
| 0 | 0 | 0.001218677 | 0 | 0.002763752 |
| 0 | 0 | 0 | 0 | 0 |
| 0.016428737 | 0.036853553 | 0.016368981 | 0.002573629 | 0 |
| 0.013874125 | 0.032748526 | 0 | 0 | 0 |
| 0 | 0 | 0 | 0 | 0 |
| 0.042227935 | 0 | 0.034423248 | 0.010255007 | 0.031451086 |
| 0 | 0 | 0 | 0 | 0 |
| 0 | 0 | 0 | 0 | 0 |
| 0.00416316 | 0 | 0.001469062 | 0 | 0 |
| 0.000372215 | 0 | 0 | 0 | 0 |
| 0 | 0 | 0 | 0 | 0 |
| 0 | 0 | 0 | 0 | 0 |
| 0 | 0 | 0 | 0 | 0 |
| 0 | 0 | 0 | 0 | 0 |
| 0 | 0 | 0 | 0 | 0.008000052 |
| 0 | 0 | 0 | 0 | 0 |
| 1.193017471 | 0.009350028 | 3.37291752 | 2.381567756 | 0.014718602 |
| 0 | 0 | 0 | 0 | 0 |
| 0 | 0 | 0 | 0 | 0 |
| 0.023080558 | 0.009831399 | 0.009693615 | 0.001872451 | 0 |
| 0.002822113 | 0 | 0.000896773 | 0.007795063 | 0.005546532 |
| 0 | 1.117017651 | 0 | 0 | 0 |
| 0 | 0 | 0 | 0 | 0 |
| 0.001562111 | 0 | 0.004421593 | 0 | 0.002552435 |
| 0.01388861 | 0.005059587 | 0.01234698 | 0.002649983 | 0.010747803 |
| 0 | 0 | 0 | 0 | 0 |

| | | | | |
|---|---|---|---|---|
| 0 | 0 | 0 | 0 | 0 |
| 0.007040784 | 0.013167554 | 0.002102012 | 0.016300979 | 0.033725933 |
| 0 | 0 | 0.013195013 | 0 | 0 |
| 0 | 0 | 0 | 0 | 0 |
| 0 | 0 | 0 | 0 | 0 |
| 0 | 0 | 0.783969375 | 0 | 0 |
| 0 | 0 | 0 | 0 | 0 |
| 0 | 0 | 0 | 0 | 0 |
| 0 | 0 | 0 | 0 | 0 |
| 0 | 0 | 0 | 0 | 0 |
| 0 | 0 | 0 | 0 | 0 |
| 0 | 0 | 0.016316736 | 0 | 0 |
| 0.170693908 | 0 | 0.116851732 | 0.666729561 | 4.843483654 |
| 0 | 0 | 0 | 0 | 0 |
| 0.003972886 | 0 | 0.017112883 | 0.031008446 | 0 |
| 0 | 0 | 0 | 0 | 0 |
| 0 | 0 | 0 | 0 | 0 |
| 0 | 0 | 0 | 0 | 0 |
| 0 | 0 | 0.006310427 | 0 | 0 |
| 0 | 0 | 0 | 0 | 0 |
| 0 | 0 | 0 | 0 | 0 |
| 0 | 0 | 0.011204134 | 0.001023622 | 0.001092528 |
| 0.002369488 | 0 | 0 | 0.024776936 | 0.024010222 |
| 0 | 0 | 0 | 0 | 0 |
| 0 | 0 | 0 | 0 | 0 |
| 0 | 0 | 0 | 0 | 0 |
| 0 | 0 | 0 | 0 | 0 |
| 0 | 0 | 0 | 0 | 0 |
| 0 | 0 | 0 | 0 | 0 |
| 0.010824698 | 0 | 0.029903761 | 0 | 0 |
| 0 | 0 | 0 | 0 | 0 |
| 0 | 0 | 0 | 0 | 0 |
| 0 | 0 | 0 | 0 | 0 |
| 0 | 0 | 0 | 0 | 0 |
| 0.0337396 | 0 | 0.833751501 | 0 | 0 |
| 0 | 0 | 0 | 0 | 0 |
| 0.002573983 | 0 | 0 | 0 | 0 |
| 0 | 0 | 0 | 0 | 0 |
| 0 | 0.305684408 | 0 | 0.007922127 | 0.002836655 |
| 0 | 0 | 0 | 0 | 0 |
| 0.327999704 | 0.218962959 | 0.223825066 | 0.290545225 | 0.037955901 |
| 0 | 0 | 0 | 0 | 0 |
| 0 | 0 | 0 | 0 | 0 |
| 0 | 0 | 0 | 0 | 1.605214776 |
| 0 | 0 | 0 | 0 | 0 |
| 0.002645083 | 0.056851523 | 0.026697449 | 0.003410496 | 0.006496138 |
| 0 | 0 | 0 | 0 | 0 |
| 0 | 0 | 0 | 0 | 0 |

| | | | | |
|---|---|---|---|---|
| 0 | 0 | 0 | 0 | 0 |
| 0 | 0 | 0.001011255 | 0 | 0 |
| 0 | 0 | 0 | 0 | 0 |
| 0 | 0 | 0 | 0 | 0.031842048 |
| 0 | 0 | 0 | 0 | 0.009513877 |
| 0 | 0 | 0 | 0 | 0 |
| 0 | 0 | 0 | 0 | 0 |
| 0 | 0 | 0 | 0 | 0 |
| 0 | 0 | 0 | 0 | 0 |
| 0.001829756 | 0 | 0 | 0.001282873 | 0.019936008 |
| 0.021027496 | 0 | 0.012167458 | 0.009200216 | 0 |
| 0 | 0 | 0 | 0 | 0 |
| 0.005026111 | 0 | 0.003412621 | 0 | 0 |
| 0 | 0 | 0 | 0 | 0 |
| 0 | 0 | 0 | 0 | 0 |
| 0 | 0 | 0 | 0 | 0 |
| 0 | 0 | 0 | 0 | 0 |
| 0 | 0 | 0 | 0 | 0 |
| 0 | 0 | 0 | 0 | 0 |
| 0.10569207 | 3.326521434 | 0.010961318 | 2.039487929 | 0.969341825 |
| 0 | 0 | 0 | 0 | 0.17108115 |
| 0 | 0 | 0 | 0 | 0 |
| 0 | 0 | 0 | 0 | 0 |
| 0 | 0 | 0 | 0 | 0 |
| 0 | 0 | 0 | 0 | 0 |
| 0.00068573 | 0 | 0 | 0 | 0 |
| 0 | 0 | 0 | 0 | 0 |
| 0 | 0 | 0 | 0 | 0 |
| 0 | 0 | 0 | 0 | 0 |
| 0 | 0 | 0 | 0 | 0 |
| 0 | 0 | 0 | 0 | 0 |
| 0 | 0 | 0 | 0 | 0 |
| 0 | 0 | 0 | 0 | 0 |
| 0 | 0 | 0 | 0 | 0 |
| 0 | 0 | 0 | 0 | 0 |
| 0 | 0 | 0.001538585 | 0 | 0 |
| 0 | 0 | 0 | 0 | 0 |
| 0 | 0 | 0 | 0 | 0 |
| 0 | 0 | 0 | 0 | 0 |
| 0 | 0 | 0.546797611 | 0 | 0 |
| 0 | 0 | 0 | 0 | 0 |
| 0 | 0 | 0 | 0 | 0 |
| 0.101064527 | 0 | 0.026456224 | 0 | 0.026140307 |
| 0 | 0 | 0 | 0 | 0 |
| 0.000541331 | 0.01641151 | 0.00434138 | 0 | 0.07500491 |
| 0 | 0 | 0.029334246 | 0 | 0.05953844 |
| 0 | 0 | 0.015282389 | 0 | 0 |
| 0 | 0 | 0 | 0 | 0 |

| | | | | |
|---|---|---|---|---|
| 0 | 0 | 0 | 0 | 0 |
| 0 | 0 | 0 | 0 | 0 |
| 0 | 0 | 0 | 0 | 0 |
| 0 | 0 | 0 | 0 | 0 |
| 0 | 0 | 0.002061563 | 0 | 0 |
| 0.006100363 | 0.014791844 | 0.024816868 | 0.01189484 | 0.043565801 |
| 0 | 0 | 0 | 0 | 0 |
| 0 | 0 | 0 | 0 | 0 |
| 0 | 0 | 0 | 0 | 0 |
| 0 | 0 | 0 | 0 | 0 |
| 0 | 0 | 0 | 0 | 0 |
| 0 | 0 | 0 | 0 | 0 |
| 0 | 0 | 0 | 0 | 0 |
| 0 | 0 | 0.024033819 | 0 | 0 |
| 0 | 0 | 0 | 0 | 0 |
| 0 | 0 | 0 | 0 | 0 |
| 0.000863687 | 0 | 0.043244531 | 0.104985286 | 0 |
| 0 | 0 | 0 | 0 | 0 |
| 0 | 0 | 0 | 0 | 0 |
| 0 | 0 | 0 | 0 | 0 |
| 0 | 0 | 0 | 0 | 0 |
| 0 | 0 | 0 | 0 | 0 |
| 0 | 0 | 0 | 0 | 0 |
| 0 | 0 | 0 | 0 | 0 |
| 0 | 0 | 0 | 0 | 0 |
| 0 | 0 | 0 | 0 | 0.002717408 |
| 0 | 0 | 0 | 0 | 0 |
| 0 | 0 | 0 | 0 | 0 |
| 0 | 0 | 0 | 0 | 0 |
| 0 | 0 | 0 | 0 | 0 |
| 0 | 0 | 0 | 0 | 0 |
| 0 | 0 | 0 | 0 | 0 |
| 0 | 0 | 0 | 0 | 0 |
| 0 | 0 | 0 | 0 | 0 |
| 0 | 0 | 0 | 0 | 0 |
| 0 | 0 | 0 | 0 | 0 |
| 0 | 0 | 0 | 0 | 0 |
| 0 | 0 | 0 | 0 | 0 |
| 0.004292022 | 0 | 1.014990799 | 0.000662788 | 1.044506366 |
| 0 | 0 | 0 | 0 | 0 |
| 0 | 0 | 0 | 0 | 0 |
| 0 | 0 | 0 | 0 | 0 |
| 0 | 0 | 0 | 0 | 0 |
| 0 | 0 | 0 | 0 | 0 |
| 0 | 0 | 0 | 0 | 0 |
| 0 | 0 | 0 | 0 | 0 |
| 0 | 0 | 0 | 0 | 0 |
| 0 | 0 | 0 | 0 | 0 |

| | | | | |
|---|---|---|---|---|
| 0 | 0 | 0 | 0 | 0 |
| 0 | 0 | 0.037737135 | 0 | 0.189865076 |
| 0 | 0 | 0 | 0 | 0 |
| 0 | 0 | 0 | 0 | 0 |
| 0 | 0 | 0 | 0 | 0 |
| 0 | 0 | 0 | 0 | 0 |
| 0 | 0 | 0 | 0 | 0.002074308 |
| 0 | 0 | 0 | 0 | 0 |
| 0 | 0 | 0 | 0 | 0 |
| 0 | 0 | 0.011401865 | 0 | 0 |
| 0.008125002 | 0.007228291 | 0.001221769 | 0 | 0.005485557 |
| 0 | 0 | 0 | 0 | 0 |
| 0.000311691 | 0 | 0.01139867 | 0.056107414 | 0.455553968 |
| 0 | 0 | 0 | 0 | 0 |
| 0 | 0 | 0 | 0 | 0 |
| 0 | 0 | 0 | 0 | 0 |
| 0 | 0 | 0 | 0 | 0 |
| 0 | 0 | 0 | 0 | 0 |
| 0 | 0 | 0 | 0 | 0 |
| 0 | 0 | 0 | 0 | 0 |
| 0 | 0 | 0 | 0 | 0.001590717 |
| 0 | 0 | 0 | 0 | 0 |
| 0 | 0 | 0 | 0 | 0 |
| 1.016109052 | 1.209765277 | 0.714612532 | 1.664253822 | 1.002477872 |
| 0 | 0 | 0 | 0 | 0 |
| 0 | 0 | 0 | 0 | 0 |
| 0 | 0 | 0 | 0 | 0 |
| 0 | 0 | 0 | 0 | 0 |
| 0 | 0 | 0 | 0 | 0 |
| 0 | 0 | 0 | 0 | 0 |
| 0 | 0 | 0 | 0 | 0 |
| 0 | 0 | 0 | 0 | 0 |
| 0.007891305 | 0 | 0.002753702 | 0 | 0.002300763 |
| 0 | 0 | 0 | 0 | 0 |

| | | | | |
|---|---|---|---|---|
| PHL018-0200_V1 | PHL018-0055_V1 | PHL018-0202_V1 | PHL018-0203_V1 | PHL018-0197_V3 |
| EYCF | EYCF | REF | EYCF | EYCF |
| 0.13497512 | 4.118229838 | 1.570632103 | 0.148722909 | 0.313878288 |
| 6.424331962 | 0.10968514 | 7.417481908 | 0 | 0.00411362 |
| 0 | 1.110639151 | 5.441638177 | 0 | 0 |
| 0 | 0 | 0 | 0 | 0 |
| 0 | 0 | 0 | 0 | 0 |
| 0 | 0 | 0 | 0 | 0 |
| 0 | 0 | 0 | 0 | 0 |
| 0 | 0 | 0 | 0 | 0 |
| 0 | 0 | 0 | 0 | 0 |
| 0 | 0 | 0 | 0 | 0 |
| 0 | 0 | 0 | 0 | 0 |
| 0 | 0 | 0 | 0 | 0 |
| 0 | 0 | 0 | 0 | 0 |
| 3.466189661 | 0.053269205 | 0.180108315 | 0 | 0.927801857 |
| 0 | 0 | 0 | 0 | 0 |
| 0 | 0 | 0 | 0 | 0 |
| 0 | 0 | 0 | 0 | 0 |
| 0 | 0 | 0 | 0 | 0 |
| 0 | 0.104138332 | 0 | 0 | 0 |
| 0 | 0 | 0 | 0 | 0 |
| 0 | 0 | 0 | 0 | 0 |
| 0 | 0 | 0 | 0 | 0 |
| 0 | 0 | 0 | 0 | 0 |
| 0 | 0 | 0 | 0 | 0 |
| 0 | 0 | 0 | 0 | 0 |
| 0 | 0 | 0 | 0 | 0 |
| 0 | 0 | 0 | 0 | 0 |
| 0 | 0 | 0 | 0 | 0 |
| 0 | 0 | 0 | 0 | 0 |
| 0 | 0 | 0 | 0 | 0 |
| 0 | 0 | 0 | 0 | 0 |
| 0 | 0 | 0 | 0 | 0 |
| 0 | 0 | 0 | 0 | 0 |
| 0 | 0 | 0 | 0 | 0 |
| 0 | 0 | 0 | 0 | 0 |
| 7.293593093 | 0.401872416 | 0 | 0 | 0 |
| 0 | 0 | 0 | 0 | 0 |
| 0 | 0 | 0 | 0 | 0 |
| 0 | 0 | 0 | 0 | 0 |
| 0 | 0 | 0 | 0 | 0 |
| 0 | 0 | 0 | 0 | 0 |
| 0.05743151 | 0.001194654 | 0.185143952 | 0.007522701 | 0.003168296 |
| 0 | 0.363956508 | 0.227157231 | 0.011620168 | 0.000801357 |
| 0.025897686 | 0.009990573 | 0.061406344 | 0.0063439 | 0.004156174 |
| 0 | 0.014123236 | 0.020372866 | 0 | 0 |
| 0 | 0 | 0 | 0 | 0 |

| | | | | |
|---|---|---|---|---|
| 0 | 0.059305027 | 0 | 0 | 0 |
| 0.026484847 | 0 | 0.052631183 | 0.064341813 | 0.290100242 |
| 0 | 0.831079073 | 0 | 0.043459248 | 0.074371116 |
| 0.751779122 | 0.115124615 | 1.141320274 | 0 | 0.103234764 |
| 0.00056119 | 0.006303702 | 0 | 0.002394255 | 0.016470133 |
| 0 | 0 | 0 | 0 | 0 |
| 0.516652199 | 0.001889595 | 0 | 0 | 0 |
| 0 | 0 | 0 | 0 | 0 |
| 0 | 0 | 0 | 0 | 0 |
| 0 | 0 | 0 | 0 | 0 |
| 0 | 0 | 0 | 0 | 0 |
| 0 | 0.383486718 | 0.0163826 | 0 | 0 |
| 0 | 0 | 0 | 0 | 0 |
| 0.022266434 | 0.008699585 | 0 | 0 | 0.313947836 |
| 0 | 0 | 0 | 0 | 0 |
| 0 | 0 | 0 | 0 | 0 |
| 0.000796302 | 0.000795081 | 0 | 0 | 0 |
| 0 | 0 | 0 | 0 | 0 |
| 0 | 0 | 0 | 0 | 0 |
| 0 | 0 | 0 | 0 | 0 |
| 0.422699362 | 0.040243958 | 0 | 0 | 0.059053349 |
| 0.011910047 | 0.015536028 | 0 | 0.033300127 | 0.053071533 |
| 0 | 0 | 0 | 0 | 0 |
| 0 | 0 | 0 | 0 | 0 |
| 0 | 0 | 0 | 0 | 0 |
| 0 | 0 | 0 | 0 | 0 |
| 0 | 0 | 0 | 0 | 0 |
| 0 | 0 | 0 | 0 | 0 |
| 0 | 0 | 0 | 0 | 0 |
| 0 | 0 | 0 | 0.081569902 | 0 |
| 0 | 0 | 0.003818336 | 0 | 0.092221193 |
| 0 | 0 | 0 | 0 | 0 |
| 0 | 0 | 0 | 0 | 0 |
| 0 | 0.000532018 | 0 | 0 | 0 |
| 0 | 0 | 0 | 0 | 0 |
| 0 | 0 | 0 | 0 | 0 |
| 0 | 0 | 0 | 0 | 0 |
| 0 | 0 | 0 | 0.003138335 | 0 |
| 0 | 0 | 0 | 0 | 0 |
| 0 | 0 | 0 | 0 | 0 |
| 0 | 0 | 0 | 0 | 0 |
| 0 | 0 | 0 | 0 | 0 |
| 0 | 0 | 0 | 0 | 0 |
| 0 | 0 | 0 | 0 | 0 |
| 0 | 0 | 0 | 0 | 0 |
| 0.010665831 | 0 | 0 | 0 | 0 |
| 0 | 0 | 0 | 0 | 0.001677048 |
| 0 | 0 | 0 | 0 | 0 |

| | | | | |
|---|---|---|---|---|
| 0 | 0 | 0 | 0 | 0 |
| 0 | 0 | 0 | 0 | 0 |
| 0 | 0 | 0 | 0 | 0 |
| 0 | 0 | 0 | 0 | 0 |
| 0 | 0 | 0 | 0 | 0.000965004 |
| 0 | 0 | 0 | 0.000848294 | 0 |
| 0 | 0 | 0 | 0 | 0 |
| 0 | 0 | 0 | 0 | 0 |
| 0 | 0 | 0 | 0.000833865 | 0 |
| 0 | 0 | 0 | 0 | 0 |
| 0 | 0 | 0 | 0 | 0 |
| 0.001724542 | 0 | 0.027067561 | 0 | 0 |
| 0 | 0 | 0 | 0 | 0 |
| 0 | 0 | 0 | 0 | 0 |
| 0 | 0 | 0 | 0 | 0 |
| 0 | 0 | 0 | 0 | 0 |
| 0 | 0 | 0 | 0 | 0 |
| 0 | 0 | 0 | 0 | 0 |
| 0 | 0.532128237 | 0.266748736 | 0 | 0 |
| 0 | 0 | 0 | 0 | 0 |
| 0 | 0 | 0 | 0 | 0 |
| 0 | 0 | 0 | 0 | 0 |
| 0 | 0 | 0 | 0 | 0 |
| 0 | 0 | 0 | 0 | 0 |
| 0 | 0 | 0 | 0 | 0 |
| 0 | 0 | 0 | 0 | 0 |
| 0 | 0 | 0 | 0 | 0 |
| 0 | 0 | 0 | 0 | 0 |
| 0 | 0 | 0 | 0 | 0 |
| 0 | 0 | 0 | 0 | 0 |
| 0 | 0.012695201 | 0 | 0 | 0 |
| 0 | 0.000719912 | 0 | 0 | 0 |
| 0 | 0 | 0 | 0 | 0 |
| 0 | 0 | 0 | 0 | 0 |
| 0 | 0 | 0 | 0 | 0 |
| 0 | 0 | 0 | 0 | 0 |
| 0 | 0 | 0 | 0 | 0 |
| 0 | 0 | 0 | 0 | 0 |
| 0 | 0 | 0 | 0 | 0 |
| 0 | 0 | 0 | 0 | 0 |
| 0 | 0 | 0 | 0 | 0 |
| 0 | 0 | 0 | 0 | 0 |
| 0 | 0 | 0.011733862 | 0 | 0 |
| 0 | 0 | 0 | 0 | 0 |
| 0 | 0 | 0 | 0 | 0 |
| 0 | 0 | 0 | 0 | 0 |
| 0 | 0 | 0 | 0 | 0 |
| 0 | 0 | 0 | 0 | 0 |

| | | | | |
|---|---|---|---|---|
| 0 | 0 | 0 | 0 | 0 |
| 0 | 0 | 0 | 0 | 0 |
| 0 | 0 | 0 | 0 | 0 |
| 0 | 0 | 0 | 0 | 0 |
| 0 | 0 | 0 | 0 | 0 |
| 0 | 0 | 0 | 0 | 0 |
| 0 | 0 | 0 | 0 | 0 |
| 0 | 0 | 0 | 0 | 0 |
| 0 | 0 | 0 | 0 | 0 |
| 0 | 0 | 0 | 0 | 0 |
| 0 | 0 | 0 | 0 | 0 |
| 0 | 0 | 0 | 0 | 0 |
| 0 | 0 | 0 | 0 | 0 |
| 0 | 0 | 0 | 0 | 0 |
| 0 | 0 | 0 | 0 | 0 |
| 0 | 0 | 0 | 0 | 0 |
| 0 | 0 | 0 | 0 | 0 |
| 0 | 0 | 0 | 0 | 0 |
| 0 | 0 | 0 | 0 | 0 |
| 0 | 0 | 0 | 0 | 0 |
| 0 | 0 | 0 | 0.019563627 | 0 |
| 0 | 0 | 0 | 0 | 0 |
| 0 | 0 | 0 | 0 | 0 |
| 0 | 0 | 0 | 0 | 0 |
| 0 | 0 | 0 | 0 | 0 |
| 0 | 0 | 0 | 0 | 0 |
| 0 | 0 | 0 | 0 | 0 |
| 0 | 0 | 0 | 0 | 0 |
| 0 | 0 | 0 | 0 | 0 |
| 0 | 0 | 0 | 0 | 0 |
| 0 | 0 | 0 | 0 | 0 |
| 0 | 0 | 0 | 0 | 0 |
| 0 | 0 | 0 | 0 | 0 |
| 0 | 0 | 0 | 0 | 0 |
| 0 | 0 | 0 | 0 | 0 |
| 0 | 0 | 0 | 0 | 0 |
| 0 | 0 | 0 | 0 | 0 |
| 0 | 0 | 0 | 0 | 0 |
| 0 | 0 | 0 | 0 | 0 |
| 0 | 0 | 0 | 0 | 0 |
| 0 | 0 | 0 | 0 | 0 |
| 0 | 0 | 0 | 0 | 0 |
| 0 | 0 | 0 | 0 | 0 |
| 0 | 0 | 0 | 0 | 0 |
| 0 | 0 | 0 | 0 | 0 |
| 0 | 0 | 0 | 0 | 0 |
| 0 | 0 | 0 | 0 | 0 |
| 0 | 0 | 0 | 0 | 0 |

| | | | | |
|---|---|---|---|---|
| 0 | 0 | 0 | 0 | 0 |
| 0 | 0 | 0 | 0 | 0 |
| 0 | 0 | 0 | 0 | 0 |
| 0 | 0 | 0 | 0 | 0 |
| 0 | 0 | 0 | 0 | 0 |
| 0 | 0 | 0 | 0 | 0 |
| 0 | 0 | 0 | 0 | 0 |
| 0 | 0 | 0 | 0 | 0 |
| 0 | 0 | 0 | 0 | 0 |
| 0 | 0 | 0 | 0 | 0 |
| 0 | 0 | 0 | 0 | 0 |
| 0 | 0 | 0 | 0 | 0 |
| 0 | 0.001390943 | 0 | 0.006434985 | 0.007377735 |
| 0 | 0 | 0 | 0 | 0 |
| 0 | 0.214976603 | 0 | 0 | 0 |
| 0 | 0 | 0 | 0 | 0 |
| 0 | 0 | 0 | 0 | 0 |
| 0 | 0 | 0 | 0 | 0 |
| 0 | 0 | 0 | 0 | 0 |
| 0 | 0 | 0 | 0 | 0 |
| 0.018605188 | 0.002229197 | 0.023704984 | 0.028074376 | 0 |
| 0 | 0 | 0 | 0 | 0 |
| 0 | 0 | 0 | 0 | 0 |
| 0 | 0 | 0.012538892 | 0 | 0 |
| 0 | 0 | 0 | 0 | 0 |
| 0 | 0 | 0 | 0 | 0 |
| 0.009968405 | 0.006707531 | 0.000800303 | 0.00350354 | 0 |
| 0 | 0 | 0 | 0 | 0 |
| 0 | 0 | 0 | 0 | 0 |
| 0.024575973 | 0 | 0 | 0.00583082 | 0 |
| 1.095640988 | 0 | 0.250498531 | 0 | 0 |
| 0 | 0 | 0 | 0 | 0.012909246 |
| 0 | 0 | 0 | 0 | 0 |
| 0 | 0.005676445 | 0 | 0 | 0 |
| 0.008715979 | 0.007735651 | 0.014753129 | 0.0319667 | 0.003846641 |
| 0 | 0 | 0 | 0 | 0 |
| 0 | 0 | 0 | 0 | 0 |
| 0 | 0 | 0 | 0 | 0 |
| 0 | 0 | 0 | 0 | 0 |
| 0 | 0 | 0 | 0 | 0 |
| 0 | 0 | 0 | 0 | 0 |
| 0 | 0 | 0 | 0 | 0 |
| 0 | 0 | 0 | 0 | 0 |
| 0 | 0 | 0 | 0 | 0 |
| 0 | 0 | 0 | 0 | 0 |
| 0 | 0.05938178 | 0 | 0 | 0 |
| 0 | 0 | 0 | 0 | 0.009949459 |
| 0.001552811 | 0.006718522 | 0 | 0.002479492 | 0.008528242 |

| | | | | |
|---|---|---|---|---|
| 0 | 0 | 0 | 0 | 0 |
| 0 | 0 | 0 | 0 | 0 |
| 0 | 0 | 0 | 0 | 0 |
| 0 | 0 | 0 | 0 | 0 |
| 0 | 0 | 0 | 0 | 0 |
| 0 | 0 | 0 | 0 | 0 |
| 0 | 0 | 0 | 0 | 0 |
| 0 | 0 | 0.002518886 | 0 | 0 |
| 0 | 0 | 0 | 0 | 0 |
| 0 | 0 | 0 | 0 | 0 |
| 0 | 0 | 0 | 0 | 0 |
| 0 | 0 | 0 | 0 | 0 |
| 0 | 0 | 0 | 0 | 0 |
| 0 | 0 | 0 | 0 | 0 |
| 0.000816089 | 0 | 0 | 0 | 0 |
| 0 | 0 | 0 | 0 | 0 |
| 0 | 0 | 0 | 0 | 0 |
| 0 | 0 | 0 | 0 | 0 |
| 0 | 0.00385434 | 0.00081001 | 0 | 0 |
| 0 | 0.017667584 | 0 | 0 | 0 |
| 0.003623632 | 0 | 0 | 0.702085218 | 0.077219735 |
| 0 | 0 | 0 | 0 | 0 |
| 0 | 0 | 0 | 0 | 0 |
| 0 | 0 | 0 | 0 | 0 |
| 0.005142223 | 0 | 0.006171927 | 0 | 0.00408497 |
| 0 | 0.003238486 | 0 | 0 | 0 |
| 0 | 0 | 0 | 0 | 0 |
| 0 | 0 | 0 | 0 | 0 |
| 0 | 0 | 0.004757275 | 0 | 0 |
| 0 | 0 | 0 | 0 | 0 |
| 0.04453447 | 0.005890814 | 0.717089796 | 0.09760753 | 0.846486992 |
| 0 | 0 | 0 | 0 | 0 |
| 0 | 0 | 0 | 0 | 0.162925449 |
| 0 | 0.023601743 | 0 | 0 | 0 |
| 0 | 0 | 0 | 0 | 0 |
| 0 | 0 | 0 | 0 | 0 |
| 0 | 0 | 0 | 0 | 0 |
| 0 | 0.002392074 | 0.00516113 | 0 | 0 |
| 0 | 0 | 0 | 0 | 0 |
| 0 | 0.022966987 | 0 | 0 | 0 |
| 0 | 0 | 0 | 0 | 0 |
| 0 | 0 | 0 | 0.01986408 | 0 |
| 0 | 0 | 0 | 0.001461991 | 0.014367241 |
| 0 | 0 | 0 | 0 | 0 |
| 0 | 0 | 0 | 0 | 0 |
| 0 | 0 | 0 | 0 | 0 |
| 0 | 0 | 0 | 0 | 0 |
| 0 | 0 | 0 | 0 | 0 |

| | | | | |
|---|---|---|---|---|
| 0 | 0 | 0 | 0 | 0 |
| 0 | 0 | 0 | 0 | 0 |
| 0.016320943 | 0.011674677 | 0.067920976 | 0.004157104 | 0.005590354 |
| 0 | 0 | 0 | 0 | 0 |
| 0.007187583 | 0 | 0 | 0 | 0 |
| 0 | 0 | 0 | 0 | 0 |
| 0 | 0 | 0 | 0 | 0 |
| 0 | 0 | 0 | 0 | 0 |
| 0 | 0 | 0 | 0 | 0 |
| 0 | 0 | 0 | 0 | 0 |
| 0 | 0 | 0 | 0 | 0 |
| 0 | 0 | 0 | 0 | 0 |
| 0 | 0 | 0 | 0 | 0 |
| 0.000807653 | 0 | 0.006462545 | 0 | 0.007604124 |
| 0.003999672 | 0.004880988 | 0.003077299 | 0.0051891 | 0.000980658 |
| 0 | 0 | 0 | 0 | 0 |
| 0 | 0 | 0.003506306 | 0.001819235 | 0.001787794 |
| 0.013747804 | 0 | 0 | 0 | 0 |
| 0 | 0 | 0 | 0 | 0 |
| 0 | 0 | 0 | 0 | 0 |
| 0 | 0 | 0 | 0 | 0 |
| 0.001828405 | 0.004335801 | 0 | 0.001026406 | 0.001210401 |
| 0 | 0 | 0 | 0 | 0 |
| 0 | 0 | 0.041544008 | 0.030232116 | 0.038466154 |
| 0 | 0 | 0 | 0 | 0 |
| 0 | 0 | 0 | 0 | 0 |
| 0 | 0 | 0 | 0 | 0 |
| 0 | 0 | 0 | 0 | 0 |
| 0 | 0 | 0 | 0 | 0 |
| 0 | 0 | 0.004883714 | 0 | 0 |
| 0 | 0 | 0 | 0 | 0 |
| 0 | 0 | 0 | 0 | 0 |
| 0 | 0 | 0 | 0 | 0 |
| 0.001054073 | 0 | 0 | 0 | 0 |
| 0 | 0 | 0 | 0 | 0 |
| 0.005021472 | 0 | 0.001264405 | 0 | 0 |
| 0 | 0 | 0 | 0 | 0 |
| 0.000861451 | 0.285562877 | 0.566552639 | 0.000773744 | 0.003041487 |
| 0 | 0 | 0 | 0 | 0 |
| 0 | 0 | 0 | 0 | 0 |
| 0 | 0 | 0 | 0 | 0 |
| 0 | 0 | 0 | 0 | 0 |
| 0 | 0 | 0 | 0 | 0 |
| 0 | 0 | 0 | 0 | 0.012593342 |
| 0 | 0 | 0 | 0 | 0 |
| 0 | 0 | 0 | 0 | 0 |
| 0 | 0 | 0 | 0 | 0 |
| 0 | 0 | 0 | 0.920953234 | 0.163017165 |

| | | | | |
|---|---|---|---|---|
| 0 | 0 | 0 | 0 | 0.139446456 |
| 0 | 0 | 0 | 0 | 0 |
| 0 | 0.001912174 | 0.021659919 | 0 | 0 |
| 0 | 0 | 0 | 0 | 0 |
| 0.003853575 | 0 | 0.003557872 | 0.004326539 | 0 |
| 0 | 0 | 0 | 0 | 0 |
| 0 | 0 | 0 | 0 | 0 |
| 0 | 0.002359589 | 0.013714638 | 0.209835042 | 0 |
| 0 | 0 | 0 | 0 | 0 |
| 0 | 0.018761756 | 0.294927821 | 0.064384384 | 0.462251728 |
| 0 | 0 | 0 | 0 | 0 |
| 0.028060938 | 0.000800511 | 0.045400144 | 0.00216034 | 0.001415336 |
| 0.003851611 | 0.003845702 | 0.01516004 | 0.187539522 | 0.000894652 |
| 0 | 0 | 0 | 0.164257819 | 0.009511223 |
| 0.004169783 | 0 | 0 | 0.056178678 | 0 |
| 0.006999151 | 0 | 0 | 0 | 0 |
| 0 | 0 | 0 | 0 | 0 |
| 0 | 0 | 0 | 0 | 0 |
| 0 | 0 | 0 | 0 | 0.249999627 |
| 0 | 0 | 0 | 0 | 0 |
| 0 | 0 | 0 | 0 | 0 |
| 0 | 0 | 0 | 0.003939247 | 0 |
| 0.004183542 | 0 | 0 | 0 | 0 |
| 0 | 0 | 0 | 0 | 0 |
| 0 | 0 | 0 | 0 | 0 |
| 0.010625079 | 0.007308269 | 0.003051928 | 0 | 0 |
| 0 | 0 | 0 | 0 | 0 |
| 0 | 0 | 0 | 0 | 0 |
| 0 | 0 | 0 | 0 | 0.003840842 |
| 0 | 0 | 0 | 0.004145742 | 0 |
| 0 | 0 | 0 | 0.035955495 | 0 |
| 0 | 0 | 1.436162405 | 0 | 0 |
| 0 | 0 | 0 | 0 | 0 |
| 0 | 0 | 0 | 0 | 0 |
| 0 | 0 | 0 | 0 | 0 |
| 0 | 0 | 0.034927228 | 0 | 0 |
| 0 | 0 | 0 | 0 | 0.115763601 |
| 0 | 0 | 0 | 0 | 0 |
| 0 | 0.0020986 | 0 | 0.000943916 | 0 |
| 0 | 0 | 0 | 0 | 0.375476356 |
| 0 | 0 | 0 | 0 | 0 |
| 0.00249567 | 0 | 0 | 0 | 0 |
| 0 | 0 | 0 | 0 | 0 |
| 0.903549064 | 0.000949645 | 0.039339847 | 2.469693221 | 0 |
| 0.057040503 | 0 | 0 | 0 | 0 |
| 0 | 0 | 0 | 0 | 0 |
| 0 | 0.015245076 | 0.001355469 | 0 | 0.008552685 |
| 0 | 0.002661065 | 0 | 0 | 0 |

| | | | | |
|---|---|---|---|---|
| 0 | 0 | 0 | 0 | 0 |
| 0.066941885 | 0 | 0 | 0.001178948 | 0 |
| 0 | 0 | 0 | 0 | 0 |
| 0 | 0.003470792 | 0 | 0 | 0 |
| 0 | 0 | 0 | 0 | 0 |
| 0.023820323 | 0.001616373 | 0 | 0 | 0.025107937 |
| 0.001285771 | 0 | 0 | 0.050352031 | 0 |
| 0.001196452 | 0 | 0 | 0.004835869 | 0 |
| 0 | 0 | 0 | 0 | 0 |
| 0 | 0 | 0 | 0 | 0 |
| 0.001341144 | 0.005356345 | 0.007924682 | 0.017346212 | 0 |
| 18.58310861 | 0 | 0 | 0 | 0 |
| 0 | 0 | 0 | 0 | 0 |
| 0.001013636 | 0 | 0 | 0.00182087 | 0 |
| 0 | 0 | 0 | 0 | 0 |
| 0 | 0 | 0 | 0 | 0 |
| 0 | 0 | 0 | 0.005723272 | 0 |
| 0 | 0 | 0 | 0 | 0 |
| 0 | 0 | 0 | 2.5882471 | 0 |
| 0 | 0 | 0 | 0.00928078 | 0.403577076 |
| 0 | 0 | 0 | 0 | 0 |
| 0.334926003 | 0 | 0.002645214 | 0 | 0 |
| 0 | 0 | 0 | 0 | 0 |
| 0 | 0 | 0 | 0 | 0 |
| 0 | 0 | 0 | 0 | 0 |
| 0 | 0 | 0 | 0 | 0 |
| 0 | 0 | 0 | 0 | 0 |
| 0 | 0 | 0 | 0 | 0 |
| 0 | 0 | 0 | 0 | 0 |
| 1.079789463 | 0.496230382 | 2.260330176 | 0 | 0.005901495 |
| 0 | 0.019911881 | 0.108568473 | 0 | 0 |
| 0.001448204 | 0 | 0 | 0.000650379 | 0 |
| 0 | 0 | 0 | 0 | 0.001168374 |
| 0.000973979 | 0 | 0 | 0 | 0 |
| 0 | 0 | 0 | 0 | 0.00343398 |
| 0 | 0 | 0 | 0 | 0.000697547 |
| 0 | 0 | 0 | 0 | 0 |
| 0.002582913 | 0 | 0 | 0 | 0 |
| 0 | 0 | 0 | 0 | 0 |
| 0 | 0 | 0 | 0 | 0 |
| 0.027082877 | 0.001040051 | 0 | 0.039107901 | 0.005884329 |
| 5.59734701 | 0.034169432 | 6.434485579 | 0 | 0.001265193 |
| 0.001183058 | 0 | 0 | 0 | 0 |
| 0 | 0 | 0 | 0 | 0 |
| 0 | 0 | 0 | 0 | 0 |
| 0 | 0 | 0 | 0 | 0 |
| 0.001093285 | 0 | 0.006863869 | 0 | 0 |
| 0 | 0.003065258 | 0 | 0 | 0 |

| | | | | |
|---|---|---|---|---|
| 0 | 0.041213955 | 0.008717947 | 0 | 0 |
| 0 | 0 | 0 | 0 | 0 |
| 0.006616878 | 0 | 0 | 0.011226033 | 0 |
| 0 | 0 | 0 | 0 | 0 |
| 0 | 0.004332555 | 0 | 0 | 0 |
| 0 | 0.006149371 | 0 | 0 | 0.001208038 |
| 0 | 0 | 0 | 0 | 0 |
| 0 | 0 | 0 | 0 | 0 |
| 0.043802644 | 0 | 0 | 0.152910293 | 0 |
| 0 | 0 | 0 | 0 | 0.034346866 |
| 0 | 0.091701076 | 0 | 0 | 0 |
| 0 | 0 | 0 | 0.019411124 | 0.009236632 |
| 0 | 0 | 0 | 0 | 0 |
| 0.03270521 | 0 | 0 | 0.039240267 | 0 |
| 0 | 0 | 0 | 0 | 0 |
| 0.001525884 | 0.075796263 | 0 | 0 | 0.010269679 |
| 0.001902694 | 0.176890123 | 0.181134375 | 0.01215271 | 0 |
| 0 | 0 | 0 | 0.028145913 | 0.163009573 |
| 0 | 0 | 0 | 0.022948146 | 0.015998105 |
| 0 | 0 | 0 | 0 | 0 |
| 0 | 0.003458371 | 0 | 0 | 0 |
| 0 | 0 | 0.00203767 | 0 | 0.006926445 |
| 0 | 0 | 0 | 0 | 0 |
| 0 | 0 | 0 | 0 | 0 |
| 0 | 0 | 0 | 0 | 0 |
| 0 | 0 | 0 | 0.004752688 | 0.006882915 |
| 0.002147098 | 0 | 0 | 0.318201743 | 0.000710687 |
| 0 | 0 | 0 | 0 | 0 |
| 0 | 0 | 0.007811185 | 0 | 0 |
| 0 | 0 | 0 | 0 | 0 |
| 0.009388955 | 0.003872097 | 0 | 0.049131613 | 0.073144383 |
| 0.003955894 | 0.005165156 | 0.00365234 | 0 | 0 |
| 0.219908361 | 0 | 0.004280027 | 0 | 0 |
| 0 | 0 | 0.00191678 | 0.004475316 | 0.002931981 |
| 0 | 0.00355086 | 0 | 0 | 0 |
| 0 | 0 | 0 | 0 | 0 |
| 0 | 0 | 0.00072552 | 0.015763153 | 0 |
| 0.007784265 | 0.0047043 | 0.00075652 | 0.013431473 | 0.224750485 |
| 0 | 0 | 3.180195273 | 0 | 0 |
| 0 | 0.02274302 | 0.011133586 | 0 | 0.070516365 |
| 0 | 0 | 0 | 0 | 0 |
| 0 | 0 | 0 | 0 | 0 |
| 0 | 0.001835289 | 0 | 0.100708896 | 0.08707056 |
| 0 | 0.003352957 | 0 | 0.088906379 | 0 |
| 0 | 0 | 0 | 0 | 0 |
| 0 | 0 | 0 | 0 | 0.00453332 |
| 0 | 0 | 0 | 0 | 0 |
| 0 | 0 | 0 | 0 | 0 |

| | | | | |
|---|---|---|---|---|
| 0 | 0 | 0 | 0 | 0 |
| 0.153243936 | 0.33444552 | 1.049925529 | 0.002757849 | 0.001478283 |
| 0 | 0 | 0 | 0 | 0 |
| 0 | 0 | 0 | 0.01032523 | 0 |
| 0 | 0 | 0 | 0.320734101 | 0 |
| 0 | 0 | 0 | 0 | 0 |
| 0.924736838 | 0 | 0.005483631 | 0 | 0 |
| 0 | 0 | 0 | 0 | 0 |
| 0 | 0 | 0 | 0 | 0 |
| 0.059635073 | 0 | 0 | 0 | 0 |
| 0.003526246 | 0.00207108 | 0 | 0.002608306 | 0.050715293 |
| 0.134400755 | 0.066634537 | 0.023105958 | 0.244209135 | 0.003272572 |
| 0 | 0 | 0 | 0 | 0 |
| 0 | 0 | 0 | 0 | 0.003213566 |
| 0 | 0.001188153 | 0 | 0.020735168 | 0 |
| 0 | 0 | 0 | 0.000844992 | 0 |
| 0.014552748 | 0.005638671 | 0.02947909 | 0.002341091 | 0.006518454 |
| 0.006767097 | 0.000844589 | 0.042693474 | 0.55285552 | 0.045793565 |
| 0 | 0 | 0 | 0 | 0 |
| 0 | 0 | 0.037020229 | 0 | 0.019716226 |
| 0.033822997 | 0.008945988 | 0.028332326 | 0.01267484 | 0 |
| 0 | 0 | 0 | 0 | 0 |
| 0 | 0 | 0 | 0 | 0 |
| 0 | 0 | 0 | 0 | 0 |
| 0.014677674 | 0 | 0 | 0 | 0 |
| 0 | 0 | 0 | 0 | 0.005076581 |
| 0.026310024 | 0 | 0 | 0.163036267 | 0 |
| 0.008570587 | 0.007606612 | 0.101549218 | 0 | 0 |
| 0 | 0 | 0 | 0 | 0 |
| 0 | 0 | 0 | 0 | 0.004816026 |
| 0.015837982 | 0 | 0 | 0.001835545 | 0 |
| 0 | 0 | 0 | 0 | 0 |
| 0 | 0 | 0 | 0 | 0.040816871 |
| 0 | 0 | 0 | 0 | 0 |
| 0 | 0 | 0 | 0 | 0 |
| 0.946464818 | 0.031782037 | 0.973907295 | 0 | 0 |
| 0 | 0 | 0 | 0 | 0 |
| 0 | 0 | 0 | 0 | 0 |
| 0.012334908 | 0 | 0.020336415 | 0 | 0 |
| 0 | 0 | 0 | 0 | 0 |
| 0 | 0 | 0 | 0 | 0 |
| 0 | 0 | 0 | 0 | 0.163999744 |
| 0.086162732 | 0 | 0.051551723 | 0.05800263 | 0.004723774 |
| 0.002583494 | 0 | 0 | 0 | 0 |
| 0 | 0.000677781 | 0 | 0 | 0 |
| 0 | 0.005152693 | 0 | 0 | 0 |
| 0 | 0 | 0 | 0 | 0 |
| 0 | 0 | 0 | 0.021254786 | 0 |

| | | | | |
|---|---|---|---|---|
| 0 | 0.006220835 | 0 | 0 | 0 |
| 0 | 0 | 0 | 0 | 0 |
| 0 | 0 | 0 | 0 | 0 |
| 0.004363144 | 0.855316394 | 0.005946597 | 0 | 0 |
| 0 | 0 | 0 | 0 | 0 |
| 0 | 0 | 0 | 0 | 0 |
| 0 | 0 | 0 | 0 | 0 |
| 0.003717429 | 0 | 0 | 0 | 0 |
| 0 | 0 | 0 | 0 | 0 |
| 0 | 0.001668684 | 0 | 0 | 0 |
| 0 | 0 | 0 | 0 | 0 |
| 0 | 0 | 0 | 0 | 0.026707523 |
| 0 | 0 | 0 | 0 | 0 |
| 0 | 0 | 0 | 0 | 0 |
| 0.008377709 | 0 | 0.130885756 | 0 | 0 |
| 0 | 0 | 0 | 0 | 0 |
| 0 | 0.087931603 | 0 | 0 | 0 |
| 0 | 0 | 0 | 0 | 0 |
| 0 | 0 | 0 | 0.002737752 | 0 |
| 0 | 0 | 0 | 0 | 0 |
| 0 | 0 | 0 | 0 | 0 |
| 0 | 0 | 0 | 0 | 0 |
| 0 | 0 | 0 | 0 | 0 |
| 0.26045144 | 0.040445488 | 0.003067911 | 0.29319931 | 0.037430638 |
| 0 | 0 | 0 | 0 | 0.023524503 |
| 0 | 0 | 0 | 0 | 0 |
| 0 | 0 | 0 | 0 | 0 |
| 0 | 0 | 0 | 0 | 0 |
| 0 | 0 | 0 | 0 | 0 |
| 0 | 0 | 0 | 0 | 0 |
| 0 | 0.079996407 | 0 | 0 | 0 |
| 0 | 0 | 0 | 0 | 0 |
| 0 | 0.006470672 | 0 | 0.003174984 | 0 |
| 0 | 0 | 0 | 0 | 0 |
| 0 | 0 | 0 | 0 | 0 |
| 0 | 0 | 0 | 0 | 0 |
| 0.015470154 | 0.247989084 | 0 | 0 | 0 |
| 0 | 0 | 0 | 0 | 0 |
| 0 | 0 | 0 | 0 | 0 |
| 0 | 0 | 0 | 0 | 0 |
| 0.001718704 | 0.010677751 | 1.550851858 | 0.003087435 | 0.134679302 |
| 0 | 0 | 0 | 0 | 0 |
| 0.108817033 | 0.313598451 | 1.312839195 | 0 | 0.016378865 |
| 0 | 0 | 0 | 0 | 0 |
| 0 | 0 | 0 | 0 | 0 |
| 0 | 0 | 0 | 0 | 0 |
| 0 | 0 | 0 | 0 | 0 |
| 0 | 0.781155078 | 0.013140827 | 0.008660053 | 0.000810513 |

| | | | | |
|---|---|---|---|---|
| 0 | 0 | 0 | 0 | 0 |
| 0 | 0 | 0 | 0 | 0 |
| 0.45531111 | 0.019555591 | 0.034133715 | 1.068737159 | 2.290502281 |
| 0 | 0 | 0 | 0 | 0 |
| 0 | 0 | 0 | 0 | 0 |
| 0.001560217 | 0.046388514 | 0.008963075 | 0 | 0 |
| 0 | 0 | 0 | 0 | 0 |
| 0 | 0 | 0 | 0 | 0 |
| 0 | 0 | 0 | 0 | 0 |
| 0 | 0 | 0 | 0 | 0 |
| 0 | 0.083272442 | 0.478760785 | 0.042306207 | 16.7223252 |
| 1.006806703 | 0.11675135 | 0.338555179 | 0 | 0 |
| 0 | 0 | 0 | 0 | 0 |
| 0.008394632 | 0.386491924 | 0.219596716 | 0.001047215 | 0.022846392 |
| 0.058986352 | 0.00862338 | 0.074140321 | 1.203208289 | 1.996965728 |
| 0 | 0 | 0 | 0 | 0 |
| 0 | 0 | 0 | 0.001072584 | 0 |
| 0 | 0 | 0 | 0 | 0 |
| 0 | 0 | 0 | 0 | 0 |
| 0.001245854 | 0.000710825 | 0 | 0 | 0.035189476 |
| 0.103403978 | 0.013922751 | 0.001337573 | 0.039037245 | 0.001438597 |
| 0 | 0.386761399 | 0.485533983 | 0.818892378 | 0.731306141 |
| 0.059866282 | 0.343617605 | 7.0281304 | 4.481745939 | 36.50826443 |
| 0 | 0 | 0 | 0 | 0 |
| 0 | 0 | 0 | 0 | 0 |
| 0.011718188 | 0 | 0 | 0 | 0 |
| 1.622299563 | 0 | 0 | 0 | 0 |
| 0 | 0 | 0 | 0 | 0 |
| 0 | 0.000687473 | 0 | 0 | 0 |
| 0 | 0 | 0 | 0 | 0 |
| 0 | 0 | 0 | 0 | 0 |
| 0.043254263 | 0.012973441 | 0.079712475 | 0 | 0 |
| 0.407191158 | 0.022599333 | 0 | 0.00298965 | 3.276436765 |
| 0.000676871 | 0 | 0 | 0 | 0 |
| 0.002343335 | 0.000731169 | 0 | 0.006051165 | 0.001163462 |
| 0.002817149 | 0.153499981 | 0.039757775 | 0.041208178 | 0 |
| 0 | 0 | 0.00412769 | 0 | 0 |
| 0 | 0 | 0 | 0 | 0 |
| 0 | 0 | 0 | 0 | 0 |
| 0 | 0 | 0 | 0 | 0 |
| 0 | 0 | 0 | 0 | 0 |
| 0 | 0 | 0 | 0 | 0 |
| 0 | 0 | 0 | 0 | 0 |
| 0 | 0 | 0 | 0 | 0 |
| 0 | 0.337267769 | 0.291315144 | 0.036275491 | 0.941243563 |
| 0 | 0 | 0 | 0 | 0 |
| 0 | 0 | 0 | 0 | 0 |
| 0.007197023 | 5.415189673 | 0.176341755 | 0.127545077 | 0 |

| | | | | |
|---|---|---|---|---|
| 0 | 0 | 0 | 0 | 0 |
| 0 | 0 | 0 | 0 | 0 |
| 0 | 0 | 0 | 0 | 0 |
| 0 | 0.010545112 | 0 | 0 | 0 |
| 0 | 0 | 0 | 0 | 0 |
| 0 | 0 | 0 | 0 | 0 |
| 0 | 0 | 0 | 0 | 0 |
| 0.002519565 | 0 | 0 | 0.038956628 | 0.297212332 |
| 0 | 0 | 0 | 0 | 0 |
| 0 | 0 | 0 | 0 | 0 |
| 0 | 0.006524702 | 0.001206657 | 0.132257313 | 0.425291634 |
| 0 | 0 | 0 | 0 | 0 |
| 0 | 0 | 0 | 0 | 0 |
| 0 | 0 | 0 | 0 | 0 |
| 0 | 0 | 0 | 0 | 0 |
| 0 | 0 | 0 | 0 | 0 |
| 0 | 0 | 0 | 0 | 0 |
| 0.001088002 | 0.035124768 | 0.020592553 | 0.03583175 | 0 |
| 0 | 0.003810368 | 0 | 0 | 0 |
| 0 | 0 | 0 | 0 | 0 |
| 0 | 0 | 0 | 0 | 0 |
| 0 | 0 | 0 | 0 | 0 |
| 0 | 0.002713673 | 0 | 0 | 0 |
| 0 | 0 | 0 | 0 | 0 |
| 0 | 0 | 0 | 0 | 0 |
| 0 | 0 | 0 | 0 | 0 |
| 0 | 0 | 0 | 0 | 0 |
| 0 | 0 | 0 | 0 | 0 |
| 0 | 0 | 0 | 0 | 0 |
| 0.44313715 | 0 | 0 | 0 | 0 |
| 0 | 0.029291896 | 0 | 0 | 0.082529686 |
| 0 | 0 | 0 | 0 | 0 |
| 0 | 0 | 0 | 0 | 0 |
| 0 | 0 | 0 | 0 | 0 |
| 0 | 0.032858124 | 0.000959475 | 0 | 0 |
| 0 | 0 | 0 | 0 | 0 |
| 0 | 0 | 0 | 0 | 0 |
| 0.001106352 | 0.016385713 | 0 | 0.010433968 | 0.031900224 |
| 0 | 0 | 0 | 0 | 0 |
| 0 | 0 | 0 | 0 | 0 |
| 0 | 0 | 0 | 0 | 0 |
| 0 | 0 | 0 | 0 | 0 |
| 0 | 0 | 0 | 0 | 0 |
| 0 | 0 | 0 | 0 | 0 |
| 0.07181571 | 0.061043774 | 0 | 0 | 0 |
| 0 | 0 | 0 | 0 | 0 |
| 0 | 0.000874673 | 0 | 0 | 0 |
| 0 | 0 | 0 | 0 | 0 |

| | | | | |
|---|---|---|---|---|
| 0 | 0 | 0 | 0 | 0 |
| 0 | 0 | 0 | 0 | 0 |
| 0 | 0 | 0 | 0 | 0 |
| 0 | 0.012453155 | 0 | 0 | 0 |
| 0 | 0 | 0 | 0 | 0 |
| 0 | 0 | 0 | 0 | 0 |
| 0 | 0 | 0 | 0 | 0 |
| 0 | 0 | 0 | 0 | 0 |
| 0.013986908 | 0.005840097 | 0.070437963 | 0.238009077 | 0.140068058 |
| 0 | 0 | 0.000756312 | 0 | 0 |
| 0 | 0 | 0 | 0 | 0 |
| 0 | 0 | 0 | 0 | 0 |
| 0 | 0 | 0 | 0 | 0 |
| 0 | 0 | 0 | 0 | 0 |
| 0 | 0.287940241 | 0 | 0 | 0 |
| 0 | 0.050295179 | 0 | 0 | 0.009342203 |
| 0 | 0.281624443 | 0 | 0 | 0.00205414 |
| 0 | 0 | 0 | 0 | 0 |
| 0 | 0 | 0 | 0 | 0 |
| 0 | 0 | 0 | 0 | 0 |
| 0 | 0 | 0 | 0 | 0 |
| 0 | 0 | 0 | 0 | 0 |
| 0 | 0 | 0 | 0 | 0 |
| 0 | 0 | 0 | 0 | 0 |
| 0 | 0.219769197 | 0.006350533 | 0 | 0 |
| 0.507374798 | 0.995180429 | 0.619904319 | 0 | 0 |
| 0 | 0.006441556 | 0 | 0 | 0 |
| 0 | 0 | 0 | 0 | 0 |
| 0 | 5.74804789 | 0 | 0 | 0 |
| 0 | 0 | 0 | 0 | 0 |
| 0 | 0 | 0 | 0 | 0 |
| 0 | 0 | 0 | 0 | 0 |
| 0 | 0 | 0 | 0 | 0.001278268 |
| 0.000798582 | 0 | 0 | 0.001721462 | 0 |
| 0.005789565 | 0.204951469 | 0.027941364 | 0 | 0 |
| 0 | 0.000928691 | 0.001889241 | 0.00618211 | 0.002955536 |
| 0 | 0 | 0 | 0 | 0 |
| 0 | 0.093749577 | 0 | 0 | 0.002781092 |
| 0 | 3.017623347 | 0 | 0 | 0 |
| 0 | 0 | 0 | 0 | 0 |
| 0 | 0 | 0 | 0 | 0 |
| 0 | 0 | 0 | 0 | 0 |
| 0.408506867 | 0.053554889 | 0.112655199 | 0 | 0 |
| 0 | 0 | 0 | 0 | 0 |
| 0 | 0 | 0 | 0 | 0 |
| 0.001057404 | 0 | 0 | 0 | 0 |
| 0 | 0 | 0 | 0 | 0 |
| 0.004492192 | 0.046815318 | 0.757175224 | 0 | 0 |

| | | | | |
|---|---|---|---|---|
| 0 | 0 | 0 | 0 | 0 |
| 0 | 0 | 0 | 0 | 0 |
| 0 | 0 | 0 | 0 | 0 |
| 0 | 0 | 0 | 0 | 0 |
| 0 | 0.001248761 | 0.008852772 | 0 | 0.328235279 |
| 0 | 0 | 0 | 0 | 0 |
| 0 | 0 | 0 | 0 | 0 |
| 0 | 0 | 0 | 0 | 0 |
| 0 | 0 | 0 | 0 | 0 |
| 0 | 0 | 0 | 0 | 0 |
| 0 | 0 | 0 | 0 | 0 |
| 0 | 1.01439036 | 0 | 0 | 0 |
| 0 | 0.019908404 | 0.582562721 | 0 | 0 |
| 0 | 0 | 0 | 0 | 0 |
| 0.392941709 | 0.000722096 | 0.092144069 | 0.04113968 | 0.052131727 |
| 0 | 0 | 0 | 0 | 0 |
| 0 | 0 | 0 | 0 | 0 |
| 0 | 0 | 0 | 0 | 0 |
| 0 | 0 | 0 | 0 | 0.045320266 |
| 0 | 0 | 0 | 0 | 0 |
| 0 | 0 | 0 | 0 | 0 |
| 0 | 0 | 0 | 0 | 0 |
| 0.028352978 | 0.124315537 | 0 | 0 | 0.273111555 |
| 0 | 0 | 0 | 0 | 0 |
| 0 | 0.002405231 | 0 | 0 | 0 |
| 0 | 0 | 0 | 0 | 0 |
| 0 | 0 | 0 | 0 | 0 |
| 0 | 0 | 0 | 0 | 0 |
| 0 | 0 | 0 | 0 | 0 |
| 0 | 0 | 0 | 0 | 0 |
| 0 | 0 | 0 | 0 | 0 |
| 0.049738401 | 0.017491051 | 0.122457975 | 0.012643695 | 0.014910217 |
| 0 | 0 | 0 | 0 | 0 |
| 0 | 0.02122141 | 0 | 0.021476344 | 0.478797922 |
| 0 | 0.001414908 | 0.002803592 | 0.004000245 | 0.003037676 |
| 0 | 0.904958855 | 0 | 0 | 0 |
| 0 | 0 | 0 | 0 | 0 |
| 0 | 0 | 0 | 0 | 0 |
| 0.001162871 | 0 | 0 | 0 | 0 |
| 0 | 0 | 0 | 0 | 0 |
| 0 | 0 | 0 | 0 | 0 |
| 0 | 0 | 0 | 0 | 0 |
| 0 | 0 | 0 | 0 | 0 |
| 0 | 0.002303731 | 0 | 0 | 0 |
| 0.005384449 | 0.019314454 | 0 | 0.013613127 | 0.032564525 |
| 0 | 0 | 0 | 0 | 0 |
| 0.177522759 | 0 | 0 | 0 | 0 |
| 0 | 0 | 0 | 0 | 0 |

| | | | | |
|---|---|---|---|---|
| 0 | 0 | 0 | 0 | 0 |
| 0 | 0 | 0 | 0 | 0 |
| 0 | 0 | 0 | 0.009077495 | 0 |
| 0 | 0 | 0 | 0 | 0 |
| 0 | 0 | 0 | 0 | 0 |
| 0 | 0 | 0 | 0 | 0 |
| 0 | 0.018135375 | 0 | 0 | 0 |
| 0 | 0 | 0 | 0 | 0 |
| 0 | 0 | 0 | 0 | 0 |
| 0 | 0 | 0 | 0 | 0 |
| 0 | 0 | 0 | 0 | 0 |
| 0 | 0 | 0 | 0 | 0.010558387 |
| 0 | 0 | 0 | 0 | 0 |
| 0 | 0 | 0 | 0 | 0 |
| 0 | 0 | 0 | 0 | 0 |
| 0 | 0 | 0 | 0 | 0 |
| 0 | 0 | 0 | 0 | 0 |
| 0 | 0 | 0 | 0 | 0 |
| 0.006391297 | 2.548195514 | 0 | 0 | 0 |
| 0 | 0 | 0 | 0.000525717 | 0 |
| 0.000937003 | 0 | 0 | 0 | 0 |
| 0 | 0 | 0 | 0 | 0 |
| 0 | 0 | 0 | 0 | 0 |
| 0 | 0 | 0 | 0 | 0 |
| 0 | 0.001305323 | 0 | 0 | 0 |
| 0.002599187 | 0.018351769 | 0.026225726 | 0.022845303 | 0.026711155 |
| 0 | 0 | 0 | 0 | 0 |
| 0.019807539 | 0.006531903 | 0 | 0 | 0 |
| 0 | 0 | 0 | 0 | 0 |
| 0 | 0.027187651 | 0.170670833 | 0 | 0 |
| 0 | 0 | 0 | 0 | 0 |
| 0 | 0 | 0 | 0 | 0 |
| 0 | 0 | 0 | 0 | 0 |
| 0 | 0 | 0 | 0 | 0 |
| 0 | 0.000777031 | 0 | 0.020620254 | 0.0437906 |
| 0 | 0 | 0 | 0 | 0 |
| 0 | 0 | 0 | 0 | 0 |
| 0 | 0.010800858 | 0 | 0.006146919 | 0.09236402 |
| 0 | 0 | 0 | 0 | 0 |
| 0 | 0 | 0 | 0 | 0 |
| 25.23562573 | 0.466802178 | 20.82221956 | 0 | 0 |
| 0.905447716 | 0.10407242 | 1.886635957 | 0 | 0.015405011 |
| 0 | 0 | 0 | 0 | 0 |
| 0 | 0 | 0 | 0 | 0 |
| 0 | 0 | 0 | 0 | 0 |
| 0 | 0 | 0 | 0 | 0 |
| 0 | 0 | 0 | 0 | 0 |
| 0 | 0.082922511 | 0.012132455 | 0 | 0.036343294 |

| | | | | |
|---|---|---|---|---|
| 0 | 0 | 0 | 0 | 0 |
| 0.006668095 | 0 | 0 | 0 | 0 |
| 0 | 0 | 0 | 0 | 0 |
| 0 | 0 | 0 | 0 | 0 |
| 1.473229572 | 0.249391769 | 2.599532497 | 0 | 0 |
| 0 | 0 | 0 | 0 | 0 |
| 0 | 0 | 0 | 0 | 0 |
| 0 | 0 | 0 | 0 | 0 |
| 0.315097861 | 0.002515478 | 0.045025145 | 53.00302039 | 2.885771437 |
| 0 | 0 | 0 | 0 | 0 |
| 0 | 0 | 0.001879858 | 0.298642388 | 0 |
| 0 | 0 | 0 | 0 | 0 |
| 0 | 0.002622669 | 0 | 0 | 0 |
| 0.094509658 | 0.012682266 | 0.018683836 | 0 | 0 |
| 0 | 0 | 0 | 0 | 0 |
| 0.739324022 | 0.016580606 | 0 | 0 | 0 |
| 0 | 0.002976145 | 0.066219844 | 0 | 0 |
| 0 | 0 | 0 | 0 | 0 |
| 0 | 0 | 0 | 0 | 0 |
| 0 | 0 | 0 | 0 | 0 |
| 0 | 0 | 0 | 0 | 0 |
| 0 | 0 | 0 | 0 | 0 |
| 0 | 0 | 0 | 0 | 0 |
| 0 | 0 | 0 | 0 | 0 |
| 0 | 0 | 0 | 0 | 0 |
| 0 | 0 | 0 | 0 | 0 |
| 0 | 0.007116671 | 2.292915362 | 0 | 0.01701506 |
| 0 | 0.007300334 | 0.00067505 | 0.03201481 | 0.007583032 |
| 0 | 0 | 0 | 0 | 0 |
| 0 | 0 | 0 | 0 | 0 |
| 0 | 0 | 0 | 0 | 0 |
| 0 | 0 | 0 | 0 | 0.021158795 |
| 0 | 0 | 0 | 0 | 0 |
| 0 | 0 | 0 | 0 | 0 |
| 0 | 0 | 0 | 0 | 0 |
| 0.001903038 | 0 | 0.003904465 | 0.386298224 | 0 |
| 0 | 0 | 0 | 0.143649973 | 0 |
| 0 | 0.215110694 | 0.096781688 | 0 | 0 |
| 0 | 0 | 0 | 0 | 0 |
| 0 | 0 | 0 | 0 | 0 |
| 7.542248943 | 0.047577416 | 0 | 0 | 0 |
| 0 | 0 | 0 | 0 | 0 |
| 0 | 0 | 0 | 0 | 0 |
| 0 | 0 | 0 | 0 | 0 |
| 0 | 0 | 0 | 0 | 0 |
| 0 | 0 | 0 | 0 | 0 |
| 0 | 0 | 0 | 0 | 0 |
| 0 | 0 | 0 | 0 | 0 |

| | | | | |
|---|---|---|---|---|
| 0 | 0.234000395 | 0 | 0 | 0 |
| 0 | 0 | 0 | 0 | 0 |
| 0 | 0 | 0 | 0 | 0 |
| 0 | 0 | 0 | 0 | 0 |
| 0 | 0 | 0 | 0 | 0 |
| 0 | 0 | 0 | 0 | 0 |
| 0.002230103 | 0 | 0.002230558 | 0.167254613 | 0 |
| 0 | 0 | 0 | 0 | 0 |
| 0 | 0 | 0 | 0 | 0 |
| 0 | 0 | 0 | 0 | 0 |
| 0 | 0 | 0 | 0 | 0 |
| 0 | 0 | 0 | 0 | 0 |
| 0 | 0.001984152 | 0 | 0 | 0 |
| 0 | 0 | 0 | 0 | 0 |
| 0 | 0 | 0 | 0 | 0 |
| 0 | 0 | 0 | 0 | 0 |
| 0 | 0.001908574 | 0 | 0 | 0 |
| 0 | 0 | 0 | 0 | 0 |
| 0 | 0 | 0 | 0 | 0 |
| 0 | 0 | 0 | 0 | 0 |
| 0.00194625 | 0.450660625 | 0 | 0.001112425 | 0 |
| 0 | 0 | 0 | 0 | 0 |
| 0 | 0.069563514 | 0 | 0 | 0 |
| 0 | 0.020618865 | 0 | 0 | 0.004288823 |
| 0 | 0 | 0 | 0 | 0 |
| 0 | 0 | 0 | 0 | 0.029455539 |
| 0 | 0 | 0 | 0 | 0 |
| 0 | 0 | 0 | 0 | 0 |
| 0 | 0 | 0 | 0 | 0 |
| 1.349432369 | 0.186530965 | 0.004150641 | 0.213111332 | 0 |
| 0 | 0 | 0 | 0 | 0 |
| 0 | 0 | 0 | 0 | 0 |
| 0 | 0 | 0 | 0 | 0 |
| 0.007968926 | 0 | 0 | 0 | 0 |
| 0 | 0 | 0 | 0 | 0 |
| 0 | 0 | 0 | 0 | 0 |
| 0 | 0.023848315 | 0 | 0.00468069 | 0.078963164 |
| 0 | 0 | 0 | 0 | 0 |
| 0 | 0 | 0 | 0 | 0 |
| 0 | 0 | 0 | 0 | 0 |
| 0 | 0 | 0 | 0 | 0 |
| 0 | 0 | 0 | 0 | 0 |
| 0 | 0 | 0 | 0 | 0 |
| 0 | 0 | 0 | 0 | 0 |
| 0 | 0 | 0 | 0 | 0 |
| 0 | 0 | 0 | 0 | 0 |
| 0 | 0 | 0 | 0 | 0 |
| 0 | 0 | 0 | 0 | 0 |

| | | | | |
|---|---|---|---|---|
| 0.00746915 | 0.000745769 | 0 | 0.446128151 | 4.656954786 |
| 0 | 0 | 0 | 0 | 0 |
| 0 | 0 | 0 | 0 | 0 |
| 0 | 0 | 0 | 0 | 0 |
| 0 | 0 | 0 | 0 | 0 |
| 0 | 0 | 0 | 0 | 0 |
| 0 | 0.010749601 | 0 | 0 | 0.003167622 |
| 4.125799834 | 0.632498196 | 4.602947725 | 0 | 0 |
| 0 | 0.429608957 | 0.302810334 | 0.016585227 | 0.001810955 |
| 0.024485594 | 0.00285227 | 0 | 0.053515431 | 0.019991615 |
| 0 | 0.002629855 | 0 | 0 | 0 |
| 0 | 0 | 0 | 0 | 0 |
| 0 | 0 | 0 | 0 | 0 |
| 0 | 2.806093465 | 0.002203694 | 1.135258665 | 0 |
| 0 | 0 | 0 | 0 | 0 |
| 0 | 0 | 0 | 0 | 0 |
| 0 | 0.068855354 | 0 | 0 | 0.067606092 |
| 0 | 0 | 0 | 0 | 0 |
| 0 | 0 | 0 | 0 | 0 |
| 0 | 0.012670558 | 0 | 0 | 0 |
| 0 | 0 | 0 | 0 | 0 |
| 0 | 0 | 0 | 0 | 0 |
| 0 | 0 | 0 | 0 | 0 |
| 0 | 0 | 0 | 0 | 0 |
| 0 | 0 | 0 | 0 | 0 |
| 0 | 0 | 0 | 0 | 0 |
| 0 | 0 | 0 | 0 | 0 |
| 0 | 0 | 0 | 0 | 0 |
| 0 | 0 | 0 | 0 | 0 |
| 0 | 0 | 0 | 0 | 0 |
| 0 | 0 | 0 | 0 | 0 |
| 0 | 0 | 0 | 0 | 0 |
| 0 | 0 | 0 | 0 | 0 |
| 0 | 0 | 0 | 0 | 0 |
| 0 | 0 | 0 | 0 | 0 |
| 0 | 0.014075258 | 0 | 0 | 0 |
| 0 | 0 | 0 | 0 | 0 |
| 0 | 0 | 0 | 0 | 0 |
| 0.002442386 | 0 | 0 | 0 | 0 |
| 0.052568686 | 0 | 0.161954922 | 0 | 0 |
| 0 | 0 | 0 | 0 | 0 |
| 0 | 0 | 3.789869781 | 0 | 0 |
| 0 | 0 | 0 | 0 | 0 |
| 0 | 0 | 0 | 0 | 0 |
| 0 | 0 | 0 | 0 | 0 |
| 0 | 0 | 0 | 0 | 0 |
| 0 | 0 | 0 | 0 | 0 |
| 0 | 0 | 0 | 0 | 0 |

| | | | | |
|---|---|---|---|---|
| 0 | 0 | 0 | 0 | 0 |
| 0 | 0 | 0 | 0 | 0 |
| 0 | 0 | 0 | 0 | 0 |
| 0 | 0 | 0 | 0 | 0 |
| 0.014137791 | 36.01652496 | 0 | 0 | 0 |
| 0 | 0 | 0 | 0 | 0 |
| 0 | 0 | 0 | 0 | 0 |
| 0.00855117 | 0.005929202 | 0.007456388 | 0.044163163 | 0.035642443 |
| 0 | 0 | 0 | 0 | 0 |
| 0 | 0 | 0 | 0 | 0 |
| 0 | 0 | 0 | 0 | 0 |
| 0 | 0 | 0 | 0 | 0 |
| 0 | 0 | 0 | 0 | 0 |
| 0 | 0 | 0 | 0 | 0 |
| 0 | 0 | 0 | 0 | 0 |
| 0 | 0 | 0 | 0 | 0 |
| 0 | 0 | 0 | 0 | 0 |
| 0 | 0 | 0 | 0 | 0 |
| 0 | 0 | 0 | 0 | 0 |
| 0 | 0 | 0 | 0 | 0 |
| 0 | 0 | 0 | 0 | 0 |
| 0 | 0 | 0 | 0 | 0 |
| 0 | 0 | 0 | 0 | 0 |
| 0 | 0.001436746 | 0 | 0.07440817 | 0.058788084 |
| 0 | 0 | 0 | 0 | 0 |
| 0 | 0 | 0 | 0 | 0 |
| 0 | 7.991594379 | 0 | 0.047959155 | 0.200635694 |
| 0 | 0 | 0 | 0 | 0 |
| 0 | 0 | 0 | 0 | 0.00468667 |
| 0 | 0 | 0 | 0 | 0 |
| 0 | 0 | 0 | 0 | 0 |
| 0 | 0 | 0 | 0 | 0 |
| 0 | 0 | 0 | 0 | 0 |
| 0 | 0 | 0 | 0 | 0 |
| 0 | 0 | 0 | 0 | 0 |
| 0 | 0 | 0 | 0 | 0 |
| 0 | 0 | 0 | 0 | 0 |
| 0 | 0 | 0 | 0 | 0 |
| 0 | 0 | 0 | 0 | 0 |
| 0 | 0 | 0 | 0 | 0 |
| 0 | 0 | 0 | 0 | 0 |
| 0 | 0 | 0 | 0 | 0 |
| 0 | 0 | 0 | 0 | 0 |
| 0 | 0 | 0 | 0 | 0 |
| 0 | 0 | 0 | 0 | 0 |
| 0 | 0 | 0 | 0 | 0 |
| 0 | 0 | 0 | 0 | 0 |
| 0 | 0 | 0 | 0 | 0 |

| | | | | |
|---|---|---|---|---|
| 0 | 0.040956852 | 0 | 0 | 0.018571538 |
| 0 | 0 | 0 | 0 | 0 |
| 0 | 0 | 0 | 0 | 0 |
| 0 | 0 | 0 | 0 | 0 |
| 0 | 0 | 0 | 0 | 0 |
| 0.002048439 | 0 | 0 | 0 | 0 |
| 0 | 0 | 0 | 0 | 0 |
| 0 | 0 | 0 | 0 | 0 |
| 0 | 0 | 0 | 0 | 0 |
| 0 | 0 | 0 | 0 | 0 |
| 0 | 0 | 0 | 0 | 0 |
| 0 | 0 | 0 | 0 | 0 |
| 0 | 0 | 0 | 0 | 0 |
| 0 | 0 | 0 | 0 | 0 |
| 0 | 0 | 0 | 0 | 0 |
| 0 | 0 | 0 | 0 | 0 |
| 0 | 0 | 0 | 0 | 0 |
| 0 | 0 | 0 | 0 | 0 |
| 0 | 0 | 0 | 0 | 0 |
| 0 | 0 | 0 | 0 | 0 |
| 0 | 0 | 0 | 3.150322786 | 1.381888138 |
| 0 | 0 | 0 | 0 | 0 |
| 0 | 0.70676404 | 0.004773801 | 0.001161033 | 0.014604386 |
| 0 | 0.018427921 | 0 | 0 | 0 |
| 0 | 0 | 0 | 0 | 0 |
| 0 | 0 | 0 | 0 | 0 |
| 0 | 0 | 0 | 0 | 0 |
| 0 | 0 | 0 | 0 | 0 |
| 0 | 0 | 0 | 0 | 0 |
| 0 | 0 | 0 | 0 | 0 |
| 0 | 0 | 0 | 0 | 0 |
| 0 | 0 | 0 | 0 | 0 |
| 0.032442812 | 0.554065977 | 0.022576758 | 2.23788645 | 0.103859315 |
| 0 | 0 | 0 | 0 | 0 |
| 0 | 0.053626146 | 2.869861431 | 0 | 0 |
| 0.00271997 | 0.004849637 | 0.002870004 | 0.001919533 | 0.17954703 |
| 0 | 0 | 0 | 0 | 0 |
| 0 | 0 | 0 | 0 | 0 |
| 0.004719113 | 0 | 0 | 0 | 0.00099176 |
| 0 | 0 | 0 | 0 | 0 |
| 0 | 0 | 0 | 0 | 0 |
| 0 | 0 | 0 | 0 | 0 |
| 0 | 0.25632717 | 0 | 0 | 0 |
| 0 | 0 | 0 | 0 | 0 |
| 0 | 0 | 0 | 0 | 0 |
| 0 | 0.010001305 | 0 | 0 | 0 |
| 0 | 0 | 0 | 0 | 0 |
| 0 | 0 | 0 | 0 | 0 |

| | | | | |
|---|---|---|---|---|
| 0 | 0 | 0 | 0 | 0 |
| 0 | 0 | 0 | 0 | 0 |
| 0 | 0 | 0 | 0 | 0 |
| 0 | 0 | 0 | 0 | 0 |
| 0 | 0 | 0 | 0 | 0 |
| 0 | 0 | 0 | 0 | 0 |
| 0 | 0 | 0 | 0 | 0 |
| 0 | 0 | 0 | 0 | 0 |
| 0 | 0 | 0 | 0 | 0 |
| 0 | 0.281137838 | 0 | 0 | 0 |
| 0 | 0 | 0 | 0 | 0 |
| 0 | 0 | 0 | 0 | 0 |
| 0 | 0 | 0 | 0 | 0 |
| 0 | 0 | 0 | 0 | 0 |
| 0 | 0 | 0 | 0 | 0 |
| 0.04462144 | 0 | 0.001171491 | 7.317441965 | 2.81262765 |
| 0 | 0 | 0 | 0 | 0 |
| 0 | 0 | 0 | 0 | 0 |
| 0 | 0 | 0 | 0 | 0 |
| 0 | 0 | 0 | 0 | 0 |
| 0.013694045 | 0 | 0 | 0 | 0 |
| 0 | 0 | 0 | 0 | 0 |
| 0 | 0 | 0 | 0 | 0 |
| 0 | 0.001687549 | 0 | 0 | 0 |
| 0 | 0 | 0 | 0 | 0 |
| 0 | 0 | 0 | 0 | 0 |
| 0 | 0 | 0 | 0 | 0 |
| 0 | 0.003121187 | 0 | 0 | 0 |
| 0 | 0 | 0 | 0 | 0 |
| 0 | 0 | 1.146774447 | 0 | 0 |
| 0 | 0.394549934 | 0 | 0 | 0 |
| 0 | 0 | 0 | 0 | 0 |
| 0 | 0 | 0 | 0 | 0 |
| 0 | 0 | 0 | 0 | 0 |
| 0 | 0 | 0 | 0 | 0 |
| 0 | 0 | 0 | 0 | 0 |
| 0 | 0 | 0 | 0 | 0 |
| 0 | 0 | 0 | 0 | 0 |
| 0 | 0 | 0 | 0 | 0 |
| 0 | 0 | 0 | 0 | 0 |
| 0 | 0 | 0 | 0 | 0 |
| 0 | 0 | 0 | 0 | 0 |
| 0 | 0 | 0 | 0 | 0 |
| 0 | 0 | 0 | 0 | 0 |
| 0 | 0 | 0 | 0 | 0 |
| 0 | 0 | 0 | 0 | 0 |
| 0.358285092 | 0.001296926 | 0.505282582 | 5.813137663 | 0.003057357 |
| 0 | 0 | 0 | 0 | 0 |

| | | | | |
|---|---|---|---|---|
| 0 | 0 | 0 | 0 | 0 |
| 0 | 0 | 0 | 0 | 0 |
| 0 | 0 | 0 | 0 | 0 |
| 0 | 0 | 0 | 0 | 0 |
| 0 | 0.168750176 | 0.039719421 | 0 | 0.097501425 |
| 0 | 0 | 0 | 0 | 0 |
| 0 | 0 | 0 | 0 | 0 |
| 0 | 0 | 0 | 0 | 0 |
| 0 | 0 | 0 | 0 | 0 |
| 0 | 0 | 0 | 0 | 0 |
| 0 | 0 | 0 | 0 | 0 |
| 0.001571785 | 0.302889112 | 0 | 0.007058785 | 0.025368839 |
| 0 | 0.102128327 | 0 | 0 | 0.004958755 |
| 0 | 0 | 0 | 0 | 0 |
| 0 | 0 | 0 | 0 | 0 |
| 0 | 0.055615849 | 0 | 0 | 0 |
| 0.010361908 | 0.010173577 | 0 | 0 | 0 |
| 0 | 0 | 0 | 0 | 0 |
| 0.079911057 | 0.017994843 | 0 | 0 | 0.475583477 |
| 0 | 0 | 0 | 0 | 0 |
| 0 | 0 | 0 | 0 | 0 |
| 0 | 0 | 0 | 0 | 0 |
| 0 | 0 | 0 | 0 | 0 |
| 0 | 0 | 0 | 0 | 0 |
| 0 | 0 | 0 | 0 | 0 |
| 0 | 0 | 0 | 0 | 0 |
| 0 | 0.001953875 | 0 | 0.037549615 | 0.039413052 |
| 0 | 0 | 0 | 0 | 0 |
| 0 | 0 | 0 | 0 | 0 |
| 0 | 0 | 0 | 0 | 0 |
| 0.001008283 | 0 | 0 | 0 | 0 |
| 0 | 0 | 0 | 0 | 0.00211481 |
| 0 | 0 | 0 | 0 | 0 |
| 0 | 0 | 0 | 0 | 0 |
| 0 | 0 | 0 | 0 | 0 |
| 0 | 0 | 0 | 0 | 0 |
| 0 | 0 | 0 | 0 | 0 |
| 0.003541952 | 0.377134288 | 0 | 0.054718972 | 0.970670497 |
| 0.020789984 | 0 | 0 | 0 | 0 |
| 0 | 0 | 0 | 0 | 0 |
| 0 | 0.161983239 | 0 | 0 | 0.003245954 |
| 0 | 0 | 0 | 0 | 0 |
| 0 | 0 | 0 | 0 | 0 |
| 0.074582177 | 0 | 0 | 0 | 0.016874993 |
| 0 | 0 | 0 | 0 | 0 |
| 0 | 0 | 0 | 0 | 0 |
| 0 | 0 | 0 | 0 | 0 |
| 0 | 0 | 0 | 0 | 0 |

| | | | | |
|---|---|---|---|---|
| 0 | 0 | 0 | 0 | 0 |
| 0 | 0.029762201 | 0 | 0 | 0 |
| 0 | 0 | 0 | 0 | 0 |
| 0 | 0.001978543 | 0 | 0.006328294 | 0.026236091 |
| 0.004901032 | 0.005097409 | 0.001319778 | 0 | 0 |
| 0 | 0 | 0.004623243 | 0 | 0 |
| 0 | 0 | 0 | 0 | 0 |
| 0.000860983 | 0.001031595 | 0 | 0 | 0.003039836 |
| 0 | 0 | 0 | 0 | 0 |
| 0 | 0 | 0 | 0 | 0 |
| 0 | 0 | 0 | 0 | 0 |
| 0 | 0 | 0 | 0 | 0 |
| 0.015139154 | 0.296396831 | 0 | 0 | 0 |
| 0 | 0 | 0 | 0 | 0 |
| 0 | 0 | 0 | 0 | 0 |
| 0 | 0 | 0 | 0 | 0 |
| 0 | 0 | 0 | 0 | 0 |
| 0 | 0 | 0 | 0 | 0 |
| 0 | 0.047963021 | 0 | 0 | 0 |
| 0 | 0 | 0 | 0 | 0 |
| 0 | 0 | 0 | 0 | 0 |
| 0 | 0 | 0 | 0 | 0 |
| 0 | 0 | 0 | 0 | 0.071289848 |
| 0 | 0.021066226 | 0 | 0 | 0 |
| 0 | 0 | 0 | 0 | 0 |
| 0 | 0 | 0 | 0 | 0 |
| 0 | 0.02764288 | 0 | 0 | 0 |
| 0 | 0 | 0 | 0 | 0 |
| 0 | 0 | 0 | 0 | 0 |
| 0 | 0 | 0 | 0 | 0 |
| 0 | 0 | 0.004865944 | 0 | 0 |
| 0 | 0 | 0 | 0 | 0 |
| 0 | 0 | 0 | 0 | 0 |
| 0 | 0 | 0 | 0 | 0 |
| 0 | 0 | 0 | 0 | 0 |
| 0 | 0 | 0 | 0.382975326 | 0.091129037 |
| 0 | 0 | 0 | 0.546005639 | 0.245614235 |
| 0 | 0 | 0 | 0 | 0 |
| 0.011438544 | 0 | 0.003042946 | 0.001741348 | 0 |
| 0 | 0 | 0 | 0 | 0 |
| 0 | 0 | 0 | 0 | 0 |
| 0 | 0 | 0 | 0 | 0 |
| 0 | 0 | 0 | 0 | 0 |
| 0 | 0 | 0 | 0 | 0 |
| 0 | 0 | 0 | 0 | 0 |
| 0.399149634 | 0.326153347 | 2.135374523 | 0 | 0 |
| 0 | 0 | 0 | 0 | 0 |
| 0 | 0 | 0 | 0 | 0 |

| | | | | |
|---|---|---|---|---|
| 0 | 0 | 0 | 0 | 0 |
| 0 | 0 | 0 | 0 | 0 |
| 0 | 0 | 0 | 0 | 0 |
| 0 | 0 | 0 | 0 | 0 |
| 0 | 0 | 0 | 0.002908518 | 0 |
| 0 | 0 | 0 | 0 | 0 |
| 0 | 0 | 0 | 0 | 0.030788436 |
| 0 | 0 | 0 | 0 | 0 |
| 0 | 0 | 0 | 0 | 0 |
| 0 | 0 | 0 | 0 | 0 |
| 0 | 0.012249287 | 0.006389432 | 0 | 0.00819088 |
| 0 | 0.000703393 | 0 | 0.001687332 | 0.004767241 |
| 0 | 0 | 0 | 0 | 0 |
| 0 | 0 | 0 | 0 | 0 |
| 0 | 0 | 0 | 0 | 0 |
| 0 | 0 | 0 | 0 | 0 |
| 0 | 0 | 0 | 0 | 0 |
| 0 | 0 | 0 | 0 | 0 |
| 0 | 0 | 0 | 0 | 0 |
| 0 | 0 | 0.178328322 | 0.001498786 | 0 |
| 0 | 0 | 0 | 0 | 0 |
| 0 | 0 | 0 | 0 | 0 |
| 0 | 0 | 0 | 0 | 0 |
| 0 | 0 | 0 | 0 | 0 |
| 0 | 0 | 0 | 0 | 0 |
| 0 | 0 | 0 | 0 | 0 |
| 0 | 0 | 0 | 0 | 0 |
| 0 | 0.003226724 | 0 | 0 | 0 |
| 0 | 0 | 0 | 0 | 0 |
| 0 | 0 | 0 | 0 | 0 |
| 0.0044799 | 0.065127269 | 0 | 0 | 0.075921545 |
| 0 | 0 | 0 | 0 | 0 |
| 0.016645184 | 0.003531675 | 0.009989149 | 0 | 0 |
| 0.001275718 | 0.001698348 | 0.001570435 | 0.000954861 | 0 |
| 0 | 0 | 0 | 0 | 0 |
| 0 | 0 | 0 | 0 | 0 |
| 0 | 0 | 0 | 0 | 0 |
| 0 | 0 | 0 | 0 | 0 |
| 0 | 0 | 0 | 0 | 0 |
| 0 | 0 | 0 | 0 | 0 |
| 0 | 0 | 0 | 0 | 0 |
| 0.363742673 | 0.043069422 | 0 | 0 | 1.203720446 |
| 0 | 0.044766669 | 0 | 0 | 0 |
| 0 | 0 | 0 | 0 | 0 |
| 0 | 0 | 0 | 0 | 0 |
| 0 | 1.036258731 | 0 | 0 | 0 |
| 0 | 0 | 0 | 0 | 0 |
| 0 | 0 | 0 | 0 | 0 |

| | | | | |
|---|---|---|---|---|
| 0 | 0 | 0 | 0 | 0 |
| 0.008301866 | 0 | 0 | 0 | 0 |
| 0.001786374 | 0.006115313 | 0.082700453 | 0.003208995 | 0 |
| 0 | 0 | 0 | 0 | 0 |
| 0 | 0.008137972 | 0 | 0.007137636 | 3.391314464 |
| 0.041056508 | 0.019011487 | 0 | 0.09887157 | 0 |
| 0 | 0 | 0 | 0 | 0 |
| 0 | 0 | 0 | 0 | 0 |
| 0 | 0 | 0 | 0 | 0 |
| 0 | 0 | 0 | 0 | 0 |
| 0 | 0 | 0 | 0 | 0 |
| 0 | 0 | 0 | 0 | 0 |
| 0 | 0 | 0 | 0 | 0 |
| 0 | 0 | 0 | 0 | 0 |
| 0 | 0 | 0 | 0 | 0 |
| 0 | 0 | 0 | 0 | 0 |
| 0 | 0 | 0 | 0 | 0 |
| 0 | 0 | 0 | 0 | 0 |
| 0 | 0 | 0 | 0 | 0 |
| 0 | 0 | 0 | 0 | 0 |
| 0 | 0 | 0 | 0 | 0 |
| 0 | 0 | 0 | 0 | 0 |
| 0 | 0 | 0 | 0 | 0 |
| 0 | 0 | 0 | 0 | 0 |
| 0 | 0 | 0 | 0 | 0 |
| 0 | 0.005703747 | 0.001562715 | 0.06727185 | 0 |
| 0 | 0 | 0 | 0 | 0 |
| 0 | 0.016393582 | 0 | 0 | 0 |
| 0 | 0 | 0 | 0 | 0 |
| 0 | 0 | 0 | 0 | 0 |
| 0 | 0 | 0 | 0 | 0 |
| 0 | 0 | 0 | 0 | 0 |
| 0 | 0 | 0 | 0 | 0 |
| 0 | 0 | 0 | 0 | 0 |
| 0 | 0 | 0 | 0 | 0 |
| 0 | 0 | 0 | 0 | 0 |
| 0 | 0 | 0 | 0 | 0 |
| 0 | 0 | 0 | 0 | 0.030300586 |
| 0 | 0 | 0 | 0 | 0 |
| 0 | 0.014956705 | 0 | 0 | 0 |
| 0 | 0 | 0 | 0 | 0 |
| 0 | 0 | 0 | 0 | 0 |
| 0.0044591 | 0.001272074 | 0 | 0.348444222 | 3.619142733 |
| 0 | 0 | 0 | 0 | 0 |
| 0 | 0 | 0 | 0 | 0 |
| 0 | 0 | 0 | 0 | 0 |
| 0 | 0.002270362 | 0 | 0.01342111 | 0 |
| 0 | 0 | 0 | 0 | 0 |

| | | | | |
|---|---|---|---|---|
| 0 | 0 | 0 | 0 | 0 |
| 0.004918596 | 0.03683287 | 0.043897963 | 0.023009473 | 0.030028488 |
| 0 | 0 | 0 | 0 | 0 |
| 0 | 0 | 0 | 0 | 0 |
| 0 | 0 | 0 | 0 | 0 |
| 0 | 0 | 0 | 0 | 0 |
| 0 | 0 | 0 | 0 | 0 |
| 0 | 0 | 0 | 0 | 0 |
| 0 | 0 | 0 | 0 | 0 |
| 0 | 0 | 0 | 0 | 0 |
| 0 | 0 | 0 | 0 | 0 |
| 0 | 0 | 0 | 0 | 0 |
| 0.018631152 | 0.136490934 | 0 | 0 | 0 |
| 0 | 0 | 0 | 0 | 0 |
| 0 | 0 | 0 | 0 | 0 |
| 0 | 0 | 0 | 0 | 0 |
| 0 | 0 | 0 | 0 | 0 |
| 0 | 0 | 0 | 0 | 0 |
| 0 | 0 | 0 | 0 | 0 |
| 0 | 0 | 0 | 0 | 0 |
| 0 | 0 | 0 | 0 | 0 |
| 0 | 0 | 0 | 0.037191265 | 0 |
| 0 | 0.007701601 | 0 | 0 | 0 |
| 0.003618708 | 4.683940097 | 0 | 0 | 0 |
| 0 | 0 | 0 | 0 | 0 |
| 0 | 0 | 0 | 0 | 0 |
| 0 | 0 | 0 | 0 | 0 |
| 0 | 0 | 0 | 0 | 0 |
| 0 | 0 | 0 | 0 | 0 |
| 0 | 0 | 0 | 0 | 0 |
| 0 | 0 | 0 | 0 | 0 |
| 0 | 0 | 0 | 0 | 0 |
| 0 | 0 | 0 | 0 | 0 |
| 0 | 0 | 0 | 0 | 0 |
| 0 | 0.064382683 | 0 | 0 | 0 |
| 0 | 0 | 0 | 0 | 0 |
| 0 | 0 | 0 | 0 | 0 |
| 0 | 0 | 0 | 0 | 0 |
| 0 | 0.02302035 | 0.006478515 | 0.001350545 | 0 |
| 0 | 0 | 0 | 0 | 0 |
| 0.166503545 | 0.001794851 | 0.220321212 | 0 | 0 |
| 0 | 0 | 0 | 0 | 0 |
| 0 | 0 | 0 | 0 | 0 |
| 0 | 0 | 0 | 0 | 0 |
| 0 | 0 | 0 | 0 | 0 |
| 0.033187509 | 0.033136592 | 1.569097884 | 4.458810576 | 3.269055455 |
| 0 | 0 | 0 | 0 | 0 |
| 0 | 0 | 0 | 0 | 0 |

| | | | | |
|---|---|---|---|---|
| 0 | 0 | 0 | 0 | 0 |
| 0 | 0 | 0 | 0 | 0 |
| 0 | 0 | 0 | 0 | 0 |
| 0 | 0 | 0 | 0 | 0 |
| 0 | 0 | 0 | 0 | 0.002927801 |
| 0 | 0 | 0 | 0 | 0 |
| 0 | 0 | 0 | 0 | 0 |
| 0 | 0 | 0 | 0 | 0 |
| 0 | 0 | 0 | 0 | 0 |
| 0.008554673 | 0 | 0.07665932 | 0 | 0.003775452 |
| 0 | 0 | 0 | 0 | 0 |
| 0 | 0 | 0 | 0 | 0 |
| 0.001475554 | 0.004174322 | 0 | 0 | 0 |
| 0 | 0 | 0 | 0 | 0 |
| 0 | 0 | 0 | 0 | 0 |
| 0 | 0 | 0 | 0 | 0 |
| 0 | 0 | 0 | 0 | 0 |
| 0 | 0 | 0 | 0 | 0 |
| 0 | 0 | 0 | 0 | 0 |
| 0.026704726 | 1.186100006 | 2.552051265 | 0 | 1.831799559 |
| 0 | 0 | 0 | 0 | 0 |
| 0 | 0 | 0 | 0 | 0 |
| 0 | 0 | 0 | 0 | 0 |
| 0 | 0 | 0 | 0 | 0 |
| 0 | 0 | 0 | 0 | 0 |
| 0 | 0 | 0 | 0 | 0 |
| 0 | 0 | 0 | 0 | 0 |
| 0 | 0 | 0 | 0 | 0 |
| 0 | 0 | 0 | 0 | 0 |
| 0 | 0 | 0.00086417 | 0 | 0 |
| 0 | 0 | 0 | 0 | 0 |
| 0 | 0 | 0 | 0 | 0 |
| 0 | 0 | 0 | 0 | 0 |
| 0 | 0 | 0 | 0 | 0 |
| 0 | 0 | 0 | 0 | 0 |
| 0 | 0 | 0 | 0 | 0 |
| 0 | 0 | 0 | 0 | 0 |
| 0 | 0 | 0 | 0 | 0 |
| 0 | 0 | 0 | 0 | 0 |
| 0 | 0 | 0 | 0 | 0 |
| 0 | 0 | 0 | 0 | 0 |
| 0 | 0 | 0 | 0 | 0 |
| 0 | 0 | 0 | 0 | 0 |
| 0 | 0 | 0 | 0 | 0 |
| 0 | 0.249409414 | 0.01727314 | 0.001292612 | 0.029397737 |
| 0 | 0 | 0 | 0 | 0 |
| 0 | 0 | 0 | 0 | 0 |
| 0 | 0 | 0.008550476 | 0 | 0 |

| | | | | |
|---|---|---|---|---|
| 0 | 0 | 0 | 0 | 0 |
| 0 | 0 | 0 | 0 | 0 |
| 0 | 0 | 0 | 0 | 0 |
| 0 | 0 | 0 | 0 | 0 |
| 0 | 0 | 0 | 0 | 0 |
| 0 | 0.001532459 | 0 | 0.005789911 | 0.022352971 |
| 0 | 0 | 0 | 0 | 0 |
| 0 | 0 | 0 | 0 | 0 |
| 0 | 0 | 0 | 0 | 0 |
| 0 | 0 | 0 | 0 | 0 |
| 0 | 0 | 0 | 0 | 0 |
| 0 | 0 | 0 | 0 | 0 |
| 0 | 0 | 0 | 0 | 0 |
| 0 | 0 | 0 | 0 | 0 |
| 0 | 0 | 0 | 0 | 0 |
| 0 | 0 | 0 | 0 | 0 |
| 0 | 0 | 0 | 0 | 0 |
| 0 | 0 | 0 | 0 | 0.000667367 |
| 0 | 0 | 0 | 0 | 0 |
| 0 | 0 | 0 | 0 | 0 |
| 0 | 0 | 0 | 0 | 0 |
| 0 | 0 | 0 | 0 | 0 |
| 0 | 0 | 0 | 0 | 0 |
| 0 | 0 | 0 | 0 | 0 |
| 0 | 0 | 0 | 0 | 0 |
| 0 | 0 | 0 | 0 | 0 |
| 0 | 0 | 0 | 0 | 0 |
| 0 | 0 | 0 | 0 | 0 |
| 0 | 0 | 0 | 0 | 0 |
| 0 | 0 | 0 | 0 | 0.031348391 |
| 0 | 0 | 0 | 0 | 0 |
| 0 | 0 | 0 | 0 | 0 |
| 0 | 0 | 0 | 0 | 0 |
| 0 | 0 | 0 | 0 | 0 |
| 0 | 0 | 0 | 0 | 0 |
| 0 | 0 | 0 | 0 | 0 |
| 0 | 0 | 0 | 0 | 0 |
| 0 | 0 | 0 | 0 | 0 |
| 0 | 0 | 0 | 0 | 0 |
| 0 | 0 | 0 | 0 | 0 |
| 0 | 0 | 0 | 0 | 0 |
| 0 | 0 | 0 | 0 | 0 |
| 0 | 0 | 0 | 0 | 0 |
| 0 | 0 | 0 | 0 | 0 |
| 0 | 0 | 0 | 0 | 0 |
| 0 | 0 | 0 | 0 | 0 |
| 0 | 0 | 0 | 0 | 0 |
| 0 | 0 | 0 | 0 | 0 |

| | | | | |
|---|---|---|---|---|
| 0 | 0 | 0 | 0 | 0 |
| 0.001890183 | 0 | 0 | 0 | 0 |
| 0 | 0 | 0 | 0 | 0 |
| 0 | 0 | 0 | 0 | 0 |
| 0 | 0 | 0 | 0 | 0 |
| 0 | 0 | 0 | 0 | 0 |
| 0 | 0 | 0 | 0 | 0 |
| 0 | 0 | 0 | 0 | 0 |
| 0 | 0 | 0 | 0 | 0 |
| 0 | 0 | 0 | 0 | 0 |
| 0.000900016 | 0.423257314 | 0.007270846 | 0 | 0 |
| 0 | 0 | 0 | 0 | 0 |
| 0 | 0 | 0 | 0 | 0 |
| 0 | 0 | 0 | 0 | 0 |
| 0 | 0 | 0 | 0 | 0 |
| 0 | 0 | 0 | 0 | 0 |
| 0 | 0 | 0 | 0 | 0 |
| 0 | 0 | 0 | 0 | 0 |
| 0 | 0 | 0 | 0 | 0 |
| 0 | 0 | 0 | 0 | 0 |
| 0 | 0 | 0 | 0 | 0 |
| 0 | 0 | 0 | 0 | 0 |
| 0 | 0 | 0 | 0 | 0 |
| 0 | 1.07552671 | 1.998301698 | 0 | 0 |
| 0 | 0 | 0 | 0 | 0 |
| 0 | 0 | 0 | 0 | 0 |
| 0 | 0 | 0 | 0 | 0 |
| 0 | 0 | 0 | 0 | 0 |
| 0 | 0 | 0 | 0 | 0 |
| 0 | 0 | 0 | 0 | 0 |
| 0 | 0 | 0 | 0 | 0 |
| 0 | 0 | 0 | 0 | 0 |
| 0.001201094 | 0.019667719 | 0 | 0 | 0 |
| 0 | 0 | 0 | 0 | 0 |

| PHL018-0271_V2 | PHL018-0205_V1 | PHL018-0207_V1 | PHL018-0214_V1 | PHL018-0267_V3 |
|---|---|---|---|---|
| REF | REF | REF | EYCF | REF |
| 0.12872009 | 0.134476259 | 0.067273725 | 0.101013024 | 0.063907759 |
| 0.050397677 | 11.08523504 | 0.151093986 | 0.478299772 | 0.234758084 |
| 0 | 0 | 2.153228489 | 10.51641644 | 0 |
| 0 | 0 | 0 | 0 | 0 |
| 0 | 0 | 0 | 0 | 0 |
| 0 | 0 | 0 | 0 | 0 |
| 0 | 0 | 0 | 0 | 0 |
| 0 | 0 | 0 | 0 | 0 |
| 0 | 0 | 0 | 0 | 0 |
| 0 | 0 | 0 | 0 | 0 |
| 0 | 0 | 0 | 0 | 0 |
| 0 | 0 | 0 | 0 | 0 |
| 0 | 0 | 0 | 0 | 0 |
| 0.024113337 | 1.077685512 | 0.399870681 | 2.816039011 | 0.186142286 |
| 0 | 0 | 0 | 0 | 0 |
| 0 | 0 | 0 | 0 | 0 |
| 0 | 0 | 0 | 0.010923655 | 0 |
| 0 | 0 | 0 | 0.004792823 | 0 |
| 0 | 0 | 0 | 0 | 0 |
| 0 | 0 | 0 | 0 | 0 |
| 0 | 0 | 0 | 0 | 0 |
| 0 | 0 | 0 | 0 | 0 |
| 0 | 0 | 0 | 0 | 0 |
| 0 | 0 | 0 | 0 | 0 |
| 0.020835356 | 0 | 0 | 0 | 0.012296733 |
| 0 | 0 | 0 | 0 | 0 |
| 0 | 0 | 0 | 0 | 0 |
| 0 | 0 | 0 | 0 | 0 |
| 0 | 0 | 0 | 0 | 0 |
| 0 | 0 | 0 | 0 | 0 |
| 0 | 0 | 0 | 0 | 0 |
| 0 | 0 | 0 | 0 | 0 |
| 0 | 0 | 0 | 0 | 0 |
| 0 | 0 | 0 | 0 | 0 |
| 0 | 0 | 0 | 0 | 0 |
| 0.646213777 | 9.690736313 | 0.005103191 | 0 | 9.647929245 |
| 0 | 0 | 0 | 0 | 0 |
| 0 | 0 | 0 | 0 | 0 |
| 0 | 0 | 0 | 0 | 0 |
| 0 | 0 | 0 | 0 | 0 |
| 0 | 0 | 0 | 0 | 0 |
| 0.083217652 | 0.01898824 | 0.00929053 | 0.139307 | 0.011405491 |
| 0.67788742 | 0.023644757 | 2.807948514 | 0.036048783 | 0.106726533 |
| 0.043524843 | 0.014940496 | 0.000551962 | 0.062197205 | 0.005861874 |
| 0.03392419 | 0.001335675 | 0 | 0 | 0.001006239 |
| 0 | 0 | 0 | 0 | 0 |

| | | | | |
|---|---|---|---|---|
| 0.012640365 | 0 | 0.036102207 | 0 | 0.032602695 |
| 0 | 0.15550941 | 0 | 0.02170667 | 0.06567339 |
| 0 | 0.011498574 | 0.011734912 | 0 | 0.004154183 |
| 0.393713671 | 1.617859583 | 0.236202657 | 0.444255529 | 1.57045594 |
| 0.003049344 | 0 | 0 | 0.001331642 | 0 |
| 0 | 0 | 0 | 0 | 0 |
| 0.006686672 | 0.013711653 | 0.002738183 | 0.1532822 | 0.012308858 |
| 0 | 0 | 0 | 0 | 0 |
| 0.005394108 | 0 | 0 | 0 | 0 |
| 0 | 0 | 0 | 0 | 0 |
| 0.00167557 | 0 | 0 | 0 | 0 |
| 0.023451774 | 0.135821985 | 0.012560636 | 1.244014201 | 0.548482022 |
| 0.000702148 | 0 | 0 | 0 | 0 |
| 0.003627867 | 0.000235912 | 0.020154729 | 0.025238653 | 0.125892312 |
| 0 | 0.016911946 | 0 | 0.276852858 | 0 |
| 0 | 0 | 0 | 0 | 0 |
| 0.310739353 | 1.636014748 | 0.004779243 | 0.214315479 | 0.000755296 |
| 0 | 0 | 0 | 0 | 0 |
| 0 | 0 | 0 | 0 | 0 |
| 0 | 0 | 0 | 0 | 0 |
| 0.001073391 | 0.253244214 | 0.005180435 | 0.51228713 | 0.001746557 |
| 0.000431918 | 0.006816881 | 0.001621308 | 0.025040047 | 0.011068964 |
| 0.168748923 | 0.000557857 | 0 | 0 | 0.126450445 |
| 0 | 0 | 0 | 0 | 0 |
| 0 | 0 | 0 | 0 | 0 |
| 0 | 0 | 0 | 0 | 0 |
| 0 | 0 | 0 | 0 | 0 |
| 0 | 0 | 0 | 0 | 0 |
| 0.002026011 | 0.001047989 | 0 | 0.452978024 | 0 |
| 0 | 0 | 0 | 0 | 0 |
| 0 | 0.000430968 | 0 | 0 | 0 |
| 0 | 0 | 0 | 0 | 0 |
| 0 | 0 | 0 | 0 | 0 |
| 0 | 0 | 0 | 0 | 0.000758095 |
| 0 | 0 | 0 | 0 | 0.000629513 |
| 0 | 0 | 0 | 0 | 0 |
| 0 | 0 | 0 | 0 | 0 |
| 0.004539134 | 0.041579265 | 0.196731677 | 0.004421897 | 0 |
| 0 | 0 | 0 | 0 | 0 |
| 0 | 0 | 0 | 0 | 0 |
| 0 | 0 | 0 | 0 | 0 |
| 0 | 0 | 0 | 0 | 0 |
| 0 | 0 | 0 | 0 | 0 |
| 0 | 0 | 0 | 0 | 0 |
| 0.001713798 | 0 | 0.001781487 | 0 | 0 |
| 0 | 0 | 0 | 0 | 0 |
| 0 | 0 | 0 | 0 | 0 |
| 0 | 0 | 0.020093358 | 0.464584552 | 0.466658503 |

| | | | | |
|---|---|---|---|---|
| 0 | 0 | 0 | 0 | 0 |
| 0 | 0 | 0 | 0.09995206 | 0 |
| 0 | 0 | 0 | 0 | 0 |
| 0 | 0 | 0 | 0 | 0 |
| 0.005735749 | 0 | 0.000738188 | 0 | 0 |
| 0 | 0.000431528 | 0 | 0 | 0 |
| 0 | 0 | 0 | 0 | 0 |
| 0 | 0 | 0 | 0 | 0 |
| 0.008627978 | 0.001244284 | 0.004805838 | 0.000352473 | 0 |
| 0 | 0 | 0 | 0 | 0 |
| 0 | 0 | 0 | 0 | 0 |
| 0 | 0.002232547 | 0.000849052 | 0.014595319 | 0.00168685 |
| 0 | 0 | 0 | 0 | 0 |
| 0 | 0 | 0 | 0 | 0 |
| 0 | 0 | 0 | 0 | 0 |
| 0 | 0 | 0 | 0 | 0 |
| 0 | 0 | 0 | 0 | 0 |
| 0 | 0 | 0 | 0 | 0 |
| 2.453523412 | 0 | 0.163752847 | 0 | 0.006823718 |
| 0 | 0 | 0 | 0 | 0 |
| 0 | 0 | 0 | 0 | 0 |
| 0 | 0 | 0 | 0 | 0 |
| 0 | 0 | 0 | 0 | 0 |
| 0 | 0 | 0 | 0 | 0 |
| 0 | 0 | 0 | 0 | 0 |
| 0 | 0 | 0 | 0 | 0 |
| 0 | 0 | 0 | 0 | 0 |
| 0 | 0 | 0 | 0.000632655 | 0 |
| 0 | 0 | 0 | 0 | 0 |
| 0 | 0 | 0 | 0 | 0 |
| 0.008370406 | 0 | 0.00908772 | 0.090131235 | 0.016378708 |
| 0 | 0.200134401 | 0.004361211 | 0 | 0.056420779 |
| 0 | 0 | 0 | 0 | 0 |
| 0 | 0 | 0 | 0 | 0 |
| 0 | 0 | 0 | 0 | 0 |
| 0 | 0 | 0 | 0 | 0 |
| 0 | 0 | 0 | 0 | 0 |
| 0 | 0 | 0 | 0 | 0 |
| 0 | 0 | 0.037609863 | 0 | 0 |
| 0 | 0 | 0 | 0 | 0 |
| 0 | 0 | 0 | 0 | 0 |
| 0.34610846 | 0 | 0 | 0 | 0 |
| 0 | 0.001650046 | 0 | 0 | 0.0023714 |
| 0 | 0 | 0 | 0 | 0 |
| 0 | 0 | 0 | 0 | 0 |
| 0 | 0 | 0 | 0 | 0 |
| 0 | 0 | 0 | 0 | 0 |
| 0 | 0 | 0 | 0 | 0 |

| | | | | |
|---|---|---|---|---|
| 0 | 0.00024653 | 0 | 0 | 0 |
| 0 | 0 | 0 | 0 | 0 |
| 0 | 0 | 0 | 0 | 0 |
| 0 | 0 | 0 | 0 | 0 |
| 0.080781842 | 0 | 0.003089719 | 0 | 0.05334904 |
| 0 | 0 | 0 | 0 | 0 |
| 0 | 0 | 0 | 0 | 0 |
| 0 | 0 | 0 | 0 | 0 |
| 0 | 0 | 0 | 0 | 0 |
| 0 | 0 | 0 | 0 | 0 |
| 0 | 0 | 0 | 0 | 0 |
| 0 | 0 | 0 | 0 | 0 |
| 0 | 0 | 0 | 0 | 0 |
| 0 | 0 | 0 | 0 | 0 |
| 0 | 0 | 0 | 0 | 0 |
| 0 | 0 | 0 | 0 | 0 |
| 0 | 0 | 0 | 0 | 0 |
| 0 | 0 | 0 | 0 | 0 |
| 0 | 0 | 0 | 0 | 0 |
| 0 | 0 | 0 | 0 | 0 |
| 0 | 0 | 0 | 0.000889195 | 0 |
| 0 | 0 | 0 | 0 | 0 |
| 0.001548436 | 0 | 0 | 0 | 0 |
| 0 | 0 | 0 | 0 | 0 |
| 0.017900381 | 0.010687384 | 0.011060142 | 0 | 0.02751545 |
| 0 | 0 | 0 | 0 | 0 |
| 0.001467512 | 0 | 0 | 0 | 0 |
| 0 | 0 | 0 | 0 | 0 |
| 0 | 0 | 0 | 0 | 0 |
| 0 | 0 | 0 | 0 | 0 |
| 0 | 0 | 0 | 0 | 0 |
| 0 | 0 | 0 | 1.092545816 | 0 |
| 0 | 0 | 0 | 0 | 0 |
| 0 | 0 | 0 | 0 | 0 |
| 0 | 0 | 0 | 0 | 0 |
| 0 | 0 | 0 | 0 | 0 |
| 0 | 0 | 0 | 0 | 0 |
| 0.021896814 | 0 | 0 | 0 | 0 |
| 0 | 0 | 0 | 0 | 0 |
| 0 | 0 | 0 | 0 | 0 |
| 0 | 0 | 0 | 0 | 0 |
| 0 | 0 | 0 | 0 | 0 |
| 0 | 0 | 0 | 0 | 0 |
| 0 | 0 | 0 | 0 | 0 |
| 0 | 0 | 0 | 0 | 0 |
| 0 | 0 | 0 | 0 | 0 |
| 0 | 0 | 0 | 0 | 0 |
| 0 | 0.003495095 | 0 | 0 | 0 |

| | | | | |
|---|---|---|---|---|
| 0.194696226 | 0 | 0.368935084 | 0 | 0 |
| 0 | 0 | 0 | 0 | 0 |
| 0 | 0 | 0 | 0 | 0 |
| 0 | 0 | 0 | 0 | 0 |
| 0 | 0 | 0 | 0 | 0 |
| 0 | 0 | 0 | 0 | 0 |
| 0 | 0 | 0 | 0 | 0 |
| 0 | 0 | 0 | 0 | 0 |
| 0 | 0 | 0 | 0 | 0 |
| 0 | 0 | 0 | 0 | 0 |
| 0 | 0 | 0 | 0 | 0 |
| 0 | 0 | 0 | 0 | 0 |
| 0.001193237 | 0.000727441 | 0.002911414 | 0.001007429 | 0.00184044 |
| 0 | 0 | 0 | 0 | 0 |
| 0.00501745 | 0 | 0.272913491 | 0.004509443 | 0 |
| 0 | 0 | 0 | 0 | 0 |
| 0 | 0 | 0 | 0 | 0 |
| 0 | 0.000437426 | 0 | 0 | 0.000851305 |
| 0 | 0 | 0 | 0 | 0 |
| 0 | 0 | 0 | 0 | 0 |
| 0.011155333 | 0.018475221 | 0.014446596 | 0.224154323 | 0.071735399 |
| 0 | 0 | 0 | 0 | 0 |
| 0 | 9.94531E-05 | 0 | 0 | 0 |
| 0 | 0 | 0 | 0.000982134 | 0 |
| 0 | 0 | 0 | 0 | 0 |
| 0 | 0 | 0 | 0 | 0 |
| 0.002273811 | 0.002046292 | 0.000731596 | 0.002742483 | 0.003391491 |
| 0 | 0 | 0 | 0 | 0 |
| 0 | 0 | 0 | 0 | 0 |
| 0 | 0 | 0 | 0.009535958 | 0 |
| 0 | 0 | 0 | 0.069848333 | 0 |
| 0 | 0 | 0 | 0 | 0 |
| 0 | 0 | 0 | 0 | 0 |
| 0.204693829 | 0 | 0.000639773 | 0 | 0.011324042 |
| 0.006532429 | 0.021645109 | 0.000408684 | 0.005055612 | 0.017050969 |
| 0 | 0 | 0 | 0 | 0 |
| 0 | 0 | 0 | 0 | 0 |
| 0 | 0 | 0 | 0 | 0 |
| 0 | 0 | 0 | 0 | 0 |
| 0 | 0 | 0 | 0.002350237 | 0 |
| 0 | 0 | 0 | 0 | 0 |
| 0 | 0.000515566 | 0 | 0 | 0 |
| 0 | 0 | 0 | 0 | 0 |
| 0 | 0 | 0 | 0 | 0 |
| 0 | 0 | 0 | 0 | 0 |
| 0 | 0 | 0 | 0 | 0 |
| 0 | 0.001317521 | 0 | 0 | 0.000932407 |
| 0.000905175 | 0.001129932 | 0.004077347 | 0 | 0.001472846 |

| | | | | |
|---|---|---|---|---|
| 0 | 0 | 0 | 0 | 0 |
| 0 | 0 | 0 | 0 | 0 |
| 0 | 0 | 0 | 0 | 0 |
| 0 | 0 | 0 | 0 | 0 |
| 0 | 0 | 0 | 0 | 0 |
| 0 | 0 | 0 | 0 | 0 |
| 0 | 0.000149623 | 0 | 0 | 0 |
| 0.002788297 | 0.000566616 | 0 | 0.004865161 | 0 |
| 0 | 0 | 0 | 0 | 0 |
| 0 | 0 | 0 | 0 | 0 |
| 0 | 0 | 0 | 0 | 0 |
| 0 | 0 | 0 | 0 | 0 |
| 0 | 0 | 0 | 0 | 0 |
| 0 | 0 | 0 | 0 | 0 |
| 0.001631041 | 0 | 0 | 0 | 0 |
| 0 | 0 | 0 | 0 | 0 |
| 0 | 0 | 0 | 0 | 0 |
| 0.006659728 | 0 | 0 | 0 | 0 |
| 0.00631239 | 0 | 0.01155134 | 0.00044412 | 0 |
| 0.179938512 | 0 | 0.007467198 | 0 | 0 |
| 0 | 0 | 0.000582544 | 0.005404757 | 0.001657138 |
| 0 | 0 | 0 | 0 | 0 |
| 0 | 0 | 0 | 0 | 0 |
| 0 | 0 | 0 | 0 | 0 |
| 0.003083183 | 0.004229146 | 0.004340048 | 0 | 0.004755481 |
| 0 | 0 | 0 | 0 | 0 |
| 0 | 0 | 0 | 0 | 0 |
| 0 | 0 | 0 | 0 | 0 |
| 0 | 0 | 0 | 0.047198927 | 0 |
| 0.001762421 | 0 | 0.001157741 | 0 | 0 |
| 0.026530892 | 0.000405804 | 0.020560518 | 0.160289327 | 0.291896821 |
| 0 | 0 | 0 | 0 | 0 |
| 0 | 0 | 0 | 0.008834805 | 0 |
| 0.00055363 | 0 | 0 | 0.001090646 | 0 |
| 0 | 0 | 0 | 0 | 0 |
| 0 | 0 | 0 | 0 | 0 |
| 0 | 0 | 0 | 0 | 0 |
| 0 | 0 | 0 | 0 | 0 |
| 0 | 0 | 0 | 0 | 0 |
| 0 | 0 | 0 | 0 | 0 |
| 0 | 0 | 0 | 0 | 0 |
| 0.030097169 | 0.025025815 | 0.004584569 | 0 | 0.021804899 |
| 0 | 0 | 0 | 0.001029969 | 0 |
| 0 | 0 | 0 | 0 | 0 |
| 0 | 0 | 0 | 0 | 0 |
| 0 | 0 | 0 | 0 | 0 |
| 0 | 0 | 0 | 0 | 0 |
| 0 | 0 | 0 | 0 | 0 |

| | | | | |
|---|---|---|---|---|
| 0 | 0 | 0 | 0 | 0 |
| 0 | 0 | 0 | 0 | 0 |
| 0.0544058 | 0.015470862 | 0 | 0.058419224 | 0.00606511 |
| 0 | 0 | 0 | 0 | 0 |
| 0.003168782 | 0 | 0 | 0 | 0 |
| 0 | 0 | 0 | 0 | 0 |
| 0 | 0 | 0 | 0 | 0 |
| 0 | 0 | 0 | 0 | 0 |
| 0 | 0 | 0 | 0 | 0 |
| 0 | 0 | 0 | 0 | 0.001426398 |
| 0 | 0 | 0 | 0 | 0 |
| 0 | 0 | 0 | 0 | 0 |
| 0 | 0 | 0 | 0 | 0 |
| 0.0272393 | 0.003690237 | 0 | 0.002044236 | 0 |
| 0.011491043 | 0.001116796 | 0.005626221 | 0.010685912 | 0.000632284 |
| 0.022109163 | 0 | 0 | 0 | 0 |
| 0.001518031 | 0.001966574 | 0.002136858 | 0.000640823 | 0.001801077 |
| 0 | 0.16589112 | 0 | 0 | 0 |
| 0 | 0 | 0 | 0 | 0 |
| 0 | 0 | 0 | 0 | 0 |
| 0 | 0 | 0 | 0 | 0 |
| 0.000513881 | 0.001775255 | 0.004115143 | 0 | 0.000812929 |
| 0 | 0 | 0 | 0 | 0 |
| 0 | 0 | 0 | 0.001345165 | 0 |
| 0 | 0.002958146 | 0 | 0 | 0 |
| 0 | 0 | 0 | 0 | 0 |
| 3.131568035 | 0 | 0 | 0 | 0 |
| 0 | 0 | 0 | 0 | 0 |
| 0 | 0 | 0 | 0 | 0 |
| 0 | 0.001158083 | 0 | 0 | 0.002822183 |
| 0 | 0 | 0.002168031 | 0 | 0 |
| 0 | 0 | 0 | 0 | 0 |
| 0 | 0 | 0 | 0 | 0 |
| 0 | 0.000120404 | 0 | 0.003001437 | 0 |
| 0 | 0 | 0 | 0 | 0 |
| 0.004105614 | 0.006465905 | 0 | 0 | 0 |
| 0 | 0 | 0 | 0 | 0 |
| 0.277624264 | 0.002449093 | 0.124247588 | 0.000908501 | 0.01082643 |
| 0 | 0 | 0 | 0 | 0 |
| 0 | 0 | 0 | 0 | 0 |
| 0.002484901 | 0 | 0 | 0 | 0 |
| 0 | 0 | 0 | 0 | 0 |
| 0 | 0 | 0 | 0 | 0 |
| 0 | 0.00038027 | 0 | 0 | 0 |
| 0 | 0 | 0 | 0 | 0 |
| 0 | 0.00026896 | 0 | 0 | 0 |
| 0 | 0.000663682 | 0 | 0 | 0 |
| 0 | 0.025479822 | 0 | 0.004841859 | 0.001237125 |

| | | | | |
|---|---|---|---|---|
| 0.127856902 | 0 | 0.026964442 | 0.12448762 | 0.913605087 |
| 0 | 0 | 0 | 0 | 0 |
| 0 | 0.000328137 | 0 | 0 | 0 |
| 0 | 0 | 0 | 0 | 0 |
| 0 | 0 | 0.003930015 | 0 | 0.019189428 |
| 0 | 0.000299066 | 0.000662922 | 0 | 0 |
| 0 | 0 | 0 | 0 | 0 |
| 0.009361929 | 0.131732296 | 0.001139748 | 0.08502259 | 0.001921301 |
| 0 | 0 | 0 | 0 | 0 |
| 0.003651178 | 0.034554303 | 0.021341525 | 0.017394813 | 0 |
| 0 | 0 | 0 | 0 | 0 |
| 0.008813006 | 0.008873141 | 0.000902228 | 0.752014366 | 0.001330795 |
| 0.008660085 | 0.038345852 | 0.008782778 | 0.022576031 | 0.003605197 |
| 0 | 0 | 0.001212616 | 0.011104367 | 0 |
| 0 | 0 | 0 | 0.000527703 | 0.000889887 |
| 0 | 0 | 0 | 0.001347912 | 0 |
| 0 | 0 | 0 | 0 | 0 |
| 0 | 0 | 0 | 0.002004705 | 0 |
| 0 | 0 | 0 | 6.871329554 | 0 |
| 0 | 0 | 0 | 0.008636033 | 0 |
| 0 | 0 | 0 | 0 | 0 |
| 0 | 0 | 0 | 0 | 0 |
| 0 | 0 | 0 | 0.000529444 | 0.002976078 |
| 0.077012635 | 0 | 0 | 0 | 0 |
| 0 | 0 | 0 | 0 | 0 |
| 0.008671101 | 0.000287685 | 0 | 0.004780967 | 0 |
| 0.002129697 | 0 | 0.001421213 | 0 | 0 |
| 0 | 0.093800241 | 0 | 0.042277353 | 0.001218701 |
| 0 | 0 | 0 | 0.029523094 | 0 |
| 0 | 0 | 0 | 0 | 0 |
| 0 | 0 | 0 | 0.001763519 | 0 |
| 0 | 0 | 0 | 0.032560576 | 0 |
| 0 | 0 | 0 | 0 | 0 |
| 0 | 0 | 0 | 0 | 0 |
| 0 | 0 | 0 | 0 | 0 |
| 0 | 0 | 0.000673601 | 0.19756237 | 0 |
| 0 | 0 | 0 | 0.148872829 | 0 |
| 0 | 0 | 0 | 0 | 0 |
| 0 | 0 | 0 | 0.000531989 | 0 |
| 0 | 0 | 0 | 0 | 0 |
| 0 | 0 | 0 | 0 | 0 |
| 0 | 0 | 0 | 1.309320603 | 0 |
| 0 | 0 | 0 | 0.002604462 | 0 |
| 0 | 0 | 0 | 0.001324027 | 0 |
| 0 | 0 | 0 | 0 | 0 |
| 0.018514476 | 0 | 0 | 0 | 0 |
| 0.003300979 | 0.001520477 | 0 | 0.003158552 | 0.005013078 |
| 0 | 0 | 0 | 0 | 0 |

| | | | | |
|---|---|---|---|---|
| 0 | 0 | 0 | 0 | 0 |
| 0 | 0.029586764 | 0 | 0.03031564 | 0 |
| 0 | 0 | 0 | 0 | 0 |
| 0.038368625 | 0 | 0 | 0 | 0 |
| 0 | 0 | 0 | 0 | 0.00433554 |
| 0 | 0 | 0.000390377 | 0.00468281 | 0 |
| 0 | 0.050523267 | 0 | 0 | 0 |
| 0 | 0.000273335 | 0 | 0.033690027 | 0 |
| 0 | 0 | 0 | 0 | 0 |
| 0 | 0 | 0 | 0 | 0 |
| 0.002814439 | 0.001879192 | 0 | 0.003224817 | 0.005342734 |
| 0 | 0.014549301 | 0 | 0 | 0 |
| 0 | 0 | 0 | 0 | 0 |
| 0.000569773 | 0.000964873 | 0.004134972 | 0.002886295 | 0.008292398 |
| 0 | 0 | 0 | 0.006003069 | 0 |
| 0 | 0 | 0 | 0 | 0 |
| 0 | 0 | 0.017926687 | 0 | 0 |
| 0 | 0 | 0 | 0.204732607 | 0 |
| 0 | 0 | 0 | 0 | 0.341335297 |
| 0.003302671 | 0 | 0 | 0 | 0 |
| 0 | 0.000873273 | 0 | 0.026261081 | 0 |
| 0.041657631 | 0.415761338 | 0.061420271 | 0 | 0.655670426 |
| 0 | 0 | 0 | 0.07653861 | 0 |
| 0 | 0 | 0 | 0 | 0 |
| 0 | 0 | 0 | 0 | 0 |
| 0 | 0 | 0 | 0 | 0 |
| 0 | 0 | 0 | 2.144582464 | 0 |
| 0 | 0 | 0 | 0.006318988 | 0 |
| 0 | 0 | 0 | 0 | 0 |
| 0.073773164 | 1.047884405 | 0.162811062 | 0.1135239 | 1.335191411 |
| 0.010952128 | 0 | 0 | 0 | 0.01088457 |
| 0 | 0.000294087 | 0 | 0.001221841 | 0 |
| 0 | 0 | 0 | 0.011056216 | 0 |
| 0 | 0 | 0 | 0.008320114 | 0 |
| 0 | 0 | 0 | 0.050973229 | 0 |
| 0 | 0 | 0 | 0 | 0 |
| 0.043754172 | 0 | 0.714107569 | 0 | 0 |
| 0.001161501 | 0.014673262 | 0.000871994 | 0.049522048 | 0 |
| 0 | 0 | 0 | 0 | 0 |
| 0 | 0 | 0 | 0.017645334 | 0 |
| 0 | 0.004981488 | 0 | 0.011468769 | 0.045645959 |
| 0.429714164 | 9.3031037 | 0 | 0 | 0.61652043 |
| 0.00070934 | 0.013261474 | 0.001065071 | 0.048060374 | 0 |
| 0 | 0 | 0 | 0 | 2.157641176 |
| 0 | 0 | 0 | 0.002974108 | 0.00986354 |
| 0 | 0 | 0 | 0 | 0 |
| 0 | 0.030771137 | 0 | 0.011345497 | 0.001555478 |
| 0 | 0 | 0 | 0 | 0 |

| | | | | |
|---|---|---|---|---|
| 0.03336277 | 0.001725746 | 0.000910801 | 0 | 0 |
| 0 | 0 | 0 | 0.012544242 | 0 |
| 0.39590944 | 3.81003965 | 0.002275232 | 0.487781309 | 0.002092044 |
| 0 | 0 | 0 | 0 | 0 |
| 0.2581387 | 0 | 0 | 0 | 0.091061142 |
| 0 | 0 | 0 | 0.005340496 | 0.061499718 |
| 0 | 0 | 0 | 0 | 0 |
| 0 | 0 | 0 | 0.000765337 | 0 |
| 0 | 0 | 0 | 0 | 0 |
| 0 | 0 | 0 | 0 | 0.171739143 |
| 0 | 0 | 0 | 0 | 0 |
| 0 | 0 | 0 | 0 | 0 |
| 0 | 0 | 0 | 0.001469469 | 0 |
| 0.001646316 | 0.000185862 | 0 | 0 | 0.024828378 |
| 0 | 0 | 0 | 0 | 0 |
| 0.10892937 | 0.080351078 | 0 | 0.010741568 | 0.079511387 |
| 0.440008175 | 0.009176546 | 0.005709788 | 0.046152114 | 0.017445545 |
| 0 | 0 | 0 | 0 | 0 |
| 0 | 0.01912365 | 0 | 0.000829072 | 0 |
| 0 | 0 | 0 | 4.643528359 | 0 |
| 0 | 0 | 0.020238632 | 0.005394996 | 0 |
| 0 | 0 | 0 | 0 | 0 |
| 0 | 0 | 0 | 0 | 0 |
| 0 | 0 | 0 | 0 | 0 |
| 0.002221265 | 0 | 0 | 0 | 0 |
| 0 | 0 | 0 | 0 | 0.007924631 |
| 0 | 0 | 0 | 0.002377585 | 0.003627568 |
| 0 | 0 | 0 | 0 | 0.01316996 |
| 0 | 0 | 0 | 0 | 0 |
| 0 | 0 | 0 | 0 | 0 |
| 0.001376767 | 0.05045288 | 0.001952364 | 0.047657574 | 0.024393207 |
| 0.002508722 | 0.012188916 | 0 | 0.002118069 | 0 |
| 0.006731629 | 0.149282865 | 0.007010045 | 0.341920328 | 0.554025059 |
| 0.090558075 | 0.001833926 | 0.001635408 | 0.013399605 | 0.001476882 |
| 0 | 0 | 0 | 0.003000442 | 0 |
| 0 | 0 | 0 | 0 | 0 |
| 0 | 0.000877672 | 0.000736926 | 0.010773603 | 0 |
| 0 | 0.003151109 | 0.008414131 | 0.003499805 | 0.016321205 |
| 0.027258575 | 1.473770752 | 0 | 0 | 0.680056321 |
| 0 | 0.001658049 | 0.002921394 | 0 | 0 |
| 0.0232394 | 0 | 0 | 0 | 0 |
| 0 | 0 | 0 | 0 | 0 |
| 0 | 0.000326607 | 0.000517123 | 0 | 0.000653795 |
| 0 | 0 | 0.013226509 | 0.247906375 | 0.041634826 |
| 0 | 0 | 0 | 0.186526809 | 0 |
| 0.003012479 | 0 | 0 | 0 | 0 |
| 0 | 0 | 0 | 0 | 0 |
| 0 | 0 | 0 | 0 | 0 |

| | | | | |
|---|---|---|---|---|
| 0 | 0 | 0 | 0.005765611 | 0 |
| 1.40480309 | 0.6100151 | 0.125647352 | 0.033735755 | 0.696579543 |
| 0 | 0 | 0 | 0 | 0 |
| 0 | 0 | 0 | 0.003423104 | 0.001683652 |
| 0 | 0 | 0 | 0 | 0.000584416 |
| 0 | 0 | 0 | 0 | 0 |
| 0 | 0 | 0 | 0.061276115 | 0 |
| 0 | 0.000946973 | 0 | 0 | 0 |
| 0 | 0.0027631 | 0.005607216 | 0.01600706 | 0 |
| 0 | 0 | 0 | 0 | 0 |
| 0 | 0.007834718 | 0.00326794 | 0.001050025 | 0 |
| 0.012041325 | 0.019857881 | 0.001043076 | 0.290520899 | 0.018902145 |
| 0.07778782 | 0 | 0 | 0 | 0 |
| 0 | 0 | 0 | 0 | 0 |
| 0 | 0.000217484 | 0 | 0.028462757 | 0.000677219 |
| 0 | 0 | 0 | 0 | 0 |
| 0.017255779 | 0.028317309 | 0.011732589 | 0 | 0.006953171 |
| 0 | 0 | 0 | 0.948645413 | 0 |
| 0 | 0 | 0 | 0.004370359 | 0 |
| 0 | 0 | 0 | 0.036262599 | 0 |
| 0.092668622 | 0.032511428 | 0.071209181 | 0.000566945 | 0.070111296 |
| 0 | 0 | 0 | 0 | 0.00733628 |
| 0 | 0 | 0 | 0 | 0 |
| 0 | 0 | 0 | 0 | 0 |
| 0 | 0.01564816 | 0 | 0.00809688 | 0 |
| 0 | 0 | 0 | 0 | 0 |
| 0.001325631 | 0 | 0 | 0.049524906 | 0.059451971 |
| 0.022838972 | 0.051052703 | 0.028264606 | 0.028019937 | 0.299765421 |
| 0 | 0 | 0 | 0 | 0 |
| 0 | 0 | 0 | 0 | 0 |
| 0.00076582 | 0 | 0 | 0 | 0.00545167 |
| 0 | 0 | 0 | 0 | 0 |
| 0 | 0 | 0 | 0 | 0 |
| 0 | 0 | 0 | 0 | 0 |
| 0.452939041 | 0 | 0.117316541 | 0 | 0 |
| 0.104787485 | 0.210179344 | 0.076175118 | 0.062719514 | 0.153920308 |
| 0 | 0 | 0 | 0 | 0 |
| 0 | 0 | 0 | 0 | 0 |
| 0.002641352 | 0.010332528 | 0.009749695 | 0.218172918 | 0.001671386 |
| 0 | 0 | 0 | 0 | 0 |
| 0 | 0 | 0 | 0.19147991 | 0 |
| 0 | 0 | 0 | 0 | 0 |
| 0 | 0.104520749 | 0.031216565 | 0.008240268 | 0.093020032 |
| 0.001678101 | 0.002150988 | 0 | 0.003269516 | 0 |
| 0 | 0 | 0 | 0 | 0 |
| 0.181518015 | 0 | 0.200456248 | 0 | 0 |
| 0 | 0 | 0 | 0.088956457 | 0 |
| 0 | 0 | 0 | 0 | 0 |

| | | | | |
|---|---|---|---|---|
| 0 | 0 | 0 | 0.037216333 | 0 |
| 0 | 0.000376073 | 0 | 0 | 0 |
| 0 | 0.000298789 | 0 | 0 | 0.001162988 |
| 0.639378026 | 0.002056845 | 0.756374636 | 0 | 0.072718608 |
| 0 | 0 | 0 | 0 | 0 |
| 0.002700624 | 0 | 0 | 0 | 0 |
| 0 | 0 | 0 | 0 | 0 |
| 0.02967227 | 0.000594484 | 0 | 0 | 0 |
| 0.529027896 | 0.103267539 | 4.683306269 | 0.027968792 | 0.23622359 |
| 0.159632094 | 0 | 0.054540763 | 0 | 0.000792592 |
| 0 | 0 | 0 | 0 | 0 |
| 0.008056506 | 0.003971676 | 0.000560037 | 0.008313513 | 0.047581044 |
| 0 | 0 | 0 | 0 | 0 |
| 0 | 0 | 0 | 0 | 3.778318622 |
| 0.028193503 | 0.028721357 | 0.06340618 | 0 | 0.038095428 |
| 0 | 0 | 0 | 0 | 0 |
| 0.001284749 | 0 | 0 | 0 | 0 |
| 2.910591144 | 0 | 0 | 0 | 0.321218432 |
| 0 | 0 | 0 | 0 | 0 |
| 0.018519637 | 0 | 0 | 0 | 0 |
| 0 | 0 | 0 | 0 | 0 |
| 0.001385162 | 0 | 0 | 0 | 0 |
| 0.000686915 | 0 | 0 | 0 | 0 |
| 0.119896518 | 0 | 0.011574245 | 0.263429192 | 0.55111029 |
| 0 | 0.000738024 | 0 | 0.004481451 | 0 |
| 0.142087746 | 0 | 0.311153759 | 0 | 0 |
| 0.131964927 | 0 | 0 | 0 | 0 |
| 1.136416582 | 0.746483011 | 2.543214672 | 0 | 0.534665692 |
| 0 | 0 | 0 | 0 | 0 |
| 0 | 0 | 0 | 0 | 0.028683468 |
| 0.97352563 | 0.007132805 | 0 | 0 | 0.135053593 |
| 0.206352056 | 0 | 0 | 0 | 0 |
| 0 | 0.014468757 | 0.013508388 | 0.010065454 | 0 |
| 0 | 0 | 0 | 0 | 0 |
| 0.019953482 | 0 | 0 | 0 | 0 |
| 0.059276296 | 0 | 0 | 0 | 0 |
| 2.193430772 | 0 | 0.942835458 | 0 | 0.020351935 |
| 0 | 0 | 0 | 0.002359086 | 0 |
| 0 | 0 | 0.000699599 | 0 | 0.007223398 |
| 0.000640737 | 0 | 0 | 0 | 0 |
| 0.09474909 | 0.100633522 | 0.022134932 | 0.316717249 | 0.005162289 |
| 0.036794807 | 0 | 0 | 0 | 0 |
| 0 | 4.795499982 | 0.478350931 | 19.52790593 | 0.416873213 |
| 0.000748086 | 0 | 0 | 0 | 0 |
| 0 | 0 | 0 | 0 | 0 |
| 0 | 0 | 0 | 0 | 0 |
| 0 | 0 | 0 | 0 | 0 |
| 0.00103232 | 0.000664303 | 0 | 0.001162092 | 0.017963765 |

| | | | | |
|---|---|---|---|---|
| 0 | 0 | 0 | 0 | 0 |
| 0.107684102 | 0 | 0.195695581 | 0 | 0 |
| 0.006927165 | 0.049268925 | 0 | 0.022140682 | 0.003600607 |
| 0 | 0 | 0 | 0 | 0 |
| 0.002569393 | 0 | 0 | 0 | 2.528439122 |
| 0.178650264 | 0.002719491 | 0.222690038 | 0 | 0.005179549 |
| 0 | 0 | 0 | 0 | 0 |
| 0 | 0 | 0 | 0 | 0 |
| 0 | 0 | 0 | 0 | 0 |
| 0 | 0 | 0 | 0.355155831 | 0 |
| 0.017326988 | 0.028300054 | 0.003160869 | 0.023105408 | 0.083153064 |
| 0.425685905 | 1.561452301 | 0.618107093 | 2.824515662 | 3.800709908 |
| 0 | 0 | 0 | 0 | 0 |
| 0.661315593 | 0.017437677 | 0.0073475 | 0 | 0.010118804 |
| 0 | 0.069939216 | 0.001033948 | 0.161740862 | 0.331509464 |
| 0.010614233 | 0 | 0 | 0 | 0 |
| 0 | 0 | 0 | 0 | 0 |
| 0 | 0 | 0 | 0 | 0 |
| 0.056800227 | 0 | 0 | 0 | 0 |
| 0.002000871 | 0.001355337 | 0 | 0.031083111 | 0.004811196 |
| 0.047775175 | 0.413824494 | 0.203077046 | 0.047554909 | 0.028728052 |
| 0 | 0.185995258 | 0.106184858 | 4.306642064 | 0 |
| 0.091531576 | 0.883958143 | 0.954147593 | 0.341800341 | 0.014844797 |
| 0 | 0 | 0 | 0 | 0 |
| 0.008646541 | 0 | 0 | 0 | 0 |
| 1.459361335 | 0.003602477 | 1.474979612 | 0 | 0.014356541 |
| 0 | 3.54005139 | 0 | 0 | 0 |
| 0.000566235 | 0.000862993 | 0.000956473 | 0 | 0.000671812 |
| 0 | 0 | 0 | 0 | 0 |
| 0 | 0 | 0 | 0 | 0 |
| 0.001946114 | 0 | 0 | 0 | 0 |
| 0.035749592 | 0.147169749 | 0.114378041 | 0 | 0 |
| 0.000665244 | 0 | 0.009239456 | 0.006318601 | 0.040095549 |
| 0.003931573 | 0 | 0 | 0 | 0 |
| 0.001975811 | 0 | 0.010053716 | 0.006023846 | 0.010418725 |
| 0.117785357 | 0.005822954 | 0.328346341 | 0.060481355 | 0.017893363 |
| 0.002330941 | 0.001095374 | 0.003281153 | 0 | 0.002074169 |
| 0 | 0 | 0 | 0 | 0 |
| 4.511592763 | 0.129681796 | 0.594101435 | 0 | 0.1832568 |
| 0 | 0 | 0 | 0 | 0 |
| 0.011420705 | 0.003751101 | 0 | 0 | 0 |
| 0 | 0 | 0 | 0 | 0 |
| 0.015949562 | 0 | 0 | 0 | 0 |
| 0 | 0 | 0 | 0 | 0 |
| 0.022530126 | 0.002027075 | 0.020013265 | 0.009293086 | 0.020078864 |
| 0.039613552 | 0 | 0 | 0 | 0 |
| 0 | 0 | 0 | 0 | 0 |
| 0.003319388 | 0.016441906 | 27.54354422 | 0.543159571 | 0.592321588 |

| | | | | |
|---|---|---|---|---|
| 0 | 0 | 0 | 0 | 0 |
| 0 | 0 | 0 | 0 | 0 |
| 0 | 0 | 0 | 0 | 0 |
| 0.005570143 | 0 | 0 | 0 | 0.00834787 |
| 0 | 0 | 0 | 0 | 0 |
| 0.026539489 | 0 | 0.057343556 | 0 | 0 |
| 0.093559931 | 0.014218616 | 0.002048161 | 0 | 0.477529908 |
| 0.001888357 | 0 | 0.159590174 | 0 | 0.091154415 |
| 0 | 0 | 0 | 0 | 0 |
| 0 | 0 | 0 | 0 | 0 |
| 0.000979526 | 0.001194309 | 0.00306407 | 0.000826996 | 0.001704508 |
| 0 | 0 | 0 | 0 | 0 |
| 0.229850619 | 0 | 0 | 0 | 0 |
| 0.027304682 | 0 | 0 | 0 | 0 |
| 0 | 0 | 0 | 0 | 0 |
| 0 | 0 | 0 | 0 | 0 |
| 0.02549783 | 0.032149768 | 0.018960124 | 0 | 0.048864276 |
| 0.035471336 | 0.000994235 | 0.014488362 | 0 | 0.004772879 |
| 0.112690802 | 0 | 0 | 0 | 0 |
| 0.035175941 | 0 | 0 | 0 | 0.017915389 |
| 0.001569637 | 0 | 0 | 0 | 0 |
| 0 | 0 | 0 | 0 | 0 |
| 0.006424262 | 0.000445776 | 0 | 0.01283213 | 0.031826938 |
| 0 | 0 | 0 | 0 | 0 |
| 0 | 0 | 0 | 0 | 0 |
| 0 | 0 | 0 | 0 | 0 |
| 0 | 0 | 0 | 0 | 0 |
| 0 | 0 | 0 | 0 | 0 |
| 0 | 0 | 0 | 0 | 0 |
| 0.663585779 | 0.088609104 | 0.675052293 | 1.428771593 | 0.151775409 |
| 0.019930049 | 0.007864891 | 0 | 0 | 0 |
| 0 | 0 | 0 | 0 | 0 |
| 0 | 0.007819903 | 0 | 0 | 0 |
| 0 | 0 | 0 | 0 | 0 |
| 0.01376006 | 0 | 0.007504109 | 0 | 0 |
| 0 | 0 | 0 | 0 | 0 |
| 0.078562866 | 0 | 0 | 0 | 0.159569037 |
| 0 | 0 | 0 | 0.007350702 | 0.000787034 |
| 0 | 0 | 0 | 0 | 0 |
| 0 | 0 | 0 | 0 | 0.01552574 |
| 0 | 0 | 0 | 0 | 0 |
| 0 | 0 | 0 | 0 | 0 |
| 0 | 0 | 0 | 0 | 0 |
| 0.000939444 | 0 | 0.002317366 | 0 | 0.031845976 |
| 0.015535278 | 0.195667495 | 0.049950932 | 0 | 0.114240707 |
| 0 | 0 | 0 | 0 | 0 |
| 0.005252436 | 0 | 0 | 0 | 0 |
| 0 | 0 | 0 | 0 | 0 |

| | | | | |
|---|---|---|---|---|
| 0 | 0 | 0 | 0 | 0 |
| 0 | 0 | 0 | 0 | 0 |
| 0 | 0 | 0 | 0 | 0 |
| 0.533781062 | 0 | 0.003302476 | 0 | 0.134783828 |
| 0 | 0 | 0 | 0 | 0 |
| 0 | 0 | 0 | 0 | 0 |
| 0 | 0 | 0 | 0 | 0 |
| 0 | 0 | 0 | 0 | 0 |
| 0.001143586 | 0.182697621 | 0.003147995 | 0.768062518 | 0.021528123 |
| 0.000460462 | 0.005801433 | 0 | 0 | 0.00058274 |
| 0.001771682 | 0 | 0 | 0 | 0 |
| 0.001278173 | 0 | 0 | 0 | 0 |
| 0 | 0 | 0 | 0 | 0 |
| 0 | 0 | 0 | 0 | 0 |
| 0.398989208 | 0.021459745 | 2.357863362 | 1.922284682 | 0.440719288 |
| 0 | 0 | 0 | 1.165702739 | 0 |
| 0 | 0 | 0 | 0 | 0 |
| 0 | 0 | 0 | 0 | 0 |
| 0 | 0 | 0 | 0 | 0 |
| 0 | 0 | 0 | 0 | 0 |
| 0.05436073 | 0 | 0.004923601 | 0 | 0.006363206 |
| 0 | 0 | 0 | 0 | 0 |
| 0 | 0 | 0 | 0 | 0 |
| 0 | 0 | 0 | 0 | 0 |
| 0.058499858 | 0.000774306 | 0.357911549 | 0.005487821 | 0.002765664 |
| 2.451658724 | 0.666622405 | 0.536233517 | 0.140014226 | 1.93180465 |
| 1.198133748 | 0 | 0 | 0 | 0 |
| 0 | 0 | 0 | 0.001879474 | 0.002245015 |
| 0.188121764 | 0.005973221 | 0 | 0 | 0 |
| 0 | 0 | 0 | 0 | 0 |
| 0 | 0 | 0 | 0 | 0 |
| 0.047489741 | 0 | 0 | 0.032051421 | 0 |
| 0 | 0 | 0 | 0 | 0.004738975 |
| 0 | 0.00046218 | 0 | 0.00485106 | 0.027382085 |
| 0.340820313 | 0.008416868 | 0.001184583 | 0.022646875 | 0.004118567 |
| 0.000836523 | 0.003399832 | 0.000732687 | 0.000941682 | 0.000661665 |
| 0 | 0 | 0 | 3.294853415 | 0 |
| 0.720241588 | 3.498286681 | 0.802274234 | 0 | 0.565431395 |
| 0.003873798 | 0 | 5.276877211 | 0 | 0 |
| 0 | 0 | 0 | 0 | 0 |
| 0 | 0 | 0 | 0 | 0 |
| 0.000948624 | 0 | 0 | 0.00213575 | 0.001500668 |
| 0.211096838 | 4.044089094 | 3.801956981 | 0.024610534 | 2.636219747 |
| 0.504492119 | 0 | 0 | 0 | 0 |
| 0 | 0 | 0 | 0 | 0 |
| 0 | 0.004965576 | 0 | 0.001895767 | 0.006393817 |
| 0 | 0 | 0 | 0 | 0 |
| 0.001893823 | 0.009321865 | 1.12162858 | 0.071063146 | 0.005192923 |

| | | | | |
|---|---|---|---|---|
| 0 | 0 | 0.023796159 | 0 | 0.004170243 |
| 0 | 0 | 0 | 0 | 0 |
| 0.018682407 | 0 | 0 | 0 | 0 |
| 0 | 0 | 0 | 0 | 0 |
| 0 | 0.000666687 | 0 | 0.000395697 | 0.001482843 |
| 0.554534131 | 0 | 0 | 0.000508067 | 0 |
| 0 | 0 | 0 | 0 | 0 |
| 0 | 0 | 0 | 0 | 0 |
| 0 | 0 | 0 | 0 | 0 |
| 0.001254445 | 0 | 0 | 0 | 0.001637178 |
| 0.000597702 | 0 | 0 | 0 | 0.002950051 |
| 0.309011458 | 0.534107774 | 1.168928893 | 1.21283549 | 0.248705492 |
| 0.007755655 | 0.336350605 | 0.060213576 | 0 | 0.104495918 |
| 0 | 0 | 0 | 0 | 0 |
| 0.079497173 | 0.073026974 | 0.010037479 | 1.376835244 | 0.005144723 |
| 0 | 0 | 0 | 0 | 0 |
| 0 | 0 | 0 | 0 | 0 |
| 0 | 0 | 0 | 0 | 0 |
| 0 | 0.006725816 | 0 | 0.010027624 | 0.007515552 |
| 0.001580377 | 0 | 0 | 0 | 0.002000051 |
| 0.058605573 | 0.052699585 | 0 | 0 | 0 |
| 0 | 0 | 0 | 0 | 0 |
| 1.329813444 | 0.074473979 | 0.068899847 | 0.050442598 | 0.427801697 |
| 0 | 0 | 0 | 0 | 0 |
| 0 | 0 | 0 | 0 | 0 |
| 0 | 0 | 0 | 0 | 0 |
| 0 | 0 | 0 | 0 | 0 |
| 0 | 0 | 0 | 0 | 0 |
| 0.006672456 | 0 | 0 | 0 | 0 |
| 0 | 0 | 0 | 0 | 0 |
| 0 | 0 | 0 | 0 | 0 |
| 0.03985677 | 0.058506088 | 0.007744614 | 0.010886882 | 0.069875435 |
| 0 | 0 | 0 | 0 | 0 |
| 0.00117128 | 0.018422608 | 0.009145088 | 0 | 0.031684524 |
| 0.001820693 | 0.003514875 | 0.000797347 | 0.000512393 | 0.001008081 |
| 0.512840173 | 0.001311634 | 0.026871651 | 0.00198161 | 0.032302809 |
| 0.003232346 | 0 | 0 | 0 | 0 |
| 0.011316533 | 0 | 0 | 0 | 0 |
| 0.001568781 | 0 | 0 | 0 | 0 |
| 0 | 0 | 0 | 0 | 0 |
| 0.081439967 | 0 | 0 | 0 | 0 |
| 0 | 0 | 0 | 0 | 0 |
| 0 | 0 | 0 | 0 | 0 |
| 0 | 0 | 0 | 0 | 0.011723865 |
| 0.007024798 | 0.059682646 | 0.00516164 | 0.007192808 | 0.036885107 |
| 0 | 0 | 0 | 0 | 0 |
| 0.162853323 | 0 | 0.203738081 | 0 | 0 |
| 0.00070683 | 0 | 0 | 0.003699937 | 0.008855857 |

| | | | | |
|---|---|---|---|---|
| 0.00357169 | 0 | 0 | 0 | 0 |
| 0.034842607 | 0 | 0 | 0 | 0 |
| 0.005874153 | 0 | 0 | 0.01265963 | 0.018340592 |
| 0 | 0 | 0 | 0 | 0 |
| 0 | 0 | 0 | 0 | 0 |
| 0.003723613 | 0 | 0 | 0 | 0 |
| 2.666263941 | 0 | 0.001021988 | 0 | 0.273762057 |
| 0.021898374 | 0.000237334 | 0.001863217 | 0.002095353 | 0.004572737 |
| 0 | 0 | 0 | 0 | 0 |
| 0 | 0 | 0 | 0 | 0 |
| 0 | 0 | 0 | 0 | 0 |
| 0 | 0 | 0 | 0 | 0 |
| 0 | 0 | 0 | 0 | 0 |
| 0.063879434 | 0 | 0.058500476 | 0 | 0 |
| 0 | 0 | 0 | 0 | 0 |
| 0 | 0 | 0 | 0 | 0 |
| 0 | 0 | 0 | 0 | 0 |
| 0.001161297 | 0 | 0 | 0 | 0 |
| 1.580578398 | 0 | 0.43476574 | 0 | 0.501696603 |
| 0 | 0 | 0 | 0 | 0.000728658 |
| 0.026966874 | 0 | 0 | 0 | 0.002133001 |
| 0 | 0 | 0 | 0 | 0 |
| 0 | 0 | 0 | 0 | 0 |
| 0 | 0 | 0 | 0 | 0 |
| 0.003429346 | 0.000895994 | 0 | 0 | 0 |
| 0 | 0.002516562 | 0 | 0.028194664 | 0 |
| 0 | 0 | 0 | 0 | 0 |
| 0.078720952 | 0.186207853 | 0.039058879 | 0.053929076 | 0.041625558 |
| 0 | 0 | 0 | 0 | 0 |
| 0.825059715 | 0.348589824 | 0.199460225 | 0.236189768 | 0.278163816 |
| 0 | 0 | 0 | 0 | 0 |
| 0 | 0 | 0 | 0 | 0 |
| 0.002732401 | 0 | 0 | 0 | 0 |
| 0 | 0 | 0 | 0 | 0 |
| 0 | 0.004652141 | 0 | 0.000738657 | 0.001245627 |
| 0 | 0 | 0 | 0 | 0 |
| 0 | 0 | 0 | 0 | 0 |
| 0 | 0.000168115 | 0.000621085 | 0 | 0 |
| 0 | 0 | 0 | 0 | 0 |
| 0 | 0 | 0 | 0 | 0 |
| 0.889060258 | 11.95283627 | 2.307400735 | 0.894093956 | 13.52280213 |
| 0 | 0 | 0.061103266 | 2.009535258 | 0.286707717 |
| 5.846173392 | 0 | 0 | 0 | 0 |
| 0 | 0 | 0 | 0 | 0 |
| 0 | 0 | 0 | 0 | 0 |
| 0 | 0 | 0 | 0 | 0 |
| 0 | 0 | 0 | 0 | 0 |
| 0.000656583 | 0.000480332 | 0.013801979 | 0 | 0.000747846 |

| | | | | |
|---|---|---|---|---|
| 0 | 0 | 0 | 0 | 0 |
| 0.003511817 | 0 | 0 | 0.005701855 | 0.003076887 |
| 0 | 0 | 0 | 0 | 0.003743365 |
| 0 | 0 | 0 | 0 | 0 |
| 1.356006481 | 2.842042633 | 2.705703509 | 0.334780005 | 3.73300411 |
| 0 | 0 | 0 | 0 | 0 |
| 0 | 0 | 0 | 0 | 0 |
| 0 | 0 | 0 | 0 | 0 |
| 0 | 0 | 0 | 1.645862229 | 0.050181721 |
| 0 | 0 | 0 | 0 | 0 |
| 0 | 0 | 0 | 0 | 0 |
| 0 | 0 | 0 | 0 | 0 |
| 0.02454252 | 0 | 0.04522558 | 0 | 0.004235436 |
| 0.005485915 | 0.015410528 | 0.009811809 | 0.001226028 | 0.03537723 |
| 4.499707018 | 0.128599186 | 0 | 0 | 0 |
| 0.232764481 | 0.554826345 | 0.084988051 | 0.064500234 | 2.409640516 |
| 0.206363946 | 0 | 0.039937261 | 0.06672114 | 0.00066263 |
| 0 | 0 | 0 | 0 | 0 |
| 0 | 0 | 0 | 0 | 0 |
| 0 | 0 | 0 | 0 | 0 |
| 0 | 0 | 0 | 0 | 0 |
| 0 | 0 | 0 | 0 | 0 |
| 0 | 0 | 0 | 0 | 0 |
| 0 | 0 | 0 | 0 | 0 |
| 0 | 0 | 0 | 0 | 0 |
| 0 | 0 | 0 | 0 | 0 |
| 0.01578312 | 0 | 0.108009454 | 0.003997622 | 0.026504465 |
| 0 | 0 | 0.000514247 | 0.002891583 | 0 |
| 0 | 0 | 0 | 0 | 0 |
| 0 | 0 | 0 | 0 | 0 |
| 0 | 0 | 0 | 0 | 0 |
| 0 | 0 | 0 | 0 | 0 |
| 0.084910514 | 0 | 0 | 0 | 0.070362139 |
| 0.003733819 | 0 | 0 | 0 | 0 |
| 0.428872491 | 0 | 0.32580041 | 0 | 0.320981323 |
| 0 | 0 | 0 | 0 | 0 |
| 0 | 0 | 0 | 0.000547033 | 0 |
| 0 | 0 | 0 | 0 | 0 |
| 0 | 0 | 0 | 0 | 0 |
| 0 | 0 | 0 | 0 | 0 |
| 0.062691815 | 0.965892711 | 0.014280004 | 0 | 9.96513206 |
| 0.009627616 | 0 | 0 | 0.042484568 | 0 |
| 0 | 0 | 0 | 0 | 0 |
| 0.417669141 | 0 | 0 | 0 | 0 |
| 0 | 0 | 0 | 0 | 0 |
| 0 | 0 | 0 | 0 | 0 |
| 0 | 0 | 0 | 0 | 0 |
| 0 | 0 | 0 | 0 | 0 |

| | | | | |
|---|---|---|---|---|
| 0.868937467 | 0 | 1.031737456 | 0 | 0.019647431 |
| 0 | 0 | 0 | 0 | 0 |
| 0 | 0 | 0 | 0 | 0 |
| 0 | 0 | 0 | 0 | 0 |
| 0.017521364 | 0 | 0.00557378 | 0 | 0 |
| 0 | 0 | 0 | 0 | 0 |
| 0.175916037 | 0.000679302 | 0.022063706 | 0.022343094 | 0.012162745 |
| 0 | 0 | 0 | 0 | 0 |
| 0 | 0 | 0.000590461 | 0 | 0 |
| 0 | 0 | 0 | 0.018629857 | 0.025660101 |
| 0.019255169 | 0 | 0 | 0 | 0 |
| 0 | 0 | 0 | 0 | 0.001849835 |
| 0 | 0 | 0 | 0 | 0 |
| 0 | 0 | 0 | 0 | 0 |
| 0 | 0 | 0 | 0 | 0 |
| 0 | 0 | 0 | 0 | 0 |
| 0 | 0 | 0 | 0 | 0 |
| 0 | 0 | 0.000402122 | 0 | 0.0005084 |
| 0 | 0 | 0 | 0 | 0 |
| 0 | 0 | 0 | 0 | 0 |
| 0.002784734 | 0.001994094 | 1.74746998 | 0.44603755 | 0.009691631 |
| 0 | 0 | 0 | 0 | 0 |
| 0.024133624 | 0 | 0.000329423 | 0.004322095 | 0.025405712 |
| 0.01313604 | 0.001480064 | 0.001952842 | 0.005490359 | 0.012097924 |
| 0.003887221 | 0 | 0.024397171 | 0 | 0 |
| 0 | 0.0016526 | 0 | 0.003368242 | 0.00131077 |
| 0 | 0 | 0 | 0 | 0 |
| 0 | 0 | 0 | 0 | 0 |
| 0 | 0 | 0 | 0 | 0 |
| 0.001310306 | 0.030506931 | 0.005059074 | 1.227167191 | 0.004797115 |
| 0 | 0 | 0 | 0 | 0 |
| 0 | 0 | 0 | 0 | 0 |
| 0 | 0 | 0 | 0 | 0 |
| 0.001250064 | 0 | 0 | 0 | 0 |
| 0.191329659 | 0 | 0.003955312 | 0 | 0 |
| 0 | 0 | 0 | 0 | 0 |
| 0 | 0 | 0.004887021 | 0.007144655 | 0 |
| 0 | 0 | 0 | 0 | 0 |
| 0 | 0 | 0 | 0 | 0 |
| 0.473073642 | 0 | 0 | 0 | 0 |
| 0 | 0 | 0 | 0 | 0 |
| 0 | 0 | 0 | 0 | 0 |
| 0 | 0 | 0 | 0 | 0 |
| 0 | 0 | 0 | 0 | 0 |
| 0 | 0 | 0 | 0 | 0 |
| 0.967679574 | 0 | 0 | 0 | 0 |
| 0 | 0 | 0 | 0 | 0 |
| 0 | 0 | 0 | 0 | 0 |

| | | | | |
|---|---|---|---|---|
| 0 | 0.019310298 | 0.001786128 | 0.00165419 | 0.00398504 |
| 0.301423015 | 0 | 0 | 0 | 0 |
| 0 | 0 | 0 | 0 | 0 |
| 0 | 0 | 0 | 0 | 0 |
| 0 | 0 | 0 | 0 | 0 |
| 0 | 0 | 0.212383218 | 0 | 0 |
| 0 | 0 | 0 | 0 | 0 |
| 0.484830163 | 1.450167845 | 0.052105744 | 0.247128719 | 0.758607674 |
| 0.912293336 | 0.03517615 | 3.35507964 | 0.052115454 | 0.138029608 |
| 0 | 0.124649359 | 0 | 0.017688707 | 0.008418943 |
| 0.001794587 | 0 | 0 | 0 | 0 |
| 0 | 0 | 0 | 0 | 0 |
| 0.011257417 | 0 | 0 | 0 | 0 |
| 0.002114141 | 0 | 1.427002436 | 0.065355958 | 0 |
| 0 | 0 | 0 | 0 | 0 |
| 0 | 0 | 0 | 0 | 0 |
| 0 | 0 | 0 | 0.640750496 | 0 |
| 0 | 0 | 3.085850762 | 0 | 0 |
| 0 | 0 | 0 | 0 | 0 |
| 0 | 0 | 0 | 0 | 0 |
| 0 | 0 | 0 | 0 | 0 |
| 0 | 0 | 0 | 0 | 0 |
| 0 | 0 | 0 | 0 | 0 |
| 0 | 0 | 0 | 0 | 0 |
| 0.032517617 | 0 | 0.004385978 | 0 | 0.302786789 |
| 0 | 0 | 0 | 0 | 0 |
| 0 | 0 | 0 | 0 | 0 |
| 0 | 0 | 0 | 0 | 0 |
| 0 | 0 | 0 | 0 | 0 |
| 0 | 0 | 0 | 0 | 0 |
| 0 | 0 | 0 | 0 | 0 |
| 0 | 0 | 0 | 0 | 0 |
| 0 | 0 | 0 | 0 | 0 |
| 0.000778959 | 0 | 0 | 0 | 0 |
| 0.020500779 | 0 | 0.003943125 | 0 | 0 |
| 0 | 0.040278849 | 0 | 0 | 0 |
| 0 | 0 | 0 | 0 | 0 |
| 0 | 0.12174631 | 0 | 0 | 0 |
| 0 | 0 | 0 | 0 | 0 |
| 0 | 0.593833427 | 0 | 0 | 0 |
| 0 | 0 | 0 | 0 | 0 |
| 0.094699091 | 0.42269429 | 0.040254765 | 3.344021041 | 0.515094742 |
| 0 | 0 | 0 | 0 | 0 |
| 0 | 0 | 0 | 0 | 0 |
| 0 | 0 | 0 | 0 | 0 |
| 0 | 0 | 0 | 0 | 0 |
| 0 | 0 | 0 | 0 | 0 |
| 0 | 0 | 0 | 0.000440125 | 0.000742201 |

| | | | | |
|---|---|---|---|---|
| 0 | 0 | 0 | 0 | 0 |
| 0.045345313 | 0.022895615 | 0 | 0 | 0 |
| 0 | 0 | 0 | 0 | 0 |
| 0 | 0 | 0 | 0 | 0 |
| 2.304266699 | 0.048411692 | 0.782363039 | 0.103326012 | 0.114485687 |
| 0 | 0 | 0 | 0 | 0 |
| 0 | 0 | 0 | 0 | 0 |
| 0.000890126 | 0.107807086 | 0.009890248 | 0.074851155 | 0.002365654 |
| 0.020974687 | 0.003178672 | 0.000667233 | 0 | 0.009954219 |
| 0 | 0 | 0 | 0 | 0 |
| 0 | 0 | 0 | 0 | 0 |
| 0.159460815 | 0 | 0 | 0 | 0 |
| 0 | 0 | 0 | 0 | 0 |
| 0 | 0 | 0 | 0 | 0 |
| 0 | 0 | 0.000470431 | 0 | 0.032355121 |
| 0 | 0.038592398 | 0.004868446 | 0.001303571 | 0.161938871 |
| 0 | 0 | 0 | 0 | 0 |
| 0 | 0 | 0 | 0 | 0 |
| 0 | 0 | 0 | 0 | 0 |
| 0 | 0 | 0 | 0 | 0 |
| 0 | 0 | 0 | 0 | 0 |
| 0 | 0 | 0 | 0 | 0 |
| 0 | 0 | 0 | 0 | 0 |
| 0 | 0.000626162 | 0.001734972 | 0 | 0.001169874 |
| 0.027826368 | 0 | 0.07502084 | 0 | 0.459409418 |
| 0 | 0 | 0 | 0 | 0 |
| 0 | 0 | 0.008251509 | 0 | 0 |
| 0.001186998 | 0 | 0 | 0.002004322 | 0 |
| 0 | 0 | 0 | 0 | 0 |
| 0 | 0 | 0 | 0 | 0 |
| 0 | 0 | 0 | 0 | 0 |
| 0 | 0 | 0 | 0 | 0 |
| 0 | 0 | 0 | 0 | 0 |
| 0 | 0 | 0 | 0 | 0 |
| 0 | 0 | 0 | 0 | 0 |
| 0 | 0 | 0 | 0 | 0 |
| 0 | 0 | 0 | 0 | 0 |
| 0.071244654 | 0 | 0.002929295 | 0 | 0.002468991 |
| 0 | 0 | 0 | 0 | 0 |
| 0 | 0 | 0 | 0 | 0 |
| 0 | 0 | 0 | 0 | 0 |
| 0.478387199 | 0 | 0 | 0 | 0.719557591 |
| 0 | 0 | 0 | 0 | 0 |
| 0 | 0 | 0 | 0 | 0.064475062 |
| 0 | 0 | 0 | 0 | 0 |
| 0 | 0 | 0 | 0 | 0 |
| 0 | 0 | 0 | 0 | 0 |
| 0 | 0 | 0 | 0 | 0.670889921 |

| | | | | |
|---|---|---|---|---|
| 0.167233484 | 0 | 0 | 0 | 0.124772032 |
| 0 | 0 | 0.00342129 | 0 | 0 |
| 0 | 0 | 0 | 0 | 0 |
| 0 | 0 | 0 | 0 | 0 |
| 0 | 0 | 0 | 0 | 0 |
| 0.001842311 | 0.003587806 | 0.366869972 | 0.010239915 | 0 |
| 0 | 0 | 0 | 0 | 0 |
| 0 | 0 | 0 | 0 | 0 |
| 0 | 0 | 0 | 0 | 0 |
| 0 | 0 | 0 | 0 | 0 |
| 0 | 0 | 0 | 0 | 0 |
| 0 | 0 | 0.000533122 | 0 | 0 |
| 0 | 0 | 0 | 0 | 0 |
| 0 | 0 | 0 | 0 | 0 |
| 0 | 0 | 0 | 0 | 0 |
| 0.914373095 | 0 | 0 | 0 | 0 |
| 0 | 0 | 0 | 0 | 0 |
| 0 | 0 | 0 | 0.090196488 | 0 |
| 0 | 0 | 0 | 0 | 0 |
| 0 | 0 | 0 | 0 | 0 |
| 0 | 0.006453686 | 0.000460725 | 0.034455398 | 0 |
| 0 | 0 | 0 | 0 | 0 |
| 0.001162566 | 0.00362246 | 0.034911737 | 0 | 0.006252978 |
| 0.014009871 | 0 | 0.049660351 | 0 | 0.245249588 |
| 0.082493438 | 0 | 0 | 0 | 0 |
| 0 | 0 | 0 | 0 | 0 |
| 0 | 0 | 0 | 0 | 0 |
| 0 | 0 | 0 | 0 | 0 |
| 0.332954996 | 0 | 0 | 0 | 0 |
| 0 | 0 | 0 | 0 | 0 |
| 0 | 0 | 0 | 0 | 0 |
| 1.458860783 | 0 | 0 | 0 | 0 |
| 0 | 0.00778044 | 0.047679767 | 0.007353618 | 0 |
| 0 | 0 | 0 | 0 | 0.005923542 |
| 0.009251775 | 2.420609726 | 0.049838783 | 0 | 0.088122728 |
| 0 | 0.002722274 | 0.001749075 | 0 | 0.004284478 |
| 0 | 0 | 0 | 0 | 0 |
| 0 | 0 | 0 | 0 | 0 |
| 0.005557935 | 0 | 0 | 0.019196452 | 0.000479582 |
| 0 | 0 | 0 | 0 | 0 |
| 0 | 0 | 0 | 0 | 0 |
| 0.016326157 | 0 | 0 | 0 | 0 |
| 0.562843433 | 0.010645266 | 0.527822027 | 0.004655551 | 0.041684245 |
| 0 | 0 | 0 | 0 | 0 |
| 0 | 0 | 0 | 0 | 0 |
| 0.028934266 | 0 | 0 | 0.014129024 | 1.766860966 |
| 0 | 0 | 0 | 0 | 0 |
| 0 | 0 | 0 | 0 | 0 |

| | | | | |
|---|---|---|---|---|
| 0 | 0 | 0 | 0 | 0 |
| 0 | 0 | 0 | 0 | 0 |
| 0 | 0 | 0 | 0 | 0 |
| 0 | 0 | 0 | 0 | 0 |
| 0 | 0 | 0 | 0 | 0 |
| 0 | 0 | 0 | 0 | 0 |
| 0 | 0 | 0 | 0 | 0 |
| 0 | 0 | 0 | 0.100292959 | 0 |
| 0.00322374 | 0 | 0 | 0 | 0.102088097 |
| 0.260646455 | 0 | 0.001330332 | 0 | 0 |
| 0 | 0 | 0 | 0 | 0 |
| 0 | 0 | 0 | 0 | 0 |
| 0 | 0 | 0 | 0 | 0 |
| 0 | 0 | 0 | 0 | 0 |
| 0 | 0 | 0 | 0 | 0.044483832 |
| 0 | 0.022352628 | 0 | 0.005754077 | 0 |
| 0 | 0 | 0 | 0 | 0 |
| 0 | 0 | 0 | 0 | 0 |
| 0 | 0 | 0 | 0 | 0 |
| 0.140394762 | 0 | 0.078303594 | 0 | 1.795831325 |
| 0.004138458 | 0 | 5.407064954 | 0 | 0.011155745 |
| 0 | 0 | 0 | 0 | 0 |
| 0 | 0 | 0 | 0 | 0 |
| 0 | 0 | 0 | 0 | 0 |
| 0 | 0 | 0.000804097 | 0 | 0 |
| 0 | 0 | 0 | 0 | 0 |
| 0 | 0 | 0 | 0 | 0 |
| 0.559317097 | 0 | 3.31668229 | 0 | 0 |
| 0 | 0 | 0 | 0 | 0 |
| 0.057330032 | 0 | 0.028118071 | 0 | 0.074529181 |
| 0.466076136 | 0.941232925 | 0.083656237 | 0.002283995 | 0.779856173 |
| 0 | 0 | 0 | 0 | 0 |
| 0 | 0 | 0 | 0 | 0 |
| 0 | 0 | 0 | 0 | 0 |
| 0 | 0 | 0 | 0 | 0 |
| 0 | 0 | 0 | 0 | 0 |
| 0 | 0 | 0 | 0 | 0 |
| 0 | 0 | 0 | 0 | 0 |
| 0 | 0 | 0.008610895 | 0 | 0 |
| 0 | 0 | 0 | 0 | 0 |
| 0.003433073 | 0 | 0 | 0 | 0 |
| 0.038971547 | 0 | 0 | 0 | 0.156389904 |
| 0.000975322 | 0 | 0 | 0 | 0.005025456 |
| 0 | 0 | 0.000508397 | 0 | 0 |
| 0 | 0 | 0 | 0 | 0 |
| 0 | 0 | 0 | 0 | 0 |
| 0 | 0.048797824 | 0 | 1.936985051 | 0 |
| 0.001714203 | 0 | 0 | 0 | 0 |

| | | | | |
|---|---|---|---|---|
| 0 | 0 | 0 | 0 | 0 |
| 0 | 0 | 0 | 0 | 0 |
| 0 | 0 | 0 | 0 | 0 |
| 0 | 0 | 0 | 0 | 0 |
| 0.382673004 | 0.117125991 | 0.006643088 | 0.287879415 | 0.09871479 |
| 0 | 0 | 0 | 0 | 0 |
| 0 | 0 | 0 | 0 | 0 |
| 0 | 0 | 0 | 0 | 0 |
| 0 | 0 | 0 | 0 | 0 |
| 0 | 0 | 0 | 0 | 0 |
| 0.009541789 | 0 | 0 | 0 | 0.005843053 |
| 0.00370234 | 0.009507097 | 0.005053677 | 0.000710414 | 0.008146395 |
| 0.001016332 | 0.000295044 | 0 | 0 | 0 |
| 0 | 0 | 0 | 0 | 0 |
| 0 | 0 | 0 | 0 | 0 |
| 0 | 0 | 0 | 0 | 2.282827672 |
| 0 | 0 | 0.008551131 | 0 | 0.004054176 |
| 0 | 0 | 0 | 0.003273118 | 0 |
| 0.001146859 | 0.437445342 | 0 | 0.030661966 | 0.009555123 |
| 0.048750263 | 0.118643029 | 0.064829193 | 0.037366173 | 0 |
| 0 | 0 | 0 | 0 | 0 |
| 0 | 0 | 0 | 0 | 0 |
| 0 | 0 | 0 | 0 | 0 |
| 0 | 0 | 0 | 0 | 0 |
| 0.002467749 | 0 | 0 | 0 | 0 |
| 0 | 0 | 0 | 0 | 0 |
| 0.003866583 | 0 | 0 | 0.002701645 | 0.013541127 |
| 0 | 0 | 0 | 0.027041007 | 0 |
| 0 | 0 | 0 | 0 | 0 |
| 0 | 0 | 0 | 0 | 0 |
| 0 | 0.000337842 | 0 | 0.001276023 | 0 |
| 0 | 0 | 0 | 0.005513015 | 0 |
| 0 | 0 | 0 | 0 | 0 |
| 0 | 0 | 0 | 0 | 0 |
| 0 | 0 | 0 | 0 | 0 |
| 0 | 0 | 0 | 0 | 0 |
| 0 | 0 | 0 | 0 | 0 |
| 0.012423588 | 0.010314272 | 0.007812341 | 0.006096179 | 0.019048671 |
| 0.22503948 | 0.016895977 | 0.030491153 | 0 | 0.115406815 |
| 0 | 0 | 0 | 0 | 0 |
| 0 | 0.000215418 | 0.020851013 | 0.00125299 | 0 |
| 0 | 0 | 0 | 0 | 0 |
| 0 | 0 | 0 | 0 | 0 |
| 0 | 0.007231413 | 0 | 0.001063513 | 0 |
| 0 | 0 | 0 | 0 | 0 |
| 0 | 0 | 0 | 0 | 0 |
| 0 | 0 | 0 | 0 | 0 |
| 0 | 0 | 0 | 0 | 0 |

| | | | | |
|---|---|---|---|---|
| 0 | 0 | 0 | 0 | 0 |
| 0.019245017 | 0 | 0 | 0 | 0 |
| 0.052074889 | 0 | 0 | 0 | 0 |
| 0.002145219 | 0.000972469 | 0 | 0.003900983 | 0 |
| 0.007040323 | 0.00108856 | 0 | 0.000516871 | 0.002760133 |
| 0 | 0.001043633 | 0 | 0 | 0.00074994 |
| 0.049161856 | 0 | 0 | 0 | 0 |
| 0 | 0 | 0.012886314 | 0 | 0.009554753 |
| 0 | 0 | 0 | 0 | 0 |
| 0 | 0 | 0 | 0 | 0 |
| 0 | 0 | 0 | 0 | 0 |
| 0 | 0 | 0 | 0 | 0 |
| 0.280035113 | 0.014119662 | 0.187315241 | 0 | 0.024981157 |
| 0 | 0 | 0 | 0 | 0 |
| 0 | 0 | 0 | 0 | 0 |
| 0 | 0 | 0 | 0 | 0 |
| 0 | 0 | 0 | 0 | 0 |
| 0 | 0 | 0 | 0 | 0 |
| 0 | 0 | 0 | 0 | 0 |
| 0 | 0 | 0 | 0 | 0 |
| 0 | 0 | 0 | 0 | 0 |
| 0 | 0 | 0 | 0 | 0 |
| 0 | 0 | 0 | 0.008062643 | 0.006725083 |
| 0.003453396 | 0.00158822 | 0.004502987 | 0 | 0.011041154 |
| 0 | 0 | 0 | 0 | 0.009290677 |
| 0.003221515 | 0 | 0 | 0 | 0 |
| 0.006033556 | 0 | 0.004750637 | 0 | 0 |
| 0.000757189 | 0 | 0 | 0 | 0 |
| 0 | 0 | 0 | 0 | 0 |
| 0 | 0 | 0 | 0 | 0 |
| 0.019063131 | 0.009143702 | 0.010185733 | 0.004349909 | 0.00668339 |
| 0 | 0 | 0 | 0 | 0 |
| 0 | 0 | 0 | 0 | 0 |
| 0 | 0 | 0 | 0.000904859 | 0 |
| 0.011374648 | 0 | 0 | 0 | 0.159486319 |
| 0 | 0 | 0 | 0 | 0 |
| 0 | 0 | 0 | 0 | 0 |
| 0 | 0 | 0 | 0 | 0 |
| 0.002324862 | 0.012106277 | 0.001309038 | 0.001104097 | 0 |
| 0 | 0 | 0 | 0 | 0 |
| 0 | 0 | 0 | 0 | 0 |
| 0 | 0 | 0 | 0 | 0 |
| 0 | 0 | 0 | 0 | 0 |
| 0 | 0 | 0 | 0 | 0 |
| 14.43314342 | 0 | 0 | 0 | 0 |
| 0.095499031 | 0.443752863 | 0.201074179 | 0.067675822 | 0.128989192 |
| 0 | 0 | 0 | 0.195779085 | 0 |
| 0 | 0 | 0 | 0 | 0 |

| | | | | |
|---|---|---|---|---|
| 0 | 0 | 0 | 0 | 0 |
| 0 | 0 | 0 | 0 | 0 |
| 0.025963959 | 0 | 0.070782929 | 0 | 0.056756063 |
| 0.018529588 | 0 | 0 | 0 | 0 |
| 0 | 0 | 0 | 0 | 0 |
| 0.004354873 | 0 | 0 | 0 | 0 |
| 0 | 0.00150071 | 0 | 0.001122296 | 0 |
| 0 | 0 | 0 | 0 | 0 |
| 0.057945683 | 0 | 0 | 0 | 0 |
| 0 | 0 | 0 | 0 | 0 |
| 0 | 0 | 0 | 0 | 0 |
| 0.000703983 | 0 | 0 | 0.001337311 | 0.000501147 |
| 0 | 0 | 0 | 0 | 0 |
| 0 | 0 | 0 | 0 | 0 |
| 0 | 0 | 0 | 0 | 0 |
| 0 | 0 | 0 | 0 | 0 |
| 0.000519837 | 0 | 0 | 0.000438889 | 0 |
| 0 | 0 | 0 | 0 | 0 |
| 0 | 0 | 0 | 0 | 0 |
| 0 | 0 | 0 | 0.011746783 | 0.012464139 |
| 0 | 0 | 0 | 0 | 0 |
| 0 | 0 | 0 | 0 | 0 |
| 0 | 0 | 0 | 0 | 0 |
| 0 | 0 | 0 | 0 | 0 |
| 0 | 0 | 0 | 0 | 0 |
| 0.004915329 | 0 | 0 | 0 | 0 |
| 0 | 0 | 0 | 0 | 0 |
| 0.043078286 | 0.048481142 | 0.004416029 | 0 | 0 |
| 0 | 0 | 0 | 0 | 0 |
| 0 | 0 | 0 | 0 | 0 |
| 0.071315069 | 0.014164579 | 0.002924009 | 0 | 0.033653668 |
| 0 | 0 | 0 | 0 | 0 |
| 0.003742556 | 0.020090737 | 0.012526646 | 0.2247389 | 0.080814899 |
| 0 | 0.004339265 | 0.029071142 | 0 | 0.004688839 |
| 0 | 0 | 0 | 0 | 0 |
| 0 | 0 | 0 | 0 | 0 |
| 0 | 0 | 0 | 0 | 0 |
| 0 | 0 | 0 | 0 | 0 |
| 0 | 0 | 0 | 0 | 0 |
| 0 | 0 | 0 | 0 | 0 |
| 0 | 0 | 0 | 0 | 0 |
| 0.002052645 | 0.45234295 | 0.017914334 | 0.030219386 | 0.260950221 |
| 0.034853507 | 0.023026564 | 0.061249992 | 0.055685446 | 0.013499718 |
| 0 | 0 | 0 | 0 | 0 |
| 0 | 0 | 0 | 0 | 0 |
| 0.246182319 | 0.00238662 | 0.068841497 | 0.004957827 | 0.016506796 |
| 0 | 0 | 0.000889349 | 0 | 0 |
| 0 | 0 | 0 | 0 | 0 |

| | | | | |
|---|---|---|---|---|
| 0 | 0 | 0 | 0 | 0 |
| 0 | 0.008684417 | 0.002581451 | 0 | 0.012122353 |
| 0.01568363 | 0.019822226 | 0.001866681 | 0.000807403 | 0.012526321 |
| 0 | 0 | 0 | 0 | 0 |
| 0.002290724 | 0 | 0.101465561 | 0 | 0.021162941 |
| 0.000445955 | 0.001752055 | 0 | 0.095885104 | 0 |
| 0 | 0 | 0 | 0 | 0 |
| 0 | 0 | 0 | 0 | 0 |
| 0 | 0 | 0 | 0 | 0 |
| 0 | 0 | 0 | 0 | 0 |
| 0 | 0 | 0 | 0 | 0 |
| 0 | 0 | 0 | 0 | 0 |
| 0 | 0 | 0 | 0 | 0 |
| 0 | 0 | 0 | 0 | 0 |
| 0 | 0 | 0 | 0 | 0 |
| 0 | 0 | 0 | 0 | 0 |
| 0.634651463 | 0 | 0 | 0 | 0 |
| 0 | 0 | 0 | 0 | 0 |
| 0 | 0 | 0 | 0.085057742 | 0 |
| 0 | 0 | 0 | 0 | 0 |
| 0 | 0 | 0 | 0 | 0 |
| 0 | 0 | 0 | 0.080143659 | 0 |
| 0 | 0 | 0 | 0 | 0 |
| 0 | 0 | 0 | 0 | 0 |
| 0.076922584 | 0 | 0.006153629 | 0 | 0.010323449 |
| 0.001268562 | 0 | 0.001547601 | 0.050472033 | 0 |
| 0 | 0 | 0 | 0 | 0 |
| 0.356859408 | 0 | 0.000692874 | 0 | 0.003503984 |
| 0 | 0 | 0 | 0 | 0 |
| 0 | 0 | 0 | 0 | 0 |
| 0.003649828 | 0 | 0 | 0 | 0 |
| 0 | 0.000395612 | 0 | 0.022893612 | 0 |
| 0 | 0 | 0 | 0 | 0 |
| 0 | 0 | 0 | 0 | 0 |
| 0 | 0 | 0 | 0 | 0 |
| 0 | 0 | 0 | 0 | 0 |
| 0 | 0 | 0 | 0 | 0 |
| 0 | 0 | 0 | 0 | 0 |
| 0.002015571 | 0 | 0.177474983 | 1.822896884 | 0 |
| 0 | 0 | 0 | 0 | 0 |
| 0 | 0 | 0 | 0 | 0 |
| 0.06020851 | 0.001153595 | 0.006328195 | 0 | 0.002612471 |
| 0 | 0.015393695 | 0.001433711 | 0.000940529 | 0.003625262 |
| 0 | 0 | 0 | 0 | 0 |
| 0 | 0 | 0 | 0 | 0 |
| 0.004943373 | 0 | 0 | 0 | 0 |
| 0 | 0 | 0 | 0.024254479 | 0.007394588 |
| 0 | 0 | 0 | 0 | 0 |

| | | | | |
|---|---|---|---|---|
| 0 | 0 | 0 | 0 | 0 |
| 0.005068767 | 0.015575371 | 0.000922512 | 0.018025644 | 0.013558542 |
| 0 | 0 | 0 | 0 | 0 |
| 0 | 0 | 0 | 0 | 0 |
| 0 | 0 | 0 | 0 | 0 |
| 0 | 0 | 0 | 0 | 0 |
| 0 | 0 | 0 | 0 | 0 |
| 0 | 0 | 0 | 0 | 0 |
| 0 | 0 | 0 | 0 | 0 |
| 0 | 0 | 0 | 0 | 0 |
| 0 | 0 | 0 | 0 | 0 |
| 0.000671164 | 0 | 0 | 0 | 0 |
| 0.317104198 | 0.009172634 | 0.173093997 | 0 | 0.077981567 |
| 0 | 0 | 0 | 0 | 0 |
| 0 | 0 | 0.007045183 | 0 | 0.016178342 |
| 0 | 0 | 0 | 0 | 0 |
| 0 | 0 | 0 | 0 | 0 |
| 0 | 0 | 0 | 0 | 0 |
| 0 | 0 | 0 | 0 | 0.005831749 |
| 0 | 0 | 0 | 0 | 0 |
| 0 | 0 | 0 | 0 | 0 |
| 0.017679768 | 0.053508929 | 0.005789306 | 0.014132739 | 0 |
| 0 | 0 | 0 | 0.005857 | 0.011248688 |
| 0 | 0 | 0 | 0 | 0 |
| 0 | 0 | 0 | 0 | 0 |
| 0 | 0 | 0 | 0 | 0 |
| 0 | 0 | 0 | 0 | 0 |
| 0 | 0 | 0 | 0 | 0 |
| 0 | 0 | 0 | 0 | 0 |
| 0.006919606 | 0 | 0 | 0 | 0 |
| 0 | 0 | 0 | 0 | 0 |
| 0 | 0 | 0 | 0 | 0 |
| 0 | 0 | 0 | 0 | 0 |
| 0 | 0 | 0 | 0 | 0 |
| 3.400172959 | 0 | 0.015769102 | 0.740059839 | 0 |
| 0.003373488 | 0 | 0 | 0 | 0 |
| 0 | 0 | 0 | 0 | 0 |
| 0 | 0 | 0 | 0 | 0 |
| 0.018218857 | 0.000496183 | 0.078259361 | 0.00190291 | 0.000713101 |
| 0 | 0 | 0 | 0 | 0 |
| 0.062984743 | 0.948616845 | 0.06220464 | 0.005971737 | 0.604542698 |
| 0 | 0 | 0 | 0 | 0 |
| 0 | 0 | 0 | 0 | 0 |
| 0 | 0 | 0 | 0 | 0 |
| 0 | 0 | 0 | 0 | 0 |
| 0.04319082 | 0.095409564 | 0.027503689 | 0.291233554 | 0 |
| 0 | 0 | 0 | 0 | 0 |
| 0 | 0 | 0 | 0 | 0 |

| | | | | |
|---|---|---|---|---|
| 0 | 0 | 0 | 0 | 0 |
| 0 | 0 | 0 | 0 | 0 |
| 0 | 0 | 0 | 0 | 0 |
| 0 | 0 | 0 | 0 | 0 |
| 0 | 0 | 0 | 0.000493859 | 0 |
| 0 | 0 | 0 | 0 | 0 |
| 0 | 0 | 0 | 0 | 0 |
| 0.002709291 | 0.000338807 | 0.000938766 | 0 | 0 |
| 0.000653098 | 0.012156923 | 0 | 0 | 0 |
| 0.04054352 | 0.011036338 | 0 | 0.101894466 | 0.000715953 |
| 0.023268814 | 0.00788084 | 0 | 0 | 0 |
| 0 | 0 | 0 | 0 | 0 |
| 0.000921579 | 0 | 0.001660495 | 0.002956675 | 0.002099352 |
| 0 | 0 | 0 | 0 | 0 |
| 0 | 0 | 0 | 0 | 0 |
| 0 | 0 | 0 | 0 | 0 |
| 0.393138554 | 0 | 0 | 0 | 0 |
| 0 | 0 | 0 | 0 | 0 |
| 0 | 0 | 0 | 0 | 0 |
| 0.753992119 | 0.079710563 | 0.641803047 | 0.207069252 | 0.051385717 |
| 0 | 0 | 0 | 0 | 0 |
| 0 | 0 | 0 | 0 | 0 |
| 0 | 0 | 0 | 0 | 0 |
| 0 | 0 | 0 | 0 | 0 |
| 0 | 0 | 0 | 0 | 0 |
| 0.006886212 | 0 | 0 | 0 | 0 |
| 0 | 0 | 0 | 0 | 0 |
| 0 | 0 | 0 | 0 | 0 |
| 0 | 0 | 0 | 0 | 0 |
| 0 | 0.015003848 | 0 | 0 | 0 |
| 0 | 0 | 0 | 0 | 0 |
| 0.090673827 | 0 | 0 | 0 | 0 |
| 0 | 0 | 0 | 0 | 0 |
| 0 | 0 | 0 | 0 | 0 |
| 0 | 0 | 0 | 0 | 0 |
| 0 | 0 | 0 | 0 | 0 |
| 0 | 0 | 0 | 0 | 0 |
| 0 | 0 | 0 | 0.012725515 | 0 |
| 0 | 0 | 0 | 0 | 0 |
| 0 | 0 | 0 | 0 | 0 |
| 0 | 0 | 0 | 0 | 0 |
| 0 | 0 | 0 | 0 | 0 |
| 0.001090945 | 0 | 0 | 0 | 0.319577142 |
| 0 | 0 | 0 | 0 | 0 |
| 0.013405441 | 0.001816098 | 0.005668287 | 0.001431008 | 0.000877516 |
| 0 | 0 | 0 | 0 | 0 |
| 0 | 0 | 0 | 0 | 0 |
| 0 | 0 | 0 | 0 | 0 |

| | | | | |
|---|---|---|---|---|
| 0 | 0 | 0 | 0 | 0 |
| 0 | 0 | 0 | 0 | 0 |
| 0 | 0 | 0 | 0 | 0 |
| 0 | 0 | 0 | 0 | 0 |
| 0 | 0 | 0 | 0 | 0 |
| 0.075575262 | 0.007807469 | 0.037738935 | 0.002816436 | 0.010699958 |
| 0 | 0 | 0 | 0 | 0 |
| 0 | 0 | 0 | 0 | 0 |
| 0 | 0 | 0 | 0 | 0 |
| 0 | 0 | 0 | 0 | 0 |
| 0 | 0 | 0 | 0 | 0 |
| 0 | 0 | 0 | 0 | 0 |
| 0 | 0 | 0 | 0 | 0 |
| 0.008236011 | 0 | 0 | 0 | 0 |
| 0 | 0 | 0 | 0 | 0 |
| 0 | 0 | 0 | 0 | 0 |
| 0.10116041 | 0 | 0 | 0 | 0 |
| 0 | 0 | 0 | 0 | 0 |
| 0 | 0 | 0 | 0 | 0 |
| 0 | 0 | 0 | 0 | 0 |
| 0 | 0 | 0.000600794 | 0 | 0 |
| 0 | 0 | 0 | 0 | 0 |
| 0 | 0 | 0 | 0 | 0 |
| 0 | 0 | 0 | 0 | 0 |
| 0 | 0 | 0 | 0 | 0 |
| 0 | 0.000146254 | 0 | 0.014431414 | 0 |
| 0 | 0 | 0 | 0 | 0 |
| 0.15984537 | 0 | 0.546941054 | 0 | 0.387287666 |
| 0 | 0 | 0 | 0 | 0 |
| 0 | 0 | 0 | 0 | 0 |
| 0 | 0 | 0 | 0 | 0 |
| 0 | 0 | 0 | 0 | 0 |
| 0 | 0 | 0 | 0 | 0 |
| 0 | 0 | 0 | 0 | 0 |
| 0 | 0 | 0 | 0 | 0 |
| 0 | 0 | 0 | 0 | 0 |
| 0 | 0 | 0 | 0 | 0 |
| 0 | 0 | 0 | 0 | 0 |
| 0.250221946 | 0 | 0 | 0.275142555 | 5.667834161 |
| 0 | 0 | 0 | 0 | 0 |
| 0 | 0 | 0 | 0 | 0 |
| 0 | 0 | 0 | 0 | 0 |
| 0 | 0 | 0 | 0 | 0 |
| 0 | 0 | 0 | 0 | 0 |
| 0 | 0 | 0 | 0 | 0 |
| 0 | 0 | 0 | 0 | 0 |
| 0 | 0 | 0 | 0 | 0 |
| 0 | 0 | 0 | 0 | 0 |

| | | | | |
|---|---|---|---|---|
| 0 | 0 | 0 | 0 | 0 |
| 0.009787759 | 0 | 0 | 0 | 0.003544944 |
| 0 | 0 | 0 | 0.001996806 | 0 |
| 0 | 0 | 0 | 0 | 0 |
| 0 | 0 | 0 | 0 | 0 |
| 0.004944921 | 0 | 0 | 0 | 0 |
| 0 | 0 | 0 | 0 | 0 |
| 0 | 0 | 0 | 0 | 0 |
| 0 | 0 | 0 | 0 | 0 |
| 0 | 0 | 0 | 0 | 0 |
| 0.096628112 | 0.0010966 | 0.007216351 | 0 | 0 |
| 0 | 0 | 0 | 0 | 0 |
| 0.029064797 | 0 | 0.033289983 | 0 | 0.000825261 |
| 0 | 0 | 0 | 0 | 0 |
| 0 | 0 | 0 | 0 | 0 |
| 0 | 0 | 0 | 0 | 0 |
| 0 | 0 | 0 | 0 | 0 |
| 0 | 0 | 0 | 0 | 0 |
| 0 | 0 | 0 | 0 | 0 |
| 0 | 0 | 0 | 0 | 0 |
| 0.814697002 | 0.028102829 | 0.03790575 | 0.004190586 | 0 |
| 0 | 0 | 0 | 0 | 0 |
| 0 | 0 | 0 | 0 | 0 |
| 0.51226773 | 6.054884712 | 0.658252373 | 0 | 3.408161657 |
| 0 | 0 | 0 | 0 | 0 |
| 0 | 0 | 0 | 0 | 0 |
| 0 | 0 | 0 | 0 | 0 |
| 0 | 0 | 0 | 0 | 0 |
| 0 | 0 | 0 | 0.019345897 | 0 |
| 0 | 0 | 0 | 0 | 0 |
| 0 | 0 | 0 | 0.003971229 | 0 |
| 0 | 0 | 0 | 0 | 0 |
| 0.012422658 | 0 | 0 | 0 | 0 |
| 0 | 0 | 0 | 0 | 0 |

| PHL018-0171_V2 | PHL018-0236_V3 | PHL018-0201_V1 | PHL018-0144_V2 | PHL018-0231_V3 |
|---|---|---|---|---|
| REF | EYCF | CTRL | REF | EYCF |
| 0.111217757 | 1.792157806 | 0.03312351 | 3.209480661 | 0.005528323 |
| 0.094797274 | 0.475327142 | 0.518735939 | 0.026423372 | 3.046528265 |
| 0 | 14.76615262 | 5.88125023 | 1.47627843 | 5.645249483 |
| 0 | 0 | 0 | 0 | 0 |
| 0 | 0 | 0 | 0 | 0 |
| 0 | 0 | 0 | 0 | 0 |
| 0 | 0 | 0 | 0 | 0 |
| 0 | 0 | 0 | 0 | 0 |
| 0 | 0 | 0 | 0 | 0 |
| 0 | 0 | 0 | 0 | 0 |
| 0 | 0 | 0 | 0 | 0 |
| 0 | 0 | 0 | 0 | 0 |
| 0 | 0 | 0 | 0 | 0 |
| 1.36974045 | 3.946888101 | 0 | 0.645640864 | 7.348041682 |
| 0 | 0 | 0 | 0.294246823 | 0 |
| 0 | 0 | 0 | 0 | 0 |
| 0 | 0 | 0 | 0 | 0 |
| 0 | 0 | 0 | 0 | 0 |
| 0 | 0 | 0 | 0 | 0.005171269 |
| 0 | 0 | 0 | 0 | 0 |
| 0 | 0 | 0 | 0 | 0 |
| 0 | 0 | 0 | 0 | 0 |
| 0 | 0 | 0 | 0 | 0 |
| 0 | 0 | 0 | 0 | 0 |
| 0.106458222 | 0 | 0 | 0 | 0 |
| 0 | 0 | 0 | 0 | 0 |
| 0 | 0 | 0 | 0 | 0 |
| 0 | 0 | 0 | 0 | 0 |
| 0 | 0 | 0 | 0 | 0 |
| 0 | 0 | 0 | 0 | 0 |
| 0 | 0 | 0 | 0 | 0 |
| 0 | 0 | 0 | 0 | 0 |
| 0 | 0 | 0 | 0 | 0 |
| 0 | 0 | 0 | 0 | 0 |
| 0 | 0 | 0 | 0 | 0 |
| 0.155971266 | 0 | 0 | 0.883235205 | 4.337732514 |
| 0 | 0 | 0 | 0 | 0 |
| 0 | 0 | 0 | 0 | 0 |
| 0 | 0 | 0 | 0 | 0 |
| 0 | 0 | 0 | 0 | 0 |
| 0 | 0 | 0 | 0 | 0 |
| 0.009251807 | 0 | 0.006177928 | 0.005020558 | 0.144402242 |
| 0.045433026 | 0.138845562 | 0.875910851 | 3.192879343 | 0.281001713 |
| 0.005505483 | 0.000992709 | 0.119225435 | 0.040133275 | 0.002887607 |
| 0.010055455 | 0 | 0.016854378 | 0.001632212 | 0 |
| 0 | 0 | 0 | 0 | 0 |

| | | | | |
|---|---|---|---|---|
| 0.054414686 | 0 | 0 | 0.046923036 | 0 |
| 0.000943746 | 0.004084069 | 0.002755062 | 0.005159715 | 0.044986019 |
| 0 | 0 | 0 | 0.069134923 | 0.006861407 |
| 0.473653744 | 0.072841768 | 0.438218249 | 0.33323001 | 0.573560636 |
| 0.000437437 | 0.002981496 | 0 | 0.002134032 | 0 |
| 0 | 0 | 0 | 0 | 0 |
| 0 | 0 | 0 | 0.007941062 | 0.198254553 |
| 0 | 0 | 0 | 0 | 0 |
| 0 | 0 | 0 | 0 | 0 |
| 0 | 0 | 0 | 0 | 0 |
| 0.000619609 | 0 | 0 | 0 | 0 |
| 0.414521805 | 0 | 0 | 0 | 0 |
| 0 | 0 | 0 | 0 | 0 |
| 0.009055449 | 0.054176744 | 0.022782515 | 0.012897618 | 0.008884457 |
| 0 | 0 | 0.08889343 | 0 | 0.046688239 |
| 0 | 0 | 0 | 0 | 0 |
| 0.066208325 | 0.088238283 | 0.001976735 | 0.386621136 | 0.405375493 |
| 0 | 0 | 0 | 0 | 0 |
| 0 | 0 | 0 | 0 | 0 |
| 0 | 0 | 0 | 0 | 0 |
| 0 | 0 | 0.045507163 | 0.025218666 | 0.159421154 |
| 0.005031146 | 0.003985112 | 0.032903444 | 0.003728005 | 0 |
| 0.046084642 | 0 | 0 | 0 | 0 |
| 0 | 0 | 0 | 0 | 0 |
| 0 | 0 | 0 | 0 | 0 |
| 0 | 0 | 0 | 0 | 0 |
| 0 | 0 | 0.351199387 | 0.001804609 | 0 |
| 0 | 0 | 0 | 0 | 0 |
| 0 | 0 | 0.002440175 | 0 | 0.003616943 |
| 0 | 0 | 0 | 0 | 0 |
| 0 | 0 | 0 | 0 | 0 |
| 0 | 0 | 0 | 0 | 0 |
| 0 | 0 | 0 | 0 | 0 |
| 0 | 0 | 0 | 0 | 0 |
| 0 | 0 | 0 | 0 | 0 |
| 0 | 0 | 0.002439091 | 0 | 0 |
| 0 | 0 | 0 | 0 | 0 |
| 0 | 0.007307483 | 0.84099913 | 10.93276098 | 0.009881877 |
| 0 | 0 | 0 | 0 | 0 |
| 0 | 0 | 0 | 0 | 0 |
| 0 | 0 | 0 | 0 | 0 |
| 0 | 0 | 0 | 0 | 0 |
| 0 | 0 | 0 | 0 | 0 |
| 0 | 0 | 0 | 0 | 0 |
| 0 | 0 | 0 | 0 | 0 |
| 0 | 0 | 0 | 0 | 0 |
| 0 | 0 | 0.059427983 | 0 | 0 |
| 0 | 0 | 0 | 0 | 0 |

| | | | | |
|---|---|---|---|---|
| 0 | 0 | 0 | 0 | 0 |
| 0 | 0 | 0 | 0 | 0 |
| 0 | 0 | 0 | 0 | 0 |
| 0 | 0 | 0 | 0 | 0 |
| 0 | 0 | 0 | 0 | 0 |
| 0 | 0 | 0 | 0 | 0 |
| 0 | 0 | 0 | 0 | 0 |
| 0 | 0 | 0 | 0 | 0 |
| 0.043535404 | 0 | 0.000460924 | 0.001558231 | 0.000643015 |
| 0 | 0 | 0 | 0 | 0 |
| 0 | 0 | 0 | 0 | 0 |
| 0.003495046 | 0.00065444 | 0 | 0 | 0.30980894 |
| 0 | 0 | 0 | 0 | 0 |
| 0 | 0 | 0 | 0 | 0 |
| 0 | 0 | 0 | 0 | 0 |
| 0 | 0 | 0 | 0.001573062 | 0 |
| 0 | 0 | 0 | 0 | 0 |
| 0 | 0 | 0 | 0 | 0 |
| 0.07907664 | 0 | 0 | 0 | 0 |
| 0.033822687 | 0 | 0 | 0 | 0 |
| 0 | 0 | 0 | 0 | 0 |
| 0 | 0 | 0 | 0 | 0 |
| 0.000633216 | 0 | 0 | 0 | 0 |
| 0 | 0 | 0 | 0 | 0 |
| 0 | 0 | 0 | 0 | 0 |
| 0 | 0 | 0 | 0 | 0 |
| 0 | 0 | 0 | 0 | 0 |
| 0 | 0 | 0 | 0 | 0 |
| 0 | 0 | 0 | 0 | 0 |
| 0 | 0 | 0 | 0 | 0 |
| 0.033536279 | 0 | 0 | 0.003694975 | 0 |
| 0.105547434 | 0.004469071 | 0.000626448 | 0 | 0 |
| 0 | 0 | 0 | 0 | 0 |
| 0 | 0 | 0 | 0 | 0 |
| 0 | 0 | 0 | 0 | 0 |
| 0 | 0 | 0 | 0 | 0 |
| 0 | 0 | 0 | 0 | 0 |
| 0 | 0 | 0 | 0 | 0 |
| 0 | 0 | 0 | 0 | 0 |
| 0 | 0 | 0 | 0 | 0 |
| 0 | 0 | 0 | 0 | 0 |
| 0.054666243 | 0 | 0 | 0 | 0 |
| 0 | 0.000316257 | 0 | 0 | 0 |
| 0 | 0 | 0 | 0 | 0 |
| 0 | 0 | 0 | 0 | 0 |
| 0 | 0 | 0 | 0 | 0 |
| 0 | 0 | 0 | 0 | 0 |
| 0 | 0 | 0 | 0 | 0 |

| | | | | |
|---|---|---|---|---|
| 0 | 0 | 0 | 0 | 0 |
| 0 | 0 | 0 | 0 | 0 |
| 0 | 0 | 0 | 0 | 0 |
| 0 | 0 | 0 | 0 | 0 |
| 0.018392986 | 0 | 0 | 0.025180997 | 0 |
| 0 | 0 | 0 | 0 | 0 |
| 0 | 0 | 0 | 0 | 0 |
| 0 | 0 | 0 | 0 | 0 |
| 0 | 0 | 0 | 0 | 0 |
| 0 | 0 | 0 | 0 | 0 |
| 0 | 0 | 0 | 0 | 0 |
| 0 | 0 | 0 | 0 | 0 |
| 0 | 0 | 0 | 0 | 0 |
| 0.003068881 | 0 | 0 | 0 | 0 |
| 0 | 0 | 0 | 0 | 0 |
| 0 | 0 | 0 | 0 | 0 |
| 0 | 0 | 0 | 0 | 0 |
| 0 | 0 | 0 | 0 | 0 |
| 0 | 0 | 0 | 0 | 0 |
| 0 | 0 | 0 | 0 | 0 |
| 0 | 0 | 0 | 0 | 0 |
| 0 | 0 | 0 | 0 | 0 |
| 0.00271983 | 0 | 0 | 0 | 0 |
| 0 | 0 | 0 | 0 | 0 |
| 0 | 0 | 0 | 0 | 0 |
| 0 | 0 | 0 | 0 | 0 |
| 0.023931782 | 0 | 0 | 0 | 0 |
| 0 | 0 | 0 | 0 | 0 |
| 0 | 0 | 0 | 0 | 0 |
| 0 | 0 | 0 | 0 | 0 |
| 0.059226701 | 0 | 0 | 0 | 0 |
| 0 | 0 | 0 | 0 | 0.400269443 |
| 0 | 0 | 0 | 0 | 0 |
| 0 | 0 | 0 | 0 | 0 |
| 0 | 0 | 0 | 0 | 0 |
| 0 | 0 | 0 | 0 | 0 |
| 0 | 0 | 0 | 0 | 0 |
| 0 | 0 | 0 | 0 | 0 |
| 0 | 0 | 0 | 0 | 0 |
| 0 | 0 | 0 | 0 | 0 |
| 0 | 0 | 0 | 0 | 0 |
| 0 | 0 | 0 | 0 | 0 |
| 0 | 0 | 0 | 0 | 0 |
| 0 | 0 | 0 | 0 | 0 |
| 0 | 0 | 0 | 0 | 0 |
| 0 | 0 | 0 | 0 | 0 |
| 0 | 0 | 0 | 0 | 0 |
| 0 | 0 | 0 | 0 | 0 |

| | | | | |
|---|---|---|---|---|
| 0 | 0 | 0 | 0 | 0 |
| 0 | 0 | 0 | 0 | 0 |
| 0 | 0 | 0 | 0 | 0 |
| 0 | 0 | 0 | 0 | 0 |
| 0 | 0 | 0 | 0 | 0 |
| 0 | 0 | 0 | 0 | 0 |
| 0 | 0 | 0 | 0 | 0 |
| 0 | 0 | 0 | 0 | 0 |
| 0 | 0 | 0 | 0 | 0 |
| 0 | 0 | 0 | 0 | 0 |
| 0 | 0 | 0 | 0 | 0 |
| 0.037766939 | 0 | 0 | 0 | 0 |
| 0.002978416 | 0 | 0 | 0.017327635 | 0 |
| 0 | 0 | 0 | 0 | 0 |
| 0 | 0 | 0 | 0 | 0 |
| 0 | 0 | 0 | 0 | 0 |
| 0 | 0 | 0 | 0 | 0 |
| 0 | 0 | 0 | 0 | 0 |
| 0 | 0 | 0 | 0 | 0 |
| 0 | 0 | 0 | 0 | 0 |
| 0.007193191 | 0.048763936 | 0.070571262 | 0.010734448 | 0.028994039 |
| 0 | 0 | 0 | 0 | 0 |
| 0 | 0 | 0 | 0.003253771 | 0 |
| 0 | 0.002748701 | 0.026778107 | 0 | 0 |
| 0 | 0 | 0 | 0 | 0 |
| 0 | 0 | 0 | 0 | 0 |
| 0.005540482 | 0.001096485 | 0.002044193 | 0 | 0.00806749 |
| 0 | 0 | 0 | 0 | 0 |
| 0 | 0 | 0.000777102 | 0 | 0 |
| 0 | 0.002815471 | 0.031424471 | 0.006323553 | 0 |
| 0 | 0.082194319 | 0 | 0 | 0.334394176 |
| 0 | 0 | 0 | 0 | 0 |
| 0 | 0 | 0 | 0 | 0 |
| 0.112169713 | 0 | 0 | 0 | 0 |
| 0.013738868 | 0.013597892 | 0 | 0.049017816 | 0.002515342 |
| 0 | 0 | 0 | 0 | 0 |
| 0 | 0 | 0 | 0 | 0 |
| 0 | 0 | 0 | 0 | 0 |
| 0 | 0 | 0 | 0 | 0 |
| 0 | 0 | 0 | 0 | 0 |
| 0 | 0 | 0 | 0 | 0 |
| 0 | 0.000367032 | 0 | 0 | 0 |
| 0.000627693 | 0 | 0 | 0 | 0 |
| 0 | 0 | 0 | 0 | 0 |
| 0 | 0 | 0 | 0 | 0 |
| 0.033924088 | 0 | 0 | 0 | 0 |
| 0 | 0 | 0 | 0 | 0 |
| 0.003765649 | 0 | 0 | 0.008506615 | 0 |

| | | | | |
|---|---|---|---|---|
| 0 | 0 | 0 | 0 | 0 |
| 0 | 0 | 0 | 0 | 0 |
| 0 | 0.000478634 | 0 | 0 | 0.007097783 |
| 0 | 0 | 0 | 0 | 0 |
| 0 | 0 | 0 | 0 | 0 |
| 0 | 0 | 0 | 0 | 0 |
| 0 | 0 | 0 | 0 | 0 |
| 0 | 0 | 0.003078434 | 0 | 0.008803906 |
| 0 | 0 | 0 | 0 | 0 |
| 0 | 0 | 0 | 0 | 0 |
| 0 | 0 | 0 | 0 | 0 |
| 0 | 0 | 0 | 0 | 0 |
| 0 | 0 | 0 | 0 | 0 |
| 0 | 0 | 0 | 0 | 0 |
| 0 | 0 | 0 | 0 | 0 |
| 0 | 0 | 0 | 0 | 0 |
| 0 | 0 | 0 | 0 | 0 |
| 0 | 0 | 0 | 0 | 0 |
| 0 | 0.001065394 | 0.00522692 | 0.014541331 | 0 |
| 0.18820791 | 0 | 0 | 0 | 0 |
| 0 | 0.062207772 | 0.042791825 | 0 | 0.007394898 |
| 0 | 0 | 0 | 0 | 0 |
| 0 | 0 | 0 | 0 | 0 |
| 0 | 0 | 0 | 0 | 0 |
| 0.002565292 | 0 | 0.005106006 | 0.003506285 | 0.015581924 |
| 0 | 0 | 0 | 0 | 0 |
| 0 | 0 | 0 | 0 | 0 |
| 0 | 0 | 0 | 0 | 0 |
| 0 | 0 | 0 | 0 | 0.04461017 |
| 0 | 0 | 0 | 0 | 0 |
| 0.026702924 | 1.287306026 | 1.125268596 | 0.50830197 | 0.054868973 |
| 0 | 0 | 0 | 0 | 0 |
| 0 | 0.04741644 | 0 | 0 | 0.520396904 |
| 0 | 0 | 0 | 0 | 0 |
| 0 | 0 | 0 | 0 | 0 |
| 0 | 0 | 0 | 0 | 0 |
| 0 | 0 | 0 | 0 | 0 |
| 0 | 0 | 0 | 0 | 0 |
| 0 | 0 | 0 | 0 | 0 |
| 0 | 0 | 0 | 0.00059826 | 0 |
| 0 | 0 | 0 | 0 | 0 |
| 0.00290338 | 0.004417172 | 0 | 0.017399866 | 0 |
| 0 | 0 | 0 | 0 | 0 |
| 0 | 0 | 0 | 0 | 0 |
| 0 | 0 | 0 | 0 | 0 |
| 0 | 0 | 0 | 0 | 0 |
| 0 | 0 | 0 | 0 | 0 |
| 0 | 0 | 0 | 0 | 0 |

| | | | | |
|---|---|---|---|---|
| 0 | 0 | 0 | 0 | 0 |
| 0 | 0 | 0 | 0 | 0 |
| 0.001367127 | 0.000739531 | 0.12589882 | 0.038458285 | 0.002108981 |
| 0 | 0 | 0 | 0 | 0 |
| 7.749895871 | 0 | 0 | 0 | 0 |
| 0 | 0 | 0 | 0 | 0 |
| 0 | 0 | 0 | 0 | 0 |
| 0 | 0 | 0 | 0 | 0 |
| 0 | 0 | 0 | 0 | 0 |
| 0 | 0 | 0 | 0 | 0 |
| 0 | 0 | 0 | 0 | 0 |
| 0 | 0 | 0 | 0 | 0 |
| 0 | 0 | 0 | 0 | 0 |
| 0 | 0.001498412 | 0.002138573 | 0.012143814 | 0 |
| 0.007066721 | 0.005059404 | 0.003971503 | 0.001048932 | 0.000961887 |
| 0.041389692 | 0 | 0 | 0 | 0 |
| 0.000947282 | 0.000896737 | 0.00314248 | 0.001593552 | 0.00613751 |
| 0 | 0 | 0 | 0 | 0 |
| 0 | 0 | 0 | 0 | 0 |
| 0 | 0 | 0 | 0 | 0 |
| 0 | 0 | 0 | 0 | 0 |
| 0.003848063 | 0 | 0.001702058 | 0.00887089 | 0 |
| 0 | 0 | 0 | 0 | 0 |
| 0 | 0 | 0 | 0 | 0 |
| 0.001371143 | 0 | 0 | 0 | 0 |
| 0 | 0 | 0 | 0 | 0 |
| 0.081255166 | 0 | 0 | 0 | 0 |
| 0 | 0 | 0 | 0 | 0 |
| 0 | 0 | 0 | 0 | 0 |
| 0 | 0 | 0 | 0 | 0.001717343 |
| 0.000873728 | 0 | 0 | 0 | 0 |
| 0 | 0 | 0 | 0 | 0 |
| 0.001474778 | 0 | 0 | 0 | 0 |
| 0 | 0 | 0 | 0 | 0 |
| 0 | 0 | 0 | 0 | 0 |
| 0.002846651 | 0 | 0 | 0 | 0.002371329 |
| 0 | 0 | 0 | 0 | 0 |
| 0.443896396 | 0 | 0.004276918 | 0.058136583 | 0.000994424 |
| 0 | 0 | 0 | 0 | 0 |
| 0 | 0 | 0 | 0 | 0 |
| 0 | 0 | 0 | 0 | 0 |
| 0 | 0 | 0 | 0 | 0 |
| 0 | 0 | 0 | 0 | 0 |
| 0 | 0 | 0.048875932 | 0 | 0.004632111 |
| 0 | 0 | 0 | 0 | 0 |
| 0.004955509 | 0 | 0 | 0 | 0 |
| 0.001552779 | 0 | 0 | 0 | 0 |
| 0 | 0.001935847 | 0 | 0 | 0 |

| | | | | |
|---|---|---|---|---|
| 0 | 0 | 0 | 0 | 0.07345082 |
| 0.166888523 | 0 | 0 | 0 | 0.033291846 |
| 0 | 0 | 0.035774305 | 0 | 0.00145079 |
| 0 | 0 | 0 | 0 | 0 |
| 0.002403033 | 0.012917683 | 0 | 0.009221884 | 0.020643399 |
| 0.016897939 | 0 | 0 | 0 | 0 |
| 0 | 0 | 0 | 0 | 0 |
| 0.015578754 | 0.005807898 | 0 | 0 | 0.002338284 |
| 0 | 0 | 0 | 0 | 0 |
| 0.015189394 | 0.290336832 | 2.483446851 | 0.021536817 | 0.246133521 |
| 0 | 0 | 0 | 0 | 0 |
| 0.008165915 | 0 | 0.000796094 | 0 | 0.047431724 |
| 0.004930027 | 0 | 0.002314819 | 0.000744286 | 0 |
| 0 | 0 | 0 | 0 | 0 |
| 0 | 0 | 0 | 0 | 0 |
| 0 | 0 | 0 | 0 | 0 |
| 0 | 0 | 0 | 0 | 0 |
| 0 | 0 | 0 | 0 | 0.000914293 |
| 0 | 0.018887198 | 0.01853246 | 0 | 0.171224874 |
| 0 | 0.096014773 | 0 | 0 | 0 |
| 0 | 0 | 0 | 0 | 0 |
| 0 | 0 | 0 | 0 | 0 |
| 0 | 0 | 0.008308155 | 0 | 0 |
| 0.064916935 | 0 | 0 | 0 | 0 |
| 0 | 0 | 0 | 0 | 0 |
| 0.004122623 | 0.000477875 | 0.011722497 | 0 | 0.000613258 |
| 0 | 0 | 0 | 0 | 0 |
| 0 | 0.003611776 | 0 | 0.137480614 | 0.049687144 |
| 0 | 0.241090448 | 0.131423502 | 0 | 0.157390476 |
| 0 | 0.002212531 | 0 | 0.003695157 | 0 |
| 0 | 0 | 0.133210308 | 0 | 0 |
| 0 | 0 | 0 | 0 | 0 |
| 0 | 0 | 0.001042535 | 0 | 0 |
| 0 | 0.032498601 | 0 | 0 | 0 |
| 0 | 0 | 0 | 0 | 0 |
| 0 | 0.022048896 | 0.20600585 | 0 | 0 |
| 0 | 0 | 0.004916801 | 0 | 1.056457921 |
| 0 | 0 | 0 | 0 | 0 |
| 0 | 0 | 0 | 0 | 0 |
| 0 | 0 | 0 | 0 | 0.011394828 |
| 0 | 0 | 0 | 0 | 0 |
| 0 | 0 | 0 | 0 | 0 |
| 0 | 0 | 0 | 0 | 0 |
| 0 | 0 | 0.069791582 | 0.003990902 | 0 |
| 0 | 0 | 0 | 0 | 0 |
| 0.004416687 | 0 | 0 | 0 | 0 |
| 0.001281702 | 0.001634258 | 0 | 0.001232072 | 0.001016846 |
| 0 | 0 | 0 | 0 | 0.001845927 |

| | | | | |
|---|---|---|---|---|
| 0 | 0 | 0 | 0 | 0 |
| 0 | 0.004649013 | 0 | 0 | 0.001704595 |
| 0 | 0 | 0.425285134 | 0 | 0 |
| 0 | 0 | 0 | 0 | 0 |
| 0 | 0 | 0 | 0 | 0 |
| 0.000576854 | 0.014861035 | 0.042942022 | 0 | 0 |
| 0 | 0 | 0 | 0 | 0 |
| 0 | 0.008626707 | 0 | 0 | 0 |
| 0 | 0 | 0 | 0 | 0 |
| 0 | 0 | 0 | 0 | 0 |
| 0 | 0.007464539 | 0 | 0 | 0.081742928 |
| 0 | 0 | 0 | 0 | 0 |
| 0 | 0 | 0 | 0 | 0 |
| 0.002054288 | 0.002949069 | 0.029817481 | 0.013823195 | 0.001535756 |
| 0 | 0.011805975 | 0 | 0 | 0 |
| 0 | 0.020528819 | 0 | 0 | 0 |
| 0.0004415 | 0.006985615 | 0 | 0 | 0 |
| 0 | 0 | 0 | 0 | 0.008967234 |
| 0 | 0 | 0 | 0 | 0 |
| 0 | 0.010071984 | 0 | 0 | 0 |
| 0 | 0 | 0 | 0 | 0 |
| 0 | 0.434176305 | 0 | 0.023743482 | 0 |
| 0.004798575 | 0 | 0.0127747 | 0 | 0 |
| 0 | 0 | 0.004730818 | 0 | 0 |
| 0 | 0 | 0 | 0 | 0 |
| 0 | 0 | 0 | 0 | 0 |
| 0 | 0 | 0 | 0 | 0.037109426 |
| 0 | 0 | 0 | 0 | 0 |
| 0 | 0 | 0 | 0 | 0 |
| 0.327984389 | 1.064891987 | 0.061963195 | 0.986404282 | 0.087792793 |
| 0.014365534 | 0.000434937 | 0 | 0 | 0 |
| 0 | 0 | 0 | 0 | 0.0390771 |
| 0 | 0 | 0 | 0.006387446 | 0 |
| 0 | 0.003449708 | 0 | 0 | 0 |
| 0 | 0.006021319 | 0 | 0 | 0 |
| 0 | 0 | 0 | 0 | 0 |
| 0 | 0 | 0 | 0.029196404 | 0 |
| 0 | 0 | 0 | 0 | 0.063955521 |
| 0 | 0.010535709 | 0 | 0 | 0.001638847 |
| 0 | 0 | 0 | 0 | 0 |
| 0 | 0.001686578 | 0.001965194 | 0 | 0.000721462 |
| 0.006703758 | 0 | 0 | 0.239548806 | 0 |
| 0.001032833 | 0 | 0 | 0 | 0.024786975 |
| 0 | 0 | 0 | 0 | 0 |
| 0 | 0.032224399 | 0.001050083 | 0 | 0 |
| 0 | 0.001002616 | 0 | 0 | 0 |
| 0.000409054 | 0.045803519 | 0.011507207 | 0 | 0.005111278 |
| 0 | 0 | 0 | 0.002576361 | 0 |

| | | | | |
|---|---|---|---|---|
| 0.010935255 | 0 | 0.056122134 | 0.001981074 | 0 |
| 0 | 0 | 0 | 0.02079607 | 0 |
| 0.004126188 | 0.203113214 | 0.001825077 | 0.022944602 | 0.879990341 |
| 0 | 0 | 0 | 0 | 0 |
| 0 | 0 | 0 | 0 | 0 |
| 0 | 0.003000834 | 0.001019241 | 0 | 0 |
| 0 | 0 | 0 | 0.000603294 | 0 |
| 0 | 0 | 0 | 0 | 0 |
| 0 | 0 | 0 | 0 | 0 |
| 0 | 0.087462602 | 0 | 0 | 0 |
| 0 | 0 | 0 | 0 | 0 |
| 0 | 0 | 0 | 0 | 0 |
| 0 | 0 | 0.000494126 | 0 | 0 |
| 0 | 0.009015087 | 0 | 0 | 0.141787134 |
| 0 | 0 | 0 | 0 | 0 |
| 0.014153843 | 0 | 0 | 0.00560238 | 0 |
| 0.102433796 | 0 | 4.760913899 | 0.028276101 | 0.023428178 |
| 0 | 0.001299642 | 0.008161485 | 0 | 0 |
| 0 | 0.005524588 | 0 | 0 | 0 |
| 0 | 1.328288605 | 0 | 0 | 0.00781112 |
| 0.000498439 | 0 | 0.014550843 | 0 | 0.014763085 |
| 0 | 0 | 0 | 0 | 0 |
| 0 | 0 | 0 | 0 | 0 |
| 0.05879268 | 0 | 0 | 0 | 0.237402922 |
| 0 | 0 | 0 | 0 | 0 |
| 0 | 0.229548744 | 0 | 0 | 0.029174702 |
| 0 | 0 | 0.002798218 | 0 | 0 |
| 0 | 0 | 0 | 0 | 0 |
| 0 | 0 | 0 | 0 | 0 |
| 0 | 0 | 0 | 0 | 0 |
| 0.001145507 | 0.090158904 | 0.009424165 | 0 | 0 |
| 0.00056927 | 0 | 0.001208626 | 0.002394118 | 0 |
| 0.001806741 | 0.007330016 | 0 | 0.181146184 | 0.00326096 |
| 0.004919548 | 0 | 0 | 0 | 0.002156896 |
| 0 | 0 | 0 | 0.001709877 | 0 |
| 0 | 0 | 0 | 0 | 0 |
| 0 | 0 | 0.007932897 | 0.000549559 | 0.001133897 |
| 0.00153289 | 0.030887674 | 0.039765939 | 0 | 0.003724393 |
| 0.070886452 | 0 | 0 | 0 | 0 |
| 0 | 0.014406547 | 0 | 0 | 0 |
| 0 | 0 | 0 | 0 | 0 |
| 0 | 0 | 0 | 0 | 0 |
| 0 | 0 | 0 | 0.005302576 | 0 |
| 0 | 0 | 0 | 0 | 0.012875396 |
| 0 | 0 | 0 | 10.21254992 | 0 |
| 0 | 0 | 0 | 0 | 0 |
| 0 | 0 | 0.000452063 | 0 | 0 |
| 0 | 0 | 0.001136499 | 0 | 0 |

| | | | | |
|---|---|---|---|---|
| 0 | 0 | 0 | 0 | 0 |
| 0.610439439 | 0.001553598 | 0.022727689 | 0.095018598 | 0.079266003 |
| 0 | 0.001275954 | 0 | 0 | 0 |
| 0 | 0 | 0 | 0 | 0 |
| 0 | 0 | 0.001359569 | 0 | 0 |
| 0 | 0 | 0 | 0 | 0 |
| 0 | 0.079573061 | 0 | 0 | 0.328873344 |
| 0 | 0 | 0 | 0 | 0.000556872 |
| 0 | 0.001034321 | 0 | 0 | 0.030971373 |
| 0 | 0.007184349 | 0 | 0 | 0 |
| 0.000517392 | 0.003568433 | 0.003604393 | 0 | 0 |
| 0.02177142 | 0.014226196 | 0.086208331 | 0.09464077 | 0.013909706 |
| 0.421730093 | 0 | 0 | 0 | 0 |
| 0 | 0 | 0 | 0 | 0 |
| 0 | 0 | 0.036393199 | 0.001872463 | 0 |
| 0 | 0 | 0.291641543 | 0 | 0 |
| 0.00975211 | 0.00230794 | 0.002480268 | 0.002506374 | 0.03196843 |
| 0 | 0.050647081 | 0.048435937 | 0 | 0.429641242 |
| 0 | 0 | 0 | 0 | 0 |
| 0 | 0 | 0.007181184 | 0 | 0.002377726 |
| 0.064112645 | 0 | 0 | 0.005404381 | 0.010084181 |
| 0 | 0.001410292 | 0 | 0 | 0 |
| 0 | 0.00159967 | 0 | 0 | 0 |
| 0 | 0 | 0 | 0 | 0 |
| 0 | 0 | 0 | 0.009080368 | 0 |
| 0 | 0 | 0 | 0 | 0 |
| 0.003860363 | 0.030987675 | 0.009000955 | 0 | 0.012633404 |
| 0.020190304 | 0.002168281 | 0 | 0.005119705 | 0.058742864 |
| 0 | 0 | 0 | 0 | 0 |
| 0 | 0 | 0.000427722 | 0.006326186 | 0 |
| 0.000955774 | 0 | 0 | 0.000803919 | 0 |
| 0 | 0 | 0 | 0 | 0 |
| 0 | 0 | 0 | 0 | 0 |
| 0 | 0 | 0.001879923 | 0 | 0 |
| 0.004398611 | 0 | 0 | 0 | 0 |
| 0.153266472 | 0.013863258 | 0.080617064 | 0.011412169 | 0.065930357 |
| 0 | 0 | 0 | 0 | 0 |
| 0 | 0 | 0 | 0 | 0 |
| 0.015200595 | 0 | 0.002478769 | 0.002002548 | 0.021104102 |
| 0 | 0 | 0 | 0 | 0 |
| 0 | 0.000559487 | 0 | 10.35658702 | 0 |
| 0 | 0 | 0.044305198 | 0 | 0 |
| 0 | 0.004874174 | 0.03170705 | 0 | 0 |
| 0 | 0 | 0.011201802 | 0 | 0 |
| 0 | 0.030568989 | 0 | 0 | 0.076597359 |
| 0.10730636 | 0 | 0 | 0 | 0 |
| 0 | 0 | 0 | 0 | 0 |
| 0 | 0.001549525 | 0.0424336 | 0 | 0 |

| | | | | |
|---|---|---|---|---|
| 0 | 0 | 0 | 0 | 0 |
| 0 | 0 | 0 | 0 | 0 |
| 0 | 0 | 0 | 0 | 0 |
| 2.521397374 | 0 | 0 | 1.634757946 | 0 |
| 0 | 0 | 0 | 0 | 0 |
| 0.000449398 | 0 | 0 | 0 | 0 |
| 0 | 0 | 0 | 0 | 0 |
| 0.004752175 | 0 | 0 | 0.001364878 | 0 |
| 1.195974033 | 0 | 0 | 0.001529881 | 0 |
| 0.084646996 | 0 | 0 | 0 | 0 |
| 0 | 0 | 0 | 0 | 0 |
| 0.04655021 | 0 | 0 | 0 | 0 |
| 0 | 0 | 0 | 0 | 0 |
| 0 | 0 | 0 | 0 | 0 |
| 0.002970633 | 0.302047282 | 0 | 0.408142129 | 0 |
| 0 | 0 | 0 | 0 | 0.20315681 |
| 0.028727929 | 0 | 0 | 0.000337166 | 0 |
| 3.488202362 | 0 | 0 | 0 | 0 |
| 0 | 0 | 0 | 0 | 0 |
| 0 | 0 | 0 | 0 | 0 |
| 0 | 0 | 0 | 0 | 0 |
| 0 | 0 | 0 | 0 | 0 |
| 0.001943207 | 0 | 0 | 0 | 0 |
| 0.117616717 | 0.269972442 | 0 | 0.012864165 | 0.013699323 |
| 0.030682357 | 0 | 0.00129544 | 0 | 0 |
| 0.23596255 | 0 | 0 | 0 | 0 |
| 0.003318843 | 0 | 0 | 0 | 0 |
| 3.3719565 | 0.017530697 | 0 | 0 | 0 |
| 0 | 0 | 0 | 0 | 0 |
| 0 | 0 | 0 | 0 | 0 |
| 0.048271725 | 0 | 0 | 0 | 0 |
| 0.108244586 | 0 | 0 | 0 | 0 |
| 0.039677393 | 0 | 0 | 0.005407724 | 0 |
| 0 | 0 | 0 | 0 | 0 |
| 0.047799913 | 0 | 0 | 0 | 0 |
| 0 | 0 | 0 | 0 | 0 |
| 0.431602365 | 0 | 0 | 0 | 0 |
| 0 | 0.006392784 | 0 | 0 | 0 |
| 0 | 0.018454101 | 0 | 0 | 0 |
| 0 | 0 | 0 | 0.000896818 | 0 |
| 0.448352321 | 0.003865036 | 0.034131985 | 0.051985102 | 0.008928007 |
| 0.012411191 | 0 | 0 | 0 | 0 |
| 0.03357121 | 11.61986083 | 0 | 0.015140032 | 0.089450336 |
| 0 | 0 | 0 | 0.055058481 | 0 |
| 0 | 0 | 0 | 0 | 0 |
| 0 | 0 | 0 | 0 | 0 |
| 0 | 0 | 0 | 0 | 0 |
| 0.000429459 | 0.012544784 | 0 | 0.026249072 | 0 |

| | | | | |
|---|---|---|---|---|
| 0 | 0 | 0 | 0 | 0 |
| 0 | 0 | 0 | 0 | 0 |
| 0.128857356 | 0.089746544 | 0.44718783 | 0.034396717 | 0.637112595 |
| 0.005191802 | 0 | 0 | 0 | 0 |
| 0 | 0 | 0 | 0 | 0 |
| 0.105292467 | 0 | 0 | 0.286148973 | 0 |
| 0 | 0 | 0 | 1.085571328 | 0 |
| 0.025963892 | 0 | 0 | 0.09362078 | 0 |
| 0 | 0 | 0 | 0 | 0 |
| 0 | 0.000432916 | 0 | 0 | 0 |
| 0.110212278 | 0.176449031 | 0.006865327 | 0.340774593 | 0.004115339 |
| 0.300734371 | 1.460871042 | 0 | 0.408061205 | 1.94562681 |
| 0 | 0 | 0 | 0 | 0 |
| 0.143519916 | 0 | 0.007177799 | 0.022626837 | 0.01029607 |
| 0.007333687 | 0.199810292 | 0.00419667 | 0.10380426 | 0.021477436 |
| 12.93513557 | 0 | 0 | 0 | 0 |
| 0 | 0 | 0 | 0 | 0 |
| 0 | 0 | 0 | 0 | 0 |
| 0 | 0 | 0 | 0 | 0 |
| 0.010987537 | 0.003241953 | 0 | 0 | 0 |
| 0.400099638 | 0.004031705 | 0.029669864 | 0.065444527 | 0.036687588 |
| 0 | 6.244707957 | 18.12950563 | 0.049641917 | 5.868646173 |
| 0.762207335 | 17.26566753 | 0.636227575 | 0.829509002 | 47.99908513 |
| 0 | 0 | 0 | 0 | 0 |
| 0.023574692 | 0 | 0 | 0 | 0 |
| 2.100710394 | 0 | 0 | 0.001252023 | 0.003569624 |
| 0 | 0 | 0 | 0 | 0 |
| 0 | 0 | 0 | 0 | 0 |
| 0.009445851 | 0 | 0 | 0 | 0 |
| 0.007548747 | 0 | 0 | 0 | 0 |
| 0.020053408 | 0 | 0 | 0 | 0 |
| 0.023028095 | 0 | 0.015957569 | 0 | 0 |
| 0.038745045 | 0.225230984 | 0.069517173 | 0.092646348 | 0.019019523 |
| 0 | 0 | 0 | 0 | 0 |
| 0 | 0.005409687 | 0.090964547 | 0.221468512 | 0.003043181 |
| 0.119081381 | 0.008959822 | 0 | 0.479276631 | 0.002264787 |
| 0 | 0.009736949 | 0 | 0.030892166 | 0 |
| 0 | 0 | 0 | 0 | 0 |
| 1.315668492 | 0 | 0 | 0.038534538 | 0 |
| 0 | 0 | 0 | 0 | 0 |
| 0.042981487 | 0 | 0 | 0 | 0 |
| 0 | 0 | 0 | 0 | 0 |
| 0 | 0 | 0 | 0 | 0 |
| 0 | 0 | 0 | 0 | 0 |
| 0.070403592 | 0.018345344 | 0.018570428 | 0.645617931 | 0 |
| 0.4396449 | 0 | 0 | 0 | 0 |
| 0.002435173 | 0 | 0 | 0 | 0 |
| 0.044879578 | 0.007423186 | 0 | 4.672749918 | 0 |

| | | | | |
|---|---|---|---|---|
| 0 | 0 | 0 | 0 | 0 |
| 0 | 0 | 0 | 0 | 0 |
| 0 | 0 | 0 | 0 | 0 |
| 0.01768431 | 0 | 0 | 0 | 0 |
| 0 | 0 | 0 | 0 | 0 |
| 0 | 0 | 0 | 0 | 0 |
| 0.008144952 | 0 | 0 | 0 | 0 |
| 0.004489039 | 1.049660285 | 0.01384936 | 0.884578629 | 0.00296042 |
| 0 | 0 | 0 | 0 | 0 |
| 0 | 0 | 0 | 0 | 0 |
| 0.005840776 | 0.002534948 | 0.069104694 | 0.025592175 | 0 |
| 0 | 0 | 0 | 0 | 0 |
| 0 | 0 | 0 | 0.000700248 | 0 |
| 0 | 0 | 0 | 0 | 0.121820564 |
| 0 | 0 | 0 | 0 | 0 |
| 0 | 0 | 0 | 0 | 0 |
| 0 | 0 | 0 | 0 | 0 |
| 0.006897699 | 0.003761814 | 0 | 0.003328891 | 0 |
| 0.037123954 | 0 | 0 | 0 | 0 |
| 0 | 0 | 0 | 0 | 0 |
| 0 | 0 | 0 | 0.020364206 | 0 |
| 0.245522753 | 0 | 0 | 0 | 0 |
| 0.021380636 | 0 | 0 | 0 | 0 |
| 0 | 0 | 0 | 0 | 0 |
| 0 | 0 | 0 | 0 | 0 |
| 0 | 0 | 0 | 0 | 0 |
| 0.112184997 | 0 | 0 | 0 | 0 |
| 0 | 0 | 0 | 0 | 0 |
| 0 | 0 | 0 | 0 | 0 |
| 1.153826841 | 0 | 0 | 0 | 0 |
| 0 | 0 | 0 | 0 | 0 |
| 0 | 0 | 0 | 0 | 0 |
| 0.495741658 | 0 | 0 | 0 | 0 |
| 0 | 0 | 0 | 0 | 0 |
| 0.007236468 | 0 | 0 | 0 | 0 |
| 0.036916138 | 0 | 0 | 0 | 0 |
| 0.038227683 | 0 | 0 | 0.02102692 | 0 |
| 0 | 0.006251034 | 0.006774455 | 0.007350362 | 0.004629588 |
| 0 | 0 | 0 | 0 | 0 |
| 0.005217045 | 0 | 0 | 0 | 0 |
| 0.292472442 | 0 | 0 | 0.708883342 | 0 |
| 0 | 0 | 0 | 0 | 0 |
| 0 | 0 | 0 | 0 | 0 |
| 0.196415646 | 0 | 0 | 0 | 0 |
| 0 | 0 | 0 | 0.00274548 | 0 |
| 0 | 0 | 0 | 0 | 0 |
| 0.00207583 | 0 | 0 | 0 | 0 |
| 0.233039948 | 0 | 0 | 0 | 0 |

| | | | | |
|---|---|---|---|---|
| 0 | 0 | 0 | 0 | 0 |
| 0 | 0 | 0 | 0 | 0 |
| 0 | 0 | 0 | 0 | 0 |
| 1.054656843 | 0 | 0 | 0 | 0 |
| 0.016163114 | 0 | 0 | 0 | 0 |
| 0 | 0 | 0 | 0 | 0 |
| 0 | 0 | 0 | 0 | 0 |
| 0 | 0 | 0 | 0 | 0 |
| 0.001902988 | 0.070256514 | 0.223729478 | 0.044551102 | 0.076839868 |
| 0.002298702 | 0 | 0 | 0 | 0 |
| 0.001020522 | 0 | 0 | 0 | 0 |
| 0 | 0 | 0 | 0 | 0 |
| 0 | 0 | 0 | 0 | 0 |
| 0.000660877 | 0 | 0 | 0 | 0 |
| 0.666788567 | 0 | 0 | 0.047639976 | 0 |
| 0 | 0.083603134 | 0.131466733 | 0.001028047 | 0.002036308 |
| 0 | 0 | 0 | 0 | 0.000402964 |
| 0 | 0 | 0 | 0 | 0 |
| 0 | 0 | 0 | 0 | 0 |
| 0 | 0 | 0 | 0 | 0 |
| 0.114468456 | 0 | 0 | 0 | 0 |
| 0 | 0 | 0 | 0 | 0 |
| 0 | 0.060103648 | 0 | 0 | 0 |
| 0.066800948 | 0 | 0 | 0 | 0 |
| 0.002424348 | 0.165693274 | 0.258645215 | 0.234660713 | 0.007767464 |
| 4.7084323 | 0 | 0 | 0 | 0 |
| 0.407990917 | 0 | 0 | 0 | 0 |
| 0 | 0 | 0 | 0 | 0 |
| 0 | 0 | 0 | 0 | 0 |
| 0.064630785 | 0 | 0 | 0 | 0 |
| 0 | 0 | 0 | 0 | 0 |
| 0.032546971 | 0 | 0 | 0 | 0 |
| 0.001715836 | 0 | 0 | 0 | 0 |
| 0.000995967 | 0.021334779 | 0 | 0 | 0 |
| 0.06843136 | 0 | 0.001567852 | 0 | 0.006379453 |
| 0.001972028 | 0.028613877 | 0 | 0.009171682 | 0 |
| 0 | 0.401940404 | 0 | 0 | 0 |
| 0.120245037 | 0 | 0 | 0 | 0 |
| 2.517244477 | 0 | 2.074134406 | 0 | 0 |
| 0 | 0 | 0 | 0 | 0 |
| 0 | 0 | 0 | 0 | 0 |
| 0 | 0 | 0 | 0 | 0 |
| 0.426889727 | 0 | 0.33028411 | 0 | 0 |
| 0 | 0 | 0 | 0 | 0 |
| 0.089521184 | 0 | 0 | 0 | 0 |
| 0.002088042 | 0.00214011 | 0 | 0 | 0 |
| 0 | 0 | 0 | 0 | 0 |
| 3.209023936 | 0 | 0.006272654 | 0.001178098 | 0 |

| | | | | |
|---|---|---|---|---|
| 0.008225068 | 0.01504272 | 0 | 0 | 0 |
| 0.248664109 | 0 | 0 | 0 | 0 |
| 0.001028665 | 0 | 0 | 0 | 0 |
| 0 | 0 | 0 | 0 | 0 |
| 0.000779906 | 0.003480517 | 0 | 0.005138611 | 0.009023335 |
| 0 | 0 | 0 | 0 | 0 |
| 0 | 0 | 0 | 0 | 0 |
| 0.001040831 | 0 | 0 | 0 | 0 |
| 0 | 0 | 0 | 0 | 0 |
| 0 | 0 | 0 | 0 | 0 |
| 0 | 0 | 0 | 0 | 0 |
| 0.382540893 | 0 | 0 | 2.40484434 | 0 |
| 0.009285901 | 0 | 0 | 0.085254046 | 0 |
| 0 | 0 | 0 | 0 | 0 |
| 0.045699345 | 0.001951621 | 0.711409729 | 0.000379328 | 1.395015815 |
| 0 | 0 | 0 | 0 | 0 |
| 0.001882332 | 0 | 0 | 0 | 0 |
| 0 | 0 | 0 | 0 | 0 |
| 0 | 0.012829415 | 0 | 0 | 0 |
| 0.002775934 | 0 | 0 | 0 | 0 |
| 0.025198852 | 0 | 0 | 0 | 0 |
| 0.146673153 | 0 | 0 | 0 | 0 |
| 0.128503927 | 0 | 0 | 0 | 0 |
| 0 | 0 | 0 | 0 | 0 |
| 0 | 0 | 0 | 0 | 0 |
| 0 | 0 | 0 | 0 | 0 |
| 0 | 0 | 0 | 0 | 0 |
| 0 | 0 | 0 | 0 | 0 |
| 0.034664041 | 0 | 0 | 0 | 0 |
| 0 | 0 | 0 | 0 | 0 |
| 0 | 0 | 0 | 0 | 0 |
| 0.099875858 | 0.001582813 | 0.003572095 | 0.003445617 | 0.026812176 |
| 0 | 0 | 0 | 0 | 0 |
| 0.019100911 | 0.012098416 | 0 | 0 | 0 |
| 0.001641105 | 0.028783152 | 0 | 0.009768725 | 0 |
| 0 | 0 | 0.002461756 | 0.183420821 | 0 |
| 0.025773406 | 0 | 0 | 0 | 0 |
| 0 | 0 | 0 | 0 | 0 |
| 0 | 0 | 0 | 0 | 0 |
| 0 | 0 | 0 | 0 | 0 |
| 0.183597999 | 0 | 0 | 0 | 0 |
| 0.010822671 | 0 | 0 | 0 | 0 |
| 0 | 0 | 0 | 0 | 0 |
| 0.10544205 | 0 | 0 | 0 | 0 |
| 0.008705053 | 0.004439818 | 0.073465239 | 0.840249479 | 0.002417173 |
| 0 | 0 | 0 | 0 | 0 |
| 0.215332512 | 0 | 0 | 0 | 0 |
| 0.000352861 | 0 | 0 | 0 | 0 |

| | | | | |
|---|---|---|---|---|
| 0 | 0 | 0 | 0 | 0 |
| 0 | 0 | 0 | 0 | 0 |
| 0 | 0.002295941 | 0 | 0 | 0 |
| 0 | 0 | 0 | 0 | 0 |
| 0 | 0.510346521 | 0 | 0 | 0.0032597 |
| 0 | 0 | 0 | 0 | 0 |
| 0 | 0 | 0 | 0 | 0 |
| 0.069843475 | 0 | 0 | 0 | 0 |
| 0 | 0 | 0 | 0 | 0 |
| 0 | 0 | 0 | 0 | 0 |
| 0 | 0 | 0 | 0 | 0 |
| 0 | 0 | 0 | 0 | 0 |
| 0 | 0 | 0 | 0 | 0 |
| 0 | 0 | 0 | 0 | 0 |
| 0 | 0 | 0 | 0 | 0 |
| 0 | 0 | 0 | 0 | 0 |
| 0 | 0 | 0 | 0 | 0 |
| 0 | 0 | 0 | 0 | 0 |
| 2.002242211 | 0 | 0 | 0.005201834 | 0 |
| 0 | 0 | 0 | 0 | 0 |
| 0.003973248 | 0 | 0 | 0 | 0 |
| 0 | 0 | 0 | 0 | 0 |
| 0 | 0 | 0 | 0 | 0 |
| 0 | 0 | 0 | 0 | 0 |
| 0.008831667 | 0 | 0 | 0.00079999 | 0 |
| 0 | 0.027805852 | 0.070882068 | 0.420771968 | 0.017520166 |
| 0 | 0 | 0 | 0 | 0 |
| 1.124570365 | 0 | 0.004601238 | 0.0244004 | 0 |
| 0 | 0 | 0 | 0 | 0 |
| 0.625724041 | 0 | 0 | 0.094563339 | 0 |
| 0 | 0 | 0 | 0 | 0 |
| 0 | 0 | 0 | 0 | 0 |
| 0.005743406 | 0 | 0 | 0 | 0 |
| 0 | 0 | 0 | 0 | 0 |
| 0.001941159 | 0.010631729 | 0 | 0.002449116 | 0 |
| 0 | 0 | 0 | 0 | 0 |
| 0 | 0 | 0 | 0 | 0 |
| 0 | 0.000670217 | 0.000767233 | 0.036590558 | 0 |
| 0.177475921 | 0 | 0 | 0 | 0 |
| 0 | 0 | 0 | 0 | 0 |
| 2.218916192 | 0.708147486 | 2.572755781 | 0.525347345 | 3.768713652 |
| 1.37674308 | 0.008421994 | 0 | 0.015002685 | 0 |
| 4.54494196 | 0 | 0 | 0 | 0 |
| 0.008731915 | 0 | 0 | 0 | 0 |
| 0 | 0 | 0 | 0 | 0 |
| 0 | 0 | 0 | 0 | 0 |
| 0.004061755 | 0 | 0 | 0 | 0 |
| 0.004698132 | 0.008067468 | 0.001391815 | 0.02959169 | 0 |

| | | | | |
|---|---|---|---|---|
| 0 | 0 | 0 | 0 | 0 |
| 0.009215307 | 0.000911874 | 0 | 0 | 0 |
| 0.002625112 | 0 | 0 | 0 | 0 |
| 0 | 0 | 0 | 0 | 0 |
| 1.709634356 | 0 | 3.718690402 | 0 | 0 |
| 0.047593625 | 0 | 0 | 0 | 0 |
| 0 | 0 | 0 | 0 | 0 |
| 0.003455461 | 0 | 0 | 0 | 0 |
| 0 | 0.223717213 | 0.047262365 | 0.000566322 | 0.717916514 |
| 0 | 0 | 0 | 0 | 0 |
| 0 | 0.001545342 | 0.162650349 | 0.006087322 | 0.058877195 |
| 0 | 0 | 0 | 0 | 0 |
| 0.043570027 | 0 | 0 | 0 | 0 |
| 0.003893189 | 0.050652345 | 0 | 0.040989372 | 0.016029216 |
| 0 | 0 | 0 | 0 | 0 |
| 0.195098434 | 0 | 0 | 0 | 0.002600102 |
| 0.815402456 | 0 | 0 | 0.018370109 | 0 |
| 0 | 0 | 0 | 0 | 0 |
| 0 | 0 | 0 | 0 | 0 |
| 0 | 0 | 0 | 0 | 0 |
| 0.004837277 | 0 | 0 | 0 | 0 |
| 0 | 0 | 0 | 0 | 0 |
| 0 | 0 | 0 | 0 | 0 |
| 0 | 0 | 0 | 0 | 0 |
| 0 | 0 | 0 | 0 | 0 |
| 0 | 0 | 0 | 0 | 0 |
| 0.426183286 | 0 | 0.009007609 | 0 | 0 |
| 0 | 0.00295961 | 0.109173099 | 0.000479372 | 0 |
| 0 | 0 | 0 | 0 | 0 |
| 0 | 0 | 0 | 0 | 0 |
| 0 | 0 | 0 | 0 | 0 |
| 0 | 0 | 0 | 0 | 0 |
| 0.001720121 | 0 | 0 | 0 | 0 |
| 0.028557171 | 0 | 0 | 0 | 0 |
| 0 | 0 | 0 | 0.933819131 | 0 |
| 0 | 0 | 0 | 0 | 0 |
| 0 | 0 | 0.00053651 | 0 | 0 |
| 0.122642069 | 0 | 0 | 0 | 0 |
| 0 | 0 | 0 | 0 | 0 |
| 0 | 0 | 0 | 0 | 0 |
| 0.204123394 | 0 | 0 | 0.905050103 | 4.682369579 |
| 0 | 0 | 0 | 0 | 0 |
| 0.026629447 | 0 | 0 | 0 | 0 |
| 0 | 0 | 0 | 0 | 0 |
| 0 | 0 | 0 | 0 | 0 |
| 0 | 0 | 0 | 0 | 0 |
| 0.007154636 | 0 | 0 | 0 | 0 |
| 0 | 0 | 0 | 0 | 0 |

| | | | | |
|---|---|---|---|---|
| 1.242780697 | 0 | 0 | 0 | 0 |
| 0 | 0 | 0 | 0 | 0 |
| 0 | 0 | 0 | 0 | 0 |
| 0 | 0 | 0 | 0 | 0 |
| 0.012848624 | 0 | 0 | 0 | 0 |
| 0.005547211 | 0 | 0 | 0 | 0 |
| 1.499941829 | 0 | 0.005351457 | 0.041427192 | 0 |
| 0.000896171 | 0 | 0 | 0 | 0 |
| 0 | 0 | 0 | 0 | 0 |
| 0.000729514 | 0 | 0 | 0 | 0 |
| 0.02009629 | 0 | 0 | 0 | 0 |
| 0.037295504 | 0 | 0 | 0 | 0 |
| 0 | 0 | 0 | 0 | 0 |
| 0 | 0 | 0 | 0 | 0 |
| 0.000797089 | 0 | 0 | 0 | 0 |
| 0 | 0 | 0 | 0 | 0 |
| 0 | 0 | 41.34334301 | 0.318493314 | 0 |
| 0 | 0.009311457 | 0 | 0.003561083 | 0 |
| 0 | 0 | 0 | 0 | 0 |
| 0 | 0 | 0 | 0 | 0 |
| 0.145197028 | 0 | 0 | 0.016890026 | 0 |
| 0 | 0 | 0 | 0 | 0 |
| 0.052085849 | 0 | 0 | 0 | 0 |
| 0.015474496 | 0.002019516 | 0 | 0.009921197 | 0.001802885 |
| 0 | 0 | 0 | 0 | 0 |
| 0 | 0.006526185 | 0 | 0 | 0 |
| 0 | 0 | 0 | 0 | 0 |
| 0 | 0 | 0 | 0 | 0 |
| 0.070058312 | 0 | 0 | 0 | 0 |
| 0 | 0.025148041 | 0.898369542 | 0.162439188 | 0.097952725 |
| 0.034931926 | 0 | 0 | 0 | 0 |
| 0 | 0 | 0 | 0 | 0 |
| 0.023781591 | 0 | 0 | 0 | 0 |
| 0 | 1.828431257 | 0 | 0 | 0 |
| 0.002422478 | 0 | 0 | 0 | 0 |
| 0.599168439 | 0 | 0 | 0 | 0 |
| 0.035340134 | 0 | 0.038917013 | 0.089745154 | 0.015603143 |
| 0 | 0 | 0 | 0 | 0 |
| 0 | 0 | 0 | 0 | 0 |
| 0 | 0 | 0 | 0 | 0 |
| 0 | 0 | 0 | 0 | 0 |
| 0.541527304 | 0 | 0 | 0 | 0 |
| 0 | 0 | 0 | 0 | 0 |
| 0 | 0 | 0 | 0 | 0 |
| 0.268599349 | 0 | 0 | 0 | 0 |
| 0 | 0 | 0 | 0 | 0 |
| 0 | 0 | 0 | 0 | 0 |
| 0 | 0 | 0 | 0 | 0 |

| | | | | |
|---|---|---|---|---|
| 0.000815089 | 0.004346143 | 0 | 0.030753449 | 0 |
| 0 | 0 | 0 | 0 | 0 |
| 0 | 0 | 0 | 0 | 0 |
| 0 | 0 | 0 | 0 | 0 |
| 0 | 0 | 0 | 0 | 0 |
| 0 | 0 | 0 | 0 | 0 |
| 0 | 0 | 0 | 0 | 0 |
| 0.531678457 | 0 | 0 | 0 | 0.131631982 |
| 0.048617384 | 0.171141302 | 1.111606484 | 3.943404763 | 0.400680662 |
| 0 | 0.114187734 | 0 | 0 | 0.043104378 |
| 0 | 0 | 0 | 0 | 0 |
| 0 | 0 | 0 | 0 | 0 |
| 0.004187365 | 0 | 0 | 0 | 0 |
| 0 | 0.029094636 | 0 | 0 | 0.575039406 |
| 0 | 0 | 0 | 0 | 0 |
| 0 | 0 | 0 | 0 | 0 |
| 0 | 0 | 0.044116605 | 0.019443565 | 0 |
| 0.193308592 | 0 | 0 | 0 | 0 |
| 0 | 0 | 0 | 0 | 0 |
| 0 | 0 | 0 | 0 | 0 |
| 0 | 0 | 0 | 0 | 0 |
| 0 | 0 | 0 | 0 | 0 |
| 0.795145727 | 0 | 0 | 0 | 0 |
| 0 | 0 | 0 | 0 | 0 |
| 0.010822213 | 0 | 0 | 0 | 0 |
| 0 | 0 | 0 | 0 | 0 |
| 0 | 0 | 0 | 0 | 0 |
| 0 | 0 | 0 | 0 | 0 |
| 0 | 0 | 0 | 0 | 0 |
| 0 | 0 | 0 | 0 | 0 |
| 0 | 0 | 0 | 0 | 0 |
| 0 | 0 | 0 | 0 | 0 |
| 0 | 0 | 0 | 0 | 0 |
| 0.008147733 | 0 | 0 | 0 | 0 |
| 0.000563874 | 0 | 0 | 0 | 0 |
| 0 | 0 | 0 | 0 | 0 |
| 0 | 0 | 0 | 0 | 0 |
| 0 | 0.006224305 | 0 | 0 | 0 |
| 0 | 0 | 0 | 0 | 0 |
| 0 | 0 | 0 | 0.000586654 | 0 |
| 0 | 0 | 0 | 0 | 0 |
| 0.096901206 | 0.183039866 | 0 | 0 | 0 |
| 0 | 0 | 0 | 0 | 0 |
| 0 | 0 | 0 | 0 | 0 |
| 0 | 0 | 0 | 0 | 0 |
| 0.00041416 | 0 | 0 | 0 | 0 |
| 0.001568286 | 0 | 0 | 0 | 0 |
| 0 | 0 | 0 | 0 | 0 |

| | | | | |
|---|---|---|---|---|
| 0 | 0 | 0 | 0 | 0 |
| 0 | 0 | 0 | 0.000265188 | 0 |
| 0 | 0 | 0 | 0 | 0 |
| 0 | 0 | 0 | 0 | 0 |
| 0 | 0.004768977 | 0.099554374 | 10.72340865 | 0 |
| 0 | 0 | 0 | 0 | 0 |
| 0.008920187 | 0 | 0 | 0 | 0.005484587 |
| 0.005184262 | 0.006970854 | 0.017099819 | 0.003862226 | 0.005963802 |
| 0.008429964 | 0 | 0 | 0 | 0 |
| 0 | 0 | 0 | 0 | 0 |
| 0 | 0 | 0 | 0 | 0 |
| 0 | 0 | 0 | 0 | 0 |
| 0 | 0 | 0 | 0 | 0 |
| 0 | 0 | 0 | 0 | 0 |
| 0 | 0 | 0 | 0 | 0 |
| 0.259499118 | 0.062333968 | 0 | 0.272836835 | 0 |
| 0 | 0 | 0 | 0 | 0 |
| 0 | 0 | 0 | 0 | 0 |
| 0 | 0 | 0 | 0 | 0 |
| 0 | 0 | 0 | 0 | 0 |
| 0 | 0 | 0 | 0 | 0 |
| 0 | 0 | 0 | 0 | 0 |
| 0 | 0 | 0 | 0 | 0 |
| 0.00551205 | 0.002184253 | 0.045416041 | 0.002048591 | 0 |
| 0 | 0 | 0 | 0.167493981 | 0 |
| 0 | 0 | 0 | 0 | 0 |
| 0.20388339 | 0.736122442 | 0 | 0.42305483 | 0.075690524 |
| 0.000617259 | 0 | 0 | 0 | 0 |
| 0 | 0 | 0 | 0 | 0 |
| 0 | 0 | 0 | 0 | 0 |
| 0 | 0 | 0 | 0 | 0 |
| 0 | 0 | 0 | 0 | 0 |
| 0 | 0 | 0 | 0 | 0 |
| 0 | 0 | 0 | 0 | 0 |
| 0 | 0 | 0 | 0 | 0 |
| 0 | 0 | 0 | 0 | 0 |
| 0 | 0 | 0 | 0 | 0 |
| 0.192862197 | 0 | 0 | 0 | 0 |
| 0 | 0 | 0 | 0 | 0 |
| 0 | 0 | 0 | 0 | 0 |
| 0 | 0 | 0 | 0 | 0 |
| 0 | 0 | 0 | 0 | 0 |
| 0 | 0 | 0 | 0 | 0 |
| 0 | 0 | 0 | 0 | 0 |
| 0 | 0 | 0 | 0 | 0 |
| 0 | 0 | 0 | 0 | 0 |
| 0 | 0 | 0 | 0 | 0 |
| 0 | 0 | 0 | 0 | 0 |

| | | | | |
|---|---|---|---|---|
| 0.329664705 | 0 | 0 | 0 | 0.00596997 |
| 0 | 0 | 0 | 0 | 0 |
| 0.00294879 | 0 | 0 | 0 | 0 |
| 0.000987286 | 0 | 0 | 0 | 0 |
| 0.001736105 | 0 | 0 | 0 | 0 |
| 0 | 0 | 0.130583597 | 0 | 0 |
| 0 | 0 | 0 | 0 | 0 |
| 0 | 0 | 0 | 0 | 0 |
| 0 | 0 | 0 | 0 | 0 |
| 0 | 0 | 0 | 0 | 0 |
| 0 | 0 | 0 | 0 | 0 |
| 0.002481531 | 0 | 0 | 0 | 0 |
| 0 | 0 | 0 | 0 | 0 |
| 0 | 0 | 0 | 0 | 0 |
| 0 | 0 | 0 | 0 | 0 |
| 0 | 0 | 0 | 0 | 0 |
| 0 | 0 | 0 | 0 | 0 |
| 0 | 0 | 0 | 0 | 0 |
| 0 | 0 | 0 | 0 | 0 |
| 0 | 0 | 0 | 0 | 0 |
| 0 | 0 | 0 | 0 | 0.00549406 |
| 0 | 0 | 0 | 0 | 0 |
| 0.055941378 | 0.118645164 | 0 | 0.214655341 | 0.00626712 |
| 0.317531408 | 0 | 0 | 0 | 0 |
| 0.007095361 | 0 | 0 | 0 | 0 |
| 0 | 0 | 0 | 0 | 0 |
| 0 | 0 | 0 | 0 | 0 |
| 0 | 0 | 0 | 0 | 0 |
| 0 | 0 | 0 | 0 | 0 |
| 0 | 0 | 0 | 0 | 0 |
| 0 | 0 | 0 | 0 | 0 |
| 0.068892878 | 0 | 0 | 0 | 0 |
| 0 | 0.182897778 | 0.051206343 | 0.371697274 | 0.010270981 |
| 0.102322952 | 0 | 0 | 0 | 0 |
| 0.009183252 | 0 | 0 | 0.087238043 | 0 |
| 0.014417132 | 0 | 0.001350407 | 0 | 0 |
| 0 | 0 | 0 | 0 | 0 |
| 0 | 0 | 0 | 0 | 0 |
| 0.014853979 | 0 | 0.013946061 | 0.452846907 | 0 |
| 0 | 0 | 0 | 0 | 0 |
| 0 | 0 | 0 | 0 | 0 |
| 0.002522706 | 0 | 0 | 0 | 0 |
| 0.102082385 | 0 | 0 | 0.055266789 | 0 |
| 0 | 0 | 0 | 0 | 0 |
| 0 | 0 | 0 | 0 | 0 |
| 0.258048199 | 0 | 0 | 0 | 0 |
| 0 | 0 | 0 | 0 | 0 |
| 0 | 0 | 0 | 0 | 0 |

| | | | | |
|---|---|---|---|---|
| 0 | 0 | 0 | 0 | 0 |
| 0 | 0 | 0 | 0 | 0 |
| 0 | 0 | 0 | 0 | 0 |
| 0 | 0 | 0 | 0 | 0 |
| 0 | 0 | 0 | 0 | 0 |
| 0 | 0 | 0 | 0 | 0 |
| 0 | 0 | 0 | 0 | 0 |
| 0 | 0.017855613 | 0 | 0 | 0 |
| 0.05864349 | 0 | 0 | 0 | 0 |
| 0 | 0 | 0 | 0 | 0 |
| 0 | 0 | 0 | 0 | 0 |
| 0 | 0 | 0 | 0 | 0 |
| 0 | 0 | 0 | 0 | 0 |
| 0 | 0 | 0 | 0 | 0 |
| 0 | 0 | 0 | 0 | 0 |
| 0 | 0.019367548 | 0.001784886 | 0 | 0.25797302 |
| 0 | 0 | 0 | 0 | 0 |
| 0 | 0 | 0 | 0 | 0 |
| 0 | 0 | 0 | 0 | 0 |
| 1.323025391 | 0 | 0 | 0 | 0 |
| 0.004407435 | 0 | 0 | 0.002316984 | 0.006692837 |
| 0 | 0 | 0 | 0 | 0 |
| 0 | 0 | 0 | 0 | 0 |
| 0.001932238 | 0 | 0 | 0 | 0 |
| 0.001782303 | 0 | 0 | 0 | 0 |
| 0.247652625 | 0 | 0 | 0 | 0 |
| 0 | 0 | 0 | 0 | 0 |
| 0 | 0 | 0 | 0 | 0 |
| 0 | 0 | 0 | 0 | 0 |
| 0.065058058 | 0 | 0 | 0 | 0 |
| 0.476321009 | 0 | 0 | 0.025423299 | 0 |
| 0 | 0 | 0 | 0 | 0 |
| 0 | 0 | 0 | 0 | 0 |
| 0 | 0 | 0 | 0 | 0 |
| 0 | 0 | 0 | 0 | 0 |
| 0 | 0 | 0 | 0 | 0 |
| 0 | 0 | 0 | 0 | 0 |
| 0 | 0 | 0 | 0 | 0 |
| 0 | 0 | 0 | 0.000508033 | 0 |
| 0 | 0 | 0 | 0 | 0 |
| 0 | 0 | 0 | 0 | 0 |
| 0.585845827 | 0 | 0 | 0 | 0 |
| 0.002434483 | 0 | 0 | 0 | 0 |
| 0.000422579 | 0.001485829 | 0 | 0 | 0 |
| 0 | 0 | 0 | 0 | 0 |
| 0 | 0 | 0 | 0 | 0 |
| 0 | 0.015225812 | 1.096627419 | 0.511425161 | 0.029051235 |
| 0 | 0 | 0 | 0 | 0 |

| | | | | |
|---|---|---|---|---|
| 0 | 0 | 0 | 0 | 0 |
| 0 | 0 | 0 | 0 | 0 |
| 0 | 0 | 0 | 0 | 0 |
| 0 | 0 | 0 | 0 | 0 |
| 0.094297947 | 0 | 0 | 0 | 0.04690744 |
| 0 | 0 | 0 | 0 | 0 |
| 0 | 0 | 0 | 0 | 0 |
| 0 | 0 | 0 | 0 | 0 |
| 0 | 0 | 0 | 0 | 0 |
| 0 | 0 | 0 | 0 | 0 |
| 0.015237806 | 0 | 0 | 0 | 0 |
| 0.012041746 | 0.03215263 | 0.013377558 | 0.194562053 | 0 |
| 0.00326166 | 0 | 0 | 0.007620673 | 0 |
| 0 | 0 | 0 | 0 | 0 |
| 0.006295609 | 0 | 0.001953971 | 0 | 0 |
| 0 | 0 | 0 | 0 | 0 |
| 0.022938444 | 0.000611676 | 0 | 0 | 0 |
| 0 | 0 | 0 | 0 | 0 |
| 0 | 0.133344667 | 0 | 0 | 0.002796822 |
| 0.811230636 | 0 | 0.004849579 | 0.022356564 | 0.01172419 |
| 0 | 0 | 0 | 0 | 0 |
| 0 | 0 | 0 | 0 | 0 |
| 0 | 0 | 0 | 0 | 0 |
| 0 | 0 | 0 | 0 | 0 |
| 0 | 0 | 0 | 0 | 0 |
| 0 | 0 | 0 | 0 | 0 |
| 0.007210744 | 0.069670148 | 0 | 0.01185025 | 0.000770093 |
| 0 | 0 | 0 | 0 | 0 |
| 0 | 0 | 0 | 0 | 0 |
| 0 | 0 | 0 | 0 | 0 |
| 0.074066811 | 0.00377528 | 0.002502955 | 0.076578 | 0 |
| 0.000407474 | 0 | 0 | 0 | 0 |
| 0 | 0 | 0 | 0 | 0 |
| 0 | 0 | 0 | 0 | 0 |
| 0 | 0 | 0 | 0 | 0 |
| 0 | 0 | 0 | 0 | 0 |
| 0 | 0 | 0 | 0 | 0 |
| 0.010425108 | 0.00767046 | 0.060281491 | 0.768083012 | 0 |
| 0.641202933 | 0 | 0 | 0 | 0 |
| 0 | 0 | 0 | 0 | 0 |
| 0.006174011 | 0.010878074 | 0.009690653 | 0.007418677 | 0 |
| 0.001359094 | 0 | 0 | 0 | 0 |
| 0 | 0 | 0 | 0 | 0 |
| 0 | 0.005527707 | 0 | 0 | 0 |
| 0 | 0 | 0 | 0 | 0 |
| 0 | 0 | 0 | 0 | 0 |
| 0 | 0 | 0 | 0 | 0 |
| 0.000761296 | 0 | 0 | 0 | 0 |

| | | | | |
|---|---|---|---|---|
| 0 | 0 | 0 | 0 | 0 |
| 0.0454617 | 0 | 0 | 0 | 0 |
| 0 | 0 | 0 | 0 | 0 |
| 0.000823791 | 0.00133686 | 0.005465631 | 0.004503886 | 0 |
| 0.000636711 | 0 | 0 | 0 | 0.001414388 |
| 0.000986084 | 0.007867804 | 0.003402048 | 0 | 0.002053578 |
| 0.031710583 | 0 | 0 | 0 | 0 |
| 0.006442758 | 0.02143358 | 0 | 0 | 0 |
| 0 | 0 | 0 | 0 | 0 |
| 0.021101297 | 0 | 0 | 0 | 0 |
| 0.018946462 | 0 | 0 | 0 | 0 |
| 0 | 0 | 0 | 0 | 0 |
| 0.15367664 | 0 | 0 | 0.160285972 | 0 |
| 0.005358103 | 0 | 0 | 0 | 0 |
| 0 | 0 | 0 | 0 | 0 |
| 0 | 0 | 0 | 0 | 0 |
| 0 | 0 | 0 | 0 | 0 |
| 0 | 0 | 0 | 0 | 0 |
| 0 | 0 | 0 | 0 | 0 |
| 0 | 0 | 0 | 0 | 0 |
| 0 | 0 | 0 | 0 | 0 |
| 0 | 0 | 0 | 0 | 0 |
| 0 | 0.012998241 | 0 | 0.015630059 | 0.007117079 |
| 0.00725891 | 0 | 0 | 0 | 0 |
| 0 | 0 | 0 | 0 | 0 |
| 0.035850181 | 0 | 0 | 0 | 0 |
| 0 | 0 | 0 | 0 | 0 |
| 0 | 0 | 0 | 0 | 0 |
| 0 | 0 | 0 | 0 | 0 |
| 0 | 0 | 0 | 0 | 0 |
| 0.014860793 | 0 | 0 | 0.007692114 | 0 |
| 0 | 0 | 0 | 0 | 0 |
| 0 | 0 | 0 | 0 | 0 |
| 0 | 0 | 0 | 0 | 0 |
| 0.055586105 | 0 | 0 | 0 | 0 |
| 0 | 0 | 0 | 0 | 0.006636455 |
| 0 | 0.089553208 | 0 | 0 | 0 |
| 0 | 0 | 0 | 0 | 0.021057047 |
| 0.002659728 | 0 | 0 | 0 | 0.000839249 |
| 0 | 0 | 0 | 0 | 0 |
| 0 | 0 | 0 | 0 | 0 |
| 0 | 0 | 0 | 0 | 0 |
| 0.002080822 | 0 | 0 | 0 | 0 |
| 0 | 0 | 0 | 0 | 0 |
| 0 | 0 | 0 | 0 | 0 |
| 0.414733369 | 0.009925506 | 0 | 0.083385159 | 0 |
| 0 | 0 | 0.014816752 | 0 | 0.029704625 |
| 0 | 0 | 0 | 0 | 0 |

| | | | | |
|---|---|---|---|---|
| 0 | 0 | 0 | 0 | 0 |
| 0 | 0 | 0 | 0 | 0 |
| 0 | 0 | 0 | 0 | 0 |
| 0 | 0 | 0 | 0 | 0 |
| 0 | 0 | 0 | 0 | 0 |
| 0.006463312 | 0 | 0 | 0 | 0 |
| 0.002212011 | 0 | 0 | 0 | 0 |
| 0 | 0 | 0 | 0 | 0 |
| 0.00134234 | 0 | 0 | 0 | 0 |
| 0 | 0 | 0 | 0 | 0 |
| 0.004707823 | 0.001830399 | 0 | 0.003464857 | 0 |
| 0.000732166 | 0.00118817 | 0.001049269 | 0.003571862 | 0 |
| 0 | 0 | 0 | 0 | 0 |
| 0 | 0 | 0 | 0 | 0 |
| 0 | 0 | 0 | 0 | 0 |
| 0 | 0 | 0 | 0 | 0 |
| 0.000865037 | 0 | 0 | 0 | 0 |
| 0 | 0 | 0 | 0 | 0 |
| 0 | 0 | 0 | 0 | 0 |
| 0 | 0 | 0.000724905 | 0 | 0 |
| 0 | 0 | 0 | 0 | 0 |
| 0 | 0 | 0 | 0 | 0 |
| 0 | 0 | 0 | 0 | 0 |
| 0 | 0 | 0 | 0 | 0 |
| 0 | 0 | 0 | 0 | 0 |
| 0 | 0 | 0 | 0 | 0 |
| 0 | 0 | 0 | 0 | 0 |
| 0.006333578 | 0 | 0 | 0 | 0 |
| 0 | 0 | 0 | 0 | 0 |
| 0 | 0 | 0.006029835 | 0 | 0 |
| 0.042686204 | 0 | 0 | 0.001879798 | 0 |
| 0 | 0 | 0 | 0 | 0 |
| 0.003503146 | 5.038241922 | 0 | 0.017679362 | 0.025759463 |
| 0.003845008 | 0 | 0.000633367 | 0.00234194 | 0.003129353 |
| 0 | 0 | 0 | 0 | 0 |
| 0 | 0 | 0 | 0 | 0 |
| 0 | 0 | 0 | 0 | 0 |
| 0 | 0 | 0 | 0 | 0 |
| 0 | 0 | 0 | 0 | 0 |
| 0 | 0 | 0 | 0 | 0 |
| 0 | 0 | 0 | 0 | 0 |
| 0 | 0.055603904 | 0.149741334 | 0.15550207 | 0.023118629 |
| 0.049415432 | 0 | 0 | 0.115504797 | 0 |
| 0 | 0 | 0 | 0 | 0 |
| 0 | 0 | 0 | 0 | 0 |
| 0.179649343 | 0 | 0.003291509 | 0.309641584 | 0 |
| 0.008254681 | 0 | 0 | 0 | 0 |
| 0 | 0 | 0 | 0 | 0 |

| | | | | |
|---|---|---|---|---|
| 0 | 0 | 0 | 0 | 0 |
| 0 | 0.012933407 | 0 | 0.000687537 | 0 |
| 0.00938906 | 0 | 0 | 0.002007791 | 0.001104705 |
| 0 | 0 | 0 | 0 | 0 |
| 0.371150812 | 0.16485662 | 0 | 0.707405352 | 0 |
| 0 | 0 | 0 | 0 | 0 |
| 0 | 0 | 0 | 0 | 0 |
| 0 | 0 | 0 | 0 | 0 |
| 0 | 0 | 0 | 0 | 0 |
| 0 | 0 | 0 | 0 | 0 |
| 0 | 0 | 0 | 0 | 0 |
| 0 | 0 | 0 | 0 | 0 |
| 0 | 0 | 0 | 0 | 0 |
| 0 | 0.053723652 | 0 | 0 | 0 |
| 0 | 0 | 0 | 0 | 0 |
| 0 | 0 | 0 | 0 | 0 |
| 0.444417311 | 0 | 0 | 0 | 0 |
| 0 | 0 | 0 | 0 | 0 |
| 0 | 0 | 0 | 0 | 0 |
| 0 | 0.015818217 | 0 | 0 | 0 |
| 0 | 0 | 0 | 0 | 0 |
| 0 | 0 | 0 | 0 | 0 |
| 0 | 0 | 0 | 0 | 0 |
| 0 | 0 | 0 | 0 | 0 |
| 0 | 0 | 0 | 0.005074419 | 0 |
| 0 | 0 | 0.008403376 | 0.003551128 | 0 |
| 0 | 0 | 0 | 0 | 0 |
| 0.014205925 | 0 | 0 | 0.00333707 | 0 |
| 0 | 0 | 0 | 0 | 0 |
| 0 | 0 | 0 | 0 | 0 |
| 0 | 0 | 0 | 0 | 0 |
| 0 | 0 | 0.065494324 | 0.016446491 | 0 |
| 0 | 0 | 0 | 0 | 0 |
| 0 | 0 | 0 | 0 | 0 |
| 0 | 0 | 0 | 0 | 0 |
| 0 | 0 | 0 | 0 | 0 |
| 0 | 0 | 0 | 0 | 0 |
| 0 | 0 | 0 | 0 | 0.021515532 |
| 0.719437026 | 0 | 0 | 7.789030059 | 0 |
| 0 | 0 | 0 | 0 | 0 |
| 0 | 0 | 0 | 0 | 0 |
| 0.013310984 | 0.036815063 | 0 | 0 | 0 |
| 0.000926876 | 0.003115738 | 0 | 0.022274651 | 0 |
| 0 | 0 | 0 | 0 | 0 |
| 0 | 0 | 0 | 0 | 0 |
| 0.000914004 | 0 | 0 | 0 | 0 |
| 0 | 0.011833991 | 0.000967644 | 0.007837447 | 0 |
| 0 | 0 | 0 | 0 | 0 |

| | | | | |
|---|---|---|---|---|
| 0 | 0 | 0 | 0 | 0 |
| 0.008179104 | 0.015036017 | 0.011904638 | 0.032570578 | 0.011000786 |
| 0.066371393 | 0 | 0 | 0 | 0 |
| 0 | 0 | 0 | 0 | 0 |
| 0 | 0 | 0 | 0 | 0 |
| 0 | 0 | 0 | 0 | 0 |
| 0 | 0 | 0 | 0 | 0 |
| 0 | 0 | 0 | 0 | 0 |
| 0 | 0 | 0 | 0 | 0 |
| 0 | 0 | 0 | 0 | 0 |
| 0 | 0 | 0 | 0 | 0 |
| 0 | 0 | 0 | 0 | 0 |
| 0.943769007 | 0 | 0 | 0 | 0 |
| 0 | 0 | 0 | 0 | 0 |
| 0.003186909 | 0.005301059 | 0 | 0.022784826 | 0 |
| 0 | 0 | 0 | 0 | 0 |
| 0 | 0 | 0 | 0 | 0 |
| 0 | 0 | 0 | 0 | 0 |
| 0.001410219 | 0 | 0 | 0 | 0 |
| 0 | 0 | 0 | 0 | 0 |
| 0 | 0 | 0 | 0 | 0 |
| 0.180276271 | 0 | 0.053699219 | 0.00816084 | 0 |
| 0.006613729 | 0.017172576 | 0 | 0 | 0 |
| 0 | 0 | 0 | 0.010032522 | 0 |
| 0 | 0 | 0 | 0 | 0 |
| 0 | 0 | 0 | 0 | 0 |
| 0 | 0 | 0 | 0 | 0 |
| 0 | 0 | 0 | 0 | 0 |
| 0 | 0 | 0 | 0 | 0 |
| 0 | 0 | 0 | 0 | 0 |
| 0 | 0 | 0 | 0 | 0 |
| 0 | 0 | 0 | 0 | 0 |
| 0 | 0 | 0 | 0 | 0 |
| 0 | 0 | 0 | 0 | 0 |
| 1.518310127 | 20.9698633 | 0 | 0.198983314 | 0 |
| 0 | 0 | 0 | 0 | 0 |
| 0 | 0 | 0 | 0 | 0 |
| 0 | 0 | 0 | 0 | 0 |
| 0.298952199 | 0.00063401 | 0.012815295 | 0.005257798 | 0.003471467 |
| 0 | 0 | 0 | 0 | 0 |
| 0.019336605 | 0.00943666 | 0.001896508 | 0.051857422 | 0.273911393 |
| 0 | 0 | 0 | 0 | 0 |
| 0 | 0 | 0 | 0 | 0 |
| 0 | 0 | 0 | 0 | 0 |
| 0 | 0 | 0 | 0 | 0 |
| 0 | 0.124184748 | 3.162157079 | 3.194953867 | 1.111221454 |
| 0 | 0 | 0 | 0 | 0 |
| 0 | 0 | 0 | 0 | 0 |

| | | | | |
|---|---|---|---|---|
| 0 | 0.008579604 | 0 | 0 | 0 |
| 0 | 0.165906656 | 0 | 0 | 0 |
| 0 | 0 | 0 | 0 | 0 |
| 0 | 0 | 0 | 0 | 0 |
| 0.002920144 | 0 | 0 | 0 | 0 |
| 0 | 0 | 0 | 0 | 0 |
| 0 | 0 | 0 | 0 | 0 |
| 0.020981431 | 0 | 0 | 0 | 0 |
| 0 | 0 | 0 | 0 | 0 |
| 0.005491467 | 0 | 0.040223592 | 0 | 0.124383501 |
| 0 | 0 | 0 | 0 | 0 |
| 0 | 0 | 0 | 0 | 0 |
| 0 | 0.001244339 | 0 | 0.004256678 | 0 |
| 0 | 0 | 0 | 0 | 0 |
| 0 | 0 | 0 | 0 | 0 |
| 0 | 0 | 0 | 0 | 0 |
| 0 | 0 | 0 | 0 | 0 |
| 0 | 0 | 0 | 0 | 0 |
| 0 | 0 | 0 | 0 | 0 |
| 0.162158833 | 0.104331185 | 1.145540874 | 1.663373233 | 0.012697121 |
| 0 | 0 | 0 | 0 | 0 |
| 0 | 0 | 0 | 0 | 0 |
| 0 | 0 | 0 | 0 | 0 |
| 0 | 0 | 0 | 0 | 0 |
| 0 | 0 | 0 | 0 | 0 |
| 0 | 0 | 0 | 0 | 0 |
| 0 | 0 | 0 | 0 | 0 |
| 0 | 0 | 0 | 0 | 0 |
| 0 | 0 | 0 | 0 | 0 |
| 0 | 0.038242831 | 0 | 0.008100456 | 0 |
| 0 | 0 | 0 | 0 | 0 |
| 0 | 0 | 0 | 0 | 0 |
| 0 | 0 | 0 | 0 | 0 |
| 0 | 0 | 0 | 0 | 0 |
| 0 | 0 | 0 | 0 | 0 |
| 0 | 0 | 0 | 0 | 0 |
| 0 | 0 | 0 | 0 | 0 |
| 0 | 0 | 0 | 0 | 0 |
| 0 | 0 | 0 | 0 | 0 |
| 0 | 0 | 0 | 0 | 0 |
| 0 | 0 | 0 | 0 | 0 |
| 0 | 0 | 0 | 0 | 0 |
| 0.000907696 | 0 | 0 | 0 | 0 |
| 0 | 0 | 0 | 0 | 0 |
| 0.006281962 | 0.011754841 | 0.001326928 | 0.105030469 | 0 |
| 0 | 0 | 0 | 0 | 0 |
| 0 | 0 | 0 | 0 | 0 |
| 0 | 0 | 0.012772083 | 0 | 0.00231631 |

| | | | | |
|---|---|---|---|---|
| 0 | 0 | 0 | 0 | 0 |
| 0 | 0 | 0 | 0 | 0 |
| 0 | 0 | 0 | 0 | 0 |
| 0 | 0 | 0 | 0 | 0 |
| 0 | 0 | 0 | 0 | 0 |
| 0.001818466 | 0.001397859 | 0.031699296 | 0 | 0 |
| 0 | 0 | 0 | 0 | 0 |
| 0 | 0 | 0 | 0 | 0 |
| 0 | 0 | 0 | 0 | 0 |
| 0 | 0 | 0 | 0 | 0 |
| 0 | 0 | 0 | 0 | 0 |
| 0 | 0 | 0 | 0 | 0 |
| 0 | 0 | 0 | 0 | 0 |
| 0 | 0 | 0 | 0 | 0 |
| 0 | 0 | 0 | 0 | 0 |
| 0 | 0 | 0 | 0 | 0 |
| 0.055725144 | 0 | 0 | 0 | 0 |
| 0 | 0 | 0 | 0 | 0 |
| 0 | 0 | 0 | 0 | 0 |
| 0 | 0 | 0 | 0 | 0 |
| 0 | 0.005132534 | 0 | 0 | 0 |
| 0 | 0 | 0 | 0 | 0 |
| 0 | 0 | 0 | 0 | 0 |
| 0 | 0 | 0 | 0 | 0 |
| 0 | 0 | 0 | 0 | 0 |
| 0.000898227 | 0 | 0.00083433 | 0 | 0.003117687 |
| 0 | 0 | 0 | 0 | 0 |
| 0.001792696 | 0 | 0 | 0 | 0 |
| 0 | 0 | 0 | 0 | 0 |
| 0 | 0 | 0 | 0 | 0 |
| 0 | 0 | 0 | 0 | 0 |
| 0 | 0 | 0 | 0 | 0 |
| 0 | 0 | 0 | 0 | 0 |
| 0 | 0 | 0 | 0 | 0 |
| 0 | 0 | 0 | 0 | 0 |
| 0 | 0 | 0 | 0 | 0 |
| 0 | 0 | 0 | 0 | 0 |
| 0 | 0 | 0 | 0 | 0 |
| 0.12415827 | 0 | 0 | 0 | 0 |
| 0 | 0 | 0 | 0 | 0 |
| 0 | 0 | 0 | 0 | 0 |
| 0 | 0 | 0 | 0 | 0 |
| 0 | 0 | 0 | 0 | 0 |
| 0 | 0 | 0 | 0 | 0 |
| 0 | 0 | 0 | 0 | 0 |
| 0 | 0 | 0 | 0 | 0 |
| 0 | 0 | 0 | 0 | 0 |
| 0 | 0 | 0 | 0 | 0 |

| | | | | |
|---|---|---|---|---|
| 0 | 0 | 0 | 0 | 0 |
| 0.257061648 | 0 | 0 | 0.032176023 | 0 |
| 0 | 0 | 0 | 0 | 0 |
| 0 | 0 | 0 | 0 | 0 |
| 0 | 0 | 0 | 0 | 0 |
| 0 | 0 | 0 | 0.000988748 | 0 |
| 0 | 0 | 0 | 0 | 0 |
| 0 | 0 | 0 | 0 | 0 |
| 0 | 0 | 0 | 0 | 0 |
| 0.01091168 | 0 | 0 | 0 | 0 |
| 0.109581512 | 0 | 0.001563936 | 0.00389455 | 0.001948016 |
| 0 | 0 | 0 | 0 | 0 |
| 0.129904472 | 0 | 0 | 0 | 0 |
| 0 | 0 | 0 | 0 | 0 |
| 0 | 0 | 0 | 0 | 0 |
| 0 | 0 | 0 | 0 | 0 |
| 0 | 0 | 0 | 0 | 0 |
| 0 | 0 | 0 | 0 | 0 |
| 0 | 0 | 0 | 0 | 0 |
| 0 | 0 | 0 | 0 | 0 |
| 0.002990514 | 0.069675955 | 0.001700679 | 0 | 0 |
| 0 | 0 | 0 | 0 | 0 |
| 0 | 0 | 0 | 0 | 0 |
| 0.470694708 | 0 | 0 | 0.909699857 | 0 |
| 0 | 0 | 0 | 0 | 0 |
| 0 | 0 | 0 | 0 | 0 |
| 0 | 0 | 0 | 0 | 0 |
| 0 | 0 | 0 | 0 | 0 |
| 0.001121475 | 0 | 0 | 0 | 0 |
| 0 | 0 | 0 | 0 | 0 |
| 0 | 0.000340233 | 0 | 0 | 0 |
| 0 | 0 | 0 | 0 | 0 |
| 0.007190251 | 0 | 0 | 0 | 0 |
| 0 | 0 | 0 | 0 | 0 |

| PHL018-0193_V1 | PHL018-0074_V1 | PHL018-0140_V3 | PHL018-0036_V1 | PHL018-0192_V1 |
|---|---|---|---|---|
| REF | CTRL | CTRL | CTRL | CTRL |
| 0.199220561 | 0.006545199 | 0.022559208 | 0.449187448 | 0.747050828 |
| 5.019501436 | 3.991266271 | 1.652093379 | 0.092155309 | 2.372011076 |
| 8.589197834 | 0.451689762 | 0 | 2.558538084 | 1.393582613 |
| 0 | 0 | 0 | 0 | 0 |
| 0 | 0 | 0 | 0 | 0 |
| 0 | 0 | 0 | 0 | 0 |
| 0 | 0 | 0 | 0 | 0 |
| 0 | 0 | 0 | 0 | 0 |
| 0 | 0 | 0 | 0 | 0 |
| 0 | 0 | 0 | 0 | 0 |
| 0 | 0 | 0 | 0 | 0 |
| 0 | 0 | 0 | 0 | 0 |
| 0 | 0 | 0 | 0 | 0 |
| 0 | 0.388218004 | 2.185199597 | 0.678255715 | 0.494593663 |
| 0 | 0 | 0 | 0 | 0 |
| 0 | 0 | 0 | 0 | 0 |
| 0 | 0 | 0 | 0 | 0 |
| 0 | 0 | 0 | 0 | 0 |
| 0 | 0 | 0 | 0 | 0 |
| 0 | 0 | 0 | 0 | 0 |
| 0 | 0 | 0 | 0 | 0 |
| 0 | 0 | 0 | 0 | 0 |
| 0 | 0 | 0 | 0 | 0 |
| 0 | 0 | 0 | 0 | 0 |
| 0 | 0 | 0 | 0 | 0 |
| 0 | 0 | 0 | 0 | 0 |
| 0 | 0 | 0 | 0 | 0 |
| 0 | 0 | 0 | 0 | 0 |
| 0 | 0 | 0 | 0 | 0 |
| 0 | 0 | 0 | 0 | 0 |
| 0 | 0 | 0 | 0 | 0 |
| 0 | 0 | 0 | 0 | 0 |
| 0 | 0 | 0 | 0 | 0 |
| 0 | 0 | 0 | 0 | 0 |
| 0 | 0 | 0 | 0 | 0 |
| 0 | 0 | 3.04108147 | 0.013817201 | 0.007312392 |
| 0 | 0 | 0 | 0 | 0 |
| 0 | 0 | 0 | 0 | 0 |
| 0 | 0 | 0 | 0 | 0 |
| 0 | 0 | 0 | 0 | 0 |
| 0 | 0 | 0 | 0 | 0 |
| 0 | 0.342244737 | 0.018679717 | 0.004452846 | 0.007743436 |
| 1.077953805 | 0.786773469 | 0.009016934 | 0.948502548 | 1.060731846 |
| 0.019757339 | 0.078996929 | 0.038971323 | 0.170818096 | 0.036552866 |
| 0.025420675 | 0.007071469 | 0 | 0.039619788 | 0 |
| 0 | 0 | 0 | 0 | 0 |

| | | | | |
|---|---|---|---|---|
| 0 | 0.016347983 | 0 | 0.043807476 | 0 |
| 0.016022124 | 0.038341369 | 0.010899664 | 0 | 0.002968043 |
| 0 | 0 | 0.00305253 | 0 | 0.031903097 |
| 0.518447999 | 0.280468287 | 0.466466436 | 0.34979985 | 0.755369642 |
| 0 | 0 | 0 | 0 | 0.007566472 |
| 0 | 0 | 0 | 0 | 0 |
| 0 | 0.001461183 | 0.013566985 | 0 | 0.037644952 |
| 0 | 0 | 0 | 0 | 0 |
| 0 | 0 | 0 | 0 | 0 |
| 0 | 0 | 0 | 0 | 0 |
| 0 | 0 | 0 | 0 | 0 |
| 0 | 0.003222481 | 0.001462781 | 0.754170637 | 0.713192988 |
| 0 | 0.000632918 | 0 | 0 | 0 |
| 0.030147199 | 0 | 0.002294114 | 0.015359708 | 0.111447862 |
| 0 | 0 | 0 | 0 | 0 |
| 0 | 0 | 0 | 0 | 0 |
| 0.23234496 | 14.00307878 | 0 | 0 | 0.037219762 |
| 0 | 0 | 0 | 0 | 0 |
| 0 | 0 | 0 | 0 | 0 |
| 0 | 0 | 0 | 0 | 0 |
| 0.093667046 | 0.456824792 | 0.190654012 | 0.003206021 | 0.085064418 |
| 0.002232087 | 0.00960352 | 0 | 0.007525353 | 0.006027713 |
| 6.056912114 | 0.001018923 | 0 | 0 | 0.002112745 |
| 0 | 0 | 0 | 0 | 0 |
| 0 | 0 | 0 | 0 | 0 |
| 0 | 0 | 0 | 0 | 0 |
| 0.000533038 | 0 | 0 | 0 | 0 |
| 0 | 0 | 0 | 0 | 0 |
| 0.003426582 | 0.120366453 | 0.000685115 | 0 | 0 |
| 0 | 0 | 0 | 0 | 0 |
| 0.038443853 | 0.438067896 | 0.241063335 | 0 | 0 |
| 0.122009812 | 0 | 0 | 0 | 0 |
| 0 | 0 | 0 | 0 | 0 |
| 0.004953052 | 0.013199026 | 0 | 0 | 0 |
| 0 | 0 | 0 | 0 | 0 |
| 0.018682144 | 0.007392271 | 0 | 0 | 0 |
| 0 | 0 | 0 | 0 | 0 |
| 1.719438306 | 0.29364984 | 0 | 0 | 0 |
| 0 | 0 | 0 | 0 | 0 |
| 0 | 0 | 0 | 0 | 0 |
| 0 | 0 | 0 | 0 | 0 |
| 0 | 0 | 0 | 0 | 0 |
| 0 | 0 | 0 | 0 | 0 |
| 0 | 0 | 0 | 0 | 0 |
| 0 | 0 | 0 | 0 | 0 |
| 0 | 0 | 0 | 0 | 0 |
| 0.074877693 | 0 | 0 | 0.00093039 | 0 |
| 0.022400961 | 0.001180054 | 0 | 0 | 0 |

| | | | | |
|---|---|---|---|---|
| 0 | 0 | 0 | 0 | 0 |
| 0 | 0 | 0 | 0 | 0 |
| 0 | 0 | 0 | 0 | 0 |
| 0 | 0 | 0 | 0 | 0 |
| 0 | 0.049190896 | 0 | 0.001284872 | 0.002144097 |
| 0 | 0 | 0 | 0 | 0 |
| 0 | 0 | 0 | 0 | 0 |
| 0 | 0 | 0 | 0 | 0.000583704 |
| 0.01280109 | 0.012794873 | 0 | 0 | 0 |
| 0 | 0 | 0 | 0 | 0 |
| 0 | 0 | 0 | 0 | 0 |
| 0 | 0.010485596 | 0 | 0 | 0.006130029 |
| 0 | 0 | 0 | 0 | 0 |
| 0 | 0 | 0 | 0 | 0 |
| 0 | 0 | 0 | 0 | 0 |
| 0 | 0 | 0 | 0 | 0 |
| 0 | 0 | 0 | 0 | 0 |
| 0 | 0 | 0 | 0 | 0 |
| 0 | 0.007257981 | 0 | 1.43395108 | 0.929117652 |
| 0 | 0 | 0 | 0 | 0 |
| 0 | 0 | 0 | 0 | 0 |
| 0 | 0 | 0 | 0 | 0 |
| 0 | 0 | 0 | 0 | 0 |
| 0 | 0.001835913 | 0 | 0 | 0 |
| 0 | 0 | 0 | 0 | 0 |
| 0 | 0 | 0 | 0 | 0 |
| 0 | 0 | 0 | 0 | 0 |
| 0 | 0.001013186 | 0 | 0 | 0 |
| 0 | 0 | 0 | 0 | 0 |
| 0 | 0 | 0 | 0 | 0 |
| 0 | 0 | 0 | 0.576173821 | 0.071605174 |
| 0.000977422 | 0.001095988 | 0 | 0 | 0.000707012 |
| 0 | 0 | 0 | 0 | 0 |
| 0 | 0 | 0 | 0 | 0 |
| 0 | 0 | 0 | 0 | 0 |
| 0 | 0 | 0 | 0 | 0 |
| 0 | 0 | 0 | 0 | 0 |
| 0 | 0 | 0 | 0 | 0 |
| 0 | 0 | 0 | 0 | 0 |
| 0 | 0 | 0 | 0 | 0 |
| 0 | 0 | 0 | 0 | 0 |
| 0 | 0 | 0 | 0 | 0 |
| 0 | 0 | 0.001161684 | 0.002856655 | 0.001021493 |
| 0 | 0 | 0 | 0 | 0 |
| 0.001204466 | 0 | 0 | 0 | 0 |
| 0 | 0 | 0 | 0 | 0 |
| 0.020812014 | 0 | 0 | 0 | 0 |
| 0 | 0 | 0 | 0 | 0 |

| | | | | |
|---|---|---|---|---|
| 0 | 0 | 0 | 0 | 0.001587238 |
| 0 | 0 | 0 | 0 | 0 |
| 0 | 0 | 0 | 0 | 0 |
| 0 | 0 | 0 | 0 | 0 |
| 0 | 0 | 0.000437393 | 0.160261032 | 0.164304928 |
| 0 | 0 | 0 | 0 | 0 |
| 0 | 0 | 0 | 0 | 0 |
| 0 | 0 | 0 | 0 | 0 |
| 0 | 0 | 0 | 0 | 0 |
| 0 | 0 | 0 | 0 | 0 |
| 0 | 0 | 0 | 0 | 0 |
| 0 | 0 | 0 | 0 | 0 |
| 0 | 0 | 0 | 0 | 0 |
| 0 | 0 | 0 | 0 | 0 |
| 0 | 0 | 0 | 0 | 0 |
| 0 | 0 | 2.269017541 | 0.182359516 | 0 |
| 0 | 0 | 0 | 0 | 0 |
| 0 | 0 | 0 | 0 | 0 |
| 0 | 0 | 0 | 0 | 0 |
| 0 | 0 | 0 | 0 | 0 |
| 0 | 0 | 0 | 0 | 0 |
| 0 | 0 | 0 | 0 | 0 |
| 0 | 0 | 0 | 0 | 0 |
| 0 | 0 | 0 | 0 | 0 |
| 0 | 0 | 0 | 0.061065014 | 0 |
| 0 | 0 | 0 | 0 | 0 |
| 0 | 0 | 0 | 0 | 0 |
| 0 | 0 | 0 | 0 | 0 |
| 0 | 0 | 0 | 0 | 0 |
| 0 | 0 | 0 | 0 | 0 |
| 0 | 0 | 0 | 0 | 0 |
| 0.000805525 | 0 | 0 | 0 | 0 |
| 0 | 0 | 0 | 0 | 0 |
| 0 | 0 | 0 | 0 | 0 |
| 0 | 0 | 0 | 0 | 0 |
| 0 | 0 | 0 | 0 | 0 |
| 0 | 0 | 0 | 0 | 0 |
| 0 | 0 | 0 | 0 | 0 |
| 0 | 0 | 0 | 0 | 0 |
| 0 | 0 | 0 | 0 | 0 |
| 0 | 0 | 0 | 0 | 0 |
| 0 | 0 | 0 | 0 | 0 |
| 0 | 0 | 0 | 0 | 0 |
| 0 | 0 | 0 | 0 | 0 |
| 0 | 0 | 0 | 0 | 0 |
| 0 | 0 | 0 | 0 | 0 |
| 0 | 0 | 0 | 0 | 0 |
| 0 | 0 | 0 | 0 | 0 |

| | | | | |
|---|---|---|---|---|
| 0 | 0 | 0 | 0 | 0 |
| 0 | 0 | 0 | 0 | 0 |
| 0 | 0 | 0 | 0 | 0 |
| 0 | 0 | 0 | 0 | 0 |
| 0 | 0 | 0 | 0 | 0 |
| 0 | 0 | 0 | 0 | 0 |
| 0 | 0 | 0 | 0 | 0 |
| 0 | 0 | 0 | 0 | 0 |
| 0 | 0 | 0 | 0 | 0 |
| 0 | 0 | 0 | 0 | 0 |
| 0 | 0 | 0 | 0 | 0 |
| 0 | 0 | 0 | 0 | 0 |
| 0.003237394 | 0.001075586 | 0 | 0.001754146 | 0.008391266 |
| 0 | 0 | 0 | 0 | 0 |
| 0.004851309 | 0 | 0 | 0 | 0 |
| 0 | 0 | 0 | 0 | 0 |
| 0 | 0 | 0 | 0 | 0 |
| 0 | 0 | 0 | 0 | 0 |
| 0 | 0 | 0 | 0 | 0 |
| 0 | 0 | 0 | 0 | 0 |
| 0.026230315 | 0.145301071 | 0.046682039 | 0.00437311 | 0.009486766 |
| 0 | 0 | 0 | 0 | 0 |
| 0 | 0 | 0 | 0 | 0 |
| 0.004508727 | 0.012320813 | 0 | 0 | 0 |
| 0 | 0 | 0 | 0 | 0 |
| 0 | 0 | 0 | 0 | 0 |
| 0.01812966 | 0.019471365 | 0.012687042 | 0.000636699 | 0.005949859 |
| 0 | 0 | 0 | 0 | 0 |
| 1.374685614 | 0 | 0 | 0 | 0 |
| 0.097342436 | 0 | 0 | 0 | 0 |
| 0.003681373 | 0.196869257 | 0 | 0 | 0 |
| 0 | 0 | 0 | 0 | 0 |
| 0 | 0 | 0 | 0 | 0 |
| 0 | 0 | 0 | 0.01158115 | 0.002675872 |
| 0.035258969 | 0.010141035 | 0.025142776 | 0.02679398 | 0.062912996 |
| 0 | 0 | 0 | 0.031073095 | 0 |
| 0 | 0 | 0 | 0 | 0 |
| 0 | 0 | 0 | 0 | 0 |
| 0 | 0 | 0 | 0 | 0 |
| 0 | 0 | 0 | 0 | 0 |
| 0 | 0 | 0 | 0 | 0 |
| 0 | 0 | 0 | 0 | 0 |
| 0 | 0 | 0 | 0 | 0 |
| 0 | 0 | 0 | 0 | 0 |
| 0 | 0 | 0 | 0 | 0 |
| 0 | 0 | 0.01494158 | 0.022575918 | 0 |
| 0.003249029 | 0 | 0 | 0 | 0.002313443 |
| 0.054529838 | 0.000932487 | 0 | 0.005238208 | 0.038573729 |

| | | | | |
|---|---|---|---|---|
| 0 | 0 | 0 | 0 | 0 |
| 0 | 0 | 0 | 0 | 0 |
| 0.000732768 | 0.001917197 | 0 | 0 | 0 |
| 0 | 0 | 0.00061142 | 0 | 0 |
| 0 | 0 | 0 | 0 | 0 |
| 0 | 0 | 0 | 0 | 0 |
| 0 | 0 | 0 | 0 | 0 |
| 0 | 0 | 0 | 0 | 0.000972815 |
| 0 | 0 | 0 | 0 | 0 |
| 0 | 0 | 0 | 0 | 0 |
| 0 | 0 | 0 | 0 | 0 |
| 0 | 0 | 0 | 0 | 0 |
| 0 | 0 | 0 | 0 | 0 |
| 0 | 0 | 0 | 0 | 0 |
| 0 | 0 | 0 | 0 | 0 |
| 0 | 0 | 0 | 0 | 0 |
| 0 | 0 | 0 | 0 | 0 |
| 0 | 0 | 0 | 0 | 0 |
| 0.005572833 | 0.004148958 | 0 | 0.004983531 | 0.005161741 |
| 0 | 0 | 0 | 0 | 0 |
| 0.000802 | 0.019934157 | 0 | 0 | 0 |
| 0 | 0 | 0 | 0 | 0 |
| 0 | 0 | 0 | 0 | 0 |
| 0 | 0 | 0 | 0 | 0 |
| 0.005178361 | 0.011811545 | 0.00107519 | 0.005539731 | 0 |
| 0 | 0 | 0 | 0 | 0 |
| 0 | 0 | 0 | 0 | 0 |
| 0 | 0 | 0 | 0 | 0 |
| 0.001330481 | 0.115868803 | 0.001539105 | 0 | 0 |
| 0 | 0 | 0 | 0 | 0 |
| 0.315789703 | 0.106074576 | 0.017243962 | 0 | 0.418031735 |
| 0 | 0 | 0 | 0 | 0 |
| 0 | 0 | 0 | 0.00880699 | 0 |
| 0 | 0.036762823 | 0 | 0 | 0 |
| 0 | 0 | 0 | 0 | 0 |
| 0 | 0 | 0 | 0 | 0 |
| 0 | 0 | 0 | 0 | 0 |
| 0 | 0 | 0 | 0 | 0 |
| 0 | 0 | 0 | 0 | 0 |
| 0 | 0 | 0 | 0 | 0 |
| 0 | 0 | 0 | 0 | 0 |
| 0 | 0 | 0.012574567 | 0.004939867 | 0.012174681 |
| 0 | 0 | 0 | 0.08193814 | 0 |
| 0 | 0 | 0 | 0 | 0 |
| 0 | 0 | 0 | 0 | 0 |
| 0 | 0 | 0 | 0 | 0 |
| 0 | 0 | 0 | 0 | 0 |
| 0 | 0 | 0 | 0 | 0 |

| | | | | |
|---|---|---|---|---|
| 0 | 0 | 0 | 0 | 0 |
| 0 | 0 | 0 | 0 | 0 |
| 0.022266427 | 0.086892302 | 0.036162906 | 0.18417592 | 0.033441003 |
| 0 | 0 | 0 | 0 | 0 |
| 0 | 0 | 0 | 0 | 0 |
| 0 | 0 | 0 | 0 | 0 |
| 0 | 0 | 0 | 0 | 0 |
| 0 | 0 | 0 | 0 | 0 |
| 0 | 0 | 0 | 0 | 0 |
| 0 | 0.002107187 | 0.000621114 | 0 | 0 |
| 0 | 0 | 0 | 0 | 0 |
| 0 | 0 | 0 | 0 | 0 |
| 0 | 0 | 0 | 0 | 0 |
| 0.00208546 | 0.00200066 | 0.003189818 | 0.003427616 | 0 |
| 0.009294879 | 0.009006998 | 0.001858431 | 0.001523334 | 0 |
| 0 | 0 | 0 | 0 | 0 |
| 0.002549608 | 0.004276111 | 0.001411677 | 0 | 0.001737844 |
| 0 | 0 | 0 | 0 | 0.020028546 |
| 0 | 0 | 0 | 0 | 0 |
| 0 | 0 | 0 | 0 | 0 |
| 0 | 0 | 0 | 0 | 0 |
| 0.058955568 | 0 | 0 | 0.003357522 | 0.035409551 |
| 0 | 0 | 0 | 0 | 0 |
| 0 | 0 | 0 | 0 | 0 |
| 0 | 0 | 0 | 0 | 0 |
| 0 | 0 | 0 | 0 | 0 |
| 0 | 0 | 3.188679254 | 0 | 0 |
| 0 | 0 | 0 | 0 | 0 |
| 0 | 0 | 0 | 0 | 0 |
| 0.002560954 | 0 | 0 | 0 | 0 |
| 0 | 0 | 0 | 0 | 0 |
| 0 | 0 | 0 | 0 | 0 |
| 0 | 0 | 0 | 0 | 0 |
| 0 | 0 | 0 | 0 | 0 |
| 0 | 0 | 0 | 0 | 0 |
| 0 | 0 | 0 | 0 | 0.016114681 |
| 0 | 0 | 0 | 0 | 0 |
| 1.118413921 | 0.050438191 | 0.006204177 | 0.007874307 | 0.172885433 |
| 0 | 0 | 0 | 0 | 0 |
| 0 | 0 | 0 | 0 | 0 |
| 0 | 0 | 0 | 0 | 0 |
| 0 | 0 | 0 | 0 | 0 |
| 0 | 0 | 0 | 0 | 0 |
| 0 | 0 | 0 | 0 | 0 |
| 0 | 0 | 0 | 0 | 0 |
| 0 | 0 | 0 | 0 | 0 |
| 0.001446682 | 0 | 0 | 0 | 0 |
| 0.002963699 | 0 | 0.001696894 | 0 | 0 |

| | | | | |
|---|---|---|---|---|
| 8.769256034 | 0.505150146 | 0 | 0.614628436 | 0 |
| 0 | 0 | 0 | 0 | 0 |
| 0 | 0 | 0 | 0 | 0 |
| 0 | 0 | 0 | 0 | 0 |
| 0.00167913 | 0 | 0.027193939 | 0.000707636 | 0.004251059 |
| 0.008822362 | 0 | 0 | 0 | 0 |
| 0 | 0 | 0 | 0 | 0 |
| 0.202415688 | 0 | 0.011294308 | 0 | 0.014234899 |
| 0 | 0 | 0 | 0 | 0 |
| 0.207825574 | 0.025154011 | 0.045109992 | 0.001703969 | 0.002729715 |
| 0.011698128 | 0 | 0 | 0 | 0 |
| 0.002484232 | 0.026540381 | 0 | 0 | 0.003406725 |
| 0.004867977 | 0.29800573 | 0.002967026 | 0.054596724 | 0 |
| 0 | 0.010191681 | 0 | 0 | 0.001207574 |
| 0 | 0 | 0 | 0 | 0 |
| 0 | 0.001850278 | 0.001696761 | 0 | 0 |
| 0.003599905 | 0.003438574 | 0 | 0 | 0 |
| 0 | 0 | 0 | 0 | 0 |
| 0.002536447 | 0.029863345 | 0.073941121 | 0 | 0 |
| 0 | 0 | 0 | 0 | 0 |
| 0 | 0.010923626 | 0 | 0 | 0 |
| 0 | 0 | 0 | 0 | 0 |
| 0 | 0 | 0 | 0 | 0 |
| 0 | 0 | 0 | 0 | 0 |
| 0 | 0 | 0 | 0 | 0 |
| 0.002743525 | 0.022969899 | 0 | 0.008093434 | 0.000926106 |
| 0 | 0 | 0 | 0 | 0 |
| 0 | 0.041858386 | 0.003383047 | 0 | 0 |
| 0.197377263 | 0.143393639 | 0 | 0 | 0 |
| 0.026157413 | 0 | 0 | 0 | 0.017865863 |
| 0 | 0.001369331 | 0 | 0 | 0 |
| 0.001089088 | 0 | 0 | 0 | 0 |
| 0 | 0 | 0 | 0 | 0 |
| 0 | 0.064461707 | 0 | 0 | 0 |
| 0 | 0 | 0 | 0 | 0 |
| 0.016136073 | 0 | 0 | 0 | 0 |
| 0 | 0.022603068 | 0.009514016 | 0 | 0 |
| 0 | 0 | 0 | 0 | 0 |
| 0 | 0 | 0 | 0 | 0 |
| 0 | 0 | 0 | 0 | 0 |
| 0 | 0 | 0 | 0 | 0 |
| 0 | 0 | 0 | 0 | 0 |
| 0 | 0 | 0 | 0 | 0 |
| 0 | 0.027115493 | 0 | 0 | 0 |
| 0 | 0 | 0 | 0 | 0 |
| 0 | 0 | 0 | 0 | 0 |
| 0 | 0 | 0 | 0 | 0.00633427 |
| 0 | 0 | 0 | 0 | 0 |

| | | | | |
|---|---|---|---|---|
| 0 | 0 | 0 | 0 | 0 |
| 0 | 0.001995204 | 0.021955952 | 0 | 0.004183039 |
| 0 | 0 | 0 | 0 | 0 |
| 0.53046865 | 0 | 0 | 0.893190639 | 0 |
| 0.011465473 | 0.002156718 | 0 | 0 | 0 |
| 0 | 0.035622319 | 0 | 0.006115321 | 0 |
| 0 | 0 | 0 | 0 | 0 |
| 0 | 0 | 0.01146598 | 0 | 0 |
| 0 | 0 | 0 | 0 | 0 |
| 0 | 0 | 0 | 0 | 0 |
| 0 | 0 | 0.02916374 | 0.002626942 | 0.004734332 |
| 0 | 0 | 0 | 0.01034058 | 0.006409213 |
| 0 | 0 | 0 | 0 | 0 |
| 0.0017667 | 0.352325789 | 0.003179127 | 0 | 0.001490919 |
| 0 | 0.088516319 | 0 | 0 | 0 |
| 0 | 0 | 0 | 0 | 0 |
| 0 | 0 | 0 | 0 | 0 |
| 0 | 0 | 0 | 0 | 0 |
| 0.006059003 | 0 | 0 | 0 | 0.005025039 |
| 0 | 0.008623132 | 0 | 0 | 0 |
| 0 | 0 | 0 | 0 | 0 |
| 0 | 0 | 0.231036704 | 0.096805168 | 0 |
| 0 | 0 | 0 | 0 | 0 |
| 0 | 0 | 0 | 0 | 0 |
| 0 | 0 | 0 | 0 | 0 |
| 0 | 0 | 0 | 0 | 0 |
| 0 | 0.207899012 | 0 | 0 | 0 |
| 0 | 0.001285047 | 0 | 0 | 0 |
| 0 | 0 | 0 | 0 | 0 |
| 0.319097696 | 0.623336519 | 1.535966229 | 0.29102422 | 0.568561909 |
| 0 | 0.009059258 | 0.006709998 | 0.003367412 | 0.151383399 |
| 0 | 0.001304504 | 0 | 0 | 0 |
| 0 | 0 | 0 | 0 | 0 |
| 0 | 0 | 0 | 0 | 0 |
| 0 | 0 | 0 | 0 | 0.01997212 |
| 0 | 0 | 0 | 0 | 0 |
| 0 | 0 | 0.111278976 | 1.669775583 | 0 |
| 0 | 0.038214738 | 0 | 0 | 0.001519644 |
| 0 | 0 | 0 | 0 | 0 |
| 0 | 0.46024182 | 0 | 0.001761965 | 0 |
| 0.007262097 | 0.032652493 | 0.003920383 | 0.004692717 | 0.005311358 |
| 0 | 1.607119197 | 7.979802211 | 1.945846977 | 0.034787018 |
| 0 | 0.038843575 | 0 | 0 | 0.001392092 |
| 0 | 0 | 0 | 0 | 0 |
| 0 | 0.002222724 | 0 | 0 | 0 |
| 0 | 0 | 0 | 0 | 0 |
| 0.012364726 | 0.002511247 | 0 | 0 | 0 |
| 0 | 0 | 0 | 0 | 0.002608956 |

| | | | | |
|---|---|---|---|---|
| 0.010867286 | 0.020231104 | 0 | 0.017966922 | 0 |
| 0 | 0 | 0 | 0 | 0 |
| 0.015946549 | 31.4840904 | 0 | 0 | 0.009372053 |
| 0 | 0.023081565 | 0.001190616 | 0 | 0 |
| 0 | 0 | 0 | 0.211598534 | 0 |
| 0 | 0 | 0 | 0 | 0.5301062 |
| 0 | 0 | 0 | 0 | 0 |
| 0 | 0 | 0 | 0 | 0 |
| 0.025185997 | 0 | 0 | 0 | 0 |
| 0.040190963 | 0.03451505 | 0 | 0 | 0 |
| 0 | 0 | 0 | 0 | 0 |
| 0 | 0 | 0 | 0 | 0 |
| 0 | 0 | 0 | 0 | 0 |
| 0 | 0 | 0.058517242 | 0.005019686 | 0 |
| 0 | 0 | 0 | 0 | 0 |
| 0 | 0.001932857 | 0.009039694 | 0.003175598 | 0.249124572 |
| 1.691050478 | 0.394053337 | 0.01281914 | 0.256946826 | 0.003938772 |
| 0 | 0.001561738 | 0 | 0 | 0 |
| 0 | 0 | 0 | 0 | 0 |
| 0 | 0 | 0.182946511 | 0 | 0 |
| 0 | 0 | 0.017455653 | 0.066278672 | 0.023774817 |
| 0 | 0.001159685 | 0 | 0 | 0 |
| 0 | 0 | 0 | 0 | 0 |
| 0 | 0 | 0 | 0 | 0.259730347 |
| 0 | 0.011613063 | 0 | 0 | 0.009881016 |
| 0 | 0.110630061 | 0 | 0 | 0 |
| 0.002702734 | 0.002357122 | 0.000561172 | 0 | 0 |
| 0 | 0.000917962 | 0 | 0 | 0 |
| 0 | 0 | 0 | 0 | 0 |
| 0 | 0.009308746 | 0 | 0 | 0.007656367 |
| 0 | 0.000551565 | 0 | 0.00839564 | 0 |
| 0.001885773 | 0.024669453 | 0.004453833 | 0.072121011 | 0 |
| 0 | 0.021922744 | 0.072696873 | 0.008228924 | 0.024433137 |
| 0 | 0.113327159 | 0.003472724 | 0 | 0 |
| 0 | 0.07548096 | 0 | 0.695716441 | 0 |
| 0 | 0 | 0 | 0 | 0 |
| 0 | 0.0077863 | 0 | 0 | 0.004366478 |
| 0 | 0.001660704 | 0.003712108 | 0.002407467 | 0.082356649 |
| 0.214190284 | 0 | 0.074879811 | 1.030138441 | 0 |
| 0 | 0 | 0.000466927 | 0 | 0 |
| 0 | 0.003161968 | 0 | 0 | 0 |
| 0 | 0.006009917 | 0 | 0 | 0 |
| 0.313013328 | 0.00206965 | 0 | 0.003600363 | 0.00510681 |
| 0 | 0.034678272 | 0 | 0.028346481 | 0 |
| 0 | 0.001902671 | 0.005452523 | 0 | 0 |
| 0 | 0.002413737 | 0 | 0 | 0 |
| 0 | 0 | 0 | 0 | 0 |
| 0 | 0 | 0 | 0 | 0 |

| | | | | |
|---|---|---|---|---|
| 0.002588026 | 0.010875037 | 0.002116856 | 0 | 0.002084764 |
| 0.039569459 | 0.1355866 | 0.220810682 | 9.322848602 | 0.001642264 |
| 0 | 0.001239009 | 0 | 0 | 0 |
| 0 | 0 | 0 | 0 | 0 |
| 0 | 0 | 0 | 0 | 0 |
| 0 | 0 | 0 | 0 | 0 |
| 0.001600302 | 0.195043988 | 0 | 0 | 0 |
| 0 | 0.00043454 | 0 | 0 | 0.001681909 |
| 0 | 0.001381011 | 0 | 0 | 0.022550365 |
| 0 | 0.004385118 | 0 | 0 | 0 |
| 0.023780635 | 0 | 0.001445696 | 0 | 0.017898933 |
| 0.126848807 | 2.105481342 | 0.0260562 | 0.045086219 | 0.022116485 |
| 0 | 0 | 0.001777682 | 10.22300699 | 0 |
| 0 | 0 | 0 | 0 | 0 |
| 0 | 0.203554014 | 0 | 0 | 0.004434079 |
| 0 | 0 | 0 | 0 | 0 |
| 0.013292154 | 0.015114172 | 0.006206794 | 0.01680515 | 0.003407776 |
| 0 | 0.030097134 | 0.018865818 | 0 | 0.083775025 |
| 0 | 0.000777672 | 0 | 0 | 0 |
| 0 | 0.020533051 | 0 | 0 | 0.003590701 |
| 0 | 0.006204345 | 0.033877249 | 0.275969367 | 0.147379971 |
| 0 | 0 | 0 | 0 | 0 |
| 0 | 0 | 0 | 0 | 0 |
| 0 | 0 | 0 | 0 | 0 |
| 0 | 0 | 0 | 0 | 0 |
| 0 | 0 | 0 | 0 | 0 |
| 0 | 0.01643024 | 0.036982715 | 0.022302657 | 0.009105512 |
| 0 | 0.058866214 | 0.042311803 | 0.143626274 | 0.029180915 |
| 0 | 0 | 0 | 0 | 0 |
| 0.018908484 | 0 | 0 | 0 | 0.01182687 |
| 0 | 0 | 0.003026707 | 0 | 0.052101747 |
| 0 | 0 | 0 | 0 | 0 |
| 0 | 0.000976273 | 0 | 0 | 0 |
| 0 | 0 | 0 | 0 | 0 |
| 0 | 0 | 0.014314931 | 0 | 0.012612851 |
| 0.097693094 | 0.011704877 | 0.219885447 | 0.086419285 | 0.20861051 |
| 0 | 0 | 0 | 0 | 0 |
| 0 | 0 | 0 | 0 | 0 |
| 0.053462905 | 0.01904736 | 0.004707905 | 0 | 0.010079418 |
| 0 | 0 | 0 | 0 | 0 |
| 0 | 0.00149404 | 0.00462402 | 0 | 0 |
| 0 | 0 | 0 | 0 | 0 |
| 0 | 0.001084655 | 0.000497331 | 0 | 0 |
| 0.001067343 | 0.001279929 | 0 | 0 | 0.014017644 |
| 0 | 0 | 0 | 0 | 0 |
| 0 | 0 | 0.210721946 | 0 | 0 |
| 0 | 0 | 0 | 0 | 0 |
| 0 | 0 | 0 | 0 | 0 |

| | | | | |
|---|---|---|---|---|
| 0 | 0 | 0 | 0 | 0 |
| 0 | 0 | 0 | 0 | 0 |
| 0 | 0 | 0 | 0 | 0 |
| 5.433031084 | 0 | 0.002027317 | 0.004273116 | 0.078233418 |
| 0 | 0 | 0 | 0 | 0 |
| 0 | 0 | 0 | 0 | 0 |
| 0 | 0 | 0 | 0 | 0 |
| 0 | 0 | 0 | 0 | 0 |
| 0 | 0.082458835 | 0.041752828 | 0 | 1.016445626 |
| 0.468361053 | 0 | 0 | 0.00909313 | 0 |
| 0 | 0 | 0 | 0 | 0 |
| 0 | 0 | 0.005549678 | 0.00955291 | 0.034159647 |
| 0 | 0 | 0 | 0 | 0 |
| 0 | 0 | 0 | 0 | 0 |
| 0 | 0.023527071 | 0.006411452 | 0 | 0.122258103 |
| 0 | 0 | 0 | 0 | 0 |
| 0 | 0 | 0 | 0.001678824 | 0 |
| 0 | 0 | 0 | 0 | 0 |
| 0 | 0 | 0 | 0 | 0 |
| 0 | 0 | 0 | 0 | 0 |
| 0 | 0 | 0 | 0 | 0 |
| 0 | 0 | 0 | 0 | 0 |
| 0 | 0 | 0 | 0 | 0 |
| 0 | 0 | 0.014005993 | 0.305441927 | 0.319162625 |
| 0 | 0 | 0 | 0 | 0.001096531 |
| 0 | 0 | 0.200943955 | 0 | 0 |
| 0 | 0 | 0 | 0.621141667 | 0 |
| 0 | 0 | 0.053239238 | 2.547303997 | 0.731642072 |
| 0 | 0 | 0 | 0 | 0 |
| 0 | 0 | 0 | 0 | 0 |
| 0 | 0 | 0 | 0.311590254 | 0.010437115 |
| 0 | 0 | 0 | 0 | 0 |
| 0 | 0 | 0 | 0 | 0.003177364 |
| 0 | 0 | 0 | 0 | 0 |
| 0 | 0 | 0 | 0 | 0 |
| 0 | 0 | 0 | 0 | 0.373449154 |
| 0 | 0 | 0.002954031 | 0.058528308 | 0 |
| 0 | 0 | 0 | 0 | 0 |
| 0 | 0.005739923 | 0 | 0 | 0.003860821 |
| 0 | 0 | 0 | 0 | 0 |
| 0.169675861 | 0.010450102 | 0.015173183 | 0 | 0.052993892 |
| 0 | 0.248527703 | 0.004508315 | 0 | 0 |
| 0 | 1.984121313 | 2.570327479 | 7.944865613 | 1.354345157 |
| 0 | 0 | 0 | 0 | 0.01794384 |
| 0 | 0 | 0 | 0 | 0 |
| 0 | 0 | 0 | 0 | 0 |
| 0 | 0 | 0 | 0 | 0 |
| 0.333251525 | 0.000465267 | 0 | 0.003597241 | 0.743972288 |

| | | | | |
|---|---|---|---|---|
| 0 | 0 | 0 | 0 | 0 |
| 0 | 0 | 0 | 0 | 0 |
| 0.214620675 | 0.318600795 | 0 | 0 | 0.026973579 |
| 0 | 0 | 0 | 0 | 0 |
| 0 | 0 | 0 | 0 | 0 |
| 0 | 0.004216207 | 0 | 0.054329731 | 0 |
| 0 | 0 | 0 | 0 | 0 |
| 0 | 0.001898989 | 0 | 0.004301408 | 0 |
| 0 | 0 | 0 | 0 | 0 |
| 0.534197806 | 0 | 0 | 0 | 0 |
| 0.121343603 | 0.014158899 | 0.003915321 | 0.590377922 | 0.024788342 |
| 0 | 2.035819902 | 2.245561873 | 0.815489828 | 0.626348172 |
| 0 | 0 | 0 | 0 | 0 |
| 0.055273714 | 0.05765768 | 0.007476016 | 0.646751574 | 0.010746767 |
| 0.1346591 | 0.3039029 | 0.000640368 | 0.003239396 | 0.053696203 |
| 0 | 0 | 0 | 0.089287873 | 0 |
| 0 | 0.000907599 | 0 | 0 | 0 |
| 0 | 0 | 0 | 0 | 0 |
| 0 | 0 | 0 | 0 | 0 |
| 0 | 0.002044067 | 0 | 0.010632716 | 0 |
| 0.162866374 | 0.62309138 | 0.177838341 | 0 | 0.097311127 |
| 13.09502932 | 0.018176487 | 1.308513255 | 0 | 0.018710901 |
| 3.923557801 | 0.271093547 | 2.440550419 | 0.08476369 | 0.971792496 |
| 0 | 0 | 0 | 0 | 0 |
| 0 | 0 | 0 | 0 | 0 |
| 0 | 0 | 0.057624202 | 1.60542033 | 2.856255666 |
| 0 | 0 | 0 | 0.973871418 | 1.740944688 |
| 0 | 0 | 0 | 0 | 0.005185812 |
| 0 | 0 | 0 | 0 | 0.002970681 |
| 0 | 0 | 0 | 0 | 0 |
| 0 | 0 | 0 | 0.006259815 | 0 |
| 0 | 0 | 0.033364122 | 0.144331378 | 0.164962148 |
| 0.192177324 | 0.002098782 | 0 | 0 | 0.000870368 |
| 0 | 0 | 0.001179396 | 0.382993367 | 0.063717429 |
| 0.177892854 | 0.017414218 | 0.001633231 | 0 | 0.106992036 |
| 0 | 0.010195769 | 0.008274601 | 0.157855382 | 0.935870403 |
| 0 | 0 | 0 | 0 | 0.052416289 |
| 0 | 0 | 0 | 0 | 0 |
| 0 | 0 | 0.001805297 | 0 | 0 |
| 0 | 0 | 0 | 0 | 0 |
| 0 | 0 | 0 | 0 | 0 |
| 0 | 0 | 0 | 0 | 0 |
| 0 | 0 | 0 | 0 | 0 |
| 0 | 0 | 0 | 0 | 0 |
| 0.505902482 | 0.000620113 | 0 | 0.550835917 | 0.067730011 |
| 0 | 0 | 0 | 0 | 0 |
| 0 | 0 | 0 | 0.118785655 | 0 |
| 0 | 0.026741916 | 0.023537083 | 0.946670577 | 0.019814504 |

| | | | | |
|---|---|---|---|---|
| 0 | 0 | 0 | 0 | 0 |
| 0 | 0 | 0 | 0 | 0 |
| 0.175354361 | 0 | 0 | 0 | 0 |
| 0 | 0.000924909 | 0.001363131 | 0.005171697 | 0.005753423 |
| 0 | 0 | 0 | 0 | 0 |
| 0 | 0 | 0 | 0.851436447 | 0 |
| 0 | 0 | 0 | 0 | 0 |
| 0.086992176 | 0.002066919 | 0 | 0.356564837 | 0.199767469 |
| 0 | 0 | 0 | 0 | 0 |
| 0 | 0 | 0 | 0 | 0 |
| 0.092297856 | 0.003826102 | 0 | 0.001066648 | 0.019650556 |
| 0 | 0 | 0 | 0 | 0 |
| 0 | 0.003092355 | 0.000531709 | 0 | 0.035159266 |
| 0 | 0 | 0 | 0.028034213 | 0 |
| 0 | 0 | 0 | 0 | 0 |
| 0 | 0 | 0 | 0 | 0 |
| 0 | 0 | 0 | 0.128991494 | 0 |
| 0 | 0 | 0.000884687 | 0 | 0 |
| 0.475059172 | 0 | 0 | 0.008222456 | 0 |
| 0 | 0.005523982 | 0 | 0 | 0 |
| 0 | 0 | 0.000449129 | 0 | 0.001737685 |
| 0 | 0 | 0 | 0 | 0 |
| 0 | 0 | 0.000728558 | 0.005937779 | 0 |
| 0 | 0 | 0 | 0 | 0 |
| 0 | 0 | 0 | 0 | 0 |
| 0 | 0 | 0 | 0 | 0 |
| 0.005328931 | 0 | 0 | 0 | 0 |
| 0 | 0 | 0 | 0 | 0 |
| 0 | 0 | 0 | 0 | 0 |
| 0 | 0.04913119 | 0.021679232 | 0.030677743 | 0 |
| 0 | 0 | 0 | 0 | 0 |
| 0 | 0 | 0 | 0 | 0 |
| 0 | 0 | 0 | 0 | 0 |
| 0 | 0 | 0 | 0 | 0 |
| 0 | 0 | 0 | 0.003731846 | 0.00135871 |
| 0 | 0 | 0 | 0 | 0 |
| 0 | 0 | 0.033159095 | 0 | 0.110894082 |
| 0.050278567 | 0.00074743 | 0 | 0 | 0.03399232 |
| 0 | 0 | 0 | 0 | 0 |
| 0 | 0.003440375 | 0 | 0 | 0 |
| 0 | 0 | 0.001205793 | 0.563881673 | 0.047500524 |
| 0 | 0 | 0 | 0 | 0 |
| 0 | 0 | 0 | 0 | 0 |
| 0 | 0 | 5.510709631 | 0 | 0.004565292 |
| 0 | 0 | 0.049907373 | 0.144153688 | 0 |
| 0 | 0 | 0 | 0 | 0 |
| 0 | 0 | 0 | 0 | 0.007387399 |
| 0 | 0 | 0 | 0 | 0 |

| | | | | |
|---|---|---|---|---|
| 0 | 0 | 0 | 0 | 0 |
| 0 | 0 | 0 | 0 | 0 |
| 0 | 0 | 0 | 0 | 0 |
| 0 | 0 | 0 | 2.355147967 | 0 |
| 0 | 0 | 0 | 0 | 0 |
| 0 | 1.030856536 | 0 | 0 | 0 |
| 0 | 0 | 0 | 0 | 0 |
| 0 | 0 | 0 | 0 | 0 |
| 0.537010698 | 0.293271233 | 0.00124071 | 0 | 0.002493675 |
| 0 | 0 | 0 | 0.000601701 | 0 |
| 0 | 0 | 0.001140617 | 0 | 0 |
| 0 | 0 | 0 | 0 | 0 |
| 0 | 0 | 0 | 0 | 0 |
| 0 | 0 | 0 | 0 | 0 |
| 0 | 0 | 0 | 0.191867784 | 0.88687097 |
| 1.643713564 | 0 | 0 | 0 | 0 |
| 0 | 0 | 0 | 0 | 0 |
| 0 | 0 | 0 | 0 | 0 |
| 0 | 0 | 0 | 0 | 0 |
| 0 | 0 | 0 | 0 | 0 |
| 0 | 0 | 0 | 0 | 0 |
| 0 | 0 | 0 | 0 | 0 |
| 0 | 0 | 0 | 0 | 0 |
| 0 | 0 | 0 | 0 | 0 |
| 1.736766378 | 0.076572347 | 0 | 0.142050464 | 0.130495832 |
| 0 | 0 | 0 | 0.036163693 | 30.24197059 |
| 0 | 0 | 0 | 0 | 0 |
| 0 | 0 | 0 | 0 | 0 |
| 0 | 0 | 0 | 0 | 0 |
| 0 | 0 | 0 | 0 | 0 |
| 0 | 0 | 0 | 0 | 0 |
| 0 | 0 | 0 | 0 | 0.060244431 |
| 0 | 0 | 0 | 0 | 0 |
| 0 | 0 | 0 | 0.011262295 | 0.008300533 |
| 0.011619734 | 0.003377954 | 0.002017573 | 0.326288711 | 0.001548301 |
| 0.03429598 | 0.000879717 | 0 | 0.010384561 | 0.02444296 |
| 0 | 0 | 0.210196603 | 0 | 0 |
| 0 | 0 | 0.704478099 | 0.023863366 | 1.958181353 |
| 0 | 0 | 0 | 5.757123853 | 0 |
| 0 | 0 | 0 | 0 | 0 |
| 0 | 0 | 0 | 0 | 0 |
| 0.036440745 | 0.001852698 | 0 | 0.001859396 | 0 |
| 0 | 0 | 0.213493077 | 0.032862645 | 0.081510216 |
| 0 | 0 | 0 | 0 | 0 |
| 0 | 0 | 0 | 0 | 0 |
| 0 | 0.000952482 | 0 | 0 | 0 |
| 0 | 0 | 0 | 0 | 0 |
| 0.356246237 | 1.518556068 | 0.014676174 | 0.151507807 | 3.220555153 |

| | | | | |
|---|---|---|---|---|
| 0 | 0.005940586 | 0 | 0.013737984 | 0.001642381 |
| 0 | 0 | 0 | 0 | 0 |
| 0 | 0 | 0 | 0 | 0 |
| 0 | 0 | 0 | 0 | 0 |
| 0.001130293 | 0 | 0.000726404 | 0.007349241 | 0 |
| 0 | 0 | 0 | 0 | 0 |
| 0 | 0 | 0 | 0 | 0 |
| 0 | 0 | 0 | 0 | 0 |
| 0 | 0 | 0 | 0 | 0 |
| 0 | 0 | 0 | 0 | 0 |
| 0 | 0 | 0 | 0 | 0 |
| 0 | 0 | 0.103605802 | 1.240954676 | 4.285437106 |
| 0.021701873 | 0 | 1.547652261 | 0.011123303 | 0 |
| 0 | 0 | 0 | 0 | 0 |
| 0.002427627 | 1.966059598 | 0.010921119 | 0.018651455 | 0.112519627 |
| 0 | 0 | 0 | 0 | 0 |
| 0 | 0.000453174 | 0 | 0 | 0 |
| 0 | 0.003805677 | 0 | 0 | 0 |
| 0 | 0 | 0 | 0.022969881 | 0 |
| 0.003629842 | 0 | 0 | 0.01973346 | 0 |
| 0 | 0 | 0.010363468 | 0 | 0 |
| 0 | 0 | 0 | 0 | 0 |
| 3.917258412 | 0.390450749 | 0.002720736 | 0.064188227 | 0.550403391 |
| 0 | 0 | 0 | 0 | 0 |
| 0 | 0 | 0 | 0 | 0 |
| 0 | 0 | 0 | 0 | 0 |
| 0 | 0 | 0 | 0 | 0 |
| 0 | 0 | 0 | 0 | 0 |
| 0 | 0 | 0.000454023 | 0 | 0 |
| 0 | 0 | 0 | 0 | 0 |
| 0 | 0 | 0 | 0 | 0 |
| 0.018901113 | 0.03360228 | 0.048401736 | 0.262861559 | 0.018404616 |
| 0 | 0 | 0.011722649 | 0 | 0 |
| 0 | 0.069787964 | 0.001524907 | 0 | 0.025438473 |
| 0.03732262 | 0 | 0 | 0.00733574 | 0.021438859 |
| 0 | 0.000881531 | 0.001091328 | 0.784391446 | 0.058857188 |
| 0 | 0 | 0 | 0 | 0.034008402 |
| 0 | 0 | 0 | 0 | 0 |
| 0 | 0 | 0 | 0.000911101 | 0.003192789 |
| 0 | 0 | 0 | 0 | 0 |
| 0 | 0 | 0 | 0.001351368 | 0 |
| 0 | 0 | 0 | 0 | 0 |
| 0 | 0 | 0 | 0 | 0 |
| 0 | 0 | 0 | 0 | 0 |
| 0 | 0.00485017 | 0 | 0.038085286 | 0.101881668 |
| 0 | 0 | 0 | 0 | 0 |
| 0 | 0 | 0 | 0 | 0 |
| 0 | 0.00050971 | 0 | 0 | 0 |

| | | | | |
|---|---|---|---|---|
| 0 | 0 | 0 | 0 | 0 |
| 0 | 0 | 0 | 0 | 0 |
| 0 | 0.000418024 | 0 | 0 | 0 |
| 0 | 0 | 0 | 0 | 0 |
| 0 | 0 | 0 | 0 | 0 |
| 0 | 0 | 0 | 0 | 0 |
| 0 | 0 | 0 | 0 | 0 |
| 0 | 0.118435409 | 0.006788056 | 0 | 0.032088707 |
| 0 | 0 | 0 | 0 | 0 |
| 0 | 0.008546631 | 0 | 0 | 0 |
| 0 | 0 | 0 | 0 | 0 |
| 0.002113336 | 0 | 0 | 0 | 0.003086735 |
| 0 | 0 | 0 | 0 | 0 |
| 0 | 0 | 0 | 0 | 0 |
| 0 | 0 | 0 | 0 | 0 |
| 0 | 0 | 0 | 0 | 0 |
| 0 | 0 | 0 | 0 | 0 |
| 0 | 0 | 0 | 0 | 0 |
| 0 | 0 | 0 | 2.837244009 | 0 |
| 0 | 0 | 0 | 0 | 0 |
| 0 | 0 | 0 | 0 | 0 |
| 0 | 0 | 0 | 0 | 0 |
| 0 | 0 | 0 | 0 | 0 |
| 0 | 0 | 0 | 0 | 0 |
| 0.001519057 | 0 | 0 | 0 | 0 |
| 0.102253277 | 0.111503436 | 0.000517586 | 0 | 0.223375189 |
| 0 | 0 | 0 | 0 | 0 |
| 0.458620181 | 0.069117709 | 0.030776806 | 0 | 0.978266456 |
| 0 | 0 | 0 | 0 | 0 |
| 0 | 1.261645364 | 0.171863407 | 1.033575747 | 0.242792076 |
| 0 | 0 | 0 | 0 | 0 |
| 0 | 0 | 0 | 0 | 0 |
| 0 | 0 | 0 | 0 | 0 |
| 0 | 0 | 0 | 0 | 0 |
| 0.025470061 | 0.000657192 | 0 | 0 | 0.002289324 |
| 0 | 0 | 0.282236883 | 0.006893948 | 0.015338793 |
| 0 | 0 | 0 | 0 | 0 |
| 0.002907207 | 0 | 0 | 0.028323399 | 0.001515332 |
| 0 | 0 | 0 | 0 | 0 |
| 0 | 0 | 0 | 0 | 0.006080913 |
| 4.067130693 | 7.84497601 | 43.79867747 | 1.012150065 | 7.995534057 |
| 1.194859268 | 2.269511512 | 0 | 0.469417197 | 1.63342038 |
| 0 | 0 | 0.057241164 | 0 | 0 |
| 0 | 0 | 0 | 0 | 0 |
| 0 | 0 | 0 | 0 | 0 |
| 0 | 0 | 0 | 0 | 0 |
| 0 | 0 | 0 | 0 | 0 |
| 0.019001462 | 0 | 0 | 0.019905084 | 0.006700486 |

| | | | | |
|---|---|---|---|---|
| 0 | 0 | 0 | 0 | 0 |
| 0 | 0.057710567 | 0 | 0.004942005 | 0 |
| 0 | 0 | 0 | 0 | 0 |
| 0 | 0 | 0 | 0 | 0 |
| 0 | 0 | 0.83030591 | 0.163830325 | 0.777385577 |
| 0 | 0 | 0 | 0 | 0 |
| 0 | 0 | 0 | 0 | 0 |
| 0 | 0 | 0 | 0 | 0 |
| 0.08126919 | 0.309281072 | 0.395579943 | 0 | 0.11275634 |
| 0 | 0 | 0 | 0 | 0 |
| 0 | 0 | 0 | 0 | 0 |
| 0 | 0 | 0 | 0 | 0 |
| 0 | 0 | 0 | 0.002058 | 0.112299447 |
| 0.006503907 | 0.006981196 | 0.145394746 | 0.001897576 | 0.033354126 |
| 0 | 0 | 1.290675057 | 0 | 0 |
| 0 | 0 | 0 | 0.242394401 | 0.160236663 |
| 0 | 0 | 0.008050185 | 0 | 0.001461409 |
| 0 | 0 | 0 | 0 | 0 |
| 0 | 0 | 0 | 0 | 0 |
| 0 | 0 | 0 | 0 | 0 |
| 0 | 0 | 0 | 0 | 0 |
| 0 | 0 | 0 | 0 | 0 |
| 0 | 0 | 0 | 0 | 0 |
| 0 | 0 | 0 | 0 | 0 |
| 0 | 0 | 0 | 0 | 0 |
| 0 | 0 | 0 | 0 | 0 |
| 0 | 0.007113467 | 0 | 0.200849068 | 0.04908291 |
| 0.045735152 | 0.016547457 | 0 | 0.006444617 | 0 |
| 0 | 0 | 0 | 0 | 0 |
| 0 | 0 | 0 | 0 | 0 |
| 0 | 0 | 0 | 0 | 0 |
| 0 | 0 | 0 | 0 | 0 |
| 0 | 0 | 0 | 0 | 0 |
| 0 | 0 | 0 | 0.034341369 | 0 |
| 0 | 0 | 0.038855875 | 0 | 0 |
| 0.039147222 | 0 | 0 | 0 | 0 |
| 0 | 0 | 0 | 0 | 0 |
| 0 | 0 | 0 | 0 | 0 |
| 0 | 0 | 0 | 0 | 0 |
| 0 | 0 | 0 | 0 | 0 |
| 0 | 0 | 0.886948203 | 0.017500241 | 0.023616394 |
| 0 | 0 | 0 | 0 | 0 |
| 0 | 0 | 0 | 0 | 0 |
| 0 | 0 | 0 | 0 | 0 |
| 0 | 0 | 0 | 0 | 0 |
| 0 | 0 | 0 | 0 | 0 |
| 0 | 0 | 0 | 0 | 0 |
| 0 | 0 | 0 | 0 | 0 |

| | | | | |
|---|---|---|---|---|
| 0 | 0.357085191 | 0 | 0.055469984 | 0 |
| 0 | 0 | 0 | 0 | 0 |
| 0 | 0 | 0 | 0 | 0 |
| 0.002405989 | 0 | 0 | 0 | 0.001421291 |
| 0.072591642 | 0 | 0 | 0.003686601 | 0 |
| 0 | 0 | 0 | 0 | 0 |
| 0.02591279 | 0.009541884 | 0.019558422 | 0 | 0 |
| 0 | 0 | 0 | 0.001231539 | 0 |
| 0 | 0.001063426 | 0 | 0 | 0 |
| 0.208884736 | 0.013304077 | 0 | 0 | 0 |
| 0 | 0 | 0 | 0 | 0 |
| 0 | 0 | 0 | 0 | 0 |
| 0 | 0 | 0 | 0 | 0 |
| 0 | 0 | 0 | 9.96565293 | 0 |
| 0 | 0 | 0 | 0 | 0 |
| 0 | 0 | 0 | 0 | 0 |
| 4.233139804 | 0 | 0 | 0 | 0 |
| 0 | 0 | 0 | 0 | 0.005255905 |
| 0 | 0 | 0 | 0 | 0 |
| 0 | 0 | 0 | 0 | 0 |
| 0.210109745 | 0.131006684 | 0.00456269 | 0 | 0.837460005 |
| 0 | 0 | 0 | 0 | 0 |
| 0 | 0.002768703 | 0 | 0.025802333 | 0.032149202 |
| 0 | 0 | 0.00108853 | 0.066791663 | 0.013783212 |
| 0 | 0 | 0 | 0 | 0 |
| 0 | 0 | 0 | 0.006616722 | 0 |
| 0 | 0 | 0 | 0 | 0 |
| 0 | 0 | 0 | 0 | 0 |
| 0 | 0 | 0 | 0 | 0 |
| 2.274242954 | 4.040584529 | 1.517213307 | 0.009539505 | 0.45527767 |
| 0 | 0 | 0 | 0 | 0 |
| 0 | 0 | 0 | 0 | 0 |
| 0 | 0 | 0 | 0 | 0 |
| 0 | 0 | 0 | 0 | 0 |
| 0 | 0 | 0 | 0 | 0 |
| 0 | 0 | 0 | 0 | 0 |
| 0.239687152 | 0.005721019 | 0 | 0 | 0 |
| 0 | 0 | 0 | 0 | 0 |
| 0 | 0 | 0 | 0 | 0 |
| 0 | 0 | 0 | 0 | 0 |
| 0 | 0 | 0 | 0 | 0 |
| 0 | 0 | 0 | 0 | 0 |
| 0 | 0 | 0 | 0 | 0 |
| 0 | 0 | 0 | 0 | 0 |
| 0 | 0 | 0 | 0 | 0 |
| 0 | 0 | 0 | 0 | 0 |
| 0 | 0 | 0 | 0 | 0 |
| 0 | 0 | 0 | 0 | 0 |

| | | | | |
|---|---|---|---|---|
| 0.301588889 | 0.003153756 | 0 | 0.003108889 | 0.018676358 |
| 0 | 0 | 0 | 0 | 0 |
| 0 | 0 | 0 | 0 | 0 |
| 0 | 0 | 0 | 0 | 0 |
| 0 | 0 | 0 | 0 | 0 |
| 0 | 0 | 0 | 0 | 0 |
| 0.003224735 | 0 | 0 | 0 | 0 |
| 0 | 0 | 0.293181091 | 0.395072588 | 0.309078422 |
| 1.450591219 | 0.963129268 | 0.01143972 | 1.189252614 | 1.330686906 |
| 0.009641782 | 0.071957625 | 0.008817274 | 0 | 0.072229964 |
| 0 | 0 | 0 | 0 | 0 |
| 0 | 0 | 0 | 0 | 0 |
| 0 | 0 | 0 | 0 | 0 |
| 0 | 0 | 0 | 0 | 0 |
| 0 | 0 | 0 | 0 | 0 |
| 0 | 0 | 0 | 0 | 0 |
| 0.069678055 | 0.055796065 | 0 | 0 | 0 |
| 0 | 0 | 0 | 0 | 0 |
| 0 | 0 | 0 | 0 | 0 |
| 0 | 0.001028776 | 0 | 0 | 0 |
| 0 | 0 | 0 | 0 | 0 |
| 0 | 0 | 0 | 0 | 0 |
| 0 | 0 | 0 | 0 | 0 |
| 0 | 0 | 0 | 0 | 0 |
| 0 | 0 | 0 | 0.025495092 | 0 |
| 0 | 0 | 0 | 0 | 0 |
| 0 | 0 | 0 | 0 | 0 |
| 0 | 0 | 0 | 0 | 0 |
| 0 | 0 | 0 | 0 | 0 |
| 0 | 0 | 0 | 0 | 0 |
| 0 | 0 | 0 | 0 | 0 |
| 0 | 0 | 0 | 0 | 0 |
| 0 | 0 | 0 | 0 | 0 |
| 0 | 0 | 0 | 0 | 0 |
| 0 | 0 | 0 | 0 | 0.001551691 |
| 0 | 0 | 0 | 0 | 0 |
| 0 | 0 | 0 | 0 | 0 |
| 0 | 0.044423986 | 0 | 0 | 0 |
| 0 | 0 | 0 | 0.002826897 | 0 |
| 0 | 0 | 0.033910312 | 0 | 0 |
| 0 | 0 | 0 | 0 | 0 |
| 0 | 0.193980776 | 0.062060635 | 0.206808979 | 0.229882794 |
| 0 | 0 | 0 | 0 | 0 |
| 0 | 0 | 0 | 0 | 0 |
| 0 | 0 | 0 | 0 | 0 |
| 0 | 0 | 0 | 0 | 0 |
| 0 | 0 | 0 | 0 | 0 |
| 0 | 0 | 0 | 0 | 0 |

| | | | | |
|---|---|---|---|---|
| 0 | 0 | 0 | 0 | 0 |
| 0.135641656 | 0.055903357 | 0 | 0.000825267 | 0.000495771 |
| 0 | 0 | 0 | 0 | 0 |
| 0 | 0 | 0 | 0 | 0 |
| 0 | 0.060013449 | 0.003612986 | 3.260750482 | 2.03326408 |
| 0 | 0 | 0 | 0 | 0 |
| 0 | 0.002070848 | 0 | 0 | 0 |
| 0.073600558 | 0.025354617 | 0 | 0.004187363 | 0.004192532 |
| 0 | 0 | 0 | 0.060623443 | 0 |
| 0 | 0 | 0 | 0 | 0 |
| 0 | 0 | 0 | 0 | 0 |
| 0 | 0 | 0 | 0 | 0 |
| 0 | 0 | 0 | 0 | 0 |
| 0 | 0 | 0 | 0 | 0 |
| 0.150125839 | 0.00779471 | 0 | 0 | 0.005082957 |
| 0 | 0 | 0 | 0 | 0.070298928 |
| 0 | 0 | 0 | 0 | 0 |
| 0 | 0 | 0 | 0 | 0 |
| 0 | 0 | 0 | 0 | 0 |
| 0 | 0 | 0 | 0 | 0 |
| 0 | 0 | 0 | 0 | 0 |
| 0 | 0 | 0 | 0 | 0 |
| 0 | 0 | 0 | 0 | 0 |
| 0.011783616 | 0.00124988 | 0 | 0.000805293 | 0.012094303 |
| 0 | 0 | 0.02982848 | 0 | 0 |
| 0 | 0 | 0 | 0 | 0 |
| 0.353471369 | 0 | 0 | 0.960621555 | 0.007743129 |
| 0 | 0 | 0 | 0 | 0.000582376 |
| 0 | 0 | 0 | 0 | 0.003014316 |
| 0 | 0 | 0 | 0 | 0 |
| 0.000730953 | 0 | 0 | 0 | 0 |
| 0 | 0 | 0 | 0 | 0 |
| 0 | 0 | 0 | 0 | 0 |
| 0 | 0 | 0 | 0 | 0 |
| 0 | 0 | 0 | 0 | 0 |
| 0 | 0 | 0 | 0 | 0 |
| 0 | 0 | 0 | 0 | 0 |
| 0 | 0 | 0 | 0 | 0 |
| 0 | 0 | 0 | 0 | 0 |
| 0 | 0 | 0 | 0 | 0 |
| 0 | 0 | 0 | 0 | 0 |
| 0 | 0 | 0 | 0 | 0 |
| 0 | 0 | 0 | 0 | 0 |
| 0 | 0 | 0 | 0 | 0 |
| 0 | 0 | 0 | 0 | 0 |
| 0 | 0 | 0 | 0 | 0 |
| 0 | 0 | 0 | 0 | 0 |
| 0 | 0 | 0 | 0 | 0 |

| | | | | |
|---|---|---|---|---|
| 0 | 0.048018477 | 0.001355537 | 0.010216509 | 0.207529745 |
| 0 | 0 | 0 | 0 | 0 |
| 0 | 0 | 0 | 0 | 0 |
| 0 | 0 | 0 | 0 | 0 |
| 0 | 0 | 0 | 0 | 0 |
| 0.040939346 | 0 | 0 | 0 | 0 |
| 0 | 0 | 0 | 0 | 0 |
| 0 | 0 | 0 | 0 | 0 |
| 0 | 0 | 0 | 0 | 0 |
| 0 | 0 | 0 | 0 | 0 |
| 0 | 0 | 0 | 0 | 0 |
| 0 | 0 | 0 | 0 | 0 |
| 0 | 0 | 0 | 0 | 0 |
| 0 | 0 | 0 | 0 | 0 |
| 0 | 0 | 0 | 0 | 0 |
| 0 | 0 | 0 | 0 | 0 |
| 0 | 0 | 0 | 0 | 0 |
| 0 | 0 | 0 | 0 | 0 |
| 0 | 0 | 0 | 0 | 0 |
| 0 | 0 | 0 | 0 | 0 |
| 0 | 0.784270867 | 0.004850893 | 0 | 0.061222372 |
| 0 | 0 | 0 | 0 | 0 |
| 0.014018577 | 0.001397252 | 0.00090093 | 0.008355392 | 0.055264356 |
| 0.321313181 | 0 | 0 | 0.025954441 | 0.050116775 |
| 0 | 0 | 0 | 0 | 0 |
| 0 | 0 | 0 | 0 | 0 |
| 0 | 0 | 0 | 0 | 0 |
| 0 | 0 | 0 | 0 | 0 |
| 0 | 0 | 0 | 0 | 0 |
| 0 | 0 | 0 | 0 | 0 |
| 0 | 0 | 0 | 0 | 0 |
| 0 | 0 | 0.027901241 | 0 | 0 |
| 0.029257442 | 0.01831929 | 0.001856178 | 0.006164986 | 0.003323705 |
| 0 | 0 | 0 | 0 | 0 |
| 0.541195474 | 0 | 1.461856069 | 0.005302453 | 0 |
| 0.001354493 | 0.035438615 | 0.010426536 | 0 | 0 |
| 0 | 0 | 0 | 0 | 0 |
| 0 | 0 | 0 | 0 | 0 |
| 0.357378231 | 0.018369755 | 0 | 0 | 0.011017724 |
| 0 | 0 | 0 | 0 | 0 |
| 0 | 0 | 0 | 0 | 0 |
| 0 | 0 | 0 | 0 | 0 |
| 0 | 0.016509228 | 0.009889491 | 0 | 0.105383109 |
| 0 | 0 | 0 | 0 | 0 |
| 0 | 0 | 0 | 0 | 0 |
| 0 | 0.0260814 | 0.002763434 | 0.001226249 | 0.000818508 |
| 0 | 0 | 0 | 0 | 0 |
| 0 | 0 | 0 | 0 | 0 |

| | | | | |
|---|---|---|---|---|
| 0 | 0 | 0 | 0 | 0 |
| 0 | 0 | 0 | 0 | 0 |
| 0 | 0 | 0 | 0 | 0 |
| 0 | 0 | 0 | 0 | 0 |
| 0 | 0 | 0 | 0 | 0 |
| 0 | 0 | 0 | 0 | 0 |
| 0 | 0 | 0 | 0 | 0 |
| 0 | 0.149855053 | 0 | 0 | 0.031242061 |
| 0 | 0 | 0 | 0 | 0 |
| 0 | 0 | 0 | 0 | 0 |
| 0 | 0 | 0 | 0 | 0 |
| 0 | 0 | 0 | 0 | 0 |
| 0 | 0 | 0 | 0 | 0 |
| 0 | 0 | 0 | 0 | 0 |
| 0 | 0 | 0 | 0 | 0 |
| 0.114999595 | 0.001571559 | 0 | 0 | 0 |
| 0 | 0 | 0 | 0 | 0 |
| 0 | 0 | 0 | 0 | 0 |
| 0 | 0 | 0 | 0 | 0 |
| 0 | 0 | 2.142001169 | 0.121727572 | 0.671543305 |
| 0 | 0 | 0.002770932 | 0 | 0 |
| 0 | 0 | 0 | 0 | 0 |
| 0 | 0 | 0 | 0 | 0 |
| 0 | 0 | 0 | 0 | 0 |
| 0 | 0 | 0 | 0 | 0 |
| 0 | 0 | 0 | 0 | 0 |
| 0 | 0 | 0 | 0 | 0 |
| 0 | 0 | 0 | 0 | 0 |
| 0 | 0 | 0 | 0 | 0 |
| 0 | 0 | 0 | 0.020392336 | 0 |
| 0.188800926 | 0.754663393 | 0.252889804 | 0.081305897 | 0.386974644 |
| 0 | 0 | 0 | 0 | 0 |
| 0 | 0 | 0 | 0 | 0 |
| 0 | 0 | 0 | 0 | 0 |
| 0 | 0 | 0 | 0 | 0 |
| 0 | 0 | 0 | 0 | 0 |
| 0 | 0 | 0 | 0 | 0 |
| 0 | 0 | 0 | 0 | 0 |
| 0 | 0 | 0 | 0.017833713 | 0 |
| 0 | 0 | 0 | 0 | 0 |
| 0 | 0 | 0 | 0 | 0 |
| 0 | 0 | 0 | 0 | 0 |
| 0 | 0 | 0 | 0 | 0 |
| 0 | 0 | 0 | 0 | 0 |
| 0 | 0 | 0 | 0 | 0 |
| 0 | 0 | 0 | 0 | 0 |
| 2.485625052 | 0 | 0.008751288 | 0 | 0 |
| 0 | 0 | 0 | 0 | 0.001380166 |

| | | | | |
|---|---|---|---|---|
| 0 | 0 | 0 | 0 | 0 |
| 0 | 0 | 0 | 0 | 0 |
| 0 | 0 | 0 | 0 | 0 |
| 0 | 0 | 0 | 0 | 0 |
| 0.014783417 | 0.119533327 | 0.004170708 | 0.11166806 | 0.14828157 |
| 0 | 0 | 0 | 0 | 0 |
| 0 | 0 | 0 | 0 | 0 |
| 0 | 0 | 0 | 0 | 0.001690062 |
| 0 | 0 | 0 | 0 | 0 |
| 0 | 0 | 0 | 0 | 0 |
| 0 | 0 | 0 | 0.004022117 | 0 |
| 0 | 0.000910172 | 0 | 0.033645862 | 0.027082019 |
| 0 | 0 | 0 | 0 | 0.004559016 |
| 0 | 0 | 0 | 0 | 0 |
| 0 | 0.041732464 | 0 | 0 | 0 |
| 0 | 0.187445719 | 0 | 0.000684037 | 0 |
| 0 | 0.001283388 | 0 | 0 | 0 |
| 0 | 0 | 0 | 0 | 0 |
| 0 | 0 | 0 | 0.004329397 | 0.031676955 |
| 0.006706772 | 0.011248356 | 0.001701989 | 0 | 0.008054404 |
| 0 | 0 | 0 | 0 | 0 |
| 0 | 0 | 0 | 0 | 0 |
| 0 | 0 | 0 | 0 | 0 |
| 0 | 0 | 0 | 0 | 0 |
| 0 | 0 | 0 | 0 | 0 |
| 0 | 0 | 0 | 0 | 0 |
| 0 | 0 | 0 | 0 | 0.024596355 |
| 0 | 0 | 0 | 0 | 0 |
| 0 | 0 | 0 | 0 | 0 |
| 0 | 0 | 0 | 0 | 0 |
| 0.00765432 | 0 | 0 | 0 | 0.004548007 |
| 0 | 0.008681845 | 0 | 0 | 0 |
| 0 | 0 | 0 | 0 | 0 |
| 0 | 0 | 0 | 0 | 0 |
| 0 | 0 | 0 | 0 | 0 |
| 0 | 0 | 0 | 0 | 0 |
| 0 | 0 | 0 | 0 | 0 |
| 0.07023939 | 0.025460167 | 0.000493726 | 0.381575497 | 0.025006678 |
| 0 | 0 | 0.041925891 | 0 | 0 |
| 0 | 0 | 0 | 0 | 0 |
| 0.00668346 | 0.002102195 | 0 | 0 | 0 |
| 0 | 0 | 0 | 0 | 0 |
| 0 | 0 | 0 | 0 | 0 |
| 0 | 0 | 0 | 0 | 0 |
| 0 | 0 | 0 | 0 | 0 |
| 0 | 0 | 0 | 0 | 0 |
| 0 | 0 | 0 | 0 | 0 |
| 0 | 0 | 0 | 0 | 0 |

| | | | | |
|---|---|---|---|---|
| 0 | 0 | 0 | 0 | 0 |
| 0 | 0 | 0 | 0.214198364 | 0 |
| 0 | 0 | 0 | 0 | 0 |
| 0.016032298 | 0.001636212 | 0 | 0.001940696 | 0.026771478 |
| 0.020406291 | 0 | 0.000711639 | 0.000599988 | 0 |
| 0.007349659 | 0.001958557 | 0 | 0 | 0 |
| 0 | 0 | 0.000595669 | 0 | 0 |
| 0 | 0 | 0 | 0.006577099 | 0 |
| 0 | 0 | 0 | 0 | 0 |
| 0 | 0 | 0 | 0 | 0 |
| 0 | 0 | 0 | 0 | 0 |
| 0 | 0 | 0 | 0 | 0 |
| 0 | 0 | 0.004895033 | 0.427989373 | 0.363168693 |
| 0 | 0 | 0 | 0.011004387 | 0 |
| 0 | 0 | 0 | 0 | 0 |
| 0 | 0 | 0 | 0 | 0 |
| 0 | 0 | 0 | 0 | 0 |
| 0 | 0 | 0 | 0 | 0 |
| 0 | 0 | 0 | 0 | 0 |
| 0 | 0 | 0 | 0 | 0 |
| 0 | 0 | 0 | 0 | 0 |
| 0 | 0 | 0 | 0 | 0 |
| 0 | 0 | 0.001289126 | 0 | 0.04735745 |
| 0 | 0 | 0.00135219 | 0 | 0 |
| 0 | 0 | 0 | 0 | 0 |
| 0 | 0 | 0 | 0 | 0 |
| 0 | 0 | 0 | 0.004807465 | 0 |
| 0 | 0 | 0 | 0 | 0 |
| 0 | 0 | 0 | 0 | 0 |
| 0 | 0 | 0 | 0 | 0 |
| 0 | 0.019660449 | 0.006707726 | 0.031194115 | 0.01235823 |
| 0 | 0 | 0 | 0 | 0 |
| 0 | 0 | 0 | 0 | 0 |
| 0 | 0 | 0 | 0 | 0 |
| 0 | 0 | 0 | 0.443086383 | 0 |
| 0 | 0 | 0 | 0 | 0 |
| 0 | 0 | 0 | 0 | 0 |
| 0 | 0 | 0 | 0 | 0 |
| 0.001051271 | 0.002881498 | 0 | 0 | 0.020341516 |
| 0 | 0 | 0 | 0 | 0 |
| 0 | 0 | 0 | 0 | 0 |
| 0 | 0 | 0 | 0 | 0 |
| 0 | 0 | 0 | 0 | 0 |
| 0 | 0 | 0 | 0 | 0 |
| 0 | 0 | 0 | 0 | 0 |
| 0 | 0 | 0.133866385 | 0.784500202 | 0.514501937 |
| 0.089955828 | 0 | 0 | 0 | 0 |
| 0 | 0 | 0 | 0 | 0 |

| | | | | |
|---|---|---|---|---|
| 0 | 0 | 0 | 0 | 0 |
| 0 | 0 | 0 | 0 | 0 |
| 0 | 0 | 0 | 0 | 0 |
| 0 | 0 | 0 | 0.004287756 | 0 |
| 0 | 0 | 0 | 0 | 0 |
| 0 | 0 | 0 | 0 | 0 |
| 0 | 0 | 0 | 0 | 0 |
| 0 | 0 | 0 | 0 | 0 |
| 0.070034807 | 0 | 0 | 0 | 0 |
| 0 | 0 | 0 | 0 | 0 |
| 0.007553927 | 0 | 0 | 0.005237295 | 0.002004967 |
| 0.004183788 | 0.001903715 | 0 | 0 | 0.005986844 |
| 0 | 0 | 0 | 0 | 0 |
| 0 | 0 | 0 | 0 | 0 |
| 0 | 0 | 0 | 0 | 0 |
| 0 | 0 | 0 | 0 | 0 |
| 0 | 0 | 0 | 0 | 0 |
| 0 | 0 | 0 | 0 | 0 |
| 0 | 0 | 0 | 0 | 0 |
| 0 | 0 | 0.000581509 | 0.005107024 | 0.003477063 |
| 0 | 0 | 0 | 0 | 0 |
| 0 | 0 | 0 | 0 | 0 |
| 0 | 0 | 0 | 0 | 0 |
| 0 | 0 | 0 | 0 | 0 |
| 0 | 0 | 0 | 0 | 0 |
| 0 | 0 | 0 | 0 | 0 |
| 0 | 0 | 0 | 0 | 0 |
| 0 | 0 | 0.343970777 | 0.008816786 | 0.012222925 |
| 0 | 0 | 0 | 0 | 0 |
| 0 | 0 | 0 | 0 | 0 |
| 0 | 0 | 0.002248092 | 0.019304834 | 0.121770571 |
| 0 | 0 | 0 | 0 | 0 |
| 0 | 0.021225234 | 0.034803516 | 0.00998481 | 0.024686799 |
| 0.002470545 | 0.00086185 | 0.003260163 | 0.016450367 | 0.001459427 |
| 0 | 0 | 0 | 0 | 0 |
| 0 | 0 | 0 | 0 | 0 |
| 0 | 0 | 0 | 0 | 0 |
| 0 | 0 | 0 | 0 | 0 |
| 0 | 0 | 0 | 0 | 0 |
| 0 | 0 | 0 | 0 | 0 |
| 0 | 0 | 0 | 0 | 0 |
| 0.11655268 | 0.012373599 | 0 | 0.000838205 | 0.332506852 |
| 0.008077021 | 0.001831485 | 0.008575282 | 0.050854288 | 0.189558679 |
| 0 | 0 | 0 | 0 | 0 |
| 0 | 0 | 0 | 0 | 0 |
| 0 | 0.009772148 | 0.001680255 | 0.085588412 | 0.06146332 |
| 0 | 0 | 0 | 0 | 0 |
| 0 | 0 | 0 | 0 | 0 |

| | | | | |
|---|---|---|---|---|
| 0 | 0 | 0 | 0 | 0 |
| 0 | 0 | 0.008374678 | 0.05135095 | 0 |
| 0 | 0.139648475 | 0.012628328 | 0.085226318 | 0 |
| 0 | 0 | 0 | 0 | 0 |
| 0 | 0.070893619 | 0.00099411 | 0 | 0 |
| 0 | 0.014605455 | 0 | 0 | 0 |
| 0 | 0 | 0 | 0 | 0 |
| 0 | 0 | 0 | 0 | 0 |
| 0 | 0 | 0 | 0 | 0 |
| 0 | 0 | 0 | 0 | 0 |
| 0 | 0 | 0 | 0.002114461 | 0 |
| 0 | 0 | 0 | 0 | 0 |
| 0 | 0 | 0 | 0 | 0 |
| 0 | 0 | 0 | 0 | 0 |
| 0 | 0 | 0 | 0 | 0 |
| 0 | 0 | 0 | 0 | 0 |
| 0 | 0 | 0 | 0.829109047 | 0 |
| 0 | 0 | 0 | 0 | 0 |
| 0 | 0 | 0 | 0 | 0 |
| 0 | 0 | 0 | 0 | 0 |
| 0 | 0 | 0 | 0 | 0 |
| 0 | 0.001525483 | 0 | 0 | 0 |
| 0 | 0 | 0 | 0 | 0 |
| 0 | 0 | 0 | 0 | 0 |
| 0 | 0 | 0 | 0 | 0.005155804 |
| 0.00098336 | 0.003430453 | 0.000884765 | 0.006837894 | 0.000622394 |
| 0 | 0 | 0 | 0 | 0 |
| 0 | 0 | 0 | 0.090650941 | 0.08090168 |
| 0 | 0 | 0 | 0 | 0 |
| 0 | 0 | 0 | 0 | 0 |
| 0 | 0 | 0 | 0 | 0 |
| 0 | 0 | 0 | 0 | 0 |
| 0 | 0 | 0 | 0 | 0 |
| 0 | 0 | 0 | 0 | 0 |
| 0 | 0 | 0 | 0 | 0 |
| 0 | 0 | 0 | 0 | 0 |
| 0 | 0 | 0 | 0 | 0 |
| 0.069498526 | 0 | 0 | 0 | 0 |
| 0 | 0 | 0 | 0 | 0 |
| 0 | 0 | 0 | 0 | 0 |
| 0 | 0 | 0 | 0 | 0 |
| 0 | 0 | 0 | 0 | 0 |
| 0.249030121 | 0.00200832 | 0 | 0.002079568 | 0.015824221 |
| 0 | 0 | 0 | 0 | 0 |
| 0 | 0 | 0 | 0 | 0 |
| 0 | 0 | 0 | 0 | 0 |
| 0 | 0 | 0 | 0.016861056 | 0.001274103 |
| 0 | 0 | 0 | 0 | 0 |

| | | | | |
|---|---|---|---|---|
| 0 | 0 | 0 | 0 | 0 |
| 0.033814856 | 0.011353262 | 0.014140942 | 0.019268424 | 0.048029556 |
| 0 | 0 | 0 | 0 | 0 |
| 0 | 0 | 0 | 0 | 0 |
| 0 | 0 | 0 | 0 | 0 |
| 0 | 0 | 0 | 0 | 0 |
| 0 | 0 | 0 | 0 | 0 |
| 0 | 0 | 0 | 0 | 0 |
| 0 | 0 | 0 | 0 | 0 |
| 0 | 0 | 0 | 0 | 0 |
| 0 | 0 | 0 | 0 | 0 |
| 0 | 0 | 0 | 0 | 0 |
| 0.022487653 | 0 | 0.022497839 | 0.224689262 | 0 |
| 0 | 0 | 0 | 0 | 0 |
| 0 | 0.005006322 | 0.000712389 | 0.00175181 | 0.023803907 |
| 0 | 0 | 0 | 0 | 0 |
| 0 | 0 | 0 | 0 | 0 |
| 0 | 0 | 0 | 0 | 0 |
| 0 | 0.002291705 | 0 | 0 | 0 |
| 0 | 0 | 0 | 0 | 0 |
| 0 | 0 | 0 | 0 | 0 |
| 0.025465933 | 0.144955021 | 0.001224339 | 0 | 0 |
| 0 | 0 | 0 | 0 | 0 |
| 1.283257431 | 0.005937195 | 0 | 0 | 0 |
| 0 | 0 | 0 | 0 | 0 |
| 0 | 0 | 0 | 0 | 0 |
| 0 | 0 | 0 | 0 | 0 |
| 0 | 0 | 0 | 0 | 0 |
| 0 | 0 | 0 | 0 | 0 |
| 0 | 0 | 0 | 0 | 0 |
| 0 | 0 | 0 | 0 | 0 |
| 0 | 0 | 0 | 0 | 0 |
| 0 | 0 | 0 | 0 | 0 |
| 0 | 0 | 0 | 0 | 0 |
| 0 | 0 | 0.001588045 | 0 | 0 |
| 0 | 0 | 0 | 0 | 0 |
| 0 | 0 | 0 | 0 | 0 |
| 0 | 0 | 0 | 0 | 0 |
| 0.188304509 | 0.010327559 | 0 | 0 | 0.001228688 |
| 0 | 0 | 0 | 0 | 0 |
| 0.071887503 | 0 | 0.025057545 | 1.187249019 | 0.028423385 |
| 0 | 0 | 0 | 0 | 0 |
| 0 | 0 | 0 | 0 | 0 |
| 0 | 0 | 0 | 0 | 0 |
| 0 | 0 | 0 | 0 | 0 |
| 2.690200289 | 1.667136061 | 0.019006584 | 0.001763714 | 0.049444937 |
| 0 | 0 | 0 | 0 | 0 |
| 0 | 0 | 0 | 0 | 0 |

| | | | | |
|---|---|---|---|---|
| 0 | 0 | 0 | 0 | 0 |
| 0 | 0 | 0 | 0 | 0 |
| 0 | 0 | 0 | 0 | 0 |
| 0 | 0 | 0 | 0 | 0 |
| 0 | 0 | 0 | 0 | 0 |
| 0 | 0 | 0 | 0 | 0 |
| 0 | 0 | 0 | 0 | 0 |
| 0.007323705 | 0 | 0 | 0 | 0 |
| 0 | 0 | 0 | 0 | 0.013500756 |
| 0.004677716 | 0.011048792 | 0 | 0 | 0.025658936 |
| 0 | 0.063640734 | 0 | 0 | 0 |
| 0 | 0 | 0 | 0 | 0 |
| 0 | 0.002325995 | 0 | 0.003371918 | 0 |
| 0 | 0 | 0 | 0 | 0 |
| 0 | 0 | 0 | 0 | 0 |
| 0 | 0 | 0 | 0 | 0 |
| 0 | 0 | 0 | 0 | 0 |
| 0 | 0 | 0 | 0 | 0 |
| 0 | 0 | 0 | 0 | 0 |
| 0.010512792 | 0.360084307 | 0.025325012 | 0.197439038 | 4.313604663 |
| 0 | 0 | 0 | 0 | 0 |
| 0 | 0 | 0 | 0 | 0 |
| 0 | 0 | 0 | 0 | 0 |
| 0 | 0 | 0 | 0 | 0 |
| 0 | 0 | 0 | 0 | 0 |
| 0 | 0 | 0 | 0 | 0 |
| 0 | 0 | 0 | 0 | 0 |
| 0 | 0 | 0 | 0 | 0 |
| 0 | 0 | 0 | 0 | 0 |
| 0 | 0.336562065 | 0 | 0 | 0.271672127 |
| 0 | 0 | 0 | 0 | 0 |
| 0 | 0 | 0 | 0 | 0.003206288 |
| 0 | 0 | 0 | 0 | 0 |
| 0 | 0 | 0 | 0 | 0 |
| 0 | 0 | 0 | 0 | 0 |
| 0 | 0 | 0.009281834 | 0 | 0 |
| 0 | 0 | 0 | 0 | 0 |
| 0 | 0 | 0 | 0 | 0 |
| 0 | 0 | 0 | 0 | 0 |
| 0 | 0 | 0 | 0 | 0 |
| 0 | 0 | 0 | 0 | 0 |
| 0 | 0 | 0 | 0 | 0 |
| 0 | 0 | 0 | 0 | 0 |
| 0 | 0 | 0 | 0 | 0 |
| 0.014651742 | 0 | 0 | 0.023155082 | 0.023586856 |
| 0 | 0 | 0 | 0 | 0 |
| 0 | 0 | 0 | 0 | 0 |
| 0 | 0 | 0 | 0 | 0 |

| | | | | |
|---|---|---|---|---|
| 0 | 0 | 0 | 0 | 0 |
| 0 | 0 | 0 | 0 | 0 |
| 0 | 0 | 0 | 0 | 0 |
| 0 | 0 | 0 | 0 | 0 |
| 0 | 0 | 0 | 0 | 0 |
| 0 | 0 | 0.000534859 | 0.005862266 | 0.004515001 |
| 0 | 0 | 0 | 0 | 0 |
| 0 | 0 | 0 | 0 | 0 |
| 0 | 0 | 0 | 0 | 0 |
| 0 | 0 | 0 | 0 | 0 |
| 0 | 0 | 0 | 0 | 0 |
| 0 | 0 | 0 | 0 | 0 |
| 0 | 0 | 0 | 0 | 0 |
| 0 | 0 | 0 | 0 | 0 |
| 0 | 0 | 0 | 0 | 0 |
| 0 | 0 | 0 | 0 | 0 |
| 0 | 0 | 0 | 0 | 0 |
| 0 | 0.003703258 | 0 | 0 | 0.017052092 |
| 0 | 0 | 0 | 0 | 0 |
| 0 | 0 | 0 | 0 | 0 |
| 0 | 0 | 0 | 0 | 0 |
| 0 | 0 | 0 | 0 | 0 |
| 0 | 0 | 0 | 0 | 0 |
| 0 | 0 | 0 | 0 | 0 |
| 0 | 0 | 0 | 0 | 0 |
| 0 | 0 | 0 | 0 | 0 |
| 0 | 0 | 0 | 0 | 0 |
| 0 | 0 | 0 | 0 | 0 |
| 0 | 0 | 0 | 0 | 0 |
| 0 | 0 | 0 | 0 | 0 |
| 0 | 0 | 0 | 0 | 0 |
| 0 | 0 | 0 | 0.001061953 | 0 |
| 0 | 0 | 0 | 0 | 0 |
| 0 | 0 | 0 | 0 | 0 |
| 0 | 0 | 0 | 0 | 0 |
| 0 | 0 | 0 | 0 | 0 |
| 0 | 0 | 0 | 0 | 0 |
| 0 | 0 | 0 | 0 | 0 |
| 0 | 0 | 0.027633038 | 0 | 3.231490216 |
| 0 | 0 | 0 | 0 | 0 |
| 0 | 0 | 0 | 0 | 0 |
| 0 | 0 | 0 | 0 | 0 |
| 0 | 0 | 0 | 0 | 0 |
| 0 | 0 | 0 | 0 | 0 |
| 0 | 0 | 0 | 0 | 0 |
| 0 | 0 | 0 | 0 | 0 |
| 0 | 0 | 0 | 0 | 0 |
| 0 | 0 | 0 | 0 | 0 |

| | | | | |
|---|---|---|---|---|
| 0 | 0 | 0 | 0 | 0 |
| 0 | 0.000696529 | 0 | 0.005216965 | 0.032519906 |
| 0 | 0 | 0 | 0 | 0 |
| 0 | 0 | 0 | 0 | 0 |
| 0 | 0 | 0 | 0 | 0 |
| 0 | 0.006845233 | 0 | 0 | 0 |
| 0 | 0 | 0 | 0 | 0 |
| 0 | 0 | 0 | 0 | 0 |
| 0 | 0 | 0 | 0 | 0 |
| 0 | 0 | 0 | 0 | 0 |
| 0.005228892 | 0.003952216 | 0.001254566 | 0.01917144 | 0.002868234 |
| 0 | 0 | 0 | 0 | 0 |
| 0 | 0 | 1.353735555 | 0.511267993 | 0 |
| 0 | 0 | 0 | 0 | 0 |
| 0 | 0 | 0 | 0 | 0 |
| 0 | 0 | 0 | 0 | 0 |
| 0 | 0 | 0 | 0 | 0 |
| 0 | 0 | 0 | 0 | 0 |
| 0 | 0 | 0 | 0 | 0 |
| 0 | 0 | 0 | 0 | 0 |
| 0 | 0 | 0.001432473 | 0 | 0.025751903 |
| 0 | 0 | 0 | 0 | 0 |
| 0 | 0 | 0 | 0 | 0 |
| 0 | 2.500119311 | 1.341453282 | 1.645052275 | 2.655292514 |
| 0 | 0 | 0 | 0 | 0 |
| 0 | 0 | 0 | 0 | 0 |
| 0 | 0 | 0 | 0 | 0 |
| 0 | 0 | 0 | 0 | 0 |
| 0 | 0 | 0 | 0 | 0 |
| 0 | 0 | 0 | 0 | 0 |
| 0 | 0 | 0 | 0 | 0 |
| 0 | 0 | 0 | 0 | 0 |
| 0 | 0 | 0 | 0 | 0 |
| 0 | 0 | 0 | 0 | 0 |

| PHL018-0271_V3 | PHL018-0196_V1 | PHL018-0069_V1 | PHL018-0186_V1 | PHL018-0233_V3 |
|---|---|---|---|---|
| REF | REF | CTRL | REF | REF |
| 0.337327774 | 0.415681986 | 6.65058264 | 0.19885299 | 0 |
| 0.131836257 | 0.228659245 | 3.560292181 | 4.31468382 | 0.039391313 |
| 0 | 6.839290929 | 0 | 0 | 0 |
| 0 | 0 | 0 | 0 | 0 |
| 0 | 0 | 0 | 0 | 0 |
| 0 | 0 | 0 | 0 | 0 |
| 0 | 0 | 0 | 0 | 0 |
| 0 | 0 | 0 | 0 | 0 |
| 0 | 0 | 0 | 0 | 0 |
| 0 | 0 | 0 | 0 | 0 |
| 0 | 0 | 0 | 0 | 0 |
| 0 | 0 | 0 | 0 | 0 |
| 0 | 0 | 0 | 0 | 0 |
| 0.154183448 | 0 | 4.093279058 | 0.967127547 | 0.110214551 |
| 0 | 0 | 0 | 0 | 0 |
| 0 | 0 | 0 | 0 | 0 |
| 0 | 0 | 0 | 0 | 0 |
| 0 | 0 | 0 | 0 | 0 |
| 0 | 0 | 0 | 0 | 0 |
| 0 | 0 | 0 | 0 | 0 |
| 0 | 0 | 0 | 0 | 0 |
| 0 | 0 | 0 | 0.057640847 | 0 |
| 0 | 0 | 0 | 0 | 0 |
| 0 | 0 | 0 | 0 | 0 |
| 0.02765882 | 0 | 0 | 0 | 0 |
| 0 | 0 | 0 | 0 | 0 |
| 0 | 0 | 0 | 0 | 0 |
| 0 | 0 | 0 | 0 | 0.064126764 |
| 0 | 0 | 0 | 0 | 0 |
| 0 | 0 | 0 | 0 | 0 |
| 0 | 0 | 0 | 0 | 0 |
| 0 | 0 | 0 | 0 | 0 |
| 0 | 0 | 0 | 0 | 0 |
| 0 | 0 | 0 | 0 | 0 |
| 0 | 0 | 0 | 0 | 0 |
| 0.000490696 | 0 | 0.05076869 | 0 | 0.105674633 |
| 0 | 0 | 0 | 0 | 0 |
| 0 | 0 | 0 | 0 | 0 |
| 0 | 0 | 0 | 0 | 0 |
| 0 | 0 | 0 | 0 | 0 |
| 0 | 0 | 0.000484604 | 0 | 0 |
| 0.064377914 | 0.029201761 | 0 | 0.213777591 | 0 |
| 0.164120504 | 0.667659849 | 0.246735274 | 0.232242363 | 18.57283634 |
| 0.011702772 | 0.13045054 | 0.015246711 | 0.001913513 | 0.000687315 |
| 0.015841434 | 0 | 0.042453177 | 0.000677686 | 0.461503373 |
| 0 | 0 | 0 | 0 | 0 |

| | | | | |
|---|---|---|---|---|
| 0 | 0 | 0 | 0 | 0 |
| 0.001228213 | 0.001497439 | 0.008787042 | 0.015192188 | 0 |
| 0.005077654 | 0 | 0.049181442 | 0.050888125 | 0 |
| 0.182708857 | 0.293514867 | 0.216787028 | 0.681103143 | 0.097499809 |
| 0.019213572 | 0 | 0 | 0 | 0 |
| 0 | 0 | 0 | 0 | 0 |
| 0 | 0.000356669 | 0 | 0 | 0 |
| 0 | 0 | 0 | 0 | 0 |
| 0 | 0 | 0 | 0 | 0 |
| 0 | 0 | 0 | 0 | 0 |
| 0.001965534 | 0 | 0 | 0 | 0 |
| 0.414588472 | 0 | 0 | 0 | 0.056703336 |
| 0 | 0 | 0 | 0.000280529 | 0 |
| 0.024277038 | 0.010408994 | 0 | 0.142956244 | 0 |
| 0 | 0.044946408 | 0 | 0 | 0 |
| 0 | 0 | 0 | 0 | 0 |
| 0.002100273 | 0.000612806 | 0.064490471 | 0 | 0.309029089 |
| 0 | 0 | 0 | 0 | 0 |
| 0 | 0 | 0 | 0 | 0 |
| 0 | 0 | 0 | 0 | 0 |
| 0.02814397 | 0.034346888 | 0.005482674 | 0.107300377 | 0 |
| 0.000584612 | 0.011974349 | 0 | 0.031883282 | 0 |
| 0 | 0 | 0 | 0 | 0.001509611 |
| 0 | 0 | 0 | 0 | 0 |
| 0 | 0 | 0 | 0 | 0 |
| 0 | 0 | 0 | 0 | 0 |
| 0 | 0.401246033 | 0 | 0 | 0 |
| 0 | 0 | 0 | 0 | 0 |
| 0.032757465 | 0.01337343 | 0 | 0.006349463 | 0 |
| 0 | 0 | 0 | 0 | 0 |
| 0 | 0 | 0 | 0.085103945 | 0 |
| 0 | 0 | 0 | 0 | 0 |
| 0 | 0 | 0 | 0 | 0 |
| 0 | 0 | 0 | 0 | 0 |
| 0 | 0 | 0 | 0 | 0 |
| 0 | 0.00935356 | 0 | 0 | 0 |
| 0 | 0 | 0 | 0 | 0 |
| 0 | 0.046018913 | 0.001040354 | 0 | 30.63131348 |
| 0 | 0 | 0 | 0 | 0 |
| 0 | 0 | 0 | 0 | 0 |
| 0 | 0 | 0 | 0 | 0 |
| 0 | 0 | 0 | 0 | 0 |
| 0 | 0 | 0 | 0 | 0 |
| 0 | 0 | 0 | 0 | 0 |
| 0 | 0 | 0 | 0 | 0 |
| 0 | 0 | 0 | 0 | 0 |
| 0 | 0.185172198 | 0 | 0 | 0 |
| 0 | 0 | 0.073331838 | 0 | 0 |

| | | | | |
|---|---|---|---|---|
| 0 | 0 | 0 | 0 | 0 |
| 0 | 0 | 0 | 0 | 0 |
| 0 | 0 | 0 | 0 | 0 |
| 0 | 0 | 0 | 0 | 0 |
| 0.012111017 | 0 | 0 | 0 | 0 |
| 0 | 0 | 0 | 0 | 0 |
| 0 | 0 | 0 | 0 | 0 |
| 0 | 0 | 0 | 0 | 0 |
| 0 | 0.00055115 | 0 | 0.152976475 | 0 |
| 0 | 0 | 0 | 0 | 0 |
| 0 | 0 | 0 | 0 | 0 |
| 0 | 0 | 0 | 0.029766428 | 0 |
| 0 | 0 | 0 | 0 | 0 |
| 0 | 0 | 0 | 0 | 0 |
| 0 | 0 | 0 | 0 | 0 |
| 0 | 0 | 0 | 0 | 0 |
| 0 | 0 | 0 | 0 | 0 |
| 0 | 0 | 0 | 0 | 0 |
| 0 | 0 | 0 | 0 | 0.002464286 |
| 0.071782032 | 0 | 0 | 0 | 0 |
| 0 | 0 | 0 | 0 | 0 |
| 0 | 0 | 0 | 0 | 0 |
| 0 | 0 | 0 | 0 | 0 |
| 0 | 0 | 0 | 0 | 0 |
| 0 | 0 | 0 | 0 | 0 |
| 0 | 0 | 0 | 0 | 0 |
| 0 | 0 | 0 | 0 | 0 |
| 0 | 0 | 0 | 0 | 0 |
| 0 | 0 | 0 | 0 | 0 |
| 0 | 0 | 0 | 0 | 0 |
| 0.007948111 | 0 | 0 | 0.919566497 | 0 |
| 0.04026502 | 0.000297252 | 0 | 0.000277586 | 0 |
| 0 | 0 | 0 | 0 | 0 |
| 0 | 0 | 0 | 0 | 0 |
| 0 | 0 | 0 | 0 | 0 |
| 0 | 0 | 0 | 0 | 0 |
| 0 | 0 | 0 | 0 | 0 |
| 0 | 0 | 0 | 0 | 0 |
| 0 | 0 | 0 | 0 | 0 |
| 0 | 0 | 0 | 0 | 0 |
| 0 | 0 | 0 | 0 | 0 |
| 0 | 0 | 0 | 0 | 0 |
| 0 | 0 | 0 | 0.000213897 | 0 |
| 0 | 0 | 0 | 0 | 0 |
| 0 | 0 | 0 | 0 | 0 |
| 0 | 0 | 0 | 0 | 0 |
| 0 | 0 | 0 | 0 | 0 |
| 0 | 0 | 0 | 0 | 0 |

| | | | | |
|---|---|---|---|---|
| 0.000738921 | 0 | 0.018434929 | 0 | 0 |
| 0 | 0 | 0 | 0 | 0 |
| 0 | 0 | 0 | 0 | 0 |
| 0 | 0 | 0 | 0 | 0 |
| 0.031321894 | 0 | 0 | 0 | 0 |
| 0 | 0 | 0 | 0 | 0 |
| 0 | 0 | 0 | 0 | 0 |
| 0 | 0 | 0 | 0 | 0 |
| 0 | 0 | 0 | 0 | 0 |
| 0 | 0 | 0 | 0 | 0 |
| 0 | 0 | 0 | 0 | 0 |
| 0 | 0 | 0 | 0 | 0 |
| 0 | 0 | 0 | 0 | 0 |
| 0 | 0 | 0 | 0 | 0 |
| 0 | 0 | 0 | 0 | 0 |
| 0 | 0 | 0 | 0 | 0 |
| 0 | 0 | 0 | 0 | 0 |
| 0 | 0 | 0 | 0 | 0 |
| 0 | 0 | 0 | 0 | 0 |
| 0 | 0 | 0 | 0 | 0 |
| 0 | 0 | 0 | 0 | 0 |
| 0 | 0 | 0 | 0 | 0 |
| 0 | 0 | 0 | 0 | 0 |
| 0 | 0 | 0 | 0 | 0 |
| 0.23233719 | 0 | 0 | 0 | 0 |
| 0 | 0 | 0 | 0 | 0 |
| 0 | 0 | 0 | 0 | 0 |
| 0 | 0 | 0 | 0 | 0 |
| 0 | 0 | 0 | 0 | 0 |
| 0 | 0 | 0 | 0 | 0 |
| 0 | 0 | 0 | 0 | 0 |
| 0 | 0 | 0 | 0 | 0.056343816 |
| 0 | 0 | 0 | 0 | 0 |
| 0 | 0 | 0 | 0 | 0 |
| 0 | 0 | 0 | 0 | 0 |
| 0 | 0 | 0 | 0 | 0 |
| 0 | 0 | 0 | 0 | 0 |
| 0 | 0 | 0 | 0 | 0 |
| 0 | 0 | 0 | 0 | 0 |
| 0 | 0 | 0 | 0 | 0 |
| 0 | 0 | 0 | 0 | 0 |
| 0 | 0 | 0 | 0 | 0 |
| 0 | 0 | 0 | 0 | 0 |
| 0 | 0 | 0 | 0 | 0 |
| 0 | 0 | 0 | 0 | 0 |
| 0 | 0 | 0 | 0 | 0 |
| 0 | 0 | 0 | 0 | 0 |
| 0 | 0 | 0 | 0 | 0 |

| | | | | |
|---|---|---|---|---|
| 0 | 0 | 0 | 0 | 0 |
| 0 | 0 | 0 | 0 | 0 |
| 0 | 0 | 0 | 0 | 0 |
| 0 | 0 | 0 | 0 | 0 |
| 0 | 0 | 0 | 0 | 0 |
| 0 | 0 | 0 | 0 | 0 |
| 0 | 0 | 0 | 0 | 0 |
| 0 | 0 | 0 | 0 | 0 |
| 0 | 0 | 0 | 0 | 0 |
| 0 | 0 | 0 | 0 | 0 |
| 0 | 0 | 0 | 0 | 0 |
| 0 | 0 | 0 | 0 | 0 |
| 0.000484522 | 0 | 0 | 0.002179354 | 0 |
| 0 | 0 | 0 | 0 | 0 |
| 0.055731964 | 0 | 0 | 0 | 0 |
| 0 | 0 | 0 | 0 | 0 |
| 0.001881975 | 0 | 0 | 0 | 0 |
| 0 | 0 | 0 | 0 | 0 |
| 0 | 0 | 0 | 0 | 0 |
| 0 | 0 | 0 | 0 | 0 |
| 0.002944307 | 0.083452873 | 0.016618964 | 0.020310669 | 0.002824391 |
| 0 | 0 | 0 | 0 | 0 |
| 0.000794907 | 0.001022992 | 0.001020748 | 0.000502797 | 0 |
| 0 | 0.003583364 | 0 | 0.002523003 | 0 |
| 0 | 0 | 0 | 0 | 0 |
| 0 | 0 | 0 | 0 | 0 |
| 0.003033691 | 0.005145987 | 0.012194901 | 0.000185399 | 0.003289718 |
| 0 | 0 | 0 | 0 | 0 |
| 0 | 0.000172078 | 0 | 0 | 0 |
| 0 | 0.022634187 | 0 | 0.00392744 | 0 |
| 0 | 0 | 0 | 0 | 0 |
| 0 | 0 | 0 | 0 | 0 |
| 0 | 0 | 0 | 0 | 0 |
| 0.00761275 | 0 | 0 | 0.098639467 | 0 |
| 0.043177445 | 0.007825428 | 0.012759557 | 0.023240531 | 0 |
| 0 | 0 | 0 | 0 | 0 |
| 0 | 0 | 0 | 0 | 0 |
| 0 | 0 | 0 | 0 | 0 |
| 0 | 0 | 0 | 0 | 0 |
| 0 | 0 | 0 | 0 | 0 |
| 0 | 0 | 0 | 0 | 0 |
| 0 | 0 | 0 | 0 | 0 |
| 0 | 0 | 0 | 0 | 0 |
| 0 | 0 | 0 | 0 | 0 |
| 0 | 0 | 0 | 0 | 0 |
| 0 | 0 | 0 | 0 | 0 |
| 0.001595553 | 0.00077812 | 0 | 0.002780972 | 0 |
| 0.003465497 | 0.000967371 | 0.018493841 | 0.003690247 | 0 |

| | | | | |
|---|---|---|---|---|
| 0 | 0 | 0 | 0 | 0 |
| 0 | 0 | 0 | 0 | 0 |
| 0 | 0 | 0 | 0.00102511 | 0 |
| 0 | 0 | 0 | 0.000187631 | 0 |
| 0 | 0 | 0 | 0 | 0 |
| 0 | 0 | 0 | 0 | 0 |
| 0 | 0 | 0 | 0 | 0 |
| 0.001811533 | 0.004662647 | 0 | 0.003522386 | 0 |
| 0 | 0 | 0 | 0 | 0 |
| 0 | 0 | 0 | 0 | 0 |
| 0 | 0 | 0 | 0 | 0 |
| 0 | 0 | 0 | 0 | 0 |
| 0 | 0 | 0 | 0 | 0 |
| 0 | 0 | 0 | 0 | 0 |
| 0.001490166 | 0 | 0 | 0 | 0 |
| 0 | 0 | 0 | 0 | 0 |
| 0 | 0 | 0 | 0 | 0 |
| 0 | 0 | 0 | 0 | 0.135848542 |
| 0.001601994 | 0.004050984 | 0 | 0.00066052 | 0.090463269 |
| 0 | 0 | 0 | 0 | 0 |
| 0 | 0.025524405 | 0 | 0.020505322 | 0 |
| 0 | 0 | 0 | 0 | 0 |
| 0 | 0 | 0 | 0 | 0 |
| 0 | 0 | 0 | 0 | 0 |
| 0.003755846 | 0.003052753 | 0.005511909 | 0.003783457 | 0 |
| 0 | 0 | 0.004680094 | 0 | 0.000728371 |
| 0 | 0 | 0 | 0 | 0 |
| 0 | 0 | 0 | 0 | 0 |
| 0.017622603 | 0 | 0 | 0.028213124 | 0 |
| 0 | 0 | 0 | 0 | 0 |
| 0.02629553 | 0.493619452 | 0 | 2.724641137 | 0.001111214 |
| 0 | 0 | 0 | 0 | 0 |
| 0 | 0 | 0 | 0 | 0 |
| 0 | 0 | 0 | 0 | 0 |
| 0 | 0 | 0 | 0 | 0 |
| 0 | 0 | 0 | 0 | 0 |
| 0 | 0 | 0 | 0 | 0 |
| 0 | 0 | 0 | 0 | 0 |
| 0 | 0 | 0 | 0 | 0 |
| 0 | 0 | 0.005433821 | 0 | 0.002330892 |
| 0 | 0 | 0 | 0 | 0 |
| 0.019601105 | 0 | 0.323700664 | 0 | 0.010534095 |
| 0.001981452 | 0 | 0 | 0 | 0 |
| 0 | 0 | 0 | 0 | 0.000490941 |
| 0 | 0 | 0 | 0 | 0 |
| 0 | 0 | 0 | 0 | 0 |
| 0 | 0 | 0 | 0 | 0 |
| 0 | 0 | 0 | 0 | 0 |

| | | | | |
|---|---|---|---|---|
| 0 | 0 | 0 | 0 | 0 |
| 0 | 0 | 0 | 0 | 0 |
| 0.004003224 | 0.126216162 | 0.007833266 | 0.001750609 | 0.002275663 |
| 0 | 0 | 0.001316551 | 0 | 0 |
| 0 | 0 | 0 | 0 | 0 |
| 0 | 0 | 0 | 0 | 0 |
| 0 | 0 | 0 | 0 | 0 |
| 0 | 0 | 0 | 0 | 0 |
| 0 | 0 | 0 | 0 | 0 |
| 0 | 0.000661316 | 0 | 0 | 0 |
| 0 | 0 | 0 | 0 | 0 |
| 0 | 0 | 0 | 0 | 0 |
| 0 | 0 | 0 | 0 | 0 |
| 0.007865396 | 0.000976708 | 0 | 0.004698653 | 0 |
| 0.001622964 | 0.000659573 | 0 | 0.003916144 | 0 |
| 0.045611916 | 0 | 0 | 0 | 0.00472268 |
| 0.00261973 | 0.002421577 | 0.001356917 | 0.00134296 | 0 |
| 0 | 0 | 0 | 0 | 0 |
| 0 | 0 | 0 | 0 | 0 |
| 0 | 0 | 0 | 0 | 0 |
| 0 | 0 | 0 | 0 | 0 |
| 0.004312408 | 0.002336752 | 0.022456657 | 0.004106212 | 0 |
| 0 | 0 | 0 | 0 | 0 |
| 0 | 0 | 0 | 0.038254013 | 0 |
| 0 | 0 | 0 | 0.012854613 | 0 |
| 0 | 0 | 0 | 0 | 0 |
| 2.078937483 | 0 | 0 | 0 | 0 |
| 0 | 0 | 0 | 0 | 0 |
| 0 | 0 | 0 | 0 | 0 |
| 0 | 0 | 0 | 0.011426799 | 0 |
| 0 | 0 | 0 | 0.005394266 | 0 |
| 0 | 0 | 0 | 0 | 0 |
| 0 | 0 | 0 | 0 | 0 |
| 0 | 0 | 0 | 0 | 0 |
| 0 | 0 | 0 | 0 | 0 |
| 0.022506038 | 0 | 0 | 0.016637478 | 0 |
| 0 | 0 | 0 | 0 | 0 |
| 0.117625079 | 0.022255913 | 0.096442169 | 0.002653215 | 0.093386024 |
| 0 | 0 | 0 | 0 | 0 |
| 0 | 0 | 0 | 0 | 0 |
| 0 | 0 | 0 | 0 | 0 |
| 0 | 0 | 0 | 0 | 0 |
| 0 | 0 | 0 | 0 | 0 |
| 0 | 0.029880478 | 0 | 0 | 0 |
| 0 | 0 | 0 | 0 | 0 |
| 0 | 0 | 0 | 0.004079277 | 0 |
| 0 | 0 | 0 | 0.004899832 | 0 |
| 0.000635097 | 0 | 0 | 0.002201986 | 0 |

| | | | | |
|---|---|---|---|---|
| 0 | 0 | 0 | 0 | 0 |
| 0 | 0 | 0 | 0 | 0 |
| 0.00276843 | 0.01871203 | 0.021169438 | 0 | 0 |
| 0 | 0 | 0 | 0 | 0 |
| 0.001905736 | 0 | 0.01054168 | 0.007170717 | 0 |
| 0 | 0 | 0 | 0.016665178 | 0 |
| 0 | 0 | 0 | 0 | 0 |
| 0.001027427 | 0 | 0.009046827 | 0.000404364 | 0 |
| 0 | 0 | 0 | 0 | 0 |
| 0.002753376 | 2.334808259 | 0.004351555 | 0.008990689 | 0 |
| 0 | 0 | 0 | 0.000215331 | 0 |
| 0.076451566 | 0.0052885 | 0 | 0.258043207 | 0 |
| 0.002714483 | 0.002473767 | 0.008148379 | 0.003052397 | 0 |
| 0 | 0 | 0 | 0.004425085 | 0 |
| 0 | 0 | 0 | 0.00060646 | 0 |
| 0 | 0 | 0 | 0 | 0 |
| 0.002155109 | 0.000729864 | 0 | 0 | 0 |
| 0 | 0 | 0 | 0 | 0 |
| 0 | 0.008639449 | 0 | 0.075132185 | 0 |
| 0 | 0 | 0.0028133 | 0 | 0 |
| 0 | 0 | 0 | 0 | 0 |
| 0 | 0 | 0 | 0 | 0 |
| 0 | 0.007174899 | 0 | 0 | 0 |
| 0.08140851 | 0 | 0 | 0 | 0 |
| 0 | 0 | 0 | 0 | 0 |
| 0.002155691 | 0.003504294 | 0 | 0.000363607 | 0.014750981 |
| 0 | 0 | 0 | 0 | 8.534495561 |
| 0 | 0 | 0 | 0 | 0 |
| 0 | 0.049114247 | 0 | 0 | 0 |
| 0.006653805 | 0 | 0 | 0 | 0 |
| 0 | 0.069774415 | 0 | 0.627132562 | 0 |
| 0 | 0 | 0 | 0 | 0 |
| 0 | 0.000593624 | 0 | 0 | 0 |
| 0 | 0 | 0 | 0 | 0 |
| 0 | 0 | 0 | 0 | 0 |
| 0 | 0.074071593 | 0 | 0 | 0 |
| 0 | 0.002295208 | 0 | 0 | 0 |
| 0 | 0 | 0 | 0 | 0 |
| 0 | 0 | 0.000563232 | 0 | 0 |
| 0 | 0 | 0 | 0.001572175 | 0 |
| 0 | 0 | 0 | 0 | 0 |
| 0 | 0 | 0 | 0 | 0 |
| 0 | 0 | 0 | 0 | 0 |
| 0 | 0.029235625 | 0.012063862 | 0.028642459 | 0 |
| 0.005595105 | 0 | 0 | 0 | 0 |
| 0 | 0 | 0 | 0 | 0 |
| 0.003038209 | 0 | 0.003147344 | 0.000552656 | 0 |
| 0 | 0 | 0 | 0 | 0 |

| | | | | |
|---|---|---|---|---|
| 0 | 0 | 0 | 0 | 0 |
| 0 | 0 | 0 | 0 | 0 |
| 0 | 0.233144398 | 0 | 0 | 0 |
| 0 | 0 | 0 | 0 | 0 |
| 0 | 0 | 0 | 0 | 0 |
| 0.020269757 | 0.01685664 | 0 | 0.164814308 | 0 |
| 0.007199912 | 0 | 0 | 0 | 0 |
| 0 | 0 | 0 | 0 | 0 |
| 0 | 0 | 0 | 0 | 0 |
| 0 | 0 | 0 | 0 | 0 |
| 0 | 0 | 0.00790659 | 0.000734336 | 0.004854947 |
| 0 | 0 | 0 | 0 | 0 |
| 0 | 0 | 0 | 0 | 0 |
| 0 | 0.043460409 | 0 | 0.038633969 | 0 |
| 0 | 0 | 0 | 0 | 0 |
| 0 | 0 | 0 | 0 | 0 |
| 0.028657111 | 0 | 0 | 0 | 0 |
| 0 | 0 | 0 | 0 | 0 |
| 0 | 0 | 0 | 0 | 0 |
| 0 | 0 | 0 | 0 | 0 |
| 0 | 0 | 0 | 0 | 0 |
| 0 | 0 | 0 | 0 | 0.027434413 |
| 0 | 0.002617271 | 0 | 0 | 0 |
| 0 | 0.001669564 | 0 | 0 | 0 |
| 0 | 0 | 0 | 0 | 0 |
| 0 | 0 | 0 | 0 | 0 |
| 0 | 0 | 0 | 0 | 0 |
| 0 | 0 | 0 | 0 | 0 |
| 0 | 0 | 0 | 0 | 0 |
| 0.245301062 | 0.031116884 | 0.090578807 | 0.501909809 | 0.029495137 |
| 0.003557752 | 0 | 0.105191726 | 0 | 0 |
| 0 | 0 | 0 | 0.010324952 | 0 |
| 0 | 0 | 0 | 0 | 0 |
| 0 | 0 | 0 | 0.000124991 | 0 |
| 0 | 0 | 0 | 0 | 0 |
| 0 | 0 | 0 | 0 | 0 |
| 0 | 0 | 0 | 0 | 0.021720037 |
| 0 | 0 | 0 | 1.20959022 | 0 |
| 0 | 0 | 0 | 0 | 0 |
| 0 | 0 | 0 | 0 | 0 |
| 0.010904996 | 0.00116807 | 0 | 0.009303823 | 0 |
| 0.009378757 | 0 | 0 | 1.344129035 | 0.119678102 |
| 0 | 0 | 0 | 1.162719073 | 0 |
| 0 | 0 | 0 | 0 | 0 |
| 0 | 0.000930101 | 0 | 0.023202026 | 0 |
| 0 | 0.000653534 | 0 | 0 | 0 |
| 0 | 0.005264471 | 0 | 0.005013464 | 0 |
| 0 | 0 | 0 | 0 | 0 |

| | | | | |
|---|---|---|---|---|
| 0.020033422 | 0.025359043 | 0.0395215 | 0.00055395 | 0.1934352 |
| 0.059387157 | 0 | 0 | 0 | 0 |
| 0.001342479 | 0.001697372 | 0.004597043 | 0 | 0.0158829 |
| 0 | 0 | 0 | 0 | 0 |
| 0 | 0 | 0 | 0 | 0 |
| 0.05886745 | 0 | 0 | 0.001912491 | 0 |
| 0 | 0 | 0 | 0 | 0 |
| 0 | 0 | 0 | 0.002431723 | 0 |
| 0 | 0 | 0 | 0.000463606 | 0 |
| 0 | 0 | 0 | 0 | 0 |
| 0 | 0 | 0 | 0 | 0 |
| 0 | 0 | 0 | 0.000467101 | 0 |
| 0 | 0.002188339 | 0 | 0 | 0 |
| 0 | 0 | 0 | 0 | 0.003135106 |
| 0 | 0 | 0 | 0 | 0 |
| 0.581975735 | 0 | 0.238863311 | 0.028850497 | 0 |
| 0.114780173 | 11.66515876 | 0.311398801 | 0.02127028 | 0.29439622 |
| 0 | 0.002880288 | 0.001376607 | 0.002461196 | 0 |
| 0.002126623 | 0 | 0 | 0.002391357 | 0 |
| 0 | 0 | 0 | 0 | 0 |
| 0.042650721 | 0.009841954 | 0 | 0 | 0 |
| 0 | 0 | 0.003942823 | 0.000335679 | 0 |
| 0 | 0 | 0 | 0 | 0 |
| 0.100140737 | 0 | 0 | 0 | 0 |
| 0.026216985 | 0 | 0 | 0 | 0 |
| 0 | 0 | 0 | 0 | 0 |
| 0.001143498 | 0.00115073 | 0 | 0.007485468 | 0 |
| 0 | 0 | 0 | 0 | 0 |
| 0 | 0 | 0 | 0 | 0 |
| 0 | 0 | 0 | 0 | 0 |
| 0.008323575 | 0.00451027 | 0 | 0.068172426 | 0 |
| 0.003519092 | 0.001505431 | 0 | 0.00312408 | 0 |
| 0.005290502 | 0 | 0.001811623 | 0.012840124 | 0.009980894 |
| 0.002021815 | 0 | 0 | 0.008241442 | 0 |
| 0 | 0 | 0 | 0 | 0 |
| 0 | 0 | 0 | 0 | 0 |
| 0.000478298 | 0.010366952 | 0 | 0.002734015 | 0 |
| 0 | 0.019052477 | 0 | 0.318195239 | 0 |
| 0.110314898 | 0 | 0 | 0 | 0.002665542 |
| 0.006728149 | 0 | 0 | 0 | 0 |
| 0 | 0 | 0 | 0 | 0 |
| 0 | 0 | 0 | 0 | 0 |
| 0.0026105 | 0 | 0 | 0.042249785 | 0 |
| 0.049541472 | 0 | 0 | 0 | 0 |
| 0.044759759 | 0 | 0 | 0.004711904 | 0 |
| 0 | 0 | 0 | 0 | 0 |
| 0 | 0.000825846 | 0 | 0 | 0 |
| 0 | 0.003225835 | 0 | 0 | 0 |

| | | | | |
|---|---|---|---|---|
| 0 | 0 | 0 | 0 | 0 |
| 0.181960056 | 0.024488429 | 0.277258991 | 0.135356866 | 0.03683298 |
| 0 | 0 | 0 | 0 | 0 |
| 0 | 0 | 0 | 0 | 0 |
| 0 | 0.000451585 | 0 | 0.011292427 | 0 |
| 0 | 0 | 0 | 0.000552676 | 0 |
| 0 | 0 | 0 | 0 | 0 |
| 0 | 0.001202123 | 0 | 0 | 0 |
| 0 | 0 | 0 | 0.001311659 | 0 |
| 0.001481533 | 0.000535196 | 0 | 0.006872094 | 0 |
| 0.016412799 | 0.003557422 | 0 | 0.02157927 | 0 |
| 0.006581974 | 0.130535883 | 0.004967694 | 0.037905401 | 0 |
| 0.007881648 | 0 | 0 | 0 | 0.010653636 |
| 0 | 0 | 0 | 0 | 0 |
| 0 | 0.061853401 | 0 | 0.003705782 | 0 |
| 0 | 0.130441882 | 0 | 0 | 0 |
| 0.023135767 | 0.000859646 | 0.007178634 | 0.003567886 | 0.0006341 |
| 0 | 0.017762039 | 0 | 0.000627197 | 0 |
| 0 | 0 | 0 | 0 | 0 |
| 0 | 0.004408183 | 0 | 0.057205624 | 0 |
| 0.004908099 | 0 | 0.007402983 | 0.005672393 | 0.09861068 |
| 0 | 0 | 0 | 0 | 0 |
| 0 | 0 | 0 | 0 | 0 |
| 0 | 0 | 0 | 0 | 0 |
| 0 | 0 | 0 | 0 | 0 |
| 0 | 0 | 0 | 0.009770844 | 0 |
| 0.019781866 | 0.010427463 | 0 | 0 | 0 |
| 0.005506395 | 0 | 0.009144215 | 0.072632154 | 0.009313231 |
| 0 | 0 | 0 | 0 | 0 |
| 0 | 0.000662989 | 0 | 0.00167066 | 0 |
| 0.005286433 | 0 | 0.002738163 | 0 | 0.000745754 |
| 0 | 0 | 0 | 0 | 0 |
| 0 | 0 | 0 | 0 | 0 |
| 0 | 0.0009515 | 0 | 0.001912852 | 0 |
| 0.924962347 | 0 | 0 | 0.043059708 | 0.007389041 |
| 0.12605504 | 0.032557862 | 0.114090798 | 0.556646773 | 0.041404886 |
| 0 | 0 | 0 | 0 | 0 |
| 0 | 0 | 0 | 0.00043632 | 0 |
| 0.030388634 | 0.006392914 | 0 | 0.161410452 | 0 |
| 0 | 0 | 0 | 0 | 0 |
| 0.016152586 | 0 | 0 | 0.003594829 | 0 |
| 0 | 0.020052082 | 0 | 0 | 0 |
| 0 | 0.052599084 | 0 | 0.001709345 | 0 |
| 0 | 0.006362129 | 0 | 0.018124279 | 0 |
| 0 | 0 | 0 | 0 | 0 |
| 0.340318017 | 0 | 0 | 0 | 0.01548128 |
| 0 | 0 | 0 | 0 | 0 |
| 0 | 0.024975159 | 0 | 0.310352279 | 0 |

| | | | | |
|---|---|---|---|---|
| 0 | 0 | 0 | 0 | 0 |
| 0 | 0 | 0 | 0 | 0 |
| 0.006865937 | 0 | 0 | 0 | 0.000572722 |
| 0.200440363 | 0 | 0.000927941 | 21.0967525 | 0.001698344 |
| 0 | 0 | 0 | 0 | 0 |
| 0 | 0 | 0 | 0 | 0 |
| 0 | 0 | 0 | 0 | 0 |
| 0.040727788 | 0 | 0 | 0 | 0 |
| 0 | 0 | 0 | 0.240750758 | 0.355278167 |
| 0.105904286 | 0 | 0 | 0 | 0.001843167 |
| 0 | 0 | 0 | 0 | 0 |
| 0.001817446 | 0 | 0 | 0 | 0 |
| 0 | 0 | 0 | 0 | 0 |
| 0 | 0 | 0 | 0 | 0 |
| 0.232962829 | 0 | 0.197501333 | 0.039350713 | 0 |
| 0 | 0 | 0 | 0 | 0 |
| 0.406781316 | 0 | 0 | 0 | 0.021143367 |
| 0 | 0 | 0 | 0 | 0.004133135 |
| 0 | 0 | 0 | 0 | 0 |
| 0 | 0 | 0 | 0 | 0 |
| 0 | 0 | 0 | 0 | 0 |
| 0 | 0 | 0 | 0 | 0 |
| 0.002231413 | 0 | 0 | 0 | 0 |
| 0.090627958 | 0 | 0 | 0.77555973 | 0 |
| 0 | 0.001967013 | 0 | 0 | 0 |
| 0 | 0 | 0 | 0 | 0 |
| 0.014712337 | 0 | 0 | 0 | 0 |
| 0 | 0 | 0.803188787 | 0 | 0.291015785 |
| 0 | 0 | 0 | 0 | 0 |
| 0 | 0 | 0 | 0 | 0 |
| 0.009423291 | 0 | 0 | 0 | 0 |
| 0.019088748 | 0 | 0 | 0 | 0 |
| 0.019632846 | 0 | 0 | 0 | 0 |
| 0 | 0 | 0 | 0 | 0 |
| 0 | 0 | 0 | 0 | 0 |
| 0 | 0 | 0 | 0 | 0.001149673 |
| 5.450235079 | 0 | 0 | 0 | 0.008661384 |
| 0 | 0 | 0 | 0 | 0 |
| 0 | 0 | 0 | 0 | 0 |
| 0 | 0 | 0 | 0 | 0 |
| 0.733903216 | 0.027145896 | 0.003582183 | 0.393385622 | 0 |
| 0 | 0 | 0 | 0 | 0 |
| 0.004322556 | 0 | 0.147539543 | 0.021951328 | 0.002541408 |
| 0 | 0 | 0 | 0 | 0 |
| 0 | 0 | 0 | 0 | 0 |
| 0 | 0 | 0 | 0 | 0 |
| 0 | 0 | 0 | 0 | 0 |
| 0.002375358 | 0.00333139 | 0.055980534 | 0.001492649 | 0.000536145 |

| | | | | |
|---|---|---|---|---|
| 0 | 0 | 0 | 0 | 0 |
| 0.25217178 | 0 | 0 | 0 | 0.001343218 |
| 0.010983411 | 0.221223394 | 0.005503965 | 0.30869185 | 0 |
| 0 | 0 | 0 | 0 | 0 |
| 0 | 0 | 0 | 0 | 0 |
| 0.020681126 | 0 | 0.205641353 | 0 | 0.000404874 |
| 0.00094565 | 0 | 0 | 0 | 0 |
| 0 | 0 | 0 | 0 | 0 |
| 0 | 0 | 0 | 0 | 0 |
| 0 | 0 | 0 | 0 | 0 |
| 0.067595777 | 0.014537151 | 0.041880364 | 0.296943623 | 0.001639992 |
| 0.351607034 | 0 | 0.180661504 | 1.322236777 | 0.098842364 |
| 0 | 0 | 0 | 0 | 0 |
| 0.145904237 | 0.014074033 | 0.807541501 | 0.012927473 | 3.384953265 |
| 0.106477805 | 0.001878783 | 0.006073208 | 0.018110835 | 0 |
| 0 | 0 | 0 | 0 | 0.522232556 |
| 0 | 0 | 0 | 0 | 0 |
| 0 | 0 | 0 | 0 | 0 |
| 0 | 0 | 0 | 0 | 0 |
| 0.068463957 | 0 | 0 | 0.140939305 | 0 |
| 0.008266812 | 0.069940119 | 0.00097056 | 0.647398124 | 0 |
| 0 | 13.78911843 | 0.178796613 | 2.060771031 | 0 |
| 0.249446088 | 0.725476827 | 0.267375859 | 1.158461145 | 0.013182739 |
| 0 | 0 | 0 | 0 | 0 |
| 0 | 0 | 0 | 0 | 0 |
| 0 | 0 | 0 | 0 | 2.515798835 |
| 0 | 0 | 0.007545426 | 0 | 0 |
| 0 | 0 | 0 | 0 | 0 |
| 0 | 0 | 0 | 0 | 0 |
| 0 | 0 | 0 | 0 | 0 |
| 0 | 0 | 0 | 0 | 0 |
| 0.045786423 | 0.011277341 | 0 | 0 | 0.009183615 |
| 0 | 0.015588711 | 0.073933455 | 0.120058698 | 0 |
| 0 | 0 | 0 | 0 | 0 |
| 0.031200249 | 0.204567257 | 0.000523292 | 0.011226952 | 0 |
| 0.222419707 | 0 | 0.394355866 | 0.1703308 | 0.018680839 |
| 0.012777685 | 0 | 0.008160571 | 0.00129714 | 0 |
| 0 | 0 | 0 | 0 | 0 |
| 0.891903846 | 0 | 0 | 0 | 0.050045087 |
| 0 | 0 | 0 | 0 | 0 |
| 0.005715938 | 0 | 0 | 0 | 0 |
| 0 | 0 | 0 | 0 | 0 |
| 0 | 0 | 0 | 0 | 0 |
| 0 | 0 | 0 | 0 | 0 |
| 0.151544166 | 0.059563047 | 0.090804521 | 0.224790953 | 0 |
| 0.048148142 | 0 | 0 | 0 | 0 |
| 0 | 0 | 0 | 0 | 0 |
| 10.51584468 | 0 | 27.40799777 | 0.042390845 | 1.941872847 |

| | | | | |
|---|---|---|---|---|
| 0 | 0 | 0 | 0 | 0 |
| 0 | 0 | 0 | 0 | 0 |
| 0 | 0 | 0 | 0 | 0 |
| 0 | 0 | 0 | 0 | 0 |
| 0 | 0 | 0 | 0 | 0 |
| 0 | 0 | 0 | 0 | 0 |
| 0.718890506 | 0 | 0 | 0 | 0 |
| 0 | 0.107315955 | 0 | 0 | 0 |
| 0 | 0 | 0 | 0 | 0 |
| 0 | 0 | 0 | 0 | 0 |
| 0.001856136 | 0.04856476 | 0.005642533 | 0.049720048 | 0 |
| 0 | 0 | 0 | 0 | 0 |
| 0 | 0 | 0 | 0 | 0.000668144 |
| 0.177169907 | 0 | 0 | 0 | 0.005534905 |
| 0 | 0 | 0 | 0 | 0 |
| 0 | 0 | 0 | 0 | 0 |
| 0 | 0 | 0 | 0 | 0 |
| 0.032780128 | 0 | 0.032395034 | 0.014210636 | 0.002964518 |
| 0.000348418 | 0 | 0 | 0 | 0 |
| 0 | 0 | 0 | 0.023476687 | 0 |
| 0 | 0 | 0 | 0 | 0 |
| 0 | 0 | 0 | 0 | 0 |
| 0.94393779 | 0 | 0.000933729 | 0.021213147 | 0 |
| 0 | 0 | 0 | 0 | 0 |
| 0 | 0 | 0 | 0 | 0 |
| 0 | 0 | 0 | 0 | 0 |
| 0 | 0 | 0 | 0 | 0 |
| 0 | 0 | 0 | 0 | 0 |
| 0 | 0 | 0 | 0 | 0 |
| 0.038531486 | 0 | 0 | 0.017728537 | 0.006650635 |
| 0 | 0 | 0 | 0 | 0 |
| 0 | 0 | 0 | 0 | 0 |
| 0 | 0 | 0 | 0 | 0 |
| 0 | 0 | 0 | 0 | 0 |
| 0.000737955 | 0 | 0 | 0 | 0 |
| 0 | 0 | 0 | 0 | 0 |
| 0.029622369 | 0 | 0 | 0 | 0.005737829 |
| 0 | 0.012771143 | 0 | 0.014387207 | 0 |
| 0 | 0 | 0 | 0 | 0 |
| 0 | 0 | 0 | 0 | 0 |
| 0 | 0 | 0 | 0 | 0 |
| 0 | 0 | 0 | 0 | 0 |
| 0 | 0 | 0 | 0 | 0 |
| 0.008828258 | 0 | 0 | 0 | 0.047780208 |
| 0 | 0 | 0 | 0 | 0.041808916 |
| 0 | 0 | 0 | 0 | 0 |
| 0 | 0 | 0 | 0 | 0 |
| 0 | 0 | 0 | 0 | 0 |

| | | | | |
|---|---|---|---|---|
| 0 | 0 | 0 | 0 | 0 |
| 0 | 0 | 0 | 0 | 0 |
| 0.025195277 | 0 | 0 | 0 | 0 |
| 0.060619556 | 0 | 0 | 0 | 0.002141829 |
| 0 | 0 | 0 | 0 | 0 |
| 0 | 0 | 0 | 0 | 0 |
| 0 | 0 | 0 | 0 | 0 |
| 0 | 0 | 0 | 0 | 0 |
| 0.010370733 | 0.090416171 | 0.007269071 | 0.254164983 | 0 |
| 0 | 0 | 0 | 0 | 0 |
| 0.000774743 | 0 | 0 | 0 | 0 |
| 0 | 0 | 0 | 0 | 0 |
| 0 | 0 | 0 | 0 | 0 |
| 0 | 0 | 0 | 0 | 0 |
| 0.553611953 | 0 | 0 | 0 | 0 |
| 0 | 0.08772194 | 0 | 0.35707282 | 0 |
| 0 | 0 | 0 | 0 | 0 |
| 0 | 0 | 0 | 0 | 0 |
| 0 | 0 | 0 | 0 | 0 |
| 0 | 0 | 0 | 0 | 0 |
| 0 | 0 | 0 | 0 | 0 |
| 0 | 0 | 0 | 0 | 0 |
| 0 | 0 | 0 | 0 | 0 |
| 0 | 0 | 0 | 0 | 0 |
| 0.011831636 | 0.102776038 | 0.016829308 | 0.014123242 | 0.705256569 |
| 5.818843147 | 0 | 0.010071317 | 1.481606766 | 0.042461326 |
| 0.206086666 | 0 | 0 | 0 | 0.148075061 |
| 0 | 0 | 0 | 0 | 0 |
| 0 | 0 | 0 | 0 | 0.00274789 |
| 0 | 0 | 0 | 0 | 0 |
| 0 | 0 | 0 | 0 | 0 |
| 0.043842232 | 0 | 0 | 0 | 0 |
| 0 | 0 | 0 | 0 | 0 |
| 0 | 0 | 0 | 0.007269431 | 0 |
| 0.035719367 | 0.000781149 | 0.184058089 | 0.003872498 | 0.979844433 |
| 0.00566127 | 0 | 0.05500043 | 0.039946746 | 0.00054307 |
| 0 | 0 | 0 | 0 | 0 |
| 0.364375451 | 0 | 0 | 0 | 0.012509681 |
| 0 | 1.753666805 | 0 | 0 | 0 |
| 0 | 0 | 0 | 0 | 0 |
| 0 | 0 | 0 | 0 | 0 |
| 0 | 0 | 0 | 0 | 0 |
| 0.133681978 | 0.153930433 | 0 | 0.625999702 | 0.750574665 |
| 0 | 0 | 0 | 0 | 0.000669158 |
| 0.007436485 | 0 | 0 | 0 | 0 |
| 0.000536334 | 0 | 0 | 0.00030155 | 0 |
| 0 | 0 | 0 | 0.176518671 | 0 |
| 0.012304004 | 0.003148369 | 0.25204279 | 0.04780033 | 0 |

| | | | | |
|---|---|---|---|---|
| 0 | 0 | 0 | 0 | 0 |
| 0 | 0 | 0 | 0 | 0 |
| 0 | 0 | 0 | 0 | 0.000642102 |
| 0 | 0 | 0 | 0 | 0 |
| 0.001649354 | 0 | 0.092910618 | 0.005100355 | 0 |
| 0 | 0 | 0 | 0 | 0.00199242 |
| 0 | 0 | 0 | 0 | 0 |
| 0 | 0 | 0 | 0 | 0 |
| 0 | 0 | 0 | 2.554300554 | 0 |
| 0 | 0 | 0 | 0 | 0 |
| 0 | 0 | 0 | 0 | 0 |
| 1.558738835 | 0 | 2.269154331 | 0 | 0.037502071 |
| 0.500191373 | 0 | 0 | 0 | 0.297971535 |
| 0 | 0 | 0 | 0 | 0 |
| 0.654558378 | 3.972281144 | 0 | 1.163295172 | 0 |
| 0 | 0 | 0 | 0 | 0 |
| 0 | 0 | 0 | 0.000430416 | 0 |
| 0 | 0 | 0 | 0 | 0 |
| 0.016718976 | 0 | 0 | 0.003796203 | 0 |
| 0 | 0 | 0 | 0 | 0 |
| 0 | 0 | 0 | 0 | 0 |
| 0 | 0 | 0 | 0 | 0 |
| 2.221072081 | 0 | 0 | 0.130320183 | 0.001199602 |
| 0 | 0 | 0 | 0 | 0 |
| 0 | 0 | 0 | 0 | 0 |
| 0 | 0 | 0 | 0 | 0 |
| 0 | 0 | 0 | 0 | 0 |
| 0 | 0 | 0 | 0 | 0 |
| 0 | 0 | 0 | 0 | 0 |
| 0 | 0 | 0 | 0 | 0 |
| 0 | 0 | 0 | 0 | 0 |
| 0.038841903 | 0.00361103 | 0.018162597 | 0.025264237 | 0.002009115 |
| 0 | 0 | 0 | 0 | 0 |
| 0 | 0 | 0 | 0 | 0 |
| 0.005421574 | 0 | 0.04611128 | 0.038518678 | 0.001181992 |
| 0.153282939 | 0.053880737 | 0.633825887 | 0 | 0.008380521 |
| 0 | 0 | 0 | 0 | 0 |
| 0 | 0 | 0 | 0 | 0.000565129 |
| 0 | 0 | 0 | 0 | 0 |
| 0 | 0 | 0 | 0 | 0 |
| 0.003254437 | 0 | 0 | 0 | 0.089628479 |
| 0.12688256 | 0 | 0 | 0 | 0 |
| 0 | 0 | 0 | 0 | 0 |
| 0 | 0 | 0 | 0 | 0 |
| 0.119318194 | 0.153710863 | 0.190959909 | 0.111002069 | 0 |
| 0 | 0 | 0 | 0 | 0 |
| 0 | 0 | 0 | 0 | 0 |
| 0.001377664 | 0 | 0 | 0 | 0 |

| | | | | |
|---|---|---|---|---|
| 0.003793119 | 0 | 0 | 0 | 0 |
| 0.007633388 | 0 | 0 | 0 | 0 |
| 0 | 0 | 0 | 0 | 0 |
| 0 | 0 | 0 | 0 | 0 |
| 0 | 0 | 0 | 0 | 0 |
| 0 | 0 | 0 | 0 | 0 |
| 0 | 0 | 0 | 0 | 0.000927938 |
| 0 | 0 | 0 | 0.305400193 | 0 |
| 0 | 0 | 0 | 0 | 0 |
| 0 | 0 | 0 | 0 | 0 |
| 0 | 0 | 0 | 0 | 0 |
| 0 | 0 | 0 | 0.000502718 | 0 |
| 0 | 0 | 0 | 0 | 0 |
| 0 | 0 | 0 | 0 | 0.003001364 |
| 0 | 0 | 0 | 0 | 0 |
| 0 | 0 | 0 | 0 | 0 |
| 0.001959997 | 0 | 0 | 0 | 0 |
| 0 | 0 | 0 | 0 | 0 |
| 0.00053657 | 0 | 0 | 0 | 0.011581131 |
| 0 | 0 | 0 | 0 | 0 |
| 0 | 0 | 0 | 0 | 0.000875343 |
| 0 | 0 | 0 | 0 | 0 |
| 0 | 0 | 0 | 0 | 0 |
| 0 | 0 | 0 | 0 | 0 |
| 0 | 0 | 0.00408717 | 0.002013245 | 0.008523653 |
| 0.00384206 | 0.063987359 | 0.061525248 | 0.129927884 | 0 |
| 0 | 0 | 0 | 0 | 0 |
| 0.028757624 | 0.0037159 | 0.015258146 | 0.645786692 | 0 |
| 0 | 0 | 0 | 0 | 0 |
| 1.520541095 | 0 | 0.035199658 | 0 | 0.012639796 |
| 0 | 0 | 0 | 0 | 0 |
| 0 | 0 | 0 | 0 | 0 |
| 0 | 0 | 0 | 0 | 0 |
| 0 | 0 | 0 | 0 | 0 |
| 0.001894702 | 0 | 0.001390289 | 0.274576495 | 0 |
| 0 | 0 | 0 | 0 | 0 |
| 0 | 0 | 0 | 0 | 0.000967785 |
| 0.001746819 | 0.001092166 | 0.002957945 | 0.002228694 | 0 |
| 0.004676355 | 0 | 0 | 0 | 0 |
| 0 | 0 | 0 | 0 | 0 |
| 3.45939254 | 1.092914164 | 2.709442695 | 18.3181276 | 0.370477913 |
| 0.413702039 | 0 | 0.054275083 | 2.910857839 | 0 |
| 1.300395854 | 0 | 0 | 0 | 0 |
| 0 | 0 | 0 | 0 | 0 |
| 0 | 0 | 0 | 0 | 0 |
| 0 | 0 | 0 | 0 | 0 |
| 0 | 0 | 0 | 0 | 0 |
| 0.011304267 | 0.002542623 | 0.008607827 | 0.006175863 | 0 |

| | | | | |
|---|---|---|---|---|
| 0 | 0 | 0 | 0 | 0 |
| 0.001425998 | 0 | 0 | 0.009528577 | 0 |
| 0.005124572 | 0 | 0 | 0 | 0 |
| 0 | 0 | 0 | 0 | 0 |
| 1.766144133 | 1.841820191 | 0 | 0.400977826 | 0.888233669 |
| 0 | 0 | 0 | 0 | 0 |
| 0 | 0 | 0 | 0 | 0 |
| 0 | 0 | 0 | 0 | 0 |
| 0 | 0.01946711 | 0.026683162 | 0.012070211 | 0 |
| 0 | 0 | 0 | 0 | 0 |
| 0 | 0.129538553 | 0 | 0 | 0 |
| 0 | 0 | 0 | 0 | 0 |
| 0.020890593 | 0 | 0 | 0 | 0.000511218 |
| 0.01598627 | 0 | 0.009999653 | 0.060080719 | 0.000754189 |
| 15.5467954 | 0 | 0 | 0 | 0.081990204 |
| 0.126881119 | 0 | 0.021776513 | 1.282904875 | 0.002888117 |
| 0.029594912 | 0 | 0 | 0.01740456 | 0 |
| 0 | 0 | 0 | 0 | 0 |
| 0 | 0 | 0 | 0 | 0 |
| 0 | 0 | 0 | 0 | 0 |
| 0 | 0 | 0 | 0 | 0 |
| 0 | 0 | 0 | 0 | 0 |
| 0 | 0 | 0 | 0 | 0 |
| 0 | 0 | 0 | 0 | 0 |
| 0 | 0 | 0 | 0 | 0 |
| 0 | 0 | 0 | 0 | 0 |
| 0.394587882 | 0.020676181 | 0 | 0 | 0.011633586 |
| 0.001001308 | 0.103209964 | 0 | 0.000469148 | 0.000640351 |
| 0 | 0 | 0 | 0 | 0 |
| 0 | 0 | 0 | 0 | 0 |
| 0 | 0 | 0 | 0 | 0 |
| 0 | 0 | 0 | 0 | 0 |
| 0.14918706 | 0 | 0 | 0 | 0 |
| 0.019826496 | 0 | 0 | 0 | 0 |
| 0 | 0 | 0 | 0 | 0.100298628 |
| 0 | 0 | 0 | 0.025941413 | 0 |
| 0 | 0.00057025 | 0 | 0 | 0 |
| 0 | 0 | 0 | 0 | 0 |
| 0 | 0 | 0 | 0 | 0 |
| 0 | 0 | 0 | 0 | 0 |
| 0 | 0 | 0.052741756 | 0 | 0.015725362 |
| 0 | 0 | 0 | 0 | 0.040101522 |
| 0 | 0 | 0 | 0 | 0 |
| 0 | 0 | 0 | 0 | 0.003134937 |
| 0 | 0 | 0 | 0 | 0 |
| 0 | 0 | 0 | 0 | 0 |
| 0 | 0 | 0 | 0 | 0 |
| 0 | 0 | 0 | 0 | 0 |

| | | | | |
|---|---|---|---|---|
| 0.292478744 | 0 | 0 | 1.160491441 | 0.012503389 |
| 0 | 0 | 0 | 0 | 0 |
| 0 | 0 | 0 | 0 | 0 |
| 0 | 0 | 0 | 0.000744034 | 0 |
| 0.012299946 | 0 | 0 | 0 | 0.000347029 |
| 0 | 0 | 0 | 0 | 0 |
| 0.028052458 | 0.037266234 | 0 | 1.097605145 | 0 |
| 0 | 0 | 0 | 0 | 0 |
| 0 | 0 | 0 | 0 | 0 |
| 0 | 0 | 0 | 0 | 0 |
| 0 | 0 | 0 | 0 | 0 |
| 0.013651096 | 0 | 0 | 0.056507487 | 0 |
| 0 | 0 | 0 | 0 | 0 |
| 0 | 0 | 0 | 0 | 0 |
| 0 | 0 | 0 | 0 | 0 |
| 0 | 0 | 0 | 0 | 0 |
| 0 | 39.0090252 | 0 | 0 | 0 |
| 0 | 0 | 0 | 0 | 0 |
| 0 | 0 | 0 | 0 | 0 |
| 0 | 0 | 0 | 0 | 0 |
| 0.004953811 | 0 | 0.002370648 | 0.01047494 | 0.025103236 |
| 0 | 0 | 0 | 0 | 0 |
| 0.004513769 | 0 | 0 | 0 | 0 |
| 0.008344251 | 0 | 0 | 0.011164569 | 0 |
| 0 | 0 | 0 | 0 | 0 |
| 0.003613745 | 0 | 0 | 0.002136895 | 0 |
| 0 | 0 | 0 | 0 | 0 |
| 0 | 0 | 0 | 0 | 0 |
| 0 | 0 | 0 | 0 | 0 |
| 0 | 1.128838562 | 0 | 0.180342505 | 0 |
| 0 | 0 | 0 | 0 | 0 |
| 0 | 0 | 0 | 0 | 0 |
| 0 | 0 | 0 | 0 | 0 |
| 0 | 0 | 0 | 0 | 0 |
| 0 | 0 | 0 | 0 | 0 |
| 0 | 0 | 0 | 0 | 0 |
| 0 | 0.008092423 | 0 | 0.134979157 | 0 |
| 0 | 0 | 0 | 0 | 0 |
| 0 | 0 | 0 | 0 | 0 |
| 0 | 0 | 0 | 0 | 0 |
| 0 | 0 | 0 | 0 | 0 |
| 0 | 0 | 0 | 0 | 0 |
| 0.012637329 | 0 | 0 | 0 | 0 |
| 0 | 0 | 0 | 0 | 0 |
| 0 | 0 | 0 | 0 | 0 |
| 0 | 0 | 0 | 0 | 0 |
| 0 | 0 | 0 | 0 | 0 |
| 0 | 0 | 0 | 0 | 0.019576176 |

| | | | | |
|---|---|---|---|---|
| 0.003295982 | 0 | 0 | 0.397435053 | 0 |
| 0 | 0 | 0 | 0 | 0.00568697 |
| 0 | 0 | 0 | 0 | 0.001425228 |
| 0 | 0 | 0 | 0 | 0 |
| 0 | 0 | 0 | 0 | 0 |
| 0 | 0 | 0 | 0 | 0 |
| 0 | 0 | 0 | 0 | 0 |
| 0.046095529 | 0 | 0.033053182 | 3.275007367 | 0.048472055 |
| 0.228884499 | 0.825254117 | 0.317573888 | 0.327940396 | 22.96404913 |
| 0 | 0.000403784 | 0.011300326 | 0.074715836 | 0 |
| 0 | 0 | 0 | 0 | 0 |
| 0 | 0 | 0 | 0 | 0 |
| 0 | 0 | 0 | 0 | 0 |
| 0 | 0 | 0 | 0 | 0 |
| 0 | 0 | 0 | 0 | 0 |
| 0 | 0 | 0 | 0 | 0 |
| 0 | 0.020766549 | 0 | 0.148966432 | 0 |
| 0 | 0 | 0 | 0 | 0.046929125 |
| 0 | 0 | 0 | 0 | 0 |
| 0 | 0 | 0 | 0 | 0 |
| 0 | 0 | 0 | 0 | 0 |
| 0 | 0 | 0 | 0 | 0 |
| 0 | 0 | 0 | 0 | 0 |
| 0 | 0 | 0 | 0 | 0 |
| 0 | 0 | 0 | 0 | 0 |
| 0 | 0 | 0 | 0 | 0 |
| 0 | 0 | 0 | 0 | 0 |
| 0 | 0 | 0 | 0 | 0 |
| 0 | 0 | 0 | 0 | 0 |
| 0 | 0 | 0 | 0 | 0 |
| 0 | 0 | 0 | 0 | 0 |
| 0 | 0 | 0 | 0 | 0 |
| 0 | 0 | 0 | 0 | 0 |
| 0 | 0 | 0 | 0 | 0 |
| 0.047470219 | 0 | 0 | 0.002552936 | 0 |
| 0 | 0 | 0 | 0 | 0 |
| 0 | 0 | 0 | 0 | 0 |
| 0 | 0 | 0 | 0.05158847 | 0 |
| 0 | 0 | 0 | 0 | 0 |
| 0 | 0 | 0 | 0.000542246 | 0 |
| 0 | 0 | 0 | 0 | 0 |
| 0.271806482 | 0 | 0 | 0 | 0.001459764 |
| 0 | 0 | 0 | 0 | 0 |
| 0 | 0 | 0 | 0 | 0 |
| 0 | 0 | 0 | 0 | 0 |
| 0 | 0 | 0 | 0 | 0 |
| 0 | 0 | 0 | 0 | 0 |
| 0 | 0 | 0 | 0 | 0 |

| | | | | |
|---|---|---|---|---|
| 0 | 0 | 0 | 0 | 0 |
| 0 | 0 | 0 | 0 | 0.002952012 |
| 0 | 0 | 0 | 0 | 0 |
| 0 | 0 | 0 | 0 | 0 |
| 5.877730846 | 1.498012923 | 35.28821343 | 0 | 0.090113338 |
| 0 | 0 | 0 | 0 | 0 |
| 0 | 0 | 0 | 0 | 0 |
| 0.008819186 | 0.00501385 | 0.002970441 | 0.118029115 | 0 |
| 0.008805631 | 0 | 0 | 0 | 0 |
| 0 | 0 | 0 | 0 | 0 |
| 0 | 0 | 0 | 0 | 0 |
| 0 | 0 | 0 | 0 | 0 |
| 0 | 0 | 0 | 0 | 0 |
| 0 | 0 | 0 | 0 | 0 |
| 0 | 0 | 0 | 0 | 0 |
| 0 | 0 | 0 | 0 | 0 |
| 0 | 0 | 0 | 0 | 0 |
| 0 | 0 | 0 | 0 | 0 |
| 0 | 0 | 0 | 0 | 0 |
| 0 | 0 | 0 | 0 | 0 |
| 0 | 0 | 0 | 0 | 0 |
| 0 | 0 | 0 | 0 | 0 |
| 0 | 0 | 0 | 0 | 0 |
| 0.001751671 | 0.037560113 | 0.001652575 | 0.045479618 | 0 |
| 0 | 0 | 0 | 0 | 0.004378278 |
| 0 | 0 | 0 | 0 | 0 |
| 0.021742808 | 0 | 0.011252036 | 0.257137669 | 0 |
| 0 | 0 | 0 | 0 | 0 |
| 0 | 0 | 0 | 0.001291066 | 0 |
| 0 | 0 | 0 | 0 | 0 |
| 0 | 0 | 0 | 0 | 0 |
| 0 | 0 | 0 | 0 | 0 |
| 0 | 0 | 0 | 0 | 0 |
| 0 | 0 | 0 | 0 | 0 |
| 0 | 0 | 0 | 0 | 0 |
| 0 | 0 | 0 | 0 | 0 |
| 0 | 0 | 0 | 0 | 0 |
| 0.004107623 | 0 | 0 | 0 | 0 |
| 0 | 0 | 0 | 0 | 0 |
| 0 | 0 | 0 | 0.00291535 | 0 |
| 0 | 0 | 0 | 0 | 0 |
| 0.996629246 | 0 | 0 | 0 | 0.052281347 |
| 0 | 0 | 0 | 0 | 0 |
| 0 | 0 | 0 | 0 | 0 |
| 0 | 0 | 0 | 0 | 0 |
| 0 | 0 | 0 | 0 | 0 |
| 0 | 0 | 0 | 0 | 0 |
| 0 | 0 | 0 | 0 | 0 |

| | | | | |
|---|---|---|---|---|
| 0.405178149 | 0 | 0 | 0 | 0.000825872 |
| 0 | 0 | 0 | 0 | 0 |
| 0 | 0 | 0 | 0 | 0 |
| 0 | 0 | 0 | 0 | 0 |
| 0 | 0 | 0 | 0 | 0 |
| 0.172931945 | 0.235380026 | 1.89306489 | 0 | 0.001315629 |
| 0 | 0 | 0 | 0 | 0 |
| 0 | 0 | 0 | 0 | 0 |
| 0 | 0 | 0 | 0 | 0 |
| 0 | 0 | 0 | 0 | 0 |
| 0 | 0 | 0 | 0 | 0 |
| 0 | 0 | 0 | 0.000421519 | 0 |
| 0 | 0 | 0 | 0 | 0 |
| 0 | 0 | 0 | 0 | 0 |
| 0 | 0 | 0 | 0 | 0 |
| 0 | 0 | 0 | 0 | 0 |
| 0 | 0 | 0 | 0 | 0 |
| 0 | 0 | 0 | 0 | 0 |
| 0 | 0 | 0 | 0 | 0 |
| 0 | 0 | 0 | 0 | 0 |
| 0.000373789 | 0 | 0 | 0.058389479 | 0 |
| 0 | 0 | 0 | 0 | 0 |
| 0.049724513 | 0.00042633 | 0.06743115 | 0.003008058 | 0 |
| 0.00950535 | 0 | 0 | 0 | 0 |
| 0 | 0 | 0 | 0 | 0 |
| 0 | 0 | 0 | 0 | 0.004396237 |
| 0 | 0 | 0 | 0 | 0 |
| 0 | 0 | 0 | 0 | 0 |
| 0 | 0 | 0 | 0 | 0.004704208 |
| 0 | 0 | 0 | 0 | 0 |
| 0 | 0 | 0 | 0 | 0 |
| 0 | 0 | 0 | 0 | 0 |
| 0.000589453 | 0.026830038 | 0.011029448 | 0.013836608 | 0.000706808 |
| 0.000496886 | 0 | 0 | 0 | 0 |
| 0.582889323 | 0 | 0 | 0 | 0.263010158 |
| 0 | 0.001858242 | 0.007115244 | 0.038229919 | 0 |
| 0 | 0 | 0 | 0 | 0 |
| 0 | 0 | 0 | 0 | 0 |
| 0.011899699 | 0.011339681 | 0 | 0.012765818 | 0 |
| 0 | 0 | 0 | 0 | 0 |
| 0 | 0 | 0 | 0 | 0 |
| 0 | 0 | 0 | 0 | 0 |
| 0.027668692 | 0 | 0.237529364 | 0.020412314 | 0.037434685 |
| 0 | 0 | 0 | 0 | 0 |
| 0 | 0 | 0 | 0 | 0 |
| 0.14047952 | 0 | 0.009506537 | 0.842824885 | 0 |
| 0 | 0 | 0 | 0 | 0 |
| 0 | 0 | 0 | 0 | 0 |

| | | | | |
|---|---|---|---|---|
| 0 | 0 | 0 | 0 | 0 |
| 0 | 0 | 0 | 0 | 0 |
| 0 | 0 | 0 | 0 | 0 |
| 0 | 0 | 0 | 0 | 0 |
| 0 | 0 | 0 | 0 | 0 |
| 0 | 0 | 0 | 0 | 0 |
| 0 | 0 | 0 | 0 | 0 |
| 0 | 0 | 0 | 0 | 0 |
| 0.108767889 | 0 | 0 | 0 | 0 |
| 0 | 0 | 0.045886256 | 0 | 0.002384437 |
| 0 | 0 | 0 | 0 | 0 |
| 0 | 0 | 0 | 0 | 0 |
| 0 | 0 | 0 | 0 | 0 |
| 0 | 0 | 0 | 0 | 0.005612815 |
| 0 | 0 | 0 | 0 | 0 |
| 0 | 0.001534448 | 0 | 0.00209874 | 0 |
| 0 | 0 | 0 | 0 | 0 |
| 0 | 0 | 0 | 0 | 0 |
| 0 | 0 | 0 | 0 | 0 |
| 0.469213842 | 0 | 0 | 0 | 0.004858722 |
| 0 | 0 | 0 | 0 | 0.14559668 |
| 0 | 0 | 0 | 0 | 0 |
| 0 | 0 | 0 | 0 | 0 |
| 0 | 0 | 0 | 0 | 0 |
| 0 | 0 | 0 | 0.000489052 | 0 |
| 0 | 0 | 0 | 0 | 0 |
| 0 | 0 | 0 | 0 | 0 |
| 0 | 0 | 0 | 0 | 0 |
| 0 | 0 | 0 | 0 | 0 |
| 0 | 0 | 0 | 0.490541226 | 0.005886723 |
| 0.062666044 | 0 | 0 | 0.11040623 | 0.0123439 |
| 0 | 0 | 0 | 0 | 0 |
| 0 | 0 | 0 | 0 | 0 |
| 0 | 0 | 0 | 0 | 0 |
| 0 | 0 | 0 | 0 | 0 |
| 0 | 0 | 0 | 0 | 0 |
| 0 | 0 | 0 | 0 | 0 |
| 0 | 0 | 0 | 0 | 0 |
| 0 | 0 | 0 | 0 | 0 |
| 0 | 0 | 0 | 0 | 0 |
| 0 | 0 | 0 | 0 | 0 |
| 0 | 0 | 0 | 0 | 0 |
| 0 | 0 | 0 | 0 | 0 |
| 0 | 0 | 0 | 0 | 0 |
| 0 | 0 | 0 | 0 | 0 |
| 0 | 0 | 0 | 0 | 0 |
| 0 | 0.518864826 | 0.005801254 | 0.22269948 | 0 |
| 0 | 0 | 0 | 0 | 0 |

| | | | | |
|---|---|---|---|---|
| 0 | 0 | 0 | 0 | 0 |
| 0 | 0 | 0 | 0 | 0 |
| 0 | 0 | 0 | 0 | 0 |
| 0 | 0 | 0 | 0 | 0.000453133 |
| 0.039277451 | 0 | 0 | 0.130591295 | 0.000283291 |
| 0 | 0 | 0 | 0 | 0 |
| 0 | 0 | 0 | 0 | 0 |
| 0 | 0 | 0 | 0 | 0 |
| 0 | 0 | 0 | 0 | 0 |
| 0 | 0 | 0 | 0 | 0 |
| 0 | 0 | 0 | 0 | 0 |
| 0.026240493 | 0.026067905 | 0.039311634 | 0.025857092 | 0 |
| 0.003773156 | 0 | 0 | 0.005071542 | 0 |
| 0 | 0 | 0 | 0 | 0 |
| 0 | 0.001072289 | 0 | 0 | 0 |
| 0.212028011 | 0 | 0 | 3.365909049 | 0.097057503 |
| 0.01513654 | 0 | 0 | 0 | 0 |
| 0 | 0 | 0 | 0 | 0 |
| 0 | 0 | 0 | 0.197595107 | 0.000595632 |
| 0.18606318 | 0.004319875 | 0 | 0.760550069 | 0 |
| 0 | 0 | 0 | 0 | 0 |
| 0 | 0 | 0 | 0 | 0 |
| 0 | 0 | 0 | 0 | 0 |
| 0 | 0 | 0 | 0 | 0 |
| 0 | 0 | 0 | 0 | 0 |
| 0 | 0 | 0 | 0 | 0 |
| 0.005955376 | 0 | 0 | 0.021459987 | 0 |
| 0 | 0 | 0 | 0 | 0 |
| 0 | 0 | 0 | 0 | 0 |
| 0 | 0 | 0 | 0 | 0 |
| 0.002332071 | 0.002926399 | 0 | 0.016458883 | 0 |
| 0 | 0 | 0 | 0 | 0 |
| 0 | 0 | 0 | 0 | 0 |
| 0 | 0 | 0 | 0 | 0 |
| 0 | 0 | 0 | 0 | 0 |
| 0 | 0 | 0 | 0 | 0 |
| 0 | 0 | 0 | 0 | 0 |
| 0.050446866 | 0.032942772 | 0.092130615 | 0.049017746 | 0.000330887 |
| 2.732342879 | 0 | 0 | 0 | 0.016218078 |
| 0 | 0 | 0 | 0 | 0 |
| 0.008953286 | 0.013155002 | 0.117496877 | 0 | 0 |
| 0 | 0 | 0 | 0 | 0 |
| 0 | 0 | 0 | 0 | 0 |
| 0.013426773 | 0 | 0 | 0.054497352 | 0 |
| 0 | 0 | 0 | 0 | 0 |
| 0 | 0 | 0 | 0 | 0 |
| 0 | 0 | 0 | 0 | 0 |
| 0 | 0 | 0 | 0 | 0 |

| | | | | |
|---|---|---|---|---|
| 0 | 0 | 0 | 0 | 0 |
| 0.034209265 | 0 | 0 | 0 | 0 |
| 0.010904238 | 0 | 0 | 0 | 0 |
| 0.000938088 | 0.003993934 | 0 | 0.008476604 | 0 |
| 0.000745767 | 0 | 0.01586958 | 0 | 0.013711709 |
| 0 | 0.001564615 | 0 | 0.006629083 | 0 |
| 0.022238381 | 0 | 0 | 0 | 0 |
| 0.000524047 | 0 | 0 | 0 | 0 |
| 0.004572509 | 0 | 0 | 0 | 0 |
| 0.009291558 | 0 | 0 | 0 | 0 |
| 0 | 0 | 0 | 0 | 0 |
| 0 | 0 | 0 | 0 | 0 |
| 0.096800964 | 0 | 0 | 0.172757338 | 0.068527544 |
| 0 | 0 | 0 | 0 | 0 |
| 0 | 0 | 0 | 0 | 0 |
| 0 | 0 | 0 | 0 | 0 |
| 0 | 0 | 0 | 0 | 0 |
| 0 | 0 | 0 | 0 | 0 |
| 0 | 0 | 0 | 0 | 0 |
| 0 | 0 | 0 | 0 | 0 |
| 0 | 0 | 0 | 0 | 0 |
| 0 | 0 | 0 | 0 | 0 |
| 0 | 0 | 0 | 0.174312307 | 0 |
| 0.004280632 | 0 | 0.017329826 | 0 | 0.000707981 |
| 0 | 0 | 0 | 0 | 0 |
| 0 | 0 | 0 | 0 | 0 |
| 0.000478043 | 0 | 0 | 0.150716136 | 0 |
| 0 | 0 | 0 | 0 | 0 |
| 0 | 0 | 0 | 0 | 0 |
| 0 | 0 | 0 | 0 | 0 |
| 0.048675883 | 0 | 0.001228101 | 0 | 0.000535168 |
| 0 | 0 | 0 | 0 | 0 |
| 0 | 0 | 0 | 0 | 0 |
| 0 | 0 | 0 | 0 | 0 |
| 0 | 0 | 0 | 0 | 0 |
| 0 | 0 | 0 | 0.014017792 | 0 |
| 0 | 0 | 0 | 0 | 0 |
| 0 | 0 | 0 | 0 | 0 |
| 0.044723274 | 0.000575479 | 0 | 0.045314682 | 0 |
| 0 | 0 | 0 | 0 | 0 |
| 0 | 0 | 0 | 0 | 0 |
| 0 | 0 | 0 | 0 | 0 |
| 0 | 0 | 0 | 0 | 0 |
| 0 | 0 | 0 | 0 | 0 |
| 3.145535472 | 0 | 0 | 0 | 0 |
| 0.295585402 | 0 | 0.038244017 | 0.589081102 | 0.057565884 |
| 0 | 0.005222737 | 0 | 0 | 0 |
| 0 | 0 | 0 | 0 | 0 |

| | | | | |
|---|---|---|---|---|
| 0 | 0 | 0 | 0 | 0 |
| 0 | 0 | 0 | 0 | 0 |
| 0.015015576 | 0 | 0 | 0 | 0.045153038 |
| 0 | 0 | 0 | 0 | 0 |
| 0 | 0 | 0 | 0 | 0 |
| 0 | 0 | 0 | 0 | 0 |
| 0 | 0 | 0 | 0 | 0 |
| 0 | 0 | 0 | 0 | 0 |
| 0 | 0 | 0 | 0 | 0 |
| 0 | 0 | 0 | 0 | 0 |
| 0.001148788 | 0.000778113 | 0 | 0.001372533 | 0 |
| 0 | 0 | 0 | 0.000843786 | 0 |
| 0 | 0 | 0 | 0 | 0 |
| 0 | 0 | 0 | 0 | 0 |
| 0 | 0 | 0 | 0 | 0 |
| 0 | 0 | 0 | 0 | 0 |
| 0 | 0 | 0 | 0 | 0 |
| 0 | 0 | 0 | 0 | 0 |
| 0 | 0 | 0 | 0 | 0 |
| 0.001904363 | 0.000619147 | 0 | 0.01206338 | 0.004262539 |
| 0 | 0 | 0 | 0 | 0 |
| 0 | 0 | 0 | 0 | 0 |
| 0 | 0 | 0 | 0 | 0 |
| 0 | 0 | 0 | 0 | 0 |
| 0 | 0 | 0 | 0 | 0 |
| 0 | 0 | 0 | 0 | 0 |
| 0 | 0 | 0 | 0.008356178 | 0 |
| 0 | 0 | 0 | 0 | 0.005795441 |
| 0 | 0 | 0 | 0.000244081 | 0 |
| 0 | 0.000296715 | 0 | 0.000153936 | 0 |
| 0.008289294 | 0 | 0.424174028 | 0.013736491 | 0.001778667 |
| 0 | 0 | 0 | 0 | 0 |
| 0.003292665 | 0 | 0.002416083 | 0.048560424 | 0.004373394 |
| 0.036494563 | 0.001192122 | 0 | 0.004111037 | 0 |
| 0 | 0 | 0 | 0 | 0 |
| 0 | 0 | 0 | 0 | 0 |
| 0 | 0 | 0 | 0 | 0 |
| 0 | 0 | 0 | 0 | 0 |
| 0 | 0 | 0 | 0 | 0 |
| 0 | 0 | 0 | 0 | 0 |
| 0 | 0 | 0 | 0 | 0 |
| 0 | 0.05425341 | 0 | 0.801904025 | 0 |
| 0.106746776 | 0 | 0.111392055 | 0.048527904 | 0.001461106 |
| 0 | 0 | 0 | 0 | 0 |
| 0 | 0 | 0 | 0 | 0 |
| 0 | 0.060031217 | 0.898251561 | 0.00046407 | 0.044339407 |
| 0 | 0 | 0 | 0.001284644 | 0 |
| 0 | 0 | 0 | 0 | 0 |

| | | | | |
|---|---|---|---|---|
| 0 | 0 | 0 | 0 | 0 |
| 0 | 0 | 0 | 0 | 0.013393649 |
| 0.001242624 | 0 | 0.006610619 | 0.003318619 | 0 |
| 0 | 0 | 0 | 0 | 0 |
| 0 | 0 | 0 | 0 | 0 |
| 0 | 0 | 0 | 0 | 0.001389666 |
| 0 | 0 | 0 | 0 | 0 |
| 0 | 0 | 0 | 0 | 0 |
| 0 | 0 | 0 | 0 | 0.002791627 |
| 0 | 0 | 0 | 0 | 0 |
| 0 | 0 | 0 | 0 | 0 |
| 0 | 0 | 0 | 0 | 0 |
| 0 | 0 | 0 | 0 | 0 |
| 0 | 0 | 0 | 0 | 0 |
| 0 | 0 | 0 | 0 | 0 |
| 0 | 0 | 0 | 0 | 0 |
| 0 | 0 | 0 | 0 | 0 |
| 0 | 0 | 0 | 0 | 0 |
| 0 | 0 | 0 | 0 | 0 |
| 0 | 0 | 0 | 0 | 0 |
| 0 | 0 | 0 | 0 | 0 |
| 0 | 0 | 0 | 0 | 0 |
| 0 | 0 | 0 | 0 | 0 |
| 0 | 0 | 0 | 0 | 0 |
| 0 | 0 | 0 | 0.000503817 | 0.005403133 |
| 0 | 0.010118116 | 0.001322913 | 0.003004768 | 0 |
| 0 | 0 | 0 | 0 | 0 |
| 2.164460647 | 0 | 0 | 0.089408451 | 0 |
| 0 | 0 | 0 | 0 | 0 |
| 0 | 0 | 0 | 0 | 0 |
| 0 | 0 | 0 | 0 | 0 |
| 0 | 0.045772382 | 0 | 0.000285723 | 0 |
| 0 | 0 | 0 | 0 | 0 |
| 0 | 0 | 0 | 0 | 0 |
| 0 | 0 | 0 | 0 | 0 |
| 0 | 0 | 0 | 0 | 0 |
| 0 | 0 | 0 | 0 | 0 |
| 0.002307364 | 0 | 0 | 0 | 0 |
| 1.057069879 | 0 | 0 | 0 | 0 |
| 0 | 0 | 0 | 0 | 0 |
| 0 | 0 | 0 | 0 | 0 |
| 0.010617459 | 0 | 0 | 0 | 0.000670068 |
| 0.002455597 | 0 | 0 | 0.316457639 | 0 |
| 0 | 0 | 0 | 0 | 0 |
| 0 | 0 | 0 | 0 | 0 |
| 0 | 0 | 0 | 0.001003187 | 0 |
| 0.001384004 | 0.001499893 | 0 | 0.068921543 | 0 |
| 0 | 0 | 0 | 0 | 0 |

| | | | | |
|---|---|---|---|---|
| 0 | 0 | 0 | 0 | 0 |
| 0.034303482 | 0.047855456 | 0.01446195 | 0.016797122 | 0 |
| 0 | 0 | 0 | 0 | 0 |
| 0 | 0 | 0 | 0 | 0 |
| 0 | 0 | 0 | 0 | 0 |
| 0 | 0 | 0 | 0 | 0 |
| 0 | 0 | 0 | 0 | 0 |
| 0 | 0 | 0 | 0 | 0 |
| 0 | 0 | 0 | 0 | 0 |
| 0 | 0 | 0 | 0 | 0 |
| 0 | 0 | 0 | 0 | 0 |
| 0 | 0 | 0 | 0 | 0 |
| 2.438244761 | 0 | 0.202608585 | 0.062498318 | 0 |
| 0 | 0 | 0 | 0 | 0 |
| 0.020996806 | 0 | 0.018260124 | 0 | 0 |
| 0 | 0 | 0 | 0 | 0 |
| 0 | 0 | 0 | 0 | 0 |
| 0 | 0 | 0 | 0 | 0 |
| 0 | 0 | 0 | 0 | 0 |
| 0 | 0 | 0 | 0 | 0 |
| 0 | 0 | 0 | 0 | 0 |
| 0.002443914 | 0.066296571 | 0 | 0.377698217 | 0 |
| 0 | 0 | 0 | 0 | 0 |
| 0 | 0.000341 | 0 | 0 | 0 |
| 0 | 0 | 0 | 0 | 0 |
| 0 | 0 | 0 | 0 | 0 |
| 0 | 0 | 0 | 0 | 0 |
| 0 | 0 | 0 | 0 | 0 |
| 0 | 0 | 0 | 0 | 0 |
| 0 | 0 | 0 | 0 | 0 |
| 0 | 0 | 0 | 0 | 0 |
| 0 | 0 | 0 | 0 | 0 |
| 0 | 0 | 0 | 0 | 0 |
| 0 | 0 | 0 | 0 | 0 |
| 3.252962398 | 0 | 0 | 0 | 0.103634504 |
| 0 | 0 | 0 | 0 | 0 |
| 0 | 0 | 0 | 0 | 0 |
| 0 | 0 | 0 | 0 | 0 |
| 0.002288009 | 0.00900919 | 0.002686223 | 0.150510752 | 0 |
| 0 | 0 | 0 | 0 | 0 |
| 0.128013792 | 0.00133397 | 0.053590331 | 0.026144701 | 0.057726525 |
| 0 | 0 | 0 | 0 | 0 |
| 0 | 0 | 0 | 0 | 0 |
| 0.448737613 | 0 | 0 | 0 | 0 |
| 0 | 0 | 0 | 0 | 0 |
| 0.026306912 | 1.595864067 | 0.486492002 | 0.033587601 | 0 |
| 0 | 0 | 0 | 0 | 0 |
| 0 | 0 | 0 | 0 | 0 |

| | | | | |
|---|---|---|---|---|
| 0 | 0 | 0 | 0 | 0 |
| 0 | 0 | 0 | 0 | 0 |
| 0 | 0 | 0 | 0 | 0 |
| 0 | 0 | 0 | 0 | 0 |
| 0 | 0.00051482 | 0 | 0 | 0 |
| 0 | 0 | 0 | 0 | 0 |
| 0 | 0 | 0 | 0 | 0 |
| 0 | 0 | 0 | 0.017081178 | 0 |
| 0 | 0 | 0 | 0 | 0 |
| 0.024043617 | 0.075183239 | 0 | 0.070777596 | 0 |
| 0.028910697 | 0 | 0 | 0 | 0 |
| 0 | 0 | 0 | 0 | 0 |
| 0 | 0 | 0 | 0.004376296 | 0 |
| 0 | 0 | 0 | 0 | 0 |
| 0 | 0 | 0 | 0 | 0 |
| 0 | 0 | 0 | 0 | 0 |
| 0 | 0 | 0 | 0 | 0.000775648 |
| 0 | 0 | 0 | 0 | 0 |
| 0 | 0 | 0 | 0 | 0 |
| 0.292041446 | 3.949259914 | 4.07274736 | 2.37589224 | 0.026711117 |
| 0 | 0 | 0 | 0 | 0 |
| 0 | 0 | 0 | 0 | 0 |
| 0 | 0 | 0 | 0 | 0 |
| 0 | 0 | 0 | 0 | 0 |
| 0 | 0 | 0 | 0 | 0 |
| 0 | 0 | 0 | 0 | 0 |
| 0 | 0 | 0 | 0 | 0 |
| 0 | 0 | 0 | 0 | 0 |
| 0 | 0 | 0 | 0 | 0 |
| 0 | 0 | 0 | 0 | 0 |
| 0 | 0 | 0 | 0 | 0 |
| 0 | 0 | 0 | 0 | 0 |
| 0 | 0 | 0 | 0 | 0 |
| 0 | 0 | 0 | 0 | 0 |
| 0 | 0 | 0 | 0 | 0 |
| 0.002069567 | 0 | 0 | 0 | 0 |
| 0 | 0 | 0 | 0 | 0 |
| 0 | 0 | 0 | 0 | 0 |
| 0 | 0 | 0 | 0 | 0 |
| 0 | 0 | 0 | 0 | 0 |
| 0 | 0 | 0 | 0 | 0 |
| 0 | 0 | 0 | 0 | 0 |
| 0.015836752 | 0 | 0 | 0 | 0 |
| 0 | 0 | 0 | 0 | 0 |
| 0.039667815 | 0.016680414 | 0.287767852 | 0.007492944 | 0 |
| 0 | 0 | 0 | 0 | 0 |
| 0 | 0 | 0 | 0 | 0 |
| 0 | 0.002850814 | 0 | 0 | 0 |

| | | | | |
|---|---|---|---|---|
| 0 | 0 | 0 | 0 | 0 |
| 0 | 0 | 0 | 0 | 0 |
| 0 | 0 | 0 | 0 | 0 |
| 0 | 0 | 0 | 0 | 0 |
| 0 | 0 | 0 | 0 | 0 |
| 0.06968997 | 0.177069575 | 0.000548385 | 0.005830131 | 0.002957244 |
| 0 | 0 | 0 | 0 | 0 |
| 0 | 0 | 0 | 0 | 0 |
| 0 | 0 | 0 | 0 | 0 |
| 0 | 0 | 0 | 0 | 0 |
| 0 | 0 | 0 | 0 | 0 |
| 0 | 0 | 0 | 0 | 0 |
| 0 | 0 | 0 | 0 | 0 |
| 0 | 0 | 0 | 0 | 0.000572872 |
| 0 | 0 | 0 | 0 | 0 |
| 0 | 0 | 0 | 0 | 0 |
| 0.011898139 | 0 | 0 | 0 | 0 |
| 0 | 0 | 0 | 0 | 0 |
| 0 | 0 | 0 | 0 | 0 |
| 0 | 0 | 0 | 0 | 0 |
| 0 | 0 | 0 | 0 | 0 |
| 0 | 0 | 0 | 0 | 0 |
| 0 | 0 | 0 | 0 | 0 |
| 0.021357767 | 0 | 0 | 0 | 0 |
| 0 | 0 | 0 | 0 | 0 |
| 0 | 0.000633428 | 0 | 0.001560962 | 0 |
| 0 | 0 | 0 | 0 | 0 |
| 0 | 0 | 0.001925937 | 0 | 0 |
| 0 | 0 | 0 | 0 | 0 |
| 0 | 0 | 0 | 0 | 0 |
| 0 | 0 | 0 | 0 | 0 |
| 0.006863872 | 0 | 0 | 0 | 0 |
| 0 | 0 | 0 | 0 | 0 |
| 0 | 0 | 0 | 0 | 0 |
| 0 | 0 | 0 | 0 | 0 |
| 0 | 0 | 0 | 0 | 0 |
| 0 | 0 | 0 | 0 | 0 |
| 0 | 0 | 0 | 0 | 0 |
| 2.583428916 | 0 | 0 | 0 | 0 |
| 0 | 0 | 0 | 0 | 0 |
| 0 | 0 | 0 | 0 | 0 |
| 0 | 0 | 0 | 0 | 0 |
| 0 | 0 | 0 | 0 | 0 |
| 0 | 0 | 0 | 0 | 0 |
| 0 | 0 | 0 | 0 | 0 |
| 0 | 0 | 0 | 0 | 0 |
| 0 | 0 | 0 | 0 | 0 |
| 0 | 0 | 0 | 0 | 0 |

| | | | | |
|---|---|---|---|---|
| 0 | 0 | 0 | 0 | 0 |
| 0.000522946 | 0 | 0 | 0.083473007 | 0 |
| 0 | 0 | 0 | 0 | 0 |
| 0 | 0 | 0 | 0 | 0 |
| 0 | 0 | 0 | 0 | 0 |
| 0 | 0 | 0 | 0 | 0 |
| 0 | 0 | 0 | 0 | 0 |
| 0 | 0 | 0 | 0 | 0 |
| 0 | 0 | 0 | 0 | 0 |
| 0 | 0 | 0 | 0 | 0 |
| 0.094496368 | 0.002745747 | 0.033966202 | 0.000269499 | 0.482929077 |
| 0 | 0 | 0 | 0 | 0 |
| 0 | 0 | 0 | 0 | 0.085635471 |
| 0 | 0 | 0 | 0 | 0 |
| 0 | 0 | 0 | 0 | 0 |
| 0 | 0 | 0 | 0 | 0 |
| 0 | 0 | 0 | 0 | 0 |
| 0 | 0 | 0 | 0 | 0 |
| 0 | 0 | 0 | 0 | 0 |
| 0 | 0 | 0 | 0 | 0 |
| 0.003891924 | 0.009038165 | 0.008975389 | 0 | 0.433609393 |
| 0.002579715 | 0 | 0 | 0 | 0 |
| 0 | 0 | 0 | 0 | 0 |
| 2.082517777 | 0 | 0.471628768 | 0.921924336 | 0.305299474 |
| 0 | 0 | 0 | 0 | 0 |
| 0 | 0 | 0 | 0 | 0 |
| 0 | 0 | 0 | 0 | 0 |
| 0 | 0 | 0 | 0 | 0 |
| 0 | 0 | 0 | 0 | 0 |
| 0 | 0 | 0 | 0 | 0 |
| 0 | 0 | 0 | 0 | 0 |
| 0 | 0 | 0 | 0 | 0 |
| 0.001169695 | 0 | 0 | 0 | 0 |
| 0 | 0 | 0 | 0 | 0 |

| PHL018-0152_V2 | PHL018-0259_V2 | PHL018-0185_V1 | PHL018-0166_V3 | PHL018-0125_V3 |
|---|---|---|---|---|
| REF | REF | REF | EYCF | CTRL |
| 0.004398397 | 0.221036824 | 0 | 0.05003837 | 0.030302348 |
| 0.466622821 | 0.157329535 | 4.413488749 | 2.569466901 | 3.669245803 |
| 1.865321925 | 3.575731353 | 0 | 0.089041383 | 0 |
| 0 | 0 | 0 | 0 | 0 |
| 0 | 0 | 0 | 0 | 0 |
| 0 | 0 | 0 | 0 | 0 |
| 0 | 0 | 0 | 0 | 0 |
| 0 | 0 | 0 | 0 | 0 |
| 0 | 0 | 0 | 0 | 0 |
| 0 | 0 | 0 | 0 | 0 |
| 0 | 0 | 0 | 0 | 0 |
| 0 | 0 | 0 | 0 | 0 |
| 0 | 0 | 0 | 0 | 0 |
| 0.334334083 | 3.206045454 | 0.792695118 | 1.82131694 | 3.941537508 |
| 0 | 0 | 0 | 0 | 0 |
| 0 | 0 | 0 | 0 | 0 |
| 0 | 0 | 0 | 0 | 0 |
| 0 | 0 | 0 | 0 | 0 |
| 0 | 0 | 0 | 0 | 0 |
| 0 | 0 | 0 | 0 | 0 |
| 0 | 0 | 0 | 0 | 0 |
| 0 | 0 | 0 | 0 | 0 |
| 0 | 0 | 0 | 0 | 0 |
| 0 | 0 | 0 | 0 | 0 |
| 0 | 0.190387526 | 0 | 0 | 0 |
| 0 | 0 | 0 | 0 | 0 |
| 0 | 0 | 0 | 0 | 0 |
| 0 | 0 | 0 | 0 | 0 |
| 0 | 0 | 0 | 0 | 0 |
| 0 | 0 | 0 | 0 | 0 |
| 0 | 0 | 0 | 0 | 0 |
| 0 | 0 | 0 | 0 | 0 |
| 0 | 0 | 0 | 0 | 0 |
| 0 | 0 | 0 | 0 | 0 |
| 0 | 0 | 0 | 0 | 0 |
| 4.08422323 | 0.837026187 | 0 | 10.86613474 | 7.305266726 |
| 0 | 0 | 0 | 0 | 0 |
| 0 | 0 | 0 | 0 | 0 |
| 0 | 0 | 0 | 0 | 0 |
| 0 | 0 | 0 | 0 | 0 |
| 0 | 0 | 0 | 0 | 0 |
| 0.052273091 | 0.007215346 | 0 | 0.008311539 | 0.07985463 |
| 0.625824474 | 0.010090431 | 0.01381256 | 0.016442507 | 0.045679544 |
| 0.947035614 | 0.01004291 | 0.002118319 | 0.001168188 | 0.009439986 |
| 0 | 0.000277721 | 0 | 0.000842222 | 0.045226282 |
| 0 | 0 | 0 | 0 | 0 |

| | | | | |
|---|---|---|---|---|
| 0.001063152 | 0.001002669 | 0 | 0 | 0.004647408 |
| 0 | 0 | 0.046793089 | 0.00100125 | 0.207233549 |
| 0.00335975 | 0 | 0.161516232 | 0.011176237 | 0.012624515 |
| 0.523071841 | 0.277564757 | 0.820186438 | 0.508651697 | 0.22850058 |
| 0 | 0.00131827 | 0 | 0.001670728 | 0.004064802 |
| 0 | 0 | 0 | 0 | 0 |
| 0.837957829 | 0.129178256 | 0 | 0.022805045 | 0.542853486 |
| 0 | 0.006440576 | 0 | 0 | 0 |
| 0 | 0 | 0 | 0 | 0 |
| 0 | 0 | 0 | 0 | 0 |
| 0 | 0 | 0 | 0.005423239 | 0.000385553 |
| 0.232121223 | 0.047869997 | 0 | 0 | 0.033759878 |
| 0 | 0.000525545 | 0.000689673 | 0 | 0 |
| 0.010355484 | 0.003068393 | 0.069367723 | 0.032856656 | 0.045879609 |
| 0 | 0 | 0.001825236 | 0 | 0 |
| 0 | 0 | 0 | 0 | 0 |
| 0.904627535 | 0.016639641 | 0.004950192 | 0.049257546 | 4.066273115 |
| 0 | 0 | 0 | 0 | 0 |
| 0 | 0 | 0 | 0 | 0 |
| 0 | 0 | 0 | 0 | 0 |
| 0.430978581 | 0.001664213 | 1.555774211 | 0.164460162 | 0.010267729 |
| 0.010026429 | 0.009024981 | 0.017535432 | 0.004765804 | 0.024647613 |
| 0 | 0.0116334 | 0 | 0 | 0 |
| 0 | 0 | 0 | 0 | 0 |
| 0 | 0 | 0 | 0 | 0 |
| 0 | 0 | 0 | 0 | 0 |
| 0 | 0 | 0 | 0 | 0.008069548 |
| 0 | 0 | 0 | 0 | 0 |
| 0 | 0.000620358 | 0.00434185 | 0.002438737 | 0.020978569 |
| 0 | 0 | 0 | 0 | 0 |
| 0 | 0 | 0 | 0 | 0 |
| 0 | 0 | 0 | 0 | 0 |
| 0 | 0 | 0 | 0 | 0 |
| 0 | 0 | 0 | 0 | 0 |
| 0 | 0 | 0 | 0 | 0 |
| 0 | 0.003382271 | 0 | 0 | 0 |
| 0 | 0 | 0 | 0 | 0 |
| 0.0169613 | 0.000522686 | 3.333119509 | 1.278323443 | 0.13979788 |
| 0 | 0 | 0 | 0 | 0 |
| 0 | 0 | 0 | 0 | 0 |
| 0 | 0 | 0 | 0 | 0 |
| 0 | 0 | 0 | 0 | 0 |
| 0 | 0 | 0 | 0 | 0 |
| 0 | 0 | 0 | 0 | 0 |
| 0 | 0.0790369 | 0 | 0 | 0 |
| 0 | 0 | 0 | 0.020399161 | 0 |
| 0.00474418 | 0.001298985 | 0 | 0 | 0 |
| 0 | 3.274015172 | 0 | 0.000963001 | 0 |

| | | | | |
|---|---|---|---|---|
| 0 | 0 | 0 | 0 | 0 |
| 0 | 0 | 0 | 0 | 0 |
| 0 | 0 | 0 | 0 | 0 |
| 0 | 0 | 0 | 0 | 0 |
| 0.114690506 | 0.1192256 | 0 | 0 | 0 |
| 0 | 0 | 0 | 0 | 0 |
| 0 | 0 | 0 | 0 | 0 |
| 0 | 0.000779163 | 0 | 0 | 0 |
| 0 | 0.000572877 | 0 | 0.018794616 | 0.045918549 |
| 0 | 0 | 0 | 0 | 0 |
| 0 | 0 | 0 | 0 | 0 |
| 0 | 0.052966038 | 0 | 0.008129086 | 0.003345849 |
| 0 | 0 | 0 | 0 | 0 |
| 0 | 0 | 0 | 0 | 0 |
| 0 | 0 | 0 | 0 | 0 |
| 0 | 0 | 0 | 0 | 0 |
| 0 | 0 | 0 | 0 | 0 |
| 0 | 0 | 0 | 0 | 0 |
| 0.092591728 | 0.187311563 | 0 | 0 | 0 |
| 0 | 0.003277854 | 0 | 0 | 0 |
| 0 | 0 | 0 | 0 | 0 |
| 0 | 0 | 0 | 0 | 0 |
| 0 | 0 | 0 | 0 | 0 |
| 0 | 0 | 0 | 0 | 0 |
| 0 | 0 | 0 | 0 | 0 |
| 0 | 0 | 0 | 0 | 0 |
| 0 | 0.000228954 | 0 | 0 | 0 |
| 0 | 0 | 0 | 0 | 0 |
| 0 | 0 | 0 | 0 | 0 |
| 0 | 0 | 0 | 0 | 0 |
| 0 | 0.039325524 | 0 | 0 | 0.000266684 |
| 0.000929214 | 0.044997244 | 0.002477 | 0.000894398 | 0.001492133 |
| 0 | 0 | 0 | 0 | 0 |
| 0 | 0 | 0 | 0 | 0 |
| 0 | 0 | 0 | 0 | 0 |
| 0 | 0 | 0 | 0 | 0 |
| 0 | 0 | 0 | 0 | 0 |
| 0 | 0 | 0 | 0 | 0 |
| 0 | 0.009083168 | 0 | 0.040714188 | 0 |
| 0 | 0 | 0 | 0 | 0 |
| 0 | 0 | 0 | 0 | 0 |
| 0 | 0 | 0 | 0 | 0 |
| 0.000805518 | 0 | 0.001840513 | 0 | 0.000862335 |
| 0 | 0 | 0 | 0 | 0 |
| 0 | 0.000934543 | 0 | 0 | 0 |
| 0 | 0 | 0 | 0 | 0 |
| 0 | 0 | 0 | 0 | 0 |
| 0 | 0 | 0 | 0 | 0 |

| | | | | |
|---|---|---|---|---|
| 0 | 0 | 0 | 0 | 0 |
| 0 | 0 | 0 | 0 | 0 |
| 0 | 0 | 0 | 0 | 0 |
| 0 | 0 | 0 | 0 | 0 |
| 0 | 0.009571224 | 0 | 0 | 0.006412495 |
| 0 | 0 | 0 | 0 | 0 |
| 0 | 0 | 0 | 0 | 0 |
| 0 | 0 | 0 | 0 | 0 |
| 0 | 0 | 0 | 0 | 0 |
| 0 | 0 | 0 | 0 | 0 |
| 0 | 0 | 0 | 0 | 0 |
| 0 | 0 | 0 | 0 | 0 |
| 0 | 0 | 0 | 0 | 0 |
| 0 | 0 | 0 | 0 | 0 |
| 0 | 0 | 0 | 0 | 0 |
| 0.902481336 | 0 | 0 | 0 | 0 |
| 0 | 0 | 0 | 0 | 0 |
| 0 | 0 | 0 | 0 | 0 |
| 0 | 0 | 0 | 0 | 0 |
| 0 | 0 | 0 | 0 | 0 |
| 0 | 0 | 0 | 0 | 0 |
| 0 | 0 | 0 | 0 | 0 |
| 0 | 0.000998007 | 0 | 0 | 0 |
| 0 | 0 | 0 | 0 | 0 |
| 0 | 0 | 0 | 0 | 0.380654323 |
| 0 | 0 | 0 | 0 | 0 |
| 0 | 0.006645354 | 0 | 0 | 0 |
| 0 | 0 | 0 | 0 | 0 |
| 0 | 0 | 0 | 0 | 0 |
| 0 | 0 | 0 | 0 | 0 |
| 0 | 0 | 0 | 0 | 0 |
| 0 | 0.00662089 | 0 | 0 | 0 |
| 0 | 0 | 0 | 0 | 0 |
| 0 | 0 | 0 | 0 | 0 |
| 0 | 0 | 0 | 0 | 0 |
| 0 | 0 | 0 | 0 | 0 |
| 0 | 0 | 0 | 0 | 0 |
| 0 | 0 | 0 | 0 | 0.000775156 |
| 0 | 0 | 0 | 0 | 0 |
| 0 | 0 | 0 | 0 | 0 |
| 0 | 0 | 0 | 0 | 0 |
| 0 | 0 | 0 | 0 | 0 |
| 0 | 0 | 0 | 0 | 0 |
| 0 | 0 | 0 | 0 | 0 |
| 0 | 0 | 0 | 0 | 0 |
| 0 | 0 | 0 | 0 | 0 |
| 0 | 0 | 0 | 0 | 0 |
| 0 | 0 | 0 | 0 | 0.015830372 |

| | | | | |
|---|---|---|---|---|
| 0 | 0 | 0 | 0 | 0 |
| 0 | 0 | 0 | 0 | 0 |
| 0 | 0 | 0 | 0 | 0 |
| 0 | 0 | 0 | 0 | 0 |
| 0 | 0 | 0 | 0 | 0 |
| 0 | 0 | 0 | 0 | 0 |
| 0 | 0 | 0 | 0 | 0 |
| 0 | 0 | 0 | 0 | 0 |
| 0 | 0 | 0 | 0 | 0 |
| 0 | 0 | 0 | 0 | 0 |
| 0 | 0 | 0 | 0 | 0 |
| 0 | 0 | 0 | 0 | 0 |
| 0 | 0 | 0.000976697 | 0 | 0 |
| 0 | 0 | 0 | 0 | 0 |
| 0 | 0 | 0 | 0 | 0.003240129 |
| 0 | 0 | 0 | 0 | 0 |
| 0 | 0 | 0 | 0 | 0 |
| 0.00524365 | 0 | 0 | 0 | 0.002600375 |
| 0 | 0 | 0 | 0 | 0 |
| 0 | 0 | 0 | 0 | 0 |
| 0.021100183 | 0.029484393 | 0.039810927 | 0.109671787 | 0.013476077 |
| 0 | 0 | 0 | 0 | 0 |
| 0 | 0 | 0.000961421 | 0 | 0 |
| 0 | 0.000870691 | 0 | 0 | 0 |
| 0 | 0 | 0 | 0 | 0 |
| 0 | 0 | 0 | 0 | 0 |
| 0.000698196 | 0.000759779 | 0.001914353 | 0.024945961 | 0.002915027 |
| 0 | 0 | 0 | 0 | 0 |
| 0 | 0 | 0 | 0 | 0 |
| 0 | 0 | 0.006144412 | 0 | 0 |
| 0 | 0 | 0.083328452 | 0 | 0 |
| 0 | 0 | 0 | 0 | 0 |
| 0 | 0 | 0 | 0 | 0 |
| 0 | 0.018274166 | 0 | 0.00451344 | 0 |
| 0.002652175 | 0.003836823 | 0.021625533 | 0.003363691 | 0.006388299 |
| 0 | 0 | 0 | 0 | 0 |
| 0 | 0 | 0 | 0 | 0 |
| 0 | 0 | 0 | 0 | 0 |
| 0 | 0 | 0 | 0 | 0 |
| 0 | 0 | 0 | 0 | 0 |
| 0 | 0 | 0 | 0 | 0 |
| 0 | 0.0392678 | 0 | 0 | 0 |
| 0 | 0 | 0 | 0 | 0 |
| 0 | 0 | 0 | 0 | 0 |
| 0 | 0 | 0 | 0 | 0 |
| 0 | 0.0028772 | 0 | 0.034313577 | 0 |
| 0 | 0 | 0.000771913 | 0 | 0 |
| 0 | 0.000992064 | 0.000677404 | 0 | 0 |

| | | | | |
|---|---|---|---|---|
| 0 | 0 | 0 | 0 | 0 |
| 0 | 0 | 0 | 0 | 0 |
| 0 | 0.00053065 | 0 | 0 | 0 |
| 0 | 0.000123029 | 0 | 0 | 0 |
| 0 | 0 | 0 | 0 | 0 |
| 0 | 0 | 0 | 0 | 0 |
| 0 | 0 | 0 | 0 | 0 |
| 0.000799096 | 0.001252194 | 0 | 0 | 0 |
| 0 | 0 | 0 | 0 | 0 |
| 0 | 0 | 0 | 0 | 0 |
| 0 | 0 | 0 | 0 | 0 |
| 0 | 0 | 0 | 0 | 0 |
| 0 | 0 | 0 | 0 | 0 |
| 0 | 0 | 0 | 0 | 0 |
| 0 | 0.000305201 | 0 | 0.006748866 | 0 |
| 0 | 0.000224148 | 0 | 0 | 0 |
| 0 | 0 | 0 | 0 | 0 |
| 0.003917665 | 0 | 0 | 0 | 0 |
| 0.003391992 | 0 | 0 | 0 | 0 |
| 0 | 0.015340514 | 0 | 0 | 0 |
| 0.008506008 | 0.001306767 | 0.060973213 | 0.002054852 | 0 |
| 0 | 0 | 0 | 0 | 0 |
| 0 | 0 | 0 | 0 | 0 |
| 0 | 0 | 0 | 0 | 0 |
| 0.002982173 | 0.000288463 | 0.004542607 | 0.01581929 | 0.002660433 |
| 0 | 0 | 0 | 0 | 0 |
| 0 | 0 | 0 | 0 | 0 |
| 0 | 0 | 0 | 0 | 0 |
| 0 | 0 | 0 | 0 | 0.028816396 |
| 0 | 0 | 0 | 0 | 0 |
| 0.106939071 | 0.034137418 | 0.749467994 | 0.007932391 | 0.019299137 |
| 0 | 0 | 0 | 0 | 0 |
| 0 | 0 | 0 | 0.027711195 | 0 |
| 0.000793321 | 0.014606956 | 0 | 0 | 0 |
| 0 | 0 | 0 | 0 | 0 |
| 0 | 0 | 0 | 0 | 0 |
| 0 | 0 | 0 | 0 | 0 |
| 0 | 0 | 0 | 0 | 0 |
| 0 | 0 | 0 | 0 | 0 |
| 0 | 0 | 0 | 0 | 0 |
| 0 | 0 | 0 | 0 | 0 |
| 0 | 0 | 0 | 0.007893236 | 0 |
| 0 | 0 | 0 | 0 | 0 |
| 0 | 0 | 0 | 0 | 0 |
| 0 | 0 | 0 | 0 | 0 |
| 0 | 0 | 0 | 0 | 0 |
| 0 | 0 | 0 | 0 | 0 |
| 0 | 0 | 0 | 0 | 0 |

| | | | | |
|---|---|---|---|---|
| 0 | 0 | 0 | 0 | 0.001594689 |
| 0 | 0 | 0 | 0 | 0 |
| 1.065495893 | 0.011512776 | 0 | 0 | 0.006553528 |
| 0 | 0 | 0 | 0 | 0 |
| 0 | 0 | 0 | 0 | 0 |
| 0 | 0 | 0 | 0 | 0 |
| 0 | 0 | 0 | 0 | 0 |
| 0 | 0 | 0 | 0 | 0 |
| 0 | 0 | 0 | 0 | 0 |
| 0 | 0.000406182 | 0 | 0 | 0 |
| 0 | 0 | 0 | 0 | 0 |
| 0 | 0 | 0 | 0 | 0 |
| 0 | 0 | 0 | 0 | 0.001316466 |
| 0 | 0.002189835 | 0.001057001 | 0.008282087 | 0.015878517 |
| 0.001575013 | 0.000560924 | 0.001526728 | 0.014774103 | 0.001034656 |
| 0 | 0.007907153 | 0 | 0 | 0 |
| 0.000815721 | 0 | 0.00149106 | 0.015075014 | 0.00275076 |
| 0 | 0 | 0 | 0 | 0.00040644 |
| 0 | 0 | 0 | 0 | 0 |
| 0 | 0 | 0 | 0 | 0 |
| 0 | 0 | 0 | 0 | 0 |
| 0 | 0.000737208 | 0 | 0 | 0 |
| 0 | 0 | 0 | 0 | 0 |
| 0 | 0 | 0.004520987 | 0 | 0 |
| 0 | 0 | 0 | 0 | 0 |
| 0 | 0 | 0 | 0 | 0 |
| 0 | 0.023342353 | 0 | 0 | 0 |
| 0 | 0 | 0 | 0 | 0 |
| 0 | 0 | 0 | 0 | 0 |
| 0.003195468 | 0 | 0 | 0 | 0 |
| 0.000902859 | 0 | 0 | 0 | 0 |
| 0 | 0 | 0 | 0 | 0 |
| 0 | 0 | 0 | 0 | 0 |
| 0 | 0 | 0 | 0 | 0 |
| 0 | 0 | 0 | 0 | 0 |
| 0.002059093 | 0 | 0 | 0 | 0.001180891 |
| 0 | 0 | 0 | 0 | 0 |
| 0.000740131 | 0.00060406 | 0.002536667 | 0.010543514 | 0.040260569 |
| 0 | 0 | 0 | 0 | 0 |
| 0 | 0 | 0 | 0 | 0 |
| 0 | 0 | 0 | 0 | 0 |
| 0 | 0 | 0 | 0 | 0 |
| 0 | 0 | 0 | 0 | 0 |
| 0 | 0 | 0 | 0 | 0 |
| 0 | 0 | 0 | 0 | 0 |
| 0.001991395 | 0 | 0 | 0 | 0 |
| 0 | 0 | 0 | 0 | 0 |
| 0 | 0 | 0 | 0 | 0 |

| | | | | |
|---|---|---|---|---|
| 0 | 0.058653952 | 0 | 0 | 0 |
| 0 | 0 | 0.270932208 | 0 | 0 |
| 0 | 0 | 0 | 0 | 0 |
| 0 | 0 | 0 | 0 | 0 |
| 0.00155197 | 0 | 0.00212764 | 0.012906605 | 0.007974899 |
| 0.008351072 | 0 | 0 | 0 | 0.002235028 |
| 0 | 0 | 0 | 0 | 0 |
| 0.014140279 | 0.001420385 | 0.032805909 | 0.002010161 | 0.002445314 |
| 0 | 0 | 0 | 0 | 0 |
| 0.016816953 | 0.011175365 | 0 | 0.070099836 | 0.001080188 |
| 0 | 0 | 0 | 0 | 0 |
| 0.007404928 | 0.003560543 | 0.008131793 | 0.060732728 | 0.024334753 |
| 0.070145946 | 0.042217736 | 0.001392831 | 0.20660159 | 0.128791902 |
| 0 | 0 | 0.015109656 | 0 | 0.060050353 |
| 0 | 0.002660753 | 0.001432495 | 0 | 0 |
| 0 | 0 | 0 | 0 | 0.003778593 |
| 0 | 0 | 0 | 0 | 0 |
| 0 | 0 | 0 | 0 | 0 |
| 0 | 0 | 0.076813471 | 0 | 0.060139179 |
| 0 | 0 | 0 | 0 | 0 |
| 0 | 0 | 0 | 0 | 0 |
| 0 | 0 | 0 | 0 | 0 |
| 0 | 0 | 0 | 0.001037907 | 0 |
| 0 | 0.320459502 | 0 | 0 | 0 |
| 0 | 0 | 0 | 0 | 0 |
| 0.003195056 | 0.011986929 | 0 | 0.001171559 | 0.016491298 |
| 0 | 0 | 0 | 0 | 0 |
| 0 | 0 | 0 | 0 | 0 |
| 0 | 0 | 0 | 0 | 0 |
| 0 | 0.000306623 | 0 | 0 | 0 |
| 0 | 0 | 0 | 0 | 0 |
| 0 | 0 | 0 | 0 | 0 |
| 0 | 0 | 0.001472223 | 0 | 0 |
| 0 | 0 | 0 | 0 | 0 |
| 0 | 0 | 0 | 0 | 0 |
| 0 | 0 | 0 | 0 | 0 |
| 0 | 0 | 0.831244259 | 0.074372589 | 0 |
| 0 | 0 | 0 | 0 | 0 |
| 0 | 0 | 0 | 0 | 0 |
| 0.003200363 | 0 | 0 | 0 | 0.001284787 |
| 0 | 0 | 0 | 0 | 0 |
| 0 | 0 | 0 | 0 | 0 |
| 0 | 0 | 0 | 0 | 0 |
| 0 | 0 | 0 | 0 | 0 |
| 0 | 0 | 0 | 0 | 0 |
| 0 | 0.051651602 | 0 | 0 | 0 |
| 0 | 0 | 0 | 0.001165541 | 0.00106339 |
| 0 | 0 | 0 | 0 | 0 |

| | | | | |
|---|---|---|---|---|
| 0 | 0 | 0 | 0 | 0 |
| 0.003171747 | 0.006949034 | 0 | 0 | 0.039557158 |
| 0 | 0 | 0 | 0 | 0 |
| 0 | 0 | 0 | 0 | 0 |
| 0 | 0 | 0 | 0 | 0 |
| 0.02190621 | 0 | 0.61856972 | 0 | 0 |
| 0 | 0 | 0.036346948 | 0 | 0 |
| 0 | 0 | 0.00381672 | 0.026417984 | 0 |
| 0 | 0 | 0 | 0 | 0 |
| 0 | 0 | 0 | 0 | 0 |
| 0.000518521 | 0 | 0.003159354 | 0.017834239 | 0.004579528 |
| 0 | 0 | 0 | 0 | 0 |
| 0 | 0 | 0 | 0 | 0 |
| 0.000489872 | 0.001634781 | 0 | 0.003017712 | 0.001704382 |
| 0.00363486 | 0 | 0.02541901 | 0.015775808 | 0 |
| 0 | 0 | 0 | 0 | 0 |
| 0 | 0 | 0 | 0 | 0 |
| 0 | 0 | 0.017670208 | 0 | 0 |
| 0 | 0 | 0 | 0.007762069 | 0 |
| 0 | 0 | 0 | 0.002563493 | 0 |
| 0.010732285 | 0 | 0 | 0 | 0 |
| 0.401359352 | 0.297937522 | 0 | 0.430746154 | 0 |
| 0 | 0.000910563 | 0 | 0 | 0 |
| 0 | 0 | 0 | 0 | 0 |
| 0 | 0 | 0 | 0 | 0 |
| 0 | 0 | 0 | 0 | 0 |
| 0 | 0 | 0.007821089 | 0.003271999 | 0 |
| 0 | 0 | 0 | 0 | 0 |
| 0 | 0 | 0 | 0 | 0 |
| 0.497130079 | 0.113769638 | 1.94144639 | 0.611525905 | 0.382234805 |
| 0 | 0.000506314 | 0 | 0.001876671 | 0 |
| 0 | 0 | 0 | 0 | 0 |
| 0 | 0 | 0 | 0 | 0 |
| 0 | 0 | 0 | 0 | 0 |
| 0 | 0 | 0 | 0.000908422 | 0.002249616 |
| 0 | 0 | 0 | 0 | 0 |
| 0 | 2.984670524 | 0 | 0 | 0 |
| 0 | 0 | 0.002281729 | 0 | 0.004677128 |
| 0.001219764 | 0 | 0 | 0 | 0 |
| 0 | 0 | 0 | 0.059512095 | 0 |
| 0 | 0 | 0 | 0.013644882 | 0.006251451 |
| 1.481973394 | 0 | 6.069853914 | 2.843468409 | 0.604764866 |
| 0 | 0 | 0.000928983 | 0 | 0.00550871 |
| 0 | 0 | 0 | 0 | 0 |
| 0 | 0 | 0.002768048 | 0 | 0 |
| 0 | 0 | 0 | 0 | 0 |
| 0 | 0.000858621 | 0.002146225 | 0 | 0.000791887 |
| 0 | 0 | 0 | 0 | 0 |

| | | | | |
|---|---|---|---|---|
| 0.000695375 | 0.004880778 | 0 | 0.002409554 | 0.040757161 |
| 0 | 0 | 0 | 0.001197493 | 0 |
| 1.907084441 | 0.047841884 | 0.001082464 | 0.008025604 | 9.390888806 |
| 0 | 0 | 0 | 0 | 0 |
| 0.078057376 | 0.054718072 | 0 | 0 | 0 |
| 0 | 0 | 0 | 0.171131497 | 0 |
| 0 | 0 | 0 | 0 | 0 |
| 0.001753593 | 0.000339247 | 0 | 0 | 0 |
| 0 | 0.000474974 | 0.002216207 | 0.003809096 | 0 |
| 0 | 0 | 0 | 0 | 0 |
| 0 | 0 | 0 | 0 | 0 |
| 0 | 0 | 0 | 0 | 0 |
| 0 | 0 | 0 | 0 | 0 |
| 0 | 0 | 0 | 0.25698155 | 0.007450159 |
| 0 | 0 | 0 | 0 | 0 |
| 0.019541961 | 0.031510589 | 0 | 0.004921294 | 0.001578894 |
| 0.010353316 | 0.015091203 | 0 | 0.03063047 | 0.407649739 |
| 0.000662046 | 0 | 0 | 0.001019584 | 0 |
| 0 | 0.000765622 | 0.015218261 | 0 | 0 |
| 0 | 0 | 0 | 0.493553442 | 0 |
| 0 | 0 | 0 | 0 | 0.027707187 |
| 0 | 0 | 0 | 0.001703481 | 0.000507489 |
| 0 | 0 | 0 | 0 | 0 |
| 0 | 0 | 0 | 0 | 0.605143656 |
| 0 | 0.031034755 | 0 | 0 | 0.000511119 |
| 0 | 0.004608723 | 0.001093429 | 0.008843889 | 0 |
| 0 | 0 | 0 | 0.00159804 | 0 |
| 0 | 0.009980482 | 0 | 0 | 0 |
| 0 | 0 | 0 | 0 | 0 |
| 0 | 0 | 0 | 0 | 0 |
| 0 | 0.004322312 | 0.034258417 | 0.014583654 | 0 |
| 0.035883263 | 0.070841724 | 0 | 0.006945501 | 0.020781489 |
| 19.71305896 | 0.017472221 | 0 | 0.083382014 | 0.01573949 |
| 0.001337782 | 0.00052408 | 0 | 0.002060251 | 0.002363033 |
| 0.048123718 | 0.02331641 | 0 | 0 | 0 |
| 0 | 0 | 0 | 0 | 0 |
| 0.001350302 | 0.001102053 | 0 | 0.001039767 | 0 |
| 0.014123986 | 0.010859392 | 0.034986056 | 0.017889207 | 0.002755553 |
| 0 | 0.015600691 | 0 | 0 | 1.993322457 |
| 0 | 0 | 0.027947097 | 0 | 0 |
| 0 | 0 | 0 | 0 | 0 |
| 0 | 0 | 0 | 0 | 0 |
| 0 | 0 | 0.014433543 | 0 | 0 |
| 0.885750465 | 0 | 0 | 0.104735484 | 0 |
| 0 | 0 | 0 | 0.038507058 | 0 |
| 0 | 0.00106476 | 0 | 0 | 0 |
| 0 | 0 | 0 | 0 | 0 |
| 0 | 0 | 0 | 0 | 0 |

| | | | | |
|---|---|---|---|---|
| 0 | 0 | 0 | 0 | 0 |
| 0.048563973 | 0.079584414 | 0.335902441 | 0.408303268 | 10.23267022 |
| 0 | 0 | 0 | 0 | 0 |
| 0 | 0 | 0 | 0 | 0 |
| 0 | 0 | 0 | 0 | 0 |
| 0 | 0 | 0 | 0 | 0 |
| 0 | 0 | 0.002027755 | 0 | 0 |
| 0 | 0 | 0.002525369 | 0 | 0.001220186 |
| 0 | 0.001684255 | 0 | 0 | 0.000661 |
| 0 | 0 | 0 | 0 | 0 |
| 0 | 0 | 0.008143956 | 0.000823376 | 0.000536581 |
| 0.346617681 | 0.239746795 | 0.011827387 | 0.024528831 | 0.028293528 |
| 0 | 0.22328814 | 0 | 0 | 0 |
| 0 | 0 | 0 | 0 | 0 |
| 0.003527251 | 0.014752683 | 0 | 0.003070342 | 0 |
| 0 | 0 | 0 | 0 | 0 |
| 0.011896758 | 0.000152307 | 0.033258798 | 0.02694354 | 0.011799427 |
| 0.007994353 | 0 | 0.071404005 | 0.037214929 | 0 |
| 0 | 0 | 0 | 0 | 0 |
| 0.004837175 | 0 | 0 | 0 | 0.000284178 |
| 0.007361135 | 0.017905633 | 0.003297905 | 0.088247024 | 0.27917295 |
| 0.001257221 | 0.039557989 | 0 | 0 | 0 |
| 0 | 0 | 0 | 0 | 0 |
| 0 | 0 | 0 | 0 | 0 |
| 0.027161275 | 0.000422243 | 0 | 0 | 0.269871651 |
| 0 | 0 | 0 | 0 | 0 |
| 1.507726507 | 0.006077291 | 0 | 0 | 0 |
| 0.11981153 | 0.036526251 | 0.110296524 | 0.034257093 | 0.04015384 |
| 0 | 0 | 0 | 0 | 0 |
| 0 | 0.000241641 | 0 | 0 | 0 |
| 0 | 0 | 0 | 0.012421632 | 0 |
| 0 | 0 | 0 | 0 | 0 |
| 0 | 0 | 0 | 0 | 0 |
| 0 | 0 | 0 | 0 | 0 |
| 0.002005264 | 0.000756474 | 0 | 0 | 0 |
| 0.026222997 | 0.028479456 | 1.428502278 | 0.090765695 | 0.425050056 |
| 0 | 0 | 0 | 0 | 0 |
| 0 | 0 | 0 | 0 | 0 |
| 0.002838686 | 0.005148448 | 0.018160965 | 0.020401358 | 0.01884125 |
| 0 | 0 | 0 | 0 | 0 |
| 0 | 0 | 0 | 0.035479712 | 0 |
| 0 | 0 | 0 | 0 | 0 |
| 0 | 0 | 0.037471221 | 0.03735332 | 0 |
| 0 | 0 | 0 | 0 | 0 |
| 0 | 0 | 0 | 0 | 0 |
| 0 | 0.116346972 | 0 | 0 | 0.051097247 |
| 0 | 0 | 0 | 0 | 0 |
| 0 | 0 | 0 | 0 | 0 |

| | | | | |
|---|---|---|---|---|
| 0 | 0 | 0 | 0 | 0 |
| 0 | 0 | 0 | 0 | 0 |
| 0 | 0.018479484 | 0 | 0 | 0.003806177 |
| 0.001338813 | 0.046300306 | 0 | 0 | 0.006127122 |
| 0 | 0 | 0 | 0 | 0 |
| 0 | 0.000126335 | 0 | 0 | 0 |
| 0 | 0 | 0 | 0 | 0 |
| 0 | 0 | 0 | 0 | 0 |
| 0.068096026 | 0.451568069 | 0 | 0.252715368 | 1.305567888 |
| 0 | 0.033542379 | 0 | 0 | 0.07436022 |
| 0 | 1.313312129 | 0 | 0 | 0 |
| 0.000897908 | 0.003824677 | 0 | 0.008099399 | 0.000411961 |
| 0 | 0 | 0 | 0 | 0 |
| 0 | 0 | 0 | 0 | 0 |
| 0.009949985 | 0.151287059 | 0 | 0.014997446 | 0.013088118 |
| 0 | 0 | 0 | 0 | 0 |
| 0 | 0.052888633 | 0.010726348 | 0 | 0 |
| 0 | 0 | 0 | 0 | 0 |
| 0 | 0 | 0 | 0 | 0 |
| 0 | 0 | 0 | 0 | 0 |
| 0 | 0 | 0 | 0 | 0 |
| 0 | 0.040019266 | 0 | 0 | 0 |
| 0 | 0.000822629 | 0 | 0 | 0 |
| 0.119295012 | 0.043338998 | 0 | 0.240249661 | 0 |
| 0 | 0.003737702 | 0 | 0 | 0 |
| 0 | 0.269538729 | 0 | 0 | 0 |
| 0 | 0.063754829 | 0 | 0 | 0 |
| 0.41276049 | 2.527925389 | 0 | 0 | 5.194093316 |
| 0 | 0 | 0 | 0 | 0 |
| 0 | 0 | 0 | 0 | 0 |
| 0.044394222 | 0.003935362 | 0 | 0 | 0.090111633 |
| 0 | 0.014652363 | 0 | 0 | 0 |
| 0 | 0 | 0 | 0.001023142 | 0.000228605 |
| 0 | 0 | 0 | 0 | 0 |
| 0 | 0.006364838 | 0 | 0 | 0 |
| 0.031614492 | 0.07773469 | 0 | 0 | 0 |
| 0 | 0.001159087 | 0 | 0.04710795 | 0.368174626 |
| 0 | 0 | 0 | 0.001233249 | 0 |
| 0 | 0.000842797 | 0 | 0 | 0 |
| 0 | 0 | 0 | 0.001979577 | 0 |
| 0.193195817 | 0.015158447 | 0.008247513 | 0.021225162 | 0.28434838 |
| 0 | 0.028935861 | 0 | 0 | 0 |
| 2.652864122 | 22.09670164 | 8.24252133 | 9.33893931 | 0.155147332 |
| 0 | 0.002403028 | 0 | 0 | 0 |
| 0 | 0 | 0 | 0 | 0 |
| 0 | 0.084241019 | 0 | 0 | 0 |
| 0 | 0 | 0 | 0 | 0 |
| 0 | 0.001931681 | 0 | 0 | 0 |

| | | | | |
|---|---|---|---|---|
| 0 | 0 | 0 | 0 | 0 |
| 0.007546818 | 0.063382714 | 0 | 0 | 0.01363353 |
| 0.003261483 | 0 | 0.709569384 | 0 | 0.017305842 |
| 0 | 0.002935733 | 0 | 0 | 0 |
| 0 | 0 | 0 | 0 | 0 |
| 0.000781952 | 0.003087635 | 0 | 0.004128841 | 0 |
| 0 | 0 | 0.579931026 | 0.079513007 | 0 |
| 0 | 0 | 0 | 0 | 0.000272677 |
| 0 | 0 | 0 | 0 | 0 |
| 0 | 0 | 0 | 0 | 0 |
| 0.00208839 | 0.003272535 | 0.076559617 | 0.041810931 | 0.007266003 |
| 1.235422311 | 0.526769626 | 0.417979987 | 2.028465603 | 1.120138012 |
| 0 | 0 | 0 | 0 | 0 |
| 0.014124286 | 0 | 0.000686645 | 0.028231464 | 0.528052486 |
| 1.416175821 | 0.00036079 | 0.188303651 | 0.191291826 | 0.071683507 |
| 0 | 0.880263405 | 0 | 0 | 0 |
| 0 | 0 | 0 | 0 | 0 |
| 0 | 0 | 0 | 0.001012552 | 0 |
| 0 | 0 | 0 | 0 | 0 |
| 0 | 0.008586327 | 0 | 0 | 0 |
| 0.025205446 | 0.024609603 | 0.334350266 | 0.103154328 | 0.243318137 |
| 0.851477487 | 0.10102074 | 0.250600395 | 0.121190421 | 0 |
| 1.237751592 | 0.958032513 | 0.123567368 | 3.388384735 | 0.076813113 |
| 0 | 0 | 0 | 0 | 0 |
| 0 | 0.008114614 | 0 | 0 | 0 |
| 0.003355965 | 0.297209778 | 0 | 0.019381332 | 0.01100257 |
| 0 | 0 | 0 | 0 | 0 |
| 0.002799273 | 0 | 0 | 0 | 0.00107491 |
| 0 | 0 | 0 | 0 | 0.000213735 |
| 0 | 0 | 0 | 0 | 0 |
| 0 | 0.003837662 | 0 | 0 | 0 |
| 0.3621159 | 0.115687226 | 0 | 0.276717598 | 0.212020486 |
| 0 | 0.000155601 | 0.01938492 | 0.053951407 | 0.005281065 |
| 0.005342958 | 0.003061369 | 0 | 0 | 0.008492183 |
| 0 | 0 | 0.067134073 | 0 | 0 |
| 0.006483228 | 0.055678632 | 0.082560023 | 0.010583567 | 0.002290371 |
| 0.000375763 | 0.006079072 | 0 | 0 | 0.0009051 |
| 0 | 0 | 0 | 0 | 0 |
| 0.503300731 | 0.848999575 | 0.026810958 | 0 | 0.056284131 |
| 0 | 0 | 0 | 0 | 0 |
| 0 | 0.006833581 | 0 | 0 | 0 |
| 0 | 0 | 0 | 0 | 0 |
| 0 | 0 | 0 | 0 | 0 |
| 0 | 0.003411468 | 0 | 0 | 0 |
| 0.001182942 | 0.004956034 | 0.238078715 | 0 | 0.00189957 |
| 0 | 0 | 0 | 0 | 0 |
| 0 | 0 | 0 | 0 | 0 |
| 0.01730178 | 0 | 0.014671856 | 0.090650021 | 0.147079244 |

| | | | | |
|---|---|---|---|---|
| 0 | 0 | 0 | 0 | 0 |
| 0 | 0 | 0 | 0 | 0 |
| 0 | 0 | 0 | 0 | 0 |
| 0 | 0.000274286 | 0 | 0.000776352 | 0 |
| 0 | 0 | 0 | 0 | 0 |
| 0 | 0.038444731 | 0 | 0 | 0 |
| 0.100446249 | 0.011411064 | 0 | 0 | 0 |
| 0.001623548 | 0.128695167 | 0 | 0.011608746 | 0.027095172 |
| 0 | 0 | 0 | 0 | 0 |
| 0 | 0 | 0 | 0 | 0 |
| 0.000561443 | 0.00051932 | 0.00085522 | 0 | 0 |
| 0 | 0 | 0 | 0 | 0 |
| 0.01314996 | 0.038275465 | 0 | 0 | 0 |
| 0 | 0 | 0 | 0 | 0.195314227 |
| 0 | 0 | 0 | 0 | 0 |
| 0 | 0 | 0 | 0 | 0 |
| 0 | 0.012813986 | 0 | 0.429572338 | 0.196799921 |
| 0 | 0.00038146 | 0 | 0.001619554 | 0.000469083 |
| 0 | 0.281272642 | 0 | 0 | 0 |
| 0 | 0 | 0 | 0 | 0 |
| 0 | 0.000903727 | 0 | 0 | 0 |
| 0 | 0 | 0 | 0 | 0 |
| 0.018321427 | 0.017503904 | 0 | 0.022614231 | 0 |
| 0 | 0 | 0 | 0 | 0 |
| 0 | 0 | 0 | 0 | 0 |
| 0 | 0 | 0 | 0 | 0 |
| 0 | 0 | 0 | 0 | 0 |
| 0 | 0 | 0 | 0 | 0 |
| 0 | 0 | 0 | 0 | 0 |
| 0 | 0.525712275 | 0.001935069 | 0.317590068 | 0 |
| 0 | 0 | 0.000841976 | 0 | 0 |
| 0 | 0 | 0 | 0 | 0 |
| 0 | 0 | 0 | 0 | 0 |
| 0 | 0 | 0 | 0 | 0 |
| 0 | 0.001165942 | 0 | 0 | 0 |
| 0 | 0 | 0 | 0 | 0 |
| 0 | 0.032507836 | 0 | 0.08529343 | 0.082155895 |
| 0.00166345 | 0 | 0.009411465 | 0 | 0.000381596 |
| 0 | 0.004074035 | 0 | 0 | 0 |
| 0 | 0.000102026 | 0 | 0.006317036 | 0.001411444 |
| 0 | 0.007594227 | 0 | 0 | 0 |
| 0 | 0.532337775 | 0 | 0 | 0 |
| 0 | 0 | 0 | 0 | 0 |
| 0.002653883 | 0 | 0 | 0 | 0 |
| 0 | 0.070530673 | 0 | 0 | 0 |
| 0 | 0 | 0 | 0 | 0 |
| 0 | 0.001990247 | 0 | 0 | 0 |
| 0 | 0 | 0 | 0 | 0 |

| | | | | |
|---|---|---|---|---|
| 0 | 0 | 0 | 0 | 0 |
| 0 | 0 | 0 | 0 | 0 |
| 0 | 0.061374381 | 0 | 0 | 0 |
| 0 | 0.268776755 | 0 | 0.022934129 | 0.010438338 |
| 0 | 0.158807768 | 0 | 0 | 0 |
| 0 | 0 | 0 | 0 | 0.280104999 |
| 0 | 0 | 0 | 0 | 0 |
| 0 | 0 | 0 | 0 | 0 |
| 0.036542931 | 0.001069941 | 0.426592109 | 0.031041203 | 0.005394414 |
| 0 | 0 | 0 | 0 | 0 |
| 0 | 0.001122061 | 0 | 0 | 0 |
| 0 | 0 | 0 | 0 | 0 |
| 0 | 0 | 0 | 0 | 0 |
| 0 | 0 | 0 | 0 | 0 |
| 0.001648655 | 3.749457067 | 0 | 0.02829184 | 1.134794433 |
| 0 | 0 | 0 | 0 | 0 |
| 0 | 0 | 0 | 0 | 0 |
| 0 | 0 | 0 | 0 | 0 |
| 0 | 0 | 0 | 0 | 0 |
| 0 | 0.752981872 | 0 | 0 | 0 |
| 0 | 0.010199274 | 0 | 0 | 0 |
| 0 | 0 | 0 | 0 | 0 |
| 0 | 0 | 0 | 0 | 0 |
| 0 | 0.001080274 | 0 | 0 | 0 |
| 0.196945567 | 0.01467924 | 0.002054781 | 0 | 0.005105887 |
| 0.406677338 | 0.384674634 | 6.701870829 | 1.135961979 | 6.487993768 |
| 0 | 0.039778599 | 0 | 0.002207669 | 0 |
| 0 | 0 | 0 | 0 | 0 |
| 0.003706564 | 0.000293345 | 0.050557698 | 0 | 0 |
| 0 | 0.071414633 | 0 | 0 | 0 |
| 0 | 0 | 0 | 0 | 0 |
| 0 | 0.047888788 | 0 | 0.001720134 | 0 |
| 0 | 0 | 0 | 0 | 0 |
| 0 | 0.000201591 | 0 | 0.002218972 | 0.000826325 |
| 0.000678301 | 0.000295252 | 0 | 0.00626771 | 0.417170321 |
| 0 | 0.000600034 | 0.006573284 | 0 | 0 |
| 0 | 0 | 0 | 0.016002578 | 0 |
| 0.443552003 | 0.24574149 | 2.987505647 | 0.003543659 | 0.26755807 |
| 22.45770087 | 0 | 0 | 0 | 0 |
| 0 | 0 | 0 | 0 | 0 |
| 0 | 0 | 0 | 0 | 0 |
| 0 | 0.000236676 | 0 | 0 | 0 |
| 0.22817817 | 0.114366159 | 0 | 0.203578279 | 0.201879982 |
| 0 | 6.407600422 | 0 | 0 | 0 |
| 0 | 0 | 0 | 0 | 0 |
| 0 | 0.000790896 | 0 | 0 | 0.001732387 |
| 0 | 0 | 0 | 0 | 0 |
| 0.414117519 | 0.017994269 | 0 | 0.005851026 | 0.030794671 |

| | | | | |
|---|---|---|---|---|
| 0 | 0.000734045 | 0 | 0 | 0 |
| 0 | 0.005373746 | 0 | 0 | 0 |
| 0 | 0 | 0 | 0 | 0 |
| 0 | 0 | 0 | 0 | 0 |
| 0 | 0 | 0.000818402 | 0 | 0 |
| 0 | 0 | 0 | 0 | 0.00093467 |
| 0 | 0.064789633 | 0 | 0 | 0 |
| 0 | 0 | 0 | 0 | 0 |
| 0 | 0 | 0 | 0 | 0 |
| 0 | 0.333407757 | 0 | 0 | 0 |
| 0 | 0 | 0 | 0 | 0 |
| 0.242133773 | 0.26308633 | 0 | 0.625186557 | 0.552385372 |
| 0.135150196 | 0.003008672 | 0 | 0.89767387 | 0.075997716 |
| 0 | 0 | 0 | 0 | 0 |
| 0.024232857 | 0.234256029 | 0.350671496 | 0.338749138 | 1.116134229 |
| 0 | 0 | 0 | 0 | 0 |
| 0 | 0 | 0 | 0 | 0 |
| 0 | 0 | 0 | 0 | 0 |
| 0 | 0.000977877 | 0 | 0 | 0 |
| 0 | 0.003252929 | 0 | 0 | 0 |
| 0 | 0.072151876 | 0 | 0.000694396 | 0 |
| 0 | 0 | 0 | 0 | 0 |
| 0.034951275 | 0.197777356 | 0 | 0.157198501 | 0.043262878 |
| 0 | 0 | 0 | 0 | 0 |
| 0 | 0 | 0 | 0 | 0 |
| 0 | 0 | 0 | 0 | 0 |
| 0 | 0 | 0 | 0 | 0 |
| 0 | 0 | 0 | 0 | 0 |
| 0 | 0 | 0 | 0 | 0 |
| 0 | 0 | 0 | 0 | 0 |
| 0 | 0 | 0 | 0 | 0 |
| 0.065108353 | 0.017065695 | 0.041144447 | 0.051842458 | 0.142260708 |
| 0 | 0.000196567 | 0 | 0 | 0 |
| 0 | 0.03379607 | 0 | 0.101582258 | 0.002183068 |
| 0 | 0.000823332 | 0.006954676 | 0.000803584 | 0 |
| 0.036155038 | 0.127364912 | 0.002817703 | 0.014243869 | 0.150275238 |
| 0 | 0.000252016 | 0 | 0 | 0 |
| 0 | 0.005483923 | 0 | 0 | 0 |
| 0 | 0.001793923 | 0 | 0 | 0 |
| 0 | 0 | 0 | 0 | 0 |
| 0.128924673 | 0.065479673 | 0 | 0 | 0 |
| 0 | 0.002269898 | 0 | 0 | 0 |
| 0 | 0 | 0 | 0 | 0 |
| 0 | 0 | 0 | 0 | 0 |
| 0.006296701 | 0.007858932 | 0.231957065 | 0.011478362 | 0.012174774 |
| 0 | 0 | 0 | 0 | 0 |
| 0 | 0.530845213 | 0 | 0 | 0.101307272 |
| 0 | 0.000423239 | 0 | 0 | 0 |

| | | | | |
|---|---|---|---|---|
| 0 | 0 | 0 | 0 | 0 |
| 0 | 0 | 0 | 0 | 0 |
| 0 | 0 | 0.000607346 | 0 | 0 |
| 0 | 0 | 0 | 0 | 0 |
| 0 | 0 | 0 | 0 | 0 |
| 0 | 0 | 0 | 0 | 0 |
| 0.004974183 | 0.112716197 | 0 | 0 | 0 |
| 0 | 0.007867456 | 0 | 0.012371334 | 0.050086411 |
| 0 | 0 | 0 | 0 | 0 |
| 0 | 0.000913972 | 0.021824475 | 0 | 0 |
| 0 | 0 | 0 | 0 | 0 |
| 0 | 0 | 0 | 0 | 0 |
| 0 | 0 | 0 | 0 | 0 |
| 0 | 0.330153368 | 0 | 0 | 0 |
| 0 | 0 | 0 | 0 | 0 |
| 0 | 0 | 0 | 0 | 0 |
| 0 | 6.995721948 | 0 | 0 | 0 |
| 0 | 0 | 0 | 0 | 0 |
| 2.191414679 | 4.330919588 | 0 | 0.025807589 | 0 |
| 0 | 0 | 0 | 0 | 0 |
| 0 | 0.00102498 | 0 | 0 | 0 |
| 0 | 0 | 0 | 0 | 0 |
| 0 | 0 | 0 | 0 | 0 |
| 0 | 0 | 0 | 0 | 0 |
| 0 | 0 | 0 | 0.001729808 | 0.004058237 |
| 0.002273017 | 0 | 0.026605667 | 0 | 0.000480265 |
| 0 | 0 | 0 | 0 | 0 |
| 0.002107735 | 0.05813116 | 0.021582469 | 0.104233248 | 0.177597711 |
| 0 | 0 | 0 | 0 | 0 |
| 0.24760559 | 0.329192925 | 0 | 0.029539267 | 0.393055401 |
| 0 | 0 | 0 | 0 | 0 |
| 0 | 0 | 0 | 0 | 0 |
| 0 | 0.015015724 | 0 | 0 | 0 |
| 0 | 0 | 0 | 0 | 0 |
| 0 | 0 | 0.005156093 | 0 | 0 |
| 0 | 0 | 0 | 0 | 0 |
| 0 | 0 | 0 | 0 | 0 |
| 0 | 0 | 0 | 0.001533562 | 0 |
| 0 | 0 | 0 | 0 | 0 |
| 0 | 0.017194624 | 0 | 0 | 0 |
| 0.433285039 | 0.387070565 | 33.68376902 | 19.26937414 | 8.386992881 |
| 0.100382794 | 0.269760824 | 0 | 0 | 1.116157327 |
| 0.36311939 | 0.070464268 | 0 | 0 | 0 |
| 0 | 0.054936188 | 0 | 0 | 0 |
| 0 | 0 | 0 | 0 | 0 |
| 0 | 0 | 0 | 0 | 0 |
| 0 | 0 | 0 | 0 | 0 |
| 0 | 0.000761732 | 0.014962078 | 0 | 0 |

| | | | | |
|---|---|---|---|---|
| 0 | 0 | 1.68095802 | 0 | 0 |
| 0 | 0.001086796 | 0 | 0 | 0 |
| 0 | 0.001328355 | 0 | 0 | 0 |
| 0 | 0 | 0 | 0 | 0 |
| 0.94561531 | 1.638937328 | 0 | 3.128886405 | 2.722233617 |
| 0 | 0 | 0 | 0 | 0 |
| 0 | 0 | 0 | 0 | 0 |
| 0 | 0 | 0 | 0 | 0 |
| 0.015306388 | 0 | 0.659193142 | 0.196259831 | 0.010520567 |
| 0 | 0 | 0 | 0 | 0 |
| 0 | 0 | 0 | 0 | 0 |
| 0 | 0 | 0 | 0 | 0 |
| 0.056419413 | 0.084652549 | 0 | 0 | 0.003080345 |
| 0.015537072 | 0.003109772 | 0.206240154 | 0.065801653 | 0.009690191 |
| 0.051894544 | 0.130187465 | 0 | 0 | 0.420417543 |
| 0.186690705 | 0.17632586 | 0 | 0 | 0.096791386 |
| 0.018966949 | 0.051808773 | 0 | 0.005546213 | 0.094648511 |
| 0 | 0 | 0 | 0 | 0 |
| 0 | 0.039018859 | 0 | 0 | 0 |
| 0 | 0 | 0 | 0 | 0 |
| 0 | 0 | 0 | 0 | 0 |
| 0 | 0 | 0 | 0 | 0 |
| 0 | 0 | 0 | 0 | 0 |
| 0 | 0 | 0 | 0 | 0 |
| 0 | 0 | 0 | 0 | 0 |
| 0 | 0 | 0 | 0 | 0 |
| 0.002609579 | 0.001113184 | 0 | 0.00450115 | 0.095626456 |
| 0.001060063 | 0 | 0 | 0 | 0 |
| 0 | 0 | 0 | 0 | 0 |
| 0 | 0 | 0 | 0 | 0 |
| 0 | 0 | 0 | 0 | 0 |
| 0 | 0 | 0 | 0 | 0 |
| 0 | 0 | 0 | 0 | 0 |
| 0 | 0.011635606 | 0 | 0.013500635 | 0 |
| 0.015333352 | 0.043732716 | 0 | 0 | 0 |
| 0.0109002 | 0 | 0.004981131 | 0 | 0 |
| 0.000417799 | 0.000636511 | 0 | 0 | 0 |
| 0 | 0 | 0 | 0.010699797 | 0 |
| 0 | 0 | 0 | 0 | 0 |
| 0 | 0 | 0 | 0 | 0 |
| 0.555524385 | 0.084005005 | 0 | 11.2257879 | 7.913733885 |
| 0 | 0 | 0 | 0 | 0.055472118 |
| 0 | 0 | 0 | 0 | 0 |
| 0 | 0.000125499 | 0 | 0 | 0 |
| 0 | 0 | 0 | 0 | 0 |
| 0 | 0 | 0 | 0 | 0 |
| 0 | 0 | 0 | 0 | 0 |
| 0 | 0 | 0 | 0 | 0 |

| | | | | |
|---|---|---|---|---|
| 0 | 0.058015852 | 0 | 0 | 0 |
| 0 | 0 | 0 | 0 | 0 |
| 0 | 0 | 0 | 0 | 0 |
| 0 | 0 | 0 | 0 | 0 |
| 0 | 0.017726695 | 0 | 0 | 0.011582632 |
| 0 | 0 | 0 | 0 | 0 |
| 0.015088763 | 0.036931176 | 0.001094477 | 0.092212124 | 0.475649345 |
| 0 | 0 | 0 | 0 | 0 |
| 0 | 0.000239151 | 0 | 0.001822433 | 0 |
| 0 | 0 | 0 | 0 | 0 |
| 0 | 0.017951966 | 0 | 0.002012801 | 0 |
| 0 | 0.003601219 | 0 | 0 | 0 |
| 0 | 0 | 0 | 0 | 0 |
| 0 | 0 | 0 | 0 | 0 |
| 0 | 0 | 0 | 0 | 0.000377898 |
| 0 | 0 | 0 | 0 | 0 |
| 0 | 0 | 0 | 0 | 0 |
| 0 | 0 | 0 | 0 | 0 |
| 0 | 0 | 0 | 0 | 0 |
| 0 | 0 | 0 | 0 | 0 |
| 0.387750258 | 0.055581937 | 0.021361061 | 0.333430525 | 0.094651588 |
| 0 | 0 | 0 | 0 | 0 |
| 0 | 0.001368453 | 0 | 0.00445433 | 0 |
| 0.000782748 | 0.000584084 | 0 | 0.001549892 | 0 |
| 0 | 0 | 0 | 0 | 0 |
| 0 | 0.000401968 | 0 | 0 | 0 |
| 0 | 0 | 0 | 0 | 0 |
| 0 | 0.001440314 | 0 | 0 | 0 |
| 0 | 0 | 0 | 0 | 0 |
| 5.840722187 | 9.995806893 | 0 | 0.120455558 | 0.01744421 |
| 0 | 0.011314599 | 0 | 0.001782788 | 0 |
| 0 | 0 | 0 | 0 | 0 |
| 0 | 0 | 0 | 0 | 0 |
| 0 | 0 | 0 | 0 | 0.003164074 |
| 0 | 0 | 0 | 0 | 0 |
| 0 | 0 | 0 | 0 | 0 |
| 0 | 0.000395872 | 0.011935252 | 0.049560317 | 0.000449359 |
| 0 | 0 | 0 | 0 | 0 |
| 0 | 0.045654909 | 0 | 0 | 0 |
| 0 | 0 | 0 | 0 | 0 |
| 0 | 0 | 0 | 0 | 0 |
| 0 | 0 | 0 | 0 | 0 |
| 0 | 0.000154882 | 0 | 0 | 0 |
| 0 | 0.00491085 | 0 | 0 | 0 |
| 0 | 0.023747873 | 0 | 0 | 0 |
| 0 | 0 | 0 | 0 | 0 |
| 0 | 0 | 0 | 0 | 0 |
| 0 | 0.002902519 | 0 | 0 | 0 |

| | | | | |
|---|---|---|---|---|
| 0 | 0.000471372 | 0.05021963 | 0.005744435 | 0 |
| 0 | 0.041128305 | 0 | 0 | 0 |
| 0 | 0 | 0 | 0 | 0 |
| 0 | 0 | 0 | 0 | 0 |
| 0 | 0 | 0 | 0 | 0 |
| 0 | 0 | 0 | 0 | 0 |
| 0 | 0 | 0 | 0 | 0 |
| 0.226087551 | 0.163078446 | 3.276397432 | 1.365536881 | 2.295955206 |
| 0.870482986 | 0.01261921 | 0.017836823 | 0.020069548 | 0.058229989 |
| 0.001051862 | 0 | 0 | 0.030778496 | 0.000527838 |
| 0 | 0 | 0 | 0 | 0 |
| 0 | 0.00017534 | 0 | 0 | 0 |
| 0 | 0.015265876 | 0 | 0 | 0 |
| 0.172958259 | 0.635575395 | 0 | 0 | 0 |
| 0 | 0 | 0 | 0 | 0 |
| 0 | 0 | 0 | 0 | 0 |
| 0 | 0 | 0.03585094 | 0 | 0 |
| 0 | 0 | 0 | 0 | 0 |
| 0 | 0 | 0 | 0 | 0 |
| 0 | 0.000616956 | 0 | 0 | 0.000262618 |
| 0 | 0 | 0 | 0 | 0 |
| 0 | 0 | 0 | 0 | 0 |
| 0 | 0 | 0 | 0 | 0 |
| 0 | 0 | 0 | 0 | 0 |
| 0 | 0.046450062 | 0 | 0 | 0.000380461 |
| 0 | 0 | 0 | 0 | 0 |
| 0 | 0 | 0 | 0 | 0 |
| 0 | 0 | 0 | 0 | 0 |
| 0 | 0 | 0 | 0 | 0 |
| 0 | 0 | 0 | 0 | 0 |
| 0 | 0 | 0 | 0 | 0.002282704 |
| 0 | 0 | 0 | 0 | 0 |
| 0 | 0 | 0 | 0 | 0 |
| 0 | 0 | 0 | 0 | 0 |
| 0 | 0 | 0 | 0 | 0 |
| 0 | 0 | 0 | 0 | 0 |
| 0 | 0 | 0 | 0 | 0 |
| 0 | 0 | 0 | 0.032627592 | 0 |
| 0 | 0.003923484 | 0 | 0 | 0 |
| 0 | 0 | 0 | 0 | 1.053660864 |
| 0 | 0 | 0 | 0 | 0 |
| 0.01127724 | 0.243874364 | 0 | 0.213181518 | 0.01651701 |
| 0 | 0.000778744 | 0 | 0 | 0 |
| 0 | 0 | 0 | 0 | 0 |
| 0 | 0.000101904 | 0 | 0 | 0 |
| 0 | 0 | 0 | 0 | 0 |
| 0 | 0 | 0 | 0 | 0 |
| 0 | 0 | 0 | 0 | 0 |

| | | | | |
|---|---|---|---|---|
| 0 | 0 | 0 | 0 | 0 |
| 0 | 0 | 0.004300952 | 0.028933435 | 0 |
| 0 | 0 | 0 | 0 | 0 |
| 0 | 0 | 0 | 0 | 0 |
| 0.080927665 | 0.19331514 | 0.090431307 | 0.268907287 | 0.115812466 |
| 0 | 0 | 0 | 0 | 0 |
| 0 | 0 | 0.019255864 | 0 | 0 |
| 0.091530055 | 0.001265862 | 0.037070796 | 0.014143234 | 0.00823378 |
| 0.007029951 | 0 | 0 | 0 | 0 |
| 0 | 0.027000153 | 0 | 0 | 0 |
| 0 | 0 | 0 | 0 | 0 |
| 0 | 0.062190244 | 0 | 0 | 0 |
| 0 | 0 | 0 | 0 | 0 |
| 0 | 0 | 0 | 0 | 0 |
| 0 | 0 | 0 | 0 | 0 |
| 0 | 0 | 0 | 0.006542038 | 0.001385586 |
| 0 | 0 | 0 | 0 | 0 |
| 0 | 0 | 0 | 0 | 0 |
| 0 | 0 | 0 | 0 | 0 |
| 0 | 0 | 0 | 0 | 0 |
| 0 | 0 | 0 | 0 | 0 |
| 0 | 0 | 0 | 0 | 0 |
| 0 | 0.001440203 | 0 | 0 | 0 |
| 0 | 0.000288289 | 0 | 0 | 0 |
| 3.496823185 | 0.571115428 | 0 | 0.260089373 | 0 |
| 0 | 0 | 0 | 0 | 0.002229414 |
| 0.00181726 | 0 | 0.978817329 | 0.005970501 | 0 |
| 0 | 0 | 0 | 0 | 0 |
| 0 | 0 | 0 | 0 | 0 |
| 0 | 0 | 0 | 0 | 0 |
| 0 | 0 | 0 | 0 | 0 |
| 0 | 0 | 0 | 0 | 0 |
| 0 | 0 | 0 | 0 | 0 |
| 0 | 0.000343231 | 0 | 0 | 0 |
| 0 | 0 | 0 | 0 | 0 |
| 0 | 0 | 0 | 0 | 0 |
| 0 | 0 | 0 | 0 | 0 |
| 0 | 0.183150304 | 0 | 0 | 0 |
| 0 | 0.007120053 | 0 | 0 | 0 |
| 0 | 0 | 0 | 0 | 0 |
| 0 | 0 | 0 | 0 | 0 |
| 0 | 0.003162672 | 0 | 0 | 0 |
| 0 | 0 | 0 | 0 | 0 |
| 0 | 0.000774259 | 0 | 0 | 0 |
| 0 | 0 | 0 | 0 | 0 |
| 0 | 0 | 0 | 0 | 0 |
| 0 | 0 | 0 | 0 | 0 |
| 0 | 0.001841402 | 0 | 0 | 0 |

| | | | | |
|---|---|---|---|---|
| 0 | 0.023977923 | 0 | 0.106329174 | 0.032382476 |
| 0 | 0 | 0 | 0 | 0 |
| 0 | 0 | 0 | 0 | 0 |
| 0 | 0.000246708 | 0 | 0 | 0 |
| 0 | 0 | 0 | 0 | 0 |
| 0 | 0 | 0 | 0 | 0.003815285 |
| 0 | 0 | 0 | 0 | 0 |
| 0 | 0 | 0 | 0 | 0 |
| 0 | 0 | 0 | 0 | 0 |
| 0 | 0 | 0 | 0 | 0 |
| 0 | 0 | 0 | 0 | 0 |
| 0 | 0 | 0 | 0 | 0 |
| 0 | 0 | 0 | 0 | 0 |
| 0 | 0 | 0 | 0 | 0 |
| 0 | 0.002102355 | 0 | 0 | 0 |
| 0 | 0.017579635 | 0 | 0 | 0 |
| 0 | 0 | 0 | 0 | 0 |
| 0 | 0 | 0 | 0 | 0 |
| 0 | 0 | 0 | 0 | 0 |
| 0 | 0 | 0 | 0 | 0 |
| 0.000527629 | 0 | 0 | 0 | 0.000847268 |
| 0 | 0 | 0 | 0 | 0 |
| 0.000666358 | 0.009716778 | 0.020300631 | 0.001795892 | 0.004815171 |
| 0 | 0.19704637 | 0 | 0 | 0 |
| 0 | 0.001791121 | 0 | 0 | 0 |
| 0 | 0 | 0 | 0 | 0 |
| 0 | 0.007633888 | 0 | 0 | 0 |
| 0 | 0 | 0 | 0 | 0 |
| 0.015142422 | 0 | 0 | 0 | 0 |
| 0 | 0 | 0 | 0 | 0 |
| 0 | 0 | 0 | 0 | 0 |
| 0 | 0 | 0 | 0 | 0 |
| 0.063184158 | 0.004719632 | 0.058460231 | 0.04324746 | 0.066137687 |
| 0 | 0.000343465 | 0 | 0 | 0 |
| 0.151830881 | 0.002551236 | 0 | 0.78830716 | 1.165605845 |
| 0 | 0 | 0.002288385 | 0.001156809 | 0 |
| 0 | 0 | 0 | 0 | 0 |
| 0 | 0 | 0 | 0 | 0 |
| 0 | 0 | 0 | 0 | 0.002790318 |
| 0 | 0 | 0 | 0 | 0 |
| 0 | 0 | 0 | 0 | 0 |
| 0 | 0 | 0 | 0 | 0 |
| 0.007958006 | 0.050817214 | 0 | 0.074577419 | 0.088501224 |
| 0 | 0 | 0 | 0 | 0 |
| 0 | 0 | 0 | 0 | 0 |
| 0.023263114 | 0.01393057 | 0 | 0.038104344 | 0 |
| 0 | 0 | 0 | 0 | 0 |
| 0 | 0 | 0 | 0 | 0 |

| | | | | |
|---|---|---|---|---|
| 0 | 0 | 0 | 0 | 0 |
| 0 | 0 | 0 | 0 | 0 |
| 0 | 0 | 0 | 0 | 0 |
| 0 | 0 | 0 | 0 | 0 |
| 0 | 0 | 0 | 0 | 0 |
| 0 | 0 | 0 | 0 | 0 |
| 0 | 0 | 0 | 0 | 0 |
| 0 | 0 | 1.503878689 | 0.001801021 | 0 |
| 0 | 0.026788791 | 0 | 0 | 0 |
| 0.006094064 | 0.001929188 | 0 | 0 | 0.005560146 |
| 0 | 0 | 0 | 0 | 0 |
| 0 | 0 | 0 | 0 | 0 |
| 0 | 0 | 0 | 0 | 0 |
| 0 | 0 | 0 | 0 | 0 |
| 0 | 0 | 0 | 0 | 0 |
| 0 | 0 | 0 | 0 | 0 |
| 0 | 0 | 0 | 0 | 0 |
| 0 | 0 | 0 | 0 | 0 |
| 0 | 0 | 0 | 0 | 0 |
| 0.10074116 | 0.211481845 | 0 | 4.064556185 | 0 |
| 0 | 0.000836341 | 0 | 0.009205848 | 0.013027057 |
| 0 | 0 | 0 | 0 | 0 |
| 0 | 0 | 0 | 0 | 0 |
| 0 | 0 | 0 | 0.000838625 | 0 |
| 0 | 0 | 0 | 0 | 0 |
| 0 | 0 | 0 | 0 | 0 |
| 0 | 0 | 0 | 0 | 0 |
| 0 | 0.008446423 | 0 | 0 | 0 |
| 0 | 0 | 0 | 0 | 0 |
| 0 | 0 | 0 | 0 | 0.062594871 |
| 0.318092733 | 1.340486379 | 0 | 0 | 0.123696766 |
| 0 | 0 | 0 | 0 | 0 |
| 0 | 0.000855077 | 0 | 0 | 0 |
| 0 | 0 | 0 | 0 | 0 |
| 0 | 0 | 0 | 0 | 0 |
| 0 | 0 | 0 | 0 | 0 |
| 0 | 0 | 0 | 0 | 0 |
| 0 | 0 | 0 | 0 | 0 |
| 0 | 0.03463847 | 0 | 0 | 0 |
| 0 | 0 | 0 | 0 | 0 |
| 0 | 0 | 0 | 0 | 0 |
| 0 | 0.917762009 | 0 | 0 | 0 |
| 0 | 0 | 0 | 0 | 0 |
| 0 | 0 | 0 | 0 | 0 |
| 0 | 0 | 0 | 0 | 0 |
| 0 | 0 | 0 | 0 | 0 |
| 0 | 0 | 0.34806181 | 0.0165423 | 0.000560018 |
| 0 | 0.000370111 | 0 | 0 | 0 |

| | | | | |
|---|---|---|---|---|
| 0 | 0 | 0 | 0 | 0 |
| 0 | 0 | 0 | 0 | 0 |
| 0 | 0 | 0 | 0 | 0 |
| 0 | 0 | 0 | 0 | 0 |
| 0.063050658 | 0.019710345 | 2.027838707 | 0.400763095 | 0.001338801 |
| 0 | 0 | 0 | 0 | 0 |
| 0 | 0 | 0 | 0 | 0 |
| 0 | 0 | 0 | 0 | 0 |
| 0 | 0 | 0 | 0 | 0 |
| 0 | 0 | 0 | 0 | 0 |
| 0.020509299 | 0.033405448 | 0 | 0 | 0.034846155 |
| 0.000868132 | 0.002645173 | 0.070086438 | 0.002005451 | 0.001510219 |
| 0 | 0.01211696 | 0.028712288 | 0 | 0.003608137 |
| 0 | 0 | 0 | 0 | 0 |
| 0 | 0 | 0 | 0 | 0 |
| 0.000787612 | 0 | 1.535143627 | 0.734015234 | 0.507796474 |
| 0 | 0 | 0 | 0 | 0 |
| 0 | 0 | 0 | 0 | 0 |
| 0 | 0.001335293 | 0 | 0.09060616 | 0 |
| 0.067806089 | 0.008157026 | 0 | 0.031063071 | 0.17940448 |
| 0 | 0 | 0 | 0 | 0 |
| 0 | 0.000272974 | 0 | 0 | 0 |
| 0 | 0 | 0 | 0 | 0 |
| 0 | 0 | 0 | 0 | 0 |
| 0 | 0 | 0 | 0 | 0 |
| 0 | 0 | 0 | 0 | 0 |
| 0 | 0.000299386 | 0 | 0.000706163 | 0 |
| 0 | 0 | 0 | 0 | 0 |
| 0 | 0 | 0 | 0 | 0 |
| 0 | 0 | 0 | 0 | 0 |
| 0 | 0.000169685 | 0 | 0 | 0 |
| 0 | 0 | 0 | 0 | 0 |
| 0 | 0 | 0 | 0 | 0 |
| 0 | 0 | 0 | 0 | 0 |
| 0 | 0.000991541 | 0 | 0 | 0 |
| 0 | 0 | 0 | 0 | 0 |
| 0 | 0 | 0 | 0 | 0 |
| 0.002008468 | 0.003795035 | 0.0604928 | 0.015184513 | 0.0013194 |
| 0 | 0.045670388 | 0 | 0 | 0 |
| 0 | 0 | 0 | 0 | 0 |
| 0 | 0 | 0 | 0 | 0.00197578 |
| 0 | 0.000458484 | 0 | 0 | 0 |
| 0 | 0 | 0 | 0 | 0 |
| 0 | 0 | 0 | 0.00208488 | 0 |
| 0 | 0 | 0 | 0 | 0 |
| 0 | 0 | 0 | 0 | 0 |
| 0 | 0 | 0 | 0 | 0 |
| 0 | 0 | 0 | 0 | 0 |

| | | | | |
|---|---|---|---|---|
| 0 | 0 | 0 | 0 | 0 |
| 0 | 0.016217702 | 0 | 0 | 0.032051197 |
| 0 | 0 | 0 | 0 | 0 |
| 0 | 0.000370536 | 0 | 0.003495942 | 0 |
| 0 | 0 | 0 | 0.007092817 | 0.00211304 |
| 0 | 0 | 0 | 0 | 0.00136354 |
| 0 | 0.000958873 | 0 | 0 | 0.000442174 |
| 0 | 0.000893524 | 0 | 0.001879714 | 0 |
| 0 | 0 | 0 | 0 | 0 |
| 0 | 0.008563525 | 0 | 0 | 0 |
| 0 | 0.040619871 | 0 | 0 | 0.00160017 |
| 0 | 0 | 0 | 0 | 0 |
| 0.002011404 | 0.048350978 | 0 | 0.003252547 | 0.037790019 |
| 0 | 0.000896042 | 0 | 0 | 0 |
| 0 | 0 | 0 | 0 | 0 |
| 0 | 0.005644357 | 0 | 0 | 0 |
| 0 | 0 | 0 | 0 | 0 |
| 0 | 0 | 0 | 0 | 0 |
| 0 | 0 | 0 | 0 | 0 |
| 0 | 0 | 0 | 0 | 0 |
| 0 | 0 | 0 | 0 | 0 |
| 0 | 0 | 0 | 0 | 0 |
| 0 | 0 | 0 | 0.005098637 | 0.006911215 |
| 0 | 0.044111718 | 0.033801377 | 0.126829226 | 0.005269689 |
| 0 | 0 | 0 | 0 | 0 |
| 0 | 0.008539827 | 0 | 0 | 0 |
| 0 | 0.000413049 | 0 | 0 | 0 |
| 0 | 0 | 0 | 0 | 0 |
| 0 | 0 | 0 | 0 | 0 |
| 0 | 0 | 0 | 0 | 0 |
| 0.0057586 | 0.010926256 | 0 | 0 | 0.018375784 |
| 0 | 0 | 0 | 0 | 0 |
| 0 | 0 | 0 | 0 | 0 |
| 0 | 0 | 0 | 0 | 0 |
| 0 | 0.008028758 | 0 | 0 | 0 |
| 0 | 0 | 0 | 0 | 0 |
| 0 | 0 | 0 | 0 | 0 |
| 0 | 0 | 0 | 0 | 0 |
| 0.001624058 | 0.000435029 | 0 | 0 | 0.00090274 |
| 0 | 0 | 0 | 0 | 0 |
| 0 | 0 | 0 | 0 | 0 |
| 0 | 0 | 0 | 0 | 0 |
| 0 | 0 | 0 | 0 | 0 |
| 0 | 0 | 0 | 0 | 0 |
| 0 | 0 | 0 | 0 | 0 |
| 0.769079514 | 0.117816415 | 0.204263818 | 0.334657812 | 0.424577161 |
| 0 | 0 | 0 | 0 | 0 |
| 0 | 0 | 0 | 0 | 0 |

| | | | | |
|---|---|---|---|---|
| 0 | 0 | 0 | 0 | 0 |
| 0 | 0 | 0 | 0 | 0 |
| 0.10383562 | 0.149775733 | 0 | 0 | 0 |
| 0 | 0.00772911 | 0 | 0 | 0 |
| 0 | 0 | 0 | 0 | 0 |
| 0 | 0 | 0 | 0 | 0 |
| 0 | 0 | 0 | 0 | 0 |
| 0 | 0 | 0 | 0 | 0 |
| 0 | 0 | 0 | 0 | 0 |
| 0 | 0 | 0 | 0 | 0 |
| 0 | 0.000220578 | 0.002470101 | 0 | 0 |
| 0 | 0 | 0 | 0 | 0 |
| 0 | 0 | 0 | 0 | 0 |
| 0 | 0 | 0 | 0 | 0 |
| 0 | 0 | 0 | 0 | 0 |
| 0 | 0 | 0 | 0 | 0 |
| 0 | 0.007781773 | 0 | 0 | 0 |
| 0 | 0 | 0 | 0 | 0 |
| 0 | 0 | 0 | 0 | 0 |
| 0 | 0 | 0.001228417 | 0 | 0 |
| 0 | 0 | 0 | 0 | 0 |
| 0 | 0 | 0 | 0 | 0 |
| 0 | 0 | 0 | 0 | 0 |
| 0 | 0 | 0 | 0 | 0 |
| 0 | 0 | 0 | 0 | 0 |
| 0 | 0 | 0 | 0 | 0 |
| 0 | 0 | 0 | 0 | 0 |
| 0.017105584 | 0.002168978 | 0 | 0 | 0 |
| 0 | 0 | 0 | 0 | 0 |
| 0 | 0 | 0 | 0 | 0 |
| 0.000346409 | 0.001105759 | 0.002814233 | 0.006046188 | 0.00139066 |
| 0 | 0 | 0 | 0 | 0 |
| 1.07820706 | 0.013743559 | 0.013274669 | 0.033655856 | 0.022821063 |
| 0.000959049 | 0 | 0.001669572 | 0.005485942 | 0.001650046 |
| 0 | 0 | 0 | 0 | 0 |
| 0 | 0 | 0 | 0 | 0 |
| 0 | 0 | 0 | 0 | 0 |
| 0 | 0 | 0 | 0 | 0 |
| 0 | 0 | 0 | 0 | 0 |
| 0 | 0 | 0 | 0 | 0 |
| 0 | 0 | 0 | 0 | 0 |
| 0 | 0 | 0.112737805 | 0.068456515 | 0.037372243 |
| 0.011421984 | 0.026145704 | 0 | 0.023798818 | 0.020607148 |
| 0 | 0 | 0 | 0 | 0 |
| 0 | 0 | 0 | 0 | 0 |
| 0.002718563 | 0.012256023 | 0 | 0.0086725 | 0.0063923 |
| 0 | 0 | 0 | 0 | 0 |
| 0 | 0 | 0 | 0 | 0 |

| | | | | |
|---|---|---|---|---|
| 0 | 0 | 0 | 0 | 0 |
| 0.010417496 | 0.006035845 | 0 | 0.012141004 | 0 |
| 0.024337472 | 0.139077456 | 0.005510724 | 0.015194973 | 0.093081798 |
| 0 | 0 | 0 | 0 | 0 |
| 0.007352762 | 0.008172759 | 0 | 0.296840714 | 0.002740929 |
| 0 | 0 | 0 | 0.02499711 | 0 |
| 0 | 0 | 0 | 0 | 0 |
| 0 | 0 | 0 | 0 | 0 |
| 0 | 0 | 0 | 0 | 0 |
| 0 | 0 | 0 | 0 | 0 |
| 0 | 0 | 0 | 0 | 0 |
| 0 | 0 | 0 | 0 | 0 |
| 0 | 0 | 0 | 0 | 0 |
| 0 | 0 | 0 | 0 | 0 |
| 0 | 0 | 0 | 0 | 0 |
| 0 | 0 | 0 | 0 | 0 |
| 0 | 0 | 0 | 0 | 0 |
| 0 | 0 | 0 | 0 | 0 |
| 0 | 0 | 0 | 0 | 0 |
| 0 | 0 | 0 | 0 | 0 |
| 0 | 0 | 0 | 0 | 0 |
| 0 | 0 | 0.001208929 | 0 | 0 |
| 0 | 0 | 0 | 0 | 0 |
| 0 | 0 | 0 | 0 | 0 |
| 0 | 0.00350996 | 0 | 0.002295846 | 0.00108812 |
| 0.030402387 | 0.0680373 | 0 | 0 | 0 |
| 0 | 0 | 0 | 0 | 0 |
| 0.003702953 | 0.010102701 | 0 | 0.000814676 | 0.000424729 |
| 0 | 0 | 0 | 0 | 0 |
| 0 | 0 | 0 | 0 | 0 |
| 0 | 0 | 0 | 0 | 0 |
| 0 | 0 | 0.008923627 | 0.000552369 | 0 |
| 0 | 0 | 0 | 0 | 0 |
| 0 | 0 | 0 | 0 | 0 |
| 0 | 0 | 0 | 0 | 0 |
| 0 | 0 | 0 | 0 | 0 |
| 0 | 0 | 0 | 0.000799816 | 0 |
| 0 | 0 | 0.034935452 | 0 | 0 |
| 0 | 0.917536128 | 0 | 4.693063378 | 1.403787727 |
| 0 | 0 | 0 | 0 | 0 |
| 0 | 0 | 0 | 0 | 0 |
| 0.006359739 | 0.008369208 | 0 | 0 | 0.001543747 |
| 0 | 0.000387125 | 0.044810329 | 0.001475028 | 0 |
| 0 | 0 | 0 | 0 | 0 |
| 0 | 0 | 0 | 0 | 0 |
| 0 | 0.000308335 | 0 | 0 | 0 |
| 0.002302481 | 0.000546669 | 0.02518852 | 0.001611789 | 0 |
| 0 | 0 | 0 | 0 | 0 |

| | | | | |
|---|---|---|---|---|
| 0 | 0 | 0 | 0 | 0 |
| 0.001584713 | 0.002241838 | 0.239983869 | 0.024202 | 0.00349901 |
| 0 | 0 | 0 | 0 | 0 |
| 0 | 0 | 0 | 0 | 0 |
| 0 | 0 | 0 | 0 | 0 |
| 0 | 0 | 0 | 0 | 0 |
| 0 | 0 | 0 | 0 | 0 |
| 0 | 0 | 0 | 0 | 0 |
| 0 | 0 | 0 | 0 | 0 |
| 0 | 0 | 0 | 0 | 0 |
| 0 | 0 | 0 | 0 | 0 |
| 0 | 0 | 0 | 0 | 0 |
| 0 | 0.087075733 | 0 | 0 | 0 |
| 0 | 0 | 0 | 0 | 0 |
| 0 | 0 | 0 | 0.002535819 | 0 |
| 0 | 0 | 0 | 0 | 0 |
| 0 | 0 | 0 | 0 | 0 |
| 0 | 0 | 0 | 0 | 0 |
| 0 | 0 | 0 | 0 | 0 |
| 0 | 0 | 0 | 0 | 0 |
| 0 | 0 | 0 | 0 | 0 |
| 0.012936587 | 0.001900482 | 0.010591816 | 0.004233636 | 0.036743291 |
| 0 | 0 | 0 | 0 | 0 |
| 0.000666231 | 0 | 0 | 0.001282537 | 0 |
| 0 | 0 | 0 | 0 | 0 |
| 0 | 0 | 0 | 0 | 0 |
| 0 | 0 | 0 | 0 | 0 |
| 0 | 0 | 0 | 0 | 0 |
| 0 | 0 | 0 | 0 | 0 |
| 0 | 0.038798556 | 0 | 0 | 0 |
| 0 | 0 | 0 | 0 | 0 |
| 0 | 0 | 0 | 0 | 0 |
| 0 | 0 | 0 | 0 | 0 |
| 0 | 0 | 0 | 0 | 0 |
| 3.400527184 | 0.00022368 | 0 | 0 | 0.000353649 |
| 0 | 0 | 0 | 0 | 0 |
| 0 | 0 | 0 | 0 | 0 |
| 0 | 0 | 0 | 0 | 0 |
| 0.036656945 | 0.006009893 | 0 | 0.009699067 | 0.028913267 |
| 0 | 0 | 0 | 0 | 0 |
| 0.036487601 | 0.214096964 | 0.515216606 | 0.01449416 | 0.015926753 |
| 0 | 0 | 0 | 0 | 0 |
| 0 | 0 | 0 | 0 | 0 |
| 0 | 0 | 0 | 0 | 0 |
| 0 | 0 | 0 | 0 | 0 |
| 0.039123387 | 0 | 1.984306298 | 2.323020454 | 0.010248896 |
| 0 | 0 | 0 | 0 | 0 |
| 0 | 0.000320833 | 0 | 0 | 0 |

| | | | | |
|---|---|---|---|---|
| 0 | 0 | 0 | 0 | 0 |
| 0 | 0 | 0 | 0 | 0 |
| 0 | 0 | 0 | 0 | 0 |
| 0 | 0.068229168 | 0 | 0 | 0 |
| 0 | 0.000136819 | 0 | 0 | 0 |
| 0 | 0 | 0 | 0 | 0 |
| 0 | 0 | 0 | 0 | 0 |
| 0.01021335 | 0 | 0 | 0 | 0.001582515 |
| 0 | 0.002346395 | 0 | 0 | 0 |
| 0.012646158 | 0.014573177 | 0.011854343 | 0.001747821 | 0.003905235 |
| 0 | 0.03576292 | 0 | 0 | 0 |
| 0 | 0 | 0 | 0 | 0 |
| 0 | 0 | 0 | 0 | 0 |
| 0 | 0 | 0 | 0 | 0 |
| 0 | 0 | 0 | 0 | 0 |
| 0 | 0 | 0 | 0 | 0 |
| 0 | 0 | 0 | 0 | 0 |
| 0 | 0 | 0 | 0 | 0 |
| 0 | 0 | 0 | 0 | 0 |
| 0.139073604 | 0.012340537 | 0.368686575 | 0.253714662 | 0.03929698 |
| 0 | 0 | 0 | 0 | 0 |
| 0 | 0 | 0 | 0 | 0 |
| 0 | 0.027571604 | 0 | 0 | 0 |
| 0 | 0 | 0 | 0 | 0 |
| 0 | 0 | 0 | 0 | 0 |
| 0 | 0.065716102 | 0 | 0 | 0 |
| 0 | 0 | 0 | 0 | 0 |
| 0 | 0 | 0 | 0 | 0 |
| 0 | 0 | 0 | 0 | 0 |
| 0 | 0 | 0 | 0.003018772 | 0.00096851 |
| 0 | 0 | 0 | 0 | 0 |
| 0 | 0.001232395 | 0 | 0 | 0 |
| 0 | 0 | 0 | 0 | 0 |
| 0 | 0 | 0 | 0 | 0 |
| 0 | 0 | 0 | 0 | 0 |
| 0.001095503 | 0 | 0 | 0 | 0 |
| 0 | 0 | 0 | 0 | 0 |
| 0 | 0 | 0 | 0 | 0 |
| 0 | 0 | 0 | 0 | 0 |
| 0 | 0.022506163 | 0 | 0 | 0 |
| 0 | 0 | 0 | 0 | 0 |
| 0 | 0 | 0 | 0 | 0 |
| 0 | 0.012401391 | 0 | 0 | 0 |
| 0 | 0 | 0 | 0 | 0 |
| 0 | 0.00305625 | 0 | 0 | 0 |
| 0 | 0.012672098 | 0 | 0 | 0 |
| 0 | 0 | 0 | 0 | 0 |
| 0 | 0 | 0 | 0 | 0 |

| | | | | |
|---|---|---|---|---|
| 0 | 0 | 0 | 0 | 0 |
| 0 | 0 | 0 | 0 | 0 |
| 0 | 0 | 0 | 0 | 0 |
| 0 | 0 | 0 | 0 | 0 |
| 0 | 0 | 0 | 0 | 0 |
| 0.020670053 | 0.012096837 | 0 | 0.001066174 | 0.000555847 |
| 0 | 0 | 0 | 0 | 0 |
| 0 | 0 | 0 | 0 | 0 |
| 0 | 0 | 0 | 0 | 0 |
| 0 | 0 | 0 | 0 | 0 |
| 0 | 0 | 0 | 0 | 0 |
| 0 | 0 | 0 | 0 | 0 |
| 0 | 0 | 0 | 0 | 0 |
| 0 | 0.007155227 | 0 | 0 | 0 |
| 0 | 0 | 0 | 0 | 0 |
| 0 | 0 | 0 | 0 | 0 |
| 0 | 0.002088737 | 0 | 0 | 0 |
| 0 | 0 | 0 | 0 | 0 |
| 0 | 0 | 0 | 0 | 0 |
| 0 | 0 | 0 | 0 | 0 |
| 0.001720096 | 0 | 0 | 0 | 0 |
| 0 | 0 | 0 | 0 | 0 |
| 0 | 0 | 0 | 0 | 0 |
| 0 | 0 | 0 | 0 | 0 |
| 0 | 0 | 0 | 0 | 0 |
| 0.000464088 | 0.000303013 | 0 | 0 | 0 |
| 0 | 0 | 0 | 0 | 0 |
| 0 | 0.262036704 | 0 | 0 | 0 |
| 0 | 0 | 0 | 0 | 0 |
| 0 | 0 | 0 | 0 | 0 |
| 0 | 0 | 0 | 0 | 0 |
| 0 | 0 | 0 | 0 | 0 |
| 0 | 0 | 0 | 0 | 0 |
| 0 | 0 | 0 | 0 | 0 |
| 0 | 0 | 0 | 0 | 0 |
| 0 | 0 | 0 | 0 | 0 |
| 0 | 0 | 0 | 0 | 0 |
| 0 | 0 | 0 | 0 | 0 |
| 0 | 0.626455894 | 0 | 2.097111131 | 0.001457166 |
| 0 | 0 | 0 | 0 | 0 |
| 0 | 0.000354142 | 0 | 0 | 0 |
| 0 | 0 | 0 | 0 | 0 |
| 0 | 0 | 0 | 0 | 0 |
| 0 | 0 | 0 | 0 | 0 |
| 0 | 0 | 0 | 0 | 0 |
| 0 | 0 | 0 | 0 | 0 |
| 0 | 0 | 0 | 0 | 0 |
| 0 | 0 | 0 | 0 | 0 |

| | | | | |
|---|---|---|---|---|
| 0 | 0 | 0 | 0 | 0 |
| 0 | 0.002144771 | 0 | 0.00255786 | 0.000622315 |
| 0 | 0 | 0 | 0 | 0 |
| 0 | 0 | 0 | 0 | 0 |
| 0 | 0 | 0 | 0 | 0 |
| 0 | 0.000660926 | 0 | 0 | 0 |
| 0 | 0 | 0 | 0 | 0 |
| 0 | 0 | 0 | 0 | 0 |
| 0 | 0 | 0 | 0 | 0 |
| 0 | 0 | 0 | 0 | 0 |
| 0.001884835 | 0.000315551 | 0 | 0.002232877 | 0.056109813 |
| 0 | 0 | 0 | 0 | 0 |
| 0.019649444 | 0.070051251 | 0 | 0 | 0.015433586 |
| 0 | 0 | 0 | 0 | 0 |
| 0 | 0 | 0 | 0 | 0 |
| 0 | 0 | 0 | 0.005231097 | 0 |
| 0 | 0 | 0 | 0 | 0 |
| 0 | 0 | 0 | 0 | 0 |
| 0 | 0 | 0 | 0 | 0 |
| 0 | 0.000472634 | 0 | 0 | 0 |
| 0.000588613 | 0 | 0 | 0.003399354 | 0.000472598 |
| 0 | 0 | 0 | 0 | 0 |
| 0 | 0 | 0 | 0 | 0 |
| 1.603008399 | 0.778625735 | 3.243957799 | 0.861139902 | 1.761946447 |
| 0 | 0 | 0 | 0 | 0 |
| 0 | 0 | 0 | 0 | 0 |
| 0 | 0 | 0 | 0 | 0 |
| 0 | 0 | 0 | 0 | 0 |
| 0 | 0 | 0 | 0 | 0 |
| 0 | 0 | 0 | 0 | 0 |
| 0 | 0.007701344 | 0 | 0 | 0 |
| 0 | 0 | 0 | 0 | 0 |
| 0 | 0.000202133 | 0 | 0 | 0 |
| 0 | 0 | 0 | 0 | 0 |

| | | | | |
|---|---|---|---|---|
| PHL018-0178_V1 | PHL018-0051_V1 | PHL018-0204_V3 | PHL018-0042_V1 | PHL018-0194_V1 |
| EYCF | EYCF | EYCF | CTRL | EYCF |
| 0.012885561 | 0.045349168 | 1.603692937 | 0.183246437 | 0.318245156 |
| 0.307717537 | 27.45294158 | 5.220028159 | 0.014173476 | 12.68541169 |
| 29.67482749 | 0 | 0 | 0 | 6.244735564 |
| 0 | 0 | 0 | 0 | 0 |
| 0 | 0 | 0 | 0 | 0 |
| 0 | 0 | 0 | 0 | 0 |
| 0 | 0 | 0 | 0 | 0 |
| 0 | 0 | 0 | 0 | 0 |
| 0 | 0 | 0 | 0 | 0 |
| 0 | 0 | 0 | 0 | 0 |
| 0 | 0 | 0 | 0 | 0 |
| 0 | 0 | 0 | 0 | 0 |
| 0 | 0 | 0 | 0 | 0 |
| 6.241069684 | 0 | 0.027967337 | 0.176438291 | 8.72696655 |
| 0 | 0 | 0 | 0 | 0 |
| 0 | 0 | 0 | 0 | 0 |
| 0 | 0.003285471 | 0 | 0 | 0 |
| 0 | 0 | 0 | 0 | 0 |
| 0 | 0 | 0 | 0 | 0 |
| 0 | 0 | 0 | 0 | 0 |
| 0 | 0 | 0 | 0 | 0 |
| 0 | 0 | 0.00406117 | 0 | 0 |
| 0 | 0 | 0 | 0 | 0 |
| 0 | 0 | 0 | 0 | 0 |
| 0 | 0 | 0 | 0.038015268 | 0 |
| 0 | 0 | 0 | 0 | 0 |
| 0 | 0 | 0 | 0 | 0 |
| 0 | 0 | 0 | 0 | 0 |
| 0 | 0 | 0 | 0 | 0 |
| 0 | 0 | 0 | 0 | 0 |
| 0 | 0 | 0 | 0 | 0 |
| 0 | 0 | 0 | 0 | 0 |
| 0 | 0 | 0 | 0 | 0 |
| 0 | 0 | 0 | 0 | 0 |
| 0 | 0 | 0 | 0 | 0 |
| 1.351290455 | 0 | 0 | 3.071217595 | 0 |
| 0 | 0 | 0 | 0 | 0 |
| 0 | 0 | 0 | 0 | 0 |
| 0 | 0 | 0 | 0 | 0 |
| 0 | 0 | 0 | 0 | 0 |
| 0 | 0 | 0 | 0 | 0 |
| 0.001261613 | 0 | 0.017285142 | 0.251616506 | 0.002760132 |
| 0.018906666 | 0.079614718 | 1.341136489 | 2.078407327 | 0.030005751 |
| 0 | 0.105017678 | 0.003792418 | 0.001420937 | 0.104270466 |
| 0 | 0.001517315 | 0 | 0.003352731 | 0 |
| 0 | 0 | 0 | 0 | 0 |

| | | | | |
|---|---|---|---|---|
| 0 | 0 | 0 | 0.020277659 | 0.006506101 |
| 0.063033817 | 0.360943184 | 0 | 0.000830373 | 0.00290937 |
| 0.001649323 | 0 | 0.001909614 | 0 | 0.03552844 |
| 0.136512316 | 2.057828466 | 0.241854001 | 0.141905191 | 0.655406451 |
| 0.000591735 | 0 | 0.006993936 | 0.001662714 | 0.00809116 |
| 0 | 0 | 0.207990569 | 0.019873283 | 0 |
| 0 | 0 | 0.00275051 | 0.003026089 | 0.022308379 |
| 0 | 0 | 0 | 0 | 0 |
| 0 | 0 | 0 | 0.013623674 | 0 |
| 0 | 0 | 0 | 0 | 0 |
| 0.001047704 | 0 | 0 | 0.0068692 | 0 |
| 0.012473322 | 0 | 0.09394947 | 0.032033823 | 0.087012963 |
| 0 | 0.001507428 | 0 | 0 | 0 |
| 0.001735357 | 0.028246626 | 0.141039729 | 0.012238249 | 0 |
| 0.060083286 | 0.005984163 | 0 | 0.002973401 | 0 |
| 0 | 0 | 0 | 0 | 0 |
| 0 | 0 | 0 | 0.01135966 | 0.788007137 |
| 0 | 0 | 0 | 0 | 0 |
| 0 | 0 | 0 | 0 | 0 |
| 0 | 0 | 0 | 0 | 0 |
| 0.094922908 | 0.613419997 | 0.012905314 | 0 | 0.072248996 |
| 0.006886805 | 0.003503044 | 0.013875727 | 0.00809463 | 0.008692395 |
| 0 | 0 | 0.035291305 | 0 | 0 |
| 0 | 0 | 0 | 0 | 0 |
| 0 | 0 | 0 | 0 | 0 |
| 0 | 0 | 0 | 0 | 0 |
| 0.007497521 | 0 | 0 | 0 | 0 |
| 0 | 0 | 0 | 0 | 0 |
| 0 | 0 | 0 | 0.016342072 | 0 |
| 0 | 0 | 0 | 0 | 0 |
| 0.048657181 | 0.847336291 | 0 | 0 | 0.176517457 |
| 0 | 0 | 0 | 0 | 0 |
| 0 | 0 | 0 | 0 | 0 |
| 0 | 0 | 0 | 0 | 0 |
| 0 | 0 | 0 | 0 | 0 |
| 0 | 0 | 0 | 0 | 0 |
| 0 | 0 | 0 | 0.001105608 | 0 |
| 0 | 0 | 0 | 0 | 0.014260599 |
| 0 | 0 | 0 | 0 | 0 |
| 0 | 0 | 0 | 0 | 0 |
| 0 | 0 | 0 | 0 | 0 |
| 0 | 0 | 0 | 0 | 0 |
| 0 | 0 | 0 | 0 | 0 |
| 0 | 0 | 0 | 0 | 0 |
| 0 | 0 | 0 | 0 | 0 |
| 0 | 0 | 0 | 0 | 0 |
| 0 | 0 | 0 | 0 | 0.012643249 |
| 0 | 0 | 0 | 0 | 0 |

| | | | | |
|---|---|---|---|---|
| 0 | 0 | 0 | 0 | 0 |
| 0 | 0 | 0 | 0 | 0 |
| 0 | 0 | 0 | 0 | 0 |
| 0 | 0 | 0 | 0 | 0 |
| 0 | 0.073180102 | 0.017573824 | 0.873439413 | 0.020370437 |
| 0 | 0 | 0 | 0.001243669 | 0 |
| 0 | 0 | 0 | 0 | 0 |
| 0 | 0 | 0 | 0.002874145 | 0 |
| 0 | 0 | 0 | 0 | 0 |
| 0 | 0 | 0 | 0 | 0 |
| 0 | 0 | 0 | 0 | 0 |
| 0 | 0 | 0 | 0.028102445 | 0 |
| 0 | 0 | 0 | 0 | 0 |
| 0 | 0 | 0 | 0 | 0 |
| 0 | 0 | 0 | 0 | 0 |
| 0 | 0 | 0 | 0 | 0 |
| 0 | 0 | 0.000724682 | 0 | 0 |
| 0 | 0 | 0 | 0 | 0 |
| 0 | 0 | 0 | 0.587479122 | 0 |
| 0 | 0 | 0.020148246 | 0 | 0 |
| 0 | 0 | 0 | 0 | 0 |
| 0 | 0.95046991 | 0 | 0 | 0 |
| 0 | 0 | 0 | 0 | 0 |
| 0 | 0 | 0 | 0 | 0 |
| 0 | 0 | 0 | 0 | 0 |
| 0 | 0 | 0 | 0 | 0 |
| 0 | 0 | 0 | 0 | 0 |
| 0 | 0 | 0 | 0 | 0 |
| 0 | 0 | 0 | 0 | 0 |
| 0 | 0 | 0.001531217 | 0 | 0 |
| 0 | 0 | 0.019752803 | 0.036992837 | 0.228977113 |
| 0 | 0 | 0.354848354 | 0.003441751 | 0 |
| 0 | 0 | 0 | 0 | 0 |
| 0 | 0 | 0 | 0 | 0 |
| 0 | 0 | 0 | 0 | 0 |
| 0 | 0 | 0 | 0 | 0 |
| 0 | 0 | 0 | 0 | 0 |
| 0 | 0 | 0 | 0 | 0 |
| 0 | 0 | 0 | 0 | 0 |
| 0 | 0 | 0 | 0 | 0 |
| 0 | 0 | 0 | 0 | 0 |
| 0 | 0 | 0 | 0 | 0 |
| 0 | 0 | 0 | 0.000548706 | 0 |
| 0 | 0 | 0 | 0 | 0 |
| 0 | 0 | 0 | 0 | 0 |
| 0 | 0 | 0 | 0 | 0 |
| 0 | 0 | 0 | 0 | 0 |
| 0 | 0 | 0 | 0 | 0 |

| | | | | |
|---|---|---|---|---|
| 0 | 0 | 0 | 0 | 0 |
| 0 | 0 | 0 | 0 | 0 |
| 0 | 0 | 0 | 0 | 0 |
| 0 | 0 | 0 | 0 | 0 |
| 0 | 0 | 0.038626869 | 0.036670945 | 0 |
| 0 | 0 | 0 | 0 | 0 |
| 0 | 0 | 0 | 0 | 0 |
| 0 | 0 | 0 | 0 | 0 |
| 0 | 0 | 0 | 0 | 0 |
| 0 | 0 | 0 | 0 | 0 |
| 0 | 0 | 0 | 0 | 0 |
| 0 | 0 | 0 | 0 | 0 |
| 0 | 0 | 0 | 0 | 0 |
| 0 | 0 | 0 | 0 | 0 |
| 0 | 0 | 0 | 0 | 0 |
| 0 | 0.00068403 | 0.626690653 | 2.942459216 | 0 |
| 0 | 0 | 0 | 0 | 0 |
| 0 | 0 | 0 | 0 | 0 |
| 0 | 0 | 0 | 0 | 0 |
| 0 | 0 | 0 | 0 | 0 |
| 0 | 0 | 0.01200897 | 0 | 0 |
| 0 | 0 | 0 | 0 | 0 |
| 0 | 0 | 0 | 0 | 0 |
| 0 | 0 | 0 | 0 | 0 |
| 0 | 0 | 0 | 0.014472845 | 0 |
| 0 | 0 | 0 | 0 | 0 |
| 0 | 0 | 0.003585677 | 0.012763022 | 0 |
| 0 | 0 | 0 | 0 | 0 |
| 0 | 0 | 0 | 0 | 0 |
| 0 | 0 | 0 | 0 | 0 |
| 0 | 0 | 0.02211358 | 0 | 0 |
| 0 | 0 | 0 | 0.47652454 | 0 |
| 0 | 0 | 0 | 0 | 0 |
| 0 | 0 | 0 | 0 | 0 |
| 0 | 0 | 0 | 0 | 0 |
| 0 | 0 | 0 | 0 | 0 |
| 0 | 0 | 0 | 0 | 0 |
| 0 | 0 | 0 | 0.005425575 | 0 |
| 0 | 0 | 0 | 0 | 0 |
| 0 | 0 | 0 | 0 | 0 |
| 0 | 0 | 0 | 0 | 0 |
| 0 | 0 | 0 | 0 | 0 |
| 0 | 0 | 0 | 0 | 0 |
| 0 | 0 | 0 | 0 | 0 |
| 0 | 0 | 0 | 0 | 0 |
| 0 | 0 | 0 | 0 | 0 |
| 0 | 0 | 0 | 0 | 0 |
| 0 | 0 | 0 | 0 | 0 |

| | | | | |
|---|---|---|---|---|
| 0 | 0 | 0.379522705 | 0 | 0 |
| 0 | 0 | 0 | 0 | 0 |
| 0 | 0 | 0 | 0 | 0 |
| 0 | 0 | 0 | 0 | 0 |
| 0 | 0 | 0 | 0 | 0 |
| 0 | 0 | 0 | 0 | 0 |
| 0 | 0 | 0 | 0 | 0 |
| 0 | 0 | 0 | 0 | 0 |
| 0 | 0 | 0 | 0 | 0 |
| 0 | 0 | 0 | 0 | 0 |
| 0 | 0 | 0 | 0 | 0 |
| 0 | 0 | 0.004107715 | 0 | 0 |
| 0 | 0 | 0.000809867 | 0.000393092 | 0.000529721 |
| 0 | 0 | 0 | 0 | 0 |
| 0 | 0 | 0 | 0 | 0 |
| 0 | 0 | 0 | 0 | 0 |
| 0 | 0 | 0 | 0 | 0 |
| 0 | 0 | 0 | 0.00047275 | 0 |
| 0 | 0 | 0 | 0 | 0 |
| 0 | 0 | 0 | 0 | 0 |
| 0.017852232 | 0.017163559 | 0.024228115 | 0.003919938 | 0.025421563 |
| 0 | 0 | 0 | 0 | 0 |
| 0 | 0 | 0 | 0 | 0 |
| 0 | 0 | 0 | 0 | 0 |
| 0 | 0 | 0 | 0 | 0 |
| 0 | 0 | 0 | 0 | 0 |
| 0.000571248 | 0 | 0.010582412 | 0 | 0.003845433 |
| 0 | 0 | 0 | 0 | 0 |
| 0 | 0 | 0 | 0 | 0 |
| 0.00061117 | 0 | 0 | 0 | 0 |
| 0.146801006 | 0.072296698 | 0 | 0 | 0 |
| 0 | 0 | 0 | 0 | 0 |
| 0 | 0 | 0 | 0 | 0 |
| 0 | 0 | 0.008097425 | 0.001646991 | 0 |
| 0.003191103 | 0.005713618 | 0.00960625 | 0.003985189 | 0.027066475 |
| 0 | 0 | 0 | 0 | 0 |
| 0 | 0 | 0 | 0 | 0 |
| 0 | 0 | 0 | 0 | 0 |
| 0 | 0 | 0 | 0.000463525 | 0 |
| 0 | 0 | 0 | 0 | 0 |
| 0 | 0 | 0 | 0 | 0 |
| 0 | 0 | 0 | 0 | 0 |
| 0 | 0 | 0 | 0.001159804 | 0 |
| 0 | 0 | 0 | 0 | 0 |
| 0 | 0 | 0 | 0 | 0 |
| 0 | 0 | 0 | 0.042041219 | 0 |
| 0 | 0 | 0.00293362 | 0 | 0 |
| 0 | 0 | 0.002984012 | 0.001135981 | 0.000918491 |

| | | | | |
|---|---|---|---|---|
| 0 | 0 | 0 | 0 | 0 |
| 0 | 0 | 0 | 0 | 0 |
| 0 | 0 | 0 | 0 | 0 |
| 0 | 0 | 0.001115608 | 0 | 0 |
| 0 | 0 | 0 | 0 | 0 |
| 0 | 0 | 0 | 0 | 0 |
| 0 | 0 | 0 | 0.005659693 | 0 |
| 0 | 0 | 0 | 0.004898982 | 0 |
| 0 | 0 | 0 | 0.006939428 | 0.002111603 |
| 0 | 0 | 0 | 0 | 0 |
| 0 | 0 | 0 | 0.000843298 | 0 |
| 0 | 0 | 0 | 0 | 0 |
| 0 | 0 | 0 | 0 | 0 |
| 0 | 0 | 0 | 0 | 0 |
| 0 | 0 | 0 | 0.007164264 | 0 |
| 0 | 0 | 0 | 0 | 0 |
| 0 | 0 | 0 | 0 | 0 |
| 0 | 0 | 0 | 0 | 0 |
| 0 | 0 | 0.010532264 | 0.013285783 | 0 |
| 0 | 0 | 0 | 0.03230326 | 0 |
| 0.025654352 | 0.003331748 | 0 | 0 | 0 |
| 0 | 0 | 0 | 0 | 0 |
| 0 | 0 | 0.001462117 | 0 | 0 |
| 0 | 0 | 0 | 0 | 0 |
| 0 | 0 | 0.003662052 | 0 | 0 |
| 0 | 0 | 0 | 0 | 0 |
| 0 | 0 | 0 | 0 | 0 |
| 0 | 0 | 0 | 0 | 0 |
| 0 | 0 | 0 | 0.040226012 | 0 |
| 0 | 0 | 0 | 0 | 0 |
| 0.017358557 | 0.062879052 | 0.248224166 | 0.026063875 | 0.279126296 |
| 0 | 0 | 0 | 0 | 0 |
| 0 | 0 | 0.029612805 | 0 | 0 |
| 0 | 0 | 0.010521182 | 0 | 0.002184676 |
| 0 | 0 | 0 | 0 | 0 |
| 0 | 0 | 0 | 0 | 0 |
| 0 | 0 | 0 | 0 | 0 |
| 0 | 0 | 0 | 0 | 0 |
| 0 | 0 | 0 | 0 | 0 |
| 0 | 0 | 0 | 0 | 0 |
| 0 | 0 | 0 | 0 | 0 |
| 0.003579744 | 0 | 0.006907811 | 0.006386487 | 0.01239301 |
| 0 | 0 | 0 | 0 | 0 |
| 0 | 0 | 0 | 0 | 0 |
| 0 | 0 | 0 | 0 | 0 |
| 0 | 0 | 0 | 0 | 0 |
| 0 | 0 | 0 | 0 | 0 |
| 0 | 0 | 0 | 0 | 0 |

| | | | | |
|---|---|---|---|---|
| 0 | 0 | 0 | 0 | 0 |
| 0 | 0 | 0 | 0 | 0 |
| 0 | 0.119938326 | 0 | 0.0011227 | 0.106768926 |
| 0 | 0 | 0 | 0 | 0 |
| 0 | 0.004308562 | 1.770024874 | 0.084208226 | 0 |
| 0 | 0 | 0 | 0 | 0 |
| 0 | 0 | 0 | 0 | 0 |
| 0 | 0 | 0 | 0 | 0 |
| 0 | 0 | 0 | 0 | 0 |
| 0 | 0 | 0 | 0 | 0 |
| 0 | 0 | 0 | 0 | 0 |
| 0 | 0 | 0 | 0 | 0 |
| 0 | 0 | 0 | 0 | 0 |
| 0 | 0.002599088 | 0.001643352 | 0.000531766 | 0.0033441 |
| 0.002694429 | 0.003813702 | 0.002486682 | 0 | 0.001380058 |
| 0 | 0 | 0 | 0.003445958 | 0 |
| 0 | 0.007061228 | 0.005151547 | 0 | 0 |
| 0 | 0 | 0 | 0 | 0 |
| 0 | 0 | 0 | 0 | 0 |
| 0 | 0 | 0 | 0.000849072 | 0 |
| 0 | 0 | 0 | 0 | 0 |
| 0 | 0 | 0.00395282 | 0 | 0 |
| 0 | 0 | 0 | 0 | 0 |
| 0 | 0 | 0.015499662 | 0 | 0 |
| 0 | 0 | 0 | 0 | 0 |
| 0 | 0 | 0 | 0 | 0 |
| 0 | 0 | 0 | 0.154136036 | 0 |
| 0 | 0 | 0 | 0 | 0 |
| 0 | 0 | 0 | 0 | 0 |
| 0 | 0 | 0 | 0 | 0 |
| 0 | 0 | 0 | 0 | 0 |
| 0 | 0 | 0 | 0 | 0 |
| 0 | 0 | 0 | 0.002802841 | 0 |
| 0 | 0 | 0 | 0.000520508 | 0 |
| 0 | 0 | 0 | 0 | 0 |
| 0 | 0 | 0.057124066 | 0.030657292 | 0 |
| 0 | 0 | 0 | 0 | 0 |
| 0 | 0 | 0.013146113 | 0.038379603 | 0 |
| 0 | 0 | 0 | 0 | 0 |
| 0 | 0 | 0 | 0 | 0 |
| 0 | 0 | 0 | 0 | 0 |
| 0 | 0 | 0 | 0 | 0 |
| 0 | 0 | 0 | 0 | 0 |
| 0 | 0.004973976 | 0 | 0 | 0 |
| 0 | 0 | 0 | 0 | 0 |
| 0 | 0 | 0 | 0 | 0 |
| 0 | 0 | 0 | 0 | 0 |
| 0 | 0.013058287 | 0 | 0 | 0 |

| | | | | |
|---|---|---|---|---|
| 0 | 0 | 0.072548553 | 0.229925891 | 0 |
| 0 | 0 | 0 | 0 | 0.052941677 |
| 0 | 0 | 0 | 0 | 0 |
| 0 | 0 | 0 | 0 | 0 |
| 0 | 0 | 0 | 0 | 0.002778015 |
| 0 | 0 | 0 | 0.008532925 | 0 |
| 0 | 0 | 0 | 0 | 0 |
| 0.000711954 | 0 | 0.002060782 | 0.00266736 | 0.038041386 |
| 0 | 0 | 0 | 0 | 0 |
| 2.659205201 | 0.009331713 | 0 | 0.03803206 | 0.061439417 |
| 0 | 0 | 0 | 0 | 0 |
| 0 | 0 | 0.00462208 | 0.213436606 | 0.002608273 |
| 0.002351249 | 0.004566494 | 0.007218259 | 0.001735111 | 0.014388856 |
| 0 | 0 | 0 | 0 | 0 |
| 0 | 0 | 0 | 0 | 0 |
| 0 | 0 | 0 | 0 | 0 |
| 0 | 0 | 0 | 0.001894141 | 0 |
| 0 | 0 | 0 | 0.001390619 | 0 |
| 0.210780536 | 1.803260453 | 0 | 0 | 0.045321035 |
| 0 | 0 | 0.0491313 | 0 | 0 |
| 0 | 0 | 0 | 0 | 0 |
| 0 | 0 | 0 | 0 | 0 |
| 0 | 0 | 0 | 0 | 0 |
| 0 | 0 | 0.082945845 | 0.029834665 | 0 |
| 0 | 0 | 0 | 0 | 0 |
| 0 | 0.069650976 | 0 | 0.00233188 | 0.008379674 |
| 0.000998746 | 0 | 0 | 0.001247278 | 0 |
| 0 | 0.00643191 | 0 | 0 | 0.003293341 |
| 0.085392619 | 0 | 0 | 0 | 0 |
| 0 | 0 | 0 | 0 | 0.004095274 |
| 0.003472976 | 0.012773623 | 0 | 0 | 0 |
| 0 | 0 | 0 | 0 | 0 |
| 0 | 0.05888679 | 0 | 0 | 0 |
| 0.019535959 | 0.001987433 | 0 | 0 | 0 |
| 0 | 0 | 0 | 0 | 0 |
| 0.012875609 | 1.148826004 | 0 | 0 | 0 |
| 0.0503774 | 0.372100532 | 0 | 0 | 0.0097367 |
| 0 | 0 | 0 | 0 | 0 |
| 0 | 0 | 0.001710653 | 0 | 0 |
| 0.769436014 | 0 | 0 | 0 | 0 |
| 0 | 0 | 0 | 0 | 0 |
| 0 | 1.224617176 | 0 | 0 | 0 |
| 0 | 0 | 0 | 0 | 0 |
| 0 | 0.962495813 | 0.029802621 | 0 | 0 |
| 0 | 0.007837987 | 0 | 0 | 0 |
| 0 | 0 | 0.015114402 | 0 | 0 |
| 0.000619213 | 0 | 0.002688507 | 0.002513226 | 0.007294551 |
| 0 | 0 | 0 | 0 | 0 |

| | | | | |
|---|---|---|---|---|
| 0 | 0 | 0 | 0 | 0 |
| 0 | 0 | 0 | 0.002808706 | 0.113548094 |
| 0 | 0 | 0 | 0 | 0 |
| 0.00116624 | 0.001016948 | 0.007072957 | 0.644791845 | 0.077946211 |
| 0 | 0.002490512 | 0 | 0 | 0 |
| 0.003048158 | 0.026296087 | 0.011764027 | 0 | 0.012516639 |
| 0 | 0 | 0 | 0 | 0 |
| 0 | 0 | 0 | 0 | 0 |
| 0 | 0 | 0 | 0.056846421 | 0 |
| 0 | 0 | 0 | 0 | 0 |
| 0.00141414 | 0 | 0.020739322 | 0 | 0.005235709 |
| 0 | 0 | 0 | 0.000662073 | 0 |
| 0 | 0.001948021 | 0 | 0 | 0 |
| 0 | 0.016581614 | 0.001546853 | 0 | 0.002248379 |
| 0.016627146 | 0 | 0 | 0 | 0.104420477 |
| 0 | 0 | 0 | 0 | 0 |
| 0 | 0 | 0 | 0 | 0 |
| 0.028603345 | 0.00687789 | 0 | 0 | 0 |
| 0 | 0 | 0 | 0 | 0 |
| 0 | 0 | 0.000772955 | 0.000900423 | 0 |
| 0 | 0 | 0 | 0 | 0 |
| 3.020400775 | 0 | 0 | 0.104316908 | 0 |
| 0 | 0 | 0 | 0 | 0 |
| 0 | 0 | 0 | 0 | 0 |
| 0 | 0 | 0 | 0 | 0 |
| 0 | 0 | 0 | 0 | 0 |
| 0.927095643 | 0.164757095 | 0.002515797 | 0 | 0.01072649 |
| 0 | 0 | 0 | 0 | 0 |
| 0 | 0 | 0 | 0 | 0 |
| 0.118086012 | 6.130880473 | 0.666226605 | 0.030325708 | 1.456349212 |
| 0 | 0 | 0.002798445 | 0.054558619 | 0.00488111 |
| 0 | 0 | 0 | 0.003178365 | 0 |
| 0 | 0.00369177 | 0 | 0 | 0 |
| 0.000385121 | 0 | 0.001486333 | 0 | 0.002808537 |
| 0.011703404 | 0 | 0.005820616 | 0 | 0 |
| 0 | 0 | 0 | 0 | 0 |
| 0.470840153 | 0 | 0 | 0.448379016 | 1.512485818 |
| 0 | 0 | 0 | 0.009353362 | 0 |
| 0.698588254 | 0 | 0 | 0 | 1.577543478 |
| 0 | 0 | 0 | 0 | 0.089389719 |
| 0.013729305 | 0 | 0.026917255 | 0.001920325 | 0 |
| 1.507995394 | 0 | 0.015676361 | 0.008375771 | 0.109531213 |
| 0 | 0 | 0 | 0.004362036 | 0 |
| 1.427604149 | 0 | 0 | 0 | 0 |
| 0.000495596 | 0.003529259 | 0.000717261 | 0 | 0.00750638 |
| 0 | 0 | 0 | 0 | 0 |
| 0.001383349 | 0 | 0 | 0 | 0 |
| 0 | 0 | 0 | 0 | 0 |

| | | | | |
|---|---|---|---|---|
| 0 | 0.018521429 | 0 | 0.001598665 | 0 |
| 0 | 0 | 0 | 0 | 0 |
| 0 | 0 | 0.003365881 | 0.015974224 | 1.770059628 |
| 0 | 0 | 0 | 0 | 0 |
| 0 | 0 | 0 | 0.204035478 | 0 |
| 0 | 0 | 0 | 0 | 0.010928863 |
| 0 | 0 | 0 | 0 | 0 |
| 0 | 0 | 0 | 0.001791779 | 0 |
| 0 | 0.092035851 | 0.07465477 | 0 | 0 |
| 0.002512082 | 0 | 0 | 0 | 0 |
| 0 | 0 | 0 | 0 | 0 |
| 0 | 0 | 0 | 0 | 0 |
| 0 | 0 | 0.004050169 | 0 | 0 |
| 0 | 0.030893633 | 0.072257135 | 0 | 0 |
| 0 | 0 | 0 | 0 | 0 |
| 0 | 0 | 0.299608712 | 0.015572145 | 0.002199998 |
| 0.003343757 | 0.021317147 | 0.01096913 | 0.019487209 | 0.000937873 |
| 0.005145877 | 0.009918958 | 0 | 0 | 0 |
| 0 | 0.001639713 | 0 | 0 | 0 |
| 0 | 0 | 0 | 0 | 0.002751702 |
| 0.007866293 | 0 | 0 | 0 | 0.018911782 |
| 0 | 0.002630509 | 0 | 0 | 0.005656994 |
| 0 | 0.276504978 | 0.028981778 | 0 | 0 |
| 0 | 0 | 0 | 0 | 0.143905802 |
| 0 | 0 | 0 | 0 | 0 |
| 0.114965558 | 0 | 0.005099963 | 0 | 0 |
| 0.00056599 | 0 | 0 | 0 | 0 |
| 0 | 0.004372641 | 0.075166209 | 0 | 0 |
| 0 | 0 | 0 | 0 | 0 |
| 0 | 0 | 0 | 0 | 0 |
| 0 | 0.007663062 | 0.001661212 | 0 | 0.034770276 |
| 0.003529483 | 0.029704181 | 0.01176415 | 0 | 0.004049861 |
| 0.030092154 | 0 | 0.030066006 | 0.007058248 | 0.023136089 |
| 0 | 0 | 0 | 0.081194643 | 0 |
| 0 | 0.010490783 | 0.003919567 | 0 | 0.009729003 |
| 0 | 0 | 0 | 0 | 0 |
| 0 | 0.011520205 | 0 | 0 | 0 |
| 0.004103975 | 0.012525172 | 0.078568953 | 0.013487431 | 0.021628601 |
| 0 | 0 | 0 | 0.005410479 | 0.168098405 |
| 0 | 0 | 0.011416328 | 0.002095197 | 0 |
| 0 | 0 | 0 | 0 | 0 |
| 0 | 0 | 0 | 0 | 0 |
| 0 | 0.007886895 | 0.002493363 | 0.000806817 | 0 |
| 0 | 0 | 0 | 0.000421144 | 0 |
| 0 | 1.095307731 | 0.030562655 | 0 | 0 |
| 0 | 0.012455788 | 0 | 0 | 0 |
| 0 | 0 | 0 | 0 | 0 |
| 0 | 0 | 0 | 0 | 0 |

| | | | | |
|---|---|---|---|---|
| 0 | 0.009774039 | 0 | 0 | 0 |
| 0.324935462 | 1.232426878 | 0.933439689 | 0.154929367 | 0.364557664 |
| 0 | 0 | 0 | 0 | 0 |
| 0 | 0 | 0 | 0 | 0 |
| 0 | 0 | 0 | 0 | 0 |
| 0 | 0 | 0 | 0 | 0 |
| 0.243835263 | 0.01412729 | 0 | 0 | 0 |
| 0 | 0 | 0 | 0 | 0.001902301 |
| 0 | 0 | 0 | 0 | 0 |
| 0 | 0.004351703 | 0 | 0 | 0 |
| 0 | 0.000890017 | 0.011606518 | 0.000682856 | 0 |
| 0.005864109 | 0.18427316 | 0.013201928 | 0.001830839 | 0.001370661 |
| 0 | 0 | 0 | 0.05632214 | 0 |
| 0 | 0 | 0 | 0 | 0 |
| 0 | 0.003574143 | 0 | 0 | 0 |
| 0.001686367 | 0 | 0 | 0 | 0 |
| 0.000687082 | 0 | 0.009943936 | 0.002145145 | 0.003468887 |
| 0.783558158 | 0.154253899 | 0.002868578 | 0 | 0.053786968 |
| 0 | 0 | 0 | 0 | 0 |
| 0.012355648 | 0.005892025 | 0.002421516 | 0 | 0.018844036 |
| 0.010037861 | 0.010572134 | 0.009343178 | 0.051175234 | 0.005365947 |
| 0 | 0 | 0 | 0 | 0 |
| 0 | 0 | 0 | 0 | 0 |
| 0 | 0 | 0 | 0 | 0 |
| 0 | 0 | 0 | 0 | 0 |
| 0 | 0.001322038 | 0 | 0 | 0 |
| 0 | 0 | 0.013720809 | 0.052987189 | 0.090628278 |
| 0.232328023 | 0.027070034 | 0.089858177 | 0.06755859 | 0.235310199 |
| 0 | 0 | 0 | 0 | 0 |
| 0 | 0 | 0.000876469 | 0.000945377 | 0 |
| 0 | 0 | 0 | 0 | 0.004532999 |
| 0 | 0 | 0 | 0 | 0 |
| 0 | 0 | 0 | 0 | 0 |
| 0 | 0 | 0 | 0 | 0 |
| 0 | 0 | 0.033348283 | 0.035651394 | 0 |
| 2.901957439 | 0.617630209 | 0.807128682 | 0.009815635 | 1.931898338 |
| 0 | 0 | 0 | 0 | 0 |
| 0 | 0 | 0 | 0 | 0 |
| 0 | 0 | 0.004780598 | 0.073866106 | 0.001302879 |
| 0 | 0 | 0 | 0 | 0 |
| 0 | 1.086094061 | 0.022498875 | 0 | 0 |
| 0 | 0 | 0 | 0 | 0 |
| 0.012414618 | 0.006123464 | 0.030489986 | 0.003171256 | 0.0014245 |
| 0.000363215 | 0 | 0.016646209 | 0.001133996 | 0 |
| 0.005606861 | 0 | 0 | 0 | 0 |
| 0.006848778 | 0 | 0 | 0.027416638 | 0 |
| 0 | 0 | 0 | 0 | 0 |
| 0.001320993 | 0.005972771 | 0 | 0 | 0 |

| | | | | |
|---|---|---|---|---|
| 0 | 0.012029922 | 0 | 0 | 0 |
| 0 | 0 | 0 | 0 | 0 |
| 0 | 0 | 0 | 0 | 0 |
| 0 | 0.110098669 | 0.992833548 | 2.093672887 | 0 |
| 0 | 0 | 0 | 0 | 0 |
| 0 | 0 | 0 | 0 | 0 |
| 0 | 0 | 0 | 0 | 0 |
| 0 | 0 | 0 | 0.000734276 | 0 |
| 0 | 0 | 0 | 14.75249738 | 0 |
| 0 | 0 | 0.197230654 | 0.02139595 | 0 |
| 0 | 0 | 0 | 0 | 0 |
| 0.001679197 | 0 | 0.004657984 | 0.010223141 | 0.003355796 |
| 0 | 0 | 0 | 0 | 0 |
| 0 | 0 | 0.067489656 | 0 | 0 |
| 0.006668573 | 0 | 0 | 0.242377414 | 0.070250526 |
| 0 | 0 | 0 | 0 | 0 |
| 0 | 0 | 0 | 0 | 0 |
| 0 | 0 | 0 | 2.770472536 | 0 |
| 0 | 0 | 0 | 0 | 0 |
| 0 | 0 | 0 | 0.012992041 | 0 |
| 0 | 0 | 0 | 0.037579133 | 0 |
| 0 | 0 | 0 | 0 | 0 |
| 0 | 0 | 0 | 0 | 0 |
| 0.258359971 | 0.471184761 | 0.20444489 | 0.029587651 | 0.675148302 |
| 0 | 0 | 0.03564695 | 0.000613556 | 0 |
| 0 | 0 | 0 | 0.058680551 | 0 |
| 0 | 0 | 0 | 0.000730037 | 0 |
| 0 | 0 | 0 | 2.130478026 | 8.637612306 |
| 0 | 0 | 0 | 0 | 0 |
| 0 | 0 | 0 | 0 | 0 |
| 0 | 0 | 0 | 0.100660543 | 0 |
| 0 | 0 | 0 | 0.00540233 | 0 |
| 0 | 0 | 0.000899064 | 0.015418987 | 0.008102195 |
| 0 | 0 | 0 | 0 | 0 |
| 0 | 0 | 0 | 0 | 0 |
| 0 | 0 | 0.031717923 | 0.356844762 | 0 |
| 0 | 0 | 5.437845907 | 0.23580537 | 0 |
| 0 | 0 | 0 | 0 | 0 |
| 0.001529538 | 0 | 0.011595342 | 0.004638947 | 0.004044971 |
| 0 | 0 | 0.041458317 | 0 | 0 |
| 0.137428978 | 0.009013347 | 0.001359979 | 0.096941151 | 0.028465271 |
| 0 | 0 | 0 | 0 | 0 |
| 18.58627218 | 0 | 0 | 0.005279181 | 3.821074354 |
| 0 | 0 | 0 | 0.004928896 | 0.007970455 |
| 0 | 0 | 0 | 0 | 0 |
| 0 | 0 | 0 | 0 | 0 |
| 0 | 0 | 0 | 0 | 0 |
| 0 | 0 | 0.092952948 | 0.081317119 | 0.003258907 |

| | | | | |
|---|---|---|---|---|
| 0 | 0 | 0 | 0 | 0 |
| 0 | 0 | 0.467670166 | 1.205694673 | 0 |
| 0.005685006 | 1.155514223 | 0.031119898 | 0.038686036 | 0.029483034 |
| 0 | 0 | 0 | 0.00391552 | 0 |
| 0 | 0 | 0 | 0 | 0 |
| 0 | 0 | 0.012522079 | 0.027736009 | 0 |
| 0 | 0 | 0 | 0 | 0.066954989 |
| 0 | 0 | 0 | 0 | 0 |
| 0 | 0 | 0 | 0 | 0 |
| 0 | 0 | 0 | 0 | 0 |
| 0.003759082 | 0.03036176 | 0.143538895 | 0.070275344 | 0.023771138 |
| 1.570836805 | 0.434742945 | 0.690091735 | 0.613224106 | 2.589893744 |
| 0 | 0 | 0 | 0 | 0.001259732 |
| 0.00122938 | 0 | 0.008065916 | 0.028096062 | 0.004758548 |
| 0.037395634 | 0.154144523 | 0.964467589 | 0.004718526 | 0.031955826 |
| 0 | 0 | 0 | 0.266767999 | 0 |
| 0 | 0 | 0 | 0 | 0 |
| 0 | 0 | 0 | 0 | 0.004526476 |
| 0 | 0 | 0 | 0.023294416 | 0 |
| 0.006568322 | 0 | 0.003440317 | 0.003281123 | 0.005369028 |
| 0.002864241 | 0.00058277 | 0.373448307 | 0.126029248 | 0.0006427 |
| 2.739256376 | 0 | 0.001164772 | 0.000879442 | 0.281040855 |
| 1.593808671 | 1.313350918 | 0.003480325 | 1.315608483 | 0.125355029 |
| 0 | 0 | 0 | 0 | 0 |
| 0 | 0 | 0 | 0 | 0 |
| 0.002516961 | 0 | 0 | 0.092298453 | 0 |
| 0 | 0.175649077 | 0 | 0 | 0 |
| 0 | 0 | 0 | 0.002984588 | 0 |
| 0 | 0 | 0 | 0 | 0 |
| 0 | 0 | 0 | 0 | 0.113607771 |
| 0 | 0 | 0.002032086 | 0.013414109 | 0 |
| 0 | 0 | 0 | 0.05448149 | 0.157605517 |
| 0.000701941 | 0.856442619 | 0 | 0.001022717 | 0.006497162 |
| 0 | 0 | 0 | 0.015932263 | 0.073568138 |
| 0.00046329 | 0 | 0.051554441 | 0.000675006 | 0.008966252 |
| 0.016655916 | 0.001510966 | 0.056707064 | 0.042661181 | 0.006070272 |
| 0 | 0 | 0 | 0.012286246 | 0.002759435 |
| 0 | 0 | 0 | 0 | 0 |
| 0 | 0 | 0.015811102 | 8.655144589 | 0.038860019 |
| 0 | 0 | 0 | 0 | 0 |
| 0 | 0 | 0 | 0.013999497 | 0 |
| 0 | 0 | 0 | 0 | 0 |
| 0 | 0 | 0 | 0 | 0 |
| 0 | 0 | 0 | 0 | 0 |
| 0.006291067 | 0 | 0.112293477 | 0.043966516 | 0.010587286 |
| 0 | 0 | 0 | 0 | 0 |
| 0 | 0 | 0 | 0 | 0 |
| 0.014009992 | 0 | 0 | 0.011481941 | 0.018911142 |

| | | | | |
|---|---|---|---|---|
| 0 | 0 | 0 | 0 | 0 |
| 0 | 0 | 0 | 0 | 0 |
| 0 | 0 | 0 | 0 | 0 |
| 0 | 0 | 0.001392829 | 0.001158943 | 0.005899981 |
| 0 | 0 | 0 | 0 | 0 |
| 0 | 0 | 0 | 0.162975237 | 0 |
| 0 | 0 | 0.147913643 | 0.011278415 | 0 |
| 0 | 0.028378518 | 0.006225182 | 0.029267775 | 0.560948714 |
| 0 | 0 | 0 | 0 | 0 |
| 0 | 0 | 0 | 0 | 0 |
| 0 | 0 | 0.00576176 | 0.001434173 | 0.02299856 |
| 0 | 0 | 0 | 0 | 0 |
| 0 | 0 | 0.001164201 | 0.047089885 | 0 |
| 0 | 0 | 0 | 0.024231649 | 0 |
| 0 | 0 | 0 | 0 | 0 |
| 0 | 0 | 0 | 0 | 0 |
| 0 | 0 | 0 | 0.027199494 | 0 |
| 0 | 0 | 0 | 0.015998474 | 0.003378666 |
| 0 | 0.07061512 | 0 | 0.036684435 | 0 |
| 0 | 0 | 0 | 0.006244944 | 0 |
| 0 | 0 | 0.002458467 | 0 | 0 |
| 0 | 0 | 0 | 0.00480062 | 0 |
| 0 | 0 | 0 | 0.007157798 | 0 |
| 0 | 0 | 0 | 0 | 0 |
| 0 | 0 | 0 | 0 | 0 |
| 0 | 0 | 0 | 0 | 0 |
| 0 | 0 | 0 | 0 | 0 |
| 0 | 0 | 0 | 0 | 0 |
| 0 | 0 | 0 | 0 | 0 |
| 0.35084649 | 0 | 0.051256119 | 0 | 0.008940207 |
| 0 | 0 | 0.001570856 | 0.046425354 | 0 |
| 0 | 0 | 0 | 0 | 0 |
| 0 | 0 | 0 | 0.152686523 | 0 |
| 0 | 0 | 0 | 0 | 0 |
| 0 | 0 | 0.015154096 | 0 | 0 |
| 0 | 0 | 0 | 0 | 0 |
| 0 | 0 | 0 | 0.012324407 | 0.176737183 |
| 0.005443989 | 0.002966937 | 0 | 0 | 0.057097232 |
| 0 | 0 | 0 | 0.022191209 | 0 |
| 0 | 0 | 0.001850319 | 0 | 0.017911887 |
| 0 | 0 | 0.207338744 | 0 | 0.11328265 |
| 0 | 0 | 0 | 0.011746552 | 0 |
| 0 | 0 | 0 | 0 | 0 |
| 0 | 0 | 0.006376143 | 0.001650584 | 0 |
| 0 | 0 | 0.36531295 | 0.042597002 | 0.50733463 |
| 0 | 0 | 0 | 0 | 0 |
| 0 | 0 | 0.00784279 | 0.004383501 | 0 |
| 0 | 0 | 0 | 0.004865925 | 0 |

| | | | | |
|---|---|---|---|---|
| 0 | 0 | 0 | 0 | 0 |
| 0 | 0 | 0 | 0.011823898 | 0 |
| 0 | 0 | 0 | 0.111968359 | 0 |
| 0 | 0.006295962 | 0.054673958 | 0.114362195 | 0 |
| 0 | 0 | 0 | 0 | 0 |
| 0 | 0 | 0 | 0 | 0 |
| 0 | 0 | 0 | 0 | 0 |
| 0 | 0 | 0 | 0 | 0 |
| 0.025876374 | 0.021005092 | 0.040360749 | 0.002846447 | 0.100181945 |
| 0 | 0 | 0.001041742 | 0.001483209 | 0 |
| 0 | 0 | 0 | 0.000478894 | 0 |
| 0 | 0 | 0 | 0 | 0 |
| 0 | 0 | 0.005433624 | 0 | 0 |
| 0 | 0 | 0 | 0 | 0 |
| 0.001348892 | 2.190708305 | 4.635667988 | 0.405977978 | 2.415183641 |
| 0 | 0.271432053 | 0 | 0 | 0.05964879 |
| 0 | 0 | 0 | 0 | 0 |
| 0 | 0 | 0 | 0 | 0 |
| 0 | 0 | 0 | 0 | 0 |
| 0 | 0 | 0 | 0.001196813 | 0 |
| 0 | 0 | 0.011869839 | 0.02419775 | 0 |
| 0 | 0 | 0 | 0 | 0 |
| 0 | 0.012098552 | 0 | 0 | 0 |
| 0 | 0 | 0 | 0 | 0 |
| 0.001103674 | 0.002245581 | 0.693387095 | 0.196508609 | 0.033963564 |
| 0.017684404 | 0 | 17.19867572 | 0.796546837 | 0.822560272 |
| 0 | 0 | 0 | 0.124940677 | 0 |
| 0 | 0 | 0.001322038 | 0 | 0 |
| 0 | 0 | 0.858654517 | 2.466895397 | 0 |
| 0 | 0 | 0 | 0.004264998 | 0 |
| 0 | 0 | 0 | 0 | 0 |
| 0 | 0 | 0.01469543 | 0 | 0.008202271 |
| 0 | 0 | 0.000736668 | 0 | 0 |
| 0 | 0 | 0 | 0.001051585 | 0 |
| 0.001248685 | 0.059563546 | 0.007764202 | 0.040544722 | 0.002568404 |
| 0 | 0.000798184 | 0.007191631 | 0.000857357 | 0.004456351 |
| 0 | 0.029035559 | 0.150189022 | 0 | 0 |
| 0 | 0 | 1.366610543 | 0.169556444 | 0 |
| 0 | 0 | 0 | 0 | 0 |
| 0 | 0 | 0 | 0 | 0 |
| 0 | 0 | 0 | 0 | 0 |
| 0 | 0 | 0 | 0.001388927 | 0 |
| 0.085370929 | 0.045538089 | 0.158015722 | 0.014895967 | 0.130364335 |
| 0 | 0 | 0.960175152 | 0 | 0 |
| 0 | 0 | 0 | 0.022289366 | 0 |
| 0 | 0.004537075 | 0.000896469 | 0 | 0.004534562 |
| 0 | 0 | 0 | 0 | 0 |
| 0.773561441 | 0 | 0.132249767 | 0.031055807 | 0 |

| | | | | |
|---|---|---|---|---|
| 0.002869879 | 0 | 0.008519982 | 0.002619099 | 0.002229112 |
| 0 | 0 | 0 | 0 | 0 |
| 0 | 0 | 0 | 0 | 0 |
| 0 | 0 | 0 | 0 | 0 |
| 0 | 0 | 0.003605118 | 0.001235187 | 0 |
| 0 | 0 | 0 | 0 | 0 |
| 0 | 0 | 0 | 0 | 0 |
| 0 | 0 | 0 | 0 | 0 |
| 0 | 0 | 0 | 0 | 0 |
| 0 | 0 | 0 | 0 | 0 |
| 0 | 0 | 0 | 0.001155165 | 0 |
| 0.080297789 | 0 | 0.755577809 | 0.335784045 | 0.861307314 |
| 0 | 0 | 0.057517472 | 0.004320607 | 0 |
| 0 | 0 | 0 | 0 | 0 |
| 0.01016758 | 0 | 0.171677813 | 0.791544091 | 0.015399992 |
| 0 | 0 | 0 | 0 | 0 |
| 0 | 0 | 0.000853048 | 0 | 0 |
| 0 | 0 | 0 | 0 | 0 |
| 0 | 0 | 0 | 0 | 0 |
| 0 | 0 | 0 | 0.122020106 | 0 |
| 0 | 0.030211593 | 0 | 0 | 0.024057588 |
| 0 | 0 | 0 | 0 | 0 |
| 0 | 0.1747703 | 1.086504383 | 0.450598199 | 0.058151076 |
| 0 | 0 | 0 | 0 | 0 |
| 0 | 0 | 0 | 0 | 0 |
| 0 | 0 | 0 | 0 | 0 |
| 0 | 0 | 0 | 0 | 0 |
| 0 | 0 | 0 | 0 | 0 |
| 0 | 0 | 0 | 0.001286711 | 0 |
| 0 | 0 | 0 | 0 | 0 |
| 0 | 0 | 0 | 0 | 0 |
| 0.020780325 | 0.004697833 | 0.04360044 | 0.0123578 | 0.00943672 |
| 0 | 0 | 0 | 0 | 0 |
| 0 | 0 | 0.037363333 | 0.000617374 | 0.057266371 |
| 0 | 0.000868624 | 0.00556079 | 0.000933019 | 0.004849627 |
| 0.002751744 | 0 | 0.074871336 | 2.443521817 | 0.000926186 |
| 0 | 0 | 0 | 0.000828212 | 0 |
| 0 | 0 | 0 | 0 | 0 |
| 0 | 0 | 0 | 0 | 0 |
| 0 | 0 | 0 | 0 | 0 |
| 0 | 0 | 0 | 0.025437931 | 0 |
| 0 | 0 | 0 | 0.01617761 | 0 |
| 0 | 0 | 0 | 0 | 0 |
| 0 | 0 | 0.005365327 | 0 | 0 |
| 0.002102784 | 0.008128978 | 0.01095587 | 0.075761497 | 0.00406961 |
| 0 | 0 | 0 | 0 | 0 |
| 0 | 0 | 0 | 0.287234721 | 0 |
| 0 | 0 | 0.00710009 | 0 | 0.002008238 |

| | | | | |
|---|---|---|---|---|
| 0 | 0 | 0 | 0 | 0 |
| 0 | 0 | 0 | 0 | 0 |
| 0.001848584 | 0 | 0.005875391 | 0.005431983 | 0.015005984 |
| 0 | 0 | 0 | 0 | 0 |
| 0 | 0 | 0 | 0 | 0 |
| 0 | 0 | 0.000722077 | 0 | 0 |
| 0 | 0 | 0 | 0.273558165 | 0 |
| 0 | 0 | 0.008124967 | 0.009362249 | 0.000691305 |
| 0 | 0 | 0 | 0 | 0 |
| 0 | 0 | 0 | 0.008581796 | 0 |
| 0 | 0 | 0 | 0 | 0 |
| 0 | 0 | 0.000640507 | 0 | 0 |
| 0 | 0 | 0 | 0 | 0 |
| 0 | 0 | 0 | 0 | 0.170959429 |
| 0 | 0 | 0.038455214 | 0 | 0 |
| 0 | 0 | 0 | 0 | 0 |
| 0 | 0 | 0 | 0 | 0 |
| 0 | 0 | 0 | 0 | 0 |
| 2.310371651 | 0 | 0 | 2.111441542 | 0 |
| 0 | 0 | 0 | 0 | 0 |
| 0 | 0 | 0 | 0.00690962 | 0.003491706 |
| 0 | 0 | 0 | 0 | 0 |
| 0 | 0 | 0 | 0 | 0 |
| 0 | 0 | 0 | 0 | 0 |
| 0 | 0 | 0.005541778 | 0.001291134 | 0 |
| 0.001076687 | 0.000597456 | 0.007932945 | 0 | 0.004612274 |
| 0 | 0 | 0 | 0 | 0 |
| 0 | 0 | 0.101681585 | 0.35522983 | 0 |
| 0 | 0 | 0 | 0.086361858 | 0 |
| 0.854121962 | 0 | 0.766369911 | 0.120040171 | 0.140604651 |
| 0 | 0 | 0 | 0 | 0 |
| 0 | 0 | 0 | 0 | 0 |
| 0 | 0 | 0 | 0.001010689 | 0 |
| 0 | 0 | 0 | 0 | 0 |
| 0 | 0 | 0 | 0.01396262 | 0.000863103 |
| 0 | 0 | 2.488390877 | 0.004377181 | 0 |
| 0 | 0 | 0 | 0.667795306 | 0 |
| 0 | 0 | 0.012802048 | 0.005814123 | 0 |
| 0 | 0 | 0 | 0 | 0 |
| 0 | 0 | 0 | 0 | 0 |
| 3.016773016 | 21.61521601 | 8.602251557 | 0.205520418 | 17.6595805 |
| 0.035442404 | 0.372177095 | 0.672257215 | 0.004468762 | 0.140512934 |
| 0 | 0 | 1.609151287 | 2.417855263 | 0 |
| 0 | 0 | 0 | 0.006068553 | 0 |
| 0 | 0 | 0 | 0.000810377 | 0 |
| 0 | 0 | 0 | 0 | 0 |
| 0 | 0 | 0 | 0.000494836 | 0 |
| 0 | 0 | 0.003921563 | 0.014304669 | 0.000621825 |

| | | | | |
|---|---|---|---|---|
| 0 | 0 | 0 | 0 | 0 |
| 0 | 0.004446341 | 0.003666956 | 0 | 0.001439097 |
| 0 | 0 | 0.025054365 | 0 | 0 |
| 0 | 0 | 0 | 0 | 0 |
| 1.199863122 | 0.087961753 | 1.904919533 | 0.055933759 | 1.293303772 |
| 0 | 0 | 1.185960326 | 0 | 0 |
| 0 | 0 | 0 | 0 | 0 |
| 0 | 0 | 0 | 0 | 0 |
| 0.744854378 | 0.254225977 | 0.024349304 | 0 | 0.132042059 |
| 0 | 0 | 0 | 0 | 0 |
| 0.001395497 | 0 | 0 | 0 | 0 |
| 0 | 0 | 0 | 0 | 0 |
| 0 | 0 | 0.140206905 | 0.021214466 | 0 |
| 0.003632024 | 0.021245861 | 0.034386417 | 0 | 0.061123627 |
| 0 | 0 | 0 | 7.843160292 | 0 |
| 0 | 0.112564292 | 0.652591195 | 0.084328151 | 0 |
| 0 | 0 | 0.002084823 | 0.215878202 | 0 |
| 0 | 0 | 0 | 0 | 0 |
| 0 | 0 | 0 | 0 | 0 |
| 0 | 0 | 0 | 0 | 0 |
| 0 | 0 | 0 | 0 | 0 |
| 0 | 0 | 0 | 0 | 0 |
| 0 | 0 | 0 | 0 | 0 |
| 0 | 0 | 0 | 0 | 0 |
| 0 | 0 | 0 | 0 | 0 |
| 0 | 0 | 0 | 0 | 0 |
| 0 | 0 | 0.023731715 | 0.043302431 | 0 |
| 0 | 0 | 0 | 0 | 0 |
| 0 | 0 | 0 | 0 | 0 |
| 0 | 0 | 0 | 0.001658099 | 0 |
| 0 | 0 | 0 | 0 | 0 |
| 0 | 0 | 0 | 0 | 0 |
| 0 | 0 | 0 | 0.01855096 | 0 |
| 0 | 0 | 0 | 0.012111219 | 0.004420203 |
| 0 | 0 | 0 | 0.703086604 | 0 |
| 0.000557394 | 0 | 0 | 0.001252976 | 0 |
| 0 | 0 | 0.001099392 | 0.024048554 | 0.000767034 |
| 0 | 0.006853812 | 0 | 0.007493375 | 0 |
| 0 | 0 | 0 | 0 | 0 |
| 0 | 0 | 0 | 0 | 0 |
| 1.42144276 | 0 | 0 | 0.346301985 | 0 |
| 0 | 0 | 0 | 0 | 0 |
| 0 | 0 | 0 | 0 | 0 |
| 0 | 0 | 0.005007241 | 0 | 0 |
| 0 | 0 | 0 | 0 | 0 |
| 0 | 0 | 0 | 0 | 0 |
| 0 | 0 | 0 | 0 | 0 |
| 0 | 0 | 0 | 0 | 0 |

| | | | | |
|---|---|---|---|---|
| 0 | 0 | 0 | 0.189092207 | 0 |
| 0 | 0 | 0 | 0.000754547 | 0 |
| 0 | 0 | 0 | 0.001009625 | 0 |
| 0 | 0 | 0 | 0 | 0 |
| 0 | 0 | 0.68510034 | 0.00404375 | 0 |
| 0 | 0 | 0 | 0 | 0 |
| 0 | 0 | 0.198711006 | 0.081981077 | 0 |
| 0 | 0 | 0 | 0 | 0 |
| 0 | 0 | 0 | 0 | 0 |
| 0 | 0.008993521 | 0 | 0.003722885 | 0 |
| 0 | 0 | 0.001604936 | 0.010535073 | 0 |
| 0 | 0.024232735 | 0.42394198 | 0.005885328 | 0 |
| 0 | 0 | 0 | 0 | 0 |
| 0 | 0 | 0 | 0 | 0 |
| 0 | 0 | 0 | 0 | 0 |
| 0 | 0 | 0 | 0 | 0 |
| 0 | 0 | 0 | 0 | 0 |
| 0 | 0 | 0.002181234 | 0 | 0 |
| 0 | 0 | 0 | 0 | 0 |
| 0 | 0 | 0 | 0 | 0 |
| 0 | 0 | 0.440649206 | 0.002679347 | 0 |
| 0 | 0 | 0 | 0 | 0 |
| 0 | 0 | 0.024222309 | 0.012463702 | 0.00259728 |
| 0.000548938 | 0.003536821 | 0.014653397 | 0.004913017 | 0.001693654 |
| 0 | 0 | 0 | 0 | 0 |
| 0 | 0 | 0 | 0 | 0 |
| 0 | 0 | 0 | 0 | 0 |
| 0 | 0 | 0 | 0 | 0 |
| 0 | 0 | 0 | 0 | 0 |
| 0 | 1.368490814 | 0 | 0 | 0 |
| 0 | 0 | 0 | 0 | 0 |
| 0 | 0 | 0 | 0 | 0 |
| 0 | 0 | 0 | 0 | 0 |
| 0 | 0 | 0.065612444 | 0 | 0 |
| 0 | 0 | 0 | 0.065405494 | 0 |
| 0 | 0 | 0 | 0 | 0 |
| 0.009845037 | 0.026785793 | 0 | 0.001572605 | 0.007898837 |
| 0 | 0 | 0 | 0 | 0 |
| 0 | 0 | 0 | 0 | 0 |
| 0 | 0 | 0 | 0 | 0 |
| 0 | 0 | 0 | 0 | 0 |
| 0 | 0 | 0 | 0 | 0 |
| 0 | 0 | 0 | 0 | 0 |
| 0 | 0 | 0 | 0 | 0 |
| 0 | 0 | 0 | 0 | 0 |
| 0 | 0 | 0 | 0 | 0 |
| 0 | 0 | 0 | 0 | 0 |
| 0 | 0 | 0 | 0 | 0 |

| | | | | |
|---|---|---|---|---|
| 0 | 0.011016629 | 0.005889094 | 0.001475325 | 0.000994054 |
| 0 | 0 | 0 | 0 | 0 |
| 0 | 0 | 0 | 0 | 0 |
| 0 | 0 | 0 | 0 | 0 |
| 0 | 0 | 0 | 0 | 0 |
| 0 | 0 | 0 | 0 | 0 |
| 0 | 0 | 0 | 0.00198478 | 0 |
| 0 | 1.341760069 | 0.56195939 | 0.051125352 | 0.268776086 |
| 0.02426056 | 0.132364348 | 1.528510039 | 2.833698766 | 0.048889929 |
| 0 | 0.029986433 | 0 | 0 | 0.465999451 |
| 0 | 0 | 0 | 0 | 0 |
| 0 | 0 | 0.025439397 | 0 | 0 |
| 0 | 0 | 0 | 0.012914558 | 0 |
| 0 | 0 | 0 | 0 | 0 |
| 0 | 0 | 0 | 0 | 0 |
| 0 | 0 | 0 | 0 | 0 |
| 0.014657884 | 0 | 0 | 0 | 0.088997806 |
| 0 | 0 | 0 | 0 | 0 |
| 0 | 0 | 0 | 0 | 0 |
| 0 | 0 | 0 | 0.049128679 | 0 |
| 0 | 0 | 0 | 0 | 0 |
| 0 | 0 | 0 | 0 | 0 |
| 0 | 0 | 0 | 0.0069636 | 0 |
| 0 | 0 | 0 | 0 | 0 |
| 0 | 0 | 0.055362481 | 0.003776574 | 0 |
| 0 | 0 | 0 | 0 | 0 |
| 0 | 0 | 0 | 0 | 0 |
| 0 | 0 | 0 | 0 | 0 |
| 0 | 0 | 0 | 0 | 0 |
| 0 | 0 | 0 | 0 | 0 |
| 0.010821593 | 0 | 0 | 0.001263035 | 0.250879555 |
| 0 | 0 | 0 | 0 | 0 |
| 0 | 0 | 0 | 0 | 0 |
| 0 | 0 | 0 | 0 | 0 |
| 0 | 0 | 0.001839891 | 0.003274496 | 0 |
| 0 | 0 | 0 | 0 | 0 |
| 0 | 0 | 0 | 0 | 0 |
| 0 | 0 | 0 | 0 | 0 |
| 0 | 0 | 0 | 0 | 0 |
| 0 | 0 | 0 | 0 | 0 |
| 0 | 0 | 0 | 0 | 0 |
| 0.237665779 | 0 | 0 | 0.092562388 | 0.499450153 |
| 0 | 0 | 0 | 0 | 0 |
| 0 | 0 | 0 | 0 | 0 |
| 0 | 0 | 0.001478479 | 0 | 0 |
| 0 | 0 | 0 | 0 | 0 |
| 0 | 0 | 0 | 0 | 0 |
| 0 | 0 | 0 | 0 | 0 |

| | | | | |
|---|---|---|---|---|
| 0 | 0 | 0 | 0 | 0 |
| 0 | 0.101418497 | 0.001714569 | 0.056812598 | 0.002542002 |
| 0 | 0 | 0 | 0 | 0 |
| 0 | 0 | 0 | 0 | 0 |
| 0.007702099 | 0 | 0.761113889 | 8.324845794 | 0.041394532 |
| 0 | 0 | 0 | 0 | 0 |
| 0 | 0 | 0 | 0 | 0.002900998 |
| 0.013023973 | 0.038219865 | 0.007249698 | 0.004066227 | 0.030558956 |
| 0 | 0 | 0 | 0.003591518 | 0 |
| 0 | 0 | 0 | 0 | 0 |
| 0 | 0 | 0 | 0 | 0 |
| 0 | 0 | 0 | 0 | 0 |
| 0 | 0 | 0 | 0 | 0 |
| 0 | 0 | 0 | 0 | 0 |
| 0 | 0 | 0.001190824 | 0 | 0.012759095 |
| 0 | 0.022098536 | 0.02137787 | 0.004747357 | 0.002741747 |
| 0 | 0 | 0 | 0 | 0 |
| 0 | 0 | 0 | 0 | 0 |
| 0 | 0 | 0 | 0 | 0 |
| 0 | 0 | 0 | 0 | 0 |
| 0 | 0 | 0 | 0 | 0 |
| 0 | 0 | 0 | 0 | 0 |
| 0 | 0 | 0 | 0 | 0 |
| 0 | 0 | 0.004182678 | 0 | 0.001094328 |
| 0.191692653 | 0 | 0.054704737 | 0.075864354 | 0 |
| 0 | 0 | 0 | 0 | 0 |
| 0 | 0.023394794 | 0.021996817 | 0.106582221 | 0 |
| 0 | 0 | 0 | 0 | 0 |
| 0 | 0 | 0 | 0 | 0 |
| 0 | 0 | 0 | 0 | 0 |
| 0 | 0 | 0 | 0 | 0 |
| 0 | 0 | 0 | 0 | 0 |
| 0 | 0 | 0 | 0 | 0 |
| 0 | 0 | 0 | 0 | 0 |
| 0 | 0 | 0 | 0 | 0 |
| 0 | 0 | 0 | 0 | 0 |
| 0 | 0 | 0 | 0 | 0 |
| 0 | 0 | 0.085724295 | 0.014980424 | 0 |
| 0 | 0 | 0 | 0 | 0 |
| 0 | 0 | 0 | 0 | 0 |
| 0 | 0 | 0 | 0 | 0 |
| 0 | 0 | 0 | 0.991326263 | 0 |
| 0 | 0 | 0 | 0 | 0 |
| 0 | 0 | 0 | 0 | 0 |
| 0 | 0 | 0 | 0 | 0 |
| 0 | 0 | 0 | 0 | 0 |
| 0 | 0 | 0 | 0 | 0 |
| 0 | 0 | 0 | 0 | 0 |

| | | | | |
|---|---|---|---|---|
| 0 | 0 | 1.038082999 | 0.074504172 | 0 |
| 0 | 0 | 0.001427549 | 0 | 0 |
| 0 | 0 | 0.001781803 | 0 | 0 |
| 0 | 0 | 0.001252789 | 0 | 0 |
| 0 | 0 | 0 | 0 | 0 |
| 0.001403956 | 0 | 0 | 0.001078967 | 0 |
| 0 | 0 | 0 | 0 | 0 |
| 0 | 0 | 0.001416923 | 0 | 0 |
| 0 | 0 | 0 | 0 | 0 |
| 0 | 0 | 0.055343786 | 0 | 0 |
| 0 | 0 | 0 | 0 | 0 |
| 0 | 0 | 0 | 0 | 0 |
| 0 | 0 | 0 | 0 | 0 |
| 0 | 0 | 0 | 0 | 0 |
| 0 | 0 | 0 | 0 | 0 |
| 0 | 0 | 0 | 0 | 0 |
| 0 | 0 | 0 | 0 | 0 |
| 0 | 0 | 0 | 0 | 0 |
| 0 | 0 | 0.002897604 | 0 | 0 |
| 0 | 0 | 0 | 0 | 0 |
| 0.000431695 | 0 | 0.042484967 | 0 | 0.011079159 |
| 0 | 0 | 0 | 0 | 0 |
| 0 | 0 | 0.004997318 | 0.200345456 | 0.000917519 |
| 0 | 0 | 0 | 0.069794594 | 0 |
| 0 | 0 | 0 | 0 | 0 |
| 0 | 0 | 0 | 0 | 0 |
| 0 | 0 | 0 | 0 | 0 |
| 0 | 0 | 0 | 0 | 0 |
| 0 | 0 | 0 | 0 | 0 |
| 0 | 0 | 0 | 0 | 0 |
| 0 | 0 | 0 | 0 | 0 |
| 0 | 0 | 0 | 0 | 0 |
| 0.003829323 | 0.176602741 | 0 | 0 | 0.118147312 |
| 0 | 0 | 0 | 0 | 0 |
| 0.00388202 | 0 | 0.940575117 | 0.009696073 | 0 |
| 0 | 0 | 0 | 0 | 0 |
| 0 | 0 | 0 | 0 | 0 |
| 0 | 0 | 0 | 0 | 0.005927783 |
| 0.002606459 | 0.002410549 | 0.003429319 | 0 | 0.00937101 |
| 0 | 0 | 0 | 0 | 0 |
| 0 | 0 | 0 | 0 | 0 |
| 0 | 0 | 0 | 0 | 0 |
| 0 | 0 | 0.007217824 | 1.625048905 | 0 |
| 0 | 0 | 0 | 0 | 0 |
| 0 | 0 | 0 | 0 | 0 |
| 0 | 0 | 0.000849215 | 0.010029986 | 0 |
| 0 | 0 | 0 | 0 | 0 |
| 0 | 0 | 0 | 0 | 0 |

| | | | | |
|---|---|---|---|---|
| 0 | 0 | 0 | 0 | 0 |
| 0 | 0 | 0 | 0 | 0 |
| 0 | 0 | 0 | 0 | 0 |
| 0 | 0 | 0 | 0 | 0 |
| 0 | 0 | 0 | 0 | 0 |
| 0 | 0 | 0 | 0 | 0 |
| 0 | 0 | 0 | 0 | 0 |
| 0 | 0 | 0.041235707 | 0 | 0 |
| 0 | 0 | 0.665549575 | 0.011714277 | 0 |
| 0.001473144 | 0 | 0 | 0.069626593 | 0.000572115 |
| 0 | 0 | 0 | 0 | 0 |
| 0 | 0 | 0 | 0 | 0 |
| 0 | 0 | 0.004116172 | 0 | 0 |
| 0 | 0 | 0 | 0 | 0 |
| 0 | 0 | 0 | 0 | 0 |
| 0 | 0.012249821 | 0 | 0 | 0.005066084 |
| 0 | 0 | 0 | 0 | 0 |
| 0 | 0 | 0 | 0 | 0 |
| 0 | 0 | 0 | 0 | 0 |
| 0 | 0 | 4.142234293 | 0.097222777 | 0 |
| 0.00209604 | 0 | 0 | 0.234132209 | 0 |
| 0 | 0 | 0 | 0 | 0 |
| 0 | 0 | 0 | 0 | 0 |
| 0 | 0 | 0 | 0 | 0 |
| 0 | 0 | 0 | 0 | 0 |
| 0 | 0 | 0 | 0.158934263 | 0 |
| 0 | 0 | 0 | 0 | 0 |
| 0 | 0 | 0 | 0 | 0 |
| 0 | 0 | 0 | 0 | 0 |
| 0 | 0 | 0 | 0.003607726 | 0 |
| 0 | 0 | 0.112963449 | 0.133991343 | 0.014945256 |
| 0 | 0 | 0 | 0 | 0 |
| 0 | 0 | 0.014399798 | 0 | 0 |
| 0 | 0 | 0 | 0.011174512 | 0 |
| 0 | 0 | 0 | 0 | 0 |
| 0 | 0 | 0 | 0 | 0 |
| 0 | 0 | 0 | 0 | 0 |
| 0 | 0 | 0 | 0 | 0 |
| 6.025501528 | 0 | 0 | 0.003316175 | 0.019078282 |
| 0 | 0 | 0 | 0 | 0 |
| 0 | 0 | 0 | 0 | 0 |
| 0 | 0 | 0.477002813 | 0.07800632 | 0 |
| 0 | 0 | 0 | 0.005630471 | 0 |
| 0 | 0 | 0 | 0 | 0 |
| 0 | 0 | 0.040058227 | 0 | 0 |
| 0 | 0 | 0.089196856 | 0 | 0 |
| 0.936018949 | 0 | 0 | 0 | 0.051669052 |
| 0 | 0 | 0 | 0.001621749 | 0 |

| | | | | |
|---|---|---|---|---|
| 0 | 0 | 0 | 0 | 0 |
| 0 | 0 | 0 | 0 | 0 |
| 0 | 0 | 0 | 0 | 0 |
| 0 | 0 | 0 | 0.001405194 | 0 |
| 0 | 0.074079404 | 0.455280998 | 0.042594151 | 0.036735182 |
| 0 | 0 | 0 | 0 | 0 |
| 0 | 0 | 0.089119858 | 0 | 0 |
| 0 | 0 | 0 | 0 | 0 |
| 0 | 0 | 0 | 0 | 0 |
| 0 | 0 | 0 | 0 | 0 |
| 0 | 0 | 0 | 0.001487194 | 0 |
| 0 | 0.002890355 | 0.00319815 | 0.033707395 | 0.001195347 |
| 0 | 0 | 0 | 0.004846832 | 0.064798944 |
| 0 | 0 | 1.042612085 | 0 | 0 |
| 0.000451073 | 0.000917772 | 0 | 0 | 0.001973698 |
| 0 | 0 | 0.140871616 | 0.005978239 | 0 |
| 0 | 0 | 0.023894778 | 0.022627511 | 0 |
| 0 | 0 | 0 | 0 | 0 |
| 0 | 0.123837916 | 0.026638229 | 0.002126953 | 0.019460151 |
| 0 | 0 | 0.00406536 | 0.042095781 | 0 |
| 0 | 0 | 0 | 0 | 0 |
| 0 | 0 | 0 | 0 | 0 |
| 0 | 0 | 0 | 0 | 0 |
| 0 | 0 | 0 | 0 | 0 |
| 0 | 0 | 0 | 0 | 0 |
| 0 | 0 | 0 | 0 | 0 |
| 0.001688225 | 0 | 0.006515518 | 0.001405553 | 0.004261692 |
| 0 | 0 | 0.003203515 | 0 | 0 |
| 0 | 0 | 0 | 0 | 0 |
| 0 | 0 | 0 | 0 | 0 |
| 0 | 0 | 0.001436105 | 0 | 0 |
| 0 | 0 | 0 | 0 | 0 |
| 0 | 0 | 0 | 0 | 0 |
| 0 | 0 | 0 | 0 | 0 |
| 0 | 0 | 0 | 0.000735011 | 0 |
| 0 | 0 | 0 | 0 | 0 |
| 0 | 0 | 0 | 0 | 0 |
| 0 | 0 | 0.002882759 | 0.116229388 | 0.022626775 |
| 0 | 0 | 0 | 0.3017035 | 0 |
| 0 | 0 | 0.022864014 | 0 | 0 |
| 0 | 0 | 0 | 0.002793765 | 0 |
| 0 | 0 | 0 | 0 | 0 |
| 0 | 0 | 0 | 0 | 0 |
| 0 | 0.002478987 | 0 | 0 | 0.000559211 |
| 0 | 0 | 0 | 0 | 0 |
| 0 | 0 | 0 | 0 | 0 |
| 0 | 0 | 0 | 0 | 0 |
| 0 | 0 | 0 | 0 | 0 |

| | | | | |
|---|---|---|---|---|
| 0 | 0 | 0 | 0 | 0 |
| 0 | 0 | 0.083121002 | 0.01844351 | 0 |
| 0 | 0 | 0 | 0 | 0 |
| 0 | 0 | 0.003135982 | 0.003189244 | 0 |
| 0 | 0.005914457 | 0 | 0.00080672 | 0.001630671 |
| 0 | 0 | 0 | 0 | 0 |
| 0 | 0 | 0 | 0.001969494 | 0 |
| 0 | 0 | 0.007883386 | 0 | 0 |
| 0 | 0 | 0 | 0 | 0 |
| 0 | 0 | 0 | 0.016249073 | 0 |
| 0 | 0 | 0 | 0 | 0 |
| 0 | 0 | 0 | 0 | 0 |
| 0.001481117 | 0 | 0.208641565 | 0.184865658 | 0.004846691 |
| 0 | 0 | 0 | 0.009428859 | 0 |
| 0 | 0 | 0 | 0 | 0 |
| 0 | 0 | 0 | 0 | 0 |
| 0 | 0 | 0 | 0 | 0 |
| 0 | 0 | 0 | 0 | 0 |
| 0 | 0 | 0 | 0 | 0 |
| 0 | 0 | 0 | 0 | 0 |
| 0 | 0 | 0 | 0 | 0 |
| 0 | 0 | 0 | 0 | 0 |
| 0.000866796 | 0.000661357 | 0 | 0 | 0.03537438 |
| 0 | 0 | 0.008388587 | 0 | 0 |
| 0 | 0 | 0 | 0 | 0 |
| 0 | 0 | 0.001275452 | 0.012263629 | 0 |
| 0 | 0 | 0 | 0.001809899 | 0 |
| 0 | 0 | 0 | 0 | 0 |
| 0 | 0 | 0 | 0 | 0 |
| 0 | 0 | 0 | 0 | 0 |
| 0.02440345 | 0 | 0.015153109 | 0.010711896 | 0.001829796 |
| 0 | 0 | 0 | 0 | 0 |
| 0 | 0 | 0 | 0 | 0 |
| 0 | 0 | 0 | 0 | 0 |
| 0 | 0 | 0 | 0.005818425 | 0 |
| 0 | 0 | 0 | 0 | 0 |
| 0 | 0 | 0 | 0 | 0 |
| 0 | 0 | 0 | 0 | 0 |
| 0 | 0 | 0.096844696 | 0.050931546 | 0 |
| 0 | 0 | 0 | 0 | 0 |
| 0 | 0 | 0 | 0 | 0 |
| 0 | 0 | 0 | 0 | 0 |
| 0 | 0 | 0 | 0 | 0 |
| 0 | 0 | 0 | 0 | 0 |
| 0 | 0 | 0 | 0 | 0 |
| 0 | 0.113811227 | 0.364430552 | 0.124438525 | 0.282106606 |
| 0.631822489 | 0 | 0 | 0 | 0 |
| 0 | 0 | 0 | 0 | 0 |

| | | | | |
|---|---|---|---|---|
| 0 | 0 | 0 | 0 | 0 |
| 0 | 0 | 0 | 0 | 0 |
| 0.407069212 | 0 | 0 | 0 | 0.775873577 |
| 0 | 0 | 0 | 0 | 0 |
| 0 | 0 | 0 | 0 | 0 |
| 0 | 0 | 0 | 0 | 0 |
| 0 | 0 | 0 | 0 | 0 |
| 0 | 0 | 0 | 0 | 0 |
| 0 | 0.000577273 | 0 | 0 | 0 |
| 0 | 0 | 0 | 0.000437038 | 0 |
| 0 | 0 | 0.000800071 | 0.001656907 | 0 |
| 0 | 0 | 0 | 0.001236884 | 0 |
| 0 | 0 | 0 | 0 | 0 |
| 0 | 0 | 0 | 0 | 0 |
| 0 | 0 | 0 | 0 | 0 |
| 0 | 0 | 0 | 0 | 0 |
| 0 | 0 | 0 | 0.007458971 | 0 |
| 0 | 0 | 0 | 0 | 0 |
| 0 | 0 | 0 | 0 | 0 |
| 0 | 0 | 0.000848825 | 0.002472012 | 0.003516284 |
| 0 | 0 | 0 | 0 | 0 |
| 0 | 0 | 0 | 0 | 0 |
| 0 | 0 | 0 | 0 | 0 |
| 0 | 0 | 0 | 0 | 0 |
| 0 | 0.001161323 | 0 | 0 | 0 |
| 0 | 0 | 0 | 0 | 0 |
| 0 | 0 | 0 | 0 | 0 |
| 0 | 0 | 0 | 0.012462629 | 0 |
| 0 | 0 | 0 | 0 | 0 |
| 0 | 0 | 0 | 0 | 0 |
| 0 | 0.006919993 | 0 | 0.07291418 | 0.002225891 |
| 0 | 0 | 0 | 0 | 0 |
| 0.014589397 | 0.045977005 | 0.010583863 | 0.017260937 | 0.043936255 |
| 0 | 0 | 0.020116942 | 0.001679886 | 0 |
| 0 | 0 | 0 | 0 | 0 |
| 0 | 0 | 0 | 0 | 0 |
| 0 | 0 | 0 | 0 | 0 |
| 0 | 0 | 0 | 0 | 0 |
| 0 | 0 | 0 | 0 | 0 |
| 0 | 0 | 0 | 0 | 0 |
| 0 | 0 | 0 | 0 | 0 |
| 0.003339059 | 0.165070647 | 0 | 0 | 0.372394498 |
| 0.003408254 | 0 | 0.078286181 | 0.053136056 | 0.037745117 |
| 0 | 0 | 0 | 0 | 0 |
| 0 | 0 | 0 | 0 | 0 |
| 0 | 0 | 0.040775484 | 1.451976203 | 0 |
| 0 | 0 | 0.001206025 | 0 | 0 |
| 0 | 0 | 0 | 0 | 0 |

| | | | | |
|---|---|---|---|---|
| 0 | 0 | 0 | 0 | 0 |
| 0.083621096 | 0 | 0 | 0.028624667 | 0 |
| 0.004843551 | 0.014782329 | 0.014798739 | 0.002016279 | 0.003169931 |
| 0 | 0 | 0 | 0 | 0 |
| 0.001718819 | 0 | 0.03068035 | 0.012342582 | 0.19163566 |
| 0.007738031 | 0.010637915 | 0 | 0 | 0.004223479 |
| 0 | 0 | 0 | 0 | 0 |
| 0 | 0 | 0 | 0 | 0 |
| 0 | 0 | 0 | 0 | 0 |
| 0 | 0 | 0 | 0 | 0 |
| 0 | 0 | 0 | 0 | 0 |
| 0 | 0 | 0 | 0 | 0 |
| 0 | 0 | 0 | 0 | 0 |
| 0 | 0 | 0.015057404 | 0 | 0 |
| 0 | 0 | 0.050517223 | 0 | 0 |
| 0 | 0 | 0 | 0 | 0 |
| 0 | 0 | 0 | 0.052074321 | 0 |
| 0 | 0 | 0 | 0 | 0 |
| 0 | 0 | 0 | 0 | 0 |
| 0 | 0 | 0 | 0 | 0 |
| 0 | 0 | 0 | 0 | 0 |
| 0 | 0.018937 | 0 | 0 | 0 |
| 0 | 0 | 0 | 0 | 0 |
| 0 | 0 | 0 | 0 | 0 |
| 0 | 0 | 0 | 0.023263768 | 0 |
| 0 | 0.010212938 | 0 | 0 | 0.004693003 |
| 0 | 0 | 0 | 0 | 0 |
| 0 | 0 | 1.252579067 | 0.029322813 | 0 |
| 0 | 0 | 0 | 0 | 0 |
| 0 | 0 | 0 | 0 | 0 |
| 0 | 0 | 0 | 0 | 0 |
| 0.004695285 | 0 | 0 | 0 | 0.001481575 |
| 0 | 0 | 0 | 0 | 0 |
| 0 | 0 | 0 | 0 | 0 |
| 0 | 0 | 0 | 0 | 0 |
| 0 | 0 | 0 | 0 | 0 |
| 0 | 0 | 0 | 0 | 0 |
| 0.000999304 | 0 | 0 | 0 | 0 |
| 0 | 0 | 0 | 0 | 1.838313081 |
| 0 | 0 | 0 | 0 | 0 |
| 0 | 0 | 0 | 0.001139968 | 0 |
| 0 | 0 | 0 | 0.070573791 | 0 |
| 0 | 0.008427611 | 0.007992916 | 0.000559221 | 0.001130386 |
| 0 | 0 | 0 | 0 | 0 |
| 0 | 0 | 0 | 0 | 0 |
| 0 | 0 | 0 | 0 | 0 |
| 0.001370064 | 0 | 0 | 0.000855498 | 0.013257728 |
| 0 | 0 | 0 | 0 | 0 |

| | | | | |
|---|---|---|---|---|
| 0 | 0 | 0 | 0 | 0 |
| 0.002917298 | 0.008353881 | 0.001667998 | 0.003643249 | 0.046549745 |
| 0 | 0 | 0 | 0 | 0 |
| 0 | 0 | 0 | 0 | 0 |
| 0 | 0 | 0 | 0 | 0 |
| 0 | 0 | 0 | 0 | 0 |
| 0 | 0 | 0 | 0 | 0 |
| 0 | 0 | 0 | 0 | 0 |
| 0 | 0 | 0 | 0 | 0 |
| 0 | 0 | 0 | 0 | 0 |
| 0 | 0 | 0 | 0 | 0 |
| 0 | 0 | 0 | 0 | 0 |
| 0 | 0 | 0.446315426 | 0.133652188 | 0 |
| 0 | 0 | 0 | 0 | 0 |
| 0.000808318 | 0 | 0.001819774 | 0 | 0.006801623 |
| 0 | 0 | 0 | 0 | 0 |
| 0 | 0 | 0 | 0 | 0 |
| 0 | 0 | 0 | 0 | 0 |
| 0 | 0 | 0.001150366 | 0 | 0 |
| 0 | 0 | 0 | 0 | 0 |
| 0 | 0 | 0 | 0 | 0 |
| 0 | 0 | 0.025020265 | 0.003139345 | 0 |
| 0.001728319 | 0 | 0 | 0 | 0.062278292 |
| 0 | 0 | 0 | 0 | 0.000917345 |
| 0 | 0 | 0 | 0 | 0 |
| 0 | 0 | 0 | 0 | 0 |
| 0 | 0 | 0 | 0 | 0 |
| 0 | 0 | 0 | 0 | 0 |
| 0 | 0 | 0 | 0 | 0 |
| 0 | 0 | 0 | 0 | 0 |
| 0 | 0 | 0 | 0 | 0 |
| 0 | 0 | 0 | 0 | 0 |
| 0 | 0 | 0 | 0 | 0 |
| 0 | 0 | 0 | 0 | 0 |
| 0 | 0 | 5.013504623 | 4.424344092 | 0.006266984 |
| 0 | 0 | 0 | 0 | 0 |
| 0 | 0 | 0 | 0 | 0 |
| 0 | 0 | 0 | 0 | 0 |
| 0.000396369 | 0 | 0 | 0.013695069 | 0 |
| 0 | 0 | 0 | 0 | 0 |
| 0.014334341 | 0.322022767 | 0.096470234 | 0.009320539 | 0.393549981 |
| 0 | 0 | 0 | 0 | 0 |
| 0 | 0 | 0 | 0 | 0 |
| 0 | 0 | 0 | 0 | 0 |
| 0 | 0 | 0 | 0 | 0 |
| 0.262996426 | 13.30031038 | 0.025911691 | 0.00457345 | 0.121891572 |
| 0 | 0 | 0 | 0 | 0 |
| 0 | 0 | 0 | 0 | 0 |

| | | | | |
|---|---|---|---|---|
| 0 | 0 | 0 | 0 | 0 |
| 0 | 0 | 0 | 0 | 0 |
| 0 | 0 | 0 | 0 | 0.006189866 |
| 0 | 0 | 0 | 0.013527075 | 0 |
| 0 | 0 | 0.001588045 | 0 | 0 |
| 0 | 0 | 0 | 0 | 0 |
| 0 | 0 | 0 | 0 | 0 |
| 0 | 0 | 0 | 0.024899349 | 0 |
| 0 | 0 | 0.030674105 | 0 | 0.005412099 |
| 0 | 0 | 0.002559765 | 0.096083257 | 0.00156268 |
| 0 | 0 | 0 | 0 | 0 |
| 0 | 0 | 0 | 0 | 0 |
| 0 | 0 | 0 | 0.002752631 | 0.001090991 |
| 0 | 0 | 0 | 0 | 0 |
| 0 | 0 | 0 | 0 | 0 |
| 0 | 0 | 0 | 0 | 0 |
| 0 | 0 | 0 | 0 | 0 |
| 0 | 0 | 0 | 0 | 0 |
| 0 | 0 | 0 | 0 | 0 |
| 0.01857981 | 0.012444545 | 0.017592639 | 0.162422966 | 0.150125777 |
| 0 | 0 | 0 | 0 | 0 |
| 0 | 0 | 0 | 0 | 0 |
| 0 | 0 | 0 | 0 | 0 |
| 0 | 0 | 0 | 0 | 0 |
| 0 | 0 | 0 | 0 | 0 |
| 0 | 0 | 0.008679863 | 0 | 0 |
| 0 | 0 | 0 | 0 | 0 |
| 0 | 0 | 0 | 0 | 0 |
| 0 | 0 | 0 | 0 | 0 |
| 0 | 0 | 0.001190305 | 0 | 0.01806256 |
| 0 | 0 | 0 | 0 | 0 |
| 0 | 0 | 0 | 0 | 0 |
| 0 | 0 | 0 | 0 | 0 |
| 0 | 0 | 0 | 0 | 0 |
| 0 | 0 | 0 | 0 | 0 |
| 0 | 0 | 0 | 0 | 0 |
| 0 | 0 | 0 | 0 | 0 |
| 0 | 0 | 0 | 0 | 0 |
| 0 | 0 | 0 | 0 | 0 |
| 0 | 0 | 0 | 0 | 0 |
| 0 | 0 | 0 | 0 | 0 |
| 0 | 0 | 0 | 0 | 0 |
| 0 | 0.026153757 | 0.056273461 | 0.004791921 | 0 |
| 0 | 0 | 0 | 0 | 0 |
| 0 | 0 | 0.017360294 | 0.054686479 | 0.002371342 |
| 0 | 0 | 0 | 0.006723997 | 0 |
| 0 | 0 | 0 | 0 | 0 |
| 0 | 0.005077547 | 0 | 0 | 0 |

| | | | | |
|---|---|---|---|---|
| 0 | 0 | 0 | 0 | 0 |
| 0 | 0 | 0 | 0 | 0 |
| 0 | 0 | 0 | 0 | 0 |
| 0 | 0 | 0 | 0 | 0 |
| 0 | 0 | 0 | 0.002410453 | 0 |
| 0.001051929 | 0 | 0.038099672 | 0.013440105 | 0.003540598 |
| 0 | 0 | 0 | 0 | 0 |
| 0 | 0 | 0 | 0 | 0 |
| 0 | 0 | 0 | 0 | 0 |
| 0 | 0 | 0 | 0 | 0 |
| 0 | 0 | 0 | 0 | 0 |
| 0 | 0 | 0 | 0 | 0 |
| 0.001156305 | 0 | 0 | 0 | 0 |
| 0 | 0 | 0 | 0.004134429 | 0 |
| 0 | 0 | 0 | 0 | 0 |
| 0 | 0 | 0 | 0 | 0 |
| 0 | 0 | 0 | 0.093648897 | 0 |
| 0 | 0 | 0 | 0 | 0 |
| 0 | 0 | 0 | 0 | 0 |
| 0 | 0 | 0 | 0 | 0 |
| 0 | 0 | 0 | 0 | 0 |
| 0 | 0 | 0 | 0 | 0 |
| 0 | 0 | 0 | 0 | 0 |
| 0 | 0 | 0 | 0.001367064 | 0 |
| 0 | 0 | 0 | 0 | 0 |
| 0 | 0 | 0 | 0.003477419 | 0 |
| 0 | 0 | 0 | 0 | 0 |
| 0 | 0 | 0 | 0.018099956 | 0 |
| 0 | 0 | 0 | 0 | 0 |
| 0 | 0 | 0 | 0 | 0 |
| 0 | 0 | 0 | 0 | 0 |
| 0 | 0 | 0 | 0 | 0 |
| 0 | 0 | 0 | 0 | 0 |
| 0 | 0 | 0 | 0 | 0 |
| 0 | 0 | 0 | 0 | 0 |
| 0 | 0 | 0 | 0 | 0 |
| 0 | 0 | 0 | 0 | 0 |
| 0 | 0 | 0 | 0 | 0 |
| 0 | 0 | 0 | 0.208833693 | 0 |
| 0 | 0 | 0 | 0 | 0 |
| 0 | 0 | 0 | 0 | 0 |
| 0 | 0 | 0 | 0 | 0 |
| 0 | 0 | 0 | 0 | 0 |
| 0 | 0 | 0 | 0 | 0 |
| 0 | 0 | 0 | 0 | 0 |
| 0 | 0 | 0 | 0 | 0 |
| 0 | 0 | 0 | 0 | 0 |
| 0 | 0 | 0 | 0 | 0 |

| | | | | |
|---|---|---|---|---|
| 0 | 0 | 0 | 0 | 0 |
| 0.005435625 | 0 | 0.016083279 | 0.006788247 | 0.007470587 |
| 0 | 0 | 0 | 0 | 0 |
| 0 | 0 | 0 | 0 | 0 |
| 0 | 0 | 0 | 0 | 0 |
| 0 | 0 | 0.000958911 | 0 | 0 |
| 0 | 0 | 0 | 0 | 0 |
| 0 | 0 | 0 | 0 | 0 |
| 0 | 0 | 0 | 0 | 0 |
| 0 | 0 | 0 | 0 | 0 |
| 0 | 0 | 0.001144553 | 0.019555077 | 0 |
| 0 | 0 | 0 | 0 | 0 |
| 0 | 0 | 0.112290443 | 0 | 0.962093651 |
| 0 | 0 | 0 | 0 | 0 |
| 0 | 0 | 0 | 0 | 0 |
| 0 | 0 | 0 | 0 | 0 |
| 0 | 0 | 0 | 0 | 0 |
| 0 | 0 | 0 | 0 | 0 |
| 0 | 0 | 0 | 0 | 0 |
| 0 | 0 | 0 | 0.000616365 | 0 |
| 0 | 0 | 0 | 0.125999818 | 0.008915195 |
| 0 | 0 | 0 | 0.002790559 | 0 |
| 0 | 0 | 0 | 0 | 0 |
| 0 | 2.232428511 | 2.991252428 | 0.130784137 | 8.883729 |
| 0 | 0 | 0 | 0 | 0 |
| 0 | 0 | 0 | 0 | 0 |
| 0 | 0 | 0 | 0 | 0 |
| 0 | 0 | 0 | 0 | 0 |
| 0 | 0 | 0 | 0 | 0 |
| 0 | 0 | 0 | 0 | 0 |
| 0 | 0 | 0 | 0 | 0 |
| 0 | 0 | 0 | 0 | 0 |
| 0 | 0 | 0 | 0.010280531 | 0 |
| 0 | 0 | 0 | 0 | 0 |

| PHL018-0265_V3 | PHL018-0273_V2 | PHL018-0187_V1 | PHL018-0189_V3 | PHL018-0234_V2 |
|---|---|---|---|---|
| REF | REF | CTRL | EYCF | CTRL |
| 0 | 0.141766163 | 1.499670912 | 0.151940338 | 0.053737539 |
| 0 | 0.242176126 | 0.065187911 | 1.848109799 | 0.25260511 |
| 0 | 0.529954219 | 0.507816461 | 9.09899097 | 0 |
| 0 | 0 | 0 | 0 | 0 |
| 0 | 0 | 0 | 0 | 0 |
| 0 | 0 | 0 | 0 | 0 |
| 0 | 0 | 0 | 0 | 0 |
| 0 | 0 | 0 | 0 | 0 |
| 0 | 0 | 0 | 0 | 0 |
| 0 | 0 | 0 | 0 | 0 |
| 0 | 0 | 0 | 0 | 0 |
| 0 | 0 | 0 | 0 | 0 |
| 0 | 0 | 0 | 0 | 0 |
| 0 | 0.377806029 | 7.541677046 | 13.03430369 | 1.568340371 |
| 0 | 0 | 0 | 0 | 0 |
| 0 | 0 | 0 | 0 | 0 |
| 0 | 0 | 0 | 0 | 0 |
| 0 | 0 | 0 | 0 | 0 |
| 0 | 0 | 0 | 0 | 0 |
| 0 | 0 | 0 | 0 | 0 |
| 0 | 0 | 0 | 0 | 0 |
| 0 | 0 | 0 | 0 | 0 |
| 0 | 0 | 0 | 0 | 0 |
| 0 | 0 | 0 | 0 | 0 |
| 0 | 0.002857667 | 0 | 0 | 0.093143861 |
| 0 | 0 | 0 | 0 | 0 |
| 0 | 0 | 0 | 0 | 0 |
| 0 | 0 | 0 | 0 | 0 |
| 0 | 0 | 0 | 0 | 0 |
| 0 | 0 | 0 | 0 | 0 |
| 0 | 0 | 0 | 0 | 0 |
| 0 | 0 | 0 | 0 | 0 |
| 0 | 0 | 0 | 0 | 0 |
| 0 | 0 | 0 | 0 | 0 |
| 0 | 0 | 0 | 0 | 0 |
| 0 | 1.243995039 | 0 | 2.542686713 | 2.707677174 |
| 0 | 0 | 0 | 0 | 0 |
| 0 | 0 | 0 | 0 | 0 |
| 0 | 0 | 0 | 0 | 0 |
| 0 | 0 | 0 | 0 | 0 |
| 0 | 0 | 0 | 0 | 0 |
| 0.160628874 | 0.166925252 | 0.000683908 | 0.002013747 | 0.002757846 |
| 4.661085421 | 2.224033385 | 0.013550163 | 0.019736821 | 2.155866797 |
| 0.262774589 | 0.009910404 | 0.000623021 | 0.056960279 | 0.005187208 |
| 0.068042333 | 0.018599817 | 0 | 0 | 0.00268498 |
| 0 | 0 | 0 | 0 | 0 |

| | | | | |
|---|---|---|---|---|
| 0 | 0.000881918 | 0 | 0 | 0 |
| 0 | 0 | 0.007288959 | 0.277887557 | 0 |
| 0.001311427 | 0.036621434 | 0.117091566 | 0 | 0 |
| 0 | 0.698409509 | 0.002314698 | 0.206325769 | 0.368592396 |
| 0 | 0 | 0.075158994 | 0 | 0.001141335 |
| 0 | 0.070233589 | 0 | 0 | 0 |
| 0 | 0.040102654 | 0.001030231 | 0 | 0.001649537 |
| 0 | 0 | 0 | 0 | 0 |
| 0 | 0.002885327 | 0 | 0 | 0 |
| 0 | 0 | 0 | 0 | 0 |
| 0 | 0.025693019 | 0.00039441 | 0 | 0 |
| 0 | 0.018663668 | 0.007149428 | 0 | 0.263867293 |
| 0.000736372 | 0.00507035 | 0.001041248 | 0 | 0 |
| 0 | 0.003881116 | 0.795184732 | 0 | 0.013126074 |
| 0 | 0.001277981 | 0 | 0 | 0.000900454 |
| 0 | 0 | 0 | 0 | 0 |
| 0.001877699 | 2.839710211 | 0.002107234 | 0.121736058 | 0 |
| 0 | 0 | 0 | 0 | 0 |
| 0 | 0 | 0 | 0 | 0 |
| 0 | 0 | 0 | 0 | 0 |
| 0 | 0 | 0.002858708 | 0.075613028 | 0.04951668 |
| 0 | 0.002772414 | 0.119257163 | 0.002155393 | 0.003516152 |
| 0 | 0.499606772 | 0 | 0 | 0 |
| 0 | 0 | 0 | 0 | 0 |
| 0 | 0 | 0 | 0 | 0 |
| 0 | 0 | 0 | 0 | 0 |
| 0 | 0 | 0 | 0 | 0 |
| 0 | 0 | 0 | 0 | 0 |
| 0.048483169 | 0.012807598 | 0 | 0 | 0 |
| 0 | 0 | 0 | 0 | 0 |
| 0 | 0 | 0 | 0 | 0 |
| 0 | 0 | 0 | 0 | 0 |
| 0 | 0 | 0 | 0 | 0 |
| 0 | 0 | 0 | 0 | 0.000963262 |
| 0 | 0.002305959 | 0 | 0 | 0 |
| 0 | 0 | 0 | 0 | 0 |
| 0 | 0 | 0 | 0 | 0 |
| 0 | 0.0102723 | 0 | 0 | 0.002842469 |
| 0 | 0 | 0 | 0 | 0 |
| 0 | 0 | 0 | 0 | 0 |
| 0 | 0 | 0 | 0 | 0 |
| 0 | 0 | 0 | 0 | 0 |
| 0 | 0 | 0 | 0 | 0 |
| 0 | 0 | 0 | 0 | 0 |
| 0 | 0 | 0 | 0 | 0 |
| 0 | 0 | 0 | 0 | 0 |
| 0 | 0 | 0 | 0 | 0 |
| 0 | 0 | 0 | 0 | 0 |

| | | | | |
|---|---|---|---|---|
| 0 | 0 | 0 | 0 | 0 |
| 0 | 0 | 0 | 0 | 0 |
| 0 | 0 | 0 | 0 | 0 |
| 0 | 0 | 0 | 0 | 0 |
| 0 | 0 | 0 | 0 | 0 |
| 0 | 0.002082458 | 0 | 0 | 0 |
| 0 | 0 | 0 | 0 | 0 |
| 0 | 0.055842091 | 0 | 0 | 0 |
| 0 | 0.005955012 | 0 | 0 | 0.002769259 |
| 0 | 0 | 0 | 0 | 0 |
| 0 | 0 | 0 | 0 | 0 |
| 0 | 0.015555716 | 0 | 0 | 0 |
| 0 | 0 | 0 | 0 | 0 |
| 0 | 0 | 0 | 0 | 0 |
| 0 | 0 | 0 | 0 | 0 |
| 0 | 0 | 0 | 0 | 0 |
| 0 | 0 | 0 | 0 | 0 |
| 0 | 0 | 0 | 0 | 0 |
| 0 | 0.695115567 | 0 | 0 | 0.056964858 |
| 0 | 0 | 0 | 0 | 0 |
| 0 | 0 | 0 | 0 | 0 |
| 0 | 0 | 0 | 0 | 0 |
| 0 | 0 | 0 | 0 | 0 |
| 0 | 0 | 0 | 0 | 0 |
| 0 | 0 | 0 | 0 | 0 |
| 0 | 0 | 0 | 0 | 0 |
| 0.017109379 | 0.000872652 | 0.001036847 | 0 | 0 |
| 0 | 0 | 0 | 0 | 0 |
| 0 | 0 | 0 | 0 | 0 |
| 0 | 0 | 0 | 0 | 0 |
| 0 | 0 | 0 | 0 | 0.010133885 |
| 0 | 0.12935197 | 0 | 0.00126407 | 0.000488797 |
| 0 | 0 | 0 | 0 | 0 |
| 0 | 0 | 0 | 0 | 0 |
| 0 | 0 | 0 | 0 | 0 |
| 0 | 0 | 0 | 0 | 0 |
| 0 | 0 | 0 | 0 | 0 |
| 0 | 0 | 0 | 0 | 0 |
| 0 | 0 | 0 | 0 | 0.105315738 |
| 0 | 0 | 0 | 0 | 0 |
| 0 | 0 | 0 | 0 | 0 |
| 0 | 0 | 0 | 0 | 0 |
| 0 | 0 | 0 | 0.000438319 | 0.0009887 |
| 0 | 0 | 0 | 0 | 0 |
| 0 | 0 | 0 | 0 | 0 |
| 0 | 0 | 0 | 0 | 0 |
| 0 | 0 | 0 | 0 | 0 |
| 0 | 0 | 0 | 0 | 0 |

| | | | | |
|---|---|---|---|---|
| 0 | 0 | 0 | 0 | 0.00150885 |
| 0 | 0 | 0 | 0 | 0 |
| 0 | 0 | 0 | 0 | 0 |
| 0 | 0 | 0 | 0 | 0 |
| 0 | 0.044531322 | 0 | 0 | 0.027121939 |
| 0 | 0 | 0 | 0 | 0 |
| 0 | 0 | 0 | 0 | 0 |
| 0 | 0 | 0 | 0 | 0 |
| 0 | 0 | 0 | 0 | 0 |
| 0 | 0 | 0 | 0 | 0 |
| 0 | 0 | 0 | 0 | 0 |
| 0 | 0 | 0 | 0 | 0 |
| 0 | 0 | 0 | 0 | 0 |
| 0 | 0 | 0 | 0 | 0 |
| 0 | 0 | 0 | 0 | 0 |
| 0 | 0 | 0 | 0 | 0 |
| 0 | 0 | 0 | 0 | 0 |
| 0 | 0 | 0 | 0 | 0 |
| 0 | 0 | 0 | 0 | 0 |
| 0 | 0 | 0 | 0 | 0 |
| 0 | 0 | 0 | 0 | 0 |
| 0 | 0 | 0 | 0 | 0 |
| 0 | 0.001472469 | 0.036156792 | 0 | 0.009026145 |
| 0 | 0 | 0 | 0 | 0 |
| 0 | 0 | 0 | 0 | 0 |
| 0 | 0 | 0 | 0 | 0 |
| 0 | 0.003767889 | 0 | 0 | 0 |
| 0 | 0 | 0 | 0 | 0 |
| 0 | 0 | 0 | 0 | 0 |
| 0 | 0 | 0 | 0 | 0 |
| 0 | 0 | 0 | 0 | 0 |
| 0 | 0.071703946 | 0 | 0 | 0 |
| 0 | 0 | 0 | 0 | 0 |
| 0 | 0 | 0 | 0 | 0 |
| 0 | 0 | 0 | 0 | 0 |
| 0 | 0 | 0 | 0 | 0 |
| 0 | 0 | 0 | 0 | 0 |
| 0 | 0 | 0 | 0 | 0 |
| 0 | 0 | 0 | 0 | 0.002511409 |
| 0 | 0 | 0 | 0 | 0 |
| 0 | 0 | 0 | 0 | 0 |
| 0 | 0 | 0 | 0 | 0 |
| 0 | 0 | 0 | 0 | 0 |
| 0 | 0 | 0 | 0 | 0 |
| 0 | 0 | 0 | 0 | 0 |
| 0 | 0 | 0 | 0 | 0 |
| 0 | 0 | 0 | 0 | 0 |
| 0 | 0 | 0 | 0 | 0 |

| | | | | |
|---|---|---|---|---|
| 0 | 0 | 0 | 0 | 0 |
| 0 | 0 | 0 | 0 | 0 |
| 0 | 0 | 0 | 0 | 0 |
| 0 | 0 | 0 | 0 | 0 |
| 0 | 0 | 0 | 0 | 0 |
| 0 | 0 | 0 | 0 | 0 |
| 0 | 0 | 0 | 0 | 0 |
| 0 | 0 | 0 | 0 | 0 |
| 0 | 0 | 0 | 0 | 0 |
| 0 | 0 | 0 | 0 | 0 |
| 0 | 0 | 0 | 0 | 0 |
| 0 | 0 | 0 | 0 | 0 |
| 0 | 0 | 0.014535233 | 0.001116485 | 0.001214235 |
| 0 | 0 | 0 | 0 | 0 |
| 0 | 0 | 0 | 0 | 0 |
| 0 | 0 | 0 | 0 | 0 |
| 0 | 0 | 0 | 0 | 0 |
| 0 | 0 | 0 | 0 | 0 |
| 0 | 0 | 0 | 0 | 0 |
| 0 | 0 | 0 | 0 | 0 |
| 0.059957658 | 0.022674694 | 0.048328496 | 0.040707402 | 0.017153589 |
| 0 | 0 | 0 | 0 | 0 |
| 0 | 0 | 0.005080336 | 0 | 0 |
| 0 | 0 | 0.000739319 | 0 | 0 |
| 0 | 0 | 0 | 0 | 0 |
| 0 | 0 | 0 | 0 | 0 |
| 0.288030313 | 0.003648807 | 0.001445117 | 0.007902344 | 0 |
| 0 | 0 | 0 | 0 | 0 |
| 0 | 0 | 0 | 0 | 0 |
| 0.002186819 | 0 | 0 | 0 | 0 |
| 0 | 0 | 0.001097552 | 0 | 0 |
| 0 | 0 | 0 | 0 | 0 |
| 0 | 0 | 0 | 0 | 0 |
| 0 | 0.340234768 | 0.0031774 | 0 | 0 |
| 0 | 0 | 0.086339466 | 0.036333915 | 0.010832773 |
| 0 | 0 | 0 | 0 | 0 |
| 0 | 0 | 0 | 0 | 0 |
| 0 | 0 | 0 | 0 | 0 |
| 0 | 0.003763144 | 0 | 0 | 0 |
| 0 | 0 | 0 | 0 | 0 |
| 0 | 0.097188717 | 0 | 0 | 0 |
| 0 | 0.003877413 | 0 | 0 | 0 |
| 0 | 0.000807077 | 0 | 0 | 0 |
| 0 | 0 | 0 | 0 | 0 |
| 0 | 0 | 0 | 0 | 0 |
| 0 | 0.023103306 | 0 | 0 | 0 |
| 0 | 0 | 0.002580555 | 0.001470653 | 0 |
| 0.002034202 | 0.00055335 | 0.01013593 | 0 | 0.001169656 |

| | | | | |
|---|---|---|---|---|
| 0 | 0 | 0 | 0 | 0 |
| 0 | 0 | 0 | 0 | 0 |
| 0 | 0 | 0 | 0 | 0 |
| 0 | 0 | 0 | 0 | 0 |
| 0 | 0 | 0 | 0 | 0 |
| 0 | 0 | 0 | 0 | 0 |
| 0 | 0.074042526 | 0 | 0 | 0 |
| 0 | 0.014715834 | 0 | 0 | 0 |
| 0 | 0 | 0.424518101 | 0 | 0 |
| 0 | 0 | 0 | 0 | 0 |
| 0 | 0 | 0 | 0 | 0 |
| 0 | 0 | 0 | 0 | 0 |
| 0 | 0 | 0 | 0 | 0 |
| 0 | 0 | 0 | 0 | 0 |
| 0 | 0.008070155 | 0 | 0 | 0 |
| 0 | 0 | 0 | 0 | 0 |
| 0 | 0 | 0 | 0 | 0 |
| 0.026834533 | 0 | 0 | 0 | 0 |
| 0.027031922 | 0.014912957 | 0 | 0 | 0.014274384 |
| 0 | 0 | 0 | 0 | 0.264041967 |
| 0 | 0.00456565 | 0.002958929 | 0.031038231 | 0 |
| 0 | 0 | 0 | 0 | 0 |
| 0 | 0 | 0 | 0 | 0 |
| 0 | 0 | 0 | 0 | 0 |
| 0 | 0.004947615 | 0.003102562 | 0.002524258 | 0.004531858 |
| 0 | 0 | 0 | 0 | 0 |
| 0 | 0 | 0 | 0 | 0 |
| 0 | 0 | 0 | 0 | 0 |
| 0.938643152 | 0.0332478 | 0 | 0 | 0 |
| 0 | 0 | 0 | 0 | 0 |
| 0.000747958 | 0.033876396 | 0.370654531 | 0.375435126 | 0.180243188 |
| 0 | 0 | 0 | 0 | 0 |
| 0 | 0 | 0 | 0.139234072 | 0 |
| 0 | 0 | 0 | 0 | 0 |
| 0 | 0 | 0 | 0 | 0 |
| 0 | 0 | 0 | 0 | 0 |
| 0 | 0 | 0 | 0 | 0 |
| 0 | 0 | 0 | 0 | 0 |
| 0 | 0 | 0 | 0 | 0 |
| 0.000597685 | 0.000457268 | 0 | 0 | 0 |
| 0 | 0 | 0 | 0 | 0 |
| 0.003202159 | 0.054441321 | 0.002895414 | 0.000680223 | 0.017360122 |
| 0 | 0 | 0 | 0 | 0 |
| 0 | 0 | 0 | 0 | 0 |
| 0 | 0 | 0 | 0 | 0 |
| 0 | 0 | 0 | 0 | 0 |
| 0 | 0 | 0 | 0 | 0 |
| 0 | 0 | 0 | 0 | 0 |

| | | | | |
|---|---|---|---|---|
| 0 | 0 | 0 | 0 | 0 |
| 0 | 0 | 0 | 0 | 0 |
| 0.236463528 | 0.008593833 | 0 | 0.057910177 | 0 |
| 0 | 0 | 0 | 0 | 0 |
| 0 | 0 | 0 | 0 | 0 |
| 0 | 0 | 0 | 0 | 0 |
| 0 | 0 | 0 | 0 | 0 |
| 0 | 0 | 0 | 0 | 0 |
| 0 | 0 | 0 | 0 | 0 |
| 0 | 0.000535899 | 0.000848976 | 0 | 0 |
| 0 | 0 | 0 | 0 | 0 |
| 0 | 0 | 0 | 0 | 0 |
| 0 | 0 | 0 | 0 | 0 |
| 0 | 0.003022007 | 0.000512945 | 0.001699147 | 0 |
| 0.002794465 | 0.04756932 | 0.001834599 | 0 | 0.000602551 |
| 0 | 0.017930144 | 0 | 0 | 0.002419165 |
| 0.037014487 | 0.002638999 | 0.002251158 | 0.00106529 | 0.001029829 |
| 0 | 0 | 0 | 0 | 0 |
| 0 | 0 | 0 | 0 | 0 |
| 0 | 0 | 0 | 0 | 0 |
| 0 | 0 | 0 | 0 | 0 |
| 0.001077855 | 0 | 0.002467615 | 0 | 0 |
| 0 | 0 | 0 | 0 | 0 |
| 0 | 0 | 0.000562554 | 0 | 0 |
| 0 | 0 | 0 | 0 | 0 |
| 0 | 0 | 0 | 0 | 0 |
| 0 | 0.224407302 | 0 | 0 | 2.709012362 |
| 0 | 0 | 0 | 0 | 0 |
| 0 | 0 | 0 | 0 | 0 |
| 0.00337811 | 0 | 0 | 0 | 0 |
| 0 | 0 | 0 | 0 | 0 |
| 0 | 0 | 0 | 0 | 0 |
| 0 | 0 | 0 | 0 | 0 |
| 0 | 0 | 0 | 0 | 0 |
| 0 | 0.007364182 | 0 | 0 | 0 |
| 0 | 0.140184811 | 0.000579848 | 0 | 0 |
| 0 | 0 | 0 | 0 | 0 |
| 0.105854377 | 0.064005577 | 0.025805459 | 0 | 0.062487951 |
| 0 | 0 | 0 | 0 | 0 |
| 0 | 0 | 0 | 0 | 0 |
| 0 | 0 | 0 | 0 | 0 |
| 0 | 0 | 0 | 0 | 0 |
| 0 | 0 | 0 | 0 | 0 |
| 0 | 0 | 0 | 0 | 0 |
| 0 | 0 | 0 | 0 | 0 |
| 0 | 0 | 0 | 0 | 0 |
| 0 | 0 | 0 | 0 | 0 |
| 0.001640293 | 0 | 0.001656725 | 0 | 0.005658959 |

| | | | | |
|---|---|---|---|---|
| 0 | 0 | 5.936804561 | 0.104839537 | 0.129951429 |
| 0 | 0 | 0 | 0 | 0 |
| 0.005080363 | 0 | 0.000456112 | 0.000503628 | 0.001460591 |
| 0 | 0 | 0 | 0 | 0 |
| 0.178328802 | 0.008690007 | 0.029522087 | 0.003546891 | 0.013062186 |
| 0 | 0 | 0 | 0 | 0 |
| 0 | 0.004084488 | 0 | 0 | 0 |
| 0.313122228 | 0.041144569 | 0.002680162 | 0.098014476 | 0.006408335 |
| 0 | 0 | 0 | 0 | 0 |
| 0.023521865 | 0.00139502 | 0.016575007 | 1.941205299 | 0.011323216 |
| 0.075670264 | 0 | 0 | 0 | 0 |
| 0.06763874 | 0.347986748 | 0.00033946 | 0 | 0 |
| 0.016092997 | 0.007314171 | 0.020792525 | 0 | 0.001649119 |
| 0 | 0 | 0 | 0.010795122 | 0 |
| 0 | 0 | 0.004170998 | 0 | 0 |
| 0 | 0 | 0 | 0 | 0 |
| 0.046963904 | 0.030696126 | 0 | 0 | 0 |
| 0 | 0 | 0 | 0 | 0 |
| 0 | 0 | 0 | 0.079908272 | 0 |
| 0 | 0 | 0 | 0 | 0 |
| 0 | 0 | 0 | 0 | 0 |
| 0 | 0 | 0.011838081 | 0 | 0 |
| 0 | 0.012746516 | 0 | 0 | 0 |
| 0 | 0.0081038 | 0 | 0 | 0.204984085 |
| 0 | 0 | 0 | 0 | 0 |
| 0.001855865 | 0.035401778 | 0 | 0 | 0 |
| 0 | 0.775909726 | 0 | 0 | 0 |
| 0 | 0 | 0 | 0 | 0 |
| 0 | 0 | 0 | 0 | 0 |
| 0 | 0 | 0 | 0 | 0 |
| 0 | 0 | 0 | 0.0007107 | 0 |
| 0 | 0 | 0.009554205 | 0 | 0 |
| 0 | 0 | 0 | 0 | 0 |
| 0 | 0 | 0 | 0 | 0 |
| 0 | 0 | 0 | 0 | 0 |
| 0 | 0 | 0 | 0 | 0 |
| 0 | 0 | 0 | 0 | 0 |
| 0 | 0 | 0 | 0 | 0 |
| 0 | 0.006066842 | 0 | 0 | 0 |
| 0 | 0 | 0 | 0 | 0 |
| 0 | 0 | 0 | 0 | 0 |
| 0 | 0 | 0 | 0 | 0 |
| 0 | 0 | 0 | 0 | 0 |
| 0 | 0 | 1.722755205 | 0 | 0 |
| 0 | 0 | 0 | 0 | 0 |
| 0 | 0 | 0 | 0 | 0 |
| 0 | 0.001647991 | 0.001118897 | 0 | 0.000796224 |
| 0 | 0 | 0 | 0 | 0 |

| | | | | |
|---|---|---|---|---|
| 0 | 0 | 0 | 0 | 0 |
| 0 | 0.010787366 | 0 | 0 | 0 |
| 0 | 0 | 0 | 0 | 0 |
| 0 | 0 | 0 | 0 | 0 |
| 0 | 0 | 0 | 0 | 0 |
| 0 | 0 | 0.65595651 | 0 | 0 |
| 0 | 0 | 0 | 0 | 0 |
| 0 | 0.001438965 | 0 | 0.005243967 | 0 |
| 0 | 0 | 0 | 0 | 0.182458247 |
| 0 | 0 | 0 | 0 | 0 |
| 0.018553027 | 0.022151665 | 0.027767634 | 0.001316704 | 0.002363909 |
| 0 | 0 | 0.002155412 | 0.002644396 | 0 |
| 0 | 0 | 0 | 0 | 0 |
| 0.150780357 | 0.002844553 | 0.003540713 | 0.001954788 | 0.00274868 |
| 0 | 0 | 0 | 0.076238811 | 0.003302639 |
| 0 | 0 | 0 | 0 | 0 |
| 0 | 0 | 0 | 0 | 0 |
| 0 | 0 | 0 | 0 | 0 |
| 0 | 0 | 0 | 0 | 0 |
| 0 | 0 | 0 | 0 | 0 |
| 0 | 0 | 0 | 0 | 0 |
| 0 | 0.266934235 | 0 | 0.002611936 | 0.339514096 |
| 0 | 0 | 0 | 0 | 0 |
| 0 | 0 | 0 | 0 | 0 |
| 0 | 0 | 0 | 0 | 0 |
| 0 | 0 | 0 | 0 | 0 |
| 0 | 0 | 0 | 0 | 0 |
| 0 | 0 | 0 | 0 | 0 |
| 0 | 0 | 0 | 0 | 0 |
| 0 | 0.679035834 | 1.455543308 | 0.122947809 | 0.377717701 |
| 0 | 0.005651565 | 0.001419418 | 0.009765419 | 0 |
| 0.002086882 | 0.027287356 | 0.000536528 | 0 | 0 |
| 0 | 0 | 0 | 0 | 0 |
| 0 | 0 | 0 | 0 | 0.001155499 |
| 0 | 0 | 0 | 0 | 0.006981486 |
| 0 | 0 | 0 | 0 | 0 |
| 0 | 0 | 0 | 0 | 0.450625712 |
| 0 | 0.034585185 | 0.001230317 | 0 | 0 |
| 0 | 0 | 0 | 0 | 0 |
| 0 | 0 | 0 | 0.554913073 | 0.01727833 |
| 0.000982493 | 0.005679291 | 0.041611944 | 0.00525943 | 0.007061606 |
| 0 | 0.139062604 | 0.051719106 | 3.618791116 | 0.368040711 |
| 0 | 0.006639996 | 0 | 0 | 0 |
| 0 | 0 | 0 | 0 | 0 |
| 0 | 0 | 0 | 0 | 0 |
| 0 | 0 | 0 | 0 | 0 |
| 0 | 0.001402547 | 0.002430231 | 0.000766687 | 0 |
| 0 | 0 | 0 | 0 | 0.008879828 |

| | | | | |
|---|---|---|---|---|
| 0.028415233 | 0.011897238 | 0.000771042 | 0 | 0.002011844 |
| 0 | 0 | 0 | 0 | 0.002687883 |
| 0.010690772 | 0.355755537 | 0.005486487 | 0.282537371 | 0 |
| 0 | 0 | 0 | 0 | 0 |
| 0 | 0 | 0 | 0 | 0 |
| 0 | 0 | 0 | 0 | 0 |
| 0 | 0 | 0 | 0 | 0 |
| 0 | 0.003394209 | 0 | 0 | 0 |
| 0 | 0 | 0 | 0 | 0 |
| 0 | 0 | 0.020090377 | 0 | 0 |
| 0 | 0 | 0 | 0 | 0 |
| 0.001251656 | 0 | 0 | 0 | 0 |
| 0 | 0 | 0 | 0 | 0 |
| 0.059854042 | 0.00058708 | 0 | 0 | 0.000992763 |
| 0 | 0 | 0 | 0 | 0 |
| 0 | 0.036091699 | 0.817615379 | 0 | 0.06788485 |
| 0.275344016 | 0.089839235 | 0.00422944 | 0.00237209 | 0.015048656 |
| 0 | 0 | 0 | 0.011527986 | 0 |
| 0 | 0 | 0.001525575 | 0 | 0 |
| 0 | 0 | 0 | 0 | 0 |
| 0 | 0 | 0.161093439 | 0 | 0.010656854 |
| 0 | 0 | 0.000622975 | 0 | 0 |
| 0 | 0 | 0 | 0 | 0.075936268 |
| 0 | 0 | 0.031495672 | 0 | 0 |
| 0 | 0 | 0 | 0 | 0 |
| 0 | 0 | 0.009462777 | 0 | 0 |
| 0 | 0 | 0.004090901 | 0 | 0 |
| 0 | 0 | 0 | 0 | 0.000460574 |
| 0 | 0 | 0.006716551 | 0 | 0 |
| 0 | 0 | 0 | 0 | 0 |
| 0 | 0.001472875 | 0.032861193 | 0 | 0.018956669 |
| 0.022722269 | 0.012443293 | 0.001449468 | 0 | 0.004744727 |
| 0 | 0.010686158 | 0.050051233 | 0.073958159 | 0.008249569 |
| 0 | 0.046275839 | 0.000527414 | 0 | 0 |
| 0 | 0 | 0 | 0 | 0 |
| 0 | 0 | 0 | 0 | 0 |
| 0 | 0 | 0 | 0 | 0.00124303 |
| 0 | 0 | 0.003642822 | 0.008331952 | 0.044994667 |
| 0 | 0.483786511 | 0 | 0 | 0.0587004 |
| 0 | 0 | 0 | 0 | 0 |
| 0 | 0 | 0 | 0 | 0 |
| 0 | 0 | 0 | 0 | 0 |
| 0 | 0 | 0.707440233 | 0.015468108 | 0 |
| 0 | 0 | 0.04722526 | 0.004261317 | 0.056588479 |
| 0 | 0 | 0 | 0 | 0 |
| 0 | 0 | 0 | 0 | 0 |
| 0 | 0 | 0 | 0 | 0 |
| 0 | 0 | 0 | 0 | 0 |

| | | | | |
|---|---|---|---|---|
| 0 | 0 | 0 | 0 | 0 |
| 0.005485014 | 1.973311226 | 0.068961463 | 0.446108008 | 0.593618596 |
| 0 | 0 | 0 | 0 | 0 |
| 0 | 0 | 0 | 0 | 0 |
| 0 | 0 | 0.000434797 | 0 | 0 |
| 0 | 0 | 0 | 0 | 0 |
| 0 | 0 | 0 | 0 | 0 |
| 0 | 0.002062894 | 0.000340422 | 0 | 0 |
| 0 | 0.008195096 | 0 | 0 | 0 |
| 0 | 0 | 0.041224018 | 0 | 0 |
| 0 | 0.000997881 | 0.052233939 | 0.007709466 | 0.001828054 |
| 0.204237364 | 0.029231981 | 0.026981188 | 0.0654341 | 0.020945681 |
| 0 | 0.087622659 | 0 | 0 | 0.210818507 |
| 0 | 0 | 0 | 0 | 0 |
| 0.011223984 | 0 | 0 | 0.000834495 | 0 |
| 0 | 0 | 0 | 0 | 0 |
| 0.003073049 | 0.008533557 | 0.01289816 | 0.015993527 | 0.029597042 |
| 0 | 0 | 0 | 0.335748096 | 0.001720345 |
| 0 | 0 | 0 | 0 | 0 |
| 0 | 0 | 0.002383791 | 0.000898776 | 0 |
| 0.001936665 | 0.485809726 | 0.001493724 | 0.02167699 | 0.0182221 |
| 0 | 0 | 0 | 0 | 0 |
| 0 | 0 | 0 | 0 | 0 |
| 0 | 0 | 0 | 0 | 0 |
| 0 | 0 | 0 | 0 | 0 |
| 0 | 0 | 0 | 0 | 0 |
| 0 | 0.000886356 | 0 | 0 | 0 |
| 0.007485052 | 0.02615125 | 0.002419212 | 0.078467658 | 0.05794081 |
| 0 | 0 | 0 | 0 | 0 |
| 0 | 0 | 0.001185493 | 0 | 0 |
| 0 | 0 | 0 | 0 | 0 |
| 0 | 0 | 0 | 0 | 0 |
| 0 | 0 | 0 | 0 | 0 |
| 0 | 0 | 0 | 0 | 0 |
| 0 | 0.07934562 | 0 | 0 | 0 |
| 0 | 0.262972089 | 0.656634996 | 0.047187033 | 0.148867531 |
| 0 | 0 | 0.000829617 | 0 | 0 |
| 0 | 0 | 0 | 0 | 0 |
| 0.001154206 | 0.110203724 | 0.003357415 | 0 | 0.002986482 |
| 0 | 0 | 0 | 0 | 0 |
| 0 | 0 | 0 | 0 | 0 |
| 0 | 0 | 0 | 0 | 0 |
| 0 | 0.024744702 | 0.136239591 | 0.006755396 | 0 |
| 0.008122583 | 0.000552383 | 0.002133029 | 0 | 0.004553683 |
| 0 | 0 | 0 | 0 | 0 |
| 0 | 0.61438548 | 0 | 0 | 0.326326558 |
| 0 | 0 | 0 | 0 | 0 |
| 0 | 0 | 0 | 0 | 0 |

| | | | | |
|---|---|---|---|---|
| 0 | 0 | 0 | 0 | 0 |
| 0 | 0.034505626 | 0 | 0 | 0 |
| 0 | 0 | 0 | 0 | 0 |
| 0 | 0.052308675 | 4.1491303 | 0 | 1.780366349 |
| 0 | 0 | 0 | 0 | 0 |
| 0 | 0 | 0 | 0 | 0 |
| 0.001012871 | 0 | 0 | 0 | 0 |
| 0 | 0.008345733 | 0.211896146 | 0 | 0.003528196 |
| 0 | 1.515276767 | 0 | 0 | 2.799948235 |
| 0 | 0.01905777 | 0 | 0 | 0.126138614 |
| 0 | 0 | 0 | 0 | 0 |
| 0 | 0.001755703 | 0.000189641 | 0 | 0 |
| 0 | 0 | 0 | 0 | 0 |
| 0 | 0 | 0 | 0 | 0 |
| 0 | 0.047020483 | 0.117369029 | 0.029951127 | 0.046215238 |
| 0 | 0 | 0 | 0 | 0 |
| 0 | 0.168368117 | 0 | 0 | 0.08221829 |
| 0 | 0 | 0 | 0 | 2.37343296 |
| 0 | 0 | 0 | 0 | 0 |
| 0 | 0 | 0 | 0 | 0 |
| 0 | 0 | 0 | 0 | 0 |
| 0 | 0 | 0 | 0 | 0 |
| 0 | 0 | 0 | 0 | 0 |
| 0 | 0 | 0.082204606 | 0.012095064 | 0.159081658 |
| 0 | 0.014196338 | 2.957605621 | 0 | 0 |
| 0 | 0.034035886 | 0 | 0 | 0.363301522 |
| 0 | 0.016358036 | 0 | 0 | 0.007967782 |
| 0 | 1.30542866 | 0 | 0 | 3.281478883 |
| 0 | 0 | 0 | 0 | 0 |
| 0 | 0 | 0 | 0 | 0 |
| 0 | 0.377367349 | 0 | 0 | 0.006822167 |
| 0 | 0 | 0 | 0 | 0.277505593 |
| 0 | 0.000472375 | 0.000795112 | 0 | 0 |
| 0 | 0 | 0 | 0 | 0 |
| 0 | 0 | 0 | 0 | 0 |
| 0 | 0 | 0 | 0 | 0 |
| 0 | 0.259802334 | 0 | 0 | 0 |
| 0 | 0 | 0 | 0 | 0 |
| 0 | 0 | 0 | 0 | 0 |
| 0 | 0 | 0 | 0 | 0.000386378 |
| 0 | 0.071199234 | 0.008307973 | 0 | 0.020972761 |
| 0 | 0.082317236 | 0 | 0 | 0 |
| 0 | 3.006877552 | 0.227482017 | 0.333193687 | 0.033079954 |
| 0 | 0 | 0 | 0 | 0.031577806 |
| 0 | 0 | 0 | 0 | 0 |
| 0 | 0 | 0 | 0 | 0 |
| 0 | 0 | 0 | 0 | 0 |
| 0.001082636 | 0.006442232 | 0.0121741 | 0.001126905 | 0.072522444 |

| | | | | |
|---|---|---|---|---|
| 0 | 0 | 0 | 0 | 0 |
| 0 | 0.049467753 | 0 | 0 | 0.008081524 |
| 9.085154173 | 0.004411147 | 2.420283994 | 0.038735237 | 0.002983732 |
| 0 | 0 | 0 | 0 | 0 |
| 0 | 0 | 0 | 0 | 0 |
| 0 | 0.009729809 | 0.005890365 | 0 | 0 |
| 0 | 0 | 0 | 0 | 0 |
| 0 | 0 | 0 | 0.004558396 | 0 |
| 0 | 0 | 0 | 0 | 0 |
| 0 | 0 | 0 | 0 | 0 |
| 0.000509482 | 0.115203653 | 1.490515593 | 0.046970723 | 0.002783021 |
| 0.001371768 | 0.393209532 | 0.095754177 | 1.825620276 | 0.602875299 |
| 0 | 0 | 0 | 0 | 0 |
| 0.961694135 | 0.176122097 | 0.009700327 | 0 | 0.083150894 |
| 0.218530563 | 0.051567769 | 0.471257997 | 0.05528261 | 0.019682743 |
| 0 | 0 | 0 | 0 | 14.07522665 |
| 0 | 0 | 0 | 0 | 0 |
| 0 | 0 | 0.020736652 | 0 | 0 |
| 0 | 0 | 0 | 0 | 0.051508642 |
| 0 | 0 | 0.022776671 | 0 | 0 |
| 0.002704466 | 0.092056443 | 0.000862598 | 0 | 0.020747696 |
| 1.355242068 | 0.001376956 | 0.055807099 | 24.56835334 | 0 |
| 0.261358879 | 0.5326688 | 0.059747928 | 14.54005674 | 0.256960298 |
| 0 | 0 | 0 | 0 | 0 |
| 0 | 0 | 0 | 0 | 0 |
| 0 | 8.64742623 | 0 | 0.002739201 | 0.003677805 |
| 0 | 0.015977207 | 0.323749682 | 0.958436477 | 0 |
| 0 | 0.025593424 | 0 | 0 | 0 |
| 0 | 0.001324944 | 0 | 0 | 0 |
| 0 | 0 | 0 | 0 | 0 |
| 0 | 0.002722571 | 0 | 0 | 0.00379146 |
| 0 | 0.162713247 | 0 | 0 | 0.433936583 |
| 0.037325256 | 0.004981775 | 0 | 0 | 0.05385516 |
| 0 | 0.013567818 | 0 | 0.000474669 | 0 |
| 0 | 0 | 0.00488337 | 0 | 0 |
| 0 | 0.043884836 | 1.054342339 | 0.034995984 | 0.082940641 |
| 0 | 0.002026096 | 0.004691182 | 0.000817878 | 0.001713082 |
| 0 | 0 | 0 | 0 | 0 |
| 0 | 0.185214677 | 0 | 0 | 2.834935167 |
| 0 | 0 | 0 | 0 | 0 |
| 0 | 0.002557245 | 0 | 0 | 0 |
| 0 | 0 | 0 | 0 | 0 |
| 0 | 0 | 0 | 0 | 0 |
| 0 | 0 | 0 | 0 | 0 |
| 0 | 0.05206958 | 0.443050615 | 0.017379703 | 0.011200783 |
| 0 | 0.469295914 | 0 | 0 | 0 |
| 0 | 0.001841827 | 0 | 0 | 0 |
| 1.233642609 | 0 | 0 | 0.031058772 | 0.259903034 |

| | | | | |
|---|---|---|---|---|
| 0 | 0 | 0 | 0 | 0 |
| 0 | 0 | 0 | 0 | 0 |
| 0 | 0 | 0 | 0 | 0 |
| 0 | 0.013486104 | 0 | 0 | 0.000662943 |
| 0 | 0 | 0 | 0 | 0 |
| 0 | 0.006232417 | 0 | 0 | 0 |
| 0 | 0.006180579 | 0 | 0 | 0 |
| 0 | 0.789456208 | 0 | 0.001262067 | 0.1034607 |
| 0 | 0 | 0 | 0 | 0 |
| 0 | 0 | 0 | 0 | 0 |
| 0.027222631 | 0.000523952 | 0.533859418 | 0.001069274 | 0.003396378 |
| 0 | 0 | 0 | 0 | 0 |
| 0 | 0.004587607 | 0 | 0 | 0 |
| 0 | 0 | 0 | 0 | 0 |
| 0 | 0 | 0 | 0 | 0 |
| 0 | 0 | 0 | 0 | 0 |
| 0 | 0.142233975 | 0 | 0 | 0.019768784 |
| 0 | 0.004507171 | 0.018368985 | 0 | 0.003687923 |
| 0 | 0 | 0 | 0 | 0 |
| 0 | 0 | 0 | 0 | 0.004545391 |
| 0 | 0.000904189 | 0 | 0 | 0 |
| 0 | 0 | 0 | 0 | 0 |
| 0 | 0.089010086 | 0.001327782 | 0 | 0 |
| 0 | 0 | 0 | 0 | 0 |
| 0 | 0 | 0 | 0 | 0 |
| 0 | 0 | 0 | 0 | 0 |
| 0 | 0 | 0 | 0 | 0 |
| 0 | 0 | 0 | 0 | 0 |
| 0 | 0 | 0 | 0 | 0 |
| 0 | 0.035302229 | 0 | 0 | 0.322540403 |
| 0 | 0.000962897 | 0.012312395 | 0 | 0 |
| 0 | 0 | 0 | 0 | 0 |
| 0 | 0 | 0 | 0 | 0 |
| 0 | 0 | 0 | 0 | 0 |
| 0 | 0 | 0.000385008 | 0 | 0 |
| 0 | 0 | 0 | 0 | 0 |
| 0 | 0.037771158 | 0 | 0.004375567 | 0 |
| 0 | 0 | 0.036596377 | 0.023534137 | 0 |
| 0 | 0 | 0 | 0 | 0 |
| 0 | 0 | 0 | 0 | 0 |
| 0 | 0 | 0.069496993 | 0 | 0 |
| 0 | 0 | 0 | 0 | 0 |
| 0 | 0 | 0 | 0 | 0 |
| 0 | 0.780616091 | 0 | 0 | 0 |
| 0 | 0.131614941 | 0 | 0 | 0.122920765 |
| 0 | 0 | 0 | 0 | 0 |
| 0 | 0.006602432 | 0 | 0 | 0 |
| 0 | 0 | 0 | 0 | 0 |

| | | | | |
|---|---|---|---|---|
| 0 | 0 | 0 | 0 | 0 |
| 0 | 0 | 0 | 0 | 0 |
| 0 | 0.00136866 | 0 | 0 | 0 |
| 0 | 0.109708406 | 0 | 0 | 0.041032998 |
| 0 | 0 | 0 | 0 | 0 |
| 0 | 0.283928159 | 0 | 0 | 0 |
| 0 | 0 | 0 | 0 | 0 |
| 0 | 0 | 0 | 0 | 0 |
| 0.099743893 | 0.001427318 | 0.339417575 | 0.073118558 | 0.024481013 |
| 0 | 0 | 0.000455227 | 0 | 0.000971839 |
| 0 | 0 | 0 | 0 | 0 |
| 0 | 0 | 0 | 0 | 0 |
| 0 | 0 | 0 | 0 | 0 |
| 0 | 0 | 0 | 0 | 0 |
| 0 | 2.096988049 | 6.288558477 | 0 | 2.486641728 |
| 0 | 0 | 0 | 0 | 0.00172737 |
| 0 | 0 | 0 | 0 | 0 |
| 0 | 0 | 0 | 0 | 0 |
| 0 | 0 | 0 | 0 | 0 |
| 0 | 0 | 0 | 0 | 0 |
| 0 | 0.001247302 | 0 | 0 | 0.019510199 |
| 0 | 0 | 0 | 0 | 0 |
| 0 | 0 | 0 | 0 | 0 |
| 0 | 0 | 0.017862044 | 0 | 0 |
| 7.354622365 | 0.321789695 | 0.001899336 | 0.028521967 | 0.082312138 |
| 0 | 12.98495502 | 0 | 0.078672726 | 0.566582439 |
| 0 | 0.038706431 | 0 | 0 | 0.023376168 |
| 0 | 0 | 0 | 0 | 0.000755098 |
| 0 | 0.489298598 | 0 | 0 | 0.13435708 |
| 0 | 0 | 0 | 0 | 0 |
| 0 | 0 | 0 | 0 | 0 |
| 0 | 0 | 0 | 0 | 0 |
| 0 | 0 | 0 | 0 | 0 |
| 0 | 0.023434958 | 0 | 0.003360124 | 0.007362753 |
| 0.065570616 | 0.24708361 | 0 | 0 | 0.00249766 |
| 0.015937546 | 0 | 0.083528276 | 0.002087243 | 0.001134992 |
| 0 | 0 | 0 | 0 | 0.295683163 |
| 0 | 2.264063502 | 0 | 0 | 0.787330775 |
| 0 | 0.701879341 | 0 | 0 | 0.000796357 |
| 0 | 0 | 0 | 0 | 0 |
| 0 | 0 | 0 | 0 | 0 |
| 0 | 0 | 0.038317421 | 0.000887606 | 0.00114408 |
| 0 | 0.526875282 | 0 | 0.785795996 | 0.998490646 |
| 0 | 0 | 0 | 0 | 1.216833774 |
| 0 | 0 | 0 | 0 | 0 |
| 0 | 0.001413037 | 0.034697443 | 0 | 0 |
| 0 | 0 | 0 | 0 | 0 |
| 0 | 0.103552346 | 0.01783135 | 0.443198779 | 0.028931032 |

| | | | | |
|---|---|---|---|---|
| 0 | 0 | 0 | 0 | 0 |
| 0 | 0 | 0 | 0 | 0 |
| 0 | 0.014255123 | 0 | 0 | 0 |
| 0 | 0 | 0 | 0 | 0 |
| 0.002949132 | 0.002172711 | 0.031110621 | 0 | 0.006359009 |
| 0 | 0.058745029 | 0 | 0 | 0 |
| 0 | 0 | 0 | 0 | 0 |
| 0 | 0 | 0 | 0 | 0 |
| 0 | 0 | 0 | 0 | 0 |
| 0 | 0 | 0 | 0 | 0 |
| 0 | 0.000383655 | 0 | 0 | 0 |
| 0 | 0.422325343 | 0.688714661 | 0 | 1.100051114 |
| 0 | 0.362034782 | 0.538112919 | 0.315228027 | 0 |
| 0 | 0 | 0 | 0 | 0 |
| 44.82701261 | 1.434388352 | 0.010104862 | 0.016736339 | 0.002614824 |
| 0 | 0 | 0 | 0 | 0 |
| 0 | 0 | 0.032431054 | 0 | 0 |
| 0 | 0 | 0 | 0 | 0 |
| 0 | 0.000677664 | 0 | 0.001037226 | 0.001718913 |
| 0 | 0 | 0 | 0 | 0 |
| 0 | 0.104915174 | 0 | 0 | 0.08787658 |
| 0 | 0.006048526 | 0 | 0 | 0 |
| 0 | 0.292073483 | 0.785854826 | 0 | 0.006685638 |
| 0 | 0 | 0 | 0 | 0 |
| 0 | 0 | 0 | 0 | 0 |
| 0 | 0 | 0 | 0 | 0 |
| 0 | 0 | 0 | 0 | 0 |
| 0 | 0 | 0 | 0 | 0 |
| 0 | 0.000696413 | 0 | 0 | 0 |
| 0 | 0 | 0 | 0 | 0 |
| 0 | 0 | 0 | 0 | 0 |
| 0.010081039 | 0.163784075 | 0.048079061 | 0.007677989 | 0.103913699 |
| 0 | 0 | 0 | 0 | 0 |
| 0 | 0.047239717 | 0.087492409 | 0.001534316 | 0 |
| 0.011297496 | 0 | 0.087556849 | 0.001419652 | 0.000960676 |
| 0 | 0.845767454 | 0 | 0 | 0.254408594 |
| 0 | 0 | 0 | 0 | 0 |
| 0 | 0.123703623 | 0 | 0 | 0 |
| 0 | 0.001678293 | 0 | 0 | 0.00078834 |
| 0 | 0 | 0 | 0 | 0 |
| 0 | 0.09044407 | 0 | 0 | 0.681927331 |
| 0 | 0 | 0 | 0 | 0 |
| 0 | 0 | 0 | 0 | 0 |
| 0 | 0 | 0 | 0 | 0.003128316 |
| 0 | 0.035976907 | 2.262063988 | 0.009369945 | 0.019468031 |
| 0 | 0 | 0 | 0 | 0 |
| 0 | 0.052333739 | 0 | 0 | 0.140025925 |
| 0 | 0 | 0 | 0 | 0.00153444 |

| | | | | |
|---|---|---|---|---|
| 0 | 0 | 0 | 0 | 0 |
| 0 | 0 | 0 | 0 | 0 |
| 0 | 0 | 0 | 0 | 0 |
| 0 | 0 | 0 | 0 | 0 |
| 0 | 0 | 0 | 0 | 0 |
| 0 | 0 | 0 | 0 | 0 |
| 0 | 0.390294066 | 0 | 0 | 0 |
| 0 | 0.004060685 | 0 | 0 | 0.001452569 |
| 0 | 0 | 0 | 0 | 0 |
| 0 | 0.001997934 | 0 | 0 | 0 |
| 0 | 0 | 0 | 0 | 0 |
| 0 | 0 | 0.001999237 | 0 | 0 |
| 0 | 0 | 0 | 0 | 0 |
| 0 | 0 | 0 | 0 | 0 |
| 0 | 0 | 0 | 0 | 0 |
| 0 | 0 | 0 | 0 | 0 |
| 0 | 0 | 0 | 0 | 0 |
| 0 | 0 | 0 | 0 | 0 |
| 0 | 0.163100953 | 0 | 0 | 0.742257537 |
| 0 | 0 | 0 | 0 | 0 |
| 0 | 0 | 0 | 0 | 0 |
| 0 | 0 | 0 | 0 | 0 |
| 0 | 0 | 0 | 0 | 0 |
| 0 | 0 | 0 | 0 | 0 |
| 0.005822898 | 0 | 0 | 0 | 0.000886271 |
| 0 | 0 | 0.047577215 | 0.005337995 | 0.001762056 |
| 0 | 0 | 0 | 0 | 0 |
| 0 | 0.128072734 | 0.012579878 | 0 | 0.003449404 |
| 0 | 0.48482914 | 0 | 0 | 0 |
| 0 | 0.2180532 | 0.256771779 | 0.047678592 | 0.401637994 |
| 0 | 0 | 0 | 0 | 0 |
| 0 | 0 | 0 | 0 | 0 |
| 0 | 0 | 0 | 0 | 0.005853652 |
| 0 | 0 | 0 | 0 | 0 |
| 0.000917536 | 0 | 0.061164063 | 0 | 0.005539576 |
| 0 | 0 | 0 | 0 | 0.238735026 |
| 0 | 0 | 0 | 0 | 0 |
| 0 | 0.001062048 | 0.018507546 | 0 | 0.001047633 |
| 0 | 0 | 0 | 0 | 0 |
| 0 | 0 | 0 | 0 | 0 |
| 0 | 4.39924347 | 36.01460166 | 8.426430461 | 4.251282 |
| 0 | 0.027598543 | 0.134962907 | 0.00124658 | 0.091257755 |
| 0 | 0 | 0 | 0 | 0.682399045 |
| 0 | 0 | 0 | 0 | 0 |
| 0 | 0 | 0 | 0 | 0 |
| 0 | 0 | 0 | 0 | 0.259468447 |
| 0 | 0 | 0 | 0 | 0 |
| 0 | 0.002950149 | 0.055082296 | 0 | 0.007008014 |

| | | | | |
|---|---|---|---|---|
| 0 | 0 | 0 | 0 | 0 |
| 0 | 0 | 0.000343374 | 0 | 0.012706193 |
| 0 | 0 | 0 | 0 | 0.003139255 |
| 0 | 0 | 0 | 0 | 0 |
| 0 | 3.311860383 | 0 | 0.549285441 | 1.118370909 |
| 0 | 0 | 0 | 0 | 0 |
| 0 | 0 | 0 | 0 | 0 |
| 0 | 0 | 0 | 0 | 0.003497502 |
| 0 | 0 | 0.00508587 | 0.646752207 | 0.004391813 |
| 0 | 0 | 0 | 0 | 0 |
| 0 | 0 | 0 | 0 | 0 |
| 0 | 0 | 0 | 0 | 0 |
| 0 | 0.024430486 | 0 | 0 | 0.057220036 |
| 0 | 0.024338543 | 0.043137696 | 0.024004548 | 0.006129753 |
| 0 | 0.628988401 | 0 | 0 | 3.268865035 |
| 0 | 0.059916097 | 0 | 0 | 0.156529778 |
| 0 | 0.003734261 | 0.000295792 | 0 | 0.028795046 |
| 0 | 0 | 0 | 0 | 0 |
| 0 | 0.046415586 | 0 | 0 | 0 |
| 0 | 0 | 0 | 0 | 0 |
| 0 | 0 | 0 | 0 | 0 |
| 0 | 0 | 0 | 0 | 0 |
| 0 | 0 | 0 | 0 | 0 |
| 0 | 0 | 0 | 0 | 0 |
| 0 | 0 | 0 | 0 | 0 |
| 0 | 0 | 0 | 0 | 0 |
| 0 | 0.009091862 | 0 | 0 | 0.012738874 |
| 0 | 0.000439678 | 0.045681482 | 0 | 0 |
| 0 | 0 | 0 | 0 | 0 |
| 0 | 0 | 0 | 0 | 0 |
| 0 | 0 | 0 | 0 | 0 |
| 0 | 0 | 0 | 0 | 0 |
| 0 | 0 | 0 | 0 | 0 |
| 0 | 0.037179246 | 0 | 0 | 0 |
| 0 | 0.542395968 | 0 | 0 | 0.347983423 |
| 0 | 0 | 0.023299702 | 0 | 0 |
| 0 | 0.135511949 | 0 | 0 | 0 |
| 0 | 0 | 0 | 0 | 0 |
| 0 | 0 | 0 | 0 | 0 |
| 0 | 0 | 0 | 0 | 0 |
| 0 | 0.806270971 | 0 | 2.659828356 | 2.862196159 |
| 0 | 0 | 0 | 0 | 0 |
| 0 | 0.006307315 | 0 | 0 | 0 |
| 0 | 0 | 0 | 0 | 0.014104742 |
| 0 | 0 | 0 | 0 | 0 |
| 0 | 0 | 0 | 0 | 0 |
| 0 | 0 | 0 | 0 | 0 |
| 0 | 0 | 0 | 0 | 0 |

| | | | | |
|---|---|---|---|---|
| 0 | 0.161431501 | 0 | 0 | 0.268234852 |
| 0 | 0 | 0 | 0 | 0 |
| 0 | 0 | 0 | 0 | 0 |
| 0 | 0 | 0.001643842 | 0 | 0 |
| 0 | 0.005242098 | 0 | 0 | 0.039084308 |
| 0 | 0 | 0 | 0 | 0 |
| 0 | 0.053791616 | 0.001003248 | 0 | 0.020409889 |
| 0 | 0 | 0 | 0 | 0 |
| 0 | 0 | 0 | 0.000919885 | 0 |
| 0 | 0 | 0.053185781 | 0 | 0 |
| 0 | 0.076614971 | 0 | 0 | 0 |
| 0 | 0 | 0.040255323 | 0 | 0 |
| 0 | 0 | 0 | 0 | 0 |
| 0 | 0 | 0 | 0 | 0 |
| 0 | 0 | 0 | 0 | 0 |
| 0 | 0 | 0 | 0 | 0 |
| 0 | 0 | 0.001011674 | 0.82729978 | 0 |
| 0 | 0 | 0 | 0 | 0 |
| 0 | 0 | 0 | 0 | 0 |
| 0 | 0 | 0 | 0 | 0 |
| 0 | 0.282307776 | 0.000955148 | 0.002605614 | 0.021110604 |
| 0 | 0.011698952 | 0 | 0 | 0 |
| 0 | 0.010327325 | 0 | 0 | 0.002778315 |
| 0 | 0.003339332 | 0.007163807 | 0.003772516 | 0.011176099 |
| 0 | 0 | 0 | 0 | 0 |
| 0 | 0 | 0 | 0.000430715 | 0 |
| 0 | 0 | 0 | 0 | 0 |
| 0 | 0 | 0 | 0 | 0 |
| 0 | 0 | 0 | 0 | 0 |
| 10.12666265 | 0.005206589 | 0.048300249 | 0.384971505 | 0.001354529 |
| 0 | 0 | 0 | 0 | 0 |
| 0 | 0 | 0 | 0 | 0 |
| 0 | 0 | 0 | 0 | 0 |
| 0 | 0 | 0 | 0 | 0 |
| 0 | 0 | 0 | 0 | 0 |
| 0 | 0 | 0 | 0 | 0 |
| 0 | 0.00417837 | 0 | 0 | 0.052845535 |
| 0 | 0 | 0 | 0 | 0 |
| 0 | 0 | 0 | 0 | 0.003119824 |
| 0 | 0 | 0 | 0 | 0 |
| 0 | 0 | 0 | 0 | 0 |
| 0 | 0 | 0 | 0 | 0 |
| 0 | 0 | 0 | 0 | 0 |
| 0 | 0.001661323 | 0 | 0 | 0 |
| 0 | 0.001647713 | 0 | 0 | 0 |
| 0 | 0 | 0 | 0 | 0 |
| 0 | 0 | 0 | 0 | 0 |
| 0 | 0 | 0 | 0 | 0 |

| | | | | |
|---|---|---|---|---|
| 0 | 0 | 0.634825505 | 0.0136185 | 0 |
| 0 | 0 | 0 | 0 | 0.113212533 |
| 0 | 0.000533776 | 0 | 0 | 0 |
| 0 | 0 | 0 | 0 | 0 |
| 0 | 0 | 0 | 0 | 0 |
| 0 | 0 | 0 | 0 | 0 |
| 0 | 0 | 0.000683762 | 0 | 0 |
| 0 | 0.231666434 | 0 | 0.944398553 | 0.377974496 |
| 5.696093893 | 2.828055236 | 0.017591052 | 0.028570223 | 3.111828132 |
| 0 | 0 | 0.325856919 | 0.001717099 | 0 |
| 0 | 0.008639374 | 0 | 0 | 0 |
| 0 | 0 | 0 | 0 | 0 |
| 0 | 0.00184191 | 0 | 0 | 0 |
| 0 | 0 | 0 | 0 | 0 |
| 0 | 0 | 0 | 0 | 0 |
| 0 | 0 | 0 | 0 | 0 |
| 0 | 0 | 0 | 0 | 0 |
| 0 | 0 | 0 | 0 | 0 |
| 0 | 0 | 0 | 0 | 0 |
| 0 | 0.000406992 | 0 | 0 | 0 |
| 0 | 0 | 0 | 0 | 0 |
| 0 | 0 | 0 | 0 | 0 |
| 0 | 0 | 0 | 0 | 0 |
| 0 | 0 | 0 | 0 | 0 |
| 0 | 0.005306578 | 0 | 0 | 0.01575351 |
| 0 | 0.000372863 | 0 | 0 | 0 |
| 0 | 0 | 0 | 0 | 0 |
| 0 | 0 | 0 | 0 | 0 |
| 0 | 0 | 0 | 0 | 0 |
| 0 | 0 | 0 | 0 | 0 |
| 0.014625772 | 0 | 0 | 0 | 0 |
| 0 | 0 | 0 | 0 | 0 |
| 0 | 0 | 0 | 0 | 0 |
| 0 | 0 | 0 | 0 | 0 |
| 0 | 0.002900082 | 0 | 0 | 0 |
| 0 | 0.03788834 | 0 | 0 | 0 |
| 0 | 0.013091112 | 0 | 0 | 0 |
| 0 | 0.001255322 | 0 | 0 | 0 |
| 0 | 0.004984751 | 0 | 0 | 0 |
| 0 | 0 | 0 | 1.147122708 | 0 |
| 0 | 0 | 0 | 0 | 0 |
| 0 | 0.055357975 | 0 | 0 | 0.764796764 |
| 0 | 0 | 0 | 0 | 0 |
| 0 | 0 | 0 | 0 | 0 |
| 0 | 0 | 0 | 0 | 0 |
| 0 | 0 | 0.00411266 | 0 | 0.001125626 |
| 0 | 0 | 0 | 0 | 0 |
| 0 | 0 | 0 | 0 | 0 |

| | | | | |
|---|---|---|---|---|
| 0 | 0 | 0 | 0 | 0 |
| 0 | 0 | 0 | 0.00709113 | 0 |
| 0 | 0 | 0 | 0 | 0 |
| 0 | 0 | 0 | 0 | 0 |
| 0 | 1.237094213 | 0 | 0 | 0.866107602 |
| 0 | 0 | 0 | 0 | 0 |
| 0 | 0 | 0 | 0 | 0 |
| 0.010081927 | 0.002666336 | 0.147841049 | 0.017823442 | 0.021738964 |
| 0 | 0.005704795 | 0 | 0 | 0 |
| 0 | 0 | 0 | 0 | 0 |
| 0 | 0 | 0 | 0 | 0 |
| 0 | 0 | 0 | 0 | 0.319030245 |
| 0 | 0 | 0 | 0 | 0 |
| 0 | 0 | 0 | 0 | 0 |
| 0 | 0 | 0 | 0.000469049 | 0 |
| 0 | 0 | 0.045204599 | 0 | 0 |
| 0 | 0 | 0 | 0 | 0 |
| 0 | 0 | 0 | 0 | 0 |
| 0 | 0 | 0 | 0 | 0 |
| 0 | 0 | 0 | 0 | 0 |
| 0 | 0 | 0 | 0 | 0 |
| 0 | 0 | 0 | 0 | 0 |
| 0 | 0 | 0 | 0 | 0 |
| 0 | 0 | 0.074416555 | 0.000576625 | 0.002926511 |
| 0 | 0 | 0 | 0 | 0.167881441 |
| 0 | 0 | 0 | 0 | 0 |
| 0 | 0.026682312 | 0.444495141 | 0 | 0.278496578 |
| 0 | 0 | 0 | 0 | 0 |
| 0 | 0 | 0 | 0 | 0 |
| 0 | 0 | 0 | 0 | 0 |
| 0 | 0 | 0 | 0 | 0 |
| 0 | 0 | 0 | 0 | 0 |
| 0 | 0 | 0 | 0 | 0 |
| 0 | 0 | 0 | 0 | 0 |
| 0 | 0 | 0 | 0 | 0 |
| 0 | 0 | 0 | 0 | 0 |
| 0 | 0 | 0 | 0 | 0 |
| 0 | 0.122443533 | 0 | 0 | 0.008365827 |
| 0 | 0.512212469 | 0 | 0 | 0.001234726 |
| 0 | 0 | 0 | 0 | 0 |
| 0 | 0 | 0 | 0 | 0 |
| 0 | 0 | 0 | 0 | 0.080038246 |
| 0 | 0 | 0 | 0 | 0 |
| 0 | 0 | 0 | 0 | 0 |
| 0 | 0 | 0 | 0 | 0 |
| 0 | 0 | 0 | 0 | 0 |
| 0 | 0 | 0 | 0 | 0 |
| 0 | 0.001830901 | 0 | 0 | 0 |

| | | | | |
|---|---|---|---|---|
| 0 | 0.017153572 | 0 | 0 | 0 |
| 0 | 0 | 0 | 0 | 0 |
| 0 | 0 | 0 | 0 | 0 |
| 0 | 0 | 0.057873545 | 0 | 0 |
| 0 | 0 | 0 | 0 | 0 |
| 0 | 1.151179326 | 0 | 0 | 0.355504514 |
| 0 | 0 | 0 | 0 | 0 |
| 0 | 0 | 0 | 0 | 0 |
| 0 | 0 | 0 | 0 | 0 |
| 0 | 0 | 0 | 0 | 0 |
| 0 | 0 | 0 | 0 | 0 |
| 0 | 0 | 0.005235278 | 0 | 0 |
| 0 | 0 | 0 | 0 | 0 |
| 0 | 0 | 0 | 0 | 0 |
| 0 | 0 | 0 | 0 | 0 |
| 0 | 0 | 0 | 0 | 0 |
| 0 | 0 | 0 | 0 | 0 |
| 0 | 0 | 0 | 0 | 0 |
| 0 | 0 | 0 | 0 | 0 |
| 0 | 0 | 0 | 0 | 0 |
| 0 | 0 | 0.01449607 | 0 | 0.075493652 |
| 0 | 0 | 0 | 0 | 0 |
| 0.001219231 | 0.058040299 | 0.016501362 | 0 | 0.113920845 |
| 0 | 0 | 0 | 0 | 0.000962458 |
| 0 | 0 | 0 | 0 | 0 |
| 0 | 0 | 0 | 0 | 0 |
| 0 | 0 | 0 | 0 | 0 |
| 0 | 0 | 0 | 0 | 0 |
| 0 | 0 | 0 | 0 | 0 |
| 0 | 0 | 0 | 0 | 0 |
| 0 | 0 | 0 | 0 | 0 |
| 0 | 0 | 0 | 0 | 0.02044229 |
| 0.084683822 | 0 | 0.043977003 | 0.210023614 | 0.032170053 |
| 0 | 0 | 0 | 0 | 0 |
| 0 | 0.392257945 | 0.544944295 | 1.204493376 | 0 |
| 0 | 0.001401982 | 0.004133584 | 0 | 0.001843937 |
| 0 | 0 | 0 | 0 | 0 |
| 0 | 0 | 0 | 0 | 0 |
| 0 | 0 | 0.174690147 | 0.001891068 | 0.003351551 |
| 0 | 0 | 0 | 0 | 0 |
| 0 | 0.178121801 | 0 | 0 | 0 |
| 0 | 0 | 0 | 0 | 0 |
| 0 | 0.175710105 | 0 | 0 | 0.306926367 |
| 0 | 0 | 0 | 0 | 0 |
| 0 | 0 | 0 | 0 | 0 |
| 0 | 0.04551085 | 0.002518151 | 0 | 0.003678219 |
| 0 | 0 | 0 | 0 | 0 |
| 0 | 0 | 0 | 0 | 0 |

| | | | | |
|---|---|---|---|---|
| 0 | 0 | 0 | 0 | 0 |
| 0 | 0 | 0 | 0 | 0 |
| 0 | 0 | 0 | 0 | 0 |
| 0 | 0 | 0 | 0 | 0 |
| 0 | 0 | 0 | 0 | 0 |
| 0 | 0 | 0 | 0 | 0 |
| 0 | 0 | 0 | 0 | 0 |
| 0 | 0 | 0 | 0 | 0 |
| 0 | 0 | 0 | 0 | 0.045065045 |
| 0 | 0.003963752 | 0 | 0 | 0.009471313 |
| 0 | 0.16084177 | 0 | 0 | 0 |
| 0 | 0 | 0 | 0 | 0 |
| 0 | 0 | 0 | 0 | 0 |
| 0 | 0.001185806 | 0 | 0 | 0 |
| 0 | 0 | 0 | 0 | 0 |
| 0 | 0 | 0.000738703 | 0 | 0 |
| 0 | 0 | 0 | 0 | 0 |
| 0 | 0 | 0 | 0 | 0 |
| 0 | 0 | 0 | 0 | 0 |
| 0 | 0.083473356 | 0 | 0 | 1.246508603 |
| 0 | 0.056492988 | 0 | 0.00309781 | 0.004342303 |
| 0 | 0 | 0 | 0 | 0 |
| 0 | 0 | 0 | 0 | 0 |
| 0 | 0.000903433 | 0 | 0 | 0 |
| 0 | 0 | 0 | 0 | 0 |
| 0 | 0 | 0 | 0 | 0 |
| 0 | 0 | 0 | 0 | 0 |
| 0 | 0 | 0 | 0 | 0 |
| 0 | 0 | 0 | 0 | 0 |
| 0 | 0.023197223 | 0 | 0 | 0 |
| 0 | 0.279486969 | 0.848360787 | 0.007232123 | 0.384526067 |
| 0 | 0 | 0 | 0 | 0 |
| 0 | 0 | 0 | 0 | 0.001771457 |
| 0 | 0 | 0 | 0 | 0 |
| 0 | 0 | 0 | 0 | 0 |
| 0 | 0 | 0 | 0 | 0 |
| 0 | 0 | 0 | 0 | 0 |
| 0 | 0 | 0 | 0 | 0 |
| 0 | 0 | 0 | 0 | 0.004071107 |
| 0 | 0 | 0 | 0 | 0 |
| 0 | 0 | 0 | 0 | 0 |
| 0 | 0.005101555 | 0 | 0 | 0 |
| 0 | 0 | 0 | 0 | 0.00063015 |
| 0 | 0 | 0.001291157 | 0 | 0 |
| 0 | 0 | 0 | 0 | 0 |
| 0 | 0 | 0 | 0 | 0 |
| 0 | 0 | 0.018080166 | 0.124336713 | 0.000733809 |
| 0 | 0 | 0 | 0 | 0 |

| | | | | |
|---|---|---|---|---|
| 0 | 0 | 0 | 0 | 0 |
| 0 | 0 | 0 | 0 | 0 |
| 0 | 0 | 0 | 0 | 0 |
| 0 | 0 | 0 | 0 | 0 |
| 0 | 0.130583415 | 0 | 0 | 0.059206658 |
| 0 | 0 | 0 | 0 | 0 |
| 0 | 0 | 0 | 0 | 0 |
| 0 | 0 | 0 | 0 | 0 |
| 0 | 0 | 0 | 0 | 0 |
| 0 | 0 | 0 | 0 | 0 |
| 0 | 0.036550758 | 0 | 0 | 0 |
| 0 | 0.008641719 | 0.403435388 | 0.001732098 | 0.023746617 |
| 0 | 0.015998532 | 0.016363518 | 0 | 0.050299065 |
| 0 | 0 | 0 | 0 | 0 |
| 0 | 0 | 0 | 0.007799881 | 0 |
| 0 | 0.005180125 | 0.007873702 | 0 | 0.001420489 |
| 0 | 0 | 0 | 0 | 0 |
| 0 | 0 | 0 | 0 | 0 |
| 0.010022984 | 0.018812733 | 0.324272608 | 0.314296225 | 0.019364234 |
| 0.009597106 | 0.047347417 | 0 | 0 | 0.054029051 |
| 0 | 0 | 0 | 0 | 0 |
| 0 | 0.001092455 | 0 | 0 | 0.05199 |
| 0 | 0 | 0 | 0 | 0 |
| 0 | 0 | 0 | 0 | 0 |
| 0 | 0 | 0 | 0 | 0 |
| 0 | 0 | 0 | 0 | 0 |
| 0 | 0 | 0.001638267 | 0 | 0.001688422 |
| 0 | 0 | 0 | 0 | 0 |
| 0 | 0 | 0 | 0 | 0 |
| 0 | 0 | 0 | 0 | 0 |
| 0 | 0 | 0.000960549 | 0 | 0 |
| 0 | 0 | 0 | 0 | 0 |
| 0 | 0 | 0 | 0 | 0 |
| 0 | 0 | 0 | 0 | 0 |
| 0 | 0 | 0 | 0 | 0 |
| 0 | 0 | 0 | 0 | 0 |
| 0 | 0 | 0 | 0 | 0 |
| 0.005011203 | 0.019311473 | 0.328473969 | 0.015201235 | 0.016039883 |
| 0 | 0.064901051 | 0 | 0 | 0 |
| 0 | 0 | 0 | 0 | 0 |
| 0 | 0.053641191 | 0.007815174 | 0 | 0.027423415 |
| 0 | 0 | 0 | 0 | 0 |
| 0 | 0 | 0 | 0 | 0 |
| 0 | 0 | 0.005737479 | 0.004419903 | 0 |
| 0 | 0 | 0 | 0 | 0 |
| 0 | 0 | 0 | 0 | 0 |
| 0 | 0 | 0 | 0 | 0 |
| 0 | 0 | 0 | 0 | 0 |

| | | | | |
|---|---|---|---|---|
| 0 | 0 | 0 | 0 | 0 |
| 0 | 0.796182618 | 0 | 0 | 0 |
| 0 | 0 | 0 | 0 | 0 |
| 0 | 0 | 0.003286122 | 0 | 0.000746315 |
| 0 | 0.003438424 | 0 | 0 | 0 |
| 0 | 0.040995116 | 0.008787631 | 0 | 0 |
| 0 | 0.004968133 | 0 | 0 | 0.000579393 |
| 0.016647788 | 0.00441813 | 0 | 0 | 0 |
| 0 | 0 | 0 | 0 | 0 |
| 0 | 0 | 0 | 0 | 0 |
| 0 | 0.011669965 | 0 | 0 | 0 |
| 0 | 0 | 0 | 0 | 0 |
| 0 | 0.04167138 | 0.187590855 | 0 | 0.043803844 |
| 0 | 0 | 0 | 0 | 0 |
| 0 | 0 | 0 | 0 | 0 |
| 0 | 0 | 0 | 0 | 0 |
| 0 | 0 | 0 | 0 | 0 |
| 0 | 0 | 0 | 0 | 0 |
| 0 | 0 | 0 | 0 | 0 |
| 0 | 0 | 0 | 0 | 0 |
| 0 | 0 | 0 | 0 | 0 |
| 0 | 0 | 0 | 0 | 0 |
| 0 | 0 | 0.05168693 | 0.006485402 | 0 |
| 0 | 0.078750464 | 0 | 0 | 0.00180846 |
| 0 | 0 | 0 | 0 | 0 |
| 0 | 0.002871999 | 0 | 0 | 0 |
| 0 | 0 | 0 | 0 | 0 |
| 0 | 0 | 0 | 0 | 0 |
| 0 | 0 | 0 | 0 | 0 |
| 0 | 0 | 0 | 0 | 0 |
| 0 | 0.007532869 | 0.010332777 | 0.001285546 | 0.015689756 |
| 0 | 0 | 0 | 0 | 0 |
| 0 | 0 | 0 | 0 | 0 |
| 0 | 0 | 0 | 0 | 0 |
| 0 | 0 | 0 | 0 | 0.061341633 |
| 0.049706672 | 0 | 0 | 0 | 0 |
| 0 | 0 | 0 | 0 | 0 |
| 0 | 0 | 0 | 0 | 0 |
| 0 | 0.273742287 | 0.001231302 | 0 | 0.001314319 |
| 0 | 0 | 0 | 0 | 0 |
| 0 | 0 | 0 | 0 | 0 |
| 0 | 0 | 0 | 0 | 0 |
| 0 | 10.64041173 | 0 | 0 | 0 |
| 0 | 0 | 0 | 0 | 0 |
| 0 | 0 | 0 | 0 | 0 |
| 0 | 0.167814017 | 0.572004855 | 0.051317077 | 0.180395833 |
| 0 | 0 | 0 | 0 | 0 |
| 0 | 0 | 0 | 0 | 0 |

| | | | | |
|---|---|---|---|---|
| 0 | 0 | 0 | 0 | 0 |
| 0 | 0 | 0 | 0 | 0 |
| 0 | 0 | 0 | 0 | 0.091568598 |
| 0 | 0.009085577 | 0 | 0 | 0 |
| 0 | 0 | 0 | 0 | 0 |
| 0 | 0 | 0 | 0 | 0 |
| 0 | 0 | 0 | 0 | 0 |
| 0 | 0 | 0 | 0 | 0 |
| 0 | 0 | 0 | 0 | 0 |
| 0 | 0.000709622 | 0 | 0 | 0 |
| 0 | 0.000756655 | 0.009689485 | 0 | 0 |
| 0 | 0 | 0.011110816 | 0 | 0 |
| 0 | 0 | 0 | 0 | 0 |
| 0 | 0 | 0 | 0 | 0 |
| 0 | 0 | 0 | 0 | 0 |
| 0 | 0 | 0 | 0 | 0 |
| 0 | 0.001668376 | 0 | 0 | 0 |
| 0 | 0 | 0 | 0 | 0 |
| 0 | 0 | 0 | 0 | 0 |
| 0 | 0 | 0.001390971 | 0 | 0 |
| 0 | 0 | 0 | 0 | 0 |
| 0 | 0 | 0 | 0 | 0 |
| 0 | 0 | 0 | 0 | 0 |
| 0 | 0 | 0 | 0 | 0 |
| 0 | 0 | 0 | 0 | 0 |
| 0 | 0 | 0 | 0 | 0 |
| 0 | 0.000889917 | 0 | 0 | 0 |
| 0 | 0.02665182 | 0 | 0 | 0 |
| 0 | 0 | 0 | 0 | 0 |
| 0 | 0 | 0 | 0 | 0 |
| 0 | 0.000574714 | 0.003584964 | 0.000628323 | 0 |
| 0 | 0 | 0 | 0 | 0 |
| 0 | 0.007673972 | 0.003766079 | 0.014007305 | 0.010155776 |
| 0.054147097 | 0.003239074 | 0.001485395 | 0.000596414 | 0.006198033 |
| 0 | 0 | 0 | 0 | 0 |
| 0 | 0 | 0 | 0 | 0 |
| 0 | 0 | 0 | 0 | 0 |
| 0 | 0 | 0 | 0 | 0 |
| 0 | 0 | 0 | 0 | 0 |
| 0 | 0 | 0 | 0 | 0 |
| 0 | 0 | 0 | 0 | 0 |
| 0 | 0.000960721 | 0.001576338 | 0.002400767 | 0.004757745 |
| 0 | 0.030096738 | 0.038540916 | 0 | 0.056549029 |
| 0 | 0 | 0 | 0 | 0 |
| 0 | 0 | 0 | 0 | 0 |
| 0 | 0.079735112 | 0 | 0 | 0.111952671 |
| 0 | 0 | 0 | 0 | 0 |
| 0 | 0 | 0 | 0 | 0 |

| | | | | |
|---|---|---|---|---|
| 0 | 0 | 0 | 0 | 0 |
| 0 | 0.008583255 | 0 | 0 | 0.013773906 |
| 0 | 0.031510752 | 0.00705033 | 0.002147535 | 0.013148308 |
| 0 | 0 | 0 | 0 | 0 |
| 0 | 0 | 0.169851043 | 0 | 0.01022202 |
| 0 | 0 | 0.043080793 | 0.011127213 | 0 |
| 0 | 0 | 0 | 0 | 0 |
| 0 | 0 | 0 | 0 | 0 |
| 0 | 0 | 0 | 0 | 0 |
| 0 | 0 | 0 | 0 | 0 |
| 0 | 0 | 0 | 0 | 0 |
| 0 | 0 | 0 | 0 | 0 |
| 0 | 0 | 0 | 0 | 0 |
| 0 | 0 | 0 | 0 | 0 |
| 0 | 0 | 0 | 0 | 0 |
| 0 | 0 | 0 | 0 | 0 |
| 0 | 0 | 0 | 0 | 0.454441495 |
| 0 | 0 | 0 | 0 | 0 |
| 0 | 0 | 0 | 0 | 0 |
| 0 | 0 | 0 | 0 | 0 |
| 0 | 0 | 0 | 0 | 0 |
| 0 | 0 | 0 | 0 | 0 |
| 0 | 0 | 0 | 0 | 0 |
| 0 | 0 | 0 | 0 | 0 |
| 0 | 0.009443378 | 0 | 0 | 0.005845766 |
| 1.794534339 | 0 | 0 | 0.001038595 | 0 |
| 0 | 0 | 0 | 0 | 0 |
| 0 | 0.007405033 | 0 | 0 | 0.283554724 |
| 0 | 0 | 0 | 0 | 0 |
| 0 | 0 | 0 | 0 | 0 |
| 0 | 0 | 0 | 0 | 0.004952075 |
| 0 | 0 | 0.000942691 | 0 | 0 |
| 0 | 0 | 0 | 0 | 0 |
| 0 | 0 | 0 | 0 | 0 |
| 0 | 0 | 0 | 0 | 0 |
| 0 | 0 | 0 | 0 | 0 |
| 0 | 0 | 0 | 0 | 0.007212264 |
| 0 | 0 | 0 | 0 | 0 |
| 0 | 0 | 0.071430618 | 0 | 1.059899446 |
| 0 | 0 | 0 | 0 | 0 |
| 0 | 0 | 0 | 0 | 0 |
| 0 | 0.00202436 | 0 | 0 | 0.019761372 |
| 0 | 0 | 0.507130165 | 0.007296402 | 0 |
| 0 | 0 | 0 | 0 | 0.006148118 |
| 0 | 0 | 0 | 0 | 0 |
| 0 | 0.005288454 | 0 | 0 | 0 |
| 0 | 0 | 0.005467082 | 0 | 0.031270454 |
| 0 | 0 | 0 | 0 | 0 |

| | | | | |
|---|---|---|---|---|
| 0 | 0 | 0 | 0 | 0 |
| 0 | 0 | 0.16068187 | 0.025869445 | 0.007224624 |
| 0 | 0.015781918 | 0 | 0 | 0 |
| 0 | 0.000355421 | 0 | 0 | 0 |
| 0 | 0 | 0 | 0 | 0 |
| 0 | 0 | 0 | 0 | 0 |
| 0 | 0 | 0 | 0 | 0 |
| 0 | 0 | 0 | 0 | 0 |
| 0 | 0 | 0 | 0 | 0 |
| 0 | 0 | 0 | 0 | 0 |
| 0 | 0 | 0 | 0 | 0 |
| 0 | 0 | 0 | 0 | 0 |
| 0 | 0.055758373 | 0.016442017 | 0 | 0.011896201 |
| 0 | 0 | 0 | 0 | 0 |
| 0 | 0 | 0.004787533 | 0 | 0.014897874 |
| 0 | 0 | 0 | 0 | 0 |
| 0 | 0 | 0 | 0 | 0 |
| 0 | 0 | 0 | 0 | 0 |
| 0 | 0 | 0 | 0 | 0.001149831 |
| 0 | 0 | 0 | 0 | 0.004153264 |
| 0 | 0 | 0 | 0 | 0 |
| 0 | 0.010865451 | 0 | 0 | 0.027560511 |
| 0 | 0 | 0.033802034 | 0 | 0 |
| 0 | 0.036475555 | 0 | 0 | 0 |
| 0 | 0 | 0 | 0 | 0 |
| 0 | 0 | 0 | 0 | 0 |
| 0 | 0 | 0 | 0 | 0 |
| 0 | 0 | 0 | 0 | 0 |
| 0 | 0 | 0 | 0 | 0 |
| 0 | 0.001298709 | 0 | 0 | 0 |
| 0 | 0 | 0 | 0 | 0 |
| 0 | 0 | 0 | 0 | 0 |
| 0 | 0 | 0 | 0 | 0 |
| 0 | 0 | 0 | 0 | 0 |
| 0 | 0.164237731 | 0 | 0 | 17.14659666 |
| 0 | 0 | 0 | 0 | 0 |
| 0 | 0 | 0 | 0 | 0 |
| 0 | 0 | 0 | 0 | 0 |
| 0 | 0.039985984 | 0.001034547 | 0 | 0.004077408 |
| 0 | 0 | 0 | 0 | 0 |
| 0 | 0.581931133 | 1.274176803 | 0.019539422 | 0.132832329 |
| 0 | 0 | 0 | 0 | 0 |
| 0 | 0 | 0 | 0 | 0 |
| 0 | 0 | 0 | 0 | 0 |
| 0 | 0 | 0 | 0 | 0 |
| 7.936301641 | 0.001856484 | 3.083452659 | 1.419495571 | 0.002092899 |
| 0 | 0.002421306 | 0 | 0 | 0 |
| 0 | 0 | 0 | 0 | 0 |

| | | | | |
|---|---|---|---|---|
| 0 | 0 | 0 | 0 | 0 |
| 0 | 0 | 0 | 0 | 0 |
| 0 | 0 | 0 | 0 | 0 |
| 0 | 0.001317839 | 0 | 0 | 0 |
| 0 | 0.001095113 | 0.295736248 | 0 | 0 |
| 0 | 0 | 0 | 0 | 0 |
| 0 | 0 | 0 | 0 | 0 |
| 0 | 0 | 0 | 0 | 0 |
| 0 | 0.000838427 | 0 | 0 | 0 |
| 0.001779892 | 0.164013216 | 0.001438179 | 0 | 0.006822861 |
| 0 | 0.054892549 | 0 | 0 | 0 |
| 0 | 0 | 0 | 0 | 0 |
| 0 | 0 | 0.010855119 | 0 | 0.000500158 |
| 0 | 0 | 0 | 0 | 0 |
| 0 | 0 | 0 | 0 | 0 |
| 0 | 0 | 0 | 0 | 0 |
| 0 | 0 | 0 | 0 | 0 |
| 0 | 0 | 0 | 0 | 0 |
| 0 | 0 | 0 | 0 | 0 |
| 0.08086357 | 0.143739456 | 0.256976655 | 0.218279344 | 0.067221551 |
| 0 | 0 | 0 | 0 | 0 |
| 0 | 0 | 0 | 0 | 0 |
| 0 | 0 | 0 | 0 | 0 |
| 0 | 0 | 0 | 0 | 0 |
| 0 | 0 | 0 | 0 | 0 |
| 0 | 0 | 0 | 0 | 0.012457588 |
| 0 | 0 | 0 | 0 | 0 |
| 0 | 0 | 0 | 0 | 0 |
| 0 | 0 | 0 | 0 | 0 |
| 0 | 0 | 0 | 0.003774202 | 0 |
| 0 | 0 | 0 | 0 | 0 |
| 0 | 0 | 0 | 0 | 0 |
| 0 | 0 | 0 | 0 | 0 |
| 0 | 0 | 0 | 0 | 0 |
| 0 | 0 | 0 | 0 | 0 |
| 0 | 0.002271886 | 0 | 0 | 0 |
| 0 | 0 | 0 | 0 | 0 |
| 0 | 0 | 0 | 0 | 0 |
| 0 | 0 | 0 | 0 | 0 |
| 0 | 0.283005013 | 0 | 0 | 0 |
| 0 | 0 | 0 | 0 | 0 |
| 0 | 0 | 0 | 0 | 0 |
| 0 | 0 | 0 | 0 | 0 |
| 0 | 0 | 0 | 0 | 0 |
| 0 | 0.005934302 | 0.118755026 | 0 | 0 |
| 0 | 0.026167098 | 0 | 0 | 0.011137512 |
| 0 | 0 | 0 | 0 | 0 |
| 0 | 0 | 0 | 0 | 0 |

| | | | | |
|---|---|---|---|---|
| 0 | 0 | 0 | 0 | 0 |
| 0 | 0 | 0 | 0 | 0 |
| 0 | 0 | 0 | 0 | 0 |
| 0 | 0 | 0 | 0 | 0 |
| 0 | 0.001565546 | 0 | 0 | 0 |
| 0.007962079 | 0.126998707 | 0.029730456 | 0.000538159 | 0.121529211 |
| 0 | 0 | 0 | 0 | 0 |
| 0 | 0 | 0 | 0 | 0 |
| 0 | 0 | 0 | 0 | 0 |
| 0 | 0 | 0 | 0 | 0 |
| 0 | 0 | 0 | 0 | 0 |
| 0 | 0 | 0 | 0 | 0 |
| 0 | 0 | 0 | 0 | 0 |
| 0 | 0 | 0 | 0 | 0.002527586 |
| 0 | 0 | 0 | 0 | 0 |
| 0 | 0 | 0 | 0 | 0 |
| 0 | 0.024928397 | 0 | 0 | 0 |
| 0 | 0 | 0 | 0 | 0.010763917 |
| 0 | 0 | 0 | 0 | 0 |
| 0 | 0 | 0 | 0 | 0 |
| 0 | 0 | 0 | 0 | 0 |
| 0 | 0 | 0 | 0 | 0 |
| 0 | 0 | 0 | 0 | 0 |
| 0 | 0 | 0 | 0 | 0 |
| 0 | 0 | 0 | 0 | 0 |
| 0 | 0.005582143 | 0 | 0 | 0 |
| 0 | 0 | 0 | 0 | 0 |
| 0 | 0 | 0 | 0 | 0.001705301 |
| 0 | 0 | 0 | 0 | 0 |
| 0 | 0 | 0 | 0 | 0 |
| 0 | 0 | 0 | 0 | 0 |
| 0 | 0 | 0 | 0 | 0 |
| 0 | 0 | 0 | 0 | 0 |
| 0 | 0 | 0 | 0 | 0 |
| 0 | 0 | 0 | 0 | 0 |
| 0 | 0 | 0 | 0 | 0 |
| 0 | 0 | 0 | 0 | 0 |
| 0 | 0 | 0 | 0 | 0 |
| 0 | 0.380601456 | 0.322665874 | 0 | 0 |
| 0 | 0 | 0 | 0 | 0 |
| 0 | 0 | 0 | 0 | 0 |
| 0 | 0 | 0 | 0 | 0 |
| 0 | 0 | 0 | 0 | 0 |
| 0 | 0 | 0 | 0 | 0 |
| 0 | 0 | 0 | 0 | 0 |
| 0 | 0 | 0 | 0 | 0 |
| 0 | 0 | 0 | 0 | 0 |
| 0 | 0 | 0 | 0 | 0 |

| | | | | |
|---|---|---|---|---|
| 0 | 0 | 0 | 0 | 0 |
| 0 | 0 | 0.146238555 | 0 | 0.003028772 |
| 0 | 0 | 0 | 0 | 0 |
| 0 | 0 | 0 | 0 | 0 |
| 0 | 0 | 0 | 0 | 0 |
| 0 | 0 | 0 | 0 | 0 |
| 0 | 0 | 0.002327735 | 0 | 0 |
| 0 | 0 | 0 | 0 | 0 |
| 0 | 0 | 0 | 0 | 0 |
| 0 | 0 | 0 | 0 | 0 |
| 0.018039202 | 0.003968961 | 0.001929168 | 0 | 0.001677896 |
| 0 | 0 | 0 | 0 | 0 |
| 0 | 0.152412353 | 0 | 0 | 0.013145008 |
| 0 | 0 | 0 | 0 | 0 |
| 0 | 0 | 0 | 0 | 0 |
| 0 | 0 | 0 | 0 | 0 |
| 0 | 0 | 0 | 0 | 0 |
| 0 | 0 | 0 | 0 | 0 |
| 0 | 0 | 0 | 0 | 0 |
| 0 | 0 | 0 | 0 | 0 |
| 0 | 0 | 0.018927232 | 0 | 0.005263706 |
| 0 | 0 | 0 | 0 | 0 |
| 0 | 0 | 0 | 0 | 0 |
| 0 | 2.042399541 | 0.064279168 | 0.072434912 | 0.862471728 |
| 0 | 0.001407326 | 0 | 0 | 0 |
| 0 | 0 | 0 | 0 | 0 |
| 0 | 0 | 0 | 0 | 0 |
| 0 | 0 | 0 | 0 | 0 |
| 0 | 0 | 0 | 0 | 0 |
| 0 | 0 | 0 | 0 | 0 |
| 0 | 0 | 0 | 0 | 0 |
| 0 | 0 | 0 | 0 | 0 |
| 0 | 0.004237341 | 0 | 0 | 0 |
| 0 | 0 | 0 | 0 | 0 |

| PHL018-0222_V1 | PHL018-0180_V1 | PHL018-0267_V2 | PHL018-0277_V3 | PHL018-0105_V3 |
|---|---|---|---|---|
| REF | CTRL | REF | REF | CTRL |
| 0.139757267 | 0.077562677 | 0.621908221 | 2.139479356 | 0 |
| 1.1031707 | 1.659817707 | 1.954602452 | 4.294863793 | 0 |
| 0 | 2.099484483 | 0 | 0 | 0.006620941 |
| 0 | 0 | 0 | 0 | 0 |
| 0 | 0 | 0 | 0 | 0 |
| 0 | 0 | 0 | 0 | 0 |
| 0 | 0 | 0 | 0 | 0 |
| 0 | 0 | 0 | 0 | 0 |
| 0 | 0 | 0 | 0 | 0 |
| 0 | 0 | 0 | 0 | 0 |
| 0 | 0 | 0 | 0 | 0 |
| 0 | 0 | 0 | 0 | 0 |
| 0 | 0 | 0 | 0 | 0 |
| 0 | 12.26982767 | 0.0735667 | 0.783642027 | 0.000847468 |
| 0 | 0 | 0 | 0 | 0 |
| 0 | 0 | 0 | 0 | 0 |
| 0 | 0 | 0 | 0 | 0 |
| 0 | 0 | 0 | 0 | 0 |
| 0 | 0 | 0 | 0 | 0 |
| 0 | 0 | 0 | 0 | 0 |
| 0 | 0 | 0 | 0 | 0 |
| 0 | 0 | 0 | 0 | 0 |
| 0 | 0 | 0 | 0 | 0 |
| 0 | 0 | 0 | 0 | 0 |
| 0 | 0 | 0 | 0 | 0 |
| 0 | 0 | 0 | 0 | 0 |
| 0 | 0 | 0 | 0 | 0 |
| 0 | 0 | 0 | 0 | 0 |
| 0 | 0 | 0 | 0 | 0 |
| 0 | 0 | 0 | 0 | 0 |
| 0 | 0 | 0 | 0 | 0 |
| 0 | 0 | 0 | 0 | 0 |
| 0 | 0 | 0 | 0 | 0 |
| 0 | 0 | 0 | 0 | 0 |
| 0 | 0 | 0 | 0 | 0 |
| 0 | 6.720415039 | 1.346003563 | 0 | 0 |
| 0 | 0 | 0 | 0 | 0 |
| 0 | 0 | 0 | 0 | 0 |
| 0 | 0 | 0 | 0 | 0 |
| 0 | 0 | 0 | 0 | 0 |
| 0 | 0 | 0 | 0 | 0 |
| 0.012704897 | 0.042698125 | 0.002937713 | 0 | 0.110048119 |
| 0.253136744 | 0.065973612 | 0.128173711 | 0.036585026 | 0.188921817 |
| 0.110226666 | 0.022585262 | 0.00154836 | 0.011661426 | 0 |
| 0.001593737 | 0.001465486 | 0.001488417 | 0 | 0 |
| 0 | 0 | 0 | 0 | 0 |

| | | | | |
|---|---|---|---|---|
| 0 | 0 | 0.0419562 | 0 | 0 |
| 0.020927445 | 0.033392131 | 0.021897054 | 0.036591759 | 0.538263827 |
| 0 | 0.003241151 | 0.042428507 | 0.033617084 | 0 |
| 0.26205342 | 0.840059018 | 0.133791254 | 1.565791092 | 0 |
| 0.002011875 | 0 | 0.017223457 | 0.016395345 | 0 |
| 0 | 0 | 0 | 0 | 0 |
| 2.470992944 | 0.011359745 | 0 | 0.013073487 | 0 |
| 0 | 0 | 0 | 0 | 0 |
| 0 | 0 | 0 | 0 | 0 |
| 0 | 0 | 0 | 0 | 0 |
| 0 | 0 | 0 | 0 | 3.036832625 |
| 0.650789623 | 0.048760086 | 0.485526682 | 0 | 0 |
| 0 | 0 | 0 | 0.000943807 | 0 |
| 0.109706373 | 0 | 0.091003889 | 0.104161377 | 0.004230517 |
| 0 | 0 | 0 | 0 | 0.007462673 |
| 0 | 0 | 0 | 0 | 0 |
| 0.001450035 | 0.001375015 | 0.027465093 | 1.367508669 | 0 |
| 0 | 0 | 0 | 0 | 0 |
| 0 | 0 | 0 | 0 | 0 |
| 0 | 0 | 0 | 0 | 0 |
| 2.671778272 | 0.186065769 | 0.002332314 | 0.776030653 | 0.000638622 |
| 0.11261438 | 0 | 0.00505342 | 0.334796197 | 0.088741369 |
| 0 | 0.004092496 | 0.016437199 | 0 | 0 |
| 0 | 0 | 0 | 0 | 0 |
| 0 | 0 | 0 | 0 | 0 |
| 0 | 0 | 0 | 0 | 0 |
| 0 | 0.027196577 | 0 | 0 | 0 |
| 0 | 0 | 0 | 0 | 0 |
| 0 | 0.019378458 | 0 | 0 | 0.015085646 |
| 0 | 0 | 0 | 0 | 0 |
| 0 | 2.350488341 | 0 | 0.006359231 | 7.512317281 |
| 0 | 0 | 0 | 0 | 0 |
| 0 | 0 | 0 | 0 | 0 |
| 0.000151605 | 0 | 0 | 0 | 0 |
| 0 | 0 | 0 | 0 | 0 |
| 0 | 0 | 0 | 0 | 0 |
| 0 | 0 | 0 | 0 | 0 |
| 0.000159064 | 0 | 0 | 0 | 0 |
| 0 | 0 | 0 | 0 | 0 |
| 0 | 0 | 0 | 0 | 0 |
| 0 | 0 | 0 | 0 | 0 |
| 0 | 0 | 0 | 0 | 0 |
| 0 | 0 | 0 | 0 | 0 |
| 0 | 0 | 0 | 0 | 0 |
| 0 | 0 | 0 | 0 | 0 |
| 0 | 0 | 0 | 0 | 0 |
| 0 | 0 | 0 | 0 | 0 |
| 0.000231927 | 0 | 0 | 0 | 0 |

| | | | | |
|---|---|---|---|---|
| 0 | 0 | 0 | 0 | 0 |
| 0 | 0 | 0 | 0 | 0 |
| 0 | 0 | 0 | 0 | 0 |
| 0 | 0 | 0 | 0 | 0 |
| 0.004852647 | 0 | 0.002300844 | 0 | 0 |
| 0 | 0 | 0 | 0 | 0 |
| 0 | 0 | 0 | 0 | 0 |
| 0 | 0 | 0 | 0 | 0 |
| 0.000190187 | 0 | 0.002767898 | 0.000561168 | 0 |
| 0 | 0 | 0 | 0 | 0 |
| 0 | 0 | 0 | 0 | 0 |
| 0 | 0 | 0 | 0 | 0 |
| 0 | 0 | 0 | 0 | 0 |
| 0 | 0 | 0 | 0 | 0 |
| 0 | 0 | 0 | 0 | 0 |
| 0 | 0 | 0 | 0 | 0 |
| 0 | 0 | 0 | 0 | 0 |
| 0 | 0 | 0 | 0 | 0 |
| 0 | 0 | 1.441697108 | 0 | 0 |
| 0 | 0 | 0 | 0 | 0 |
| 0 | 0 | 0 | 0 | 0 |
| 0 | 0 | 0 | 0 | 0 |
| 0.000242692 | 0 | 0 | 0 | 0 |
| 0 | 0 | 0 | 0 | 0 |
| 0 | 0 | 0 | 0 | 0 |
| 0 | 0 | 0 | 0 | 0 |
| 0.000185802 | 0 | 0 | 0 | 0 |
| 0 | 0 | 0.008986433 | 0 | 0 |
| 0 | 0 | 0 | 0 | 0 |
| 0 | 0 | 0 | 0 | 0 |
| 0.155167982 | 0 | 0.654194392 | 0 | 0 |
| 0.004431191 | 0 | 0.01454174 | 0 | 0 |
| 0 | 0 | 0 | 0 | 0 |
| 0 | 0 | 0 | 0 | 0 |
| 0 | 0 | 0 | 0 | 0 |
| 0 | 0 | 0 | 0 | 0 |
| 0 | 0 | 0 | 0 | 0 |
| 0 | 0 | 0 | 0 | 0 |
| 0 | 0 | 0 | 0 | 0 |
| 0 | 0 | 0 | 0 | 0 |
| 0 | 0 | 0 | 0 | 0 |
| 0 | 0 | 0 | 0 | 0 |
| 0.02941453 | 0 | 0.001644256 | 0 | 0 |
| 0 | 0 | 0 | 0 | 0 |
| 0 | 0 | 0 | 0 | 0 |
| 0 | 0 | 0 | 0 | 0 |
| 0.05835755 | 0 | 0.013024533 | 0 | 0 |
| 0 | 0 | 0 | 0 | 0 |

| | | | | |
|---|---|---|---|---|
| 0 | 0 | 0 | 0 | 0 |
| 0 | 0 | 0 | 0 | 0 |
| 0 | 0 | 0 | 0 | 0 |
| 0 | 0 | 0 | 0 | 0 |
| 0.001044564 | 0 | 0.088736148 | 0 | 0 |
| 0 | 0 | 0 | 0 | 0 |
| 0 | 0 | 0 | 0 | 0 |
| 0 | 0 | 0 | 0 | 0 |
| 0 | 0 | 0 | 0 | 0 |
| 0 | 0 | 0 | 0 | 0 |
| 0 | 0 | 0 | 0 | 0 |
| 0 | 0 | 0 | 0 | 0 |
| 0 | 0 | 0 | 0 | 0 |
| 0 | 0 | 0 | 0 | 0 |
| 0 | 0 | 0 | 0 | 0 |
| 1.332168136 | 0 | 4.430293166 | 0 | 0 |
| 0 | 0 | 0 | 0 | 0 |
| 0 | 0 | 0 | 0 | 0 |
| 0 | 0 | 0 | 0 | 0 |
| 0 | 0 | 0 | 0 | 0 |
| 0 | 0 | 0 | 0 | 0 |
| 0 | 0 | 0 | 0 | 0 |
| 0.000470268 | 0 | 0 | 0 | 0 |
| 0 | 0 | 0 | 0 | 0 |
| 0 | 0 | 0.015197828 | 0 | 0 |
| 0 | 0 | 0 | 0 | 0 |
| 0 | 0 | 0 | 0 | 0 |
| 0 | 0 | 0 | 0 | 0 |
| 0 | 0 | 0 | 0 | 0 |
| 0 | 0 | 0 | 0 | 0 |
| 0 | 0 | 0 | 0 | 0 |
| 0.000355044 | 0.848960838 | 0 | 0 | 0 |
| 0 | 0 | 0 | 0 | 0 |
| 0 | 0 | 0 | 0 | 0 |
| 0 | 0 | 0 | 0 | 0 |
| 0 | 0 | 0 | 0 | 0 |
| 0 | 0 | 0 | 0 | 0 |
| 0 | 0 | 0 | 0 | 0 |
| 0 | 0 | 0 | 0 | 0 |
| 0 | 0 | 0 | 0 | 0 |
| 0 | 0 | 0 | 0 | 0 |
| 0 | 0 | 0 | 0 | 0 |
| 0 | 0 | 0 | 0 | 0 |
| 0 | 0 | 0 | 0 | 0 |
| 0 | 0 | 0 | 0 | 0 |
| 0 | 0 | 0 | 0 | 0 |
| 0 | 0 | 0 | 0 | 0 |
| 0 | 0 | 0 | 0.180582821 | 0 |

| | | | | |
|---|---|---|---|---|
| 0 | 0 | 0 | 0 | 0 |
| 0 | 0 | 0 | 0 | 0 |
| 0 | 0 | 0 | 0 | 0 |
| 0 | 0 | 0 | 0 | 0 |
| 0 | 0 | 0 | 0 | 0 |
| 0 | 0 | 0 | 0 | 0 |
| 0 | 0 | 0 | 0 | 0 |
| 0 | 0 | 0 | 0 | 0 |
| 0 | 0 | 0 | 0 | 0 |
| 0 | 0 | 0 | 0 | 0 |
| 0 | 0 | 0 | 0 | 0 |
| 0 | 0 | 0 | 0 | 0 |
| 0.00159679 | 0 | 0.002094121 | 0.011427881 | 0.001449431 |
| 0 | 0 | 0 | 0 | 0 |
| 0 | 0 | 0 | 0 | 0 |
| 0 | 0 | 0 | 0 | 0 |
| 0 | 0 | 0 | 0 | 0 |
| 0.003595586 | 0 | 0 | 0 | 0 |
| 0 | 0 | 0 | 0 | 0 |
| 0 | 0 | 0 | 0 | 0 |
| 0.08385129 | 0.007496184 | 0.017293474 | 0.054355647 | 0.006194488 |
| 0 | 0 | 0 | 0 | 0 |
| 0 | 0 | 0 | 0.001480149 | 0 |
| 0.001324849 | 0.001786746 | 0 | 0 | 0 |
| 0 | 0 | 0 | 0 | 0 |
| 0 | 0 | 0 | 0 | 0 |
| 0.003995428 | 0.00947959 | 0.001140148 | 0 | 0 |
| 0 | 0 | 0 | 0 | 0 |
| 0 | 0 | 0 | 0 | 0 |
| 0.136630422 | 0 | 0 | 0 | 0 |
| 0 | 0 | 0 | 0.036342778 | 0.033938085 |
| 0 | 0 | 0 | 0 | 0 |
| 0 | 0 | 0 | 0 | 0 |
| 0 | 0 | 0.01595275 | 0 | 0 |
| 0.005001161 | 0.004319998 | 0.022079506 | 0.110734298 | 0.004282368 |
| 0 | 0 | 0 | 0 | 0 |
| 0 | 0 | 0 | 0 | 0 |
| 0 | 0 | 0 | 0 | 0 |
| 0 | 0 | 0 | 0 | 0 |
| 0 | 0 | 0 | 0 | 0 |
| 0 | 0 | 0 | 0 | 0 |
| 0 | 0 | 0 | 0 | 0 |
| 0 | 0 | 0 | 0 | 0 |
| 0 | 0 | 0 | 0 | 0 |
| 0 | 0 | 0 | 0 | 0 |
| 0.035456583 | 0 | 0 | 0 | 0 |
| 0.001566301 | 0 | 0.008275235 | 0.005809929 | 0 |
| 0.006185389 | 0 | 0.004538784 | 0.015295767 | 0.002872217 |

| | | | | |
|---|---|---|---|---|
| 0 | 0 | 0 | 0 | 0 |
| 0 | 0 | 0 | 0 | 0 |
| 0 | 0.005081751 | 0 | 0 | 0 |
| 0 | 0 | 0 | 0 | 0 |
| 0 | 0 | 0 | 0 | 0 |
| 0 | 0 | 0 | 0 | 0 |
| 0 | 0 | 0 | 0 | 0 |
| 0.000254046 | 0 | 0 | 0 | 0.005161082 |
| 0 | 0 | 0 | 0.00068503 | 0 |
| 0 | 0 | 0 | 0 | 0 |
| 0 | 0 | 0 | 0 | 0 |
| 0 | 0 | 0 | 0 | 0 |
| 0 | 0 | 0 | 0 | 0 |
| 0 | 0 | 0 | 0.012249013 | 0 |
| 0 | 0 | 0 | 0 | 0 |
| 0 | 0 | 0 | 0 | 0 |
| 0 | 0 | 0 | 0 | 0 |
| 0 | 0 | 0 | 0 | 0 |
| 0.000659005 | 0 | 0 | 0 | 0 |
| 0 | 0 | 0 | 0 | 0 |
| 0.166847559 | 0 | 0 | 0.071706978 | 0.076481396 |
| 0 | 0 | 0 | 0 | 0 |
| 0 | 0 | 0 | 0 | 0 |
| 0 | 0 | 0 | 0 | 0 |
| 0.006671696 | 0.012727041 | 0.002254567 | 0.007252563 | 0 |
| 0 | 0 | 0 | 0 | 0 |
| 0 | 0 | 0 | 0 | 0 |
| 0 | 0 | 0 | 0 | 0 |
| 0 | 0 | 0 | 0 | 0 |
| 0 | 0 | 0 | 0 | 0 |
| 2.13908886 | 0.001862248 | 0.237313449 | 1.850398916 | 0.559595158 |
| 0 | 0 | 0 | 0 | 0 |
| 0 | 0 | 0 | 0 | 0 |
| 0.003068558 | 0 | 0 | 0 | 0 |
| 0 | 0 | 0 | 0 | 0 |
| 0 | 0 | 0 | 0 | 0 |
| 0 | 0 | 0 | 0 | 0 |
| 0 | 0 | 0 | 0 | 0 |
| 0 | 0 | 0 | 0 | 0 |
| 0 | 0 | 0 | 0 | 0 |
| 0 | 0 | 0 | 0 | 0 |
| 0 | 0.00178659 | 0.011397614 | 0.17198561 | 0 |
| 0 | 0 | 0 | 0.617876109 | 0.004403242 |
| 0 | 0 | 0 | 0 | 0 |
| 0 | 0 | 0 | 0 | 0 |
| 0 | 0 | 0 | 0 | 0 |
| 0 | 0 | 0 | 0 | 0 |
| 0 | 0 | 0 | 0 | 0 |

| | | | | |
|---|---|---|---|---|
| 0 | 0 | 0 | 0 | 0 |
| 0 | 0 | 0 | 0 | 0 |
| 0.109910551 | 0.01079616 | 0.001922451 | 0.005889716 | 0 |
| 0 | 0 | 0 | 0 | 0 |
| 0 | 0 | 0 | 0 | 0 |
| 0 | 0 | 0 | 0 | 0 |
| 0 | 0 | 0 | 0 | 0 |
| 0 | 0 | 0 | 0 | 0 |
| 0 | 0 | 0 | 0 | 0 |
| 0 | 0 | 0 | 0 | 0 |
| 0 | 0 | 0 | 0 | 0 |
| 0 | 0 | 0 | 0 | 0 |
| 0 | 0 | 0 | 0 | 0 |
| 0.011581798 | 0.002634273 | 0 | 0.063193475 | 0 |
| 0.010090328 | 0.002557938 | 0 | 0.00470093 | 0 |
| 0 | 0.002875541 | 0 | 0 | 0 |
| 0.006742616 | 0.007286352 | 0.001332066 | 0.014283436 | 0 |
| 0.000201168 | 0 | 0 | 0 | 0 |
| 0 | 0 | 0 | 0 | 0 |
| 0 | 0 | 0 | 0 | 0 |
| 0 | 0 | 0 | 0 | 0 |
| 0.005384351 | 0 | 0.002204538 | 0.014966095 | 0 |
| 0 | 0 | 0 | 0 | 0 |
| 0.000846794 | 0 | 0 | 0 | 0 |
| 0 | 0 | 0 | 0 | 0 |
| 0 | 0 | 0 | 0 | 0 |
| 0 | 0 | 0 | 0 | 0 |
| 0 | 0 | 0 | 0 | 0 |
| 0 | 0 | 0 | 0 | 0 |
| 0 | 0 | 0.000869696 | 0 | 0 |
| 0 | 0 | 0 | 0 | 0 |
| 0 | 0 | 0 | 0 | 0 |
| 0 | 0 | 0 | 0 | 0 |
| 0.000944717 | 0 | 0 | 0 | 0 |
| 0 | 0 | 0 | 0 | 0 |
| 0 | 0 | 0 | 0 | 0.540032901 |
| 0 | 0 | 0 | 0 | 0 |
| 0.000686292 | 0.004131972 | 0.006798535 | 0.021406889 | 0 |
| 0 | 0 | 0 | 0 | 0 |
| 0 | 0 | 0 | 0 | 0 |
| 0 | 0 | 0 | 0 | 0 |
| 0 | 0 | 0 | 0 | 0 |
| 0 | 0 | 0 | 0 | 0 |
| 0 | 0 | 0 | 0 | 0 |
| 0 | 0 | 0 | 0 | 0 |
| 0 | 0 | 0 | 0 | 0 |
| 0 | 0 | 0 | 0 | 0 |
| 0.008015837 | 0.001401359 | 0.042698604 | 0.006307081 | 0.161036635 |

| | | | | |
|---|---|---|---|---|
| 0 | 0 | 0.090277895 | 0 | 0 |
| 0 | 0 | 0 | 0.016475204 | 0 |
| 0.00106255 | 0.015294502 | 0 | 0.001929331 | 0 |
| 0 | 0 | 0 | 0 | 0 |
| 0.002837035 | 0.074859292 | 0.007814261 | 0.003154277 | 0 |
| 0 | 0 | 0 | 0 | 0 |
| 0 | 0 | 0 | 0 | 0 |
| 0.057633754 | 0.004274998 | 0.002664342 | 0.021766958 | 0.03161324 |
| 0 | 0 | 0 | 0 | 0 |
| 0.000772257 | 0.241346892 | 0.006407788 | 0 | 0.002534346 |
| 0 | 0 | 0.004689951 | 0.014262715 | 0 |
| 0.000729969 | 0.21658265 | 0 | 0.001884619 | 0.004726974 |
| 0.010831906 | 0.085059817 | 0.007821387 | 0.007147752 | 0.003585577 |
| 0.002785611 | 0 | 0 | 0 | 0 |
| 0.00096099 | 0 | 0.001279746 | 0.00441078 | 0 |
| 0.00114291 | 0.000700625 | 0 | 0 | 0 |
| 0 | 0 | 0 | 0 | 0 |
| 0 | 0 | 0 | 0 | 0 |
| 0 | 0.890884648 | 0 | 0 | 0 |
| 0 | 0 | 0 | 0 | 0.61497091 |
| 0 | 0 | 0 | 0 | 0 |
| 0 | 0 | 0 | 0 | 0 |
| 0 | 0.026006146 | 0 | 0 | 0 |
| 0 | 0 | 0 | 0 | 0 |
| 0 | 0 | 0 | 0 | 0 |
| 0.0010077 | 0.008290059 | 0 | 0 | 0 |
| 0 | 0 | 0 | 0 | 0.073584493 |
| 0 | 0.004601633 | 0 | 0.078815021 | 0 |
| 0 | 0 | 0 | 0 | 0 |
| 0 | 0 | 0 | 0 | 0 |
| 0.000259516 | 0 | 0 | 0.193984759 | 0.016080242 |
| 0 | 0 | 0 | 0 | 0 |
| 0 | 0 | 0 | 0 | 0 |
| 0 | 0 | 0 | 0 | 0 |
| 0 | 0 | 0 | 0 | 0 |
| 0 | 0 | 0 | 0 | 0 |
| 0 | 0.40610092 | 0.002581758 | 2.647648496 | 0.019215465 |
| 0 | 0 | 0 | 0 | 0 |
| 0 | 0 | 0 | 0 | 0 |
| 0 | 0 | 0 | 0.21339851 | 0 |
| 0 | 0 | 0 | 0 | 0 |
| 0.058368627 | 0 | 0 | 0.317627226 | 0 |
| 0 | 0 | 0 | 0 | 0 |
| 0.660966834 | 0 | 0 | 0 | 0.223248521 |
| 0 | 0 | 0 | 0 | 0 |
| 0 | 0 | 0 | 0 | 0 |
| 0.001804543 | 0 | 0.002574752 | 0.002662246 | 0.001222009 |
| 0 | 0.499169478 | 0 | 0 | 0 |

| | | | | |
|---|---|---|---|---|
| 0 | 0 | 0 | 0 | 0 |
| 0.098090214 | 0 | 0 | 0 | 0 |
| 0 | 0 | 0 | 0 | 0 |
| 0 | 0 | 0 | 0 | 0 |
| 0 | 0 | 0.003562822 | 0 | 0 |
| 0.036282541 | 0 | 0.023929562 | 0 | 0.057267567 |
| 0 | 0 | 0 | 0.332378553 | 0 |
| 0.000357442 | 0.001721644 | 0 | 0 | 0 |
| 0 | 0 | 0 | 0 | 0 |
| 0 | 0.003472885 | 0 | 0 | 0 |
| 0.00402958 | 0.035200459 | 0.008232197 | 0.004323525 | 0 |
| 0.017896098 | 0 | 0 | 0 | 0.008631491 |
| 0.00163868 | 0 | 0 | 0 | 0 |
| 0.001643908 | 0.002479586 | 0.003333157 | 0.011488075 | 0.002109276 |
| 0 | 0 | 0.001349022 | 0 | 0 |
| 0 | 0 | 0 | 0 | 0 |
| 0 | 0 | 0 | 0 | 0 |
| 0 | 0 | 0 | 0.257631189 | 0 |
| 0 | 0 | 0.035239819 | 0.06548393 | 0 |
| 0.013307545 | 0 | 0 | 0 | 0.001897193 |
| 0 | 0 | 0 | 0 | 0 |
| 0.002641191 | 0 | 0.056527425 | 0 | 0 |
| 0 | 0 | 0 | 0 | 0 |
| 0 | 0 | 0 | 0 | 0 |
| 0 | 0 | 0 | 0 | 0 |
| 0 | 0 | 0 | 0 | 0 |
| 0 | 0.009805219 | 0 | 0.026849848 | 0 |
| 0 | 0 | 0 | 0 | 0 |
| 0 | 0 | 0 | 0 | 0 |
| 10.39118972 | 0.856471901 | 0.277182584 | 0.11413176 | 0 |
| 0.006808961 | 0 | 0.004371812 | 0.027884291 | 0 |
| 0 | 0 | 0.001058256 | 0 | 0 |
| 0 | 0 | 0 | 0.001866926 | 0 |
| 0.001122341 | 0 | 0.00192165 | 0 | 0 |
| 0 | 0 | 0.003210818 | 0 | 0 |
| 0 | 0 | 0 | 0 | 0 |
| 0 | 0 | 0 | 0 | 0 |
| 0 | 0 | 0 | 0 | 0 |
| 0 | 0 | 0.008536561 | 0 | 0 |
| 0 | 0 | 0.004732773 | 0 | 0 |
| 0.07223258 | 0.01462916 | 0.007672609 | 0.185740808 | 0.054189209 |
| 1.008161659 | 10.91369239 | 0.169996818 | 0 | 0 |
| 0 | 0 | 0 | 0 | 0 |
| 0 | 0 | 0 | 0 | 0 |
| 0.004011921 | 0 | 0.016279836 | 0 | 0 |
| 0 | 0 | 0 | 0 | 0 |
| 0.001231826 | 0.008558164 | 0 | 0.016080482 | 0 |
| 0 | 0 | 0 | 0 | 0 |

| | | | | |
|---|---|---|---|---|
| 0.000184226 | 0.001242275 | 0 | 0 | 0 |
| 0 | 0 | 0 | 0 | 0 |
| 0.036084461 | 0.006559193 | 0.018696096 | 0.098508762 | 0 |
| 0 | 0 | 0 | 0 | 0 |
| 0 | 0 | 0.08894158 | 0 | 0 |
| 0 | 0 | 0.025002652 | 0.009191922 | 0.017799851 |
| 0 | 0 | 0.001008599 | 0.000965623 | 0 |
| 0.007226808 | 0 | 0 | 0 | 0 |
| 0.008865397 | 0 | 0 | 0 | 0.029130077 |
| 0 | 0 | 0.080778511 | 0 | 0 |
| 0 | 0 | 0 | 0 | 0 |
| 0 | 0 | 0 | 0 | 0 |
| 0 | 0 | 0 | 0.002673738 | 0 |
| 0.000374996 | 0.016155414 | 0.017335624 | 0 | 0 |
| 0 | 0 | 0 | 0 | 0 |
| 0.007293758 | 0.014381724 | 0.24368759 | 0.882360454 | 0 |
| 0.007362551 | 0.00425895 | 0.012976774 | 0.004472582 | 0.002419582 |
| 0.011093817 | 0 | 0 | 0.008021638 | 0.00454317 |
| 0.001677565 | 0 | 0 | 0.037178707 | 0 |
| 0 | 0 | 0 | 0 | 0 |
| 0 | 0.101215505 | 0 | 0.040799158 | 0 |
| 0 | 0.001411474 | 0 | 0.006423192 | 0 |
| 0 | 0 | 0 | 0 | 0 |
| 0.281707645 | 0 | 0 | 0 | 0 |
| 0 | 0 | 0 | 0 | 0 |
| 0.609982426 | 0 | 0.004395758 | 0.351546348 | 0.285414801 |
| 0.005589764 | 0.001235833 | 0.002118101 | 0.002433419 | 0.027365871 |
| 0 | 0 | 0.000595745 | 0 | 0 |
| 0.000586753 | 0 | 0 | 0 | 0 |
| 0.001254541 | 0 | 0 | 0.003238946 | 0 |
| 0.059792746 | 0.007048859 | 0.014139331 | 0 | 0 |
| 0.007584483 | 0.026622759 | 0 | 0.012568916 | 0 |
| 0.280548502 | 0.011340103 | 0.042428606 | 0.016053705 | 0.00361745 |
| 0 | 0 | 0.001456391 | 0 | 0.003456106 |
| 0 | 0 | 0 | 0 | 0 |
| 0 | 0 | 0 | 0 | 0 |
| 0.002101702 | 0.005879958 | 0 | 0.001319423 | 0 |
| 0.040146498 | 0.044569107 | 0.074725643 | 0.126504828 | 0.005115247 |
| 0 | 0 | 0.23538982 | 0.208802887 | 0 |
| 0 | 0 | 0 | 0 | 0 |
| 0 | 0 | 0 | 0 | 0 |
| 0 | 0 | 0 | 0 | 0 |
| 0.016526421 | 0 | 0.01396901 | 0.157707914 | 0.001274974 |
| 1.848226407 | 0 | 0.029026065 | 0 | 0 |
| 0 | 0 | 0 | 0.177801721 | 0 |
| 0 | 0 | 0 | 0 | 0 |
| 0 | 0 | 0 | 0 | 0 |
| 0 | 0 | 0 | 0 | 0 |

| | | | | |
|---|---|---|---|---|
| 0 | 0 | 0 | 0 | 0 |
| 0.534990959 | 1.786907059 | 0.554153015 | 0.437836724 | 0.040659342 |
| 0 | 0 | 0 | 0 | 0 |
| 0.005367954 | 0 | 0 | 0 | 0 |
| 0 | 0 | 0 | 0.005517513 | 0 |
| 0 | 0 | 0 | 0 | 0 |
| 0 | 0 | 0 | 0.002081213 | 0 |
| 0.0019399 | 0.000617035 | 0 | 0 | 0 |
| 0.000479838 | 0 | 0 | 0.008494872 | 0 |
| 0.030472555 | 0 | 0.003557351 | 0.287719769 | 0 |
| 0.011898021 | 0 | 0.00309211 | 0.051405714 | 0 |
| 0.237617823 | 0.054951437 | 0.003251143 | 0.018053153 | 0.05497053 |
| 0 | 0 | 0 | 0 | 0 |
| 0 | 0 | 0 | 0 | 0 |
| 0.000121888 | 0.000410959 | 0 | 0 | 0 |
| 0 | 0 | 0 | 0 | 0 |
| 0.035671146 | 0.115518173 | 0.006856698 | 0.062800736 | 0 |
| 0.029747562 | 0.192966139 | 0 | 0.264740061 | 0.590549447 |
| 0 | 0 | 0 | 0 | 0 |
| 0 | 0 | 0 | 0 | 0 |
| 0.029099905 | 0.155356815 | 0.017088216 | 0.008349289 | 0 |
| 0.000333076 | 0 | 0 | 0.001684759 | 0 |
| 0 | 0 | 0 | 0 | 0 |
| 0 | 0 | 0 | 0 | 0 |
| 0.010601037 | 0 | 0 | 0 | 0 |
| 0.001823446 | 0 | 0 | 0.014899198 | 0 |
| 0.23872977 | 0 | 0.018224032 | 0 | 0 |
| 0.889012903 | 0.034531604 | 0.02359005 | 0.165009576 | 0 |
| 0 | 0 | 0 | 0 | 0 |
| 0.00215707 | 0 | 0 | 0.00665396 | 0 |
| 0.000130828 | 0.004705072 | 0 | 0 | 0 |
| 0 | 0 | 0 | 0 | 0 |
| 0 | 0 | 0 | 0 | 0 |
| 0 | 0 | 0 | 0 | 0 |
| 0 | 0 | 0 | 1.820419851 | 0 |
| 0.127939948 | 0.13497476 | 0.078082922 | 0.14247583 | 0 |
| 0.000445999 | 0 | 0 | 0.017859523 | 0 |
| 0 | 0 | 0 | 0 | 0 |
| 0.002155891 | 0.061700347 | 0.005150613 | 0.032545574 | 0.051946574 |
| 0 | 0 | 0 | 0 | 0 |
| 0 | 0 | 0 | 0.049756676 | 0 |
| 0 | 0 | 0 | 0 | 0 |
| 0 | 0.00318304 | 0.015955662 | 0.113759758 | 0.004825775 |
| 0.001146707 | 0 | 0.00090617 | 0.005899398 | 0 |
| 0 | 0 | 0 | 0 | 0 |
| 0 | 0 | 0.012639517 | 0 | 0 |
| 0 | 0 | 0 | 0 | 0 |
| 0.000712909 | 0 | 0 | 0.060300666 | 0.004344939 |

| | | | | |
|---|---|---|---|---|
| 0.000723263 | 0 | 0 | 0 | 0 |
| 0 | 0 | 0 | 0 | 0 |
| 0 | 0 | 0 | 0 | 0 |
| 0 | 0 | 2.029405377 | 0 | 0 |
| 0 | 0 | 0 | 0 | 0 |
| 0 | 0 | 0 | 0 | 0 |
| 0 | 0 | 0 | 0 | 0 |
| 0 | 0.004707317 | 0 | 0.030521995 | 0 |
| 0.576039278 | 0 | 4.514313942 | 0.222869637 | 0 |
| 0 | 0 | 0.083532985 | 0 | 0 |
| 0 | 0 | 0 | 0 | 0 |
| 0.008325923 | 0 | 0.046432116 | 0 | 0 |
| 0 | 0 | 0 | 0 | 0 |
| 0 | 0 | 23.13201898 | 0 | 0 |
| 0.021761487 | 0 | 0.431558035 | 0.841177801 | 0.001025481 |
| 0 | 0 | 0 | 0 | 0 |
| 0 | 0 | 0.001315256 | 0 | 0 |
| 0 | 0 | 0 | 0.565757658 | 0 |
| 0 | 0 | 0 | 0 | 0 |
| 0 | 0 | 0 | 0 | 0 |
| 0 | 0 | 0 | 0 | 0 |
| 0 | 0 | 0 | 0 | 0 |
| 0 | 0 | 0 | 0 | 0 |
| 0.089736335 | 0.002732287 | 0.519903751 | 1.746950018 | 0.000987801 |
| 0.010976902 | 0 | 0 | 0.464516599 | 0 |
| 0 | 0 | 0 | 0 | 0 |
| 0.000757149 | 0 | 0 | 0 | 0 |
| 0 | 0 | 1.361896315 | 0 | 0 |
| 0 | 0 | 0 | 0 | 0 |
| 0 | 0 | 0 | 0 | 0 |
| 0 | 0.000779728 | 0.010521338 | 0 | 0 |
| 0 | 0 | 0 | 0 | 0 |
| 0.016393922 | 0 | 0 | 0 | 0 |
| 0 | 0 | 0 | 0 | 0 |
| 0 | 0 | 0 | 0 | 0 |
| 0.030897463 | 0 | 0 | 0.160016038 | 0 |
| 0 | 0.094654575 | 0.905545978 | 0 | 0 |
| 0.000141435 | 0 | 0 | 0 | 0 |
| 0.002157977 | 0 | 0.020533645 | 0.01022949 | 0 |
| 0 | 0 | 0 | 0 | 0 |
| 0.013822806 | 0.091671164 | 0.624108782 | 0.101579277 | 0.101134124 |
| 0.000169887 | 0 | 0 | 0 | 0 |
| 0 | 0.24051171 | 0.031940574 | 0.025156046 | 0.014062957 |
| 0 | 0 | 0 | 0 | 0 |
| 0 | 0 | 0 | 0 | 0 |
| 0 | 0 | 0 | 0 | 0 |
| 0 | 0 | 0 | 0 | 0 |
| 0.000783802 | 0.001057067 | 0.058176107 | 0.027520991 | 0 |

| | | | | |
|---|---|---|---|---|
| 0 | 0 | 0 | 0 | 0 |
| 0 | 0 | 0 | 0.173610586 | 0 |
| 2.467436905 | 0.116151605 | 0.007718805 | 2.310972515 | 0.002976545 |
| 0 | 0 | 0 | 0 | 0 |
| 0 | 0 | 0.075154407 | 0 | 0 |
| 8.87842E-05 | 0 | 0.023258207 | 0.022179861 | 0 |
| 0 | 0 | 0 | 0 | 0 |
| 0 | 0 | 0 | 0.001061914 | 0 |
| 0 | 0 | 0 | 0 | 0 |
| 0 | 0 | 0 | 0 | 0 |
| 0.026004092 | 0 | 0.484076545 | 0.094032326 | 0 |
| 16.61371015 | 2.576725924 | 0.318826941 | 2.945363969 | 0.007263322 |
| 0 | 0 | 0 | 0 | 0 |
| 0.007842186 | 0.066840027 | 0.010428255 | 0.00634272 | 0.000970466 |
| 1.293113942 | 0.18742104 | 0.017402538 | 2.098178017 | 8.001351088 |
| 0 | 0 | 0 | 0 | 0 |
| 0 | 0 | 0 | 0 | 0 |
| 0 | 0 | 0 | 0 | 0 |
| 0 | 0 | 0 | 0 | 0 |
| 0.061162443 | 0 | 0.049629366 | 0 | 0.00759232 |
| 0.030049633 | 0.001459276 | 0.113222567 | 1.143882198 | 0 |
| 0 | 0.438991559 | 0 | 0 | 0.052810089 |
| 0.001460011 | 0.99849518 | 0.270278411 | 0.028604539 | 0.459150765 |
| 0 | 0 | 0 | 0 | 0 |
| 0 | 0 | 0 | 0 | 0 |
| 0 | 0.002747878 | 0.003425165 | 0 | 0 |
| 1.595111603 | 0 | 0 | 0 | 1.944828858 |
| 0.016154272 | 0 | 0 | 0.00323474 | 0 |
| 0 | 0 | 0.002535795 | 0 | 0 |
| 0 | 0 | 0 | 0 | 0 |
| 0.006512858 | 0 | 0 | 0 | 0 |
| 0.007705203 | 0 | 0 | 0.994656231 | 0 |
| 3.651190366 | 0 | 0.005642903 | 0.559993028 | 0 |
| 0 | 0 | 0 | 0 | 0 |
| 0.018852003 | 0.000927289 | 0.076542556 | 0.174841831 | 0 |
| 0.018240976 | 0.009960666 | 0.062463652 | 0.122442262 | 0 |
| 0.013605308 | 0 | 0.018749417 | 0.038968929 | 0 |
| 0 | 0 | 0 | 0 | 0 |
| 0.031556151 | 0 | 4.258305011 | 0 | 0 |
| 0 | 0 | 0 | 0 | 0 |
| 0 | 0 | 0 | 0 | 0 |
| 0 | 0 | 0 | 0 | 0 |
| 0 | 0 | 0.026090704 | 0 | 0 |
| 0 | 0 | 0 | 0 | 0 |
| 0.004230867 | 0.004754934 | 0.512714494 | 0.106804227 | 0.012654095 |
| 0 | 0 | 0 | 0 | 0 |
| 0 | 0 | 0 | 0 | 0 |
| 0.654203568 | 0 | 0.842027307 | 0.195204673 | 0.043776849 |

| | | | | |
|---|---|---|---|---|
| 0 | 0 | 0 | 0 | 0 |
| 0 | 0 | 0 | 0 | 0 |
| 0 | 0 | 0 | 0 | 0 |
| 0.00063438 | 0 | 0.000514503 | 0.007487218 | 0 |
| 0 | 0 | 0 | 0 | 0 |
| 0 | 0 | 0 | 0 | 0 |
| 0 | 0 | 0.114721915 | 0 | 0 |
| 0.107869864 | 0.007354684 | 0.244135073 | 3.76117548 | 0.001872487 |
| 0 | 0 | 0 | 0 | 0 |
| 0 | 0 | 0 | 0 | 0 |
| 0.071917479 | 0 | 0.016999594 | 0.056615916 | 0.094960195 |
| 0 | 0 | 0 | 0 | 0 |
| 0 | 0 | 0 | 0.195020296 | 0 |
| 0 | 0 | 0 | 0 | 0 |
| 0 | 0 | 0 | 0 | 0 |
| 0 | 0 | 0 | 0 | 0 |
| 0.017921411 | 0 | 0 | 0 | 0 |
| 0.004953023 | 0 | 0.019876087 | 0.010412764 | 0 |
| 0 | 0 | 0 | 0 | 0 |
| 0 | 0 | 0.551371125 | 0 | 0 |
| 0 | 0 | 0 | 0 | 0 |
| 0 | 0 | 0 | 0 | 0 |
| 0 | 0 | 0.0071497 | 0 | 0 |
| 0 | 0 | 0 | 0 | 0 |
| 0 | 0 | 0 | 0 | 0 |
| 0 | 0 | 0 | 0 | 0 |
| 0 | 0 | 0 | 0 | 0 |
| 0 | 0 | 0 | 0.2921079 | 0 |
| 0 | 0 | 0 | 0 | 0 |
| 0.001869752 | 0 | 0.79944148 | 8.258574712 | 0 |
| 0 | 0 | 0 | 0 | 0 |
| 0 | 0 | 0 | 0 | 0 |
| 0.001099701 | 0 | 0 | 0 | 0 |
| 0 | 0 | 0 | 0 | 0 |
| 0 | 0 | 0 | 0 | 0 |
| 0 | 0 | 0 | 0 | 0 |
| 0 | 0 | 0.068528245 | 0 | 0 |
| 0.003431156 | 0.004139185 | 0.002101976 | 0.04383962 | 0 |
| 0 | 0 | 0 | 0 | 0 |
| 0.005464562 | 0 | 0.005103442 | 0 | 0 |
| 0 | 0 | 0 | 0 | 0 |
| 0 | 0 | 0 | 0 | 0 |
| 0 | 0 | 0 | 0 | 0 |
| 0 | 0 | 0 | 0.052495247 | 0 |
| 0 | 0 | 0 | 0 | 0 |
| 0 | 0 | 0 | 0 | 0 |
| 0.021983732 | 0 | 0 | 0 | 0 |
| 0 | 0 | 0 | 0 | 0 |

| | | | | |
|---|---|---|---|---|
| 0 | 0 | 0 | 0 | 0 |
| 0 | 0 | 0 | 0 | 0 |
| 0 | 0 | 0 | 0 | 0 |
| 0 | 0 | 0.457413738 | 0 | 0 |
| 0 | 0 | 0 | 0 | 0 |
| 0 | 0 | 0 | 0 | 0 |
| 0 | 0 | 0 | 3.565319341 | 0 |
| 0 | 0 | 0 | 0 | 0 |
| 1.253278188 | 0.045522649 | 0.010480915 | 0.293343943 | 6.058414212 |
| 0 | 0 | 0 | 0 | 0 |
| 0.000310424 | 0 | 0 | 0 | 0 |
| 0 | 0 | 0 | 0 | 0 |
| 0 | 0 | 0 | 0 | 0 |
| 0.001356932 | 0 | 0 | 0 | 0 |
| 0 | 0 | 1.045166065 | 4.996531641 | 0 |
| 0 | 0 | 0.099169514 | 0.117289494 | 0 |
| 0 | 0 | 0 | 0.000625196 | 0 |
| 0 | 0 | 0 | 0 | 0 |
| 0 | 0 | 0 | 0 | 0 |
| 0 | 0 | 0 | 0 | 0 |
| 0 | 0 | 0.059679934 | 0 | 0 |
| 0 | 0 | 0 | 0 | 0 |
| 0 | 0 | 0 | 0 | 0 |
| 1.115854078 | 0 | 0.001570825 | 0 | 0 |
| 0.014550317 | 0 | 0.015472132 | 0.00241023 | 0.253280403 |
| 0 | 0.000973455 | 2.537217309 | 0 | 0 |
| 0 | 0.042639486 | 0.010924208 | 0 | 0 |
| 0.000475372 | 0 | 0 | 0 | 0.002317783 |
| 0 | 0 | 0 | 0 | 0 |
| 0 | 0 | 0 | 0 | 0 |
| 0 | 0 | 0 | 0 | 0 |
| 0.00177546 | 0 | 0 | 0 | 0 |
| 0.000370842 | 0 | 0.000507959 | 0 | 0 |
| 0.023775932 | 0 | 0.036833871 | 0.006436087 | 0.006813253 |
| 0.005930195 | 0.019691315 | 0.001846101 | 0 | 0 |
| 0.041093604 | 0 | 0.006361728 | 0.086955847 | 0.007935484 |
| 0 | 0 | 0 | 0.900454628 | 0 |
| 0 | 0 | 0.363410961 | 0 | 0 |
| 0 | 0.000676136 | 0 | 0 | 0 |
| 0 | 0 | 0 | 0 | 0 |
| 0 | 0 | 0 | 0 | 0 |
| 0 | 0 | 0 | 0 | 0 |
| 0 | 3.070293426 | 2.045404535 | 0.021255641 | 0 |
| 0 | 0 | 0 | 0 | 0 |
| 0 | 0 | 0 | 0 | 0 |
| 0.004482785 | 0 | 0 | 0.075810675 | 0 |
| 0 | 0 | 0 | 0 | 0 |
| 0.016056648 | 0.001939223 | 0.05982558 | 0.013859483 | 0 |

| | | | | |
|---|---|---|---|---|
| 0.001036508 | 0 | 0.013401959 | 0.300770506 | 0.03463551 |
| 0 | 0 | 0 | 0 | 0 |
| 0 | 0 | 0 | 0 | 0 |
| 0 | 0 | 0 | 0 | 0 |
| 0.000533775 | 0 | 0.007676911 | 0 | 0 |
| 0 | 0 | 0 | 0 | 0 |
| 0 | 0 | 0 | 0 | 0 |
| 0 | 0 | 0.012879806 | 0 | 0 |
| 0 | 0 | 0 | 0 | 0 |
| 0 | 0 | 0.003852695 | 0 | 0 |
| 0 | 0 | 0.001538477 | 0 | 0 |
| 0.003545576 | 0.46887318 | 0.57013646 | 0 | 0 |
| 0 | 0 | 0.004205059 | 0 | 0 |
| 0 | 0 | 0 | 0 | 0 |
| 0.023828179 | 16.92325375 | 0.001691114 | 0.008500044 | 8.539663085 |
| 0 | 0 | 0 | 0 | 0 |
| 0.010850227 | 0 | 0 | 0.007771383 | 0.001570333 |
| 0 | 0 | 0 | 0 | 0 |
| 0.003534993 | 0 | 0.00092641 | 0.021925046 | 0.003517483 |
| 0 | 0 | 0.013148675 | 0 | 0 |
| 0 | 0 | 0 | 0 | 0 |
| 0 | 0 | 0 | 0 | 0 |
| 0 | 0.009105807 | 0.545889267 | 0.455980971 | 0 |
| 0 | 0 | 0 | 0 | 0 |
| 0 | 0 | 0 | 0 | 0 |
| 0 | 0 | 0 | 0 | 0 |
| 0 | 0 | 0 | 0 | 0 |
| 0 | 0 | 0 | 0 | 0 |
| 0 | 0 | 0 | 0 | 0 |
| 0 | 0 | 0 | 0 | 0 |
| 0 | 0 | 0 | 0 | 0 |
| 0.063282673 | 0.14116235 | 0.021121645 | 0.051053125 | 0.001301895 |
| 0 | 0 | 0 | 0 | 0 |
| 0.185901139 | 0 | 0 | 0.093676731 | 0 |
| 0.040365627 | 0 | 0.004615436 | 0.089123279 | 0.007793278 |
| 0.030339593 | 0.002853991 | 0.38811376 | 0.120937593 | 0 |
| 0.00024542 | 0 | 0 | 0 | 0 |
| 0 | 0 | 0 | 0 | 0 |
| 0 | 0 | 0 | 0 | 0 |
| 0 | 0 | 0 | 0 | 0 |
| 0 | 0 | 0 | 0 | 0 |
| 0 | 0 | 0 | 0 | 0 |
| 0 | 0 | 0 | 0 | 0 |
| 0 | 0 | 0.01329538 | 0 | 0 |
| 0.08303498 | 0 | 0.11209284 | 0.257559849 | 0.168482542 |
| 0 | 0 | 0 | 0 | 0 |
| 0 | 0 | 0 | 0 | 0 |
| 0.001674409 | 0 | 0.005127321 | 0.029453087 | 0 |

| | | | | |
|---|---|---|---|---|
| 0 | 0 | 0 | 0 | 0 |
| 0 | 0 | 0 | 0 | 0 |
| 0 | 0 | 0.059684206 | 0.031375649 | 0 |
| 0 | 0 | 0 | 0 | 0 |
| 0 | 0 | 0.006052841 | 0 | 0 |
| 0 | 0 | 0 | 0 | 0 |
| 0 | 0 | 0.209241785 | 0.355211854 | 0 |
| 0 | 0.004148317 | 0.006661454 | 0.117151831 | 0 |
| 0 | 0 | 0 | 0 | 0 |
| 0 | 0 | 0 | 0 | 0 |
| 0 | 0 | 0 | 0 | 0 |
| 0 | 0 | 0 | 0.00211417 | 0.000873391 |
| 0 | 0 | 0 | 0 | 0 |
| 0 | 0 | 0 | 0 | 0 |
| 0.010546589 | 0 | 0 | 0 | 0 |
| 0 | 0 | 0 | 0 | 0 |
| 0 | 0 | 0 | 0 | 0.000773177 |
| 0 | 0 | 0 | 0 | 0 |
| 0 | 0.259540271 | 2.154806099 | 0 | 0 |
| 0 | 0 | 0 | 0 | 0 |
| 0 | 0 | 0.002136272 | 0 | 0 |
| 0 | 0 | 0 | 0 | 0 |
| 0 | 0 | 0 | 0 | 0 |
| 0 | 0 | 0 | 0 | 0 |
| 0.000185984 | 0.001128715 | 0 | 0 | 0 |
| 0.00183827 | 0.000748024 | 0 | 0.017768296 | 0 |
| 0 | 0 | 0 | 0 | 0 |
| 0.002357704 | 0.003974621 | 0.107719179 | 0.02343293 | 0 |
| 0 | 0 | 0 | 0 | 0 |
| 0 | 0 | 0.659066143 | 0 | 0 |
| 0 | 0 | 0 | 0 | 0 |
| 0 | 0 | 0 | 0 | 0 |
| 0 | 0 | 0 | 0 | 0 |
| 0 | 0 | 0 | 0 | 0 |
| 0.005048479 | 0 | 0.006312324 | 0.006664027 | 0.004655801 |
| 0 | 0 | 0 | 0 | 0 |
| 0 | 0 | 0 | 0 | 0 |
| 0.010816398 | 0 | 0.004258866 | 0.00889614 | 0.007809609 |
| 0 | 0 | 0 | 0 | 0 |
| 0 | 0 | 0 | 0 | 0 |
| 6.438843833 | 0.897526342 | 2.640483159 | 3.06689255 | 0.047975592 |
| 0.064886125 | 1.921866574 | 0.22956205 | 0.728043235 | 0 |
| 0 | 0 | 0 | 0.671653445 | 0 |
| 0 | 0 | 0 | 0 | 0 |
| 0 | 0 | 0 | 0 | 0 |
| 0 | 0 | 0 | 0 | 0 |
| 0 | 0 | 0 | 0 | 0 |
| 0.003679068 | 0 | 0.034876196 | 0.006883957 | 0 |

| | | | | |
|---|---|---|---|---|
| 0 | 0 | 0 | 0 | 0 |
| 0.005722487 | 0 | 0.009955956 | 0.041394967 | 0 |
| 0 | 0 | 0 | 0.034351806 | 0 |
| 0 | 0 | 0 | 0 | 0 |
| 0 | 1.944101544 | 2.108553227 | 1.525472855 | 0 |
| 0 | 0 | 0 | 0 | 0 |
| 0 | 0 | 0 | 0 | 0 |
| 0 | 0 | 0 | 0 | 0 |
| 0.365607024 | 1.325487386 | 0.010414671 | 1.72333512 | 55.3816161 |
| 0 | 0 | 0 | 0 | 0 |
| 0 | 0 | 0 | 0 | 0 |
| 0 | 0 | 0 | 0 | 0 |
| 0 | 0.084564707 | 0.020422694 | 0 | 0 |
| 0.479395803 | 0.00421307 | 0.009438957 | 0.007590884 | 0 |
| 0 | 0 | 0 | 0.286363424 | 0 |
| 0 | 0.938938931 | 0.302850603 | 0 | 0.001838064 |
| 0 | 0 | 0.010291621 | 0 | 0 |
| 0 | 0 | 0 | 0 | 0 |
| 0 | 0 | 0 | 0 | 0 |
| 0 | 0 | 0 | 0 | 0 |
| 0 | 0 | 0 | 0 | 0 |
| 0 | 0 | 0 | 0 | 0 |
| 0 | 0 | 0 | 0 | 0 |
| 0 | 0 | 0 | 0 | 0 |
| 0 | 0 | 0 | 0 | 0 |
| 0 | 0 | 0 | 0 | 0 |
| 0 | 0 | 0.135229143 | 0 | 0 |
| 0.000218434 | 0.000841681 | 0 | 0.001289022 | 0 |
| 0 | 0 | 0 | 0 | 0 |
| 0 | 0 | 0 | 0 | 0 |
| 0 | 0 | 0 | 0 | 0 |
| 7.81968E-05 | 0 | 0 | 0 | 0 |
| 0 | 0 | 0.036282463 | 0 | 0 |
| 0.007719785 | 0 | 0 | 0 | 0 |
| 0 | 0.043828411 | 0.105125076 | 0 | 0 |
| 0 | 0 | 0 | 0.002343205 | 0 |
| 0 | 0 | 0 | 0.005225537 | 0 |
| 0 | 0.001608954 | 0 | 0 | 0 |
| 0 | 0 | 0 | 0 | 0 |
| 0 | 0 | 0 | 0 | 0 |
| 0 | 0.615315209 | 1.425878921 | 0 | 0 |
| 0 | 0 | 0 | 0 | 0 |
| 0 | 0 | 0 | 0 | 0 |
| 0 | 0 | 0 | 0 | 0 |
| 0 | 0 | 0 | 0 | 0 |
| 0 | 0 | 0 | 0 | 0 |
| 0 | 0 | 0 | 0 | 0 |
| 0 | 0 | 0 | 0 | 0 |

| | | | | |
|---|---|---|---|---|
| 0 | 0 | 0.32870523 | 0 | 0 |
| 0 | 0 | 0 | 0 | 0 |
| 0 | 0 | 0 | 0 | 0 |
| 0 | 0 | 0 | 0 | 0 |
| 0 | 0 | 0.014071965 | 0 | 0 |
| 0 | 0 | 0 | 0 | 0 |
| 0 | 0 | 0.097777105 | 0.171682313 | 0 |
| 0 | 0 | 0 | 0 | 0 |
| 0 | 0 | 0 | 0 | 0 |
| 0 | 0 | 0.082067276 | 0 | 0 |
| 0 | 0 | 0 | 0 | 0 |
| 0 | 0 | 0 | 0 | 0 |
| 0 | 0 | 0 | 0 | 0 |
| 0 | 0 | 0 | 0 | 0 |
| 0 | 0 | 0.01393075 | 0 | 0 |
| 0 | 0 | 0 | 0 | 0 |
| 0 | 0.019437412 | 0 | 0 | 0 |
| 0 | 0 | 0.002976743 | 0 | 0 |
| 0 | 0 | 0 | 0 | 0 |
| 0 | 0 | 0 | 0 | 0 |
| 0.050200583 | 0.147055505 | 0.277871422 | 0 | 0 |
| 0 | 0 | 0 | 0 | 0 |
| 0.000932846 | 0 | 0.016257211 | 0.042663105 | 0 |
| 0.001629368 | 0 | 0.022216712 | 0.016957779 | 0.000902768 |
| 0 | 0 | 0 | 0 | 0 |
| 0.005417355 | 0 | 0 | 0.012787575 | 0.001065063 |
| 0 | 0 | 0 | 0 | 0 |
| 0 | 0 | 0 | 0 | 0 |
| 0 | 0 | 0 | 0 | 0 |
| 0.002771413 | 0.006756506 | 0 | 0.002516105 | 0.001559153 |
| 0 | 0 | 0 | 0 | 0 |
| 0 | 0 | 0 | 0 | 0 |
| 0 | 0 | 0 | 0 | 0 |
| 0 | 0 | 0 | 0 | 0 |
| 0 | 0 | 0 | 0 | 0 |
| 0 | 0 | 0 | 0 | 0 |
| 0.005337847 | 0 | 0 | 0.018593595 | 0.000677758 |
| 0 | 0 | 0 | 0 | 0 |
| 0 | 0 | 0 | 0 | 0 |
| 0 | 0 | 0 | 0 | 0 |
| 0 | 0 | 0 | 0 | 0 |
| 0 | 0 | 0 | 0 | 0 |
| 0 | 0 | 0 | 0 | 0 |
| 0 | 0 | 0 | 0 | 0 |
| 0.004174808 | 0 | 0 | 0 | 0 |
| 0 | 0 | 0 | 0 | 0 |
| 0 | 0 | 0 | 0 | 0 |
| 0 | 0 | 0 | 0 | 0 |

| | | | | |
|---|---|---|---|---|
| 0.10347406 | 0 | 0.018666306 | 0.171184809 | 0.00291423 |
| 0 | 0 | 0 | 0 | 0 |
| 0 | 0 | 0 | 0 | 0 |
| 0 | 0 | 0 | 0 | 0 |
| 0 | 0 | 0 | 0 | 0 |
| 0 | 0 | 0 | 0.032756462 | 0 |
| 0 | 0 | 0 | 0 | 0 |
| 0.000163654 | 0.741258246 | 0.79656909 | 0.165917369 | 0 |
| 0.314528665 | 0 | 0.217176646 | 0.050492028 | 0.204014749 |
| 0.326009763 | 0.000352337 | 0.052246183 | 0.741871854 | 0 |
| 0 | 0 | 0 | 0 | 0 |
| 0 | 0 | 0 | 0 | 0 |
| 0 | 0 | 0 | 0 | 0 |
| 0 | 0 | 0 | 0 | 0 |
| 0.002223092 | 0 | 0 | 0 | 0 |
| 0 | 0 | 0 | 0 | 0 |
| 0 | 0 | 0 | 0 | 0.155128895 |
| 0 | 0 | 0 | 0 | 0 |
| 0 | 0 | 0 | 0 | 0 |
| 0.000346621 | 0 | 0 | 0 | 0 |
| 0 | 0 | 0 | 0 | 0 |
| 0 | 0 | 0 | 0 | 0 |
| 0 | 0 | 0 | 0 | 0 |
| 0 | 0 | 0 | 0 | 0 |
| 0 | 0 | 0.021021715 | 0 | 0 |
| 0 | 0 | 0 | 0 | 0 |
| 0 | 0 | 0 | 0 | 0 |
| 0 | 0 | 0 | 0 | 0 |
| 0 | 0 | 0 | 0 | 0 |
| 0 | 0 | 0 | 0 | 0 |
| 0 | 0 | 0 | 0 | 0 |
| 0 | 0 | 0 | 0 | 0 |
| 0 | 0 | 0 | 0 | 0 |
| 0 | 0 | 0 | 0 | 0 |
| 0 | 0 | 0 | 0.074015467 | 0 |
| 0 | 0 | 0 | 0 | 0 |
| 0 | 0 | 0 | 0 | 0 |
| 0 | 0 | 0 | 0 | 0 |
| 0 | 0 | 0 | 0 | 0 |
| 0.099130621 | 0 | 0 | 0.184229697 | 0 |
| 0 | 0 | 0 | 0 | 0 |
| 0.057915166 | 0 | 0.975759329 | 0 | 0 |
| 0 | 0 | 0 | 0 | 0 |
| 0 | 0 | 0 | 0 | 0 |
| 0.000620228 | 0 | 0 | 0.003294085 | 0 |
| 0 | 0 | 0 | 0 | 0 |
| 0 | 0 | 0 | 0 | 0 |
| 0 | 0 | 0 | 0 | 0 |

| | | | | |
|---|---|---|---|---|
| 0 | 0 | 0 | 0 | 0 |
| 0.002215348 | 0 | 0 | 0 | 0 |
| 0 | 0 | 0 | 0 | 0 |
| 0 | 0 | 0 | 0 | 0 |
| 1.455642272 | 0.110669664 | 5.455391372 | 0.125573675 | 0 |
| 0 | 0 | 0 | 0 | 0 |
| 0 | 0 | 0 | 0 | 0 |
| 0.093914655 | 0.008613342 | 0.01145587 | 0.028476239 | 0.412230815 |
| 0 | 0 | 0.000623905 | 0 | 0 |
| 0 | 0 | 0 | 0 | 0 |
| 0 | 0 | 0 | 0 | 0 |
| 0 | 0 | 0 | 0 | 0 |
| 0 | 0 | 0 | 0 | 0 |
| 0 | 0 | 0 | 0 | 0 |
| 22.41945202 | 0 | 0.012756617 | 0 | 0 |
| 0.050362054 | 0 | 0.353345713 | 0 | 0 |
| 0 | 0 | 0 | 0 | 0 |
| 0 | 0 | 0 | 0 | 0 |
| 0 | 0 | 0 | 0 | 0 |
| 0 | 0 | 0 | 0 | 0 |
| 0 | 0 | 0 | 0 | 0 |
| 0 | 0 | 0 | 0 | 0 |
| 0 | 0 | 0 | 0 | 0 |
| 0.035443935 | 0 | 0.010094368 | 0.010354554 | 0.00192492 |
| 0 | 0 | 0.158292915 | 0 | 0 |
| 0 | 0 | 0 | 0 | 0 |
| 0.002664012 | 0.001839685 | 0.001154056 | 0.011553907 | 0.003325498 |
| 0 | 0 | 0 | 0.003390493 | 0 |
| 0 | 0 | 0 | 0 | 0 |
| 0 | 0 | 0 | 0 | 0 |
| 0 | 0 | 0 | 0 | 0 |
| 0 | 0 | 0 | 0 | 0 |
| 0 | 0 | 0 | 0 | 0 |
| 0 | 0 | 0 | 0 | 0 |
| 0 | 0 | 0 | 0 | 0 |
| 0 | 0 | 0 | 0 | 0 |
| 0 | 0 | 0 | 0 | 0 |
| 0.092397678 | 0 | 0 | 0 | 0 |
| 0 | 0 | 0 | 0 | 0 |
| 0 | 0 | 0 | 0 | 0 |
| 0 | 0 | 0 | 0 | 0 |
| 0 | 0 | 2.113212182 | 0 | 0 |
| 0 | 0 | 0 | 0 | 0 |
| 0 | 0 | 0 | 0 | 0 |
| 0 | 0 | 0 | 0 | 0 |
| 0 | 0 | 0 | 0 | 0 |
| 0 | 0 | 0 | 0 | 0 |
| 0 | 0 | 0.039713583 | 0 | 0 |

| | | | | |
|---|---|---|---|---|
| 0 | 0 | 0.390549325 | 0.001068733 | 0 |
| 0 | 0 | 0 | 0 | 0 |
| 0 | 0 | 0 | 0.094571783 | 0 |
| 0 | 0 | 0 | 0.009924418 | 0 |
| 0 | 0 | 0 | 0 | 0 |
| 0 | 0 | 0.070053536 | 0.091832363 | 0.099105428 |
| 0 | 0 | 0 | 0 | 0 |
| 0 | 0 | 0 | 0 | 0 |
| 0 | 0 | 0 | 0 | 0 |
| 0 | 0 | 0 | 0 | 0 |
| 0 | 0 | 0 | 0 | 0 |
| 0.005823028 | 0 | 0 | 0.000954525 | 0.000788653 |
| 0 | 0 | 0 | 0 | 0 |
| 0 | 0 | 0 | 0 | 0 |
| 0 | 0 | 0 | 0 | 0 |
| 0 | 0 | 0 | 0 | 0 |
| 0 | 0 | 0 | 0 | 0 |
| 0 | 0 | 0 | 0.003028483 | 0 |
| 0 | 0 | 0 | 0 | 0 |
| 0 | 0 | 0 | 0 | 0 |
| 0.178365946 | 0.003299101 | 0 | 0.010517495 | 0 |
| 0 | 0 | 0 | 0 | 0 |
| 0.031512133 | 0 | 0.070050078 | 0.005469401 | 0.001075943 |
| 0 | 0 | 1.429792885 | 0 | 0 |
| 0 | 0 | 0 | 0 | 0 |
| 0 | 0 | 0 | 0 | 0 |
| 0 | 0 | 0 | 0 | 0 |
| 0 | 0 | 0 | 0 | 0 |
| 0 | 0 | 0 | 0 | 0 |
| 0 | 0 | 0 | 0 | 0 |
| 0 | 0 | 0 | 0 | 0 |
| 0 | 0 | 0 | 0 | 0 |
| 1.571315141 | 0.027452888 | 0.002547635 | 0.025366447 | 0.032999423 |
| 0 | 0 | 0.012312661 | 0 | 0 |
| 0 | 0 | 0.008355775 | 0 | 0 |
| 0 | 0.094493269 | 0 | 0.005088878 | 0 |
| 0 | 0 | 0 | 0 | 0 |
| 0 | 0 | 0 | 0 | 0 |
| 0.000191815 | 0.001422794 | 0.045913386 | 0.056823427 | 0 |
| 0 | 0 | 0 | 0 | 0 |
| 0 | 0 | 0 | 0 | 0 |
| 0 | 0 | 0 | 0 | 0 |
| 0.054188309 | 0.001966158 | 0.457372166 | 0.032030586 | 0 |
| 0 | 0 | 0 | 0 | 0 |
| 0 | 0 | 0 | 0 | 0 |
| 0 | 0 | 0.137241621 | 0 | 0 |
| 0 | 0 | 0 | 0 | 0 |
| 0 | 0 | 0 | 0 | 0 |

| | | | | |
|---|---|---|---|---|
| 0 | 0 | 0 | 0 | 0 |
| 0 | 0 | 0 | 0 | 0 |
| 0 | 0 | 0 | 0 | 0 |
| 0 | 0 | 0 | 0 | 0 |
| 0 | 0 | 0 | 0 | 0 |
| 0 | 0 | 0 | 0 | 0 |
| 0 | 0 | 0 | 0 | 0 |
| 0 | 0 | 0 | 0 | 0 |
| 0 | 0 | 0.000857218 | 1.046086963 | 0 |
| 0 | 0 | 0.111201119 | 0 | 0 |
| 0 | 0 | 0 | 0 | 0 |
| 0 | 0 | 0 | 0 | 0 |
| 0 | 0 | 0 | 0 | 0 |
| 0 | 0 | 0 | 0 | 0 |
| 0 | 0 | 0.01294675 | 0 | 0 |
| 0.05155385 | 0.006329546 | 0 | 0.00724357 | 0.204628179 |
| 0 | 0 | 0 | 0 | 0 |
| 0 | 0 | 0 | 0 | 0 |
| 0 | 0 | 0 | 0 | 0 |
| 0 | 0 | 0 | 3.932322943 | 0 |
| 0 | 0 | 0.001549431 | 0 | 0 |
| 0 | 0 | 0 | 0 | 0 |
| 0 | 0 | 0 | 0 | 0 |
| 0.001538841 | 0 | 0 | 0 | 0 |
| 0.003757061 | 0 | 0 | 0 | 0.001486884 |
| 0 | 0 | 0 | 0 | 0 |
| 0 | 0 | 0 | 0 | 0 |
| 0 | 0 | 0 | 0 | 0 |
| 0 | 0 | 0 | 0 | 0 |
| 0 | 0 | 0.019988212 | 0 | 0 |
| 0 | 0 | 0.192620719 | 9.347758886 | 0 |
| 0 | 0 | 0 | 0 | 0 |
| 0 | 0 | 0 | 0 | 0 |
| 0 | 0 | 0 | 0 | 0 |
| 0 | 0 | 0 | 0 | 0 |
| 0 | 0 | 0 | 0 | 0 |
| 0 | 0 | 0 | 0 | 0 |
| 0 | 0 | 0 | 0 | 0 |
| 0 | 0 | 0 | 0 | 0 |
| 0 | 0 | 0 | 0 | 0 |
| 0 | 0 | 0 | 0 | 0 |
| 0.003745524 | 0 | 0 | 0 | 0 |
| 0 | 0 | 0 | 0 | 0 |
| 0.000308498 | 0 | 0.000990382 | 0.002730765 | 0 |
| 0.009157937 | 0 | 0 | 0 | 0 |
| 0 | 0 | 0 | 0 | 0 |
| 0.007724116 | 0.668261833 | 0 | 0.009378034 | 0 |
| 0 | 0 | 0 | 0.035448926 | 0 |

| | | | | |
|---|---|---|---|---|
| 0 | 0 | 0 | 0 | 0 |
| 0 | 0 | 0 | 0 | 0 |
| 0 | 0 | 0 | 0 | 0 |
| 0 | 0 | 0 | 0 | 0 |
| 0 | 0.005213038 | 0.067364439 | 1.22068933 | 0 |
| 0 | 0 | 0 | 0 | 0 |
| 0 | 0 | 0 | 0 | 0 |
| 0 | 0 | 0 | 0 | 0 |
| 0 | 0 | 0 | 0 | 0 |
| 0 | 0 | 0 | 0 | 0 |
| 0 | 0 | 0.039433567 | 0 | 0 |
| 0.024916111 | 0 | 0.029337542 | 0.043431915 | 0.020558866 |
| 0.001058281 | 0 | 0 | 0.00448869 | 0 |
| 0 | 0 | 0 | 0 | 0 |
| 0 | 0 | 0 | 0 | 0 |
| 0 | 0 | 2.172238949 | 0 | 0 |
| 0.002889258 | 0 | 0.033164613 | 0.012352631 | 0 |
| 0 | 0 | 0 | 0 | 0 |
| 0.14518591 | 0 | 0.043981836 | 4.745476119 | 0.075890545 |
| 0 | 0.000766673 | 0.105509721 | 0.002012825 | 0 |
| 0 | 0 | 0 | 0 | 0 |
| 0 | 0 | 0 | 0 | 0 |
| 0 | 0 | 0 | 0 | 0 |
| 0 | 0 | 0 | 0 | 0 |
| 0 | 0 | 0 | 0 | 0 |
| 0 | 0 | 0 | 0 | 0 |
| 0.012694596 | 0.003174247 | 0.029795216 | 0.170437038 | 0.001110564 |
| 0 | 0 | 0 | 0 | 0 |
| 0 | 0 | 0 | 0 | 0 |
| 0 | 0 | 0 | 0 | 0 |
| 0.002031123 | 0 | 0.002121954 | 0.030066773 | 0 |
| 0 | 0 | 0 | 0 | 0 |
| 0 | 0 | 0 | 0 | 0 |
| 0 | 0 | 0 | 0 | 0 |
| 0 | 0 | 0 | 0 | 0 |
| 0 | 0 | 0 | 0 | 0 |
| 0 | 0 | 0 | 0 | 0 |
| 0.219396464 | 0.005871422 | 0.195049422 | 0.167993227 | 0.140038567 |
| 0 | 0 | 0.205615139 | 0 | 0 |
| 0 | 0 | 0.004926735 | 0 | 0 |
| 0.000941694 | 0 | 0.002232484 | 0.002279849 | 0.004120528 |
| 0 | 0 | 0 | 0 | 0 |
| 0 | 0 | 0 | 0 | 0 |
| 0.004770129 | 0 | 0 | 0.288267999 | 0.0019673 |
| 0 | 0 | 0 | 0 | 0 |
| 0 | 0 | 0 | 0 | 0 |
| 0 | 0 | 0 | 0 | 0 |
| 0 | 0 | 0 | 0 | 0 |

| | | | | |
|---|---|---|---|---|
| 0 | 0 | 0 | 0 | 0 |
| 0 | 0 | 0 | 0 | 0 |
| 0 | 0 | 0 | 0 | 0 |
| 0.002217653 | 0 | 0.004826721 | 0.007763374 | 0 |
| 0.000906403 | 0.009285633 | 0 | 0 | 0 |
| 0.001574731 | 0 | 0 | 0 | 0 |
| 0 | 0 | 0 | 0 | 0 |
| 0.008671984 | 0 | 0.11928727 | 0 | 0 |
| 0 | 0 | 0 | 0 | 0 |
| 0 | 0 | 0 | 0 | 0 |
| 0.060689298 | 0 | 0 | 0 | 0 |
| 0 | 0 | 0 | 0 | 0 |
| 0.008126461 | 0.028836531 | 0.260852547 | 0.139308047 | 0 |
| 0 | 0 | 0 | 0 | 0 |
| 0 | 0 | 0 | 0 | 0 |
| 0 | 0 | 0 | 0 | 0 |
| 0 | 0 | 0 | 0 | 0 |
| 0 | 0 | 0 | 0 | 0 |
| 0 | 0 | 0 | 0 | 0 |
| 0 | 0 | 0 | 0 | 0 |
| 0 | 0 | 0 | 0 | 0 |
| 0 | 0 | 0 | 0 | 0 |
| 0.011507613 | 0 | 0.045593453 | 0.001863351 | 0 |
| 0 | 0 | 0.009569498 | 0 | 0 |
| 0 | 0 | 0 | 0 | 0 |
| 0 | 0 | 0 | 0 | 0 |
| 0 | 0 | 0 | 0 | 0 |
| 0 | 0 | 0 | 0 | 0 |
| 0 | 0 | 0 | 0 | 0 |
| 0 | 0 | 0 | 0 | 0 |
| 0 | 0 | 0.016878521 | 0 | 0 |
| 0 | 0 | 0 | 0 | 0 |
| 0 | 0 | 0 | 0 | 0 |
| 0 | 0 | 0 | 0 | 0 |
| 0 | 0 | 0.093157935 | 0 | 0 |
| 0.002412725 | 0 | 0 | 0 | 0 |
| 0 | 0 | 0.003202177 | 0.031393114 | 0.026849675 |
| 0 | 0 | 0 | 0 | 0 |
| 0 | 0 | 0 | 0.001757818 | 1.096930989 |
| 0 | 0 | 0 | 0 | 0 |
| 0 | 0 | 0 | 0 | 0 |
| 0 | 0 | 0 | 0 | 0 |
| 0 | 0 | 0 | 0 | 0 |
| 0 | 0 | 0 | 0 | 0 |
| 0 | 0 | 0 | 0 | 0 |
| 0.044000348 | 0.22542386 | 0.428834677 | 0.036602614 | 0 |
| 0 | 0 | 0 | 0 | 0 |
| 0 | 0 | 0 | 0 | 0 |

| | | | | |
|---|---|---|---|---|
| 0 | 0 | 0 | 0 | 0 |
| 0 | 0 | 0 | 0 | 0 |
| 0 | 0 | 0.03206534 | 0 | 0 |
| 0 | 0 | 0 | 0 | 0 |
| 0 | 0 | 0 | 0 | 0 |
| 0 | 0 | 0 | 0 | 0 |
| 0 | 0 | 0 | 0.04725072 | 0 |
| 0 | 0 | 0 | 0 | 0 |
| 0 | 0 | 0.007393082 | 0 | 0 |
| 0 | 0 | 0 | 0 | 0 |
| 0.000295357 | 0 | 0.006068454 | 0.000792256 | 0 |
| 0.000400881 | 0 | 0.001029571 | 0.012774683 | 0 |
| 0 | 0 | 0 | 0 | 0 |
| 0 | 0 | 0 | 0 | 0 |
| 0 | 0 | 0 | 0 | 0 |
| 0 | 0 | 0 | 0 | 0 |
| 0 | 0 | 0 | 0 | 0 |
| 0 | 0 | 0 | 0 | 0 |
| 0 | 0 | 0 | 0 | 0 |
| 0.001424343 | 0 | 0.000731619 | 0 | 0 |
| 0 | 0 | 0 | 0 | 0 |
| 0 | 0 | 0 | 0 | 0 |
| 0 | 0 | 0 | 0 | 0 |
| 0 | 0 | 0 | 0 | 0 |
| 0 | 0 | 0 | 0 | 0 |
| 0 | 0 | 0 | 0 | 0 |
| 0 | 0 | 0 | 0 | 0 |
| 0 | 0.016873782 | 0 | 0 | 0 |
| 0 | 0 | 0 | 0 | 0 |
| 0 | 0 | 0 | 0 | 0 |
| 0.005751193 | 0 | 0.013042136 | 0 | 0 |
| 0 | 0 | 0 | 0 | 0 |
| 0.296909917 | 1.322254768 | 0.0074804 | 0.043179584 | 0 |
| 0.001016326 | 0.04368977 | 0.008017043 | 0.13301715 | 0 |
| 0 | 0 | 0 | 0 | 0 |
| 0 | 0 | 0 | 0 | 0 |
| 0 | 0 | 0 | 0 | 0 |
| 0 | 0 | 0 | 0 | 0 |
| 0 | 0 | 0 | 0 | 0 |
| 0 | 0 | 0 | 0 | 0 |
| 0 | 0 | 0 | 0 | 0 |
| 0 | 0.257838142 | 0.543508286 | 0.002241771 | 0.032235563 |
| 0 | 0.019084834 | 0.050835869 | 0 | 0 |
| 0 | 0 | 0 | 0 | 0 |
| 0 | 0 | 0 | 0 | 0 |
| 0.030455473 | 0.004336307 | 0.170176689 | 0.017000924 | 0 |
| 0.013457807 | 0 | 0 | 0.005971226 | 0.002672356 |
| 0 | 0 | 0 | 0 | 0 |

| | | | | |
|---|---|---|---|---|
| 0 | 0 | 0 | 0 | 0 |
| 0 | 0 | 0 | 0 | 0 |
| 0.00470511 | 0.0176264 | 0.007832215 | 0.007455639 | 0.003717266 |
| 0 | 0 | 0 | 0 | 0 |
| 0.213755179 | 0 | 0.612142701 | 0 | 0.010176199 |
| 0.001489828 | 0.03653169 | 0 | 0 | 0.024350901 |
| 0 | 0 | 0 | 0 | 0 |
| 0 | 0 | 0 | 0 | 0 |
| 0 | 0 | 0 | 0 | 0 |
| 0 | 0 | 0 | 0 | 0 |
| 0 | 0 | 0 | 0 | 0 |
| 0 | 0 | 0 | 0 | 0 |
| 0 | 0 | 0 | 0 | 0 |
| 0 | 0 | 0 | 0 | 0 |
| 0 | 0 | 0 | 0 | 0 |
| 0 | 0 | 0 | 0 | 0 |
| 0 | 0 | 0 | 0 | 0 |
| 0 | 0 | 0 | 0 | 0 |
| 0 | 0 | 0 | 0 | 0 |
| 0 | 0 | 0 | 0 | 0 |
| 0 | 0 | 0 | 0 | 0 |
| 0 | 0 | 0 | 0 | 0 |
| 0 | 0 | 0 | 0 | 0 |
| 0 | 0 | 0.121287401 | 0 | 0 |
| 0.000646279 | 0 | 0.012725269 | 0.012500869 | 0 |
| 0.002781159 | 0.006819609 | 0 | 0 | 0.003522124 |
| 0 | 0 | 0 | 0 | 0 |
| 0 | 0.000708778 | 0.011517891 | 0 | 0 |
| 0 | 0 | 0 | 0 | 0 |
| 0 | 0 | 0 | 0 | 0 |
| 0 | 0 | 0 | 0 | 0 |
| 0.000253393 | 0 | 0 | 0 | 0 |
| 0 | 0 | 0 | 0 | 0 |
| 0 | 0 | 0 | 0 | 0 |
| 0 | 0 | 0 | 0 | 0 |
| 0 | 0 | 0 | 0 | 0 |
| 0.132802036 | 0 | 0 | 0 | 0 |
| 0 | 0 | 0 | 0.036492952 | 0 |
| 0 | 0 | 0 | 0 | 0 |
| 0 | 0 | 0 | 0 | 0 |
| 0 | 0 | 0 | 0 | 0 |
| 0.000274284 | 0 | 0.054543495 | 0 | 0 |
| 0.078044757 | 0 | 0.016012853 | 0.142467166 | 0.002209273 |
| 0 | 0 | 0 | 0 | 0 |
| 0 | 0 | 0 | 0 | 0 |
| 0 | 0 | 0 | 0 | 0 |
| 0.01902084 | 0.00224364 | 0.010824049 | 0.029452282 | 0 |
| 0 | 0 | 0 | 0 | 0 |

| | | | | |
|---|---|---|---|---|
| 0 | 0 | 0 | 0 | 0 |
| 0.005213002 | 0.0027131 | 0.009704339 | 0.099721688 | 0.00447786 |
| 0 | 0 | 0 | 0 | 0 |
| 0 | 0 | 0 | 0 | 0 |
| 0 | 0 | 0 | 0 | 0 |
| 0 | 0 | 0 | 0 | 0 |
| 0 | 0 | 0 | 0 | 0 |
| 0 | 0 | 0 | 0 | 0 |
| 0 | 0 | 0 | 0 | 0 |
| 0 | 0 | 0 | 0 | 0 |
| 0 | 0 | 0 | 0 | 0 |
| 0 | 0 | 0 | 0 | 0 |
| 0.000147936 | 0 | 0.468654443 | 0 | 0 |
| 0 | 0 | 0 | 0 | 0 |
| 0.006368959 | 0 | 0.055345601 | 0 | 0.002392807 |
| 0 | 0 | 0 | 0 | 0 |
| 0 | 0 | 0 | 0 | 0 |
| 0 | 0 | 0 | 0 | 0 |
| 0 | 0 | 0.05651683 | 0 | 0 |
| 0 | 0 | 0 | 0 | 0 |
| 0 | 0 | 0 | 0 | 0 |
| 0.00068546 | 0 | 0.030147626 | 0.039186325 | 0 |
| 0 | 0 | 0.028577015 | 0 | 0 |
| 0 | 0.004463287 | 0 | 0 | 0 |
| 0.000404893 | 0 | 0 | 0 | 0 |
| 0 | 0 | 0 | 0 | 0 |
| 0 | 0 | 0 | 0 | 0 |
| 0 | 0 | 0 | 0 | 0 |
| 0 | 0 | 0 | 0 | 0 |
| 0 | 0 | 0 | 0 | 0 |
| 0 | 0 | 0 | 0 | 0 |
| 0 | 0 | 0 | 0 | 0 |
| 0 | 0 | 0 | 0 | 0 |
| 0 | 0 | 0 | 0 | 0 |
| 0 | 1.652186913 | 0 | 0 | 0 |
| 0 | 0 | 0 | 0 | 0 |
| 0 | 0 | 0 | 0 | 0 |
| 0 | 0 | 0 | 0 | 0 |
| 0 | 0 | 0.064387396 | 0 | 0 |
| 0 | 0 | 0 | 0 | 0 |
| 0.079366556 | 0.016554778 | 0.071918474 | 0.016751342 | 0 |
| 0 | 0 | 0 | 0 | 0 |
| 0 | 0 | 0 | 0 | 0 |
| 0 | 0 | 0 | 0 | 0 |
| 0 | 0 | 0 | 0 | 0 |
| 1.022005186 | 0.932896834 | 0.046923523 | 1.361978619 | 0.645362194 |
| 0 | 0 | 0 | 0 | 0 |
| 0 | 0 | 0 | 0 | 0 |

| | | | | |
|---|---|---|---|---|
| 0 | 0 | 0 | 0 | 0 |
| 0 | 0 | 0 | 0 | 0 |
| 0 | 0 | 0 | 0 | 0 |
| 0 | 0 | 0 | 0 | 0 |
| 0.001065906 | 0 | 0 | 0.040689296 | 0 |
| 0 | 0 | 0 | 0 | 0 |
| 0 | 0 | 0 | 0 | 0 |
| 0 | 0 | 0.000731505 | 0 | 0.002893178 |
| 0.02171925 | 0 | 0 | 0.101833531 | 0 |
| 0.000987925 | 0 | 0 | 0 | 0.04110035 |
| 0 | 0 | 0 | 0 | 0 |
| 0 | 0 | 0 | 0 | 0 |
| 0.00117553 | 0 | 0.020270947 | 0.031216702 | 0 |
| 0 | 0 | 0 | 0 | 0 |
| 0 | 0 | 0 | 0 | 0 |
| 0 | 0 | 0 | 0 | 0 |
| 0 | 0 | 0 | 0 | 0 |
| 0 | 0 | 0 | 0 | 0 |
| 0 | 0 | 0 | 0 | 0 |
| 0.157879878 | 0.002771782 | 1.490047847 | 0.693744641 | 0.01844738 |
| 0 | 0 | 0 | 0 | 0 |
| 0 | 0 | 0 | 0 | 0 |
| 0 | 0 | 0 | 0 | 0 |
| 0 | 0 | 0 | 0 | 0 |
| 0 | 0 | 0 | 0 | 0 |
| 0 | 0 | 0 | 0 | 0 |
| 0 | 0 | 0 | 0 | 0 |
| 0 | 0 | 0 | 0 | 0 |
| 0 | 0 | 0 | 0 | 0 |
| 0.039387758 | 0 | 0 | 0.107731175 | 0 |
| 0 | 0 | 0 | 0 | 0 |
| 0 | 0 | 0 | 0 | 0 |
| 0 | 0 | 0 | 0 | 0 |
| 0 | 0 | 0 | 0 | 0 |
| 0 | 0 | 0 | 0 | 0 |
| 0 | 0 | 0 | 0 | 0 |
| 0 | 0 | 0 | 0 | 0 |
| 0 | 0 | 0 | 0 | 0 |
| 0 | 0 | 0 | 0 | 0 |
| 0 | 0 | 0 | 0 | 0.000965155 |
| 0 | 0 | 0 | 0 | 0 |
| 0 | 0 | 0 | 0 | 0 |
| 0 | 0 | 0.027123497 | 0 | 0 |
| 0 | 0 | 0 | 0 | 0 |
| 0.008177698 | 0.001775018 | 0.054804887 | 0.04100919 | 0 |
| 0 | 0 | 0 | 0 | 0 |
| 0 | 0 | 0 | 0 | 0 |
| 0 | 0 | 0 | 0 | 0.286930962 |

| | | | | |
|---|---|---|---|---|
| 0 | 0 | 0 | 0 | 0 |
| 0 | 0 | 0 | 0 | 0 |
| 0 | 0 | 0 | 0 | 0 |
| 0 | 0 | 0 | 0 | 0 |
| 0 | 0 | 0 | 0 | 0 |
| 0.002620159 | 0 | 0.232832842 | 0.066950886 | 0.000798449 |
| 0 | 0 | 0 | 0 | 0 |
| 0.000561787 | 0 | 0 | 0 | 0 |
| 0 | 0 | 0 | 0 | 0 |
| 0 | 0 | 0 | 0 | 0 |
| 0 | 0 | 0 | 0 | 0 |
| 0 | 0 | 0 | 0 | 0 |
| 0 | 0 | 0 | 0 | 0 |
| 0 | 0 | 0 | 0 | 0 |
| 0 | 0 | 0 | 0 | 0 |
| 0 | 0 | 0 | 0 | 0 |
| 0 | 0 | 0 | 0 | 0 |
| 0.000838985 | 0 | 0 | 0 | 0 |
| 0 | 0 | 0 | 0 | 0 |
| 0 | 0 | 0 | 0 | 0 |
| 0 | 0 | 0 | 0 | 0 |
| 0 | 0 | 0 | 0 | 0 |
| 0 | 0 | 0 | 0 | 0 |
| 0 | 0 | 0 | 0 | 0 |
| 0 | 0 | 0 | 0 | 0 |
| 0.001024592 | 0 | 0 | 0 | 0 |
| 0 | 0 | 0 | 0 | 0 |
| 0 | 0 | 0.999058543 | 0 | 0 |
| 0 | 0 | 0 | 0 | 0 |
| 0 | 0 | 0 | 0 | 0 |
| 0 | 0 | 0 | 0 | 0 |
| 0 | 0 | 0 | 0 | 0 |
| 0 | 0 | 0 | 0 | 0 |
| 0 | 0 | 0 | 0 | 0 |
| 0 | 0 | 0 | 0 | 0 |
| 0 | 0 | 0 | 0 | 0 |
| 0 | 0 | 0 | 0 | 0 |
| 0 | 0 | 0 | 0 | 0 |
| 0 | 0 | 0 | 0 | 0 |
| 0 | 0 | 0 | 0 | 0 |
| 0 | 0 | 0 | 0 | 0 |
| 0 | 0 | 0 | 0 | 0 |
| 0 | 0 | 0 | 0 | 0 |
| 0 | 0 | 0 | 0 | 0 |
| 0 | 0 | 0 | 0 | 0 |
| 0 | 0 | 0 | 0 | 0 |
| 0 | 0 | 0 | 0 | 0 |

| | | | | |
|---|---|---|---|---|
| 0 | 0 | 0 | 0 | 0 |
| 0 | 0 | 0.331042349 | 0 | 0 |
| 0 | 0 | 0 | 0 | 0 |
| 0 | 0 | 0 | 0 | 0 |
| 0 | 0 | 0 | 0 | 0 |
| 0 | 0 | 0 | 0.004747724 | 0 |
| 0 | 0 | 0 | 0 | 0 |
| 0 | 0 | 0 | 0 | 0 |
| 0 | 0 | 0 | 0 | 0 |
| 0 | 0 | 0 | 0 | 0 |
| 0.000768232 | 0.000518034 | 0.00078921 | 0.000680024 | 0 |
| 0 | 0 | 0 | 0 | 0 |
| 0 | 0 | 0 | 0 | 0 |
| 0 | 0 | 0 | 0 | 0 |
| 0 | 0 | 0 | 0 | 0 |
| 0 | 0 | 0 | 0 | 0 |
| 0 | 0 | 0 | 0 | 0 |
| 0 | 0 | 0 | 0 | 0 |
| 0 | 0 | 0 | 0 | 0 |
| 0 | 0 | 0 | 0 | 0 |
| 0.011383749 | 0 | 0 | 0.239263605 | 0 |
| 0 | 0 | 0 | 0 | 0 |
| 0 | 0 | 0 | 0 | 0 |
| 2.232944748 | 15.39606213 | 0.337912228 | 0 | 0 |
| 0 | 0 | 0 | 0 | 0 |
| 0 | 0 | 0 | 0 | 0 |
| 0 | 0 | 0 | 0 | 0 |
| 0 | 0 | 0 | 0 | 0 |
| 0 | 0 | 0 | 0 | 0 |
| 0 | 0 | 0 | 0 | 0 |
| 0 | 0 | 0 | 0 | 0 |
| 0 | 0 | 0 | 0 | 0 |
| 0 | 0 | 0 | 0 | 0 |
| 0 | 0 | 0 | 0 | 0 |

| PHL018-0177_V1 | PHL018-0239_V2 | PHL018-0199_V1 | PHL018-0071_V1 | PHL018-0202_V3 |
|---|---|---|---|---|
| CTRL | REF | CTRL | CTRL | REF |
| 0.025909964 | 0.124654019 | 0.002540354 | 0.007945299 | 0.103843278 |
| 5.427021151 | 0.462314221 | 2.354045367 | 0.229331802 | 2.474082089 |
| 12.72605646 | 0 | 0.046008268 | 0 | 0 |
| 0 | 0 | 0 | 0 | 0 |
| 0 | 0 | 0 | 0 | 0 |
| 0 | 0 | 0 | 0 | 0 |
| 0.055121181 | 0 | 0 | 0 | 0 |
| 0 | 0 | 0 | 0 | 0 |
| 0 | 0 | 0 | 0 | 0 |
| 0 | 0 | 0 | 0 | 0 |
| 0 | 0 | 0 | 0 | 0 |
| 0 | 0 | 0 | 0 | 0 |
| 0 | 0 | 0 | 0 | 0 |
| 1.060825016 | 0.954326691 | 1.394088027 | 0.115541874 | 0.086619273 |
| 0.004158814 | 0 | 0 | 0 | 0 |
| 0 | 0 | 0 | 0 | 0 |
| 0 | 0 | 0 | 0 | 0 |
| 0.007107573 | 0 | 0 | 0 | 0 |
| 0 | 0 | 0 | 0 | 0 |
| 0 | 0 | 0 | 0 | 0 |
| 0 | 0 | 0 | 0 | 0 |
| 0 | 0 | 0 | 0 | 0 |
| 0 | 0 | 0 | 0 | 0 |
| 0 | 0 | 0 | 0 | 0 |
| 0 | 0.000564397 | 0 | 0.003354967 | 0 |
| 0 | 0 | 0 | 0 | 0 |
| 0 | 0 | 0 | 0 | 0 |
| 0 | 0 | 0 | 0 | 0 |
| 0 | 0 | 0 | 0 | 0 |
| 0 | 0 | 0 | 0.051961401 | 0 |
| 0 | 0 | 0 | 0 | 0 |
| 0 | 0 | 0 | 0 | 0 |
| 0 | 0 | 0 | 0 | 0 |
| 0 | 0 | 0 | 0 | 0 |
| 0 | 0 | 0 | 0 | 0 |
| 0 | 0.820889882 | 2.038352705 | 0.111092693 | 6.252243632 |
| 0 | 0 | 0 | 0 | 0 |
| 0 | 0 | 0 | 0 | 0 |
| 0 | 0 | 0 | 0 | 0 |
| 0 | 0 | 0 | 0 | 0 |
| 0 | 0 | 0 | 0 | 0 |
| 0 | 0.003789448 | 0.038651642 | 0 | 0.014383591 |
| 0.059507704 | 0.593230243 | 18.35413578 | 0.366416758 | 0.506614968 |
| 0.05062858 | 0.07456504 | 0.005658847 | 0.034063796 | 0.00238123 |
| 0 | 0 | 0.01240268 | 0.005473948 | 0.001471528 |
| 0 | 0 | 0 | 0 | 0 |

| | | | | |
|---|---|---|---|---|
| 0 | 0.001161209 | 0.003172528 | 0 | 0 |
| 0 | 0.011783307 | 0 | 0 | 0.330778788 |
| 0.014638786 | 0.000990799 | 0 | 0 | 0.005183115 |
| 0.645693577 | 1.145765319 | 0.217313723 | 0.119556389 | 0.621678938 |
| 0 | 0.012330478 | 0 | 0 | 0.003973205 |
| 0 | 0 | 0 | 0 | 0 |
| 0.001116262 | 0.001712516 | 0.449471847 | 0 | 0.002968836 |
| 0 | 0 | 0 | 0 | 0.005789081 |
| 0 | 0 | 0 | 0.001176194 | 0 |
| 0 | 0 | 0 | 0 | 0 |
| 0 | 0 | 0 | 0 | 0.000574271 |
| 0 | 0.035652695 | 0.060535061 | 0.025942669 | 4.967017138 |
| 0 | 0.00092723 | 0 | 0.002572158 | 0 |
| 0.003397586 | 0.044152197 | 0 | 0 | 0.340749412 |
| 0 | 0 | 0 | 0 | 0.022723017 |
| 0 | 0 | 0 | 0 | 0 |
| 0.01972686 | 0.0006305 | 0.248970682 | 0 | 0.009971605 |
| 0 | 0 | 0 | 0 | 0 |
| 0 | 0 | 0 | 0 | 0 |
| 0 | 0 | 0 | 0 | 0 |
| 0.093978822 | 0.035477449 | 0 | 0 | 0.14733784 |
| 0.019233673 | 0.000760499 | 0 | 0 | 0.020317388 |
| 0 | 0.029342808 | 0.368396383 | 0.162254466 | 0 |
| 0 | 0 | 0 | 0 | 0 |
| 0 | 0 | 0 | 0 | 0 |
| 0 | 0 | 0 | 0 | 0 |
| 0 | 0 | 0 | 0 | 0.002783897 |
| 0 | 0 | 0 | 0 | 0 |
| 0 | 0 | 0.002551726 | 0 | 0.000852191 |
| 0 | 0 | 0 | 0 | 0 |
| 0.002920829 | 0 | 0 | 0 | 0.523212245 |
| 0 | 0 | 0 | 0 | 0 |
| 0 | 0 | 0 | 0 | 0 |
| 0 | 0 | 0 | 0 | 0 |
| 0 | 0 | 0 | 0 | 0 |
| 0 | 0 | 0.002061085 | 0 | 0 |
| 0 | 0 | 0 | 0 | 0 |
| 0.003847068 | 0 | 14.6869727 | 0 | 0 |
| 0 | 0 | 0 | 0 | 0 |
| 0 | 0 | 0 | 0 | 0 |
| 0 | 0 | 0 | 0 | 0 |
| 0 | 0 | 0 | 0 | 0 |
| 0 | 0 | 0 | 0 | 0 |
| 0 | 0 | 0 | 0 | 0 |
| 0 | 0 | 0 | 0.000367947 | 0 |
| 0 | 0 | 0 | 0 | 0 |
| 0 | 0 | 0 | 0 | 0 |
| 0.003004985 | 0 | 0 | 0.007003245 | 0 |

| | | | | |
|---|---|---|---|---|
| 0 | 0 | 0 | 0 | 0 |
| 0.036232794 | 0 | 0 | 0 | 0 |
| 0 | 0 | 0 | 0 | 0 |
| 0 | 0 | 0 | 0 | 0 |
| 0 | 0 | 0.00378404 | 0 | 0 |
| 0.001218592 | 0 | 0 | 0 | 0 |
| 0 | 0 | 0 | 0 | 0 |
| 0 | 0 | 0 | 0.000518834 | 0 |
| 0 | 0.092473332 | 0 | 0 | 0.004507162 |
| 0 | 0 | 0 | 0 | 0 |
| 0 | 0 | 0 | 0 | 0 |
| 0 | 0 | 0.002238345 | 0 | 0.000747532 |
| 0 | 0 | 0 | 0 | 0 |
| 0 | 0 | 0 | 0 | 0 |
| 0 | 0 | 0 | 0 | 0 |
| 0 | 0 | 0 | 0 | 0 |
| 0 | 0 | 0 | 0 | 0 |
| 0 | 0 | 0 | 0 | 0 |
| 0.164558612 | 0 | 0.073040837 | 1.153060832 | 0 |
| 0 | 0 | 0 | 0.000653138 | 0 |
| 0 | 0 | 0 | 0 | 0 |
| 0 | 0 | 0 | 0 | 0 |
| 0 | 0 | 0 | 0 | 0 |
| 0 | 0 | 0 | 0 | 0 |
| 0 | 0 | 0 | 0 | 0 |
| 0 | 0 | 0 | 0 | 0 |
| 0 | 0 | 0 | 0 | 0 |
| 0.001032024 | 0 | 0 | 0 | 0 |
| 0 | 0 | 0 | 0 | 0 |
| 0 | 0 | 0 | 0 | 0 |
| 0.037244545 | 0.030337971 | 0.001982328 | 0.006110145 | 0 |
| 0 | 0.007707029 | 0.001455745 | 0.00179105 | 0.003750453 |
| 0 | 0 | 0 | 0 | 0 |
| 0 | 0 | 0 | 0 | 0 |
| 0 | 0 | 0 | 0 | 0 |
| 0 | 0 | 0 | 0 | 0 |
| 0 | 0 | 0 | 0 | 0 |
| 0 | 0 | 0 | 0 | 0 |
| 0 | 0 | 0 | 0 | 0 |
| 0 | 0 | 0 | 0 | 0 |
| 0 | 0 | 0 | 0 | 0 |
| 0 | 0 | 0 | 0 | 0 |
| 0 | 0.003299299 | 0 | 0 | 0.000481659 |
| 0 | 0 | 0 | 0 | 0 |
| 0 | 0 | 0 | 0 | 0 |
| 0 | 0 | 0 | 0 | 0 |
| 0 | 0 | 0 | 0 | 0 |
| 0 | 0 | 0 | 0 | 0 |

| | | | | |
|---|---|---|---|---|
| 0 | 0 | 0 | 0 | 0 |
| 0 | 0 | 0 | 0 | 0 |
| 0 | 0 | 0 | 0 | 0 |
| 0 | 0 | 0 | 0 | 0 |
| 0 | 0 | 0.159830683 | 0.049885008 | 0 |
| 0 | 0 | 0 | 0 | 0 |
| 0 | 0 | 0 | 0 | 0 |
| 0 | 0 | 0 | 0 | 0 |
| 0 | 0 | 0 | 0 | 0 |
| 0 | 0 | 0 | 0 | 0 |
| 0 | 0 | 0 | 0 | 0 |
| 0 | 0 | 0 | 0 | 0 |
| 0 | 0 | 0 | 0 | 0 |
| 0 | 0 | 0 | 0 | 0 |
| 0 | 0 | 0 | 0 | 0 |
| 0 | 4.332063028 | 0 | 0 | 0 |
| 0 | 0 | 0 | 0 | 0 |
| 0 | 0 | 0 | 0 | 0 |
| 0 | 0 | 0 | 0 | 0 |
| 0 | 0 | 0 | 0 | 0 |
| 0 | 0 | 0 | 0 | 0 |
| 0 | 0 | 0 | 0 | 0 |
| 0 | 0 | 0 | 0 | 0 |
| 0 | 0 | 0 | 0 | 0 |
| 0 | 0 | 0 | 0.0019896 | 0 |
| 0 | 0 | 0 | 0 | 0 |
| 0 | 0 | 0 | 0.001023979 | 0 |
| 0 | 0 | 0 | 0 | 0 |
| 0 | 0 | 0 | 0 | 0 |
| 0 | 0 | 0 | 0 | 0 |
| 0 | 0 | 0 | 0 | 0 |
| 0.013739138 | 0 | 0 | 0 | 0 |
| 0 | 0 | 0 | 0 | 0 |
| 0 | 0 | 0 | 0 | 0 |
| 0 | 0 | 0 | 0 | 0 |
| 0 | 0 | 0 | 0 | 0 |
| 0 | 0 | 0 | 0 | 0 |
| 0 | 0 | 0 | 0 | 0 |
| 0 | 0 | 0 | 0 | 0 |
| 0 | 0 | 0 | 0 | 0 |
| 0 | 0 | 0 | 0 | 0 |
| 0 | 0 | 0 | 0 | 0 |
| 0 | 0 | 0 | 0 | 0 |
| 0 | 0 | 0 | 0 | 0 |
| 0 | 0 | 0 | 0 | 0 |
| 0 | 0 | 0 | 0 | 0 |
| 0 | 0 | 0 | 0 | 0 |
| 0 | 0 | 0 | 0 | 0 |

| | | | | |
|---|---|---|---|---|
| 0 | 0 | 0 | 0.002224807 | 0 |
| 0 | 0 | 0 | 0 | 0 |
| 0 | 0 | 0 | 0 | 0 |
| 0 | 0 | 0 | 0 | 0 |
| 0 | 0 | 0 | 0 | 0 |
| 0 | 0 | 0 | 0 | 0 |
| 0 | 0 | 0 | 0 | 0 |
| 0 | 0 | 0 | 0 | 0 |
| 0 | 0 | 0 | 0 | 0 |
| 0 | 0 | 0 | 0 | 0 |
| 0 | 0 | 0 | 0 | 0 |
| 0 | 0 | 0 | 0 | 0 |
| 0.00102711 | 0 | 0 | 0 | 0.00092016 |
| 0 | 0 | 0 | 0 | 0 |
| 0 | 0 | 0 | 0 | 0 |
| 0 | 0 | 0 | 0 | 0 |
| 0 | 0 | 0 | 0 | 0 |
| 0 | 0 | 0 | 0 | 0 |
| 0 | 0 | 0 | 0 | 0 |
| 0 | 0 | 0 | 0 | 0 |
| 0.039945338 | 0.050675669 | 0 | 0.004670283 | 0.021936089 |
| 0 | 0 | 0 | 0 | 0 |
| 0 | 0 | 0 | 0 | 0 |
| 0.013617965 | 0 | 0 | 0 | 0.012783048 |
| 0 | 0 | 0 | 0 | 0 |
| 0 | 0 | 0 | 0 | 0 |
| 0.001342109 | 0.004632747 | 0.00650043 | 0.001813242 | 0.01077113 |
| 0 | 0 | 0 | 0 | 0 |
| 0.018849198 | 0 | 0 | 0 | 0 |
| 0 | 0 | 0 | 0 | 0.01313185 |
| 0 | 0 | 0 | 0 | 0 |
| 0 | 0 | 0 | 0 | 0 |
| 0 | 0 | 0 | 0 | 0 |
| 0.00258205 | 0 | 0 | 0.017481896 | 0 |
| 0.006747551 | 0.007092877 | 0 | 0 | 0.032603527 |
| 0 | 0 | 0 | 0 | 0 |
| 0 | 0 | 0 | 0 | 0 |
| 0 | 0 | 0 | 0 | 0 |
| 0 | 0 | 0 | 0 | 0 |
| 0 | 0 | 0 | 0 | 0 |
| 0 | 0 | 0 | 0 | 0 |
| 0 | 0 | 0 | 0 | 0 |
| 0 | 0 | 0 | 0 | 0 |
| 0 | 0 | 0 | 0 | 0 |
| 0 | 0 | 0 | 0 | 0 |
| 0.123967189 | 0 | 0 | 0.000482201 | 0 |
| 0 | 0 | 0 | 0 | 0.009393386 |
| 0.00249329 | 0 | 0 | 0 | 0 |

| | | | | |
|---|---|---|---|---|
| 0 | 0 | 0 | 0 | 0 |
| 0 | 0 | 0 | 0 | 0 |
| 0 | 0 | 0 | 0.002638367 | 0 |
| 0 | 0 | 0 | 0.000458769 | 0 |
| 0 | 0 | 0 | 0 | 0 |
| 0 | 0 | 0 | 0 | 0 |
| 0 | 0 | 0 | 0 | 0 |
| 0 | 0 | 0.001716886 | 0 | 0.001720147 |
| 0 | 0.000942197 | 0 | 0 | 0 |
| 0 | 0 | 0 | 0 | 0 |
| 0 | 0 | 0 | 0 | 0 |
| 0 | 0 | 0 | 0 | 0 |
| 0 | 0 | 0 | 0 | 0 |
| 0 | 0 | 0 | 0 | 0 |
| 0.000842377 | 0 | 0 | 0 | 0 |
| 0 | 0 | 0 | 0 | 0 |
| 0 | 0 | 0 | 0 | 0 |
| 0 | 0 | 0.159442038 | 0 | 0 |
| 0 | 0.001667179 | 0.10668556 | 0.002202284 | 0.001926827 |
| 0 | 0 | 0 | 0 | 0 |
| 0.190758117 | 0.01311605 | 0 | 0 | 0.002692673 |
| 0 | 0 | 0 | 0 | 0 |
| 0 | 0 | 0 | 0 | 0 |
| 0 | 0 | 0 | 0 | 0 |
| 0 | 0.010993137 | 0.005339423 | 0.001075669 | 0.006835554 |
| 0 | 0 | 0 | 0.002740013 | 0 |
| 0 | 0 | 0 | 0 | 0 |
| 0 | 0 | 0 | 0 | 0 |
| 0 | 0 | 0.00736015 | 0 | 0.008508611 |
| 0 | 0 | 0 | 0 | 0 |
| 0.185644014 | 0.019589835 | 0.003293613 | 0 | 1.51804962 |
| 0 | 0 | 0 | 0 | 0 |
| 0 | 0 | 0 | 0 | 0 |
| 0.000762482 | 0 | 0.00255672 | 0 | 0 |
| 0 | 0 | 0 | 0 | 0 |
| 0 | 0 | 0 | 0 | 0 |
| 0 | 0 | 0 | 0 | 0 |
| 0 | 0 | 0 | 0 | 0 |
| 0 | 0 | 0 | 0 | 0 |
| 0 | 0 | 0 | 0.008052642 | 0 |
| 0 | 0 | 0 | 0 | 0 |
| 0.010012339 | 0.005683365 | 0 | 0.003449401 | 0.013118303 |
| 0.001176101 | 0 | 0 | 0 | 0 |
| 0 | 0 | 0 | 0 | 0 |
| 0 | 0 | 0 | 0 | 0 |
| 0 | 0 | 0 | 0 | 0 |
| 0 | 0 | 0 | 0 | 0 |
| 0 | 0 | 0 | 0 | 0 |

| | | | | |
|---|---|---|---|---|
| 0 | 0 | 0 | 0 | 0 |
| 0 | 0 | 0 | 0 | 0 |
| 0.044002486 | 0.078114647 | 0.003091463 | 0.027643824 | 0 |
| 0 | 0 | 0 | 0 | 0 |
| 0 | 0 | 0 | 0 | 0 |
| 0 | 0 | 0 | 0 | 0 |
| 0 | 0 | 0 | 0 | 0 |
| 0 | 0 | 0 | 0 | 0 |
| 0 | 0 | 0 | 0 | 0 |
| 0 | 0 | 0 | 0.023768192 | 0 |
| 0 | 0 | 0 | 0 | 0 |
| 0 | 0 | 0 | 0 | 0 |
| 0 | 0 | 0 | 0 | 0 |
| 0.001667338 | 0.005755386 | 0 | 0 | 0.001711557 |
| 0 | 0.013019389 | 0 | 0 | 0.005008544 |
| 0 | 0 | 0.027995822 | 0.106247307 | 0 |
| 0 | 0.006615352 | 0.001460505 | 0.001059229 | 0.003219213 |
| 0 | 0 | 0 | 0 | 0 |
| 0 | 0 | 0 | 0 | 0 |
| 0 | 0 | 0 | 0 | 0 |
| 0 | 0 | 0 | 0 | 0 |
| 0.003066865 | 0 | 0 | 0 | 0 |
| 0 | 0 | 0 | 0 | 0 |
| 0 | 0 | 0 | 0 | 0 |
| 0 | 0.00087049 | 0 | 0 | 0 |
| 0 | 0 | 0 | 0 | 0 |
| 0 | 0 | 0.049802346 | 0.200019958 | 0 |
| 0 | 0 | 0 | 0 | 0 |
| 0 | 0 | 0 | 0 | 0 |
| 0 | 0.004711761 | 0 | 0 | 0.0021018 |
| 0 | 0 | 0 | 0 | 0 |
| 0 | 0 | 0 | 0 | 0 |
| 0 | 0 | 0 | 0 | 0 |
| 0 | 0 | 0 | 0 | 0 |
| 0 | 0 | 0 | 0 | 0 |
| 0 | 0 | 0 | 0 | 0.078623005 |
| 0 | 0 | 0 | 0 | 0 |
| 0 | 0.032740003 | 0.01424553 | 0.036190468 | 0.003485166 |
| 0 | 0 | 0 | 0 | 0 |
| 0 | 0 | 0 | 0 | 0 |
| 0 | 0 | 0 | 0 | 0 |
| 0 | 0 | 0 | 0 | 0 |
| 0 | 0 | 0 | 0 | 0 |
| 0.003313574 | 0 | 0 | 0 | 0 |
| 0 | 0 | 0 | 0 | 0 |
| 0 | 0 | 0 | 0 | 0 |
| 0 | 0 | 0 | 0 | 0.000899475 |
| 0.007539305 | 0 | 0 | 0 | 0.039399832 |

| | | | | |
|---|---|---|---|---|
| 2.286091639 | 0.155226501 | 0 | 0.318245015 | 0 |
| 0 | 0 | 0 | 0 | 0 |
| 0 | 0 | 0 | 0 | 0 |
| 0 | 0 | 0 | 0 | 0 |
| 0 | 0.014111804 | 0.000833616 | 0.000881678 | 0.022689578 |
| 0 | 0.036381588 | 0 | 0 | 0 |
| 0 | 0 | 0 | 0 | 0 |
| 0.004878675 | 0.036888519 | 0.002726491 | 0.006179326 | 0.012097394 |
| 0 | 0 | 0 | 0 | 0 |
| 0.200423874 | 0.052348849 | 0 | 0 | 0.03921716 |
| 0 | 0 | 0 | 0 | 0 |
| 0 | 0 | 0.000924986 | 0 | 0.10271397 |
| 0.009416077 | 0.01861889 | 0.000701635 | 0.000954112 | 0.022260626 |
| 0.001906746 | 0.003900316 | 0 | 0 | 0.315127545 |
| 0 | 0 | 0 | 0 | 0.007832345 |
| 0 | 0 | 0 | 0 | 0 |
| 0 | 0 | 0 | 0 | 0 |
| 0 | 0 | 0 | 0 | 0 |
| 0.376176471 | 0 | 0 | 0 | 0 |
| 0 | 0 | 0 | 0 | 0 |
| 0 | 0 | 0 | 0 | 0 |
| 0 | 0 | 0 | 0 | 0 |
| 0 | 0 | 0 | 0 | 0 |
| 0 | 0 | 0 | 0.004492745 | 0 |
| 0 | 0 | 0 | 0 | 0 |
| 0 | 0 | 0.005175761 | 0.012841653 | 0.000545852 |
| 0.000977703 | 0 | 0 | 0 | 0 |
| 0 | 0 | 0 | 0 | 0 |
| 0.478104311 | 0 | 0 | 0 | 0 |
| 0.001003022 | 0 | 0 | 0 | 0 |
| 0 | 0 | 0 | 0 | 0.000878593 |
| 0 | 0 | 0 | 0 | 0 |
| 0.038395679 | 0 | 0 | 0 | 0 |
| 0.062154091 | 0 | 0 | 0 | 0 |
| 0 | 0 | 0 | 0 | 0 |
| 0.000988574 | 0 | 0 | 0 | 0 |
| 0.003315358 | 0 | 0 | 0 | 0.012530271 |
| 0 | 0 | 0 | 0 | 0 |
| 0 | 0 | 0 | 0 | 0 |
| 0 | 0 | 0 | 0 | 0 |
| 0 | 0 | 0 | 0 | 0.233072027 |
| 0.003720975 | 0 | 0 | 0 | 0 |
| 0.002683294 | 0 | 0 | 0 | 0 |
| 0 | 0 | 0 | 0 | 0 |
| 0 | 0 | 0 | 0 | 0 |
| 0 | 0 | 0 | 0 | 0 |
| 0.003636998 | 0.001627414 | 0 | 0 | 0.001866727 |
| 0 | 0 | 0 | 0 | 0 |

| | | | | |
|---|---|---|---|---|
| 0.030364965 | 0 | 0 | 0 | 0 |
| 0 | 0 | 0.007950382 | 0 | 0.007206864 |
| 0 | 0 | 0 | 0 | 0 |
| 0 | 0 | 0 | 0.001322089 | 0 |
| 0 | 0 | 0 | 0 | 0 |
| 0.319638954 | 0.088260106 | 0 | 0 | 0.05186055 |
| 0.005574189 | 0 | 0 | 0 | 0 |
| 0 | 0 | 0 | 0 | 0 |
| 0 | 0.149193991 | 0 | 0 | 0 |
| 0 | 0 | 0 | 0 | 0 |
| 0 | 0.009769452 | 0.004332458 | 0.000981909 | 0.035190562 |
| 0 | 0.001990479 | 0 | 0 | 0 |
| 0 | 0 | 0 | 0 | 0 |
| 0 | 0.003410971 | 0 | 0.001192704 | 0.003368565 |
| 0.346179999 | 0 | 0 | 0 | 0 |
| 0 | 0 | 0 | 0 | 0 |
| 0 | 0 | 0 | 0 | 0 |
| 0.026982466 | 0 | 0 | 0 | 0 |
| 0 | 0 | 0 | 0 | 0 |
| 0.026350375 | 0 | 0 | 0 | 0 |
| 0 | 0 | 0 | 0 | 0 |
| 0.216773925 | 0.191916234 | 0.116353095 | 0.020526876 | 0.097034675 |
| 0.043934779 | 0 | 0 | 0 | 0 |
| 0 | 0 | 0 | 0 | 0 |
| 0 | 0 | 0 | 0 | 0 |
| 0 | 0 | 0 | 0 | 0 |
| 0.070619646 | 0 | 0 | 0 | 0.310931804 |
| 0 | 0 | 0 | 0 | 0 |
| 0 | 0 | 0 | 0 | 0 |
| 0.118473507 | 0.272634481 | 0.477826068 | 0.104700981 | 0.050449504 |
| 0 | 0 | 0 | 0.010159387 | 0 |
| 0 | 0 | 0 | 0 | 0 |
| 0 | 0 | 0 | 0 | 0 |
| 0 | 0 | 0 | 0 | 0 |
| 0.003070898 | 0 | 0 | 0 | 0 |
| 0 | 0 | 0 | 0 | 0 |
| 0 | 0 | 0.021615861 | 0.030388148 | 0 |
| 0 | 0 | 0 | 0 | 0 |
| 0 | 0 | 0 | 0 | 0 |
| 0.156300697 | 0.023741482 | 0 | 0 | 0 |
| 0.032471107 | 0.050145506 | 0 | 0 | 0.001324461 |
| 0 | 2.701979072 | 2.866931219 | 0.098103798 | 0 |
| 0 | 0 | 0 | 0 | 0 |
| 0 | 0 | 0 | 0 | 0 |
| 0 | 0 | 0 | 0 | 0 |
| 0 | 0 | 0 | 0 | 0 |
| 0.006771011 | 0 | 0 | 0 | 0.001769239 |
| 0 | 0 | 0 | 0 | 0 |

| | | | | |
|---|---|---|---|---|
| 0.001670858 | 0 | 0.012139064 | 0.00253957 | 0.001247397 |
| 0 | 0 | 0 | 0 | 0 |
| 0.012142253 | 0.00174638 | 0 | 0 | 0.004674103 |
| 0 | 0 | 0 | 0 | 0 |
| 0.186810999 | 0 | 0 | 0.159035317 | 0 |
| 0.027076984 | 0.117938041 | 0 | 0 | 0 |
| 0 | 0 | 0 | 0 | 0 |
| 0 | 0 | 0 | 0 | 0.009611792 |
| 0 | 0 | 0 | 0 | 0 |
| 0.786655623 | 0 | 0 | 0 | 0 |
| 0 | 0 | 0 | 0 | 0 |
| 0 | 0 | 0 | 0 | 0 |
| 0 | 0 | 0 | 0 | 0 |
| 0 | 0.003478497 | 0 | 0 | 0 |
| 0 | 0 | 0 | 0 | 0 |
| 0.248855544 | 0.034734925 | 0 | 0.072615754 | 0.009700837 |
| 0.008947031 | 0.000669566 | 0.037318032 | 0.009508079 | 0.028591519 |
| 0.408563975 | 0.000610122 | 0 | 0 | 0.006977206 |
| 0 | 0 | 0 | 0 | 0 |
| 0.245915897 | 0 | 0 | 0 | 0 |
| 0.086906205 | 0.03016741 | 0 | 0 | 0 |
| 0.001265621 | 0 | 0 | 0 | 0.002692858 |
| 0 | 0 | 0 | 0 | 0 |
| 0.06142638 | 0 | 0 | 0 | 0 |
| 0.020394794 | 0 | 0 | 0 | 0 |
| 0.006324268 | 0 | 0 | 0 | 0 |
| 0.002770324 | 0.004250097 | 0 | 0 | 0 |
| 0 | 0 | 0 | 0 | 0 |
| 0 | 0 | 0 | 0 | 0 |
| 0 | 0 | 0 | 0 | 0 |
| 0.015590494 | 0.003878624 | 0 | 0 | 0.021705623 |
| 0.002826914 | 0.002650337 | 0 | 0.002705317 | 0.007034874 |
| 0.384301257 | 0.042210917 | 0.004679818 | 0.001363674 | 0.004353798 |
| 0 | 0.000986288 | 0.008143771 | 0 | 0.008399216 |
| 0 | 0.006570284 | 0.020173073 | 0 | 0 |
| 0.015691026 | 0 | 0 | 0 | 0 |
| 0.002433396 | 0.001451799 | 0 | 0 | 0.001211118 |
| 0.075063891 | 0.026600197 | 0 | 0 | 0.053750789 |
| 0 | 0.536906853 | 0.194705956 | 0.030798224 | 0 |
| 0.00397624 | 0 | 0 | 0 | 0.009195908 |
| 0 | 0 | 0 | 0 | 0 |
| 0 | 0 | 0 | 0 | 0 |
| 0.001264878 | 0 | 0 | 0 | 0 |
| 0.115377567 | 0 | 0 | 0 | 0.463213849 |
| 0 | 0 | 0 | 0 | 0 |
| 0 | 0 | 0 | 0 | 0 |
| 0.001174835 | 0 | 0 | 0 | 0 |
| 0.001933246 | 0 | 0 | 0 | 0 |

| | | | | |
|---|---|---|---|---|
| 0 | 0 | 0 | 0 | 0 |
| 0.16999325 | 0.45572894 | 0.312380277 | 0.116926083 | 0.877939264 |
| 0.001183168 | 0 | 0 | 0 | 0 |
| 0.002093987 | 0 | 0 | 0 | 0 |
| 0 | 0 | 0 | 0 | 0 |
| 0 | 0 | 0 | 0 | 0 |
| 0 | 0 | 0 | 0 | 0 |
| 0 | 0 | 0 | 0 | 0.000371747 |
| 0 | 0 | 0 | 0 | 0 |
| 0 | 0.001606059 | 0 | 0 | 0 |
| 0.005138586 | 0.001149658 | 0 | 0 | 0.004195915 |
| 0.376006166 | 0.016145942 | 0.008555091 | 0.056067328 | 0.039821012 |
| 0 | 0 | 0 | 0.129999592 | 0 |
| 0.027872066 | 0 | 0 | 0 | 0 |
| 0.004421914 | 0 | 0 | 0.005102944 | 0.000412653 |
| 0 | 0 | 0 | 0 | 0 |
| 0 | 0.040759191 | 0.009773177 | 0.00454358 | 0.037534997 |
| 0.040164182 | 0 | 0 | 0 | 0.027866487 |
| 0 | 0 | 0 | 0 | 0 |
| 0.122087077 | 0 | 0 | 0 | 0 |
| 0 | 0.017026029 | 0.080370459 | 0.020147981 | 0.041038302 |
| 0 | 0 | 0 | 0 | 0 |
| 0 | 0 | 0 | 0 | 0 |
| 0 | 0 | 0 | 0 | 0 |
| 0 | 0 | 0 | 0 | 0 |
| 0 | 0 | 0 | 0 | 0.086425945 |
| 0.276139392 | 0.057944106 | 0.001785555 | 0 | 0 |
| 0.330766761 | 0.09198879 | 0.021149482 | 0.001867529 | 0.036325115 |
| 0 | 0 | 0 | 0 | 0 |
| 0 | 0 | 0 | 0 | 0.003319439 |
| 0 | 0 | 0.001768318 | 0 | 0 |
| 0 | 0 | 0 | 0 | 0 |
| 0.003878254 | 0 | 0 | 0 | 0 |
| 0 | 0 | 0 | 0 | 0 |
| 0 | 0 | 0.11441148 | 0.091678063 | 0.139449026 |
| 0.502408645 | 0.130801375 | 0.070658752 | 0.026549719 | 0.027944586 |
| 0 | 0 | 0 | 0 | 0 |
| 0 | 0 | 0 | 0 | 0 |
| 0.002122039 | 0.032322759 | 0.001694172 | 0 | 0.044641354 |
| 0 | 0 | 0 | 0 | 0 |
| 0 | 0 | 0 | 0 | 0 |
| 0 | 0 | 0 | 0 | 0 |
| 0.02264885 | 0.075002173 | 0 | 0 | 0.072996253 |
| 0.01564471 | 0.008591395 | 0 | 0 | 0 |
| 0.090155223 | 0.006091451 | 0 | 0 | 0 |
| 0 | 1.679524619 | 0.274700947 | 0.135777044 | 0 |
| 0 | 0 | 0 | 0 | 0 |
| 0.002011582 | 0 | 0 | 0 | 0 |

| | | | | |
|---|---|---|---|---|
| 0 | 0 | 0 | 0 | 0 |
| 0 | 0 | 0 | 0 | 0 |
| 0 | 0 | 0 | 0 | 0 |
| 0.019515981 | 0.575284806 | 0.031641359 | 0.043353195 | 0 |
| 0 | 0 | 0 | 0 | 0 |
| 0 | 0 | 0 | 0 | 0 |
| 0 | 0 | 0 | 0 | 0 |
| 0 | 0.000588681 | 0 | 0.020121065 | 0 |
| 0 | 0.044945776 | 0.116745146 | 12.65744611 | 0.093106671 |
| 0 | 0.0799842 | 0.004499026 | 0.023694965 | 0 |
| 0 | 0 | 0 | 0 | 0 |
| 0 | 0.000630466 | 0 | 0.01186772 | 0.015416802 |
| 0 | 0 | 0 | 0 | 0 |
| 0 | 0.916823311 | 0 | 0 | 0 |
| 0 | 0.042141217 | 0.010613143 | 0.005154354 | 0.021551421 |
| 0 | 0 | 0 | 0 | 0 |
| 0 | 0.002714544 | 0.071691704 | 0.124495019 | 0 |
| 0 | 0 | 0 | 3.708003495 | 0 |
| 0 | 0 | 0 | 0 | 0 |
| 0 | 0 | 0 | 0 | 0 |
| 0 | 0 | 0 | 0.199829921 | 0 |
| 0 | 0 | 0 | 0.000579911 | 0 |
| 0 | 0 | 0 | 0.000671028 | 0 |
| 0.40963134 | 0.149967819 | 0 | 0 | 0.064473468 |
| 0.015967456 | 0 | 0 | 0 | 0 |
| 0 | 0 | 0.020609377 | 0.033148422 | 0 |
| 0.002746816 | 0.015978203 | 0 | 0 | 0 |
| 0 | 0 | 0.095997803 | 0.156364811 | 0 |
| 0 | 0 | 0 | 0 | 0 |
| 0 | 0 | 0 | 0 | 0 |
| 0.004794208 | 0.005363052 | 0.001339643 | 0.002327731 | 0 |
| 0.001882097 | 0.061502103 | 0 | 0.062743277 | 0 |
| 0.060356317 | 0 | 0 | 0 | 0.009108404 |
| 0 | 0 | 0 | 0 | 0 |
| 0 | 0 | 0 | 0.000560652 | 0 |
| 0 | 0 | 0.018363193 | 0.039215673 | 0.035752949 |
| 0 | 5.351986216 | 0.541070807 | 0.346551341 | 0 |
| 0 | 0 | 0 | 0 | 0 |
| 0 | 0 | 0 | 0 | 0 |
| 0 | 0 | 0 | 0.00037257 | 0.011693738 |
| 0.103881426 | 0.220607799 | 0 | 0 | 0.385635014 |
| 0 | 0.294165238 | 0 | 0.017000447 | 0 |
| 6.844796483 | 0.532013102 | 2.705947179 | 0.056427669 | 0.599124164 |
| 0 | 0 | 0 | 0 | 0 |
| 0 | 0 | 0 | 0 | 0 |
| 0 | 0 | 0 | 0 | 0 |
| 0 | 0 | 0 | 0 | 0 |
| 0 | 0.002726483 | 0 | 0 | 0.000796069 |

| | | | | |
|---|---|---|---|---|
| 0 | 0 | 0 | 0 | 0 |
| 0 | 0 | 0.007721244 | 8.460544412 | 0.084490626 |
| 0.01862646 | 0.047568275 | 0.000761675 | 0 | 0.691134183 |
| 0 | 0 | 0 | 0.008086033 | 0 |
| 0 | 0 | 0 | 0 | 0 |
| 0 | 0 | 0.001200037 | 0.02538449 | 0 |
| 0.054754922 | 0 | 0 | 0 | 0 |
| 0 | 0 | 0 | 0 | 0 |
| 0 | 0 | 0 | 0 | 0 |
| 0 | 0 | 0 | 0 | 0 |
| 0.023194385 | 0.002908283 | 0.005484088 | 0.001807876 | 0.0004995 |
| 1.781235462 | 1.881312072 | 0.339389119 | 0.120778911 | 0.433727902 |
| 0 | 0 | 0 | 0 | 0 |
| 0 | 0.00443116 | 0.16276291 | 0.050241697 | 0.020080772 |
| 0.310880992 | 0.161792463 | 0.000848023 | 0 | 0.002230287 |
| 0 | 0 | 0.414352161 | 1.456137208 | 0 |
| 0 | 0 | 0 | 0 | 0 |
| 0 | 0 | 0 | 0 | 0 |
| 0 | 0 | 0 | 0.013851411 | 0 |
| 0.273180039 | 0.004932241 | 0 | 0 | 0 |
| 0.032898987 | 1.242873811 | 0 | 0 | 0.104559046 |
| 3.138984595 | 0.005892309 | 0 | 0 | 0.201046053 |
| 3.561138424 | 1.166340244 | 0.006314881 | 0 | 3.198037212 |
| 0 | 0 | 0 | 0 | 0 |
| 0 | 0 | 0 | 0.000278458 | 0 |
| 0 | 0 | 1.684182036 | 0.296647164 | 0.011162248 |
| 0 | 0.59835262 | 0 | 0 | 0 |
| 0 | 0 | 0 | 0.000987747 | 0 |
| 0 | 0 | 0 | 0 | 0 |
| 0 | 0 | 0 | 0 | 0 |
| 0 | 0 | 0 | 0.000940106 | 0 |
| 0 | 0.537139458 | 0.067063812 | 0.010997183 | 0.204142627 |
| 0.062530749 | 0.008257853 | 0 | 0 | 0.003590999 |
| 0 | 0.010986695 | 0.000585691 | 0.006283079 | 0.008606446 |
| 0.004686459 | 0.001043672 | 0 | 0 | 0 |
| 0.008723681 | 0.013224123 | 0.030876946 | 0.061771453 | 0 |
| 0 | 0.002770335 | 0 | 0 | 0.001011091 |
| 0 | 0 | 0 | 0 | 0 |
| 0 | 0.008762892 | 0.031513853 | 0.162672203 | 0.090910499 |
| 0 | 0 | 0 | 0 | 0 |
| 0 | 0 | 0.005110659 | 0.004447799 | 0 |
| 0 | 0 | 0 | 0 | 0 |
| 0 | 0 | 0 | 0 | 0 |
| 0 | 0 | 0 | 0 | 0 |
| 0.034819288 | 0.04069948 | 0.011119469 | 0.003240153 | 0.00066313 |
| 0 | 0 | 0 | 0 | 0 |
| 0 | 0 | 0 | 0.174669598 | 0 |
| 0.035715627 | 0.076929633 | 0 | 0 | 0.688727919 |

| | | | | |
|---|---|---|---|---|
| 0 | 0 | 0 | 0 | 0 |
| 0 | 0 | 0 | 0 | 0 |
| 0 | 0 | 0 | 0 | 0 |
| 0 | 0 | 0 | 0.001329644 | 0 |
| 0 | 0 | 0 | 0 | 0 |
| 0 | 0 | 0 | 0.150766372 | 0 |
| 0 | 0.335178781 | 0 | 0 | 0 |
| 0 | 0.020662008 | 0.018894693 | 0.001788212 | 0.002808383 |
| 0 | 0 | 0 | 0 | 0 |
| 0 | 0 | 0 | 0 | 0 |
| 0.001349044 | 0.003621867 | 0 | 0 | 0.027822328 |
| 0 | 0 | 0 | 0 | 0 |
| 0 | 0 | 0 | 0.012666972 | 0.00121252 |
| 0 | 0 | 0 | 0 | 0 |
| 0 | 0 | 0 | 0 | 0 |
| 0 | 0 | 0 | 0 | 0 |
| 0 | 0.020780091 | 0.776226177 | 0.013555432 | 0 |
| 0 | 0.001866504 | 0.01171568 | 0.029871501 | 0.012157143 |
| 0 | 0 | 0 | 0 | 0 |
| 0 | 0 | 0 | 0 | 0 |
| 0 | 0 | 0 | 0.002274726 | 0.001303528 |
| 0 | 0 | 0 | 0 | 0 |
| 0 | 0.002648548 | 0 | 0.071722048 | 0.003020758 |
| 0 | 0 | 0 | 0 | 0 |
| 0 | 0 | 0 | 0 | 0 |
| 0 | 0 | 0 | 0 | 0 |
| 0 | 0 | 0 | 0 | 0 |
| 0 | 0 | 0 | 0 | 0 |
| 0 | 0 | 0 | 0 | 0 |
| 4.095221515 | 0.217667203 | 0.013141548 | 0.005269483 | 0 |
| 0 | 0 | 0 | 0 | 0 |
| 0 | 0 | 0 | 0 | 0 |
| 0 | 0 | 0 | 0.00217979 | 0 |
| 0 | 0 | 0 | 0 | 0 |
| 0 | 0 | 0 | 0.023822079 | 0 |
| 0 | 0 | 0 | 0 | 0 |
| 0 | 0 | 0.051650244 | 0.017783367 | 0 |
| 0.001903316 | 0 | 0 | 0 | 0.007459937 |
| 0 | 0 | 0 | 0 | 0 |
| 0 | 0 | 0 | 0 | 0 |
| 0 | 0.080545983 | 0 | 0 | 0.000599937 |
| 0 | 0 | 0 | 0 | 0 |
| 0 | 0.018269217 | 0 | 0 | 0 |
| 0 | 0.136252586 | 0.073340463 | 0.352384279 | 0 |
| 0 | 0.247012312 | 0.011111792 | 0.017519159 | 0.089426196 |
| 0 | 0 | 0 | 0.000524185 | 0 |
| 0 | 0.001387233 | 0 | 0.008612676 | 0 |
| 0 | 0 | 0 | 0 | 0 |

| | | | | |
|---|---|---|---|---|
| 0 | 0 | 0 | 0 | 0 |
| 0 | 0 | 0 | 0 | 0 |
| 0 | 0 | 0 | 0 | 0 |
| 0 | 0.093235072 | 0.04528475 | 0.083353722 | 0 |
| 0 | 0 | 0 | 0 | 0 |
| 0 | 0 | 0.587119609 | 0.180543689 | 0 |
| 0 | 0 | 0 | 0 | 0 |
| 0 | 0 | 0 | 0 | 0 |
| 0.272736509 | 0.029398073 | 0 | 0 | 0.028219884 |
| 0 | 0 | 0 | 0.003534242 | 0 |
| 0 | 0.000719881 | 0 | 0 | 0.000945848 |
| 0 | 0 | 0 | 0 | 0 |
| 0 | 0 | 0 | 0 | 0 |
| 0 | 0 | 0 | 0 | 0 |
| 0 | 0.009116114 | 0.154338466 | 0.189232316 | 0 |
| 0.070001099 | 0 | 0 | 0 | 0 |
| 0 | 0 | 0 | 0 | 0 |
| 0 | 0 | 0 | 0 | 0 |
| 0 | 0 | 0 | 0 | 0 |
| 0 | 0 | 0 | 0 | 0 |
| 0 | 0.002001559 | 0 | 0 | 0 |
| 0 | 0 | 0 | 0 | 0 |
| 0 | 0 | 0 | 0 | 0 |
| 0.008135926 | 0 | 0 | 0 | 0 |
| 0.005865137 | 0.145388879 | 0.517889892 | 0.132466837 | 0.064470292 |
| 1.394835936 | 12.21576216 | 0.118374645 | 6.753843127 | 0 |
| 0.00068889 | 0 | 0.07622843 | 0.27688823 | 0 |
| 0 | 0 | 0 | 0 | 0 |
| 0 | 0 | 0.031673754 | 2.218239467 | 0.018587005 |
| 0 | 0.052571993 | 0 | 0.17154406 | 0 |
| 0 | 0 | 0 | 0 | 0 |
| 0 | 0 | 0 | 0 | 0 |
| 0 | 0 | 0 | 0 | 0 |
| 0.000659444 | 0 | 0 | 0 | 0 |
| 0 | 0 | 0.378912344 | 0.024634526 | 0 |
| 0.008064662 | 0 | 0 | 0.001361959 | 0.03429682 |
| 1.934983213 | 0 | 0 | 0 | 0 |
| 0.992597351 | 6.900974812 | 0.466751598 | 0.162466835 | 0 |
| 2.070040935 | 0 | 0 | 1.446720598 | 0 |
| 0 | 0 | 0 | 0 | 0 |
| 0 | 0 | 0 | 0 | 0 |
| 0.010016382 | 0.000501086 | 0 | 0.003309585 | 0 |
| 0.046443556 | 1.457836984 | 0.108220807 | 0.044667402 | 0.21068209 |
| 0 | 0 | 0 | 0.247331013 | 0 |
| 0 | 0 | 0 | 0.001996687 | 0 |
| 0.001455287 | 0.000837236 | 0 | 0.000737306 | 0 |
| 0 | 0 | 0 | 0 | 0 |
| 0.00550631 | 7.266640294 | 0 | 0.00102779 | 0 |

| | | | | |
|---|---|---|---|---|
| 0 | 0 | 0 | 0 | 0 |
| 0 | 0 | 0 | 0.00098228 | 0 |
| 0 | 0 | 0 | 0 | 0 |
| 0 | 0 | 0 | 0 | 0 |
| 0 | 0 | 0 | 0 | 0 |
| 0 | 0 | 0 | 0.032541989 | 0 |
| 0 | 0 | 0 | 0.02846416 | 0 |
| 0 | 0 | 0 | 0 | 0 |
| 0 | 0 | 0 | 0 | 0 |
| 0 | 0 | 0 | 0 | 0 |
| 0 | 0 | 0 | 0.000834111 | 0 |
| 0.419827035 | 0.553481014 | 1.550848639 | 0.166741528 | 0 |
| 0.002084178 | 0.649281261 | 0.000873571 | 0.008975383 | 0 |
| 0 | 0 | 0 | 0 | 0 |
| 0.002737171 | 0 | 0.077597128 | 0 | 0.168585756 |
| 0 | 0 | 0 | 0 | 0 |
| 0 | 0 | 0 | 0 | 0 |
| 0 | 0 | 0 | 0 | 0 |
| 0.001744815 | 0.000501902 | 0 | 0.001104991 | 0 |
| 0 | 0 | 0 | 0.31158317 | 0 |
| 0 | 0.016621153 | 0.012916766 | 0.02662158 | 0 |
| 0 | 0 | 0 | 0 | 0 |
| 0.134667264 | 1.662235274 | 0.044563405 | 2.192421912 | 0 |
| 0 | 0 | 0 | 0 | 0 |
| 0 | 0 | 0 | 0 | 0 |
| 0 | 0 | 0 | 0 | 0 |
| 0 | 0 | 0 | 5.975109539 | 0 |
| 0 | 0 | 0 | 0 | 0 |
| 0 | 0 | 0 | 0 | 0 |
| 0 | 0 | 0 | 0 | 0 |
| 0 | 0 | 0 | 0 | 0 |
| 0.041330759 | 0.097836022 | 0.033203468 | 0.011451455 | 0.15241133 |
| 0 | 0 | 0.204830453 | 0.000732992 | 0 |
| 0 | 0.005073329 | 0.004868182 | 0.000817279 | 0.001535487 |
| 0.006268838 | 0 | 0.000700679 | 0.001376285 | 0.036270519 |
| 0.002963141 | 0.004545907 | 0.014452169 | 0.938823841 | 0.029864211 |
| 0 | 0.001138269 | 0 | 0.074616788 | 0 |
| 0 | 0 | 0 | 0.050313511 | 0 |
| 0 | 0.000552446 | 0.00375656 | 0.003283924 | 0 |
| 0 | 0 | 0 | 0 | 0 |
| 0 | 0.032229796 | 0 | 0.017408598 | 0 |
| 0 | 0 | 0 | 0.002549509 | 0 |
| 0 | 0 | 0 | 0 | 0 |
| 0 | 0.01161362 | 0 | 0 | 0 |
| 0.001440935 | 0.011526763 | 0.005061755 | 0 | 0.00495611 |
| 0 | 0 | 0 | 0 | 0 |
| 0 | 0 | 0.005226766 | 0.009915503 | 0 |
| 0.001557561 | 0 | 0 | 0 | 0.001962248 |

| | | | | |
|---|---|---|---|---|
| 0 | 0 | 0 | 0 | 2.272927939 |
| 0 | 0 | 0 | 0.012099444 | 0 |
| 0.029379943 | 0 | 0 | 0 | 0 |
| 0 | 0 | 0 | 0 | 0 |
| 0.002375004 | 0 | 0 | 0 | 0 |
| 0 | 0 | 0 | 0 | 0 |
| 0 | 0 | 0 | 4.235086079 | 0 |
| 0.009852057 | 0.174125776 | 0 | 0.020169506 | 0 |
| 0 | 0 | 0 | 0 | 0 |
| 0 | 0 | 0 | 0.002706486 | 0 |
| 0 | 0 | 0 | 0 | 0 |
| 0 | 0 | 0 | 0 | 0 |
| 0 | 0 | 0 | 0 | 0 |
| 0 | 0 | 0.003443601 | 0.020291925 | 0.126601036 |
| 0 | 0 | 0 | 0 | 0 |
| 0 | 0 | 0 | 0 | 0 |
| 0 | 0.663899254 | 0 | 0 | 0 |
| 0 | 0 | 0 | 0 | 0 |
| 4.428066384 | 0 | 0 | 0 | 1.125490136 |
| 0 | 0 | 0 | 0 | 0 |
| 0 | 0 | 0 | 0.001176035 | 0 |
| 0 | 0 | 0 | 0 | 0 |
| 0 | 0 | 0 | 0 | 0 |
| 0 | 0 | 0 | 0 | 0 |
| 0 | 0 | 0 | 0 | 0 |
| 0.005940732 | 0.001763997 | 0 | 0 | 0.01437836 |
| 0 | 0 | 0 | 0 | 0 |
| 0.093599444 | 9.636597714 | 0 | 0 | 0.003360849 |
| 0 | 0 | 0 | 0.000704058 | 0 |
| 0 | 0.377498036 | 0.396884825 | 0.152039501 | 0 |
| 0 | 0 | 0 | 0 | 0 |
| 0 | 0 | 0 | 0 | 0 |
| 0 | 0 | 0 | 0 | 0 |
| 0 | 0 | 0 | 0 | 0 |
| 0 | 0 | 0 | 0 | 0.006184476 |
| 0 | 0 | 0.3176318 | 0 | 0 |
| 0 | 0 | 0 | 0.536955897 | 0 |
| 0 | 0.002272371 | 0.002801706 | 0 | 0 |
| 0 | 0 | 0 | 0 | 0 |
| 0 | 0 | 0 | 0 | 0 |
| 6.114805442 | 2.738054323 | 1.259880744 | 0.447993765 | 0.407162604 |
| 1.293079512 | 0.72126986 | 0 | 0.006253777 | 0 |
| 0 | 0.157689796 | 0 | 0 | 0.066322006 |
| 0 | 0 | 0 | 0 | 0 |
| 0 | 0 | 0 | 0.00128733 | 0 |
| 0 | 0 | 0 | 0 | 0 |
| 0 | 0 | 0 | 0 | 0 |
| 0.001085128 | 0 | 0.001212867 | 0 | 0 |

| | | | | |
|---|---|---|---|---|
| 0 | 0 | 0 | 0 | 0 |
| 0.004464574 | 0 | 0 | 0 | 0 |
| 0 | 0 | 0 | 0 | 0 |
| 0 | 0 | 0 | 0.000723155 | 0 |
| 1.461604205 | 2.746935881 | 0.357233301 | 0.248207836 | 7.524014671 |
| 0 | 0 | 0 | 0.053830201 | 0 |
| 0 | 0 | 0 | 0 | 0 |
| 0 | 0.008623708 | 0 | 0.000999264 | 0 |
| 0.189836149 | 0.002564713 | 0 | 0 | 1.025697481 |
| 0.013080479 | 0 | 0 | 0 | 0 |
| 0 | 0 | 0 | 0 | 0 |
| 0 | 0 | 0 | 0 | 0 |
| 0 | 0.380183714 | 0.045255149 | 0.005952513 | 0 |
| 0.015333083 | 0.005795589 | 0.006954575 | 0.004053051 | 0 |
| 0 | 0 | 0.733893064 | 2.404306478 | 0.063590256 |
| 2.40666804 | 1.01522736 | 0.045043543 | 0.013857936 | 0.000306306 |
| 0.006538054 | 0.32598692 | 0 | 0 | 0.009259645 |
| 0 | 0 | 0 | 0 | 0 |
| 0 | 0 | 0 | 0 | 0 |
| 0 | 0 | 0 | 0 | 0 |
| 0 | 0 | 0 | 0 | 0 |
| 0 | 0 | 0 | 0 | 0 |
| 0 | 0 | 0 | 0 | 0 |
| 0 | 0 | 0 | 0 | 0 |
| 0 | 0 | 0 | 0.000348844 | 0 |
| 0 | 0 | 0 | 0 | 0 |
| 0 | 0.111074894 | 0.04578867 | 0.014062646 | 0 |
| 0 | 0 | 0 | 0 | 0.005282196 |
| 0 | 0 | 0 | 0 | 0 |
| 0 | 0 | 0 | 0 | 0 |
| 0 | 0 | 0 | 0 | 0 |
| 0 | 0 | 0 | 0 | 0 |
| 0 | 0.135569474 | 0.002955149 | 0 | 0 |
| 0 | 0 | 0.000663199 | 0.0020041 | 0 |
| 0 | 0.010048549 | 0.020132605 | 12.50080969 | 0.010740318 |
| 0 | 0.002343905 | 0 | 0 | 0.000855457 |
| 0.000669263 | 0 | 0 | 0 | 0 |
| 0 | 0.005533277 | 0 | 0 | 0 |
| 0 | 0 | 0 | 0 | 0 |
| 0 | 0 | 0 | 0 | 0 |
| 0 | 0.891346318 | 0.201676145 | 0.011376196 | 6.697289015 |
| 0 | 0 | 0 | 1.611022262 | 0.025143615 |
| 0 | 0 | 0 | 0 | 0 |
| 0 | 0 | 0 | 0.139341305 | 0 |
| 0 | 0 | 0 | 0 | 0 |
| 0 | 0 | 0 | 0 | 0 |
| 0 | 0 | 0 | 0 | 0 |
| 0 | 0 | 0 | 0 | 0 |

| | | | | |
|---|---|---|---|---|
| 0 | 0 | 0.00369443 | 0.265321478 | 0 |
| 0 | 0 | 0 | 0 | 0 |
| 0 | 0.003035371 | 0 | 0.000320769 | 0 |
| 0 | 0 | 0 | 0 | 0 |
| 0 | 0.004235436 | 0.002742897 | 0.005076816 | 0 |
| 0 | 0 | 0 | 0 | 0 |
| 0.016880876 | 1.221906385 | 0 | 0 | 0.094949099 |
| 0 | 0 | 0 | 0 | 0 |
| 0 | 0 | 0 | 0 | 0 |
| 0 | 0 | 0 | 0 | 0 |
| 0 | 0 | 0 | 0.005657091 | 0 |
| 0 | 0 | 0 | 0.000509955 | 0 |
| 0 | 0 | 0 | 0 | 0 |
| 0 | 0 | 0 | 0 | 0 |
| 0 | 0 | 0 | 0 | 0 |
| 0 | 0 | 0 | 0 | 0 |
| 0.088459697 | 0.01538726 | 0 | 0 | 0 |
| 0.004794988 | 0 | 0 | 0 | 0 |
| 0 | 0 | 0 | 0 | 0 |
| 0 | 0 | 0 | 0 | 0 |
| 0.011140496 | 0.087277242 | 0 | 0.039972163 | 0 |
| 0 | 0 | 0 | 0 | 0 |
| 0 | 0.002472334 | 0.002701856 | 0.008164659 | 0 |
| 0.001612114 | 0.00178622 | 0.005005247 | 0.004809815 | 0.001203541 |
| 0 | 0 | 0 | 0 | 0 |
| 0.000845303 | 0 | 0 | 0.001177727 | 0 |
| 0 | 0 | 0 | 0.603117354 | 0 |
| 0 | 0 | 0 | 0 | 0 |
| 0 | 0 | 0 | 0 | 0 |
| 0.598614294 | 0 | 0 | 0.132571488 | 0.003031311 |
| 0 | 0.025862442 | 0 | 0.000427042 | 0 |
| 0 | 0 | 0 | 0 | 0 |
| 0 | 0 | 0 | 0 | 0 |
| 0 | 0 | 0 | 0 | 0 |
| 0 | 0 | 0 | 0.003579434 | 0 |
| 0 | 0 | 0 | 0.041483739 | 0 |
| 0.038774538 | 0.014269768 | 0 | 0 | 0.004141339 |
| 0 | 0 | 0 | 0 | 0 |
| 0 | 0 | 0 | 0 | 0 |
| 0 | 0 | 0.038871561 | 0 | 0.010496999 |
| 0 | 0 | 0 | 0 | 0 |
| 0 | 0 | 0 | 0.009342626 | 0 |
| 0 | 0 | 0 | 0 | 0 |
| 0 | 0 | 0 | 0.001083573 | 0 |
| 0 | 0 | 0 | 0 | 0 |
| 0 | 0 | 0 | 0 | 0 |
| 0 | 0 | 0 | 0 | 0 |
| 0 | 0 | 0 | 4.110507078 | 0 |

| | | | | |
|---|---|---|---|---|
| 0.002505667 | 0.000739245 | 0 | 0 | 0.055579344 |
| 0 | 0 | 0 | 1.330375856 | 0 |
| 0 | 0.080823728 | 0 | 0 | 0 |
| 0 | 0 | 0 | 0 | 0 |
| 0 | 0 | 0 | 0 | 0 |
| 0 | 0 | 0 | 0 | 0 |
| 0 | 0 | 0 | 0 | 0 |
| 2.122917223 | 0.354622185 | 0.41696379 | 0.057352193 | 0 |
| 0.098628039 | 0.842186063 | 22.94275966 | 0.420596959 | 0.765680355 |
| 0.004633624 | 0 | 0 | 0 | 0.150950426 |
| 0 | 0 | 0 | 0.001878301 | 0 |
| 0 | 0 | 0 | 0 | 0 |
| 0 | 0.05414761 | 0 | 0 | 0 |
| 0.053306207 | 0 | 0 | 0 | 0 |
| 0 | 0 | 0 | 0 | 0 |
| 0 | 0 | 0 | 0.019078604 | 0 |
| 0.274319917 | 0 | 0 | 0 | 0 |
| 0 | 0 | 0 | 0.030275047 | 0 |
| 0 | 0 | 0 | 0 | 0 |
| 0 | 0 | 0 | 0 | 0.000391162 |
| 0 | 0 | 0 | 0 | 0 |
| 0 | 0 | 0 | 0 | 0 |
| 0 | 0 | 0 | 0.003341668 | 0 |
| 0 | 0 | 0 | 0 | 0 |
| 0 | 0.202858571 | 0 | 0.002615081 | 0 |
| 0 | 0 | 0 | 0 | 0 |
| 0 | 0 | 0 | 0 | 0 |
| 0 | 0 | 0 | 0 | 0 |
| 0 | 0 | 0 | 0 | 0 |
| 0 | 0 | 0 | 0 | 0 |
| 0 | 0 | 0 | 0 | 0 |
| 0 | 0 | 0 | 0 | 0 |
| 0 | 0 | 0 | 0 | 0 |
| 0 | 0 | 0 | 0.003882363 | 0 |
| 0 | 0 | 0.006520298 | 0.003546628 | 0.006402028 |
| 0 | 0 | 0 | 0 | 0 |
| 0 | 0 | 0 | 0 | 0 |
| 0 | 0 | 0 | 0 | 0 |
| 0 | 0.000527412 | 0.007172665 | 0.003367344 | 0 |
| 0 | 0 | 0 | 0 | 1.168645074 |
| 0 | 0 | 0 | 0 | 0 |
| 0 | 0.211205592 | 0.091415553 | 0.077945598 | 4.103061699 |
| 0 | 0 | 0 | 0 | 0 |
| 0 | 0 | 0 | 0 | 0 |
| 0 | 0 | 0 | 0 | 0 |
| 0 | 0 | 0 | 0 | 0 |
| 0 | 0 | 0 | 0 | 0 |
| 0 | 0 | 0 | 0 | 0 |

| | | | | |
|---|---|---|---|---|
| 0 | 0 | 0 | 0 | 0 |
| 0 | 0.00040032 | 0 | 0 | 0 |
| 0 | 0 | 0 | 0 | 0 |
| 0 | 0 | 0 | 0 | 0 |
| 0.146904792 | 0.098881074 | 0.007611833 | 0.694618394 | 0.194334213 |
| 0 | 0 | 0.001242215 | 0 | 0 |
| 0.003374961 | 0 | 0 | 0 | 0 |
| 0.113520096 | 0.004889938 | 0 | 0 | 0.001510117 |
| 0 | 0 | 0.001368125 | 0.002852665 | 0 |
| 0 | 0 | 0.00634125 | 0.027625839 | 0 |
| 0 | 0 | 0 | 0 | 0 |
| 0 | 0 | 0 | 0 | 0 |
| 0 | 0 | 0 | 0 | 0 |
| 0 | 0 | 0 | 0 | 0 |
| 0.050917218 | 0.002515559 | 0 | 0 | 0 |
| 0 | 0 | 0.002376783 | 0 | 0.0044054 |
| 0 | 0 | 0 | 0 | 0 |
| 0 | 0 | 0 | 0 | 0 |
| 0 | 0 | 0 | 0 | 0 |
| 0 | 0 | 0 | 0 | 0 |
| 0 | 0 | 0 | 0 | 0 |
| 0 | 0 | 0 | 0 | 0 |
| 0 | 0 | 0 | 0 | 0 |
| 0 | 0.000976578 | 0 | 0 | 0.005108722 |
| 0 | 0.263415721 | 0.010761507 | 0.009373375 | 0.384450368 |
| 0 | 0 | 0 | 0 | 0 |
| 0 | 0.012802413 | 0 | 0 | 0.026513753 |
| 0.001021739 | 0 | 0 | 0 | 0.000686511 |
| 0 | 0 | 0 | 0 | 0 |
| 0 | 0 | 0 | 0 | 0 |
| 0 | 0 | 0 | 0 | 0 |
| 0 | 0 | 0 | 0 | 0 |
| 0 | 0 | 0 | 0 | 0 |
| 0 | 0 | 0 | 0 | 0 |
| 0 | 0 | 0 | 0 | 0 |
| 0 | 0 | 0 | 0 | 0 |
| 0 | 0 | 0 | 0 | 0 |
| 0 | 0 | 0 | 0.006856835 | 0 |
| 0 | 0 | 5.001636973 | 0 | 0 |
| 0 | 0 | 0 | 0 | 0 |
| 0 | 0 | 0 | 0 | 0 |
| 0 | 0 | 0 | 1.031699235 | 0.002172292 |
| 0 | 0 | 0 | 0 | 0 |
| 0 | 0 | 0 | 0.002285688 | 0 |
| 0 | 0 | 0 | 0 | 0 |
| 0 | 0 | 0 | 0 | 0 |
| 0 | 0 | 0 | 0 | 0 |
| 0 | 0 | 0 | 0.005771859 | 0 |

| | | | | |
|---|---|---|---|---|
| 0 | 0.193053046 | 0.009383485 | 0.01608876 | 0 |
| 0 | 0 | 0 | 0 | 0 |
| 0 | 0 | 0 | 0 | 0 |
| 0.000544746 | 0 | 0 | 0 | 0 |
| 0 | 0 | 0 | 0 | 0 |
| 0.084154029 | 0 | 0 | 0.004392127 | 0.001894253 |
| 0 | 0 | 0 | 0 | 0 |
| 0 | 0 | 0 | 0 | 0 |
| 0 | 0 | 0 | 0 | 0 |
| 0 | 0 | 0 | 0 | 0 |
| 0 | 0 | 0 | 0 | 0 |
| 0 | 0 | 0 | 0 | 0 |
| 0 | 0 | 0 | 0.002325673 | 0 |
| 0 | 0 | 0 | 0 | 0 |
| 0 | 0 | 0 | 0.00183892 | 0 |
| 0 | 0 | 0 | 0 | 0 |
| 0 | 0 | 0 | 0 | 0 |
| 0 | 0 | 0 | 0 | 0 |
| 0 | 0 | 0 | 0 | 0 |
| 0 | 0 | 0 | 0 | 0 |
| 0.034864398 | 0.032416528 | 0 | 0 | 0.054186359 |
| 0 | 0 | 0 | 0 | 0 |
| 0.007205105 | 0.000818794 | 0 | 0.002298395 | 0.001344762 |
| 0 | 0.161121688 | 0 | 0.022149785 | 0 |
| 0 | 0 | 0 | 0.000364311 | 0 |
| 0 | 0 | 0 | 0 | 0 |
| 0 | 0 | 0 | 0 | 0 |
| 0 | 0 | 0 | 0 | 0 |
| 0 | 0 | 0 | 0 | 0 |
| 0 | 0 | 0 | 0 | 0 |
| 0 | 0 | 0 | 0 | 0 |
| 0.001878262 | 0 | 0.454962948 | 0 | 0 |
| 0.001749365 | 0.008051373 | 0 | 0 | 0.008815546 |
| 0 | 0 | 0 | 0 | 0 |
| 0.073098453 | 4.381851034 | 0 | 0.036694812 | 0 |
| 0 | 0.00430726 | 0 | 0 | 0.00258261 |
| 0 | 0 | 0 | 0 | 0 |
| 0 | 0 | 0 | 0 | 0 |
| 0.006030914 | 0 | 0 | 0 | 0 |
| 0 | 0 | 0 | 0 | 0 |
| 0 | 0 | 0 | 0 | 0 |
| 0 | 0.08476038 | 0 | 0.010415386 | 0 |
| 0 | 0.003953006 | 0.013945229 | 1.417025719 | 0.008656388 |
| 0 | 0 | 0 | 0 | 0 |
| 0 | 0 | 0 | 0 | 0 |
| 0 | 0.030732804 | 0.001685315 | 0.188797521 | 0 |
| 0 | 0 | 0 | 0 | 0 |
| 0 | 0 | 0 | 0 | 0 |

| | | | | |
|---|---|---|---|---|
| 0 | 0 | 0 | 0 | 0 |
| 0 | 0 | 0 | 0 | 0 |
| 0 | 0 | 0 | 0 | 0 |
| 0 | 0 | 0 | 0 | 0 |
| 0 | 0 | 0 | 0 | 0 |
| 0 | 0 | 0 | 0 | 0 |
| 0 | 0 | 0 | 0 | 0 |
| 0.003954796 | 0 | 0 | 0 | 0 |
| 0 | 0.01253923 | 0 | 0.007975197 | 0 |
| 0 | 0 | 0.008679282 | 0.022480851 | 0.00832309 |
| 0 | 0 | 0 | 0 | 0 |
| 0 | 0 | 0 | 0 | 0 |
| 0 | 0 | 0 | 0 | 0 |
| 0 | 0.542319387 | 0 | 0 | 0 |
| 0.002689425 | 0 | 0 | 0 | 0 |
| 0.010477823 | 0 | 0 | 0 | 0.004033388 |
| 0 | 0 | 0 | 0 | 0 |
| 0 | 0 | 0 | 0 | 0 |
| 0 | 0 | 0 | 0 | 0 |
| 0 | 1.05015534 | 0 | 0.004351836 | 0 |
| 0 | 0 | 0.003312717 | 0.243604093 | 0.009574064 |
| 0 | 0 | 0 | 0 | 0 |
| 0 | 0 | 0 | 0 | 0 |
| 0.005815282 | 0 | 0 | 0 | 0 |
| 0 | 0 | 0 | 0 | 0 |
| 0 | 0 | 0 | 0 | 0 |
| 0 | 0 | 0 | 0 | 0 |
| 0 | 0 | 0 | 0 | 0 |
| 0 | 0 | 0 | 0 | 0 |
| 0 | 0 | 0.149446819 | 0.064646544 | 0 |
| 0.35554637 | 0.185495198 | 0.268304858 | 0.032269497 | 0.091094916 |
| 0 | 0 | 0 | 0 | 0 |
| 0 | 0 | 0 | 0 | 0 |
| 0 | 0 | 0 | 0.002218918 | 0 |
| 0 | 0 | 0 | 0 | 0 |
| 0 | 0 | 0 | 0 | 0 |
| 0 | 0 | 0 | 0 | 0 |
| 0 | 0 | 0 | 0 | 0 |
| 0 | 0 | 0 | 0 | 0 |
| 0 | 0 | 0 | 0 | 0 |
| 0 | 0 | 0 | 0 | 0 |
| 0 | 0.308004131 | 0 | 0.001032645 | 0 |
| 0 | 0 | 0 | 0.000583325 | 0 |
| 0 | 0 | 0 | 0 | 0 |
| 0 | 0 | 0 | 0 | 0 |
| 0 | 0 | 0 | 0 | 0 |
| 0.484460897 | 0 | 0 | 0 | 0.007382063 |
| 0 | 0 | 0 | 0.00064406 | 0 |

| | | | | |
|---|---|---|---|---|
| 0 | 0 | 0 | 0 | 0 |
| 0 | 0 | 0 | 0 | 0 |
| 0 | 0 | 0 | 0 | 0 |
| 0 | 0 | 0 | 0.001116115 | 0.008185895 |
| 0.092318471 | 0.167113781 | 1.31174979 | 0.117141955 | 0 |
| 0 | 0 | 0 | 0 | 0 |
| 0 | 0 | 0 | 0 | 0 |
| 0 | 0 | 0 | 0 | 0 |
| 0 | 0 | 0 | 0 | 0 |
| 0 | 0 | 0 | 0 | 0 |
| 0 | 0.059344365 | 0.008054887 | 0.00161079 | 0 |
| 0 | 0.002311242 | 0.001295287 | 0.004227324 | 0.000778648 |
| 0.002199584 | 0 | 0 | 0.000709161 | 0 |
| 0 | 0 | 0 | 0 | 0 |
| 0.002296159 | 0.00379363 | 0 | 0 | 0 |
| 0 | 0.100649837 | 0.247788264 | 0.404069352 | 0 |
| 0.005526102 | 0.004238939 | 0.000597738 | 0 | 0.009082907 |
| 0.002184259 | 0 | 0 | 0 | 0 |
| 0 | 0 | 0 | 0 | 0 |
| 0.011686642 | 0.344343819 | 0 | 0 | 0.07108134 |
| 0 | 0 | 0 | 0 | 0 |
| 0 | 0 | 0 | 0.02972298 | 0 |
| 0 | 0 | 0 | 0 | 0 |
| 0 | 0 | 0 | 0 | 0 |
| 0 | 0 | 0 | 0 | 0 |
| 0 | 0 | 0 | 0 | 0 |
| 0.008446901 | 0.001971995 | 0 | 0 | 0.009356366 |
| 0 | 0 | 0 | 0 | 0 |
| 0 | 0 | 0 | 0 | 0 |
| 0 | 0 | 0 | 0 | 0 |
| 0.001040761 | 0 | 0 | 0 | 0.001048938 |
| 0.017086983 | 0 | 0 | 0 | 0 |
| 0 | 0 | 0 | 0 | 0 |
| 0 | 0 | 0 | 0 | 0 |
| 0 | 0 | 0 | 0 | 0 |
| 0 | 0 | 0 | 0.00482782 | 0 |
| 0 | 0 | 0 | 0 | 0 |
| 0.008920747 | 0.020528668 | 0.007028652 | 0.00237094 | 0.012118794 |
| 0 | 0.005936822 | 0.799790409 | 0.706611465 | 0 |
| 0.008815139 | 0 | 0 | 0 | 0 |
| 0.003503914 | 0 | 0 | 0.00029587 | 0 |
| 0 | 0 | 0 | 0 | 0 |
| 0 | 0 | 0 | 0 | 0 |
| 0 | 0.005988474 | 0 | 0 | 0 |
| 0 | 0 | 0 | 0 | 0 |
| 0 | 0 | 0 | 0 | 0 |
| 0 | 0 | 0 | 0 | 0 |
| 0 | 0 | 0 | 0 | 0 |

| | | | | |
|---|---|---|---|---|
| 0 | 0 | 0 | 0 | 0 |
| 0 | 0 | 0.005800731 | 0.017372544 | 0 |
| 0 | 0 | 0 | 0 | 0 |
| 0.002727218 | 0 | 0 | 0 | 0.00432657 |
| 0 | 0 | 0.006361228 | 0.000533967 | 0 |
| 0.003536541 | 0 | 0 | 0 | 0.001218573 |
| 0 | 0 | 0 | 0.008045107 | 0 |
| 0 | 0 | 0 | 0 | 0 |
| 0 | 0.003540598 | 0 | 0 | 0 |
| 0 | 0 | 0 | 0.004789962 | 0 |
| 0 | 0.001008372 | 0 | 0.003996069 | 0 |
| 0 | 0 | 0 | 0 | 0 |
| 0.001772112 | 0.006673146 | 0.019627152 | 0.102351764 | 0.148385254 |
| 0 | 0 | 0 | 0 | 0 |
| 0 | 0 | 0 | 0 | 0 |
| 0 | 0 | 0 | 0.108297821 | 0 |
| 0 | 0 | 0 | 0 | 0 |
| 0 | 0 | 0 | 0 | 0 |
| 0 | 0 | 0 | 0 | 0 |
| 0 | 0 | 0 | 0 | 0 |
| 0 | 0 | 0 | 0 | 0 |
| 0 | 0 | 0 | 0 | 0 |
| 0 | 0 | 0 | 0 | 0 |
| 0.028036322 | 0.018625719 | 0.006155423 | 0.001141418 | 0 |
| 0 | 0 | 0 | 0 | 0 |
| 0 | 0 | 0 | 0.040367854 | 0 |
| 0 | 0 | 0 | 0.002464394 | 0 |
| 0 | 0 | 0 | 0 | 0 |
| 0 | 0 | 0 | 0 | 0 |
| 0 | 0 | 0 | 0 | 0 |
| 0 | 0.014877626 | 0.01427603 | 0.002396684 | 0 |
| 0 | 0 | 0 | 0 | 0 |
| 0 | 0 | 0 | 0 | 0 |
| 0 | 0 | 0 | 0 | 0 |
| 0.083483992 | 0 | 0 | 0.016576937 | 0 |
| 0 | 0 | 0 | 0 | 0 |
| 0 | 0 | 0 | 0 | 0 |
| 0.111458517 | 0 | 0 | 0 | 0 |
| 0 | 0 | 0 | 0 | 0.1282978 |
| 0 | 0 | 0 | 0 | 0 |
| 0 | 0 | 0 | 0 | 0 |
| 0 | 0 | 0 | 0 | 0 |
| 0 | 0 | 0 | 0.219526839 | 0 |
| 0 | 0 | 0 | 0 | 0 |
| 0 | 0 | 0 | 3.967663525 | 0 |
| 0 | 0.382895212 | 0.285160378 | 0.256552761 | 0.11238794 |
| 0 | 0 | 0 | 0 | 0 |
| 0 | 0 | 0 | 0.002982704 | 0 |

| | | | | |
|---|---|---|---|---|
| 0 | 0 | 0 | 0.01266459 | 0 |
| 0 | 0 | 0 | 0 | 0 |
| 0 | 0 | 0 | 0.01097985 | 0.002325788 |
| 0 | 0 | 0.013964803 | 0.006060612 | 0 |
| 0 | 0 | 0 | 0 | 0 |
| 0 | 0 | 0 | 0 | 0 |
| 0 | 0 | 0 | 0 | 0 |
| 0 | 0 | 0 | 0 | 0 |
| 0 | 0 | 0 | 0 | 0 |
| 0 | 0 | 0 | 0 | 0 |
| 0 | 0 | 0 | 0 | 0 |
| 0 | 0 | 0 | 0 | 0.00081431 |
| 0 | 0 | 0 | 0 | 0 |
| 0 | 0 | 0 | 0 | 0 |
| 0 | 0 | 0 | 0 | 0 |
| 0 | 0 | 0 | 0 | 0 |
| 0 | 0 | 0 | 0 | 0 |
| 0 | 0 | 0 | 0 | 0 |
| 0 | 0 | 0 | 0 | 0 |
| 0 | 0.000825771 | 0 | 0 | 0.001808294 |
| 0 | 0 | 0 | 0 | 0 |
| 0 | 0 | 0 | 0 | 0 |
| 0 | 0 | 0 | 0 | 0 |
| 0 | 0 | 0 | 0 | 0 |
| 0 | 0 | 0 | 0 | 0 |
| 0 | 0 | 0 | 0 | 0 |
| 0 | 0 | 0 | 0 | 0 |
| 0 | 0 | 0.011185381 | 0.000845019 | 0 |
| 0 | 0 | 0 | 0 | 0 |
| 0 | 0 | 0 | 0 | 0 |
| 0.007768663 | 0.002128269 | 0.000620227 | 0.05819529 | 0.000724972 |
| 0 | 0 | 0 | 0 | 0 |
| 0.215196421 | 0.02191075 | 0.008641757 | 0.002284998 | 0.008537917 |
| 0 | 0.004040375 | 0.001471823 | 0.000333574 | 0.00454674 |
| 0 | 0 | 0 | 0 | 0 |
| 0 | 0 | 0 | 0 | 0 |
| 0 | 0 | 0 | 0 | 0 |
| 0 | 0 | 0 | 0 | 0 |
| 0 | 0 | 0 | 0 | 0 |
| 0 | 0 | 0 | 0 | 0 |
| 0 | 0 | 0 | 0 | 0 |
| 0.028093196 | 0.025344511 | 0 | 0 | 0 |
| 0.022171193 | 0.089658188 | 0.082764195 | 0.015813175 | 0 |
| 0 | 0 | 0 | 0 | 0 |
| 0 | 0 | 0 | 0 | 0 |
| 0.003732689 | 0.002386046 | 0.002781396 | 1.567903177 | 0.008185868 |
| 0 | 0 | 0 | 0 | 0 |
| 0 | 0 | 0 | 0 | 0 |

| | | | | |
|---|---|---|---|---|
| 0 | 0 | 0 | 0 | 0 |
| 0.751390759 | 0.004497514 | 0.007561611 | 0 | 0.001767727 |
| 0.005531746 | 0.006465933 | 0.005299656 | 0.011076952 | 0.008112074 |
| 0 | 0 | 0 | 0 | 0 |
| 0.155430463 | 0.039688537 | 0 | 0.003516453 | 0 |
| 0 | 0 | 0 | 0 | 0 |
| 0 | 0 | 0 | 0 | 0 |
| 0 | 0 | 0 | 0 | 0 |
| 0 | 0 | 0 | 0 | 0 |
| 0 | 0 | 0 | 0 | 0 |
| 0 | 0 | 0 | 0 | 0 |
| 0 | 0 | 0 | 0 | 0 |
| 0 | 0 | 0 | 0 | 0 |
| 0.047381399 | 0 | 0 | 0 | 0 |
| 0 | 0 | 0 | 0 | 0 |
| 0 | 0 | 0 | 0 | 0 |
| 0 | 0 | 0.006694918 | 0 | 0 |
| 0 | 0 | 0 | 0 | 0 |
| 0 | 0 | 0 | 0 | 0 |
| 0 | 0 | 0 | 0 | 0 |
| 0 | 0 | 0 | 0 | 0 |
| 0 | 0 | 0 | 0 | 0 |
| 0 | 0 | 0 | 0.001056331 | 0 |
| 0 | 0 | 0 | 0 | 0 |
| 0 | 0 | 0 | 0.179389678 | 0 |
| 0.007425248 | 0 | 0 | 0 | 0 |
| 0 | 0 | 0 | 0 | 0 |
| 0 | 0.01105011 | 0.762443159 | 1.803562877 | 0 |
| 0 | 0 | 0 | 0 | 0 |
| 0 | 0 | 0 | 0 | 0 |
| 0 | 0 | 0 | 0.111477041 | 0 |
| 0 | 0 | 0 | 0 | 0 |
| 0 | 0 | 0 | 0 | 0 |
| 0 | 0 | 0 | 0 | 0 |
| 0.001034486 | 0 | 0 | 0 | 0 |
| 0 | 0 | 0 | 0 | 0 |
| 0 | 0 | 0 | 0 | 0 |
| 0.07869465 | 0.034351218 | 0 | 0 | 0 |
| 0 | 0 | 0 | 0.005525126 | 0 |
| 0 | 0 | 0 | 0 | 0 |
| 0 | 0 | 0 | 0 | 0 |
| 0 | 0 | 0 | 0.001440391 | 0.001857182 |
| 0.001315067 | 0.000672504 | 0 | 0 | 0.046511691 |
| 0 | 0 | 0 | 0 | 0 |
| 0 | 0 | 0 | 0 | 0 |
| 0 | 0 | 0 | 0 | 0 |
| 0.002011795 | 0.002057599 | 0.00224862 | 0.000679504 | 0 |
| 0 | 0 | 0 | 0 | 0 |

| | | | | |
|---|---|---|---|---|
| 0 | 0 | 0 | 0 | 0 |
| 0.006134746 | 0.004219007 | 0 | 0.000428704 | 0.018477765 |
| 0 | 0 | 0 | 0 | 0 |
| 0 | 0 | 0 | 0 | 0 |
| 0 | 0 | 0 | 0 | 0 |
| 0 | 0 | 0 | 0 | 0 |
| 0 | 0 | 0 | 0 | 0 |
| 0 | 0 | 0 | 0 | 0 |
| 0 | 0 | 0 | 0 | 0 |
| 0 | 0 | 0 | 0 | 0 |
| 0 | 0 | 0 | 0 | 0 |
| 0 | 0 | 0 | 0 | 0 |
| 0 | 0.051631501 | 0.316178772 | 0.115787078 | 0 |
| 0.00397257 | 0 | 0 | 0 | 0 |
| 0.002637622 | 0.008497675 | 0 | 0.001336324 | 0.006793548 |
| 0 | 0 | 0 | 0 | 0 |
| 0 | 0 | 0 | 0 | 0 |
| 0 | 0 | 0 | 0 | 0 |
| 0.007878311 | 0 | 0 | 0 | 0 |
| 0 | 0 | 0 | 0 | 0 |
| 0 | 0 | 0 | 0 | 0 |
| 0.007512023 | 0.394617609 | 0 | 0 | 0.008992426 |
| 0.003781901 | 0 | 0 | 0 | 0 |
| 0.008270959 | 0 | 0 | 0 | 0.001195117 |
| 0 | 0 | 0 | 0 | 0 |
| 0 | 0 | 0 | 0 | 0 |
| 0 | 0 | 0 | 0 | 0 |
| 0 | 0 | 0 | 0 | 0 |
| 0 | 0 | 0 | 0 | 0 |
| 0 | 0 | 0 | 0 | 0 |
| 0 | 0 | 0 | 0 | 0 |
| 0 | 0 | 0 | 0 | 0 |
| 0 | 0 | 0 | 0 | 0 |
| 0 | 0 | 0 | 0 | 0 |
| 16.99480248 | 0.00108245 | 4.853210387 | 0.088771521 | 41.216201 |
| 0 | 0 | 0 | 0 | 0 |
| 0 | 0 | 0 | 0 | 0 |
| 0 | 0 | 0 | 0 | 0 |
| 0.002069425 | 0.013889799 | 0 | 0 | 0.040120438 |
| 0 | 0 | 0 | 0 | 0 |
| 0.109939139 | 0.549756978 | 0.066625322 | 0.330201871 | 0.006623239 |
| 0 | 0 | 0 | 0 | 0 |
| 0 | 0 | 0 | 0 | 0 |
| 0 | 0 | 0 | 0 | 0 |
| 0 | 0 | 0 | 0 | 0 |
| 0.086836321 | 0.003055506 | 0.024338831 | 0 | 0.507476831 |
| 0 | 0 | 0 | 0 | 0 |
| 0 | 0 | 0 | 0 | 0 |

| | | | | |
|---|---|---|---|---|
| 0.001419125 | 0 | 0 | 0 | 0 |
| 0 | 0 | 0 | 0 | 0 |
| 0 | 0 | 0 | 0 | 0 |
| 0 | 0 | 0 | 0 | 0 |
| 0.003222448 | 0 | 0 | 0 | 0 |
| 0 | 0 | 0 | 0 | 0 |
| 0 | 0 | 0 | 0 | 0 |
| 0 | 0.022677652 | 0 | 0 | 0.000482137 |
| 0 | 0 | 0 | 0 | 0.002719624 |
| 0 | 0 | 0.017997704 | 0 | 0.026466156 |
| 0 | 0.023111577 | 0 | 0.002220331 | 0.008281684 |
| 0 | 0 | 0 | 0 | 0 |
| 0 | 0.001557761 | 0 | 0 | 0 |
| 0 | 0 | 0 | 0 | 0 |
| 0 | 0 | 0 | 0 | 0 |
| 0 | 0 | 0 | 0 | 0 |
| 0 | 0 | 0 | 0 | 0 |
| 0 | 0 | 0 | 0 | 0 |
| 0 | 0 | 0 | 0 | 0 |
| 0.122008693 | 0.039342428 | 0.186879189 | 0.055404061 | 0.006831515 |
| 0 | 0 | 0 | 0 | 0 |
| 0 | 0 | 0 | 0 | 0 |
| 0 | 0 | 0 | 0 | 0 |
| 0 | 0 | 0 | 0 | 0 |
| 0 | 0 | 0 | 0 | 0 |
| 0 | 0 | 0 | 0.008237076 | 0 |
| 0 | 0 | 0 | 0 | 0 |
| 0 | 0 | 0 | 0 | 0 |
| 0 | 0 | 0 | 0 | 0 |
| 0 | 0.001852762 | 0 | 0 | 0 |
| 0 | 0 | 0 | 0 | 0 |
| 0 | 0 | 0 | 0.001068734 | 0 |
| 0 | 0 | 0 | 0 | 0 |
| 0 | 0 | 0 | 0 | 0 |
| 0 | 0 | 0 | 0 | 0 |
| 0 | 0 | 0 | 0 | 0 |
| 0 | 0 | 0 | 0 | 0 |
| 0 | 0 | 0 | 0 | 0 |
| 0 | 0 | 0 | 0 | 0 |
| 0 | 0 | 0 | 0.048026264 | 0 |
| 0 | 0 | 0 | 0 | 0 |
| 0 | 0 | 0 | 0 | 0 |
| 0.058972943 | 0.021898029 | 0 | 0.000666071 | 0 |
| 0 | 0 | 0 | 0 | 0 |
| 0.001273278 | 0.002116185 | 0 | 0.00096764 | 0 |
| 0 | 0.009492382 | 0 | 0.007546676 | 0 |
| 0 | 0 | 0 | 0.000351678 | 0 |
| 0 | 0 | 0 | 0 | 0 |

| | | | | |
|---|---|---|---|---|
| 0 | 0 | 0 | 0 | 0 |
| 0 | 0 | 0 | 0 | 0 |
| 0 | 0 | 0 | 0 | 0 |
| 0 | 0 | 0 | 0 | 0 |
| 0 | 0 | 0 | 0.008041196 | 0 |
| 0.001742678 | 0 | 0 | 0.004173756 | 0.003104694 |
| 0 | 0 | 0 | 0 | 0 |
| 0 | 0 | 0 | 0 | 0 |
| 0 | 0 | 0 | 0 | 0 |
| 0 | 0 | 0 | 0 | 0 |
| 0 | 0 | 0 | 0 | 0 |
| 0 | 0 | 0 | 0 | 0 |
| 0 | 0 | 0 | 0 | 0 |
| 0 | 0 | 0 | 0.013135551 | 0 |
| 0 | 0 | 0 | 0 | 0 |
| 0 | 0 | 0 | 0 | 0 |
| 0 | 0 | 0 | 0.000876243 | 0 |
| 0 | 0.000748318 | 0 | 0 | 0 |
| 0 | 0 | 0 | 0 | 0 |
| 0 | 0 | 0 | 0 | 0 |
| 0 | 0 | 0 | 0 | 0 |
| 0 | 0 | 0 | 0 | 0 |
| 0 | 0 | 0 | 0 | 0 |
| 0 | 0 | 0 | 0 | 0 |
| 0 | 0 | 0 | 0 | 0 |
| 0 | 0 | 0 | 0 | 0.000693752 |
| 0 | 0 | 0 | 0 | 0 |
| 0 | 0 | 0 | 0 | 0 |
| 0 | 0 | 0 | 0 | 0 |
| 0.007446494 | 0 | 0 | 0 | 0 |
| 0 | 0 | 0 | 0 | 0 |
| 0 | 0 | 0 | 0.012248471 | 0 |
| 0 | 0 | 0 | 0 | 0 |
| 0 | 0 | 0 | 0 | 0 |
| 0 | 0 | 0 | 0 | 0 |
| 0 | 0 | 0 | 0 | 0 |
| 0 | 0 | 0 | 0 | 0 |
| 0 | 0 | 0 | 0 | 0 |
| 0.987780356 | 0 | 0 | 0.163266164 | 0 |
| 0 | 0 | 0 | 0 | 0 |
| 0 | 0 | 0 | 0 | 0 |
| 0 | 0 | 0 | 0 | 0 |
| 0 | 0 | 0 | 0 | 0 |
| 0 | 0 | 0 | 0 | 0 |
| 0 | 0 | 0 | 0 | 0 |
| 0 | 0 | 0 | 0 | 0 |
| 0 | 0 | 0 | 0 | 0 |
| 0 | 0 | 0 | 0 | 0 |

| | | | | |
|---|---|---|---|---|
| 0 | 0 | 0 | 0 | 0 |
| 0 | 0 | 0 | 0 | 0 |
| 0 | 0 | 0 | 0 | 0 |
| 0 | 0 | 0 | 0 | 0 |
| 0 | 0 | 0 | 0 | 0 |
| 0 | 0 | 0 | 0.001355512 | 0 |
| 0 | 0 | 0 | 0 | 0 |
| 0 | 0 | 0 | 0 | 0 |
| 0 | 0 | 0 | 0 | 0 |
| 0 | 0 | 0.002153403 | 0 | 0 |
| 0 | 0 | 0.037121656 | 0.006001067 | 0.001820595 |
| 0 | 0 | 0 | 0 | 0 |
| 0 | 0.263096051 | 0 | 0.003986747 | 0.237637352 |
| 0 | 0 | 0 | 0 | 0 |
| 0 | 0 | 0 | 0 | 0 |
| 0 | 0 | 0 | 0 | 0 |
| 0 | 0 | 0 | 0 | 0 |
| 0 | 0 | 0 | 0 | 0 |
| 0 | 0 | 0 | 0 | 0 |
| 0 | 0 | 0 | 0 | 0 |
| 0 | 0 | 0 | 0 | 0.008051105 |
| 0 | 0 | 0 | 0 | 0 |
| 0 | 0 | 0 | 0 | 0 |
| 0 | 3.837273629 | 0.591262693 | 0.530475458 | 1.21199613 |
| 0 | 0 | 0 | 0 | 0 |
| 0 | 0 | 0 | 0 | 0 |
| 0 | 0 | 0 | 0 | 0 |
| 0 | 0 | 0 | 0 | 0 |
| 0 | 0 | 0 | 0 | 0 |
| 0 | 0 | 0 | 0 | 0 |
| 0.003239049 | 0 | 0 | 0 | 0 |
| 0 | 0 | 0 | 0 | 0 |
| 0 | 0 | 0 | 0.014698085 | 0 |
| 0 | 0 | 0 | 0 | 0 |

| PHL018-0252_V3 | PHL018-0213_V1 | PHL018-0121_V2 | PHL018-0249_V3 | PHL018-0116_V2 |
|---|---|---|---|---|
| REF | REF | EYCF | REF | CTRL |
| 0.126228743 | 0.008445553 | 0.186055706 | 0.041092258 | 0.22114521 |
| 5.906607084 | 0.111584467 | 0.066062896 | 0.563208615 | 2.65536983 |
| 0 | 0.010258459 | 5.161817114 | 0 | 0 |
| 0 | 0 | 0 | 0 | 0 |
| 0 | 0 | 0 | 0 | 0 |
| 0 | 0 | 0 | 0 | 0 |
| 0 | 0 | 0 | 0 | 0 |
| 0 | 0 | 0 | 0 | 0 |
| 0 | 0 | 0 | 0 | 0 |
| 0 | 0 | 0 | 0 | 0 |
| 0 | 0 | 0 | 0 | 0 |
| 0 | 0 | 0 | 0 | 0 |
| 0 | 0 | 0 | 0 | 0 |
| 11.44320625 | 0.211052202 | 6.236684794 | 0.76167807 | 3.347623421 |
| 0 | 0 | 0 | 0 | 0 |
| 0 | 0 | 0 | 0 | 0 |
| 0 | 0 | 0 | 0 | 0 |
| 0 | 0 | 0.000403494 | 0 | 0 |
| 0 | 0 | 0 | 0 | 0 |
| 0 | 0 | 0 | 0 | 0 |
| 0 | 0 | 0 | 0 | 0 |
| 0.024108423 | 0 | 0 | 0 | 0 |
| 0 | 0 | 0 | 0 | 0 |
| 0 | 0 | 0 | 0 | 0 |
| 0 | 0.009361284 | 0 | 0 | 0 |
| 0 | 0 | 0 | 0 | 0 |
| 0 | 0 | 0 | 0 | 0 |
| 0 | 0 | 0 | 0 | 0 |
| 0 | 0 | 0 | 0 | 0 |
| 0 | 0.001921767 | 0 | 0 | 0 |
| 0 | 0 | 0 | 0 | 0 |
| 0 | 0 | 0 | 0 | 0 |
| 0 | 0 | 0 | 0 | 0 |
| 0 | 0 | 0 | 0 | 0 |
| 0 | 0 | 0 | 0 | 0 |
| 0 | 1.55098864 | 0.001535341 | 5.059621927 | 5.25643185 |
| 0 | 0 | 0 | 0 | 0 |
| 0 | 0 | 0 | 0 | 0 |
| 0 | 0 | 0 | 0 | 0 |
| 0 | 0 | 0 | 0 | 0 |
| 0 | 0 | 0 | 0 | 0 |
| 0 | 0.008180877 | 0.036738783 | 0 | 0.00108554 |
| 0.168920799 | 3.881623849 | 0.000931008 | 1.009613068 | 0.038507829 |
| 0 | 0.07149003 | 0.000574833 | 0.00104988 | 0.120507023 |
| 0 | 0.02311714 | 0 | 0.001135389 | 0 |
| 0 | 0 | 0 | 0 | 0 |

| | | | | |
|---|---|---|---|---|
| 0 | 0.005502915 | 0 | 0.004904964 | 0.005322307 |
| 0.1035882 | 0 | 0.000886837 | 0.003543154 | 0 |
| 0.043987891 | 0.001391216 | 0.007821533 | 0.003069112 | 0.014191404 |
| 0.580600357 | 0.292707431 | 0.137944482 | 1.310356511 | 0.339831005 |
| 0 | 0 | 0.004603864 | 0 | 0 |
| 0 | 0.002983854 | 0.021151693 | 0 | 0 |
| 0 | 0 | 0.307660191 | 0 | 0.000981147 |
| 0 | 0 | 0 | 0 | 0 |
| 0 | 0 | 0 | 0 | 0 |
| 0 | 0 | 0 | 0 | 0 |
| 0 | 0 | 0.000727807 | 0 | 0.000676113 |
| 0.218582954 | 0.031500365 | 0 | 0.495312814 | 0 |
| 0 | 0 | 0 | 0 | 0.001274965 |
| 0.02572579 | 0 | 0.010211272 | 0.008418384 | 0.01932334 |
| 0 | 0 | 0 | 0 | 0 |
| 0 | 0 | 0 | 0 | 0 |
| 0 | 0.557743229 | 0 | 0.152692515 | 0.000722461 |
| 0 | 0 | 0 | 0 | 0 |
| 0 | 0 | 0 | 0 | 0 |
| 0 | 0 | 0 | 0 | 0 |
| 0.140673677 | 0 | 0.016904599 | 0.013275032 | 0.063483966 |
| 0.044264271 | 0 | 0.003827236 | 0.023899966 | 0.047928204 |
| 0 | 0.058029093 | 0 | 0.026518357 | 0 |
| 0 | 0 | 0 | 0 | 0 |
| 0 | 0 | 0 | 0 | 0 |
| 0 | 0 | 0 | 0 | 0 |
| 0 | 0 | 0 | 0 | 0 |
| 0 | 0 | 0 | 0 | 0 |
| 0 | 0 | 0 | 0 | 0 |
| 0 | 0 | 0 | 0 | 0 |
| 0.0857511 | 0 | 0.045917791 | 0 | 0 |
| 0 | 0 | 0 | 0 | 0 |
| 0 | 0 | 0 | 0 | 0 |
| 0 | 0 | 0 | 0 | 0 |
| 0 | 0 | 0 | 0 | 0 |
| 0 | 0 | 0 | 0 | 0 |
| 0 | 0 | 0 | 0 | 0 |
| 0 | 29.0079899 | 0 | 7.916392287 | 0 |
| 0 | 0 | 0 | 0 | 0 |
| 0 | 0 | 0 | 0 | 0 |
| 0 | 0 | 0 | 0 | 0 |
| 0 | 0 | 0 | 0 | 0 |
| 0 | 0 | 0 | 0 | 0 |
| 0 | 0 | 0 | 0 | 0 |
| 0 | 0 | 0 | 0 | 0 |
| 0 | 0 | 0 | 0 | 0 |
| 0 | 0 | 0 | 0 | 0 |
| 0 | 0 | 0 | 0.001557852 | 0 |

| | | | | |
|---|---|---|---|---|
| 0 | 0 | 0 | 0 | 0 |
| 0.001874406 | 0 | 0 | 0 | 0 |
| 0 | 0 | 0 | 0 | 0 |
| 0 | 0 | 0 | 0 | 0 |
| 0 | 0 | 0 | 0 | 0 |
| 0 | 0 | 0 | 0 | 0 |
| 0 | 0 | 0 | 0 | 0 |
| 0 | 0 | 0 | 0 | 0 |
| 0.071768198 | 0 | 0 | 0 | 0.000673838 |
| 0 | 0 | 0 | 0 | 0 |
| 0 | 0 | 0 | 0 | 0 |
| 0 | 0 | 0.003789566 | 0 | 0 |
| 0 | 0 | 0 | 0 | 0 |
| 0 | 0 | 0 | 0 | 0 |
| 0 | 0 | 0 | 0 | 0 |
| 0 | 0 | 0 | 0 | 0 |
| 0 | 0 | 0 | 0 | 0 |
| 0 | 0 | 0 | 0 | 0 |
| 0 | 1.361471306 | 0.025856111 | 0.212507257 | 0.098689422 |
| 0 | 0 | 0 | 0 | 0 |
| 0 | 0 | 0 | 0 | 0 |
| 0 | 0 | 0 | 0 | 0 |
| 0 | 0 | 0 | 0 | 0 |
| 0 | 0 | 0 | 0 | 0 |
| 0 | 0 | 0 | 0 | 0 |
| 0 | 0 | 0 | 0 | 0 |
| 0 | 0 | 0 | 0 | 0 |
| 0 | 0 | 0.002489973 | 0 | 0 |
| 0 | 0 | 0 | 0 | 0 |
| 0 | 0 | 0 | 0 | 0 |
| 0 | 0.002005764 | 0.003121195 | 0.105506668 | 0.257214404 |
| 0 | 0.015781655 | 0.004541917 | 0.003978897 | 0 |
| 0 | 0 | 0 | 0 | 0 |
| 0 | 0 | 0 | 0 | 0 |
| 0 | 0 | 0 | 0 | 0 |
| 0 | 0 | 0 | 0 | 0 |
| 0 | 0 | 0 | 0 | 0 |
| 0 | 0 | 0 | 0 | 0 |
| 0 | 0.055158224 | 0 | 0.455895422 | 0 |
| 0 | 0 | 0 | 0 | 0 |
| 0 | 0 | 0 | 0 | 0 |
| 0 | 0 | 0 | 0 | 0 |
| 0.245461696 | 0 | 0 | 0.002647895 | 0 |
| 0 | 0 | 0 | 0 | 0 |
| 0 | 0 | 0 | 0 | 0 |
| 0 | 0 | 0 | 0 | 0 |
| 0 | 0 | 0 | 0 | 0 |
| 0 | 0 | 0 | 0 | 0 |

| | | | | |
|---|---|---|---|---|
| 0 | 0 | 0 | 0 | 0 |
| 0 | 0 | 0 | 0 | 0 |
| 0 | 0 | 0 | 0 | 0 |
| 0 | 0 | 0 | 0 | 0 |
| 0 | 0.012951222 | 0 | 0.080072912 | 0.005693708 |
| 0 | 0 | 0 | 0 | 0 |
| 0 | 0 | 0 | 0 | 0 |
| 0 | 0 | 0 | 0 | 0 |
| 0 | 0 | 0 | 0 | 0 |
| 0 | 0 | 0 | 0 | 0 |
| 0 | 0 | 0 | 0 | 0 |
| 0 | 0 | 0 | 0 | 0 |
| 0 | 0 | 0 | 0 | 0 |
| 0 | 0 | 0 | 0 | 0 |
| 0 | 0 | 0 | 0 | 0 |
| 0 | 0 | 0 | 0 | 0 |
| 0 | 0 | 0 | 0 | 0 |
| 0 | 0 | 0 | 0 | 0 |
| 0 | 0 | 0 | 0 | 0 |
| 0 | 0 | 0 | 0 | 0 |
| 0 | 0 | 0 | 0 | 0 |
| 0 | 0 | 0 | 0 | 0 |
| 0 | 0 | 0 | 0 | 0 |
| 0 | 0 | 0 | 0 | 0 |
| 0 | 0.005815892 | 0 | 0 | 0 |
| 0 | 0 | 0 | 0 | 0 |
| 0 | 0 | 0.000688428 | 0 | 0 |
| 0 | 0 | 0 | 0 | 0 |
| 0 | 0 | 0 | 0 | 0 |
| 0 | 0 | 0 | 0 | 0 |
| 0.036879476 | 0 | 0 | 0 | 0 |
| 0.002125609 | 0 | 0.000609348 | 0.474242382 | 0 |
| 0 | 0 | 0 | 0 | 0 |
| 0 | 0 | 0 | 0 | 0 |
| 0 | 0 | 0 | 0 | 0 |
| 0 | 0 | 0 | 0 | 0 |
| 0 | 0 | 0 | 0 | 0 |
| 0 | 0.020405145 | 0 | 0 | 0 |
| 0 | 0 | 0 | 0 | 0 |
| 0 | 0 | 0 | 0 | 0 |
| 0 | 0 | 0 | 0 | 0 |
| 0 | 0 | 0 | 0 | 0 |
| 0 | 0 | 0 | 0 | 0 |
| 0 | 0 | 0 | 0 | 0 |
| 0 | 0 | 0 | 0 | 0 |
| 0 | 0 | 0 | 0 | 0 |
| 0 | 0 | 0 | 0 | 0 |
| 0 | 0 | 0 | 0 | 0 |

| | | | | |
|---|---|---|---|---|
| 0 | 0.001995373 | 0 | 0 | 0 |
| 0 | 0 | 0 | 0 | 0 |
| 0 | 0 | 0 | 0 | 0 |
| 0 | 0 | 0 | 0 | 0 |
| 0 | 0 | 0 | 0 | 0 |
| 0 | 0 | 0 | 0 | 0 |
| 0 | 0 | 0 | 0 | 0 |
| 0 | 0 | 0 | 0 | 0 |
| 0 | 0 | 0 | 0 | 0 |
| 0 | 0 | 0 | 0 | 0 |
| 0 | 0 | 0 | 0 | 0 |
| 0 | 0 | 0 | 0 | 0 |
| 0 | 0 | 0 | 0.001064954 | 0 |
| 0 | 0 | 0 | 0 | 0 |
| 0 | 0 | 0 | 0 | 0 |
| 0 | 0 | 0 | 0 | 0 |
| 0 | 0 | 0 | 0.001316152 | 0 |
| 0 | 0 | 0 | 0 | 0 |
| 0 | 0 | 0 | 0 | 0 |
| 0 | 0 | 0 | 0 | 0 |
| 0.062578114 | 0.010859482 | 0.003270693 | 0.047540087 | 0.100266803 |
| 0 | 0 | 0 | 0 | 0 |
| 0 | 0 | 0 | 0 | 0 |
| 0 | 0 | 0 | 0 | 0 |
| 0 | 0 | 0 | 0 | 0 |
| 0 | 0 | 0 | 0 | 0 |
| 0.003422919 | 0.004878751 | 0 | 0.018844028 | 0 |
| 0 | 0 | 0 | 0 | 0 |
| 0 | 0.001015103 | 0 | 0 | 0 |
| 0 | 0 | 0.001766171 | 0 | 0 |
| 0.001806389 | 0 | 0 | 0 | 0 |
| 0 | 0 | 0 | 0 | 0 |
| 0 | 0 | 0 | 0 | 0 |
| 0 | 0.1035316 | 0 | 0.006845077 | 0.008046454 |
| 0.002249539 | 0 | 0.000993107 | 0.004048706 | 0.031630994 |
| 0 | 0 | 0 | 0 | 0 |
| 0 | 0 | 0 | 0 | 0 |
| 0 | 0 | 0 | 0 | 0 |
| 0 | 0 | 0.001650147 | 0.000470913 | 0 |
| 0 | 0 | 0 | 0.005382948 | 0 |
| 0 | 0 | 0 | 0 | 0 |
| 0 | 0 | 0 | 0 | 0 |
| 0 | 0 | 0 | 0 | 0 |
| 0 | 0 | 0 | 0 | 0 |
| 0 | 0 | 0 | 0 | 0 |
| 0 | 0 | 0 | 0.004471579 | 0 |
| 0 | 0 | 0 | 0 | 0.001426998 |
| 0.000801541 | 0 | 0 | 0.002423583 | 0.001252283 |

| | | | | |
|---|---|---|---|---|
| 0 | 0 | 0 | 0 | 0 |
| 0 | 0 | 0 | 0 | 0 |
| 0 | 0 | 0 | 0 | 0 |
| 0 | 0 | 0 | 0 | 0 |
| 0 | 0 | 0 | 0 | 0 |
| 0 | 0 | 0 | 0 | 0 |
| 0 | 0 | 0 | 0 | 0 |
| 0 | 0 | 0 | 0 | 0 |
| 0 | 0 | 0 | 0 | 0 |
| 0 | 0 | 0 | 0 | 0 |
| 0 | 0 | 0 | 0 | 0 |
| 0 | 0 | 0 | 0 | 0 |
| 0 | 0 | 0 | 0 | 0 |
| 0 | 0 | 0 | 0 | 0 |
| 0 | 0 | 0 | 0 | 0 |
| 0 | 0 | 0 | 0 | 0 |
| 0 | 0 | 0 | 0 | 0 |
| 0 | 0 | 0 | 0 | 0 |
| 0 | 0.024287299 | 0 | 0.003286357 | 0 |
| 0.076786165 | 1.372828129 | 0 | 0 | 0 |
| 0.024770409 | 0 | 0.002730066 | 0 | 0 |
| 0 | 0 | 0 | 0 | 0 |
| 0 | 0 | 0 | 0 | 0 |
| 0 | 0 | 0 | 0 | 0 |
| 0.002388915 | 0.003430188 | 0 | 0 | 0.010730387 |
| 0 | 0.014354615 | 0 | 0 | 0 |
| 0 | 0 | 0 | 0 | 0 |
| 0 | 0 | 0 | 0 | 0 |
| 0 | 0 | 0.003738266 | 0 | 0 |
| 0 | 0 | 0 | 0 | 0 |
| 4.02398358 | 0 | 0.051300636 | 0.048502928 | 0.04230417 |
| 0 | 0 | 0 | 0 | 0 |
| 0 | 0 | 0.008597556 | 0 | 0 |
| 0 | 0 | 0 | 0 | 0 |
| 0 | 0 | 0 | 0 | 0 |
| 0 | 0 | 0 | 0 | 0 |
| 0 | 0 | 0 | 0 | 0 |
| 0 | 0 | 0 | 0.00267087 | 0 |
| 0 | 0 | 0 | 0 | 0 |
| 0 | 0.320035979 | 0 | 0 | 0 |
| 0 | 0 | 0 | 0 | 0 |
| 0 | 0.026043522 | 0.028760857 | 0.00116789 | 0 |
| 0 | 0 | 0 | 0 | 0 |
| 0 | 0 | 0 | 0 | 0 |
| 0 | 0 | 0 | 0 | 0 |
| 0 | 0 | 0 | 0 | 0 |
| 0 | 0 | 0 | 0 | 0 |
| 0 | 0 | 0 | 0 | 0 |

| | | | | |
|---|---|---|---|---|
| 0 | 0 | 0 | 0 | 0 |
| 0 | 0 | 0 | 0 | 0 |
| 0 | 0.061544622 | 0 | 0.00081471 | 0.137245693 |
| 0 | 0 | 0 | 0 | 0 |
| 3.05097828 | 0 | 0.002973032 | 0.077631652 | 0 |
| 0 | 0 | 0 | 0 | 0 |
| 0 | 0 | 0 | 0 | 0 |
| 0 | 0 | 0 | 0 | 0 |
| 0 | 0 | 0 | 0 | 0 |
| 0 | 0 | 0 | 0 | 0.000606395 |
| 0 | 0 | 0 | 0 | 0 |
| 0 | 0 | 0 | 0 | 0 |
| 0 | 0 | 0 | 0 | 0 |
| 0.015758834 | 0 | 0.000631027 | 0.003421526 | 0 |
| 0.003716244 | 0 | 0 | 0.001337695 | 0.009878355 |
| 0 | 0.127351533 | 0 | 0.006206107 | 0.003237577 |
| 0 | 0.002955542 | 0 | 0.000846768 | 0.002297041 |
| 0 | 0.002783949 | 0 | 0 | 0 |
| 0 | 0 | 0 | 0 | 0 |
| 0 | 0 | 0 | 0 | 0 |
| 0 | 0 | 0 | 0 | 0 |
| 0 | 0 | 0 | 0.001834536 | 0.000829429 |
| 0 | 0 | 0 | 0 | 0 |
| 0.030286272 | 0 | 0 | 0 | 0 |
| 0.002042992 | 0 | 0 | 0 | 0 |
| 0 | 0 | 0 | 0 | 0 |
| 0 | 0 | 0.000705905 | 0.037335139 | 0 |
| 0 | 0 | 0 | 0 | 0 |
| 0 | 0 | 0 | 0 | 0 |
| 0.00276456 | 0 | 0 | 0 | 0 |
| 0 | 0 | 0 | 0 | 0 |
| 0 | 0 | 0 | 0 | 0 |
| 0 | 0 | 0 | 0 | 0 |
| 0 | 0 | 0.00154417 | 0 | 0 |
| 0 | 0 | 0 | 0 | 0 |
| 0 | 0 | 0 | 0 | 0 |
| 0 | 0 | 0 | 0 | 0 |
| 0.001867614 | 0.497065473 | 0 | 0 | 0.006773595 |
| 0 | 0 | 0 | 0 | 0 |
| 0 | 0 | 0 | 0 | 0 |
| 0 | 0 | 0 | 0.004712725 | 0 |
| 0 | 0 | 0 | 0 | 0 |
| 0 | 0 | 0 | 0 | 0 |
| 0 | 0 | 0 | 0 | 0 |
| 0 | 0 | 0 | 0 | 0 |
| 0 | 0 | 0 | 0 | 0 |
| 0 | 0 | 0 | 0 | 0 |
| 0.025853131 | 0 | 0 | 0 | 0 |

| | | | | |
|---|---|---|---|---|
| 0 | 0 | 0 | 0 | 0 |
| 0 | 0 | 0 | 0 | 0.193085391 |
| 0 | 0.003792581 | 0.001122222 | 0 | 0 |
| 0 | 0 | 0 | 0 | 0 |
| 0.004923187 | 0 | 0 | 0.003060976 | 0.003933263 |
| 0 | 0 | 0 | 0 | 0 |
| 0 | 0 | 0 | 0 | 0 |
| 0.001882072 | 0.018297659 | 0.010682758 | 0.061875434 | 0.001531482 |
| 0 | 0 | 0 | 0 | 0 |
| 0.110573866 | 0 | 0.260592482 | 0 | 0.007576951 |
| 0 | 0 | 0 | 0 | 0 |
| 0.013905313 | 0 | 0.017382825 | 0 | 0 |
| 0 | 0.005408985 | 0.000712729 | 0.000813584 | 0.011402948 |
| 0.002002399 | 0 | 0 | 0 | 0 |
| 0 | 0 | 0 | 0 | 0 |
| 0 | 0 | 0.001069925 | 0 | 0 |
| 0 | 0 | 0 | 0 | 0 |
| 0 | 0 | 0.02722828 | 0 | 0 |
| 0.817476475 | 0 | 0.036071989 | 0 | 0 |
| 0 | 0 | 0 | 0 | 0 |
| 0 | 0 | 0 | 0 | 0 |
| 0 | 0 | 0 | 0 | 0 |
| 0 | 0 | 0 | 0 | 0 |
| 0 | 0.014748233 | 0 | 0.022310094 | 0 |
| 0 | 0 | 0 | 0 | 0 |
| 0 | 0.0124033 | 0 | 0 | 0 |
| 0 | 4.334139887 | 0 | 2.758602696 | 0 |
| 0.541197555 | 0 | 0.003011638 | 0 | 0 |
| 0 | 0 | 0.288983533 | 0 | 0 |
| 0 | 0 | 0 | 0 | 0 |
| 0.00117704 | 0 | 0 | 0 | 0 |
| 0 | 0 | 0 | 0 | 0 |
| 0 | 0 | 0 | 0 | 0 |
| 0.525619796 | 0 | 0.023884976 | 0 | 0 |
| 0 | 0 | 0 | 0 | 0 |
| 0.008453644 | 0 | 0 | 0 | 0 |
| 0.003730367 | 0 | 0.004234759 | 0 | 0 |
| 0 | 0 | 0 | 0 | 0 |
| 0 | 0 | 0 | 0 | 0 |
| 0.640283873 | 0 | 0.016879991 | 0 | 0 |
| 0 | 0 | 0 | 0 | 0 |
| 0.323088886 | 0 | 0 | 0 | 0 |
| 0 | 0 | 0 | 0 | 0 |
| 0 | 0 | 0 | 0 | 0 |
| 0.1403802 | 0 | 0 | 0 | 0 |
| 0 | 0 | 0 | 0 | 0 |
| 0 | 0 | 0 | 0 | 0 |
| 0 | 0 | 0.00140556 | 0 | 0 |

| | | | | |
|---|---|---|---|---|
| 0.014955676 | 0 | 0 | 0 | 0 |
| 0 | 0 | 0.112103923 | 0 | 0 |
| 0 | 0 | 0 | 0 | 0 |
| 0 | 0 | 0 | 0 | 0 |
| 0 | 0 | 0 | 0 | 0 |
| 0.003652912 | 0 | 0.000406552 | 0 | 0 |
| 0.003583974 | 0 | 0 | 0 | 0.013065271 |
| 0 | 0 | 0 | 0 | 0 |
| 0 | 0 | 0 | 0 | 0 |
| 0 | 0 | 0 | 0 | 0 |
| 0.009221189 | 0.009169935 | 0 | 0.008073845 | 0.000973424 |
| 0 | 0 | 0 | 0 | 0 |
| 0 | 0 | 0 | 0 | 0 |
| 0.005415391 | 0 | 0.001039453 | 0.002712094 | 0.028968723 |
| 0.349728816 | 0 | 0.036209877 | 0 | 0 |
| 0 | 0 | 0 | 0 | 0 |
| 0 | 0 | 0 | 0 | 0 |
| 0 | 0 | 0 | 0 | 0 |
| 0.256366074 | 0 | 0 | 0 | 0 |
| 0 | 0 | 0 | 0 | 0 |
| 0 | 0 | 0 | 0 | 0 |
| 0 | 0.041568143 | 0 | 0.517249159 | 0 |
| 0 | 0 | 0 | 0 | 0 |
| 0 | 0 | 0 | 0 | 0 |
| 0 | 0 | 0 | 0 | 0 |
| 0 | 0 | 0 | 0 | 0 |
| 0.157515206 | 0 | 0.033167228 | 0 | 0 |
| 0 | 0 | 0 | 0 | 0 |
| 0 | 0 | 0 | 0 | 0 |
| 1.304219804 | 0.161055077 | 0.405500499 | 0.251690052 | 0.552007043 |
| 0.001597353 | 0.007884258 | 0 | 0 | 0 |
| 0 | 0 | 0 | 0 | 0 |
| 0 | 0 | 0 | 0 | 0 |
| 0.00067872 | 0 | 0 | 0 | 0 |
| 0 | 0 | 0.008046168 | 0 | 0 |
| 0 | 0 | 0 | 0 | 0 |
| 0 | 0.30814681 | 0 | 0.232303432 | 0 |
| 0 | 0 | 0.000756771 | 0 | 0 |
| 0 | 0 | 0 | 0 | 0 |
| 1.687757082 | 0 | 0 | 0 | 0 |
| 0.000967835 | 0 | 0.00305194 | 0 | 0.185798253 |
| 6.655035685 | 0.2016889 | 0 | 1.754706562 | 2.660258775 |
| 0 | 0 | 0.000831901 | 0 | 0 |
| 0 | 0 | 0 | 0 | 0 |
| 0.003930366 | 0 | 0 | 0 | 0 |
| 0 | 0 | 0 | 0 | 0 |
| 0.061151894 | 0 | 0.001665679 | 0.002632683 | 0 |
| 0 | 0 | 0 | 0 | 0 |

| | | | | |
|---|---|---|---|---|
| 0 | 0.020021043 | 0 | 0 | 0 |
| 0 | 0 | 0.005454799 | 0 | 0 |
| 0 | 0.00367823 | 0 | 0.004180152 | 0.003001648 |
| 0.002040716 | 0 | 0 | 0 | 0 |
| 0 | 0 | 0.000635675 | 0.06832988 | 0 |
| 0.225716437 | 0 | 0 | 0 | 0.013037999 |
| 0 | 0 | 0 | 0 | 0 |
| 0 | 0 | 0 | 0 | 0 |
| 0 | 0 | 0 | 0.003322644 | 0 |
| 0 | 0 | 0 | 0 | 0 |
| 0 | 0 | 0 | 0 | 0 |
| 0 | 0 | 0 | 0 | 0 |
| 0 | 0 | 0 | 0 | 0 |
| 0.0442208 | 0 | 0 | 0 | 0 |
| 0 | 0 | 0 | 0 | 0 |
| 0 | 0.013995591 | 0.035206802 | 0.083822278 | 0.005364508 |
| 0.000589288 | 0.104005287 | 0.001238828 | 0.009757488 | 0.002877096 |
| 0.000954616 | 0 | 0 | 0 | 0 |
| 0.00060869 | 0 | 0 | 0 | 0 |
| 0 | 0 | 0 | 0 | 0 |
| 0.097537373 | 0 | 0.027634572 | 0 | 0 |
| 0 | 0 | 0 | 0.001640316 | 0.000667456 |
| 0 | 0 | 0.054130639 | 0 | 0 |
| 0 | 0 | 0 | 0 | 0 |
| 0 | 0 | 0.00277873 | 0 | 0 |
| 0 | 0 | 0.015829789 | 0 | 0 |
| 0 | 0 | 0 | 0 | 0.013635986 |
| 0.002367167 | 0 | 0 | 0 | 0 |
| 0 | 0 | 0 | 0 | 0 |
| 0 | 0 | 0 | 0 | 0 |
| 0.28863829 | 0 | 0.000837225 | 0.023892415 | 0.020555076 |
| 0 | 0.001776137 | 0 | 0 | 0.021810474 |
| 0.013998553 | 0.008214023 | 2.096942746 | 0.034497095 | 0.021817989 |
| 0 | 0 | 0 | 0 | 0 |
| 0 | 0 | 0 | 0.000792942 | 0 |
| 0 | 0 | 0 | 0 | 0 |
| 0 | 0 | 0 | 0 | 0.003564753 |
| 0.001903329 | 0 | 0.010323262 | 0.001096192 | 0.00910682 |
| 0.088228969 | 0.067378031 | 0 | 0.046314899 | 0.210963243 |
| 0 | 0 | 0.003631301 | 0.002688746 | 0 |
| 0 | 0 | 0 | 0 | 0 |
| 0 | 0 | 0 | 0 | 0 |
| 0.004269638 | 0 | 0.000897583 | 0 | 0.001389718 |
| 0.679894224 | 0.000666688 | 0.029142068 | 0 | 0.302785175 |
| 0 | 0 | 0.008435028 | 0 | 0 |
| 0 | 0 | 0 | 0 | 0 |
| 0 | 0 | 0 | 0 | 0 |
| 0 | 0 | 0 | 0 | 0 |

| | | | | |
|---|---|---|---|---|
| 0.003668607 | 0 | 0 | 0 | 0 |
| 0.05161119 | 0.566976761 | 0 | 0.334028748 | 0.259580154 |
| 0 | 0 | 0 | 0 | 0 |
| 0 | 0 | 0 | 0 | 0 |
| 0 | 0 | 0 | 0 | 0 |
| 0 | 0 | 0 | 0 | 0 |
| 0 | 0 | 0 | 0 | 0 |
| 0 | 0 | 0.005276738 | 0 | 0 |
| 0 | 0 | 0 | 0 | 0 |
| 0.007161729 | 0 | 0 | 0 | 0 |
| 0.005010913 | 0 | 0 | 0 | 0.002446491 |
| 0.001722438 | 0.010562661 | 0.005612529 | 0.00537339 | 0.331054134 |
| 0 | 0.101593533 | 0 | 0 | 0 |
| 0 | 0 | 0 | 0 | 0 |
| 0 | 0 | 0.000836767 | 0 | 0.004534454 |
| 0 | 0 | 0 | 0 | 0 |
| 0.016750515 | 0.028615697 | 0 | 0.002469913 | 0.012020893 |
| 0.433841716 | 0 | 0.032879764 | 0 | 0 |
| 0 | 0 | 0 | 0 | 0 |
| 0.066515838 | 0 | 0.009977825 | 0 | 0.009468474 |
| 0.002289326 | 0.019050761 | 0 | 0.257706957 | 0.033328492 |
| 0 | 0 | 0 | 0 | 0.008092089 |
| 0 | 0 | 0 | 0 | 0 |
| 0 | 0 | 0 | 0 | 0 |
| 0 | 0 | 0 | 0 | 0 |
| 0 | 0 | 0 | 0 | 0 |
| 0.189402213 | 0 | 0.048583638 | 0 | 0 |
| 0.370291198 | 0.011315141 | 0.310540133 | 0.029460653 | 0.054214877 |
| 0 | 0 | 0 | 0 | 0 |
| 0.001334104 | 0 | 0 | 0.000960445 | 0 |
| 0 | 0.005751074 | 0 | 0.001537847 | 0 |
| 0 | 0 | 0 | 0 | 0 |
| 0 | 0 | 0.013379374 | 0 | 0 |
| 0 | 0 | 0 | 0 | 0 |
| 0 | 0.00813047 | 0 | 0.168747332 | 0.030304235 |
| 0.21339103 | 0.106166026 | 0.024791068 | 0.31452478 | 0.217142369 |
| 0 | 0 | 0 | 0 | 0 |
| 0 | 0 | 0 | 0 | 0 |
| 0.033563835 | 0 | 0.006195461 | 0 | 0.002398092 |
| 0 | 0 | 0 | 0 | 0 |
| 0 | 0 | 0.003023754 | 0 | 0 |
| 0 | 0 | 0 | 0 | 0 |
| 0.00331499 | 0 | 0 | 0 | 0.006473945 |
| 0.004160728 | 0.000430838 | 0.003128688 | 0 | 0.008594399 |
| 0.277936878 | 0 | 0 | 0 | 0 |
| 0 | 0.231326966 | 0 | 0.199379494 | 0.115437213 |
| 0 | 0 | 0 | 0 | 0 |
| 0 | 0 | 0 | 0 | 0 |

| | | | | |
|---|---|---|---|---|
| 0.219514717 | 0 | 0 | 0 | 0 |
| 0 | 0 | 0 | 0 | 0 |
| 0 | 0 | 0 | 0 | 0 |
| 0 | 0.283426595 | 0.000486995 | 0.017789052 | 0.669183615 |
| 0 | 0 | 0 | 0 | 0 |
| 0 | 0.00330503 | 0 | 0 | 0 |
| 0 | 0 | 0 | 0 | 0 |
| 0 | 0.005114506 | 0 | 0.003232575 | 0 |
| 0 | 0.06024129 | 0.317514082 | 0.175143805 | 2.355219552 |
| 0 | 0.030038418 | 0 | 0.204824318 | 0.028640718 |
| 0 | 0 | 0.002187873 | 0 | 0 |
| 0 | 0.005975497 | 0.004199344 | 0.000399465 | 0.000903028 |
| 0 | 0 | 0 | 0 | 0 |
| 0 | 0 | 0.000438415 | 0 | 0 |
| 0 | 0.009998963 | 0.126772581 | 0.058895541 | 0.01758627 |
| 0 | 0 | 0 | 0 | 0 |
| 0 | 0.178310759 | 0 | 0.024795829 | 11.98193633 |
| 0 | 0.220361596 | 0.017095097 | 0 | 0 |
| 0 | 0 | 0 | 0 | 0 |
| 0 | 0.004743361 | 0.000520851 | 0 | 0 |
| 0 | 0 | 0 | 0 | 0 |
| 0 | 0.000462318 | 0 | 0 | 0 |
| 0 | 0 | 0 | 0 | 0 |
| 0.504573057 | 0 | 0.077515347 | 0.256350178 | 0.157341815 |
| 0 | 0 | 0.004805362 | 0 | 0 |
| 0 | 0.025145349 | 0 | 0 | 0 |
| 0 | 0.058431043 | 0.031447012 | 0.299932302 | 0 |
| 0 | 0.423562238 | 0 | 2.478852933 | 0 |
| 0 | 0 | 0 | 0 | 0 |
| 0 | 0 | 0 | 0 | 0 |
| 0 | 0.185410365 | 0 | 0.01190933 | 0 |
| 0 | 0.017940411 | 0 | 0 | 0 |
| 0 | 0 | 0.160618059 | 0.089949105 | 0 |
| 0 | 0 | 0 | 0 | 0 |
| 0 | 0 | 0.006386873 | 0 | 0 |
| 0 | 0.175814903 | 0 | 0.084404272 | 0.036771315 |
| 0 | 0.484171648 | 0.036385125 | 0.646822012 | 1.53103512 |
| 0 | 0 | 0 | 0 | 0 |
| 0.025608065 | 0 | 0 | 0 | 0.023313294 |
| 0 | 0 | 0.053603055 | 0 | 0 |
| 0.147803059 | 0 | 0.007609427 | 0.002043813 | 0.192317242 |
| 0 | 0 | 0.043249474 | 0 | 0 |
| 0.437416279 | 0.034192694 | 49.90573032 | 0.045971331 | 7.715628435 |
| 0 | 0 | 0 | 0 | 0 |
| 0 | 0 | 0 | 0 | 0 |
| 0 | 0 | 0 | 0 | 0 |
| 0 | 0 | 0 | 0 | 0 |
| 0.001919671 | 0 | 0.000807124 | 0 | 0 |

| | | | | |
|---|---|---|---|---|
| 0 | 0 | 0 | 0 | 0 |
| 0 | 0.044642999 | 0.032476314 | 0.128702431 | 0 |
| 0.879216962 | 0 | 0.042167708 | 0.700528833 | 0.573614408 |
| 0 | 0.036893019 | 0 | 0 | 0 |
| 0 | 0 | 0 | 3.008897876 | 0 |
| 0 | 0.023850827 | 0.004876051 | 0.002551101 | 0 |
| 0 | 0 | 0 | 0 | 0 |
| 0 | 0 | 0 | 0 | 0 |
| 0 | 0 | 0 | 0 | 0 |
| 0 | 0 | 0 | 0 | 0 |
| 0.33706321 | 0.004323835 | 0.261321408 | 0.016331339 | 0.056455982 |
| 0.832762578 | 0.23461504 | 1.518675068 | 0.822362362 | 1.216930731 |
| 0 | 0 | 0 | 0 | 0 |
| 0.001083301 | 0.386732755 | 0 | 0.005771164 | 0 |
| 0.319447973 | 0 | 0.05351646 | 0 | 0.564007877 |
| 0 | 4.134200386 | 0 | 0 | 0.00361376 |
| 0 | 0 | 0 | 0 | 0 |
| 0 | 0 | 0 | 0 | 0 |
| 0 | 0.004513782 | 0 | 0.010604298 | 0 |
| 0 | 0 | 0.002398731 | 0.01250032 | 0 |
| 1.653659664 | 0 | 0.011672888 | 0 | 0.060955334 |
| 0.280301625 | 0 | 4.748541024 | 0.156227639 | 0.007617338 |
| 4.155906653 | 0.024531252 | 13.0209093 | 0 | 0.633777252 |
| 0 | 0 | 0 | 0 | 0 |
| 0 | 0.001035966 | 0 | 0 | 0 |
| 0 | 0.017732492 | 0 | 5.43485893 | 0.007875232 |
| 0 | 0 | 0.000761526 | 0.000434643 | 0 |
| 0 | 0 | 0 | 0 | 0 |
| 0 | 0 | 0 | 0 | 0 |
| 0 | 0 | 0 | 0 | 0 |
| 0 | 0.001498946 | 0 | 0 | 0 |
| 0.910371499 | 0.069709871 | 0.030455551 | 0.466471008 | 0 |
| 1.07789807 | 0 | 0.004551083 | 0.379241388 | 0 |
| 0 | 0.003104173 | 0.021021638 | 0.013809202 | 0 |
| 0.074707906 | 0 | 0.00042911 | 0.014401053 | 0 |
| 0.017574763 | 0.050000583 | 0.01473928 | 0.011171781 | 0.016430885 |
| 0 | 0 | 0.003758817 | 0.002730452 | 0.000529067 |
| 0 | 0 | 0 | 0 | 0 |
| 0.008313924 | 4.32017679 | 0.007589091 | 0.066153746 | 0 |
| 0 | 0 | 0 | 0 | 0 |
| 0 | 0.011228625 | 0 | 0.10915865 | 0 |
| 0 | 0 | 0 | 0 | 0 |
| 0 | 0.103346125 | 0 | 0.051799031 | 0 |
| 0 | 0 | 0 | 0 | 0 |
| 0.128780655 | 0.008610401 | 0.027969304 | 0.024712818 | 0.015406438 |
| 0 | 0.002070495 | 0 | 0.179146588 | 0 |
| 0 | 0.002298493 | 0 | 0 | 0 |
| 0 | 0.037622199 | 0 | 0 | 0.478673202 |

| | | | | |
|---|---|---|---|---|
| 0 | 0 | 0 | 0 | 0 |
| 0 | 0 | 0 | 0 | 0 |
| 0 | 0 | 0 | 0 | 0 |
| 0 | 0.001141562 | 0.00126067 | 0.001046591 | 0 |
| 0 | 0 | 0 | 0 | 0 |
| 0 | 0.245141473 | 0 | 0.561653393 | 0 |
| 0 | 0 | 0 | 3.967822244 | 0 |
| 0 | 0.012005066 | 0.047878207 | 0.001805724 | 0.038207627 |
| 0 | 0 | 0 | 0 | 0 |
| 0 | 0 | 0 | 0 | 0 |
| 0.009107487 | 0 | 0.004339799 | 0.003059765 | 0 |
| 0 | 0 | 0 | 0 | 0 |
| 0.037922309 | 0.038167122 | 0.027828191 | 0.007654467 | 0 |
| 0 | 0.015653842 | 0 | 0.454311423 | 0 |
| 0 | 0 | 0 | 0 | 0 |
| 0 | 0 | 0 | 0 | 0 |
| 0 | 0.093883065 | 0 | 0.00823572 | 0 |
| 0 | 0.010886495 | 0 | 0.000970356 | 0 |
| 0.208492739 | 0.038503141 | 0 | 0 | 0 |
| 0.003038891 | 0 | 0 | 0.001093876 | 0 |
| 0 | 0.007522506 | 0 | 0.000862085 | 0 |
| 0 | 0.031411566 | 0.000534068 | 0 | 0 |
| 0 | 0.013248647 | 0 | 0.030346122 | 0.003983244 |
| 0 | 0 | 0 | 0 | 0 |
| 0 | 0 | 0 | 0 | 0 |
| 0 | 0 | 0 | 0 | 0 |
| 0 | 0 | 0 | 0 | 0 |
| 0 | 0 | 0 | 0 | 0 |
| 0 | 0 | 0 | 0 | 0 |
| 0.554407665 | 0.394706307 | 0 | 0.215388328 | 0.048293115 |
| 0 | 0 | 0 | 0 | 0 |
| 0 | 0 | 0 | 0 | 0 |
| 0 | 0.011848472 | 0.000555109 | 0 | 0 |
| 0 | 0 | 0 | 0 | 0 |
| 0 | 0.004043813 | 0 | 0 | 0 |
| 0 | 0 | 0 | 0 | 0 |
| 0 | 0.029004905 | 0 | 0.134703337 | 0.082790803 |
| 0.008394949 | 0 | 0 | 0.002590145 | 0.017398514 |
| 0 | 0 | 0 | 0 | 0 |
| 0 | 0 | 0 | 0 | 0 |
| 0 | 0 | 0.025141763 | 0 | 0.181762713 |
| 0 | 0 | 0 | 0 | 0 |
| 0 | 0 | 0 | 0.002224705 | 0 |
| 0 | 0.150803968 | 0 | 0.008863569 | 0.998814642 |
| 0 | 0.024790809 | 0.189556337 | 0.058121389 | 0 |
| 0 | 0 | 0 | 0 | 0 |
| 0 | 0.000730448 | 0.010061272 | 0 | 0 |
| 0 | 0 | 0.002598399 | 0 | 0 |

| | | | | |
|---|---|---|---|---|
| 0 | 0 | 0 | 0 | 0 |
| 0 | 0 | 0 | 0 | 0 |
| 0 | 0.004803774 | 0 | 0 | 0 |
| 0 | 0.331568746 | 0.00365427 | 0.502036698 | 0 |
| 0 | 0 | 0 | 0 | 0 |
| 0 | 0.303469928 | 0.000371707 | 0 | 0 |
| 0 | 0 | 0 | 0 | 0 |
| 0 | 0 | 0 | 0 | 0 |
| 0.908521496 | 0 | 0.101631393 | 0.040145195 | 0.030917223 |
| 0 | 0.000683048 | 0 | 0 | 0 |
| 0 | 0.000454865 | 0 | 0 | 0 |
| 0 | 0.001551304 | 0 | 0.002903749 | 0 |
| 0 | 0 | 0 | 0 | 0 |
| 0 | 0 | 0 | 0 | 0 |
| 0 | 0.843749884 | 0.523560294 | 0.281892834 | 1.990328837 |
| 0 | 0 | 0 | 0 | 0 |
| 0 | 0 | 0.002363748 | 0 | 0 |
| 0 | 0 | 0 | 0 | 0 |
| 0 | 0 | 0 | 0 | 0 |
| 0 | 0 | 0 | 0 | 0 |
| 0 | 0.000810709 | 0.000569734 | 0.099244182 | 0 |
| 0 | 0 | 0.002606723 | 0 | 0 |
| 0 | 0 | 0 | 0 | 0 |
| 0.061084565 | 0 | 0 | 0 | 0 |
| 0.006113056 | 0.204976778 | 0 | 0.829768844 | 0.00488388 |
| 0.812415172 | 8.890216664 | 0.845689295 | 2.783892049 | 10.7148562 |
| 0 | 0.445378026 | 0 | 0.012350947 | 0.284990884 |
| 0 | 0 | 0 | 0 | 0 |
| 0 | 0 | 0.004289946 | 0.005071891 | 0.34372758 |
| 0 | 0 | 0 | 0 | 0 |
| 0 | 0 | 0 | 0 | 0 |
| 0 | 0 | 0 | 0 | 0 |
| 0 | 0 | 0 | 0 | 0 |
| 0 | 0 | 0.000935909 | 0.001602519 | 0 |
| 0 | 0.250372091 | 0 | 0.000880148 | 0 |
| 0.033876935 | 0 | 0.000654039 | 0.014931794 | 0.000675095 |
| 0 | 0 | 0.048596442 | 0 | 0 |
| 0 | 0.243981418 | 0.039387923 | 1.086664132 | 2.655468086 |
| 0 | 0.036437266 | 0 | 0 | 0.049877921 |
| 0 | 0 | 0 | 0 | 0 |
| 0 | 0.07815732 | 0 | 0 | 0 |
| 0 | 0 | 0 | 0 | 0 |
| 6.869851185 | 0.055534363 | 0.076714101 | 0.896950551 | 5.26648884 |
| 0 | 0.442773704 | 0 | 0 | 0 |
| 0 | 0 | 0 | 0 | 0 |
| 0 | 0 | 0.00072289 | 0.000589417 | 0 |
| 0 | 0 | 0 | 0 | 0 |
| 0.052955882 | 0 | 0.015958591 | 0.41260162 | 0.207602508 |

| | | | | |
|---|---|---|---|---|
| 0.021203592 | 0 | 0 | 0 | 0.016525668 |
| 0 | 0 | 0 | 0 | 0 |
| 0 | 0.001069819 | 0 | 0 | 0 |
| 0 | 0 | 0 | 0 | 0 |
| 0.008328048 | 0.000869044 | 0.004275111 | 0 | 0.002269409 |
| 0 | 0 | 0 | 0 | 0 |
| 0 | 0 | 0 | 0 | 0 |
| 0 | 0 | 0 | 0 | 0 |
| 0 | 0 | 0 | 0 | 0 |
| 0 | 0 | 0 | 0 | 0 |
| 0 | 0 | 0 | 0 | 0 |
| 0 | 0.231705417 | 0.008902314 | 0.59595871 | 1.322853549 |
| 0 | 0.031567809 | 0.041263385 | 0.095386555 | 0 |
| 0 | 0 | 0 | 0 | 0 |
| 0.611705057 | 0 | 0.121485225 | 0 | 0 |
| 0 | 0 | 0 | 0 | 0 |
| 0 | 0 | 0 | 0 | 0 |
| 0 | 0 | 0 | 0 | 0 |
| 0.001963227 | 0.000704738 | 0.003219204 | 0.013709606 | 0 |
| 0 | 0.106080742 | 0 | 0.006268769 | 0 |
| 0 | 0.047697727 | 0 | 0 | 0 |
| 0 | 0 | 0 | 0 | 0 |
| 0.626142655 | 0.372439867 | 0 | 0.101835773 | 0.191810176 |
| 0 | 0 | 0 | 0 | 0 |
| 0 | 0 | 0 | 0 | 0 |
| 0 | 0 | 0 | 0 | 0 |
| 0 | 0 | 0 | 0 | 0 |
| 0 | 0 | 0 | 0 | 0 |
| 0 | 0 | 0 | 0 | 0 |
| 0 | 0 | 0 | 0 | 0 |
| 0 | 0 | 0 | 0 | 0 |
| 0.011044807 | 0.040169131 | 0 | 0.024063295 | 0.038882299 |
| 0 | 0 | 0 | 0.037033172 | 0 |
| 0.039495798 | 0.004039623 | 0.078206797 | 0.001881643 | 0.09783357 |
| 0.038183258 | 0 | 0.000474506 | 0.014759993 | 0 |
| 0 | 0.283756279 | 0.058569852 | 0.057780516 | 0 |
| 0 | 0.001798069 | 0 | 0 | 0 |
| 0 | 0.017271144 | 0 | 0 | 0 |
| 0 | 0.001357489 | 0 | 0 | 0 |
| 0 | 0 | 0.000400995 | 0 | 0 |
| 0 | 0.149427705 | 0 | 0.013152429 | 0 |
| 0 | 0.000813009 | 0 | 0 | 0 |
| 0 | 0 | 0 | 0 | 0 |
| 0 | 0 | 0 | 0 | 0 |
| 0.042061351 | 0.005154852 | 0.01460737 | 0.007336734 | 0.052831998 |
| 0 | 0 | 0 | 0 | 0 |
| 0 | 0.101154147 | 0.000294967 | 0 | 0 |
| 0 | 0 | 0.002210553 | 0 | 0 |

| | | | | |
|---|---|---|---|---|
| 0 | 0 | 0 | 0.377620179 | 0 |
| 0 | 0.040927628 | 0 | 0 | 0 |
| 0.021846801 | 0 | 0.001208614 | 0 | 0 |
| 0 | 0 | 0 | 0 | 0 |
| 0.01030051 | 0 | 0 | 0 | 0 |
| 0 | 0 | 0 | 0 | 0 |
| 0 | 0.359694666 | 0 | 0.417029196 | 0 |
| 0.342966094 | 0.002923558 | 0.015295068 | 0.129512315 | 0.003887957 |
| 0 | 0 | 0 | 0 | 0 |
| 0 | 0.004155511 | 0 | 0 | 0 |
| 0 | 0 | 0 | 0 | 0 |
| 0.004679703 | 0 | 0 | 0 | 0 |
| 0 | 0 | 0 | 0 | 0 |
| 0 | 0.07300463 | 0 | 0.275076629 | 0 |
| 0 | 0 | 0 | 0 | 0 |
| 0 | 0.001489813 | 0 | 0 | 0 |
| 0 | 0.036145279 | 3.023211756 | 0 | 0 |
| 0 | 0 | 0 | 0 | 0 |
| 0 | 0.341623246 | 0 | 0 | 0 |
| 0 | 0 | 0 | 0 | 0 |
| 0 | 0.0023439 | 0 | 0.002005642 | 0 |
| 0 | 0 | 0 | 0 | 0 |
| 0 | 0 | 0 | 0 | 0 |
| 0 | 0 | 0 | 0 | 0 |
| 0 | 0.000908407 | 0 | 0 | 0 |
| 0.009832507 | 0 | 0.005004398 | 0 | 0.000673762 |
| 0 | 0 | 0 | 0 | 0 |
| 0.07648602 | 0 | 0.00234267 | 0.004619023 | 0.089689082 |
| 0 | 0.021970502 | 0.001127008 | 0 | 0 |
| 0.582839941 | 0.196162857 | 0.011496221 | 0.177273646 | 1.364268668 |
| 0 | 0 | 0 | 0 | 0 |
| 0 | 0 | 0.000298223 | 0 | 0 |
| 0.001426272 | 0.024479404 | 0 | 0 | 0 |
| 0 | 0 | 0 | 0 | 0 |
| 2.014856392 | 0 | 0.000912052 | 0.056350226 | 0 |
| 0 | 0 | 0.000802053 | 0 | 0 |
| 0 | 0.536504601 | 0 | 0 | 0 |
| 0 | 0.000552239 | 0.003104736 | 0.002215046 | 0.002002929 |
| 0 | 0 | 0 | 0 | 0 |
| 0 | 0 | 0 | 0 | 0 |
| 6.273317698 | 1.62468716 | 0.766894768 | 3.095867664 | 3.488460189 |
| 0.12071984 | 0.021694301 | 0.000284085 | 0.041400357 | 0.303053916 |
| 0 | 0 | 0.210176091 | 0.027023989 | 0 |
| 0 | 0 | 0.083300078 | 0 | 0 |
| 0 | 0 | 0 | 0 | 0 |
| 0 | 0 | 0 | 0 | 0 |
| 0 | 0 | 0 | 0 | 0 |
| 0.006023411 | 0.001899246 | 0.001540057 | 0.001523585 | 0.006835433 |

| | | | | |
|---|---|---|---|---|
| 0 | 0 | 0 | 0 | 0.023412965 |
| 0.002009376 | 0 | 0.004012991 | 0 | 0 |
| 0 | 0 | 0 | 0 | 0 |
| 0 | 0 | 0 | 0 | 0 |
| 5.152892808 | 0.378796379 | 0.621781497 | 4.209237007 | 3.160981753 |
| 0 | 0 | 0.000541449 | 0 | 0 |
| 0 | 0 | 0 | 0 | 0 |
| 0 | 0 | 0 | 0 | 0 |
| 1.269725222 | 0 | 0.114834312 | 0.002166678 | 0.012734755 |
| 0 | 0 | 0 | 0 | 0 |
| 0 | 0 | 0 | 0 | 0 |
| 0 | 0 | 0 | 0 | 0 |
| 0 | 0.025260514 | 0.02442196 | 0.091949903 | 0.136950303 |
| 0.020802947 | 0.007718931 | 0.02245516 | 0.012528338 | 0.016797583 |
| 0 | 1.21025006 | 0.040279212 | 0.162064413 | 0 |
| 4.687233337 | 0.027104051 | 0.140994103 | 0.141447755 | 0.236892034 |
| 0.001211654 | 0 | 0 | 0 | 0.004056475 |
| 0 | 0 | 0 | 0 | 0 |
| 0 | 0 | 0 | 0 | 0 |
| 0 | 0 | 0 | 0 | 0 |
| 0 | 0 | 0 | 0 | 0 |
| 0 | 0 | 0 | 0 | 0 |
| 0 | 0 | 0 | 0 | 0 |
| 0 | 0 | 0 | 0 | 0 |
| 0 | 0 | 0 | 0 | 0 |
| 0 | 0 | 0 | 0 | 0 |
| 0.004966893 | 0.033768211 | 0 | 0.021021094 | 0.0130803 |
| 0.001698359 | 0 | 0 | 0 | 0 |
| 0 | 0 | 0 | 0 | 0 |
| 0 | 0 | 0 | 0.002045497 | 0 |
| 0 | 0 | 0 | 0 | 0 |
| 0 | 0 | 0 | 0 | 0 |
| 0 | 0 | 0.569202166 | 0 | 0 |
| 0 | 0.005911534 | 0.001122809 | 0.005767618 | 0 |
| 0 | 0.027211209 | 0 | 0.055330391 | 0 |
| 0 | 0 | 0 | 0 | 0.005371551 |
| 0.000602432 | 0 | 0.000506584 | 0 | 0 |
| 0 | 0 | 0 | 0.004140298 | 0 |
| 0 | 0 | 0 | 0 | 0 |
| 0 | 0 | 0 | 0 | 0 |
| 0 | 0.165839539 | 0.003338543 | 0.507897832 | 5.743499003 |
| 0 | 0 | 0 | 0 | 0 |
| 0 | 0 | 0 | 0.021949675 | 0 |
| 0 | 0.323464253 | 0 | 0 | 0 |
| 0 | 0 | 0 | 0 | 0 |
| 0 | 0 | 0 | 0 | 0 |
| 0 | 0 | 0 | 0 | 0 |
| 0 | 0 | 0 | 0 | 0 |

| | | | | |
|---|---|---|---|---|
| 0 | 0.155865648 | 0 | 0.023427563 | 0.127286164 |
| 0 | 0 | 0 | 0 | 0 |
| 0 | 0 | 0 | 0.001128288 | 0 |
| 0 | 0 | 0 | 0 | 0 |
| 0 | 0.021640921 | 0 | 0.0539367 | 0 |
| 0 | 0 | 0 | 0.001286559 | 0 |
| 0.003108107 | 0 | 0 | 0 | 0.038442809 |
| 0 | 0 | 0 | 0 | 0 |
| 0.002193822 | 0 | 0 | 0 | 0 |
| 0 | 0 | 0.022736521 | 0 | 0 |
| 0 | 0.002442893 | 0 | 0 | 0 |
| 0 | 0 | 0 | 0 | 0 |
| 0 | 0 | 0 | 0 | 0 |
| 0 | 0 | 0 | 0 | 0 |
| 0 | 0 | 0 | 0 | 0 |
| 0 | 0 | 0 | 0 | 0 |
| 0 | 0 | 0 | 0 | 0 |
| 0 | 0 | 0 | 0 | 0 |
| 0 | 0 | 0 | 0 | 0 |
| 0 | 0 | 0 | 0 | 0 |
| 0.020220455 | 0 | 0 | 0.465470831 | 0.009792024 |
| 0 | 0 | 0 | 0 | 0 |
| 0 | 0.001789987 | 0.003773797 | 0.058612332 | 0.003009994 |
| 0 | 0.000578791 | 0.002237133 | 0.002437623 | 0.001574422 |
| 0 | 0 | 0 | 0 | 0 |
| 0.00285335 | 0 | 0.001199688 | 0.005203913 | 0 |
| 0 | 0 | 0 | 0 | 0 |
| 0 | 0 | 0 | 0.021029806 | 0 |
| 0 | 0 | 0.039205613 | 0 | 0 |
| 0 | 0.001874285 | 0.031465823 | 0.037756102 | 1.167197567 |
| 0 | 0 | 0 | 0 | 0 |
| 0 | 0 | 0 | 0 | 0 |
| 0 | 0.005210864 | 0 | 0 | 0 |
| 0 | 0 | 0 | 0 | 0 |
| 0 | 0.04056743 | 0 | 0 | 0 |
| 0 | 0 | 0.001379318 | 0 | 0 |
| 0.186617944 | 0 | 0.005980179 | 0.46768156 | 0 |
| 0 | 0 | 0 | 0 | 0 |
| 0 | 0 | 0 | 0 | 0 |
| 0 | 0 | 0 | 0 | 0 |
| 0 | 0 | 0 | 0 | 0 |
| 0 | 0 | 0.005744923 | 0 | 0 |
| 0 | 0 | 0 | 0 | 0 |
| 0 | 0 | 0.002306898 | 0 | 0 |
| 0 | 0 | 0.000361263 | 0 | 0.14114008 |
| 0 | 0 | 0 | 0 | 0 |
| 0 | 0 | 0 | 0 | 0 |
| 0 | 0.045731798 | 0 | 0.002491899 | 0 |

| | | | | |
|---|---|---|---|---|
| 0.033311354 | 0 | 0.002188395 | 0 | 0.002541202 |
| 0 | 0 | 0.042547019 | 0 | 0 |
| 0 | 0 | 0 | 0.02968702 | 0 |
| 0.003834085 | 0 | 0 | 0 | 0 |
| 0 | 0 | 0 | 0 | 0 |
| 0 | 0 | 0 | 0 | 0 |
| 0 | 0 | 0 | 0 | 0.000520948 |
| 1.80493304 | 0.162426467 | 0.000337048 | 0.58006306 | 0.989597684 |
| 0.238287898 | 5.496672669 | 0.000515252 | 1.323955506 | 0.057172768 |
| 0.532361133 | 0 | 0.010521958 | 0 | 0 |
| 0 | 0.004392438 | 0 | 0 | 0 |
| 0 | 0 | 0.000523308 | 0 | 0 |
| 0 | 0.001915498 | 0 | 0 | 0 |
| 0 | 0 | 0.087606397 | 0 | 0 |
| 0 | 0 | 0 | 0 | 0 |
| 0 | 0 | 0 | 0 | 0 |
| 0.314800191 | 0 | 0 | 0.004558129 | 0 |
| 0 | 0.086357892 | 0 | 0.662570031 | 0 |
| 0 | 0 | 0 | 0 | 0 |
| 0 | 0 | 0 | 0.012675989 | 0.005065845 |
| 0 | 0 | 0 | 0 | 0 |
| 0 | 0 | 0 | 0 | 0 |
| 0 | 0.151758998 | 0 | 0 | 0 |
| 0 | 0 | 0 | 0 | 0 |
| 0 | 0.002943245 | 0 | 0.012100571 | 0 |
| 0 | 0 | 0 | 0 | 0 |
| 0 | 0 | 0 | 0 | 0 |
| 0 | 0 | 0 | 0 | 0 |
| 0 | 0 | 0 | 0 | 0 |
| 0 | 0 | 0 | 0 | 0 |
| 0 | 0.003838916 | 0 | 0 | 0 |
| 0 | 0 | 0 | 0 | 0 |
| 0 | 0 | 0 | 0 | 0 |
| 0 | 0 | 0 | 0 | 0 |
| 0 | 0 | 0 | 0.007711862 | 0.037738184 |
| 0 | 0 | 0 | 0 | 0 |
| 0 | 0 | 0 | 0 | 0 |
| 0.058187693 | 0 | 0 | 0 | 0.029626666 |
| 0 | 0 | 0 | 0 | 0 |
| 0 | 0 | 0 | 0.001246926 | 0 |
| 0 | 0 | 0 | 0 | 0 |
| 0.286083714 | 0.107602286 | 0 | 1.497012906 | 0 |
| 0 | 0 | 0 | 0 | 0 |
| 0 | 0 | 0 | 0 | 0 |
| 0 | 0 | 0.001703155 | 0 | 0 |
| 0 | 0 | 0 | 0 | 0 |
| 0 | 0 | 0 | 0 | 0 |
| 0 | 0 | 0.001528431 | 0 | 0 |

| | | | | |
|---|---|---|---|---|
| 0 | 0 | 0 | 0 | 0 |
| 0 | 0 | 0 | 0.041935803 | 0 |
| 0 | 0 | 0 | 0 | 0 |
| 0 | 0 | 0 | 0 | 0 |
| 0 | 0.66349613 | 0.13614187 | 0.113008737 | 0 |
| 0 | 0 | 0.000504743 | 0.01037101 | 0.005470421 |
| 0 | 0 | 0 | 0 | 0.016130059 |
| 0.145220787 | 0 | 0.005706783 | 0.047189138 | 0.01293037 |
| 0 | 0 | 0 | 0.001427776 | 0.003012449 |
| 0 | 0.028389212 | 0 | 0.135111976 | 0 |
| 0 | 0 | 0 | 0 | 0 |
| 0 | 0 | 0 | 0 | 0 |
| 0 | 0 | 0 | 0 | 0 |
| 0 | 0 | 0 | 0 | 0 |
| 0.009011182 | 0 | 0 | 0 | 0 |
| 0.009570585 | 0 | 0.011709676 | 0 | 0 |
| 0 | 0 | 0 | 0 | 0 |
| 0 | 0 | 0 | 0 | 0 |
| 0 | 0 | 0 | 0 | 0 |
| 0 | 0 | 0 | 0 | 0 |
| 0 | 0 | 0 | 0 | 0 |
| 0 | 0 | 0 | 0 | 0 |
| 0 | 0 | 0 | 0 | 0 |
| 0.004201954 | 0 | 0.003011434 | 0.00151253 | 0.001119017 |
| 0 | 0.025163032 | 0 | 0.039026107 | 0 |
| 0 | 0 | 0.001317429 | 0 | 0 |
| 0.760597103 | 0 | 0.001762751 | 0 | 0.000511734 |
| 0 | 0 | 0 | 0 | 0 |
| 0 | 0 | 0 | 0 | 0 |
| 0 | 0 | 0 | 0 | 0 |
| 0 | 0 | 0 | 0 | 0 |
| 0 | 0 | 0 | 0 | 0 |
| 0 | 0 | 0 | 0 | 0 |
| 0 | 0 | 0 | 0 | 0 |
| 0 | 0 | 0 | 0 | 0 |
| 0 | 0 | 0 | 0 | 0 |
| 0 | 0 | 0 | 0 | 0 |
| 0 | 0.003376317 | 0 | 0 | 0 |
| 0 | 0.031322733 | 0 | 0 | 0 |
| 0 | 0 | 0 | 0 | 0 |
| 0 | 0 | 0 | 0 | 0 |
| 0 | 0.098997811 | 0 | 0.020556617 | 0 |
| 0 | 0 | 0 | 0 | 0 |
| 0 | 0 | 0 | 0 | 0 |
| 0 | 0 | 0 | 0 | 0 |
| 0 | 0 | 0 | 0 | 0 |
| 0 | 0 | 0 | 0 | 0 |
| 0 | 0 | 0 | 0 | 0 |

| | | | | |
|---|---|---|---|---|
| 0 | 0.031992056 | 0.007666932 | 0.008397042 | 0.020693396 |
| 0 | 0 | 0 | 0 | 0 |
| 0 | 0 | 0 | 0 | 0 |
| 0 | 0 | 0 | 0 | 0 |
| 0 | 0 | 0 | 0 | 0 |
| 0 | 0.041676379 | 0 | 0.086734003 | 0 |
| 0 | 0 | 0 | 0 | 0 |
| 0 | 0 | 0 | 0 | 0 |
| 0 | 0 | 0 | 0 | 0 |
| 0 | 0 | 0 | 0 | 0 |
| 0 | 0 | 0 | 0 | 0 |
| 0 | 0 | 0 | 0 | 0 |
| 0 | 0.009751069 | 0 | 0 | 0 |
| 0 | 0 | 0 | 0 | 0 |
| 0 | 0 | 0 | 0 | 0 |
| 0 | 0 | 0 | 0 | 0 |
| 0 | 0 | 0 | 0 | 0 |
| 0 | 0 | 0 | 0 | 0 |
| 0 | 0 | 0 | 0 | 0 |
| 0 | 0 | 0 | 0 | 0 |
| 0.113168698 | 0 | 0.005757782 | 0 | 0.002228678 |
| 0 | 0 | 0 | 0 | 0 |
| 0.005284578 | 0.013580801 | 0.000908957 | 0.00449618 | 0.107191583 |
| 0 | 0.023281319 | 0 | 0.794380006 | 0 |
| 0 | 0.015683559 | 0 | 0 | 0 |
| 0 | 0 | 0 | 0 | 0 |
| 0 | 0 | 0 | 0 | 0 |
| 0 | 0 | 0 | 0 | 0 |
| 0 | 0 | 0 | 0 | 0 |
| 0 | 0 | 0 | 0 | 0 |
| 0 | 0 | 0 | 0 | 0 |
| 0 | 0 | 0 | 0.414880022 | 0 |
| 0.025419791 | 0 | 0.076894886 | 0.001457534 | 0.019549712 |
| 0 | 0 | 0 | 0 | 0 |
| 0 | 0.034490708 | 0.038959609 | 0.057220464 | 0 |
| 0.088452878 | 0 | 0 | 0.003638787 | 0.164457754 |
| 0 | 0 | 0 | 0 | 0 |
| 0 | 0 | 0 | 0 | 0 |
| 0.064100187 | 0 | 0.003292044 | 0 | 0.002854341 |
| 0 | 0 | 0 | 0 | 0 |
| 0 | 0 | 0 | 0 | 0 |
| 0 | 0 | 0 | 0.021336043 | 0 |
| 0 | 0.025634791 | 0.03356664 | 0.024079641 | 0.412429448 |
| 0 | 0 | 0 | 0 | 0 |
| 0 | 0 | 0 | 0 | 0 |
| 0.001163358 | 0.023490542 | 0 | 0.074260255 | 0.000567989 |
| 0 | 0 | 0 | 0 | 0 |
| 0 | 0 | 0 | 0 | 0 |

| | | | | |
|---|---|---|---|---|
| 0 | 0 | 0 | 0 | 0 |
| 0 | 0 | 0 | 0 | 0 |
| 0 | 0 | 0 | 0 | 0 |
| 0 | 0 | 0 | 0 | 0 |
| 0 | 0 | 0 | 0 | 0 |
| 0 | 0 | 0 | 0 | 0 |
| 0 | 0 | 0.000401646 | 0 | 0 |
| 0 | 0 | 0.020678751 | 0 | 0.002927241 |
| 0 | 0.004564718 | 0 | 0 | 0 |
| 0 | 0.101439774 | 0 | 0.004025644 | 0 |
| 0 | 0 | 0 | 0 | 0 |
| 0 | 0 | 0 | 0 | 0 |
| 0 | 0 | 0 | 0 | 0 |
| 0 | 0 | 0.000541645 | 0.153336011 | 0 |
| 0 | 0 | 0 | 0 | 0 |
| 0.002161393 | 0 | 0.000867448 | 0 | 0 |
| 0 | 0 | 0 | 0 | 0 |
| 0 | 0 | 0 | 0 | 0 |
| 0 | 0 | 0 | 0 | 0 |
| 0 | 0.010481951 | 0 | 2.211600406 | 0.01734871 |
| 0 | 0 | 0 | 0.014330929 | 0.009017564 |
| 0 | 0 | 0 | 0 | 0 |
| 0 | 0 | 0 | 0 | 0 |
| 0 | 0 | 0 | 0 | 0 |
| 0 | 0 | 0 | 0 | 0 |
| 0 | 0 | 0 | 0.087418659 | 0 |
| 0 | 0 | 0 | 0 | 0 |
| 0 | 0 | 0 | 0 | 0 |
| 0 | 0 | 0 | 0 | 0 |
| 0 | 0.009320634 | 0 | 0.098228075 | 0 |
| 0 | 0.067647047 | 0.011179884 | 0.315598349 | 0.264479696 |
| 0 | 0 | 0 | 0 | 0 |
| 0 | 0 | 0 | 0 | 0 |
| 0 | 0 | 0 | 0 | 0 |
| 0 | 0 | 0 | 0 | 0 |
| 0 | 0 | 0 | 0 | 0 |
| 0 | 0 | 0 | 0 | 0 |
| 0 | 0 | 0 | 0 | 0 |
| 0 | 0.003876654 | 0 | 0.009977508 | 0 |
| 0 | 0 | 0 | 0 | 0 |
| 0 | 0 | 0 | 0 | 0 |
| 0 | 0 | 0.001157092 | 0 | 0 |
| 0 | 0.003565304 | 0 | 0 | 0 |
| 0.027704114 | 0 | 0 | 0 | 0 |
| 0 | 0 | 0 | 0 | 0 |
| 0 | 0 | 0 | 0 | 0 |
| 1.180724803 | 0 | 0.003932574 | 0 | 0.013748821 |
| 0 | 0 | 0 | 0 | 0 |

| | | | | |
|---|---|---|---|---|
| 0 | 0 | 0 | 0 | 0 |
| 0 | 0 | 0 | 0 | 0 |
| 0 | 0 | 0 | 0 | 0 |
| 0 | 0.000647121 | 0 | 0 | 0 |
| 0.007964353 | 0.058527707 | 0.009571954 | 0.013414639 | 0.011151801 |
| 0 | 0 | 0 | 0 | 0 |
| 0 | 0 | 0 | 0 | 0 |
| 0 | 0 | 0 | 0 | 0 |
| 0 | 0 | 0 | 0 | 0 |
| 0 | 0 | 0 | 0 | 0 |
| 0 | 0.006506392 | 0 | 0.002747085 | 0 |
| 0.030251226 | 0.0052424 | 0.004342037 | 0.001201566 | 0.013038024 |
| 0.005579826 | 0 | 0.000794624 | 0 | 0.026012224 |
| 0 | 0 | 0 | 0 | 0 |
| 0 | 0 | 0.011731681 | 0 | 0 |
| 0.05126744 | 0 | 0.000511598 | 0.102665879 | 0.82600447 |
| 0 | 0 | 0.001517974 | 0 | 0 |
| 0 | 0 | 0 | 0 | 0 |
| 0.001421777 | 0 | 0.023413232 | 0 | 0.004164947 |
| 0.074668818 | 0 | 0 | 0 | 0.010272064 |
| 0 | 0 | 0 | 0 | 0 |
| 0 | 0.045768607 | 0 | 0 | 0 |
| 0 | 0 | 0 | 0 | 0 |
| 0 | 0 | 0 | 0 | 0 |
| 0 | 0 | 0 | 0 | 0 |
| 0 | 0 | 0 | 0 | 0 |
| 0.000826458 | 0 | 0.009069317 | 0 | 0 |
| 0 | 0 | 0 | 0 | 0 |
| 0 | 0 | 0 | 0 | 0 |
| 0 | 0 | 0 | 0 | 0 |
| 0.0044968 | 0 | 0.002245177 | 0 | 0 |
| 0.007285602 | 0 | 0 | 0 | 0 |
| 0 | 0 | 0 | 0 | 0 |
| 0 | 0 | 0 | 0 | 0 |
| 0 | 0 | 0 | 0 | 0 |
| 0 | 0 | 0 | 0 | 0 |
| 0 | 0 | 0 | 0 | 0 |
| 0.007240111 | 0.009450825 | 0.068990326 | 0.002558757 | 0.047174295 |
| 0 | 0.124615178 | 0 | 0.040807492 | 0 |
| 0 | 0 | 0 | 0 | 0 |
| 0 | 0.001179371 | 0 | 0.006244245 | 0.000769948 |
| 0 | 0 | 0 | 0 | 0 |
| 0 | 0 | 0 | 0 | 0 |
| 0.070663496 | 0 | 0 | 0 | 0 |
| 0 | 0 | 0 | 0 | 0 |
| 0 | 0 | 0 | 0 | 0 |
| 0 | 0 | 0 | 0 | 0 |
| 0 | 0 | 0 | 0 | 0 |

| | | | | |
|---|---|---|---|---|
| 0 | 0 | 0 | 0 | 0 |
| 0 | 0.013175996 | 0 | 0.001921792 | 0.687989036 |
| 0 | 0.026040823 | 0 | 0 | 0 |
| 0.003273179 | 0 | 0.001290192 | 0.000736381 | 0.001598073 |
| 0 | 0.003746071 | 0 | 0 | 0 |
| 0.002448761 | 0 | 0 | 0 | 0 |
| 0 | 0 | 0 | 0 | 0 |
| 0 | 0 | 0 | 0 | 0 |
| 0 | 0 | 0.000733689 | 0.103133433 | 0 |
| 0 | 0.002333627 | 0 | 0 | 0 |
| 0 | 0.007281721 | 0.02985096 | 0 | 0 |
| 0 | 0 | 0 | 0 | 0 |
| 0 | 0.090056067 | 0.001097477 | 0.043638388 | 0.00792966 |
| 0 | 0 | 0 | 0 | 0 |
| 0 | 0 | 0 | 0 | 0 |
| 0 | 0 | 0 | 0 | 0 |
| 0 | 0 | 0 | 0 | 0 |
| 0 | 0 | 0 | 0 | 0 |
| 0 | 0 | 0 | 0 | 0 |
| 0 | 0 | 0 | 0 | 0 |
| 0 | 0 | 0 | 0 | 0 |
| 0 | 0 | 0 | 0 | 0 |
| 0.061867775 | 0 | 0.001445125 | 0.007423282 | 0 |
| 0 | 0.062686356 | 0 | 0.015086198 | 0.074808318 |
| 0 | 0.002856369 | 0 | 0 | 0 |
| 0 | 0 | 0 | 0 | 0 |
| 0 | 0.001637224 | 0 | 0 | 0 |
| 0 | 0 | 0.000296006 | 0 | 0 |
| 0 | 0 | 0 | 0 | 0 |
| 0 | 0 | 0 | 0 | 0 |
| 0.01894884 | 0.004012445 | 0.001208481 | 0.002299146 | 0.030768843 |
| 0.008067521 | 0 | 0.018482839 | 0 | 0 |
| 0 | 0 | 0 | 0 | 0 |
| 0 | 0 | 0 | 0 | 0 |
| 0 | 0 | 0.003152063 | 0.015677427 | 0 |
| 0.000667706 | 0 | 0 | 0 | 0 |
| 0.025560544 | 0 | 0 | 0 | 0 |
| 0 | 0 | 0 | 0 | 0 |
| 0.000720541 | 0 | 0.000454426 | 0 | 0 |
| 0 | 0 | 0 | 0 | 0 |
| 0 | 0 | 0 | 0 | 0 |
| 0 | 0 | 0 | 0 | 0 |
| 0 | 0 | 0.001303564 | 0 | 0 |
| 0 | 0 | 0 | 0 | 0 |
| 0 | 11.82117541 | 0 | 0 | 0 |
| 0.097470869 | 0.215765737 | 0.053475905 | 0.056022685 | 0.012690256 |
| 0 | 0 | 0 | 0 | 0 |
| 0 | 0 | 0 | 0 | 0 |

| | | | | |
|---|---|---|---|---|
| 0 | 0 | 0 | 0 | 0 |
| 0 | 0 | 0 | 0 | 0 |
| 0 | 0.021299878 | 0 | 0.197084232 | 0 |
| 0 | 0 | 0 | 0 | 0 |
| 0 | 0 | 0 | 0 | 0 |
| 0 | 0 | 0 | 0 | 0 |
| 0 | 0 | 0 | 0 | 0 |
| 0 | 0 | 0 | 0 | 0 |
| 0.000666692 | 0.001047033 | 0 | 0 | 0 |
| 0 | 0 | 0 | 0 | 0 |
| 0.002045931 | 0 | 0.000645157 | 0.00305101 | 0.000428096 |
| 0.001745475 | 0 | 0 | 0 | 0.001278297 |
| 0 | 0 | 0 | 0 | 0 |
| 0 | 0 | 0 | 0 | 0 |
| 0 | 0 | 0 | 0 | 0 |
| 0 | 0 | 0 | 0 | 0 |
| 0 | 0 | 0 | 0 | 0 |
| 0 | 0 | 0 | 0 | 0 |
| 0 | 0 | 0 | 0 | 0 |
| 0 | 0.002434938 | 0 | 0 | 0.010976085 |
| 0 | 0 | 0 | 0 | 0 |
| 0 | 0 | 0 | 0 | 0 |
| 0 | 0 | 0 | 0 | 0 |
| 0 | 0 | 0 | 0 | 0 |
| 0.007711955 | 0 | 0 | 0 | 0 |
| 0 | 0 | 0 | 0 | 0 |
| 0 | 0 | 0 | 0 | 0 |
| 0 | 0.006736676 | 0 | 0.054659593 | 0 |
| 0 | 0 | 0 | 0 | 0 |
| 0 | 0 | 0 | 0 | 0 |
| 0 | 0.004333146 | 0.001890102 | 0.008270656 | 0.000487738 |
| 0 | 0 | 0 | 0 | 0 |
| 0.04929679 | 0.005898676 | 0.030236727 | 0.007237099 | 0.017366932 |
| 0.000592659 | 0.00230475 | 0 | 0.000426665 | 0.015046472 |
| 0 | 0 | 0 | 0 | 0 |
| 0 | 0 | 0 | 0 | 0 |
| 0 | 0 | 0 | 0 | 0 |
| 0 | 0 | 0 | 0 | 0 |
| 0 | 0 | 0 | 0 | 0 |
| 0 | 0 | 0 | 0 | 0 |
| 0 | 0 | 0 | 0 | 0 |
| 0.159202265 | 0 | 0 | 0 | 0.057771671 |
| 0 | 0.054251875 | 0.002077383 | 0.047845358 | 0.064517002 |
| 0 | 0 | 0 | 0 | 0 |
| 0 | 0 | 0 | 0 | 0 |
| 0 | 0.064828893 | 0.005650753 | 0.047773029 | 0.542164726 |
| 0 | 0 | 0 | 0 | 0 |
| 0 | 0 | 0 | 0 | 0 |

| | | | | |
|---|---|---|---|---|
| 0 | 0 | 0 | 0 | 0 |
| 0 | 0.002003835 | 0 | 0.01417511 | 0 |
| 0.001422674 | 0.004043011 | 0.012262318 | 0.003584727 | 0.007872086 |
| 0 | 0 | 0 | 0 | 0 |
| 0 | 0.009684448 | 0 | 0.00463409 | 0.557744081 |
| 0 | 0 | 0.001162241 | 0 | 0 |
| 0 | 0 | 0 | 0 | 0 |
| 0 | 0 | 0 | 0 | 0 |
| 0 | 0 | 0 | 0 | 0 |
| 0 | 0 | 0 | 0 | 0 |
| 0 | 0 | 0 | 0 | 0 |
| 0 | 0 | 0 | 0 | 0 |
| 0 | 0 | 0 | 0 | 0 |
| 0.032286447 | 0 | 0.033308552 | 0 | 0 |
| 0 | 0 | 0 | 0 | 0 |
| 0 | 0 | 0 | 0 | 0 |
| 0 | 0 | 0 | 0 | 0 |
| 0 | 0 | 0 | 0 | 0 |
| 0 | 0 | 0 | 0 | 0 |
| 0 | 0 | 0 | 0 | 0 |
| 0 | 0 | 0 | 0 | 0 |
| 0 | 0 | 0.001443456 | 0 | 0 |
| 0 | 0 | 0 | 0 | 0 |
| 0 | 0 | 0 | 0 | 0 |
| 0 | 0.014730988 | 0 | 0.016178414 | 0.001526519 |
| 0 | 0.001587753 | 0.009546376 | 0 | 0.004414971 |
| 0 | 0 | 0 | 0 | 0 |
| 0.002860367 | 0.177975836 | 0.000360792 | 0.000549128 | 0.039661301 |
| 0 | 0 | 0 | 0 | 0 |
| 0 | 0 | 0 | 0 | 0 |
| 0 | 0 | 0 | 0 | 0 |
| 0.001034343 | 0 | 0 | 0 | 0 |
| 0 | 0 | 0 | 0 | 0 |
| 0 | 0 | 0 | 0 | 0 |
| 0 | 0 | 0 | 0 | 0 |
| 0 | 0 | 0 | 0 | 0 |
| 0.102892147 | 0 | 0 | 0 | 0 |
| 0 | 0 | 0 | 0 | 0 |
| 0 | 0.158090497 | 0.597597541 | 0 | 1.587344828 |
| 0 | 0 | 0 | 0 | 0 |
| 0 | 0 | 0 | 0 | 0 |
| 0 | 0.052056777 | 0.102890972 | 0 | 0 |
| 0.019531832 | 0 | 0.001617543 | 0 | 0.00096324 |
| 0 | 0 | 0 | 0 | 0 |
| 0 | 0 | 0 | 0 | 0 |
| 0 | 0 | 0 | 0 | 0 |
| 0.012072679 | 0 | 0.017321667 | 0.00325925 | 0.0085467 |
| 0 | 0 | 0 | 0 | 0 |

| | | | | |
|---|---|---|---|---|
| 0 | 0 | 0 | 0 | 0 |
| 0.006283819 | 0 | 0.001801381 | 0.003015887 | 0.035514026 |
| 0 | 0 | 0 | 0.048926033 | 0 |
| 0 | 0 | 0 | 0 | 0 |
| 0 | 0 | 0 | 0 | 0 |
| 0 | 0 | 0 | 0 | 0 |
| 0 | 0 | 0 | 0 | 0 |
| 0 | 0 | 0 | 0 | 0 |
| 0 | 0 | 0 | 0 | 0 |
| 0.007599076 | 0 | 0 | 0 | 0 |
| 0 | 0 | 0 | 0 | 0 |
| 0 | 0 | 0 | 0 | 0 |
| 0 | 0.024025361 | 0.011425317 | 0.026373952 | 0.047369083 |
| 0 | 0 | 0 | 0 | 0 |
| 0.105178604 | 0 | 0.013326568 | 0 | 0 |
| 0 | 0 | 0 | 0 | 0 |
| 0 | 0 | 0 | 0 | 0 |
| 0 | 0 | 0 | 0 | 0 |
| 0 | 0 | 0 | 0 | 0 |
| 0 | 0 | 0 | 0 | 0 |
| 0 | 0 | 0 | 0 | 0 |
| 0.056426802 | 0 | 0 | 0 | 0.003415216 |
| 0.010033575 | 0 | 0.017853748 | 0.004901556 | 0 |
| 0 | 0 | 0 | 0 | 0 |
| 0 | 0 | 0 | 0 | 0 |
| 0 | 0 | 0 | 0 | 0 |
| 0 | 0 | 0 | 0 | 0 |
| 0 | 0 | 0 | 0 | 0 |
| 0 | 0 | 0 | 0 | 0 |
| 0 | 0 | 0 | 0 | 0 |
| 0 | 0 | 0 | 0 | 0 |
| 0 | 0 | 0 | 0 | 0 |
| 0 | 0 | 0 | 0 | 0 |
| 0 | 0 | 0 | 0 | 0 |
| 1.496748088 | 0.044077247 | 0.00106813 | 21.67659191 | 1.087356275 |
| 0 | 0 | 0 | 0 | 0 |
| 0 | 0 | 0 | 0 | 0 |
| 0 | 0 | 0 | 0 | 0 |
| 0.032598589 | 0 | 0 | 0.002179199 | 0.009776784 |
| 0 | 0 | 0 | 0 | 0 |
| 0.072654809 | 0.260433651 | 0.014352127 | 0.116033964 | 0.109475322 |
| 0 | 0 | 0 | 0 | 0 |
| 0 | 0 | 0 | 0 | 0 |
| 0 | 0 | 0 | 0 | 0 |
| 0 | 0 | 0 | 0 | 0 |
| 1.458841685 | 0 | 0.005125643 | 0.192393017 | 0.422941022 |
| 0 | 0 | 0 | 0.053024648 | 0 |
| 0 | 0 | 0 | 0 | 0 |

| | | | | |
|---|---|---|---|---|
| 0 | 0 | 0.013292917 | 0 | 0 |
| 0 | 0 | 0 | 0 | 0 |
| 0 | 0 | 0 | 0 | 0 |
| 0 | 0 | 0 | 0 | 0 |
| 0 | 0 | 0 | 0 | 0 |
| 0 | 0 | 0 | 0 | 0 |
| 0 | 0 | 0 | 0 | 0 |
| 0 | 0 | 0 | 0 | 0 |
| 0 | 0 | 0.042688506 | 0 | 0 |
| 0 | 0 | 0.005602638 | 0 | 0 |
| 0 | 0.007080385 | 0 | 0.048989334 | 0 |
| 0 | 0 | 0 | 0 | 0 |
| 0.002970483 | 0 | 0 | 0.003948005 | 0.002008089 |
| 0 | 0 | 0 | 0 | 0 |
| 0 | 0 | 0 | 0 | 0 |
| 0 | 0 | 0 | 0 | 0 |
| 0 | 0 | 0 | 0 | 0 |
| 0 | 0 | 0 | 0 | 0 |
| 0 | 0 | 0 | 0 | 0 |
| 0.298358395 | 0.301701993 | 0.259226324 | 0.040703865 | 0.49807243 |
| 0 | 0 | 0 | 0 | 0 |
| 0 | 0 | 0 | 0 | 0 |
| 0 | 0 | 0 | 0 | 0 |
| 0 | 0 | 0 | 0 | 0 |
| 0 | 0 | 0 | 0 | 0 |
| 0 | 0.011491857 | 0 | 0 | 0 |
| 0 | 0 | 0 | 0 | 0 |
| 0 | 0 | 0 | 0 | 0 |
| 0 | 0 | 0 | 0 | 0 |
| 0 | 0 | 0.002468139 | 0 | 0 |
| 0 | 0 | 0 | 0 | 0 |
| 0 | 0.005623319 | 0 | 0 | 0 |
| 0 | 0 | 0 | 0 | 0 |
| 0 | 0 | 0 | 0 | 0 |
| 0 | 0 | 0 | 0 | 0 |
| 0 | 0 | 0 | 0 | 0 |
| 0 | 0 | 0 | 0 | 0 |
| 0 | 0 | 0 | 0 | 0 |
| 0 | 0 | 0 | 0 | 0 |
| 0 | 0.043895355 | 2.349742537 | 0 | 0 |
| 0 | 0 | 0 | 0 | 0 |
| 0 | 0 | 0 | 0 | 0 |
| 0 | 0.00075858 | 0 | 0.01922977 | 0 |
| 0 | 0 | 0 | 0 | 0 |
| 0.003820439 | 0 | 0.002168505 | 0.00151272 | 0.002238315 |
| 0 | 0 | 0 | 0 | 0 |
| 0 | 0 | 0 | 0 | 0 |
| 0 | 0 | 0 | 0 | 0 |

| | | | | |
|---|---|---|---|---|
| 0 | 0 | 0 | 0 | 0 |
| 0 | 0 | 0 | 0 | 0 |
| 0 | 0 | 0 | 0 | 0 |
| 0 | 0 | 0 | 0 | 0 |
| 0 | 0.001628093 | 0 | 0 | 0 |
| 0.131767294 | 0.005375045 | 0.016998167 | 0.004722546 | 0.006266211 |
| 0 | 0 | 0 | 0 | 0 |
| 0 | 0 | 0 | 0 | 0 |
| 0 | 0 | 0 | 0 | 0 |
| 0 | 0 | 0 | 0 | 0 |
| 0 | 0 | 0 | 0 | 0 |
| 0 | 0 | 0 | 0 | 0 |
| 0 | 0 | 0 | 0 | 0 |
| 0 | 0.049741742 | 0 | 0 | 0 |
| 0 | 0 | 0 | 0 | 0 |
| 0 | 0 | 0 | 0 | 0 |
| 0 | 0.014436904 | 0 | 0 | 0 |
| 0 | 0 | 0 | 0 | 0 |
| 0 | 0 | 0 | 0 | 0 |
| 0 | 0 | 0 | 0 | 0 |
| 0 | 0 | 0 | 0 | 0 |
| 0 | 0 | 0 | 0 | 0 |
| 0 | 0 | 0 | 0 | 0 |
| 0 | 0 | 0 | 0.003888786 | 0 |
| 0 | 0 | 0 | 0 | 0 |
| 0 | 0 | 0 | 0 | 0 |
| 0 | 0 | 0 | 0 | 0 |
| 0 | 0.002546929 | 0 | 0 | 0 |
| 0 | 0 | 0 | 0 | 0 |
| 0.008507532 | 0 | 0 | 0 | 0 |
| 0 | 0 | 0 | 0 | 0 |
| 0 | 0 | 0 | 0 | 0 |
| 0 | 0 | 0 | 0 | 0 |
| 0 | 0 | 0 | 0 | 0 |
| 0 | 0 | 0 | 0 | 0 |
| 0 | 0 | 0 | 0 | 0 |
| 0 | 0 | 0 | 0 | 0 |
| 0 | 0 | 0 | 0 | 0 |
| 0 | 0.021136212 | 0 | 0.548085549 | 0.388604309 |
| 0 | 0 | 0 | 0 | 0 |
| 0 | 0 | 0 | 0 | 0 |
| 0 | 0 | 0 | 0 | 0 |
| 0 | 0 | 0 | 0 | 0 |
| 0 | 0 | 0 | 0 | 0 |
| 0 | 0 | 0 | 0 | 0 |
| 0 | 0 | 0 | 0 | 0 |
| 0 | 0 | 0 | 0 | 0 |
| 0 | 0 | 0 | 0 | 0 |

| | | | | |
|---|---|---|---|---|
| 0 | 0 | 0 | 0 | 0 |
| 0.043267351 | 0 | 0 | 0 | 0 |
| 0 | 0 | 0 | 0 | 0 |
| 0 | 0 | 0 | 0 | 0 |
| 0 | 0 | 0 | 0 | 0 |
| 0 | 0 | 0.001104631 | 0 | 0 |
| 0 | 0 | 0 | 0 | 0 |
| 0 | 0 | 0 | 0 | 0 |
| 0 | 0 | 0 | 0 | 0 |
| 0 | 0.004279906 | 0 | 0 | 0 |
| 0 | 0.024765172 | 0 | 0.000451517 | 0 |
| 0 | 0 | 0 | 0 | 0 |
| 0.075143709 | 0.019484492 | 0 | 0 | 0.509717242 |
| 0 | 0 | 0 | 0 | 0 |
| 0 | 0 | 0 | 0 | 0 |
| 0 | 0 | 0 | 0 | 0 |
| 0 | 0 | 0 | 0 | 0 |
| 0 | 0 | 0 | 0 | 0 |
| 0 | 0 | 0 | 0 | 0 |
| 0 | 0 | 0.001097128 | 0 | 0 |
| 0 | 0.005522118 | 0 | 0.000611017 | 0.008494762 |
| 0 | 0 | 0 | 0 | 0 |
| 0 | 0 | 0 | 0 | 0 |
| 2.419681277 | 0.296397377 | 0.243437441 | 0.817837956 | 2.668932652 |
| 0 | 0 | 0 | 0 | 0 |
| 0 | 0 | 0 | 0 | 0 |
| 0 | 0 | 0 | 0 | 0 |
| 0 | 0 | 0.265736799 | 0 | 0 |
| 0 | 0 | 0 | 0 | 0 |
| 0 | 0 | 0 | 0 | 0 |
| 0 | 0 | 0 | 0 | 0 |
| 0 | 0 | 0 | 0 | 0 |
| 0 | 0.001001507 | 0 | 0.00337434 | 0 |
| 0 | 0 | 0 | 0 | 0 |

| | | | | |
|---|---|---|---|---|
| PHL018-0189_V1 | PHL018-0258_V2 | PHL018-0158_V2 | PHL018-0176_V1 | PHL018-0200_V3 |
| EYCF | REF | EYCF | REF | EYCF |
| 0.048784969 | 0.333724773 | 0.078359157 | 0.013332405 | 0.071595609 |
| 8.667085493 | 0.882826699 | 0.656978261 | 5.724456555 | 1.534830663 |
| 0 | 0 | 4.797418206 | 3.558917803 | 1.910324502 |
| 0 | 0 | 0 | 0 | 0 |
| 0 | 0 | 0 | 0 | 0 |
| 0 | 0 | 0 | 0 | 0 |
| 0 | 0 | 0 | 0 | 0 |
| 0 | 0 | 0 | 0 | 0 |
| 0 | 0 | 0 | 0 | 0 |
| 0 | 0 | 0 | 0 | 0 |
| 0 | 0 | 0 | 0 | 0 |
| 0 | 0 | 0 | 0 | 0 |
| 0 | 0 | 0 | 0 | 0 |
| 0 | 1.132977762 | 10.14058046 | 0.130753517 | 5.648428784 |
| 0 | 0 | 0 | 0 | 0 |
| 0 | 0 | 0 | 0 | 0 |
| 0 | 0 | 0 | 0 | 0 |
| 0 | 0 | 0 | 0 | 0 |
| 0 | 0 | 0 | 0 | 0 |
| 0 | 0 | 0 | 0 | 0 |
| 0 | 0 | 0 | 0 | 0 |
| 0 | 0.020900811 | 0 | 0 | 0 |
| 0 | 0 | 0 | 0 | 0 |
| 0 | 0 | 0 | 0 | 0 |
| 0 | 0 | 0 | 0 | 0 |
| 0 | 0 | 0 | 0 | 0 |
| 0 | 0 | 0 | 0 | 0 |
| 0 | 0 | 0 | 0 | 0 |
| 0 | 0 | 0 | 0 | 0 |
| 0 | 0 | 0 | 0 | 0 |
| 0 | 0 | 0 | 0 | 0 |
| 0 | 0 | 0 | 0 | 0 |
| 0 | 0 | 0 | 0 | 0 |
| 0 | 0 | 0 | 0 | 0 |
| 0 | 0 | 0 | 0 | 0 |
| 0 | 0 | 0 | 0.172554912 | 3.194679589 |
| 0 | 0 | 0 | 0 | 0 |
| 0 | 0 | 0 | 0 | 0 |
| 0 | 0 | 0 | 0 | 0 |
| 0 | 0 | 0 | 0 | 0 |
| 0 | 0 | 0 | 0 | 0 |
| 0 | 0.014966809 | 0.00378932 | 0 | 0.031458102 |
| 0.172385236 | 1.838846993 | 0.008890727 | 0.00159804 | 0.003891864 |
| 0.105172915 | 0.015128536 | 0 | 0.017957554 | 0.001993564 |
| 0.004515031 | 0 | 0 | 0 | 0 |
| 0 | 0 | 0 | 0 | 0 |

| | | | | |
|---|---|---|---|---|
| 0 | 0 | 0 | 0 | 0 |
| 0.806791079 | 0.237289149 | 0.009039518 | 0 | 0 |
| 0 | 0 | 0.003824008 | 0 | 0 |
| 0.239978578 | 0.444686074 | 0.347656319 | 0.718727507 | 0.334771996 |
| 0 | 0.021877048 | 0.003390913 | 0 | 0.001247387 |
| 0 | 0 | 0 | 0 | 0 |
| 0.001269078 | 0 | 0 | 0.014466651 | 0 |
| 0 | 0 | 0 | 0 | 0 |
| 0 | 0 | 0 | 0 | 0 |
| 0 | 0 | 0 | 0 | 0 |
| 0 | 0 | 0 | 0 | 0 |
| 0.196499195 | 1.681953173 | 0 | 0.02203099 | 0.05125508 |
| 0.000879531 | 0 | 0 | 0 | 0 |
| 0 | 0.110980409 | 0.218332941 | 0.001363232 | 0.008607441 |
| 0 | 0 | 0 | 0 | 0 |
| 0 | 0 | 0 | 0 | 0 |
| 0.004485491 | 0 | 1.244586553 | 0.713151687 | 0 |
| 0 | 0 | 0 | 0 | 0 |
| 0 | 0 | 0 | 0 | 0 |
| 0 | 0 | 0 | 0 | 0 |
| 0.060889347 | 0.063807249 | 0.038707557 | 0.11005542 | 0.050544517 |
| 0.007394119 | 0.008887568 | 0.013448372 | 0.004598514 | 0.001219958 |
| 0 | 0 | 0 | 0 | 0 |
| 0 | 0 | 0 | 0 | 0 |
| 0 | 0 | 0 | 0 | 0 |
| 0 | 0 | 0 | 0 | 0 |
| 0 | 0.011201627 | 0 | 0 | 0 |
| 0 | 0 | 0 | 0 | 0 |
| 0 | 0 | 0 | 0 | 0 |
| 0 | 0 | 0 | 0 | 0 |
| 0 | 0 | 0.000725642 | 0 | 0 |
| 0 | 0 | 0 | 0 | 0 |
| 0 | 0 | 0 | 0 | 0 |
| 0 | 0 | 0 | 0 | 0 |
| 0 | 0.002084992 | 0 | 0 | 0 |
| 0 | 0 | 0 | 0 | 0 |
| 0 | 0 | 0 | 0 | 0 |
| 0 | 0 | 0.04271029 | 0.016728545 | 0 |
| 0 | 0 | 0 | 0 | 0 |
| 0 | 0 | 0 | 0 | 0 |
| 0 | 0 | 0 | 0 | 0 |
| 0 | 0 | 0 | 0 | 0 |
| 0 | 0 | 0 | 0 | 0 |
| 0 | 0 | 0 | 0 | 0 |
| 0 | 0 | 0 | 0 | 0 |
| 0 | 0 | 0 | 0 | 0 |
| 0 | 0.073050264 | 0 | 0 | 0 |
| 0 | 0 | 0 | 0.008745388 | 0 |

| | | | | |
|---|---|---|---|---|
| 0 | 0 | 0 | 0 | 0 |
| 0 | 0 | 0.000504398 | 0 | 0 |
| 0 | 0 | 0 | 0 | 0 |
| 0 | 0 | 0 | 0 | 0 |
| 0 | 0 | 0 | 0 | 0 |
| 0 | 0 | 0 | 0 | 0 |
| 0 | 0 | 0 | 0 | 0 |
| 0 | 0 | 0 | 0 | 0 |
| 0 | 0 | 0 | 0 | 0 |
| 0 | 0 | 0 | 0 | 0 |
| 0 | 0 | 0 | 0 | 0 |
| 0 | 0.002770399 | 0 | 0 | 0 |
| 0 | 0 | 0 | 0 | 0 |
| 0 | 0 | 0 | 0 | 0 |
| 0 | 0 | 0 | 0 | 0 |
| 0 | 0 | 0 | 0 | 0 |
| 0 | 0 | 0 | 0 | 0 |
| 0 | 0 | 0 | 0 | 0 |
| 0.010086017 | 0.00608163 | 0 | 0 | 0 |
| 0 | 0 | 0 | 0 | 0 |
| 0 | 0 | 0 | 0 | 0 |
| 0 | 0 | 0 | 0 | 0 |
| 0 | 0 | 0 | 0 | 0 |
| 0 | 0 | 0 | 0 | 0 |
| 0 | 0 | 0 | 0 | 0 |
| 0 | 0 | 0 | 0 | 0 |
| 0 | 0 | 0 | 0 | 0 |
| 0 | 0 | 0.002291514 | 0 | 0 |
| 0 | 0 | 0 | 0 | 0 |
| 0 | 0 | 0 | 0 | 0 |
| 0 | 0 | 0.004868803 | 0.004355058 | 0.010257126 |
| 0.003046067 | 0 | 0.003004587 | 0 | 0.007784278 |
| 0 | 0 | 0 | 0 | 0 |
| 0 | 0 | 0 | 0 | 0 |
| 0 | 0 | 0 | 0 | 0 |
| 0 | 0 | 0 | 0 | 0 |
| 0 | 0 | 0 | 0 | 0 |
| 0 | 0 | 0 | 0 | 0 |
| 0 | 0 | 0 | 0.079842341 | 0 |
| 0 | 0 | 0 | 0 | 0 |
| 0 | 0 | 0 | 0 | 0 |
| 0 | 0 | 0 | 0 | 0 |
| 0 | 0 | 0 | 0.001173523 | 0.000793888 |
| 0 | 0 | 0 | 0 | 0 |
| 0 | 0 | 0 | 0 | 0 |
| 0 | 0 | 0 | 0 | 0 |
| 0 | 0 | 0 | 0 | 0 |
| 0 | 0 | 0 | 0 | 0 |

| | | | | |
|---|---|---|---|---|
| 0 | 0.002600339 | 0 | 0 | 0 |
| 0 | 0 | 0 | 0 | 0 |
| 0 | 0 | 0 | 0 | 0 |
| 0 | 0 | 0 | 0 | 0 |
| 0.019295259 | 0 | 0 | 0 | 0.00797098 |
| 0 | 0 | 0 | 0 | 0 |
| 0 | 0 | 0 | 0 | 0 |
| 0 | 0 | 0 | 0 | 0 |
| 0 | 0 | 0 | 0 | 0 |
| 0 | 0 | 0 | 0 | 0 |
| 0 | 0 | 0 | 0 | 0 |
| 0 | 0 | 0 | 0 | 0 |
| 0 | 0 | 0 | 0 | 0 |
| 0 | 0 | 0 | 0 | 0 |
| 0 | 0 | 0 | 0 | 0 |
| 0 | 0 | 0 | 0 | 6.638415607 |
| 0 | 0 | 0 | 0 | 0 |
| 0 | 0 | 0 | 0 | 0 |
| 0 | 0 | 0 | 0 | 0 |
| 0 | 0 | 0 | 0 | 0 |
| 0 | 0 | 0 | 0 | 0 |
| 0 | 0 | 0 | 0 | 0 |
| 0 | 0 | 0 | 0 | 0 |
| 0 | 0 | 0 | 0 | 0 |
| 0 | 0 | 0 | 0 | 0 |
| 0 | 0 | 0 | 0 | 0 |
| 0 | 0 | 0 | 0 | 0 |
| 0 | 0 | 0 | 0 | 0 |
| 0 | 0 | 0 | 0 | 0 |
| 0 | 0 | 0 | 0 | 0 |
| 0.054342859 | 0 | 0 | 0 | 0 |
| 0 | 0 | 0 | 0 | 0 |
| 0 | 0 | 0 | 0 | 0 |
| 0 | 0 | 0 | 0 | 0 |
| 0 | 0 | 0 | 0 | 0 |
| 0 | 0 | 0 | 0 | 0 |
| 0 | 0 | 0 | 0 | 0 |
| 0 | 0 | 0 | 0 | 0 |
| 0 | 0 | 0 | 0 | 0 |
| 0 | 0 | 0 | 0 | 0 |
| 0 | 0 | 0 | 0 | 0 |
| 0 | 0 | 0 | 0 | 0 |
| 0 | 0 | 0 | 0 | 0 |
| 0 | 0 | 0 | 0 | 0 |
| 0 | 0 | 0 | 0 | 0 |
| 0 | 0 | 0 | 0 | 0 |
| 0 | 0 | 0 | 0 | 0 |
| 0 | 0 | 0 | 0 | 0 |

| | | | | |
|---|---|---|---|---|
| 0 | 0 | 0 | 0 | 0 |
| 0 | 0 | 0 | 0 | 0 |
| 0 | 0 | 0 | 0 | 0 |
| 0 | 0 | 0 | 0 | 0 |
| 0 | 0 | 0 | 0 | 0 |
| 0 | 0 | 0 | 0 | 0 |
| 0 | 0 | 0 | 0 | 0 |
| 0 | 0 | 0 | 0 | 0 |
| 0 | 0 | 0 | 0 | 0 |
| 0 | 0 | 0 | 0 | 0 |
| 0 | 0 | 0 | 0 | 0 |
| 0 | 0 | 0 | 0 | 0 |
| 0 | 0.007843383 | 0.001161034 | 0 | 0 |
| 0 | 0 | 0 | 0 | 0 |
| 0 | 0.007030947 | 0 | 0 | 0 |
| 0 | 0 | 0 | 0 | 0 |
| 0 | 0 | 0 | 0 | 0 |
| 0 | 0 | 0 | 0 | 0 |
| 0 | 0 | 0 | 0 | 0 |
| 0 | 0 | 0 | 0 | 0 |
| 0.051352568 | 0.049096681 | 0.004341709 | 0.026722742 | 0.005198881 |
| 0 | 0 | 0 | 0 | 0 |
| 0 | 0.000699339 | 0 | 0 | 0 |
| 0.002550019 | 0 | 0.004204134 | 0 | 0 |
| 0 | 0 | 0 | 0 | 0 |
| 0 | 0 | 0 | 0 | 0 |
| 0.018717009 | 0 | 0.000722431 | 0 | 0.000412869 |
| 0 | 0 | 0 | 0 | 0 |
| 0 | 0 | 0 | 0 | 0 |
| 0 | 0.014103619 | 0 | 0 | 0 |
| 0.750971608 | 0.027576723 | 0.004140812 | 0 | 0 |
| 0 | 0 | 0 | 0 | 0 |
| 0 | 0 | 0 | 0 | 0 |
| 0.001494452 | 0.027082293 | 0 | 0.003522433 | 0 |
| 0.00522784 | 0.031944918 | 0.004439203 | 0 | 0.010763039 |
| 0 | 0 | 0 | 0 | 0 |
| 0 | 0 | 0 | 0 | 0 |
| 0 | 0 | 0 | 0 | 0 |
| 0 | 0 | 0 | 0 | 0 |
| 0 | 0 | 0 | 0 | 0 |
| 0 | 0 | 0 | 0 | 0 |
| 0 | 0 | 0 | 0 | 0 |
| 0 | 0 | 0 | 0 | 0 |
| 0 | 0 | 0 | 0 | 0 |
| 0 | 0 | 0 | 0 | 0 |
| 0 | 0 | 0 | 0 | 0 |
| 0 | 0.035486237 | 0.006991238 | 0 | 0 |
| 0 | 0.010199818 | 0 | 0 | 0 |

| | | | | |
|---|---|---|---|---|
| 0 | 0 | 0 | 0 | 0 |
| 0 | 0 | 0 | 0 | 0 |
| 0 | 0 | 0 | 0.001776045 | 0 |
| 0 | 0 | 0 | 0 | 0 |
| 0 | 0 | 0 | 0 | 0 |
| 0 | 0 | 0 | 0 | 0 |
| 0 | 0 | 0 | 0 | 0 |
| 0 | 0 | 0 | 0 | 0 |
| 0 | 0 | 0 | 0 | 0 |
| 0 | 0 | 0 | 0 | 0 |
| 0 | 0 | 0 | 0 | 0 |
| 0 | 0 | 0 | 0 | 0 |
| 0 | 0 | 0 | 0 | 0 |
| 0 | 0 | 0 | 0 | 0 |
| 0 | 0 | 0 | 0 | 0 |
| 0 | 0 | 0 | 0 | 0 |
| 0 | 0 | 0 | 0 | 0 |
| 0 | 0 | 0 | 0 | 0 |
| 0 | 0.007817445 | 0 | 0 | 0 |
| 0 | 0 | 0 | 0 | 0 |
| 0 | 0.561718411 | 0.040986264 | 0.035353681 | 0 |
| 0 | 0 | 0 | 0 | 0 |
| 0 | 0 | 0 | 0 | 0 |
| 0 | 0 | 0 | 0 | 0 |
| 0.018827657 | 0.006168027 | 0 | 0.002896394 | 0 |
| 0 | 0 | 0 | 0 | 0 |
| 0 | 0 | 0 | 0 | 0 |
| 0 | 0 | 0 | 0 | 0 |
| 0 | 0 | 0 | 0 | 0 |
| 0 | 0 | 0 | 0 | 0 |
| 0.045204506 | 2.261154877 | 0.690300119 | 0.046095197 | 0.482617335 |
| 0 | 0 | 0 | 0 | 0 |
| 0 | 0 | 0.017947702 | 0 | 0.00646235 |
| 0 | 0 | 0 | 0 | 0 |
| 0 | 0 | 0 | 0 | 0 |
| 0 | 0 | 0 | 0 | 0 |
| 0 | 0 | 0 | 0 | 0 |
| 0 | 0 | 0 | 0 | 0 |
| 0 | 0 | 0 | 0 | 0 |
| 0 | 0 | 0 | 0 | 0 |
| 0 | 0 | 0 | 0 | 0 |
| 0.002367669 | 0.042716662 | 0.006952387 | 0.001912237 | 0 |
| 0 | 0.000418376 | 0.003934092 | 0 | 0 |
| 0 | 0 | 0 | 0 | 0 |
| 0 | 0 | 0 | 0 | 0 |
| 0 | 0 | 0 | 0 | 0 |
| 0 | 0 | 0 | 0 | 0 |
| 0 | 0 | 0 | 0 | 0 |

| | | | | |
|---|---|---|---|---|
| 0 | 0 | 0 | 0 | 0 |
| 0 | 0 | 0 | 0 | 0 |
| 0.100167156 | 0.020035909 | 0 | 0.018522987 | 0 |
| 0 | 0 | 0 | 0 | 0 |
| 4.098905003 | 0 | 0 | 0 | 0 |
| 0 | 0 | 0 | 0 | 0 |
| 0 | 0 | 0 | 0 | 0 |
| 0 | 0 | 0 | 0 | 0 |
| 0 | 0 | 0 | 0 | 0 |
| 0 | 0.001336175 | 0 | 0.000941166 | 0 |
| 0 | 0 | 0 | 0 | 0 |
| 0 | 0 | 0 | 0 | 0 |
| 0 | 0 | 0 | 0 | 0 |
| 0.003538445 | 0 | 0 | 0.006065554 | 0.001538755 |
| 0.007927135 | 0.011232367 | 0 | 0 | 0 |
| 0.00201009 | 0 | 0 | 0 | 0 |
| 0.0159253 | 0.003253799 | 0.000633027 | 0.006060769 | 0 |
| 0 | 0 | 0 | 0 | 0 |
| 0 | 0 | 0 | 0 | 0 |
| 0 | 0 | 0 | 0 | 0 |
| 0 | 0 | 0 | 0 | 0 |
| 0 | 0.008420121 | 0 | 0 | 0 |
| 0 | 0 | 0 | 0 | 0 |
| 0 | 0.003035607 | 0 | 0 | 0 |
| 0 | 0 | 0 | 0 | 0 |
| 0 | 0 | 0 | 0 | 0 |
| 0 | 0 | 0 | 0 | 0 |
| 0 | 0 | 0 | 0 | 0 |
| 0 | 0 | 0 | 0 | 0 |
| 0 | 0 | 0 | 0 | 0 |
| 0 | 0 | 0 | 0 | 0 |
| 0 | 0 | 0 | 0 | 0 |
| 0 | 0 | 0 | 0 | 0 |
| 0 | 0 | 0 | 0 | 0 |
| 0 | 0 | 0 | 0 | 0 |
| 0.013285591 | 0 | 0 | 0 | 0 |
| 0 | 0 | 0 | 0 | 0 |
| 0.004582885 | 0.007134681 | 0 | 0 | 0.001823612 |
| 0 | 0 | 0 | 0 | 0 |
| 0 | 0 | 0 | 0 | 0 |
| 0 | 0 | 0 | 0 | 0 |
| 0 | 0 | 0 | 0 | 0 |
| 0 | 0 | 0 | 0 | 0 |
| 0 | 0 | 0 | 0 | 0 |
| 0 | 0 | 0 | 0 | 0 |
| 0 | 0 | 0 | 0 | 0 |
| 0 | 0 | 0 | 0 | 0 |
| 0 | 0.001191985 | 0 | 0 | 0 |

| | | | | |
|---|---|---|---|---|
| 0 | 0.807031633 | 0.150969244 | 0 | 1.278965465 |
| 0 | 0 | 0 | 0 | 0 |
| 0 | 0 | 0 | 0 | 0 |
| 0 | 0 | 0 | 0 | 0 |
| 0.00090445 | 0.004333417 | 0 | 0 | 0.002141386 |
| 0 | 0 | 0 | 0 | 0 |
| 0 | 0 | 0 | 0 | 0 |
| 0.052613114 | 0.002024746 | 0.002532307 | 0 | 0 |
| 0 | 0 | 0 | 0 | 0 |
| 0.024501269 | 0.161887657 | 0.40630607 | 0 | 1.710009194 |
| 0 | 0 | 0.002884286 | 0 | 0 |
| 0 | 0 | 0.001336389 | 0.002634254 | 0 |
| 0.015605706 | 0.003647333 | 0.001351598 | 0.01798358 | 0.001416135 |
| 0.021388746 | 0.056448702 | 0.080056399 | 0 | 0 |
| 0 | 0.000893143 | 0 | 0 | 0 |
| 0 | 0 | 0 | 0.004713646 | 0 |
| 0 | 0 | 0 | 0 | 0 |
| 0 | 0 | 0 | 0 | 0 |
| 0 | 0 | 0.027562446 | 0 | 0 |
| 0 | 0.1099616 | 0.075684955 | 0 | 0 |
| 0 | 0 | 0 | 0 | 0 |
| 0 | 0 | 0 | 0 | 0 |
| 0.000589137 | 0.005734974 | 0 | 0 | 0 |
| 0 | 0 | 0 | 0 | 0 |
| 0 | 0 | 0 | 0 | 0 |
| 0.006428334 | 0 | 0 | 0.002992327 | 0 |
| 0 | 0 | 0 | 0 | 0 |
| 0.004556963 | 2.104324041 | 0 | 0 | 0 |
| 0 | 0 | 0.174882681 | 0 | 0 |
| 0 | 0.007076658 | 0.000607396 | 0 | 0.001542784 |
| 0.211457912 | 0 | 0 | 0 | 0 |
| 0 | 0 | 0 | 0 | 0 |
| 0 | 0 | 0 | 0 | 0 |
| 0 | 0.095508267 | 0.11040591 | 0 | 0 |
| 0 | 0 | 0 | 0 | 0 |
| 0 | 0 | 0 | 0 | 0 |
| 0 | 0.002522554 | 0.001561518 | 0 | 0 |
| 0 | 0 | 0 | 0 | 0 |
| 0.000591968 | 0 | 0 | 0 | 0 |
| 0 | 0.015318971 | 0.003932348 | 0.002098108 | 0 |
| 0 | 0 | 0 | 0 | 0 |
| 0 | 0 | 0.011324247 | 0 | 0 |
| 0 | 0 | 0 | 0 | 0 |
| 0 | 0 | 0.133804035 | 0.015803633 | 0 |
| 0 | 0 | 0 | 0 | 0 |
| 0 | 0 | 0 | 0 | 0 |
| 0.002756601 | 0.008553411 | 0 | 0 | 0 |
| 0 | 0 | 0 | 0 | 0 |

| | | | | |
|---|---|---|---|---|
| 0 | 0 | 0 | 0 | 0 |
| 0 | 0.016306589 | 0.00082046 | 0.025876325 | 0.000833587 |
| 0 | 0 | 0 | 0 | 0 |
| 0 | 0 | 0 | 0 | 0 |
| 0 | 0.145709787 | 0.005079391 | 0.001634672 | 0 |
| 0 | 0.043591364 | 0.042403502 | 0 | 0 |
| 0 | 0 | 0 | 0 | 0 |
| 0 | 0 | 0 | 0 | 0 |
| 0 | 0 | 0 | 0 | 0 |
| 0 | 0 | 0 | 0 | 0 |
| 0.006043623 | 0.00344718 | 0.014754277 | 0 | 0.001533102 |
| 0 | 0 | 0 | 0 | 0 |
| 0 | 0.001333642 | 0 | 0 | 0 |
| 0.020221895 | 0.005210756 | 0.003801572 | 0.160219465 | 0 |
| 0 | 0.24099027 | 0.005342372 | 0 | 0 |
| 0 | 0 | 0 | 0 | 0 |
| 0 | 0 | 0 | 0 | 0 |
| 0 | 0 | 0.012742874 | 0 | 0 |
| 0 | 0 | 0 | 0 | 0 |
| 0 | 0 | 0.004052541 | 0 | 0 |
| 0 | 0 | 0 | 0.164171422 | 0 |
| 0.324386255 | 0 | 0 | 0.160166491 | 0.265468395 |
| 0 | 0 | 0.001402921 | 0 | 0 |
| 0 | 0 | 0 | 0 | 0 |
| 0 | 0 | 0 | 0 | 0 |
| 0 | 0 | 0 | 0 | 0 |
| 0 | 0 | 0.021067545 | 0 | 0 |
| 0 | 0 | 0 | 0 | 0 |
| 0 | 0 | 0 | 0 | 0 |
| 0.419159487 | 0.074469301 | 0.32071734 | 0.277738044 | 0.61025825 |
| 0.006940174 | 0.001178361 | 0 | 0 | 0 |
| 0 | 0 | 0 | 0 | 0 |
| 0 | 0 | 0.003750887 | 0 | 0 |
| 0 | 0 | 0 | 0 | 0.005721558 |
| 0 | 0 | 0 | 0 | 0 |
| 0 | 0 | 0 | 0 | 0 |
| 0 | 0 | 0 | 0 | 0 |
| 0 | 0 | 0 | 0 | 0 |
| 0 | 0 | 0 | 0 | 0 |
| 0 | 1.334608055 | 0.019742246 | 0.003800057 | 0 |
| 0 | 0.007496665 | 0.043754287 | 0 | 0.021697938 |
| 2.752253134 | 0 | 0.094524317 | 6.541393476 | 2.547359755 |
| 0 | 0 | 0 | 0 | 0 |
| 0 | 0 | 0 | 0 | 0 |
| 0 | 0.006845842 | 0.007051017 | 0 | 0 |
| 0 | 0 | 0.002595152 | 0 | 0 |
| 0.004926694 | 0.001873402 | 0.000911176 | 0.001539502 | 0 |
| 0 | 0 | 0 | 0 | 0 |

| | | | | |
|---|---|---|---|---|
| 0.002532797 | 0 | 0 | 0.001899488 | 0 |
| 0 | 0.049921144 | 0 | 0 | 0 |
| 0.013114296 | 0 | 0.102634659 | 1.681293076 | 0 |
| 0 | 0 | 0 | 0 | 0 |
| 0 | 0 | 0 | 0 | 0 |
| 0 | 0 | 0 | 0 | 0 |
| 0 | 0 | 0 | 0 | 0 |
| 0 | 0 | 0 | 0 | 0 |
| 0 | 0.014702106 | 0.02295763 | 0 | 0 |
| 0 | 0 | 0 | 0 | 0 |
| 0 | 0 | 0 | 0 | 0 |
| 0 | 0 | 0 | 0 | 0 |
| 0 | 0 | 0 | 0 | 0 |
| 0 | 0.000836449 | 0.015459462 | 0 | 0.014105118 |
| 0 | 0 | 0 | 0 | 0 |
| 0.067328689 | 0.020100368 | 0.121607848 | 0 | 0.022045812 |
| 0.099436284 | 0.003622628 | 0 | 0.002083873 | 0.00147687 |
| 0 | 1.380263917 | 0.003639203 | 0.00434027 | 0 |
| 0 | 0.004677376 | 0 | 0 | 0 |
| 0 | 0 | 0 | 0 | 0 |
| 0 | 0 | 0 | 0.135065456 | 0.016074659 |
| 0.004374207 | 0 | 0 | 0 | 0 |
| 0 | 0 | 1.213610171 | 0 | 0 |
| 0 | 0.381297613 | 0.000972489 | 0 | 0 |
| 0 | 0 | 0 | 0 | 0.002384116 |
| 0 | 0.015032348 | 0.066614601 | 0 | 0 |
| 0.018645513 | 0.006898433 | 0.003578907 | 0 | 0.002215789 |
| 0 | 0 | 0 | 0 | 0 |
| 0 | 0 | 0 | 0 | 0 |
| 0 | 0.001574057 | 0 | 0 | 0 |
| 0 | 0.000524624 | 0.001530984 | 0 | 0.00777739 |
| 0.060564479 | 0 | 0 | 0 | 0.000773009 |
| 0.035632972 | 0.01961076 | 0.112043477 | 0.293750937 | 0.435717337 |
| 0 | 0.012451197 | 0 | 0 | 0 |
| 0 | 0 | 0 | 0 | 0 |
| 0 | 0 | 0 | 0 | 0 |
| 0.017705769 | 0 | 0 | 0.039097967 | 0 |
| 0 | 0.073889626 | 0.038242886 | 0.002596111 | 0.012228834 |
| 0 | 0 | 0 | 1.375501111 | 0 |
| 0 | 0 | 0 | 0 | 0.000531826 |
| 0 | 0 | 0 | 0 | 0.002353131 |
| 0 | 0 | 0 | 0 | 0 |
| 0.01207953 | 0.002362272 | 0.009446922 | 0 | 0 |
| 0 | 0.308676841 | 0 | 0 | 0 |
| 0 | 0.023057385 | 0.573245373 | 0.183309244 | 0 |
| 0 | 0 | 0 | 0 | 0 |
| 0 | 0 | 0 | 0 | 0 |
| 0 | 0 | 0 | 0 | 0 |

| | | | | |
|---|---|---|---|---|
| 0 | 0 | 0 | 0 | 0 |
| 2.634696615 | 0.280528074 | 0.024194292 | 0.013757065 | 0.018436033 |
| 0 | 0 | 0 | 0 | 0 |
| 0 | 0 | 0 | 0.001190257 | 0 |
| 0 | 0 | 0 | 0 | 0 |
| 0 | 0 | 0 | 0 | 0 |
| 0.008242777 | 0.021018649 | 0.002367419 | 0 | 0 |
| 0 | 0 | 0 | 0 | 0 |
| 0 | 0 | 0 | 0 | 0 |
| 0 | 0 | 0.021976899 | 0 | 0.003864549 |
| 0.003115763 | 0.357212867 | 0.024721131 | 0.001168343 | 0.001449037 |
| 0.769828789 | 0.139505211 | 0.001287511 | 1.515349197 | 0.001373516 |
| 0 | 0 | 0 | 0 | 0 |
| 0 | 0 | 0 | 0 | 0 |
| 0.014970097 | 0 | 0 | 0.019437653 | 0 |
| 0 | 0 | 0 | 0 | 0 |
| 0.062194336 | 0.015445987 | 0.000905126 | 0.001835136 | 0.000551764 |
| 0 | 0.171762399 | 0.018094663 | 0.007940867 | 0.001969731 |
| 0 | 0 | 0 | 0 | 0 |
| 0 | 0.018039881 | 0.000457782 | 0.000773458 | 0 |
| 0.06434834 | 0.015736798 | 0 | 0 | 0.015363214 |
| 0 | 0 | 0 | 0 | 0 |
| 0 | 0 | 0 | 0 | 0 |
| 0 | 0 | 0 | 0 | 0 |
| 0 | 0 | 0 | 0.011129066 | 0 |
| 0 | 0.040829412 | 0 | 0 | 0 |
| 0 | 0.364646779 | 0.003501036 | 0.035180299 | 0.060657039 |
| 0.038443004 | 0.009178844 | 0.111311384 | 0.040228713 | 0.063954577 |
| 0 | 0 | 0 | 0 | 0 |
| 0 | 0.001230204 | 0 | 0 | 0 |
| 0.000959288 | 0 | 0.006387016 | 0 | 0 |
| 0 | 0 | 0 | 0 | 0 |
| 0 | 0 | 0 | 0 | 0 |
| 0.006771128 | 0 | 0 | 0 | 0 |
| 0 | 0 | 0.007953664 | 0 | 0 |
| 0.928275304 | 0.048675221 | 0.270996465 | 0.835172272 | 0.49641668 |
| 0 | 0 | 0.000466577 | 0 | 0 |
| 0 | 0 | 0 | 0 | 0 |
| 0.009374461 | 0 | 0 | 0.005789772 | 0.005968426 |
| 0 | 0 | 0 | 0 | 0 |
| 0 | 0.027473915 | 0.579047144 | 0.196188043 | 0 |
| 0 | 0 | 0 | 0 | 0 |
| 0.014235477 | 0 | 0.00223014 | 0.000753599 | 0 |
| 0 | 0.00059026 | 0 | 0 | 0 |
| 0 | 0 | 0 | 0 | 0 |
| 0 | 0 | 0 | 0.616431794 | 0 |
| 0 | 0 | 0 | 0 | 0 |
| 0.104808421 | 0.001192637 | 0 | 0 | 0 |

| | | | | |
|---|---|---|---|---|
| 0 | 0 | 0 | 0 | 0 |
| 0 | 0 | 0 | 0 | 0 |
| 0 | 0 | 0 | 0 | 0 |
| 0 | 0 | 0 | 0.031012226 | 0.014514301 |
| 0 | 0 | 0 | 0 | 0 |
| 0 | 0 | 0 | 0 | 0 |
| 0 | 0 | 0 | 0 | 0 |
| 0.064041211 | 0 | 0 | 0 | 0.002242794 |
| 0 | 0 | 0 | 0 | 0.210030338 |
| 0 | 0 | 0 | 0 | 0 |
| 0 | 0 | 0 | 0 | 0 |
| 0 | 0.001876094 | 0 | 0.022705252 | 0.002528411 |
| 0 | 0 | 0 | 0 | 0 |
| 0 | 0 | 0 | 0 | 0 |
| 0 | 0 | 0.034500409 | 0.01457278 | 0.03661722 |
| 0 | 0 | 0 | 0 | 0 |
| 0 | 0 | 0 | 0 | 0 |
| 0 | 0 | 0 | 0 | 0 |
| 0 | 0 | 0 | 0 | 0 |
| 0 | 0 | 0 | 0 | 0 |
| 0 | 0 | 0 | 0 | 0 |
| 0 | 0 | 0 | 0 | 0 |
| 0 | 0 | 0 | 0 | 0 |
| 0.386383048 | 0.069276382 | 0.311622844 | 0.36987231 | 0 |
| 0 | 0.002714596 | 0.026250904 | 0 | 0 |
| 0 | 0 | 0 | 0 | 0 |
| 0 | 0 | 0 | 0 | 0 |
| 0 | 0 | 0.08912822 | 1.313044344 | 0 |
| 0 | 0 | 0 | 0 | 0 |
| 0 | 0 | 0 | 0 | 0 |
| 0 | 0 | 0 | 0 | 0 |
| 0 | 0 | 0 | 0 | 0 |
| 0 | 0.004290519 | 0 | 0 | 0.010008534 |
| 0 | 0 | 0 | 0 | 0 |
| 0 | 0 | 0 | 0 | 0 |
| 0 | 0 | 0 | 0 | 0.016881605 |
| 0 | 0 | 0 | 0 | 0.014131369 |
| 0 | 0.002839309 | 0 | 0 | 0 |
| 0 | 0 | 0 | 0 | 0.005000945 |
| 0 | 0 | 0 | 0.001003198 | 0 |
| 0.010039854 | 0 | 0.000795788 | 0.001613454 | 0.01515974 |
| 0 | 0 | 0 | 0 | 0 |
| 0 | 14.35259981 | 0.217468133 | 0.569047671 | 0.180259331 |
| 0 | 0 | 0 | 0 | 0 |
| 0 | 0 | 0 | 0 | 0 |
| 0 | 0 | 0 | 0 | 0 |
| 0 | 0 | 0 | 0 | 0 |
| 0 | 0.564683435 | 0.132397327 | 0 | 0.00072895 |

| | | | | |
|---|---|---|---|---|
| 0 | 0 | 0 | 0 | 0 |
| 0 | 0 | 0 | 0 | 0 |
| 0.008883764 | 1.559456063 | 0.286115238 | 0.03873509 | 0.15401128 |
| 0 | 0 | 0 | 0 | 0 |
| 0 | 0 | 0 | 0 | 0 |
| 0 | 0 | 0 | 0.00122056 | 0.000660567 |
| 0 | 0 | 0 | 0 | 0 |
| 0 | 0 | 0 | 0 | 0 |
| 0 | 0 | 0 | 0 | 0 |
| 0 | 0 | 0 | 0 | 0 |
| 0.009939343 | 0.253241148 | 0.011704768 | 0.006084958 | 0.007546867 |
| 0.898419876 | 3.975605927 | 1.778243213 | 2.806511505 | 2.568910039 |
| 0 | 0 | 0 | 0 | 0 |
| 0.030210577 | 0.00779168 | 0 | 0 | 0.014364617 |
| 0.005693001 | 0.072264758 | 0.254993655 | 0.391759177 | 0.028124624 |
| 0 | 0 | 0 | 0 | 0 |
| 0 | 0 | 0 | 0 | 0 |
| 0 | 0 | 0 | 0 | 0 |
| 0 | 0 | 0 | 0 | 0 |
| 0 | 0.047423875 | 0 | 0.000751861 | 0.002147564 |
| 0.509188688 | 0 | 0 | 0.110926982 | 0.191601681 |
| 0 | 0 | 33.57674955 | 0.163098283 | 2.942291925 |
| 0.28936754 | 3.646128638 | 1.04606961 | 0 | 19.85482648 |
| 0 | 0 | 0 | 0 | 0 |
| 0 | 0 | 0 | 0 | 0 |
| 0 | 0 | 0 | 0 | 0.005512516 |
| 0.0965881 | 0 | 0 | 0 | 0 |
| 0 | 0 | 0 | 0 | 0.000359851 |
| 0 | 0 | 0.002524898 | 0 | 0 |
| 0 | 0 | 0.049734449 | 0 | 0 |
| 0 | 0 | 0 | 0 | 0 |
| 0 | 0 | 0 | 0.137705481 | 0.178063713 |
| 0 | 0 | 0 | 0.000937412 | 0 |
| 0 | 0 | 0 | 0 | 0.009779368 |
| 0 | 0.034131128 | 0.001098564 | 0.000618703 | 0 |
| 0.010579146 | 0.072582387 | 0 | 0.014734382 | 0.017251971 |
| 0 | 0 | 0.002106031 | 0.001642294 | 0.001481348 |
| 0 | 0 | 0 | 0 | 0 |
| 0 | 0 | 0 | 0.007626359 | 0.056676756 |
| 0 | 0 | 0 | 0 | 0 |
| 0 | 0 | 0 | 0 | 0 |
| 0 | 0 | 0 | 0 | 0 |
| 0 | 0 | 0 | 0 | 0 |
| 0 | 0 | 0 | 0 | 0 |
| 0.048041824 | 1.060896831 | 0 | 0.008078313 | 0.016030597 |
| 0 | 0 | 0 | 0.043457535 | 0 |
| 0 | 0 | 0 | 0 | 0 |
| 0.00207898 | 0 | 0 | 0 | 0.082095548 |

| | | | | |
|---|---|---|---|---|
| 0 | 0 | 0 | 0 | 0 |
| 0 | 0 | 0 | 0 | 0 |
| 0 | 0 | 0 | 0 | 0 |
| 0 | 0 | 0 | 0 | 0.001863111 |
| 0 | 0 | 0 | 0 | 0 |
| 0 | 0 | 0 | 0 | 0 |
| 0 | 0 | 0 | 0 | 0.035899629 |
| 0.010475385 | 0.328122888 | 0 | 0.007349238 | 0.002171572 |
| 0 | 0 | 0 | 0 | 0 |
| 0 | 0 | 0 | 0 | 0 |
| 0 | 0.020528928 | 0.000544623 | 0 | 0 |
| 0 | 0 | 0 | 0 | 0 |
| 0 | 0 | 0 | 0 | 0.003512548 |
| 0 | 0 | 0 | 0 | 0 |
| 0 | 0 | 0 | 0 | 0 |
| 0 | 0 | 0 | 0 | 0 |
| 0 | 0 | 0 | 0 | 0 |
| 0 | 0 | 0 | 0.003830157 | 0.00264868 |
| 0 | 0 | 0 | 0 | 0 |
| 0 | 0 | 0 | 0 | 0 |
| 0 | 0 | 0 | 0.001361117 | 0 |
| 0 | 0 | 0 | 0 | 0 |
| 0 | 0 | 0 | 0 | 0.033723901 |
| 0 | 0 | 0 | 0 | 0 |
| 0 | 0 | 0 | 0 | 0 |
| 0 | 0 | 0 | 0 | 0 |
| 0 | 0 | 0 | 0 | 0 |
| 0 | 0 | 0 | 0 | 0 |
| 0 | 0 | 0 | 0 | 0 |
| 0.025500268 | 0 | 0.714000845 | 0 | 0.037142978 |
| 0 | 0.091427309 | 0.011009752 | 0 | 0.004358142 |
| 0 | 0 | 0 | 0 | 0 |
| 0 | 0 | 0 | 0.01307275 | 0 |
| 0 | 0 | 0 | 0 | 0 |
| 0 | 0 | 0.000721763 | 0 | 0 |
| 0 | 0 | 0 | 0 | 0 |
| 0.357175894 | 0 | 0 | 0 | 0 |
| 0.079803853 | 0.009794927 | 0.004456653 | 0.004154406 | 0.002224943 |
| 0 | 0 | 0 | 0 | 0 |
| 0 | 0 | 0 | 0 | 0.001963555 |
| 0 | 0 | 0 | 0 | 0.009119286 |
| 0 | 0 | 0 | 0 | 0 |
| 0 | 0 | 0 | 0 | 0 |
| 0.000672442 | 0 | 0 | 2.373752123 | 0 |
| 0 | 0 | 0 | 0 | 0.027125558 |
| 0 | 0 | 0 | 0 | 0 |
| 0 | 0 | 0 | 0 | 0.000556334 |
| 0 | 0 | 0 | 0 | 0 |

| | | | | |
|---|---|---|---|---|
| 0 | 0 | 0 | 0 | 0 |
| 0 | 0 | 0 | 0 | 0 |
| 0 | 0 | 0 | 0 | 0 |
| 0 | 0 | 0 | 0 | 0 |
| 0 | 0 | 0 | 0 | 0 |
| 0 | 0 | 0 | 0 | 0.126896508 |
| 0 | 0.251870375 | 0 | 0 | 0 |
| 0 | 0 | 0 | 0 | 0 |
| 0.083084315 | 0.248824126 | 0.008477887 | 0.009310632 | 0.021479978 |
| 0 | 0 | 0 | 0 | 0.000650291 |
| 0 | 0 | 0 | 0 | 0 |
| 0 | 0 | 0 | 0 | 0 |
| 0 | 0 | 0 | 0 | 0 |
| 0 | 0.008323457 | 0 | 0 | 0 |
| 0 | 0.715872645 | 0 | 0 | 0 |
| 0 | 0.017968474 | 0 | 0 | 0 |
| 0 | 0.069653338 | 0 | 0 | 0 |
| 0 | 0 | 0 | 0 | 0 |
| 0 | 0 | 0 | 0 | 0 |
| 0 | 0 | 0 | 0 | 0 |
| 0 | 0 | 0 | 0 | 0 |
| 0 | 0 | 0 | 0 | 0 |
| 0 | 0 | 0 | 0 | 0 |
| 0 | 0 | 0 | 0 | 0 |
| 0.070190221 | 2.417492264 | 0 | 0.001684467 | 0.011775265 |
| 0.665541522 | 0 | 0.005638145 | 9.414152958 | 0.043141636 |
| 0.038011217 | 0 | 0 | 0.013783486 | 0 |
| 0 | 0 | 0 | 0 | 0.000471578 |
| 0 | 0 | 0 | 0.015831592 | 0 |
| 0 | 0 | 0 | 0 | 0 |
| 0 | 0 | 0 | 0 | 0 |
| 0 | 0 | 0 | 0.007051241 | 0.008194895 |
| 0 | 0 | 0 | 0 | 0 |
| 0.001199555 | 0.07129423 | 0 | 0 | 0 |
| 0.019270751 | 0 | 0 | 0 | 0.003008285 |
| 0.001397137 | 0.005418942 | 0.004961201 | 0 | 0 |
| 0 | 0 | 0.0057598 | 0 | 0.028162513 |
| 2.093092758 | 0 | 0.005076861 | 0.168005596 | 0.509049664 |
| 0 | 0 | 0 | 0 | 0 |
| 0 | 0 | 0 | 0 | 0 |
| 0 | 0 | 0 | 0 | 0 |
| 0 | 0.002711084 | 0.002109766 | 0 | 0.006430559 |
| 4.76205308 | 0 | 0.005084677 | 0 | 3.249107868 |
| 0 | 0 | 0 | 0 | 0 |
| 0 | 0 | 0 | 0 | 0 |
| 0 | 0 | 0 | 0.00893386 | 0.003581484 |
| 0 | 0 | 0 | 0 | 0 |
| 1.169522355 | 0 | 0 | 0.026488656 | 0.945190397 |

| | | | | |
|---|---|---|---|---|
| 0 | 0 | 0.124935182 | 0 | 0.001329616 |
| 0 | 0 | 0 | 0 | 0 |
| 0 | 0 | 0 | 0 | 0 |
| 0 | 0 | 0 | 0 | 0 |
| 0 | 0.00250029 | 0 | 0 | 0 |
| 0 | 0 | 0 | 0 | 0.008158637 |
| 0 | 0 | 0 | 0 | 0 |
| 0 | 0 | 0 | 0 | 0 |
| 0 | 0 | 0 | 0 | 0 |
| 0 | 0 | 0 | 0 | 0 |
| 0 | 0 | 0 | 0 | 0 |
| 0.23668721 | 0 | 0.008334188 | 0 | 0 |
| 0.948511352 | 0 | 0 | 0.336923497 | 0 |
| 0 | 0 | 0 | 0 | 0 |
| 0.004300064 | 0.102651259 | 0.011853898 | 0.01018377 | 0.009032661 |
| 0 | 0 | 0 | 0 | 0 |
| 0 | 0.071408778 | 0 | 0 | 0 |
| 0 | 0 | 0 | 0 | 0 |
| 0 | 0.025706703 | 0 | 0 | 0.004294018 |
| 0 | 0 | 0 | 0 | 0 |
| 0 | 0 | 0 | 0.011577574 | 0 |
| 0 | 0 | 0 | 0 | 0 |
| 1.428126873 | 0 | 0 | 0.131926258 | 0 |
| 0 | 0 | 0 | 0.004822766 | 0 |
| 0 | 0 | 0 | 0 | 0 |
| 0 | 0 | 0 | 0 | 0 |
| 0 | 0 | 0 | 0 | 0 |
| 0 | 0 | 0 | 0 | 0 |
| 0 | 0 | 0 | 0 | 0 |
| 0 | 0 | 0 | 0 | 0 |
| 0 | 0 | 0 | 0 | 0 |
| 0.135680505 | 0.080673065 | 0.008342828 | 0.040525588 | 0.002383961 |
| 0 | 0 | 0 | 0.112500103 | 0 |
| 0.041521149 | 0.080204056 | 0.054964619 | 0.005501599 | 0.080391538 |
| 0.001900544 | 0.00485649 | 0.003880553 | 0 | 0.000514257 |
| 0 | 0 | 0 | 0 | 0 |
| 0 | 0 | 0 | 0 | 0 |
| 0 | 0 | 0 | 0 | 0 |
| 0 | 0 | 0 | 0.003274976 | 0 |
| 0 | 0 | 0 | 0 | 0 |
| 0.157393068 | 0 | 0 | 0 | 0 |
| 0 | 0 | 0 | 0 | 0 |
| 0 | 0 | 0 | 0 | 0 |
| 0 | 0 | 0 | 0 | 0 |
| 0.002995562 | 0.206230147 | 0.007645466 | 0 | 0.014564599 |
| 0 | 0 | 0 | 0 | 0 |
| 0 | 0 | 0 | 0 | 3.185804783 |
| 0 | 0.007272209 | 0 | 0 | 0.001557231 |

| | | | | |
|---|---|---|---|---|
| 0 | 0 | 0 | 0 | 0 |
| 0 | 0 | 0 | 0 | 0 |
| 0 | 0.014843961 | 0.001375187 | 0 | 0.005632417 |
| 0 | 0 | 0 | 0 | 0 |
| 0 | 0 | 0 | 0 | 0 |
| 0 | 0 | 0 | 0 | 0 |
| 0 | 0 | 0 | 0 | 0.059549292 |
| 0 | 0 | 0 | 0 | 0 |
| 0 | 0 | 0 | 0 | 0 |
| 0 | 0 | 0 | 0 | 0 |
| 0 | 0 | 0 | 0 | 0 |
| 0 | 0 | 0 | 0 | 0 |
| 0 | 0 | 0 | 0 | 0 |
| 0 | 0 | 0 | 0 | 0 |
| 0 | 0 | 0 | 0 | 0 |
| 0 | 0 | 0 | 0 | 0 |
| 0 | 0 | 0 | 0 | 0.072205849 |
| 0 | 0 | 0 | 0 | 0 |
| 0 | 0 | 0.021490449 | 0 | 1.407610931 |
| 0 | 0 | 0 | 0 | 0 |
| 0 | 0 | 0 | 0.001319434 | 0.00083309 |
| 0 | 0 | 0 | 0 | 0 |
| 0 | 0 | 0 | 0 | 0 |
| 0 | 0 | 0 | 0 | 0 |
| 0 | 0 | 0 | 0 | 0 |
| 0.001568675 | 0.040243658 | 0 | 0.002091445 | 0.000707431 |
| 0 | 0 | 0 | 0 | 0 |
| 0.002047227 | 0 | 0 | 0.00179122 | 0.137332761 |
| 0 | 0 | 0 | 0 | 0 |
| 1.954571039 | 0.129254254 | 0 | 0 | 0.067129872 |
| 0 | 0 | 0 | 0 | 0 |
| 0 | 0 | 0 | 0 | 0 |
| 0 | 0 | 0 | 0 | 0 |
| 0 | 0 | 0 | 0 | 0 |
| 0 | 0.031671321 | 0.000486447 | 0 | 0 |
| 0 | 0 | 0 | 0 | 0 |
| 0 | 0 | 0 | 0 | 0 |
| 0 | 0.018536273 | 0.000551974 | 0 | 0 |
| 0 | 0 | 0 | 0 | 0 |
| 0 | 0 | 0 | 0 | 0 |
| 12.65682683 | 6.743209921 | 6.330216028 | 26.68766463 | 16.80360427 |
| 0.764027968 | 0.053028445 | 0.099395735 | 0.003640907 | 0.188938177 |
| 0 | 0 | 0 | 0 | 0 |
| 0 | 0 | 0 | 0 | 0 |
| 0 | 0 | 0 | 0 | 0 |
| 0 | 0 | 0 | 0 | 0 |
| 0 | 0 | 0 | 0 | 0 |
| 0.001151436 | 0.040681558 | 0.025846564 | 0 | 0.000556358 |

| | | | | |
|---|---|---|---|---|
| 0 | 0 | 0 | 0 | 0 |
| 0 | 0.018683267 | 0.006158183 | 0 | 0.00114452 |
| 0 | 0 | 0 | 0 | 0 |
| 0 | 0 | 0 | 0 | 0 |
| 6.713456493 | 0 | 0.000820104 | 0 | 12.95810392 |
| 0 | 0 | 0 | 0 | 0 |
| 0 | 0 | 0 | 0 | 0 |
| 0 | 0 | 0 | 0 | 0 |
| 0 | 2.007879816 | 0.134230428 | 0.037249757 | 0.004457046 |
| 0 | 0 | 0 | 0 | 0 |
| 0 | 0 | 0.000678777 | 0 | 0 |
| 0 | 0 | 0 | 0 | 0 |
| 0 | 0 | 0 | 0.041672789 | 0.031805943 |
| 0.031529497 | 0.021949569 | 0.009330043 | 0.079122313 | 0.050039842 |
| 0 | 0 | 0 | 0 | 0 |
| 0.442658399 | 0.027093166 | 0 | 0.112192208 | 0.015258135 |
| 0 | 0 | 0 | 0.001049319 | 0 |
| 0 | 0 | 0 | 0 | 0 |
| 0 | 0 | 0 | 0 | 0 |
| 0 | 0 | 0 | 0 | 0 |
| 0 | 0 | 0 | 0 | 0 |
| 0 | 0 | 0 | 0 | 0 |
| 0 | 0 | 0 | 0 | 0 |
| 0 | 0 | 0 | 0 | 0 |
| 0 | 0 | 0 | 0 | 0 |
| 0 | 0 | 0 | 0 | 0 |
| 0.404836679 | 0 | 0 | 0.02668847 | 0 |
| 0 | 0 | 0 | 0.000772178 | 0.00058042 |
| 0 | 0 | 0 | 0 | 0 |
| 0 | 0 | 0 | 0 | 0 |
| 0 | 0 | 0 | 0 | 0 |
| 0 | 0 | 0 | 0 | 0 |
| 0 | 0 | 0 | 0 | 0 |
| 0 | 0 | 0 | 0 | 0 |
| 0 | 0 | 0 | 0 | 0.032814542 |
| 0 | 0.000724657 | 0 | 0.002382 | 0 |
| 0 | 0.000555515 | 0 | 0 | 0 |
| 0.000749805 | 0 | 0 | 0 | 0 |
| 0 | 0 | 0 | 0 | 0 |
| 0 | 0 | 0 | 0 | 0 |
| 0 | 0 | 0 | 0.165194218 | 3.373502415 |
| 0 | 0 | 0 | 0 | 0 |
| 0 | 0 | 0 | 0 | 0 |
| 0 | 0 | 0 | 0 | 0 |
| 0 | 0 | 0 | 0 | 0 |
| 0 | 0 | 0 | 0 | 0 |
| 0 | 0 | 0 | 2.909726543 | 0 |
| 0 | 0 | 0 | 0 | 0 |

| | | | | |
|---|---|---|---|---|
| 0.022171205 | 0 | 0 | 0 | 0 |
| 0 | 0 | 0 | 0 | 0 |
| 0 | 0 | 0 | 0 | 0 |
| 0 | 0 | 0 | 0 | 0 |
| 0 | 0 | 0 | 0.001673884 | 0 |
| 0 | 0 | 0 | 0 | 0 |
| 0.009944873 | 0 | 0 | 0.003663688 | 0 |
| 0 | 0 | 0 | 0 | 0 |
| 0 | 0.000899097 | 0 | 0 | 0 |
| 0 | 0.207646661 | 0 | 0 | 0 |
| 0 | 0 | 0 | 0 | 0 |
| 0 | 0 | 0 | 0 | 0 |
| 0 | 0 | 0 | 0 | 0 |
| 0 | 0 | 0 | 0 | 0 |
| 0 | 0 | 0 | 0 | 0 |
| 0 | 0 | 0 | 0 | 0 |
| 0 | 0 | 0.078858686 | 0 | 0 |
| 0 | 0 | 0 | 0 | 0 |
| 0 | 0 | 0 | 0 | 0 |
| 0 | 0 | 0 | 0 | 0 |
| 0.29351027 | 0 | 0 | 0.018436755 | 0.012921918 |
| 0 | 0 | 0 | 0 | 0 |
| 0 | 0 | 0 | 0 | 0.000867564 |
| 0.002280833 | 0.004311714 | 0.002603309 | 0 | 0.00231434 |
| 0 | 0 | 0 | 0 | 0 |
| 0 | 0.017891692 | 0 | 0 | 0.000910133 |
| 0 | 0 | 0 | 0 | 0 |
| 0 | 0 | 0 | 0 | 0 |
| 0 | 0 | 0 | 0 | 0 |
| 1.912145347 | 0 | 0 | 0.730465067 | 0.224279068 |
| 0 | 0 | 0 | 0 | 0 |
| 0 | 0 | 0 | 0 | 0 |
| 0 | 0 | 0 | 0 | 0 |
| 0 | 7.917300543 | 4.457551002 | 0.05976042 | 0 |
| 0 | 0 | 0 | 0 | 0 |
| 0 | 0 | 0 | 0 | 0 |
| 0 | 0 | 0 | 0 | 0 |
| 0 | 0 | 0 | 0 | 0 |
| 0 | 0 | 0 | 0 | 0 |
| 0 | 0 | 0 | 0 | 0 |
| 0 | 0.049404219 | 0 | 0 | 0 |
| 0 | 0 | 0 | 0 | 0 |
| 0 | 0 | 0 | 0 | 0 |
| 0 | 0 | 0 | 0 | 0 |
| 0 | 0 | 0 | 0 | 0 |
| 0 | 0 | 0 | 0 | 0 |
| 0 | 0 | 0 | 0 | 0 |
| 0 | 0 | 0 | 0 | 0 |

| | | | | |
|---|---|---|---|---|
| 0.011745372 | 0.417080231 | 0.00933751 | 0 | 0.009012556 |
| 0 | 0 | 0 | 0 | 0 |
| 0 | 0 | 0 | 0 | 0 |
| 0 | 0 | 0 | 0 | 0 |
| 0 | 0 | 0 | 0 | 0 |
| 0 | 0 | 0 | 0 | 0 |
| 0 | 0 | 0 | 0 | 0 |
| 8.614507792 | 0 | 0.070148634 | 0.500004427 | 0.098748407 |
| 0.178032442 | 2.370322499 | 0.012946685 | 0.004127251 | 0.010423781 |
| 0 | 0.65435976 | 0.035712259 | 0.01723961 | 0.028249389 |
| 0 | 0 | 0 | 0 | 0 |
| 0 | 0 | 0 | 0 | 0 |
| 0 | 0 | 0 | 0 | 0 |
| 0 | 0 | 0 | 0 | 0 |
| 0 | 0 | 0 | 0 | 0 |
| 0 | 0 | 0 | 0 | 0 |
| 0 | 0 | 0 | 0 | 0 |
| 0 | 0 | 0 | 0 | 0 |
| 0 | 0 | 0 | 0 | 0 |
| 0 | 0.0004349 | 0 | 0 | 0.003331088 |
| 0 | 0 | 0 | 0 | 0 |
| 0 | 0 | 0 | 0 | 0 |
| 0 | 0 | 0 | 0 | 0 |
| 0 | 0 | 0 | 0 | 0 |
| 0 | 0 | 0 | 0 | 0 |
| 0 | 0 | 0 | 0 | 0 |
| 0 | 0 | 0 | 0 | 0 |
| 0 | 0 | 0 | 0 | 0 |
| 0 | 0 | 0 | 0 | 0 |
| 0 | 0 | 0 | 0 | 0 |
| 0 | 0 | 0 | 0 | 0 |
| 0 | 0 | 0 | 0 | 0 |
| 0 | 0 | 0 | 0 | 0 |
| 0 | 0 | 0 | 0 | 0 |
| 0 | 0 | 0 | 0 | 0 |
| 0.020503986 | 0 | 0 | 0 | 0 |
| 0 | 0 | 0 | 0 | 0 |
| 0 | 0 | 0 | 0 | 0 |
| 0 | 0 | 0 | 0.004689858 | 0.002961175 |
| 3.826907766 | 0 | 0.043656907 | 0 | 0 |
| 0 | 0 | 0 | 0 | 0 |
| 0 | 0 | 0.059305137 | 0 | 0.15596819 |
| 0 | 0 | 0 | 0 | 0 |
| 0 | 0 | 0 | 0 | 0 |
| 0 | 0 | 0 | 0 | 0 |
| 0 | 0 | 0 | 0 | 0 |
| 0 | 0 | 0 | 0 | 0 |
| 0 | 0 | 0 | 0 | 0 |

| | | | | |
|---|---|---|---|---|
| 0 | 0 | 0 | 0 | 0 |
| 0.005695905 | 0 | 0.006180175 | 0 | 0 |
| 0 | 0 | 0 | 0 | 0 |
| 0 | 0 | 0 | 0 | 0 |
| 0 | 0 | 0 | 0 | 0.024242003 |
| 0 | 0 | 0 | 0 | 0 |
| 0 | 0 | 0 | 0 | 0 |
| 0.051958876 | 0.05128507 | 0.002375598 | 0.001672399 | 0.006184863 |
| 0 | 0 | 0 | 0 | 0 |
| 0 | 0 | 0 | 0 | 0 |
| 0 | 0 | 0 | 0 | 0 |
| 0 | 0 | 0 | 0 | 0 |
| 0 | 0 | 0 | 0 | 0 |
| 0 | 0 | 0 | 0 | 0 |
| 0 | 0.200285025 | 0 | 0 | 0 |
| 0.001547246 | 0.010060762 | 0 | 0 | 0.008634869 |
| 0 | 0 | 0 | 0 | 0 |
| 0 | 0 | 0 | 0 | 0 |
| 0 | 0 | 0 | 0 | 0 |
| 0 | 0 | 0 | 0 | 0 |
| 0 | 0 | 0 | 0 | 0 |
| 0 | 0 | 0 | 0 | 0 |
| 0 | 0 | 0 | 0 | 0 |
| 0 | 0.022719871 | 0 | 0 | 0 |
| 0 | 0 | 0 | 0 | 0 |
| 0 | 0 | 0 | 0 | 0 |
| 0 | 0 | 0.002193729 | 0.007677701 | 0 |
| 0 | 0 | 0 | 0 | 0 |
| 0 | 0 | 0 | 0 | 0 |
| 0 | 0 | 0 | 0 | 0 |
| 0 | 0 | 0 | 0 | 0 |
| 0 | 0.13748623 | 0 | 0 | 0 |
| 0 | 0 | 0 | 0 | 0 |
| 0 | 0 | 0 | 0 | 0 |
| 0 | 0 | 0 | 0 | 0 |
| 0 | 0 | 0 | 0 | 0 |
| 0 | 0 | 0 | 0 | 0 |
| 0 | 0 | 0 | 0 | 0.040409548 |
| 0 | 0 | 0 | 0 | 0 |
| 0 | 0 | 0 | 0 | 0 |
| 0 | 0 | 0 | 0 | 0 |
| 0 | 0 | 0 | 0 | 0 |
| 0 | 0 | 0 | 0 | 0 |
| 0 | 0 | 0 | 0 | 0.011394969 |
| 0 | 0 | 0 | 0 | 0 |
| 0 | 0 | 0 | 0 | 0 |
| 0 | 0 | 0 | 0 | 0 |
| 0 | 0 | 0 | 0 | 0 |

| | | | | |
|---|---|---|---|---|
| 0 | 0 | 0.053785709 | 0 | 0 |
| 0 | 0 | 0 | 0 | 0 |
| 0 | 0 | 0 | 0 | 0 |
| 0 | 0 | 0 | 0 | 0 |
| 0 | 0 | 0 | 0 | 0 |
| 0.003076974 | 0 | 0 | 0 | 0 |
| 0 | 0 | 0 | 0 | 0 |
| 0 | 0 | 0 | 0 | 0 |
| 0 | 0 | 0 | 0 | 0 |
| 0 | 0 | 0 | 0 | 0 |
| 0 | 0 | 0 | 0 | 0 |
| 0 | 0.062994649 | 0 | 0 | 0 |
| 0 | 0 | 0 | 0 | 0 |
| 0 | 0 | 0 | 0 | 0 |
| 0 | 0 | 0 | 0 | 0 |
| 0 | 0 | 0 | 0 | 0 |
| 0 | 0 | 0 | 0 | 0 |
| 0 | 0 | 3.62138066 | 0 | 0 |
| 0 | 0 | 0 | 0 | 0 |
| 0 | 0 | 0 | 0 | 0 |
| 0 | 0.101899724 | 0.00136486 | 0.001441273 | 0 |
| 0 | 0 | 0 | 0 | 0 |
| 0 | 0.078632763 | 0.021115556 | 0 | 0.008045019 |
| 0 | 0 | 0 | 0 | 0 |
| 0 | 0 | 0 | 0 | 0 |
| 0 | 0 | 0 | 0 | 0.008808485 |
| 0 | 0 | 0 | 0 | 0 |
| 0 | 0 | 0 | 0 | 0 |
| 0 | 0 | 0 | 0 | 0 |
| 0 | 0 | 0 | 0 | 0 |
| 0 | 0 | 0 | 0 | 0 |
| 0 | 0 | 0 | 0 | 0 |
| 0.009546491 | 3.609775682 | 0 | 0.020114651 | 0 |
| 0 | 0 | 0 | 0 | 0 |
| 4.094183224 | 0 | 0 | 2.076756773 | 0 |
| 0 | 0.000832288 | 0 | 0.025169273 | 0.001603993 |
| 0 | 0 | 0 | 0 | 0 |
| 0 | 0 | 0 | 0 | 0 |
| 0 | 0.028495098 | 0 | 0.002215062 | 0 |
| 0 | 0 | 0 | 0 | 0 |
| 0 | 0 | 0 | 0 | 0 |
| 0 | 0 | 0 | 0.002629211 | 0 |
| 0 | 0 | 0 | 0 | 0.019190572 |
| 0 | 0 | 0 | 0 | 0 |
| 0 | 0 | 0 | 0 | 0 |
| 0.006856951 | 0 | 0.044001898 | 0 | 0 |
| 0 | 0 | 0 | 0 | 0 |
| 0 | 0 | 0 | 0 | 0 |

| | | | | |
|---|---|---|---|---|
| 0 | 0 | 0 | 0 | 0 |
| 0 | 0 | 0 | 0 | 0 |
| 0 | 0 | 0 | 0 | 0 |
| 0 | 0 | 0 | 0 | 0 |
| 0 | 0 | 0 | 0 | 0 |
| 0 | 0 | 0 | 0 | 0 |
| 0 | 0 | 0 | 0 | 0 |
| 0 | 0 | 0 | 0.003691407 | 0 |
| 0 | 0 | 0 | 0 | 0.00533913 |
| 0 | 0 | 0 | 0 | 0 |
| 0 | 0 | 0 | 0 | 0 |
| 0 | 0.003267286 | 0 | 0 | 0 |
| 0 | 0 | 0 | 0 | 0 |
| 0 | 0 | 0 | 0 | 0 |
| 0 | 0 | 0 | 0 | 0 |
| 0 | 0.023192014 | 0.015862523 | 0.023525292 | 0.018399451 |
| 0 | 0 | 0 | 0 | 0 |
| 0 | 0 | 0 | 0 | 0 |
| 0 | 0 | 0 | 0 | 0 |
| 2.630623497 | 0 | 0 | 0.056345857 | 0 |
| 0 | 0 | 0 | 0 | 0.00561078 |
| 0 | 0 | 0 | 0 | 0 |
| 0 | 0 | 0 | 0 | 0 |
| 0 | 0 | 0 | 0 | 0 |
| 0 | 0.036487105 | 0 | 0 | 0 |
| 0 | 0 | 0 | 0 | 0 |
| 0 | 0 | 0 | 0 | 0 |
| 0 | 0 | 0 | 0 | 0 |
| 0 | 0 | 0 | 0 | 0 |
| 0.0072019 | 0 | 0 | 0 | 0 |
| 0.147326672 | 0 | 0 | 11.83062849 | 0.026197765 |
| 0 | 0 | 0 | 0 | 0 |
| 0 | 0 | 0 | 0 | 0 |
| 0 | 0 | 0 | 0 | 0 |
| 0 | 0 | 0 | 0 | 0 |
| 0 | 0 | 0 | 0 | 0 |
| 0 | 0 | 0 | 0 | 0 |
| 0 | 0 | 0 | 0 | 0 |
| 0 | 0 | 0 | 0 | 0 |
| 0 | 0 | 0 | 0 | 0 |
| 0 | 0 | 0 | 0 | 0 |
| 0 | 0 | 0 | 0 | 0 |
| 0 | 0 | 0 | 0 | 0 |
| 0 | 0 | 0.001506084 | 0 | 0 |
| 0 | 0 | 0 | 0 | 0 |
| 0 | 0 | 0 | 0 | 0 |
| 0 | 12.67703127 | 0.302574577 | 0.049079871 | 0 |
| 0 | 0 | 0 | 0 | 0 |

| | | | | |
|---|---|---|---|---|
| 0 | 0 | 0 | 0 | 0 |
| 0 | 0 | 0 | 0 | 0 |
| 0 | 0 | 0 | 0 | 0 |
| 0 | 0 | 0 | 0 | 0 |
| 0.125416763 | 0.57436327 | 0.479858718 | 0.019130259 | 0 |
| 0 | 0 | 0 | 0 | 0 |
| 0 | 0 | 0 | 0 | 0 |
| 0 | 0 | 0 | 0 | 0 |
| 0 | 0 | 0 | 0 | 0 |
| 0 | 0 | 0 | 0 | 0 |
| 0.011566758 | 0 | 0 | 0 | 0.001173668 |
| 0.001054013 | 0.039438134 | 0.001871388 | 0 | 0.002138994 |
| 0 | 0 | 0 | 0 | 0 |
| 0 | 0 | 0 | 0 | 0 |
| 0.012369764 | 0.026536023 | 0 | 0 | 0 |
| 1.78764448 | 5.027578852 | 0.292509107 | 0 | 0 |
| 0 | 0 | 0 | 0 | 0.001974168 |
| 0 | 0 | 0 | 0 | 0 |
| 0.039266745 | 0.043555958 | 0.00892726 | 0.105822517 | 0.026562399 |
| 0.002292579 | 0 | 0 | 0 | 0 |
| 0 | 0 | 0 | 0 | 0 |
| 0 | 0 | 0 | 0 | 0 |
| 0 | 0 | 0 | 0 | 0 |
| 0 | 0 | 0 | 0 | 0 |
| 0 | 0 | 0 | 0 | 0 |
| 0 | 0 | 0 | 0 | 0 |
| 0 | 0.013565263 | 0.006671938 | 0 | 0.003615297 |
| 0 | 0 | 0 | 0 | 0 |
| 0 | 0 | 0 | 0 | 0 |
| 0 | 0 | 0 | 0 | 0 |
| 0 | 0 | 0 | 0 | 0 |
| 0 | 0 | 0.007442787 | 0 | 0 |
| 0 | 0 | 0 | 0 | 0 |
| 0 | 0 | 0 | 0 | 0 |
| 0 | 0 | 0 | 0 | 0 |
| 0 | 0 | 0 | 0 | 0 |
| 0 | 0 | 0 | 0 | 0 |
| 0.002926214 | 0.32228091 | 0.005077381 | 0.004389067 | 0.002069444 |
| 0 | 0 | 0 | 0 | 0 |
| 0 | 0 | 0 | 0 | 0 |
| 0 | 0 | 0 | 0 | 0 |
| 0 | 0 | 0 | 0 | 0 |
| 0 | 0 | 0 | 0 | 0 |
| 0.00059171 | 0.070920256 | 0.459976252 | 0 | 0.010273552 |
| 0 | 0 | 0 | 0 | 0 |
| 0 | 0 | 0 | 0 | 0 |
| 0 | 0 | 0 | 0 | 0 |
| 0 | 0 | 0 | 0 | 0 |

| | | | | |
|---|---|---|---|---|
| 0 | 0 | 0 | 0 | 0 |
| 11.34573192 | 0 | 0 | 0 | 0 |
| 0 | 0 | 0 | 0 | 0 |
| 0.001653639 | 0.004904679 | 0 | 0 | 0 |
| 0.008243763 | 0 | 0 | 0 | 0 |
| 0 | 0.004064494 | 0.00109826 | 0 | 0 |
| 0.001404139 | 0 | 0 | 0 | 0 |
| 0 | 0 | 0 | 0 | 0 |
| 0 | 0 | 0 | 0 | 0 |
| 0 | 0 | 0 | 0 | 0 |
| 0 | 0 | 0 | 0 | 0 |
| 0 | 0 | 0 | 0 | 0 |
| 0 | 0 | 0.005723361 | 0.027032224 | 0.002378835 |
| 0 | 0 | 0 | 0 | 0 |
| 0 | 0 | 0 | 0 | 0 |
| 0 | 0 | 0 | 0 | 0 |
| 0 | 0 | 0 | 0 | 0 |
| 0 | 0 | 0 | 0 | 0 |
| 0 | 0 | 0 | 0 | 0 |
| 0 | 0 | 0 | 0 | 0 |
| 0 | 0 | 0 | 0 | 0 |
| 0 | 0 | 0 | 0 | 0 |
| 0.004823479 | 0.050710687 | 0.014045012 | 0.003472703 | 0.018533232 |
| 0.003415123 | 0 | 0 | 0.006146864 | 0 |
| 0 | 0 | 0 | 0 | 0 |
| 0 | 0 | 0 | 0 | 0 |
| 0 | 0 | 0 | 0 | 0 |
| 0 | 0 | 0 | 0 | 0 |
| 0 | 0 | 0 | 0 | 0 |
| 0 | 0 | 0 | 0 | 0 |
| 0.058299234 | 0 | 0 | 0 | 0 |
| 0 | 0.007894687 | 0 | 0 | 0 |
| 0 | 0 | 0 | 0 | 0 |
| 0 | 0 | 0 | 0 | 0 |
| 0 | 0 | 0 | 0 | 0 |
| 0 | 0.00164188 | 0 | 0 | 0 |
| 0 | 0.058403275 | 0.001623194 | 0 | 0.017934653 |
| 0 | 0 | 0 | 0 | 0 |
| 0.026209698 | 0 | 0 | 0 | 0 |
| 0 | 0 | 0 | 0 | 0 |
| 0 | 0 | 0 | 0 | 0 |
| 0 | 0 | 0 | 0 | 0 |
| 0 | 0 | 0 | 0 | 0 |
| 0 | 0 | 0 | 0 | 0 |
| 0 | 0 | 0 | 0 | 0 |
| 0.211865627 | 0.049397383 | 0 | 0.175343446 | 0.043331432 |
| 0 | 0 | 0.001184409 | 0 | 0 |
| 0 | 0 | 0 | 0 | 0 |

| | | | | |
|---|---|---|---|---|
| 0 | 0 | 0 | 0 | 0 |
| 0 | 0 | 0 | 0 | 0 |
| 0 | 0 | 0 | 0 | 0 |
| 0 | 0 | 0 | 0 | 0 |
| 0 | 0 | 0 | 0 | 0 |
| 0 | 0 | 0 | 0 | 0 |
| 0 | 0 | 0 | 0 | 0 |
| 0 | 0 | 0 | 0 | 0 |
| 0 | 0 | 0 | 0 | 0 |
| 0 | 0 | 0 | 0 | 0 |
| 0 | 0.011319564 | 0 | 0 | 0 |
| 0 | 0.000603577 | 0 | 0 | 0 |
| 0 | 0 | 0 | 0 | 0 |
| 0 | 0 | 0 | 0 | 0 |
| 0 | 0 | 0 | 0 | 0 |
| 0 | 0 | 0 | 0 | 0 |
| 0 | 0 | 0 | 0 | 0 |
| 0 | 0 | 0 | 0 | 0 |
| 0 | 0 | 0 | 0 | 0 |
| 0 | 0 | 0 | 0 | 0 |
| 0 | 0 | 0 | 0 | 0 |
| 0 | 0 | 0 | 0 | 0 |
| 0 | 0 | 0 | 0 | 0 |
| 0 | 0 | 0 | 0 | 0 |
| 0 | 0 | 0 | 0 | 0 |
| 0 | 0 | 0 | 0 | 0 |
| 0 | 0 | 0 | 0 | 0 |
| 0 | 0 | 0 | 0 | 0 |
| 0 | 0 | 0 | 0 | 0 |
| 0 | 0 | 0 | 0 | 0 |
| 0.001177625 | 0.005834172 | 0 | 0 | 0.005576318 |
| 0 | 0 | 0 | 0 | 0 |
| 0.007618053 | 0.38344792 | 0.193522488 | 0.103130726 | 0.03052346 |
| 0.001996109 | 0.003279014 | 0.000708814 | 0 | 0 |
| 0 | 0 | 0 | 0 | 0 |
| 0 | 0 | 0 | 0 | 0 |
| 0 | 0 | 0 | 0 | 0 |
| 0 | 0 | 0 | 0 | 0 |
| 0 | 0 | 0 | 0 | 0 |
| 0 | 0 | 0 | 0 | 0 |
| 0 | 0 | 0 | 0 | 0 |
| 0.066851234 | 0.08594098 | 0 | 0.015185275 | 0 |
| 0.009201567 | 0 | 0 | 0 | 0.003575774 |
| 0 | 0 | 0 | 0 | 0 |
| 0 | 0 | 0 | 0 | 0 |
| 0 | 0 | 0 | 0 | 0.003253449 |
| 0 | 0.152348963 | 0 | 0 | 0 |
| 0 | 0 | 0 | 0 | 0 |

| | | | | |
|---|---|---|---|---|
| 0 | 0 | 0 | 0 | 0 |
| 0.005537793 | 0 | 0 | 0.009844393 | 0.004856052 |
| 0.033301963 | 0.002842399 | 0 | 0 | 0 |
| 0 | 0 | 0 | 0 | 0 |
| 0 | 0.552715021 | 0.059777024 | 0 | 0.005693756 |
| 0.006330986 | 0.068313272 | 0.001322423 | 0 | 0 |
| 0 | 0 | 0 | 0 | 0 |
| 0 | 0 | 0 | 0 | 0 |
| 0 | 0 | 0 | 0 | 0 |
| 0 | 0 | 0 | 0 | 0 |
| 0 | 0 | 0 | 0 | 0 |
| 0 | 0 | 0 | 0 | 0 |
| 0 | 0 | 0 | 0 | 0 |
| 0 | 0 | 0 | 0 | 0 |
| 0 | 0 | 0 | 0 | 0 |
| 0 | 0 | 0.001179811 | 0 | 0 |
| 0 | 0 | 0 | 0 | 0 |
| 0 | 0 | 0 | 0 | 0 |
| 0 | 0 | 0 | 0 | 0 |
| 0 | 0 | 0 | 0 | 0 |
| 0 | 0 | 0 | 0 | 0 |
| 0 | 0 | 0.000855414 | 0 | 0 |
| 0 | 0 | 0 | 0 | 0 |
| 0 | 0 | 0 | 0 | 0 |
| 0 | 0 | 0 | 0 | 0.005609808 |
| 0.003575334 | 0 | 0 | 0.043199441 | 0 |
| 0 | 0 | 0 | 0 | 0 |
| 0 | 0 | 0 | 0 | 0 |
| 0 | 0 | 0 | 0 | 0 |
| 0 | 0 | 0 | 0 | 0 |
| 0 | 0 | 0 | 0 | 0 |
| 0 | 0 | 0 | 0 | 0 |
| 0 | 0 | 0 | 0 | 0 |
| 0 | 0 | 0 | 0 | 0 |
| 0 | 0 | 0 | 0 | 0 |
| 0 | 0 | 0 | 0 | 0 |
| 0 | 0.000552425 | 0 | 0.003404738 | 0 |
| 0 | 0.025261744 | 0 | 0.000889684 | 0 |
| 0 | 0 | 0 | 0.086287736 | 0 |
| 0 | 0 | 0 | 0 | 0 |
| 0 | 0 | 0 | 0 | 0 |
| 0 | 0 | 0 | 0 | 0 |
| 0.007375822 | 0.320008225 | 0.008317531 | 0 | 0.008765253 |
| 0 | 0 | 0 | 0 | 0 |
| 0 | 0 | 0 | 0 | 0 |
| 0 | 0 | 0 | 0 | 0 |
| 0 | 0.012524018 | 0.005685278 | 0 | 0.005776237 |
| 0 | 0 | 0 | 0 | 0 |

| | | | | |
|---|---|---|---|---|
| 0 | 0 | 0 | 0 | 0 |
| 0.005194867 | 0.014222707 | 0.004440909 | 0.008946195 | 0.006247328 |
| 0 | 0 | 0 | 0 | 0 |
| 0 | 0 | 0 | 0 | 0 |
| 0 | 0 | 0 | 0 | 0 |
| 0 | 0 | 0 | 0 | 0 |
| 0 | 0 | 0 | 0 | 0 |
| 0 | 0 | 0 | 0 | 0 |
| 0 | 0 | 0 | 0 | 0 |
| 0 | 0 | 0 | 0 | 0 |
| 0 | 0 | 0 | 0 | 0 |
| 0 | 0 | 0 | 0 | 0 |
| 0 | 0 | 0 | 0.010677169 | 0.031093734 |
| 0 | 0 | 0 | 0 | 0 |
| 0.028307833 | 0 | 0 | 0 | 0 |
| 0 | 0 | 0 | 0 | 0 |
| 0 | 0 | 0 | 0 | 0 |
| 0 | 0 | 0 | 0 | 0 |
| 0 | 0 | 0 | 0 | 0 |
| 0 | 0 | 0 | 0 | 0 |
| 0 | 0 | 0 | 0 | 0 |
| 0.019554607 | 0 | 0 | 0 | 0 |
| 0 | 0 | 0.00289287 | 0.001900784 | 0 |
| 0.008493257 | 0 | 0 | 0 | 0 |
| 0 | 0 | 0 | 0 | 0 |
| 0 | 0 | 0 | 0 | 0 |
| 0 | 0 | 0 | 0 | 0 |
| 0 | 0 | 0 | 0 | 0 |
| 0 | 0 | 0 | 0 | 0 |
| 0 | 0 | 0 | 0 | 0 |
| 0 | 0 | 0 | 0 | 0 |
| 0 | 0 | 0 | 0 | 0 |
| 0 | 0 | 0 | 0 | 0 |
| 0 | 0 | 0 | 0 | 0 |
| 0 | 0 | 20.20390055 | 0.001604229 | 0 |
| 0 | 0 | 0 | 0 | 0 |
| 0 | 0 | 0 | 0 | 0 |
| 0 | 0 | 0 | 0 | 0 |
| 0.033408736 | 0 | 0 | 0.008116439 | 0 |
| 0 | 0 | 0 | 0 | 0 |
| 0.377184268 | 0.057178017 | 0.144387065 | 0.592322077 | 0.11530279 |
| 0 | 0 | 0 | 0 | 0 |
| 0 | 0 | 0 | 0 | 0 |
| 0 | 0 | 0 | 0 | 0 |
| 0 | 0 | 0 | 0 | 0 |
| 0.068110621 | 3.004113284 | 0.085765687 | 1.452338347 | 0.037251432 |
| 0 | 0 | 0 | 0 | 0 |
| 0 | 0 | 0 | 0 | 0 |

| | | | | |
|---|---|---|---|---|
| 0 | 0 | 0 | 0 | 0 |
| 0 | 0 | 0 | 0 | 0 |
| 0 | 0 | 0.002824121 | 0 | 0 |
| 0 | 0 | 0 | 0 | 0 |
| 0 | 0.006519724 | 0.002927105 | 0 | 0 |
| 0 | 0 | 0 | 0 | 0 |
| 0 | 0 | 0 | 0 | 0 |
| 0 | 0 | 0 | 0 | 0 |
| 0 | 0 | 0 | 0 | 0 |
| 0 | 0.000862287 | 0.001258185 | 0 | 0.001438104 |
| 0 | 0 | 0 | 0 | 0 |
| 0 | 0 | 0 | 0 | 0 |
| 0 | 0.008638833 | 0.00122977 | 0 | 0.000624722 |
| 0 | 0 | 0 | 0 | 0 |
| 0 | 0 | 0 | 0 | 0 |
| 0 | 0 | 0 | 0 | 0 |
| 0 | 0 | 0 | 0 | 0 |
| 0 | 0 | 0 | 0 | 0 |
| 0 | 0 | 0 | 0 | 0 |
| 0.091652421 | 0.120963789 | 0.101431238 | 0.04530983 | 0.930822612 |
| 0 | 0 | 0 | 0 | 0 |
| 0 | 0 | 0 | 0 | 0 |
| 0 | 0 | 0 | 0 | 0 |
| 0 | 0 | 0 | 0 | 0 |
| 0 | 0 | 0 | 0 | 0 |
| 0 | 0 | 0 | 0 | 0 |
| 0 | 0 | 0 | 0 | 0 |
| 0 | 0 | 0 | 0 | 0 |
| 0 | 0 | 0 | 0 | 0 |
| 0 | 0.103249313 | 0 | 0 | 0 |
| 0 | 0 | 0 | 0 | 0 |
| 0 | 0 | 0 | 0 | 0 |
| 0 | 0 | 0 | 0 | 0 |
| 0 | 0 | 0 | 0 | 0 |
| 0 | 0 | 0 | 0 | 0 |
| 0 | 0 | 0 | 0 | 0 |
| 0 | 0 | 0 | 0 | 0 |
| 0 | 0 | 0 | 0 | 0 |
| 0 | 0 | 0 | 0 | 0 |
| 0 | 0 | 0 | 0.05126986 | 0 |
| 0 | 0 | 0 | 0 | 0 |
| 0 | 0 | 0 | 0 | 0 |
| 0 | 0.019174627 | 0 | 0 | 0 |
| 0 | 0 | 0 | 0 | 0 |
| 0 | 0.036990486 | 0.002570178 | 0 | 0.000652825 |
| 0 | 0 | 0 | 0 | 0 |
| 0 | 0 | 0 | 0 | 0 |
| 0 | 0 | 0 | 0 | 0 |

| | | | | |
|---|---|---|---|---|
| 0 | 0 | 0 | 0 | 0 |
| 0 | 0 | 0 | 0 | 0 |
| 0 | 0 | 0 | 0 | 0 |
| 0 | 0 | 0 | 0 | 0 |
| 0 | 0 | 0 | 0 | 0 |
| 0 | 0.020645377 | 0.00370956 | 0 | 0.005360949 |
| 0 | 0 | 0 | 0 | 0 |
| 0 | 0.003132742 | 0 | 0 | 0 |
| 0 | 0 | 0 | 0 | 0 |
| 0 | 0 | 0 | 0 | 0 |
| 0 | 0 | 0 | 0 | 0 |
| 0 | 0 | 0 | 0 | 0 |
| 0 | 0 | 0 | 0 | 0 |
| 0 | 0 | 0 | 0 | 0 |
| 0 | 0 | 0 | 0 | 0 |
| 0 | 0 | 0 | 0 | 0 |
| 0 | 0 | 0 | 0 | 0 |
| 0 | 0 | 0 | 0 | 0 |
| 0 | 0 | 0 | 0 | 0 |
| 0 | 0 | 0 | 0 | 0 |
| 0 | 0 | 0 | 0.002004743 | 0 |
| 0 | 0 | 0 | 0 | 0 |
| 0 | 0 | 0 | 0 | 0 |
| 0 | 0 | 0 | 0 | 0 |
| 0 | 0 | 0 | 0 | 0 |
| 0 | 0 | 0 | 0 | 0 |
| 0 | 0 | 0 | 0 | 0 |
| 0 | 0 | 0 | 0 | 0 |
| 0 | 0 | 0 | 0 | 0 |
| 0 | 0 | 0 | 0 | 0 |
| 0 | 0 | 0 | 0 | 0 |
| 0 | 0 | 0 | 0 | 0 |
| 0 | 0 | 0 | 0 | 0 |
| 0 | 0 | 0 | 0 | 0 |
| 0 | 0 | 0 | 0 | 0 |
| 0 | 0 | 0 | 0 | 0 |
| 0 | 0 | 0 | 0 | 0 |
| 0 | 0 | 0 | 0 | 0 |
| 0 | 0 | 0 | 2.95041058 | 0 |
| 0 | 0 | 0 | 0 | 0 |
| 0 | 0 | 0 | 0 | 0 |
| 0 | 0 | 0 | 0 | 0 |
| 0 | 0 | 0 | 0 | 0 |
| 0 | 0 | 0 | 0 | 0 |
| 0 | 0 | 0 | 0 | 0 |
| 0 | 0 | 0 | 0 | 0 |
| 0 | 0 | 0 | 0 | 0 |
| 0 | 0 | 0 | 0 | 0 |

| | | | | |
|---|---|---|---|---|
| 0 | 0 | 0 | 0 | 0 |
| 0 | 0 | 0 | 0 | 0 |
| 0 | 0 | 0 | 0 | 0 |
| 0 | 0 | 0 | 0 | 0 |
| 0 | 0 | 0 | 0 | 0 |
| 0 | 0 | 0 | 0 | 0 |
| 0 | 0 | 0 | 0 | 0 |
| 0 | 0 | 0 | 0 | 0 |
| 0 | 0 | 0 | 0 | 0 |
| 0 | 0 | 0 | 0 | 0 |
| 0.003802276 | 0 | 0 | 0 | 0.000428682 |
| 0 | 0 | 0 | 0 | 0 |
| 0 | 0 | 0 | 0.605354436 | 0 |
| 0 | 0 | 0 | 0 | 0 |
| 0 | 0 | 0 | 0 | 0 |
| 0 | 0 | 0 | 0 | 0 |
| 0 | 0 | 0 | 0 | 0 |
| 0 | 0 | 0 | 0 | 0 |
| 0 | 0 | 0 | 0 | 0 |
| 0 | 0 | 0 | 0 | 0 |
| 0.001286361 | 0 | 0 | 0.001607857 | 0 |
| 0 | 0 | 0 | 0 | 0 |
| 0 | 0 | 0 | 0 | 0 |
| 1.635549928 | 1.127450522 | 0.045880926 | 3.499827264 | 0 |
| 0 | 0.001315849 | 0 | 0 | 0 |
| 0 | 0 | 0 | 0 | 0 |
| 0 | 0 | 0 | 0 | 0 |
| 0 | 0 | 0 | 0 | 0 |
| 0 | 0 | 0 | 0 | 0 |
| 0 | 0 | 0 | 0 | 0 |
| 0 | 0 | 0 | 0 | 0 |
| 0 | 0 | 0 | 0 | 0 |
| 0 | 0 | 0 | 0 | 0.000457669 |
| 0 | 0 | 0 | 0 | 0 |

| PHL018-0260_V3 | PHL018-0224_V1 | PHL018-0141_V2 | PHL018-0195_V1 | PHL018-0197_V1 |
|---|---|---|---|---|
| REF | EYCF | EYCF | CTRL | EYCF |
| 1.742331684 | 0.026143863 | 0.456977523 | 3.54221375 | 0.012357453 |
| 0.165470233 | 0 | 0.132038607 | 0.070025513 | 0.014992178 |
| 0.000485143 | 2.228932411 | 2.456881537 | 1.014076804 | 0 |
| 0 | 0 | 0 | 0 | 0 |
| 0 | 0 | 0 | 0 | 0 |
| 0 | 0 | 0 | 0 | 0 |
| 0 | 0 | 0 | 0.004079272 | 0 |
| 0 | 0 | 0 | 0 | 0 |
| 0 | 0 | 0 | 0 | 0 |
| 0 | 0 | 0 | 0 | 0 |
| 0 | 0 | 0 | 0 | 0 |
| 0 | 0 | 0 | 0 | 0 |
| 0 | 0 | 0 | 0 | 0 |
| 0.001164326 | 62.47713278 | 21.46570845 | 0.066297458 | 35.28707786 |
| 0 | 0 | 0 | 0 | 0 |
| 0 | 0 | 0 | 0 | 0 |
| 0 | 0.000554083 | 0 | 0 | 0 |
| 0 | 0 | 0 | 0 | 0 |
| 0 | 0 | 0 | 0 | 0 |
| 0 | 0 | 0 | 0 | 0 |
| 0.002791131 | 0 | 0 | 0 | 0 |
| 0.000634024 | 0 | 0 | 0.006125232 | 0 |
| 0 | 0 | 0 | 0 | 0 |
| 0 | 0 | 0 | 0 | 0 |
| 0.001120839 | 0 | 0 | 0.432982563 | 0 |
| 0 | 0 | 0 | 0 | 0 |
| 0 | 0 | 0 | 0 | 0 |
| 0 | 0 | 0 | 0 | 0 |
| 0 | 0 | 0 | 0 | 0 |
| 0 | 0 | 0 | 0 | 0 |
| 0 | 0 | 0 | 0 | 0 |
| 0 | 0 | 0 | 0 | 0 |
| 0 | 0 | 0 | 0 | 0 |
| 0 | 0 | 0 | 0 | 0 |
| 0 | 0 | 0 | 0 | 0 |
| 5.131253937 | 0 | 0 | 0 | 0 |
| 0 | 0 | 0 | 0 | 0 |
| 0 | 0 | 0 | 0 | 0 |
| 0 | 0 | 0 | 0 | 0 |
| 0 | 0 | 0 | 0 | 0 |
| 0 | 0 | 0 | 0 | 0 |
| 0 | 0.001111119 | 0.041125612 | 0.038654659 | 0.017657911 |
| 0.002984912 | 0.002785262 | 0.305918963 | 0.153682644 | 0.046942457 |
| 0 | 0.011777662 | 0.086091873 | 0 | 0.039761578 |
| 0 | 0 | 0.039980956 | 0.001420652 | 0 |
| 0 | 0 | 0 | 0 | 0 |

| | | | | |
|---|---|---|---|---|
| 0.001467487 | 0 | 0 | 0.036363627 | 0 |
| 0 | 0.020077651 | 0 | 0 | 0.021496314 |
| 0.053662738 | 0.019782763 | 0.022197007 | 0.094085292 | 0.008886961 |
| 0.316471876 | 1.249203866 | 0.200740575 | 0.423586901 | 0.440593444 |
| 0.004893422 | 0 | 0 | 0.024424115 | 0 |
| 0 | 0 | 0 | 0.00807365 | 0 |
| 0 | 0.000896667 | 0 | 0 | 0 |
| 0 | 0 | 0 | 0 | 0 |
| 0.013888899 | 0 | 0 | 0 | 0 |
| 0 | 0 | 0 | 0 | 0 |
| 0 | 0 | 0 | 0 | 0 |
| 0.099822246 | 0 | 0 | 0.129668634 | 0 |
| 0 | 0 | 0 | 0 | 0 |
| 0.017602844 | 0.00417407 | 0.011654914 | 0.053639703 | 0.014287399 |
| 0 | 0 | 0 | 0 | 0 |
| 0 | 0 | 0 | 0 | 0 |
| 0 | 0.000264102 | 0 | 0 | 0 |
| 0 | 0 | 0 | 0 | 0 |
| 0 | 0 | 0 | 0 | 0 |
| 0 | 0 | 0 | 0 | 0 |
| 0.001052875 | 0.109046227 | 0.00553312 | 0.002054888 | 0.109296529 |
| 0.019441418 | 0.001337934 | 0 | 0.022348178 | 0.017369328 |
| 0.002710328 | 0 | 0.319161921 | 0.269776035 | 0 |
| 0 | 0 | 0 | 0 | 0 |
| 0 | 0 | 0 | 0 | 0 |
| 0 | 0 | 0 | 0 | 0 |
| 0 | 0.003042933 | 0 | 0 | 0 |
| 0 | 0 | 0 | 0 | 0 |
| 0 | 0 | 0.00128632 | 0.009255685 | 0 |
| 0 | 0 | 0 | 0 | 0 |
| 0 | 0.044181343 | 0.057916324 | 0 | 0.418059349 |
| 0 | 0 | 0 | 0.000625511 | 0 |
| 0 | 0 | 0 | 0 | 0 |
| 0 | 0 | 0 | 0 | 0 |
| 0 | 0 | 0 | 0 | 0 |
| 0 | 0 | 0 | 0 | 0 |
| 0 | 0 | 0 | 0 | 0 |
| 0 | 0 | 0 | 0 | 0 |
| 0 | 0 | 0 | 0 | 0 |
| 0 | 0 | 0 | 0 | 0 |
| 0.002786105 | 0 | 0 | 0 | 0 |
| 0 | 0 | 0 | 0 | 0 |
| 0 | 0 | 0 | 0 | 0 |
| 0 | 0 | 0 | 0 | 0 |
| 0.107213083 | 0 | 0 | 0.530870952 | 0 |
| 0 | 0 | 0 | 0 | 0 |
| 0 | 0 | 0 | 0 | 0 |
| 0 | 0 | 0 | 0 | 0.396962402 |

| | | | | |
|---|---|---|---|---|
| 0 | 0 | 0 | 0 | 0 |
| 0 | 0 | 0 | 0 | 0 |
| 0 | 0 | 0 | 0 | 0 |
| 0 | 0 | 0 | 0 | 0 |
| 0.002500498 | 0 | 0 | 0 | 0.001136254 |
| 0 | 0 | 0 | 0 | 0 |
| 0 | 0 | 0 | 0 | 0 |
| 0.00567275 | 0 | 0 | 0.006642878 | 0 |
| 0.106061369 | 0 | 0 | 0.009168883 | 0 |
| 0 | 0 | 0 | 0 | 0 |
| 0 | 0 | 0 | 0 | 0 |
| 0 | 0 | 0.015608779 | 0.004690969 | 0 |
| 0 | 0 | 0 | 0 | 0 |
| 0 | 0 | 0 | 0 | 0 |
| 0 | 0 | 0 | 0 | 0 |
| 0.00034297 | 0 | 0 | 0 | 0 |
| 0 | 0 | 0 | 0 | 0 |
| 0 | 0 | 0 | 0 | 0 |
| 3.261538338 | 0 | 0 | 3.451067523 | 0 |
| 0.034175615 | 0 | 0 | 0 | 0 |
| 0 | 0 | 0 | 0 | 0 |
| 0 | 0 | 0 | 0 | 0 |
| 0.002786957 | 0 | 0 | 0.000849888 | 0 |
| 0 | 0 | 0 | 0 | 0 |
| 0 | 0 | 0 | 0 | 0 |
| 0 | 0 | 0 | 0 | 0 |
| 0 | 0 | 0 | 0 | 0 |
| 0 | 0 | 0 | 0 | 0 |
| 0 | 0 | 0 | 0 | 0 |
| 0.00749939 | 0 | 0 | 0 | 0 |
| 0.008350769 | 0 | 0 | 0.056372368 | 0 |
| 0.002679739 | 0 | 0.009120557 | 0.000754331 | 0 |
| 0 | 0 | 0 | 0 | 0 |
| 0 | 0 | 0 | 0 | 0 |
| 0 | 0 | 0 | 0 | 0 |
| 0 | 0 | 0 | 0 | 0 |
| 0 | 0 | 0 | 0 | 0 |
| 0 | 0 | 0 | 0 | 0 |
| 0 | 0 | 5.626084019 | 0.032987773 | 0 |
| 0 | 0 | 0 | 0 | 0 |
| 0 | 0 | 0 | 0 | 0 |
| 0.652613039 | 0 | 0 | 0 | 0 |
| 0.001072161 | 0 | 0 | 0 | 0 |
| 0 | 0 | 0 | 0 | 0 |
| 0.001016065 | 0 | 0 | 0.002416837 | 0 |
| 0 | 0 | 0 | 0 | 0 |
| 0 | 0 | 0 | 0 | 0 |
| 0 | 0 | 0 | 0 | 0 |

| | | | | |
|---|---|---|---|---|
| 0 | 0 | 0 | 0.010330153 | 0 |
| 0 | 0 | 0 | 0 | 0 |
| 0 | 0 | 0 | 0 | 0 |
| 0 | 0 | 0 | 0 | 0 |
| 0.028258035 | 0 | 0 | 0 | 0 |
| 0 | 0 | 0 | 0 | 0 |
| 0 | 0 | 0 | 0 | 0 |
| 0 | 0 | 0 | 0 | 0 |
| 0 | 0 | 0 | 0 | 0 |
| 0 | 0 | 0 | 0 | 0 |
| 0 | 0 | 0 | 0 | 0 |
| 0 | 0 | 0 | 0 | 0 |
| 0 | 0 | 0 | 0 | 0 |
| 0 | 0 | 0 | 0 | 0 |
| 0 | 0 | 0 | 0 | 0 |
| 0 | 0 | 0 | 0 | 0 |
| 0 | 0 | 0 | 0 | 0 |
| 0 | 0 | 0 | 0 | 0 |
| 0 | 0 | 0 | 0 | 0 |
| 0 | 0 | 0 | 0 | 0 |
| 0 | 0 | 0 | 0 | 0 |
| 0 | 0 | 0 | 0 | 0 |
| 0.000656289 | 0 | 0 | 0 | 0 |
| 0 | 0 | 0 | 0 | 0 |
| 0.495014789 | 0 | 0 | 0.042456299 | 0 |
| 0 | 0 | 0 | 0 | 0 |
| 0.116548503 | 0 | 0 | 0.073254811 | 0 |
| 0 | 0 | 0 | 0 | 0 |
| 0 | 0 | 0 | 0 | 0 |
| 0 | 0 | 0 | 0 | 0 |
| 0.0582245 | 0 | 0 | 0 | 0 |
| 0 | 0 | 0 | 0.001218469 | 0 |
| 0 | 0 | 0 | 0 | 0 |
| 0 | 0 | 0 | 0 | 0 |
| 0 | 0 | 0 | 0 | 0 |
| 0 | 0 | 0 | 0 | 0 |
| 0 | 0 | 0 | 0 | 0 |
| 0 | 0 | 0 | 0 | 0 |
| 0 | 0 | 0 | 0 | 0 |
| 0 | 0 | 0 | 0 | 0 |
| 0 | 0 | 0 | 0 | 0 |
| 0 | 0 | 0 | 0 | 0 |
| 0 | 0 | 0 | 0 | 0 |
| 0 | 0 | 0 | 0 | 0 |
| 0 | 0 | 0 | 0 | 0 |
| 0 | 0 | 0 | 0 | 0 |
| 0 | 0 | 0 | 0 | 0 |
| 0 | 0 | 0 | 0 | 0 |

| | | | | |
|---|---|---|---|---|
| 0 | 0 | 0 | 0.159116811 | 0 |
| 0 | 0 | 0 | 0 | 0 |
| 0 | 0 | 0 | 0 | 0 |
| 0 | 0 | 0 | 0 | 0 |
| 0 | 0 | 0 | 0 | 0 |
| 0 | 0 | 0 | 0 | 0 |
| 0 | 0 | 0 | 0 | 0 |
| 0 | 0 | 0 | 0 | 0 |
| 0 | 0 | 0 | 0 | 0 |
| 0 | 0 | 0 | 0 | 0 |
| 0 | 0 | 0 | 0 | 0 |
| 0.043748862 | 0 | 0 | 0 | 0 |
| 0.000682751 | 0 | 0 | 0.002165345 | 0 |
| 0 | 0 | 0 | 0 | 0 |
| 0.000617399 | 0 | 0 | 0 | 0 |
| 0 | 0 | 0 | 0 | 0 |
| 0 | 0 | 0 | 0 | 0 |
| 0 | 0 | 0 | 0 | 0 |
| 0 | 0 | 0 | 0 | 0 |
| 0 | 0 | 0 | 0 | 0 |
| 0.002978688 | 0.011847556 | 0.040577119 | 0.005294421 | 0.046794132 |
| 0 | 0 | 0 | 0 | 0 |
| 0.000466717 | 0 | 0 | 0 | 0 |
| 0.001331217 | 0 | 0 | 0 | 0 |
| 0 | 0 | 0 | 0 | 0 |
| 0 | 0 | 0 | 0 | 0 |
| 0 | 0.006827865 | 0.026277847 | 0 | 0.017567276 |
| 0 | 0 | 0 | 0 | 0 |
| 0 | 0 | 0 | 0 | 0 |
| 0 | 0 | 0 | 0 | 0 |
| 0 | 0 | 0 | 0 | 0 |
| 0 | 0 | 0 | 0 | 0.006258878 |
| 0 | 0 | 0 | 0 | 0 |
| 0.259795498 | 0 | 0 | 0.030262578 | 0 |
| 0.001799658 | 0 | 0.010560655 | 0.014387243 | 0.0057874 |
| 0 | 0 | 0 | 0 | 0 |
| 0 | 0 | 0 | 0 | 0 |
| 0 | 0 | 0 | 0 | 0 |
| 0 | 0 | 0 | 0 | 0 |
| 0 | 0 | 0 | 0 | 0 |
| 0 | 0 | 0 | 0 | 0 |
| 0 | 0 | 0 | 0 | 0 |
| 0 | 0 | 0 | 0 | 0 |
| 0 | 0 | 0 | 0 | 0 |
| 0 | 0 | 0 | 0 | 0 |
| 0.092131973 | 0 | 0 | 0.318917763 | 0 |
| 0 | 0 | 0 | 0 | 0 |
| 0 | 0 | 0.001655678 | 0.004476548 | 0.002869049 |

| | | | | |
|---|---|---|---|---|
| 0 | 0 | 0 | 0 | 0 |
| 0 | 0 | 0 | 0 | 0 |
| 0 | 0 | 0 | 0 | 0 |
| 0 | 0 | 0 | 0 | 0 |
| 0 | 0 | 0 | 0 | 0 |
| 0 | 0 | 0 | 0 | 0 |
| 0 | 0 | 0 | 0 | 0 |
| 0 | 0 | 0.004976504 | 0 | 0 |
| 0 | 0 | 0 | 0.051789738 | 0 |
| 0 | 0 | 0 | 0 | 0 |
| 0 | 0 | 0.001117359 | 0 | 0 |
| 0 | 0 | 0 | 0 | 0 |
| 0 | 0 | 0 | 0 | 0 |
| 0 | 0.539860352 | 0 | 0 | 0 |
| 0 | 0 | 0 | 0 | 0 |
| 0 | 0 | 0 | 0 | 0 |
| 0 | 0 | 0 | 0 | 0 |
| 0 | 0 | 0 | 0 | 0 |
| 0.000300987 | 0 | 0.002602257 | 0.001321728 | 0 |
| 0.09623203 | 0 | 0 | 0 | 0 |
| 0.005771831 | 0.105330503 | 0.010160984 | 0 | 0.003497047 |
| 0 | 0 | 0 | 0 | 0 |
| 0 | 0 | 0 | 0 | 0 |
| 0 | 0 | 0 | 0 | 0 |
| 0.001470123 | 0.00119383 | 0.031401911 | 0.001291153 | 0.008016487 |
| 0 | 0 | 0 | 0 | 0 |
| 0 | 0 | 0 | 0 | 0 |
| 0 | 0 | 0 | 0 | 0 |
| 0 | 0 | 0 | 0.008009079 | 0 |
| 0.000840357 | 0 | 0 | 0 | 0 |
| 0.057892851 | 0.004165986 | 0.016270978 | 0.095732729 | 0.029867533 |
| 0 | 0 | 0 | 0 | 0 |
| 0 | 0 | 0.170111308 | 0 | 0 |
| 0 | 0 | 0 | 0 | 0 |
| 0 | 0 | 0 | 0 | 0 |
| 0 | 0 | 0 | 0 | 0 |
| 0 | 0 | 0 | 0 | 0 |
| 0 | 0 | 0 | 0 | 0 |
| 0 | 0 | 0 | 0 | 0 |
| 0 | 0 | 0.034329143 | 0.000477323 | 0 |
| 0 | 0 | 0 | 0 | 0 |
| 0.000998326 | 0 | 0.001184683 | 0.027927419 | 0 |
| 0.000558421 | 0 | 0 | 0 | 0 |
| 0 | 0 | 0 | 0 | 0 |
| 0 | 0 | 0 | 0 | 0 |
| 0 | 0 | 0 | 0 | 0 |
| 0 | 0 | 0 | 0 | 0 |
| 0 | 0 | 0 | 0 | 0 |

| | | | | |
|---|---|---|---|---|
| 0 | 0 | 0 | 0 | 0 |
| 0 | 0 | 0 | 0 | 0 |
| 0 | 0.014542412 | 0.090316383 | 0 | 0.041519898 |
| 0 | 0 | 0 | 0 | 0 |
| 5.11265362 | 0 | 0 | 0 | 0 |
| 0 | 0 | 0 | 0 | 0 |
| 0 | 0 | 0 | 0 | 0 |
| 0 | 0 | 0 | 0 | 0 |
| 0 | 0 | 0 | 0 | 0 |
| 0 | 0 | 0.000777437 | 0 | 0 |
| 0 | 0 | 0 | 0 | 0 |
| 0 | 0 | 0 | 0 | 0 |
| 0 | 0 | 0 | 0 | 0 |
| 0 | 0.000625022 | 0 | 0.009914297 | 0.017907116 |
| 0.000635265 | 0.001105444 | 0.012987748 | 0.001115857 | 0 |
| 0.036591253 | 0 | 0 | 0.016128081 | 0 |
| 0 | 0.001175584 | 0.020761898 | 0.00063571 | 0.003683851 |
| 0.001684455 | 0 | 0 | 0 | 0 |
| 0 | 0 | 0 | 0 | 0 |
| 0 | 0 | 0 | 0 | 0 |
| 0 | 0 | 0 | 0 | 0 |
| 0.000653409 | 0 | 0.000598151 | 0.003825763 | 0.0007126 |
| 0 | 0 | 0 | 0 | 0 |
| 0 | 0 | 0 | 0 | 0 |
| 0 | 0 | 0 | 0 | 0 |
| 0 | 0 | 0 | 0 | 0 |
| 0.953991091 | 0 | 0 | 0 | 0 |
| 0 | 0 | 0 | 0 | 0 |
| 0 | 0 | 0 | 0 | 0 |
| 0 | 0 | 0 | 0 | 0 |
| 0 | 0 | 0 | 0 | 0 |
| 0 | 0 | 0 | 0 | 0 |
| 0 | 0 | 0 | 0 | 0 |
| 0 | 0 | 0 | 0 | 0 |
| 0 | 0 | 0 | 0 | 0 |
| 0 | 0 | 0 | 0.001910354 | 0 |
| 0 | 0 | 0 | 0 | 0 |
| 0.000492565 | 0 | 0.397050404 | 0.191065332 | 0 |
| 0 | 0 | 0 | 0 | 0 |
| 0 | 0 | 0 | 0 | 0 |
| 0 | 0 | 0 | 0 | 0 |
| 0 | 0 | 0 | 0 | 0 |
| 0 | 0 | 0 | 0 | 0 |
| 0 | 0.000354895 | 0 | 0 | 0 |
| 0 | 0 | 0 | 0 | 0 |
| 0 | 0 | 0 | 0 | 0 |
| 0 | 0 | 0 | 0 | 0 |
| 0 | 0.006690596 | 0 | 0.005822089 | 0 |

| | | | | |
|---|---|---|---|---|
| 0 | 0.064792665 | 0 | 0.501845448 | 0.384863975 |
| 0 | 0 | 0 | 0 | 0 |
| 0 | 0 | 0 | 0 | 0.001243988 |
| 0 | 0 | 0 | 0 | 0 |
| 0.007023392 | 0.000718919 | 0.007984258 | 0.002418971 | 0 |
| 0 | 0 | 0 | 0.001314784 | 0 |
| 0 | 0 | 0 | 0 | 0 |
| 0.000772143 | 0.014891905 | 0.036814788 | 0 | 0.031227493 |
| 0 | 0 | 0 | 0 | 0 |
| 0.002785528 | 0.078824411 | 0.171211446 | 0.005824814 | 0.137065366 |
| 0 | 0 | 0 | 0 | 0 |
| 0 | 0 | 0.008626244 | 0.006262914 | 0 |
| 0 | 0.013312128 | 0.00760439 | 0.000848329 | 0.010534181 |
| 0 | 0 | 0 | 0.001030714 | 0 |
| 0 | 0 | 0 | 0.005234923 | 0 |
| 0 | 0 | 0 | 0 | 0.001897626 |
| 0 | 0 | 0 | 0.001666947 | 0 |
| 0.001509579 | 0 | 0 | 0 | 0 |
| 0 | 0 | 0 | 0 | 0.210989779 |
| 0 | 0 | 0 | 0 | 0 |
| 0 | 0 | 0 | 0 | 0 |
| 0 | 0 | 0 | 0 | 0 |
| 0 | 0 | 0 | 0.00052522 | 0 |
| 0.25408635 | 0 | 0 | 0 | 0 |
| 0 | 0 | 0 | 0 | 0 |
| 0 | 0 | 0.060970328 | 0 | 0 |
| 0 | 0 | 0 | 0 | 0 |
| 0 | 0 | 0 | 0 | 0 |
| 0.004285022 | 0.006334014 | 0 | 0 | 0 |
| 0 | 0.000483422 | 0 | 0 | 0 |
| 0 | 0 | 0 | 0 | 0 |
| 0 | 0 | 0 | 0 | 0 |
| 0 | 0 | 0 | 0 | 0 |
| 0 | 0.003379666 | 0.002424466 | 0 | 0 |
| 0 | 0 | 0 | 0 | 0 |
| 0 | 0 | 0 | 0 | 0 |
| 0 | 0.117178482 | 0.024751041 | 0 | 0.890168868 |
| 0 | 0 | 0 | 0 | 0 |
| 0 | 0 | 0 | 0 | 0 |
| 0 | 0 | 0.010182039 | 0 | 0.252928508 |
| 0 | 0 | 0 | 0 | 0 |
| 0 | 0 | 0.008224901 | 0 | 0 |
| 0 | 0 | 0 | 0 | 0 |
| 0 | 0 | 0 | 0.000477623 | 0 |
| 0 | 0 | 0 | 0 | 0 |
| 0.21197249 | 0 | 0 | 0 | 0 |
| 0 | 0 | 0 | 0.004423552 | 0 |
| 0.002590623 | 0 | 0 | 0.002676758 | 0 |

| | | | | |
|---|---|---|---|---|
| 0 | 0 | 0 | 0 | 0 |
| 0 | 0 | 0 | 0 | 0.012959652 |
| 0 | 0 | 0 | 0 | 0 |
| 0 | 0 | 0 | 0.003967341 | 0 |
| 0 | 0 | 0 | 0 | 0 |
| 0.001983511 | 0.004295284 | 0.001210511 | 0.002709842 | 0.003124609 |
| 0 | 0 | 0 | 0.017218259 | 0 |
| 0 | 0 | 0 | 0 | 0 |
| 0 | 0 | 0 | 0 | 0 |
| 0 | 0 | 0 | 0 | 0 |
| 0 | 0.01281036 | 0.058265551 | 0.002918463 | 0.033731271 |
| 0 | 0 | 0 | 0 | 0 |
| 0 | 0 | 0 | 0 | 0 |
| 0 | 0 | 0.002431767 | 0 | 0 |
| 0 | 0.038097548 | 1.123782583 | 0 | 0 |
| 0 | 0.000428541 | 0 | 0 | 0 |
| 0 | 0 | 0 | 0 | 0 |
| 0 | 0 | 0 | 0 | 0 |
| 0 | 0 | 0 | 0 | 0 |
| 0 | 0 | 0 | 0 | 0 |
| 0 | 0 | 0 | 0 | 0 |
| 0.058811422 | 0 | 0 | 0 | 0 |
| 0 | 0 | 0 | 0.006239269 | 0 |
| 0 | 0 | 0 | 0 | 0 |
| 0 | 0 | 0 | 0 | 0 |
| 0 | 0 | 0 | 0 | 0 |
| 0 | 0.185171724 | 0 | 0 | 0.005425666 |
| 0 | 0 | 0 | 0 | 0 |
| 0 | 0 | 0 | 0 | 0 |
| 0.040565824 | 0.017000693 | 0.078094104 | 0.045389192 | 2.730484323 |
| 0.002850703 | 0 | 0 | 0.023453856 | 0 |
| 0 | 0 | 0.001684518 | 0.005858441 | 0 |
| 0 | 0 | 0 | 0 | 0 |
| 0.000556906 | 0 | 0 | 0.000815183 | 0 |
| 0 | 0.026109905 | 0.00199646 | 0 | 0 |
| 0 | 0 | 0 | 0 | 0 |
| 0 | 0 | 0.115834663 | 0 | 0 |
| 0 | 0 | 0.002703947 | 0.013619336 | 0 |
| 0 | 0 | 0.301492223 | 0 | 0 |
| 0 | 0 | 1.198830633 | 0 | 0 |
| 0 | 0 | 0.003816611 | 0.012205484 | 0 |
| 0.014547636 | 0 | 0.421872524 | 0.077229633 | 7.806664869 |
| 0 | 0 | 0.001444911 | 0.004752835 | 0.000983644 |
| 0 | 0 | 0 | 0 | 0 |
| 0 | 0.000467655 | 0 | 0 | 0 |
| 0 | 0 | 0 | 0 | 0 |
| 0 | 0.00029008 | 0.000763011 | 0 | 0.002045258 |
| 0 | 0 | 0 | 0 | 0 |

| | | | | |
|---|---|---|---|---|
| 0 | 0 | 0.041893518 | 0.001129003 | 0 |
| 0 | 0 | 0 | 0 | 0 |
| 0 | 0.001341117 | 0 | 0 | 0.001528208 |
| 0 | 0 | 0 | 0 | 0 |
| 0.312204727 | 0 | 0 | 0.441106799 | 0 |
| 0 | 0 | 0.06944905 | 0 | 0.001659487 |
| 0 | 0 | 0 | 0 | 0 |
| 0 | 0 | 0 | 0 | 0 |
| 0.000387305 | 0 | 0 | 0.003590523 | 0.001877286 |
| 0 | 0 | 0 | 0 | 0.017882557 |
| 0 | 0 | 0 | 0 | 0 |
| 0 | 0 | 0 | 0 | 0 |
| 0 | 0 | 0 | 0 | 0 |
| 0.002511988 | 0.000323792 | 0 | 0 | 0.024605141 |
| 0 | 0 | 0 | 0 | 0 |
| 0.170569416 | 0 | 0 | 0.16171359 | 0 |
| 0 | 0.09283417 | 0.216152938 | 0.011678199 | 0.000878878 |
| 0.014392879 | 0.009285573 | 0 | 0 | 0.010944969 |
| 0 | 0 | 0 | 0.000731073 | 0 |
| 0.002579372 | 0 | 2.301222158 | 0 | 0 |
| 0 | 0.137233435 | 0.052995255 | 0.00602094 | 0 |
| 0 | 0 | 0 | 0 | 0.001784041 |
| 0 | 0 | 0 | 0 | 0 |
| 0.000934065 | 0 | 0.111362893 | 0.089066915 | 0 |
| 0.006552567 | 0 | 0 | 0.037373571 | 0 |
| 0 | 0 | 0.04081646 | 0 | 0.000868325 |
| 0 | 0 | 0 | 0.002021668 | 0 |
| 0.000776928 | 0.001239293 | 0 | 0 | 0 |
| 0 | 0 | 0 | 0 | 0 |
| 0 | 0 | 0 | 0 | 0 |
| 0.001750587 | 0.000880028 | 0 | 0.037924334 | 0 |
| 0.002783904 | 0 | 0.062915382 | 0.001528122 | 0 |
| 0.0125906 | 0 | 0.037408374 | 0 | 0 |
| 0 | 0 | 0 | 0.004170254 | 0.00906227 |
| 0 | 0 | 0 | 0 | 0 |
| 0 | 0 | 0 | 0 | 0 |
| 0 | 0.003648752 | 0.00251363 | 0 | 0.002450112 |
| 0.009604632 | 0.014471285 | 0.008756635 | 0.016630729 | 0.020225515 |
| 0.063866881 | 0 | 0 | 0.043431686 | 0 |
| 0 | 0 | 0.000730555 | 0 | 0 |
| 0.037354849 | 0 | 0 | 0 | 0 |
| 0 | 0 | 0 | 0 | 0 |
| 0.001401338 | 0 | 0 | 0.003846066 | 0 |
| 0.018835432 | 0.002174467 | 0.005580108 | 0 | 0 |
| 0 | 0 | 0.000818979 | 0 | 0 |
| 0 | 0 | 0 | 0 | 0.001856628 |
| 0 | 0 | 0 | 0 | 0 |
| 0 | 0 | 0 | 0 | 0 |

| | | | | |
|---|---|---|---|---|
| 0 | 0 | 0 | 0 | 0.019886597 |
| 0.049956059 | 0.00101835 | 2.961821008 | 0.106999554 | 0.10443725 |
| 0 | 0 | 0 | 0 | 0 |
| 0 | 0 | 0 | 0 | 0 |
| 0 | 0 | 0 | 0 | 0 |
| 0 | 0 | 0 | 0 | 0 |
| 0 | 0 | 0 | 0 | 0 |
| 0 | 0 | 0 | 0 | 0 |
| 0 | 0 | 0 | 0 | 0 |
| 0 | 0 | 0 | 0 | 0 |
| 0.001897652 | 0 | 0 | 0 | 0.003664836 |
| 0.001413303 | 0.004918664 | 0.020484869 | 0.00206875 | 0.217070658 |
| 0.05339951 | 0 | 0 | 0 | 0 |
| 0 | 0 | 0 | 0 | 0 |
| 0 | 0 | 0.00062287 | 0 | 0.000742048 |
| 0 | 0 | 0 | 0 | 0 |
| 0.001614531 | 0.007131796 | 0.128661065 | 0.005090188 | 0.067496831 |
| 0 | 2.47218157 | 0.901513552 | 0 | 0.097876758 |
| 0 | 0 | 0 | 0 | 0 |
| 0 | 0 | 0.006229314 | 0.001838888 | 0.007801791 |
| 0.029201346 | 0.002377271 | 0.053932602 | 0.005624215 | 0.002793562 |
| 0 | 0 | 0 | 0 | 0 |
| 0 | 0 | 0 | 0 | 0 |
| 0.013898725 | 0 | 0 | 0 | 0 |
| 0 | 0 | 0 | 0 | 0 |
| 0 | 0 | 0 | 0 | 0 |
| 0.05688787 | 0 | 0 | 0.020355066 | 0 |
| 0.010209447 | 0 | 0.186505471 | 0.019128674 | 0 |
| 0 | 0 | 0 | 0 | 0 |
| 0 | 0 | 0 | 0.001442104 | 0 |
| 0 | 0 | 0.12568817 | 0 | 0 |
| 0 | 0 | 0 | 0 | 0 |
| 0 | 0 | 0 | 0 | 0 |
| 0 | 0 | 0 | 0 | 0 |
| 0 | 0 | 0 | 0 | 0 |
| 0.027646209 | 0.84843832 | 2.096713328 | 0.078911964 | 0.335006645 |
| 0 | 0 | 0 | 0 | 0 |
| 0 | 0 | 0 | 0 | 0 |
| 0.003694385 | 0 | 0.003586921 | 0.017943825 | 0.002441848 |
| 0 | 0 | 0 | 0 | 0 |
| 0 | 0 | 0.000964634 | 0 | 0 |
| 0 | 0 | 0 | 0 | 0 |
| 0.078744913 | 0.014199631 | 0.021476256 | 0.002836818 | 0.001668618 |
| 0.000886322 | 0.000514105 | 0.005258857 | 0 | 0.010740126 |
| 0 | 0.030168549 | 0.256616972 | 0 | 0 |
| 0.09869786 | 0 | 0 | 0 | 0 |
| 0 | 0 | 0 | 0 | 0 |
| 0 | 0 | 0 | 0.000932044 | 0 |

| | | | | |
|---|---|---|---|---|
| 0 | 0 | 0 | 0 | 0 |
| 0 | 0 | 0 | 0 | 0 |
| 0.009534724 | 0 | 0 | 0.000547319 | 0 |
| 0.157765206 | 0 | 0.003806332 | 3.978705877 | 0 |
| 0.008459968 | 0 | 0 | 0 | 0 |
| 0.002163355 | 0 | 0 | 0.006634892 | 0 |
| 0.034538361 | 0.000587657 | 0 | 0.297135617 | 0 |
| 0.030608219 | 0 | 0 | 0.040914192 | 0 |
| 0 | 0 | 0.000978675 | 11.36171309 | 0 |
| 0.091524729 | 0 | 0 | 0 | 0 |
| 4.945629452 | 0 | 0 | 0 | 0 |
| 0.002048802 | 0 | 0 | 0.010996234 | 0 |
| 0 | 0 | 0 | 0 | 0 |
| 1.091871407 | 0 | 0 | 0 | 0 |
| 0.300658595 | 0.012367286 | 0.021352488 | 2.78575502 | 0 |
| 0 | 0 | 0 | 0 | 0 |
| 0.002817933 | 0 | 0 | 0.124999507 | 0 |
| 0 | 0 | 0.064374485 | 7.934633196 | 0 |
| 0 | 0 | 0 | 0 | 0 |
| 0.743746811 | 0 | 0 | 0 | 0 |
| 0 | 0 | 0 | 0 | 0 |
| 0 | 0 | 0 | 0 | 0 |
| 0.01635053 | 0 | 0 | 0 | 0 |
| 0 | 0.014956683 | 0.016702807 | 0.314930208 | 0 |
| 0.113387042 | 0 | 0 | 0 | 0 |
| 0.278524315 | 0 | 0 | 0 | 0 |
| 0.003043152 | 0 | 0 | 0.00853774 | 0 |
| 0 | 0 | 0 | 0 | 0 |
| 0 | 0 | 0 | 0 | 0 |
| 0 | 0 | 0 | 0 | 0 |
| 0.068605909 | 0 | 0 | 0.007774702 | 0 |
| 0 | 0 | 0 | 0.12330772 | 0 |
| 0.239511286 | 0 | 0.000642452 | 0.26343506 | 0 |
| 0 | 0 | 0 | 0 | 0 |
| 0.081528549 | 0 | 0 | 0 | 0 |
| 0 | 0 | 0 | 0 | 0 |
| 0.35879225 | 0 | 0 | 1.985289342 | 0 |
| 0 | 0 | 0 | 0 | 0 |
| 0.001658846 | 0 | 0 | 0.015956537 | 0 |
| 0 | 0 | 0 | 0.064278293 | 0 |
| 0.059946495 | 0.001203388 | 0 | 0.097337773 | 0 |
| 0.255390988 | 0 | 0 | 0 | 0 |
| 0.005239576 | 1.21248178 | 1.480909872 | 0.001917383 | 0.781894926 |
| 0.034623914 | 0 | 0 | 0 | 0 |
| 0 | 0 | 0 | 0 | 0 |
| 0.476730329 | 0 | 0 | 0 | 0 |
| 0 | 0 | 0 | 0 | 0 |
| 0.012601104 | 0 | 0.003404557 | 1.460045778 | 0 |

| | | | | |
|---|---|---|---|---|
| 0 | 0 | 0 | 0 | 0 |
| 0 | 0 | 0.004013897 | 0 | 0 |
| 0 | 0.162467466 | 0.119411263 | 0 | 3.11254721 |
| 0 | 0 | 0 | 0 | 0 |
| 0 | 0 | 0 | 0 | 0 |
| 0.005947393 | 0 | 0 | 0.163085117 | 0 |
| 0 | 0 | 0 | 0 | 0 |
| 0.008839773 | 0 | 0 | 0 | 0 |
| 0 | 0 | 0 | 0.002350498 | 0 |
| 0 | 0 | 0 | 0 | 0 |
| 0.327509133 | 0.007739235 | 0.016398617 | 0.99225046 | 0.00179644 |
| 0.233175848 | 0 | 6.31233983 | 0.091699169 | 0 |
| 0 | 0 | 0 | 0 | 0 |
| 0.000666656 | 0 | 0.648114999 | 0.008392148 | 0.000727047 |
| 0.012293208 | 0.081179098 | 0 | 0.101660795 | 1.421275094 |
| 0.481055472 | 0 | 0 | 0 | 0 |
| 0 | 0 | 0.005469293 | 0 | 0 |
| 0 | 0 | 0 | 0 | 0 |
| 0.019566433 | 0 | 0 | 0 | 0 |
| 0.004782996 | 0.000944458 | 0 | 0.017577489 | 0 |
| 0.247061149 | 0.003483555 | 0.010268846 | 0.246145955 | 0.147180363 |
| 0.000654633 | 7.625135777 | 1.282773722 | 0 | 0.998913373 |
| 0.109929293 | 2.353359966 | 14.81228014 | 0.084750348 | 9.427449784 |
| 0.015053128 | 0 | 0 | 0 | 0 |
| 0.029024816 | 0 | 0 | 0.034166771 | 0 |
| 0.007692921 | 0 | 0 | 0 | 0 |
| 0.013251526 | 0 | 0 | 0 | 0.171532407 |
| 0 | 0 | 0 | 0 | 0 |
| 0.037006948 | 0 | 0 | 0.269234624 | 0 |
| 0.005654335 | 0 | 0 | 0 | 0 |
| 0.017816563 | 0 | 0 | 0.009361814 | 0 |
| 0.030792988 | 0.006804295 | 0.005667599 | 0.009873316 | 0 |
| 0.131955801 | 0.004710161 | 0 | 0.000742896 | 0.636291243 |
| 0.345419862 | 0 | 0 | 0.636621714 | 0 |
| 0.003265968 | 0 | 0 | 0.013606389 | 0.003653151 |
| 0.005637814 | 0 | 0 | 0.0835351 | 0 |
| 0.011225534 | 0.00038681 | 0.001865317 | 0.12087796 | 0 |
| 0 | 0 | 0 | 0 | 0 |
| 0 | 0 | 0.005341006 | 0 | 0.011746969 |
| 0 | 0 | 0 | 0 | 0 |
| 0.039512404 | 0 | 0 | 0.018834116 | 0 |
| 0 | 0 | 0 | 0 | 0 |
| 0.151200046 | 0 | 0 | 0 | 0 |
| 0 | 0 | 0 | 0 | 0 |
| 0.132653245 | 0.000989397 | 0.023622345 | 0.419461415 | 0.005246848 |
| 0.114193217 | 0 | 0 | 0 | 0 |
| 0.214094476 | 0 | 0 | 0 | 0 |
| 0.100821199 | 0 | 14.87028637 | 0.28403742 | 0 |

| | | | | |
|---|---|---|---|---|
| 0 | 0 | 0 | 0 | 0 |
| 0 | 0 | 0 | 0 | 0 |
| 0 | 0.004721054 | 0 | 0.000661879 | 0 |
| 0.003914039 | 0 | 0 | 0.005074479 | 0 |
| 0 | 0 | 0 | 0 | 0 |
| 0.065383939 | 0 | 0.000753455 | 0.025589336 | 0 |
| 0.033791293 | 0 | 0 | 0 | 0 |
| 0.234928835 | 0.009430791 | 0.03626742 | 0.611545969 | 0.012905909 |
| 0 | 0 | 0 | 0 | 0 |
| 0 | 0 | 0 | 0 | 0 |
| 0.009590248 | 0.001155898 | 0 | 0.008386284 | 0.029203702 |
| 0 | 0 | 0 | 0 | 0 |
| 0.250601722 | 0 | 0 | 0.035437221 | 0 |
| 0.002338181 | 0 | 0 | 0 | 0 |
| 0 | 0 | 0 | 0 | 0 |
| 0 | 0 | 0 | 0 | 0 |
| 0.007225251 | 0 | 0 | 0 | 0 |
| 0.001866311 | 0 | 0 | 0.001973002 | 0 |
| 0.034585647 | 0 | 0 | 0.383603276 | 0 |
| 0.01094017 | 0 | 0 | 0.051600301 | 0 |
| 0.00865881 | 0 | 0 | 0 | 0 |
| 1.433798824 | 0 | 0 | 0 | 0 |
| 0.007411489 | 0 | 0 | 0.001399834 | 0 |
| 0 | 0 | 0 | 0 | 0 |
| 0 | 0 | 0 | 0 | 0 |
| 0 | 0 | 0 | 0 | 0 |
| 0 | 0 | 0 | 0 | 0 |
| 0 | 0 | 0 | 0 | 0 |
| 0.001225722 | 0 | 0 | 0 | 0 |
| 0.040956507 | 0 | 0.300019221 | 0.235036786 | 0 |
| 0.014616296 | 0 | 0 | 0.042071825 | 0 |
| 0 | 0 | 0 | 0 | 0 |
| 0.964913535 | 0 | 0.000495852 | 0.037898979 | 0 |
| 0.001221763 | 0 | 0 | 0 | 0 |
| 0.00059421 | 0 | 0 | 0.105099229 | 0 |
| 0 | 0 | 0 | 0 | 0 |
| 0.242511113 | 0 | 0.001632969 | 0.021149987 | 0 |
| 0 | 0.000305778 | 0.004825814 | 0 | 0.009965233 |
| 0.021290571 | 0 | 0 | 0 | 0 |
| 0 | 0 | 0 | 0.002587768 | 0 |
| 0.673855976 | 0 | 0 | 1.797388097 | 0 |
| 0 | 0 | 0 | 0 | 0 |
| 0 | 0 | 0 | 0 | 0 |
| 0.088770026 | 0 | 0 | 0 | 0 |
| 0 | 0 | 0 | 0.018049622 | 0 |
| 0.006973028 | 0 | 0 | 0 | 0 |
| 0.043377357 | 0 | 0 | 0 | 0 |
| 5.412472234 | 0 | 0 | 0 | 0 |

| | | | | |
|---|---|---|---|---|
| 0 | 0 | 0 | 0 | 0 |
| 0.151314731 | 0 | 0 | 0 | 0 |
| 0 | 0 | 0 | 0 | 0 |
| 0.06365886 | 0 | 0 | 0.38255234 | 0 |
| 0.948782142 | 0 | 0 | 0 | 0 |
| 0.210275936 | 0 | 0 | 0 | 0 |
| 0 | 0 | 0 | 0 | 0 |
| 0.003745552 | 0 | 0 | 0 | 0 |
| 0.001017864 | 1.089298532 | 0.711483982 | 0.007662439 | 0.674235247 |
| 0 | 0 | 0 | 0 | 0 |
| 0.008629685 | 0 | 0 | 0 | 0 |
| 0.11914359 | 0 | 0 | 0 | 0 |
| 0 | 0 | 0 | 0 | 0 |
| 0 | 0 | 0 | 0.002069699 | 0 |
| 0.086835254 | 0 | 0 | 0.277004082 | 0 |
| 0 | 0 | 0 | 0 | 0 |
| 0 | 0 | 0 | 0.002207485 | 0 |
| 0 | 0 | 0 | 0 | 0 |
| 0 | 0 | 0 | 0 | 0 |
| 0 | 0 | 0 | 0 | 0 |
| 0.008116592 | 0 | 0 | 0.053219557 | 0 |
| 4.735416334 | 0 | 0 | 0 | 0 |
| 0 | 0 | 0 | 0 | 0 |
| 0 | 0 | 0 | 0 | 0 |
| 0.000683988 | 0.001487786 | 0.032350773 | 0.025030029 | 0.0077703 |
| 1.283411656 | 0 | 0.00516396 | 4.552518521 | 0.00087886 |
| 0.177700597 | 0 | 0.00834517 | 0.89968829 | 0 |
| 0.000955313 | 0 | 0 | 0.002330596 | 0 |
| 0 | 0 | 0 | 0 | 0 |
| 0 | 0 | 0 | 0 | 0 |
| 0 | 0 | 0 | 0 | 0 |
| 0 | 0 | 0 | 0.037397187 | 0 |
| 0 | 0 | 0 | 0 | 0 |
| 0.002009115 | 0 | 0 | 0.013233966 | 0 |
| 0 | 0.004974983 | 1.48031939 | 0.001101283 | 0.001367525 |
| 0 | 0 | 0 | 0.013856825 | 0.000580005 |
| 0 | 0 | 0 | 0 | 0 |
| 1.153489486 | 0 | 0.004031424 | 1.34333506 | 10.75824146 |
| 0 | 0 | 0 | 0 | 0 |
| 0 | 0 | 0 | 0 | 0 |
| 0 | 0 | 0 | 0 | 0 |
| 0 | 0 | 0 | 0 | 0.001753946 |
| 0.063808845 | 0 | 0.014858174 | 0.952737175 | 0 |
| 0 | 0 | 0 | 0 | 0 |
| 0 | 0 | 0 | 0 | 0 |
| 0.000806144 | 0 | 0 | 0.028025218 | 0 |
| 0 | 0 | 0 | 0.066671845 | 0 |
| 0.034836212 | 0 | 0.020819217 | 0.108094025 | 0.00262617 |

| | | | | |
|---|---|---|---|---|
| 0.047525477 | 0 | 0 | 0.008235813 | 0 |
| 0.23907011 | 0 | 0 | 0 | 0 |
| 0 | 0 | 0 | 0 | 0 |
| 0 | 0 | 0 | 0 | 0 |
| 0.007747135 | 0 | 0.043097334 | 0.01517818 | 0 |
| 0 | 0 | 0 | 0 | 0 |
| 0.038048176 | 0 | 0 | 0 | 0 |
| 0.00458098 | 0 | 0 | 0.000977885 | 0 |
| 0 | 0 | 0 | 0 | 0 |
| 0.00203369 | 0 | 0 | 0 | 0 |
| 0.00072959 | 0 | 0 | 0 | 0 |
| 0.084447231 | 0 | 0 | 3.854643699 | 0 |
| 0.033480892 | 0 | 0.079926342 | 0.158854507 | 0 |
| 0 | 0 | 0 | 0 | 0 |
| 0.000964876 | 0.029422653 | 0.194320959 | 0.051248337 | 0.14957425 |
| 0 | 0 | 0 | 0 | 0 |
| 0.001917744 | 0 | 0 | 0.009263547 | 0 |
| 0 | 0 | 0 | 0 | 0 |
| 0.047118142 | 0 | 0 | 0.039790439 | 0 |
| 0 | 0 | 0 | 0.006275021 | 0 |
| 0.024405944 | 0 | 0 | 0.04114194 | 0 |
| 0 | 0 | 0 | 0 | 0 |
| 0.542038417 | 0 | 0 | 1.747964155 | 0 |
| 0.197171273 | 0 | 0 | 0 | 0 |
| 0 | 0 | 0 | 0 | 0 |
| 0.007919064 | 0 | 0 | 0 | 0 |
| 0 | 0 | 0 | 0 | 0 |
| 0 | 0 | 0 | 0 | 0 |
| 0.006952872 | 0 | 0 | 0 | 0 |
| 0 | 0 | 0 | 0 | 0 |
| 0.165355265 | 0 | 0 | 0 | 0 |
| 0.0078701 | 0.017533753 | 0.094423033 | 0.012567285 | 0.021132459 |
| 0 | 0 | 0 | 0 | 0 |
| 0.169870176 | 0 | 0.001636027 | 0.189529172 | 0 |
| 0 | 0 | 0 | 0.012368741 | 0 |
| 0.104602427 | 0 | 0.019351603 | 0.164253354 | 0 |
| 0.000616499 | 0 | 0 | 0.010227337 | 0 |
| 0 | 0 | 0 | 0 | 0 |
| 0 | 0 | 0 | 0 | 0 |
| 0.543879469 | 0 | 0 | 0 | 0 |
| 0.062328764 | 0 | 0 | 0.023097452 | 0 |
| 0.050287264 | 0 | 0 | 0 | 0 |
| 0 | 0 | 0 | 0 | 0 |
| 0.006030919 | 0 | 0 | 0.055174213 | 0 |
| 0.071723428 | 0.000595269 | 0.011482292 | 0.517421831 | 0 |
| 0.099340768 | 0 | 0 | 0 | 0 |
| 0.213179433 | 0 | 0 | 0.388722783 | 0 |
| 0.001941297 | 0 | 0 | 0.001420805 | 0 |

| | | | | |
|---|---|---|---|---|
| 0.007126622 | 0 | 0 | 0 | 0 |
| 0 | 0 | 0 | 0 | 0 |
| 0 | 0.000547251 | 0 | 0 | 0 |
| 0 | 0 | 0 | 0 | 0 |
| 0 | 0.000901862 | 0.009488848 | 0 | 0 |
| 0.028103518 | 0 | 0 | 0 | 0 |
| 0 | 0 | 0.015449169 | 0 | 0 |
| 0.04588727 | 0 | 0 | 0.104176203 | 0 |
| 0 | 0 | 0 | 0 | 0 |
| 0.013809486 | 0 | 0 | 0 | 0 |
| 0 | 0 | 0 | 0 | 0.005178366 |
| 0 | 0 | 0 | 0 | 0 |
| 0 | 0 | 0 | 0 | 0 |
| 0.35574202 | 0 | 0 | 0 | 0 |
| 0 | 0 | 0 | 0 | 0 |
| 0 | 0 | 0 | 0 | 0 |
| 0 | 0 | 0 | 0.066964491 | 0 |
| 0 | 0 | 0 | 0 | 0 |
| 0.140255224 | 0 | 0.007498299 | 0.038736095 | 0 |
| 0 | 0 | 0 | 0.001469645 | 0 |
| 0.006536324 | 0 | 0 | 0 | 0 |
| 0 | 0 | 0 | 0 | 0 |
| 0 | 0 | 0 | 0 | 0 |
| 0 | 0 | 0 | 0 | 0 |
| 0 | 0 | 0.003611555 | 0.005288555 | 0 |
| 0 | 0.006834804 | 0.069806198 | 0.013674069 | 0.043607379 |
| 0 | 0 | 0 | 0 | 0 |
| 0.193679118 | 0 | 0.179138777 | 0.143575899 | 0 |
| 0.397102618 | 0 | 0 | 0 | 0 |
| 0.062277529 | 0 | 0.032459547 | 0.317078053 | 0 |
| 0 | 0 | 0 | 0 | 0 |
| 0.241915821 | 0 | 0 | 0 | 0 |
| 0.002852585 | 0 | 0 | 0 | 0 |
| 0 | 0 | 0 | 0 | 0 |
| 0.000556222 | 0 | 0 | 0.008141806 | 0.005863882 |
| 1.410282463 | 0 | 0 | 0.492761417 | 0 |
| 0 | 0 | 0 | 0 | 0 |
| 0.005301651 | 0.000439313 | 0 | 0.168511429 | 0 |
| 0.491064943 | 0 | 0 | 0 | 0 |
| 0 | 0 | 0 | 0 | 0 |
| 0.376105991 | 0.654939018 | 1.642617243 | 1.802108617 | 13.83631501 |
| 0.655953129 | 0.180620128 | 0.384727686 | 1.233646551 | 0.001007711 |
| 0 | 0 | 0 | 0 | 0 |
| 0 | 0 | 0 | 0 | 0 |
| 0 | 0 | 0 | 0 | 0 |
| 0 | 0 | 0 | 0 | 0 |
| 0.00530004 | 0 | 0 | 0 | 0 |
| 0.009317014 | 0 | 0 | 0.017890657 | 0 |

| | | | | |
|---|---|---|---|---|
| 0 | 0 | 0 | 0 | 0 |
| 0.008037622 | 0 | 0 | 0.013876931 | 0 |
| 0.001684934 | 0 | 0 | 0.008103736 | 0 |
| 0 | 0 | 0 | 0 | 0 |
| 0.476465105 | 0 | 0.456830505 | 1.23125217 | 0 |
| 1.276658022 | 0 | 0 | 0 | 0 |
| 0 | 0 | 0 | 0 | 0 |
| 0 | 0 | 0 | 0 | 0 |
| 0 | 2.427200279 | 1.349154876 | 0 | 2.496606044 |
| 0.002373632 | 0 | 0 | 0 | 0 |
| 0 | 0.001350585 | 0 | 0 | 0 |
| 0 | 0 | 0 | 0 | 0 |
| 0.026333446 | 0 | 0 | 0.037080461 | 0 |
| 0 | 0.000856814 | 0.0018781 | 0.002522582 | 0.0069361 |
| 0 | 0 | 0 | 0 | 0 |
| 0.172528203 | 0 | 0 | 0.204191929 | 0 |
| 0.085855639 | 0 | 0.002925368 | 0.015280322 | 0 |
| 0 | 0 | 0 | 0 | 0 |
| 0 | 0 | 0 | 0 | 0 |
| 0 | 0 | 0 | 0 | 0 |
| 0 | 0 | 0 | 0 | 0 |
| 0 | 0 | 0 | 0 | 0 |
| 0 | 0 | 0 | 0 | 0 |
| 0 | 0 | 0 | 0 | 0 |
| 0.001144241 | 0 | 0 | 0 | 0 |
| 0 | 0 | 0 | 0 | 0 |
| 0.002778722 | 0 | 0 | 0.045148237 | 0 |
| 0 | 0 | 0 | 0 | 0.003609484 |
| 0 | 0 | 0 | 0 | 0 |
| 0 | 0 | 0 | 0.000451663 | 0 |
| 0 | 0 | 0 | 0 | 0 |
| 0 | 0 | 0 | 0.000638957 | 0 |
| 0 | 0 | 0 | 0.046668851 | 0 |
| 0.018296616 | 0 | 0 | 0.029027226 | 0 |
| 0.005053658 | 0 | 0 | 0 | 0 |
| 0 | 0 | 0 | 0 | 0 |
| 0 | 0.004014103 | 0 | 0 | 0 |
| 0.022605048 | 0 | 0 | 0 | 0 |
| 0 | 0 | 0 | 0 | 0 |
| 0 | 0 | 0 | 0 | 0 |
| 5.278589463 | 0 | 0 | 0 | 0 |
| 0 | 0 | 0 | 0 | 0 |
| 0.016449607 | 0 | 0 | 0 | 0 |
| 0 | 0 | 0 | 0 | 0 |
| 0 | 0 | 0 | 0 | 0 |
| 0 | 0 | 0 | 0.000540779 | 0 |
| 0 | 0 | 0.001000919 | 0 | 0 |
| 0 | 0 | 0 | 0 | 0 |

| | | | | |
|---|---|---|---|---|
| 0.123933252 | 0 | 0 | 0.258965279 | 0 |
| 0 | 0 | 0 | 0 | 0 |
| 0 | 0 | 0 | 0 | 0 |
| 0 | 0 | 0 | 0 | 0 |
| 0.091305162 | 0 | 0 | 0 | 0 |
| 0.024057362 | 0 | 0 | 0 | 0 |
| 0.118906765 | 0 | 0.001134877 | 0.499758124 | 0.002510897 |
| 0.000924444 | 0 | 0 | 0.003307754 | 0 |
| 0 | 0 | 0 | 0.007575367 | 0 |
| 0.019955948 | 0 | 0.000510287 | 0.000815947 | 0.002026415 |
| 0.019071225 | 0 | 0 | 0.027915875 | 0 |
| 0.240125445 | 0 | 0 | 0.025164602 | 0 |
| 0 | 0 | 0 | 0 | 0 |
| 0 | 0 | 0 | 0 | 0 |
| 0 | 0 | 0 | 0 | 0 |
| 0 | 0 | 0 | 0 | 0 |
| 0 | 0.031698536 | 0 | 0 | 0 |
| 0 | 0 | 0 | 0 | 0 |
| 0 | 0 | 0 | 0 | 0 |
| 0 | 0 | 0 | 0 | 0 |
| 0.00303501 | 0 | 0.002160932 | 0.077300448 | 0 |
| 0 | 0 | 0 | 0 | 0 |
| 0.217036407 | 0 | 0 | 0.037894278 | 0 |
| 0.008731727 | 0.000287772 | 0 | 0.025707711 | 0.005771339 |
| 0 | 0 | 0 | 0 | 0 |
| 0.018261706 | 0 | 0 | 0.01148059 | 0 |
| 0 | 0 | 0 | 0 | 0 |
| 0 | 0 | 0 | 0 | 0 |
| 0 | 0 | 0 | 0 | 0 |
| 0 | 0.080432001 | 0.03507916 | 0 | 0.167943196 |
| 0 | 0 | 0 | 0 | 0 |
| 0 | 0 | 0 | 0 | 0 |
| 0 | 0 | 0 | 0 | 0 |
| 0 | 0 | 0.009377056 | 0 | 0 |
| 0 | 0 | 0 | 0 | 0 |
| 2.076186039 | 0 | 0 | 0 | 0 |
| 0.171582402 | 0.003024646 | 0 | 0 | 0.025952048 |
| 0 | 0 | 0 | 0 | 0 |
| 0 | 0 | 0 | 0 | 0 |
| 0 | 0 | 0 | 0 | 0 |
| 0 | 0 | 0 | 0 | 0 |
| 0 | 0 | 0 | 0 | 0 |
| 0 | 0 | 0 | 0 | 0 |
| 0.01208435 | 0 | 0 | 0.005896234 | 0 |
| 0.269200448 | 0 | 0 | 0 | 0 |
| 0 | 0 | 0 | 0 | 0 |
| 0.007900166 | 0 | 0 | 0 | 0 |
| 0 | 0 | 0 | 0 | 0 |

| | | | | |
|---|---|---|---|---|
| 0.008221192 | 0 | 0 | 0.007032816 | 0.002134744 |
| 0 | 0 | 0 | 0 | 0 |
| 0.080571353 | 0 | 0 | 0.008914989 | 0 |
| 0 | 0 | 0 | 0 | 0 |
| 0 | 0 | 0 | 0 | 0 |
| 0 | 0 | 0 | 0 | 0 |
| 0 | 0 | 0 | 0 | 0 |
| 0.175513399 | 0 | 0.361282536 | 0.90296002 | 0 |
| 0.005530456 | 0.003985579 | 0.367177176 | 0.190853102 | 0.067625274 |
| 0 | 0.008526944 | 0 | 0.001024675 | 0.00551372 |
| 0.003285868 | 0 | 0 | 0 | 0 |
| 0.002144645 | 0 | 0 | 0 | 0 |
| 0.028594378 | 0 | 0 | 0 | 0 |
| 0 | 0 | 0.05319191 | 0 | 0 |
| 0 | 0 | 0.00093373 | 0 | 0 |
| 0 | 0 | 0 | 0 | 0 |
| 0 | 0 | 0 | 0 | 0.198300601 |
| 0 | 0 | 0 | 0 | 0 |
| 0 | 0 | 0 | 0 | 0 |
| 0.04934059 | 0 | 0.00118086 | 0.145862632 | 0 |
| 0 | 0 | 0 | 0 | 0 |
| 0 | 0 | 0 | 0 | 0 |
| 0.00075593 | 0 | 0 | 0 | 0 |
| 0 | 0 | 0 | 0 | 0 |
| 0.017996368 | 0 | 0 | 0 | 0 |
| 0 | 0 | 0 | 0 | 0 |
| 0 | 0 | 0 | 0 | 0 |
| 0 | 0 | 0 | 0 | 0 |
| 0 | 0 | 0 | 0 | 0 |
| 0 | 0 | 0 | 0 | 0 |
| 0 | 0 | 0.010208391 | 0 | 0 |
| 0 | 0 | 0 | 0 | 0 |
| 0 | 0 | 0 | 0 | 0 |
| 0.012649618 | 0 | 0 | 0 | 0 |
| 0 | 0 | 0 | 0 | 0 |
| 0 | 0 | 0 | 0 | 0 |
| 0 | 0 | 0 | 0 | 0 |
| 0 | 0 | 0 | 0 | 0 |
| 0 | 0 | 0 | 0 | 0 |
| 0 | 0 | 0 | 0 | 0 |
| 0 | 0 | 0 | 0 | 0 |
| 0.074996191 | 0 | 0.006375477 | 0.872744275 | 0 |
| 0 | 0 | 0 | 0 | 0 |
| 0 | 0 | 0 | 0 | 0 |
| 0 | 0 | 0 | 0 | 0 |
| 0.000284817 | 0 | 0 | 0 | 0 |
| 0 | 0 | 0 | 0 | 0 |
| 0 | 0 | 0 | 0 | 0 |

| | | | | |
|---|---|---|---|---|
| 0 | 0 | 0 | 0 | 0 |
| 0.000770908 | 0 | 0.004851774 | 0.016926464 | 0 |
| 0 | 0 | 0 | 0 | 0 |
| 0 | 0 | 0 | 0 | 0 |
| 5.722776704 | 0 | 0.290661072 | 7.685396093 | 0 |
| 0.663539949 | 0 | 0 | 0.061278831 | 0 |
| 0 | 0 | 0 | 0 | 0 |
| 0.001629811 | 0.083113007 | 0.053462579 | 0.010536696 | 0.036289615 |
| 0.001627306 | 0 | 0 | 0 | 0 |
| 0 | 0 | 0 | 0 | 0 |
| 0 | 0 | 0 | 0 | 0 |
| 0 | 0 | 0 | 0 | 0 |
| 0 | 0 | 0 | 0 | 0 |
| 0 | 0 | 0 | 0 | 0 |
| 0 | 0 | 0 | 0.004198564 | 0 |
| 0.046999628 | 0 | 0 | 0.119144017 | 0 |
| 0 | 0 | 0 | 0 | 0 |
| 0.001662393 | 0 | 0 | 0 | 0 |
| 0.094865991 | 0 | 0 | 0 | 0 |
| 0 | 0 | 0 | 0 | 0 |
| 0 | 0 | 0 | 0 | 0 |
| 0 | 0 | 0 | 0 | 0 |
| 0.011450162 | 0 | 0 | 0 | 0 |
| 0.001410467 | 0.00177262 | 0 | 0.007398144 | 0 |
| 0 | 0 | 0 | 0 | 0 |
| 0 | 0 | 0 | 0.00062723 | 0 |
| 0.335193343 | 0.006547405 | 0 | 0.587736336 | 0.009379282 |
| 0.001471559 | 0 | 0 | 0.002154023 | 0 |
| 0 | 0 | 0 | 0 | 0 |
| 0 | 0 | 0 | 0 | 0 |
| 0 | 0 | 0 | 0.000592322 | 0 |
| 0.006922876 | 0 | 0 | 0 | 0 |
| 0.181988744 | 0 | 0 | 0 | 0 |
| 0 | 0 | 0 | 0 | 0 |
| 0 | 0 | 0 | 0 | 0 |
| 0 | 0 | 0 | 0 | 0 |
| 0 | 0 | 0 | 0 | 0 |
| 0.219398085 | 0 | 0 | 0.789314498 | 0 |
| 0 | 0 | 0.066535074 | 0 | 0 |
| 0 | 0 | 0 | 0 | 0 |
| 0 | 0 | 0 | 0 | 0 |
| 0 | 0 | 0 | 0 | 0 |
| 0 | 0 | 0 | 0 | 0 |
| 0 | 0 | 0 | 0 | 0 |
| 0 | 0 | 0 | 0 | 0 |
| 0 | 0 | 0 | 0 | 0 |
| 0 | 0 | 0 | 0 | 0 |
| 0 | 0 | 0 | 0 | 0 |

| | | | | |
|---|---|---|---|---|
| 0.040438801 | 0 | 0 | 0.51971523 | 0 |
| 0 | 0 | 0 | 0 | 0 |
| 0 | 0 | 0 | 0 | 0 |
| 0.00321875 | 0 | 0 | 0 | 0 |
| 0 | 0 | 0 | 0 | 0 |
| 0 | 0 | 0 | 0 | 0 |
| 0 | 0 | 0 | 0 | 0 |
| 0.007849727 | 0 | 0 | 0 | 0 |
| 0 | 0 | 0 | 0 | 0 |
| 0 | 0 | 0 | 0 | 0 |
| 0 | 0 | 0 | 0 | 0 |
| 0.000541761 | 0 | 0 | 0.010943574 | 0 |
| 0 | 0 | 0 | 0 | 0 |
| 0 | 0 | 0 | 0 | 0 |
| 0 | 0 | 0 | 0 | 0 |
| 0 | 0 | 0 | 0 | 0 |
| 0 | 0 | 0 | 0 | 0 |
| 0 | 0 | 0 | 0 | 0 |
| 0 | 0 | 0 | 0 | 0 |
| 0 | 0 | 0 | 0 | 0 |
| 0 | 0 | 0.047859863 | 0 | 0.040209927 |
| 0 | 0 | 0 | 0 | 0 |
| 0.067111438 | 0 | 0 | 0.014280956 | 0 |
| 0.024353082 | 0 | 0 | 0.656504379 | 0 |
| 0.007435385 | 0 | 0 | 0 | 0 |
| 0 | 0 | 0 | 0 | 0 |
| 0 | 0 | 0 | 0 | 0 |
| 0 | 0 | 0 | 0 | 0 |
| 0 | 0 | 4.438440905 | 0 | 0 |
| 0 | 0 | 0 | 0 | 0 |
| 0 | 0 | 0 | 0 | 0 |
| 0.165133987 | 0 | 0 | 0 | 0 |
| 0 | 0.011000888 | 0 | 0 | 0.071210182 |
| 0 | 0 | 0 | 0 | 0 |
| 0.043588032 | 0 | 0.087096319 | 0.167119906 | 0 |
| 0.001333061 | 0.000257745 | 0 | 0 | 0.032710941 |
| 0 | 0 | 0 | 0 | 0 |
| 0 | 0 | 0 | 0 | 0 |
| 0 | 0.010210691 | 0.001372291 | 0 | 0.005838793 |
| 0 | 0 | 0 | 0 | 0 |
| 0 | 0 | 0 | 0 | 0 |
| 0 | 0 | 0 | 0 | 0 |
| 0.065356536 | 0 | 0 | 1.151521553 | 0 |
| 0 | 0 | 0 | 0 | 0 |
| 0 | 0 | 0 | 0 | 0 |
| 0.631324859 | 0 | 0 | 0.142695358 | 0 |
| 0 | 0 | 0 | 0 | 0 |
| 0 | 0 | 0 | 0 | 0 |

| | | | | |
|---|---|---|---|---|
| 0 | 0 | 0 | 0 | 0 |
| 0 | 0 | 0 | 0 | 0 |
| 0 | 0 | 0 | 0 | 0 |
| 0 | 0 | 0 | 0 | 0 |
| 0 | 0 | 0 | 0 | 0 |
| 0 | 0 | 0 | 0 | 0 |
| 0.883180772 | 0 | 0 | 0 | 0 |
| 0 | 2.240207344 | 0.007212616 | 0 | 0 |
| 0.031413626 | 0 | 0 | 0.170838168 | 0 |
| 0 | 0 | 0.00168758 | 0.042095553 | 0 |
| 0 | 0 | 0 | 0 | 0 |
| 0.002616579 | 0 | 0 | 0 | 0 |
| 0 | 0 | 0 | 0 | 0 |
| 0.566046716 | 0 | 0 | 0.053844916 | 0 |
| 0.087202093 | 0 | 0 | 0.00043614 | 0 |
| 0 | 1.056000748 | 0.00092244 | 0 | 0.407046559 |
| 0 | 0 | 0 | 0 | 0 |
| 0 | 0 | 0 | 0 | 0 |
| 0 | 0 | 0 | 0 | 0 |
| 0.046062874 | 0 | 0.014673628 | 0.394179404 | 0 |
| 0.008208925 | 0 | 0 | 0 | 0 |
| 0 | 0 | 0 | 0 | 0 |
| 0 | 0 | 0 | 0 | 0 |
| 0.005476257 | 0 | 0 | 0.024283698 | 0 |
| 0 | 0 | 0 | 0.006698065 | 0 |
| 0 | 0 | 0 | 0 | 0 |
| 0.000899242 | 0 | 0 | 0 | 0 |
| 0 | 0 | 0 | 0 | 0 |
| 0 | 0 | 0 | 0 | 0 |
| 0 | 0 | 0 | 0.005870212 | 0 |
| 0.019361051 | 0 | 0.001949307 | 0.156805704 | 0 |
| 0 | 0 | 0 | 0 | 0 |
| 0.349620915 | 0 | 0 | 0 | 0 |
| 0 | 0 | 0 | 0 | 0 |
| 0 | 0 | 0 | 0 | 0 |
| 0 | 0 | 0 | 0 | 0 |
| 0 | 0 | 0 | 0 | 0 |
| 0 | 0 | 0 | 0 | 0 |
| 0 | 0 | 0 | 0 | 0 |
| 0 | 0 | 0 | 0 | 0 |
| 0 | 0 | 0 | 0 | 0 |
| 0.815179775 | 0 | 0 | 0 | 0 |
| 0.005208595 | 0 | 0 | 0.00280072 | 0 |
| 0 | 0 | 0 | 0 | 0 |
| 0 | 0 | 0 | 0 | 0 |
| 0 | 0 | 0 | 0 | 0 |
| 0 | 8.435490637 | 0.169028884 | 0 | 0.036674172 |
| 0.006941326 | 0 | 0 | 0 | 0 |

| | | | | |
|---|---|---|---|---|
| 0 | 0 | 0 | 0 | 0 |
| 0 | 0 | 0 | 0 | 0 |
| 0 | 0 | 0 | 0 | 0 |
| 0 | 0 | 0 | 0 | 0 |
| 0.259024441 | 0 | 0 | 0.058070771 | 0.025213202 |
| 0 | 0 | 0 | 0 | 0 |
| 0 | 0 | 0 | 0 | 0 |
| 0 | 0 | 0 | 0 | 0 |
| 0 | 0 | 0 | 0 | 0 |
| 0 | 0 | 0 | 0 | 0 |
| 0.017259565 | 0 | 0 | 0.015983361 | 0 |
| 0.030171428 | 0 | 0.001175313 | 0.098852216 | 0 |
| 0.149757533 | 0 | 0 | 0.049938485 | 0 |
| 0 | 0 | 0 | 0 | 0 |
| 0 | 0 | 0 | 0.003437215 | 0 |
| 0.740019204 | 0 | 0 | 0.081255924 | 0 |
| 0 | 0 | 0.000753296 | 0.013731489 | 0 |
| 0 | 0 | 0 | 0 | 0 |
| 0 | 0 | 0 | 0.001138425 | 0.109204232 |
| 0.170474628 | 0 | 0.002711353 | 0.194537388 | 0 |
| 0 | 0 | 0 | 0 | 0 |
| 0 | 0 | 0 | 0 | 0 |
| 0 | 0 | 0 | 0 | 0 |
| 0 | 0 | 0 | 0 | 0 |
| 0 | 0 | 0 | 0 | 0 |
| 0 | 0 | 0 | 0 | 0 |
| 0.001220633 | 0.000826023 | 0.001862341 | 0.007444686 | 0.003512905 |
| 0 | 0 | 0 | 0 | 0 |
| 0 | 0 | 0 | 0 | 0 |
| 0 | 0 | 0 | 0 | 0 |
| 0.001153043 | 0 | 0 | 0.003544356 | 0 |
| 0.002739925 | 0 | 0 | 0 | 0 |
| 0.004680488 | 0 | 0 | 0 | 0 |
| 0 | 0 | 0 | 0 | 0 |
| 0 | 0 | 0 | 0.001712953 | 0 |
| 0 | 0 | 0 | 0 | 0 |
| 0 | 0 | 0 | 0 | 0 |
| 0.171820998 | 0.005779647 | 0.004202316 | 0.328582746 | 0.007509566 |
| 0.074539872 | 0 | 0 | 0.208805024 | 0 |
| 0.010280157 | 0 | 0 | 0 | 0 |
| 0.007278617 | 0.000422191 | 0 | 0 | 0 |
| 0.002284694 | 0 | 0 | 0 | 0 |
| 0 | 0 | 0 | 0 | 0 |
| 0 | 0 | 0.000733118 | 0 | 0.002620173 |
| 0 | 0 | 0 | 0 | 0 |
| 0.000751758 | 0 | 0 | 0.003458402 | 0 |
| 0 | 0 | 0 | 0 | 0 |
| 0.001279771 | 0 | 0 | 0 | 0 |

| | | | | |
|---|---|---|---|---|
| 0 | 0 | 0 | 0 | 0 |
| 0.093021822 | 0 | 0 | 0 | 0 |
| 0 | 0 | 0 | 0 | 0 |
| 0 | 0 | 0 | 0.009766777 | 0.000915318 |
| 0 | 0.000541826 | 0.100832345 | 0.000512747 | 0.000636707 |
| 0 | 0 | 0 | 0 | 0 |
| 0.008796234 | 0 | 0.001491175 | 0.03290446 | 0 |
| 0 | 0 | 0 | 0 | 0 |
| 0.016977378 | 0 | 0 | 0 | 0 |
| 0.011783642 | 0 | 0 | 0 | 0 |
| 0.049725203 | 0 | 0 | 0 | 0 |
| 0.000968403 | 0 | 0 | 0 | 0 |
| 0.061040606 | 0 | 0 | 0.259165427 | 0 |
| 0 | 0 | 0 | 0 | 0 |
| 0.001434167 | 0 | 0 | 0 | 0 |
| 0 | 0 | 0 | 0 | 0 |
| 0 | 0 | 0 | 0 | 0 |
| 0 | 0 | 0 | 0 | 0 |
| 0 | 0 | 0 | 0 | 0 |
| 0 | 0 | 0 | 0 | 0 |
| 0 | 0 | 0 | 0 | 0 |
| 0 | 0 | 0 | 0 | 0 |
| 0 | 0.001567693 | 0 | 0 | 0.005126168 |
| 0.01982911 | 0 | 0.000634692 | 0.00487137 | 0 |
| 0 | 0 | 0 | 0 | 0 |
| 0 | 0 | 0 | 0.001199184 | 0 |
| 0.002357671 | 0 | 0 | 0 | 0 |
| 0.001925563 | 0 | 0 | 0 | 0 |
| 0.159094046 | 0 | 0 | 0 | 0 |
| 0 | 0 | 0 | 0 | 0 |
| 0.001179202 | 0 | 0.000999516 | 0.007863252 | 0 |
| 0.001621127 | 0 | 0 | 0.029513643 | 0 |
| 0 | 0 | 0 | 0 | 0 |
| 0 | 0 | 0 | 0 | 0 |
| 7.213054672 | 0 | 0 | 0 | 0 |
| 0 | 0 | 0 | 0 | 0.018920804 |
| 0 | 0.003229723 | 0 | 0 | 0 |
| 0 | 0 | 0 | 0.020882852 | 0 |
| 0 | 0 | 0 | 0.004381149 | 0 |
| 0 | 0 | 0 | 0 | 0 |
| 0 | 0 | 0 | 0 | 0 |
| 0.026877275 | 0 | 0 | 0 | 0 |
| 1.2805034 | 0 | 0 | 0 | 0 |
| 0 | 0 | 0 | 0 | 0 |
| 0 | 0 | 0 | 0 | 0 |
| 0.338733023 | 0 | 0.012499894 | 0.444288431 | 0 |
| 0 | 0 | 0 | 0 | 0 |
| 0 | 0 | 0 | 0 | 0 |

| | | | | |
|---|---|---|---|---|
| 0 | 0 | 0 | 0 | 0 |
| 0 | 0 | 0 | 0 | 0 |
| 0 | 0 | 0.03388499 | 0 | 0 |
| 0.020910156 | 0 | 0 | 0 | 0 |
| 0 | 0 | 0 | 0 | 0 |
| 0 | 0 | 0 | 0.024841778 | 0 |
| 0.006648195 | 0 | 0 | 0 | 0 |
| 0 | 0 | 0 | 0 | 0 |
| 0.005333614 | 0 | 0 | 0 | 0 |
| 0 | 0 | 0 | 0 | 0 |
| 0.005036211 | 0 | 0 | 0.01895619 | 0 |
| 0.000402808 | 0 | 0 | 0.003930785 | 0 |
| 0 | 0 | 0 | 0 | 0 |
| 0 | 0 | 0 | 0 | 0 |
| 0 | 0 | 0 | 0 | 0 |
| 0 | 0 | 0 | 0 | 0 |
| 0 | 0 | 0 | 0 | 0 |
| 0 | 0 | 0 | 0 | 0 |
| 0 | 0 | 0 | 0 | 0 |
| 0.000596329 | 0 | 0 | 0.000872888 | 0 |
| 0 | 0 | 0 | 0 | 0 |
| 0 | 0 | 0 | 0 | 0 |
| 0 | 0 | 0 | 0 | 0 |
| 0 | 0 | 0 | 0 | 0 |
| 0 | 0 | 0 | 0 | 0 |
| 0 | 0 | 0 | 0 | 0 |
| 0 | 0 | 0 | 0 | 0 |
| 0 | 0 | 0 | 0 | 0 |
| 0 | 0 | 0 | 0 | 0 |
| 0 | 0 | 0 | 0 | 0 |
| 0.006250158 | 0 | 0.00062531 | 0.106935861 | 0.02104499 |
| 0 | 0 | 0 | 0 | 0 |
| 0.003688019 | 0 | 0.28043631 | 0.001741422 | 0 |
| 0.000607864 | 0 | 0.000927429 | 0.004804771 | 0 |
| 0 | 0 | 0 | 0 | 0 |
| 0 | 0 | 0 | 0 | 0 |
| 0 | 0 | 0 | 0 | 0 |
| 0 | 0 | 0 | 0 | 0 |
| 0 | 0 | 0 | 0 | 0 |
| 0 | 0 | 0 | 0 | 0 |
| 0 | 0 | 0 | 0 | 0 |
| 0.160513677 | 0 | 0 | 0.105586488 | 0.006760221 |
| 0.007869732 | 0 | 0 | 0.234054745 | 0 |
| 0 | 0 | 0 | 0 | 0 |
| 0 | 0 | 0 | 0 | 0 |
| 0.186264653 | 0 | 0.006572325 | 0.212115991 | 0 |
| 0.000677819 | 0 | 0 | 0.012236773 | 0 |
| 0 | 0 | 0 | 0 | 0 |

| | | | | |
|---|---|---|---|---|
| 0 | 0 | 0 | 0 | 0 |
| 0.273195666 | 0 | 0 | 0 | 0 |
| 0 | 0.000423192 | 0.029832263 | 0.002135894 | 0.00132613 |
| 0 | 0 | 0 | 0 | 0 |
| 0.315387244 | 0 | 0 | 3.039379774 | 0.009529662 |
| 1.860802112 | 0 | 0 | 1.786529499 | 0 |
| 0 | 0 | 0 | 0 | 0 |
| 0 | 0 | 0 | 0 | 0 |
| 0 | 0 | 0 | 0 | 0 |
| 0 | 0 | 0 | 0 | 0 |
| 0 | 0 | 0 | 0 | 0 |
| 0 | 0 | 0 | 0 | 0 |
| 0 | 0 | 0 | 0 | 0 |
| 0 | 0 | 0 | 0 | 0 |
| 0 | 0 | 0 | 0 | 0 |
| 0 | 0 | 0 | 0 | 0 |
| 0 | 0 | 0 | 0 | 0 |
| 0 | 0 | 0 | 0 | 0 |
| 0 | 0 | 0 | 0 | 0 |
| 0 | 0 | 0 | 0 | 0 |
| 0.006044573 | 0 | 0 | 0.000449891 | 0 |
| 0 | 0.001565883 | 0 | 0 | 0 |
| 0.001979923 | 0 | 0 | 0 | 0 |
| 0 | 0 | 0 | 0 | 0 |
| 0.011544621 | 0 | 0 | 0.011793858 | 0 |
| 0 | 0 | 0.002399462 | 0 | 0.001319339 |
| 0 | 0 | 0 | 0 | 0 |
| 0.203367767 | 0 | 0 | 0.02095637 | 0 |
| 0 | 0 | 0 | 0 | 0 |
| 0 | 0 | 0 | 0 | 0 |
| 0 | 0 | 0 | 0 | 0 |
| 0 | 0 | 0 | 0 | 0 |
| 0 | 0 | 0 | 0 | 0 |
| 0 | 0 | 0 | 0 | 0 |
| 0 | 0 | 0 | 0 | 0 |
| 0 | 0 | 0 | 0 | 0 |
| 0 | 0 | 0 | 0.003237893 | 0 |
| 0 | 0.023678878 | 0.002388463 | 0 | 0.00678534 |
| 0.148717862 | 0 | 0.006032832 | 1.675430916 | 0 |
| 0 | 0 | 0 | 0 | 0 |
| 0 | 0 | 0 | 0 | 0 |
| 0 | 0 | 0.544236858 | 0 | 0 |
| 0.007163295 | 0 | 0 | 0.005509284 | 0.001324101 |
| 0 | 0 | 0 | 0 | 0 |
| 0 | 0 | 0 | 0 | 0 |
| 0.003072959 | 0 | 0 | 0 | 0 |
| 0 | 0 | 0 | 0.009243752 | 0.002700821 |
| 0 | 0 | 0 | 0 | 0 |

| | | | | |
|---|---|---|---|---|
| 0 | 0 | 0 | 0 | 0 |
| 0.001640555 | 0.013730133 | 0.006615135 | 0.019039625 | 0.035144372 |
| 0.033485815 | 0 | 0 | 0 | 0 |
| 0 | 0 | 0 | 0 | 0 |
| 0 | 0 | 0 | 0 | 0 |
| 0 | 0 | 0 | 0 | 0 |
| 0 | 0 | 0 | 0 | 0 |
| 0 | 0 | 0 | 0 | 0 |
| 0 | 0 | 0 | 0 | 0 |
| 0 | 0 | 0 | 0 | 0 |
| 0 | 0 | 0 | 0 | 0 |
| 0 | 0 | 0 | 0 | 0 |
| 0.019571821 | 0 | 0.006614796 | 0.141067314 | 0 |
| 0 | 0 | 0 | 0 | 0 |
| 0 | 0 | 0.003789657 | 0 | 0 |
| 0 | 0 | 0 | 0 | 0 |
| 0 | 0 | 0 | 0 | 0 |
| 0.002504681 | 0 | 0 | 0 | 0 |
| 0.001616343 | 0 | 0 | 0.052287567 | 0 |
| 0 | 0 | 0 | 0 | 0 |
| 0 | 0 | 0 | 0 | 0 |
| 0.150521517 | 0.002829845 | 0 | 0.107538932 | 0.004173036 |
| 0 | 0 | 0 | 0.010167962 | 0 |
| 0.000443384 | 0 | 0 | 0.096270006 | 0 |
| 0 | 0 | 0 | 0 | 0 |
| 0 | 0 | 0 | 0 | 0 |
| 0 | 0 | 0 | 0 | 0 |
| 0 | 0 | 0 | 0 | 0 |
| 0 | 0 | 0 | 0 | 0 |
| 0.147350315 | 0 | 0 | 0 | 0 |
| 0 | 0 | 0 | 0 | 0 |
| 0 | 0 | 0 | 0 | 0 |
| 0 | 0 | 0 | 0 | 0 |
| 0 | 0 | 0 | 0 | 0 |
| 2.842609296 | 0 | 0.000596318 | 0.047866174 | 0 |
| 0 | 0 | 0 | 0 | 0 |
| 0 | 0 | 0 | 0 | 0 |
| 0 | 0 | 0 | 0 | 0 |
| 0.005158534 | 0 | 0.004153858 | 0.028106143 | 0 |
| 0 | 0 | 0 | 0 | 0 |
| 0.030706906 | 0.005514809 | 0 | 0.066951577 | 0.271131185 |
| 0 | 0 | 0 | 0 | 0 |
| 0 | 0 | 0 | 0 | 0 |
| 0 | 0 | 0 | 0 | 0 |
| 0 | 0 | 0 | 0 | 0 |
| 0.013827564 | 0.897282126 | 0.004488716 | 0.005167753 | 2.897851136 |
| 0 | 0 | 0 | 0 | 0 |
| 0 | 0 | 0 | 0 | 0 |

| | | | | |
|---|---|---|---|---|
| 0.003668525 | 0 | 0 | 0 | 0 |
| 0.00047919 | 0 | 0 | 0 | 0 |
| 0 | 0 | 0 | 0 | 0 |
| 0 | 0 | 0 | 0 | 0 |
| 0.011044989 | 0 | 0 | 0.000489919 | 0 |
| 0 | 0 | 0 | 0 | 0 |
| 0 | 0 | 0 | 0 | 0 |
| 0 | 0 | 0 | 0.001396403 | 0 |
| 0 | 0 | 0 | 0 | 0 |
| 0 | 0 | 0.054655139 | 0.004843484 | 0.003137962 |
| 0 | 0 | 0 | 0 | 0 |
| 0 | 0 | 0 | 0 | 0 |
| 0 | 0 | 0 | 0.008850705 | 0 |
| 0 | 0 | 0 | 0 | 0 |
| 0 | 0 | 0 | 0 | 0 |
| 0 | 0 | 0 | 0 | 0 |
| 0 | 0 | 0 | 0 | 0 |
| 0 | 0 | 0 | 0 | 0 |
| 0 | 0 | 0 | 0 | 0 |
| 0.031289685 | 0.020617696 | 0.133860817 | 0.12915849 | 0.110789663 |
| 0 | 0 | 0 | 0 | 0 |
| 0 | 0 | 0 | 0 | 0 |
| 0 | 0 | 0 | 0 | 0 |
| 0 | 0 | 0 | 0 | 0 |
| 0 | 0 | 0 | 0 | 0 |
| 0 | 0 | 0 | 0 | 0 |
| 0 | 0 | 0 | 0 | 0 |
| 0 | 0 | 0 | 0 | 0 |
| 0 | 0 | 0 | 0 | 0 |
| 0.000535187 | 0 | 0.040010668 | 0.002741866 | 0 |
| 0 | 0 | 0 | 0 | 0 |
| 0 | 0 | 0 | 0.004618182 | 0 |
| 0 | 0 | 0 | 0 | 0 |
| 0 | 0 | 0 | 0.007453225 | 0 |
| 0 | 0 | 0 | 0 | 0 |
| 0 | 0 | 0 | 0.001185764 | 0 |
| 0 | 0 | 0 | 0 | 0 |
| 0 | 0 | 0 | 0 | 0 |
| 0 | 0 | 0 | 0 | 0 |
| 0.000397805 | 0 | 0 | 0 | 0 |
| 0 | 0 | 0 | 0 | 0 |
| 0 | 0 | 0 | 0 | 0 |
| 0.055867804 | 0 | 0 | 0.053909244 | 0 |
| 0 | 0 | 0 | 0 | 0 |
| 0.051253408 | 0 | 0.004483844 | 0.091542032 | 0 |
| 0 | 0 | 0 | 0 | 0 |
| 0 | 0 | 0 | 0 | 0 |
| 0 | 0 | 0 | 0 | 0 |

| | | | | |
|---|---|---|---|---|
| 0 | 0 | 0 | 0 | 0 |
| 0 | 0 | 0 | 0 | 0 |
| 0 | 0 | 0 | 0 | 0 |
| 0 | 0 | 0 | 0 | 0 |
| 0 | 0 | 0 | 0 | 0 |
| 0.007196189 | 0.000661748 | 0.000401684 | 0.030701476 | 0.000478541 |
| 0 | 0 | 0 | 0 | 0 |
| 0 | 0 | 0 | 0.000612058 | 0 |
| 0 | 0 | 0 | 0 | 0 |
| 0 | 0 | 0 | 0 | 0 |
| 0 | 0 | 0 | 0 | 0 |
| 0 | 0 | 0 | 0 | 0 |
| 0 | 0 | 0 | 0 | 0 |
| 0 | 0 | 0 | 0 | 0 |
| 0 | 0 | 0 | 0 | 0 |
| 0 | 0 | 0 | 0 | 0 |
| 0 | 0 | 0 | 0.015581872 | 0 |
| 0 | 0 | 0 | 0 | 0 |
| 0 | 0 | 0 | 0 | 0 |
| 0 | 0 | 0 | 0 | 0 |
| 0 | 0 | 0 | 0 | 0 |
| 0 | 0 | 0 | 0 | 0 |
| 0 | 0 | 0 | 0 | 0 |
| 0 | 0 | 0 | 0.00590851 | 0 |
| 0 | 0 | 0 | 0 | 0 |
| 0 | 0 | 0.000628301 | 0 | 0 |
| 0 | 0 | 0 | 0 | 0 |
| 0.091024323 | 0 | 0 | 0 | 0 |
| 0 | 0 | 0 | 0 | 0 |
| 0 | 0 | 0 | 0 | 0.031144081 |
| 0 | 0 | 0 | 0 | 0 |
| 0 | 0 | 0 | 0 | 0 |
| 0 | 0 | 0 | 0 | 0 |
| 0 | 0 | 0 | 0 | 0 |
| 0 | 0 | 0 | 0 | 0 |
| 0 | 0 | 0 | 0 | 0 |
| 0 | 0 | 0 | 0 | 0 |
| 0 | 0 | 0 | 0 | 0 |
| 0.327040154 | 0 | 0 | 0 | 0 |
| 0 | 0 | 0 | 0 | 0 |
| 0 | 0 | 0 | 0 | 0 |
| 0 | 0 | 0 | 0 | 0 |
| 0 | 0 | 0 | 0 | 0 |
| 0 | 0 | 0 | 0 | 0 |
| 0 | 0.000212004 | 0 | 0 | 0 |
| 0 | 0 | 0 | 0 | 0 |
| 0 | 0 | 0 | 0 | 0 |
| 0 | 0 | 0 | 0 | 0 |

| | | | | |
|---|---|---|---|---|
| 0 | 0 | 0 | 0 | 0 |
| 0.254965496 | 0 | 0 | 0 | 0 |
| 0 | 0 | 0 | 0 | 0 |
| 0 | 0 | 0 | 0 | 0 |
| 0 | 0 | 0 | 0 | 0 |
| 0.005119874 | 0 | 0 | 0 | 0 |
| 0 | 0 | 0 | 0 | 0 |
| 0 | 0 | 0 | 0 | 0 |
| 0 | 0 | 0 | 0.006821648 | 0 |
| 0 | 0 | 0 | 0 | 0 |
| 0 | 0 | 0.057512886 | 0.003954715 | 0 |
| 0.017693027 | 0 | 0 | 0 | 0 |
| 0.085189394 | 0 | 0 | 0.154935761 | 0 |
| 0 | 0 | 0 | 0 | 0 |
| 0 | 0 | 0 | 0 | 0 |
| 0 | 0 | 0 | 0 | 0 |
| 0 | 0 | 0 | 0 | 0 |
| 0 | 0 | 0 | 0 | 0 |
| 0 | 0 | 0 | 0 | 0 |
| 0.003425751 | 0 | 0 | 0 | 0 |
| 0 | 0 | 0 | 0 | 0 |
| 0 | 0 | 0 | 0 | 0 |
| 0 | 0 | 0 | 0 | 0 |
| 0.150355123 | 0 | 0.17691819 | 0.357203073 | 0 |
| 0 | 0 | 0 | 0 | 0 |
| 0 | 0 | 0 | 0 | 0 |
| 0 | 0 | 0 | 0 | 0 |
| 0 | 0 | 0 | 0 | 0 |
| 0 | 0 | 0 | 0 | 0 |
| 0 | 0 | 0 | 0 | 0 |
| 0 | 0 | 0 | 0.000562791 | 0 |
| 0 | 0 | 0 | 0 | 0 |
| 0.000412061 | 0 | 0 | 0.001608431 | 0 |
| 0 | 0 | 0 | 0 | 0 |

| PHL018-0182_V3 | PHL018-0265_V2 | PHL018-0191_V1 | PHL018-0163_V3 | PHL018-0227_V2 |
|---|---|---|---|---|
| EYCF | REF | EYCF | CTRL | REF |
| 0.086159301 | 0.002747648 | 0.024357834 | 0.112414117 | 0.01062117 |
| 2.040359346 | 2.435363492 | 2.985555621 | 2.456027165 | 0.01582485 |
| 11.94364684 | 26.62549444 | 4.738916287 | 0 | 0 |
| 0 | 0 | 0 | 0 | 0 |
| 0 | 0 | 0 | 0 | 0 |
| 0 | 0 | 0 | 0 | 0 |
| 0 | 0 | 0 | 0 | 0 |
| 0 | 0 | 0 | 0 | 0 |
| 0 | 0 | 0 | 0 | 0 |
| 0 | 0 | 0 | 0 | 0 |
| 0 | 0 | 0 | 0 | 0 |
| 0 | 0 | 0 | 0 | 0 |
| 0 | 0 | 0 | 0 | 0 |
| 39.28016585 | 10.08826709 | 6.498631369 | 0.843719922 | 0 |
| 0 | 0 | 0 | 0 | 0 |
| 0 | 0 | 0 | 0 | 0 |
| 0 | 0.001042364 | 0 | 0 | 0 |
| 0 | 0 | 0 | 0 | 0 |
| 0 | 0 | 0 | 0 | 0 |
| 0 | 0 | 0 | 0 | 0 |
| 0 | 0 | 0 | 0 | 0 |
| 0 | 0 | 0 | 0 | 0 |
| 0 | 0 | 0 | 0 | 0 |
| 0 | 0 | 0 | 0 | 0 |
| 0 | 0 | 0 | 0 | 0 |
| 0 | 0 | 0 | 0 | 0 |
| 0 | 0 | 0 | 0 | 0 |
| 0 | 0 | 0 | 0 | 0 |
| 0 | 0 | 0 | 0 | 0 |
| 0 | 0 | 0 | 0 | 0 |
| 0 | 0 | 0 | 0 | 0 |
| 0 | 0 | 0 | 0 | 0 |
| 0 | 0 | 0 | 0 | 0 |
| 0 | 0 | 0 | 0 | 0 |
| 0 | 0 | 0 | 0 | 0 |
| 0 | 6.121606969 | 0 | 7.791751216 | 0.002536176 |
| 0 | 0 | 0 | 0 | 0 |
| 0 | 0 | 0 | 0 | 0 |
| 0 | 0 | 0 | 0 | 0 |
| 0 | 0 | 0 | 0 | 0 |
| 0 | 0 | 0 | 0 | 0 |
| 0.080582694 | 0 | 0.005255442 | 0.009035123 | 1.342750185 |
| 0 | 0 | 0 | 5.746917979 | 0.149231183 |
| 0 | 0.005875789 | 0.004787566 | 0.014010173 | 0.05657714 |
| 0 | 0.001765098 | 0 | 0.017647445 | 0.002966554 |
| 0 | 0 | 0 | 0 | 0 |

| | | | | |
|---|---|---|---|---|
| 0 | 0 | 0 | 0.001203762 | 0.010188505 |
| 0 | 0.100565023 | 0 | 0.000828143 | 0.006083553 |
| 0.011658383 | 0.00143139 | 0.002374991 | 0.007018568 | 0 |
| 0.793394586 | 0 | 0.2788 | 0.777648049 | 0.006775037 |
| 0 | 0 | 0 | 0 | 0 |
| 0 | 0 | 0 | 0 | 0 |
| 0 | 0.001012108 | 0.001979186 | 0 | 0.774532357 |
| 0 | 0 | 0 | 0 | 0 |
| 0 | 0 | 0 | 0 | 0 |
| 0 | 0 | 0 | 0 | 0 |
| 0.001171096 | 0 | 0 | 0 | 0.004428247 |
| 0 | 0 | 0.45193008 | 0.04244044 | 0 |
| 0 | 0 | 0 | 0 | 0 |
| 0.022038007 | 0.087343192 | 0.032443415 | 0.108863083 | 0.001421255 |
| 0 | 0 | 0 | 0 | 0.001392838 |
| 0 | 0 | 0 | 0 | 0 |
| 0.458182118 | 0 | 0 | 0.001742946 | 0.003896782 |
| 0 | 0 | 0 | 0 | 0 |
| 0 | 0 | 0 | 0 | 0 |
| 0 | 0 | 0 | 0 | 0 |
| 0.15212589 | 0.021573763 | 0.174242649 | 0.078761021 | 0 |
| 0.002846278 | 0 | 0.002656301 | 0.066538264 | 0 |
| 0 | 0 | 0 | 0.103103168 | 0 |
| 0 | 0 | 0 | 0 | 0 |
| 0 | 0 | 0 | 0 | 0 |
| 0 | 0 | 0 | 0 | 0 |
| 0 | 0 | 0 | 0 | 0 |
| 0 | 0 | 0 | 0 | 0 |
| 0 | 0 | 0 | 0 | 0.060301437 |
| 0 | 0 | 0.001028016 | 0 | 0 |
| 0.009822037 | 0 | 0 | 0 | 0.608064282 |
| 0 | 0 | 0 | 0 | 0 |
| 0 | 0 | 0 | 0 | 0 |
| 0.001794305 | 0 | 0 | 0 | 0 |
| 0 | 0 | 0 | 0 | 0.001623917 |
| 0 | 0 | 0 | 0 | 0 |
| 0 | 0 | 0 | 0 | 0 |
| 0.006275293 | 0 | 0 | 0.075331158 | 0 |
| 0 | 0 | 0 | 0 | 0 |
| 0 | 0 | 0 | 0 | 0 |
| 0 | 0 | 0 | 0 | 0 |
| 0 | 0 | 0 | 0 | 0 |
| 0 | 0 | 0 | 0 | 0 |
| 0 | 0 | 0 | 0 | 0 |
| 0 | 0 | 0 | 0 | 0 |
| 0 | 0 | 0 | 0 | 0 |
| 0 | 0.000444549 | 0 | 0.001559506 | 0 |
| 0 | 0 | 0 | 0 | 0.007530179 |

| | | | | |
|---|---|---|---|---|
| 0 | 0 | 0 | 0 | 0 |
| 0.016575876 | 0.008433281 | 0 | 0 | 0 |
| 0 | 0 | 0 | 0 | 0 |
| 0 | 0 | 0 | 0 | 0 |
| 0 | 0 | 0.001956423 | 0 | 0 |
| 0 | 0 | 0 | 0 | 0.002723539 |
| 0 | 0 | 0 | 0 | 0 |
| 0 | 0 | 0 | 0 | 0 |
| 0 | 0 | 0 | 0 | 0.003894127 |
| 0 | 0 | 0 | 0 | 0 |
| 0 | 0 | 0 | 0 | 0 |
| 0 | 0 | 0.001227406 | 0 | 0.268999783 |
| 0 | 0 | 0 | 0 | 0 |
| 0 | 0 | 0 | 0 | 0 |
| 0 | 0 | 0 | 0 | 0 |
| 0 | 0 | 0 | 0 | 0 |
| 0 | 0 | 0 | 0 | 0 |
| 0 | 0 | 0 | 0 | 0 |
| 0 | 0 | 0.14215946 | 0.13888543 | 0 |
| 0 | 0 | 0 | 0 | 0 |
| 0 | 0 | 0 | 0 | 0 |
| 0 | 0 | 0 | 0 | 0 |
| 0 | 0 | 0 | 0 | 0 |
| 0 | 0 | 0 | 0 | 0 |
| 0 | 0 | 0 | 0 | 0 |
| 0 | 0 | 0 | 0 | 0 |
| 0 | 0 | 0 | 0 | 0 |
| 0 | 0 | 0 | 0 | 0 |
| 0 | 0 | 0 | 0 | 0 |
| 0 | 0 | 0 | 0 | 0 |
| 0 | 0 | 0.219841614 | 0 | 0 |
| 0 | 0 | 0 | 0.120137744 | 0 |
| 0 | 0 | 0 | 0 | 0 |
| 0 | 0 | 0 | 0 | 0 |
| 0 | 0 | 0 | 0 | 0 |
| 0 | 0 | 0 | 0 | 0 |
| 0 | 0 | 0 | 0 | 0 |
| 0 | 0 | 0 | 0 | 0 |
| 0 | 0 | 0 | 0.022513112 | 0 |
| 0 | 0 | 0 | 0 | 0 |
| 0 | 0 | 0 | 0 | 0 |
| 0 | 0 | 0 | 0 | 0 |
| 0 | 0 | 0 | 0.005130304 | 0 |
| 0 | 0 | 0 | 0 | 0 |
| 0 | 0 | 0 | 0 | 0 |
| 0 | 0 | 0 | 0 | 0 |
| 0 | 0 | 0 | 0 | 0 |
| 0 | 0 | 0 | 0 | 0 |

| | | | | |
|---|---|---|---|---|
| 0 | 0 | 0 | 0 | 0 |
| 0 | 0 | 0 | 0 | 0 |
| 0 | 0 | 0 | 0 | 0 |
| 0 | 0 | 0 | 0 | 0 |
| 0 | 0 | 0.013654909 | 0.100960575 | 0 |
| 0 | 0 | 0 | 0 | 0 |
| 0 | 0 | 0 | 0 | 0 |
| 0 | 0 | 0 | 0 | 0 |
| 0 | 0 | 0 | 0 | 0 |
| 0 | 0 | 0 | 0 | 0 |
| 0 | 0 | 0 | 0 | 0 |
| 0 | 0 | 0 | 0 | 0 |
| 0 | 0 | 0 | 0 | 0 |
| 0 | 0 | 0 | 0 | 0 |
| 0 | 0 | 0 | 0 | 0 |
| 0 | 0 | 0 | 0 | 0 |
| 0 | 0 | 0 | 0 | 0 |
| 0 | 0 | 0 | 0 | 0 |
| 0 | 0 | 0 | 0 | 0 |
| 0 | 0 | 0 | 0 | 0 |
| 0 | 0 | 0 | 0 | 0 |
| 0 | 0 | 0 | 0 | 0 |
| 0 | 0 | 0 | 0 | 0 |
| 0 | 0 | 0 | 0 | 0 |
| 0 | 0 | 0 | 0.004163646 | 0 |
| 0 | 0 | 0 | 0 | 0 |
| 0 | 0 | 0 | 0 | 0 |
| 0 | 0 | 0 | 0 | 0 |
| 0 | 0 | 0 | 0 | 0 |
| 0 | 0 | 0 | 0 | 0 |
| 0 | 0 | 0 | 0 | 0 |
| 0 | 0 | 0 | 0 | 0.005452204 |
| 0 | 0 | 0 | 0 | 0 |
| 0 | 0 | 0 | 0 | 0 |
| 0 | 0 | 0 | 0 | 0 |
| 0 | 0 | 0 | 0 | 0 |
| 0 | 0 | 0 | 0 | 0 |
| 0 | 0 | 0 | 0 | 0 |
| 0 | 0 | 0 | 0 | 0 |
| 0 | 0 | 0 | 0 | 0 |
| 0 | 0 | 0 | 0 | 0 |
| 0 | 0 | 0 | 0 | 0 |
| 0 | 0 | 0 | 0 | 0 |
| 0 | 0 | 0 | 0 | 0 |
| 0 | 0 | 0 | 0 | 0 |
| 0 | 0 | 0 | 0 | 0 |
| 0 | 0 | 0 | 0 | 0 |
| 0 | 0 | 0 | 0 | 0 |

| | | | | |
|---|---|---|---|---|
| 0 | 0 | 0 | 0 | 0 |
| 0 | 0 | 0 | 0 | 0 |
| 0 | 0 | 0 | 0 | 0 |
| 0 | 0 | 0 | 0 | 0 |
| 0 | 0 | 0 | 0 | 0 |
| 0 | 0 | 0 | 0 | 0 |
| 0 | 0 | 0 | 0 | 0 |
| 0 | 0 | 0 | 0 | 0 |
| 0 | 0 | 0 | 0 | 0 |
| 0 | 0 | 0 | 0 | 0 |
| 0 | 0 | 0 | 0 | 0 |
| 0 | 0 | 0 | 0 | 0 |
| 0 | 0 | 0.000485631 | 0.002776927 | 0 |
| 0 | 0 | 0 | 0 | 0 |
| 0 | 0 | 0 | 0 | 0 |
| 0 | 0 | 0 | 0 | 0 |
| 0 | 0 | 0 | 0 | 0 |
| 0 | 0 | 0 | 0 | 0 |
| 0 | 0 | 0 | 0 | 0 |
| 0 | 0 | 0 | 0 | 0 |
| 0.01954751 | 0.021823773 | 0.024213666 | 0.058030308 | 0.00897456 |
| 0 | 0 | 0 | 0 | 0 |
| 0 | 0 | 0 | 0 | 0 |
| 0 | 0 | 0 | 0.003503439 | 0 |
| 0 | 0 | 0 | 0 | 0 |
| 0 | 0 | 0 | 0 | 0 |
| 0.002335181 | 0.010275888 | 0 | 0.017253484 | 0.010453144 |
| 0 | 0 | 0 | 0 | 0 |
| 0 | 0 | 0 | 0 | 0 |
| 0 | 0.027774357 | 0 | 0 | 0 |
| 0.072161157 | 0.48399665 | 0.049198693 | 0 | 0.041906562 |
| 0 | 0 | 0 | 0 | 0 |
| 0 | 0 | 0 | 0 | 0 |
| 0 | 0 | 0 | 0.002239864 | 0 |
| 0.001956716 | 0.005287126 | 0.106147302 | 0.073527979 | 0 |
| 0 | 0 | 0 | 0 | 0 |
| 0 | 0 | 0 | 0 | 0 |
| 0 | 0 | 0 | 0 | 0 |
| 0 | 0 | 0 | 0 | 0.000738243 |
| 0 | 0 | 0 | 0 | 0.002596543 |
| 0 | 0 | 0 | 0 | 0 |
| 0 | 0 | 0 | 0 | 0 |
| 0 | 0 | 0 | 0 | 0.003694372 |
| 0 | 0 | 0 | 0 | 0 |
| 0 | 0 | 0 | 0 | 0 |
| 0 | 0 | 0 | 0 | 0 |
| 0 | 0 | 0.001279361 | 0.010327946 | 0 |
| 0 | 0 | 0.003368168 | 0 | 0 |

| | | | | |
|---|---|---|---|---|
| 0 | 0 | 0 | 0 | 0 |
| 0 | 0 | 0 | 0 | 0 |
| 0 | 0 | 0 | 0.006987924 | 0 |
| 0 | 0 | 0 | 0.00198014 | 0 |
| 0 | 0 | 0 | 0 | 0 |
| 0 | 0 | 0 | 0 | 0 |
| 0 | 0 | 0 | 0 | 0.006953679 |
| 0 | 0 | 0 | 0 | 0.088948114 |
| 0 | 0.004454757 | 0 | 0 | 0 |
| 0 | 0 | 0 | 0 | 0 |
| 0 | 0 | 0 | 0 | 0.00313389 |
| 0 | 0 | 0 | 0 | 0 |
| 0 | 0 | 0 | 0 | 0 |
| 0 | 0 | 0 | 0 | 0 |
| 0.0018321 | 0 | 0 | 0 | 0.003423094 |
| 0 | 0 | 0 | 0 | 0 |
| 0 | 0 | 0 | 0 | 0 |
| 0 | 0 | 0 | 0 | 0 |
| 0 | 0 | 0 | 0.043206848 | 0.000735996 |
| 0 | 0 | 0 | 0 | 0 |
| 0.003312093 | 0 | 0.013474188 | 0 | 0.436438871 |
| 0 | 0 | 0 | 0 | 0 |
| 0 | 0 | 0 | 0 | 0 |
| 0 | 0 | 0 | 0 | 0 |
| 0.004848548 | 0.001604201 | 0.009201954 | 0 | 0.010245325 |
| 0 | 0 | 0 | 0 | 0 |
| 0 | 0 | 0 | 0 | 0 |
| 0 | 0 | 0 | 0 | 0 |
| 0.00462704 | 0 | 0 | 0 | 0.111152041 |
| 0 | 0 | 0 | 0 | 0 |
| 0.609870536 | 0.00795597 | 0.042416761 | 0.326954169 | 0.097591179 |
| 0 | 0 | 0 | 0 | 0 |
| 0.030441425 | 0 | 0 | 0 | 0 |
| 0 | 0 | 0 | 0 | 0 |
| 0 | 0 | 0 | 0 | 0 |
| 0 | 0 | 0 | 0 | 0 |
| 0 | 0 | 0 | 0 | 0 |
| 0 | 0 | 0 | 0 | 0 |
| 0 | 0 | 0 | 0 | 0 |
| 0 | 0 | 0.000618517 | 0.000624141 | 0 |
| 0 | 0 | 0 | 0 | 0 |
| 0 | 0.002057708 | 0 | 0.027228903 | 0 |
| 0 | 0 | 0 | 0 | 0 |
| 0 | 0 | 0 | 0 | 0 |
| 0 | 0 | 0 | 0 | 0 |
| 0 | 0 | 0 | 0 | 0 |
| 0 | 0 | 0 | 0 | 0 |
| 0 | 0 | 0 | 0 | 0 |

| | | | | |
|---|---|---|---|---|
| 0 | 0 | 0 | 0 | 0 |
| 0 | 0 | 0 | 0 | 0 |
| 0 | 0.004711618 | 0 | 0.014196001 | 0.047001266 |
| 0 | 0 | 0 | 0 | 0 |
| 0 | 0 | 0 | 0 | 0 |
| 0 | 0 | 0 | 0 | 0 |
| 0 | 0 | 0 | 0 | 0 |
| 0 | 0 | 0 | 0 | 0 |
| 0 | 0 | 0 | 0 | 0 |
| 0 | 0 | 0 | 0.006948945 | 0 |
| 0 | 0 | 0 | 0 | 0 |
| 0 | 0 | 0 | 0 | 0 |
| 0 | 0 | 0 | 0 | 0 |
| 0 | 0 | 0.00153288 | 0.000662922 | 0 |
| 0.001795833 | 0.001247764 | 0.001988159 | 0.002735775 | 0.001398056 |
| 0 | 0 | 0.000774031 | 0 | 0 |
| 0.001364127 | 0.002369525 | 0.002471268 | 0 | 0.016726068 |
| 0 | 0 | 0 | 0 | 0 |
| 0 | 0 | 0 | 0 | 0 |
| 0 | 0 | 0 | 0 | 0 |
| 0 | 0 | 0 | 0 | 0 |
| 0.000461782 | 0 | 0.002416754 | 0 | 0 |
| 0 | 0 | 0 | 0 | 0 |
| 0 | 0 | 0 | 0 | 0 |
| 0 | 0 | 0 | 0 | 0 |
| 0 | 0 | 0 | 0 | 0 |
| 0 | 0 | 0 | 0 | 0 |
| 0 | 0 | 0 | 0 | 0 |
| 0 | 0 | 0 | 0 | 0 |
| 0 | 0 | 0 | 0.001356785 | 0.002773416 |
| 0 | 0 | 0 | 0 | 0 |
| 0 | 0 | 0 | 0 | 0.003824145 |
| 0 | 0 | 0 | 0 | 0.003968004 |
| 0 | 0 | 0 | 0 | 0 |
| 0 | 0 | 0 | 0 | 0 |
| 0 | 0 | 0 | 0.00374693 | 0.023456097 |
| 0 | 0 | 0 | 0 | 0 |
| 0 | 0.000895813 | 0.007007079 | 0.014612958 | 0.442637126 |
| 0 | 0 | 0 | 0 | 0 |
| 0 | 0 | 0 | 0 | 0 |
| 0 | 0 | 0 | 0 | 0 |
| 0 | 0 | 0 | 0 | 0 |
| 0 | 0 | 0 | 0 | 0 |
| 0 | 0.004896053 | 0 | 0 | 0 |
| 0 | 0 | 0 | 0 | 0 |
| 0 | 0 | 0 | 0 | 0 |
| 0 | 0 | 0 | 0 | 0.001631981 |
| 0 | 0.010850537 | 0.001980377 | 0 | 0.037566594 |

| | | | | |
|---|---|---|---|---|
| 0 | 0 | 0 | 0 | 0 |
| 0 | 0 | 0 | 0 | 0 |
| 0 | 0.002240441 | 0 | 0 | 0 |
| 0 | 0 | 0 | 0 | 0 |
| 0.003244193 | 0.007513664 | 0.004114051 | 0.001581507 | 0 |
| 0 | 0 | 0 | 0 | 0 |
| 0 | 0 | 0 | 0 | 0 |
| 0.002046352 | 0.018957697 | 0 | 0.030204306 | 0.004248226 |
| 0 | 0 | 0 | 0 | 0 |
| 7.682012826 | 0.851080293 | 0.050098684 | 0.010853439 | 0.042327811 |
| 0.001059449 | 0.001766678 | 0 | 0 | 0.008907629 |
| 0.011879266 | 0 | 0.001956419 | 0 | 0.386734797 |
| 0 | 0.002150184 | 0.041717265 | 0.01447589 | 0.173460355 |
| 0 | 0 | 0 | 0.004295948 | 0 |
| 0 | 0.002081332 | 0 | 0 | 0 |
| 0 | 0 | 0.000742583 | 0 | 0.021061181 |
| 0 | 0 | 0 | 0 | 0.001856457 |
| 0.002844958 | 0 | 0 | 0 | 0 |
| 0.511841714 | 0.482553611 | 0.005859451 | 0 | 0 |
| 0 | 0 | 0 | 0 | 0 |
| 0 | 0 | 0 | 0 | 0 |
| 0 | 0 | 0 | 0 | 0 |
| 0 | 0 | 0 | 0 | 0.00394828 |
| 0 | 0 | 0 | 0 | 0 |
| 0 | 0 | 0 | 0 | 0 |
| 0 | 0.005008833 | 0 | 0.010852878 | 0 |
| 0 | 0 | 0 | 0 | 0 |
| 0 | 0.046318777 | 0 | 0.013358506 | 0 |
| 0.108596809 | 0.015234133 | 0 | 0 | 0 |
| 0 | 0 | 0 | 0.03110597 | 0 |
| 0 | 0.020076271 | 0 | 0 | 0 |
| 0 | 0.021754769 | 0 | 0 | 0 |
| 0 | 0.020026897 | 0 | 0 | 0 |
| 0.145578494 | 0.016761913 | 0 | 0 | 0.013599939 |
| 0 | 0 | 0 | 0 | 0 |
| 0.002741336 | 0.013594393 | 0 | 0 | 0 |
| 0.011717274 | 0.022211097 | 0 | 0 | 0 |
| 0 | 0 | 0 | 0 | 0 |
| 0 | 0 | 0 | 0 | 0 |
| 0.06489246 | 0 | 0 | 0 | 0 |
| 0 | 0 | 0 | 0 | 0 |
| 0.26893041 | 0 | 0 | 0 | 0 |
| 0 | 0 | 0 | 0 | 0 |
| 0 | 0 | 0 | 0 | 0.100932243 |
| 0 | 0 | 0 | 0 | 0 |
| 0 | 0 | 0 | 0 | 0 |
| 0 | 0 | 0 | 0.001205037 | 0 |
| 0 | 0 | 0 | 0 | 0 |

| | | | | |
|---|---|---|---|---|
| 0 | 0 | 0 | 0 | 0 |
| 0.013702261 | 0 | 0.001868411 | 0 | 0.005161552 |
| 0 | 0 | 0 | 0 | 0 |
| 0 | 0 | 0 | 0 | 0 |
| 0 | 0 | 0.001416386 | 0 | 0 |
| 0.003270864 | 0 | 0.002821683 | 0 | 0 |
| 0 | 0 | 0 | 0.033982616 | 0 |
| 0.002820305 | 0 | 0 | 0 | 0 |
| 0 | 0 | 0 | 0 | 0 |
| 0 | 0 | 0 | 0 | 0.001265417 |
| 0.018426308 | 0.002008281 | 0.001527249 | 0.018273455 | 0 |
| 0 | 0 | 0 | 0 | 0 |
| 0 | 0 | 0 | 0 | 0 |
| 0.002560037 | 0 | 0 | 0.001455987 | 0.002125854 |
| 0.122434704 | 0 | 0 | 0 | 0 |
| 0 | 0 | 0 | 0 | 0 |
| 0 | 0 | 0 | 0 | 0 |
| 0 | 0 | 0 | 0 | 0 |
| 0 | 0 | 0 | 0 | 0.000848492 |
| 0 | 0 | 0.024349023 | 0 | 0 |
| 0 | 0 | 0 | 0 | 0 |
| 0 | 0 | 0 | 1.863913034 | 0 |
| 0.041460947 | 0 | 0.008802084 | 0 | 0 |
| 0 | 0 | 0 | 0 | 0 |
| 0 | 0 | 0 | 0 | 0 |
| 0 | 0 | 0 | 0 | 0 |
| 0.35566706 | 0 | 0 | 0 | 0 |
| 0 | 0 | 0 | 0 | 0 |
| 0 | 0 | 0 | 0 | 0 |
| 0.183621232 | 0.227353155 | 0.345944196 | 0.588421569 | 1.413131598 |
| 0 | 0 | 0 | 0 | 0 |
| 0 | 0 | 0 | 0 | 0.047836746 |
| 0 | 0 | 0 | 0 | 0 |
| 0 | 0 | 0 | 0 | 0.002042681 |
| 0.001233037 | 0.028557572 | 0 | 0 | 0 |
| 0 | 0 | 0 | 0 | 0 |
| 0 | 0 | 0 | 0.336317132 | 0 |
| 0 | 0 | 0 | 0 | 0.042523673 |
| 0 | 0 | 0 | 0 | 0 |
| 0.569283617 | 0 | 0 | 0 | 0 |
| 0.002244937 | 0.012998384 | 0.013725968 | 0 | 0 |
| 2.048998675 | 0 | 2.530331768 | 3.654557685 | 0 |
| 0 | 0 | 0 | 0 | 0.00868411 |
| 0.00372391 | 0 | 0 | 0 | 0 |
| 0.000633102 | 0 | 0 | 0 | 0 |
| 0 | 0 | 0 | 0 | 0 |
| 0 | 0 | 0.007114266 | 0.005204737 | 0 |
| 0 | 0 | 0 | 0 | 0 |

| | | | | |
|---|---|---|---|---|
| 0 | 0.000673314 | 0.001975007 | 0.013064985 | 0.004809389 |
| 0 | 0 | 0.039822206 | 0.005942743 | 0 |
| 1.000713363 | 0.000917442 | 0.000897033 | 0.004525943 | 0.024670722 |
| 0 | 0 | 0 | 0 | 0 |
| 0 | 0 | 0 | 0 | 0 |
| 0 | 0 | 0 | 0 | 0.002152769 |
| 0 | 0 | 0 | 0 | 0 |
| 0 | 0 | 0 | 0 | 0.312640256 |
| 0 | 0 | 0.013774199 | 0 | 0 |
| 0 | 0 | 0 | 0 | 0 |
| 0 | 0 | 0 | 0 | 0 |
| 0 | 0.001277441 | 0 | 0 | 0 |
| 0 | 0 | 0 | 0 | 0 |
| 0.00969833 | 0.010050647 | 0 | 0 | 0 |
| 0 | 0 | 0 | 0 | 0 |
| 0 | 0 | 0.17391765 | 0.165009659 | 0 |
| 0 | 0.005144331 | 0.006190638 | 0.506347758 | 0.010197777 |
| 0 | 0 | 0.000835709 | 0 | 0.044980727 |
| 0 | 0 | 0.008525948 | 0 | 0 |
| 0 | 0 | 0 | 0 | 0 |
| 0.003947757 | 0 | 0 | 0 | 0 |
| 0 | 0.001071028 | 0.00139627 | 0.006038421 | 0 |
| 0 | 0 | 0 | 0 | 0 |
| 0.035206823 | 0 | 0 | 0 | 0 |
| 0 | 0 | 0 | 0 | 0 |
| 0 | 0 | 0 | 0 | 0.002920385 |
| 0 | 0.001339645 | 0.001309844 | 0 | 0 |
| 0.002402204 | 0 | 0 | 0 | 0 |
| 0 | 0 | 0 | 0 | 0 |
| 0 | 0 | 0 | 0 | 0 |
| 0.036153303 | 0 | 0.051134813 | 0 | 0 |
| 0 | 0.002468213 | 0 | 0.006493994 | 0.011487489 |
| 0.002146473 | 0 | 0.003063725 | 0.011890694 | 0 |
| 0 | 0 | 0 | 0 | 0.876477477 |
| 0.001796364 | 0 | 0 | 0.004865039 | 0.001864626 |
| 0 | 0 | 0 | 0 | 0 |
| 0 | 0 | 0.002343693 | 0 | 0.01428322 |
| 0.086089737 | 0 | 0.005665255 | 0 | 0.007088743 |
| 0 | 0 | 0 | 0.123906949 | 0 |
| 0 | 0 | 0 | 0 | 0 |
| 0 | 0 | 0 | 0 | 0 |
| 0 | 0.004050341 | 0 | 0 | 0.000756367 |
| 0 | 0 | 0.001245938 | 0 | 0.013171193 |
| 0 | 0.015564602 | 0.025103791 | 0 | 0 |
| 0.184199503 | 0 | 0 | 0 | 0 |
| 0 | 0 | 0 | 0 | 0 |
| 0 | 0 | 0 | 0 | 0 |
| 0 | 0 | 0 | 0 | 0 |

| | | | | |
|---|---|---|---|---|
| 0 | 0 | 0 | 0 | 0 |
| 0.02030334 | 0.275173092 | 0.680462514 | 0.785625659 | 0.207236136 |
| 0 | 0 | 0 | 0 | 0 |
| 0.001252419 | 0 | 0 | 0 | 0 |
| 0 | 0 | 0 | 0 | 0 |
| 0 | 0 | 0 | 0 | 0 |
| 0.061393349 | 0.007250439 | 0.062804612 | 0 | 0.003249499 |
| 0 | 0 | 0 | 0 | 0.000674488 |
| 0 | 0 | 0 | 0 | 0.123792266 |
| 0 | 0 | 0 | 0 | 0 |
| 0.0022352 | 0 | 0.008267349 | 0.060270411 | 0 |
| 0 | 0.002506073 | 0.005528937 | 0.013440813 | 0.59614451 |
| 0 | 0 | 0 | 0 | 0 |
| 0 | 0 | 0 | 0 | 0 |
| 0 | 0 | 0 | 0 | 0.076368057 |
| 0 | 0 | 0 | 0 | 0 |
| 0.004827441 | 0.012332455 | 0.027031376 | 0.002139384 | 0.00341651 |
| 0.2483886 | 0.010203685 | 0.050456872 | 0.002082912 | 0.041689964 |
| 0.001085377 | 0 | 0 | 0 | 0 |
| 0.01319426 | 0 | 0 | 0 | 0.005183863 |
| 0.001206863 | 0.039271672 | 0.001639775 | 0.177418816 | 0.007751225 |
| 0.003191212 | 0 | 0 | 0 | 0 |
| 0 | 0 | 0 | 0 | 0 |
| 0 | 0 | 0 | 0 | 0 |
| 0 | 0 | 0 | 0 | 0.005919775 |
| 0 | 0 | 0 | 0 | 0 |
| 0.069836089 | 0 | 0.005574969 | 0 | 0.012983255 |
| 0.007215283 | 0.966704094 | 0.156661803 | 0.015437338 | 0 |
| 0 | 0 | 0 | 0 | 0 |
| 0 | 0 | 0 | 0 | 0 |
| 0 | 0 | 0 | 0.003145116 | 0 |
| 0.003319748 | 0 | 0 | 0 | 0 |
| 0 | 0.00072131 | 0 | 0 | 0 |
| 0 | 0 | 0 | 0 | 0 |
| 0 | 0 | 0 | 0.167390778 | 0 |
| 0.101219241 | 9.479914016 | 0.199920718 | 0.139563268 | 0.545467622 |
| 0 | 0 | 0 | 0 | 0 |
| 0 | 0 | 0 | 0 | 0 |
| 0.003560344 | 0 | 0.00358331 | 0 | 0.1693833 |
| 0 | 0 | 0 | 0 | 0 |
| 0.075215847 | 0 | 0 | 0 | 0 |
| 0 | 0 | 0 | 0 | 0 |
| 0.007569125 | 0 | 0 | 0 | 0.001683589 |
| 0.001391971 | 0 | 0 | 0 | 0 |
| 0.007010121 | 0 | 0 | 0 | 0 |
| 0 | 0 | 0 | 0 | 0 |
| 0 | 0 | 0 | 0 | 0 |
| 0 | 0.004863702 | 0 | 0 | 0 |

| | | | | |
|---|---|---|---|---|
| 0 | 0 | 0 | 0 | 0 |
| 0 | 0 | 0 | 0 | 0 |
| 0 | 0 | 0 | 0 | 0 |
| 0 | 0 | 0 | 0.044680579 | 0 |
| 0 | 0 | 0 | 0 | 0 |
| 0 | 0 | 0 | 0 | 0 |
| 0 | 0.000603013 | 0.021880667 | 0 | 0 |
| 0 | 0 | 0 | 0.001525632 | 0 |
| 0 | 0 | 1.531922677 | 0.042909605 | 0 |
| 0 | 0 | 0 | 0 | 0 |
| 0 | 0 | 0 | 0 | 0 |
| 0 | 0 | 0.005505287 | 0.005391948 | 0 |
| 0 | 0 | 0 | 0 | 0 |
| 0 | 0 | 0 | 0 | 0 |
| 0.005199789 | 0 | 0.039817854 | 0 | 0 |
| 0 | 0 | 0 | 0 | 0 |
| 0 | 0 | 0 | 0.088289879 | 0 |
| 0 | 0 | 0 | 0.015318091 | 0 |
| 0 | 0 | 0 | 0 | 0 |
| 0 | 0 | 0 | 0 | 0 |
| 0 | 0 | 0 | 0 | 0 |
| 0 | 0 | 0 | 0 | 0 |
| 0 | 0 | 0 | 0 | 0 |
| 0.198963785 | 0 | 0.19138787 | 0 | 0.008794154 |
| 0 | 0 | 0 | 0.009331597 | 0 |
| 0 | 0 | 0.961959197 | 0 | 0 |
| 0 | 0 | 0 | 0 | 0 |
| 0 | 0 | 0 | 0.939131015 | 0.074951979 |
| 0 | 0 | 0 | 0 | 0 |
| 0 | 0 | 0 | 0 | 0 |
| 0 | 0 | 0.002046383 | 0.067826913 | 0 |
| 0 | 0 | 0 | 0.131253286 | 0 |
| 0 | 0 | 0.015215063 | 0 | 0 |
| 0 | 0 | 0 | 0 | 0 |
| 0 | 0 | 0 | 0 | 0 |
| 0 | 0 | 0.082484153 | 0.024916354 | 0 |
| 0 | 0.033055955 | 0 | 0.335363675 | 0 |
| 0 | 0 | 0 | 0 | 0 |
| 0 | 0 | 0 | 0 | 0 |
| 0 | 0 | 0.078810748 | 0 | 0 |
| 0.000643075 | 0.007983095 | 0.034639367 | 0.010972203 | 0.765568666 |
| 0 | 0 | 0 | 0 | 0 |
| 0.813722231 | 0.255998123 | 1.268799837 | 1.966854241 | 0 |
| 0 | 0 | 0.029363521 | 0.005734934 | 0 |
| 0 | 0 | 0 | 0 | 0 |
| 0 | 0 | 0 | 0 | 0 |
| 0 | 0 | 0 | 0 | 0 |
| 0.001236879 | 0.00515638 | 0 | 0.04842558 | 0.002648003 |

| | | | | |
|---|---|---|---|---|
| 0 | 0 | 0 | 0 | 0 |
| 0 | 0 | 0 | 0.272598895 | 0 |
| 0.05720946 | 0.32719731 | 0.118856338 | 0 | 8.491672735 |
| 0 | 0 | 0 | 0 | 0 |
| 0 | 0 | 0 | 0 | 0 |
| 0 | 0 | 0 | 0.001138334 | 0 |
| 0 | 0 | 0 | 0 | 0 |
| 0 | 0 | 0 | 0 | 0 |
| 0 | 0 | 0 | 0 | 0 |
| 0 | 0 | 0 | 0 | 0 |
| 0.012659975 | 0.007144874 | 0.033216137 | 0.036178304 | 0.014727039 |
| 0.804628495 | 0.118812142 | 2.775440218 | 1.021364304 | 0.001830102 |
| 0 | 0 | 0 | 0 | 0 |
| 0 | 0.014112658 | 0.017070576 | 0.234461998 | 0.029587332 |
| 0.062746174 | 0.393602848 | 0.129141776 | 0 | 0.06917708 |
| 0 | 0 | 0.171893736 | 0 | 0 |
| 0 | 0 | 0 | 0 | 0 |
| 0 | 0 | 0 | 0 | 0 |
| 0 | 0 | 0 | 0 | 0 |
| 0.001318544 | 0 | 0.005066918 | 0 | 0 |
| 0 | 0 | 0.027398109 | 0 | 0.015761608 |
| 20.55833529 | 3.954211821 | 2.607066636 | 0.015975384 | 0 |
| 0.756163313 | 11.01025003 | 0.739336769 | 0.366430803 | 1.663063931 |
| 0 | 0 | 0 | 0 | 0 |
| 0 | 0 | 0 | 0 | 0 |
| 0 | 0.005686616 | 0 | 3.497849837 | 0 |
| 0 | 0 | 0 | 0 | 0 |
| 0 | 0 | 0 | 0 | 0 |
| 0 | 0 | 0 | 0 | 0 |
| 0 | 0 | 0 | 0 | 0 |
| 0 | 0 | 0 | 0 | 0 |
| 0 | 0 | 0.051453703 | 0.285287668 | 0.009681072 |
| 0.005679089 | 0.000553809 | 0 | 0 | 0.06259445 |
| 0 | 0 | 0 | 0 | 0 |
| 0.005523765 | 0.001462084 | 0 | 0.00396703 | 0.001689384 |
| 0 | 0 | 0.112609941 | 0.010735749 | 0 |
| 0 | 0 | 0.00126488 | 0 | 0 |
| 0 | 0 | 0 | 0 | 0 |
| 0 | 0 | 0 | 0.037688984 | 0 |
| 0 | 0 | 0 | 0 | 0 |
| 0 | 0 | 0 | 0.023851574 | 0 |
| 0 | 0 | 0 | 0 | 0 |
| 0 | 0 | 0 | 0 | 0 |
| 0 | 0 | 0 | 0 | 0 |
| 0.001545492 | 0 | 0.013252527 | 0.010171027 | 0.004331393 |
| 0 | 0 | 0 | 0 | 0 |
| 0 | 0 | 0 | 0 | 0 |
| 0 | 0.024524819 | 0.052461654 | 0.049303359 | 9.643030219 |

| | | | | |
|---|---|---|---|---|
| 0 | 0 | 0 | 0 | 0 |
| 0 | 0 | 0 | 0 | 0 |
| 0 | 0 | 0 | 0 | 0 |
| 0 | 0 | 0.00572708 | 0 | 0 |
| 0 | 0 | 0 | 0 | 0 |
| 0 | 0 | 0 | 0 | 0 |
| 0 | 0 | 0 | 0 | 0 |
| 0 | 0 | 0.010100739 | 0.022305487 | 0 |
| 0 | 0 | 0 | 0 | 0 |
| 0 | 0 | 0 | 0 | 0 |
| 0 | 0.000679539 | 0.00212615 | 0.00071516 | 0.020100653 |
| 0 | 0 | 0 | 0 | 0 |
| 0 | 0 | 0 | 0 | 0 |
| 0 | 0 | 0 | 0.669462726 | 0 |
| 0 | 0 | 0 | 0 | 0 |
| 0 | 0 | 0 | 0 | 0 |
| 0 | 0 | 0 | 0 | 0 |
| 0 | 0 | 0.001032483 | 0 | 0 |
| 0 | 0 | 0 | 0 | 0 |
| 0 | 0 | 0 | 0 | 0 |
| 0 | 0 | 0 | 0 | 0 |
| 0 | 0 | 0 | 0 | 0 |
| 0 | 0 | 0 | 0 | 0 |
| 0 | 0 | 0 | 0 | 0 |
| 0 | 0 | 0 | 0 | 0 |
| 0 | 0 | 0 | 0 | 0 |
| 0 | 0 | 0 | 0 | 0 |
| 0 | 0 | 0 | 0 | 0 |
| 0 | 0 | 0 | 0 | 0 |
| 0.000737641 | 0 | 0.052205518 | 0.000719183 | 0 |
| 0 | 0 | 0 | 0 | 0 |
| 0 | 0 | 0 | 0 | 0 |
| 0 | 0 | 0 | 0 | 0 |
| 0 | 0 | 0 | 0 | 0 |
| 0 | 0 | 0 | 0 | 0 |
| 0 | 0 | 0 | 0 | 0 |
| 0 | 0 | 0 | 0 | 0 |
| 0.001986987 | 0.004832027 | 0.007799235 | 0.024821226 | 0 |
| 0 | 0 | 0 | 0 | 0 |
| 0.000367473 | 0 | 0 | 0 | 0 |
| 0 | 0 | 0 | 0 | 0 |
| 0 | 0 | 0 | 0 | 0 |
| 0 | 0 | 0 | 0 | 0 |
| 0 | 0 | 0.002767416 | 0.899209874 | 0 |
| 0 | 0 | 0.330395084 | 0.237847532 | 0 |
| 0 | 0 | 0 | 0 | 0 |
| 0 | 0 | 0 | 0 | 0 |
| 0 | 0 | 0 | 0 | 0 |

| | | | | |
|---|---|---|---|---|
| 0 | 0 | 0 | 0 | 0 |
| 0 | 0 | 0 | 0 | 0 |
| 0 | 0 | 0 | 0.007005498 | 0 |
| 0 | 0 | 0 | 0 | 0 |
| 0 | 0 | 0 | 0 | 0 |
| 0 | 0 | 0 | 0.14643527 | 0 |
| 0 | 0 | 0 | 0 | 0 |
| 0 | 0 | 0 | 0 | 0 |
| 0.094200779 | 1.340292034 | 0.147694301 | 0 | 3.635830122 |
| 0 | 0 | 0 | 0 | 0 |
| 0 | 0 | 0 | 0 | 0 |
| 0 | 0 | 0 | 0 | 0 |
| 0 | 0 | 0.002051478 | 0 | 0 |
| 0 | 0 | 0 | 0 | 0 |
| 0 | 0.823716646 | 0.053811768 | 0.42825803 | 0.009007024 |
| 0.004752207 | 0 | 0.070786351 | 0 | 3.422122577 |
| 0 | 0 | 0 | 0 | 0 |
| 0 | 0 | 0 | 0 | 0 |
| 0 | 0 | 0 | 0 | 0 |
| 0 | 0 | 0 | 0 | 0 |
| 0 | 0 | 0.018347735 | 0.014843562 | 0 |
| 0 | 0 | 0 | 0 | 0 |
| 0 | 0 | 0 | 0 | 0 |
| 0 | 0 | 0 | 0 | 0 |
| 0 | 0 | 0 | 9.362846274 | 0.009094422 |
| 0 | 0 | 8.074034273 | 2.482986007 | 0 |
| 0 | 0 | 0.014295298 | 0 | 0 |
| 0 | 0 | 0 | 0 | 0 |
| 0 | 0 | 0.096554315 | 0.059661112 | 0 |
| 0 | 0 | 0 | 0 | 0 |
| 0 | 0 | 0 | 0 | 0 |
| 0 | 0 | 0.003603129 | 0 | 0 |
| 0 | 0 | 0 | 0 | 0 |
| 0 | 0 | 0.002208537 | 0 | 0.002344761 |
| 0 | 0.004925864 | 0.0059936 | 0.119233845 | 0 |
| 0.004009138 | 0.000812464 | 0.006203811 | 0.011604301 | 0 |
| 0.170664408 | 0.320044686 | 0 | 0 | 0.003824821 |
| 0.055738801 | 0 | 0 | 2.464102907 | 0.715868494 |
| 0 | 0 | 0.10338549 | 0 | 0 |
| 0 | 0 | 0 | 0 | 0 |
| 0 | 0 | 0 | 0 | 0 |
| 0 | 0 | 0 | 0 | 0 |
| 0.043143643 | 0.177956668 | 0 | 0.572538654 | 0 |
| 0 | 0 | 0 | 0 | 0 |
| 0 | 0 | 0 | 0 | 0 |
| 0 | 0 | 0 | 0 | 0 |
| 0 | 0 | 0 | 0 | 0 |
| 0 | 0.099500343 | 0.482780602 | 0.366414511 | 0 |

| | | | | |
|---|---|---|---|---|
| 0 | 0 | 0 | 0 | 0 |
| 0 | 0 | 0 | 0 | 0 |
| 0 | 0 | 0 | 0 | 0 |
| 0 | 0 | 0 | 0 | 0 |
| 0 | 0.013655957 | 0 | 0.001026558 | 0 |
| 0 | 0 | 0 | 0.00115332 | 0 |
| 0 | 0 | 0 | 0 | 0 |
| 0 | 0 | 0 | 0 | 0 |
| 0 | 0 | 0 | 0 | 0 |
| 0 | 0 | 0 | 0 | 0 |
| 0 | 0 | 0 | 0 | 0 |
| 0.149591214 | 0 | 0 | 0.396820238 | 0 |
| 0 | 0 | 0.08581416 | 1.190154704 | 0 |
| 0 | 0 | 0 | 0 | 0 |
| 0.127937991 | 0 | 0.011961258 | 0.00395738 | 3.625277346 |
| 0 | 0 | 0 | 0 | 0 |
| 0 | 0 | 0.003410157 | 0 | 0 |
| 0 | 0 | 0 | 0 | 0 |
| 0 | 0 | 0.001203082 | 0 | 0.010192243 |
| 0 | 0 | 0 | 0 | 0 |
| 0 | 0 | 0 | 0 | 0 |
| 0 | 0 | 0 | 0 | 0 |
| 0 | 0.168186588 | 0.082891078 | 0.237611697 | 0.011203255 |
| 0 | 0 | 0 | 0 | 0 |
| 0 | 0 | 0 | 0 | 0 |
| 0 | 0 | 0 | 0 | 0 |
| 0 | 0 | 0 | 0 | 0 |
| 0 | 0 | 0 | 0 | 0 |
| 0 | 0 | 0 | 0 | 0 |
| 0 | 0 | 0 | 0 | 0 |
| 0 | 0 | 0 | 0 | 0 |
| 0.001685456 | 0.007416794 | 0.022645985 | 0.030041235 | 0.079055669 |
| 0 | 0 | 0 | 0.680875817 | 0 |
| 0 | 0 | 0 | 0 | 0 |
| 0.003135868 | 0 | 0.007080633 | 0.008806639 | 0.001061423 |
| 0 | 0 | 0.009764617 | 0.06447548 | 0.080182531 |
| 0 | 0 | 0 | 0.000884986 | 0 |
| 0 | 0 | 0 | 0.039722588 | 0 |
| 0 | 0 | 0.000945884 | 0 | 0 |
| 0 | 0 | 0 | 0 | 0 |
| 0 | 0 | 0 | 0 | 0 |
| 0 | 0 | 0 | 0 | 0 |
| 0 | 0 | 0 | 0 | 0 |
| 0 | 0 | 0 | 0 | 0 |
| 0.019475038 | 0 | 0.019789912 | 0 | 0.001463851 |
| 0 | 0 | 0 | 0 | 0 |
| 0 | 0 | 0.00887132 | 0 | 0 |
| 0 | 0 | 0.000920543 | 0 | 0 |

| | | | | |
|---|---|---|---|---|
| 0 | 0 | 0 | 0 | 0 |
| 0 | 0 | 0 | 0 | 0 |
| 0.002222572 | 0 | 0.006207424 | 0 | 0 |
| 0 | 0 | 0 | 0 | 0 |
| 0 | 0 | 0 | 0 | 0 |
| 0 | 0 | 0 | 0 | 0 |
| 0 | 0 | 0 | 0.024226125 | 1.048746594 |
| 0 | 0 | 0.003010386 | 0 | 0 |
| 0 | 0 | 0 | 0 | 0 |
| 0 | 0 | 0 | 0 | 0 |
| 0 | 0 | 0 | 0 | 0 |
| 0.000565353 | 0 | 0 | 0 | 0 |
| 0 | 0 | 0 | 0 | 0 |
| 0 | 0 | 0 | 0 | 0 |
| 0 | 0 | 0 | 0 | 0 |
| 0 | 0 | 0 | 0 | 0 |
| 0 | 0 | 0 | 0 | 0 |
| 0 | 0 | 0 | 0 | 0 |
| 0.417238748 | 0 | 42.56685027 | 0 | 0 |
| 0 | 0 | 0 | 0 | 0 |
| 0 | 0 | 0 | 0 | 0 |
| 0 | 0 | 0 | 0 | 0 |
| 0 | 0 | 0 | 0 | 0 |
| 0 | 0 | 0 | 0 | 0 |
| 0 | 0 | 0.002304005 | 0.00143074 | 0 |
| 0.001000305 | 0.009730312 | 0.025219272 | 0.001981026 | 0 |
| 0 | 0 | 0 | 0 | 0 |
| 0 | 0 | 0.027493084 | 0.034604232 | 0 |
| 0 | 0 | 0 | 0 | 0 |
| 0 | 0 | 0.78620422 | 0.547503317 | 0 |
| 0 | 0 | 0 | 0 | 0 |
| 0 | 0 | 0 | 0 | 0 |
| 0 | 0 | 0 | 0 | 0 |
| 0 | 0 | 0 | 0 | 0 |
| 0.001965483 | 0 | 0.000633012 | 0.000638767 | 0.015913331 |
| 0 | 0 | 0 | 0 | 0 |
| 0 | 0 | 0 | 0.015089179 | 0 |
| 0 | 0 | 0 | 0 | 0.007639487 |
| 0 | 0 | 0 | 0 | 0 |
| 0 | 0 | 0 | 0 | 0 |
| 0.43612045 | 8.40978899 | 1.943647361 | 3.454975162 | 16.77857882 |
| 0 | 0 | 0.096218891 | 0 | 0.003897552 |
| 0 | 0 | 0.568990111 | 0.106020935 | 0 |
| 0 | 0 | 0 | 0 | 0 |
| 0 | 0 | 0 | 0 | 0 |
| 0 | 0 | 0 | 0 | 0 |
| 0 | 0 | 0 | 0 | 0 |
| 0.000472013 | 0.000765239 | 0.013254104 | 0.003595325 | 0 |

| | | | | |
|---|---|---|---|---|
| 0 | 0 | 0 | 0 | 0 |
| 0 | 0 | 0 | 0 | 0 |
| 0 | 0 | 0 | 0 | 0 |
| 0 | 0 | 0 | 0 | 0 |
| 0.025183533 | 1.776894897 | 0 | 4.655986366 | 0 |
| 0 | 0 | 0 | 0 | 0 |
| 0 | 0 | 0 | 0 | 0 |
| 0 | 0 | 0 | 0 | 0 |
| 0.23184998 | 0.430364116 | 0.085336411 | 0.056571189 | 0.037552051 |
| 0 | 0 | 0 | 0 | 0 |
| 0.008776296 | 0.000889272 | 0.003809192 | 0 | 0 |
| 0 | 0 | 0 | 0 | 0 |
| 0 | 0 | 0.017804738 | 0.041107619 | 0 |
| 0.001884898 | 0.010342852 | 0.003502258 | 0.006361381 | 0.050568604 |
| 0 | 0 | 0 | 0 | 0 |
| 0 | 0 | 0.040091293 | 0.498084834 | 0 |
| 0 | 0.000697413 | 0.001060731 | 0.001682019 | 0 |
| 0 | 0 | 0 | 0 | 0 |
| 0 | 0 | 0 | 0 | 0 |
| 0 | 0 | 0 | 0 | 0 |
| 0 | 0 | 0 | 0 | 0 |
| 0 | 0 | 0 | 0 | 0 |
| 0 | 0 | 0 | 0 | 0 |
| 0 | 0 | 0 | 0 | 0 |
| 0 | 0 | 0 | 0 | 0 |
| 0 | 0 | 0 | 0 | 0 |
| 0 | 0 | 0.009091732 | 0.125649294 | 0 |
| 0 | 0 | 0 | 0.001650363 | 0.003450192 |
| 0 | 0 | 0 | 0 | 0 |
| 0 | 0 | 0 | 0 | 0 |
| 0 | 0 | 0 | 0 | 0 |
| 0 | 0 | 0 | 0 | 0 |
| 0 | 0 | 0 | 0 | 0 |
| 0 | 0 | 0 | 0 | 0 |
| 0 | 0 | 0 | 0.104353833 | 0 |
| 0 | 0.001978947 | 0 | 0.150994613 | 0.011308277 |
| 0 | 0 | 0 | 0 | 0.044193962 |
| 0 | 0 | 0 | 0 | 0 |
| 0 | 0 | 0 | 0 | 0 |
| 0 | 0 | 0 | 0 | 0 |
| 0 | 6.317134022 | 0 | 0.895951789 | 0 |
| 0 | 0 | 0 | 0.109515023 | 0 |
| 0 | 0 | 0 | 0 | 0 |
| 0 | 0 | 0 | 0.027360477 | 0 |
| 0 | 0 | 0 | 0 | 0 |
| 0 | 0 | 0 | 0 | 0 |
| 0 | 0 | 0 | 0 | 0 |
| 0 | 0 | 0 | 0 | 0 |

| | | | | |
|---|---|---|---|---|
| 0 | 0 | 0 | 0.003778258 | 0 |
| 0 | 0 | 0 | 0 | 0 |
| 0 | 0 | 0 | 0 | 0 |
| 0 | 0 | 0 | 0.001699579 | 0 |
| 0 | 0 | 0 | 0 | 0.001038768 |
| 0 | 0 | 0 | 0 | 0 |
| 0 | 0 | 0.015872283 | 0 | 0.256070907 |
| 0 | 0 | 0 | 0 | 0 |
| 0 | 0 | 0.014297508 | 0 | 0 |
| 0.008666888 | 0.006690939 | 0 | 0 | 0 |
| 0 | 0 | 0 | 0 | 0 |
| 0 | 0 | 0 | 0 | 0 |
| 0 | 0 | 0 | 0 | 0 |
| 0 | 0 | 0 | 0 | 0 |
| 0 | 0 | 0 | 0 | 0 |
| 0 | 0 | 0 | 0 | 0 |
| 0 | 0 | 0.017621378 | 0 | 0 |
| 0 | 0 | 0 | 0 | 0 |
| 0 | 0 | 0 | 0 | 0 |
| 0 | 0 | 0 | 0 | 0 |
| 0 | 0 | 0.018421392 | 4.061814937 | 0 |
| 0 | 0 | 0 | 0 | 0 |
| 0 | 0 | 0 | 0.02354695 | 0 |
| 0.002571225 | 0.003139934 | 0.004516906 | 0.007691581 | 0 |
| 0 | 0 | 0 | 0 | 0 |
| 0 | 0 | 0 | 0 | 0.002790927 |
| 0 | 0 | 0 | 0 | 0 |
| 0 | 0 | 0 | 0 | 0 |
| 0 | 0 | 0 | 0 | 0 |
| 0 | 0 | 0 | 0.002049998 | 9.207397389 |
| 0 | 0 | 0 | 0 | 0 |
| 0 | 0 | 0 | 0 | 0 |
| 0 | 0 | 0 | 0 | 0 |
| 0 | 0 | 0 | 0 | 0.976925674 |
| 0 | 0 | 0 | 0 | 0 |
| 0 | 0 | 0 | 0 | 0 |
| 0.000731197 | 0.001490261 | 0 | 0 | 0.058517562 |
| 0 | 0 | 0 | 0 | 0 |
| 0 | 0 | 0 | 0 | 0 |
| 0 | 0 | 0 | 0.037809102 | 0 |
| 0 | 0 | 0 | 0 | 0 |
| 0 | 0 | 0 | 0 | 0 |
| 0 | 0 | 0 | 0 | 0 |
| 0 | 0 | 0 | 0 | 0 |
| 0 | 0 | 0 | 0 | 0 |
| 0 | 0 | 0 | 0 | 0 |
| 0 | 0 | 0 | 0 | 0 |
| 0 | 0 | 0 | 0.093601799 | 0 |

| | | | | |
|---|---|---|---|---|
| 0.003269771 | 0 | 0.002126404 | 0 | 0.072645111 |
| 0 | 0 | 0 | 0.032124049 | 0 |
| 0.000747269 | 0 | 0 | 0 | 0 |
| 0 | 0 | 0 | 0 | 0 |
| 0 | 0 | 0 | 0 | 0 |
| 0 | 0 | 0 | 0 | 0 |
| 0 | 0 | 0 | 0 | 0 |
| 0.068179337 | 0 | 0.347462359 | 0.239675252 | 0.198236791 |
| 0 | 0 | 0 | 7.324800764 | 0.163203656 |
| 0.051946223 | 0 | 0.005975007 | 0.037180873 | 0 |
| 0 | 0 | 0 | 0 | 0 |
| 0 | 0 | 0 | 0 | 0 |
| 0 | 0 | 0 | 0 | 0 |
| 0 | 0 | 0.037240978 | 0 | 0 |
| 0 | 0 | 0 | 0 | 0 |
| 0 | 0 | 0 | 0 | 0 |
| 0.025884269 | 0 | 0 | 0 | 0 |
| 0 | 0 | 0 | 0 | 0 |
| 0 | 0 | 0 | 0 | 0 |
| 0 | 0 | 0 | 0 | 0 |
| 0 | 0 | 0 | 0 | 0 |
| 0 | 0 | 0 | 0 | 0 |
| 0 | 0 | 0 | 0 | 0 |
| 0 | 0 | 0 | 0 | 0 |
| 0 | 0 | 0 | 0.004345882 | 0 |
| 0 | 0 | 0 | 0 | 0 |
| 0 | 0 | 0 | 0 | 0 |
| 0 | 0 | 0 | 0 | 0 |
| 0.002409182 | 0.004201223 | 0 | 0 | 0 |
| 0 | 0 | 0 | 0 | 0 |
| 0 | 0.001555971 | 0 | 0 | 0 |
| 0 | 0 | 0 | 0 | 0 |
| 0 | 0 | 0 | 0 | 0 |
| 0 | 0 | 0 | 0 | 0 |
| 0 | 0 | 0 | 0.010761969 | 0 |
| 0 | 0 | 0 | 0 | 0 |
| 0 | 0 | 0 | 0 | 0 |
| 0 | 0 | 0.032747057 | 0 | 0 |
| 0 | 0 | 0 | 0 | 0 |
| 0 | 0.282167332 | 0 | 0 | 0.017397598 |
| 0 | 0 | 0 | 0 | 0 |
| 0 | 0 | 0.171607038 | 0 | 0 |
| 0 | 0 | 0 | 0 | 0 |
| 0 | 0 | 0 | 0 | 0 |
| 0 | 0 | 0 | 0 | 0 |
| 0 | 0 | 0 | 0 | 0 |
| 0 | 0 | 0 | 0 | 0 |
| 0 | 0 | 0.000424324 | 0 | 0 |

| | | | | |
|---|---|---|---|---|
| 0 | 0 | 0 | 0 | 0 |
| 0.149258133 | 0 | 0 | 0.002351612 | 0.001590535 |
| 0 | 0 | 0 | 0 | 0 |
| 0 | 0 | 0 | 0 | 0 |
| 0 | 0 | 0.427023728 | 0.225510145 | 0.516166753 |
| 0 | 0 | 0 | 0 | 0 |
| 0 | 0 | 0 | 0 | 0 |
| 0.006878984 | 0.096332832 | 0.015456798 | 0 | 0.321566326 |
| 0 | 0 | 0 | 0 | 0 |
| 0 | 0 | 0 | 0.028822822 | 0 |
| 0 | 0 | 0 | 0 | 0 |
| 0 | 0 | 0 | 0 | 0 |
| 0 | 0 | 0 | 0 | 0 |
| 0 | 0 | 0 | 0 | 0 |
| 0 | 0 | 0 | 0 | 0.314394911 |
| 0 | 0 | 0 | 0 | 0 |
| 0 | 0 | 0 | 0 | 0 |
| 0 | 0 | 0 | 0 | 0 |
| 0 | 0 | 0 | 0 | 0 |
| 0 | 0 | 0 | 0 | 0 |
| 0 | 0 | 0 | 0 | 0 |
| 0 | 0 | 0 | 0 | 0 |
| 0 | 0 | 0 | 0 | 0 |
| 0 | 0 | 0 | 0 | 0.005389024 |
| 0 | 0 | 0 | 0.097728269 | 0 |
| 0 | 0 | 0 | 0 | 0 |
| 0.025831484 | 0 | 0.002446881 | 0 | 0.033319186 |
| 0 | 0 | 0.001610304 | 0 | 0 |
| 0 | 0 | 0 | 0 | 0 |
| 0 | 0 | 0 | 0 | 0 |
| 0 | 0 | 0 | 0 | 0 |
| 0 | 0 | 0 | 0 | 0 |
| 0 | 0 | 0 | 0 | 0 |
| 0 | 0 | 0 | 0 | 0 |
| 0 | 0 | 0 | 0 | 0 |
| 0 | 0 | 0 | 0 | 0 |
| 0 | 0 | 0 | 0 | 0 |
| 0 | 0 | 0 | 0 | 0 |
| 0 | 0 | 0 | 0 | 0 |
| 0 | 0 | 0 | 0 | 0 |
| 0 | 0 | 0 | 0 | 0 |
| 0 | 0 | 0 | 0 | 0 |
| 0 | 0 | 0 | 0 | 0 |
| 0 | 0 | 0 | 0 | 0 |
| 0 | 0 | 0 | 0 | 0 |
| 0 | 0 | 0 | 0 | 0 |
| 0 | 0 | 0 | 0 | 0 |
| 0 | 0 | 0 | 0 | 0 |

| | | | | |
|---|---|---|---|---|
| 0.002262517 | 0 | 0.134037963 | 0.02263952 | 0 |
| 0 | 0 | 0 | 0 | 0 |
| 0 | 0 | 0.000427378 | 0 | 0 |
| 0 | 0 | 0 | 0 | 0 |
| 0 | 0 | 0 | 0 | 0 |
| 0 | 0 | 0 | 0.056998057 | 0 |
| 0 | 0 | 0 | 0 | 0 |
| 0 | 0 | 0 | 0 | 0 |
| 0 | 0 | 0 | 0 | 0 |
| 0 | 0 | 0 | 0 | 0 |
| 0 | 0 | 0 | 0 | 0 |
| 0 | 0 | 0.004624154 | 0 | 0 |
| 0 | 0 | 0 | 0 | 0 |
| 0 | 0 | 0 | 0 | 0 |
| 0 | 0 | 0 | 0 | 0 |
| 0 | 0 | 0 | 0 | 0 |
| 0 | 0 | 0 | 0 | 0 |
| 0 | 0 | 0 | 0 | 0 |
| 0 | 0 | 0 | 0 | 0 |
| 0 | 0 | 0 | 0 | 0 |
| 0 | 0.002426728 | 0.01315416 | 0.041333467 | 0.117204268 |
| 0 | 0 | 0 | 0 | 0 |
| 0 | 0.009516949 | 0.141103195 | 0.006153795 | 0 |
| 0 | 0 | 0 | 0 | 0 |
| 0 | 0 | 0 | 0 | 0 |
| 0 | 0 | 0 | 0 | 0 |
| 0 | 0 | 0 | 0 | 0 |
| 0 | 0 | 0 | 0 | 0 |
| 0 | 0 | 0 | 0 | 0 |
| 0 | 0 | 0 | 0 | 0 |
| 0 | 0 | 0 | 0 | 0 |
| 0 | 0 | 0 | 0 | 0 |
| 0.004239551 | 3.029221033 | 0.014573099 | 0 | 0.145475408 |
| 0 | 0 | 0 | 0 | 0 |
| 0 | 0 | 0.603335403 | 1.365536298 | 0 |
| 0.000523394 | 0 | 0 | 0 | 0.086585716 |
| 0 | 0 | 0 | 0 | 0 |
| 0 | 0 | 0 | 0 | 0 |
| 0 | 0.00462693 | 0 | 0 | 0.005891181 |
| 0 | 0 | 0 | 0 | 0 |
| 0 | 0 | 0 | 0 | 0 |
| 0 | 0 | 0 | 0 | 0 |
| 0 | 0 | 0.183383684 | 0.007980054 | 0.021492294 |
| 0 | 0 | 0 | 0 | 0 |
| 0 | 0 | 0 | 0 | 0 |
| 0 | 0 | 0 | 0.005652407 | 0 |
| 0 | 0 | 0 | 0 | 0 |
| 0 | 0 | 0 | 0 | 0 |

| | | | | |
|---|---|---|---|---|
| 0 | 0 | 0 | 0 | 0 |
| 0 | 0 | 0 | 0 | 0 |
| 0 | 0 | 0 | 0 | 0 |
| 0 | 0 | 0 | 0 | 0 |
| 0 | 0 | 0 | 0 | 0 |
| 0 | 0 | 0 | 0 | 0 |
| 0 | 0 | 0 | 0 | 0 |
| 0 | 0 | 0 | 0 | 0 |
| 0 | 0 | 0 | 0 | 0 |
| 0 | 0 | 0 | 0.092621401 | 0 |
| 0 | 0 | 0 | 0 | 0 |
| 0 | 0 | 0 | 0 | 0 |
| 0 | 0 | 0 | 0 | 0 |
| 0.002614641 | 0 | 0 | 0 | 0 |
| 0 | 0 | 0 | 0.024237379 | 0 |
| 0 | 0 | 0.008772792 | 0 | 0.025280593 |
| 0 | 0 | 0 | 0 | 0 |
| 0 | 0 | 0 | 0 | 0 |
| 0 | 0 | 0 | 0 | 0 |
| 0 | 0 | 0.458837637 | 0.019909692 | 3.353094364 |
| 0 | 0.00633922 | 0 | 0.01015232 | 0 |
| 0 | 0 | 0 | 0 | 0 |
| 0 | 0 | 0 | 0 | 0 |
| 0 | 0 | 0 | 0 | 0 |
| 0 | 0 | 0 | 0 | 0 |
| 0 | 0 | 0 | 0 | 0.063846799 |
| 0 | 0 | 0 | 0 | 0 |
| 0 | 0 | 0 | 0 | 0 |
| 0 | 0 | 0 | 0 | 0 |
| 0 | 0 | 0 | 0 | 0 |
| 0 | 0 | 0 | 0.238284993 | 0 |
| 0 | 0 | 0 | 0 | 0 |
| 0 | 0 | 0 | 0 | 0 |
| 0 | 0 | 0 | 0 | 0 |
| 0 | 0 | 0 | 0 | 0 |
| 0 | 0 | 0 | 0 | 0 |
| 0 | 0 | 0 | 0 | 0 |
| 0 | 0 | 0 | 0 | 0 |
| 0 | 0 | 0 | 0.004770098 | 0 |
| 0 | 0 | 0 | 0 | 0 |
| 0 | 0 | 0 | 0 | 0 |
| 0 | 0 | 0 | 0 | 0 |
| 0 | 0 | 0 | 0 | 0 |
| 0 | 0 | 0 | 0 | 0 |
| 0 | 0 | 0 | 0 | 0 |
| 0 | 0 | 0 | 0 | 0 |
| 0.088501823 | 0.952670129 | 0.20215251 | 0 | 0 |
| 0 | 0 | 0 | 0 | 0 |

| | | | | |
|---|---|---|---|---|
| 0 | 0 | 0 | 0 | 0 |
| 0 | 0 | 0 | 0 | 0 |
| 0 | 0 | 0 | 0 | 0 |
| 0 | 0 | 0 | 0 | 0 |
| 0 | 0.001917283 | 0.170229783 | 0.019912383 | 0 |
| 0 | 0 | 0 | 0 | 0 |
| 0 | 0 | 0 | 0.015617248 | 0 |
| 0 | 0 | 0 | 0 | 0 |
| 0 | 0 | 0 | 0 | 0 |
| 0 | 0 | 0 | 0 | 0 |
| 0 | 0 | 0 | 0 | 0 |
| 0.005141701 | 0 | 0.011689114 | 0 | 0 |
| 0.001435174 | 0 | 0 | 0 | 0 |
| 0 | 0 | 0 | 0 | 0 |
| 0 | 0 | 0 | 0 | 0 |
| 0 | 0 | 0 | 1.727313022 | 0 |
| 0.000348933 | 0 | 0 | 0 | 0 |
| 0 | 0 | 0 | 0 | 0 |
| 0.03240621 | 0.032037151 | 0.077723409 | 0.257200361 | 0.020325215 |
| 0 | 0 | 0.001805749 | 0.0043732 | 0 |
| 0 | 0 | 0 | 0 | 0 |
| 0 | 0 | 0 | 0 | 0 |
| 0 | 0 | 0 | 0 | 0 |
| 0 | 0 | 0 | 0 | 0 |
| 0 | 0 | 0 | 0 | 0 |
| 0 | 0 | 0 | 0 | 0 |
| 0.005631207 | 0 | 0.011865641 | 0 | 0 |
| 0 | 0 | 0 | 0 | 0 |
| 0 | 0 | 0 | 0 | 0 |
| 0 | 0 | 0 | 0 | 0 |
| 0.0009507 | 0 | 0 | 0.002648315 | 0.008881433 |
| 0 | 0 | 0 | 0 | 0 |
| 0 | 0 | 0 | 0 | 0 |
| 0 | 0 | 0 | 0 | 0 |
| 0 | 0 | 0 | 0 | 0 |
| 0 | 0 | 0 | 0 | 0 |
| 0 | 0 | 0 | 0 | 0 |
| 0.000477096 | 0.00521546 | 0.019936793 | 0 | 0.070569587 |
| 0 | 0 | 0 | 0 | 0 |
| 0 | 0 | 0 | 0 | 0 |
| 0 | 0 | 0 | 0.004179393 | 0 |
| 0 | 0 | 0 | 0 | 0 |
| 0 | 0 | 0 | 0 | 0 |
| 0 | 0.00157299 | 0.001367109 | 0.002414193 | 0.004626444 |
| 0 | 0 | 0 | 0 | 0 |
| 0 | 0 | 0 | 0 | 0 |
| 0 | 0 | 0 | 0 | 0 |
| 0 | 0 | 0 | 0 | 0 |

| | | | | |
|---|---|---|---|---|
| 0 | 0 | 0 | 0 | 0 |
| 0 | 0 | 0 | 0 | 0 |
| 0 | 0 | 0 | 0 | 0 |
| 0 | 0.001099 | 0.003044565 | 0.001987924 | 0 |
| 0 | 0.014652511 | 0 | 0.001005691 | 0 |
| 0 | 0 | 0 | 0 | 0 |
| 0 | 0 | 0 | 0 | 0 |
| 0.001507651 | 0 | 0 | 0 | 0 |
| 0 | 0 | 0 | 0 | 0 |
| 0 | 0 | 0 | 0 | 0 |
| 0 | 0 | 0 | 0 | 0 |
| 0 | 0 | 0 | 0 | 0 |
| 0 | 0 | 0.006982309 | 0.033563587 | 0.000654653 |
| 0 | 0 | 0 | 0 | 0 |
| 0 | 0 | 0 | 0 | 0 |
| 0 | 0 | 0 | 0 | 0 |
| 0 | 0 | 0 | 0 | 0 |
| 0 | 0 | 0 | 0 | 0 |
| 0 | 0 | 0 | 0 | 0 |
| 0 | 0 | 0 | 0 | 0 |
| 0 | 0 | 0 | 0 | 0 |
| 0 | 0 | 0 | 0 | 0 |
| 0.008166283 | 0 | 0 | 0 | 0 |
| 0 | 0 | 0 | 0.059318177 | 0.007120551 |
| 0 | 0 | 0 | 0 | 0 |
| 0 | 0 | 0 | 0.002058049 | 0 |
| 0 | 0 | 0 | 0 | 0 |
| 0 | 0 | 0 | 0 | 0 |
| 0 | 0 | 0 | 0 | 0 |
| 0 | 0 | 0 | 0 | 0 |
| 0 | 0 | 0.019757189 | 0.037804723 | 0 |
| 0 | 0 | 0.052032886 | 0.000581784 | 0 |
| 0 | 0 | 0 | 0 | 0 |
| 0 | 0 | 0 | 0 | 0 |
| 0 | 0 | 0 | 0 | 0 |
| 0 | 0.021522089 | 0 | 0 | 0 |
| 0 | 0 | 0 | 0 | 0 |
| 0 | 0 | 0 | 0 | 0 |
| 0 | 0 | 0 | 0.003500891 | 0.041473452 |
| 0 | 0 | 0 | 0 | 0 |
| 0 | 0 | 0 | 0 | 0 |
| 0 | 0 | 0 | 0 | 0 |
| 0 | 0 | 0 | 0 | 0 |
| 0 | 0 | 0 | 0 | 0 |
| 0 | 0 | 0 | 0 | 0 |
| 0 | 0 | 0.178429787 | 0.622061628 | 0.001252101 |
| 0 | 0.000760019 | 0 | 0 | 0 |
| 0 | 0 | 0 | 0 | 0 |

| | | | | |
|---|---|---|---|---|
| 0 | 0 | 0 | 0 | 0 |
| 0 | 0 | 0 | 0 | 0 |
| 0 | 0 | 0 | 0.180230458 | 0 |
| 0 | 0 | 0 | 0 | 0 |
| 0 | 0 | 0 | 0 | 0 |
| 0 | 0 | 0 | 0 | 0 |
| 0 | 0 | 0 | 0.016498948 | 0 |
| 0 | 0 | 0 | 0 | 0 |
| 0 | 0.001343088 | 0 | 0 | 0 |
| 0 | 0 | 0 | 0 | 0.034570902 |
| 0 | 0 | 0.00051174 | 0.001290982 | 0 |
| 0 | 0 | 0 | 0.00096372 | 0.001477462 |
| 0 | 0 | 0 | 0 | 0 |
| 0 | 0 | 0 | 0 | 0 |
| 0 | 0 | 0 | 0 | 0 |
| 0 | 0 | 0 | 0 | 0 |
| 0 | 0 | 0 | 0 | 0.00654595 |
| 0 | 0 | 0 | 0 | 0 |
| 0 | 0 | 0 | 0 | 0 |
| 0 | 0 | 0 | 0.000856032 | 0 |
| 0 | 0 | 0 | 0 | 0 |
| 0 | 0 | 0 | 0 | 0 |
| 0 | 0 | 0 | 0 | 0 |
| 0 | 0 | 0 | 0 | 0 |
| 0 | 0 | 0 | 0 | 0 |
| 0 | 0 | 0 | 0 | 0 |
| 0 | 0 | 0 | 0 | 0 |
| 0 | 0 | 0 | 0.048882961 | 0 |
| 0 | 0 | 0 | 0 | 0 |
| 0 | 0 | 0 | 0 | 0 |
| 0 | 0 | 0.005684594 | 0.020003428 | 0 |
| 0 | 0 | 0 | 0 | 0 |
| 3.096734822 | 0.020511321 | 0.045187299 | 0.015028599 | 0 |
| 0.004582328 | 0 | 0.041852875 | 0.001396145 | 0 |
| 0 | 0 | 0 | 0 | 0 |
| 0 | 0 | 0 | 0 | 0 |
| 0 | 0 | 0 | 0 | 0 |
| 0 | 0 | 0 | 0 | 0 |
| 0 | 0 | 0 | 0 | 0 |
| 0 | 0 | 0 | 0 | 0 |
| 0 | 0 | 0 | 0 | 0 |
| 0.009683835 | 0.009505244 | 0.037314414 | 0 | 0.002333458 |
| 0.00674151 | 0 | 0 | 0.016945545 | 0 |
| 0 | 0 | 0 | 0 | 0 |
| 0 | 0 | 0 | 0 | 0 |
| 0 | 0 | 0.013481589 | 0.009646587 | 0.03002042 |
| 0 | 0 | 0.006910422 | 0 | 0 |
| 0 | 0 | 0 | 0 | 0 |

| | | | | |
|---|---|---|---|---|
| 0 | 0 | 0 | 0 | 0 |
| 0 | 0 | 0 | 0.219846642 | 0 |
| 0 | 0.002069929 | 0.007887954 | 0.013196295 | 0.004281693 |
| 0 | 0 | 0 | 0 | 0 |
| 0 | 0 | 0.351702035 | 0 | 0 |
| 0.025513957 | 0 | 0.091152088 | 0 | 0 |
| 0 | 0 | 0 | 0 | 0 |
| 0 | 0 | 0 | 0 | 0 |
| 0 | 0 | 0 | 0 | 0 |
| 0 | 0 | 0 | 0 | 0 |
| 0 | 0 | 0 | 0 | 0 |
| 0 | 0 | 0 | 0 | 0 |
| 0 | 0 | 0 | 0 | 0 |
| 0 | 0 | 0.035243844 | 0 | 0 |
| 0 | 0 | 0 | 0 | 0 |
| 0 | 0 | 0 | 0 | 0 |
| 0 | 0 | 0 | 0 | 0 |
| 0 | 0 | 0 | 0 | 0 |
| 0 | 0 | 0 | 0 | 0 |
| 0 | 0 | 0 | 0 | 0 |
| 0 | 0 | 0 | 0 | 0 |
| 0 | 0 | 0 | 0 | 0 |
| 0 | 0 | 0 | 0 | 0 |
| 0 | 0 | 0 | 0 | 0 |
| 0 | 0 | 0.001026439 | 0 | 0.038815886 |
| 0 | 0 | 0 | 0 | 0.149535567 |
| 0 | 0 | 0 | 0 | 0 |
| 0 | 0 | 0.004340408 | 0.001684566 | 0 |
| 0 | 0 | 0 | 0 | 0 |
| 0 | 0 | 0 | 0 | 0 |
| 0 | 0 | 0 | 0 | 0 |
| 0 | 0 | 0 | 0 | 0 |
| 0 | 0 | 0 | 0 | 0 |
| 0 | 0 | 0 | 0 | 0 |
| 0 | 0 | 0 | 0 | 0 |
| 0 | 0 | 0 | 0 | 0 |
| 0 | 0 | 0 | 0 | 0 |
| 0 | 0 | 0 | 0 | 0 |
| 0 | 0 | 3.264360527 | 0 | 0 |
| 0 | 0 | 0 | 0 | 0 |
| 0 | 0 | 0 | 0 | 0 |
| 0 | 0 | 0 | 0 | 0 |
| 0.001716098 | 0 | 0.001209017 | 0 | 0.067467069 |
| 0 | 0 | 0 | 0 | 0 |
| 0 | 0 | 0 | 0 | 0 |
| 0 | 0 | 0 | 0 | 0 |
| 0.002625293 | 0 | 0.025365391 | 0.01413113 | 0 |
| 0 | 0 | 0 | 0 | 0 |

| | | | | |
|---|---|---|---|---|
| 0 | 0 | 0 | 0 | 0 |
| 0.002208425 | 0.037082226 | 0.079516043 | 0.045418298 | 0.00236398 |
| 0 | 0 | 0 | 0 | 0 |
| 0 | 0 | 0 | 0 | 0 |
| 0 | 0 | 0 | 0 | 0 |
| 0 | 0 | 0 | 0 | 0 |
| 0 | 0 | 0 | 0 | 0 |
| 0 | 0 | 0 | 0 | 0 |
| 0 | 0 | 0 | 0 | 0 |
| 0 | 0 | 0 | 0 | 0 |
| 0 | 0 | 0 | 0 | 0 |
| 0 | 0 | 0 | 0 | 0 |
| 0 | 0 | 0 | 0 | 0 |
| 0 | 0 | 0 | 0 | 0 |
| 0.00103259 | 0 | 0.007066905 | 0.002516879 | 0 |
| 0 | 0 | 0 | 0 | 0 |
| 0 | 0 | 0 | 0 | 0 |
| 0 | 0 | 0 | 0 | 0 |
| 0 | 0 | 0 | 0 | 0 |
| 0 | 0 | 0 | 0 | 0 |
| 0 | 0 | 0 | 0 | 0 |
| 0 | 0 | 0 | 0 | 0.004210479 |
| 0 | 0 | 0.003607632 | 0 | 0 |
| 0 | 0 | 0 | 0 | 0 |
| 0 | 0 | 0 | 0 | 0 |
| 0 | 0 | 0 | 0 | 0 |
| 0 | 0 | 0 | 0 | 0 |
| 0 | 0 | 0 | 0 | 0 |
| 0 | 0 | 0 | 0 | 0 |
| 0 | 0 | 0 | 0 | 0 |
| 0 | 0 | 0 | 0 | 0 |
| 0 | 0 | 0 | 0 | 0 |
| 0 | 0 | 0 | 0 | 0 |
| 0 | 0 | 0 | 0 | 0 |
| 0 | 0 | 0.005374697 | 9.505635372 | 0 |
| 0 | 0 | 0 | 0 | 0 |
| 0 | 0 | 0 | 0 | 0 |
| 0 | 0 | 0 | 0 | 0 |
| 0 | 0 | 0.002242281 | 0.009667763 | 0.013139602 |
| 0 | 0 | 0 | 0 | 0 |
| 0.010290749 | 0.063509896 | 0.012428557 | 0.073036118 | 0.337419029 |
| 0 | 0 | 0 | 0 | 0 |
| 0 | 0 | 0 | 0 | 0 |
| 0 | 0 | 0 | 0 | 0 |
| 0 | 0 | 0 | 0 | 0 |
| 0.261461034 | 0.481874075 | 0.408271184 | 0.201240353 | 16.15186727 |
| 0 | 0 | 0 | 0 | 0 |
| 0 | 0 | 0 | 0 | 0 |

| | | | | |
|---|---|---|---|---|
| 0 | 0 | 0 | 0 | 0 |
| 0 | 0 | 0 | 0 | 0 |
| 0 | 0 | 0 | 0 | 0 |
| 0 | 0 | 0 | 0 | 0 |
| 0 | 0 | 0 | 0.001441375 | 0 |
| 0 | 0 | 0 | 0 | 0 |
| 0 | 0 | 0 | 0 | 0 |
| 0 | 0 | 0 | 0 | 0 |
| 0 | 0 | 0 | 0 | 0 |
| 0.014234315 | 0 | 0 | 0 | 1.524749416 |
| 0 | 0 | 0 | 0 | 0 |
| 0 | 0 | 0 | 0 | 0 |
| 0 | 0 | 0.002800518 | 0 | 0 |
| 0 | 0 | 0 | 0 | 0 |
| 0 | 0 | 0 | 0 | 0 |
| 0 | 0 | 0 | 0 | 0 |
| 0 | 0 | 0 | 0 | 0 |
| 0 | 0 | 0 | 0 | 0 |
| 0 | 0 | 0 | 0 | 0 |
| 0.148158697 | 0.057633989 | 1.568060476 | 0.116064136 | 4.689866265 |
| 0 | 0 | 0 | 0 | 0 |
| 0 | 0 | 0 | 0 | 0 |
| 0 | 0 | 0 | 0 | 0 |
| 0 | 0 | 0 | 0 | 0 |
| 0 | 0 | 0 | 0 | 0 |
| 0 | 0 | 0 | 0 | 0 |
| 0 | 0 | 0 | 0 | 0 |
| 0 | 0 | 0 | 0 | 0 |
| 0 | 0 | 0 | 0 | 0 |
| 0.006303842 | 0 | 0 | 0 | 0 |
| 0 | 0 | 0 | 0 | 0 |
| 0 | 0 | 0 | 0 | 0 |
| 0 | 0 | 0 | 0 | 0 |
| 0 | 0 | 0 | 0 | 0 |
| 0 | 0 | 0 | 0 | 0 |
| 0 | 0 | 0 | 0.010465799 | 0 |
| 0 | 0 | 0 | 0 | 0 |
| 0 | 0 | 0 | 0 | 0 |
| 0 | 0 | 0 | 0 | 0 |
| 0 | 0 | 0 | 0 | 0 |
| 0 | 0 | 0 | 0 | 0 |
| 0 | 0 | 0 | 0 | 0 |
| 0 | 0 | 0 | 0 | 0 |
| 0 | 0 | 0 | 0 | 0 |
| 0 | 0 | 0.003511795 | 0.002531231 | 0.003449409 |
| 0 | 0 | 0 | 0 | 0 |
| 0 | 0 | 0 | 0 | 0 |
| 0 | 0.007704429 | 0 | 0 | 0 |

| | | | | |
|---|---|---|---|---|
| 0 | 0 | 0 | 0 | 0 |
| 0 | 0 | 0 | 0 | 0 |
| 0 | 0 | 0 | 0 | 0 |
| 0 | 0 | 0 | 0 | 0 |
| 0 | 0 | 0 | 0 | 0 |
| 0.001137054 | 0.003734723 | 0.002247164 | 0.014235459 | 0.000643779 |
| 0 | 0 | 0 | 0 | 0 |
| 0 | 0 | 0 | 0 | 0 |
| 0 | 0 | 0 | 0 | 0 |
| 0 | 0 | 0 | 0 | 0 |
| 0 | 0 | 0 | 0 | 0 |
| 0 | 0 | 0 | 0 | 0 |
| 0 | 0 | 0 | 0 | 0 |
| 0 | 0 | 0 | 0 | 0 |
| 0 | 0 | 0 | 0 | 0 |
| 0 | 0 | 0 | 0 | 0 |
| 0 | 0 | 0 | 0 | 0 |
| 0 | 0 | 0 | 0 | 0 |
| 0 | 0 | 0 | 0 | 0 |
| 0 | 0 | 0 | 0 | 0 |
| 0 | 0 | 0 | 0 | 0 |
| 0 | 0 | 0 | 0 | 0 |
| 0 | 0 | 0 | 0 | 0 |
| 0 | 0 | 0 | 0 | 0 |
| 0 | 0 | 0 | 0 | 0 |
| 0 | 0 | 0 | 0 | 0.14777417 |
| 0 | 0 | 0 | 0 | 0 |
| 0 | 0 | 0.011399662 | 0 | 0 |
| 0 | 0 | 0 | 0 | 0 |
| 0 | 0 | 0 | 0 | 0 |
| 0 | 0 | 0 | 0 | 0 |
| 0 | 0 | 0 | 0 | 0 |
| 0 | 0 | 0 | 0 | 0 |
| 0 | 0 | 0 | 0 | 0 |
| 0 | 0 | 0 | 0 | 0 |
| 0 | 0 | 0 | 0 | 0 |
| 0 | 0 | 0 | 0 | 0 |
| 0 | 0 | 0 | 0 | 0 |
| 0 | 0 | 0 | 1.096668689 | 0 |
| 0 | 0 | 0 | 0 | 0 |
| 0 | 0 | 0 | 0 | 0 |
| 0 | 0 | 0 | 0 | 0 |
| 0 | 0 | 0 | 0 | 0 |
| 0 | 0 | 0 | 0 | 0 |
| 0 | 0 | 0 | 0 | 0 |
| 0 | 0 | 0 | 0 | 0 |
| 0 | 0 | 0 | 0 | 0 |
| 0 | 0 | 0 | 0 | 0 |

| | | | | |
|---|---|---|---|---|
| 0 | 0 | 0 | 0 | 0 |
| 0 | 0 | 0 | 0 | 0 |
| 0 | 0 | 0 | 0 | 0 |
| 0 | 0 | 0 | 0 | 0 |
| 0 | 0 | 0 | 0 | 0 |
| 0 | 0 | 0 | 0 | 0 |
| 0 | 0 | 0 | 0 | 0 |
| 0 | 0 | 0 | 0 | 0 |
| 0 | 0 | 0 | 0 | 0 |
| 0 | 0 | 0 | 0 | 0 |
| 0 | 0 | 0.01061512 | 0.063161686 | 0.001651618 |
| 0 | 0 | 0 | 0 | 0 |
| 0 | 0 | 0 | 0.184862847 | 0 |
| 0 | 0 | 0 | 0 | 0 |
| 0 | 0 | 0 | 0 | 0 |
| 0 | 0 | 0 | 0 | 0 |
| 0 | 0 | 0 | 0 | 0 |
| 0 | 0 | 0 | 0 | 0 |
| 0 | 0 | 0 | 0 | 0 |
| 0 | 0 | 0 | 0 | 0 |
| 0.001384223 | 0 | 0 | 0.007497697 | 0 |
| 0 | 0 | 0 | 0 | 0 |
| 0 | 0 | 0 | 0 | 0 |
| 0.969080064 | 0 | 3.014319819 | 2.059131347 | 0.014878293 |
| 0 | 0 | 0 | 0 | 0 |
| 0 | 0 | 0 | 0 | 0 |
| 0 | 0 | 0 | 0 | 0 |
| 0 | 0 | 0 | 0 | 0 |
| 0 | 0 | 0 | 0 | 0 |
| 0 | 0 | 0 | 0 | 0 |
| 0.001660528 | 0 | 0 | 0 | 0 |
| 0 | 0 | 0 | 0 | 0 |
| 0 | 0 | 0 | 0 | 0 |
| 0 | 0 | 0 | 0 | 0 |

| PHL018-0147_V3 | PHL018-0197_V2 | PHL018-0212_V1 | PHL018-0175_V2 | PHL018-0188_V1 |
|---|---|---|---|---|
| CTRL | EYCF | REF | CTRL | CTRL |
| 0.062130345 | 0.146284865 | 0.007519634 | 0.084743939 | 0.018149111 |
| 0 | 3.42655318 | 1.214486231 | 0 | 0.903646301 |
| 0.007301873 | 0 | 0 | 0.438434285 | 0 |
| 0 | 0 | 0 | 0 | 0 |
| 0 | 0 | 0 | 0 | 0 |
| 0 | 0 | 0 | 0 | 0 |
| 0 | 0 | 0 | 0 | 0 |
| 0 | 0 | 0 | 0 | 0 |
| 0 | 0 | 0 | 0 | 0 |
| 0 | 0 | 0 | 0 | 0 |
| 0 | 0 | 0 | 0 | 0 |
| 0 | 0 | 0 | 0 | 0 |
| 0 | 0 | 0 | 0 | 0 |
| 0.433538819 | 39.46932606 | 1.315653457 | 0 | 2.987151904 |
| 0 | 0 | 0 | 0 | 0 |
| 0 | 0 | 0 | 0 | 0 |
| 0 | 0 | 0 | 0 | 0 |
| 0 | 0 | 0 | 0 | 0 |
| 0 | 0 | 0 | 0 | 0 |
| 0 | 0 | 0 | 0 | 0 |
| 0 | 0 | 0 | 0 | 0 |
| 0 | 0 | 0 | 0 | 0 |
| 0 | 0 | 0 | 0 | 0 |
| 0 | 0 | 0 | 0 | 0 |
| 0 | 0 | 0.003783096 | 0 | 0 |
| 0 | 0 | 0 | 0 | 0 |
| 0 | 0 | 0 | 0 | 0 |
| 0 | 0 | 0 | 0 | 0 |
| 0 | 0 | 0 | 0 | 0 |
| 0 | 0 | 0 | 0 | 0 |
| 0 | 0 | 0 | 0 | 0 |
| 0 | 0 | 0 | 0 | 0 |
| 0 | 0 | 0 | 0 | 0 |
| 0 | 0 | 0 | 0 | 0 |
| 0 | 0 | 0 | 0 | 0 |
| 0 | 0 | 0.589337202 | 0 | 0 |
| 0 | 0 | 0 | 0 | 0 |
| 0 | 0 | 0 | 0 | 0 |
| 0 | 0 | 0 | 0 | 0 |
| 0 | 0 | 0 | 0 | 0 |
| 0 | 0 | 0 | 0 | 0 |
| 0.736134235 | 0.018691561 | 0.001766976 | 0.298724895 | 0.117269797 |
| 0.033975505 | 0.962221328 | 0.557883692 | 0.00090523 | 0.004251872 |
| 0.000716306 | 0.01886124 | 0 | 0 | 0.129959706 |
| 0.005902069 | 0 | 0 | 0 | 0 |
| 0 | 0 | 0 | 0 | 0 |

| | | | | |
|---|---|---|---|---|
| 0 | 0 | 0.018612629 | 0 | 0.013874078 |
| 0.010787859 | 0.038971387 | 0 | 0.019113924 | 0.092046349 |
| 0 | 0.16630616 | 0 | 0 | 0.000937666 |
| 0.002160126 | 1.23953274 | 1.635340878 | 0 | 0.424915036 |
| 0 | 0 | 0.000388484 | 0 | 0 |
| 0 | 0 | 0.003159345 | 0 | 0 |
| 3.367326058 | 0 | 0.028197965 | 0 | 0.001215509 |
| 0 | 0 | 0.000697751 | 0 | 0 |
| 0 | 0 | 0 | 0 | 0 |
| 0 | 0 | 0 | 0 | 0 |
| 0.036924935 | 0 | 0 | 0 | 0.002419772 |
| 0.058216017 | 0 | 0.365772962 | 0 | 0.039459528 |
| 0.000610792 | 0 | 0 | 0 | 0 |
| 0.041505716 | 0.228185772 | 0 | 0.130698198 | 0.003336953 |
| 0.486605545 | 0 | 0 | 0 | 0 |
| 0 | 0 | 0 | 0 | 0 |
| 0.068529127 | 0.00592373 | 0.021314627 | 0.004234526 | 8.322606641 |
| 0 | 0 | 0 | 0 | 0 |
| 0 | 0 | 0 | 0 | 0 |
| 0 | 0 | 0 | 0 | 0 |
| 0.252588042 | 0.287309352 | 0 | 8.574920242 | 0.461003406 |
| 0.295467569 | 0.004287068 | 0.007092242 | 0.016472076 | 0 |
| 0 | 0 | 0.131253806 | 0 | 0 |
| 0 | 0 | 0 | 0 | 0 |
| 0.007856472 | 0 | 0 | 0 | 0 |
| 0 | 0 | 0 | 0 | 0 |
| 0 | 0 | 0 | 0 | 0 |
| 0 | 0 | 0 | 0 | 0 |
| 0.707142217 | 0 | 0 | 0 | 0 |
| 0 | 0 | 0 | 0 | 0 |
| 0 | 0.156007566 | 0 | 1.365560057 | 0.002609657 |
| 0 | 0 | 0 | 0 | 0 |
| 0 | 0 | 0 | 0 | 0 |
| 0 | 0 | 0 | 0 | 0 |
| 0 | 0 | 0 | 0 | 0 |
| 0 | 0 | 0 | 0 | 0 |
| 0 | 0 | 0 | 0 | 0 |
| 0.003037347 | 0 | 1.036095682 | 0 | 0.189382729 |
| 0 | 0 | 0 | 0 | 0 |
| 0 | 0 | 0 | 0 | 0 |
| 0 | 0 | 0 | 0 | 0 |
| 0 | 0 | 0 | 0 | 0 |
| 0 | 0 | 0 | 0 | 0 |
| 0 | 0 | 0 | 0 | 0 |
| 0 | 0 | 0.003247053 | 0 | 0 |
| 0 | 0 | 0 | 0 | 0.006248415 |
| 0.000867104 | 0 | 0 | 0 | 0 |
| 0.007490784 | 0 | 0 | 0 | 0 |

| | | | | |
|---|---|---|---|---|
| 0 | 0 | 0 | 0 | 0 |
| 0 | 0 | 0 | 0 | 0 |
| 0 | 0 | 0 | 0 | 0 |
| 0 | 0 | 0 | 0 | 0 |
| 0 | 0.070381745 | 0 | 0 | 0 |
| 0 | 0 | 0 | 0 | 0 |
| 0 | 0 | 0 | 0 | 0 |
| 0 | 0 | 0 | 0 | 0 |
| 0 | 0 | 0 | 0.016476955 | 0 |
| 0 | 0 | 0 | 0 | 0 |
| 0 | 0 | 0 | 0 | 0 |
| 0.013154756 | 0 | 0 | 0.009457241 | 0.00242295 |
| 0 | 0 | 0 | 0 | 0 |
| 0 | 0 | 0 | 0 | 0 |
| 0 | 0 | 0 | 0 | 0 |
| 0 | 0 | 0 | 0 | 0 |
| 0 | 0 | 0 | 0 | 0 |
| 0 | 0 | 0 | 0 | 0 |
| 0 | 0 | 0.095442406 | 0 | 0 |
| 0 | 0 | 0 | 0 | 0 |
| 0 | 0 | 0.017316087 | 0 | 0 |
| 0 | 0 | 0 | 0 | 0 |
| 0 | 0 | 0 | 0 | 0 |
| 0 | 0 | 0 | 0 | 0 |
| 0 | 0 | 0 | 0 | 0 |
| 0 | 0 | 0 | 0 | 0 |
| 0.03987834 | 0 | 0 | 0 | 0 |
| 0 | 0 | 0 | 0 | 0 |
| 0 | 0 | 0 | 0 | 0 |
| 0 | 0 | 0 | 0 | 0 |
| 0 | 0 | 0.074537404 | 0 | 0 |
| 0.003008496 | 0 | 0 | 0 | 0 |
| 0 | 0 | 0 | 0 | 0 |
| 0 | 0 | 0 | 0 | 0 |
| 0 | 0 | 0 | 0 | 0 |
| 0 | 0 | 0 | 0 | 0 |
| 0 | 0 | 0 | 0 | 0 |
| 0 | 0 | 0 | 0 | 0 |
| 0 | 0 | 0.277202731 | 0 | 0 |
| 0 | 0 | 0 | 0 | 0 |
| 0 | 0 | 0 | 0 | 0 |
| 0 | 0 | 0 | 0 | 0 |
| 0.008041362 | 0 | 0 | 0 | 0 |
| 0 | 0 | 0 | 0 | 0 |
| 0 | 0 | 0 | 0 | 0 |
| 0 | 0 | 0 | 0 | 0 |
| 0 | 0 | 0 | 0 | 0 |
| 0 | 0 | 0 | 0 | 0 |

| | | | | |
|---|---|---|---|---|
| 0 | 0 | 0 | 0 | 0 |
| 0 | 0 | 0 | 0 | 0 |
| 0 | 0 | 0 | 0 | 0 |
| 0 | 0 | 0 | 0 | 0 |
| 0 | 0 | 0.007819771 | 0 | 0 |
| 0 | 0 | 0 | 0 | 0 |
| 0 | 0 | 0 | 0 | 0 |
| 0 | 0 | 0 | 0 | 0 |
| 0 | 0 | 0 | 0 | 0 |
| 0 | 0 | 0 | 0 | 0 |
| 0 | 0 | 0 | 0 | 0 |
| 0 | 0 | 0 | 0 | 0 |
| 0 | 0 | 0 | 0 | 0 |
| 0 | 0 | 0 | 0 | 0 |
| 0 | 0 | 0 | 0 | 0 |
| 0 | 0 | 2.106904289 | 0 | 0 |
| 0 | 0 | 0 | 0 | 0 |
| 0 | 0 | 0 | 0 | 0 |
| 0 | 0 | 0 | 0 | 0 |
| 0 | 0 | 0 | 0 | 0 |
| 0 | 0 | 0 | 0 | 0 |
| 0 | 0 | 0.000985751 | 0 | 0 |
| 0 | 0 | 0 | 0 | 0 |
| 0 | 0 | 0 | 0 | 0 |
| 0.016756457 | 0 | 0.093056232 | 0 | 0 |
| 0 | 0 | 0 | 0 | 0 |
| 0 | 0 | 0.000542183 | 0 | 0 |
| 0 | 0 | 0 | 0 | 0 |
| 0 | 0 | 0 | 0 | 0 |
| 0 | 0 | 0 | 0 | 0 |
| 0 | 0 | 0.010249324 | 0 | 0 |
| 0 | 0 | 0 | 0 | 14.79159791 |
| 0 | 0 | 0 | 0 | 0 |
| 0 | 0 | 0 | 0 | 0 |
| 0 | 0 | 0 | 0 | 0 |
| 0 | 0 | 0 | 0 | 0 |
| 0 | 0 | 0 | 0 | 0 |
| 0 | 0 | 0 | 0 | 0 |
| 0 | 0 | 0 | 0 | 0 |
| 0 | 0 | 0 | 0 | 0 |
| 0 | 0 | 0 | 0 | 0 |
| 0 | 0 | 0 | 0 | 0 |
| 0 | 0 | 0 | 0 | 0 |
| 0 | 0 | 0 | 0 | 0 |
| 0 | 0 | 0 | 0 | 0 |
| 0 | 0 | 0 | 0 | 0 |
| 0 | 0 | 0 | 0 | 0 |
| 0 | 0 | 0 | 0 | 0 |

| | | | | |
|---|---|---|---|---|
| 0 | 0 | 0.87540562 | 0 | 0 |
| 0 | 0 | 0 | 0 | 0 |
| 0 | 0 | 0 | 0 | 0 |
| 0 | 0 | 0 | 0 | 0 |
| 0 | 0 | 0 | 0 | 0 |
| 0 | 0 | 0 | 0 | 0 |
| 0 | 0 | 0 | 0 | 0 |
| 0 | 0 | 0 | 0 | 0 |
| 0 | 0 | 0 | 0 | 0 |
| 0 | 0 | 0 | 0 | 0 |
| 0 | 0 | 0 | 0 | 0 |
| 0 | 0 | 0 | 0 | 0 |
| 0.001764576 | 0 | 0 | 0 | 0.000596496 |
| 0 | 0 | 0 | 0 | 0 |
| 0 | 0 | 0 | 0 | 0 |
| 0 | 0 | 0 | 0 | 0 |
| 0 | 0 | 0 | 0 | 0 |
| 0 | 0 | 0 | 0 | 0.004304234 |
| 0 | 0 | 0 | 0 | 0 |
| 0 | 0 | 0 | 0 | 0 |
| 0.011062471 | 0.045034812 | 0.050487413 | 0.055652337 | 0.039872138 |
| 0 | 0 | 0 | 0 | 0 |
| 0.003235544 | 0 | 0 | 0 | 0 |
| 0 | 0 | 0 | 0 | 0 |
| 0 | 0 | 0 | 0 | 0 |
| 0 | 0 | 0 | 0 | 0 |
| 0.001808427 | 0.007812361 | 0.004200383 | 0.001080354 | 0.007144798 |
| 0 | 0 | 0 | 0 | 0 |
| 0 | 0 | 0 | 0 | 0 |
| 0 | 0 | 0 | 0.00577927 | 0 |
| 0.175312092 | 0 | 0 | 0.090892819 | 0 |
| 0 | 0.011076764 | 0 | 0 | 0 |
| 0 | 0 | 0 | 0 | 0 |
| 0 | 0 | 0.022039122 | 0 | 0 |
| 0.005051111 | 0.003740687 | 0.000893872 | 0.045142269 | 0.011247987 |
| 0 | 0 | 0 | 0 | 0 |
| 0 | 0 | 0 | 0 | 0 |
| 0 | 0 | 0 | 0 | 0 |
| 0 | 0 | 0 | 0 | 0 |
| 0.001452915 | 0 | 0 | 0 | 0 |
| 0 | 0 | 0 | 0 | 0 |
| 0.00542288 | 0 | 0 | 0 | 0 |
| 0 | 0 | 0 | 0 | 0 |
| 0 | 0 | 0 | 0 | 0 |
| 0 | 0 | 0 | 0 | 0 |
| 0 | 0 | 1.421914603 | 0 | 0 |
| 0.001093802 | 0 | 0 | 0 | 0 |
| 0.009088885 | 0 | 0 | 0 | 0.000387853 |

| | | | | |
|---|---|---|---|---|
| 0 | 0 | 0 | 0 | 0 |
| 0 | 0 | 0 | 0 | 0 |
| 0.002960281 | 0 | 0 | 0 | 0.000708823 |
| 0 | 0 | 0 | 0 | 0 |
| 0 | 0 | 0 | 0 | 0 |
| 0 | 0 | 0 | 0 | 0 |
| 0 | 0 | 0 | 0 | 0.00165631 |
| 0.168945175 | 0 | 0 | 0.005687813 | 0 |
| 0 | 0 | 0 | 0 | 0 |
| 0 | 0 | 0 | 0 | 0 |
| 0.00367419 | 0 | 0 | 0 | 0 |
| 0 | 0 | 0 | 0 | 0 |
| 0 | 0 | 0 | 0 | 0 |
| 0 | 0 | 0 | 0 | 0 |
| 0 | 0 | 0 | 0 | 0.006115149 |
| 0 | 0 | 0 | 0 | 0 |
| 0 | 0 | 0 | 0 | 0 |
| 0 | 0 | 0 | 0 | 0 |
| 0 | 0.003765298 | 0 | 0 | 0 |
| 0 | 0 | 0.443383164 | 0 | 0 |
| 1.17354308 | 0.006220701 | 0 | 0.052647089 | 0.033359074 |
| 0 | 0 | 0 | 0 | 0 |
| 0 | 0 | 0 | 0 | 0 |
| 0 | 0 | 0 | 0 | 0 |
| 0.004559456 | 0.014124271 | 0 | 0.021534176 | 0.006550421 |
| 0 | 0 | 0 | 0 | 0 |
| 0 | 0 | 0 | 0 | 0 |
| 0 | 0 | 0 | 0 | 0 |
| 0.143972052 | 0 | 0 | 0.022262909 | 0.006986593 |
| 0 | 0 | 0 | 0 | 0 |
| 0.736442857 | 0.671899513 | 0.179879686 | 0.221451884 | 0.158154447 |
| 0 | 0 | 0 | 0 | 0 |
| 0 | 0.042656312 | 0 | 0 | 0 |
| 0 | 0 | 0 | 0 | 0 |
| 0 | 0 | 0 | 0 | 0 |
| 0 | 0 | 0 | 0 | 0 |
| 0 | 0 | 0 | 0 | 0 |
| 0 | 0 | 0 | 0 | 0 |
| 0 | 0 | 0 | 0 | 0 |
| 0 | 0 | 0 | 0 | 0 |
| 0 | 0 | 0 | 0 | 0 |
| 0.00252959 | 0 | 0.0008953 | 0 | 0 |
| 0 | 0 | 0 | 0 | 0 |
| 0 | 0 | 0 | 0 | 0 |
| 0 | 0 | 0 | 0 | 0 |
| 0 | 0 | 0 | 0 | 0 |
| 0 | 0 | 0 | 0 | 0 |
| 0 | 0 | 0 | 0 | 0 |

| | | | | |
|---|---|---|---|---|
| 0 | 0 | 0 | 0 | 0 |
| 0 | 0 | 0 | 0 | 0 |
| 0 | 0.015890927 | 0 | 0 | 0.135073952 |
| 0 | 0 | 0 | 0 | 0 |
| 0 | 0 | 1.49365693 | 0 | 0 |
| 0 | 0 | 0 | 0 | 0 |
| 0 | 0 | 0 | 0 | 0 |
| 0 | 0 | 0 | 0 | 0 |
| 0 | 0 | 0 | 0 | 0 |
| 0 | 0 | 0 | 0 | 0 |
| 0 | 0 | 0 | 0 | 0 |
| 0 | 0 | 0 | 0 | 0 |
| 0 | 0 | 0 | 0 | 0.002542239 |
| 0.000561664 | 0.001386501 | 0 | 0 | 0.001210387 |
| 0.000347685 | 0.003433125 | 0 | 0.005095747 | 0.008158633 |
| 0 | 0 | 0.003732869 | 0 | 0 |
| 0.000792313 | 0.011083257 | 0 | 0.004796382 | 0.001517719 |
| 0 | 0 | 0 | 0 | 0.002207476 |
| 0 | 0 | 0 | 0 | 0 |
| 0 | 0 | 0 | 0 | 0 |
| 0 | 0 | 0 | 0 | 0 |
| 0.01156292 | 0 | 0 | 0 | 0.001027551 |
| 0 | 0 | 0 | 0 | 0 |
| 0.139151016 | 0 | 0 | 0.024022332 | 0 |
| 0 | 0 | 0 | 0 | 0 |
| 0 | 0 | 0 | 0 | 0 |
| 0 | 0 | 0.411771427 | 0 | 0 |
| 0 | 0 | 0 | 0 | 0 |
| 0 | 0 | 0 | 0 | 0 |
| 0.017070733 | 0 | 0 | 0.03164485 | 0 |
| 0 | 0 | 0 | 0 | 0 |
| 0 | 0 | 0 | 0 | 0 |
| 0 | 0 | 0 | 0 | 0 |
| 0.053602924 | 0 | 0 | 0 | 0 |
| 0 | 0 | 0 | 0 | 0 |
| 0.03476198 | 0 | 0 | 0 | 0.002736515 |
| 0 | 0 | 0 | 0 | 0 |
| 0.002096769 | 0.020950449 | 0 | 0 | 0.003657864 |
| 0 | 0 | 0 | 0 | 0 |
| 0 | 0 | 0 | 0 | 0 |
| 0 | 0.001422937 | 0 | 0 | 0 |
| 0.001759164 | 0 | 0 | 0 | 0 |
| 0 | 0 | 0 | 0 | 0 |
| 0 | 0.016838887 | 0 | 0 | 0 |
| 0 | 0 | 0 | 0 | 0 |
| 0.007138301 | 0 | 0 | 0 | 0 |
| 0.00373392 | 0 | 0 | 0.001241401 | 0 |
| 0.017052357 | 0.007463603 | 0 | 0.074213888 | 0 |

| | | | | |
|---|---|---|---|---|
| 0 | 1.417264002 | 0 | 0 | 0 |
| 0 | 0 | 0 | 0 | 0 |
| 0 | 0 | 0 | 0 | 0 |
| 0 | 0 | 0 | 0 | 0 |
| 0.004271063 | 0.016952075 | 0.002000728 | 0.005763459 | 0 |
| 0.011675538 | 0 | 0 | 0.010572572 | 0 |
| 0 | 0 | 0 | 0 | 0 |
| 0 | 0 | 0.056089217 | 0 | 0.175057477 |
| 0 | 0 | 0 | 0 | 0 |
| 0.132308372 | 0.094682778 | 0.002890619 | 0.251890192 | 0.034418339 |
| 0 | 0 | 0 | 0.00094108 | 0 |
| 0.366869854 | 0 | 0 | 0.039170428 | 0.009812471 |
| 0.007083972 | 0.006264068 | 0 | 0.002021235 | 0.165115473 |
| 0 | 0 | 0 | 0 | 0.016840893 |
| 0 | 0 | 0 | 0.000623638 | 0 |
| 0.00476159 | 0 | 0 | 0 | 0 |
| 0 | 0 | 0 | 0 | 0 |
| 0 | 0 | 0 | 0 | 0 |
| 0 | 0.931138696 | 0 | 0.149201107 | 0.906100751 |
| 0 | 0 | 0 | 0 | 0 |
| 0 | 0 | 0 | 0 | 0 |
| 0.010039551 | 0 | 0 | 0 | 0 |
| 0.001418309 | 0 | 0 | 0.000834262 | 0 |
| 0 | 0 | 0.001399323 | 0 | 0 |
| 0 | 0 | 0 | 0 | 0 |
| 0 | 0 | 0 | 0 | 0.0021231 |
| 0 | 0 | 13.22170425 | 0 | 0 |
| 0 | 0.475213544 | 0 | 0 | 0.017971922 |
| 0 | 0 | 0 | 0 | 0 |
| 0 | 0 | 0 | 0 | 0 |
| 0 | 0 | 0 | 0.001515726 | 0 |
| 0 | 0 | 0 | 0 | 0 |
| 0 | 0 | 0 | 0 | 0 |
| 0 | 0 | 0 | 0 | 0 |
| 0 | 0 | 0 | 0 | 0 |
| 0 | 0 | 0 | 0 | 0 |
| 0.2988893 | 0.181268624 | 0 | 0.146514594 | 14.87492329 |
| 0 | 0 | 0 | 0 | 0 |
| 0 | 0 | 0 | 0 | 0 |
| 0 | 0.515194915 | 0 | 0.121573611 | 0 |
| 0 | 0 | 0 | 0 | 0 |
| 0 | 0 | 0 | 0 | 0 |
| 0 | 0 | 0 | 0 | 0 |
| 0 | 0 | 0 | 0 | 0 |
| 0 | 0 | 0 | 0 | 0.002296047 |
| 0 | 0 | 0.003986196 | 0 | 0 |
| 0.030935538 | 0.001764234 | 0 | 0.004977028 | 0 |
| 0 | 0 | 0 | 0 | 0 |

| | | | | |
|---|---|---|---|---|
| 0 | 0 | 0 | 0 | 0 |
| 0.464163205 | 0.002534985 | 0 | 0 | 0 |
| 0 | 0 | 0 | 0 | 0 |
| 0 | 0 | 0 | 0 | 0 |
| 0 | 0 | 0 | 0 | 0 |
| 0 | 0.001588055 | 0.014728592 | 0 | 0 |
| 0 | 0 | 0 | 0 | 0 |
| 0 | 0 | 0.001104325 | 0 | 0 |
| 0 | 0 | 3.301598334 | 0 | 0 |
| 0 | 0 | 0 | 0 | 0 |
| 0.001538903 | 0.0679192 | 0.000990301 | 0.005883776 | 0.001406931 |
| 0 | 0.029777951 | 0 | 0 | 0 |
| 0 | 0 | 0 | 0 | 0.018078944 |
| 0.001453878 | 0.003262711 | 0.000935586 | 0.002021344 | 0.000569656 |
| 0 | 0 | 0 | 0 | 0 |
| 0 | 0 | 0 | 0 | 0 |
| 0 | 0 | 0 | 0 | 0 |
| 0 | 0 | 0 | 0 | 0 |
| 0.019694564 | 0 | 0 | 0.172381715 | 0 |
| 0 | 0 | 0 | 0 | 0 |
| 0.004557074 | 0 | 0 | 0 | 0 |
| 0.02166784 | 0 | 0.311252289 | 0 | 0 |
| 0 | 0 | 0.006436743 | 0 | 0 |
| 0 | 0 | 0 | 0 | 0 |
| 0 | 0 | 0 | 0 | 0 |
| 0 | 0 | 0 | 0 | 0 |
| 0.00229293 | 0.001179214 | 0 | 0.001095836 | 0.183650351 |
| 0 | 0 | 0 | 0 | 0 |
| 0.001687456 | 0 | 0 | 0 | 0 |
| 0.098610938 | 1.515080375 | 0.34627183 | 1.075891024 | 5.291930507 |
| 0 | 0 | 0 | 0 | 0 |
| 0.031976102 | 0 | 0 | 0 | 0 |
| 0 | 0.00136343 | 0 | 0.00131982 | 0 |
| 0 | 0 | 0 | 0 | 0 |
| 0.038792697 | 0.001964349 | 0 | 0 | 0 |
| 0.000549579 | 0 | 0 | 0 | 0 |
| 0 | 0 | 0.143357459 | 0 | 0 |
| 0.066146153 | 0.003990686 | 0 | 0 | 0 |
| 0 | 0 | 0 | 0 | 0 |
| 0 | 0 | 0 | 0 | 0 |
| 0.052590834 | 0 | 0 | 0.002700368 | 0.009054175 |
| 0 | 1.662963041 | 1.563631731 | 9.364574148 | 0 |
| 0.041218835 | 0.002030959 | 0 | 0 | 0 |
| 0 | 0 | 0 | 0 | 0 |
| 0 | 0 | 0 | 0 | 0.000528289 |
| 0.000885862 | 0 | 0 | 0 | 0 |
| 0.000456181 | 0 | 0 | 0 | 0 |
| 0 | 0 | 0 | 0 | 0 |

| | | | | |
|---|---|---|---|---|
| 0.00379924 | 0 | 0 | 0 | 0 |
| 0 | 0 | 0 | 0 | 0 |
| 0.203427304 | 0.011674748 | 0 | 0.008796703 | 19.55011507 |
| 0 | 0 | 0 | 0 | 0 |
| 0 | 0 | 0.03587896 | 0 | 0 |
| 0 | 0.000587381 | 0 | 0 | 0 |
| 0.002499644 | 0 | 0 | 0 | 0 |
| 0.063714947 | 0 | 0 | 0 | 0 |
| 0 | 0 | 0.005418433 | 0 | 0.020302488 |
| 0 | 0 | 0 | 0 | 0 |
| 0 | 0 | 0 | 0 | 0 |
| 0 | 0 | 0 | 0.001360942 | 0 |
| 0 | 0 | 0 | 0 | 0 |
| 0 | 0.139734344 | 0 | 0 | 0 |
| 0.00057243 | 0 | 0 | 0 | 0 |
| 0 | 0.003765517 | 0.002640733 | 0 | 0.00057169 |
| 0.227477719 | 0.009073223 | 0 | 0 | 0.014098919 |
| 0.071940965 | 0.014330695 | 0 | 0.106283271 | 0.001924678 |
| 0 | 0 | 0 | 0.015894465 | 0.003763503 |
| 0 | 0 | 0 | 0 | 0 |
| 0.002640352 | 0 | 0 | 0 | 0 |
| 0 | 0.002104893 | 0 | 0 | 0.000551259 |
| 0 | 0 | 0 | 0 | 0 |
| 0 | 0 | 0 | 0 | 0.016088046 |
| 0 | 0 | 0.014954304 | 0 | 0 |
| 4.296943925 | 0 | 0 | 0.001388408 | 0 |
| 0.00237971 | 0 | 0 | 0 | 0.001206653 |
| 0 | 0 | 0 | 0 | 0 |
| 0 | 0 | 0 | 0 | 0 |
| 0 | 0 | 0 | 0 | 0 |
| 0.06153193 | 0 | 0.035794375 | 0 | 0 |
| 0 | 0.016716544 | 0 | 0 | 0.019381603 |
| 0.093390284 | 0.012434696 | 0.059100884 | 0.106310431 | 0.051888044 |
| 0.701671721 | 0.022809768 | 0 | 0 | 0.000933397 |
| 0 | 0 | 0 | 0 | 0 |
| 0 | 0 | 0 | 0 | 0 |
| 0 | 0 | 0 | 0.002611738 | 0.001177666 |
| 0.38752062 | 0.097587047 | 0 | 0 | 0.011972864 |
| 0 | 0 | 0.525596287 | 0 | 0 |
| 0 | 0.003450265 | 0 | 0 | 0 |
| 0 | 0 | 0 | 0 | 0 |
| 0 | 0 | 0.000665753 | 0 | 0 |
| 0.052888496 | 0.001840701 | 0 | 0 | 0 |
| 0.198669181 | 0 | 0 | 0 | 0 |
| 0 | 0.620470881 | 0 | 0.664899155 | 0.104111005 |
| 0 | 0.003258421 | 0 | 0 | 0 |
| 0 | 0 | 0 | 0 | 0 |
| 0 | 0 | 0 | 0 | 0 |

| | | | | |
|---|---|---|---|---|
| 3.547028682 | 0 | 0 | 0 | 0 |
| 0.012374941 | 0.559212656 | 0 | 0 | 0.130516366 |
| 0 | 0 | 0 | 0 | 0 |
| 0 | 0 | 0 | 0.002022709 | 0 |
| 0 | 0 | 0 | 0 | 0 |
| 0.001990972 | 0 | 0 | 0 | 0 |
| 0.168898047 | 0 | 0 | 0 | 0 |
| 0.000447306 | 0 | 0 | 0 | 0.002249206 |
| 0.001732554 | 0 | 0 | 0 | 0.000957348 |
| 0 | 0 | 0 | 0.006067416 | 0.002735876 |
| 0.010818746 | 0.006231562 | 0 | 0.003929573 | 0.000621718 |
| 0.001933779 | 0.018298973 | 0 | 0 | 0.055795468 |
| 0 | 0 | 0.049438106 | 0 | 0 |
| 0 | 0.002604208 | 0 | 0 | 0 |
| 0 | 0 | 0 | 0 | 0.008738453 |
| 0 | 0 | 0 | 0 | 0 |
| 0.004191652 | 0.057982348 | 0.001804324 | 0.078614913 | 0.008951634 |
| 0.166475685 | 0.026138465 | 0 | 0 | 0.221845111 |
| 0 | 0 | 0 | 0 | 0 |
| 0.004297286 | 0.012258244 | 0 | 0 | 0 |
| 0.001051456 | 0.024609926 | 0.012611499 | 0 | 0 |
| 0.002180613 | 0 | 0 | 0 | 0 |
| 0 | 0 | 0 | 0 | 0 |
| 0 | 0 | 0.000641026 | 0 | 0 |
| 0.063402797 | 0 | 0 | 0 | 0 |
| 0 | 0 | 0 | 0 | 0 |
| 0.191423664 | 0 | 0.01244818 | 0 | 0.070906734 |
| 0.032408718 | 0.405224776 | 0.047683657 | 0.467155297 | 1.097116221 |
| 0 | 0 | 0 | 0 | 0 |
| 0.000898678 | 0 | 0 | 0 | 0 |
| 0 | 0 | 0 | 0 | 0.000574247 |
| 0 | 0 | 0 | 0 | 0 |
| 0 | 0.002976944 | 0 | 0 | 0 |
| 0 | 0 | 0 | 0 | 0 |
| 0 | 0 | 0.004978699 | 0 | 0 |
| 0.036986853 | 0.203705118 | 0.237270569 | 0.941286524 | 0.447746336 |
| 0 | 0 | 0 | 0 | 0 |
| 0 | 0 | 0 | 0 | 0 |
| 0.133725466 | 0 | 0 | 0.009956201 | 0.001980608 |
| 0 | 0 | 0 | 0 | 0 |
| 0 | 0.605221148 | 0 | 0.654892069 | 0.025133025 |
| 0 | 0.002923601 | 0 | 0 | 0 |
| 0.364730138 | 0.002909317 | 0.001975856 | 0.017609038 | 0.012297861 |
| 0.007725522 | 0.004435091 | 0 | 0 | 0.001935873 |
| 0 | 0 | 0 | 0 | 0 |
| 0 | 0 | 0.46022795 | 0 | 0 |
| 0 | 0 | 0 | 0 | 0 |
| 0.001016441 | 0 | 0 | 0 | 0 |

| | | | | |
|---|---|---|---|---|
| 0 | 0 | 0 | 0 | 0 |
| 0 | 0 | 0 | 0 | 0 |
| 0 | 0 | 0 | 0 | 0 |
| 0 | 0 | 0.064243149 | 0 | 0 |
| 0 | 0 | 0 | 0 | 0 |
| 0 | 0 | 0.005654007 | 0 | 0 |
| 0 | 0 | 0 | 0 | 0 |
| 0 | 0 | 0 | 0 | 0 |
| 0.177759327 | 0 | 0.015738984 | 0 | 0.011600585 |
| 0.007070244 | 0 | 0.180136931 | 0 | 0 |
| 0 | 0 | 0 | 0 | 0 |
| 0 | 0.002221279 | 0.012861579 | 0 | 0 |
| 0 | 0 | 0 | 0 | 0 |
| 0 | 0 | 0 | 0 | 0 |
| 0.003855501 | 0 | 0.002577542 | 0 | 0.003692715 |
| 0 | 0 | 0 | 0 | 0 |
| 0 | 0 | 0.009888757 | 0 | 0 |
| 0 | 0 | 0 | 0 | 0 |
| 0 | 0 | 0 | 0 | 0 |
| 0 | 0 | 0.000984492 | 0 | 0 |
| 0 | 0 | 0 | 0 | 0 |
| 0 | 0 | 0.012793964 | 0 | 0 |
| 0 | 0 | 0 | 0 | 0 |
| 0.240986679 | 0.042918961 | 0 | 0 | 0.143229771 |
| 0 | 0.000799879 | 0 | 0 | 0 |
| 0 | 0 | 0.202109894 | 0 | 0 |
| 0 | 0 | 0.144096665 | 0 | 0 |
| 0 | 0 | 1.135886918 | 0 | 0 |
| 0 | 0 | 0 | 0 | 0 |
| 0 | 0 | 0 | 0 | 0 |
| 0 | 0 | 0.002679353 | 0 | 0 |
| 0 | 0 | 0.00050489 | 0 | 0 |
| 0.002611895 | 0 | 0 | 0 | 0 |
| 0 | 0 | 0 | 0 | 0 |
| 0 | 0 | 0.00362826 | 0 | 0 |
| 0.005306162 | 0 | 0.005300848 | 0 | 0 |
| 0 | 0 | 0.093889006 | 0 | 0 |
| 0 | 0 | 0 | 0 | 0 |
| 0 | 0 | 0 | 0 | 0 |
| 0 | 0 | 0 | 0 | 0 |
| 1.443743524 | 0.000819585 | 0.001586364 | 0.266381699 | 0.000715481 |
| 0 | 0 | 0.045007553 | 0 | 0 |
| 2.933019477 | 0.853214266 | 9.752225181 | 0 | 0.036563883 |
| 0 | 0 | 0 | 0 | 0 |
| 0 | 0 | 0 | 0 | 0 |
| 0 | 0 | 0 | 0 | 0 |
| 0 | 0 | 0 | 0 | 0 |
| 0 | 0 | 0 | 0.011215759 | 0 |

| | | | | |
|---|---|---|---|---|
| 0 | 0 | 0 | 0 | 0 |
| 0 | 0 | 0.01177519 | 0 | 0 |
| 1.925640961 | 1.392132052 | 0.001015593 | 1.343946388 | 0.34694683 |
| 0 | 0 | 0 | 0 | 0 |
| 0 | 0 | 0.008003748 | 0 | 0 |
| 0 | 0 | 0 | 0 | 0 |
| 0 | 0 | 0 | 0 | 0 |
| 0 | 0 | 0 | 0 | 0 |
| 0 | 0 | 0 | 0 | 0 |
| 0 | 0 | 0 | 0 | 0 |
| 0.003155383 | 0.057306578 | 0.000997132 | 0.001723451 | 0.000809506 |
| 0.715366052 | 1.424122599 | 1.575592306 | 5.291352581 | 4.435292749 |
| 0 | 0 | 0 | 0 | 0 |
| 0.022621706 | 0.008406469 | 0 | 0 | 0.005241913 |
| 0.208362374 | 3.187704129 | 0.003901001 | 6.575975055 | 0.023132641 |
| 0 | 0 | 0 | 0 | 0 |
| 0 | 0 | 0 | 0 | 0 |
| 0 | 0 | 0 | 0 | 0 |
| 0 | 0 | 0 | 0 | 0 |
| 0.001299603 | 0.000916612 | 0 | 0 | 0 |
| 0.039764588 | 0.027202222 | 0 | 0.018236883 | 0.001628365 |
| 0.022193818 | 1.330276971 | 0 | 18.85072216 | 0.78282999 |
| 31.13761034 | 7.184981427 | 0.247549492 | 7.451509593 | 1.070212828 |
| 0 | 0 | 0 | 0 | 0 |
| 0 | 0 | 0.005569949 | 0 | 0 |
| 0 | 0 | 0.373548722 | 0 | 0 |
| 0 | 2.509666528 | 0 | 0.01187791 | 0 |
| 0 | 0 | 0.011505979 | 0 | 0 |
| 0 | 0 | 0 | 0 | 0 |
| 0 | 0 | 0 | 0 | 0 |
| 0 | 0 | 0.008111868 | 0 | 0 |
| 0 | 0 | 0.245696374 | 0 | 0 |
| 0 | 0.493889706 | 0 | 1.323317837 | 1.09576422 |
| 0 | 0 | 0 | 0 | 0.001648386 |
| 0.000381942 | 0.00892561 | 0 | 0.004088844 | 0.002743619 |
| 0 | 0 | 0.006500572 | 0 | 0.013871999 |
| 0 | 0 | 0 | 0 | 0.006408773 |
| 0 | 0 | 0 | 0 | 0 |
| 0 | 0.009903846 | 0.025863738 | 0 | 0.010233873 |
| 0 | 0 | 0 | 0 | 0 |
| 0 | 0 | 0.008994984 | 0 | 0 |
| 0 | 0 | 0 | 0 | 0 |
| 0 | 0 | 0 | 0 | 0 |
| 0 | 0 | 0 | 0 | 0 |
| 0.353196433 | 0.155211984 | 0 | 0.007550503 | 0.004470711 |
| 0 | 0 | 0.059589832 | 0 | 0 |
| 0 | 0 | 0.01229744 | 0 | 0 |
| 4.238857877 | 0 | 0 | 0.004967984 | 0.33975267 |

| | | | | |
|---|---|---|---|---|
| 0 | 0 | 0 | 0 | 0 |
| 0 | 0 | 0 | 0 | 0 |
| 0 | 0 | 0 | 0 | 0 |
| 0 | 0 | 0.000722081 | 0 | 0 |
| 0 | 0 | 0 | 0 | 0 |
| 0 | 0 | 0.020567034 | 0 | 0 |
| 0 | 0 | 0.635792864 | 0 | 0 |
| 0.12193263 | 0.352824572 | 0.001328891 | 0.011484328 | 0.026970995 |
| 0 | 0 | 0 | 0 | 0 |
| 0 | 0 | 0 | 0 | 0 |
| 0.034537671 | 0.00822667 | 0 | 0.017374979 | 0.004080521 |
| 0 | 0 | 0 | 0 | 0 |
| 0 | 0 | 0.001584329 | 0 | 0.000428738 |
| 0 | 0 | 1.183614948 | 0 | 0 |
| 0 | 0 | 0 | 0 | 0 |
| 0 | 0 | 0 | 0 | 0 |
| 0 | 0 | 0.306984267 | 0 | 0 |
| 0 | 0 | 0.001338968 | 0 | 0 |
| 0 | 0 | 0 | 0 | 0 |
| 0 | 0 | 0 | 0 | 0 |
| 0 | 0 | 0 | 0 | 0 |
| 0 | 0 | 0 | 0 | 0 |
| 0 | 0 | 0.006560872 | 0 | 0 |
| 0 | 0 | 0 | 0 | 0 |
| 0 | 0 | 0 | 0 | 0 |
| 0 | 0 | 0 | 0 | 0 |
| 0 | 0 | 0 | 0 | 0 |
| 0 | 0 | 0 | 0 | 0 |
| 0 | 0 | 0 | 0 | 0 |
| 0 | 0 | 0.008491674 | 0 | 0 |
| 0 | 0.007068457 | 0 | 0 | 0 |
| 0 | 0 | 0 | 0 | 0 |
| 0 | 0 | 0 | 0 | 0 |
| 0 | 0 | 0 | 0 | 0 |
| 0 | 0 | 0.000799333 | 0 | 0 |
| 0 | 0 | 0 | 0 | 0 |
| 0.005776452 | 0 | 0.178962659 | 0 | 0 |
| 0 | 0.007913659 | 0 | 0.004780173 | 0.002348878 |
| 0 | 0 | 0.001333247 | 0 | 0 |
| 0.002181785 | 0 | 0 | 0 | 0 |
| 0 | 0 | 0 | 0 | 0 |
| 0 | 0 | 0 | 0 | 0 |
| 0 | 0 | 0.023599175 | 0 | 0 |
| 0 | 0 | 0.385429236 | 0 | 0 |
| 1.544472332 | 0 | 0.029761019 | 0 | 0 |
| 0 | 0 | 0.007092922 | 0 | 0 |
| 0 | 0 | 0 | 0 | 0 |
| 0 | 0 | 0 | 0 | 0 |

| | | | | |
|---|---|---|---|---|
| 0 | 0 | 0 | 0 | 0 |
| 0 | 0 | 0 | 0 | 0 |
| 0 | 0 | 0.002166388 | 0 | 0 |
| 0 | 0 | 0.00846465 | 0 | 0 |
| 0 | 0 | 0.01931466 | 0 | 0 |
| 0 | 0 | 0.212239243 | 0 | 0 |
| 0 | 0 | 0 | 0 | 0 |
| 0 | 0 | 0 | 0 | 0 |
| 2.982792923 | 0.932950761 | 0.011032111 | 10.62482693 | 0.319757254 |
| 0 | 0 | 0 | 0 | 0 |
| 0 | 0 | 0.001007017 | 0 | 0 |
| 0 | 0 | 0 | 0 | 0 |
| 0 | 0 | 0 | 0 | 0 |
| 0 | 0 | 0 | 0 | 0 |
| 0 | 0 | 0.520715548 | 0 | 0 |
| 0 | 0 | 0 | 0 | 0.292709473 |
| 0 | 0 | 0 | 0 | 0 |
| 0 | 0 | 0 | 0 | 0 |
| 0 | 0 | 0 | 0 | 0 |
| 0 | 0 | 0 | 0 | 0 |
| 0 | 0 | 0.003769112 | 0 | 0 |
| 0 | 0 | 0 | 0 | 0 |
| 0 | 0 | 0 | 0 | 0 |
| 0 | 0 | 0 | 0 | 0 |
| 0 | 0.040814843 | 0.016285818 | 0 | 0 |
| 0 | 0 | 2.591818938 | 0 | 0 |
| 0 | 0 | 0.042093516 | 0 | 0 |
| 0.00048412 | 0 | 0 | 0 | 0 |
| 0 | 0 | 0.001206649 | 0 | 0 |
| 0 | 0 | 0.05547142 | 0 | 0 |
| 0 | 0 | 0 | 0 | 0 |
| 0 | 0 | 0.001244358 | 0 | 0 |
| 0 | 0 | 0 | 0 | 0 |
| 0.123122374 | 0 | 0 | 0.004777477 | 0 |
| 0.00288241 | 0.00203297 | 0 | 0 | 0.032142547 |
| 0.046862837 | 0.000798368 | 0 | 0.007790146 | 0.00069696 |
| 0 | 0 | 0 | 0 | 0 |
| 0.486763695 | 1.237573708 | 4.835143661 | 0 | 0 |
| 0 | 0 | 0.008890812 | 0 | 0 |
| 0 | 0 | 0 | 0.001594318 | 0 |
| 0 | 0 | 0.294272063 | 0 | 0 |
| 0 | 0.004707853 | 0 | 0 | 0 |
| 0.007159629 | 0.477633119 | 0.722663193 | 0 | 0 |
| 0 | 0 | 0 | 0 | 0 |
| 0 | 0 | 0 | 0 | 0 |
| 0.00128686 | 0 | 0 | 0 | 0 |
| 0 | 0 | 0 | 0 | 0 |
| 0 | 0 | 0.002073147 | 0.019595866 | 0.00126229 |

| | | | | |
|---|---|---|---|---|
| 0 | 0 | 0 | 0 | 0.000470645 |
| 0 | 0 | 0.029819299 | 0 | 0 |
| 0 | 0 | 0 | 0 | 0 |
| 0 | 0 | 0 | 0 | 0 |
| 0 | 0.001431364 | 0 | 0 | 0 |
| 0 | 0 | 0 | 0 | 0 |
| 0 | 0 | 0 | 0 | 0 |
| 0 | 0 | 0 | 0 | 0 |
| 0 | 0 | 0 | 0 | 0 |
| 0 | 0 | 0.00135177 | 0 | 0 |
| 0 | 0 | 0 | 0 | 0 |
| 0 | 0 | 0.046149436 | 0.088585245 | 0 |
| 0 | 0 | 0.007687619 | 0 | 0 |
| 0 | 0 | 0 | 0 | 0 |
| 4.743703788 | 0.200506977 | 0.037158548 | 0.678305954 | 0.25614518 |
| 0 | 0 | 0 | 0 | 0 |
| 0 | 0 | 0 | 0 | 0 |
| 0 | 0 | 0 | 0 | 0 |
| 0.212696938 | 0.003264588 | 0 | 0 | 0 |
| 0 | 0 | 0.009731369 | 0 | 0 |
| 0 | 0 | 0.004843908 | 0 | 0 |
| 0 | 0 | 0 | 0 | 0 |
| 0 | 0.658681393 | 0.009861084 | 0 | 0.228466713 |
| 0 | 0 | 0 | 0 | 0 |
| 0 | 0 | 0.005240965 | 0 | 0 |
| 0 | 0 | 0 | 0 | 0 |
| 0 | 0 | 0 | 0 | 0 |
| 0 | 0 | 0 | 0 | 0 |
| 0 | 0 | 0 | 0 | 0 |
| 0 | 0 | 0 | 0 | 0 |
| 0 | 0 | 0 | 0 | 0 |
| 0.00228421 | 0.076525248 | 0 | 0.002807148 | 0.027659615 |
| 0 | 0 | 0.001940547 | 0 | 0 |
| 0.003504966 | 0.007914425 | 0 | 0 | 0 |
| 0.045507987 | 0 | 0 | 0.003835119 | 0.00045508 |
| 0.055126338 | 0 | 0 | 0 | 0.003715471 |
| 0 | 0 | 0.000663453 | 0 | 0 |
| 0 | 0 | 0.000893371 | 0 | 0 |
| 0 | 0 | 0 | 0 | 0 |
| 0 | 0 | 0 | 0 | 0 |
| 0 | 0 | 0.58585226 | 0 | 0 |
| 0 | 0 | 0.007019641 | 0 | 0 |
| 0 | 0 | 0 | 0 | 0 |
| 0 | 0 | 0 | 0 | 0 |
| 0.650293989 | 0.008729973 | 0 | 0 | 0.000747166 |
| 0 | 0 | 0 | 0 | 0 |
| 0 | 0 | 0 | 0 | 0 |
| 0 | 0 | 0 | 0 | 0 |

| | | | | |
|---|---|---|---|---|
| 0 | 0 | 0 | 0 | 0 |
| 0 | 0 | 0 | 0 | 0 |
| 0.013016726 | 0 | 0.002094101 | 0 | 0 |
| 0 | 0 | 0 | 0 | 0 |
| 0 | 0 | 0 | 0 | 0 |
| 0 | 0 | 0 | 0 | 0 |
| 0 | 0 | 0.12845931 | 0 | 0 |
| 0 | 0.008861417 | 0.014922513 | 0 | 0 |
| 0 | 0 | 0 | 0 | 0 |
| 0 | 0 | 0 | 0 | 0 |
| 0 | 0 | 0 | 0 | 0 |
| 0 | 0 | 0 | 0 | 0 |
| 0 | 0 | 0 | 0 | 0 |
| 0 | 0 | 0.727306764 | 0 | 0 |
| 0 | 0 | 0 | 0 | 0 |
| 0 | 0 | 0.031516813 | 0 | 0 |
| 0 | 0 | 0 | 0 | 0 |
| 0 | 0 | 0 | 0 | 0 |
| 0 | 0 | 6.459385124 | 0 | 0.087525843 |
| 0 | 0 | 0 | 0 | 0 |
| 0 | 0 | 0 | 0 | 0 |
| 0 | 0 | 0 | 0 | 0 |
| 0 | 0 | 0 | 0 | 0 |
| 0 | 0 | 0 | 0 | 0 |
| 0.013296355 | 0 | 0 | 0 | 0 |
| 0 | 0.08844387 | 0 | 0.002961813 | 0.019337209 |
| 0 | 0 | 0 | 0 | 0 |
| 0.00037912 | 0.002027741 | 0.00083864 | 0 | 0.00054467 |
| 0 | 0 | 0.000710075 | 0 | 0 |
| 0.002910984 | 0 | 0.100218844 | 0 | 0 |
| 0 | 0 | 0 | 0 | 0 |
| 0 | 0 | 0 | 0 | 0 |
| 0 | 0 | 0.00920953 | 0 | 0 |
| 0 | 0 | 0 | 0 | 0 |
| 0.0916318 | 0.002504965 | 0 | 0.000969937 | 0 |
| 0 | 0 | 0 | 0 | 0 |
| 0 | 0 | 0.009874807 | 0 | 0 |
| 0.260686572 | 0.001894934 | 0 | 0.00176095 | 0 |
| 0 | 0 | 0 | 0 | 0 |
| 0 | 0 | 0 | 0 | 0 |
| 1.543790749 | 7.997699358 | 2.972717288 | 8.991621135 | 3.361121475 |
| 0.200685227 | 0.12581002 | 0.021627807 | 0 | 0.153301319 |
| 0 | 0 | 0 | 0 | 0 |
| 0 | 0 | 0 | 0 | 0 |
| 0 | 0 | 0 | 0 | 0 |
| 0 | 0 | 0 | 0 | 0 |
| 0 | 0 | 0.002312311 | 0 | 0 |
| 0.02622745 | 0.006316484 | 0 | 0 | 0 |

| | | | | |
|---|---|---|---|---|
| 0 | 0 | 0 | 0 | 0 |
| 0 | 0 | 0 | 0 | 0 |
| 0 | 0 | 0 | 0 | 0 |
| 0 | 0 | 0 | 0 | 0 |
| 0.007099706 | 1.289183845 | 9.083488657 | 0 | 0 |
| 0 | 0 | 0 | 0 | 0 |
| 0 | 0 | 0 | 0 | 0 |
| 0 | 0 | 0.002469119 | 0 | 0 |
| 3.897840249 | 1.462301324 | 0.008470932 | 0 | 1.192956936 |
| 0 | 0 | 0 | 0 | 0 |
| 0 | 0 | 0 | 0.000609043 | 0.000762848 |
| 0 | 0 | 0 | 0 | 0 |
| 0 | 0 | 0.004363998 | 0 | 0 |
| 0.006400292 | 0 | 0.019672004 | 0.031554195 | 0.244718432 |
| 0.008519216 | 0 | 0.004606576 | 0 | 0 |
| 0 | 0 | 0.21344242 | 0 | 0 |
| 0.013991899 | 0 | 0 | 0.002043251 | 0 |
| 0 | 0 | 0 | 0 | 0 |
| 0 | 0 | 0 | 0 | 0 |
| 0 | 0 | 0 | 0 | 0 |
| 0 | 0 | 0 | 0 | 0 |
| 0 | 0 | 0 | 0 | 0 |
| 0 | 0 | 0 | 0 | 0 |
| 0 | 0 | 0 | 0 | 0 |
| 0 | 0 | 0.002001006 | 0 | 0 |
| 0 | 0 | 0 | 0 | 0 |
| 0 | 0.10664772 | 0.142041864 | 0 | 0 |
| 0.00104871 | 0 | 0 | 0.002733814 | 0.001232712 |
| 0 | 0 | 0 | 0 | 0 |
| 0 | 0 | 0.000996186 | 0 | 0 |
| 0 | 0 | 0 | 0 | 0 |
| 0 | 0 | 0 | 0 | 0 |
| 0 | 0 | 0 | 0 | 0 |
| 0 | 0 | 0 | 0 | 0 |
| 0 | 0 | 0.007111955 | 0 | 0 |
| 0 | 0 | 0 | 0 | 0 |
| 0 | 0 | 4.791613551 | 0 | 0.008583262 |
| 0 | 0 | 0 | 0 | 0 |
| 0 | 0 | 0 | 0 | 0 |
| 0 | 0 | 0 | 0 | 0 |
| 0 | 0 | 0.04684979 | 0 | 0 |
| 0 | 0 | 0 | 0 | 0 |
| 0 | 0 | 0.001452153 | 0 | 0 |
| 0 | 0 | 0 | 0 | 0 |
| 0 | 0 | 0 | 0 | 0 |
| 0 | 0 | 0 | 0 | 0 |
| 0 | 0 | 0.088320079 | 0 | 0 |
| 0 | 0 | 0 | 0 | 0 |

| | | | | |
|---|---|---|---|---|
| 0 | 0 | 0.002586165 | 0 | 0 |
| 0 | 0 | 0 | 0 | 0 |
| 0 | 0 | 0 | 0 | 0 |
| 0 | 0 | 0 | 0 | 0 |
| 0.001033334 | 0 | 0.464109828 | 0 | 0 |
| 0 | 0 | 0 | 0 | 0 |
| 0.00630043 | 0.002552283 | 0 | 0.031315431 | 0.003063626 |
| 0 | 0 | 0 | 0 | 0 |
| 0 | 0 | 0 | 0 | 0 |
| 0 | 0.00050208 | 0.018176471 | 0 | 0 |
| 0 | 0 | 0 | 0 | 0 |
| 0 | 0 | 0.002850153 | 0 | 0 |
| 0 | 0 | 0 | 0 | 0 |
| 0 | 0 | 0 | 0 | 0 |
| 0 | 0 | 0 | 0 | 0 |
| 0 | 0 | 0 | 0 | 0 |
| 0 | 0 | 0 | 5.798750021 | 0.025782062 |
| 0 | 0 | 0 | 0 | 0 |
| 0 | 0 | 0 | 0 | 0 |
| 0 | 0 | 0 | 0 | 0 |
| 0 | 0.000759349 | 0 | 0 | 0 |
| 0 | 0 | 0 | 0 | 0 |
| 0 | 0 | 0.001441026 | 0 | 0 |
| 0.002534288 | 0.006851847 | 0.002082235 | 0.007440141 | 0.000650166 |
| 0 | 0 | 0 | 0 | 0 |
| 0.065136881 | 0.000878656 | 0 | 0 | 0 |
| 0 | 0 | 0 | 0 | 0 |
| 0 | 0 | 0 | 0 | 0 |
| 0 | 0 | 0 | 0 | 0 |
| 0 | 0 | 0 | 0 | 1.262127264 |
| 0 | 0 | 0.064517577 | 0 | 0 |
| 0 | 0 | 0 | 0 | 0 |
| 0 | 0 | 0 | 0 | 0 |
| 0.001449893 | 0.000795365 | 0 | 0 | 0 |
| 0 | 0 | 0 | 0 | 0 |
| 0 | 0 | 0 | 0 | 0 |
| 0 | 0.015283079 | 0 | 0.007577535 | 0.005531975 |
| 0 | 0 | 0 | 0 | 0 |
| 0 | 0 | 0 | 0 | 0 |
| 0 | 0 | 0.006802693 | 0 | 0 |
| 0 | 0 | 0 | 0 | 0 |
| 0 | 0 | 0.000804357 | 0 | 0 |
| 0 | 0 | 0 | 0 | 0 |
| 0 | 0 | 0.024096976 | 0 | 0 |
| 0 | 0 | 0.001138075 | 0 | 0 |
| 0 | 0 | 0 | 0 | 0 |
| 0 | 0 | 0 | 0 | 0 |
| 0 | 0 | 0.000573084 | 0 | 0 |

| | | | | |
|---|---|---|---|---|
| 0.187772285 | 0.024202116 | 0 | 0.006144029 | 0 |
| 0 | 0 | 0.003432589 | 0 | 0 |
| 0 | 0 | 0.056940409 | 0 | 0 |
| 0 | 0 | 0 | 0 | 0 |
| 0 | 0 | 0 | 0 | 0 |
| 0 | 0 | 0 | 0 | 0 |
| 0.002470729 | 0 | 0 | 0 | 0 |
| 0 | 0 | 1.00109198 | 0.11680337 | 1.522002858 |
| 0.027058306 | 1.181931446 | 0.756402559 | 0.00175345 | 0.003074762 |
| 0.000744973 | 0.082667838 | 0 | 0.009155246 | 0.00198766 |
| 0 | 0 | 0.057462039 | 0 | 0 |
| 0 | 0 | 0 | 0 | 0 |
| 0 | 0 | 0.116129636 | 0 | 0 |
| 0 | 0 | 0 | 0 | 0 |
| 0.002512117 | 0 | 0 | 0 | 0 |
| 0 | 0 | 0 | 0 | 0 |
| 0.000995165 | 0.120257541 | 0 | 0.053539996 | 0 |
| 0 | 0 | 0.574866121 | 0 | 0 |
| 0 | 0 | 0 | 0 | 0 |
| 0 | 0 | 0 | 0 | 0 |
| 0 | 0 | 0 | 0 | 0 |
| 0 | 0 | 0 | 0 | 0 |
| 0 | 0 | 0 | 0 | 0 |
| 0 | 0 | 0 | 0 | 0 |
| 0.007977831 | 0 | 0.018869255 | 0 | 0 |
| 0 | 0 | 0.015166035 | 0 | 0 |
| 0 | 0 | 0 | 0 | 0 |
| 0 | 0 | 0 | 0 | 0 |
| 0 | 0 | 0 | 0 | 0 |
| 0 | 0 | 0 | 0 | 0 |
| 0 | 0 | 0 | 0 | 0 |
| 0 | 0 | 0 | 0 | 0 |
| 0 | 0 | 0.03938615 | 0 | 0 |
| 0 | 0 | 0 | 0 | 0 |
| 0 | 0 | 0 | 0 | 0 |
| 0 | 0 | 0.074769887 | 0 | 0 |
| 0 | 0 | 0 | 0 | 0 |
| 0 | 0 | 0.012179992 | 0 | 0 |
| 0.000463228 | 0 | 0 | 0 | 0 |
| 0 | 0 | 0 | 0 | 0 |
| 0 | 0 | 0 | 0 | 0 |
| 0.009027713 | 0 | 0.717319345 | 0 | 0 |
| 0 | 0 | 0 | 0 | 0 |
| 0 | 0 | 0 | 0 | 0 |
| 0 | 0 | 0 | 0 | 0 |
| 0.014202648 | 0 | 0 | 0 | 0 |
| 0 | 0 | 0 | 0 | 0 |
| 0 | 0 | 0 | 0 | 0 |

| | | | | |
|---|---|---|---|---|
| 0 | 0 | 0 | 0 | 0 |
| 0 | 0.004629079 | 0.162553937 | 0 | 0 |
| 0 | 0 | 0 | 0 | 0 |
| 0 | 0 | 0 | 0 | 0 |
| 0.589294214 | 0 | 0.032840442 | 0 | 0.006003143 |
| 0 | 0 | 0 | 0 | 0 |
| 0 | 0 | 0 | 0 | 0 |
| 4.881762086 | 0.057495962 | 0.021705009 | 1.360870381 | 0.085968551 |
| 0 | 0 | 0 | 0 | 0 |
| 0 | 0 | 0.005495887 | 0 | 0 |
| 0 | 0 | 0 | 0 | 0 |
| 0 | 0 | 0.065350805 | 0 | 0 |
| 0 | 0 | 0.002132167 | 0 | 0 |
| 0 | 0 | 0 | 0 | 0 |
| 0 | 0 | 0 | 0 | 0.013281565 |
| 0.000859591 | 0 | 0 | 0.026086779 | 0 |
| 0 | 0 | 0 | 0 | 0 |
| 0 | 0 | 0 | 0 | 0 |
| 0 | 0 | 0 | 0 | 0 |
| 0 | 0 | 0 | 0 | 0 |
| 0 | 0 | 0 | 0 | 0 |
| 0 | 0 | 0 | 0 | 0 |
| 0 | 0 | 0 | 0 | 0 |
| 0.002894848 | 0.001764469 | 0 | 0.003484382 | 0.000462104 |
| 0.000953927 | 0 | 0.074614976 | 0 | 0 |
| 0 | 0 | 0 | 0 | 0 |
| 0 | 0.902279472 | 0.024468494 | 0.006373726 | 0 |
| 0 | 0 | 0 | 0 | 0 |
| 0 | 0 | 0 | 0 | 0 |
| 0 | 0 | 0 | 0 | 0 |
| 0 | 0 | 0 | 0 | 0 |
| 0 | 0 | 0 | 0 | 0 |
| 0 | 0 | 0 | 0 | 0 |
| 0 | 0 | 0 | 0 | 0 |
| 0 | 0 | 0 | 0 | 0 |
| 0 | 0 | 0 | 0 | 0 |
| 0 | 0 | 0 | 0 | 0 |
| 0 | 0 | 0.04538254 | 0 | 0 |
| 0 | 0 | 0.0397157 | 0 | 0 |
| 0 | 0 | 0.000916088 | 0 | 0 |
| 0 | 0 | 0 | 0 | 0 |
| 0.078760459 | 0 | 0.002448822 | 0 | 0 |
| 0 | 0 | 0 | 0 | 0 |
| 0 | 0 | 0 | 0 | 0 |
| 0 | 0 | 0 | 0 | 0 |
| 0 | 0 | 0 | 0 | 0 |
| 0 | 0 | 0 | 0 | 0 |
| 0 | 0 | 0.000878196 | 0 | 0 |

| | | | | |
|---|---|---|---|---|
| 0 | 0 | 0.002937512 | 0 | 0 |
| 0 | 0 | 0.006043992 | 0 | 0 |
| 0 | 0 | 0 | 0 | 0 |
| 0 | 0 | 0 | 0 | 0 |
| 0 | 0 | 0 | 0 | 0 |
| 0.001282087 | 0 | 0 | 0 | 0 |
| 0 | 0 | 0 | 0 | 0 |
| 0 | 0 | 0 | 0 | 0 |
| 0 | 0 | 0 | 0 | 0 |
| 0 | 0 | 0.00445949 | 0 | 0 |
| 0 | 0 | 0 | 0 | 0 |
| 0 | 0 | 0 | 0 | 0 |
| 0 | 0 | 0.013986416 | 0 | 0 |
| 0 | 0 | 0 | 0 | 0 |
| 0 | 0 | 0 | 0 | 0 |
| 0 | 0 | 0 | 0 | 0 |
| 0 | 0 | 0 | 0 | 0 |
| 0 | 0 | 0 | 0 | 0.12802213 |
| 0 | 0 | 0 | 0 | 0 |
| 0 | 0 | 0 | 0 | 0 |
| 0 | 0.46527817 | 0 | 0.000816427 | 0.111515016 |
| 0 | 0 | 0 | 0 | 0 |
| 0.144414414 | 0.001331448 | 0 | 0 | 0 |
| 0 | 0 | 0.001310393 | 0 | 0 |
| 0 | 0 | 0.124954778 | 0 | 0 |
| 0 | 0 | 0.004023504 | 0 | 0 |
| 0 | 0 | 0 | 0 | 0 |
| 0 | 0 | 0 | 0 | 0 |
| 0 | 0 | 0.002685416 | 0 | 0 |
| 0 | 0 | 0 | 0 | 0 |
| 0 | 0 | 0 | 0 | 0 |
| 0 | 0 | 0 | 0 | 0 |
| 0.052657834 | 0.042394453 | 0 | 0.034279156 | 7.646998391 |
| 0 | 0 | 0 | 0 | 0 |
| 0 | 0 | 0.009994555 | 0 | 0 |
| 0.011450587 | 0.090052338 | 0 | 0.000774862 | 0 |
| 0 | 0 | 0 | 0 | 0 |
| 0 | 0 | 0.000499996 | 0 | 0 |
| 0 | 0.060566979 | 0 | 0 | 0.037382082 |
| 0 | 0 | 0 | 0 | 0 |
| 0 | 0 | 0 | 0 | 0 |
| 0 | 0 | 0.015941243 | 0 | 0 |
| 0.018912948 | 0 | 0.027891153 | 0 | 0.000984479 |
| 0 | 0 | 0 | 0 | 0 |
| 0 | 0 | 0 | 0 | 0 |
| 0 | 0 | 0 | 0 | 0 |
| 0 | 0 | 0 | 0 | 0 |
| 0 | 0 | 0 | 0 | 0 |

| | | | | |
|---|---|---|---|---|
| 0 | 0 | 0 | 0 | 0 |
| 0 | 0 | 0 | 0 | 0 |
| 0 | 0 | 0 | 0 | 0 |
| 0 | 0 | 0 | 0 | 0 |
| 0 | 0 | 0 | 0 | 0 |
| 0 | 0 | 0 | 0 | 0 |
| 0 | 0 | 0 | 0 | 0 |
| 0 | 0 | 0 | 0 | 0 |
| 0 | 0 | 0.013354029 | 0 | 0 |
| 0 | 0 | 0.096218436 | 0 | 0 |
| 0 | 0 | 0 | 0 | 0 |
| 0 | 0 | 0 | 0 | 0 |
| 0 | 0 | 0 | 0 | 0 |
| 0 | 0 | 0.382899246 | 0 | 0 |
| 0 | 0 | 0 | 0 | 0 |
| 0.008272478 | 0.011435808 | 0 | 0.504793053 | 0 |
| 0 | 0 | 0 | 0 | 0 |
| 0 | 0 | 0 | 0 | 0 |
| 0 | 0 | 0 | 0 | 0 |
| 0 | 0 | 0.031050035 | 0 | 0 |
| 0 | 0 | 0 | 0 | 0 |
| 0 | 0 | 0 | 0 | 0 |
| 0 | 0 | 0 | 0 | 0 |
| 0 | 0 | 0.063180065 | 0 | 0 |
| 0 | 0 | 0 | 0 | 0 |
| 0 | 0 | 0 | 0 | 0 |
| 0 | 0 | 0 | 0 | 0 |
| 0 | 0 | 0 | 0 | 0 |
| 0 | 0 | 0 | 0 | 0 |
| 0 | 0 | 0.042281102 | 0 | 0 |
| 0 | 0 | 0.423904483 | 0 | 0 |
| 0 | 0 | 0 | 0 | 0 |
| 0 | 0 | 0 | 0 | 0 |
| 0 | 0 | 0.295104152 | 0 | 0 |
| 0 | 0 | 0 | 0 | 0 |
| 0 | 0 | 0 | 0 | 0 |
| 0 | 0 | 0 | 0 | 0 |
| 0 | 0 | 0 | 0 | 0 |
| 0 | 0 | 0.002324416 | 0 | 0 |
| 0 | 0 | 0 | 0 | 0 |
| 0 | 0 | 0 | 0 | 0 |
| 0 | 0 | 0.458883511 | 0 | 0 |
| 0 | 0 | 0 | 0 | 0 |
| 0 | 0 | 0 | 0 | 0 |
| 0 | 0 | 0 | 0 | 0 |
| 0 | 0 | 0 | 0 | 0 |
| 0 | 2.025640013 | 0 | 0.960763351 | 0.281448961 |
| 0 | 0 | 0 | 0 | 0 |

| | | | | |
|---|---|---|---|---|
| 0 | 0 | 0 | 0 | 0 |
| 0 | 0 | 0 | 0 | 0 |
| 0 | 0 | 0 | 0 | 0 |
| 0 | 0 | 0.028294834 | 0 | 0 |
| 0 | 0.05275256 | 0.008508859 | 0 | 0 |
| 0 | 0 | 0 | 0 | 0 |
| 0 | 0 | 0.0175618 | 0 | 0 |
| 0 | 0 | 0 | 0 | 0 |
| 0 | 0 | 0 | 0 | 0 |
| 0 | 0 | 0 | 0 | 0 |
| 0 | 0 | 0 | 0 | 0 |
| 0.140653774 | 0.029873906 | 0 | 0 | 0 |
| 0 | 0.003824478 | 0 | 0 | 0.000435482 |
| 0 | 0 | 0 | 0 | 0 |
| 0.009192643 | 0 | 0 | 0 | 0.024490221 |
| 0 | 0 | 0.523954298 | 0 | 0 |
| 0.001891564 | 0 | 0.392922726 | 0 | 0 |
| 0 | 0 | 0 | 0 | 0 |
| 0.113332149 | 0.054581543 | 0.003452498 | 0 | 0.003057664 |
| 0.010420933 | 0.000952766 | 0.001024525 | 0.094294976 | 0.010147291 |
| 0 | 0 | 0 | 0 | 0 |
| 0 | 0 | 0 | 0 | 0 |
| 0 | 0 | 0 | 0 | 0 |
| 0 | 0 | 0 | 0 | 0 |
| 0 | 0 | 0 | 0 | 0 |
| 0 | 0 | 0 | 0 | 0 |
| 0.008629131 | 0.004428269 | 0 | 0 | 0 |
| 0 | 0 | 0 | 0 | 0 |
| 0 | 0 | 0 | 0 | 0 |
| 0 | 0 | 0 | 0 | 0 |
| 0.00936086 | 0 | 0 | 0.002010669 | 0 |
| 0 | 0 | 0 | 0 | 0 |
| 0 | 0 | 0 | 0 | 0 |
| 0 | 0 | 0 | 0 | 0 |
| 0 | 0 | 0.024214086 | 0 | 0 |
| 0 | 0 | 0 | 0 | 0 |
| 0 | 0 | 0.00090164 | 0 | 0 |
| 1.428467175 | 0.034780347 | 0.000653844 | 0.005226759 | 0.000636976 |
| 0 | 0 | 0.007864409 | 0 | 0 |
| 0 | 0 | 0 | 0 | 0 |
| 0.000368856 | 0.011897747 | 0 | 0 | 0 |
| 0 | 0 | 0.000838193 | 0 | 0 |
| 0 | 0 | 0 | 0 | 0 |
| 0.002081966 | 0.002704974 | 0 | 0 | 0.091307012 |
| 0 | 0 | 0 | 0 | 0 |
| 0 | 0 | 0.00104016 | 0 | 0 |
| 0 | 0 | 0 | 0 | 0 |
| 0 | 0 | 0 | 0 | 0 |

| | | | | |
|---|---|---|---|---|
| 0 | 0 | 0 | 0 | 0 |
| 0 | 0 | 0 | 0 | 0 |
| 0 | 0 | 0.001340729 | 0 | 0 |
| 0.003100608 | 0 | 0 | 0.001756256 | 0.000659932 |
| 0.000319529 | 0.006134929 | 0 | 0 | 0.001224153 |
| 0 | 0 | 0 | 0 | 0 |
| 0 | 0 | 0.002760967 | 0 | 0 |
| 0 | 0 | 0 | 0 | 0 |
| 0 | 0 | 0 | 0 | 0 |
| 0 | 0 | 0.001643847 | 0 | 0 |
| 0 | 0 | 0.003022665 | 0 | 0 |
| 0 | 0 | 0 | 0 | 0 |
| 0 | 0 | 0.225735879 | 0 | 0 |
| 0 | 0 | 0 | 0 | 0 |
| 0 | 0 | 0 | 0 | 0 |
| 0 | 0 | 0 | 0 | 0 |
| 0 | 0 | 0 | 0 | 0 |
| 0 | 0 | 0 | 0 | 0 |
| 0 | 0 | 0 | 0 | 0 |
| 0 | 0 | 0 | 0 | 0 |
| 0 | 0 | 0 | 0 | 0 |
| 0 | 0 | 0 | 0 | 0 |
| 0 | 0.018875071 | 0 | 0 | 0.000769979 |
| 0.010624949 | 0 | 0.410520995 | 0 | 0 |
| 0 | 0 | 0 | 0 | 0 |
| 0 | 0 | 0 | 0 | 0 |
| 0 | 0 | 0.055094254 | 0 | 0 |
| 0 | 0 | 0 | 0 | 0 |
| 0 | 0 | 0 | 0 | 0 |
| 0 | 0 | 0 | 0 | 0 |
| 0 | 0 | 0.002538024 | 0 | 0 |
| 0 | 0 | 0 | 0 | 0 |
| 0 | 0 | 0 | 0 | 0 |
| 0 | 0 | 0 | 0 | 0 |
| 0 | 0 | 0 | 0 | 0 |
| 0 | 0 | 0 | 0.006687598 | 0 |
| 0 | 0 | 0 | 0 | 0.019154487 |
| 0 | 0 | 0 | 0 | 0 |
| 0.102534484 | 0.002828999 | 0 | 0 | 0.00566569 |
| 0 | 0 | 0 | 0 | 0 |
| 0 | 0 | 0 | 0 | 0 |
| 0 | 0 | 0 | 0 | 0 |
| 0 | 0 | 0 | 0 | 0 |
| 0 | 0 | 0 | 0 | 0 |
| 0 | 0 | 0 | 0 | 0 |
| 0.036832693 | 0 | 0 | 0 | 0 |
| 0.074651335 | 0 | 0 | 0 | 0 |
| 0 | 0 | 0 | 0 | 0 |

| | | | | |
|---|---|---|---|---|
| 0 | 0 | 0.048490373 | 0 | 0 |
| 0 | 0 | 0 | 0 | 0 |
| 0 | 0 | 0.381442314 | 0 | 0 |
| 0 | 0 | 0 | 0 | 0 |
| 0 | 0 | 0 | 0 | 0 |
| 0 | 0 | 0 | 0 | 0 |
| 0 | 0.001860702 | 0 | 0 | 0 |
| 0 | 0 | 0 | 0 | 0 |
| 0 | 0 | 0 | 0 | 0 |
| 0 | 0 | 0 | 0 | 0 |
| 0.007465135 | 0.001890063 | 0 | 0 | 0 |
| 0.000612388 | 0 | 0 | 0 | 0 |
| 0 | 0 | 0 | 0 | 0 |
| 0 | 0 | 0 | 0 | 0 |
| 0 | 0 | 0 | 0 | 0 |
| 0 | 0 | 0 | 0 | 0 |
| 0.000723523 | 0 | 0 | 0 | 0 |
| 0 | 0 | 0 | 0 | 0 |
| 0 | 0 | 0 | 0 | 0 |
| 0.002067044 | 0 | 0 | 0 | 0.00062519 |
| 0 | 0 | 0 | 0 | 0 |
| 0 | 0 | 0 | 0 | 0 |
| 0 | 0 | 0 | 0 | 0 |
| 0 | 0 | 0 | 0 | 0 |
| 0 | 0 | 0 | 0 | 0 |
| 0 | 0 | 0 | 0 | 0 |
| 0 | 0 | 0 | 0 | 0 |
| 0 | 0 | 0.081388994 | 0 | 0 |
| 0 | 0 | 0 | 0 | 0 |
| 0.005044207 | 0 | 0 | 0 | 0 |
| 0 | 0.004306768 | 0 | 0.00178672 | 0.000671379 |
| 0 | 0 | 0 | 0 | 0 |
| 0.004557862 | 0.075187543 | 0.043978036 | 0.051449172 | 0.023230273 |
| 0.006542876 | 0.001277517 | 0 | 0.044519554 | 0 |
| 0 | 0 | 0 | 0 | 0 |
| 0 | 0 | 0 | 0 | 0 |
| 0 | 0 | 0 | 0 | 0 |
| 0 | 0 | 0 | 0 | 0 |
| 0 | 0 | 0 | 0 | 0 |
| 0 | 0 | 0 | 0 | 0 |
| 0 | 0 | 0 | 0 | 0 |
| 2.763894546 | 0.033205418 | 0.001263942 | 0.338443461 | 0.083371594 |
| 0 | 0.000715992 | 0.013473577 | 0.003326835 | 0 |
| 0 | 0 | 0 | 0 | 0 |
| 0 | 0 | 0 | 0 | 0 |
| 0.008015956 | 0 | 0.002225172 | 0 | 0 |
| 0 | 0 | 0 | 0 | 0 |
| 0 | 0 | 0 | 0 | 0 |

| | | | | |
|---|---|---|---|---|
| 0 | 0 | 0 | 0 | 0 |
| 0.001595267 | 0 | 0.005646144 | 0 | 0 |
| 0 | 0.012266692 | 0 | 0 | 0.005354288 |
| 0 | 0 | 0 | 0 | 0 |
| 0 | 0.070484435 | 0 | 0 | 0.045499716 |
| 0 | 0 | 0 | 0.067436363 | 0 |
| 0 | 0 | 0 | 0 | 0 |
| 0 | 0 | 0 | 0 | 0 |
| 0 | 0 | 0 | 0 | 0 |
| 0 | 0 | 0 | 0 | 0 |
| 0 | 0 | 0 | 0 | 0 |
| 0 | 0 | 0 | 0 | 0 |
| 0 | 0 | 0 | 0 | 0 |
| 0 | 0 | 0 | 0 | 0 |
| 0 | 0 | 0 | 0 | 0 |
| 0 | 0 | 0 | 0 | 0 |
| 0 | 0 | 0 | 0 | 0 |
| 0 | 0 | 0 | 0 | 0 |
| 0 | 0 | 0 | 0 | 0 |
| 0 | 0 | 0 | 0 | 0 |
| 0 | 0 | 0 | 0 | 0 |
| 0 | 0 | 0 | 0.014122613 | 0 |
| 0 | 0 | 0 | 0 | 0 |
| 0 | 0 | 0 | 0 | 0 |
| 0 | 0 | 0.00310597 | 0 | 0 |
| 0 | 0 | 0 | 0 | 0.003329293 |
| 0 | 0 | 0 | 0 | 0 |
| 0 | 0 | 0 | 0 | 0 |
| 0 | 0 | 0 | 0 | 0 |
| 0 | 0 | 0 | 0 | 0 |
| 0 | 0 | 0 | 0 | 0 |
| 0 | 0 | 0 | 0 | 0 |
| 0 | 0 | 0 | 0 | 0 |
| 0 | 0 | 0 | 0 | 0 |
| 0 | 0 | 0 | 0 | 0.00060078 |
| 0 | 0 | 0 | 0 | 0 |
| 0 | 0 | 0 | 0 | 0 |
| 0 | 0.195053864 | 0 | 0 | 0 |
| 0.004207985 | 0 | 0.874630145 | 0 | 0 |
| 0 | 0 | 0 | 0 | 0 |
| 0 | 0 | 0 | 0 | 0 |
| 0 | 0 | 0 | 0 | 0 |
| 0.150619118 | 0.017497031 | 0 | 0.006139797 | 0 |
| 0 | 0 | 0 | 0 | 0 |
| 0 | 0 | 0 | 0 | 0 |
| 0 | 0 | 0 | 0 | 0 |
| 0 | 0.001115293 | 0 | 0.034008009 | 0.002190664 |
| 0 | 0 | 0 | 0 | 0 |

| | | | | |
|---|---|---|---|---|
| 0 | 0 | 0 | 0 | 0 |
| 0.01293387 | 0.027090422 | 0 | 0.030446981 | 0.004760592 |
| 0 | 0 | 0.00314008 | 0 | 0 |
| 0 | 0 | 0 | 0 | 0 |
| 0 | 0 | 0 | 0 | 0 |
| 0 | 0 | 0 | 0 | 0 |
| 0 | 0 | 0 | 0 | 0 |
| 0 | 0 | 0 | 0 | 0 |
| 0 | 0 | 0 | 0 | 0 |
| 0 | 0 | 0 | 0 | 0 |
| 0 | 0 | 0 | 0 | 0 |
| 0 | 0 | 0 | 0 | 0 |
| 0 | 0 | 0.327334423 | 0 | 0 |
| 0 | 0 | 0 | 0 | 0 |
| 0.050645479 | 0 | 0 | 0 | 0.001914755 |
| 0 | 0 | 0 | 0 | 0 |
| 0 | 0 | 0 | 0 | 0 |
| 0 | 0 | 0 | 0 | 0 |
| 0 | 0 | 0 | 0 | 0 |
| 0 | 0 | 0 | 0 | 0 |
| 0 | 0 | 0 | 0 | 0 |
| 0 | 0 | 0 | 0.006755882 | 0 |
| 0 | 0 | 0 | 0 | 0 |
| 0 | 0 | 0 | 0 | 0 |
| 0 | 0 | 0 | 0 | 0 |
| 0 | 0 | 0 | 0 | 0 |
| 0 | 0 | 0 | 0 | 0 |
| 0 | 0 | 0 | 0 | 0 |
| 0 | 0 | 0 | 0 | 0 |
| 0 | 0 | 0 | 0 | 0 |
| 0 | 0 | 0 | 0 | 0 |
| 0 | 0 | 0 | 0 | 0 |
| 0 | 0 | 0 | 0 | 0 |
| 0 | 0 | 0 | 0 | 0 |
| 0 | 0.245642936 | 0.014300792 | 0 | 0.002048803 |
| 0 | 0 | 0 | 0 | 0 |
| 0 | 0 | 0 | 0 | 0 |
| 0 | 0 | 0 | 0 | 0 |
| 0.001307086 | 0.000860431 | 0 | 0.004497711 | 0.010515947 |
| 0 | 0 | 0 | 0 | 0 |
| 0.001640766 | 0.151597839 | 0.265263537 | 0.084240554 | 0.046639563 |
| 0 | 0 | 0 | 0 | 0 |
| 0 | 0 | 0 | 0 | 0 |
| 0 | 0 | 0 | 0 | 0 |
| 0 | 0 | 0 | 0 | 0 |
| 4.44253798 | 2.792344325 | 0 | 1.572437954 | 1.280807918 |
| 0 | 0 | 0 | 0 | 0 |
| 0 | 0 | 0 | 0 | 0 |

| | | | | |
|---|---|---|---|---|
| 0 | 0 | 0 | 0 | 0 |
| 0 | 0 | 0 | 0 | 0 |
| 0 | 0 | 0 | 0 | 0 |
| 0 | 0 | 0.01156752 | 0 | 0 |
| 0 | 0 | 0 | 0 | 0 |
| 0 | 0 | 0 | 0 | 0 |
| 0 | 0 | 0 | 0 | 0 |
| 0.027701321 | 0 | 0 | 0.017467168 | 0 |
| 0 | 0 | 0 | 0 | 0 |
| 0.277816872 | 0.002159681 | 0 | 0.124934373 | 0.011689223 |
| 0 | 0 | 0.005388332 | 0 | 0 |
| 0 | 0 | 0 | 0 | 0 |
| 0.012057158 | 0 | 0 | 0 | 0 |
| 0 | 0 | 0 | 0 | 0 |
| 0 | 0 | 0 | 0 | 0 |
| 0 | 0 | 0 | 0 | 0 |
| 0 | 0 | 0 | 0 | 0 |
| 0 | 0 | 0 | 0 | 0 |
| 0 | 0 | 0 | 0 | 0 |
| 1.090297279 | 0.267361205 | 0.003030549 | 0.024444121 | 0.016894096 |
| 0 | 0 | 0 | 0 | 0 |
| 0 | 0 | 0 | 0 | 0 |
| 0 | 0 | 0 | 0 | 0 |
| 0 | 0 | 0 | 0 | 0 |
| 0 | 0 | 0 | 0 | 0 |
| 0 | 0 | 0 | 0 | 0 |
| 0 | 0 | 0.001115899 | 0 | 0 |
| 0 | 0 | 0 | 0 | 0 |
| 0 | 0 | 0 | 0 | 0 |
| 0 | 0 | 0 | 0 | 0 |
| 0 | 0 | 0 | 0 | 0 |
| 0 | 0 | 0 | 0 | 0 |
| 0 | 0 | 0 | 0 | 0 |
| 0 | 0 | 0 | 0 | 0 |
| 0 | 0 | 0 | 0 | 0 |
| 0 | 0 | 0 | 0 | 0 |
| 0 | 0 | 0.47447919 | 0 | 0 |
| 0 | 0 | 0.021562227 | 0 | 0 |
| 0 | 0 | 0 | 0 | 0 |
| 0 | 0 | 0 | 0 | 0 |
| 0 | 0 | 0 | 0 | 0 |
| 0 | 0 | 0 | 0 | 0 |
| 0 | 0 | 0 | 0 | 0 |
| 0 | 0 | 0 | 0 | 0 |
| 0.009757937 | 0.0074117 | 0 | 0 | 0 |
| 0 | 0 | 0.004098323 | 0 | 0 |
| 0 | 0 | 0 | 0 | 0 |
| 0.003542244 | 0 | 0 | 0 | 0 |

| | | | | |
|---|---|---|---|---|
| 0 | 0 | 0 | 0 | 0 |
| 0 | 0 | 0 | 0 | 0 |
| 0 | 0 | 0 | 0 | 0 |
| 0 | 0 | 0 | 0 | 0 |
| 0 | 0 | 0 | 0 | 0 |
| 0.028578358 | 0.001449151 | 0.009349782 | 0.000918195 | 0 |
| 0 | 0 | 0 | 0 | 0 |
| 0 | 0 | 0 | 0 | 0 |
| 0 | 0 | 0 | 0 | 0 |
| 0 | 0 | 0 | 0 | 0 |
| 0 | 0 | 0 | 0 | 0 |
| 0 | 0 | 0 | 0 | 0 |
| 0 | 0 | 0 | 0 | 0 |
| 0 | 0 | 0 | 0 | 0 |
| 0 | 0 | 0 | 0 | 0 |
| 0 | 0 | 0 | 0 | 0 |
| 0 | 0 | 0.002749387 | 0 | 0 |
| 0 | 0 | 0 | 0 | 0.000991463 |
| 0 | 0 | 0 | 0 | 0 |
| 0 | 0 | 0 | 0 | 0 |
| 0.000464093 | 0 | 0 | 0 | 0 |
| 0 | 0 | 0 | 0 | 0 |
| 0 | 0 | 0 | 0 | 0 |
| 0 | 0 | 0 | 0 | 0 |
| 0 | 0 | 0 | 0 | 0 |
| 0.015025663 | 0 | 0 | 0.001196843 | 0 |
| 0 | 0 | 0 | 0 | 0 |
| 0 | 0 | 0.688904603 | 0 | 0 |
| 0 | 0 | 0 | 0 | 0 |
| 0 | 0 | 0 | 0 | 0 |
| 0 | 0 | 0 | 0 | 0 |
| 0 | 0 | 0 | 0 | 0 |
| 0 | 0 | 0 | 0 | 0 |
| 0 | 0 | 0 | 0 | 0 |
| 0 | 0 | 0 | 0 | 0 |
| 0 | 0 | 0 | 0 | 0 |
| 0 | 0 | 0 | 0 | 0 |
| 0 | 0 | 0 | 0 | 0 |
| 0 | 0 | 0.003999411 | 0 | 0 |
| 0 | 0 | 0 | 0 | 0 |
| 0 | 0 | 0 | 0 | 0 |
| 0 | 0 | 0 | 0 | 0 |
| 0 | 0 | 0 | 0 | 0 |
| 0 | 0 | 0 | 0 | 0 |
| 0 | 0 | 0 | 0 | 0 |
| 0 | 0 | 0 | 0 | 0 |
| 0 | 0 | 0 | 0 | 0 |
| 0 | 0 | 0 | 0 | 0 |

| | | | | |
|---|---|---|---|---|
| 0 | 0 | 0 | 0 | 0 |
| 0 | 0 | 0 | 0 | 0 |
| 0 | 0 | 0 | 0 | 0 |
| 0 | 0 | 0 | 0 | 0 |
| 0 | 0 | 0 | 0 | 0 |
| 0 | 0 | 0 | 0 | 0 |
| 0 | 0 | 0 | 0 | 0 |
| 0 | 0 | 0 | 0 | 0 |
| 0 | 0 | 0 | 0 | 0 |
| 0 | 0 | 0 | 0 | 0 |
| 0.0010562 | 0.002510724 | 0 | 0 | 0.001011605 |
| 0 | 0 | 0 | 0 | 0 |
| 0 | 0 | 0.143482036 | 0 | 0 |
| 0 | 0 | 0 | 0 | 0 |
| 0 | 0 | 0 | 0 | 0 |
| 0 | 0 | 0 | 0 | 0 |
| 0 | 0 | 0 | 0 | 0 |
| 0 | 0 | 0 | 0 | 0 |
| 0 | 0 | 0 | 0 | 0 |
| 0 | 0 | 0 | 0 | 0 |
| 0.043593829 | 0 | 0 | 0.013889564 | 0.011293903 |
| 0 | 0 | 0 | 0 | 0 |
| 0 | 0 | 0.001265678 | 0 | 0 |
| 0.015787152 | 0 | 1.197309526 | 0.780529163 | 0 |
| 0 | 0 | 0 | 0 | 0 |
| 0 | 0 | 0 | 0 | 0 |
| 0 | 0 | 0 | 0 | 0 |
| 0 | 0 | 0 | 0 | 0 |
| 0 | 0 | 0 | 0 | 0 |
| 0 | 0 | 0 | 0 | 0 |
| 0 | 0 | 0.00662022 | 0 | 0 |
| 0 | 0 | 0 | 0 | 0 |
| 0 | 0 | 0 | 0 | 0 |
| 0 | 0 | 0 | 0 | 0 |

| PHL018-0178_V2 | PHL018-0259_V3 | PHL018-0184_V1 | PHL018-0276_V2 | PHL018-0203_V3 |
|---|---|---|---|---|
| EYCF | REF | EYCF | REF | EYCF |
| 0.102820977 | 0.02557801 | 0.347245036 | 0.420649433 | 0.124937238 |
| 0.002566149 | 0.105314044 | 3.578813107 | 4.868257987 | 4.869120383 |
| 0.111221846 | 0.342171785 | 7.960151191 | 3.192504045 | 0 |
| 0 | 0 | 0 | 0 | 0 |
| 0 | 0 | 0 | 0 | 0 |
| 0 | 0 | 0 | 0 | 0 |
| 0 | 0 | 0 | 0 | 0 |
| 0 | 0 | 0 | 0 | 0 |
| 0 | 0 | 0 | 0 | 0 |
| 0 | 0 | 0 | 0 | 0 |
| 0 | 0 | 0 | 0 | 0 |
| 0 | 0 | 0 | 0 | 0 |
| 0 | 0 | 0 | 0 | 0 |
| 4.73805777 | 1.093311341 | 2.311880397 | 0.475843292 | 0 |
| 0 | 0 | 0 | 0 | 0 |
| 0 | 0 | 0 | 0 | 0 |
| 0 | 0 | 0 | 0 | 0.039437128 |
| 0 | 0 | 0 | 0 | 0.001690081 |
| 0 | 0 | 0 | 0 | 0 |
| 0 | 0 | 0 | 0 | 0 |
| 0 | 0 | 0 | 0 | 0 |
| 0 | 0 | 0 | 0 | 0.003650178 |
| 0 | 0 | 0 | 0 | 0 |
| 0 | 0 | 0 | 0 | 0 |
| 0 | 0.336354559 | 0 | 0 | 0 |
| 0 | 0 | 0 | 0 | 0 |
| 0 | 0 | 0 | 0 | 0.00060624 |
| 0 | 0 | 0 | 0 | 0 |
| 0 | 0 | 0 | 0 | 0 |
| 0 | 0 | 0 | 0 | 0 |
| 0 | 0 | 0 | 0 | 0 |
| 0 | 0 | 0 | 0 | 0 |
| 0 | 0 | 0 | 0 | 0 |
| 0 | 0 | 0 | 0 | 0 |
| 0 | 0 | 0 | 0 | 0 |
| 0.266394304 | 4.310251114 | 0 | 9.766870692 | 0 |
| 0 | 0 | 0 | 0 | 0 |
| 0 | 0 | 0 | 0 | 0 |
| 0 | 0 | 0 | 0 | 0 |
| 0 | 0 | 0 | 0 | 0 |
| 0 | 0 | 0 | 0 | 0 |
| 0 | 0 | 0.071112768 | 0.104965116 | 0 |
| 0.039064378 | 0.080406955 | 1.075616668 | 2.596344828 | 0.001179721 |
| 0 | 0.195581057 | 0.003533553 | 0.022497488 | 0.002634366 |
| 0 | 0 | 0.000955338 | 0.001889696 | 0 |
| 0 | 0 | 0 | 0 | 0 |

| | | | | |
|---|---|---|---|---|
| 0 | 0.005588168 | 0 | 0 | 0 |
| 0 | 0 | 0.402898111 | 0.19853513 | 0.01726206 |
| 0.002504323 | 0 | 0 | 0 | 0.023216028 |
| 0.025565508 | 0.201449652 | 1.052675166 | 1.194195191 | 0.71977599 |
| 0.000311975 | 0.001040272 | 0 | 0.002967654 | 0.003524563 |
| 0 | 0 | 0 | 0 | 0 |
| 0 | 0.006013894 | 0.087037877 | 0 | 0 |
| 0 | 0.01339034 | 0 | 0 | 0 |
| 0 | 0 | 0 | 0 | 0 |
| 0 | 0 | 0 | 0 | 0 |
| 0 | 0.00102326 | 0.008668181 | 0 | 0 |
| 0.012388691 | 0.569031534 | 3.99712509 | 0.023790368 | 0 |
| 0 | 0 | 0 | 0 | 0 |
| 0.031946694 | 0.01978012 | 0 | 0.015520121 | 0.053002612 |
| 0.013350174 | 0 | 0 | 0 | 0 |
| 0 | 0 | 0 | 0 | 0 |
| 0 | 0 | 0.055275558 | 0.173757615 | 0.001264669 |
| 0 | 0 | 0 | 0 | 0 |
| 0 | 0 | 0 | 0 | 0 |
| 0 | 0 | 0 | 0 | 0 |
| 0.03967226 | 0 | 0.334174533 | 0.473549673 | 0.207901029 |
| 0 | 0.017883587 | 0.03762493 | 0.005560386 | 0.003244993 |
| 0 | 0.020946074 | 0 | 0 | 0 |
| 0 | 0 | 0 | 0 | 0 |
| 0 | 0 | 0 | 0.025744961 | 0 |
| 0 | 0 | 0 | 0 | 0 |
| 0.455979806 | 0 | 0 | 0 | 0 |
| 0 | 0 | 0 | 0 | 0 |
| 0 | 0 | 0.420473644 | 0.004377441 | 0 |
| 0 | 0 | 0 | 0 | 0 |
| 0.090124498 | 0 | 0 | 0.005573546 | 0.696637774 |
| 0 | 0 | 0 | 0 | 0 |
| 0 | 0 | 0 | 0 | 0 |
| 0 | 0 | 0 | 0 | 0 |
| 0 | 0 | 0 | 0 | 0 |
| 0 | 0 | 0 | 0 | 0 |
| 0 | 0 | 0 | 0 | 0 |
| 0 | 0 | 0.002884288 | 0 | 0 |
| 0 | 0 | 0 | 0 | 0 |
| 0 | 0 | 0 | 0 | 0 |
| 0 | 0 | 0 | 0 | 0 |
| 0 | 0 | 0 | 0 | 0 |
| 0 | 0 | 0 | 0 | 0 |
| 0 | 0 | 0 | 0 | 0 |
| 0 | 1.916205561 | 0 | 0 | 0 |
| 0 | 0 | 0 | 0 | 0 |
| 0 | 0.000978328 | 0.060953618 | 0 | 0 |
| 0 | 0 | 0 | 0 | 0 |

| | | | | |
|---|---|---|---|---|
| 0 | 0 | 0 | 0 | 0 |
| 0 | 0 | 0 | 0 | 0.130021196 |
| 0 | 0 | 0 | 0 | 0 |
| 0 | 0 | 0 | 0 | 0 |
| 0 | 0.035308075 | 0 | 0 | 0 |
| 0 | 0 | 0 | 0 | 0 |
| 0 | 0 | 0 | 0 | 0 |
| 0 | 0 | 0 | 0 | 0 |
| 0 | 0 | 0 | 0.004134268 | 0 |
| 0 | 0 | 0 | 0 | 0 |
| 0 | 0 | 0 | 0 | 0 |
| 0 | 0.00621593 | 0 | 0.34894572 | 0 |
| 0 | 0 | 0 | 0 | 0 |
| 0 | 0 | 0 | 0 | 0 |
| 0 | 0 | 0 | 0 | 0 |
| 0 | 0 | 0 | 0 | 0 |
| 0 | 0 | 0 | 0 | 0 |
| 0 | 0 | 0 | 0 | 0 |
| 0 | 0.016595662 | 0 | 0.261208385 | 0 |
| 0 | 0.010436416 | 0 | 0 | 0 |
| 0 | 0 | 0 | 0 | 0 |
| 0 | 0 | 0 | 0 | 0 |
| 0 | 0 | 0 | 0 | 0 |
| 0 | 0 | 0 | 0 | 0 |
| 0 | 0 | 0 | 0 | 0 |
| 0 | 0 | 0 | 0 | 0 |
| 0 | 0 | 0 | 0 | 0 |
| 0 | 0 | 0 | 0 | 0.001082651 |
| 0 | 0 | 0 | 0 | 0 |
| 0 | 0 | 0 | 0 | 0.103281405 |
| 0 | 0.017977645 | 0 | 0 | 0.01265176 |
| 0 | 0.01410796 | 0 | 0.000602029 | 0 |
| 0 | 0 | 0 | 0 | 0 |
| 0 | 0 | 0 | 0 | 0 |
| 0 | 0 | 0 | 0 | 0 |
| 0 | 0 | 0 | 0 | 0 |
| 0 | 0 | 0 | 0 | 0 |
| 0 | 0 | 0 | 0 | 0 |
| 0 | 0.001519133 | 0 | 0 | 0 |
| 0 | 0 | 0 | 0 | 0 |
| 0 | 0 | 0 | 0 | 0 |
| 0 | 0 | 0 | 0 | 0 |
| 0 | 0 | 0 | 0 | 0 |
| 0 | 0 | 0 | 0 | 0 |
| 0 | 0 | 0 | 0 | 0 |
| 0 | 0 | 0 | 0 | 0 |
| 0 | 0 | 0 | 0 | 0 |
| 0 | 0 | 0 | 0 | 0 |

| | | | | |
|---|---|---|---|---|
| 0 | 0 | 0 | 0 | 0 |
| 0 | 0 | 0 | 0 | 0 |
| 0 | 0 | 0 | 0 | 0 |
| 0 | 0 | 0 | 0 | 0 |
| 0 | 0.060492117 | 0 | 0.001397328 | 0 |
| 0 | 0 | 0 | 0 | 0 |
| 0 | 0 | 0 | 0 | 0 |
| 0 | 0 | 0 | 0 | 0 |
| 0 | 0 | 0 | 0 | 0 |
| 0 | 0 | 0 | 0 | 0 |
| 0 | 0 | 0 | 0 | 0 |
| 0 | 0 | 0 | 0 | 0 |
| 0 | 0 | 0 | 0 | 0 |
| 0 | 0 | 0 | 0 | 0.000799361 |
| 0 | 0 | 0 | 0 | 0 |
| 0 | 0.56265781 | 0 | 0 | 0.091423077 |
| 0 | 0 | 0 | 0 | 0 |
| 0 | 0 | 0 | 0 | 0 |
| 0 | 0 | 0 | 0 | 0 |
| 0 | 0 | 0 | 0 | 0 |
| 0 | 0 | 0 | 0 | 0.023494464 |
| 0 | 0 | 0 | 0 | 0 |
| 0 | 0 | 0 | 0 | 0 |
| 0 | 0 | 0 | 0 | 0 |
| 0 | 0 | 0 | 0 | 0 |
| 0 | 0 | 0 | 0 | 0 |
| 0 | 0.142442043 | 0 | 0 | 0 |
| 0 | 0 | 0 | 0 | 0 |
| 0 | 0 | 0 | 0 | 0 |
| 0 | 0 | 0 | 0 | 0 |
| 0 | 0 | 0 | 0 | 0.019034257 |
| 0 | 0 | 0.207185206 | 0 | 0 |
| 0 | 0 | 0 | 0 | 0 |
| 0 | 0 | 0 | 0 | 0 |
| 0 | 0 | 0 | 0 | 0 |
| 0 | 0 | 0 | 0 | 0 |
| 0 | 0 | 0 | 0 | 0 |
| 0 | 0 | 0 | 0 | 0 |
| 0 | 0 | 0 | 0 | 0 |
| 0 | 0 | 0 | 0 | 0 |
| 0 | 0 | 0 | 0 | 0 |
| 0 | 0 | 0 | 0 | 0 |
| 0 | 0 | 0 | 0 | 0 |
| 0 | 0 | 0 | 0 | 0 |
| 0 | 0 | 0 | 0 | 0 |
| 0 | 0 | 0 | 0 | 0 |
| 0 | 0 | 0 | 0 | 0 |
| 0 | 0 | 0 | 0.007380599 | 0 |

| | | | | |
|---|---|---|---|---|
| 0 | 0 | 0 | 0 | 0 |
| 0 | 0 | 0 | 0 | 0 |
| 0 | 0 | 0 | 0 | 0 |
| 0 | 0 | 0 | 0 | 0 |
| 0 | 0 | 0 | 0 | 0 |
| 0 | 0 | 0 | 0 | 0 |
| 0 | 0 | 0 | 0 | 0 |
| 0 | 0 | 0 | 0 | 0 |
| 0 | 0 | 0 | 0 | 0 |
| 0 | 0 | 0 | 0 | 0 |
| 0 | 0 | 0 | 0 | 0 |
| 0 | 0 | 0 | 0 | 0 |
| 0 | 0 | 0.006496522 | 0 | 0 |
| 0 | 0 | 0 | 0 | 0 |
| 0 | 0 | 0 | 0 | 0 |
| 0 | 0 | 0 | 0 | 0 |
| 0 | 0 | 0 | 0 | 0 |
| 0 | 0 | 0 | 0 | 0 |
| 0 | 0 | 0 | 0 | 0 |
| 0 | 0 | 0 | 0 | 0 |
| 0.002978762 | 0.098712992 | 0.020105272 | 0.041011818 | 0.028044045 |
| 0 | 0 | 0 | 0 | 0 |
| 0.000290409 | 0 | 0 | 0 | 0 |
| 0 | 0 | 0 | 0 | 0 |
| 0 | 0 | 0 | 0 | 0 |
| 0 | 0 | 0 | 0 | 0 |
| 0 | 0.00589164 | 0.000975736 | 0.001447534 | 0 |
| 0 | 0 | 0 | 0 | 0 |
| 0 | 0 | 0 | 0 | 0 |
| 0.000618665 | 0 | 0.308219006 | 0 | 0 |
| 0.106829942 | 0 | 0.006322183 | 0.070463859 | 0 |
| 0 | 0 | 0 | 0 | 0 |
| 0 | 0 | 0 | 0 | 0 |
| 0 | 0.003559682 | 0 | 0.017721932 | 0 |
| 0.007752564 | 0.001196794 | 0.129725484 | 0.016172409 | 0.001048675 |
| 0 | 0 | 0 | 0 | 0 |
| 0 | 0 | 0 | 0 | 0 |
| 0 | 0 | 0 | 0 | 0 |
| 0 | 0 | 0.001056627 | 0 | 0 |
| 0 | 0 | 0 | 0 | 0 |
| 0 | 0 | 0 | 0 | 0 |
| 0 | 0 | 0 | 0 | 0 |
| 0 | 0 | 0 | 0 | 0 |
| 0 | 0 | 0 | 0 | 0 |
| 0 | 0.008045668 | 0 | 0 | 0 |
| 0 | 0.027347473 | 0 | 0 | 0 |
| 0 | 0 | 0 | 0 | 0.001021893 |
| 0 | 0.000710723 | 0.004855351 | 0 | 0.000298926 |

| | | | | |
|---|---|---|---|---|
| 0 | 0 | 0 | 0 | 0 |
| 0 | 0 | 0 | 0 | 0 |
| 0 | 0 | 0 | 0 | 0 |
| 0 | 0 | 0.0017281 | 0 | 0 |
| 0 | 0 | 0 | 0 | 0 |
| 0 | 0 | 0 | 0 | 0 |
| 0 | 0 | 0 | 0.027342572 | 0 |
| 0 | 0 | 0 | 0.004694054 | 0 |
| 0 | 0 | 0 | 0 | 0 |
| 0 | 0 | 0 | 0 | 0 |
| 0 | 0 | 0 | 0 | 0 |
| 0 | 0 | 0 | 0 | 0 |
| 0 | 0 | 0 | 0 | 0 |
| 0 | 0 | 0 | 0 | 0 |
| 0 | 0 | 0.008573913 | 0 | 0 |
| 0 | 0 | 0 | 0 | 0 |
| 0 | 0 | 0 | 0 | 0 |
| 0 | 0 | 0.005790843 | 0 | 0 |
| 0 | 0 | 0.005530403 | 0.012892817 | 0 |
| 0 | 0 | 0 | 0 | 0 |
| 0.102333316 | 0.001705927 | 0.058270771 | 0 | 0.031054406 |
| 0 | 0 | 0 | 0 | 0 |
| 0 | 0 | 0 | 0 | 0 |
| 0 | 0 | 0 | 0 | 0 |
| 0 | 0.002541889 | 0 | 0.004579844 | 0.001113649 |
| 0 | 0.001233307 | 0 | 0 | 0 |
| 0 | 0 | 0 | 0 | 0 |
| 0 | 0 | 0 | 0 | 0 |
| 0 | 0 | 0.036181651 | 0.024857851 | 0 |
| 0 | 0 | 0 | 0 | 0 |
| 0.175375677 | 0.05142907 | 9.11965775 | 0.062080875 | 0.070151562 |
| 0 | 0 | 0 | 0 | 0 |
| 0.003157325 | 0 | 0 | 0 | 0.065190347 |
| 0 | 0.012171534 | 0 | 0 | 0 |
| 0 | 0 | 0 | 0 | 0 |
| 0 | 0 | 0 | 0 | 0 |
| 0 | 0 | 0 | 0 | 0 |
| 0 | 0 | 0 | 0 | 0 |
| 0 | 0 | 0 | 0 | 0 |
| 0 | 0 | 0 | 0 | 0 |
| 0 | 0 | 0 | 0 | 0 |
| 0.003537365 | 0 | 0.072063512 | 0.026781092 | 0.003865289 |
| 0 | 0 | 0 | 0 | 0.003454628 |
| 0 | 0 | 0 | 0 | 0 |
| 0 | 0 | 0 | 0 | 0 |
| 0 | 0 | 0 | 0 | 0 |
| 0 | 0 | 0 | 0 | 0 |
| 0 | 0 | 0 | 0 | 0 |

| | | | | |
|---|---|---|---|---|
| 0 | 0.00190454 | 0 | 0 | 0 |
| 0 | 0 | 0 | 0 | 0 |
| 0 | 0.188447527 | 0 | 0.026305827 | 0.002532264 |
| 0 | 0 | 0 | 0 | 0 |
| 0 | 0 | 0.00095185 | 0 | 2.804464344 |
| 0 | 0 | 0 | 0 | 0 |
| 0 | 0 | 0 | 0 | 0 |
| 0 | 0 | 0 | 0 | 0 |
| 0 | 0 | 0 | 0 | 0 |
| 0 | 0 | 0 | 0 | 0 |
| 0 | 0 | 0 | 0 | 0 |
| 0 | 0 | 0 | 0 | 0 |
| 0 | 0 | 0 | 0 | 0 |
| 0 | 0.013086096 | 0.004242642 | 0 | 0.001282696 |
| 0 | 0.002929047 | 0 | 0.001731648 | 0.000866207 |
| 0 | 0.004507901 | 0 | 0 | 0 |
| 0 | 0.002503039 | 0.001709968 | 0.001691193 | 0.001096628 |
| 0 | 0 | 0 | 0 | 0 |
| 0 | 0 | 0 | 0 | 0 |
| 0 | 0 | 0 | 0 | 0 |
| 0 | 0 | 0 | 0 | 0 |
| 0 | 0.001318059 | 0.005402646 | 0.000763332 | 0 |
| 0 | 0 | 0 | 0 | 0 |
| 0 | 0 | 0 | 0 | 0.018722565 |
| 0 | 0 | 0 | 0 | 0.000793657 |
| 0 | 0 | 0 | 0 | 0 |
| 0 | 0.0711268 | 0 | 0 | 0 |
| 0 | 0 | 0 | 0 | 0 |
| 0 | 0 | 0 | 0 | 0 |
| 0 | 0 | 0.002604992 | 0 | 0 |
| 0 | 0 | 0 | 0 | 0 |
| 0 | 0 | 0 | 0 | 0 |
| 0 | 0 | 0 | 0 | 0 |
| 0 | 0 | 0 | 0 | 0 |
| 0 | 0 | 0 | 0 | 0 |
| 0 | 0 | 0.004110863 | 0.302388406 | 0 |
| 0 | 0 | 0 | 0 | 0 |
| 0 | 0.019398917 | 0.00387878 | 0.002237778 | 0.00062188 |
| 0 | 0 | 0 | 0 | 0 |
| 0 | 0 | 0 | 0 | 0 |
| 0 | 0 | 0 | 0 | 0 |
| 0 | 0 | 0 | 0 | 0 |
| 0 | 0 | 0 | 0 | 0 |
| 0 | 0 | 0 | 0 | 0 |
| 0 | 0 | 0 | 0 | 0 |
| 0 | 0 | 0 | 0 | 0 |
| 0 | 0 | 0 | 0 | 0 |
| 0 | 0 | 0 | 0 | 0.027569047 |

| | | | | |
|---|---|---|---|---|
| 0 | 1.138540136 | 0 | 0 | 0 |
| 0 | 0 | 0 | 0 | 0 |
| 0 | 0 | 0.001347345 | 0 | 0 |
| 0 | 0 | 0 | 0 | 0 |
| 0.000535566 | 0.005555924 | 0.008404467 | 0.016892509 | 0 |
| 0 | 0 | 0 | 0 | 0 |
| 0 | 0 | 0 | 0 | 0 |
| 0 | 0.032542087 | 0 | 0.013154761 | 0.002778345 |
| 0 | 0 | 0 | 0 | 0 |
| 18.06351087 | 0.007453741 | 0.078731271 | 0 | 9.772149724 |
| 0 | 0 | 0 | 0 | 0 |
| 0.001901656 | 0 | 0.003910758 | 0.202614152 | 0 |
| 0 | 0.012692781 | 0.034684629 | 0.005190706 | 0.000263413 |
| 0 | 0 | 0.014208929 | 0.020907997 | 0.121083796 |
| 0 | 0.002576494 | 0.005632476 | 0 | 0 |
| 0 | 0 | 0 | 0 | 0 |
| 0 | 0 | 0 | 0 | 0 |
| 0 | 0 | 0 | 0 | 0.006975579 |
| 1.252349031 | 0 | 0 | 0 | 0.669559395 |
| 0 | 0 | 0 | 0 | 0 |
| 0 | 0 | 0 | 0 | 0 |
| 0 | 0 | 0 | 0 | 0 |
| 0 | 0 | 0 | 0.00419176 | 0 |
| 0 | 0.064255314 | 0 | 0 | 0 |
| 0 | 0 | 0 | 0 | 0 |
| 0 | 0.001945244 | 0 | 0 | 0 |
| 0 | 0 | 0 | 0 | 0 |
| 0 | 0 | 0 | 0.00858261 | 0 |
| 0.053326036 | 0 | 0 | 0 | 1.142065765 |
| 0 | 0 | 0 | 0 | 0 |
| 0.000901427 | 0 | 0.00370757 | 0 | 0.058052968 |
| 0 | 0 | 0 | 0 | 0 |
| 0.01427496 | 0 | 0 | 0 | 0.335097085 |
| 0.003158591 | 0 | 0 | 0.044347167 | 0 |
| 0 | 0 | 0 | 0 | 0 |
| 0 | 0 | 0 | 0 | 0 |
| 0.052018888 | 0 | 0 | 0 | 0 |
| 0 | 0 | 0 | 0 | 0 |
| 0 | 0 | 0 | 0 | 0.000728306 |
| 0.81748161 | 0 | 0 | 0 | 0 |
| 0 | 0 | 0 | 0 | 0 |
| 0 | 0 | 0 | 0.005556828 | 0 |
| 0 | 0 | 0 | 0 | 0.064743281 |
| 0 | 0 | 0.002141227 | 0.004447206 | 0 |
| 0 | 0 | 0 | 0 | 0 |
| 0 | 1.332843275 | 0 | 0 | 0 |
| 0 | 0 | 0 | 0.005230254 | 0.004324136 |
| 0 | 0 | 0 | 0.016615758 | 0.180776289 |

| | | | | |
|---|---|---|---|---|
| 0 | 0 | 0 | 0 | 0.008414409 |
| 0 | 0 | 0 | 0 | 0 |
| 0 | 0 | 0 | 0 | 0 |
| 0 | 0 | 0 | 0 | 0 |
| 0 | 0 | 0 | 0 | 0 |
| 0.000514256 | 0 | 0.322021431 | 0.06874347 | 0.010217342 |
| 0 | 0 | 0 | 0 | 0 |
| 0 | 0 | 0 | 0 | 0 |
| 0 | 2.067912484 | 0 | 0 | 0 |
| 0 | 0 | 0 | 0 | 0 |
| 0.00035787 | 0 | 0.010265709 | 0.005972348 | 0.000929443 |
| 0 | 0 | 0.078668341 | 0 | 0 |
| 0 | 0 | 0 | 0 | 0 |
| 0 | 0.027558176 | 0.003137758 | 0.001692712 | 0 |
| 0.012999587 | 0 | 0 | 0 | 0 |
| 0 | 0 | 0 | 0 | 0.143385705 |
| 0 | 0 | 0 | 0 | 0 |
| 0.009102996 | 0 | 0 | 0.001647283 | 0.100299787 |
| 0 | 0 | 0 | 0 | 0 |
| 0 | 0 | 0 | 0 | 0.022706764 |
| 0 | 0 | 0 | 0 | 0 |
| 0.089065747 | 0.845737446 | 0 | 0.334595134 | 0 |
| 0 | 0.00059435 | 0 | 0.031590627 | 0 |
| 0 | 0 | 0 | 0 | 0 |
| 0 | 0 | 0 | 0 | 0 |
| 0 | 0 | 0 | 0 | 0 |
| 0.961315197 | 0 | 0 | 0 | 0 |
| 0.002855059 | 0 | 0 | 0 | 0.020082331 |
| 0 | 0 | 0 | 0 | 0 |
| 0.02015362 | 0.113839083 | 0.303135446 | 1.07889184 | 0.332714665 |
| 0 | 0.002124549 | 0 | 0 | 0 |
| 0 | 0 | 0 | 0 | 0 |
| 0 | 0 | 0 | 0 | 0 |
| 0 | 0 | 0 | 0 | 0 |
| 0 | 0 | 0 | 0.001528672 | 0 |
| 0 | 0 | 0 | 0 | 0 |
| 0.027653951 | 1.127923816 | 0 | 0.370010223 | 0 |
| 0 | 0 | 0 | 0 | 0 |
| 0.869993074 | 0 | 0 | 0 | 0 |
| 0 | 0 | 0 | 0 | 0 |
| 0.00750474 | 0 | 0.205428269 | 0.013684005 | 0.008572399 |
| 0.017211715 | 0.003837947 | 1.996691935 | 6.196982938 | 3.835782979 |
| 0 | 0 | 0 | 0 | 0 |
| 0 | 0 | 0 | 0 | 0 |
| 0 | 0 | 0 | 0 | 0 |
| 0 | 0 | 0 | 0 | 0 |
| 0.003111808 | 0.00054043 | 0.008368491 | 0.010467472 | 0.049627479 |
| 0 | 0 | 0 | 0 | 0 |

| | | | | |
|---|---|---|---|---|
| 0 | 0.002889465 | 0 | 0 | 0 |
| 0 | 0 | 0 | 0 | 0 |
| 0 | 0 | 0.006206925 | 0.009003535 | 0.001114567 |
| 0 | 0 | 0 | 0.016301266 | 0 |
| 0 | 0 | 0 | 0.00100642 | 0 |
| 0 | 0 | 0 | 0 | 0 |
| 0 | 0 | 0.000863157 | 0 | 0 |
| 0 | 0 | 0 | 0.002019797 | 0 |
| 0 | 0.004278394 | 0 | 0 | 0 |
| 0.01141945 | 0 | 0 | 0 | 0.366788635 |
| 0 | 0 | 0 | 0 | 0 |
| 0 | 0 | 0 | 0 | 0 |
| 0 | 0 | 0 | 0 | 0 |
| 0 | 0.000603237 | 0.002197894 | 0 | 0.004545775 |
| 0 | 0 | 0 | 0 | 0 |
| 0 | 0.000471419 | 0 | 0.109994526 | 0.003882905 |
| 0 | 0.00348346 | 0.002855698 | 0.012003457 | 0.015658489 |
| 0.014735932 | 0 | 0.002023911 | 0 | 0.000556271 |
| 0 | 0 | 0 | 0 | 0.004067155 |
| 0 | 0 | 0 | 0 | 0 |
| 0.009360979 | 0 | 0 | 0.006229082 | 0.019387938 |
| 0 | 0 | 0 | 0.004368121 | 0 |
| 0 | 0 | 0 | 0 | 0 |
| 0 | 0 | 0 | 0 | 0 |
| 0 | 0.017580401 | 0 | 0 | 0 |
| 0.475360456 | 0.004359402 | 0 | 0 | 0 |
| 0.000358082 | 0 | 0.01238655 | 0 | 0.016507357 |
| 0 | 0 | 0 | 0 | 0 |
| 0 | 0 | 0 | 0 | 0 |
| 0.001225585 | 0 | 0 | 0 | 0 |
| 0 | 0.004203914 | 0.071223908 | 0.025904432 | 0.014675567 |
| 0 | 0.020306766 | 0.042131907 | 0.003726523 | 0 |
| 0.033759845 | 0.013128559 | 0.002935264 | 0.006773748 | 0 |
| 0 | 0 | 0.007478256 | 0.055933353 | 0 |
| 0 | 0.003174073 | 0 | 0 | 0 |
| 0 | 0 | 0 | 0 | 0.001899319 |
| 0 | 0.01359554 | 0 | 0 | 0 |
| 0.058305988 | 0 | 0 | 0.065224359 | 0.01712453 |
| 0 | 0.009528285 | 0.42550339 | 2.88279675 | 0.585245634 |
| 0.000819347 | 0 | 0 | 0 | 0 |
| 0 | 0 | 0 | 0 | 0 |
| 0 | 0 | 0 | 0 | 0.059880449 |
| 0 | 0 | 0.008621139 | 0.002046355 | 0.001857699 |
| 0 | 0 | 0 | 0.036317457 | 0 |
| 0.126504886 | 0 | 0 | 0 | 1.405698649 |
| 0 | 0 | 0 | 0 | 0 |
| 0 | 0.00337571 | 0 | 0 | 0.045552013 |
| 0 | 0 | 0 | 0 | 0.000450685 |

| | | | | |
|---|---|---|---|---|
| 0 | 0 | 0 | 0 | 0 |
| 0.001241395 | 0.020007121 | 1.303329913 | 0.403983922 | 0.176518881 |
| 0 | 0 | 0 | 0 | 0 |
| 0 | 0 | 0 | 0 | 0 |
| 0 | 0 | 0 | 0 | 0.011979798 |
| 0 | 0 | 0 | 0 | 0 |
| 0.093598875 | 0 | 0 | 0.005462321 | 0 |
| 0 | 0 | 0 | 0 | 0 |
| 0 | 0 | 0 | 0.008818642 | 0 |
| 0 | 0 | 0 | 0 | 0 |
| 0 | 0 | 0 | 0 | 0.005031287 |
| 0 | 0.204909798 | 0.010433701 | 0.012038996 | 0.104384114 |
| 0 | 0.306148272 | 0 | 0 | 0 |
| 0 | 0 | 0 | 0 | 0 |
| 0 | 0.066665686 | 0.002679006 | 0 | 0 |
| 0.013723315 | 0 | 0 | 0 | 0.000896471 |
| 0 | 0.003399995 | 0.007579431 | 0.017168737 | 0.000846721 |
| 17.08784834 | 0 | 0.079665359 | 0.005336418 | 0.298605189 |
| 0 | 0 | 0 | 0 | 0 |
| 0.010178357 | 0 | 0.069867118 | 0.034855744 | 0 |
| 0.001494258 | 0.04004509 | 0.015380471 | 0.047380378 | 0.003298689 |
| 0 | 0 | 0 | 0 | 0 |
| 0 | 0 | 0 | 0 | 0 |
| 0 | 0 | 0 | 0 | 0 |
| 0 | 0 | 0.002118202 | 0 | 0 |
| 0 | 0 | 0 | 0.001829437 | 0 |
| 0 | 0.002185797 | 0 | 0.192482305 | 0.143262733 |
| 0.187934117 | 0.01882928 | 0 | 0.108402968 | 0 |
| 0 | 0 | 0 | 0 | 0 |
| 0 | 0 | 0 | 0 | 0 |
| 0 | 0 | 0 | 0 | 0 |
| 0 | 0 | 0 | 0 | 0 |
| 0 | 0 | 0 | 0 | 0.003379989 |
| 0 | 0 | 0 | 0 | 0.000312398 |
| 0 | 0.005298548 | 0 | 0 | 0 |
| 0.477907289 | 0.12016678 | 0.374179568 | 0.095097061 | 0.02530561 |
| 0 | 0 | 0 | 0 | 0 |
| 0 | 0 | 0 | 0 | 0 |
| 0.004048997 | 0.002419586 | 0.00264473 | 0.118033043 | 0 |
| 0 | 0 | 0 | 0 | 0 |
| 0.132447718 | 0 | 0 | 0 | 0.312908503 |
| 0 | 0 | 0 | 0 | 0 |
| 0.00420482 | 0 | 0 | 0.028399987 | 0.056637446 |
| 0 | 0.000709481 | 0 | 0 | 0 |
| 0.104556155 | 0 | 0 | 0 | 0 |
| 0 | 0.761774907 | 0 | 0 | 0 |
| 0 | 0 | 0 | 0 | 0 |
| 0 | 0 | 0.003447215 | 0 | 0.032075986 |

| | | | | |
|---|---|---|---|---|
| 0 | 0 | 0 | 0.189498571 | 0 |
| 0 | 0 | 0 | 0 | 0 |
| 0 | 0.060609682 | 0 | 0 | 0 |
| 0 | 0.004279319 | 0 | 0 | 0.037077022 |
| 0 | 0 | 0 | 0 | 0 |
| 0 | 0 | 0 | 0 | 0 |
| 0 | 0 | 0 | 0.001075966 | 0 |
| 0 | 0.001071927 | 0.002301495 | 0 | 0 |
| 0.008182281 | 0.367663528 | 0.135877738 | 2.640756161 | 0 |
| 0 | 0.063489338 | 0 | 0 | 0 |
| 0 | 0.00963826 | 0 | 0 | 0 |
| 0 | 0.004100048 | 0 | 0.003545882 | 0.005690702 |
| 0 | 0 | 0 | 0 | 0 |
| 0 | 0.721587286 | 0 | 0 | 0 |
| 0.267676176 | 0.025036937 | 0 | 0.021762652 | 0 |
| 0 | 0 | 0 | 0 | 0 |
| 0 | 0.004236777 | 0.035697382 | 0 | 0 |
| 0 | 0 | 0 | 0 | 0 |
| 0 | 0 | 0 | 0 | 0.110680588 |
| 0 | 0.001318123 | 0 | 0 | 0 |
| 0 | 0.077794617 | 0 | 0 | 0 |
| 0 | 0.093642207 | 0 | 0 | 0 |
| 0 | 0.015705874 | 0 | 0 | 0 |
| 0.095564344 | 0.059054029 | 0.725820463 | 0.181268392 | 0.111303882 |
| 0 | 0.001381929 | 0 | 0.003319852 | 0.201009057 |
| 0 | 1.140327616 | 0 | 0 | 0 |
| 0 | 0.579518241 | 0 | 0 | 0 |
| 0 | 1.966856229 | 0 | 0 | 0 |
| 0 | 0 | 0 | 0 | 0 |
| 0 | 0 | 0 | 0 | 0.007772208 |
| 0 | 0 | 0 | 0.062480534 | 0 |
| 0 | 2.061168659 | 0 | 0.022938319 | 0 |
| 0 | 0.001456127 | 0 | 0.010986215 | 0.001467299 |
| 0 | 0 | 0 | 0 | 0 |
| 0 | 0.578964973 | 0 | 0 | 0 |
| 0 | 0.484910186 | 0 | 0 | 0 |
| 0 | 0.130769958 | 0 | 0 | 0 |
| 0 | 0 | 0 | 0 | 0 |
| 0.010137647 | 0.004951054 | 0 | 0 | 0.114082278 |
| 0 | 0 | 0 | 0 | 0 |
| 0.003736891 | 0.048457799 | 0.147679419 | 0 | 0.106950903 |
| 0 | 0.058893587 | 0 | 0 | 0 |
| 15.43448527 | 1.737892772 | 0.178448963 | 0.448620264 | 0 |
| 0 | 0.015761376 | 0 | 0 | 0 |
| 0 | 0 | 0 | 0 | 0 |
| 0 | 0 | 0 | 0 | 0 |
| 0 | 0 | 0 | 0 | 0 |
| 0 | 0 | 0.05297403 | 0.000766718 | 0.011335396 |

| | | | | |
|---|---|---|---|---|
| 0 | 0 | 0 | 0 | 0 |
| 0 | 0.154382752 | 0 | 0 | 0 |
| 0.046585836 | 0 | 0.899453402 | 0.238135311 | 0.749199811 |
| 0 | 0.013522523 | 0 | 0 | 0 |
| 0 | 0.030403857 | 0 | 0 | 0 |
| 0 | 0.002427984 | 0 | 0 | 0 |
| 0 | 0 | 0 | 0 | 0 |
| 0 | 0 | 0 | 0.01193068 | 0 |
| 0 | 0 | 0 | 0 | 0 |
| 0 | 0 | 0 | 0 | 0 |
| 0.068128092 | 0.003204112 | 0.206730426 | 0.159960308 | 0.081606481 |
| 1.341727628 | 0.502921884 | 0.824665888 | 0.506680605 | 0 |
| 0 | 0 | 0 | 0 | 0 |
| 0 | 0 | 0.007874538 | 0.025441048 | 0.001414017 |
| 0.054836308 | 0 | 0.497262409 | 0.034368894 | 0.18441655 |
| 0 | 0.25942359 | 0 | 0 | 0 |
| 0 | 0 | 0 | 0 | 0.00607757 |
| 0 | 0 | 0 | 0 | 0 |
| 0 | 0.158642395 | 0 | 0 | 0 |
| 0.002928668 | 0.007624775 | 0 | 0 | 0.001695975 |
| 0.045826067 | 0.04968208 | 0.114766252 | 0.019354509 | 0.023923699 |
| 1.322573716 | 0.001572057 | 0.386411218 | 0 | 45.15599157 |
| 9.180279956 | 0.577203013 | 0.52717202 | 0.144342169 | 9.443861028 |
| 0 | 0.047601595 | 0 | 0 | 0 |
| 0 | 0.026613154 | 0 | 0 | 0 |
| 0 | 0.300888228 | 0 | 0.006764481 | 0 |
| 0 | 0.001070668 | 7.623005261 | 0 | 0 |
| 0 | 0 | 0 | 0 | 0 |
| 0 | 0 | 0 | 0.000919843 | 0 |
| 0 | 0 | 0 | 0 | 0 |
| 0 | 0.012506461 | 0 | 0 | 0 |
| 0 | 0.126907587 | 0 | 0.184244076 | 0 |
| 0.109740324 | 0.003656341 | 0 | 0.008893538 | 0 |
| 0 | 0.023142416 | 0.091240234 | 0.008477514 | 0 |
| 0.000195405 | 0 | 0.020277955 | 0.00260882 | 0.003489028 |
| 0.004564058 | 0.00862064 | 0 | 0.00179218 | 0.005055186 |
| 0.008558293 | 0.001281142 | 0 | 0.001298416 | 0 |
| 0 | 0 | 0 | 0 | 0 |
| 0.00178029 | 0.05315542 | 0.181037259 | 0.663243337 | 0 |
| 0 | 0 | 0 | 0 | 0 |
| 0 | 0.030655589 | 0 | 0 | 0 |
| 0 | 0 | 0 | 0 | 0 |
| 0 | 0 | 0 | 0 | 0 |
| 0 | 0.00334014 | 0 | 0 | 0 |
| 0.033780107 | 0.015502513 | 0.204063626 | 0.029890246 | 0.057350474 |
| 0 | 0 | 0 | 0 | 0.183455492 |
| 0 | 0 | 0 | 0 | 0 |
| 0.000738635 | 0 | 0.049854481 | 0.015100294 | 0 |

| | | | | |
|---|---|---|---|---|
| 0 | 0 | 0 | 0 | 0 |
| 0 | 0 | 0 | 0 | 0 |
| 0 | 0 | 0 | 0 | 0 |
| 0 | 0 | 0 | 0.001959641 | 0 |
| 0 | 0 | 0 | 0 | 0 |
| 0 | 0.267399991 | 0 | 0 | 0 |
| 0 | 1.269252558 | 0 | 0 | 0 |
| 0 | 0.072207265 | 0.184147608 | 0.098175561 | 0.04209385 |
| 0 | 0 | 0 | 0 | 0 |
| 0 | 0 | 0 | 0 | 0 |
| 0.001065609 | 0 | 0.135837801 | 0 | 0.010000908 |
| 0 | 0 | 0 | 0 | 0 |
| 0 | 0.023569306 | 0 | 0 | 0 |
| 0 | 0 | 0.180985312 | 0 | 0 |
| 0 | 0 | 0 | 0 | 0 |
| 0 | 0 | 0 | 0 | 0 |
| 0 | 0.022823169 | 0 | 0 | 0 |
| 0 | 0.001792727 | 0.008981232 | 0 | 0.004712563 |
| 0 | 0.009589324 | 0 | 0 | 0 |
| 0 | 0 | 0 | 0 | 0 |
| 0 | 0.003450843 | 0 | 0 | 0 |
| 0 | 0 | 0 | 0 | 0 |
| 0 | 0 | 0 | 0.037705586 | 0.001811083 |
| 0 | 0 | 0 | 0 | 0 |
| 0 | 0 | 0 | 0 | 0 |
| 0 | 0 | 0 | 0 | 0 |
| 0 | 0 | 0 | 0 | 0 |
| 0 | 0 | 0 | 0 | 0 |
| 0 | 0 | 0 | 0 | 0 |
| 0 | 1.211110897 | 0 | 0.097302713 | 0 |
| 0 | 0 | 0 | 0 | 0.005796157 |
| 0 | 0 | 0 | 0 | 0 |
| 0 | 0 | 0 | 0 | 0 |
| 0 | 0 | 0 | 0 | 0 |
| 0 | 0 | 0 | 0 | 0 |
| 0 | 0 | 0 | 0 | 0 |
| 0 | 0.021461518 | 0 | 0 | 0 |
| 0.004346275 | 0 | 0 | 0.002258108 | 0 |
| 0 | 0.008594769 | 0 | 0 | 0 |
| 0 | 0 | 0 | 0 | 0 |
| 0 | 0.012827927 | 0.266020392 | 0 | 0 |
| 0 | 1.262086564 | 0 | 0 | 0 |
| 0 | 0 | 0 | 0 | 0 |
| 0 | 0 | 0 | 0.028108862 | 0 |
| 0 | 0.141274453 | 0 | 0 | 0 |
| 0 | 0 | 0 | 0 | 0 |
| 0 | 0.022373216 | 0 | 0 | 0 |
| 0 | 0.035517417 | 0 | 0 | 0 |

| | | | | |
|---|---|---|---|---|
| 0 | 0 | 0 | 0 | 0 |
| 0 | 0 | 0 | 0 | 0 |
| 0 | 0.112330328 | 0 | 0 | 0 |
| 0 | 0.107741052 | 0 | 0 | 0 |
| 0 | 0.593216096 | 0 | 0 | 0 |
| 0 | 0 | 0 | 0 | 0 |
| 0 | 0 | 0 | 0 | 0 |
| 0 | 0 | 0 | 0 | 0 |
| 0.04037629 | 0 | 0.786075414 | 0.034823619 | 0.012116552 |
| 0 | 0.005061625 | 0 | 0 | 0 |
| 0 | 0.016179389 | 0 | 0 | 0 |
| 0 | 0 | 0 | 0 | 0 |
| 0 | 0 | 0 | 0 | 0 |
| 0 | 0 | 0 | 0 | 0 |
| 0 | 1.375237112 | 0.013782933 | 1.062247335 | 0 |
| 0 | 0 | 0 | 0 | 0.034722478 |
| 0 | 0 | 0 | 0 | 0.037900919 |
| 0 | 0 | 0 | 0 | 0 |
| 0 | 0 | 0 | 0 | 0 |
| 0 | 1.289777294 | 0 | 0 | 0 |
| 0 | 0.456580355 | 0 | 0.001082425 | 0 |
| 0 | 0.025450851 | 0 | 0 | 0 |
| 0 | 0 | 0 | 0 | 0.767710283 |
| 0 | 0 | 0 | 0 | 0 |
| 0.001862014 | 0.005174037 | 0.176397235 | 0.133175885 | 0 |
| 0 | 0.409878834 | 0 | 0.009414287 | 0 |
| 0 | 0.053334348 | 0 | 0 | 0 |
| 0 | 0.000611765 | 0 | 0 | 0 |
| 0 | 0 | 0 | 0.071315706 | 0 |
| 0 | 0.003409599 | 0 | 0 | 0 |
| 0 | 0 | 0 | 0 | 0 |
| 0 | 0.303697871 | 0 | 0 | 0 |
| 0 | 0 | 0 | 0 | 0 |
| 0 | 0 | 0.009348831 | 0.003200601 | 0.001937023 |
| 0 | 0 | 0.001184917 | 0 | 0.000911886 |
| 0.001406424 | 0 | 0.042507826 | 0.000621297 | 0.000322296 |
| 0.014599283 | 0 | 0 | 0 | 0 |
| 0 | 0.477457488 | 0 | 0 | 3.189377599 |
| 0 | 0 | 0.006347047 | 0 | 0 |
| 0 | 0 | 0 | 0 | 0 |
| 0 | 0 | 0 | 0 | 0 |
| 0 | 0.004171094 | 0 | 0 | 0 |
| 0 | 0.137595189 | 0 | 5.625562405 | 0 |
| 0 | 2.966687693 | 0 | 0 | 0 |
| 0 | 2.95241262 | 0 | 0 | 0 |
| 0 | 0.00174231 | 0 | 0.011968218 | 0.002213682 |
| 0 | 0 | 0 | 0 | 0 |
| 0 | 0.075175457 | 0.10776987 | 0 | 0 |

| | | | | |
|---|---|---|---|---|
| 0.006610901 | 0.002414824 | 0 | 0.047782266 | 0.063569453 |
| 0 | 0.714465916 | 0 | 0 | 0 |
| 0 | 0 | 0 | 0 | 0 |
| 0 | 0 | 0 | 0 | 0 |
| 0.001019734 | 0 | 0.004223496 | 0.002784748 | 0.000812577 |
| 0 | 0 | 0 | 0 | 0 |
| 0 | 0.03650247 | 0 | 0 | 0 |
| 0 | 0 | 0 | 0 | 0 |
| 0 | 0 | 0 | 0 | 0 |
| 0 | 0.369385267 | 0 | 0 | 0 |
| 0 | 0.001182641 | 0 | 0 | 0 |
| 0.007098533 | 0.497321266 | 0 | 0 | 0 |
| 0 | 0.008941266 | 0 | 1.119732452 | 0 |
| 0 | 0 | 0 | 0 | 0 |
| 0.006968721 | 0 | 0.988563017 | 1.178725375 | 0.01263436 |
| 0 | 0 | 0 | 0 | 0 |
| 0 | 0 | 0 | 0 | 0.006464653 |
| 0 | 0 | 0 | 0 | 0 |
| 0 | 0.002959334 | 0 | 0.000705703 | 0.000366082 |
| 0 | 0.547805622 | 0 | 0.000782511 | 0 |
| 0 | 0.037615585 | 0 | 0 | 0 |
| 0 | 0 | 0 | 0 | 0 |
| 0 | 0.155387714 | 0.007169452 | 6.50494605 | 0 |
| 0 | 0 | 0 | 0 | 0 |
| 0 | 0 | 0 | 0 | 0 |
| 0 | 0 | 0 | 0 | 0 |
| 0 | 0 | 0 | 0 | 0 |
| 0 | 0 | 0 | 0 | 0 |
| 0 | 0 | 0 | 0 | 0 |
| 0 | 0 | 0 | 0 | 0 |
| 0 | 0 | 0 | 0 | 0 |
| 0 | 0.033503675 | 0.075003057 | 0.067214316 | 0.020053204 |
| 0 | 1.155898604 | 0 | 0 | 0 |
| 0 | 0.057681235 | 0.056821059 | 0.024879861 | 0.025722495 |
| 0.002250797 | 0 | 0.050816719 | 0.000901502 | 0.000438424 |
| 0.010793766 | 0.053750842 | 0.009694144 | 0.007553948 | 0 |
| 0 | 0.00222072 | 0 | 0 | 0 |
| 0 | 0.013237098 | 0 | 0 | 0 |
| 0 | 0 | 0 | 0 | 0 |
| 0 | 0 | 0 | 0 | 0 |
| 0 | 0.441361456 | 0 | 0 | 1.90355459 |
| 0 | 0.060568154 | 0 | 0.002605191 | 0 |
| 0 | 0 | 0 | 0 | 0 |
| 0 | 0 | 0 | 0 | 0 |
| 0.053302941 | 0.014786808 | 0.094956029 | 0.05062 | 0.033255662 |
| 0 | 0 | 0 | 0 | 0 |
| 0 | 0.004700041 | 0 | 0 | 0 |
| 0 | 0 | 0 | 0.0018899 | 0.002532652 |

| | | | | |
|---|---|---|---|---|
| 0 | 0 | 0 | 0 | 0 |
| 0 | 0 | 0 | 0 | 0 |
| 0.006329238 | 0 | 0 | 0 | 0 |
| 0 | 0 | 0 | 0 | 0 |
| 0.004093124 | 0 | 0 | 0 | 0 |
| 0 | 0 | 0 | 0 | 0.034661866 |
| 0 | 0.07257541 | 0.042168872 | 0 | 0 |
| 0 | 0 | 0 | 0 | 0 |
| 0 | 0 | 0 | 0 | 0 |
| 0 | 0.007580009 | 0 | 0 | 0 |
| 0 | 0 | 0 | 0 | 0.025549507 |
| 0 | 0 | 0.037560296 | 0 | 0 |
| 0 | 0 | 0 | 0 | 0 |
| 0 | 0.292899504 | 0 | 0 | 0 |
| 0 | 0 | 0 | 0 | 0.004577174 |
| 0 | 0 | 0 | 0 | 0 |
| 0 | 0.008775247 | 0 | 0 | 0 |
| 0 | 0.03815479 | 0 | 0 | 0 |
| 0.000490072 | 2.726100177 | 0.082363302 | 0.007360749 | 0 |
| 0 | 0 | 0 | 0 | 0 |
| 0 | 0.001080745 | 0.002320429 | 0 | 0 |
| 0 | 0 | 0 | 0 | 0 |
| 0 | 0 | 0 | 0 | 0 |
| 0 | 0 | 0 | 0 | 0 |
| 0 | 0 | 0 | 0 | 0 |
| 0.006357692 | 0 | 0.093625088 | 0.00124014 | 0.005468218 |
| 0 | 0 | 0 | 0 | 0 |
| 0 | 0.227263756 | 0.015341575 | 0 | 0 |
| 0 | 0 | 0 | 0 | 0 |
| 0 | 0.789484473 | 1.266205621 | 4.449431275 | 0 |
| 0 | 0 | 0 | 0 | 0 |
| 0 | 0 | 0 | 0 | 0 |
| 0 | 0 | 0 | 0 | 0 |
| 0 | 0 | 0 | 0 | 0 |
| 0.000519153 | 0 | 0.16359517 | 0 | 0.001432589 |
| 0 | 0 | 0 | 0 | 0 |
| 0 | 0 | 0 | 0 | 0 |
| 0 | 0.002182552 | 0.006958116 | 0.001228878 | 0.000478108 |
| 0 | 0 | 0 | 0 | 0 |
| 0 | 0.010431031 | 0 | 0 | 0 |
| 0.503486827 | 1.108937007 | 13.56666516 | 9.770504653 | 0.024487678 |
| 0.073378305 | 0.021434977 | 0.27176781 | 0.160118878 | 0.055576426 |
| 0 | 0.899305027 | 0 | 0 | 0 |
| 0 | 0.242117108 | 0 | 0 | 0 |
| 0 | 1.801101767 | 0 | 0 | 0 |
| 0 | 0 | 0 | 0 | 0 |
| 0 | 0 | 0 | 0 | 0 |
| 0.002103894 | 0.001154796 | 0.027414503 | 0.001170367 | 0.003415079 |

| | | | | |
|---|---|---|---|---|
| 0 | 0 | 0.126501737 | 0 | 0 |
| 0 | 0.001039328 | 0 | 0.046347022 | 0.001717316 |
| 0 | 0 | 0 | 0 | 0 |
| 0 | 0 | 0 | 0 | 0 |
| 0 | 2.467334211 | 0 | 3.910366296 | 0 |
| 0 | 0 | 0 | 0 | 0 |
| 0 | 0 | 0 | 0 | 0 |
| 0 | 0 | 0 | 0 | 0 |
| 16.16306991 | 0 | 0.212085341 | 0.288290199 | 0.27319622 |
| 0 | 0 | 0 | 0 | 0 |
| 0.236340511 | 0 | 0.044005238 | 0 | 0 |
| 0 | 0 | 0 | 0 | 0 |
| 0 | 0.094207752 | 0 | 0.120090749 | 0.120598955 |
| 0 | 0.003192556 | 0.040469963 | 0.009826646 | 0 |
| 0 | 0.760120461 | 0 | 0.293726646 | 0 |
| 0 | 0.082086711 | 0 | 0.204935337 | 0.682162841 |
| 0 | 0.027626623 | 0.017405386 | 0 | 0 |
| 0 | 0 | 0 | 0 | 0 |
| 0 | 0.095061514 | 0 | 0 | 0 |
| 0 | 0 | 0 | 0 | 0 |
| 0 | 0.002939797 | 0 | 0 | 0 |
| 0 | 0 | 0 | 0 | 0 |
| 0 | 0 | 0 | 0 | 0 |
| 0 | 0 | 0 | 0 | 0 |
| 0 | 0 | 0 | 0 | 0 |
| 0 | 0 | 0 | 0 | 0 |
| 0 | 0.106499682 | 0 | 0.00198378 | 0 |
| 0 | 0 | 0.096294183 | 0.001220986 | 0.000870903 |
| 0 | 0 | 0 | 0 | 0 |
| 0 | 0.001185582 | 0 | 0 | 0 |
| 0 | 0 | 0 | 0 | 0 |
| 0 | 0 | 0 | 0.002039796 | 0 |
| 0 | 0.877374955 | 0 | 0 | 0 |
| 0 | 0.051936384 | 0 | 0.009172752 | 0 |
| 0 | 0.015496757 | 0 | 0 | 0 |
| 0 | 0 | 0.012376931 | 0 | 0 |
| 0 | 0 | 0.001216415 | 0 | 0 |
| 0 | 0 | 0 | 0 | 0 |
| 0 | 0 | 0 | 0 | 0 |
| 0 | 0.62881529 | 0 | 0 | 0 |
| 0.273765176 | 0.398548435 | 0 | 0.905581523 | 0 |
| 0 | 0 | 0 | 0 | 0 |
| 0 | 0 | 0 | 0 | 0 |
| 0 | 0 | 1.008824881 | 0 | 0 |
| 0 | 0 | 0 | 0 | 0 |
| 0 | 0.00399236 | 0 | 0 | 0 |
| 0 | 0 | 0 | 0 | 0 |
| 0 | 0 | 0 | 0 | 0 |

| | | | | |
|---|---|---|---|---|
| 0 | 0.044024271 | 0 | 0 | 0 |
| 0 | 0 | 0 | 0 | 0 |
| 0 | 0 | 0 | 0 | 0 |
| 0 | 0 | 0 | 0 | 0 |
| 0 | 0.119642584 | 0 | 0 | 0 |
| 0 | 0 | 0 | 0 | 0 |
| 0.00041325 | 0.08436253 | 0.002510322 | 0 | 0 |
| 0 | 0 | 0 | 0 | 0 |
| 0 | 0.001188765 | 0 | 0 | 0 |
| 0 | 0 | 0 | 0 | 0.019086322 |
| 0 | 0.012625463 | 0 | 0 | 0 |
| 0 | 0.00227623 | 0 | 0 | 0 |
| 0 | 0 | 0 | 0 | 0 |
| 0 | 0 | 0 | 0 | 0 |
| 0 | 0 | 0 | 0 | 0 |
| 0 | 0 | 0 | 0 | 0 |
| 0 | 0 | 0 | 0 | 0 |
| 0 | 0 | 0 | 0 | 0 |
| 0 | 0 | 0 | 0 | 0 |
| 0 | 0 | 0 | 0 | 0 |
| 0 | 0.01224446 | 3.661230078 | 0 | 0 |
| 0 | 0.016829283 | 0 | 0 | 0 |
| 0 | 0.036336488 | 0 | 0 | 0 |
| 0 | 0 | 0.024808071 | 0.008886939 | 0.00120263 |
| 0 | 0 | 0 | 0 | 0 |
| 0 | 0.002698723 | 0 | 0.000911704 | 0 |
| 0 | 0 | 0 | 0 | 0 |
| 0 | 0.018303819 | 0 | 0 | 0 |
| 0 | 0 | 0 | 0 | 0 |
| 0 | 2.818768551 | 0.014056941 | 0 | 0.331785413 |
| 0 | 0.023411874 | 0 | 0 | 0 |
| 0 | 0 | 0 | 0 | 0 |
| 0 | 0 | 0 | 0 | 0 |
| 5.652171852 | 0 | 0 | 0 | 0 |
| 0 | 0 | 0 | 0 | 0 |
| 0 | 0.876154478 | 0 | 0 | 0 |
| 0.01351945 | 0.005366702 | 0 | 0.006526876 | 0 |
| 0 | 0 | 0 | 0 | 0 |
| 0 | 0.018957125 | 0 | 0 | 0 |
| 0 | 0 | 0 | 0 | 0 |
| 0 | 0 | 0 | 0 | 0.007123455 |
| 0 | 0 | 0 | 0 | 0 |
| 0 | 0 | 0 | 0 | 0 |
| 0 | 0.101569104 | 0 | 0 | 0 |
| 0 | 0.126471541 | 0 | 0 | 0 |
| 0 | 0 | 0 | 0 | 0 |
| 0 | 0 | 0 | 0 | 0 |
| 0 | 0.031032814 | 0 | 0 | 0 |

| | | | | |
|---|---|---|---|---|
| 0 | 0.005076677 | 1.030360446 | 0.01205727 | 0.00501453 |
| 0 | 0.678288936 | 0 | 0 | 0 |
| 0 | 0 | 0 | 0 | 0 |
| 0 | 0 | 0 | 0 | 0 |
| 0 | 0 | 0 | 0 | 0 |
| 0 | 0 | 0 | 0 | 0 |
| 0 | 0 | 0 | 0 | 0 |
| 0 | 0.206450554 | 0 | 1.978039718 | 0 |
| 0.06022201 | 0.129188027 | 1.46751831 | 3.363879292 | 0.001777336 |
| 0 | 0 | 0.280216617 | 0.124485775 | 0 |
| 0 | 0 | 0 | 0 | 0 |
| 0 | 0 | 0 | 0 | 0 |
| 0 | 0.033921235 | 0 | 0 | 0.007654004 |
| 0.004921545 | 0.531652026 | 0 | 0 | 0.144173296 |
| 0 | 0 | 0 | 0 | 0 |
| 0 | 0 | 0 | 0 | 0 |
| 0 | 0 | 0 | 0 | 0.000590312 |
| 0 | 0 | 0 | 0 | 0 |
| 0 | 0 | 0 | 0 | 0 |
| 0 | 0.024673397 | 0 | 0.001695327 | 0 |
| 0 | 0 | 0 | 0 | 0 |
| 0 | 0 | 0 | 0 | 0 |
| 0 | 0 | 0 | 0 | 0 |
| 0 | 0 | 0 | 0 | 0 |
| 0 | 0.12007878 | 0 | 0 | 0 |
| 0 | 0 | 0 | 0 | 0 |
| 0 | 0 | 0 | 0 | 0 |
| 0 | 0 | 0 | 0 | 0 |
| 0 | 0 | 0 | 0 | 0 |
| 0 | 0 | 0 | 0 | 0 |
| 0.089750161 | 0 | 0 | 0 | 0 |
| 0 | 0 | 0 | 0 | 0 |
| 0 | 0 | 0 | 0 | 0 |
| 0 | 0 | 0 | 0 | 0 |
| 0 | 0 | 0 | 0 | 0 |
| 0 | 0 | 0 | 0 | 0 |
| 0 | 0.014909973 | 0 | 0 | 0 |
| 0 | 0 | 0 | 0 | 0.054444866 |
| 0 | 0 | 0 | 0 | 0 |
| 0 | 0 | 1.48543819 | 0.83137839 | 0 |
| 0 | 0 | 0 | 0 | 0 |
| 0.152415396 | 0.945436557 | 0 | 1.30177316 | 0 |
| 0 | 0.004757761 | 0 | 0 | 0 |
| 0 | 0 | 0 | 0 | 0 |
| 0 | 0 | 0 | 0 | 0 |
| 0 | 0 | 0 | 0.007763746 | 0 |
| 0 | 0 | 0 | 0 | 0 |
| 0 | 0 | 0 | 0 | 0 |

| | | | | |
|---|---|---|---|---|
| 0 | 0 | 0 | 0 | 0 |
| 0 | 0 | 0 | 0 | 0 |
| 0 | 0 | 0 | 0 | 0 |
| 0 | 0.029544509 | 0 | 0 | 0 |
| 0.008278594 | 0.104245902 | 0.240039964 | 0.515993758 | 0 |
| 0 | 0.743174451 | 0 | 0 | 0 |
| 0 | 0 | 0 | 0 | 0 |
| 0.008239803 | 0 | 0.053475851 | 0.01375106 | 0.006070192 |
| 0 | 0 | 0 | 0 | 0 |
| 0 | 0.103520338 | 0 | 0 | 0 |
| 0 | 0 | 0 | 0 | 0 |
| 0 | 0.461974273 | 0 | 0 | 0 |
| 0 | 0 | 0 | 0 | 0 |
| 0 | 0 | 0 | 0 | 0 |
| 0 | 0 | 0 | 0.002233906 | 0 |
| 0 | 0 | 0 | 0 | 0 |
| 0 | 0 | 0 | 0 | 0 |
| 0 | 0 | 0 | 0 | 0 |
| 0 | 0 | 0 | 0 | 0 |
| 0 | 0 | 0 | 0 | 0 |
| 0 | 0 | 0 | 0 | 0 |
| 0 | 0 | 0 | 0 | 0 |
| 0 | 0.002961192 | 0 | 0 | 0 |
| 0.000365686 | 0 | 0.007867421 | 0 | 0.00338345 |
| 0.04790893 | 1.565115979 | 0 | 0 | 0 |
| 0 | 0 | 0 | 0 | 0 |
| 0.000334461 | 0 | 0 | 0 | 0.016612841 |
| 0 | 0 | 0 | 0 | 0 |
| 0 | 0 | 0.004404006 | 0 | 0 |
| 0 | 0 | 0 | 0 | 0 |
| 0 | 0 | 0 | 0 | 0 |
| 0 | 0 | 0 | 0.001408272 | 0 |
| 0 | 0 | 0 | 0 | 0 |
| 0 | 0 | 0 | 0 | 0 |
| 0 | 0 | 0 | 0 | 0 |
| 0 | 0 | 0 | 0 | 0.004095141 |
| 0 | 0 | 0 | 0 | 0 |
| 0 | 0.193167852 | 0 | 0 | 0 |
| 0 | 0.016130638 | 0 | 0 | 0 |
| 0 | 0 | 0 | 0 | 0 |
| 0 | 0 | 0 | 0 | 0 |
| 0 | 0.038615756 | 0 | 0 | 0 |
| 0 | 0 | 0 | 0 | 0 |
| 0 | 0.037524503 | 0 | 0 | 0 |
| 0 | 0 | 0 | 0 | 0 |
| 0 | 0 | 0 | 0 | 0 |
| 0 | 0 | 0 | 0 | 0 |
| 0 | 0.015207083 | 0 | 0 | 0 |

| | | | | |
|---|---|---|---|---|
| 0 | 0.620830779 | 0 | 0 | 0 |
| 0 | 0 | 0 | 0 | 0 |
| 0 | 0 | 0 | 0 | 0 |
| 0 | 0 | 0 | 0 | 0.007391005 |
| 0 | 0 | 0 | 0 | 0 |
| 0.005921554 | 0 | 0 | 0 | 0 |
| 0 | 0 | 0 | 0 | 0 |
| 0 | 0 | 0 | 0 | 0 |
| 0 | 0 | 0 | 0 | 0 |
| 0 | 0 | 0 | 0 | 0 |
| 0 | 0 | 0 | 0 | 0 |
| 0 | 0 | 0 | 0 | 0.000902869 |
| 0 | 0 | 0 | 0 | 0 |
| 0 | 0 | 0 | 0 | 0 |
| 0 | 0.0362888 | 0 | 0 | 0 |
| 0 | 0 | 0 | 0 | 0 |
| 0 | 0 | 0 | 0 | 0 |
| 0 | 0 | 0 | 0 | 0 |
| 0 | 0 | 0 | 0 | 0 |
| 0 | 0 | 0 | 0 | 0 |
| 0.003368452 | 0 | 1.717145189 | 0.006836921 | 0 |
| 0 | 0 | 0 | 0 | 0 |
| 0.005288894 | 0.003833846 | 0.01425964 | 0.009497986 | 0.03280974 |
| 0 | 0.046395933 | 0 | 0.072442099 | 0 |
| 0 | 0.004017504 | 0 | 0 | 0 |
| 0 | 0 | 0 | 0 | 0 |
| 0 | 0.007268091 | 0 | 0 | 0 |
| 0 | 0 | 0 | 0 | 0 |
| 0 | 0 | 0 | 0 | 0 |
| 0 | 0 | 0 | 0 | 0 |
| 0 | 0 | 0 | 0 | 0 |
| 0 | 0.010708137 | 0 | 0 | 0 |
| 0.001399769 | 0.010372213 | 0.167880038 | 0.012398847 | 0.012688971 |
| 0 | 0.024212411 | 0 | 0 | 0 |
| 0.011268777 | 0.008742632 | 0 | 1.196350019 | 0 |
| 0 | 0 | 0 | 0.000648884 | 0.001346429 |
| 0 | 0 | 0 | 0 | 0 |
| 0 | 0 | 0 | 0 | 0 |
| 0.009894081 | 0 | 0.031366598 | 0 | 0 |
| 0 | 0 | 0 | 0 | 0 |
| 0 | 0.280520911 | 0 | 0 | 0 |
| 0 | 0 | 0 | 0 | 0 |
| 0.00093488 | 0.058233591 | 0.097312237 | 0.299847572 | 0 |
| 0 | 0 | 0 | 0 | 0 |
| 0 | 0 | 0 | 0 | 0 |
| 0 | 0.053468381 | 0 | 0.003949489 | 0 |
| 0 | 0 | 0 | 0 | 0 |
| 0 | 0 | 0 | 0 | 0 |

| | | | | |
|---|---|---|---|---|
| 0 | 0 | 0 | 0 | 0 |
| 0 | 0 | 0 | 0 | 0 |
| 0 | 0 | 0 | 0 | 0.000694793 |
| 0 | 0 | 0 | 0 | 0 |
| 0 | 0 | 0 | 0 | 0 |
| 0 | 0 | 0 | 0 | 0 |
| 0 | 0 | 0 | 0 | 0 |
| 0 | 0 | 0.049937767 | 0.006959423 | 0 |
| 0 | 0.010147845 | 0 | 0 | 0 |
| 0 | 0.00070832 | 0 | 0 | 0 |
| 0 | 0 | 0 | 0 | 0 |
| 0 | 0 | 0 | 0 | 0.022051004 |
| 0 | 0 | 0 | 0 | 0 |
| 0 | 0 | 0 | 0 | 0 |
| 0 | 0 | 0 | 0.001547032 | 0.010758777 |
| 0.026051941 | 0 | 0.002380487 | 0 | 0.23119468 |
| 0 | 0 | 0 | 0 | 0 |
| 0 | 0 | 0 | 0 | 0 |
| 0 | 0 | 0 | 0 | 0 |
| 0 | 1.128437761 | 0 | 1.90608825 | 0 |
| 0 | 0 | 0 | 0 | 0 |
| 0 | 0.003647433 | 0 | 0 | 0 |
| 0 | 0 | 0 | 0 | 0 |
| 0 | 0 | 0 | 0 | 0 |
| 0 | 0 | 0 | 0 | 0 |
| 0 | 0 | 0 | 0 | 0 |
| 0 | 0 | 0 | 0 | 0 |
| 0 | 0.02043133 | 0 | 0 | 0 |
| 0 | 0 | 0 | 0 | 0 |
| 0 | 0 | 0 | 0 | 0 |
| 0 | 0.244413397 | 0 | 1.16554272 | 0 |
| 0 | 0 | 0 | 0 | 0 |
| 0 | 0.002511596 | 0 | 0 | 0 |
| 0 | 0 | 0 | 0 | 0 |
| 0 | 0 | 0 | 0 | 0 |
| 0 | 0 | 0 | 0 | 0 |
| 0 | 0 | 0 | 0 | 0 |
| 0 | 0 | 0 | 0 | 0 |
| 0.320401235 | 0.012288926 | 0 | 0.004313289 | 0 |
| 0 | 0 | 0 | 0 | 0 |
| 0 | 0 | 0 | 0 | 0 |
| 0 | 3.075038672 | 0 | 0 | 0 |
| 0 | 0.107365334 | 0 | 0 | 0 |
| 0 | 0 | 0 | 0 | 0.002152487 |
| 0 | 0 | 0 | 0 | 0 |
| 0 | 0 | 0 | 0 | 0 |
| 0.087132164 | 0 | 1.58032126 | 0.005302287 | 0 |
| 0 | 0 | 0 | 0 | 0 |

| | | | | |
|---|---|---|---|---|
| 0 | 0 | 0 | 0 | 0 |
| 0 | 0 | 0 | 0 | 0 |
| 0 | 0 | 0 | 0 | 0 |
| 0 | 0.003676466 | 0 | 0 | 0 |
| 0 | 0 | 0.082288256 | 0.001080328 | 0 |
| 0 | 0 | 0 | 0 | 0 |
| 0 | 0 | 0 | 0 | 0 |
| 0 | 0 | 0 | 0 | 0 |
| 0 | 0 | 0 | 0 | 0 |
| 0 | 0 | 0 | 0 | 0.000716317 |
| 0 | 0.013195584 | 0 | 0 | 0 |
| 0.010085924 | 0.004254782 | 0.015418056 | 0.008499313 | 0.011670883 |
| 0 | 0 | 0 | 0 | 0.001426446 |
| 0 | 0 | 0 | 0 | 0 |
| 0.004413845 | 0 | 0 | 0.000571506 | 0 |
| 0 | 0 | 0 | 0 | 0 |
| 0.001843312 | 0 | 0 | 0.000961321 | 0 |
| 0 | 0 | 0 | 0 | 0.004073617 |
| 0 | 0.000700379 | 0 | 0.098239502 | 0 |
| 0 | 0.001828964 | 0 | 0.0009886 | 0.004134725 |
| 0 | 0 | 0 | 0 | 0 |
| 0 | 0.014913477 | 0 | 0 | 0 |
| 0 | 0 | 0 | 0 | 0 |
| 0 | 0 | 0 | 0 | 0 |
| 0 | 0 | 0 | 0 | 0 |
| 0 | 0 | 0 | 0 | 0 |
| 0.014082833 | 0 | 0 | 0.002376632 | 0.019109524 |
| 0 | 0 | 0 | 0 | 0.030687467 |
| 0 | 0 | 0 | 0 | 0 |
| 0 | 0 | 0 | 0 | 0.001250437 |
| 0.000717467 | 0 | 0.012938741 | 0.001796024 | 0 |
| 0 | 0 | 0 | 0 | 0 |
| 0 | 0 | 0 | 0 | 0.004012089 |
| 0 | 0 | 0 | 0 | 0 |
| 0 | 0.003985428 | 0 | 0 | 0 |
| 0 | 0 | 0 | 0 | 0 |
| 0 | 0 | 0 | 0 | 0 |
| 0.001575222 | 0.062213451 | 0.336663153 | 0.041482815 | 0.012027798 |
| 0 | 0.004352216 | 0 | 0 | 0 |
| 0 | 0 | 0 | 0 | 0 |
| 0.000503226 | 0 | 0 | 0 | 0 |
| 0 | 0.006284578 | 0 | 0 | 0 |
| 0 | 0 | 0 | 0 | 0 |
| 0 | 0 | 0 | 0.003508389 | 0 |
| 0 | 0 | 0 | 0 | 0 |
| 0 | 0 | 0 | 0 | 0 |
| 0 | 0 | 0 | 0 | 0 |
| 0 | 0.004358473 | 0 | 0 | 0 |

| | | | | |
|---|---|---|---|---|
| 0 | 0 | 0 | 0 | 0 |
| 0 | 0.017456621 | 0 | 0.014593397 | 0 |
| 0 | 0 | 0 | 0 | 0 |
| 0.000293758 | 0 | 0.011400722 | 0 | 0.001621236 |
| 0 | 0 | 0 | 0 | 0 |
| 0.000820472 | 0 | 0.005340031 | 0 | 0 |
| 0 | 0.013554282 | 0 | 0 | 0 |
| 0 | 0 | 0 | 0 | 0 |
| 0 | 0 | 0 | 0 | 0 |
| 0 | 0.016978568 | 0 | 0 | 0 |
| 0 | 0.021883815 | 0 | 0 | 0 |
| 0 | 0 | 0 | 0 | 0.000825283 |
| 0 | 0.011958238 | 0.089423632 | 0.002432583 | 0 |
| 0 | 0.007805009 | 0 | 0 | 0 |
| 0 | 0 | 0 | 0 | 0 |
| 0 | 0.00379784 | 0 | 0 | 0 |
| 0 | 0 | 0 | 0 | 0 |
| 0 | 0 | 0 | 0 | 0 |
| 0 | 0 | 0 | 0 | 0 |
| 0 | 0 | 0 | 0 | 0 |
| 0 | 0 | 0 | 0 | 0 |
| 0 | 0 | 0 | 0 | 0 |
| 0 | 0 | 0 | 0.032946699 | 0.000534095 |
| 0 | 0.062936017 | 0 | 0.034558434 | 0 |
| 0 | 0 | 0 | 0 | 0 |
| 0 | 0.005164317 | 0 | 0 | 0 |
| 0 | 0 | 0 | 0 | 0 |
| 0 | 0 | 0 | 0 | 0 |
| 0 | 0.054413682 | 0 | 0 | 0 |
| 0 | 0 | 0 | 0 | 0 |
| 0 | 0.023031754 | 0.032500346 | 0.097706021 | 0 |
| 0 | 0 | 0 | 0.002170035 | 0 |
| 0 | 0 | 0 | 0 | 0 |
| 0 | 0 | 0 | 0 | 0 |
| 0 | 0.002848914 | 0 | 0 | 0 |
| 0 | 0 | 0 | 0 | 0.008196689 |
| 0 | 0 | 0 | 0 | 0 |
| 0 | 0 | 0 | 0 | 0 |
| 0 | 0 | 0.009165849 | 0.60542644 | 0 |
| 0 | 0 | 0 | 0 | 0 |
| 0 | 0 | 0 | 0 | 0 |
| 0 | 0 | 0 | 0 | 0 |
| 0 | 0 | 0 | 0 | 0 |
| 0 | 0 | 0 | 0 | 0 |
| 0 | 0 | 0 | 0 | 0 |
| 0 | 0.00660499 | 0.496922673 | 0.184964597 | 0.050610171 |
| 0.184099078 | 0 | 0 | 0 | 0.073953471 |
| 0 | 0 | 0 | 0 | 0 |

| | | | | |
|---|---|---|---|---|
| 0 | 0 | 0 | 0 | 0 |
| 0 | 0 | 0 | 0 | 0 |
| 0.180212527 | 0.036901294 | 0 | 0 | 0 |
| 0 | 0.002546078 | 0 | 0 | 0 |
| 0 | 0 | 0 | 0 | 0.002992203 |
| 0 | 0.000517523 | 0 | 0 | 0 |
| 0 | 0 | 0 | 0.004607315 | 0.264867082 |
| 0 | 0 | 0 | 0 | 0 |
| 0 | 0 | 0 | 0 | 0 |
| 0 | 0 | 0 | 0 | 0 |
| 0 | 0.001295798 | 0.00318684 | 0 | 0.000817507 |
| 0 | 0 | 0.002378981 | 0 | 0 |
| 0 | 0 | 0 | 0 | 0 |
| 0 | 0 | 0 | 0 | 0 |
| 0 | 0 | 0 | 0 | 0 |
| 0 | 0 | 0 | 0 | 0 |
| 0 | 0 | 0 | 0 | 0 |
| 0 | 0 | 0 | 0 | 0 |
| 0 | 0 | 0 | 0 | 0 |
| 0 | 0 | 0 | 0 | 0.000632424 |
| 0 | 0 | 0 | 0 | 0 |
| 0 | 0 | 0 | 0 | 0 |
| 0 | 0 | 0 | 0 | 0 |
| 0 | 0 | 0 | 0 | 0 |
| 0 | 0 | 0 | 0 | 0 |
| 0 | 0 | 0 | 0 | 0 |
| 0 | 0 | 0 | 0 | 0 |
| 0 | 0.012171244 | 0 | 0.021329882 | 0 |
| 0 | 0 | 0 | 0 | 0 |
| 0 | 0 | 0 | 0 | 0 |
| 0 | 0.000590531 | 0.07846619 | 0.002194476 | 0 |
| 0 | 0 | 0 | 0 | 0 |
| 0.012214388 | 0.010284999 | 2.928547269 | 0 | 0 |
| 0 | 0 | 0.014120826 | 0.001893665 | 0.000276282 |
| 0 | 0 | 0 | 0 | 0 |
| 0 | 0 | 0 | 0 | 0 |
| 0 | 0 | 0 | 0 | 0 |
| 0 | 0 | 0 | 0 | 0 |
| 0 | 0 | 0 | 0 | 0 |
| 0 | 0 | 0 | 0 | 0 |
| 0 | 0 | 0 | 0 | 0 |
| 0.00182703 | 0 | 0.362241189 | 0.06765088 | 0.640362063 |
| 0 | 0.032814886 | 0 | 0 | 0 |
| 0 | 0 | 0 | 0 | 0 |
| 0 | 0.854367712 | 0 | 0 | 0 |
| 0 | 0.007696406 | 0.008480404 | 0.030194244 | 0 |
| 0 | 0 | 0 | 0 | 0.015746165 |
| 0 | 0 | 0 | 0 | 0 |

| | | | | |
|---|---|---|---|---|
| 0 | 0 | 0 | 0 | 0 |
| 0.002526182 | 0.019978806 | 0 | 0.009485638 | 0 |
| 0 | 0.031957459 | 0.042227585 | 0.004545734 | 0.002210709 |
| 0 | 0 | 0 | 0 | 0 |
| 0.004168502 | 0.044821733 | 0.014450012 | 0.011342343 | 0.007237094 |
| 0 | 0 | 0 | 0.009053266 | 0.148193147 |
| 0 | 0 | 0 | 0 | 0 |
| 0 | 0 | 0 | 0 | 0 |
| 0 | 0 | 0 | 0 | 0 |
| 0 | 0 | 0 | 0 | 0 |
| 0 | 0 | 0 | 0 | 0 |
| 0 | 0 | 0 | 0 | 0 |
| 0 | 0 | 0 | 0 | 0 |
| 0 | 0 | 0 | 0 | 0 |
| 0 | 0 | 0 | 0 | 0 |
| 0 | 0 | 0 | 0 | 0 |
| 0 | 0 | 0 | 0 | 0 |
| 0 | 0 | 0 | 0 | 0 |
| 0 | 0 | 0 | 0 | 0 |
| 0 | 0 | 0 | 0 | 0.115306613 |
| 0 | 0 | 0 | 0 | 0 |
| 0 | 0 | 0 | 0 | 0.007913257 |
| 0 | 0 | 0 | 0 | 0 |
| 0 | 0 | 0 | 0 | 0 |
| 0 | 0.002252544 | 0.003787915 | 0.00491705 | 0 |
| 0 | 0.137703448 | 0 | 0.001413267 | 0 |
| 0 | 0 | 0 | 0 | 0 |
| 0 | 0.00067634 | 0 | 0.077685394 | 0 |
| 0 | 0 | 0 | 0 | 0 |
| 0 | 0 | 0 | 0 | 0 |
| 0 | 0 | 0 | 0 | 0 |
| 0.000742635 | 0 | 0.00229737 | 0.000619676 | 0.001928733 |
| 0 | 0 | 0 | 0 | 0 |
| 0 | 0 | 0 | 0 | 0 |
| 0 | 0 | 0 | 0 | 0 |
| 0 | 0 | 0 | 0 | 0 |
| 0 | 0 | 0 | 0 | 0 |
| 0 | 0 | 0 | 0.006096086 | 0 |
| 0 | 0.005222518 | 0 | 0.174506467 | 1.337820919 |
| 0 | 0 | 0 | 0 | 0 |
| 0 | 0 | 0 | 0 | 0 |
| 0 | 0 | 0.001291832 | 0.009454593 | 0 |
| 0 | 0.002798996 | 0.820552556 | 0.009219393 | 0.004690568 |
| 0 | 0 | 0 | 0 | 0 |
| 0 | 0 | 0 | 0 | 0 |
| 0 | 0 | 0 | 0 | 0 |
| 0.001300186 | 0.013113369 | 0 | 0.001084913 | 0.002532583 |
| 0 | 0 | 0 | 0 | 0 |

| | | | | |
|---|---|---|---|---|
| 0 | 0 | 0 | 0 | 0 |
| 0.008294122 | 0 | 0.127111877 | 0.023272327 | 0.001153986 |
| 0 | 0 | 0 | 0 | 0 |
| 0 | 0 | 0 | 0 | 0 |
| 0 | 0 | 0 | 0 | 0 |
| 0 | 0 | 0 | 0 | 0 |
| 0 | 0 | 0 | 0 | 0 |
| 0 | 0 | 0 | 0 | 0 |
| 0 | 0 | 0 | 0 | 0 |
| 0 | 0 | 0 | 0 | 0 |
| 0 | 0 | 0 | 0 | 0 |
| 0 | 0 | 0 | 0 | 0 |
| 0 | 0.077461408 | 0 | 0 | 0 |
| 0 | 0 | 0 | 0 | 0.000777924 |
| 0 | 0 | 0 | 0.011948211 | 0 |
| 0 | 0 | 0 | 0 | 0 |
| 0 | 0 | 0 | 0 | 0 |
| 0 | 0 | 0 | 0 | 0 |
| 0 | 0 | 0 | 0 | 0 |
| 0 | 0 | 0 | 0 | 0 |
| 0 | 0 | 0 | 0 | 0 |
| 0 | 0 | 0.020056412 | 0.003073213 | 0.000760879 |
| 0.004888345 | 0 | 0 | 0 | 0 |
| 0 | 0 | 0 | 0 | 0 |
| 0 | 0 | 0 | 0 | 0.069935202 |
| 0 | 0 | 0 | 0 | 0 |
| 0 | 0 | 0 | 0 | 0.000414944 |
| 0 | 0 | 0 | 0 | 0 |
| 0 | 0 | 0 | 0 | 0 |
| 0 | 0.003202681 | 0 | 0 | 0 |
| 0 | 0 | 0 | 0 | 0 |
| 0 | 0 | 0 | 0 | 0 |
| 0 | 0 | 0 | 0 | 0 |
| 0 | 0 | 0 | 0 | 0 |
| 0 | 10.04530546 | 0 | 3.176890807 | 0 |
| 0 | 0.21121867 | 0 | 0 | 0 |
| 0 | 0 | 0 | 0 | 0 |
| 0 | 0 | 0 | 0 | 0 |
| 0 | 0.004542218 | 0.077011969 | 0.000836993 | 0 |
| 0 | 0 | 0 | 0 | 0 |
| 0 | 0.169202147 | 0.014670264 | 0.09105765 | 0.066960867 |
| 0 | 0 | 0 | 0 | 0 |
| 0 | 0 | 0 | 0 | 0 |
| 0 | 0 | 0 | 0 | 0 |
| 0 | 0 | 0 | 0 | 0 |
| 0.122309963 | 0 | 3.385933452 | 0.713744481 | 0.420100942 |
| 0 | 0 | 0 | 0 | 0 |
| 0 | 0 | 0 | 0 | 0 |

| | | | | |
|---|---|---|---|---|
| 0 | 0 | 0 | 0 | 0 |
| 0 | 0 | 0 | 0 | 0.012886359 |
| 0 | 0 | 0 | 0 | 0 |
| 0 | 0.104435524 | 0 | 0 | 0 |
| 0 | 0 | 0 | 0 | 0.038368994 |
| 0 | 0 | 0 | 0 | 0 |
| 0 | 0 | 0 | 0 | 0 |
| 0 | 0 | 0 | 0 | 0.000361329 |
| 0 | 0.012204599 | 0 | 0 | 0 |
| 0 | 0 | 0.003681905 | 0.094118188 | 0 |
| 0 | 0.089633045 | 0 | 0.028655725 | 0 |
| 0 | 0 | 0 | 0 | 0 |
| 0 | 0 | 0 | 0.001642725 | 0.00074564 |
| 0 | 0 | 0 | 0 | 0 |
| 0 | 0 | 0 | 0 | 0 |
| 0 | 0 | 0 | 0 | 0 |
| 0 | 0 | 0 | 0 | 0 |
| 0 | 0 | 0 | 0 | 0 |
| 0 | 0 | 0 | 0 | 0 |
| 0.000486741 | 0.004508403 | 1.922132151 | 0.151087929 | 0.122779378 |
| 0 | 0 | 0 | 0 | 0 |
| 0 | 0 | 0 | 0 | 0 |
| 0 | 0.159561629 | 0 | 0 | 0 |
| 0 | 0 | 0 | 0 | 0 |
| 0 | 0 | 0 | 0 | 0 |
| 0 | 0.029017027 | 0 | 0 | 0 |
| 0 | 0 | 0 | 0 | 0 |
| 0 | 0 | 0 | 0 | 0 |
| 0 | 0 | 0 | 0 | 0 |
| 0 | 0 | 0 | 0.013025439 | 0 |
| 0 | 0 | 0 | 0 | 0 |
| 0 | 0 | 0 | 0 | 0 |
| 0 | 0 | 0 | 0 | 0 |
| 0 | 0 | 0 | 0 | 0 |
| 0 | 0 | 0 | 0 | 0 |
| 0 | 0 | 0 | 0 | 0 |
| 0 | 0 | 0 | 0 | 0 |
| 0 | 0 | 0 | 0 | 0 |
| 0 | 0 | 0 | 0 | 0 |
| 0 | 0.175202143 | 0 | 0 | 0 |
| 0 | 0 | 0 | 0 | 0 |
| 0 | 0 | 0 | 0 | 0 |
| 0 | 0.021136251 | 0 | 0.029574477 | 0 |
| 0 | 0 | 0 | 0 | 0 |
| 0.00155436 | 0.00101627 | 0.01272831 | 0.008239781 | 0.011309267 |
| 0 | 0.005487191 | 0 | 0 | 0 |
| 0 | 0 | 0 | 0 | 0 |
| 0 | 0 | 0 | 0 | 0 |

| | | | | |
|---|---|---|---|---|
| 0 | 0 | 0 | 0 | 0 |
| 0 | 0 | 0 | 0 | 0 |
| 0 | 0 | 0 | 0 | 0 |
| 0 | 0 | 0 | 0 | 0 |
| 0 | 0 | 0 | 0 | 0 |
| 0 | 0.010874484 | 0.016240086 | 0.011960894 | 0.000332394 |
| 0 | 0 | 0 | 0 | 0 |
| 0 | 0 | 0 | 0 | 0 |
| 0 | 0 | 0 | 0 | 0 |
| 0 | 0 | 0 | 0 | 0 |
| 0 | 0 | 0 | 0 | 0 |
| 0 | 0 | 0 | 0 | 0 |
| 0 | 0 | 0 | 0 | 0 |
| 0 | 0.035728687 | 0 | 0 | 0 |
| 0 | 0 | 0 | 0 | 0 |
| 0 | 0 | 0 | 0 | 0 |
| 0 | 0.140496001 | 0 | 0 | 0 |
| 0 | 0 | 0 | 0 | 0 |
| 0 | 0 | 0 | 0 | 0 |
| 0 | 0 | 0 | 0 | 0 |
| 0 | 0 | 0 | 0 | 0 |
| 0 | 0 | 0 | 0 | 0 |
| 0 | 0 | 0 | 0 | 0 |
| 0 | 0 | 0 | 0 | 0 |
| 0 | 0 | 0 | 0 | 0 |
| 0 | 0 | 0 | 0.002672692 | 0 |
| 0 | 0 | 0 | 0 | 0 |
| 0 | 0.004736911 | 0 | 0 | 0.078447435 |
| 0 | 0 | 0 | 0 | 0 |
| 0 | 0 | 0 | 0 | 0.006730157 |
| 0 | 0 | 0 | 0 | 0 |
| 0 | 0 | 0 | 0 | 0 |
| 0 | 0 | 0 | 0 | 0 |
| 0 | 0 | 0 | 0 | 0 |
| 0 | 0 | 0 | 0 | 0 |
| 0 | 0 | 0 | 0 | 0 |
| 0 | 0 | 0 | 0 | 0 |
| 0 | 0 | 0 | 0.03089021 | 0 |
| 0 | 2.96153868 | 0 | 0 | 0 |
| 0 | 0 | 0 | 0 | 0 |
| 0 | 0.001702164 | 0 | 0 | 0 |
| 0 | 0 | 0 | 0 | 0 |
| 0 | 0 | 0 | 0 | 0 |
| 0 | 0 | 0 | 0 | 0.596356844 |
| 0 | 0 | 0 | 0 | 0 |
| 0 | 0 | 0 | 0 | 0 |
| 0 | 0 | 0 | 0 | 0 |
| 0 | 0 | 0 | 0 | 0 |

| | | | | |
|---|---|---|---|---|
| 0 | 0 | 0 | 0 | 0 |
| 0 | 0 | 0 | 0.011669461 | 0 |
| 0 | 0 | 0 | 0 | 0 |
| 0 | 0 | 0 | 0 | 0 |
| 0 | 0 | 0 | 0 | 0 |
| 0 | 0 | 0 | 0 | 0 |
| 0 | 0 | 0 | 0 | 0 |
| 0 | 0 | 0 | 0 | 0 |
| 0 | 0 | 0 | 0 | 0 |
| 0 | 0 | 0 | 0 | 0 |
| 0 | 0.000556117 | 0 | 0.001753469 | 0 |
| 0 | 0 | 0 | 0 | 0 |
| 0 | 0.007099875 | 0 | 0 | 0 |
| 0 | 0 | 0 | 0 | 0 |
| 0 | 0 | 0 | 0 | 0 |
| 0 | 0 | 0 | 0 | 0 |
| 0 | 0 | 0 | 0 | 0 |
| 0 | 0 | 0 | 0 | 0 |
| 0 | 2.102486371 | 0 | 0 | 0 |
| 0 | 0 | 0 | 0 | 0 |
| 0 | 0 | 0.012853025 | 0.002288142 | 0 |
| 0 | 0 | 0 | 0 | 0 |
| 0 | 0.000753156 | 0 | 0 | 0 |
| 0 | 0.235187475 | 6.485568934 | 1.457636313 | 0 |
| 0 | 0 | 0 | 0 | 0 |
| 0 | 0 | 0 | 0 | 0 |
| 0 | 0 | 0 | 0 | 0 |
| 0 | 0 | 0 | 0 | 0 |
| 0 | 0 | 0 | 0 | 0 |
| 0 | 0 | 0 | 0 | 0 |
| 0 | 0.00572449 | 0 | 0 | 0.002330018 |
| 0 | 0 | 0 | 0 | 0 |
| 0 | 0 | 0 | 0 | 0 |
| 0 | 0 | 0 | 0 | 0 |

| PHL018-0190_V1 | PHL018-0142_V2 | PHL018-0221_V1 | PHL018-0251_V3 | PHL018-0135_V2 |
|---|---|---|---|---|
| REF | EYCF | CTRL | REF | EYCF |
| 0.530628353 | 0.477479605 | 0.092023891 | 0.151591181 | 0.822831117 |
| 0.175255333 | 0.966203486 | 2.766313477 | 31.73746306 | 2.962670344 |
| 0 | 10.0351169 | 4.526508215 | 0 | 0 |
| 0 | 0 | 0 | 0 | 0 |
| 0 | 0 | 0 | 0 | 0 |
| 0 | 0 | 0 | 0 | 0 |
| 0 | 0 | 0 | 0 | 0 |
| 0 | 0 | 0 | 0 | 0 |
| 0 | 0 | 0 | 0 | 0 |
| 0 | 0 | 0 | 0 | 0 |
| 0 | 0 | 0 | 0 | 0 |
| 0 | 0 | 0 | 0 | 0 |
| 0 | 0 | 0 | 0 | 0 |
| 2.668557077 | 2.457718079 | 0.425519127 | 0 | 9.458125788 |
| 0 | 0 | 0 | 0 | 0 |
| 0 | 0 | 0 | 0 | 0 |
| 0 | 0 | 0.007222907 | 0 | 0 |
| 0 | 0 | 0 | 0 | 0 |
| 0 | 0 | 0 | 0 | 0 |
| 0 | 0 | 0 | 0 | 0 |
| 0 | 0 | 0 | 0 | 0 |
| 0 | 0 | 0 | 0 | 0 |
| 0 | 0 | 0 | 0 | 0 |
| 0 | 0 | 0 | 0 | 0 |
| 0 | 0.053521453 | 0 | 0 | 0 |
| 0 | 0 | 0 | 0 | 0 |
| 0 | 0 | 0 | 0 | 0 |
| 0 | 0 | 0 | 0 | 0 |
| 0 | 0 | 0 | 0 | 0 |
| 0 | 0 | 0 | 0 | 0 |
| 0 | 0 | 0 | 0 | 0 |
| 0 | 0 | 0 | 0 | 0 |
| 0 | 0 | 0 | 0 | 0 |
| 0 | 0 | 0 | 0 | 0 |
| 0 | 0 | 0 | 0 | 0 |
| 0.004753015 | 0 | 0 | 0 | 0 |
| 0 | 0 | 0 | 0 | 0 |
| 0 | 0 | 0 | 0 | 0 |
| 0 | 0 | 0 | 0 | 0 |
| 0 | 0 | 0 | 0 | 0 |
| 0 | 0.002586345 | 0 | 0 | 0 |
| 0 | 0 | 0.001839277 | 0.005547877 | 0.110521886 |
| 0.002459293 | 0.041522994 | 1.031018177 | 0.896615028 | 0.110885433 |
| 0.009253569 | 0.009470035 | 0.182842372 | 0.108770142 | 0.003020474 |
| 0 | 0 | 0 | 0.001140646 | 0.002322832 |
| 0 | 0 | 0 | 0 | 0 |

| | | | | |
|---|---|---|---|---|
| 0.196764128 | 0 | 0 | 0 | 0 |
| 0.002250747 | 0.00105217 | 0.009451285 | 0.085429416 | 0.001553303 |
| 0.007576922 | 0 | 0.013024435 | 0.003363624 | 0.11409101 |
| 0.378745667 | 2.291390682 | 0.689656044 | 2.093029957 | 0.181760812 |
| 0 | 0.004849059 | 0 | 0 | 0.012527546 |
| 0 | 0 | 0 | 0 | 0 |
| 0 | 0.027227253 | 0 | 0 | 0 |
| 0.009516043 | 0 | 0 | 0 | 0 |
| 0 | 0 | 0 | 0 | 0 |
| 0 | 0 | 0 | 0 | 0 |
| 0 | 0 | 0 | 0 | 0 |
| 0.459436874 | 0.175094946 | 0.084690253 | 0.022309577 | 0.172718451 |
| 0 | 0 | 0 | 0 | 0 |
| 0.01431533 | 0.16903241 | 0.148075768 | 0.175457733 | 0.415928658 |
| 0 | 0 | 0 | 0 | 0 |
| 0 | 0 | 0 | 0 | 0 |
| 0 | 0.065167863 | 0.157092534 | 0 | 0 |
| 0 | 0 | 0 | 0 | 0 |
| 0 | 0 | 0 | 0 | 0 |
| 0 | 0 | 0 | 0 | 0 |
| 0.047727962 | 0.020848601 | 0.421055001 | 1.047258651 | 0.083995892 |
| 0.108846403 | 0.027702354 | 0.006275073 | 0.023623357 | 0.005520477 |
| 0 | 0 | 0 | 0 | 0 |
| 0 | 0 | 0 | 0 | 0 |
| 0 | 0 | 0 | 0 | 0 |
| 0 | 0 | 0 | 0 | 0 |
| 0 | 0 | 0 | 0 | 0 |
| 0 | 0 | 0 | 0 | 0 |
| 0 | 0 | 0 | 0 | 0.003026694 |
| 0 | 0 | 0 | 0 | 0 |
| 0 | 0 | 0.070685248 | 1.155845398 | 0 |
| 0 | 0 | 0 | 0 | 0 |
| 0 | 0 | 0 | 0 | 0 |
| 0 | 0 | 0 | 0 | 0 |
| 0 | 0 | 0 | 0 | 0 |
| 0 | 0 | 0 | 0 | 0 |
| 0 | 0 | 0 | 0 | 0 |
| 0.000445403 | 0.00152691 | 0 | 0 | 0 |
| 0 | 0 | 0 | 0 | 0 |
| 0 | 0 | 0 | 0 | 0 |
| 0 | 0 | 0 | 0 | 0 |
| 0 | 0 | 0 | 0 | 0 |
| 0 | 0 | 0 | 0 | 0 |
| 0 | 0 | 0 | 0 | 0 |
| 0 | 0 | 0 | 0 | 0 |
| 0 | 0 | 0 | 0 | 0 |
| 0 | 0 | 0 | 0 | 0.002437568 |
| 0 | 0 | 0 | 0 | 0 |

| | | | | |
|---|---|---|---|---|
| 0 | 0 | 0 | 0 | 0 |
| 0 | 0 | 0.004845538 | 0 | 0 |
| 0 | 0 | 0 | 0 | 0 |
| 0 | 0 | 0 | 0 | 0 |
| 0 | 0 | 0.033402486 | 0 | 0.006059323 |
| 0 | 0 | 0 | 0 | 0 |
| 0 | 0 | 0 | 0 | 0 |
| 0 | 0 | 0 | 0 | 0 |
| 0 | 0 | 0.013022663 | 0.006176363 | 0.000762282 |
| 0 | 0 | 0 | 0 | 0 |
| 0 | 0 | 0 | 0 | 0 |
| 0 | 0 | 0.002071103 | 0 | 0 |
| 0 | 0 | 0 | 0 | 0 |
| 0 | 0 | 0 | 0 | 0 |
| 0 | 0 | 0 | 0 | 0 |
| 0 | 0 | 0 | 0 | 0 |
| 0 | 0 | 0 | 0 | 0 |
| 0 | 0 | 0 | 0 | 0 |
| 0 | 0 | 0 | 0 | 0 |
| 0 | 0 | 0 | 0 | 0 |
| 0 | 0 | 0 | 0 | 0 |
| 0 | 0 | 0 | 0 | 0 |
| 0 | 0 | 0 | 0 | 0 |
| 0 | 0 | 0 | 0 | 0 |
| 0 | 0 | 0 | 0 | 0 |
| 0 | 0 | 0 | 0 | 0 |
| 0 | 0 | 0 | 0 | 0 |
| 0 | 0 | 0 | 0 | 0.00153925 |
| 0 | 0 | 0 | 0 | 0 |
| 0 | 0 | 0 | 0 | 0 |
| 0 | 0.13883977 | 0.003631746 | 0 | 0.117271667 |
| 0.028837026 | 0 | 0.000865913 | 0.028162934 | 0.023310605 |
| 0 | 0 | 0 | 0 | 0 |
| 0 | 0 | 0 | 0 | 0 |
| 0 | 0 | 0 | 0 | 0 |
| 0 | 0 | 0 | 0 | 0 |
| 0 | 0 | 0 | 0 | 0 |
| 0 | 0 | 0 | 0 | 0 |
| 0 | 0 | 0 | 0 | 0 |
| 0 | 0 | 0 | 0 | 0 |
| 0 | 0 | 0 | 0 | 0 |
| 0 | 0 | 0 | 0 | 0 |
| 0.001726309 | 0.000372465 | 0 | 0 | 0.019672939 |
| 0 | 0 | 0 | 0 | 0 |
| 0 | 0 | 0 | 0 | 0 |
| 0 | 0 | 0 | 0 | 0 |
| 0 | 0 | 0 | 0 | 0 |
| 0 | 0 | 0 | 0 | 0 |

| | | | | |
|---|---|---|---|---|
| 0 | 0 | 0.001166382 | 0 | 0 |
| 0 | 0 | 0 | 0 | 0 |
| 0 | 0 | 0 | 0 | 0 |
| 0 | 0 | 0 | 0 | 0 |
| 0.228394246 | 0 | 0 | 0 | 0.002737438 |
| 0 | 0 | 0 | 0 | 0 |
| 0 | 0 | 0 | 0 | 0 |
| 0 | 0 | 0 | 0 | 0 |
| 0 | 0 | 0 | 0 | 0 |
| 0 | 0 | 0 | 0 | 0 |
| 0 | 0 | 0 | 0 | 0 |
| 0 | 0 | 0 | 0 | 0 |
| 0 | 0 | 0 | 0 | 0 |
| 0 | 0 | 0 | 0 | 0 |
| 0 | 0 | 0 | 0 | 0 |
| 0 | 0 | 0 | 0 | 0 |
| 0 | 0 | 0 | 0 | 0 |
| 0 | 0 | 0 | 0 | 0 |
| 0 | 0 | 0 | 0 | 0 |
| 0 | 0 | 0 | 0 | 0 |
| 0 | 0 | 0 | 0 | 0 |
| 0 | 0 | 0 | 0 | 0 |
| 0 | 0 | 0 | 0 | 0 |
| 0 | 0 | 0 | 0 | 0 |
| 0.013740927 | 0 | 0 | 0 | 0 |
| 0 | 0 | 0 | 0 | 0 |
| 0.021340753 | 0 | 0 | 0 | 0 |
| 0 | 0 | 0 | 0 | 0 |
| 0 | 0 | 0 | 0 | 0 |
| 0 | 0 | 0 | 0 | 0 |
| 0 | 0 | 0 | 0 | 0.047770372 |
| 0 | 0.00123934 | 0 | 0 | 0 |
| 0 | 0 | 0 | 0 | 0 |
| 0 | 0 | 0 | 0 | 0 |
| 0 | 0 | 0 | 0 | 0 |
| 0 | 0 | 0 | 0 | 0 |
| 0 | 0 | 0 | 0 | 0 |
| 0 | 0 | 0 | 0 | 0 |
| 0 | 0 | 0 | 0 | 0 |
| 0 | 0 | 0 | 0 | 0 |
| 0 | 0 | 0 | 0 | 0 |
| 0 | 0 | 0 | 0 | 0 |
| 0 | 0 | 0 | 0 | 0 |
| 0 | 0 | 0 | 0 | 0 |
| 0 | 0 | 0 | 0 | 0 |
| 0 | 0 | 0 | 0 | 0 |
| 0 | 0 | 0 | 0 | 0 |
| 0 | 0 | 0 | 0.089700301 | 0 |

| | | | | |
|---|---|---|---|---|
| 0 | 0 | 0 | 0 | 0 |
| 0 | 0 | 0 | 0 | 0 |
| 0 | 0 | 0 | 0 | 0 |
| 0 | 0 | 0 | 0 | 0 |
| 0 | 0 | 0 | 0 | 0 |
| 0 | 0 | 0 | 0 | 0 |
| 0 | 0 | 0 | 0 | 0 |
| 0 | 0 | 0 | 0 | 0 |
| 0 | 0 | 0 | 0 | 0 |
| 0 | 0 | 0 | 0 | 0 |
| 0 | 0 | 0 | 0 | 0 |
| 0 | 0 | 0 | 0 | 0 |
| 0.014840708 | 0 | 0.002485648 | 0.003009051 | 0.00102128 |
| 0 | 0 | 0 | 0 | 0 |
| 0 | 0 | 0 | 0 | 0 |
| 0 | 0 | 0 | 0 | 0 |
| 0 | 0 | 0 | 0 | 0 |
| 0 | 0 | 0 | 0 | 0 |
| 0 | 0 | 0 | 0 | 0 |
| 0 | 0 | 0 | 0 | 0 |
| 0.014704365 | 0.005986974 | 0.013227686 | 0.070515061 | 0.027879417 |
| 0 | 0 | 0 | 0 | 0 |
| 0 | 0 | 0 | 0 | 0.002848368 |
| 0 | 0.00208107 | 0 | 0 | 0.001194765 |
| 0 | 0 | 0 | 0 | 0 |
| 0 | 0 | 0 | 0 | 0 |
| 0 | 0.006715072 | 0.001041012 | 0.007645323 | 0.001112076 |
| 0 | 0 | 0 | 0 | 0 |
| 0 | 0 | 0 | 0 | 0 |
| 0 | 0.001519767 | 0.01492443 | 0 | 0 |
| 0 | 0 | 0.051678091 | 0 | 0 |
| 0 | 0 | 0 | 0 | 0 |
| 0 | 0 | 0 | 0 | 0 |
| 0 | 0 | 0 | 0 | 0 |
| 0.022375472 | 0.006348131 | 0.040474456 | 0.008053555 | 0.056407431 |
| 0 | 0 | 0 | 0 | 0 |
| 0 | 0 | 0 | 0 | 0 |
| 0 | 0 | 0 | 0 | 0 |
| 0 | 0 | 0 | 0 | 0 |
| 0 | 0 | 0 | 0 | 0 |
| 0 | 0 | 0 | 0 | 0 |
| 0 | 0 | 0 | 0 | 0 |
| 0 | 0 | 0 | 0 | 0 |
| 0 | 0 | 0 | 0 | 0 |
| 0 | 0 | 0 | 0 | 0 |
| 0 | 0 | 0 | 0 | 0 |
| 0.003174531 | 0.00140591 | 0 | 0 | 0.001883347 |
| 0.062571817 | 0.001850666 | 0.048900813 | 0.003478291 | 0 |

| | | | | |
|---|---|---|---|---|
| 0 | 0 | 0 | 0 | 0 |
| 0 | 0 | 0 | 0 | 0 |
| 0 | 0 | 0 | 0 | 0 |
| 0 | 0 | 0 | 0.00110534 | 0 |
| 0 | 0 | 0 | 0 | 0 |
| 0 | 0 | 0 | 0 | 0 |
| 0 | 0 | 0 | 0 | 0 |
| 0 | 0 | 0.000953164 | 0 | 0 |
| 0 | 0 | 0 | 0 | 0.170985643 |
| 0 | 0 | 0 | 0 | 0 |
| 0 | 0 | 0 | 0 | 0 |
| 0 | 0 | 0 | 0 | 0 |
| 0 | 0 | 0 | 0 | 0 |
| 0 | 0 | 0 | 0 | 0 |
| 0 | 0 | 0 | 0 | 0 |
| 0 | 0 | 0 | 0 | 0 |
| 0 | 0 | 0 | 0 | 0 |
| 0 | 0 | 0 | 0.007542658 | 0 |
| 0 | 0 | 0 | 0.005482969 | 0.000900453 |
| 0 | 0 | 0 | 0 | 0 |
| 0 | 0.020051136 | 0.011687781 | 0.102012372 | 0 |
| 0 | 0 | 0 | 0 | 0 |
| 0 | 0 | 0 | 0 | 0 |
| 0 | 0 | 0 | 0 | 0 |
| 0 | 0 | 0.001976191 | 0 | 0 |
| 0 | 0 | 0 | 0 | 0 |
| 0 | 0 | 0 | 0 | 0 |
| 0 | 0 | 0 | 0 | 0 |
| 0 | 0 | 0 | 0 | 0 |
| 0 | 0 | 0 | 0 | 0 |
| 1.749429508 | 0.309728844 | 0.388010875 | 0.614657127 | 0.622919217 |
| 0 | 0 | 0 | 0 | 0 |
| 0 | 0.031729327 | 0 | 0 | 0.026959919 |
| 0 | 0 | 0 | 0 | 0.004801645 |
| 0 | 0 | 0 | 0 | 0 |
| 0 | 0 | 0 | 0 | 0 |
| 0 | 0 | 0 | 0 | 0 |
| 0 | 0 | 0 | 0 | 0 |
| 0 | 0 | 0 | 0 | 0 |
| 0 | 0 | 0 | 0 | 0 |
| 0 | 0 | 0 | 0 | 0 |
| 0 | 0.006011456 | 0 | 0.006062036 | 0 |
| 0 | 0 | 0.001407468 | 0 | 0 |
| 0 | 0 | 0 | 0 | 0 |
| 0 | 0 | 0 | 0 | 0 |
| 0 | 0 | 0 | 0 | 0 |
| 0 | 0 | 0 | 0 | 0 |
| 0 | 0 | 0 | 0 | 0 |

| | | | | |
|---|---|---|---|---|
| 0 | 0 | 0 | 0 | 0 |
| 0 | 0 | 0 | 0 | 0 |
| 0.008539328 | 0.01146772 | 0.197060979 | 0.123508949 | 0.00175011 |
| 0 | 0 | 0 | 0 | 0 |
| 0 | 0.020659856 | 0 | 0 | 5.350492758 |
| 0 | 0 | 0 | 0 | 0 |
| 0 | 0 | 0 | 0 | 0 |
| 0 | 0 | 0 | 0 | 0 |
| 0 | 0 | 0 | 0 | 0 |
| 0 | 0 | 0 | 0 | 0 |
| 0 | 0 | 0 | 0 | 0 |
| 0 | 0 | 0 | 0 | 0 |
| 0 | 0 | 0 | 0 | 0 |
| 0.004118181 | 0 | 0.009570255 | 0 | 0.033986433 |
| 0.001133007 | 0.001721372 | 0.003202294 | 0.007839348 | 0 |
| 0 | 0 | 0 | 0 | 0 |
| 0.001452329 | 0 | 0.001945988 | 0.001786447 | 0 |
| 0 | 0 | 0 | 0 | 0 |
| 0 | 0 | 0 | 0 | 0 |
| 0 | 0 | 0 | 0 | 0 |
| 0 | 0 | 0 | 0 | 0 |
| 0.053898264 | 0.001225757 | 0.046990968 | 0.005529091 | 0 |
| 0 | 0 | 0 | 0 | 0 |
| 0 | 0 | 0 | 0.003571397 | 0.001818214 |
| 0 | 0 | 0 | 0 | 0 |
| 0 | 0 | 0 | 0 | 0 |
| 0 | 7.814530161 | 0 | 0 | 0 |
| 0 | 0 | 0 | 0 | 0 |
| 0 | 0 | 0 | 0 | 0 |
| 0 | 0 | 0.011117058 | 0 | 0 |
| 0 | 0 | 0.00235579 | 0 | 0 |
| 0 | 0 | 0 | 0 | 0 |
| 0 | 0 | 0 | 0 | 0 |
| 0 | 0.002041425 | 0 | 0 | 0 |
| 0 | 0 | 0 | 0 | 0 |
| 0 | 0 | 0.010964693 | 0 | 0 |
| 0 | 0 | 0 | 0 | 0 |
| 0.008968005 | 0 | 0.096283694 | 0.002315577 | 0.00117887 |
| 0 | 0 | 0 | 0 | 0 |
| 0 | 0 | 0 | 0 | 0 |
| 0 | 0 | 0 | 0 | 0 |
| 0 | 0 | 0 | 0 | 0 |
| 0 | 0 | 0 | 0 | 0 |
| 0 | 0 | 0.023988419 | 0 | 0 |
| 0 | 0 | 0 | 0 | 0 |
| 0 | 0 | 0 | 0 | 0 |
| 0 | 0 | 0 | 0 | 0 |
| 0 | 0.005388851 | 0.006014976 | 0.027696851 | 0 |

| | | | | |
|---|---|---|---|---|
| 0 | 0 | 0.005287156 | 0 | 0 |
| 0 | 0 | 0 | 0 | 0 |
| 0 | 0 | 0 | 0 | 0 |
| 0 | 0 | 0 | 0 | 0 |
| 0.003530711 | 0.011768936 | 0 | 0.005341048 | 0 |
| 0 | 0 | 0 | 0 | 0 |
| 0 | 0 | 0 | 0 | 0 |
| 0 | 0 | 0.009406352 | 0.015653914 | 0.029105912 |
| 0 | 0 | 0 | 0 | 0 |
| 0.225114315 | 0.025505343 | 0 | 0.011574986 | 0 |
| 0 | 0 | 0 | 0.050740768 | 0 |
| 0 | 0.003503605 | 0.011040772 | 0 | 0.004662927 |
| 0 | 0 | 0.003505737 | 0.00286073 | 0 |
| 0 | 0.004036222 | 0.172152723 | 0.147720157 | 0 |
| 0 | 0.000496962 | 0 | 0.002101568 | 0 |
| 0 | 0 | 0 | 0 | 0 |
| 0 | 0 | 0 | 0 | 0 |
| 0 | 0 | 0 | 0 | 0 |
| 0 | 0 | 0.216752973 | 0.017162916 | 0.011762309 |
| 0 | 0 | 0 | 0 | 0 |
| 0 | 0 | 0 | 0 | 0 |
| 0 | 0 | 0 | 0 | 0 |
| 0 | 0 | 0 | 0 | 0 |
| 0 | 0 | 0 | 0 | 0 |
| 0 | 0 | 0 | 0 | 0 |
| 0 | 0 | 0 | 0 | 0.000969341 |
| 0 | 0 | 0 | 0 | 0 |
| 0.001273897 | 0 | 0.263625423 | 0.300473584 | 0.033993868 |
| 0 | 0 | 0.03692952 | 0 | 0.004962325 |
| 0.007896645 | 0.003474343 | 0.002800795 | 0 | 0 |
| 0 | 0 | 0 | 0 | 0 |
| 0 | 0 | 0 | 0 | 0 |
| 0 | 0 | 0 | 0 | 0 |
| 0 | 0.020425321 | 0.107397224 | 0 | 0.001584649 |
| 0 | 0 | 0 | 0 | 0 |
| 0 | 0 | 0 | 0 | 0 |
| 0 | 0 | 0.158665937 | 1.229204517 | 0 |
| 0 | 0 | 0 | 0 | 0 |
| 0 | 0 | 0 | 0 | 0 |
| 0 | 0 | 0 | 0.373472422 | 0.003313369 |
| 0 | 0 | 0 | 0 | 0 |
| 0 | 0 | 0.004529097 | 0 | 0 |
| 0 | 0 | 0 | 0 | 0.015119031 |
| 0 | 0.021083883 | 0 | 0.193659805 | 0.009566454 |
| 0 | 0 | 0 | 0 | 0 |
| 0 | 0 | 0 | 0 | 0 |
| 0.032563631 | 0 | 0.01184842 | 0.003551684 | 0.014766784 |
| 0 | 0 | 0 | 0 | 0 |

| | | | | |
|---|---|---|---|---|
| 0 | 0 | 0 | 0 | 0.017090164 |
| 0 | 0.01114608 | 0 | 0 | 0 |
| 0 | 0 | 0 | 0 | 0 |
| 0 | 0 | 0 | 0 | 0 |
| 0 | 0 | 0 | 0 | 0 |
| 0.00884407 | 0.000689065 | 0.028662774 | 0 | 0.013766869 |
| 0 | 0 | 0.005188372 | 0 | 0 |
| 0 | 0 | 0 | 0 | 0 |
| 0 | 4.587618756 | 0 | 0 | 0 |
| 0 | 0 | 0 | 0 | 0 |
| 0 | 0.005274713 | 0.001546232 | 0.026767052 | 0 |
| 0 | 0.005393027 | 0 | 0 | 0.042830722 |
| 0 | 0 | 0 | 0.073744525 | 0 |
| 0.019058761 | 0.001510086 | 0 | 0.00510872 | 0 |
| 0 | 0.07563281 | 0.011085521 | 0 | 0 |
| 0 | 0 | 0 | 0 | 0 |
| 0 | 0 | 0 | 0 | 0 |
| 0 | 0.001469557 | 0.042174035 | 0.006960254 | 0 |
| 0 | 0.013983112 | 0 | 0.036159024 | 0 |
| 0 | 0 | 0 | 0.012559798 | 0 |
| 0 | 0 | 0 | 0 | 0 |
| 0.09258334 | 0 | 0.165159552 | 0 | 0 |
| 0 | 0 | 0 | 0 | 0.063183432 |
| 0 | 0 | 0 | 0 | 0 |
| 0 | 0 | 0 | 0 | 0 |
| 0 | 0 | 0 | 0 | 0 |
| 0 | 0.003765866 | 0.011879224 | 0.011632331 | 0.002820033 |
| 0 | 0 | 0 | 0 | 0 |
| 0 | 0 | 0 | 0 | 0 |
| 0.428512164 | 0.581257135 | 7.938968431 | 1.219097482 | 0.216540517 |
| 0.018518182 | 0.001434263 | 0 | 0 | 0.010234011 |
| 0 | 0 | 0 | 0 | 0 |
| 0 | 0.001104318 | 0 | 0 | 0 |
| 0 | 0 | 0 | 0 | 0 |
| 0 | 0.0058641 | 0 | 0 | 0.000939529 |
| 0 | 0 | 0 | 0 | 0 |
| 2.005840945 | 0 | 0 | 0 | 0 |
| 0 | 0 | 0 | 0 | 0 |
| 0 | 0 | 0 | 0 | 0.179294682 |
| 0 | 0.462071165 | 0 | 0 | 0 |
| 0 | 0.001986328 | 0.012209542 | 0.031499429 | 0.006200778 |
| 2.96631515 | 6.585702909 | 0 | 3.819988865 | 0 |
| 0 | 0 | 0 | 0 | 0 |
| 0 | 0.001198156 | 0 | 0 | 0 |
| 0 | 0.000700213 | 0.004741524 | 0 | 0 |
| 0 | 0.003711109 | 0 | 0 | 0 |
| 0.000696826 | 0.001433296 | 0 | 0.00110203 | 0.001346516 |
| 0 | 0 | 0.032838031 | 0 | 0 |

| | | | | |
|---|---|---|---|---|
| 0 | 0 | 0 | 0 | 0.001384479 |
| 0 | 0.03615952 | 0 | 0 | 0 |
| 0 | 0.100109739 | 0.295966606 | 0.001482179 | 0 |
| 0 | 0 | 0 | 0 | 0 |
| 0 | 0 | 0 | 0 | 0 |
| 0 | 0 | 0.023449609 | 0 | 0 |
| 0 | 0 | 0 | 0 | 0 |
| 0 | 0 | 0 | 0 | 0 |
| 0 | 0 | 0 | 0 | 0.030589283 |
| 0 | 0 | 0.000762981 | 0 | 0 |
| 0 | 0 | 0 | 0 | 0 |
| 0 | 0 | 0 | 0 | 0 |
| 0 | 0 | 0 | 0.107344863 | 0 |
| 0 | 0.029670293 | 0 | 0 | 0 |
| 0 | 0 | 0 | 0 | 0 |
| 0.156215884 | 0.025687361 | 0.04544659 | 0.001922611 | 0.050898111 |
| 0.001212717 | 0.003275512 | 0.014218131 | 0.010868173 | 0.190292975 |
| 0 | 0.000612249 | 0 | 0.321566131 | 0 |
| 0 | 0.004164132 | 0 | 0.002201177 | 0 |
| 0 | 0 | 0 | 0 | 0 |
| 0 | 0.005874572 | 0.015103065 | 0.292739598 | 0 |
| 0 | 0 | 0.024738439 | 0.002966239 | 0 |
| 0 | 0 | 0.022852238 | 0 | 0.237877299 |
| 0 | 0.43959227 | 0 | 0 | 0 |
| 0 | 0.158362526 | 0.026956099 | 0.095598667 | 0 |
| 0 | 0 | 0 | 0.002807236 | 0 |
| 0 | 0 | 0 | 0.001893738 | 0 |
| 0 | 0 | 0 | 0 | 0 |
| 0 | 0 | 0 | 0 | 0 |
| 0 | 0 | 0 | 0 | 0.0009428 |
| 0 | 0.050111359 | 0.004706399 | 0.172821876 | 0.017387387 |
| 0 | 0 | 0.019880548 | 0.002147135 | 0 |
| 0.048382888 | 0.084083049 | 0.001670203 | 0.010513866 | 0.012489528 |
| 0 | 0 | 0 | 0 | 0 |
| 0 | 0 | 0 | 0 | 0 |
| 0 | 0 | 0 | 0 | 0 |
| 0.003005139 | 0 | 0 | 0 | 0 |
| 0 | 0.006958063 | 0.044479565 | 0.08631182 | 0.056556571 |
| 0 | 0 | 0 | 0 | 0 |
| 0 | 0.003593007 | 0.058089994 | 0 | 0 |
| 0.107765296 | 0 | 0 | 0 | 0 |
| 0.176842313 | 0 | 0 | 0 | 0 |
| 0.303432089 | 0 | 0.008535639 | 0 | 0.004716371 |
| 0 | 0.028587506 | 0.018743617 | 0.709876478 | 0.125062539 |
| 0 | 0 | 0.027659871 | 0.013563528 | 0 |
| 0 | 0 | 0 | 0 | 0.000916748 |
| 0 | 0 | 0 | 0 | 0.002102701 |
| 0 | 0 | 0 | 0 | 0 |

| | | | | |
|---|---|---|---|---|
| 0 | 0 | 0.050774458 | 0 | 0 |
| 0.005693135 | 0 | 0.19952714 | 0.667117743 | 0.002864889 |
| 0 | 0 | 0 | 0 | 0 |
| 0 | 0.001208885 | 0 | 0 | 0 |
| 0 | 0 | 0 | 0 | 0 |
| 0 | 0 | 0 | 0 | 0 |
| 0 | 0 | 0.000744309 | 0 | 0 |
| 0 | 0.000638824 | 0 | 0 | 0.003080751 |
| 0 | 0 | 0 | 0 | 0 |
| 0 | 0.001208743 | 0 | 0.086283323 | 0.011242036 |
| 0.015071589 | 0.009888563 | 0.004982207 | 0.00271811 | 0.003619173 |
| 0.098659907 | 0.002025278 | 0.01335805 | 0.023669333 | 0.011098832 |
| 0 | 0.958367107 | 0 | 0 | 0 |
| 0 | 0 | 0 | 0 | 0.052936613 |
| 0.003488891 | 0 | 0 | 0 | 0.004641084 |
| 0 | 0 | 0 | 0 | 0 |
| 0.000830638 | 0.002200385 | 0.003547621 | 0.005035679 | 0 |
| 0 | 0 | 0.04036412 | 0.35299838 | 0.071480053 |
| 0 | 0 | 0 | 0 | 0 |
| 0.012603256 | 0.019202627 | 0.003518168 | 0.057396953 | 0.065770825 |
| 0.137769468 | 0 | 0.006025752 | 0.009257186 | 0 |
| 0 | 0 | 0 | 0 | 0 |
| 0 | 0 | 0 | 0 | 0 |
| 0 | 0 | 0 | 0 | 0 |
| 0 | 0.003588328 | 0 | 0 | 0 |
| 0 | 0 | 0 | 0 | 0 |
| 0.039879641 | 0.056125911 | 0.030375848 | 0.01827461 | 0.018607367 |
| 0.11866028 | 0.205426361 | 0.010979117 | 0.048475181 | 0.090896636 |
| 0 | 0 | 0 | 0 | 0 |
| 0.004209767 | 0.005989455 | 0.001931327 | 0 | 0.005305289 |
| 0 | 0 | 0 | 0.004377408 | 0 |
| 0 | 0 | 0 | 0 | 0 |
| 0 | 0 | 0 | 0 | 0.003559609 |
| 0 | 0 | 0 | 0 | 0 |
| 0.203767048 | 0 | 0 | 0 | 0 |
| 0.039291935 | 0.166632784 | 1.149343237 | 0.433357047 | 0.339516899 |
| 0 | 0 | 0 | 0 | 0 |
| 0 | 0 | 0 | 0 | 0 |
| 0 | 0.000700045 | 0.011286641 | 0.006512823 | 0.00572713 |
| 0 | 0 | 0 | 0 | 0 |
| 0 | 0 | 0.019122549 | 0.011517431 | 0 |
| 0 | 0 | 0 | 0 | 0 |
| 0 | 0.009780019 | 0.039248756 | 0.006833071 | 0 |
| 0 | 0 | 0 | 0.006423591 | 0 |
| 0 | 0 | 0 | 0 | 0 |
| 0 | 0 | 0 | 0 | 0 |
| 0 | 0 | 0 | 0 | 0 |
| 0 | 0 | 0 | 0 | 0 |

| | | | | |
|---|---|---|---|---|
| 0 | 0 | 0.027295962 | 0 | 0 |
| 0 | 0 | 0 | 0 | 0 |
| 0 | 0 | 0 | 0 | 0 |
| 0 | 0 | 0.122365458 | 0 | 0.165050946 |
| 0 | 0 | 0 | 0 | 0 |
| 0 | 0 | 0 | 0 | 0 |
| 0 | 0.006143234 | 0 | 0 | 0 |
| 0 | 0.00232601 | 0.019643708 | 0 | 0 |
| 1.801293782 | 0 | 4.430950935 | 0.169622172 | 4.488637709 |
| 0 | 0 | 0 | 0 | 0 |
| 0 | 0 | 0 | 0 | 0 |
| 0 | 0 | 0.003442588 | 0 | 0.007559499 |
| 0 | 0 | 0 | 0 | 0 |
| 0 | 0 | 0 | 0 | 0 |
| 0.419217756 | 0.443144221 | 0.181339824 | 0.041064519 | 0 |
| 0 | 0 | 0 | 0 | 0 |
| 0 | 0 | 0 | 0 | 0 |
| 0 | 0 | 0 | 0 | 0 |
| 0 | 0 | 0 | 0 | 0 |
| 0 | 0 | 0 | 0 | 0 |
| 0 | 0 | 0 | 0 | 0 |
| 0 | 0 | 0 | 0 | 0 |
| 0.025119059 | 0 | 0 | 0 | 0 |
| 0.298725649 | 0.511535993 | 0.157953978 | 0.51151206 | 0.217782255 |
| 0 | 0.027099312 | 0.055688741 | 0 | 0.035770689 |
| 0 | 0 | 0 | 0 | 0 |
| 0 | 0 | 0 | 0 | 0 |
| 3.136457775 | 0.152978248 | 0.078107887 | 0 | 0.209159655 |
| 0 | 0 | 0 | 0 | 0 |
| 0 | 0 | 0 | 0 | 0 |
| 0 | 0 | 0 | 0 | 0 |
| 0 | 0 | 0 | 0 | 0 |
| 0 | 0 | 0 | 0 | 0 |
| 0 | 0 | 0 | 0 | 0 |
| 0 | 0 | 0 | 0 | 0 |
| 0.329544801 | 0.095336603 | 0 | 0.020344201 | 0 |
| 0 | 0 | 0 | 0 | 0 |
| 0 | 0 | 0 | 0 | 0.013817688 |
| 0 | 0 | 0 | 0 | 0 |
| 0 | 0 | 0 | 0.005601375 | 0 |
| 0.161152515 | 0.041422803 | 0.605467177 | 0.028681696 | 0.033319945 |
| 0 | 0 | 0 | 0 | 0 |
| 1.160079357 | 17.60234216 | 1.966819564 | 0 | 6.940674724 |
| 0.016752881 | 0 | 0 | 0 | 0 |
| 0 | 0 | 0 | 0 | 0 |
| 0 | 0 | 0 | 0 | 0 |
| 0 | 0 | 0 | 0 | 0 |
| 0.003950561 | 0.002188789 | 0 | 0 | 0.005890352 |

| | | | | |
|---|---|---|---|---|
| 0 | 0 | 0 | 0 | 0 |
| 0.005065362 | 0 | 0 | 0 | 0 |
| 0.022021046 | 0.04340655 | 0.166606547 | 0.88507615 | 0.030943191 |
| 0 | 0 | 0 | 0 | 0 |
| 0 | 0 | 0 | 0 | 0 |
| 0.021767033 | 0.008367718 | 0.035309895 | 0 | 0.036118965 |
| 0 | 0 | 0 | 0 | 0 |
| 0 | 0 | 0 | 0 | 0 |
| 0 | 0 | 0 | 0 | 0 |
| 0 | 0 | 0 | 0 | 0 |
| 0.060455506 | 0.310940626 | 0 | 0.070128842 | 1.152250044 |
| 2.0178168 | 1.034453588 | 1.04164323 | 3.039302051 | 2.396662152 |
| 0 | 0 | 0 | 0 | 0 |
| 0 | 0.000416869 | 0.021059356 | 0.015043161 | 0.003111281 |
| 0.030685862 | 0.129213543 | 0.044666828 | 0.351377396 | 0.103730381 |
| 0 | 0 | 0 | 0 | 0 |
| 0 | 0 | 0 | 0 | 0 |
| 0 | 0 | 0 | 0 | 0 |
| 0 | 0 | 0 | 0 | 0 |
| 0 | 0.008590804 | 0.024965498 | 0 | 0.02612174 |
| 0.001385074 | 0.003777015 | 0.167550418 | 0.286407291 | 0.087356611 |
| 13.93872822 | 3.493470203 | 0 | 0.693392523 | 24.61487219 |
| 21.39096303 | 3.38910807 | 0.164335857 | 1.130695446 | 0.048101829 |
| 0 | 0 | 0 | 0 | 0 |
| 0 | 0 | 0 | 0 | 0 |
| 2.422104963 | 0 | 0 | 0 | 0 |
| 0 | 2.915972576 | 0 | 0 | 6.05487643 |
| 0.00060192 | 0 | 0.001088798 | 0 | 0.009498844 |
| 0 | 0 | 0 | 0 | 0 |
| 0 | 0 | 0 | 0 | 0 |
| 0 | 0 | 0 | 0 | 0 |
| 0.102647843 | 0.124293309 | 0 | 0 | 0.090367529 |
| 0.000990036 | 0 | 0.023451631 | 0.554720142 | 0.357568047 |
| 0.083802651 | 0.014016531 | 0 | 0.013304569 | 0 |
| 0 | 0.135277509 | 0.113414219 | 0.038826667 | 0.044644463 |
| 0.344363094 | 0.05300094 | 0.289415424 | 0.016835261 | 0.150558782 |
| 0.019451045 | 0.021776603 | 0.009524475 | 0.084644063 | 0 |
| 0 | 0 | 0 | 0 | 0 |
| 4.850250439 | 0 | 0.015157776 | 0.0227863 | 0 |
| 0 | 0 | 0 | 0 | 0 |
| 0 | 0 | 0 | 0 | 0 |
| 0 | 0 | 0 | 0 | 0 |
| 0 | 0 | 0 | 0 | 0 |
| 0 | 0 | 0 | 0 | 0 |
| 0.075177433 | 0.081910616 | 0.03395257 | 0.016422988 | 0.069714375 |
| 0 | 0 | 0 | 0 | 0 |
| 0 | 0 | 0 | 0 | 0 |
| 0.055045861 | 0 | 0.016222862 | 0.007812636 | 0 |

| | | | | |
|---|---|---|---|---|
| 0 | 0 | 0 | 0 | 0 |
| 0 | 0 | 0 | 0 | 0 |
| 0 | 0 | 0 | 0 | 0 |
| 0.016579309 | 0 | 0 | 0 | 0 |
| 0 | 0 | 0 | 0 | 0 |
| 0 | 0 | 0 | 0 | 0 |
| 0 | 0 | 0 | 0 | 0 |
| 0.096697587 | 0.070674456 | 0 | 0.006893521 | 0 |
| 0 | 0 | 0 | 0 | 0 |
| 0 | 0 | 0 | 0 | 0 |
| 0.032209555 | 0.02557125 | 0.01778866 | 0.034691517 | 0.012296052 |
| 0 | 0 | 0 | 0 | 0 |
| 0.147209494 | 0.0019321 | 0 | 0.003075963 | 0 |
| 0 | 0 | 0 | 0 | 0 |
| 0 | 0 | 0 | 0 | 0 |
| 0 | 0 | 0 | 0 | 0 |
| 0 | 0 | 0 | 0 | 0.179741312 |
| 0.10529008 | 0.000864466 | 0 | 0 | 0.007630609 |
| 0 | 0 | 0 | 0 | 0 |
| 0 | 0 | 0 | 0 | 0 |
| 0.105863601 | 0 | 0 | 0.001786282 | 0 |
| 0 | 0 | 0 | 0 | 0 |
| 0 | 0 | 0 | 0 | 0 |
| 0 | 0 | 0 | 0 | 0 |
| 0 | 0 | 0 | 0 | 0 |
| 0 | 0 | 0 | 0 | 0 |
| 0 | 0 | 0 | 0 | 0 |
| 0 | 0 | 0 | 0 | 0 |
| 0 | 0 | 0 | 0 | 0 |
| 0.009354221 | 7.533215636 | 0 | 0 | 0 |
| 0 | 0 | 0 | 0 | 0.004225968 |
| 0 | 0 | 0 | 0 | 0 |
| 0 | 0 | 0 | 0 | 0 |
| 0 | 0 | 0 | 0 | 0 |
| 0.002207885 | 0 | 0.019968954 | 0 | 0 |
| 0 | 0 | 0 | 0 | 0 |
| 0.040365108 | 0.419595322 | 0 | 0 | 0 |
| 0.000705154 | 0.008241057 | 0.004960418 | 0.012453085 | 0.004352781 |
| 0 | 0 | 0 | 0 | 0 |
| 0 | 0 | 0 | 0 | 0 |
| 0 | 0 | 0 | 0 | 0 |
| 0 | 0 | 0 | 0 | 0 |
| 0 | 0 | 0 | 0 | 0 |
| 0 | 0 | 0 | 0 | 0.616048867 |
| 0.245012669 | 0 | 0.042243775 | 0 | 0 |
| 0 | 0 | 0 | 0 | 0 |
| 0.072138209 | 0 | 0 | 0 | 0.01960043 |
| 0 | 0 | 0 | 0 | 0 |

| | | | | |
|---|---|---|---|---|
| 0 | 0 | 0 | 0 | 0 |
| 0 | 0 | 0 | 0 | 0 |
| 0 | 0 | 0 | 0 | 0 |
| 0.016249598 | 0 | 0 | 0 | 0 |
| 0 | 0 | 0 | 0 | 0 |
| 0 | 0 | 0 | 0 | 0 |
| 0 | 0 | 0 | 0 | 0.019042483 |
| 0 | 0 | 0 | 0 | 0 |
| 0.020098851 | 0.038037526 | 0.596165628 | 0.077158806 | 0.189492671 |
| 0 | 0 | 0 | 0 | 0 |
| 0.003129236 | 0 | 0 | 0 | 0 |
| 0 | 0 | 0 | 0 | 0 |
| 0 | 0 | 0.000712688 | 0 | 0 |
| 0 | 0 | 0.000509112 | 0 | 0 |
| 0 | 0 | 2.802466095 | 0 | 2.69160212 |
| 0 | 0.003036777 | 1.016320193 | 0 | 0 |
| 0 | 0 | 0 | 0 | 0.000955413 |
| 0 | 0 | 0 | 0 | 0 |
| 0 | 0 | 0 | 0 | 0 |
| 0 | 0 | 0 | 0 | 0 |
| 0 | 0.05411459 | 0 | 0 | 0 |
| 0 | 0 | 0 | 0 | 0 |
| 0 | 0 | 0 | 0 | 0 |
| 0 | 0 | 0 | 0 | 0 |
| 0 | 0.008554145 | 0.029307184 | 0.027130533 | 0.009208181 |
| 0 | 0.126314658 | 0 | 0 | 0 |
| 0 | 0 | 0 | 0 | 0 |
| 0 | 0.001327489 | 0 | 0 | 0.000762126 |
| 0 | 0 | 0.277580973 | 0 | 0 |
| 0 | 0 | 0 | 0 | 0 |
| 0 | 0 | 0 | 0 | 0 |
| 0 | 0 | 0 | 0 | 0 |
| 0 | 0 | 0 | 0 | 0 |
| 0 | 0 | 0.024552028 | 0.001931926 | 0 |
| 0 | 0.001097743 | 0.002781212 | 0.001061068 | 0.002160777 |
| 0.016599184 | 0.029154356 | 0.006970291 | 0.003562721 | 0.008591668 |
| 0 | 1.780021949 | 0.056622978 | 0 | 2.557059257 |
| 0.129596865 | 0.467017746 | 0 | 0 | 1.299566883 |
| 0 | 0 | 0 | 0 | 0 |
| 0 | 0 | 0 | 0 | 0 |
| 0 | 0 | 0 | 0 | 0 |
| 0 | 0 | 0 | 0 | 0 |
| 0.651166452 | 0.258429468 | 0.385152984 | 0 | 0.267426309 |
| 0 | 0 | 0 | 0 | 0 |
| 0 | 0 | 0 | 0 | 0 |
| 0 | 0.002310426 | 0.001523879 | 0 | 0.198423577 |
| 0 | 0 | 0 | 0 | 0 |
| 0.019684371 | 0.004851945 | 0.261114967 | 0 | 0.001728966 |

| | | | | |
|---|---|---|---|---|
| 0 | 0 | 0.036005946 | 0 | 0 |
| 0 | 0 | 0 | 0 | 0 |
| 0 | 0 | 0 | 0 | 0 |
| 0 | 0 | 0 | 0 | 0 |
| 0.000797144 | 0.655358921 | 0 | 0 | 0.027170361 |
| 0 | 0 | 0 | 0 | 0 |
| 0 | 0 | 0 | 0 | 0 |
| 0 | 0 | 0 | 0 | 0 |
| 0 | 0 | 0 | 0 | 0 |
| 0 | 0 | 0 | 0 | 0 |
| 0.000609955 | 0 | 0 | 0 | 0 |
| 0 | 3.186008132 | 3.468141571 | 0 | 0.745873179 |
| 0.097324474 | 0 | 0 | 0 | 0 |
| 0 | 0 | 0 | 0 | 0 |
| 0 | 0.001580202 | 0.039142208 | 0.007289903 | 0.600986643 |
| 0 | 0 | 0 | 0 | 0 |
| 0 | 0.000599596 | 0.007572579 | 0.001014236 | 0 |
| 0 | 0 | 0 | 0 | 0 |
| 0.000538693 | 0 | 0 | 0 | 0 |
| 0 | 0 | 0 | 0 | 0 |
| 0.150744033 | 0 | 0 | 0 | 0 |
| 0 | 0 | 0 | 0 | 0 |
| 0.118118158 | 0 | 0.716941187 | 0.595204698 | 0.331277978 |
| 0 | 0 | 0 | 0 | 0 |
| 0 | 0 | 0 | 0 | 0 |
| 0 | 0 | 0 | 0 | 0 |
| 0 | 0 | 0 | 0 | 0 |
| 0 | 0 | 0 | 0 | 0 |
| 0 | 0 | 0 | 0 | 0 |
| 0 | 0 | 0 | 0 | 0 |
| 0 | 0 | 0 | 0 | 0 |
| 0.008174648 | 0.004473892 | 0.033661294 | 0.128651497 | 0 |
| 0 | 0 | 0 | 0 | 0 |
| 0.22242387 | 0 | 0.084083012 | 0 | 0.395220648 |
| 0.015096403 | 0.023162096 | 0.002528474 | 0.00550961 | 0.008518787 |
| 0 | 0.003110286 | 0.031592173 | 0.008417843 | 0 |
| 0 | 0 | 0 | 0 | 0 |
| 0.083899572 | 0 | 0 | 0 | 0 |
| 0.00192706 | 0 | 0 | 0 | 0 |
| 0 | 0 | 0 | 0 | 0 |
| 0 | 0 | 0 | 0 | 0 |
| 0 | 0 | 0 | 0 | 0 |
| 0 | 0 | 0 | 0 | 0 |
| 0 | 0 | 0 | 0 | 0 |
| 0.108803235 | 0.032799417 | 0.045984011 | 0.04181199 | 0.797226559 |
| 0 | 0 | 0 | 0 | 0 |
| 0 | 0 | 0 | 0 | 0 |
| 0 | 0 | 0.000724877 | 0 | 0.002516672 |

| | | | | |
|---|---|---|---|---|
| 0 | 0 | 0 | 0 | 0 |
| 0 | 0 | 0 | 0 | 0 |
| 0 | 0 | 0.013673205 | 0.002079041 | 0.057367936 |
| 0 | 0 | 0 | 0 | 0 |
| 0 | 0 | 0 | 0 | 0 |
| 0 | 0 | 0 | 0 | 0 |
| 0.004630671 | 0 | 0 | 0 | 0 |
| 0.004717764 | 0 | 0 | 0 | 0.03418653 |
| 0 | 0 | 0 | 0 | 0 |
| 0 | 0 | 0 | 0 | 0 |
| 0 | 0 | 0 | 0 | 0 |
| 0 | 0 | 0 | 0.000634612 | 0.010123294 |
| 0 | 0 | 0 | 0 | 0 |
| 0.09099827 | 0 | 0 | 0 | 0 |
| 0 | 0 | 0 | 0 | 0 |
| 0 | 0 | 0 | 0 | 0 |
| 0 | 0 | 0 | 0 | 0 |
| 0 | 0 | 0 | 0 | 0 |
| 1.721873124 | 0 | 0 | 0 | 0 |
| 0 | 0 | 0 | 0 | 0 |
| 0 | 0 | 0 | 0 | 0 |
| 0 | 0 | 0 | 0 | 0 |
| 0 | 0 | 0 | 0 | 0 |
| 0 | 0 | 0 | 0 | 0 |
| 0 | 0 | 0 | 0 | 0 |
| 0 | 0.00188078 | 0.017658868 | 0.027884048 | 0.014100615 |
| 0 | 0 | 0 | 0 | 0 |
| 0.023512054 | 0 | 0.878891599 | 0.001831741 | 0.00317066 |
| 0 | 0 | 0 | 0 | 0 |
| 0.145740362 | 0.014613699 | 0 | 0 | 0.162995269 |
| 0 | 0 | 0 | 0 | 0 |
| 0 | 0 | 0 | 0 | 0 |
| 0 | 0 | 0 | 0 | 0 |
| 0 | 0 | 0 | 0 | 0 |
| 0.063614059 | 0.003014381 | 0.00579462 | 0.006667824 | 0.010383538 |
| 0.373630703 | 0 | 0 | 0 | 0 |
| 0.563859836 | 0 | 0 | 0 | 0 |
| 0.002954873 | 0.001183998 | 0.007847816 | 0.001780242 | 0.001812658 |
| 0 | 0 | 0 | 0 | 0 |
| 0 | 0 | 0 | 0 | 0 |
| 1.030542822 | 5.010357757 | 33.59102547 | 19.62029939 | 3.87225961 |
| 0 | 0 | 0.080269653 | 0.417169279 | 0.009785692 |
| 0 | 0 | 0 | 0 | 0 |
| 0 | 0 | 0 | 0 | 0 |
| 0 | 0 | 0 | 0 | 0 |
| 0 | 0 | 0 | 0 | 0 |
| 0 | 0 | 0 | 0 | 0 |
| 0.012284122 | 0.001044096 | 0.002525052 | 0.003532246 | 0.018342472 |

| | | | | |
|---|---|---|---|---|
| 0 | 0 | 0 | 0 | 0 |
| 0 | 0.015464721 | 0.002250928 | 0.001816603 | 0.010543182 |
| 0 | 0 | 0 | 0 | 0 |
| 0 | 0 | 0 | 0 | 0 |
| 4.394322551 | 3.371299138 | 2.796826168 | 0 | 1.079292904 |
| 0 | 0 | 0 | 0 | 0 |
| 0 | 0 | 0 | 0 | 0 |
| 0 | 0 | 0 | 0 | 0 |
| 0.00160575 | 0.051687541 | 1.107379632 | 3.29118527 | 0.187465835 |
| 0 | 0 | 0 | 0 | 0 |
| 0 | 0 | 0 | 0 | 0 |
| 0 | 0 | 0 | 0 | 0 |
| 0.032925442 | 0 | 0 | 0 | 0.065421008 |
| 0.002058228 | 0.011417768 | 0.323287369 | 0.013237363 | 0.015246057 |
| 3.323448014 | 0 | 0 | 0 | 0 |
| 0.05922644 | 0.114116425 | 0 | 0 | 0.214678147 |
| 0 | 0 | 0.042240689 | 0 | 0.013575506 |
| 0 | 0 | 0 | 0 | 0 |
| 0 | 0 | 0 | 0 | 0 |
| 0 | 0 | 0 | 0 | 0 |
| 0 | 0 | 0 | 0 | 0 |
| 0 | 0 | 0 | 0 | 0 |
| 0 | 0 | 0 | 0 | 0 |
| 0 | 0 | 0 | 0 | 0 |
| 0 | 0 | 0 | 0 | 0 |
| 0 | 0 | 0 | 0 | 0 |
| 0 | 0 | 0.104275892 | 0 | 0 |
| 0.006407709 | 0.000544627 | 0.001580557 | 0 | 0.000750423 |
| 0 | 0 | 0 | 0 | 0 |
| 0 | 0 | 0 | 0 | 0 |
| 0 | 0 | 0 | 0 | 0 |
| 0 | 0 | 0 | 0 | 0 |
| 0 | 0 | 0 | 0 | 0 |
| 0.011602072 | 0 | 0 | 0 | 0.05821165 |
| 0.063266376 | 0 | 0 | 0 | 0 |
| 0 | 0.082028636 | 0 | 0.00490222 | 0 |
| 0 | 0.000515165 | 0 | 0 | 0 |
| 0 | 0 | 0 | 0 | 0 |
| 0 | 0 | 0 | 0 | 0 |
| 0 | 0 | 0 | 0 | 0 |
| 0.016340885 | 0 | 0 | 0 | 0 |
| 0 | 0 | 0 | 0 | 0 |
| 0 | 0 | 0 | 0 | 0 |
| 0.016825683 | 0 | 0 | 0 | 0 |
| 0 | 0 | 0 | 0 | 0 |
| 0 | 0 | 0 | 0 | 0 |
| 0 | 0 | 0 | 0 | 0 |
| 0 | 0 | 0 | 0 | 0 |

| | | | | |
|---|---|---|---|---|
| 0 | 0 | 0 | 0 | 0 |
| 0 | 0 | 0 | 0 | 0 |
| 0 | 0 | 0 | 0 | 0 |
| 0 | 0 | 0 | 0 | 0 |
| 0 | 0 | 0 | 0 | 0 |
| 0 | 0 | 0 | 0 | 0 |
| 0.003908842 | 0 | 0.131234804 | 0.00112397 | 0.325401392 |
| 0 | 0 | 0 | 0 | 0 |
| 0 | 0 | 0 | 0 | 0 |
| 0 | 0 | 0 | 0 | 0.06583838 |
| 0 | 0 | 0 | 0 | 0 |
| 0 | 0 | 0.00078073 | 0 | 0 |
| 0 | 0 | 0 | 0 | 0 |
| 0 | 0 | 0 | 0 | 0 |
| 0 | 0 | 0 | 0 | 0 |
| 0 | 0 | 0 | 0 | 0 |
| 0 | 0 | 0 | 0 | 0 |
| 0 | 0.002235858 | 0 | 0 | 0 |
| 0.014702469 | 0 | 0 | 0 | 0 |
| 0 | 0 | 0 | 0 | 0 |
| 0.004059747 | 0.000527176 | 0.229486863 | 0 | 0.000907973 |
| 0 | 0 | 0 | 0 | 0 |
| 0.008458231 | 0.002325892 | 0 | 0 | 0.011216695 |
| 0 | 0.002275028 | 0.014338415 | 0.004373052 | 0.017810716 |
| 0 | 0 | 0 | 0 | 0 |
| 0 | 0 | 0 | 0 | 0 |
| 0 | 0 | 0 | 0 | 0 |
| 0 | 0 | 0 | 0 | 0 |
| 0 | 0 | 0 | 0 | 0 |
| 0.467849607 | 0.043756544 | 0 | 0.496206861 | 0.025377459 |
| 0 | 0 | 0 | 0 | 0 |
| 0 | 0 | 0 | 0 | 0 |
| 0 | 0 | 0 | 0 | 0 |
| 0.0615403 | 0 | 0 | 0 | 0 |
| 0 | 0 | 0 | 0 | 0 |
| 0 | 0 | 0 | 0 | 0 |
| 0.001799143 | 0 | 0.032335651 | 0.020574044 | 0.012257201 |
| 0 | 0 | 0 | 0 | 0 |
| 0 | 0 | 0 | 0 | 0 |
| 0 | 0 | 0 | 0 | 0 |
| 0 | 0 | 0 | 0 | 0 |
| 0 | 0 | 0 | 0 | 0 |
| 0 | 0 | 0 | 0 | 0 |
| 0 | 0.002222503 | 0 | 0 | 0 |
| 0 | 0 | 0 | 0 | 0 |
| 0 | 0 | 0 | 0 | 0 |
| 0 | 0 | 0 | 0 | 0 |
| 0.001714862 | 0 | 0 | 0 | 0 |

| | | | | |
|---|---|---|---|---|
| 0.317413031 | 0.009680772 | 0.018657895 | 0 | 0.013798773 |
| 0 | 0 | 0 | 0 | 0 |
| 0 | 0 | 0 | 0 | 0 |
| 0 | 0 | 0 | 0 | 0 |
| 0 | 0 | 0 | 0 | 0 |
| 0.009160636 | 0 | 0 | 0 | 0 |
| 0 | 0 | 0 | 0 | 0.002799293 |
| 0.219351722 | 0.041016636 | 1.154420026 | 0 | 0.320752798 |
| 0.002054931 | 0.052572679 | 1.472766393 | 1.166409952 | 0.159436127 |
| 0.000650264 | 0.07854914 | 0.047441984 | 0.201565153 | 0.24104687 |
| 0 | 0 | 0 | 0 | 0 |
| 0 | 0 | 0 | 0 | 0 |
| 0 | 0 | 0 | 0 | 0 |
| 0.001936202 | 0 | 0 | 0 | 0 |
| 0 | 0 | 0 | 0 | 0.187665095 |
| 0 | 0 | 0 | 0 | 0 |
| 0 | 0 | 0.234382476 | 0 | 0.073576062 |
| 0 | 0 | 0 | 0 | 0 |
| 0 | 0 | 0 | 0 | 0 |
| 0 | 0 | 0 | 0 | 0 |
| 0 | 0 | 0 | 0 | 0 |
| 0 | 0 | 0 | 0 | 0 |
| 0 | 0 | 0 | 0 | 0 |
| 0 | 0 | 0 | 0 | 0 |
| 0.028497209 | 0 | 0 | 0 | 0 |
| 0 | 0 | 0 | 0 | 0 |
| 0 | 0 | 0 | 0 | 0 |
| 0 | 0 | 0 | 0 | 0 |
| 0 | 0 | 0 | 0 | 0 |
| 0 | 0 | 0 | 0 | 0 |
| 0 | 0 | 0 | 0 | 0 |
| 0 | 0 | 0 | 0 | 0 |
| 0 | 0 | 0 | 0 | 0 |
| 0 | 0 | 0 | 0 | 0 |
| 0.01239126 | 0 | 0 | 0 | 0 |
| 0 | 0 | 0 | 0 | 0 |
| 0 | 0 | 0 | 0 | 0 |
| 0 | 0 | 0 | 0 | 0 |
| 0.008632607 | 0 | 0 | 0 | 0 |
| 0 | 0 | 0 | 0.367541891 | 0 |
| 0 | 0 | 0 | 0 | 0 |
| 0.453170817 | 0 | 0 | 0 | 0 |
| 0 | 0 | 0 | 0 | 0 |
| 0 | 0 | 0 | 0 | 0 |
| 0 | 0.001082503 | 0 | 0 | 0 |
| 0 | 0 | 0 | 0 | 0 |
| 0 | 0 | 0 | 0 | 0 |
| 0 | 0.002590537 | 0 | 0 | 0 |

| | | | | |
|---|---|---|---|---|
| 0 | 0 | 0 | 0 | 0 |
| 0 | 0.007933888 | 0 | 0 | 0.06676726 |
| 0 | 0 | 0 | 0 | 0 |
| 0 | 0 | 0 | 0 | 0 |
| 0.02432965 | 0.121598362 | 2.05376828 | 0.086692826 | 0 |
| 0 | 0 | 0 | 0 | 0 |
| 0 | 0 | 0 | 0 | 0 |
| 0.0211954 | 0.035245357 | 0.159521164 | 0.026816436 | 0.029011273 |
| 0.018139561 | 0 | 0 | 0 | 0 |
| 0 | 0 | 0 | 0 | 0 |
| 0 | 0 | 0 | 0 | 0 |
| 0.3589829 | 0 | 0 | 0 | 0 |
| 0 | 0 | 0 | 0 | 0 |
| 0 | 0 | 0 | 0 | 0 |
| 0 | 0 | 0 | 0 | 0.007208087 |
| 0 | 0 | 0.026522322 | 0.027410854 | 0.666667682 |
| 0 | 0 | 0 | 0 | 0 |
| 0 | 0 | 0 | 0 | 0 |
| 0 | 0 | 0 | 0 | 0 |
| 0 | 0 | 0 | 0 | 0 |
| 0 | 0 | 0 | 0 | 0 |
| 0 | 0 | 0 | 0 | 0 |
| 0 | 0 | 0 | 0 | 0 |
| 0.001048164 | 0.016904697 | 0.009479995 | 0.040198561 | 0.005696516 |
| 0.371979776 | 0 | 0 | 0 | 0 |
| 0 | 0 | 0 | 0 | 0 |
| 0.00419416 | 0.110133499 | 0.016979791 | 0 | 0.12002551 |
| 0.001387984 | 0 | 0 | 0 | 0.001625504 |
| 0 | 0 | 0 | 0.001408462 | 0.008891473 |
| 0 | 0 | 0 | 0 | 0 |
| 0 | 0 | 0 | 0 | 0 |
| 0 | 0 | 0 | 0 | 0 |
| 0 | 0 | 0 | 0 | 0 |
| 0 | 0 | 0 | 0 | 0 |
| 0 | 0 | 0 | 0 | 0 |
| 0 | 0 | 0 | 0 | 0 |
| 0 | 0 | 0 | 0 | 0 |
| 0.001843439 | 0.017350155 | 0 | 0 | 0 |
| 0 | 0 | 0 | 0 | 0 |
| 0 | 0 | 0 | 0 | 0 |
| 0 | 0 | 0 | 0 | 0 |
| 0.610877193 | 0 | 0 | 0 | 0 |
| 0 | 0 | 0 | 0 | 0 |
| 0.003958673 | 0 | 0 | 0 | 0 |
| 0 | 0 | 0 | 0 | 0 |
| 0 | 0 | 0 | 0 | 0 |
| 0 | 0 | 0 | 0 | 0 |
| 0 | 0 | 0 | 0 | 0 |

| | | | | |
|---|---|---|---|---|
| 0 | 0 | 0 | 0 | 0 |
| 0 | 0 | 0 | 0 | 0 |
| 0 | 0 | 0 | 0 | 0 |
| 0 | 0.000419318 | 0.001183098 | 0 | 0 |
| 0 | 0 | 0 | 0 | 0 |
| 0.155367657 | 0 | 0 | 0.003923166 | 0 |
| 0 | 0 | 0 | 0 | 0 |
| 0 | 0 | 0 | 0 | 0 |
| 0 | 0 | 0 | 0 | 0 |
| 0 | 0 | 0 | 0 | 0 |
| 0 | 0 | 0 | 0 | 0 |
| 0 | 0 | 0.003459205 | 0 | 0 |
| 0 | 0 | 0 | 0 | 0 |
| 0 | 0 | 0 | 0 | 0 |
| 0 | 0 | 0 | 0 | 0 |
| 0 | 0 | 0 | 0 | 0 |
| 0 | 0 | 0 | 0 | 0 |
| 0 | 0 | 0.008183296 | 0 | 0 |
| 0 | 0 | 0.000576087 | 0 | 0 |
| 0 | 0 | 0 | 0 | 0 |
| 0 | 0 | 0.313498823 | 0.362338102 | 0.144969079 |
| 0 | 0 | 0 | 0 | 0 |
| 0.070030507 | 0.002773064 | 0.023596604 | 0.040131789 | 0.279669565 |
| 0 | 0 | 0 | 0 | 0 |
| 0 | 0 | 0 | 0 | 0 |
| 0.3667475 | 0 | 0 | 0 | 0 |
| 0 | 0 | 0 | 0 | 0 |
| 0 | 0 | 0 | 0 | 0 |
| 0.062478584 | 0 | 0 | 0 | 0 |
| 0 | 0 | 0 | 0 | 0 |
| 0 | 0 | 0 | 0 | 0 |
| 0 | 0 | 0 | 0 | 0 |
| 0.001543136 | 0 | 1.437541031 | 0.604992788 | 0 |
| 0 | 0 | 0 | 0 | 0 |
| 0.130164917 | 0 | 0 | 0 | 0 |
| 0 | 0 | 0.00149329 | 0 | 0.003788667 |
| 0 | 0 | 0 | 0 | 0 |
| 0.473360869 | 0 | 0 | 0 | 0 |
| 0 | 0.002410422 | 0.00734071 | 0.254378744 | 0.023986738 |
| 0 | 0 | 0 | 0 | 0 |
| 0 | 0 | 0 | 0 | 0 |
| 0 | 0 | 0 | 0 | 0 |
| 0.194609282 | 0 | 0.433466241 | 0.011389256 | 0.962251332 |
| 0 | 0 | 0 | 0 | 0 |
| 0 | 0 | 0 | 0 | 0 |
| 0 | 0 | 0 | 0 | 0 |
| 0 | 0 | 0 | 0 | 0 |
| 0 | 0 | 0 | 0 | 0 |

| | | | | |
|---|---|---|---|---|
| 0 | 0 | 0 | 0 | 0 |
| 0 | 0 | 0 | 0 | 0 |
| 0 | 0 | 0 | 0 | 0 |
| 0 | 0 | 0 | 0 | 0 |
| 0 | 0 | 0 | 0 | 0 |
| 0 | 0 | 0 | 0 | 0 |
| 0 | 0 | 0 | 0 | 0 |
| 0 | 0 | 0.587614194 | 0 | 0 |
| 0 | 0 | 0 | 0 | 0 |
| 0.080279187 | 0 | 0.034899533 | 0 | 0 |
| 0 | 0 | 0 | 0 | 0 |
| 0 | 0 | 0 | 0 | 0 |
| 0 | 0 | 0 | 0 | 0 |
| 0 | 0 | 0 | 0 | 0 |
| 0 | 0 | 0 | 0.030115352 | 0 |
| 0 | 0 | 0.001219075 | 0 | 0.006294423 |
| 0 | 0 | 0 | 0 | 0 |
| 0 | 0 | 0 | 0 | 0 |
| 0 | 0 | 0 | 0 | 0 |
| 0.396438141 | 0.193502066 | 0 | 0 | 0.029256974 |
| 0.005349512 | 0 | 0 | 0 | 0 |
| 0 | 0 | 0 | 0 | 0 |
| 0 | 0 | 0 | 0 | 0 |
| 0 | 0 | 0 | 0 | 0 |
| 0 | 0 | 0.001647613 | 0 | 0 |
| 0 | 0 | 0 | 0 | 0 |
| 0 | 0 | 0 | 0 | 0 |
| 0 | 0 | 0 | 0 | 0 |
| 0 | 0 | 0 | 0 | 0 |
| 0 | 0 | 0 | 0 | 0 |
| 0.309922634 | 0.418511798 | 0 | 0 | 0.073034312 |
| 0 | 0 | 0 | 0 | 0 |
| 0 | 0 | 0 | 0 | 0 |
| 0 | 0 | 0 | 0 | 0 |
| 0 | 0 | 0 | 0 | 0 |
| 0 | 0 | 0 | 0 | 0 |
| 0 | 0 | 0 | 0 | 0 |
| 0 | 0 | 0 | 0 | 0 |
| 0.020372452 | 0 | 0 | 0 | 0 |
| 0 | 0 | 0 | 0 | 0 |
| 0 | 0 | 0 | 0 | 0 |
| 0 | 0 | 0 | 0 | 0 |
| 0 | 0 | 0 | 0 | 0 |
| 0 | 0 | 0 | 0 | 0 |
| 0 | 0 | 0 | 0 | 0 |
| 0 | 0 | 0 | 0 | 0 |
| 0 | 0.038056801 | 0.060525978 | 1.360153276 | 0 |
| 0.001233513 | 0 | 0 | 0 | 0 |

| | | | | |
|---|---|---|---|---|
| 0 | 0 | 0 | 0 | 0 |
| 0 | 0 | 0 | 0 | 0 |
| 0 | 0 | 0 | 0 | 0 |
| 0.001978604 | 0 | 0 | 0 | 0 |
| 0.077621184 | 0.271687312 | 0.684166072 | 0 | 0.361700217 |
| 0 | 0 | 0 | 0 | 0 |
| 0 | 0 | 0 | 0 | 0 |
| 0 | 0 | 0 | 0 | 0 |
| 0 | 0 | 0 | 0 | 0 |
| 0 | 0 | 0 | 0 | 0 |
| 0 | 0 | 0 | 0 | 0.033509931 |
| 0.019320547 | 0.009500206 | 0.018984357 | 0.012448522 | 0.148645451 |
| 0 | 0.003347778 | 0.001488974 | 0 | 0.007555435 |
| 0 | 0 | 0 | 0 | 0 |
| 0 | 0 | 0 | 0 | 0 |
| 0.15914803 | 0 | 0 | 0 | 0.454127805 |
| 0.019923131 | 0.004013586 | 0.002654768 | 0 | 0.005671987 |
| 0 | 0 | 0 | 0 | 0 |
| 0.029909464 | 0.018845232 | 0 | 0.107628837 | 0.162812386 |
| 0 | 0 | 0.149302469 | 0.003804169 | 0.014354504 |
| 0 | 0 | 0 | 0 | 0 |
| 0 | 0 | 0 | 0 | 0 |
| 0 | 0 | 0 | 0 | 0 |
| 0 | 0 | 0 | 0 | 0 |
| 0 | 0 | 0 | 0 | 0 |
| 0 | 0 | 0 | 0 | 0 |
| 0 | 0.061168267 | 0.028372462 | 0.004303701 | 0.014241713 |
| 0 | 0 | 0 | 0 | 0.001385068 |
| 0 | 0 | 0 | 0 | 0 |
| 0 | 0 | 0 | 0 | 0 |
| 0.023520909 | 0.001201689 | 0.007943546 | 0.001422887 | 0.007657923 |
| 0 | 0 | 0 | 0 | 0.069301328 |
| 0 | 0 | 0 | 0 | 0 |
| 0 | 0 | 0 | 0 | 0 |
| 0 | 0 | 0 | 0 | 0 |
| 0 | 0 | 0 | 0 | 0 |
| 0 | 0 | 0 | 0 | 0 |
| 0.017699023 | 0.015112463 | 0.04873083 | 0.045842438 | 0.028064466 |
| 0 | 0 | 0 | 0 | 0 |
| 0 | 0.002678468 | 0 | 0 | 0 |
| 0.008023318 | 0 | 0 | 0 | 0.007984228 |
| 0 | 0 | 0 | 0 | 0 |
| 0 | 0 | 0 | 0 | 0 |
| 0 | 0 | 0 | 0.001058855 | 0.00301878 |
| 0 | 0 | 0 | 0 | 0 |
| 0 | 0 | 0 | 0 | 0 |
| 0 | 0 | 0 | 0 | 0 |
| 0 | 0 | 0 | 0 | 0 |

| | | | | |
|---|---|---|---|---|
| 0 | 0 | 0 | 0 | 0 |
| 0 | 0 | 0 | 0 | 0 |
| 0 | 0 | 0 | 0 | 0 |
| 0.031434634 | 0.004854574 | 0.002749991 | 0.002441308 | 0.01288076 |
| 0 | 0 | 0 | 0 | 0 |
| 0 | 0 | 0 | 0 | 0 |
| 0.013890635 | 0 | 0 | 0 | 0 |
| 0 | 0 | 0.005955871 | 0 | 0 |
| 0 | 0 | 0 | 0 | 0 |
| 0 | 0 | 0 | 0 | 0 |
| 0 | 0 | 0 | 0 | 0 |
| 0 | 0 | 0 | 0 | 0 |
| 0.026261789 | 0.003348204 | 0.331390629 | 0 | 0.070642365 |
| 0 | 0 | 0 | 0 | 0 |
| 0 | 0 | 0 | 0 | 0 |
| 0 | 0 | 0 | 0 | 0 |
| 0 | 0 | 0 | 0 | 0 |
| 0 | 0 | 0 | 0 | 0 |
| 0 | 0 | 0 | 0 | 0 |
| 0 | 0 | 0 | 0 | 0 |
| 0 | 0 | 0 | 0 | 0 |
| 0 | 0 | 0 | 0 | 0 |
| 0 | 0.016777982 | 0 | 0.005593239 | 0.223162947 |
| 0.003709677 | 0 | 0 | 0 | 0.010453949 |
| 0 | 0 | 0 | 0 | 0 |
| 0 | 0 | 0 | 0 | 0 |
| 0 | 0 | 0 | 0 | 0 |
| 0 | 0 | 0 | 0 | 0 |
| 0 | 0 | 0 | 0 | 0 |
| 0 | 0 | 0 | 0 | 0 |
| 0.005257821 | 0 | 0 | 0 | 0.00329259 |
| 0.001468243 | 0 | 0 | 0 | 0 |
| 0 | 0 | 0 | 0 | 0 |
| 0 | 0 | 0 | 0 | 0 |
| 0 | 0 | 0 | 0 | 0 |
| 0 | 0 | 0 | 0.013038804 | 0.003441984 |
| 0 | 0.004352237 | 0 | 0.019631875 | 0 |
| 0 | 0 | 0 | 0 | 0 |
| 0 | 0.000808716 | 0.030920387 | 0.000977122 | 0 |
| 0 | 0 | 0 | 0 | 0 |
| 0 | 0 | 0 | 0 | 0 |
| 0 | 0 | 0 | 0 | 0 |
| 0 | 0 | 0 | 0 | 0 |
| 0 | 0 | 0 | 0 | 0 |
| 0 | 0 | 0 | 0 | 0 |
| 0.299524263 | 0.02337907 | 0 | 0 | 0.157564787 |
| 0 | 0 | 0 | 0 | 0 |
| 0 | 0 | 0 | 0 | 0 |

| | | | | |
|---|---|---|---|---|
| 0 | 0 | 0 | 0 | 0 |
| 0 | 0 | 0 | 0 | 0 |
| 0.243328239 | 0 | 0 | 0 | 0 |
| 0 | 0 | 0 | 0 | 0 |
| 0 | 0 | 0 | 0 | 0 |
| 0 | 0 | 0 | 0 | 0 |
| 0 | 0 | 0.001325363 | 0 | 0 |
| 0 | 0 | 0 | 0 | 0 |
| 0 | 0 | 0 | 0 | 0 |
| 0 | 0 | 0 | 0 | 0 |
| 0.01042574 | 0.000562359 | 0.001208902 | 0 | 0.004197137 |
| 0.011225238 | 0 | 0.000902447 | 0 | 0.001687091 |
| 0 | 0 | 0 | 0 | 0 |
| 0 | 0 | 0 | 0 | 0 |
| 0 | 0 | 0 | 0 | 0 |
| 0 | 0 | 0 | 0 | 0 |
| 0 | 0 | 0 | 0 | 0 |
| 0 | 0 | 0 | 0 | 0 |
| 0 | 0 | 0 | 0 | 0 |
| 0.021271265 | 0.001615867 | 0 | 0.00126152 | 0 |
| 0 | 0 | 0 | 0 | 0 |
| 0 | 0 | 0 | 0 | 0 |
| 0 | 0 | 0 | 0 | 0 |
| 0 | 0 | 0 | 0 | 0 |
| 0 | 0 | 0 | 0 | 0 |
| 0 | 0 | 0 | 0 | 0 |
| 0 | 0 | 0 | 0 | 0 |
| 0.013241391 | 0 | 0 | 0 | 0 |
| 0 | 0 | 0 | 0 | 0 |
| 0 | 0 | 0 | 0 | 0 |
| 0.000342642 | 0.003096748 | 0.004476306 | 0.00270944 | 0.000919592 |
| 0 | 0 | 0 | 0 | 0 |
| 0.054636938 | 1.339933622 | 0.031384582 | 0.211826344 | 0.045272164 |
| 0.005149648 | 0 | 0.000612834 | 0.008358494 | 0 |
| 0 | 0 | 0 | 0 | 0 |
| 0 | 0 | 0 | 0 | 0 |
| 0 | 0 | 0 | 0 | 0 |
| 0 | 0 | 0 | 0 | 0 |
| 0 | 0 | 0 | 0 | 0 |
| 0 | 0 | 0 | 0 | 0 |
| 0 | 0 | 0 | 0 | 0 |
| 0.123283311 | 0.028560965 | 0.523138975 | 1.699022938 | 0.010013985 |
| 0 | 0.141790881 | 0.147862343 | 0 | 0.038525867 |
| 0 | 0 | 0 | 0 | 0 |
| 0 | 0 | 0 | 0 | 0 |
| 0.012868775 | 0 | 0.057036936 | 0.006075218 | 0 |
| 0 | 0.000941888 | 0.012907931 | 0.001194926 | 0 |
| 0 | 0 | 0 | 0 | 0 |

| | | | | |
|---|---|---|---|---|
| 0 | 0 | 0 | 0 | 0 |
| 0.00208869 | 0 | 0.005247473 | 0 | 0 |
| 0 | 0 | 0.005639235 | 0.007717124 | 0 |
| 0 | 0 | 0 | 0 | 0 |
| 0.031428879 | 0.469301576 | 0.897203456 | 0.116867901 | 1.596301913 |
| 0 | 0.048577259 | 0.000952796 | 0 | 0.321636652 |
| 0 | 0 | 0 | 0 | 0 |
| 0 | 0 | 0 | 0 | 0 |
| 0 | 0 | 0 | 0 | 0 |
| 0 | 0 | 0 | 0 | 0 |
| 0 | 0 | 0 | 0 | 0 |
| 0 | 0 | 0 | 0 | 0 |
| 0 | 0 | 0 | 0 | 0 |
| 0 | 0 | 0 | 0 | 0 |
| 0 | 0 | 0 | 0 | 0 |
| 0 | 0 | 0 | 0 | 0 |
| 0 | 0 | 0 | 0 | 0 |
| 0 | 0 | 0 | 0 | 0 |
| 0 | 0 | 0 | 0 | 0 |
| 0 | 0 | 0 | 0 | 0 |
| 0 | 0 | 0 | 0 | 0 |
| 0 | 0 | 0.12286948 | 0 | 0 |
| 0 | 0 | 0 | 0 | 0 |
| 0 | 0 | 0 | 0 | 0 |
| 0.002010747 | 0 | 0.004041324 | 0 | 0.009066123 |
| 0.001618211 | 0 | 0.002032738 | 0.00405207 | 0 |
| 0 | 0 | 0 | 0 | 0 |
| 0.01582802 | 0.814769629 | 0 | 0 | 0 |
| 0 | 0 | 0 | 0 | 0 |
| 0 | 0 | 0 | 0 | 0 |
| 0.00747752 | 0 | 0 | 0 | 0 |
| 0 | 0 | 0 | 0 | 0 |
| 0 | 0 | 0 | 0 | 0 |
| 0 | 0 | 0 | 0 | 0 |
| 0 | 0 | 0 | 0 | 0 |
| 0 | 0 | 0 | 0 | 0 |
| 0 | 0 | 0.001393819 | 0 | 0.001985289 |
| 0 | 0.009914585 | 0 | 0 | 0.034800909 |
| 4.804555909 | 1.035723202 | 0 | 0.023816431 | 0 |
| 0 | 0 | 0 | 0 | 0 |
| 0 | 0 | 0 | 0 | 0 |
| 0.05281157 | 0 | 1.625678883 | 0 | 0 |
| 0.260254201 | 0.009363521 | 0.017340633 | 0 | 0.011986362 |
| 0 | 0 | 0 | 0 | 0 |
| 0 | 0 | 0 | 0 | 0 |
| 0 | 0 | 0 | 0 | 0 |
| 0.002691521 | 0.010452907 | 0.004681372 | 0.037000044 | 0.013335846 |
| 0 | 0 | 0 | 0 | 0 |

| | | | | |
|---|---|---|---|---|
| 0 | 0 | 0 | 0 | 0 |
| 0.081378268 | 0.005007181 | 0.028156845 | 0.004338196 | 0.042909832 |
| 0 | 0 | 0 | 0 | 0 |
| 0 | 0 | 0 | 0 | 0 |
| 0 | 0 | 0 | 0 | 0 |
| 0 | 0 | 0 | 0 | 0 |
| 0 | 0 | 0 | 0 | 0 |
| 0 | 0 | 0 | 0 | 0 |
| 0 | 0 | 0 | 0 | 0 |
| 0 | 0 | 0 | 0 | 0 |
| 0 | 0 | 0 | 0 | 0 |
| 0 | 0 | 0 | 0 | 0 |
| 0 | 0 | 0 | 0 | 0 |
| 0 | 0 | 0 | 0 | 0 |
| 0 | 0.017054594 | 0.024796095 | 0.010303003 | 0 |
| 0 | 0 | 0 | 0 | 0 |
| 0 | 0 | 0 | 0 | 0 |
| 0 | 0 | 0 | 0 | 0 |
| 0 | 0.00235835 | 0 | 0 | 0.00551253 |
| 0.008297685 | 0 | 0 | 0.002573097 | 0 |
| 0 | 0 | 0 | 0 | 0 |
| 0 | 0 | 0.05907101 | 0.072093327 | 0.004378607 |
| 0 | 0 | 0.005743381 | 0 | 0.06610451 |
| 0 | 0 | 0.0221021 | 0 | 0 |
| 0 | 0 | 0 | 0 | 0 |
| 0 | 0 | 0 | 0 | 0 |
| 0 | 0 | 0 | 0 | 0 |
| 0 | 0 | 0 | 0 | 0 |
| 0 | 0 | 0 | 0 | 0 |
| 0 | 0 | 0 | 0 | 0 |
| 0 | 0 | 0 | 0 | 0 |
| 0 | 0 | 0 | 0 | 0 |
| 0 | 0 | 0 | 0 | 0 |
| 0 | 0 | 0 | 0 | 0 |
| 0 | 0 | 0 | 0 | 0 |
| 0 | 0 | 0 | 0 | 0 |
| 0 | 0 | 0 | 0 | 0 |
| 0 | 0 | 0 | 0 | 0 |
| 0 | 0.004928155 | 0.021910405 | 0.006315255 | 0.005401408 |
| 0 | 0 | 0 | 0 | 0 |
| 0.118440795 | 0.09359701 | 0.166447679 | 0.251866292 | 0.067341829 |
| 0 | 0 | 0 | 0 | 0 |
| 0 | 0 | 0 | 0 | 0 |
| 0 | 0 | 0 | 0 | 0 |
| 0 | 0 | 0 | 0 | 0 |
| 0.109206806 | 0.009505088 | 0.136934476 | 9.057455693 | 0.010033797 |
| 0 | 0 | 0 | 0 | 0 |
| 0 | 0 | 0 | 0 | 0 |

| | | | | |
|---|---|---|---|---|
| 0 | 0 | 0 | 0 | 0 |
| 0 | 0 | 0 | 0 | 0 |
| 0 | 0 | 0 | 0 | 0 |
| 0 | 0 | 0 | 0 | 0 |
| 0 | 0.002557993 | 0.006373738 | 0 | 0.004806237 |
| 0 | 0 | 0 | 0 | 0 |
| 0 | 0 | 0 | 0 | 0 |
| 0 | 0 | 0 | 0 | 0 |
| 0 | 0.014435921 | 0 | 0 | 0 |
| 0 | 0 | 0.009427731 | 0 | 0.001032955 |
| 0.032642073 | 0 | 0 | 0 | 0 |
| 0 | 0 | 0 | 0 | 0 |
| 0.007523772 | 0 | 0.008978536 | 0.011155174 | 0 |
| 0 | 0 | 0 | 0 | 0 |
| 0 | 0 | 0 | 0 | 0 |
| 0 | 0 | 0 | 0 | 0 |
| 0 | 0 | 0 | 0 | 0 |
| 0 | 0 | 0 | 0 | 0 |
| 0 | 0 | 0 | 0 | 0 |
| 1.6166921 | 0.078002587 | 0.899467844 | 0.094434798 | 2.619756208 |
| 0 | 0 | 0 | 0 | 0 |
| 0 | 0 | 0 | 0 | 0 |
| 0 | 0 | 0 | 0 | 0 |
| 0 | 0 | 0 | 0 | 0 |
| 0 | 0 | 0 | 0 | 0 |
| 0 | 0 | 0 | 0 | 0 |
| 0 | 0 | 0 | 0 | 0 |
| 0 | 0 | 0 | 0 | 0 |
| 0 | 0 | 0 | 0 | 0 |
| 0 | 0.004183246 | 0 | 0.00117935 | 0.016331206 |
| 0 | 0 | 0 | 0 | 0 |
| 0 | 0 | 0 | 0 | 0 |
| 0 | 0 | 0 | 0 | 0 |
| 0 | 0 | 0 | 0 | 0 |
| 0 | 0 | 0 | 0 | 0 |
| 0 | 0 | 0 | 0 | 0 |
| 0 | 0 | 0 | 0 | 0 |
| 0 | 0 | 0 | 0 | 0 |
| 0 | 0 | 0 | 0 | 0 |
| 0 | 0 | 0 | 0 | 0 |
| 0 | 0 | 0 | 0 | 0 |
| 0 | 0 | 0 | 0 | 0 |
| 0 | 0 | 0 | 0 | 0 |
| 0 | 0 | 0 | 0 | 0 |
| 0.004324221 | 0.006615713 | 0.017382176 | 0.002694057 | 0.011183456 |
| 0 | 0 | 0 | 0 | 0 |
| 0 | 0 | 0 | 0 | 0 |
| 0 | 0 | 0 | 0 | 0 |

| | | | | |
|---|---|---|---|---|
| 0 | 0 | 0 | 0 | 0 |
| 0 | 0 | 0 | 0 | 0 |
| 0 | 0 | 0 | 0 | 0 |
| 0 | 0 | 0 | 0 | 0 |
| 0 | 0 | 0 | 0 | 0 |
| 0.170311945 | 0.010426539 | 0.020939304 | 0.029085306 | 0.000472578 |
| 0 | 0 | 0 | 0 | 0 |
| 0 | 0 | 0 | 0 | 0 |
| 0 | 0 | 0 | 0 | 0 |
| 0 | 0 | 0 | 0 | 0 |
| 0 | 0 | 0 | 0 | 0 |
| 0 | 0 | 0 | 0 | 0 |
| 0 | 0 | 0 | 0 | 0 |
| 0.025139519 | 0 | 0 | 0 | 0 |
| 0 | 0 | 0 | 0 | 0 |
| 0 | 0 | 0 | 0 | 0 |
| 0.242778514 | 0 | 0 | 0 | 0 |
| 0 | 0 | 0 | 0 | 0 |
| 0.001577361 | 0 | 0 | 0 | 0 |
| 0 | 0 | 0 | 0 | 0 |
| 0 | 0 | 0 | 0 | 0 |
| 0 | 0 | 0 | 0 | 0 |
| 0 | 0 | 0 | 0 | 0 |
| 0 | 0 | 0 | 0 | 0 |
| 0 | 0 | 0 | 0 | 0 |
| 0 | 0 | 0 | 0 | 0 |
| 0 | 0 | 0 | 0 | 0 |
| 0 | 0 | 0 | 0.011591265 | 0 |
| 0 | 0 | 0 | 0 | 0 |
| 0 | 0 | 0 | 0 | 0 |
| 0 | 0 | 0 | 0 | 0 |
| 0 | 0 | 0 | 0 | 0 |
| 0 | 0 | 0 | 0 | 0 |
| 0 | 0 | 0 | 0 | 0 |
| 0 | 0 | 0 | 0 | 0 |
| 0 | 0 | 0 | 0 | 0 |
| 0 | 0 | 0 | 0 | 0 |
| 0 | 0 | 0 | 0 | 0 |
| 3.692325354 | 0 | 0 | 0 | 0.865656228 |
| 0 | 0 | 0 | 0 | 0 |
| 0 | 0 | 0 | 0 | 0 |
| 0 | 0 | 0 | 0 | 0 |
| 0 | 0 | 0 | 0 | 0 |
| 0 | 0 | 0 | 0 | 0 |
| 0 | 0 | 0 | 0 | 0 |
| 0 | 0 | 0 | 0 | 0 |
| 0 | 0 | 0 | 0 | 0 |
| 0 | 0 | 0 | 0 | 0 |

| | | | | |
|---|---|---|---|---|
| 0 | 0 | 0 | 0 | 0 |
| 0 | 0 | 0.001981119 | 0 | 0 |
| 0 | 0 | 0 | 0.001060299 | 0 |
| 0 | 0 | 0 | 0 | 0 |
| 0 | 0 | 0 | 0 | 0 |
| 0 | 0 | 0 | 0 | 0 |
| 0 | 0 | 0 | 0 | 0.013730211 |
| 0 | 0 | 0 | 0 | 0 |
| 0 | 0 | 0 | 0 | 0 |
| 0 | 0 | 0 | 0 | 0 |
| 0 | 0 | 0.001513235 | 0 | 0 |
| 0 | 0 | 0 | 0 | 0 |
| 0 | 0 | 0.232149937 | 0 | 0 |
| 0 | 0 | 0 | 0 | 0 |
| 0 | 0 | 0 | 0 | 0 |
| 0 | 0 | 0 | 0 | 0 |
| 0 | 0 | 0 | 0 | 0 |
| 0 | 0 | 0 | 0 | 0 |
| 0 | 0 | 0 | 0 | 0 |
| 0 | 0 | 0 | 0 | 0 |
| 0.000727765 | 0.001496935 | 0.001316437 | 0.007135959 | 0.000937535 |
| 0 | 0 | 0 | 0 | 0 |
| 0 | 0 | 0 | 0 | 0 |
| 0.726350172 | 0.035849012 | 3.040777082 | 0.826455445 | 1.153330504 |
| 0 | 0 | 0 | 0 | 0 |
| 0 | 0 | 0 | 0 | 0 |
| 0 | 0 | 0 | 0 | 0 |
| 0 | 0 | 0 | 0.001850675 | 0 |
| 0 | 0 | 0.014137146 | 0 | 0 |
| 0 | 0 | 0 | 0 | 0 |
| 0 | 0 | 0 | 0 | 0.003450712 |
| 0 | 0 | 0 | 0 | 0 |
| 0 | 0 | 0 | 0 | 0 |
| 0 | 0 | 0 | 0 | 0 |

| PHL018-0183_V1 | PHL018-0204_V1 | PHL018-0207_V3 | PHL018-0181_V1 | PHL018-0180_V3 |
|---|---|---|---|---|
| REF | EYCF | REF | CTRL | CTRL |
| 0.073521771 | 0.012697283 | 0.041083969 | 0.025955689 | 0.02462628 |
| 1.679518095 | 5.841214461 | 0.18601602 | 1.592063131 | 8.569248406 |
| 0.045142585 | 3.839162134 | 1.577694498 | 0.849613772 | 0 |
| 0 | 0 | 0 | 0 | 0 |
| 0 | 0 | 0 | 0 | 0 |
| 0 | 0 | 0 | 0 | 0 |
| 0 | 0 | 0 | 0 | 0 |
| 0 | 0 | 0 | 0 | 0 |
| 0 | 0 | 0 | 0 | 0 |
| 0 | 0 | 0 | 0 | 0 |
| 0 | 0 | 0 | 0 | 0 |
| 0 | 0 | 0 | 0 | 0 |
| 0 | 0 | 0 | 0 | 0 |
| 1.127675749 | 5.819189069 | 0.856353021 | 1.168302069 | 0 |
| 0 | 0 | 0 | 0 | 0 |
| 0 | 0 | 0 | 0 | 0 |
| 0 | 0.017245745 | 0 | 0 | 0 |
| 0 | 0 | 0 | 0 | 0 |
| 0 | 0 | 0 | 0 | 0 |
| 0 | 0 | 0 | 0 | 0 |
| 0 | 0 | 0 | 0 | 0 |
| 0 | 0 | 0 | 0 | 0 |
| 0 | 0 | 0 | 0 | 0 |
| 0 | 0 | 0 | 0 | 0 |
| 0 | 0 | 0.029718629 | 0 | 0 |
| 0 | 0 | 0 | 0 | 0 |
| 0 | 0 | 0 | 0 | 0 |
| 0 | 0 | 0 | 0 | 0 |
| 0 | 0 | 0 | 0 | 0 |
| 0 | 0 | 0 | 0 | 0 |
| 0 | 0 | 0 | 0 | 0 |
| 0 | 0 | 0 | 0 | 0 |
| 0 | 0 | 0 | 0 | 0 |
| 0 | 0 | 0 | 0 | 0 |
| 0 | 0 | 0 | 0 | 0 |
| 0 | 0 | 0.779299667 | 6.240639575 | 0 |
| 0 | 0 | 0 | 0 | 0 |
| 0 | 0 | 0 | 0 | 0 |
| 0 | 0 | 0 | 0 | 0 |
| 0 | 0 | 0 | 0 | 0 |
| 0 | 0 | 0 | 0 | 0 |
| 0 | 0.157754779 | 0.066974488 | 0.020753316 | 0.030550904 |
| 0.066209333 | 0.706130135 | 0.207579156 | 6.771770423 | 0.143448923 |
| 0.025376251 | 0.150988983 | 0.013466887 | 0.055175308 | 0.074936826 |
| 0 | 0.001007009 | 0 | 0.019294224 | 0 |
| 0 | 0 | 0 | 0 | 0 |

| | | | | |
|---|---|---|---|---|
| 0 | 0 | 0.003721812 | 0 | 0 |
| 0.004164875 | 0.06572367 | 0.001463124 | 0.001699063 | 0.019694257 |
| 0.001147889 | 0 | 0.060689809 | 0.002809692 | 0 |
| 0.989148209 | 0 | 0.293889235 | 0.305339374 | 0.547453486 |
| 0.006177497 | 0 | 0.001966708 | 0 | 0 |
| 0 | 0 | 0 | 0 | 0 |
| 0 | 0 | 0 | 0.140909257 | 0 |
| 0 | 0 | 0 | 0 | 0 |
| 0 | 0 | 0 | 0 | 0 |
| 0 | 0 | 0 | 0 | 0 |
| 0 | 0 | 0.001200752 | 0 | 0 |
| 0.022052876 | 0 | 0.156050646 | 1.61037013 | 0 |
| 0 | 0 | 0.001358575 | 0 | 0 |
| 0.006749285 | 0.07857063 | 0.006364343 | 0.01402047 | 0 |
| 0 | 0 | 0 | 0 | 0 |
| 0 | 0 | 0 | 0 | 0 |
| 0 | 0.000566905 | 0.005003953 | 3.227269382 | 2.181694342 |
| 0 | 0 | 0 | 0 | 0 |
| 0 | 0 | 0 | 0 | 0 |
| 0 | 0 | 0 | 0 | 0 |
| 0.558916971 | 0.045008268 | 0.003857072 | 0.059778089 | 0.109232405 |
| 0.020617167 | 0.046224374 | 0.00287856 | 0 | 0.002634904 |
| 0 | 0 | 0 | 2.018806212 | 0 |
| 0 | 0 | 0 | 0 | 0 |
| 0 | 0 | 0 | 0 | 0 |
| 0 | 0 | 0 | 0 | 0 |
| 0 | 0 | 0 | 0 | 0.010406557 |
| 0 | 0 | 0 | 0 | 0 |
| 0 | 0.002974212 | 0.00356372 | 0.003931474 | 0.001555756 |
| 0 | 0 | 0 | 0 | 0 |
| 0.003793752 | 0 | 0 | 0 | 0.125167104 |
| 0 | 0 | 0 | 0 | 0 |
| 0 | 0 | 0 | 0 | 0 |
| 0 | 0 | 0 | 0 | 0 |
| 0 | 0 | 0 | 0 | 0 |
| 0 | 0 | 0 | 0 | 0 |
| 0 | 0 | 0 | 0 | 0 |
| 0 | 0 | 0.003040163 | 0.348725928 | 0 |
| 0 | 0 | 0 | 0 | 0 |
| 0 | 0 | 0 | 0 | 0 |
| 0 | 0 | 0 | 0 | 0 |
| 0 | 0 | 0 | 0 | 0 |
| 0 | 0 | 0 | 0 | 0 |
| 0 | 0 | 0 | 0 | 0 |
| 0 | 0 | 0.030779256 | 0 | 0 |
| 0 | 0 | 0 | 0.036738871 | 0 |
| 0 | 0 | 0 | 0 | 0 |
| 0 | 0.000552681 | 0 | 0 | 0 |

| | | | | |
|---|---|---|---|---|
| 0 | 0 | 0 | 0 | 0 |
| 0 | 0.135434125 | 0 | 0 | 0 |
| 0 | 0 | 0 | 0 | 0 |
| 0 | 0 | 0 | 0 | 0 |
| 0 | 0 | 0 | 0 | 0 |
| 0 | 0 | 0 | 0 | 0 |
| 0 | 0 | 0 | 0 | 0 |
| 0 | 0 | 0 | 0 | 0 |
| 0.001021954 | 0 | 0.029259595 | 0.00140706 | 0 |
| 0 | 0 | 0 | 0 | 0 |
| 0 | 0 | 0 | 0 | 0 |
| 0 | 0.074124771 | 0.000729413 | 0.048281039 | 0 |
| 0 | 0 | 0 | 0 | 0 |
| 0 | 0 | 0 | 0 | 0 |
| 0 | 0 | 0 | 0 | 0 |
| 0 | 0 | 0 | 0 | 0 |
| 0 | 0 | 0 | 0 | 0 |
| 0 | 0 | 0 | 0 | 0 |
| 0.017422377 | 0 | 0.334123473 | 0 | 0 |
| 0 | 0 | 0 | 0 | 0 |
| 0 | 0 | 0 | 0 | 0 |
| 0 | 0 | 0 | 0 | 0 |
| 0.000745191 | 0 | 0 | 0 | 0 |
| 0 | 0 | 0 | 0 | 0 |
| 0 | 0 | 0 | 0 | 0 |
| 0 | 0 | 0 | 0 | 0 |
| 0 | 0 | 0 | 0 | 0 |
| 0 | 0 | 0 | 0 | 0 |
| 0 | 0 | 0 | 0 | 0 |
| 0 | 0 | 0 | 0 | 0 |
| 0.188822098 | 0 | 0.010797168 | 0 | 0 |
| 0 | 0 | 0.045221548 | 0.030810131 | 0 |
| 0 | 0 | 0 | 0 | 0 |
| 0 | 0 | 0 | 0 | 0 |
| 0 | 0 | 0 | 0 | 0 |
| 0 | 0 | 0 | 0 | 0 |
| 0 | 0 | 0 | 0 | 0 |
| 0 | 0 | 0 | 0 | 0 |
| 0 | 0 | 0.085678043 | 0.025229346 | 0 |
| 0 | 0 | 0 | 0 | 0 |
| 0 | 0 | 0 | 0 | 0 |
| 0 | 0 | 0 | 0 | 0 |
| 0 | 0 | 0 | 0 | 0.001099143 |
| 0 | 0 | 0 | 0 | 0 |
| 0 | 0 | 0 | 0 | 0 |
| 0 | 0 | 0 | 0 | 0 |
| 0 | 0 | 0 | 0 | 0 |
| 0 | 0 | 0 | 0 | 0 |

| | | | | |
|---|---|---|---|---|
| 0 | 0 | 0 | 0 | 0 |
| 0 | 0 | 0 | 0 | 0 |
| 0 | 0 | 0 | 0 | 0 |
| 0 | 0 | 0 | 0 | 0 |
| 0 | 0 | 0.006522121 | 0 | 0 |
| 0 | 0 | 0 | 0 | 0 |
| 0 | 0 | 0 | 0 | 0 |
| 0 | 0 | 0 | 0 | 0 |
| 0 | 0 | 0 | 0 | 0 |
| 0 | 0 | 0 | 0 | 0 |
| 0 | 0 | 0 | 0 | 0 |
| 0 | 0 | 0 | 0 | 0 |
| 0 | 0 | 0 | 0 | 0 |
| 0 | 0 | 0 | 0 | 0 |
| 0 | 0 | 0 | 0 | 0 |
| 0 | 0 | 0 | 0 | 0 |
| 0 | 0 | 0 | 0 | 0 |
| 0 | 0 | 0 | 0 | 0 |
| 0 | 0 | 0 | 0 | 0 |
| 0 | 0 | 0 | 0 | 0 |
| 0 | 0 | 0 | 0 | 0 |
| 0 | 0 | 0 | 0 | 0 |
| 0 | 0 | 0 | 0 | 0 |
| 0 | 0 | 0 | 0 | 0 |
| 0 | 0 | 0.004393247 | 0 | 0 |
| 0 | 0 | 0 | 0 | 0 |
| 0 | 0 | 0 | 0 | 0 |
| 0 | 0 | 0 | 0 | 0 |
| 0 | 0 | 0 | 0 | 0 |
| 0 | 0 | 0 | 0 | 0 |
| 0 | 0 | 0.004339813 | 0 | 0 |
| 0 | 0.002517053 | 0 | 0 | 0 |
| 0 | 0 | 0 | 0 | 0 |
| 0 | 0 | 0 | 0 | 0 |
| 0 | 0 | 0 | 0 | 0 |
| 0 | 0 | 0 | 0 | 0 |
| 0 | 0 | 0 | 0 | 0 |
| 0 | 0 | 0 | 0 | 0 |
| 0 | 0 | 0 | 0 | 0 |
| 0 | 0 | 0 | 0 | 0 |
| 0 | 0 | 0 | 0 | 0 |
| 0 | 0 | 0 | 0 | 0 |
| 0 | 0 | 0 | 0 | 0 |
| 0 | 0 | 0 | 0 | 0 |
| 0 | 0 | 0 | 0 | 0 |
| 0 | 0 | 0 | 0 | 0 |
| 0 | 0 | 0 | 0 | 0 |
| 0 | 0 | 0 | 0 | 0 |

| | | | | |
|---|---|---|---|---|
| 0 | 0 | 0.743403524 | 0 | 0 |
| 0 | 0 | 0 | 0 | 0 |
| 0 | 0 | 0 | 0 | 0 |
| 0 | 0 | 0 | 0 | 0 |
| 0 | 0 | 0 | 0 | 0 |
| 0 | 0 | 0 | 0 | 0 |
| 0 | 0 | 0 | 0 | 0 |
| 0 | 0 | 0 | 0 | 0 |
| 0 | 0 | 0 | 0 | 0 |
| 0 | 0 | 0 | 0 | 0 |
| 0 | 0 | 0 | 0 | 0 |
| 0 | 0 | 0 | 0 | 0 |
| 0.001643017 | 0 | 0.003270762 | 0.002513511 | 0 |
| 0 | 0 | 0 | 0 | 0 |
| 0 | 0 | 0 | 0 | 0 |
| 0 | 0 | 0 | 0 | 0 |
| 0 | 0 | 0 | 0 | 0 |
| 0 | 0 | 0 | 0.001007619 | 0 |
| 0 | 0 | 0 | 0 | 0 |
| 0 | 0 | 0 | 0 | 0 |
| 0.095574803 | 0.149673544 | 0.008094097 | 0.02412493 | 0.064388395 |
| 0 | 0 | 0 | 0 | 0 |
| 0 | 0 | 0 | 0 | 0 |
| 0 | 0.002486221 | 0 | 0 | 0 |
| 0 | 0 | 0 | 0 | 0 |
| 0 | 0 | 0 | 0 | 0 |
| 0.00974059 | 0.015890502 | 0 | 0.002919437 | 0.017148605 |
| 0 | 0 | 0 | 0 | 0 |
| 0 | 0 | 0 | 0 | 0 |
| 0.008719871 | 0 | 0 | 0.002537812 | 0 |
| 0.824136739 | 0 | 0 | 0 | 0.200572506 |
| 0 | 0 | 0 | 0 | 0 |
| 0 | 0 | 0 | 0 | 0 |
| 0 | 0 | 0 | 0 | 0 |
| 0.007106978 | 0.009652377 | 0.011937289 | 0 | 0.005109088 |
| 0 | 0 | 0 | 0.001453863 | 0 |
| 0 | 0 | 0 | 0 | 0 |
| 0 | 0 | 0 | 0 | 0 |
| 0 | 0 | 0 | 0 | 0 |
| 0 | 0 | 0 | 0 | 0 |
| 0 | 0 | 0 | 0 | 0 |
| 0 | 0 | 0 | 0 | 0 |
| 0 | 0 | 0 | 0 | 0 |
| 0 | 0 | 0 | 0 | 0 |
| 0 | 0 | 0 | 0.005873419 | 0 |
| 0 | 0 | 0 | 0 | 0 |
| 0 | 0 | 0 | 0 | 0 |
| 0.000791348 | 0.002579454 | 0.001251005 | 0.002614928 | 0.001092262 |

| | | | | |
|---|---|---|---|---|
| 0 | 0 | 0 | 0 | 0 |
| 0 | 0 | 0 | 0 | 0 |
| 0 | 0.000897924 | 0 | 0 | 0.004657737 |
| 0 | 0 | 0 | 0 | 0 |
| 0 | 0 | 0 | 0 | 0 |
| 0 | 0 | 0 | 0 | 0 |
| 0 | 0 | 0.002255667 | 0 | 0 |
| 0 | 0 | 0.001198895 | 0.023807045 | 0 |
| 0 | 0 | 0 | 0 | 0.00197301 |
| 0 | 0 | 0 | 0 | 0 |
| 0 | 0 | 0 | 0.159789226 | 0 |
| 0 | 0 | 0 | 0 | 0 |
| 0 | 0 | 0 | 0 | 0 |
| 0 | 0 | 0 | 0 | 0 |
| 0 | 0 | 0.000920463 | 0 | 0 |
| 0 | 0 | 0 | 0 | 0 |
| 0 | 0 | 0 | 0 | 0 |
| 0 | 0 | 0 | 0 | 0 |
| 0 | 0.003372799 | 0 | 0.033389801 | 0.000925686 |
| 0 | 0 | 0.738211721 | 0 | 0 |
| 0.008310085 | 0.138016327 | 0 | 0 | 0 |
| 0 | 0 | 0 | 0 | 0 |
| 0 | 0 | 0 | 0 | 0 |
| 0 | 0 | 0 | 0 | 0 |
| 0.002830241 | 0.00878607 | 0.003728499 | 0.012123282 | 0.032011247 |
| 0 | 0 | 0 | 0.001260457 | 0 |
| 0 | 0 | 0 | 0 | 0 |
| 0 | 0 | 0 | 0 | 0 |
| 0 | 0.003318934 | 0.015655906 | 0 | 0.007507755 |
| 0 | 0 | 0 | 0 | 0 |
| 0.105098919 | 0.063274665 | 0.11922773 | 0.082207052 | 0.003815344 |
| 0 | 0 | 0 | 0 | 0 |
| 0 | 0 | 0 | 0 | 0 |
| 0 | 0 | 0 | 0 | 0 |
| 0 | 0 | 0 | 0 | 0 |
| 0 | 0 | 0 | 0 | 0 |
| 0 | 0 | 0 | 0 | 0 |
| 0 | 0 | 0 | 0 | 0 |
| 0 | 0 | 0 | 0 | 0 |
| 0 | 0 | 0 | 0.004001628 | 0 |
| 0 | 0 | 0 | 0 | 0 |
| 0.022956615 | 0 | 0.001687956 | 0.004410338 | 0.0020469 |
| 0 | 0 | 0 | 0 | 0 |
| 0 | 0 | 0 | 0 | 0 |
| 0 | 0 | 0 | 0 | 0 |
| 0 | 0 | 0 | 0 | 0 |
| 0 | 0 | 0 | 0 | 0 |
| 0 | 0 | 0 | 0 | 0 |

| | | | | |
|---|---|---|---|---|
| 0 | 0 | 0 | 0 | 0 |
| 0 | 0 | 0 | 0 | 0 |
| 0.032401102 | 0.1297193 | 0.013423543 | 0.055994373 | 0.062713348 |
| 0 | 0 | 0 | 0 | 0 |
| 0.004072345 | 0 | 0.451053547 | 0 | 0 |
| 0 | 0 | 0 | 0 | 0 |
| 0 | 0 | 0 | 0 | 0 |
| 0 | 0 | 0 | 0 | 0 |
| 0 | 0 | 0 | 0 | 0 |
| 0 | 0 | 0 | 0 | 0.002820843 |
| 0 | 0 | 0 | 0 | 0 |
| 0 | 0 | 0 | 0 | 0 |
| 0 | 0 | 0 | 0 | 0 |
| 0 | 0.026066045 | 0 | 0.001133405 | 0 |
| 0.001222994 | 0.00885875 | 0.002148195 | 0.005425767 | 0.01266034 |
| 0 | 0 | 0.014259545 | 0 | 0 |
| 0.001161244 | 0.003965402 | 0.002447676 | 0.009379852 | 0.010685423 |
| 0 | 0.001957473 | 0 | 0 | 0 |
| 0 | 0 | 0 | 0 | 0 |
| 0 | 0 | 0 | 0 | 0 |
| 0 | 0 | 0 | 0 | 0 |
| 0 | 0.001301682 | 0.001104776 | 0.00288659 | 0 |
| 0 | 0 | 0 | 0 | 0 |
| 0 | 0.001261191 | 0 | 0 | 0 |
| 0 | 0 | 0 | 0 | 0 |
| 0 | 0 | 0 | 0 | 0 |
| 0 | 0 | 0 | 3.651534848 | 0 |
| 0 | 0 | 0 | 0 | 0 |
| 0 | 0 | 0 | 0 | 0 |
| 0 | 0 | 0 | 0 | 0 |
| 0 | 0 | 0 | 0 | 0 |
| 0 | 0 | 0 | 0 | 0 |
| 0 | 0 | 0 | 0 | 0 |
| 0 | 0 | 0 | 0 | 0 |
| 0 | 0 | 0 | 0 | 0 |
| 0 | 0.003899888 | 0.035968129 | 0 | 0 |
| 0 | 0 | 0 | 0 | 0 |
| 0 | 0 | 0.110904164 | 0.329304461 | 0 |
| 0 | 0 | 0 | 0 | 0 |
| 0 | 0 | 0 | 0 | 0 |
| 0 | 0 | 0 | 0 | 0 |
| 0 | 0 | 0 | 0 | 0 |
| 0 | 0 | 0 | 0 | 0 |
| 0 | 0.003555053 | 0 | 0 | 0.001881721 |
| 0 | 0 | 0 | 0 | 0 |
| 0 | 0 | 0 | 0 | 0 |
| 0 | 0 | 0.000902749 | 0 | 0 |
| 0.003190543 | 0.002723758 | 0 | 0 | 0.007339611 |

| | | | | |
|---|---|---|---|---|
| 0 | 0 | 0.007758587 | 0 | 0 |
| 0 | 0 | 0 | 0 | 0 |
| 0 | 0 | 0 | 0 | 0.002273246 |
| 0 | 0 | 0 | 0 | 0 |
| 0 | 0.01920412 | 0.001746333 | 0.018048667 | 0.08513112 |
| 0 | 0 | 0.00303739 | 0 | 0 |
| 0 | 0 | 0 | 0 | 0 |
| 0.232576808 | 0.080035363 | 0 | 0.075044755 | 0.002493463 |
| 0 | 0 | 0 | 0 | 0 |
| 0.078524102 | 1.403639745 | 0.026912768 | 0.01904457 | 2.082487812 |
| 0 | 0.001343881 | 0 | 0.002649039 | 0 |
| 0 | 0 | 0.103087884 | 0.021152028 | 0.00225581 |
| 0.021198981 | 0.003607957 | 0.001273867 | 0.064178258 | 0.022244459 |
| 0 | 0 | 0 | 0 | 0 |
| 0.003633778 | 0 | 0 | 0 | 0 |
| 0 | 0 | 0 | 0.011530318 | 0 |
| 0 | 0 | 0.001818505 | 0 | 0 |
| 0 | 0 | 0 | 0 | 0 |
| 0.056768953 | 1.017523725 | 0 | 0 | 0 |
| 0 | 0 | 0 | 0 | 0 |
| 0 | 0 | 0 | 0.130598197 | 0 |
| 0 | 0 | 0 | 0 | 0 |
| 0 | 0 | 0 | 0.003170282 | 0 |
| 0 | 0 | 0 | 0 | 0 |
| 0 | 0 | 0 | 0 | 0 |
| 0 | 0.005547102 | 0 | 0.007355847 | 0 |
| 0 | 0 | 0 | 0 | 0 |
| 0 | 0 | 0 | 0 | 0.002711365 |
| 0 | 0 | 0 | 0 | 0 |
| 0.027187113 | 0 | 0 | 0 | 0 |
| 0 | 0 | 0 | 0 | 0 |
| 0 | 0 | 0 | 0 | 0 |
| 0 | 0 | 0 | 0 | 0 |
| 0 | 0 | 0 | 0 | 0 |
| 0 | 0 | 0 | 0 | 0 |
| 0 | 0.686938895 | 0 | 0 | 0 |
| 0.026026013 | 0 | 0 | 0.006310296 | 0.00169446 |
| 0 | 0 | 0 | 0 | 0 |
| 0 | 0 | 0 | 0 | 0 |
| 0 | 0 | 0 | 0 | 0 |
| 0 | 0 | 0 | 0 | 0 |
| 0.504161428 | 0 | 0 | 0 | 0 |
| 0 | 0 | 0 | 0 | 0 |
| 0.00488582 | 0 | 0 | 0 | 0 |
| 0 | 0 | 0 | 0 | 0 |
| 0 | 0 | 0 | 0 | 0 |
| 0.002693484 | 0.001254229 | 0.002129002 | 0 | 0.002168657 |
| 0 | 0.011004819 | 0 | 0 | 0 |

| | | | | |
|---|---|---|---|---|
| 0.028258071 | 0 | 0 | 0 | 0 |
| 0 | 0 | 0 | 0.019340918 | 0 |
| 0 | 0 | 0 | 0 | 0 |
| 0 | 0 | 0 | 0 | 0 |
| 0 | 0.003719022 | 0 | 0 | 0 |
| 0.099071473 | 0 | 0.000782529 | 0 | 0 |
| 0 | 0 | 0 | 0 | 0 |
| 0 | 0 | 0 | 0 | 0 |
| 0 | 0 | 0 | 0 | 0 |
| 0 | 0 | 0 | 0 | 0 |
| 0.017223632 | 0.140545103 | 0 | 0.011969955 | 0.01584868 |
| 0 | 0 | 0 | 0.009101862 | 0 |
| 0 | 0 | 0 | 0 | 0 |
| 0.001394745 | 0.004149373 | 0.002082394 | 0.00256044 | 0.004010635 |
| 0.003763283 | 2.114543429 | 0 | 0.006908548 | 0 |
| 0 | 0.008830863 | 0 | 0 | 0 |
| 0 | 0 | 0.028747909 | 0 | 0 |
| 0.035063889 | 0 | 0 | 0 | 0 |
| 0 | 0.000768064 | 0 | 0.007494287 | 0 |
| 0.003484738 | 0 | 0 | 0 | 0 |
| 0 | 0 | 0 | 0.003218564 | 0 |
| 0.449682197 | 0 | 0.051657696 | 0.366922928 | 0 |
| 0.040368154 | 0 | 0 | 0 | 0 |
| 0 | 0 | 0 | 0 | 0 |
| 0 | 0 | 0 | 0 | 0 |
| 0 | 0 | 0 | 0 | 0 |
| 0.201384288 | 0 | 0 | 0 | 0.026961257 |
| 0 | 0 | 0 | 0 | 0 |
| 0 | 0 | 0 | 0 | 0 |
| 1.036512614 | 0.67775886 | 0.089595551 | 0.136674272 | 1.114219145 |
| 0.002996374 | 0 | 0.000332409 | 0 | 0 |
| 0 | 0 | 0 | 0 | 0 |
| 0 | 0 | 0 | 0 | 0 |
| 0 | 0 | 0 | 0 | 0 |
| 0 | 0.144350944 | 0 | 0 | 0 |
| 0 | 0 | 0 | 0 | 0 |
| 0 | 0 | 1.352315652 | 0 | 0 |
| 0 | 0 | 0.000374561 | 0 | 0 |
| 0 | 0 | 0 | 0 | 0 |
| 0.002200453 | 9.762758721 | 0 | 0.037618231 | 0 |
| 0 | 0.040193264 | 0 | 0.005086988 | 0 |
| 13.42119771 | 0 | 1.021986265 | 0.620046569 | 2.073283797 |
| 0 | 0 | 0 | 0 | 0 |
| 0 | 0 | 0 | 0.225374424 | 0 |
| 0 | 0 | 0 | 0 | 0 |
| 0 | 0 | 0 | 0 | 0 |
| 0 | 0.000622667 | 0.001585429 | 0.009389555 | 0.003691333 |
| 0 | 0 | 0 | 0.063588736 | 0 |

| | | | | |
|---|---|---|---|---|
| 0 | 0.000960334 | 0.000782461 | 0.010449338 | 0 |
| 0 | 0 | 0 | 0 | 0 |
| 0 | 0.001570234 | 0.004086959 | 7.49910527 | 0.127219637 |
| 0 | 0 | 0 | 0 | 0 |
| 0.010780387 | 0 | 0 | 0 | 0 |
| 0 | 0.321212594 | 0 | 0.049552403 | 0 |
| 0 | 0 | 0 | 0 | 0 |
| 0 | 0 | 0.00087698 | 0.003055197 | 0 |
| 0 | 0.072012709 | 0 | 0.000760449 | 0 |
| 0 | 0 | 0 | 0 | 0.00139651 |
| 0 | 0 | 0 | 0 | 0 |
| 0 | 0 | 0 | 0 | 0.003360377 |
| 0 | 0 | 0 | 0 | 0 |
| 0 | 0.00173758 | 0 | 0.001027527 | 0.031932545 |
| 0 | 0 | 0 | 0 | 0 |
| 0.072785722 | 0 | 0.024524774 | 0 | 0 |
| 0 | 0.019114487 | 0.017066116 | 0.166258761 | 0.001338374 |
| 0 | 0.004388666 | 0 | 0 | 0 |
| 0.003004744 | 0.009483253 | 0 | 0 | 0 |
| 0.05405412 | 0 | 0 | 0.075016975 | 0 |
| 0.036905685 | 0 | 0.000643957 | 0 | 0 |
| 0 | 0.004888273 | 0.001896604 | 0.000825917 | 0.004657331 |
| 0.066217691 | 0 | 0 | 0 | 0 |
| 0 | 0 | 0 | 0 | 0 |
| 0 | 0 | 0.024354692 | 0 | 0 |
| 0 | 0 | 0 | 0.003282904 | 0 |
| 0 | 0.003057133 | 0 | 0 | 0 |
| 0 | 0 | 0 | 0 | 0 |
| 0 | 0 | 0 | 0 | 0 |
| 0 | 0.009482515 | 0 | 0.015275709 | 0 |
| 0.039318783 | 0 | 0.011444828 | 0.002062301 | 0 |
| 0.018702406 | 0.004332753 | 0 | 0.002818417 | 0.007705703 |
| 0.059800362 | 0.015470123 | 0.004341637 | 0.073918624 | 0 |
| 0 | 0.001478013 | 0.007225536 | 0 | 0.002190511 |
| 0 | 0 | 0 | 0 | 0.002814241 |
| 0 | 0 | 0 | 0 | 0 |
| 0.000961132 | 0 | 0.000886322 | 0.001984976 | 0.005251155 |
| 0.000939562 | 0.071605774 | 0.020497278 | 0.010693921 | 0 |
| 0 | 0 | 0.028688195 | 0 | 0 |
| 0 | 0 | 0.002266866 | 0 | 0 |
| 0 | 0 | 0 | 0 | 0 |
| 0 | 0 | 0 | 0 | 0 |
| 0.001405113 | 0.001526689 | 0 | 0 | 0 |
| 1.319905408 | 0 | 0.043905171 | 0.002827522 | 0 |
| 0.150416976 | 0 | 0 | 0 | 0 |
| 0 | 0 | 0 | 0 | 0 |
| 0 | 0 | 0 | 0 | 0 |
| 0 | 0 | 0 | 0 | 0 |

| | | | | |
|---|---|---|---|---|
| 0 | 0 | 0 | 0 | 0 |
| 1.238023133 | 0.737651898 | 0.023747367 | 0.367350987 | 0.516583161 |
| 0 | 0 | 0 | 0 | 0 |
| 0 | 0 | 0 | 0 | 0 |
| 0 | 0 | 0 | 0 | 0 |
| 0 | 0 | 0 | 0 | 0 |
| 0.80232507 | 0 | 0 | 0 | 0.002452201 |
| 0 | 0 | 0 | 0.000541582 | 0 |
| 0 | 0 | 0.007534457 | 0.002151501 | 0 |
| 0 | 0 | 0 | 0 | 0 |
| 0.001141659 | 0.002067399 | 0 | 0 | 0 |
| 0.037695145 | 0.146933714 | 0.016727164 | 0.028096194 | 0.253624819 |
| 0 | 0 | 0.051403013 | 0 | 0 |
| 0 | 0 | 0 | 0 | 0 |
| 0.000873274 | 0.000677738 | 0 | 0 | 0 |
| 0 | 0 | 0 | 0 | 0 |
| 0.013947282 | 0.017009653 | 0.018478841 | 0.036760174 | 0.119630089 |
| 0.025606349 | 0.073669842 | 0 | 0.018518125 | 0.063963069 |
| 0 | 0 | 0 | 0 | 0 |
| 0.046019337 | 0.056961635 | 0 | 0.000924977 | 0 |
| 0.008835374 | 0.040982723 | 0.019056347 | 0.045264493 | 0.026233596 |
| 0 | 0 | 0 | 0.007275234 | 0 |
| 0 | 0 | 0 | 0 | 0 |
| 0 | 0 | 0 | 0 | 0 |
| 0 | 0 | 0 | 0.012041692 | 0 |
| 0 | 0 | 0 | 0 | 0 |
| 0.075243592 | 0 | 0 | 0 | 0 |
| 0.23869844 | 0.004745682 | 0.044190728 | 0.032072983 | 0 |
| 0 | 0 | 0 | 0 | 0 |
| 0 | 0 | 0 | 0 | 0 |
| 0 | 0.002909783 | 0 | 0.016131721 | 0 |
| 0 | 0 | 0 | 0 | 0 |
| 0 | 0.00205758 | 0 | 0 | 0 |
| 0 | 0 | 0 | 0 | 0 |
| 0 | 0 | 0.10579986 | 0 | 0 |
| 0.674167203 | 19.08437992 | 0.036031723 | 0.016623743 | 0.562446225 |
| 0 | 0 | 0 | 0 | 0 |
| 0 | 0 | 0 | 0 | 0 |
| 0.005657535 | 0.004390748 | 0.041311547 | 0.014342534 | 0.026649237 |
| 0 | 0 | 0 | 0 | 0 |
| 0.146400703 | 0 | 0 | 0 | 0 |
| 0 | 0 | 0 | 0 | 0 |
| 0 | 0.001778005 | 0.002328241 | 0.026285828 | 0.000752891 |
| 0 | 0.004291575 | 0.000749291 | 0 | 0.000726902 |
| 0 | 4.046086696 | 0 | 0 | 0 |
| 0.471761283 | 0 | 0.396032005 | 0 | 0 |
| 0.000694574 | 0 | 0 | 0 | 0 |
| 0 | 0 | 0 | 0 | 0 |

| | | | | |
|---|---|---|---|---|
| 0.001600276 | 0 | 0 | 0 | 0 |
| 0 | 0 | 0 | 0 | 0 |
| 0 | 0 | 0 | 0 | 0 |
| 0.215748623 | 0 | 0.426032555 | 0 | 0 |
| 0 | 0 | 0 | 0 | 0 |
| 0 | 0 | 0 | 0 | 0 |
| 0 | 0 | 0 | 0 | 0 |
| 0 | 0 | 0 | 0 | 0 |
| 0.217343481 | 0 | 8.972882668 | 0.893013971 | 0 |
| 0 | 0 | 0.080157029 | 0 | 0 |
| 0 | 0 | 0 | 0 | 0 |
| 0 | 0 | 0.001828268 | 0 | 0 |
| 0 | 0 | 0 | 0 | 0 |
| 0 | 0 | 0 | 0 | 0 |
| 0.031945689 | 0 | 0.658424847 | 0.002351339 | 0 |
| 0 | 0 | 0 | 0 | 0 |
| 0 | 0 | 0 | 0 | 0 |
| 0 | 0 | 0 | 0 | 0 |
| 0 | 0 | 0 | 0 | 0 |
| 0 | 0 | 0 | 0 | 0 |
| 0 | 0 | 0 | 0 | 0 |
| 0 | 0 | 0 | 0 | 0 |
| 0 | 0 | 0 | 0 | 0 |
| 0.245739641 | 0.484167783 | 0.099509851 | 0.044099758 | 0 |
| 0.001538696 | 0 | 0 | 0 | 0 |
| 0.310285637 | 0 | 0.125901969 | 0 | 0 |
| 0 | 0 | 0 | 0 | 0 |
| 0 | 0 | 2.020937414 | 0.443332046 | 0 |
| 0 | 0 | 0 | 0 | 0 |
| 0 | 0 | 0 | 0 | 0 |
| 0 | 0 | 0.009915958 | 0 | 0 |
| 0 | 0 | 0.00220346 | 0 | 0 |
| 0.014456682 | 0 | 0.049908369 | 0.000620076 | 0 |
| 0 | 0 | 0 | 0 | 0 |
| 0 | 0 | 0 | 0 | 0 |
| 0 | 0 | 0 | 0 | 0 |
| 0.732882663 | 0 | 2.213595606 | 0 | 0 |
| 0 | 0.021233436 | 0 | 0 | 0 |
| 0.003991957 | 0 | 0 | 0 | 0 |
| 0 | 0 | 0 | 0 | 0 |
| 0.145222676 | 0.007069594 | 0.156466052 | 0.013989079 | 0 |
| 0 | 0 | 0.051310726 | 0 | 0 |
| 2.345458253 | 0.08448227 | 0.161123211 | 2.960122913 | 0 |
| 0.025179504 | 0 | 0 | 0 | 0 |
| 0 | 0 | 0 | 0 | 0 |
| 0 | 0 | 0 | 0 | 0 |
| 0 | 0 | 0 | 0 | 0 |
| 0.035799317 | 0 | 0 | 0 | 0 |

| | | | | |
|---|---|---|---|---|
| 0 | 0 | 0 | 0 | 0 |
| 0.042398449 | 0 | 0.234741801 | 0 | 0 |
| 0.014130812 | 0.665212331 | 0.001595625 | 0.008894065 | 0.942932164 |
| 0 | 0 | 0 | 0 | 0 |
| 0 | 0 | 0 | 0 | 0 |
| 0 | 0 | 0.124356424 | 0.087288081 | 0 |
| 0 | 0 | 0 | 0 | 0 |
| 0 | 0.007504286 | 0.001103979 | 0 | 0 |
| 0 | 0 | 0 | 0 | 0 |
| 0 | 0 | 0 | 0 | 0 |
| 0.010306364 | 0.003537864 | 0.037076695 | 0.018738262 | 0 |
| 6.393801494 | 0.269064126 | 0.828526481 | 1.853347338 | 0 |
| 0 | 0 | 0 | 0 | 0 |
| 0.001711238 | 0.005312289 | 0.02434696 | 0.295885501 | 0 |
| 0.036072946 | 0.807691799 | 0 | 0.004332273 | 1.363017816 |
| 0 | 0 | 0 | 0 | 0 |
| 0 | 0 | 0 | 0 | 0 |
| 0 | 0 | 0 | 0 | 0 |
| 0 | 0 | 0 | 0 | 0 |
| 0.010122372 | 0 | 0.004731772 | 0 | 0 |
| 0.263632772 | 0.09050463 | 0.338944299 | 0.011435988 | 1.378596524 |
| 0.397338901 | 0.689825336 | 0 | 0.036310616 | 2.818049169 |
| 1.96130704 | 2.507768819 | 1.283375576 | 0.825239767 | 24.73175643 |
| 0 | 0 | 0 | 0 | 0 |
| 0 | 0 | 0.012526082 | 0 | 0 |
| 0 | 0 | 0.222169524 | 0.008222246 | 0 |
| 0 | 0 | 0 | 1.007610759 | 0.008455721 |
| 0.005024551 | 0 | 0.000730398 | 0 | 0 |
| 0.067581472 | 0 | 0 | 0 | 0 |
| 0 | 0 | 0 | 0 | 0 |
| 0 | 0 | 0.014386149 | 0 | 0 |
| 0.379842772 | 0 | 0.072063729 | 0.003598816 | 0 |
| 0 | 0.136808382 | 0.014266116 | 0.044094615 | 1.084034151 |
| 0.016298845 | 0 | 0.047115146 | 0 | 0 |
| 0.014509734 | 0.044939081 | 0 | 0.006289206 | 0 |
| 0.005353048 | 0.005873516 | 0.153003847 | 0.584537206 | 0 |
| 0.00267464 | 0 | 0.026214937 | 0 | 0 |
| 0 | 0 | 0 | 0 | 0 |
| 0.086186019 | 0 | 5.275635867 | 0 | 0 |
| 0 | 0 | 0 | 0 | 0 |
| 0 | 0 | 0.049248801 | 0 | 0 |
| 0 | 0 | 0 | 0 | 0 |
| 0 | 0 | 0 | 0 | 0 |
| 0 | 0 | 0 | 0 | 0 |
| 0.013682597 | 0.008005021 | 0.0405982 | 0.004057555 | 0.014769418 |
| 0 | 0 | 0 | 0 | 0 |
| 0 | 0 | 0 | 0 | 0 |
| 0.260270379 | 1.214715924 | 0.154544438 | 19.73843462 | 0 |

| | | | | |
|---|---|---|---|---|
| 0 | 0 | 0 | 0 | 0 |
| 0 | 0 | 0 | 0 | 0 |
| 0 | 0 | 0 | 0 | 0 |
| 0 | 0 | 0.002079882 | 0 | 0 |
| 0 | 0 | 0 | 0 | 0 |
| 0 | 0 | 0.296498657 | 0 | 0 |
| 0 | 0 | 0.008797791 | 0 | 0 |
| 0 | 0.007687433 | 0.008699402 | 0.021972366 | 0.045573091 |
| 0 | 0 | 0 | 0 | 0 |
| 0 | 0 | 0 | 0 | 0 |
| 0.001398703 | 0.003256552 | 0.002527024 | 0.003484746 | 0.001378977 |
| 0 | 0 | 0 | 0 | 0 |
| 0 | 0 | 0 | 0 | 0 |
| 0 | 0 | 0.023091255 | 0 | 0 |
| 0 | 0 | 0 | 0 | 0 |
| 0 | 0 | 0 | 0 | 0 |
| 0 | 0 | 0.058184337 | 0 | 0 |
| 0 | 0 | 0.000788884 | 0 | 0 |
| 0 | 0 | 0 | 0 | 0 |
| 0 | 0 | 0 | 0 | 0 |
| 0 | 0 | 0 | 0 | 0 |
| 0 | 0 | 0 | 0 | 0 |
| 0 | 0 | 0.001111643 | 0 | 0 |
| 0 | 0 | 0 | 0 | 0 |
| 0 | 0 | 0 | 0 | 0 |
| 0 | 0 | 0 | 0 | 0 |
| 0 | 0 | 0 | 0 | 0 |
| 0 | 0 | 0 | 0 | 0 |
| 0 | 0 | 0 | 0 | 0 |
| 0 | 0 | 0.949046559 | 0.001844393 | 0 |
| 0.007554066 | 0 | 0 | 0 | 0 |
| 0 | 0 | 0 | 0 | 0 |
| 0 | 0 | 0 | 0 | 0 |
| 0 | 0 | 0 | 0 | 0 |
| 0 | 0 | 0 | 0.046084444 | 0 |
| 0 | 0 | 0 | 0 | 0 |
| 0 | 0 | 0.015381956 | 0 | 0 |
| 0.03010811 | 0.018903281 | 0 | 0 | 0.003112876 |
| 0 | 0 | 0 | 0 | 0 |
| 0 | 0 | 0 | 0 | 0 |
| 0.097125288 | 0 | 0 | 0 | 0 |
| 0 | 0 | 0 | 0 | 0 |
| 0 | 0 | 0.000502403 | 0 | 0 |
| 0 | 0 | 0 | 0 | 0 |
| 0.43598598 | 0 | 0.022771923 | 0 | 0 |
| 0 | 0 | 0 | 0 | 0 |
| 0 | 0 | 0.006690541 | 0 | 0 |
| 0 | 0 | 0 | 0 | 0 |

| | | | | |
|---|---|---|---|---|
| 0 | 0 | 0 | 0 | 0 |
| 0 | 0 | 0 | 0 | 0 |
| 0 | 0 | 0.01000473 | 0 | 0 |
| 0 | 0 | 0.210568181 | 0.000617744 | 0 |
| 0 | 0 | 0 | 0 | 0 |
| 0 | 0 | 0 | 0 | 0 |
| 0 | 0 | 0 | 0 | 0 |
| 0 | 0 | 0 | 0 | 0 |
| 0.041057548 | 0.179598719 | 0.000983424 | 0.008779184 | 0.663295709 |
| 0 | 0 | 0 | 0 | 0 |
| 0 | 0 | 0.004922252 | 0 | 0 |
| 0 | 0 | 0 | 0 | 0 |
| 0 | 0 | 0 | 0 | 0 |
| 0 | 0 | 0 | 0 | 0 |
| 0 | 0 | 1.088782054 | 0 | 0 |
| 1.844712697 | 0.068234097 | 0.00047372 | 0.003135631 | 0 |
| 0 | 0 | 0 | 0 | 0 |
| 0 | 0 | 0 | 0 | 0 |
| 0 | 0 | 0 | 0 | 0 |
| 0 | 0 | 0 | 0 | 0 |
| 0 | 0 | 0.414334786 | 0 | 0 |
| 0 | 0 | 0 | 0 | 0 |
| 0 | 0 | 0 | 0 | 0 |
| 0 | 0 | 0 | 0 | 0 |
| 0.0913527 | 0.05194912 | 0.010263749 | 0.349003943 | 0.030922999 |
| 3.676086133 | 0 | 1.790237065 | 0 | 0 |
| 0 | 0 | 0.612199025 | 0 | 0 |
| 0 | 0 | 0 | 0 | 0 |
| 0.031179849 | 0 | 0 | 0 | 0 |
| 0 | 0 | 0 | 0 | 0 |
| 0 | 0 | 0 | 0 | 0 |
| 0.012013669 | 0 | 0 | 0 | 0 |
| 0 | 0 | 0 | 0 | 0 |
| 0.002051156 | 0.001478173 | 0 | 0.001748252 | 0 |
| 0 | 0.027915285 | 0 | 0.063815624 | 0 |
| 0.005119302 | 0.000529738 | 0 | 0.004542317 | 0 |
| 0.619869453 | 0 | 0 | 0.519347743 | 0 |
| 7.781234786 | 0 | 0.757053125 | 0 | 0 |
| 0 | 0 | 4.440878031 | 0 | 0 |
| 0 | 0 | 0 | 0 | 0 |
| 0.299166567 | 0 | 0 | 0 | 0 |
| 0 | 0 | 0 | 0 | 0 |
| 0 | 0 | 0.263328279 | 0.071275291 | 0 |
| 0 | 0 | 0 | 0 | 0 |
| 0 | 0 | 0 | 0 | 0 |
| 0.002424948 | 0 | 0 | 0 | 0 |
| 0 | 0 | 0 | 0 | 0 |
| 2.332362562 | 0.88986888 | 0.469033538 | 0.43686934 | 0 |

| | | | | |
|---|---|---|---|---|
| 0.002688763 | 0 | 0.024288807 | 0 | 0 |
| 0 | 0 | 0 | 0 | 0 |
| 0 | 0 | 0 | 0 | 0 |
| 0 | 0 | 0 | 0 | 0 |
| 0 | 0.007568288 | 0.001209116 | 0 | 0 |
| 0 | 0 | 0 | 0 | 0 |
| 0 | 0 | 0 | 0 | 0 |
| 0 | 0 | 0 | 0 | 0 |
| 0 | 0 | 0 | 0 | 0 |
| 0 | 0 | 0 | 0 | 0 |
| 0 | 0 | 0 | 0 | 0 |
| 0.211510451 | 0 | 0.386702068 | 1.668917524 | 0 |
| 0.011344715 | 0 | 0.000366007 | 0.367224105 | 0.088945163 |
| 0 | 0 | 0 | 0 | 0 |
| 0.001105686 | 0.082035337 | 0.700220102 | 0.791213412 | 0.250285291 |
| 0 | 0 | 0 | 0 | 0 |
| 0 | 0 | 0.002350811 | 0 | 0 |
| 0 | 0 | 0 | 0 | 0 |
| 0 | 0 | 0 | 0 | 0 |
| 0 | 0 | 0.006115688 | 0 | 0 |
| 0 | 0 | 0.022915749 | 0.003093168 | 0 |
| 0 | 0 | 0 | 0 | 0 |
| 0.127970747 | 0.099462886 | 0.087893087 | 0.454628066 | 0 |
| 0 | 0 | 0 | 0 | 0 |
| 0 | 0 | 0 | 0 | 0 |
| 0 | 0 | 0 | 0 | 0 |
| 0 | 0 | 0 | 0 | 0 |
| 0 | 0 | 0 | 0 | 0 |
| 0 | 0 | 0 | 0 | 0 |
| 0 | 0 | 0.04118627 | 0 | 0 |
| 0 | 0 | 0 | 0 | 0 |
| 0.109904329 | 0.046767756 | 0.007711817 | 0.033451046 | 0.068652718 |
| 0 | 0 | 0 | 0 | 0 |
| 0.074976499 | 0.001446365 | 0.03293673 | 0 | 0 |
| 0.004271164 | 0.001008853 | 0 | 0.004431816 | 0 |
| 0.03159993 | 0 | 0.343629006 | 0.012744715 | 0 |
| 0 | 0 | 0.000521184 | 0 | 0 |
| 0 | 0 | 0 | 0 | 0 |
| 0.002133451 | 0.001324597 | 0 | 0 | 0 |
| 0 | 0 | 0 | 0 | 0 |
| 0 | 0 | 0.05144173 | 0 | 0 |
| 0 | 0 | 0 | 0 | 0 |
| 0 | 0 | 0 | 0 | 0 |
| 0 | 0 | 0 | 0 | 0 |
| 0.017927714 | 0.000709872 | 0.02660593 | 0.003022465 | 0 |
| 0 | 0 | 0 | 0 | 0 |
| 0 | 0 | 0.113469184 | 0 | 0 |
| 0 | 0 | 0.001732335 | 0 | 0 |

| | | | | |
|---|---|---|---|---|
| 0 | 0 | 0 | 0 | 0 |
| 0 | 0 | 0 | 0 | 0 |
| 0 | 0 | 0 | 0 | 0 |
| 0 | 0 | 0 | 0 | 0 |
| 0 | 0.131342157 | 0 | 0 | 0 |
| 0.023438611 | 0 | 0 | 0 | 0 |
| 0 | 0 | 0 | 0 | 0 |
| 0.020905922 | 0 | 0.000564944 | 0 | 0 |
| 0 | 0 | 0 | 0 | 0 |
| 0 | 0 | 0.000555926 | 0 | 0 |
| 0 | 0 | 0 | 0 | 0 |
| 0 | 0 | 0 | 0 | 0 |
| 0 | 0 | 0 | 0 | 0 |
| 1.08074907 | 0 | 0.063242395 | 0 | 0 |
| 0 | 0 | 0 | 0 | 0 |
| 0 | 0 | 0 | 0 | 0 |
| 7.179429695 | 0 | 0 | 0 | 0 |
| 0 | 0 | 0 | 0 | 0 |
| 1.107476093 | 0 | 3.216008516 | 0.146598449 | 0 |
| 0 | 0 | 0 | 0 | 0 |
| 0 | 0 | 0.000362345 | 0 | 0 |
| 0 | 0 | 0 | 0 | 0 |
| 0 | 0 | 0 | 0 | 0 |
| 0 | 0 | 0 | 0 | 0 |
| 0 | 0 | 0 | 0 | 0 |
| 0.004598272 | 0.103226961 | 0 | 0.025324207 | 0.006856103 |
| 0 | 0 | 0 | 0 | 0 |
| 0.495087306 | 0.018241295 | 0.115159176 | 0.001224067 | 0.001342152 |
| 0 | 0 | 0 | 0 | 0 |
| 0 | 0 | 0.08551379 | 0.035031523 | 0 |
| 0 | 0 | 0.004186478 | 0 | 0 |
| 0 | 0 | 0 | 0 | 0 |
| 0 | 0 | 0.017437414 | 0 | 0 |
| 0 | 0 | 0 | 0 | 0 |
| 0.003033999 | 0.000554036 | 0 | 0.006552643 | 0 |
| 0.119458198 | 0 | 0 | 0 | 0 |
| 0 | 0 | 0 | 0 | 0 |
| 0.000810047 | 0 | 0.002347705 | 0.0011153 | 0.000698795 |
| 0 | 0 | 0 | 0 | 0 |
| 0 | 0 | 0 | 0 | 0 |
| 7.792035752 | 17.71546078 | 0.998644663 | 3.63080265 | 27.81866149 |
| 0.373267183 | 0.8092406 | 0.384482175 | 0 | 0.021995415 |
| 0.068378753 | 0 | 1.180538379 | 0 | 0 |
| 0 | 0 | 0 | 0 | 0 |
| 0 | 0 | 0 | 0 | 0 |
| 0 | 0 | 0 | 0 | 0 |
| 0 | 0 | 0 | 0 | 0 |
| 0.009482749 | 0 | 0.010671447 | 0 | 0.000554603 |

| | | | | |
|---|---|---|---|---|
| 0 | 0 | 0 | 0 | 0 |
| 0.020499492 | 0 | 0.001655181 | 0 | 0 |
| 0.004827066 | 0 | 0.000712216 | 0 | 0 |
| 0 | 0 | 0 | 0 | 0 |
| 0 | 0 | 0.468758064 | 0.873277297 | 0 |
| 0 | 0 | 0 | 0 | 0 |
| 0 | 0 | 0 | 0 | 0 |
| 0 | 0 | 0 | 0 | 0 |
| 0.761324727 | 5.211748769 | 0 | 0.101215506 | 1.243845993 |
| 0 | 0 | 0 | 0 | 0 |
| 0 | 0 | 0 | 0 | 0 |
| 0 | 0 | 0 | 0 | 0 |
| 0 | 0 | 0.046640426 | 0 | 0 |
| 0.028042798 | 0.019311444 | 0.002705683 | 0.013776425 | 0.097219894 |
| 0 | 0 | 2.291880588 | 0 | 0 |
| 0 | 0 | 0.114258183 | 0 | 0 |
| 0.008031933 | 0 | 0.235215493 | 0 | 0 |
| 0 | 0 | 0 | 0 | 0 |
| 0 | 0 | 0 | 0 | 0 |
| 0 | 0 | 0 | 0 | 0 |
| 0 | 0 | 0 | 0 | 0 |
| 0 | 0 | 0 | 0 | 0 |
| 0 | 0 | 0 | 0 | 0 |
| 0 | 0 | 0 | 0 | 0 |
| 0 | 0 | 0 | 0 | 0 |
| 0 | 0 | 0 | 0 | 0 |
| 0.013522399 | 0 | 0.012841824 | 0.092749423 | 0 |
| 0.002012112 | 0 | 0 | 0.002462526 | 0 |
| 0 | 0 | 0 | 0 | 0 |
| 0 | 0 | 0 | 0 | 0 |
| 0 | 0 | 0 | 0 | 0 |
| 0 | 0 | 0 | 0 | 0 |
| 0 | 0 | 0 | 0 | 0 |
| 0 | 0 | 0.008243341 | 0.004517423 | 0 |
| 0.056337678 | 0 | 0.415511465 | 0 | 0 |
| 0 | 0.016859921 | 0.002964113 | 0.034539655 | 0.029895752 |
| 0 | 0.00061546 | 0.001044718 | 0 | 0 |
| 0 | 0.00312725 | 0 | 0 | 0 |
| 0 | 0 | 0 | 0 | 0 |
| 0 | 0 | 0 | 0 | 0 |
| 0 | 0 | 0.090130998 | 6.556403331 | 0 |
| 0 | 0 | 0 | 0 | 0 |
| 0 | 0 | 0 | 0 | 0 |
| 0 | 0 | 0 | 0 | 0 |
| 0 | 0 | 0 | 0 | 0 |
| 0 | 0 | 0 | 0 | 0 |
| 4.660161723 | 0 | 0 | 0 | 0 |
| 0 | 0 | 0 | 0 | 0 |

| | | | | |
|---|---|---|---|---|
| 0 | 0 | 0.155852544 | 0 | 0 |
| 0 | 0 | 0 | 0 | 0 |
| 0 | 0 | 0 | 0 | 0 |
| 0.004938544 | 0 | 0 | 0 | 0 |
| 0 | 0 | 0.01063022 | 0 | 0 |
| 0 | 0 | 0 | 0 | 0 |
| 0.003920963 | 0 | 0.69018621 | 0 | 0 |
| 0 | 0 | 0 | 0 | 0 |
| 0 | 0 | 0.001775411 | 0 | 0 |
| 0.008585172 | 0 | 0 | 0 | 0 |
| 0 | 0 | 0.108392421 | 0 | 0 |
| 0 | 0 | 0 | 0 | 0 |
| 0 | 0 | 0 | 0 | 0 |
| 0 | 0 | 0 | 0 | 0 |
| 0 | 0 | 0 | 0 | 0 |
| 0 | 0 | 0 | 0 | 0 |
| 0 | 0 | 0 | 0 | 8.937371879 |
| 0 | 0 | 0 | 0 | 0 |
| 0 | 0 | 0 | 0 | 0 |
| 0 | 0 | 0 | 0 | 0 |
| 0.425559204 | 0.005038468 | 0.07295362 | 0.012365035 | 0 |
| 0 | 0 | 0 | 0 | 0 |
| 0 | 0 | 0.013489877 | 0.000492961 | 0 |
| 0.005889896 | 0.001111884 | 0.003774757 | 0.012712037 | 0 |
| 0 | 0 | 0.003108811 | 0 | 0 |
| 0 | 0 | 0 | 0 | 0 |
| 0 | 0 | 0 | 0 | 0 |
| 0 | 0 | 0 | 0 | 0 |
| 0 | 0 | 0 | 0 | 0 |
| 0.636457965 | 0.820934627 | 0.100928607 | 0 | 0.288292392 |
| 0 | 0 | 0 | 0 | 0 |
| 0 | 0 | 0 | 0 | 0 |
| 0 | 0 | 0 | 0 | 0 |
| 0 | 0 | 0 | 0 | 0 |
| 0 | 0 | 0 | 0 | 0 |
| 0 | 0 | 0 | 0 | 0 |
| 0 | 0 | 0.013854715 | 0.048083808 | 0.005040275 |
| 0 | 0 | 0 | 0 | 0 |
| 0 | 0 | 0 | 0 | 0 |
| 0 | 0 | 0 | 0 | 0 |
| 0 | 0 | 0 | 0 | 0 |
| 0 | 0 | 0 | 0 | 0 |
| 0 | 0 | 0 | 0 | 0 |
| 0 | 0 | 0 | 0 | 0 |
| 0 | 0 | 0.006456884 | 0 | 0 |
| 0 | 0 | 0 | 0 | 0 |
| 0 | 0 | 0 | 0 | 0 |
| 0 | 0 | 0 | 0 | 0 |

| | | | | |
|---|---|---|---|---|
| 0.002569353 | 0.030442424 | 0.000812354 | 0.011791899 | 0 |
| 0.037038805 | 0 | 0 | 0 | 0 |
| 0 | 0 | 0 | 0 | 0 |
| 0 | 0 | 0 | 0 | 0 |
| 0 | 0 | 0 | 0 | 0 |
| 0 | 0 | 0.177787013 | 0 | 0 |
| 0 | 0 | 0 | 0 | 0 |
| 2.349180153 | 0 | 0.059870162 | 0.17434966 | 0 |
| 0.086843624 | 0.919789013 | 0.260122913 | 7.830360421 | 0.176892663 |
| 0.042863359 | 0.029198329 | 0 | 0.046559677 | 0.002368219 |
| 0 | 0 | 0 | 0 | 0 |
| 0 | 0 | 0 | 0 | 0 |
| 0 | 0 | 0 | 0 | 0 |
| 0.010748484 | 0 | 1.35336411 | 0.002009722 | 0 |
| 0 | 0 | 0 | 0 | 0 |
| 0 | 0 | 0 | 0 | 0 |
| 0 | 0.002328616 | 0 | 0 | 0 |
| 0 | 0 | 0.919362035 | 0 | 0 |
| 0 | 0 | 0 | 0 | 0 |
| 0 | 0.001686405 | 0.040730756 | 0 | 0 |
| 0 | 0 | 0.072032882 | 0 | 0 |
| 0 | 0 | 0 | 0 | 0 |
| 0 | 0 | 0 | 0 | 0 |
| 0 | 0 | 0 | 0 | 0 |
| 0 | 0 | 0.009739813 | 0 | 0 |
| 0 | 0 | 0 | 0 | 0 |
| 0 | 0 | 0 | 0 | 0 |
| 0 | 0 | 0 | 0 | 0 |
| 0 | 0 | 0 | 0 | 0 |
| 0 | 0 | 0 | 0 | 0 |
| 0.009326475 | 0 | 0 | 0 | 0 |
| 0 | 0 | 0 | 0 | 0 |
| 0 | 0 | 0 | 0 | 0 |
| 0 | 0 | 0 | 0 | 0 |
| 0 | 0 | 0.003824603 | 0 | 0 |
| 0 | 0 | 0 | 0 | 0 |
| 0 | 0 | 0 | 0 | 0 |
| 0 | 0 | 0 | 0 | 0 |
| 0.006517678 | 0.004267936 | 0 | 0 | 0 |
| 0 | 0 | 0 | 0.003453122 | 0 |
| 0 | 0 | 0 | 0 | 0 |
| 0.99764469 | 0 | 0.275358734 | 0.39236174 | 0 |
| 0 | 0 | 0 | 0 | 0 |
| 0 | 0 | 0 | 0 | 0 |
| 0 | 0 | 0 | 0 | 0 |
| 0.013555751 | 0 | 0 | 0 | 0 |
| 0 | 0 | 0 | 0 | 0 |
| 0 | 0 | 0 | 0 | 0 |

| | | | | |
|---|---|---|---|---|
| 0 | 0 | 0 | 0 | 0 |
| 0 | 0 | 0 | 0.004966596 | 0.011558262 |
| 0 | 0 | 0 | 0 | 0 |
| 0 | 0 | 0 | 0 | 0 |
| 0.233440065 | 0 | 3.951632339 | 0.339462341 | 0 |
| 0 | 0 | 0 | 0 | 0 |
| 0 | 0 | 0 | 0 | 0 |
| 0.00980521 | 0.043290794 | 0.000803735 | 0.002800028 | 0.046114826 |
| 0 | 0 | 0.010088568 | 0 | 0 |
| 0 | 0 | 0 | 0 | 0 |
| 0 | 0 | 0 | 0 | 0 |
| 0 | 0 | 0 | 0 | 0 |
| 0 | 0 | 0 | 0 | 0 |
| 0 | 0 | 0 | 0 | 0 |
| 0 | 0.022142132 | 0 | 0 | 0 |
| 0.005102391 | 0 | 0.003584949 | 0.008673012 | 0 |
| 0 | 0 | 0 | 0 | 0 |
| 0 | 0 | 0 | 0 | 0 |
| 0 | 0 | 0 | 0 | 0 |
| 0 | 0 | 0 | 0 | 0 |
| 0 | 0 | 0 | 0 | 0 |
| 0 | 0 | 0 | 0 | 0 |
| 0 | 0 | 0 | 0 | 0 |
| 0 | 0 | 0.000794933 | 0 | 0 |
| 0 | 0 | 0.18875207 | 0.305838759 | 0 |
| 0 | 0 | 0 | 0 | 0 |
| 0.034838472 | 0.006692509 | 0.069524789 | 0.006807163 | 0.008728483 |
| 0.00127122 | 0 | 0 | 0 | 0 |
| 0 | 0 | 0 | 0 | 0 |
| 0 | 0 | 0 | 0 | 0 |
| 0 | 0 | 0 | 0 | 0 |
| 0 | 0 | 0 | 0 | 0 |
| 0 | 0 | 0 | 0 | 0 |
| 0 | 0 | 0 | 0 | 0 |
| 0 | 0 | 0 | 0 | 0 |
| 0 | 0 | 0 | 0 | 0 |
| 0 | 0 | 0 | 0 | 0 |
| 0 | 0 | 0.120607135 | 0 | 0 |
| 0 | 0 | 0 | 0 | 0 |
| 0 | 0 | 0 | 0 | 0 |
| 0 | 0 | 0 | 0 | 0 |
| 0 | 0 | 0 | 0 | 0 |
| 0 | 0 | 0 | 0 | 0 |
| 0 | 0 | 0 | 0 | 0 |
| 0 | 0 | 0 | 0 | 0 |
| 0 | 0 | 0 | 0 | 0 |
| 0 | 0 | 0 | 0 | 0 |
| 0 | 0 | 0.005702999 | 0 | 0 |

| | | | | |
|---|---|---|---|---|
| 0.005027921 | 0 | 0.066236739 | 0.180732129 | 0 |
| 0 | 0 | 0.007687141 | 0 | 0 |
| 0 | 0 | 0 | 0 | 0 |
| 0 | 0 | 0 | 0 | 0 |
| 0 | 0 | 0 | 0 | 0 |
| 0 | 0 | 2.751814458 | 0 | 0 |
| 0 | 0 | 0 | 0 | 0 |
| 0 | 0 | 0 | 0 | 0 |
| 0 | 0 | 0 | 0 | 0 |
| 0.059422316 | 0 | 0 | 0 | 0 |
| 0 | 0 | 0.160376398 | 0 | 0 |
| 0 | 0 | 0.001465603 | 0 | 0 |
| 0 | 0 | 0 | 0 | 0 |
| 0 | 0 | 0 | 0 | 0 |
| 0 | 0 | 0 | 0 | 0 |
| 0 | 0 | 0 | 0 | 0 |
| 0 | 0 | 0 | 0 | 0 |
| 0 | 0 | 0 | 0 | 0 |
| 0 | 0 | 0 | 0 | 0 |
| 0 | 0 | 0 | 0 | 0 |
| 0.006947884 | 0.30793677 | 0 | 0 | 0.020086865 |
| 0 | 0 | 0 | 0 | 0 |
| 0.014229197 | 0 | 0.041989329 | 0.001523762 | 0 |
| 0 | 0 | 0.385337622 | 0 | 0 |
| 0.014271799 | 0 | 0 | 0 | 0 |
| 0 | 0 | 0 | 0 | 0 |
| 0 | 0 | 0 | 0 | 0 |
| 0 | 0 | 0 | 0 | 0 |
| 0 | 0 | 0 | 0 | 0 |
| 0 | 0 | 0 | 0 | 0 |
| 0 | 0 | 0 | 0 | 0 |
| 0 | 0 | 0 | 0 | 0 |
| 0.013103491 | 0.024648033 | 0.015214192 | 0.010804405 | 0.275890753 |
| 0 | 0 | 0 | 0 | 0 |
| 0.35619953 | 0 | 0.004710815 | 0.408461401 | 0.088887569 |
| 0 | 0.023790142 | 0 | 0.000817932 | 0.030338735 |
| 0 | 0 | 0 | 0 | 0 |
| 0 | 0 | 0 | 0 | 0 |
| 0 | 0.01151877 | 0 | 0.018164506 | 0.046235482 |
| 0 | 0 | 0 | 0 | 0 |
| 0 | 0 | 0 | 0 | 0 |
| 0 | 0 | 0 | 0 | 0 |
| 0.026032266 | 0 | 0.946142857 | 0.22036241 | 0 |
| 0 | 0 | 0 | 0 | 0 |
| 0 | 0 | 0 | 0 | 0 |
| 0.052259618 | 0 | 0.031774934 | 0 | 0 |
| 0 | 0 | 0 | 0 | 0 |
| 0 | 0 | 0 | 0 | 0 |

| | | | | |
|---|---|---|---|---|
| 0 | 0 | 0 | 0 | 0 |
| 0 | 0 | 0 | 0 | 0 |
| 0 | 0 | 0 | 0 | 0 |
| 0 | 0 | 0 | 0 | 0 |
| 0 | 0 | 0 | 0 | 0 |
| 0 | 0 | 0 | 0 | 0 |
| 0 | 0 | 0 | 0 | 0 |
| 0.023677349 | 2.293949852 | 0 | 0 | 0 |
| 0.111196697 | 0 | 0.021169896 | 0 | 0 |
| 0.001774511 | 0 | 0.137145196 | 0 | 0 |
| 0 | 0 | 0 | 0 | 0 |
| 0 | 0 | 0 | 0 | 0 |
| 0 | 0 | 0 | 0 | 0 |
| 0 | 0 | 0 | 0 | 0 |
| 0 | 0 | 0 | 0 | 0 |
| 0.003297854 | 0.121648017 | 0 | 0 | 0.014503546 |
| 0 | 0 | 0 | 0 | 0 |
| 0 | 0 | 0 | 0 | 0 |
| 0 | 0 | 0 | 0 | 0 |
| 0 | 0 | 0.337291218 | 0 | 0 |
| 0 | 0 | 3.374544735 | 0.004277393 | 0 |
| 0 | 0 | 0 | 0 | 0 |
| 0 | 0 | 0 | 0 | 0 |
| 0 | 0 | 0 | 0 | 0 |
| 0 | 0 | 0.000690793 | 0 | 0 |
| 0 | 0 | 0 | 0 | 0 |
| 0 | 0 | 0 | 0 | 0 |
| 0 | 0 | 0.485751713 | 0 | 0 |
| 0 | 0 | 0 | 0 | 0 |
| 0 | 0 | 0.045451416 | 0 | 0 |
| 5.082516809 | 0 | 0.280328171 | 0 | 0 |
| 0 | 0 | 0 | 0 | 0 |
| 0 | 0 | 0 | 0 | 0 |
| 0 | 0 | 0 | 0 | 0 |
| 0 | 0 | 0 | 0 | 0 |
| 0 | 0 | 0 | 0 | 0 |
| 0 | 0 | 0 | 0 | 0 |
| 0 | 0 | 0 | 0 | 0 |
| 0 | 0 | 0.016386972 | 0 | 0 |
| 0 | 0 | 0 | 0 | 0 |
| 0 | 0 | 0 | 0 | 0 |
| 0 | 0 | 0.369496904 | 0 | 0 |
| 0 | 0 | 0 | 0 | 0 |
| 0 | 0 | 0 | 0 | 0 |
| 0 | 0 | 0 | 0 | 0 |
| 0 | 0 | 0 | 0 | 0 |
| 0.04090904 | 1.128015406 | 0 | 0.01731672 | 3.558068159 |
| 0.000484176 | 0 | 0.001275685 | 0 | 0 |

| | | | | |
|---|---|---|---|---|
| 0 | 0 | 0 | 0 | 0 |
| 0 | 0 | 0 | 0 | 0 |
| 0 | 0 | 0 | 0 | 0 |
| 0 | 0 | 0 | 0 | 0 |
| 0.242864908 | 0 | 0.014697533 | 0.046164245 | 0 |
| 0 | 0 | 0 | 0 | 0 |
| 0 | 0 | 0.003096112 | 0 | 0 |
| 0 | 0 | 0 | 0 | 0 |
| 0 | 0 | 0 | 0 | 0 |
| 0 | 0 | 0 | 0 | 0 |
| 0 | 0 | 0.006749636 | 0 | 0 |
| 0.002883663 | 0 | 0.010528311 | 0.001512504 | 0 |
| 0.002132464 | 0 | 0.001966483 | 0 | 0 |
| 0 | 0 | 0 | 0 | 0 |
| 0.002040584 | 0 | 0 | 0 | 0 |
| 0 | 0 | 0 | 0.13849589 | 0 |
| 0 | 0.001475375 | 0.020202062 | 0 | 0 |
| 0 | 0 | 0 | 0 | 0 |
| 0.490045637 | 0.002178783 | 0.001643731 | 0.018181262 | 0 |
| 0.008349415 | 0.002686781 | 0.341355084 | 0.007663832 | 0 |
| 0 | 0 | 0 | 0 | 0 |
| 0 | 0 | 0 | 0 | 0 |
| 0 | 0 | 0 | 0 | 0 |
| 0 | 0 | 0 | 0 | 0 |
| 0 | 0 | 0 | 0 | 0 |
| 0 | 0 | 0 | 0 | 0 |
| 0.012402401 | 0.001139845 | 0.002407802 | 0 | 0 |
| 0.035280975 | 0 | 0 | 0 | 0 |
| 0 | 0 | 0 | 0 | 0 |
| 0 | 0 | 0 | 0 | 0 |
| 0 | 0 | 0.002436937 | 0 | 0 |
| 0.009990205 | 0 | 0 | 0 | 0 |
| 0 | 0 | 0 | 0 | 0 |
| 0 | 0 | 0 | 0 | 0 |
| 0 | 0 | 0 | 0 | 0 |
| 0 | 0 | 0 | 0 | 0 |
| 0 | 0 | 0 | 0 | 0 |
| 0.010787036 | 0.002118139 | 0.026640608 | 0.006918978 | 0.036325255 |
| 0.018133166 | 0 | 0.222902 | 0 | 0 |
| 0 | 0 | 0 | 0 | 0 |
| 0 | 0 | 0.123681256 | 0 | 0 |
| 0 | 0 | 0 | 0 | 0 |
| 0 | 0 | 0 | 0 | 0 |
| 0.011755954 | 0.130423465 | 0 | 0 | 0.009975138 |
| 0 | 0 | 0 | 0 | 0 |
| 0 | 0 | 0.000635532 | 0 | 0 |
| 0 | 0 | 0 | 0 | 0 |
| 0 | 0 | 0 | 0 | 0 |

| | | | | |
|---|---|---|---|---|
| 0 | 0 | 0 | 0 | 0 |
| 0 | 0 | 0 | 0 | 0 |
| 0 | 0 | 0 | 0 | 0 |
| 0.00100986 | 0.000940489 | 0.000957865 | 0.000926939 | 0.001858487 |
| 0 | 0.004506823 | 0 | 0.005330271 | 0 |
| 0 | 0.002251544 | 0 | 0.002441016 | 0 |
| 0 | 0 | 0.00309845 | 0 | 0 |
| 0 | 0 | 0.009988442 | 0.014208952 | 0 |
| 0 | 0 | 0 | 0 | 0 |
| 0 | 0 | 0 | 0 | 0 |
| 0 | 0 | 0.004573103 | 0 | 0 |
| 0 | 0 | 0 | 0 | 0 |
| 0.014603312 | 0 | 0.128631131 | 0.06347289 | 0 |
| 0 | 0 | 0 | 0 | 0 |
| 0 | 0 | 0 | 0 | 0 |
| 0 | 0 | 0 | 0 | 0 |
| 0 | 0 | 0 | 0 | 0 |
| 0 | 0 | 0 | 0 | 0 |
| 0 | 0 | 0 | 0 | 0 |
| 0 | 0 | 0 | 0 | 0 |
| 0 | 0 | 0 | 0 | 0 |
| 0 | 0 | 0 | 0 | 0 |
| 0.006598254 | 0.007608088 | 0 | 0.003287793 | 0.011058839 |
| 0.013570176 | 0 | 0.009495354 | 0 | 0 |
| 0 | 0 | 0 | 0 | 0 |
| 0 | 0 | 0.036360646 | 0 | 0 |
| 0 | 0 | 0.003511758 | 0 | 0 |
| 0 | 0 | 0 | 0 | 0 |
| 0 | 0 | 0 | 0 | 0 |
| 0 | 0 | 0 | 0 | 0 |
| 0 | 0 | 0.002658373 | 0 | 0 |
| 0.000487648 | 0 | 0 | 0 | 0 |
| 0 | 0 | 0 | 0 | 0 |
| 0 | 0 | 0 | 0 | 0 |
| 0 | 0 | 0 | 0 | 0 |
| 0 | 0 | 0 | 0 | 0.13926286 |
| 0 | 0 | 0 | 0 | 0 |
| 0 | 0 | 0 | 0 | 0 |
| 0 | 0.001242206 | 0.067100168 | 0 | 0 |
| 0 | 0 | 0 | 0 | 0 |
| 0 | 0 | 0 | 0 | 0 |
| 0 | 0 | 0 | 0 | 0 |
| 0 | 0 | 0 | 0 | 0 |
| 0 | 0 | 0 | 0 | 0 |
| 0 | 0 | 0 | 0 | 0 |
| 0.063528301 | 0.402564946 | 0.228644939 | 0.101830387 | 0 |
| 0 | 0 | 0 | 0 | 0 |
| 0 | 0 | 0 | 0 | 0 |

| | | | | |
|---|---|---|---|---|
| 0 | 0 | 0 | 0 | 0 |
| 0 | 0 | 0 | 0 | 0 |
| 0.244598357 | 0 | 0.073579755 | 0 | 0 |
| 0 | 0 | 0 | 0 | 0 |
| 0 | 0 | 0 | 0 | 0 |
| 0 | 0 | 0 | 0 | 0 |
| 0 | 0 | 0 | 0 | 0 |
| 0 | 0 | 0 | 0 | 0 |
| 0 | 0.007151644 | 0 | 0 | 0 |
| 0 | 0 | 0 | 0 | 0 |
| 0 | 0 | 0.001140423 | 0 | 0 |
| 0 | 0 | 0.000340531 | 0 | 0 |
| 0 | 0 | 0 | 0 | 0 |
| 0 | 0 | 0 | 0 | 0 |
| 0 | 0 | 0 | 0 | 0 |
| 0 | 0 | 0 | 0 | 0 |
| 0 | 0 | 0 | 0 | 0 |
| 0 | 0 | 0 | 0 | 0 |
| 0 | 0 | 0 | 0 | 0 |
| 0.001913393 | 0 | 0 | 0 | 0 |
| 0 | 0 | 0 | 0 | 0 |
| 0 | 0 | 0 | 0 | 0 |
| 0 | 0 | 0 | 0 | 0 |
| 0 | 0 | 0 | 0 | 0 |
| 0 | 0 | 0 | 0 | 0 |
| 0 | 0 | 0 | 0 | 0 |
| 0 | 0 | 0 | 0 | 0 |
| 0 | 0 | 0.006136986 | 0 | 0 |
| 0 | 0 | 0 | 0 | 0 |
| 0 | 0 | 0 | 0 | 0 |
| 0.004109506 | 0 | 0.003031714 | 0.023688591 | 0 |
| 0 | 0 | 0 | 0 | 0 |
| 0.11852543 | 0.036290858 | 0.017801792 | 0.033461335 | 0 |
| 0.003510729 | 0.001059581 | 0.007913817 | 0.002864407 | 0.016601023 |
| 0 | 0 | 0.106082731 | 0 | 0 |
| 0 | 0 | 0 | 0 | 0 |
| 0 | 0 | 0 | 0 | 0 |
| 0 | 0 | 0 | 0 | 0 |
| 0 | 0 | 0 | 0 | 0 |
| 0 | 0 | 0 | 0 | 0 |
| 0 | 0 | 0 | 0 | 0 |
| 0.001570206 | 0.01169874 | 0.001572102 | 0.252511232 | 0 |
| 0.022955445 | 0 | 0.039514914 | 0.09393949 | 0 |
| 0 | 0 | 0 | 0 | 0 |
| 0 | 0 | 0 | 0 | 0 |
| 0.012716015 | 0 | 0.239332284 | 0.006597152 | 0 |
| 0 | 0 | 0.002960623 | 0 | 0 |
| 0.001544781 | 0 | 0 | 0 | 0 |

| | | | | |
|---|---|---|---|---|
| 0 | 0 | 0 | 0 | 0 |
| 0.016433396 | 0 | 0.002534517 | 0.012692656 | 0 |
| 0.011470746 | 0.005268716 | 0 | 0.013322239 | 0.00403892 |
| 0 | 0 | 0 | 0 | 0 |
| 0.066541765 | 0 | 0.034276311 | 0.008749927 | 0 |
| 0 | 0.004942108 | 0 | 0.005678085 | 0 |
| 0 | 0 | 0 | 0 | 0 |
| 0 | 0 | 0 | 0 | 0 |
| 0 | 0 | 0 | 0 | 0 |
| 0 | 0 | 0 | 0 | 0 |
| 0 | 0 | 0 | 0 | 0 |
| 0 | 0 | 0 | 0 | 0 |
| 0 | 0 | 0 | 0 | 0 |
| 0 | 0 | 0 | 0.003070334 | 0 |
| 0 | 0 | 0 | 0 | 0 |
| 0 | 0 | 0 | 0 | 0 |
| 0 | 0 | 0 | 0 | 0 |
| 0 | 0 | 0 | 0 | 0 |
| 0 | 0 | 0 | 0 | 0 |
| 0 | 0 | 0 | 0 | 0 |
| 0 | 0 | 0 | 0 | 0 |
| 0 | 0 | 0 | 0 | 0 |
| 0 | 0 | 0 | 0 | 0 |
| 0 | 0 | 0 | 0 | 0 |
| 0.002315145 | 0 | 0.015147943 | 0.002302132 | 0 |
| 0.00116449 | 0.002409993 | 0.001738618 | 0 | 0.002232354 |
| 0 | 0 | 0 | 0 | 0 |
| 0 | 0 | 0.008829427 | 0.006566676 | 0 |
| 0 | 0 | 0 | 0 | 0 |
| 0 | 0 | 0 | 0 | 0 |
| 0 | 0 | 0 | 0 | 0 |
| 0 | 0 | 0 | 0 | 0 |
| 0 | 0 | 0 | 0 | 0 |
| 0 | 0 | 0 | 0 | 0 |
| 0 | 0 | 0 | 0 | 0 |
| 0 | 0 | 0 | 0 | 0 |
| 0 | 0 | 0 | 0.000950069 | 0 |
| 0.109926277 | 0.033885111 | 0 | 0 | 0 |
| 1.941492369 | 0 | 2.918698729 | 0 | 0 |
| 0 | 0 | 0 | 0 | 0 |
| 0 | 0 | 0 | 0 | 0 |
| 0 | 0 | 0.053588343 | 0 | 0 |
| 0.001947822 | 0.0195007 | 0.001129048 | 0.007866687 | 0 |
| 0 | 0 | 0 | 0 | 0 |
| 0 | 0 | 0 | 0 | 0 |
| 0 | 0 | 0 | 0 | 0 |
| 0.008641381 | 0 | 0.001570202 | 0.00218809 | 0 |
| 0 | 0 | 0 | 0 | 0 |

| | | | | |
|---|---|---|---|---|
| 0 | 0 | 0 | 0 | 0 |
| 0.014851773 | 0.007003315 | 0.006835508 | 0.01294203 | 0.008865708 |
| 0 | 0 | 0 | 0 | 0 |
| 0 | 0 | 0 | 0 | 0 |
| 0 | 0 | 0 | 0 | 0 |
| 0 | 0 | 0 | 0 | 0 |
| 0 | 0 | 0 | 0 | 0 |
| 0 | 0 | 0 | 0 | 0 |
| 0 | 0 | 0 | 0 | 0 |
| 0 | 0 | 0 | 0 | 0 |
| 0 | 0 | 0 | 0 | 0 |
| 0 | 0 | 0 | 0 | 0 |
| 0 | 0 | 0.014242034 | 0 | 0 |
| 0 | 0 | 0 | 0 | 0 |
| 0 | 0.00109151 | 0.004323184 | 0 | 0 |
| 0 | 0 | 0 | 0 | 0 |
| 0 | 0 | 0 | 0 | 0 |
| 0 | 0 | 0 | 0 | 0 |
| 0 | 0 | 0.002049669 | 0 | 0 |
| 0 | 0 | 0 | 0 | 0 |
| 0 | 0 | 0 | 0 | 0 |
| 0.014963215 | 0.014113992 | 0.066718267 | 0.011621554 | 0 |
| 0 | 0 | 0 | 0.001136573 | 0 |
| 0 | 0 | 0 | 0 | 0.002999976 |
| 0 | 0 | 0 | 0 | 0 |
| 0 | 0 | 0 | 0 | 0 |
| 0 | 0 | 0 | 0 | 0 |
| 0 | 0 | 0 | 0 | 0 |
| 0 | 0 | 0 | 0 | 0 |
| 0 | 0 | 0 | 0 | 0 |
| 0 | 0 | 0 | 0 | 0 |
| 0 | 0 | 0 | 0 | 0 |
| 0 | 0 | 0 | 0 | 0 |
| 0 | 0 | 0 | 0 | 0 |
| 0 | 0 | 17.31914421 | 0 | 0 |
| 0 | 0 | 0 | 0 | 0 |
| 0 | 0 | 0 | 0 | 0 |
| 0 | 0 | 0 | 0 | 0 |
| 0.00551726 | 0 | 0.0993337 | 0.066890041 | 0 |
| 0 | 0 | 0 | 0 | 0 |
| 0.145522888 | 0.147972003 | 0.048117255 | 0 | 0.282368822 |
| 0 | 0 | 0 | 0 | 0 |
| 0 | 0 | 0 | 0 | 0 |
| 0 | 0 | 0 | 0 | 0 |
| 0 | 0 | 0 | 0 | 0 |
| 0.896788177 | 1.867259696 | 0.069889689 | 0.54414653 | 3.762252009 |
| 0 | 0 | 0 | 0 | 0 |
| 0 | 0 | 0.000491483 | 0 | 0 |

| | | | | |
|---|---|---|---|---|
| 0 | 0 | 0 | 0.005595126 | 0 |
| 0 | 0 | 0 | 0 | 0 |
| 0 | 0 | 0 | 0 | 0 |
| 0 | 0 | 0.029197648 | 0 | 0 |
| 0 | 0 | 0 | 0 | 0 |
| 0 | 0 | 0 | 0 | 0 |
| 0 | 0 | 0 | 0 | 0 |
| 0 | 0 | 0.001209729 | 0 | 0 |
| 0.001119102 | 0 | 0 | 0 | 0 |
| 0 | 0 | 0.027730085 | 0.29320534 | 0.001327377 |
| 0 | 0 | 0.027577837 | 0 | 0 |
| 0 | 0 | 0 | 0 | 0 |
| 0 | 0 | 0.000832135 | 0 | 0 |
| 0 | 0 | 0 | 0 | 0 |
| 0 | 0 | 0 | 0 | 0 |
| 0 | 0 | 0 | 0 | 0 |
| 0 | 0 | 0 | 0 | 0 |
| 0 | 0 | 0 | 0 | 0 |
| 0 | 0 | 0 | 0 | 0 |
| 0.169469707 | 0.044568383 | 0.136810261 | 2.500111519 | 0.00692866 |
| 0 | 0 | 0 | 0 | 0 |
| 0 | 0 | 0 | 0 | 0 |
| 0 | 0 | 0 | 0 | 0 |
| 0 | 0 | 0 | 0 | 0 |
| 0 | 0 | 0 | 0 | 0 |
| 0 | 0 | 0 | 0 | 0 |
| 0 | 0 | 0 | 0 | 0 |
| 0 | 0 | 0 | 0 | 0 |
| 0 | 0 | 0 | 0 | 0 |
| 0 | 0.386985177 | 0 | 0.038223072 | 0 |
| 0 | 0 | 0 | 0 | 0 |
| 0 | 0 | 0 | 0 | 0 |
| 0 | 0 | 0 | 0 | 0 |
| 0 | 0 | 0 | 0 | 0 |
| 0 | 0 | 0 | 0 | 0 |
| 0 | 0 | 0 | 0.00190864 | 0 |
| 0 | 0 | 0 | 0 | 0 |
| 0 | 0 | 0 | 0 | 0 |
| 0 | 0 | 0 | 0 | 0 |
| 0 | 0 | 0 | 0 | 0 |
| 0 | 0 | 0 | 0 | 0 |
| 0 | 0 | 0 | 0 | 0 |
| 0 | 0 | 0.000439761 | 0 | 0 |
| 0 | 0 | 0 | 0 | 0 |
| 0.002451704 | 0.000439094 | 0.02077024 | 0 | 0 |
| 0 | 0 | 0 | 0 | 0 |
| 0 | 0 | 0 | 0 | 0 |
| 0 | 0 | 0 | 0 | 0 |

| | | | | |
|---|---|---|---|---|
| 0 | 0 | 0 | 0 | 0 |
| 0 | 0 | 0 | 0 | 0 |
| 0 | 0 | 0.098525442 | 0 | 0 |
| 0 | 0 | 0 | 0 | 0 |
| 0 | 0 | 0 | 0 | 0 |
| 0.010700171 | 0.001201935 | 0.09503789 | 0.025587761 | 0.004048513 |
| 0 | 0 | 0.002044506 | 0 | 0 |
| 0 | 0 | 0 | 0 | 0 |
| 0 | 0 | 0 | 0 | 0 |
| 0 | 0 | 0 | 0 | 0 |
| 0 | 0 | 0 | 0 | 0 |
| 0 | 0 | 0 | 0 | 0 |
| 0 | 0 | 0 | 0 | 0 |
| 0 | 0 | 0.00208682 | 0 | 0 |
| 0 | 0 | 0 | 0 | 0 |
| 0 | 0 | 0 | 0 | 0 |
| 0 | 0 | 0 | 0 | 0 |
| 0 | 0 | 0 | 0 | 0 |
| 0 | 0 | 0 | 0 | 0 |
| 0 | 0 | 0 | 0 | 0 |
| 0 | 0 | 0 | 0 | 0.001126608 |
| 0 | 0 | 0 | 0 | 0 |
| 0 | 0 | 0 | 0 | 0 |
| 0 | 0 | 0 | 0 | 0 |
| 0 | 0 | 0 | 0 | 0 |
| 0 | 0 | 0.000464185 | 0.12573055 | 0 |
| 0 | 0 | 0 | 0 | 0 |
| 0 | 0 | 0.660166431 | 0 | 0 |
| 0 | 0 | 0 | 0 | 0 |
| 0 | 0 | 0 | 0 | 0 |
| 0 | 0 | 0 | 0 | 0 |
| 0 | 0 | 0 | 0 | 0 |
| 0 | 0 | 0 | 0 | 0 |
| 0 | 0 | 0 | 0 | 0 |
| 0 | 0 | 0 | 0 | 0 |
| 0 | 0 | 0 | 0 | 0 |
| 0 | 0 | 0 | 0 | 0 |
| 0 | 0 | 0 | 0 | 0 |
| 0 | 0 | 0 | 0 | 0 |
| 0 | 0 | 0 | 0 | 0 |
| 0 | 0 | 0 | 0 | 0 |
| 0 | 0 | 0 | 0 | 0 |
| 0 | 0 | 0 | 0 | 0 |
| 0 | 0 | 0 | 0 | 0 |
| 0 | 0 | 0 | 0 | 0 |
| 0 | 0 | 0 | 0 | 0 |
| 0 | 0 | 0 | 0 | 0 |

| | | | | |
|---|---|---|---|---|
| 0 | 0 | 0 | 0 | 0 |
| 0 | 0 | 0.010299703 | 0 | 0 |
| 0 | 0 | 0 | 0 | 0 |
| 0 | 0 | 0 | 0 | 0 |
| 0 | 0 | 0 | 0 | 0 |
| 0 | 0 | 0 | 0 | 0 |
| 0 | 0 | 0 | 0 | 0 |
| 0 | 0 | 0 | 0 | 0 |
| 0 | 0 | 0 | 0 | 0 |
| 0 | 0 | 0 | 0 | 0 |
| 0 | 0.000800927 | 0.002719082 | 0.013830075 | 0 |
| 0 | 0 | 0 | 0 | 0 |
| 0 | 0 | 0.029800788 | 0 | 0 |
| 0 | 0 | 0 | 0 | 0 |
| 0 | 0 | 0 | 0 | 0 |
| 0 | 0 | 0 | 0.000924673 | 0 |
| 0 | 0 | 0 | 0 | 0 |
| 0 | 0 | 0 | 0 | 0 |
| 0 | 0 | 0 | 0 | 0 |
| 0 | 0 | 0 | 0 | 0 |
| 0.005237114 | 0 | 0.02351268 | 0.004037953 | 0 |
| 0 | 0 | 0.005560879 | 0 | 0 |
| 0 | 0 | 0.000441898 | 0 | 0 |
| 0.510372078 | 0 | 0.315304127 | 0.641928828 | 0 |
| 0 | 0 | 0 | 0 | 0 |
| 0 | 0 | 0 | 0 | 0 |
| 0 | 0 | 0 | 0 | 0 |
| 0 | 0 | 0 | 0 | 0 |
| 0 | 0 | 0 | 0 | 0 |
| 0 | 0 | 0 | 0 | 0 |
| 0.029402083 | 0 | 0.002708648 | 0 | 0 |
| 0 | 0 | 0 | 0 | 0 |
| 0 | 0 | 0 | 0 | 0 |
| 0 | 0 | 0 | 0 | 0 |

| PHL018-0198_V1 | PHL018-0121_V3 | PHL018-0220_V1 | PHL018-0228_V3 | PHL018-0096_V1 |
|---|---|---|---|---|
| REF | EYCF | REF | REF | EYCF |
| 0.344184497 | 0.001015761 | 0.011464805 | 0.001310049 | 0.643582211 |
| 1.20015427 | 0.002559457 | 5.420506191 | 0.116062673 | 12.6823206 |
| 2.755960335 | 0 | 0 | 0 | 3.163840283 |
| 0 | 0 | 0 | 0 | 0 |
| 0 | 0 | 0 | 0 | 0 |
| 0 | 0 | 0 | 0 | 0 |
| 0 | 0 | 0 | 0 | 0 |
| 0 | 0 | 0 | 0 | 0 |
| 0 | 0 | 0 | 0 | 0 |
| 0 | 0 | 0 | 0 | 0 |
| 0 | 0 | 0 | 0 | 0 |
| 0 | 0 | 0 | 0 | 0 |
| 0 | 0 | 0 | 0 | 0 |
| 0 | 0.001221191 | 8.203412599 | 0.373273981 | 1.761014421 |
| 0 | 0 | 0 | 0 | 0 |
| 0 | 0 | 0 | 0 | 0 |
| 0 | 0 | 0 | 0 | 0 |
| 0 | 0 | 0 | 0 | 0 |
| 0 | 0 | 0 | 0 | 0 |
| 0 | 0 | 0 | 0 | 0 |
| 0 | 0 | 0 | 0 | 0 |
| 0 | 0 | 0 | 0 | 0 |
| 0 | 0 | 0 | 0 | 0 |
| 0 | 0 | 0 | 0 | 0 |
| 0 | 0 | 0 | 0 | 0 |
| 0 | 0 | 0 | 0 | 0 |
| 0 | 0 | 0 | 0 | 0 |
| 0 | 0 | 0 | 0 | 0 |
| 0 | 0 | 0 | 0 | 0 |
| 0 | 0 | 0 | 0 | 0 |
| 0 | 0 | 0 | 0 | 0 |
| 0 | 0 | 0 | 0 | 0 |
| 0 | 0 | 0 | 0 | 0 |
| 0 | 0 | 0 | 0 | 0 |
| 0 | 0 | 0 | 0 | 0 |
| 0 | 0 | 0 | 0.462749556 | 0 |
| 0 | 0 | 0 | 0 | 0 |
| 0 | 0 | 0 | 0 | 0 |
| 0 | 0 | 0 | 0 | 0 |
| 0 | 0 | 0 | 0 | 0 |
| 0 | 0 | 0 | 0 | 0 |
| 0.115403316 | 0.050228337 | 0.679816339 | 0.004555995 | 0.04512509 |
| 0.031930913 | 0.037798657 | 0.067147758 | 1.298147349 | 0.003533068 |
| 0.065538595 | 0.026790273 | 0.534921729 | 0.043890361 | 0.001396112 |
| 0.005404795 | 0.09370308 | 0 | 0.002861371 | 0.00163564 |
| 0 | 0 | 0 | 0 | 0 |

| | | | | |
|---|---|---|---|---|
| 0 | 0 | 0 | 0.001636059 | 0.029269883 |
| 0.002774572 | 0 | 0 | 0.057027726 | 0.006057803 |
| 0.003743031 | 0.000481056 | 0.001151741 | 0 | 0 |
| 0.131683269 | 0.00252196 | 1.588714807 | 1.315046133 | 0.554428443 |
| 0 | 0 | 0 | 0 | 0.003057449 |
| 0 | 0 | 0 | 0 | 0 |
| 0.007512995 | 0.003741599 | 7.77179988 | 0.230664643 | 0.003126253 |
| 0 | 0 | 0 | 0.00213642 | 0 |
| 0 | 0 | 0 | 0 | 0 |
| 0 | 0 | 0 | 0 | 0 |
| 0 | 0 | 0 | 0 | 0 |
| 0.155860001 | 0 | 0 | 0.129070834 | 0.621236556 |
| 0 | 0 | 0.001847736 | 0 | 0 |
| 0.02466095 | 0 | 0.001069253 | 0.049189793 | 0.282879476 |
| 0 | 0.010294067 | 0 | 0 | 0 |
| 0 | 0 | 0 | 0 | 0 |
| 0.092663425 | 26.78657025 | 0.014867727 | 1.660739843 | 8.511483028 |
| 0 | 0 | 0 | 0 | 0 |
| 0 | 0 | 0 | 0 | 0 |
| 0 | 0 | 0 | 0 | 0 |
| 0.024637629 | 0 | 0.030829333 | 0.060012605 | 0.090539394 |
| 0.002224263 | 0 | 0.020004385 | 0.009143365 | 0.003502829 |
| 0 | 0 | 0 | 0 | 0 |
| 0 | 0 | 0 | 0 | 0 |
| 0 | 0 | 0 | 0 | 0 |
| 0 | 0 | 0 | 0 | 0 |
| 0 | 0 | 0 | 0 | 0 |
| 0 | 0 | 0 | 0 | 0 |
| 0.005833224 | 0.419006633 | 0.598877393 | 0 | 0.000874367 |
| 0 | 0 | 0 | 0 | 0 |
| 0 | 0 | 0.162791535 | 0 | 0.094967768 |
| 0 | 0 | 0 | 0 | 0 |
| 0 | 0 | 0 | 0 | 0 |
| 0 | 0 | 0 | 0 | 0 |
| 0 | 0 | 0 | 0 | 0 |
| 0 | 0.003296502 | 0 | 0 | 0 |
| 0 | 0 | 0 | 0 | 0 |
| 0.000674288 | 0.040296829 | 0 | 41.19843559 | 0.019683759 |
| 0 | 0 | 0 | 0 | 0 |
| 0 | 0 | 0 | 0 | 0 |
| 0 | 0 | 0 | 0 | 0 |
| 0 | 0 | 0 | 0 | 0 |
| 0 | 0 | 0 | 0 | 0 |
| 0 | 0 | 0 | 0 | 0 |
| 0 | 0 | 0 | 0 | 0 |
| 0 | 0 | 0 | 0 | 0 |
| 0 | 0 | 0.008806175 | 0 | 0 |
| 0.577986316 | 0 | 0 | 0.596181798 | 0 |

| | | | | |
|---|---|---|---|---|
| 0 | 0 | 0 | 0 | 0 |
| 0 | 0 | 0 | 0 | 0 |
| 0 | 0 | 0 | 0 | 0 |
| 0 | 0 | 0 | 0 | 0 |
| 0 | 0.014424409 | 0.310502828 | 0 | 0 |
| 0 | 0 | 0 | 0 | 0 |
| 0 | 0 | 0 | 0 | 0 |
| 0 | 0 | 0 | 0 | 0 |
| 0.027196541 | 0.000685247 | 0 | 0 | 0 |
| 0 | 0 | 0 | 0 | 0 |
| 0 | 0 | 0 | 0 | 0 |
| 0.005865646 | 0.065136347 | 0.00357135 | 0 | 0.008628582 |
| 0 | 0 | 0 | 0 | 0 |
| 0 | 0 | 0 | 0 | 0 |
| 0 | 0 | 0 | 0 | 0 |
| 0.000463016 | 0 | 0 | 0 | 0 |
| 0 | 0 | 0 | 0 | 0 |
| 0 | 0 | 0 | 0 | 0 |
| 0 | 0 | 0.077642181 | 0 | 0 |
| 0 | 0 | 0 | 0 | 0 |
| 0 | 0 | 0 | 0 | 0 |
| 0 | 0 | 0 | 0 | 0 |
| 0 | 0 | 0 | 0 | 0 |
| 0 | 0 | 0 | 0 | 0 |
| 0 | 0 | 0 | 0 | 0 |
| 0 | 0 | 0 | 0 | 0 |
| 0 | 0.005020834 | 0 | 0 | 0 |
| 0 | 0 | 0 | 0 | 0 |
| 0 | 0 | 0 | 0 | 0 |
| 0 | 0 | 0 | 0 | 0 |
| 0.004177902 | 0 | 0.014775108 | 0.01247179 | 0 |
| 0.000939211 | 0.00640288 | 0.000568821 | 0 | 0.015793038 |
| 0 | 0 | 0 | 0 | 0 |
| 0 | 0 | 0 | 0 | 0 |
| 0 | 0 | 0 | 0 | 0 |
| 0 | 0 | 0 | 0 | 0 |
| 0 | 0 | 0 | 0 | 0 |
| 0 | 0 | 0 | 0 | 0 |
| 0 | 0 | 0 | 0 | 0 |
| 0 | 0 | 0 | 0 | 0 |
| 0 | 0 | 0 | 0 | 0 |
| 0 | 0 | 0 | 0 | 0 |
| 0.001085579 | 0 | 0 | 0.000867715 | 0.000370645 |
| 0 | 0 | 0 | 0 | 0 |
| 0 | 0 | 0 | 0 | 0 |
| 0 | 0 | 0 | 0 | 0 |
| 0 | 0 | 0 | 0 | 0 |
| 0 | 0 | 0 | 0 | 0 |

| | | | | |
|---|---|---|---|---|
| 0 | 0 | 0 | 0 | 0 |
| 0 | 0 | 0 | 0 | 0 |
| 0 | 0 | 0 | 0 | 0 |
| 0 | 0 | 0 | 0 | 0 |
| 0 | 0 | 0 | 0.023056308 | 0 |
| 0 | 0 | 0 | 0 | 0 |
| 0 | 0 | 0 | 0 | 0 |
| 0 | 0 | 0 | 0 | 0 |
| 0 | 0 | 0 | 0 | 0 |
| 0 | 0 | 0 | 0 | 0 |
| 0 | 0 | 0 | 0 | 0 |
| 0 | 0 | 0 | 0 | 0 |
| 0 | 0 | 0 | 0 | 0 |
| 0 | 0 | 0 | 0 | 0 |
| 0 | 0 | 0 | 0 | 0 |
| 0 | 0 | 0 | 0 | 0 |
| 0 | 0 | 0 | 0 | 0 |
| 0 | 0 | 0 | 0 | 0 |
| 0 | 0 | 0 | 0 | 0 |
| 0 | 0 | 0 | 0 | 0 |
| 0 | 0 | 0 | 0 | 0 |
| 0 | 0 | 0 | 0 | 0 |
| 0 | 0 | 0 | 0 | 0 |
| 0 | 0 | 0 | 0 | 0 |
| 0 | 0 | 0 | 0.002137807 | 0.027448678 |
| 0 | 0 | 0 | 0 | 0 |
| 0 | 0 | 0 | 0 | 0 |
| 0 | 0 | 0 | 0 | 0 |
| 0 | 0 | 0 | 0 | 0 |
| 0 | 0 | 0 | 0 | 0 |
| 0 | 0 | 0 | 0 | 0.010612806 |
| 0 | 0.000671595 | 0 | 0 | 0 |
| 0 | 0 | 0 | 0 | 0 |
| 0 | 0 | 0 | 0 | 0 |
| 0 | 0 | 0 | 0 | 0 |
| 0 | 0 | 0 | 0 | 0 |
| 0 | 0 | 0 | 0 | 0 |
| 0 | 0 | 0 | 0 | 0 |
| 0 | 0 | 0 | 0 | 0 |
| 0 | 0 | 0 | 0 | 0 |
| 0 | 0 | 0 | 0 | 0 |
| 0 | 0 | 0 | 0 | 0 |
| 0 | 0 | 0 | 0 | 0 |
| 0 | 0 | 0 | 0 | 0 |
| 0 | 0 | 0 | 0 | 0 |
| 0 | 0 | 0 | 0 | 0 |
| 0 | 0 | 0 | 0 | 0 |
| 0 | 0 | 0 | 0 | 0 |

| | | | | |
|---|---|---|---|---|
| 0 | 0 | 0 | 0 | 0 |
| 0 | 0 | 0 | 0 | 0 |
| 0 | 0 | 0 | 0 | 0 |
| 0 | 0 | 0 | 0 | 0 |
| 0 | 0 | 0 | 0 | 0 |
| 0 | 0 | 0 | 0 | 0 |
| 0 | 0 | 0 | 0 | 0 |
| 0 | 0 | 0 | 0 | 0 |
| 0 | 0 | 0 | 0 | 0 |
| 0 | 0 | 0 | 0 | 0 |
| 0 | 0 | 0 | 0 | 0 |
| 0 | 0 | 0 | 0 | 0 |
| 0.002419535 | 0 | 0 | 0 | 0.000531059 |
| 0 | 0 | 0 | 0 | 0 |
| 0.001625326 | 0 | 0 | 0 | 0.000864408 |
| 0 | 0 | 0 | 0 | 0 |
| 0 | 0 | 0 | 0 | 0 |
| 0 | 0 | 0 | 0 | 0 |
| 0 | 0 | 0 | 0 | 0 |
| 0 | 0 | 0 | 0 | 0 |
| 0.016587811 | 0.13320612 | 0.100103189 | 0.014390361 | 0.014894289 |
| 0 | 0 | 0 | 0 | 0 |
| 0 | 0 | 0 | 0 | 0 |
| 0 | 0.003580082 | 0 | 0 | 0 |
| 0 | 0 | 0 | 0 | 0 |
| 0 | 0 | 0 | 0 | 0 |
| 0.000752755 | 0.003998768 | 0.001880572 | 0.053506953 | 0.005493586 |
| 0 | 0 | 0 | 0 | 0 |
| 0 | 0 | 0 | 0 | 0 |
| 0 | 0 | 0 | 0 | 0 |
| 0 | 0 | 0.230406751 | 0 | 0.214723601 |
| 0 | 0 | 0 | 0 | 0 |
| 0 | 0 | 0 | 0 | 0 |
| 0 | 0 | 0 | 0 | 0 |
| 0.007709224 | 0 | 0.000764017 | 0.002736917 | 0.030257513 |
| 0 | 0 | 0 | 0 | 0 |
| 0 | 0 | 0 | 0 | 0 |
| 0 | 0 | 0 | 0 | 0 |
| 0 | 0 | 0 | 0 | 0 |
| 0 | 0 | 0 | 0 | 0 |
| 0 | 0 | 0 | 0 | 0 |
| 0 | 0 | 0 | 0 | 0 |
| 0 | 0 | 0 | 0 | 0 |
| 0 | 0 | 0 | 0 | 0 |
| 0 | 0 | 0.001100625 | 0 | 0 |
| 0 | 0 | 0 | 0 | 0 |
| 0 | 0 | 0 | 0 | 0.014689914 |
| 0.010787828 | 0 | 0 | 0 | 0.0056016 |

| | | | | |
|---|---|---|---|---|
| 0 | 0 | 0 | 0 | 0 |
| 0 | 0 | 0 | 0 | 0 |
| 0 | 0 | 0 | 0 | 0 |
| 0 | 0 | 0.000692079 | 0 | 0 |
| 0 | 0 | 0 | 0 | 0 |
| 0 | 0 | 0 | 0 | 0 |
| 0 | 0 | 0.004650186 | 0 | 0 |
| 0.001292308 | 0.011899292 | 0.056156592 | 0 | 0.001103069 |
| 0.000688921 | 0 | 0 | 0 | 0 |
| 0 | 0 | 0 | 0 | 0 |
| 0 | 0 | 0 | 0 | 0 |
| 0 | 0 | 0 | 0 | 0 |
| 0 | 0 | 0 | 0 | 0 |
| 0 | 0 | 0 | 0 | 0 |
| 0 | 0.000627458 | 0 | 0 | 0 |
| 0 | 0 | 0 | 0 | 0 |
| 0 | 0 | 0 | 0 | 0 |
| 0 | 0 | 0 | 0 | 0 |
| 0 | 0 | 0 | 0.006577017 | 0 |
| 0 | 0 | 0 | 0 | 0 |
| 0 | 0 | 0.016335618 | 0 | 0 |
| 0 | 0 | 0 | 0 | 0 |
| 0 | 0 | 0 | 0 | 0 |
| 0 | 0 | 0 | 0 | 0 |
| 0.002679339 | 0.038666681 | 0.014604371 | 0.010096227 | 0.005031376 |
| 0 | 0.001035873 | 0.003936633 | 0 | 0 |
| 0 | 0 | 0 | 0 | 0 |
| 0 | 0 | 0 | 0 | 0 |
| 0.038780139 | 0.19480123 | 0.072976537 | 0 | 0.004801515 |
| 0 | 0.0012204 | 0.003014629 | 0 | 0 |
| 0.048811084 | 0 | 0.016666058 | 0.255907581 | 1.02765684 |
| 0 | 0 | 0 | 0 | 0 |
| 0 | 0.14355654 | 0 | 0 | 0 |
| 0 | 0.086555174 | 0.00854716 | 0 | 0 |
| 0 | 0 | 0 | 0 | 0 |
| 0 | 0 | 0 | 0 | 0 |
| 0 | 0 | 0 | 0 | 0 |
| 0 | 0 | 0 | 0 | 0 |
| 0 | 0 | 0 | 0 | 0 |
| 0 | 0 | 0 | 0 | 0 |
| 0 | 0 | 0 | 0 | 0 |
| 0.000561566 | 0.161984628 | 0.001530475 | 0.007733319 | 0 |
| 0 | 0 | 0 | 0 | 0 |
| 0 | 0 | 0 | 0 | 0 |
| 0 | 0 | 0 | 0 | 0 |
| 0 | 0 | 0 | 0 | 0 |
| 0 | 0 | 0 | 0 | 0 |
| 0 | 0 | 0 | 0 | 0 |

| | | | | |
|---|---|---|---|---|
| 0 | 0 | 0 | 0 | 0 |
| 0 | 0 | 0 | 0 | 0 |
| 0.067693347 | 0.028543195 | 0.570187227 | 0.037972224 | 0 |
| 0 | 0 | 0 | 0 | 0 |
| 0 | 0 | 0 | 0 | 0.786189646 |
| 0 | 0 | 0 | 0 | 0 |
| 0 | 0 | 0 | 0 | 0 |
| 0 | 0 | 0 | 0 | 0 |
| 0 | 0 | 0 | 0 | 0 |
| 0.009415756 | 0.000308331 | 0.000703052 | 0 | 0 |
| 0 | 0 | 0 | 0 | 0 |
| 0 | 0 | 0 | 0 | 0 |
| 0 | 0.002738945 | 0 | 0 | 0 |
| 0 | 0.006954888 | 0.012106205 | 0 | 0.001197338 |
| 0.000900501 | 0.006662905 | 0.056446529 | 0.002115272 | 0.00385416 |
| 0 | 0 | 0 | 0 | 0 |
| 0.003811006 | 0.027096827 | 0.00858875 | 0.244330213 | 0.001576425 |
| 0 | 0 | 0.016578626 | 0 | 0 |
| 0 | 0 | 0 | 0 | 0 |
| 0 | 0 | 0 | 0 | 0 |
| 0 | 0 | 0 | 0 | 0 |
| 0.011776245 | 0 | 0 | 0 | 0.002846124 |
| 0 | 0 | 0 | 0 | 0 |
| 0 | 0 | 0 | 0 | 0.001575765 |
| 0.000636489 | 0 | 0 | 0 | 0 |
| 0 | 0 | 0 | 0 | 0 |
| 0 | 0 | 0 | 0 | 0 |
| 0 | 0 | 0 | 0 | 0 |
| 0 | 0 | 0 | 0 | 0 |
| 0.003253771 | 0 | 0 | 0 | 0 |
| 0.00162235 | 0.008887538 | 0 | 0 | 0 |
| 0 | 0 | 0 | 0 | 0 |
| 0 | 0 | 0 | 0 | 0 |
| 0 | 0 | 0.949655739 | 0 | 0 |
| 0 | 0 | 0 | 0 | 0 |
| 0.003964277 | 0 | 0.00504192 | 0 | 0.075728692 |
| 0 | 0 | 0 | 0 | 0 |
| 0.171562836 | 0.005298679 | 0.009061471 | 0 | 0.019922648 |
| 0 | 0 | 0 | 0 | 0 |
| 0 | 0 | 0 | 0 | 0 |
| 0 | 0.003823746 | 0 | 0 | 0 |
| 0 | 0 | 0 | 0 | 0 |
| 0 | 0 | 0 | 0 | 0 |
| 0 | 0 | 0 | 0 | 0.002538158 |
| 0 | 0 | 0 | 0 | 0 |
| 0.002555961 | 0 | 0 | 0 | 0 |
| 0 | 0 | 0 | 0 | 0 |
| 0 | 0 | 0 | 0.005380355 | 0.000618752 |

| | | | | |
|---|---|---|---|---|
| 0 | 0 | 0 | 0 | 0 |
| 0 | 0 | 0 | 0 | 0 |
| 0 | 0.00419649 | 0.004721432 | 0 | 0 |
| 0 | 0 | 0 | 0 | 0 |
| 0.003485926 | 0 | 0.005130233 | 0.003725731 | 0.002785033 |
| 0.001818918 | 0 | 0 | 0 | 0 |
| 0 | 0 | 0 | 0 | 0 |
| 0.002540872 | 0.046722363 | 0.046875649 | 0.077332162 | 0.002252214 |
| 0 | 0 | 0 | 0 | 0 |
| 0.05922815 | 0.068865585 | 0.160320431 | 0.000621101 | 0.002321406 |
| 0 | 0 | 0 | 0 | 0 |
| 0.014853145 | 0.012081959 | 0.011595916 | 0.000636015 | 0.052534958 |
| 0.05527694 | 0.000841646 | 0.031505364 | 0.009045745 | 0.060766178 |
| 0.005168983 | 0 | 0 | 0 | 0.057508937 |
| 0 | 0 | 0 | 0 | 0.000649075 |
| 0 | 0.002386519 | 0.108753988 | 0 | 0 |
| 0 | 0 | 0 | 0 | 0 |
| 0 | 0 | 0 | 0 | 0 |
| 0 | 0 | 1.018037251 | 0.002532593 | 0.222954463 |
| 0 | 0 | 0.000828236 | 0 | 0 |
| 0 | 0 | 0 | 0 | 0 |
| 0 | 0 | 0 | 0 | 0 |
| 0 | 0 | 0 | 0 | 0 |
| 0 | 0 | 0 | 0 | 0 |
| 0 | 0 | 0 | 0 | 0 |
| 0.001366954 | 0.053589929 | 0 | 0.001685758 | 0 |
| 0 | 0 | 0 | 4.680094987 | 0 |
| 0.000661211 | 0 | 0.001351533 | 0.019875861 | 0.430866671 |
| 0 | 0 | 0 | 0 | 0 |
| 0.000843854 | 0 | 0 | 0 | 0 |
| 0 | 0 | 0.00279833 | 0 | 0 |
| 0 | 0 | 0 | 0 | 0 |
| 0 | 0 | 0 | 0 | 0 |
| 0 | 0 | 0.037272396 | 0 | 0.29774118 |
| 0 | 0 | 0 | 0 | 0 |
| 0 | 0 | 0 | 0 | 0 |
| 0 | 0 | 0 | 0.001592483 | 0.989819326 |
| 0 | 0 | 0 | 0 | 0 |
| 0 | 0 | 0 | 0 | 0 |
| 0 | 0 | 0 | 0 | 0 |
| 0 | 0 | 0 | 0 | 0 |
| 0 | 0 | 0 | 0 | 0 |
| 0 | 0 | 0 | 0 | 0 |
| 0 | 0 | 0 | 0 | 0 |
| 0 | 0 | 0 | 0 | 0 |
| 0 | 0 | 0 | 0 | 0 |
| 0 | 0 | 0 | 0.009783075 | 0 |
| 0 | 0 | 0 | 0 | 0 |

| | | | | |
|---|---|---|---|---|
| 0 | 0 | 0 | 0 | 0 |
| 0 | 0.602489178 | 0.46701818 | 0.021476066 | 0 |
| 0 | 0 | 0 | 0 | 0 |
| 0 | 0 | 0 | 0 | 0 |
| 0 | 0 | 0.00206061 | 0 | 0 |
| 0.003347224 | 0 | 0.002554277 | 0.02655596 | 0.048341653 |
| 0 | 0 | 0 | 0.0379437 | 0 |
| 0 | 0.002345755 | 0.07503983 | 0 | 0 |
| 0 | 0 | 0 | 0 | 0 |
| 0 | 0 | 0 | 0 | 0 |
| 0 | 0 | 0.006136671 | 0.008936916 | 0.018371276 |
| 0 | 0 | 0 | 0 | 0 |
| 0 | 0 | 0 | 0 | 0.02953751 |
| 0.011296584 | 0.000935213 | 0.008596468 | 0.001960017 | 0.002028653 |
| 0.021672682 | 0 | 0 | 0 | 0.008780707 |
| 0 | 0 | 0 | 0 | 0 |
| 0 | 0 | 0 | 0 | 0 |
| 0 | 0 | 0 | 0 | 0.000548391 |
| 0 | 0 | 0.000851127 | 0 | 0 |
| 0 | 0 | 0 | 0 | 0 |
| 0 | 0 | 0.021324927 | 0 | 0 |
| 0 | 0 | 0 | 0.245977266 | 0 |
| 0.001531417 | 0 | 0 | 0 | 0 |
| 0 | 0 | 0 | 0 | 0 |
| 0 | 0 | 0 | 0 | 0 |
| 0 | 0 | 0 | 0 | 0 |
| 0 | 0 | 0.004335278 | 0 | 0.014786183 |
| 0 | 0 | 0 | 0 | 0 |
| 0 | 0 | 0 | 0 | 0 |
| 0.08881338 | 0.020391042 | 0.168203655 | 0.140996722 | 0.129185579 |
| 0 | 0 | 0 | 0.003784575 | 0 |
| 0.003912649 | 0 | 0 | 0 | 0 |
| 0 | 0 | 0 | 0.001417598 | 0 |
| 0 | 0 | 0 | 0 | 0.001082936 |
| 0.001722388 | 0 | 0 | 0 | 0 |
| 0 | 0 | 0 | 0 | 0 |
| 0 | 0 | 0 | 0 | 0 |
| 0.004486063 | 0 | 0 | 0 | 0 |
| 0.01189833 | 0 | 0 | 0 | 0 |
| 0.098080702 | 0 | 0 | 0 | 0 |
| 0.002774045 | 0 | 0.032049077 | 0 | 0.01714101 |
| 0.540434696 | 0.000247965 | 0 | 1.78899533 | 2.633716591 |
| 0.001232857 | 0 | 0 | 0 | 0 |
| 0 | 0 | 0 | 0 | 2.284298549 |
| 0 | 0 | 0 | 0 | 0.030937523 |
| 0 | 0 | 0 | 0 | 0 |
| 0.000632948 | 0.012205269 | 0.011557605 | 0 | 0.001944943 |
| 0 | 0 | 0 | 0 | 0 |

| | | | | |
|---|---|---|---|---|
| 0.003592376 | 0.274675812 | 0 | 0.002728745 | 0.002039773 |
| 0 | 0 | 0 | 0 | 0 |
| 0.009151312 | 1.315275748 | 0.083232317 | 1.356021507 | 0.397773588 |
| 0 | 0 | 0 | 0 | 0 |
| 0 | 0 | 0 | 0 | 0 |
| 0.124267525 | 0 | 0.049847454 | 0 | 0.255651004 |
| 0.000443935 | 0 | 0 | 0 | 0 |
| 0.009803226 | 0 | 0 | 0 | 0 |
| 0 | 0 | 0.037584652 | 0 | 0 |
| 0 | 0 | 0 | 0 | 0 |
| 0 | 0 | 0 | 0 | 0 |
| 0 | 0 | 0 | 0 | 0 |
| 0 | 0 | 0 | 0 | 0 |
| 0 | 0 | 0.00231059 | 0 | 0 |
| 0 | 0 | 0 | 0 | 0 |
| 0.001545944 | 0 | 0.001655212 | 0.019064925 | 0.637490547 |
| 0.058259617 | 1.548143327 | 0.006504598 | 0.03207457 | 0.030087741 |
| 0.001040929 | 0 | 0 | 0 | 0 |
| 0 | 0 | 0 | 0 | 0 |
| 0.007018817 | 0 | 0 | 0.005200165 | 0 |
| 0.005553053 | 0 | 0 | 0 | 0.040052066 |
| 0 | 0 | 0 | 0 | 0.000436253 |
| 0 | 0 | 0 | 0 | 0 |
| 0 | 0 | 0.749508589 | 0 | 0 |
| 0 | 0 | 0 | 0 | 0 |
| 0 | 0 | 0 | 0 | 0 |
| 0 | 0 | 0.006352031 | 0 | 0.001790467 |
| 0 | 0 | 0.002411903 | 0 | 0 |
| 0 | 0 | 0 | 0 | 0 |
| 0 | 0 | 0 | 0 | 0.012869059 |
| 0.003781322 | 0 | 0.032043652 | 0 | 0.025417376 |
| 0.004051944 | 0 | 0.005761586 | 0.007785098 | 0.004330764 |
| 0.199274028 | 2.224487583 | 0.005257234 | 0.006722934 | 0.005154341 |
| 0.030048308 | 0.004214022 | 0.011137407 | 0 | 0.063156062 |
| 0 | 0.090232187 | 0.029926684 | 0.003526496 | 0 |
| 0 | 0 | 0 | 0 | 0 |
| 0 | 0.117332722 | 0.002066479 | 0 | 0 |
| 0.006285563 | 0 | 0.001292866 | 0.004509517 | 0.042182103 |
| 0 | 0 | 0 | 0 | 0 |
| 0 | 0 | 0 | 0.001594374 | 0 |
| 0.000338708 | 0 | 0 | 0 | 0 |
| 0 | 0 | 0 | 0 | 0 |
| 0 | 0 | 0 | 0.025517745 | 0.008447435 |
| 0.000462892 | 0 | 0.037468079 | 0 | 0.048930219 |
| 0 | 0 | 0 | 0 | 0.004731909 |
| 0 | 0 | 0 | 0 | 0 |
| 0 | 0 | 0 | 0 | 0 |
| 0 | 0 | 0 | 0 | 0 |

| | | | | |
|---|---|---|---|---|
| 0 | 0 | 0 | 0 | 0.004052401 |
| 0.215253128 | 0.007597337 | 1.997320331 | 0.237653859 | 0.341146492 |
| 0 | 0 | 0.094226863 | 0 | 0 |
| 0 | 0 | 0 | 0.004928496 | 0 |
| 0 | 0 | 0.001728304 | 0 | 0 |
| 0 | 0 | 0 | 0 | 0 |
| 0 | 0 | 0.011204849 | 0 | 0.209369654 |
| 0 | 0.013945989 | 0.001353167 | 0.00038269 | 0.00143033 |
| 0 | 0 | 0.07519147 | 0 | 0 |
| 0 | 0 | 0 | 0 | 0 |
| 0.002882101 | 0 | 0.000818207 | 0.001851182 | 0.143083218 |
| 0.169572366 | 0.555445257 | 0.053380919 | 0.019853199 | 0.00769509 |
| 0 | 0 | 0 | 0 | 0 |
| 0 | 0 | 0 | 0 | 0 |
| 0 | 0.038536703 | 0 | 0.0024072 | 0 |
| 0 | 0 | 0 | 0 | 0 |
| 0.002263482 | 0.00941882 | 0.09441724 | 0.012793804 | 0.019030477 |
| 0.023832976 | 0 | 0.052051886 | 0.026841414 | 0.036366571 |
| 0 | 0 | 0 | 0 | 0 |
| 0.003497975 | 0 | 0 | 0 | 0.036073262 |
| 0.001426468 | 0.001343312 | 0.032691613 | 0.249079925 | 0.005180262 |
| 0.008552324 | 0 | 0 | 0.009452423 | 0 |
| 0 | 0 | 0 | 0 | 0 |
| 0 | 0 | 0 | 0 | 0 |
| 0 | 0 | 0.026127918 | 0.021496025 | 0 |
| 0 | 0 | 0 | 0 | 0 |
| 0 | 0 | 0.013256148 | 0 | 0 |
| 0.055545314 | 0.070947813 | 0.068365809 | 0.011048339 | 0.037270394 |
| 0 | 0 | 0 | 0 | 0 |
| 0 | 0 | 0 | 0 | 0.000638591 |
| 0 | 0.009898875 | 0 | 0.002279784 | 0.005112507 |
| 0 | 0 | 0 | 0 | 0 |
| 0 | 0 | 0 | 0 | 0 |
| 0 | 0 | 0 | 0 | 0 |
| 0 | 0 | 0 | 0 | 0 |
| 0.099372302 | 0.000743498 | 0.399563877 | 0.107637123 | 0.741703133 |
| 0 | 0 | 0 | 0 | 0 |
| 0 | 0 | 0 | 0 | 0 |
| 0.008841466 | 0.026961131 | 0.016450324 | 0 | 0.034613517 |
| 0 | 0 | 0 | 0 | 0 |
| 0 | 0 | 0 | 0 | 0.003278515 |
| 0 | 0 | 0 | 0 | 0 |
| 0.014045769 | 0 | 0.012243936 | 0 | 0.00983801 |
| 0.003988508 | 0.006594663 | 0.004619815 | 0.002561831 | 0.001072401 |
| 0 | 0 | 0 | 0 | 0.060280233 |
| 0 | 0 | 0 | 0.761777614 | 0 |
| 0 | 0 | 0 | 0 | 0 |
| 0 | 0 | 0.001317807 | 0 | 0.001361999 |

| | | | | |
|---|---|---|---|---|
| 0.004328436 | 0 | 0 | 0 | 0.000443352 |
| 0 | 0 | 0 | 0 | 0 |
| 0 | 0 | 0 | 0 | 0.000363545 |
| 0.158055617 | 0 | 0 | 0.005068255 | 0.395399206 |
| 0 | 0 | 0 | 0 | 0 |
| 0 | 0 | 0 | 0 | 0 |
| 0 | 0 | 0 | 0 | 0 |
| 0 | 0 | 0 | 0.012385827 | 1.053575732 |
| 0 | 6.627401471 | 0 | 0.324919838 | 0 |
| 0 | 0 | 0 | 0.010827263 | 0 |
| 0 | 0 | 0 | 0 | 0 |
| 0 | 0 | 0 | 0.001065936 | 0 |
| 0 | 0 | 0 | 0 | 0 |
| 0 | 0 | 0 | 0 | 0 |
| 0.042035056 | 0 | 0.004535786 | 0.007916521 | 0.368578053 |
| 0 | 0 | 0 | 0 | 0 |
| 0 | 0 | 0 | 1.118439246 | 0 |
| 0 | 0.000593742 | 0 | 0.221049914 | 0 |
| 0 | 0 | 0 | 0 | 0 |
| 0 | 0 | 0 | 0 | 0 |
| 0 | 0 | 0 | 0 | 0 |
| 0 | 0 | 0 | 0 | 0 |
| 0 | 0 | 0 | 0 | 0 |
| 0.017222218 | 0 | 0.935991385 | 0.003106742 | 0.318370454 |
| 0 | 0 | 0.003234753 | 0 | 0 |
| 0 | 0 | 0.082434569 | 0 | 0 |
| 0 | 0 | 0.037788771 | 0 | 0 |
| 0 | 0 | 3.400103428 | 1.159568115 | 0 |
| 0 | 0 | 0 | 0 | 0 |
| 0 | 0 | 0 | 0 | 0 |
| 0 | 0 | 0.005648956 | 0.004260229 | 0 |
| 0 | 0 | 0 | 0 | 0 |
| 0.000341081 | 0 | 0.000348589 | 0.001664989 | 0.023188892 |
| 0 | 0 | 0 | 0 | 0 |
| 0 | 0 | 0 | 0 | 0 |
| 0 | 0 | 0.004789683 | 0.005271856 | 0 |
| 0 | 0 | 0 | 0 | 0 |
| 0 | 0 | 0 | 0 | 0 |
| 0.004079045 | 0 | 0 | 0 | 0 |
| 0 | 0 | 0 | 0 | 0.002471427 |
| 0.330128236 | 0.009250097 | 0.120073282 | 0.018292739 | 0.002547963 |
| 0 | 0 | 0 | 0 | 0 |
| 0.021220594 | 0.005072744 | 7.81282223 | 0.089276849 | 3.997209838 |
| 0 | 0 | 0 | 0.002177503 | 0.107281077 |
| 0 | 0 | 0 | 0 | 0 |
| 0 | 0 | 0 | 0 | 0 |
| 0 | 0 | 0 | 0 | 0 |
| 0.002790989 | 0 | 0.00923643 | 0.000546335 | 0.00214406 |

| | | | | |
|---|---|---|---|---|
| 0 | 0 | 0 | 0 | 0 |
| 0 | 0 | 0 | 0.011198843 | 0 |
| 0.046757159 | 0.050556326 | 0.05555552 | 0.002618617 | 0.855210504 |
| 0 | 0 | 0 | 0 | 0 |
| 0.001167502 | 0 | 0 | 0 | 0 |
| 0.000401455 | 0.000479835 | 0 | 0 | 0 |
| 0 | 0 | 0 | 0 | 0 |
| 0 | 0 | 0 | 0 | 0 |
| 0 | 0 | 0 | 0 | 0 |
| 0 | 0 | 0 | 0 | 0 |
| 0.004503162 | 0.001694448 | 0.002386373 | 0.006684646 | 0 |
| 1.299695875 | 0.206732981 | 0.839719287 | 0.519526428 | 0.741346173 |
| 0 | 0 | 0 | 0 | 0 |
| 0.100439925 | 0.418345885 | 0.018580184 | 0.022059241 | 0.022193518 |
| 0.09287419 | 0 | 0.011017665 | 0.066582191 | 0.134357386 |
| 0 | 0 | 0 | 0.044500117 | 0 |
| 0 | 0 | 0 | 0 | 0 |
| 0 | 0 | 0 | 0 | 0 |
| 0 | 0 | 0 | 0 | 0 |
| 0.001339513 | 0 | 0.00617805 | 0 | 0.006490772 |
| 0.148351476 | 0.024477506 | 0.407455462 | 0 | 0.211096674 |
| 0.010052859 | 38.84220851 | 0 | 0 | 0.001527565 |
| 2.408506499 | 3.483878766 | 7.645753613 | 0.013433292 | 0.319656487 |
| 0 | 0 | 0 | 0 | 0 |
| 0 | 0 | 0 | 0 | 0 |
| 0 | 0 | 0 | 5.495322676 | 0 |
| 0 | 0 | 0.086804538 | 0 | 0 |
| 0 | 0 | 0 | 0 | 0 |
| 0.000398618 | 0 | 0 | 0 | 0 |
| 0 | 0 | 0 | 0 | 0 |
| 0 | 0 | 0 | 0 | 0 |
| 0 | 0 | 0.394245456 | 0.210762059 | 0 |
| 0.012332973 | 0 | 0.010153593 | 0 | 0.012237626 |
| 0 | 0 | 0 | 0.004429892 | 0 |
| 0.029846357 | 0 | 0.004390636 | 0.000522829 | 0.062596636 |
| 0.520277201 | 0.028776759 | 0.006191215 | 0.004429735 | 0.049311318 |
| 0.001237883 | 0 | 0 | 0.000346951 | 0.012448829 |
| 0 | 0 | 0 | 0 | 0 |
| 2.445269322 | 0 | 0.020062998 | 0.008826275 | 0.11059099 |
| 0 | 0 | 0 | 0 | 0 |
| 0 | 0 | 0 | 0 | 0 |
| 0 | 0 | 0 | 0 | 0 |
| 0 | 0 | 0 | 0 | 0 |
| 0 | 0 | 0 | 0 | 0 |
| 0.013949461 | 0.004446124 | 0.005250024 | 0.00054612 | 0.008062582 |
| 0 | 0 | 0 | 0 | 0 |
| 0 | 0 | 0 | 0 | 0 |
| 36.21701676 | 0.003320101 | 0.083857181 | 0 | 1.072040517 |

| | | | | |
|---|---|---|---|---|
| 0 | 0 | 0 | 0 | 0 |
| 0 | 0 | 0 | 0 | 0 |
| 0 | 0 | 0 | 0 | 0 |
| 0 | 0 | 0 | 0 | 0 |
| 0 | 0 | 0 | 0 | 0 |
| 0 | 0 | 0 | 0 | 0 |
| 0 | 0 | 0 | 0 | 0 |
| 0.077414202 | 0 | 0.005821178 | 0.001927367 | 0 |
| 0 | 0 | 0 | 0 | 0 |
| 0 | 0 | 0 | 0 | 0 |
| 0.020429396 | 0 | 0 | 0.001036789 | 0.018503454 |
| 0 | 0 | 0 | 0 | 0 |
| 0 | 0 | 0 | 0.001361685 | 0 |
| 0 | 0 | 0 | 0 | 0 |
| 0 | 0 | 0 | 0 | 0 |
| 0 | 0 | 0 | 0 | 0 |
| 0 | 0 | 0 | 0 | 0 |
| 0.040942792 | 0.001338434 | 0.001573623 | 0 | 0 |
| 0 | 0 | 0 | 0 | 0 |
| 0.001041435 | 0 | 0 | 0 | 0.015346206 |
| 0 | 0 | 0 | 0 | 0 |
| 0 | 0 | 0 | 0 | 0 |
| 0 | 0 | 0 | 0.008831481 | 0 |
| 0 | 0 | 0 | 0 | 0 |
| 0 | 0 | 0 | 0 | 0 |
| 0 | 0 | 0 | 0 | 0 |
| 0 | 0 | 0 | 0 | 0 |
| 0 | 0 | 0 | 0 | 0 |
| 0 | 0 | 0 | 0 | 0 |
| 0 | 0 | 0.092799587 | 0.03101798 | 0 |
| 0 | 0 | 0.003383667 | 0 | 0 |
| 0 | 0 | 0 | 0 | 0 |
| 0 | 0 | 0 | 0 | 0 |
| 0 | 0 | 0 | 0 | 0 |
| 0 | 0 | 0 | 0 | 0 |
| 0 | 0 | 0 | 0 | 0 |
| 0 | 0 | 0 | 0.045661371 | 0 |
| 0.002348546 | 0 | 0.194492584 | 0 | 0.005002478 |
| 0 | 0 | 0 | 0 | 0 |
| 0 | 0 | 0.001865279 | 0.000541047 | 0 |
| 0 | 0 | 0 | 0 | 0 |
| 0 | 0 | 0 | 0 | 0 |
| 0 | 0 | 0 | 0 | 0 |
| 0 | 0 | 0 | 0 | 0 |
| 0 | 0.000869293 | 0 | 0.084833373 | 0 |
| 0 | 0 | 0 | 0 | 0 |
| 0 | 0 | 0 | 0 | 0 |
| 0 | 0 | 0 | 0 | 0 |

| | | | | |
|---|---|---|---|---|
| 0 | 0 | 0 | 0 | 0 |
| 0 | 0 | 0 | 0 | 0 |
| 0 | 0 | 0 | 0 | 0 |
| 0 | 0 | 0 | 0 | 0 |
| 0 | 0 | 0 | 0 | 0 |
| 0 | 0 | 0 | 0 | 0 |
| 0 | 0 | 0.000710541 | 0 | 0 |
| 0 | 0 | 0 | 0 | 0 |
| 0.106741012 | 0 | 0.089479461 | 0.00060522 | 0.044901697 |
| 0 | 0 | 0 | 0 | 0 |
| 0 | 0 | 0 | 0 | 0 |
| 0 | 0 | 0 | 0 | 0 |
| 0 | 0 | 0 | 0 | 0 |
| 0 | 0 | 0 | 0 | 0 |
| 0.100300896 | 0 | 5.543262141 | 0 | 0 |
| 0.19483395 | 0 | 0.88694905 | 0 | 0.690483475 |
| 0 | 0 | 0 | 0.00193846 | 0 |
| 0 | 0 | 0 | 0 | 0 |
| 0 | 0 | 0 | 0 | 0 |
| 0 | 0 | 0 | 0 | 0 |
| 0 | 0 | 0 | 0 | 0 |
| 0 | 0 | 0 | 0 | 0 |
| 0 | 0 | 0 | 0 | 0 |
| 0 | 0 | 0 | 0 | 0 |
| 0.037051337 | 0.001241643 | 0.001887449 | 0.061385981 | 0 |
| 12.93794428 | 0 | 3.877329681 | 0.243510272 | 0 |
| 0 | 0 | 0 | 0 | 0 |
| 0 | 0 | 0 | 0 | 0 |
| 0 | 0 | 0 | 0.005911357 | 0 |
| 0 | 0 | 0 | 0 | 0 |
| 0 | 0 | 0 | 0 | 0 |
| 0 | 0 | 0 | 0 | 0 |
| 0 | 0 | 0 | 0 | 0 |
| 0.000832197 | 0 | 0.000378007 | 0.000665184 | 0.058744611 |
| 0.056219103 | 0.300345654 | 0.005605528 | 0.017536203 | 0.004564583 |
| 0.035109131 | 0 | 0.00044027 | 0.002766957 | 0.033920737 |
| 0 | 0 | 0.00121159 | 0.422831117 | 0.205506499 |
| 0.32043764 | 0 | 0 | 0.893195811 | 0 |
| 0 | 0 | 0 | 0 | 0 |
| 0 | 0 | 0 | 0 | 0 |
| 0 | 0 | 0 | 0 | 0 |
| 0 | 0 | 0 | 0 | 0 |
| 1.454181704 | 0 | 0.695491867 | 8.659467501 | 0.044092154 |
| 0 | 0 | 0 | 0 | 0 |
| 0 | 0 | 0 | 0 | 0 |
| 0 | 0 | 0 | 0.006605789 | 0 |
| 0 | 0 | 0 | 0 | 0 |
| 2.100726729 | 0 | 0.537351665 | 0 | 0.011737603 |

| | | | | |
|---|---|---|---|---|
| 0.002302984 | 0 | 0 | 0.007598489 | 0 |
| 0 | 0 | 0 | 0 | 0 |
| 0 | 0 | 0 | 0 | 0 |
| 0 | 0 | 0 | 0 | 0 |
| 0 | 0 | 0 | 0 | 0 |
| 0 | 0 | 0 | 0 | 0 |
| 0 | 0 | 0 | 0 | 0 |
| 0 | 0 | 0 | 0 | 0 |
| 0 | 0 | 0 | 0 | 0 |
| 0 | 0 | 0 | 0 | 0 |
| 0 | 0 | 0 | 0 | 0 |
| 1.65007285 | 0.000709708 | 2.01762192 | 0.071052146 | 2.795800527 |
| 0.782620148 | 0.007800984 | 0.32296577 | 0.331865969 | 0 |
| 0 | 0 | 0 | 0 | 0 |
| 0.361192805 | 8.113678689 | 2.404854701 | 0.013195402 | 0.109894921 |
| 0 | 0 | 0 | 0 | 0 |
| 0 | 0 | 0 | 0 | 0 |
| 0 | 0 | 0 | 0 | 0 |
| 0 | 0 | 0 | 0.00314286 | 0 |
| 0 | 0 | 0 | 0 | 0 |
| 0 | 0 | 0 | 0 | 0 |
| 0 | 0 | 0 | 0 | 0 |
| 0.782553036 | 0 | 0.001782961 | 0.022736668 | 0 |
| 0 | 0 | 0 | 0 | 0 |
| 0 | 0 | 0 | 0 | 0 |
| 0 | 0 | 0 | 0 | 0 |
| 0 | 0 | 0 | 0 | 0 |
| 0 | 0 | 0 | 0 | 0 |
| 0 | 0 | 0 | 0 | 0 |
| 0 | 0 | 0 | 0 | 0 |
| 0 | 0 | 0 | 0 | 0 |
| 0.064654269 | 0.026841455 | 0.337112518 | 0.0214036 | 0.038676995 |
| 0 | 0 | 0 | 0 | 0 |
| 0 | 0 | 0.003575441 | 0.003440315 | 0 |
| 0.036097914 | 0 | 0.00079854 | 0.002047577 | 0.033042766 |
| 0.032785733 | 0.027448848 | 0.063000178 | 0 | 0 |
| 0 | 0 | 0 | 0 | 0 |
| 0 | 0 | 0 | 0 | 0 |
| 0 | 0 | 0 | 0 | 0 |
| 0 | 0 | 0 | 0 | 0 |
| 0 | 0 | 0 | 0 | 0 |
| 0 | 0 | 0 | 0 | 0 |
| 0 | 0 | 0 | 0 | 0 |
| 0 | 0 | 0 | 0 | 0 |
| 0.002770914 | 0 | 0.001101298 | 0.006288509 | 0.28124641 |
| 0 | 0 | 0 | 0 | 0 |
| 0 | 0 | 0 | 0 | 0 |
| 0 | 0 | 0 | 0 | 0 |

| | | | | |
|---|---|---|---|---|
| 0 | 0 | 0 | 0 | 0 |
| 0 | 0 | 0 | 0 | 0 |
| 0 | 0 | 0 | 0 | 0 |
| 0 | 0 | 0 | 0 | 0 |
| 0 | 0 | 0 | 0 | 0.001581485 |
| 0 | 0 | 0 | 0 | 0 |
| 0 | 0 | 0 | 0.142781732 | 1.806151226 |
| 0 | 0 | 0 | 0 | 0 |
| 0 | 0 | 0 | 0 | 0 |
| 0 | 0 | 0 | 0 | 0 |
| 0 | 0 | 0 | 0 | 0 |
| 0 | 0 | 0 | 0 | 0.000373336 |
| 0 | 0 | 0 | 0 | 0 |
| 0 | 0 | 0 | 0.18330717 | 0 |
| 0 | 0 | 0 | 0 | 0 |
| 0 | 0 | 0 | 0 | 0 |
| 0 | 0 | 0 | 0 | 0 |
| 0 | 0 | 0 | 0 | 0 |
| 9.223664997 | 0 | 0 | 0.002097999 | 0 |
| 0 | 0 | 0 | 0 | 0 |
| 0 | 0 | 0 | 0 | 0 |
| 0 | 0 | 0 | 0 | 0 |
| 0 | 0 | 0 | 0 | 0 |
| 0 | 0 | 0 | 0 | 0 |
| 0 | 0.009548947 | 0 | 0 | 0.009690512 |
| 0.30321239 | 0 | 0.001611134 | 0 | 0.158699726 |
| 0 | 0 | 0 | 0 | 0 |
| 0.127719436 | 0.001005967 | 0.001863704 | 0 | 0.000727377 |
| 0 | 0 | 0 | 0 | 0 |
| 0 | 0 | 0 | 0.764449915 | 0.198050852 |
| 0 | 0 | 0 | 0 | 0 |
| 0 | 0 | 0 | 0 | 0 |
| 0 | 0 | 0 | 0 | 0 |
| 0 | 0 | 0 | 0 | 0 |
| 0.012277389 | 0 | 0 | 0.000578775 | 0.044475574 |
| 0 | 0 | 0 | 0 | 0 |
| 0 | 0 | 0 | 0 | 0 |
| 0 | 0 | 0 | 0 | 0.002061871 |
| 0 | 0 | 0 | 0 | 0 |
| 0 | 0 | 0 | 0 | 0 |
| 2.760516408 | 0.005179139 | 10.06729236 | 1.803981569 | 25.8910449 |
| 0.950915014 | 0 | 0.624440437 | 0 | 0.310196876 |
| 0 | 0.004287477 | 0 | 0 | 0 |
| 0 | 0 | 0 | 0 | 0 |
| 0 | 0 | 0 | 0 | 0 |
| 0 | 0 | 0 | 0 | 0 |
| 0 | 0 | 0 | 0 | 0 |
| 0.000405747 | 0 | 0 | 0 | 0.00654566 |

| | | | | |
|---|---|---|---|---|
| 0 | 0 | 0.05056829 | 0 | 0 |
| 0 | 0 | 0 | 0.003431187 | 0 |
| 0 | 0 | 0 | 0 | 0 |
| 0 | 0 | 0 | 0 | 0 |
| 1.18000695 | 0.001740104 | 0.473025657 | 8.15628591 | 0.117286574 |
| 0 | 0 | 0 | 0 | 0 |
| 0 | 0 | 0 | 0 | 0 |
| 0 | 0 | 0 | 0 | 0 |
| 0.026149737 | 0 | 0.18782294 | 0.076445479 | 0.495648296 |
| 0 | 0 | 0 | 0 | 0 |
| 0 | 0 | 0 | 0 | 0 |
| 0 | 0 | 0 | 0 | 0 |
| 0 | 0 | 0 | 0 | 0 |
| 0.005733288 | 0 | 0.028533197 | 0.008197585 | 0.069033543 |
| 0 | 0.016221237 | 0 | 0 | 0.294308979 |
| 0.177755653 | 0 | 0.356647451 | 0.06558707 | 0 |
| 0.004422003 | 0.020282025 | 0.001910617 | 0 | 0 |
| 0 | 0 | 0 | 0 | 0 |
| 0 | 0 | 0 | 0 | 0 |
| 0 | 0 | 0 | 0 | 0 |
| 0 | 0 | 0 | 0 | 0 |
| 0 | 0 | 0 | 0 | 0 |
| 0 | 0 | 0 | 0 | 0 |
| 0 | 0 | 0 | 0 | 0 |
| 0 | 0 | 0 | 0 | 0 |
| 0 | 0 | 0 | 0 | 0 |
| 0.002063232 | 0 | 0 | 0 | 0 |
| 0.010159096 | 0 | 0 | 0.000870029 | 0.021299238 |
| 0 | 0 | 0 | 0 | 0 |
| 0 | 0 | 0 | 0 | 0 |
| 0 | 0 | 0 | 0 | 0 |
| 0 | 0 | 0 | 0 | 0 |
| 0.002737667 | 0 | 0 | 0.005744146 | 0 |
| 0 | 0 | 0 | 0 | 0 |
| 0 | 0 | 0 | 0.1329473 | 0.248248118 |
| 0.001468996 | 0 | 0 | 0 | 0.024450715 |
| 0 | 0 | 0 | 0 | 0 |
| 0 | 0 | 0 | 0 | 0 |
| 0 | 0 | 0 | 0 | 0 |
| 0 | 0 | 0 | 0 | 0 |
| 0 | 0 | 0 | 0.492745405 | 0 |
| 0 | 0 | 0 | 0.020278993 | 0 |
| 0 | 0 | 0 | 0 | 0 |
| 0 | 0 | 0 | 0 | 0 |
| 0 | 0 | 0 | 0 | 0 |
| 0 | 0 | 0 | 0 | 0 |
| 0 | 0 | 0 | 0 | 0 |
| 0 | 0 | 0 | 0 | 0 |

| | | | | |
|---|---|---|---|---|
| 0 | 0 | 0 | 0 | 0.047976494 |
| 0 | 0 | 0 | 0 | 0 |
| 0 | 0 | 0 | 0 | 0 |
| 0 | 0 | 0.010128041 | 0 | 0 |
| 0 | 0 | 0 | 0.00212175 | 0 |
| 0 | 0 | 0 | 0 | 0 |
| 0.483193348 | 0.015517458 | 0.002932264 | 0 | 0 |
| 0 | 0 | 0 | 0 | 0 |
| 0 | 0 | 0 | 0 | 0 |
| 0 | 0 | 0.009998372 | 0 | 0 |
| 0 | 0 | 0 | 0 | 0 |
| 0 | 0 | 0 | 0 | 0 |
| 0 | 0 | 0 | 0 | 0 |
| 0 | 0 | 0 | 0 | 0 |
| 0 | 0 | 0 | 0 | 0 |
| 0 | 0 | 0 | 0 | 0 |
| 0 | 0 | 0 | 0 | 0 |
| 0.017998198 | 0 | 0.001973345 | 0 | 0.096148847 |
| 0 | 0 | 0 | 0 | 0 |
| 0 | 0 | 0 | 0 | 0 |
| 0.125787832 | 0 | 0.046202284 | 0 | 0 |
| 0 | 0 | 0 | 0 | 0 |
| 0 | 0 | 0 | 0 | 0 |
| 0.005324695 | 0 | 0.00246426 | 0.002168194 | 0.004630726 |
| 0 | 0 | 0 | 0 | 0 |
| 0 | 0 | 0 | 0.001218087 | 0 |
| 0 | 0 | 0 | 0 | 0 |
| 0 | 0 | 0 | 0 | 0 |
| 0 | 0 | 0 | 0 | 0 |
| 0.075044316 | 1.153316859 | 0.011231053 | 0.335829527 | 0 |
| 0 | 0 | 0 | 0 | 0 |
| 0 | 0 | 0 | 0 | 0 |
| 0 | 0 | 0 | 0 | 0 |
| 0 | 0 | 0 | 0 | 0 |
| 0 | 0 | 0 | 0 | 0 |
| 0 | 0 | 0 | 0 | 0 |
| 0.010056714 | 0 | 0.017301457 | 0 | 0.001255418 |
| 0 | 0 | 0 | 0 | 0 |
| 0 | 0 | 0 | 0 | 0 |
| 0 | 0 | 0 | 0 | 0 |
| 0 | 0 | 0 | 0 | 0 |
| 0 | 0 | 0 | 0 | 0 |
| 0 | 0 | 0 | 0 | 0 |
| 0 | 0 | 0 | 0 | 0 |
| 0 | 0 | 0.003566728 | 0 | 0 |
| 0 | 0 | 0 | 0 | 0 |
| 0 | 0 | 0 | 0 | 0 |
| 0 | 0 | 0 | 0 | 0 |

| | | | | |
|---|---|---|---|---|
| 0.025080344 | 0 | 0 | 0.020664192 | 0.1685856 |
| 0 | 0.000816957 | 0 | 0 | 0 |
| 0 | 0 | 0 | 0.002244488 | 0 |
| 0 | 0 | 0 | 0 | 0 |
| 0 | 0 | 0 | 0 | 0 |
| 0 | 0 | 0 | 0 | 0 |
| 0 | 0 | 0 | 0 | 0 |
| 1.152950051 | 0 | 1.566720254 | 0.039697174 | 0 |
| 0.045645783 | 0.031233775 | 0.06428185 | 1.607303752 | 0.000651774 |
| 0.00129944 | 0 | 0.018673119 | 0.02173085 | 0.676081253 |
| 0 | 0 | 0 | 0 | 0 |
| 0 | 0 | 0 | 0 | 0 |
| 0 | 0 | 0 | 0 | 0 |
| 0 | 0.028928663 | 0 | 0 | 0 |
| 0 | 0 | 0 | 0 | 0 |
| 0 | 0 | 0 | 0 | 0 |
| 0 | 0 | 0 | 0 | 0 |
| 0 | 0 | 0 | 0 | 0 |
| 0 | 0 | 0 | 0 | 0 |
| 0 | 0 | 0 | 0 | 0.017834579 |
| 0 | 0 | 0 | 0 | 0 |
| 0 | 0 | 0 | 0 | 0 |
| 0 | 0 | 0 | 0 | 0 |
| 0 | 0 | 0 | 0 | 0 |
| 0 | 0 | 0 | 0 | 0 |
| 0 | 0 | 0 | 0 | 0 |
| 0 | 0 | 0 | 0 | 0 |
| 0 | 0 | 0 | 0 | 0 |
| 0 | 0 | 0 | 0 | 0 |
| 0 | 0 | 0 | 0 | 0 |
| 0 | 0 | 0 | 0 | 0 |
| 0 | 0 | 0 | 0 | 0 |
| 0 | 0 | 0 | 0 | 0 |
| 0 | 0 | 0 | 0 | 0 |
| 0 | 0 | 0 | 0 | 0 |
| 0 | 0 | 0 | 0 | 0 |
| 0 | 0 | 0 | 0 | 0 |
| 0 | 0 | 0 | 0 | 0 |
| 0 | 0 | 0 | 0 | 0 |
| 0 | 0 | 0 | 0.310105494 | 0 |
| 0 | 0 | 0 | 0 | 0 |
| 0.018371313 | 0 | 0 | 0.171292379 | 0.186975776 |
| 0 | 0 | 0 | 0 | 0 |
| 0 | 0 | 0 | 0 | 0 |
| 0 | 0 | 0 | 0 | 0 |
| 0 | 0 | 0 | 0 | 0 |
| 0 | 0 | 0 | 0 | 0 |
| 0 | 0 | 0 | 0 | 0 |

| | | | | |
|---|---|---|---|---|
| 0 | 0 | 0 | 0 | 0 |
| 0 | 0.059553411 | 0.449525281 | 0 | 0 |
| 0 | 0 | 0 | 0 | 0 |
| 0 | 0 | 0 | 0 | 0 |
| 0.835410225 | 0.058480489 | 0.836698147 | 0.016122131 | 0 |
| 0 | 0 | 0 | 0 | 0 |
| 0 | 0 | 0 | 0 | 0 |
| 0.010313798 | 0 | 0.108312946 | 0.001130596 | 0.028312034 |
| 0 | 0 | 0 | 0 | 0 |
| 0 | 0 | 0 | 0 | 0 |
| 0 | 0 | 0 | 0 | 0 |
| 0 | 0 | 0 | 0 | 0 |
| 0 | 0 | 0 | 0 | 0 |
| 0 | 0 | 0 | 0 | 0 |
| 0.000290422 | 0 | 0.001451096 | 0 | 0.079573956 |
| 0.010972631 | 0 | 0.001300194 | 0.001103127 | 0.041504968 |
| 0 | 0 | 0 | 0 | 0 |
| 0 | 0 | 0 | 0 | 0 |
| 0 | 0 | 0 | 0 | 0 |
| 0 | 0 | 0 | 0 | 0 |
| 0 | 0 | 0 | 0 | 0 |
| 0 | 0 | 0 | 0 | 0 |
| 0 | 0 | 0 | 0 | 0 |
| 0.01190099 | 0 | 0 | 0 | 0.001097093 |
| 0 | 0 | 0 | 0 | 0 |
| 0 | 0 | 0 | 0 | 0 |
| 0.043539224 | 0.007719245 | 0.065263229 | 0 | 0.034283368 |
| 0 | 0 | 0 | 0 | 0.000352188 |
| 0 | 0 | 0 | 0 | 0 |
| 0 | 0 | 0 | 0 | 0 |
| 0 | 0 | 0 | 0 | 0 |
| 0 | 0 | 0 | 0 | 0 |
| 0 | 0 | 0 | 0 | 0 |
| 0 | 0 | 0 | 0 | 0 |
| 0 | 0 | 0 | 0 | 0 |
| 0 | 0 | 0 | 0 | 0 |
| 0 | 0 | 0 | 0 | 0 |
| 0 | 0 | 0 | 0 | 0 |
| 0 | 0 | 0 | 0 | 0 |
| 0 | 0 | 0 | 0 | 0 |
| 0 | 0 | 0 | 0 | 0 |
| 0 | 0 | 0 | 0.051170324 | 0 |
| 0 | 0 | 0 | 0 | 0 |
| 0 | 0 | 0 | 0 | 0 |
| 0 | 0 | 0 | 0 | 0 |
| 0 | 0 | 0 | 0 | 0 |
| 0 | 0 | 0 | 0 | 0 |
| 0 | 0 | 0 | 0 | 0 |

| | | | | |
|---|---|---|---|---|
| 0 | 0 | 0.146601224 | 0.02461589 | 0 |
| 0 | 0 | 0 | 0 | 0 |
| 0 | 0 | 0 | 0 | 0 |
| 0 | 0 | 0 | 0 | 0 |
| 0 | 0 | 0 | 0 | 0 |
| 0 | 0 | 0 | 0 | 0.149410186 |
| 0 | 0 | 0 | 0 | 0 |
| 0 | 0 | 0 | 0 | 0 |
| 0 | 0 | 0 | 0 | 0 |
| 0 | 0 | 0 | 0 | 0 |
| 0 | 0 | 0 | 0 | 0 |
| 0 | 0 | 0 | 0 | 0 |
| 0 | 0 | 0 | 0 | 0 |
| 0 | 0 | 0 | 0 | 0 |
| 0 | 0 | 0 | 0 | 0 |
| 0 | 0 | 0 | 0 | 0 |
| 0 | 0 | 0 | 0 | 0 |
| 0 | 0 | 0.000790534 | 0 | 0 |
| 0 | 0 | 0 | 0 | 0 |
| 0 | 0 | 0 | 0 | 0 |
| 0.028917026 | 0 | 0.008720728 | 0.004384563 | 0.33002797 |
| 0 | 0 | 0 | 0 | 0 |
| 0.003143123 | 0 | 0 | 0 | 0.088994262 |
| 0 | 0 | 0 | 0 | 0 |
| 0 | 0 | 0 | 0 | 0 |
| 0 | 0 | 0 | 0 | 0 |
| 0 | 0 | 0 | 0 | 0 |
| 0 | 0 | 0 | 0 | 0 |
| 0 | 0 | 0 | 0 | 0 |
| 0 | 0 | 0 | 0 | 0 |
| 0 | 0 | 0 | 0 | 0 |
| 0 | 0 | 0 | 0 | 0 |
| 0.327011267 | 0.002345478 | 0.029796672 | 0.389506452 | 0.017013121 |
| 0 | 0 | 0 | 0 | 0 |
| 0.710524782 | 0.006593774 | 0.305996735 | 0.363873728 | 0 |
| 0.00089983 | 0 | 0 | 0 | 0 |
| 0 | 0 | 0 | 0 | 0 |
| 0 | 0 | 0 | 0 | 0 |
| 0.108893217 | 0 | 0.003013835 | 0.002674024 | 0.071459562 |
| 0 | 0 | 0 | 0 | 0 |
| 0 | 0 | 0 | 0 | 0 |
| 0 | 0 | 0 | 0 | 0 |
| 0.006693509 | 0.583177365 | 0.003316778 | 0.041272911 | 0.01706212 |
| 0 | 0 | 0 | 0 | 0 |
| 0 | 0 | 0 | 0 | 0 |
| 0 | 0 | 0 | 0.050532519 | 0 |
| 0 | 0 | 0 | 0 | 0 |
| 0 | 0 | 0 | 0 | 0 |

| | | | | |
|---|---|---|---|---|
| 0 | 0 | 0 | 0 | 0 |
| 0 | 0 | 0 | 0 | 0 |
| 0 | 0 | 0 | 0 | 0 |
| 0 | 0 | 0 | 0 | 0 |
| 0 | 0 | 0 | 0 | 0 |
| 0 | 0 | 0 | 0 | 0 |
| 0 | 0 | 0 | 0 | 0 |
| 0 | 0 | 0 | 0 | 0.428510189 |
| 0 | 0 | 0 | 0 | 0 |
| 0 | 0 | 0 | 0.003069166 | 0 |
| 0 | 0 | 0 | 0 | 0 |
| 0 | 0 | 0 | 0 | 0 |
| 0 | 0 | 0 | 0 | 0 |
| 0 | 0 | 0 | 0.008139279 | 0 |
| 0 | 0 | 0 | 0 | 0.023175761 |
| 0 | 0 | 0.002335709 | 0 | 0.053610421 |
| 0 | 0 | 0 | 0 | 0 |
| 0 | 0 | 0 | 0 | 0 |
| 0 | 0 | 0 | 0 | 0 |
| 0 | 0 | 0 | 0 | 0 |
| 0 | 0 | 0 | 0.025888124 | 0 |
| 0 | 0 | 0 | 0 | 0 |
| 0 | 0 | 0 | 0 | 0 |
| 0 | 0 | 0 | 0 | 0 |
| 0 | 0 | 0 | 0 | 0 |
| 0 | 0 | 0 | 0 | 0 |
| 0 | 0 | 0 | 0 | 0 |
| 0 | 0 | 0 | 0 | 0 |
| 0 | 0 | 0 | 0 | 0 |
| 0 | 0 | 0 | 0 | 0 |
| 0.07144775 | 0 | 0 | 0.023418528 | 0 |
| 0 | 0 | 0 | 0 | 0 |
| 0 | 0 | 0 | 0 | 0 |
| 0 | 0 | 0 | 0 | 0 |
| 0 | 0 | 0 | 0 | 0 |
| 0 | 0 | 0 | 0 | 0 |
| 0 | 0 | 0 | 0 | 0 |
| 0 | 0 | 0 | 0 | 0 |
| 0 | 0 | 0 | 0 | 0 |
| 0 | 0 | 0 | 0 | 0 |
| 0 | 0 | 0 | 0 | 0 |
| 0 | 0 | 0 | 0 | 0 |
| 0 | 0 | 0 | 0 | 0 |
| 0 | 0 | 0 | 0 | 0 |
| 0 | 0 | 0 | 0 | 0 |
| 0 | 0 | 0 | 0 | 0 |
| 0.165940738 | 0 | 0.147732596 | 0.000515211 | 0.055650851 |
| 0 | 0 | 0 | 0 | 0 |

| | | | | |
|---|---|---|---|---|
| 0 | 0 | 0 | 0 | 0 |
| 0 | 0 | 0 | 0 | 0 |
| 0 | 0 | 0 | 0 | 0 |
| 0 | 0 | 0 | 0 | 0 |
| 0.003464422 | 0 | 0.743479949 | 0.003560334 | 0 |
| 0 | 0 | 0 | 0 | 0 |
| 0 | 0 | 0 | 0 | 0 |
| 0 | 0 | 0 | 0 | 0 |
| 0 | 0 | 0 | 0 | 0 |
| 0 | 0 | 0 | 0 | 0 |
| 0 | 0 | 0 | 0 | 0 |
| 0.003509887 | 0 | 0.002834292 | 0.001736732 | 0.052728119 |
| 0.006392758 | 0 | 0 | 0 | 0 |
| 0 | 0 | 0 | 0 | 0 |
| 0.017039335 | 0 | 0.004994835 | 0 | 0 |
| 0.003639247 | 0 | 0.701167807 | 0.160300513 | 0.035334531 |
| 0.001299769 | 0 | 0 | 0 | 0 |
| 0 | 0 | 0 | 0 | 0 |
| 0.024805264 | 0 | 0.099929744 | 0 | 0.101554286 |
| 0.298947458 | 0.063290386 | 0.007355795 | 0 | 0.002863269 |
| 0 | 0 | 0 | 0 | 0 |
| 0 | 0 | 0 | 0 | 0 |
| 0 | 0 | 0 | 0 | 0 |
| 0 | 0 | 0 | 0 | 0 |
| 0 | 0 | 0 | 0 | 0 |
| 0 | 0 | 0 | 0 | 0 |
| 0.002059848 | 0 | 0.005052459 | 0.001799343 | 0.002294468 |
| 0 | 0 | 0 | 0 | 0 |
| 0 | 0 | 0 | 0 | 0 |
| 0 | 0 | 0 | 0 | 0 |
| 0.002685187 | 0 | 0 | 0 | 0.008609877 |
| 0 | 0 | 0 | 0 | 0 |
| 0 | 0 | 0 | 0 | 0 |
| 0 | 0 | 0 | 0 | 0 |
| 0 | 0 | 0 | 0 | 0 |
| 0 | 0 | 0 | 0 | 0 |
| 0 | 0 | 0 | 0 | 0 |
| 0.00328093 | 0 | 0.00312961 | 0.005226232 | 0.064397175 |
| 0 | 0 | 0 | 0 | 0 |
| 0 | 0 | 0 | 0 | 0 |
| 0 | 0 | 0 | 0 | 0.006290522 |
| 0 | 0 | 0 | 0 | 0 |
| 0 | 0 | 0 | 0 | 0 |
| 0.001094671 | 0 | 0.001988921 | 0 | 0.001308121 |
| 0 | 0 | 0 | 0 | 0 |
| 0 | 0 | 0 | 0 | 0 |
| 0 | 0 | 0 | 0 | 0 |
| 0 | 0 | 0 | 0 | 0 |

| | | | | |
|---|---|---|---|---|
| 0 | 0 | 0 | 0 | 0 |
| 0 | 0 | 0 | 0 | 0 |
| 0 | 0 | 0 | 0 | 0 |
| 0.002931784 | 0 | 0 | 0 | 0.007344198 |
| 0 | 0.00876108 | 0.001449934 | 0.000485994 | 0.001998077 |
| 0 | 0.000729485 | 0 | 0 | 0 |
| 0 | 0 | 0 | 0 | 0 |
| 0.004286743 | 0 | 0.000407545 | 0 | 0 |
| 0 | 0 | 0 | 0 | 0 |
| 0 | 0 | 0 | 0 | 0 |
| 0 | 0 | 0 | 0 | 0 |
| 0 | 0 | 0 | 0 | 0 |
| 0.210442592 | 0 | 0.000738772 | 0.016993203 | 0.050602342 |
| 0 | 0 | 0 | 0 | 0 |
| 0 | 0 | 0 | 0 | 0 |
| 0 | 0 | 0 | 0 | 0 |
| 0 | 0 | 0 | 0 | 0 |
| 0 | 0 | 0 | 0 | 0 |
| 0 | 0 | 0 | 0 | 0 |
| 0 | 0 | 0 | 0 | 0 |
| 0 | 0 | 0 | 0 | 0 |
| 0 | 0 | 0 | 0 | 0 |
| 0.0005949 | 0 | 0.001297057 | 0.001222739 | 0.055480652 |
| 0.001965607 | 0 | 0 | 0 | 0 |
| 0 | 0 | 0 | 0 | 0 |
| 0 | 0 | 0 | 0 | 0 |
| 0 | 0 | 0 | 0 | 0 |
| 0 | 0 | 0 | 0 | 0 |
| 0 | 0 | 0 | 0 | 0 |
| 0 | 0 | 0 | 0 | 0 |
| 0 | 0 | 0 | 0.014315165 | 0.003363101 |
| 0 | 0 | 0 | 0 | 0.028844201 |
| 0 | 0 | 0 | 0 | 0 |
| 0 | 0 | 0 | 0 | 0 |
| 0 | 0 | 0 | 0 | 0 |
| 0 | 0 | 0 | 0 | 0.002450333 |
| 0.014940047 | 0 | 0 | 0 | 0 |
| 0 | 0 | 0.028285438 | 0 | 0 |
| 0.005948794 | 0 | 0.001376546 | 0 | 0.165982654 |
| 0 | 0 | 0 | 0 | 0 |
| 0 | 0 | 0 | 0 | 0 |
| 0 | 0 | 0 | 0 | 0 |
| 0 | 0 | 0 | 0 | 0 |
| 0 | 0 | 0 | 0 | 0 |
| 0 | 0 | 0 | 0 | 0 |
| 0.133712754 | 0.007396839 | 0.016417582 | 0.013294912 | 0.291391859 |
| 0 | 0 | 0 | 0 | 0 |
| 0 | 0 | 0 | 0 | 0 |

| | | | | |
|---|---|---|---|---|
| 0 | 0 | 0 | 0 | 0 |
| 0 | 0 | 0 | 0 | 0 |
| 0 | 0 | 0 | 0 | 0 |
| 0 | 0 | 0 | 0 | 0 |
| 0 | 0 | 0 | 0 | 0 |
| 0 | 0 | 0 | 0 | 0 |
| 0 | 0 | 0 | 0 | 0 |
| 0 | 0 | 0 | 0 | 0 |
| 0 | 0 | 0.004811647 | 0 | 0 |
| 0 | 0 | 0 | 0 | 0 |
| 0 | 0 | 0 | 0 | 0 |
| 0 | 0 | 0 | 0 | 0.002819816 |
| 0 | 0 | 0 | 0 | 0 |
| 0 | 0 | 0 | 0 | 0 |
| 0 | 0 | 0 | 0 | 0 |
| 0 | 0 | 0 | 0 | 0 |
| 0 | 0 | 0 | 0 | 0 |
| 0 | 0 | 0 | 0 | 0 |
| 0 | 0 | 0 | 0 | 0 |
| 0 | 0 | 0.00724044 | 0 | 0 |
| 0 | 0 | 0 | 0 | 0 |
| 0 | 0 | 0 | 0 | 0 |
| 0 | 0 | 0 | 0 | 0 |
| 0 | 0 | 0 | 0 | 0 |
| 0 | 0 | 0 | 0 | 0 |
| 0 | 0 | 0 | 0 | 0 |
| 0 | 0 | 0 | 0 | 0 |
| 0 | 0 | 0 | 0.010712466 | 0 |
| 0 | 0 | 0 | 0 | 0 |
| 0 | 0.007159104 | 0 | 0 | 0 |
| 0.056229197 | 0.001818651 | 0.005654807 | 0.000959544 | 0.043036336 |
| 0 | 0 | 0 | 0 | 0 |
| 0.068660626 | 0.018524834 | 0.006073371 | 0.01596918 | 0.011659566 |
| 0.005662335 | 0 | 0.007380496 | 0 | 0.0047281 |
| 0 | 0 | 0 | 0 | 0 |
| 0 | 0 | 0 | 0 | 0 |
| 0 | 0 | 0 | 0 | 0 |
| 0 | 0 | 0 | 0 | 0 |
| 0 | 0 | 0 | 0 | 0 |
| 0 | 0 | 0 | 0 | 0 |
| 0 | 0 | 0 | 0 | 0 |
| 0.232585535 | 0 | 0.076568098 | 0 | 0.935545883 |
| 0.09175527 | 0 | 0.077323317 | 0.010049916 | 0.122981529 |
| 0 | 0 | 0 | 0 | 0 |
| 0 | 0 | 0 | 0 | 0 |
| 0.017620878 | 0.030583907 | 0 | 0.014522827 | 0.031093749 |
| 0 | 0 | 0 | 0 | 0 |
| 0 | 0 | 0 | 0 | 0 |

| | | | | |
|---|---|---|---|---|
| 0 | 0 | 0 | 0 | 0 |
| 0 | 0 | 0 | 0.00571925 | 0 |
| 0.029844198 | 0.002354521 | 0.024159335 | 0.003795846 | 0.006128882 |
| 0 | 0 | 0 | 0.086954895 | 0 |
| 0.013448137 | 0 | 0 | 0 | 0.009424744 |
| 0.001607585 | 0 | 0 | 0 | 0 |
| 0 | 0 | 0 | 0 | 0 |
| 0 | 0 | 0 | 0 | 0 |
| 0 | 0 | 0 | 0 | 0 |
| 0 | 0 | 0 | 0 | 0 |
| 0 | 0 | 0 | 0 | 0 |
| 0 | 0 | 0 | 0 | 0 |
| 0 | 0 | 0 | 0 | 0 |
| 0 | 0 | 0 | 0 | 0 |
| 0 | 0 | 0 | 0 | 0 |
| 0 | 0 | 0 | 0 | 0 |
| 0 | 0 | 0 | 0 | 0 |
| 0 | 0 | 0 | 0 | 0 |
| 0 | 0 | 0 | 0 | 0 |
| 0 | 0 | 0 | 0 | 0 |
| 0 | 0 | 0 | 0 | 0 |
| 0 | 0 | 0 | 0 | 0 |
| 0 | 0 | 0 | 0 | 0 |
| 0 | 0 | 0 | 0 | 0 |
| 0 | 0.012224877 | 0 | 0.005380694 | 0.094567072 |
| 0.005879479 | 0.199304669 | 0.014187692 | 0.002643485 | 0 |
| 0 | 0 | 0 | 0 | 0 |
| 0 | 0 | 0 | 0.05543714 | 0 |
| 0 | 0 | 0 | 0 | 0 |
| 0 | 0 | 0 | 0 | 0 |
| 0 | 0 | 0 | 0 | 0 |
| 0 | 0 | 0 | 0 | 0.002393012 |
| 0 | 0 | 0 | 0 | 0 |
| 0 | 0 | 0 | 0 | 0 |
| 0 | 0 | 0 | 0 | 0 |
| 0 | 0 | 0 | 0 | 0 |
| 0 | 0 | 0 | 0 | 0.065453015 |
| 0 | 0 | 0.000830744 | 0 | 0.780610972 |
| 0 | 0 | 0 | 3.065126867 | 0 |
| 0 | 0 | 0 | 0 | 0 |
| 0 | 0 | 0 | 0 | 0 |
| 0.025114668 | 0 | 0 | 0 | 0.001429133 |
| 0.017702097 | 0 | 0 | 0.017434171 | 0.135611214 |
| 0 | 0 | 0 | 0 | 0 |
| 0 | 0 | 0 | 0 | 0 |
| 0 | 0 | 0 | 0 | 0 |
| 0.001128364 | 0 | 0 | 0 | 0.009679476 |
| 0 | 0 | 0 | 0 | 0 |

| | | | | |
|---|---|---|---|---|
| 0 | 0 | 0 | 0 | 0 |
| 0.005101908 | 0 | 0.002101855 | 0.002316742 | 0.021966463 |
| 0 | 0 | 0 | 0 | 0 |
| 0 | 0 | 0 | 0 | 0 |
| 0 | 0 | 0 | 0 | 0 |
| 0 | 0 | 0 | 0 | 0 |
| 0 | 0 | 0 | 0 | 0 |
| 0 | 0 | 0 | 0 | 0 |
| 0 | 0 | 0 | 0 | 0 |
| 0 | 0 | 0 | 0 | 0 |
| 0 | 0 | 0 | 0 | 0 |
| 0 | 0 | 0 | 0 | 0 |
| 0.109744714 | 0 | 0.010938322 | 0 | 0 |
| 0 | 0 | 0 | 0 | 0 |
| 0.001923187 | 0 | 0 | 0.05959697 | 0.06523324 |
| 0 | 0 | 0 | 0 | 0 |
| 0 | 0 | 0 | 0 | 0 |
| 0 | 0 | 0 | 0 | 0 |
| 0 | 0 | 0 | 0 | 0 |
| 0 | 0 | 0 | 0 | 0 |
| 0 | 0 | 0 | 0 | 0 |
| 0.058259786 | 0.017480972 | 0.010096942 | 0 | 0 |
| 0 | 0 | 0 | 0 | 0.036963947 |
| 0 | 0 | 0 | 0 | 0 |
| 0 | 0 | 0 | 0 | 0 |
| 0 | 0 | 0 | 0 | 0 |
| 0 | 0 | 0 | 0 | 0 |
| 0 | 0 | 0 | 0 | 0 |
| 0 | 0 | 0 | 0 | 0 |
| 0 | 0 | 0 | 0 | 0 |
| 0 | 0 | 0 | 0 | 0 |
| 0 | 0 | 0 | 0 | 0 |
| 0 | 0 | 0 | 0 | 0 |
| 0 | 0 | 0 | 0 | 0 |
| 0 | 0 | 0 | 0 | 0.00070935 |
| 0 | 0 | 0 | 0 | 0 |
| 0 | 0 | 0 | 0 | 0 |
| 0 | 0 | 0 | 0 | 0 |
| 0.188031327 | 0.152025023 | 0 | 0 | 0.002117667 |
| 0 | 0 | 0 | 0 | 0 |
| 0.043059359 | 0 | 0.201910304 | 0.133823886 | 0.585520363 |
| 0 | 0 | 0 | 0 | 0 |
| 0 | 0 | 0 | 0 | 0 |
| 0 | 0 | 0 | 0 | 0 |
| 0 | 0 | 0 | 0 | 0 |
| 1.347037839 | 0.001068134 | 2.7186741 | 0.072400301 | 0.594750003 |
| 0 | 0 | 0 | 0 | 0 |
| 0 | 0 | 0 | 0 | 0 |

| | | | | |
|---|---|---|---|---|
| 0 | 0 | 0 | 0 | 0 |
| 0 | 0 | 0 | 0 | 0 |
| 0 | 0 | 0 | 0 | 0 |
| 0 | 0 | 0 | 0 | 0 |
| 0 | 0 | 0 | 0 | 0 |
| 0 | 0 | 0 | 0 | 0 |
| 0 | 0 | 0 | 0 | 0 |
| 0.000965916 | 0.00064139 | 0 | 0 | 0 |
| 0 | 0 | 0 | 0 | 0 |
| 0.024617588 | 0.15878483 | 0.896611033 | 0.00149699 | 0.016673394 |
| 0 | 0 | 0 | 0 | 0 |
| 0 | 0 | 0 | 0 | 0 |
| 0.001139012 | 0 | 0 | 0 | 0.005249988 |
| 0 | 0 | 0 | 0 | 0 |
| 0 | 0 | 0 | 0 | 0 |
| 0 | 0 | 0 | 0 | 0 |
| 0 | 0 | 0 | 0 | 0 |
| 0 | 0 | 0 | 0 | 0 |
| 0 | 0 | 0 | 0 | 0 |
| 4.105512619 | 1.788118198 | 0.151791223 | 0.021488521 | 0.807236563 |
| 0 | 0 | 0 | 0 | 0 |
| 0 | 0 | 0 | 0 | 0 |
| 0 | 0 | 0 | 0 | 0 |
| 0 | 0 | 0 | 0 | 0 |
| 0 | 0 | 0 | 0 | 0 |
| 0 | 0 | 0 | 0 | 0 |
| 0 | 0 | 0 | 0 | 0 |
| 0 | 0 | 0 | 0 | 0 |
| 0 | 0 | 0 | 0 | 0 |
| 0 | 0 | 0 | 0 | 0.488296721 |
| 0 | 0 | 0 | 0 | 0 |
| 0 | 0 | 0 | 0 | 0 |
| 0 | 0 | 0 | 0 | 0 |
| 0 | 0 | 0 | 0 | 0 |
| 0 | 0 | 0 | 0 | 0 |
| 0.007053837 | 0 | 0.002458922 | 0 | 0 |
| 0 | 0 | 0 | 0 | 0 |
| 0 | 0 | 0 | 0 | 0 |
| 0 | 0 | 0 | 0 | 0 |
| 0 | 0 | 0 | 0 | 0 |
| 0 | 0 | 0 | 0 | 0 |
| 0 | 0 | 0 | 0 | 0 |
| 0 | 0 | 0 | 0 | 0 |
| 0 | 0 | 0 | 0 | 0 |
| 0.005832218 | 0 | 0.004703605 | 0 | 0.005394716 |
| 0 | 0 | 0 | 0 | 0 |
| 0 | 0 | 0 | 0 | 0 |
| 0 | 0 | 0 | 0 | 0 |

| | | | | |
|---|---|---|---|---|
| 0 | 0 | 0 | 0 | 0 |
| 0 | 0 | 0 | 0 | 0 |
| 0 | 0 | 0 | 0 | 0 |
| 0 | 0 | 0 | 0 | 0 |
| 0 | 0 | 0 | 0 | 0 |
| 0.006219968 | 0 | 0.001453003 | 0.001552383 | 0.027918517 |
| 0 | 0 | 0 | 0 | 0 |
| 0 | 0 | 0 | 0 | 0 |
| 0 | 0 | 0 | 0 | 0 |
| 0 | 0 | 0 | 0 | 0 |
| 0 | 0 | 0 | 0 | 0 |
| 0 | 0 | 0 | 0 | 0 |
| 0 | 0 | 0 | 0 | 0 |
| 0 | 0 | 0 | 0 | 0.172191099 |
| 0 | 0 | 0 | 0 | 0 |
| 0 | 0 | 0 | 0 | 0 |
| 0 | 0 | 0 | 0 | 0 |
| 0 | 0 | 0 | 0 | 0.000252199 |
| 0 | 0 | 0 | 0 | 0 |
| 0 | 0 | 0 | 0 | 0 |
| 0 | 0 | 0 | 0 | 0 |
| 0 | 0 | 0 | 0 | 0 |
| 0 | 0 | 0 | 0 | 0 |
| 0 | 0 | 0 | 0 | 0 |
| 0 | 0 | 0 | 0 | 0 |
| 0.001945815 | 0.002547214 | 0.012689469 | 0 | 0 |
| 0 | 0 | 0 | 0 | 0 |
| 0.005201099 | 0 | 0 | 0 | 0 |
| 0 | 0 | 0 | 0 | 0 |
| 0 | 0 | 0 | 0 | 0 |
| 0 | 0 | 0 | 0 | 0 |
| 0 | 0 | 0.003079585 | 0 | 0 |
| 0 | 0 | 0 | 0 | 0 |
| 0 | 0 | 0 | 0 | 0 |
| 0 | 0 | 0 | 0 | 0 |
| 0 | 0 | 0 | 0 | 0 |
| 0 | 0 | 0 | 0 | 0 |
| 0 | 0 | 0 | 0 | 0 |
| 0 | 0 | 0 | 0.232667522 | 4.569959109 |
| 0 | 0 | 0 | 0 | 0 |
| 0 | 0 | 0 | 0 | 0 |
| 0 | 0 | 0 | 0 | 0 |
| 0 | 0 | 0 | 0 | 0 |
| 0 | 0 | 0 | 0 | 0 |
| 0 | 0 | 0 | 0 | 0 |
| 0 | 0 | 0 | 0 | 0 |
| 0 | 0 | 0 | 0 | 0 |
| 0 | 0 | 0 | 0 | 0 |

| | | | | |
|---|---|---|---|---|
| 0 | 0 | 0 | 0 | 0 |
| 0.003282913 | 0 | 0 | 0 | 0 |
| 0 | 0 | 0 | 0 | 0 |
| 0 | 0 | 0 | 0 | 0 |
| 0 | 0 | 0 | 0 | 0 |
| 0 | 0 | 0 | 0 | 0 |
| 0 | 0 | 0 | 0 | 0 |
| 0 | 0 | 0 | 0 | 0 |
| 0 | 0 | 0 | 0 | 0 |
| 0 | 0 | 0 | 0 | 0 |
| 0.017585651 | 0.094456139 | 0.00142007 | 0.003614502 | 0.001200839 |
| 0 | 0 | 0 | 0 | 0 |
| 0 | 0 | 0 | 0 | 0 |
| 0 | 0 | 0 | 0 | 0 |
| 0 | 0 | 0 | 0 | 0 |
| 0 | 0 | 0 | 0 | 0 |
| 0 | 0 | 0 | 0 | 0 |
| 0 | 0 | 0 | 0 | 0 |
| 0 | 0 | 0 | 0 | 0 |
| 0 | 0 | 0 | 0 | 0 |
| 0 | 1.652077963 | 0.007746912 | 0.001494575 | 0 |
| 0 | 0 | 0 | 0 | 0 |
| 0 | 0 | 0 | 0 | 0 |
| 0.52560029 | 0.000785437 | 5.426252783 | 1.884508558 | 1.427962509 |
| 0 | 0 | 0 | 0 | 0 |
| 0 | 0 | 0 | 0 | 0 |
| 0 | 0 | 0 | 0 | 0 |
| 0 | 0 | 0 | 0 | 0 |
| 0 | 0 | 0 | 0 | 0 |
| 0 | 0 | 0 | 0 | 0 |
| 0 | 0 | 0 | 0.004667486 | 0 |
| 0 | 0 | 0 | 0 | 0 |
| 0 | 0 | 0 | 0 | 0 |
| 0 | 0 | 0 | 0 | 0 |

| PHL018-0099_V3 | PHL018-0119_V2 | PHL018-0089_V1 | PHL018-0214_V3 | PHL018-0098_V3 |
|---|---|---|---|---|
| EYCF | REF | CTRL | EYCF | CTRL |
| 0.034195999 | 0.011168576 | 0.225118044 | 0.0361148 | 0.026258155 |
| 0.721683404 | 9.873733096 | 5.128470414 | 0.079937255 | 0.768274563 |
| 1.234345881 | 0 | 4.857978003 | 4.343926579 | 0 |
| 0 | 0 | 0 | 0 | 0 |
| 0 | 0 | 0 | 0 | 0 |
| 0 | 0 | 0 | 0 | 0 |
| 0 | 0 | 0 | 0 | 0 |
| 0 | 0 | 0 | 0 | 0 |
| 0 | 0 | 0 | 0 | 0 |
| 0 | 0 | 0 | 0 | 0 |
| 0 | 0 | 0 | 0 | 0 |
| 0 | 0 | 0 | 0 | 0 |
| 0 | 0 | 0 | 0 | 0 |
| 1.088298826 | 1.092438451 | 7.118439151 | 4.546663624 | 0.292113684 |
| 0 | 0 | 0 | 0 | 0 |
| 0 | 0 | 0 | 0 | 0 |
| 0.003857325 | 0 | 0 | 0 | 0 |
| 0 | 0 | 0 | 0 | 0 |
| 0 | 0 | 0 | 0 | 0 |
| 0 | 0 | 0 | 0 | 0 |
| 0 | 0 | 0 | 0 | 0 |
| 0.001455285 | 0 | 0 | 0 | 0 |
| 0 | 0 | 0 | 0 | 0 |
| 0 | 0 | 0 | 0 | 0 |
| 0 | 0 | 0 | 0 | 0 |
| 0 | 0 | 0 | 0 | 0 |
| 0 | 0 | 0 | 0 | 0 |
| 0 | 0 | 0 | 0 | 0 |
| 0 | 0 | 0 | 0 | 0 |
| 0 | 0 | 0 | 0 | 0 |
| 0 | 0 | 0 | 0 | 0 |
| 0 | 0 | 0 | 0 | 0 |
| 0 | 0 | 0 | 0 | 0 |
| 0 | 0 | 0 | 0 | 0 |
| 0 | 0 | 0 | 0 | 0 |
| 0 | 0.259357983 | 10.51584946 | 0 | 12.49854319 |
| 0 | 0 | 0 | 0 | 0 |
| 0 | 0 | 0 | 0 | 0 |
| 0 | 0 | 0 | 0 | 0 |
| 0 | 0 | 0 | 0 | 0 |
| 0 | 0 | 0 | 0 | 0 |
| 0 | 0.081141028 | 0.0617024 | 0 | 0.36722385 |
| 0.005461188 | 0.171428095 | 0.020603282 | 0 | 0.476248917 |
| 0.166653868 | 0.111512105 | 0.008981534 | 0.001352079 | 0.085141823 |
| 0 | 0.001572544 | 0 | 0 | 0.000822845 |
| 0 | 0 | 0 | 0 | 0 |

| | | | | |
|---|---|---|---|---|
| 0.000400993 | 0 | 0.011046169 | 0 | 0.003199276 |
| 0 | 0.001577366 | 0.003552944 | 0.003940133 | 0.087599091 |
| 0 | 0.006810939 | 0 | 0 | 0.002062498 |
| 0.440276749 | 1.902973999 | 0.857492705 | 0.12634487 | 1.692357482 |
| 0.011559282 | 0 | 0.011582707 | 0 | 0.000897759 |
| 0.09268238 | 0 | 0 | 0 | 0 |
| 0.018135431 | 0.059925265 | 0.00074048 | 0 | 17.94172523 |
| 0 | 0 | 0 | 0 | 0 |
| 0 | 0 | 0 | 0 | 0 |
| 0 | 0 | 0 | 0 | 0 |
| 0.000905595 | 0.006386237 | 0 | 0 | 0.005780145 |
| 2.259793726 | 0.01153385 | 1.10539652 | 0.2680616 | 0 |
| 0 | 0 | 0 | 0 | 0.000544989 |
| 0.011435109 | 0.00565046 | 0.016894111 | 0.023395501 | 0.012389752 |
| 0 | 0 | 0 | 0.005736287 | 0 |
| 0 | 0 | 0 | 0 | 0 |
| 0.003677162 | 0.048874723 | 0.883300977 | 0 | 0.426478864 |
| 0 | 0 | 0 | 0 | 0 |
| 0 | 0 | 0 | 0 | 0 |
| 0 | 0 | 0 | 0 | 0 |
| 0.089506582 | 0.25830802 | 0.144095835 | 0.245714403 | 0.03582489 |
| 0.013165947 | 0.003870807 | 0.032244581 | 0.004412813 | 0.002570199 |
| 0 | 0 | 0 | 0 | 0 |
| 0 | 0 | 0 | 0 | 0 |
| 0 | 0 | 0 | 0 | 0 |
| 0.000649135 | 0 | 0 | 0 | 0 |
| 0 | 0 | 1.272871776 | 0.244484115 | 0 |
| 0 | 0 | 0 | 0 | 0 |
| 0 | 0.002902827 | 0 | 0 | 0.034881684 |
| 0 | 0 | 0 | 0.000414752 | 0 |
| 0.216317822 | 0.001663675 | 0 | 0.21600883 | 0.002025961 |
| 0 | 0 | 0.009649853 | 0 | 0 |
| 0 | 0 | 0 | 0 | 0 |
| 0 | 0 | 0 | 0 | 0 |
| 0 | 0 | 0 | 0 | 0 |
| 0 | 0 | 0 | 0 | 0.00506625 |
| 0 | 0 | 0 | 0 | 0 |
| 0 | 0.000647415 | 0.020051254 | 0 | 0.007046308 |
| 0 | 0 | 0 | 0 | 0 |
| 0 | 0 | 0 | 0 | 0 |
| 0 | 0 | 0 | 0 | 0 |
| 0 | 0 | 0 | 0 | 0 |
| 0 | 0 | 0 | 0 | 0 |
| 0 | 0 | 0 | 0 | 0 |
| 0.089006069 | 0 | 0 | 0 | 0 |
| 0 | 0 | 0 | 0 | 0.006430192 |
| 0 | 0 | 0 | 0 | 0 |
| 0.059433875 | 0 | 0 | 0.000624172 | 0.000451606 |

| | | | | |
|---|---|---|---|---|
| 0 | 0 | 0 | 0 | 0 |
| 0.003530186 | 0 | 0 | 0 | 0 |
| 0 | 0 | 0 | 0 | 0 |
| 0 | 0 | 0 | 0 | 0 |
| 0.015783424 | 0 | 0.948795829 | 0 | 0.016408556 |
| 0 | 0 | 0 | 0 | 0 |
| 0 | 0 | 0 | 0 | 0 |
| 0.006365696 | 0 | 0 | 0 | 0 |
| 0 | 0 | 0 | 0 | 0.001188143 |
| 0 | 0 | 0 | 0 | 0 |
| 0 | 0 | 0 | 0 | 0 |
| 0 | 0.009286607 | 0.000674763 | 0 | 0.511259626 |
| 0 | 0 | 0 | 0 | 0 |
| 0 | 0 | 0 | 0 | 0 |
| 0 | 0 | 0 | 0 | 0 |
| 0 | 0 | 0 | 0 | 0.000581552 |
| 0 | 0 | 0 | 0 | 0 |
| 0 | 0 | 0 | 0 | 0 |
| 0.018464062 | 0 | 0 | 0 | 0 |
| 0 | 0 | 0 | 0 | 0 |
| 0 | 0 | 0 | 0 | 0 |
| 0 | 0 | 0 | 0 | 0 |
| 0.002073083 | 0 | 0 | 0 | 0 |
| 0.00184933 | 0 | 0 | 0 | 0 |
| 0 | 0 | 0 | 0 | 0 |
| 0 | 0 | 0 | 0 | 0 |
| 0 | 0 | 0 | 0 | 0 |
| 0.008747921 | 0 | 0.0005868 | 0.002411594 | 0 |
| 0 | 0 | 0 | 0 | 0 |
| 0 | 0 | 0 | 0 | 0 |
| 0.577409374 | 0 | 0.059362615 | 0.01326196 | 0.012993768 |
| 0 | 0 | 0.000493699 | 0 | 0.012792727 |
| 0 | 0 | 0 | 0 | 0 |
| 0 | 0 | 0 | 0 | 0 |
| 0 | 0 | 0 | 0 | 0 |
| 0 | 0 | 0 | 0 | 0 |
| 0 | 0 | 0 | 0 | 0 |
| 0 | 0 | 0 | 0 | 0 |
| 0 | 0 | 0 | 0 | 0 |
| 0 | 0 | 0 | 0 | 0 |
| 0 | 0 | 0 | 0 | 0 |
| 0 | 0 | 0 | 0 | 0 |
| 0 | 0 | 0 | 0.003733129 | 0 |
| 0 | 0 | 0 | 0 | 0 |
| 0 | 0 | 0 | 0 | 0 |
| 0 | 0 | 0 | 0 | 0 |
| 0 | 0 | 0 | 0 | 0 |
| 0 | 0 | 0 | 0 | 0 |

| | | | | |
|---|---|---|---|---|
| 0 | 0 | 0 | 0 | 0 |
| 0 | 0 | 0 | 0 | 0 |
| 0 | 0 | 0 | 0 | 0 |
| 0 | 0 | 0 | 0 | 0 |
| 0.033402874 | 0 | 0.000920805 | 0 | 0.001277743 |
| 0 | 0 | 0 | 0 | 0 |
| 0 | 0 | 0 | 0 | 0 |
| 0 | 0 | 0 | 0 | 0 |
| 0 | 0 | 0 | 0 | 0 |
| 0 | 0 | 0 | 0 | 0 |
| 0 | 0 | 0 | 0 | 0 |
| 0 | 0 | 0 | 0 | 0 |
| 0 | 0 | 0 | 0 | 0 |
| 0 | 0 | 0 | 0 | 0 |
| 0 | 0 | 0 | 0 | 0 |
| 0 | 0 | 0 | 0 | 0 |
| 0 | 0 | 0 | 0 | 0 |
| 0 | 0 | 0 | 0 | 0 |
| 0 | 0 | 0 | 0 | 0 |
| 0 | 0 | 0 | 0 | 0 |
| 0 | 0 | 0 | 0 | 0 |
| 0 | 0 | 0 | 0 | 0 |
| 0 | 0 | 0 | 0 | 0 |
| 0 | 0 | 0 | 0 | 0 |
| 4.105307725 | 0 | 0 | 0 | 0 |
| 0 | 0 | 0 | 0 | 0 |
| 0 | 0 | 0 | 0 | 0 |
| 0 | 0 | 0 | 0 | 0 |
| 0 | 0 | 0 | 0 | 0 |
| 0 | 0 | 0 | 0 | 0 |
| 0 | 0 | 0 | 0 | 0 |
| 0 | 0.031791804 | 0 | 0 | 0.017421695 |
| 0 | 0 | 0 | 0 | 0 |
| 0 | 0 | 0 | 0 | 0 |
| 0 | 0 | 0 | 0 | 0 |
| 0 | 0 | 0 | 0 | 0 |
| 0 | 0 | 0 | 0 | 0 |
| 0 | 0 | 0 | 0 | 0 |
| 0 | 0 | 0 | 0 | 0 |
| 0 | 0 | 0 | 0 | 0 |
| 0 | 0 | 0 | 0 | 0 |
| 0 | 0 | 0 | 0 | 0 |
| 0 | 0 | 0 | 0 | 0 |
| 0 | 0 | 0 | 0 | 0 |
| 0 | 0 | 0 | 0 | 0 |
| 0 | 0 | 0 | 0 | 0 |
| 0 | 0 | 0 | 0 | 0 |
| 0.005577405 | 0 | 0 | 0 | 0 |

| | | | | |
|---|---|---|---|---|
| 0 | 0 | 0 | 0 | 0 |
| 0 | 0 | 0 | 0 | 0 |
| 0 | 0 | 0 | 0 | 0 |
| 0 | 0 | 0 | 0 | 0 |
| 0 | 0 | 0 | 0 | 0 |
| 0 | 0 | 0 | 0 | 0 |
| 0 | 0 | 0 | 0 | 0 |
| 0 | 0 | 0 | 0 | 0 |
| 0 | 0 | 0 | 0 | 0 |
| 0 | 0 | 0 | 0 | 0 |
| 0 | 0 | 0 | 0 | 0 |
| 0 | 0 | 0 | 0 | 0 |
| 0 | 0 | 0 | 0 | 0 |
| 0 | 0 | 0 | 0 | 0 |
| 0 | 0 | 0 | 0 | 0 |
| 0 | 0 | 0 | 0 | 0 |
| 0 | 0 | 0 | 0 | 0 |
| 0 | 0 | 0 | 0 | 0 |
| 0 | 0 | 0 | 0 | 0 |
| 0 | 0 | 0 | 0 | 0 |
| 0.052362914 | 0.015513043 | 0.029264274 | 0.081546661 | 0.034200955 |
| 0 | 0 | 0 | 0 | 0 |
| 0 | 0 | 0.000766495 | 0 | 0 |
| 0 | 0.001887315 | 0.001366422 | 0 | 0.000902904 |
| 0 | 0 | 0 | 0 | 0 |
| 0 | 0 | 0 | 0 | 0 |
| 0.001264038 | 0.024091814 | 0.00025437 | 0.001991231 | 0.004286115 |
| 0 | 0 | 0 | 0 | 0 |
| 0 | 0 | 0 | 0 | 0 |
| 0 | 0 | 0.00698511 | 0.004580333 | 0 |
| 0 | 0 | 0 | 0.158793516 | 0 |
| 0 | 0 | 0 | 0 | 0 |
| 0 | 0 | 0 | 0 | 0 |
| 0 | 0 | 0 | 0 | 0 |
| 0.00370711 | 0.001345815 | 0.044144566 | 0.110900269 | 0.004788613 |
| 0 | 0 | 0 | 0 | 0 |
| 0 | 0 | 0 | 0 | 0 |
| 0 | 0 | 0 | 0 | 0 |
| 0 | 0 | 0.000918194 | 0 | 0 |
| 0 | 0 | 0 | 0 | 0 |
| 0 | 0 | 0 | 0 | 0 |
| 0 | 0 | 0.000567646 | 0 | 0 |
| 0 | 0 | 0 | 0 | 0 |
| 0 | 0 | 0 | 0 | 0 |
| 0 | 0 | 0 | 0 | 0 |
| 0 | 0 | 0 | 0.021075464 | 0 |
| 0 | 0 | 0.001641093 | 0.002529173 | 0 |
| 0.000628997 | 0.000959064 | 0.003645418 | 0.00047561 | 0 |

| | | | | |
|---|---|---|---|---|
| 0 | 0 | 0 | 0 | 0 |
| 0 | 0 | 0 | 0 | 0 |
| 0 | 0 | 0.000740246 | 0 | 0 |
| 0 | 0 | 0 | 0 | 0 |
| 0 | 0 | 0 | 0 | 0 |
| 0 | 0 | 0 | 0 | 0 |
| 0 | 0 | 0.002114121 | 0 | 0.003428925 |
| 0 | 0 | 0.000291131 | 0 | 0.034915786 |
| 0 | 0 | 0 | 0 | 0 |
| 0 | 0 | 0 | 0 | 0.001676299 |
| 0 | 0 | 0 | 0 | 0.00074508 |
| 0 | 0 | 0 | 0 | 0 |
| 0 | 0 | 0 | 0 | 0 |
| 0 | 0 | 0 | 0 | 0 |
| 0 | 0.008694726 | 0 | 0 | 0.005855112 |
| 0 | 0.001903798 | 0 | 0 | 0 |
| 0 | 0 | 0 | 0 | 0 |
| 0 | 0 | 0 | 0 | 0 |
| 0 | 0 | 0 | 0 | 0.002245608 |
| 0.132242271 | 0 | 0 | 0 | 0 |
| 0.002642084 | 0.000899224 | 0.228269382 | 0.081338463 | 0 |
| 0 | 0 | 0 | 0 | 0 |
| 0 | 0 | 0 | 0 | 0 |
| 0 | 0 | 0 | 0 | 0 |
| 0.002999462 | 0.024296375 | 0.001408402 | 0.001417509 | 0.003290491 |
| 0 | 0 | 0 | 0 | 0 |
| 0 | 0 | 0 | 0 | 0 |
| 0 | 0 | 0 | 0 | 0 |
| 0 | 0.009888311 | 0.003072133 | 0 | 0.11889799 |
| 0 | 0 | 0 | 0 | 0 |
| 0.042462377 | 0.019968207 | 0.352149756 | 0.405978345 | 0.328263217 |
| 0 | 0 | 0 | 0 | 0 |
| 0.002588168 | 0 | 0 | 0.054252921 | 0 |
| 0 | 0 | 0 | 0 | 0 |
| 0 | 0 | 0 | 0 | 0 |
| 0 | 0 | 0 | 0 | 0 |
| 0 | 0 | 0 | 0 | 0 |
| 0 | 0 | 0 | 0 | 0 |
| 0 | 0 | 0 | 0 | 0 |
| 0 | 0 | 0 | 0 | 0 |
| 0 | 0 | 0 | 0 | 0 |
| 0.000565795 | 0.023589382 | 0.003415757 | 0.013725915 | 0.000677119 |
| 0 | 0 | 0 | 0 | 0 |
| 0 | 0 | 0 | 0 | 0 |
| 0 | 0 | 0 | 0 | 0 |
| 0 | 0 | 0 | 0 | 0 |
| 0 | 0 | 0 | 0 | 0 |
| 0 | 0 | 0 | 0 | 0 |

| | | | | |
|---|---|---|---|---|
| 0 | 0 | 0 | 0 | 0 |
| 0 | 0 | 0 | 0 | 0 |
| 0.180454041 | 0.128467691 | 0.008959339 | 0.000671499 | 0.094659458 |
| 0 | 0 | 0 | 0 | 0 |
| 0.000924821 | 0 | 0 | 0 | 0 |
| 0 | 0 | 0 | 0 | 0 |
| 0 | 0 | 0 | 0 | 0 |
| 0 | 0 | 0 | 0 | 0 |
| 0 | 0 | 0 | 0 | 0 |
| 0 | 0 | 0.00052303 | 0 | 0 |
| 0 | 0 | 0 | 0 | 0 |
| 0 | 0 | 0 | 0 | 0 |
| 0 | 0 | 0 | 0 | 0.001233514 |
| 0.02944404 | 0.001122372 | 0.03591995 | 0.001979007 | 0.003602041 |
| 0.008910807 | 0.012197215 | 0.003825447 | 0.002041764 | 0.006850856 |
| 0 | 0 | 0 | 0 | 0 |
| 0.001661411 | 0.013018035 | 0.002773753 | 0.000814243 | 0.003682045 |
| 0 | 0 | 0 | 0 | 0.001942233 |
| 0 | 0 | 0 | 0 | 0 |
| 0 | 0 | 0 | 0 | 0 |
| 0 | 0 | 0 | 0 | 0 |
| 0 | 0 | 0.004471263 | 0 | 0 |
| 0 | 0 | 0 | 0 | 0 |
| 0 | 0 | 0.002079443 | 0 | 0 |
| 0 | 0 | 0 | 0 | 0 |
| 0 | 0 | 0 | 0 | 0 |
| 0 | 0 | 0 | 0 | 0 |
| 0 | 0 | 0 | 0 | 0 |
| 0 | 0 | 0 | 0 | 0.000883941 |
| 0 | 0 | 0 | 0 | 0 |
| 0 | 0 | 0 | 0 | 0 |
| 0 | 0 | 0 | 0 | 0 |
| 0 | 0 | 0 | 0 | 0 |
| 0 | 0 | 0 | 0.003753151 | 0 |
| 0 | 0 | 0 | 0 | 0 |
| 0 | 0.005709428 | 0.0012503 | 0 | 0.245638944 |
| 0 | 0 | 0 | 0 | 0 |
| 0 | 0.003790919 | 0 | 0 | 0.007349859 |
| 0 | 0 | 0 | 0 | 0 |
| 0 | 0 | 0 | 0 | 0 |
| 0.000906534 | 0 | 0 | 0 | 0 |
| 0 | 0 | 0 | 0 | 0 |
| 0 | 0 | 0 | 0 | 0 |
| 0 | 0 | 0 | 0.000983242 | 0 |
| 0 | 0 | 0 | 0 | 0 |
| 0 | 0 | 0 | 0 | 0 |
| 0 | 0 | 0 | 0 | 0 |
| 0 | 0.008458494 | 0 | 0 | 0 |

| | | | | |
|---|---|---|---|---|
| 3.299380004 | 0 | 0 | 0 | 0 |
| 0 | 0 | 0 | 0 | 0.001542712 |
| 0.00890178 | 0.001164351 | 0 | 0.000439934 | 0 |
| 0 | 0 | 0 | 0 | 0 |
| 0.001580479 | 0.016400293 | 0.001319318 | 0 | 0.007145475 |
| 0 | 0 | 0 | 0 | 0 |
| 0 | 0 | 0 | 0 | 0 |
| 0.069907847 | 0.001407467 | 0.000924657 | 0.007134881 | 0.090732778 |
| 0 | 0 | 0 | 0 | 0 |
| 0.004820617 | 0.017650259 | 0.022056632 | 2.048589856 | 0.004048495 |
| 0 | 0 | 0 | 0 | 0 |
| 0 | 0.0300822 | 0.031056105 | 0.001350608 | 11.3266444 |
| 0.018032298 | 0.011268354 | 0.04513139 | 0.000651944 | 0.01344345 |
| 0.00359166 | 0 | 0.067820169 | 0.105803047 | 0 |
| 0 | 0 | 0 | 0 | 0 |
| 0 | 0 | 0.00059534 | 0.001537921 | 0 |
| 0 | 0.002152699 | 0 | 0 | 0.006179196 |
| 0 | 0 | 0 | 0.000363888 | 0 |
| 0 | 0.236615276 | 0.053937088 | 0.145321086 | 0 |
| 0.004401443 | 0 | 0 | 0.039430868 | 0 |
| 0 | 0 | 0 | 0 | 0 |
| 0 | 0 | 0.001239353 | 0 | 0 |
| 0 | 0 | 0 | 0 | 0.004258918 |
| 0 | 0 | 0 | 0 | 0 |
| 0 | 0 | 0 | 0 | 0 |
| 0 | 0.00639832 | 0.002032451 | 0.001084784 | 0.002472346 |
| 0.006215601 | 0.011627182 | 0 | 0 | 0.001469346 |
| 0 | 0.040075493 | 0.014858505 | 0.020726533 | 0 |
| 0 | 0 | 0 | 0.237903957 | 0 |
| 0 | 0 | 0.023002495 | 0.001004496 | 0 |
| 0 | 0 | 0 | 0.000465613 | 0.081210108 |
| 0 | 0 | 0 | 0.003875608 | 0 |
| 0 | 0 | 0 | 0 | 0 |
| 0 | 0 | 0.231614721 | 0.10469996 | 0 |
| 0 | 0 | 0 | 0 | 0 |
| 0 | 0 | 0 | 0 | 0 |
| 0.001951564 | 0.0042775 | 0 | 0.053369598 | 0.013701882 |
| 0 | 0 | 0 | 0 | 0 |
| 0 | 0 | 0 | 0 | 0 |
| 0 | 0 | 0 | 0.002510014 | 0 |
| 0 | 0.000922943 | 0 | 0 | 0 |
| 0 | 0 | 0 | 0 | 0 |
| 0 | 0 | 0 | 0 | 0 |
| 0.00083217 | 0 | 0 | 0.000611758 | 0 |
| 0 | 0 | 0 | 0 | 0 |
| 0 | 0 | 0 | 0 | 0 |
| 0 | 0.001020105 | 0.000919096 | 0.009982712 | 0.000683235 |
| 0.013019659 | 0 | 0 | 0 | 0 |

| | | | | |
|---|---|---|---|---|
| 0.040469762 | 0 | 0 | 0 | 0 |
| 0 | 0.008892299 | 0.035950377 | 0.052465132 | 0.491543659 |
| 0 | 0 | 0 | 0 | 0 |
| 0.00668195 | 0 | 0 | 0 | 0 |
| 0 | 0 | 0 | 0.000400008 | 0 |
| 0.036253615 | 0 | 0 | 0.018912728 | 0.001210793 |
| 0 | 0 | 0 | 0 | 0 |
| 0 | 0 | 0 | 0 | 0.000696005 |
| 0 | 0 | 0 | 0 | 0 |
| 0 | 0 | 0 | 0 | 0 |
| 0 | 0.110520221 | 0 | 0.00616168 | 0.002496562 |
| 0 | 0 | 0 | 0 | 0 |
| 0 | 0 | 0.007714574 | 0 | 0 |
| 0.006097314 | 0.002993317 | 0.000495758 | 0.041912966 | 0.000737071 |
| 0.001121706 | 0.004632128 | 0.033207116 | 0 | 0.001044097 |
| 0 | 0 | 0 | 0.00356184 | 0 |
| 0.138735004 | 0 | 0 | 0 | 0 |
| 0.065809804 | 0.001542166 | 0 | 0.003965508 | 0.000573831 |
| 0 | 0 | 0.020367608 | 0 | 0 |
| 0.186464159 | 0 | 0 | 0.002233997 | 0.008485885 |
| 0 | 0 | 0 | 0 | 0 |
| 0.360142244 | 0 | 0 | 0 | 0.000500003 |
| 0.190764611 | 0 | 0 | 0.021985852 | 0 |
| 0 | 0 | 0 | 0 | 0 |
| 0 | 0 | 0 | 0 | 0 |
| 0 | 0 | 0 | 0 | 0 |
| 0.012621195 | 0 | 0 | 0.555183381 | 0.02669261 |
| 0 | 0 | 0 | 0 | 0 |
| 0 | 0 | 0 | 0 | 0 |
| 0.777281622 | 1.078827539 | 3.68948694 | 0.801564305 | 0.079516813 |
| 0 | 0.010750912 | 0 | 0.007582564 | 0.000700062 |
| 0 | 0 | 0.00056664 | 0 | 0 |
| 0 | 0 | 0 | 0 | 0 |
| 0.001242767 | 0 | 0.012480682 | 0.003356096 | 0 |
| 0.000667445 | 0 | 0.017998117 | 0 | 0.001331283 |
| 0 | 0 | 0 | 0 | 0 |
| 0 | 0 | 0 | 0 | 0 |
| 0 | 0.004666216 | 0.000303186 | 0 | 0 |
| 0 | 0 | 0 | 0 | 0 |
| 0 | 0 | 0 | 0 | 0 |
| 0.034025296 | 0 | 0.023475863 | 0.008709969 | 0.028237384 |
| 6.338354808 | 1.791736108 | 0 | 0 | 0 |
| 0 | 0.005260992 | 0 | 0 | 0 |
| 0 | 0 | 0.066673167 | 0.048381744 | 0 |
| 0.002570244 | 0 | 0.008091721 | 0 | 0 |
| 0 | 0 | 0 | 0.00728305 | 0 |
| 0 | 0.002886681 | 0 | 0.001205504 | 0 |
| 0 | 0 | 0 | 0 | 0 |

| | | | | |
|---|---|---|---|---|
| 0 | 0.001312198 | 0.002744547 | 0 | 0.000784706 |
| 0 | 0 | 0 | 0.000633599 | 0 |
| 0.010721191 | 0.010472398 | 1.97525729 | 0 | 0.32696795 |
| 0 | 0.000847836 | 0 | 0 | 0 |
| 0 | 0 | 0 | 0 | 0 |
| 0 | 0.007290768 | 0 | 0 | 0 |
| 0 | 0 | 0 | 0 | 0 |
| 0 | 0 | 0 | 0 | 0.005452874 |
| 0.076386924 | 0 | 0 | 0 | 0.000525383 |
| 0 | 0 | 0.165615274 | 0 | 0 |
| 0 | 0 | 0 | 0 | 0 |
| 0 | 0 | 0 | 0 | 0 |
| 0 | 0 | 0 | 0 | 0 |
| 0.000889784 | 0.268796182 | 0.003342395 | 0 | 0 |
| 0 | 0 | 0 | 0 | 0 |
| 0.042972682 | 0 | 0.069554478 | 0 | 0.027516951 |
| 0 | 0.009841989 | 0.002812438 | 0.002136844 | 0.003566494 |
| 0.00074912 | 0 | 0.022813516 | 0.005074356 | 0 |
| 0 | 0 | 0.00649361 | 0.000501636 | 0 |
| 0 | 0 | 0 | 10.74526268 | 0.043955485 |
| 0 | 0.107930759 | 0.147304088 | 0.001346516 | 0.01859916 |
| 0 | 0.001363128 | 0.004029878 | 0.017350396 | 0 |
| 0 | 0 | 0.010713344 | 0 | 0 |
| 0 | 0.326253887 | 0 | 0 | 0 |
| 0 | 0 | 0.150056149 | 0 | 0 |
| 0 | 0 | 0.020376902 | 0 | 0 |
| 0 | 0 | 0.001575182 | 0.001479677 | 0.003434801 |
| 0 | 0 | 0 | 0 | 0 |
| 0 | 0 | 0 | 0 | 0 |
| 0 | 0 | 0 | 0 | 0 |
| 0.155963574 | 0 | 0.053746744 | 0.008345901 | 0 |
| 0.10250517 | 0.017549354 | 0.022026765 | 0 | 0.01115216 |
| 0.026406596 | 0.762993546 | 0.083889834 | 0.539511026 | 0.003708 |
| 0 | 0.028562262 | 0 | 0 | 0.258691569 |
| 0.001944751 | 0 | 0.009856341 | 0.000544632 | 0.001206797 |
| 0 | 0 | 0 | 0 | 0 |
| 0 | 0.002366064 | 0.001127384 | 0.000601721 | 0.002590404 |
| 0.092603659 | 0.00113869 | 0.000801515 | 0.016564199 | 0.006792442 |
| 0 | 1.891747093 | 0 | 0 | 0.048954092 |
| 0.004274118 | 0 | 0 | 0 | 0 |
| 0 | 0 | 0 | 0 | 0 |
| 0 | 0 | 0 | 0 | 0 |
| 0.00536085 | 0 | 0.00910985 | 0 | 0 |
| 0.058880206 | 0 | 0.157233698 | 0.180878645 | 0.091069987 |
| 0 | 0 | 0 | 2.462634742 | 0 |
| 0.009280478 | 0 | 0 | 0 | 0 |
| 0.006970918 | 0 | 0 | 0 | 0 |
| 0.001213859 | 0 | 0 | 0.000286828 | 0 |

| | | | | |
|---|---|---|---|---|
| 0 | 0 | 0 | 0 | 0 |
| 0.474412011 | 0.380144501 | 0.066949768 | 0.00192365 | 0.940170479 |
| 0 | 0 | 0 | 0 | 0.007468143 |
| 0 | 0 | 0 | 0 | 0 |
| 0 | 0 | 0 | 0 | 0.002708113 |
| 0 | 0 | 0 | 0 | 0 |
| 0 | 0 | 0 | 0.142370116 | 0 |
| 0 | 0 | 0 | 0.001477566 | 0.000748342 |
| 0.000931543 | 0 | 0.001199746 | 0 | 0.013823891 |
| 0 | 0 | 0 | 0.0195703 | 0 |
| 0.000907439 | 0 | 0.004544952 | 0.008100398 | 0.001930638 |
| 0.02523091 | 0.145124533 | 0.485131258 | 0.083750656 | 0.086811383 |
| 0 | 0 | 0 | 0 | 0 |
| 0 | 0 | 0 | 0 | 0 |
| 0 | 0.000826839 | 0.001769295 | 0.012246467 | 0 |
| 0 | 0 | 0 | 0.003198464 | 0 |
| 0.015520251 | 0.061576085 | 0.001359777 | 0.000299375 | 0.031588378 |
| 0.088196403 | 0.011363204 | 0.003199387 | 0.526334344 | 0.062493263 |
| 0 | 0 | 0 | 0 | 0 |
| 0.063811479 | 0.002671588 | 0.02113334 | 0 | 0.020343184 |
| 0.017475168 | 0.834377604 | 0.051884125 | 0 | 0.068017825 |
| 0.001422573 | 0 | 0 | 0 | 0 |
| 0 | 0 | 0 | 0 | 0 |
| 0 | 0 | 0 | 0 | 0 |
| 0.020854928 | 0.002353514 | 0 | 0 | 0.049478688 |
| 0 | 0 | 0.000642962 | 0 | 0 |
| 0.219560312 | 0.038559349 | 0 | 0.041545247 | 0 |
| 0.04999229 | 0.007609351 | 0.235950514 | 0.025813319 | 0.027698432 |
| 0 | 0 | 0 | 0 | 0 |
| 0 | 0 | 0 | 0 | 0 |
| 0 | 0 | 0 | 0 | 0.001040215 |
| 0 | 0 | 0 | 0 | 0 |
| 0.006111174 | 0 | 0 | 0 | 0 |
| 0 | 0 | 0 | 0 | 0 |
| 0 | 0.045840526 | 0 | 0.048515703 | 0 |
| 0.434208521 | 0.06607165 | 0.224653905 | 0.012773041 | 0.061175716 |
| 0 | 0 | 0 | 0 | 0 |
| 0 | 0 | 0 | 0 | 0 |
| 0.001927221 | 0.013060154 | 0.00942276 | 0.001214375 | 4.282415016 |
| 0 | 0 | 0 | 0 | 0 |
| 0 | 0 | 0 | 1.617020724 | 0 |
| 0.013783146 | 0 | 0 | 0.004772836 | 0 |
| 0.001951039 | 0.004462277 | 0 | 0.002704643 | 0 |
| 0.004144122 | 0.007659101 | 0 | 0 | 0 |
| 0 | 0 | 0 | 0 | 0 |
| 0 | 0 | 0 | 0 | 0 |
| 0 | 0 | 0 | 0.001855325 | 0 |
| 0.000811958 | 0 | 0 | 0.000511629 | 0.027046096 |

| | | | | |
|---|---|---|---|---|
| 0 | 0 | 0 | 0 | 0 |
| 0 | 0 | 0 | 0 | 0 |
| 0.068659404 | 0 | 0 | 0 | 0 |
| 0.250260766 | 0 | 0 | 0 | 0 |
| 0 | 0 | 0 | 0 | 0 |
| 0 | 0 | 0 | 0 | 0 |
| 0 | 0 | 0 | 0 | 0 |
| 0 | 0 | 0 | 0 | 0.001405637 |
| 0.077870107 | 0.373066075 | 0.027126733 | 0.022059664 | 0 |
| 0 | 0 | 0 | 0 | 0.006913459 |
| 0 | 0 | 0 | 0 | 0 |
| 0 | 0.001936441 | 0.009813924 | 0 | 0.000810606 |
| 0 | 0 | 0 | 0 | 0 |
| 0 | 0 | 0 | 0 | 0 |
| 0.057730399 | 0.000456559 | 0.158048817 | 0.010264026 | 0.006020225 |
| 0 | 0 | 0 | 0 | 0 |
| 0.019997808 | 0 | 0 | 0 | 0.671481504 |
| 0 | 0 | 0 | 0 | 0 |
| 0 | 0 | 0 | 0 | 0 |
| 0 | 0 | 0 | 0 | 0 |
| 0 | 0 | 0 | 0 | 0 |
| 0 | 0 | 0 | 0 | 0 |
| 0 | 0 | 0 | 0 | 0 |
| 1.333581756 | 0.013633278 | 0.109893736 | 0.112245253 | 0.031480392 |
| 0.018888878 | 0 | 0.043754006 | 0 | 0 |
| 0 | 0 | 0 | 0 | 0 |
| 0 | 0 | 0 | 0.000407533 | 0 |
| 0 | 0.605258843 | 0 | 0 | 0.19742312 |
| 0 | 0 | 0 | 0 | 0 |
| 0 | 0 | 0 | 0 | 0 |
| 0.000423312 | 0 | 0 | 0 | 0 |
| 0 | 0 | 0 | 0 | 0 |
| 0.195558145 | 0 | 0.285033086 | 0.004601466 | 0 |
| 0 | 0 | 0 | 0 | 0 |
| 0 | 0 | 0 | 0 | 0 |
| 0 | 0 | 0 | 0 | 0 |
| 0.278747353 | 0 | 0 | 0 | 0 |
| 0 | 0 | 0 | 0 | 0 |
| 0.079777726 | 0 | 0 | 0 | 0.000723292 |
| 0.004155588 | 0 | 0.000947756 | 0 | 0 |
| 0.004177171 | 0.014861351 | 0.005753432 | 0.000263211 | 0.056767082 |
| 0 | 0 | 0 | 0 | 0 |
| 0.023050737 | 4.182897935 | 18.64717948 | 17.02195965 | 0.208028706 |
| 0 | 0 | 0 | 0 | 0.013161273 |
| 0 | 0 | 0 | 0 | 0 |
| 2.906507052 | 0 | 0 | 0 | 0 |
| 0 | 0 | 0 | 0 | 0 |
| 0.029793872 | 0 | 0.001616794 | 0 | 0 |

| | | | | |
|---|---|---|---|---|
| 0 | 0 | 0 | 0 | 0 |
| 0.039037211 | 0 | 0 | 0 | 0 |
| 0.009145871 | 0.115598245 | 0.190635168 | 0.287440773 | 0.038916918 |
| 0.031740062 | 0 | 0 | 0 | 0 |
| 0 | 0 | 0.746908403 | 0 | 0 |
| 0.000884796 | 0 | 0 | 0 | 0.000907616 |
| 0 | 0 | 0 | 0 | 0.004131651 |
| 0.00678897 | 0 | 0 | 0 | 0 |
| 0 | 0 | 0 | 0 | 0 |
| 0.296073252 | 0 | 0 | 0 | 0 |
| 0.053562665 | 0.001050898 | 0.135262821 | 0.007445019 | 0.007289973 |
| 1.550324107 | 0.354902349 | 4.386008757 | 3.063759827 | 0.374851152 |
| 0 | 0 | 0 | 0 | 0 |
| 0 | 0.004374663 | 0.000912382 | 0 | 0.006239849 |
| 0.180474792 | 0.003575084 | 0.781400675 | 0.039341977 | 1.131499744 |
| 0 | 0 | 0 | 0 | 0 |
| 0 | 0 | 0.001168106 | 0 | 0 |
| 0 | 0 | 0 | 0 | 0 |
| 0.194248283 | 0 | 0 | 0 | 0 |
| 0.011679231 | 0 | 0.547007297 | 0.005887427 | 0.001449487 |
| 0.046345834 | 0.159677767 | 0.013320994 | 0.000416002 | 0.15127692 |
| 0 | 0.049056585 | 0.245558755 | 35.3167738 | 0 |
| 0.288339755 | 0.299737456 | 1.40021618 | 0.447146945 | 0.067064275 |
| 0 | 0 | 0 | 0 | 0 |
| 0 | 0 | 0 | 0 | 0 |
| 0 | 23.33149381 | 0.016641804 | 0 | 0.018178007 |
| 0.027952776 | 0 | 0.016356267 | 0 | 0 |
| 0 | 0 | 0 | 0 | 0 |
| 0 | 0 | 0 | 0 | 0 |
| 0 | 0 | 0 | 0 | 0 |
| 0 | 0 | 0 | 0 | 0 |
| 0 | 0 | 0 | 0 | 0.222679166 |
| 0.047244099 | 0.041630126 | 0.143085994 | 0.046285172 | 0.003227152 |
| 0 | 0 | 0 | 0 | 0.007382859 |
| 0.000854293 | 0.000508822 | 0.003724821 | 0.00141305 | 0.001277976 |
| 0.001027028 | 0 | 0.015510495 | 0.005824332 | 0 |
| 0.003684924 | 0 | 0.006997103 | 0 | 0 |
| 0 | 0 | 0 | 0 | 0 |
| 0 | 0.001963385 | 0.082367857 | 0 | 0.009725637 |
| 0 | 0 | 0 | 0 | 0 |
| 0 | 0 | 0 | 0 | 0 |
| 0 | 0 | 0 | 0 | 0 |
| 0 | 0 | 0 | 0 | 0 |
| 0 | 0 | 0 | 0 | 0 |
| 0.004182889 | 0.002072807 | 0.00395061 | 0.00063257 | 0.01174718 |
| 0.003491687 | 0 | 0 | 0 | 0 |
| 0 | 0 | 0 | 0 | 0 |
| 0.212928826 | 0.076742181 | 0.059568942 | 0.153627952 | 0.007890286 |

| | | | | |
|---|---|---|---|---|
| 0 | 0 | 0 | 0 | 0 |
| 0 | 0 | 0 | 0 | 0 |
| 0 | 0 | 0 | 0 | 0 |
| 0.008698744 | 0 | 0.011784811 | 0 | 0.000568868 |
| 0 | 0 | 0 | 0 | 0 |
| 0 | 0 | 0 | 0 | 0 |
| 0 | 0 | 0 | 0 | 0 |
| 0.020339098 | 0 | 0.046052471 | 0 | 0.001361 |
| 0 | 0 | 0 | 0 | 0 |
| 0 | 0 | 0 | 0 | 0 |
| 0.000635285 | 0 | 0.003068219 | 0.00050038 | 0 |
| 0 | 0 | 0 | 0 | 0 |
| 0 | 0 | 0 | 0.001139117 | 0 |
| 0 | 0 | 0 | 0 | 0 |
| 0 | 0 | 0 | 0 | 0 |
| 0 | 0 | 0 | 0 | 0 |
| 0 | 0 | 0 | 0 | 0.041560534 |
| 0.001454366 | 0 | 0 | 0 | 0 |
| 0 | 0 | 0 | 0 | 0 |
| 0.0042925 | 0 | 0 | 0 | 0.00085618 |
| 0 | 0 | 0 | 0 | 0 |
| 0 | 0 | 0 | 0 | 0 |
| 0.003658429 | 0 | 0 | 0 | 0.013742839 |
| 0 | 0 | 0 | 0 | 0 |
| 0 | 0 | 0 | 0 | 0 |
| 0 | 0 | 0 | 0 | 0 |
| 0 | 0 | 0 | 0 | 0 |
| 0 | 0 | 0 | 0 | 0 |
| 0 | 0 | 0 | 0 | 0 |
| 0 | 0.002739659 | 0.016970128 | 0 | 0 |
| 0.009256618 | 0 | 0 | 0.004808081 | 0 |
| 0 | 0 | 0 | 0 | 0 |
| 0 | 0 | 0 | 0 | 0 |
| 0 | 0 | 0 | 0 | 0 |
| 0 | 0 | 0 | 0 | 0 |
| 0 | 0 | 0 | 0 | 0 |
| 0 | 0 | 0 | 0 | 0 |
| 0.00107556 | 0.004356149 | 0.010172763 | 0.01626549 | 0.000965388 |
| 0 | 0 | 0 | 0 | 0 |
| 0.005834801 | 0 | 0 | 0 | 0.002645014 |
| 0 | 0 | 0 | 0.00341784 | 0 |
| 0 | 0 | 0 | 0 | 0 |
| 0 | 0 | 0 | 0 | 0 |
| 0 | 0 | 0 | 0 | 0 |
| 0 | 0 | 0.903768581 | 0 | 0.091714374 |
| 0 | 0 | 0 | 0 | 0 |
| 0.000894217 | 0 | 0 | 0 | 0 |
| 0 | 0 | 0 | 0 | 0 |

| | | | | |
|---|---|---|---|---|
| 0 | 0 | 0 | 0 | 0 |
| 0 | 0 | 0 | 0 | 0 |
| 0 | 0 | 0 | 0 | 0 |
| 0 | 0 | 0 | 0 | 0 |
| 0 | 0 | 0 | 0 | 0 |
| 0 | 0 | 0 | 0 | 0 |
| 0 | 0 | 0 | 0 | 0 |
| 0 | 0 | 0 | 0 | 0 |
| 0.059705912 | 0.025268342 | 0.63234228 | 0.408230712 | 0.005917471 |
| 0.005226186 | 0 | 0 | 0 | 0 |
| 0 | 0 | 0 | 0 | 0 |
| 0 | 0 | 0 | 0 | 0 |
| 0 | 0 | 0.004934101 | 0 | 0 |
| 0 | 0 | 0 | 0 | 0 |
| 2.390890505 | 0 | 0 | 0 | 1.216945019 |
| 0.385142342 | 0 | 0.058821089 | 0.082125616 | 0 |
| 0 | 0 | 0 | 0 | 0 |
| 0 | 0 | 0 | 0 | 0 |
| 0 | 0 | 0 | 0 | 0 |
| 0 | 0 | 0 | 0 | 0 |
| 0 | 0 | 0 | 0 | 0 |
| 0 | 0 | 0 | 0 | 0 |
| 0 | 0 | 0 | 0 | 0 |
| 0 | 0 | 0 | 0 | 0 |
| 0.001090255 | 0 | 0.002691295 | 0 | 0.019484575 |
| 10.56117162 | 0.013905814 | 0 | 0 | 6.38938866 |
| 0 | 0 | 0 | 0 | 0.012294201 |
| 0.004602052 | 0 | 0.001888518 | 0 | 0.000755936 |
| 0.032599271 | 0 | 0.031713828 | 0 | 0 |
| 0 | 0 | 0 | 0 | 0 |
| 0 | 0 | 0 | 0 | 0 |
| 0.056872575 | 0 | 0 | 0 | 0.000784261 |
| 0 | 0 | 0 | 0 | 0 |
| 0 | 0 | 0.00581179 | 0.000220138 | 0.01663104 |
| 0 | 0.030353797 | 0.000926709 | 0.001209058 | 0.003674105 |
| 0 | 0.006721299 | 0.004803854 | 0.0089739 | 0 |
| 1.252882229 | 0 | 0 | 0 | 0 |
| 0.720134614 | 1.982613111 | 0.202674604 | 0.015344385 | 0.783378202 |
| 0 | 0 | 0 | 0 | 0 |
| 0 | 0 | 0 | 0 | 0 |
| 0 | 0 | 0 | 0 | 0 |
| 0.021533509 | 0 | 0 | 0 | 0 |
| 5.076473341 | 1.932865224 | 0 | 0 | 0.116527062 |
| 0 | 0 | 0 | 0 | 0 |
| 0 | 0 | 0 | 0 | 0 |
| 0.001156471 | 0 | 0 | 0 | 0 |
| 0 | 0 | 0 | 0 | 0.016478588 |
| 0.183011461 | 0.008583664 | 0.013731753 | 0.005933622 | 0.014046039 |

| | | | | |
|---|---|---|---|---|
| 0.061977147 | 0 | 0 | 0 | 0 |
| 0 | 0 | 0 | 0 | 0 |
| 0 | 0 | 0 | 0 | 0 |
| 0 | 0 | 0 | 0 | 0 |
| 0.039059842 | 0 | 0.003017688 | 0 | 0 |
| 0 | 0 | 0.020577538 | 0.000276668 | 0 |
| 0 | 0 | 0 | 0 | 0 |
| 0 | 0 | 0 | 0 | 0 |
| 0 | 0 | 0 | 0 | 0 |
| 0.02440762 | 0 | 0 | 0 | 0 |
| 0 | 0 | 0 | 0 | 0 |
| 1.817604081 | 0.056120599 | 0.107372469 | 0 | 0.743479074 |
| 0.156237868 | 0 | 0.32440623 | 0 | 0 |
| 0 | 0 | 0 | 0 | 0 |
| 0.000703077 | 0.545054291 | 0.129694295 | 0.015616489 | 4.9583026 |
| 0 | 0 | 0 | 0 | 0 |
| 0 | 0 | 0 | 0 | 0 |
| 0 | 0 | 0 | 0 | 0 |
| 0 | 0 | 0 | 0.000485383 | 0 |
| 0.014862142 | 0 | 0 | 0 | 0 |
| 0 | 0 | 0 | 0 | 0 |
| 0 | 0 | 0 | 0 | 0 |
| 0.081642925 | 0.037117151 | 0 | 0 | 0.241786795 |
| 0 | 0 | 0 | 0 | 0 |
| 0 | 0 | 0 | 0 | 0 |
| 0 | 0 | 0 | 0 | 0 |
| 0 | 0 | 0 | 0 | 0 |
| 0 | 0 | 0 | 0 | 0 |
| 0 | 0 | 0 | 0 | 0 |
| 0 | 0 | 0 | 0 | 0 |
| 0 | 0 | 0 | 0 | 0 |
| 0.109937077 | 0.251701003 | 0.017135642 | 0.003305569 | 0.23784136 |
| 0 | 0 | 0 | 0 | 0 |
| 0.011052902 | 0 | 0.117036607 | 0.001148804 | 0.023727924 |
| 0 | 0.005204518 | 0.004752542 | 0.007626666 | 0 |
| 0.019218288 | 0.001087987 | 0.009496235 | 0 | 0.020950146 |
| 0 | 0 | 0 | 0 | 0 |
| 0 | 0 | 0 | 0 | 0 |
| 0 | 0 | 0.001455992 | 0 | 0 |
| 0 | 0 | 0 | 0 | 0 |
| 0 | 0 | 0 | 0 | 0 |
| 0 | 0 | 0 | 0 | 0 |
| 0 | 0 | 0 | 0 | 0 |
| 0 | 0 | 0 | 0 | 0 |
| 0.00334432 | 0.0004157 | 0.082476487 | 0.004855983 | 0.003364284 |
| 0 | 0 | 0 | 0 | 0 |
| 5.449909403 | 0 | 1.481430759 | 0 | 0 |
| 0 | 0 | 0 | 0.000866583 | 0.000329173 |

| | | | | |
|---|---|---|---|---|
| 0 | 0 | 0 | 0 | 0 |
| 0 | 0 | 0 | 0 | 0 |
| 0.02601667 | 0 | 0 | 0.007675593 | 0.004799326 |
| 0 | 0 | 0 | 0 | 0 |
| 0 | 0 | 0 | 0 | 0 |
| 0 | 0 | 0 | 0 | 0 |
| 0 | 0 | 0 | 0 | 0 |
| 0 | 0 | 0 | 0 | 0.017676807 |
| 0 | 0 | 0 | 0 | 0 |
| 0 | 0 | 0 | 0 | 0 |
| 0 | 0 | 0 | 0 | 0 |
| 0 | 0 | 0 | 0 | 0 |
| 0 | 0 | 0 | 0 | 0 |
| 0.014266713 | 0 | 0 | 0 | 0 |
| 0.00148173 | 0 | 0 | 0 | 0 |
| 0 | 0 | 0 | 0 | 0 |
| 0 | 0 | 0 | 0 | 0 |
| 0 | 0 | 0 | 0 | 0 |
| 0.39915819 | 0.049615538 | 0.195488031 | 0 | 0 |
| 0 | 0 | 0 | 0 | 0 |
| 0 | 0.0010417 | 0 | 0 | 0 |
| 0 | 0 | 0 | 0 | 0 |
| 0 | 0 | 0 | 0 | 0 |
| 0 | 0 | 0 | 0 | 0 |
| 0 | 0 | 0 | 0 | 0 |
| 0 | 0.011610041 | 0.020557659 | 0.056295963 | 0 |
| 0 | 0 | 0 | 0 | 0 |
| 0.016032152 | 0.009343668 | 0.000213306 | 0 | 0.017865197 |
| 0 | 0 | 0 | 0 | 0 |
| 0.093102647 | 0 | 0 | 0 | 1.052131202 |
| 0 | 0 | 0 | 0 | 0 |
| 0 | 0 | 0 | 0 | 0 |
| 0.002938016 | 0 | 0 | 0 | 0 |
| 0 | 0 | 0 | 0 | 0 |
| 0.000425568 | 0 | 0.006470559 | 0 | 0 |
| 0 | 0 | 0 | 0.000808512 | 0 |
| 0 | 0 | 0 | 0 | 0 |
| 0.003702195 | 0.000613579 | 0 | 0 | 0 |
| 0 | 0 | 0 | 0 | 0 |
| 0 | 0 | 0 | 0 | 0 |
| 13.27051238 | 18.80891754 | 2.46033409 | 0.570421622 | 0.894623332 |
| 0.44880344 | 0.544024208 | 0 | 0.298390174 | 0.112432283 |
| 0 | 0.099239362 | 0 | 0 | 0 |
| 0 | 0 | 0 | 0 | 0 |
| 0 | 0 | 0 | 0 | 0 |
| 0.00316999 | 0 | 0 | 0 | 0 |
| 0 | 0 | 0 | 0 | 0 |
| 0 | 0 | 0.000959769 | 0 | 0.001121168 |

| | | | | |
|---|---|---|---|---|
| 0 | 0 | 0 | 0 | 0.018643862 |
| 0.005913097 | 0 | 0 | 0.002401164 | 0.000524188 |
| 0.007366575 | 0 | 0 | 0 | 0 |
| 0 | 0 | 0 | 0 | 0 |
| 2.930260187 | 7.561665473 | 0 | 0 | 1.752855828 |
| 0 | 0 | 0 | 0 | 0 |
| 0 | 0 | 0 | 0 | 0 |
| 0 | 0 | 0 | 0 | 0 |
| 0.094338865 | 0.236446302 | 0.094490788 | 0.572536056 | 0.212297242 |
| 0.002070965 | 0 | 0 | 0 | 0 |
| 0 | 0 | 0 | 0.002057998 | 0 |
| 0 | 0 | 0 | 0 | 0 |
| 0.58758152 | 0 | 0 | 0 | 0.00173175 |
| 0.040341054 | 0.056992582 | 0.171753875 | 0.039959201 | 0 |
| 0 | 0 | 0 | 0 | 0 |
| 2.065044947 | 0.10446014 | 0 | 0 | 0.004727312 |
| 0.000950827 | 0.004659991 | 0.000291567 | 0 | 0.064806592 |
| 0 | 0 | 0 | 0 | 0 |
| 0 | 0 | 0 | 0 | 0 |
| 0 | 0 | 0 | 0 | 0 |
| 0 | 0 | 0 | 0 | 0 |
| 0 | 0 | 0 | 0 | 0 |
| 0 | 0 | 0 | 0 | 0 |
| 0 | 0 | 0 | 0 | 0 |
| 0 | 0 | 0 | 0 | 0 |
| 0 | 0 | 0 | 0 | 0 |
| 0.000590559 | 0 | 0 | 0 | 0 |
| 0 | 0 | 0.002360158 | 0 | 0 |
| 0 | 0 | 0 | 0 | 0 |
| 0 | 0 | 0 | 0 | 0 |
| 0 | 0 | 0 | 0 | 0 |
| 0.000556632 | 0 | 0 | 0 | 0 |
| 0 | 0 | 0 | 0 | 0 |
| 0 | 0 | 0 | 0 | 0 |
| 0.030899566 | 0 | 0 | 0 | 0 |
| 0 | 0.003673049 | 0.059651023 | 0 | 0.06248644 |
| 0 | 0.007658768 | 0 | 0.000893662 | 0 |
| 0 | 0 | 0 | 0 | 0 |
| 0 | 0 | 0 | 0 | 0 |
| 0 | 0 | 0 | 0 | 0 |
| 0 | 0.26883272 | 10.99008606 | 0 | 13.62185459 |
| 0 | 0 | 0 | 0.023292413 | 0 |
| 0 | 0 | 0 | 0 | 0 |
| 0 | 0 | 0 | 0 | 0 |
| 0 | 0 | 0 | 0 | 0 |
| 0 | 0 | 0 | 0 | 0 |
| 0 | 0 | 0 | 0 | 0 |
| 0 | 0 | 0 | 0 | 0 |

| | | | | |
|---|---|---|---|---|
| 0.025682756 | 0 | 0 | 0 | 0 |
| 0 | 0 | 0 | 0 | 0 |
| 0 | 0 | 0 | 0 | 0 |
| 0.00513317 | 0 | 0 | 0 | 0 |
| 0 | 0 | 0 | 0 | 0.000922008 |
| 0 | 0 | 0 | 0 | 0 |
| 0.00257454 | 0.001033036 | 0 | 0 | 0.195136225 |
| 0 | 0 | 0 | 0 | 0 |
| 0 | 0 | 0 | 0 | 0 |
| 0 | 0 | 0 | 0.049358484 | 0 |
| 0 | 0 | 0 | 0 | 0 |
| 0 | 0 | 0 | 0 | 0 |
| 0 | 0 | 0 | 0 | 0 |
| 0 | 0 | 0 | 0 | 0 |
| 0 | 0 | 0 | 0 | 0 |
| 0 | 0 | 0 | 0 | 0 |
| 0 | 0 | 0 | 0 | 0 |
| 0.000521084 | 0 | 0.017057405 | 0 | 0 |
| 0 | 0 | 0 | 0 | 0 |
| 0 | 0 | 0 | 0 | 0 |
| 0.066179851 | 0.023235363 | 0 | 0.000243866 | 0 |
| 0 | 0 | 0 | 0 | 0 |
| 0 | 0 | 0 | 0 | 0 |
| 0.000632641 | 0 | 0.004073929 | 0.000877003 | 0 |
| 0 | 0 | 0 | 0 | 0 |
| 0 | 0 | 0 | 0.000376242 | 0 |
| 0 | 0 | 0 | 0 | 0 |
| 0 | 0 | 0 | 0 | 0 |
| 0 | 0 | 0 | 0.19793683 | 0 |
| 0.074480385 | 2.691594564 | 0.122934689 | 0.097534629 | 0.579849087 |
| 0 | 0 | 0 | 0 | 0 |
| 0 | 0 | 0 | 0 | 0 |
| 0 | 0 | 0 | 0 | 0 |
| 0 | 0 | 0 | 0 | 0 |
| 0 | 0 | 0 | 0 | 0 |
| 0 | 0 | 0 | 0 | 0 |
| 0.051295502 | 0.152835048 | 0.123105613 | 0.037808965 | 0.003410458 |
| 0 | 0 | 0 | 0 | 0 |
| 0 | 0 | 0 | 0 | 0 |
| 0 | 0 | 0 | 0 | 0 |
| 0 | 0 | 0 | 0 | 0 |
| 0 | 0 | 0 | 0 | 0 |
| 0 | 0 | 0 | 0 | 0 |
| 0.009819877 | 0 | 0 | 0 | 0 |
| 0 | 0 | 0 | 0 | 0 |
| 0 | 0 | 0 | 0 | 0 |
| 0 | 0 | 0 | 0 | 0 |
| 0 | 0 | 0 | 0 | 0 |

| | | | | |
|---|---|---|---|---|
| 0.0108919 | 0 | 0.045444282 | 0.002745269 | 0.003258739 |
| 0 | 0 | 0 | 0 | 0 |
| 0.013429338 | 0 | 0 | 0 | 0 |
| 0 | 0 | 0 | 0 | 0 |
| 0 | 0 | 0 | 0 | 0 |
| 0 | 0 | 0 | 0 | 0 |
| 0 | 0 | 0 | 0 | 0 |
| 0.785083079 | 1.159142757 | 0 | 0 | 1.017291974 |
| 0.012608649 | 0.252532607 | 0.030734527 | 0 | 0.584312646 |
| 0.000446327 | 0.009499175 | 0.220032206 | 0.035998497 | 0.002255278 |
| 0 | 0 | 0 | 0 | 0 |
| 0 | 0 | 0 | 0 | 0 |
| 0 | 0 | 0 | 0 | 0 |
| 0.011391137 | 0 | 0.101455984 | 0.13109364 | 0 |
| 0 | 0 | 0 | 0 | 0 |
| 0 | 0 | 0 | 0 | 0 |
| 0.009340805 | 0 | 0 | 0 | 0 |
| 0 | 0 | 0 | 0 | 0 |
| 0 | 0 | 0 | 0 | 0 |
| 0.01640799 | 0 | 0.114399116 | 0 | 0 |
| 0 | 0 | 0 | 0 | 0 |
| 0 | 0 | 0 | 0 | 0 |
| 0 | 0 | 0 | 0 | 0 |
| 0 | 0 | 0 | 0 | 0 |
| 0.009222297 | 0.026672297 | 0 | 0 | 0 |
| 0 | 0 | 0 | 0 | 0 |
| 0 | 0 | 0 | 0 | 0 |
| 0 | 0 | 0 | 0 | 0 |
| 0.000920538 | 0 | 0 | 0.001836814 | 0 |
| 0 | 0 | 0 | 0 | 0 |
| 0 | 0 | 0 | 0 | 0 |
| 0 | 0 | 0 | 0 | 0 |
| 0 | 0 | 0 | 0 | 0 |
| 0 | 0 | 0 | 0 | 0 |
| 0 | 0.002984433 | 0 | 0.002492651 | 0 |
| 0 | 0 | 0 | 0 | 0.000924 |
| 0 | 0 | 0 | 0 | 0 |
| 0.008480063 | 0 | 0 | 0 | 0 |
| 0 | 0 | 0 | 0 | 0 |
| 0 | 0.001683849 | 0 | 0 | 0.449272708 |
| 0 | 0 | 0 | 0 | 0 |
| 1.184758183 | 0.086458546 | 0 | 0.075280988 | 0.063210249 |
| 0 | 0 | 0 | 0 | 0 |
| 0 | 0 | 0 | 0 | 0 |
| 0 | 0 | 0 | 0 | 0 |
| 0.000847447 | 0 | 0 | 0 | 0 |
| 0 | 0 | 0 | 0 | 0 |
| 0 | 0 | 0 | 0 | 0 |

| | | | | |
|---|---|---|---|---|
| 0 | 0 | 0 | 0 | 0 |
| 0.09093135 | 0 | 0.01008901 | 0 | 0.00647054 |
| 0 | 0 | 0 | 0 | 0 |
| 0 | 0 | 0 | 0 | 0 |
| 0.052228355 | 0.038147662 | 0.241827814 | 0.009310061 | 0.224660313 |
| 0 | 0 | 0 | 0 | 0 |
| 0 | 0 | 0 | 0 | 0.009629022 |
| 0.031693966 | 0.004951377 | 0.051674405 | 0.018552213 | 0.000690892 |
| 0 | 0 | 0 | 0 | 0 |
| 0 | 0 | 0 | 0 | 0 |
| 0 | 0 | 0 | 0 | 0 |
| 0 | 0 | 0 | 0 | 0 |
| 0 | 0 | 0 | 0 | 0 |
| 0 | 0 | 0 | 0 | 0 |
| 0.034015793 | 0 | 0.001439376 | 0 | 0 |
| 0 | 0 | 0 | 0 | 0 |
| 0 | 0 | 0 | 0 | 0 |
| 0 | 0 | 0 | 0 | 0 |
| 0 | 0 | 0 | 0 | 0 |
| 0 | 0 | 0 | 0 | 0 |
| 0 | 0 | 0 | 0 | 0 |
| 0 | 0 | 0 | 0 | 0 |
| 0 | 0 | 0 | 0 | 0 |
| 0 | 0 | 0.000723884 | 0.000566662 | 0 |
| 0 | 0 | 0 | 0 | 0 |
| 0 | 0 | 0 | 0 | 0 |
| 0.076904492 | 0.004572315 | 0.003751753 | 0 | 0 |
| 0 | 0 | 0 | 0 | 0 |
| 0 | 0 | 0 | 0 | 0 |
| 0 | 0 | 0 | 0 | 0 |
| 0 | 0 | 0 | 0 | 0 |
| 0 | 0 | 0 | 0 | 0 |
| 0 | 0 | 0 | 0 | 0 |
| 0 | 0 | 0 | 0 | 0 |
| 0 | 0 | 0 | 0 | 0 |
| 0 | 0 | 0 | 0 | 0 |
| 0 | 0 | 0 | 0 | 0 |
| 0 | 0 | 0 | 0 | 0 |
| 0 | 0 | 0 | 0 | 0 |
| 0 | 0 | 0 | 0 | 0.000384912 |
| 0 | 0 | 0 | 0 | 0 |
| 0 | 0 | 0 | 0.018182394 | 0 |
| 0 | 0 | 0 | 0 | 0 |
| 0 | 0 | 0 | 0 | 0 |
| 0 | 0 | 0 | 0 | 0 |
| 0 | 0 | 0 | 0 | 0 |
| 0 | 0 | 0 | 0 | 0 |
| 0 | 0 | 0 | 0 | 0 |

| | | | | |
|---|---|---|---|---|
| 0.003609657 | 0 | 0 | 0 | 0.009771159 |
| 0 | 0 | 0 | 0 | 0 |
| 0 | 0 | 0 | 0 | 0 |
| 0 | 0 | 0.001376614 | 0 | 0 |
| 0 | 0 | 0 | 0 | 0 |
| 0 | 0 | 0 | 0.000282337 | 0.004528181 |
| 0 | 0 | 0 | 0 | 0 |
| 0 | 0 | 0 | 0 | 0 |
| 0 | 0 | 0 | 0 | 0 |
| 0 | 0 | 0 | 0 | 0 |
| 0 | 0 | 0 | 0 | 0 |
| 0 | 0 | 0 | 0 | 0 |
| 0 | 0 | 0 | 0 | 0 |
| 0 | 0 | 0 | 0 | 0 |
| 0 | 0 | 0 | 0 | 0 |
| 0 | 0 | 0 | 0 | 0 |
| 0 | 0 | 0 | 0 | 0 |
| 0 | 0 | 0 | 0 | 0 |
| 0 | 0 | 0.005044126 | 0 | 0 |
| 0 | 0 | 0 | 0 | 0 |
| 0.001194048 | 0.009529849 | 0.013295824 | 0.020784785 | 0.004048792 |
| 0 | 0 | 0 | 0 | 0 |
| 0 | 0 | 0.018106666 | 0 | 0.002255881 |
| 1.593954694 | 0 | 0 | 0 | 0 |
| 0 | 0 | 0 | 0 | 0 |
| 0 | 0 | 0 | 0 | 0 |
| 0 | 0 | 0 | 0 | 0 |
| 0 | 0 | 0 | 0 | 0 |
| 0 | 0 | 0 | 0 | 0 |
| 0 | 0 | 0 | 0 | 0 |
| 0 | 0 | 0 | 0 | 0 |
| 0 | 0 | 0 | 0 | 0 |
| 0.00176529 | 0.029439703 | 0.564311543 | 0.175749674 | 0.0091547 |
| 0 | 0 | 0 | 0 | 0 |
| 0.147798499 | 0 | 0.290137009 | 0 | 0 |
| 0 | 0 | 0 | 0 | 0.001356233 |
| 0 | 0 | 0 | 0 | 0 |
| 0 | 0 | 0 | 0 | 0 |
| 0.000819244 | 0.002342146 | 0.002637789 | 0.009601692 | 0.001307249 |
| 0 | 0 | 0 | 0 | 0 |
| 0 | 0 | 0 | 0 | 0 |
| 0 | 0 | 0 | 0 | 0 |
| 0.010920542 | 0.043088422 | 0.034236454 | 0.005553266 | 0 |
| 0 | 0 | 0 | 0 | 0 |
| 0 | 0 | 0 | 0 | 0 |
| 0.548669001 | 0 | 0 | 0 | 0.000364184 |
| 0 | 0 | 0 | 0 | 0 |
| 0 | 0 | 0 | 0 | 0 |

| | | | | |
|---|---|---|---|---|
| 0 | 0 | 0 | 0 | 0 |
| 0 | 0 | 0 | 0 | 0 |
| 0 | 0 | 0 | 0 | 0 |
| 0 | 0 | 0 | 0 | 0 |
| 0 | 0 | 0 | 0 | 0 |
| 0 | 0 | 0 | 0 | 0 |
| 0 | 0 | 0 | 0 | 0 |
| 0 | 0.260893431 | 1.546922034 | 0.103577956 | 0.042230028 |
| 0.238351336 | 0 | 0 | 0 | 0 |
| 0.020686699 | 0 | 0 | 0 | 0 |
| 0 | 0 | 0 | 0 | 0 |
| 0 | 0 | 0 | 0 | 0 |
| 0 | 0 | 0 | 0 | 0 |
| 0.077774672 | 0 | 0 | 0 | 0 |
| 0.287240065 | 0 | 0 | 0 | 0 |
| 0.002775467 | 0 | 0.04517846 | 0.003206261 | 0.014393423 |
| 0 | 0 | 0 | 0 | 0 |
| 0 | 0 | 0 | 0 | 0 |
| 0 | 0 | 0 | 0 | 0 |
| 0 | 0 | 0 | 0 | 0 |
| 0 | 0.031461224 | 0.011493951 | 0 | 0.014133504 |
| 0 | 0 | 0 | 0 | 0 |
| 0 | 0 | 0 | 0 | 0 |
| 0 | 0 | 0 | 0.00155302 | 0 |
| 0 | 0 | 0 | 0 | 0 |
| 0 | 0 | 0 | 0 | 0 |
| 0 | 0 | 0 | 0 | 0 |
| 0 | 0 | 0 | 0 | 0 |
| 0 | 0 | 0 | 0 | 0 |
| 0 | 0 | 0 | 0 | 0.265203166 |
| 2.419243478 | 0.080973181 | 0 | 0 | 0 |
| 0 | 0 | 0 | 0 | 0 |
| 0 | 0 | 0 | 0 | 0 |
| 0 | 0 | 0 | 0 | 0 |
| 0 | 0 | 0 | 0 | 0 |
| 0 | 0 | 0 | 0 | 0 |
| 0 | 0 | 0 | 0 | 0 |
| 0 | 0 | 0 | 0 | 0 |
| 0 | 0 | 0 | 0 | 0 |
| 0 | 0 | 0 | 0 | 0 |
| 0 | 0 | 0 | 0 | 0 |
| 0 | 0 | 0 | 0 | 0 |
| 0 | 0 | 0 | 0 | 0 |
| 0.002141097 | 0 | 0 | 0 | 0 |
| 0.001541425 | 0 | 0 | 0 | 0 |
| 0 | 0 | 0 | 0 | 0 |
| 0.020677807 | 0.001955491 | 1.133690001 | 2.325208221 | 0.002014966 |
| 0 | 0 | 0 | 0 | 0 |

| | | | | |
|---|---|---|---|---|
| 0 | 0 | 0 | 0 | 0 |
| 0 | 0 | 0 | 0 | 0 |
| 0 | 0 | 0 | 0 | 0 |
| 0 | 0 | 0.010476231 | 0 | 0 |
| 0.054480262 | 0.001348521 | 0.026577963 | 0.13204013 | 0 |
| 0 | 0 | 0 | 0 | 0 |
| 0 | 0 | 0 | 0 | 0 |
| 0 | 0 | 0 | 0 | 0 |
| 0 | 0 | 0 | 0 | 0 |
| 0 | 0 | 0 | 0 | 0 |
| 0 | 0 | 0 | 0 | 0 |
| 0.002455776 | 0 | 0.016516979 | 0.000722133 | 0 |
| 0.000564991 | 0 | 0 | 0 | 0 |
| 0 | 0 | 0 | 0 | 0 |
| 0 | 0 | 0.030932108 | 0 | 0 |
| 0.063911774 | 0 | 0 | 0 | 1.266146851 |
| 0.005162675 | 0 | 0.000608147 | 0.000555406 | 0 |
| 0 | 0 | 0 | 0 | 0 |
| 0.090125125 | 0.00697016 | 0.121807775 | 0.100299276 | 0.075070254 |
| 0 | 0 | 0 | 0 | 0.057855676 |
| 0 | 0 | 0 | 0 | 0 |
| 0 | 0 | 0 | 0 | 0 |
| 0 | 0 | 0 | 0 | 0 |
| 0 | 0 | 0 | 0 | 0 |
| 0 | 0 | 0 | 0 | 0 |
| 0 | 0 | 0 | 0 | 0 |
| 0.015046354 | 0 | 0.057773221 | 0.003677961 | 0.000517438 |
| 0.135458303 | 0 | 0 | 0 | 0 |
| 0 | 0 | 0 | 0 | 0 |
| 0 | 0 | 0 | 0 | 0 |
| 0.00147033 | 0 | 0.016648403 | 0.001111776 | 0 |
| 0.031762562 | 0 | 0 | 0.002301626 | 0 |
| 0 | 0 | 0 | 0 | 0 |
| 0 | 0 | 0 | 0 | 0 |
| 0 | 0 | 0 | 0 | 0 |
| 0 | 0 | 0 | 0 | 0 |
| 0 | 0 | 0 | 0 | 0 |
| 0.052890187 | 0.002657958 | 0.013359721 | 0.001301836 | 0.018873623 |
| 0 | 0 | 0 | 0 | 0.005452229 |
| 0.016713966 | 0 | 0 | 0.004130101 | 0 |
| 0 | 0 | 0 | 0 | 0 |
| 0 | 0 | 0 | 0 | 0 |
| 0 | 0 | 0 | 0 | 0 |
| 0.001685008 | 0 | 0.003370297 | 0.039091783 | 0 |
| 0 | 0 | 0 | 0 | 0 |
| 0 | 0 | 0 | 0 | 0 |
| 0 | 0 | 0 | 0 | 0 |
| 0 | 0 | 0 | 0 | 0 |

| | | | | |
|---|---|---|---|---|
| 0 | 0 | 0 | 0 | 0 |
| 0 | 0 | 0 | 0 | 0 |
| 0 | 0 | 0 | 0 | 0 |
| 0 | 0 | 0.00111993 | 0.000809252 | 0 |
| 0 | 0.012202714 | 0 | 0 | 0.000475175 |
| 0 | 0 | 0 | 0.000363255 | 0.000766573 |
| 0 | 0 | 0 | 0 | 0 |
| 0.007658734 | 0 | 0 | 0 | 0 |
| 0 | 0 | 0 | 0 | 0.003121534 |
| 0 | 0 | 0 | 0 | 0 |
| 0 | 0 | 0 | 0 | 0 |
| 0 | 0 | 0 | 0 | 0 |
| 0.005917464 | 0.0204962 | 0.006228847 | 0 | 0.001634254 |
| 0 | 0 | 0 | 0 | 0 |
| 0 | 0 | 0 | 0 | 0 |
| 0 | 0 | 0 | 0 | 0 |
| 0 | 0 | 0.048644372 | 0 | 0.173863218 |
| 0 | 0 | 0 | 0 | 0 |
| 0 | 0 | 0 | 0 | 0 |
| 0 | 0 | 0 | 0 | 0 |
| 0 | 0 | 0 | 0 | 0 |
| 0 | 0 | 0 | 0 | 0 |
| 0.004345503 | 0.000713989 | 0.121662354 | 0.008308948 | 0.002630143 |
| 0.010078865 | 0.05156291 | 0 | 0 | 0 |
| 0 | 0 | 0 | 0 | 0 |
| 0 | 0 | 0 | 0 | 0 |
| 0 | 0.002455756 | 0 | 0 | 0.014163473 |
| 0 | 0 | 0 | 0 | 0 |
| 0 | 0 | 0 | 0 | 0 |
| 0 | 0 | 0 | 0 | 0 |
| 0.002673223 | 0 | 0 | 0 | 0.022527686 |
| 0 | 0 | 0 | 0 | 0 |
| 0 | 0 | 0 | 0 | 0 |
| 0 | 0 | 0 | 0 | 0 |
| 0 | 0 | 0 | 0 | 0 |
| 0 | 0 | 0 | 0 | 0 |
| 0 | 0 | 0 | 0 | 0 |
| 0.123523939 | 0 | 0 | 0 | 0 |
| 0 | 0.014144569 | 0.001441285 | 0 | 0.471537246 |
| 0 | 0 | 0 | 0 | 0 |
| 0 | 0 | 0 | 0 | 0 |
| 0 | 0 | 0 | 0 | 0 |
| 0 | 0 | 0.10993318 | 0 | 0 |
| 0 | 0 | 0 | 0 | 0 |
| 0 | 0 | 0 | 0 | 0 |
| 0.450101967 | 0.021334088 | 0.17679887 | 0 | 0.02163184 |
| 0 | 0 | 0 | 0.102694287 | 0 |
| 0 | 0 | 0 | 0 | 0 |

| | | | | |
|---|---|---|---|---|
| 0 | 0 | 0 | 0 | 0 |
| 0 | 0 | 0 | 0 | 0 |
| 0 | 0 | 0 | 0 | 0 |
| 0 | 0 | 0 | 0 | 0 |
| 0 | 0 | 0 | 0 | 0 |
| 0.002404568 | 0 | 0 | 0 | 0 |
| 0 | 0 | 0 | 0 | 0 |
| 0 | 0 | 0 | 0 | 0 |
| 0 | 0 | 0 | 0 | 0 |
| 0 | 0 | 0 | 0 | 0.000514849 |
| 0 | 0 | 0 | 0 | 0 |
| 0 | 0 | 0 | 0 | 0 |
| 0 | 0 | 0 | 0 | 0 |
| 0 | 0 | 0 | 0 | 0 |
| 0 | 0 | 0 | 0 | 0 |
| 0 | 0 | 0 | 0 | 0 |
| 0 | 0 | 0.000361847 | 0 | 0 |
| 0 | 0 | 0 | 0 | 0 |
| 0 | 0 | 0 | 0 | 0 |
| 0.001216677 | 0.002173981 | 0 | 0.000383324 | 0 |
| 0 | 0 | 0 | 0 | 0 |
| 0 | 0 | 0 | 0 | 0 |
| 0 | 0 | 0 | 0 | 0 |
| 0 | 0 | 0 | 0 | 0 |
| 0 | 0 | 0 | 0 | 0 |
| 0 | 0 | 0 | 0 | 0 |
| 0 | 0 | 0 | 0 | 0 |
| 0 | 0.089659039 | 0 | 0 | 0 |
| 0 | 0 | 0 | 0 | 0 |
| 0 | 0 | 0 | 0 | 0 |
| 0 | 0.00224121 | 0.007993018 | 0.000905617 | 0.001250912 |
| 0 | 0 | 0 | 0 | 0 |
| 0.022755811 | 0.052045454 | 0.118206913 | 3.571897402 | 0.005083525 |
| 0.001395238 | 0.010932433 | 0 | 0 | 0.020531874 |
| 0 | 0 | 0 | 0 | 0 |
| 0 | 0 | 0 | 0 | 0 |
| 0 | 0 | 0 | 0 | 0 |
| 0 | 0 | 0 | 0 | 0 |
| 0 | 0 | 0 | 0 | 0 |
| 0 | 0 | 0 | 0 | 0 |
| 0 | 0 | 0 | 0 | 0 |
| 0.051420345 | 0.002021929 | 0.170652396 | 0.01840242 | 0.019018917 |
| 0.0776974 | 0 | 0.007588287 | 0 | 0.032511351 |
| 0 | 0 | 0 | 0 | 0 |
| 0 | 0 | 0 | 0 | 0 |
| 0.004833893 | 0.003559611 | 0.004480568 | 0 | 0 |
| 0 | 0 | 0 | 0 | 0 |
| 0 | 0 | 0 | 0 | 0 |

| | | | | |
|---|---|---|---|---|
| 0 | 0 | 0 | 0 | 0 |
| 0.004778767 | 0 | 0 | 0 | 0.001906341 |
| 0.009303507 | 0.011880391 | 0.019470922 | 0 | 0.012321666 |
| 0 | 0 | 0 | 0 | 0 |
| 1.144716873 | 0 | 0.060432401 | 0.002246767 | 0.099686439 |
| 0.264501306 | 0 | 0 | 0 | 0 |
| 0 | 0 | 0 | 0 | 0 |
| 0 | 0 | 0 | 0 | 0 |
| 0 | 0 | 0 | 0 | 0 |
| 0 | 0 | 0 | 0 | 0 |
| 0 | 0 | 0 | 0 | 0 |
| 0 | 0 | 0 | 0 | 0 |
| 0 | 0 | 0 | 0 | 0 |
| 0 | 0 | 0 | 0 | 0 |
| 0 | 0 | 0 | 0 | 0 |
| 0 | 0 | 0 | 0 | 0 |
| 0 | 0 | 0 | 0 | 0 |
| 0 | 0 | 0 | 0 | 0 |
| 0 | 0 | 0 | 0 | 0 |
| 0 | 0 | 0 | 0 | 0 |
| 0.021164021 | 0 | 0 | 0 | 0 |
| 0.040710763 | 0 | 0 | 0.001037419 | 0 |
| 0.01367571 | 0 | 0 | 0 | 0 |
| 0 | 0 | 0 | 0 | 0 |
| 0 | 0 | 0 | 0 | 0 |
| 0.009255865 | 0.004851309 | 0.100912434 | 0.001944093 | 0.004184652 |
| 0 | 0 | 0 | 0 | 0 |
| 0 | 0 | 0 | 0 | 0 |
| 0 | 0 | 0 | 0 | 0 |
| 0 | 0 | 0 | 0 | 0 |
| 0 | 0 | 0 | 0 | 0 |
| 0 | 0 | 0.001379319 | 0 | 0 |
| 0 | 0 | 0 | 0 | 0 |
| 0 | 0 | 0 | 0 | 0 |
| 0 | 0 | 0 | 0 | 0 |
| 0 | 0 | 0 | 0 | 0 |
| 0 | 0.006720144 | 0 | 0 | 0 |
| 0.026872522 | 0 | 0 | 0 | 0 |
| 0 | 1.218741565 | 2.996195268 | 1.763760042 | 0 |
| 0 | 0 | 0 | 0 | 0 |
| 0 | 0 | 0 | 0 | 0 |
| 0 | 0 | 0.000898068 | 0 | 0.000534083 |
| 0.008824808 | 0 | 0.032235715 | 0.002048665 | 0.001852833 |
| 0 | 0 | 0 | 0 | 0 |
| 0 | 0 | 0 | 0 | 0 |
| 0 | 0 | 0 | 0 | 0 |
| 0.007815922 | 0 | 0.060117734 | 0.015222562 | 0 |
| 0 | 0 | 0 | 0 | 0 |

| | | | | |
|---|---|---|---|---|
| 0.074867008 | 0 | 0 | 0 | 0 |
| 0.005379412 | 0 | 0.033678815 | 0.067134097 | 0.002265169 |
| 0 | 0 | 0 | 0 | 0 |
| 0 | 0 | 0.061803546 | 0 | 0 |
| 0 | 0 | 0 | 0 | 0 |
| 0 | 0 | 0 | 0 | 0 |
| 0 | 0 | 0 | 0 | 0 |
| 0 | 0 | 0 | 0 | 0 |
| 0 | 0 | 0 | 0 | 0 |
| 0 | 0 | 0 | 0 | 0 |
| 0 | 0 | 0 | 0 | 0 |
| 0 | 0 | 0 | 0 | 0 |
| 0.099829936 | 0.004666429 | 0 | 0 | 0.029489962 |
| 0 | 0 | 0 | 0 | 0 |
| 0 | 0.00088776 | 0.013497559 | 0 | 0.005054054 |
| 0 | 0 | 0 | 0 | 0 |
| 0 | 0 | 0 | 0 | 0 |
| 0 | 0 | 0 | 0 | 0 |
| 0 | 0 | 0 | 0 | 0 |
| 0 | 0 | 0 | 0 | 0 |
| 0 | 0 | 0 | 0 | 0 |
| 0.000548926 | 0.001918071 | 0 | 0 | 0.06846682 |
| 0.000703008 | 0 | 0.024068896 | 0.004606965 | 0.00661846 |
| 0 | 0 | 0 | 0 | 0 |
| 0 | 0 | 0 | 0 | 0 |
| 0 | 0 | 0 | 0 | 0 |
| 0 | 0 | 0 | 0 | 0 |
| 0 | 0 | 0 | 0 | 0 |
| 0 | 0 | 0 | 0 | 0 |
| 0 | 0 | 0 | 0 | 0 |
| 0 | 0 | 0 | 0 | 0 |
| 0 | 0 | 0 | 0 | 0 |
| 0 | 0 | 0 | 0 | 0 |
| 0 | 0 | 0 | 0 | 0 |
| 0.021430959 | 0 | 0 | 0 | 0 |
| 0 | 0 | 0 | 0 | 0 |
| 0 | 0 | 0 | 0 | 0 |
| 0 | 0 | 0 | 0 | 0 |
| 0 | 0.003308462 | 0 | 0 | 0.007434952 |
| 0 | 0 | 0 | 0 | 0 |
| 0.077330305 | 0.037471252 | 0.04475079 | 0.011768846 | 0.049279208 |
| 0 | 0 | 0 | 0 | 0 |
| 0 | 0 | 0 | 0 | 0 |
| 0 | 0 | 0 | 0 | 0 |
| 0 | 0 | 0 | 0 | 0 |
| 0.013318207 | 2.474017276 | 0.471094952 | 0.067490837 | 0.016761777 |
| 0 | 0 | 0 | 0 | 0 |
| 0 | 0 | 0 | 0 | 0 |

| | | | | |
|---|---|---|---|---|
| 0 | 0 | 0 | 0 | 0 |
| 0 | 0 | 0 | 0 | 0 |
| 0 | 0 | 0 | 0 | 0 |
| 0 | 0 | 0 | 0 | 0 |
| 0 | 0 | 0.003206448 | 0 | 0 |
| 0 | 0 | 0 | 0 | 0 |
| 0 | 0 | 0 | 0 | 0 |
| 0 | 0 | 0 | 0 | 0 |
| 0 | 0 | 0 | 0.000400354 | 0 |
| 0 | 0.008041999 | 0.006497497 | 0 | 0.748371761 |
| 0 | 0 | 0 | 0 | 0 |
| 0 | 0 | 0 | 0 | 0 |
| 0.001613797 | 0 | 0.001732024 | 0.002033762 | 0.001144487 |
| 0 | 0 | 0 | 0 | 0 |
| 0 | 0 | 0 | 0 | 0 |
| 0 | 0 | 0 | 0 | 0 |
| 0 | 0 | 0 | 0 | 0 |
| 0 | 0 | 0 | 0 | 0 |
| 0 | 0 | 0 | 0 | 0 |
| 0.21976812 | 0.031331213 | 0.310722877 | 0.001508493 | 0.008276729 |
| 0 | 0 | 0 | 0 | 0 |
| 0 | 0 | 0 | 0 | 0 |
| 0 | 0 | 0 | 0 | 0 |
| 0 | 0 | 0 | 0 | 0 |
| 0 | 0 | 0 | 0 | 0 |
| 0 | 0 | 0 | 0 | 0 |
| 0 | 0 | 0 | 0 | 0 |
| 0 | 0 | 0 | 0 | 0 |
| 0 | 0 | 0 | 0 | 0 |
| 0 | 0 | 0 | 0 | 0.002586326 |
| 0 | 0 | 0 | 0 | 0 |
| 0 | 0 | 0 | 0 | 0 |
| 0 | 0 | 0 | 0 | 0 |
| 0 | 0 | 0 | 0 | 0 |
| 0 | 0 | 0 | 0 | 0 |
| 0 | 0 | 0 | 0 | 0 |
| 0 | 0 | 0 | 0 | 0 |
| 0 | 0 | 0 | 0 | 0 |
| 0 | 0 | 0 | 0 | 0 |
| 0 | 0 | 0 | 0 | 0 |
| 0 | 0 | 0 | 0 | 0 |
| 0 | 0 | 0 | 0 | 0 |
| 0.090610305 | 0 | 0 | 0 | 0 |
| 0 | 0 | 0 | 0 | 0 |
| 0.001798817 | 0 | 0.003056783 | 0.000283366 | 0 |
| 0.572062442 | 0 | 0 | 0 | 0 |
| 0 | 0 | 0 | 0 | 0 |
| 0 | 0 | 0 | 0 | 0 |

| | | | | |
|---|---|---|---|---|
| 0 | 0 | 0 | 0 | 0 |
| 0 | 0 | 0 | 0 | 0 |
| 0 | 0 | 0 | 0 | 0 |
| 0 | 0 | 0 | 0 | 0 |
| 0 | 0 | 0 | 0 | 0 |
| 0.019248049 | 0 | 0.008647607 | 0.000282059 | 0 |
| 0 | 0 | 0 | 0 | 0 |
| 0 | 0 | 0 | 0 | 0 |
| 0 | 0 | 0 | 0 | 0 |
| 0 | 0 | 0 | 0 | 0 |
| 0 | 0 | 0 | 0 | 0 |
| 0 | 0 | 0 | 0 | 0 |
| 0 | 0 | 0 | 0 | 0 |
| 0 | 0 | 0 | 0 | 0 |
| 0 | 0 | 0 | 0 | 0 |
| 0 | 0 | 0 | 0 | 0 |
| 0 | 0 | 0 | 0 | 0 |
| 0 | 0 | 0 | 0 | 0 |
| 0 | 0 | 0 | 0 | 0 |
| 0 | 0 | 0 | 0 | 0 |
| 0 | 0 | 0 | 0 | 0 |
| 0 | 0 | 0 | 0 | 0 |
| 0 | 0 | 0 | 0 | 0 |
| 0 | 0 | 0 | 0 | 0 |
| 0 | 0 | 0 | 0 | 0 |
| 0 | 0 | 0.001221125 | 0 | 0.036170625 |
| 0 | 0 | 0 | 0 | 0 |
| 1.432819624 | 0 | 0 | 0 | 0 |
| 0 | 0 | 0 | 0 | 0 |
| 0 | 0 | 0 | 0 | 0 |
| 0 | 0 | 0 | 0 | 0 |
| 0 | 0 | 0 | 0 | 0 |
| 0 | 0 | 0 | 0 | 0 |
| 0 | 0 | 0 | 0 | 0 |
| 0 | 0 | 0 | 0 | 0 |
| 0 | 0 | 0 | 0 | 0 |
| 0 | 0 | 0 | 0 | 0 |
| 0 | 0 | 0 | 0 | 0 |
| 0.064766523 | 0 | 0 | 0 | 0 |
| 0 | 0 | 0 | 0 | 0 |
| 0 | 0 | 0 | 0 | 0 |
| 0 | 0 | 0 | 0 | 0 |
| 0 | 0 | 0 | 0 | 0 |
| 0 | 0 | 0 | 0 | 0 |
| 0 | 0 | 0 | 0 | 0 |
| 0 | 0 | 0 | 0 | 0 |
| 0 | 0 | 0 | 0 | 0 |
| 0 | 0 | 0 | 0 | 0 |

| | | | | |
|---|---|---|---|---|
| 0 | 0 | 0 | 0 | 0 |
| 0.021173874 | 0.000955175 | 0 | 0 | 0.334267794 |
| 0 | 0 | 0 | 0 | 0.001101433 |
| 0 | 0 | 0 | 0 | 0 |
| 0 | 0 | 0 | 0 | 0 |
| 0 | 0 | 0 | 0 | 0 |
| 0 | 0 | 0 | 0 | 0 |
| 0 | 0 | 0 | 0 | 0 |
| 0 | 0 | 0 | 0 | 0 |
| 0 | 0 | 0 | 0 | 0 |
| 0 | 0.002657792 | 0 | 0 | 0.000654452 |
| 0 | 0 | 0 | 0 | 0 |
| 0 | 0 | 0 | 0 | 0.002699405 |
| 0 | 0 | 0 | 0 | 0 |
| 0 | 0 | 0 | 0 | 0 |
| 0 | 0 | 0 | 0 | 0 |
| 0 | 0 | 0 | 0 | 0 |
| 0 | 0 | 0 | 0 | 0 |
| 0 | 0 | 0 | 0 | 0 |
| 0 | 0 | 0 | 0 | 0 |
| 0 | 0.084257175 | 0.000536117 | 0.01238045 | 0.001062766 |
| 0 | 0 | 0 | 0 | 0 |
| 0 | 0 | 0 | 0 | 0 |
| 4.049481966 | 5.86505481 | 1.076692741 | 0 | 1.747034097 |
| 0 | 0 | 0 | 0 | 0 |
| 0 | 0 | 0 | 0 | 0 |
| 0 | 0 | 0 | 0 | 0 |
| 0 | 0 | 0 | 0 | 0 |
| 0 | 0 | 0 | 0 | 0 |
| 0 | 0 | 0 | 0 | 0 |
| 0 | 0 | 0 | 0 | 0 |
| 0 | 0 | 0 | 0 | 0 |
| 0 | 0 | 0 | 0 | 0 |
| 0 | 0 | 0 | 0 | 0 |

| PHL018-0105_V1 | PHL018-0081_V1 | PHL018-0219_V3 | PHL018-0082_V1 | PHL018-0233_V2 |
|---|---|---|---|---|
| CTRL | EYCF | REF | REF | REF |
| 0.521101573 | 0.058453441 | 0.368692124 | 0.041768412 | 0.013765964 |
| 2.28207058 | 7.220825646 | 0.102805937 | 1.357524653 | 0.057960333 |
| 6.82548166 | 2.574894716 | 2.260168322 | 2.367292451 | 0 |
| 0 | 0 | 0 | 0 | 0 |
| 0 | 0 | 0 | 0 | 0 |
| 0 | 0 | 0 | 0 | 0 |
| 0 | 0 | 0.423320323 | 0 | 0 |
| 0 | 0 | 0 | 0 | 0 |
| 0 | 0 | 0.268329165 | 0 | 0 |
| 0 | 0 | 0 | 0 | 0 |
| 0 | 0 | 0 | 0 | 0 |
| 0 | 0 | 0 | 0 | 0 |
| 0 | 0 | 0 | 0 | 0 |
| 0 | 1.294085624 | 0.190552307 | 0.837211092 | 0.090552829 |
| 0 | 0 | 0 | 0 | 0 |
| 0 | 0 | 0 | 0 | 0 |
| 0.0017186 | 0 | 0 | 0 | 0 |
| 0 | 0 | 0 | 0 | 0 |
| 0 | 0 | 0 | 0 | 0 |
| 0 | 0 | 0 | 0 | 0 |
| 0 | 0 | 0 | 0 | 0 |
| 0 | 0 | 0 | 0 | 0 |
| 0 | 0 | 0 | 0 | 0 |
| 0 | 0 | 0 | 0 | 0 |
| 0 | 0.040350948 | 0 | 0 | 0 |
| 0 | 0 | 0 | 0 | 0 |
| 0 | 0 | 0 | 0 | 0 |
| 0 | 0 | 0 | 0 | 0.094872528 |
| 0 | 0 | 0 | 0 | 0 |
| 0 | 0 | 0 | 0 | 0 |
| 0 | 0 | 0 | 0 | 0 |
| 0 | 0 | 0 | 0 | 0 |
| 0 | 0 | 0 | 0 | 0 |
| 0 | 0 | 0 | 0 | 0 |
| 0 | 0 | 0 | 0 | 0 |
| 3.969615275 | 0.296456035 | 0.885766982 | 0 | 0.053450822 |
| 0 | 0 | 0 | 0 | 0 |
| 0 | 0 | 0 | 0 | 0 |
| 0 | 0 | 0 | 0 | 0 |
| 0 | 0 | 0 | 0 | 0 |
| 0 | 0 | 0 | 0 | 0 |
| 0.085455623 | 0.003593135 | 2.919660587 | 0.10396978 | 0.010617282 |
| 0.475314435 | 0.110011696 | 0.596813445 | 0.086818131 | 8.760529413 |
| 0.031831087 | 0.045249398 | 0.00221355 | 0.032118809 | 0 |
| 0.007732846 | 0.004077902 | 0 | 0.004557793 | 0.005705355 |
| 0 | 0 | 0 | 0 | 0 |

| | | | | |
|---|---|---|---|---|
| 0 | 0 | 0 | 0 | 0 |
| 0.132111689 | 0.001514965 | 0.00284584 | 0.008520859 | 0 |
| 0 | 0 | 0.076681493 | 0.00292653 | 0 |
| 0.088812884 | 0.695998093 | 0.328738311 | 0.454850035 | 0.238882041 |
| 0 | 0.00488734 | 0.004934909 | 0 | 0 |
| 0 | 0.012615399 | 0 | 0 | 0 |
| 0.006198086 | 0.024844118 | 0.005023993 | 0 | 0 |
| 0 | 0 | 0 | 0 | 0 |
| 0 | 0 | 0 | 0 | 0 |
| 0 | 0 | 0 | 0 | 0 |
| 0 | 0 | 0.009067294 | 0 | 0 |
| 0.182368269 | 0.448985548 | 0 | 0.248295898 | 0.258964434 |
| 0.00598901 | 0.000844027 | 0 | 0.000608614 | 0 |
| 0.136922188 | 0.015699332 | 0.016062614 | 0.038238274 | 0 |
| 0 | 0 | 0.001586738 | 0 | 0 |
| 0 | 0 | 0 | 0 | 0 |
| 0.015915207 | 0.000637692 | 0.038931611 | 0 | 0.36401338 |
| 0 | 0 | 0 | 0 | 0 |
| 0 | 0 | 0 | 0 | 0 |
| 0 | 0 | 0 | 0 | 0 |
| 0.098140169 | 0.025559901 | 0.005159864 | 0.064649729 | 0.055497734 |
| 0.029641929 | 0.014883534 | 0.003824691 | 0.002995056 | 0.003293005 |
| 0 | 0.104043953 | 0 | 0 | 0.002824433 |
| 0 | 0 | 0 | 0 | 0 |
| 0 | 0 | 0 | 0 | 0 |
| 0 | 0 | 0 | 0 | 0 |
| 0.002629232 | 0 | 0 | 0 | 0 |
| 0 | 0 | 0 | 0 | 0 |
| 0.002816959 | 0.025829925 | 0.05219079 | 0.028481132 | 0 |
| 0 | 0 | 0 | 0 | 0 |
| 0 | 0 | 0.001733519 | 0.166405824 | 0 |
| 0 | 0 | 0 | 0 | 0 |
| 0 | 0 | 0 | 0 | 0 |
| 0 | 0 | 0.000353628 | 0 | 0 |
| 0 | 0 | 0 | 0 | 0 |
| 0 | 0 | 0 | 0 | 0 |
| 0 | 0 | 0 | 0 | 0 |
| 0.004313275 | 0 | 0.004792435 | 0 | 37.69500566 |
| 0 | 0 | 0 | 0 | 0 |
| 0 | 0 | 0 | 0 | 0 |
| 0 | 0 | 0 | 0 | 0 |
| 0 | 0 | 0 | 0 | 0 |
| 0 | 0 | 0 | 0 | 0 |
| 0 | 0 | 0 | 0 | 0 |
| 0 | 0.116210388 | 0 | 0 | 0 |
| 0 | 0 | 0 | 0 | 0 |
| 0 | 0 | 0 | 0 | 0 |
| 0 | 0 | 0 | 0.732487809 | 0 |

| | | | | |
|---|---|---|---|---|
| 0 | 0 | 0 | 0 | 0 |
| 0 | 0 | 0 | 0 | 0 |
| 0 | 0 | 0 | 0 | 0 |
| 0 | 0 | 0 | 0 | 0 |
| 0 | 0.009849618 | 0 | 0.005000093 | 0 |
| 0 | 0 | 0.010812401 | 0 | 0 |
| 0 | 0 | 0 | 0 | 0 |
| 0 | 0.001906801 | 0.232033148 | 0 | 0 |
| 0 | 0 | 0.011213821 | 0.001447478 | 0 |
| 0 | 0 | 0 | 0 | 0 |
| 0 | 0 | 0 | 0 | 0 |
| 0.003041237 | 0 | 0.001001463 | 0.003136921 | 0.000652119 |
| 0 | 0 | 0 | 0 | 0 |
| 0 | 0 | 0 | 0 | 0 |
| 0 | 0 | 0 | 0 | 0 |
| 0 | 0 | 0 | 0 | 0 |
| 0 | 0 | 0 | 0 | 0 |
| 0 | 0 | 0 | 0 | 0 |
| 0 | 0.004148088 | 1.571792949 | 0 | 0.394254587 |
| 0 | 0 | 0 | 0 | 0 |
| 0 | 0 | 0 | 0 | 0 |
| 0 | 0 | 0 | 0 | 0 |
| 0 | 0 | 0 | 0 | 0 |
| 0 | 0 | 0 | 0 | 0 |
| 0 | 0 | 0 | 0 | 0 |
| 0 | 0 | 0 | 0 | 0 |
| 0 | 0 | 0 | 0 | 0 |
| 0.003305971 | 0 | 0 | 0 | 0 |
| 0 | 0 | 0 | 0 | 0 |
| 0 | 0 | 0 | 0 | 0 |
| 0 | 0.012383702 | 0.096927489 | 0 | 0.029800587 |
| 0 | 0.296099799 | 0.006340382 | 0 | 0 |
| 0 | 0 | 0 | 0 | 0 |
| 0 | 0 | 0 | 0 | 0 |
| 0 | 0 | 0 | 0 | 0 |
| 0 | 0 | 0 | 0 | 0 |
| 0 | 0 | 0 | 0 | 0 |
| 0 | 0 | 0 | 0 | 0 |
| 0 | 0 | 0.102336565 | 0 | 0 |
| 0 | 0 | 0 | 0 | 0 |
| 0 | 0 | 0 | 0 | 0 |
| 0 | 0 | 0 | 0 | 0 |
| 0 | 0.001376486 | 0 | 0 | 0.001260543 |
| 0 | 0 | 0 | 0 | 0 |
| 0 | 0 | 0 | 0 | 0 |
| 0 | 0 | 0 | 0 | 0 |
| 0 | 0 | 0 | 0.00034056 | 0 |
| 0 | 0 | 0 | 0 | 0 |

| | | | | |
|---|---|---|---|---|
| 0 | 0 | 0 | 0 | 0 |
| 0 | 0 | 0 | 0 | 0 |
| 0 | 0 | 0 | 0 | 0 |
| 0 | 0 | 0 | 0 | 0 |
| 0 | 0.002826925 | 0 | 0 | 0 |
| 0 | 0 | 0 | 0 | 0 |
| 0 | 0 | 0 | 0 | 0 |
| 0 | 0 | 0 | 0 | 0 |
| 0 | 0 | 0 | 0 | 0 |
| 0 | 0 | 0 | 0 | 0 |
| 0 | 0 | 0 | 0 | 0 |
| 0 | 0 | 0 | 0 | 0 |
| 0 | 0 | 0 | 0 | 0 |
| 0 | 0 | 0 | 0 | 0 |
| 0 | 0 | 0 | 0 | 0 |
| 0 | 0 | 0.680800568 | 0 | 0 |
| 0 | 0 | 0 | 0 | 0 |
| 0 | 0 | 0 | 0 | 0 |
| 0 | 0 | 0 | 0 | 0 |
| 0 | 0 | 0 | 0 | 0 |
| 0 | 0 | 0 | 0 | 0 |
| 0 | 0 | 0 | 0 | 0 |
| 0 | 0 | 0 | 0 | 0 |
| 0 | 0 | 0 | 0 | 0 |
| 0 | 0.054332999 | 0 | 0 | 0 |
| 0 | 0 | 0 | 0 | 0 |
| 0 | 0.023285354 | 0 | 0 | 0 |
| 0 | 0 | 0 | 0 | 0 |
| 0 | 0 | 0 | 0 | 0 |
| 0 | 0 | 0 | 0 | 0 |
| 0 | 0 | 0 | 0 | 0 |
| 0 | 0 | 0 | 0 | 0.211394185 |
| 0 | 0 | 0 | 0 | 0 |
| 0 | 0 | 0 | 0 | 0 |
| 0 | 0 | 0 | 0 | 0 |
| 0 | 0 | 0 | 0 | 0 |
| 0 | 0 | 0 | 0 | 0 |
| 0 | 0 | 0 | 0 | 0 |
| 0 | 0 | 0 | 0 | 0 |
| 0 | 0 | 0 | 0 | 0 |
| 0 | 0 | 0 | 0 | 0 |
| 0 | 0 | 0 | 0 | 0 |
| 0 | 0 | 0 | 0 | 0 |
| 0 | 0 | 0 | 0 | 0 |
| 0 | 0 | 0 | 0 | 0 |
| 0 | 0 | 0 | 0 | 0 |
| 0 | 0 | 0 | 0 | 0 |
| 0 | 0 | 0 | 0 | 0 |

| | | | | |
|---|---|---|---|---|
| 0 | 0.367624433 | 0 | 0 | 0 |
| 0 | 0 | 0 | 0 | 0 |
| 0 | 0 | 0 | 0 | 0 |
| 0 | 0 | 0 | 0 | 0 |
| 0 | 0 | 0 | 0 | 0 |
| 0 | 0 | 0 | 0 | 0 |
| 0 | 0 | 0 | 0 | 0 |
| 0 | 0 | 0 | 0 | 0 |
| 0 | 0 | 0 | 0 | 0 |
| 0 | 0 | 0 | 0 | 0 |
| 0 | 0 | 0 | 0 | 0 |
| 0 | 0 | 0.030414024 | 0 | 0 |
| 0.002632183 | 0.000717173 | 0.002245332 | 0 | 0 |
| 0 | 0 | 0 | 0 | 0 |
| 0 | 0 | 0.122827903 | 0 | 0 |
| 0 | 0 | 0 | 0 | 0 |
| 0 | 0 | 0 | 0 | 0 |
| 0 | 0 | 0 | 0 | 0 |
| 0 | 0 | 0 | 0 | 0 |
| 0 | 0 | 0 | 0 | 0 |
| 0.033138439 | 0.027936283 | 0.020497246 | 0.017211389 | 0.005253077 |
| 0 | 0 | 0 | 0 | 0 |
| 0.002591013 | 0 | 0 | 0 | 0 |
| 0.007185062 | 0 | 0 | 0 | 0 |
| 0 | 0 | 0 | 0 | 0 |
| 0 | 0 | 0 | 0 | 0 |
| 0.006687788 | 0.000911088 | 0.002876413 | 0.000563118 | 0.002840749 |
| 0 | 0 | 0 | 0 | 0 |
| 0 | 0 | 0 | 0 | 0 |
| 0 | 0 | 0 | 0 | 0 |
| 1.831788243 | 0 | 0 | 0.003480267 | 0 |
| 0 | 0 | 0 | 0 | 0 |
| 0 | 0 | 0 | 0 | 0 |
| 0 | 0 | 0 | 0 | 0 |
| 0.164594261 | 0.003344538 | 0.063308682 | 0.00402648 | 0 |
| 0 | 0 | 0 | 0 | 0 |
| 0 | 0 | 0 | 0 | 0 |
| 0 | 0 | 0 | 0 | 0 |
| 0 | 0 | 0 | 0 | 0 |
| 0 | 0 | 0 | 0 | 0 |
| 0 | 0 | 0 | 0 | 0 |
| 0.001066019 | 0 | 0 | 0.001507966 | 0 |
| 0 | 0 | 0 | 0 | 0 |
| 0 | 0 | 0 | 0 | 0 |
| 0 | 0 | 0 | 0 | 0 |
| 0 | 0 | 0 | 0 | 0 |
| 0.004854018 | 0 | 0 | 0.002043566 | 0 |
| 0.013083413 | 0.005492201 | 0.009504022 | 0.003945395 | 0 |

| | | | | |
|---|---|---|---|---|
| 0 | 0 | 0 | 0 | 0 |
| 0 | 0 | 0 | 0 | 0 |
| 0.001390157 | 0 | 0 | 0 | 0 |
| 0.013730016 | 0 | 0 | 0.000569899 | 0 |
| 0 | 0 | 0 | 0 | 0 |
| 0 | 0 | 0 | 0 | 0 |
| 0.003248385 | 0 | 0 | 0.004978013 | 0 |
| 0.000874774 | 0 | 0.000823024 | 0.006831673 | 0 |
| 0 | 0 | 0 | 0 | 0 |
| 0 | 0 | 0 | 0 | 0 |
| 0 | 0 | 0 | 0 | 0 |
| 0 | 0 | 0 | 0 | 0 |
| 0 | 0 | 0 | 0 | 0 |
| 0 | 0 | 0 | 0 | 0 |
| 0 | 0 | 0.005416154 | 0 | 0 |
| 0 | 0 | 0 | 0 | 0 |
| 0 | 0 | 0 | 0 | 0 |
| 0 | 0 | 0 | 0 | 0 |
| 0 | 0.000526937 | 0.001746782 | 0.000455959 | 0.046003317 |
| 0 | 0 | 0 | 0 | 0 |
| 0.072347244 | 0 | 0.001889566 | 0.074522552 | 0.000782995 |
| 0 | 0 | 0 | 0 | 0 |
| 0 | 0 | 0 | 0 | 0 |
| 0.00527613 | 0 | 0 | 0 | 0 |
| 0 | 0 | 0.003412748 | 0.004543198 | 0 |
| 0 | 0 | 0 | 0 | 0 |
| 0 | 0 | 0 | 0 | 0 |
| 0 | 0 | 0 | 0 | 0 |
| 0.019471596 | 0.00618951 | 0.197038526 | 0.150472203 | 0 |
| 0 | 0 | 0 | 0 | 0 |
| 1.096997251 | 0.101848045 | 0.54287545 | 0.127891243 | 0.0095956 |
| 0 | 0 | 0 | 0 | 0 |
| 0 | 0 | 0 | 0 | 0 |
| 0.001954019 | 0.001183107 | 0 | 0 | 0 |
| 0 | 0 | 0 | 0 | 0 |
| 0.001120275 | 0 | 0 | 0 | 0 |
| 0 | 0 | 0 | 0 | 0 |
| 0 | 0 | 0 | 0 | 0 |
| 0 | 0 | 0 | 0 | 0 |
| 0 | 0.000685064 | 0.000883155 | 0 | 0 |
| 0 | 0 | 0 | 0 | 0 |
| 0 | 0 | 0.003347523 | 0.003125506 | 0.001173734 |
| 0 | 0 | 0 | 0 | 0 |
| 0 | 0 | 0 | 0 | 0 |
| 0 | 0 | 0 | 0 | 0 |
| 0 | 0 | 0 | 0 | 0 |
| 0 | 0 | 0 | 0 | 0 |
| 0 | 0 | 0 | 0 | 0 |

| | | | | |
|---|---|---|---|---|
| 0 | 0 | 0 | 0 | 0 |
| 0 | 0 | 0 | 0 | 0 |
| 0.029426435 | 0.044038398 | 0 | 0.035068087 | 0 |
| 0 | 0 | 0 | 0 | 0 |
| 0 | 0 | 0 | 0 | 0 |
| 0 | 0 | 0 | 0 | 0 |
| 0 | 0 | 0 | 0 | 0 |
| 0 | 0 | 0 | 0 | 0 |
| 0 | 0 | 0 | 0 | 0 |
| 0.001571573 | 0.000401434 | 0 | 0 | 0 |
| 0 | 0 | 0 | 0 | 0 |
| 0 | 0 | 0 | 0 | 0 |
| 0 | 0 | 0 | 0 | 0 |
| 0.001661684 | 0.001455261 | 0.001250706 | 0.000979408 | 0 |
| 0.001959289 | 0.001734961 | 0.004276645 | 0 | 0 |
| 0 | 0 | 0.002210414 | 0 | 0.008156292 |
| 0.002232434 | 0.001064448 | 0.001008177 | 0.001578975 | 0 |
| 0 | 0 | 0.008863343 | 0 | 0 |
| 0 | 0 | 0 | 0 | 0 |
| 0 | 0 | 0 | 0 | 0 |
| 0 | 0 | 0 | 0 | 0 |
| 0.016122008 | 0.004941737 | 0.012437977 | 0.00427609 | 0.000921858 |
| 0 | 0 | 0 | 0 | 0 |
| 0 | 0 | 0 | 0.011968876 | 0 |
| 0 | 0 | 0.0017025 | 0.002666402 | 0 |
| 0 | 0 | 0 | 0 | 0 |
| 0 | 0 | 0 | 0 | 0 |
| 0 | 0 | 0 | 0 | 0 |
| 0.001071871 | 0 | 0 | 0 | 0 |
| 0 | 0 | 0 | 0.006529038 | 0 |
| 0 | 0 | 0 | 0 | 0 |
| 0 | 0 | 0 | 0 | 0 |
| 0 | 0 | 0 | 0 | 0 |
| 0 | 0 | 0 | 0 | 0 |
| 0 | 0 | 0 | 0 | 0 |
| 0.057560036 | 0.02166008 | 0.156632651 | 0.018030752 | 0 |
| 0 | 0 | 0 | 0 | 0 |
| 0.015698116 | 0.003449322 | 0.016579897 | 0.001193881 | 0.003764214 |
| 0 | 0 | 0 | 0 | 0 |
| 0 | 0 | 0 | 0 | 0 |
| 0.001827163 | 0 | 0.001650309 | 0 | 0 |
| 0 | 0 | 0 | 0 | 0 |
| 0 | 0 | 0 | 0 | 0 |
| 0 | 0 | 0 | 0.004078214 | 0 |
| 0 | 0 | 0 | 0 | 0 |
| 0 | 0 | 0 | 0.00137668 | 0 |
| 0 | 0 | 0 | 0.001132358 | 0 |
| 0.001533416 | 0 | 0 | 0.024764282 | 0 |

| | | | | |
|---|---|---|---|---|
| 0 | 0.006334979 | 0 | 1.297605292 | 0 |
| 0 | 0 | 0 | 0 | 0 |
| 0 | 0 | 0 | 0 | 0 |
| 0 | 0 | 0 | 0 | 0 |
| 0 | 0.000578627 | 0.00223782 | 0 | 0.001700057 |
| 0 | 0 | 0 | 0.001632829 | 0 |
| 0 | 0 | 0 | 0 | 0 |
| 0.163724795 | 0 | 0.005377368 | 0.062637614 | 0.006808584 |
| 0 | 0 | 0 | 0 | 0 |
| 0.083763693 | 0.17430392 | 0.019629934 | 0.001085075 | 0 |
| 0.000832233 | 0.000944804 | 0 | 0 | 0 |
| 0.070929355 | 0.001284096 | 0.168496101 | 0.359867108 | 0.007815065 |
| 0.053623688 | 0.001095786 | 0.039150305 | 0.001580309 | 0 |
| 0 | 0 | 0.000817309 | 0 | 0 |
| 0 | 0.000626106 | 0 | 0 | 0 |
| 0 | 0 | 0 | 0 | 0 |
| 0 | 0 | 0.001762415 | 0 | 0 |
| 0 | 0 | 0 | 0 | 0 |
| 0.011548734 | 0 | 0 | 0.081172325 | 0 |
| 0 | 0 | 0 | 0 | 0 |
| 0.077622942 | 0 | 0 | 0.001089321 | 0 |
| 0 | 0 | 0 | 0 | 0 |
| 0 | 0 | 0 | 0 | 0 |
| 0 | 0.20083007 | 0 | 0 | 0 |
| 0 | 0 | 0 | 0 | 0 |
| 0.001873738 | 0.017017489 | 0 | 0.000736263 | 0 |
| 0 | 0 | 0 | 0 | 1.883186655 |
| 0 | 0 | 0.000947478 | 0.124054906 | 0 |
| 0 | 0 | 0 | 0 | 0 |
| 0.01049602 | 0 | 0 | 0 | 0 |
| 0.00804249 | 0 | 0 | 0 | 0 |
| 0 | 0 | 0 | 0 | 0 |
| 0.321112539 | 0 | 0 | 0 | 0 |
| 0.006806957 | 0 | 0 | 0.00176531 | 0 |
| 0 | 0 | 0 | 0 | 0 |
| 0.193596044 | 0 | 0 | 0.134887483 | 0 |
| 0.854349643 | 0 | 0 | 0.568243348 | 0 |
| 0 | 0.014395031 | 0 | 0 | 0 |
| 0 | 0 | 0 | 0 | 0 |
| 0.391729877 | 0 | 0 | 0.008517958 | 0 |
| 0 | 0 | 0 | 0.073726991 | 0 |
| 0 | 0 | 0 | 0 | 0 |
| 0 | 0 | 0 | 0 | 0 |
| 0.285975705 | 0 | 0 | 0.089567032 | 0 |
| 0 | 0 | 0 | 0 | 0 |
| 0 | 0.021477405 | 0 | 0 | 0 |
| 0.001553427 | 0 | 0 | 0.000610401 | 0 |
| 0 | 0 | 0 | 0 | 0 |

| | | | | |
|---|---|---|---|---|
| 0 | 0 | 0.017497467 | 0 | 0 |
| 0 | 0.049863511 | 0.001306689 | 0.034449347 | 0 |
| 0 | 0 | 0 | 0.001664247 | 0 |
| 0 | 0 | 0.275792123 | 0 | 0 |
| 0 | 0.001917392 | 0 | 0 | 0 |
| 0.135402012 | 0 | 0.004911496 | 0.007572039 | 0 |
| 0 | 0 | 0 | 0 | 0 |
| 0 | 0 | 0 | 0 | 0 |
| 0 | 0 | 0 | 0 | 0 |
| 0 | 0 | 0 | 0 | 0 |
| 0 | 0 | 0.005696508 | 0 | 0 |
| 0.004211635 | 0 | 0 | 0 | 0 |
| 0 | 0 | 0 | 0.066740236 | 0 |
| 0.001340664 | 0.020851498 | 0.001177263 | 0.000790197 | 0.001022125 |
| 0.04386053 | 0 | 0.052071526 | 0 | 0 |
| 0 | 0 | 0 | 0 | 0 |
| 0 | 0 | 0 | 0 | 0 |
| 0 | 0 | 0 | 0 | 0 |
| 0 | 0.000647977 | 0 | 0 | 0.001087894 |
| 0.074629515 | 0 | 0 | 0 | 0 |
| 0 | 0 | 0 | 0 | 0 |
| 0 | 0.433123884 | 0 | 0 | 0.051425208 |
| 0 | 0 | 0 | 0 | 0 |
| 0 | 0 | 0 | 0 | 0 |
| 0 | 0 | 0 | 0 | 0 |
| 0 | 0 | 0 | 0 | 0 |
| 0.033675876 | 0 | 0 | 0.010138593 | 0 |
| 0 | 0 | 0 | 0 | 0 |
| 0 | 0 | 0 | 0 | 0 |
| 0.45820349 | 0.563759479 | 0.39270207 | 14.32925676 | 0.114988424 |
| 0 | 0.00474978 | 0.010040542 | 0 | 0 |
| 0 | 0 | 0 | 0 | 0 |
| 0 | 0 | 0 | 0.001375873 | 0 |
| 0.013955627 | 0 | 0 | 0 | 0 |
| 0 | 0 | 0 | 0.114060119 | 0 |
| 0 | 0 | 0 | 0 | 0 |
| 0 | 0.858485515 | 0.473047987 | 0.441277987 | 0.132248683 |
| 0 | 0 | 0 | 0 | 0.002083634 |
| 0 | 0 | 0.116296455 | 0 | 0 |
| 0 | 0 | 0 | 0 | 0 |
| 0.051067296 | 0 | 0.000553051 | 0 | 0 |
| 0.050871402 | 1.102928253 | 0.642242063 | 0 | 3.531973595 |
| 0 | 0 | 0 | 0 | 0.001192965 |
| 0 | 0 | 0 | 5.266548854 | 0 |
| 0.009324805 | 0 | 0 | 0 | 0 |
| 0 | 0 | 0 | 0 | 0 |
| 0.005976844 | 0 | 0 | 0 | 0.001763905 |
| 0 | 0 | 0 | 0 | 0 |

| | | | | |
|---|---|---|---|---|
| 0.00261673 | 0.009398147 | 0 | 0.028883429 | 0 |
| 0 | 0 | 0 | 0 | 0 |
| 0.032413597 | 0.001987091 | 0.077094083 | 0 | 0.025206423 |
| 0 | 0 | 0 | 0.456588165 | 0 |
| 0 | 0.042893483 | 0 | 0 | 0 |
| 0 | 0 | 0 | 0 | 0 |
| 0 | 0 | 0 | 0 | 0 |
| 0 | 0 | 0 | 0 | 0 |
| 0.052957424 | 0.003525872 | 0 | 0 | 0 |
| 0.17482467 | 0 | 0.083273339 | 0 | 0 |
| 0 | 0 | 0 | 0 | 0 |
| 0 | 0 | 0 | 0 | 0 |
| 0 | 0 | 0 | 0.001724162 | 0 |
| 0 | 0 | 0.006965206 | 0.000634226 | 0 |
| 0 | 0 | 0 | 0 | 0 |
| 0 | 0.097947242 | 0.004810259 | 0.027557396 | 0.004423653 |
| 0.145960307 | 0.054091689 | 0.002665835 | 0.083393079 | 0.049031583 |
| 0.0079269 | 0 | 0 | 0.209580188 | 0 |
| 0 | 0 | 0 | 0.028939897 | 0 |
| 0 | 0 | 0 | 0 | 0 |
| 0 | 0 | 0 | 0 | 0 |
| 0 | 0 | 0 | 0.001529354 | 0 |
| 0 | 0 | 0 | 0 | 0 |
| 1.331158608 | 0 | 0.341513585 | 0 | 0 |
| 0 | 0.031566528 | 0.001573567 | 0 | 0 |
| 0.268893091 | 0 | 0.004805578 | 0.001157899 | 0 |
| 0.002603162 | 0 | 0 | 0 | 0 |
| 0 | 0 | 0 | 0 | 0 |
| 0 | 0 | 0 | 0 | 0 |
| 0.005264813 | 0 | 0 | 0 | 0 |
| 0.065869923 | 0.034815477 | 0.021131844 | 0 | 0.01687699 |
| 0.002683169 | 0.002924259 | 0.001817595 | 0.000948887 | 0 |
| 0 | 0.020012086 | 0.012691041 | 0.020177466 | 0.01460803 |
| 0.009519095 | 0 | 0.849852274 | 0.022658276 | 0.002512168 |
| 0 | 0.002135969 | 0 | 0.005544769 | 0 |
| 0 | 0 | 0 | 0 | 0 |
| 0.013626997 | 0.000629298 | 0.000521526 | 0.028315661 | 0 |
| 0.136011493 | 0.002337664 | 0.059003408 | 0 | 0 |
| 0.40576556 | 0.037747777 | 0 | 0 | 0.020102023 |
| 0 | 0 | 0.002556567 | 0 | 0 |
| 0 | 0 | 0 | 0 | 0 |
| 0 | 0 | 0 | 0 | 0 |
| 0.050513586 | 0.001839986 | 0 | 0.001114499 | 0 |
| 0 | 0 | 0.105066447 | 0.061000501 | 0 |
| 1.997144281 | 0 | 0 | 0.024720069 | 0 |
| 0 | 0.004425899 | 0 | 0.000928421 | 0 |
| 0 | 0 | 0 | 0 | 0 |
| 0 | 0 | 0 | 0 | 0 |

| | | | | |
|---|---|---|---|---|
| 0.001606852 | 0 | 0 | 0 | 0 |
| 0.168842469 | 0.053145289 | 0.162466531 | 0 | 0.012103225 |
| 0 | 0 | 0 | 0 | 0 |
| 0 | 0 | 0 | 0 | 0.001193284 |
| 0 | 0 | 0 | 0 | 0 |
| 0 | 0 | 0 | 0 | 0 |
| 1.676317488 | 0 | 0 | 0 | 0 |
| 0 | 0.000643867 | 0 | 0 | 0 |
| 0 | 0 | 0.002374169 | 0 | 0 |
| 0 | 0 | 0 | 0.006325102 | 0 |
| 0.015180581 | 0 | 0.003854562 | 0.010241159 | 0 |
| 0.107610877 | 0.030804566 | 0.01004759 | 0.033559895 | 0 |
| 0 | 0 | 0 | 0 | 0.55653037 |
| 0.03718535 | 0 | 0 | 0 | 0 |
| 0 | 0.011149572 | 0 | 0.004576491 | 0 |
| 0 | 0 | 0 | 0 | 0 |
| 0.002872818 | 0.001695931 | 0.009946555 | 0.01038532 | 0.000876096 |
| 0.22301301 | 0 | 0.002806957 | 0.011576575 | 0 |
| 0 | 0 | 0 | 0 | 0 |
| 0 | 0.003189048 | 0.013487915 | 0.0453888 | 0 |
| 0.016985574 | 0.039687315 | 0.007284246 | 0.001979001 | 0.062791612 |
| 0.000737296 | 0 | 0 | 0 | 0 |
| 0 | 0 | 0 | 0 | 0 |
| 0 | 0 | 0 | 0 | 0 |
| 0 | 0 | 0 | 0 | 0 |
| 0 | 0.006908764 | 0 | 0 | 0 |
| 0 | 0 | 0 | 0 | 0 |
| 0 | 0.054049845 | 0.212670137 | 0.04454228 | 0.039130679 |
| 0 | 0 | 0 | 0 | 0 |
| 0.001266066 | 0 | 0 | 0.004477368 | 0.001158301 |
| 0.00608161 | 0 | 0 | 0 | 0 |
| 0 | 0 | 0 | 0 | 0 |
| 0 | 0 | 0 | 0 | 0 |
| 0 | 0 | 0 | 0 | 0 |
| 0 | 0.064171557 | 0 | 0 | 0 |
| 0.342132531 | 0.178354269 | 0.1553642 | 0.223433213 | 0.156017882 |
| 0 | 0 | 0 | 0 | 0 |
| 0 | 0 | 0 | 0 | 0 |
| 0.030039399 | 0.006350146 | 0.063541518 | 0.143627665 | 0.003553772 |
| 0 | 0 | 0 | 0 | 0 |
| 1.997437127 | 0 | 0 | 0.026243174 | 0 |
| 0 | 0 | 0 | 0 | 0 |
| 0.032560368 | 0 | 0.002959823 | 0 | 0 |
| 0.023691217 | 0 | 0 | 0.000626579 | 0 |
| 0 | 0 | 0.007808931 | 0 | 0 |
| 0 | 0.532370802 | 0 | 0 | 0.040733401 |
| 0 | 0 | 0 | 0.059026975 | 0 |
| 0.002454809 | 0 | 0 | 0 | 0 |

| | | | | |
|---|---|---|---|---|
| 0 | 0 | 0 | 0 | 0 |
| 0 | 0 | 0 | 0 | 0 |
| 0 | 0.000785524 | 0 | 0 | 0.011704561 |
| 0 | 0.00698816 | 1.559401126 | 0 | 0.010475433 |
| 0 | 0 | 0 | 0 | 0 |
| 0 | 0 | 0 | 0 | 0 |
| 0 | 0 | 0 | 0 | 0 |
| 0 | 0 | 0 | 0 | 0 |
| 0 | 5.681011007 | 1.752187131 | 0.415352796 | 0.462222873 |
| 0 | 0.0036805 | 0.083433476 | 0 | 0.03333036 |
| 0 | 0 | 0 | 0 | 0 |
| 0 | 0.000956487 | 0.001849592 | 0 | 0 |
| 0 | 0 | 0 | 0 | 0 |
| 0 | 0 | 0 | 0 | 0 |
| 0.20756202 | 0.088795743 | 0.214770516 | 0 | 0 |
| 0 | 0 | 0 | 0 | 0 |
| 0 | 0.049033119 | 0.087599991 | 0 | 0.130289372 |
| 0 | 0 | 0 | 0 | 0.23836993 |
| 0 | 0 | 0 | 0 | 0 |
| 0 | 0 | 0 | 0 | 0 |
| 0 | 0 | 0 | 0 | 0 |
| 0 | 0 | 0 | 0 | 0 |
| 0 | 0 | 0 | 0 | 0 |
| 0 | 0 | 0.057548082 | 0.187990114 | 0 |
| 0 | 0.026753548 | 0 | 0 | 0 |
| 0 | 0 | 1.063036861 | 0 | 0.047066181 |
| 0 | 0 | 0 | 0 | 0 |
| 0 | 1.565945646 | 0 | 0 | 0.47068704 |
| 0 | 0 | 0 | 0 | 0 |
| 0 | 0 | 0 | 0 | 0 |
| 0 | 0 | 0.00128437 | 0 | 0.04078542 |
| 0 | 0 | 0 | 0 | 0 |
| 0 | 0.003096179 | 0 | 0 | 0 |
| 0 | 0 | 0 | 0 | 0 |
| 0 | 0.001609755 | 0 | 0 | 0 |
| 0 | 0.055978136 | 0 | 0 | 0.036732476 |
| 0 | 0.286129502 | 0.964404617 | 0 | 0.066245313 |
| 0 | 0 | 0 | 0 | 0 |
| 0 | 0 | 0.001925425 | 0 | 0 |
| 0 | 0 | 0 | 0 | 0 |
| 0.243654105 | 0.001720459 | 0.006210235 | 0.369102177 | 0 |
| 0 | 0 | 0 | 0 | 0 |
| 0.00585798 | 8.409713064 | 0.027975281 | 1.203337328 | 0.002772083 |
| 0 | 0 | 0 | 0 | 0 |
| 0 | 0 | 0 | 0 | 0 |
| 0 | 0 | 0 | 0 | 0 |
| 0 | 0 | 0 | 0 | 0 |
| 0.823237771 | 0.002068189 | 0.013864341 | 0 | 0.000925947 |

| | | | | |
|---|---|---|---|---|
| 0 | 0.036073417 | 0 | 0 | 0 |
| 0 | 0.096721196 | 0 | 0 | 0.048715823 |
| 0.600946192 | 0.003833013 | 0 | 0.079715429 | 0.001331437 |
| 0 | 0 | 0 | 0 | 0.00215883 |
| 0 | 0 | 0 | 0 | 0 |
| 0 | 0.0030195 | 0 | 0 | 0 |
| 0 | 0 | 0 | 0 | 0 |
| 0 | 0 | 0 | 0 | 0.010451997 |
| 0 | 0 | 0 | 0 | 0 |
| 0 | 0 | 0 | 0 | 0 |
| 0.906847768 | 0.039709718 | 0.048324133 | 0.011229041 | 0.001307235 |
| 0 | 1.688317635 | 0.649785697 | 0.970005072 | 0.134139667 |
| 0 | 0 | 0 | 0 | 0 |
| 0.024467677 | 0.017506677 | 0.000928547 | 0.000605942 | 0.043970606 |
| 0.109045612 | 0.005959843 | 0.031830274 | 0.371403763 | 0 |
| 0 | 0.173575294 | 0 | 0 | 2.071158943 |
| 0 | 0 | 0 | 0 | 0 |
| 0 | 0 | 0 | 0 | 0 |
| 0 | 0.019623966 | 0.830034293 | 0 | 0 |
| 0.506581626 | 0.073544853 | 0.065084001 | 0 | 0 |
| 0.016287257 | 0 | 0.084971623 | 0.257595214 | 0 |
| 0.103811553 | 0 | 0 | 0.108193334 | 0 |
| 3.471590221 | 4.400248074 | 2.766224947 | 0.25637245 | 0.097968008 |
| 0 | 0 | 0 | 0 | 0 |
| 0 | 0 | 0 | 0 | 0 |
| 0.008419106 | 1.279238584 | 0 | 0 | 1.393887782 |
| 1.002663399 | 0 | 0 | 0 | 0 |
| 0 | 0 | 0 | 0 | 0 |
| 0 | 0 | 0 | 0 | 0.014719061 |
| 0 | 0 | 0.132165115 | 0 | 0 |
| 0 | 0.009957243 | 0 | 0 | 0 |
| 0 | 0.054011807 | 0 | 0 | 0.225323081 |
| 0.110353979 | 0 | 0.073487958 | 0.01960528 | 0 |
| 0 | 0.010265076 | 0.041777506 | 0.003517775 | 0.003753968 |
| 0.010202065 | 0.000263894 | 0.013219245 | 0.008524997 | 0 |
| 0.064229861 | 0.013415267 | 0.09334957 | 0 | 0.039161632 |
| 0.005656051 | 0.004669897 | 0.006966263 | 0 | 0 |
| 0 | 0 | 0 | 0 | 0 |
| 0 | 0.262774289 | 0 | 0.010402156 | 1.065656263 |
| 0 | 0.534730577 | 0 | 0 | 0 |
| 0 | 0.034161393 | 0 | 0 | 0 |
| 0 | 0 | 0 | 0 | 0 |
| 0 | 0 | 0 | 0 | 0 |
| 0 | 0 | 0 | 0 | 0 |
| 1.087776647 | 0.010612535 | 0 | 0.005605232 | 0.004627917 |
| 0 | 0 | 0.003766798 | 0 | 0 |
| 0 | 0 | 0 | 0 | 0 |
| 3.596957108 | 0.005818917 | 0.133557205 | 0.700168792 | 0.065594754 |

| | | | | |
|---|---|---|---|---|
| 0 | 0 | 0 | 0 | 0 |
| 0 | 0.038767239 | 0 | 0 | 0 |
| 0 | 0 | 0 | 0 | 0 |
| 0 | 0.003641502 | 0.008177455 | 0.000914805 | 0.001577742 |
| 0 | 0 | 0 | 0 | 0 |
| 0 | 0 | 0 | 0 | 0 |
| 0 | 0 | 0 | 0 | 0.075570552 |
| 0 | 0.000756641 | 0.406574793 | 0.092502971 | 0 |
| 0 | 0 | 0 | 0 | 0 |
| 0 | 0 | 0 | 0 | 0 |
| 0.046864357 | 0.000915796 | 0.018070476 | 0.002377318 | 0.001757187 |
| 0 | 0 | 0 | 0 | 0 |
| 0 | 0.010996816 | 0.001265769 | 0 | 0.003654066 |
| 0 | 0.00971262 | 0 | 0 | 0.027177672 |
| 0 | 0 | 0 | 0 | 0 |
| 0 | 0 | 0 | 0 | 0 |
| 0 | 1.00074699 | 0 | 0 | 0 |
| 0 | 0.000544557 | 0 | 0 | 0.000731409 |
| 0 | 0 | 0 | 0 | 0 |
| 0 | 0 | 0.004449821 | 0 | 0 |
| 0.006676394 | 0.001741665 | 0.001496852 | 0 | 0 |
| 0 | 0 | 0 | 0 | 0 |
| 0 | 0.010798761 | 0.001665005 | 0 | 0.016865242 |
| 0 | 0 | 0 | 0 | 0 |
| 0 | 0 | 0 | 0 | 0 |
| 0 | 0 | 0 | 0 | 0 |
| 0 | 0 | 0 | 0 | 0 |
| 0 | 0 | 0 | 0 | 0 |
| 0 | 0 | 0 | 0 | 0 |
| 0 | 0.027891553 | 0.675712197 | 0 | 0 |
| 0 | 0 | 0 | 0 | 0 |
| 0 | 0 | 0 | 0 | 0 |
| 0 | 0 | 0 | 0 | 0 |
| 0 | 0 | 0 | 0 | 0 |
| 0.008857897 | 0.002392646 | 0 | 0 | 0 |
| 0 | 0 | 0 | 0 | 0 |
| 0 | 0.156833768 | 0 | 0 | 0.015257486 |
| 0.086171698 | 0 | 0 | 0.01542872 | 0 |
| 0 | 0 | 0 | 0 | 0 |
| 0 | 0.00240319 | 0 | 0 | 0 |
| 0 | 0 | 0 | 0 | 0 |
| 0 | 0 | 0 | 0 | 0 |
| 0 | 0 | 0 | 0 | 0 |
| 0 | 0.018175888 | 0 | 0 | 0 |
| 0 | 0.126885228 | 0 | 0 | 0.042436845 |
| 0 | 0 | 0 | 0 | 0 |
| 0 | 0.001157523 | 0.001511607 | 0 | 0 |
| 0 | 0 | 0 | 0 | 0 |

| | | | | |
|---|---|---|---|---|
| 0 | 0 | 0 | 0 | 0 |
| 0 | 0 | 0 | 0 | 0 |
| 0 | 0.048955298 | 0 | 0 | 0 |
| 0 | 0.119856266 | 0.014363909 | 0 | 0.136679512 |
| 0 | 0 | 0 | 0 | 0 |
| 0 | 0.373939038 | 0 | 0 | 0.013752324 |
| 0 | 0 | 0 | 0 | 0 |
| 0 | 0 | 0 | 0 | 0 |
| 0.147098919 | 0.023674743 | 0.010970483 | 1.737458732 | 0.00256437 |
| 0 | 0 | 0 | 0 | 0 |
| 0 | 0.002730351 | 0 | 0 | 0 |
| 0 | 0 | 0 | 0 | 0 |
| 0 | 0 | 0 | 0 | 0 |
| 0 | 0 | 0 | 0 | 0 |
| 0 | 0.203391202 | 0 | 0 | 0.010319815 |
| 0 | 0 | 0.008455256 | 0 | 0 |
| 0 | 0 | 0 | 0 | 0 |
| 0 | 0 | 0 | 0 | 0 |
| 0 | 0 | 0 | 0 | 0 |
| 0 | 0 | 0 | 0 | 0 |
| 0 | 0 | 0 | 0 | 0 |
| 0 | 0 | 0 | 0 | 0 |
| 0 | 0 | 0 | 0 | 0 |
| 0 | 0 | 0.043037547 | 0 | 0 |
| 0.032698222 | 0.014189839 | 0.011313163 | 0.001709662 | 1.857022658 |
| 0 | 1.413974034 | 2.327367892 | 17.63165996 | 0.145812732 |
| 0 | 0 | 0.006939223 | 0 | 0.131679873 |
| 0 | 0 | 0 | 0 | 0 |
| 0 | 0 | 0.169013597 | 0 | 0.018191986 |
| 0 | 0 | 0 | 0 | 0 |
| 0 | 0 | 0 | 0 | 0 |
| 0 | 0.041874395 | 0 | 0 | 0 |
| 0 | 0 | 0 | 0 | 0 |
| 0.043094053 | 0 | 0 | 0.001992152 | 0 |
| 0.006729269 | 0.049314247 | 0 | 0.00752225 | 0.018044874 |
| 0.056753273 | 0.000893825 | 0.003827218 | 0.00396382 | 0 |
| 0 | 0 | 0 | 0 | 0 |
| 0 | 0.843760982 | 0.297771564 | 0.118555113 | 0.158858907 |
| 0 | 0 | 6.907996618 | 0 | 0 |
| 0 | 0 | 0 | 0 | 0 |
| 0 | 0 | 0.283502816 | 0 | 0 |
| 0 | 0 | 0 | 0.003727564 | 0 |
| 0.016138479 | 0.288409794 | 0.109660495 | 0.011097486 | 0.458067766 |
| 0 | 0.685476348 | 0 | 0 | 0.095342579 |
| 0 | 0.131322328 | 0 | 0 | 0 |
| 0 | 0.017994233 | 0 | 0 | 0 |
| 0 | 0 | 0 | 0 | 0 |
| 0.002723188 | 0 | 0.004285678 | 0.009046735 | 0.001415566 |

| | | | | |
|---|---|---|---|---|
| 0 | 0 | 0.001528443 | 0 | 0 |
| 0 | 0.028203423 | 0 | 0 | 0 |
| 0 | 0.018384273 | 0 | 0 | 0 |
| 0 | 0 | 0 | 0 | 0 |
| 0.016174252 | 0.000751174 | 0.089782763 | 0 | 0 |
| 0 | 0 | 0 | 0 | 0.00101206 |
| 0 | 0 | 0 | 0 | 0 |
| 0 | 0 | 0 | 0 | 0 |
| 0 | 0 | 0 | 0 | 0 |
| 0 | 0 | 0 | 0 | 0 |
| 0 | 0 | 0 | 0 | 0 |
| 0 | 0.665863474 | 1.567650093 | 0.481026534 | 0.167737096 |
| 0 | 0.033652993 | 0.155947783 | 0.009444316 | 0.010180236 |
| 0 | 0 | 0 | 0 | 0 |
| 0.777554522 | 0.327946722 | 7.190767917 | 7.422190376 | 0.021877854 |
| 0 | 0 | 0 | 0 | 0 |
| 0 | 0 | 0 | 0 | 0 |
| 0 | 0 | 0.025849713 | 0 | 0 |
| 0 | 0.001903604 | 0 | 0 | 0 |
| 0 | 0 | 0 | 0 | 0.004252594 |
| 0 | 0.129758087 | 0 | 0 | 0 |
| 0 | 0 | 0 | 0 | 0 |
| 0.602181268 | 0.013944168 | 0.404430695 | 0 | 0.303306958 |
| 0 | 0 | 0 | 0 | 0 |
| 0 | 0 | 0 | 0 | 0 |
| 0 | 0 | 0 | 0 | 0 |
| 0 | 0 | 0 | 0 | 0 |
| 0 | 0 | 0 | 0 | 0 |
| 0 | 0.010955122 | 0 | 0 | 0 |
| 0 | 0 | 0 | 0 | 0 |
| 0 | 0 | 0 | 0 | 0 |
| 0.089093054 | 0.008830537 | 0.033009974 | 0.004714705 | 0 |
| 0 | 0 | 0 | 0 | 0 |
| 0 | 0.084007825 | 0.088990325 | 0.01291401 | 0 |
| 0.047124597 | 0.000607941 | 0.002418371 | 0.003997998 | 0 |
| 0 | 0.975405567 | 0.038103679 | 0.012070985 | 0.040525957 |
| 0 | 0 | 0 | 0 | 0 |
| 0 | 0 | 0 | 0 | 0.007222436 |
| 0 | 0 | 0 | 0 | 0 |
| 0 | 0 | 0 | 0 | 0 |
| 0 | 0 | 0 | 0 | 0.076700614 |
| 0 | 0.0050363 | 0 | 0 | 0 |
| 0 | 0 | 0 | 0 | 0 |
| 0 | 0 | 0.00404693 | 0 | 0 |
| 0.485325626 | 0.054498365 | 0.112234592 | 0.009638786 | 0.003820787 |
| 0 | 0 | 0 | 0 | 0 |
| 0 | 0.381341492 | 0 | 0 | 0 |
| 0 | 0.004644778 | 0.006634615 | 0 | 0 |

| | | | | |
|---|---|---|---|---|
| 0 | 0 | 0 | 0 | 0 |
| 0 | 0 | 0 | 0 | 0 |
| 0.021460102 | 0 | 0.003011475 | 0.011469614 | 0 |
| 0 | 0 | 0 | 0 | 0 |
| 0 | 0 | 0 | 0 | 0 |
| 0 | 0 | 0 | 0 | 0 |
| 0 | 1.020543957 | 0 | 0 | 0.051511902 |
| 1.262912666 | 0 | 0.040980264 | 0.166933937 | 0 |
| 0 | 0 | 0 | 0 | 0 |
| 0 | 0 | 0 | 0 | 0 |
| 0 | 0 | 0 | 0 | 0 |
| 0 | 0 | 0 | 0 | 0 |
| 0 | 0 | 0 | 0 | 0 |
| 0 | 0.190623287 | 0.240790938 | 0.115048928 | 0.098319233 |
| 0 | 0 | 0 | 0 | 0 |
| 0 | 0 | 0 | 0 | 0 |
| 0 | 0 | 0 | 0 | 0 |
| 0 | 0 | 0 | 0 | 0 |
| 8.831475993 | 2.363709301 | 26.66054909 | 0 | 0.016889869 |
| 0 | 0 | 0 | 0 | 0 |
| 0 | 0 | 0 | 0 | 0.00075588 |
| 0 | 0 | 0 | 0 | 0 |
| 0 | 0 | 0 | 0 | 0 |
| 0 | 0 | 0 | 0 | 0 |
| 0 | 0.000392599 | 0.000578425 | 0 | 0 |
| 0.056477477 | 0 | 0.002710726 | 0.017367765 | 0 |
| 0 | 0 | 0 | 0 | 0 |
| 0.085243696 | 0 | 0 | 1.579356075 | 0 |
| 0 | 0 | 0 | 0 | 0 |
| 0 | 0.603790102 | 0 | 0 | 0.043147421 |
| 0 | 0 | 0 | 0 | 0 |
| 0 | 0 | 0 | 0 | 0 |
| 0 | 0 | 0 | 0 | 0 |
| 0 | 0 | 0 | 0 | 0 |
| 0.170006487 | 0 | 0.0120083 | 0.000808904 | 0 |
| 0 | 0 | 0 | 0 | 0 |
| 0 | 0 | 0 | 0 | 0.419371903 |
| 0.003309212 | 0 | 0.002637272 | 0 | 0 |
| 0 | 0 | 0 | 0 | 0 |
| 0 | 0 | 0 | 0 | 0 |
| 19.64009368 | 9.65237642 | 6.916358413 | 10.20317761 | 1.633645969 |
| 0.377032875 | 0.198001046 | 0.243564086 | 0.885711686 | 0.077891909 |
| 0 | 0.284231632 | 0 | 0 | 0 |
| 0 | 0 | 0 | 0 | 0 |
| 0 | 0 | 0 | 0 | 0 |
| 0 | 0 | 0 | 0 | 0 |
| 0 | 0 | 0 | 0 | 0 |
| 0.035378794 | 0.00326225 | 0.009884011 | 0.000364236 | 0 |

| | | | | |
|---|---|---|---|---|
| 0 | 0 | 0 | 0 | 0 |
| 0.002224717 | 0.004004943 | 0.003947002 | 0 | 0 |
| 0 | 0.000884941 | 0 | 0 | 0 |
| 0 | 0 | 0 | 0 | 0 |
| 0.322622924 | 3.363635 | 0.355482782 | 0.144726669 | 0.827042024 |
| 0 | 0 | 0 | 0 | 0 |
| 0 | 0 | 0 | 0 | 0 |
| 0 | 0 | 0.000634068 | 0 | 0 |
| 1.305570163 | 0.006376845 | 0.003582894 | 0.14178176 | 0 |
| 0 | 0 | 0 | 0 | 0 |
| 0 | 0 | 0 | 0 | 0 |
| 0 | 0 | 0 | 0 | 0 |
| 0 | 0.083614338 | 0.040327556 | 0 | 0.002295536 |
| 0.073273274 | 0.01861953 | 0.005572238 | 0.702528595 | 0.003473388 |
| 0 | 0.751303096 | 0 | 0 | 0.032712338 |
| 0.157951566 | 0.50123779 | 0.264386493 | 0 | 0.010332111 |
| 0 | 0 | 0 | 0.000774556 | 0 |
| 0 | 0 | 0 | 0 | 0 |
| 0 | 0 | 0 | 0 | 0 |
| 0 | 0 | 0 | 0 | 0 |
| 0 | 0 | 0 | 0 | 0 |
| 0 | 0 | 0 | 0 | 0 |
| 0 | 0 | 0 | 0 | 0 |
| 0 | 0 | 0 | 0 | 0 |
| 0 | 0 | 0 | 0 | 0 |
| 0 | 0 | 0 | 0 | 0 |
| 0 | 0.002043172 | 0.039348254 | 0 | 0.011924374 |
| 0.002095268 | 0.000768497 | 0.000363935 | 0 | 0 |
| 0 | 0 | 0 | 0 | 0 |
| 0 | 0 | 0 | 0 | 0 |
| 0 | 0 | 0 | 0 | 0 |
| 0 | 0.001925793 | 0 | 0 | 0 |
| 0 | 0 | 0 | 0 | 0 |
| 0 | 0.004833539 | 0 | 0 | 0 |
| 0 | 0.043284261 | 0 | 0 | 0.047532795 |
| 0.002983126 | 0 | 0 | 0.000439569 | 0 |
| 0 | 0 | 0 | 0.004155953 | 0 |
| 0 | 0.000359769 | 0 | 0 | 0 |
| 0 | 0 | 0 | 0 | 0 |
| 0 | 0 | 0 | 0 | 0 |
| 4.08765052 | 0.316058959 | 0.94219812 | 0 | 0.009923277 |
| 0 | 0 | 0 | 0.009613569 | 0.010362677 |
| 0 | 0 | 0 | 0 | 0 |
| 0 | 0 | 0 | 0 | 0 |
| 0 | 0 | 0 | 0 | 0 |
| 0 | 0 | 0 | 0 | 0 |
| 0 | 0 | 0 | 0 | 0 |
| 0 | 0 | 0 | 0 | 0 |

| | | | | |
|---|---|---|---|---|
| 0 | 0 | 0 | 0 | 0.053278518 |
| 0 | 0 | 0 | 0 | 0 |
| 0 | 0 | 0 | 0 | 0 |
| 0.055138865 | 0 | 0 | 0 | 0 |
| 0 | 0.016778035 | 0.015909852 | 0 | 0.08530549 |
| 0 | 0 | 0 | 0 | 0 |
| 0 | 0 | 0 | 0.175396651 | 0 |
| 0 | 0 | 0 | 0 | 0 |
| 0 | 0 | 0 | 0 | 0 |
| 0 | 0 | 0.010093329 | 0 | 0 |
| 0 | 0.003248552 | 0 | 0 | 0 |
| 0 | 0 | 0 | 0 | 0 |
| 0 | 0 | 0 | 0 | 0 |
| 0 | 0 | 0 | 0 | 0 |
| 0 | 0 | 0 | 0 | 0 |
| 0 | 0 | 0 | 0 | 0 |
| 0 | 0 | 0 | 0 | 0 |
| 0.083811524 | 0 | 0 | 0 | 0 |
| 0 | 0 | 0 | 0 | 0 |
| 0 | 0 | 0 | 0 | 0 |
| 0.001560101 | 0 | 0 | 0.012045896 | 0.010169568 |
| 0 | 0 | 0 | 0 | 0 |
| 0 | 0.033053982 | 0.004015089 | 0 | 0.003738999 |
| 0 | 0.000833814 | 0.008407383 | 0.003156563 | 0 |
| 0 | 0 | 0 | 0 | 0 |
| 0 | 0.000860749 | 0 | 0 | 0 |
| 0 | 0 | 0 | 0 | 0 |
| 0 | 0 | 0 | 0 | 0 |
| 0 | 0 | 0 | 0 | 0 |
| 0.119581118 | 2.245000586 | 0.002154829 | 0.164328055 | 0 |
| 0 | 0.000490453 | 0 | 0 | 0 |
| 0 | 0 | 0 | 0 | 0 |
| 0 | 0 | 0 | 0 | 0 |
| 0 | 0.118654204 | 1.207803299 | 0 | 0 |
| 0 | 0 | 0 | 0 | 0 |
| 0 | 0.001413757 | 0 | 0 | 0 |
| 0.140914687 | 0 | 0.092660824 | 0.021328263 | 0 |
| 0 | 0 | 0 | 0 | 0 |
| 0 | 0.001922803 | 0 | 0 | 0 |
| 0 | 0 | 0 | 0 | 0 |
| 0 | 0 | 0 | 0 | 0 |
| 0 | 0 | 0 | 0 | 0 |
| 0 | 0 | 0 | 0 | 0 |
| 0 | 0 | 0 | 0 | 0 |
| 0 | 0 | 0 | 0 | 0.038336392 |
| 0 | 0 | 0 | 0 | 0 |
| 0 | 0 | 0 | 0 | 0 |
| 0 | 0 | 0 | 0 | 0.008973989 |

| | | | | |
|---|---|---|---|---|
| 0.044619749 | 0.009757196 | 0.003717798 | 0.066378764 | 0.001694632 |
| 0 | 0 | 0 | 0 | 0.005535848 |
| 0 | 0 | 0 | 0 | 0.039159199 |
| 0 | 0 | 0 | 0 | 0 |
| 0 | 0 | 0 | 0 | 0 |
| 0 | 0 | 0 | 0 | 0 |
| 0.00316435 | 0 | 0 | 0 | 0 |
| 0 | 0.431944248 | 0.435049357 | 0 | 0.100920238 |
| 0.611728861 | 0.13132502 | 0.836636663 | 0.10327748 | 11.1849183 |
| 0.00755658 | 0.006372756 | 0 | 0.169248228 | 0.048558257 |
| 0 | 0.015100447 | 0 | 0.010702032 | 0 |
| 0 | 0 | 0 | 0 | 0 |
| 0 | 0 | 0 | 0 | 0 |
| 1.779159916 | 0 | 0.00057603 | 0 | 0.001531623 |
| 0 | 0 | 0 | 0 | 0 |
| 0 | 0 | 0 | 0 | 0 |
| 0 | 0 | 0.002894392 | 0 | 0 |
| 0 | 0 | 0 | 0 | 0.154956773 |
| 0 | 0 | 0 | 0 | 0 |
| 0 | 0 | 0.060750787 | 0 | 0 |
| 0 | 0 | 0 | 0 | 0 |
| 0 | 0 | 0 | 0 | 0 |
| 0 | 0 | 3.223746223 | 0 | 0 |
| 0 | 0 | 0 | 0 | 0 |
| 0 | 0.025352143 | 0 | 0 | 0.011716206 |
| 0 | 0.00744816 | 0 | 0 | 0 |
| 0 | 0 | 0 | 0 | 0 |
| 0 | 0 | 0 | 0 | 0 |
| 0 | 0 | 0 | 0 | 0 |
| 0 | 0 | 0 | 0 | 0 |
| 0 | 0 | 0.511417412 | 0 | 0 |
| 0 | 0 | 0 | 0 | 0 |
| 0 | 0 | 0 | 0 | 0 |
| 0 | 0 | 0 | 0 | 0 |
| 0 | 0.002579732 | 0 | 0 | 0 |
| 0 | 0 | 0 | 0 | 0 |
| 0 | 0 | 0 | 0 | 0 |
| 0.029779461 | 0 | 0 | 0 | 0 |
| 0 | 0 | 0 | 0 | 0 |
| 0.277951001 | 0 | 0 | 0.016372936 | 0 |
| 0 | 0 | 0 | 0 | 0 |
| 0 | 1.50797241 | 0 | 0 | 0.190826529 |
| 0 | 0 | 0 | 0 | 0 |
| 0 | 0 | 0 | 0 | 0 |
| 0 | 0 | 0 | 0 | 0 |
| 0 | 0.003290946 | 0.000991763 | 0 | 0 |
| 0 | 0 | 0 | 0 | 0 |
| 0 | 0 | 0 | 0 | 0 |

| | | | | |
|---|---|---|---|---|
| 0 | 0 | 0 | 0 | 0 |
| 0.006687112 | 0.000910996 | 0 | 0 | 0 |
| 0 | 0 | 0 | 0 | 0 |
| 0 | 0 | 0 | 0 | 0 |
| 0 | 1.875170455 | 0.933437517 | 0.097232772 | 0.762943385 |
| 0 | 0 | 0 | 0 | 0 |
| 0 | 0 | 0 | 0 | 0 |
| 0.034139509 | 0.036522254 | 0.003310516 | 0.181839256 | 0 |
| 0 | 0.065382608 | 0 | 0 | 0 |
| 0 | 0 | 0 | 0 | 0 |
| 0 | 0 | 0 | 0 | 0 |
| 0 | 0.096347246 | 0 | 0 | 0 |
| 0 | 0 | 0 | 0 | 0 |
| 0 | 0 | 0 | 0 | 0 |
| 0 | 0 | 0 | 0.000434516 | 0 |
| 0.135874161 | 0 | 0 | 0 | 0 |
| 0 | 0 | 0 | 0 | 0 |
| 0 | 0 | 0 | 0 | 0 |
| 0 | 0 | 0 | 0 | 0 |
| 0 | 0 | 0 | 0 | 0 |
| 0 | 0 | 0 | 0 | 0 |
| 0 | 0 | 0 | 0 | 0 |
| 0 | 0 | 0 | 0 | 0 |
| 0.007431548 | 0.000987719 | 0.002455696 | 0 | 0 |
| 0 | 0.668431861 | 0 | 0 | 0.073401809 |
| 0 | 0 | 0.000373023 | 0 | 0 |
| 0 | 0 | 3.219285288 | 0 | 0 |
| 0 | 0.001189039 | 0 | 0 | 0 |
| 0 | 0 | 0 | 0 | 0 |
| 0 | 0 | 0 | 0 | 0 |
| 0 | 0 | 0 | 0 | 0 |
| 0 | 0 | 0 | 0 | 0 |
| 0 | 0 | 0 | 0 | 0 |
| 0 | 0 | 0 | 0 | 0 |
| 0 | 0 | 0 | 0 | 0 |
| 0 | 0 | 0 | 0 | 0 |
| 0 | 0 | 0 | 0 | 0 |
| 0 | 0.067863938 | 0 | 0 | 0 |
| 0 | 0 | 0 | 0 | 0 |
| 0 | 0 | 0 | 0 | 0 |
| 0 | 0 | 0 | 0 | 0 |
| 0 | 0 | 0 | 0 | 0.044588819 |
| 0 | 0 | 0 | 0 | 0 |
| 0 | 0 | 0 | 0 | 0 |
| 0 | 0 | 0 | 0 | 0 |
| 0 | 0 | 0 | 0 | 0 |
| 0 | 0 | 0 | 0 | 0 |
| 0 | 0 | 0 | 0 | 0 |

| | | | | |
|---|---|---|---|---|
| 0 | 0.028672379 | 0 | 0 | 0.067393637 |
| 0 | 0 | 0 | 0 | 0 |
| 0 | 0 | 0 | 0 | 0 |
| 0 | 0.0007396 | 0 | 0 | 0 |
| 0 | 0 | 0 | 0 | 0 |
| 0.012101649 | 0 | 0 | 0.001490435 | 0.153886703 |
| 0 | 0 | 0 | 0 | 0 |
| 0 | 0.006333489 | 0 | 0 | 0 |
| 0 | 0 | 0 | 0 | 0 |
| 0 | 0 | 0 | 0 | 0 |
| 0 | 0 | 0 | 0 | 0 |
| 0 | 0 | 0 | 0 | 0 |
| 0 | 0 | 0 | 0 | 0 |
| 0 | 0 | 0 | 0 | 0 |
| 0 | 0.001000411 | 0 | 0 | 0 |
| 0 | 0 | 0 | 0 | 0 |
| 0 | 0 | 0 | 0 | 0 |
| 0 | 0 | 0 | 14.33468988 | 0 |
| 0 | 0 | 0 | 0 | 0 |
| 0 | 0 | 0 | 0 | 0 |
| 0.196203016 | 0 | 0.054342906 | 0.014681501 | 0 |
| 0 | 0 | 0 | 0 | 0 |
| 0.033281838 | 0.050464473 | 0.138291733 | 0.000671799 | 0 |
| 0 | 0.001989625 | 0 | 0 | 0.003340399 |
| 0 | 0 | 0 | 0 | 0 |
| 0 | 0 | 0 | 0 | 0.016783441 |
| 0 | 0 | 0 | 0 | 0 |
| 0 | 0 | 0 | 0 | 0 |
| 0.006314852 | 0 | 0.096545772 | 0 | 0.021890716 |
| 0 | 0 | 0 | 0 | 0 |
| 0 | 0 | 0 | 0 | 0 |
| 0 | 0 | 0 | 0 | 0 |
| 0.036078379 | 0 | 0.004017048 | 3.014952519 | 0.00122069 |
| 0 | 0 | 0 | 0 | 0 |
| 0 | 0.036680512 | 0.142579392 | 0.252229241 | 0.006114648 |
| 0.00119917 | 0.003734054 | 0.074527619 | 0.003735938 | 0 |
| 0 | 0 | 0 | 0 | 0 |
| 0 | 0 | 0 | 0 | 0 |
| 0.046762714 | 0 | 0.054883558 | 0.003036521 | 0 |
| 0 | 0 | 0 | 0 | 0 |
| 0 | 0 | 0 | 0 | 0 |
| 0 | 0 | 0 | 0 | 0 |
| 0 | 0.339312545 | 0.163751712 | 0.028268261 | 0.051668184 |
| 0 | 0 | 0 | 0 | 0 |
| 0 | 0 | 0 | 0 | 0 |
| 0 | 0.031835416 | 0.018973881 | 0 | 0.007154559 |
| 0 | 0 | 0 | 0 | 0 |
| 0 | 0 | 0 | 0 | 0 |

| | | | | |
|---|---|---|---|---|
| 0 | 0 | 0 | 0 | 0 |
| 0 | 0 | 0 | 0 | 0 |
| 0 | 0 | 0 | 0 | 0 |
| 0 | 0 | 0 | 0 | 0 |
| 0 | 0 | 0 | 0 | 0 |
| 0 | 0.004542592 | 0 | 0 | 0 |
| 0 | 0 | 0 | 0 | 0 |
| 1.652536543 | 0 | 0 | 0.014462478 | 0 |
| 0 | 0.019023413 | 0 | 0 | 0.004140186 |
| 0 | 0 | 0.001854435 | 0 | 0.012137369 |
| 0 | 0 | 0 | 0 | 0 |
| 0 | 0 | 0 | 0 | 0 |
| 0 | 0 | 0 | 0 | 0 |
| 0 | 0 | 0 | 0 | 0.256880321 |
| 0 | 0.003338444 | 0 | 0 | 0 |
| 0.084284204 | 0 | 0 | 0.000769345 | 0 |
| 0 | 0 | 0 | 0 | 0 |
| 0 | 0 | 0 | 0 | 0 |
| 0 | 0 | 0 | 0 | 0 |
| 0.474286921 | 0.808724377 | 0 | 0 | 0.221028916 |
| 0.008179674 | 0.004908359 | 0.001172691 | 0 | 0.677740931 |
| 0 | 0.0014318 | 0 | 0 | 0 |
| 0 | 0 | 0 | 0 | 0 |
| 0 | 0 | 0 | 0 | 0 |
| 0 | 0 | 0 | 0 | 0 |
| 0 | 0 | 0 | 0 | 0 |
| 0 | 0 | 0 | 0 | 0 |
| 0 | 0 | 0 | 0 | 0 |
| 0 | 0 | 0 | 0 | 0 |
| 0 | 0 | 0.004654806 | 0 | 0.200680798 |
| 0 | 0.118409842 | 0.730524029 | 0 | 0.039690539 |
| 0 | 0 | 0 | 0 | 0 |
| 0 | 0 | 0 | 0 | 0 |
| 0 | 0 | 0 | 0 | 0 |
| 0 | 0 | 0 | 0 | 0 |
| 0 | 0 | 0 | 0 | 0 |
| 0 | 0 | 0 | 0 | 0 |
| 0 | 0 | 0 | 0 | 0 |
| 0 | 0 | 0.007199631 | 0.006544011 | 0 |
| 0 | 0 | 0 | 0 | 0 |
| 0 | 0 | 0 | 0 | 0 |
| 0 | 0.226390675 | 0 | 0 | 0 |
| 0 | 0.034390378 | 0 | 0 | 0 |
| 0 | 0 | 0 | 0 | 0 |
| 0 | 0 | 0 | 0 | 0 |
| 0 | 0 | 0 | 0 | 0 |
| 2.450803906 | 0 | 0.002586167 | 0.05906795 | 0 |
| 0 | 0 | 0 | 0 | 0 |

| | | | | |
|---|---|---|---|---|
| 0 | 0 | 0 | 0 | 0 |
| 0 | 0 | 0 | 0 | 0 |
| 0 | 0 | 0 | 0 | 0 |
| 0 | 0.00081572 | 0 | 0 | 0 |
| 0 | 0.073144942 | 0.101325817 | 0.013448585 | 0.059526359 |
| 0 | 0 | 0 | 0 | 0 |
| 0 | 0 | 0 | 0 | 0 |
| 0 | 0 | 0 | 0 | 0 |
| 0 | 0 | 0 | 0 | 0 |
| 0 | 0 | 0 | 0 | 0 |
| 0 | 0.050072609 | 0 | 0 | 0 |
| 0.069890228 | 0.012811893 | 0.134268236 | 0.00186714 | 0.002717053 |
| 0 | 0 | 0.014399602 | 0.000805436 | 0 |
| 0 | 0 | 0 | 0 | 0 |
| 0 | 0 | 0.004996846 | 0 | 0 |
| 0 | 0.173776791 | 0 | 0 | 0.020248564 |
| 0 | 0.07758623 | 0.002406917 | 0 | 0 |
| 0 | 0 | 0 | 0 | 0 |
| 0.237621049 | 0.003369904 | 0.088579258 | 0 | 0 |
| 0.001370236 | 0 | 0.003683361 | 0.307936584 | 0 |
| 0 | 0 | 0 | 0 | 0 |
| 0 | 0 | 0 | 0 | 0 |
| 0 | 0 | 0 | 0 | 0 |
| 0 | 0 | 0 | 0 | 0 |
| 0 | 0 | 0 | 0 | 0 |
| 0 | 0 | 0 | 0 | 0 |
| 0.102273957 | 0.002564346 | 0.034121023 | 0 | 0 |
| 0 | 0 | 0 | 0 | 0 |
| 0 | 0 | 0 | 0 | 0 |
| 0 | 0 | 0 | 0 | 0 |
| 0.002844979 | 0.000847823 | 0.010974373 | 0 | 0 |
| 0.018245397 | 0 | 0.02934489 | 0 | 0 |
| 0 | 0 | 0 | 0 | 0 |
| 0 | 0 | 0 | 0 | 0 |
| 0 | 0 | 0 | 0 | 0 |
| 0 | 0 | 0 | 0 | 0 |
| 0 | 0 | 0 | 0 | 0 |
| 0.030106922 | 0.001446592 | 0.103148444 | 0.011412906 | 0.001285781 |
| 0 | 0.086588308 | 0 | 0 | 0 |
| 0 | 0 | 0.002088134 | 0 | 0 |
| 0.007233493 | 0 | 0.056885223 | 0 | 0.009555864 |
| 0 | 0 | 0 | 0 | 0 |
| 0 | 0 | 0 | 0 | 0 |
| 0.007595165 | 0 | 0.00111545 | 0 | 0 |
| 0 | 0 | 0 | 0 | 0 |
| 0 | 0 | 0 | 0 | 0 |
| 0 | 0 | 0 | 0 | 0 |
| 0 | 0 | 0 | 0 | 0 |

| | | | | |
|---|---|---|---|---|
| 0 | 0 | 0 | 0 | 0 |
| 0 | 0 | 0 | 0 | 0 |
| 0 | 0 | 0 | 0 | 0 |
| 0.010677736 | 0.000661202 | 0.005991107 | 0.004462686 | 0.002442217 |
| 0.002700935 | 0 | 0 | 0.00148582 | 0 |
| 0.005809694 | 0 | 0.001399297 | 0 | 0 |
| 0 | 0 | 0 | 0 | 0 |
| 0 | 0 | 0 | 0 | 0.000868193 |
| 0 | 0 | 0 | 0 | 0 |
| 0 | 0.002444985 | 0 | 0 | 0 |
| 0 | 0 | 0 | 0 | 0 |
| 0 | 0 | 0 | 0 | 0 |
| 0.168032059 | 0.004686994 | 0.078100636 | 0 | 0.056657063 |
| 0 | 0 | 0 | 0 | 0 |
| 0 | 0 | 0 | 0 | 0 |
| 0 | 0 | 0 | 0 | 0 |
| 0 | 0 | 0 | 0 | 0 |
| 0 | 0 | 0 | 0 | 0 |
| 0 | 0 | 0 | 0 | 0 |
| 0 | 0 | 0 | 0 | 0 |
| 0 | 0 | 0 | 0 | 0 |
| 0 | 0 | 0 | 0 | 0 |
| 0 | 0.007899765 | 0.016503588 | 0.026274629 | 0 |
| 0 | 0.006263215 | 0.009656926 | 0.00390713 | 0.00709176 |
| 0 | 0 | 0 | 0 | 0 |
| 0 | 0.004195171 | 0 | 0 | 0 |
| 0 | 0 | 0 | 0 | 0.004751863 |
| 0 | 0 | 0 | 0 | 0 |
| 0 | 0 | 0 | 0 | 0 |
| 0 | 0 | 0 | 0 | 0 |
| 0 | 0.016102496 | 0 | 0 | 0.002310652 |
| 0 | 0 | 0 | 0 | 0 |
| 0 | 0 | 0 | 0 | 0 |
| 0 | 0 | 0 | 0 | 0 |
| 0 | 0 | 0 | 0 | 0 |
| 0 | 0.003309073 | 0 | 0 | 0 |
| 0.056885857 | 0 | 0 | 0 | 0 |
| 0 | 0 | 0 | 0 | 0 |
| 0.099662992 | 0.033186249 | 0.292978084 | 0.029119133 | 0 |
| 0 | 0 | 0 | 0 | 0 |
| 0 | 0 | 0 | 0 | 0 |
| 0 | 0 | 0 | 0 | 0 |
| 0 | 0 | 0 | 0 | 0 |
| 0 | 0 | 0 | 0 | 0 |
| 0 | 0 | 0 | 0 | 0 |
| 0.124536553 | 0.963445296 | 0.042679035 | 0.090955526 | 0.16908111 |
| 0 | 0 | 0 | 0 | 0 |
| 0 | 0 | 0 | 0 | 0 |

| | | | | |
|---|---|---|---|---|
| 0 | 0 | 0 | 0 | 0 |
| 0 | 0 | 0 | 0 | 0 |
| 0 | 0.127048079 | 0.084998582 | 0 | 0.056104288 |
| 0 | 0 | 0 | 0 | 0 |
| 0 | 0 | 0 | 0 | 0 |
| 0 | 0 | 0.020740621 | 0 | 0 |
| 0 | 0 | 0 | 0 | 0 |
| 0 | 0 | 0 | 0 | 0 |
| 0 | 0 | 0.001904841 | 0.002703617 | 0 |
| 0 | 0 | 0 | 0 | 0 |
| 0.006795568 | 0.001983795 | 0 | 0 | 0 |
| 0.001035286 | 0 | 0.001090926 | 0.000610205 | 0 |
| 0 | 0 | 0 | 0 | 0 |
| 0 | 0 | 0 | 0 | 0 |
| 0 | 0 | 0 | 0 | 0 |
| 0 | 0 | 0 | 0 | 0 |
| 0.001630888 | 0 | 0 | 0 | 0 |
| 0 | 0 | 0 | 0 | 0 |
| 0 | 0 | 0 | 0 | 0 |
| 0 | 0.015785121 | 0.020903217 | 0 | 0.001682653 |
| 0 | 0 | 0 | 0 | 0 |
| 0 | 0 | 0 | 0 | 0 |
| 0 | 0 | 0 | 0 | 0 |
| 0 | 0 | 0 | 0 | 0 |
| 0 | 0 | 0 | 0 | 0 |
| 0 | 0 | 0 | 0 | 0 |
| 0 | 0 | 0 | 0 | 0 |
| 0 | 0.003604695 | 0 | 0 | 0.002094907 |
| 0 | 0 | 0 | 0 | 0 |
| 0 | 0 | 0 | 0 | 0 |
| 0.010428428 | 0.009578839 | 0.024855812 | 0.004656506 | 0 |
| 0 | 0 | 0 | 0 | 0 |
| 0 | 0.017995158 | 0.015051455 | 0.011354023 | 0.019512066 |
| 0.002812168 | 0 | 0.013123199 | 0 | 0 |
| 0 | 0 | 0 | 0 | 0 |
| 0 | 0 | 0 | 0 | 0 |
| 0 | 0 | 0 | 0 | 0 |
| 0 | 0 | 0 | 0 | 0 |
| 0 | 0 | 0 | 0 | 0 |
| 0 | 0 | 0 | 0 | 0 |
| 0 | 0 | 0 | 0 | 0 |
| 0.80220472 | 0.000514044 | 0.126780621 | 0.000533763 | 0 |
| 0 | 0.035821497 | 0.079164874 | 0.024818782 | 0.010514166 |
| 0 | 0 | 0 | 0 | 0 |
| 0 | 0 | 0 | 0 | 0 |
| 0 | 0.104253086 | 0.027799695 | 0.007360911 | 0.049531465 |
| 0 | 0 | 0 | 0 | 0 |
| 0 | 0 | 0 | 0 | 0 |

| | | | | |
|---|---|---|---|---|
| 0 | 0 | 0 | 0 | 0 |
| 0 | 0.017189293 | 0 | 0 | 0.024232963 |
| 0.013726212 | 0 | 0.007452128 | 0 | 0 |
| 0 | 0 | 0 | 0 | 0 |
| 0.29483443 | 0.002814786 | 0.48872494 | 0 | 0.004931238 |
| 0 | 0 | 0.019744961 | 0 | 0.000800006 |
| 0 | 0 | 0 | 0 | 0 |
| 0 | 0 | 0 | 0 | 0 |
| 0 | 0 | 0 | 0 | 0.006917476 |
| 0 | 0 | 0 | 0 | 0 |
| 0 | 0 | 0 | 0 | 0 |
| 0 | 0 | 0 | 0 | 0 |
| 0 | 0 | 0 | 0 | 0 |
| 0 | 0 | 0 | 0 | 0 |
| 0 | 0 | 0 | 0 | 0 |
| 0 | 0 | 0 | 0 | 0 |
| 0 | 0 | 0 | 0 | 0 |
| 0 | 0 | 0 | 0 | 0 |
| 0 | 0 | 0 | 0 | 0 |
| 0 | 0 | 0 | 0 | 0 |
| 0 | 0 | 0 | 0 | 0 |
| 0.027150371 | 0 | 0.000817415 | 0 | 0 |
| 0 | 0 | 0 | 0 | 0 |
| 0 | 0 | 0 | 0 | 0 |
| 0 | 0.044843394 | 0.004466619 | 0 | 0.280580249 |
| 0.000559669 | 0.069636743 | 0 | 0.018912704 | 0 |
| 0 | 0 | 0 | 0 | 0 |
| 0 | 0.006533127 | 0.017843307 | 0 | 0.00124172 |
| 0 | 0 | 0 | 0 | 0 |
| 0 | 0 | 0 | 0 | 0 |
| 0 | 0 | 0 | 0 | 0 |
| 0 | 0 | 0.015268931 | 0 | 0 |
| 0 | 0 | 0 | 0 | 0 |
| 0 | 0 | 0 | 0 | 0 |
| 0 | 0 | 0 | 0 | 0 |
| 0 | 0 | 0 | 0 | 0 |
| 0 | 0 | 0 | 0 | 0 |
| 0 | 0 | 0 | 0 | 0 |
| 0 | 0 | 0 | 0 | 0.017626849 |
| 0 | 0 | 0 | 0 | 0 |
| 0 | 0 | 0 | 0 | 0 |
| 0 | 0.002067841 | 0 | 0 | 0.014349763 |
| 0.035386399 | 0.008927308 | 0.002113842 | 0.051866584 | 0 |
| 0 | 0 | 0 | 0 | 0 |
| 0 | 0 | 0 | 0 | 0 |
| 0 | 0 | 0 | 0 | 0 |
| 0.053561416 | 0 | 0.002587012 | 0.003545234 | 0.00327556 |
| 0 | 0 | 0 | 0 | 0 |

| | | | | |
|---|---|---|---|---|
| 0 | 0 | 0 | 0 | 0 |
| 0.043911954 | 0.001230905 | 0.044476539 | 0.023645293 | 0.002066577 |
| 0 | 0 | 0 | 0 | 0 |
| 0 | 0 | 0 | 0 | 0 |
| 0 | 0 | 0 | 0 | 0 |
| 0 | 0 | 0 | 0 | 0 |
| 0 | 0 | 0 | 0 | 0 |
| 0 | 0 | 0 | 0 | 0 |
| 0 | 0 | 0 | 0 | 0 |
| 0 | 0 | 0 | 0 | 0 |
| 0 | 0 | 0 | 0 | 0 |
| 0 | 0 | 0 | 0 | 0 |
| 0 | 0.022380216 | 0.014952987 | 0 | 0 |
| 0 | 0 | 0 | 0 | 0 |
| 0 | 0.010743269 | 0.006105197 | 0.011553802 | 0 |
| 0 | 0 | 0 | 0 | 0 |
| 0 | 0 | 0 | 0 | 0 |
| 0 | 0 | 0 | 0 | 0 |
| 0 | 0 | 0 | 0 | 0 |
| 0 | 0 | 0 | 0 | 0 |
| 0 | 0 | 0 | 0 | 0 |
| 0.014604301 | 0 | 0.022650745 | 0.02699743 | 0 |
| 0.005611105 | 0 | 0.004479291 | 0 | 0 |
| 2.953772714 | 0 | 0 | 0 | 0 |
| 0 | 0 | 0 | 0 | 0 |
| 0 | 0.133297174 | 0 | 0 | 0 |
| 0 | 0 | 0 | 0 | 0 |
| 0 | 0 | 0 | 0 | 0 |
| 0 | 0 | 0 | 0 | 0 |
| 0 | 0 | 0 | 0 | 0 |
| 0 | 0 | 0 | 0 | 0 |
| 0 | 0 | 0 | 0 | 0 |
| 0 | 0 | 0 | 0 | 0 |
| 0 | 0 | 0 | 0 | 0 |
| 0 | 6.772146829 | 0 | 0 | 14.86857652 |
| 0 | 0 | 0 | 0 | 0 |
| 0 | 0 | 0 | 0 | 0 |
| 0 | 0 | 0 | 0 | 0 |
| 0.006408192 | 0 | 0.001995839 | 0.022141227 | 0.001516226 |
| 0 | 0 | 0 | 0 | 0 |
| 0.018029899 | 0.013495837 | 0.373266907 | 0.189766914 | 0.113744574 |
| 0 | 0 | 0 | 0 | 0 |
| 0 | 0 | 0 | 0 | 0 |
| 0 | 0 | 0 | 0 | 0 |
| 0 | 0 | 0 | 0 | 0 |
| 1.554350397 | 0.051300027 | 0.067613512 | 0.265671317 | 0.00415075 |
| 0 | 0 | 0 | 0 | 0 |
| 0 | 0 | 0 | 0 | 0 |

| | | | | |
|---|---|---|---|---|
| 0 | 0 | 0.00167281 | 0 | 0 |
| 0 | 0 | 0.00139049 | 0 | 0 |
| 0 | 0 | 0.009789282 | 0 | 0 |
| 0 | 0 | 0 | 0 | 0 |
| 0 | 0.002109426 | 0 | 0 | 0 |
| 0 | 0 | 0 | 0 | 0 |
| 0 | 0 | 0 | 0 | 0 |
| 0 | 0 | 0 | 0.001878707 | 0 |
| 0 | 0.002721561 | 0 | 0 | 0 |
| 0.019967038 | 0.002115666 | 0.012523887 | 0.111148929 | 0.005328016 |
| 0 | 0.066141274 | 0 | 0 | 0 |
| 0 | 0 | 0 | 0 | 0 |
| 0.002168455 | 0.002806416 | 0.001795353 | 0.001533723 | 0 |
| 0 | 0 | 0 | 0 | 0 |
| 0 | 0 | 0 | 0 | 0 |
| 0 | 0 | 0 | 0 | 0 |
| 0 | 0 | 0 | 0 | 0.007463378 |
| 0 | 0 | 0 | 0 | 0 |
| 0 | 0 | 0 | 0 | 0 |
| 1.028731733 | 0.042858845 | 0.438575915 | 0.450490193 | 0.00419376 |
| 0 | 0 | 0 | 0 | 0 |
| 0 | 0 | 0 | 0 | 0 |
| 0 | 0 | 0 | 0 | 0 |
| 0 | 0 | 0 | 0 | 0 |
| 0 | 0 | 0 | 0 | 0 |
| 0 | 0.00709514 | 0 | 0 | 0.00154416 |
| 0 | 0 | 0 | 0 | 0 |
| 0 | 0 | 0 | 0 | 0 |
| 0 | 0 | 0 | 0 | 0 |
| 0 | 0 | 0 | 0.011674698 | 0 |
| 0 | 0.000897297 | 0 | 0 | 0 |
| 0 | 0 | 0 | 0 | 0 |
| 0 | 0 | 0 | 0 | 0 |
| 0 | 0 | 0 | 0 | 0 |
| 0 | 0 | 0 | 0 | 0 |
| 0 | 0 | 0.000752205 | 0 | 0 |
| 0 | 0 | 0 | 0 | 0 |
| 0 | 0.028305875 | 0 | 0 | 0 |
| 0 | 8.123744843 | 0 | 0 | 0 |
| 0 | 0 | 0 | 0 | 0 |
| 0 | 0 | 0 | 0 | 0 |
| 0 | 0 | 0 | 0 | 0 |
| 0 | 0.031582525 | 0 | 0 | 0 |
| 0 | 0 | 0 | 0 | 0 |
| 0.026466887 | 0.010866273 | 0.009005352 | 0.000747934 | 0 |
| 0 | 0 | 0 | 0 | 0 |
| 0 | 0 | 0 | 0 | 0 |
| 0 | 0 | 0 | 0 | 0 |

| | | | | |
|---|---|---|---|---|
| 0 | 0 | 0 | 0 | 0 |
| 0 | 0 | 0 | 0 | 0 |
| 0 | 0 | 0 | 0 | 0 |
| 0 | 0 | 0 | 0 | 0 |
| 0 | 0 | 0 | 0 | 0 |
| 0.019487895 | 0.00285767 | 0.130586174 | 0.000478596 | 0.014238536 |
| 0 | 0 | 0 | 0 | 0 |
| 0 | 0 | 0 | 0 | 0 |
| 0.055293049 | 0 | 0 | 0 | 0 |
| 0 | 0 | 0 | 0 | 0 |
| 0 | 0 | 0 | 0 | 0 |
| 0 | 0 | 0 | 0 | 0 |
| 0 | 0 | 0 | 0 | 0 |
| 0 | 0 | 0 | 0 | 0 |
| 0 | 0 | 0 | 0 | 0 |
| 0 | 0 | 0 | 0 | 0 |
| 0 | 0.001229989 | 0 | 0 | 0 |
| 0 | 0 | 0 | 0 | 0 |
| 0 | 0 | 0 | 0 | 0 |
| 0 | 0 | 0 | 0 | 0 |
| 0 | 0 | 0 | 0 | 0 |
| 0 | 0 | 0 | 0 | 0 |
| 0 | 0 | 0 | 0 | 0 |
| 0 | 0 | 0 | 0 | 0 |
| 0 | 0 | 0 | 0 | 0 |
| 0 | 0 | 0 | 0.000998139 | 0 |
| 0 | 0 | 0 | 0 | 0 |
| 0 | 0.001726654 | 0.045194276 | 0 | 0 |
| 0 | 0 | 0 | 0 | 0 |
| 0 | 0 | 0.009207318 | 0 | 0 |
| 0 | 0 | 0 | 0 | 0 |
| 0 | 0 | 0 | 0 | 0 |
| 0 | 0 | 0 | 0 | 0 |
| 0 | 0 | 0 | 0 | 0 |
| 0 | 0 | 0 | 0 | 0 |
| 0 | 0 | 0 | 0 | 0 |
| 0 | 0 | 0 | 0 | 0 |
| 0 | 0 | 0 | 0 | 0 |
| 0 | 0.00234224 | 0 | 0 | 0 |
| 0 | 0 | 0 | 0 | 0 |
| 0 | 0 | 0 | 0 | 0 |
| 0 | 0 | 0 | 0 | 0 |
| 0 | 0 | 0 | 0 | 0 |
| 0 | 0 | 0 | 0 | 0 |
| 0 | 0 | 0 | 0 | 0 |
| 0 | 0 | 0 | 0 | 0 |
| 0 | 0 | 0 | 0 | 0 |
| 0 | 0 | 0 | 0 | 0 |

| | | | | |
|---|---|---|---|---|
| 0 | 0 | 0 | 0 | 0 |
| 0 | 0.052015918 | 0.007298683 | 0 | 0 |
| 0 | 0 | 0 | 0 | 0 |
| 0 | 0 | 0 | 0 | 0 |
| 0 | 0 | 0 | 0 | 0 |
| 0 | 0 | 0 | 0 | 0 |
| 0 | 0 | 0 | 0 | 0 |
| 0 | 0 | 0 | 0 | 0 |
| 0 | 0 | 0 | 0 | 0 |
| 0 | 0 | 0 | 0 | 0 |
| 0.002380774 | 0.001081123 | 0 | 0 | 0.003403324 |
| 0 | 0 | 0 | 0 | 0 |
| 0 | 0.001891216 | 0 | 0 | 0.037433693 |
| 0 | 0 | 0 | 0 | 0 |
| 0 | 0 | 0 | 0 | 0 |
| 0 | 0 | 0 | 0 | 0 |
| 0 | 0 | 0 | 0 | 0 |
| 0 | 0 | 0 | 0 | 0 |
| 0 | 0 | 0 | 0 | 0 |
| 0 | 0 | 0 | 0 | 0 |
| 0 | 0 | 0.00772955 | 0.00462868 | 0 |
| 0 | 0 | 0 | 0 | 0 |
| 0 | 0 | 0 | 0 | 0 |
| 0.757809273 | 1.77115069 | 0.081859957 | 1.121806754 | 0.436082315 |
| 0 | 0 | 0 | 0 | 0 |
| 0 | 0 | 0 | 0 | 0 |
| 0 | 0 | 0 | 0 | 0 |
| 0 | 0 | 0.025762458 | 0 | 0 |
| 0 | 0 | 0 | 0 | 0 |
| 0 | 0 | 0 | 0 | 0 |
| 0 | 0.001884705 | 0 | 0 | 0 |
| 0 | 0 | 0 | 0 | 0 |
| 0 | 0 | 0 | 0 | 0 |
| 0 | 0 | 0 | 0 | 0 |

| PHL018-0106_V1 | PHL018-0171_V3 | PHL018-0107_V1 | PHL018-0097_V1 | PHL018-0211_V3 |
|---|---|---|---|---|
| CTRL | REF | EYCF | CTRL | REF |
| 0.108912998 | 0.809614349 | 0.121762666 | 0.506741887 | 0.780751509 |
| 4.838741803 | 0.116254341 | 19.36374105 | 5.132986608 | 3.574773053 |
| 7.604540857 | 0 | 12.26305565 | 0 | 1.688709563 |
| 0 | 0 | 0 | 0 | 0 |
| 0 | 0 | 0 | 0 | 0 |
| 0 | 0 | 0 | 0 | 0 |
| 0 | 0 | 0 | 0 | 0 |
| 0 | 0 | 0 | 0 | 0 |
| 0 | 0 | 0 | 0 | 0 |
| 0 | 0 | 0 | 0 | 0 |
| 0 | 0 | 0 | 0 | 0 |
| 0 | 0 | 0 | 0 | 0 |
| 0 | 0 | 0 | 0 | 0 |
| 6.355950974 | 0.56838656 | 5.232194475 | 0.181569389 | 0 |
| 0 | 0 | 0 | 0 | 0 |
| 0 | 0 | 0 | 0 | 0 |
| 0.001751685 | 0 | 0 | 0 | 0 |
| 0 | 0 | 0 | 0 | 0 |
| 0 | 0 | 0 | 0 | 0 |
| 0 | 0 | 0 | 0 | 0 |
| 0 | 0.001814647 | 0 | 0 | 0 |
| 0.000523191 | 0 | 0 | 0 | 0 |
| 0 | 0 | 0 | 0 | 0 |
| 0 | 0 | 0 | 0 | 0 |
| 0 | 0.110044809 | 0 | 0 | 0 |
| 0 | 0 | 0 | 0 | 0 |
| 0 | 0 | 0 | 0 | 0 |
| 0 | 0 | 0 | 0 | 0 |
| 0 | 0 | 0 | 0 | 0 |
| 0 | 0 | 0 | 0 | 0 |
| 0 | 0 | 0 | 0 | 0 |
| 0 | 0 | 0 | 0 | 0 |
| 0 | 0 | 0 | 0 | 0 |
| 0 | 0 | 0 | 0 | 0 |
| 0 | 0 | 0 | 0 | 0 |
| 0 | 0.074174886 | 0 | 0 | 0 |
| 0 | 0 | 0 | 0 | 0 |
| 0 | 0 | 0 | 0 | 0 |
| 0 | 0 | 0 | 0 | 0 |
| 0 | 0 | 0 | 0 | 0 |
| 0 | 0 | 0 | 0 | 0 |
| 0 | 0.005032466 | 0.001370286 | 0.017991944 | 0.063282316 |
| 0.056044917 | 1.540235586 | 0.008726555 | 1.226556949 | 0.180853159 |
| 0.001079993 | 0.00600145 | 0.000962969 | 0.067993619 | 0.003802989 |
| 0.0075639 | 0 | 0 | 0 | 0.006658708 |
| 0 | 0 | 0 | 0 | 0 |

| | | | | |
|---|---|---|---|---|
| 0.002450745 | 0.026023097 | 0 | 0.008183819 | 0.065992817 |
| 0.004760529 | 0 | 0.031569944 | 0.026669189 | 0.051337594 |
| 0.019680879 | 0 | 0.002559133 | 0 | 0.242558628 |
| 0 | 0.817690896 | 1.250162158 | 0.509317516 | 0.22994298 |
| 0 | 0.007107961 | 0 | 0.004285312 | 0 |
| 0 | 0 | 0 | 0 | 0 |
| 0.005102519 | 0.000904534 | 0.04578067 | 0.145783845 | 0 |
| 0 | 0 | 0 | 0 | 0 |
| 0 | 0.000429992 | 0 | 0 | 0 |
| 0 | 0 | 0 | 0 | 0 |
| 0 | 0.005342732 | 0 | 0 | 0 |
| 0.314785466 | 0.092105159 | 0 | 0.115659734 | 0.443145051 |
| 0 | 0 | 0 | 0 | 0 |
| 0.021621061 | 0.001480683 | 0.040389383 | 0.048222289 | 0.322901191 |
| 0 | 0 | 0 | 0 | 0 |
| 0 | 0 | 0 | 0 | 0 |
| 1.371380385 | 0 | 0 | 0.01715057 | 7.115137863 |
| 0 | 0 | 0 | 0 | 0 |
| 0 | 0 | 0 | 0 | 0 |
| 0 | 0 | 0 | 0 | 0 |
| 0.231685891 | 0 | 0.416247143 | 0.397313434 | 0.017563173 |
| 0.015257346 | 0.091447186 | 0.016500051 | 0.001579716 | 0.006516252 |
| 0.002033215 | 0.03552831 | 0 | 0 | 0 |
| 0 | 0 | 0 | 0 | 0 |
| 0 | 0 | 0 | 0 | 0 |
| 0 | 0 | 0 | 0 | 0 |
| 0 | 0 | 0 | 0 | 0 |
| 0 | 0 | 0 | 0 | 0 |
| 0.00251229 | 0 | 0.001206191 | 0.002598317 | 0 |
| 0 | 0 | 0 | 0 | 0 |
| 0.012153131 | 0 | 0 | 0.266197423 | 0.004972651 |
| 0 | 0 | 0 | 0 | 0 |
| 0 | 0 | 0 | 0 | 0 |
| 0 | 0 | 0 | 0 | 0 |
| 0 | 0 | 0 | 0 | 0 |
| 0 | 0 | 0 | 0 | 0 |
| 0 | 0 | 0 | 0 | 0 |
| 0.002564514 | 0.000668008 | 0 | 0.002736531 | 6.830242961 |
| 0 | 0 | 0 | 0 | 0 |
| 0 | 0 | 0 | 0 | 0 |
| 0 | 0 | 0 | 0 | 0 |
| 0 | 0 | 0 | 0 | 0 |
| 0 | 0 | 0 | 0 | 0 |
| 0 | 0 | 0 | 0 | 0 |
| 0 | 0.050778966 | 0 | 0 | 0 |
| 0 | 0 | 0 | 0 | 0 |
| 0 | 0 | 0 | 0 | 0 |
| 0 | 0 | 0 | 0 | 0 |

| | | | | |
|---|---|---|---|---|
| 0 | 0 | 0 | 0 | 0 |
| 0.036011859 | 0.001832005 | 0 | 0 | 0 |
| 0 | 0 | 0 | 0 | 0 |
| 0 | 0 | 0 | 0 | 0 |
| 0 | 0.000313526 | 0 | 0 | 0 |
| 0 | 0 | 0 | 0 | 0 |
| 0 | 0 | 0 | 0 | 0 |
| 0 | 0 | 0 | 0 | 0 |
| 0.010659027 | 0.005502243 | 0 | 0 | 0 |
| 0 | 0 | 0 | 0 | 0 |
| 0 | 0 | 0 | 0 | 0 |
| 0 | 0 | 0.0024688 | 0 | 0 |
| 0 | 0 | 0 | 0 | 0 |
| 0 | 0 | 0 | 0 | 0 |
| 0 | 0 | 0 | 0 | 0 |
| 0 | 0 | 0 | 0 | 0 |
| 0 | 0 | 0 | 0 | 0 |
| 0 | 0 | 0 | 0 | 0 |
| 0.279249959 | 0.183020878 | 0 | 0 | 0.869108868 |
| 0 | 0.166885534 | 0 | 0 | 0 |
| 0 | 0 | 0 | 0 | 0 |
| 0 | 0 | 0 | 0 | 0 |
| 0 | 0.001553086 | 0 | 0 | 0 |
| 0 | 0 | 0 | 0 | 0 |
| 0 | 0 | 0 | 0 | 0 |
| 0 | 0 | 0 | 0 | 0 |
| 0 | 0 | 0 | 0 | 0 |
| 0 | 0 | 0 | 0 | 0.000692151 |
| 0 | 0 | 0 | 0 | 0 |
| 0 | 0 | 0 | 0 | 0 |
| 0 | 0.004681001 | 0.041323415 | 0.047233448 | 0.008802102 |
| 0.001984495 | 0.033083135 | 0 | 0.002752889 | 0.00287008 |
| 0 | 0 | 0 | 0 | 0 |
| 0 | 0 | 0 | 0 | 0 |
| 0 | 0 | 0 | 0 | 0 |
| 0 | 0 | 0 | 0 | 0 |
| 0 | 0 | 0 | 0 | 0 |
| 0 | 0 | 0 | 0 | 0 |
| 0 | 0 | 0 | 0 | 0 |
| 0 | 0 | 0 | 0 | 0 |
| 0 | 0 | 0 | 0 | 0 |
| 0 | 0.356303235 | 0 | 0 | 0 |
| 0 | 0.002091188 | 0.120327124 | 0.005874285 | 0 |
| 0 | 0 | 0 | 0 | 0 |
| 0 | 0 | 0 | 0 | 0 |
| 0 | 0 | 0 | 0 | 0 |
| 0 | 0 | 0 | 0 | 0 |
| 0 | 0 | 0 | 0 | 0 |

| | | | | |
|---|---|---|---|---|
| 0 | 0 | 0 | 0 | 0.00210108 |
| 0 | 0 | 0 | 0 | 0 |
| 0 | 0 | 0 | 0 | 0 |
| 0 | 0 | 0 | 0 | 0 |
| 0 | 0.010948147 | 0 | 0 | 0 |
| 0 | 0 | 0 | 0 | 0 |
| 0 | 0 | 0 | 0 | 0 |
| 0 | 0 | 0 | 0 | 0 |
| 0 | 0 | 0 | 0 | 0 |
| 0 | 0 | 0 | 0 | 0 |
| 0 | 0 | 0 | 0 | 0 |
| 0 | 0 | 0 | 0 | 0 |
| 0 | 0 | 0 | 0 | 0 |
| 0 | 0 | 0 | 0 | 0 |
| 0 | 0 | 0 | 0 | 0 |
| 0 | 0 | 0 | 0 | 0 |
| 0 | 0 | 0 | 0 | 0 |
| 0 | 0 | 0 | 0 | 0 |
| 0 | 0 | 0 | 0 | 0 |
| 0 | 0 | 0 | 0 | 0 |
| 0 | 0 | 0 | 0 | 0 |
| 0 | 0 | 0 | 0 | 0 |
| 0 | 0.00307213 | 0 | 0 | 0 |
| 0 | 0 | 0 | 0 | 0 |
| 0 | 0.060410962 | 0 | 0 | 0 |
| 0 | 0 | 0 | 0 | 0 |
| 0 | 0.028824583 | 0 | 0 | 0 |
| 0 | 0 | 0 | 0 | 0 |
| 0 | 0 | 0 | 0 | 0 |
| 0 | 0 | 0 | 0 | 0 |
| 0 | 0.06933511 | 0 | 0 | 0 |
| 0 | 0 | 0 | 0 | 0 |
| 0 | 0 | 0 | 0 | 0 |
| 0 | 0 | 0 | 0 | 0 |
| 0 | 0 | 0 | 0 | 0 |
| 0 | 0 | 0 | 0 | 0 |
| 0 | 0 | 0 | 0 | 0 |
| 0 | 0 | 0 | 0 | 0 |
| 0.00940171 | 0.009577427 | 0 | 0 | 0 |
| 0 | 0 | 0 | 0 | 0 |
| 0 | 0 | 0 | 0 | 0 |
| 0 | 0 | 0 | 0 | 0 |
| 0 | 0 | 0 | 0 | 0 |
| 0 | 0 | 0 | 0 | 0 |
| 0 | 0 | 0 | 0 | 0 |
| 0 | 0 | 0 | 0 | 0 |
| 0 | 0 | 0 | 0 | 0 |
| 0 | 0 | 0 | 0 | 0 |

| | | | | |
|---|---|---|---|---|
| 0 | 0 | 0 | 0 | 0 |
| 0 | 0 | 0 | 0 | 0 |
| 0 | 0 | 0 | 0 | 0 |
| 0 | 0 | 0 | 0 | 0 |
| 0 | 0 | 0 | 0 | 0 |
| 0 | 0 | 0 | 0 | 0 |
| 0 | 0 | 0 | 0 | 0 |
| 0 | 0 | 0 | 0 | 0 |
| 0 | 0 | 0 | 0 | 0 |
| 0 | 0 | 0 | 0 | 0 |
| 0 | 0 | 0 | 0 | 0 |
| 0 | 0.039175083 | 0 | 0 | 0 |
| 0 | 0.001426786 | 0.001627993 | 0 | 0.002342109 |
| 0 | 0 | 0 | 0 | 0 |
| 0 | 0.004954426 | 0 | 0 | 0 |
| 0 | 0 | 0 | 0 | 0 |
| 0 | 0 | 0 | 0 | 0 |
| 0 | 0 | 0 | 0 | 0 |
| 0 | 0 | 0 | 0 | 0 |
| 0 | 0 | 0 | 0 | 0 |
| 0.031602659 | 0.006224741 | 0.009131866 | 0.059014256 | 0.006697572 |
| 0 | 0 | 0.004653184 | 0 | 0 |
| 0 | 0 | 0 | 0 | 0 |
| 0 | 0 | 0 | 0.007293629 | 0 |
| 0 | 0 | 0 | 0 | 0 |
| 0 | 0 | 0 | 0 | 0 |
| 0.003749094 | 0.003107265 | 0 | 0.002036653 | 0.007650997 |
| 0 | 0 | 0 | 0 | 0 |
| 0 | 0 | 0 | 0 | 0 |
| 0 | 0 | 0.00398289 | 0 | 0 |
| 0 | 0 | 0.658954783 | 0.323055062 | 0 |
| 0 | 0 | 0 | 0 | 0 |
| 0 | 0 | 0 | 0 | 0 |
| 0.04363439 | 0.015434083 | 0 | 0 | 0 |
| 0.006854125 | 0.013307631 | 0.021390042 | 0.071675932 | 0 |
| 0 | 0 | 0 | 0 | 0 |
| 0 | 0 | 0 | 0 | 0 |
| 0 | 0 | 0 | 0 | 0 |
| 0 | 0 | 0 | 0 | 0 |
| 0 | 0 | 0 | 0 | 0 |
| 0 | 0 | 0 | 0 | 0 |
| 0 | 0 | 0 | 0 | 0 |
| 0 | 0 | 0 | 0 | 0 |
| 0 | 0 | 0 | 0 | 0 |
| 0 | 0 | 0 | 0 | 0 |
| 0 | 0.011458371 | 0.035074175 | 0 | 0 |
| 0 | 0 | 0.002787745 | 0.001231841 | 0.004536861 |
| 0 | 0.000659713 | 0.00127026 | 0 | 0.005494318 |

| | | | | |
|---|---|---|---|---|
| 0 | 0 | 0 | 0 | 0 |
| 0 | 0 | 0 | 0 | 0 |
| 0.002479608 | 0 | 0.001805591 | 0 | 0.0032743 |
| 0.000689861 | 0 | 0 | 0 | 0 |
| 0 | 0 | 0 | 0 | 0 |
| 0 | 0 | 0 | 0 | 0 |
| 0 | 0 | 0 | 0 | 0 |
| 0 | 0 | 0 | 0 | 0 |
| 0 | 0 | 0 | 0 | 0 |
| 0 | 0 | 0 | 0 | 0 |
| 0 | 0 | 0 | 0 | 0 |
| 0 | 0 | 0 | 0 | 0 |
| 0 | 0 | 0 | 0 | 0 |
| 0 | 0 | 0 | 0 | 0 |
| 0 | 0.003900559 | 0 | 0 | 0 |
| 0 | 0 | 0 | 0 | 0 |
| 0 | 0 | 0 | 0 | 0 |
| 0 | 0 | 0 | 0 | 0 |
| 0.000551938 | 0.008470415 | 0 | 0.004534986 | 0 |
| 0 | 0 | 0 | 0 | 0 |
| 0.028902823 | 0 | 0.078341527 | 0.00516921 | 0 |
| 0 | 0 | 0 | 0 | 0 |
| 0 | 0 | 0 | 0 | 0 |
| 0 | 0 | 0 | 0 | 0 |
| 0 | 0.001474658 | 0.004206541 | 0.004228711 | 0.002847875 |
| 0 | 0 | 0 | 0 | 0 |
| 0 | 0 | 0 | 0 | 0 |
| 0 | 0 | 0 | 0 | 0 |
| 0 | 0 | 0 | 0 | 0 |
| 0 | 0 | 0 | 0 | 0 |
| 0.064205694 | 0.035657896 | 0.795876131 | 0.615748675 | 1.224775682 |
| 0 | 0 | 0 | 0 | 0 |
| 0 | 0 | 0 | 0.003058093 | 0 |
| 0 | 0 | 0 | 0 | 0 |
| 0 | 0 | 0 | 0 | 0 |
| 0 | 0 | 0 | 0 | 0 |
| 0 | 0 | 0 | 0 | 0 |
| 0 | 0 | 0 | 0 | 0 |
| 0 | 0 | 0 | 0 | 0 |
| 0 | 0 | 0 | 0 | 0 |
| 0 | 0 | 0 | 0 | 0 |
| 0 | 0.028465867 | 0.00587182 | 0.017320863 | 0 |
| 0 | 0.001011371 | 0 | 0.004015654 | 0 |
| 0 | 0 | 0 | 0 | 0 |
| 0 | 0 | 0 | 0 | 0 |
| 0 | 0 | 0 | 0 | 0 |
| 0 | 0 | 0 | 0 | 0 |
| 0 | 0 | 0 | 0 | 0 |

| | | | | |
|---|---|---|---|---|
| 0 | 0 | 0 | 0 | 0 |
| 0 | 0 | 0 | 0 | 0 |
| 0 | 0.000582143 | 0.001394899 | 0.054659159 | 0.002866813 |
| 0 | 0 | 0 | 0 | 0 |
| 0 | 9.123200668 | 0 | 0 | 0 |
| 0 | 0 | 0 | 0 | 0 |
| 0 | 0 | 0 | 0 | 0 |
| 0 | 0 | 0 | 0 | 0 |
| 0 | 0 | 0 | 0 | 0 |
| 0.000876 | 0 | 0 | 0 | 0.000616933 |
| 0 | 0 | 0 | 0 | 0 |
| 0 | 0 | 0 | 0 | 0 |
| 0 | 0 | 0 | 0 | 0 |
| 0.019900655 | 0.001286746 | 0.002202307 | 0.014232308 | 0 |
| 0.007164264 | 0.005310203 | 0.015814132 | 0.007048144 | 0 |
| 0 | 0.060263897 | 0 | 0 | 0 |
| 0 | 0.00060505 | 0.002278237 | 0.006840991 | 0.003330164 |
| 0 | 0 | 0 | 0.009921169 | 0 |
| 0 | 0 | 0 | 0 | 0 |
| 0 | 0 | 0 | 0 | 0 |
| 0 | 0 | 0 | 0 | 0 |
| 0 | 0.000327712 | 0.000934817 | 0 | 0.007119919 |
| 0 | 0 | 0 | 0 | 0 |
| 0 | 0 | 0.010723933 | 0.003746104 | 0 |
| 0.008357367 | 0.000525468 | 0 | 0 | 0 |
| 0 | 0 | 0 | 0 | 0 |
| 0 | 0.552741558 | 0 | 0 | 0 |
| 0 | 0 | 0 | 0 | 0 |
| 0 | 0 | 0 | 0 | 0 |
| 0.006239514 | 0 | 0 | 0 | 0 |
| 0 | 0.000837104 | 0 | 0 | 0 |
| 0 | 0 | 0 | 0 | 0 |
| 0 | 0 | 0 | 0 | 0 |
| 0 | 0 | 0.001940114 | 0 | 0 |
| 0 | 0 | 0 | 0.002839463 | 0 |
| 0.417497547 | 0 | 0 | 0 | 0.009480684 |
| 0 | 0 | 0 | 0 | 0 |
| 0.013097037 | 0.115423553 | 0 | 0 | 0.002584238 |
| 0 | 0 | 0 | 0 | 0 |
| 0 | 0 | 0 | 0 | 0 |
| 0 | 0 | 0 | 0 | 0 |
| 0 | 0 | 0 | 0 | 0 |
| 0 | 0 | 0 | 0 | 0 |
| 0 | 0 | 0.002626045 | 0.007961561 | 0.043458845 |
| 0 | 0 | 0 | 0 | 0 |
| 0.020136487 | 0.002743168 | 0 | 0 | 0 |
| 0.002202938 | 0.000446308 | 0 | 0 | 0 |
| 0 | 0 | 0.024184463 | 0.00858068 | 0 |

| | | | | |
|---|---|---|---|---|
| 3.571652555 | 0.008329052 | 0 | 0.111789233 | 0 |
| 0 | 0.289840976 | 0 | 0 | 0 |
| 0 | 0.000572089 | 0 | 0 | 0.002154406 |
| 0 | 0 | 0 | 0 | 0 |
| 0.002272798 | 0.002302306 | 0.00728987 | 0 | 0 |
| 0.002161836 | 0.004317237 | 0 | 0 | 0 |
| 0 | 0 | 0 | 0 | 0 |
| 0 | 0.013715552 | 0.064624203 | 0.047989876 | 0.000934857 |
| 0 | 0 | 0 | 0 | 0 |
| 0.079356108 | 0.0039916 | 0.315968455 | 0.017169393 | 0.00891309 |
| 0 | 0 | 0 | 0 | 0 |
| 0.020385457 | 0 | 0.025796994 | 0.002511667 | 0.012580613 |
| 0.00382592 | 0.00455382 | 0.002984939 | 0.024291151 | 0.00238571 |
| 0.013558032 | 0 | 0 | 0 | 0 |
| 0 | 0 | 0.00289939 | 0 | 0 |
| 0 | 0 | 0 | 0.003217504 | 0 |
| 0 | 0 | 0 | 0 | 0 |
| 0 | 0 | 0 | 0 | 0 |
| 0.021886811 | 0 | 0.157767394 | 0.308888982 | 0.400311864 |
| 0 | 0 | 0 | 0 | 0 |
| 0 | 0 | 0 | 0 | 0 |
| 0 | 0 | 0 | 0 | 0 |
| 0 | 0 | 0.006160144 | 0 | 0.000868851 |
| 0 | 0.09114006 | 0 | 0 | 0 |
| 0 | 0.000485754 | 0 | 0 | 0 |
| 0 | 0 | 0 | 0 | 0.040536978 |
| 0.004469153 | 0.348743471 | 0 | 0 | 0 |
| 0.00538881 | 0 | 1.443005288 | 0.762773145 | 0.20932533 |
| 0 | 0 | 0 | 0 | 0 |
| 0 | 0 | 0.001391087 | 0.007705571 | 0.001513577 |
| 0.010360496 | 0 | 0 | 0 | 0 |
| 0 | 0 | 0 | 0 | 0.744571338 |
| 0 | 0 | 0 | 0 | 0 |
| 0 | 0 | 0.004546876 | 0.363853412 | 0.110732771 |
| 0 | 0 | 0 | 0 | 0 |
| 0 | 0 | 0 | 0 | 0 |
| 0 | 0 | 0 | 0.037732955 | 0.137117276 |
| 0 | 0 | 0 | 0 | 0 |
| 0 | 0 | 0 | 0 | 0 |
| 0 | 0 | 1.010234963 | 0 | 0 |
| 0 | 0 | 0 | 0.031209192 | 0 |
| 0 | 0 | 0 | 0.588004926 | 0 |
| 0 | 0 | 0 | 0 | 0 |
| 0 | 0 | 0 | 0.11866203 | 0.019094406 |
| 0 | 0 | 0 | 0 | 0.023823241 |
| 0 | 0.006109953 | 0 | 0 | 0 |
| 0 | 0.000421021 | 0.006004921 | 0.001724734 | 0.001829429 |
| 0 | 0 | 0 | 0 | 0 |

| | | | | |
|---|---|---|---|---|
| 0 | 0 | 0 | 0 | 0 |
| 0 | 0 | 0.046707995 | 0 | 0 |
| 0 | 0 | 0 | 0 | 0 |
| 0 | 0 | 0 | 0 | 0 |
| 0.002510454 | 0 | 0 | 0.003068495 | 0 |
| 0.023096701 | 0.00044214 | 0 | 0.00733556 | 0.018731661 |
| 0 | 0 | 0 | 0 | 0.087942878 |
| 0 | 0 | 0 | 0 | 0 |
| 0 | 0 | 0 | 0 | 0 |
| 0 | 0 | 0 | 0 | 0 |
| 0.002953039 | 0.000769212 | 0.002413639 | 0 | 0 |
| 0.00256998 | 0 | 0 | 0 | 0.054994293 |
| 0.008772532 | 0 | 0.012733452 | 0 | 0 |
| 0.003985547 | 0.001211187 | 0.000829194 | 0 | 0.008245177 |
| 0.264139062 | 0 | 0.303926177 | 0 | 0.010650176 |
| 0 | 0 | 0 | 0 | 0 |
| 0 | 0.003722348 | 0 | 0 | 0 |
| 0 | 0 | 0 | 0 | 0 |
| 0 | 0 | 0.024049485 | 0 | 0 |
| 0 | 0.005955413 | 0 | 0.01374544 | 0 |
| 0 | 0 | 0 | 0 | 0 |
| 0 | 0 | 0 | 0 | 0.106311233 |
| 0 | 0.007126026 | 0 | 0 | 0 |
| 0 | 0 | 0 | 0 | 0.096474305 |
| 0 | 0 | 0 | 0 | 0 |
| 0 | 0 | 0 | 0 | 0 |
| 0 | 0 | 0.011687861 | 0.033892679 | 0.002282542 |
| 0 | 0 | 0 | 0 | 0 |
| 0 | 0 | 0 | 0 | 0 |
| 0.135825973 | 0.110249054 | 0.091764605 | 0.96655574 | 1.463912932 |
| 0.001487132 | 0.029087982 | 0 | 0.008684577 | 0 |
| 0 | 0 | 0 | 0 | 0 |
| 0 | 0 | 0 | 0 | 0.041602777 |
| 0 | 0.00081466 | 0 | 0 | 0 |
| 0 | 0 | 0.062715137 | 0 | 0.021704708 |
| 0 | 0 | 0 | 0 | 0 |
| 0 | 0 | 0 | 0 | 0.136282643 |
| 0 | 0 | 0 | 0.001213749 | 0 |
| 0 | 0 | 2.074101062 | 0 | 0 |
| 1.421378978 | 0 | 0 | 0 | 0 |
| 0.077326661 | 0 | 0.024540767 | 0.018355737 | 0 |
| 2.611416748 | 0.001712736 | 0 | 0 | 0.581485407 |
| 0 | 0 | 0 | 0.00305765 | 0 |
| 0 | 0 | 0 | 0 | 0.025402509 |
| 0 | 0 | 0 | 0.044173286 | 0.006019527 |
| 0 | 0 | 0 | 0 | 0 |
| 0.000687796 | 0 | 0.004292883 | 0.002568755 | 0.003178798 |
| 0 | 0 | 0 | 0 | 0 |

| | | | | |
|---|---|---|---|---|
| 0.005516058 | 0.001160517 | 0 | 0.000950824 | 0.079375892 |
| 0.001897861 | 0 | 0 | 0 | 0 |
| 0.071402493 | 0 | 0 | 0.033252971 | 0.351696996 |
| 0 | 0 | 0 | 0 | 0 |
| 0.181562591 | 0.240406384 | 0 | 0 | 0 |
| 0 | 0 | 0.204325259 | 0 | 0 |
| 0 | 0 | 0 | 0 | 0 |
| 0 | 0 | 0 | 0 | 0 |
| 0.043856237 | 0.000431666 | 0 | 0 | 0 |
| 0 | 0 | 0 | 0.019280802 | 0 |
| 0 | 0 | 0 | 0 | 0 |
| 0 | 0 | 0 | 0 | 0 |
| 0 | 0 | 0.024153522 | 0 | 0 |
| 0 | 0 | 0 | 0 | 0 |
| 0 | 0 | 0 | 0 | 0 |
| 0 | 0.081591288 | 0 | 0.01814996 | 0 |
| 0.383871734 | 0.014348415 | 0.001037652 | 0.136102487 | 0.032490722 |
| 0.002221863 | 0 | 0.217471963 | 0.012673516 | 0 |
| 0 | 0 | 0 | 0 | 0 |
| 0 | 0 | 0 | 0 | 0 |
| 0 | 0 | 0.028334304 | 0.046012035 | 0.009960216 |
| 0.000771366 | 0 | 0.010632 | 0 | 0.002206929 |
| 0 | 0 | 0 | 0 | 0 |
| 0 | 0.064470804 | 0.23557836 | 0 | 0 |
| 0 | 0 | 0.119445353 | 0 | 0 |
| 0 | 0 | 0.002733848 | 0 | 0 |
| 0.006120623 | 0 | 0 | 0.002522383 | 0 |
| 0 | 0 | 0 | 0 | 0 |
| 0 | 0 | 0 | 0 | 0 |
| 0 | 0 | 0 | 0 | 0.092208511 |
| 0.043176511 | 0.011121248 | 0.082649237 | 0.010550456 | 0.02882502 |
| 0.03063001 | 0.002181634 | 0.008463587 | 0.004289846 | 0.000842639 |
| 0.001367058 | 0.003185056 | 0.142919498 | 0 | 0 |
| 0 | 0 | 0 | 0.001951169 | 0 |
| 0 | 0 | 0 | 0.009400307 | 0 |
| 0 | 0 | 0 | 0 | 0 |
| 0 | 0 | 0.001285662 | 0.015386174 | 0.026656238 |
| 0.001127633 | 0.002447728 | 0.009049029 | 0.015401858 | 0.023399036 |
| 0 | 0.055932255 | 0 | 0 | 0 |
| 0 | 0 | 0 | 0.000983796 | 0 |
| 0.004294011 | 0 | 0 | 0 | 0 |
| 0 | 0 | 0 | 0 | 0 |
| 0 | 0.000585692 | 0.00100243 | 0 | 0.025873776 |
| 0 | 0 | 0.122309814 | 0.061054541 | 0.069631563 |
| 0.002706903 | 0 | 0 | 0 | 0.025779305 |
| 0 | 0 | 0 | 0 | 0 |
| 0 | 0 | 0 | 0 | 0 |
| 0 | 0 | 0 | 0 | 0 |

| | | | | |
|---|---|---|---|---|
| 0 | 0 | 0 | 0.011774732 | 0.000630782 |
| 1.031459589 | 0.739911938 | 0.273781653 | 0.028857104 | 0.363092889 |
| 0 | 0 | 0 | 0 | 0 |
| 0.001994119 | 0 | 0 | 0 | 0 |
| 0 | 0 | 0 | 0 | 0 |
| 0 | 0 | 0 | 0 | 0 |
| 0 | 0 | 0.029785655 | 0.323999404 | 0 |
| 0 | 0 | 0.002104686 | 0 | 0.010241437 |
| 0 | 0 | 0 | 0 | 0 |
| 0 | 0 | 0.029535793 | 0.014297654 | 0.057853769 |
| 0.00114182 | 0.000495705 | 0.000509048 | 0.004142588 | 0.029724464 |
| 0.016278826 | 0.007235986 | 0.016428579 | 0.297273004 | 0.431204206 |
| 0 | 0.096588158 | 0.021025336 | 0 | 0 |
| 0 | 0 | 0 | 0 | 0 |
| 0 | 0 | 0 | 0.019851188 | 0.084521348 |
| 0 | 0 | 0 | 0 | 0 |
| 0.001708071 | 0.009135706 | 0.030916739 | 0.001531019 | 0 |
| 0.70710103 | 0 | 0.095261055 | 0.009606125 | 0.001561569 |
| 0 | 0 | 0 | 0 | 0 |
| 0 | 0 | 0.09226211 | 0.200037093 | 0.001774502 |
| 0.006869665 | 0.049889629 | 0.003481472 | 0.001842011 | 0.005737423 |
| 0 | 0 | 0 | 0.00327441 | 0 |
| 0 | 0 | 0 | 0 | 0 |
| 0 | 0.193190707 | 0 | 0 | 0 |
| 0 | 0 | 0.001231479 | 0 | 0 |
| 0 | 0 | 0 | 0 | 0 |
| 0.094408247 | 0.002007785 | 0 | 0.023116571 | 0 |
| 0 | 0.025716138 | 0.099128852 | 0.08166765 | 0.09756854 |
| 0 | 0 | 0 | 0.001363334 | 0 |
| 0 | 0 | 0 | 0 | 0 |
| 0 | 0 | 0 | 0 | 0 |
| 0 | 0 | 0 | 0 | 0 |
| 0 | 0 | 0 | 0 | 0.004801926 |
| 0 | 0 | 0 | 0 | 0 |
| 0 | 0.021914031 | 0 | 0 | 0.297297422 |
| 0.467357588 | 0.289447402 | 0.30851161 | 0.813685273 | 0.273355329 |
| 0 | 0 | 0 | 0 | 0.001550226 |
| 0 | 0 | 0 | 0 | 0.003177518 |
| 0.070440476 | 0.005193708 | 0.006246463 | 0.002415148 | 0.006302725 |
| 0 | 0 | 0 | 0 | 0 |
| 0.00492741 | 0 | 0 | 0 | 0.025005758 |
| 0 | 0 | 0 | 0 | 0 |
| 0.001823692 | 0 | 0.047281533 | 0.020118828 | 0 |
| 0.005823987 | 0 | 0.000563573 | 0.542263102 | 0 |
| 0 | 0 | 0 | 0 | 0 |
| 0 | 0.122732243 | 0 | 0 | 0 |
| 0 | 0 | 0 | 0 | 0 |
| 0.002627169 | 0.000665319 | 0 | 0 | 0 |

| | | | | |
|---|---|---|---|---|
| 0.006663313 | 0 | 0.070334558 | 0 | 0 |
| 0 | 0 | 0 | 0 | 0 |
| 0 | 0 | 0 | 0 | 0 |
| 0 | 3.174173166 | 0 | 0 | 0 |
| 0 | 0 | 0 | 0 | 0 |
| 0 | 0.001894469 | 0 | 0 | 0 |
| 0.00050668 | 0 | 0 | 0 | 0 |
| 0 | 0.001687934 | 0 | 0.057210152 | 0 |
| 0 | 1.934796378 | 0 | 0.189517528 | 1.000137719 |
| 0 | 0.318626559 | 0 | 0 | 0 |
| 0 | 0.090178874 | 0 | 0 | 0 |
| 0 | 0.022549249 | 0 | 0 | 0 |
| 0 | 0 | 0 | 0 | 0 |
| 0 | 0.136243382 | 0 | 0 | 0 |
| 0.014984729 | 0.012719186 | 0 | 0 | 0 |
| 0 | 0 | 0 | 0 | 0 |
| 0 | 0.051104571 | 0 | 0 | 0 |
| 0 | 10.83492951 | 0 | 0 | 0 |
| 0 | 0 | 0 | 0 | 0 |
| 0 | 0 | 0 | 0 | 0 |
| 0 | 0 | 0 | 0 | 0.002178983 |
| 0 | 0.000353339 | 0 | 0 | 0 |
| 0 | 0.004731048 | 0 | 0 | 0 |
| 0 | 0.086670393 | 0.422624163 | 0.449853826 | 0.28601171 |
| 0 | 0.086318109 | 0.00060985 | 0 | 0 |
| 0 | 0.270643552 | 0 | 0.004061745 | 0 |
| 0 | 0.008055312 | 0.001451255 | 0 | 0 |
| 0 | 4.60686708 | 0 | 0 | 0 |
| 0 | 0 | 0 | 0 | 0 |
| 0 | 0 | 0 | 0 | 0 |
| 0.069092138 | 0.004994821 | 0 | 0 | 0 |
| 0 | 0.101790074 | 0 | 0 | 0 |
| 0 | 0.033016085 | 0 | 0.03201064 | 0 |
| 0 | 0 | 0 | 0 | 0 |
| 0 | 0.031512984 | 0 | 0 | 0 |
| 0 | 0 | 0.060000831 | 0 | 0 |
| 0 | 1.213236017 | 0 | 0 | 0.005281463 |
| 0 | 0 | 0 | 0 | 0 |
| 0 | 0.001188545 | 0 | 0.310881799 | 0 |
| 0 | 0.000953427 | 0 | 0 | 0 |
| 0.098964586 | 0.074662254 | 0.052020799 | 0.050477776 | 0.010708376 |
| 0 | 0.107414979 | 0 | 0 | 0 |
| 0.152588745 | 0.012701367 | 0.170724501 | 1.474025959 | 0 |
| 0 | 0 | 0.010886946 | 0.025406472 | 0 |
| 0 | 0 | 0 | 0 | 0 |
| 0 | 0.368594484 | 0 | 0 | 0 |
| 0 | 0 | 0 | 0 | 0 |
| 0.000722105 | 0.005047212 | 0.012957696 | 0.058252875 | 0.004449821 |

| | | | | |
|---|---|---|---|---|
| 0 | 0 | 0 | 0 | 0 |
| 0 | 0 | 0.013002419 | 0 | 0 |
| 0.164055923 | 0.00115699 | 0.910723707 | 0.273780705 | 0.690731534 |
| 0 | 0 | 0 | 0 | 0 |
| 0 | 0 | 0 | 0 | 0 |
| 0 | 0.175077845 | 0 | 0 | 0 |
| 0 | 0 | 0 | 0 | 0 |
| 0 | 0.069189789 | 0 | 0 | 0 |
| 0 | 0 | 0 | 0 | 0 |
| 0.222302757 | 0 | 0 | 0 | 0 |
| 0.072891002 | 0.624570992 | 0.013432864 | 0.14239764 | 0.427786283 |
| 0.775269638 | 0.435425284 | 1.144674859 | 0.981172589 | 0.082962131 |
| 0 | 0 | 0 | 0 | 0 |
| 0.012102606 | 0.010587952 | 0.003815077 | 0 | 0.088139748 |
| 0.570938531 | 0.052433645 | 0.076212932 | 0.178529971 | 0.009668077 |
| 0 | 3.485147513 | 0 | 0 | 0 |
| 0 | 0 | 0 | 0 | 0 |
| 0 | 0 | 0 | 0 | 0 |
| 0 | 0.006653149 | 0 | 0 | 0 |
| 0 | 0.00459359 | 0.028682019 | 0.004390841 | 0 |
| 1.210190353 | 0.055481525 | 0.000740705 | 0.03361378 | 0.000376099 |
| 0.537646135 | 0 | 1.015083713 | 0.031607645 | 0 |
| 2.005523285 | 0.238228112 | 2.810768971 | 0.413139269 | 1.261083559 |
| 0 | 0 | 0 | 0 | 0 |
| 0 | 0.031642341 | 0 | 0 | 0 |
| 0 | 1.625786264 | 0 | 0 | 0 |
| 0.159426643 | 0.002795131 | 0 | 0 | 12.01546675 |
| 0 | 0 | 0 | 0 | 0.001961323 |
| 0 | 0.084838829 | 0 | 0 | 0 |
| 0 | 0.074319827 | 0 | 0 | 0 |
| 0 | 0.014415597 | 0 | 0 | 0 |
| 0 | 0.027455976 | 0 | 0 | 0.304647032 |
| 0.290195276 | 0.00095454 | 0.00181525 | 0.368873817 | 0.006912813 |
| 0 | 0.213520391 | 0.008582466 | 0 | 0 |
| 0.000691037 | 0 | 0.003594263 | 0.008430815 | 0.032850304 |
| 0.008070265 | 0.043471981 | 0.016460991 | 0.002363964 | 0 |
| 0.000917149 | 0.000464528 | 0 | 0 | 0 |
| 0 | 0 | 0 | 0 | 0 |
| 0 | 2.810172401 | 0.010787103 | 0.197238299 | 0.121444788 |
| 0 | 0 | 0 | 0 | 0 |
| 0 | 0.048441905 | 0 | 0 | 0 |
| 0 | 0 | 0 | 0 | 0 |
| 0 | 0 | 0 | 0 | 0 |
| 0 | 0 | 0 | 0 | 0 |
| 0.028692394 | 0.067123483 | 0.026468368 | 0.00242625 | 0.497072137 |
| 0 | 0.302555327 | 0 | 0 | 0 |
| 0 | 0.375491667 | 0 | 0 | 0 |
| 0.123205984 | 0 | 0.035098249 | 0 | 0 |

| | | | | |
|---|---|---|---|---|
| 0 | 0 | 0 | 0 | 0 |
| 0 | 0 | 0 | 0 | 0 |
| 0 | 0 | 0 | 0 | 0 |
| 0 | 0.010002239 | 0 | 0.001608353 | 0 |
| 0 | 0.004983559 | 0 | 0 | 0 |
| 0 | 0.056801247 | 0 | 0 | 0.13225599 |
| 0 | 0.395447821 | 0 | 0 | 0 |
| 0.01089743 | 0.010322098 | 0 | 0.106558278 | 0 |
| 0 | 0 | 0 | 0 | 0 |
| 0 | 0 | 0 | 0 | 0 |
| 0.005310113 | 0.002290436 | 0.003385587 | 0.009468192 | 0.005579406 |
| 0 | 0 | 0 | 0 | 0 |
| 0 | 0 | 0 | 0.019044922 | 0 |
| 0 | 0 | 0 | 0 | 0 |
| 0 | 0 | 0 | 0 | 0 |
| 0 | 0 | 0 | 0 | 0 |
| 0 | 0.015486192 | 0 | 0 | 0 |
| 0 | 0.001733396 | 0 | 0 | 0 |
| 0 | 0.004195182 | 0 | 0 | 0 |
| 0 | 0 | 0 | 0 | 0 |
| 0 | 0 | 0 | 0 | 0 |
| 0 | 0.007695839 | 0 | 0 | 0 |
| 0.001479651 | 0.022982537 | 0 | 0 | 0 |
| 0 | 0 | 0 | 0 | 0 |
| 0 | 0 | 0 | 0 | 0 |
| 0 | 0 | 0 | 0 | 0 |
| 0 | 0.001095999 | 0 | 0 | 0 |
| 0 | 0 | 0 | 0 | 0 |
| 0 | 0 | 0 | 0 | 0 |
| 0 | 0.333353142 | 0.012364187 | 0 | 0 |
| 0 | 0 | 0 | 0 | 0.068291404 |
| 0 | 0 | 0 | 0 | 0 |
| 0 | 0.172054837 | 0 | 0 | 0 |
| 0 | 0 | 0 | 0 | 0 |
| 0.001225841 | 0.004221972 | 0 | 0 | 0 |
| 0 | 0.026880245 | 0 | 0 | 0 |
| 0 | 0.027585477 | 0 | 0 | 0.245021428 |
| 0.00739517 | 0 | 0.008899557 | 0.014730252 | 0.003574211 |
| 0 | 0 | 0 | 0 | 0 |
| 0 | 0.000695425 | 0 | 0 | 0.006673087 |
| 0 | 0.409063467 | 0 | 0 | 0 |
| 0 | 0.003436318 | 0 | 0 | 0 |
| 0 | 0.001092878 | 0 | 0 | 0 |
| 0 | 0.130946839 | 0 | 0 | 0 |
| 0 | 0.13289622 | 0 | 0.128642996 | 0.100157481 |
| 0 | 0 | 0 | 0 | 0 |
| 0 | 0.003433329 | 0 | 0 | 0 |
| 0 | 0.085128786 | 0 | 0 | 0 |

| | | | | |
|---|---|---|---|---|
| 0 | 0 | 0 | 0 | 0 |
| 0 | 0 | 0 | 0 | 0 |
| 0 | 0 | 0 | 0 | 0 |
| 0 | 0.420178602 | 0 | 0 | 0 |
| 0 | 0.003068709 | 0 | 0 | 0 |
| 0 | 0 | 0 | 0 | 0 |
| 0 | 0 | 0 | 0 | 0 |
| 0 | 0 | 0 | 0 | 0 |
| 0.132389282 | 0.001458577 | 1.512611394 | 0.489961535 | 0.049646443 |
| 0 | 0.001076704 | 0 | 0 | 0 |
| 0 | 0.000434553 | 0 | 0 | 0 |
| 0 | 0.001037423 | 0 | 0 | 0 |
| 0.000933334 | 0.00829634 | 0 | 0 | 0 |
| 0 | 0.002110585 | 0 | 0 | 0 |
| 0 | 0.596934076 | 0 | 2.853077722 | 0.578300293 |
| 1.229982008 | 0 | 0 | 0.177991254 | 0 |
| 0 | 0 | 0 | 0 | 0 |
| 0 | 0 | 0 | 0 | 0 |
| 0 | 0 | 0 | 0 | 0 |
| 0 | 0.232404268 | 0 | 0 | 0 |
| 0 | 0.074631596 | 0 | 0 | 0 |
| 0 | 0.008977196 | 0 | 0 | 0 |
| 0 | 0 | 0 | 0 | 0 |
| 0 | 0.038614582 | 0 | 0 | 0 |
| 0 | 0.048598622 | 0 | 0.22344573 | 0.006417968 |
| 5.77718946 | 4.538723464 | 0 | 21.82364466 | 0 |
| 0 | 0.414999307 | 0 | 0 | 0 |
| 0.001751811 | 0 | 0 | 0 | 0.002698792 |
| 0 | 0 | 0 | 0 | 0 |
| 0 | 0.516874102 | 0 | 0 | 0 |
| 0 | 0 | 0 | 0 | 0 |
| 0 | 0.024670016 | 0 | 0 | 0 |
| 0 | 0.00027687 | 0 | 0 | 0 |
| 0 | 0.002977166 | 0 | 0.031522285 | 0 |
| 0.013451553 | 0.006792134 | 0 | 0.006801505 | 0.021861729 |
| 0.001609151 | 0 | 0.005630463 | 0.003059534 | 0.002961938 |
| 0 | 0 | 0.066427945 | 0 | 0 |
| 0 | 0.850256501 | 0.640416054 | 0 | 0 |
| 0 | 2.152618881 | 0 | 0 | 0 |
| 0 | 0 | 0 | 0 | 0 |
| 0 | 0 | 0 | 0 | 0 |
| 0.009621618 | 0 | 0 | 0.00123912 | 0 |
| 0.152525986 | 0.1699771 | 0 | 0.064505442 | 0.698640892 |
| 0 | 0 | 0 | 0 | 0 |
| 0 | 0.00109492 | 0 | 0 | 0 |
| 0 | 0.00353776 | 0.006487488 | 0 | 0.009394217 |
| 0 | 0 | 0 | 0 | 0 |
| 0.019208497 | 0.283414098 | 0.215434554 | 0 | 0 |

| | | | | |
|---|---|---|---|---|
| 0 | 0.015310302 | 0 | 0.204145736 | 0 |
| 0 | 0.054942433 | 0 | 0 | 0 |
| 0 | 0.148664233 | 0 | 0 | 0 |
| 0 | 0 | 0 | 0 | 0 |
| 0.001311356 | 0.009663976 | 0 | 0.001469282 | 0.009668291 |
| 0 | 0 | 0 | 0 | 0 |
| 0 | 0.168540682 | 0 | 0 | 0 |
| 0 | 0.002712392 | 0 | 0 | 0 |
| 0 | 0 | 0 | 0 | 0 |
| 0 | 0 | 0 | 0 | 0 |
| 0 | 0 | 0 | 0 | 0 |
| 4.778749009 | 0.32849978 | 1.244033812 | 0.286915599 | 0 |
| 0 | 0.004704679 | 0 | 0.375674657 | 0 |
| 0 | 0 | 0 | 0 | 0 |
| 0.296871224 | 0.001497866 | 0.369953861 | 0.049843934 | 0.533535316 |
| 0 | 0 | 0 | 0 | 0 |
| 0 | 0.019717515 | 0 | 0 | 0 |
| 0 | 0 | 0 | 0 | 0 |
| 0.00132928 | 0.000605942 | 0.003802661 | 0.006205686 | 0.802758269 |
| 0 | 0 | 0 | 0 | 0 |
| 0 | 0.038879 | 0 | 0 | 0 |
| 0 | 0.301481871 | 0 | 0 | 0 |
| 0.005008609 | 0.046055517 | 0 | 0.451115785 | 0 |
| 0 | 0 | 0 | 0 | 0 |
| 0 | 0 | 0 | 0 | 0 |
| 0 | 0 | 0 | 0 | 0 |
| 0 | 0 | 0 | 0 | 0 |
| 0 | 0 | 0 | 0 | 0 |
| 0 | 0.023455301 | 0 | 0 | 0 |
| 0 | 0 | 0 | 0 | 0 |
| 0 | 0 | 0 | 0 | 0 |
| 0.050921462 | 0.039920427 | 0.067557385 | 0.040424672 | 0.019490241 |
| 0 | 0 | 0 | 0 | 0 |
| 0.006636387 | 0.045788017 | 0 | 0 | 0.165528807 |
| 0.001114374 | 0 | 0.004802521 | 0.002378243 | 0.002242317 |
| 0 | 0 | 0.001921355 | 0.013183127 | 0 |
| 0 | 0.031864803 | 0 | 0 | 0 |
| 0 | 0 | 0 | 0 | 0 |
| 0 | 0.000666963 | 0 | 0 | 0 |
| 0 | 0 | 0 | 0 | 0 |
| 0 | 0.25564043 | 0 | 0 | 0 |
| 0 | 0.028054614 | 0 | 0 | 0 |
| 0 | 0 | 0 | 0 | 0 |
| 0 | 0.013627178 | 0 | 0 | 0.042327687 |
| 0.004234256 | 0.056141244 | 0.023981098 | 0.028582254 | 0.075931192 |
| 0 | 0 | 0 | 0 | 0 |
| 0 | 0.149727615 | 0.423308458 | 0 | 0 |
| 0 | 0.00045076 | 0 | 0 | 0 |

| | | | | |
|---|---|---|---|---|
| 0 | 0 | 0 | 0 | 0 |
| 0 | 0 | 0 | 0 | 0 |
| 0 | 0 | 0 | 0 | 0.035267942 |
| 0 | 0 | 0 | 0 | 0 |
| 0 | 0 | 0 | 0 | 0 |
| 0 | 0 | 0 | 0 | 0 |
| 0 | 0 | 0 | 0.100821765 | 0 |
| 0.029103924 | 0.008099731 | 0 | 0.356066275 | 0 |
| 0 | 0 | 0 | 0 | 0 |
| 0 | 0 | 0 | 0 | 0 |
| 0 | 0 | 0 | 0 | 0 |
| 0 | 0 | 0.009613675 | 0 | 0 |
| 0 | 0 | 0 | 0 | 0 |
| 0 | 0 | 0 | 0 | 0.023303197 |
| 0 | 0 | 0 | 0 | 0 |
| 0 | 0 | 0 | 0 | 0 |
| 0 | 0 | 0 | 0 | 0 |
| 0 | 0 | 0 | 0 | 0 |
| 4.354099222 | 0.683214641 | 1.775289345 | 0 | 0 |
| 0 | 0 | 0 | 0 | 0 |
| 0.000884215 | 0.011823173 | 0 | 0 | 0 |
| 0 | 0 | 0 | 0 | 0 |
| 0 | 0 | 0 | 0 | 0 |
| 0 | 0 | 0 | 0 | 0 |
| 0.015078286 | 0.000937271 | 0 | 0 | 0 |
| 0.005985897 | 0 | 0.031741691 | 0.028571957 | 0.035087484 |
| 0 | 0 | 0 | 0 | 0 |
| 0.267513484 | 0.036913239 | 0 | 0 | 0 |
| 0 | 0 | 0 | 0 | 0 |
| 0 | 0.487114402 | 0.990603491 | 0 | 1.613750513 |
| 0 | 0 | 0 | 0 | 0 |
| 0 | 0 | 0 | 0 | 0 |
| 0 | 0 | 0 | 0 | 0 |
| 0 | 0 | 0 | 0 | 0 |
| 0 | 0 | 0.00238732 | 0 | 0.000404061 |
| 0 | 0 | 0 | 1.679638053 | 0 |
| 0 | 0 | 0 | 0 | 0 |
| 0 | 0.003165479 | 0.012099794 | 0.003630915 | 0.006724526 |
| 0 | 0.035763599 | 0 | 0 | 0 |
| 0 | 0 | 0 | 0 | 0 |
| 14.95969825 | 1.962604192 | 9.127451548 | 24.84491069 | 11.47871895 |
| 2.001389228 | 0.470071498 | 0.752721677 | 1.660774432 | 0.258089514 |
| 0 | 5.455042993 | 0 | 0 | 0 |
| 0 | 0.025330687 | 0 | 0 | 0 |
| 0 | 0 | 0 | 0 | 0 |
| 0.249126601 | 0.924915784 | 0 | 0 | 0 |
| 0 | 0.003232939 | 0 | 0 | 0 |
| 0.012676068 | 0.009546738 | 0.003869892 | 0.008439251 | 0.018679345 |

| | | | | |
|---|---|---|---|---|
| 0 | 0 | 0 | 0 | 0 |
| 0.001417216 | 0.006718679 | 0 | 0.003387499 | 0.011478045 |
| 0.014566004 | 0 | 0 | 0 | 0 |
| 0 | 0 | 0 | 0 | 0 |
| 1.295105253 | 0.85229047 | 0 | 1.122865379 | 1.935314376 |
| 0 | 0 | 0 | 0 | 0 |
| 0 | 0 | 0 | 0 | 0 |
| 0 | 0.025115053 | 0 | 0 | 0 |
| 1.229744544 | 0.000516059 | 0.770343191 | 0.147561769 | 0.052197937 |
| 0 | 0 | 0 | 0 | 0 |
| 0 | 0 | 0.002998094 | 0 | 0 |
| 0 | 0 | 0 | 0 | 0 |
| 0 | 0.050887587 | 0 | 0 | 0 |
| 0.01557526 | 0.005659311 | 0.011975531 | 0.10521172 | 0.0140094 |
| 0 | 0 | 0 | 0 | 3.984949305 |
| 0 | 0.251535377 | 0 | 0 | 0 |
| 0.011573476 | 0.239787465 | 0 | 0.001531998 | 0 |
| 0 | 0 | 0 | 0 | 0 |
| 0 | 0 | 0 | 0 | 0 |
| 0 | 0 | 0 | 0 | 0 |
| 0 | 0.03448079 | 0 | 0 | 0 |
| 0 | 0 | 0 | 0 | 0 |
| 0 | 0 | 0 | 0 | 0 |
| 0 | 0 | 0 | 0 | 0 |
| 0 | 0 | 0 | 0 | 0 |
| 0 | 0 | 0 | 0 | 0 |
| 0 | 0.036544518 | 0 | 0 | 0.004934279 |
| 0.001437426 | 0 | 0 | 0.01009268 | 0 |
| 0 | 0 | 0 | 0 | 0 |
| 0 | 0 | 0 | 0 | 0 |
| 0 | 0 | 0 | 0 | 0 |
| 0 | 0 | 0 | 0 | 0 |
| 0 | 0.063283957 | 0 | 0 | 0 |
| 0 | 0.028756782 | 0 | 0 | 0 |
| 0 | 0 | 0 | 0 | 0.225761108 |
| 0.004822129 | 0.001414889 | 0 | 0 | 0 |
| 0 | 0.001549486 | 0 | 0 | 0 |
| 0 | 0.003531005 | 0 | 0 | 0 |
| 0 | 0 | 0 | 0 | 0 |
| 0 | 0 | 0 | 0 | 0 |
| 0 | 0.105581563 | 0 | 0 | 0 |
| 0 | 0 | 0 | 0 | 0 |
| 0 | 0.05467889 | 0 | 0 | 0 |
| 0 | 0 | 0 | 0 | 0 |
| 0 | 0 | 0 | 0 | 0 |
| 0 | 0 | 0 | 0 | 0 |
| 0 | 0.056263694 | 0 | 0 | 0 |
| 0 | 0 | 0 | 0 | 0 |

| | | | | |
|---|---|---|---|---|
| 0 | 0.824848357 | 0 | 0 | 0 |
| 0 | 0 | 0 | 0 | 0 |
| 0 | 0.001099373 | 0 | 0 | 0 |
| 0 | 0 | 0 | 0 | 0 |
| 0 | 0.04895614 | 0 | 0 | 0 |
| 0 | 0.633396278 | 0 | 0 | 0 |
| 0.066495279 | 0.215221785 | 0 | 0.005239791 | 0 |
| 0 | 0 | 0 | 0 | 0 |
| 0 | 0 | 0 | 0 | 0 |
| 0 | 0.004286803 | 0 | 0 | 0 |
| 0 | 0.068290842 | 0 | 0 | 0 |
| 0.001022442 | 0.052899203 | 0 | 0 | 0 |
| 0 | 0 | 0 | 0 | 0 |
| 0 | 0 | 0 | 0 | 0 |
| 0 | 0.001745549 | 0 | 0 | 0 |
| 0 | 0 | 0 | 0 | 0 |
| 0 | 0 | 0 | 0 | 0 |
| 0.00084301 | 0 | 0 | 0 | 0 |
| 0 | 0 | 0 | 0 | 0 |
| 0 | 0 | 0 | 0 | 0 |
| 0.004313239 | 0.485154471 | 0 | 0.001662858 | 0 |
| 0 | 0 | 0 | 0 | 0 |
| 0 | 0.05923018 | 0 | 0 | 0.009322002 |
| 0.002456365 | 0.00489357 | 0 | 0.00224252 | 0.030874344 |
| 0 | 0 | 0 | 0 | 0 |
| 0 | 0 | 0.001674781 | 0.003367216 | 0.376436693 |
| 0 | 0 | 0 | 0 | 0 |
| 0 | 0 | 0 | 0 | 0 |
| 0 | 0.109243187 | 0.090955657 | 0 | 0 |
| 0 | 0.004894285 | 0.002043103 | 0.740567462 | 0.389026051 |
| 0 | 0.013035816 | 0 | 0 | 0 |
| 0 | 0 | 0 | 0 | 0 |
| 0 | 0.02412503 | 0 | 0 | 0 |
| 0 | 0 | 0.050786744 | 5.251093845 | 0 |
| 0 | 0.103447454 | 0 | 0 | 0 |
| 0 | 0.446417224 | 0 | 0 | 0 |
| 0.043371192 | 0.001868072 | 0.012611431 | 0.246160438 | 0.027508247 |
| 0 | 0 | 0 | 0 | 0 |
| 0 | 0 | 0 | 0 | 0 |
| 0 | 0 | 0 | 0 | 0 |
| 0 | 0 | 0 | 0 | 0 |
| 0 | 0.118604219 | 0 | 0 | 0 |
| 0 | 0 | 0 | 0 | 0 |
| 0 | 0.004555513 | 0 | 0 | 0 |
| 0 | 0.391314876 | 0 | 0 | 0 |
| 0 | 0 | 0 | 0 | 0 |
| 0 | 0 | 0 | 0 | 0 |
| 0 | 0 | 0 | 0 | 0 |

| | | | | |
|---|---|---|---|---|
| 0.005873636 | 0.001249478 | 0.009165089 | 0.006361629 | 0.130560737 |
| 0 | 0 | 0 | 0 | 0 |
| 0 | 0 | 0 | 0 | 0 |
| 0 | 0 | 0 | 0 | 0 |
| 0 | 0 | 0 | 0 | 0 |
| 0 | 0 | 0 | 0 | 0 |
| 0 | 0 | 0 | 0 | 0.000695626 |
| 0 | 0.364718974 | 0 | 0.705901624 | 0 |
| 0.0537039 | 1.883493434 | 0.008152057 | 1.536394607 | 0.251602981 |
| 0.015243645 | 0 | 0.218495876 | 0.220054976 | 0.316839031 |
| 0 | 0 | 0.001371129 | 0 | 0 |
| 0 | 0 | 0 | 0 | 0 |
| 0 | 0.020692675 | 0 | 0 | 0 |
| 0 | 0 | 0.063723668 | 0 | 0 |
| 0 | 0 | 0 | 0 | 0 |
| 0 | 0 | 0 | 0 | 0 |
| 0 | 0 | 0 | 0.044189819 | 0 |
| 0 | 0.576242748 | 0 | 0 | 0 |
| 0 | 0 | 0 | 0 | 0 |
| 0 | 0 | 0 | 0 | 0 |
| 0 | 0 | 0 | 0 | 0 |
| 0 | 0 | 0 | 0 | 0 |
| 0 | 0.064241612 | 0 | 0 | 0 |
| 0 | 0 | 0 | 0 | 0 |
| 0 | 0.010755209 | 0 | 0 | 0 |
| 0 | 0 | 0 | 0 | 0 |
| 0 | 0 | 0 | 0 | 0 |
| 0 | 0 | 0 | 0 | 0 |
| 0 | 0 | 0 | 0 | 0 |
| 0 | 0 | 0 | 0 | 0 |
| 0 | 0 | 0 | 0 | 0 |
| 0 | 0 | 0 | 0 | 0 |
| 0 | 0 | 0 | 0 | 0 |
| 0 | 0.014264072 | 0 | 0 | 0 |
| 0 | 0.000864382 | 0 | 0 | 0.016119209 |
| 0 | 0 | 0 | 0 | 0 |
| 0 | 0 | 0 | 0 | 0 |
| 0 | 0 | 0 | 0 | 0.040257483 |
| 0 | 0 | 0 | 0 | 0 |
| 0 | 0 | 0.003049876 | 0 | 0.224159856 |
| 0 | 0 | 0 | 0 | 0 |
| 0 | 0.237705754 | 0 | 0.816298577 | 0 |
| 0 | 0 | 0 | 0 | 0 |
| 0 | 0 | 0 | 0 | 0 |
| 0 | 0 | 0 | 0 | 0 |
| 0 | 0 | 0 | 0 | 0 |
| 0 | 0 | 0 | 0 | 0 |
| 0 | 0 | 0 | 0 | 0 |

| | | | | |
|---|---|---|---|---|
| 0 | 0 | 0 | 0 | 0 |
| 0 | 0 | 0 | 0.031480072 | 0 |
| 0 | 0 | 0 | 0 | 0 |
| 0 | 0 | 0 | 0 | 0 |
| 0 | 0 | 0.143216591 | 0.343804081 | 0 |
| 0 | 0 | 0 | 0 | 0 |
| 0 | 0.022171502 | 0 | 0 | 0 |
| 0.018118849 | 0.0026112 | 0.149230801 | 0.134223093 | 0.002631013 |
| 0 | 0.006687999 | 0 | 0 | 0 |
| 0 | 0.114421259 | 0 | 0 | 0 |
| 0 | 0 | 0 | 0 | 0 |
| 0 | 0 | 0 | 0 | 0 |
| 0 | 0 | 0 | 0 | 0 |
| 0 | 0 | 0 | 0 | 0 |
| 0 | 0.000239764 | 0 | 1.39571628 | 0 |
| 0.024624536 | 0.523927418 | 0.011291137 | 0.049371684 | 0 |
| 0 | 0 | 0 | 0 | 0 |
| 0 | 0 | 0 | 0 | 0 |
| 0 | 0 | 0 | 0 | 0 |
| 0 | 0 | 0 | 0 | 0 |
| 0 | 0 | 0 | 0 | 0 |
| 0 | 0 | 0 | 0 | 0 |
| 0 | 0 | 0 | 0 | 0 |
| 0.003071413 | 0.002652783 | 0.00050448 | 0.001931959 | 0.002561541 |
| 0 | 0 | 0 | 0 | 0 |
| 0 | 0 | 0 | 0 | 0 |
| 0.00473121 | 0.081774332 | 0 | 2.898977399 | 0 |
| 0 | 0.001324703 | 0 | 0 | 0 |
| 0 | 0 | 0.001600249 | 0 | 0 |
| 0 | 0 | 0 | 0 | 0 |
| 0 | 0 | 0 | 0 | 0 |
| 0 | 0 | 0 | 0 | 0 |
| 0 | 0 | 0 | 0 | 0 |
| 0 | 0 | 0 | 0 | 0 |
| 0 | 0 | 0 | 0 | 0 |
| 0 | 0 | 0 | 0.001234122 | 0 |
| 0 | 0 | 0 | 0 | 0 |
| 0 | 0.323200379 | 0 | 0 | 0 |
| 0 | 0 | 0 | 0 | 0 |
| 0 | 0 | 0 | 0 | 0 |
| 0 | 0 | 0 | 0 | 0 |
| 0 | 0 | 0 | 0 | 1.091299164 |
| 0 | 0 | 0 | 0 | 0 |
| 0 | 0 | 0 | 0 | 0 |
| 0 | 0 | 0 | 0 | 0 |
| 0 | 0 | 0 | 0 | 0 |
| 0 | 0 | 0 | 0 | 0 |
| 0 | 0 | 0 | 0 | 0 |

| | | | | |
|---|---|---|---|---|
| 0 | 0.478396253 | 0 | 0 | 0 |
| 0 | 0 | 0 | 0 | 0 |
| 0 | 0.002511277 | 0 | 0 | 0 |
| 0 | 0.002438325 | 0 | 0 | 0 |
| 0 | 0.000626196 | 0 | 0 | 0 |
| 0 | 0 | 0.010556936 | 0.011791752 | 0.11685044 |
| 0 | 0 | 0 | 0 | 0 |
| 0 | 0.000950959 | 0 | 0 | 0 |
| 0 | 0.146646312 | 0 | 0 | 0 |
| 0 | 0 | 0 | 0 | 0 |
| 0 | 0 | 0 | 0 | 0 |
| 0 | 0.019201256 | 0 | 0 | 0 |
| 0 | 0 | 0 | 0 | 0 |
| 0 | 0 | 0 | 0 | 0 |
| 0 | 0 | 0 | 0 | 0 |
| 0 | 0 | 0 | 0 | 0 |
| 0 | 0 | 0 | 0 | 0 |
| 0 | 0 | 0 | 0 | 0 |
| 0.002239748 | 0.003367417 | 0 | 0 | 0 |
| 0 | 0 | 0 | 0 | 0 |
| 0.006439102 | 0 | 0.021099602 | 0.089460551 | 1.12605105 |
| 0 | 0 | 0 | 0 | 0 |
| 0.006708997 | 0.11553369 | 0.007190531 | 0.040090444 | 0.01485477 |
| 0 | 0.08832605 | 0 | 0 | 0 |
| 0 | 0.007769084 | 0 | 0 | 0 |
| 0 | 0 | 0 | 0 | 0 |
| 0 | 0 | 0 | 0 | 0 |
| 0 | 0 | 0 | 0 | 0 |
| 0 | 0.017970702 | 0 | 0 | 0 |
| 0 | 0 | 0 | 0 | 0 |
| 0 | 0 | 0 | 0 | 0 |
| 0 | 0.07578942 | 0 | 0 | 0 |
| 5.85399373 | 0 | 0.441070597 | 0.038113392 | 0.045086443 |
| 0 | 0.085372436 | 0 | 0 | 0 |
| 0 | 0.009833419 | 0 | 0.375777991 | 0 |
| 0.060348845 | 0.008078756 | 0.001748248 | 0.000684725 | 0 |
| 0 | 0 | 0 | 0 | 0 |
| 0 | 0 | 0 | 0 | 0 |
| 0.004241194 | 0.029858988 | 0.365269281 | 0.006599943 | 0 |
| 0 | 0 | 0 | 0 | 0 |
| 0 | 0 | 0 | 0 | 0 |
| 0 | 0.028706086 | 0 | 0 | 0 |
| 0 | 0.160002047 | 0 | 0.018212855 | 0.100412085 |
| 0 | 0 | 0 | 0 | 0 |
| 0 | 0 | 0 | 0 | 0 |
| 0 | 1.066126025 | 0 | 0 | 0 |
| 0 | 0 | 0 | 0 | 0 |
| 0 | 0 | 0 | 0 | 0 |

| | | | | |
|---|---|---|---|---|
| 0 | 0 | 0 | 0 | 0 |
| 0 | 0 | 0 | 0 | 0 |
| 0 | 0 | 0 | 0 | 0 |
| 0 | 0 | 0 | 0 | 0 |
| 0 | 0 | 0 | 0 | 0 |
| 0 | 0 | 0 | 0 | 0 |
| 0 | 0 | 0 | 0 | 0 |
| 0 | 0 | 0 | 0 | 0.558116136 |
| 0.488770758 | 0.001588609 | 0 | 0 | 0 |
| 0 | 0 | 0.003516568 | 0 | 0.004166322 |
| 0 | 0 | 0 | 0 | 0 |
| 0 | 0 | 0 | 0 | 0 |
| 0 | 0 | 0 | 0 | 0 |
| 0 | 0 | 0 | 0 | 0 |
| 0 | 0 | 0 | 0 | 0 |
| 0 | 0 | 0 | 0.014657917 | 0 |
| 0 | 0 | 0 | 0 | 0 |
| 0 | 0 | 0 | 0 | 0 |
| 0 | 0 | 0 | 0 | 0 |
| 0 | 1.155039413 | 0 | 0 | 0 |
| 0 | 0 | 0 | 0 | 0 |
| 0 | 0 | 0 | 0 | 0 |
| 0 | 0 | 0 | 0 | 0 |
| 0 | 0 | 0.002304338 | 0 | 0 |
| 0 | 0.008333066 | 0 | 0 | 0 |
| 0 | 0.325989354 | 0 | 0 | 0 |
| 0 | 0 | 0 | 0 | 0 |
| 0 | 0 | 0 | 0 | 0 |
| 0 | 0 | 0 | 0 | 0 |
| 0 | 0.017704114 | 0 | 0 | 0 |
| 0 | 0.173341078 | 0.17252363 | 0.131048978 | 0 |
| 0 | 0 | 0 | 0 | 0 |
| 0 | 0.000520387 | 0 | 0 | 0 |
| 0 | 0 | 0 | 0 | 0 |
| 0 | 0 | 0 | 0 | 0 |
| 0 | 0 | 0 | 0 | 0 |
| 0 | 0 | 0 | 0 | 0 |
| 0 | 0 | 0 | 0 | 0 |
| 0 | 0 | 0 | 0 | 0 |
| 0 | 0 | 0 | 0 | 0 |
| 0 | 0 | 0 | 0 | 0 |
| 0 | 0.328436777 | 0 | 0 | 0 |
| 0 | 0.013683639 | 0 | 0 | 0 |
| 0 | 0 | 0 | 0.008226428 | 0 |
| 0 | 0 | 0 | 0 | 0 |
| 0 | 0 | 0 | 0 | 0 |
| 0.393599365 | 0 | 0.573963105 | 0.009918693 | 0.366728549 |
| 0 | 0 | 0 | 0 | 0 |

| | | | | |
|---|---|---|---|---|
| 0 | 0 | 0 | 0 | 0 |
| 0 | 0 | 0 | 0 | 0 |
| 0 | 0 | 0 | 0 | 0 |
| 0 | 0 | 0 | 0 | 0 |
| 0.773656886 | 0.089823527 | 1.204484495 | 0.636310409 | 0.538318406 |
| 0 | 0 | 0 | 0 | 0 |
| 0 | 0 | 0 | 0 | 0 |
| 0 | 0 | 0 | 0 | 0 |
| 0 | 0 | 0 | 0 | 0 |
| 0 | 0 | 0 | 0 | 0 |
| 0 | 0.023947417 | 0 | 0 | 0 |
| 0.00123603 | 0.015990816 | 0.005510495 | 0.091270483 | 0.023938441 |
| 0.000761702 | 0.007314681 | 0.002852501 | 0 | 0.008314781 |
| 0 | 0 | 0 | 0 | 0 |
| 0.009419411 | 0.006297524 | 0 | 0.006834833 | 0 |
| 0 | 0 | 0 | 0.441239347 | 0.501410542 |
| 0 | 0.010647985 | 0 | 0.004057678 | 0 |
| 0 | 0 | 0 | 0 | 0 |
| 0.47277782 | 0.001381488 | 0 | 0.333967658 | 0 |
| 0.228346534 | 0.093585521 | 0 | 0.002086412 | 0 |
| 0 | 0 | 0 | 0 | 0 |
| 0 | 0 | 0 | 0 | 0 |
| 0 | 0 | 0 | 0 | 0 |
| 0 | 0 | 0 | 0 | 0 |
| 0 | 0 | 0 | 0 | 0 |
| 0 | 0 | 0 | 0 | 0 |
| 0.008113993 | 0.002465801 | 0 | 0.004806819 | 0.02721718 |
| 0 | 0 | 0 | 0 | 0 |
| 0 | 0 | 0 | 0 | 0 |
| 0 | 0 | 0 | 0 | 0 |
| 0 | 0.000385533 | 0.000824815 | 0.001895229 | 0.003908864 |
| 0 | 0.000520526 | 0 | 0 | 0 |
| 0 | 0 | 0 | 0 | 0 |
| 0 | 0 | 0 | 0 | 0 |
| 0 | 0.009870961 | 0 | 0 | 0 |
| 0 | 0 | 0 | 0 | 0 |
| 0 | 0 | 0 | 0 | 0 |
| 0.005570684 | 0.064398007 | 0.024106837 | 0.034786289 | 0.107300074 |
| 0 | 0.220725666 | 0 | 0 | 0 |
| 0 | 0 | 0 | 0 | 0 |
| 0 | 0.003380125 | 0.000347111 | 0.057824494 | 0.004308302 |
| 0 | 0.013802533 | 0 | 0 | 0 |
| 0 | 0 | 0 | 0 | 0 |
| 0.106396757 | 0 | 0 | 0.005429858 | 0 |
| 0 | 0 | 0 | 0 | 0 |
| 0.001181607 | 0.000359084 | 0 | 0 | 0 |
| 0 | 0 | 0 | 0 | 0 |
| 0 | 0.001069765 | 0 | 0 | 0 |

| | | | | |
|---|---|---|---|---|
| 0 | 0 | 0 | 0 | 0 |
| 0 | 0.121356319 | 0 | 0 | 0 |
| 0 | 0 | 0 | 0 | 0 |
| 0.001385146 | 0.000947113 | 0.00072045 | 0 | 0.002591187 |
| 0.002087639 | 0.000390414 | 0 | 0 | 0.005937527 |
| 0.002072538 | 0 | 0 | 0.002167324 | 0 |
| 0 | 0.003370036 | 0 | 0 | 0 |
| 0 | 0.007736451 | 0 | 0 | 0 |
| 0 | 0.004445516 | 0 | 0 | 0 |
| 0 | 0.04219688 | 0 | 0 | 0 |
| 0 | 0.024956685 | 0 | 0 | 0.01095759 |
| 0 | 0 | 0 | 0 | 0 |
| 0 | 0.175152834 | 0 | 0.006234016 | 0 |
| 0 | 0.006949983 | 0 | 0 | 0 |
| 0 | 0 | 0 | 0 | 0 |
| 0 | 0 | 0 | 0 | 0 |
| 0 | 0 | 0 | 0 | 0 |
| 0 | 0 | 0 | 0 | 0 |
| 0.00556382 | 0 | 0 | 0 | 0 |
| 0 | 0 | 0 | 0 | 0 |
| 0 | 0 | 0 | 0 | 0 |
| 0 | 0 | 0 | 0 | 0 |
| 0.013575466 | 0 | 0.021687204 | 0.084501964 | 0 |
| 0 | 0.076732882 | 0.005554776 | 0 | 0 |
| 0 | 0 | 0 | 0 | 0 |
| 0 | 0.120954445 | 0 | 0 | 0.009422889 |
| 0 | 0 | 0 | 0 | 0 |
| 0 | 0 | 0 | 0 | 0 |
| 0 | 0 | 0 | 0 | 0 |
| 0 | 0 | 0 | 0 | 0 |
| 0 | 0.008542728 | 0.028679952 | 0 | 0.052031534 |
| 0 | 0 | 0 | 0 | 0 |
| 0 | 0 | 0 | 0 | 0 |
| 0 | 0 | 0 | 0 | 0 |
| 0 | 0.058122639 | 0 | 0 | 0 |
| 0.022039701 | 0 | 0 | 0.006472484 | 0 |
| 0.24773188 | 0 | 0 | 0.114115285 | 0 |
| 0 | 0 | 0 | 0 | 0 |
| 0.919178474 | 0.00027799 | 0.00507508 | 0 | 0.017179429 |
| 0 | 0 | 0 | 0 | 0 |
| 0 | 0 | 0 | 0 | 0 |
| 0 | 0 | 0 | 0 | 0 |
| 0 | 1.803916659 | 0 | 0 | 0 |
| 0 | 0 | 0 | 0 | 0 |
| 0 | 0 | 0 | 0 | 0 |
| 0.375455609 | 0.174226261 | 0.110933116 | 0 | 0 |
| 0 | 0 | 0 | 0 | 0 |
| 0 | 0.046241634 | 0 | 0 | 0 |

| | | | | |
|---|---|---|---|---|
| 0 | 0 | 0 | 0 | 0 |
| 0 | 0 | 0 | 0 | 0 |
| 0 | 0 | 0 | 0 | 0 |
| 0 | 0.000492363 | 0 | 0 | 0 |
| 0 | 0 | 0 | 0 | 0 |
| 0 | 0.02484463 | 0 | 0 | 0 |
| 0 | 0.030894553 | 0 | 0 | 0 |
| 0 | 0 | 0 | 0 | 0 |
| 0 | 0.002057718 | 0 | 0 | 0 |
| 0 | 0 | 0 | 0 | 0 |
| 0.000494742 | 0.005938809 | 0.000385992 | 0.008869178 | 0 |
| 0 | 0.000673417 | 0 | 0 | 0 |
| 0 | 0 | 0 | 0 | 0 |
| 0 | 0 | 0 | 0 | 0 |
| 0 | 0 | 0 | 0 | 0 |
| 0 | 0 | 0 | 0 | 0 |
| 0 | 0 | 0 | 0 | 0 |
| 0 | 0 | 0 | 0 | 0 |
| 0 | 0 | 0 | 0 | 0 |
| 0 | 0 | 0 | 0 | 0.000721993 |
| 0 | 0 | 0 | 0 | 0 |
| 0 | 0 | 0 | 0 | 0 |
| 0 | 0 | 0 | 0 | 0 |
| 0 | 0 | 0 | 0 | 0 |
| 0 | 0 | 0 | 0 | 0 |
| 0 | 0 | 0 | 0 | 0 |
| 0 | 0 | 0 | 0 | 0 |
| 0 | 0.002537568 | 0 | 0 | 0 |
| 0 | 0 | 0 | 0 | 0 |
| 0 | 0 | 0 | 0 | 0 |
| 0.001127338 | 0.019270813 | 0 | 0.00147362 | 0.002357013 |
| 0 | 0 | 0 | 0 | 0 |
| 2.966908085 | 0.006364551 | 0.081358084 | 0.022976591 | 0.002160578 |
| 0.049324337 | 0.001321101 | 0.003652556 | 0 | 0 |
| 0 | 0 | 0 | 0 | 0 |
| 0 | 0 | 0 | 0 | 0 |
| 0 | 0 | 0 | 0 | 0 |
| 0 | 0 | 0 | 0 | 0 |
| 0 | 0 | 0 | 0 | 0 |
| 0 | 0 | 0 | 0 | 0 |
| 0 | 0 | 0 | 0 | 0 |
| 0.037690263 | 0 | 0.063291875 | 0 | 0.293996455 |
| 0.21613961 | 0.077843685 | 0.051689219 | 0.016169113 | 0 |
| 0 | 0 | 0 | 0 | 0 |
| 0 | 0 | 0 | 0 | 0 |
| 0 | 0.253353514 | 0.003697748 | 0.00672643 | 0.090957471 |
| 0.000932216 | 0.038622658 | 0 | 0 | 0 |
| 0 | 0 | 0 | 0 | 0 |

| | | | | |
|---|---|---|---|---|
| 0 | 0 | 0 | 0 | 0 |
| 0 | 0 | 0 | 0 | 0.043708912 |
| 0.011037552 | 0.010489672 | 0.004592732 | 0.001499006 | 0.003003331 |
| 0 | 0 | 0 | 0 | 0 |
| 0 | 0.515385319 | 0 | 0 | 0 |
| 0.020900289 | 0 | 0 | 0.070135401 | 0 |
| 0 | 0 | 0 | 0 | 0 |
| 0 | 0 | 0 | 0 | 0 |
| 0 | 0 | 0 | 0 | 0 |
| 0 | 0 | 0 | 0 | 0 |
| 0 | 0 | 0 | 0 | 0 |
| 0 | 0 | 0 | 0 | 0 |
| 0 | 0 | 0 | 0 | 0 |
| 0 | 0 | 0 | 0 | 0 |
| 0 | 0 | 0 | 0 | 0 |
| 0 | 0 | 0 | 0 | 0 |
| 0 | 0.007087921 | 0 | 0 | 0 |
| 0 | 0 | 0 | 0 | 0 |
| 0 | 0 | 0 | 0 | 0 |
| 0 | 0 | 0 | 0 | 0 |
| 0 | 0 | 0 | 0 | 0 |
| 0.004358504 | 0 | 0 | 0 | 0 |
| 0 | 0.001930862 | 0 | 0 | 0 |
| 0 | 0 | 0 | 0 | 0 |
| 0 | 0 | 0 | 0.001729548 | 0 |
| 0 | 0 | 0 | 0 | 0.073527036 |
| 0 | 0 | 0 | 0 | 0 |
| 0 | 0.642413839 | 0 | 0 | 0 |
| 0 | 0 | 0 | 0 | 0 |
| 0 | 0 | 0 | 0 | 0 |
| 0 | 0 | 0 | 0 | 0 |
| 0 | 0 | 0.001669554 | 0 | 0.001027551 |
| 0 | 0 | 0 | 0 | 0 |
| 0 | 0 | 0 | 0 | 0 |
| 0 | 0 | 0 | 0 | 0 |
| 0 | 0 | 0 | 0 | 0 |
| 0 | 0 | 0.007779861 | 0.024996471 | 0 |
| 0.590423018 | 0 | 0.013782631 | 0.042060649 | 0 |
| 6.647061506 | 0.561248513 | 1.338358445 | 1.13962532 | 0 |
| 0 | 0 | 0 | 0 | 0 |
| 0 | 0 | 0 | 0 | 0 |
| 0 | 0.024683292 | 0 | 0.004044657 | 0.001112268 |
| 0.005844291 | 0.001319351 | 0.008337645 | 0.00249452 | 0.110394614 |
| 0 | 0 | 0 | 0 | 0 |
| 0 | 0 | 0 | 0 | 0 |
| 0.000960521 | 0 | 0 | 0 | 0 |
| 0 | 0 | 0.001594371 | 0.006487418 | 0.033731456 |
| 0 | 0 | 0 | 0 | 0 |

| | | | | |
|---|---|---|---|---|
| 0 | 0 | 0 | 0 | 0 |
| 0.004512568 | 0.01126463 | 0.014250267 | 0.032984474 | 0.137052514 |
| 0 | 0.132172905 | 0 | 0 | 0 |
| 0 | 0 | 0 | 0 | 0 |
| 0 | 0 | 0 | 0 | 0 |
| 0 | 0 | 0 | 0 | 0 |
| 0 | 0 | 0 | 0 | 0 |
| 0 | 0 | 0 | 0 | 0 |
| 0 | 0 | 0 | 0 | 0 |
| 0 | 0 | 0 | 0 | 0 |
| 0 | 0 | 0 | 0 | 0 |
| 0 | 0 | 0 | 0 | 0 |
| 0 | 0.341906651 | 0 | 0 | 0.025345915 |
| 0 | 0 | 0 | 0 | 0 |
| 0 | 0 | 0.009615588 | 0.000900584 | 0 |
| 0 | 0 | 0 | 0 | 0 |
| 0 | 0 | 0 | 0 | 0 |
| 0 | 0 | 0 | 0 | 0 |
| 0 | 0 | 0 | 0 | 0 |
| 0 | 0 | 0 | 0 | 0 |
| 0 | 0 | 0 | 0 | 0 |
| 0.108342384 | 0.038022328 | 0 | 0.001768889 | 0 |
| 0 | 0.000737339 | 0 | 0 | 0 |
| 0.314856516 | 0 | 0 | 0 | 0 |
| 0 | 0 | 0 | 0 | 0 |
| 0 | 0 | 0 | 0 | 0 |
| 0 | 0 | 0 | 0 | 0 |
| 0 | 0 | 0 | 0 | 0 |
| 0 | 0 | 0 | 0 | 0 |
| 0 | 0 | 0 | 0 | 0 |
| 0 | 0 | 0 | 0 | 0 |
| 0 | 0 | 0 | 0 | 0 |
| 0 | 0 | 0 | 0 | 0 |
| 0 | 0 | 0 | 0 | 0 |
| 0 | 1.293437433 | 0 | 0 | 16.43809029 |
| 0 | 0 | 0 | 0 | 0 |
| 0 | 0 | 0 | 0 | 0 |
| 0 | 0 | 0 | 0 | 0 |
| 0.000788291 | 0.045276405 | 0 | 0.055348759 | 0 |
| 0 | 0 | 0 | 0 | 0 |
| 0.473030852 | 0.044247803 | 0.18510108 | 0.160497391 | 0.12739993 |
| 0 | 0 | 0 | 0 | 0 |
| 0 | 0 | 0 | 0 | 0 |
| 0 | 0 | 0 | 0 | 0 |
| 0 | 0 | 0 | 0 | 0 |
| 0.398553837 | 0 | 7.037613677 | 1.060843173 | 0.891926936 |
| 0 | 0 | 0 | 0 | 0 |
| 0 | 0 | 0 | 0 | 0 |

| | | | | |
|---|---|---|---|---|
| 0 | 0 | 0 | 0 | 0 |
| 0 | 0.000600835 | 0 | 0 | 0 |
| 0 | 0.002858092 | 0 | 0 | 0 |
| 0 | 0 | 0 | 0 | 0 |
| 0 | 0.011937033 | 0 | 0 | 0 |
| 0 | 0 | 0 | 0 | 0 |
| 0 | 0 | 0 | 0 | 0 |
| 0.001530692 | 0.004651694 | 0 | 0 | 0 |
| 0.010415967 | 0 | 0 | 0 | 0 |
| 0.025524076 | 0 | 0.010085439 | 0.001182343 | 0.01463133 |
| 0 | 0 | 0 | 0 | 0 |
| 0 | 0 | 0 | 0 | 0 |
| 0.00232071 | 0.000705252 | 0.002414123 | 0.001444551 | 0.006980198 |
| 0 | 0 | 0 | 0 | 0 |
| 0 | 0 | 0 | 0 | 0 |
| 0 | 0 | 0 | 0 | 0 |
| 0 | 0 | 0 | 0 | 0 |
| 0 | 0 | 0 | 0 | 0 |
| 0 | 0 | 0 | 0 | 0 |
| 0.250969905 | 0.116652053 | 0.128387181 | 0.296494561 | 3.313277375 |
| 0 | 0 | 0 | 0 | 0 |
| 0 | 0 | 0 | 0 | 0 |
| 0 | 0 | 0 | 0 | 0 |
| 0 | 0 | 0 | 0 | 0 |
| 0 | 0 | 0 | 0 | 0 |
| 0 | 0 | 0 | 0 | 0 |
| 0 | 0 | 0 | 0 | 0 |
| 0 | 0 | 0 | 0 | 0 |
| 0 | 0 | 0 | 0 | 0 |
| 0 | 0 | 0 | 0.055529411 | 0.004211782 |
| 0 | 0 | 0 | 0 | 0 |
| 0 | 0.045908048 | 0 | 0 | 0 |
| 0 | 0 | 0 | 0 | 0 |
| 0 | 0 | 0 | 0 | 0 |
| 0 | 0 | 0 | 0 | 0 |
| 0 | 0 | 0 | 0.002995879 | 0 |
| 0 | 0 | 0 | 0 | 0 |
| 0 | 0 | 0 | 0 | 0 |
| 0 | 0 | 0 | 0 | 0 |
| 0 | 0 | 0 | 0 | 0 |
| 0 | 0 | 0 | 0 | 0 |
| 0 | 0 | 0 | 0 | 0 |
| 0 | 0.017740825 | 0 | 0 | 0 |
| 0 | 0 | 0 | 0 | 0 |
| 0.002101757 | 0.016901336 | 0.003027263 | 0.008211858 | 0 |
| 0 | 0 | 0 | 0 | 0 |
| 0 | 0 | 0 | 0 | 0 |
| 0 | 0 | 0 | 0 | 0 |

| | | | | |
|---|---|---|---|---|
| 0 | 0 | 0 | 0 | 0 |
| 0 | 0 | 0 | 0 | 0 |
| 0 | 0 | 0 | 0 | 0 |
| 0 | 0 | 0 | 0 | 0 |
| 0 | 0 | 0 | 0 | 0 |
| 0.000482783 | 0.007115686 | 0.026742969 | 0.006130471 | 0.111670519 |
| 0 | 0 | 0 | 0 | 0 |
| 0 | 0.001135951 | 0 | 0 | 0 |
| 0 | 0 | 0 | 0 | 0 |
| 0 | 0 | 0 | 0 | 0 |
| 0 | 0 | 0 | 0 | 0 |
| 0 | 0 | 0 | 0 | 0 |
| 0 | 0 | 0 | 0.00093694 | 0 |
| 0 | 0 | 0 | 0 | 0 |
| 0 | 0 | 0 | 0 | 0 |
| 0 | 0 | 0 | 0 | 0 |
| 0 | 0.063533316 | 0 | 0 | 0 |
| 0 | 0 | 0.028451231 | 0 | 0.000654273 |
| 0 | 0 | 0 | 0 | 0 |
| 0 | 0 | 0 | 0 | 0 |
| 0 | 0 | 0 | 0 | 0 |
| 0 | 0 | 0 | 0 | 0 |
| 0 | 0 | 0 | 0 | 0 |
| 0 | 0 | 0 | 0 | 0 |
| 0 | 0 | 0 | 0 | 0 |
| 0 | 0 | 0 | 0 | 0 |
| 0 | 0 | 0 | 0 | 0 |
| 0 | 0.005582046 | 0 | 0 | 0.001741401 |
| 0 | 0 | 0 | 0 | 0 |
| 0 | 0 | 0 | 0 | 0 |
| 0 | 0 | 0 | 0 | 0 |
| 0 | 0 | 0 | 0 | 0 |
| 0 | 0 | 0 | 0 | 0 |
| 0 | 0 | 0 | 0 | 0 |
| 0 | 0 | 0 | 0 | 0 |
| 0 | 0 | 0 | 0 | 0 |
| 0 | 0 | 0 | 0 | 0 |
| 0 | 0 | 0 | 0 | 0 |
| 0 | 0.561038276 | 0 | 0 | 0 |
| 0 | 0 | 0 | 0 | 0 |
| 0 | 0 | 0 | 0 | 0 |
| 0 | 0 | 0 | 0 | 0 |
| 0 | 0 | 0 | 0 | 0 |
| 0 | 0 | 0 | 0 | 0 |
| 0 | 0 | 0 | 0 | 0 |
| 0 | 0 | 0 | 0 | 0 |
| 0 | 0 | 0 | 0 | 0 |
| 0 | 0 | 0 | 0 | 0 |

| | | | | |
|---|---|---|---|---|
| 0 | 0 | 0 | 0 | 0 |
| 0.603213675 | 0.045253481 | 0 | 0 | 0 |
| 0 | 0 | 0 | 0 | 0 |
| 0 | 0 | 0 | 0 | 0 |
| 0 | 0 | 0 | 0 | 0 |
| 0 | 0 | 0 | 0 | 0 |
| 0 | 0 | 0 | 0 | 0 |
| 0 | 0 | 0 | 0 | 0 |
| 0 | 0 | 0 | 0 | 0 |
| 0 | 0.006021677 | 0 | 0 | 0 |
| 0.003361861 | 0.014303135 | 0 | 0 | 0.001713417 |
| 0 | 0 | 0 | 0 | 0 |
| 0 | 0.080794602 | 0 | 0 | 0 |
| 0 | 0 | 0 | 0 | 0 |
| 0 | 0 | 0 | 0 | 0 |
| 0 | 0 | 0 | 0 | 0 |
| 0 | 0 | 0 | 0 | 0 |
| 0 | 0 | 0 | 0 | 0 |
| 0 | 0 | 0 | 0 | 0 |
| 0 | 0 | 0 | 0 | 0 |
| 0 | 0.003492769 | 0 | 0.093898325 | 0 |
| 0 | 0 | 0 | 0 | 0 |
| 0 | 0 | 0 | 0 | 0 |
| 0 | 0.58455893 | 13.54725485 | 0.789410014 | 0.896052867 |
| 0 | 0 | 0 | 0 | 0 |
| 0 | 0 | 0 | 0 | 0 |
| 0 | 0 | 0 | 0 | 0 |
| 0 | 0 | 0 | 0 | 0 |
| 0 | 0.001328432 | 0 | 0 | 0 |
| 0 | 0 | 0 | 0 | 0 |
| 0 | 0 | 0 | 0 | 0 |
| 0 | 0 | 0 | 0 | 0 |
| 0.000944524 | 0.001033329 | 0 | 0 | 0 |
| 0 | 0 | 0 | 0 | 0 |

| PHL018-0218_V2 | PHL018-0090_V3 | PHL018-0111_V1 | PHL018-0072_V2 | PHL018-0080_V3 |
|---|---|---|---|---|
| REF | CTRL | EYCF | CTRL | CTRL |
| 1.417458567 | 0.348899688 | 3.619200129 | 0.021636256 | 0.097326149 |
| 0.10609881 | 0.872044563 | 0 | 0.442282224 | 0.612252245 |
| 0.558391507 | 2.304260541 | 0 | 0.004856686 | 0 |
| 0 | 0 | 0 | 0 | 0 |
| 0 | 0 | 0 | 0 | 0 |
| 0 | 0 | 0 | 0 | 0 |
| 0.084690944 | 0 | 0 | 0 | 0 |
| 0 | 0 | 0 | 0 | 0 |
| 0 | 0 | 0 | 0 | 0 |
| 0 | 0 | 0 | 0 | 0 |
| 0 | 0 | 0 | 0 | 0 |
| 0 | 0 | 0 | 0 | 0 |
| 0 | 0 | 0 | 0 | 0 |
| 0.164146371 | 0.826866567 | 1.656155935 | 0.584736046 | 0.567477914 |
| 0 | 0 | 0 | 0 | 0 |
| 0 | 0 | 0 | 0 | 0 |
| 0 | 0 | 0.0027196 | 0 | 0 |
| 0 | 0 | 0 | 0 | 0 |
| 0 | 0 | 0 | 0 | 0 |
| 0 | 0 | 0 | 0 | 0 |
| 0 | 0 | 0 | 0 | 0 |
| 0 | 0 | 0 | 0 | 0 |
| 0 | 0 | 0 | 0 | 0 |
| 0 | 0 | 0 | 0 | 0 |
| 1.985307445 | 0 | 0 | 0.00170008 | 0 |
| 0 | 0 | 0 | 0 | 0 |
| 0 | 0 | 0 | 0 | 0 |
| 0 | 0 | 0 | 0 | 0 |
| 0 | 0 | 0 | 0 | 0 |
| 0 | 0.213658068 | 0 | 0 | 0 |
| 0 | 0 | 0 | 0 | 0 |
| 0 | 0 | 0 | 0 | 0 |
| 0 | 0 | 0 | 0 | 0 |
| 0 | 0 | 0 | 0 | 0 |
| 0 | 0 | 0 | 0 | 0 |
| 0.642270959 | 1.626648147 | 0 | 0 | 0 |
| 0 | 0 | 0 | 0 | 0 |
| 0 | 0 | 0 | 0 | 0 |
| 0 | 0 | 0 | 0 | 0 |
| 0 | 0 | 0 | 0 | 0 |
| 0 | 0 | 0 | 0 | 0 |
| 0 | 0 | 0.05249929 | 0 | 0.000543355 |
| 0.00308074 | 3.837967362 | 0.952242934 | 0.224543846 | 0.007215119 |
| 0.043051854 | 0.006435437 | 0.001916293 | 0.001497368 | 0.091621127 |
| 0.003199883 | 0 | 0 | 0.002159099 | 0 |
| 0 | 0 | 0 | 0 | 0 |

| | | | | |
|---|---|---|---|---|
| 0 | 0.009096865 | 0 | 0 | 0.006243804 |
| 0.003043267 | 0.008061544 | 0.092292991 | 0 | 0.004963694 |
| 0.007077042 | 0.104544245 | 0 | 0 | 0.009313285 |
| 0.089069116 | 0.600522762 | 0.230141142 | 1.562578836 | 2.404402846 |
| 0.00655924 | 0 | 0 | 0 | 0 |
| 0.20179619 | 0 | 0 | 0 | 0 |
| 0 | 0 | 0.338004514 | 0 | 0.056272179 |
| 0 | 0 | 0 | 0 | 0 |
| 0 | 0 | 0 | 0 | 0 |
| 0 | 0 | 0 | 0 | 0 |
| 0 | 0 | 0 | 0 | 0 |
| 0.521258975 | 1.001506991 | 0 | 0.011649438 | 0.090689385 |
| 0 | 0 | 0 | 0 | 0.001772694 |
| 0.109210973 | 0.084028957 | 0.230284833 | 0 | 0.016266693 |
| 0 | 0 | 0 | 0 | 0 |
| 0 | 0 | 0 | 0 | 0 |
| 0 | 0 | 0 | 0 | 0 |
| 0 | 0 | 0 | 0 | 0 |
| 0 | 0 | 0 | 0 | 0 |
| 0 | 0 | 0 | 0 | 0 |
| 0.026844936 | 0.064013364 | 0.036278127 | 0.003903758 | 4.158471247 |
| 0.003041956 | 0.020195669 | 0.02958743 | 0 | 0.051760115 |
| 0 | 0.153669799 | 0.001780699 | 0.004316554 | 0 |
| 0 | 0 | 0 | 0 | 0 |
| 0 | 0 | 0 | 0 | 0 |
| 0 | 0 | 0 | 0 | 0 |
| 0 | 0 | 0 | 0 | 0 |
| 0 | 0 | 0 | 0 | 0 |
| 0 | 0 | 0.001292469 | 0 | 0 |
| 0 | 0 | 0 | 0 | 0 |
| 0 | 0 | 0.775509515 | 0 | 0.685017908 |
| 0 | 0 | 0.280757065 | 0 | 0 |
| 0 | 0 | 0 | 0 | 0 |
| 0 | 0 | 0 | 0 | 0 |
| 0 | 0 | 0 | 0 | 0 |
| 0 | 0 | 0.00117445 | 0 | 0 |
| 0 | 0 | 0 | 0 | 0 |
| 0 | 0 | 0 | 0 | 0.26974666 |
| 0 | 0 | 0 | 0 | 0 |
| 0 | 0 | 0 | 0 | 0 |
| 0 | 0 | 0 | 0 | 0 |
| 0 | 0 | 0 | 0 | 0 |
| 0 | 0 | 0 | 0 | 0 |
| 0 | 0 | 0 | 0 | 0 |
| 0 | 0 | 0 | 0.000671227 | 0 |
| 0 | 0 | 0 | 0 | 0 |
| 0 | 0 | 0 | 0 | 0 |
| 0 | 0 | 0 | 0 | 0 |

| | | | | |
|---|---|---|---|---|
| 0 | 0 | 0 | 0 | 0 |
| 0 | 0 | 0.032890857 | 0 | 0.01847535 |
| 0 | 0 | 0 | 0 | 0 |
| 0 | 0 | 0 | 0 | 0 |
| 0 | 0 | 0.053392286 | 0 | 0.036243583 |
| 0 | 0 | 0 | 0 | 0 |
| 0 | 0 | 0 | 0 | 0 |
| 0 | 0 | 0 | 0 | 0 |
| 0 | 0.000468497 | 0.000976537 | 0 | 0.000843206 |
| 0 | 0 | 0 | 0 | 0 |
| 0 | 0 | 0 | 0 | 0 |
| 0 | 0 | 0.000971777 | 0 | 0 |
| 0 | 0 | 0 | 0 | 0 |
| 0 | 0 | 0 | 0 | 0 |
| 0 | 0 | 0 | 0 | 0 |
| 0 | 0 | 0 | 0 | 0 |
| 0 | 0 | 0 | 0 | 0 |
| 0 | 0 | 0 | 0 | 0 |
| 0.118625096 | 0.020653197 | 0.311773346 | 0.015007777 | 0 |
| 0 | 0 | 0 | 0 | 0 |
| 0 | 0 | 0 | 0 | 0 |
| 0 | 0 | 0 | 0 | 0 |
| 0 | 0.000512429 | 0 | 0 | 0 |
| 0 | 0 | 0 | 0 | 0 |
| 0 | 0 | 0 | 0 | 0 |
| 0 | 0 | 0 | 0 | 0 |
| 0 | 0 | 0 | 0 | 0 |
| 0 | 0.002312492 | 0 | 0 | 0 |
| 0 | 0 | 0 | 0 | 0 |
| 0 | 0 | 0 | 0 | 0 |
| 0.028633787 | 0.330207788 | 1.30681916 | 0 | 0 |
| 0 | 0.044116967 | 0.000677155 | 0 | 0.00054572 |
| 0 | 0 | 0 | 0 | 0 |
| 0 | 0 | 0 | 0 | 0 |
| 0 | 0 | 0 | 0 | 0 |
| 0 | 0 | 0 | 0 | 0 |
| 0 | 0 | 0 | 0 | 0 |
| 0 | 0 | 0 | 0 | 0 |
| 0.036846739 | 0 | 0 | 0 | 0 |
| 0 | 0 | 0 | 0 | 0 |
| 0 | 0 | 0 | 0 | 0 |
| 0 | 0 | 0 | 0 | 0 |
| 0 | 0 | 0.003287271 | 0 | 0.009776868 |
| 0 | 0 | 0 | 0 | 0 |
| 0 | 0 | 0 | 0 | 0 |
| 0 | 0 | 0 | 0 | 0 |
| 0 | 0 | 0 | 0 | 0.000661294 |
| 0 | 0 | 0 | 0 | 0 |

| | | | | |
|---|---|---|---|---|
| 0.002746349 | 0.042203601 | 0.054423441 | 0 | 0 |
| 0 | 0 | 0 | 0 | 0 |
| 0 | 0 | 0 | 0 | 0 |
| 0 | 0 | 0 | 0 | 0 |
| 0.082223352 | 0 | 0 | 0.244120431 | 0.001187468 |
| 0 | 0 | 0 | 0 | 0 |
| 0 | 0 | 0 | 0 | 0 |
| 0 | 0 | 0 | 0 | 0 |
| 0 | 0 | 0 | 0 | 0 |
| 0 | 0 | 0 | 0 | 0 |
| 0 | 0 | 0 | 0 | 0 |
| 0 | 0 | 0 | 0 | 0 |
| 0 | 0 | 0 | 0 | 0 |
| 0 | 0 | 0.004390859 | 0 | 0 |
| 0 | 0 | 0 | 0 | 0 |
| 0 | 0 | 0 | 0 | 6.50390498 |
| 0 | 0 | 0 | 0 | 0 |
| 0 | 0 | 0 | 0 | 0 |
| 0 | 0 | 0 | 0 | 0 |
| 0 | 0 | 0 | 0 | 0 |
| 0 | 0 | 0 | 0 | 0 |
| 0 | 0 | 0 | 0 | 0 |
| 0 | 0.004247579 | 0.001897215 | 0 | 0 |
| 0 | 0 | 0 | 0 | 0 |
| 0.032035629 | 0.104704917 | 0 | 0 | 0 |
| 0 | 0 | 0 | 0 | 0 |
| 0 | 0 | 0 | 0.002428388 | 0 |
| 0 | 0 | 0 | 0 | 0 |
| 0 | 0 | 0 | 0 | 0 |
| 0 | 0 | 0 | 0 | 0 |
| 0 | 0 | 0 | 0 | 0 |
| 0 | 0 | 0 | 0 | 0 |
| 0 | 0 | 0 | 0 | 0 |
| 0 | 0 | 0 | 0 | 0 |
| 0 | 0 | 0 | 0 | 0 |
| 0 | 0 | 0 | 0 | 0 |
| 0 | 0 | 0 | 0 | 0 |
| 0 | 0 | 0 | 0 | 0 |
| 0 | 0 | 0 | 0 | 0 |
| 0 | 0 | 0 | 0 | 0 |
| 0 | 0 | 0 | 0 | 0 |
| 0 | 0 | 0 | 0 | 0 |
| 0 | 0 | 0 | 0 | 0 |
| 0 | 0 | 0 | 0 | 0 |
| 0 | 0 | 0 | 0 | 0 |
| 0 | 0 | 0 | 0 | 0 |
| 0 | 0 | 0 | 0 | 0 |
| 0 | 0 | 0 | 0 | 0 |

| | | | | |
|---|---|---|---|---|
| 0 | 0 | 0 | 0.02942484 | 0 |
| 0 | 0 | 0 | 0 | 0 |
| 0 | 0 | 0 | 0 | 0 |
| 0 | 0 | 0 | 0 | 0 |
| 0 | 0 | 0 | 0 | 0 |
| 0 | 0 | 0 | 0 | 0 |
| 0 | 0 | 0 | 0 | 0 |
| 0 | 0 | 0 | 0 | 0 |
| 0 | 0 | 0 | 0 | 0 |
| 0 | 0 | 0 | 0 | 0 |
| 0 | 0 | 0 | 0 | 0 |
| 0 | 0 | 0 | 0 | 0 |
| 0.009304259 | 0 | 0.027811395 | 0 | 0.000753133 |
| 0 | 0 | 0 | 0 | 0 |
| 0 | 0 | 0 | 0 | 0 |
| 0 | 0 | 0 | 0 | 0.001215925 |
| 0 | 0 | 0 | 0 | 0 |
| 0 | 0 | 0 | 0 | 0 |
| 0 | 0 | 0 | 0 | 0 |
| 0 | 0 | 0 | 0 | 0 |
| 0.013468435 | 0.015022137 | 0.029634766 | 0 | 0.132087079 |
| 0 | 0 | 0 | 0 | 0 |
| 0.003200631 | 0 | 0.001103887 | 0 | 0 |
| 0 | 0 | 0 | 0 | 0 |
| 0 | 0 | 0 | 0 | 0 |
| 0 | 0 | 0 | 0 | 0 |
| 0.000490231 | 0.002733918 | 0.001139718 | 0.001033688 | 0.004428484 |
| 0 | 0 | 0 | 0 | 0 |
| 0 | 0 | 0 | 0 | 0 |
| 0 | 0 | 0 | 0 | 0.041062384 |
| 0 | 0 | 0.170253519 | 0 | 0 |
| 0 | 0 | 0 | 0 | 0 |
| 0 | 0 | 0 | 0 | 0 |
| 0.161363285 | 0.02276023 | 0 | 0 | 0 |
| 0.044364113 | 0.070252085 | 1.407582941 | 0.001616826 | 0.172252281 |
| 0 | 0 | 0 | 0 | 0 |
| 0 | 0 | 0 | 0 | 0 |
| 0 | 0 | 0 | 0 | 0 |
| 0 | 0 | 0 | 0 | 0 |
| 0 | 0 | 0 | 0 | 0 |
| 0 | 0 | 0 | 0 | 0 |
| 0 | 0 | 0.003633372 | 0 | 0 |
| 0 | 0 | 0 | 0 | 0 |
| 0 | 0 | 0 | 0 | 0 |
| 0.008766784 | 0.002346746 | 0 | 0 | 0 |
| 0.267690453 | 0 | 0 | 0 | 0 |
| 0 | 0 | 0.00118173 | 0 | 0.001190447 |
| 0.039681461 | 0.00682025 | 0.039018813 | 0 | 0.001175282 |

| | | | | |
|---|---|---|---|---|
| 0 | 0 | 0 | 0 | 0 |
| 0 | 0 | 0 | 0 | 0 |
| 0 | 0 | 0 | 0 | 0 |
| 0 | 0 | 0 | 0 | 0 |
| 0 | 0 | 0 | 0 | 0 |
| 0 | 0 | 0 | 0 | 0 |
| 0 | 0 | 0 | 0 | 0 |
| 0 | 0 | 0 | 0 | 0 |
| 0.063837809 | 0 | 0 | 0 | 0 |
| 0 | 0 | 0 | 0 | 0 |
| 0 | 0 | 0 | 0 | 0 |
| 0 | 0 | 0 | 0 | 0 |
| 0 | 0 | 0 | 0 | 0 |
| 0 | 0 | 0 | 0 | 0 |
| 0 | 0 | 0 | 0 | 0 |
| 0 | 0 | 0 | 0 | 0 |
| 0 | 0 | 0 | 0 | 0 |
| 0 | 0 | 0 | 0 | 0 |
| 0 | 0.02803975 | 0.004218671 | 0.002343548 | 0 |
| 0 | 0 | 0 | 0 | 0 |
| 0.001561414 | 0 | 0.04492206 | 0 | 0 |
| 0 | 0 | 0 | 0 | 0 |
| 0 | 0 | 0 | 0 | 0 |
| 0 | 0 | 0 | 0 | 0 |
| 0 | 0 | 0.002318108 | 0 | 0.034638919 |
| 0 | 0 | 0 | 0.008092747 | 0 |
| 0 | 0 | 0 | 0 | 0 |
| 0 | 0 | 0 | 0 | 0 |
| 0 | 0 | 0 | 0 | 0 |
| 0 | 0 | 0 | 0 | 0 |
| 0.498421831 | 0.227137719 | 1.23573764 | 0 | 0.292127084 |
| 0 | 0 | 0 | 0 | 0 |
| 0 | 0 | 0 | 0.03386427 | 0 |
| 0 | 0 | 0.001479998 | 0 | 0.004472747 |
| 0 | 0 | 0 | 0 | 0 |
| 0 | 0 | 0 | 0 | 0 |
| 0 | 0 | 0 | 0 | 0 |
| 0 | 0 | 0 | 0 | 0 |
| 0 | 0 | 0 | 0 | 0 |
| 0 | 0 | 0 | 0.002357656 | 0 |
| 0 | 0 | 0 | 0 | 0 |
| 0.010532735 | 0.015990032 | 0.370076156 | 0.047009125 | 0.044049554 |
| 0 | 0 | 0 | 0 | 0 |
| 0 | 0 | 0 | 0 | 0 |
| 0 | 0 | 0 | 0 | 0 |
| 0 | 0 | 0 | 0 | 0 |
| 0 | 0 | 0 | 0 | 0 |
| 0 | 0 | 0 | 0 | 0 |

| | | | | |
|---|---|---|---|---|
| 0 | 0 | 0 | 0 | 0 |
| 0 | 0 | 0 | 0 | 0 |
| 0.044452651 | 0.00655612 | 0.000915107 | 0.001394355 | 0.107488612 |
| 0 | 0 | 0 | 0 | 0 |
| 0 | 0 | 0 | 0 | 0 |
| 0 | 0 | 0 | 0 | 0 |
| 0 | 0 | 0 | 0 | 0 |
| 0 | 0 | 0 | 0 | 0 |
| 0 | 0 | 0 | 0 | 0 |
| 0 | 0 | 0 | 0 | 0 |
| 0 | 0 | 0 | 0 | 0 |
| 0 | 0 | 0 | 0 | 0 |
| 0 | 0 | 0 | 0 | 0 |
| 0 | 0.009283548 | 0 | 0 | 0.001426347 |
| 0.002345804 | 0.018875554 | 0.002003386 | 0 | 0.005718133 |
| 0.03175245 | 0 | 0 | 0 | 0 |
| 0 | 0.03172815 | 0 | 0.001449222 | 0.007665059 |
| 0 | 0 | 0 | 0 | 0 |
| 0 | 0 | 0 | 0 | 0 |
| 0 | 0 | 0 | 0 | 0 |
| 0 | 0 | 0 | 0 | 0 |
| 0.040155986 | 0.005574513 | 0.026787902 | 0 | 0.002767744 |
| 0 | 0 | 0 | 0 | 0 |
| 0 | 0 | 0.007986031 | 0 | 0 |
| 0 | 0 | 0 | 0 | 0 |
| 0 | 0 | 0 | 0 | 0 |
| 4.397073617 | 0 | 0 | 0 | 0 |
| 0 | 0 | 0 | 0 | 0 |
| 0 | 0 | 0 | 0 | 0 |
| 0 | 0 | 0.001738735 | 0 | 0 |
| 0 | 0 | 0 | 0 | 0 |
| 0 | 0 | 0 | 0 | 0 |
| 0 | 0 | 0 | 0 | 0 |
| 0 | 0 | 0 | 0 | 0 |
| 0 | 0 | 0 | 0 | 0 |
| 0 | 0 | 0 | 0 | 0 |
| 0 | 0 | 0 | 0 | 0 |
| 0.012775358 | 0.00772833 | 0.005177881 | 0.01287534 | 0.001304019 |
| 0 | 0 | 0 | 0 | 0 |
| 0 | 0 | 0 | 0 | 0 |
| 0 | 0 | 0 | 0 | 0 |
| 0 | 0 | 0 | 0 | 0 |
| 0 | 0 | 0 | 0 | 0 |
| 0 | 0 | 0 | 0 | 0 |
| 0 | 0 | 0 | 0 | 0 |
| 0 | 0 | 0 | 0 | 0 |
| 0 | 0 | 0 | 0 | 0 |
| 0.003671835 | 0 | 0.015679281 | 0 | 0.025008657 |

| | | | | |
|---|---|---|---|---|
| 0 | 1.257766394 | 0 | 0 | 0 |
| 0 | 0 | 0 | 0 | 0 |
| 0 | 0 | 0 | 0 | 0 |
| 0 | 0 | 0 | 0 | 0 |
| 0.000726467 | 0.002430813 | 0.002352441 | 0 | 0.001458335 |
| 0 | 0 | 0 | 0 | 0 |
| 0 | 0 | 0 | 0 | 0 |
| 0.002545757 | 0 | 0.009131439 | 0.004186973 | 0.02708535 |
| 0 | 0 | 0 | 0 | 0 |
| 0.006035124 | 0 | 0.016209507 | 0.055770875 | 0.163290785 |
| 0 | 0 | 0 | 0 | 0 |
| 0 | 0 | 0.002409493 | 0 | 0 |
| 0.001528624 | 0.005626381 | 0.003198451 | 0 | 0.011507302 |
| 0 | 0.028742348 | 0 | 0 | 0 |
| 0 | 0 | 0.000626578 | 0 | 0 |
| 0 | 0 | 0.00137183 | 0 | 0 |
| 0 | 0 | 0 | 0 | 0 |
| 0 | 0 | 0.001388522 | 0 | 0 |
| 0 | 0 | 0.142195933 | 0 | 0.344481573 |
| 0 | 0.007505337 | 0 | 0 | 0 |
| 0 | 0 | 0 | 0 | 0 |
| 0 | 0 | 0 | 0 | 0 |
| 0 | 0 | 0 | 0 | 0 |
| 0 | 0 | 0 | 0 | 0 |
| 0 | 0 | 0 | 0 | 0 |
| 0.002848731 | 0 | 0 | 0 | 0.000893538 |
| 0 | 0 | 0 | 0 | 0 |
| 0 | 0 | 0.101470205 | 0 | 0 |
| 0.001276052 | 0 | 0.017436578 | 0 | 0 |
| 0 | 0 | 0 | 0.000695272 | 0 |
| 0 | 0.002130923 | 0 | 0.002900509 | 0 |
| 0 | 0 | 0 | 0 | 0 |
| 0 | 0 | 0 | 0 | 0 |
| 0 | 0 | 1.154922586 | 0 | 0.30359806 |
| 0 | 0 | 0 | 0 | 0 |
| 0 | 0 | 0 | 0 | 0 |
| 0 | 0 | 0.294611473 | 0 | 0 |
| 0 | 0 | 0 | 0 | 0 |
| 0 | 0 | 0 | 0 | 0 |
| 0 | 0 | 0 | 0 | 0 |
| 0 | 0 | 0.151893658 | 0 | 0 |
| 0 | 0 | 0 | 0 | 1.19420902 |
| 0 | 0 | 0 | 0 | 0 |
| 0 | 0 | 0.021009081 | 0 | 0.005327007 |
| 0 | 0 | 0 | 0 | 0 |
| 0.001302653 | 0 | 0 | 0.005108933 | 0 |
| 0.002435553 | 0 | 0.005735853 | 0 | 0.00533369 |
| 0 | 0 | 0 | 0 | 0 |

| | | | | |
|---|---|---|---|---|
| 0 | 0 | 0 | 0 | 0 |
| 0 | 0 | 0 | 0 | 0.009189522 |
| 0 | 0.001346643 | 0 | 0 | 0 |
| 0 | 0 | 0 | 0 | 0 |
| 0 | 0 | 0 | 0 | 0 |
| 0.017962198 | 0.035983929 | 0.033535077 | 0 | 0.373007278 |
| 0 | 0 | 0 | 0 | 0 |
| 0 | 0 | 0 | 0 | 0 |
| 0 | 0 | 0 | 0 | 0 |
| 0 | 0 | 0 | 0 | 0 |
| 0.000809053 | 0.008121463 | 0 | 0 | 0.004060306 |
| 0 | 0.119883669 | 0 | 0 | 0 |
| 0 | 0 | 0 | 0 | 0 |
| 0 | 0.00767275 | 0 | 0 | 0.005370363 |
| 0 | 0 | 0.014833643 | 0.001223058 | 0.113528481 |
| 0 | 0 | 0.112030627 | 0 | 0 |
| 0 | 0 | 0 | 0 | 0 |
| 0 | 0 | 0.454502413 | 0 | 0 |
| 0 | 0 | 0 | 0 | 0 |
| 0 | 0 | 0 | 0.001594605 | 0 |
| 0 | 0 | 0 | 0 | 0 |
| 0 | 0.329163362 | 0 | 0.187777143 | 0 |
| 0 | 0 | 0 | 0 | 0 |
| 0 | 0 | 0 | 0 | 0 |
| 0 | 0 | 0 | 0 | 0 |
| 0 | 0 | 0 | 0 | 0 |
| 0 | 0 | 0.251441147 | 0 | 0 |
| 0 | 0 | 0 | 0 | 0 |
| 0 | 0 | 0 | 0 | 0 |
| 1.140268164 | 0.277729651 | 0.008093263 | 0.060595699 | 0.168400865 |
| 0.001037106 | 0 | 0 | 0 | 0.001431322 |
| 0 | 0 | 0 | 0 | 0 |
| 0 | 0 | 0 | 0 | 0 |
| 0 | 0 | 0 | 0 | 0 |
| 0.00138056 | 0.004426981 | 0.017538204 | 0 | 0.030312018 |
| 0 | 0 | 0 | 0 | 0 |
| 0 | 0 | 0 | 0.460062808 | 0 |
| 0 | 0 | 0 | 0 | 0 |
| 0 | 0 | 0.529067 | 0.02579822 | 1.396077741 |
| 0 | 0 | 0 | 0 | 0 |
| 0.017751773 | 0.164003729 | 0.017061198 | 0 | 0 |
| 1.027365928 | 0.105636903 | 0 | 0.001709318 | 7.763453731 |
| 0 | 0 | 0 | 0 | 0 |
| 0 | 0 | 0 | 0 | 0 |
| 0 | 0.005139021 | 0.529174524 | 0 | 0.034684757 |
| 0 | 0 | 0 | 0 | 0 |
| 0 | 0 | 0 | 0 | 0.000827479 |
| 0 | 0 | 0 | 0 | 0 |

| | | | | |
|---|---|---|---|---|
| 0.002644691 | 0 | 0 | 0 | 0 |
| 0 | 0.004929533 | 0.038078461 | 0 | 0 |
| 0 | 0 | 0 | 0 | 0.016693804 |
| 0 | 0 | 0 | 0 | 0 |
| 0.202323067 | 0.271586103 | 0 | 0 | 0 |
| 0 | 0.003069588 | 0 | 0 | 0 |
| 0 | 0 | 0 | 0 | 0 |
| 0 | 0 | 0.004089317 | 0 | 0 |
| 0 | 0.0024687 | 0 | 0 | 0 |
| 0 | 0 | 0 | 0 | 0 |
| 0 | 0 | 0 | 0 | 0 |
| 0 | 0 | 0.00410207 | 0 | 0 |
| 0 | 0 | 0 | 0 | 0 |
| 0 | 0.090732075 | 0 | 0 | 0.002586209 |
| 0 | 0 | 0 | 0 | 0 |
| 0.343029731 | 0.08037032 | 0.117510581 | 0.025474825 | 0 |
| 0.065839766 | 0.005334259 | 0.001111875 | 0.01230291 | 0.003360231 |
| 0 | 0 | 0.002508786 | 0 | 0 |
| 0 | 0 | 0.005742415 | 0 | 0 |
| 0 | 0.010908008 | 0 | 0 | 0 |
| 0.01727847 | 0 | 0.236415785 | 0 | 0.046784977 |
| 0 | 0 | 0.007185568 | 0 | 0.000742418 |
| 0 | 0 | 0 | 0.272561277 | 0 |
| 0.30408963 | 0.112467511 | 0 | 0.001113186 | 1.479823188 |
| 0.045889484 | 0 | 0 | 0.001130976 | 0 |
| 0.006930183 | 0 | 0 | 0.003188246 | 0 |
| 0 | 0.022121489 | 0.000806591 | 0 | 0 |
| 0.024334545 | 0.143708416 | 0.002297005 | 0 | 0 |
| 0 | 0 | 0 | 0 | 0 |
| 0 | 0 | 0 | 0 | 0 |
| 0.015083335 | 0.036908325 | 0.060574261 | 0 | 0 |
| 0.007801751 | 0 | 0.005944376 | 0 | 0.016582778 |
| 0.004369601 | 0.079440872 | 0 | 0.009397893 | 0 |
| 0 | 0 | 0 | 0 | 0 |
| 0.00111738 | 0 | 0.002003979 | 0 | 0 |
| 0 | 0 | 0 | 0 | 0 |
| 0 | 0 | 0 | 0.001249462 | 0.001784298 |
| 0.063332949 | 0.09372584 | 0.181767188 | 0.016611336 | 0.027132855 |
| 0.092948767 | 0.049659796 | 0 | 0.023228351 | 0 |
| 0 | 0 | 0 | 0 | 0.003549421 |
| 0 | 0 | 0 | 0 | 0 |
| 0 | 0 | 0 | 0 | 0 |
| 0.04204392 | 0.013913621 | 0.001150858 | 0 | 0 |
| 0.005908584 | 0.008607596 | 0 | 0 | 0.188083175 |
| 0 | 0.051585393 | 0 | 0 | 3.62280998 |
| 0 | 0 | 0.006375411 | 0 | 0.002028139 |
| 0 | 0 | 0 | 0 | 0 |
| 0 | 0 | 0 | 0 | 0 |

| | | | | |
|---|---|---|---|---|
| 0 | 0 | 0.015973776 | 0 | 0 |
| 0.026731696 | 0.099776052 | 0.237823952 | 0 | 0.504931305 |
| 0 | 0 | 0 | 0 | 0 |
| 0 | 0 | 0 | 0 | 0 |
| 0 | 0.008061117 | 0 | 0 | 0 |
| 0 | 0 | 0 | 0 | 0 |
| 0 | 0 | 0.183542464 | 0 | 0 |
| 0 | 0 | 0 | 0 | 0 |
| 0.001156095 | 0.00300874 | 0.057210812 | 0 | 0 |
| 0 | 0 | 0.009753623 | 0 | 0.005219835 |
| 0 | 0.000523373 | 0.007480601 | 0 | 0.006279819 |
| 0.010717219 | 0 | 0.03064206 | 0.005895139 | 0.220987496 |
| 0.005917923 | 0 | 0 | 0 | 0 |
| 0 | 0 | 0 | 0 | 0 |
| 0.004845567 | 0 | 0 | 0 | 0 |
| 0 | 0 | 0 | 0 | 0 |
| 0 | 0.002557553 | 0.01207629 | 0 | 0.063128415 |
| 0 | 0 | 0.205703625 | 0 | 0.04502949 |
| 0 | 0 | 0 | 0 | 0 |
| 0.001794831 | 0.003541789 | 0.217211182 | 0 | 0.094093946 |
| 0.001182347 | 0.014317763 | 0.001851226 | 0.035472751 | 0.021022302 |
| 0 | 0 | 0 | 0 | 0 |
| 0 | 0 | 0 | 0 | 0 |
| 0 | 0 | 0 | 0 | 0 |
| 0 | 0 | 0.473347056 | 0 | 0.079188275 |
| 0.001858707 | 0 | 0.007099164 | 0 | 0 |
| 0.03851218 | 0.160662477 | 0 | 0 | 0 |
| 0.007539963 | 0.212646893 | 0.005366127 | 0.025891939 | 0 |
| 0 | 0 | 0 | 0.008192687 | 0 |
| 0.006712948 | 0 | 0.007551607 | 0 | 0.002390869 |
| 0 | 0 | 0 | 0 | 0.016627618 |
| 0 | 0 | 0 | 0 | 0 |
| 0.001743516 | 0 | 0 | 0 | 0 |
| 0 | 0 | 0 | 0 | 0 |
| 0 | 0.095794278 | 1.563723255 | 0 | 0.04082607 |
| 0.295020729 | 0.373786322 | 0.016347329 | 0.014606854 | 0.175352817 |
| 0 | 0 | 0.000420617 | 0 | 0 |
| 0 | 0 | 0 | 0 | 0 |
| 0 | 0.005434202 | 0.003530516 | 0 | 0.00088914 |
| 0 | 0 | 0 | 0 | 0 |
| 0 | 0.054249835 | 0 | 0 | 2.736694931 |
| 0 | 0 | 0 | 0 | 0 |
| 0 | 0.021305273 | 0.115400334 | 0.003573907 | 0.009586369 |
| 0 | 0.001738296 | 0.00129404 | 0 | 0 |
| 0 | 0 | 0.936057798 | 0 | 0 |
| 1.605564182 | 0 | 0 | 0.083016347 | 0 |
| 0 | 0 | 0 | 0 | 0 |
| 0 | 0.00257567 | 0 | 0.001062387 | 0 |

| | | | | |
|---|---|---|---|---|
| 0 | 0 | 0.055424113 | 0 | 0 |
| 0 | 0 | 0 | 0 | 0 |
| 0 | 0.002062494 | 0 | 0 | 0 |
| 0.866397624 | 3.210429584 | 0 | 0.011892774 | 0 |
| 0 | 0 | 0 | 0 | 0 |
| 0 | 0 | 0 | 0 | 0 |
| 0 | 0 | 0 | 0 | 0 |
| 0.201269966 | 0 | 0 | 0.015959043 | 0 |
| 0 | 0.589142976 | 0 | 7.222438074 | 0.19868756 |
| 0.000420079 | 0.001249438 | 0 | 0 | 0 |
| 0 | 0 | 0 | 0 | 0 |
| 0.003602613 | 0.001171977 | 0 | 0.101981274 | 0.016272029 |
| 0 | 0 | 0 | 0 | 0 |
| 0.477855378 | 0 | 0 | 0 | 0 |
| 2.119171761 | 0.226029335 | 1.530041668 | 0.102177272 | 0.009946459 |
| 0 | 0 | 0 | 0 | 0 |
| 0.035062394 | 0 | 0 | 0.001226515 | 0 |
| 0 | 0 | 0 | 0 | 0.012339149 |
| 0 | 0 | 0.00839712 | 0 | 0 |
| 0 | 0 | 0 | 0 | 0 |
| 0 | 0 | 0 | 0 | 0 |
| 0 | 0 | 0 | 0 | 0 |
| 0.222265604 | 0 | 0 | 0 | 0 |
| 0.17301222 | 0.202419609 | 0.574097587 | 0 | 1.074028919 |
| 0 | 0.026334473 | 0 | 0 | 0 |
| 0.277371356 | 0 | 0 | 0.011361203 | 0 |
| 0.00418053 | 0 | 0 | 0 | 0 |
| 0.076527111 | 4.218198196 | 0 | 0.55097644 | 3.921109259 |
| 0 | 0 | 0 | 0 | 0 |
| 0 | 0 | 0 | 0 | 0 |
| 0.034585746 | 0.053387211 | 0 | 0 | 0 |
| 0 | 0 | 0 | 0 | 0 |
| 0.034985325 | 0.183337652 | 0 | 0.005339477 | 0 |
| 0 | 0 | 0 | 0 | 0 |
| 0 | 0 | 0 | 0.001022767 | 0 |
| 0 | 0 | 0.487218819 | 0.76704925 | 0.044212897 |
| 0.614280872 | 0.183584254 | 0 | 0.060651935 | 0 |
| 0.032798681 | 0 | 0 | 0 | 0 |
| 0.049691744 | 0 | 0 | 0 | 0 |
| 0.007735963 | 0 | 1.013905433 | 0 | 0 |
| 0.00273605 | 0.101990269 | 0.188675675 | 0 | 0.019368091 |
| 0 | 0 | 0 | 0 | 0 |
| 0.247721822 | 0.023121231 | 0.071086136 | 4.847703254 | 1.45561799 |
| 0.380851356 | 0.008394978 | 0.000749937 | 0 | 0 |
| 0 | 0 | 0 | 0 | 0 |
| 0 | 0 | 0 | 0 | 0 |
| 0 | 0 | 0 | 0 | 0 |
| 0.55498322 | 0.267798035 | 0.02276722 | 0.001533042 | 0.000521254 |

| | | | | |
|---|---|---|---|---|
| 0 | 0 | 0 | 0 | 0 |
| 0 | 0.204218474 | 0 | 0.155426642 | 0.017095569 |
| 0.04082204 | 0.01092009 | 0.452537047 | 0.006088319 | 0.176220678 |
| 0.03497846 | 0 | 0 | 0 | 0 |
| 0 | 0 | 0 | 0 | 0 |
| 0.047191255 | 0.010497875 | 0 | 0.005788448 | 0.001705726 |
| 0 | 0.097070291 | 0 | 0 | 0 |
| 0 | 0 | 0 | 0 | 0 |
| 0 | 0 | 0 | 0 | 0 |
| 0 | 0.081891285 | 0 | 0 | 0 |
| 0.936014739 | 0.055243455 | 1.601922626 | 0.005359272 | 0.001962391 |
| 0.12414549 | 0.944640154 | 0.179932642 | 0.583663336 | 0.000440308 |
| 0 | 0 | 0 | 0 | 0 |
| 0.011781099 | 0.000784487 | 0.002277582 | 0.016684456 | 0.003176838 |
| 0.054318081 | 0.127968558 | 0.194116927 | 0.00052592 | 0.25980459 |
| 0.009844962 | 2.643176183 | 0 | 0 | 0 |
| 0 | 0 | 0 | 0 | 0 |
| 0 | 0 | 0 | 0 | 0 |
| 0 | 0 | 0 | 0 | 0 |
| 0.41913421 | 0.007185168 | 1.993250965 | 0 | 0 |
| 0 | 0.114841517 | 0.111433719 | 0 | 0.010691021 |
| 0.009928299 | 0 | 0.188095896 | 0.117051372 | 0.169697409 |
| 0.076872738 | 1.585051758 | 1.287953632 | 0.478116246 | 5.552824995 |
| 0 | 0 | 0 | 0 | 0 |
| 0 | 0 | 0 | 0 | 0 |
| 0 | 0.002555178 | 0 | 0.774679569 | 0 |
| 0 | 0.011294509 | 0 | 0 | 0 |
| 0.000569705 | 0.002223988 | 0 | 0 | 0 |
| 0 | 0 | 0 | 0 | 0 |
| 0 | 0 | 0 | 0 | 0 |
| 0.083292027 | 0 | 0 | 0 | 0 |
| 0.069616791 | 0 | 0.19447593 | 0.039633854 | 0.218943573 |
| 0 | 0.018663311 | 4.54401797 | 0 | 0.07457078 |
| 0.762933369 | 0.116134915 | 0.273602502 | 0.011139019 | 0.010886508 |
| 0.053011178 | 0.031657305 | 0.210174163 | 0.004750561 | 0.036469828 |
| 0.175560572 | 0.096298713 | 0.138940507 | 0.275476984 | 0.008224301 |
| 2.023782973 | 0.007193361 | 0.148819053 | 0.005377772 | 0 |
| 0 | 0 | 0 | 0 | 0 |
| 0 | 0.025746155 | 0 | 0.1033361 | 0.007128958 |
| 0 | 0 | 0 | 0 | 0 |
| 0 | 0 | 0 | 0 | 0 |
| 0 | 0 | 0 | 0 | 0 |
| 0 | 0 | 0 | 0 | 0 |
| 0 | 0 | 0 | 0 | 0 |
| 1.045159634 | 0.077007527 | 0.072585108 | 0.116684371 | 0.00868415 |
| 0 | 0 | 0 | 0 | 0 |
| 0 | 0 | 0 | 0 | 0 |
| 0.061158808 | 0.307079469 | 0.579210377 | 3.642155081 | 0.659115784 |

| | | | | |
|---|---|---|---|---|
| 0 | 0 | 0 | 0 | 0 |
| 0 | 0 | 0 | 0.010192135 | 0 |
| 0 | 0 | 0 | 0 | 0 |
| 0.004276949 | 0.024180653 | 0 | 0.121653185 | 0 |
| 0 | 0 | 0 | 0 | 0 |
| 0.072551643 | 0.142810508 | 0 | 0 | 0 |
| 0 | 0.002701348 | 0 | 0 | 0 |
| 0.7647215 | 0.846278804 | 0.496108688 | 0 | 0 |
| 0 | 0 | 0 | 0 | 0 |
| 0 | 0 | 0 | 0 | 0 |
| 0.017082499 | 0.056060127 | 0.485573912 | 0 | 0.000989193 |
| 0 | 0 | 0 | 0 | 0 |
| 0.06486188 | 0 | 0.399651316 | 0 | 0.007650687 |
| 0 | 0 | 0 | 0 | 0 |
| 0 | 0 | 0 | 0 | 0 |
| 0 | 0 | 0 | 0 | 0 |
| 0 | 0 | 0 | 0 | 2.463610505 |
| 0.006153227 | 0.002135173 | 0 | 0.016261404 | 0.002882181 |
| 0 | 0 | 0 | 0 | 0 |
| 0.187685985 | 0 | 0.372288449 | 0.002028151 | 0 |
| 0 | 0 | 0.033649131 | 0 | 0.00377992 |
| 0 | 0 | 0 | 0 | 0 |
| 0.158122504 | 0.096885535 | 0 | 0.00279229 | 0 |
| 0 | 0 | 0 | 0 | 0 |
| 0 | 0 | 0 | 0 | 0 |
| 0 | 0 | 0 | 0 | 0 |
| 0 | 0 | 0 | 0 | 0 |
| 0 | 0 | 0 | 0 | 0 |
| 0 | 0 | 0 | 0 | 0 |
| 0.639406861 | 3.59089605 | 0 | 0.256621051 | 2.676890884 |
| 0 | 0 | 0 | 0 | 0 |
| 0 | 0 | 0 | 0 | 0 |
| 0 | 0 | 0 | 0 | 0 |
| 0 | 0 | 0 | 0 | 0 |
| 0 | 0 | 0 | 0 | 0 |
| 0 | 0 | 0 | 0 | 0 |
| 0.042580423 | 0.04211347 | 0 | 0.22723762 | 0 |
| 0 | 0.002326269 | 0.582143146 | 0.001935025 | 0.056243281 |
| 0 | 0 | 0 | 0 | 0 |
| 0 | 0.003212304 | 0 | 0 | 0.005781541 |
| 0 | 0.071298777 | 0 | 0.00907766 | 0 |
| 0 | 0 | 0 | 0 | 0 |
| 0 | 0 | 0 | 0.000793235 | 0 |
| 0 | 0.008283302 | 0 | 0 | 0 |
| 0 | 0 | 0 | 0 | 0 |
| 0 | 0 | 0 | 0 | 0 |
| 0.010569249 | 0 | 0 | 0 | 0 |
| 0 | 0 | 0 | 0 | 0 |

| | | | | |
|---|---|---|---|---|
| 0 | 0 | 0 | 0 | 0 |
| 0 | 0 | 0 | 0 | 0 |
| 0 | 0 | 0 | 0 | 0 |
| 0.318524665 | 0 | 0 | 0.015048298 | 0 |
| 0 | 0 | 0 | 0 | 0 |
| 0 | 0 | 0 | 0.832695101 | 0 |
| 0 | 0 | 0 | 0 | 0 |
| 0 | 0 | 0 | 0 | 0 |
| 0.006136501 | 0.012833251 | 1.593134683 | 0 | 3.380694208 |
| 0 | 0 | 0 | 0.002930605 | 0 |
| 0 | 0 | 0 | 0 | 0 |
| 0 | 0 | 0 | 0 | 0 |
| 0 | 0 | 0 | 0 | 0 |
| 0 | 0 | 0.002388789 | 0 | 0 |
| 0.202853187 | 0.986633803 | 0 | 0.000697387 | 0.009682414 |
| 0.00177359 | 0 | 0.695961714 | 0 | 7.292521131 |
| 0.020824526 | 0.039537786 | 0 | 0 | 0 |
| 0 | 0 | 0 | 0 | 0 |
| 0 | 0 | 0 | 0 | 0 |
| 0 | 0 | 0 | 0 | 0 |
| 0.035191765 | 0.04170466 | 0 | 0 | 0 |
| 0 | 0 | 0 | 0 | 0 |
| 0 | 0 | 0 | 0 | 0 |
| 0 | 0 | 0 | 0 | 0 |
| 0 | 0.217318151 | 0.055498898 | 0.014265187 | 0.001584446 |
| 5.734905582 | 2.009434106 | 0 | 0 | 7.780004836 |
| 0.392039801 | 0 | 0 | 0 | 0 |
| 0 | 0 | 0 | 0 | 0 |
| 0 | 0.027268261 | 0 | 0.24519662 | 0.070648181 |
| 0 | 0 | 0 | 0 | 0 |
| 0 | 0 | 0 | 0 | 0 |
| 0.118507932 | 0.01619839 | 0 | 0 | 0 |
| 0 | 0 | 0 | 0 | 0 |
| 0.268454819 | 0.005776226 | 0.3530397 | 0 | 0 |
| 0.001587541 | 0 | 0 | 0.033390777 | 0.006373774 |
| 0.050498972 | 0.115308709 | 0.546902138 | 0 | 0.000703982 |
| 0 | 0.58706549 | 0 | 0 | 0.323723981 |
| 0.003643077 | 0.652960649 | 0 | 0.062522285 | 0 |
| 0 | 0.409225181 | 0 | 0 | 0 |
| 0 | 0 | 0 | 0 | 0 |
| 0 | 0 | 0 | 0.020304532 | 0 |
| 0 | 0.0053227 | 0 | 0 | 0 |
| 0.073991636 | 0.076754592 | 0 | 0.120286474 | 0 |
| 0 | 0 | 0 | 0.096215615 | 0 |
| 0 | 0 | 0 | 0 | 0 |
| 0.013820859 | 0.00044467 | 0.005296408 | 0.000504386 | 0 |
| 0 | 0 | 0 | 0 | 0 |
| 0.068172 | 0.040851768 | 0 | 0.003749885 | 0.052700343 |

| | | | | |
|---|---|---|---|---|
| 0.030785737 | 0 | 0 | 0 | 0 |
| 0 | 0 | 0 | 0 | 0 |
| 0 | 0 | 0 | 0 | 0 |
| 0 | 0 | 0 | 0 | 0 |
| 0.007073254 | 0 | 0.028190254 | 0 | 0 |
| 0 | 0.003714185 | 0 | 0.231522073 | 0.184288702 |
| 0 | 0.09406822 | 0 | 0 | 0 |
| 0 | 0.001263436 | 0 | 0 | 0 |
| 0 | 0 | 0 | 0 | 0 |
| 0 | 0 | 0 | 0 | 0 |
| 0 | 0 | 0 | 0 | 0 |
| 0.102233276 | 1.67190759 | 0 | 0.272255621 | 0 |
| 0.104486541 | 0 | 0 | 0 | 0.001146175 |
| 0 | 0 | 0 | 0 | 0 |
| 0 | 0 | 0.041658147 | 0 | 0.026274016 |
| 0 | 0 | 0 | 0.001598455 | 0 |
| 0 | 0.001410439 | 0.002267942 | 0 | 0 |
| 0 | 0 | 0 | 0 | 0 |
| 0 | 0.003021106 | 0.009366434 | 0 | 0.017431735 |
| 1.196786643 | 0 | 0 | 0 | 0 |
| 0 | 0 | 0.105672151 | 0 | 0 |
| 0 | 0 | 0 | 0 | 0 |
| 1.85945241 | 0.838430036 | 0.284650599 | 0.274409582 | 0 |
| 0 | 0 | 0 | 0 | 0 |
| 0 | 0 | 0 | 0 | 0 |
| 0 | 0 | 0 | 0 | 0 |
| 0 | 0 | 0 | 0 | 0 |
| 0 | 0 | 0 | 0 | 0 |
| 0.006713346 | 0 | 0 | 0 | 0 |
| 0 | 0 | 0 | 0 | 0 |
| 0 | 0 | 0 | 0 | 0 |
| 0.008491982 | 0.004998903 | 0.133497956 | 0.001193731 | 0.071503135 |
| 0 | 0 | 0 | 0 | 0 |
| 0.175472347 | 0.215036348 | 0 | 0.172946496 | 0.013366079 |
| 0.044117082 | 0.103009886 | 0.493411796 | 0 | 0.001072553 |
| 0.09682061 | 0.047190628 | 0.035685669 | 0.066474299 | 0.022912491 |
| 0 | 0 | 0 | 0 | 0 |
| 0 | 0 | 0 | 0 | 0.017288836 |
| 0 | 0.000782434 | 0 | 0.908365919 | 0 |
| 0 | 0 | 0 | 0 | 0 |
| 0 | 0 | 0 | 0 | 0 |
| 0 | 0 | 0 | 0 | 0 |
| 0 | 0 | 0 | 0 | 0 |
| 0.218101966 | 0.030494375 | 0 | 0 | 0 |
| 0.363016718 | 0.150954638 | 0.271798079 | 0.009512598 | 0.003018776 |
| 0 | 0 | 0 | 0 | 0 |
| 0 | 0 | 0 | 0 | 0 |
| 0.013512072 | 0.014277608 | 0.010770396 | 0 | 0.00642425 |

| | | | | |
|---|---|---|---|---|
| 0 | 0 | 0 | 0 | 0.290219806 |
| 0 | 0 | 0 | 0 | 0 |
| 0.047903331 | 0.012143046 | 0.11854853 | 0 | 0 |
| 0 | 0 | 0 | 0 | 0 |
| 0.014037988 | 0 | 0.031431463 | 0 | 0 |
| 0 | 0 | 0 | 0 | 0 |
| 0 | 0 | 0 | 0.034872324 | 0.2007103 |
| 0.014247731 | 0 | 0 | 0 | 0 |
| 0 | 0 | 0 | 0 | 0 |
| 0 | 0 | 0 | 0 | 0 |
| 0 | 0 | 0 | 0 | 0 |
| 0 | 0 | 0.010091106 | 0 | 0 |
| 0 | 0 | 0 | 0 | 0 |
| 0 | 0 | 0 | 0 | 0 |
| 0 | 0 | 0 | 0 | 0 |
| 0 | 0 | 0 | 0 | 0 |
| 0.143452981 | 0 | 0 | 0 | 0 |
| 0 | 0 | 0 | 0 | 0 |
| 0.590802674 | 1.687522947 | 0 | 0.001471772 | 0 |
| 0 | 0 | 0 | 0.00265235 | 0 |
| 0 | 0 | 0 | 0.003754411 | 0 |
| 0 | 0 | 0 | 0 | 0 |
| 0 | 0 | 0 | 0 | 0 |
| 0 | 0 | 0 | 0 | 0 |
| 0 | 0 | 0 | 0 | 0 |
| 0 | 0.014834045 | 0.366299116 | 0 | 0.00674488 |
| 0 | 0 | 0 | 0 | 0 |
| 0.004659022 | 0.054868728 | 0.000409597 | 0 | 0.02214386 |
| 0 | 0 | 0 | 0 | 0 |
| 0.462054075 | 0.860017333 | 0 | 0.028235944 | 0 |
| 0 | 0 | 0 | 0 | 0 |
| 0 | 0 | 0 | 0 | 0 |
| 0 | 0 | 0 | 0 | 0 |
| 0 | 0 | 0 | 0 | 0 |
| 0.053108695 | 0.193250909 | 0.042537287 | 0 | 0.002061563 |
| 0 | 0.021379138 | 0 | 0 | 0 |
| 0 | 0 | 0 | 0 | 0.005474358 |
| 0.002164134 | 0.007984062 | 0.114282516 | 0 | 0.001336726 |
| 0 | 0 | 0 | 0 | 0 |
| 0.000681956 | 0 | 0 | 0 | 0 |
| 4.689783357 | 5.540875015 | 0.390425095 | 0.079563909 | 5.006294506 |
| 1.541140088 | 0.146788809 | 0.118987247 | 0 | 0.375250737 |
| 0 | 0 | 0 | 0 | 0.139826568 |
| 0 | 0 | 0 | 0 | 0 |
| 0 | 0 | 0 | 0 | 0 |
| 0 | 0 | 0 | 0 | 0 |
| 0 | 0 | 0 | 0 | 0 |
| 0.02999153 | 0 | 0.051471789 | 0.004847419 | 0 |

| | | | | |
|---|---|---|---|---|
| 0 | 0 | 0 | 0 | 0 |
| 0.014948741 | 0.010458649 | 0 | 0 | 0.016641151 |
| 0.051742974 | 0.005487949 | 0 | 0 | 0 |
| 0 | 0 | 0 | 0 | 0 |
| 0.661505516 | 0.667193106 | 0 | 2.050865104 | 0 |
| 0 | 0 | 0 | 0 | 0 |
| 0 | 0 | 0 | 0 | 0 |
| 0 | 0 | 0 | 0 | 0 |
| 0 | 0.006205397 | 1.199804031 | 0.001201736 | 0.038272697 |
| 0 | 0 | 0 | 0 | 0 |
| 0 | 0 | 0 | 0 | 0 |
| 0 | 0 | 0 | 0 | 0 |
| 0.008979245 | 0.020913843 | 0 | 0.009355322 | 0 |
| 0.007305266 | 0.013217854 | 0 | 0 | 0.005377112 |
| 0 | 0 | 0 | 0 | 0.002747244 |
| 0.053019334 | 0.405479895 | 0 | 0.003370707 | 0 |
| 0 | 0 | 0 | 0 | 0.010857131 |
| 0 | 0 | 0 | 0 | 0 |
| 0 | 0 | 0 | 0 | 0 |
| 0 | 0 | 0 | 0 | 0 |
| 0 | 0 | 0 | 0 | 0 |
| 0 | 0 | 0 | 0 | 0 |
| 0 | 0 | 0 | 0 | 0 |
| 0 | 0 | 0 | 0 | 0 |
| 0 | 0 | 0 | 0 | 0 |
| 0 | 0 | 0 | 0 | 0 |
| 0.154767398 | 0.008719617 | 0 | 0 | 0 |
| 0.012129532 | 0.001383625 | 0.033784401 | 0 | 0 |
| 0 | 0 | 0 | 0 | 0 |
| 0 | 0 | 0 | 0 | 0 |
| 0 | 0 | 0 | 0 | 0 |
| 0 | 0 | 0 | 0 | 0 |
| 0.120643246 | 0 | 0 | 0 | 0 |
| 0 | 0.01337599 | 0 | 0 | 0 |
| 0 | 0.168495002 | 0 | 1.972411988 | 0.031903969 |
| 0 | 0 | 0.001270944 | 0 | 0 |
| 0 | 0 | 0 | 0 | 0 |
| 0 | 0.003022778 | 0 | 0 | 0 |
| 0 | 0 | 0 | 0 | 0 |
| 0 | 0 | 0 | 0 | 0 |
| 0.691102154 | 1.666022782 | 0 | 0.002470595 | 0 |
| 0 | 0.218962147 | 0 | 13.58388848 | 0.00077781 |
| 0 | 0 | 0 | 0 | 0 |
| 0 | 0 | 0 | 0.182694544 | 0.09245199 |
| 0 | 0 | 0 | 0 | 0 |
| 0 | 0 | 0 | 0.00138691 | 0 |
| 0 | 0 | 0 | 0 | 0 |
| 0 | 0 | 0 | 0 | 0 |

| | | | | |
|---|---|---|---|---|
| 1.069096499 | 0.1331464 | 0 | 0 | 0 |
| 0 | 0 | 0.001435064 | 0 | 0 |
| 0 | 0 | 0 | 0 | 0 |
| 0 | 0 | 0.013068181 | 0 | 0.001723933 |
| 0.000597584 | 0.004499017 | 0 | 0 | 0 |
| 0.548602584 | 0 | 0 | 0 | 0 |
| 0 | 0.017818654 | 0 | 0 | 0.049652572 |
| 0.001761003 | 0.001661974 | 0.001079758 | 0 | 0 |
| 0 | 0 | 0 | 0 | 0 |
| 0 | 0.0883897 | 0.108991085 | 0 | 0 |
| 0.036537119 | 0 | 0 | 0.001074993 | 0 |
| 0.192776177 | 0 | 0 | 0 | 0 |
| 0 | 0 | 0 | 0 | 0 |
| 0 | 0 | 0 | 0 | 0 |
| 0 | 0 | 0 | 0 | 0 |
| 0 | 0 | 0 | 0 | 0 |
| 0 | 0 | 0 | 0 | 0 |
| 0 | 0.001352429 | 0 | 0 | 0 |
| 1.760770861 | 0 | 0 | 0 | 0 |
| 0 | 0 | 0 | 0 | 0 |
| 0.012141133 | 0.0751491 | 0 | 0 | 0.035621406 |
| 0 | 0 | 0 | 0 | 0 |
| 0 | 0 | 0 | 0.064976628 | 0.006499145 |
| 0.012823971 | 0.007443569 | 0.044590496 | 0.01970077 | 0.00354627 |
| 0 | 0 | 0 | 0.000593764 | 0 |
| 0 | 0.000516573 | 0.002307314 | 0 | 0.003254068 |
| 0 | 0 | 0 | 0.001199036 | 0 |
| 0 | 0 | 0 | 0 | 0 |
| 0 | 0 | 0 | 0 | 0.092936184 |
| 0.454637511 | 0.001050298 | 0.039781724 | 0.021682491 | 0.555948578 |
| 0 | 0 | 0 | 0 | 0 |
| 0 | 0 | 0 | 0 | 0 |
| 0 | 0 | 0 | 0 | 0 |
| 0 | 0 | 0 | 0.000530401 | 0 |
| 0 | 0 | 0 | 0 | 0 |
| 0 | 0 | 0 | 0 | 0 |
| 0 | 0.022280149 | 4.230901483 | 0 | 0.020707411 |
| 0 | 0 | 0 | 0 | 0 |
| 0 | 0 | 0 | 0 | 0 |
| 0 | 0 | 0 | 0 | 0 |
| 0 | 0 | 0 | 0 | 0 |
| 0 | 0 | 0 | 0 | 0 |
| 0 | 0 | 0 | 0 | 0 |
| 0 | 0 | 0 | 0 | 0 |
| 0 | 0 | 0 | 0 | 0 |
| 0 | 0 | 0 | 0 | 0 |
| 0 | 0 | 0 | 0 | 0 |
| 0 | 0.003133245 | 0 | 0 | 0 |

| | | | | |
|---|---|---|---|---|
| 0.067164754 | 0.019474187 | 0.11616442 | 0 | 0.019644883 |
| 0 | 0 | 0 | 0.292617512 | 0.027879666 |
| 0 | 0 | 0 | 0 | 0 |
| 0 | 0 | 0 | 0 | 0 |
| 0 | 0 | 0 | 0 | 0 |
| 0 | 0 | 0 | 0 | 0 |
| 0.00216491 | 0 | 0.002049194 | 0 | 0 |
| 0.920663587 | 0.206728583 | 0 | 0.055421762 | 0.00072557 |
| 0.002210171 | 5.574653905 | 1.263590102 | 0.281296095 | 0.014641362 |
| 0 | 0.02625713 | 1.607726774 | 0.025695375 | 0.118144707 |
| 0.022750363 | 0 | 0 | 0 | 0 |
| 0 | 0 | 0 | 0 | 0 |
| 0 | 0 | 0 | 0 | 0 |
| 0 | 0 | 0 | 0.321066908 | 0 |
| 0 | 0 | 0 | 0 | 0 |
| 0 | 0 | 0 | 0 | 0 |
| 0 | 0.032782833 | 0.003686278 | 0 | 0.010108892 |
| 0 | 0 | 0 | 0 | 0 |
| 0 | 0 | 0 | 0 | 0 |
| 0 | 0.064322979 | 0 | 0 | 0 |
| 0 | 0 | 0 | 0 | 0 |
| 0 | 0 | 0 | 0 | 0 |
| 0 | 0 | 0 | 0 | 0 |
| 0 | 0 | 0 | 0 | 0 |
| 0 | 0.020781358 | 0 | 0.001683725 | 0 |
| 0 | 0 | 0 | 0 | 0 |
| 0 | 0 | 0 | 0 | 0 |
| 0 | 0 | 0 | 0 | 0 |
| 0 | 0 | 0 | 0 | 0 |
| 0.545253143 | 0 | 0 | 0 | 0 |
| 0.203239672 | 0.07115171 | 0 | 0 | 0 |
| 0 | 0 | 0 | 0 | 0 |
| 0 | 0 | 0 | 0 | 0 |
| 0 | 0 | 0 | 0 | 0 |
| 0 | 0.005133537 | 0.427505103 | 0 | 0.0025665 |
| 0 | 0 | 0 | 0.200402036 | 0 |
| 0 | 0 | 0 | 0 | 0 |
| 0 | 0 | 0 | 0.006187894 | 0 |
| 0 | 0 | 0 | 0.030714344 | 0 |
| 0 | 0 | 0 | 0 | 0 |
| 0 | 0 | 0 | 0 | 0 |
| 0 | 0.143473605 | 0 | 0.010824943 | 0 |
| 0 | 0 | 0 | 0 | 0 |
| 0 | 0 | 0 | 0 | 0 |
| 0 | 0 | 0 | 0 | 0 |
| 0 | 0.105854147 | 0 | 0 | 0 |
| 0 | 0 | 0 | 0 | 0 |
| 0 | 0 | 0 | 0 | 0 |

| | | | | |
|---|---|---|---|---|
| 0 | 0 | 0 | 0 | 0 |
| 0 | 0.001559188 | 0.003038941 | 0 | 0.011735207 |
| 0 | 0 | 0 | 0 | 0 |
| 0 | 0 | 0 | 0 | 0 |
| 4.114392277 | 1.272504761 | 1.286698558 | 0.717987458 | 0.053859299 |
| 0 | 0 | 0 | 0 | 0 |
| 0 | 0 | 0 | 0 | 0 |
| 0.012179902 | 0.024572797 | 0.175788662 | 0 | 0.254391695 |
| 0.007511318 | 0.010572004 | 0 | 0 | 0 |
| 0 | 0 | 0 | 0 | 0 |
| 0 | 0 | 0 | 0 | 0 |
| 0 | 0 | 0 | 1.689331847 | 0.0530514 |
| 0 | 0 | 0 | 0 | 0 |
| 0 | 0 | 0 | 0 | 0 |
| 0 | 0 | 0.121990147 | 0 | 0 |
| 0 | 0.051286941 | 0.474716155 | 0 | 0 |
| 0 | 0 | 0 | 0 | 0 |
| 0 | 0 | 0 | 0 | 0 |
| 0 | 0 | 0 | 0 | 0 |
| 0 | 0 | 0 | 0 | 0 |
| 0 | 0 | 0 | 0 | 0 |
| 0 | 0 | 0 | 0 | 0 |
| 0 | 0 | 0 | 0 | 0 |
| 0.01906627 | 0.010373528 | 0.122631172 | 0.000784441 | 0 |
| 0.151707317 | 0 | 0 | 0.0914292 | 0 |
| 0 | 0 | 0 | 0.002323574 | 0 |
| 1.003054856 | 0.132354371 | 0.491583418 | 0.008788892 | 0.001280715 |
| 0 | 0 | 0 | 0 | 0.00299678 |
| 0 | 0 | 0.00251958 | 0 | 0 |
| 0 | 0 | 0 | 0 | 0 |
| 0 | 0 | 0 | 0 | 0 |
| 0 | 0 | 0 | 0 | 0 |
| 0 | 0 | 0 | 0 | 0 |
| 0 | 0 | 0 | 0 | 0 |
| 0 | 0 | 0 | 0 | 0 |
| 0 | 0 | 0.041673937 | 0 | 0 |
| 0 | 0 | 0 | 0 | 0 |
| 1.677312448 | 0 | 0 | 0.004782686 | 0 |
| 0.714173714 | 0 | 0 | 0.002389148 | 0 |
| 0 | 0 | 0 | 0 | 0 |
| 0 | 0 | 0 | 0 | 0 |
| 0 | 0 | 0 | 14.2213371 | 0.007697604 |
| 0 | 0 | 0 | 0 | 0 |
| 0 | 0 | 0 | 0 | 0 |
| 0 | 0 | 0 | 0 | 0 |
| 0 | 0 | 0 | 0 | 0 |
| 0 | 0 | 0 | 0 | 0 |
| 0 | 0 | 0 | 0 | 0 |

| | | | | |
|---|---|---|---|---|
| 3.050249029 | 0.094726451 | 0 | 0.006747098 | 0 |
| 0 | 0 | 0 | 0 | 0 |
| 0 | 0 | 0 | 0 | 0 |
| 0 | 0.000739777 | 0 | 0 | 0 |
| 0 | 0.005647307 | 0 | 0 | 0 |
| 0.00664207 | 20.58850143 | 22.19274064 | 0 | 0 |
| 0 | 0 | 0 | 0 | 0 |
| 0 | 0 | 0 | 0 | 0 |
| 0.003069492 | 0.00106987 | 0 | 0 | 0 |
| 0 | 0 | 0 | 0 | 0 |
| 0 | 0 | 0 | 0 | 0 |
| 0 | 0.001275032 | 0.04399412 | 0 | 0 |
| 0 | 0.01620799 | 0 | 0 | 0.000373991 |
| 0 | 0 | 0 | 0 | 0 |
| 0 | 0 | 0 | 0 | 0 |
| 0 | 0 | 0 | 0 | 0 |
| 0 | 0 | 0 | 0 | 0 |
| 0 | 0 | 0 | 0 | 0 |
| 0 | 0 | 0 | 0 | 0 |
| 0.000803994 | 0 | 0.002860978 | 0 | 0 |
| 0 | 0 | 0.091358201 | 0 | 0.017817665 |
| 0 | 0 | 0 | 0 | 0 |
| 0.026123085 | 0 | 0.679064183 | 0.004439478 | 0.001369712 |
| 0.083325376 | 0 | 0 | 0 | 0 |
| 0 | 0 | 0 | 0 | 0 |
| 0 | 0 | 0 | 9.755291947 | 0 |
| 0 | 0 | 0 | 0 | 0 |
| 0 | 0 | 0 | 0 | 0 |
| 0.001687634 | 0 | 0 | 10.21858753 | 0 |
| 0 | 0 | 0 | 0 | 0 |
| 0 | 0 | 0 | 0 | 0 |
| 0 | 0 | 0 | 0 | 0 |
| 0.039799904 | 0.003868955 | 0.095682169 | 0.001385851 | 1.311866044 |
| 0 | 0 | 0 | 0 | 0 |
| 0.603254346 | 0 | 0 | 0 | 0.000983482 |
| 0 | 0 | 0.001605685 | 0.106760393 | 0 |
| 0 | 0 | 0 | 0 | 0 |
| 0 | 0 | 0 | 0 | 0 |
| 0.010167266 | 0.004347057 | 0.386472373 | 0 | 0.065822669 |
| 0 | 0 | 0 | 0 | 0 |
| 0 | 0 | 0 | 0 | 0 |
| 0 | 0 | 0 | 0 | 0 |
| 0 | 0.06983617 | 0 | 0.750032564 | 0.023865558 |
| 0 | 0 | 0 | 0 | 0 |
| 0 | 0 | 0 | 0 | 0 |
| 0.03477654 | 0.015796148 | 0 | 0 | 0 |
| 0 | 0 | 0 | 0 | 0 |
| 0 | 0 | 0 | 0 | 0 |

| | | | | |
|---|---|---|---|---|
| 0 | 0 | 0 | 0 | 0 |
| 0 | 0 | 0 | 0 | 0 |
| 0 | 0 | 0 | 0 | 0 |
| 0 | 0 | 0 | 0 | 0 |
| 0 | 0 | 0 | 0 | 0 |
| 0 | 0 | 0 | 0 | 0 |
| 0.975956595 | 0 | 0 | 0 | 0 |
| 0 | 0.006784037 | 0.727992248 | 0 | 4.927492963 |
| 1.327474532 | 0.183349686 | 0 | 0 | 0 |
| 0.01264216 | 0.001626986 | 0 | 0 | 0.005856535 |
| 0 | 0 | 0 | 0 | 0 |
| 0 | 0 | 0 | 0 | 0 |
| 0 | 0 | 0 | 0 | 0 |
| 0 | 0 | 0 | 0 | 0 |
| 0 | 0 | 0 | 0 | 0 |
| 0 | 0 | 0.29203723 | 0 | 0.014565519 |
| 0 | 0 | 0 | 0 | 0 |
| 0 | 0 | 0 | 0 | 0 |
| 0 | 0 | 0 | 0 | 0 |
| 0.326302048 | 1.778768141 | 0 | 0.003378224 | 0 |
| 0.001498975 | 0 | 0 | 0 | 0 |
| 0 | 0 | 0 | 0 | 0 |
| 0 | 0 | 0 | 0 | 0 |
| 0.47665786 | 0 | 0 | 0 | 0 |
| 0 | 0 | 0.005153812 | 0 | 0 |
| 0 | 0 | 0 | 0 | 0 |
| 0 | 0 | 0 | 0 | 0 |
| 0 | 0 | 0 | 0 | 0 |
| 0 | 0 | 0 | 0 | 0 |
| 0 | 0 | 0 | 0 | 0 |
| 0.037327951 | 0.137190004 | 0 | 0.004099411 | 0 |
| 0 | 0 | 0 | 0 | 0 |
| 0.014121399 | 0 | 0 | 0 | 0 |
| 0.483880783 | 0 | 0 | 0 | 0 |
| 0 | 0 | 0 | 0 | 0 |
| 0 | 0 | 0 | 0 | 0 |
| 0 | 0 | 0 | 0 | 0 |
| 0 | 0 | 0 | 0 | 0 |
| 0 | 0 | 0 | 0.008501162 | 0 |
| 0 | 0 | 0 | 0 | 0 |
| 0 | 0 | 0 | 0 | 0 |
| 10.31828477 | 0 | 0 | 0.010674871 | 0 |
| 0.008691509 | 0 | 0 | 0 | 0 |
| 0.001533009 | 0 | 0 | 0 | 0 |
| 0 | 0 | 0 | 0 | 0 |
| 0 | 0 | 0 | 0 | 0 |
| 0 | 0 | 0.122965738 | 0 | 0.217105118 |
| 0 | 0 | 0 | 0 | 0 |

| | | | | |
|---|---|---|---|---|
| 0 | 0 | 0 | 0 | 0 |
| 0 | 0 | 0 | 0 | 0 |
| 0 | 0 | 0 | 0 | 0 |
| 0 | 0 | 0 | 0 | 0 |
| 1.649345976 | 0.009385778 | 0 | 0.003240154 | 0 |
| 0 | 0 | 0 | 0 | 0 |
| 0 | 0 | 0 | 0 | 0 |
| 0 | 0 | 0 | 0 | 0 |
| 0 | 0 | 0 | 0 | 0 |
| 0 | 0 | 0 | 0 | 0 |
| 0 | 0 | 0 | 0 | 0 |
| 0.108872437 | 0.2088703 | 0.086545418 | 0.002570562 | 0 |
| 0.001314721 | 0.050671754 | 0.450475678 | 0.008501363 | 0 |
| 0 | 0 | 0 | 0 | 0 |
| 0.079355468 | 0 | 0 | 0 | 0 |
| 0.817538485 | 0.12556461 | 0 | 0 | 0.224406706 |
| 0 | 0.037764894 | 0.015570456 | 0 | 0 |
| 0 | 0 | 0 | 0 | 0 |
| 0.007307953 | 0 | 0.292917053 | 0 | 0.404461177 |
| 0 | 0.003174141 | 0.001667948 | 0 | 0 |
| 0 | 0 | 0 | 0 | 0 |
| 0 | 0 | 0 | 0 | 0 |
| 0 | 0 | 0 | 0 | 0 |
| 0 | 0 | 0 | 0 | 0 |
| 0 | 0 | 0 | 0 | 0 |
| 0 | 0 | 0 | 0 | 0 |
| 0.019987982 | 0.020797545 | 0.99256137 | 0.001018295 | 0.012522102 |
| 0 | 0 | 0.000506963 | 0 | 0 |
| 0 | 0 | 0 | 0 | 0 |
| 0 | 0 | 0 | 0 | 0 |
| 0.011860935 | 0.002883287 | 0.255750513 | 0.000577146 | 0.000763143 |
| 0 | 0 | 0.078551757 | 0 | 0 |
| 0 | 0 | 0 | 0 | 0 |
| 0 | 0 | 0 | 0 | 0 |
| 0 | 0 | 0 | 0 | 0 |
| 0 | 0 | 0 | 0 | 0 |
| 0 | 0 | 0 | 0 | 0 |
| 0.149355667 | 0.130360629 | 0.19788536 | 0.004054857 | 0.013511286 |
| 0 | 0 | 0 | 0 | 0 |
| 0 | 0.013798319 | 0 | 0 | 0 |
| 0.011092225 | 1.044585248 | 0.798927521 | 0 | 0 |
| 0 | 0 | 0 | 0 | 0 |
| 0 | 0 | 0 | 0 | 0 |
| 0 | 0 | 0.010575797 | 0 | 0.013834052 |
| 0 | 0 | 0 | 0 | 0 |
| 0 | 0 | 0 | 0 | 0 |
| 0 | 0 | 0 | 0 | 0 |
| 0 | 0 | 0 | 0 | 0 |

| | | | | |
|---|---|---|---|---|
| 0 | 0 | 0 | 0 | 0 |
| 0.004739185 | 0 | 0 | 0 | 0 |
| 0.004563939 | 0 | 0 | 0 | 0 |
| 0.043831429 | 0.001666629 | 0.032257897 | 0 | 0.002499681 |
| 0 | 0 | 0 | 0 | 0 |
| 0 | 0 | 0.002574196 | 0 | 0 |
| 0 | 0.00089852 | 0 | 0.003669066 | 0 |
| 0 | 0 | 0 | 0.00312126 | 0 |
| 0.031696358 | 0 | 0 | 0 | 0 |
| 0 | 0 | 0 | 0 | 0 |
| 0 | 0 | 0 | 0 | 0 |
| 0 | 0 | 0 | 0 | 0 |
| 0.814301124 | 0.168153312 | 0 | 0.064322845 | 0.023980009 |
| 0 | 0 | 0 | 0 | 0 |
| 0 | 0 | 0 | 0 | 0 |
| 0 | 0 | 0 | 0 | 0 |
| 0 | 0 | 0 | 0 | 0 |
| 0 | 0 | 0 | 0 | 0 |
| 0 | 0 | 0 | 0 | 0 |
| 0 | 0 | 0 | 0 | 0 |
| 0 | 0 | 0 | 0 | 0 |
| 0 | 0 | 0 | 0 | 0 |
| 0.030839303 | 0.05773146 | 1.011221311 | 0.042349182 | 0.038264689 |
| 0.291131511 | 0 | 0 | 0 | 0 |
| 0 | 0 | 0 | 0 | 0 |
| 0 | 0 | 0 | 0 | 0 |
| 0 | 0 | 0 | 0 | 0 |
| 0 | 0 | 0.002024167 | 0 | 0 |
| 0 | 0 | 0 | 0 | 0 |
| 0 | 0 | 0 | 0 | 0 |
| 0.014168976 | 0.016959845 | 0 | 0.00153025 | 0 |
| 0 | 0.008644077 | 0.014245561 | 0 | 0 |
| 0 | 0 | 0 | 0.011184072 | 0 |
| 0 | 0 | 0 | 0 | 0 |
| 0 | 0.008078837 | 0 | 0 | 0 |
| 0 | 0 | 0.001619788 | 0 | 0 |
| 0 | 0 | 0 | 0 | 0 |
| 0 | 0 | 0 | 0 | 0 |
| 0 | 0 | 0.001165307 | 0 | 0 |
| 0 | 0 | 0 | 0 | 0 |
| 0 | 0 | 0 | 0 | 0 |
| 0 | 0 | 0 | 0 | 0 |
| 0 | 0 | 0 | 0 | 0 |
| 0 | 0 | 0 | 0 | 0 |
| 0 | 0 | 0 | 0 | 0 |
| 0.470902993 | 0.277031138 | 0.000640798 | 0.380628878 | 0.00077463 |
| 0 | 0 | 0.001220272 | 0 | 0 |
| 0 | 0 | 0 | 0 | 0 |

| | | | | |
|---|---|---|---|---|
| 0 | 0 | 0 | 0 | 0 |
| 0 | 0 | 0 | 0 | 0 |
| 0 | 0 | 0 | 0.212150679 | 0 |
| 0.341790948 | 0 | 0 | 0 | 0 |
| 0 | 0 | 0 | 0 | 0 |
| 0.002960514 | 0.012283573 | 0 | 0 | 0 |
| 0 | 0 | 0 | 0 | 0 |
| 0 | 0 | 0 | 0 | 0 |
| 0.020560836 | 0 | 0 | 0 | 0 |
| 0 | 0 | 0 | 0 | 0 |
| 0.007590578 | 0 | 0.005790428 | 0.001200396 | 0 |
| 0.02248844 | 0 | 0.025935384 | 0 | 0 |
| 0 | 0 | 0 | 0 | 0 |
| 0 | 0 | 0 | 0 | 0 |
| 0 | 0 | 0 | 0 | 0 |
| 0 | 0 | 0 | 0 | 0 |
| 0 | 0 | 0.00156337 | 0 | 0 |
| 0 | 0 | 0 | 0 | 0 |
| 0 | 0 | 0 | 0 | 0 |
| 0 | 0.007368151 | 0 | 0.589810706 | 0.002052341 |
| 0 | 0 | 0 | 0 | 0 |
| 0 | 0 | 0 | 0 | 0 |
| 0 | 0 | 0 | 0 | 0 |
| 0 | 0 | 0 | 0 | 0 |
| 0 | 0 | 0.768672648 | 0 | 0 |
| 0 | 0 | 0 | 0 | 0 |
| 0 | 0 | 0 | 0 | 0 |
| 0 | 0 | 0 | 0.00286283 | 0 |
| 0 | 0 | 0 | 0 | 0 |
| 0 | 0 | 0 | 0 | 0 |
| 0 | 0.003466431 | 0.011174754 | 0.346182384 | 0.008137725 |
| 0 | 0 | 0 | 0 | 0 |
| 0.002353432 | 0.005949822 | 0.00250121 | 0.014093167 | 0 |
| 0 | 0.003218862 | 0.004153444 | 0 | 0.002252967 |
| 0 | 0 | 0 | 0 | 0 |
| 0 | 0 | 0 | 0 | 0 |
| 0 | 0 | 0 | 0 | 0 |
| 0 | 0 | 0 | 0 | 0 |
| 0 | 0 | 0 | 0 | 0 |
| 0 | 0 | 0 | 0 | 0 |
| 0 | 0 | 0 | 0 | 0 |
| 0.167670367 | 0.233657795 | 1.991361314 | 0 | 0 |
| 0.036954118 | 0.067175103 | 0 | 0.153188264 | 0 |
| 0 | 0 | 0 | 0 | 0 |
| 0 | 0 | 0 | 0 | 0 |
| 0.074534538 | 0.065897913 | 0.054565797 | 0.182268982 | 0.009351455 |
| 0 | 0.004154283 | 0.001039105 | 0 | 0 |
| 0 | 0 | 0 | 0 | 0 |

| | | | | |
|---|---|---|---|---|
| 0 | 0 | 0 | 0 | 0 |
| 0 | 0.008084837 | 0 | 0.002709487 | 0 |
| 0 | 0 | 0.006519057 | 0 | 0.012554836 |
| 0 | 0 | 0 | 0 | 0 |
| 0.210040648 | 0.680875946 | 2.1382462 | 0.012635371 | 0 |
| 0.078670019 | 0 | 0.131882302 | 0.015515906 | 0 |
| 0 | 0 | 0 | 0 | 0 |
| 0 | 0 | 0 | 0 | 0 |
| 0 | 0 | 0 | 0 | 0 |
| 0 | 0 | 0 | 0 | 0 |
| 0.002306397 | 0 | 0 | 0 | 0 |
| 0 | 0 | 0 | 0 | 0 |
| 0 | 0 | 0 | 0.151207873 | 0 |
| 0 | 0.00595341 | 0 | 0 | 0.000487046 |
| 0 | 0 | 0 | 0 | 0 |
| 0 | 0 | 0 | 0 | 0 |
| 3.174714601 | 0 | 0 | 0 | 0 |
| 0 | 0 | 0 | 0 | 0 |
| 0 | 0 | 0 | 0 | 0 |
| 0 | 0 | 0 | 0 | 0 |
| 0 | 0.000602792 | 0 | 0 | 0 |
| 0 | 0 | 0 | 0 | 0 |
| 0 | 0 | 0 | 0 | 0 |
| 0 | 0 | 0 | 0 | 0 |
| 0 | 0 | 0 | 0.024277981 | 0.00827756 |
| 0.001276336 | 0 | 0 | 0 | 0.003683109 |
| 0 | 0 | 0 | 0 | 0 |
| 0.48517685 | 0.012600854 | 0 | 0 | 0 |
| 0 | 0 | 0 | 0 | 0 |
| 0 | 0 | 0 | 0 | 0 |
| 0.010015077 | 0 | 0 | 0 | 0 |
| 0 | 0 | 0.005715432 | 0 | 0 |
| 0 | 0 | 0 | 0 | 0 |
| 0 | 0 | 0 | 0 | 0 |
| 0 | 0 | 0 | 0 | 0 |
| 0 | 0 | 0 | 0 | 0 |
| 0 | 0 | 0 | 0 | 0 |
| 0 | 0 | 0.560780913 | 0 | 0 |
| 1.848741559 | 6.321903665 | 0 | 0 | 5.097088465 |
| 0 | 0 | 0 | 0 | 0 |
| 0 | 0 | 0 | 0 | 0 |
| 0 | 0 | 0 | 0 | 0 |
| 0.048035358 | 0.013572754 | 0.098752971 | 0 | 0.015107035 |
| 0 | 0 | 0 | 0 | 0 |
| 0 | 0 | 0 | 0 | 0 |
| 0.001657889 | 0 | 0 | 0 | 0 |
| 0.008328266 | 0.034697163 | 0.015863856 | 0 | 0.132272492 |
| 0 | 0 | 0 | 0 | 0 |

| | | | | |
|---|---|---|---|---|
| 0 | 0 | 0 | 0 | 0 |
| 0.039253233 | 0.053951142 | 0.558789737 | 0.002541703 | 0.094309815 |
| 0 | 0 | 0 | 0 | 0 |
| 0 | 0 | 0 | 0 | 0 |
| 0 | 0 | 0 | 0 | 0 |
| 0 | 0 | 0 | 0 | 0 |
| 0 | 0 | 0 | 0 | 0 |
| 0 | 0 | 0 | 0 | 0 |
| 0 | 0 | 0 | 0 | 0 |
| 0 | 0 | 0 | 0 | 0 |
| 0 | 0 | 0 | 0 | 0 |
| 0 | 0 | 0 | 0 | 0 |
| 0.185253389 | 0.012207943 | 0 | 0.001240064 | 0 |
| 0 | 0 | 0 | 0 | 0 |
| 0.019076156 | 0.007092253 | 0.097722191 | 0.000975114 | 0.005028525 |
| 0 | 0 | 0 | 0 | 0 |
| 0 | 0 | 0 | 0 | 0 |
| 0 | 0 | 0 | 0 | 0 |
| 0.006608063 | 0.011649925 | 0 | 0 | 0 |
| 0 | 0 | 0 | 0 | 0 |
| 0 | 0 | 0 | 0 | 0 |
| 0 | 0.008653674 | 0.053359069 | 0 | 0.001139633 |
| 0.022393419 | 0.036816299 | 0 | 0 | 0.034882627 |
| 0.003313494 | 0 | 0 | 0 | 0 |
| 0 | 0 | 0 | 0 | 0 |
| 0 | 0 | 0 | 0 | 0 |
| 0 | 0 | 0 | 0 | 0 |
| 0 | 0 | 0 | 0 | 0 |
| 0 | 0 | 0 | 0 | 0 |
| 0 | 0 | 0 | 0 | 0 |
| 0 | 0 | 0 | 0 | 0 |
| 0 | 0 | 0 | 0 | 0 |
| 0 | 0 | 0 | 0 | 0 |
| 0 | 0 | 0 | 0 | 0 |
| 0 | 0 | 0 | 1.204016539 | 0 |
| 0 | 0 | 0 | 0 | 0 |
| 0 | 0 | 0 | 0 | 0 |
| 0 | 0 | 0 | 0 | 0 |
| 0.003590512 | 0.001264645 | 0 | 0 | 0 |
| 0 | 0 | 0 | 0 | 0 |
| 0.095394979 | 0.095060413 | 0.014181303 | 0 | 0.002551056 |
| 0 | 0 | 0 | 0 | 0 |
| 0 | 0 | 0 | 0 | 0 |
| 0 | 0 | 0 | 0 | 0 |
| 0 | 0 | 0 | 0 | 0 |
| 0.000969989 | 0.138048618 | 2.273707687 | 0.055713756 | 1.703402055 |
| 0 | 0 | 0 | 0 | 0 |
| 0 | 0 | 0 | 0 | 0 |

| | | | | |
|---|---|---|---|---|
| 0 | 0 | 0.00327104 | 0 | 0 |
| 0 | 0 | 0 | 0 | 0 |
| 0 | 0 | 0 | 0 | 0 |
| 0 | 0 | 0 | 0 | 0 |
| 0 | 0.00229748 | 0 | 0 | 0 |
| 0 | 0 | 0 | 0 | 0 |
| 0 | 0 | 0 | 0 | 0 |
| 0 | 0 | 0 | 0 | 0 |
| 0 | 0 | 0 | 0 | 0 |
| 0 | 0 | 0.001134246 | 0 | 0 |
| 0 | 0 | 0 | 0.006835107 | 0 |
| 0 | 0 | 0 | 0 | 0 |
| 0.01483562 | 0.001241028 | 0.056540139 | 0 | 0.001489077 |
| 0 | 0 | 0 | 0 | 0 |
| 0 | 0 | 0 | 0 | 0 |
| 0 | 0 | 0 | 0 | 0 |
| 0 | 0 | 0 | 0 | 0 |
| 0 | 0 | 0 | 0 | 0 |
| 0 | 0 | 0 | 0 | 0 |
| 0.127590882 | 0.057439325 | 2.626588246 | 9.10596943 | 0.152750509 |
| 0.052105404 | 0 | 0 | 0 | 0 |
| 0 | 0 | 0 | 0 | 0 |
| 0 | 0 | 0 | 0 | 0 |
| 0 | 0 | 0 | 0 | 0 |
| 0 | 0 | 0 | 0 | 0 |
| 0 | 0 | 0 | 0.001252205 | 0 |
| 0 | 0 | 0 | 0 | 0 |
| 0 | 0 | 0 | 0 | 0 |
| 0 | 0 | 0 | 0 | 0 |
| 0 | 0.011021153 | 0.866392849 | 0 | 0.001593964 |
| 0 | 0 | 0 | 0 | 0 |
| 0 | 0 | 0 | 0 | 0 |
| 0 | 0 | 0 | 0 | 0 |
| 0 | 0 | 0 | 0 | 0 |
| 0 | 0 | 0 | 0 | 0 |
| 0 | 0 | 0 | 0 | 0 |
| 0 | 0 | 0 | 0 | 0 |
| 0 | 0 | 0 | 0 | 0 |
| 0 | 0 | 0 | 0 | 0 |
| 0.742868656 | 0 | 0 | 0 | 0 |
| 0 | 0 | 0 | 0 | 0 |
| 0 | 0 | 0 | 0 | 0 |
| 0.347262932 | 0 | 0 | 0 | 0 |
| 0 | 0 | 0 | 0 | 0 |
| 0.056275822 | 0.016599731 | 0.041397085 | 0 | 0.002334093 |
| 0 | 0 | 0 | 0 | 0 |
| 0 | 0 | 0 | 0 | 0 |
| 0 | 0 | 0 | 0 | 0 |

| | | | | |
|---|---|---|---|---|
| 0 | 0 | 0 | 0 | 0 |
| 0 | 0 | 0 | 0 | 0 |
| 0 | 0 | 0 | 0 | 0 |
| 0 | 0 | 0 | 0 | 0 |
| 0 | 0 | 0 | 0.001862747 | 0 |
| 0.127772304 | 0.005937998 | 0.090753237 | 0.110548893 | 0.006737631 |
| 0 | 0 | 0 | 0 | 0 |
| 0 | 0 | 0 | 0 | 0 |
| 0 | 0 | 0 | 0 | 0 |
| 0 | 0 | 0 | 0 | 0 |
| 0 | 0 | 0 | 0 | 0 |
| 0 | 0 | 0 | 0 | 0 |
| 0 | 0 | 0 | 0 | 0 |
| 0 | 0 | 0 | 0 | 0 |
| 0 | 0 | 0 | 0 | 0 |
| 0 | 0 | 0 | 0 | 0 |
| 0 | 0 | 0 | 0 | 0 |
| 0 | 0 | 0 | 0 | 0 |
| 0 | 0 | 0 | 0 | 0 |
| 0 | 0 | 0 | 0 | 0 |
| 0 | 0 | 0 | 0 | 0.000673466 |
| 0 | 0 | 0 | 0 | 0 |
| 0 | 0 | 0 | 0 | 0 |
| 0 | 0 | 0 | 0 | 0 |
| 0 | 0 | 0 | 0 | 0 |
| 0 | 0 | 0 | 0 | 0 |
| 0 | 0 | 0 | 0 | 0 |
| 0 | 0 | 0 | 0 | 0 |
| 0 | 0 | 0 | 0 | 0 |
| 0 | 0 | 0.093394088 | 0 | 0 |
| 0 | 0 | 0 | 0 | 0 |
| 0 | 0 | 0 | 0 | 0 |
| 0 | 0 | 0 | 0 | 0 |
| 0 | 0 | 0 | 0 | 0 |
| 0 | 0 | 0 | 0 | 0 |
| 0 | 0 | 0 | 0 | 0 |
| 0 | 0 | 0 | 0 | 0 |
| 0 | 0.042220816 | 0 | 0 | 0.023763991 |
| 1.44126278 | 0 | 0 | 0 | 0 |
| 0 | 0 | 0 | 0 | 0 |
| 0 | 0 | 0 | 0 | 0 |
| 0 | 0 | 0 | 0 | 0 |
| 0 | 0 | 0 | 0 | 0 |
| 0 | 0 | 0 | 0 | 0 |
| 0 | 0 | 0 | 0 | 0 |
| 0 | 0 | 0 | 0 | 0 |
| 0 | 0 | 0 | 0 | 0 |
| 0 | 0.001897452 | 0 | 0 | 0 |

| | | | | |
|---|---|---|---|---|
| 0 | 0 | 0 | 0 | 0 |
| 0.319144547 | 0.004624743 | 0 | 0 | 0 |
| 0 | 0 | 0 | 0 | 0 |
| 0 | 0 | 0 | 0 | 0 |
| 0 | 0 | 0 | 0 | 0 |
| 0.002132239 | 0 | 0.006019402 | 0 | 0 |
| 0.001243689 | 0 | 0 | 0 | 0 |
| 0 | 0 | 0 | 0 | 0 |
| 0 | 0 | 0 | 0 | 0 |
| 0 | 0 | 0 | 0 | 0 |
| 0.001187682 | 0 | 0 | 0 | 0 |
| 0 | 0 | 0 | 0 | 0 |
| 0.129593601 | 0 | 0 | 0 | 0.004139333 |
| 0.064791176 | 0 | 0 | 0 | 0 |
| 0 | 0 | 0 | 0 | 0 |
| 0.019377816 | 0 | 0 | 0 | 0 |
| 0 | 0 | 0 | 0 | 0 |
| 0 | 0 | 0 | 0 | 0 |
| 0 | 0 | 0 | 0 | 0 |
| 0.000705919 | 0 | 0.003938788 | 0 | 0 |
| 0 | 0.091616879 | 0.111011136 | 0.012200295 | 0.039062728 |
| 0 | 0 | 0 | 0 | 0 |
| 0 | 0 | 0 | 0 | 0 |
| 0.926106449 | 0.143357949 | 0 | 1.655336283 | 0.091924023 |
| 0 | 0 | 0 | 0 | 0 |
| 0 | 0 | 0 | 0 | 0 |
| 0 | 0 | 0 | 0 | 0 |
| 0 | 0 | 0 | 0 | 1.176604732 |
| 0 | 0 | 0 | 0 | 0 |
| 0 | 0 | 0 | 0 | 0 |
| 0 | 0 | 0.005220523 | 0 | 0 |
| 0 | 0 | 0 | 0 | 0 |
| 0 | 0 | 0 | 0 | 0 |
| 0 | 0 | 0 | 0 | 0 |

| PHL018-0098_V1 | PHL018-0100_V3 | PHL018-0078_V3 | PHL018-0112_V1 | PHL018-0217_V2 |
|---|---|---|---|---|
| CTRL | EYCF | CTRL | CTRL | REF |
| 0.026762596 | 0.116053475 | 0.002575475 | 0.020176737 | 0.074708169 |
| 8.535715867 | 0 | 0.299580486 | 0.148145929 | 1.533802572 |
| 0 | 0.010965005 | 0 | 0 | 0 |
| 0 | 0 | 0 | 0 | 0 |
| 0 | 0 | 0 | 0 | 0 |
| 0 | 0 | 0 | 0 | 0 |
| 0 | 0 | 0 | 0 | 0 |
| 0 | 0 | 0 | 0 | 0 |
| 0 | 0 | 0 | 0 | 0 |
| 0 | 0 | 0 | 0 | 0 |
| 0 | 0 | 0 | 0 | 0 |
| 0 | 0 | 0 | 0 | 0 |
| 0 | 0 | 0 | 0 | 0 |
| 1.226539125 | 47.23951663 | 0.229519192 | 1.19529126 | 1.960927894 |
| 0 | 0 | 0 | 0 | 0 |
| 0 | 0 | 0 | 0 | 0 |
| 0 | 0 | 0 | 0 | 0 |
| 0 | 0 | 0 | 0 | 0 |
| 0 | 0 | 0 | 0 | 0 |
| 0 | 0 | 0 | 0 | 0 |
| 0 | 0 | 0 | 0 | 0 |
| 0 | 0 | 0 | 0 | 0 |
| 0 | 0 | 0 | 0 | 0 |
| 0 | 0 | 0 | 0 | 0 |
| 0 | 0 | 0.034392668 | 0.169087301 | 0.065564547 |
| 0 | 0 | 0 | 0 | 0 |
| 0 | 0 | 0 | 0 | 0 |
| 0 | 0 | 0 | 0 | 0 |
| 0 | 0 | 0 | 0 | 0 |
| 0 | 0 | 0 | 0.029196057 | 0 |
| 0 | 0 | 0 | 0 | 0 |
| 0 | 0 | 0 | 0 | 0 |
| 0 | 0 | 0 | 0 | 0 |
| 0 | 0 | 0 | 0 | 0 |
| 0 | 0 | 0 | 0 | 0 |
| 0.021861199 | 0 | 1.161468871 | 0.1261665 | 0.927674927 |
| 0 | 0 | 0 | 0 | 0 |
| 0 | 0 | 0 | 0 | 0 |
| 0 | 0 | 0 | 0 | 0 |
| 0 | 0 | 0 | 0 | 0 |
| 0 | 0 | 0 | 0 | 0 |
| 0.006258868 | 0.004850992 | 0.234607191 | 0.019633103 | 0.005036731 |
| 0.173204968 | 0.530507618 | 3.570489702 | 0.02793264 | 0.533717566 |
| 0.012677809 | 0.012727074 | 0 | 0.012229315 | 0.007928628 |
| 0 | 0.004587885 | 0.054147066 | 0.001657879 | 0.023579577 |
| 0 | 0 | 0 | 0 | 0 |

| | | | | |
|---|---|---|---|---|
| 0 | 0 | 0.013450357 | 0.00488329 | 0 |
| 0.007955545 | 0 | 0 | 0 | 0 |
| 0.004492244 | 0.002113922 | 0.00055442 | 0.005444434 | 0.016329774 |
| 0.800574235 | 0.903714018 | 0.337867001 | 2.204866062 | 0.285086371 |
| 0.001726823 | 0 | 0.001044286 | 0.003512991 | 0 |
| 0 | 0 | 0 | 0.013511063 | 0 |
| 0.127074004 | 0.001096123 | 0.2227977 | 0 | 0 |
| 0 | 0 | 0 | 0 | 0 |
| 0 | 0 | 0 | 0.010304784 | 0 |
| 0 | 0 | 0 | 0 | 0 |
| 0 | 0 | 0.00396209 | 0 | 0 |
| 0 | 0 | 0.063705132 | 0.044420562 | 0.278382369 |
| 0 | 0 | 0 | 0 | 0 |
| 0.022193051 | 0 | 0.031397429 | 0.010934223 | 0.015486249 |
| 0 | 0 | 0 | 0 | 0 |
| 0 | 0 | 0 | 0 | 0 |
| 0 | 0 | 0.399237292 | 0.04072663 | 0.067489076 |
| 0 | 0 | 0 | 0 | 0 |
| 0 | 0 | 0 | 0 | 0 |
| 0 | 0 | 0 | 0 | 0 |
| 0.228216576 | 0 | 0.011856846 | 0.008829082 | 0.00258382 |
| 0.004581012 | 0 | 0.019711538 | 0.023333822 | 0.002264225 |
| 0 | 0 | 0.003092939 | 0.02582724 | 0.004172188 |
| 0 | 0 | 0 | 0 | 0 |
| 0 | 0 | 0 | 0 | 0 |
| 0 | 0 | 0 | 0 | 0 |
| 0 | 0.116243979 | 0 | 0 | 0 |
| 0 | 0 | 0 | 0 | 0 |
| 0 | 0 | 1.291934202 | 0.001309242 | 0 |
| 0 | 0 | 0 | 0 | 0 |
| 0.003683788 | 0 | 0 | 0 | 0 |
| 0 | 0 | 0 | 0 | 0 |
| 0 | 0 | 0 | 0 | 0 |
| 0 | 0 | 0 | 0 | 0 |
| 0 | 0 | 0 | 0 | 0 |
| 0 | 0 | 0.362114689 | 0 | 0 |
| 0 | 0 | 0 | 0 | 0 |
| 0 | 0 | 2.684861315 | 0 | 0 |
| 0 | 0 | 0 | 0 | 0 |
| 0 | 0 | 0 | 0 | 0 |
| 0 | 0 | 0 | 0 | 0 |
| 0 | 0 | 0 | 0 | 0 |
| 0 | 0 | 0 | 0 | 0 |
| 0 | 0 | 0 | 0 | 0 |
| 0 | 0 | 0.053025182 | 0 | 0 |
| 0.018199931 | 0 | 0 | 0 | 0 |
| 0 | 0 | 0.016541416 | 0 | 0 |
| 0 | 0 | 0.041893799 | 0 | 0 |

| | | | | |
|---|---|---|---|---|
| 0 | 0 | 0 | 0 | 0 |
| 0 | 0 | 0 | 0 | 0 |
| 0 | 0 | 0 | 0 | 0 |
| 0 | 0 | 0 | 0 | 0 |
| 2.024695607 | 0 | 0.027435759 | 0 | 0.12462984 |
| 0 | 0.005983031 | 0.001129805 | 0 | 0 |
| 0 | 0 | 0 | 0 | 0 |
| 0 | 0 | 0 | 0.003382751 | 0 |
| 0 | 0 | 0 | 0.011079142 | 0.002657526 |
| 0 | 0 | 0 | 0 | 0 |
| 0 | 0 | 0 | 0 | 0 |
| 0 | 0 | 0.280223121 | 0 | 0.000725961 |
| 0 | 0 | 0 | 0 | 0 |
| 0 | 0 | 0 | 0 | 0 |
| 0 | 0 | 0 | 0 | 0 |
| 0 | 0 | 0 | 0 | 0 |
| 0 | 0 | 0 | 0 | 0 |
| 0 | 0 | 0 | 0 | 0 |
| 0.05091525 | 0 | 0.991524005 | 0.077677227 | 0.217403102 |
| 0 | 0 | 0 | 0 | 0 |
| 0 | 0 | 0 | 0 | 0 |
| 0 | 0 | 0 | 0 | 0 |
| 0 | 0 | 0 | 0 | 0 |
| 0 | 0 | 0 | 0 | 0 |
| 0 | 0 | 0 | 0 | 0 |
| 0 | 0 | 0 | 0 | 0 |
| 0 | 0.643694657 | 0 | 0 | 0 |
| 0 | 0 | 0 | 0 | 0 |
| 0 | 0 | 0 | 0 | 0 |
| 0 | 0 | 0 | 0 | 0 |
| 0.003172224 | 0 | 0.018361668 | 0.017483858 | 0.088820423 |
| 0.001247977 | 0 | 0.001629208 | 0.001344446 | 0.091359246 |
| 0 | 0 | 0 | 0 | 0 |
| 0 | 0 | 0 | 0 | 0 |
| 0 | 0 | 0 | 0 | 0 |
| 0 | 0 | 0 | 0 | 0 |
| 0 | 0 | 0 | 0 | 0 |
| 0 | 0 | 0 | 0 | 0 |
| 0 | 0 | 0.001102898 | 0.128197953 | 0 |
| 0 | 0 | 0 | 0 | 0 |
| 0 | 0 | 0 | 0 | 0 |
| 0 | 0 | 0 | 0 | 0 |
| 0 | 0 | 0 | 0.000443991 | 0 |
| 0 | 0 | 0 | 0 | 0 |
| 0 | 0 | 0 | 0.000710034 | 0.003366207 |
| 0 | 0 | 0 | 0 | 0 |
| 0 | 0 | 0 | 0 | 0 |
| 0.000474177 | 0 | 0 | 0 | 0 |

| | | | | |
|---|---|---|---|---|
| 0 | 0 | 0 | 0 | 0 |
| 0 | 0 | 0 | 0 | 0 |
| 0 | 0 | 0 | 0 | 0 |
| 0 | 0 | 0 | 0 | 0 |
| 0 | 0 | 0.021187204 | 0.13214771 | 0.00541567 |
| 0 | 0 | 0 | 0 | 0 |
| 0 | 0 | 0 | 0 | 0 |
| 0 | 0 | 0 | 0 | 0 |
| 0 | 0 | 0 | 0 | 0 |
| 0 | 0 | 0 | 0 | 0 |
| 0 | 0 | 0 | 0 | 0 |
| 0 | 0 | 0 | 0 | 0 |
| 0 | 0 | 0 | 0 | 0 |
| 0 | 0 | 0 | 0 | 0 |
| 0 | 0 | 0 | 0 | 0 |
| 0.630022626 | 0 | 1.226395151 | 1.038091658 | 1.119376899 |
| 0 | 0 | 0 | 0 | 0 |
| 0 | 0 | 0 | 0 | 0 |
| 0 | 0 | 0 | 0 | 0 |
| 0 | 0 | 0 | 0 | 0 |
| 0 | 0 | 0 | 0 | 0 |
| 0 | 0 | 0 | 0 | 0 |
| 0 | 0 | 0 | 0.002445972 | 0 |
| 0 | 0 | 0 | 0 | 0 |
| 0 | 0 | 0.063828567 | 0 | 0 |
| 0 | 0 | 0 | 0 | 0 |
| 0.002410022 | 0 | 0.008536471 | 0.015021553 | 0.033464853 |
| 0 | 0 | 0 | 0 | 0 |
| 0 | 0 | 0 | 0 | 0 |
| 0 | 0 | 0 | 0 | 0 |
| 0 | 0 | 0 | 0 | 0 |
| 0 | 0 | 0.230767512 | 0 | 0 |
| 0 | 0 | 0 | 0 | 0 |
| 0 | 0 | 0 | 0 | 0 |
| 0 | 0 | 0 | 0 | 0 |
| 0 | 0 | 0 | 0 | 0 |
| 0 | 0 | 0 | 0 | 0 |
| 0 | 0 | 0 | 0.127580538 | 0 |
| 0 | 0 | 0 | 0 | 0 |
| 0 | 0 | 0 | 0 | 0 |
| 0 | 0 | 0 | 0 | 0 |
| 0 | 0 | 0 | 0 | 0 |
| 0 | 0 | 0 | 0 | 0 |
| 0 | 0 | 0 | 0 | 0 |
| 0 | 0 | 0 | 0 | 0 |
| 0 | 0 | 0 | 0 | 0 |
| 0 | 0 | 0 | 0 | 0 |
| 0 | 0 | 0 | 0 | 0 |

| | | | | |
|---|---|---|---|---|
| 0 | 0 | 0 | 5.465214424 | 0 |
| 0 | 0 | 0 | 0 | 0 |
| 0 | 0 | 0 | 0 | 0 |
| 0 | 0 | 0 | 0 | 0 |
| 0 | 0 | 0 | 0 | 0 |
| 0 | 0 | 0 | 0 | 0 |
| 0 | 0 | 0 | 0 | 0 |
| 0 | 0 | 0 | 0 | 0 |
| 0 | 0 | 0 | 0 | 0 |
| 0 | 0 | 0 | 0 | 0 |
| 0 | 0 | 0 | 0 | 0 |
| 0 | 0 | 0 | 0 | 0.001529702 |
| 0 | 0.003832599 | 0.00063485 | 0 | 0.001675514 |
| 0 | 0 | 0 | 0 | 0 |
| 0 | 0 | 0 | 0 | 0.007818098 |
| 0 | 0 | 0 | 0 | 0 |
| 0 | 0 | 0 | 0 | 0 |
| 0 | 0 | 0 | 0 | 0 |
| 0 | 0 | 0 | 0 | 0 |
| 0 | 0 | 0 | 0 | 0 |
| 0.028911015 | 0.026024039 | 0.013255016 | 0.017643441 | 0.012217953 |
| 0 | 0 | 0 | 0 | 0 |
| 0 | 0 | 0 | 0 | 0 |
| 0 | 0 | 0 | 0 | 0 |
| 0 | 0 | 0 | 0 | 0 |
| 0 | 0 | 0 | 0 | 0 |
| 0.009502122 | 0.006809122 | 0.002765161 | 0.000577254 | 0.002554263 |
| 0 | 0 | 0 | 0 | 0 |
| 0 | 0 | 0 | 0 | 0 |
| 0 | 0.01809494 | 0 | 0 | 0 |
| 0.00515144 | 0 | 0 | 0 | 0 |
| 0 | 0 | 0 | 0 | 0 |
| 0 | 0 | 0 | 0 | 0 |
| 0.004023546 | 0 | 0 | 0.013811389 | 0.022974936 |
| 0.00558744 | 0.022331435 | 0.001802118 | 0.005546402 | 0.010599535 |
| 0 | 0 | 0 | 0 | 0 |
| 0 | 0 | 0 | 0 | 0 |
| 0 | 0 | 0 | 0 | 0 |
| 0 | 0 | 0 | 0 | 0 |
| 0 | 0 | 0.036532442 | 0 | 0 |
| 0 | 0 | 0.002151963 | 0 | 0 |
| 0 | 0 | 0.301462001 | 0 | 0 |
| 0 | 0 | 0.002274478 | 0 | 0 |
| 0 | 0 | 0 | 0 | 0 |
| 0 | 0 | 0 | 0 | 0 |
| 0.018974136 | 0 | 0 | 0.31844327 | 0 |
| 0.000806628 | 0 | 0 | 0 | 0 |
| 0 | 0 | 0 | 0.001011109 | 0 |

| | | | | |
|---|---|---|---|---|
| 0 | 0 | 0 | 0 | 0 |
| 0 | 0 | 0 | 0 | 0 |
| 0 | 0 | 0 | 0 | 0 |
| 0.000674845 | 0 | 0 | 0 | 0 |
| 0 | 0 | 0 | 0 | 0 |
| 0 | 0 | 0 | 0 | 0 |
| 0 | 0 | 0.005876023 | 0.002224374 | 0 |
| 0.00076318 | 0 | 0.041010132 | 0.002114164 | 0 |
| 0 | 0 | 0 | 0 | 0 |
| 0 | 0 | 0.000824956 | 0 | 0 |
| 0 | 0 | 0.001929411 | 0 | 0 |
| 0 | 0 | 0 | 0 | 0 |
| 0 | 0 | 0 | 0 | 0 |
| 0 | 0 | 0 | 0 | 0 |
| 0 | 0 | 0 | 0 | 0 |
| 0 | 0 | 0 | 0 | 0 |
| 0 | 0 | 0 | 0 | 0 |
| 0 | 0 | 0 | 0 | 0 |
| 0 | 0.003734865 | 0.018937889 | 0 | 0.004136394 |
| 0 | 0 | 0 | 0.06106121 | 0.003893668 |
| 0 | 0.041975671 | 0 | 0 | 0.000653743 |
| 0 | 0 | 0 | 0 | 0 |
| 0 | 0 | 0 | 0 | 0 |
| 0 | 0 | 0 | 0 | 0 |
| 0 | 0.015636289 | 0 | 0.003287467 | 0.001298799 |
| 0 | 0 | 0 | 0 | 0.001470392 |
| 0 | 0 | 0 | 0 | 0 |
| 0 | 0 | 0 | 0 | 0 |
| 0 | 0 | 0.062486952 | 0 | 0 |
| 0 | 0 | 0 | 0 | 0 |
| 0.026499453 | 0 | 0.076092119 | 0.162939187 | 0.058645043 |
| 0 | 0 | 0 | 0 | 0 |
| 0 | 0.265286718 | 0 | 0 | 0 |
| 0.000757666 | 0 | 0 | 0 | 0 |
| 0 | 0 | 0 | 0 | 0 |
| 0 | 0 | 0 | 0 | 0 |
| 0 | 0 | 0 | 0 | 0 |
| 0 | 0 | 0 | 0 | 0 |
| 0 | 0 | 0 | 0 | 0 |
| 0 | 0.000770738 | 0.012128529 | 0 | 0.004801495 |
| 0 | 0 | 0 | 0 | 0 |
| 0.013132802 | 0.039130145 | 0.000515713 | 0.011472262 | 0.060922182 |
| 0 | 0 | 0 | 0 | 0 |
| 0 | 0 | 0 | 0 | 0 |
| 0 | 0 | 0 | 0 | 0 |
| 0 | 0 | 0 | 0 | 0 |
| 0 | 0 | 0 | 0 | 0 |
| 0 | 0 | 0 | 0 | 0 |

| | | | | |
|---|---|---|---|---|
| 0 | 0 | 0 | 0 | 0 |
| 0 | 0 | 0 | 0 | 0 |
| 0.014616486 | 0 | 0 | 0.001297776 | 0.0051272 |
| 0 | 0 | 0 | 0 | 0 |
| 0 | 0 | 0 | 0.999768359 | 0 |
| 0 | 0 | 0 | 0 | 0 |
| 0 | 0 | 0 | 0 | 0 |
| 0 | 0 | 0 | 0 | 0 |
| 0 | 0 | 0 | 0 | 0 |
| 0 | 0 | 0 | 0 | 0.000750288 |
| 0 | 0 | 0 | 0 | 0 |
| 0 | 0 | 0 | 0 | 0 |
| 0 | 0 | 0 | 0 | 0 |
| 0.001242604 | 0 | 0.001574482 | 0.013769056 | 0.001133296 |
| 0.000854673 | 0.007657594 | 0.005316255 | 0.002012473 | 0.003180318 |
| 0.03806566 | 0 | 0 | 0.000506969 | 0.005407863 |
| 0 | 0.00975163 | 0 | 0.004046528 | 0.002984213 |
| 0.003928136 | 0 | 0 | 0 | 0 |
| 0 | 0 | 0 | 0 | 0 |
| 0 | 0 | 0 | 0 | 0 |
| 0 | 0 | 0 | 0 | 0 |
| 0 | 0 | 0 | 0.001948192 | 0.003271156 |
| 0 | 0 | 0 | 0 | 0 |
| 0 | 0.751279241 | 0 | 0 | 0 |
| 0 | 0 | 0 | 0 | 0 |
| 0 | 0 | 0 | 0 | 0 |
| 0 | 0 | 0 | 0 | 0 |
| 0 | 0 | 0 | 0 | 0 |
| 0 | 0 | 0 | 0 | 0 |
| 0 | 0 | 0 | 0 | 0 |
| 0 | 0 | 0.003352234 | 0 | 0.000884731 |
| 0 | 0 | 0 | 0 | 0 |
| 0 | 0 | 0.013076923 | 0 | 0 |
| 0.001013581 | 0 | 0 | 0 | 0 |
| 0 | 0 | 0 | 0 | 0 |
| 0 | 0 | 0.018931037 | 0.022861014 | 0 |
| 0 | 0 | 0 | 0 | 0 |
| 0.001104478 | 0.075964606 | 0.508082169 | 0.031514156 | 0.332174091 |
| 0 | 0 | 0 | 0 | 0 |
| 0 | 0 | 0 | 0 | 0 |
| 0 | 0 | 0 | 0 | 0 |
| 0 | 0 | 0 | 0 | 0 |
| 0 | 0 | 0 | 0 | 0 |
| 0 | 0 | 0 | 0 | 0 |
| 0 | 0 | 0 | 0 | 0 |
| 0 | 0 | 0 | 0 | 0.048320682 |
| 0 | 0 | 0 | 0 | 0.014937177 |
| 0 | 0 | 0 | 0.001297091 | 0 |

| | | | | |
|---|---|---|---|---|
| 0.122766509 | 0 | 0 | 0.349249789 | 2.760761948 |
| 0.017170611 | 0.017696 | 0 | 0 | 0 |
| 0 | 0.049498947 | 0.000509103 | 0.002168115 | 0 |
| 0 | 0 | 0 | 0 | 0 |
| 0.008708024 | 0.002441211 | 0.000640258 | 0.006543999 | 0.001013873 |
| 0 | 0 | 0 | 0 | 0.009919273 |
| 0 | 0 | 0 | 0 | 0 |
| 0.02623037 | 0 | 0.000538478 | 0.002158319 | 0.001989633 |
| 0 | 0 | 0 | 0 | 0 |
| 0.003479908 | 0 | 0 | 0.033369392 | 0.007324135 |
| 0 | 0 | 0 | 0 | 0 |
| 0.003906109 | 0 | 0.395712252 | 0.112193668 | 0.011475004 |
| 0.005925845 | 0.019108795 | 0.010454468 | 0.007991895 | 0.002389388 |
| 0 | 0.249957916 | 0 | 0 | 0 |
| 0 | 0 | 0 | 0 | 0 |
| 0.003979645 | 0 | 0.002912269 | 0 | 0 |
| 0 | 0 | 0.291669898 | 0 | 0 |
| 0 | 0 | 0 | 0 | 0 |
| 0.366999279 | 0 | 0 | 0 | 0 |
| 0 | 0 | 0 | 0 | 0 |
| 0 | 0 | 0 | 0 | 0 |
| 0 | 0 | 0 | 0 | 0 |
| 0 | 0 | 0.053271024 | 0 | 0 |
| 0 | 0 | 0 | 0 | 0 |
| 0 | 0 | 0 | 0 | 0 |
| 0.008536807 | 0 | 0.019959893 | 0 | 0.001789089 |
| 0 | 0 | 0.201269291 | 0 | 0 |
| 0 | 0 | 0.010474573 | 0 | 0 |
| 0 | 0 | 0 | 0 | 0 |
| 0 | 0 | 0 | 0 | 0 |
| 0 | 0 | 0 | 0 | 0 |
| 0 | 0 | 0 | 0 | 0 |
| 0 | 0 | 0 | 0 | 0 |
| 0.006413692 | 0 | 0 | 0 | 0 |
| 0 | 0 | 0 | 0 | 0 |
| 0 | 0 | 0 | 0 | 0 |
| 0.00205901 | 0 | 0 | 0 | 0 |
| 0 | 0 | 0 | 0 | 0 |
| 0 | 0 | 0 | 0 | 0 |
| 0 | 0 | 0 | 0 | 0 |
| 0 | 0 | 0 | 0 | 0 |
| 0 | 0 | 0 | 0 | 0 |
| 0 | 0 | 0 | 0 | 0 |
| 0.000438993 | 0 | 0 | 0 | 0 |
| 0 | 0 | 0 | 0 | 0 |
| 0 | 0 | 0 | 0 | 0 |
| 0 | 0 | 0 | 0.000938585 | 0.005932996 |
| 0 | 0 | 0 | 0 | 0 |

| | | | | |
|---|---|---|---|---|
| 0 | 0 | 0 | 0 | 0 |
| 0.008835146 | 0 | 0.001570188 | 0 | 0 |
| 0 | 0 | 0 | 0 | 0 |
| 0.005105049 | 0 | 0 | 0.115156401 | 0 |
| 0 | 0 | 0 | 0 | 0 |
| 0.038071685 | 0 | 0.001598436 | 0.004558704 | 0.001168238 |
| 0 | 0 | 0 | 0 | 0 |
| 0 | 0 | 0.001431262 | 0 | 0 |
| 0 | 0 | 0 | 0.80518641 | 0 |
| 0 | 0 | 0 | 0 | 0 |
| 0.022078372 | 0.017399906 | 0.002852179 | 0.004144123 | 0.007226452 |
| 0 | 0 | 0 | 0 | 0 |
| 0 | 0 | 0 | 0 | 0 |
| 0.000779758 | 0.003852787 | 0.000538919 | 0.001755071 | 0.001066748 |
| 0 | 0 | 0 | 0 | 0 |
| 0 | 0 | 0 | 0 | 0 |
| 0 | 0 | 0 | 0 | 0 |
| 0 | 0 | 0 | 0 | 0 |
| 0 | 0 | 0 | 0.001293233 | 0 |
| 0 | 0.001078123 | 0 | 0 | 0 |
| 0 | 0 | 0.082163839 | 0 | 0 |
| 0.088600937 | 0 | 0.102820459 | 0.210030337 | 0 |
| 0 | 0 | 0 | 0.007174369 | 0.002429503 |
| 0 | 0 | 0 | 0 | 0 |
| 0 | 0 | 0 | 0 | 0 |
| 0 | 0 | 0 | 0 | 0 |
| 0 | 0 | 0 | 0 | 0 |
| 0 | 0 | 0 | 0 | 0 |
| 0 | 0 | 0 | 0 | 0 |
| 0.190874725 | 0 | 0.668093829 | 0.345911277 | 0.493481468 |
| 0.001587012 | 0.011675016 | 0.005392803 | 0.00183181 | 0.000578963 |
| 0 | 0 | 0.030028651 | 0 | 0 |
| 0 | 0.026028536 | 0 | 0 | 0 |
| 0 | 0 | 0 | 0.000518894 | 0 |
| 0.012499413 | 0 | 0 | 0 | 0 |
| 0 | 0 | 0 | 0 | 0 |
| 0 | 0 | 0.149562837 | 0 | 0 |
| 0 | 0 | 0.04078568 | 0 | 0 |
| 0.016225968 | 0 | 0 | 0 | 0 |
| 0.001153317 | 0 | 0 | 0 | 0 |
| 0.004647585 | 0 | 0.020048017 | 0.000998903 | 0 |
| 0 | 0 | 1.03710049 | 0.182510702 | 0.038013823 |
| 0 | 0 | 0.03748812 | 0 | 0 |
| 0 | 0 | 0 | 0 | 0 |
| 0 | 0 | 0 | 0 | 0 |
| 0 | 0 | 0 | 0 | 0.002859911 |
| 0.000504618 | 0.000886513 | 0.001162532 | 0.000349473 | 0 |
| 0 | 0 | 0 | 0 | 0.003963147 |

| | | | | |
|---|---|---|---|---|
| 0 | 0.003007973 | 0.05436261 | 0.001581035 | 0.009994065 |
| 0 | 0 | 0 | 0 | 0 |
| 0.002827864 | 0.001490398 | 0.584181675 | 0.005091939 | 0.007427825 |
| 0.055568659 | 0 | 0 | 0 | 0 |
| 0 | 0 | 0.325386655 | 0 | 0.068194204 |
| 0 | 0 | 0 | 0 | 0 |
| 0 | 0 | 0 | 0 | 0 |
| 0 | 0 | 0.333745536 | 0 | 0 |
| 0.086845337 | 0 | 0.000960351 | 0 | 0 |
| 0 | 0 | 0 | 0 | 0 |
| 0 | 0 | 0 | 0 | 0 |
| 0 | 0 | 0 | 0 | 0 |
| 0 | 0 | 0 | 0.000530233 | 0 |
| 0.004693859 | 0.002226467 | 0 | 0 | 0 |
| 0 | 0 | 0 | 0 | 0 |
| 0.011151237 | 0 | 0.011763348 | 0.01392135 | 0.031153229 |
| 0.004228415 | 0.02271401 | 1.671056497 | 0.020611363 | 0.12121135 |
| 0 | 0.00416553 | 0 | 0 | 0 |
| 0.00083993 | 0 | 0 | 0 | 0 |
| 0.016917403 | 0 | 0 | 0 | 0 |
| 0 | 0 | 0 | 0.000567785 | 0.001121592 |
| 0.002452371 | 0 | 0 | 0 | 0 |
| 0 | 0 | 0 | 0 | 0 |
| 0 | 0 | 0 | 0.057564702 | 0.061497882 |
| 0 | 0 | 0.012185687 | 0.006000048 | 0 |
| 0 | 0.003669643 | 0 | 0 | 0 |
| 0.000619385 | 0 | 0.002283091 | 0 | 0 |
| 0 | 0 | 0.000632117 | 0 | 0 |
| 0 | 0 | 0 | 0 | 0 |
| 0 | 0 | 0 | 0 | 0 |
| 0 | 0 | 0.001302213 | 0.001522358 | 0.005498943 |
| 0.002340881 | 0 | 0.003559307 | 0.003404473 | 0.002818148 |
| 0.020951047 | 0 | 0.04806135 | 0.041059234 | 0.028784041 |
| 0 | 0 | 0.240405832 | 0.005309042 | 0.001048737 |
| 0.022342055 | 0 | 0.008193395 | 0 | 0.001247553 |
| 0 | 0 | 0 | 0 | 0 |
| 0 | 0.002123993 | 0 | 0.000837302 | 0 |
| 0.054524038 | 0 | 0.018623938 | 0.02182696 | 0.011037832 |
| 0.100675346 | 0 | 0.074839334 | 0.325751139 | 0.181930248 |
| 0 | 0 | 0 | 0 | 0 |
| 0 | 0 | 0 | 0.001246751 | 0 |
| 0 | 0 | 0 | 0 | 0 |
| 0 | 0.001552571 | 0.001303021 | 0 | 0 |
| 0.011440272 | 0 | 0 | 0.000937125 | 0.000942417 |
| 0.018535866 | 0 | 0 | 0 | 0 |
| 0.000515338 | 0 | 0.002493181 | 0 | 0 |
| 0 | 0 | 0 | 0 | 0.000766599 |
| 0 | 0 | 0 | 0 | 0 |

| | | | | |
|---|---|---|---|---|
| 0 | 0 | 0 | 0 | 0 |
| 0.335620004 | 0.619890366 | 0.603716863 | 0.332366864 | 0.387291492 |
| 0.005819684 | 0 | 0.012797886 | 0 | 0 |
| 0 | 0 | 0.000539283 | 0 | 0 |
| 0 | 0 | 0 | 0 | 0 |
| 0 | 0 | 0 | 0 | 0 |
| 0 | 0 | 0 | 0 | 0 |
| 0 | 0.000869267 | 0 | 0 | 0 |
| 0 | 0.050763071 | 0.284477778 | 0 | 0 |
| 0.000936232 | 0 | 0 | 0.000648388 | 0 |
| 0.001276532 | 0 | 0.000992539 | 0 | 0 |
| 0.00190143 | 0.009394967 | 0.010184637 | 0.007407202 | 0.007543628 |
| 0 | 0 | 0 | 0.000940739 | 0.442527181 |
| 0 | 0 | 0 | 0 | 0 |
| 0 | 0 | 0 | 0 | 0 |
| 0 | 0 | 0 | 0 | 0 |
| 0.000501267 | 0.054598845 | 0.000577406 | 0.007984515 | 0.010789269 |
| 0.070277135 | 0 | 0 | 0 | 0 |
| 0 | 0 | 0 | 0 | 0 |
| 0.037746889 | 0 | 0 | 0 | 0 |
| 0.81348032 | 0.036552962 | 0.085387777 | 0.015752095 | 0.024704509 |
| 0 | 0 | 0 | 0 | 0 |
| 0 | 0 | 0 | 0 | 0 |
| 0 | 0 | 0 | 0 | 0 |
| 0 | 0 | 0 | 0 | 0 |
| 0 | 0 | 0 | 0 | 0 |
| 0 | 0 | 0.058538964 | 0.036985643 | 0.002233702 |
| 0.057506329 | 0 | 0.028352902 | 0.097028376 | 0.01603496 |
| 0 | 0 | 0 | 0 | 0 |
| 0 | 0 | 0 | 0 | 0 |
| 0 | 0.068873371 | 0.001765602 | 0 | 0.003226036 |
| 0 | 0 | 0 | 0 | 0 |
| 0 | 0 | 0 | 0 | 0 |
| 0 | 0 | 0 | 0 | 0 |
| 0 | 0.008201003 | 0.001103017 | 0.000884219 | 0.421121868 |
| 0.300036937 | 0 | 0.539195282 | 0.195555068 | 0.436246768 |
| 0 | 0 | 0 | 0 | 0 |
| 0 | 0 | 0 | 0 | 0 |
| 0.005648128 | 0.001190716 | 0.160360897 | 0.050694503 | 0.017802825 |
| 0 | 0 | 0 | 0 | 0 |
| 0.022399532 | 0 | 0 | 0 | 0 |
| 0 | 0 | 0 | 0 | 0 |
| 0.000411691 | 0 | 0 | 0 | 0 |
| 0.007287134 | 0 | 0 | 0 | 0.001740072 |
| 0 | 0 | 0 | 0 | 0 |
| 0.38768079 | 0 | 0.406735406 | 0.283562487 | 0 |
| 0 | 0 | 0 | 0 | 0 |
| 0 | 0 | 0 | 0 | 0 |

| | | | | |
|---|---|---|---|---|
| 0.053679863 | 0 | 0 | 0 | 0 |
| 0 | 0 | 0 | 0 | 0.324160299 |
| 0 | 0 | 0.00130379 | 0 | 0 |
| 0 | 0 | 1.307344215 | 0.172455469 | 2.884851082 |
| 0 | 0 | 0 | 0 | 0 |
| 0.000554388 | 0 | 0.000766315 | 0.000479927 | 0.018809063 |
| 0 | 0 | 0 | 0 | 0 |
| 0 | 0 | 0.001976442 | 0.000495122 | 0.019874042 |
| 0 | 0 | 0.311345998 | 0.168064478 | 10.67396396 |
| 0.632391085 | 0 | 0.233886298 | 0.115649047 | 0.130855977 |
| 0 | 0 | 0 | 0 | 0 |
| 0.004134626 | 0 | 0.026988336 | 0.003181596 | 0.004357503 |
| 0 | 0 | 0 | 0 | 0 |
| 0 | 0 | 0 | 0 | 0 |
| 0.013394881 | 1.579261197 | 0.013013159 | 0.071412539 | 0.002351115 |
| 0 | 0 | 0 | 0 | 0 |
| 0 | 0 | 0.004442997 | 0 | 0 |
| 0 | 0 | 0 | 0.462727377 | 0 |
| 0 | 0 | 0 | 0 | 0 |
| 0 | 0 | 0.001512239 | 0.002841252 | 0 |
| 0.008393902 | 0 | 0.046856877 | 0 | 0 |
| 0 | 0 | 0 | 0.369333271 | 0.008402445 |
| 0 | 0 | 0 | 0 | 0 |
| 0.034569507 | 0 | 0.416010831 | 0 | 0.112703419 |
| 0 | 0 | 0 | 0 | 0 |
| 0.114810052 | 0 | 0.072507135 | 0.494466819 | 1.247956032 |
| 0 | 0 | 0 | 0.007206736 | 0 |
| 0 | 0 | 1.905597625 | 2.379095766 | 0.351645871 |
| 0 | 0 | 0 | 0 | 0 |
| 0 | 0 | 0 | 0 | 0 |
| 0.006103778 | 0 | 0.069863147 | 0.005051988 | 0.141913891 |
| 0.002805313 | 0 | 2.069822007 | 0 | 0 |
| 0 | 0 | 0.008927087 | 0 | 0.000620017 |
| 0 | 0 | 0 | 0 | 0 |
| 0 | 0 | 0 | 0 | 0 |
| 0 | 0.002993709 | 0 | 0.320302434 | 0.231536436 |
| 2.821118188 | 0 | 0.717828644 | 0.581218658 | 5.095116831 |
| 0 | 0 | 0 | 0 | 0 |
| 0.001753541 | 0 | 0 | 0.00077281 | 0.002442548 |
| 0 | 0 | 0.000984815 | 0 | 0 |
| 0.058174367 | 0 | 0.051070248 | 0.061450406 | 0.061256665 |
| 0 | 0 | 0 | 0.225323121 | 0 |
| 0.116664619 | 0.48065689 | 1.150515891 | 0.026118383 | 0.061719592 |
| 0.003327309 | 0.017199017 | 0.001149812 | 0.002481585 | 0.000980415 |
| 0 | 0 | 0 | 0 | 0 |
| 0 | 0 | 0 | 0 | 0 |
| 0 | 0 | 0 | 0 | 0 |
| 0 | 0 | 0 | 0 | 0 |

| | | | | |
|---|---|---|---|---|
| 0 | 0 | 0 | 0 | 0 |
| 0 | 0 | 0.093068612 | 0.023621174 | 0.142271375 |
| 0.216857674 | 0.762620393 | 0 | 0.024854948 | 0.007225737 |
| 0 | 0 | 0.034147538 | 0.00861772 | 0.004005475 |
| 0 | 0 | 0 | 0 | 0.089305609 |
| 0.000533434 | 0 | 0.015115682 | 0.006926805 | 0.152228716 |
| 0.034660155 | 0 | 0 | 0 | 0 |
| 0 | 0.000712274 | 0 | 0 | 0 |
| 0 | 0 | 0 | 0 | 0.014028875 |
| 0 | 0 | 0 | 0 | 0 |
| 0.014460322 | 0.077306689 | 0.007926315 | 0.000690655 | 0.008185838 |
| 0.991700046 | 0 | 0.542401805 | 0.770586261 | 0.314929569 |
| 0 | 0 | 0 | 0 | 0 |
| 0.009148444 | 0.043961616 | 0.70493231 | 0.017019585 | 0.293108731 |
| 0.149840867 | 0.211399484 | 0.006936594 | 0.093199309 | 0.032488944 |
| 0 | 0 | 0 | 0.001178907 | 0 |
| 0 | 0 | 0 | 0 | 0 |
| 0 | 0 | 0 | 0 | 0 |
| 0 | 0 | 0 | 0 | 0.003743718 |
| 0 | 0 | 0.005961441 | 0.009671608 | 0.004195629 |
| 0.077316472 | 0.040381772 | 0.006835982 | 0.107669966 | 0.165729752 |
| 0 | 0.189540286 | 0.003246424 | 0 | 0 |
| 0.803958005 | 0 | 0.306286418 | 1.62642126 | 0.87124323 |
| 0 | 0 | 0 | 0 | 0 |
| 0 | 0 | 0 | 0.010023183 | 0 |
| 20.32103303 | 0 | 0.983563676 | 0 | 0.022654692 |
| 0 | 0 | 0 | 0 | 0 |
| 0 | 0 | 0.008334843 | 0.005031264 | 0.001431167 |
| 0 | 0 | 0.025990959 | 0 | 0.006608413 |
| 0 | 0 | 0 | 0 | 0 |
| 0 | 0 | 0 | 0.00851306 | 0 |
| 0.29959877 | 0 | 0.110076333 | 0.450861174 | 0.042318038 |
| 0.053942027 | 4.989351551 | 0.004365213 | 0.004374146 | 0.015693194 |
| 0.002863827 | 0 | 0.002878973 | 0.010367482 | 0.0474417 |
| 0 | 0 | 0 | 0.00023408 | 0.002712732 |
| 0 | 0.218032405 | 0.089867463 | 0.024013698 | 0.059125797 |
| 0 | 0.060485699 | 0.000620077 | 0.002692498 | 0 |
| 0 | 0 | 0 | 0 | 0 |
| 0.025359959 | 0 | 2.038035438 | 0.473073227 | 0 |
| 0 | 0 | 0 | 0 | 0 |
| 0 | 0 | 0 | 0.024449218 | 0.08891029 |
| 0 | 0 | 0 | 0 | 0 |
| 0 | 0 | 0 | 0 | 0.000829481 |
| 0 | 0 | 0 | 0 | 0 |
| 0.00511929 | 0.005814808 | 0.004392156 | 0.006846224 | 0.022217822 |
| 0 | 0 | 0 | 0 | 0.028969471 |
| 0 | 0 | 0 | 0 | 0 |
| 0.011711706 | 0.83160924 | 0 | 0.008995776 | 0 |

| | | | | |
|---|---|---|---|---|
| 0 | 0 | 0 | 0 | 0 |
| 0 | 0 | 0 | 0 | 0 |
| 0 | 0 | 0 | 0 | 0 |
| 0.004062243 | 0 | 0.001247806 | 0.002292325 | 0 |
| 0 | 0 | 0 | 0 | 0 |
| 0 | 0 | 0 | 0.377949831 | 0.175823252 |
| 0 | 0 | 0.079917405 | 0.264883135 | 0 |
| 0.001107555 | 0.09266651 | 0.000956839 | 0 | 0.003030382 |
| 0 | 0 | 0 | 0 | 0 |
| 0 | 0 | 0 | 0 | 0 |
| 0.000837827 | 0.074625022 | 0.001389725 | 0.003017233 | 0.001650511 |
| 0 | 0 | 0 | 0 | 0 |
| 0 | 0.00154651 | 0.004664449 | 0.062082843 | 0 |
| 0 | 0 | 0.032216738 | 0 | 0.026603653 |
| 0 | 0 | 0 | 0 | 0 |
| 0 | 0 | 0 | 0 | 0 |
| 0 | 0 | 0.132643986 | 0.051353688 | 0 |
| 0 | 0.094472026 | 0.008869677 | 0.0085014 | 0.000763341 |
| 0 | 0 | 0 | 0.114769621 | 0.506842386 |
| 0 | 0 | 0.030691737 | 0.076668552 | 0 |
| 0 | 0 | 0.001970002 | 0.010556788 | 0.005560977 |
| 0 | 0 | 0 | 0 | 0 |
| 0 | 0 | 0.081475361 | 0.006905607 | 0.023762081 |
| 0 | 0 | 0 | 0 | 0.003090764 |
| 0 | 0 | 0 | 0 | 0 |
| 0 | 0 | 0 | 0 | 0 |
| 0 | 0 | 0 | 0 | 0 |
| 0 | 0 | 0 | 0 | 0 |
| 0 | 0 | 0 | 0 | 0 |
| 0 | 0 | 0.015605888 | 1.326997719 | 1.683770323 |
| 0 | 0 | 0 | 0 | 0.003322124 |
| 0 | 0 | 0 | 0 | 0 |
| 0 | 0 | 0 | 0 | 0 |
| 0 | 0 | 0 | 0 | 0 |
| 0.004530152 | 0.003335578 | 0 | 0 | 0.010353427 |
| 0 | 0 | 0 | 0 | 0 |
| 0 | 0 | 0.063858939 | 0.033989291 | 0.032963482 |
| 0.000851082 | 0 | 0.009215341 | 0 | 0 |
| 0 | 0 | 0 | 0.046771684 | 0 |
| 0 | 0 | 0.006091901 | 0 | 0 |
| 0 | 0 | 0.002207558 | 0.004700662 | 0.004224034 |
| 0 | 0 | 0 | 0 | 0 |
| 0 | 0 | 0 | 0 | 0 |
| 0 | 0 | 0.000761633 | 0.608263731 | 0.011759227 |
| 0.296361012 | 0 | 0.074138019 | 0.166649945 | 0.111583208 |
| 0 | 0 | 0 | 0 | 0 |
| 0 | 0 | 0 | 0 | 0.303716548 |
| 0 | 0 | 0 | 0 | 0 |

| | | | | |
|---|---|---|---|---|
| 0 | 0 | 0 | 0 | 0 |
| 0 | 0 | 0 | 0 | 0 |
| 0 | 0 | 0 | 0.142664112 | 0 |
| 0.057143945 | 0 | 0.002145366 | 0.142079341 | 0.177275603 |
| 0 | 0 | 0 | 0.010515017 | 0 |
| 0 | 0 | 0 | 0.974462512 | 0 |
| 0 | 0 | 0 | 0 | 0 |
| 0 | 0 | 0 | 0 | 0 |
| 0.121682272 | 0.246421426 | 0 | 0.000541936 | 0 |
| 0.002678189 | 0 | 0.001524347 | 0 | 0 |
| 0 | 0 | 0.010634524 | 0.003003135 | 0.002143294 |
| 0 | 0 | 0 | 0 | 0 |
| 0 | 0 | 0 | 0 | 0 |
| 0 | 0 | 0 | 0 | 0 |
| 4.43448392 | 0 | 0.991748849 | 1.612127174 | 1.161351989 |
| 0 | 0 | 0 | 0 | 0 |
| 0 | 0 | 0 | 0 | 0 |
| 0 | 0 | 0 | 0 | 0 |
| 0 | 0 | 0 | 0 | 0 |
| 0 | 0 | 0 | 0 | 0 |
| 0 | 0 | 0.042284644 | 0.105565349 | 0.004747727 |
| 0 | 0 | 0 | 0 | 0 |
| 0 | 0 | 0 | 0 | 0 |
| 0 | 0 | 0 | 0 | 0 |
| 0.008282019 | 0.981811982 | 0.154707039 | 0.00971885 | 0.061938506 |
| 0.598688204 | 0 | 0.78633727 | 2.497641504 | 10.39275956 |
| 1.426577001 | 0 | 0 | 0.023703847 | 0.026596087 |
| 0 | 0 | 0 | 0 | 0 |
| 0 | 0 | 0.127890582 | 0 | 0 |
| 0 | 0 | 0 | 0 | 0.548857008 |
| 0 | 0 | 0 | 0 | 0 |
| 0 | 0 | 0 | 0.029653264 | 0 |
| 0 | 0 | 0 | 0 | 0 |
| 0.005528912 | 0 | 0.000424581 | 0 | 0.001613617 |
| 0.002226865 | 0.004001069 | 0.363919235 | 0.002383425 | 0.03345569 |
| 0 | 0.012632943 | 0 | 0.000396422 | 0.0018794 |
| 0 | 0 | 0 | 0 | 0 |
| 8.017302474 | 0 | 1.094562112 | 0.412154049 | 0.345205909 |
| 0.051327731 | 0 | 2.646468275 | 4.245367506 | 0 |
| 0 | 0 | 0 | 0 | 0 |
| 0 | 0 | 0 | 0 | 0 |
| 0.000811395 | 0 | 0 | 0.001910568 | 0.008525019 |
| 0.085847156 | 0 | 0.26368302 | 1.774801345 | 0.062921724 |
| 0 | 0 | 0 | 0.654843911 | 0 |
| 0 | 0 | 0 | 1.589577859 | 0 |
| 0 | 0 | 0.000374793 | 0.001126679 | 0.003264242 |
| 0 | 0 | 0 | 0 | 0.017055747 |
| 6.102984665 | 0 | 0.117925254 | 0.549698944 | 0.577945623 |

| | | | | |
|---|---|---|---|---|
| 0.002254805 | 0.000848838 | 0 | 0 | 0.001938945 |
| 0 | 0 | 0 | 0.332326796 | 0 |
| 0 | 0 | 0 | 0 | 0 |
| 0 | 0 | 0 | 0 | 0 |
| 0.00112246 | 0.001901512 | 0 | 0 | 0 |
| 0 | 0.007121051 | 0.26680618 | 0.041065294 | 0 |
| 0 | 0 | 0 | 0 | 0 |
| 0.002439584 | 0 | 0 | 0 | 0 |
| 0 | 0 | 0 | 0 | 0 |
| 0 | 0 | 0 | 0 | 0.003346774 |
| 0 | 0 | 0 | 0 | 0 |
| 0.243647986 | 0 | 0.491685618 | 0.313810561 | 0.028981052 |
| 0 | 0 | 0.005770136 | 0.068442078 | 0.501272523 |
| 0 | 0 | 0 | 0 | 0 |
| 0.089941509 | 0.013683277 | 0.484349468 | 0.234643383 | 0.018063434 |
| 0 | 0 | 0 | 0 | 0 |
| 0 | 0 | 0 | 0 | 0 |
| 0 | 0 | 0 | 0 | 0 |
| 0 | 0 | 0 | 0 | 0.001423149 |
| 0 | 0 | 0.022172829 | 0.096237275 | 0.087384261 |
| 0 | 0 | 0 | 0 | 0.052136992 |
| 0 | 0 | 0 | 0 | 0 |
| 0.924756958 | 0 | 0.713412468 | 0.098293586 | 2.446073596 |
| 0.46952709 | 0 | 0 | 0 | 0 |
| 0 | 0 | 0 | 0 | 0 |
| 0 | 0 | 0 | 0 | 0 |
| 0 | 0 | 0 | 0 | 0 |
| 0 | 0 | 0 | 0 | 0 |
| 0 | 0 | 0 | 0 | 0 |
| 0 | 0 | 0 | 0 | 0 |
| 0 | 0 | 0 | 0 | 0 |
| 0.022139124 | 0.042487488 | 0.06020668 | 0.036997886 | 0.03107753 |
| 0 | 0 | 0 | 0 | 0 |
| 0.007694139 | 0.012513846 | 0.011300105 | 0.010657157 | 0.028288531 |
| 0 | 0.007386882 | 0.000430525 | 0 | 0.00187482 |
| 0.021882416 | 0.004761293 | 0.038433687 | 0.017518247 | 1.903614948 |
| 0 | 0 | 0 | 0 | 0 |
| 0 | 0 | 0 | 0 | 0.246097412 |
| 0.001252383 | 0 | 0 | 0 | 0 |
| 0 | 0 | 0 | 0 | 0 |
| 0 | 0 | 0.011004269 | 0.101905903 | 0.017280461 |
| 0 | 0 | 0 | 0.031790639 | 0.00935822 |
| 0 | 0 | 0 | 0 | 0 |
| 0 | 0 | 0 | 0 | 0.047453311 |
| 0.002761391 | 0.070746852 | 0.000954248 | 0.004781006 | 0.009234428 |
| 0 | 0 | 0 | 0 | 0 |
| 0 | 0 | 0.382796797 | 0.317072662 | 0 |
| 0 | 0 | 0 | 0 | 0 |

| | | | | |
|---|---|---|---|---|
| 0 | 0 | 0 | 0 | 0 |
| 0 | 0 | 0 | 0 | 0 |
| 0 | 0 | 0 | 0 | 0 |
| 0 | 0 | 0 | 0 | 0 |
| 0 | 0 | 0 | 0 | 0 |
| 0 | 0 | 0 | 0 | 0.001045721 |
| 0 | 0 | 0.089446426 | 0.069550532 | 0.000764598 |
| 0.017082243 | 0 | 0.291425554 | 0.017953018 | 0.001639955 |
| 0 | 0 | 0 | 0 | 0 |
| 0 | 0 | 0.000611458 | 0 | 0 |
| 0 | 0 | 0 | 0 | 0 |
| 0 | 0 | 0 | 0 | 0 |
| 0 | 0 | 0 | 0 | 0 |
| 0 | 0 | 0 | 0.158132716 | 0.256459276 |
| 0 | 0 | 0 | 0 | 0 |
| 0 | 0 | 0 | 0 | 0 |
| 0 | 0 | 0 | 0.002639342 | 1.339087571 |
| 0 | 0 | 0 | 0 | 0 |
| 0 | 0 | 1.935953987 | 2.647218236 | 0.268490752 |
| 0 | 0 | 0 | 0 | 0 |
| 0 | 0 | 0.000697446 | 0 | 0 |
| 0 | 0 | 0 | 0 | 0 |
| 0 | 0 | 0 | 0 | 0 |
| 0 | 0 | 0 | 0 | 0 |
| 0 | 0.014134323 | 0.006139751 | 0.006964885 | 0.005870205 |
| 0.004855863 | 0.069249866 | 0 | 0 | 0 |
| 0 | 0 | 0 | 0 | 0 |
| 12.59245183 | 0.000736759 | 0.108015785 | 0.118111744 | 0.166304252 |
| 0 | 0 | 0 | 0 | 0.214387848 |
| 0 | 0 | 1.027780247 | 0.968883394 | 0.517021581 |
| 0 | 0 | 0 | 0 | 0 |
| 0 | 0 | 0 | 0 | 0 |
| 0 | 0 | 0 | 0.002657878 | 0 |
| 0 | 0 | 0 | 0 | 0 |
| 0 | 0.000986 | 0.001344714 | 0.003627793 | 0 |
| 0 | 0 | 0 | 0 | 0 |
| 0 | 0 | 0 | 0 | 0 |
| 0 | 0.012754558 | 0 | 0 | 0.000929327 |
| 0 | 0 | 0 | 0 | 0 |
| 0 | 0.002619017 | 0 | 0.006401164 | 0 |
| 6.360463573 | 0 | 4.30001508 | 6.289126238 | 6.51216921 |
| 0.438331792 | 0 | 0.01073975 | 0.297109784 | 0.515374019 |
| 0 | 0 | 0.623819035 | 0 | 0 |
| 0.003408823 | 0 | 0 | 0.316816885 | 0 |
| 0 | 0 | 0 | 0 | 0 |
| 0 | 0 | 0 | 0 | 0 |
| 0 | 0 | 0 | 0.000667335 | 0 |
| 0.000808705 | 0.004794974 | 0 | 0.000466724 | 0.000885079 |

| | | | | |
|---|---|---|---|---|
| 0.103741132 | 0 | 0 | 0 | 0 |
| 0 | 0 | 0.001724702 | 0.009697297 | 0.005007072 |
| 0 | 0 | 0 | 0 | 0.001063269 |
| 0 | 0 | 0 | 0 | 0 |
| 0.408132539 | 0 | 2.23148856 | 3.145435388 | 1.736446282 |
| 0 | 0 | 0 | 0 | 0 |
| 0 | 0 | 0 | 0 | 0 |
| 0 | 0 | 0 | 0 | 0.025900483 |
| 0.324485148 | 0 | 0.000765404 | 0 | 0 |
| 0 | 0 | 0 | 0 | 0 |
| 0 | 0.089154606 | 0 | 0 | 0 |
| 0 | 0 | 0 | 0 | 0 |
| 0 | 0 | 0.095988486 | 0.095811745 | 0.057645528 |
| 0.025338506 | 0 | 0.008126648 | 0.009404913 | 0.017037635 |
| 0 | 0 | 0 | 0 | 7.570879616 |
| 0.178843497 | 0 | 0.136003263 | 0.242069213 | 0.217597287 |
| 0 | 0 | 0.000396189 | 0.007741493 | 0.00250952 |
| 0 | 0 | 0 | 0 | 0 |
| 0 | 0 | 0 | 0 | 0 |
| 0 | 0 | 0 | 0.080570085 | 0.00757727 |
| 0 | 0 | 0 | 0 | 0 |
| 0 | 0 | 0 | 0.004768068 | 0 |
| 0 | 0 | 0 | 0 | 0 |
| 0 | 0 | 0 | 0 | 0 |
| 0 | 0 | 0 | 0 | 0 |
| 0 | 0 | 0 | 0 | 0 |
| 0.090844044 | 0 | 0.090431916 | 0.079656636 | 0.169387912 |
| 0.000703068 | 0.053543769 | 0 | 0 | 0.000615573 |
| 0 | 0 | 0 | 0 | 0 |
| 0 | 0 | 0 | 0 | 0 |
| 0 | 0 | 0 | 0 | 0 |
| 0 | 0 | 0 | 0 | 0 |
| 0 | 0 | 0 | 0.005983279 | 0 |
| 0.005159022 | 0 | 0.025570343 | 0.025520587 | 0.04629916 |
| 0 | 0 | 0.143262191 | 0.064231667 | 0.179351896 |
| 0.020007278 | 0 | 0.000337262 | 0.026022331 | 0 |
| 0 | 0 | 0.008847879 | 0 | 0 |
| 0.01535979 | 0 | 0.000849256 | 0.002021111 | 0 |
| 0 | 0 | 0 | 0 | 0 |
| 0 | 0 | 0 | 0 | 0 |
| 0.178498988 | 0 | 0.197304704 | 0.179496662 | 1.011012982 |
| 0 | 0.449982447 | 0.001274878 | 0 | 0 |
| 0 | 0 | 0 | 0.034245851 | 0 |
| 0 | 0 | 0.117159583 | 0.004886673 | 0 |
| 0 | 0 | 0 | 0 | 0 |
| 0 | 0 | 0 | 0.001635072 | 0 |
| 0 | 0 | 0 | 0 | 0 |
| 0 | 0 | 0 | 0 | 0 |

| | | | | |
|---|---|---|---|---|
| 0 | 0 | 0.029474584 | 0.016420125 | 4.048178962 |
| 0 | 0 | 0 | 0 | 0 |
| 0 | 0 | 0.007908204 | 0 | 0 |
| 0 | 0 | 0 | 0 | 0 |
| 0.08001375 | 0 | 0.076893864 | 0.004749733 | 0.005337614 |
| 0 | 0 | 0 | 0 | 0.145942621 |
| 0.019442872 | 0.00131857 | 0.006916447 | 0.020593785 | 0.278818184 |
| 0 | 0 | 0 | 0 | 0 |
| 0 | 0 | 0 | 0 | 0 |
| 0 | 0 | 0 | 0 | 0 |
| 0 | 0 | 0 | 0.006483462 | 0.026942763 |
| 0 | 0 | 0 | 0.003982904 | 0 |
| 0 | 0 | 0 | 0 | 0 |
| 0 | 0 | 0 | 0 | 0 |
| 0 | 0 | 0 | 0 | 0 |
| 0 | 0 | 0 | 0 | 0 |
| 0 | 0 | 0 | 0 | 0 |
| 0 | 0 | 0 | 0 | 0 |
| 0 | 0 | 0 | 0 | 0 |
| 0 | 0 | 0 | 0 | 0 |
| 0.14536342 | 0 | 0.064343323 | 0.250264781 | 0.058691065 |
| 0 | 0 | 0 | 0 | 0 |
| 0 | 0 | 0.012554716 | 0.020794062 | 0.006572187 |
| 0.005072781 | 0.019524016 | 0.003875032 | 0.002958447 | 0.002191521 |
| 0 | 0 | 0 | 0 | 0 |
| 0 | 0 | 0 | 0 | 0 |
| 0 | 0 | 0 | 0 | 0 |
| 0 | 0 | 0 | 0 | 0 |
| 0 | 0 | 0 | 0 | 0.047863538 |
| 0.059175888 | 0 | 0.004780354 | 0 | 0.001682193 |
| 0 | 0 | 0 | 0 | 0 |
| 0 | 0 | 0 | 0 | 0 |
| 0 | 0 | 0 | 0 | 0 |
| 0.352417719 | 0 | 0 | 0 | 0 |
| 0 | 0 | 0 | 0 | 0 |
| 0 | 0 | 0 | 0 | 0 |
| 0.057794409 | 0.005942335 | 0.005772225 | 0.003586103 | 0.01809826 |
| 0 | 0 | 0 | 0 | 0 |
| 0 | 0 | 0 | 0.002984747 | 0 |
| 0 | 0 | 0 | 0 | 0 |
| 0 | 0 | 0 | 0 | 0 |
| 0 | 0 | 0 | 0 | 0 |
| 0 | 0 | 0 | 0 | 0 |
| 0 | 0 | 0 | 0.006623244 | 0 |
| 0 | 0 | 0 | 0 | 0 |
| 0 | 0 | 0 | 0 | 0 |
| 0 | 0 | 0 | 0.000485336 | 0 |
| 0 | 0 | 0 | 0.00132314 | 0 |

| | | | | |
|---|---|---|---|---|
| 0.000430934 | 0.002838991 | 0.001489169 | 0 | 0 |
| 0 | 0 | 0.126167355 | 0 | 0 |
| 0 | 0 | 0 | 0 | 0 |
| 0 | 0 | 0 | 0 | 0 |
| 0 | 0 | 0 | 0 | 0 |
| 0 | 0 | 0.02229804 | 0 | 0 |
| 0 | 0 | 0 | 0 | 0 |
| 2.355717878 | 0 | 0.325583502 | 0.642795814 | 1.347852411 |
| 0.198415076 | 0.726067246 | 4.645183816 | 0.043253683 | 0.614938836 |
| 0.035003556 | 0.214885196 | 0.029182539 | 0 | 0 |
| 0 | 0 | 0 | 0.012629041 | 0.00645098 |
| 0 | 0 | 0 | 0 | 0 |
| 0 | 0 | 0 | 0.036904272 | 0.014579989 |
| 0 | 0.210917336 | 0 | 0 | 0 |
| 0 | 0 | 0 | 0 | 0 |
| 0 | 0 | 0 | 0 | 0 |
| 0 | 0 | 0 | 0 | 0 |
| 0 | 0 | 0 | 0 | 0 |
| 0 | 0 | 0 | 0 | 0 |
| 0 | 0 | 0.019700934 | 0 | 0 |
| 0 | 0 | 0 | 0 | 0 |
| 0 | 0 | 0 | 0 | 0 |
| 0 | 0 | 0 | 0 | 0 |
| 0 | 0 | 0 | 0 | 0 |
| 0 | 0 | 0.005317268 | 0.058374575 | 0.18846151 |
| 0 | 0 | 0 | 0 | 0 |
| 0 | 0 | 0 | 0 | 0 |
| 0.000601742 | 0 | 0 | 0 | 0 |
| 0 | 0 | 0 | 0 | 0 |
| 0 | 0 | 0 | 0 | 0 |
| 0 | 0 | 0.000509955 | 0.001328596 | 0 |
| 0 | 0 | 0 | 0 | 0 |
| 0 | 0 | 0 | 0 | 0 |
| 0 | 0 | 0 | 0.000316408 | 0 |
| 0 | 0 | 0 | 0 | 0.020174471 |
| 0.020466686 | 0 | 0 | 0.033531527 | 0.002507413 |
| 0 | 0 | 0 | 0 | 0 |
| 0 | 0 | 0 | 0 | 0 |
| 0.001708046 | 0 | 0.00141659 | 0.001419489 | 0 |
| 0.007887123 | 0 | 0 | 0 | 0 |
| 0 | 0 | 0 | 0 | 0 |
| 0 | 0 | 0.08445851 | 1.752872949 | 0.125647391 |
| 0 | 0 | 0 | 0 | 0 |
| 0 | 0 | 0 | 0 | 0 |
| 0 | 0 | 0 | 0 | 0 |
| 0 | 0 | 0 | 0.001857637 | 0.008387514 |
| 0 | 0 | 0 | 0 | 0 |
| 0 | 0 | 0 | 0 | 0 |

| | | | | |
|---|---|---|---|---|
| 0 | 0 | 0 | 0 | 0 |
| 0.003241137 | 0 | 0.008333047 | 0.004130157 | 0 |
| 0 | 0 | 0 | 0 | 0 |
| 0 | 0 | 0 | 0 | 0 |
| 0.658345785 | 0.409835101 | 0.145070892 | 0.199096091 | 1.421399971 |
| 0 | 0 | 0 | 0.004244798 | 0 |
| 0.026567131 | 0.003314069 | 0 | 0 | 0 |
| 0.014252648 | 0.013241808 | 0 | 0.00088583 | 0 |
| 0 | 0 | 0.001765324 | 0 | 0 |
| 0 | 0 | 0 | 0.013990368 | 0 |
| 0 | 0 | 0 | 0 | 0 |
| 0 | 0 | 0 | 0 | 0 |
| 0 | 0 | 0 | 0 | 0 |
| 0 | 0 | 0 | 0 | 0 |
| 0 | 0 | 0 | 0 | 0 |
| 0 | 0 | 0.005147874 | 0.004060875 | 0.017691254 |
| 0 | 0 | 0 | 0 | 0 |
| 0 | 0 | 0 | 0 | 0 |
| 0 | 0 | 0 | 0 | 0.012793904 |
| 0 | 0 | 0 | 0 | 0 |
| 0 | 0 | 0 | 0 | 0 |
| 0 | 0 | 0 | 0.000624266 | 0 |
| 0 | 0 | 0 | 0 | 0 |
| 0.000790673 | 0.006875817 | 0 | 0 | 0.000692275 |
| 0 | 0 | 0.120188213 | 0.471213289 | 0.336760059 |
| 0 | 0 | 0 | 0 | 0 |
| 0 | 0 | 0 | 0.043070914 | 0.002374365 |
| 0.000913757 | 0 | 0 | 0.000421882 | 0 |
| 0 | 0 | 0 | 0 | 0 |
| 0 | 0 | 0 | 0 | 0 |
| 0 | 0 | 0 | 0 | 0 |
| 0 | 0 | 0 | 0 | 0 |
| 0 | 0 | 0 | 0 | 0 |
| 0 | 0 | 0 | 0 | 0 |
| 0 | 0 | 0 | 0 | 0 |
| 0 | 0 | 0 | 0 | 0 |
| 0 | 0 | 0 | 0 | 0 |
| 0 | 0 | 0 | 0.055060596 | 0.024675176 |
| 0 | 0 | 0 | 0 | 0 |
| 0.005599053 | 0 | 0 | 0 | 0 |
| 0 | 0 | 0 | 0 | 0 |
| 0 | 0 | 0 | 0 | 0 |
| 0 | 0 | 0 | 0 | 0 |
| 0 | 0 | 0 | 0.020098723 | 0 |
| 0 | 0 | 0 | 0 | 0 |
| 0 | 0 | 0 | 0 | 0 |
| 0 | 0 | 0 | 0 | 0 |
| 0 | 0 | 0 | 0 | 0 |

| | | | | |
|---|---|---|---|---|
| 0 | 0 | 0.048318109 | 0.019404473 | 0.104499041 |
| 0 | 0 | 0 | 0.012708477 | 0 |
| 0.000943105 | 0 | 0 | 0 | 0.003391422 |
| 0 | 0 | 0 | 0 | 0 |
| 0 | 0 | 0 | 0 | 0 |
| 0.020170237 | 0.025122917 | 0.007079098 | 0 | 0.265591178 |
| 0 | 0 | 0 | 0 | 0 |
| 0 | 0 | 0 | 0 | 0 |
| 0 | 0 | 0 | 0 | 0 |
| 0 | 0 | 0 | 0 | 0 |
| 0 | 0 | 0 | 0 | 0 |
| 0 | 0 | 0 | 0 | 0 |
| 0 | 0 | 0 | 0 | 0 |
| 0 | 0 | 0 | 0 | 0 |
| 0 | 0 | 0 | 0 | 0 |
| 0 | 0 | 0 | 0 | 0 |
| 0 | 0 | 0 | 0 | 0 |
| 0 | 0 | 0 | 0 | 0 |
| 0 | 0 | 0 | 0 | 0 |
| 0 | 0 | 0 | 0 | 0 |
| 0.033856798 | 1.828791151 | 0.014039791 | 0 | 0 |
| 0 | 0 | 0 | 0 | 0 |
| 0.002585408 | 0.039306161 | 0.002199223 | 0.009365823 | 0.001958935 |
| 0 | 0 | 1.965918811 | 0.27317233 | 0.007570106 |
| 0 | 0 | 0 | 0 | 0 |
| 0 | 0 | 0 | 0.078538427 | 0 |
| 0 | 0 | 0 | 0 | 0 |
| 0 | 0 | 0 | 0 | 0 |
| 0 | 0 | 0 | 0 | 0 |
| 0 | 0 | 0 | 0 | 0 |
| 0 | 0 | 0 | 0 | 0 |
| 0 | 0 | 0 | 0 | 0 |
| 0.067421772 | 0 | 0 | 0.007352208 | 0 |
| 0 | 0 | 0 | 0 | 0 |
| 0 | 0 | 0.007138794 | 0.115377457 | 0.528943457 |
| 0.020624956 | 0 | 0 | 0 | 0 |
| 0 | 0 | 0 | 0 | 0 |
| 0 | 0 | 0 | 0.043674527 | 0 |
| 0.016768365 | 0.002277732 | 0 | 0.004070514 | 0 |
| 0 | 0 | 0 | 0 | 0 |
| 0 | 0 | 0 | 0 | 0 |
| 0.007061995 | 0 | 0.166608953 | 0 | 0 |
| 0 | 0.001864346 | 0.050153674 | 0.021278437 | 0.826280108 |
| 0 | 0 | 0 | 0 | 0 |
| 0 | 0 | 0 | 0 | 0 |
| 0.000963189 | 0 | 0.169530177 | 0.104060843 | 0.00263538 |
| 0 | 0 | 0 | 0 | 0 |
| 0 | 0 | 0 | 0 | 0 |

| | | | | |
|---|---|---|---|---|
| 0 | 0 | 0 | 0 | 0 |
| 0 | 0 | 0 | 0 | 0 |
| 0 | 0 | 0 | 0 | 0 |
| 0 | 0 | 0 | 0 | 0 |
| 0 | 0 | 0 | 0 | 0 |
| 0 | 0 | 0 | 0 | 0 |
| 0 | 0 | 0 | 0 | 0 |
| 0.022337892 | 0 | 0 | 0 | 0 |
| 0 | 0 | 0 | 0.017811653 | 0.019752857 |
| 0.001157419 | 0.000871438 | 0.119075921 | 0.011794548 | 0.033477756 |
| 0 | 0 | 0 | 0 | 0 |
| 0 | 0 | 0 | 0 | 0 |
| 0 | 0 | 0 | 0 | 0 |
| 0 | 0 | 0 | 0 | 0 |
| 0 | 0 | 0 | 0 | 0 |
| 0 | 0.025721953 | 0 | 0 | 0 |
| 0 | 0 | 0 | 0 | 0 |
| 0 | 0 | 0 | 0 | 0 |
| 0 | 0 | 0 | 0 | 0 |
| 0 | 0 | 0.113714304 | 0.276661073 | 0.940303893 |
| 0.018690059 | 0 | 0 | 2.561461523 | 0.001487647 |
| 0 | 0 | 0 | 0 | 0 |
| 0 | 0 | 0 | 0 | 0 |
| 0.001083478 | 0 | 0 | 0.018909143 | 0 |
| 0 | 0 | 0 | 0 | 0 |
| 0 | 0 | 0 | 0.776042102 | 0 |
| 0 | 0 | 0 | 0 | 0 |
| 0 | 0 | 0 | 0 | 0 |
| 0 | 0 | 0 | 0 | 0 |
| 2.838502655 | 0 | 0.014100193 | 0.106726855 | 0.069198924 |
| 0 | 0.033623947 | 0.061318815 | 0.296599222 | 0.30374704 |
| 0 | 0 | 0 | 0 | 0 |
| 0 | 0 | 0 | 0.000928083 | 0 |
| 0 | 0 | 0 | 0 | 0 |
| 0 | 0 | 0 | 0 | 0 |
| 0 | 0 | 0 | 0 | 0 |
| 0 | 0 | 0 | 0 | 0 |
| 0 | 0 | 0 | 0 | 0 |
| 0 | 0 | 0 | 0 | 0.000489283 |
| 0 | 0 | 0 | 0 | 0 |
| 0 | 0 | 0.00329492 | 0 | 0 |
| 0 | 0 | 0 | 0 | 0 |
| 0 | 0 | 0 | 0.00168372 | 0.012051807 |
| 0.003822884 | 0 | 0 | 0 | 0 |
| 0 | 0 | 0 | 0 | 0 |
| 0 | 0 | 0 | 0 | 0 |
| 0.036304891 | 0 | 0 | 0 | 0 |
| 0 | 0 | 0.001590197 | 0.00037493 | 0 |

| | | | | |
|---|---|---|---|---|
| 0 | 0 | 0 | 0 | 0 |
| 0 | 0 | 0 | 0 | 0 |
| 0 | 0 | 0 | 0 | 0 |
| 0 | 0 | 0 | 0 | 0 |
| 0 | 0 | 0.107484585 | 0.001723273 | 0.456015603 |
| 0 | 0 | 0 | 0.027597798 | 0 |
| 0 | 0 | 0 | 0 | 0 |
| 0 | 0 | 0 | 0 | 0 |
| 0 | 0 | 0 | 0 | 0 |
| 0 | 0 | 0 | 0 | 0 |
| 0.018323799 | 0 | 0 | 0 | 0 |
| 0 | 0.010241633 | 0 | 0.002990642 | 0.004158987 |
| 0 | 0 | 0 | 0.0045411 | 0 |
| 0 | 0 | 0 | 0 | 0 |
| 0 | 0 | 0 | 0.016227034 | 0.002160981 |
| 1.28279123 | 0 | 0.003249258 | 0.0025117 | 0 |
| 0 | 0 | 0.038563744 | 0.055295807 | 0.000436194 |
| 0 | 0 | 0 | 0 | 0 |
| 0.015695248 | 0.049632147 | 0.001627133 | 0.013949511 | 0.008588752 |
| 0.012979034 | 0 | 0.0969026 | 0.017740725 | 0.445020536 |
| 0 | 0 | 0 | 0 | 0 |
| 0 | 0 | 0 | 0 | 0 |
| 0 | 0 | 0 | 0 | 0 |
| 0 | 0 | 0 | 0 | 0 |
| 0 | 0 | 0 | 0 | 0 |
| 0 | 0 | 0 | 0 | 0 |
| 0 | 0 | 0.002080822 | 0.002748515 | 0.001947081 |
| 0 | 0 | 0 | 0 | 0 |
| 0 | 0 | 0 | 0 | 0 |
| 0 | 0 | 0 | 0 | 0 |
| 0 | 0.003474745 | 0 | 0 | 0.009337809 |
| 0 | 0 | 0 | 0 | 0 |
| 0 | 0 | 0 | 0 | 0 |
| 0 | 0 | 0 | 0 | 0 |
| 0 | 0 | 0 | 0 | 0 |
| 0 | 0 | 0 | 0 | 0 |
| 0 | 0 | 0 | 0 | 0 |
| 0.00643032 | 0.315639928 | 0.000602607 | 0.00083028 | 0.025764768 |
| 0 | 0 | 0.122312124 | 0.02614665 | 0.017359991 |
| 0 | 0 | 0 | 0 | 0 |
| 0.001088056 | 0.002007073 | 0 | 0 | 0.01369434 |
| 0 | 0 | 0 | 0 | 0 |
| 0 | 0 | 0 | 0 | 0 |
| 0 | 0 | 0 | 0.002014691 | 0 |
| 0 | 0 | 0 | 0 | 0 |
| 0 | 0 | 0 | 0 | 0 |
| 0 | 0 | 0 | 0 | 0 |
| 0 | 0 | 0 | 0.002818443 | 0 |

| | | | | |
|---|---|---|---|---|
| 0 | 0 | 0 | 0 | 0 |
| 0 | 0 | 0 | 0.005835564 | 0.002418876 |
| 0 | 0 | 0.00051486 | 0 | 0 |
| 0.002201867 | 0.00490969 | 0.000585304 | 0.001173003 | 0 |
| 0 | 0 | 0.006297174 | 0 | 0.001719278 |
| 0 | 0 | 0 | 0 | 0 |
| 0.00230111 | 0 | 0.066568755 | 0.008195834 | 0 |
| 0 | 0 | 0 | 0 | 0.004494239 |
| 0 | 0 | 0 | 0 | 0 |
| 0 | 0 | 0 | 0.023043042 | 0 |
| 0 | 0 | 0 | 0 | 0 |
| 0 | 0 | 0 | 0 | 0 |
| 0.006483381 | 0.315914443 | 0.227032151 | 0.058620324 | 0.184618824 |
| 0 | 0 | 0 | 0 | 0 |
| 0 | 0 | 0 | 0 | 0 |
| 0 | 0 | 0 | 0.037310862 | 0 |
| 0.091966258 | 0 | 0 | 0 | 0 |
| 0 | 0 | 0 | 0 | 0 |
| 0 | 0 | 0 | 0 | 0 |
| 0 | 0 | 0 | 0 | 0 |
| 0 | 0 | 0 | 0 | 0 |
| 0 | 0 | 0 | 0 | 0 |
| 0.02608564 | 0.001249834 | 0 | 0.00854011 | 0 |
| 0.005410161 | 0 | 0.006704696 | 0.242767377 | 0.008167029 |
| 0 | 0 | 0 | 0 | 1.473944613 |
| 0 | 0 | 0 | 0.007156175 | 0 |
| 0.186390622 | 0 | 0 | 0 | 0.006447115 |
| 0 | 0 | 0 | 0 | 0 |
| 0 | 0 | 0 | 0 | 0 |
| 0 | 0 | 0 | 0 | 0 |
| 0 | 0 | 0.026802597 | 0.026857445 | 0.021993149 |
| 0 | 0 | 0 | 0 | 0 |
| 0 | 0 | 0 | 0 | 0 |
| 0 | 0 | 0 | 0 | 0 |
| 0 | 0 | 0.243194195 | 0.013817052 | 0.129555159 |
| 0.016031759 | 0 | 0 | 0 | 0 |
| 0.002996468 | 0 | 0 | 0 | 0 |
| 0 | 0 | 0 | 0 | 0 |
| 0 | 0 | 0.037829118 | 0.038836119 | 0.000816128 |
| 0 | 0 | 0 | 0.000320194 | 0 |
| 0 | 0 | 0 | 0 | 0 |
| 0 | 0 | 0 | 0 | 0 |
| 0 | 0 | 0 | 0 | 0 |
| 0 | 0 | 0 | 0 | 0 |
| 0 | 0 | 0 | 0 | 0 |
| 0 | 0.10667597 | 0.581778876 | 0.172755242 | 0.113596816 |
| 0 | 0 | 0 | 0 | 0 |
| 0 | 0 | 0 | 0 | 0 |

| | | | | |
|---|---|---|---|---|
| 0 | 0 | 0 | 0 | 0 |
| 0 | 0 | 0 | 0 | 0 |
| 0 | 0 | 0 | 0 | 0 |
| 0 | 0 | 0.204563456 | 0 | 0 |
| 0 | 0 | 0 | 0 | 0 |
| 0 | 0 | 0 | 0 | 0 |
| 0 | 0 | 0 | 0 | 0 |
| 0 | 0 | 0 | 0 | 0 |
| 0 | 0 | 0.022698842 | 0.001146821 | 0.00105719 |
| 0 | 0 | 0 | 0 | 0 |
| 0 | 0.001115945 | 0 | 0 | 0 |
| 0 | 0 | 0 | 0 | 0 |
| 0 | 0 | 0 | 0 | 0 |
| 0 | 0 | 0 | 0 | 0 |
| 0 | 0 | 0 | 0 | 0 |
| 0 | 0 | 0 | 0 | 0 |
| 0 | 0 | 0.070925897 | 0 | 0 |
| 0 | 0 | 0 | 0 | 0 |
| 0 | 0 | 0 | 0 | 0 |
| 0 | 0 | 0 | 0 | 0 |
| 0 | 0 | 0 | 0 | 0 |
| 0 | 0 | 0 | 0 | 0 |
| 0 | 0 | 0 | 0 | 0 |
| 0 | 0 | 0 | 0 | 0 |
| 0 | 0 | 0 | 0 | 0 |
| 0 | 0 | 0.00038743 | 0 | 0 |
| 0 | 0 | 0 | 0 | 0 |
| 0.012607728 | 0 | 0.011045478 | 0 | 0 |
| 0 | 0 | 0 | 0 | 0 |
| 0 | 0.229791139 | 0 | 0 | 0 |
| 0.001516348 | 0 | 0.016482224 | 0.006109948 | 0.001659552 |
| 0 | 0 | 0 | 0 | 0 |
| 0.022248018 | 0 | 0.012610865 | 0.012525824 | 0.005780725 |
| 0 | 0.074997056 | 0.00135652 | 0.017444293 | 0.00537025 |
| 0 | 0 | 0 | 0 | 0 |
| 0 | 0 | 0 | 0 | 0 |
| 0 | 0 | 0 | 0 | 0 |
| 0 | 0 | 0 | 0 | 0 |
| 0 | 0 | 0 | 0 | 0 |
| 0 | 0 | 0 | 0 | 0 |
| 0 | 0 | 0 | 0 | 0 |
| 0.062810407 | 0 | 0 | 0 | 0.00158525 |
| 0.033046713 | 0 | 0.027274679 | 0.021442217 | 0.037747778 |
| 0 | 0 | 0 | 0 | 0 |
| 0 | 0 | 0 | 0 | 0 |
| 0 | 0 | 0.019386466 | 0.019426139 | 0.366868112 |
| 0 | 0 | 0 | 0 | 0 |
| 0 | 0 | 0 | 0 | 0 |

| | | | | |
|---|---|---|---|---|
| 0 | 0 | 0 | 0 | 0 |
| 0.383013762 | 0 | 0.003136154 | 0.007099883 | 0 |
| 0.043974426 | 0.01396785 | 0.010990076 | 0.010604693 | 0.004941649 |
| 0 | 0 | 0 | 0 | 0 |
| 0.001880971 | 0.001445715 | 0.010833391 | 0.007924559 | 0.006004279 |
| 0 | 0 | 0 | 0 | 0 |
| 0 | 0 | 0 | 0 | 0 |
| 0 | 0 | 0 | 0 | 0 |
| 0 | 0 | 0 | 0 | 0.20319549 |
| 0 | 0 | 0 | 0 | 0 |
| 0 | 0 | 0 | 0 | 0 |
| 0 | 0 | 0 | 0 | 0 |
| 0 | 0 | 0 | 0 | 0 |
| 0 | 0 | 0 | 0 | 0.001625316 |
| 0 | 0 | 0 | 0 | 0 |
| 0 | 0 | 0 | 0 | 0 |
| 0 | 0 | 0 | 0 | 0 |
| 0 | 0 | 0 | 0 | 0 |
| 0 | 0 | 0 | 0 | 0 |
| 0 | 0 | 0 | 0 | 0 |
| 0 | 0 | 0 | 0 | 0 |
| 0 | 0 | 0 | 0 | 0.002938033 |
| 0 | 0 | 0 | 0 | 0 |
| 0 | 0 | 0 | 0 | 0 |
| 0 | 0 | 0.273733771 | 0.159780701 | 0.013988542 |
| 0.000488273 | 0 | 0 | 0 | 0.000890642 |
| 0 | 0 | 0 | 0 | 0 |
| 0.012314682 | 0 | 0.026406294 | 0.122789061 | 1.031042376 |
| 0 | 0 | 0 | 0 | 0 |
| 0 | 0 | 0 | 0 | 0 |
| 0 | 0 | 0 | 0 | 0 |
| 0 | 0.001504476 | 0 | 0 | 0 |
| 0 | 0 | 0 | 0 | 0 |
| 0 | 0 | 0 | 0 | 0 |
| 0 | 0 | 0 | 0 | 0 |
| 0 | 0 | 0 | 0 | 0 |
| 0 | 0 | 0 | 0 | 0 |
| 0.005994425 | 0 | 0 | 0 | 0 |
| 0 | 0 | 0 | 0.435656784 | 0.568630635 |
| 0 | 0 | 0 | 0 | 0 |
| 0 | 0 | 0 | 0 | 0 |
| 0 | 0 | 0.0176946 | 0.005502665 | 0 |
| 0 | 0.004089245 | 0.00124183 | 0 | 0 |
| 0 | 0 | 0 | 0 | 0 |
| 0 | 0 | 0 | 0 | 0 |
| 0 | 0 | 0 | 0 | 0 |
| 0 | 0.004938745 | 0 | 0.00121141 | 0 |
| 0 | 0 | 0 | 0 | 0 |

| | | | | |
|---|---|---|---|---|
| 0 | 0 | 0 | 0 | 0 |
| 0.00567558 | 0.016410375 | 0.00108961 | 0.004258177 | 0.011042811 |
| 0 | 0 | 0 | 0.07651167 | 0 |
| 0 | 0 | 0 | 0 | 0 |
| 0 | 0 | 0 | 0 | 0 |
| 0 | 0 | 0 | 0 | 0 |
| 0 | 0 | 0 | 0 | 0 |
| 0 | 0 | 0 | 0 | 0 |
| 0 | 0 | 0 | 0 | 0 |
| 0 | 0 | 0 | 0 | 0 |
| 0 | 0 | 0 | 0 | 0 |
| 0 | 0 | 0 | 0 | 0 |
| 0 | 0 | 0.112877855 | 0.300200116 | 0.338341269 |
| 0 | 0 | 0 | 0 | 0 |
| 0.003734869 | 0 | 0.005162604 | 0.001633632 | 0.013123312 |
| 0 | 0 | 0 | 0 | 0 |
| 0 | 0 | 0 | 0 | 0 |
| 0 | 0 | 0 | 0 | 0 |
| 0 | 0 | 0 | 0.004969856 | 0 |
| 0 | 0 | 0 | 0 | 0 |
| 0 | 0 | 0 | 0 | 0 |
| 0.001930493 | 0 | 0.000667116 | 0.031686028 | 0.051129952 |
| 0 | 0 | 0.005536343 | 0.003492979 | 0.00649408 |
| 0 | 0 | 0 | 0 | 0 |
| 0 | 0 | 0 | 0 | 0 |
| 0 | 0 | 0 | 0 | 0 |
| 0 | 0 | 0 | 0 | 0 |
| 0 | 0 | 0 | 0 | 0 |
| 0 | 0 | 0 | 0 | 0 |
| 0 | 0 | 0 | 0 | 0 |
| 0 | 0 | 0 | 0 | 0 |
| 0 | 0 | 0 | 0 | 0 |
| 0 | 0 | 0 | 0 | 0 |
| 0 | 0 | 0 | 0 | 0 |
| 0.029972535 | 0 | 26.01028777 | 25.92197573 | 0 |
| 0 | 0 | 0 | 0 | 0 |
| 0 | 0 | 0 | 0 | 0 |
| 0 | 0 | 0 | 0 | 0 |
| 0 | 0.034799503 | 0.124179023 | 0.06555437 | 0.304879736 |
| 0 | 0 | 0 | 0 | 0 |
| 0.044251006 | 0 | 0.055551992 | 0.222662695 | 0.154370827 |
| 0 | 0 | 0 | 0 | 0 |
| 0 | 0 | 0 | 0 | 0 |
| 0 | 0 | 0 | 0 | 0 |
| 0 | 0 | 0 | 0 | 0 |
| 0.093610378 | 27.63439308 | 0.020927521 | 0.030564624 | 0.001732785 |
| 0 | 0 | 0 | 0 | 0 |
| 0 | 0 | 0 | 0 | 0 |

| | | | | |
|---|---|---|---|---|
| 0 | 0 | 0 | 0 | 0 |
| 0 | 0 | 0 | 0 | 0.000881971 |
| 0 | 0 | 0 | 0 | 0 |
| 0 | 0 | 0 | 0.10562464 | 0 |
| 0 | 0 | 0 | 0 | 0 |
| 0 | 0 | 0 | 0 | 0 |
| 0 | 0 | 0 | 0 | 0 |
| 0 | 0 | 0 | 0 | 0.007959808 |
| 0 | 0.002473089 | 0 | 0.001021344 | 0.001614034 |
| 0.012774572 | 0 | 0.617358767 | 0.020509033 | 0.002330167 |
| 0 | 0 | 0 | 0 | 0 |
| 0 | 0 | 0 | 0 | 0 |
| 0 | 0 | 0.001569013 | 0.004716671 | 0 |
| 0 | 0 | 0 | 0 | 0 |
| 0 | 0 | 0 | 0 | 0 |
| 0 | 0 | 0 | 0 | 0 |
| 0 | 0 | 0 | 0 | 0 |
| 0 | 0 | 0 | 0 | 0 |
| 0 | 0 | 0 | 0 | 0 |
| 0.068533138 | 0.021520958 | 0.157459007 | 0.009445883 | 0.325453389 |
| 0 | 0 | 0 | 0 | 0 |
| 0 | 0 | 0 | 0 | 0 |
| 0 | 0 | 0 | 0 | 0 |
| 0 | 0 | 0 | 0 | 0 |
| 0 | 0 | 0 | 0 | 0 |
| 0 | 0 | 0 | 0.005128077 | 0 |
| 0 | 0 | 0 | 0 | 0 |
| 0 | 0 | 0 | 0 | 0 |
| 0 | 0 | 0 | 0 | 0 |
| 0 | 0 | 0 | 0.001121978 | 0 |
| 0 | 0 | 0 | 0 | 0 |
| 0 | 0 | 0 | 0.00076213 | 0 |
| 0 | 0 | 0 | 0 | 0 |
| 0 | 0 | 0 | 0 | 0 |
| 0 | 0 | 0 | 0 | 0 |
| 0 | 0 | 0 | 0 | 0.000954229 |
| 0 | 0 | 0 | 0 | 0 |
| 0 | 0 | 0 | 0 | 0 |
| 0 | 0 | 0 | 0 | 0 |
| 0 | 0 | 0 | 0 | 0 |
| 0 | 0 | 0 | 0 | 0 |
| 0 | 0 | 0 | 0 | 0 |
| 0 | 0 | 0 | 0.041294601 | 0.009038081 |
| 0 | 0 | 0 | 0 | 0 |
| 0 | 0 | 0 | 0.000985769 | 0.002250176 |
| 0 | 0 | 0 | 0 | 0 |
| 0 | 0 | 0 | 0 | 0 |
| 0 | 0 | 0 | 0 | 0 |

| | | | | |
|---|---|---|---|---|
| 0 | 0 | 0 | 0 | 0 |
| 0 | 0 | 0 | 0 | 0 |
| 0 | 0 | 0 | 0 | 0 |
| 0 | 0 | 0 | 0 | 0 |
| 0 | 0 | 0 | 0 | 0 |
| 0 | 0.000777835 | 0.013872226 | 0.006705002 | 0.003618126 |
| 0 | 0 | 0 | 0 | 0 |
| 0 | 0 | 0 | 0 | 0 |
| 0 | 0 | 0 | 0 | 0 |
| 0 | 0 | 0 | 0 | 0 |
| 0 | 0 | 0 | 0 | 0 |
| 0 | 0 | 0 | 0 | 0 |
| 0 | 0 | 0 | 0 | 0 |
| 0 | 0 | 0 | 0.010454412 | 0.015860301 |
| 0 | 0 | 0 | 0 | 0 |
| 0 | 0 | 0 | 0 | 0 |
| 0 | 0 | 0 | 0.110193859 | 0 |
| 0 | 0.000957945 | 0 | 0.000906318 | 0 |
| 0 | 0 | 0 | 0.002527065 | 0 |
| 0 | 0 | 0 | 0 | 0 |
| 0 | 0 | 0.001513852 | 0 | 0 |
| 0 | 0 | 0 | 0 | 0 |
| 0 | 0 | 0 | 0 | 0 |
| 0 | 0 | 0 | 0 | 0.033038863 |
| 0 | 0 | 0 | 0 | 0 |
| 0.004801649 | 0 | 0.303651201 | 0 | 0 |
| 0 | 0 | 0 | 0 | 0 |
| 0 | 0 | 0 | 0.671215546 | 0.439749837 |
| 0 | 0 | 0 | 0 | 0 |
| 0 | 0 | 0 | 0 | 0 |
| 0 | 0 | 0 | 0 | 0 |
| 0 | 0 | 0 | 0 | 0 |
| 0 | 0 | 0 | 0 | 0 |
| 0 | 0 | 0 | 0 | 0 |
| 0 | 0 | 0 | 0 | 0 |
| 0 | 0 | 0 | 0 | 0 |
| 0 | 0 | 0 | 0 | 0 |
| 0 | 0 | 0 | 0 | 0 |
| 0 | 0 | 1.545932617 | 0 | 0.01222106 |
| 0 | 0 | 0 | 0 | 0 |
| 0 | 0 | 0 | 0 | 0 |
| 0 | 0 | 0 | 0 | 0 |
| 0 | 0 | 0 | 0 | 0 |
| 0 | 0 | 0 | 0 | 0 |
| 0 | 0 | 0.033645591 | 0 | 0 |
| 0 | 0 | 0 | 0 | 0 |
| 0 | 0 | 0 | 0 | 0 |
| 0 | 0 | 0 | 0.000881404 | 0 |

| | | | | |
|---|---|---|---|---|
| 0 | 0 | 0 | 0 | 0 |
| 0 | 0 | 0.006577867 | 0.02517521 | 0 |
| 0 | 0 | 0 | 0.001905358 | 0 |
| 0 | 0 | 0 | 0 | 0 |
| 0 | 0 | 0 | 0 | 0 |
| 0 | 0 | 0 | 0.001506445 | 0 |
| 0 | 0 | 0 | 0 | 0 |
| 0 | 0 | 0 | 0 | 0 |
| 0 | 0 | 0 | 0 | 0 |
| 0 | 0 | 0 | 0 | 0.014732131 |
| 0.001038531 | 0.00615766 | 0.632950227 | 0.002157705 | 0.009092879 |
| 0 | 0 | 0 | 0 | 0 |
| 0 | 0 | 0.061149631 | 0.091117521 | 0.096972269 |
| 0 | 0 | 0 | 0 | 0 |
| 0 | 0 | 0 | 0 | 0 |
| 0 | 0 | 0 | 0 | 0 |
| 0 | 0 | 0 | 0 | 0 |
| 0 | 0 | 0 | 0 | 0 |
| 0 | 0 | 0 | 0 | 0 |
| 0 | 0 | 0 | 0.00079798 | 0 |
| 0.002283768 | 3.515083399 | 0.000364245 | 0.011558022 | 0.000576795 |
| 0 | 0 | 0 | 0 | 0 |
| 0 | 0 | 0 | 0.001217586 | 0 |
| 2.117732723 | 0 | 1.066551135 | 1.627692341 | 0.832197955 |
| 0 | 0 | 0 | 0 | 0 |
| 0 | 0 | 0 | 0.003295011 | 0 |
| 0 | 0 | 0 | 0 | 0 |
| 0 | 0 | 0 | 0.06846956 | 0.015899172 |
| 0 | 0 | 0 | 0 | 0 |
| 0 | 0 | 0 | 0 | 0 |
| 0 | 0 | 0 | 0.003224139 | 0 |
| 0 | 0 | 0 | 0 | 0 |
| 0 | 0 | 0.006641279 | 0 | 0 |
| 0 | 0 | 0 | 0 | 0 |

| | | | | |
|---|---|---|---|---|
| PHL018-0113_V1 | PHL018-0097_V3 | PHL018-0108_V1 | PHL018-0209_V2 | PHL018-0099_V1 |
| EYCF | CTRL | CTRL | REF | EYCF |
| 0.329736304 | 0.061973869 | 0.391566103 | 0.010439364 | 0.113280829 |
| 20.49740917 | 0.198280867 | 0.409208677 | 0.063249046 | 1.245796673 |
| 6.678420851 | 0 | 0 | 0 | 1.279974531 |
| 0 | 0 | 0 | 0 | 0 |
| 0 | 0 | 0 | 0 | 0 |
| 0 | 0 | 0 | 0 | 0 |
| 0 | 0 | 0 | 0 | 0 |
| 0 | 0 | 0 | 0 | 0 |
| 0 | 0 | 0 | 0 | 0 |
| 0 | 0 | 0 | 0 | 0 |
| 0 | 0 | 0 | 0 | 0 |
| 0 | 0 | 0 | 0 | 0 |
| 0 | 0 | 0 | 0 | 0 |
| 11.42559318 | 0 | 18.57077122 | 0.098170522 | 0.20197037 |
| 0 | 0 | 0 | 0 | 0 |
| 0 | 0 | 0 | 0 | 0 |
| 0 | 0 | 0 | 0 | 0 |
| 0 | 0 | 0.001690853 | 0 | 0 |
| 0 | 0 | 0 | 0 | 0 |
| 0 | 0 | 0 | 0 | 0 |
| 0 | 0 | 0 | 0 | 0 |
| 0 | 0 | 0 | 0 | 0 |
| 0 | 0 | 0 | 0 | 0 |
| 0 | 0 | 0 | 0 | 0 |
| 0 | 0 | 0 | 0 | 0 |
| 0 | 0 | 0 | 0 | 0 |
| 0 | 0 | 0 | 0 | 0 |
| 0 | 0 | 0 | 0.102828488 | 0 |
| 0 | 0 | 0 | 0 | 0 |
| 0 | 0 | 0 | 0 | 0.402969626 |
| 0 | 0 | 0 | 0 | 0 |
| 0 | 0 | 0 | 0 | 0 |
| 0 | 0 | 0 | 0 | 0 |
| 0 | 0 | 0 | 0 | 0 |
| 0 | 0 | 0 | 0 | 0 |
| 0 | 0 | 0 | 0.068821758 | 1.273046646 |
| 0 | 0 | 0 | 0 | 0 |
| 0 | 0 | 0 | 0 | 0 |
| 0 | 0 | 0 | 0 | 0 |
| 0 | 0 | 0 | 0 | 0 |
| 0 | 0 | 0 | 0 | 0 |
| 0.035489625 | 0.003799377 | 0.056587252 | 0.008031013 | 0 |
| 0.094770007 | 0.043407762 | 0.029595018 | 8.165160581 | 0.007772377 |
| 0.008505301 | 0.035822699 | 0.001011718 | 0 | 0.019115635 |
| 0.020655231 | 0 | 0.005001691 | 0.006480228 | 0.006227713 |
| 0 | 0 | 0 | 0 | 0 |

| | | | | |
|---|---|---|---|---|
| 0 | 0 | 0 | 0 | 0.597422084 |
| 0.108196965 | 0.030400127 | 0.06354902 | 0 | 0.002673532 |
| 0 | 0.005117491 | 0.072671337 | 0 | 0.104492022 |
| 0.859538405 | 0.818011847 | 0.322062184 | 0.299080517 | 0.698515601 |
| 0 | 0.01377018 | 0.014262761 | 0 | 0 |
| 0 | 0 | 0 | 0 | 0 |
| 0.00082248 | 0.007024103 | 0 | 0 | 0 |
| 0 | 0 | 0 | 0 | 0 |
| 0 | 0 | 0 | 0 | 0 |
| 0 | 0 | 0 | 0 | 0 |
| 0 | 0.000717088 | 0 | 0 | 0 |
| 0.162089299 | 0.294258248 | 1.054588156 | 0.199214169 | 0.342239286 |
| 0 | 0 | 0 | 0 | 0 |
| 0.032544148 | 0.010759555 | 0.114678885 | 0 | 0.170020916 |
| 0 | 0 | 0 | 0 | 0 |
| 0 | 0 | 0 | 0 | 0 |
| 0.001211256 | 0 | 0 | 0.33709614 | 0.001731333 |
| 0 | 0 | 0 | 0 | 0 |
| 0 | 0 | 0 | 0 | 0 |
| 0 | 0 | 0 | 0 | 0 |
| 0.254573021 | 0.134219592 | 0.176193694 | 0.080108647 | 0.145212453 |
| 0 | 0.17315498 | 0.012877684 | 0.003069968 | 0.024824749 |
| 0 | 0 | 0 | 0.005050213 | 0.483446563 |
| 0 | 0 | 0 | 0 | 0 |
| 0 | 0 | 0.005313852 | 0 | 0 |
| 0 | 0 | 0 | 0 | 0 |
| 0.002791173 | 0 | 0 | 0 | 0 |
| 0 | 0 | 0 | 0 | 0 |
| 0.020746331 | 0 | 0.003982793 | 0 | 0 |
| 0 | 0 | 0 | 0 | 0 |
| 0.009270642 | 0.1616442 | 0.109839651 | 0 | 0 |
| 0 | 0 | 0 | 0 | 0 |
| 0 | 0 | 0 | 0 | 0 |
| 0 | 0 | 0 | 0 | 0 |
| 0 | 0 | 0 | 0 | 0 |
| 0 | 0 | 0 | 0 | 0 |
| 0 | 0 | 0 | 0 | 0 |
| 0 | 0 | 0 | 36.12145541 | 0 |
| 0 | 0 | 0 | 0 | 0 |
| 0 | 0 | 0 | 0 | 0 |
| 0 | 0 | 0 | 0 | 0 |
| 0 | 0 | 0 | 0 | 0 |
| 0 | 0 | 0 | 0 | 0 |
| 0 | 0 | 0 | 0 | 0 |
| 0 | 0 | 0 | 0 | 0 |
| 0 | 0 | 0 | 0 | 0 |
| 0 | 0 | 0 | 0 | 0 |
| 0.000885648 | 0 | 0.009865202 | 0 | 0.001740639 |

| | | | | |
|---|---|---|---|---|
| 0 | 0 | 0 | 0 | 0 |
| 0 | 0 | 0 | 0 | 0 |
| 0 | 0 | 0 | 0 | 0 |
| 0 | 0 | 0 | 0 | 0 |
| 0 | 0 | 0 | 0 | 0 |
| 0 | 0 | 0 | 0 | 0 |
| 0 | 0 | 0 | 0 | 0 |
| 0 | 0.00572798 | 0 | 0 | 0 |
| 0.01228587 | 0.004154046 | 0.003419585 | 0 | 0 |
| 0 | 0 | 0 | 0 | 0 |
| 0 | 0 | 0 | 0 | 0 |
| 0.00147555 | 0 | 0.000476409 | 0.00068901 | 0 |
| 0 | 0 | 0 | 0 | 0 |
| 0 | 0 | 0 | 0 | 0 |
| 0 | 0 | 0 | 0 | 0 |
| 0 | 0 | 0 | 0 | 0 |
| 0 | 0 | 0 | 0 | 0 |
| 0 | 0 | 0 | 0 | 0 |
| 0 | 0 | 0 | 0.8656156 | 0.520947706 |
| 0 | 0 | 0 | 0 | 0 |
| 0 | 0 | 0 | 0 | 0 |
| 0 | 0 | 0 | 0 | 0 |
| 0 | 0 | 0 | 0 | 0 |
| 0 | 0 | 0 | 0 | 0 |
| 0 | 0 | 0 | 0 | 0 |
| 0 | 0 | 0 | 0 | 0 |
| 0 | 0 | 0 | 0 | 0 |
| 0 | 0 | 0.001288943 | 0 | 0.003668322 |
| 0 | 0 | 0 | 0 | 0 |
| 0 | 0 | 0 | 0 | 0 |
| 0 | 0 | 0 | 0.033408603 | 0.07557879 |
| 0 | 0.003902638 | 0.00185904 | 0 | 0 |
| 0 | 0 | 0 | 0 | 0 |
| 0 | 0 | 0 | 0 | 0 |
| 0 | 0 | 0 | 0 | 0 |
| 0 | 0 | 0 | 0 | 0 |
| 0 | 0 | 0 | 0 | 0 |
| 0 | 0 | 0 | 0 | 0 |
| 0 | 0 | 0 | 0 | 0 |
| 0 | 0 | 0 | 0 | 0 |
| 0 | 0 | 0 | 0 | 0 |
| 0 | 0 | 0 | 0 | 0 |
| 0 | 0.013532496 | 0 | 0 | 0 |
| 0 | 0 | 0 | 0 | 0 |
| 0 | 0 | 0 | 0 | 0.002626034 |
| 0 | 0 | 0 | 0 | 0 |
| 0 | 2.080982961 | 0 | 0 | 0 |
| 0 | 0 | 0 | 0 | 0 |

| | | | | |
|---|---|---|---|---|
| 0.001231088 | 0 | 0 | 0 | 0 |
| 0 | 0 | 0.02050099 | 0 | 0 |
| 0 | 0 | 0 | 0 | 0 |
| 0 | 0 | 0 | 0 | 0 |
| 0 | 0 | 0 | 0 | 0 |
| 0 | 0 | 0 | 0 | 0 |
| 0 | 0 | 0 | 0 | 0 |
| 0 | 0 | 0 | 0 | 0 |
| 0 | 0 | 0 | 0 | 0 |
| 0 | 0 | 0 | 0 | 0 |
| 0 | 0 | 0 | 0 | 0 |
| 0 | 0 | 0.004716352 | 0 | 0 |
| 0 | 0 | 0 | 0 | 0 |
| 0 | 0 | 0 | 0 | 0 |
| 0 | 0 | 0 | 0 | 0 |
| 0 | 0 | 0 | 0 | 0 |
| 0 | 0 | 0 | 0 | 0 |
| 0 | 0 | 0 | 0 | 0 |
| 0 | 0 | 0 | 0 | 0 |
| 0 | 0 | 0 | 0 | 0 |
| 0 | 0 | 0 | 0 | 0 |
| 0 | 0 | 0 | 0 | 0 |
| 0 | 0 | 0 | 0.000611439 | 0 |
| 0 | 0 | 0 | 0 | 0 |
| 0 | 0 | 0.030673699 | 0 | 0.057385662 |
| 0 | 0 | 0 | 0 | 0 |
| 0 | 0 | 0 | 0 | 0 |
| 0 | 0 | 0 | 0 | 0 |
| 0 | 0 | 0 | 0 | 0 |
| 0 | 0 | 0 | 0 | 0 |
| 0 | 0 | 0.008180278 | 0 | 0.005490025 |
| 0 | 0 | 0 | 0.215487062 | 0 |
| 0 | 0 | 0 | 0 | 0 |
| 0 | 0 | 0 | 0 | 0 |
| 0 | 0 | 0 | 0 | 0 |
| 0 | 0 | 0 | 0 | 0 |
| 0 | 0 | 0 | 0 | 0 |
| 0 | 0 | 0 | 0 | 0 |
| 0 | 0 | 0 | 0 | 0 |
| 0 | 0 | 0 | 0 | 0 |
| 0 | 0 | 0 | 0 | 0 |
| 0 | 0 | 0 | 0 | 0 |
| 0 | 0 | 0 | 0 | 0 |
| 0 | 0 | 0 | 0 | 0 |
| 0 | 0 | 0 | 0 | 0 |
| 0 | 0 | 0 | 0 | 0 |
| 0 | 0 | 0 | 0.002683888 | 0 |
| 0 | 0 | 0 | 0 | 0 |

| | | | | |
|---|---|---|---|---|
| 0 | 0 | 0 | 0 | 0 |
| 0 | 0 | 0 | 0 | 0 |
| 0 | 0 | 0 | 0 | 0 |
| 0 | 0 | 0 | 0 | 0 |
| 0 | 0 | 0 | 0 | 0 |
| 0 | 0 | 0 | 0 | 0 |
| 0 | 0 | 0 | 0 | 0 |
| 0 | 0 | 0 | 0 | 0 |
| 0 | 0 | 0 | 0 | 0 |
| 0 | 0 | 0 | 0 | 0 |
| 0 | 0 | 0 | 0 | 0 |
| 0 | 0 | 0 | 0 | 0 |
| 0 | 0.002585244 | 0 | 0 | 0.004786676 |
| 0 | 0.000961144 | 0 | 0 | 0 |
| 0 | 0 | 0 | 0 | 0 |
| 0 | 0 | 0 | 0 | 0 |
| 0 | 0 | 0 | 0 | 0 |
| 0 | 0.000518188 | 0 | 0 | 0 |
| 0 | 0 | 0 | 0 | 0 |
| 0 | 0 | 0 | 0 | 0 |
| 0.029998374 | 0.081637254 | 0.013705934 | 0.003964467 | 0.011073648 |
| 0 | 0 | 0 | 0 | 0 |
| 0.000620795 | 0 | 0 | 0 | 0 |
| 0.015936251 | 0 | 0.001715104 | 0 | 0 |
| 0 | 0 | 0 | 0 | 0 |
| 0 | 0 | 0 | 0 | 0 |
| 0.027029592 | 0.001876722 | 0.002234964 | 0.004502184 | 0.001855201 |
| 0 | 0 | 0 | 0 | 0 |
| 0 | 0 | 0 | 0 | 0 |
| 0 | 0 | 0 | 0 | 0 |
| 0.001807214 | 0.002650261 | 0.183481765 | 0 | 0 |
| 0 | 0 | 0 | 0 | 0 |
| 0 | 0 | 0 | 0 | 0 |
| 0 | 0.011816436 | 0 | 0 | 0.006303997 |
| 0.002209646 | 0.034246841 | 0.289650502 | 0.000902823 | 0.272807019 |
| 0 | 0 | 0 | 0 | 0 |
| 0 | 0 | 0 | 0 | 0 |
| 0 | 0 | 0 | 0 | 0 |
| 0 | 0 | 0 | 0 | 0 |
| 0 | 0 | 0 | 0 | 0 |
| 0 | 0 | 0 | 0 | 0 |
| 0 | 0 | 0.000534373 | 0 | 0 |
| 0 | 0 | 0 | 0 | 0 |
| 0 | 0 | 0 | 0 | 0 |
| 0 | 0 | 0 | 0 | 0 |
| 0 | 0 | 0 | 0 | 0 |
| 0 | 0 | 0.002703573 | 0 | 0 |
| 0.005380066 | 0 | 0.003304638 | 0.000459554 | 0.174645142 |

| | | | | |
|---|---|---|---|---|
| 0 | 0 | 0 | 0 | 0 |
| 0 | 0 | 0 | 0 | 0 |
| 0 | 0 | 0.001161427 | 0 | 0 |
| 0 | 0 | 0 | 0 | 0 |
| 0 | 0 | 0 | 0 | 0 |
| 0 | 0 | 0 | 0 | 0 |
| 0.109833474 | 0 | 0 | 0 | 0 |
| 0.007545311 | 0 | 0 | 0 | 0 |
| 0 | 0 | 0.004758773 | 0 | 0 |
| 0 | 0 | 0 | 0 | 0 |
| 0 | 0 | 0 | 0 | 0 |
| 0 | 0 | 0 | 0 | 0 |
| 0 | 0 | 0 | 0 | 0 |
| 0 | 0 | 0 | 0 | 0 |
| 0 | 0 | 0 | 0 | 0 |
| 0 | 0 | 0 | 0 | 0 |
| 0 | 0 | 0 | 0 | 0 |
| 0 | 0 | 0 | 0 | 0 |
| 0 | 0 | 0 | 0.040099798 | 0 |
| 0 | 0 | 0 | 0 | 1.706983349 |
| 0.021457069 | 0.258338443 | 0.139573111 | 0.003447045 | 0.002532393 |
| 0 | 0 | 0 | 0 | 0 |
| 0 | 0 | 0 | 0 | 0 |
| 0 | 0 | 0 | 0 | 0 |
| 0.011341662 | 0.002597764 | 0 | 0 | 0.001048153 |
| 0.00075902 | 0 | 0 | 0 | 0 |
| 0 | 0 | 0 | 0 | 0 |
| 0 | 0 | 0 | 0 | 0 |
| 0 | 0 | 0 | 0 | 0 |
| 0 | 0 | 0 | 0 | 0 |
| 0 | 0.734552063 | 0.675634675 | 0.013382743 | 0.292034298 |
| 0 | 0 | 0 | 0 | 0 |
| 0 | 0.00102471 | 0.025800915 | 0 | 0 |
| 0 | 0 | 0 | 0 | 0 |
| 0 | 0 | 0 | 0 | 0 |
| 0 | 0 | 0 | 0 | 0 |
| 0 | 0 | 0 | 0 | 0 |
| 0 | 0 | 0 | 0 | 0 |
| 0 | 0 | 0 | 0 | 0 |
| 0.000722908 | 0 | 0 | 0 | 0 |
| 0 | 0 | 0 | 0 | 0 |
| 0.003098446 | 0.013860624 | 0.047875817 | 0.002790303 | 0.017952564 |
| 0 | 0 | 0 | 0 | 0 |
| 0 | 0 | 0 | 0 | 0 |
| 0 | 0 | 0 | 0 | 0 |
| 0 | 0 | 0 | 0 | 0 |
| 0 | 0 | 0 | 0 | 0 |
| 0 | 0 | 0 | 0 | 0 |

| | | | | |
|---|---|---|---|---|
| 0 | 0 | 0 | 0 | 0 |
| 0 | 0 | 0 | 0 | 0 |
| 0 | 0.03533599 | 0.000897253 | 0 | 0.023237539 |
| 0 | 0 | 0 | 0 | 0 |
| 0 | 0 | 0.481679788 | 0 | 0.512513941 |
| 0 | 0 | 0 | 0 | 0 |
| 0 | 0 | 0 | 0 | 0 |
| 0 | 0 | 0 | 0 | 0 |
| 0 | 0 | 0 | 0 | 0 |
| 0 | 0 | 0 | 0 | 0 |
| 0 | 0 | 0 | 0 | 0 |
| 0 | 0 | 0 | 0 | 0 |
| 0 | 0 | 0 | 0 | 0 |
| 0 | 0.023120723 | 0.002181579 | 0 | 0 |
| 0.005069918 | 0.004650511 | 0 | 0 | 0.000634094 |
| 0.002894942 | 0 | 0 | 0.008871171 | 0 |
| 0.009242747 | 0.004933403 | 0 | 0 | 0.001135347 |
| 0 | 0 | 0 | 0 | 0 |
| 0 | 0 | 0 | 0 | 0 |
| 0 | 0 | 0 | 0 | 0 |
| 0 | 0 | 0 | 0 | 0 |
| 0.00451943 | 0 | 0.002188772 | 0.001339263 | 0.183456058 |
| 0 | 0 | 0 | 0 | 0 |
| 0 | 0 | 0.002871081 | 0 | 0 |
| 0 | 0 | 0 | 0 | 0 |
| 0 | 0 | 0 | 0 | 0 |
| 9.921205742 | 0 | 0 | 0 | 0 |
| 0 | 0 | 0 | 0 | 0 |
| 0 | 0 | 0 | 0 | 0 |
| 0 | 0 | 0.001217722 | 0 | 0.000539098 |
| 0 | 0 | 0.001032179 | 0 | 0 |
| 0 | 0 | 0 | 0 | 0 |
| 0 | 0 | 0 | 0 | 0 |
| 0 | 0.001141071 | 0 | 0 | 0 |
| 0 | 0 | 0 | 0 | 0 |
| 0 | 0 | 0.002354025 | 0 | 0 |
| 0 | 0 | 0 | 0 | 0 |
| 0.225817485 | 0 | 0.002749965 | 0.003365292 | 0.001521796 |
| 0 | 0 | 0 | 0 | 0 |
| 0 | 0 | 0 | 0 | 0 |
| 0 | 0 | 0 | 0 | 0 |
| 0 | 0 | 0 | 0 | 0 |
| 0 | 0 | 0 | 0 | 0 |
| 0 | 0 | 0.004992449 | 0 | 0 |
| 0 | 0 | 0 | 0 | 0 |
| 0 | 0 | 0 | 0 | 0 |
| 0 | 0 | 0 | 0 | 0 |
| 0.003967915 | 0.022089035 | 0 | 0 | 0.005529829 |

| | | | | |
|---|---|---|---|---|
| 0.382087595 | 0 | 0 | 0 | 0 |
| 0 | 0.000637966 | 0 | 0 | 0 |
| 0.004187549 | 0 | 0 | 0 | 0 |
| 0 | 0 | 0 | 0 | 0 |
| 0.016669134 | 0 | 0.001064559 | 0.001796233 | 0.000294557 |
| 0 | 0 | 0 | 0 | 0 |
| 0 | 0 | 0 | 0 | 0 |
| 0.082677781 | 0.151790273 | 0 | 0.004136412 | 0.000688143 |
| 0 | 0 | 0 | 0 | 0 |
| 0 | 1.097434817 | 0.015636911 | 0.003151275 | 0 |
| 0 | 0 | 0 | 0 | 0 |
| 0.010772501 | 0.001928695 | 0.003346851 | 0.006833527 | 0 |
| 0.013104955 | 0.022968847 | 0 | 0 | 0.002809775 |
| 0 | 0 | 0.01202052 | 0 | 0.022215184 |
| 0.003488462 | 0.001504646 | 0 | 0 | 0.002039851 |
| 0 | 0 | 0 | 0 | 0 |
| 0 | 0 | 0 | 0 | 0 |
| 0 | 0 | 0 | 0 | 0 |
| 0.006972902 | 0.008979706 | 1.108860289 | 0 | 0 |
| 0 | 0 | 0 | 0 | 0.097152614 |
| 0 | 0 | 0 | 0 | 0 |
| 0 | 0 | 0 | 0 | 0 |
| 0 | 0 | 0.00061638 | 0 | 0 |
| 0 | 0 | 0 | 0 | 0 |
| 0 | 0 | 0 | 0 | 0 |
| 0.002873211 | 0 | 0 | 0.000377339 | 0.001058805 |
| 0 | 0 | 0 | 1.85466332 | 0 |
| 0.001085784 | 0 | 0.459380993 | 0 | 0 |
| 0 | 0 | 0 | 0 | 0 |
| 0 | 0 | 0.006800488 | 0 | 0 |
| 0 | 0.001828488 | 0 | 0 | 0 |
| 0 | 0 | 0 | 0 | 0 |
| 0 | 0 | 0 | 0 | 0 |
| 0 | 0 | 0 | 0 | 0 |
| 0 | 0 | 0 | 0 | 0 |
| 0 | 0 | 0.17056172 | 0 | 0 |
| 0 | 0.11165008 | 0.122840922 | 0 | 0 |
| 0 | 0 | 0 | 0 | 0 |
| 0 | 0 | 0 | 0.00089573 | 0 |
| 0 | 0.002329828 | 0 | 0 | 0 |
| 0 | 3.708114021 | 0 | 0 | 0 |
| 0 | 0 | 0 | 0 | 0 |
| 0 | 0 | 0 | 0 | 0 |
| 0 | 0.019493832 | 0.033491192 | 0 | 0 |
| 0 | 0 | 0 | 0 | 0 |
| 0 | 0 | 0 | 0 | 0 |
| 0 | 0.001271438 | 0.002595666 | 0 | 0.000598504 |
| 0 | 0 | 0 | 0 | 0 |

| | | | | |
|---|---|---|---|---|
| 0 | 0 | 0 | 0 | 0 |
| 0 | 0.001894561 | 0 | 0 | 0 |
| 0 | 0 | 0 | 0 | 0 |
| 0 | 0 | 0 | 0 | 0 |
| 0 | 0 | 0 | 0 | 0 |
| 0 | 0 | 0.028962357 | 0 | 0.02602096 |
| 0 | 0 | 0 | 0 | 0 |
| 0 | 0 | 0 | 0 | 0 |
| 0 | 0 | 0 | 0 | 0 |
| 0 | 0 | 0 | 0 | 0 |
| 0.004488031 | 0.00096789 | 0.004149533 | 0.002429098 | 0.009731935 |
| 0.001338371 | 0.009797044 | 0.019260096 | 0 | 0 |
| 0 | 0 | 0 | 0 | 0 |
| 0.002698224 | 0.002926125 | 0.000746718 | 0.001214942 | 0 |
| 0 | 0.274791139 | 0 | 0 | 0 |
| 0 | 0 | 0 | 0 | 0 |
| 0 | 0 | 0 | 0 | 0 |
| 0 | 0.011212394 | 0 | 0 | 0 |
| 0.017231067 | 0 | 0 | 0.002586237 | 0 |
| 0 | 0.016559091 | 0.066492065 | 0 | 0 |
| 0 | 0 | 0 | 0 | 0 |
| 0 | 0 | 0 | 0.074328283 | 0.050106661 |
| 0 | 0 | 0 | 0 | 0 |
| 0 | 0 | 0 | 0 | 0 |
| 0 | 0 | 0 | 0 | 0 |
| 0 | 0 | 0 | 0 | 0 |
| 0 | 0.68609512 | 0.002833741 | 0 | 0 |
| 0 | 0 | 0 | 0 | 0 |
| 0 | 0 | 0 | 0 | 0 |
| 0.478276248 | 0.250643484 | 0.088383491 | 0.137323324 | 0.206089561 |
| 0.00418407 | 0 | 0 | 0 | 0 |
| 0 | 0 | 0 | 0 | 0 |
| 0 | 0 | 0 | 0 | 0 |
| 0 | 0.011598026 | 0 | 0 | 0 |
| 0 | 0 | 0.047710544 | 0 | 0 |
| 0 | 0 | 0 | 0 | 0 |
| 0 | 0 | 0 | 0.143491107 | 0 |
| 0 | 0 | 0 | 0.000550377 | 0 |
| 0 | 0 | 0 | 0 | 0 |
| 0 | 2.645239022 | 0 | 0 | 0 |
| 0.004594912 | 0.369709596 | 0.034837892 | 0 | 0.022081502 |
| 0 | 2.608768278 | 0 | 3.549003419 | 0 |
| 0 | 0 | 0.001220137 | 0.000882317 | 0 |
| 0 | 0 | 0 | 0 | 0 |
| 0 | 0.006444882 | 0.037394603 | 0 | 0 |
| 0 | 0 | 0 | 0 | 0 |
| 0.01496697 | 0.000631211 | 0.001127551 | 0.002795539 | 0 |
| 0 | 0 | 0.000603083 | 0 | 0 |

| | | | | |
|---|---|---|---|---|
| 0.011080038 | 0 | 0.003776138 | 0.002874355 | 0.002089667 |
| 0 | 0 | 0 | 0 | 0.001194684 |
| 0 | 0.005305928 | 0 | 0.020561779 | 0.004345927 |
| 0 | 0 | 0 | 0 | 0 |
| 0 | 0.127741821 | 0 | 0 | 0 |
| 0 | 0 | 0 | 0 | 0 |
| 0 | 0 | 0 | 0 | 0 |
| 0.002299718 | 0 | 0 | 0.000805392 | 0 |
| 0 | 0 | 0.012474824 | 0 | 0.007087501 |
| 0 | 0 | 0.015990524 | 0 | 0 |
| 0 | 0 | 0 | 0 | 0 |
| 0 | 0 | 0 | 0 | 0 |
| 0 | 0 | 0 | 0 | 0 |
| 0.001485012 | 0.001056852 | 0 | 0 | 0.00119398 |
| 0 | 0 | 0 | 0 | 0 |
| 0.01900337 | 0.022024308 | 0 | 0.003556234 | 0.105826267 |
| 0.064958434 | 0.006408058 | 0.057468164 | 0.041669558 | 0.057226644 |
| 0.042195902 | 0.569827412 | 0.005487333 | 0 | 0 |
| 0 | 0.001575962 | 0.039573516 | 0 | 0.012730188 |
| 0 | 0 | 0 | 0 | 0 |
| 0 | 0.221512492 | 0 | 0 | 0 |
| 0 | 0.002123718 | 0.00560017 | 0 | 0.001399577 |
| 0 | 0 | 0 | 0 | 0 |
| 0 | 0 | 0 | 0 | 0.129766985 |
| 0 | 0 | 0.050507359 | 0 | 0 |
| 0 | 0.004421731 | 0 | 0 | 0 |
| 0 | 0.10866144 | 0.001977135 | 0 | 0.002735304 |
| 0 | 0 | 0 | 0 | 0.001384814 |
| 0 | 0 | 0 | 0 | 0 |
| 0 | 0 | 0 | 0 | 0 |
| 0.005277961 | 0.218302346 | 0.089614911 | 0.017614294 | 0.033782306 |
| 0.035178129 | 0.019545289 | 0 | 0 | 0.001984838 |
| 0 | 0.021746234 | 0.053573283 | 0.025312474 | 0.090121481 |
| 0.001263175 | 0 | 0.003058794 | 0.002654286 | 0 |
| 0 | 0.000855519 | 0 | 0 | 0 |
| 0 | 0 | 0 | 0 | 0 |
| 0.001394528 | 0.005482142 | 0 | 0 | 0 |
| 0 | 0.035333175 | 0.076660614 | 0 | 0.061880816 |
| 0 | 0 | 0 | 0.036803863 | 0.276188596 |
| 0 | 0.001813085 | 0 | 0 | 0 |
| 0 | 0 | 0 | 0 | 0 |
| 0 | 0 | 0 | 0 | 0 |
| 0 | 0 | 0.009252911 | 0.000652786 | 0.08942044 |
| 0 | 0.118059755 | 0 | 0 | 0.075098591 |
| 0 | 0 | 0.922793235 | 0 | 0 |
| 0 | 0 | 0 | 0 | 0 |
| 0 | 0 | 0 | 0 | 0 |
| 0 | 0 | 0 | 0 | 0 |

| | | | | |
|---|---|---|---|---|
| 0 | 0.007233403 | 0 | 0 | 0 |
| 3.322833606 | 0.050160407 | 0.127901581 | 0.015464471 | 0.34388183 |
| 0 | 0 | 0 | 0 | 0 |
| 0 | 0 | 0 | 0.000450282 | 0 |
| 0 | 0 | 0 | 0 | 0.001004725 |
| 0 | 0 | 0 | 0 | 0 |
| 0 | 0 | 0.159035724 | 0 | 0 |
| 0 | 0.001856794 | 0 | 0 | 0 |
| 0 | 0.000922043 | 0 | 0 | 0.005000061 |
| 0 | 0 | 0.066943284 | 0 | 0 |
| 0 | 0.000898185 | 0.009168314 | 0 | 0.007356763 |
| 0.078250025 | 0.913989324 | 0.001138039 | 0 | 0.008187091 |
| 0 | 0 | 0 | 0.435419116 | 0 |
| 0 | 0 | 0 | 0 | 0 |
| 0.002896122 | 0.003091664 | 0 | 0.000507131 | 0 |
| 0 | 0 | 0 | 0 | 0 |
| 0.054018905 | 0.001724303 | 0.000640037 | 0 | 0.006198293 |
| 0.01586904 | 0.045378146 | 0.541094891 | 0 | 0 |
| 0 | 0 | 0 | 0 | 0 |
| 0 | 0.074788643 | 0.091987203 | 0 | 0 |
| 0.140715841 | 0 | 0.001980118 | 0.088895996 | 0.018080238 |
| 0 | 0 | 0 | 0 | 0 |
| 0 | 0 | 0 | 0 | 0 |
| 0 | 0 | 0 | 0 | 0 |
| 0.001717401 | 0 | 0 | 0 | 0 |
| 0 | 0 | 0 | 0 | 0 |
| 0 | 0.244446488 | 0.019707622 | 0 | 0.021271107 |
| 0.077858198 | 0.394828841 | 0.27569901 | 0.047939188 | 0.065220058 |
| 0 | 0 | 0 | 0 | 0 |
| 0 | 0 | 0 | 0.000917871 | 0.003980361 |
| 0 | 0.000737427 | 0 | 0 | 0 |
| 0 | 0 | 0 | 0 | 0 |
| 0 | 0 | 0.006174429 | 0 | 0 |
| 0 | 0 | 0 | 0 | 0 |
| 0 | 0 | 0 | 0 | 0.480896326 |
| 0.499195136 | 0.104391866 | 0.027604443 | 0.173859034 | 0.079336385 |
| 0 | 0.000538697 | 0 | 0 | 0 |
| 0 | 0 | 0 | 0 | 0 |
| 0.012731785 | 0.007206366 | 0.002379874 | 0.003441915 | 0.004070702 |
| 0 | 0 | 0 | 0 | 0 |
| 0 | 0 | 0.846612396 | 0 | 0 |
| 0 | 0 | 0 | 0 | 0 |
| 0.001763776 | 0 | 0.008279182 | 0 | 0.093231822 |
| 0.000392975 | 0 | 0.000761276 | 0 | 0.002948962 |
| 0 | 0 | 0 | 0 | 0 |
| 0 | 0 | 0 | 0.069580888 | 0 |
| 0 | 0 | 0 | 0 | 0 |
| 0 | 0 | 0 | 0 | 0 |

| | | | | |
|---|---|---|---|---|
| 0 | 0 | 0 | 0 | 0 |
| 0 | 0 | 0 | 0 | 0 |
| 0 | 0 | 0 | 0.012366711 | 0.000733114 |
| 0.015959874 | 0 | 0 | 0.014609824 | 1.865520436 |
| 0 | 0 | 0 | 0 | 0 |
| 0 | 0 | 0 | 0 | 0 |
| 0 | 0 | 0 | 0 | 0 |
| 0 | 0 | 0 | 0 | 0.00068196 |
| 0 | 0 | 0 | 0.397653634 | 0.609628611 |
| 0 | 0 | 0 | 0.034919161 | 0 |
| 0 | 0 | 0 | 0 | 0 |
| 0.004541963 | 0.00416623 | 0.00601247 | 0 | 0 |
| 0 | 0 | 0 | 0 | 0 |
| 0 | 0 | 0 | 0 | 0 |
| 0 | 0 | 0.436491499 | 0 | 0.028661747 |
| 0 | 0 | 0 | 0 | 0 |
| 0 | 0 | 0 | 0.130925484 | 0 |
| 0 | 0 | 0 | 0.280308657 | 0 |
| 0 | 0 | 0 | 0 | 0 |
| 0 | 0 | 0 | 0 | 0 |
| 0 | 0 | 0 | 0 | 0 |
| 0 | 0 | 0 | 0 | 0 |
| 0 | 0 | 0 | 0 | 0 |
| 0 | 1.199848518 | 0.180560573 | 0 | 0.146880377 |
| 0 | 0 | 0 | 0 | 0.025680853 |
| 0 | 0 | 0 | 0.056277457 | 0 |
| 0 | 0 | 0 | 0 | 0 |
| 0 | 0 | 0 | 0.488740341 | 1.540887323 |
| 0 | 0 | 0 | 0 | 0 |
| 0 | 0 | 0 | 0 | 0 |
| 0.068146845 | 0 | 0 | 0.047525716 | 0.030768433 |
| 0 | 0 | 0 | 0 | 0 |
| 0 | 0.023597568 | 0.048174926 | 0 | 0.047014205 |
| 0 | 0 | 0 | 0 | 0 |
| 0 | 0 | 0 | 0 | 0 |
| 0 | 0 | 0 | 0.026135903 | 0 |
| 0.473211862 | 0 | 0 | 0.094829146 | 0 |
| 0 | 0 | 0 | 0 | 0 |
| 0 | 0.178117994 | 0 | 0 | 0 |
| 0 | 0 | 0 | 0 | 0 |
| 0.045314894 | 0.088204208 | 0.017866393 | 0 | 0.101439432 |
| 0 | 0 | 0 | 0 | 0 |
| 0.277902056 | 0 | 0.36030951 | 0.000854264 | 0.312767943 |
| 0 | 0.013446544 | 0.012745293 | 0 | 0.000745999 |
| 0 | 0 | 0.036450497 | 0 | 0 |
| 0 | 0 | 0 | 0 | 0 |
| 0 | 0 | 0 | 0 | 0 |
| 0.581576601 | 0 | 0.003382276 | 0 | 0.069159643 |

| | | | | |
|---|---|---|---|---|
| 0 | 0 | 0 | 0 | 0 |
| 0 | 0 | 0 | 0.045009949 | 0.030352201 |
| 0.037323333 | 0.325417175 | 0.266840756 | 0.00035169 | 0.006100431 |
| 0 | 0 | 0 | 0.002280959 | 0 |
| 0 | 0 | 0 | 0 | 0 |
| 0.022809533 | 0 | 0.001021662 | 0 | 0 |
| 0 | 0 | 0 | 0 | 0.006062269 |
| 0 | 0 | 0 | 0.012259921 | 0 |
| 0 | 0 | 0 | 0 | 0 |
| 0 | 0 | 0 | 0 | 0.006417866 |
| 0 | 0.038514877 | 0.009390904 | 0.003683168 | 0.065973032 |
| 0.168277039 | 5.924216984 | 3.195453919 | 0.164764212 | 0.261347338 |
| 0 | 0 | 0 | 0 | 0 |
| 0.249883711 | 0.000841434 | 0.048098525 | 0.047203427 | 0.052093499 |
| 1.190312387 | 1.700540193 | 0.259232744 | 0 | 0.064199426 |
| 0 | 0 | 0 | 3.175339604 | 0 |
| 0 | 0 | 0 | 0 | 0 |
| 0 | 0 | 0 | 0 | 0 |
| 0.013527612 | 0 | 0 | 0 | 0 |
| 0 | 0.041102668 | 0 | 0 | 0.024016023 |
| 0.198606841 | 0.200482863 | 0.047892793 | 0 | 0.027905946 |
| 2.192255572 | 10.73461186 | 3.348485821 | 0 | 0.004045024 |
| 1.563414903 | 24.99860436 | 0.742675252 | 0.096220797 | 0.068614067 |
| 0 | 0 | 0 | 0 | 0 |
| 0 | 0 | 0 | 0 | 0 |
| 0 | 0 | 0 | 2.085191044 | 0.002300085 |
| 0.254016197 | 3.570385435 | 1.126971711 | 0 | 0 |
| 0 | 0 | 0 | 0.000402466 | 0 |
| 0 | 0 | 0 | 0.011810525 | 0 |
| 0 | 0 | 0 | 0 | 0 |
| 0 | 0 | 0 | 0 | 0 |
| 0 | 0 | 0 | 0.246904327 | 0 |
| 0.006244416 | 2.087808638 | 3.023369172 | 0 | 0 |
| 0 | 0 | 0.042134465 | 0.002343745 | 0.030284092 |
| 0 | 0.001796859 | 0.037006931 | 0 | 0.186521637 |
| 0.330416882 | 0.004792327 | 0.021346115 | 0.052203729 | 0.422959232 |
| 0 | 0 | 0.019331302 | 0 | 0.001743329 |
| 0 | 0 | 0 | 0 | 0 |
| 0 | 0 | 0 | 0.847843762 | 0.918571506 |
| 0 | 0 | 0 | 0 | 0 |
| 0 | 0 | 0 | 0 | 0 |
| 0 | 0 | 0 | 0 | 0 |
| 0 | 0 | 0 | 0 | 0 |
| 0 | 0 | 0 | 0 | 0 |
| 0.062655052 | 0.001159265 | 0 | 0.005867673 | 0.080358316 |
| 0 | 0 | 0 | 0 | 0 |
| 0 | 0 | 0 | 0 | 0 |
| 0.840212388 | 1.156868165 | 0.241438113 | 0.065176736 | 3.933109104 |

| | | | | |
|---|---|---|---|---|
| 0 | 0 | 0 | 0 | 0 |
| 0 | 0 | 0 | 0 | 0 |
| 0 | 0 | 0 | 0 | 0 |
| 0 | 0 | 0 | 0.002708871 | 0.048954931 |
| 0 | 0 | 0 | 0 | 0 |
| 0 | 0 | 0 | 0 | 0 |
| 0 | 0 | 0 | 0.081670772 | 0.000523743 |
| 0.103614663 | 0.046887329 | 0.027045952 | 0 | 0.077035455 |
| 0 | 0 | 0 | 0 | 0 |
| 0 | 0 | 0 | 0 | 0 |
| 0.021370945 | 0.001886419 | 0.001765117 | 0.001392446 | 0.088621801 |
| 0 | 0 | 0 | 0 | 0 |
| 0 | 0.009772603 | 0 | 0.00741677 | 0 |
| 0 | 0 | 0 | 0.032576862 | 0 |
| 0 | 0 | 0 | 0 | 0 |
| 0 | 0 | 0 | 0 | 0 |
| 0 | 0.184962066 | 0 | 0 | 0 |
| 0.002206614 | 0 | 0 | 0.001062581 | 0.077619648 |
| 0 | 0 | 0 | 0 | 0.229549733 |
| 0 | 0 | 0 | 0 | 0 |
| 0 | 0 | 0 | 0 | 0 |
| 0 | 0 | 0 | 0 | 0 |
| 0 | 0 | 0 | 0.012696281 | 0.342021875 |
| 0 | 0 | 0 | 0 | 0 |
| 0 | 0 | 0 | 0 | 0 |
| 0 | 0 | 0 | 0 | 0 |
| 0 | 0 | 0 | 0 | 0 |
| 0 | 0 | 0 | 0 | 0 |
| 0 | 0 | 0 | 0 | 0 |
| 0 | 0 | 0 | 0.012835883 | 1.341448028 |
| 0 | 0 | 0 | 0 | 0 |
| 0 | 0 | 0 | 0 | 0 |
| 0 | 0 | 0.036847037 | 0 | 0 |
| 0 | 0 | 0 | 0 | 0 |
| 0.003424973 | 0 | 0 | 0 | 0.001200352 |
| 0 | 0 | 0 | 0 | 0 |
| 0.088028088 | 0 | 0 | 0.030994824 | 0.024609588 |
| 0 | 0.007851361 | 0.027982335 | 0 | 0.102983237 |
| 0 | 0 | 0 | 0 | 0 |
| 0 | 0 | 0 | 0 | 0 |
| 0 | 0 | 0.174476761 | 0 | 0 |
| 0 | 0 | 0 | 0 | 0 |
| 0 | 0 | 0 | 0 | 0 |
| 0 | 0 | 0 | 0.000476953 | 0.008686907 |
| 0 | 0.667879882 | 0 | 0.045060654 | 0 |
| 0 | 0 | 0 | 0 | 0 |
| 0 | 0 | 0 | 0 | 0 |
| 0 | 0 | 0 | 0 | 0 |

| | | | | |
|---|---|---|---|---|
| 0 | 0 | 0 | 0 | 0 |
| 0 | 0 | 0 | 0 | 0 |
| 0 | 0 | 0 | 0 | 0 |
| 0 | 0.007412694 | 0 | 0.102983745 | 0 |
| 0 | 0 | 0 | 0 | 0 |
| 0 | 0 | 0 | 0.016811242 | 0 |
| 0 | 0 | 0 | 0 | 0 |
| 0 | 0 | 0 | 0 | 0 |
| 0.357621591 | 0.364859915 | 0.407092704 | 0.001761137 | 0.044579099 |
| 0 | 0 | 0 | 0 | 0 |
| 0 | 0 | 0 | 0 | 0 |
| 0 | 0 | 0 | 0 | 0 |
| 0 | 0 | 0 | 0 | 0 |
| 0 | 0 | 0 | 0 | 0.001296134 |
| 0 | 0 | 0 | 0.010611566 | 0.706350316 |
| 0 | 0.167338375 | 0.782444917 | 0 | 0.001598063 |
| 0 | 0 | 0 | 0 | 0.021377839 |
| 0 | 0 | 0 | 0 | 0 |
| 0 | 0 | 0 | 0 | 0 |
| 0 | 0 | 0 | 0 | 0 |
| 0 | 0 | 0 | 0 | 0.044947428 |
| 0 | 0 | 0 | 0 | 0 |
| 0 | 0 | 0 | 0 | 0 |
| 0 | 0 | 0 | 0 | 0 |
| 0.002274488 | 0.002589929 | 0.011015403 | 1.698577183 | 0 |
| 5.632190349 | 0 | 0 | 0.207367575 | 2.045639023 |
| 0.143984906 | 0 | 0 | 0.216159946 | 0 |
| 0 | 0.00522499 | 0.000547022 | 0 | 0.000908145 |
| 0 | 0 | 0.074733552 | 0.016714037 | 0.070775219 |
| 0 | 0 | 0 | 0 | 0 |
| 0 | 0 | 0 | 0 | 0 |
| 0 | 0.008756624 | 0 | 0 | 0.053468296 |
| 0 | 0 | 0.001386897 | 0 | 0 |
| 0.028910397 | 0.004841159 | 0.000705952 | 0 | 0.024416554 |
| 0.028821299 | 0.002279057 | 0.000969321 | 0.016261928 | 0.012552 |
| 0.040321887 | 0.010471612 | 0.003014848 | 0.000396387 | 0.107610438 |
| 0 | 0 | 1.733384878 | 0 | 0 |
| 0 | 0 | 0 | 0.174000217 | 0.120055053 |
| 0 | 0 | 0 | 0 | 0 |
| 0 | 0 | 0 | 0 | 0 |
| 0 | 0 | 0 | 0 | 0 |
| 0.001283518 | 0 | 0 | 0 | 0 |
| 0 | 0.034122998 | 0.22247156 | 0.609107181 | 0.063629084 |
| 0 | 0 | 0 | 0.078960976 | 0 |
| 0 | 0 | 0 | 0 | 0 |
| 0 | 0.016788603 | 0.03764978 | 0 | 0.002801937 |
| 0 | 0 | 0 | 0 | 0 |
| 0.001067668 | 0.041537747 | 0.00703221 | 0.001196518 | 0.482436007 |

| | | | | |
|---|---|---|---|---|
| 0 | 0.107882764 | 0 | 0 | 0 |
| 0 | 0 | 0 | 0 | 0 |
| 0 | 0 | 0 | 0 | 0 |
| 0 | 0 | 0 | 0 | 0 |
| 0 | 0.001053037 | 0.010134759 | 0 | 0.000509859 |
| 0 | 0 | 0 | 0 | 0.007855784 |
| 0 | 0 | 0 | 0 | 0 |
| 0 | 0 | 0 | 0 | 0 |
| 0 | 0 | 0 | 0 | 0 |
| 0 | 0 | 0 | 0 | 0 |
| 0 | 0 | 0 | 0 | 0 |
| 0.290029274 | 0.222749691 | 3.013945968 | 0.201402852 | 1.314312665 |
| 0.858047812 | 0.301092385 | 0 | 0.011831767 | 0 |
| 0 | 0 | 0 | 0 | 0 |
| 0.431410047 | 0.017571654 | 0.077606221 | 0.021446073 | 0 |
| 0 | 0 | 0 | 0 | 0 |
| 0 | 0 | 0 | 0 | 0.006289549 |
| 0 | 0 | 0.011975205 | 0 | 0 |
| 0 | 0.003964741 | 0 | 0 | 0.001636621 |
| 0 | 0 | 0 | 0.002246586 | 0 |
| 0 | 0 | 0 | 0 | 0 |
| 0 | 0 | 0 | 0 | 0 |
| 0.122820242 | 0.003113792 | 0 | 0.309301868 | 3.722180831 |
| 0 | 0 | 0 | 0 | 0 |
| 0 | 0 | 0 | 0 | 0 |
| 0 | 0 | 0 | 0 | 0 |
| 0 | 0 | 0 | 0 | 0 |
| 0 | 0 | 0 | 0 | 0 |
| 0 | 0 | 0 | 0 | 0 |
| 0 | 0 | 0 | 0 | 0 |
| 0 | 0 | 0 | 0 | 0 |
| 0.34069481 | 0.057117077 | 0.004608921 | 0 | 0.038640358 |
| 0 | 0 | 0 | 0 | 0 |
| 0 | 0 | 0.040294406 | 0 | 0.101935614 |
| 0.035874084 | 0.008765952 | 0.002684381 | 0.000431368 | 0.097878116 |
| 0 | 0.007156904 | 0 | 0.031418836 | 0.067131143 |
| 0 | 0 | 0 | 0 | 0 |
| 0 | 0 | 0 | 0.005671707 | 0 |
| 0 | 0 | 0 | 0 | 0.002844372 |
| 0 | 0 | 0 | 0 | 0 |
| 0 | 0 | 0 | 0.077823857 | 0 |
| 0 | 0 | 0 | 0 | 0 |
| 0 | 0 | 0 | 0 | 0 |
| 0 | 0.005709034 | 0 | 0 | 0 |
| 0.231906437 | 0.008491419 | 0.025709293 | 0.003399525 | 0.066827182 |
| 0 | 0 | 0 | 0 | 0 |
| 0 | 0 | 0 | 0 | 0 |
| 0 | 0 | 0.003890623 | 0 | 0.078892856 |

| | | | | |
|---|---|---|---|---|
| 0 | 0 | 0 | 0 | 0 |
| 0 | 0 | 0 | 0 | 0 |
| 0 | 0.00342359 | 0 | 0 | 0.074497328 |
| 0 | 0 | 0 | 0 | 0 |
| 0 | 0 | 0 | 0 | 0 |
| 0 | 0 | 0 | 0 | 0.018228562 |
| 0 | 0 | 0 | 0.045475973 | 0.002776697 |
| 0 | 0.10922798 | 0 | 0.001348951 | 0 |
| 0 | 0 | 0 | 0 | 0 |
| 0 | 0 | 0 | 0 | 0 |
| 0 | 0 | 0 | 0 | 0 |
| 0.005586285 | 0 | 0.000927584 | 0 | 0.000684417 |
| 0 | 0 | 0 | 0 | 0 |
| 0 | 0 | 0 | 0.080472709 | 0 |
| 0 | 0 | 0 | 0 | 0 |
| 0 | 0 | 0 | 0 | 0 |
| 0 | 0 | 0 | 0 | 0 |
| 0 | 0 | 0 | 0 | 0 |
| 0 | 9.495302715 | 0 | 0.01491027 | 0.607982385 |
| 0 | 0 | 0 | 0 | 0.000775484 |
| 0 | 0.009196669 | 0 | 0.000598981 | 0 |
| 0 | 0 | 0 | 0 | 0 |
| 0 | 0 | 0 | 0 | 0 |
| 0 | 0 | 0 | 0 | 0 |
| 0.001657148 | 0 | 0.00947022 | 0 | 0.001954159 |
| 0.008048449 | 0.000803918 | 0.016959218 | 0 | 0.130027898 |
| 0 | 0 | 0 | 0 | 0.000312527 |
| 0 | 0.008262193 | 0.047791075 | 0 | 0.036670179 |
| 0 | 0 | 0 | 0 | 0 |
| 0.405182783 | 0.290740236 | 0 | 0.054868199 | 0.751317309 |
| 0 | 0 | 0 | 0 | 0 |
| 0 | 0 | 0 | 0 | 0 |
| 0 | 0 | 0 | 0 | 0.049750028 |
| 0 | 0 | 0 | 0 | 0 |
| 0.002663456 | 0.000702047 | 0.017342239 | 0 | 0.020304346 |
| 0 | 0 | 0 | 0 | 0.000956545 |
| 0 | 0 | 0 | 0.357367039 | 0 |
| 0.000839512 | 0 | 0 | 0 | 0.002159952 |
| 0 | 0 | 0 | 0 | 0 |
| 0 | 0 | 0 | 0 | 0 |
| 4.598344268 | 1.94513793 | 3.965722829 | 1.776473058 | 2.087679689 |
| 2.10868597 | 0.040702299 | 0 | 0.087291407 | 0.113180011 |
| 0 | 0 | 0 | 0 | 0 |
| 0 | 0 | 0 | 0 | 0 |
| 0 | 0 | 0 | 0 | 0 |
| 0 | 0 | 0 | 0 | 0 |
| 0 | 0 | 0 | 0 | 0 |
| 0.001465823 | 0.002276062 | 0.000387219 | 0.000560019 | 0.002357104 |

| | | | | |
|---|---|---|---|---|
| 0 | 0.00390501 | 0 | 0 | 0 |
| 0 | 0.016387827 | 0.014338325 | 0 | 0.02542026 |
| 0 | 0 | 0 | 0 | 0.000600653 |
| 0 | 0 | 0 | 0 | 0 |
| 0 | 0.703405101 | 0.734076579 | 1.120112677 | 1.123547302 |
| 0 | 0 | 0 | 0 | 0 |
| 0 | 0 | 0 | 0 | 0 |
| 0 | 0 | 0 | 0 | 0 |
| 0.05419788 | 0.350390246 | 5.600272993 | 0 | 0.007922949 |
| 0 | 0 | 0 | 0 | 0 |
| 0 | 0 | 0.003299862 | 0 | 0 |
| 0 | 0 | 0 | 0 | 0 |
| 0.030232407 | 0 | 0 | 0.003824668 | 0.128926035 |
| 0.014408612 | 0.005903996 | 0.010942973 | 0.002867098 | 0.003510553 |
| 0 | 0 | 0 | 0.02791622 | 0 |
| 0.434920478 | 0 | 0 | 0.015810275 | 0.134746334 |
| 0 | 0.05176216 | 0.003842674 | 0 | 0.016632167 |
| 0 | 0 | 0 | 0 | 0 |
| 0 | 0 | 0 | 0 | 0 |
| 0 | 0 | 0 | 0 | 0 |
| 0 | 0 | 0 | 0 | 0 |
| 0 | 0 | 0 | 0 | 0 |
| 0 | 0 | 0 | 0 | 0 |
| 0 | 0 | 0 | 0 | 0 |
| 0 | 0 | 0 | 0 | 0 |
| 0 | 0 | 0 | 0 | 0 |
| 0 | 0 | 0 | 0.023903504 | 0.147265054 |
| 0.020713983 | 0.002770254 | 0 | 0.000876361 | 0.005216166 |
| 0 | 0 | 0 | 0 | 0 |
| 0 | 0 | 0 | 0 | 0 |
| 0 | 0 | 0 | 0 | 0 |
| 0 | 0.000440764 | 0 | 0 | 0 |
| 0 | 0 | 0 | 0 | 0 |
| 0 | 0 | 0 | 0 | 0.053117074 |
| 0 | 0 | 0 | 0.075212003 | 0.219974093 |
| 0.004342067 | 0.00350981 | 0 | 0 | 0.389280572 |
| 0 | 0.002183664 | 0 | 0 | 0 |
| 0 | 0 | 0 | 0 | 0.036303295 |
| 0 | 0 | 0 | 0 | 0 |
| 0 | 0 | 0 | 0 | 0 |
| 0 | 0 | 0 | 0.010760569 | 1.356298811 |
| 0 | 0 | 0 | 0.0095803 | 0.417126762 |
| 0 | 0 | 0 | 0 | 0 |
| 0 | 0 | 0 | 0 | 0 |
| 0 | 0 | 0 | 0 | 0 |
| 0 | 0 | 0 | 0 | 0.000460952 |
| 0 | 0 | 0 | 0 | 0 |
| 0 | 0 | 0 | 0 | 0 |

| | | | | |
|---|---|---|---|---|
| 0 | 0 | 0 | 0.062902703 | 0 |
| 0 | 0 | 0 | 0 | 0 |
| 0 | 0 | 0 | 0 | 0 |
| 0 | 0 | 0 | 0 | 0.002954446 |
| 0 | 0 | 0 | 0.073139406 | 0 |
| 0 | 0 | 0 | 0 | 0 |
| 0 | 0.115745776 | 0.027380928 | 0 | 0.019167609 |
| 0 | 0 | 0 | 0 | 0.0004394 |
| 0 | 0 | 0 | 0 | 0 |
| 0 | 0 | 0.13544804 | 0 | 0.006676818 |
| 0 | 0 | 0 | 0 | 0 |
| 0 | 0 | 0 | 0 | 0 |
| 0 | 0 | 0 | 0 | 0 |
| 0 | 0 | 0 | 0 | 0 |
| 0 | 0 | 0 | 0 | 0 |
| 0 | 0 | 0 | 0 | 0 |
| 0 | 0 | 0 | 0 | 0 |
| 0 | 0 | 0.002500769 | 0 | 0 |
| 0 | 0 | 0 | 0 | 0 |
| 0 | 0 | 0 | 0 | 0 |
| 0.039158565 | 0.009321367 | 0 | 0.009142574 | 0.281519407 |
| 0 | 0 | 0 | 0 | 0 |
| 0 | 0 | 0.045142368 | 0.002287144 | 0 |
| 0.00197972 | 0.003757133 | 0.022499447 | 0 | 0.061759343 |
| 0 | 0 | 0 | 0 | 0 |
| 0 | 0.002216286 | 0 | 0 | 0.000417309 |
| 0 | 0 | 0 | 0 | 0 |
| 0 | 0 | 0 | 0 | 0 |
| 0 | 0 | 0 | 0 | 0 |
| 0.032481799 | 2.483076186 | 0 | 0 | 0.001730888 |
| 0 | 0 | 0 | 0 | 0 |
| 0 | 0 | 0 | 0 | 0 |
| 0 | 0 | 0 | 0 | 0 |
| 0 | 0 | 0 | 0 | 0 |
| 0 | 0 | 0 | 0 | 0 |
| 0 | 0 | 0 | 0 | 0 |
| 0 | 0.002350574 | 0.220102427 | 0 | 0 |
| 0 | 0 | 0 | 0 | 0 |
| 0 | 0 | 0 | 0 | 0 |
| 0 | 0 | 0 | 0 | 0 |
| 0 | 0 | 0 | 0 | 0 |
| 0 | 0 | 0 | 0 | 0 |
| 0 | 0 | 0 | 0 | 0 |
| 0 | 0 | 0.004426524 | 0 | 0 |
| 0 | 0 | 0 | 0.065355618 | 0 |
| 0 | 0 | 0 | 0 | 0 |
| 0 | 0 | 0 | 0 | 0 |
| 0 | 0 | 0 | 0.007717634 | 0.006412285 |

| | | | | |
|---|---|---|---|---|
| 0 | 0.018462161 | 0.026411132 | 0.000994723 | 0.087617397 |
| 0 | 0 | 0 | 0.002830172 | 0 |
| 0 | 0 | 0 | 0.043620557 | 0 |
| 0 | 0 | 0 | 0 | 0 |
| 0 | 0 | 0 | 0 | 0 |
| 0 | 0 | 0 | 0 | 0 |
| 0 | 0 | 0 | 0 | 0 |
| 6.886006751 | 0.455508672 | 0.270476428 | 0.114596066 | 0.680078513 |
| 0.110789417 | 0.057746742 | 0.029796671 | 10.37746666 | 0.009439413 |
| 0.058816124 | 0.257554983 | 0.682883633 | 0.128915419 | 1.849235437 |
| 0 | 0 | 0 | 0 | 0 |
| 0 | 0 | 0 | 0 | 0 |
| 0 | 0 | 0 | 0 | 0 |
| 0 | 0 | 0 | 0.001387088 | 0 |
| 0 | 0 | 0.004993684 | 0 | 0 |
| 0 | 0 | 0 | 0 | 0 |
| 0 | 0 | 0 | 0 | 0 |
| 0 | 0 | 0 | 0.21285874 | 0 |
| 0 | 0 | 0 | 0 | 0 |
| 0 | 0 | 0 | 0 | 0 |
| 0 | 0 | 0 | 0 | 0 |
| 0 | 0 | 0 | 0 | 0 |
| 0 | 0 | 0 | 0 | 0 |
| 0 | 0 | 0 | 0 | 0 |
| 0 | 0 | 0 | 0.013554238 | 0.017327658 |
| 0 | 0 | 0 | 0 | 0 |
| 0 | 0 | 0 | 0 | 0 |
| 0 | 0 | 0 | 0 | 0 |
| 0 | 0 | 0 | 0 | 0 |
| 0 | 0 | 0 | 0 | 0 |
| 0 | 0 | 0 | 0 | 0.01673547 |
| 0 | 0 | 0 | 0 | 0 |
| 0 | 0 | 0 | 0 | 0 |
| 0 | 0 | 0 | 0 | 0 |
| 0 | 0 | 0 | 0 | 0.03004332 |
| 0 | 0 | 0 | 0 | 0 |
| 0 | 0 | 0 | 0 | 0 |
| 0.255840268 | 0 | 0 | 0 | 0 |
| 0 | 0 | 0 | 0 | 0 |
| 0 | 0.019641815 | 0 | 0 | 0 |
| 0 | 0 | 0 | 0 | 0 |
| 0 | 0.598621307 | 0.163494197 | 0.158695335 | 0.530623065 |
| 0 | 0 | 0 | 0 | 0 |
| 0 | 0 | 0 | 0 | 0 |
| 0 | 0 | 0.000535284 | 0 | 0 |
| 0 | 0 | 0 | 0 | 0 |
| 0 | 0 | 0 | 0 | 0 |
| 0 | 0 | 0 | 0 | 0 |

| | | | | |
|---|---|---|---|---|
| 0 | 0 | 0 | 0 | 0 |
| 0 | 0.028217479 | 0 | 0 | 0.000755746 |
| 0 | 0 | 0 | 0 | 0 |
| 0 | 0 | 0 | 0 | 0 |
| 0 | 0.198450374 | 0.083145816 | 0.586942555 | 2.706053581 |
| 0 | 0 | 0 | 0 | 0 |
| 0 | 0 | 0 | 0 | 0 |
| 0.049318913 | 0.045256163 | 0.042346058 | 0.000759216 | 0.082018207 |
| 0 | 0 | 0 | 0 | 0.012182302 |
| 0 | 0 | 0 | 0 | 0 |
| 0 | 0 | 0 | 0 | 0 |
| 0 | 0 | 0 | 0 | 0 |
| 0 | 0 | 0 | 0 | 0 |
| 0 | 0 | 0 | 0 | 0.002831063 |
| 0 | 0 | 0.010716865 | 0 | 0.004021886 |
| 0 | 0 | 0.04112752 | 0 | 0.04316737 |
| 0 | 0 | 0 | 0 | 0 |
| 0 | 0 | 0 | 0 | 0 |
| 0 | 0 | 0 | 0 | 0 |
| 0 | 0 | 0 | 0 | 0 |
| 0 | 0 | 0 | 0 | 0 |
| 0 | 0 | 0 | 0 | 0 |
| 0 | 0 | 0 | 0 | 0 |
| 0 | 0.00074177 | 0.000908604 | 0 | 0.066035706 |
| 0 | 0 | 0 | 0.071611063 | 0 |
| 0 | 0 | 0 | 0 | 0 |
| 0.181314882 | 0.529992093 | 0.136287514 | 0 | 0 |
| 0 | 0 | 0.002625116 | 0 | 0.002340471 |
| 0.002125412 | 0 | 0.000617605 | 0 | 0 |
| 0 | 0 | 0 | 0 | 0 |
| 0 | 0 | 0 | 0 | 0 |
| 0 | 0 | 0 | 0 | 0 |
| 0 | 0 | 0 | 0 | 0 |
| 0 | 0 | 0 | 0 | 0 |
| 0 | 0 | 0 | 0 | 0 |
| 0 | 0 | 0 | 0 | 0 |
| 0 | 0 | 0 | 0 | 0 |
| 0 | 0 | 0 | 0 | 0 |
| 0 | 0 | 0 | 0 | 0 |
| 0 | 0 | 0 | 0.00026436 | 0 |
| 0 | 0 | 0 | 0 | 0 |
| 0 | 0 | 0 | 0.029915673 | 0 |
| 0 | 0 | 0 | 0 | 0 |
| 0 | 0 | 0 | 0 | 0 |
| 0 | 0 | 0 | 0 | 0 |
| 0 | 0 | 0 | 0 | 0 |
| 0 | 0 | 0 | 0 | 0 |
| 0 | 0 | 0 | 0 | 0 |

| | | | | |
|---|---|---|---|---|
| 0 | 0 | 0 | 0.078081961 | 0.153888829 |
| 0 | 0 | 0 | 0 | 0 |
| 0 | 0 | 0 | 0 | 0 |
| 0 | 0 | 0 | 0 | 0.000358574 |
| 0 | 0 | 0 | 0 | 0 |
| 0.003583264 | 0 | 0 | 0.08500635 | 0.037578502 |
| 0 | 0 | 0 | 0 | 0 |
| 0 | 0 | 0 | 0 | 0 |
| 0 | 0 | 0 | 0 | 0 |
| 0 | 0 | 0 | 0 | 0 |
| 0 | 0 | 0 | 0 | 0 |
| 0 | 0 | 0 | 0 | 0.007570673 |
| 0 | 0 | 0 | 0 | 0.006714608 |
| 0 | 0 | 0 | 0 | 0 |
| 0 | 0 | 0 | 0 | 0 |
| 0 | 0.001765835 | 0 | 0 | 0 |
| 0 | 0 | 0 | 0 | 0 |
| 0 | 0 | 0 | 0 | 0 |
| 0 | 0 | 0 | 0 | 0 |
| 0 | 0 | 0 | 0 | 0 |
| 0 | 0.002216009 | 0.089274164 | 0 | 0.041809122 |
| 0 | 0 | 0 | 0 | 0 |
| 0 | 0.007463088 | 0.006856222 | 0.000964044 | 0.001580895 |
| 0 | 0 | 0 | 0.001764686 | 0 |
| 0 | 0 | 0 | 0 | 0 |
| 0 | 0 | 0 | 0.019738421 | 0 |
| 0 | 0 | 0 | 0 | 0 |
| 0 | 0 | 0 | 0 | 0 |
| 0 | 0 | 0 | 0.015975782 | 0 |
| 0 | 0 | 0 | 0 | 0 |
| 0 | 0 | 0 | 0 | 0 |
| 0 | 0 | 0 | 0 | 0 |
| 0.05616184 | 0.026558774 | 5.899310343 | 0.000644873 | 0.011547909 |
| 0 | 0 | 0 | 0 | 0 |
| 0.793571975 | 2.719516685 | 0 | 0.013959436 | 0 |
| 0.03679283 | 0.003785738 | 0.194790936 | 0 | 0.002692891 |
| 0 | 0 | 0 | 0 | 0 |
| 0 | 0 | 0 | 0 | 0 |
| 0 | 0.008919788 | 0.254773511 | 0 | 0.009619095 |
| 0 | 0 | 0 | 0 | 0 |
| 0 | 0 | 0 | 0 | 0 |
| 0 | 0 | 0 | 0 | 0 |
| 0 | 0 | 0.005226431 | 0.048432153 | 0.08762419 |
| 0 | 0 | 0 | 0 | 0 |
| 0 | 0 | 0 | 0 | 0 |
| 0 | 0 | 0 | 0.00478015 | 0.004934169 |
| 0 | 0 | 0 | 0 | 0 |
| 0 | 0 | 0 | 0 | 0 |

| | | | | |
|---|---|---|---|---|
| 0 | 0 | 0 | 0 | 0 |
| 0 | 0 | 0 | 0 | 0 |
| 0 | 0 | 0.227829239 | 0 | 0 |
| 0 | 0 | 0 | 0 | 0 |
| 0 | 0 | 0 | 0 | 0 |
| 0 | 0 | 0 | 0 | 0 |
| 0 | 0 | 0 | 0 | 0 |
| 0.182045647 | 0.00058212 | 0 | 0 | 0.002137366 |
| 0 | 0 | 0 | 0.005103472 | 0.018969664 |
| 0 | 0 | 0 | 0.004465501 | 0.182694174 |
| 0 | 0 | 0 | 0 | 0 |
| 0 | 0 | 0 | 0 | 0 |
| 0 | 0 | 0 | 0 | 0 |
| 0 | 0 | 0 | 0.293669164 | 0 |
| 0 | 0 | 0 | 0 | 0.00545248 |
| 0 | 0.003357643 | 0.333231838 | 0 | 0.006724493 |
| 0 | 0 | 0 | 0 | 0 |
| 0 | 0 | 0 | 0 | 0 |
| 0 | 0 | 0 | 0 | 0 |
| 0 | 0 | 0 | 0.187694097 | 2.565961733 |
| 0 | 0 | 0 | 0.720553172 | 0.003151461 |
| 0 | 0 | 0 | 0 | 0 |
| 0 | 0 | 0 | 0 | 0 |
| 0 | 0 | 0 | 0 | 0 |
| 0 | 0 | 0 | 0 | 0.00279641 |
| 0 | 0 | 0 | 0 | 0 |
| 0 | 0 | 0 | 0 | 0 |
| 0 | 0 | 0 | 0 | 0 |
| 0 | 0 | 0 | 0 | 0 |
| 0 | 0 | 0 | 0.161477211 | 0 |
| 0 | 0.141411727 | 0 | 0.042759979 | 0.266266932 |
| 0 | 0 | 0 | 0 | 0 |
| 0 | 0 | 0 | 0 | 0 |
| 0 | 0 | 0 | 0 | 0 |
| 0 | 0 | 0 | 0 | 0 |
| 0 | 0 | 0 | 0 | 0 |
| 0 | 0 | 0 | 0 | 0 |
| 0 | 0 | 0 | 0 | 0 |
| 0 | 0 | 0 | 0.001341538 | 0 |
| 0 | 0 | 0 | 0 | 0 |
| 0 | 0 | 0 | 0 | 0 |
| 0 | 0 | 0 | 0 | 0 |
| 0 | 0 | 0 | 0 | 0 |
| 0 | 0.002608322 | 0 | 0 | 0 |
| 0 | 0 | 0 | 0 | 0 |
| 0 | 0 | 0 | 0 | 0 |
| 0.314643837 | 0.040621396 | 0.002551675 | 0 | 0 |
| 0 | 0 | 0 | 0 | 0 |

| | | | | |
|---|---|---|---|---|
| 0 | 0 | 0 | 0 | 0 |
| 0 | 0 | 0 | 0 | 0 |
| 0 | 0 | 0 | 0 | 0 |
| 0 | 0 | 0 | 0 | 0 |
| 0.153510168 | 0 | 0.036457825 | 0.086672939 | 0.015362238 |
| 0 | 0 | 0 | 0 | 0 |
| 0 | 0 | 0 | 0 | 0 |
| 0 | 0 | 0 | 0 | 0 |
| 0 | 0 | 0 | 0 | 0 |
| 0 | 0 | 0 | 0 | 0 |
| 0 | 0 | 0 | 0 | 0 |
| 0.04474291 | 0.019769957 | 0.011413499 | 0.001794226 | 0.015469834 |
| 0 | 0 | 0.003425034 | 0 | 0.00710764 |
| 0 | 0 | 0 | 0 | 0 |
| 0 | 0.00049397 | 0.063532363 | 0 | 0 |
| 0 | 0 | 0 | 0.032928257 | 0 |
| 0 | 0.001121713 | 0.005470557 | 0 | 0.044494813 |
| 0 | 0.000848425 | 0 | 0 | 0 |
| 0 | 1.16016913 | 0.290082913 | 0 | 0 |
| 0.000675365 | 0.007850507 | 0.002780189 | 0 | 0.000814511 |
| 0 | 0 | 0 | 0 | 0 |
| 0 | 0 | 0 | 0 | 0 |
| 0 | 0 | 0 | 0 | 0 |
| 0 | 0 | 0 | 0 | 0 |
| 0 | 0 | 0 | 0 | 0 |
| 0 | 0 | 0 | 0 | 0 |
| 0 | 0.049429057 | 0.049013647 | 0 | 0.033505541 |
| 0 | 0 | 0 | 0 | 0.0061433 |
| 0 | 0 | 0 | 0 | 0 |
| 0 | 0 | 0 | 0 | 0 |
| 0 | 0.002183004 | 0.006090752 | 0 | 0.011509179 |
| 0 | 0 | 0 | 0 | 0 |
| 0 | 0 | 0 | 0 | 0 |
| 0 | 0 | 0 | 0 | 0 |
| 0 | 0 | 0 | 0 | 0 |
| 0 | 0 | 0 | 0 | 0 |
| 0 | 0 | 0 | 0 | 0 |
| 0.022412682 | 0.007157337 | 0.011272001 | 0.00067926 | 0.025470947 |
| 0 | 0 | 0 | 0.001634021 | 0 |
| 0 | 0.019949534 | 0 | 0 | 0 |
| 0.002259023 | 0 | 0.002709077 | 0.005048229 | 0.001902792 |
| 0 | 0 | 0 | 0 | 0 |
| 0 | 0 | 0 | 0 | 0 |
| 0 | 0.02259143 | 0.010833763 | 0 | 0 |
| 0 | 0 | 0 | 0 | 0 |
| 0 | 0 | 0 | 0 | 0 |
| 0 | 0 | 0 | 0 | 0 |
| 0 | 0 | 0 | 0 | 0 |

| | | | | |
|---|---|---|---|---|
| 0 | 0 | 0 | 0 | 0 |
| 0 | 0 | 0 | 0 | 0 |
| 0 | 0 | 0 | 0 | 0 |
| 0 | 0 | 0 | 0.004105147 | 0.016156459 |
| 0.00559122 | 0 | 0 | 0.000326354 | 0.000332997 |
| 0 | 0.097003528 | 0.000776607 | 0.000561587 | 0 |
| 0 | 0 | 0 | 0 | 0 |
| 0 | 0 | 0.028922385 | 0.000917309 | 0 |
| 0 | 0 | 0 | 0 | 0 |
| 0 | 0 | 0 | 0 | 0 |
| 0 | 0 | 0 | 0 | 0 |
| 0 | 0 | 0 | 0 | 0 |
| 0 | 0 | 0 | 0.057866856 | 0.661009076 |
| 0 | 0 | 0 | 0 | 0 |
| 0 | 0 | 0 | 0 | 0 |
| 0 | 0 | 0 | 0 | 0 |
| 0 | 0 | 0 | 0 | 0 |
| 0 | 0 | 0 | 0 | 0 |
| 0 | 0 | 0 | 0 | 0 |
| 0 | 0 | 0 | 0 | 0 |
| 0 | 0 | 0 | 0 | 0 |
| 0 | 0 | 0 | 0 | 0 |
| 0 | 0.024323958 | 0.042390862 | 0 | 0.225453589 |
| 0 | 0 | 0 | 0.008268088 | 0 |
| 0 | 0 | 0 | 0 | 0 |
| 0 | 0 | 0 | 0 | 0 |
| 0 | 0 | 0 | 0.006798845 | 0 |
| 0 | 0 | 0 | 0 | 0 |
| 0 | 0 | 0 | 0 | 0 |
| 0 | 0 | 0 | 0 | 0 |
| 0.032938391 | 0.010087728 | 0 | 0.004150329 | 0.022419599 |
| 0 | 0 | 0 | 0 | 0.027233871 |
| 0 | 0 | 0 | 0 | 0 |
| 0 | 0 | 0 | 0 | 0 |
| 0 | 0 | 0 | 0 | 0 |
| 0 | 0 | 0.012881945 | 0 | 0 |
| 0 | 0.035314952 | 0.113524628 | 0 | 0 |
| 0 | 0 | 0.043248388 | 0 | 0 |
| 0.001179605 | 0 | 0.001428214 | 0 | 0 |
| 0 | 0 | 0 | 0 | 0 |
| 0 | 0 | 0 | 0 | 0 |
| 0 | 0 | 0 | 0 | 0 |
| 0 | 0 | 0 | 0 | 0 |
| 0 | 0 | 0 | 0 | 0 |
| 0 | 0 | 0 | 0 | 0 |
| 0.231831386 | 0 | 0 | 0.225079877 | 0.39942765 |
| 0 | 0 | 0 | 0 | 0 |
| 0 | 0 | 0 | 0 | 0 |

| | | | | |
|---|---|---|---|---|
| 0 | 0 | 0 | 0 | 0 |
| 0 | 0 | 0 | 0 | 0 |
| 0 | 0 | 0 | 0.060955903 | 0 |
| 0 | 0 | 0 | 0 | 0 |
| 0 | 0 | 0 | 0 | 0 |
| 0 | 0 | 0 | 0 | 0.015621005 |
| 0 | 0 | 0 | 0 | 0 |
| 0 | 0 | 0 | 0 | 0 |
| 0 | 0 | 0.001057182 | 0 | 0 |
| 0 | 0 | 0 | 0 | 0 |
| 0.001674707 | 0 | 0 | 0 | 0.000384714 |
| 0 | 0 | 0.001729889 | 0.000500374 | 0.004020656 |
| 0 | 0 | 0 | 0 | 0 |
| 0 | 0 | 0 | 0 | 0 |
| 0 | 0 | 0 | 0 | 0 |
| 0 | 0 | 0 | 0 | 0 |
| 0 | 0 | 0 | 0 | 0 |
| 0 | 0 | 0 | 0 | 0 |
| 0 | 0 | 0 | 0 | 0 |
| 0 | 0 | 0.002458541 | 0.003000111 | 0.002380919 |
| 0 | 0 | 0 | 0 | 0 |
| 0 | 0 | 0 | 0 | 0 |
| 0 | 0 | 0 | 0 | 0 |
| 0 | 0 | 0 | 0 | 0 |
| 0 | 0 | 0 | 0 | 0 |
| 0 | 0 | 0 | 0 | 0 |
| 0 | 0 | 0 | 0 | 0 |
| 0 | 0 | 0 | 0.006271357 | 0 |
| 0 | 0 | 0 | 0 | 0 |
| 0 | 0 | 0 | 0 | 0 |
| 0.007495814 | 0.002457156 | 0.000660044 | 0.000668216 | 0.008620136 |
| 0 | 0 | 0 | 0 | 0 |
| 0 | 0.12598282 | 0.074185418 | 0.023830209 | 0.002629445 |
| 0.022962521 | 0 | 0.00422904 | 0 | 0.001213487 |
| 0 | 0 | 0 | 0 | 0 |
| 0 | 0 | 0 | 0 | 0 |
| 0 | 0 | 0 | 0 | 0 |
| 0 | 0 | 0 | 0 | 0 |
| 0 | 0 | 0 | 0 | 0 |
| 0 | 0 | 0 | 0 | 0 |
| 0 | 0 | 0 | 0 | 0 |
| 0.165596149 | 0.237679181 | 0.560759996 | 0 | 0.68832287 |
| 0.01792212 | 0.014748911 | 0.134420579 | 0.008887179 | 0.053048308 |
| 0 | 0 | 0 | 0 | 0 |
| 0 | 0 | 0 | 0 | 0 |
| 0 | 0 | 0 | 0.047517586 | 0.110319435 |
| 0 | 0 | 0 | 0 | 0.015867177 |
| 0 | 0 | 0 | 0 | 0 |

| | | | | |
|---|---|---|---|---|
| 0 | 0 | 0 | 0 | 0 |
| 0 | 0 | 0 | 0.024323676 | 0 |
| 0.021155756 | 0.003867624 | 0.002819941 | 0.000407837 | 0.003641213 |
| 0 | 0 | 0 | 0 | 0 |
| 0 | 0 | 0.630594601 | 0.006295667 | 0.441914692 |
| 0 | 0 | 0.189507419 | 0.002007502 | 0 |
| 0 | 0 | 0 | 0 | 0 |
| 0 | 0 | 0 | 0 | 0 |
| 0 | 0 | 0 | 0.004097364 | 0 |
| 0 | 0 | 0 | 0 | 0 |
| 0 | 0 | 0 | 0 | 0 |
| 0 | 0 | 0 | 0 | 0 |
| 0 | 0 | 0 | 0 | 0 |
| 0 | 0 | 0 | 0 | 0.002710759 |
| 0 | 0 | 0 | 0 | 0 |
| 0 | 0 | 0 | 0 | 0 |
| 0 | 0 | 0 | 0 | 0 |
| 0 | 0 | 0 | 0 | 0 |
| 0 | 0 | 0 | 0 | 0 |
| 0 | 0 | 0 | 0 | 0 |
| 0 | 0 | 0 | 0 | 0 |
| 0 | 0 | 0 | 0 | 0 |
| 0 | 0 | 0 | 0 | 0 |
| 0 | 0 | 0.704146717 | 0 | 0 |
| 0 | 0 | 0 | 0.250741618 | 0.001286088 |
| 0 | 0.000610765 | 0 | 0 | 0.000776264 |
| 0 | 0 | 0 | 0 | 0 |
| 0 | 0 | 0 | 0.000983975 | 0.129767607 |
| 0 | 0 | 0 | 0 | 0 |
| 0 | 0 | 0 | 0 | 0 |
| 0 | 0 | 0 | 0.000471674 | 0 |
| 0.001270001 | 0 | 0 | 0 | 0.007034291 |
| 0 | 0 | 0 | 0 | 0 |
| 0 | 0 | 0 | 0 | 0 |
| 0 | 0 | 0 | 0 | 0 |
| 0 | 0 | 0 | 0 | 0 |
| 0 | 0 | 0.003324897 | 0 | 0 |
| 0 | 0.09362663 | 0.11139532 | 0 | 0 |
| 0 | 0.676661012 | 0 | 0.062932863 | 1.349197347 |
| 0 | 0 | 0 | 0 | 0 |
| 0 | 0 | 0 | 0 | 0 |
| 0 | 0 | 0 | 0.012960686 | 0.010292665 |
| 0 | 0.01394505 | 0.025966332 | 0 | 0.074790338 |
| 0 | 0 | 0 | 0 | 0 |
| 0 | 0 | 0 | 0 | 0 |
| 0 | 0 | 0 | 0 | 0 |
| 0 | 0.007970645 | 0.017708024 | 0.001903475 | 0.041846074 |
| 0 | 0 | 0 | 0 | 0 |

| | | | | |
|---|---|---|---|---|
| 0 | 0 | 0 | 0 | 0 |
| 0.01698964 | 0.019671264 | 0.223593669 | 0.001310092 | 0.077030835 |
| 0 | 0 | 0 | 0 | 0 |
| 0 | 0 | 0 | 0 | 0 |
| 0 | 0 | 0 | 0 | 0 |
| 0 | 0 | 0 | 0 | 0 |
| 0 | 0 | 0 | 0 | 0 |
| 0 | 0 | 0 | 0 | 0 |
| 0 | 0 | 0 | 0 | 0 |
| 0 | 0 | 0 | 0 | 0 |
| 0 | 0 | 0 | 0 | 0 |
| 0 | 0 | 0 | 0 | 0 |
| 0 | 0.004065045 | 0 | 0 | 0.00900835 |
| 0 | 0 | 0 | 0 | 0 |
| 0 | 0 | 0.036707349 | 0 | 0.014479796 |
| 0 | 0 | 0 | 0 | 0 |
| 0 | 0 | 0 | 0 | 0 |
| 0 | 0 | 0 | 0 | 0 |
| 0 | 0 | 0 | 0 | 0.006914419 |
| 0 | 0 | 0 | 0 | 0 |
| 0 | 0 | 0 | 0 | 0 |
| 0.04332553 | 0.012134329 | 0.028285003 | 0 | 0.011151173 |
| 0 | 0 | 0 | 0 | 0.085373185 |
| 0.094164838 | 0.071445516 | 0 | 0 | 0 |
| 0 | 0 | 0 | 0 | 0 |
| 0 | 0 | 0 | 0 | 0 |
| 0 | 0 | 0 | 0 | 0 |
| 0 | 0 | 0 | 0 | 0 |
| 0 | 0 | 0 | 0 | 0 |
| 0 | 0 | 0 | 0 | 0 |
| 0 | 0 | 0 | 0 | 0 |
| 0 | 0 | 0 | 0 | 0 |
| 0 | 0 | 0 | 0 | 0 |
| 0 | 0 | 0 | 0 | 0 |
| 0.744113691 | 0 | 21.83037012 | 14.56718939 | 39.2664352 |
| 0 | 0 | 0 | 0 | 0 |
| 0 | 0.050022166 | 0 | 0 | 0 |
| 0 | 0 | 0 | 0 | 0 |
| 0.003430767 | 0 | 0.050953565 | 0.000934501 | 0.023804039 |
| 0 | 0 | 0 | 0 | 0 |
| 0.123045455 | 0.015405636 | 0 | 0.114792805 | 0.036274738 |
| 0 | 0 | 0 | 0 | 0 |
| 0 | 0 | 0 | 0 | 0 |
| 0 | 0 | 0 | 0 | 0 |
| 0 | 0 | 0 | 0 | 0 |
| 4.667584317 | 0.407354321 | 4.515360539 | 0.001233441 | 0.429269972 |
| 0 | 0 | 0 | 0 | 0 |
| 0 | 0 | 0 | 0 | 0 |

| | | | | |
|---|---|---|---|---|
| 0 | 0 | 0 | 0 | 0 |
| 0 | 0 | 0.024849347 | 0 | 0 |
| 0 | 0 | 0 | 0 | 0 |
| 0 | 0 | 0 | 0 | 0 |
| 0 | 0 | 0 | 0 | 0.002863542 |
| 0 | 0 | 0 | 0 | 0 |
| 0 | 0 | 0 | 0 | 0 |
| 0 | 0 | 0 | 0 | 0 |
| 0 | 0 | 0.006676181 | 0 | 0 |
| 0.018370608 | 0.008534402 | 0 | 0.0058975 | 0 |
| 0 | 0 | 0 | 0 | 0 |
| 0 | 0 | 0 | 0 | 0 |
| 0 | 0.000709928 | 0.007246669 | 0 | 0.007519153 |
| 0 | 0 | 0 | 0 | 0 |
| 0 | 0 | 0 | 0 | 0 |
| 0 | 0 | 0 | 0 | 0 |
| 0 | 0 | 0 | 0.006773524 | 0 |
| 0 | 0 | 0 | 0 | 0 |
| 0 | 0 | 0 | 0 | 0 |
| 0.11070768 | 0.58891818 | 1.091992931 | 0.003731376 | 0.360506819 |
| 0 | 0 | 0 | 0 | 0 |
| 0 | 0 | 0 | 0 | 0 |
| 0 | 0 | 0 | 0 | 0 |
| 0 | 0 | 0 | 0 | 0 |
| 0 | 0 | 0 | 0 | 0 |
| 0 | 0 | 0 | 0.000815758 | 0 |
| 0 | 0 | 0 | 0 | 0 |
| 0 | 0 | 0 | 0 | 0 |
| 0 | 0 | 0 | 0 | 0 |
| 0 | 0.008781459 | 0.090671937 | 0 | 0 |
| 0 | 0 | 0 | 0 | 0 |
| 0 | 0 | 0 | 0 | 0 |
| 0 | 0 | 0 | 0 | 0 |
| 0 | 0 | 0 | 0 | 0 |
| 0 | 0 | 0 | 0 | 0 |
| 0.005172038 | 0 | 0 | 0 | 0 |
| 0 | 0 | 0 | 0 | 0 |
| 0 | 0 | 0 | 0 | 0 |
| 0 | 0 | 0 | 0 | 0 |
| 0 | 0 | 0 | 0 | 0 |
| 0 | 0 | 0 | 0 | 0 |
| 0 | 0 | 0 | 0 | 0 |
| 0 | 0 | 0 | 0 | 0.00927189 |
| 0 | 0 | 0 | 0 | 0 |
| 0 | 0.002077218 | 0.003937779 | 0 | 0.018271058 |
| 0 | 0 | 0 | 0 | 0 |
| 0 | 0 | 0 | 0 | 0 |
| 0 | 0 | 0 | 0 | 0 |

| | | | | |
|---|---|---|---|---|
| 0 | 0 | 0 | 0 | 0 |
| 0 | 0 | 0 | 0 | 0 |
| 0 | 0 | 0 | 0 | 0 |
| 0 | 0 | 0 | 0 | 0 |
| 0 | 0 | 0 | 0 | 0 |
| 0 | 0.004652173 | 0.009158311 | 0.015044038 | 0.003441298 |
| 0 | 0 | 0 | 0 | 0 |
| 0 | 0 | 0 | 0 | 0.00126701 |
| 0 | 0 | 0 | 0 | 0 |
| 0 | 0 | 0 | 0 | 0 |
| 0 | 0 | 0 | 0 | 0 |
| 0 | 0 | 0 | 0 | 0 |
| 0 | 0 | 0 | 0 | 0 |
| 0 | 0 | 0 | 0 | 0 |
| 0 | 0 | 0 | 0 | 0 |
| 0 | 0 | 0 | 0 | 0 |
| 0 | 0 | 0 | 0 | 0 |
| 0 | 0 | 0 | 0 | 0.000308229 |
| 0 | 0 | 0 | 0 | 0 |
| 0 | 0 | 0 | 0 | 0 |
| 0 | 0 | 0 | 0 | 0 |
| 0 | 0 | 0 | 0 | 0 |
| 0 | 0 | 0 | 0 | 0 |
| 0 | 0 | 0 | 0 | 0 |
| 0 | 0 | 0 | 0 | 0 |
| 0.002008444 | 0 | 0 | 0.000511552 | 0 |
| 0 | 0 | 0 | 0 | 0 |
| 0 | 0 | 0 | 0 | 0 |
| 0 | 0 | 0 | 0 | 0 |
| 0 | 0 | 0 | 0 | 0 |
| 0 | 0 | 0 | 0 | 0 |
| 0 | 0 | 0 | 0 | 0 |
| 0 | 0 | 0 | 0 | 0 |
| 0 | 0 | 0 | 0 | 0 |
| 0 | 0 | 0 | 0 | 0 |
| 0 | 0 | 0 | 0 | 0 |
| 0 | 0 | 0 | 0 | 0 |
| 0 | 0 | 0 | 0 | 0 |
| 0 | 0 | 0 | 0 | 0 |
| 0 | 0 | 0 | 0 | 0 |
| 0 | 0 | 0 | 0 | 0 |
| 0 | 0 | 0 | 0 | 0 |
| 0 | 0 | 0 | 0 | 0 |
| 0 | 0 | 0 | 0 | 0 |
| 0 | 0 | 0 | 0 | 0 |
| 0 | 0 | 0 | 0 | 0 |
| 0 | 0 | 0 | 0 | 0 |
| 0 | 0 | 0 | 0 | 0 |
| 0 | 0 | 0 | 0 | 0.000429195 |

| | | | | |
|---|---|---|---|---|
| 0 | 0 | 0 | 0 | 0 |
| 0.016205434 | 0 | 0 | 0 | 0.213936074 |
| 0 | 0 | 0 | 0 | 0 |
| 0 | 0 | 0 | 0 | 0 |
| 0 | 0 | 0 | 0 | 0 |
| 0 | 0 | 0 | 0 | 0 |
| 0 | 0 | 0 | 0 | 0 |
| 0 | 0 | 0 | 0 | 0 |
| 0 | 0 | 0 | 0 | 0 |
| 0 | 0 | 0 | 0 | 0 |
| 0.010267612 | 0 | 0.00381235 | 0.002756824 | 0.001651075 |
| 0 | 0 | 0 | 0 | 0 |
| 0.979610772 | 0.001554845 | 0 | 0.101615291 | 0 |
| 0 | 0 | 0 | 0 | 0 |
| 0 | 0 | 0 | 0 | 0 |
| 0 | 0 | 0 | 0 | 0 |
| 0 | 0 | 0 | 0 | 0 |
| 0 | 0 | 0 | 0 | 0 |
| 0 | 0 | 0 | 0 | 0 |
| 0 | 0 | 0 | 0 | 0 |
| 0.002952619 | 0.002472138 | 0 | 0.001094874 | 0.012474964 |
| 0 | 0 | 0 | 0 | 0 |
| 0 | 0 | 0 | 0 | 0 |
| 1.957621933 | 0 | 0 | 0.819014513 | 0.845249503 |
| 0 | 0 | 0 | 0 | 0 |
| 0 | 0 | 0 | 0 | 0 |
| 0 | 0 | 0 | 0 | 0 |
| 0 | 0 | 0 | 0 | 0 |
| 0 | 0 | 0 | 0 | 0 |
| 0 | 0 | 0 | 0 | 0 |
| 0 | 0 | 0.011393165 | 0 | 0 |
| 0 | 0 | 0 | 0 | 0 |
| 0 | 0 | 0 | 0 | 0 |
| 0 | 0 | 0 | 0 | 0 |

| PHL018-0090_V1 | PHL018-0130_V3 | PHL018-0216_V2 | PHL018-0091_V1 | PHL018-0088_V3 |
|---|---|---|---|---|
| CTRL | REF | CTRL | EYCF | EYCF |
| 1.221050971 | 0.196038393 | 0.041930767 | 0.746444703 | 0.014157831 |
| 0.268869767 | 0.293157589 | 0.277155728 | 2.760854409 | 2.349533032 |
| 0.800429908 | 0 | 3.541054531 | 0.437628783 | 3.056845105 |
| 0 | 0 | 0 | 0 | 0 |
| 0 | 0 | 0 | 0 | 0 |
| 0 | 0 | 0 | 0 | 0 |
| 3.418461341 | 0 | 0 | 0 | 0 |
| 0 | 0 | 0 | 0 | 0 |
| 0 | 0 | 0 | 0 | 0 |
| 0 | 0 | 0 | 0 | 0 |
| 0 | 0 | 0 | 0 | 0 |
| 0 | 0 | 0 | 0 | 0 |
| 0 | 0 | 0 | 0 | 0 |
| 0.005603778 | 0.052994386 | 1.270221652 | 1.301085976 | 3.121690238 |
| 0 | 0 | 0 | 0 | 0 |
| 0 | 0 | 0 | 0 | 0 |
| 0.000934449 | 0 | 0.001390134 | 0 | 0 |
| 0 | 0 | 0.001309633 | 0 | 0 |
| 0 | 0 | 0 | 0 | 0 |
| 0 | 0 | 0 | 0 | 0 |
| 0 | 0 | 0 | 0 | 0 |
| 0.001190826 | 0 | 0 | 0 | 0 |
| 0 | 0 | 0 | 0 | 0 |
| 0 | 0 | 0 | 0 | 0 |
| 0 | 0.446003687 | 0 | 0 | 0.003497806 |
| 0 | 0 | 0 | 0 | 0 |
| 0 | 0 | 0 | 0 | 0 |
| 0 | 0 | 0 | 0 | 0 |
| 0 | 0 | 0 | 0 | 0 |
| 0 | 0 | 0 | 0 | 0 |
| 0 | 0 | 0 | 0 | 0 |
| 0 | 0 | 0 | 0 | 0 |
| 0 | 0 | 0 | 0 | 0 |
| 0 | 0 | 0 | 0 | 0 |
| 0 | 0 | 0 | 0 | 0 |
| 0 | 0 | 2.427922951 | 1.192352261 | 2.166380102 |
| 0 | 0 | 0 | 0 | 0 |
| 0 | 0 | 0 | 0 | 0 |
| 0 | 0 | 0 | 0 | 0 |
| 0 | 0 | 0 | 0 | 0 |
| 0 | 0 | 0 | 0 | 0 |
| 0.004336687 | 0.000524437 | 0 | 0.003682254 | 0.053934057 |
| 0.931128169 | 0.065784103 | 0.199438856 | 1.247385892 | 0.005837599 |
| 0.057020402 | 0.001827387 | 0 | 0.027635375 | 0.025495769 |
| 0 | 0.001704976 | 0 | 0.010380639 | 0.004020964 |
| 0 | 0 | 0 | 0 | 0 |

| | | | | |
|---|---|---|---|---|
| 0.016980394 | 0 | 0 | 0.353046771 | 0.00099262 |
| 0.022517241 | 0.00082919 | 0.003957848 | 0.005672744 | 0 |
| 0 | 0.04399291 | 0 | 0.296731247 | 0.001270427 |
| 0.180690154 | 0.46707871 | 0.291253394 | 0.238250283 | 0.461173498 |
| 0.049532592 | 0.020831175 | 0 | 0 | 0.000474788 |
| 0.156332699 | 0 | 0 | 0 | 0 |
| 0.00072586 | 0 | 0.002313914 | 0.007360322 | 0.905779855 |
| 0 | 0 | 0 | 0 | 0 |
| 0 | 0.071718087 | 0 | 0 | 0 |
| 0 | 0 | 0 | 0 | 0 |
| 0 | 0 | 0 | 0 | 0 |
| 0 | 0 | 0.139892614 | 0.465234559 | 0.006069474 |
| 0 | 0 | 0 | 0 | 0.000475567 |
| 0.006497977 | 0.066832531 | 0.030519219 | 0 | 0.010877042 |
| 0 | 0 | 0 | 0 | 0 |
| 0 | 0 | 0 | 0 | 0 |
| 0 | 0 | 0.00530082 | 0.077289986 | 0 |
| 0 | 0 | 0 | 0 | 0 |
| 0 | 0 | 0 | 0 | 0 |
| 0 | 0 | 0 | 0 | 0 |
| 0.051497708 | 0 | 0.063769207 | 0.04020136 | 0.064530801 |
| 0.006446844 | 0.059675954 | 0.011371773 | 0.017280916 | 0.000780105 |
| 0 | 0 | 0 | 0 | 2.00136734 |
| 0 | 0 | 0 | 0 | 0 |
| 0 | 0 | 0 | 0 | 0 |
| 0.129971137 | 0 | 0 | 0 | 0 |
| 0 | 0 | 0 | 0.005430006 | 0 |
| 0 | 0 | 0 | 0 | 0 |
| 0 | 0 | 0 | 0 | 0.002827609 |
| 0.005816902 | 0 | 0 | 0 | 0 |
| 0.059754781 | 0 | 0.032136061 | 0 | 0 |
| 0 | 0 | 0 | 0 | 0 |
| 0 | 0 | 0 | 0 | 0 |
| 0.001549783 | 0.000681185 | 0 | 0.00336375 | 0 |
| 0 | 0 | 0 | 0 | 0 |
| 0 | 0 | 0 | 0 | 0 |
| 0 | 0 | 0 | 0 | 0 |
| 0 | 0.007785124 | 0 | 5.471993445 | 0 |
| 0 | 0 | 0 | 0 | 0 |
| 0 | 0 | 0 | 0 | 0 |
| 0 | 0 | 0 | 0 | 0 |
| 0 | 0 | 0 | 0 | 0 |
| 0 | 0 | 0 | 0 | 0 |
| 0 | 0 | 0 | 0 | 0 |
| 0.485356146 | 0 | 0 | 0 | 0 |
| 0 | 0 | 0 | 1.105018315 | 0 |
| 0 | 0 | 0 | 0 | 0 |
| 0.00065134 | 0 | 0 | 0 | 0 |

| | | | | |
|---|---|---|---|---|
| 0 | 0 | 0 | 0 | 0 |
| 0.000947845 | 0 | 0.002014374 | 0 | 0 |
| 0 | 0 | 0 | 0 | 0 |
| 0 | 0 | 0 | 0 | 0 |
| 0.013208741 | 0 | 0 | 0.003364638 | 0 |
| 0 | 0 | 0 | 0 | 0 |
| 0 | 0 | 0 | 0 | 0 |
| 0.185789142 | 0 | 0 | 0.025365644 | 0 |
| 0.000623138 | 0.010071371 | 0.017878082 | 0.00637365 | 0.001508065 |
| 0 | 0 | 0 | 0 | 0 |
| 0 | 0 | 0 | 0 | 0 |
| 0 | 0 | 0 | 0 | 0.001167222 |
| 0 | 0 | 0 | 0 | 0 |
| 0 | 0 | 0 | 0 | 0 |
| 0 | 0 | 0 | 0 | 0 |
| 0 | 0 | 0 | 0 | 0 |
| 0 | 0 | 0 | 0 | 0 |
| 0 | 0 | 0 | 0 | 0 |
| 0.607009954 | 0.076835651 | 0.03653343 | 0 | 0.015321904 |
| 0 | 0 | 0 | 0 | 0 |
| 0 | 0 | 0 | 0 | 0.011904171 |
| 0 | 0 | 0 | 0 | 0 |
| 0.003544173 | 0 | 0 | 0 | 0.000687284 |
| 0.022374693 | 0 | 0 | 0 | 0 |
| 0 | 0 | 0 | 0 | 0 |
| 0 | 0 | 0 | 0 | 0 |
| 0 | 0 | 0 | 0 | 0 |
| 0.014931606 | 0 | 0.000891373 | 0 | 0 |
| 0 | 0 | 0 | 0 | 0 |
| 0 | 0 | 0.001840233 | 0 | 0 |
| 0.720100183 | 0.029183064 | 0.050121757 | 0 | 0.001860692 |
| 0 | 0 | 0 | 0.002133623 | 0.000732011 |
| 0 | 0 | 0 | 0 | 0 |
| 0 | 0 | 0 | 0 | 0 |
| 0 | 0 | 0 | 0 | 0 |
| 0 | 0 | 0 | 0 | 0 |
| 0 | 0 | 0 | 0 | 0 |
| 0 | 0 | 0 | 0 | 0 |
| 0 | 0 | 0.246587419 | 0 | 0.181898772 |
| 0 | 0 | 0 | 0 | 0 |
| 0 | 0 | 0 | 0 | 0 |
| 0 | 0 | 0 | 0 | 0 |
| 0 | 0.006049995 | 0 | 0 | 0 |
| 0 | 0 | 0 | 0 | 0 |
| 0 | 0 | 0 | 0 | 0 |
| 0 | 0 | 0 | 0 | 0 |
| 0 | 0 | 0 | 0 | 0 |
| 0 | 0 | 0 | 0 | 0 |

| | | | | |
|---|---|---|---|---|
| 0.003531019 | 0.001219435 | 0 | 0.003831976 | 0 |
| 0 | 0 | 0 | 0 | 0 |
| 0 | 0 | 0 | 0 | 0 |
| 0 | 0 | 0 | 0 | 0 |
| 0.129210691 | 0.004212007 | 0 | 0 | 0.011893137 |
| 0 | 0 | 0 | 0 | 0 |
| 0 | 0 | 0 | 0 | 0 |
| 0 | 0 | 0 | 0 | 0 |
| 0 | 0 | 0 | 0 | 0 |
| 0 | 0 | 0 | 0 | 0 |
| 0 | 0 | 0 | 0 | 0 |
| 0 | 0 | 0 | 0 | 0 |
| 0 | 0 | 0 | 0 | 0 |
| 0 | 0 | 0 | 0 | 0 |
| 0 | 0 | 0 | 0 | 0 |
| 0 | 0 | 0 | 0 | 2.520681553 |
| 0 | 0 | 0 | 0 | 0 |
| 0 | 0 | 0 | 0 | 0 |
| 0 | 0 | 0 | 0 | 0 |
| 0 | 0 | 0 | 0 | 0 |
| 0 | 0 | 0 | 0 | 0 |
| 0 | 0 | 0 | 0 | 0 |
| 0 | 0.0115711 | 0 | 0 | 0 |
| 0 | 0 | 0 | 0 | 0 |
| 0.281463686 | 0 | 0 | 0 | 0 |
| 0 | 0 | 0 | 0 | 0 |
| 0 | 0.038835257 | 0 | 0 | 0 |
| 0 | 0 | 0 | 0 | 0 |
| 0 | 0 | 0 | 0 | 0 |
| 0 | 0 | 0 | 0 | 0 |
| 0 | 0 | 0 | 0 | 0 |
| 0 | 0.000341843 | 0 | 0 | 1.337818202 |
| 0 | 0 | 0 | 0 | 0 |
| 0 | 0 | 0 | 0 | 0 |
| 0 | 0 | 0 | 0 | 0 |
| 0 | 0 | 0 | 0 | 0 |
| 0 | 0 | 0 | 0 | 0 |
| 0 | 0 | 0 | 0 | 0 |
| 0 | 0 | 0 | 0 | 0 |
| 0 | 0 | 0 | 0 | 0 |
| 0 | 0 | 0 | 0 | 0 |
| 0 | 0 | 0 | 0 | 0 |
| 0 | 0 | 0 | 0 | 0 |
| 0 | 0 | 0 | 0 | 0 |
| 0 | 0 | 0 | 0 | 0 |
| 0 | 0 | 0 | 0 | 0 |
| 0 | 0 | 0 | 0 | 0 |
| 0 | 0 | 0 | 0 | 0 |

| | | | | |
|---|---|---|---|---|
| 0 | 0 | 0 | 0 | 0 |
| 0 | 0 | 0 | 0 | 0 |
| 0 | 0 | 0 | 0 | 0 |
| 0 | 0 | 0 | 0 | 0 |
| 0 | 0 | 0 | 0 | 0 |
| 0 | 0 | 0 | 0 | 0 |
| 0 | 0 | 0 | 0 | 0 |
| 0 | 0 | 0 | 0 | 0 |
| 0 | 0 | 0 | 0 | 0 |
| 0 | 0 | 0 | 0 | 0 |
| 0 | 0 | 0 | 0 | 0 |
| 0 | 0 | 0 | 0 | 0 |
| 0.006812457 | 0.022788674 | 0.001703287 | 0.008480318 | 0 |
| 0 | 0 | 0 | 0 | 0 |
| 0 | 0 | 0 | 0.011758504 | 0 |
| 0 | 0 | 0 | 0 | 0 |
| 0 | 0 | 0 | 0 | 0 |
| 0 | 0 | 0 | 0 | 0 |
| 0 | 0 | 0 | 0 | 0 |
| 0 | 0 | 0 | 0 | 0 |
| 0.029721202 | 0.028746017 | 0.053078953 | 0.02202244 | 0.01561477 |
| 0 | 0 | 0 | 0 | 0 |
| 0 | 0.000626099 | 0 | 0.004637598 | 0 |
| 0.000781342 | 0 | 0 | 0 | 0 |
| 0 | 0 | 0 | 0 | 0 |
| 0 | 0 | 0 | 0 | 0 |
| 0.000727266 | 0.003561916 | 0.000463679 | 0.002308563 | 0.01628061 |
| 0 | 0 | 0 | 0 | 0 |
| 0 | 0 | 0 | 0 | 0 |
| 0.002178654 | 0 | 0 | 0 | 0 |
| 0 | 0 | 0.002311046 | 0 | 0 |
| 0 | 0 | 0 | 0 | 0 |
| 0 | 0 | 0 | 0 | 0 |
| 0 | 0 | 0.011677881 | 0.033916044 | 0.002000905 |
| 0.572345657 | 0.025999457 | 0.011569559 | 0.022926357 | 0.004096694 |
| 0 | 0 | 0 | 0 | 0 |
| 0 | 0 | 0 | 0 | 0 |
| 0 | 0 | 0 | 0 | 0 |
| 0 | 0 | 0 | 0 | 0 |
| 0 | 0 | 0 | 0 | 0 |
| 0 | 0 | 0 | 0 | 0 |
| 0 | 0 | 0 | 0 | 0 |
| 0 | 0 | 0 | 0 | 0 |
| 0 | 0 | 0 | 0 | 0 |
| 0 | 0.014523049 | 0 | 0 | 0 |
| 0 | 5.953160128 | 0 | 0 | 0 |
| 0.000527852 | 0 | 0.000747866 | 0 | 0 |
| 0.010654159 | 0.058892304 | 0.001230564 | 0.007760522 | 0 |

| | | | | |
|---|---|---|---|---|
| 0 | 0 | 0 | 0 | 0 |
| 0 | 0 | 0 | 0 | 0 |
| 0 | 0 | 0 | 0 | 0 |
| 0 | 0 | 0 | 0 | 0.001039068 |
| 0 | 0 | 0 | 0 | 0 |
| 0 | 0 | 0 | 0 | 0 |
| 0 | 0 | 0 | 0 | 0 |
| 0 | 0 | 0 | 0 | 0 |
| 0.000912814 | 0.009313894 | 0 | 0 | 0 |
| 0 | 0 | 0 | 0 | 0 |
| 0 | 0 | 0 | 0 | 0 |
| 0 | 0 | 0 | 0 | 0 |
| 0 | 0 | 0 | 0 | 0 |
| 0 | 0 | 0 | 0 | 0 |
| 0 | 0 | 0 | 0 | 0 |
| 0 | 0 | 0 | 0 | 0 |
| 0 | 0 | 0 | 0 | 0 |
| 0 | 0 | 0 | 0 | 0 |
| 0.00471096 | 0 | 0.000876033 | 0.005192368 | 0 |
| 0.325667177 | 0.032169083 | 0 | 0 | 0.006260356 |
| 0 | 0.012053955 | 0.11039403 | 0 | 0 |
| 0 | 0 | 0 | 0 | 0 |
| 0 | 0 | 0 | 0 | 0 |
| 0 | 0 | 0 | 0 | 0 |
| 0.001380595 | 0.000845214 | 0.001833788 | 0.004869364 | 0 |
| 0 | 0 | 0 | 0 | 0 |
| 0 | 0 | 0 | 0 | 0 |
| 0 | 0 | 0 | 0 | 0 |
| 0 | 0 | 0 | 0 | 0.003321408 |
| 0 | 0 | 0 | 0.001391722 | 0 |
| 1.857888515 | 0.2751784 | 0.362833761 | 0.172785452 | 0.030271097 |
| 0 | 0 | 0 | 0 | 0 |
| 0 | 0 | 0 | 0 | 0.023064348 |
| 0 | 0 | 0 | 0.00495473 | 0 |
| 0 | 0 | 0 | 0 | 0 |
| 0 | 0 | 0 | 0 | 0 |
| 0 | 0 | 0 | 0 | 0 |
| 0 | 0 | 0 | 0 | 0 |
| 0 | 0 | 0 | 0 | 0 |
| 0 | 0 | 0 | 0 | 0 |
| 0 | 0 | 0 | 0 | 0 |
| 0.052996426 | 0.002976113 | 0.010100619 | 0 | 0.01378688 |
| 0.002731324 | 0 | 0 | 0 | 0 |
| 0 | 0 | 0 | 0 | 0 |
| 0 | 0 | 0 | 0 | 0 |
| 0 | 0 | 0 | 0 | 0 |
| 0 | 0 | 0 | 0 | 0 |
| 0 | 0 | 0 | 0 | 0 |

| | | | | |
|---|---|---|---|---|
| 0 | 0 | 0 | 0 | 0 |
| 0 | 0 | 0 | 0 | 0 |
| 0.060823128 | 0.001601571 | 0.001042437 | 0.032870528 | 0.037261365 |
| 0 | 0 | 0 | 0 | 0 |
| 0 | 0 | 0 | 0 | 0 |
| 0 | 0 | 0 | 0 | 0 |
| 0 | 0 | 0 | 0 | 0 |
| 0 | 0 | 0 | 0 | 0 |
| 0 | 0 | 0 | 0 | 0 |
| 0 | 0 | 0 | 0 | 0 |
| 0 | 0 | 0 | 0 | 0 |
| 0 | 0 | 0 | 0 | 0 |
| 0 | 0 | 0 | 0 | 0 |
| 0.006505213 | 0.013865198 | 0.007104503 | 0 | 0 |
| 0.000745723 | 0.005234978 | 0.011093746 | 0.001841117 | 0.00135355 |
| 0 | 0 | 0 | 0 | 0.01111018 |
| 0 | 0 | 0.001625185 | 0 | 0 |
| 0 | 0 | 0.03459876 | 0.001859759 | 0 |
| 0 | 0 | 0 | 0 | 0 |
| 0 | 0 | 0 | 0 | 0 |
| 0 | 0 | 0 | 0 | 0 |
| 0.01595406 | 0.066742774 | 0.000815044 | 0.003516878 | 0 |
| 0 | 0 | 0 | 0 | 0 |
| 0 | 0 | 0 | 0 | 0 |
| 0 | 0 | 0 | 0 | 0 |
| 0 | 0 | 0 | 0 | 0 |
| 0 | 0 | 0 | 0 | 0 |
| 0 | 0 | 0 | 0 | 0 |
| 0 | 0 | 0 | 0 | 0 |
| 0 | 0 | 0.003065317 | 0 | 0 |
| 0 | 0 | 0 | 0 | 0.003556243 |
| 0 | 0 | 0 | 0 | 0 |
| 0 | 0 | 0 | 0 | 0 |
| 0 | 0 | 0 | 0 | 0 |
| 0 | 0 | 0 | 0 | 0 |
| 0 | 0 | 0 | 0 | 0 |
| 0 | 0 | 0 | 0 | 0 |
| 0.002120104 | 0.016991335 | 0.001228821 | 0.008837185 | 0.013993346 |
| 0 | 0 | 0 | 0 | 0 |
| 0 | 0 | 0 | 0 | 0 |
| 0 | 0 | 0 | 0 | 0 |
| 0 | 0 | 0 | 0 | 0 |
| 0 | 0 | 0 | 0 | 0 |
| 0 | 0 | 0 | 0 | 0 |
| 0 | 0 | 0 | 0 | 0 |
| 0 | 0 | 0 | 0 | 0 |
| 0 | 0 | 0 | 0 | 0 |
| 0.089412336 | 0 | 0 | 0 | 0 |

| | | | | |
|---|---|---|---|---|
| 3.685523855 | 0.863945413 | 0 | 0 | 0.074182338 |
| 0 | 0 | 0 | 0 | 0 |
| 0 | 0 | 0 | 0 | 0 |
| 0 | 0 | 0 | 0 | 0 |
| 0 | 0.040247434 | 0.001717798 | 0 | 0.009128759 |
| 0 | 0 | 0.00112041 | 0 | 0 |
| 0 | 0 | 0 | 0 | 0 |
| 0.018581225 | 0.000924847 | 0.003611808 | 0.004795322 | 0.023306936 |
| 0 | 0 | 0 | 0 | 0 |
| 0 | 0.002478474 | 0.008686461 | 0.008237737 | 0.339383237 |
| 0 | 0 | 0 | 0 | 0 |
| 0.017040873 | 0 | 0 | 0.140135444 | 0.000904405 |
| 0.003129479 | 0.001888132 | 0.005060403 | 0.036952251 | 0.002881297 |
| 0.003099698 | 0.002024177 | 0.009442121 | 0.00655959 | 0 |
| 0.040442377 | 0 | 0 | 0 | 0 |
| 0 | 0 | 0 | 0 | 0 |
| 0 | 0 | 0 | 0 | 0 |
| 0.005847798 | 0 | 0.00828522 | 0 | 0 |
| 0 | 0 | 0 | 0 | 0 |
| 0.002818644 | 0 | 0 | 0 | 0 |
| 0 | 0 | 0 | 0 | 0 |
| 0 | 0 | 0 | 0 | 0 |
| 0 | 0 | 0 | 0.000742791 | 0 |
| 0 | 0 | 0 | 0 | 0 |
| 0 | 0 | 0 | 0 | 0 |
| 0 | 0.00090555 | 0 | 0.003633243 | 0.028605699 |
| 0 | 0 | 0 | 0 | 0 |
| 0 | 0 | 0 | 0.012842832 | 0 |
| 0.004705565 | 0 | 0.455015443 | 0 | 0 |
| 0.001467506 | 0 | 0.000693059 | 0 | 0 |
| 0 | 0 | 0.014998499 | 0 | 0 |
| 0 | 0 | 0 | 0 | 0 |
| 0 | 0 | 0.002567447 | 0 | 0 |
| 0 | 0 | 0.146018616 | 0 | 0 |
| 0.563002734 | 0 | 0 | 0 | 0 |
| 0 | 0 | 0 | 0 | 0 |
| 0.002694803 | 0 | 0.00458163 | 0 | 0 |
| 0 | 0 | 0 | 0 | 0 |
| 0 | 0.000921257 | 0 | 0 | 0 |
| 0 | 0 | 0.008678689 | 0 | 0 |
| 0 | 0 | 0.007768471 | 0 | 0 |
| 0 | 0 | 0.169098227 | 0 | 0 |
| 0 | 0 | 0 | 0 | 0 |
| 0.004596381 | 0.003335052 | 0 | 0 | 0 |
| 0 | 0 | 0 | 0 | 0 |
| 0 | 0 | 0 | 0 | 0.000701533 |
| 0 | 0.001286997 | 0.001047108 | 0.001390223 | 0.001987343 |
| 0 | 0.004380646 | 0.015777157 | 0 | 0 |

| | | | | |
|---|---|---|---|---|
| 0.029245107 | 0 | 0.057599604 | 0 | 0 |
| 0 | 0 | 0 | 0 | 0 |
| 0.007701911 | 0 | 0 | 0 | 0 |
| 0 | 0 | 0 | 0 | 0 |
| 0 | 0.083638071 | 0 | 0 | 0 |
| 0.009624041 | 0.005701872 | 0.053112236 | 0 | 0.001565274 |
| 0 | 0.015498188 | 0 | 0 | 0 |
| 0.001003831 | 0 | 0 | 0 | 0 |
| 0 | 0 | 0 | 0 | 0.035404816 |
| 0 | 0 | 0 | 0 | 0 |
| 0 | 0.015871707 | 0.006121863 | 0.007937372 | 0.026868718 |
| 0 | 0 | 0 | 0 | 0 |
| 0 | 0 | 0 | 0 | 0 |
| 0.000680358 | 0.004859423 | 0 | 0 | 0.003087269 |
| 0.0013768 | 0.003231081 | 0.053033686 | 0 | 0 |
| 0 | 0 | 0.004239216 | 0 | 0 |
| 0.21265888 | 0.091643457 | 0 | 0 | 0 |
| 0.020690569 | 0 | 0 | 0 | 0 |
| 0 | 0 | 0 | 0 | 0 |
| 0.259058515 | 0.146276676 | 0.028286396 | 0 | 0 |
| 0 | 0 | 0 | 0 | 0 |
| 0 | 0 | 0 | 0 | 0.291843967 |
| 0 | 0 | 0 | 0 | 0 |
| 0 | 0 | 0 | 0 | 0 |
| 0 | 0 | 0 | 0 | 0 |
| 0 | 0 | 0 | 0 | 0 |
| 0.002028642 | 0 | 0.078779248 | 0 | 0 |
| 0 | 0 | 0 | 0 | 0 |
| 0 | 0 | 0 | 0 | 0 |
| 0.128896152 | 0.238311757 | 0.83901634 | 1.919593027 | 0.454288841 |
| 0 | 0 | 0 | 0.005290855 | 0.003956215 |
| 0 | 0 | 0 | 0 | 0 |
| 0 | 0 | 0 | 0 | 0 |
| 0 | 0.000533634 | 0 | 0 | 0 |
| 0.001382456 | 0 | 0 | 0 | 0 |
| 0 | 0 | 0.060310099 | 0 | 0 |
| 0 | 0 | 0 | 0.030854971 | 0.431335778 |
| 0 | 0 | 0 | 0 | 0.000699278 |
| 0 | 0 | 0 | 0 | 0 |
| 0 | 0 | 0 | 0 | 0 |
| 0.005593283 | 0.018034456 | 0.004309012 | 0 | 0.002538071 |
| 0.747425128 | 0.220875856 | 0 | 0.852143073 | 0.210402332 |
| 0 | 0 | 0 | 0 | 0 |
| 0 | 0 | 0 | 0 | 0 |
| 0.02066366 | 0 | 0.000670454 | 0 | 0 |
| 0 | 0 | 0.095295376 | 0 | 0 |
| 0 | 0 | 0 | 0.00155291 | 0 |
| 0 | 0 | 0 | 0 | 0 |

| | | | | |
|---|---|---|---|---|
| 0 | 0.002069389 | 0 | 0.005428752 | 0.002556393 |
| 0 | 0.025973333 | 0 | 0 | 0 |
| 0 | 0 | 0 | 0.015229303 | 0 |
| 0 | 0 | 0 | 0 | 0 |
| 0 | 0.027637476 | 0.378860867 | 0 | 0 |
| 0 | 0 | 0.053687845 | 0 | 0.00077194 |
| 0 | 0 | 0 | 0 | 0 |
| 0 | 0 | 0 | 0 | 0 |
| 0.035614087 | 0.003958613 | 0 | 0 | 0.001375374 |
| 0.14907046 | 0 | 0.403466145 | 0 | 0 |
| 0 | 0 | 0 | 0 | 0 |
| 0 | 0 | 0 | 0 | 0 |
| 0 | 0 | 0 | 0 | 0 |
| 0 | 0.033965685 | 0 | 0.016467147 | 0 |
| 0 | 0 | 0 | 0 | 0 |
| 0.143129348 | 0.09426112 | 0 | 0.173164267 | 0.00438925 |
| 0.004398893 | 0.038223887 | 0.007689974 | 0.13087583 | 0.011160942 |
| 0 | 0 | 0.015022108 | 0 | 0 |
| 0 | 0.001555367 | 0 | 0 | 0 |
| 0 | 0 | 0 | 0 | 0 |
| 0 | 0 | 0 | 0.00725361 | 0 |
| 0 | 0 | 0.001399211 | 0.004353991 | 0 |
| 0 | 0 | 0 | 0 | 0 |
| 0 | 0 | 0.194076469 | 0 | 0 |
| 0 | 0 | 0 | 0 | 0.000936114 |
| 0 | 0.042418784 | 0 | 0 | 0 |
| 0 | 0.001323424 | 0 | 0 | 0 |
| 0 | 0.0024195 | 0 | 0 | 0.002758982 |
| 0 | 0 | 0 | 0 | 0 |
| 0 | 0 | 0 | 0 | 0 |
| 0.018220758 | 0.027398075 | 0 | 0.00144958 | 0 |
| 0.006944414 | 0.002167401 | 0.001953314 | 0 | 0 |
| 0.005639665 | 0 | 0.015911788 | 0.029836687 | 0.080401414 |
| 0.001393481 | 0.000909977 | 0 | 0 | 0.040187557 |
| 0 | 0 | 0 | 0 | 0 |
| 0 | 0 | 0 | 0 | 0 |
| 0 | 0 | 0.003175987 | 0.005270864 | 0 |
| 0.093497129 | 0.00953998 | 0.023084443 | 0 | 0.034246077 |
| 0 | 0 | 0 | 0 | 0.232053341 |
| 0.003484922 | 0 | 0.01098986 | 0 | 0.004421 |
| 0 | 0 | 0 | 0 | 0 |
| 0 | 0 | 0 | 0 | 0 |
| 0.012954357 | 0.004531878 | 0 | 0.015955265 | 0 |
| 0.03574169 | 0.06010272 | 0 | 0 | 0 |
| 0 | 0.212427171 | 0.010713066 | 0 | 0 |
| 0 | 0 | 0 | 0.001057266 | 0 |
| 0.003666949 | 0 | 0 | 0 | 0 |
| 0.000698395 | 0 | 0 | 0 | 0 |

| | | | | |
|---|---|---|---|---|
| 0 | 0 | 0 | 0 | 0 |
| 0.10229607 | 0.198203115 | 0.003384451 | 0.121918244 | 0.344789225 |
| 0 | 0 | 0 | 0 | 0 |
| 0 | 0 | 0.000723442 | 0 | 0 |
| 0 | 0 | 0 | 0 | 0 |
| 0 | 0 | 0 | 0 | 0 |
| 0 | 0 | 0.004420317 | 0 | 0 |
| 0 | 0 | 0 | 0 | 0 |
| 0.010290509 | 0 | 0 | 0 | 0 |
| 0 | 0 | 0 | 0 | 0 |
| 0.007378956 | 0.000340941 | 0.003106785 | 0.006138116 | 0 |
| 0.001866424 | 0.010495392 | 0.462764512 | 0.096548995 | 0.001672947 |
| 0 | 8.877227698 | 0 | 0 | 0.004647778 |
| 0.001832771 | 0 | 0 | 0 | 0 |
| 0 | 0 | 0.010354453 | 0.006197594 | 0.000644331 |
| 0 | 0 | 0 | 0 | 0 |
| 0.007581042 | 0.000952039 | 0 | 0.004884895 | 0.002352177 |
| 0.099737184 | 0.004325573 | 0.203717754 | 0 | 0.000572522 |
| 0 | 0 | 0 | 0 | 0 |
| 0.055547211 | 0.000401257 | 0.071822325 | 0 | 0 |
| 0.004510366 | 0.006749823 | 0.00846719 | 0.003711885 | 0.017283029 |
| 0 | 0 | 0 | 0 | 0.00301838 |
| 0 | 0 | 0 | 0 | 0 |
| 0 | 0 | 0 | 0 | 0 |
| 0 | 0 | 0 | 0 | 0.030567066 |
| 0 | 0 | 0 | 0 | 0 |
| 0.035318853 | 0.108900154 | 0 | 0 | 0.012869231 |
| 0.018536231 | 0 | 0.026318922 | 0.021416725 | 0.056719268 |
| 0 | 0 | 0 | 0.017033193 | 0 |
| 0 | 0 | 0 | 0 | 0 |
| 0 | 0 | 0 | 0.000907112 | 0 |
| 0 | 0 | 0 | 0 | 0 |
| 0.005237733 | 0 | 0 | 0 | 0 |
| 0 | 0 | 0 | 0 | 0 |
| 0 | 0 | 0 | 2.728262951 | 0 |
| 0.109800128 | 0.166451351 | 0.619006856 | 0.136878341 | 0.332706398 |
| 0 | 0 | 0.003992852 | 0 | 0 |
| 0 | 0 | 0 | 0 | 0 |
| 0 | 0.012390016 | 0.005236667 | 0.04588724 | 0.021269156 |
| 0 | 0 | 0 | 0 | 0 |
| 0 | 0.10884719 | 0.012618376 | 0 | 0 |
| 0.004458029 | 0 | 0 | 0 | 0 |
| 0.013171049 | 0.000977388 | 0.023029219 | 0 | 0 |
| 0.000924829 | 0 | 0.021988595 | 0.042608261 | 0 |
| 0 | 0 | 0.589539419 | 0 | 0 |
| 0 | 0 | 0 | 0 | 0.102858659 |
| 0 | 0 | 0 | 0 | 0 |
| 0.006353387 | 0 | 0.006949721 | 0 | 0 |

| | | | | |
|---|---|---|---|---|
| 0 | 0 | 0 | 0 | 0 |
| 0 | 0 | 0 | 0 | 0 |
| 0.005925489 | 0 | 0 | 0 | 0.111757412 |
| 2.375903271 | 0.391136147 | 2.487310053 | 0 | 0.007453084 |
| 0 | 0 | 0 | 0 | 0 |
| 0 | 0.229011968 | 0 | 0 | 0.001097484 |
| 0.004108436 | 0.003794902 | 0 | 0 | 0 |
| 0 | 0 | 0 | 0 | 0.000880624 |
| 0.349084172 | 0 | 0.050408088 | 14.59432981 | 1.316012212 |
| 0 | 0.007121818 | 0 | 0 | 0.150945851 |
| 0 | 0 | 0 | 0 | 0 |
| 0 | 0 | 0 | 0 | 0.003637794 |
| 0 | 0 | 0 | 0 | 0 |
| 0 | 0 | 13.6136328 | 0 | 0.001519156 |
| 0.623619703 | 0.084871384 | 0.02917311 | 0 | 0.010673483 |
| 0 | 0 | 0 | 0 | 0 |
| 0.004746126 | 0.00363937 | 0 | 0 | 0 |
| 0 | 0 | 0 | 0 | 0 |
| 0 | 0 | 0 | 0 | 0 |
| 0 | 0 | 0 | 0 | 0 |
| 0 | 0 | 0 | 0 | 0 |
| 0 | 0.475819344 | 0 | 0 | 0 |
| 0 | 0.110173088 | 0 | 0 | 0 |
| 0.426420266 | 0.19306861 | 0 | 0.033713265 | 0.026452928 |
| 0.698037052 | 0 | 0 | 0 | 0 |
| 0 | 0.990401484 | 2.174681532 | 0 | 0 |
| 0 | 0.179090084 | 0 | 0 | 0 |
| 0 | 0.122715159 | 0.480018513 | 1.251306273 | 5.940991826 |
| 0 | 0 | 0 | 0 | 0 |
| 0 | 0 | 0 | 0 | 0 |
| 0.009092647 | 0 | 0 | 0 | 0 |
| 0 | 0.003896122 | 0 | 0 | 0.010983563 |
| 1.684468801 | 0.005003221 | 0.003886812 | 0.058054933 | 0.000299064 |
| 0.001788745 | 0 | 0 | 0 | 0 |
| 0 | 0 | 0 | 0 | 0 |
| 0 | 0 | 0 | 0 | 0.011105942 |
| 0 | 0.136544312 | 0 | 2.791137531 | 2.144766246 |
| 0 | 0 | 0 | 0 | 0 |
| 0 | 0.094463303 | 0 | 0.100813451 | 0 |
| 0.114094794 | 0.005464653 | 0 | 0 | 0 |
| 0.013074169 | 0.504417596 | 0.055843256 | 0.003616679 | 0.030245049 |
| 0 | 0.090872162 | 0 | 0 | 0 |
| 0 | 0.000376547 | 0.017319583 | 2.250674767 | 17.00462338 |
| 0 | 0 | 0 | 0.0031506 | 0 |
| 0 | 0 | 0 | 0 | 0 |
| 0 | 0.336678629 | 0 | 0 | 0 |
| 0 | 0 | 0 | 0 | 0 |
| 0.695847031 | 0.305007666 | 0 | 0.386670259 | 0 |

| | | | | |
|---|---|---|---|---|
| 0 | 0 | 0 | 0 | 0 |
| 0.183892079 | 0 | 0.05474327 | 0 | 0 |
| 0.255682015 | 0.009886809 | 0.006278213 | 0.152642951 | 0.018618198 |
| 0 | 0 | 0 | 0 | 0 |
| 0 | 0 | 0 | 0.02410882 | 0 |
| 0.009541391 | 0 | 0 | 0.19247605 | 0.000938669 |
| 0 | 0 | 0 | 0 | 0 |
| 0.024291154 | 0.004235189 | 0 | 0 | 0 |
| 0 | 0 | 0 | 0 | 0 |
| 0.752997068 | 0 | 0.608404546 | 0 | 0 |
| 1.682554059 | 0.588226151 | 0.067959113 | 0.136313666 | 0.001023667 |
| 0.443053652 | 0.001806154 | 1.712638054 | 0.597471181 | 1.61250767 |
| 0 | 0 | 0 | 0 | 0 |
| 0.00140863 | 0.036922535 | 0 | 0.078111858 | 0.051766983 |
| 0.084364155 | 0.488295547 | 0 | 0.168134709 | 0.029227672 |
| 0 | 0 | 0 | 0 | 3.770654696 |
| 0.004809183 | 0 | 0 | 0 | 0 |
| 0 | 0 | 0 | 0 | 0 |
| 1.125460412 | 0 | 0 | 0 | 0 |
| 0.006689793 | 0.033447058 | 0.001015518 | 0 | 0.00204785 |
| 0.017260161 | 0.571601732 | 0.057476386 | 0.002786608 | 0.490889253 |
| 0 | 0 | 0.00653256 | 0.379675866 | 0 |
| 0.013547264 | 0.488254764 | 0.198364106 | 0.151037383 | 6.063311117 |
| 0 | 0 | 0 | 0 | 0 |
| 0 | 0 | 0 | 0 | 0 |
| 0 | 0 | 0.003869323 | 0 | 0.003965613 |
| 0.040270253 | 0 | 0 | 0 | 0 |
| 0 | 0 | 0 | 0 | 0.009204289 |
| 0 | 0 | 0 | 0 | 0.006617451 |
| 0 | 0 | 0 | 0 | 0 |
| 0 | 0.001422671 | 0 | 0 | 0.000811143 |
| 0 | 0.346768256 | 0 | 0.018415401 | 0.068965918 |
| 1.066276866 | 0.000729471 | 0.039408501 | 0 | 0 |
| 0 | 0 | 0 | 0 | 0.001947041 |
| 0.104595105 | 0.00048146 | 0 | 0 | 0 |
| 0.051053902 | 0.053250469 | 0.000861115 | 1.134233719 | 0.038951813 |
| 0.041750156 | 0.005005473 | 0.00055455 | 0 | 0.000657813 |
| 0 | 0 | 0 | 0 | 0 |
| 0 | 0 | 0 | 0 | 0.858716977 |
| 0 | 0 | 0 | 0 | 0 |
| 0 | 0.0239788 | 0 | 0 | 0 |
| 0 | 0 | 0 | 0 | 0 |
| 0 | 0 | 0 | 0 | 0 |
| 0 | 0 | 0 | 0 | 0 |
| 0.231652524 | 0.002137358 | 0.001309339 | 1.711219712 | 0.000776576 |
| 0 | 0 | 0 | 0 | 0 |
| 0 | 0 | 0 | 0 | 0 |
| 2.050066197 | 0 | 0.148860406 | 2.578609997 | 0.122153808 |

| | | | | |
|---|---|---|---|---|
| 0 | 0 | 0 | 0 | 0 |
| 0 | 0 | 0 | 0 | 0 |
| 0 | 0 | 0 | 0 | 0 |
| 0 | 0.002464601 | 0 | 0.002083522 | 0 |
| 0 | 0 | 0 | 0 | 0 |
| 0 | 2.441238812 | 0 | 0 | 0 |
| 0 | 0 | 0 | 0 | 0.196952806 |
| 0.463373816 | 0.019424877 | 0 | 0.542146555 | 0.022290945 |
| 0 | 0 | 0 | 0 | 0 |
| 0 | 0 | 0 | 0 | 0 |
| 0.038013243 | 0.043799263 | 0.006991124 | 0.029650733 | 0 |
| 0 | 0 | 0.003947417 | 0 | 0 |
| 0 | 0 | 0 | 0 | 0 |
| 0 | 0 | 0 | 0 | 0.60110255 |
| 0 | 0 | 0 | 0 | 0 |
| 0 | 0 | 0 | 0 | 0 |
| 0 | 0.021923854 | 0 | 0 | 0.045833348 |
| 0.010710669 | 0.00337794 | 0 | 0.034986261 | 0.004786555 |
| 0 | 0.003659033 | 0 | 0 | 0 |
| 0 | 0.002060761 | 0 | 0 | 0 |
| 0.002054501 | 0 | 0 | 0 | 0 |
| 0 | 0 | 0 | 0 | 0 |
| 0 | 0 | 0 | 0 | 0 |
| 0.492264584 | 0 | 0 | 0 | 0 |
| 0 | 0 | 0 | 0 | 0 |
| 0 | 0 | 0 | 0 | 0 |
| 0 | 0 | 0 | 0 | 0 |
| 0 | 0 | 0 | 0 | 0 |
| 0 | 0 | 0 | 0 | 0 |
| 0 | 2.519122208 | 0 | 0.023854015 | 0 |
| 0.030860882 | 0 | 0.003181411 | 0 | 0 |
| 0 | 0 | 0 | 0 | 0 |
| 0 | 0 | 0 | 0 | 0.001410576 |
| 0 | 0 | 0 | 0 | 0 |
| 0 | 0 | 0 | 0.043668369 | 0 |
| 0 | 0 | 0 | 0 | 0 |
| 0 | 0.035724023 | 0 | 0 | 0.113917611 |
| 0.456444968 | 0 | 0.000526054 | 0.085994318 | 0 |
| 0 | 0 | 0 | 0 | 0 |
| 0 | 0.004783069 | 0 | 0 | 0 |
| 0 | 0.072418154 | 0.03899195 | 0 | 0 |
| 0 | 0 | 0 | 0 | 0 |
| 0 | 0 | 0 | 0 | 0 |
| 0 | 0.001275417 | 0.006768907 | 0 | 0.01502851 |
| 0 | 0 | 0 | 0 | 0 |
| 0 | 0 | 0 | 0 | 0 |
| 0 | 0.01843054 | 0 | 0 | 0 |
| 0 | 0 | 0 | 0 | 0 |

| | | | | |
|---|---|---|---|---|
| 0 | 0 | 0 | 0 | 0 |
| 0 | 0 | 0 | 0 | 0 |
| 0 | 0 | 0 | 0 | 0 |
| 0 | 0.207616624 | 0.120613664 | 0 | 0.018372949 |
| 0 | 0 | 0 | 0 | 0 |
| 0 | 0 | 0 | 0 | 0 |
| 0 | 0 | 0 | 0 | 0 |
| 0 | 0 | 0 | 0 | 0 |
| 0.041819656 | 0.010449964 | 0.014147572 | 0.031606739 | 0.000688492 |
| 0 | 0 | 0 | 0 | 0 |
| 0 | 0 | 0 | 0 | 0 |
| 0 | 0 | 0 | 0 | 0 |
| 0.002788206 | 0 | 0 | 0 | 0 |
| 0 | 0 | 0 | 0.001254459 | 0 |
| 0.348856747 | 0.479213888 | 0 | 0.203771979 | 3.374073736 |
| 0.357046883 | 0.00236253 | 0 | 0.181425438 | 0 |
| 0 | 0 | 0 | 0 | 0 |
| 0 | 0 | 0 | 0 | 0 |
| 0 | 0 | 0 | 0 | 0 |
| 0 | 0 | 0 | 0 | 0 |
| 0 | 0.045812178 | 0 | 0 | 0.000789676 |
| 0 | 0 | 0 | 0 | 0 |
| 0 | 0 | 0 | 0 | 0 |
| 0 | 0 | 0 | 0 | 0.007032236 |
| 0.044963405 | 0.009011831 | 0.012371208 | 0.064071631 | 0 |
| 0.564749788 | 0.256828465 | 3.280386397 | 4.018449477 | 1.519383835 |
| 0 | 0 | 0 | 0 | 0.290600781 |
| 0.006479311 | 0.002542756 | 0 | 0 | 0 |
| 0.06598611 | 0 | 0 | 0 | 0 |
| 0 | 0 | 0 | 0 | 0 |
| 0 | 0 | 0 | 0 | 0 |
| 0.122807355 | 0 | 0 | 0 | 0.000651771 |
| 0 | 0.000396726 | 0 | 0 | 0 |
| 0 | 0.000437536 | 0 | 0.029303019 | 0 |
| 0 | 0.017157924 | 0 | 0.078185884 | 0.014783634 |
| 0.072543696 | 0.001223049 | 0.00145945 | 0.006385539 | 0 |
| 0 | 0 | 0 | 0 | 0 |
| 0.24997277 | 0.273099479 | 4.435139759 | 0.780461181 | 0.122408514 |
| 0 | 0 | 0 | 0.000834274 | 0.177618768 |
| 0.001431004 | 0 | 0 | 0 | 0 |
| 0 | 0 | 0 | 0 | 0 |
| 0.017699039 | 0.004276422 | 0 | 0 | 0 |
| 1.39689457 | 0.008635146 | 0 | 0.721546031 | 0.194936542 |
| 0 | 0 | 0 | 0 | 0 |
| 0 | 0 | 0 | 0 | 0 |
| 0.018926278 | 0.012842099 | 0.002513901 | 0 | 0.003578417 |
| 0 | 0 | 0 | 0 | 0 |
| 0.016018159 | 1.789316776 | 0.02282819 | 0.396802158 | 0.238295433 |

| | | | | |
|---|---|---|---|---|
| 0 | 0.062745841 | 0 | 0.070876214 | 0 |
| 0 | 0 | 0 | 0 | 0 |
| 0 | 0 | 0 | 0 | 0 |
| 0 | 0 | 0 | 0 | 0 |
| 0.40979781 | 0.008679666 | 0.00163537 | 0.002960789 | 0 |
| 0 | 0 | 0 | 0 | 0 |
| 0 | 0 | 0 | 0 | 0 |
| 0 | 0 | 0 | 0 | 0 |
| 0 | 0 | 0 | 0 | 0 |
| 0.016008963 | 0 | 0 | 0 | 0 |
| 0 | 0 | 0 | 0 | 0 |
| 0 | 0 | 0 | 1.050040747 | 0.176364218 |
| 0.097070248 | 0 | 0 | 0.002389998 | 0.02408658 |
| 0 | 0 | 0 | 0 | 0 |
| 0.002427097 | 0.007660604 | 0 | 0.009987113 | 0.066080818 |
| 0 | 0 | 0 | 0 | 0 |
| 0.007203825 | 0 | 0 | 0 | 0 |
| 0 | 0 | 0 | 0 | 0 |
| 0.012622228 | 0.002662644 | 0 | 0.015006289 | 0 |
| 0 | 0.009884278 | 0 | 0 | 0 |
| 0 | 0 | 0 | 0 | 0.034691286 |
| 0 | 0 | 0 | 0 | 0 |
| 0.440187635 | 0.249900484 | 0.300547807 | 4.509040483 | 0.022944754 |
| 0 | 0 | 0 | 0 | 0 |
| 0 | 0 | 0 | 0 | 0 |
| 0 | 0 | 0 | 0 | 0 |
| 0 | 0 | 0 | 0 | 0 |
| 0 | 0 | 0 | 0 | 0 |
| 0 | 0 | 0 | 0 | 0.001469681 |
| 0 | 0 | 0 | 0 | 0 |
| 0 | 0 | 0 | 0 | 0 |
| 0.011548159 | 0.056559187 | 0.015840955 | 0.023327416 | 0.009315952 |
| 0 | 0 | 0 | 0 | 0 |
| 0.065769575 | 0.00222623 | 0 | 0.115059877 | 0.001269297 |
| 0.069841824 | 0.000887323 | 0.001299475 | 0.006709444 | 0 |
| 0.143459768 | 0 | 0.007631379 | 1.196690552 | 0.004239181 |
| 0 | 0 | 0 | 0 | 0.000608132 |
| 0 | 0 | 0 | 0 | 0 |
| 0 | 0 | 0 | 0.015691613 | 0 |
| 0 | 0 | 0 | 0 | 0 |
| 0 | 0.049801645 | 0 | 0 | 0 |
| 0 | 0 | 0 | 0 | 0 |
| 0 | 0 | 0 | 0 | 0 |
| 0 | 0.016433744 | 0 | 0 | 0 |
| 0.289928777 | 0.114070288 | 0.031433887 | 0.772364298 | 0.002834501 |
| 0 | 0 | 0 | 0 | 0 |
| 0.412534852 | 0.043391523 | 0 | 0.000993047 | 0.343764207 |
| 0 | 0.002686918 | 0 | 0 | 0 |

| | | | | |
|---|---|---|---|---|
| 0 | 0 | 0 | 0 | 0 |
| 0 | 0 | 0 | 0 | 0.009366418 |
| 0.029487511 | 0.00113005 | 0.004707404 | 0 | 0 |
| 0 | 0 | 0 | 0 | 0 |
| 0 | 0 | 0.025809371 | 0 | 0 |
| 0 | 0.001166605 | 0.007846356 | 0 | 0 |
| 0 | 0 | 0 | 0 | 0 |
| 0.060141647 | 0.256774724 | 0.096971888 | 0 | 0.003032284 |
| 0 | 0 | 0 | 0 | 0 |
| 0 | 0 | 0 | 0 | 0.005578564 |
| 0 | 0 | 0.431371668 | 0 | 0 |
| 0 | 0 | 0 | 0 | 0 |
| 0 | 0 | 0 | 0 | 0 |
| 0 | 0 | 0 | 0 | 0 |
| 0 | 0 | 0 | 0 | 0 |
| 0 | 0 | 0 | 0 | 0 |
| 0 | 0.000407148 | 0 | 0 | 0 |
| 0 | 0 | 0 | 0 | 0.001165258 |
| 0.686675408 | 0.216383195 | 0 | 0 | 0.733600116 |
| 0 | 0 | 0 | 0 | 0 |
| 0 | 0 | 0 | 0 | 0 |
| 0 | 0 | 0 | 0 | 0 |
| 0 | 0 | 0 | 0 | 0 |
| 0 | 0 | 0 | 0 | 0 |
| 0 | 0.000716274 | 0 | 0 | 0.001474733 |
| 0.020561219 | 0.002441267 | 0.045021171 | 0.088561709 | 0.000753947 |
| 0 | 0 | 0 | 0 | 0.003761846 |
| 0.019393513 | 0.643775188 | 0.127664029 | 0.005807633 | 0.710290269 |
| 0 | 0 | 0 | 0 | 0 |
| 0 | 0.115087656 | 0.223761231 | 0.44602499 | 0.795682449 |
| 0 | 0 | 0 | 0 | 0 |
| 0 | 0 | 0 | 0 | 0 |
| 0.007469916 | 0.00063078 | 0 | 0 | 0 |
| 0 | 0 | 0 | 0 | 0 |
| 0.022069235 | 0.001172553 | 0.023728464 | 0.02579257 | 0 |
| 0 | 0 | 0 | 0 | 0 |
| 0 | 0 | 0 | 0 | 0 |
| 0.061790313 | 0.000362865 | 0 | 0.100866667 | 0 |
| 0 | 0 | 0 | 0 | 0.0052001 |
| 0 | 0 | 0 | 0 | 0 |
| 2.843187111 | 1.849947177 | 25.01302545 | 1.191021822 | 9.058459036 |
| 0.34991493 | 0.598527311 | 0.270279996 | 0.593356805 | 0.534739981 |
| 0 | 0 | 0 | 1.471897681 | 0 |
| 0 | 0.860719864 | 0 | 0 | 0 |
| 0 | 0 | 0 | 0 | 0 |
| 0 | 0 | 0 | 0 | 0 |
| 0 | 0 | 0 | 0 | 0 |
| 0.009290592 | 0.000479983 | 0 | 0.062425035 | 0.00059294 |

| | | | | |
|---|---|---|---|---|
| 0 | 0 | 0 | 0 | 0 |
| 0.040885765 | 0.056281991 | 0.00527002 | 0 | 0 |
| 0 | 0.002652432 | 0 | 0 | 0 |
| 0 | 0 | 0 | 0 | 0 |
| 0.79906329 | 0.088237874 | 0 | 4.48350004 | 1.622731093 |
| 0 | 0 | 0 | 0 | 0 |
| 0 | 0 | 0 | 0 | 0 |
| 0 | 0 | 0 | 0 | 0 |
| 0.10592885 | 0.003549411 | 0.312911347 | 0.000852022 | 0.016199097 |
| 0.029916617 | 0 | 0 | 0 | 0 |
| 0 | 0 | 0 | 0 | 0 |
| 0 | 0 | 0 | 0 | 0 |
| 0.124824242 | 0.203111467 | 0 | 0 | 0.136181764 |
| 0.010981979 | 0.002005188 | 0.030248664 | 0.103714018 | 0.032639204 |
| 0 | 0 | 0 | 0 | 0.028405697 |
| 0.25197279 | 0.446495751 | 4.766674304 | 0.116473179 | 0.435949198 |
| 0.001000337 | 0.019699388 | 0 | 0.003969217 | 0.010465473 |
| 0 | 0 | 0 | 0 | 0 |
| 0 | 0 | 0 | 0 | 0 |
| 0 | 0 | 0 | 0 | 0 |
| 0 | 0 | 0 | 0 | 0 |
| 0 | 0 | 0 | 0 | 0 |
| 0 | 0 | 0 | 0 | 0 |
| 0 | 0 | 0 | 0 | 0 |
| 0 | 0 | 0 | 0 | 0 |
| 0 | 0 | 0 | 0 | 0.013279985 |
| 0 | 0.131000182 | 0.006470046 | 0 | 0.047803245 |
| 0.012882323 | 0.003505194 | 0.005605912 | 0.001947258 | 0 |
| 0 | 0 | 0 | 0 | 0 |
| 0 | 0 | 0 | 0 | 0 |
| 0 | 0 | 0 | 0 | 0 |
| 0.001024828 | 0.000669238 | 0 | 0 | 0 |
| 0 | 0 | 0 | 0 | 0 |
| 0 | 0.018161932 | 0 | 0 | 0.001296884 |
| 0.199418925 | 0 | 0.041378676 | 0 | 0 |
| 0.270935718 | 0.11040593 | 0 | 0.044050316 | 0.011162962 |
| 0 | 0 | 0 | 0 | 0 |
| 0 | 0.020347677 | 0 | 0 | 0.000405423 |
| 0 | 0 | 0 | 0 | 0 |
| 0 | 0 | 0 | 0 | 0 |
| 0 | 0 | 2.591223405 | 0.42669785 | 2.364687993 |
| 0 | 0 | 0 | 0 | 0 |
| 0 | 0 | 0 | 0 | 0 |
| 0 | 0 | 0 | 0 | 0 |
| 0 | 0 | 0 | 0 | 0 |
| 0 | 0 | 0 | 0 | 0 |
| 0 | 0 | 0 | 0 | 0 |
| 0 | 0 | 0 | 0 | 0 |

| | | | | |
|---|---|---|---|---|
| 0.153004988 | 0.106473189 | 0 | 0 | 0 |
| 0.002564042 | 0 | 0 | 0 | 0 |
| 0 | 0 | 0 | 0 | 0.007472692 |
| 0.024321909 | 0 | 0 | 0.003676435 | 0 |
| 0 | 0.017801876 | 0 | 0 | 0.006436481 |
| 0 | 0 | 0 | 0 | 1.778432439 |
| 0 | 0.04755049 | 0.33057374 | 0.003959565 | 0.217982699 |
| 0 | 0 | 0 | 0 | 0 |
| 0 | 0.000718693 | 0 | 0 | 0 |
| 0.117652593 | 0.202543967 | 0.007509457 | 0 | 0 |
| 0 | 0.006050268 | 0 | 0 | 0.002135462 |
| 0 | 0.028227121 | 0 | 0 | 0.00033 |
| 0 | 0 | 0 | 0 | 0 |
| 0 | 0 | 0 | 0 | 0 |
| 0 | 0 | 0 | 0 | 0.001977483 |
| 0 | 0 | 0 | 0 | 0 |
| 0 | 0 | 0 | 0.001885497 | 0 |
| 0.055404216 | 0.003034602 | 0 | 0 | 0 |
| 0 | 0 | 0 | 0 | 0 |
| 0 | 0 | 0 | 0 | 0 |
| 0.028383061 | 0.005040851 | 0.563702845 | 0.013612902 | 0.046703609 |
| 0 | 0 | 0 | 0 | 0 |
| 0 | 0.15290112 | 0 | 0.00854699 | 0.002641739 |
| 0.023411866 | 0.002186789 | 0.002475386 | 0.049092358 | 0.004580671 |
| 0 | 0 | 0 | 0 | 0 |
| 0.005496664 | 0.000785193 | 0 | 0.006058363 | 0 |
| 0 | 0 | 0 | 0 | 0 |
| 0 | 0 | 0 | 0 | 0 |
| 0 | 0 | 0 | 0 | 0 |
| 0.004861491 | 0 | 0.739430212 | 0.126824987 | 0 |
| 0 | 0 | 0 | 0 | 0 |
| 0 | 0 | 0 | 0 | 0 |
| 0 | 0 | 0 | 0 | 0 |
| 0 | 4.229741954 | 0 | 0 | 0.038507879 |
| 0 | 0 | 0 | 0 | 0 |
| 0 | 0 | 0 | 0 | 0 |
| 1.287054868 | 0.000594835 | 0 | 0 | 0 |
| 0 | 0 | 0 | 0 | 0 |
| 0 | 0 | 0 | 0 | 0 |
| 0 | 0 | 0 | 0 | 0 |
| 0 | 0 | 0 | 0 | 0 |
| 0 | 0 | 0 | 0 | 0 |
| 0 | 0 | 0 | 0 | 0 |
| 0.023781636 | 0 | 0 | 0 | 0 |
| 0 | 0 | 0 | 0 | 0.007232709 |
| 0 | 0 | 0 | 0 | 0 |
| 0 | 0 | 0 | 0 | 0 |
| 0 | 0 | 0 | 0 | 0 |

| | | | | |
|---|---|---|---|---|
| 0.101143994 | 0.00634304 | 0 | 0.017018972 | 0 |
| 0 | 0 | 0 | 0 | 0 |
| 0 | 0 | 0 | 0 | 0 |
| 0 | 0 | 0 | 0 | 0 |
| 0 | 0.006077551 | 0 | 0 | 0 |
| 0 | 0 | 0 | 0 | 0 |
| 0 | 0 | 0 | 0.002378797 | 0 |
| 0.298429789 | 1.618237615 | 0.276090008 | 1.149014144 | 0.916807375 |
| 1.270872435 | 0.083986657 | 0.275313917 | 1.500137791 | 0.006546481 |
| 0.002739143 | 0 | 0.012370197 | 0.331371293 | 0 |
| 0 | 0.078320383 | 0 | 0 | 0 |
| 0 | 0 | 0 | 0 | 0.003263975 |
| 0 | 0 | 0 | 0 | 0.001554836 |
| 0.327840771 | 0 | 0 | 0.273535123 | 0.009399223 |
| 0 | 0.001075103 | 0 | 0.002956121 | 0 |
| 0 | 0 | 0 | 0 | 0 |
| 0.004024964 | 0.000746704 | 0 | 0 | 0.015127868 |
| 0 | 0 | 0 | 0 | 0 |
| 0 | 0 | 0 | 0 | 0 |
| 0.305952657 | 0 | 0 | 0 | 0 |
| 0 | 0 | 0 | 0 | 0 |
| 0 | 0 | 0 | 0 | 0 |
| 0 | 0 | 0 | 0 | 0 |
| 0 | 0 | 0 | 0 | 0 |
| 0 | 0.085496458 | 0 | 0 | 0.004977234 |
| 0 | 0 | 0 | 0 | 0 |
| 0 | 0 | 0 | 0 | 0 |
| 0 | 0 | 0 | 0 | 0 |
| 0.005508176 | 0 | 0 | 0 | 0 |
| 0 | 0 | 0 | 0 | 0 |
| 0 | 0 | 0 | 0 | 0 |
| 0 | 0 | 0 | 0 | 0 |
| 0 | 0 | 0 | 0 | 0 |
| 0 | 0.008704354 | 0 | 0 | 0 |
| 0 | 0 | 0 | 0.106497878 | 0 |
| 0 | 0 | 0 | 0 | 0 |
| 0 | 0 | 0 | 0 | 0 |
| 0 | 0 | 0 | 0 | 0 |
| 0 | 0 | 0 | 0.041262224 | 0 |
| 0 | 0 | 0 | 0 | 0 |
| 0 | 0 | 0 | 0 | 0 |
| 0.605798043 | 1.941123898 | 0 | 0.767610493 | 0.128475103 |
| 0 | 0 | 0 | 0 | 0 |
| 0 | 0 | 0 | 0 | 0 |
| 0 | 0 | 0 | 0 | 0 |
| 0 | 0 | 0 | 0 | 0 |
| 0 | 0 | 0 | 0 | 0 |
| 0 | 0 | 0 | 0 | 0 |

| | | | | |
|---|---|---|---|---|
| 0 | 0 | 0 | 0 | 0 |
| 0.032465098 | 0 | 0 | 0.003191764 | 0 |
| 0 | 0 | 0 | 0 | 0 |
| 0 | 0 | 0 | 0 | 0 |
| 0.314730435 | 0 | 0.005713964 | 0.590728122 | 0.219127512 |
| 0 | 0 | 0 | 0 | 0 |
| 0 | 0 | 0 | 0 | 0 |
| 0.067759053 | 0.001561704 | 0.02930876 | 0.059043721 | 0.001607692 |
| 0 | 0.00129942 | 0 | 0 | 0 |
| 0 | 0 | 0 | 0 | 0 |
| 0 | 0 | 0 | 0 | 0 |
| 0 | 0 | 0 | 0 | 0 |
| 0 | 0 | 0 | 0 | 0 |
| 0 | 0 | 0 | 0 | 0 |
| 0.0506181 | 0 | 0 | 0 | 0 |
| 0 | 0.152376131 | 0 | 0 | 0.000418302 |
| 0 | 0 | 0 | 0 | 0 |
| 0 | 0 | 0 | 0 | 0 |
| 0 | 0 | 0 | 0 | 0 |
| 0 | 0 | 0 | 0 | 0 |
| 0 | 0 | 0 | 0 | 0 |
| 0 | 0 | 0 | 0 | 0 |
| 0 | 0 | 0 | 0 | 0 |
| 0.025387619 | 0.004279834 | 0.00483828 | 0.002189892 | 0 |
| 0 | 0.271660213 | 0 | 0.074038819 | 0.160306684 |
| 0 | 0 | 0 | 0 | 0.004184612 |
| 20.00870377 | 0.015348544 | 0.07589814 | 0.010459613 | 0.004581089 |
| 0.002391824 | 0.001518532 | 0 | 0 | 0 |
| 0 | 0 | 0 | 0 | 0 |
| 0 | 0 | 0 | 0 | 0 |
| 0 | 0 | 0 | 0 | 0 |
| 0 | 0 | 0 | 0 | 0 |
| 0 | 0 | 0 | 0 | 0 |
| 0 | 0 | 0 | 0 | 0 |
| 0 | 0 | 0 | 0 | 0 |
| 0.179378953 | 0 | 0 | 0 | 0 |
| 0 | 0 | 0 | 0 | 0 |
| 0 | 0.06011096 | 0 | 0 | 0.022055324 |
| 0 | 0 | 0 | 0 | 0 |
| 0 | 0 | 0 | 0 | 0.002127245 |
| 0 | 0 | 0 | 0 | 0 |
| 0 | 0 | 0 | 0 | 0 |
| 0 | 0 | 0 | 0 | 0 |
| 0 | 0 | 0.115831951 | 0 | 0 |
| 0 | 0 | 0 | 0 | 0 |
| 0 | 0 | 0 | 0 | 0 |
| 0 | 0 | 0 | 0 | 0 |
| 0 | 0 | 0 | 0 | 0 |

| | | | | |
|---|---|---|---|---|
| 0 | 0 | 0 | 0.002929981 | 0.14827112 |
| 0 | 0 | 0 | 0 | 0 |
| 0.022100259 | 0.142976597 | 0 | 0 | 0 |
| 0.01912823 | 0 | 0 | 0 | 0 |
| 0 | 0 | 0 | 0 | 0 |
| 0 | 0 | 0 | 0.722554769 | 0 |
| 0 | 0 | 0 | 0 | 0 |
| 0 | 0.001090102 | 0 | 0 | 0 |
| 0 | 0 | 0 | 0 | 0 |
| 0 | 0 | 0 | 0 | 0 |
| 0 | 0 | 0 | 0 | 0 |
| 0.004960489 | 0 | 0 | 0.007850621 | 0 |
| 0 | 0 | 0 | 0 | 0 |
| 0 | 0 | 0 | 0 | 0 |
| 0 | 0 | 0 | 0 | 0 |
| 0 | 0 | 0 | 0 | 0 |
| 0 | 0 | 0 | 0 | 0 |
| 0 | 0 | 0 | 0 | 0 |
| 0 | 0 | 0 | 0 | 0 |
| 0 | 0 | 0 | 0 | 0 |
| 0.01401474 | 0 | 0.020878238 | 0 | 0 |
| 0 | 0 | 0 | 0 | 0 |
| 0.007461594 | 0.002266326 | 0.001475116 | 0.321925171 | 0.001224858 |
| 0 | 0.02566896 | 0 | 0 | 0.001121054 |
| 0 | 0 | 0 | 0 | 0 |
| 0 | 0 | 0 | 0 | 0 |
| 0 | 0 | 0 | 0 | 0 |
| 0 | 0 | 0 | 0 | 0 |
| 0 | 0 | 0.005267551 | 0 | 0.370966924 |
| 0 | 0 | 0 | 0 | 0 |
| 0 | 0 | 0 | 0 | 0 |
| 0 | 0 | 0 | 0 | 0.060172146 |
| 0.007637776 | 0.015918035 | 0.004316999 | 0.327560326 | 0.00589924 |
| 0 | 0 | 0 | 0 | 0 |
| 0.090995814 | 0 | 0 | 0.048192666 | 0.036204999 |
| 0 | 0.011389754 | 0 | 0 | 0 |
| 0 | 0 | 0 | 0 | 0 |
| 0 | 0 | 0 | 0 | 0 |
| 0.008748315 | 0 | 0.019072883 | 0.218115389 | 0 |
| 0 | 0 | 0 | 0 | 0 |
| 0 | 0 | 0 | 0 | 0 |
| 0 | 0 | 0 | 0 | 0.135747 |
| 0.040741239 | 0 | 0.006559387 | 1.610498484 | 0.108486724 |
| 0 | 0 | 0 | 0 | 0 |
| 0 | 0 | 0 | 0 | 0 |
| 0.064431084 | 0.007660402 | 0.001041858 | 0.003705142 | 0.001271173 |
| 0 | 0 | 0 | 0 | 0 |
| 0 | 0 | 0 | 0 | 0 |

| | | | | |
|---|---|---|---|---|
| 0 | 0 | 0 | 0 | 0 |
| 0 | 0 | 0 | 0 | 0 |
| 0 | 0 | 0 | 0 | 0 |
| 0 | 0 | 0.000970509 | 0 | 0 |
| 0 | 0 | 0 | 0 | 0 |
| 0 | 0 | 0 | 0 | 0 |
| 0 | 0 | 0 | 0 | 0 |
| 0.001082797 | 0.160753076 | 0 | 0 | 0 |
| 0.000787382 | 0.061058797 | 0 | 0 | 0.001323301 |
| 0.079058894 | 0 | 0.003219307 | 0 | 0.028658992 |
| 0 | 0 | 0 | 0 | 0 |
| 0 | 0 | 0.005167752 | 0 | 0 |
| 0 | 0 | 0 | 0 | 0 |
| 0 | 0 | 0 | 0 | 0 |
| 0.010259768 | 0.053104122 | 0.009043794 | 0 | 0 |
| 0.023988505 | 0 | 0 | 0 | 0 |
| 0 | 0 | 0 | 0 | 0 |
| 0 | 0 | 0 | 0 | 0 |
| 0 | 0 | 0 | 0 | 0 |
| 0 | 0.559243038 | 4.017350105 | 0 | 1.126097111 |
| 0 | 0 | 0.005513615 | 0.001307198 | 0.003438332 |
| 0 | 0 | 0 | 0 | 0 |
| 0 | 0 | 0 | 0 | 0 |
| 0 | 0.343551417 | 0 | 0 | 0 |
| 0 | 0 | 0 | 0 | 0 |
| 0 | 0 | 1.756215915 | 0 | 0 |
| 0 | 0 | 0 | 0 | 0 |
| 0 | 0 | 0 | 0 | 0 |
| 0 | 0 | 0 | 0 | 0 |
| 0 | 0.014890678 | 0 | 0 | 0 |
| 0 | 0.230346063 | 0.164149173 | 0.146485182 | 0.457661142 |
| 0 | 0 | 0 | 0 | 0 |
| 0 | 0.003698483 | 0 | 0 | 0 |
| 0 | 0 | 0 | 0 | 0 |
| 0 | 0 | 0 | 0 | 0 |
| 0 | 0 | 0 | 0 | 0 |
| 0 | 0 | 0 | 0 | 0 |
| 0 | 0 | 0 | 0 | 0 |
| 0 | 0 | 0 | 0 | 0.004851214 |
| 0 | 0 | 0 | 0 | 0 |
| 0 | 0.004428854 | 0 | 0 | 0 |
| 0 | 0 | 0 | 0 | 0.003563953 |
| 0 | 0 | 0 | 0 | 0.001006611 |
| 0 | 0.006311828 | 0 | 0.002941132 | 0 |
| 0 | 0 | 0 | 0 | 0 |
| 0 | 0 | 0.029806104 | 0 | 0 |
| 0.057977434 | 0 | 0.085076701 | 0.016912432 | 0 |
| 0 | 0.002506273 | 0 | 0 | 0 |

| | | | | |
|---|---|---|---|---|
| 0 | 0 | 0 | 0 | 0 |
| 0 | 0 | 0 | 0 | 0 |
| 0 | 0 | 0 | 0 | 0 |
| 0 | 0 | 0 | 0 | 0 |
| 0.147743451 | 0.141157857 | 0.176257017 | 0.05972838 | 0.055717601 |
| 0 | 0 | 0 | 0 | 0 |
| 0 | 0 | 0 | 0 | 0 |
| 0 | 0 | 0 | 0 | 0 |
| 0 | 0 | 0 | 0 | 0 |
| 0 | 0 | 0 | 0 | 0 |
| 0 | 0 | 0 | 0 | 0 |
| 0.677906713 | 0.02275943 | 0.013452491 | 0.14538307 | 0 |
| 0.003900818 | 0.001698219 | 0 | 0.008254922 | 0 |
| 0 | 0 | 0.000956088 | 0 | 0.010585133 |
| 0 | 0.050064015 | 0 | 0 | 0 |
| 0 | 0.02917891 | 0 | 0 | 0.067364241 |
| 0.020980818 | 0.00293322 | 0 | 0 | 0.006662592 |
| 0.010415787 | 0 | 0.000670782 | 0 | 0 |
| 0.07988448 | 0.172894632 | 0.064391989 | 0 | 0 |
| 0.006854315 | 0.102291931 | 0.111943422 | 0.001576643 | 0.020434764 |
| 0 | 0 | 0 | 0 | 0 |
| 0 | 0 | 0 | 0 | 0 |
| 0 | 0 | 0 | 0 | 0 |
| 0 | 0 | 0 | 0 | 0 |
| 0 | 0 | 0 | 0 | 0 |
| 0 | 0 | 0 | 0 | 0 |
| 0.131346613 | 0.000389875 | 0.008247313 | 0.001474009 | 0.000963254 |
| 0.015096577 | 0.001109073 | 0 | 0 | 0 |
| 0 | 0 | 0 | 0 | 0 |
| 0 | 0 | 0 | 0 | 0 |
| 0.006496943 | 0.002209719 | 0.001150617 | 0 | 0 |
| 0.069443571 | 0.019213367 | 0.023302507 | 0 | 0 |
| 0 | 0 | 0 | 0 | 0 |
| 0 | 0 | 0 | 0 | 0 |
| 0 | 0 | 0 | 0 | 0 |
| 0 | 0 | 0 | 0 | 0 |
| 0 | 0 | 0 | 0 | 0 |
| 0.337207151 | 0.010091148 | 0.004345097 | 0.206103778 | 0.000287676 |
| 0 | 0 | 0 | 0 | 0.260895623 |
| 0.007843964 | 0 | 0.007533608 | 0 | 0 |
| 0.338065463 | 0 | 0 | 0.023271247 | 0 |
| 0 | 0 | 0 | 0 | 0 |
| 0 | 0 | 0 | 0 | 0 |
| 0.002397237 | 0.003107885 | 0.001348582 | 0 | 0 |
| 0 | 0 | 0 | 0 | 0 |
| 0 | 0 | 0 | 0 | 0 |
| 0 | 0 | 0 | 0 | 0 |
| 0 | 0 | 0 | 0 | 0 |

| | | | | |
|---|---|---|---|---|
| 0 | 0 | 0 | 0 | 0 |
| 0 | 0 | 0 | 0 | 0.007292138 |
| 0 | 0.009682898 | 0 | 0 | 0 |
| 0.007241379 | 0.002653915 | 0.001256285 | 0.000781848 | 0 |
| 0 | 0.000559424 | 0 | 0.010152125 | 0.011748301 |
| 0.000530693 | 0.000433194 | 0 | 0 | 0 |
| 0 | 0 | 0.007467166 | 0 | 0.000578457 |
| 0.317265162 | 0 | 0 | 0 | 0 |
| 0 | 0 | 0 | 0 | 0 |
| 0 | 0 | 0 | 0 | 0 |
| 0 | 0 | 0 | 0 | 0.022534439 |
| 0 | 0 | 0 | 0 | 0 |
| 0.17033573 | 0.011715072 | 0 | 0 | 0.015528412 |
| 0 | 0 | 0 | 0 | 0 |
| 0 | 0.007716864 | 0 | 0 | 0 |
| 0 | 0 | 0 | 0 | 0 |
| 0 | 0 | 0 | 0 | 0 |
| 0 | 0 | 0 | 0 | 0 |
| 0 | 0 | 0 | 0 | 0 |
| 0 | 0 | 0 | 0 | 0 |
| 0 | 0 | 0 | 0 | 0 |
| 0 | 0 | 0 | 0 | 0 |
| 0.028829777 | 0 | 0.006302842 | 0 | 0 |
| 0 | 0.000664353 | 0.006572736 | 0.269832054 | 0.31941625 |
| 0 | 0 | 0 | 0 | 0 |
| 0 | 0 | 0 | 0 | 0.000606094 |
| 0 | 0 | 0 | 0 | 0.070434709 |
| 0 | 0 | 0 | 0 | 0 |
| 0 | 0 | 0 | 0 | 0 |
| 0 | 0 | 0 | 0 | 0 |
| 0 | 0.002636493 | 0.014545581 | 0.018715147 | 0.064363661 |
| 0.003211318 | 0 | 0.003538755 | 0.011326333 | 0 |
| 0 | 0 | 0 | 0 | 0 |
| 0 | 0 | 0 | 0 | 0 |
| 0 | 0.008420481 | 0 | 0 | 0 |
| 0 | 0 | 0 | 0 | 0 |
| 0 | 0.010937553 | 0.004167267 | 0 | 0 |
| 0.017872956 | 0 | 0 | 0 | 0 |
| 0 | 0 | 0.001521037 | 0 | 0.002099513 |
| 0 | 0 | 0 | 0 | 0 |
| 0 | 0 | 0 | 0 | 0 |
| 0 | 0 | 0 | 0 | 0 |
| 0 | 0 | 0 | 0 | 0 |
| 0 | 0 | 0 | 0 | 0 |
| 0 | 0 | 0 | 0 | 3.632292762 |
| 0.095944316 | 0.043551219 | 0.125674886 | 0.427840816 | 0.215317128 |
| 0 | 0 | 0.172020399 | 0 | 0 |
| 0 | 0 | 0 | 0 | 0 |

| | | | | |
|---|---|---|---|---|
| 0 | 0 | 0 | 0 | 0 |
| 0 | 0 | 0 | 0 | 0 |
| 0 | 0 | 0 | 0 | 0 |
| 0 | 0.523908586 | 0 | 0 | 0 |
| 0 | 0 | 0.000769861 | 0 | 0 |
| 0 | 0.000258126 | 0 | 0 | 0 |
| 0 | 0 | 0 | 0 | 0 |
| 0 | 0 | 0 | 0 | 0 |
| 0.001204039 | 0.067815496 | 0 | 0 | 0.001456956 |
| 0 | 0 | 0 | 0 | 0 |
| 0.00200582 | 0 | 0 | 0.025505567 | 0 |
| 0.000788078 | 0.001801218 | 0.001172385 | 0.001389777 | 0 |
| 0 | 0 | 0 | 0 | 0 |
| 0 | 0 | 0 | 0 | 0 |
| 0 | 0 | 0 | 0 | 0 |
| 0 | 0 | 0 | 0 | 0 |
| 0 | 0 | 0 | 0 | 0 |
| 0 | 0 | 0 | 0 | 0 |
| 0 | 0 | 0 | 0 | 0 |
| 0.006020141 | 0.000571409 | 0 | 0 | 0 |
| 0 | 0 | 0 | 0 | 0 |
| 0 | 0 | 0 | 0 | 0 |
| 0 | 0 | 0 | 0 | 0 |
| 0 | 0 | 0 | 0 | 0 |
| 0 | 0 | 0.001801703 | 0 | 0 |
| 0.131081599 | 0 | 0 | 0 | 0 |
| 0 | 0 | 0 | 0 | 0 |
| 0 | 0 | 0 | 0.00601111 | 0 |
| 0.001367797 | 0 | 0 | 0 | 0 |
| 0 | 0 | 0 | 0 | 0 |
| 0.018402426 | 0.004319918 | 0.002683959 | 0.004878514 | 0.000606426 |
| 0 | 0 | 0 | 0 | 0 |
| 0.005865476 | 0 | 0.040364151 | 0.014037987 | 0.131389336 |
| 0 | 0.00745552 | 0 | 0 | 0 |
| 0 | 0 | 0 | 0 | 0 |
| 0 | 0 | 0 | 0 | 0 |
| 0 | 0 | 0 | 0 | 0 |
| 0 | 0 | 0 | 0 | 0 |
| 0 | 0 | 0 | 0 | 0 |
| 0 | 0 | 0 | 0 | 0 |
| 0 | 0 | 0 | 0 | 0 |
| 0.374548683 | 0.020538747 | 0.257233873 | 0 | 0.007414727 |
| 0 | 0.001713835 | 0.007585455 | 0.098242088 | 0.036132912 |
| 0 | 0 | 0 | 0 | 0 |
| 0 | 0 | 0 | 0 | 0 |
| 0.026901647 | 0 | 0.005158357 | 0.599256201 | 0.011421937 |
| 0 | 0 | 0 | 0 | 0 |
| 0 | 0 | 0 | 0 | 0 |

| | | | | |
|---|---|---|---|---|
| 0 | 0 | 0 | 0 | 0 |
| 0 | 0 | 0 | 0 | 0.006210425 |
| 0.003939648 | 0.004054775 | 0.010556758 | 0.003322755 | 0.003281767 |
| 0 | 0 | 0 | 0 | 0 |
| 2.316270049 | 0.11822613 | 0 | 0.090445224 | 0.000965386 |
| 1.580087547 | 0.569860913 | 0.02065346 | 0 | 0 |
| 0 | 0 | 0 | 0 | 0 |
| 0 | 0 | 0 | 0 | 0 |
| 0 | 0 | 0 | 0 | 0 |
| 0 | 0 | 0 | 0 | 0 |
| 0 | 0 | 0 | 0 | 0 |
| 0 | 0 | 0 | 0 | 0 |
| 0 | 0 | 0 | 0 | 0 |
| 0 | 0 | 0.019735285 | 0 | 0 |
| 0 | 0 | 0 | 0 | 0 |
| 0 | 0 | 0 | 0 | 0 |
| 0 | 0.217255846 | 0 | 0 | 0.00101826 |
| 0 | 0 | 0 | 0 | 0 |
| 0 | 0 | 0.026889473 | 0 | 0 |
| 0 | 0 | 0 | 0 | 0 |
| 0.014551958 | 0 | 0 | 0 | 0 |
| 0.013089313 | 0 | 0.012005494 | 0 | 0 |
| 0 | 0 | 0 | 0 | 0 |
| 0 | 0 | 0 | 0 | 0 |
| 0.000705831 | 0 | 0 | 0.011451563 | 0.000711747 |
| 0.002414169 | 0 | 0.051004481 | 0.001753044 | 0 |
| 0 | 0 | 0 | 0 | 0 |
| 0 | 0.147079771 | 0.014637907 | 0.001943444 | 0.000416727 |
| 0 | 0 | 0 | 0 | 0 |
| 0 | 0 | 0 | 0 | 0 |
| 0 | 0 | 0 | 0 | 0 |
| 0.011457154 | 0 | 0.001587973 | 0 | 0 |
| 0 | 0 | 0 | 0 | 0 |
| 0 | 0 | 0 | 0 | 0 |
| 0 | 0 | 0 | 0 | 0 |
| 0 | 0 | 0 | 0 | 0 |
| 0 | 0 | 0.010117113 | 0 | 0 |
| 0 | 0 | 0.0033046 | 0 | 0 |
| 0 | 0.003367876 | 0 | 0 | 0.084899403 |
| 0 | 0 | 0 | 0 | 0 |
| 0 | 0 | 0 | 0 | 0 |
| 0 | 0 | 0 | 0 | 0.009165649 |
| 0.074254198 | 0.004071838 | 0 | 0.013069611 | 0 |
| 0 | 0 | 0 | 0 | 0 |
| 0 | 0 | 0 | 0 | 0 |
| 0 | 0 | 0 | 0 | 0 |
| 0.036193223 | 0.002135697 | 0 | 0.001922495 | 0 |
| 0 | 0 | 0 | 0 | 0 |

| | | | | |
|---|---|---|---|---|
| 0.016356519 | 0 | 0 | 0 | 0 |
| 0.429317896 | 0.001347428 | 0.006869989 | 0.066467878 | 0.003745187 |
| 0 | 0 | 0 | 0 | 0 |
| 0 | 0 | 0 | 0 | 0 |
| 0 | 0 | 0 | 0 | 0 |
| 0 | 0 | 0 | 0 | 0 |
| 0 | 0 | 0 | 0 | 0 |
| 0 | 0 | 0 | 0 | 0 |
| 0 | 0 | 0 | 0 | 0 |
| 0 | 0 | 0 | 0 | 0 |
| 0 | 0 | 0 | 0 | 0 |
| 0 | 0 | 0 | 0 | 0 |
| 0.057291849 | 0 | 0 | 0 | 0.006744953 |
| 0 | 0 | 0 | 0 | 0 |
| 0 | 0.006440153 | 0.002004771 | 0.047789479 | 0.010204123 |
| 0 | 0 | 0 | 0 | 0 |
| 0 | 0 | 0 | 0 | 0 |
| 0 | 0 | 0 | 0 | 0 |
| 0 | 0.02152831 | 0 | 0 | 0 |
| 0 | 0 | 0.001092385 | 0 | 0 |
| 0 | 0 | 0 | 0 | 0 |
| 0.001852841 | 0.025986426 | 0.09880015 | 0.002376356 | 0.013927845 |
| 0.005565596 | 0 | 0.002063039 | 0.00205429 | 0 |
| 0 | 0.000849711 | 0.01308917 | 0.001835727 | 0 |
| 0 | 0 | 0 | 0 | 0 |
| 0 | 0 | 0 | 0 | 0 |
| 0 | 0 | 0 | 0 | 0 |
| 0 | 0 | 0 | 0 | 0 |
| 0 | 0 | 0 | 0 | 0 |
| 0 | 0.002573597 | 0 | 0 | 0 |
| 0 | 0 | 0 | 0 | 0 |
| 0 | 0 | 0 | 0 | 0 |
| 0 | 0 | 0 | 0 | 0 |
| 0 | 0 | 0 | 0 | 0 |
| 16.15842297 | 42.2322955 | 9.216217562 | 0 | 0.027604657 |
| 0 | 0 | 0 | 0 | 0 |
| 0 | 0 | 0 | 0 | 0 |
| 0 | 0 | 0 | 0 | 0 |
| 0 | 0.261702779 | 0.004289735 | 0.005636064 | 0.010685906 |
| 0 | 0 | 0 | 0 | 0 |
| 0.020209959 | 0.022898801 | 0.560279849 | 0.013830567 | 0.135051094 |
| 0 | 0 | 0 | 0 | 0 |
| 0 | 0 | 0 | 0 | 0 |
| 0 | 0 | 0 | 0 | 0 |
| 0 | 0 | 0 | 0 | 0 |
| 0.038334807 | 0 | 0.157985199 | 0.405012073 | 0.049974299 |
| 0 | 0 | 0 | 0 | 0 |
| 0 | 0 | 0 | 0 | 0 |

| | | | | |
|---|---|---|---|---|
| 0 | 0 | 0 | 0 | 0 |
| 0 | 0 | 0.00089659 | 0 | 0 |
| 0 | 0 | 0 | 0 | 0 |
| 0 | 0 | 0 | 0 | 0 |
| 0.053433389 | 0 | 0 | 0 | 0 |
| 0 | 0 | 0 | 0 | 0 |
| 0.110980552 | 0 | 0 | 0 | 0 |
| 0 | 0 | 0.000594981 | 0 | 0 |
| 0 | 0 | 0 | 0 | 0.001297546 |
| 0 | 0 | 0 | 0.038716781 | 0.003916807 |
| 0 | 0 | 0 | 0 | 0 |
| 0 | 0 | 0 | 0 | 0 |
| 0.007427997 | 0.00242533 | 0.003157219 | 0.001164381 | 0 |
| 0 | 0 | 0 | 0 | 0 |
| 0 | 0 | 0 | 0 | 0 |
| 0 | 0 | 0 | 0 | 0 |
| 0 | 0 | 0 | 0 | 0 |
| 0 | 0 | 0 | 0 | 0 |
| 0 | 0 | 0 | 0 | 0 |
| 0.438705885 | 0.002758358 | 0.28913295 | 11.22504857 | 0.076446546 |
| 0 | 0 | 0 | 0 | 0 |
| 0 | 0 | 0 | 0 | 0 |
| 0 | 0 | 0 | 0 | 0 |
| 0 | 0 | 0 | 0 | 0 |
| 0 | 0 | 0 | 0 | 0 |
| 0 | 0 | 0 | 0 | 0 |
| 0 | 0 | 0 | 0 | 0 |
| 0 | 0 | 0 | 0 | 0 |
| 0 | 0 | 0 | 0 | 0 |
| 0 | 0 | 0 | 0 | 0 |
| 0 | 0 | 0 | 0 | 0 |
| 0 | 0 | 0 | 0.008708345 | 0 |
| 0 | 0 | 0 | 0 | 0 |
| 0 | 0 | 0 | 0 | 0 |
| 0 | 0 | 0 | 0 | 0 |
| 0.001141115 | 0 | 0 | 0 | 0 |
| 0 | 0 | 0 | 0 | 0 |
| 0 | 0 | 0 | 0 | 0 |
| 0 | 0 | 0 | 0 | 0 |
| 0 | 0 | 0 | 0 | 0 |
| 0 | 0 | 0 | 0 | 0 |
| 0 | 0 | 0 | 0 | 0 |
| 0.01038075 | 1.072458511 | 0 | 0 | 0.006280648 |
| 0 | 0 | 0 | 0 | 0 |
| 0.052989517 | 0 | 0 | 0.050860549 | 0 |
| 0.024140419 | 0 | 0 | 0 | 0 |
| 0 | 0 | 0 | 0 | 0 |
| 0 | 0 | 0 | 0 | 0 |

| | | | | |
|---|---|---|---|---|
| 0 | 0 | 0 | 0 | 0 |
| 0 | 0 | 0 | 0 | 0 |
| 0 | 0 | 0 | 0 | 0 |
| 0 | 0 | 0 | 0 | 0 |
| 0 | 0 | 0 | 0 | 0 |
| 0.115070589 | 0.0370842 | 0 | 0.095559294 | 0.006128973 |
| 0 | 0 | 0 | 0 | 0 |
| 0 | 0 | 0 | 0 | 0 |
| 0 | 0 | 0 | 0 | 0 |
| 0 | 0 | 0 | 0 | 0 |
| 0 | 0 | 0.2193072 | 0 | 0 |
| 0 | 0 | 0 | 0 | 0 |
| 0 | 0 | 0 | 0 | 0.0005398 |
| 0 | 0 | 0 | 0 | 0 |
| 0 | 0 | 0 | 0 | 0 |
| 0 | 0 | 0 | 0 | 0 |
| 0 | 0.124748425 | 0 | 0 | 0.003276177 |
| 0 | 0 | 0.025749613 | 0 | 0 |
| 0 | 0 | 0 | 0 | 0 |
| 0 | 0 | 0 | 0 | 0 |
| 0 | 0 | 0 | 0 | 0 |
| 0 | 0 | 0 | 0 | 0 |
| 0 | 0 | 0 | 0 | 0 |
| 0 | 0 | 0 | 0 | 0 |
| 0 | 0 | 0 | 0 | 0 |
| 0 | 0 | 0 | 0 | 0 |
| 0 | 0 | 0 | 0 | 0 |
| 1.13146635 | 0 | 0 | 0 | 0 |
| 0 | 0 | 0 | 0 | 0 |
| 0.033150123 | 0 | 0.06372267 | 0 | 0 |
| 0 | 0 | 0 | 0 | 0 |
| 0 | 0 | 0 | 0 | 0 |
| 0 | 0 | 0 | 0 | 0 |
| 0 | 0 | 0 | 0 | 0 |
| 0 | 0 | 0 | 0 | 0 |
| 0 | 0 | 0 | 0 | 0 |
| 0 | 0 | 0 | 0 | 0 |
| 0.233776697 | 0 | 0.000390586 | 0 | 0 |
| 0 | 0.920773023 | 0.098740071 | 0 | 0.270985532 |
| 0 | 0 | 0 | 0 | 0 |
| 0 | 0 | 0 | 0 | 0 |
| 0 | 0 | 0 | 0 | 0 |
| 0 | 0 | 0 | 0 | 0 |
| 0 | 0 | 0 | 0 | 0 |
| 0 | 0 | 0 | 0 | 0 |
| 0 | 0 | 0 | 0 | 0 |
| 0 | 0 | 0 | 0 | 0 |
| 0 | 0 | 0 | 0 | 0 |

| | | | | |
|---|---|---|---|---|
| 0 | 0 | 0 | 0 | 0 |
| 0 | 0 | 0 | 0.013017348 | 0.011862909 |
| 0 | 0 | 0 | 0 | 0 |
| 0 | 0 | 0 | 0 | 0 |
| 0 | 0 | 0 | 0 | 0 |
| 0 | 0 | 0 | 0 | 0 |
| 0 | 0 | 0 | 0 | 0 |
| 0 | 0 | 0 | 0 | 0 |
| 0 | 0 | 0 | 0 | 0 |
| 0 | 0 | 0 | 0 | 0.000473735 |
| 0 | 0.002588834 | 0 | 0.006658276 | 0.004568688 |
| 0 | 0 | 0 | 0 | 0 |
| 0 | 0.21029359 | 0 | 0.000784665 | 0 |
| 0 | 0 | 0 | 0 | 0 |
| 0 | 0 | 0 | 0 | 0 |
| 0 | 0 | 0 | 0 | 0 |
| 0 | 0 | 0 | 0 | 0 |
| 0 | 0 | 0 | 0 | 0 |
| 0 | 0 | 0 | 0 | 0 |
| 0 | 0 | 0 | 0 | 0 |
| 0 | 0 | 0 | 0.024868517 | 0 |
| 0 | 0 | 0 | 0 | 0 |
| 0 | 0 | 0 | 0 | 0 |
| 0.487465653 | 0 | 0 | 1.443866238 | 2.184002175 |
| 0 | 0 | 0 | 0 | 0 |
| 0 | 0 | 0 | 0 | 0 |
| 0 | 0 | 0 | 0 | 0 |
| 0 | 0 | 0.588517309 | 0 | 0 |
| 0 | 0.010570065 | 0 | 0 | 0 |
| 0 | 0 | 0 | 0 | 0 |
| 0.000902667 | 0 | 0.001598634 | 0 | 0 |
| 0 | 0 | 0 | 0 | 0 |
| 0.001289889 | 0 | 0 | 0 | 0 |
| 0 | 0 | 0 | 0 | 0 |

| PHL018-0058_V3 | PHL018-0115_V1 | PHL018-0213_V2 | PHL018-0100_V1 | PHL018-0084_V3 |
|---|---|---|---|---|
| EYCF | EYCF | REF | EYCF | EYCF |
| 0.024853333 | 0.361051438 | 0.01618771 | 0.505818291 | 0.232547881 |
| 3.208936129 | 1.954172416 | 0.39503395 | 0.583534806 | 0.196611638 |
| 6.294952208 | 1.66907677 | 0.305981463 | 0.95272259 | 0 |
| 0 | 0 | 0 | 0 | 0 |
| 0 | 0 | 0 | 0 | 0 |
| 0 | 0 | 0 | 0 | 0 |
| 0 | 0 | 0 | 0 | 0 |
| 0 | 0 | 0 | 0 | 0 |
| 0 | 0 | 0 | 0 | 0 |
| 0 | 0 | 0 | 0 | 0 |
| 0 | 0 | 0 | 0 | 0 |
| 0 | 0 | 0 | 0 | 0 |
| 0 | 0 | 0 | 0 | 0 |
| 6.366030718 | 0.122766399 | 0.772564266 | 0.169650477 | 0.012314569 |
| 0 | 0 | 0 | 0 | 0 |
| 0 | 0 | 0 | 0 | 0 |
| 0 | 0 | 0 | 0 | 0 |
| 0 | 0 | 0 | 0 | 0 |
| 0 | 0 | 0 | 0 | 0 |
| 0 | 0 | 0 | 0 | 0 |
| 0 | 0 | 0 | 0 | 0 |
| 0 | 0 | 0 | 0 | 0 |
| 0 | 0 | 0 | 0 | 0 |
| 0 | 0 | 0 | 0 | 0 |
| 0 | 0 | 0.03513495 | 0 | 0.00044441 |
| 0 | 0 | 0 | 0 | 0 |
| 0 | 0 | 0 | 0 | 0 |
| 0 | 0 | 0 | 0 | 0 |
| 0 | 0 | 0 | 0 | 0 |
| 0 | 0 | 0 | 0 | 0 |
| 0 | 0 | 0 | 0 | 0 |
| 0 | 0 | 0 | 0 | 0 |
| 0 | 0 | 0 | 0 | 0 |
| 0 | 0 | 0 | 0 | 0 |
| 0 | 0 | 0 | 0 | 0 |
| 2.839712722 | 0.011153161 | 1.228825762 | 1.176369747 | 0.171771535 |
| 0 | 0 | 0 | 0 | 0 |
| 0 | 0 | 0 | 0 | 0 |
| 0 | 0 | 0 | 0 | 0 |
| 0 | 0 | 0 | 0 | 0 |
| 0 | 0 | 0 | 0 | 0 |
| 0.00798898 | 0.107796127 | 0 | 0 | 0.016709473 |
| 1.646351447 | 0.199638889 | 1.926016351 | 0.032038052 | 4.867117288 |
| 0.004158712 | 0.324375357 | 0.001748337 | 0.000832896 | 0 |
| 0 | 0 | 0.00929611 | 0.000750611 | 0 |
| 0 | 0 | 0 | 0 | 0 |

| | | | | |
|---|---|---|---|---|
| 0.011560379 | 0 | 0.00673869 | 0.016051327 | 0 |
| 0.009222933 | 0 | 0 | 0.000936958 | 0.044346874 |
| 0 | 0.002834664 | 0 | 0.012727379 | 0.034327098 |
| 0.374330102 | 2.075052018 | 0.714511715 | 0.616602423 | 0.222989362 |
| 0.00044608 | 0 | 0 | 0.004615889 | 0.031139074 |
| 0 | 0 | 0.015857766 | 0 | 0 |
| 0.018768116 | 0.586888468 | 0.000451728 | 0.003156269 | 0 |
| 0 | 0 | 0 | 0 | 0 |
| 0 | 0 | 0 | 0.007519093 | 0 |
| 0 | 0 | 0 | 0 | 0 |
| 0 | 0 | 0 | 0.001186364 | 0 |
| 0.022951497 | 0 | 0.256949576 | 0.262435185 | 0.558489095 |
| 0 | 0 | 0 | 0 | 0 |
| 0.001744269 | 0.011654421 | 0.00097054 | 0.043358973 | 0.043698561 |
| 0 | 0 | 0 | 0 | 0 |
| 0 | 0 | 0 | 0 | 0 |
| 6.102645521 | 0 | 0.435519623 | 0 | 0 |
| 0 | 0 | 0 | 0 | 0 |
| 0 | 0 | 0 | 0 | 0 |
| 0 | 0 | 0 | 0 | 0 |
| 0.101807238 | 0.100706016 | 0 | 0.009445702 | 0.27960669 |
| 0.002341323 | 0.097350565 | 0.001069892 | 0 | 0.006706803 |
| 0 | 0 | 0.063480472 | 0.009375486 | 0.861321885 |
| 0 | 0 | 0 | 0 | 0 |
| 0 | 0 | 0 | 0 | 0 |
| 0 | 0.001512574 | 0 | 0 | 0 |
| 0 | 0 | 0 | 0 | 0 |
| 0 | 0 | 0 | 0 | 0 |
| 0 | 0 | 0 | 0 | 0.000766066 |
| 0 | 0 | 0 | 0 | 0 |
| 0 | 0 | 0 | 0 | 0 |
| 0 | 0 | 0 | 0 | 0 |
| 0 | 0 | 0 | 0 | 0 |
| 0 | 0 | 0 | 0.001036761 | 0 |
| 0 | 0 | 0 | 0 | 0 |
| 0 | 0 | 0 | 0 | 0 |
| 0 | 0 | 0 | 0 | 0 |
| 0.134425255 | 0 | 23.21892495 | 0 | 0 |
| 0 | 0 | 0 | 0 | 0 |
| 0 | 0 | 0 | 0 | 0 |
| 0 | 0 | 0 | 0 | 0 |
| 0 | 0 | 0 | 0 | 0 |
| 0 | 0 | 0 | 0 | 0 |
| 0 | 0 | 0 | 0 | 0 |
| 0 | 0 | 0 | 0.232325187 | 0 |
| 0 | 0 | 0 | 0 | 0 |
| 0.000755132 | 0 | 0 | 0 | 0 |
| 0.026534677 | 0 | 0 | 0 | 0 |

| | | | | |
|---|---|---|---|---|
| 0 | 0 | 0 | 0 | 0 |
| 0 | 0 | 0 | 0 | 0 |
| 0 | 0 | 0 | 0 | 0 |
| 0 | 0 | 0 | 0 | 0 |
| 0.345064319 | 0.005307008 | 0 | 0.28509029 | 0 |
| 0 | 0 | 0 | 0 | 0 |
| 0 | 0 | 0 | 0 | 0 |
| 0 | 0 | 0 | 0 | 0.173685517 |
| 0.002066284 | 0 | 0 | 0.007291288 | 0.000926093 |
| 0 | 0 | 0 | 0 | 0 |
| 0 | 0 | 0 | 0 | 0 |
| 0 | 0.000837124 | 0 | 0 | 0.000940779 |
| 0 | 0 | 0 | 0 | 0 |
| 0 | 0 | 0 | 0 | 0 |
| 0 | 0 | 0 | 0 | 0 |
| 0 | 0 | 0 | 0 | 0 |
| 0 | 0 | 0 | 0 | 0 |
| 0 | 0 | 0 | 0 | 0 |
| 0 | 0.274160493 | 0.977879028 | 0.167365674 | 0.003229389 |
| 0 | 0 | 0 | 0 | 0 |
| 0 | 0 | 0 | 0 | 0 |
| 0 | 0 | 0 | 0 | 0 |
| 0 | 0 | 0 | 0 | 0 |
| 0 | 0 | 0 | 0 | 0 |
| 0 | 0 | 0 | 0 | 0 |
| 0 | 0 | 0 | 0 | 0 |
| 0.001586069 | 0 | 0.003182402 | 0 | 0 |
| 0 | 0 | 0 | 0.003183362 | 0 |
| 0 | 0 | 0 | 0 | 0 |
| 0 | 0 | 0 | 0 | 0 |
| 0 | 0.067391049 | 0.001052654 | 0.096739481 | 0.044670052 |
| 0.004202916 | 0.088023855 | 0.012950728 | 0.009565329 | 0.000337142 |
| 0 | 0 | 0 | 0 | 0 |
| 0 | 0 | 0 | 0 | 0 |
| 0 | 0 | 0 | 0 | 0 |
| 0 | 0 | 0 | 0 | 0 |
| 0 | 0 | 0 | 0 | 0 |
| 0 | 0 | 0 | 0 | 0 |
| 0 | 0.058627156 | 0.071117971 | 0.032779313 | 0 |
| 0 | 0 | 0 | 0 | 0 |
| 0 | 0 | 0 | 0 | 0 |
| 0 | 0 | 0 | 0 | 0 |
| 0.002208139 | 0.010518024 | 0.000348116 | 0.005196324 | 0 |
| 0 | 0 | 0 | 0 | 0 |
| 0 | 0 | 0 | 0.004597024 | 0 |
| 0 | 0 | 0 | 0 | 0 |
| 0 | 0 | 0 | 0 | 0 |
| 0 | 0 | 0 | 0 | 0 |

| | | | | |
|---|---|---|---|---|
| 0 | 0 | 0 | 0.001288446 | 0 |
| 0 | 0 | 0 | 0 | 0 |
| 0 | 0 | 0 | 0 | 0.053901878 |
| 0 | 0 | 0 | 0 | 0 |
| 0 | 0.039754402 | 0.001747619 | 0.090665092 | 0 |
| 0 | 0 | 0 | 0 | 0 |
| 0 | 0 | 0 | 0 | 0 |
| 0 | 0 | 0 | 0 | 0 |
| 0 | 0 | 0 | 0.000327418 | 0 |
| 0 | 0 | 0 | 0 | 0 |
| 0 | 0 | 0 | 0 | 0 |
| 0 | 0 | 0 | 0 | 0 |
| 0 | 0 | 0 | 0 | 0 |
| 0 | 0 | 0 | 0 | 0 |
| 0 | 0 | 0 | 0 | 0 |
| 0 | 0 | 0 | 0.9398146 | 0 |
| 0 | 0 | 0 | 0 | 0 |
| 0 | 0 | 0 | 0 | 0 |
| 0 | 0 | 0 | 0 | 0 |
| 0 | 0 | 0 | 0 | 0 |
| 0 | 0 | 0 | 0 | 0 |
| 0 | 0 | 0 | 0 | 0 |
| 0 | 0 | 0 | 0.002801779 | 0 |
| 0 | 0 | 0 | 0 | 0 |
| 0 | 0 | 0.008593428 | 0 | 0 |
| 0 | 0 | 0 | 0 | 0 |
| 0 | 0 | 0.001853917 | 0 | 0 |
| 0 | 0 | 0 | 0 | 0 |
| 0 | 0 | 0 | 0 | 0 |
| 0 | 0 | 0 | 0 | 0 |
| 0 | 0 | 0 | 0 | 0 |
| 0.000881683 | 0 | 0.000579161 | 0.015892309 | 0 |
| 0 | 0 | 0 | 0 | 0 |
| 0 | 0 | 0 | 0 | 0 |
| 0 | 0 | 0 | 0 | 0 |
| 0 | 0 | 0 | 0 | 0 |
| 0 | 0 | 0 | 0 | 0 |
| 0 | 0 | 0.0132123 | 0.327035903 | 0 |
| 0 | 0 | 0 | 0 | 0 |
| 0 | 0 | 0 | 0 | 0 |
| 0 | 0 | 0 | 0 | 0 |
| 0 | 0 | 0 | 0 | 0 |
| 0 | 0 | 0 | 0 | 0 |
| 0 | 0 | 0 | 0 | 0 |
| 0 | 0 | 0 | 0 | 0 |
| 0 | 0 | 0 | 0 | 0 |
| 0 | 0 | 0 | 0 | 0 |
| 0 | 0 | 0 | 0 | 0 |

| | | | | |
|---|---|---|---|---|
| 0 | 0 | 0.243902542 | 0 | 0.002652346 |
| 0 | 0 | 0 | 0 | 0 |
| 0 | 0 | 0 | 0 | 0 |
| 0 | 0 | 0 | 0 | 0 |
| 0 | 0 | 0 | 0 | 0 |
| 0 | 0 | 0 | 0 | 0 |
| 0 | 0 | 0 | 0 | 0 |
| 0 | 0 | 0 | 0 | 0 |
| 0 | 0 | 0 | 0 | 0 |
| 0 | 0 | 0 | 0 | 0 |
| 0 | 0 | 0 | 0 | 0 |
| 0 | 0 | 0 | 0 | 0 |
| 0 | 0 | 0 | 0.00961022 | 0.004342622 |
| 0 | 0 | 0 | 0 | 0 |
| 0 | 0 | 0 | 0 | 0 |
| 0 | 0 | 0 | 0 | 0 |
| 0 | 0 | 0 | 0 | 0 |
| 0 | 0 | 0 | 0 | 0 |
| 0 | 0 | 0 | 0 | 0 |
| 0 | 0 | 0 | 0 | 0 |
| 0.045352755 | 0.018423851 | 0.004559371 | 0.020338315 | 0.00811966 |
| 0 | 0 | 0 | 0 | 0 |
| 0 | 0 | 0 | 0.000779663 | 0 |
| 0.001028126 | 0 | 0 | 0 | 0 |
| 0 | 0 | 0 | 0 | 0 |
| 0 | 0 | 0 | 0 | 0 |
| 0.00160771 | 0.005259616 | 0.001569023 | 0.001724935 | 0 |
| 0 | 0 | 0 | 0 | 0 |
| 0 | 0 | 0 | 0 | 0 |
| 0 | 0.001800699 | 0 | 0 | 0 |
| 0 | 0 | 0 | 0 | 0 |
| 0 | 0 | 0 | 0 | 0 |
| 0 | 0 | 0 | 0 | 0 |
| 0 | 0.004415495 | 0.003166369 | 0.021962832 | 0 |
| 0.011931868 | 0.088378721 | 0 | 0.004496716 | 0.120001465 |
| 0 | 0 | 0 | 0 | 0 |
| 0 | 0 | 0 | 0 | 0 |
| 0 | 0 | 0 | 0 | 0 |
| 0 | 0 | 0 | 0 | 0 |
| 0 | 0 | 0 | 0 | 0 |
| 0 | 0 | 0 | 0 | 0 |
| 0 | 0.000938976 | 0 | 0 | 0 |
| 0 | 0 | 0 | 0 | 0 |
| 0 | 0 | 0 | 0 | 0 |
| 0 | 0 | 0.003626058 | 0 | 0 |
| 0 | 0 | 0 | 0 | 0 |
| 0 | 0 | 0 | 0 | 0.043963626 |
| 0.000457149 | 0.001340023 | 0 | 0.002380472 | 0.008497845 |

| | | | | |
|---|---|---|---|---|
| 0 | 0 | 0 | 0 | 0 |
| 0 | 0 | 0 | 0 | 0 |
| 0 | 0 | 0 | 0 | 0 |
| 0 | 0 | 0.00048859 | 0 | 0 |
| 0 | 0 | 0 | 0 | 0 |
| 0 | 0 | 0 | 0 | 0 |
| 0 | 0 | 0 | 0 | 0 |
| 0 | 0 | 0 | 0 | 0 |
| 0 | 0 | 0 | 0 | 0.038790342 |
| 0 | 0 | 0 | 0 | 0 |
| 0 | 0 | 0 | 0 | 0 |
| 0 | 0 | 0 | 0 | 0 |
| 0 | 0 | 0 | 0 | 0 |
| 0 | 0 | 0 | 0 | 0 |
| 0 | 0 | 0 | 0.000721772 | 0 |
| 0 | 0 | 0 | 0 | 0 |
| 0 | 0 | 0 | 0 | 0 |
| 0 | 0 | 0 | 0 | 0 |
| 0.011901888 | 0 | 0.008404479 | 0 | 0.025270383 |
| 0 | 0 | 0.476069829 | 1.326313341 | 0 |
| 0.175427808 | 0.003517955 | 0 | 0 | 0.008391221 |
| 0 | 0 | 0 | 0 | 0 |
| 0 | 0 | 0 | 0 | 0 |
| 0 | 0 | 0 | 0 | 0 |
| 0.001907483 | 0.004992258 | 0 | 0.002183004 | 0 |
| 0 | 0 | 0 | 0 | 0 |
| 0 | 0 | 0 | 0 | 0 |
| 0 | 0 | 0 | 0 | 0 |
| 0 | 0 | 0 | 0 | 0 |
| 0 | 0 | 0 | 0 | 0 |
| 0.170442903 | 0.783600823 | 0.001271979 | 0.043124294 | 1.773655426 |
| 0 | 0 | 0 | 0 | 0 |
| 0 | 0 | 0 | 0 | 0.002213073 |
| 0.001565787 | 0 | 0 | 0.004311886 | 0 |
| 0 | 0 | 0 | 0 | 0 |
| 0 | 0 | 0 | 0 | 0 |
| 0 | 0 | 0 | 0 | 0 |
| 0 | 0 | 0 | 0 | 0 |
| 0 | 0 | 0 | 0 | 0 |
| 0 | 0 | 0 | 0 | 0 |
| 0 | 0 | 0 | 0 | 0 |
| 0.005757005 | 0 | 0.002268999 | 0.02192755 | 0.053822339 |
| 0 | 0.000948146 | 0 | 0 | 0 |
| 0 | 0 | 0 | 0 | 0 |
| 0 | 0 | 0 | 0 | 0 |
| 0 | 0 | 0 | 0 | 0 |
| 0 | 0 | 0 | 0 | 0 |
| 0 | 0 | 0 | 0 | 0 |

| | | | | |
|---|---|---|---|---|
| 0 | 0 | 0 | 0 | 0 |
| 0 | 0 | 0 | 0 | 0 |
| 0.005292564 | 0.293486695 | 0.002170744 | 0 | 0 |
| 0 | 0 | 0 | 0 | 0 |
| 0 | 0 | 0 | 0 | 0 |
| 0 | 0 | 0 | 0 | 0 |
| 0 | 0 | 0 | 0 | 0 |
| 0 | 0 | 0 | 0 | 0 |
| 0 | 0 | 0 | 0 | 0 |
| 0.000354186 | 0 | 0 | 0 | 0 |
| 0 | 0 | 0 | 0 | 0 |
| 0 | 0 | 0 | 0 | 0 |
| 0 | 0 | 0 | 0 | 0 |
| 0.023967612 | 0 | 0 | 0.003428693 | 0.001342763 |
| 0.005652036 | 0.003020133 | 0.001361328 | 0.003065766 | 0 |
| 0 | 0 | 0.056576782 | 0.002651094 | 0 |
| 0.000402499 | 0 | 0 | 0.001477877 | 0 |
| 0 | 0 | 0 | 0 | 0 |
| 0 | 0 | 0 | 0 | 0 |
| 0 | 0 | 0 | 0 | 0 |
| 0 | 0 | 0 | 0 | 0 |
| 0.000484456 | 0.000665656 | 0 | 0.003395895 | 0.009261933 |
| 0 | 0 | 0 | 0 | 0 |
| 0.00131428 | 0 | 0 | 0 | 0 |
| 0 | 0 | 0 | 0 | 0 |
| 0 | 0 | 0 | 0 | 0 |
| 0 | 0 | 0 | 0.825600173 | 0 |
| 0 | 0 | 0 | 0.00054781 | 0 |
| 0 | 0 | 0 | 0 | 0 |
| 0 | 0 | 0 | 0 | 0 |
| 0 | 0 | 0 | 0 | 0 |
| 0 | 0 | 0 | 0 | 0 |
| 0 | 0 | 0 | 0 | 0 |
| 0.001955017 | 0 | 0 | 0 | 0 |
| 0 | 0 | 0 | 0 | 0 |
| 0 | 0.00487503 | 0 | 0 | 0.002276848 |
| 0 | 0 | 0 | 0 | 0 |
| 0 | 0.001115104 | 0.059173763 | 0.002895186 | 0 |
| 0 | 0 | 0 | 0 | 0 |
| 0 | 0 | 0 | 0 | 0 |
| 0.003294304 | 0 | 0 | 0 | 0 |
| 0 | 0 | 0 | 0 | 0 |
| 0 | 0 | 0 | 0 | 0 |
| 0 | 0 | 0 | 0 | 0 |
| 0 | 0 | 0 | 0 | 0 |
| 0 | 0 | 0 | 0 | 0 |
| 0 | 0 | 0 | 0 | 0 |
| 0.002027439 | 0 | 0 | 0.000553705 | 0 |

| | | | | |
|---|---|---|---|---|
| 0 | 0 | 0 | 0.124718339 | 0.001858411 |
| 0 | 0 | 0 | 0 | 0.050152614 |
| 0.00050743 | 0.001394446 | 0 | 0.000889234 | 0 |
| 0 | 0 | 0 | 0 | 0 |
| 0.001276308 | 0.015432417 | 0.001073129 | 0.001405886 | 0 |
| 0 | 0 | 0 | 0 | 0 |
| 0 | 0 | 0 | 0 | 0 |
| 0.018248077 | 0.144212863 | 0.002632397 | 0.005911978 | 0.020837522 |
| 0 | 0 | 0 | 0 | 0 |
| 0.010695136 | 0 | 0.002325659 | 0.004985675 | 0 |
| 0 | 0.001527191 | 0 | 0 | 0.001373033 |
| 0.00056648 | 0.036063998 | 0 | 0 | 0.006331436 |
| 0.231498329 | 0.006297743 | 0 | 0.003549903 | 0.000379154 |
| 0.005873367 | 0 | 0 | 0 | 0 |
| 0 | 0 | 0 | 0 | 0.000779903 |
| 0 | 0 | 0 | 0 | 0 |
| 0 | 0 | 0 | 0 | 0 |
| 0 | 0 | 0 | 0 | 0 |
| 0 | 0 | 0 | 0 | 0 |
| 0 | 0 | 0.000730892 | 0 | 0 |
| 0 | 0 | 0 | 0 | 0 |
| 0 | 0 | 0 | 0 | 0 |
| 0 | 0 | 0 | 0 | 0 |
| 0 | 0 | 0.037433876 | 0 | 0 |
| 0 | 0 | 0 | 0 | 0 |
| 0 | 0 | 0.001578044 | 0 | 0 |
| 0 | 0 | 2.896553127 | 0.007371966 | 0.000787377 |
| 0.08442903 | 0 | 0 | 0 | 0 |
| 0 | 0 | 0 | 0 | 0 |
| 0 | 0.002594304 | 0 | 0.000515651 | 0 |
| 0 | 0 | 0 | 0 | 0.00126368 |
| 0 | 0 | 0 | 0 | 0 |
| 0 | 0 | 0 | 0 | 0 |
| 0 | 0 | 0 | 0 | 0 |
| 0 | 0 | 0 | 0 | 0 |
| 0.000888158 | 0 | 0 | 0 | 0 |
| 0 | 0 | 0 | 0 | 0.025531575 |
| 0 | 0 | 0 | 0 | 0 |
| 0 | 0 | 0 | 0 | 0 |
| 0 | 0 | 0 | 0 | 0 |
| 0 | 0 | 0 | 0 | 0 |
| 0 | 0 | 0 | 0 | 0.000518647 |
| 0 | 0 | 0 | 0 | 0 |
| 0 | 0 | 0 | 0 | 0 |
| 0 | 0 | 0 | 0 | 0 |
| 0 | 0 | 0 | 0 | 0 |
| 0.001555983 | 0.008266796 | 0 | 0 | 0.022333594 |
| 0 | 0 | 0 | 0 | 0 |

| | | | | |
|---|---|---|---|---|
| 0 | 0 | 0 | 0 | 0 |
| 0.071817389 | 0.001274307 | 0 | 0 | 0 |
| 0 | 0 | 0 | 0 | 0 |
| 0 | 0 | 0 | 0 | 0 |
| 0 | 0 | 0 | 0 | 0.003980633 |
| 0 | 0 | 0.00103043 | 0 | 0.021771738 |
| 0 | 0.021470312 | 0 | 0 | 0.002885838 |
| 0 | 0 | 0 | 0 | 0 |
| 0 | 0 | 0.146453376 | 0 | 0 |
| 0 | 0 | 0 | 0 | 0 |
| 0.00511705 | 0.001041625 | 0.002539642 | 0.000427012 | 0 |
| 0 | 0 | 0 | 0 | 0.004858676 |
| 0 | 0 | 0 | 0 | 0 |
| 0 | 0.005412414 | 0 | 0.000941313 | 0 |
| 0.017799533 | 0 | 0 | 0 | 0 |
| 0 | 0 | 0 | 0 | 0 |
| 0 | 0 | 0 | 0 | 0 |
| 0 | 0 | 0 | 0 | 0 |
| 0 | 0.102644887 | 0 | 0 | 0 |
| 0.005556095 | 0 | 0 | 0.444862175 | 0.009094645 |
| 0 | 0 | 0 | 0 | 0.001872498 |
| 0 | 0.210560264 | 0.358076194 | 0.206921767 | 0.262041576 |
| 0 | 0 | 0.000803593 | 0 | 0.097450833 |
| 0 | 0 | 0 | 0 | 0 |
| 0 | 0 | 0 | 0 | 0 |
| 0 | 0 | 0 | 0 | 0 |
| 0 | 0 | 0 | 0 | 0.100211849 |
| 0 | 0 | 0 | 0 | 0 |
| 0 | 0 | 0 | 0 | 0 |
| 12.16312365 | 1.210574652 | 0.079440783 | 0.209724601 | 0.259600425 |
| 0 | 0.003838797 | 0.024607782 | 0.000821065 | 0 |
| 0 | 0.003046268 | 0 | 0 | 0 |
| 0 | 0 | 0 | 0 | 0 |
| 0.001161298 | 0 | 0 | 0 | 0.001366273 |
| 0.003759472 | 0 | 0 | 0 | 0 |
| 0 | 0 | 0 | 0 | 0 |
| 0 | 0 | 1.031697754 | 0.150863058 | 0 |
| 0 | 0.007021244 | 0 | 0 | 0.000805166 |
| 0 | 0.130479018 | 0 | 0 | 0 |
| 0 | 0 | 0 | 0 | 0 |
| 0 | 0.007483407 | 0 | 0 | 0.00233792 |
| 0 | 0 | 0.281808533 | 2.13899373 | 1.117507954 |
| 0 | 0.001378267 | 0 | 0 | 0 |
| 0 | 0 | 0 | 0 | 0 |
| 0 | 0.002509686 | 0 | 0 | 0 |
| 0 | 0 | 0 | 0 | 0 |
| 0.000926969 | 0.00615613 | 0.001826725 | 0 | 0 |
| 0 | 0 | 0 | 0.000543033 | 0.002807185 |

| | | | | |
|---|---|---|---|---|
| 0 | 0 | 0.004357485 | 0 | 0 |
| 0 | 0 | 0 | 0 | 0 |
| 13.97545231 | 0.006067024 | 0.002661596 | 0.000975359 | 0 |
| 0 | 0 | 0 | 0 | 0 |
| 0 | 0 | 0.046421463 | 0 | 0 |
| 0 | 0 | 0 | 0 | 0.002275409 |
| 0 | 0 | 0 | 0 | 0 |
| 0 | 0.003522688 | 0 | 0 | 0 |
| 0.171816481 | 0.02367383 | 0 | 0 | 0 |
| 0 | 0 | 0 | 0 | 0 |
| 0 | 0 | 0 | 0 | 0 |
| 0 | 0 | 0 | 0 | 0 |
| 0 | 0 | 0 | 0 | 0 |
| 0.002328072 | 0 | 0 | 0 | 0 |
| 0 | 0 | 0 | 0 | 0 |
| 0.021832179 | 0.083698207 | 0.003718072 | 0.134818489 | 0.053868573 |
| 0.007842169 | 0.00862029 | 0.07571041 | 0.010882077 | 0 |
| 0.012794649 | 0 | 0 | 0 | 0 |
| 0 | 0.004240052 | 0 | 0 | 0.001497592 |
| 0 | 0 | 0 | 0 | 0 |
| 0 | 0 | 0 | 0.001272481 | 0.015614518 |
| 0.001559401 | 0 | 0 | 0 | 0.013606946 |
| 0 | 0 | 0 | 0 | 0 |
| 0 | 0 | 0 | 0 | 0 |
| 0 | 0.003452783 | 0 | 0 | 0 |
| 0.016676662 | 0 | 0 | 0 | 0.043407487 |
| 0 | 0 | 0 | 0 | 0.009872328 |
| 0 | 0.057053208 | 0 | 0 | 0 |
| 0 | 0 | 0 | 0 | 0 |
| 0 | 0 | 0 | 0 | 0 |
| 0 | 0.063409686 | 0.00068207 | 0 | 0.024305163 |
| 0.008869024 | 0.001772551 | 0 | 0 | 0 |
| 0.050206449 | 0.012657813 | 0.02081651 | 0.009993716 | 0.001083819 |
| 0.000660099 | 0.021767856 | 0 | 0 | 0 |
| 0.001727522 | 0 | 0 | 0.008964121 | 0 |
| 0 | 0 | 0 | 0 | 0 |
| 0 | 0.001525805 | 0 | 0 | 0 |
| 0.002822404 | 0.002585375 | 0 | 0.007391909 | 0.074712902 |
| 0 | 0 | 0.032291631 | 0.067590541 | 0.160054388 |
| 0 | 0 | 0 | 0 | 0 |
| 0 | 0 | 0 | 0 | 0 |
| 0 | 0 | 0 | 0 | 0 |
| 0.007792431 | 0.008030266 | 0 | 0 | 0.035716369 |
| 0 | 0 | 0.024135874 | 1.005389239 | 0 |
| 0 | 0 | 0 | 0 | 0 |
| 0.261751961 | 0 | 0 | 0.000473987 | 0 |
| 0 | 0 | 0 | 0 | 0 |
| 0 | 0 | 0 | 0 | 0 |

| | | | | |
|---|---|---|---|---|
| 0 | 0 | 0 | 0 | 0 |
| 0.076325989 | 1.003213919 | 0.016168232 | 0.53842977 | 0.02801835 |
| 0 | 0 | 0 | 0 | 0 |
| 0 | 0.001148863 | 0 | 0.00843268 | 0.000737782 |
| 0 | 0 | 0 | 0 | 0.009902264 |
| 0 | 0 | 0 | 0 | 0 |
| 0 | 0 | 0 | 0 | 0 |
| 0 | 0 | 0 | 0 | 0.000401013 |
| 0 | 0.00198457 | 0 | 0.010034036 | 0 |
| 0 | 0.001969249 | 0 | 0 | 0 |
| 0.000329759 | 0.002215147 | 0 | 0.001320866 | 0.606515979 |
| 0.017355219 | 0.054292171 | 0.001376641 | 0.005574489 | 0.002889412 |
| 0 | 0 | 0.225978029 | 0.0150966 | 0 |
| 0 | 0 | 0 | 0 | 0 |
| 0 | 0 | 0 | 0 | 0 |
| 0 | 0 | 0 | 0 | 0 |
| 0.002532239 | 0.028256684 | 0.001330703 | 0.003803626 | 0.001218761 |
| 0.010160424 | 0.000821218 | 0 | 0 | 0.205499787 |
| 0 | 0 | 0 | 0 | 0 |
| 0 | 0 | 0 | 0 | 0 |
| 0.018042243 | 0.330105755 | 0.015968472 | 0.024010445 | 0.0005586 |
| 0 | 0 | 0 | 0 | 0 |
| 0 | 0 | 0 | 0 | 0 |
| 0 | 0 | 0 | 0 | 0 |
| 0 | 0 | 0 | 0 | 0 |
| 0 | 0.002791849 | 0 | 0 | 0 |
| 0 | 0.076413565 | 0.001354519 | 0.013726943 | 0.051690565 |
| 0.150129777 | 0.016410153 | 0.034872419 | 0.042574709 | 0.006233947 |
| 0 | 0 | 0 | 0 | 0 |
| 0 | 0 | 0 | 0.0013715 | 0.004654991 |
| 0 | 0 | 0 | 0 | 0 |
| 0 | 0 | 0 | 0 | 0 |
| 0 | 0 | 0 | 0 | 0 |
| 0 | 0 | 0 | 0 | 0 |
| 0 | 0.410680607 | 0 | 0.062752255 | 0.318962475 |
| 0.665106661 | 0.981841448 | 0.058346263 | 0.133847261 | 0.090684778 |
| 0 | 0.001449338 | 0 | 0 | 0.001512456 |
| 0 | 0 | 0 | 0 | 0 |
| 0.007937223 | 0.013971045 | 0 | 0.003428649 | 0.003662034 |
| 0 | 0 | 0 | 0 | 0 |
| 0 | 0 | 0 | 0 | 0 |
| 0 | 0 | 0 | 0 | 0 |
| 0 | 0.001212318 | 0 | 0 | 0.016905252 |
| 0.00063889 | 0 | 0 | 0 | 0 |
| 0 | 0 | 0 | 0 | 0 |
| 0 | 0 | 0.09157846 | 0.22503023 | 0 |
| 0 | 0 | 0 | 0 | 0 |
| 0.015343207 | 0 | 0 | 0 | 0.000520713 |

| | | | | |
|---|---|---|---|---|
| 0 | 0 | 0 | 0 | 0 |
| 0 | 0 | 0 | 0.001216728 | 0 |
| 0 | 0 | 0 | 0 | 0 |
| 0 | 0.27775405 | 0.081812174 | 0.396252529 | 0 |
| 0 | 0 | 0 | 0 | 0 |
| 0 | 0 | 0 | 0 | 0 |
| 0 | 0.000781905 | 0 | 0 | 0 |
| 0 | 0.112420864 | 0 | 0.001479509 | 0 |
| 0.04244645 | 0 | 0.305658209 | 0.206930658 | 2.930423584 |
| 0 | 0 | 0.014272355 | 0.247335085 | 0 |
| 0 | 0 | 0 | 0 | 0 |
| 0.00348113 | 0 | 0.000665218 | 0.007605727 | 0.000496433 |
| 0 | 0 | 0 | 0 | 0 |
| 0 | 0 | 0 | 0 | 0 |
| 0.014014988 | 0.064748139 | 0.023604403 | 0.68559477 | 0.577207267 |
| 0 | 0 | 0 | 0 | 0 |
| 0 | 0 | 0.099172041 | 0.24037465 | 0 |
| 0 | 0.018742904 | 0.46785266 | 1.403199015 | 0 |
| 0 | 0 | 0 | 0 | 0 |
| 0 | 0 | 0 | 0 | 0 |
| 0 | 0 | 0 | 0 | 0 |
| 0 | 0 | 0.001955753 | 0 | 0 |
| 0 | 0 | 0 | 0.001459081 | 0 |
| 0 | 0.228276878 | 0 | 0 | 0.254618919 |
| 0.003259227 | 0.004523505 | 0 | 0.001879127 | 0.032767534 |
| 0 | 0 | 0.025568022 | 1.510271286 | 0.487754328 |
| 0 | 0 | 0.009139649 | 0 | 0 |
| 0 | 0 | 1.939391876 | 1.554669485 | 0.027043531 |
| 0 | 0 | 0 | 0 | 0 |
| 0 | 0 | 0 | 0 | 0 |
| 0.000922978 | 0 | 0.023281392 | 0.000410781 | 0 |
| 0 | 0 | 0.012897197 | 0 | 0 |
| 0.000390252 | 0.034603901 | 0 | 0 | 0.001193748 |
| 0 | 0 | 0 | 0 | 0 |
| 0 | 0 | 0 | 0 | 0 |
| 0 | 0.090961544 | 0.103360923 | 1.223652994 | 0 |
| 0 | 0 | 0.285512994 | 0.278330416 | 0 |
| 0 | 0 | 0 | 0 | 0 |
| 0 | 0 | 0.000461661 | 0.276454873 | 0.006201436 |
| 0 | 0 | 0 | 0.004687868 | 0.003553332 |
| 0.020037132 | 0.021320756 | 0.000957237 | 0.158087259 | 0.001666834 |
| 0 | 0 | 0 | 0.088893787 | 0 |
| 0.30167648 | 0.174145082 | 0.149919932 | 0.003525453 | 0 |
| 0 | 0.046836677 | 0 | 0.217077155 | 0.007778733 |
| 0 | 0 | 0 | 0 | 0.005595078 |
| 0 | 0 | 0 | 0 | 0 |
| 0 | 0 | 0 | 0 | 0 |
| 0 | 0.002897304 | 0 | 0.008771032 | 0.001574356 |

| | | | | |
|---|---|---|---|---|
| 0 | 0 | 0 | 0 | 0 |
| 0 | 0.024770934 | 0.075945528 | 0.090005888 | 0 |
| 0.389615316 | 0.245906078 | 0 | 0 | 0.04898033 |
| 0 | 0 | 0.004637036 | 0.33741694 | 0 |
| 11.48850847 | 0 | 0 | 0 | 0 |
| 0.000918659 | 0 | 0.014675885 | 0.064027334 | 0.001405049 |
| 0 | 0 | 0 | 0 | 0.211854396 |
| 0 | 0 | 0 | 0.004847777 | 0.008433776 |
| 0 | 0 | 0 | 0 | 0 |
| 0 | 0 | 0 | 0 | 0 |
| 0.037440389 | 0.014683346 | 0.008543924 | 0.193538312 | 0.828306438 |
| 2.603933046 | 0.608267419 | 0.573919797 | 1.435487181 | 0.032642312 |
| 0 | 0 | 0 | 0 | 0 |
| 0.001359263 | 0.013356644 | 0.189743806 | 0.016705005 | 0 |
| 2.420510687 | 0.231205248 | 0.000511723 | 0.000682587 | 0.222256242 |
| 0 | 0 | 3.506682727 | 2.304248268 | 0 |
| 0 | 0 | 0 | 0 | 0.008096545 |
| 0 | 0 | 0 | 0 | 0 |
| 0 | 0 | 0.004953151 | 0.172767204 | 0 |
| 0 | 0.0138231 | 0.002775528 | 0.04438956 | 0.013426425 |
| 0.325801221 | 0.010900314 | 0 | 0.070055521 | 0.008693559 |
| 0.129025741 | 0 | 0 | 0 | 0 |
| 0.28805799 | 0.012620426 | 0.299512549 | 0.471625289 | 0.00085312 |
| 0 | 0 | 0 | 0 | 0 |
| 0 | 0 | 0.000889675 | 0.067342665 | 0 |
| 0.0044848 | 6.000443461 | 0.030940326 | 0.002303087 | 0 |
| 0.011248305 | 0.030069863 | 0 | 0.002758508 | 1.243426954 |
| 0 | 0 | 0 | 0 | 0 |
| 0 | 0.086898458 | 0 | 0 | 0.061213582 |
| 0 | 0 | 0 | 0 | 0.008929058 |
| 0 | 0 | 0.002781152 | 0.076739456 | 0 |
| 0 | 0.258762117 | 0.106360363 | 0.130007464 | 0.251337871 |
| 0.067732412 | 0 | 0.001359481 | 0.007654015 | 0.308512572 |
| 0 | 0.014949745 | 0.002167666 | 0.098776264 | 0.073849913 |
| 0 | 0 | 0 | 0 | 0.027301801 |
| 0.00981028 | 0.004688557 | 0.281638818 | 0.071436735 | 0.008530931 |
| 0.000515032 | 0.003019388 | 0.000974347 | 0.02516844 | 0.022662098 |
| 0 | 0 | 0 | 0 | 0 |
| 0.017719144 | 0.03840591 | 2.387282466 | 2.645313528 | 0 |
| 0 | 0 | 0 | 0 | 0 |
| 0 | 0 | 0.014309426 | 0 | 0 |
| 0 | 0 | 0 | 0 | 0 |
| 0 | 0 | 0 | 0 | 0 |
| 0 | 0 | 0 | 0 | 0 |
| 0.001216034 | 0.02272376 | 0.036424781 | 0.149048963 | 0.102006797 |
| 0 | 0 | 0.001580546 | 0 | 0 |
| 0 | 0 | 0 | 0 | 0 |
| 0.428912504 | 0.012361853 | 0.46112026 | 0.010869885 | 0.340151122 |

| | | | | |
|---|---|---|---|---|
| 0 | 0 | 0 | 0 | 0 |
| 0 | 0 | 0 | 0 | 0 |
| 0 | 0 | 0 | 0 | 0 |
| 0.000414568 | 0.004367152 | 0.000326786 | 0.002802217 | 0 |
| 0 | 0 | 0 | 0 | 0 |
| 0 | 0 | 0.043773357 | 0.334569828 | 0 |
| 0 | 0 | 0 | 0 | 0 |
| 0.015068375 | 0.103609222 | 0.035282426 | 0.306369546 | 1.361130835 |
| 0 | 0 | 0 | 0 | 0 |
| 0 | 0 | 0 | 0 | 0 |
| 0.000577149 | 0.001268831 | 0 | 0.009940732 | 0.160791709 |
| 0 | 0 | 0 | 0 | 0 |
| 0 | 0.025737249 | 0.040683486 | 0.089366665 | 0.298218385 |
| 0 | 0 | 0.006211437 | 0 | 0 |
| 0 | 0 | 0 | 0 | 0 |
| 0 | 0 | 0 | 0 | 0 |
| 0 | 0 | 0.019940496 | 0.066023775 | 0 |
| 0 | 0 | 0.0141392 | 0.001347163 | 0.000678321 |
| 0 | 0 | 0 | 1.144386606 | 0 |
| 0 | 0 | 0 | 0.135723732 | 0.089007238 |
| 0 | 0.003910062 | 0.000897255 | 0.04599318 | 0 |
| 0 | 0 | 0 | 0 | 0 |
| 0 | 0 | 0.006869912 | 0.022188329 | 0 |
| 0 | 0 | 0 | 0 | 0 |
| 0 | 0 | 0 | 0 | 0 |
| 0 | 0 | 0 | 0 | 0 |
| 0 | 0 | 0 | 0 | 0 |
| 0 | 0 | 0 | 0 | 0 |
| 0 | 0 | 0 | 0 | 0 |
| 0.031109201 | 0.16226057 | 0.044652071 | 0.003138483 | 0 |
| 0 | 0 | 0 | 0.063727947 | 0 |
| 0 | 0 | 0 | 0 | 0 |
| 0 | 0 | 0.007808615 | 0 | 0 |
| 0 | 0 | 0 | 0 | 0 |
| 0 | 0 | 0.000675262 | 0 | 0 |
| 0 | 0 | 0 | 0 | 0 |
| 0 | 0 | 0.042830655 | 0.044947246 | 0 |
| 0.002149694 | 0.005370437 | 0 | 0 | 0.017703886 |
| 0 | 0 | 0 | 0 | 0 |
| 0 | 0 | 0 | 0 | 0.008277506 |
| 0 | 0.0016796 | 0 | 0 | 0.464558245 |
| 0 | 0 | 0 | 0 | 0 |
| 0 | 0 | 0.000578866 | 0.090451864 | 0 |
| 0 | 0 | 0.085071021 | 0.134267292 | 0 |
| 0.179854016 | 0 | 0.038248765 | 0.162548647 | 0 |
| 0 | 0 | 0 | 0 | 0 |
| 0 | 0.001530844 | 0.000975799 | 0.001022701 | 0 |
| 0 | 0 | 0 | 0 | 0 |

| | | | | |
|---|---|---|---|---|
| 0 | 0 | 0 | 0 | 0 |
| 0 | 0 | 0 | 0 | 0 |
| 0 | 0 | 0.000594196 | 0 | 0 |
| 0 | 0 | 0.120337454 | 2.501989519 | 0.000686111 |
| 0 | 0 | 0 | 0 | 0 |
| 0 | 0 | 0.183888646 | 1.594699509 | 0 |
| 0 | 0 | 0 | 0 | 0 |
| 0 | 0 | 0 | 0 | 0 |
| 0.223571887 | 0.091843439 | 0 | 0.008771746 | 0.063578331 |
| 0 | 0 | 0 | 0 | 0 |
| 0 | 0 | 0 | 0 | 0 |
| 0 | 0 | 0.000345396 | 0.019251656 | 0 |
| 0 | 0 | 0 | 0 | 0.023216188 |
| 0 | 0.001886308 | 0 | 0.005905109 | 0 |
| 0 | 6.306300518 | 1.188330682 | 2.723643772 | 0.008089585 |
| 0 | 0.012368573 | 0 | 0.004888455 | 0.002199573 |
| 0 | 0 | 0 | 0 | 0 |
| 0 | 0 | 0 | 0 | 0 |
| 0 | 0 | 0 | 0 | 0 |
| 0 | 0 | 0 | 0 | 0 |
| 0 | 0 | 0.007256229 | 0.19853484 | 0 |
| 0 | 0 | 0 | 0 | 0 |
| 0 | 0 | 0 | 0 | 0 |
| 0 | 0 | 0 | 0 | 0 |
| 0.078763318 | 0.156781843 | 0.416902898 | 0.002063065 | 0.1569906 |
| 0 | 4.554431573 | 3.287704549 | 2.411360047 | 0.957939986 |
| 0 | 0 | 0.231944292 | 0.025145073 | 0 |
| 0 | 0.000720902 | 0 | 0 | 0 |
| 0.066644788 | 0.169563343 | 0 | 0 | 0 |
| 0 | 0 | 0 | 0 | 0 |
| 0 | 0 | 0 | 0 | 0 |
| 0 | 0.015332234 | 0 | 0 | 0.000960597 |
| 0.000690686 | 0 | 0 | 0 | 0 |
| 0.002793026 | 0.009768683 | 0 | 0.019493545 | 0.036146117 |
| 0.001115642 | 0.000766462 | 0.041918673 | 0.00111719 | 0 |
| 0.000887202 | 0.004334366 | 0 | 0 | 0.204352439 |
| 0 | 0 | 0 | 0 | 0 |
| 2.081749422 | 3.440248993 | 0.664500311 | 1.653040368 | 0.015846002 |
| 0 | 0 | 0.431543628 | 7.264273222 | 0 |
| 0 | 0 | 0 | 0 | 0 |
| 0 | 0 | 0.301495 | 0 | 0 |
| 0 | 0 | 0 | 0 | 0 |
| 0 | 0.725072539 | 0.085939024 | 0.064614903 | 0.077023897 |
| 0 | 0 | 0.336919624 | 0 | 0 |
| 0 | 0 | 0 | 0 | 0 |
| 0.007284441 | 0.040549396 | 0 | 0 | 0.000988868 |
| 0 | 0 | 0 | 0 | 0 |
| 0.048056554 | 0.008449795 | 0.000938227 | 0.001191906 | 0 |

| | | | | |
|---|---|---|---|---|
| 0 | 0.001422811 | 0 | 0.176539013 | 0.002349532 |
| 0 | 0 | 0 | 0 | 0 |
| 0 | 0 | 0 | 0 | 0 |
| 0 | 0 | 0 | 0 | 0 |
| 0.000773223 | 0 | 0 | 0.007079272 | 0.014425259 |
| 0.032124265 | 0 | 0 | 0.013314908 | 0 |
| 0 | 0 | 0 | 0 | 0 |
| 0 | 0 | 0 | 0 | 0 |
| 0 | 0 | 0 | 0 | 0 |
| 0 | 0 | 0 | 0 | 0 |
| 0 | 0 | 0 | 0 | 0 |
| 0.022419493 | 0 | 0.256302155 | 0.257569459 | 0.119773996 |
| 0.194603633 | 0 | 0.002670841 | 0.007500287 | 0 |
| 0 | 0 | 0 | 0 | 0 |
| 0.037685504 | 0.455204255 | 0 | 0.030190216 | 0.14608732 |
| 0 | 0 | 0 | 0 | 0 |
| 0 | 0.00618668 | 0 | 0.043961094 | 0.003345674 |
| 0 | 0 | 0 | 0 | 0 |
| 0 | 0.000615401 | 0.001647544 | 0.006839655 | 0.044394208 |
| 0 | 0 | 0 | 0.036552806 | 0 |
| 0 | 0 | 0.028644318 | 0 | 0.00032719 |
| 0 | 0 | 0 | 0 | 0 |
| 0 | 0.134838055 | 0.094518733 | 0.753048164 | 0 |
| 0 | 0 | 0 | 0 | 0 |
| 0 | 0 | 0 | 0 | 0 |
| 0 | 0 | 0 | 0 | 0 |
| 0 | 0 | 0 | 0 | 0 |
| 0 | 0 | 0 | 0 | 0 |
| 0 | 0.001897283 | 0 | 0 | 0 |
| 0 | 0 | 0 | 0 | 0 |
| 0 | 0 | 0 | 0 | 0 |
| 0.018068953 | 0.092323968 | 0.003136068 | 0.025896018 | 0.0085813 |
| 0 | 0 | 0 | 0 | 0 |
| 0.005383036 | 0.050675121 | 0.005483539 | 0.093048354 | 0.043357865 |
| 0.001072776 | 0.004913413 | 0 | 0.000751984 | 0.203454766 |
| 0.004563695 | 0.010387736 | 0.123400465 | 0.0605182 | 0.027659576 |
| 0 | 0 | 0 | 0.001430385 | 0 |
| 0 | 0 | 0.024689598 | 0 | 0 |
| 0.00061623 | 0 | 0 | 0.003702511 | 0 |
| 0 | 0 | 0 | 0 | 0 |
| 0 | 0 | 0.086648956 | 0.655429412 | 0 |
| 0 | 0 | 0 | 0.112898205 | 0 |
| 0 | 0 | 0 | 0 | 0 |
| 0 | 0 | 0 | 0 | 0 |
| 0.002219262 | 0.137268198 | 0.009663067 | 0.158738573 | 0.112644878 |
| 0 | 0 | 0 | 0 | 0 |
| 0 | 0 | 0.066724423 | 0.47885372 | 0 |
| 0 | 0 | 0 | 0 | 0.014699489 |

| | | | | |
|---|---|---|---|---|
| 0 | 0 | 0 | 0 | 0 |
| 0 | 0 | 0.002408585 | 0 | 0 |
| 0 | 0.014216991 | 0 | 0.012805682 | 0.001800267 |
| 0 | 0 | 0 | 0 | 0 |
| 0 | 0 | 0 | 0 | 0 |
| 0 | 0 | 0 | 0 | 0 |
| 0 | 0 | 0.128600474 | 0 | 0.000566199 |
| 0 | 0 | 0 | 0.040726027 | 0 |
| 0 | 0 | 0 | 0 | 0 |
| 0 | 0 | 0.000960804 | 0.017088229 | 0 |
| 0 | 0 | 0 | 0 | 0 |
| 0 | 0 | 0 | 0 | 0.005233509 |
| 0 | 0 | 0 | 0 | 0 |
| 0.395512543 | 0 | 0.169285658 | 0.215761336 | 0 |
| 0 | 0 | 0 | 0 | 0 |
| 0 | 0 | 0 | 0 | 0 |
| 0 | 0 | 0.009381314 | 6.910555667 | 0 |
| 0 | 0 | 0 | 0 | 0 |
| 0 | 0.065258491 | 2.463277481 | 4.156328865 | 0 |
| 0 | 0 | 0 | 0 | 0 |
| 0 | 0 | 0.014716626 | 0 | 0 |
| 0 | 0 | 0 | 0 | 0 |
| 0 | 0 | 0 | 0 | 0 |
| 0 | 0 | 0 | 0 | 0 |
| 0 | 0 | 0 | 0 | 0 |
| 0.005214315 | 0 | 0 | 0 | 0.00914413 |
| 0 | 0 | 0 | 0 | 0 |
| 0.0041408 | 0.036871982 | 0 | 0.032015169 | 0 |
| 0 | 0 | 0.251619122 | 0 | 0 |
| 0 | 0.170433971 | 0.215990635 | 1.147432259 | 0 |
| 0 | 0 | 0 | 0 | 0 |
| 0 | 0 | 0 | 0 | 0 |
| 0 | 0 | 0.01090061 | 0.078912862 | 0 |
| 0 | 0 | 0 | 0 | 0 |
| 0.001030996 | 0 | 0.000650152 | 0.002477822 | 0.12251022 |
| 0 | 0 | 0 | 0.069104818 | 0 |
| 0 | 0 | 0.172044445 | 0 | 0 |
| 0 | 0 | 0.000983642 | 0.001640101 | 0.004954927 |
| 0 | 0 | 0 | 0 | 0 |
| 0 | 0 | 0 | 0.10667682 | 0 |
| 8.308932253 | 29.40365405 | 0.765638095 | 1.255249455 | 3.094513885 |
| 0.542758681 | 0.285221865 | 0.062912678 | 0.637155249 | 0 |
| 0 | 0 | 0 | 0 | 0 |
| 0 | 0 | 0 | 0 | 0 |
| 0 | 0 | 0 | 0 | 0 |
| 0 | 0 | 0 | 0 | 0 |
| 0 | 0 | 0 | 0.009970559 | 0 |
| 0 | 0 | 0.001951682 | 0.010227449 | 0.015183834 |

| | | | | |
|---|---|---|---|---|
| 0 | 0 | 0 | 0 | 0 |
| 0 | 0.021870377 | 0 | 0.000956343 | 0.008651602 |
| 0 | 0.000817388 | 0 | 0.000670173 | 0 |
| 0 | 0 | 0 | 0 | 0 |
| 0 | 1.914609876 | 0.546403758 | 0.644883006 | 0.652955637 |
| 0 | 0 | 0 | 0 | 0 |
| 0 | 0 | 0 | 0 | 0 |
| 0 | 0 | 0 | 0 | 0 |
| 0.018881491 | 0.099232476 | 0 | 0.000477467 | 0.551427681 |
| 0 | 0 | 0 | 0 | 0 |
| 0 | 0 | 0 | 0 | 0 |
| 0 | 0 | 0 | 0 | 0 |
| 0 | 0 | 0.022236793 | 0.124147855 | 0 |
| 0.596543476 | 0.093425488 | 0.004796118 | 0.004798159 | 0.006308342 |
| 0 | 0.057262906 | 0.091476736 | 0 | 0 |
| 0 | 1.236404458 | 0.01377481 | 1.09711021 | 0 |
| 0.000789774 | 0.001266033 | 0 | 0.001087445 | 0 |
| 0 | 0 | 0 | 0 | 0 |
| 0 | 0 | 0 | 0 | 0 |
| 0 | 0 | 0 | 0 | 0 |
| 0 | 0 | 0 | 0 | 0 |
| 0 | 0 | 0 | 0 | 0 |
| 0 | 0 | 0 | 0 | 0 |
| 0 | 0 | 0 | 0 | 0 |
| 0 | 0 | 0 | 0 | 0 |
| 0 | 0 | 0 | 0 | 0 |
| 0 | 0 | 0.087514698 | 0.001719233 | 0 |
| 0 | 0.000709832 | 0 | 0.000581989 | 0.002279215 |
| 0 | 0 | 0 | 0 | 0 |
| 0 | 0 | 0 | 0 | 0 |
| 0 | 0 | 0 | 0 | 0 |
| 0 | 0.000444696 | 0 | 0.000972279 | 0 |
| 0 | 0 | 0 | 0.007423474 | 0 |
| 0 | 0 | 0.007683731 | 0.024098137 | 0 |
| 0 | 0 | 0.139851241 | 0 | 0 |
| 0.049302557 | 0 | 0 | 0 | 0.010150827 |
| 0 | 0 | 0 | 0.060555649 | 0 |
| 0 | 0 | 0 | 0 | 0 |
| 0 | 0 | 0 | 0 | 0 |
| 0 | 0 | 0 | 0 | 0 |
| 3.061140945 | 0.031930061 | 0.165725031 | 1.252210231 | 0.193101992 |
| 0 | 0 | 0 | 0.518159848 | 0 |
| 0 | 0 | 0 | 0.024092272 | 0 |
| 0 | 0 | 0 | 0.054966093 | 0 |
| 0 | 0 | 0 | 0 | 0 |
| 0 | 0 | 0 | 0 | 0 |
| 0 | 0 | 0 | 0.556604326 | 0 |
| 0 | 0 | 0 | 0 | 0 |

| | | | | |
|---|---|---|---|---|
| 0 | 0 | 0.07245329 | 0.049915803 | 0 |
| 0 | 0 | 0 | 0 | 0 |
| 0 | 0 | 0 | 0 | 0 |
| 0 | 0 | 0 | 0 | 0.000387286 |
| 0 | 0 | 0.014785991 | 0.068652173 | 0 |
| 0 | 0 | 0 | 0 | 0 |
| 0.008370942 | 0.006856041 | 0 | 0.090531554 | 0.003591791 |
| 0 | 0 | 0 | 0 | 0 |
| 0 | 0 | 0 | 0 | 0.020854267 |
| 0 | 0 | 0 | 0.020072519 | 0.003889933 |
| 0 | 0 | 0 | 0.014709666 | 0 |
| 0 | 0 | 0 | 0 | 0 |
| 0 | 0 | 0 | 0 | 0 |
| 0 | 0 | 0 | 0 | 0 |
| 0 | 0 | 0 | 0 | 0 |
| 0 | 0 | 0 | 0 | 0 |
| 0 | 0 | 0 | 0 | 0 |
| 0.003787191 | 0 | 0 | 0 | 0.001336695 |
| 0 | 0 | 0 | 0 | 0 |
| 0 | 0 | 0 | 0 | 0 |
| 0.044817297 | 0.001030631 | 0.005025675 | 0.008637887 | 0 |
| 0 | 0 | 0 | 0 | 0 |
| 0 | 0.001136783 | 0.000434769 | 0.038731597 | 0 |
| 0.002023108 | 0.000673893 | 0 | 0.000828783 | 0.005193159 |
| 0 | 0 | 0 | 0.00066055 | 0 |
| 0 | 0 | 0.000342077 | 0.002607407 | 0.017359152 |
| 0 | 0 | 0 | 0 | 0 |
| 0 | 0 | 0 | 0 | 0 |
| 0 | 0 | 0 | 0 | 0 |
| 0.215864837 | 0.16269899 | 0 | 0.002518161 | 0 |
| 0 | 0 | 0 | 0.064739127 | 0 |
| 0 | 0 | 0 | 0 | 0 |
| 0 | 0 | 0.002498874 | 0 | 0 |
| 0 | 0 | 0 | 0 | 0 |
| 0 | 0 | 0 | 0 | 0 |
| 0 | 0 | 0 | 1.691892511 | 0 |
| 0.057992809 | 0 | 0.000362803 | 0.008411402 | 0.284016152 |
| 0 | 0 | 0 | 0 | 0 |
| 0 | 0 | 0 | 0 | 0 |
| 0 | 0 | 0 | 0 | 0 |
| 0 | 0 | 0 | 0 | 0 |
| 0 | 0 | 0 | 0 | 0 |
| 0 | 0 | 0 | 0 | 0 |
| 0 | 0 | 0.005654635 | 0.003958279 | 0.000775081 |
| 0 | 0.008977936 | 0 | 0.035441726 | 0 |
| 0 | 1.979526962 | 0 | 0 | 0 |
| 0 | 0 | 0 | 0 | 0 |
| 0 | 0 | 0.026511937 | 0 | 0 |

| | | | | |
|---|---|---|---|---|
| 0.007718228 | 0.010333138 | 0 | 0.0008918 | 0.154252716 |
| 0 | 0 | 0 | 0 | 0 |
| 0 | 0 | 0 | 0 | 0 |
| 0 | 0 | 0 | 0 | 0 |
| 0 | 0 | 0 | 0 | 0 |
| 0 | 0 | 0 | 0 | 0 |
| 0 | 0 | 0 | 0 | 0.001968912 |
| 0 | 0.730400815 | 0.173523251 | 0.910488098 | 0.082478526 |
| 2.534037012 | 0.26121791 | 2.542170132 | 0.047282602 | 6.489987615 |
| 0.010921004 | 0 | 0 | 0.000433115 | 0.376426414 |
| 0 | 0 | 0.02173761 | 0.479663421 | 0 |
| 0 | 0.0006936 | 0 | 0 | 0 |
| 0 | 0 | 0.005511472 | 0.029538306 | 0 |
| 0 | 0 | 0 | 0 | 0 |
| 0 | 0 | 0 | 0 | 0.00789636 |
| 0 | 0 | 0 | 0 | 0 |
| 0 | 0 | 0 | 0 | 0 |
| 0 | 0 | 0.206867787 | 0 | 0 |
| 0 | 0 | 0 | 0 | 0 |
| 0 | 0.0072277 | 0 | 0.316410494 | 0.003692114 |
| 0 | 0 | 0 | 0 | 0 |
| 0 | 0 | 0 | 0 | 0 |
| 0 | 0 | 0 | 0 | 0 |
| 0 | 0 | 0 | 0 | 0 |
| 0 | 0 | 0.006061613 | 0.071412298 | 0 |
| 0 | 0 | 0 | 0 | 0 |
| 0 | 0 | 0 | 0 | 0 |
| 0 | 0 | 0 | 0 | 0 |
| 0 | 0 | 0 | 0 | 0 |
| 0 | 0 | 0 | 0 | 0 |
| 0 | 0.009125624 | 0.000747888 | 0.023515055 | 0.308085961 |
| 0 | 0 | 0 | 0 | 0 |
| 0 | 0 | 0 | 0 | 0 |
| 0 | 0 | 0 | 0.017018673 | 0 |
| 0 | 0.018435375 | 0 | 0.004558519 | 0.011416087 |
| 0 | 0.001542059 | 0 | 0 | 0 |
| 0 | 0 | 0 | 0 | 0 |
| 0 | 0 | 0 | 0 | 0 |
| 0 | 0.002586718 | 0 | 0.008601182 | 0.000415287 |
| 0 | 0 | 0 | 0 | 0 |
| 0 | 0 | 0 | 0 | 0 |
| 0.037911999 | 0.399559916 | 0.045522817 | 2.292264569 | 0.151556569 |
| 0 | 0 | 0 | 0 | 0 |
| 0 | 0 | 0 | 0 | 0 |
| 0 | 0 | 0 | 0 | 0 |
| 0 | 0 | 0 | 0 | 0 |
| 0 | 0 | 0 | 0 | 0 |
| 0 | 0 | 0 | 0 | 0 |

| | | | | |
|---|---|---|---|---|
| 0 | 0 | 0 | 0 | 0 |
| 0.0250956 | 0 | 0.001595683 | 0.021821419 | 0.007985439 |
| 0 | 0 | 0 | 0 | 0 |
| 0 | 0 | 0 | 0 | 0 |
| 0.070299218 | 0.427091385 | 0.631632421 | 2.634687767 | 0.794755075 |
| 0 | 0 | 0 | 0.027768088 | 0 |
| 0 | 0 | 0 | 0 | 0.007899097 |
| 0.017496438 | 0.197167506 | 0 | 0.01562998 | 0.005035095 |
| 0 | 0.009210033 | 0 | 0.002768799 | 0 |
| 0 | 0 | 0.065815874 | 0 | 0 |
| 0 | 0 | 0 | 0 | 0 |
| 0 | 0.08286432 | 0 | 0 | 0 |
| 0 | 0 | 0 | 0 | 0 |
| 0 | 0 | 0 | 0 | 0 |
| 0 | 0 | 0 | 0 | 0 |
| 0.00152837 | 0 | 0 | 0 | 0.007192542 |
| 0 | 0 | 0 | 0 | 0 |
| 0 | 0 | 0 | 0 | 0 |
| 0 | 0 | 0 | 0 | 0 |
| 0 | 0 | 0 | 0 | 0 |
| 0 | 0 | 0 | 0 | 0 |
| 0 | 0 | 0 | 0 | 0 |
| 0 | 0 | 0 | 0 | 0 |
| 0 | 0 | 0 | 0.000654506 | 0.00858676 |
| 0 | 0 | 0.106040305 | 0.150531536 | 0 |
| 0 | 0 | 0 | 0 | 0 |
| 0.002291532 | 0 | 0.005968696 | 0 | 28.75390013 |
| 0 | 0 | 0 | 0 | 0 |
| 0 | 0 | 0 | 0 | 0.002787667 |
| 0 | 0 | 0 | 0 | 0 |
| 0 | 0 | 0 | 0 | 0 |
| 0 | 0.001052631 | 0 | 0 | 0 |
| 0 | 0 | 0 | 0 | 0 |
| 0 | 0 | 0 | 0 | 0 |
| 0 | 0 | 0 | 0 | 0 |
| 0 | 0 | 0 | 0 | 0 |
| 0 | 0 | 0 | 0 | 0 |
| 0 | 0 | 0.002792148 | 0 | 0 |
| 0 | 0 | 0.138591658 | 0.020296524 | 0 |
| 0 | 0 | 0 | 0 | 0 |
| 0 | 0 | 0 | 0 | 0 |
| 0 | 0 | 0.124492641 | 0 | 0 |
| 0 | 0 | 0 | 0 | 0 |
| 0 | 0 | 0 | 0 | 0 |
| 0 | 0 | 0 | 0 | 0 |
| 0 | 0 | 0 | 0 | 0 |
| 0 | 0 | 0 | 0 | 0 |
| 0 | 0 | 0 | 0.022619422 | 0 |

| | | | | |
|---|---|---|---|---|
| 0 | 0.025404655 | 0.052092993 | 0.055075341 | 0 |
| 0 | 0 | 0.000521006 | 0 | 0 |
| 0 | 0 | 0 | 0 | 0 |
| 0 | 0 | 0 | 0 | 0.001096756 |
| 0 | 0 | 0 | 0 | 0 |
| 0 | 0.197676797 | 0.222130053 | 0.043263325 | 0.000638556 |
| 0 | 0 | 0 | 0 | 0 |
| 0 | 0 | 0 | 0 | 0 |
| 0 | 0 | 0 | 0 | 0 |
| 0 | 0 | 0 | 0 | 0 |
| 0 | 0 | 0 | 0 | 0 |
| 0 | 0.007726807 | 0 | 0.02976528 | 0.002481016 |
| 0 | 0 | 0.007614006 | 0.000262212 | 0 |
| 0 | 0 | 0 | 0 | 0 |
| 0 | 0 | 0 | 0 | 0 |
| 0 | 0 | 0 | 0 | 0 |
| 0 | 0 | 0 | 0 | 0 |
| 0 | 0 | 0 | 0 | 0 |
| 0.00023831 | 0 | 0 | 0 | 0.026074725 |
| 0 | 0 | 0 | 0.001131578 | 0 |
| 0.000867823 | 0 | 0 | 0 | 0 |
| 0 | 0 | 0 | 0 | 0 |
| 0.003835991 | 0.003011863 | 0.031677342 | 0.083960098 | 0.179555512 |
| 0 | 0 | 0 | 3.51740093 | 0 |
| 0 | 0 | 0.037505833 | 0 | 0 |
| 0 | 0 | 0 | 0 | 0 |
| 0 | 0 | 0 | 0 | 0 |
| 0 | 0 | 0 | 0 | 0 |
| 0 | 0 | 0 | 0 | 0 |
| 0 | 0 | 0 | 0 | 0 |
| 0 | 0 | 0 | 0 | 0 |
| 0 | 0 | 0 | 0 | 0 |
| 1.725886899 | 0.010342191 | 0 | 0 | 0.025207855 |
| 0 | 0 | 0 | 0 | 0 |
| 0.057496477 | 0 | 0.006633957 | 0.008389349 | 0 |
| 0 | 0 | 0 | 0 | 0 |
| 0 | 0 | 0 | 0 | 0 |
| 0 | 0 | 0 | 0 | 0 |
| 0.000952671 | 0 | 0 | 0.012521146 | 0.110821314 |
| 0 | 0 | 0 | 0 | 0 |
| 0 | 0 | 0 | 0 | 0 |
| 0 | 0 | 0 | 0 | 0 |
| 0.072128756 | 0 | 0.037903987 | 0.046572233 | 0.377118595 |
| 0 | 0 | 0 | 0 | 0 |
| 0 | 0 | 0 | 0 | 0 |
| 0 | 0.899318606 | 0.002092055 | 0.199438804 | 0 |
| 0 | 0 | 0 | 0 | 0 |
| 0 | 0 | 0 | 0 | 0 |

| | | | | |
|---|---|---|---|---|
| 0 | 0 | 0 | 0 | 0 |
| 0 | 0 | 0 | 0 | 0 |
| 0 | 0 | 0 | 0 | 0 |
| 0 | 0 | 0 | 0 | 0 |
| 0 | 0 | 0 | 0.003366814 | 0 |
| 0 | 0 | 0 | 0 | 0 |
| 0 | 0 | 0 | 0 | 0 |
| 0.775800736 | 0 | 0 | 0 | 0 |
| 0 | 0.01385632 | 0.0014156 | 0.005913811 | 0 |
| 0 | 0.040899413 | 0.002274504 | 0.131167535 | 0 |
| 0 | 0 | 0 | 0 | 0 |
| 0 | 0 | 0 | 0 | 0 |
| 0 | 0 | 0 | 0 | 0 |
| 0 | 0 | 0 | 0 | 0 |
| 0 | 0.030522793 | 0 | 0 | 0 |
| 0 | 0.003284939 | 0 | 0 | 0 |
| 0 | 0 | 0 | 0 | 0 |
| 0 | 0 | 0 | 0 | 0 |
| 0 | 0 | 0 | 0 | 0 |
| 0 | 0 | 0.050784622 | 0.694892065 | 0 |
| 0.005852236 | 0 | 0.000615075 | 0 | 0 |
| 0 | 0 | 0 | 0 | 0 |
| 0 | 0 | 0 | 0 | 0 |
| 0 | 0 | 0 | 0 | 0 |
| 0 | 0.004439683 | 0 | 0.01941375 | 0 |
| 0 | 0 | 0 | 0 | 0 |
| 0 | 0 | 0 | 0 | 0 |
| 0 | 0 | 0 | 0.123195311 | 0 |
| 0 | 0 | 0 | 0 | 0 |
| 0 | 0 | 0.005004957 | 0.008025296 | 0 |
| 0 | 0.315048918 | 0.03800481 | 1.704901486 | 0.039756621 |
| 0 | 0 | 0 | 0 | 0 |
| 0 | 0 | 0 | 0 | 0 |
| 0 | 0 | 1.88343197 | 0.014039254 | 0 |
| 0 | 0 | 0 | 0 | 0 |
| 0 | 0 | 0 | 0 | 0 |
| 0 | 0 | 0 | 0 | 0 |
| 0 | 0 | 0 | 0 | 0 |
| 0 | 0 | 0.010573363 | 0.00246714 | 0 |
| 0 | 0 | 0 | 0 | 0 |
| 0 | 0 | 0 | 0.001715815 | 0 |
| 0 | 0 | 0 | 0 | 0 |
| 0 | 0 | 0.000517702 | 0 | 0 |
| 0 | 0 | 0 | 0.006233149 | 0.000422491 |
| 0 | 0 | 0 | 0 | 0 |
| 0 | 0 | 0 | 0 | 0.003615726 |
| 0.003441627 | 0 | 0 | 0 | 0.035766986 |
| 0 | 0 | 0 | 0.001026992 | 0.000877518 |

| | | | | |
|---|---|---|---|---|
| 0 | 0 | 0 | 0 | 0 |
| 0 | 0 | 0 | 0 | 0 |
| 0 | 0 | 0 | 0 | 0 |
| 0.00123379 | 0 | 0.000756423 | 0 | 0 |
| 0.011227365 | 0.24698448 | 0.011529832 | 0.224257959 | 0 |
| 0 | 0 | 0 | 0 | 0 |
| 0 | 0 | 0 | 0 | 0 |
| 0 | 0 | 0 | 0 | 0 |
| 0 | 0 | 0 | 0 | 0 |
| 0 | 0 | 0 | 0 | 0 |
| 0 | 0 | 0.005718312 | 0.030619609 | 0 |
| 0.000713934 | 0.027685032 | 0.003376581 | 0.035984524 | 0.854646523 |
| 0 | 0 | 0 | 0.004420397 | 0.001811658 |
| 0 | 0 | 0 | 0 | 0 |
| 0.004533897 | 0 | 0 | 0 | 0 |
| 0 | 0 | 0 | 0.229794738 | 0 |
| 0 | 0.012574651 | 0 | 0.086053349 | 0.002261067 |
| 0 | 0 | 0 | 0 | 0 |
| 0.164070594 | 0.139645055 | 0.000852255 | 0 | 0.184125767 |
| 0.004705683 | 0.000862099 | 0.007542215 | 0.005065625 | 0.001107252 |
| 0 | 0 | 0 | 0 | 0 |
| 0 | 0 | 0.007588533 | 0.003201968 | 0 |
| 0 | 0 | 0 | 0 | 0 |
| 0 | 0 | 0 | 0 | 0 |
| 0 | 0 | 0 | 0 | 0 |
| 0 | 0 | 0 | 0 | 0 |
| 0.000754176 | 0.006562973 | 0 | 0.004436928 | 0.16137623 |
| 0 | 0 | 0 | 0 | 0 |
| 0 | 0 | 0 | 0 | 0 |
| 0 | 0 | 0 | 0.000450577 | 0 |
| 0 | 0.000783103 | 0.002920143 | 0 | 1.795967468 |
| 0 | 0 | 0 | 0.006935025 | 0 |
| 0 | 0 | 0 | 0 | 0 |
| 0 | 0 | 0 | 0 | 0 |
| 0 | 0 | 0 | 0.180088408 | 0 |
| 0 | 0 | 0 | 0 | 0 |
| 0 | 0 | 0 | 0 | 0 |
| 0.004654855 | 0.006052037 | 0.004813387 | 0.085783158 | 0.207575592 |
| 0 | 0 | 0.041190997 | 0.062124765 | 0 |
| 0 | 0 | 0 | 0.046749465 | 0 |
| 0 | 0.010161349 | 0.011895116 | 0.00202652 | 0.829262125 |
| 0 | 0 | 0 | 0 | 0 |
| 0 | 0 | 0 | 0 | 0 |
| 0.005789244 | 0.003467383 | 0 | 0 | 0.002750627 |
| 0 | 0 | 0 | 0 | 0 |
| 0 | 0 | 0 | 0.0012957 | 0 |
| 0 | 0 | 0 | 0 | 0.034469833 |
| 0 | 0 | 0 | 0 | 0 |

| | | | | |
|---|---|---|---|---|
| 0 | 0 | 0 | 0 | 0 |
| 0 | 0 | 0.001300126 | 0.134641712 | 0.000559757 |
| 0 | 0 | 0.044900424 | 0.003982569 | 0 |
| 0.00175014 | 0 | 0.000367883 | 0.003154622 | 0.049005386 |
| 0 | 0.000991271 | 0 | 0 | 0 |
| 0.001117296 | 0.001023464 | 0 | 0 | 0 |
| 0 | 0 | 0 | 0.089005074 | 0 |
| 0 | 0 | 0 | 0 | 0 |
| 0 | 0 | 0 | 0 | 0 |
| 0 | 0 | 0.002408956 | 0 | 0 |
| 0 | 0 | 0.009119616 | 0 | 0 |
| 0 | 0 | 0 | 0 | 0 |
| 0.004218051 | 0 | 0.11048254 | 0.237449495 | 0 |
| 0 | 0 | 0 | 0 | 0 |
| 0 | 0 | 0 | 0 | 0 |
| 0 | 0 | 0 | 0 | 0 |
| 0.25494902 | 0 | 0 | 0 | 0 |
| 0 | 0 | 0 | 0 | 0 |
| 0 | 0 | 0 | 0 | 0 |
| 0 | 0 | 0 | 0 | 0 |
| 0 | 0 | 0 | 0 | 0 |
| 0 | 0 | 0 | 0 | 0 |
| 0.009148096 | 0 | 0 | 0 | 0.066626763 |
| 0.004497945 | 0.03649322 | 0.037278775 | 0.011453494 | 0 |
| 0 | 0 | 0 | 0 | 0 |
| 0 | 0 | 0 | 0.008933713 | 0 |
| 0 | 0 | 0.004921095 | 0 | 0 |
| 0 | 0 | 0 | 0 | 0 |
| 0 | 0 | 0 | 0 | 0 |
| 0 | 0 | 0 | 0 | 0 |
| 0 | 0.002892027 | 0.004977321 | 0.020914858 | 0 |
| 0 | 0 | 0 | 0.001316455 | 0.001436195 |
| 0 | 0 | 0 | 0 | 0 |
| 0 | 0 | 0 | 0 | 0 |
| 0 | 0 | 0.001497955 | 0.179729103 | 0 |
| 0 | 0 | 0 | 0 | 0 |
| 0 | 0.007090539 | 0 | 0 | 0 |
| 0 | 0 | 0 | 0 | 0.005882414 |
| 0.001335592 | 0.009410982 | 0 | 0 | 0.002054818 |
| 0 | 0 | 0 | 0 | 0 |
| 0 | 0 | 0 | 0 | 0 |
| 0 | 0 | 0 | 0 | 0 |
| 0 | 0 | 0 | 0 | 0 |
| 0 | 0 | 0 | 0 | 0 |
| 0 | 0 | 0.628996832 | 0 | 0.000699764 |
| 0.176445863 | 0.030636404 | 0.238172959 | 0.301333468 | 0.116450064 |
| 0 | 0 | 0 | 0 | 0 |
| 0 | 0 | 0 | 0 | 0 |

| | | | | |
|---|---|---|---|---|
| 0 | 0 | 0 | 0 | 0 |
| 0 | 0 | 0 | 0 | 0 |
| 0 | 0 | 0.045409989 | 0.133140471 | 0 |
| 0 | 0 | 0 | 0.17629478 | 0 |
| 0 | 0 | 0 | 0 | 0 |
| 0 | 0.001219696 | 0 | 0 | 0 |
| 0 | 0 | 0 | 0.000836609 | 0 |
| 0 | 0 | 0 | 0 | 0 |
| 0 | 0 | 0 | 0.004664393 | 0 |
| 0 | 0 | 0 | 0 | 0 |
| 0 | 0 | 0.000525597 | 0.003505468 | 0.004608787 |
| 0 | 0 | 0 | 0.003862951 | 0.007612948 |
| 0 | 0 | 0 | 0 | 0 |
| 0 | 0 | 0 | 0 | 0 |
| 0 | 0 | 0 | 0 | 0 |
| 0 | 0 | 0 | 0 | 0 |
| 0 | 0 | 0 | 0 | 0 |
| 0 | 0 | 0 | 0 | 0 |
| 0 | 0 | 0 | 0 | 0 |
| 0.001768539 | 0 | 0.000348516 | 0 | 0 |
| 0 | 0 | 0 | 0 | 0 |
| 0 | 0 | 0 | 0 | 0 |
| 0 | 0 | 0 | 0 | 0 |
| 0 | 0 | 0 | 0 | 0 |
| 0 | 0 | 0 | 0 | 0 |
| 0 | 0 | 0 | 0.000644487 | 0 |
| 0 | 0 | 0 | 0 | 0 |
| 0 | 0.019945125 | 0.002057021 | 0 | 0 |
| 0 | 0 | 0 | 0 | 0 |
| 0 | 0 | 0 | 0 | 0 |
| 0.010825425 | 0.002261611 | 0.005589006 | 0.000665641 | 0 |
| 0 | 0 | 0 | 0 | 0 |
| 0.077621684 | 0.007473887 | 0.019352333 | 0.02561535 | 0.000648596 |
| 0 | 0.000619257 | 0 | 0.001805249 | 0 |
| 0 | 0 | 0 | 0 | 0 |
| 0 | 0 | 0 | 0 | 0 |
| 0 | 0 | 0 | 0 | 0 |
| 0 | 0 | 0 | 0 | 0 |
| 0 | 0 | 0 | 0 | 0 |
| 0 | 0 | 0 | 0 | 0 |
| 0 | 0 | 0 | 0 | 0 |
| 0 | 0.104195519 | 0 | 0.07957069 | 1.816461194 |
| 0.003094233 | 0 | 0.083043839 | 0.021689668 | 0.004810504 |
| 0 | 0 | 0 | 0 | 0 |
| 0 | 0 | 0 | 0 | 0 |
| 0.002075998 | 0.00702149 | 0.07703761 | 0.052291768 | 0.03250302 |
| 0 | 0.012467781 | 0 | 0.059970733 | 0.004188073 |
| 0 | 0 | 0 | 0 | 0 |

| | | | | |
|---|---|---|---|---|
| 0 | 0 | 0 | 0 | 0 |
| 0 | 0.004454076 | 0.007665684 | 0.254588146 | 0.011032728 |
| 0.007843575 | 0.032208002 | 0.000710661 | 0.005416864 | 0 |
| 0 | 0 | 0 | 0 | 0 |
| 0.002699447 | 0.002967293 | 0.007490058 | 0.017516676 | 0.42798617 |
| 0 | 0 | 0 | 0 | 0 |
| 0 | 0 | 0 | 0 | 0 |
| 0 | 0 | 0 | 0 | 0 |
| 0 | 0 | 0 | 0 | 0 |
| 0 | 0 | 0 | 0 | 0 |
| 0 | 0 | 0 | 0 | 0 |
| 0 | 0 | 0 | 0 | 0 |
| 0 | 0 | 0 | 0 | 0 |
| 0 | 0 | 0 | 0 | 3.776956255 |
| 0 | 0 | 0 | 0 | 0 |
| 0 | 0 | 0 | 0 | 0 |
| 0 | 0 | 0 | 0 | 0 |
| 0 | 0 | 0 | 0 | 0 |
| 0 | 0 | 0 | 0 | 0 |
| 0 | 0 | 0 | 0 | 0 |
| 0 | 0 | 0 | 0 | 0 |
| 0 | 0.000797157 | 0 | 0 | 0.003839413 |
| 0 | 0 | 0 | 0.001837507 | 0 |
| 0 | 0 | 0 | 0 | 0 |
| 0 | 0 | 0.005973983 | 0 | 0.007998547 |
| 0.001233297 | 0.00718915 | 0 | 0 | 0 |
| 0 | 0 | 0 | 0 | 0 |
| 0 | 0 | 0.029376635 | 0.11054279 | 0 |
| 0 | 0 | 0 | 0 | 0 |
| 0 | 0 | 0 | 0 | 0 |
| 0 | 0 | 0 | 0 | 0 |
| 0 | 0 | 0 | 0 | 0.077849241 |
| 0 | 0 | 0 | 0 | 0 |
| 0 | 0 | 0 | 0 | 0 |
| 0 | 0 | 0 | 0 | 0 |
| 0 | 0 | 0 | 0 | 0 |
| 0 | 0 | 0 | 0 | 0.430125459 |
| 0 | 0 | 0 | 0.000447035 | 0.000787816 |
| 0 | 3.080084613 | 0.005777282 | 2.070920808 | 0.000958092 |
| 0 | 0 | 0 | 0 | 0 |
| 0 | 0 | 0 | 0 | 0 |
| 0 | 0 | 0.030552552 | 0.029597247 | 0 |
| 0.006638833 | 0.008864262 | 0 | 0 | 0.120866083 |
| 0 | 0 | 0 | 0 | 0 |
| 0 | 0 | 0 | 0 | 0 |
| 0 | 0 | 0 | 0 | 0 |
| 0.001893513 | 0.006937974 | 0 | 0.00741218 | 0.016404182 |
| 0 | 0 | 0 | 0 | 0 |

| | | | | |
|---|---|---|---|---|
| 0 | 0 | 0 | 0 | 0 |
| 0.006407575 | 0.065459389 | 0 | 0.007612751 | 0.175303147 |
| 0 | 0 | 0 | 0.047196059 | 0 |
| 0 | 0 | 0 | 0 | 0 |
| 0 | 0 | 0 | 0 | 0 |
| 0 | 0 | 0 | 0 | 0 |
| 0 | 0 | 0 | 0 | 0 |
| 0 | 0 | 0 | 0 | 0 |
| 0 | 0 | 0 | 0 | 0 |
| 0 | 0 | 0 | 0 | 0 |
| 0 | 0 | 0 | 0 | 0 |
| 0 | 0 | 0 | 0 | 0 |
| 0 | 0.025869769 | 0.021959927 | 0.200264103 | 0 |
| 0 | 0 | 0 | 0 | 0 |
| 0.004603992 | 0 | 0 | 0 | 0.02931348 |
| 0 | 0 | 0 | 0 | 0 |
| 0 | 0 | 0 | 0 | 0 |
| 0 | 0 | 0 | 0 | 0 |
| 0 | 0 | 0 | 0 | 0 |
| 0 | 0 | 0 | 0 | 0 |
| 0 | 0 | 0 | 0 | 0 |
| 0.001462833 | 0 | 0 | 0.025302155 | 0 |
| 0 | 0 | 0 | 0.01811235 | 0 |
| 0 | 0 | 0 | 0 | 0 |
| 0 | 0 | 0 | 0 | 0 |
| 0 | 0 | 0 | 0 | 0 |
| 0 | 0 | 0 | 0 | 0 |
| 0 | 0 | 0 | 0 | 0 |
| 0 | 0.014787094 | 0 | 0 | 0 |
| 0 | 0 | 0 | 0 | 0 |
| 0 | 0 | 0 | 0 | 0 |
| 0 | 0 | 0 | 0 | 0 |
| 0 | 0 | 0 | 0 | 0 |
| 0 | 0 | 0 | 0 | 0 |
| 0 | 9.00084586 | 28.89667608 | 0 | 17.76150807 |
| 0 | 0 | 0 | 0 | 0 |
| 0 | 0 | 0 | 0 | 0 |
| 0 | 0 | 0 | 0 | 0 |
| 0.001195213 | 0.001459783 | 0 | 0.005319424 | 0.018905232 |
| 0 | 0 | 0 | 0 | 0 |
| 0.128599956 | 0.490178807 | 0.070207964 | 0.077982422 | 0.021574052 |
| 0 | 0 | 0 | 0 | 0 |
| 0 | 0 | 0 | 0 | 0 |
| 0 | 0 | 0 | 0 | 0 |
| 0 | 0 | 0 | 0 | 0 |
| 0.777363151 | 0.085669306 | 0 | 0 | 0.60436816 |
| 0 | 0 | 0 | 0 | 0 |
| 0 | 0 | 0 | 0 | 0 |

| | | | | |
|---|---|---|---|---|
| 0 | 0 | 0 | 0 | 0 |
| 0 | 0 | 0 | 0 | 0 |
| 0 | 0 | 0 | 0 | 0 |
| 0 | 0 | 0 | 0 | 0 |
| 0 | 0.00246256 | 0 | 0.000931866 | 0.003892718 |
| 0 | 0 | 0 | 0 | 0 |
| 0 | 0 | 0 | 0 | 0 |
| 0 | 0 | 0 | 0 | 0 |
| 0 | 0 | 0 | 0 | 0 |
| 0.000399998 | 0.014411948 | 0 | 0 | 0.000784333 |
| 0 | 0 | 0.015961434 | 0 | 0 |
| 0 | 0 | 0 | 0 | 0 |
| 0 | 0.003103798 | 0 | 0.002740544 | 0.005366333 |
| 0 | 0 | 0 | 0 | 0 |
| 0 | 0 | 0 | 0 | 0 |
| 0 | 0 | 0 | 0 | 0 |
| 0 | 0 | 0 | 0 | 0 |
| 0 | 0 | 0 | 0 | 0 |
| 0 | 0 | 0 | 0 | 0 |
| 0.173412853 | 0.061839714 | 0.039499496 | 0.010221736 | 0.012145742 |
| 0 | 0 | 0 | 0 | 0 |
| 0 | 0 | 0 | 0 | 0 |
| 0 | 0 | 0 | 0 | 0 |
| 0 | 0 | 0 | 0 | 0 |
| 0 | 0 | 0 | 0 | 0 |
| 0 | 0 | 0.010721944 | 0 | 0 |
| 0 | 0 | 0 | 0 | 0 |
| 0 | 0 | 0 | 0 | 0 |
| 0 | 0 | 0 | 0 | 0 |
| 0 | 0 | 0 | 0 | 0 |
| 0 | 0 | 0 | 0 | 0 |
| 0 | 0 | 0 | 0 | 0 |
| 0 | 0 | 0 | 0 | 0 |
| 0 | 0 | 0 | 0 | 0 |
| 0 | 0 | 0 | 0 | 0 |
| 0 | 0 | 0 | 0 | 0 |
| 0 | 0 | 0 | 0 | 0 |
| 0 | 0 | 0 | 0 | 0 |
| 0 | 0 | 0 | 0 | 0 |
| 0 | 0 | 0 | 0 | 0 |
| 0 | 0 | 0 | 0 | 0 |
| 0 | 0 | 0 | 0 | 0 |
| 0 | 0 | 0 | 0.205272795 | 0 |
| 0 | 0 | 0 | 0 | 0 |
| 0 | 0.006387039 | 0.003664136 | 0.025965341 | 0.017303856 |
| 0 | 0 | 0 | 0.056078332 | 0 |
| 0 | 0 | 0 | 0 | 0 |
| 0 | 0 | 0 | 0 | 0 |

| | | | | |
|---|---|---|---|---|
| 0 | 0 | 0 | 0 | 0 |
| 0 | 0 | 0 | 0 | 0 |
| 0 | 0 | 0 | 0 | 0 |
| 0 | 0 | 0 | 0 | 0 |
| 0 | 0 | 0 | 0 | 0 |
| 0.002765327 | 0.001639059 | 0.002649938 | 0.053265653 | 0.040189336 |
| 0 | 0 | 0 | 0 | 0 |
| 0 | 0.000887449 | 0 | 0.002231355 | 0 |
| 0 | 0 | 0 | 0 | 0 |
| 0 | 0 | 0 | 0 | 0 |
| 0 | 0 | 0 | 0 | 0 |
| 0 | 0 | 0 | 0 | 0 |
| 0 | 0 | 0 | 0 | 0 |
| 0 | 0 | 0 | 0 | 0 |
| 0 | 0 | 0 | 0 | 0 |
| 0 | 0 | 0 | 0 | 0 |
| 0 | 0 | 0.033802587 | 0 | 0 |
| 0 | 0 | 0 | 0 | 0 |
| 0 | 0 | 0 | 0 | 0 |
| 0 | 0 | 0 | 0 | 0 |
| 0 | 0 | 0 | 0 | 0 |
| 0 | 0 | 0 | 0 | 0 |
| 0 | 0 | 0 | 0 | 0 |
| 0 | 0 | 0 | 0 | 0 |
| 0 | 0 | 0 | 0 | 0 |
| 0 | 0 | 0 | 0 | 0 |
| 0 | 0 | 0 | 0 | 0 |
| 0 | 0.013257164 | 0.011107903 | 0 | 0 |
| 0 | 0 | 0 | 0 | 0 |
| 0 | 0 | 0 | 0 | 0 |
| 0 | 0 | 0 | 0 | 0 |
| 0 | 0 | 0 | 0 | 0 |
| 0 | 0 | 0 | 0 | 0 |
| 0 | 0 | 0 | 0 | 0 |
| 0 | 0 | 0 | 0 | 0 |
| 0 | 0 | 0 | 0 | 0 |
| 0 | 0 | 0 | 0 | 0 |
| 0 | 0 | 0 | 0 | 0 |
| 0 | 0 | 0 | 0.454929418 | 0 |
| 0 | 0 | 0 | 0 | 0 |
| 0 | 0 | 0 | 0 | 0 |
| 0 | 0 | 0 | 0 | 0 |
| 0 | 0 | 0 | 0 | 0 |
| 0 | 0 | 0 | 0 | 0 |
| 0 | 0 | 0 | 0 | 0 |
| 0 | 0 | 0 | 0 | 0 |
| 0 | 0 | 0 | 0 | 0 |
| 0 | 0 | 0 | 0 | 0 |

| | | | | |
|---|---|---|---|---|
| 0 | 0 | 0 | 0 | 0 |
| 0 | 0 | 0 | 0 | 0.009213303 |
| 0 | 0 | 0 | 0 | 0 |
| 0 | 0 | 0 | 0 | 0 |
| 0 | 0 | 0 | 0 | 0 |
| 0 | 0 | 0 | 0 | 0.004994908 |
| 0 | 0 | 0 | 0 | 0.005598341 |
| 0 | 0 | 0 | 0 | 0 |
| 0 | 0 | 0 | 0 | 0 |
| 0 | 0 | 0.004548003 | 0 | 0 |
| 0.000715407 | 0.000873769 | 0.008521529 | 0.001193999 | 0 |
| 0 | 0 | 0 | 0 | 0 |
| 0 | 0.4727058 | 0.040059076 | 0.356789169 | 0 |
| 0 | 0 | 0 | 0 | 0 |
| 0 | 0 | 0 | 0 | 0 |
| 0 | 0 | 0 | 0 | 0 |
| 0 | 0 | 0 | 0 | 0 |
| 0 | 0 | 0 | 0 | 0 |
| 0 | 0 | 0 | 0 | 0 |
| 0 | 0 | 0.000625666 | 0.00069548 | 0.002684767 |
| 0.008108066 | 0.004212408 | 0.004197231 | 0 | 0.093968079 |
| 0 | 0 | 0 | 0 | 0 |
| 0 | 0 | 0 | 0 | 0 |
| 1.62243838 | 6.568632498 | 0.54451682 | 2.718622751 | 0.503503439 |
| 0 | 0 | 0 | 0 | 0 |
| 0 | 0 | 0 | 0 | 0 |
| 0 | 0 | 0 | 0 | 0 |
| 0 | 0 | 0 | 0 | 0.000981136 |
| 0 | 0 | 0 | 0 | 0 |
| 0 | 0 | 0 | 0 | 0 |
| 0 | 0 | 0 | 0 | 0 |
| 0 | 0 | 0 | 0 | 0 |
| 0 | 0 | 0 | 0 | 0 |
| 0 | 0 | 0 | 0 | 0 |

| PHL018-0093_V1 | PHL018-0121_V1 | PHL018-0232_V2 | PHL018-0095_V3 | PHL018-0109_V1 |
|---|---|---|---|---|
| CTRL | EYCF | REF | CTRL | REF |
| 2.531744097 | 0.603057017 | 0.004211056 | 0.050383159 | 0.265017831 |
| 2.249928479 | 1.384990061 | 13.60644493 | 3.008837724 | 0.546289619 |
| 0 | 0.148637672 | 0 | 0 | 0 |
| 0 | 0 | 0 | 0 | 0 |
| 0 | 0 | 0 | 0 | 0 |
| 0 | 0 | 0 | 0 | 0 |
| 0 | 0 | 0 | 0 | 0 |
| 0 | 0 | 0 | 0 | 0 |
| 0 | 0 | 0 | 0 | 0 |
| 0 | 0 | 0 | 0 | 0 |
| 0 | 0 | 0 | 0 | 0 |
| 0 | 0 | 0 | 0 | 0 |
| 0 | 0 | 0 | 0 | 0 |
| 3.660881099 | 0.061203248 | 1.152290317 | 2.517899284 | 0.695218193 |
| 0 | 0 | 0 | 0 | 0 |
| 0 | 0 | 0 | 0 | 0 |
| 0 | 0 | 0 | 0 | 0 |
| 0 | 0.000596425 | 0 | 0 | 0 |
| 0 | 0 | 0 | 0 | 0 |
| 0 | 0 | 0 | 0 | 0 |
| 0 | 0 | 0 | 0 | 0 |
| 0 | 0 | 0 | 0 | 0.022511523 |
| 0 | 0 | 0 | 0 | 0 |
| 0 | 0 | 0 | 0 | 0 |
| 0 | 0.010033084 | 0 | 0 | 0.457075602 |
| 0 | 0 | 0 | 0 | 0 |
| 0 | 0 | 0 | 0 | 0 |
| 0 | 0 | 0 | 0 | 0 |
| 0 | 0 | 0 | 0 | 0 |
| 0 | 0 | 0 | 0 | 0 |
| 0 | 0 | 0 | 0 | 0 |
| 0 | 0 | 0 | 0 | 0 |
| 0 | 0 | 0 | 0 | 0 |
| 0 | 0 | 0 | 0 | 0 |
| 0 | 0 | 0 | 0 | 0 |
| 0 | 0 | 0.280493321 | 0 | 0 |
| 0 | 0 | 0 | 0 | 0 |
| 0 | 0 | 0 | 0 | 0 |
| 0 | 0 | 0 | 0 | 0 |
| 0 | 0 | 0 | 0 | 0 |
| 0 | 0 | 0 | 0 | 0.007047708 |
| 0.314433776 | 1.503142143 | 0.028163292 | 0.141783465 | 0.011060963 |
| 0.011112999 | 0.074313177 | 0.076297693 | 2.517734581 | 1.324358335 |
| 0.036071923 | 0.011725716 | 0.073456334 | 0.039688064 | 0.012269303 |
| 0.003674843 | 0 | 0 | 0.001250872 | 0 |
| 0 | 0 | 0 | 0 | 0 |

| | | | | |
|---|---|---|---|---|
| 0 | 0.002223026 | 0 | 0 | 0 |
| 0.006049004 | 0.023377346 | 0 | 0.02007532 | 0 |
| 0 | 0.005058115 | 0.001994322 | 0 | 0 |
| 0.204104834 | 1.568515898 | 1.553310215 | 0.970196103 | 0.718892053 |
| 0 | 0.008506507 | 0 | 0 | 0 |
| 0 | 0 | 0 | 0 | 0 |
| 0.002701482 | 28.59677005 | 0.058665673 | 0.031379693 | 0 |
| 0 | 0 | 0 | 0 | 0 |
| 0 | 0 | 0 | 0 | 0 |
| 0 | 0 | 0 | 0 | 0 |
| 0 | 0 | 0.01388672 | 0 | 0.000900545 |
| 0 | 0.064452718 | 0.005894502 | 0 | 0.296565935 |
| 0 | 0 | 0 | 0.001812301 | 0 |
| 0 | 0.004695849 | 0.001709065 | 0.080047218 | 0.033166406 |
| 0 | 0 | 0 | 0.011452379 | 0 |
| 0 | 0 | 0 | 0 | 0 |
| 0 | 0.015633957 | 0.038853868 | 1.183039024 | 0.00457082 |
| 0 | 0 | 0 | 0 | 0 |
| 0 | 0 | 0 | 0 | 0 |
| 0 | 0 | 0 | 0 | 0 |
| 0.739254781 | 0.426914924 | 0.29527367 | 0.076443362 | 0.134252881 |
| 0.000767797 | 0.062728717 | 0.001695616 | 0 | 0.098193936 |
| 0 | 0 | 0 | 0 | 0 |
| 0 | 0 | 0 | 0 | 0 |
| 0 | 0 | 0 | 0 | 0 |
| 0 | 0 | 0 | 0 | 0 |
| 0 | 0 | 0 | 0 | 0 |
| 0 | 0 | 0 | 0 | 0 |
| 0.00171891 | 0.006811516 | 0.000723061 | 0.046724082 | 0.005122738 |
| 0 | 0 | 0 | 0 | 0 |
| 0 | 0.579186553 | 0.00192132 | 0 | 0 |
| 0 | 0 | 0 | 0 | 0 |
| 0 | 0 | 0 | 0 | 0 |
| 0 | 0 | 0 | 0 | 0 |
| 0 | 0 | 0 | 0.000733648 | 0 |
| 0 | 0 | 0 | 0 | 0 |
| 0 | 0 | 0 | 0 | 0 |
| 0 | 0 | 0 | 0 | 0 |
| 0 | 0 | 0 | 0 | 0 |
| 0 | 0 | 0 | 0 | 0 |
| 0 | 0 | 0 | 0 | 0 |
| 0 | 0 | 0 | 0 | 0 |
| 0 | 0 | 0 | 0 | 0 |
| 0 | 0 | 0 | 0 | 0 |
| 0 | 0 | 0 | 0 | 0.4959541 |
| 0 | 0 | 0 | 0 | 0 |
| 0 | 0 | 0 | 0 | 0.000861001 |
| 0 | 0 | 0 | 0 | 0 |

| | | | | |
|---|---|---|---|---|
| 0 | 0 | 0 | 0 | 0 |
| 0 | 0 | 0 | 0 | 0 |
| 0 | 0 | 0 | 0 | 0 |
| 0 | 0 | 0 | 0 | 0 |
| 0 | 0 | 0 | 0 | 0.025366311 |
| 0 | 0 | 0 | 0 | 0 |
| 0 | 0 | 0 | 0 | 0 |
| 0.454234692 | 0 | 0 | 0 | 0 |
| 0.000742136 | 0.001447454 | 0 | 0.038997276 | 0.002019407 |
| 0 | 0 | 0 | 0 | 0 |
| 0 | 0 | 0 | 0 | 0 |
| 0.003231018 | 0.002614057 | 0.002854175 | 0.015965418 | 0 |
| 0 | 0 | 0 | 0 | 0 |
| 0 | 0 | 0 | 0 | 0 |
| 0 | 0 | 0 | 0 | 0 |
| 0 | 0 | 0 | 0 | 0 |
| 0 | 0 | 0 | 0 | 0 |
| 0 | 0 | 0 | 0 | 0 |
| 0 | 0 | 0 | 0 | 0.1003253 |
| 0 | 0 | 0 | 0 | 0 |
| 0 | 0 | 0 | 0 | 0 |
| 0 | 0 | 0 | 0 | 0 |
| 0 | 0 | 0 | 0 | 0 |
| 0 | 0 | 0 | 0 | 0 |
| 0 | 0 | 0 | 0 | 0 |
| 0 | 0 | 0 | 0 | 0 |
| 0 | 0 | 0 | 0.001503867 | 0 |
| 0 | 0.001082516 | 0 | 0 | 0 |
| 0 | 0 | 0 | 0 | 0 |
| 0 | 0 | 0 | 0 | 0 |
| 0 | 0.011013102 | 0 | 0 | 0.037062559 |
| 0 | 0.065575071 | 0.00066295 | 0.000797018 | 0.00555455 |
| 0 | 0 | 0 | 0 | 0 |
| 0 | 0 | 0 | 0 | 0 |
| 0 | 0 | 0 | 0 | 0 |
| 0 | 0 | 0 | 0 | 0 |
| 0 | 0 | 0 | 0 | 0 |
| 0 | 0 | 0 | 0 | 0 |
| 0 | 0 | 0 | 0 | 0 |
| 0 | 0 | 0 | 0 | 0 |
| 0 | 0 | 0 | 0 | 0 |
| 0 | 0 | 0 | 0 | 0 |
| 0 | 0 | 0 | 0 | 0.00830847 |
| 0 | 0 | 0 | 0 | 0 |
| 0 | 0 | 0 | 0 | 0.001409225 |
| 0 | 0 | 0 | 0 | 0 |
| 0 | 0 | 0 | 0 | 0 |
| 0 | 0 | 0 | 0 | 0 |

| | | | | |
|---|---|---|---|---|
| 0 | 0 | 0 | 0 | 0 |
| 0 | 0 | 0 | 0 | 0 |
| 0 | 0 | 0 | 0 | 0 |
| 0 | 0 | 0 | 0 | 0 |
| 0 | 0 | 0 | 0 | 0 |
| 0 | 0 | 0 | 0 | 0 |
| 0 | 0 | 0 | 0 | 0 |
| 0 | 0 | 0 | 0 | 0 |
| 0 | 0 | 0 | 0 | 0 |
| 0 | 0 | 0 | 0 | 0 |
| 0 | 0 | 0 | 0 | 0 |
| 0 | 0 | 0 | 0 | 0 |
| 0 | 0 | 0 | 0 | 0 |
| 0 | 0 | 0 | 0 | 0 |
| 0 | 0 | 0 | 0 | 0 |
| 0 | 0 | 0 | 0 | 0 |
| 0 | 0 | 0 | 0 | 0 |
| 0 | 0 | 0 | 0 | 0 |
| 0 | 0 | 0 | 0 | 0 |
| 0 | 0 | 0 | 0 | 0 |
| 0 | 0 | 0 | 0 | 0 |
| 0 | 0 | 0 | 0 | 0 |
| 0 | 0 | 0 | 0 | 0 |
| 0 | 0 | 0 | 0 | 0 |
| 0 | 0 | 0 | 0 | 0 |
| 0 | 0 | 0 | 0 | 0 |
| 0 | 0.001526402 | 0 | 0 | 0 |
| 0 | 0 | 0 | 0 | 0 |
| 0 | 0 | 0 | 0 | 0 |
| 0 | 0 | 0 | 0 | 0 |
| 0 | 0 | 0 | 0 | 0 |
| 0 | 0 | 0.049565588 | 0.003218553 | 0 |
| 0 | 0 | 0 | 0 | 0 |
| 0 | 0 | 0 | 0 | 0 |
| 0 | 0 | 0 | 0 | 0 |
| 0 | 0 | 0 | 0 | 0 |
| 0 | 0 | 0 | 0 | 0 |
| 0 | 0 | 0 | 0 | 0 |
| 0 | 0 | 0 | 0 | 0 |
| 0 | 0 | 0 | 0 | 0 |
| 0 | 0 | 0 | 0 | 0 |
| 0 | 0 | 0 | 0 | 0 |
| 0 | 0 | 0 | 0 | 0 |
| 0 | 0 | 0 | 0 | 0 |
| 0 | 0 | 0 | 0 | 0 |
| 0 | 0 | 0 | 0 | 0 |
| 0 | 0 | 0 | 0 | 0 |
| 0.014013145 | 0 | 0 | 0 | 0 |

| | | | | |
|---|---|---|---|---|
| 0 | 0 | 0 | 0 | 2.119936488 |
| 0 | 0 | 0 | 0 | 0 |
| 0 | 0 | 0 | 0 | 0 |
| 0 | 0 | 0 | 0 | 0 |
| 0 | 0 | 0 | 0 | 0 |
| 0 | 0 | 0 | 0 | 0 |
| 0 | 0 | 0 | 0 | 0 |
| 0 | 0 | 0 | 0 | 0 |
| 0 | 0 | 0 | 0 | 0 |
| 0 | 0 | 0 | 0 | 0 |
| 0 | 0 | 0 | 0 | 0 |
| 0 | 0 | 0 | 0 | 0.022139296 |
| 0.008749744 | 0.000689511 | 0 | 0 | 0.001623321 |
| 0 | 0 | 0 | 0 | 0 |
| 0 | 0 | 0 | 0 | 0.038949304 |
| 0 | 0 | 0 | 0 | 0 |
| 0 | 0 | 0 | 0 | 0 |
| 0 | 0 | 0 | 0 | 0 |
| 0 | 0 | 0 | 0 | 0 |
| 0 | 0 | 0 | 0 | 0 |
| 0.0204499 | 0.11280682 | 0.017791061 | 0.0143964 | 0.027654237 |
| 0 | 0 | 0 | 0 | 0 |
| 0 | 0 | 0 | 0 | 0 |
| 0 | 0 | 0 | 0 | 0 |
| 0 | 0 | 0 | 0 | 0 |
| 0 | 0 | 0 | 0 | 0 |
| 0.00151576 | 0.000750812 | 0.022316204 | 0.016528701 | 0 |
| 0 | 0 | 0 | 0 | 0 |
| 0 | 0 | 0 | 0 | 0 |
| 0 | 0.142180938 | 0 | 0 | 0 |
| 0.583660306 | 0 | 0 | 0 | 0 |
| 0 | 0 | 0 | 0 | 0 |
| 0 | 0 | 0 | 0 | 0 |
| 0 | 0 | 0 | 0 | 0.091716409 |
| 0.087576322 | 0.067945697 | 0.000890447 | 0.005620239 | 0.043666716 |
| 0 | 0 | 0 | 0 | 0 |
| 0 | 0 | 0 | 0 | 0 |
| 0 | 0 | 0 | 0 | 0 |
| 0 | 0.001829375 | 0 | 0.002334648 | 0 |
| 0 | 0 | 0 | 0 | 0 |
| 0 | 0 | 0 | 0.001883873 | 0 |
| 0 | 0 | 0 | 0.147005239 | 0 |
| 0 | 0 | 0 | 0.043394803 | 0 |
| 0 | 0 | 0 | 0 | 0 |
| 0 | 0 | 0 | 0 | 0 |
| 0 | 0 | 0 | 0 | 0 |
| 0.001571633 | 0.001135295 | 0 | 0.002028408 | 0 |
| 0.007930458 | 0.001344998 | 0 | 0 | 0.007349497 |

| | | | | |
|---|---|---|---|---|
| 0 | 0 | 0 | 0 | 0 |
| 0 | 0 | 0 | 0 | 0 |
| 0 | 0 | 0 | 0.01882187 | 0 |
| 0 | 0 | 0 | 0 | 0 |
| 0 | 0 | 0 | 0 | 0 |
| 0 | 0 | 0 | 0 | 0 |
| 0 | 0 | 0 | 0.005972746 | 0 |
| 0.00223047 | 0.001288973 | 0 | 0.001919151 | 0 |
| 0 | 0 | 0 | 0 | 0 |
| 0 | 0 | 0 | 0 | 0 |
| 0 | 0.000739601 | 0 | 0 | 0 |
| 0 | 0 | 0 | 0 | 0 |
| 0 | 0 | 0 | 0 | 0 |
| 0 | 0 | 0 | 0 | 0 |
| 0 | 0 | 0.014807186 | 0 | 0.000739642 |
| 0 | 0 | 0.001511558 | 0 | 0 |
| 0 | 0 | 0 | 0 | 0 |
| 0 | 0 | 0 | 0 | 0 |
| 0 | 0 | 0 | 0.012607508 | 0.004929917 |
| 0 | 0 | 0 | 0 | 0.660257615 |
| 0 | 0.004932216 | 0.001903884 | 0 | 0.004926281 |
| 0 | 0 | 0 | 0 | 0 |
| 0 | 0 | 0 | 0 | 0 |
| 0 | 0 | 0 | 0 | 0 |
| 0.008221202 | 0.001781616 | 0.021181824 | 0.009663214 | 0.002330261 |
| 0 | 0 | 0 | 0 | 0 |
| 0 | 0 | 0 | 0 | 0 |
| 0 | 0 | 0 | 0 | 0 |
| 0.008335963 | 0.022740471 | 0 | 0.065908358 | 0 |
| 0 | 0 | 0 | 0 | 0 |
| 0.267951299 | 0.133526883 | 0.002799851 | 0.162202151 | 0.070375965 |
| 0 | 0 | 0 | 0 | 0 |
| 0 | 0 | 0 | 0 | 0 |
| 0 | 0 | 0 | 0 | 0.004016945 |
| 0 | 0 | 0 | 0 | 0 |
| 0 | 0 | 0 | 0 | 0 |
| 0 | 0 | 0 | 0 | 0 |
| 0 | 0 | 0 | 0 | 0 |
| 0 | 0 | 0 | 0 | 0 |
| 0 | 0 | 0 | 0 | 0 |
| 0 | 0 | 0 | 0 | 0 |
| 0.058929784 | 0.016131372 | 0.009703539 | 0 | 0 |
| 0 | 0 | 0 | 0 | 0.002507902 |
| 0 | 0 | 0 | 0 | 0 |
| 0 | 0 | 0 | 0 | 0 |
| 0 | 0 | 0 | 0 | 0 |
| 0 | 0 | 0 | 0 | 0 |
| 0 | 0 | 0 | 0 | 0 |

| | | | | |
|---|---|---|---|---|
| 0 | 0 | 0 | 0 | 0 |
| 0 | 0 | 0 | 0 | 0 |
| 0.037484836 | 0.012870741 | 0.104104866 | 0.011578724 | 0.01501285 |
| 0 | 0 | 0 | 0 | 0 |
| 0 | 0 | 0 | 0 | 0 |
| 0 | 0 | 0 | 0 | 0 |
| 0 | 0 | 0 | 0 | 0 |
| 0 | 0 | 0 | 0 | 0 |
| 0 | 0 | 0 | 0 | 0 |
| 0 | 0 | 0 | 0.002709022 | 0.00181729 |
| 0 | 0 | 0 | 0 | 0 |
| 0 | 0 | 0 | 0 | 0 |
| 0 | 0 | 0 | 0 | 0 |
| 0 | 0.003731015 | 0.001386201 | 0.001487975 | 0.002927984 |
| 0.002664389 | 0.001924665 | 0.008662679 | 0 | 0.000805556 |
| 0 | 0 | 0 | 0 | 0 |
| 0.00632465 | 0.00263158 | 0.00763555 | 0.016232366 | 0.001147323 |
| 0 | 0 | 0 | 0 | 0 |
| 0 | 0 | 0 | 0 | 0 |
| 0 | 0 | 0 | 0 | 0 |
| 0 | 0 | 0 | 0 | 0 |
| 0.004110735 | 0.001187783 | 0 | 0 | 0.007664221 |
| 0 | 0 | 0 | 0 | 0 |
| 0 | 0 | 0 | 0 | 0 |
| 0 | 0 | 0 | 0 | 0 |
| 0 | 0 | 0 | 0 | 0 |
| 0 | 0 | 0 | 0 | 4.090632806 |
| 0 | 0 | 0 | 0 | 0 |
| 0 | 0 | 0 | 0 | 0 |
| 0 | 0.003722634 | 0 | 0.013887022 | 0.001498158 |
| 0 | 0 | 0 | 0.008518534 | 0 |
| 0 | 0 | 0 | 0 | 0 |
| 0 | 0 | 0 | 0.059605653 | 0 |
| 0 | 0.000913007 | 0 | 0 | 0 |
| 0 | 0 | 0 | 0 | 0 |
| 0.003193014 | 0 | 0.001678931 | 0.05601227 | 0 |
| 0 | 0 | 0 | 0 | 0 |
| 0.070584408 | 0 | 0.001267316 | 0 | 0.055433538 |
| 0 | 0 | 0 | 0 | 0 |
| 0 | 0 | 0 | 0 | 0 |
| 0 | 0 | 0 | 0 | 0 |
| 0 | 0 | 0 | 0 | 0 |
| 0 | 0 | 0 | 0 | 0 |
| 0 | 0 | 0 | 0 | 0 |
| 0 | 0 | 0 | 0 | 0 |
| 0 | 0 | 0 | 0.002440123 | 0 |
| 0 | 0 | 0 | 0.005161037 | 0 |
| 0 | 0 | 0.002558396 | 0 | 0 |

| | | | | |
|---|---|---|---|---|
| 0 | 0 | 0 | 0 | 0.056941263 |
| 0 | 0 | 0 | 0.002198623 | 0 |
| 0 | 0 | 0 | 0.00132311 | 0 |
| 0 | 0 | 0 | 0 | 0 |
| 0.008423185 | 0.001668926 | 0.015767335 | 0.003727294 | 0 |
| 0 | 0 | 0 | 0.010418899 | 0 |
| 0 | 0 | 0 | 0 | 0 |
| 0.002698731 | 0.103809418 | 0 | 1.455803949 | 0.002753794 |
| 0 | 0 | 0 | 0 | 0 |
| 0.00417246 | 0.022605341 | 0.017917154 | 0.017694006 | 0.016084238 |
| 0 | 0 | 0 | 0 | 0 |
| 0 | 0.056712949 | 0.003735003 | 0.235446919 | 0.012837266 |
| 0.011951028 | 0.048637565 | 0.007902948 | 0.002689006 | 0.009921249 |
| 0 | 0.004266739 | 0 | 0 | 0 |
| 0 | 0.000541761 | 0 | 0 | 0 |
| 0 | 0.012915667 | 0 | 0 | 0 |
| 0 | 0 | 0.000488337 | 0 | 0 |
| 0 | 0 | 0 | 0 | 0 |
| 0.215287141 | 0.302334855 | 0.150051466 | 0 | 0 |
| 0 | 0 | 0 | 0 | 0.034491859 |
| 0 | 0 | 0 | 0 | 0 |
| 0 | 0 | 0 | 0 | 0 |
| 0.002508186 | 0.00181183 | 0 | 0.005318189 | 0 |
| 0 | 0 | 0 | 0 | 0.490532925 |
| 0 | 0 | 0 | 0 | 0 |
| 0 | 0.007362502 | 0.001736777 | 0.010179023 | 0 |
| 0 | 0 | 0.011612116 | 0 | 0 |
| 0.169756817 | 0.007914055 | 0.036544288 | 0.013256135 | 0 |
| 0 | 0 | 0 | 0 | 0.024539191 |
| 0 | 0 | 0 | 0 | 0 |
| 0 | 0 | 0 | 0 | 0 |
| 0 | 0 | 0 | 0 | 0 |
| 0 | 0 | 0 | 0 | 0 |
| 0 | 0.133198398 | 0 | 0 | 0 |
| 0 | 0 | 0 | 0 | 0 |
| 0 | 0 | 0 | 0 | 0 |
| 0.186681084 | 0.019706189 | 0.0053158 | 0 | 0 |
| 0 | 0 | 0 | 0 | 0 |
| 0 | 0 | 0 | 0 | 0.000761974 |
| 0 | 0.047751384 | 0 | 0 | 0 |
| 0 | 0 | 0.001954105 | 0 | 0 |
| 0 | 0 | 0 | 0 | 0 |
| 0 | 0 | 0 | 0 | 0 |
| 0 | 0 | 0 | 0 | 0 |
| 0 | 0 | 0 | 0 | 0 |
| 0 | 0 | 0 | 0 | 0.018631492 |
| 0.00997554 | 0.002543293 | 0 | 0 | 0 |
| 0 | 0 | 0 | 0 | 0 |

| | | | | |
|---|---|---|---|---|
| 0 | 0 | 0 | 0 | 0 |
| 0 | 0.417819175 | 0.015447997 | 0.006891967 | 0 |
| 0 | 0 | 0 | 0 | 0 |
| 0 | 0 | 0 | 0 | 0 |
| 0 | 0.002513778 | 0 | 0 | 0.004997596 |
| 0 | 0.026040962 | 0.000680448 | 0 | 0.017606518 |
| 0 | 0 | 0 | 0 | 0.007691153 |
| 0 | 0 | 0 | 0 | 0 |
| 0 | 0 | 0 | 0 | 0 |
| 0 | 0 | 0 | 0 | 0 |
| 0.010988886 | 0 | 0.094704593 | 0 | 0.001458614 |
| 0 | 0 | 0 | 0 | 0 |
| 0 | 0.000674132 | 0 | 0 | 0 |
| 0.001266067 | 0 | 0.002796004 | 0.007563237 | 0 |
| 0.001281032 | 0 | 0.006412522 | 0.022674478 | 0 |
| 0 | 0 | 0 | 0 | 0 |
| 0 | 0 | 0 | 0 | 0 |
| 0.010103117 | 0 | 0.001451181 | 0 | 0 |
| 0 | 0.013082669 | 0 | 0 | 0 |
| 0 | 0 | 0 | 0 | 0.302705464 |
| 0 | 0 | 0 | 0 | 0 |
| 0 | 0 | 0 | 0 | 0 |
| 0 | 0.044157613 | 0 | 0 | 0.164331789 |
| 0 | 0 | 0 | 0 | 0 |
| 0 | 0 | 0 | 0 | 0 |
| 0 | 0 | 0 | 0 | 0 |
| 0.008511059 | 0.629486492 | 0.004850574 | 0 | 0 |
| 0 | 0 | 0 | 0 | 0 |
| 0 | 0 | 0 | 0 | 0 |
| 1.26156509 | 0.536009284 | 1.351091596 | 4.512461906 | 0.324866656 |
| 0 | 0 | 0.004046649 | 0 | 0 |
| 0 | 0 | 0 | 0.028548555 | 0 |
| 0 | 0 | 0 | 0 | 0 |
| 0 | 0 | 0 | 0 | 0 |
| 0 | 0.133008423 | 0 | 0 | 0 |
| 0 | 0 | 0 | 0 | 0 |
| 0 | 0 | 0 | 0 | 0 |
| 0 | 0 | 0.00246989 | 0.074234365 | 0 |
| 0 | 0 | 0 | 0 | 0 |
| 0 | 0 | 0 | 0 | 0 |
| 0.028310973 | 0.009383353 | 0 | 0 | 0.013783454 |
| 8.682077533 | 0 | 2.387580166 | 0 | 0.808531799 |
| 0 | 0 | 0.002262579 | 0.057802959 | 0 |
| 0 | 0.003918497 | 0 | 0 | 0 |
| 0 | 0.005088899 | 0 | 0 | 0 |
| 0 | 0 | 0 | 0 | 0 |
| 0 | 0.014583359 | 0.001125864 | 0.021511732 | 0.000594523 |
| 0 | 0 | 0 | 0 | 0 |

| | | | | |
|---|---|---|---|---|
| 0.001617467 | 0.006543061 | 0 | 0.012525376 | 0.001222569 |
| 0 | 0.008212331 | 0 | 0 | 0.065307118 |
| 0 | 0.448318356 | 0.006489576 | 0.073549713 | 0.009661896 |
| 0 | 0 | 0.000538525 | 0 | 0 |
| 0 | 0 | 0 | 0 | 0 |
| 0 | 0 | 0.005201395 | 0 | 0.004134733 |
| 0 | 0 | 0 | 0 | 0 |
| 0 | 0 | 0 | 0 | 0 |
| 0 | 0 | 0 | 0 | 0.020463615 |
| 0 | 0 | 0 | 0 | 0 |
| 0 | 0 | 0 | 0 | 0 |
| 0 | 0 | 0 | 0 | 0.001443252 |
| 0 | 0 | 0 | 0 | 0 |
| 0.001219401 | 0 | 0.230336753 | 0.004586515 | 0 |
| 0 | 0 | 0 | 0 | 0 |
| 0 | 0.059144971 | 0 | 0.006325205 | 0.015385277 |
| 0.054501761 | 0.003845466 | 0.002021595 | 0.04743995 | 0.01571173 |
| 0 | 0 | 0 | 0 | 0 |
| 0 | 0 | 0 | 0 | 0.002572895 |
| 0 | 0 | 0 | 1.266591843 | 0 |
| 0 | 0.219010261 | 0.155218905 | 0.033280913 | 0 |
| 0.021808177 | 0 | 0.000721522 | 0 | 0 |
| 0 | 0.126727578 | 0 | 0 | 0 |
| 0 | 0.015362535 | 0.332849785 | 0.757655497 | 0 |
| 0 | 0 | 0 | 0 | 0 |
| 0 | 0.016162069 | 0 | 0 | 0 |
| 0.0013409 | 0 | 0 | 0 | 0 |
| 0 | 0 | 0 | 0 | 0 |
| 0 | 0 | 0 | 0 | 0 |
| 0 | 0 | 0 | 0 | 0 |
| 0 | 0.018563164 | 0 | 0 | 0.038662462 |
| 0.013682905 | 0.00500793 | 0.014101533 | 0.005718368 | 0.004964295 |
| 0.029530077 | 0.322984569 | 0.687312319 | 0 | 0.049892738 |
| 0.004148969 | 0.013307017 | 0.009162657 | 0.054160108 | 0 |
| 0 | 0.002310282 | 0 | 0 | 0.014504297 |
| 0 | 0 | 0 | 0 | 0 |
| 0.001570425 | 0.01520126 | 0.002890129 | 0.158962856 | 0 |
| 0.014533051 | 0.037083742 | 0 | 0 | 0.01503844 |
| 0 | 0 | 0.502034614 | 0 | 0.051582471 |
| 0 | 0.000580282 | 0 | 0.00351764 | 0 |
| 0 | 0 | 0 | 0 | 0.004065191 |
| 0 | 0 | 0 | 0 | 0 |
| 0 | 0.005572403 | 0 | 0.002709142 | 0 |
| 0.007669749 | 0.225354537 | 0 | 0 | 0.474212019 |
| 0 | 0.135253354 | 0 | 0 | 0 |
| 0 | 0 | 0 | 0 | 0 |
| 0 | 0 | 0 | 0 | 0 |
| 0 | 0 | 0 | 0 | 0 |

| | | | | |
|---|---|---|---|---|
| 0.003338375 | 0 | 0 | 0 | 0 |
| 0.538031404 | 0.012814158 | 0.375123217 | 0.056774142 | 0.594514686 |
| 0 | 0 | 0 | 0 | 0 |
| 0 | 0 | 0 | 0 | 0.017160338 |
| 0 | 0 | 0 | 0 | 0 |
| 0 | 0 | 0 | 0 | 0 |
| 0.012127636 | 0 | 0 | 0 | 0 |
| 0 | 0.002414229 | 0 | 0 | 0 |
| 0.004340539 | 0 | 0 | 0.001977198 | 0.000926348 |
| 0 | 0.013352767 | 0 | 0 | 0 |
| 0.008497907 | 0.00754599 | 0 | 0.002980772 | 0 |
| 0.00895313 | 0.871364992 | 0.138405767 | 1.53041494 | 0.005040443 |
| 0 | 0 | 0 | 0 | 0 |
| 0 | 0 | 0 | 0 | 0 |
| 0 | 0.017110015 | 0 | 0.185998848 | 0 |
| 0 | 0 | 0 | 0 | 0 |
| 0.032555616 | 0.000940683 | 0.062150448 | 0.001920799 | 0.006693186 |
| 0.059448107 | 0.675532999 | 0.006844308 | 0 | 0 |
| 0 | 0 | 0 | 0 | 0 |
| 0.083242345 | 0.156685451 | 0.000942738 | 0 | 0 |
| 0.024396419 | 0.000970082 | 0.81705952 | 0.008913729 | 0.022128166 |
| 0 | 0 | 0 | 0 | 0 |
| 0 | 0 | 0 | 0 | 0 |
| 0 | 0 | 0 | 0 | 0 |
| 0.0037606 | 0.078562075 | 0.000830498 | 0 | 0 |
| 0 | 0.005337554 | 0 | 0 | 0 |
| 0 | 0.175992052 | 0.086844033 | 0 | 0.043482829 |
| 0.227658816 | 0.330143448 | 0.016110921 | 0 | 0.191819883 |
| 0 | 0 | 0 | 0 | 0 |
| 0 | 0 | 0 | 0.00126976 | 0 |
| 0 | 0.01836494 | 0.000939513 | 0.002541397 | 0 |
| 0 | 0 | 0 | 0 | 0 |
| 0 | 0 | 0 | 0 | 0 |
| 0 | 0 | 0 | 0 | 0 |
| 0 | 0 | 0.012208301 | 0 | 0 |
| 0.027377565 | 0.090390853 | 0.074983623 | 3.490098537 | 0.278996393 |
| 0 | 0 | 0 | 0 | 0 |
| 0 | 0 | 0 | 0 | 0 |
| 0.05839883 | 0.012464819 | 0.005400713 | 0.12693611 | 0.008517661 |
| 0 | 0 | 0 | 0 | 0 |
| 0 | 0.030009935 | 0 | 0 | 0 |
| 0 | 0 | 0 | 0 | 0 |
| 0.013368973 | 0 | 0.000787314 | 0.009070933 | 0.001616799 |
| 0 | 0.014172387 | 0.010895296 | 0.002665422 | 0.007024452 |
| 0.051098454 | 0 | 0 | 0 | 0 |
| 0 | 0 | 0 | 0 | 0.351434284 |
| 0 | 0 | 0 | 0 | 0 |
| 0 | 0.000723428 | 0 | 0 | 0 |

| | | | | |
|---|---|---|---|---|
| 0.008217767 | 0.014031112 | 0 | 0 | 0.005646759 |
| 0 | 0 | 0 | 0 | 0 |
| 0 | 0 | 0 | 0 | 0 |
| 0 | 0.020875723 | 0 | 0.014928475 | 1.539777776 |
| 0 | 0 | 0 | 0 | 0 |
| 0 | 0 | 0 | 0 | 0.002122769 |
| 0 | 0 | 0 | 0 | 0 |
| 0 | 0 | 0 | 0 | 0.014487549 |
| 0 | 0.08026287 | 0.153651037 | 0 | 0.780662386 |
| 0 | 0 | 0 | 0 | 0.30512647 |
| 0 | 0 | 0 | 0 | 0 |
| 0 | 0.004655449 | 0.000439279 | 0 | 0.004149599 |
| 0 | 0 | 0 | 0 | 0 |
| 0 | 0 | 0 | 0 | 2.670756065 |
| 0.245720459 | 0.040266582 | 0.000362494 | 0 | 0 |
| 0 | 0 | 0 | 0 | 0 |
| 0 | 0 | 0 | 0 | 0.063309861 |
| 0 | 0.015049999 | 0 | 0 | 0 |
| 0 | 0 | 0 | 0 | 0 |
| 0 | 0 | 0 | 0 | 0.021589757 |
| 0 | 0 | 0 | 0 | 0 |
| 0 | 0 | 0 | 0 | 0 |
| 0 | 0 | 0 | 0 | 0.003322677 |
| 0.216295495 | 0.469007944 | 0.005470402 | 0 | 0.073999776 |
| 0.052703542 | 0.199369694 | 0 | 0 | 0 |
| 0 | 0 | 0 | 0 | 0.02489435 |
| 0 | 0.019400109 | 0 | 0 | 0 |
| 0 | 0 | 0.40354972 | 0 | 2.99721976 |
| 0 | 0 | 0 | 0 | 0 |
| 0 | 0 | 0 | 0 | 0 |
| 0 | 0 | 0 | 0 | 0.014382507 |
| 0 | 0 | 0 | 0 | 2.714494464 |
| 0 | 0.160362218 | 0 | 0 | 0.008142863 |
| 0 | 0 | 0 | 0 | 0 |
| 0 | 0.046429976 | 0 | 0 | 0 |
| 0 | 0 | 0 | 0 | 0 |
| 0 | 0.013766904 | 0 | 0 | 0.270432072 |
| 0 | 0 | 0 | 0 | 0 |
| 0 | 0 | 0 | 0 | 0.004507547 |
| 0 | 0.013328984 | 0 | 0 | 0 |
| 0.001335737 | 0.037548077 | 0.010254297 | 0.34725322 | 0.216347739 |
| 0 | 0 | 0 | 0 | 0 |
| 0.127961559 | 0.381580187 | 4.351791147 | 0.125351713 | 0.446401353 |
| 0 | 0 | 0 | 0.004413417 | 0.002261606 |
| 0 | 0 | 0 | 0 | 0 |
| 0 | 0 | 0 | 0 | 0 |
| 0 | 0 | 0 | 0 | 0 |
| 0.01330445 | 0.005368729 | 0 | 0 | 0.427771033 |

| | | | | |
|---|---|---|---|---|
| 0 | 0 | 0 | 0 | 0 |
| 0.173033883 | 0.038403914 | 0 | 0 | 0 |
| 0.786338813 | 0.02763879 | 0.091943395 | 3.771006603 | 0.001196693 |
| 0 | 0 | 0 | 0 | 0.033417172 |
| 0 | 0 | 0 | 0 | 0 |
| 0 | 0.002102199 | 0 | 0.008431735 | 0.016497402 |
| 0 | 0 | 0 | 0 | 0 |
| 0 | 0 | 0 | 0 | 0.007961215 |
| 0 | 0 | 0 | 0 | 0 |
| 0 | 0 | 0 | 0 | 0 |
| 0.688709286 | 0.319086519 | 0 | 0.037497496 | 0.162925067 |
| 6.318296786 | 6.541166935 | 0.348607768 | 0.274159792 | 1.162937356 |
| 0 | 0 | 0 | 0 | 0 |
| 0.02376642 | 0 | 0.002058282 | 0 | 0.005917537 |
| 0.214830931 | 0.106163591 | 0.004730848 | 0.987198703 | 0.172706639 |
| 0 | 0 | 0 | 0 | 0.759302012 |
| 0 | 0 | 0 | 0 | 0 |
| 0 | 0 | 0 | 0 | 0 |
| 0 | 0 | 0 | 0 | 0 |
| 0 | 0.079855035 | 0 | 0 | 0.140982266 |
| 2.554331357 | 0.03560658 | 0.172369277 | 1.071569112 | 0.190225331 |
| 0.224416512 | 0.00826394 | 0.052774751 | 0 | 0 |
| 1.538913385 | 0.428169128 | 0.3381956 | 0.440924869 | 0.670632402 |
| 0 | 0 | 0 | 0 | 0 |
| 0 | 0 | 0 | 0 | 0 |
| 0 | 0 | 23.00782924 | 0 | 0 |
| 0 | 0 | 0 | 0 | 0.013152485 |
| 0 | 0 | 0 | 0 | 0.002054201 |
| 0 | 0 | 0 | 0 | 0 |
| 0 | 0 | 0 | 0 | 0 |
| 0 | 0 | 0 | 0 | 0.106264531 |
| 0 | 0.137274982 | 0 | 0 | 0.085377566 |
| 3.485835452 | 0.075152302 | 0.026279227 | 0.244556512 | 0.003016736 |
| 0.035508306 | 0.029021153 | 0 | 0 | 0 |
| 0.027073883 | 0.043512272 | 0 | 0.002914128 | 0.001460128 |
| 0.42204491 | 0.013246424 | 0 | 0 | 0.03428652 |
| 0.010099981 | 0.0020204 | 0 | 0 | 0 |
| 0 | 0 | 0 | 0 | 0 |
| 0 | 0.043443566 | 0.002078492 | 0 | 3.289803054 |
| 0 | 0 | 0 | 0 | 0 |
| 0 | 0 | 0 | 0 | 0 |
| 0 | 0 | 0 | 0 | 0 |
| 0 | 0 | 0 | 0 | 0.056969313 |
| 0 | 0 | 0 | 0 | 0 |
| 0.059387912 | 0.12144463 | 0.002025534 | 0.025366214 | 0.047626986 |
| 0 | 0 | 0 | 0 | 0 |
| 0 | 0 | 0 | 0 | 0 |
| 0.63865737 | 0.009111008 | 0.024433475 | 0.007955632 | 0.058705553 |

| | | | | |
|---|---|---|---|---|
| 0 | 0 | 0 | 0 | 0 |
| 0 | 0 | 0 | 0 | 0 |
| 0 | 0 | 0 | 0 | 0 |
| 0 | 0.000847028 | 0 | 0 | 0.001063553 |
| 0 | 0 | 0 | 0 | 0 |
| 0 | 0 | 0 | 0 | 0 |
| 0 | 0 | 0 | 0 | 0.01804802 |
| 0 | 0.289164325 | 0 | 0 | 0 |
| 0 | 0 | 0 | 0 | 0 |
| 0 | 0 | 0 | 0 | 0 |
| 0.058549466 | 0.002264075 | 0 | 0.005297253 | 0.00730452 |
| 0 | 0 | 0 | 0 | 0 |
| 0 | 0.028248834 | 0 | 0 | 0 |
| 0 | 0 | 0 | 0 | 0 |
| 0 | 0 | 0 | 0 | 0 |
| 0 | 0 | 0 | 0 | 0 |
| 0 | 0 | 0 | 0 | 0.2585643 |
| 0 | 0 | 0 | 0 | 0 |
| 0 | 0 | 0 | 0 | 0.059317339 |
| 0 | 0 | 0 | 0 | 0 |
| 0 | 0 | 0 | 0 | 0.001314086 |
| 0 | 0 | 0 | 0 | 0.180625262 |
| 0 | 0 | 0 | 0 | 0.058360015 |
| 0 | 0 | 0 | 0 | 0 |
| 0 | 0 | 0 | 0 | 0 |
| 0 | 0 | 0 | 0 | 0 |
| 0 | 0 | 0 | 0 | 0 |
| 0 | 0 | 0 | 0 | 0 |
| 0 | 0 | 0 | 0 | 0 |
| 0 | 0.057489452 | 0 | 0 | 0.073456084 |
| 0.003257139 | 0 | 0 | 0 | 0 |
| 0 | 0 | 0 | 0 | 0 |
| 0 | 0 | 0 | 0 | 0 |
| 0 | 0 | 0 | 0 | 0 |
| 0 | 0 | 0 | 0 | 0 |
| 0 | 0 | 0 | 0 | 0 |
| 0 | 0 | 0 | 0 | 0.351034703 |
| 0.018609212 | 0.025554383 | 0.003119568 | 0.052179982 | 0 |
| 0 | 0 | 0 | 0 | 0 |
| 0 | 0 | 0 | 0 | 0 |
| 0 | 0 | 0 | 0 | 0 |
| 0 | 0 | 0 | 0 | 1.714941651 |
| 0 | 0 | 0 | 0 | 0 |
| 0 | 0 | 0 | 0 | 0.004544187 |
| 0 | 0.029560801 | 0 | 0 | 0 |
| 0 | 0 | 0 | 0 | 0 |
| 0 | 0 | 0 | 0 | 0 |
| 0 | 0 | 0 | 0 | 0 |

| | | | | |
|---|---|---|---|---|
| 0 | 0 | 0 | 0 | 0 |
| 0 | 0 | 0 | 0 | 0 |
| 0 | 0 | 0 | 0 | 0 |
| 0 | 0.008261214 | 0 | 0 | 0.196821601 |
| 0 | 0 | 0 | 0 | 2.560370839 |
| 0 | 0 | 0 | 0 | 0 |
| 0 | 0 | 0 | 0 | 0 |
| 0 | 0 | 0 | 0 | 0 |
| 0.358296508 | 0.270716148 | 0.017957865 | 0.087988347 | 0.03411179 |
| 0 | 0.0060311 | 0 | 0 | 0 |
| 0 | 0 | 0 | 0 | 0 |
| 0 | 0 | 0 | 0 | 0 |
| 0 | 0 | 0 | 0 | 0 |
| 0 | 0.001070964 | 0 | 0 | 0.004642538 |
| 0 | 1.704038789 | 0 | 1.818944592 | 5.264633806 |
| 2.399755116 | 0 | 0 | 0 | 0.765989068 |
| 0 | 0.001209453 | 0 | 0 | 0 |
| 0 | 0 | 0 | 0 | 0 |
| 0 | 0 | 0 | 0 | 0 |
| 0 | 0 | 0 | 0 | 0 |
| 0 | 0 | 0 | 0 | 0.00793075 |
| 0 | 0 | 0 | 0 | 0 |
| 0 | 0 | 0 | 0 | 0 |
| 0 | 0 | 0 | 0 | 0 |
| 0.001792964 | 0.005439743 | 0 | 0.100496563 | 0.074537065 |
| 6.340103192 | 2.38902381 | 0.006634845 | 0 | 1.668986472 |
| 0 | 0 | 0 | 0 | 0 |
| 0 | 0.000964772 | 0 | 0 | 0.000504749 |
| 0 | 0.030347113 | 0 | 0 | 0 |
| 0 | 0 | 0 | 0 | 0.045989 |
| 0 | 0 | 0 | 0 | 0 |
| 0 | 0 | 0 | 0 | 0.084484016 |
| 0 | 0 | 0 | 0 | 0 |
| 0.002074709 | 0.000968391 | 0 | 0.007415158 | 0.067890015 |
| 0.007625805 | 0.006610351 | 0.010840161 | 0.011636036 | 0.006439776 |
| 0.00706344 | 0.04559927 | 0.003557673 | 0.006991477 | 0.002866163 |
| 0 | 0 | 0 | 0 | 0 |
| 0 | 0 | 2.369082536 | 0.987287574 | 1.096375854 |
| 0 | 0 | 0 | 0 | 0 |
| 0 | 0 | 0 | 0 | 0.001932277 |
| 0 | 0 | 0 | 0 | 0.000779381 |
| 0 | 0 | 0 | 0 | 0 |
| 0.058421912 | 0.626865275 | 1.22512899 | 0 | 0.058519549 |
| 0 | 0 | 0 | 0 | 0 |
| 0 | 0 | 0 | 0 | 0 |
| 0 | 0 | 0 | 0 | 0.000479176 |
| 0 | 0 | 0 | 0 | 0 |
| 2.166650189 | 0 | 0.011978178 | 2.319910974 | 0.018066767 |

| | | | | |
|---|---|---|---|---|
| 0 | 0 | 0 | 0 | 0.215368773 |
| 0 | 0 | 0 | 0 | 0.593952872 |
| 0 | 0 | 0 | 0 | 0 |
| 0 | 0 | 0 | 0 | 0 |
| 0.007289998 | 0.005235958 | 0 | 0 | 0.004723008 |
| 0 | 0 | 0 | 0 | 0 |
| 0 | 0 | 0 | 0 | 0.066776452 |
| 0 | 0 | 0 | 0 | 0 |
| 0 | 0 | 0 | 0 | 0 |
| 0 | 0 | 0 | 0 | 0 |
| 0 | 0 | 0 | 0 | 0 |
| 0 | 0.124427158 | 0.017823226 | 0 | 2.029445006 |
| 0 | 1.14204336 | 0 | 0 | 0.021044943 |
| 0 | 0 | 0 | 0 | 0 |
| 0.261839109 | 0.081225686 | 0.201793527 | 2.844126034 | 0.006991563 |
| 0 | 0 | 0 | 0 | 0 |
| 0 | 0.00755326 | 0 | 0 | 0.043924767 |
| 0 | 0 | 0 | 0 | 0 |
| 0 | 0.015190499 | 0 | 0 | 0.582933608 |
| 0 | 0 | 0 | 0 | 0.057545685 |
| 0 | 0 | 0 | 0 | 0.447735342 |
| 0 | 0 | 0 | 0 | 0 |
| 0.00800661 | 0 | 0.008160895 | 0.171015857 | 0.283039443 |
| 0 | 0 | 0 | 0 | 0.075151455 |
| 0 | 0 | 0 | 0 | 0 |
| 0 | 0 | 0 | 0 | 0 |
| 0 | 0 | 0 | 0 | 0 |
| 0 | 0 | 0 | 0 | 0 |
| 0 | 0 | 0 | 0 | 0 |
| 0 | 0 | 0 | 0 | 0 |
| 0 | 0 | 0 | 0 | 0 |
| 0.054388664 | 0.031159882 | 0.265076739 | 0.020747594 | 0.028068142 |
| 0 | 0 | 0 | 0 | 0.850482964 |
| 0 | 0.032081432 | 0 | 0 | 0.174075559 |
| 0.006473088 | 0.040329998 | 0.002829275 | 0.003132903 | 0.003669534 |
| 0 | 0.180383587 | 0 | 0 | 0.084438956 |
| 0 | 0 | 0 | 0 | 0 |
| 0 | 0 | 0 | 0 | 0.210710939 |
| 0 | 0 | 0 | 0 | 0 |
| 0 | 0 | 0 | 0 | 0.002067367 |
| 0.007410909 | 0 | 0 | 0 | 0.118180038 |
| 0 | 0 | 0 | 0 | 0 |
| 0 | 0 | 0 | 0 | 0 |
| 0 | 0 | 0 | 0 | 0 |
| 0.240518913 | 0.153388833 | 0 | 0.010360893 | 0.167187199 |
| 0 | 0 | 0 | 0 | 0 |
| 0 | 0 | 0 | 0 | 0 |
| 0 | 0.003104155 | 0 | 0 | 0.002222356 |

| | | | | |
|---|---|---|---|---|
| 0 | 0 | 0 | 0 | 0 |
| 0 | 0 | 0 | 0 | 0 |
| 0 | 0.020008956 | 0 | 0 | 0 |
| 0 | 0 | 0 | 0 | 0 |
| 0.001881092 | 0 | 0 | 0 | 0.03488226 |
| 0 | 0 | 0 | 0 | 0 |
| 0 | 0 | 0 | 0 | 0 |
| 1.468345487 | 0.005736464 | 0 | 0 | 0.005825857 |
| 0 | 0 | 0 | 0 | 0 |
| 0 | 0 | 0 | 0 | 0 |
| 0 | 0 | 0 | 0 | 0.21005747 |
| 0.005452115 | 0 | 0 | 0 | 0 |
| 0 | 0 | 0 | 0 | 0 |
| 0 | 0 | 0 | 0 | 0 |
| 0 | 0 | 0 | 0 | 0 |
| 0 | 0 | 0 | 0 | 0 |
| 0 | 0 | 0 | 0 | 0 |
| 0 | 0 | 0 | 0 | 0 |
| 0 | 0 | 0.054956004 | 0 | 3.595400549 |
| 0 | 0 | 0 | 0 | 0 |
| 0 | 0 | 0 | 0 | 0 |
| 0 | 0 | 0 | 0 | 0 |
| 0 | 0 | 0 | 0 | 0 |
| 0 | 0 | 0 | 0 | 0 |
| 0 | 0 | 0 | 0 | 0 |
| 0.1760517 | 0.002572959 | 0.004643152 | 0.013339659 | 0 |
| 0 | 0 | 0 | 0 | 0 |
| 11.44772286 | 0 | 0.002272361 | 0.211521339 | 0.210483782 |
| 0 | 0 | 0 | 0 | 0 |
| 0 | 0.284635254 | 0 | 0 | 1.140585124 |
| 0 | 0 | 0 | 0 | 0 |
| 0 | 0 | 0 | 0 | 0 |
| 0 | 0 | 0 | 0 | 0.013216905 |
| 0 | 0 | 0 | 0 | 0 |
| 0.002138466 | 0.003201858 | 0 | 0.010538119 | 0 |
| 0 | 0 | 0 | 0 | 0 |
| 0 | 0 | 0 | 0 | 0 |
| 0.008161973 | 0.002294634 | 0 | 0 | 0.015206398 |
| 0 | 0 | 0 | 0 | 0 |
| 0 | 0 | 0 | 0 | 0 |
| 17.80189906 | 16.83049049 | 22.50748428 | 37.62459226 | 2.700607028 |
| 0.085743602 | 0 | 0.577582595 | 0 | 0.81037717 |
| 0 | 0.039429548 | 0.0422741 | 0 | 0 |
| 0 | 0.030091642 | 0 | 0 | 0 |
| 0 | 0 | 0 | 0 | 0 |
| 0 | 0 | 0 | 0 | 0 |
| 0 | 0 | 0 | 0 | 0.002951696 |
| 0.012255285 | 0.006374021 | 0 | 0 | 0.025884054 |

| | | | | |
|---|---|---|---|---|
| 0 | 0 | 0 | 0 | 0 |
| 0 | 0.000936601 | 0 | 0 | 0.049001018 |
| 0 | 0 | 0 | 0 | 0 |
| 0 | 0 | 0 | 0 | 0 |
| 0 | 4.956534729 | 4.76387772 | 0 | 0.536140904 |
| 0 | 0 | 0 | 0 | 0 |
| 0 | 0 | 0 | 0 | 0 |
| 0 | 0 | 0 | 0 | 0 |
| 0.909499738 | 1.760395921 | 0.22396704 | 0 | 0 |
| 0 | 0 | 0 | 0 | 0 |
| 0 | 0 | 0 | 0 | 0 |
| 0 | 0 | 0 | 0 | 0 |
| 0 | 0.266953116 | 0 | 0 | 0.128713379 |
| 0.014628003 | 0.068435429 | 0.065401268 | 0.128983085 | 0.009755857 |
| 0 | 0.031243109 | 0 | 0 | 0.536017116 |
| 0.049250201 | 0.298385166 | 0.114872498 | 0 | 0.778943525 |
| 0 | 0.016620397 | 0.007262752 | 0.16309772 | 0.060446019 |
| 0 | 0 | 0 | 0 | 0 |
| 0 | 0 | 0 | 0 | 0 |
| 0 | 0 | 0 | 0 | 0 |
| 0 | 0 | 0 | 0 | 0.006186877 |
| 0 | 0 | 0 | 0 | 0 |
| 0 | 0 | 0 | 0 | 0 |
| 0 | 0 | 0 | 0 | 0 |
| 0 | 0 | 0 | 0 | 0 |
| 0 | 0 | 0 | 0 | 0 |
| 0 | 0.004489979 | 0 | 0 | 0.016296629 |
| 0.005662076 | 0.001583262 | 0 | 0.001212335 | 0 |
| 0 | 0 | 0 | 0 | 0 |
| 0 | 0 | 0 | 0 | 0 |
| 0 | 0 | 0 | 0 | 0 |
| 0 | 0 | 0 | 0 | 0.000553528 |
| 0 | 0 | 0 | 0 | 0.496257928 |
| 0 | 0.005145012 | 0 | 0 | 0 |
| 0 | 0 | 0 | 0 | 0 |
| 0 | 0 | 0.002644103 | 0 | 0.000766565 |
| 0 | 0 | 0.01383091 | 0 | 0 |
| 0 | 0 | 0 | 0 | 0.00506698 |
| 0 | 0 | 0 | 0 | 0 |
| 0 | 0 | 0 | 0 | 0 |
| 0 | 0 | 0.306690385 | 0 | 0 |
| 0 | 0 | 0 | 0 | 0 |
| 0 | 0 | 0 | 0 | 0 |
| 0 | 0 | 0 | 0 | 0 |
| 0 | 0 | 0 | 0 | 0 |
| 0 | 0 | 0 | 0 | 0 |
| 0 | 0 | 0 | 0 | 0 |
| 0 | 0 | 0 | 0 | 0 |

| | | | | |
|---|---|---|---|---|
| 0 | 0 | 0 | 0 | 0.227824236 |
| 0 | 0 | 0 | 0 | 0 |
| 0 | 0 | 0 | 0 | 0 |
| 0 | 0 | 0 | 0 | 0 |
| 0 | 0 | 0 | 0 | 0.050815666 |
| 0 | 0 | 0 | 0 | 0 |
| 0 | 0.019638427 | 0.001640398 | 0.125518174 | 0.525848053 |
| 0 | 0 | 0 | 0 | 0 |
| 0 | 0 | 0 | 0 | 0 |
| 0 | 0 | 0 | 0 | 0 |
| 0 | 0 | 0 | 0 | 0.007148844 |
| 0 | 0 | 0 | 0 | 0.000589192 |
| 0 | 0 | 0 | 0 | 0 |
| 0 | 0 | 0 | 0 | 0 |
| 0 | 0 | 0 | 0 | 0 |
| 0 | 0 | 0 | 0 | 0 |
| 0 | 0 | 0 | 0 | 0 |
| 0 | 0 | 0 | 0 | 0 |
| 0 | 0 | 0 | 0 | 0 |
| 0 | 0 | 0 | 0 | 0 |
| 1.137853769 | 0 | 0.028241655 | 0.061803754 | 0.024053651 |
| 0 | 0 | 0 | 0 | 0 |
| 0 | 0.024341417 | 0 | 0 | 0.098695666 |
| 0.0029478 | 0.00195403 | 0 | 0.001150953 | 0.001415503 |
| 0 | 0 | 0 | 0 | 0 |
| 0 | 0.005142628 | 0 | 0 | 0.196129367 |
| 0 | 0 | 0 | 0 | 0 |
| 0 | 0 | 0 | 0 | 0 |
| 0 | 0.011389374 | 0 | 0 | 0 |
| 0.001497379 | 2.085650339 | 2.003209828 | 2.054877694 | 0.002716322 |
| 0 | 0 | 0 | 0 | 0 |
| 0 | 0 | 0 | 0 | 0 |
| 0 | 0 | 0 | 0 | 0 |
| 0 | 0 | 0 | 9.208781829 | 0 |
| 0 | 0 | 0 | 0 | 0 |
| 0 | 0 | 0 | 0 | 10.01995801 |
| 0.089824872 | 0.080308767 | 0.136559645 | 0 | 0.004329505 |
| 0 | 0 | 0 | 0 | 0 |
| 0 | 0 | 0 | 0 | 0 |
| 0 | 0 | 0 | 0 | 0 |
| 0 | 0 | 0 | 0 | 0 |
| 0 | 0 | 0 | 0 | 0 |
| 0 | 0 | 0 | 0 | 0 |
| 0 | 0.011845065 | 0 | 0 | 0 |
| 0 | 0 | 0 | 0 | 0 |
| 0 | 0 | 0 | 0 | 0 |
| 0 | 0 | 0 | 0 | 0 |
| 0 | 0 | 0 | 0 | 0 |

| | | | | |
|---|---|---|---|---|
| 0 | 0.00436695 | 0 | 0.029723196 | 0.006261764 |
| 0 | 0.005982306 | 0 | 0 | 0 |
| 0 | 0 | 0 | 0 | 0 |
| 0 | 0 | 0 | 0 | 0 |
| 0 | 0 | 0 | 0 | 0.001005351 |
| 0 | 0 | 0 | 0 | 0 |
| 0 | 0 | 0 | 0 | 0.001040811 |
| 0 | 0 | 1.434553877 | 0.004874563 | 0.636338471 |
| 0.012007771 | 0.101041648 | 0.115127689 | 3.395772692 | 1.826559094 |
| 0.516654775 | 0.093848465 | 0.002251361 | 0 | 0.01276034 |
| 0 | 0 | 0 | 0 | 0 |
| 0 | 0 | 0 | 0 | 0 |
| 0 | 0 | 0 | 0 | 0 |
| 0 | 0 | 0 | 0 | 0 |
| 0 | 0 | 0 | 0 | 0 |
| 0 | 0 | 0 | 0 | 0 |
| 0.156608851 | 0 | 0 | 0 | 0 |
| 0 | 0 | 0 | 0 | 0 |
| 0 | 0 | 0 | 0 | 0 |
| 0 | 0 | 0 | 0 | 0 |
| 0 | 0 | 0 | 0 | 0 |
| 0 | 0 | 0 | 0 | 0 |
| 0 | 0 | 0 | 0 | 0 |
| 0 | 0 | 0 | 0 | 0 |
| 0 | 0 | 0.024774115 | 0 | 0.059184088 |
| 0 | 0 | 0 | 0 | 0 |
| 0 | 0 | 0 | 0 | 0 |
| 0 | 0 | 0 | 0 | 0 |
| 0 | 0 | 0 | 0 | 0 |
| 0 | 0 | 0 | 0 | 0 |
| 0 | 0 | 0 | 0 | 0 |
| 0 | 0 | 0 | 0 | 0 |
| 0 | 0 | 0 | 0 | 0 |
| 0 | 0 | 0 | 0 | 0 |
| 0 | 0 | 0.000665137 | 0 | 0 |
| 0 | 0 | 0 | 0 | 0 |
| 0 | 0 | 0 | 0 | 0 |
| 0 | 0 | 0 | 0 | 0 |
| 0 | 0 | 0 | 0 | 0 |
| 0 | 0 | 0.000822723 | 0 | 0 |
| 0 | 0 | 0 | 0 | 0 |
| 0 | 0 | 0.114585963 | 0 | 0 |
| 0 | 0 | 0 | 0 | 0 |
| 0 | 0 | 0 | 0 | 0 |
| 0 | 0 | 0 | 0 | 0 |
| 0 | 0.05508258 | 0 | 0 | 0 |
| 0 | 0 | 0 | 0 | 0 |
| 0 | 0 | 0 | 0 | 0 |

| | | | | |
|---|---|---|---|---|
| 0 | 0 | 0 | 0 | 0 |
| 0 | 0 | 0 | 0 | 0.016801804 |
| 0 | 0 | 0 | 0 | 0 |
| 0 | 0 | 0 | 0 | 0 |
| 0 | 0.325940622 | 0.019932079 | 0 | 0.128988509 |
| 0 | 0 | 0 | 0 | 0 |
| 0 | 0 | 0 | 0.018071402 | 0 |
| 0.19154068 | 0.098756922 | 0.002446107 | 0.002363129 | 0.065445592 |
| 0 | 0 | 0 | 0 | 0 |
| 0 | 0 | 0 | 0 | 0 |
| 0 | 0 | 0 | 0 | 0 |
| 0 | 0 | 0 | 0 | 0 |
| 0 | 0 | 0 | 0 | 0 |
| 0 | 0 | 0 | 0 | 0 |
| 0 | 0 | 0 | 0 | 0 |
| 0 | 0.016238029 | 0 | 0 | 0 |
| 0 | 0 | 0 | 0 | 0 |
| 0 | 0 | 0 | 0 | 0 |
| 0 | 0 | 0 | 0 | 0.015127321 |
| 0 | 0 | 0 | 0 | 0 |
| 0 | 0 | 0 | 0 | 0 |
| 0 | 0 | 0 | 0 | 0.006637499 |
| 0 | 0 | 0 | 0 | 0.032575866 |
| 0.017459515 | 0 | 0 | 0.007725908 | 0.004657711 |
| 0 | 0 | 0 | 0 | 0 |
| 0 | 0 | 0 | 0 | 0 |
| 0.774388383 | 1.707652554 | 0.001244668 | 0.030758837 | 0.756577007 |
| 0 | 0 | 0 | 0 | 0 |
| 0 | 0 | 0 | 0 | 0 |
| 0 | 0 | 0 | 0 | 0 |
| 0 | 0 | 0 | 0 | 0 |
| 0 | 0 | 0 | 0 | 0.008516595 |
| 0 | 0 | 0 | 0 | 0 |
| 0 | 0 | 0 | 0 | 0 |
| 0 | 0 | 0 | 0 | 0 |
| 0 | 0 | 0 | 0 | 0 |
| 0 | 0 | 0 | 0 | 0 |
| 0 | 0 | 0 | 0 | 0 |
| 0 | 0 | 0 | 0 | 0.156199032 |
| 0 | 0 | 0 | 0 | 0 |
| 0 | 0 | 0 | 0 | 0 |
| 0 | 0 | 0 | 0 | 0 |
| 0 | 0 | 0 | 0 | 0 |
| 0 | 0 | 0 | 0 | 0 |
| 0 | 0 | 0 | 0 | 0 |
| 0 | 0 | 0 | 0 | 0 |
| 0 | 0 | 0 | 0 | 0 |
| 0 | 0 | 0 | 0 | 0 |

| | | | | |
|---|---|---|---|---|
| 0 | 0.017765059 | 0 | 0 | 0 |
| 0 | 0 | 0 | 0 | 0.005086969 |
| 0 | 0 | 0 | 0 | 0 |
| 0.001406238 | 0.002742712 | 0 | 0 | 0 |
| 0 | 0 | 0 | 0 | 0 |
| 0 | 0 | 0 | 0 | 0.021721648 |
| 0 | 0 | 0 | 0 | 0 |
| 0 | 0 | 0 | 0 | 0 |
| 0 | 0 | 0 | 0 | 0 |
| 0 | 0 | 0 | 0 | 0 |
| 0 | 0 | 0 | 0 | 0 |
| 0 | 0.004431713 | 0 | 0 | 0.030399162 |
| 0 | 0 | 0 | 0 | 0 |
| 0 | 0 | 0 | 0 | 0 |
| 0 | 0 | 0 | 0 | 0 |
| 0 | 0 | 0 | 0 | 0 |
| 0 | 0 | 0 | 0 | 0 |
| 0 | 0 | 0 | 0 | 0 |
| 0 | 0 | 0 | 0 | 0 |
| 0 | 0 | 0 | 0 | 0 |
| 0.465754419 | 0.002127718 | 0.001656329 | 0.015387222 | 0.204268124 |
| 0 | 0 | 0 | 0 | 0 |
| 0.283127997 | 0.003135011 | 0 | 0 | 0.399030632 |
| 0 | 0 | 0 | 0 | 1.64099512 |
| 0 | 0 | 0 | 0 | 0 |
| 0 | 0 | 0 | 0 | 0 |
| 0 | 0 | 0 | 0 | 0 |
| 0 | 0 | 0 | 0 | 0 |
| 0 | 0 | 0 | 0 | 0 |
| 0 | 0 | 0 | 0 | 0 |
| 0 | 0 | 0 | 0 | 0 |
| 0 | 0 | 0 | 0 | 0.016256326 |
| 0.075963992 | 0.013421372 | 0.024220162 | 0.014719674 | 0.002413738 |
| 0 | 0 | 0 | 0 | 0 |
| 0 | 1.100928452 | 0 | 0 | 0.027282885 |
| 0.251402885 | 0 | 0 | 0 | 0 |
| 0 | 0 | 0 | 0 | 0 |
| 0 | 0 | 0 | 0 | 0 |
| 0.405754808 | 0.012457311 | 0.002148864 | 0 | 0 |
| 0 | 0 | 0 | 0 | 0 |
| 0 | 0 | 0 | 0 | 0 |
| 0 | 0 | 0 | 0 | 0 |
| 0 | 0.009331555 | 0.015655839 | 0 | 0.083114366 |
| 0 | 0 | 0 | 0 | 0 |
| 0 | 0 | 0 | 0 | 0 |
| 0 | 0 | 0 | 0 | 0.163221295 |
| 0 | 0 | 0 | 0 | 0 |
| 0 | 0 | 0 | 0 | 0 |

| | | | | |
|---|---|---|---|---|
| 0 | 0 | 0 | 0 | 0 |
| 0 | 0 | 0 | 0 | 0 |
| 0 | 0 | 0 | 0 | 0 |
| 0 | 0 | 0 | 0 | 0 |
| 0 | 0 | 0 | 0 | 0 |
| 0 | 0 | 0 | 0 | 0 |
| 0 | 0 | 0 | 0 | 0 |
| 0.112434715 | 0.079297755 | 0.365334456 | 1.135710837 | 0 |
| 0 | 0 | 0 | 0 | 0.007619581 |
| 0.002147728 | 0.006516076 | 0 | 0 | 0.105778748 |
| 0 | 0 | 0 | 0 | 0 |
| 0 | 0 | 0 | 0 | 0 |
| 0 | 0 | 0 | 0 | 0 |
| 0 | 0 | 0 | 0 | 0 |
| 0 | 0 | 0 | 0 | 0 |
| 0 | 0.027476136 | 0 | 0 | 0 |
| 0 | 0 | 0 | 0 | 0 |
| 0 | 0 | 0 | 0 | 0 |
| 0 | 0 | 0 | 0 | 0 |
| 0 | 0 | 0 | 0 | 1.025234976 |
| 0 | 0 | 0.027131948 | 0 | 0 |
| 0 | 0 | 0 | 0 | 0 |
| 0 | 0 | 0 | 0 | 0 |
| 0 | 0 | 0 | 0 | 0 |
| 0 | 0.002228084 | 0 | 0 | 0.013729206 |
| 0 | 0 | 0 | 0 | 0 |
| 0 | 0 | 0 | 0 | 0 |
| 0 | 0 | 0 | 0 | 0 |
| 0 | 0 | 0 | 0 | 0 |
| 0 | 0 | 0 | 0 | 2.322779571 |
| 0 | 0 | 0.09153619 | 0 | 0.189896648 |
| 0 | 0 | 0 | 0 | 0 |
| 0 | 0 | 0 | 0 | 0 |
| 0 | 0 | 0 | 0 | 0 |
| 0 | 0 | 0 | 0 | 0 |
| 0 | 0 | 0 | 0 | 0 |
| 0 | 0 | 0 | 0 | 0 |
| 0 | 0 | 0 | 0 | 0 |
| 0 | 0 | 0 | 0 | 0 |
| 0 | 0 | 0 | 0 | 0 |
| 0 | 0 | 0 | 0 | 0 |
| 0 | 0 | 0 | 0 | 0 |
| 0 | 0 | 0 | 0 | 0 |
| 0 | 0 | 0 | 0 | 0.006551251 |
| 0 | 0 | 0 | 0 | 0 |
| 0 | 0 | 0 | 0 | 0 |
| 0.28294559 | 0.02906469 | 0 | 0.107926746 | 0 |
| 0 | 0 | 0 | 0 | 0 |

| | | | | |
|---|---|---|---|---|
| 0 | 0 | 0 | 0 | 0 |
| 0 | 0 | 0 | 0 | 0 |
| 0 | 0 | 0 | 0 | 0 |
| 0 | 0 | 0 | 0 | 0 |
| 0 | 0.041325683 | 0 | 0.02020203 | 0.17531122 |
| 0 | 0 | 0 | 0 | 0 |
| 0 | 0 | 0 | 0 | 0 |
| 0 | 0 | 0 | 0 | 0 |
| 0 | 0 | 0 | 0 | 0 |
| 0 | 0 | 0 | 0 | 0 |
| 0 | 0 | 0 | 0 | 0.034802064 |
| 0.077406721 | 0.083825405 | 0 | 0.002730295 | 0.062273077 |
| 0 | 0 | 0 | 0 | 0 |
| 0 | 0 | 0 | 0 | 0 |
| 0 | 0.007262552 | 0 | 0 | 0 |
| 0.019541467 | 0 | 0 | 0 | 1.076927839 |
| 0 | 0.00239338 | 0 | 0 | 0 |
| 0 | 0 | 0 | 0 | 0 |
| 0.007135473 | 0.55373264 | 0.00462738 | 0 | 0.083982595 |
| 0.018633507 | 0.001922889 | 0 | 0.56392674 | 0.180479166 |
| 0 | 0 | 0 | 0 | 0 |
| 0 | 0 | 0 | 0 | 0 |
| 0 | 0 | 0 | 0 | 0 |
| 0 | 0 | 0 | 0 | 0 |
| 0 | 0 | 0 | 0 | 0 |
| 0 | 0 | 0 | 0 | 0 |
| 0.017953811 | 0.022343032 | 0 | 0 | 0.015317159 |
| 0 | 0 | 0 | 0 | 0 |
| 0 | 0 | 0 | 0 | 0 |
| 0 | 0 | 0 | 0 | 0 |
| 0.001209005 | 0 | 0 | 0 | 0.002558729 |
| 0 | 0 | 0 | 0 | 0 |
| 0 | 0 | 0 | 0 | 0 |
| 0 | 0 | 0 | 0 | 0 |
| 0 | 0 | 0 | 0 | 0 |
| 0 | 0 | 0 | 0 | 0 |
| 0 | 0 | 0 | 0 | 0 |
| 0.063989036 | 0.082588773 | 0.000625286 | 0.009866544 | 0.183107439 |
| 0 | 0 | 0 | 0 | 0 |
| 0 | 0 | 0 | 0 | 0 |
| 0 | 0 | 0 | 0 | 0.025381801 |
| 0 | 0 | 0 | 0 | 0 |
| 0 | 0 | 0 | 0 | 0 |
| 0 | 0.010554507 | 0 | 0 | 0.009139695 |
| 0 | 0 | 0 | 0 | 0 |
| 0 | 0 | 0 | 0 | 0 |
| 0 | 0 | 0 | 0 | 0 |
| 0 | 0 | 0 | 0 | 0.002766188 |

| | | | | |
|---|---|---|---|---|
| 0 | 0 | 0 | 0 | 0 |
| 0 | 0 | 0 | 0 | 0.014321556 |
| 0 | 0 | 0 | 0 | 0 |
| 0.00858023 | 0.001716389 | 0 | 0.004867676 | 0.002793712 |
| 0 | 0 | 0.00450632 | 0.060948336 | 0 |
| 0 | 0 | 0 | 0.000873996 | 0 |
| 0 | 0 | 0 | 0 | 0 |
| 0 | 0 | 0 | 0 | 0.735071122 |
| 0 | 0 | 0 | 0 | 0 |
| 0 | 0 | 0 | 0 | 0 |
| 0 | 0.06240426 | 0 | 0 | 0 |
| 0 | 0 | 0 | 0 | 0 |
| 0 | 0.019196474 | 0.003329295 | 0.001035149 | 0.019379127 |
| 0 | 0 | 0 | 0 | 0 |
| 0 | 0 | 0 | 0 | 0 |
| 0 | 0 | 0 | 0 | 0 |
| 0 | 0 | 0 | 0 | 0 |
| 0 | 0 | 0 | 0 | 0 |
| 0 | 0 | 0 | 0 | 0 |
| 0 | 0 | 0 | 0 | 0 |
| 0 | 0 | 0 | 0 | 0 |
| 0 | 0 | 0 | 0 | 0 |
| 0.094668394 | 0.031329652 | 0.000604678 | 0.008178317 | 0 |
| 0 | 0 | 0.057083255 | 0.009114546 | 0.022638899 |
| 0 | 0 | 0 | 0 | 0 |
| 0 | 0 | 0 | 0 | 0 |
| 0 | 0 | 0 | 0 | 0 |
| 0 | 0 | 0 | 0 | 0 |
| 0 | 0 | 0 | 0 | 0 |
| 0 | 0 | 0 | 0 | 0 |
| 0 | 0.017466211 | 0 | 0 | 0.022844899 |
| 0 | 0.005065023 | 0 | 0 | 0 |
| 0 | 0 | 0 | 0 | 0 |
| 0 | 0 | 0 | 0 | 0 |
| 0 | 0 | 0 | 0 | 0.00208938 |
| 0 | 0 | 0 | 0 | 0 |
| 0 | 0 | 0 | 0 | 0 |
| 0 | 0 | 0 | 0 | 0 |
| 0.003099582 | 0 | 0.003993018 | 0.118512914 | 0 |
| 0 | 0 | 0 | 0 | 0 |
| 0 | 0 | 0 | 0 | 0 |
| 0 | 0 | 0 | 0 | 0 |
| 0 | 0 | 0 | 0 | 0 |
| 0 | 0 | 0 | 0 | 0 |
| 0 | 0 | 0 | 0 | 0 |
| 0.340208265 | 0.38133893 | 0.012547128 | 0.171586483 | 0.150114581 |
| 0 | 0 | 0 | 0 | 0 |
| 0 | 0 | 0 | 0 | 0 |

| | | | | |
|---|---|---|---|---|
| 0 | 0 | 0 | 0 | 0 |
| 0 | 0 | 0 | 0 | 0 |
| 0 | 0 | 0 | 0 | 0 |
| 0 | 0 | 0 | 0 | 0 |
| 0 | 0 | 0 | 0 | 0 |
| 0 | 0 | 0 | 0 | 0.002704286 |
| 0 | 0 | 0 | 0 | 0.002143304 |
| 0 | 0 | 0 | 0 | 0 |
| 0 | 0 | 0 | 0 | 0 |
| 0 | 0 | 0 | 0 | 0 |
| 0.004872021 | 0.001226108 | 0 | 0 | 0.001995698 |
| 0.001642503 | 0 | 0 | 0 | 0 |
| 0 | 0 | 0 | 0 | 0 |
| 0 | 0 | 0 | 0 | 0 |
| 0 | 0 | 0 | 0 | 0 |
| 0 | 0 | 0 | 0 | 0 |
| 0 | 0 | 0 | 0.140307783 | 0 |
| 0 | 0 | 0 | 0 | 0 |
| 0 | 0 | 0 | 0 | 0 |
| 0 | 0 | 0 | 0 | 0.005671368 |
| 0 | 0 | 0 | 0 | 0 |
| 0 | 0 | 0 | 0 | 0 |
| 0 | 0 | 0 | 0 | 0 |
| 0 | 0 | 0 | 0 | 0 |
| 0 | 0 | 0 | 0 | 0 |
| 0 | 0 | 0 | 0 | 0.00374251 |
| 0 | 0 | 0 | 0 | 0 |
| 0 | 0 | 0.088633016 | 0 | 0 |
| 0 | 0 | 0 | 0 | 0 |
| 0 | 0 | 0 | 0 | 0 |
| 0.012892147 | 0.000931285 | 0.000659057 | 0 | 0 |
| 0 | 0 | 0 | 0 | 0 |
| 0.207904101 | 0.230681009 | 0.047138284 | 0.042779269 | 0.008297267 |
| 0.00977295 | 0 | 0.01172976 | 0 | 0.001541619 |
| 0 | 0 | 0 | 0 | 0 |
| 0 | 0 | 0 | 0 | 0 |
| 0 | 0 | 0 | 0 | 0 |
| 0 | 0 | 0 | 0 | 0 |
| 0 | 0 | 0 | 0 | 0 |
| 0 | 0 | 0 | 0 | 0 |
| 0 | 0 | 0 | 0 | 0 |
| 0.001026245 | 0.102747631 | 0.000881369 | 0 | 0.245583964 |
| 0 | 0.005780114 | 0 | 0 | 0.09450111 |
| 0 | 0 | 0 | 0 | 0 |
| 0 | 0 | 0 | 0 | 0 |
| 0 | 0.00965777 | 0 | 0 | 0.003004335 |
| 0 | 0.010610225 | 0 | 0 | 0.064821932 |
| 0 | 0 | 0 | 0 | 0.004960353 |

| | | | | |
|---|---|---|---|---|
| 0 | 0 | 0 | 0 | 0 |
| 0 | 0 | 0 | 0 | 0 |
| 0.017084942 | 0.138594199 | 0.00807174 | 0 | 0.009714206 |
| 0 | 0 | 0 | 0 | 0 |
| 0 | 0 | 0 | 0 | 0 |
| 0 | 0 | 0 | 0 | 0.010246376 |
| 0 | 0 | 0 | 0 | 0 |
| 0 | 0 | 0 | 0 | 0 |
| 0 | 0 | 0 | 0 | 0 |
| 0 | 0 | 0 | 0 | 0 |
| 0 | 0 | 0 | 0 | 0 |
| 0 | 0 | 0 | 0 | 0 |
| 0 | 0 | 0 | 0 | 0 |
| 0 | 0.013191282 | 0 | 0 | 0.039075561 |
| 0 | 0 | 0 | 0 | 0 |
| 0 | 0 | 0 | 0 | 0 |
| 0 | 0 | 0 | 0 | 0 |
| 0 | 0 | 0 | 0 | 0 |
| 0 | 0 | 0 | 0 | 0 |
| 0 | 0 | 0 | 0 | 0 |
| 0 | 0 | 0 | 0 | 0 |
| 0 | 0 | 0 | 0 | 0 |
| 0 | 0 | 0 | 0 | 0 |
| 0 | 0 | 0 | 0 | 0 |
| 0 | 0.000910854 | 0 | 0 | 0 |
| 0 | 0.199386379 | 0.002801304 | 0.503534419 | 0 |
| 0 | 0 | 0 | 0 | 0 |
| 0 | 0 | 0 | 0 | 0.015438765 |
| 0 | 0 | 0 | 0 | 0 |
| 0 | 0 | 0 | 0 | 0 |
| 0 | 0 | 0 | 0 | 0 |
| 0 | 0 | 0 | 0.006152854 | 0 |
| 0 | 0 | 0 | 0 | 0 |
| 0 | 0 | 0 | 0 | 0 |
| 0 | 0 | 0 | 0 | 0 |
| 0 | 0 | 0 | 0 | 0 |
| 0 | 0 | 0.003319924 | 0 | 0.042220878 |
| 0 | 0 | 0 | 0 | 0 |
| 0 | 3.30923646 | 0.09283279 | 0 | 1.778765741 |
| 0 | 0 | 0 | 0 | 0 |
| 0 | 0 | 0 | 0 | 0 |
| 0.11192204 | 0 | 0 | 0 | 0.046389531 |
| 0 | 0.001839209 | 0 | 0.020655343 | 0.004811181 |
| 0 | 0 | 0 | 0 | 0 |
| 0 | 0 | 0 | 0 | 0 |
| 0 | 0 | 0 | 0 | 0.001549826 |
| 0 | 0.028417669 | 0 | 0 | 0.010598639 |
| 0 | 0 | 0 | 0 | 0 |

| | | | | |
|---|---|---|---|---|
| 0 | 0 | 0 | 0 | 0 |
| 0.071264552 | 0.036567928 | 0.000904494 | 0.004983958 | 0.026375578 |
| 0 | 0 | 0 | 0 | 0 |
| 0 | 0 | 0 | 0 | 0 |
| 0 | 0 | 0 | 0 | 0 |
| 0 | 0 | 0 | 0 | 0 |
| 0 | 0 | 0 | 0 | 0 |
| 0 | 0 | 0 | 0 | 0 |
| 0 | 0 | 0 | 0 | 0 |
| 0 | 0 | 0 | 0 | 0 |
| 0 | 0 | 0 | 0 | 0 |
| 0 | 0 | 0 | 0 | 0 |
| 0 | 0.032463148 | 0.002390325 | 0 | 0 |
| 0 | 0 | 0 | 0 | 0 |
| 0 | 0.007525338 | 0.000751845 | 0 | 0.015516809 |
| 0 | 0 | 0 | 0 | 0 |
| 0 | 0 | 0 | 0 | 0 |
| 0 | 0 | 0 | 0 | 0 |
| 0 | 0 | 0 | 0 | 0.003843047 |
| 0 | 0 | 0 | 0 | 0 |
| 0 | 0 | 0 | 0 | 0 |
| 0.004513643 | 0.001304202 | 0.00203052 | 0.027740356 | 0.131007596 |
| 0 | 0.018707214 | 0 | 0 | 0.018875365 |
| 0 | 0 | 0 | 0.012857596 | 0 |
| 0 | 0 | 0 | 0 | 0 |
| 0 | 0 | 0 | 0 | 0 |
| 0 | 0 | 0 | 0 | 0 |
| 0 | 0 | 0 | 0 | 0 |
| 0 | 0 | 0 | 0 | 0 |
| 0 | 0 | 0 | 0 | 0 |
| 0 | 0 | 0 | 0 | 0 |
| 0 | 0 | 0 | 0 | 0 |
| 0 | 0 | 0 | 0 | 0 |
| 0 | 0 | 0 | 0 | 0 |
| 0 | 0 | 0 | 0 | 2.320717368 |
| 0 | 0 | 0 | 0 | 0 |
| 0 | 0 | 0 | 0 | 0 |
| 0 | 0 | 0 | 0 | 0 |
| 0.007762776 | 0.004124277 | 0.004424117 | 0.147929223 | 0.035205165 |
| 0 | 0 | 0 | 0 | 0 |
| 0.240019998 | 0.331064681 | 0.032891433 | 0.678838885 | 0.388771998 |
| 0 | 0 | 0 | 0 | 0 |
| 0 | 0 | 0 | 0 | 0 |
| 0 | 0 | 0 | 0 | 0 |
| 0 | 0 | 0 | 0 | 0 |
| 7.520548155 | 0.123674793 | 2.240015587 | 1.20721451 | 0.001399007 |
| 0 | 0 | 0 | 0 | 0 |
| 0 | 0 | 0 | 0 | 0 |

| | | | | |
|---|---|---|---|---|
| 0 | 0.120076839 | 0 | 0 | 0 |
| 0 | 0 | 0 | 0 | 0 |
| 0 | 0 | 0 | 0 | 0 |
| 0 | 0 | 0 | 0 | 0 |
| 0.002924509 | 0.02873091 | 0 | 0 | 0.007604128 |
| 0 | 0 | 0 | 0 | 0 |
| 0 | 0 | 0 | 0 | 0 |
| 0.004723536 | 0 | 0 | 0.022438743 | 0 |
| 0 | 0 | 0 | 0 | 0 |
| 0.059082221 | 0.024190829 | 0.002776133 | 0.050897611 | 0 |
| 0 | 0 | 0 | 0 | 0 |
| 0 | 0 | 0 | 0 | 0 |
| 0 | 0.003621229 | 0 | 0 | 0.00445777 |
| 0 | 0 | 0 | 0 | 0 |
| 0 | 0 | 0 | 0 | 0 |
| 0 | 0 | 0 | 0 | 0 |
| 0 | 0 | 0 | 0 | 0 |
| 0 | 0 | 0 | 0 | 0 |
| 0 | 0 | 0 | 0 | 0 |
| 0.73468767 | 0.117739765 | 0.039608451 | 0.15897011 | 0.239849256 |
| 0 | 0 | 0 | 0 | 0 |
| 0 | 0 | 0 | 0 | 0 |
| 0 | 0 | 0 | 0 | 0 |
| 0 | 0 | 0 | 0 | 0 |
| 0 | 0 | 0 | 0 | 0 |
| 0 | 0 | 0 | 0 | 0 |
| 0 | 0 | 0 | 0 | 0 |
| 0 | 0 | 0 | 0 | 0 |
| 0 | 0 | 0 | 0 | 0 |
| 0 | 0.025335288 | 0 | 0 | 0.00254494 |
| 0 | 0 | 0 | 0 | 0 |
| 0 | 0 | 0 | 0 | 0 |
| 0 | 0 | 0 | 0 | 0 |
| 0 | 0 | 0 | 0 | 0 |
| 0 | 0 | 0 | 0 | 0 |
| 0 | 0.006135726 | 0 | 0 | 0 |
| 0 | 0 | 0 | 0 | 0 |
| 0 | 0 | 0 | 0 | 0 |
| 0 | 0 | 0 | 0 | 0 |
| 0 | 0 | 0 | 0 | 0 |
| 0 | 0 | 0 | 0 | 0 |
| 0 | 0 | 0 | 0 | 0 |
| 0 | 0 | 0 | 0 | 0.324371412 |
| 0 | 0 | 0 | 0 | 0 |
| 0.024651825 | 0.012465356 | 0 | 0 | 0.010061921 |
| 0 | 0 | 0 | 0 | 0 |
| 0 | 0 | 0 | 0 | 0 |
| 0 | 0 | 0 | 0 | 0 |

| | | | | |
|---|---|---|---|---|
| 0 | 0 | 0 | 0 | 0 |
| 0 | 0 | 0 | 0 | 0 |
| 0 | 0 | 0 | 0 | 0 |
| 0 | 0 | 0 | 0 | 0 |
| 0 | 0 | 0 | 0 | 0 |
| 0.00107354 | 0.007976462 | 0 | 0 | 0.112952438 |
| 0 | 0 | 0 | 0 | 0 |
| 0 | 0 | 0 | 0 | 0.000994174 |
| 0 | 0 | 0 | 0 | 0 |
| 0 | 0 | 0 | 0 | 0 |
| 0 | 0 | 0 | 0 | 0 |
| 0 | 0 | 0 | 0 | 0 |
| 0 | 0 | 0 | 0 | 0 |
| 0 | 0 | 0 | 0 | 0 |
| 0 | 0 | 0 | 0 | 0 |
| 0 | 0 | 0 | 0 | 0 |
| 0 | 0 | 0 | 0 | 0 |
| 0 | 0 | 0 | 0 | 0 |
| 0 | 0 | 0 | 0 | 0 |
| 0 | 0 | 0 | 0 | 0 |
| 0 | 0 | 0 | 0 | 0 |
| 0 | 0 | 0 | 0 | 0 |
| 0 | 0 | 0 | 0 | 0 |
| 0 | 0 | 0 | 0 | 0 |
| 0 | 0 | 0 | 0 | 0 |
| 0 | 0 | 0 | 0.010880399 | 0 |
| 0 | 0 | 0 | 0 | 0 |
| 0 | 0 | 0 | 0 | 0.536900215 |
| 0 | 0 | 0 | 0 | 0 |
| 0 | 0 | 0 | 0 | 0 |
| 0 | 0 | 0 | 0 | 0 |
| 0 | 0 | 0 | 0 | 0 |
| 0 | 0 | 0 | 0 | 0 |
| 0 | 0 | 0 | 0 | 0 |
| 0 | 0 | 0 | 0 | 0 |
| 0 | 0 | 0 | 0 | 0 |
| 0 | 0 | 0 | 0 | 0 |
| 0 | 0 | 0 | 0 | 0 |
| 0 | 0.017827529 | 0 | 0 | 0.107211331 |
| 0 | 0 | 0 | 0 | 0 |
| 0 | 0 | 0 | 0 | 0 |
| 0 | 0 | 0 | 0 | 0 |
| 0 | 0 | 0 | 0 | 0 |
| 0 | 0 | 0 | 0 | 0 |
| 0 | 0 | 0 | 0 | 0 |
| 0 | 0 | 0 | 0 | 0 |
| 0 | 0 | 0 | 0 | 0 |
| 0 | 0 | 0 | 0 | 0 |

| | | | | |
|---|---|---|---|---|
| 0 | 0 | 0 | 0 | 0 |
| 0 | 0 | 0 | 0 | 0.325492683 |
| 0 | 0 | 0 | 0 | 0 |
| 0 | 0 | 0 | 0 | 0 |
| 0 | 0 | 0 | 0 | 0 |
| 0 | 0 | 0 | 0 | 0 |
| 0 | 0 | 0 | 0 | 0 |
| 0 | 0 | 0 | 0 | 0 |
| 0 | 0 | 0 | 0 | 0 |
| 0 | 0 | 0 | 0 | 0 |
| 0.000899321 | 0 | 0.001158545 | 0 | 0 |
| 0 | 0 | 0 | 0 | 0 |
| 0 | 0 | 0 | 0 | 0 |
| 0 | 0 | 0 | 0 | 0 |
| 0 | 0 | 0 | 0 | 0 |
| 0 | 0 | 0 | 0 | 0 |
| 0 | 0 | 0 | 0 | 0 |
| 0 | 0 | 0 | 0 | 0 |
| 0 | 0 | 0 | 0 | 0 |
| 0 | 0.000486516 | 0 | 0 | 0 |
| 0 | 0.02096717 | 0.0176378 | 0.026253416 | 0 |
| 0 | 0 | 0 | 0 | 0 |
| 0 | 0 | 0 | 0 | 0 |
| 0.323055442 | 3.009294891 | 5.912783117 | 0 | 0 |
| 0 | 0 | 0 | 0 | 0 |
| 0 | 0 | 0 | 0 | 0 |
| 0 | 0 | 0 | 0 | 0 |
| 0 | 0.556370721 | 0 | 0 | 0 |
| 0 | 0 | 0 | 0 | 0 |
| 0 | 0 | 0 | 0 | 0 |
| 0 | 0 | 0 | 0 | 0 |
| 0 | 0 | 0 | 0 | 0 |
| 0 | 0 | 0 | 0 | 0 |
| 0 | 0 | 0 | 0 | 0 |

| PHL018-0077_V3 | PHL018-0094_V1 | PHL018-0215_V2 | PHL018-0116_V1 | PHL018-0103_V1 |
|---|---|---|---|---|
| EYCF | REF | CTRL | CTRL | CTRL |
| 0.012510684 | 0.846011264 | 0.287599473 | 0.467589848 | 1.19585743 |
| 0.290416259 | 5.131702267 | 4.911512364 | 3.176933558 | 0.668215399 |
| 4.170174487 | 0 | 2.918820499 | 0 | 2.413132945 |
| 0 | 0 | 0 | 0 | 0 |
| 0 | 0 | 0 | 0 | 0 |
| 0 | 0 | 0.072277499 | 0 | 0 |
| 0 | 0 | 0 | 0 | 0 |
| 0 | 0 | 0 | 0 | 0 |
| 0 | 0 | 0 | 0 | 0 |
| 0 | 0 | 0 | 0 | 0 |
| 0 | 0 | 0 | 0 | 0 |
| 0 | 0 | 0 | 0 | 0 |
| 0 | 0 | 0 | 0 | 0 |
| 2.443806091 | 0.918897174 | 0.175350226 | 0.033179717 | 0.471374549 |
| 0 | 0 | 0 | 0 | 0 |
| 0 | 0 | 0 | 0 | 0 |
| 0 | 0.008911201 | 0 | 0 | 0 |
| 0 | 0 | 0 | 0 | 0 |
| 0 | 0 | 0 | 0 | 0 |
| 0 | 0 | 0 | 0 | 0 |
| 0 | 0 | 0 | 0 | 0 |
| 0 | 0 | 0 | 0 | 0 |
| 0 | 0 | 0 | 0 | 0 |
| 0 | 0 | 0 | 0 | 0 |
| 0.032613954 | 0 | 0.089531053 | 0 | 0 |
| 0 | 0 | 0 | 0 | 0 |
| 0 | 0 | 0 | 0 | 0 |
| 0 | 0 | 0 | 0 | 0 |
| 0 | 0 | 0 | 0 | 0 |
| 0 | 0 | 0 | 0 | 0 |
| 0 | 0 | 0 | 0 | 0 |
| 0 | 0 | 0 | 0 | 0 |
| 0 | 0 | 0 | 0 | 0 |
| 0 | 0 | 0 | 0 | 0 |
| 0 | 0 | 0 | 0 | 0 |
| 0.23457622 | 0 | 1.345012829 | 0 | 1.995792478 |
| 0 | 0 | 0 | 0 | 0 |
| 0 | 0 | 0 | 0 | 0 |
| 0 | 0 | 0 | 0 | 0 |
| 0 | 0 | 0 | 0 | 0 |
| 0 | 0 | 0 | 0 | 0 |
| 0 | 0 | 0.041806643 | 0.003591322 | 0.025185351 |
| 0 | 0.011583162 | 0.521356043 | 0.036982963 | 1.585436126 |
| 0 | 0.008994297 | 0.002839453 | 0 | 0.075300998 |
| 0 | 0.002202307 | 0.001316022 | 0.000758156 | 0.003074509 |
| 0 | 0 | 0 | 0 | 0 |

| | | | | |
|---|---|---|---|---|
| 0 | 0 | 0.013455234 | 0 | 0.009437284 |
| 0 | 0.027490527 | 0.003259401 | 0.028661663 | 0.000865667 |
| 0.000498944 | 0 | 0 | 0 | 0 |
| 0.094523184 | 0.618889787 | 0.378989149 | 6.34503439 | 0.235379453 |
| 0.001258651 | 0.022814563 | 0.09166143 | 0 | 0 |
| 0 | 0 | 0 | 0 | 0 |
| 0.013420846 | 0.001403115 | 0.456956589 | 2.795871607 | 0 |
| 0 | 0 | 0 | 0 | 0 |
| 0 | 0 | 0 | 0 | 0 |
| 0 | 0 | 0 | 0 | 0 |
| 0 | 0 | 0 | 0 | 0.001420865 |
| 0.440136318 | 0 | 0.231755915 | 0 | 0.520356284 |
| 0 | 0 | 0 | 0 | 0 |
| 0.022002429 | 0.365269025 | 0.220273613 | 0.31112995 | 0.004430003 |
| 0 | 0 | 0 | 0 | 0 |
| 0 | 0 | 0 | 0 | 0 |
| 0 | 0 | 0.18329592 | 3.622200599 | 0 |
| 0 | 0 | 0 | 0 | 0 |
| 0 | 0 | 0 | 0 | 0 |
| 0 | 0 | 0 | 0 | 0 |
| 0.002080245 | 0.985205527 | 0.04674263 | 0.347082202 | 0.015447791 |
| 0.007697697 | 0.041872286 | 0.08866717 | 0.084017413 | 0.015602756 |
| 0 | 0.551648307 | 0.010691215 | 0 | 0 |
| 0 | 0 | 0 | 0 | 0 |
| 0 | 0 | 0 | 0 | 0 |
| 0 | 0 | 0 | 0 | 0 |
| 0.000548736 | 0 | 0 | 0 | 0 |
| 0 | 0 | 0 | 0 | 0 |
| 0 | 0 | 0.004572804 | 0.005400472 | 0.00238963 |
| 0 | 0 | 0 | 0 | 0 |
| 0 | 1.228133898 | 0 | 0 | 0 |
| 0 | 0 | 0 | 0 | 0 |
| 0 | 0 | 0 | 0 | 0 |
| 0 | 0 | 0 | 0 | 0.00660354 |
| 0 | 0 | 0 | 0 | 0 |
| 0 | 0 | 0 | 0 | 0 |
| 0 | 0 | 0 | 0 | 0 |
| 0 | 0 | 0.005899659 | 0.093514805 | 0 |
| 0 | 0 | 0 | 0 | 0 |
| 0 | 0 | 0 | 0 | 0 |
| 0 | 0 | 0 | 0 | 0 |
| 0 | 0 | 0 | 0 | 0 |
| 0 | 0 | 0 | 0 | 0 |
| 0 | 0 | 0 | 0 | 0 |
| 0 | 0.154733019 | 0 | 0 | 0 |
| 0 | 0 | 0 | 0 | 0 |
| 0 | 0 | 0.000491032 | 0 | 0 |
| 0.003482313 | 0 | 0 | 0 | 0.002109245 |

| | | | | |
|---|---|---|---|---|
| 0 | 0 | 0 | 0 | 0 |
| 0 | 0.033154449 | 0 | 0 | 0 |
| 0 | 0 | 0 | 0 | 0 |
| 0 | 0 | 0 | 0 | 0 |
| 0.000523104 | 0 | 0 | 0 | 0 |
| 0 | 0 | 0 | 0 | 0.00108044 |
| 0 | 0 | 0 | 0 | 0 |
| 0 | 0 | 0 | 0 | 0 |
| 0 | 0.016863686 | 0.126012372 | 0.015326287 | 0 |
| 0 | 0 | 0 | 0 | 0 |
| 0 | 0 | 0 | 0 | 0 |
| 0 | 0 | 0.000557113 | 0.001270965 | 0.003452494 |
| 0 | 0 | 0 | 0 | 0 |
| 0 | 0 | 0 | 0 | 0 |
| 0 | 0 | 0 | 0 | 0 |
| 0 | 0 | 0 | 0.004822491 | 0 |
| 0 | 0 | 0 | 0 | 0 |
| 0 | 0 | 0 | 0 | 0 |
| 0.05610789 | 0.006969546 | 0.166094514 | 0.012750579 | 0 |
| 0 | 0 | 0 | 0 | 0 |
| 0 | 0 | 0 | 0 | 0 |
| 0 | 0 | 0 | 0 | 0 |
| 0 | 0.003162008 | 0.001364643 | 0 | 0 |
| 0 | 0 | 0 | 0 | 0 |
| 0 | 0 | 0 | 0 | 0 |
| 0 | 0 | 0 | 0 | 0 |
| 0 | 0 | 0 | 0 | 0 |
| 0 | 0 | 0 | 0 | 0 |
| 0 | 0 | 0 | 0 | 0 |
| 0 | 0 | 0 | 0.014290331 | 0 |
| 0.032987082 | 0.279258111 | 0.145915636 | 0 | 0.016609347 |
| 0 | 0.007951714 | 0.006335556 | 0.076808751 | 0 |
| 0 | 0 | 0 | 0 | 0 |
| 0 | 0 | 0 | 0 | 0 |
| 0 | 0 | 0 | 0 | 0 |
| 0 | 0 | 0 | 0 | 0 |
| 0 | 0 | 0 | 0 | 0 |
| 0 | 0 | 0 | 0 | 0 |
| 0 | 0 | 0 | 0 | 0 |
| 0 | 0 | 0 | 0 | 0 |
| 0 | 0 | 0 | 0 | 0 |
| 0 | 0 | 0 | 0 | 0 |
| 0 | 0.004865783 | 0.000861518 | 0.001675071 | 0.001906759 |
| 0 | 0 | 0 | 0 | 0 |
| 0 | 0 | 0.000688873 | 0 | 0 |
| 0 | 0 | 0 | 0 | 0 |
| 0 | 0 | 0.049676754 | 0.282387583 | 0 |
| 0 | 0 | 0 | 0 | 0 |

| | | | | |
|---|---|---|---|---|
| 0 | 0 | 0 | 0 | 0 |
| 0 | 0 | 0 | 0 | 0 |
| 0 | 0 | 0 | 0 | 0 |
| 0 | 0 | 0 | 0 | 0 |
| 0 | 0 | 0.029437082 | 0 | 0 |
| 0 | 0 | 0 | 0 | 0 |
| 0 | 0 | 0 | 0 | 0 |
| 0 | 0 | 0 | 0 | 0 |
| 0 | 0 | 0 | 0 | 0 |
| 0 | 0 | 0 | 0 | 0 |
| 0 | 0 | 0 | 0 | 0 |
| 0 | 0 | 0 | 0 | 0 |
| 0 | 0 | 0 | 0 | 0 |
| 0 | 0 | 0 | 0 | 0 |
| 0 | 0 | 0 | 0 | 0 |
| 0 | 0 | 0 | 0 | 0 |
| 0 | 0 | 0 | 0 | 0 |
| 0 | 0 | 0 | 0 | 0 |
| 0 | 0 | 0 | 0 | 0 |
| 0 | 0 | 0 | 0 | 0 |
| 0 | 0 | 0 | 0 | 0 |
| 0 | 0 | 0 | 0 | 0 |
| 0 | 0 | 0 | 0.001230409 | 0 |
| 0 | 0 | 0 | 0 | 0 |
| 0 | 0 | 0 | 0 | 0 |
| 0 | 0 | 0 | 0 | 0 |
| 0 | 0 | 0.051818239 | 0 | 0 |
| 0 | 0 | 0 | 0 | 0 |
| 0 | 0 | 0 | 0 | 0 |
| 0 | 0 | 0 | 0 | 0 |
| 0 | 0.00102181 | 0.007449293 | 0 | 0 |
| 0 | 0 | 0 | 0.000334418 | 0 |
| 0 | 0 | 0 | 0 | 0 |
| 0 | 0 | 0 | 0 | 0 |
| 0 | 0 | 0 | 0 | 0 |
| 0 | 0 | 0 | 0 | 0 |
| 0 | 0 | 0 | 0 | 0 |
| 0 | 0 | 0 | 0 | 0.049134463 |
| 0 | 0 | 0 | 0 | 0 |
| 0 | 0 | 0 | 0 | 0 |
| 0 | 0 | 0 | 0 | 0 |
| 0 | 0 | 0 | 0 | 0 |
| 0 | 0 | 0 | 0 | 0 |
| 0 | 0 | 0 | 0 | 0 |
| 0 | 0 | 0 | 0 | 0 |
| 0 | 0 | 0 | 0 | 0 |
| 0 | 0 | 0 | 0 | 0 |
| 0 | 0.155930921 | 0 | 0 | 0 |

| | | | | |
|---|---|---|---|---|
| 0 | 0 | 0 | 0 | 0 |
| 0 | 0 | 0 | 0 | 0 |
| 0 | 0 | 0 | 0 | 0 |
| 0 | 0 | 0 | 0 | 0 |
| 0 | 0 | 0 | 0 | 0 |
| 0 | 0 | 0 | 0 | 0 |
| 0 | 0 | 0 | 0 | 0 |
| 0 | 0 | 0 | 0 | 0 |
| 0 | 0 | 0 | 0 | 0 |
| 0 | 0 | 0 | 0 | 0 |
| 0 | 0 | 0 | 0 | 0 |
| 0 | 0 | 0 | 0.002596948 | 0 |
| 0.000714157 | 0 | 0.00041146 | 0.004586739 | 0.002162833 |
| 0 | 0 | 0 | 0 | 0 |
| 0.003745632 | 0 | 0 | 0 | 0 |
| 0 | 0 | 0 | 0 | 0 |
| 0 | 0 | 0 | 0 | 0 |
| 0 | 0 | 0 | 0 | 0 |
| 0 | 0 | 0 | 0 | 0 |
| 0 | 0 | 0 | 0 | 0 |
| 0.018471678 | 0.016736773 | 0.02558028 | 0.010570542 | 0.018730006 |
| 0 | 0 | 0 | 0 | 0 |
| 0 | 0 | 0 | 0.000874999 | 0.001587415 |
| 0 | 0 | 0 | 0.001663841 | 0 |
| 0 | 0 | 0 | 0 | 0 |
| 0 | 0 | 0 | 0 | 0 |
| 0 | 0.001686998 | 0.003024274 | 0.007782157 | 0.00173535 |
| 0 | 0 | 0 | 0 | 0 |
| 0 | 0 | 0 | 0 | 0 |
| 0 | 0 | 0 | 0 | 0 |
| 0 | 0.101085912 | 0 | 0 | 0 |
| 0 | 0 | 0 | 0 | 0 |
| 0 | 0 | 0 | 0 | 0 |
| 0.002720179 | 0.167196248 | 0 | 0.003656637 | 0.00351203 |
| 0.004923306 | 0.037695589 | 0.057940771 | 0.004801454 | 0.08018321 |
| 0 | 0 | 0 | 0 | 0 |
| 0 | 0 | 0 | 0 | 0 |
| 0 | 0 | 0 | 0 | 0 |
| 0 | 0 | 0 | 0 | 0 |
| 0 | 0 | 0 | 0 | 0 |
| 0 | 0 | 0 | 0 | 0 |
| 0 | 0 | 0 | 0 | 0 |
| 0 | 0 | 0 | 0 | 0 |
| 0 | 0 | 0 | 0 | 0 |
| 0 | 0 | 0 | 0 | 0 |
| 0 | 0 | 0 | 0 | 0 |
| 0 | 0 | 0 | 0.000562021 | 0 |
| 0 | 0.010894397 | 0.000624257 | 0.022194459 | 0.000789509 |

| | | | | |
|---|---|---|---|---|
| 0 | 0 | 0 | 0 | 0 |
| 0 | 0 | 0 | 0 | 0 |
| 0 | 0 | 0 | 0.000845035 | 0 |
| 0 | 0 | 0 | 0 | 0 |
| 0 | 0 | 0 | 0 | 0 |
| 0 | 0 | 0 | 0 | 0 |
| 0 | 0 | 0 | 0 | 0.001123856 |
| 0 | 0 | 0 | 0 | 0.003262935 |
| 0 | 0 | 0 | 0 | 0 |
| 0 | 0 | 0 | 0 | 0 |
| 0 | 0 | 0 | 0 | 0.000976822 |
| 0 | 0 | 0 | 0 | 0 |
| 0 | 0 | 0 | 0 | 0 |
| 0 | 0 | 0 | 0 | 0 |
| 0 | 0 | 0 | 0 | 0.001089195 |
| 0 | 0 | 0 | 0 | 0 |
| 0 | 0 | 0 | 0 | 0 |
| 0 | 0 | 0 | 0 | 0 |
| 0 | 0 | 0.003174328 | 0 | 0.00792889 |
| 0 | 0.637266547 | 0.079820075 | 0 | 0.004715593 |
| 0 | 0.003134357 | 0.000668922 | 0.289670289 | 0 |
| 0 | 0 | 0 | 0 | 0 |
| 0 | 0 | 0 | 0 | 0 |
| 0 | 0 | 0 | 0 | 0 |
| 0 | 0.007116643 | 0.012757974 | 0.004409895 | 0.001470659 |
| 0 | 0 | 0 | 0 | 0 |
| 0 | 0 | 0 | 0 | 0 |
| 0 | 0 | 0 | 0 | 0 |
| 0 | 0 | 0.011414175 | 0 | 0.004771803 |
| 0 | 0 | 0 | 0 | 0 |
| 0.104265373 | 0.91249323 | 0.550291099 | 0.402016815 | 0.108534242 |
| 0 | 0 | 0 | 0 | 0 |
| 0.000566139 | 0 | 0 | 0.00088788 | 0 |
| 0 | 0.005324563 | 0 | 0 | 0 |
| 0 | 0 | 0 | 0 | 0 |
| 0 | 0 | 0 | 0 | 0 |
| 0 | 0 | 0 | 0 | 0 |
| 0 | 0 | 0 | 0 | 0 |
| 0 | 0 | 0 | 0 | 0 |
| 0 | 0 | 0 | 0 | 0.002283473 |
| 0 | 0 | 0 | 0 | 0 |
| 0 | 0.016444749 | 0.018450334 | 0 | 0.013204896 |
| 0 | 0.000563174 | 0 | 0.00287905 | 0 |
| 0 | 0 | 0 | 0 | 0 |
| 0 | 0 | 0 | 0 | 0 |
| 0 | 0 | 0 | 0 | 0 |
| 0 | 0 | 0 | 0 | 0 |
| 0 | 0 | 0 | 0 | 0 |

| | | | | |
|---|---|---|---|---|
| 0 | 0 | 0 | 0 | 0 |
| 0 | 0 | 0 | 0 | 0 |
| 0 | 0.00210705 | 0 | 0 | 0.062143613 |
| 0 | 0 | 0 | 0 | 0 |
| 0 | 0.074787941 | 0.526781672 | 0 | 0 |
| 0 | 0 | 0 | 0 | 0 |
| 0 | 0 | 0 | 0 | 0 |
| 0 | 0 | 0 | 0 | 0 |
| 0 | 0 | 0 | 0 | 0 |
| 0 | 0 | 0 | 0 | 0 |
| 0 | 0 | 0 | 0 | 0 |
| 0 | 0 | 0 | 0 | 0 |
| 0 | 0 | 0 | 0 | 0 |
| 0 | 0.001164335 | 0.001252379 | 0 | 0.001385918 |
| 0.000797384 | 0.006534843 | 0.004364405 | 0.00166739 | 0.001906489 |
| 0 | 0 | 0.002213371 | 0 | 0.003674067 |
| 0 | 0 | 0.004056798 | 0.001899842 | 0.002606728 |
| 0 | 0 | 0 | 0.000315801 | 0 |
| 0 | 0 | 0 | 0 | 0 |
| 0 | 0 | 0 | 0 | 0 |
| 0 | 0 | 0 | 0 | 0 |
| 0 | 0.005733039 | 0 | 0.027195237 | 0.000522918 |
| 0 | 0 | 0 | 0 | 0 |
| 0 | 0 | 0 | 0 | 0 |
| 0 | 0 | 0.002273037 | 0 | 0 |
| 0 | 0 | 0 | 0 | 0 |
| 0 | 0 | 6.370023725 | 0 | 0 |
| 0 | 0 | 0 | 0 | 0 |
| 0 | 0 | 0 | 0 | 0 |
| 0 | 0 | 0.005980829 | 0.000708796 | 0 |
| 0 | 0 | 0.000724219 | 0 | 0 |
| 0 | 0 | 0 | 0 | 0 |
| 0 | 0 | 0 | 0 | 0 |
| 0 | 0 | 0.004358632 | 0 | 0 |
| 0 | 0 | 0 | 0 | 0 |
| 0 | 0 | 0.001179772 | 0 | 0.037339253 |
| 0 | 0 | 0 | 0 | 0 |
| 0 | 0.00422243 | 0.003116866 | 0 | 0.05338064 |
| 0 | 0 | 0 | 0 | 0 |
| 0 | 0 | 0 | 0 | 0 |
| 0.001239352 | 0 | 0 | 0 | 0 |
| 0 | 0 | 0.000871671 | 0 | 0 |
| 0 | 0 | 0 | 0 | 0 |
| 0 | 0 | 0 | 0 | 0 |
| 0 | 0 | 0 | 0 | 0 |
| 0 | 0 | 0.001369181 | 0 | 0 |
| 0 | 0 | 0.000804421 | 0 | 0 |
| 0 | 0 | 0 | 0 | 0 |

| | | | | |
|---|---|---|---|---|
| 0 | 1.47699306 | 0 | 0.002212191 | 0 |
| 0 | 0 | 0 | 0 | 0 |
| 0 | 0 | 0.000742412 | 0 | 0 |
| 0 | 0 | 0 | 0 | 0 |
| 0.003601207 | 0.043540606 | 0.002987753 | 0.009681942 | 0.005372787 |
| 0 | 0 | 0.001420942 | 0 | 0 |
| 0 | 0 | 0 | 0 | 0 |
| 0.017162805 | 0.001168072 | 0.003838994 | 0.00778092 | 0.02317275 |
| 0 | 0 | 0.001210221 | 0 | 0 |
| 0.195423296 | 0.062906739 | 0.020863834 | 0 | 0.009553862 |
| 0 | 0 | 0 | 0.001180409 | 0 |
| 0.000639348 | 0 | 0.032047228 | 0.003294565 | 0.180073318 |
| 0.001454904 | 0.008416555 | 0.00513422 | 0.079643461 | 0.002550954 |
| 0 | 0.018641258 | 0.00334181 | 1.742750516 | 0 |
| 0 | 0.009377588 | 0 | 0.001117483 | 0 |
| 0 | 0 | 0 | 0 | 0 |
| 0 | 0 | 0 | 0 | 0 |
| 0 | 0 | 0 | 0 | 0 |
| 0 | 0.338454272 | 0 | 0 | 0 |
| 0.002982515 | 0 | 0.035678794 | 0 | 0 |
| 0 | 0 | 0 | 0 | 0 |
| 0 | 0 | 0 | 0 | 0 |
| 0 | 0.003980532 | 0 | 0 | 0 |
| 0 | 0 | 0.11754307 | 0 | 0 |
| 0 | 0 | 0 | 0 | 0 |
| 0 | 0 | 0 | 0 | 0.014991112 |
| 0 | 0 | 0 | 0 | 0 |
| 0 | 0.008891042 | 0 | 0 | 0 |
| 0.022410644 | 0 | 0 | 0 | 0 |
| 0 | 0 | 0 | 0 | 0.002640139 |
| 0 | 0.021038607 | 0 | 0 | 0 |
| 0 | 0 | 0 | 0 | 0 |
| 0 | 0 | 0 | 0 | 0 |
| 0 | 0.00828004 | 0 | 0 | 0 |
| 0 | 0 | 0 | 0 | 0 |
| 0 | 0 | 0 | 0 | 0 |
| 0 | 0.115064453 | 0 | 0 | 0 |
| 0 | 0 | 0 | 0 | 0 |
| 0 | 0 | 0 | 0 | 0.011541409 |
| 0 | 0 | 0.000513428 | 0 | 0 |
| 0 | 0 | 0 | 0 | 0 |
| 0 | 0.107215235 | 0 | 0 | 0 |
| 0 | 0 | 0 | 0 | 0 |
| 0 | 0 | 0 | 1.937685811 | 0.000761636 |
| 0 | 0 | 0 | 0 | 0 |
| 0 | 0 | 0.000535745 | 0 | 0 |
| 0 | 0.003809681 | 0.002124764 | 0.002518085 | 0.001511565 |
| 0 | 0 | 0 | 0 | 0 |

| | | | | |
|---|---|---|---|---|
| 0 | 0.003360707 | 0 | 0 | 0 |
| 0.002158861 | 0 | 0.015095496 | 0.016005399 | 0 |
| 0.006384088 | 0 | 0 | 0 | 0 |
| 0 | 0 | 0 | 0 | 0 |
| 0 | 0 | 0 | 0 | 0 |
| 0.014384937 | 0.130425452 | 0.0039447 | 0.122220676 | 0.03571621 |
| 0 | 0 | 0 | 0 | 0 |
| 0 | 0 | 0.036641495 | 0.001122253 | 0 |
| 1.154208975 | 0 | 0 | 0 | 0 |
| 0 | 0 | 0 | 0 | 0 |
| 0.009304597 | 0.005104255 | 0.017884824 | 0.004744349 | 0.019791844 |
| 0 | 0 | 0 | 0.038267159 | 0 |
| 0 | 0 | 0 | 0 | 0 |
| 0.000909364 | 0.004164665 | 0.003143569 | 0.004753873 | 0.001014639 |
| 0 | 0 | 0 | 0 | 0 |
| 0 | 0 | 0 | 0 | 0 |
| 0 | 0 | 0 | 0 | 0 |
| 0 | 0.354308064 | 0 | 0 | 0 |
| 0 | 0 | 0 | 0 | 0 |
| 0 | 0 | 0 | 0 | 0 |
| 0 | 0 | 0.018911121 | 0 | 0 |
| 0.177524505 | 1.715419307 | 0.296030696 | 0 | 0.168304601 |
| 0 | 0 | 0 | 0 | 0 |
| 0 | 0 | 0 | 0 | 0 |
| 0 | 0 | 0 | 0 | 0 |
| 0 | 0 | 0 | 0 | 0 |
| 0 | 0.003564947 | 0 | 0 | 0 |
| 0 | 0 | 0 | 0 | 0 |
| 0 | 0 | 0 | 0 | 0 |
| 0.12773976 | 0.481293736 | 0.348657887 | 0.153698174 | 2.189016031 |
| 0 | 0.003122804 | 0.009508116 | 0.004054442 | 0.028222416 |
| 0.001154869 | 0 | 0.05713913 | 0 | 0 |
| 0 | 0.001831938 | 0 | 0 | 0 |
| 0 | 0 | 0 | 0 | 0 |
| 0 | 0.015440178 | 0 | 0 | 0 |
| 0 | 0 | 0 | 0 | 0 |
| 1.45104635 | 0 | 0 | 0 | 0.233127985 |
| 0.002008248 | 0 | 0.056606369 | 0 | 0 |
| 0 | 0 | 0 | 0 | 0.014849114 |
| 0 | 0 | 0 | 0 | 0 |
| 0.004360976 | 0.09172201 | 0.011952645 | 0.059627974 | 0.00464737 |
| 0.265008328 | 1.897419673 | 0.183739475 | 0 | 0.028900909 |
| 0 | 0 | 0.005381189 | 0 | 0 |
| 0 | 0 | 15.92974474 | 0 | 0 |
| 0 | 0.002642581 | 0 | 0 | 0 |
| 0.003216765 | 0 | 0 | 0 | 0 |
| 0 | 0 | 0.000791136 | 0.002343964 | 0.000375211 |
| 0 | 0.015224581 | 0 | 0 | 0 |

| | | | | |
|---|---|---|---|---|
| 0 | 0 | 0 | 0.000867617 | 0.002777687 |
| 0 | 0 | 0.019958968 | 0.012935579 | 0 |
| 0 | 0 | 0.410605695 | 8.120302136 | 0 |
| 0 | 0 | 0 | 0 | 0 |
| 0 | 0 | 0 | 0 | 0.031955773 |
| 0 | 0.074778592 | 0 | 0 | 0 |
| 0 | 0 | 0 | 0 | 0 |
| 0 | 0 | 0 | 0 | 0.006399405 |
| 0.002430722 | 0 | 0.003384435 | 0 | 0 |
| 0 | 0 | 0 | 0 | 0 |
| 0 | 0 | 0 | 0 | 0 |
| 0 | 0 | 0 | 0 | 0 |
| 0 | 0 | 0 | 0.056012397 | 0 |
| 0.00408728 | 0 | 0.009209257 | 0.001177367 | 0 |
| 0 | 0 | 0 | 0 | 0 |
| 0.004334902 | 0.049824485 | 0.019125943 | 0.088737791 | 0.051931358 |
| 0 | 0.03181852 | 0.015626151 | 0.000793198 | 0.025515262 |
| 0 | 0.004665652 | 0 | 0.056858595 | 0 |
| 0 | 0.000755543 | 0.010497056 | 0.071163269 | 0 |
| 0 | 0 | 0 | 0 | 0.004193008 |
| 0 | 0.007835708 | 0 | 0.004569854 | 0.010210828 |
| 0 | 0.006575533 | 0.005830588 | 0.004468906 | 0.003226145 |
| 0 | 0 | 0 | 0 | 0 |
| 0 | 0 | 0 | 0 | 0 |
| 0 | 0 | 0 | 0.00254167 | 0 |
| 0 | 0 | 0.003310826 | 0.001492713 | 0 |
| 0 | 0.006732311 | 0.015537082 | 0.002445503 | 0 |
| 0 | 0.006170373 | 0.213506428 | 0 | 0 |
| 0 | 0 | 0 | 0 | 0 |
| 0 | 0.005164648 | 0 | 0 | 0 |
| 0.034913202 | 0.016948646 | 0.03175525 | 0 | 0.007004899 |
| 0 | 0.001579272 | 0.016357777 | 0.009949209 | 0.001392459 |
| 0.576003576 | 0.024560114 | 0.004642493 | 0.004192964 | 0.129763609 |
| 0 | 0 | 0.013950143 | 0.001335317 | 0.001662513 |
| 0 | 0 | 0.001991371 | 0 | 0 |
| 0 | 0 | 0 | 0 | 0 |
| 0 | 0 | 0.001353914 | 0.000701987 | 0.003895525 |
| 0.001102658 | 0.074951222 | 0.057070591 | 0 | 0.005858618 |
| 0 | 1.854913868 | 0.650255905 | 0.463044809 | 0 |
| 0 | 0 | 0 | 0 | 0 |
| 0 | 0.00303631 | 0 | 0.004703695 | 0 |
| 0 | 0 | 0 | 0 | 0 |
| 0 | 0 | 0.011876017 | 0.297204852 | 0 |
| 0.215572621 | 0 | 0.259038359 | 0.386210461 | 0.133816913 |
| 0 | 0.033293058 | 0 | 0 | 0 |
| 0 | 0 | 0 | 0 | 0 |
| 0 | 0 | 0 | 0 | 0 |
| 0 | 0 | 0 | 0 | 0 |

| | | | | |
|---|---|---|---|---|
| 0 | 0 | 0 | 0 | 0 |
| 0.005175316 | 0.677731558 | 0.316113415 | 1.280983486 | 0.000478987 |
| 0 | 0 | 0 | 0 | 0 |
| 0 | 0 | 0.006640905 | 0 | 0 |
| 0 | 0 | 0 | 0 | 0 |
| 0 | 0 | 0 | 0 | 0 |
| 0 | 0 | 0 | 0 | 0 |
| 0.000897623 | 0 | 0.000886567 | 0.001723774 | 0.003679122 |
| 0 | 0.001768172 | 0 | 0.000410874 | 0 |
| 0 | 0 | 0 | 0.003913941 | 0 |
| 0 | 0.007894428 | 0.002573146 | 0.005225392 | 0 |
| 0 | 0.021379955 | 0.010220732 | 0.018878812 | 0.158170932 |
| 0 | 0 | 0 | 0 | 0 |
| 0 | 0.035953019 | 0 | 0 | 0 |
| 0 | 0 | 0 | 0 | 0.014157728 |
| 0 | 0 | 0 | 0 | 0 |
| 0 | 0.019915005 | 0.041876333 | 0.017812259 | 0.009069519 |
| 0.001517742 | 0.059509363 | 0.005975367 | 0.021611481 | 0 |
| 0 | 0 | 0 | 0 | 0 |
| 0.000547523 | 0.091062727 | 0 | 0 | 0 |
| 0.021434794 | 0.012593685 | 0.067266816 | 0.010805255 | 0.002818701 |
| 0 | 0 | 0 | 0 | 0 |
| 0 | 0 | 0 | 0 | 0 |
| 0 | 0 | 0 | 0 | 0 |
| 0 | 0 | 0 | 0.007060217 | 0 |
| 0 | 0.001776733 | 0 | 0.017835699 | 0.001879875 |
| 0 | 0 | 0.031083705 | 0 | 0 |
| 0.025345 | 0.025946751 | 0.01378211 | 0.003062504 | 0.002859687 |
| 0 | 0 | 0 | 0 | 0 |
| 0 | 0.001241978 | 0 | 0.000461762 | 0 |
| 0 | 0 | 0.054663582 | 0.015882669 | 0.000584466 |
| 0 | 0 | 0 | 0 | 0 |
| 0 | 0.004749875 | 0 | 0 | 0 |
| 0 | 0 | 0 | 0 | 0 |
| 0.007320774 | 0 | 0 | 0.131547587 | 0 |
| 0.002650808 | 0.302514034 | 0.271574286 | 0.424150806 | 0.120765792 |
| 0 | 0 | 0 | 0 | 0 |
| 0 | 0 | 0 | 0 | 0 |
| 0.001229558 | 0.005334703 | 0.013358537 | 0.004879841 | 0.059635679 |
| 0 | 0 | 0 | 0 | 0 |
| 0 | 0.007013922 | 0 | 0 | 0 |
| 0 | 0 | 0 | 0 | 0 |
| 0.019738232 | 0.100760393 | 0.056891462 | 0.008222959 | 0 |
| 0 | 0.010875352 | 0 | 0.003600295 | 0 |
| 0 | 0.012721917 | 0 | 0 | 0 |
| 0.050341768 | 0 | 0.651309354 | 0 | 0 |
| 0 | 0 | 0 | 0 | 0 |
| 0 | 0.008669177 | 0 | 0.001342984 | 0 |

| | | | | |
|---|---|---|---|---|
| 0 | 0 | 0 | 0 | 0 |
| 0 | 0 | 0 | 0 | 0 |
| 0 | 0 | 0 | 0 | 0 |
| 0 | 0.770798155 | 0.47735518 | 0 | 0.003802957 |
| 0 | 0 | 0 | 0 | 0 |
| 0 | 0 | 0 | 0 | 0 |
| 0 | 0 | 0.011323119 | 0.00025901 | 0 |
| 0 | 0 | 0.03026306 | 0 | 0 |
| 0.032340193 | 0.293632851 | 0 | 0.020684602 | 3.745392847 |
| 0 | 0 | 0.01574098 | 0.019806546 | 0 |
| 0 | 0 | 0 | 0 | 0 |
| 0.002857393 | 0.004649033 | 0.006585126 | 0 | 0.001594093 |
| 0 | 0 | 0 | 0 | 0 |
| 0 | 0 | 0 | 0 | 0 |
| 0.061207877 | 0.117507321 | 0.090935643 | 0.037155651 | 0 |
| 0 | 0 | 0 | 0 | 0 |
| 0 | 0 | 0 | 0 | 0 |
| 0.00333569 | 0 | 0 | 0 | 0 |
| 0 | 0 | 0 | 0 | 0 |
| 0 | 0 | 0 | 0.073558665 | 0 |
| 0 | 0 | 0 | 0 | 0 |
| 0 | 0 | 0 | 0 | 0 |
| 0 | 0 | 0 | 0 | 0 |
| 0.0459936 | 0.584699227 | 0 | 0 | 0.107976331 |
| 0 | 0.011123523 | 0 | 0.02237661 | 0 |
| 0 | 0 | 4.169546804 | 0.005456501 | 0 |
| 0 | 0 | 0 | 0 | 0 |
| 0.199386587 | 0 | 0 | 0 | 0.850917044 |
| 0 | 0 | 0 | 0 | 0 |
| 0 | 0 | 0 | 0.120391852 | 0 |
| 0 | 0 | 0.008002332 | 0 | 0.055347518 |
| 0 | 0.737956493 | 0.094057517 | 0 | 0 |
| 0.088794945 | 0.158230228 | 0 | 0 | 0.048105066 |
| 0 | 0 | 0 | 0 | 0 |
| 0 | 0.003245619 | 0.060400606 | 0 | 0 |
| 0 | 0 | 0 | 0.153999402 | 0 |
| 0.001901197 | 0 | 0.158171619 | 0.00238533 | 0.511777499 |
| 0 | 0 | 0 | 0 | 0 |
| 0 | 0 | 0 | 0.031433234 | 0 |
| 0 | 0 | 0.00994245 | 0 | 0 |
| 0.01747488 | 0.24430429 | 0.259796956 | 0.033572971 | 0.030257256 |
| 0 | 0 | 0 | 0 | 0 |
| 7.16518877 | 0.022589804 | 0.658011167 | 0 | 4.779449239 |
| 0 | 0 | 0.026311943 | 0 | 0 |
| 0 | 0 | 0.032971906 | 0 | 0 |
| 0 | 0 | 0 | 0 | 0 |
| 0 | 0 | 0 | 0 | 0 |
| 0 | 0.019062509 | 0.061345771 | 0.003937421 | 0.695151744 |

| | | | | |
|---|---|---|---|---|
| 0 | 0 | 0 | 0 | 0 |
| 0 | 0 | 0 | 0 | 0.029667383 |
| 0.001711021 | 0.062244338 | 0.000682478 | 1.01993048 | 0.002895121 |
| 0 | 0 | 0.005164082 | 0 | 0 |
| 0 | 0 | 0 | 0 | 0 |
| 0.001658925 | 0 | 0.002688149 | 0 | 0.048290603 |
| 0 | 0 | 0 | 0 | 0 |
| 0 | 0 | 0 | 0 | 0 |
| 0 | 0 | 0 | 0.002007014 | 0 |
| 0 | 0 | 0 | 0 | 0 |
| 0.044711692 | 0.5341305 | 0.06254008 | 0.100868676 | 0.794539377 |
| 0.835041596 | 1.872894788 | 0.75323414 | 0.227101609 | 0.455208073 |
| 0 | 0 | 0 | 0 | 0 |
| 0.000976251 | 0.013513825 | 0.005544298 | 0.018497721 | 0.039213686 |
| 0.016375591 | 0.801884867 | 0.483072657 | 0.63695202 | 0.029220153 |
| 0 | 0 | 0.017728478 | 0 | 0 |
| 0 | 0 | 0 | 0 | 0 |
| 0 | 0 | 0 | 0 | 0 |
| 0 | 0 | 0 | 0 | 0 |
| 0.001277364 | 0.089289752 | 0.008923405 | 0 | 0.027079591 |
| 0.04180735 | 0.204729804 | 0.206675885 | 0.198301894 | 0.018334353 |
| 6.601747959 | 0 | 0.019725749 | 0 | 2.038070424 |
| 1.991384688 | 1.947925933 | 0.308552642 | 0.004217251 | 0.856860999 |
| 0 | 0 | 0 | 0 | 0 |
| 0 | 0 | 0 | 0 | 0 |
| 0.374954374 | 0 | 1.285632033 | 0 | 0.004964984 |
| 0 | 0.028289842 | 0 | 6.425987934 | 0 |
| 0 | 0 | 0 | 0 | 0 |
| 0 | 0.065630917 | 0 | 0 | 0 |
| 0 | 0 | 0 | 0 | 0 |
| 0.055788866 | 0 | 0 | 0 | 0 |
| 0 | 0 | 0.211642807 | 0.110696194 | 0.074223489 |
| 0 | 0.109598023 | 0.229049808 | 0.438047638 | 0.015612001 |
| 0 | 0.196574224 | 0 | 0 | 0.056429468 |
| 0 | 0.026730136 | 0.001135521 | 0.000942004 | 0 |
| 0.019135645 | 0.101996142 | 0.006864601 | 0.087038756 | 0.459131415 |
| 0.002906411 | 0.005380307 | 0.004219798 | 0.001458607 | 0 |
| 0 | 0 | 0 | 0 | 0 |
| 0.024786743 | 1.260775006 | 0 | 0 | 0.251195945 |
| 0 | 0 | 0 | 0 | 0 |
| 0.000953768 | 0 | 0.006136212 | 0 | 0 |
| 0 | 0 | 0 | 0 | 0 |
| 0 | 0 | 0.004073954 | 0 | 0 |
| 0 | 0 | 0 | 0 | 0 |
| 0.025115923 | 0.091106817 | 0.034159848 | 0.129637847 | 0.042133745 |
| 0 | 0 | 0 | 0 | 0 |
| 0 | 0 | 0 | 0 | 0 |
| 0 | 0.379501206 | 0.230210534 | 0.817056297 | 0.092782988 |

| | | | | |
|---|---|---|---|---|
| 0 | 0 | 0 | 0 | 0 |
| 0 | 0 | 0 | 0 | 0 |
| 0 | 0 | 0 | 0 | 0 |
| 0.001169738 | 0 | 0.00151637 | 0 | 0.002573021 |
| 0 | 0 | 0 | 0 | 0 |
| 0 | 0 | 0 | 0 | 0 |
| 0 | 0 | 0 | 1.85109049 | 0 |
| 0.011517163 | 0.135276281 | 0.024278824 | 0.069452421 | 0 |
| 0 | 0 | 0 | 0 | 0 |
| 0 | 0 | 0 | 0 | 0 |
| 0 | 0.0190297 | 0.002476957 | 0.044482434 | 0.005606734 |
| 0 | 0 | 0 | 0 | 0 |
| 0 | 0 | 0 | 0 | 0 |
| 0 | 0 | 0 | 0 | 0 |
| 0 | 0 | 0 | 0 | 0 |
| 0 | 0 | 0 | 0 | 0 |
| 0 | 0 | 0.471687951 | 0.672344797 | 0 |
| 0.001409893 | 0.018351381 | 0.001031005 | 0 | 0.003941426 |
| 0 | 0.009407689 | 0.0701234 | 0 | 0 |
| 0 | 0 | 0 | 0 | 0 |
| 0 | 0 | 0 | 0 | 0 |
| 0 | 0 | 0 | 0 | 0 |
| 0.050265633 | 0 | 0.002917657 | 0.681081301 | 0 |
| 0 | 0 | 0 | 0 | 0 |
| 0 | 0 | 0 | 0 | 0 |
| 0 | 0 | 0 | 0 | 0 |
| 0 | 0 | 0 | 0 | 0 |
| 0 | 0 | 0 | 0 | 0 |
| 0 | 0 | 0 | 0 | 0 |
| 0.005502446 | 0.817006238 | 0.048345911 | 0 | 1.075406306 |
| 0 | 0 | 0 | 0 | 0 |
| 0 | 0 | 0 | 0 | 0 |
| 0 | 0 | 0 | 0.11333019 | 0 |
| 0 | 0 | 0 | 0 | 0 |
| 0 | 0 | 0 | 0 | 0 |
| 0 | 0 | 0 | 0 | 0 |
| 0.15776017 | 0 | 0.349194457 | 0.410347195 | 0 |
| 0 | 0.01929887 | 0.00066716 | 0.031231051 | 0 |
| 0 | 0 | 0 | 0 | 0 |
| 0.004024731 | 0.005663406 | 0 | 0.010430666 | 0 |
| 0.223603467 | 0 | 0.139768126 | 0 | 0 |
| 0 | 0 | 0.395426341 | 0 | 0 |
| 0 | 0 | 0 | 0.112975762 | 0 |
| 0 | 0 | 0.020454847 | 0 | 0 |
| 0.072379799 | 0 | 0 | 0 | 0.05868304 |
| 0 | 0 | 0 | 0 | 0 |
| 0 | 0 | 0 | 0 | 0 |
| 0 | 0 | 0 | 0 | 0 |

| | | | | |
|---|---|---|---|---|
| 0 | 0 | 0 | 0 | 0 |
| 0 | 0 | 0 | 0 | 0 |
| 0.34418102 | 0 | 0 | 0 | 0 |
| 0.108382317 | 0 | 0.637178009 | 0.000589856 | 0.38786262 |
| 0 | 0 | 0 | 0 | 0 |
| 0 | 0 | 2.621917708 | 2.237511038 | 0 |
| 0 | 0 | 0 | 0 | 0 |
| 0 | 0 | 0 | 0 | 0 |
| 0 | 0.534307014 | 0.001183017 | 0.209502997 | 0.010036893 |
| 0 | 0 | 0 | 0 | 0.000702841 |
| 0 | 0 | 0 | 0 | 0 |
| 0 | 0 | 0 | 0 | 0 |
| 0 | 0 | 0.000613467 | 0 | 0 |
| 0 | 0 | 0 | 0 | 0 |
| 0.002841841 | 0.025924499 | 1.134002317 | 0.014203244 | 1.24666043 |
| 0 | 0.795467415 | 0 | 0 | 0 |
| 0 | 0 | 0 | 0 | 0 |
| 0 | 0 | 0 | 0 | 0 |
| 0 | 0 | 0 | 0 | 0 |
| 0.035332849 | 0 | 0 | 0 | 0 |
| 0.002907516 | 0.275341065 | 0.002010192 | 0 | 0 |
| 0 | 0 | 0 | 0 | 0 |
| 0 | 0 | 0 | 0 | 0 |
| 0 | 0 | 0 | 0 | 0 |
| 0 | 0 | 0.056266099 | 0.001122047 | 0.055252195 |
| 0.001685856 | 2.605652758 | 0.752954075 | 0.006194376 | 21.87287097 |
| 0 | 0 | 0.053354024 | 0 | 0.177001898 |
| 0 | 0 | 0 | 0.00099501 | 0 |
| 0 | 0.193818171 | 0 | 0.106647662 | 0.058027284 |
| 0 | 0 | 0.837518478 | 0 | 0 |
| 0 | 0 | 0 | 0 | 0 |
| 0 | 0 | 0.093376556 | 0 | 0 |
| 0.020267792 | 0 | 0 | 0 | 0 |
| 0 | 0.029149834 | 0.002063854 | 0.14647269 | 0 |
| 0 | 0 | 0.003944672 | 0.006065255 | 0.022277998 |
| 0.000778808 | 0.009010742 | 0.003173012 | 0.081560306 | 0.010002009 |
| 0.013892673 | 0.011623546 | 0 | 0 | 0 |
| 0 | 0.015988791 | 0.534717556 | 0.899340234 | 0.387840651 |
| 0 | 0 | 0 | 0 | 0.24578802 |
| 0 | 0 | 0 | 0 | 0 |
| 0.560476252 | 0 | 0 | 0 | 0 |
| 0 | 0.012590349 | 0 | 0 | 0 |
| 0.001520364 | 0.013192864 | 0.197352673 | 0.202333266 | 0.278738389 |
| 0 | 0 | 0 | 0.521833542 | 0 |
| 0 | 0 | 0 | 0 | 0 |
| 0.002529678 | 0.009603615 | 0.008562633 | 0.004534072 | 0 |
| 0 | 0 | 0 | 0 | 0 |
| 0.007556402 | 0.021613877 | 0.042955584 | 0.046349643 | 0.762500651 |

| | | | | |
|---|---|---|---|---|
| 0 | 0 | 0 | 0.017057055 | 0 |
| 0 | 0 | 0 | 0 | 0 |
| 0 | 0 | 0 | 0 | 0 |
| 0 | 0 | 0 | 0 | 0 |
| 0.000748015 | 0.693800028 | 0.000969677 | 0.00446902 | 0.011803785 |
| 0 | 0.000868138 | 0 | 0 | 0 |
| 0 | 0 | 0 | 0 | 0 |
| 0 | 0 | 0 | 0 | 0 |
| 0 | 0 | 0 | 0 | 0 |
| 0 | 0 | 0 | 0 | 0 |
| 0 | 0 | 0 | 0 | 0 |
| 0.099833258 | 0.600652308 | 0.142836001 | 1.204933793 | 5.736255573 |
| 0 | 0.549058344 | 0.557440672 | 0 | 0.003464803 |
| 0 | 0 | 0 | 0 | 0 |
| 0.001585979 | 0 | 0.558846544 | 0.080756576 | 0.173189418 |
| 0 | 0 | 0 | 0 | 0 |
| 0 | 0.001305493 | 0 | 0 | 0 |
| 0 | 0 | 0 | 0 | 0 |
| 0.008340629 | 0 | 0.004477794 | 0.002378304 | 0.000604299 |
| 0 | 0 | 0.001937623 | 0 | 0 |
| 0 | 0.05371624 | 0.033274407 | 0 | 0 |
| 0 | 0 | 0 | 0 | 0 |
| 0.001843211 | 0.654346417 | 3.290278543 | 0.010021168 | 0.020095874 |
| 0 | 0 | 0 | 0 | 0 |
| 0 | 0 | 0 | 0 | 0 |
| 0 | 0 | 0 | 0 | 0 |
| 0 | 0 | 0 | 0 | 0 |
| 0 | 0 | 0 | 0 | 0 |
| 0 | 0 | 0 | 0 | 0 |
| 0 | 0 | 0 | 0 | 0 |
| 0 | 0 | 0 | 0 | 0 |
| 0 | 0.127985327 | 0.058531933 | 0.106469638 | 0.03184974 |
| 0 | 0 | 0 | 0 | 0 |
| 0.012898685 | 0.02919848 | 0.009370219 | 0.156136847 | 0 |
| 0.00193723 | 0.009455733 | 0.003243758 | 0.082790211 | 0.009958335 |
| 0.056658059 | 0.030473966 | 0.045458004 | 0.007413514 | 0.016270787 |
| 0 | 0 | 0 | 0 | 0 |
| 0 | 0 | 0 | 0 | 0 |
| 0.009597865 | 0.001005856 | 0 | 0 | 0.003192742 |
| 0 | 0 | 0 | 0 | 0 |
| 0 | 0 | 0.01150053 | 0 | 0.007004677 |
| 0.714479514 | 0 | 0 | 0 | 0 |
| 0 | 0 | 0 | 0 | 0 |
| 0 | 0 | 0 | 0 | 0 |
| 0.006440736 | 0.03122198 | 0.029274228 | 0.124312439 | 0.321447235 |
| 0 | 0 | 0 | 0 | 0 |
| 0 | 0 | 0.218553874 | 0 | 0 |
| 0 | 0.005547148 | 0.000682455 | 0.008795552 | 0 |

| | | | | |
|---|---|---|---|---|
| 0 | 0 | 4.210606533 | 0 | 0 |
| 0 | 0 | 0 | 0 | 0 |
| 0.00061679 | 0.030864099 | 0.004904001 | 0 | 0 |
| 0 | 0 | 0 | 0 | 0 |
| 0 | 0.000723715 | 0 | 0 | 0 |
| 0 | 0 | 0 | 0 | 0 |
| 0.070746555 | 0 | 0 | 0 | 0 |
| 0 | 0.006065533 | 0.000604092 | 0.000626427 | 0 |
| 0 | 0 | 0 | 0 | 0 |
| 0 | 0 | 0 | 0 | 0 |
| 0 | 0 | 0 | 0 | 0 |
| 0 | 0 | 0 | 0 | 0 |
| 0 | 0 | 0 | 0 | 0 |
| 0.131605664 | 0 | 0 | 0 | 0 |
| 0 | 0 | 0 | 0 | 0 |
| 0 | 0 | 0 | 0 | 0 |
| 0 | 0 | 0 | 0 | 0 |
| 0 | 0 | 0 | 0 | 0 |
| 1.31324372 | 1.341666602 | 0.340491689 | 0 | 0.020926196 |
| 0 | 0 | 0.000226901 | 0 | 0 |
| 0 | 0 | 0 | 0 | 0.001607879 |
| 0 | 0 | 0 | 0 | 0 |
| 0 | 0 | 0 | 0 | 0 |
| 0 | 0 | 0 | 0 | 0 |
| 0 | 0 | 0.002477683 | 0.001985362 | 0.000498522 |
| 0 | 0.064074733 | 0 | 0 | 0.002442466 |
| 0 | 0 | 0 | 0 | 0 |
| 0.006521079 | 0.342661135 | 0.33973643 | 0.062920749 | 0.107996766 |
| 0 | 0 | 0 | 0 | 0 |
| 0.137668617 | 0 | 1.475445615 | 0 | 0.253167258 |
| 0 | 0 | 0 | 0 | 0 |
| 0 | 0 | 0 | 0 | 0 |
| 0 | 0.018091028 | 0.00198359 | 0 | 0 |
| 0 | 0 | 0 | 0 | 0 |
| 0 | 0.007572897 | 0.000603373 | 0.023150242 | 0.051636213 |
| 0 | 0 | 0 | 0 | 0 |
| 0 | 0 | 0 | 0 | 0 |
| 0 | 0.003246252 | 0.000570543 | 0.003668158 | 0.025255163 |
| 0 | 0 | 0 | 0 | 0 |
| 0 | 0 | 0 | 0 | 0 |
| 0.084532645 | 6.822143996 | 5.448451165 | 3.230003753 | 1.520453364 |
| 0.116464616 | 0.16633899 | 1.175740857 | 0.764736161 | 3.651900948 |
| 0.005672069 | 0.182930663 | 0 | 0 | 0 |
| 0 | 0 | 0 | 0 | 0 |
| 0 | 0 | 0 | 0 | 0 |
| 0 | 0 | 0 | 0 | 0 |
| 0.001048834 | 0 | 0.000647447 | 0 | 0 |
| 0.010164754 | 0.004698151 | 0.001992385 | 0.007888557 | 0.668063001 |

| | | | | |
|---|---|---|---|---|
| 0 | 0 | 0 | 0 | 0 |
| 0.007006121 | 0.052377371 | 0.003260301 | 0 | 0 |
| 0 | 0 | 0 | 0 | 0 |
| 0 | 0 | 0 | 0 | 0 |
| 0.029524308 | 0 | 3.161979862 | 1.670431038 | 0.493336659 |
| 0 | 0 | 0 | 0 | 0 |
| 0 | 0 | 0 | 0 | 0 |
| 0 | 0 | 0 | 0.025808935 | 0 |
| 0.001722041 | 0.406104433 | 0.017765698 | 1.475541129 | 0.00051466 |
| 0 | 0 | 0 | 0 | 0 |
| 0 | 0 | 0 | 0 | 0 |
| 0 | 0 | 0 | 0 | 0 |
| 0.013510548 | 0.236745856 | 0.167538676 | 0 | 0.018129625 |
| 0.007081717 | 0.003724312 | 0.012240353 | 0.011885193 | 0.117722846 |
| 0 | 0 | 0 | 0 | 0 |
| 0.002113166 | 0.560038234 | 0.221432159 | 0 | 0.105696895 |
| 0.003119774 | 0.129879405 | 0.019836166 | 0.00339498 | 0 |
| 0 | 0 | 0 | 0 | 0 |
| 0 | 0 | 0 | 0 | 0 |
| 0 | 0 | 0 | 0 | 0 |
| 0 | 0 | 0 | 0 | 0 |
| 0 | 0 | 0 | 0 | 0 |
| 0 | 0 | 0 | 0 | 0 |
| 0 | 0 | 0 | 0 | 0 |
| 0 | 0 | 0.013791545 | 0 | 0 |
| 0 | 0 | 0 | 0 | 0 |
| 0.002034579 | 0.158754219 | 0.004605145 | 0 | 0.032430247 |
| 0 | 0.010277021 | 0.001228238 | 0.002449328 | 0 |
| 0 | 0 | 0 | 0 | 0 |
| 0 | 0 | 0 | 0 | 0 |
| 0 | 0 | 0 | 0 | 0 |
| 0 | 0 | 0.000394598 | 0 | 0 |
| 0 | 0 | 0.00478655 | 0 | 0 |
| 0 | 0 | 0.116174023 | 0 | 0 |
| 0 | 0.646100192 | 0 | 0.074460535 | 0 |
| 0 | 0.107178318 | 0.208750702 | 2.634008335 | 0.000483789 |
| 0 | 0.011964412 | 0 | 0 | 0 |
| 0 | 0 | 0 | 0 | 0 |
| 0 | 0 | 0 | 0 | 0 |
| 0 | 0 | 0 | 0 | 0 |
| 0.191904583 | 0 | 1.466665576 | 0 | 0.186642943 |
| 0 | 0.030667171 | 0 | 0 | 0 |
| 0 | 0 | 0 | 0 | 0 |
| 0 | 0 | 0 | 0 | 0 |
| 0 | 0 | 0 | 0 | 0 |
| 0 | 0 | 0 | 0 | 0 |
| 0 | 0 | 0 | 0 | 0 |
| 0 | 0 | 0 | 0 | 0 |

| | | | | |
|---|---|---|---|---|
| 0 | 0 | 0.020999654 | 0 | 0 |
| 0 | 0 | 0 | 0 | 0 |
| 0 | 0 | 0 | 0 | 0 |
| 0 | 0 | 0 | 0 | 0 |
| 0 | 0 | 0.004505777 | 0.003398089 | 0.000453293 |
| 0 | 0 | 0 | 0 | 0 |
| 0.014115996 | 0.110595226 | 0.357911824 | 0.048509848 | 0 |
| 0 | 0 | 0 | 0 | 0 |
| 0 | 0 | 0 | 0 | 0 |
| 0 | 0.054474509 | 0.02307871 | 0 | 0 |
| 0.008895946 | 0 | 0 | 0 | 0 |
| 0 | 0 | 0.015372818 | 0 | 0 |
| 0 | 0 | 0 | 0 | 0 |
| 0 | 0 | 0 | 0 | 0 |
| 0 | 0 | 0 | 0 | 0 |
| 0 | 0 | 0 | 0 | 0 |
| 0 | 0 | 0 | 0 | 0.066877759 |
| 0 | 0 | 0 | 0 | 0 |
| 0 | 0 | 0 | 0 | 0 |
| 0 | 0 | 0 | 0 | 0 |
| 0 | 0.00321388 | 0.091635402 | 0.001422508 | 0.078433007 |
| 0 | 0 | 0 | 0 | 0 |
| 0.000700318 | 0.018230904 | 0.029051074 | 0.014434992 | 0 |
| 0.003009864 | 0.004953394 | 0.003587849 | 0.000837114 | 0.007246005 |
| 0 | 0 | 0 | 0 | 0 |
| 0.005020326 | 0 | 0.001693142 | 0.00164601 | 0 |
| 0 | 0 | 0 | 0 | 0 |
| 0 | 0 | 0 | 0 | 0 |
| 0.092636979 | 0 | 0 | 0 | 0 |
| 0 | 0 | 0.001032751 | 0.004283744 | 0.317861084 |
| 0 | 0 | 0 | 0 | 0 |
| 0 | 0 | 0 | 0 | 0 |
| 0 | 0 | 0 | 0 | 0 |
| 0 | 0 | 0 | 0 | 0 |
| 0 | 0 | 0 | 0 | 0 |
| 0 | 0 | 0 | 0 | 0 |
| 0 | 0.104990009 | 0.206621356 | 0.016526372 | 0.017512257 |
| 0 | 0 | 0 | 0 | 0 |
| 0 | 0 | 0 | 0 | 0 |
| 0 | 0 | 0 | 0 | 0 |
| 0 | 0 | 0 | 0 | 0 |
| 0.311291704 | 0 | 0 | 0.00326966 | 0 |
| 0 | 0 | 0 | 0 | 0 |
| 0 | 0 | 0 | 0 | 0 |
| 0.001351994 | 0 | 0 | 0 | 0 |
| 0 | 0 | 0 | 0 | 0 |
| 0 | 0 | 0 | 0 | 0 |
| 0 | 0 | 0 | 0 | 0 |

| | | | | |
|---|---|---|---|---|
| 0.0005584 | 0.001292128 | 0.011002862 | 0.320872192 | 0 |
| 0 | 0 | 0 | 0 | 0 |
| 0 | 0 | 0.013993408 | 0.151292891 | 0 |
| 0 | 0 | 0 | 0 | 0 |
| 0 | 0 | 0 | 0 | 0 |
| 0 | 0 | 0 | 0 | 0 |
| 0 | 0 | 0 | 0 | 0.003859108 |
| 0.026259457 | 2.449628971 | 1.821169132 | 0.375501751 | 0.17886741 |
| 0 | 0.012676181 | 0.689765287 | 0.052470827 | 2.004096032 |
| 0.00305094 | 0.118252012 | 0.001054676 | 0.699293318 | 0 |
| 0 | 0 | 0.004826809 | 0 | 0 |
| 0 | 0 | 0 | 0 | 0 |
| 0 | 0 | 0 | 0 | 0 |
| 0.007656818 | 0 | 0 | 0 | 0.278885782 |
| 0 | 0 | 0 | 0 | 0 |
| 0 | 0 | 0 | 0 | 0 |
| 0 | 0 | 0 | 0 | 0 |
| 0.119120324 | 0 | 0 | 0 | 0 |
| 0 | 0 | 0 | 0 | 0 |
| 0.061021488 | 0 | 0 | 0 | 0 |
| 0 | 0 | 0 | 0 | 0 |
| 0 | 0 | 0 | 0 | 0 |
| 0 | 0 | 0 | 0 | 0 |
| 0 | 0 | 0 | 0 | 0 |
| 0 | 0 | 0.152724162 | 0 | 0 |
| 0 | 0 | 0 | 0 | 0 |
| 0 | 0 | 0 | 0 | 0 |
| 0 | 0 | 0 | 0 | 0 |
| 0 | 0 | 0 | 0 | 0 |
| 0 | 0 | 0 | 0 | 0 |
| 0 | 0 | 0 | 0 | 0 |
| 0 | 0 | 0 | 0 | 0 |
| 0 | 0 | 0 | 0 | 0 |
| 0 | 0 | 0 | 0 | 0 |
| 0 | 0 | 0 | 0.007391179 | 0 |
| 0 | 0 | 0 | 0 | 0 |
| 0 | 0 | 0 | 0 | 0 |
| 0 | 0 | 0 | 0.032367889 | 0 |
| 0.011818866 | 0 | 0.000573827 | 0 | 0.005207109 |
| 0 | 0.070624675 | 0 | 0.060748974 | 0 |
| 0 | 0 | 0 | 0 | 0 |
| 0.083967073 | 0 | 0 | 0.877839189 | 0 |
| 0 | 0 | 0 | 0 | 0 |
| 0 | 0 | 0 | 0 | 0 |
| 0 | 0 | 0 | 0 | 0 |
| 0 | 0 | 0.00291797 | 0 | 0.041123618 |
| 0 | 0 | 0 | 0 | 0 |
| 0.016640041 | 0 | 0 | 0.00298248 | 0 |

| | | | | |
|---|---|---|---|---|
| 0 | 0 | 0 | 0 | 0 |
| 0.00181433 | 0.006268336 | 0 | 0 | 0.000481993 |
| 0 | 0 | 0 | 0 | 0 |
| 0 | 0 | 0 | 0 | 0 |
| 0.16220709 | 0.040966675 | 0.396799383 | 0.410965358 | 0.553671202 |
| 0 | 0 | 0 | 0.012580847 | 0 |
| 0 | 0 | 0 | 0 | 0 |
| 0 | 0.263399784 | 0.000613879 | 0.39442262 | 0.009374828 |
| 0 | 0 | 0.001961393 | 0 | 0 |
| 0 | 0 | 0 | 0 | 0 |
| 0 | 0 | 0 | 0 | 0 |
| 0 | 0 | 0 | 0 | 0 |
| 0 | 0 | 0 | 0 | 0 |
| 0 | 0 | 0 | 0 | 0 |
| 0 | 0.004049826 | 0.016508093 | 0 | 0 |
| 0.045218788 | 0.048825147 | 0.160895061 | 0.04990964 | 0 |
| 0 | 0 | 0 | 0 | 0 |
| 0 | 0 | 0 | 0 | 0 |
| 0 | 0 | 0 | 0 | 0 |
| 0 | 0 | 0 | 0 | 0 |
| 0 | 0 | 0 | 0 | 0 |
| 0 | 0 | 0 | 0 | 0 |
| 0 | 0 | 0 | 0 | 0 |
| 0 | 0.013157848 | 0.000531262 | 0.040326194 | 0.000470327 |
| 0.374432119 | 0.682474583 | 0.014180216 | 0 | 0 |
| 0 | 0 | 0 | 0 | 0 |
| 0 | 0.045697986 | 0.052282732 | 0 | 1.412885095 |
| 0.000710423 | 0 | 0 | 0 | 0 |
| 0 | 0 | 0 | 0 | 0 |
| 0 | 0 | 0 | 0 | 0 |
| 0 | 0 | 0 | 0 | 0 |
| 0 | 0 | 0 | 0 | 0 |
| 0 | 0 | 0 | 0 | 0 |
| 0 | 0 | 0 | 0 | 0 |
| 0 | 0 | 0 | 0 | 0 |
| 0 | 0 | 0 | 0 | 0 |
| 0 | 0 | 0 | 0 | 0 |
| 0 | 0 | 0.017341463 | 0.161223681 | 0 |
| 0 | 0 | 0 | 0.002684595 | 0 |
| 0 | 0 | 0 | 0 | 0 |
| 0 | 0 | 0 | 0 | 0 |
| 0.042578892 | 0 | 0 | 0 | 0 |
| 0 | 0 | 0 | 0 | 0 |
| 0 | 0 | 0 | 0 | 0 |
| 0 | 0 | 0 | 0 | 0 |
| 0 | 0 | 0 | 0 | 0 |
| 0 | 0 | 0 | 0 | 0 |
| 0 | 0 | 0 | 0 | 0 |

| | | | | |
|---|---|---|---|---|
| 0.001586211 | 0.005658632 | 0.091389225 | 0.010045432 | 0.154330272 |
| 0 | 0 | 0 | 0 | 0 |
| 0 | 0 | 0 | 0 | 0 |
| 0 | 0.001369466 | 0 | 0 | 0 |
| 0 | 0 | 0 | 0 | 0 |
| 0 | 0 | 0 | 0.00636808 | 0.018629939 |
| 0 | 0 | 0 | 0 | 0 |
| 0 | 0 | 0 | 0.015783076 | 0 |
| 0 | 0 | 0 | 0 | 0 |
| 0 | 0 | 0 | 0 | 0 |
| 0 | 0 | 0 | 0 | 0 |
| 0 | 0.002294758 | 0 | 0 | 0 |
| 0 | 0 | 0 | 0 | 0 |
| 0 | 0 | 0 | 0 | 0 |
| 0 | 0 | 0 | 0 | 0 |
| 0 | 0 | 0 | 0 | 0 |
| 0 | 0 | 0 | 0 | 0 |
| 0 | 0 | 0 | 0 | 0 |
| 0 | 0 | 0.000309927 | 0 | 0 |
| 0 | 0 | 0 | 0 | 0 |
| 0 | 0.075429886 | 0.005184599 | 0 | 0 |
| 0 | 0 | 0 | 0 | 0 |
| 0.036800098 | 0.043606052 | 0.013095542 | 0.008314123 | 0.893321613 |
| 0.092269964 | 0 | 0.224915757 | 0 | 0 |
| 0 | 0 | 0 | 0 | 0 |
| 0 | 0 | 0 | 0 | 0 |
| 0 | 0 | 0 | 0 | 0 |
| 0 | 0 | 0 | 0 | 0 |
| 0.001338538 | 0 | 0 | 0 | 0 |
| 0 | 0 | 0 | 0 | 0 |
| 0 | 0 | 0 | 0 | 0 |
| 0 | 0 | 0 | 0 | 0 |
| 0.03026385 | 0.872443465 | 0.006757671 | 0.054373221 | 0.444954497 |
| 0 | 0.056914185 | 0 | 0 | 0 |
| 0 | 0.544179534 | 0.681820289 | 0 | 0.025022707 |
| 0.000697191 | 0.003865167 | 0.001506319 | 0.202020354 | 0 |
| 0 | 0 | 0 | 0 | 0 |
| 0 | 0 | 0 | 0 | 0 |
| 0 | 0.277765082 | 0.001277684 | 0 | 0 |
| 0 | 0 | 0 | 0 | 0 |
| 0 | 0 | 0 | 0 | 0 |
| 0 | 0 | 0 | 0 | 0 |
| 0.007700646 | 0.032729075 | 0.004481985 | 0.007746168 | 0.354106069 |
| 0 | 0 | 0 | 0 | 0 |
| 0 | 0 | 0 | 0 | 0 |
| 0.000374427 | 0.053790163 | 0.249918199 | 0.008836224 | 0.00238728 |
| 0 | 0 | 0 | 0 | 0 |
| 0 | 0 | 0 | 0 | 0 |

| | | | | |
|---|---|---|---|---|
| 0 | 0 | 0 | 0 | 0 |
| 0 | 0 | 0 | 0 | 0 |
| 0 | 0 | 0 | 0 | 0 |
| 0 | 0 | 0 | 0 | 0 |
| 0 | 0 | 0 | 0 | 0 |
| 0 | 0 | 0 | 0 | 0 |
| 0 | 0 | 0.817859385 | 0 | 0 |
| 0 | 0 | 0 | 0 | 0 |
| 0.012278134 | 0 | 0.002526437 | 0 | 0 |
| 0 | 0.004276092 | 0.000555488 | 0 | 0.010327264 |
| 0 | 0 | 0 | 0 | 0 |
| 0 | 0 | 0 | 0 | 0 |
| 0 | 0 | 0 | 0 | 0 |
| 0 | 0 | 0.074054461 | 0.995132111 | 0 |
| 0 | 0 | 0.155143238 | 0 | 0 |
| 0 | 0 | 0 | 0.001813587 | 0 |
| 0 | 0 | 0 | 0 | 0 |
| 0 | 0 | 0 | 0 | 0 |
| 0 | 0 | 0 | 0 | 0 |
| 0 | 0 | 1.926500303 | 0 | 0.044560558 |
| 0.000528402 | 0 | 0.002739939 | 0 | 0 |
| 0 | 0 | 0 | 0 | 0 |
| 0 | 0 | 0 | 0 | 0 |
| 0 | 0 | 0 | 0 | 0 |
| 0 | 0 | 0 | 0 | 0 |
| 0 | 0 | 0 | 0 | 0 |
| 0 | 0 | 0 | 0 | 0 |
| 0 | 0 | 0 | 0 | 0 |
| 0 | 0 | 0 | 0 | 0 |
| 0 | 0 | 0.383596668 | 0 | 0 |
| 0.018298438 | 6.170850788 | 0.094883494 | 0 | 1.008518117 |
| 0 | 0 | 0 | 0 | 0 |
| 0 | 0 | 0 | 0 | 0 |
| 0 | 0 | 0 | 0 | 0 |
| 0 | 0 | 0 | 0 | 0 |
| 0 | 0 | 0 | 0 | 0 |
| 0 | 0 | 0 | 0 | 0 |
| 0 | 0 | 0 | 0 | 0 |
| 0.013671464 | 0 | 0 | 0 | 0 |
| 0 | 0 | 0 | 0 | 0 |
| 0 | 0 | 0.000417817 | 0 | 0 |
| 0.702953596 | 0 | 0.43921575 | 0 | 0 |
| 0 | 0 | 0 | 0 | 0 |
| 0 | 0 | 0 | 0 | 0 |
| 0 | 0 | 0 | 0 | 0 |
| 0 | 0 | 0 | 0 | 0 |
| 0 | 1.027097601 | 0 | 0 | 0 |
| 0 | 0 | 0 | 0 | 0 |

| | | | | |
|---|---|---|---|---|
| 0 | 0 | 0 | 0 | 0 |
| 0 | 0 | 0 | 0 | 0 |
| 0 | 0 | 0 | 0 | 0 |
| 0 | 0 | 0 | 0 | 0 |
| 0.005707041 | 0.030916575 | 0.690501146 | 0.219923977 | 0.283818894 |
| 0 | 0 | 0 | 0 | 0 |
| 0 | 0 | 0 | 0 | 0 |
| 0 | 0 | 0 | 0 | 0 |
| 0 | 0 | 0 | 0 | 0 |
| 0 | 0 | 0 | 0 | 0 |
| 0 | 0 | 0.045639164 | 0 | 0 |
| 0.00389455 | 0.011459126 | 0.007892119 | 0.0271995 | 0.180709263 |
| 0.001969664 | 0 | 0 | 0.028138812 | 0 |
| 0 | 0 | 0 | 0 | 0 |
| 0 | 0 | 0 | 0 | 0 |
| 0 | 0.060263166 | 0.187366372 | 0.014693718 | 0 |
| 0.000516443 | 0.009859089 | 0.045614087 | 0 | 0 |
| 0 | 0 | 0 | 0 | 0 |
| 0.097824647 | 0.306486306 | 0.126901223 | 0.385119815 | 0 |
| 0.01035642 | 1.155023541 | 0.875173592 | 0.213585944 | 0 |
| 0 | 0 | 0 | 0 | 0 |
| 0 | 0 | 0 | 0 | 0 |
| 0 | 0 | 0 | 0 | 0 |
| 0 | 0 | 0 | 0 | 0 |
| 0 | 0 | 0 | 0 | 0 |
| 0 | 0 | 0 | 0 | 0 |
| 0.005426318 | 0 | 0 | 0.040715153 | 0.028695324 |
| 0 | 0 | 0 | 0 | 0 |
| 0 | 0 | 0 | 0 | 0 |
| 0 | 0 | 0 | 0 | 0 |
| 0 | 0.006628274 | 0 | 0.007566027 | 0 |
| 0 | 0 | 0 | 0 | 0 |
| 0 | 0 | 0 | 0 | 0 |
| 0 | 0 | 0 | 0 | 0 |
| 0 | 0 | 0 | 0 | 0 |
| 0 | 0 | 0 | 0 | 0 |
| 0 | 0 | 0 | 0 | 0 |
| 0.016354008 | 0.031617443 | 0.02138332 | 0.049207925 | 0.412083131 |
| 0 | 0.180270693 | 0.035320618 | 0 | 0 |
| 0 | 0.012441169 | 0.012777891 | 0 | 0 |
| 0 | 0.000489371 | 0.001827692 | 0.000227432 | 0.016342407 |
| 0 | 0 | 0 | 0 | 0 |
| 0 | 0 | 0 | 0 | 0 |
| 0.001507829 | 0.003271023 | 0.003257749 | 0.005517726 | 0.017424724 |
| 0 | 0 | 0 | 0 | 0 |
| 0 | 0.005198782 | 0 | 0 | 0.028147497 |
| 0 | 0 | 0 | 0 | 0 |
| 0 | 0 | 0 | 0 | 0 |

| | | | | |
|---|---|---|---|---|
| 0 | 0 | 0 | 0 | 0 |
| 0 | 0 | 0.704458056 | 0 | 0.07806 |
| 0 | 0 | 0 | 0 | 0 |
| 0 | 0.006475199 | 0.000910436 | 0.004366455 | 0 |
| 0 | 0 | 0.001372174 | 0.001094545 | 0.004672265 |
| 0 | 0 | 0.001089798 | 0.002542708 | 0 |
| 0.001150107 | 0 | 0 | 0 | 0 |
| 0 | 0 | 0 | 0 | 0 |
| 0 | 0 | 0 | 0 | 0 |
| 0 | 0 | 0 | 0 | 0 |
| 0 | 0.062160891 | 0.057363445 | 0 | 0.061919002 |
| 0 | 0 | 0 | 0 | 0 |
| 0.001586876 | 0 | 0.019441801 | 0 | 0.015176442 |
| 0 | 0 | 0 | 0 | 0 |
| 0 | 0 | 0 | 0 | 0 |
| 0 | 0 | 0 | 0 | 0 |
| 0 | 0 | 0 | 0 | 0 |
| 0 | 0 | 0 | 0 | 0 |
| 0 | 0 | 0 | 0 | 0 |
| 0 | 0 | 0 | 0 | 0 |
| 0 | 0 | 0 | 0 | 0 |
| 0 | 0 | 0 | 0 | 0 |
| 0 | 0.035730578 | 0.004673927 | 0.030732734 | 0 |
| 0.010298094 | 0 | 0 | 0.010268778 | 0 |
| 0 | 0 | 0 | 0 | 0 |
| 0 | 0 | 0.000833149 | 0 | 0 |
| 0 | 0 | 0 | 0 | 0.010557231 |
| 0 | 0 | 0 | 0 | 0 |
| 0 | 0 | 0 | 0 | 0 |
| 0 | 0 | 0 | 0 | 0 |
| 0 | 0 | 0.029136507 | 0 | 0.006946721 |
| 0.009220355 | 0 | 0 | 0 | 0 |
| 0 | 0 | 0 | 0 | 0 |
| 0 | 0 | 0 | 0 | 0 |
| 0 | 0.019442075 | 0.085694469 | 0 | 0 |
| 0 | 0 | 0 | 0 | 0 |
| 0 | 0.004211591 | 0 | 0 | 0 |
| 0 | 0 | 0.113251303 | 0 | 0 |
| 0 | 0.001173873 | 0.001002094 | 0.000727401 | 0.074521091 |
| 0 | 0 | 0 | 0 | 0 |
| 0 | 0 | 0 | 0 | 0 |
| 0 | 0 | 0 | 0 | 0 |
| 0 | 0 | 0 | 0 | 0 |
| 0 | 0 | 0 | 0 | 0 |
| 0.138419974 | 0 | 0 | 0 | 0 |
| 0.000612118 | 0.199628258 | 0.315199501 | 0.061530762 | 0.239141186 |
| 0 | 0 | 0 | 0 | 0 |
| 0 | 0 | 0 | 0 | 0 |

| | | | | |
|---|---|---|---|---|
| 0 | 0 | 0 | 0 | 0 |
| 0 | 0 | 0 | 0 | 0 |
| 0.167952501 | 0.30752136 | 0.31860272 | 0 | 0 |
| 0 | 0 | 0 | 0 | 0 |
| 0 | 0 | 0 | 0 | 0 |
| 0 | 0 | 0 | 0 | 0 |
| 0 | 0.018750631 | 0 | 0 | 0 |
| 0 | 0 | 0 | 0 | 0 |
| 0 | 0.000775818 | 0 | 0 | 0 |
| 0 | 0 | 0 | 0 | 0 |
| 0.001975452 | 0.001088372 | 0.000325187 | 0 | 0.016733515 |
| 0 | 0.001015589 | 0 | 0.00390178 | 0 |
| 0 | 0 | 0 | 0 | 0 |
| 0 | 0 | 0 | 0 | 0 |
| 0 | 0 | 0 | 0 | 0 |
| 0 | 0 | 0 | 0 | 0 |
| 0 | 0 | 0 | 0 | 0 |
| 0 | 0 | 0 | 0 | 0 |
| 0 | 0 | 0 | 0 | 0 |
| 0 | 0 | 0 | 0.050198011 | 0 |
| 0 | 0 | 0 | 0 | 0 |
| 0 | 0 | 0 | 0 | 0 |
| 0 | 0 | 0 | 0 | 0 |
| 0 | 0 | 0 | 0 | 0 |
| 0 | 0 | 0 | 0 | 0 |
| 0 | 0 | 0 | 0 | 0 |
| 0 | 0 | 0 | 0 | 0 |
| 0.001242531 | 0 | 0.003818037 | 0 | 0 |
| 0 | 0.007755741 | 0 | 0 | 0 |
| 0 | 0 | 0 | 0 | 0 |
| 0.008359631 | 0 | 0.002222947 | 0.007203556 | 0.041102086 |
| 0 | 0 | 0 | 0 | 0 |
| 0.11299225 | 0.043553025 | 0.009894094 | 0.004237791 | 0.01440042 |
| 0.00178031 | 0.018207191 | 0.008791905 | 0.009003013 | 0.001945873 |
| 0 | 0 | 0 | 0 | 0 |
| 0 | 0 | 0 | 0 | 0 |
| 0 | 0 | 0 | 0 | 0 |
| 0 | 0 | 0 | 0 | 0 |
| 0 | 0 | 0 | 0 | 0 |
| 0 | 0 | 0 | 0 | 0 |
| 0 | 0 | 0 | 0 | 0 |
| 0 | 0.311815737 | 0.570846411 | 0.411669549 | 0.186518317 |
| 0.005113652 | 0.025433527 | 0.018000692 | 0.054992769 | 0.257410207 |
| 0 | 0 | 0 | 0 | 0 |
| 0 | 0 | 0 | 0 | 0 |
| 0.008651219 | 0.030236687 | 0 | 0.020351657 | 0.042058414 |
| 0 | 0.004443317 | 0 | 0 | 0 |
| 0 | 0 | 0 | 0 | 0 |

| | | | | |
|---|---|---|---|---|
| 0 | 0 | 0 | 0 | 0 |
| 0.002743946 | 0.02758985 | 0.009937209 | 0 | 0.003249052 |
| 0 | 0.012802848 | 0.006859101 | 0.017234637 | 0 |
| 0 | 0 | 0 | 0 | 0 |
| 0 | 0 | 0.297214229 | 0.494960165 | 0.531586879 |
| 0 | 0.026591798 | 0.303864148 | 0 | 0 |
| 0 | 0 | 0 | 0 | 0 |
| 0 | 0 | 0 | 0 | 0 |
| 0 | 0 | 0 | 0 | 0 |
| 0 | 0 | 0 | 0 | 0 |
| 0 | 0 | 0 | 0 | 0 |
| 0 | 0 | 0 | 0 | 0 |
| 0 | 0 | 0 | 0 | 0 |
| 0.00410525 | 0 | 0 | 0 | 0 |
| 0 | 0 | 0 | 0 | 0 |
| 0 | 0 | 0 | 0 | 0 |
| 0 | 0 | 0 | 0 | 0 |
| 0 | 0 | 0 | 0 | 0 |
| 0 | 0 | 0 | 0 | 0 |
| 0 | 0 | 0 | 0 | 0 |
| 0 | 0 | 0 | 0 | 0 |
| 0 | 0 | 0 | 0 | 0 |
| 0 | 0 | 0 | 0 | 0 |
| 0 | 0 | 0 | 0 | 0 |
| 0.003899433 | 0.001637278 | 0 | 0 | 0.006736817 |
| 0 | 0 | 0 | 0 | 0 |
| 0 | 0 | 0 | 0 | 0 |
| 0.028477566 | 0.001597708 | 0.00318245 | 0.116054102 | 0.02300901 |
| 0 | 0 | 0 | 0 | 0 |
| 0 | 0 | 0 | 0 | 0 |
| 0 | 0 | 0 | 0 | 0 |
| 0 | 0 | 0 | 0.00188949 | 0.013584221 |
| 0 | 0 | 0 | 0 | 0 |
| 0 | 0 | 0 | 0 | 0 |
| 0 | 0 | 0 | 0 | 0 |
| 0 | 0 | 0 | 0 | 0 |
| 0 | 0 | 0 | 0 | 0 |
| 0 | 0.327902535 | 0 | 0 | 0 |
| 0 | 0 | 0 | 0 | 1.204240793 |
| 0 | 0 | 0 | 0 | 0 |
| 0 | 0 | 0 | 0 | 0 |
| 0 | 0.013698871 | 0 | 0 | 0.002625745 |
| 0.000507986 | 0.001285672 | 0.011524095 | 0.252091976 | 0 |
| 0 | 0 | 0 | 0 | 0 |
| 0 | 0 | 0 | 0 | 0 |
| 0 | 0.002324332 | 0 | 0 | 0 |
| 0.00174852 | 0.014048765 | 0.000503703 | 0.025419911 | 0.013192082 |
| 0 | 0 | 0 | 0 | 0 |

| | | | | |
|---|---|---|---|---|
| 0 | 0 | 0 | 0 | 0 |
| 0.003309465 | 0.102107277 | 0.060274238 | 0.17015262 | 0.054744178 |
| 0 | 0 | 0 | 0 | 0 |
| 0 | 0 | 0 | 0 | 0 |
| 0 | 0 | 0 | 0 | 0 |
| 0 | 0 | 0 | 0 | 0 |
| 0 | 0 | 0 | 0 | 0 |
| 0 | 0 | 0 | 0 | 0 |
| 0 | 0 | 0 | 0 | 0 |
| 0 | 0 | 0 | 0 | 0 |
| 0 | 0 | 0 | 0 | 0 |
| 0 | 0 | 0 | 0 | 0 |
| 0 | 0 | 0.00179164 | 0 | 0 |
| 0 | 0 | 0 | 0 | 0 |
| 0.001528298 | 0.031165011 | 0.018226888 | 0.010135214 | 0 |
| 0 | 0 | 0 | 0 | 0 |
| 0 | 0 | 0 | 0 | 0 |
| 0 | 0 | 0 | 0 | 0 |
| 0 | 0 | 0 | 0 | 0 |
| 0 | 0 | 0 | 0 | 0 |
| 0 | 0 | 0 | 0 | 0 |
| 0.001200727 | 0.293475098 | 0.015176292 | 0.022059285 | 0 |
| 0.009457272 | 0.024018986 | 0.027908434 | 0 | 0 |
| 0 | 0.00134147 | 0.050902606 | 0 | 0.001034939 |
| 0 | 0 | 0 | 0 | 0 |
| 0 | 0 | 0 | 0 | 0 |
| 0 | 0 | 0 | 0 | 0 |
| 0 | 0 | 0 | 0 | 0 |
| 0 | 0 | 0 | 0.001523915 | 0 |
| 0 | 0 | 0 | 0 | 0 |
| 0 | 0 | 0 | 0 | 0 |
| 0 | 0 | 0 | 0 | 0 |
| 0 | 0 | 0 | 0 | 0 |
| 0 | 0 | 0 | 0 | 0 |
| 61.68901514 | 23.34847492 | 0 | 19.20694165 | 8.642759257 |
| 0 | 0 | 0 | 0 | 0 |
| 0 | 0 | 0 | 0 | 0 |
| 0 | 0 | 0 | 0 | 0 |
| 0.002548027 | 0.001734144 | 0.084714599 | 0 | 0.027092177 |
| 0 | 0 | 0 | 0 | 0 |
| 0.000537563 | 0 | 0.056213449 | 0.144404862 | 0.030718101 |
| 0 | 0 | 0 | 0 | 0 |
| 0 | 0 | 0 | 0 | 0 |
| 0 | 0 | 0 | 0 | 0 |
| 0 | 0 | 0 | 0 | 0 |
| 0.001692551 | 3.638816627 | 0.046408761 | 6.074739389 | 0.007799235 |
| 0 | 0 | 0 | 0 | 0 |
| 0 | 0 | 0 | 0 | 0 |

| | | | | |
|---|---|---|---|---|
| 0 | 0 | 0 | 0 | 0 |
| 0 | 0 | 0 | 0 | 0 |
| 0 | 0 | 0 | 0 | 0 |
| 0 | 0 | 0 | 0.009360197 | 0 |
| 0 | 0.00135018 | 0 | 0.002405373 | 0 |
| 0 | 0 | 0 | 0 | 0 |
| 0 | 0 | 0 | 0 | 0 |
| 0 | 0 | 0.001581016 | 0 | 0 |
| 0.001250822 | 0 | 0 | 0 | 0 |
| 0 | 0.00174108 | 0.004941943 | 0.000404579 | 0.045190312 |
| 0.142518234 | 0 | 0 | 0.016071844 | 0 |
| 0 | 0 | 0 | 0 | 0 |
| 0 | 0.002978079 | 0.002415166 | 0.01014967 | 0 |
| 0 | 0 | 0 | 0 | 0 |
| 0 | 0 | 0 | 0 | 0 |
| 0 | 0 | 0 | 0 | 0 |
| 0 | 0 | 0 | 0 | 0 |
| 0 | 0 | 0 | 0 | 0 |
| 0 | 0 | 0 | 0 | 0 |
| 0.026837766 | 0.216600109 | 0.171973483 | 0.182203667 | 5.121985626 |
| 0 | 0 | 0 | 0 | 0 |
| 0 | 0 | 0 | 0 | 0 |
| 0 | 0 | 0 | 0 | 0 |
| 0 | 0 | 0 | 0 | 0 |
| 0 | 0 | 0 | 0 | 0 |
| 0 | 0 | 0 | 0.006800768 | 0 |
| 0 | 0 | 0 | 0 | 0 |
| 0 | 0 | 0 | 0 | 0 |
| 0 | 0 | 0 | 0 | 0 |
| 0 | 0.124477837 | 0 | 0 | 0 |
| 0 | 0 | 0 | 0 | 0 |
| 0 | 0 | 0 | 0 | 0 |
| 0 | 0 | 0 | 0 | 0 |
| 0 | 0 | 0 | 0 | 0 |
| 0 | 0 | 0 | 0 | 0 |
| 0 | 0 | 0.004100816 | 0 | 0 |
| 0 | 0 | 0 | 0 | 0 |
| 0 | 0 | 0 | 0 | 0 |
| 0 | 0 | 0 | 0 | 0 |
| 0 | 0 | 0 | 0 | 0 |
| 0 | 0 | 0 | 0 | 0 |
| 0 | 0 | 0 | 0 | 0 |
| 0 | 0.011488984 | 0.043167483 | 0 | 0 |
| 0 | 0 | 0 | 0 | 0 |
| 0.001598494 | 0.010492035 | 0.004038095 | 0.007162656 | 0.014581972 |
| 0 | 0 | 0.026966954 | 0 | 0 |
| 0 | 0 | 0 | 0 | 0 |
| 0 | 0 | 0 | 0 | 0 |

| | | | | |
|---|---|---|---|---|
| 0 | 0 | 0 | 0 | 0 |
| 0 | 0 | 0 | 0 | 0 |
| 0 | 0 | 0 | 0 | 0 |
| 0 | 0 | 0 | 0 | 0 |
| 0 | 0 | 0 | 0 | 0 |
| 0.005691303 | 0.011417174 | 0.02403743 | 0 | 0.143800666 |
| 0 | 0 | 0 | 0 | 0 |
| 0 | 0 | 0 | 0 | 0 |
| 0 | 0 | 0 | 0 | 0 |
| 0 | 0 | 0 | 0 | 0 |
| 0 | 0 | 0 | 0 | 0 |
| 0 | 0 | 0 | 0 | 0 |
| 0 | 0 | 0 | 0 | 0 |
| 0 | 0 | 0 | 0 | 0 |
| 0 | 0 | 0 | 0 | 0 |
| 0 | 0 | 0 | 0 | 0 |
| 0.014141062 | 0 | 0.006158624 | 0 | 0 |
| 0 | 0 | 0 | 0 | 0 |
| 0 | 0 | 0 | 0 | 0 |
| 0 | 0 | 0 | 0 | 0 |
| 0 | 0 | 0 | 0 | 0 |
| 0 | 0 | 0 | 0 | 0 |
| 0 | 0 | 0 | 0 | 0 |
| 0 | 0 | 0 | 0 | 0 |
| 0 | 0 | 0 | 0 | 0 |
| 0 | 0 | 0.000992702 | 0 | 0.000961232 |
| 0 | 0 | 0 | 0 | 0 |
| 0 | 0 | 0.003250488 | 0 | 0 |
| 0 | 0 | 0 | 0 | 0 |
| 0 | 0.14280095 | 0 | 0 | 0 |
| 0 | 0 | 0 | 0 | 0 |
| 0 | 0 | 0.003138613 | 0 | 0 |
| 0 | 0 | 0 | 0 | 0 |
| 0 | 0 | 0 | 0 | 0 |
| 0 | 0 | 0 | 0 | 0 |
| 0 | 0 | 0 | 0 | 0 |
| 0 | 0 | 0 | 0 | 0 |
| 0 | 0 | 0 | 0 | 0 |
| 0.017752072 | 0 | 0.891612286 | 0 | 3.925664837 |
| 0 | 0 | 0 | 0 | 0 |
| 0 | 0 | 0 | 0 | 0 |
| 0 | 0 | 0 | 0 | 0 |
| 0 | 0 | 0 | 0 | 0 |
| 0 | 0 | 0 | 0 | 0 |
| 0 | 0 | 0 | 0 | 0 |
| 0 | 0 | 0 | 0 | 0 |
| 0 | 0 | 0 | 0 | 0 |
| 0 | 0 | 0 | 0 | 0 |

| | | | | |
|---|---|---|---|---|
| 0 | 0 | 0 | 0 | 0 |
| 0.004419202 | 0 | 0.266986773 | 0 | 0.003145229 |
| 0 | 0 | 0.027946097 | 0 | 0 |
| 0 | 0 | 0 | 0 | 0 |
| 0 | 0 | 0 | 0 | 0 |
| 0 | 0 | 0 | 0 | 0 |
| 0 | 0 | 0 | 0 | 0 |
| 0 | 0.138244254 | 0 | 0 | 0 |
| 0 | 0 | 0 | 0 | 0 |
| 0 | 0 | 0 | 0 | 0 |
| 0 | 0 | 0.0006978 | 0.000844201 | 0.016602416 |
| 0 | 0 | 0 | 0 | 0 |
| 0.048667456 | 0 | 0.097053782 | 0.010925855 | 0.105980497 |
| 0 | 0 | 0 | 0 | 0 |
| 0 | 0 | 0 | 0 | 0 |
| 0 | 0 | 0 | 0 | 0 |
| 0 | 0 | 0 | 0 | 0 |
| 0 | 0 | 0 | 0 | 0 |
| 0 | 0 | 0 | 0 | 0 |
| 0 | 0 | 0 | 0 | 0 |
| 0 | 0.092049396 | 0.064566553 | 0.007344112 | 0.003265592 |
| 0 | 0 | 0 | 0 | 0 |
| 0 | 0 | 0 | 0 | 0 |
| 0.286595879 | 0.752555105 | 0.859115094 | 1.141636189 | 0.172700911 |
| 0 | 0 | 0 | 0 | 0 |
| 0 | 0 | 0 | 0 | 0 |
| 0 | 0 | 0 | 0 | 0 |
| 0 | 0 | 0 | 0 | 0 |
| 0 | 0 | 0 | 0 | 0 |
| 0 | 0 | 0 | 0 | 0 |
| 0 | 0 | 0 | 0 | 0 |
| 0 | 0 | 0 | 0 | 0 |
| 0 | 0 | 0 | 0 | 0 |
| 0 | 0 | 0 | 0 | 0 |

| PHL018-0122_V1 | PHL018-0221_V3 | PHL018-0110_V1 | PHL018-0169_V2 | PHL018-0084_V1 |
|---|---|---|---|---|
| CTRL | CTRL | EYCF | REF | EYCF |
| 0.032201821 | 0.01133769 | 0 | 0 | 0.472868525 |
| 0.473642369 | 3.079922588 | 1.852372187 | 0.849244175 | 2.129543059 |
| 10.64749704 | 0.715401338 | 3.032874596 | 2.478489948 | 0 |
| 0 | 0 | 0 | 0 | 0 |
| 0 | 0 | 0 | 0 | 0 |
| 0 | 0 | 0 | 0 | 0 |
| 0 | 0 | 1.443683228 | 0 | 0 |
| 0 | 0 | 0 | 0 | 0 |
| 0 | 0 | 0 | 0 | 0 |
| 0 | 0 | 0 | 0 | 0 |
| 0 | 0 | 0 | 0 | 0 |
| 0 | 0 | 0 | 0 | 0 |
| 0 | 0 | 0 | 0 | 0 |
| 53.46632135 | 2.904244951 | 4.494103941 | 0.055346638 | 0 |
| 0 | 0 | 0 | 0 | 0 |
| 0 | 0 | 0 | 0 | 0 |
| 0.000680572 | 0 | 0.000488523 | 0 | 0 |
| 0 | 0 | 0 | 0 | 0 |
| 0 | 0 | 0 | 0 | 0 |
| 0 | 0 | 0 | 0 | 0 |
| 0 | 0 | 0 | 0 | 0 |
| 0 | 0 | 0 | 0 | 0 |
| 0 | 0 | 0 | 0 | 0 |
| 0 | 0 | 0 | 0 | 0 |
| 0 | 0 | 0 | 0 | 0.034842937 |
| 0 | 0 | 0 | 0 | 0 |
| 0 | 0 | 0 | 0 | 0 |
| 0 | 0 | 0 | 0 | 0 |
| 0 | 0 | 0 | 0 | 0 |
| 0 | 0 | 0 | 0 | 0 |
| 0 | 0 | 0 | 0 | 0 |
| 0 | 0 | 0 | 0 | 0 |
| 0 | 0 | 0 | 0 | 0 |
| 0 | 0 | 0 | 0 | 0 |
| 0 | 0 | 0 | 0 | 0 |
| 0 | 0 | 0 | 1.431551793 | 4.357041546 |
| 0 | 0 | 0 | 0 | 0 |
| 0 | 0 | 0 | 0 | 0 |
| 0 | 0 | 0 | 0 | 0 |
| 0 | 0 | 0 | 0 | 0 |
| 0 | 0 | 0 | 0 | 0 |
| 0 | 0.010011374 | 0 | 0.05128248 | 0.007798026 |
| 0.000369848 | 0.028779175 | 2.247170757 | 0.228176804 | 0.023451787 |
| 0 | 0.156079851 | 0.021915652 | 0.001481269 | 0 |
| 0 | 0 | 0.033586214 | 0.01199382 | 0 |
| 0 | 0 | 0 | 0 | 0 |

| | | | | |
|---|---|---|---|---|
| 0 | 0 | 0 | 0 | 0 |
| 0.069462052 | 0.004058724 | 0.086740017 | 0.00117193 | 0.005813906 |
| 0 | 0.005369437 | 0 | 0 | 0 |
| 0.089757376 | 0.493363047 | 0 | 0.223500572 | 1.78083982 |
| 0.000228614 | 0.010023382 | 0 | 0 | 0.004014341 |
| 0 | 0 | 0 | 0 | 0.029060451 |
| 0 | 0 | 0 | 0 | 0.00386787 |
| 0 | 0 | 0 | 0 | 0 |
| 0 | 0 | 0 | 0 | 0 |
| 0 | 0 | 0 | 0 | 0 |
| 0 | 0 | 0 | 0.006347723 | 0.00296152 |
| 0 | 0.193171673 | 0 | 1.102776226 | 0.777951547 |
| 0 | 0 | 0.00136976 | 0 | 0 |
| 0.023268457 | 0.253730082 | 0.03736808 | 0.030136394 | 0.072636791 |
| 0 | 0 | 0 | 0 | 0 |
| 0 | 0 | 0 | 0 | 0 |
| 0 | 3.297618453 | 0 | 1.4837514 | 0.063923595 |
| 0 | 0 | 0 | 0 | 0 |
| 0 | 0 | 0 | 0 | 0 |
| 0 | 0 | 0 | 0 | 0 |
| 0.125938488 | 0.215176419 | 0.023452619 | 0.123577178 | 0 |
| 0.002425919 | 0.013353247 | 0.010223443 | 0.000669386 | 0.007328665 |
| 0 | 0 | 0 | 0 | 0.028598583 |
| 0 | 0 | 0 | 0 | 0 |
| 0 | 0 | 0 | 0 | 0 |
| 0 | 0 | 0 | 0 | 0 |
| 0.000934397 | 0 | 0.003219468 | 0 | 0 |
| 0 | 0 | 0 | 0 | 0 |
| 0 | 0 | 0 | 0.01070423 | 0 |
| 0.000882611 | 0 | 0 | 0 | 0 |
| 0.069252869 | 0.173302587 | 0 | 0 | 0 |
| 0 | 0 | 0 | 0 | 0 |
| 0 | 0 | 0 | 0 | 0 |
| 0 | 0 | 0.003843333 | 0 | 0 |
| 0 | 0 | 0 | 0 | 0 |
| 0 | 0 | 0 | 0 | 0 |
| 0 | 0 | 0 | 0 | 0 |
| 0 | 0.077812597 | 0 | 0.007981727 | 0 |
| 0 | 0 | 0 | 0 | 0 |
| 0 | 0 | 0 | 0 | 0 |
| 0 | 0 | 0 | 0 | 0 |
| 0 | 0 | 0 | 0 | 0 |
| 0 | 0 | 0 | 0 | 0 |
| 0 | 0 | 0 | 0 | 0 |
| 0 | 0 | 0 | 0 | 0.004054315 |
| 0 | 0 | 0 | 0 | 0 |
| 0 | 0 | 0 | 0.000413795 | 0 |
| 0.025300262 | 0 | 0 | 0 | 0 |

| | | | | |
|---|---|---|---|---|
| 0 | 0 | 0 | 0 | 0 |
| 0.03443425 | 0 | 0.005427198 | 0 | 0 |
| 0 | 0 | 0 | 0 | 0 |
| 0 | 0 | 0 | 0 | 0 |
| 0 | 0.013651383 | 0 | 0 | 0 |
| 0 | 0 | 0 | 0.00095989 | 0 |
| 0 | 0 | 0 | 0 | 0 |
| 0 | 0 | 0 | 0 | 0.235457424 |
| 0 | 0.017448298 | 0 | 0.031164515 | 0 |
| 0 | 0 | 0 | 0 | 0 |
| 0 | 0 | 0 | 0 | 0 |
| 0 | 0.00110998 | 0 | 0.000625976 | 0.006853391 |
| 0 | 0 | 0 | 0 | 0 |
| 0 | 0 | 0 | 0 | 0 |
| 0 | 0 | 0 | 0 | 0 |
| 0.002524969 | 0 | 0 | 0 | 0 |
| 0 | 0 | 0 | 0 | 0 |
| 0 | 0 | 0 | 0 | 0 |
| 0 | 0 | 6.25481078 | 4.914861548 | 0.001524799 |
| 0 | 0 | 0 | 0 | 0 |
| 0 | 0 | 0 | 0 | 0.000559166 |
| 0 | 0 | 0 | 0 | 0 |
| 0 | 0 | 0 | 0 | 0 |
| 0 | 0 | 0 | 0 | 0 |
| 0 | 0 | 0 | 0 | 0 |
| 0 | 0 | 0 | 0 | 0 |
| 0 | 0 | 0 | 0 | 0 |
| 0 | 0 | 0 | 0 | 0 |
| 0 | 0 | 0 | 0 | 0 |
| 0 | 0 | 0 | 0 | 0 |
| 0.006019575 | 0 | 0 | 0 | 0.02307933 |
| 0 | 0.013574175 | 0 | 0.001674981 | 0.012321015 |
| 0 | 0 | 0 | 0 | 0 |
| 0 | 0 | 0 | 0 | 0 |
| 0 | 0 | 0 | 0 | 0 |
| 0 | 0 | 0 | 0 | 0 |
| 0 | 0 | 0 | 0 | 0 |
| 0 | 0 | 0 | 0 | 0 |
| 0 | 0 | 0 | 0.269730504 | 0 |
| 0 | 0 | 0 | 0 | 0 |
| 0 | 0 | 0 | 0 | 0 |
| 0 | 0 | 0 | 0 | 0 |
| 0.002546236 | 0.001743289 | 0 | 0.000726005 | 0.000662379 |
| 0 | 0 | 0 | 0 | 0 |
| 0 | 0 | 0 | 0 | 0 |
| 0 | 0 | 0 | 0 | 0 |
| 0 | 0 | 0 | 0 | 0 |
| 0 | 0 | 0 | 0 | 0 |

| | | | | |
|---|---|---|---|---|
| 0 | 0.008074292 | 0 | 0 | 0.001608175 |
| 0 | 0 | 0 | 0 | 0 |
| 0 | 0 | 0 | 0 | 0.060101151 |
| 0 | 0 | 0 | 0 | 0 |
| 0 | 0 | 0 | 0 | 0.075193043 |
| 0 | 0 | 0 | 0 | 0 |
| 0 | 0 | 0 | 0 | 0 |
| 0 | 0 | 0 | 0 | 0 |
| 0 | 0 | 0 | 0 | 0 |
| 0 | 0 | 0 | 0 | 0 |
| 0 | 0 | 0 | 0 | 0 |
| 0 | 0 | 0 | 0 | 0 |
| 0 | 0 | 0 | 0 | 0 |
| 0 | 0 | 0 | 0 | 0 |
| 0 | 0 | 0 | 0 | 0 |
| 0 | 0 | 0 | 0 | 0 |
| 0 | 0 | 0 | 0 | 0 |
| 0 | 0 | 0 | 0 | 0 |
| 0 | 0 | 0 | 0 | 0 |
| 0 | 0 | 0 | 0 | 0 |
| 0 | 0 | 0 | 0 | 0 |
| 0 | 0 | 0 | 0 | 0 |
| 0 | 0 | 0 | 0 | 0 |
| 0 | 0 | 0 | 0 | 0 |
| 0 | 0 | 0 | 0.113177456 | 0 |
| 0 | 0 | 0 | 0 | 0 |
| 0 | 0 | 0 | 0 | 0.011672024 |
| 0 | 0 | 0 | 0 | 0 |
| 0 | 0 | 0 | 0 | 0 |
| 0 | 0 | 0 | 0 | 0 |
| 0.015688303 | 0 | 0 | 0 | 0 |
| 0 | 0.000669302 | 0 | 0 | 0.005124297 |
| 0 | 0 | 0 | 0 | 0 |
| 0 | 0 | 0 | 0 | 0 |
| 0 | 0 | 0 | 0 | 0 |
| 0 | 0 | 0 | 0 | 0 |
| 0 | 0 | 0 | 0 | 0 |
| 0 | 0 | 0 | 0 | 0 |
| 0 | 0 | 0 | 0 | 0 |
| 0 | 0 | 0 | 0 | 0 |
| 0 | 0 | 0 | 0 | 0 |
| 0 | 0 | 0 | 0 | 0 |
| 0 | 0 | 0 | 0 | 0 |
| 0 | 0 | 0 | 0 | 0 |
| 0 | 0 | 0 | 0 | 0 |
| 0 | 0 | 0 | 0 | 0 |
| 0 | 0 | 0 | 0 | 0 |
| 0 | 0 | 0 | 0 | 0 |

| | | | | |
|---|---|---|---|---|
| 0 | 0 | 0 | 0 | 0.218306478 |
| 0 | 0 | 0 | 0 | 0 |
| 0.001007197 | 0 | 0 | 0 | 0 |
| 0 | 0 | 0 | 0 | 0 |
| 0 | 0 | 0 | 0 | 0 |
| 0 | 0 | 0 | 0 | 0 |
| 0 | 0 | 0 | 0 | 0 |
| 0 | 0 | 0 | 0 | 0 |
| 0 | 0 | 0 | 0 | 0 |
| 0 | 0 | 0 | 0 | 0 |
| 0 | 0 | 0 | 0 | 0 |
| 0 | 0 | 0 | 0 | 0 |
| 0 | 0.001665187 | 0.0064014 | 0 | 0.001423582 |
| 0 | 0 | 0 | 0 | 0 |
| 0 | 0 | 0 | 0 | 0 |
| 0 | 0 | 0 | 0 | 0 |
| 0 | 0 | 0 | 0 | 0 |
| 0 | 0 | 0 | 0 | 0 |
| 0 | 0 | 0 | 0 | 0 |
| 0 | 0 | 0 | 0 | 0 |
| 0.021979853 | 0.055563985 | 0.006310969 | 0.024852241 | 0.013604535 |
| 0 | 0 | 0 | 0 | 0 |
| 0 | 0.000525366 | 0 | 0 | 0 |
| 0 | 0 | 0 | 0 | 0 |
| 0 | 0 | 0 | 0 | 0 |
| 0 | 0 | 0 | 0 | 0 |
| 0.001059356 | 0.000697396 | 0.001647575 | 0 | 0.002239097 |
| 0 | 0 | 0 | 0 | 0 |
| 0 | 0 | 0 | 0 | 0 |
| 0.005572494 | 0.015221112 | 0.019932244 | 0.000403952 | 0 |
| 0.017951962 | 0 | 0.002274027 | 0.136621977 | 0 |
| 0 | 0 | 0 | 0 | 0 |
| 0 | 0 | 0 | 0 | 0 |
| 0 | 0 | 0.002393921 | 0.008980817 | 0.048801013 |
| 0.001972588 | 0.046437764 | 0 | 0.08028852 | 0.004041055 |
| 0 | 0 | 0 | 0 | 0.004117135 |
| 0 | 0 | 0 | 0 | 0 |
| 0 | 0 | 0 | 0 | 0 |
| 0 | 0 | 0 | 0 | 0.000746069 |
| 0 | 0 | 0 | 0 | 0 |
| 0 | 0 | 0 | 0 | 0 |
| 0 | 0 | 0 | 0 | 0 |
| 0 | 0 | 0 | 0.002192231 | 0 |
| 0 | 0 | 0 | 0 | 0 |
| 0 | 0 | 0 | 0 | 0 |
| 0 | 0 | 0 | 0 | 0 |
| 0 | 0.000674896 | 0 | 0 | 0 |
| 0 | 0.000777348 | 0 | 0.001202433 | 0.000685658 |

| | | | | |
|---|---|---|---|---|
| 0 | 0 | 0 | 0 | 0 |
| 0 | 0 | 0 | 0 | 0 |
| 0 | 0.000608849 | 0.000922043 | 0 | 0 |
| 0 | 0 | 0 | 0 | 0 |
| 0 | 0 | 0 | 0 | 0 |
| 0 | 0 | 0 | 0 | 0 |
| 0 | 0 | 0 | 0.024533624 | 0 |
| 0 | 0.007023982 | 0 | 0.009506857 | 0 |
| 0 | 0 | 0 | 0 | 0.000540454 |
| 0 | 0 | 0 | 0 | 0 |
| 0 | 0 | 0 | 0 | 0 |
| 0 | 0 | 0 | 0 | 0 |
| 0 | 0 | 0 | 0 | 0 |
| 0 | 0 | 0 | 0 | 0 |
| 0 | 0.000700354 | 0 | 0.009637177 | 0.001513478 |
| 0 | 0 | 0 | 0 | 0 |
| 0 | 0 | 0 | 0 | 0 |
| 0 | 0 | 0 | 0 | 0.001858559 |
| 0 | 0 | 0.015085005 | 0.000509528 | 0.02705563 |
| 0 | 0 | 0 | 0 | 0.173401507 |
| 0.000738086 | 0.005497578 | 0 | 0 | 0 |
| 0 | 0 | 0 | 0 | 0 |
| 0 | 0 | 0 | 0 | 0 |
| 0 | 0 | 0 | 0 | 0 |
| 0.001256883 | 0.001323891 | 0.007819126 | 0.009855269 | 0 |
| 0 | 0 | 0 | 0 | 0 |
| 0 | 0 | 0 | 0 | 0 |
| 0 | 0 | 0 | 0 | 0 |
| 0 | 0 | 0 | 0 | 0 |
| 0 | 0 | 0 | 0 | 0 |
| 0.177467552 | 1.324426359 | 0.254554394 | 0.144574545 | 0.236126727 |
| 0 | 0 | 0 | 0 | 0 |
| 0 | 0 | 0 | 0 | 0 |
| 0 | 0 | 0 | 0.000429007 | 0 |
| 0 | 0 | 0 | 0 | 0 |
| 0 | 0 | 0 | 0 | 0 |
| 0 | 0 | 0 | 0 | 0 |
| 0 | 0 | 0 | 0 | 0 |
| 0 | 0 | 0 | 0 | 0 |
| 0 | 0 | 0 | 0 | 0 |
| 0 | 0 | 0 | 0 | 0 |
| 0.001185443 | 0.005868621 | 0 | 0.00664741 | 0.001541909 |
| 0 | 0.000628594 | 0 | 0 | 0 |
| 0 | 0 | 0 | 0 | 0 |
| 0 | 0 | 0 | 0 | 0 |
| 0 | 0 | 0 | 0 | 0 |
| 0 | 0 | 0 | 0 | 0 |
| 0 | 0 | 0 | 0 | 0 |

| | | | | |
|---|---|---|---|---|
| 0 | 0 | 0 | 0 | 0 |
| 0 | 0 | 0 | 0 | 0 |
| 0 | 0.158198722 | 0.021512031 | 0 | 0.001161672 |
| 0 | 0 | 0 | 0 | 0 |
| 1.998983578 | 0 | 0 | 0 | 0 |
| 0 | 0 | 0 | 0 | 0 |
| 0 | 0 | 0 | 0 | 0 |
| 0 | 0 | 0 | 0 | 0 |
| 0 | 0 | 0 | 0 | 0 |
| 0 | 0 | 0 | 0 | 0 |
| 0 | 0 | 0 | 0 | 0 |
| 0 | 0 | 0 | 0 | 0 |
| 0 | 0 | 0 | 0 | 0 |
| 0 | 0 | 0.000472345 | 0 | 0.00171181 |
| 0.007694201 | 0.003146414 | 0.001949293 | 0.000645246 | 0.001236265 |
| 0 | 0 | 0 | 0 | 0 |
| 0.000309418 | 0.001792529 | 0.003405602 | 0.007204963 | 0.001006153 |
| 0.000320028 | 0 | 0 | 0 | 0 |
| 0 | 0 | 0 | 0 | 0 |
| 0 | 0 | 0 | 0 | 0 |
| 0 | 0 | 0 | 0 | 0 |
| 0 | 0.000735519 | 0 | 0.000796411 | 0.000726615 |
| 0 | 0 | 0 | 0 | 0 |
| 0 | 0 | 0.132614842 | 0 | 0 |
| 0 | 0 | 0 | 0.002341165 | 0 |
| 0 | 0 | 0 | 0 | 0 |
| 0.412633349 | 0 | 0 | 0 | 0.166120398 |
| 0 | 0 | 0 | 0 | 0 |
| 0 | 0 | 0 | 0 | 0 |
| 0 | 0.00212787 | 0 | 0.001728024 | 0 |
| 0 | 0 | 0 | 0 | 0 |
| 0 | 0 | 0 | 0 | 0 |
| 0 | 0 | 0 | 0.003571148 | 0 |
| 0 | 0 | 0 | 0 | 0 |
| 0 | 0 | 0 | 0 | 0 |
| 0 | 0.633767389 | 0 | 0.02969341 | 0 |
| 0 | 0 | 0 | 0 | 0 |
| 0 | 0 | 0.272559768 | 0.374060766 | 0 |
| 0 | 0 | 0 | 0 | 0 |
| 0 | 0 | 0 | 0 | 0 |
| 0 | 0 | 0 | 0 | 0 |
| 0 | 0 | 0 | 0 | 0 |
| 0 | 0 | 0 | 0 | 0 |
| 0 | 0.018366095 | 0.04338932 | 0.003521584 | 0 |
| 0 | 0 | 0 | 0 | 0 |
| 0 | 0 | 0 | 0 | 0 |
| 0 | 0 | 0 | 0 | 0 |
| 0.008643027 | 0.008059112 | 0 | 0 | 0.245757563 |

| | | | | |
|---|---|---|---|---|
| 0 | 0 | 6.227575099 | 0 | 2.992820161 |
| 0 | 0 | 0 | 0 | 0 |
| 0 | 0 | 0 | 0.000417089 | 0 |
| 0 | 0 | 0 | 0 | 0 |
| 0.000981153 | 0 | 0.001549425 | 0.007973011 | 0.000765712 |
| 0 | 0 | 0 | 0.001824662 | 0 |
| 0 | 0 | 0 | 0 | 0 |
| 0.003850844 | 0.025206032 | 0 | 0.007058482 | 0 |
| 0 | 0 | 0 | 0 | 0 |
| 0.184140123 | 0 | 0.011599931 | 0.00242511 | 0 |
| 0 | 0 | 0 | 0 | 0 |
| 0 | 0.023393389 | 0 | 0.093901369 | 0.00063723 |
| 0.008258127 | 0.070978864 | 0.003912341 | 0.014127773 | 0.004994733 |
| 0.004640527 | 0.164868428 | 0 | 0.06633617 | 0 |
| 0 | 0.000670957 | 0.002194775 | 0 | 0 |
| 0 | 0 | 0 | 0.000706938 | 0 |
| 0 | 0 | 0 | 0.00462679 | 0 |
| 0 | 0 | 0 | 0 | 0 |
| 0.222626483 | 0.078610025 | 0.114706645 | 0.038603568 | 0 |
| 0 | 0 | 0 | 0 | 0 |
| 0 | 0 | 0 | 0 | 0 |
| 0 | 0 | 0 | 0 | 0 |
| 0 | 0 | 0 | 0 | 0 |
| 0 | 0 | 0 | 0 | 0.943763097 |
| 0 | 0 | 0.007875487 | 0 | 0 |
| 0 | 0.000607884 | 0.008561411 | 0.003016796 | 0 |
| 0 | 0 | 0 | 0 | 0.029109633 |
| 0 | 0.192045198 | 0 | 0 | 0 |
| 0 | 0.009024596 | 0 | 0 | 0 |
| 0 | 0 | 0 | 0 | 0 |
| 0 | 0 | 0 | 0.001177159 | 0 |
| 0 | 0 | 0 | 0 | 0 |
| 0 | 0 | 0.035263224 | 0 | 0 |
| 0.104912005 | 0.178279 | 0 | 0.088951184 | 0 |
| 0 | 0 | 0 | 0 | 0 |
| 0 | 0.028972083 | 0 | 0.049503303 | 0 |
| 0.048630422 | 0.063397132 | 0 | 0.000621791 | 0 |
| 0 | 0 | 0 | 0 | 0 |
| 0 | 0 | 0 | 0 | 0 |
| 0 | 0.038040659 | 0 | 0 | 0 |
| 0 | 0 | 0 | 0 | 0 |
| 0.306243508 | 0 | 0 | 0 | 0 |
| 0 | 0 | 0 | 0 | 0 |
| 0 | 0 | 0.039938025 | 0 | 0 |
| 0 | 0 | 0 | 0 | 0 |
| 0 | 0 | 0 | 0 | 0 |
| 0 | 0.002645833 | 0.004808228 | 0.010061176 | 0 |
| 0 | 0 | 0 | 0 | 0 |

| | | | | |
|---|---|---|---|---|
| 0 | 0 | 0 | 0 | 0 |
| 0 | 0.0057026 | 0 | 0 | 0 |
| 0 | 0 | 0 | 0 | 0 |
| 0 | 0 | 0 | 0 | 0 |
| 0.002009858 | 0 | 0 | 0 | 0.005536009 |
| 0.020443859 | 0.006847146 | 0.223301423 | 0 | 0.067274939 |
| 0 | 0 | 0 | 0 | 0 |
| 0 | 0 | 0 | 0 | 0 |
| 0 | 0 | 0 | 0 | 1.067791824 |
| 0 | 0 | 0 | 0 | 0 |
| 0 | 0.007078326 | 0.001725568 | 0 | 0.001279141 |
| 0.008192817 | 0 | 0 | 0 | 0 |
| 0 | 0 | 0.001668966 | 0 | 0 |
| 0.000825857 | 0 | 0.024157041 | 0 | 0.000805645 |
| 0.009714069 | 0.001980378 | 0 | 0.002382589 | 0.004687218 |
| 0.044320454 | 0 | 0 | 0 | 0 |
| 0 | 0 | 0 | 0 | 0 |
| 0.006329077 | 0.045078219 | 0 | 0 | 0 |
| 0 | 0 | 0.001419651 | 0 | 0.000643114 |
| 0 | 0 | 0 | 0 | 0.066908359 |
| 0 | 0 | 0 | 0 | 0 |
| 0 | 0.241019007 | 0 | 0 | 0.437900514 |
| 0 | 0 | 0 | 0 | 0.053134069 |
| 0 | 0 | 0 | 0.004669316 | 0 |
| 0 | 0 | 0 | 0 | 0 |
| 0 | 0 | 0 | 0 | 0 |
| 0.018132815 | 0.004067492 | 0 | 0.006542519 | 0 |
| 0 | 0 | 0 | 0 | 0 |
| 0 | 0 | 0 | 0 | 0 |
| 0.085342605 | 13.08915449 | 0.102100041 | 0.123393914 | 0.139642305 |
| 0.000770383 | 0 | 0 | 0 | 0.000683208 |
| 0 | 0 | 0 | 0 | 0 |
| 0 | 0 | 0 | 0 | 0 |
| 0.000694353 | 0 | 0 | 0.001555556 | 0 |
| 0.030671993 | 0 | 0 | 0.002791106 | 0 |
| 0 | 0 | 0 | 0 | 0 |
| 0 | 0 | 3.891924491 | 0 | 0 |
| 0 | 0 | 0 | 0 | 0 |
| 0 | 0 | 0 | 0.044602593 | 0.009886846 |
| 0 | 0 | 0 | 0.013931391 | 0 |
| 0.01256047 | 0.017699733 | 0 | 0.002903798 | 0 |
| 0 | 0 | 0 | 0.788966651 | 0.195525569 |
| 0 | 0 | 0 | 0 | 0 |
| 0 | 0 | 0 | 0 | 0 |
| 0 | 0.006806666 | 0 | 0 | 0 |
| 0.33707231 | 0 | 0 | 0 | 0 |
| 0.019774672 | 0 | 0 | 0.000507957 | 0.001390322 |
| 0 | 0 | 0 | 0 | 0 |

| | | | | |
|---|---|---|---|---|
| 0 | 0 | 0.013726915 | 0.008617594 | 0 |
| 0 | 0 | 0 | 0 | 0.004521434 |
| 0 | 7.485911743 | 0.01074941 | 0.372118834 | 0.094956737 |
| 0 | 0 | 0 | 0 | 0 |
| 0 | 0 | 0 | 0.01984564 | 0.141258629 |
| 0.023230924 | 0.017324356 | 0 | 0.011392941 | 0 |
| 0 | 0 | 0 | 0 | 0 |
| 0 | 0.001167722 | 0 | 0 | 0 |
| 0 | 0 | 0 | 0 | 0 |
| 0 | 0 | 0 | 0 | 0 |
| 0 | 0 | 0 | 0 | 0 |
| 0 | 0 | 0 | 0 | 0 |
| 0 | 0.018363525 | 0 | 0.001156035 | 0 |
| 0 | 0 | 0.001427401 | 0 | 0 |
| 0 | 0 | 0 | 0 | 0 |
| 0 | 0.238777 | 0 | 0.213238652 | 0.017736961 |
| 0 | 0.009933232 | 0.525100761 | 0.188836689 | 0 |
| 0.003348355 | 0 | 0 | 0 | 0 |
| 0.000266876 | 0 | 0 | 0 | 0 |
| 0.905754902 | 0 | 0 | 0.010643346 | 0.005378166 |
| 0.028871501 | 0.016634436 | 0 | 0.015319121 | 0 |
| 0 | 0 | 0.009178652 | 0.022074977 | 0 |
| 0.028279951 | 0 | 0 | 0 | 0 |
| 0 | 0 | 0 | 0.204657015 | 0 |
| 0 | 0.037261436 | 0 | 0 | 0 |
| 0 | 0 | 0 | 0 | 0 |
| 0.000546669 | 0.002418417 | 0 | 0 | 0 |
| 0 | 0 | 0 | 0 | 0.003914872 |
| 0 | 0 | 0 | 0 | 0 |
| 0 | 0.003843065 | 0 | 0 | 0.001095156 |
| 0 | 0 | 0 | 0 | 0.007624636 |
| 0 | 0.004896466 | 0.002313549 | 0.017142602 | 0 |
| 0.00047212 | 0 | 0.00169447 | 0.045096041 | 0.004835942 |
| 0 | 0.002405246 | 0 | 0.004340621 | 0.00165009 |
| 0 | 0.000953741 | 0 | 0.005335615 | 0 |
| 0 | 0 | 0 | 0 | 0 |
| 0 | 0.008092526 | 0.010570234 | 0.000608506 | 0 |
| 0.001669002 | 0.084646769 | 0.100994003 | 0.092915614 | 0.128787083 |
| 0 | 0 | 0 | 0 | 0.097002506 |
| 0.000955205 | 0.021128733 | 0 | 0 | 0 |
| 0 | 0 | 0 | 0.00962702 | 0 |
| 0 | 0 | 0 | 0 | 0 |
| 0 | 0.095237601 | 0.132403178 | 0 | 0 |
| 0.129504425 | 0.224786111 | 0 | 0.039470087 | 0 |
| 0.04703957 | 0 | 0 | 0 | 0 |
| 0 | 0 | 0 | 0 | 0 |
| 0.000278197 | 0 | 0 | 0 | 0.001447405 |
| 0 | 0 | 0 | 0 | 0 |

| | | | | |
|---|---|---|---|---|
| 0.005938981 | 0.09696187 | 0 | 0 | 0 |
| 0.013986492 | 0.456876334 | 0.389670363 | 1.038812377 | 0.284198576 |
| 0 | 0 | 0 | 0 | 0 |
| 0 | 0 | 0 | 0 | 0 |
| 0 | 0 | 0 | 0 | 0 |
| 0 | 0 | 0 | 0 | 0 |
| 0.035662047 | 0 | 0 | 0.138126538 | 0 |
| 0 | 0.005519924 | 0 | 0 | 0.000681638 |
| 0.000728653 | 0.001315714 | 0 | 0 | 0 |
| 0 | 0 | 0 | 0 | 0 |
| 0.001013999 | 0.003070668 | 0.007035997 | 0.001566068 | 0 |
| 0.000629325 | 0.050710043 | 0.340610325 | 0.012201831 | 0.015470873 |
| 0 | 0 | 0 | 0.004899701 | 0.004210397 |
| 0.007292143 | 0 | 0 | 0 | 0 |
| 0 | 0 | 0.017259362 | 0 | 0 |
| 0 | 0 | 0 | 0.00185767 | 0 |
| 0.002742993 | 0.001537816 | 0.009019087 | 0.029392045 | 0.002934817 |
| 0.481394948 | 0.009437041 | 0 | 0 | 0 |
| 0 | 0 | 0 | 0 | 0.00054895 |
| 0.052210667 | 0.007070678 | 0 | 0.009676393 | 0 |
| 0 | 0.006055162 | 0.004977975 | 0.024326544 | 0.019405438 |
| 0 | 0 | 0 | 0 | 0 |
| 0 | 0 | 0 | 0 | 0 |
| 0 | 0 | 0 | 0 | 0.494404322 |
| 0 | 0 | 0 | 0 | 0 |
| 0 | 0 | 0 | 0 | 0 |
| 0 | 0.019495582 | 0 | 0.030431705 | 0.008962028 |
| 0.008922543 | 0.006588976 | 0.016290258 | 0.020463196 | 0.130751883 |
| 0 | 0 | 0 | 0 | 0 |
| 0 | 0 | 0.004198689 | 0 | 0 |
| 0 | 0 | 0 | 0 | 0 |
| 0 | 0 | 0 | 0 | 0 |
| 0.002119286 | 0 | 0 | 0 | 0 |
| 0 | 0 | 0 | 0 | 0 |
| 0 | 0 | 0 | 2.896535864 | 0.10223375 |
| 4.554718399 | 0.914418735 | 0.154582135 | 0.180417847 | 0.051559554 |
| 0 | 0 | 0 | 0 | 0.006377697 |
| 0 | 0 | 0 | 0 | 0 |
| 0 | 0.063513676 | 0 | 0.043664677 | 0 |
| 0.001033218 | 0 | 0 | 0 | 0 |
| 0.025075127 | 0 | 0 | 0 | 0 |
| 0 | 0 | 0 | 0 | 0 |
| 0.00218015 | 0.159139105 | 0.00459049 | 0.046103723 | 0.000992504 |
| 0.000771794 | 0.005875345 | 0 | 0.000600166 | 0 |
| 0.211272136 | 0 | 0 | 0 | 0 |
| 0 | 0 | 0 | 0 | 0.010324946 |
| 0 | 0 | 0 | 0 | 0 |
| 0 | 0 | 0 | 0 | 0 |

| | | | | |
|---|---|---|---|---|
| 0.00365486 | 0.00802023 | 0 | 0 | 0 |
| 0 | 0 | 0 | 0 | 0 |
| 0 | 0 | 0 | 0.002088822 | 0 |
| 0 | 0.707821126 | 12.25762809 | 5.197555561 | 0.007596262 |
| 0 | 0 | 0 | 0 | 0 |
| 0 | 0 | 0.126441874 | 0 | 0 |
| 0 | 0 | 0 | 0 | 0 |
| 0 | 0.009809975 | 0 | 0.00140333 | 0 |
| 0 | 0.066429434 | 0 | 1.343977993 | 0.556031374 |
| 0 | 0 | 0 | 0 | 0.196591325 |
| 0 | 0 | 0 | 0 | 0 |
| 0 | 0.014734442 | 0 | 0 | 0.000632873 |
| 0 | 0 | 0 | 0 | 0 |
| 0 | 0 | 0 | 0 | 0 |
| 0.001806806 | 0.059525742 | 0 | 0 | 0.26843553 |
| 0 | 0 | 0 | 0 | 0 |
| 0 | 0 | 0 | 0 | 0 |
| 0 | 0 | 0 | 0 | 0 |
| 0 | 0 | 0 | 0 | 0 |
| 0 | 0 | 0 | 0 | 0 |
| 0 | 0 | 0 | 0 | 0 |
| 0 | 0 | 0 | 0 | 0 |
| 0 | 0 | 0 | 0 | 0 |
| 0.241144509 | 0.082697868 | 0 | 0.094102367 | 0.280329509 |
| 0.002885129 | 0.036107446 | 0 | 0.002273065 | 0.024293752 |
| 0 | 0 | 0 | 0 | 0.591886732 |
| 0 | 0 | 0 | 0 | 0.286943078 |
| 0 | 2.44773647 | 0 | 2.610612041 | 3.348095013 |
| 0 | 0 | 0 | 0 | 0 |
| 0 | 0 | 0 | 0 | 0 |
| 0 | 0 | 0 | 0.027536371 | 0.003691561 |
| 0 | 0 | 0 | 0 | 0.002318757 |
| 0.000600007 | 0 | 0 | 0.001112015 | 0.003160742 |
| 0 | 0 | 0 | 0 | 0 |
| 0 | 0 | 0 | 0 | 0 |
| 0 | 0 | 0 | 0 | 0 |
| 0 | 0 | 0 | 1.026581775 | 0.179215353 |
| 0 | 0 | 0 | 0 | 0.001316974 |
| 0 | 0 | 0 | 0 | 0 |
| 0 | 0 | 0 | 0 | 0.000452993 |
| 0.012680574 | 0.063792156 | 0.066452228 | 0.192868728 | 0.002276731 |
| 0 | 0 | 0 | 0 | 0.529014179 |
| 0.77627379 | 4.287549549 | 0.179800043 | 0 | 0 |
| 0 | 0 | 0 | 0 | 0.00462777 |
| 0 | 0 | 0 | 0 | 0 |
| 0 | 0 | 0 | 0 | 0 |
| 0 | 0 | 0 | 0 | 0 |
| 0 | 0 | 0 | 0 | 0.004926405 |

| | | | | |
|---|---|---|---|---|
| 0 | 0 | 0 | 0 | 0 |
| 0 | 0 | 0 | 0.262479142 | 0 |
| 0.862144594 | 0.141499399 | 0.05534141 | 0.034763246 | 0 |
| 0 | 0 | 0 | 0 | 0 |
| 0 | 0 | 0 | 0 | 0 |
| 0 | 0.003347381 | 0 | 0 | 0.002204572 |
| 0 | 0 | 0 | 0 | 0.144654591 |
| 0 | 0 | 0 | 0 | 0 |
| 0 | 0 | 0 | 0 | 0 |
| 0 | 0 | 0 | 0 | 0 |
| 0.034767274 | 0 | 0 | 0.015057945 | 0.794588226 |
| 0.501623502 | 0.58199356 | 0.424601241 | 0.799150694 | 0.588603565 |
| 0 | 0 | 0 | 0 | 0 |
| 0 | 0.010355942 | 0.506494875 | 0.17957519 | 0.000926683 |
| 0.116778754 | 0.026493289 | 0.039548346 | 0.008644407 | 0.009201293 |
| 0 | 0 | 0 | 0 | 0.005100467 |
| 0 | 0 | 0 | 0 | 0 |
| 0 | 0 | 0 | 0 | 0 |
| 0 | 0 | 0 | 0 | 0.105719664 |
| 0.081494506 | 0 | 0 | 0 | 0.100436009 |
| 0.016893878 | 0.164346932 | 0.00720185 | 0.612107618 | 0.018135666 |
| 5.910134153 | 0.027019539 | 3.680653141 | 0.0150715 | 0 |
| 4.184412181 | 0.191560007 | 0.12004097 | 0.597223484 | 0.109574905 |
| 0 | 0 | 0 | 0 | 0 |
| 0 | 0 | 0 | 0 | 0.000564277 |
| 0 | 0 | 0 | 0.002142492 | 0 |
| 0.386029085 | 0 | 0 | 0 | 0 |
| 0 | 0.001580384 | 0 | 0 | 0 |
| 0 | 0 | 0 | 0 | 0 |
| 0 | 0 | 0 | 0 | 0.149335838 |
| 0 | 0 | 0 | 0 | 0.002222574 |
| 0 | 0 | 0 | 0 | 0.271089761 |
| 0.001265561 | 0.019709822 | 0.058659007 | 0.070107556 | 0 |
| 0 | 0 | 0 | 0 | 0.131536682 |
| 0.002565521 | 0.057313813 | 0.021070919 | 0 | 0.015947652 |
| 0.00065579 | 0.099079674 | 0 | 0.075144365 | 0.013434543 |
| 0 | 0.003377986 | 0 | 0 | 0.009424198 |
| 0 | 0 | 0 | 0 | 0 |
| 0 | 0 | 0 | 0 | 0 |
| 0 | 0 | 0 | 0 | 0 |
| 0 | 0 | 0 | 0 | 0.003802433 |
| 0 | 0 | 0 | 0 | 0 |
| 0 | 0 | 0 | 0 | 0 |
| 0 | 0 | 0 | 0 | 0 |
| 0.008039426 | 0.018314576 | 0 | 0.003598332 | 0.079156283 |
| 0 | 0 | 0 | 0 | 0 |
| 0 | 0 | 0 | 0 | 0 |
| 0.018267844 | 0.018350932 | 0 | 4.137208255 | 0.001100047 |

| | | | | |
|---|---|---|---|---|
| 0 | 0 | 0 | 0 | 0 |
| 0 | 0 | 0 | 0 | 0 |
| 0 | 0 | 0.605174665 | 0 | 0 |
| 0 | 0 | 0 | 0 | 0.00186538 |
| 0 | 0 | 0 | 0 | 0 |
| 0 | 0 | 0 | 0 | 0 |
| 0 | 0 | 0 | 0.054840967 | 0.651303078 |
| 0.003225841 | 0 | 0 | 0 | 0.569730748 |
| 0 | 0 | 0.001565593 | 0 | 0 |
| 0 | 0 | 0 | 0 | 0 |
| 0 | 0.005047195 | 0.131722741 | 0.002403607 | 0.001558155 |
| 0 | 0 | 0 | 0 | 0 |
| 0 | 0 | 0 | 0 | 0.009398416 |
| 0 | 0 | 0 | 0 | 0 |
| 0 | 0 | 0 | 0 | 0 |
| 0 | 0 | 0 | 0 | 0 |
| 0 | 0 | 0 | 0.014018946 | 0.156574425 |
| 0 | 0 | 0 | 0 | 0.001008877 |
| 0 | 0 | 0.011940474 | 0.002992066 | 0.090691538 |
| 0 | 0 | 0 | 0 | 0.093973513 |
| 0 | 0 | 0 | 0 | 0 |
| 0 | 0 | 0 | 0 | 0 |
| 0 | 0 | 0 | 0 | 0.013791796 |
| 0 | 0 | 0 | 1.813910507 | 0 |
| 0 | 0 | 0 | 0 | 0 |
| 0 | 0 | 0 | 0 | 0 |
| 0 | 0 | 0 | 0 | 0 |
| 0 | 0 | 0 | 0 | 0 |
| 0 | 0 | 0 | 0 | 0 |
| 0 | 0 | 0 | 0.948022801 | 0.052056538 |
| 0 | 0 | 0 | 0 | 0 |
| 0 | 0 | 0 | 0 | 0 |
| 0 | 0 | 0 | 0 | 0 |
| 0 | 0 | 0 | 0 | 0 |
| 0 | 0.002341082 | 0 | 0 | 0 |
| 0 | 0 | 0 | 0 | 0 |
| 0 | 0 | 0 | 0 | 0 |
| 0.000300466 | 0.011749481 | 0.218589809 | 0.039623017 | 0 |
| 0 | 0 | 0 | 0 | 0 |
| 0 | 0 | 0 | 0 | 0.000722774 |
| 0.003382934 | 0 | 0 | 0 | 0.113167369 |
| 0 | 0 | 0 | 0 | 0 |
| 0 | 0 | 0 | 0 | 0 |
| 0 | 0 | 0 | 0 | 0.041416098 |
| 0 | 0.17572934 | 0.127960413 | 0 | 0 |
| 0 | 0 | 0 | 0 | 0 |
| 0 | 0 | 0 | 0 | 0.004456092 |
| 0 | 0 | 0 | 0 | 0.090459195 |

| | | | | |
|---|---|---|---|---|
| 0 | 0 | 0 | 0 | 0 |
| 0 | 0 | 0 | 0 | 0 |
| 0 | 0 | 0 | 0 | 0 |
| 0 | 0 | 0 | 0 | 0.015598502 |
| 0 | 0 | 0 | 0 | 0 |
| 0 | 0 | 0 | 0 | 0 |
| 0 | 0 | 0 | 0 | 0 |
| 0 | 0 | 0 | 0 | 0 |
| 0.274534942 | 0.153206156 | 0.132219279 | 0.015212476 | 0.017989158 |
| 0 | 0 | 0 | 0 | 0 |
| 0 | 0 | 0 | 0 | 0 |
| 0 | 0 | 0 | 0 | 0.000369676 |
| 0 | 0.000572933 | 0 | 0 | 0.003537484 |
| 0 | 0 | 0 | 0 | 0 |
| 1.25333751 | 2.165025159 | 0 | 0 | 2.43443403 |
| 0.117373517 | 1.627514146 | 0.675164108 | 0.005919255 | 0.001246269 |
| 0 | 0 | 0 | 0 | 0 |
| 0 | 0 | 0 | 0 | 0 |
| 0 | 0 | 0 | 0 | 0 |
| 0 | 0 | 0 | 0 | 0 |
| 0 | 0 | 0 | 0 | 0.196437922 |
| 0 | 0 | 0 | 0 | 0 |
| 0 | 0 | 0 | 0 | 0 |
| 0 | 0 | 0 | 0 | 0.162882122 |
| 0 | 0 | 1.269074454 | 0.013547342 | 0.004278486 |
| 0 | 0 | 22.28614538 | 2.060054716 | 24.77033865 |
| 0 | 0 | 0 | 0 | 0.002829055 |
| 0 | 0 | 0 | 0 | 0 |
| 0 | 0.061929631 | 0 | 0.420470897 | 0 |
| 0 | 0 | 0 | 0 | 0 |
| 0 | 0 | 0 | 0 | 0 |
| 0 | 0 | 0 | 0.093733125 | 0.001224608 |
| 0 | 0 | 0 | 0 | 0 |
| 0 | 0.022101874 | 0 | 0 | 0.011044101 |
| 0 | 0.001185667 | 0.018314886 | 0.153600621 | 0 |
| 0.000606249 | 0.001436782 | 0.117170396 | 0 | 0.000887118 |
| 0.838346903 | 0 | 0 | 0 | 0 |
| 0 | 0 | 0 | 0.911038186 | 7.197948661 |
| 0 | 0 | 0 | 0 | 0.053591946 |
| 0 | 0 | 0 | 0 | 0 |
| 0 | 0 | 0 | 0 | 0 |
| 0 | 0 | 0.003577802 | 0 | 0.013916314 |
| 0.148396074 | 0.766539305 | 0 | 0.146896086 | 0.135167231 |
| 0 | 0 | 0 | 0 | 0 |
| 0 | 0 | 0 | 0 | 0 |
| 0.003446068 | 0.000567154 | 0 | 0 | 0 |
| 0 | 0 | 0 | 0 | 0 |
| 0 | 0.002168507 | 0.245478531 | 0.063103427 | 0.004239882 |

| | | | | |
|---|---|---|---|---|
| 0 | 0.004446911 | 0 | 0 | 0 |
| 0 | 0 | 0 | 0 | 0 |
| 0 | 0 | 0 | 0 | 0 |
| 0 | 0 | 0 | 0 | 0 |
| 0 | 0 | 0 | 0.000363178 | 0.015904791 |
| 0 | 0 | 0 | 0 | 0 |
| 0 | 0 | 0 | 0 | 0 |
| 0 | 0 | 0 | 0 | 0 |
| 0 | 0 | 0 | 0 | 0 |
| 0 | 0 | 0 | 0 | 0.033257899 |
| 0 | 0 | 0 | 0 | 0 |
| 0 | 2.247752617 | 0 | 0 | 0.109590344 |
| 0 | 0 | 0 | 0 | 0.201405669 |
| 0 | 0 | 0 | 0 | 0 |
| 0 | 0.267807457 | 0 | 2.473751984 | 0.041577854 |
| 0 | 0 | 0 | 0 | 0 |
| 0 | 0.000863494 | 0 | 0.000681757 | 0.001999314 |
| 0 | 0 | 0 | 0 | 0 |
| 0 | 0.001359978 | 0 | 0 | 0.014946603 |
| 0 | 0 | 0 | 0.009933132 | 0.01806314 |
| 0 | 0 | 0 | 0 | 0.035315751 |
| 0 | 0 | 0 | 0 | 0 |
| 0.003096806 | 0.925014775 | 2.960819517 | 5.15760663 | 0.10565394 |
| 0 | 0 | 0 | 0 | 0 |
| 0 | 0 | 0 | 0 | 0 |
| 0 | 0 | 0 | 0 | 0 |
| 0 | 0 | 0 | 0 | 0 |
| 0 | 0 | 0 | 0 | 0 |
| 0 | 0 | 0 | 0 | 0 |
| 0 | 0 | 0 | 0 | 0 |
| 0 | 0 | 0 | 0 | 0 |
| 0.005097385 | 0.041677954 | 0.036406739 | 0.054866253 | 0.074340881 |
| 0 | 0 | 0 | 0 | 0 |
| 0.007194048 | 0.041944937 | 0 | 0 | 0.018756029 |
| 0.000247406 | 0.003908946 | 0.104068664 | 0 | 0.000804505 |
| 0 | 0.022004095 | 0 | 0.022431393 | 0.016801099 |
| 0 | 0 | 0 | 0 | 0 |
| 0 | 0 | 0 | 0 | 0 |
| 0 | 0 | 0 | 0 | 0 |
| 0 | 0 | 0 | 0 | 0 |
| 0 | 0 | 0 | 0 | 0.037562193 |
| 0 | 0 | 0 | 0 | 0.00061997 |
| 0 | 0 | 0 | 0 | 0 |
| 0 | 0 | 0 | 0 | 0 |
| 0.001543561 | 0.02002366 | 0 | 0.000463277 | 0.059280374 |
| 0 | 0 | 0 | 0 | 0 |
| 0 | 0 | 0 | 0 | 0 |
| 0.000691547 | 0.001699636 | 0 | 0 | 0.001349245 |

| | | | | |
|---|---|---|---|---|
| 0 | 0 | 0 | 0 | 0 |
| 0.003787694 | 0 | 0 | 0 | 0.002234184 |
| 0.001596429 | 0 | 0 | 0.004671662 | 0.015081787 |
| 0 | 0 | 0 | 0 | 0 |
| 0.005964771 | 0 | 0 | 0 | 0 |
| 0 | 0 | 0 | 0 | 0.04752693 |
| 0 | 0 | 0 | 0 | 0 |
| 0 | 0 | 0 | 0.03835001 | 0.010992141 |
| 0 | 0 | 0 | 0 | 0 |
| 0 | 0 | 0 | 0 | 0 |
| 0 | 0 | 0 | 0 | 0 |
| 0 | 0 | 0.035469826 | 0 | 0 |
| 0 | 0 | 0 | 0 | 0 |
| 0 | 0 | 0 | 0 | 0 |
| 0 | 0 | 0 | 0 | 0 |
| 0 | 0 | 0 | 0 | 0 |
| 0 | 0 | 0 | 0 | 0 |
| 0 | 0 | 0 | 0 | 0 |
| 0 | 0 | 0.509122281 | 1.142359737 | 1.283099831 |
| 0 | 0 | 0 | 0 | 0 |
| 0 | 0 | 0 | 0 | 0 |
| 0 | 0 | 0 | 0 | 0 |
| 0 | 0 | 0 | 0 | 0 |
| 0 | 0 | 0 | 0 | 0 |
| 0 | 0 | 0.008155441 | 0.001012342 | 0 |
| 0.007184986 | 0.020672691 | 0.265800004 | 0.002048646 | 0 |
| 0 | 0 | 0 | 0 | 0 |
| 0.000444169 | 0.029357501 | 0.002444364 | 0.990788035 | 0.03249738 |
| 0 | 0 | 0 | 0 | 0 |
| 0 | 0 | 0 | 0.513906394 | 0.167491389 |
| 0 | 0 | 0 | 0 | 0 |
| 0 | 0 | 0 | 0 | 0 |
| 0 | 0 | 0 | 0 | 0.175689856 |
| 0 | 0 | 0 | 0 | 0 |
| 0.004517652 | 0.001753132 | 0.38174358 | 0.000338977 | 0.100512572 |
| 0 | 0 | 0 | 0 | 0 |
| 0 | 0 | 0 | 0 | 0.084434413 |
| 0 | 0.005399505 | 0 | 0.008205643 | 0.002281047 |
| 0 | 0 | 0 | 0 | 0 |
| 0 | 0 | 0 | 0 | 0 |
| 3.283879045 | 26.39712089 | 1.409320003 | 1.227316624 | 1.613029077 |
| 0.021197828 | 0 | 0 | 0.324073107 | 0.476773972 |
| 0 | 0 | 0 | 0 | 0 |
| 0 | 0 | 0 | 0 | 0.143838158 |
| 0 | 0 | 0 | 0 | 0 |
| 0 | 0 | 0 | 0 | 0 |
| 0 | 0 | 0 | 0 | 0 |
| 0.006352485 | 0.001127723 | 0 | 0 | 0.009051824 |

| | | | | |
|---|---|---|---|---|
| 0 | 0 | 0 | 0 | 0 |
| 0 | 0 | 0 | 0.00188398 | 0.004297176 |
| 0 | 0 | 0 | 0 | 0.004182383 |
| 0 | 0 | 0 | 0 | 0 |
| 0.471411677 | 0.840767968 | 0 | 1.087533707 | 1.071346176 |
| 0 | 0 | 0 | 0 | 0 |
| 0 | 0 | 0 | 0 | 0 |
| 0 | 0 | 0 | 0 | 0.000759354 |
| 0.485299712 | 0.234544733 | 0.00105243 | 0.020693224 | 0 |
| 0 | 0 | 0 | 0 | 0 |
| 0 | 0 | 0.390814856 | 0 | 0 |
| 0 | 0 | 0 | 0 | 0 |
| 0 | 0 | 0 | 0.134142847 | 0.208771946 |
| 0.003508039 | 0.567007292 | 0.005036231 | 0.006751637 | 0.003079966 |
| 0 | 0 | 0 | 0 | 0 |
| 0 | 0 | 0.04132695 | 0.095827441 | 0.093092501 |
| 0 | 0.052279177 | 0 | 0.020989703 | 0.000444206 |
| 0 | 0 | 0 | 0 | 0 |
| 0 | 0 | 0 | 0 | 0 |
| 0 | 0 | 0 | 0 | 0 |
| 0 | 0 | 0 | 0 | 0 |
| 0 | 0 | 0 | 0 | 0 |
| 0 | 0 | 0 | 0 | 0 |
| 0 | 0 | 0 | 0 | 0 |
| 0 | 0 | 0 | 0 | 0.471597075 |
| 0 | 0 | 0 | 0 | 0 |
| 0 | 0.088519726 | 0 | 0.289952806 | 0.016866467 |
| 0 | 0.001058847 | 0.0446848 | 0 | 0 |
| 0 | 0 | 0 | 0 | 0 |
| 0 | 0 | 0 | 0 | 0 |
| 0 | 0 | 0 | 0 | 0 |
| 0 | 0 | 0 | 0 | 0 |
| 0 | 0 | 0 | 0 | 0 |
| 0 | 0 | 0 | 0 | 0.001979824 |
| 0 | 0 | 0 | 0.401036995 | 0 |
| 0 | 0.00108877 | 0.008285406 | 0 | 0.38519568 |
| 0 | 0 | 0 | 0 | 0 |
| 0 | 0 | 0.126815168 | 0.092652643 | 0.035548167 |
| 0 | 0 | 0 | 0 | 0 |
| 0 | 0 | 0 | 0 | 0 |
| 0 | 0 | 0 | 1.523421467 | 4.728770885 |
| 0 | 0 | 0 | 0 | 0 |
| 0 | 0 | 0 | 0 | 0 |
| 0 | 0 | 0 | 0 | 0.123223979 |
| 0 | 0 | 0 | 0 | 0 |
| 0 | 0 | 0 | 0 | 0 |
| 0 | 0 | 0 | 0 | 0 |
| 0 | 0 | 0 | 0 | 0 |

| | | | | |
|---|---|---|---|---|
| 0 | 0 | 0 | 0 | 0.008589781 |
| 0 | 0 | 0 | 0 | 0 |
| 0 | 0 | 0 | 0 | 0 |
| 0 | 0.001599289 | 0.00593382 | 0.000721538 | 0 |
| 0 | 0 | 0 | 0 | 0.212422824 |
| 0 | 0 | 0 | 0 | 0 |
| 0 | 0.069615818 | 0 | 0.261301587 | 0.033086698 |
| 0 | 0 | 0 | 0 | 0 |
| 0 | 0 | 0 | 0 | 0.003753301 |
| 0.071327258 | 0 | 0 | 0 | 0.05454952 |
| 0 | 0 | 0 | 0 | 0 |
| 0 | 0 | 0.472012519 | 0 | 0.016405079 |
| 0 | 0 | 0 | 0 | 0 |
| 0 | 0 | 0 | 0 | 0 |
| 0 | 0 | 0 | 0 | 0 |
| 0 | 0 | 0 | 0 | 0 |
| 0 | 0 | 1.327713216 | 0 | 0 |
| 0.003639219 | 0 | 0 | 0 | 0 |
| 0 | 0 | 0 | 0 | 0 |
| 0 | 0 | 0 | 0 | 0 |
| 0.000720773 | 0.003758039 | 0.003311235 | 0.017879645 | 0.000984387 |
| 0 | 0 | 0 | 0 | 0 |
| 0 | 0 | 0 | 0.013940838 | 0.004498212 |
| 0.001484556 | 0.003685869 | 0.0009134 | 0.05774879 | 0.000827557 |
| 0 | 0 | 0 | 0 | 0 |
| 0 | 0 | 0 | 0 | 0.004718932 |
| 0 | 0 | 0 | 0 | 0 |
| 0 | 0 | 0 | 0 | 0 |
| 0 | 0 | 0 | 0 | 0 |
| 0 | 0.431137976 | 1.012239767 | 0.01871154 | 0 |
| 0 | 0 | 0 | 0 | 0.036086034 |
| 0 | 0 | 0 | 0 | 0 |
| 0 | 0 | 0 | 0 | 0 |
| 0 | 0 | 0 | 0 | 0 |
| 0 | 0 | 0 | 0 | 0 |
| 0 | 0 | 0 | 0 | 0 |
| 0.002742119 | 0.030187484 | 0 | 0.074780812 | 0 |
| 0 | 0 | 0 | 0 | 0 |
| 0 | 0 | 0 | 0 | 0 |
| 0 | 0 | 0 | 0 | 0 |
| 0 | 0 | 0 | 0 | 0 |
| 0 | 0 | 0 | 0 | 0 |
| 0 | 0 | 0 | 0 | 0 |
| 0 | 0 | 0 | 0 | 0.00543458 |
| 0 | 0 | 0 | 0 | 0.015026833 |
| 0 | 0 | 0 | 0 | 0 |
| 0 | 0 | 0 | 0 | 0 |
| 0 | 0 | 0 | 0 | 0 |

| | | | | |
|---|---|---|---|---|
| 0.002358119 | 0.076558249 | 0.904878235 | 0.001735141 | 0.002968267 |
| 0 | 0 | 0 | 0 | 0 |
| 0 | 0 | 0 | 0 | 0.072666306 |
| 0 | 0 | 0 | 0 | 0 |
| 0 | 0 | 0 | 0 | 0 |
| 0 | 0 | 0 | 0 | 0 |
| 0 | 0 | 0 | 0 | 0.001483211 |
| 0 | 0.297032184 | 2.208503585 | 0.713975389 | 0.084574778 |
| 0.001074606 | 0.005848132 | 2.571906331 | 0.272867159 | 0.035409475 |
| 0.003324927 | 0.489737235 | 0.112173753 | 0.013272449 | 0 |
| 0 | 0 | 0 | 0 | 0.001141876 |
| 0 | 0 | 0 | 0 | 0.002839193 |
| 0 | 0 | 0 | 0 | 0.012078739 |
| 0 | 0 | 1.006073614 | 2.645011227 | 0 |
| 0 | 0 | 0 | 0 | 0 |
| 0 | 0 | 0 | 0 | 0 |
| 0.079948302 | 0.168596576 | 0.054199707 | 0 | 0 |
| 0 | 0 | 0 | 0 | 0 |
| 0 | 0 | 0 | 0 | 0 |
| 0 | 0 | 0 | 0 | 0.041420355 |
| 0 | 0 | 0 | 0 | 0 |
| 0 | 0 | 0 | 0 | 0 |
| 0 | 0 | 0 | 0 | 0 |
| 0 | 0 | 0 | 0 | 0 |
| 0 | 0 | 0 | 0 | 0.038225026 |
| 0 | 0 | 0 | 0 | 0.009117126 |
| 0 | 0 | 0 | 0 | 0 |
| 0 | 0 | 0 | 0 | 0 |
| 0.003771666 | 0 | 0 | 0 | 0 |
| 0 | 0 | 0 | 0 | 0 |
| 0 | 0 | 0 | 0 | 0.037812363 |
| 0 | 0 | 0 | 0 | 0 |
| 0 | 0 | 0 | 0 | 0 |
| 0 | 0 | 0 | 0 | 0 |
| 0 | 0 | 0 | 0.121705414 | 0.003773181 |
| 0 | 0 | 0 | 0 | 0 |
| 0 | 0 | 0 | 0 | 0 |
| 0 | 0 | 0 | 0.010521604 | 0 |
| 0 | 0 | 0 | 0 | 0 |
| 0 | 0 | 0.346143152 | 0 | 0 |
| 0 | 0 | 0 | 0 | 0 |
| 0 | 0 | 0 | 0 | 0.276058751 |
| 0 | 0 | 0 | 0 | 0 |
| 0 | 0 | 0 | 0 | 0 |
| 0 | 0 | 0 | 0 | 0 |
| 0 | 0 | 0 | 0 | 0 |
| 0 | 0 | 0 | 0 | 0 |
| 0 | 0 | 0 | 0 | 0 |

| | | | | |
|---|---|---|---|---|
| 0 | 0 | 0 | 0 | 0 |
| 0 | 0 | 0.011827548 | 0.002642694 | 0 |
| 0 | 0 | 0 | 0 | 0 |
| 0 | 0 | 0 | 0 | 0 |
| 0 | 1.383596121 | 0 | 0.906334947 | 0.521201111 |
| 0 | 0 | 0 | 0 | 0 |
| 0 | 0 | 0 | 0 | 0 |
| 0.039479891 | 0.075372221 | 0.007501547 | 0.040143912 | 0.005474984 |
| 0 | 0 | 0 | 0 | 0 |
| 0 | 0 | 0 | 0 | 0 |
| 0 | 0 | 0 | 0 | 0 |
| 0 | 0 | 0 | 0 | 0 |
| 0 | 0 | 0 | 0 | 0 |
| 0 | 0 | 0 | 0 | 0 |
| 0.017302163 | 0 | 0 | 0 | 0 |
| 0 | 0.045082596 | 2.195907398 | 0 | 0 |
| 0 | 0 | 0 | 0 | 0 |
| 0 | 0 | 0 | 0 | 0 |
| 0 | 0 | 0 | 0 | 0 |
| 0 | 0 | 0 | 0 | 0 |
| 0 | 0 | 0 | 0 | 0 |
| 0 | 0 | 0 | 0 | 0 |
| 0 | 0 | 0 | 0 | 0 |
| 0.000251225 | 0.00172002 | 0.119500351 | 0 | 0.002940919 |
| 0 | 0 | 0 | 0 | 0.168188839 |
| 0 | 0 | 0 | 0 | 0 |
| 0.004518883 | 0.025412479 | 0 | 0.746326006 | 0 |
| 0 | 0.000891951 | 0 | 0 | 0 |
| 0 | 0 | 0.029418584 | 0 | 0 |
| 0 | 0 | 0 | 0 | 0 |
| 0 | 0 | 0.004690769 | 0 | 0 |
| 0 | 0 | 0 | 0 | 0 |
| 0 | 0 | 0 | 0 | 0 |
| 0 | 0 | 0 | 0 | 0 |
| 0 | 0 | 0 | 0 | 0 |
| 0 | 0 | 0 | 0 | 0 |
| 0 | 0 | 0 | 0 | 0 |
| 0 | 0 | 0 | 0 | 0.001992288 |
| 0 | 0 | 0 | 0 | 0 |
| 0 | 0 | 0 | 0 | 0 |
| 0 | 0 | 0 | 0 | 0 |
| 0 | 0 | 0 | 0.234978989 | 0 |
| 0 | 0 | 0 | 0 | 0 |
| 0 | 0 | 0 | 0 | 0 |
| 0 | 0 | 0 | 0 | 0 |
| 0 | 0 | 0 | 0 | 0 |
| 0 | 0 | 0 | 0 | 0 |
| 0 | 0 | 0 | 0 | 0.003932375 |

| | | | | |
|---|---|---|---|---|
| 0 | 0 | 0 | 0 | 0.054946998 |
| 0 | 0 | 0 | 0 | 0.002416403 |
| 0 | 0 | 0 | 0.001729166 | 0 |
| 0 | 0.001245485 | 0 | 0 | 0 |
| 0 | 0 | 0 | 0 | 0 |
| 0 | 0 | 0 | 0 | 0.281989431 |
| 0 | 0 | 0 | 0 | 0 |
| 0 | 0 | 0 | 0 | 0 |
| 0 | 0 | 0 | 0 | 0 |
| 0 | 0 | 0 | 0 | 0 |
| 0 | 0 | 0 | 0 | 0 |
| 0 | 0 | 0 | 0 | 0.001355529 |
| 0 | 0 | 0 | 0 | 0.004451 |
| 0 | 0 | 0 | 0 | 0 |
| 0 | 0 | 0 | 0 | 0 |
| 0 | 0 | 0 | 0 | 0 |
| 0 | 0 | 0 | 0 | 0 |
| 0.044494924 | 0 | 0 | 0 | 0 |
| 0 | 0.000675382 | 0 | 0 | 0.001310582 |
| 0 | 0 | 0 | 0 | 0 |
| 0.004002796 | 0.097631299 | 0 | 0.032931608 | 0.04034995 |
| 0 | 0 | 0 | 0 | 0 |
| 0.091836862 | 0.007155132 | 0 | 0 | 0.093287936 |
| 0 | 0 | 0 | 1.263446089 | 1.758041309 |
| 0 | 0 | 0 | 0 | 0 |
| 0 | 0 | 0 | 0 | 0 |
| 0 | 0 | 0 | 0 | 0 |
| 0 | 0 | 0 | 0 | 0 |
| 0 | 0 | 0 | 0 | 0 |
| 0 | 0 | 0 | 0 | 0 |
| 0 | 0 | 0 | 0 | 0 |
| 0 | 0 | 0 | 0 | 0 |
| 0.008482129 | 2.645238394 | 0.00892048 | 0.003093428 | 0 |
| 0 | 0 | 0 | 0 | 0 |
| 0 | 0 | 0 | 0 | 0.161801072 |
| 0 | 0.005001927 | 0.105783938 | 0.001692509 | 0 |
| 0 | 0 | 0 | 0 | 0 |
| 0 | 0 | 0 | 0 | 0 |
| 0.002380166 | 0.0072319 | 0 | 0.055988912 | 0.008930441 |
| 0 | 0 | 0 | 0 | 0 |
| 0 | 0 | 0 | 0 | 0 |
| 0 | 0 | 0 | 0 | 0 |
| 0 | 0.00980926 | 0 | 0.127883518 | 0.06098339 |
| 0 | 0 | 0 | 0 | 0 |
| 0 | 0 | 0 | 0 | 0 |
| 0 | 0 | 0 | 0.004605423 | 0.049924137 |
| 0 | 0 | 0 | 0 | 0 |
| 0 | 0 | 0 | 0 | 0 |

| | | | | |
|---|---|---|---|---|
| 0 | 0 | 0 | 0 | 0 |
| 0 | 0 | 0 | 0 | 0 |
| 0.24249989 | 0 | 0 | 0 | 0 |
| 0 | 0 | 0 | 0 | 0 |
| 0 | 0 | 0 | 0 | 0 |
| 0 | 0 | 0 | 0 | 0 |
| 0 | 0 | 0 | 0 | 0 |
| 0.515311462 | 0.145365434 | 0 | 0 | 0 |
| 0 | 0 | 0 | 0 | 0.067131203 |
| 0 | 0.003043539 | 0.051161496 | 0 | 0 |
| 0 | 0 | 0 | 0 | 0 |
| 0 | 0 | 0 | 0 | 0 |
| 0 | 0 | 0 | 0 | 0 |
| 0 | 0 | 0 | 0 | 0.484146902 |
| 0 | 0 | 0 | 0 | 0 |
| 0.004221336 | 0.006125126 | 0.006926011 | 0.004544295 | 0 |
| 0 | 0 | 0 | 0 | 0 |
| 0 | 0 | 0 | 0 | 0 |
| 0 | 0 | 0 | 0 | 0 |
| 0 | 0 | 0 | 0 | 0.245985777 |
| 0 | 0 | 0 | 0.003078612 | 0.004564311 |
| 0 | 0 | 0 | 0 | 0 |
| 0 | 0 | 0 | 0 | 0 |
| 0 | 0 | 0 | 0 | 0 |
| 0 | 0 | 0 | 0 | 0 |
| 0 | 0 | 0 | 0 | 0 |
| 0 | 0 | 0 | 0 | 0 |
| 0 | 0 | 0 | 0 | 0.003147101 |
| 0 | 0 | 0 | 0 | 0 |
| 0 | 0 | 0 | 0.084267464 | 0.033621524 |
| 0 | 0 | 0 | 0.038831403 | 1.377194408 |
| 0 | 0 | 0 | 0 | 0 |
| 0 | 0 | 0 | 0 | 0 |
| 0 | 0 | 0 | 0 | 0 |
| 0 | 0 | 0 | 0 | 0 |
| 0 | 0 | 0 | 0 | 0 |
| 0 | 0 | 0 | 0 | 0 |
| 0 | 0 | 0 | 0 | 0 |
| 0 | 0 | 0.047696326 | 0 | 0 |
| 0 | 0 | 0 | 0 | 0 |
| 0 | 0 | 0 | 0 | 0 |
| 0 | 0 | 0 | 0 | 0.212397752 |
| 0 | 0 | 0 | 0 | 0.001920865 |
| 0 | 0 | 0 | 0 | 0 |
| 0 | 0 | 0 | 0 | 0 |
| 0 | 0 | 0 | 0 | 0.008541088 |
| 0.012170336 | 0.233392502 | 0.403376428 | 0.038598709 | 0 |
| 0 | 0 | 0 | 0 | 0.006852016 |

| | | | | |
|---|---|---|---|---|
| 0 | 0 | 0 | 0 | 0 |
| 0 | 0 | 0 | 0 | 0 |
| 0 | 0 | 0 | 0 | 0 |
| 0 | 0 | 0 | 0 | 0 |
| 0 | 1.255403735 | 2.025378899 | 0.173486352 | 0.017867812 |
| 0 | 0 | 0 | 0 | 0 |
| 0 | 0 | 0 | 0 | 0 |
| 0 | 0 | 0 | 0 | 0 |
| 0 | 0 | 0 | 0 | 0 |
| 0 | 0 | 0 | 0 | 0 |
| 0 | 0 | 0 | 0 | 0.028724608 |
| 0.005396844 | 0.00867137 | 0 | 0.025429241 | 0.014277328 |
| 0 | 0.000997494 | 0 | 0 | 0 |
| 0 | 0 | 0 | 0 | 0 |
| 0 | 0 | 0 | 0 | 0 |
| 0 | 0.461348607 | 0 | 0 | 1.336897398 |
| 0.010623269 | 0.011004366 | 0 | 0.012737842 | 0.048590746 |
| 0 | 0 | 0 | 0 | 0 |
| 0.019255134 | 0.109324086 | 0 | 0.0097757 | 0.009324379 |
| 0.000452182 | 0.170178218 | 0 | 0.263533246 | 0.000529339 |
| 0 | 0 | 0.047948373 | 0 | 0 |
| 0 | 0 | 0 | 0 | 0 |
| 0 | 0 | 0 | 0 | 0 |
| 0 | 0 | 0 | 0 | 0 |
| 0 | 0 | 0 | 0 | 0 |
| 0 | 0 | 0 | 0 | 0 |
| 0.00159436 | 0.005267081 | 0 | 0.012398117 | 0.020502209 |
| 0 | 0 | 0 | 0 | 0 |
| 0 | 0 | 0 | 0 | 0 |
| 0 | 0 | 0 | 0 | 0 |
| 0.001232243 | 0 | 0 | 0.000468464 | 0 |
| 0 | 0 | 0 | 0.004427461 | 0 |
| 0 | 0 | 0 | 0 | 0.000856065 |
| 0 | 0 | 0 | 0 | 0 |
| 0 | 0 | 0 | 0 | 0 |
| 0 | 0 | 0 | 0 | 0 |
| 0 | 0 | 0 | 0 | 0 |
| 0.00196234 | 0.055443115 | 0 | 0.021064265 | 0.036146774 |
| 0 | 0 | 0 | 0 | 0.011918972 |
| 0 | 0 | 0 | 0 | 0.004465589 |
| 0 | 0.000455182 | 0 | 0 | 0.011466621 |
| 0 | 0 | 0 | 0 | 0 |
| 0 | 0 | 0 | 0 | 0 |
| 0.022936874 | 0.006220222 | 0 | 0.000488057 | 0 |
| 0 | 0 | 0 | 0 | 0 |
| 0 | 0 | 0 | 0.001636222 | 0 |
| 0 | 0 | 0 | 0 | 0.122654238 |
| 0 | 0 | 0 | 0 | 0 |

| | | | | |
|---|---|---|---|---|
| 0 | 0 | 0 | 0 | 0 |
| 0 | 0 | 0 | 0 | 0.077354373 |
| 0 | 0 | 0 | 0 | 0 |
| 0 | 0.001417135 | 0.087046349 | 0.000383614 | 0.001399979 |
| 0 | 0.001182933 | 0.019228091 | 0.00094879 | 0 |
| 0 | 0.003223009 | 0 | 0.001071441 | 0 |
| 0 | 0 | 0 | 0 | 0 |
| 0 | 0.003103302 | 0 | 0 | 0 |
| 0 | 0 | 0 | 0 | 0.000568658 |
| 0 | 0 | 0 | 0 | 0 |
| 0 | 0 | 0 | 0 | 0.019810625 |
| 0 | 0 | 0 | 0 | 0 |
| 0 | 0.045305169 | 0 | 0 | 0.035725981 |
| 0 | 0 | 0 | 0 | 0 |
| 0 | 0 | 0 | 0 | 0 |
| 0 | 0 | 0 | 0 | 0 |
| 0 | 0 | 0 | 0 | 0 |
| 0 | 0 | 0 | 0 | 0 |
| 0 | 0 | 0 | 0 | 0 |
| 0 | 0 | 0 | 0 | 0 |
| 0 | 0 | 0 | 0 | 0 |
| 0 | 0 | 0 | 0 | 0 |
| 0.007534855 | 0.005555591 | 0 | 0.003341961 | 0 |
| 0 | 0 | 0 | 0.000985907 | 0.016769317 |
| 0 | 0 | 0 | 0 | 0 |
| 0 | 0 | 0 | 0 | 0.001281135 |
| 0 | 0 | 0 | 0 | 0 |
| 0 | 0 | 0 | 0 | 0 |
| 0 | 0 | 0 | 0 | 0 |
| 0 | 0 | 0 | 0 | 0 |
| 0 | 0 | 0 | 0.044449093 | 0.005099559 |
| 0 | 0 | 0 | 0 | 0.01239392 |
| 0 | 0 | 0 | 0 | 0 |
| 0 | 0 | 0 | 0 | 0 |
| 0 | 0 | 0 | 0 | 0.001068839 |
| 0 | 0 | 0 | 0 | 0 |
| 0 | 0 | 0 | 0 | 0 |
| 0 | 0 | 0 | 0.001095932 | 0 |
| 0.000315916 | 1.216272975 | 0 | 0.053933414 | 0 |
| 0 | 0 | 0 | 0 | 0 |
| 0 | 0 | 0 | 0 | 0 |
| 0 | 0 | 0 | 0 | 0 |
| 0 | 0 | 0 | 0 | 0 |
| 0 | 0 | 0 | 0 | 0 |
| 1.354784727 | 0 | 0 | 0 | 0.619644357 |
| 0 | 0 | 0 | 0 | 0.172045499 |
| 0 | 0 | 0 | 0 | 0 |
| 0 | 0 | 0 | 0 | 0 |

| | | | | |
|---|---|---|---|---|
| 0 | 0 | 0 | 0 | 0 |
| 0 | 0 | 0 | 0 | 0 |
| 0 | 0 | 0 | 0 | 0 |
| 0 | 0 | 0 | 0 | 0.000614072 |
| 0.002110658 | 0 | 0 | 0 | 0 |
| 0 | 0 | 0 | 0 | 0 |
| 0 | 0 | 0 | 0 | 0 |
| 0 | 0 | 0 | 0 | 0 |
| 0.001023072 | 0 | 0 | 0.001041811 | 0.000712881 |
| 0 | 0 | 0 | 0 | 0 |
| 0 | 0 | 0 | 0 | 0.00075006 |
| 0 | 0 | 0.014969396 | 0 | 0 |
| 0 | 0 | 0 | 0 | 0 |
| 0 | 0 | 0 | 0 | 0 |
| 0 | 0 | 0 | 0 | 0 |
| 0 | 0 | 0 | 0 | 0 |
| 0 | 0 | 0 | 0.011413273 | 0.000918797 |
| 0 | 0 | 0 | 0 | 0 |
| 0 | 0 | 0 | 0 | 0 |
| 0.000509832 | 0 | 0 | 0.001574816 | 0 |
| 0 | 0 | 0 | 0 | 0 |
| 0 | 0 | 0 | 0 | 0 |
| 0 | 0 | 0 | 0 | 0 |
| 0 | 0 | 0 | 0 | 0 |
| 0 | 0 | 0 | 0 | 0 |
| 0 | 0 | 0 | 0 | 0 |
| 0 | 0 | 0 | 0 | 0 |
| 0 | 0 | 0 | 0 | 0 |
| 0 | 0 | 0 | 0 | 0 |
| 0 | 0 | 0 | 0 | 0 |
| 0 | 0.001499384 | 0 | 0.001040715 | 0.000569705 |
| 0 | 0 | 0 | 0 | 0 |
| 0.932954177 | 0.014998851 | 0.004380615 | 0.021025751 | 0.011906741 |
| 0 | 0 | 0.013180846 | 0.003581006 | 0.001520925 |
| 0 | 0 | 0 | 0 | 0 |
| 0 | 0 | 0 | 0 | 0 |
| 0 | 0 | 0 | 0 | 0 |
| 0 | 0 | 0 | 0 | 0 |
| 0 | 0 | 0 | 0 | 0 |
| 0 | 0 | 0 | 0 | 0 |
| 0 | 0 | 0 | 0 | 0 |
| 0.007530985 | 0.449287039 | 0.014115246 | 0.011034736 | 0.590046932 |
| 0 | 0.09704268 | 0 | 0.007387825 | 0.005138148 |
| 0 | 0 | 0 | 0 | 0 |
| 0 | 0 | 0 | 0 | 0 |
| 0 | 0 | 0 | 0.34687994 | 0.017484634 |
| 0 | 0.001017328 | 0 | 0.000917959 | 0.003297697 |
| 0 | 0 | 0 | 0 | 0 |

| | | | | |
|---|---|---|---|---|
| 0 | 0 | 0 | 0 | 0.004354733 |
| 0 | 0.006890164 | 0 | 0 | 0.016843242 |
| 0.000727721 | 0.023159795 | 0.009402618 | 0.014376383 | 0.006490618 |
| 0 | 0 | 0 | 0 | 0 |
| 0 | 0.511480343 | 0 | 0 | 0.56440076 |
| 0.009695845 | 0.000893616 | 0 | 0 | 0 |
| 0 | 0 | 0 | 0 | 0 |
| 0 | 0 | 0 | 0 | 0 |
| 0 | 0 | 0 | 0 | 0 |
| 0 | 0 | 0 | 0 | 0 |
| 0 | 0 | 0 | 0 | 0 |
| 0 | 0 | 0 | 0 | 0 |
| 0 | 0 | 0 | 0 | 0 |
| 0 | 0 | 0 | 0 | 0 |
| 0 | 0 | 0 | 0 | 0 |
| 0 | 0 | 0 | 0 | 0 |
| 0 | 0 | 0 | 0 | 0.225313488 |
| 0 | 0 | 0 | 0 | 0 |
| 0 | 0 | 0 | 0 | 0 |
| 0 | 0 | 0 | 0 | 0 |
| 0 | 0 | 0 | 0 | 0 |
| 0 | 0 | 0 | 0 | 0.008320892 |
| 0 | 0 | 0 | 0 | 0 |
| 0 | 0 | 0 | 0 | 0 |
| 0 | 0 | 0 | 0.002809358 | 0.002340267 |
| 0 | 0.002859723 | 0.109754151 | 0 | 0 |
| 0 | 0 | 0 | 0 | 0 |
| 0 | 0 | 0 | 0.229329228 | 0.003425562 |
| 0 | 0 | 0 | 0 | 0 |
| 0 | 0 | 0 | 0 | 0 |
| 0 | 0 | 0 | 0 | 0 |
| 0 | 0.001671872 | 0 | 0.00043102 | 0 |
| 0 | 0 | 0 | 0 | 0 |
| 0 | 0 | 0 | 0 | 0 |
| 0 | 0 | 0 | 0 | 0 |
| 0 | 0 | 0 | 0 | 0 |
| 0 | 0 | 0 | 0 | 0.000512469 |
| 0.007549937 | 0 | 0 | 0 | 0 |
| 0 | 0 | 0 | 0 | 2.342064041 |
| 0 | 0 | 0 | 0 | 0 |
| 0 | 0 | 0 | 0 | 0 |
| 0 | 0.096173116 | 0 | 0 | 0 |
| 0.002681528 | 0.069564404 | 0.780241419 | 0.001356514 | 0.001518911 |
| 0 | 0 | 0 | 0 | 0 |
| 0 | 0 | 0 | 0 | 0 |
| 0 | 0 | 0 | 0 | 0 |
| 0.00251426 | 0.004599682 | 0.00474939 | 0.000754619 | 0 |
| 0 | 0 | 0 | 0 | 0 |

| | | | | |
|---|---|---|---|---|
| 0 | 0 | 0 | 0 | 0 |
| 0.005259726 | 0.044189188 | 0.395528528 | 0.06843864 | 0.002117557 |
| 0 | 0 | 0 | 0 | 0 |
| 0 | 0 | 0 | 0 | 0 |
| 0 | 0 | 0 | 0 | 0 |
| 0 | 0 | 0 | 0 | 0 |
| 0 | 0 | 0 | 0 | 0 |
| 0 | 0 | 0 | 0 | 0 |
| 0 | 0 | 0 | 0 | 0 |
| 0 | 0 | 0 | 0 | 0 |
| 0 | 0 | 0 | 0 | 0 |
| 0 | 0 | 0 | 0 | 0 |
| 0 | 0 | 0 | 0.0182437 | 0.00809284 |
| 0 | 0 | 0 | 0 | 0 |
| 0.001769642 | 0.034867519 | 0 | 0.032649022 | 0.018888114 |
| 0 | 0 | 0 | 0 | 0 |
| 0 | 0 | 0 | 0 | 0 |
| 0 | 0 | 0 | 0 | 0 |
| 0 | 0 | 0 | 0 | 0.042689005 |
| 0 | 0 | 0 | 0 | 0 |
| 0 | 0 | 0 | 0 | 0 |
| 0.001329004 | 0.026166557 | 0.01130095 | 0.121259901 | 0 |
| 0.002435243 | 0.058458827 | 0 | 0.0212787 | 0 |
| 0 | 0 | 0.250333969 | 0 | 0 |
| 0 | 0 | 0 | 0 | 0 |
| 0 | 0 | 0 | 0 | 0 |
| 0 | 0 | 0 | 0 | 0 |
| 0 | 0 | 0 | 0 | 0 |
| 0 | 0 | 0 | 0 | 0 |
| 0 | 0 | 0 | 0 | 0 |
| 0 | 0 | 0 | 0 | 0 |
| 0 | 0 | 0 | 0 | 0 |
| 0 | 0 | 0 | 0 | 0 |
| 0 | 0 | 0 | 0 | 0 |
| 0 | 0 | 0 | 25.93517848 | 8.66802705 |
| 0 | 0 | 0 | 0 | 0.088543698 |
| 0 | 0 | 0 | 0 | 0 |
| 0 | 0 | 0 | 0 | 0 |
| 0 | 0.014355631 | 0.02432949 | 1.320230443 | 0.007768145 |
| 0 | 0 | 0 | 0 | 0 |
| 0.026637349 | 0.177311579 | 0.011695783 | 0.079213213 | 0.044112741 |
| 0 | 0 | 0 | 0 | 0 |
| 0 | 0 | 0 | 0 | 0 |
| 0 | 0 | 0 | 0 | 0 |
| 0 | 0 | 0 | 0 | 0 |
| 0.472246636 | 0.023513357 | 0.499374321 | 0.18452515 | 0 |
| 0 | 0 | 0 | 0 | 0 |
| 0 | 0 | 0 | 0 | 0.001010156 |

| | | | | |
|---|---|---|---|---|
| 0 | 0 | 0 | 0 | 0 |
| 0 | 0 | 0 | 0 | 0 |
| 0 | 0 | 0 | 0 | 0.011248275 |
| 0 | 0 | 0 | 0 | 0 |
| 0 | 0.003767557 | 0 | 0.00079323 | 0.002791461 |
| 0 | 0 | 0 | 0 | 0 |
| 0 | 0 | 0 | 0 | 0 |
| 0 | 0 | 0 | 0.003552877 | 0 |
| 0 | 0 | 0 | 0 | 0 |
| 0 | 0.048583311 | 0 | 0.119531252 | 0.003199678 |
| 0 | 0 | 0 | 0 | 0.014974422 |
| 0 | 0 | 0 | 0 | 0 |
| 0 | 0.002374305 | 0.021430097 | 0.005427391 | 0.001954635 |
| 0 | 0 | 0 | 0 | 0 |
| 0 | 0 | 0 | 0 | 0 |
| 0 | 0 | 0 | 0 | 0 |
| 0 | 0 | 0 | 0 | 0 |
| 0 | 0 | 0 | 0 | 0 |
| 0 | 0 | 0 | 0 | 0 |
| 0.010254604 | 0.777412193 | 0 | 0.391927274 | 0.009568697 |
| 0 | 0 | 0 | 0 | 0 |
| 0 | 0 | 0 | 0 | 0 |
| 0 | 0 | 0 | 0 | 0 |
| 0 | 0 | 0 | 0 | 0 |
| 0 | 0 | 0 | 0 | 0 |
| 0 | 0 | 0 | 0 | 0 |
| 0 | 0 | 0 | 0 | 0 |
| 0 | 0 | 0 | 0 | 0 |
| 0 | 0 | 0 | 0 | 0 |
| 0 | 0.138561237 | 0 | 0 | 0.015064663 |
| 0 | 0 | 0 | 0 | 0 |
| 0 | 0 | 0 | 0 | 0 |
| 0 | 0 | 0 | 0 | 0 |
| 0 | 0 | 0 | 0 | 0 |
| 0 | 0 | 0 | 0 | 0 |
| 0 | 0 | 0 | 0 | 0 |
| 0 | 0 | 0 | 0 | 0.01811992 |
| 0 | 0 | 0 | 0 | 0 |
| 0 | 0 | 0 | 0 | 0 |
| 0 | 0 | 0 | 0 | 0 |
| 0 | 0 | 0 | 0 | 0 |
| 0 | 0 | 0 | 0 | 0 |
| 0 | 0 | 0 | 0 | 0.034852449 |
| 0 | 0 | 0 | 0 | 0 |
| 0.000586265 | 0.009527462 | 0 | 0 | 0.010131107 |
| 0 | 0 | 0 | 0 | 0 |
| 0 | 0 | 0 | 0 | 0.003919593 |
| 0 | 0 | 0 | 0 | 0 |

| | | | | |
|---|---|---|---|---|
| 0 | 0 | 0 | 0 | 0 |
| 0 | 0 | 0 | 0 | 0 |
| 0 | 0 | 0 | 0 | 0 |
| 0 | 0 | 0 | 0 | 0 |
| 0 | 0 | 0 | 0 | 0 |
| 0.000187573 | 0.04583689 | 0 | 0 | 0.004513561 |
| 0 | 0 | 0 | 0 | 0 |
| 0 | 0 | 0 | 0 | 0 |
| 0 | 0 | 0 | 0 | 0 |
| 0 | 0 | 0 | 0 | 0 |
| 0 | 0 | 0 | 0 | 0 |
| 0 | 0 | 0 | 0 | 0 |
| 0 | 0 | 0 | 0 | 0 |
| 0 | 0 | 0 | 0 | 0 |
| 0 | 0 | 0 | 0 | 0 |
| 0 | 0 | 0 | 0 | 0 |
| 0 | 0 | 0 | 0 | 0.00192362 |
| 0 | 0 | 0 | 0.000878219 | 0 |
| 0 | 0 | 0 | 0 | 0 |
| 0 | 0 | 0 | 0 | 0 |
| 0 | 0 | 0 | 0 | 0 |
| 0 | 0 | 0 | 0 | 0 |
| 0 | 0 | 0 | 0 | 0 |
| 0 | 0 | 0 | 0 | 0 |
| 0 | 0 | 0 | 0 | 0 |
| 0 | 0.001699704 | 0 | 0.002091385 | 0.005342678 |
| 0 | 0 | 0 | 0 | 0 |
| 0 | 0 | 0 | 0 | 0 |
| 0 | 0 | 0 | 0 | 0 |
| 0 | 0 | 0 | 0 | 0 |
| 0 | 0 | 0 | 0 | 0 |
| 0 | 0 | 0 | 0.00100759 | 0 |
| 0 | 0 | 0 | 0 | 0 |
| 0 | 0 | 0 | 0 | 0 |
| 0 | 0 | 0 | 0 | 0 |
| 0 | 0 | 0 | 0 | 0 |
| 0 | 0 | 0 | 0 | 0 |
| 0 | 0 | 0 | 0 | 0 |
| 0 | 0 | 0 | 1.310596202 | 0.007060085 |
| 0 | 0 | 0 | 0 | 0 |
| 0 | 0 | 0 | 0 | 0 |
| 0 | 0 | 0 | 0 | 0 |
| 0 | 0 | 0 | 0 | 0 |
| 0 | 0 | 0 | 0 | 0 |
| 0 | 0 | 0 | 0 | 0 |
| 0 | 0 | 0 | 0 | 0 |
| 0 | 0 | 0 | 0 | 0 |
| 0 | 0 | 0 | 0 | 0 |

| | | | | |
|---|---|---|---|---|
| 0 | 0 | 0 | 0 | 0 |
| 0 | 0.015483907 | 0 | 0 | 0.069243809 |
| 0 | 0 | 0 | 0 | 0 |
| 0 | 0 | 0 | 0 | 0 |
| 0 | 0 | 0 | 0 | 0 |
| 0 | 0 | 0 | 0 | 0 |
| 0 | 0 | 0 | 0 | 0 |
| 0 | 0 | 0 | 0 | 0 |
| 0 | 0 | 0 | 0 | 0.00165718 |
| 0 | 0 | 0 | 0 | 0 |
| 0 | 0.000579284 | 0.110930755 | 0.003397562 | 0 |
| 0 | 0 | 0 | 0 | 0 |
| 0 | 0.736329467 | 0 | 0 | 0 |
| 0 | 0 | 0 | 0 | 0 |
| 0 | 0 | 0 | 0 | 0 |
| 0 | 0 | 0 | 0 | 0 |
| 0 | 0 | 0 | 0 | 0 |
| 0 | 0 | 0 | 0 | 0 |
| 0 | 0 | 0 | 0 | 0 |
| 0 | 0 | 0 | 0 | 0 |
| 0 | 0.004850494 | 0 | 0.001724161 | 0 |
| 0 | 0 | 0 | 0 | 0 |
| 0 | 0 | 0 | 0 | 0 |
| 0 | 2.532650357 | 0 | 0.095915425 | 0.719623046 |
| 0 | 0 | 0 | 0 | 0 |
| 0 | 0 | 0 | 0 | 0 |
| 0 | 0 | 0 | 0 | 0 |
| 0 | 0 | 0 | 0 | 0.130082425 |
| 0 | 0.04893228 | 0 | 0 | 0 |
| 0 | 0 | 0 | 0 | 0 |
| 0.001278326 | 0 | 0 | 0 | 0 |
| 0 | 0 | 0 | 0 | 0 |
| 0 | 0 | 0 | 0 | 0 |
| 0 | 0 | 0 | 0 | 0 |

| PHL018-0221_V2 | PHL018-0144_V3 | PHL018-0117_V1 | PHL018-0242_V2 | PHL018-0083_V3 |
|---|---|---|---|---|
| CTRL | REF | EYCF | REF | REF |
| 0 | 1.132914703 | 0.000417017 | 1.613087141 | 0.021176819 |
| 9.523401486 | 0.879166539 | 0 | 0.148228347 | 2.293500558 |
| 0.55549569 | 4.559622269 | 0 | 0.596881606 | 0 |
| 0 | 0 | 0 | 0 | 0 |
| 0 | 0 | 0 | 0 | 0 |
| 0 | 0 | 0 | 0 | 0 |
| 0 | 0 | 0 | 0.050868671 | 0 |
| 0 | 0 | 0 | 0 | 0 |
| 0 | 0 | 0 | 0 | 0 |
| 0 | 0 | 0 | 0 | 0 |
| 0 | 0 | 0 | 0 | 0 |
| 0 | 0 | 0 | 0 | 0 |
| 0 | 0 | 0 | 0 | 0 |
| 3.112704795 | 7.198283706 | 0.000462789 | 0.133065799 | 0.080295844 |
| 0 | 0 | 0 | 0 | 0 |
| 0 | 0 | 0 | 0 | 0 |
| 0 | 0 | 0 | 0 | 0 |
| 0 | 0 | 0 | 0 | 0 |
| 0 | 0 | 0 | 0 | 0 |
| 0 | 0 | 0 | 0 | 0 |
| 0 | 0 | 0 | 0 | 0 |
| 0 | 0 | 0 | 0 | 0 |
| 0 | 0 | 0 | 0 | 0 |
| 0 | 0 | 0 | 0 | 0 |
| 0 | 0 | 0 | 1.730522521 | 0 |
| 0 | 0 | 0 | 0 | 0 |
| 0 | 0 | 0 | 0 | 0 |
| 0 | 0 | 0 | 0 | 0 |
| 0 | 0 | 0 | 0 | 0 |
| 0 | 0 | 0 | 0 | 0 |
| 0 | 0 | 0 | 0 | 0 |
| 0 | 0 | 0 | 0 | 0 |
| 0 | 0 | 0 | 0 | 0 |
| 0 | 0 | 0 | 0 | 0 |
| 0 | 0 | 0 | 0 | 0 |
| 0 | 0 | 0.000381713 | 1.029223863 | 0.013867048 |
| 0 | 0 | 0 | 0 | 0 |
| 0 | 0 | 0 | 0 | 0 |
| 0 | 0 | 0 | 0 | 0 |
| 0 | 0 | 0 | 0 | 0 |
| 0 | 0 | 0 | 0 | 0 |
| 0.458505654 | 0.005361749 | 0.68244321 | 0 | 0.01310071 |
| 0.026071496 | 2.163593094 | 13.3758177 | 0.001765697 | 0.335834171 |
| 0.067452336 | 0.074302581 | 0.035522633 | 0.040773314 | 0.132375003 |
| 0 | 0.001391417 | 0.005786487 | 0.000943191 | 0 |
| 0 | 0 | 0 | 0 | 0 |

| | | | | |
|---|---|---|---|---|
| 0 | 0.161395898 | 0 | 0 | 0 |
| 0 | 0.044110481 | 0.04289876 | 0.001962247 | 0.410539527 |
| 0 | 0.070436789 | 0 | 0.004172029 | 0.001080064 |
| 0.257026601 | 0.060929059 | 0 | 0.086899074 | 0.910623189 |
| 0.003133579 | 0 | 0 | 0.008575511 | 0 |
| 0 | 0 | 0 | 0.239480319 | 0 |
| 0.149236958 | 0 | 0.092575539 | 0 | 0.013400979 |
| 0 | 0 | 0 | 0 | 0 |
| 0 | 0 | 0 | 0 | 0 |
| 0 | 0 | 0 | 0 | 0 |
| 0 | 0.00081451 | 0.000470459 | 0 | 0 |
| 0.007305456 | 0.787129944 | 0 | 0.520773783 | 0.165575282 |
| 0 | 0 | 0.001064589 | 0 | 0 |
| 0.147190044 | 0.114806431 | 0 | 0.111893166 | 0.021056893 |
| 0 | 0 | 0 | 0 | 0 |
| 0 | 0 | 0 | 0 | 0 |
| 3.198445267 | 0.006382533 | 18.61221094 | 0 | 0.005498433 |
| 0 | 0 | 0 | 0 | 0 |
| 0 | 0 | 0 | 0 | 0 |
| 0 | 0 | 0 | 0 | 0 |
| 0.159125063 | 0.061496104 | 0 | 0.02774013 | 0.829631436 |
| 0.002604974 | 0.009868094 | 0 | 0.00469669 | 0.005258328 |
| 0 | 0 | 0 | 0 | 0 |
| 0 | 0 | 0 | 0 | 0 |
| 0 | 0 | 0 | 0 | 0 |
| 0 | 0 | 0 | 0 | 0 |
| 0 | 0 | 0 | 0 | 0 |
| 0 | 0 | 0 | 0 | 0 |
| 0.013526091 | 0 | 0.009867034 | 0 | 0 |
| 0 | 0.003058726 | 0 | 0 | 0 |
| 0.125014156 | 0 | 0 | 0 | 0.009017915 |
| 0 | 0 | 0 | 0 | 0 |
| 0 | 0 | 0 | 0 | 0 |
| 0 | 0 | 0 | 0 | 0 |
| 0 | 0 | 0 | 0 | 0 |
| 0 | 0 | 0.001691707 | 0 | 0 |
| 0 | 0 | 0 | 0 | 0 |
| 0.074882261 | 0.264527231 | 0.198789124 | 0 | 0.19473489 |
| 0 | 0 | 0 | 0 | 0 |
| 0 | 0 | 0 | 0 | 0 |
| 0 | 0 | 0 | 0 | 0 |
| 0 | 0 | 0 | 0 | 0 |
| 0 | 0 | 0 | 0 | 0 |
| 0 | 0 | 0 | 0 | 0 |
| 0 | 0 | 0 | 0 | 0 |
| 0 | 1.517286791 | 0 | 0 | 0 |
| 0 | 0 | 0.010255458 | 0 | 0 |
| 0 | 0 | 0 | 0 | 0 |

| | | | | |
|---|---|---|---|---|
| 0 | 0 | 0 | 0 | 0 |
| 0.013839329 | 0 | 0 | 0 | 0 |
| 0 | 0 | 0 | 0 | 0 |
| 0 | 0 | 0 | 0 | 0 |
| 0.01063962 | 0 | 0.667723394 | 0 | 0 |
| 0 | 0 | 0 | 0 | 0 |
| 0 | 0 | 0 | 0 | 0 |
| 0 | 0 | 0 | 0 | 0 |
| 0.010812209 | 0.000507356 | 0 | 0 | 0.001648406 |
| 0 | 0 | 0 | 0 | 0 |
| 0 | 0 | 0 | 0 | 0 |
| 0 | 0 | 0.000816533 | 0 | 0.003189612 |
| 0 | 0 | 0 | 0 | 0 |
| 0 | 0 | 0 | 0 | 0 |
| 0 | 0 | 0 | 0 | 0 |
| 0 | 0 | 0 | 0 | 0 |
| 0 | 0 | 0 | 0 | 0 |
| 0 | 0 | 0 | 0 | 0 |
| 0 | 0 | 0 | 0.121527095 | 0 |
| 0 | 0 | 0 | 0 | 0 |
| 0 | 0 | 0 | 0 | 0 |
| 0 | 0 | 0 | 0 | 0 |
| 0 | 0 | 0 | 0 | 0 |
| 0 | 0 | 0 | 0 | 0 |
| 0 | 0 | 0 | 0 | 0 |
| 0 | 0 | 0 | 0 | 0 |
| 0 | 0 | 0.000515321 | 0 | 0 |
| 0 | 0 | 0 | 0 | 0 |
| 0 | 0 | 0 | 0 | 0 |
| 0 | 0 | 0 | 0 | 0 |
| 0 | 0.100753011 | 0 | 0.026542196 | 0 |
| 0 | 0.000886569 | 0 | 0 | 0.000533421 |
| 0 | 0 | 0 | 0 | 0 |
| 0 | 0 | 0 | 0 | 0 |
| 0 | 0 | 0 | 0 | 0 |
| 0 | 0 | 0 | 0 | 0 |
| 0 | 0 | 0 | 0 | 0 |
| 0 | 0 | 0 | 0 | 0 |
| 0 | 0 | 0 | 0.039447497 | 0 |
| 0 | 0 | 0 | 0 | 0 |
| 0 | 0 | 0 | 0 | 0 |
| 0 | 0 | 0 | 0 | 0 |
| 0.005152038 | 0 | 0 | 0 | 0.00462413 |
| 0 | 0 | 0 | 0 | 0 |
| 0 | 0 | 0 | 0 | 0 |
| 0 | 0 | 0 | 0 | 0 |
| 0 | 0 | 0 | 0 | 0 |
| 0 | 0 | 0 | 0 | 0 |

| | | | | |
|---|---|---|---|---|
| 0.001129801 | 0 | 0 | 0.005396722 | 0 |
| 0 | 0 | 0 | 0 | 0 |
| 0 | 0 | 0 | 0 | 0 |
| 0 | 0 | 0 | 0 | 0 |
| 0 | 0 | 0 | 0.060328429 | 0 |
| 0 | 0 | 0 | 0 | 0 |
| 0 | 0 | 0 | 0 | 0 |
| 0 | 0 | 0 | 0 | 0 |
| 0 | 0 | 0 | 0 | 0 |
| 0 | 0 | 0 | 0 | 0 |
| 0 | 0 | 0 | 0 | 0 |
| 0 | 0 | 0 | 0 | 0 |
| 0 | 0 | 0 | 0 | 0 |
| 0 | 0 | 0 | 0 | 0 |
| 0 | 0 | 0 | 0 | 0 |
| 0 | 0 | 0 | 0 | 0 |
| 0 | 0 | 0 | 0 | 0 |
| 0 | 0 | 0 | 0 | 0 |
| 0 | 0 | 0 | 0 | 0 |
| 0 | 0 | 0 | 0 | 0 |
| 0 | 0 | 0 | 0 | 0 |
| 0 | 0 | 0 | 0 | 0 |
| 0 | 0.021703114 | 0 | 0 | 0 |
| 0 | 0 | 0 | 0 | 0 |
| 0 | 0.054750404 | 0 | 0.032180538 | 0 |
| 0 | 0 | 0 | 0 | 0 |
| 0 | 0 | 0 | 0 | 0 |
| 0 | 0 | 0 | 0 | 0 |
| 0 | 0 | 0 | 0 | 0 |
| 0 | 0 | 0 | 0 | 0 |
| 0 | 0 | 0 | 0 | 0 |
| 0 | 0 | 0 | 0 | 0.023694942 |
| 0 | 0 | 0 | 0 | 0 |
| 0 | 0 | 0 | 0 | 0 |
| 0 | 0 | 0 | 0 | 0 |
| 0 | 0 | 0 | 0 | 0 |
| 0 | 0 | 0 | 0 | 0 |
| 0 | 0 | 0 | 0 | 0 |
| 0 | 0 | 0 | 0 | 0 |
| 0 | 0 | 0 | 0 | 0 |
| 0 | 0 | 0 | 0 | 0 |
| 0 | 0 | 0 | 0 | 0 |
| 0 | 0 | 0 | 0 | 0 |
| 0 | 0 | 0 | 0 | 0 |
| 0 | 0 | 0 | 0 | 0 |
| 0 | 0 | 0 | 0 | 0 |
| 0 | 0 | 0 | 0 | 0 |
| 0 | 0 | 0 | 0 | 0 |

| | | | | |
|---|---|---|---|---|
| 0 | 0 | 0 | 0 | 0 |
| 0 | 0 | 0 | 0 | 0 |
| 0 | 0 | 0 | 0 | 0 |
| 0 | 0 | 0 | 0 | 0 |
| 0 | 0 | 0 | 0 | 0 |
| 0 | 0 | 0 | 0 | 0 |
| 0 | 0 | 0 | 0 | 0 |
| 0 | 0 | 0 | 0 | 0 |
| 0 | 0 | 0 | 0 | 0 |
| 0 | 0 | 0 | 0 | 0 |
| 0 | 0 | 0 | 0 | 0 |
| 0 | 0 | 0 | 0 | 0 |
| 0.000952494 | 0.001957647 | 0 | 0.004865733 | 0.001619553 |
| 0 | 0 | 0 | 0 | 0 |
| 0 | 0 | 0 | 0 | 0 |
| 0 | 0 | 0 | 0 | 0 |
| 0 | 0 | 0 | 0 | 0 |
| 0 | 0 | 0 | 0 | 0 |
| 0 | 0 | 0 | 0 | 0 |
| 0 | 0 | 0 | 0 | 0 |
| 0.036805984 | 0.011591044 | 0.026216048 | 0.022744381 | 0.06909743 |
| 0 | 0 | 0 | 0 | 0 |
| 0 | 0.015969345 | 0 | 0 | 0 |
| 0.002166683 | 0 | 0.000734894 | 0 | 0.002009494 |
| 0 | 0 | 0 | 0 | 0 |
| 0 | 0 | 0 | 0 | 0 |
| 0.011408918 | 0 | 0.013954266 | 0.00096333 | 0.009458973 |
| 0 | 0 | 0 | 0 | 0 |
| 0 | 0 | 0 | 0 | 0 |
| 0 | 0 | 0 | 0 | 0.000514577 |
| 0.001033887 | 0 | 0.004500305 | 0 | 0 |
| 0 | 0 | 0 | 0 | 0 |
| 0 | 0 | 0 | 0 | 0 |
| 0 | 0 | 0 | 0.172359417 | 0 |
| 0.002896928 | 0.130988305 | 0 | 0.033095289 | 0.001253822 |
| 0 | 0 | 0 | 0 | 0 |
| 0 | 0 | 0 | 0 | 0 |
| 0 | 0 | 0 | 0 | 0 |
| 0 | 0 | 0 | 0 | 0 |
| 0 | 0 | 0 | 0 | 0 |
| 0 | 0 | 0 | 0 | 0 |
| 0 | 0 | 0 | 0 | 0 |
| 0 | 0 | 0 | 0 | 0 |
| 0 | 0 | 0 | 0 | 0 |
| 0 | 0 | 0 | 0.002584079 | 0 |
| 0 | 0 | 0 | 0.31177284 | 0 |
| 0 | 0.001002807 | 0 | 0 | 0 |
| 0.003578343 | 0.221672717 | 0 | 0.036814981 | 0.000765863 |

| | | | | |
|---|---|---|---|---|
| 0 | 0 | 0 | 0 | 0 |
| 0 | 0 | 0 | 0 | 0 |
| 0 | 0 | 0.006449576 | 0 | 0.000933107 |
| 0 | 0 | 0 | 0 | 0 |
| 0 | 0 | 0 | 0 | 0 |
| 0 | 0 | 0 | 0 | 0 |
| 0 | 0 | 0.001767557 | 0 | 0 |
| 0.024136922 | 0 | 0.002442603 | 0 | 0 |
| 0 | 0.046450504 | 0 | 0.062344495 | 0 |
| 0 | 0 | 0 | 0 | 0 |
| 0 | 0 | 0 | 0 | 0 |
| 0 | 0 | 0 | 0 | 0 |
| 0 | 0 | 0 | 0 | 0 |
| 0 | 0 | 0 | 0 | 0 |
| 0 | 0 | 0 | 0 | 0 |
| 0 | 0 | 0 | 0 | 0 |
| 0 | 0 | 0 | 0 | 0 |
| 0 | 0 | 0 | 0 | 0 |
| 0 | 0.008438406 | 0.05882035 | 0 | 0.001817382 |
| 0 | 1.045901183 | 0 | 0 | 0 |
| 0.009084509 | 0.021217343 | 0 | 0.002588845 | 0 |
| 0 | 0 | 0 | 0 | 0 |
| 0 | 0 | 0 | 0 | 0 |
| 0 | 0 | 0 | 0 | 0 |
| 0.006152829 | 0.002810182 | 0.031164567 | 0 | 0.011793337 |
| 0 | 0 | 0 | 0 | 0 |
| 0 | 0 | 0 | 0 | 0 |
| 0 | 0 | 0 | 0 | 0 |
| 0.099232304 | 0 | 0.037454818 | 0 | 0 |
| 0 | 0 | 0 | 0 | 0 |
| 0.100652984 | 0.844123728 | 0 | 0.36506366 | 0.123904541 |
| 0 | 0 | 0 | 0 | 0 |
| 0 | 0 | 0 | 0 | 0 |
| 0 | 0 | 0.009979612 | 0 | 0 |
| 0 | 0 | 0 | 0 | 0 |
| 0 | 0 | 0 | 0 | 0 |
| 0 | 0 | 0 | 0 | 0 |
| 0 | 0 | 0 | 0 | 0 |
| 0 | 0 | 0 | 0 | 0 |
| 0 | 0 | 0 | 0 | 0 |
| 0 | 0 | 0 | 0 | 0 |
| 0 | 0.059582233 | 0.00073483 | 0.005174346 | 0.001722274 |
| 0 | 0 | 0 | 0 | 0 |
| 0 | 0 | 0 | 0 | 0 |
| 0 | 0 | 0 | 0 | 0 |
| 0 | 0 | 0 | 0 | 0 |
| 0 | 0 | 0 | 0 | 0 |
| 0 | 0 | 0 | 0 | 0 |

| | | | | |
|---|---|---|---|---|
| 0 | 0 | 0 | 0 | 0 |
| 0 | 0 | 0 | 0 | 0 |
| 0.077919746 | 0.092121328 | 0.046780907 | 0.046563572 | 0.153724684 |
| 0 | 0 | 0 | 0 | 0 |
| 0 | 0.005545348 | 0 | 0 | 0.045980679 |
| 0 | 0 | 0 | 0 | 0 |
| 0 | 0 | 0 | 0 | 0 |
| 0 | 0 | 0 | 0 | 0 |
| 0 | 0 | 0 | 0 | 0 |
| 0 | 0 | 0.000337558 | 0 | 0 |
| 0 | 0 | 0 | 0 | 0 |
| 0 | 0 | 0 | 0 | 0 |
| 0 | 0 | 0 | 0 | 0 |
| 0 | 0.00529651 | 0.003875052 | 0 | 0 |
| 0.002481491 | 0 | 0.004629188 | 0.001728611 | 0.001808293 |
| 0 | 0 | 0 | 0.050497849 | 0 |
| 0.006058791 | 0.000691808 | 0.032798076 | 0 | 0.003184235 |
| 0 | 0 | 0 | 0 | 0 |
| 0 | 0 | 0 | 0 | 0 |
| 0 | 0 | 0 | 0 | 0 |
| 0 | 0 | 0 | 0 | 0 |
| 0.004010864 | 0.248303331 | 0 | 0.039369655 | 0.000507257 |
| 0 | 0 | 0 | 0 | 0 |
| 0 | 0 | 0 | 0 | 0.009436366 |
| 0 | 0 | 0 | 0 | 0 |
| 0 | 0 | 0 | 0 | 0 |
| 0 | 0 | 0 | 3.539063587 | 0 |
| 0 | 0 | 0 | 0 | 0 |
| 0 | 0 | 0 | 0 | 0 |
| 0.005538035 | 0 | 0 | 0 | 0 |
| 0 | 0 | 0.01605447 | 0 | 0.001036584 |
| 0 | 0 | 0 | 0 | 0 |
| 0 | 0 | 0 | 0 | 0 |
| 0.001681641 | 0 | 0 | 0 | 0 |
| 0 | 0 | 0 | 0 | 0 |
| 0.011652553 | 0 | 0.000864567 | 0 | 0.001013173 |
| 0 | 0 | 0 | 0 | 0 |
| 0 | 0.029188099 | 0.004894541 | 0.007658926 | 0.00594828 |
| 0 | 0 | 0 | 0 | 0 |
| 0 | 0 | 0 | 0 | 0 |
| 0 | 0 | 0 | 0 | 0 |
| 0 | 0 | 0 | 0 | 0 |
| 0 | 0 | 0 | 0 | 0 |
| 0 | 0 | 0 | 0 | 0 |
| 0 | 0 | 0 | 0 | 0 |
| 0.002113021 | 0 | 0.00217397 | 0 | 0 |
| 0.001241442 | 0 | 0 | 0 | 0 |
| 0.004439111 | 0 | 0 | 0 | 0 |

| | | | | |
|---|---|---|---|---|
| 0 | 6.6738672 | 0 | 0 | 0 |
| 0 | 0 | 0 | 0 | 0 |
| 0 | 0 | 0 | 0 | 0 |
| 0 | 0 | 0 | 0 | 0 |
| 0.002305461 | 0.006449463 | 0 | 0.001873653 | 0.005345511 |
| 0.00156636 | 0 | 0.00728912 | 0 | 0.002324357 |
| 0 | 0 | 0 | 0 | 0 |
| 0.012926444 | 0 | 0.214834808 | 0.006003058 | 0.133873326 |
| 0 | 0 | 0 | 0 | 0 |
| 0.002220594 | 0.01064923 | 0.107685896 | 0.007347634 | 0.018278014 |
| 0 | 0.001547402 | 0 | 0 | 0 |
| 0.076744646 | 0.010515478 | 0.046868829 | 0 | 0.073944966 |
| 0.068400907 | 0.00077548 | 0.178986921 | 0.001351722 | 0.010048187 |
| 0.046415959 | 0 | 0 | 0 | 0.054437083 |
| 0 | 0 | 0 | 0 | 0 |
| 0 | 0 | 0.000691606 | 0 | 0 |
| 0 | 0 | 0.000586762 | 0 | 0 |
| 0 | 0 | 0 | 0 | 0 |
| 0.011753656 | 0 | 0 | 0 | 0 |
| 0 | 0 | 0 | 0 | 0 |
| 0 | 0 | 0 | 0 | 0 |
| 0 | 0 | 0 | 0 | 0 |
| 0 | 0 | 0 | 0 | 0.002630796 |
| 0 | 0 | 0 | 0 | 0 |
| 0 | 0 | 0 | 0 | 0 |
| 0.003201852 | 0.000387101 | 0.094086137 | 0.002361618 | 0 |
| 0 | 0.001897987 | 0 | 0 | 0 |
| 0 | 0 | 0 | 0 | 0 |
| 0.00944903 | 0 | 0 | 0.002955279 | 0 |
| 0 | 0.000398278 | 0 | 0 | 0 |
| 0 | 0.004846108 | 0 | 0 | 0.046110585 |
| 0 | 0 | 0 | 0 | 0 |
| 0 | 0 | 0 | 0 | 0 |
| 0.161585705 | 0 | 0 | 0 | 0 |
| 0 | 0 | 0 | 0 | 0 |
| 0 | 0 | 0 | 0 | 0 |
| 0.052736354 | 0 | 0 | 0 | 0 |
| 0 | 0 | 0 | 0 | 0 |
| 0 | 0 | 0 | 0 | 0 |
| 0.001782809 | 0 | 0 | 0 | 0 |
| 0 | 0.27874576 | 0 | 0 | 0 |
| 0.181601944 | 0 | 0 | 0 | 0 |
| 0.005875298 | 0 | 0 | 0 | 0 |
| 0 | 0 | 0 | 0 | 0 |
| 0 | 0 | 0 | 0 | 0 |
| 0 | 0 | 0 | 0.001343886 | 0 |
| 0.000936884 | 0.017811466 | 0.000667321 | 0.003915811 | 0 |
| 0 | 0 | 0 | 0 | 0 |

| | | | | |
|---|---|---|---|---|
| 0 | 0 | 0 | 0 | 0 |
| 0.312539416 | 0 | 0.00198874 | 0 | 0 |
| 0 | 0 | 0 | 0 | 0 |
| 0 | 0 | 0 | 0 | 0.005377182 |
| 0 | 0 | 0 | 0 | 0 |
| 0.010330733 | 0.014028687 | 0 | 0.014906847 | 0 |
| 0 | 0.021501501 | 0 | 0 | 0.011795315 |
| 0.003340381 | 0 | 0 | 0 | 0 |
| 0 | 0 | 0 | 0 | 0 |
| 0 | 0 | 0 | 0 | 0 |
| 0.099920579 | 0.002052187 | 0 | 0.002682843 | 0 |
| 0 | 0 | 0 | 0 | 0.002789754 |
| 0 | 0 | 0 | 0 | 0 |
| 0.001212847 | 0.013156167 | 0.015453905 | 0 | 0.001874763 |
| 0.001431714 | 0 | 0.002233796 | 0 | 0.012329998 |
| 0 | 0 | 0 | 0 | 0 |
| 0 | 0 | 0 | 0 | 0 |
| 0.062752415 | 0 | 0 | 0 | 0 |
| 0.001290889 | 0.021667362 | 0 | 0 | 0.036914834 |
| 0 | 0 | 0 | 0 | 0 |
| 0 | 0 | 0.001965019 | 0 | 0 |
| 0.231513596 | 0.133713688 | 0 | 0 | 0 |
| 0 | 0 | 0 | 0 | 0 |
| 0 | 0 | 0 | 0 | 0 |
| 0 | 0 | 0 | 0 | 0 |
| 0 | 0 | 0 | 0 | 0 |
| 0.005917714 | 0 | 0 | 0 | 0.002439287 |
| 0 | 0 | 0 | 0 | 0 |
| 0 | 0 | 0 | 0 | 0 |
| 8.441491766 | 0.151025621 | 0 | 2.526758964 | 0.531190171 |
| 0 | 0.04960651 | 0 | 0.000573178 | 0.000890314 |
| 0 | 0.000527621 | 0 | 0 | 0.001428541 |
| 0 | 0 | 0 | 0 | 0 |
| 0 | 0.003193628 | 0 | 0 | 0 |
| 0 | 0 | 0 | 0.001780323 | 0 |
| 0 | 0 | 0 | 0 | 0 |
| 0 | 0 | 0 | 0 | 0 |
| 0.00144225 | 0.001270387 | 0 | 0 | 0.002484147 |
| 0 | 0 | 0 | 0 | 0 |
| 0 | 0 | 0 | 0 | 0 |
| 0.005317826 | 0 | 0 | 0.013428479 | 0.001541259 |
| 0 | 0.136495276 | 0 | 0.973306865 | 0 |
| 0.001132452 | 0 | 0 | 0 | 0.001225344 |
| 0 | 0 | 0 | 0 | 0 |
| 0 | 0 | 0 | 0 | 0 |
| 0 | 0 | 0 | 0 | 0 |
| 0.00156978 | 0.000716966 | 0.004969413 | 0 | 0.007117711 |
| 0 | 0.057936993 | 0 | 0 | 0 |

| | | | | |
|---|---|---|---|---|
| 0 | 0.001032051 | 0.064379923 | 0 | 0 |
| 0 | 0 | 0 | 0 | 0 |
| 7.348719997 | 0 | 17.64254186 | 0 | 0 |
| 0 | 0 | 0 | 0 | 0 |
| 0 | 0 | 0 | 0.104283211 | 0 |
| 0.012736921 | 0 | 0 | 0 | 0 |
| 0 | 0 | 0 | 0 | 0 |
| 0.005788828 | 0 | 0 | 0 | 0 |
| 0 | 0 | 0 | 0 | 0 |
| 0.014358725 | 0 | 0 | 0 | 0 |
| 0 | 0 | 0 | 0 | 0 |
| 0 | 0 | 0 | 0 | 0 |
| 0 | 0 | 0 | 0 | 0 |
| 0 | 0 | 0.001664083 | 0 | 0.001083397 |
| 0 | 0 | 0 | 0 | 0 |
| 0.095852796 | 0.753517601 | 0 | 0.218967288 | 0.001411093 |
| 0.003035509 | 0.035122353 | 0.559108961 | 0.024434705 | 0.00156405 |
| 0 | 0.001965171 | 0 | 0 | 0 |
| 0 | 0 | 0 | 0 | 0.005492853 |
| 0 | 0 | 0 | 0 | 0 |
| 0.017215225 | 0.019802387 | 0 | 0.009179184 | 0.163605238 |
| 0 | 0.01299931 | 0.000650209 | 0 | 0 |
| 0.035519821 | 0 | 0 | 0 | 0 |
| 0 | 0.128172758 | 0.003182283 | 0.328601867 | 0 |
| 0.023799085 | 0.033257146 | 0 | 0.056432738 | 0 |
| 0 | 0 | 0 | 0.00619008 | 0 |
| 0 | 0 | 0 | 0 | 0 |
| 0.000948393 | 0 | 0 | 0.015666907 | 0.001256556 |
| 0 | 0 | 0 | 0 | 0 |
| 0.001831874 | 0.00301202 | 0 | 0 | 0 |
| 0.014164831 | 0 | 0 | 0.015424665 | 0 |
| 0.003641039 | 0.003325944 | 0.025242693 | 0.007439974 | 0.152185111 |
| 0 | 0.628757956 | 0 | 0.009015823 | 0.705641176 |
| 0.004305779 | 0 | 0.004875595 | 0 | 0.009522731 |
| 0.002364024 | 0 | 0.001571583 | 0 | 0 |
| 0 | 0 | 0 | 0 | 0 |
| 0.000835784 | 0.002147219 | 0.173036697 | 0 | 0.004263324 |
| 0.02402061 | 0.022613564 | 0 | 0.047580355 | 0.071228859 |
| 0 | 0 | 0 | 0.100626211 | 0.109542774 |
| 0.002404813 | 0 | 0 | 0 | 0 |
| 0 | 0 | 0 | 0 | 0 |
| 0 | 0 | 0 | 0 | 0 |
| 0.008308671 | 0.00351579 | 0 | 0.046302694 | 0.010198977 |
| 0.032399768 | 0 | 0 | 0.007464009 | 0.304987842 |
| 0 | 0 | 0 | 0 | 0 |
| 0 | 0 | 0.004652067 | 0 | 0.037501014 |
| 0 | 0 | 0 | 0 | 0 |
| 0 | 0 | 0 | 0 | 0 |

| | | | | |
|---|---|---|---|---|
| 0.011144703 | 0 | 0 | 0 | 0 |
| 0.580699215 | 0.065136208 | 0.03531406 | 0.025127157 | 0.25214522 |
| 0 | 0 | 0 | 0 | 0 |
| 0 | 0 | 0 | 0 | 0 |
| 0 | 0 | 0 | 0 | 0 |
| 0 | 0 | 0 | 0 | 0 |
| 0 | 0 | 0 | 0 | 0 |
| 0.001881296 | 0 | 0 | 0 | 0 |
| 0 | 0 | 0 | 0 | 0.005104088 |
| 0 | 0 | 0 | 0 | 0 |
| 0.000661845 | 0.000906854 | 0 | 0 | 0.001074202 |
| 0.020456119 | 0.090671411 | 1.029525879 | 0.009270936 | 0.140576164 |
| 0 | 0 | 0 | 0.004360887 | 0 |
| 0 | 0 | 0 | 0 | 0 |
| 0 | 0.000390188 | 0.17326572 | 0.006347865 | 0.003169317 |
| 0 | 0 | 0 | 0 | 0 |
| 0.028068418 | 0.002374017 | 0.009461463 | 0.000965556 | 0.06170574 |
| 0.011920643 | 0.002619532 | 0 | 0 | 0.177727589 |
| 0 | 0 | 0 | 0 | 0 |
| 0.001460499 | 0 | 0 | 0.002645204 | 0.201420448 |
| 0.015366193 | 0.000856874 | 0.006957276 | 0.004231862 | 0.395393663 |
| 0 | 0 | 0 | 0 | 0 |
| 0 | 0 | 0 | 0 | 0 |
| 0 | 0 | 0 | 0 | 0 |
| 0.020114079 | 0 | 0 | 0 | 0 |
| 0 | 0 | 0 | 0 | 0 |
| 0.014109024 | 0.228359839 | 0 | 0.028993702 | 0.000981408 |
| 0.003228418 | 0.003252608 | 0 | 0.007408214 | 0.108495799 |
| 0 | 0 | 0.062683428 | 0 | 0 |
| 0 | 0.002354044 | 0 | 0.006382886 | 0.000849814 |
| 0.013041298 | 0 | 0 | 0 | 0 |
| 0 | 0 | 0 | 0 | 0 |
| 0 | 0 | 0 | 0.00224838 | 0 |
| 0 | 0 | 0 | 0 | 0 |
| 0 | 0 | 0 | 0 | 0 |
| 1.397473858 | 0.133877834 | 0 | 0.669885279 | 0.322233935 |
| 0 | 0 | 0 | 0 | 0 |
| 0 | 0 | 0 | 0 | 0 |
| 0.038654785 | 0.004654449 | 0.062519165 | 0 | 0.033894977 |
| 0 | 0 | 0 | 0 | 0 |
| 0 | 0 | 0 | 0 | 0 |
| 0 | 0 | 0 | 0 | 0 |
| 0.022625707 | 0 | 0.001418998 | 0 | 0.019136586 |
| 0 | 0 | 0.004175287 | 0 | 0.027268047 |
| 0 | 0 | 0 | 0 | 0.070308574 |
| 0 | 0 | 0 | 1.856622409 | 0 |
| 0 | 0.001655164 | 0 | 0 | 0 |
| 0 | 0.002890735 | 0 | 0 | 0.007208804 |

| | | | | |
|---|---|---|---|---|
| 0.002584351 | 0 | 0 | 0 | 0 |
| 0 | 0 | 0 | 0 | 0 |
| 0 | 0 | 0 | 0 | 0 |
| 0.079055366 | 0.001816703 | 0 | 0.900055924 | 0.004611324 |
| 0 | 0 | 0 | 0 | 0 |
| 0 | 0 | 0 | 0 | 0 |
| 0 | 0 | 0 | 0 | 0 |
| 0.001927475 | 0.000304732 | 0 | 0.168971468 | 0 |
| 0.202779468 | 0.004046308 | 0 | 0 | 0 |
| 0 | 0 | 0 | 0.001444587 | 0 |
| 0 | 0 | 0 | 0 | 0 |
| 0.003810996 | 0 | 0 | 0 | 0 |
| 0 | 0 | 0 | 0 | 0 |
| 0 | 0 | 0 | 0.409362303 | 0 |
| 0.005896581 | 0.195073574 | 0 | 3.117919469 | 0.048246932 |
| 0 | 0 | 0 | 0 | 0 |
| 0 | 0.001170991 | 0 | 0.04262559 | 0 |
| 0 | 0 | 0 | 0 | 0 |
| 0 | 0 | 0 | 0 | 0 |
| 0 | 0 | 0 | 0 | 0 |
| 0 | 0 | 0 | 0 | 0 |
| 0 | 0 | 0 | 0 | 0 |
| 0 | 0 | 0 | 0.197602979 | 0 |
| 0.015651332 | 0.132822542 | 0 | 0.264127881 | 1.357232814 |
| 0.000743347 | 0 | 0 | 0 | 0 |
| 0 | 0 | 0 | 0.435043366 | 0 |
| 0 | 0 | 0 | 0.001478694 | 0 |
| 0.9244379 | 0 | 0 | 0.076099471 | 0 |
| 0 | 0 | 0 | 0 | 0 |
| 0 | 0 | 0 | 0 | 0 |
| 0 | 0 | 0 | 0.046025457 | 0 |
| 0 | 0 | 0 | 0 | 0 |
| 0 | 0 | 0 | 0.034865067 | 0 |
| 0 | 0 | 0 | 0 | 0 |
| 0 | 0 | 0 | 0 | 0 |
| 0 | 0 | 0 | 0 | 0.002670683 |
| 0 | 0 | 0 | 0.564530208 | 0 |
| 0 | 0 | 0 | 0.012429864 | 0 |
| 0 | 0 | 0 | 0.026484422 | 0 |
| 0 | 0 | 0 | 0.006967389 | 0 |
| 0.05438254 | 0.046962881 | 0.169408693 | 0.004881281 | 0.158375614 |
| 0 | 0 | 0 | 0 | 0 |
| 3.68149995 | 0.164631397 | 0.007375856 | 0.21080504 | 0.545567348 |
| 0 | 0 | 0 | 0.396936761 | 0 |
| 0 | 0 | 0 | 0 | 0 |
| 0 | 0 | 0 | 0 | 0 |
| 0 | 0 | 0 | 0 | 0 |
| 0 | 0.008154578 | 0 | 0.6543939 | 0 |

| | | | | |
|---|---|---|---|---|
| 0 | 0 | 0 | 0 | 0 |
| 0 | 0 | 0 | 0 | 0 |
| 0.008601546 | 0.561867914 | 0.215892595 | 0.020298934 | 0.012047685 |
| 0 | 0 | 0 | 0.026171994 | 0 |
| 0 | 0 | 0 | 0 | 0 |
| 0.00082971 | 0 | 0 | 0.02061454 | 0 |
| 0 | 0 | 0 | 0 | 0 |
| 0 | 0 | 0 | 0 | 0 |
| 0 | 0 | 0 | 0 | 0 |
| 0 | 0 | 0 | 0 | 0 |
| 0 | 0.631474547 | 0 | 1.218848723 | 0 |
| 0.375881012 | 0.133313981 | 0.035036444 | 0.125855339 | 1.833582149 |
| 0 | 0 | 0 | 0 | 0 |
| 0.007626332 | 0.015546858 | 0.003621373 | 0.002850619 | 0.003105244 |
| 0.264726214 | 0.115184373 | 0.004037056 | 0.039005626 | 1.419153996 |
| 0 | 0 | 0 | 0.006885819 | 0 |
| 0 | 0 | 0 | 0 | 0 |
| 0 | 0 | 0 | 0 | 0 |
| 0 | 0 | 0 | 0 | 0 |
| 0 | 0.001458963 | 0 | 0.443853365 | 0 |
| 0.019212571 | 0.034341104 | 1.612801576 | 0 | 0.413046349 |
| 0.003348646 | 1.95721563 | 0.00556538 | 0.011239245 | 0.128886907 |
| 0.054303774 | 0.792298501 | 0.327705173 | 0.081739144 | 0.291808321 |
| 0 | 0 | 0 | 0 | 0 |
| 0 | 0 | 0 | 0 | 0 |
| 0 | 0 | 0 | 0 | 9.192198545 |
| 0 | 0 | 0 | 0 | 0.316315523 |
| 0.002862623 | 0.007638239 | 0 | 0.001049531 | 0 |
| 0 | 0 | 0 | 0 | 0 |
| 0 | 0 | 0 | 0 | 0 |
| 0 | 0 | 0 | 0.099003078 | 0 |
| 0 | 0.185087255 | 0 | 0.106400534 | 0.276871519 |
| 0.033631681 | 0.044262789 | 0.066822024 | 0 | 0 |
| 0 | 0 | 0 | 0.464505167 | 0 |
| 0.008878931 | 0.271810168 | 0 | 0.037761544 | 0.007584667 |
| 0.031300343 | 0.030131169 | 0 | 0.095937072 | 0.017864347 |
| 0 | 0.009454236 | 0 | 3.003800912 | 0.002876119 |
| 0 | 0 | 0 | 0 | 0 |
| 0 | 1.0718132 | 0 | 0 | 0.018117575 |
| 0 | 0 | 0 | 0 | 0 |
| 0 | 0 | 0 | 0 | 0 |
| 0 | 0 | 0 | 0 | 0 |
| 0 | 0 | 0 | 0 | 0 |
| 0 | 0 | 0 | 0 | 0 |
| 0.001281341 | 0.236139047 | 0 | 1.078241907 | 0.017146652 |
| 0 | 0 | 0 | 0 | 0 |
| 0 | 0 | 0 | 0 | 0 |
| 0.006624201 | 0 | 0 | 0.036915451 | 0.026827177 |

| | | | | |
|---|---|---|---|---|
| 0 | 0 | 0 | 0 | 0 |
| 0 | 0 | 0 | 0 | 0 |
| 0 | 0 | 0 | 0 | 0 |
| 0 | 0 | 0 | 0.000652068 | 0 |
| 0 | 0 | 0 | 0 | 0 |
| 0 | 0 | 0 | 0.035897913 | 0 |
| 0 | 0 | 0 | 0.002141671 | 0 |
| 0 | 0.35062325 | 0 | 0.661822741 | 0 |
| 0 | 0 | 0 | 0 | 0 |
| 0 | 0 | 0 | 0 | 0 |
| 0.002085068 | 0.717449022 | 0 | 0.025176012 | 0.011602801 |
| 0 | 0 | 0 | 0 | 0 |
| 0 | 0 | 0 | 0.047478452 | 0 |
| 0 | 0 | 0 | 0 | 0 |
| 0 | 0 | 0 | 0 | 0 |
| 0 | 0 | 0 | 0 | 0 |
| 0 | 0 | 0 | 0.001260291 | 0 |
| 0 | 0 | 0 | 0.002418283 | 0 |
| 0 | 0 | 0 | 0 | 0 |
| 0 | 0 | 0 | 0.131216947 | 0 |
| 0 | 0.001232563 | 0 | 0 | 0 |
| 0 | 0 | 0 | 0 | 0 |
| 0 | 0.203940166 | 0 | 0.093401606 | 0 |
| 0 | 0 | 0 | 0 | 0 |
| 0 | 0 | 0 | 0 | 0 |
| 0 | 0 | 0 | 0 | 0 |
| 0 | 0 | 0 | 0 | 0 |
| 0 | 0 | 0 | 0 | 0 |
| 0 | 0 | 0 | 0 | 0 |
| 0 | 0 | 0 | 1.049951208 | 0.065308761 |
| 0 | 0 | 0 | 0 | 0 |
| 0 | 0 | 0 | 0 | 0 |
| 0 | 0 | 0 | 0 | 0 |
| 0 | 0 | 0 | 0 | 0 |
| 0.001105296 | 0.000378617 | 0 | 0 | 0.002306496 |
| 0 | 0 | 0 | 0 | 0 |
| 0 | 0 | 0 | 0.062234527 | 0 |
| 0 | 0.01985144 | 0 | 0 | 0.102585061 |
| 0 | 0 | 0 | 0 | 0 |
| 0 | 0 | 0 | 0 | 0.00538214 |
| 0 | 0.45977725 | 0 | 0 | 0 |
| 0 | 0 | 0 | 0 | 0 |
| 0 | 0 | 0 | 0 | 0 |
| 0 | 0.00420791 | 0 | 0 | 0 |
| 0.236327617 | 0 | 0 | 0 | 0 |
| 0 | 0 | 0 | 0 | 0 |
| 0 | 0.080427624 | 0 | 0.0091514 | 0 |
| 0 | 0 | 0 | 0 | 0 |

| | | | | |
|---|---|---|---|---|
| 0 | 0 | 0 | 0 | 0 |
| 0 | 0 | 0 | 0 | 0 |
| 0 | 0 | 0 | 0 | 0 |
| 0 | 0 | 0 | 0.31778309 | 0 |
| 0 | 0 | 0 | 0 | 0 |
| 0 | 0 | 0 | 0 | 0 |
| 0 | 0 | 0 | 0 | 0 |
| 0 | 0 | 0 | 0 | 0 |
| 0.004462872 | 0.183449524 | 0.025141417 | 0.001695734 | 0.283904568 |
| 0 | 0 | 0 | 0 | 0 |
| 0 | 0.002782439 | 0 | 0 | 0 |
| 0 | 0 | 0 | 0 | 0 |
| 0 | 0 | 0 | 0 | 0 |
| 0 | 0 | 0 | 0 | 0 |
| 3.797687965 | 0.025866972 | 0 | 0.173244156 | 0 |
| 0.286586159 | 0 | 0 | 0.001306949 | 0 |
| 0 | 0 | 0 | 0.028794341 | 0 |
| 0 | 0 | 0 | 0 | 0 |
| 0 | 0 | 0 | 0 | 0 |
| 0 | 0 | 0 | 0 | 0 |
| 0 | 0 | 0 | 0.029174211 | 0 |
| 0 | 0 | 0 | 0 | 0 |
| 0 | 0 | 0 | 0 | 0 |
| 0 | 0 | 0 | 0 | 0.12073358 |
| 0.01240486 | 0.089561224 | 17.11145363 | 0 | 0.020133595 |
| 0 | 0 | 0 | 4.104947937 | 4.156325948 |
| 0 | 0 | 0 | 0.410160065 | 0 |
| 0 | 0 | 0 | 0 | 0 |
| 0.011888221 | 0 | 0 | 0 | 0.01489444 |
| 0 | 0 | 0 | 0 | 0 |
| 0 | 0 | 0 | 0 | 0 |
| 0 | 0 | 0 | 0.110444095 | 0 |
| 0 | 0 | 0 | 0 | 0 |
| 0.001911055 | 0 | 0 | 0.145371937 | 0 |
| 0.020992103 | 0.002732502 | 0.011263981 | 0.002632165 | 0 |
| 0.00118711 | 0.391900856 | 0.003100353 | 0.004548186 | 0.007294041 |
| 0.046246182 | 0 | 0 | 0 | 0 |
| 0 | 0.034093273 | 0 | 0 | 0.285781732 |
| 0 | 0 | 0 | 0 | 0 |
| 0 | 0 | 0 | 0 | 0 |
| 0 | 0 | 0 | 0 | 0 |
| 0 | 0.034930965 | 0 | 0 | 0 |
| 0.089018588 | 0.014447758 | 0 | 0.079667277 | 4.635610223 |
| 0 | 0 | 0 | 0 | 0 |
| 0 | 0 | 0 | 0 | 0 |
| 0 | 0.002600383 | 0 | 0.010967945 | 0.011734281 |
| 0 | 0 | 0 | 0 | 0 |
| 0.002239603 | 0.166322591 | 0.018610867 | 0.087366258 | 0.171778162 |

| | | | | |
|---|---|---|---|---|
| 0 | 0 | 0 | 0.019079383 | 0 |
| 0 | 0 | 0 | 0 | 0 |
| 0 | 0 | 0 | 0 | 0 |
| 0 | 0 | 0 | 0 | 0 |
| 0 | 0.014922784 | 0 | 0.013436009 | 0 |
| 0 | 0 | 0 | 0 | 0 |
| 0 | 0.00181372 | 0 | 0 | 0 |
| 0 | 0 | 0 | 0 | 0 |
| 0 | 0 | 0 | 0 | 0 |
| 0 | 0 | 0 | 0 | 0 |
| 0 | 0 | 0 | 0 | 0 |
| 1.239007971 | 2.582982711 | 0.000218526 | 0.108568144 | 0 |
| 0 | 0.003034467 | 0 | 0.104624255 | 1.856410622 |
| 0 | 0 | 0 | 0 | 0 |
| 5.642003271 | 0.017126457 | 0.344613786 | 0 | 0.102192558 |
| 0 | 0 | 0 | 0 | 0 |
| 0 | 0 | 0 | 0 | 0 |
| 0 | 0 | 0 | 0 | 0 |
| 0 | 0 | 0 | 0 | 0.011567687 |
| 0 | 0 | 0 | 0.587937305 | 0 |
| 0 | 0 | 0 | 0 | 0 |
| 0 | 0 | 0 | 0 | 0 |
| 0.108331251 | 0 | 0 | 1.687374269 | 0.03024801 |
| 0 | 0 | 0 | 0 | 0 |
| 0 | 0 | 0 | 0 | 0 |
| 0 | 0 | 0 | 0 | 0 |
| 0 | 0 | 0 | 0 | 0 |
| 0 | 0 | 0 | 0 | 0 |
| 0 | 0 | 0 | 0.001930549 | 0 |
| 0 | 0 | 0 | 0 | 0 |
| 0 | 0 | 0 | 0 | 0 |
| 0.040424291 | 0.012992476 | 0.074293781 | 0.004867098 | 0.130757699 |
| 0 | 0 | 0 | 0 | 0 |
| 0.001869933 | 0.534552955 | 0 | 0.183926999 | 0.116890017 |
| 0 | 0.35402238 | 0.003987408 | 0.005174547 | 0.007488438 |
| 0 | 0.102257145 | 0 | 0.065256048 | 0.002509911 |
| 0 | 0 | 0 | 0 | 0 |
| 0 | 0 | 0 | 0 | 0 |
| 0 | 0 | 0 | 0 | 0 |
| 0 | 0 | 0 | 0 | 0 |
| 0 | 0 | 0 | 0 | 0 |
| 0 | 0 | 0 | 0 | 0 |
| 0 | 0 | 0 | 0 | 0 |
| 0 | 0.003872779 | 0 | 0.251471363 | 0 |
| 0.001749858 | 7.50580965 | 0 | 0.338647796 | 0 |
| 0 | 0 | 0 | 0 | 0 |
| 0 | 0 | 0 | 0 | 0 |
| 0 | 0 | 0 | 0.00754634 | 0 |

| | | | | |
|---|---|---|---|---|
| 0 | 0 | 0 | 0 | 0 |
| 0 | 0 | 0 | 0 | 0 |
| 0.002138844 | 0 | 0 | 0.040113318 | 0.002441447 |
| 0 | 0 | 0 | 0 | 0 |
| 0 | 0 | 0 | 0.015807359 | 0.126274977 |
| 0 | 0 | 0 | 0 | 0 |
| 0 | 0 | 0 | 0 | 0 |
| 0 | 0 | 0 | 0.020781684 | 0 |
| 0 | 0 | 0 | 0 | 0 |
| 0 | 0 | 0 | 0 | 0 |
| 0 | 0 | 0 | 0 | 0 |
| 0 | 0.004931503 | 0 | 0 | 0 |
| 0 | 0 | 0 | 0 | 0 |
| 0 | 0 | 0 | 0 | 0 |
| 0 | 0 | 0 | 0 | 0 |
| 0 | 0 | 0 | 0 | 0 |
| 0 | 0 | 0 | 0.104883911 | 0 |
| 0 | 0 | 0 | 0 | 0 |
| 0 | 1.941751105 | 0 | 0.988284366 | 0.017283074 |
| 0 | 0 | 0 | 0 | 0 |
| 0 | 0 | 0 | 0 | 0 |
| 0 | 0 | 0 | 0 | 0 |
| 0 | 0 | 0 | 0 | 0 |
| 0 | 0 | 0 | 0 | 0 |
| 0 | 0 | 0.001155452 | 0 | 0 |
| 0.005479539 | 0.023234252 | 0.00021097 | 0 | 0.004944657 |
| 0 | 0 | 0 | 0 | 0 |
| 0.002899123 | 0.007448159 | 0.299215357 | 0.007068373 | 0.09397348 |
| 0 | 0 | 0 | 0 | 0 |
| 0 | 0.446327322 | 0 | 0.493012919 | 4.540584375 |
| 0 | 0 | 0 | 0 | 0 |
| 0 | 0 | 0 | 0 | 0 |
| 0 | 0.031773032 | 0 | 0 | 0 |
| 0 | 0 | 0 | 0 | 0 |
| 0.001086365 | 0.167565678 | 0 | 0.031351664 | 0 |
| 0 | 5.443629504 | 0 | 0 | 0 |
| 0 | 0 | 0 | 0 | 0 |
| 0.000704404 | 0.120766562 | 0 | 0.004416201 | 0 |
| 0 | 0 | 0 | 0 | 0 |
| 0 | 0 | 0 | 0 | 0 |
| 38.23584602 | 3.770531371 | 0 | 10.61471923 | 30.51385758 |
| 0 | 0.317574833 | 0 | 1.423097332 | 1.270030234 |
| 0 | 0 | 0 | 0 | 0 |
| 0 | 0 | 0 | 0 | 0 |
| 0 | 0 | 0 | 0 | 0 |
| 0 | 0 | 0 | 0 | 0 |
| 0 | 0 | 0 | 0 | 0 |
| 0 | 0.026044347 | 0 | 0.032518071 | 0.000648119 |

| | | | | |
|---|---|---|---|---|
| 0 | 0 | 0 | 0 | 0 |
| 0 | 0.022750749 | 0 | 0.014620772 | 0 |
| 0 | 0 | 0 | 0.076492186 | 0 |
| 0 | 0 | 0 | 0 | 0 |
| 0.091313954 | 0.004839789 | 0 | 0.683665201 | 3.075993243 |
| 0 | 0 | 0 | 0 | 0 |
| 0 | 0 | 0 | 0 | 0 |
| 0 | 0 | 0 | 0 | 0 |
| 0.266852574 | 0.139548555 | 0 | 0 | 0.907429749 |
| 0 | 0 | 0 | 0 | 0 |
| 0 | 0 | 0.004087564 | 0 | 0 |
| 0 | 0 | 0 | 0 | 0 |
| 0 | 0 | 0 | 0.014206572 | 0.232415788 |
| 0.318729558 | 0.013882808 | 0 | 0.013876727 | 0.02166081 |
| 0 | 0 | 0 | 0 | 0 |
| 0 | 0 | 0 | 0.076961373 | 0.40489731 |
| 0.101200048 | 0.006006709 | 0.006633137 | 0 | 0 |
| 0 | 0 | 0 | 0 | 0 |
| 0 | 0 | 0 | 0 | 0 |
| 0 | 0 | 0 | 0 | 0 |
| 0 | 0 | 0 | 0 | 0 |
| 0 | 0 | 0 | 0 | 0 |
| 0 | 0 | 0 | 0 | 0 |
| 0 | 0 | 0 | 0 | 0 |
| 0 | 0 | 0 | 0 | 0 |
| 0 | 0 | 0 | 0 | 0 |
| 0.043417319 | 0 | 0 | 0.056888632 | 0 |
| 0 | 0.004694951 | 0 | 0.007109924 | 0.002028453 |
| 0 | 0 | 0 | 0 | 0 |
| 0 | 0 | 0 | 0 | 0 |
| 0 | 0 | 0 | 0 | 0 |
| 0 | 0 | 0 | 0 | 0 |
| 0 | 0 | 0 | 0.076201192 | 0 |
| 0 | 0 | 0 | 0 | 0 |
| 0 | 1.588237602 | 0 | 0 | 0 |
| 0 | 0.008666623 | 0 | 0 | 0.030504486 |
| 0 | 0 | 0.000654919 | 0 | 0 |
| 0 | 0 | 0 | 0 | 0 |
| 0 | 0.000613409 | 0 | 0 | 0 |
| 0 | 0 | 0 | 0 | 0 |
| 0 | 0 | 0.000490467 | 1.145460968 | 0.021266545 |
| 0 | 0 | 0 | 0 | 0 |
| 0 | 0 | 0 | 0 | 0 |
| 0 | 0 | 0 | 0 | 0 |
| 0 | 0 | 0 | 0 | 0 |
| 0 | 0 | 0 | 0 | 0 |
| 0 | 0 | 0 | 0 | 0 |
| 0 | 0 | 0 | 0 | 0 |

| | | | | |
|---|---|---|---|---|
| 0 | 0 | 0 | 0.968639228 | 0 |
| 0 | 0 | 0 | 0 | 0 |
| 0 | 0 | 0 | 0 | 0 |
| 0.000825861 | 0.000339476 | 0 | 0 | 0.002757407 |
| 0 | 0 | 0 | 0 | 0 |
| 0 | 0 | 0 | 0.480412337 | 0 |
| 0.285962132 | 0.028944903 | 0.025552915 | 0 | 0.080568511 |
| 0 | 0 | 0 | 0.001996422 | 0 |
| 0 | 0 | 0 | 0 | 0 |
| 0 | 0 | 0 | 0 | 0 |
| 0 | 0 | 0 | 0.032559275 | 0 |
| 0 | 0 | 0 | 0.299644676 | 0 |
| 0 | 0 | 0 | 0 | 0 |
| 0 | 0 | 0 | 0 | 0 |
| 0 | 0 | 0 | 0 | 0 |
| 0 | 0 | 0 | 0 | 0 |
| 0 | 0 | 0.087488846 | 0 | 0 |
| 0 | 0.071765623 | 0 | 0 | 0 |
| 0 | 0 | 0 | 1.010754045 | 0 |
| 0 | 0 | 0 | 0 | 0 |
| 0.002681589 | 0.012376549 | 0.227193497 | 0.004522527 | 0.003926914 |
| 0 | 0 | 0 | 0 | 0 |
| 0 | 0 | 0 | 0 | 0.00158817 |
| 0.000692129 | 0 | 0 | 0.013114173 | 0.00038515 |
| 0 | 0 | 0 | 0 | 0 |
| 0 | 0 | 0 | 0 | 0.004089515 |
| 0 | 0 | 0 | 0 | 0 |
| 0 | 0 | 0 | 0 | 0 |
| 0 | 0 | 0 | 0 | 0 |
| 0.170937025 | 0.091721061 | 4.590531543 | 0.508220153 | 0.461934381 |
| 0 | 0 | 0 | 0 | 0 |
| 0 | 0 | 0 | 0 | 0 |
| 0 | 0 | 0 | 0 | 0 |
| 0 | 3.416207322 | 0 | 0 | 0 |
| 0 | 0 | 0 | 0 | 0 |
| 0 | 0 | 0 | 0 | 0 |
| 0.033255674 | 0 | 0.030678271 | 0 | 0 |
| 0 | 0 | 0 | 0 | 0 |
| 0 | 0 | 0 | 0 | 0 |
| 0 | 0 | 0 | 0 | 0 |
| 0 | 0 | 0 | 0 | 0 |
| 0 | 0 | 0 | 0 | 0 |
| 0 | 0 | 0 | 0 | 0 |
| 0 | 0 | 0 | 0 | 0 |
| 0 | 0 | 0 | 0 | 0 |
| 0 | 0 | 0 | 0 | 0 |
| 0 | 0 | 0 | 0 | 0 |
| 0 | 0 | 0 | 0 | 0 |

| | | | | |
|---|---|---|---|---|
| 0.01027763 | 0.206846701 | 0 | 0.053124953 | 0.009670154 |
| 0 | 0 | 0 | 0 | 0 |
| 0 | 0 | 0 | 0 | 0 |
| 0 | 0 | 0 | 0 | 0 |
| 0 | 0 | 0 | 0 | 0 |
| 0 | 0 | 0 | 0 | 0 |
| 0 | 0 | 0 | 0.002233435 | 0 |
| 0.105680976 | 0.046311834 | 0 | 1.667528612 | 1.290210711 |
| 0.020106906 | 2.656680875 | 17.05445867 | 0.003583061 | 0.393129844 |
| 0.002278716 | 0.303752692 | 0 | 0 | 0.019624422 |
| 0 | 0 | 0 | 0.020756204 | 0 |
| 0 | 0 | 0 | 0 | 0 |
| 0 | 0 | 0 | 0 | 0 |
| 0 | 0 | 0 | 0 | 0 |
| 0 | 0 | 0 | 0 | 0 |
| 0 | 0 | 0 | 0 | 0 |
| 0.141111175 | 0 | 0 | 0 | 0 |
| 0 | 0 | 0 | 0 | 0 |
| 0 | 0 | 0 | 0 | 0 |
| 0 | 0.042996972 | 0 | 0 | 0 |
| 0 | 0 | 0 | 0 | 0 |
| 0 | 0 | 0 | 0 | 0 |
| 0 | 0 | 0 | 0 | 0 |
| 0 | 0 | 0 | 0 | 0 |
| 0 | 0 | 0 | 0 | 0.056689165 |
| 0 | 0 | 0 | 0 | 0 |
| 0 | 0 | 0 | 0 | 0 |
| 0 | 0 | 0 | 0 | 0 |
| 0 | 0 | 0 | 0 | 0 |
| 0 | 0 | 0 | 0.320076678 | 0 |
| 0 | 0.728599321 | 0 | 0.192510955 | 0 |
| 0 | 0 | 0 | 0 | 0 |
| 0 | 0 | 0 | 0 | 0 |
| 0 | 0 | 0 | 0 | 0 |
| 0 | 0 | 0 | 0 | 0 |
| 0 | 0 | 0 | 0 | 0 |
| 0 | 0 | 0 | 0 | 0 |
| 0 | 0 | 0 | 0 | 0 |
| 0 | 0 | 0 | 0 | 0 |
| 0 | 0 | 0.15661371 | 0 | 1.977382605 |
| 0 | 0 | 0 | 0 | 0 |
| 0 | 1.070976108 | 0 | 0 | 0 |
| 0 | 0 | 0 | 0 | 0 |
| 0 | 0 | 0 | 0 | 0 |
| 0 | 0 | 0 | 0 | 0 |
| 0 | 0 | 0 | 0 | 0 |
| 0 | 0 | 0 | 0 | 0 |
| 0 | 0 | 0 | 0 | 0 |

| | | | | |
|---|---|---|---|---|
| 0 | 0 | 0 | 0 | 0 |
| 0 | 0 | 0 | 0 | 0.004239186 |
| 0 | 0 | 0 | 0 | 0 |
| 0 | 0 | 0 | 0 | 0 |
| 0.072552155 | 6.705428731 | 0.004551879 | 2.242351224 | 0.048984734 |
| 0 | 0 | 0 | 0 | 0 |
| 0 | 0 | 0 | 0 | 0 |
| 0.00795802 | 0.371514968 | 0.066580184 | 0.004751633 | 0.108425725 |
| 0 | 0 | 0 | 0 | 0 |
| 0 | 0 | 0 | 0 | 0 |
| 0 | 0 | 0 | 0 | 0 |
| 0 | 0 | 0 | 0 | 0 |
| 0 | 0 | 0 | 0 | 0 |
| 0 | 0 | 0 | 0 | 0 |
| 0 | 0 | 0 | 0 | 0 |
| 0.000492995 | 0 | 0 | 0 | 0.015393386 |
| 0 | 0 | 0 | 0 | 0 |
| 0 | 0 | 0 | 0 | 0 |
| 0 | 0 | 0 | 0 | 0 |
| 0 | 0 | 0 | 0 | 0 |
| 0 | 0 | 0 | 0 | 0 |
| 0 | 0 | 0 | 0 | 0 |
| 0 | 0 | 0 | 0 | 0 |
| 0.000491929 | 0.816146368 | 0 | 0.02078919 | 0.00532281 |
| 0 | 0 | 0.00083133 | 0.151631073 | 0 |
| 0 | 0 | 0 | 0 | 0 |
| 0.003149477 | 0.01582307 | 0.001780396 | 0.809665224 | 0.011683953 |
| 0 | 0 | 0 | 0 | 0 |
| 0 | 0.003054421 | 0 | 0 | 0 |
| 0 | 0 | 0 | 0 | 0 |
| 0 | 0 | 0 | 0 | 0 |
| 0 | 0 | 0 | 0 | 0 |
| 0 | 0 | 0 | 0 | 0 |
| 0 | 0 | 0 | 0 | 0 |
| 0 | 0 | 0 | 0 | 0 |
| 0 | 0 | 0 | 0 | 0 |
| 0 | 0 | 0 | 0 | 0 |
| 0 | 0 | 0 | 1.401003363 | 0.000998548 |
| 0 | 0 | 0 | 0.485282547 | 0 |
| 0 | 0 | 0 | 0 | 0 |
| 0 | 0 | 0 | 0 | 0 |
| 0 | 0 | 0 | 0 | 0 |
| 0 | 0 | 0 | 0 | 0 |
| 0 | 0 | 0 | 0 | 0 |
| 0 | 0 | 0 | 0 | 0 |
| 0 | 0 | 0 | 0 | 0 |
| 0 | 0 | 0 | 0 | 0 |
| 0 | 0 | 0 | 0 | 0 |

| | | | | |
|---|---|---|---|---|
| 0 | 0 | 0 | 1.811885722 | 0.539446249 |
| 0 | 0 | 0 | 0 | 0 |
| 0 | 0 | 0 | 0 | 0 |
| 0 | 0 | 0 | 0 | 0 |
| 0 | 0 | 0 | 0 | 0 |
| 0 | 0 | 0.091092255 | 0.005918939 | 0 |
| 0 | 0 | 0 | 0 | 0 |
| 0 | 0 | 0 | 0 | 0 |
| 0 | 0 | 0 | 0.005371995 | 0 |
| 0 | 0 | 0 | 0 | 0 |
| 0 | 0 | 0 | 0 | 0 |
| 0 | 0 | 0 | 0 | 0 |
| 0 | 0 | 0 | 0 | 0 |
| 0 | 0 | 0 | 0 | 0 |
| 0 | 0 | 0 | 0 | 0 |
| 0 | 0 | 0 | 0 | 0 |
| 0 | 0 | 0 | 0 | 0 |
| 0.022780791 | 0 | 0 | 0 | 0 |
| 0 | 0 | 0 | 0 | 0 |
| 0 | 0 | 0 | 0.001523466 | 0 |
| 0.147451759 | 1.421306751 | 0 | 0 | 0.303191977 |
| 0 | 0 | 0 | 0 | 0 |
| 0 | 0.15442287 | 0 | 0.025570852 | 0.002868951 |
| 0 | 0 | 0 | 0.038655676 | 0 |
| 0 | 0 | 0 | 0 | 0 |
| 0 | 0 | 0 | 0 | 0 |
| 0 | 0 | 0 | 0 | 0 |
| 0 | 0 | 0 | 0 | 0 |
| 0 | 0 | 0 | 0 | 0 |
| 0 | 0 | 0 | 0 | 0 |
| 0 | 0 | 0 | 0 | 0 |
| 0 | 0 | 0 | 0 | 0 |
| 1.575735492 | 0.038502623 | 0.012655608 | 0.06807405 | 0.081856658 |
| 0 | 0 | 0 | 0 | 0 |
| 0 | 0.004734077 | 0 | 0.500372688 | 1.975668817 |
| 0.00061991 | 0 | 0 | 0 | 0 |
| 0 | 0 | 0 | 0 | 0 |
| 0 | 0 | 0 | 0 | 0 |
| 0.009142058 | 0 | 0 | 0.003246623 | 0.003990033 |
| 0 | 0 | 0 | 0 | 0 |
| 0 | 0 | 0 | 0 | 0 |
| 0 | 0 | 0 | 0 | 0 |
| 0.019493173 | 0.047100377 | 0 | 0.011680845 | 0 |
| 0 | 0 | 0 | 0 | 0 |
| 0 | 0 | 0 | 0 | 0 |
| 0 | 0.003535311 | 0 | 0.020176642 | 0.043691083 |
| 0 | 0 | 0 | 0 | 0 |
| 0 | 0 | 0 | 0 | 0 |

| | | | | |
|---|---|---|---|---|
| 0 | 0 | 0 | 0 | 0 |
| 0 | 0 | 0 | 0 | 0 |
| 0 | 0 | 0 | 0 | 0 |
| 0 | 0 | 0 | 0 | 0 |
| 0 | 0 | 0 | 0 | 0 |
| 0 | 0 | 0 | 0 | 0 |
| 0 | 0 | 0 | 0.78363964 | 0 |
| 0.109059668 | 0.004959044 | 0.09387369 | 0 | 0.799185412 |
| 0 | 0 | 0 | 1.95707024 | 0 |
| 0 | 0.015739947 | 0 | 0.003105314 | 0 |
| 0 | 0 | 0 | 0 | 0 |
| 0 | 0 | 0 | 0 | 0 |
| 0 | 0 | 0 | 0 | 0 |
| 0 | 0 | 0 | 0 | 0 |
| 0 | 0 | 0 | 0 | 0 |
| 0.000674767 | 0.004507226 | 0.003043932 | 0 | 0 |
| 0 | 0 | 0 | 0 | 0 |
| 0 | 0 | 0 | 0 | 0 |
| 0 | 0 | 0 | 0 | 0 |
| 0 | 0.032801227 | 0 | 0.340801913 | 0 |
| 0 | 0 | 0 | 0 | 0.037256241 |
| 0 | 0 | 0 | 0 | 0 |
| 0 | 0 | 0 | 0 | 0 |
| 0 | 0 | 0 | 0.513095728 | 0 |
| 0 | 0 | 0 | 0 | 0 |
| 0 | 0 | 0 | 0 | 0 |
| 0 | 0 | 0 | 0 | 0 |
| 0 | 0 | 0 | 0 | 0 |
| 0 | 0 | 0 | 0 | 0 |
| 0 | 0 | 0 | 0 | 0 |
| 0 | 0.017753144 | 0 | 0.041069145 | 0 |
| 0 | 0 | 0 | 0 | 0 |
| 0 | 0 | 0 | 0.015003981 | 0 |
| 0 | 0 | 0 | 0.458505649 | 0 |
| 0 | 0 | 0 | 0 | 0 |
| 0 | 0 | 0 | 0 | 0 |
| 0 | 0 | 0 | 0 | 0 |
| 0 | 0 | 0 | 0 | 0 |
| 0 | 0 | 0 | 0 | 0 |
| 0 | 0 | 0 | 0 | 0 |
| 0 | 0 | 0 | 0 | 0 |
| 0 | 0 | 0 | 9.325057188 | 0 |
| 0 | 0 | 0 | 0.009297187 | 0 |
| 0 | 0 | 0 | 0 | 0 |
| 0 | 0 | 0 | 0 | 0 |
| 0 | 0 | 0 | 0 | 0 |
| 0.022794883 | 0.037858583 | 0.053874882 | 0 | 0 |
| 0 | 0 | 0 | 0 | 0 |

| | | | | |
|---|---|---|---|---|
| 0 | 0 | 0 | 0 | 0 |
| 0 | 0 | 0 | 0 | 0 |
| 0 | 0 | 0 | 0 | 0 |
| 0 | 0 | 0 | 0 | 0 |
| 0.04360579 | 0.039242925 | 0 | 1.183221349 | 0.188810386 |
| 0 | 0 | 0 | 0 | 0 |
| 0 | 0 | 0 | 0 | 0 |
| 0 | 0.000706426 | 0 | 0 | 0 |
| 0 | 0 | 0 | 0 | 0 |
| 0 | 0 | 0 | 0 | 0 |
| 0 | 0 | 0 | 0 | 0 |
| 0.001432906 | 1.183289297 | 0 | 0.106055072 | 0 |
| 0 | 0.006563786 | 0 | 0 | 0 |
| 0 | 0 | 0 | 0 | 0 |
| 0 | 0 | 0 | 0.060093066 | 0 |
| 0.111985927 | 0 | 0 | 1.147828051 | 0 |
| 0 | 0 | 0 | 0 | 0 |
| 0 | 0 | 0 | 0 | 0 |
| 0.05628463 | 0.280143129 | 0 | 0.00774712 | 0.009182396 |
| 0.41615146 | 0.010190929 | 0.015220176 | 0 | 0.509632082 |
| 0 | 0 | 0 | 0 | 0 |
| 0 | 0 | 0 | 0 | 0 |
| 0 | 0 | 0 | 0 | 0 |
| 0 | 0 | 0 | 0 | 0 |
| 0 | 0 | 0 | 0 | 0 |
| 0 | 0 | 0 | 0 | 0 |
| 0.000851441 | 0.070387096 | 0 | 0.008501326 | 0.003685132 |
| 0 | 0 | 0 | 0 | 0 |
| 0 | 0 | 0 | 0 | 0 |
| 0 | 0 | 0 | 0 | 0 |
| 0 | 0.001102037 | 0 | 0.00649917 | 0.001641078 |
| 0 | 0 | 0 | 0 | 0 |
| 0 | 0 | 0 | 0 | 0 |
| 0 | 0 | 0 | 0 | 0 |
| 0 | 0 | 0 | 0 | 0 |
| 0 | 0 | 0 | 0 | 0 |
| 0 | 0 | 0 | 0 | 0 |
| 0.008250082 | 0.332699145 | 0 | 0.198862742 | 0.061422067 |
| 0 | 0 | 0 | 0 | 0 |
| 0 | 0 | 0 | 0 | 0 |
| 0 | 0 | 0.005089643 | 0 | 0.00136032 |
| 0 | 0 | 0 | 0 | 0 |
| 0 | 0 | 0 | 0 | 0 |
| 0.00351932 | 0.023766252 | 0 | 0 | 0.656063958 |
| 0 | 0 | 0 | 0 | 0 |
| 0 | 0 | 0 | 0 | 0 |
| 0 | 0 | 0 | 0 | 0 |
| 0 | 0 | 0 | 0 | 0 |

| | | | | |
|---|---|---|---|---|
| 0 | 0 | 0 | 0 | 0 |
| 0 | 0 | 0 | 0.003991178 | 3.586837111 |
| 0 | 0 | 0 | 0.006457235 | 0 |
| 0.002810105 | 0.007580434 | 0 | 0.015170816 | 0 |
| 0.003095014 | 0 | 0.000541458 | 0 | 0 |
| 0 | 0 | 0 | 0 | 0 |
| 0 | 0 | 0 | 0 | 0 |
| 0.006593236 | 0 | 0 | 0 | 0 |
| 0 | 0 | 0 | 0.030811209 | 0 |
| 0 | 0 | 0 | 0 | 0 |
| 0 | 0 | 0 | 0 | 0 |
| 0 | 0 | 0 | 0 | 0 |
| 0.007594381 | 0.221164805 | 0 | 0.293826288 | 0.006812492 |
| 0 | 0 | 0 | 0 | 0 |
| 0 | 0 | 0 | 0 | 0 |
| 0 | 0 | 0 | 0 | 0 |
| 0 | 0 | 0 | 0 | 0 |
| 0 | 0 | 0 | 0 | 0 |
| 0 | 0 | 0 | 0 | 0 |
| 0 | 0 | 0 | 0 | 0 |
| 0 | 0 | 0 | 0 | 0 |
| 0 | 0 | 0 | 0 | 0 |
| 0.001311482 | 0.019991403 | 0 | 0.032203727 | 0 |
| 0 | 0.000662656 | 0 | 0.38675291 | 0 |
| 0 | 0 | 0 | 0 | 0 |
| 0 | 0 | 0 | 0 | 0 |
| 0 | 0 | 0 | 0 | 0 |
| 0 | 0 | 0 | 0 | 0 |
| 0 | 0 | 0 | 0 | 0 |
| 0 | 0 | 0 | 0 | 0 |
| 0 | 0.010143351 | 0 | 0.012732983 | 0.101546211 |
| 0 | 0.000484192 | 0 | 0 | 0 |
| 0 | 0 | 0 | 0 | 0 |
| 0 | 0 | 0 | 0 | 0 |
| 0 | 0 | 0 | 0 | 0 |
| 0 | 0.002618169 | 0 | 0 | 0.000886091 |
| 0 | 0.026608809 | 0 | 0 | 0 |
| 0.062384809 | 0 | 0 | 0 | 0 |
| 0.011753441 | 0 | 0.000587488 | 0 | 0.002868619 |
| 0 | 0 | 0 | 0 | 0 |
| 0 | 0 | 0 | 0 | 0 |
| 0 | 0 | 0 | 0 | 0 |
| 0 | 0 | 0 | 0 | 0 |
| 0 | 0 | 0 | 0 | 0 |
| 0 | 0 | 0 | 0 | 0 |
| 0 | 0.268097275 | 0 | 0.351081741 | 0.458342195 |
| 0 | 0 | 0 | 0 | 0 |
| 0 | 0 | 0 | 0 | 0 |

| | | | | |
|---|---|---|---|---|
| 0 | 0 | 0 | 0 | 0 |
| 0 | 0 | 0 | 0 | 0 |
| 0 | 0 | 0 | 0 | 0 |
| 0 | 0 | 0 | 0.334864639 | 0 |
| 0 | 0 | 0 | 0 | 0 |
| 0 | 0 | 0 | 0.002268851 | 0 |
| 0 | 0 | 0 | 0 | 0 |
| 0 | 0 | 0 | 0 | 0 |
| 0 | 0 | 0 | 0.010565951 | 0 |
| 0 | 0 | 0 | 0 | 0 |
| 0 | 0.003610073 | 0 | 0.006817027 | 0 |
| 0 | 0.008726424 | 0 | 0.016963072 | 0.000868636 |
| 0 | 0 | 0 | 0 | 0 |
| 0 | 0 | 0 | 0 | 0 |
| 0 | 0 | 0 | 0 | 0 |
| 0 | 0 | 0 | 0 | 0 |
| 0 | 0 | 0 | 0 | 0 |
| 0 | 0 | 0 | 0 | 0 |
| 0 | 0 | 0 | 0 | 0 |
| 0 | 0.018352378 | 0 | 0 | 0 |
| 0 | 0 | 0 | 0 | 0 |
| 0 | 0 | 0 | 0 | 0 |
| 0 | 0 | 0 | 0 | 0 |
| 0 | 0 | 0 | 0 | 0 |
| 0 | 0 | 0 | 0 | 0 |
| 0 | 0.01035401 | 0 | 0 | 0 |
| 0 | 0 | 0 | 0 | 0 |
| 0 | 0 | 0 | 0 | 0.020151469 |
| 0 | 0 | 0 | 0 | 0 |
| 0 | 0.000491649 | 0.00136308 | 0 | 0 |
| 0.000857654 | 0.010478418 | 0 | 0 | 0.005302888 |
| 0 | 0 | 0 | 0 | 0 |
| 0.010290181 | 0.037977705 | 0.001655039 | 0.001849848 | 0.049257539 |
| 0 | 0 | 0 | 0 | 0.002202194 |
| 0 | 0 | 0 | 0 | 0 |
| 0 | 0 | 0 | 0 | 0 |
| 0 | 0 | 0 | 0 | 0 |
| 0 | 0 | 0 | 0 | 0 |
| 0 | 0 | 0 | 0 | 0 |
| 0 | 0 | 0 | 0 | 0 |
| 0 | 0 | 0 | 0 | 0 |
| 0.022802606 | 0 | 0.003630894 | 0.175583783 | 0.060912233 |
| 0.049904199 | 0.140403354 | 0 | 0.030823515 | 0 |
| 0 | 0 | 0 | 0 | 0 |
| 0 | 0 | 0 | 0 | 0 |
| 0.001923068 | 0.214420597 | 0 | 0.036839349 | 0 |
| 0 | 0 | 0 | 0 | 0 |
| 0 | 0 | 0 | 0 | 0 |

| | | | | |
|---|---|---|---|---|
| 0 | 0 | 0 | 0 | 0 |
| 0.010281982 | 0.003939941 | 0 | 0 | 0 |
| 0.018321012 | 0.001673552 | 0.146445864 | 0 | 0.019446238 |
| 0 | 0 | 0 | 0 | 0 |
| 0.009589759 | 0 | 0 | 0.139266124 | 0 |
| 0 | 0 | 0 | 0.021160712 | 0 |
| 0 | 0 | 0 | 0 | 0 |
| 0 | 0 | 0 | 0 | 0 |
| 0 | 0 | 0 | 0 | 0 |
| 0 | 0 | 0 | 0 | 0 |
| 0 | 0 | 0 | 0 | 0 |
| 0 | 0 | 0 | 0 | 0 |
| 0 | 0 | 0 | 0 | 0 |
| 0 | 0 | 0 | 0 | 0 |
| 0 | 0 | 0 | 0 | 0 |
| 0 | 0 | 0 | 0 | 0 |
| 0 | 0 | 0 | 2.41908793 | 0 |
| 0 | 0 | 0 | 0 | 0 |
| 0 | 0 | 0 | 0 | 0 |
| 0 | 0 | 0 | 0 | 0 |
| 0 | 0 | 0 | 0 | 0 |
| 0.002619935 | 0 | 0 | 0 | 0 |
| 0 | 0 | 0 | 0 | 0 |
| 0 | 0 | 0 | 0 | 0 |
| 0 | 0.005976715 | 0 | 0 | 0 |
| 0.001856473 | 0.003699959 | 0.655070721 | 0 | 0.004852317 |
| 0 | 0 | 0 | 0 | 0 |
| 0 | 0 | 0 | 0.324565956 | 0 |
| 0 | 0 | 0 | 0 | 0 |
| 0 | 0 | 0 | 0 | 0 |
| 0 | 0 | 0 | 0 | 0 |
| 0 | 0.001216742 | 0 | 0 | 0 |
| 0 | 0 | 0 | 0 | 0 |
| 0 | 0 | 0 | 0 | 0 |
| 0 | 0 | 0 | 0 | 0 |
| 0 | 0 | 0 | 0 | 0 |
| 0 | 0 | 0 | 0 | 0 |
| 0.002183956 | 0 | 0 | 0 | 0 |
| 0 | 1.109714612 | 0 | 1.752037539 | 0 |
| 0 | 0 | 0 | 0 | 0 |
| 0 | 0 | 0 | 0 | 0 |
| 0.023618579 | 0.012919694 | 0 | 0 | 0 |
| 0.011179042 | 0.163385771 | 0 | 0.045072193 | 0.008325837 |
| 0 | 0 | 0 | 0 | 0 |
| 0 | 0 | 0 | 0 | 0 |
| 0 | 0 | 0 | 0.001085947 | 0 |
| 0 | 0.004438002 | 0 | 0.006859063 | 0.003364465 |
| 0 | 0 | 0 | 0 | 0 |

| | | | | |
|---|---|---|---|---|
| 0 | 0 | 0 | 0 | 0 |
| 0.000817396 | 0.105726901 | 0.000698655 | 0.031886385 | 0.00697448 |
| 0 | 0 | 0 | 0 | 0 |
| 0 | 0 | 0 | 0 | 0 |
| 0 | 0 | 0 | 0 | 0 |
| 0 | 0 | 0 | 0 | 0 |
| 0 | 0 | 0 | 0 | 0 |
| 0 | 0 | 0 | 0 | 0 |
| 0 | 0 | 0 | 0 | 0 |
| 0 | 0 | 0 | 0 | 0 |
| 0 | 0 | 0 | 0 | 0 |
| 0 | 0 | 0 | 0 | 0 |
| 0 | 0.012549626 | 0 | 0.245577638 | 0 |
| 0 | 0 | 0 | 0 | 0 |
| 0.001223004 | 0.019969331 | 0 | 0.009371415 | 0.004348063 |
| 0 | 0 | 0 | 0 | 0 |
| 0 | 0 | 0 | 0 | 0 |
| 0 | 0 | 0 | 0 | 0 |
| 0 | 0 | 0 | 0.011309685 | 0.001254804 |
| 0 | 0 | 0 | 0 | 0.003210475 |
| 0 | 0 | 0 | 0 | 0 |
| 0.068461912 | 0.004251435 | 0.00076045 | 0 | 0.080018723 |
| 0.000461468 | 0.063967617 | 0 | 0.028502757 | 0 |
| 0 | 0.012148077 | 0 | 0.001436292 | 0 |
| 0 | 0 | 0 | 0 | 0 |
| 0 | 0 | 0 | 0 | 0 |
| 0 | 0 | 0 | 0 | 0 |
| 0 | 0 | 0 | 0 | 0 |
| 0 | 0 | 0 | 0 | 0 |
| 0 | 0 | 0 | 0 | 0 |
| 0 | 0 | 0 | 0 | 0 |
| 0 | 0 | 0 | 0 | 0 |
| 0 | 0 | 0 | 0 | 0 |
| 0 | 0 | 0 | 0 | 0 |
| 0 | 0 | 0 | 0 | 0.093049202 |
| 0 | 0 | 0 | 0 | 0 |
| 0 | 0 | 0 | 0 | 0 |
| 0 | 0 | 0 | 0 | 0 |
| 0.004597815 | 0.022460448 | 0.948206997 | 0.00167105 | 0.048575394 |
| 0 | 0 | 0 | 0 | 0 |
| 0.140883626 | 0.093203172 | 0 | 0.166562984 | 0.593302185 |
| 0 | 0 | 0 | 0 | 0 |
| 0 | 0 | 0 | 0 | 0 |
| 0 | 0 | 0 | 0 | 0 |
| 0 | 0 | 0 | 0 | 0 |
| 0.002462628 | 3.075225326 | 0.045898245 | 0 | 1.508181255 |
| 0 | 0 | 0 | 0 | 0 |
| 0 | 0 | 0 | 0 | 0 |

| | | | | |
|---|---|---|---|---|
| 0 | 0 | 0 | 0 | 0 |
| 0 | 0 | 0 | 0 | 0 |
| 0 | 0 | 0 | 0 | 0 |
| 0 | 0 | 0 | 0 | 0 |
| 0 | 0 | 0 | 0 | 0 |
| 0 | 0 | 0 | 0 | 0 |
| 0 | 0 | 0 | 0 | 0 |
| 0.002218128 | 0 | 0.008636901 | 0 | 0.001440045 |
| 0 | 0 | 0 | 0 | 0 |
| 0.583849517 | 0.000687509 | 0.035453382 | 0 | 0.010982486 |
| 0 | 0 | 0 | 0 | 0 |
| 0 | 0 | 0 | 0 | 0 |
| 0 | 0.022981777 | 0 | 0.004372917 | 0.00054582 |
| 0 | 0 | 0 | 0 | 0 |
| 0 | 0 | 0 | 0 | 0 |
| 0 | 0 | 0 | 0 | 0 |
| 0 | 0 | 0 | 0 | 0 |
| 0 | 0 | 0 | 0 | 0 |
| 0 | 0 | 0 | 0 | 0 |
| 0.027762472 | 6.957229684 | 0.000580386 | 0.060329172 | 0.062670533 |
| 0 | 0 | 0 | 0.047364027 | 0 |
| 0 | 0 | 0 | 0 | 0 |
| 0 | 0 | 0 | 0 | 0 |
| 0 | 0 | 0 | 0 | 0 |
| 0 | 0 | 0 | 0 | 0 |
| 0 | 0 | 0 | 0 | 0 |
| 0 | 0 | 0 | 0 | 0 |
| 0 | 0 | 0 | 0 | 0 |
| 0 | 0 | 0 | 0 | 0 |
| 0.072049767 | 0.079540186 | 0 | 0 | 0 |
| 0 | 0 | 0 | 0 | 0 |
| 0 | 0 | 0 | 0 | 0 |
| 0 | 0 | 0 | 0 | 0 |
| 0 | 0 | 0 | 0 | 0 |
| 0 | 0 | 0 | 0 | 0 |
| 0 | 0 | 0 | 0 | 0.012996861 |
| 0 | 0 | 0 | 0 | 0 |
| 0 | 0 | 0 | 0 | 0 |
| 0 | 0 | 0 | 0 | 0 |
| 0 | 0 | 0 | 0.567518966 | 0 |
| 0 | 0 | 0 | 0 | 0 |
| 0 | 0 | 0 | 0 | 0 |
| 0 | 0 | 0 | 0.36053976 | 0 |
| 0 | 0 | 0 | 0 | 0 |
| 0 | 0.05325561 | 0 | 0.05232223 | 0 |
| 0 | 0 | 0 | 0 | 0 |
| 0 | 0 | 0 | 0 | 0 |
| 0 | 0 | 0 | 0 | 0 |

| | | | | |
|---|---|---|---|---|
| 0 | 0 | 0 | 0 | 0 |
| 0 | 0 | 0 | 0 | 0 |
| 0 | 0 | 0 | 0 | 0 |
| 0 | 0 | 0 | 0 | 0 |
| 0 | 0 | 0 | 0 | 0 |
| 0.009182264 | 0.02793082 | 0 | 0.080850281 | 0.07233015 |
| 0 | 0 | 0 | 0 | 0 |
| 0 | 0 | 0 | 0 | 0 |
| 0 | 0 | 0 | 0 | 0 |
| 0 | 0 | 0 | 0 | 0 |
| 0 | 0 | 0 | 0 | 0 |
| 0 | 0 | 0 | 0 | 0 |
| 0 | 0 | 0 | 0 | 0 |
| 0 | 0 | 0 | 0 | 0 |
| 0 | 0 | 0 | 0 | 0 |
| 0 | 0 | 0 | 0 | 0 |
| 0 | 0 | 0 | 0 | 0 |
| 0 | 0 | 0 | 0 | 0 |
| 0 | 0 | 0 | 0 | 0 |
| 0 | 0 | 0 | 0 | 0 |
| 0 | 0 | 0 | 0 | 0 |
| 0 | 0 | 0 | 0 | 0 |
| 0 | 0 | 0 | 0 | 0 |
| 0 | 0 | 0 | 0 | 0 |
| 0 | 0 | 0 | 0 | 0 |
| 0.013213593 | 0 | 0 | 0 | 0 |
| 0 | 0 | 0 | 0 | 0 |
| 0 | 0.087685132 | 0 | 0 | 0 |
| 0 | 0 | 0 | 0 | 0 |
| 0 | 0 | 0 | 0 | 0 |
| 0 | 0 | 0 | 0 | 0 |
| 0 | 0 | 0 | 0 | 0 |
| 0 | 0 | 0 | 0 | 0 |
| 0 | 0 | 0 | 0 | 0 |
| 0 | 0 | 0 | 0 | 0 |
| 0 | 0 | 0 | 0 | 0 |
| 0 | 0 | 0 | 0 | 0 |
| 0 | 0 | 0 | 0 | 0 |
| 0 | 0 | 0 | 0.956358884 | 0 |
| 0 | 0 | 0 | 0 | 0 |
| 0 | 0 | 0 | 0 | 0 |
| 0 | 0 | 0 | 0 | 0 |
| 0 | 0 | 0 | 0 | 0 |
| 0 | 0 | 0 | 0 | 0 |
| 0 | 0 | 0 | 0 | 0 |
| 0 | 0 | 0 | 0 | 0 |
| 0 | 0 | 0 | 0 | 0 |
| 0 | 0 | 0 | 0 | 0 |

| | | | | |
|---|---|---|---|---|
| 0 | 0 | 0 | 0 | 0 |
| 0.002330198 | 0.094845381 | 0 | 0.053852722 | 0 |
| 0 | 0 | 0 | 0 | 0 |
| 0 | 0 | 0 | 0 | 0 |
| 0 | 0 | 0 | 0 | 0 |
| 0 | 0 | 0 | 0 | 0 |
| 0 | 0 | 0 | 0 | 0 |
| 0 | 0 | 0 | 0 | 0 |
| 0 | 0 | 0 | 0 | 0 |
| 0 | 0 | 0 | 0 | 0 |
| 0 | 0.014386657 | 0 | 0 | 0.000832309 |
| 0 | 0 | 0 | 0 | 0 |
| 0.422902154 | 0 | 0 | 0.155631337 | 0 |
| 0 | 0 | 0 | 0.074828776 | 0 |
| 0 | 0 | 0 | 0 | 0 |
| 0 | 0 | 0 | 0.02880292 | 0 |
| 0 | 0 | 0 | 0 | 0 |
| 0 | 0 | 0 | 0 | 0 |
| 0 | 0 | 0 | 0 | 0 |
| 0 | 0 | 0 | 0 | 0 |
| 0.001457315 | 0.003868798 | 0.685087572 | 0 | 0.00253423 |
| 0 | 0 | 0 | 0 | 0 |
| 0 | 0 | 0 | 0 | 0 |
| 0.57494755 | 0.060996681 | 0 | 1.275113199 | 3.622647418 |
| 0 | 0 | 0 | 0 | 0 |
| 0 | 0 | 0 | 0 | 0 |
| 0 | 0 | 0 | 0 | 0 |
| 0 | 0 | 0 | 0 | 0 |
| 0.001043335 | 0 | 0 | 0 | 0 |
| 0 | 0 | 0 | 0 | 0 |
| 0 | 0 | 0 | 0.002740065 | 0 |
| 0 | 0 | 0 | 0 | 0 |
| 0 | 0 | 0 | 0 | 0 |
| 0 | 0 | 0 | 0 | 0 |

| PHL018-0083_V1 | PHL018-0229_V2 | PHL018-0067_V3 | PHL018-0095_V1 | PHL018-0063_V3 |
|---|---|---|---|---|
| REF | REF | EYCF | CTRL | EYCF |
| 0.085427124 | 0.002453468 | 0.032604648 | 0.025778041 | 1.870707417 |
| 3.16121743 | 1.025302436 | 0.446876071 | 0.043496371 | 1.874157928 |
| 2.504614412 | 0 | 30.06932998 | 0.780859322 | 0.639854817 |
| 0 | 0 | 0 | 0 | 0 |
| 0 | 0 | 0 | 0 | 0 |
| 0 | 0 | 0 | 0 | 0 |
| 0 | 0 | 0 | 0 | 0 |
| 0 | 0 | 0 | 0 | 0 |
| 0 | 0 | 0 | 0 | 0 |
| 0 | 0 | 0 | 0 | 0 |
| 0 | 0 | 0 | 0 | 0 |
| 0 | 0 | 0 | 0 | 0 |
| 0 | 0 | 0 | 0 | 0 |
| 4.7125193 | 0.377499951 | 12.48091556 | 0.02368724 | 2.082220925 |
| 0 | 0 | 0 | 0 | 0 |
| 0 | 0 | 0 | 0 | 0 |
| 0 | 0 | 0 | 0 | 0 |
| 0 | 0 | 0 | 0 | 0 |
| 0 | 0 | 0 | 0 | 0 |
| 0 | 0 | 0 | 0 | 0 |
| 0 | 0 | 0 | 0 | 0 |
| 0 | 0 | 0.001220779 | 0 | 0 |
| 0 | 0 | 0 | 0 | 0 |
| 0 | 0 | 0 | 0 | 0 |
| 0 | 0 | 0 | 0.024128967 | 0 |
| 0 | 0 | 0 | 0 | 0 |
| 0 | 0 | 0 | 0 | 0 |
| 0 | 0 | 0 | 0 | 0 |
| 0 | 0 | 0 | 0 | 0 |
| 0 | 0.003210117 | 0 | 0 | 0 |
| 0 | 0 | 0 | 0 | 0 |
| 0 | 0 | 0 | 0 | 0 |
| 0 | 0 | 0 | 0 | 0 |
| 0 | 0 | 0 | 0 | 0 |
| 0 | 0 | 0 | 0 | 0 |
| 0 | 2.367596304 | 1.341223998 | 0.288350504 | 3.454496621 |
| 0 | 0 | 0 | 0 | 0 |
| 0 | 0 | 0 | 0 | 0 |
| 0 | 0 | 0 | 0 | 0 |
| 0 | 0 | 0 | 0 | 0 |
| 0 | 0 | 0 | 0 | 0 |
| 0.00067179 | 0.094990672 | 0 | 0.005549361 | 0.012361387 |
| 0.014018061 | 1.604999999 | 0.030485588 | 4.33372973 | 1.500896796 |
| 0.107292773 | 0.00751479 | 0.013283916 | 0.000513686 | 0.001876815 |
| 0 | 0.015958172 | 0 | 0 | 0.005232054 |
| 0 | 0 | 0 | 0 | 0 |

| | | | | |
|---|---|---|---|---|
| 0.011836836 | 0 | 0 | 0.007611121 | 0.008768348 |
| 0.075201618 | 0 | 0.07817188 | 0 | 0.016407845 |
| 0.014754431 | 0.001016699 | 0 | 0 | 0.004788209 |
| 1.216535535 | 0.674222262 | 0.53393287 | 0.152864713 | 1.2031407 |
| 0 | 0.001465576 | 0.001621822 | 0 | 0 |
| 0 | 0 | 0 | 0 | 0.00079379 |
| 0.001062576 | 0.000753121 | 0 | 0 | 0.016896893 |
| 0 | 0 | 0 | 0.002262044 | 0 |
| 0 | 0 | 0 | 0 | 0 |
| 0 | 0 | 0 | 0 | 0 |
| 0 | 0 | 0 | 0 | 0 |
| 0.036958723 | 0.249670517 | 0.205330809 | 0.066226785 | 0.04599476 |
| 0 | 0 | 0 | 0 | 0 |
| 0.037070778 | 0.019012493 | 0.086758339 | 0 | 0 |
| 0 | 0 | 0 | 0 | 0 |
| 0 | 0 | 0 | 0 | 0 |
| 0.082862117 | 11.78818882 | 0 | 0.120428665 | 0.040457388 |
| 0 | 0 | 0 | 0 | 0 |
| 0 | 0 | 0 | 0 | 0 |
| 0 | 0 | 0 | 0 | 0 |
| 0.219641122 | 0.000569939 | 0.443630345 | 0.020662238 | 0.717771336 |
| 0.004745686 | 0.000668896 | 0.025061331 | 0.015016904 | 0.032097135 |
| 0 | 0 | 0 | 0 | 0 |
| 0 | 0 | 0 | 0 | 0 |
| 0 | 0 | 0 | 0 | 0 |
| 0 | 0 | 0 | 0 | 0 |
| 0 | 0 | 0 | 0 | 0 |
| 0 | 0 | 0 | 0 | 0 |
| 0 | 0.086940344 | 0 | 0 | 0 |
| 0 | 0 | 0 | 0 | 0 |
| 0.075894462 | 0 | 0 | 0 | 0 |
| 0 | 0 | 0 | 0 | 0 |
| 0 | 0 | 0 | 0 | 0 |
| 0 | 0 | 0 | 0 | 0 |
| 0 | 0 | 0 | 0 | 0 |
| 0 | 0 | 0 | 0 | 0 |
| 0 | 0 | 0 | 0 | 0 |
| 2.252157922 | 11.66761652 | 0 | 6.219833282 | 0 |
| 0 | 0 | 0 | 0 | 0 |
| 0 | 0 | 0 | 0 | 0 |
| 0 | 0 | 0 | 0 | 0 |
| 0 | 0 | 0 | 0 | 0 |
| 0 | 0 | 0 | 0 | 0 |
| 0 | 0 | 0 | 0 | 0 |
| 0 | 0 | 0 | 0 | 0 |
| 0 | 0 | 0 | 0 | 0 |
| 0 | 0.025967297 | 0 | 0 | 0 |
| 0.098181935 | 0 | 0 | 0 | 0.000371319 |

| | | | | |
|---|---|---|---|---|
| 0 | 0 | 0 | 0 | 0 |
| 0 | 0 | 0 | 0 | 0 |
| 0 | 0 | 0 | 0 | 0 |
| 0 | 0 | 0 | 0 | 0 |
| 0 | 0.009593371 | 0 | 0.002453563 | 0 |
| 0 | 0 | 0 | 0 | 0 |
| 0 | 0 | 0 | 0 | 0 |
| 0 | 0 | 0 | 0 | 0 |
| 0 | 0 | 0 | 0 | 0 |
| 0 | 0 | 0 | 0 | 0 |
| 0 | 0 | 0 | 0 | 0 |
| 0 | 0 | 0 | 0 | 0.001099808 |
| 0 | 0 | 0 | 0 | 0 |
| 0 | 0 | 0 | 0 | 0 |
| 0 | 0 | 0 | 0 | 0 |
| 0 | 0 | 0 | 0 | 0 |
| 0 | 0 | 0 | 0 | 0 |
| 0 | 0 | 0 | 0 | 0 |
| 0 | 0.000400811 | 0 | 0 | 0 |
| 0 | 0 | 0 | 0 | 0 |
| 0 | 0 | 0 | 0 | 0 |
| 0 | 0 | 0 | 0 | 0 |
| 0 | 0 | 0 | 0 | 0 |
| 0 | 0 | 0 | 0 | 0 |
| 0 | 0 | 0 | 0 | 0 |
| 0 | 0 | 0 | 0 | 0 |
| 0 | 0 | 0 | 0 | 0 |
| 0 | 0 | 0 | 0 | 0 |
| 0 | 0 | 0 | 0 | 0 |
| 0 | 0 | 0 | 0 | 0 |
| 0 | 0.000837606 | 0.009079876 | 0.01285337 | 0.003068145 |
| 0 | 0.002636165 | 0 | 0.001483273 | 0 |
| 0 | 0 | 0 | 0 | 0 |
| 0 | 0 | 0 | 0 | 0 |
| 0 | 0 | 0 | 0 | 0 |
| 0 | 0 | 0 | 0 | 0 |
| 0 | 0 | 0 | 0 | 0 |
| 0 | 0 | 0 | 0 | 0 |
| 0 | 0 | 0 | 0.273289498 | 0 |
| 0 | 0 | 0 | 0 | 0 |
| 0 | 0 | 0 | 0 | 0 |
| 0 | 0 | 0 | 0 | 0 |
| 0 | 0 | 0.012845075 | 0 | 0.002657405 |
| 0 | 0 | 0 | 0 | 0 |
| 0 | 0 | 0 | 0 | 0 |
| 0 | 0 | 0 | 0 | 0 |
| 0 | 0 | 0 | 0 | 0 |
| 0 | 0 | 0 | 0 | 0 |

| | | | | |
|---|---|---|---|---|
| 0 | 0 | 0 | 0 | 0 |
| 0 | 0 | 0 | 0 | 0 |
| 0 | 0 | 0 | 0 | 0 |
| 0 | 0 | 0 | 0 | 0 |
| 0 | 0.169573257 | 0 | 0.019658739 | 0 |
| 0 | 0 | 0 | 0 | 0 |
| 0 | 0 | 0 | 0 | 0 |
| 0 | 0 | 0 | 0 | 0 |
| 0 | 0 | 0 | 0 | 0 |
| 0 | 0 | 0 | 0 | 0 |
| 0 | 0 | 0 | 0 | 0 |
| 0 | 0 | 0 | 0 | 0 |
| 0 | 0 | 0 | 0 | 0 |
| 0 | 0 | 0 | 0 | 0 |
| 0 | 0 | 0 | 0 | 0 |
| 0 | 1.027320164 | 0 | 0 | 0 |
| 0 | 0 | 0 | 0 | 0 |
| 0 | 0 | 0 | 0 | 0 |
| 0 | 0 | 0 | 0 | 0 |
| 0 | 0 | 0 | 0 | 0 |
| 0 | 0 | 0 | 0 | 0 |
| 0 | 0 | 0 | 0 | 0 |
| 0 | 0 | 0 | 0 | 0 |
| 0 | 0 | 0 | 0 | 0 |
| 0 | 0 | 0 | 0 | 0.037472558 |
| 0 | 0 | 0 | 0 | 0 |
| 0 | 0 | 0 | 0 | 0 |
| 0 | 0 | 0 | 0 | 0 |
| 0 | 0 | 0 | 0 | 0 |
| 0 | 0 | 0 | 0 | 0 |
| 0 | 0 | 0 | 0 | 0 |
| 0 | 0.08241195 | 0 | 0 | 0.001989506 |
| 0 | 0 | 0 | 0 | 0 |
| 0 | 0 | 0 | 0 | 0 |
| 0 | 0 | 0 | 0 | 0 |
| 0 | 0 | 0 | 0 | 0 |
| 0 | 0 | 0 | 0 | 0 |
| 0 | 0 | 0 | 0 | 0 |
| 0 | 0 | 0 | 0 | 0 |
| 0 | 0 | 0 | 0 | 0 |
| 0 | 0 | 0 | 0 | 0 |
| 0 | 0 | 0 | 0 | 0 |
| 0 | 0 | 0 | 0 | 0 |
| 0 | 0 | 0 | 0 | 0 |
| 0 | 0 | 0 | 0 | 0 |
| 0 | 0 | 0 | 0 | 0 |
| 0 | 0 | 0 | 0 | 0 |
| 0 | 0 | 0 | 0 | 0 |

| | | | | |
|---|---|---|---|---|
| 0 | 0 | 0 | 0 | 0 |
| 0 | 0 | 0 | 0 | 0 |
| 0 | 0 | 0 | 0 | 0 |
| 0 | 0 | 0 | 0 | 0 |
| 0 | 0 | 0 | 0 | 0 |
| 0 | 0 | 0 | 0 | 0 |
| 0 | 0 | 0 | 0 | 0 |
| 0 | 0 | 0 | 0 | 0 |
| 0 | 0 | 0 | 0 | 0 |
| 0 | 0 | 0 | 0 | 0 |
| 0 | 0 | 0 | 0 | 0 |
| 0 | 0 | 0 | 0 | 0 |
| 0 | 0 | 0.00032865 | 0 | 0.002157597 |
| 0 | 0 | 0 | 0 | 0 |
| 0 | 0 | 0 | 0 | 0 |
| 0 | 0 | 0 | 0 | 0 |
| 0 | 0 | 0 | 0 | 0 |
| 0 | 0 | 0 | 0 | 0 |
| 0 | 0 | 0 | 0 | 0 |
| 0 | 0 | 0 | 0 | 0 |
| 0.105506671 | 0.005960175 | 0.021712204 | 0.030062848 | 0.023730519 |
| 0 | 0 | 0 | 0 | 0 |
| 0 | 0 | 0 | 0 | 0 |
| 0.001089329 | 0 | 0.002563184 | 0 | 0 |
| 0 | 0 | 0 | 0 | 0 |
| 0 | 0 | 0 | 0 | 0 |
| 0.004623554 | 0 | 0.000715736 | 0.001377936 | 0 |
| 0 | 0 | 0 | 0 | 0 |
| 0 | 0 | 0 | 0 | 0 |
| 0 | 0 | 0 | 0 | 0 |
| 0.63178291 | 0 | 0.005469918 | 0 | 0 |
| 0 | 0 | 0 | 0 | 0 |
| 0 | 0 | 0 | 0 | 0 |
| 0 | 0.007126608 | 0 | 0 | 0.071787966 |
| 0.009787475 | 0.006238534 | 0.008129756 | 0 | 0.037674431 |
| 0 | 0 | 0 | 0 | 0 |
| 0 | 0 | 0 | 0 | 0 |
| 0 | 0 | 0 | 0 | 0 |
| 0 | 0 | 0 | 0 | 0 |
| 0 | 0 | 0 | 0 | 0 |
| 0 | 0 | 0 | 0 | 0 |
| 0 | 0 | 0 | 0 | 0 |
| 0 | 0 | 0 | 0 | 0 |
| 0 | 0 | 0 | 0 | 0 |
| 0 | 0 | 0 | 0 | 0 |
| 0 | 0 | 0 | 0 | 0 |
| 0 | 0.000912794 | 0.000721505 | 0 | 0.020228517 |
| 0.005327987 | 0 | 0.001234677 | 0.015294548 | 0.002750801 |

| | | | | |
|---|---|---|---|---|
| 0 | 0 | 0 | 0 | 0 |
| 0 | 0 | 0 | 0 | 0 |
| 0 | 0 | 0.001041436 | 0.000471758 | 0 |
| 0 | 0.000610932 | 0 | 0 | 0 |
| 0 | 0 | 0 | 0 | 0 |
| 0 | 0 | 0 | 0 | 0 |
| 0 | 0 | 0 | 0 | 0 |
| 0 | 0.000518177 | 0 | 0 | 0 |
| 0 | 0 | 0.005708219 | 0 | 0 |
| 0 | 0 | 0 | 0 | 0 |
| 0 | 0 | 0 | 0 | 0 |
| 0 | 0 | 0 | 0 | 0 |
| 0 | 0 | 0 | 0 | 0 |
| 0 | 0 | 0 | 0 | 0 |
| 0 | 0 | 0 | 0 | 0 |
| 0 | 0 | 0 | 0 | 0 |
| 0 | 0 | 0 | 0 | 0 |
| 0 | 0 | 0 | 0 | 0 |
| 0 | 0.007331831 | 0 | 0.023101974 | 0.006378406 |
| 0 | 0.05209204 | 0 | 0 | 0 |
| 0 | 0 | 0.006269065 | 0 | 0 |
| 0 | 0 | 0 | 0 | 0 |
| 0 | 0 | 0 | 0 | 0 |
| 0 | 0 | 0 | 0 | 0 |
| 0.002021033 | 0.005371669 | 0 | 0 | 0.000983819 |
| 0 | 0 | 0 | 0 | 0 |
| 0 | 0 | 0 | 0 | 0 |
| 0 | 0 | 0 | 0 | 0 |
| 0 | 0.005468013 | 0 | 0 | 0 |
| 0 | 0 | 0 | 0 | 0 |
| 0.101300889 | 0.036183468 | 0.247034847 | 0.105497206 | 0.822669099 |
| 0 | 0 | 0 | 0 | 0 |
| 0 | 0 | 0.07711778 | 0 | 0.0436154 |
| 0 | 0 | 0 | 0.00055259 | 0 |
| 0 | 0 | 0 | 0 | 0 |
| 0 | 0 | 0 | 0 | 0 |
| 0 | 0 | 0 | 0 | 0 |
| 0 | 0 | 0 | 0 | 0 |
| 0 | 0 | 0 | 0 | 0 |
| 0 | 0 | 0 | 0 | 0.000606176 |
| 0 | 0 | 0 | 0 | 0 |
| 0 | 0.001891437 | 0.011319746 | 0 | 0.015712444 |
| 0 | 0 | 0 | 0 | 0 |
| 0 | 0 | 0 | 0 | 0 |
| 0 | 0 | 0 | 0 | 0 |
| 0 | 0 | 0 | 0 | 0 |
| 0 | 0 | 0 | 0 | 0 |
| 0 | 0 | 0 | 0 | 0 |

| | | | | |
|---|---|---|---|---|
| 0 | 0 | 0 | 0 | 0 |
| 0 | 0 | 0 | 0 | 0 |
| 0.136908062 | 0.007125033 | 0.015018358 | 0 | 0 |
| 0 | 0 | 0 | 0 | 0 |
| 0 | 0 | 0 | 0 | 0 |
| 0 | 0 | 0 | 0 | 0 |
| 0 | 0 | 0 | 0 | 0 |
| 0 | 0 | 0 | 0 | 0 |
| 0 | 0 | 0 | 0 | 0 |
| 0 | 0 | 0.00049056 | 0.000583322 | 0 |
| 0 | 0 | 0 | 0 | 0 |
| 0 | 0 | 0 | 0 | 0 |
| 0 | 0 | 0 | 0 | 0 |
| 0 | 0 | 0.00207475 | 0 | 0.003262124 |
| 0.001996165 | 0 | 0.001467801 | 0.004238717 | 0.000991958 |
| 0 | 0.055170062 | 0 | 0 | 0 |
| 0.001847991 | 0.000528957 | 0 | 0.001704578 | 0.000968783 |
| 0.000551353 | 0 | 0 | 0 | 0 |
| 0 | 0 | 0 | 0 | 0 |
| 0 | 0 | 0 | 0 | 0 |
| 0 | 0 | 0 | 0 | 0 |
| 0.004363007 | 0 | 0.000880673 | 0.015216503 | 0.001749069 |
| 0 | 0 | 0 | 0 | 0 |
| 0 | 0 | 0 | 0 | 0 |
| 0 | 0 | 0 | 0 | 0 |
| 0 | 0 | 0 | 0 | 0 |
| 0 | 0 | 0 | 0 | 0 |
| 0 | 0 | 0 | 0 | 0 |
| 0 | 0 | 0 | 0 | 0 |
| 0 | 0 | 0 | 0 | 0 |
| 0 | 0 | 0 | 0 | 0 |
| 0 | 0 | 0 | 0 | 0 |
| 0 | 0 | 0 | 0 | 0 |
| 0 | 0 | 0 | 0 | 0.001848615 |
| 0 | 0 | 0 | 0 | 0 |
| 0 | 0 | 0 | 0 | 0 |
| 0 | 0 | 0 | 0 | 0 |
| 0 | 0.00339958 | 0 | 0.000537021 | 0.005310683 |
| 0 | 0 | 0 | 0 | 0 |
| 0 | 0 | 0 | 0 | 0 |
| 0 | 0 | 0 | 0 | 0 |
| 0 | 0 | 0 | 0 | 0 |
| 0 | 0 | 0 | 0 | 0 |
| 0 | 0 | 0 | 0 | 0 |
| 0 | 0 | 0 | 0 | 0 |
| 0 | 0 | 0 | 0 | 0 |
| 0 | 0 | 0 | 0 | 0 |
| 0.006834958 | 0 | 0 | 0 | 0.00243994 |

| | | | | |
|---|---|---|---|---|
| 0 | 0 | 0.919004796 | 0.028130596 | 0 |
| 0 | 0 | 0 | 0 | 0 |
| 0 | 0.000666855 | 0 | 0 | 0 |
| 0 | 0 | 0 | 0 | 0 |
| 0 | 0.001341841 | 0.002519019 | 0 | 0.001689584 |
| 0 | 0 | 0 | 0 | 0 |
| 0 | 0 | 0 | 0 | 0 |
| 0.289257191 | 0.032445266 | 0.005017679 | 0 | 0.001291813 |
| 0 | 0 | 0 | 0 | 0 |
| 0.066818564 | 0 | 0.124507034 | 0 | 0.126936279 |
| 0 | 0 | 0 | 0 | 0.003410902 |
| 0 | 0.012469665 | 0 | 0.000699718 | 0.001193038 |
| 0.003756014 | 0.010446905 | 0.000669536 | 0.021154625 | 0.004007686 |
| 0 | 0 | 0.020676006 | 0 | 0 |
| 0 | 0 | 0.001147671 | 0 | 0.001595541 |
| 0 | 0 | 0 | 0 | 0 |
| 0 | 0 | 0 | 0 | 0 |
| 0 | 0 | 0 | 0 | 0 |
| 0 | 0 | 0 | 0 | 0.001044097 |
| 0 | 0 | 0 | 0 | 0 |
| 0 | 0 | 0 | 0 | 0 |
| 0 | 0 | 0 | 0 | 0 |
| 0 | 0 | 0 | 0 | 0 |
| 0 | 0 | 0 | 0 | 0 |
| 0 | 0 | 0 | 0 | 0 |
| 0.002916533 | 0.012332413 | 0 | 0.000264943 | 0 |
| 0 | 0.548378584 | 0 | 0.513120105 | 0 |
| 0.735518691 | 0 | 0 | 0 | 0.016752392 |
| 0 | 0 | 0 | 0 | 0 |
| 0.000818385 | 0 | 0.00267452 | 0.013357082 | 0.083660196 |
| 0.035279106 | 0 | 0 | 0 | 0 |
| 0 | 0 | 0 | 0 | 0 |
| 0 | 0 | 0 | 0 | 0 |
| 0 | 0 | 0 | 0 | 0 |
| 0 | 0 | 0 | 0 | 0 |
| 0 | 0 | 0 | 0 | 0 |
| 1.092394756 | 0 | 0.051715598 | 0 | 0 |
| 0 | 0 | 0 | 0 | 0 |
| 0 | 0 | 0 | 0 | 0 |
| 0.726566373 | 0 | 0 | 0 | 0 |
| 0 | 0 | 0 | 0 | 0.008809879 |
| 0 | 0 | 0 | 0 | 0 |
| 0 | 0 | 0 | 0 | 0 |
| 0.114813005 | 0 | 0.004816704 | 0 | 0 |
| 0 | 0 | 0 | 0 | 0 |
| 0 | 0 | 0 | 0 | 0 |
| 0.004616104 | 0 | 0 | 0 | 0.004681405 |
| 0 | 0 | 0 | 0 | 0 |

| | | | | |
|---|---|---|---|---|
| 0 | 0 | 0 | 0 | 0 |
| 0 | 0.006627243 | 0 | 0 | 0.008998693 |
| 0 | 0 | 0 | 0 | 0 |
| 0 | 0 | 0 | 0 | 0 |
| 0 | 0 | 0.001917074 | 0 | 0 |
| 0.285405667 | 0 | 0.01005707 | 0 | 0 |
| 0.004909046 | 0 | 0 | 0 | 0 |
| 0 | 0.001249838 | 0 | 0 | 0.00343361 |
| 0 | 0.211524366 | 0 | 0 | 0 |
| 0 | 0 | 0 | 0 | 0 |
| 0 | 0.000560394 | 0.001771824 | 0 | 0.00102636 |
| 0.009739219 | 0 | 0 | 0 | 0 |
| 0.033065191 | 0 | 0 | 0 | 0 |
| 0.001280528 | 0 | 0.001813424 | 0.025402079 | 0.000484827 |
| 0.011804942 | 0 | 0.039378912 | 0 | 0 |
| 0 | 0 | 0 | 0 | 0 |
| 0 | 0 | 0 | 0 | 0 |
| 0 | 0 | 0 | 0 | 0 |
| 0 | 0 | 0 | 0 | 0.014276651 |
| 0.013565332 | 0 | 0 | 0 | 0 |
| 0 | 0.005133975 | 0 | 0 | 0 |
| 0 | 0.113530809 | 0 | 0.046080283 | 0.006212358 |
| 0 | 0 | 0 | 0 | 0 |
| 0 | 0 | 0 | 0 | 0 |
| 0 | 0 | 0 | 0 | 0 |
| 0 | 0 | 0 | 0 | 0 |
| 0.020517933 | 0 | 0.152004879 | 0 | 0 |
| 0 | 0 | 0 | 0 | 0 |
| 0 | 0 | 0 | 0 | 0 |
| 7.22681354 | 0.3140363 | 0.53080222 | 1.825861548 | 17.4982084 |
| 0.001640947 | 0.013409304 | 0 | 0 | 0.005153024 |
| 0 | 0 | 0 | 0 | 0 |
| 0 | 0 | 0 | 0 | 0 |
| 0 | 0 | 0 | 0 | 0.001242289 |
| 0.012463775 | 0 | 0 | 0 | 0 |
| 0 | 0 | 0 | 0 | 0 |
| 0 | 0.127263747 | 0 | 0.005992208 | 0.814925004 |
| 0 | 0 | 0 | 0.004154231 | 0.001647225 |
| 0 | 0 | 0.0308136 | 0 | 0 |
| 0.071045695 | 0 | 0.17611617 | 0 | 0 |
| 0.020118907 | 0 | 0.016284475 | 0 | 0 |
| 5.435791821 | 1.211719991 | 0 | 0.003752939 | 0.17928227 |
| 0 | 0 | 0 | 0.004602053 | 0 |
| 0 | 0 | 0 | 0 | 0 |
| 0.003166769 | 0 | 0 | 0 | 0 |
| 0 | 0 | 0 | 0 | 0 |
| 0.003314758 | 0 | 0.006258942 | 0 | 0.001952247 |
| 0 | 0 | 0 | 0 | 0 |

| | | | | |
|---|---|---|---|---|
| 0 | 0.019915566 | 0 | 0.011200953 | 0.004473384 |
| 0 | 0 | 0 | 0 | 0 |
| 0.006398332 | 1.037330515 | 0 | 0.001512466 | 0.076660483 |
| 0 | 0 | 0 | 0 | 0 |
| 0 | 0.001385031 | 0 | 0 | 0 |
| 0 | 0 | 0 | 0 | 0 |
| 0 | 0 | 0 | 0 | 0 |
| 0 | 0 | 0 | 0 | 0 |
| 0 | 0 | 0 | 0.000337809 | 0 |
| 0 | 0 | 0 | 0 | 0 |
| 0 | 0 | 0 | 0 | 0 |
| 0 | 0 | 0 | 0 | 0 |
| 0 | 0 | 0 | 0 | 0 |
| 0 | 0 | 0.001746584 | 0 | 0 |
| 0 | 0 | 0 | 0 | 0 |
| 0.036946655 | 0.000929815 | 0.017744029 | 0.007062835 | 0.003527546 |
| 0.008071603 | 1.47037647 | 0.000581876 | 0.006088768 | 0.077928696 |
| 0 | 0 | 0.000565565 | 0 | 0 |
| 0 | 0 | 0 | 0 | 0 |
| 0 | 0 | 0 | 0 | 0 |
| 0.022439374 | 0 | 0.005573302 | 0 | 0 |
| 0 | 0 | 0 | 0 | 0 |
| 0 | 0 | 0 | 0 | 0 |
| 0 | 0 | 0.178178984 | 0 | 0 |
| 0 | 0 | 0 | 0 | 0 |
| 0 | 0 | 0 | 0 | 0 |
| 0.001054835 | 0 | 0.000886435 | 0 | 0 |
| 0 | 0 | 0 | 0 | 0.003466179 |
| 0 | 0 | 0 | 0 | 0 |
| 0 | 0 | 0 | 0 | 0 |
| 0 | 0 | 0.036403106 | 0.010688083 | 0.216338421 |
| 0.00358794 | 0.001483428 | 0.000502524 | 0.001934918 | 0.00155251 |
| 4.070487975 | 0.02360766 | 0.008293489 | 2.360424464 | 0.049519694 |
| 0 | 0.001445815 | 0 | 0 | 0 |
| 0 | 0.004540555 | 0 | 0 | 0.003591004 |
| 0 | 0 | 0 | 0 | 0 |
| 0.002500201 | 0.0158704 | 0 | 0.002351392 | 0 |
| 0.041980863 | 0 | 0.014659236 | 0.002681726 | 0.048728835 |
| 0 | 0.064646307 | 0 | 0.000455354 | 0 |
| 0.001692673 | 0 | 0.002950258 | 0 | 0 |
| 0 | 0 | 0 | 0 | 0 |
| 0 | 0 | 0 | 0 | 0 |
| 0.005848216 | 0 | 0.003710023 | 0 | 0.010354736 |
| 0 | 0 | 0 | 0.052813934 | 0.019988249 |
| 0.278131157 | 0 | 0 | 0 | 0.715046419 |
| 0 | 0 | 0 | 0.000334091 | 0 |
| 0 | 0 | 0 | 0 | 0 |
| 0 | 0 | 0 | 0 | 0 |

| | | | | |
|---|---|---|---|---|
| 0 | 0 | 0 | 0 | 0 |
| 0.186501533 | 0.137046258 | 0.108172683 | 0.006368734 | 0.085624731 |
| 0 | 0 | 0 | 0 | 0 |
| 0 | 0 | 0 | 0 | 0 |
| 0 | 0 | 0 | 0 | 0 |
| 0 | 0 | 0 | 0 | 0 |
| 0.030012949 | 0 | 0.005401712 | 0 | 0 |
| 0 | 0 | 0 | 0 | 0.002597937 |
| 0 | 0 | 0 | 0 | 0 |
| 0.01822212 | 0 | 0 | 0 | 0 |
| 0.001863409 | 0.001588998 | 0.002397822 | 0 | 0.010582709 |
| 0.107727986 | 0.003012365 | 0.004592088 | 0.070841093 | 0.001576325 |
| 0 | 0.564442555 | 0 | 0 | 0 |
| 0 | 0 | 0 | 0 | 0.036375711 |
| 0.025656343 | 0 | 0 | 0.00836776 | 0 |
| 0 | 0 | 0 | 0 | 0 |
| 0.000487814 | 0.010588523 | 0.003826081 | 0 | 0.004986731 |
| 0.498526539 | 0 | 0.207828784 | 0 | 0.010069835 |
| 0 | 0 | 0 | 0 | 0 |
| 0 | 0 | 0.063999441 | 0 | 0.001401177 |
| 0.000880355 | 0.193274056 | 0.011836965 | 0.003937724 | 0.028855573 |
| 0.000938966 | 0 | 0 | 0 | 0.000711012 |
| 0 | 0 | 0 | 0 | 0 |
| 0 | 0 | 0 | 0 | 0 |
| 0 | 0 | 0 | 0 | 0.000720041 |
| 0 | 0 | 0 | 0 | 0 |
| 0 | 0 | 0 | 0 | 0.007472972 |
| 0.01283228 | 0.021719334 | 0.031452366 | 0.012733754 | 0.20436021 |
| 0 | 0 | 0 | 0 | 0 |
| 0.005320815 | 0 | 0 | 0 | 0.001281979 |
| 0 | 0 | 0 | 0 | 0 |
| 0 | 0 | 0 | 0 | 0 |
| 0 | 0 | 0 | 0 | 0 |
| 0 | 0 | 0 | 0 | 0 |
| 0 | 0.20834024 | 0.028893177 | 0.000232795 | 0 |
| 0.248398111 | 0.155733324 | 0.159915556 | 0.06239275 | 1.021413944 |
| 0 | 0 | 0 | 0 | 0 |
| 0 | 0 | 0 | 0 | 0 |
| 0 | 0.004295098 | 0 | 0 | 0 |
| 0 | 0 | 0 | 0 | 0 |
| 0.264063942 | 0 | 0 | 0 | 0.678850773 |
| 0 | 0 | 0 | 0 | 0 |
| 0.031750903 | 0 | 0.005204538 | 0.009941935 | 0 |
| 0.000580219 | 0 | 0 | 0 | 0 |
| 0 | 0 | 0.011832935 | 0 | 0 |
| 0 | 0.783350323 | 0 | 0 | 0 |
| 0.003259331 | 0 | 0 | 0 | 0 |
| 0.005002021 | 0 | 0 | 0 | 0 |

| | | | | |
|---|---|---|---|---|
| 0 | 0 | 0 | 0 | 0 |
| 0 | 0 | 0 | 0 | 0 |
| 0 | 0 | 0 | 0 | 0 |
| 0 | 0.008970963 | 0.009835869 | 0.001631969 | 0.105471843 |
| 0 | 0 | 0 | 0 | 0 |
| 0 | 0 | 0 | 0 | 0.000574498 |
| 0 | 0 | 0 | 0 | 0 |
| 0 | 0.009319922 | 0 | 0.000347612 | 0 |
| 0 | 0.625168424 | 0.067311902 | 7.271805673 | 0.489362479 |
| 0 | 0.055607565 | 0 | 0 | 0 |
| 0 | 0 | 0 | 0 | 0 |
| 0.012182755 | 0.008595147 | 0 | 0 | 0.007744039 |
| 0 | 0 | 0 | 0 | 0 |
| 0 | 0 | 0 | 0 | 0 |
| 0.072494008 | 0.013842278 | 0.018808424 | 0 | 0.029018705 |
| 0 | 0 | 0 | 0 | 0 |
| 0 | 0.176083679 | 0 | 0.004488181 | 0 |
| 0 | 0.151031121 | 0 | 0 | 1.285388405 |
| 0 | 0 | 0 | 0 | 0 |
| 0 | 0 | 0 | 0 | 0 |
| 0 | 0 | 0 | 0 | 0 |
| 0 | 0 | 0 | 0 | 0 |
| 0 | 0 | 0 | 0 | 0 |
| 0.24325113 | 0.161436476 | 0.194761325 | 0.003314342 | 0.516262594 |
| 0 | 0 | 0 | 0 | 0 |
| 0 | 0.003344168 | 0 | 0 | 0.220003967 |
| 0 | 0 | 0 | 0 | 0 |
| 0 | 0.288592753 | 0.106623485 | 0.439590762 | 2.872264921 |
| 0 | 0 | 0 | 0 | 0 |
| 0 | 0 | 0 | 0 | 0 |
| 0 | 0.003773654 | 0 | 0.008107133 | 0 |
| 0 | 0 | 0 | 0 | 0 |
| 0 | 0 | 0 | 0 | 0 |
| 0 | 0 | 0 | 0 | 0 |
| 0 | 0 | 0 | 0 | 0 |
| 0 | 0.009256092 | 0 | 0.037438436 | 0.024382299 |
| 0 | 0.514182188 | 0 | 0.229869893 | 0 |
| 0.041981401 | 0 | 0 | 0 | 0 |
| 0 | 0 | 0.001711081 | 0 | 0 |
| 0 | 0 | 0 | 0 | 0 |
| 0.10046682 | 0 | 0.013981192 | 0.000214285 | 0.008403319 |
| 0 | 0.261389419 | 0 | 0 | 0 |
| 0.09518302 | 0.006222071 | 0.252717302 | 0.193053104 | 11.74239379 |
| 0 | 0 | 0 | 0 | 0 |
| 0 | 0 | 0 | 0 | 0 |
| 0 | 0 | 0 | 0 | 0 |
| 0 | 0 | 0 | 0 | 0 |
| 0 | 0 | 0.002021894 | 0 | 0.014201016 |

| | | | | |
|---|---|---|---|---|
| 0 | 0 | 0 | 0 | 0 |
| 0 | 0.020827614 | 0 | 0.028630556 | 0.004534804 |
| 0.640650263 | 0 | 0.250756438 | 0.074738768 | 0.022314547 |
| 0 | 0.003644551 | 0 | 0 | 0 |
| 0 | 0 | 0 | 0 | 0 |
| 0.003698712 | 0.001086556 | 0.000286284 | 0.02982721 | 0.010392344 |
| 0 | 0 | 0 | 0 | 0 |
| 0 | 0 | 0 | 0 | 0 |
| 0 | 0 | 0 | 0 | 0 |
| 0 | 0 | 0 | 0 | 0 |
| 0.016741115 | 0.000601877 | 0.101095943 | 0 | 0.074958848 |
| 0.225266388 | 0.345850259 | 1.820100035 | 0.125841307 | 2.660205615 |
| 0 | 0 | 0 | 0 | 0 |
| 0.003011282 | 0.180255798 | 0 | 0.000610534 | 0.08610352 |
| 0.989956186 | 0 | 0.105098385 | 0.036290483 | 0.009492371 |
| 0 | 1.002300194 | 0 | 0 | 0 |
| 0 | 0 | 0 | 0 | 0 |
| 0 | 0 | 0 | 0 | 0 |
| 0 | 0.051566946 | 0 | 0 | 0 |
| 0 | 0 | 0.002645252 | 0 | 0 |
| 0.01098119 | 0 | 0.009968622 | 0.00027094 | 0.00092392 |
| 0.652462692 | 0 | 10.60467177 | 0.015844524 | 0.366653827 |
| 1.390232096 | 0.042406395 | 7.983114926 | 0.073190143 | 5.391540867 |
| 0 | 0 | 0 | 0 | 0 |
| 0 | 0 | 0 | 0 | 0 |
| 0 | 4.491257839 | 0.002269626 | 0.058359008 | 0.003183008 |
| 0.943460016 | 0 | 0.02062668 | 0 | 0 |
| 0 | 0 | 0 | 0 | 0 |
| 0 | 0 | 0 | 0 | 0 |
| 0 | 0 | 0 | 0 | 0 |
| 0 | 0.000834612 | 0 | 0 | 0 |
| 0 | 0.171212077 | 0.149166622 | 0 | 0.232236883 |
| 0.139541949 | 0 | 0 | 0.011619906 | 0 |
| 0 | 0.003653213 | 0 | 0.004936886 | 0.0166192 |
| 0.021133527 | 0 | 0.01088384 | 0 | 0.058002828 |
| 0.053327138 | 0.200534237 | 0.011519911 | 0.29955542 | 0.190825295 |
| 0.002182766 | 0 | 0.024182166 | 0 | 0.001239641 |
| 0 | 0 | 0 | 0 | 0 |
| 0 | 0.017382634 | 0 | 4.893472614 | 0.267904889 |
| 0 | 0 | 0 | 0 | 0 |
| 0 | 0.005429449 | 0 | 0 | 0 |
| 0 | 0 | 0 | 0 | 0 |
| 0 | 0 | 0 | 0 | 0 |
| 0 | 0 | 0 | 0 | 0 |
| 0.023062765 | 0.003675605 | 0.036758758 | 0.002403279 | 0.025317635 |
| 0 | 0.002346874 | 0 | 0 | 0 |
| 0 | 0 | 0 | 0 | 0 |
| 0.191320759 | 0 | 0.014170869 | 0.317855882 | 0.208837019 |

| | | | | |
|---|---|---|---|---|
| 0 | 0 | 0 | 0 | 0 |
| 0 | 0 | 0 | 0 | 0 |
| 0 | 0 | 0 | 0 | 0 |
| 0 | 0.002043067 | 0 | 0 | 0.015341667 |
| 0 | 0 | 0 | 0 | 0 |
| 0 | 0.876950042 | 0 | 0 | 0 |
| 0 | 2.064090857 | 0 | 0 | 0 |
| 0 | 0.005263976 | 0.009312352 | 0 | 0.004017064 |
| 0 | 0 | 0 | 0 | 0 |
| 0 | 0 | 0 | 0 | 0 |
| 0.011374011 | 0 | 0.001678681 | 0.009288025 | 0.036951412 |
| 0 | 0 | 0 | 0 | 0 |
| 0 | 0 | 0 | 0.042302708 | 0.037705615 |
| 0 | 0.427820053 | 0 | 0 | 0 |
| 0 | 0 | 0 | 0 | 0 |
| 0 | 0 | 0 | 0 | 0 |
| 0 | 0.30698778 | 0 | 0 | 0 |
| 0.009774044 | 0.008460976 | 0.00089837 | 0.025570088 | 0.00254416 |
| 0 | 0 | 0 | 0 | 0 |
| 0 | 0 | 0 | 0 | 0 |
| 0 | 0 | 0 | 0.001446176 | 0.002568506 |
| 0 | 0 | 0 | 0 | 0 |
| 0 | 0.049211287 | 0.00230168 | 0 | 0 |
| 0 | 0 | 0 | 0 | 0 |
| 0 | 0 | 0 | 0 | 0 |
| 0 | 0 | 0 | 0 | 0 |
| 0 | 0 | 0 | 0 | 0 |
| 0 | 0 | 0 | 0 | 0 |
| 0 | 0 | 0 | 0 | 0 |
| 0 | 0.055832937 | 0 | 0 | 0 |
| 0 | 0 | 0 | 0 | 0 |
| 0 | 0 | 0 | 0 | 0 |
| 0 | 0 | 0 | 0 | 0 |
| 0 | 0 | 0 | 0 | 0 |
| 0 | 0.000603105 | 0 | 0 | 0 |
| 0 | 0 | 0 | 0 | 0 |
| 0 | 0.072240097 | 0 | 0 | 0.061975608 |
| 0.174344694 | 0 | 0.005379625 | 0.011172992 | 0.026929037 |
| 0 | 0 | 0 | 0 | 0 |
| 0 | 0 | 0.001501745 | 0 | 0 |
| 0 | 0 | 0.026475145 | 0 | 0.133722774 |
| 0 | 0 | 0 | 0 | 0 |
| 0 | 0 | 0 | 0 | 0 |
| 0 | 0.023195003 | 0 | 0 | 0 |
| 0 | 0.050305045 | 0 | 0.046048447 | 0.608615277 |
| 0 | 0 | 0 | 0 | 0 |
| 0 | 0.001830209 | 0 | 0 | 0 |
| 0 | 0 | 0 | 0 | 0 |

| | | | | |
|---|---|---|---|---|
| 0 | 0 | 0 | 0 | 0 |
| 0 | 0 | 0 | 0 | 0 |
| 0 | 0.001634563 | 0 | 0 | 0 |
| 0 | 0.295447304 | 0 | 0 | 0 |
| 0 | 0 | 0 | 0 | 0 |
| 0 | 0 | 0 | 0 | 0 |
| 0 | 0 | 0.267525697 | 0 | 0 |
| 0 | 0 | 0 | 0 | 0 |
| 0.191189521 | 0 | 0.009239255 | 0.018167899 | 0.020110547 |
| 0 | 0 | 0 | 0 | 0 |
| 0 | 0 | 0 | 0 | 0 |
| 0 | 0.022997745 | 0 | 0 | 0 |
| 0 | 0 | 0 | 0 | 0 |
| 0 | 0 | 0 | 0 | 0 |
| 0 | 0.209742529 | 4.084745366 | 0.202401121 | 0.049183318 |
| 0 | 0 | 0.127421858 | 0 | 0.267745823 |
| 0 | 0 | 0 | 0 | 0 |
| 0 | 0 | 0 | 0 | 0 |
| 0 | 0 | 0 | 0 | 0 |
| 0 | 0 | 0 | 0 | 0 |
| 0 | 0.006635615 | 0 | 0 | 0 |
| 0 | 0 | 0 | 0 | 0 |
| 0 | 0 | 0 | 0 | 0 |
| 0 | 0 | 0 | 0 | 0 |
| 0.012089624 | 0.070811183 | 0.045435875 | 0.155371156 | 0.084870943 |
| 0 | 3.320046356 | 0 | 0.021718805 | 0.043143048 |
| 0 | 0.112321973 | 0 | 0 | 0 |
| 0 | 0 | 0.000510945 | 0 | 0 |
| 0.202881226 | 0.316101211 | 0 | 0.343068811 | 0.119055847 |
| 0 | 0.003132768 | 0 | 0 | 0 |
| 0 | 0 | 0 | 0 | 0 |
| 0 | 0 | 0 | 0 | 0 |
| 0 | 0 | 0 | 0 | 0 |
| 0.020266662 | 0 | 0 | 0 | 0 |
| 0.001034298 | 0.048199897 | 0 | 0 | 0.007887953 |
| 0.002611155 | 0.001295494 | 0.010752055 | 0.076467732 | 0.004982653 |
| 0 | 0 | 0.145513797 | 0 | 0 |
| 0 | 0.921733143 | 4.49168102 | 0.017441905 | 1.095114588 |
| 0 | 0 | 0.393930588 | 4.354673745 | 0.131251149 |
| 0 | 0 | 0 | 0 | 0 |
| 0 | 0 | 0 | 0 | 0.089802957 |
| 0 | 0 | 0.003948183 | 0 | 0 |
| 0.15129126 | 0.581902604 | 0.074303966 | 0 | 0.08316566 |
| 0 | 0 | 0 | 0 | 0.731137711 |
| 0 | 0 | 0 | 0 | 0 |
| 0 | 0 | 0.00058207 | 0 | 0.025850021 |
| 0 | 0 | 0 | 0 | 0 |
| 0 | 0 | 0.001081858 | 0.000420059 | 0.000537159 |

| | | | | |
|---|---|---|---|---|
| 0 | 0 | 0.054973763 | 0 | 0 |
| 0 | 0 | 0 | 0 | 0 |
| 0 | 0 | 0 | 0 | 0 |
| 0 | 0 | 0 | 0 | 0 |
| 0.001053324 | 0 | 0.001606412 | 0 | 0.000664673 |
| 0 | 0.001677501 | 0 | 0 | 0 |
| 0 | 0 | 0 | 0 | 0.001269934 |
| 0 | 0 | 0 | 0 | 0 |
| 0 | 0 | 0 | 0 | 0 |
| 0 | 0 | 0 | 0 | 0 |
| 0 | 0 | 0 | 0 | 0 |
| 0 | 0.033748849 | 0.24940238 | 0.173708384 | 0.804522786 |
| 3.927210638 | 0.568086707 | 0 | 0.000472018 | 0.344294542 |
| 0 | 0 | 0 | 0 | 0 |
| 0.004872676 | 0.012759135 | 0.000928908 | 0.002885379 | 0.026750565 |
| 0 | 0 | 0 | 0 | 0 |
| 0 | 0 | 0 | 0 | 0 |
| 0 | 0 | 0 | 0 | 0 |
| 0.00272862 | 0 | 0 | 0.014937049 | 0 |
| 0 | 0.002773804 | 0 | 0 | 0 |
| 0 | 0 | 0 | 0 | 0 |
| 0 | 0 | 0 | 0 | 0 |
| 0.0155732 | 0.920736299 | 0 | 0 | 0 |
| 0 | 0 | 0 | 0 | 0 |
| 0 | 0 | 0 | 0 | 0 |
| 0 | 0 | 0 | 0 | 0 |
| 0 | 0 | 0 | 0 | 0 |
| 0 | 0 | 0 | 0 | 0 |
| 0 | 0 | 0 | 0 | 0 |
| 0 | 0 | 0 | 0 | 0 |
| 0 | 0 | 0 | 0 | 0 |
| 0.029155955 | 0.010892606 | 0.020147193 | 0.001521074 | 0.008977396 |
| 0 | 0 | 0 | 0 | 0 |
| 0 | 0.004135736 | 0.037077753 | 0 | 0.00249164 |
| 0.002386937 | 0.00197375 | 0.006909121 | 0.070267969 | 0.006584313 |
| 0.008791573 | 0.007996697 | 0 | 0.085508108 | 0.002662895 |
| 0 | 0 | 0 | 0 | 0 |
| 0 | 0 | 0 | 0.034899537 | 0 |
| 0 | 0 | 0 | 0 | 0.002224825 |
| 0 | 0 | 0 | 0 | 0 |
| 0 | 0.296569687 | 0 | 0 | 0 |
| 0 | 0 | 0 | 0 | 0 |
| 0 | 0 | 0 | 0 | 0 |
| 0 | 0 | 0 | 0 | 0 |
| 0.001119701 | 0 | 0.035897091 | 0.000969691 | 0.041206499 |
| 0 | 0 | 0 | 0 | 0 |
| 2.361727966 | 0.024071475 | 3.824420505 | 0 | 1.814729096 |
| 0 | 0 | 0.000830635 | 0 | 0.020569598 |

| | | | | |
|---|---|---|---|---|
| 0 | 0 | 0 | 0 | 0 |
| 0 | 0 | 0 | 0.044725083 | 0 |
| 0 | 0 | 0.002497812 | 0 | 0 |
| 0 | 0 | 0 | 0 | 0 |
| 0 | 0 | 0 | 0 | 0 |
| 0 | 0 | 0 | 0 | 0 |
| 0 | 0.023084653 | 0 | 0.003991293 | 0 |
| 0 | 0 | 0 | 0 | 0 |
| 0 | 0 | 0 | 0 | 0 |
| 0 | 0 | 0 | 0 | 0.001589134 |
| 0 | 0 | 0 | 0 | 0 |
| 0.001296118 | 0 | 0 | 0 | 0.014454203 |
| 0 | 0 | 0 | 0 | 0 |
| 0 | 0.025521068 | 0 | 0.006140263 | 0.032253919 |
| 0 | 0 | 0 | 0 | 0 |
| 0 | 0 | 0 | 0 | 0 |
| 0 | 0 | 0 | 0 | 0 |
| 0 | 0 | 0 | 0 | 0 |
| 0.02660443 | 0 | 0.128718324 | 2.332536116 | 0.001405511 |
| 0 | 0 | 0 | 0 | 0 |
| 0 | 0.012920318 | 0 | 0 | 0 |
| 0 | 0 | 0 | 0 | 0 |
| 0 | 0 | 0 | 0 | 0 |
| 0 | 0 | 0 | 0 | 0 |
| 0 | 0 | 0 | 0 | 0 |
| 0.003335677 | 0 | 0.01359237 | 0.002638794 | 0.015392037 |
| 0 | 0 | 0 | 0 | 0 |
| 0.009488803 | 0 | 0.001900604 | 0 | 0.000579452 |
| 0 | 0 | 0 | 0 | 0 |
| 0 | 1.30361362 | 0 | 0.023659009 | 0.136549314 |
| 0 | 0 | 0 | 0 | 0 |
| 0 | 0 | 0 | 0 | 0 |
| 0 | 0.002004429 | 0 | 0 | 0 |
| 0 | 0 | 0 | 0 | 0 |
| 0.001529318 | 0 | 0.003105827 | 0.001018792 | 0.009802018 |
| 0 | 0 | 0 | 0 | 0 |
| 0 | 0 | 0 | 0 | 0 |
| 0.001983232 | 0 | 0 | 0 | 0.001126321 |
| 0 | 0 | 0 | 0 | 0 |
| 0 | 0 | 0 | 0 | 0 |
| 26.98911641 | 2.785219239 | 6.105243716 | 1.361756054 | 11.29551773 |
| 0.033934296 | 0.027572488 | 0 | 0.008039087 | 0.000412236 |
| 0 | 0.653852228 | 0 | 0 | 0 |
| 0 | 0 | 0 | 0 | 0 |
| 0 | 0 | 0 | 0 | 0 |
| 0 | 0 | 0 | 0 | 0 |
| 0 | 0 | 0 | 0 | 0 |
| 0.001869128 | 0 | 0.049960258 | 0.005308334 | 0.004134337 |

| | | | | |
|---|---|---|---|---|
| 0 | 0 | 0 | 0 | 0 |
| 0 | 0 | 0.005357076 | 0 | 0.006551159 |
| 0 | 0 | 0 | 0 | 0 |
| 0 | 0 | 0 | 0 | 0 |
| 0.488033224 | 7.88984496 | 0.79905258 | 0 | 0.043928008 |
| 0 | 0.720190997 | 0 | 0 | 0 |
| 0 | 0 | 0 | 0 | 0 |
| 0 | 0 | 0 | 0 | 0 |
| 6.283741668 | 0 | 0.228627834 | 0 | 0.070694107 |
| 0 | 0 | 0 | 0 | 0 |
| 0 | 0 | 0 | 0.003960173 | 0 |
| 0 | 0 | 0 | 0 | 0 |
| 0.046210579 | 0.040121954 | 0 | 0 | 0 |
| 0.102743673 | 0.006446845 | 0 | 0.090911315 | 0.787661669 |
| 0 | 0.385645925 | 0 | 4.012858651 | 0.091435348 |
| 0 | 0.093009704 | 0 | 0 | 0.00202794 |
| 0 | 0 | 0 | 0 | 0 |
| 0 | 0 | 0 | 0 | 0 |
| 0 | 0 | 0 | 0 | 0 |
| 0 | 0 | 0 | 0 | 0 |
| 0 | 0 | 0 | 0 | 0 |
| 0 | 0 | 0 | 0 | 0 |
| 0 | 0 | 0 | 0 | 0 |
| 0 | 0 | 0 | 0 | 0 |
| 0 | 0 | 0 | 0 | 0 |
| 0 | 0 | 0 | 0 | 0 |
| 0 | 0.174070053 | 0 | 0 | 0 |
| 0.004413087 | 0 | 0.000503099 | 0.001367388 | 0.006761152 |
| 0 | 0 | 0 | 0 | 0 |
| 0 | 0 | 0 | 0 | 0 |
| 0 | 0 | 0 | 0 | 0 |
| 0 | 0 | 0 | 0 | 0 |
| 0 | 0 | 0 | 0 | 0 |
| 0 | 0.006805479 | 0 | 0 | 0 |
| 0 | 0.013572939 | 0 | 1.39363447 | 0.434161634 |
| 0.000593608 | 0 | 0 | 0 | 0.008495496 |
| 0 | 0.033413897 | 0 | 0 | 0 |
| 0 | 0 | 0 | 0 | 0 |
| 0 | 0 | 0 | 0 | 0 |
| 0 | 0 | 0 | 0 | 0 |
| 0 | 0.389196022 | 1.365681224 | 0.027218769 | 0.354423695 |
| 0 | 0.09268024 | 0 | 0 | 0 |
| 0 | 0 | 0 | 0 | 0.002866724 |
| 0 | 0.0006232 | 0 | 0.000251035 | 0.0048509 |
| 0 | 0 | 0 | 0 | 0 |
| 0 | 0 | 0 | 0 | 0 |
| 0 | 0 | 0 | 0 | 0 |
| 0 | 0 | 0 | 0 | 0 |

| | | | | |
|---|---|---|---|---|
| 0 | 0.059514323 | 0 | 0 | 0 |
| 0 | 0 | 0 | 0 | 0 |
| 0 | 0 | 0 | 0 | 0 |
| 0.003720307 | 0 | 0 | 0 | 0.000792316 |
| 0 | 0.061949937 | 0 | 0 | 0 |
| 0 | 0 | 0 | 0 | 0 |
| 0 | 0 | 0 | 0 | 0 |
| 0 | 0 | 0 | 0 | 0 |
| 0 | 0 | 0 | 0 | 0 |
| 0 | 0 | 0 | 0 | 0 |
| 0 | 0 | 0 | 0 | 0 |
| 0 | 0 | 0 | 0 | 0 |
| 0 | 0 | 0 | 0 | 0 |
| 0 | 0 | 0 | 0 | 0 |
| 0 | 0 | 0 | 0 | 0 |
| 0 | 0 | 0 | 0 | 0 |
| 0 | 0 | 0 | 0.001549102 | 0.432150616 |
| 0 | 0 | 0 | 0 | 0 |
| 0 | 0 | 0 | 0 | 0 |
| 0 | 0 | 0 | 0 | 0 |
| 0 | 0 | 0.000292187 | 0 | 0.001918219 |
| 0 | 0 | 0 | 0 | 0 |
| 0.00504037 | 0.002718174 | 0.00075199 | 0 | 0 |
| 0 | 0.01111833 | 0.003725488 | 0.003310298 | 0.003762762 |
| 0 | 0 | 0 | 0.023279539 | 0 |
| 0.001034549 | 0 | 0 | 0.007198202 | 0 |
| 0 | 0 | 0 | 0 | 0 |
| 0 | 0 | 0 | 0 | 0 |
| 0 | 0 | 0 | 0.049399289 | 0 |
| 0.072358632 | 0.028351163 | 0 | 0.227471739 | 0.001114955 |
| 0 | 0 | 0 | 0 | 0 |
| 0 | 0 | 0 | 0 | 0 |
| 0 | 0 | 0 | 0 | 0 |
| 0 | 0 | 0 | 0 | 0 |
| 0 | 0 | 0 | 0 | 0 |
| 0 | 0 | 0 | 0 | 0 |
| 0.002194466 | 0 | 0 | 0.01291342 | 0 |
| 0 | 0 | 0 | 0 | 0 |
| 0 | 0 | 0 | 0 | 0.002296867 |
| 0 | 0 | 0 | 0 | 0 |
| 0 | 0 | 0 | 0 | 0 |
| 0 | 0 | 0 | 0 | 0 |
| 0 | 0 | 0 | 0 | 0 |
| 0 | 0 | 0 | 0 | 0 |
| 0 | 0 | 0 | 0 | 0 |
| 0 | 0 | 0 | 0 | 0 |
| 0 | 0 | 0 | 0 | 0 |
| 0 | 0 | 0 | 0.028023106 | 0 |

| | | | | |
|---|---|---|---|---|
| 0.020339367 | 0 | 0.011923484 | 0 | 0.084192579 |
| 0 | 0.138745985 | 0 | 0.154734072 | 0 |
| 0 | 0.003245342 | 0 | 0 | 0 |
| 0 | 0 | 0 | 0 | 0 |
| 0 | 0 | 0 | 0 | 0 |
| 0 | 0 | 0 | 0 | 0 |
| 0.00241793 | 0 | 0 | 0 | 0 |
| 0 | 0.232729034 | 0.399260548 | 0.023306752 | 0.108211279 |
| 0.016785673 | 2.095270765 | 0.045498511 | 5.808924683 | 2.009396969 |
| 0.02932877 | 0 | 0.043468469 | 0 | 0.412247239 |
| 0 | 0.001334022 | 0 | 0 | 0 |
| 0 | 0 | 0 | 0 | 0 |
| 0 | 0 | 0 | 0 | 0 |
| 0 | 0 | 0 | 13.35990262 | 0 |
| 0 | 0 | 0 | 0 | 0 |
| 0 | 0 | 0 | 0 | 0 |
| 0 | 0 | 0 | 0 | 0.00173831 |
| 0 | 0 | 0 | 0 | 0 |
| 0 | 0 | 0 | 0 | 0 |
| 0 | 0 | 0.010245377 | 0 | 0.000431623 |
| 0 | 0 | 0 | 0 | 0 |
| 0 | 0 | 0 | 0 | 0 |
| 0 | 0 | 0 | 0 | 0 |
| 0 | 0 | 0 | 0 | 0 |
| 0 | 0.004199419 | 0 | 0 | 0 |
| 0 | 0 | 0 | 0 | 0 |
| 0 | 0 | 0 | 0 | 0 |
| 0 | 0 | 0 | 0 | 0 |
| 0 | 0 | 0 | 0 | 0 |
| 0 | 0 | 0 | 0 | 0 |
| 0 | 0 | 0 | 0 | 0 |
| 0 | 0 | 0 | 0 | 0 |
| 0 | 0 | 0 | 0 | 0 |
| 0 | 0 | 0 | 0 | 0 |
| 0 | 0.010075662 | 0.001119961 | 0 | 0 |
| 0 | 0.012306143 | 0 | 0 | 0 |
| 0 | 0 | 0 | 0 | 0 |
| 0.0350366 | 0 | 0 | 0 | 0 |
| 0 | 0.000463885 | 0 | 0 | 0.008071229 |
| 0.198235977 | 0 | 0 | 0 | 0 |
| 0 | 0 | 0 | 0 | 0 |
| 0 | 0.092414083 | 0.109465303 | 0.025176904 | 0.279272504 |
| 0 | 0 | 0 | 0 | 0 |
| 0 | 0 | 0 | 0 | 0 |
| 0 | 0 | 0.001999925 | 0 | 0 |
| 0 | 0 | 0 | 0 | 0.003917934 |
| 0 | 0 | 0 | 0 | 0 |
| 0 | 0 | 0 | 0 | 0 |

| | | | | |
|---|---|---|---|---|
| 0 | 0 | 0 | 0 | 0 |
| 0 | 0.001056304 | 0 | 0.000189109 | 0 |
| 0 | 0 | 0 | 0 | 0 |
| 0 | 0 | 0 | 0 | 0 |
| 0.258246602 | 0.111421037 | 0 | 0.18252516 | 0.096924421 |
| 0 | 0.474042874 | 0 | 0 | 0 |
| 0 | 0 | 0 | 0 | 0 |
| 0.038009521 | 0 | 0.000784528 | 0.039003169 | 0.002575225 |
| 0 | 0.007102162 | 0 | 0 | 0 |
| 0 | 0 | 0 | 0 | 0 |
| 0 | 0 | 0 | 0 | 0 |
| 0 | 0 | 0 | 0.0307305 | 0.083969856 |
| 0 | 0 | 0 | 0 | 0 |
| 0 | 0 | 0 | 0 | 0 |
| 0 | 0 | 0 | 0 | 0 |
| 0.005089391 | 0 | 0 | 0 | 0 |
| 0 | 0 | 0 | 0 | 0 |
| 0 | 0 | 0 | 0 | 0 |
| 0 | 0 | 0 | 0 | 0 |
| 0 | 0 | 0 | 0 | 0 |
| 0 | 0 | 0 | 0 | 0 |
| 0 | 0 | 0 | 0 | 0 |
| 0 | 0 | 0 | 0 | 0 |
| 0.001731269 | 0 | 0.001018416 | 0.000230665 | 0.050210062 |
| 0 | 0.013628652 | 0.128666901 | 1.731983379 | 0.229196374 |
| 0 | 0 | 0 | 0 | 0 |
| 0.268446192 | 0.193389996 | 0.067789384 | 0.458015106 | 0 |
| 0 | 0 | 0 | 0 | 0 |
| 0.000470718 | 0 | 0 | 0 | 0.009802145 |
| 0 | 0 | 0 | 0 | 0 |
| 0 | 0 | 0 | 0 | 0 |
| 0 | 0 | 0 | 0 | 0 |
| 0 | 0 | 0 | 0 | 0 |
| 0 | 0 | 0 | 0 | 0 |
| 0 | 0 | 0 | 0 | 0 |
| 0 | 0 | 0 | 0 | 0 |
| 0 | 0 | 0 | 0 | 0 |
| 0 | 0.140189178 | 0 | 0 | 0 |
| 0 | 0.849510025 | 0 | 0 | 0 |
| 0 | 0 | 0 | 0 | 0 |
| 0 | 0 | 0 | 0 | 0 |
| 0 | 0.149660661 | 0 | 0.59533139 | 0 |
| 0 | 0 | 0 | 0 | 0 |
| 0 | 0 | 0 | 0 | 0 |
| 0 | 0 | 0 | 0 | 0 |
| 0 | 0 | 0 | 0 | 0 |
| 0 | 0 | 0 | 0 | 0 |
| 0 | 0 | 0 | 0 | 0 |

| | | | | |
|---|---|---|---|---|
| 0 | 0.266951145 | 0.005255731 | 0 | 0 |
| 0 | 0 | 0 | 0 | 0 |
| 0 | 0 | 0 | 0 | 0 |
| 0 | 0 | 0 | 0 | 0 |
| 0 | 0 | 0 | 0 | 0 |
| 0.017942056 | 0.012268422 | 0 | 0 | 0.077859674 |
| 0 | 0 | 0 | 0 | 0 |
| 0 | 0 | 0 | 0 | 0 |
| 0 | 0 | 0 | 0 | 0 |
| 0 | 0 | 0 | 0 | 0 |
| 0 | 0 | 0 | 0 | 0 |
| 0 | 0 | 0 | 0 | 0 |
| 0 | 0 | 0 | 0 | 0 |
| 0 | 0 | 0 | 0 | 0 |
| 0 | 0 | 0 | 0 | 0 |
| 0 | 0 | 0 | 0 | 0 |
| 0 | 0 | 0 | 0 | 0 |
| 0 | 0 | 0 | 0 | 0 |
| 0 | 0 | 0 | 0 | 0 |
| 0 | 0 | 0 | 0 | 0 |
| 0 | 0 | 0.019719756 | 0 | 0 |
| 0 | 0 | 0 | 0 | 0 |
| 0.016983147 | 0.003240764 | 0.034724151 | 0.039066231 | 0.037591171 |
| 0 | 0.002471764 | 0 | 0 | 0 |
| 0 | 0 | 0 | 0 | 0 |
| 0 | 0 | 0 | 0.258727265 | 0.12105667 |
| 0 | 0 | 0 | 0 | 0 |
| 0 | 0 | 0 | 0 | 0 |
| 0 | 0 | 0 | 0.100452353 | 0 |
| 0 | 0 | 0 | 0 | 0 |
| 0 | 0 | 0 | 0 | 0 |
| 0 | 0.072231983 | 0 | 0 | 0 |
| 6.640530016 | 0 | 0.481388173 | 0.381429584 | 2.595521256 |
| 0 | 0 | 0 | 0 | 0 |
| 3.64396314 | 1.926121778 | 0.004053262 | 0.002592114 | 2.748312844 |
| 0.000872671 | 0 | 0 | 0 | 0 |
| 0 | 0 | 0 | 0 | 0 |
| 0 | 0 | 0 | 0.041941798 | 0.123332001 |
| 0.006813328 | 0 | 0.00160812 | 0.000420265 | 0 |
| 0 | 0 | 0 | 0 | 0 |
| 0 | 0.101625144 | 0 | 0 | 0 |
| 0 | 0 | 0 | 0 | 0 |
| 0.003393276 | 0.063498502 | 0.006075054 | 1.173206379 | 0.05563501 |
| 0 | 0 | 0 | 0 | 0 |
| 0 | 0 | 0 | 0 | 0 |
| 0 | 0.020345923 | 0.045030037 | 0.003746587 | 0.012243717 |
| 0 | 0 | 0 | 0 | 0 |
| 0 | 0 | 0 | 0 | 0 |

| | | | | |
|---|---|---|---|---|
| 0 | 0 | 0 | 0 | 0 |
| 0 | 0 | 0 | 0.013492114 | 0 |
| 0 | 0 | 0 | 0 | 0 |
| 0 | 0 | 0 | 0 | 0 |
| 0 | 0 | 0 | 0 | 0 |
| 0 | 0 | 0 | 0 | 0 |
| 0 | 0 | 0 | 0 | 0 |
| 0 | 0 | 0 | 0 | 0 |
| 0 | 0 | 0 | 0 | 0 |
| 0 | 0.185461275 | 0 | 0.271894706 | 0 |
| 0 | 0 | 0 | 0 | 0 |
| 0 | 0 | 0 | 0 | 0 |
| 0 | 0 | 0 | 0 | 0 |
| 0 | 0.025877546 | 0 | 0 | 0.002475839 |
| 0 | 0 | 0 | 0 | 0 |
| 0 | 0 | 0 | 0 | 0 |
| 0 | 0 | 0 | 0 | 0 |
| 0 | 0 | 0 | 0 | 0 |
| 0 | 0 | 0 | 0 | 0 |
| 0 | 8.556246163 | 0 | 0.009867459 | 0 |
| 0 | 0.008306172 | 0.00255325 | 0 | 0.208188959 |
| 0 | 0 | 0 | 0 | 0 |
| 0 | 0 | 0 | 0 | 0 |
| 0 | 0 | 0 | 0 | 0 |
| 0 | 0 | 0 | 0 | 0 |
| 0 | 0 | 0 | 0 | 0 |
| 0 | 0 | 0 | 0 | 0 |
| 0 | 0 | 0 | 5.220910828 | 0 |
| 0 | 0 | 0 | 0 | 0 |
| 0 | 0.492565913 | 0 | 0.046974831 | 0 |
| 0.017178068 | 0.064399013 | 0.340238529 | 0.012407933 | 0.000548079 |
| 0 | 0 | 0 | 0 | 0 |
| 0 | 0 | 0 | 0 | 0 |
| 0 | 0 | 0 | 0 | 0 |
| 0 | 0 | 0 | 0 | 0 |
| 0 | 0 | 0 | 0 | 0 |
| 0 | 0 | 0 | 0 | 0 |
| 0 | 0 | 0 | 0 | 0 |
| 0 | 0 | 0 | 0 | 0.021249096 |
| 0 | 0 | 0 | 0 | 0 |
| 0 | 0 | 0 | 0 | 0 |
| 0 | 0 | 0 | 0 | 0 |
| 0 | 0.000517868 | 0 | 0 | 0 |
| 0 | 0 | 0.002611222 | 0 | 0 |
| 0 | 0 | 0 | 0 | 0 |
| 0 | 0 | 0 | 0 | 0 |
| 0.286250426 | 0 | 0.658053391 | 0 | 0 |
| 0 | 0 | 0 | 0 | 0.006283356 |

| | | | | |
|---|---|---|---|---|
| 0 | 0 | 0 | 0 | 0 |
| 0 | 0 | 0 | 0 | 0 |
| 0 | 0 | 0 | 0 | 0 |
| 0 | 0.001756543 | 0 | 0 | 0 |
| 0 | 0.027031685 | 0.009080901 | 0 | 0.000825141 |
| 0 | 0 | 0 | 0 | 0 |
| 0 | 0 | 0 | 0 | 0 |
| 0 | 0 | 0 | 0 | 0 |
| 0 | 0 | 0 | 0 | 0 |
| 0 | 0 | 0 | 0 | 0 |
| 0 | 0.012441307 | 0 | 0 | 0 |
| 0.009203277 | 0 | 0.012731112 | 0.015369458 | 0.007875915 |
| 0 | 0 | 0 | 0.001159342 | 0.002100251 |
| 0 | 0 | 0 | 0 | 0 |
| 0 | 0 | 0 | 0 | 0 |
| 0 | 0.319570863 | 0 | 0 | 0 |
| 0 | 0.064344197 | 0.000665458 | 0 | 0 |
| 0 | 0 | 0 | 0 | 0 |
| 0.48867333 | 0 | 0.501845248 | 0.022894114 | 0.13738168 |
| 0 | 0 | 0 | 0 | 0 |
| 0 | 0 | 0 | 0 | 0 |
| 0 | 0.013826392 | 0 | 0 | 0 |
| 0 | 0 | 0 | 0 | 0 |
| 0 | 0 | 0 | 0 | 0 |
| 0 | 0 | 0 | 0 | 0 |
| 0 | 0 | 0 | 0 | 0 |
| 0 | 0 | 0.003329536 | 0 | 0 |
| 0 | 0 | 0 | 0 | 0 |
| 0 | 0 | 0 | 0 | 0 |
| 0 | 0 | 0 | 0 | 0 |
| 0.000792565 | 0 | 0.00111006 | 0 | 0.000514417 |
| 0 | 0 | 0 | 0 | 0 |
| 0 | 0 | 0 | 0 | 0 |
| 0 | 0 | 0 | 0 | 0 |
| 0 | 0 | 0 | 0 | 0 |
| 0 | 0 | 0 | 0 | 0 |
| 0 | 0 | 0 | 0 | 0 |
| 0.041126119 | 0 | 0.010840554 | 0.003020577 | 0.022317361 |
| 0 | 0.064440851 | 0 | 0 | 0 |
| 0 | 0 | 0 | 0 | 0 |
| 0.017788773 | 0.004334043 | 0 | 0 | 0.004510105 |
| 0 | 0 | 0 | 0 | 0 |
| 0 | 0 | 0 | 0 | 0 |
| 0.040577907 | 0 | 0.018041195 | 0.000785813 | 0.004287458 |
| 0 | 0 | 0 | 0 | 0 |
| 0 | 0 | 0 | 0 | 0 |
| 0 | 0 | 0 | 0 | 0 |
| 0 | 0 | 0 | 0 | 0 |

| | | | | |
|---|---|---|---|---|
| 0 | 0 | 0 | 0 | 0 |
| 0 | 0.055398066 | 0 | 0 | 0 |
| 0 | 0 | 0 | 0 | 0 |
| 0.010013349 | 0 | 0 | 0 | 0.010250284 |
| 0.000458628 | 0 | 0 | 0 | 0 |
| 0 | 0 | 0.002901554 | 0 | 0 |
| 0 | 0.012201476 | 0 | 0 | 0 |
| 0 | 0 | 0 | 0 | 0 |
| 0 | 0 | 0 | 0 | 0 |
| 0 | 0 | 0 | 0 | 0 |
| 0 | 0 | 0 | 0 | 0.080058615 |
| 0 | 0 | 0 | 0 | 0 |
| 0.11847618 | 0.511069069 | 0.018213764 | 0.005487443 | 0.073754965 |
| 0 | 0 | 0 | 0 | 0 |
| 0 | 0 | 0 | 0 | 0 |
| 0 | 0 | 0 | 0 | 0 |
| 0 | 0 | 0 | 0 | 0 |
| 0 | 0 | 0 | 0 | 0 |
| 0 | 0 | 0 | 0 | 0 |
| 0 | 0 | 0 | 0 | 0 |
| 0 | 0 | 0 | 0 | 0 |
| 0 | 0 | 0 | 0 | 0 |
| 0 | 0 | 0.033938675 | 0 | 0.009043404 |
| 0 | 0.006080005 | 0.103859898 | 0 | 0.004639795 |
| 0 | 0 | 0 | 0 | 0 |
| 0 | 0.001995619 | 0 | 0 | 0 |
| 0 | 0 | 0 | 0 | 0 |
| 0 | 0 | 0 | 0 | 0 |
| 0 | 0 | 0 | 0 | 0 |
| 0 | 0 | 0 | 0 | 0 |
| 0 | 0.042767581 | 0 | 0.001799884 | 0.002776576 |
| 0 | 0 | 0 | 0 | 0 |
| 0 | 0 | 0 | 0 | 0 |
| 0 | 0 | 0 | 0 | 0 |
| 0 | 0 | 0 | 0 | 0 |
| 0 | 0 | 0 | 0 | 0.001680428 |
| 0 | 0 | 0 | 0 | 0 |
| 0 | 0 | 0 | 0 | 0 |
| 0 | 0 | 0 | 0 | 0 |
| 0 | 0 | 0 | 0 | 0 |
| 0 | 0 | 0 | 0 | 0 |
| 0 | 0 | 0 | 0 | 0 |
| 0 | 0 | 0 | 0 | 0 |
| 0 | 0 | 0 | 0 | 0 |
| 0 | 0 | 0 | 18.95507925 | 0 |
| 0.378783329 | 0.30553288 | 0.068263457 | 0.066417563 | 0.707854302 |
| 0 | 0 | 0 | 0 | 0 |
| 0 | 0 | 0 | 0 | 0 |

| | | | | |
|---|---|---|---|---|
| 0 | 0 | 0 | 0 | 0 |
| 0 | 0 | 0 | 0 | 0 |
| 0 | 0 | 0 | 0.029056449 | 0.076990922 |
| 0 | 0.020087282 | 0 | 0 | 0 |
| 0 | 0 | 0 | 0 | 0 |
| 0 | 0 | 0 | 0 | 0 |
| 0 | 0 | 0 | 0 | 0 |
| 0 | 0 | 0 | 0 | 0 |
| 0 | 0 | 0 | 0 | 0.000457603 |
| 0 | 0 | 0 | 0 | 0 |
| 0.0018545 | 0 | 0.000952379 | 0.000647125 | 0.000902753 |
| 0.001318464 | 0 | 0 | 0 | 0.001048299 |
| 0 | 0 | 0 | 0 | 0 |
| 0 | 0 | 0 | 0 | 0 |
| 0 | 0 | 0 | 0 | 0 |
| 0 | 0 | 0 | 0 | 0 |
| 0 | 0 | 0 | 0 | 0 |
| 0 | 0 | 0 | 0 | 0 |
| 0 | 0 | 0 | 0 | 0 |
| 0.064646704 | 0 | 0.000803739 | 0.003978921 | 0.001463251 |
| 0 | 0 | 0 | 0 | 0 |
| 0 | 0 | 0 | 0 | 0 |
| 0 | 0 | 0 | 0 | 0 |
| 0 | 0 | 0 | 0 | 0 |
| 0 | 0 | 0 | 0 | 0 |
| 0 | 0 | 0 | 0 | 0 |
| 0 | 0 | 0 | 0 | 0 |
| 0 | 0.032311993 | 0 | 0.002503865 | 0 |
| 0 | 0 | 0 | 0 | 0 |
| 0 | 0 | 0 | 0 | 0 |
| 0.013683327 | 0.016722462 | 0.00049321 | 0.002748049 | 0.001942766 |
| 0 | 0 | 0 | 0 | 0 |
| 0.232708601 | 0.011012332 | 0.032527519 | 0.140938416 | 0.048963017 |
| 0 | 0 | 0.000819286 | 0 | 0 |
| 0 | 0 | 0 | 0 | 0 |
| 0 | 0 | 0 | 0 | 0 |
| 0 | 0 | 0 | 0 | 0 |
| 0 | 0 | 0 | 0 | 0 |
| 0 | 0 | 0 | 0 | 0 |
| 0 | 0 | 0 | 0 | 0 |
| 0 | 0 | 0 | 0 | 0 |
| 0.004805403 | 0 | 0.078961125 | 0.000256099 | 0.00174662 |
| 0 | 0.016846444 | 0.00964265 | 0.009360018 | 0.040961612 |
| 0 | 0 | 0 | 0 | 0 |
| 0 | 0 | 0 | 0 | 0 |
| 0.007783128 | 0.108709813 | 0 | 0.107981588 | 0.022485416 |
| 0 | 0 | 0 | 0 | 0 |
| 0 | 0 | 0 | 0 | 0 |

| | | | | |
|---|---|---|---|---|
| 0 | 0 | 0 | 0 | 0 |
| 0 | 0.003803639 | 0 | 0.001470879 | 0 |
| 0.025647898 | 0.003198997 | 0.001966692 | 0.000668166 | 0.001464738 |
| 0 | 0 | 0 | 0 | 0 |
| 0.001144057 | 0.006527516 | 0.166452714 | 0.014328251 | 0.005457773 |
| 0.091762105 | 0 | 0 | 0 | 0 |
| 0 | 0 | 0 | 0 | 0 |
| 0 | 0 | 0 | 0 | 0 |
| 0 | 0 | 0 | 0 | 0 |
| 0 | 0 | 0 | 0 | 0 |
| 0 | 0 | 0 | 0 | 0 |
| 0 | 0 | 0 | 0 | 0 |
| 0 | 0 | 0 | 0 | 0 |
| 0 | 0 | 0 | 0 | 0 |
| 0 | 0 | 0 | 0 | 0 |
| 0 | 0 | 0 | 0 | 0 |
| 0 | 0 | 0 | 0 | 0 |
| 0 | 0 | 0 | 0 | 0 |
| 0 | 0 | 0 | 0 | 0 |
| 0 | 0 | 0 | 0 | 0 |
| 0 | 0 | 0 | 0 | 0 |
| 0 | 0 | 0 | 0 | 0 |
| 0 | 0 | 0 | 0 | 0 |
| 0 | 0 | 0 | 0 | 0 |
| 0 | 0.001757616 | 0 | 0.012271919 | 0.001743661 |
| 0.005702024 | 0 | 0.000815259 | 0.006568312 | 0 |
| 0 | 0 | 0 | 0 | 0 |
| 0 | 0.002143922 | 0 | 0.012512688 | 0.536895209 |
| 0 | 0 | 0 | 0 | 0 |
| 0 | 0 | 0 | 0 | 0 |
| 0 | 0 | 0 | 0.006292412 | 0 |
| 0 | 0 | 0 | 0 | 0 |
| 0 | 0 | 0 | 0 | 0 |
| 0 | 0 | 0 | 0 | 0 |
| 0 | 0 | 0 | 0 | 0 |
| 0 | 0 | 0 | 0 | 0 |
| 0.000452522 | 0 | 0 | 0 | 0 |
| 0 | 0 | 0 | 0 | 0 |
| 0 | 0.161334041 | 0 | 0.120276109 | 0 |
| 0 | 0 | 0 | 0 | 0 |
| 0 | 0 | 0 | 0 | 0 |
| 0 | 0 | 0 | 0.050394342 | 0.003659439 |
| 0.013233521 | 0 | 0.010870384 | 0 | 0.063510272 |
| 0 | 0 | 0 | 0 | 0 |
| 0 | 0 | 0 | 0 | 0 |
| 0 | 0 | 0 | 0 | 0.000778959 |
| 0 | 0 | 0.008761811 | 0.003644999 | 0.044125192 |
| 0 | 0.00185082 | 0 | 0 | 0 |

| | | | | |
|---|---|---|---|---|
| 0 | 0 | 0 | 0 | 0 |
| 0.019216347 | 0.011988815 | 0.004851012 | 0.012648125 | 0.034504599 |
| 0 | 0 | 0 | 0 | 0.005733976 |
| 0 | 0 | 0 | 0 | 0 |
| 0 | 0 | 0 | 0 | 0 |
| 0 | 0 | 0 | 0 | 0 |
| 0 | 0 | 0 | 0 | 0 |
| 0 | 0 | 0 | 0 | 0 |
| 0 | 0 | 0 | 0 | 0 |
| 0 | 0 | 0 | 0 | 0 |
| 0 | 0 | 0 | 0 | 0 |
| 0 | 0 | 0 | 0 | 0 |
| 0 | 0.008147083 | 0 | 0.299006037 | 0 |
| 0 | 0 | 0 | 0 | 0 |
| 0.006169313 | 0.007652078 | 0.004782519 | 0.068815878 | 0.059970109 |
| 0 | 0 | 0 | 0 | 0 |
| 0 | 0 | 0 | 0 | 0 |
| 0 | 0 | 0 | 0 | 0 |
| 0 | 0 | 0 | 0 | 0.001622506 |
| 0 | 0 | 0 | 0 | 0 |
| 0 | 0 | 0 | 0 | 0 |
| 0 | 0 | 0 | 0 | 0 |
| 0 | 0 | 0.004882921 | 0.193950529 | 0.005657035 |
| 0 | 0 | 0 | 0 | 0 |
| 0 | 0 | 0 | 0 | 0 |
| 0 | 0 | 0 | 0 | 0 |
| 0 | 0 | 0 | 0 | 0 |
| 0 | 0 | 0 | 0 | 0 |
| 0 | 0 | 0 | 0 | 0 |
| 0 | 0 | 0 | 0 | 0 |
| 0 | 0 | 0 | 0 | 0 |
| 0 | 0 | 0 | 0 | 0 |
| 0 | 0 | 0 | 0 | 0 |
| 0 | 0 | 0 | 0 | 0 |
| 0 | 0 | 0 | 1.727916064 | 0 |
| 0 | 0 | 0 | 0 | 0 |
| 0 | 0 | 0 | 0 | 0 |
| 0 | 0 | 0 | 0 | 0 |
| 0.674267484 | 0.023560896 | 0.018209534 | 0 | 0.000479463 |
| 0 | 0 | 0 | 0 | 0 |
| 0.048824829 | 0.122840633 | 0.03438619 | 0.019246559 | 0.199331345 |
| 0 | 0 | 0 | 0 | 0 |
| 0 | 0 | 0 | 0 | 0 |
| 0 | 0 | 0 | 0 | 0 |
| 0 | 0 | 0 | 0 | 0 |
| 0.947285418 | 0 | 0.05919639 | 0.010777432 | 0.533554365 |
| 0 | 0 | 0 | 0 | 0 |
| 0 | 0 | 0 | 0 | 0 |

| | | | | |
|---|---|---|---|---|
| 0 | 0 | 0 | 0 | 0 |
| 0 | 0 | 0 | 0 | 0 |
| 0 | 0 | 0 | 0 | 0 |
| 0 | 0 | 0 | 0 | 0 |
| 0 | 0 | 0 | 0 | 0 |
| 0 | 0 | 0 | 0 | 0 |
| 0 | 0 | 0 | 0 | 0 |
| 0 | 0 | 0 | 0 | 0 |
| 0 | 0 | 0.026766302 | 0.003650485 | 0.004112398 |
| 0 | 0.010513392 | 0 | 0 | 0.007541636 |
| 0 | 0 | 0 | 0 | 0 |
| 0 | 0 | 0 | 0 | 0 |
| 0.00538509 | 0 | 0.000406124 | 0 | 0.006900805 |
| 0 | 0 | 0 | 0 | 0 |
| 0 | 0 | 0 | 0 | 0 |
| 0 | 0 | 0 | 0 | 0 |
| 0 | 0 | 0 | 0 | 0 |
| 0 | 0 | 0 | 0 | 0 |
| 0 | 0 | 0 | 0 | 0 |
| 0.191478601 | 0.012499979 | 0.079043494 | 0.264367871 | 0.670616898 |
| 0 | 0 | 0 | 0 | 0 |
| 0 | 0 | 0 | 0 | 0 |
| 0 | 0 | 0 | 0 | 0 |
| 0 | 0 | 0 | 0 | 0 |
| 0 | 0 | 0 | 0 | 0 |
| 0 | 0 | 0 | 0 | 0.007952262 |
| 0 | 0 | 0 | 0 | 0 |
| 0 | 0 | 0 | 0 | 0 |
| 0 | 0 | 0 | 0 | 0 |
| 0 | 0 | 0 | 0 | 0 |
| 0 | 0 | 0 | 0 | 0 |
| 0 | 0 | 0 | 0 | 0 |
| 0 | 0 | 0 | 0 | 0 |
| 0 | 0 | 0 | 0 | 0 |
| 0 | 0 | 0 | 0 | 0 |
| 0.026727382 | 0.005327869 | 0 | 0 | 0 |
| 0 | 0 | 0 | 0 | 0 |
| 0 | 0 | 0 | 0 | 0 |
| 0 | 0 | 0 | 0 | 0 |
| 0 | 0 | 0 | 0 | 0 |
| 0 | 0 | 0 | 0 | 0 |
| 0 | 0 | 0 | 0 | 0 |
| 0 | 0 | 0 | 0 | 0 |
| 0 | 0 | 0 | 0 | 0 |
| 0.002654946 | 0.00071588 | 0 | 0.000999676 | 0.014815797 |
| 0 | 0.016698033 | 0 | 0 | 0 |
| 0 | 0 | 0 | 0 | 0 |
| 0 | 0 | 0 | 0 | 0 |

| | | | | |
|---|---|---|---|---|
| 0 | 0 | 0 | 0 | 0 |
| 0 | 0 | 0 | 0 | 0 |
| 0 | 0 | 0 | 0 | 0 |
| 0 | 0 | 0 | 0 | 0 |
| 0 | 0 | 0 | 0 | 0 |
| 0 | 0 | 0.002027688 | 0.074916702 | 0.002838553 |
| 0 | 0 | 0 | 0 | 0 |
| 0 | 0 | 0 | 0 | 0 |
| 0 | 0 | 0 | 0 | 0 |
| 0 | 0 | 0 | 0 | 0 |
| 0 | 0 | 0 | 0 | 0 |
| 0 | 0 | 0 | 0 | 0 |
| 0 | 0 | 0 | 0 | 0 |
| 0 | 0 | 0 | 0 | 0.086975789 |
| 0 | 0 | 0 | 0 | 0 |
| 0 | 0 | 0 | 0 | 0 |
| 0 | 0 | 0 | 0 | 0 |
| 0 | 0 | 0 | 0 | 0 |
| 0 | 0 | 0 | 0 | 0.01657193 |
| 0 | 0 | 0 | 0 | 0 |
| 0 | 0 | 0 | 0 | 0 |
| 0 | 0 | 0 | 0 | 0 |
| 0 | 0 | 0 | 0 | 0 |
| 0 | 0 | 0 | 0 | 0 |
| 0 | 0 | 0 | 0 | 0 |
| 0 | 0 | 0 | 0 | 0 |
| 0 | 0 | 0 | 0 | 0 |
| 0 | 0 | 0 | 0 | 0 |
| 0 | 0 | 0 | 0 | 0 |
| 0 | 0 | 0 | 0 | 0 |
| 0 | 0 | 0 | 0 | 0 |
| 0 | 0 | 0 | 0 | 0 |
| 0 | 0 | 0 | 0 | 0 |
| 0 | 0 | 0 | 0 | 0 |
| 0 | 0 | 0 | 0 | 0 |
| 0 | 0 | 0 | 0 | 0 |
| 0 | 0 | 0 | 0 | 0 |
| 0 | 0 | 0 | 0 | 0 |
| 0 | 0 | 2.538469314 | 0 | 2.305747553 |
| 0 | 0 | 0 | 0 | 0 |
| 0 | 0 | 0 | 0 | 0 |
| 0 | 0 | 0 | 0 | 0 |
| 0 | 0 | 0 | 0 | 0 |
| 0 | 0 | 0 | 0 | 0 |
| 0 | 0 | 0 | 0 | 0 |
| 0 | 0 | 0 | 0 | 0 |
| 0 | 0 | 0 | 0 | 0 |
| 0 | 0 | 0 | 0 | 0 |

| | | | | |
|---|---|---|---|---|
| 0 | 0 | 0 | 0 | 0 |
| 0 | 0.002273693 | 0.0028377 | 0 | 0.010081895 |
| 0 | 0 | 0 | 0 | 0 |
| 0 | 0 | 0 | 0 | 0 |
| 0 | 0 | 0 | 0 | 0 |
| 0 | 0 | 0 | 0.0009695 | 0 |
| 0 | 0 | 0 | 0 | 0 |
| 0 | 0 | 0 | 0 | 0 |
| 0 | 0 | 0 | 0 | 0 |
| 0 | 0 | 0 | 0 | 0 |
| 0 | 0.143376705 | 0 | 0 | 0.019658671 |
| 0 | 0 | 0 | 0 | 0 |
| 0 | 0.043280407 | 0 | 0 | 0 |
| 0 | 0 | 0 | 0 | 0 |
| 0 | 0 | 0 | 0 | 0 |
| 0 | 0 | 0 | 0 | 0 |
| 0 | 0 | 0 | 0 | 0 |
| 0 | 0 | 0 | 0 | 0 |
| 0 | 0 | 0 | 0 | 0 |
| 0 | 0 | 0 | 0 | 0 |
| 0.040004585 | 0 | 0.004399798 | 0.000597917 | 0.002184566 |
| 0 | 0 | 0 | 0 | 0 |
| 0 | 0 | 0 | 0 | 0 |
| 0 | 7.149727417 | 0 | 0.425191999 | 0.450010426 |
| 0 | 0 | 0 | 0 | 0 |
| 0 | 0 | 0 | 0 | 0 |
| 0 | 0 | 0 | 0 | 0 |
| 0 | 0 | 0 | 0 | 0 |
| 0 | 0 | 0 | 0 | 0 |
| 0 | 0 | 0 | 0 | 0 |
| 0 | 0 | 0 | 0 | 0 |
| 0 | 0 | 0 | 0 | 0 |
| 0.00080925 | 0 | 0 | 0 | 0.000765985 |
| 0 | 0 | 0 | 0 | 0 |

| PHL018-0207_V2 | PHL018-0093_V3 | PHL018-0114_V1 | PHL018-0116_V3 | PHL018-0220_V2 |
|---|---|---|---|---|
| REF | CTRL | EYCF | CTRL | REF |
| 0.060524049 | 0.1597104 | 0.059008088 | 0.089114614 | 0 |
| 0.072026468 | 6.380522859 | 0.190577513 | 1.219858572 | 0.641625587 |
| 0.204772818 | 0 | 0 | 0 | 3.954368465 |
| 0 | 0 | 0 | 0 | 0 |
| 0 | 0 | 0 | 0 | 0 |
| 0 | 0 | 0 | 0 | 0 |
| 0 | 0 | 0 | 0 | 0 |
| 0 | 0 | 0 | 0 | 0 |
| 0 | 0 | 0 | 0 | 0 |
| 0 | 0 | 0 | 0 | 0 |
| 0 | 0 | 0 | 0 | 0 |
| 0 | 0 | 0 | 0 | 0 |
| 0 | 0 | 0 | 0 | 0 |
| 0.092501113 | 10.49803593 | 0.59324129 | 0.099982053 | 0.040181724 |
| 0 | 0 | 0 | 0 | 0 |
| 0 | 0 | 0 | 0 | 0 |
| 0 | 0 | 0 | 0 | 0 |
| 0 | 0 | 0 | 0 | 0 |
| 0 | 0 | 0 | 0 | 0 |
| 0 | 0 | 0 | 0 | 0 |
| 0 | 0 | 0.000530484 | 0 | 0 |
| 0 | 0 | 0 | 0.001391551 | 0 |
| 0 | 0 | 0 | 0 | 0 |
| 0 | 0 | 0 | 0 | 0 |
| 0.185410605 | 0 | 0.018242903 | 0.007827297 | 0 |
| 0 | 0 | 0 | 0 | 0 |
| 0 | 0 | 0 | 0 | 0 |
| 0 | 0 | 0 | 0 | 0 |
| 0 | 0 | 0 | 0 | 0 |
| 0 | 0 | 0 | 0 | 0 |
| 0 | 0 | 0 | 0 | 0 |
| 0 | 0 | 0 | 0 | 0 |
| 0 | 0 | 0 | 0 | 0 |
| 0 | 0 | 0 | 0 | 0 |
| 0 | 0 | 0 | 0 | 0 |
| 0.189784771 | 0 | 0.366260756 | 0.043630683 | 1.414555762 |
| 0 | 0 | 0 | 0 | 0 |
| 0 | 0 | 0 | 0 | 0 |
| 0 | 0 | 0 | 0 | 0 |
| 0 | 0 | 0 | 0 | 0 |
| 0 | 0 | 0 | 0 | 0 |
| 0.001849151 | 0 | 0.007333534 | 0.004129216 | 0.0784461 |
| 0.398635824 | 1.59668107 | 4.836182748 | 0.584450355 | 0.355411116 |
| 0.002057529 | 0.003064031 | 0.013432375 | 0.005693702 | 0.001248855 |
| 0.051060393 | 0.001227255 | 0.047591259 | 0.003328344 | 0.018727918 |
| 0 | 0 | 0 | 0 | 0 |

| | | | | |
|---|---|---|---|---|
| 0.001011856 | 0 | 0 | 0.003594672 | 0 |
| 0.004315939 | 0.175515798 | 0 | 0.00276976 | 0.004013963 |
| 0.089559656 | 0 | 0.044468416 | 0 | 0 |
| 0.270019172 | 0.290342632 | 0.381524941 | 0.949967028 | 0.269321979 |
| 0.000774382 | 0 | 0 | 0 | 0 |
| 0 | 0 | 0.015014499 | 0 | 0 |
| 0 | 0.001641985 | 0 | 0.007421841 | 0 |
| 0 | 0 | 0 | 0 | 0 |
| 0 | 0 | 0 | 0 | 0 |
| 0 | 0 | 0 | 0 | 0 |
| 0.004524608 | 0.004094953 | 0.01360559 | 0 | 0.00252992 |
| 0.177029836 | 0 | 0.111469961 | 0.295261235 | 1.60192913 |
| 0 | 0 | 0 | 0 | 0 |
| 0.015709977 | 0 | 0.002651237 | 0.012301097 | 0.042397126 |
| 0 | 0 | 0 | 0 | 0 |
| 0 | 0 | 0 | 0 | 0 |
| 0.012306694 | 0.228225449 | 0.127036721 | 0.049757737 | 0.562127303 |
| 0 | 0 | 0 | 0 | 0 |
| 0 | 0 | 0 | 0 | 0 |
| 0 | 0 | 0 | 0 | 0 |
| 0.002710303 | 2.065386499 | 0 | 0.103024919 | 0.17151898 |
| 0.051000244 | 0.022667017 | 0 | 0.002410725 | 0.001956442 |
| 0 | 0 | 0.137117385 | 0.052557246 | 0.010286145 |
| 0 | 0 | 0 | 0 | 0 |
| 0 | 0 | 0 | 0 | 0 |
| 0 | 0 | 0 | 0 | 0 |
| 0 | 0.006567295 | 0 | 0 | 0 |
| 0 | 0 | 0 | 0 | 0 |
| 0 | 0 | 0.025798406 | 0 | 0.012983559 |
| 0 | 0 | 0 | 0 | 0 |
| 0 | 0 | 0 | 0 | 0 |
| 0 | 0 | 0 | 0 | 0 |
| 0 | 0 | 0 | 0 | 0 |
| 0 | 0 | 0 | 0 | 0 |
| 0 | 0 | 0 | 0 | 0 |
| 0 | 0 | 0 | 0 | 0 |
| 0 | 0 | 0 | 0 | 0 |
| 0 | 0 | 12.63250839 | 0 | 0.001631019 |
| 0 | 0 | 0 | 0 | 0 |
| 0 | 0 | 0 | 0 | 0 |
| 0 | 0 | 0 | 0 | 0 |
| 0 | 0 | 0 | 0 | 0 |
| 0 | 0 | 0 | 0 | 0 |
| 0 | 0 | 0 | 0 | 0 |
| 0 | 0 | 0 | 0 | 0 |
| 0 | 0 | 0 | 0 | 0 |
| 0 | 0 | 0 | 0 | 0 |
| 0 | 0 | 0 | 0 | 0 |

| | | | | |
|---|---|---|---|---|
| 0 | 0 | 0 | 0 | 0 |
| 0 | 0 | 0 | 0 | 0 |
| 0 | 0 | 0 | 0 | 0 |
| 0 | 0 | 0 | 0 | 0 |
| 0.017379267 | 0 | 0.000427722 | 0 | 0 |
| 0 | 0 | 0 | 0 | 0.000901771 |
| 0 | 0 | 0 | 0 | 0 |
| 0 | 0 | 0 | 0 | 0 |
| 0 | 0.001933187 | 0 | 0.018204337 | 0.028838577 |
| 0 | 0 | 0 | 0 | 0 |
| 0 | 0 | 0 | 0 | 0 |
| 0 | 0.00074813 | 0 | 0 | 0.00150939 |
| 0 | 0 | 0 | 0 | 0 |
| 0 | 0 | 0 | 0 | 0 |
| 0 | 0 | 0 | 0 | 0 |
| 0 | 0 | 0 | 0 | 0 |
| 0 | 0 | 0 | 0 | 0 |
| 0 | 0 | 0 | 0 | 0 |
| 0.239947467 | 0 | 3.782637946 | 0 | 5.905820837 |
| 0 | 0 | 0 | 0 | 0 |
| 0 | 0 | 0 | 0 | 0 |
| 0 | 0 | 0 | 0 | 0 |
| 0 | 0 | 0 | 0 | 0 |
| 0 | 0 | 0 | 0 | 0 |
| 0 | 0 | 0 | 0 | 0 |
| 0 | 0 | 0 | 0 | 0 |
| 0 | 0 | 0 | 0 | 0 |
| 0 | 0 | 0 | 0 | 0 |
| 0 | 0 | 0 | 0 | 0 |
| 0 | 0 | 0 | 0 | 0 |
| 0.065627544 | 0 | 0.006834094 | 0.015902399 | 0 |
| 0.028852909 | 0 | 0.005759155 | 0.036193632 | 0 |
| 0 | 0 | 0 | 0 | 0 |
| 0 | 0 | 0 | 0 | 0 |
| 0 | 0 | 0 | 0 | 0 |
| 0 | 0 | 0 | 0 | 0 |
| 0 | 0 | 0 | 0 | 0 |
| 0 | 0 | 0 | 0 | 0 |
| 0.070607319 | 0 | 0 | 0 | 0.26438163 |
| 0 | 0 | 0 | 0 | 0 |
| 0 | 0 | 0 | 0 | 0 |
| 0 | 0 | 0 | 0 | 0 |
| 0 | 0 | 0 | 0.010948772 | 0 |
| 0 | 0 | 0 | 0 | 0 |
| 0 | 0 | 0 | 0 | 0 |
| 0 | 0 | 0 | 0 | 0 |
| 0 | 0 | 0 | 0 | 0 |
| 0 | 0 | 0 | 0 | 0 |

| | | | | |
|---|---|---|---|---|
| 0 | 0 | 0 | 0 | 0 |
| 0 | 0 | 0 | 0 | 0 |
| 0 | 0 | 0 | 0 | 0 |
| 0 | 0 | 0 | 0 | 0 |
| 0.075252978 | 0 | 0.118565527 | 0.033471364 | 0 |
| 0 | 0 | 0 | 0 | 0 |
| 0 | 0 | 0 | 0 | 0 |
| 0 | 0 | 0 | 0 | 0 |
| 0 | 0 | 0 | 0 | 0 |
| 0 | 0 | 0 | 0 | 0 |
| 0 | 0 | 0 | 0 | 0 |
| 0 | 0 | 0 | 0 | 0 |
| 0 | 0 | 0 | 0 | 0 |
| 0 | 0 | 0 | 0 | 0 |
| 0 | 0 | 0 | 0 | 0 |
| 0 | 0 | 2.664677395 | 0 | 0 |
| 0 | 0 | 0 | 0 | 0 |
| 0 | 0 | 0 | 0 | 0 |
| 0 | 0 | 0 | 0 | 0 |
| 0 | 0 | 0 | 0 | 0 |
| 0 | 0 | 0 | 0 | 0 |
| 0 | 0 | 0 | 0 | 0 |
| 0 | 0 | 0 | 0 | 0 |
| 0 | 0 | 0 | 0 | 0 |
| 0.002916559 | 0 | 0.085170715 | 0.000828898 | 0.076553094 |
| 0 | 0 | 0 | 0 | 0 |
| 0 | 0 | 0 | 0 | 0 |
| 0 | 0 | 0 | 0 | 0 |
| 0 | 0 | 0 | 0 | 0 |
| 0 | 0 | 0 | 0 | 0 |
| 0.006389944 | 0 | 0 | 0.105256745 | 0 |
| 0 | 0.014074698 | 0 | 0 | 0 |
| 0 | 0 | 0 | 0 | 0 |
| 0 | 0 | 0 | 0 | 0 |
| 0 | 0 | 0 | 0 | 0 |
| 0 | 0 | 0 | 0 | 0 |
| 0 | 0 | 0 | 0 | 0 |
| 0 | 0 | 0 | 0 | 0 |
| 0 | 0 | 0 | 0 | 0 |
| 0 | 0 | 0 | 0 | 0 |
| 0 | 0 | 0 | 0 | 0 |
| 0 | 0 | 0 | 0 | 0 |
| 0 | 0 | 0 | 0 | 0 |
| 0 | 0 | 0 | 0 | 0 |
| 0 | 0 | 0 | 0 | 0 |
| 0 | 0 | 0 | 0 | 0 |
| 0 | 0 | 0 | 0 | 0 |
| 0 | 0.070793297 | 0 | 0 | 0 |

| | | | | |
|---|---|---|---|---|
| 2.048192783 | 0 | 0 | 0 | 0 |
| 0 | 0 | 0 | 0 | 0 |
| 0 | 0 | 0 | 0 | 0 |
| 0 | 0 | 0 | 0 | 0 |
| 0 | 0 | 0 | 0 | 0 |
| 0 | 0 | 0 | 0 | 0 |
| 0 | 0 | 0 | 0 | 0 |
| 0 | 0 | 0 | 0 | 0 |
| 0 | 0 | 0 | 0 | 0 |
| 0 | 0 | 0 | 0 | 0 |
| 0 | 0 | 0 | 0 | 0 |
| 0 | 0 | 0 | 0.004116935 | 0 |
| 0.002416609 | 0 | 0 | 0 | 0 |
| 0 | 0 | 0 | 0 | 0 |
| 0 | 0 | 0 | 0 | 0 |
| 0 | 0 | 0 | 0 | 0 |
| 0 | 0 | 0 | 0 | 0 |
| 0 | 0 | 0 | 0 | 0 |
| 0 | 0 | 0 | 0 | 0 |
| 0 | 0 | 0 | 0 | 0 |
| 0.003696936 | 0.028410599 | 0.005337811 | 0.003210426 | 0.019422406 |
| 0 | 0 | 0 | 0 | 0 |
| 0 | 0 | 0 | 0 | 0 |
| 0 | 0 | 0 | 0.009739139 | 0 |
| 0 | 0 | 0 | 0 | 0 |
| 0 | 0 | 0 | 0 | 0 |
| 0 | 0.001128113 | 0.003673818 | 0.004759177 | 0 |
| 0 | 0 | 0 | 0 | 0 |
| 0 | 0 | 0 | 0 | 0 |
| 0 | 0.003218537 | 0 | 0 | 0.002361294 |
| 0 | 0.10533168 | 0 | 0 | 0.202130335 |
| 0 | 0 | 0 | 0 | 0 |
| 0 | 0 | 0 | 0 | 0 |
| 0 | 0 | 0.191279946 | 0.011236985 | 0.011580262 |
| 0.000801808 | 0.014494273 | 0.008682661 | 0.006646419 | 0.171219364 |
| 0 | 0 | 0 | 0 | 0 |
| 0 | 0 | 0 | 0 | 0 |
| 0 | 0 | 0 | 0 | 0 |
| 0 | 0 | 0 | 0 | 0 |
| 0 | 0 | 0 | 0 | 0 |
| 0 | 0 | 0 | 0 | 0 |
| 0 | 0 | 0 | 0 | 0 |
| 0 | 0 | 0 | 0 | 0.004485124 |
| 0 | 0 | 0 | 0 | 0 |
| 0 | 0 | 0 | 0 | 0 |
| 0 | 0 | 0.006162553 | 0 | 0 |
| 0.000578763 | 0 | 0 | 0 | 0 |
| 0.001238012 | 0.00284422 | 0 | 0 | 0 |

| | | | | |
|---|---|---|---|---|
| 0 | 0 | 0 | 0 | 0 |
| 0 | 0 | 0 | 0 | 0 |
| 0 | 0 | 0 | 0.002473166 | 0 |
| 0 | 0.000951414 | 0 | 0 | 0 |
| 0 | 0 | 0 | 0 | 0 |
| 0 | 0 | 0 | 0 | 0 |
| 0 | 0 | 0 | 0 | 0.049245356 |
| 0 | 0 | 0 | 0 | 0.01547179 |
| 0 | 0 | 0.002881951 | 0 | 0 |
| 0 | 0 | 0 | 0 | 0 |
| 0 | 0 | 0 | 0 | 0 |
| 0 | 0 | 0 | 0 | 0 |
| 0 | 0 | 0 | 0 | 0 |
| 0 | 0 | 0 | 0 | 0 |
| 0 | 0 | 0.008868759 | 0 | 0.00069263 |
| 0 | 0 | 0 | 0 | 0 |
| 0 | 0 | 0 | 0 | 0 |
| 0 | 0 | 0 | 0 | 0 |
| 0.001936999 | 0.009591098 | 0.028145287 | 0.002752517 | 0.000781839 |
| 0.592552205 | 0 | 0 | 0 | 0 |
| 0 | 0.293062524 | 0 | 0 | 0 |
| 0 | 0 | 0 | 0 | 0 |
| 0 | 0 | 0 | 0 | 0 |
| 0 | 0 | 0 | 0 | 0 |
| 0 | 0.005353845 | 0 | 0.005243246 | 0.001963936 |
| 0.00165254 | 0 | 0 | 0 | 0 |
| 0 | 0 | 0 | 0 | 0 |
| 0 | 0 | 0 | 0 | 0 |
| 0 | 0 | 0.006079565 | 0 | 0.076530338 |
| 0 | 0 | 0 | 0 | 0 |
| 0.061137658 | 0.364597066 | 0.002978298 | 0.060754831 | 0.169360737 |
| 0 | 0 | 0 | 0 | 0 |
| 0 | 0 | 0 | 0 | 0 |
| 0 | 0 | 0 | 0 | 0 |
| 0 | 0 | 0 | 0 | 0 |
| 0 | 0 | 0 | 0 | 0 |
| 0 | 0 | 0 | 0 | 0 |
| 0 | 0 | 0 | 0 | 0 |
| 0 | 0 | 0 | 0 | 0 |
| 0.001259134 | 0 | 0 | 0 | 0 |
| 0 | 0 | 0 | 0 | 0 |
| 0.00139202 | 0.011108986 | 0.017962287 | 0.006542915 | 0.022351184 |
| 0 | 0 | 0 | 0 | 0.000310831 |
| 0 | 0 | 0 | 0 | 0 |
| 0 | 0 | 0 | 0 | 0 |
| 0 | 0 | 0 | 0 | 0 |
| 0 | 0 | 0 | 0 | 0 |
| 0 | 0 | 0 | 0 | 0 |

| | | | | |
|---|---|---|---|---|
| 0 | 0 | 0 | 0 | 0 |
| 0 | 0 | 0 | 0 | 0 |
| 0 | 0 | 0.016836538 | 0 | 0 |
| 0 | 0 | 0 | 0 | 0 |
| 0.577699829 | 0 | 0 | 9.244885076 | 0 |
| 0 | 0 | 0 | 0 | 0 |
| 0 | 0 | 0 | 0 | 0 |
| 0 | 0 | 0 | 0 | 0 |
| 0 | 0 | 0 | 0 | 0 |
| 0 | 0 | 0 | 0.023590535 | 0 |
| 0 | 0 | 0 | 0 | 0 |
| 0 | 0 | 0 | 0 | 0 |
| 0 | 0 | 0 | 0 | 0 |
| 0 | 0 | 0 | 0.001478113 | 0 |
| 0.001717059 | 0.001542324 | 0 | 0.005577092 | 0 |
| 0.028756871 | 0 | 0.009307217 | 0.933903897 | 0 |
| 0.002934653 | 0.00175734 | 0 | 0.005361691 | 0.003867839 |
| 0 | 0 | 0 | 0 | 0 |
| 0 | 0 | 0 | 0 | 0 |
| 0 | 0 | 0 | 0 | 0 |
| 0 | 0 | 0 | 0 | 0 |
| 0.001135352 | 0.001388079 | 0 | 0 | 0 |
| 0 | 0 | 0 | 0 | 0 |
| 0 | 0 | 0 | 0 | 0 |
| 0 | 0 | 0 | 0.00546129 | 0.011896822 |
| 0 | 0 | 0 | 0 | 0 |
| 0 | 0 | 0 | 0 | 0 |
| 0 | 0 | 0 | 0 | 0 |
| 0 | 0 | 0 | 0 | 0 |
| 0 | 0.011473539 | 0 | 0 | 0.010311573 |
| 0 | 0 | 0 | 0 | 0 |
| 0 | 0 | 0 | 0 | 0 |
| 0 | 0 | 0 | 0 | 0.004817325 |
| 0 | 0 | 0 | 0 | 0 |
| 0 | 0.00209706 | 0 | 0 | 0 |
| 0 | 0.003564631 | 0 | 0 | 0.025570911 |
| 0 | 0 | 0 | 0 | 0 |
| 0.173868633 | 0 | 0.020595787 | 0.034684539 | 0.697680398 |
| 0 | 0 | 0 | 0 | 0 |
| 0 | 0 | 0 | 0 | 0 |
| 0 | 0 | 0 | 0 | 0 |
| 0 | 0 | 0 | 0 | 0 |
| 0 | 0 | 0 | 0 | 0 |
| 0 | 0 | 0 | 0 | 0.00317862 |
| 0 | 0 | 0 | 0 | 0 |
| 0 | 0 | 0 | 0 | 0 |
| 0 | 0 | 0 | 0 | 0.000990647 |
| 0 | 0.003621248 | 0 | 0 | 0 |

| | | | | |
|---|---|---|---|---|
| 0.000885923 | 0 | 0 | 1.815896506 | 0 |
| 0 | 0 | 0 | 0 | 0 |
| 0 | 0 | 0 | 0 | 0 |
| 0 | 0 | 0 | 0 | 0 |
| 0.003456389 | 0.005851073 | 0.00172748 | 0.015301529 | 0.003526118 |
| 0 | 0 | 0 | 0 | 0.0049997 |
| 0 | 0 | 0 | 0 | 0 |
| 0 | 0 | 0.002641574 | 0.011651016 | 0.004512836 |
| 0 | 0 | 0 | 0 | 0 |
| 0.02227973 | 0 | 0 | 0.000818792 | 0.003100985 |
| 0 | 0 | 0 | 0 | 0 |
| 0.000442527 | 0.003014324 | 0.00104554 | 0.015720999 | 0.179129203 |
| 0.00156647 | 0.026910012 | 0 | 0.005246972 | 0.006322808 |
| 0 | 0 | 0 | 0 | 0.0680029 |
| 0 | 0 | 0 | 0 | 0 |
| 0 | 0 | 0 | 0 | 0 |
| 0 | 0 | 0 | 0 | 0.00662059 |
| 0 | 0 | 0 | 0 | 0 |
| 0 | 0 | 0 | 0 | 0.051689766 |
| 0 | 0.666076893 | 0 | 0 | 0 |
| 0.063464776 | 0 | 0 | 0 | 0 |
| 0 | 0.002536815 | 0 | 0 | 0 |
| 0 | 0 | 0 | 0 | 0 |
| 0 | 0 | 0.004942682 | 0.011891103 | 0 |
| 0 | 0 | 0 | 0 | 0 |
| 0.003909726 | 0 | 0.022631507 | 0 | 0.002705312 |
| 0.005576636 | 0.001095748 | 1.334244482 | 0 | 0 |
| 0 | 0 | 0 | 0 | 0 |
| 0 | 0 | 0 | 0 | 0 |
| 0 | 0 | 0 | 0 | 0 |
| 0 | 0 | 0 | 0 | 0 |
| 0 | 0 | 0 | 0 | 0 |
| 0 | 0 | 0 | 0 | 0 |
| 0 | 0 | 0 | 0 | 0.094544574 |
| 0 | 0 | 0 | 0 | 0 |
| 0 | 0 | 0 | 0 | 0.060962904 |
| 0 | 0.066881593 | 0 | 0 | 0 |
| 0 | 0 | 0 | 0 | 0 |
| 0 | 0 | 0 | 0 | 0.001471683 |
| 0 | 0 | 0 | 0 | 0 |
| 0 | 0 | 0 | 0 | 0 |
| 0 | 0 | 0 | 0 | 0 |
| 0 | 0 | 0 | 0 | 0 |
| 0 | 0.027231762 | 0 | 0 | 0 |
| 0 | 0 | 0 | 0 | 0 |
| 0 | 0 | 0.001464322 | 0 | 0 |
| 0 | 0 | 0 | 0.001151514 | 0.015699921 |
| 0 | 0 | 0 | 0 | 0 |

| | | | | |
|---|---|---|---|---|
| 0 | 0 | 0 | 0 | 0 |
| 0 | 0 | 0 | 0.005147588 | 0 |
| 0 | 0 | 0 | 0 | 0 |
| 0.002093097 | 0 | 0 | 0 | 0 |
| 0 | 0 | 0 | 0 | 0 |
| 0.000638243 | 0.067218512 | 0 | 0.001269739 | 0 |
| 0 | 0 | 0 | 0 | 0 |
| 0 | 0 | 0 | 0 | 0 |
| 0 | 0 | 0.044193973 | 0 | 0 |
| 0 | 0 | 0 | 0 | 0 |
| 0 | 0.006748963 | 0.00052469 | 0.00946725 | 0 |
| 0 | 0.093352934 | 0 | 0 | 0 |
| 0 | 0 | 0 | 0 | 0 |
| 0.001818322 | 0.002858243 | 0.00132187 | 0.008347903 | 0 |
| 0 | 0 | 0 | 0 | 0.002937806 |
| 0 | 0 | 0 | 0 | 0 |
| 0.108736584 | 0 | 0 | 0 | 0 |
| 0 | 0 | 0 | 0 | 0 |
| 0 | 0 | 0 | 0 | 0 |
| 0 | 0 | 0 | 0 | 0 |
| 0 | 0 | 0 | 0 | 0 |
| 0.031627855 | 0 | 0.05922023 | 0.344944095 | 0 |
| 0 | 0 | 0 | 0 | 0 |
| 0 | 0 | 0 | 0 | 0.004549065 |
| 0 | 0 | 0 | 0 | 0 |
| 0 | 0 | 0 | 0 | 0 |
| 0 | 0.036950715 | 0 | 0 | 0.005072016 |
| 0 | 0 | 0 | 0 | 0 |
| 0 | 0 | 0 | 0 | 0 |
| 0.07442281 | 1.240117362 | 0.155491105 | 0.423899618 | 0.143754937 |
| 0.001043805 | 0 | 0.011994478 | 0.002427167 | 0 |
| 0 | 0 | 0 | 0 | 0.001382758 |
| 0 | 0 | 0 | 0 | 0 |
| 0 | 0 | 0 | 0 | 0.004029841 |
| 0 | 0 | 0 | 0 | 0.005098548 |
| 0 | 0 | 0 | 0 | 0 |
| 0.126194377 | 0 | 0 | 0 | 0 |
| 0 | 0 | 0 | 0 | 0.000986475 |
| 0 | 0 | 0 | 0 | 0.071435048 |
| 0 | 0 | 0 | 0 | 0.013169675 |
| 0 | 0 | 0 | 0.016667006 | 0.001989151 |
| 2.061279196 | 0 | 1.274128323 | 1.878078119 | 0.665483362 |
| 0 | 0 | 0 | 0 | 0.00060245 |
| 0 | 0 | 0 | 0 | 0 |
| 0 | 0 | 0 | 0 | 0 |
| 0 | 0 | 0 | 0 | 0 |
| 0.000482757 | 0 | 0 | 0.003601545 | 0 |
| 0 | 0 | 0 | 0 | 0 |

| | | | | |
|---|---|---|---|---|
| 0.043779661 | 0 | 0.034478806 | 0.001481234 | 0.010132038 |
| 0 | 0 | 0 | 0 | 0 |
| 0.001420313 | 0.01722297 | 0 | 0.098608123 | 0.252480118 |
| 0 | 0 | 0 | 0 | 0 |
| 0 | 0 | 0.53073975 | 0.032616208 | 0.042121699 |
| 0 | 0 | 0 | 0 | 0.017568887 |
| 0 | 0 | 0 | 0 | 0 |
| 0 | 0 | 0 | 0.001422997 | 0.001154843 |
| 0 | 0 | 0 | 0.002833509 | 0 |
| 0 | 0 | 0 | 0 | 0 |
| 0 | 0 | 0 | 0 | 0 |
| 0 | 0 | 0 | 0 | 0 |
| 0 | 0 | 0 | 0.001821463 | 0.001478221 |
| 0 | 0.015035025 | 0.003023724 | 0.002105769 | 0 |
| 0 | 0 | 0 | 0 | 0 |
| 0.019160565 | 0 | 0.173741838 | 0.005236071 | 0.315546197 |
| 0.273753559 | 0.002751385 | 0.087809658 | 0.00099491 | 0.280176846 |
| 0 | 0.667673593 | 0 | 0 | 0 |
| 0 | 0 | 0 | 0.002055333 | 0 |
| 0 | 0 | 0 | 0 | 0.009152853 |
| 0.023676977 | 0 | 0 | 0 | 0.012719832 |
| 0 | 0.001063821 | 0.001439077 | 0.002308087 | 0.03075079 |
| 0 | 0 | 0 | 0.06030634 | 0 |
| 0 | 0 | 0 | 0 | 0.138299575 |
| 0.015159009 | 0 | 0 | 0 | 0 |
| 0 | 0.012081548 | 0 | 0 | 0 |
| 0 | 0 | 0 | 0 | 0.00170839 |
| 0 | 0 | 0 | 0 | 0 |
| 0 | 0 | 0 | 0 | 0 |
| 0 | 0 | 0 | 0 | 0 |
| 0.01498361 | 0 | 0 | 0 | 0 |
| 0.002519405 | 0.00475234 | 0 | 0.002625427 | 0.005810958 |
| 0.004637707 | 0.01483152 | 0.013602193 | 0.011817056 | 0.057356876 |
| 0 | 0 | 0.001082959 | 0 | 0.009514881 |
| 0 | 0 | 0 | 0 | 0.002641024 |
| 0 | 0 | 0 | 0 | 0 |
| 0 | 0 | 0 | 0 | 0.000500205 |
| 0.021369836 | 0 | 0.008682081 | 0.010443675 | 0.0894506 |
| 0.012347229 | 0 | 0.506248436 | 0.116016301 | 0 |
| 0.000277334 | 0 | 0 | 0 | 0 |
| 0 | 0 | 0 | 0 | 0.016385827 |
| 0 | 0 | 0 | 0 | 0 |
| 0 | 0.004784385 | 0 | 0 | 0 |
| 0.067168482 | 0 | 0 | 0.029223847 | 0.047739017 |
| 0 | 0 | 0 | 0.512975357 | 0 |
| 0 | 0 | 0 | 0 | 0 |
| 0 | 0 | 0 | 0 | 0 |
| 0 | 0 | 0 | 0 | 0 |

| | | | | |
|---|---|---|---|---|
| 0 | 0 | 0 | 0 | 0 |
| 0.367455474 | 0.746409672 | 0.337258452 | 0.843778864 | 0.47884489 |
| 0 | 0 | 0 | 0 | 0 |
| 0 | 0 | 0.004078517 | 0 | 0 |
| 0 | 0 | 0 | 0 | 0 |
| 0 | 0 | 0 | 0 | 0 |
| 0 | 0.002016462 | 0 | 0 | 0.187060131 |
| 0 | 0 | 0 | 0.001177161 | 0 |
| 0.0052184 | 0 | 0 | 0 | 0.000813252 |
| 0 | 0 | 0 | 0 | 0 |
| 0.00286226 | 0 | 0 | 0 | 0.004489187 |
| 0.007333078 | 0.010186611 | 0.005842341 | 0.019386902 | 0.010620167 |
| 0.011370829 | 0 | 0.024946437 | 0 | 0 |
| 0 | 0 | 0 | 0 | 0 |
| 0 | 0 | 0 | 0 | 0 |
| 0 | 0 | 0 | 0 | 0.001077913 |
| 0.003596639 | 0.025064297 | 0 | 0.035946669 | 0.019494606 |
| 0 | 0.20035893 | 0 | 0 | 0.001076881 |
| 0 | 0 | 0 | 0 | 0 |
| 0 | 0 | 0 | 0 | 0.007458879 |
| 0.03103232 | 0.024098527 | 0.008178989 | 0.144415211 | 0.016111602 |
| 0 | 0 | 0 | 0 | 0 |
| 0 | 0 | 0 | 0 | 0 |
| 0 | 0 | 0 | 0 | 0 |
| 0 | 0.000979597 | 0 | 0 | 0 |
| 0 | 0 | 0 | 0.004833314 | 0 |
| 0 | 0 | 0.002018268 | 0 | 0.028709762 |
| 0.015243036 | 0.120836358 | 0.006054094 | 0.027636038 | 0.015002665 |
| 0 | 0 | 0 | 0 | 0 |
| 0 | 0.002740729 | 0 | 0 | 0 |
| 0 | 0 | 0 | 0 | 0 |
| 0 | 0 | 0 | 0 | 0 |
| 0 | 0 | 0 | 0 | 0 |
| 0 | 0 | 0 | 0 | 0 |
| 0.046834887 | 0 | 0.406636383 | 0.011553275 | 4.473741297 |
| 0.033804182 | 1.022891221 | 0.216746392 | 0.182664972 | 0.178250763 |
| 0 | 0 | 0 | 0 | 0 |
| 0 | 0 | 0 | 0 | 0 |
| 0.005835721 | 0.026500474 | 0 | 0.021422766 | 0.080385962 |
| 0 | 0 | 0 | 0 | 0 |
| 0 | 0 | 0 | 0.508228233 | 0 |
| 0 | 0 | 0 | 0 | 0 |
| 0.00049232 | 0.00541719 | 0.017913049 | 0.002658471 | 0.087776237 |
| 0.001235847 | 0.000747171 | 0 | 0.006079035 | 0 |
| 0 | 0.110529178 | 0 | 0 | 0 |
| 0.258421925 | 0 | 0.252834785 | 0 | 0 |
| 0 | 0 | 0 | 0 | 0 |
| 0 | 0 | 0 | 0 | 0 |

| | | | | |
|---|---|---|---|---|
| 0 | 0 | 0 | 0 | 0 |
| 0 | 0 | 0 | 0 | 0 |
| 0 | 0 | 0 | 0 | 0.00052032 |
| 0.411469924 | 0 | 0.320125647 | 0.065347749 | 6.560408254 |
| 0 | 0 | 0 | 0 | 0 |
| 0 | 0 | 0.228022487 | 0 | 0 |
| 0 | 0 | 0 | 0 | 0 |
| 0 | 0 | 0.00387828 | 0.003207313 | 0.004259321 |
| 6.583917874 | 0 | 1.254211171 | 0.001407853 | 3.180440105 |
| 0.203145564 | 0 | 0.056181426 | 0.60720618 | 0 |
| 0 | 0 | 0 | 0 | 0 |
| 0.011866528 | 0 | 0.009475319 | 0.001249081 | 0 |
| 0 | 0 | 0 | 0 | 0 |
| 0 | 0 | 0 | 0 | 0 |
| 1.03000371 | 0.030642712 | 0.065337329 | 0.027185887 | 0 |
| 0 | 0 | 0 | 0 | 0 |
| 0 | 0 | 0.28194709 | 1.389557211 | 0 |
| 0 | 0 | 0.657389112 | 0 | 0 |
| 0 | 0 | 0 | 0 | 0 |
| 0 | 0 | 0 | 0.011572928 | 0 |
| 0 | 0 | 0 | 0 | 0 |
| 0 | 0 | 0 | 0 | 0 |
| 0 | 0 | 0 | 0.003306802 | 0 |
| 0.173452898 | 0.119577304 | 0 | 0 | 0.106462597 |
| 0 | 0 | 0 | 0.003508376 | 0.015066687 |
| 0.207692498 | 0 | 0.036696537 | 0.31954177 | 0 |
| 0 | 0 | 0 | 0 | 0 |
| 1.625355384 | 0 | 1.362413929 | 3.052377832 | 1.405037359 |
| 0 | 0 | 0 | 0 | 0 |
| 0 | 0 | 0 | 0 | 0 |
| 0.006302232 | 0 | 1.197889915 | 0 | 0.027470525 |
| 0.000503209 | 0 | 0 | 0 | 0 |
| 0.143324756 | 0 | 0.013445223 | 0 | 0.012187988 |
| 0 | 0 | 0 | 0 | 0 |
| 0 | 0 | 0 | 0 | 0 |
| 0 | 0 | 0 | 0.001673111 | 0 |
| 0.984893442 | 0 | 0.945572382 | 0.999634352 | 1.185999712 |
| 0 | 0 | 0 | 0 | 0 |
| 0.003317024 | 0 | 0 | 0.009102051 | 0 |
| 0 | 0 | 0 | 0 | 0 |
| 0.01971086 | 0.113827968 | 0.005478829 | 0.029507389 | 0.160091947 |
| 0.050259026 | 0 | 0 | 0.101860095 | 0 |
| 0.206740905 | 0.237125023 | 0.00717003 | 0.002695259 | 0 |
| 0 | 0 | 0 | 0 | 0 |
| 0 | 0 | 0 | 0 | 0 |
| 0 | 0 | 0 | 4.068693968 | 0 |
| 0 | 0 | 0 | 0 | 0 |
| 0 | 0 | 0.004191206 | 0.000900286 | 0 |

| | | | | |
|---|---|---|---|---|
| 0 | 0 | 0 | 0 | 0 |
| 0.611234305 | 0.170386678 | 0.793253332 | 0 | 0.122076155 |
| 0.138713665 | 0.733374382 | 0.001147927 | 0 | 0.010085685 |
| 0 | 0 | 0 | 0 | 0 |
| 0 | 0 | 0 | 0 | 0 |
| 0.013970139 | 0 | 0.050527344 | 0.000543887 | 0 |
| 0 | 0 | 0 | 0 | 0 |
| 0.001551497 | 0 | 0 | 0 | 0 |
| 0 | 0 | 0 | 0 | 0 |
| 0 | 0 | 0 | 0 | 0 |
| 0.126651609 | 0.070485913 | 0.025922394 | 0.010168059 | 0.06945401 |
| 0.528848958 | 3.568668683 | 0.563547488 | 0.553781027 | 0.806521 |
| 0 | 0 | 0 | 0 | 0 |
| 0.834540739 | 0.003034758 | 0.975832626 | 0.015911986 | 0.253816819 |
| 0 | 0.896655232 | 0.002516183 | 0.030411297 | 0.018481139 |
| 0 | 0 | 4.093994018 | 0 | 0 |
| 0 | 0 | 0 | 0 | 0 |
| 0 | 0 | 0 | 0 | 0 |
| 0 | 0 | 0.035297181 | 0 | 0 |
| 0.002455923 | 0 | 0 | 0.00083758 | 0 |
| 0.010595279 | 0.926543768 | 0 | 0.639742939 | 0.481263311 |
| 0.003896916 | 0 | 0 | 0 | 0.014089547 |
| 0.568539157 | 5.510863447 | 0.089669916 | 0.066006291 | 0.526968605 |
| 0 | 0 | 0 | 0 | 0 |
| 0.001175594 | 0 | 0.000347191 | 0 | 0 |
| 0.136904681 | 0 | 0 | 3.512012794 | 0 |
| 0 | 4.329587583 | 0 | 0 | 0 |
| 0.005838097 | 0 | 0.001108401 | 0 | 0.000721364 |
| 0 | 0 | 0.024153567 | 0 | 0 |
| 0 | 0 | 0 | 0 | 0 |
| 0.026680085 | 0 | 0.001172156 | 0 | 0 |
| 0.031989944 | 0 | 0.023298671 | 0.25303831 | 0 |
| 0.023638775 | 0.960719236 | 0 | 0 | 0.343801667 |
| 0.054826396 | 0 | 0.036411337 | 0.011016161 | 0 |
| 0 | 0 | 0 | 0 | 0 |
| 0.030432507 | 0.014665428 | 0.03450754 | 0 | 0.058920273 |
| 0.045490846 | 0.00151785 | 0.002661635 | 0 | 0 |
| 0 | 0 | 0 | 0 | 0 |
| 0.743566698 | 0 | 0 | 0 | 0 |
| 0 | 0 | 0 | 0 | 0 |
| 0.044988259 | 0 | 0.004505853 | 0.004447254 | 0 |
| 0 | 0 | 0 | 0 | 0 |
| 0 | 0 | 0 | 0.021791726 | 0 |
| 0 | 0 | 0.004566124 | 0 | 0 |
| 0.10221456 | 0.031457529 | 0.116110645 | 0.001979851 | 0.017674387 |
| 0 | 0 | 0.064070081 | 0 | 0 |
| 0 | 0 | 0 | 0 | 0 |
| 0.03865498 | 0.284837989 | 0.482813947 | 0.001954016 | 2.123556442 |

| | | | | |
|---|---|---|---|---|
| 0 | 0 | 0 | 0 | 0 |
| 0 | 0 | 0 | 0 | 0 |
| 0 | 0 | 0 | 0 | 0 |
| 0.000539759 | 0 | 0.010967286 | 0.001840821 | 0 |
| 0 | 0 | 0 | 0 | 0 |
| 0.344090339 | 0 | 0 | 0 | 0 |
| 0.080366888 | 0 | 0.041047494 | 2.232166007 | 0.078234783 |
| 0.000596011 | 0 | 0.044826762 | 0.001835046 | 0 |
| 0 | 0 | 0 | 0 | 0 |
| 0 | 0 | 0 | 0 | 0 |
| 0.00336643 | 0.028348569 | 0.001420308 | 0 | 0.002911727 |
| 0 | 0 | 0 | 0 | 0 |
| 0 | 0 | 0.001741028 | 0 | 0 |
| 0.076023201 | 0 | 0.22021858 | 1.51155352 | 0 |
| 0 | 0 | 0 | 0 | 0 |
| 0 | 0 | 0 | 0 | 0 |
| 0.036840714 | 0 | 0.008978852 | 0.105338212 | 0.012374618 |
| 0.000900796 | 0 | 0.009458995 | 0 | 0 |
| 0 | 0 | 0 | 0 | 0.009716702 |
| 0 | 0 | 0 | 0.000897862 | 0 |
| 0 | 0 | 0.011765001 | 0 | 0 |
| 0 | 0 | 0 | 0 | 0 |
| 0.011654876 | 0 | 0.064887991 | 0.072970429 | 0 |
| 0 | 0 | 0 | 0 | 1.895662563 |
| 0 | 0 | 0 | 0 | 0 |
| 0 | 0 | 0 | 0 | 0 |
| 0 | 0 | 0 | 0 | 0 |
| 0 | 0 | 0 | 0 | 0 |
| 0 | 0 | 0 | 0 | 0 |
| 0.366707736 | 0 | 0.068986781 | 0.264415479 | 0.705394536 |
| 0 | 0 | 0.001342381 | 0 | 0 |
| 0 | 0 | 0 | 0 | 0 |
| 0 | 0 | 0.051391805 | 0.002823473 | 0 |
| 0 | 0 | 0 | 0 | 0 |
| 0.000478004 | 0 | 0.000451744 | 0 | 0 |
| 0 | 0 | 0 | 0 | 0 |
| 0 | 0 | 0.064891335 | 0.068269974 | 0 |
| 0 | 0.020957869 | 0 | 0 | 0.105870223 |
| 0 | 0 | 0 | 0 | 0 |
| 0 | 0 | 0 | 0.000713258 | 0 |
| 0 | 0 | 0.008234911 | 0.001085564 | 0 |
| 0 | 0 | 0 | 0 | 0 |
| 0 | 0 | 0 | 0.011738928 | 0 |
| 0.013639512 | 0 | 0.034210425 | 0 | 0 |
| 0.036403344 | 0 | 0 | 0 | 0 |
| 0 | 0 | 0 | 0 | 0 |
| 0.001740685 | 0 | 0.009481933 | 0 | 0 |
| 0 | 0 | 0 | 0 | 0 |

| | | | | |
|---|---|---|---|---|
| 0 | 0 | 0 | 0 | 0 |
| 0 | 0 | 0 | 0 | 0 |
| 0.109529248 | 0 | 0 | 0 | 0 |
| 0.059932774 | 0 | 0.280809534 | 0.030210823 | 0 |
| 0 | 0 | 0 | 0 | 0 |
| 0 | 0 | 0.511637196 | 0.407603937 | 0 |
| 0 | 0 | 0 | 0 | 0 |
| 0 | 0 | 0 | 0 | 0 |
| 0.004351371 | 0.586031229 | 0.002155653 | 0.003191427 | 0.011978871 |
| 0 | 0 | 0 | 0 | 0 |
| 0.002007333 | 0 | 0.002964152 | 0 | 0 |
| 0 | 0 | 0.000707641 | 0 | 0 |
| 0 | 0 | 0 | 0 | 0 |
| 0 | 0 | 0 | 0 | 0.000562172 |
| 0.988336878 | 0 | 0.341638136 | 0.10277496 | 0 |
| 0 | 0.001360454 | 0 | 0 | 0.00573909 |
| 0 | 0 | 0 | 0 | 0 |
| 0 | 0 | 0 | 0 | 0 |
| 0 | 0 | 0 | 0 | 0 |
| 0 | 0 | 0 | 0 | 0 |
| 0.151121413 | 0 | 0.013186446 | 0 | 0 |
| 0 | 0 | 0 | 0 | 0 |
| 0 | 0 | 0 | 0 | 0 |
| 0 | 0 | 0 | 0 | 0 |
| 0.553140727 | 0.041515424 | 0.157169161 | 0.033308122 | 0.03464594 |
| 3.186181541 | 0 | 10.13112848 | 12.06270445 | 2.495793513 |
| 1.316560412 | 0 | 0.542509911 | 0.015529433 | 0 |
| 0 | 0 | 0 | 0 | 0 |
| 0.001443161 | 0 | 0.667448195 | 0.018661936 | 0.61596147 |
| 0 | 0 | 0 | 0 | 0 |
| 0 | 0 | 0 | 0 | 0 |
| 0 | 0 | 0 | 0.026737738 | 0.125536643 |
| 0 | 0 | 0 | 0 | 0 |
| 0 | 0 | 0.000833136 | 0 | 0 |
| 0.045464558 | 0.00068498 | 0.346446388 | 0.003921776 | 0.193477456 |
| 0.000616063 | 0.003227988 | 0.000849068 | 0 | 0.001184113 |
| 0 | 0 | 0 | 0 | 0 |
| 0.615642393 | 0 | 1.687802129 | 0.221263921 | 1.004272358 |
| 3.65609857 | 0 | 0 | 0 | 0 |
| 0 | 0 | 0 | 0 | 0 |
| 0 | 0 | 0 | 0 | 0 |
| 0 | 0 | 0 | 0.009100243 | 0 |
| 1.665620947 | 0.002646662 | 0.447697365 | 0.233876979 | 0.253558174 |
| 0 | 0 | 0 | 0.074609705 | 0 |
| 0 | 0 | 0 | 0 | 0 |
| 0 | 0 | 0 | 0.000967596 | 0 |
| 0 | 0 | 0 | 0 | 0 |
| 0.847523262 | 0.069425154 | 0.001647621 | 0.137413338 | 0.055580678 |

| | | | | |
|---|---|---|---|---|
| 0.055353361 | 0 | 0 | 0.008046466 | 0 |
| 0 | 0 | 0 | 0 | 0 |
| 0 | 0 | 0.028682906 | 0 | 0 |
| 0 | 0 | 0 | 0 | 0 |
| 0.002070965 | 0.000542563 | 0.000407749 | 0 | 0 |
| 0 | 0 | 4.084930817 | 0 | 0 |
| 0 | 3.130039416 | 0 | 0 | 0 |
| 0 | 0 | 0 | 0 | 0 |
| 0 | 0 | 0 | 0 | 0 |
| 0.001616726 | 0 | 0 | 0 | 0 |
| 0.000968398 | 0 | 0.000311999 | 0 | 0 |
| 0.056044194 | 0 | 0.654292381 | 0.19246332 | 0 |
| 0.005433081 | 0 | 0.018267353 | 0.479662268 | 0 |
| 0 | 0 | 0 | 0 | 0 |
| 0.003060369 | 0.002091579 | 0.238924444 | 0.032143667 | 3.285508626 |
| 0 | 0 | 0 | 0 | 0 |
| 0.001573556 | 0 | 0.001640195 | 0 | 0.002668658 |
| 0 | 0 | 0 | 0 | 0 |
| 0.000466505 | 0 | 0.001515514 | 0 | 0.001479478 |
| 0.005560752 | 0 | 0 | 0 | 0.01118526 |
| 0.032539299 | 0 | 0.022470649 | 0.04171118 | 0 |
| 0 | 0 | 0 | 0 | 0 |
| 1.634479104 | 0 | 0.165314608 | 0.457177572 | 8.400719134 |
| 0 | 0 | 0 | 0 | 0 |
| 0 | 0 | 0 | 0 | 0 |
| 0 | 0 | 0 | 0 | 0 |
| 0 | 0 | 0 | 0 | 0 |
| 0 | 0 | 0 | 0 | 0 |
| 0 | 0 | 0 | 0 | 0 |
| 0 | 0 | 0 | 0 | 0 |
| 0 | 0 | 0 | 0 | 0 |
| 0.030734071 | 0.072466891 | 0.016521755 | 0.074098193 | 0.030067463 |
| 0 | 0 | 0 | 0 | 0 |
| 0.096179473 | 0 | 0.021501119 | 0.019857434 | 0 |
| 0 | 0.003512859 | 0.000659999 | 0 | 0.001159751 |
| 0.422482101 | 0 | 0.409754583 | 0.001842211 | 0.02857225 |
| 0.000694307 | 0 | 0 | 0 | 0 |
| 0 | 0 | 0 | 0 | 0 |
| 0.000641855 | 0 | 0.001061539 | 0 | 0 |
| 0 | 0 | 0 | 0 | 0 |
| 0.18945105 | 0 | 0.267665245 | 0.109444277 | 0 |
| 0 | 0 | 0 | 0 | 0 |
| 0 | 0 | 0 | 0 | 0 |
| 0 | 0 | 0 | 0.135339098 | 0 |
| 0.038085442 | 0.009170374 | 0.016514206 | 0.014859599 | 0.003681295 |
| 0 | 0 | 0 | 0 | 0 |
| 0.271820284 | 0 | 0.106562352 | 0 | 0 |
| 0.001145209 | 0 | 0 | 0 | 0 |

| | | | | |
|---|---|---|---|---|
| 0 | 0 | 0 | 0 | 0 |
| 0 | 0 | 0 | 0 | 0 |
| 0 | 0 | 0 | 0.001294193 | 0.009190229 |
| 0 | 0 | 0 | 0 | 0 |
| 0 | 0.004900049 | 0 | 0 | 0 |
| 0 | 0 | 0 | 0 | 0 |
| 0 | 0 | 0.443824705 | 0 | 0.001300682 |
| 0.031609297 | 0 | 0.014606173 | 0.011000201 | 0.058523361 |
| 0 | 0 | 0 | 0 | 0 |
| 0 | 0 | 0 | 0 | 0 |
| 0 | 0 | 0 | 0 | 0 |
| 0 | 0 | 0 | 0 | 0 |
| 0 | 0 | 0 | 0 | 0 |
| 0.052078337 | 0 | 0 | 0 | 0 |
| 0 | 0 | 0 | 0 | 0 |
| 0 | 0 | 0 | 0 | 0 |
| 0 | 0 | 0 | 0.65066529 | 0 |
| 0 | 0 | 0 | 0 | 0 |
| 1.183611643 | 0 | 0 | 0 | 0.62455587 |
| 0 | 0 | 0 | 0 | 0 |
| 0 | 0 | 0.003665799 | 0 | 0 |
| 0 | 0 | 0 | 0 | 0 |
| 0 | 0 | 0 | 0 | 0 |
| 0 | 0 | 0 | 0 | 0 |
| 0.006975364 | 0 | 0.00284145 | 0.001367192 | 0.00097086 |
| 0 | 0.030283153 | 0 | 0 | 0.001654485 |
| 0 | 0 | 0 | 0 | 0 |
| 0.109228829 | 0.452816371 | 0.002488283 | 0.156641807 | 0.936135443 |
| 0 | 0 | 0 | 0 | 0 |
| 0.124249905 | 0 | 0.657146928 | 1.693454162 | 0.880086832 |
| 0 | 0 | 0 | 0 | 0 |
| 0 | 0 | 0 | 0 | 0 |
| 0.014615607 | 0 | 0 | 0 | 0 |
| 0 | 0 | 0 | 0 | 0 |
| 0.071949106 | 0.004895273 | 0 | 0 | 0 |
| 0 | 0 | 0 | 0 | 0 |
| 0 | 0 | 0 | 0 | 0 |
| 0.00194964 | 0.003639287 | 0.002303167 | 0 | 0.018127747 |
| 0 | 0 | 0 | 0 | 0 |
| 0 | 0 | 0 | 0 | 0 |
| 0.402678632 | 35.45827969 | 1.552450647 | 2.782923792 | 1.208116056 |
| 0.174825271 | 1.215192899 | 0.020863368 | 0.160465783 | 1.208973068 |
| 4.936951382 | 0 | 3.176111189 | 0.070367872 | 0 |
| 0 | 0 | 0.004276266 | 0 | 0 |
| 0 | 0 | 0 | 0 | 0 |
| 0 | 0 | 0 | 0 | 0 |
| 0 | 0 | 0 | 0 | 0 |
| 0.007930127 | 0.003648428 | 0.004912537 | 0 | 0 |

| | | | | |
|---|---|---|---|---|
| 0 | 0 | 0 | 0 | 0 |
| 0.013130443 | 0 | 0.000822573 | 0.009470673 | 0.002867908 |
| 0 | 0 | 0.001921438 | 0.002971669 | 0 |
| 0 | 0 | 0 | 0 | 0 |
| 1.100024033 | 0 | 0.929063311 | 1.187675623 | 2.028368996 |
| 0 | 0 | 0 | 0 | 0 |
| 0 | 0 | 0 | 0 | 0 |
| 0 | 0 | 0 | 0.043013033 | 0 |
| 0 | 1.642824901 | 0 | 0.001270304 | 0.023367627 |
| 0 | 0 | 0 | 0 | 0 |
| 0 | 0 | 0 | 0 | 0 |
| 0 | 0 | 0 | 0 | 0 |
| 0.023003897 | 0 | 0.069308137 | 0.085293639 | 0.144683462 |
| 0.002733033 | 0.104226833 | 0.004491983 | 0.007091971 | 0.011237007 |
| 5.512376522 | 0 | 0 | 0.189855968 | 0 |
| 0.160987373 | 0 | 0.176471505 | 0.332549712 | 0.126957521 |
| 0.003598938 | 0.004040867 | 0 | 0.015926978 | 0.006877871 |
| 0 | 0 | 0 | 0 | 0 |
| 0 | 0 | 0 | 0 | 0 |
| 0 | 0 | 0 | 0 | 0 |
| 0 | 0 | 0 | 0 | 0 |
| 0 | 0 | 0 | 0 | 0 |
| 0 | 0 | 0 | 0 | 0 |
| 0 | 0 | 0 | 0 | 0 |
| 0 | 0 | 0.002174752 | 0 | 0 |
| 0 | 0 | 0 | 0 | 0 |
| 0.067148504 | 0 | 0.110695196 | 0.065561171 | 0.275725898 |
| 0 | 0 | 0 | 0 | 0 |
| 0 | 0 | 0 | 0 | 0 |
| 0 | 0 | 0 | 0 | 0 |
| 0 | 0 | 0 | 0 | 0 |
| 0 | 0 | 0 | 0 | 0 |
| 0 | 0 | 0 | 0 | 0 |
| 0.012268311 | 0 | 0.02161444 | 0 | 0 |
| 0.532747412 | 0 | 0.899916234 | 0.128870143 | 0.535044978 |
| 0.067808868 | 0 | 0.031710952 | 0 | 0 |
| 0 | 0.002062687 | 0.001127388 | 0 | 0 |
| 0 | 0 | 0 | 0.001722686 | 0.119470412 |
| 0 | 0 | 0 | 0 | 0 |
| 0 | 0 | 0 | 0 | 0 |
| 0.032161607 | 0 | 0.429410511 | 0.015437231 | 1.546078395 |
| 0 | 0 | 0.021330884 | 0 | 0 |
| 0 | 0 | 0.005568943 | 0 | 0 |
| 0.019016312 | 0 | 0 | 0 | 0 |
| 0 | 0 | 0 | 0 | 0 |
| 0 | 0 | 0 | 0 | 0 |
| 0 | 0 | 0 | 0 | 0 |
| 0 | 0 | 0 | 0 | 0 |

| | | | | |
|---|---|---|---|---|
| 0.146699952 | 0 | 0.536476198 | 0.027627727 | 0 |
| 0 | 0 | 0 | 0 | 0 |
| 0 | 0 | 0 | 0 | 0 |
| 0 | 0 | 0 | 0 | 0.004085932 |
| 0.026900968 | 0 | 0.056711143 | 0.062623659 | 0 |
| 0 | 0 | 0 | 0 | 0 |
| 0.008308718 | 0.042729041 | 0 | 0.382338356 | 0.166678555 |
| 0 | 0 | 0 | 0 | 0 |
| 0.00246169 | 0 | 0 | 0 | 0 |
| 0 | 0 | 0 | 0 | 0 |
| 0.008831769 | 0 | 0.006024823 | 0.001414099 | 0 |
| 0 | 0 | 0 | 0 | 0 |
| 0 | 0 | 0 | 0 | 0 |
| 0 | 0 | 0 | 0 | 0 |
| 0 | 0 | 0 | 0 | 0 |
| 0 | 0 | 0 | 0 | 0 |
| 0 | 0 | 0 | 0 | 0 |
| 0 | 0 | 0 | 0 | 0 |
| 0 | 0 | 0 | 0 | 0 |
| 0 | 0 | 0 | 0 | 0 |
| 2.652998797 | 0.099475161 | 0.003922486 | 0.232448508 | 0.030858987 |
| 0 | 0 | 0 | 0 | 0 |
| 0.017665639 | 0 | 0.01437921 | 0.019898917 | 0.028695726 |
| 0.004118684 | 0 | 0.006110192 | 0 | 0.08030529 |
| 0.005725536 | 0 | 0 | 0 | 0 |
| 0 | 0 | 0.000533974 | 0 | 0 |
| 0 | 0 | 0 | 0 | 0 |
| 0 | 0 | 0 | 0 | 0 |
| 0 | 0 | 0 | 0 | 0 |
| 0.598289347 | 0.011918251 | 0.007816888 | 0.0035261 | 0.010492651 |
| 0 | 0 | 0 | 0 | 0 |
| 0 | 0 | 0 | 0 | 0 |
| 0 | 0 | 0 | 0 | 0 |
| 0 | 0 | 0 | 0 | 0 |
| 0 | 0 | 0 | 0 | 0 |
| 0 | 0 | 0 | 0 | 0 |
| 0.02109356 | 0.03692509 | 0 | 0 | 0.337937059 |
| 0 | 0 | 0 | 0 | 0 |
| 0 | 0 | 0 | 0 | 0 |
| 0 | 0 | 0 | 0 | 0 |
| 0 | 0 | 0 | 0 | 0 |
| 0 | 0 | 0 | 0.003246735 | 0 |
| 0 | 0 | 0 | 0 | 0 |
| 0 | 0 | 0.010267869 | 0 | 0 |
| 0.002041718 | 0 | 0 | 0 | 0 |
| 0 | 0 | 0 | 0 | 0 |
| 0 | 0 | 0 | 0 | 0 |
| 0 | 0 | 0 | 0 | 0 |

| | | | | |
|---|---|---|---|---|
| 0.001442928 | 0.022519549 | 0 | 0 | 0.004278966 |
| 0 | 0 | 0.748259547 | 0 | 0 |
| 0 | 0 | 0.205031399 | 0.128798781 | 0 |
| 0 | 0 | 0 | 0 | 0 |
| 0 | 0 | 0 | 0 | 0 |
| 0.474942943 | 0 | 0.57274019 | 0 | 0 |
| 0 | 0 | 0 | 0 | 0 |
| 0.089951908 | 0 | 0.229027698 | 1.448767482 | 0.649022176 |
| 0.49358469 | 2.156725245 | 6.620732652 | 0.751280496 | 0.438063749 |
| 0 | 0.080374617 | 0.00146352 | 0.00128034 | 0.019482549 |
| 0 | 0 | 0 | 0 | 0 |
| 0 | 0 | 0.001397529 | 0 | 0 |
| 0 | 0 | 0 | 0 | 0 |
| 0.819087128 | 0 | 0 | 0 | 1.305926639 |
| 0 | 0 | 0 | 0 | 0 |
| 0 | 0 | 0 | 0 | 0 |
| 0 | 0 | 0 | 0 | 0 |
| 2.139828565 | 0 | 0.09211885 | 0 | 0 |
| 0 | 0 | 0 | 0 | 0 |
| 0 | 0 | 0.032656487 | 0 | 0 |
| 0.043125344 | 0 | 0 | 0 | 0 |
| 0 | 0 | 0 | 0 | 0 |
| 0 | 0 | 0 | 0 | 0 |
| 0 | 0 | 0 | 0 | 0 |
| 0.009010867 | 0 | 0.005849847 | 0.013150705 | 0 |
| 0 | 0 | 0 | 0 | 0 |
| 0 | 0 | 0 | 0 | 0 |
| 0 | 0 | 0 | 0 | 0 |
| 0 | 0 | 0 | 0 | 0 |
| 0 | 0 | 0 | 0 | 0 |
| 0 | 0 | 0.019387798 | 0 | 0 |
| 0 | 0 | 0 | 0 | 0 |
| 0 | 0 | 0 | 0 | 0 |
| 0 | 0 | 0 | 0 | 0 |
| 0.002121196 | 0 | 0.021225868 | 0 | 0.174690917 |
| 0 | 0 | 0 | 0 | 0 |
| 0 | 0 | 0 | 0 | 0 |
| 0 | 0 | 0 | 0.015437676 | 0.006976129 |
| 0 | 0 | 0.001158212 | 0 | 0 |
| 0.000617355 | 0 | 0 | 0.003362884 | 0 |
| 0 | 0 | 0 | 0 | 0 |
| 0.138846548 | 0 | 0.080883166 | 0.227623542 | 0 |
| 0 | 0 | 0 | 0 | 0 |
| 0 | 0 | 0 | 0 | 0 |
| 0 | 0 | 0 | 0 | 0 |
| 0 | 0 | 0 | 0 | 0 |
| 0 | 0 | 0 | 0 | 0 |
| 0 | 0 | 0 | 0 | 0 |

| | | | | |
|---|---|---|---|---|
| 0 | 0 | 0 | 0 | 0 |
| 0 | 0.000438666 | 0 | 0 | 0.003861954 |
| 0 | 0 | 0 | 0 | 0 |
| 0 | 0 | 0 | 0 | 0 |
| 5.752967357 | 0 | 2.21442598 | 0.038811067 | 0.964420469 |
| 0 | 0 | 0.000421234 | 0.265512233 | 0 |
| 0 | 0 | 0 | 0 | 0 |
| 0.003015479 | 0.053995555 | 0.003252511 | 0.002235683 | 0.027366973 |
| 0 | 0 | 0.024588305 | 0.002418267 | 0 |
| 0 | 0 | 0 | 0 | 0 |
| 0 | 0 | 0 | 0 | 0 |
| 0 | 0 | 0 | 0 | 0 |
| 0 | 0 | 0 | 0 | 0 |
| 0 | 0 | 0 | 0 | 0 |
| 0.000369181 | 0 | 0 | 0 | 0 |
| 0.026494148 | 0 | 0 | 0.004847478 | 0 |
| 0 | 0 | 0 | 0 | 0 |
| 0 | 0 | 0 | 0 | 0 |
| 0 | 0 | 0 | 0 | 0 |
| 0 | 0 | 0 | 0 | 0 |
| 0 | 0 | 0 | 0 | 0 |
| 0 | 0 | 0 | 0.018611084 | 0 |
| 0 | 0 | 0 | 0 | 0 |
| 0.002155797 | 0.002853661 | 0 | 0 | 0.001177651 |
| 0.1024645 | 0 | 0 | 0.132836971 | 0 |
| 0 | 0 | 0 | 0 | 0 |
| 0.026877666 | 0.08515116 | 0.078949382 | 0 | 0.666243144 |
| 0 | 0 | 0 | 0 | 0 |
| 0 | 0 | 0 | 0 | 0 |
| 0 | 0 | 0 | 0 | 0 |
| 0 | 0 | 0 | 0 | 0 |
| 0 | 0 | 0 | 0 | 0 |
| 0 | 0 | 0 | 0 | 0 |
| 0 | 0 | 0 | 0 | 0 |
| 0 | 0 | 0 | 0 | 0 |
| 0 | 0 | 0 | 0 | 0 |
| 0 | 0 | 0 | 0 | 0 |
| 0.083970989 | 0 | 0.024767922 | 0.040833501 | 0 |
| 0 | 0 | 0 | 8.578784233 | 0 |
| 0 | 0 | 0 | 0 | 0 |
| 0 | 0 | 0 | 0 | 0 |
| 0 | 0 | 0 | 0 | 0.352820451 |
| 0 | 0 | 0 | 0 | 0 |
| 0 | 0 | 0 | 0 | 0 |
| 0 | 0 | 0 | 0 | 0 |
| 0 | 0 | 0 | 0 | 0 |
| 0 | 0 | 0 | 0 | 0 |
| 0.036131303 | 0 | 0 | 0 | 0 |

| | | | | |
|---|---|---|---|---|
| 0.044794374 | 0 | 0.081277621 | 0.378871618 | 0 |
| 0.014973643 | 0 | 0 | 0 | 0 |
| 0 | 0 | 0 | 0 | 0.002787079 |
| 0 | 0 | 0 | 0 | 0 |
| 0 | 0 | 0 | 0 | 0 |
| 1.084520295 | 0 | 0.097103304 | 0 | 0 |
| 0 | 0 | 0 | 0 | 0 |
| 0 | 0 | 0 | 0 | 0 |
| 0 | 0 | 0 | 0 | 0 |
| 0 | 0 | 0 | 0 | 0 |
| 0.32394942 | 0 | 0.472397411 | 0 | 0 |
| 0.001359732 | 0.000822069 | 0.001729853 | 0 | 0.001568097 |
| 0 | 0 | 0 | 0 | 0 |
| 0 | 0 | 0 | 0 | 0 |
| 0 | 0 | 0.045011818 | 0 | 0 |
| 0 | 0 | 0 | 0 | 0 |
| 0 | 0 | 0 | 0 | 0 |
| 0 | 0 | 0 | 0 | 0 |
| 0 | 0 | 0 | 0 | 0 |
| 0 | 0 | 0 | 0 | 0 |
| 0 | 0.855183812 | 0 | 0 | 0.012509095 |
| 0 | 0 | 0 | 0 | 0 |
| 0.05522353 | 0.059216865 | 0.015002868 | 0.001013874 | 0 |
| 0.467559589 | 0 | 0 | 0 | 1.387181607 |
| 0 | 0 | 0 | 0 | 0 |
| 0 | 0 | 0 | 0 | 0 |
| 0 | 0 | 0 | 0 | 0 |
| 0 | 0 | 0 | 0 | 0 |
| 0.001852948 | 0 | 0 | 0 | 0.00341903 |
| 0 | 0 | 0 | 0 | 0 |
| 0 | 0 | 0 | 0 | 0 |
| 0 | 0 | 0 | 0 | 0 |
| 0.064946438 | 0.078983287 | 0 | 0.003987851 | 0.006780965 |
| 0 | 0 | 0 | 0 | 0 |
| 0.009801916 | 0 | 0.041657213 | 0.490393254 | 0 |
| 0 | 0.13721264 | 0 | 0.054249035 | 0.002102375 |
| 0 | 0 | 0 | 0 | 0 |
| 0 | 0 | 0 | 0 | 0 |
| 0 | 0.05244777 | 0 | 0 | 0.086540936 |
| 0 | 0 | 0 | 0 | 0 |
| 0 | 0 | 0 | 0 | 0 |
| 0 | 0 | 0.034697648 | 0 | 0 |
| 0.796676266 | 0 | 0.180585957 | 0.001236585 | 0.293708518 |
| 0 | 0 | 0 | 0 | 0 |
| 0 | 0 | 0 | 0 | 0 |
| 0.008293162 | 0 | 0.018097157 | 0.003764576 | 0.006110338 |
| 0 | 0 | 0 | 0 | 0 |
| 0 | 0 | 0 | 0 | 0 |

| | | | | |
|---|---|---|---|---|
| 0 | 0 | 0 | 0 | 0 |
| 0 | 0 | 0 | 0 | 0 |
| 0 | 0 | 0 | 0 | 0 |
| 0 | 0 | 0 | 0 | 0 |
| 0 | 0 | 0 | 0 | 0 |
| 0 | 0 | 0 | 0 | 0 |
| 0 | 0 | 0 | 0 | 0 |
| 0 | 0 | 0 | 0 | 0 |
| 0.003453299 | 0 | 0.041304783 | 0.07728859 | 0 |
| 0.040455222 | 0 | 0.002943138 | 0 | 0 |
| 0 | 0 | 0 | 0 | 0 |
| 0 | 0 | 0 | 0 | 0 |
| 0 | 0 | 0 | 0 | 0 |
| 0 | 0 | 1.313660667 | 0.815766055 | 0 |
| 0 | 0 | 0 | 0 | 0 |
| 0 | 0.17962943 | 0 | 0 | 0.007807541 |
| 0 | 0 | 0 | 0 | 0 |
| 0 | 0 | 0 | 0 | 0 |
| 0 | 0 | 0 | 0 | 0 |
| 1.358593156 | 0 | 0.023666666 | 3.32596941 | 0 |
| 5.305719895 | 0 | 0 | 0 | 0.001405935 |
| 0 | 0 | 0 | 0 | 0 |
| 0 | 0 | 0 | 0 | 0 |
| 0 | 0 | 0 | 0 | 0 |
| 0 | 0 | 0 | 0 | 0.001273529 |
| 0 | 0 | 0 | 0 | 0 |
| 0 | 0 | 0 | 0 | 0 |
| 0.849476119 | 0 | 0 | 0 | 0 |
| 0 | 0 | 0 | 0 | 0 |
| 0.013791405 | 0 | 0.231091923 | 0 | 0.088732052 |
| 0.196099137 | 0 | 0.277047832 | 0.091943318 | 0.050984818 |
| 0 | 0 | 0 | 0 | 0 |
| 0.00192306 | 0 | 0 | 0 | 0 |
| 0 | 0 | 0 | 0 | 0 |
| 0 | 0 | 0 | 0 | 0 |
| 0 | 0 | 0 | 0 | 0 |
| 0 | 0 | 0 | 0 | 0 |
| 0 | 0 | 0 | 0 | 0 |
| 0.001924628 | 0 | 0 | 0 | 0 |
| 0 | 0 | 0 | 0 | 0 |
| 0 | 0 | 0.004567993 | 0 | 0 |
| 1.963766885 | 0 | 0 | 0 | 0 |
| 0 | 0 | 0.010303554 | 0 | 0 |
| 0 | 0 | 0 | 0 | 0 |
| 0 | 0 | 0 | 0 | 0 |
| 0 | 0 | 0 | 0 | 0 |
| 0 | 0.465629983 | 0 | 0 | 0.072070505 |
| 0.000388442 | 0 | 0 | 0 | 0 |

| | | | | |
|---|---|---|---|---|
| 0 | 0 | 0 | 0 | 0 |
| 0 | 0 | 0 | 0 | 0 |
| 0 | 0 | 0 | 0 | 0 |
| 0 | 0 | 0 | 0 | 0 |
| 0.031154843 | 0 | 0.020247549 | 0.023214099 | 0.258709504 |
| 0 | 0 | 0 | 0 | 0 |
| 0.000519902 | 0 | 0 | 0 | 0 |
| 0 | 0 | 0 | 0 | 0 |
| 0 | 0 | 0 | 0 | 0 |
| 0 | 0 | 0 | 0 | 0 |
| 0.008613876 | 0 | 0.016870449 | 0 | 0 |
| 0.021317141 | 0.007403079 | 0.006734873 | 0.002993988 | 0.024869641 |
| 0.002138526 | 0 | 0.001515782 | 0 | 0 |
| 0 | 0 | 0 | 0 | 0 |
| 0 | 0 | 0 | 0 | 0 |
| 0 | 0 | 0.029943801 | 0 | 0 |
| 0.022114741 | 0 | 0 | 0.001760913 | 0.026273113 |
| 0 | 0 | 0 | 0 | 0 |
| 0.000469229 | 0 | 0 | 0.013068961 | 0.009199264 |
| 0.008209771 | 0.176818301 | 0.014764798 | 0.266756865 | 0.285024304 |
| 0 | 0 | 0 | 0 | 0 |
| 0 | 0 | 0 | 0 | 0 |
| 0 | 0 | 0 | 0 | 0 |
| 0 | 0 | 0 | 0 | 0 |
| 0 | 0 | 0 | 0 | 0 |
| 0 | 0 | 0 | 0 | 0 |
| 0.000589152 | 0 | 0 | 0 | 0.019816823 |
| 0 | 0 | 0 | 0 | 0 |
| 0 | 0 | 0 | 0 | 0 |
| 0 | 0 | 0 | 0 | 0 |
| 0 | 0 | 0 | 0 | 0.001411986 |
| 0 | 0 | 0 | 0 | 0.004436681 |
| 0 | 0 | 0 | 0 | 0 |
| 0 | 0 | 0 | 0 | 0 |
| 0 | 0 | 0 | 0 | 0 |
| 0 | 0 | 0 | 0 | 0 |
| 0 | 0 | 0 | 0 | 0 |
| 0.035893909 | 0.033189455 | 0.014041799 | 0.000666745 | 0.051043948 |
| 0.194419665 | 0 | 0.103558729 | 0 | 0 |
| 0 | 0 | 0 | 0 | 0 |
| 0.052384329 | 0 | 0.010605906 | 0 | 0 |
| 0 | 0 | 0 | 0 | 0 |
| 0 | 0 | 0 | 0 | 0 |
| 0 | 0 | 0 | 0 | 0 |
| 0 | 0 | 0 | 0 | 0 |
| 0 | 0 | 0 | 0 | 0.003706245 |
| 0 | 0 | 0 | 0 | 0 |
| 0 | 0 | 0 | 0 | 0 |

| | | | | |
|---|---|---|---|---|
| 0 | 0 | 0 | 0 | 0 |
| 0 | 0 | 0.009132608 | 0.019858611 | 0 |
| 0 | 0 | 0 | 0 | 0 |
| 0.001336805 | 0.002101339 | 0 | 0 | 0.001541657 |
| 0 | 0.000531534 | 0.029027474 | 0 | 0.004029611 |
| 0 | 0 | 0.000859237 | 0.003100729 | 0 |
| 0.006368403 | 0 | 0.01086682 | 0 | 0 |
| 0.018058583 | 0 | 0 | 0 | 0 |
| 0 | 0 | 0 | 0 | 0 |
| 0 | 0 | 0 | 0.000997787 | 0 |
| 0 | 0 | 0.008185427 | 0 | 0 |
| 0 | 0 | 0 | 0 | 0 |
| 0.063920351 | 0.009614393 | 0.166185265 | 0.013588103 | 0 |
| 0 | 0 | 0 | 0 | 0 |
| 0 | 0 | 0 | 0 | 0 |
| 0 | 0 | 0 | 0 | 0 |
| 0 | 0 | 0 | 0 | 0 |
| 0 | 0 | 0 | 0 | 0 |
| 0 | 0 | 0 | 0 | 0 |
| 0 | 0 | 0 | 0 | 0 |
| 0 | 0 | 0 | 0 | 0 |
| 0 | 0 | 0 | 0 | 0 |
| 0 | 0 | 0 | 0.00805964 | 0.013081718 |
| 0.008432964 | 0 | 0.05344751 | 0 | 0 |
| 0 | 0 | 0 | 0 | 0 |
| 0.054567608 | 0 | 0.001167796 | 0 | 0 |
| 0.004009942 | 0 | 0.026758006 | 0.015554582 | 0 |
| 0 | 0 | 0 | 0 | 0 |
| 0 | 0 | 0 | 0 | 0 |
| 0 | 0 | 0 | 0 | 0 |
| 0.011889019 | 0 | 0.016136636 | 0.040439081 | 0.077889546 |
| 0 | 0 | 0 | 0 | 0 |
| 0 | 0 | 0 | 0 | 0 |
| 0 | 0 | 0 | 0 | 0 |
| 0 | 0 | 0.568451425 | 0.026971892 | 0 |
| 0 | 0 | 0 | 0 | 0 |
| 0 | 0 | 0 | 0 | 0 |
| 0 | 0 | 0 | 0 | 0 |
| 0 | 0.006223779 | 0 | 0.001216507 | 0.060223182 |
| 0 | 0 | 0 | 0 | 0 |
| 0 | 0 | 0 | 0 | 0 |
| 0 | 0 | 0 | 0 | 0 |
| 0 | 0 | 0 | 0 | 0 |
| 0 | 0 | 0 | 0 | 0 |
| 0 | 0 | 0 | 0 | 0 |
| 0.069671792 | 0.129053573 | 1.182750128 | 0.02368095 | 0 |
| 0 | 0 | 0 | 0 | 0 |
| 0 | 0 | 0 | 0 | 0 |

| | | | | |
|---|---|---|---|---|
| 0 | 0 | 0 | 0 | 0 |
| 0 | 0 | 0 | 0 | 0 |
| 0.045388986 | 0 | 0.023158997 | 0 | 0 |
| 0 | 0 | 0.288997034 | 0 | 0 |
| 0 | 0 | 0 | 0 | 0 |
| 0 | 0 | 0 | 0 | 0 |
| 0 | 0 | 0 | 0 | 0 |
| 0 | 0 | 0 | 0 | 0 |
| 0 | 0.007937599 | 0 | 0 | 0.002854634 |
| 0 | 0 | 0 | 0 | 0 |
| 0.002517598 | 0.000818781 | 0.00328178 | 0.000493457 | 0 |
| 0 | 0 | 0 | 0 | 0 |
| 0 | 0 | 0 | 0 | 0 |
| 0 | 0 | 0 | 0 | 0 |
| 0 | 0 | 0 | 0 | 0 |
| 0 | 0 | 0 | 0 | 0 |
| 0 | 0 | 0 | 0 | 0.015746953 |
| 0 | 0 | 0 | 0 | 0 |
| 0 | 0 | 0 | 0 | 0 |
| 0 | 0 | 0.000952045 | 0.001472419 | 0.005178125 |
| 0 | 0 | 0 | 0 | 0 |
| 0 | 0 | 0 | 0 | 0 |
| 0 | 0 | 0 | 0 | 0 |
| 0 | 0 | 0 | 0 | 0 |
| 0 | 0 | 0 | 0 | 0 |
| 0 | 0 | 0 | 0 | 0 |
| 0 | 0 | 0 | 0 | 0 |
| 0.004077143 | 0 | 0 | 0.00597476 | 0 |
| 0 | 0 | 0 | 0 | 0 |
| 0 | 0 | 0 | 0 | 0 |
| 0.00197817 | 0.005441643 | 0.00852958 | 0 | 0.003193828 |
| 0 | 0 | 0 | 0 | 0 |
| 0.0184897 | 0.117159357 | 0.012481389 | 0.007670125 | 0.015363202 |
| 0.019950681 | 0.01033059 | 0 | 0.023180575 | 0.002436133 |
| 0.024157992 | 0 | 0 | 0 | 0 |
| 0 | 0 | 0 | 0 | 0 |
| 0 | 0 | 0 | 0 | 0 |
| 0 | 0 | 0 | 0 | 0 |
| 0 | 0 | 0 | 0 | 0 |
| 0 | 0 | 0 | 0 | 0 |
| 0 | 0 | 0 | 0 | 0 |
| 0 | 0 | 0 | 0 | 0.016779583 |
| 0.022560285 | 0 | 0.041472426 | 0.017828533 | 0.009955648 |
| 0 | 0 | 0 | 0 | 0 |
| 0 | 0 | 0 | 0 | 0 |
| 0.249166169 | 0 | 0.161124728 | 0.015363537 | 0.723550245 |
| 0.000872419 | 0.001714207 | 0.003478323 | 0 | 0.005156305 |
| 0 | 0 | 0 | 0 | 0 |

| | | | | |
|---|---|---|---|---|
| 0 | 0 | 0 | 0 | 0 |
| 0.000602931 | 0 | 0.002706588 | 0.006854221 | 0 |
| 0.005775153 | 0.023248622 | 0.005657566 | 0.012209843 | 0.006660138 |
| 0 | 0 | 0 | 0 | 0 |
| 0 | 0 | 0.009566035 | 0.001438373 | 0 |
| 0 | 0 | 0 | 0 | 0 |
| 0 | 0 | 0 | 0 | 0 |
| 0 | 0 | 0 | 0 | 0 |
| 0 | 0 | 0 | 0 | 0 |
| 0 | 0 | 0 | 0 | 0 |
| 0 | 0 | 0 | 0 | 0 |
| 0 | 0 | 0 | 0 | 0 |
| 0 | 0 | 0 | 0 | 0 |
| 0 | 0 | 0 | 0 | 0 |
| 0 | 0 | 0 | 0 | 0 |
| 0 | 0 | 0 | 0 | 0 |
| 0 | 0 | 0 | 0 | 0 |
| 0 | 0 | 0 | 0 | 0 |
| 0 | 0 | 0 | 0 | 0 |
| 0 | 0 | 0 | 0 | 0 |
| 0 | 0 | 0 | 0 | 0 |
| 0 | 0 | 0 | 0 | 0 |
| 0 | 0 | 0.005832735 | 0 | 0 |
| 0 | 0 | 0 | 0 | 0 |
| 0.017296858 | 0 | 2.367518896 | 0 | 0.00348076 |
| 0 | 0 | 0.001241602 | 0 | 0 |
| 0.002774057 | 0 | 0 | 0 | 0 |
| 0.02922643 | 0 | 0 | 0.034448509 | 0.348155315 |
| 0 | 0 | 0 | 0 | 0 |
| 0 | 0 | 0 | 0 | 0 |
| 0 | 0 | 0.002020672 | 0 | 0 |
| 0 | 0 | 0 | 0 | 0.001062922 |
| 0 | 0 | 0 | 0 | 0 |
| 0 | 0 | 0 | 0 | 0 |
| 0 | 0 | 0 | 0 | 0 |
| 0 | 0 | 0 | 0 | 0 |
| 0 | 0 | 0 | 0 | 0.00092345 |
| 0 | 0 | 0 | 0 | 0 |
| 1.205528385 | 0 | 0.007264673 | 1.007508905 | 0 |
| 0 | 0 | 0 | 0 | 0 |
| 0 | 0 | 0 | 0 | 0 |
| 0.009248251 | 0.034053451 | 0 | 0 | 0 |
| 0.000937612 | 0.019712667 | 0 | 0 | 0.004190007 |
| 0 | 0 | 0 | 0 | 0 |
| 0 | 0 | 0 | 0 | 0 |
| 0 | 0 | 0.001115002 | 0 | 0 |
| 0.008068294 | 0 | 0.000635421 | 0 | 0.001860936 |
| 0 | 0 | 0 | 0 | 0 |

| | | | | |
|---|---|---|---|---|
| 0 | 0 | 0 | 0 | 0 |
| 0 | 0.009601903 | 0.008418736 | 0.007715731 | 0.156413483 |
| 0 | 0 | 0.008873102 | 0 | 0 |
| 0 | 0 | 0 | 0 | 0 |
| 0 | 0 | 0 | 0 | 0 |
| 0 | 0 | 0 | 0 | 0 |
| 0 | 0 | 0 | 0 | 0 |
| 0 | 0 | 0 | 0 | 0 |
| 0 | 0 | 0 | 0 | 0 |
| 0 | 0 | 0 | 0 | 0 |
| 0 | 0 | 0 | 0 | 0 |
| 0 | 0 | 0 | 0 | 0 |
| 0.019610583 | 0 | 0.316138811 | 0 | 0.019857698 |
| 0 | 0 | 0 | 0 | 0 |
| 0.011565464 | 0 | 0 | 0.003808057 | 0.071080488 |
| 0 | 0 | 0 | 0 | 0 |
| 0 | 0 | 0 | 0 | 0 |
| 0 | 0 | 0 | 0 | 0 |
| 0.000936175 | 0 | 0 | 0 | 0 |
| 0 | 0 | 0 | 0 | 0 |
| 0 | 0 | 0 | 0 | 0 |
| 0 | 0.016329961 | 0 | 0.366895267 | 0.120763906 |
| 0 | 0 | 0 | 0.00272249 | 0.021480828 |
| 0 | 0 | 0.002359554 | 0 | 0 |
| 0 | 0 | 0 | 0 | 0 |
| 0 | 0 | 0 | 0 | 0 |
| 0 | 0 | 0 | 0 | 0 |
| 0 | 0 | 0 | 0 | 0 |
| 0 | 0 | 0 | 0.002548598 | 0 |
| 0 | 0 | 0 | 0 | 0 |
| 0 | 0 | 0 | 0 | 0 |
| 0 | 0 | 0 | 0 | 0 |
| 0 | 0 | 0 | 0 | 0 |
| 0 | 0 | 0 | 0 | 0 |
| 11.51365432 | 0 | 0.000891409 | 12.32241982 | 13.81101412 |
| 0 | 0 | 0 | 0 | 0 |
| 0 | 0 | 0 | 0 | 0 |
| 0 | 0 | 0 | 0 | 0 |
| 0.011757506 | 0.151985362 | 0.652806049 | 0.105749707 | 1.499012583 |
| 0 | 0 | 0 | 0 | 0 |
| 0.33809326 | 0.214647714 | 0.068861996 | 0.128304721 | 0.080097404 |
| 0 | 0 | 0 | 0 | 0 |
| 0 | 0 | 0 | 0 | 0 |
| 0 | 0 | 0 | 0 | 0 |
| 0 | 0 | 0 | 0 | 0 |
| 0.061628282 | 5.878749082 | 0.003690491 | 0.312768226 | 0.398430148 |
| 0 | 0 | 0 | 0 | 0 |
| 0 | 0 | 0 | 0 | 0 |

| | | | | |
|---|---|---|---|---|
| 0 | 0 | 0 | 0 | 0 |
| 0 | 0 | 0 | 0 | 0 |
| 0 | 0 | 0 | 0 | 0 |
| 0.031411874 | 0 | 0 | 0.002094875 | 0 |
| 0 | 0 | 0 | 0 | 0.001490403 |
| 0 | 0 | 0 | 0 | 0 |
| 0 | 0 | 0 | 0 | 0 |
| 0 | 0 | 0 | 0 | 0.006726089 |
| 0 | 0 | 0 | 0 | 0 |
| 0.00263866 | 0 | 0 | 0 | 0.266984003 |
| 0.022262753 | 0 | 0 | 0.034549675 | 0 |
| 0 | 0 | 0 | 0 | 0 |
| 0.002579068 | 0.013869183 | 0 | 0 | 0.012836388 |
| 0 | 0 | 0 | 0 | 0 |
| 0 | 0 | 0 | 0 | 0 |
| 0 | 0 | 0 | 0 | 0 |
| 0 | 0 | 0 | 0 | 0 |
| 0 | 0 | 0 | 0 | 0 |
| 0 | 0 | 0 | 0 | 0 |
| 0.025734355 | 0.059633682 | 0.014415382 | 0.001716858 | 0.525145412 |
| 0 | 0 | 0.156927847 | 0 | 0 |
| 0 | 0 | 0 | 0 | 0 |
| 0 | 0 | 0 | 0 | 0 |
| 0 | 0 | 0 | 0 | 0 |
| 0 | 0 | 0 | 0 | 0 |
| 0 | 0 | 0 | 0 | 0 |
| 0 | 0 | 0 | 0 | 0 |
| 0 | 0 | 0 | 0 | 0 |
| 0 | 0 | 0 | 0 | 0 |
| 0 | 0 | 0 | 0 | 0 |
| 0 | 0 | 0 | 0 | 0 |
| 0 | 0 | 0 | 0 | 0 |
| 0 | 0 | 0 | 0 | 0 |
| 0 | 0 | 0 | 0 | 0 |
| 0 | 0 | 0 | 0 | 0 |
| 0 | 0 | 0 | 0.004222622 | 0 |
| 0 | 0 | 0 | 0 | 0 |
| 0 | 0 | 0 | 0 | 0 |
| 0 | 0 | 0 | 0 | 0 |
| 0 | 0 | 0 | 0 | 0 |
| 0 | 0 | 0 | 0 | 0 |
| 0 | 0 | 0 | 0 | 0 |
| 0 | 0 | 0 | 0 | 0 |
| 0 | 0 | 0 | 0 | 0 |
| 0.039149648 | 0.002675644 | 0.01099243 | 0 | 0.005496393 |
| 0 | 0 | 0.029241677 | 0 | 0 |
| 0 | 0 | 0 | 0 | 0 |
| 0 | 0 | 0 | 0 | 0 |

| | | | | |
|---|---|---|---|---|
| 0 | 0 | 0 | 0 | 0 |
| 0 | 0 | 0 | 0 | 0 |
| 0.021340668 | 0 | 0.000411039 | 0 | 0 |
| 0 | 0 | 0 | 0 | 0 |
| 0 | 0 | 0 | 0 | 0 |
| 0.039223124 | 0.007324065 | 0.001801378 | 0 | 0 |
| 0.00162403 | 0 | 0 | 0 | 0 |
| 0 | 0 | 0 | 0 | 0 |
| 0 | 0 | 0 | 0 | 0 |
| 0 | 0 | 0 | 0 | 0 |
| 0 | 0 | 0 | 0 | 0 |
| 0 | 0 | 0 | 0 | 0 |
| 0 | 0 | 0 | 0 | 0 |
| 0.059463178 | 0 | 0 | 0 | 0 |
| 0 | 0 | 0 | 0 | 0 |
| 0 | 0 | 0 | 0 | 0 |
| 0 | 0 | 0 | 0 | 0 |
| 0 | 0 | 0 | 0 | 0 |
| 0 | 0 | 0 | 0 | 0 |
| 0 | 0 | 0 | 0 | 0 |
| 0 | 0.000772015 | 0 | 0 | 0 |
| 0 | 0 | 0 | 0 | 0 |
| 0 | 0 | 0 | 0 | 0 |
| 0 | 0 | 0.020307723 | 0 | 0 |
| 0 | 0 | 0 | 0 | 0 |
| 0 | 0 | 0 | 0 | 0.00382036 |
| 0 | 0 | 0 | 0 | 0 |
| 1.307505241 | 0 | 0 | 0 | 0 |
| 0 | 0 | 0 | 0 | 0 |
| 0 | 0 | 0 | 0 | 0 |
| 0 | 0 | 0 | 0 | 0 |
| 0 | 0 | 0 | 0 | 0 |
| 0 | 0 | 0 | 0 | 0 |
| 0 | 0 | 0 | 0 | 0 |
| 0 | 0 | 0 | 0 | 0 |
| 0 | 0 | 0 | 0 | 0 |
| 0 | 0 | 0 | 0 | 0 |
| 0 | 0 | 0 | 0 | 0 |
| 0 | 0 | 0 | 0 | 1.541673351 |
| 0 | 0 | 0 | 0 | 0 |
| 0 | 0 | 0 | 0 | 0 |
| 0 | 0 | 0 | 0 | 0 |
| 0 | 0 | 0 | 0 | 0 |
| 0 | 0 | 0 | 0 | 0 |
| 0 | 0 | 0 | 0 | 0 |
| 0 | 0 | 0 | 0 | 0 |
| 0 | 0 | 0 | 0 | 0 |
| 0 | 0 | 0 | 0 | 0 |

| | | | | |
|---|---|---|---|---|
| 0 | 0 | 0 | 0 | 0 |
| 0 | 0 | 0.013557294 | 0 | 0 |
| 0 | 0 | 0 | 0 | 0 |
| 0 | 0 | 0 | 0 | 0 |
| 0 | 0 | 0 | 0 | 0 |
| 0 | 0 | 0 | 0 | 0 |
| 0 | 0 | 0 | 0 | 0 |
| 0 | 0 | 0 | 0 | 0 |
| 0 | 0 | 0 | 0 | 0 |
| 0 | 0 | 0 | 0 | 0 |
| 0.423496819 | 0 | 0.178549006 | 0.000882402 | 0.007447643 |
| 0 | 0 | 0 | 0 | 0 |
| 0.013446699 | 0 | 0.169115639 | 0 | 0 |
| 0 | 0 | 0 | 0 | 0 |
| 0 | 0 | 0 | 0 | 0 |
| 0 | 0 | 1.092718094 | 0 | 0 |
| 0 | 0 | 0 | 0 | 0 |
| 0 | 0 | 0 | 0 | 0 |
| 0 | 0 | 0 | 0 | 0 |
| 0 | 0 | 0 | 0 | 0 |
| 0.050671308 | 0.019417371 | 0 | 0.001074702 | 0.006105273 |
| 0.000779795 | 0 | 0 | 0 | 0 |
| 0 | 0 | 0 | 0 | 0 |
| 0.182418484 | 0.042018853 | 0.985831063 | 3.383814983 | 0.163890513 |
| 0 | 0 | 0 | 0 | 0 |
| 0 | 0 | 0 | 0 | 0 |
| 0 | 0 | 0 | 0 | 0 |
| 0 | 0 | 0 | 0 | 0 |
| 0 | 0 | 0 | 0 | 0 |
| 0 | 0 | 0 | 0 | 0 |
| 0 | 0 | 0 | 0 | 0 |
| 0 | 0 | 0 | 0 | 0 |
| 0.001856267 | 0 | 0.021771996 | 0 | 0 |
| 0 | 0 | 0 | 0 | 0 |

| PHL018-0062_V3 | PHL018-0224_V2 | PHL018-0087_V1 | PHL018-0082_V3 | PHL018-0214_V2 |
|---|---|---|---|---|
| EYCF | EYCF | EYCF | REF | EYCF |
| 0.072866382 | 0.057002149 | 0.022054413 | 0.082279371 | 0.015704106 |
| 1.011279172 | 0 | 11.88636102 | 1.106664154 | 0 |
| 0 | 3.678952853 | 9.573259682 | 0 | 6.483149821 |
| 0 | 0 | 0 | 0 | 0 |
| 0 | 0 | 0 | 0 | 0 |
| 0 | 0 | 0 | 0 | 0 |
| 0 | 0 | 0 | 0 | 0 |
| 0 | 0 | 0 | 0 | 0 |
| 0 | 0 | 0 | 0 | 0 |
| 0 | 0 | 0 | 0 | 0 |
| 0 | 0 | 0 | 0 | 0 |
| 0 | 0 | 0 | 0 | 0 |
| 0 | 0 | 0 | 0 | 0 |
| 4.853694006 | 25.21802742 | 5.145672123 | 0 | 3.362818862 |
| 0 | 0 | 0 | 0 | 0 |
| 0 | 0 | 0 | 0 | 0 |
| 0 | 0 | 0 | 0.031135156 | 0 |
| 0 | 0 | 0 | 0 | 0 |
| 0 | 0 | 0 | 0 | 0 |
| 0 | 0 | 0 | 0 | 0 |
| 0 | 0 | 0 | 0 | 0 |
| 0 | 0 | 0 | 0 | 0 |
| 0 | 0 | 0 | 0 | 0 |
| 0 | 0 | 0 | 0 | 0 |
| 0 | 0 | 0 | 0 | 0 |
| 0 | 0 | 0 | 0 | 0 |
| 0 | 0 | 0 | 0 | 0 |
| 0 | 0 | 0 | 0 | 0 |
| 0 | 0 | 0 | 0 | 0 |
| 0 | 0 | 0 | 0 | 0 |
| 0 | 0 | 0 | 0 | 0 |
| 0 | 0 | 0 | 0 | 0 |
| 0 | 0 | 0 | 0 | 0 |
| 0 | 0 | 0 | 0 | 0 |
| 0 | 0 | 0 | 0 | 0 |
| 0 | 0 | 0 | 0 | 0 |
| 0 | 0 | 0 | 0 | 0 |
| 0 | 0 | 0 | 0 | 0 |
| 0 | 0 | 0 | 0 | 0 |
| 0 | 0 | 0 | 0 | 0 |
| 0 | 0 | 0.029028543 | 0 | 0.016333003 |
| 0.071206909 | 0.000814471 | 0.02298645 | 0.343754453 | 6.906144744 |
| 0.009963945 | 0.001307486 | 0.137558462 | 0 | 0.006018442 |
| 0 | 0 | 0 | 0 | 0 |
| 0 | 0 | 0 | 0 | 0 |

| | | | | |
|---|---|---|---|---|
| 0 | 0 | 0 | 0 | 0 |
| 0.032025245 | 0.00400845 | 0.199870136 | 0.00232068 | 0 |
| 0 | 0.048645809 | 0 | 0.018302605 | 0 |
| 0.456221839 | 0.093841437 | 0.926080388 | 0 | 0 |
| 0 | 0.03092609 | 0.003747321 | 0 | 0 |
| 0 | 0 | 0 | 0 | 0 |
| 0 | 0 | 0.009929132 | 0 | 0 |
| 0 | 0 | 0 | 0 | 0 |
| 0 | 0 | 0 | 0 | 0 |
| 0 | 0 | 0 | 0 | 0 |
| 0 | 0 | 0 | 0 | 0 |
| 0.030945047 | 0 | 0.030577051 | 0 | 0 |
| 0 | 0 | 0 | 0 | 0.001077702 |
| 0.003172016 | 0.240200996 | 0.023272141 | 0.099072912 | 0 |
| 0 | 0 | 0 | 0 | 0 |
| 0 | 0 | 0 | 0 | 0 |
| 0 | 0.000952492 | 0.243708363 | 0 | 0.017709774 |
| 0 | 0 | 0 | 0 | 0 |
| 0 | 0 | 0 | 0 | 0 |
| 0 | 0 | 0 | 0 | 0 |
| 0.132062674 | 0.048141605 | 0.250696537 | 0.201704778 | 0 |
| 0.003147055 | 0.003348169 | 0.038054107 | 0.008700928 | 0 |
| 0 | 0 | 0 | 0 | 0 |
| 0 | 0 | 0 | 0 | 0 |
| 0 | 0 | 0 | 0 | 0 |
| 0 | 0 | 0 | 0 | 0 |
| 0 | 0 | 0 | 0.101132501 | 0.838358889 |
| 0 | 0 | 0 | 0 | 0 |
| 0 | 0 | 0.001093982 | 0 | 0.000942318 |
| 0 | 0.001018109 | 0 | 0 | 0 |
| 1.362731984 | 0 | 0.033671971 | 0.260496204 | 0 |
| 0 | 0 | 0 | 0 | 0 |
| 0 | 0 | 0 | 0 | 0 |
| 0 | 0 | 0 | 0 | 0 |
| 0 | 0 | 0 | 0 | 0 |
| 0 | 0 | 0 | 0 | 0 |
| 0 | 0 | 0 | 0 | 0 |
| 0 | 0 | 0.011406532 | 0 | 1.279060992 |
| 0 | 0 | 0 | 0 | 0 |
| 0 | 0 | 0 | 0 | 0 |
| 0 | 0 | 0 | 0 | 0 |
| 0 | 0 | 0 | 0 | 0 |
| 0 | 0 | 0 | 0 | 0 |
| 0 | 0 | 0 | 0 | 0 |
| 0 | 0 | 0 | 0 | 0 |
| 0 | 0 | 0 | 0 | 0 |
| 0 | 0 | 0 | 0 | 0.002732048 |
| 0 | 0 | 0 | 0 | 0.003423317 |

| | | | | |
|---|---|---|---|---|
| 0 | 0 | 0 | 0 | 0 |
| 0 | 0.002068333 | 0 | 0 | 0 |
| 0 | 0 | 0 | 0 | 0 |
| 0 | 0 | 0 | 0 | 0 |
| 0 | 0 | 0 | 0 | 0 |
| 0 | 0 | 0 | 0 | 0 |
| 0 | 0 | 0 | 0 | 0 |
| 0 | 0 | 0 | 0 | 0 |
| 0 | 0 | 0 | 0 | 0 |
| 0 | 0 | 0 | 0 | 0 |
| 0 | 0 | 0 | 0 | 0 |
| 0 | 0 | 0.002399073 | 0 | 0 |
| 0 | 0 | 0 | 0 | 0 |
| 0 | 0 | 0 | 0 | 0 |
| 0 | 0 | 0 | 0 | 0 |
| 0 | 0 | 0 | 0 | 0 |
| 0 | 0 | 0 | 0 | 0 |
| 0 | 0 | 0 | 0 | 0 |
| 0 | 0 | 0 | 0 | 0 |
| 0 | 0 | 0 | 0 | 0 |
| 0 | 0 | 0 | 0 | 0 |
| 0 | 0 | 0 | 0 | 0 |
| 0 | 0 | 0 | 0 | 0 |
| 0 | 0 | 0 | 0 | 0 |
| 0 | 0 | 0 | 0 | 0 |
| 0 | 0 | 0 | 0 | 0 |
| 0 | 0 | 0 | 0 | 0 |
| 0 | 0.00179389 | 0 | 0 | 0 |
| 0 | 0 | 0 | 0 | 0 |
| 0 | 0 | 0 | 0 | 0 |
| 0.008585332 | 0 | 0.007648819 | 0 | 0 |
| 0 | 0 | 0.004413353 | 0 | 0 |
| 0 | 0 | 0 | 0 | 0 |
| 0 | 0 | 0 | 0 | 0 |
| 0 | 0 | 0 | 0 | 0 |
| 0 | 0 | 0 | 0 | 0 |
| 0 | 0 | 0 | 0 | 0 |
| 0 | 0 | 0 | 0 | 0 |
| 0 | 0 | 0 | 0 | 0 |
| 0 | 0 | 0 | 0 | 0 |
| 0 | 0 | 0 | 0 | 0 |
| 0 | 0 | 0 | 0 | 0 |
| 0 | 0 | 0 | 0.029637699 | 0 |
| 0 | 0 | 0 | 0 | 0 |
| 0 | 0 | 0 | 0 | 0 |
| 0 | 0 | 0 | 0 | 0 |
| 0 | 0 | 0 | 0 | 0 |
| 0 | 0 | 0 | 0 | 0 |

| | | | | |
|---|---|---|---|---|
| 0 | 0 | 0 | 0 | 0 |
| 0 | 0 | 0 | 0 | 0 |
| 0 | 0 | 0 | 0 | 0 |
| 0 | 0 | 0 | 0 | 0 |
| 0 | 0 | 0 | 0 | 0 |
| 0 | 0 | 0 | 0 | 0 |
| 0 | 0 | 0 | 0 | 0 |
| 0 | 0 | 0 | 0 | 0 |
| 0 | 0 | 0 | 0 | 0 |
| 0 | 0 | 0 | 0 | 0 |
| 0 | 0 | 0 | 0 | 0 |
| 0 | 0 | 0 | 0 | 0 |
| 0 | 0 | 0 | 0 | 0 |
| 0 | 0 | 0 | 0 | 0 |
| 0 | 0 | 0 | 0 | 0 |
| 0 | 0 | 1.377481453 | 0 | 0 |
| 0 | 0 | 0 | 0 | 0 |
| 0 | 0 | 0 | 0 | 0 |
| 0 | 0 | 0 | 0 | 0 |
| 0 | 0 | 0 | 0 | 0 |
| 0 | 0 | 0 | 0 | 0 |
| 0 | 0 | 0 | 0 | 0 |
| 0 | 0 | 0 | 0 | 0 |
| 0 | 0 | 0 | 0 | 0 |
| 0 | 0 | 0 | 0 | 0 |
| 0 | 0 | 0 | 0 | 0 |
| 0 | 0 | 0 | 0 | 0 |
| 0 | 0 | 0 | 0 | 0 |
| 0 | 0 | 0 | 0 | 0 |
| 0 | 0 | 0 | 0 | 0 |
| 0 | 0 | 0 | 0 | 0 |
| 0 | 0.002132296 | 0 | 0 | 0 |
| 0 | 0 | 0 | 0 | 0 |
| 0 | 0 | 0 | 0 | 0 |
| 0 | 0 | 0 | 0 | 0 |
| 0 | 0 | 0 | 0 | 0 |
| 0 | 0 | 0 | 0 | 0 |
| 0 | 0 | 0 | 0 | 0 |
| 0 | 0 | 0 | 0 | 0 |
| 0 | 0 | 0 | 0 | 0 |
| 0 | 0 | 0 | 0 | 0 |
| 0 | 0 | 0 | 0 | 0 |
| 0 | 0 | 0 | 0 | 0 |
| 0 | 0 | 0 | 0 | 0 |
| 0 | 0 | 0 | 0 | 0 |
| 0 | 0 | 0 | 0 | 0 |
| 0 | 0 | 0 | 0 | 0 |
| 0 | 0 | 0 | 0 | 0 |

| | | | | |
|---|---|---|---|---|
| 0 | 0 | 0 | 0 | 0 |
| 0 | 0 | 0 | 0 | 0 |
| 0 | 0 | 0 | 0 | 0 |
| 0 | 0 | 0 | 0 | 0 |
| 0 | 0 | 0 | 0 | 0 |
| 0 | 0 | 0 | 0 | 0 |
| 0 | 0 | 0 | 0 | 0 |
| 0 | 0 | 0 | 0 | 0 |
| 0 | 0 | 0 | 0 | 0 |
| 0 | 0 | 0 | 0 | 0 |
| 0 | 0 | 0 | 0 | 0 |
| 0 | 0 | 0 | 0 | 0 |
| 0 | 0.00030606 | 0.001107409 | 0.000563381 | 0 |
| 0 | 0 | 0 | 0 | 0 |
| 0 | 0 | 0 | 0 | 0 |
| 0 | 0 | 0 | 0 | 0 |
| 0 | 0 | 0 | 0 | 0 |
| 0 | 0 | 0 | 0 | 0 |
| 0 | 0 | 0 | 0 | 0 |
| 0 | 0 | 0 | 0 | 0 |
| 0.008229632 | 0.014497204 | 0.035614122 | 0.018697614 | 0.029677986 |
| 0 | 0 | 0 | 0 | 0 |
| 0 | 0 | 0 | 0 | 0 |
| 0 | 0 | 0 | 0 | 0 |
| 0 | 0 | 0 | 0 | 0 |
| 0 | 0 | 0 | 0 | 0 |
| 0 | 0.000777629 | 0.007235172 | 0.002147137 | 0 |
| 0 | 0 | 0 | 0 | 0 |
| 0 | 0 | 0 | 0 | 0 |
| 0 | 0.026147739 | 0 | 0.026253586 | 0 |
| 0.046182998 | 0 | 0.580825528 | 0 | 0 |
| 0 | 0 | 0 | 0 | 0 |
| 0 | 0 | 0 | 0 | 0 |
| 0 | 0 | 0 | 0 | 0 |
| 0 | 0 | 0.006736176 | 0.001713474 | 0 |
| 0 | 0 | 0 | 0 | 0 |
| 0 | 0 | 0 | 0 | 0 |
| 0 | 0 | 0 | 0 | 0 |
| 0 | 0 | 0 | 0 | 0 |
| 0 | 0 | 0 | 0 | 0 |
| 0 | 0 | 0 | 0 | 0 |
| 0 | 0 | 0 | 0 | 0 |
| 0 | 0 | 0 | 0 | 0 |
| 0 | 0 | 0 | 0 | 0 |
| 0 | 0 | 0 | 0 | 0 |
| 0 | 0 | 0 | 0 | 0 |
| 0.000858911 | 0 | 0 | 0 | 0 |
| 0 | 0 | 0 | 0.001221068 | 0 |

| | | | | |
|---|---|---|---|---|
| 0 | 0 | 0 | 0 | 0 |
| 0 | 0.000733243 | 0 | 0 | 0 |
| 0.001291427 | 0 | 0 | 0 | 0 |
| 0 | 0 | 0 | 0 | 0 |
| 0 | 0 | 0 | 0 | 0 |
| 0 | 0 | 0 | 0 | 0 |
| 0 | 0 | 0 | 0 | 0 |
| 0 | 0 | 0 | 0 | 0.007608278 |
| 0 | 0 | 0 | 0 | 0 |
| 0 | 0 | 0 | 0 | 0 |
| 0 | 0 | 0 | 0 | 0 |
| 0 | 0 | 0 | 0 | 0 |
| 0 | 0 | 0 | 0 | 0 |
| 0 | 0.502792942 | 0 | 0 | 0.031275922 |
| 0 | 0 | 0 | 0 | 0 |
| 0 | 0 | 0 | 0 | 0 |
| 0 | 0 | 0 | 0 | 0 |
| 0 | 0 | 0 | 0 | 0.036224536 |
| 0 | 0 | 0 | 0.002731999 | 0.021799481 |
| 0 | 0 | 0 | 0 | 0 |
| 0.007420561 | 0.020168161 | 0.14834847 | 0.32752723 | 0 |
| 0 | 0 | 0 | 0 | 0 |
| 0 | 0 | 0 | 0 | 0 |
| 0 | 0 | 0 | 0 | 0 |
| 0 | 0.001581644 | 0.007439675 | 0 | 0.001577422 |
| 0 | 0 | 0 | 0 | 0 |
| 0 | 0 | 0 | 0 | 0 |
| 0 | 0 | 0 | 0 | 0 |
| 0 | 0 | 0 | 0 | 0 |
| 0 | 0 | 0 | 0 | 0 |
| 0.05206903 | 0.272397078 | 0.023722376 | 0.679659622 | 0 |
| 0 | 0 | 0 | 0 | 0 |
| 0.037993341 | 0.012131246 | 0 | 0.065652109 | 0.089834065 |
| 0 | 0 | 0 | 0 | 0.080253802 |
| 0 | 0 | 0 | 0 | 0 |
| 0 | 0 | 0 | 0 | 0 |
| 0 | 0 | 0 | 0 | 0 |
| 0 | 0 | 0 | 0 | 0 |
| 0 | 0 | 0 | 0 | 0 |
| 0 | 0 | 0 | 0 | 0 |
| 0 | 0 | 0 | 0 | 0 |
| 0 | 0 | 0.002159019 | 0 | 0 |
| 0 | 0 | 0.000905749 | 0 | 0 |
| 0 | 0 | 0 | 0 | 0 |
| 0 | 0 | 0 | 0 | 0 |
| 0 | 0 | 0 | 0 | 0 |
| 0 | 0 | 0 | 0 | 0 |
| 0 | 0 | 0 | 0 | 0 |

| | | | | |
|---|---|---|---|---|
| 0 | 0 | 0 | 0 | 0 |
| 0 | 0 | 0 | 0 | 0 |
| 0.000798147 | 0 | 0.145038726 | 0 | 0.008219779 |
| 0 | 0 | 0 | 0 | 0 |
| 0 | 0 | 0 | 0 | 0 |
| 0 | 0 | 0 | 0 | 0 |
| 0 | 0 | 0 | 0 | 0 |
| 0 | 0 | 0 | 0 | 0 |
| 0 | 0 | 0 | 0 | 0 |
| 0.000912474 | 0 | 0 | 0 | 0 |
| 0 | 0 | 0 | 0 | 0 |
| 0 | 0 | 0 | 0 | 0 |
| 0 | 0 | 0 | 0 | 0 |
| 0 | 0.00041403 | 0.002996149 | 0 | 0.000619387 |
| 0.002002153 | 0.000797364 | 0.001978343 | 0.001572591 | 0.007497934 |
| 0.012861412 | 0 | 0 | 0 | 0 |
| 0 | 0 | 0.004508289 | 0 | 0.000970821 |
| 0 | 0 | 0 | 0 | 0 |
| 0 | 0 | 0 | 0 | 0 |
| 0 | 0 | 0 | 0 | 0 |
| 0 | 0 | 0 | 0 | 0 |
| 0 | 0 | 0.000763068 | 0.001132255 | 0 |
| 0 | 0 | 0 | 0 | 0 |
| 0 | 0 | 0.003943101 | 0 | 0 |
| 0 | 0 | 0 | 0 | 0 |
| 0 | 0 | 0 | 0 | 0 |
| 0 | 0 | 0 | 0 | 0 |
| 0 | 0 | 0 | 0 | 0 |
| 0 | 0 | 0 | 0 | 0 |
| 0 | 0 | 0 | 0 | 0 |
| 0 | 0 | 0 | 0 | 0 |
| 0 | 0 | 0 | 0 | 0 |
| 0 | 0 | 0 | 0 | 0 |
| 0.001726844 | 0 | 0 | 0 | 0 |
| 0 | 0 | 0 | 0 | 0 |
| 0 | 0 | 0.002540201 | 0 | 0 |
| 0 | 0 | 0 | 0 | 0 |
| 0 | 0 | 0 | 0 | 0 |
| 0 | 0 | 0 | 0 | 0 |
| 0 | 0 | 0 | 0 | 0 |
| 0 | 0 | 0 | 0 | 0 |
| 0 | 0 | 0 | 0 | 0 |
| 0 | 0 | 0 | 0 | 0 |
| 0 | 0 | 0 | 0.005553566 | 0 |
| 0 | 0 | 0 | 0 | 0 |
| 0 | 0 | 0 | 0 | 0 |
| 0 | 0 | 0 | 0 | 0 |
| 0 | 0.004100887 | 0 | 0.004430774 | 0 |

| | | | | |
|---|---|---|---|---|
| 0.172044512 | 0.188305008 | 0 | 0 | 0 |
| 0 | 0 | 0.005656842 | 0 | 0 |
| 0.004313994 | 0 | 0.001065673 | 0 | 0 |
| 0 | 0 | 0 | 0 | 0 |
| 0.0155856 | 0.006173344 | 0.004824764 | 0.00613635 | 0 |
| 0 | 0 | 0 | 0 | 0 |
| 0 | 0 | 0 | 0 | 0 |
| 0 | 0.002942122 | 0.039074931 | 0 | 0 |
| 0 | 0 | 0 | 0 | 0 |
| 0.01824228 | 0.138603489 | 0.010456108 | 2.21716282 | 0.006671501 |
| 0 | 0.000806406 | 0 | 0 | 0 |
| 0 | 0 | 0.002676795 | 0 | 0.035251562 |
| 0.000498153 | 0 | 0.007670567 | 0.000860803 | 0 |
| 0 | 0 | 0 | 0 | 0 |
| 0 | 0.004168259 | 0 | 0.007377637 | 0 |
| 0 | 0 | 0.010837453 | 0 | 0.357764044 |
| 0 | 0 | 0 | 0 | 0 |
| 0 | 0.004330915 | 0 | 0.000747389 | 0 |
| 0 | 0.00167855 | 0 | 0.183765819 | 0 |
| 0 | 0 | 0.1615287 | 0 | 0 |
| 0 | 0 | 0 | 0 | 0 |
| 0 | 0 | 0 | 0 | 0 |
| 0 | 0 | 0 | 0 | 0 |
| 0 | 0 | 0 | 0 | 0 |
| 0 | 0 | 0 | 0 | 0 |
| 0 | 0 | 0 | 0 | 0 |
| 0 | 0 | 0 | 0 | 0 |
| 0 | 0 | 0 | 0 | 0 |
| 0 | 0.45431545 | 0 | 0 | 0 |
| 0 | 0 | 0.016221571 | 0.001547349 | 0 |
| 0 | 0 | 0.010714816 | 0.327958505 | 0 |
| 0 | 0 | 0 | 0 | 0 |
| 0 | 0.037983546 | 0 | 0 | 0 |
| 0 | 0.036566634 | 0 | 0 | 0 |
| 0 | 0 | 0 | 0 | 0 |
| 0 | 0 | 0 | 0 | 0 |
| 0.002850207 | 0.012074516 | 0 | 0 | 0 |
| 0 | 0 | 0 | 0 | 0 |
| 0 | 0 | 0 | 0 | 0 |
| 0 | 0.007001628 | 0 | 0 | 0 |
| 0 | 0 | 0 | 0 | 0 |
| 0 | 0 | 0 | 0 | 0 |
| 0 | 0.016879845 | 0 | 0 | 0 |
| 0 | 0 | 0 | 0.035809937 | 0 |
| 0 | 0.012082995 | 0 | 0 | 0 |
| 0 | 0 | 0 | 0 | 0 |
| 0.000962068 | 0 | 0 | 0 | 0 |
| 0 | 0 | 0 | 0 | 0 |

| | | | | |
|---|---|---|---|---|
| 0 | 0 | 0 | 0 | 0 |
| 0 | 0 | 0.114306743 | 0 | 0.003774818 |
| 0 | 0 | 0 | 0 | 0 |
| 0 | 0 | 0 | 0 | 0 |
| 0 | 0 | 0 | 0 | 0 |
| 0.019133058 | 0.017664579 | 0.021104934 | 0 | 0 |
| 0 | 0 | 0 | 0 | 0 |
| 0 | 0 | 0 | 0 | 0.004128999 |
| 0 | 0 | 0 | 0 | 0 |
| 0 | 0 | 0 | 0 | 0 |
| 0.002197141 | 0.008387679 | 0.016418285 | 0.044990156 | 0 |
| 0 | 0.002319688 | 0 | 0 | 0 |
| 0 | 0 | 0 | 0 | 0 |
| 0 | 0 | 0.003666277 | 0.001076062 | 0.005247143 |
| 0.015122048 | 0.018401794 | 0 | 0 | 0 |
| 0 | 0.051268066 | 0 | 0 | 0 |
| 0 | 0 | 0 | 0 | 0 |
| 0 | 0 | 0 | 0 | 0 |
| 0 | 0 | 0 | 0 | 0 |
| 0 | 0 | 0 | 0 | 0 |
| 0 | 0 | 0 | 0 | 0 |
| 0 | 0 | 0.370829103 | 0 | 0 |
| 0 | 0 | 0 | 0 | 0 |
| 0 | 0 | 0 | 0 | 0 |
| 0 | 0 | 0 | 0 | 0 |
| 0 | 0 | 0 | 0 | 0 |
| 0.025432468 | 0.010423073 | 0 | 0 | 0 |
| 0 | 0 | 0 | 0 | 0 |
| 0 | 0 | 0 | 0 | 0 |
| 1.053048188 | 0.360645593 | 2.512548704 | 0 | 0 |
| 0.002393992 | 0 | 0 | 0 | 0 |
| 0 | 0 | 0 | 0 | 0 |
| 0.001951693 | 0 | 0 | 0 | 0 |
| 0 | 0 | 0 | 0 | 0 |
| 0 | 0.003754135 | 0 | 0 | 0 |
| 0 | 0 | 0 | 0 | 0 |
| 0 | 0 | 0 | 0 | 0 |
| 0 | 0 | 0.002587093 | 0 | 0 |
| 0 | 0 | 0 | 0 | 0 |
| 0 | 0 | 0 | 0 | 0 |
| 0.068430294 | 0 | 0.035937088 | 0.014154237 | 0 |
| 12.09992989 | 4.581555584 | 0 | 0 | 0 |
| 0 | 0 | 0.003686581 | 0 | 0 |
| 0 | 0.003092157 | 0 | 0 | 0 |
| 0 | 0.001084251 | 0 | 0 | 0 |
| 0 | 0.005804544 | 0 | 0.224980603 | 0 |
| 0.002149302 | 0.003026453 | 0.001216727 | 0.043948667 | 0 |
| 0 | 0 | 0 | 0 | 0 |

| | | | | |
|---|---|---|---|---|
| 0 | 0 | 0 | 0 | 0 |
| 0 | 0 | 0 | 0 | 0 |
| 0 | 0 | 0.506069361 | 0 | 0.001691488 |
| 0 | 0 | 0 | 0 | 0 |
| 0 | 0 | 0 | 0 | 0 |
| 0 | 0.004092686 | 0 | 0 | 0 |
| 0 | 0 | 0 | 0 | 0 |
| 0 | 0 | 0.002019099 | 0 | 0 |
| 0 | 0 | 0.026635705 | 0.004154662 | 0 |
| 0 | 0.001899804 | 0 | 0 | 0 |
| 0 | 0 | 0 | 0 | 0 |
| 0 | 0.000583092 | 0 | 0 | 0 |
| 0 | 0 | 0 | 0 | 0 |
| 0.036985923 | 0.012136417 | 0.004074395 | 0.21177096 | 0 |
| 0 | 0 | 0 | 0 | 0 |
| 0.001249897 | 0 | 0.002759527 | 0 | 0 |
| 0.001039035 | 0.01799041 | 0.001176403 | 0.041744031 | 0.00129704 |
| 0.015779839 | 0.001185053 | 0.014006981 | 0.022541053 | 0 |
| 0 | 0 | 0 | 0.001081824 | 0 |
| 0 | 0 | 0 | 0 | 0 |
| 0.025753104 | 0.039746105 | 0 | 0 | 0 |
| 0.001205227 | 0.000502842 | 0 | 0 | 0 |
| 0 | 0 | 0 | 0 | 0 |
| 0 | 0 | 0 | 0 | 0 |
| 0 | 0 | 0 | 0 | 0 |
| 0 | 0 | 0.579279209 | 1.221614745 | 0 |
| 0.003737346 | 0 | 0.001194762 | 0.002469272 | 0 |
| 0 | 0.004788804 | 0 | 0 | 0 |
| 0 | 0 | 0 | 0 | 0 |
| 0 | 0 | 0 | 0 | 0 |
| 0 | 0.035261107 | 0 | 0.037268589 | 0 |
| 0 | 0 | 0.009482418 | 0 | 0.004433962 |
| 0.022783676 | 0 | 0.021926465 | 0 | 0 |
| 0 | 0 | 0 | 0 | 0 |
| 0 | 0 | 0 | 0 | 0.009772926 |
| 0 | 0 | 0 | 0 | 0 |
| 0 | 0 | 0.001166061 | 0 | 0.002008809 |
| 0.026679871 | 0.035074144 | 0.012538813 | 0.004784227 | 0 |
| 0.034403961 | 0 | 0 | 0 | 0 |
| 0 | 0 | 0 | 0.002844796 | 0 |
| 0 | 0 | 0 | 0 | 0 |
| 0 | 0 | 0 | 0 | 0 |
| 0 | 0.000471138 | 0 | 0.006504367 | 0 |
| 0 | 0.038364359 | 0.009788089 | 0.050248402 | 0 |
| 0 | 0.98704682 | 0.003482603 | 0 | 0 |
| 0 | 0 | 0 | 0 | 0 |
| 0 | 0 | 0 | 0 | 0 |
| 0 | 0 | 0 | 0 | 0 |

| | | | | |
|---|---|---|---|---|
| 0 | 0 | 0 | 0 | 0 |
| 0.094202138 | 0.009873398 | 0.939093512 | 0 | 0.0271864 |
| 0 | 0 | 0 | 0 | 0 |
| 0 | 0 | 0 | 0 | 0 |
| 0 | 0 | 0 | 0 | 0 |
| 0 | 0 | 0 | 0.001870316 | 0 |
| 0 | 0 | 0.005173043 | 0 | 0 |
| 0 | 0 | 0 | 0 | 0 |
| 0 | 0 | 0 | 0 | 0 |
| 0 | 0 | 0 | 0 | 0 |
| 0.002548634 | 0 | 0 | 0.001761606 | 0 |
| 0.006833274 | 0 | 0.009971723 | 0.034986156 | 0.209460311 |
| 0 | 0 | 0 | 0 | 0 |
| 0 | 0 | 0 | 0 | 0 |
| 0 | 0 | 0 | 0 | 0 |
| 0 | 0 | 0 | 0 | 0 |
| 0.009607637 | 0.004453863 | 0.051971879 | 0.006148852 | 0 |
| 0.008776621 | 1.580001965 | 0.112967462 | 3.331112644 | 0 |
| 0 | 0 | 0 | 0 | 0 |
| 0.004649248 | 0.019522634 | 0.099233239 | 0.037188971 | 0 |
| 0.001284357 | 0 | 0.041131905 | 0 | 0 |
| 0 | 0 | 0 | 0 | 0 |
| 0 | 0 | 0 | 0 | 0 |
| 0 | 0 | 0 | 0 | 0 |
| 0 | 0 | 0 | 0 | 0 |
| 0 | 0 | 0 | 0 | 0 |
| 0 | 0 | 0.114170001 | 0 | 0 |
| 0.135533136 | 0 | 0.178842858 | 0 | 0 |
| 0 | 0 | 0 | 0 | 0 |
| 0 | 0 | 0 | 0 | 0 |
| 0 | 0 | 0 | 0 | 0 |
| 0 | 0 | 0 | 0 | 0 |
| 0 | 0.009037757 | 0 | 0 | 0 |
| 0 | 0 | 0 | 0 | 0 |
| 0.010196348 | 0 | 0 | 0 | 0 |
| 0.836995841 | 0.602510319 | 0.331379499 | 0 | 0 |
| 0 | 0 | 0 | 0 | 0 |
| 0 | 0 | 0 | 0 | 0 |
| 0 | 0 | 0.000980545 | 0 | 0.010360518 |
| 0 | 0 | 0 | 0 | 0 |
| 0 | 0.879549374 | 0.004922378 | 0 | 0 |
| 0 | 0.008206489 | 0 | 0 | 0 |
| 0.009498134 | 0.02450833 | 0.002382393 | 0.026891543 | 0 |
| 0 | 0.0011478 | 0 | 0 | 0.001717104 |
| 0 | 0.075281138 | 0 | 0 | 0 |
| 0.003352614 | 0 | 0.002970668 | 0 | 0 |
| 0 | 0 | 0 | 0 | 0 |
| 0 | 0 | 0.004957363 | 0.088466972 | 0 |

| | | | | |
|---|---|---|---|---|
| 0 | 0 | 0 | 0 | 0 |
| 0 | 0 | 0 | 0 | 0 |
| 0 | 0 | 0 | 0 | 0 |
| 0.001872085 | 0 | 0.310770142 | 0 | 0 |
| 0 | 0 | 0 | 0 | 0 |
| 0 | 0 | 0 | 0 | 0 |
| 0 | 0.004128708 | 0.007887693 | 0 | 0 |
| 0 | 0 | 0.026064115 | 0 | 0 |
| 0 | 0 | 0 | 0 | 0.013249184 |
| 0.196979336 | 0 | 0 | 0 | 0 |
| 0 | 0 | 0 | 0 | 0 |
| 0 | 0 | 0.005760068 | 0 | 0 |
| 0 | 0 | 0 | 0 | 0 |
| 0 | 0 | 0 | 0 | 0 |
| 0.059878054 | 0.024926046 | 0 | 0 | 0 |
| 0 | 0 | 0 | 0 | 0 |
| 0 | 0 | 0 | 0 | 0 |
| 0 | 0 | 0 | 0 | 0 |
| 0 | 0 | 0 | 0 | 0 |
| 0 | 0 | 0 | 0 | 0 |
| 0 | 0 | 0 | 0 | 0 |
| 0 | 0 | 0 | 0 | 0 |
| 0 | 0 | 0 | 0 | 0 |
| 0.051067635 | 0.215199265 | 0.531994813 | 0.475489265 | 0 |
| 0.131139849 | 0 | 0.002903867 | 0 | 0 |
| 0 | 0 | 0 | 0 | 0 |
| 0 | 0 | 0.054788858 | 0 | 0 |
| 0 | 0 | 0 | 0 | 0 |
| 0 | 0 | 0 | 0 | 0 |
| 0 | 0 | 0 | 0 | 0 |
| 0.001743742 | 0 | 0 | 0 | 0 |
| 0 | 0 | 0 | 0 | 0 |
| 0.023646789 | 0 | 0 | 0 | 0 |
| 0 | 0 | 0 | 0 | 0 |
| 0 | 0 | 0 | 0 | 0 |
| 0 | 0 | 0 | 0 | 0 |
| 0.219764767 | 0 | 0 | 0 | 0 |
| 0 | 0 | 0 | 0 | 0 |
| 0 | 0 | 0.001844994 | 0.077582674 | 0 |
| 0 | 0 | 0.000792866 | 0 | 0 |
| 0.068358513 | 0.002936882 | 0.049306741 | 0.000810912 | 0 |
| 0 | 0 | 0 | 0 | 0 |
| 33.39212775 | 0.460593136 | 0.147108858 | 0 | 40.56777237 |
| 0 | 0 | 0 | 0 | 0 |
| 0 | 0 | 0.00296928 | 0 | 0 |
| 0 | 0 | 0 | 0 | 0 |
| 0 | 0 | 0 | 0 | 0 |
| 0 | 0.001176823 | 0 | 0.000649873 | 0 |

| | | | | |
|---|---|---|---|---|
| 0 | 0 | 0 | 0 | 0 |
| 0 | 0 | 0 | 0 | 0 |
| 0.200629834 | 0.496258914 | 0.123190008 | 2.43381276 | 0.019408082 |
| 0 | 0 | 0 | 0 | 0 |
| 0 | 0 | 0 | 0 | 0 |
| 0 | 0 | 0.003086897 | 0 | 0 |
| 0 | 0 | 0 | 0 | 0 |
| 0 | 0 | 0 | 0 | 0 |
| 0 | 0 | 0 | 0 | 0 |
| 0 | 0 | 0.006284528 | 0 | 0 |
| 0.071855404 | 0.017832543 | 0.006011468 | 0.007645653 | 0.002319775 |
| 3.004963688 | 0 | 2.524974616 | 0 | 0 |
| 0 | 0 | 0 | 0 | 0 |
| 0 | 0.002629835 | 0.007525873 | 0 | 0 |
| 2.646171376 | 0.13216327 | 0.195098809 | 2.878266214 | 0 |
| 0 | 0 | 0 | 0 | 0 |
| 0 | 0 | 0 | 0 | 0 |
| 0 | 0 | 0.00227108 | 0 | 0 |
| 0 | 0 | 0 | 0 | 0 |
| 0 | 0.022535793 | 0 | 0 | 0 |
| 0.074169224 | 0.004827376 | 0.036075629 | 0 | 0 |
| 5.685823484 | 22.17748276 | 0.071565472 | 34.2577462 | 18.75089891 |
| 0.677007943 | 1.093420723 | 0.258127143 | 40.2689949 | 0.175512232 |
| 0 | 0 | 0 | 0 | 0 |
| 0 | 0 | 0 | 0 | 0 |
| 0 | 0 | 0 | 0 | 0 |
| 0 | 1.065592929 | 0 | 0 | 0 |
| 0.002166 | 0 | 0 | 0 | 0 |
| 0 | 0 | 0.002145558 | 0 | 0 |
| 0 | 0 | 0 | 0 | 0 |
| 0 | 0 | 0 | 0 | 0 |
| 0.205162799 | 0.042506636 | 0 | 0 | 0 |
| 0.042715228 | 0.013650549 | 0.007902636 | 0.00329796 | 0 |
| 0.032435057 | 0 | 0 | 0 | 0 |
| 0.00383899 | 0.006982387 | 0 | 0.005078954 | 0 |
| 0.002637269 | 0 | 0.045460851 | 0 | 0 |
| 0.001433006 | 0.001494688 | 0 | 0 | 0 |
| 0 | 0 | 0 | 0 | 0 |
| 0 | 0 | 0 | 0 | 0 |
| 0 | 0 | 0 | 0 | 0 |
| 0 | 0 | 0 | 0 | 0 |
| 0 | 0 | 0 | 0 | 0 |
| 0 | 0 | 0 | 0 | 0 |
| 0 | 0 | 0 | 0 | 0 |
| 0.0206766 | 0.002940901 | 0 | 0 | 0.000879915 |
| 0 | 0 | 0 | 0 | 0 |
| 0 | 0 | 0 | 0 | 0 |
| 0 | 0 | 0.003080628 | 0 | 0.000636851 |

| | | | | |
|---|---|---|---|---|
| 0 | 0 | 0 | 0 | 0 |
| 0 | 0 | 0 | 0 | 0 |
| 0 | 0.001722934 | 0 | 0 | 0 |
| 0 | 0 | 0 | 0 | 0 |
| 0 | 0 | 0 | 0 | 0 |
| 0 | 0 | 0 | 0 | 0 |
| 0 | 0 | 0 | 0 | 0 |
| 0 | 0.009687092 | 0.014220525 | 0.030313654 | 0 |
| 0 | 0 | 0 | 0 | 0 |
| 0 | 0 | 0 | 0 | 0 |
| 0 | 0.004801548 | 0 | 0.009866202 | 0.001169341 |
| 0 | 0 | 0 | 0 | 0 |
| 0 | 0 | 0 | 0 | 0 |
| 0 | 0 | 0 | 0 | 0 |
| 0 | 0 | 0 | 0 | 0 |
| 0 | 0 | 0 | 0 | 0 |
| 0 | 0 | 0 | 0 | 0 |
| 0 | 0 | 0.000807232 | 0 | 0 |
| 0 | 0 | 0 | 0 | 0 |
| 0 | 0 | 0 | 0 | 0 |
| 0 | 0 | 0 | 0 | 0 |
| 0 | 0 | 0 | 0 | 0 |
| 0 | 0 | 2.682454018 | 0 | 0 |
| 0 | 0 | 0 | 0 | 0 |
| 0 | 0 | 0 | 0 | 0 |
| 0 | 0 | 0 | 0 | 0 |
| 0 | 0 | 0 | 0 | 0 |
| 0 | 0 | 0 | 0 | 0 |
| 0 | 0 | 0 | 0 | 0 |
| 0.089176672 | 0 | 0.057045038 | 0 | 0 |
| 0 | 0 | 0 | 0 | 0 |
| 0 | 0 | 0 | 0 | 0 |
| 0 | 0 | 0 | 0 | 0 |
| 0 | 0 | 0 | 0 | 0 |
| 0 | 0 | 0 | 0 | 0 |
| 0 | 0 | 0 | 0 | 0 |
| 0 | 0 | 0 | 0 | 0 |
| 0.001963533 | 0.000283576 | 0.003488597 | 0.002218477 | 0 |
| 0 | 0 | 0 | 0 | 0 |
| 0 | 0 | 0 | 0 | 0 |
| 0 | 0 | 0 | 0 | 0 |
| 0 | 0 | 0 | 0 | 0 |
| 0 | 0 | 0 | 0 | 0 |
| 0.099594817 | 0 | 0 | 0 | 0 |
| 0 | 0 | 0 | 0 | 0 |
| 0 | 0 | 0 | 0 | 0 |
| 0 | 0 | 0 | 0 | 0 |
| 0 | 0 | 0 | 0 | 0 |

| | | | | |
|---|---|---|---|---|
| 0 | 0 | 0 | 0 | 0 |
| 0 | 0 | 0 | 0 | 0 |
| 0 | 0 | 0 | 0 | 0 |
| 0 | 0 | 0.001428878 | 0 | 0 |
| 0 | 0 | 0 | 0 | 0 |
| 0 | 0 | 0 | 0 | 0 |
| 0 | 0 | 0 | 0 | 0 |
| 0 | 0 | 0 | 0 | 0 |
| 0.32892262 | 0.835387788 | 0.170661701 | 0.682661884 | 0 |
| 0 | 0 | 0 | 0 | 0 |
| 0 | 0 | 0 | 0 | 0 |
| 0 | 0 | 0 | 0 | 0 |
| 0 | 0 | 0.002476638 | 0 | 0 |
| 0 | 0 | 0 | 0 | 0 |
| 0 | 0 | 0 | 0 | 0 |
| 0 | 0 | 0 | 0.429878196 | 0 |
| 0 | 0 | 0 | 0 | 0 |
| 0 | 0 | 0 | 0 | 0 |
| 0 | 0 | 0 | 0 | 0 |
| 0 | 0 | 0 | 0 | 0 |
| 0 | 0 | 0 | 0 | 0 |
| 0 | 0 | 0 | 0 | 0 |
| 0 | 0 | 0 | 0 | 0 |
| 0 | 0 | 0 | 0 | 0 |
| 0.002449675 | 0 | 0 | 0.01926021 | 7.894578606 |
| 0 | 0 | 0 | 0 | 0 |
| 0.026422883 | 0 | 0 | 0 | 0 |
| 0 | 0.000761319 | 0 | 0 | 0 |
| 0 | 0 | 0.022405766 | 0 | 0 |
| 0 | 0 | 0 | 0 | 0 |
| 0 | 0 | 0 | 0 | 0 |
| 0 | 0 | 0 | 0 | 0 |
| 0 | 0 | 0 | 0 | 0 |
| 0.001177455 | 0 | 0 | 0 | 0 |
| 0 | 0.00121415 | 0.002928755 | 0 | 0 |
| 0 | 0.00152579 | 0.001863247 | 0.004739521 | 0 |
| 0 | 1.521824898 | 0 | 0 | 0 |
| 0.493297481 | 0 | 1.741351027 | 0 | 0 |
| 0 | 0 | 0 | 0 | 0 |
| 0 | 0 | 0 | 0 | 0 |
| 0 | 0 | 0 | 0 | 0 |
| 0.00190077 | 0.00097327 | 0 | 0 | 0 |
| 8.972065034 | 0.346373823 | 0.746590767 | 0.206892677 | 0 |
| 0 | 0 | 0 | 0 | 0 |
| 0 | 0 | 0 | 0 | 0 |
| 0 | 0 | 0 | 0 | 0 |
| 0 | 0 | 0 | 0 | 0 |
| 0 | 0.001007495 | 0.321211565 | 0 | 0 |

| | | | | |
|---|---|---|---|---|
| 0 | 0 | 0.003029048 | 0.058231697 | 0 |
| 0 | 0 | 0 | 0 | 0 |
| 0 | 0 | 0 | 0 | 0 |
| 0 | 0 | 0 | 0 | 0 |
| 0.001024469 | 0 | 0 | 0 | 0 |
| 0 | 0 | 0 | 0 | 0 |
| 0 | 0 | 0 | 0 | 0 |
| 0 | 0 | 0 | 0 | 0 |
| 0 | 0 | 0 | 0 | 0 |
| 0 | 0 | 0 | 0 | 0 |
| 0 | 0 | 0 | 0 | 0 |
| 0.005040735 | 0 | 1.049546103 | 0 | 0 |
| 0 | 0 | 3.217311283 | 0 | 0 |
| 0 | 0 | 0 | 0 | 0 |
| 0.000592399 | 0.007414782 | 0.403235952 | 0 | 0.748003161 |
| 0 | 0 | 0 | 0 | 0 |
| 0 | 0 | 0 | 0 | 0 |
| 0 | 0 | 0 | 0 | 0 |
| 0 | 0 | 0 | 0 | 0 |
| 0 | 0 | 0 | 0 | 0 |
| 0 | 0 | 0 | 0 | 0 |
| 0 | 0 | 0 | 0 | 0 |
| 0 | 0 | 0.026295243 | 0 | 0 |
| 0 | 0 | 0 | 0 | 0 |
| 0 | 0 | 0 | 0 | 0 |
| 0 | 0 | 0 | 0 | 0 |
| 0 | 0 | 0 | 0 | 0 |
| 0 | 0 | 0 | 0 | 0 |
| 0 | 0 | 0 | 0 | 0 |
| 0 | 0 | 0 | 0 | 0 |
| 0 | 0 | 0 | 0 | 0 |
| 0.021267938 | 0.005612668 | 0.041776849 | 0.004649195 | 0.007676826 |
| 0 | 0 | 0 | 0 | 0 |
| 0.004992515 | 0 | 0.008174473 | 0 | 0 |
| 0 | 0.001349109 | 0.001577084 | 0.00229235 | 0 |
| 0 | 0 | 0.008422647 | 0 | 0 |
| 0 | 0 | 0 | 0 | 0 |
| 0 | 0 | 0 | 0 | 0 |
| 0 | 0 | 0 | 0 | 0 |
| 0 | 0 | 0 | 0 | 0 |
| 0 | 0 | 0 | 0 | 0 |
| 0 | 0 | 0 | 0 | 0 |
| 0 | 0 | 0 | 0 | 0 |
| 0 | 0 | 0 | 0 | 0 |
| 0.01029131 | 0.00378256 | 0.00266329 | 0 | 0.001376439 |
| 0 | 0 | 0 | 0 | 0 |
| 0 | 0 | 0 | 0 | 0 |
| 0 | 0 | 0.000629749 | 0 | 0 |

| | | | | |
|---|---|---|---|---|
| 0 | 0 | 0 | 0 | 0 |
| 0 | 0 | 0 | 0 | 0 |
| 0 | 0.008881556 | 0.003672678 | 0.001605682 | 0 |
| 0 | 0 | 0 | 0 | 0 |
| 0 | 0.002787937 | 0 | 0 | 0 |
| 0 | 0 | 0 | 0 | 0 |
| 0 | 0 | 0 | 0 | 0 |
| 0.032549542 | 0 | 0 | 0 | 0 |
| 0 | 0 | 0 | 0 | 0 |
| 0 | 0 | 0 | 0 | 0 |
| 0 | 0 | 0 | 0 | 0 |
| 0 | 0.006239673 | 0 | 0.014258119 | 0 |
| 0 | 0 | 0 | 0 | 0 |
| 0 | 0 | 0 | 0 | 0 |
| 0 | 0 | 0 | 0 | 0 |
| 0 | 0 | 0 | 0 | 0 |
| 0 | 0 | 0.455689589 | 0 | 0 |
| 0 | 0 | 0 | 0 | 0 |
| 0 | 0 | 0 | 0 | 0 |
| 0 | 0 | 0 | 0 | 0 |
| 0 | 0 | 0 | 0 | 0 |
| 0 | 0 | 0 | 0 | 0 |
| 0 | 0 | 0 | 0 | 0 |
| 0 | 0 | 0 | 0 | 0 |
| 0.0024987 | 0 | 0 | 0 | 0 |
| 0.003801913 | 0.005995932 | 0.00289266 | 0 | 0 |
| 0 | 0 | 0 | 0 | 0 |
| 0.006400664 | 0 | 0.001617908 | 0 | 0 |
| 0 | 0 | 0 | 0 | 0 |
| 0 | 0 | 2.602933923 | 0 | 0 |
| 0 | 0 | 0 | 0 | 0 |
| 0 | 0 | 0 | 0 | 0 |
| 0 | 0 | 0 | 0 | 0 |
| 0 | 0 | 0 | 0 | 0 |
| 0.002071768 | 0.001196832 | 0 | 0.008399218 | 0 |
| 0 | 0 | 0 | 0 | 0 |
| 0 | 0 | 0 | 0 | 0 |
| 0 | 0 | 0.00343967 | 0 | 0 |
| 0 | 0 | 0 | 0 | 0 |
| 0 | 0 | 0 | 0 | 0 |
| 3.585396179 | 0.464327036 | 12.57080656 | 0 | 0 |
| 1.013570577 | 0 | 1.787136748 | 0 | 0 |
| 0 | 0 | 0 | 0 | 0 |
| 0 | 0 | 0 | 0 | 0 |
| 0 | 0 | 0 | 0 | 0 |
| 0 | 0 | 0 | 0 | 0 |
| 0 | 0 | 0 | 0 | 0 |
| 0 | 0 | 0 | 0 | 0 |

| | | | | |
|---|---|---|---|---|
| 0 | 0 | 0 | 0 | 0 |
| 0 | 0.000646703 | 0.000802269 | 0.001020361 | 0 |
| 0 | 0 | 0 | 0 | 0 |
| 0 | 0 | 0 | 0 | 0 |
| 6.045382511 | 2.638541472 | 3.573910197 | 0.705693962 | 0 |
| 0 | 0 | 0 | 0 | 0 |
| 0 | 0 | 0 | 0 | 0 |
| 0 | 0 | 0 | 0 | 0 |
| 0.394897709 | 1.404504747 | 0.248536873 | 3.218568196 | 0 |
| 0 | 0 | 0 | 0 | 0 |
| 0 | 0.007306395 | 0 | 0.031413677 | 0 |
| 0 | 0 | 0 | 0 | 0 |
| 0 | 0 | 0 | 0 | 0 |
| 0.012344182 | 0 | 0.017250588 | 0 | 0 |
| 0.025272334 | 0 | 0 | 0 | 0 |
| 0.019390846 | 0 | 0 | 0 | 0 |
| 0 | 0 | 0.003731927 | 0 | 0 |
| 0 | 0 | 0 | 0 | 0 |
| 0 | 0 | 0 | 0 | 0 |
| 0 | 0 | 0 | 0 | 0 |
| 0 | 0 | 0 | 0 | 0 |
| 0 | 0 | 0 | 0 | 0 |
| 0 | 0 | 0 | 0 | 0 |
| 0 | 0 | 0 | 0 | 0 |
| 0 | 0 | 0 | 0 | 0 |
| 0 | 0 | 0 | 0 | 0 |
| 0 | 0 | 0 | 0 | 0 |
| 0.001647005 | 0.000562221 | 0.001220564 | 0.000776184 | 0 |
| 0 | 0 | 0 | 0 | 0 |
| 0 | 0 | 0 | 0 | 0 |
| 0 | 0 | 0 | 0 | 0 |
| 0 | 0 | 0 | 0 | 0 |
| 0 | 0 | 0 | 0 | 0 |
| 0 | 0 | 0 | 0 | 0 |
| 0 | 0 | 0 | 0 | 0 |
| 0 | 0 | 0 | 0 | 0 |
| 0 | 0 | 0 | 0 | 0 |
| 0 | 0 | 0 | 0 | 0 |
| 0 | 0 | 0 | 0 | 0 |
| 0 | 0 | 0 | 0 | 0 |
| 0 | 0 | 0 | 0 | 0 |
| 0 | 0 | 0 | 0 | 0.004728665 |
| 0 | 0 | 0 | 0 | 0 |
| 0 | 0 | 0 | 0 | 0 |
| 0 | 0 | 0 | 0 | 0 |
| 0 | 0 | 0 | 0 | 0 |
| 0 | 0 | 0 | 0 | 0 |
| 0 | 0 | 0 | 0 | 0 |

| | | | | |
|---|---|---|---|---|
| 0.004754196 | 0 | 0 | 0 | 0 |
| 0 | 0 | 0 | 0 | 0 |
| 0 | 0 | 0 | 0 | 0 |
| 0 | 0 | 0 | 0 | 0 |
| 0.023369127 | 0 | 0 | 0 | 0 |
| 0 | 0 | 0 | 0 | 0 |
| 0 | 0 | 0.010961733 | 0 | 0 |
| 0 | 0 | 0 | 0 | 0 |
| 0 | 0 | 0 | 0 | 0 |
| 0 | 0 | 0 | 0 | 0 |
| 0 | 0 | 0 | 0 | 0 |
| 0 | 0 | 0 | 0 | 0 |
| 0 | 0 | 0 | 0 | 0 |
| 0 | 0 | 0 | 0 | 0 |
| 0 | 0 | 0 | 0 | 0 |
| 0 | 0 | 0 | 0 | 0 |
| 0 | 8.674249099 | 0 | 0 | 0 |
| 0 | 0 | 0.001193044 | 0 | 0 |
| 0 | 0 | 0 | 0 | 0 |
| 0 | 0 | 0 | 0 | 0 |
| 0.001594231 | 0 | 0.085064716 | 0 | 0 |
| 0 | 0 | 0 | 0 | 0 |
| 0 | 0 | 0.00304066 | 0 | 0 |
| 0.004264405 | 0.001957104 | 0.002124404 | 0.002824725 | 0 |
| 0 | 0 | 0 | 0 | 0 |
| 0 | 0 | 0 | 0 | 0 |
| 0 | 0 | 0 | 0 | 0 |
| 0 | 0 | 0 | 0 | 0 |
| 0 | 0 | 0 | 0 | 0 |
| 0 | 0.008194128 | 0.006392986 | 0.012373085 | 1.563901422 |
| 0 | 0 | 0 | 0 | 0 |
| 0 | 0 | 0 | 0 | 0 |
| 0 | 0 | 0 | 0 | 0 |
| 0 | 0 | 0 | 0 | 0 |
| 0 | 0 | 0 | 0 | 0 |
| 0 | 0 | 0 | 0 | 0 |
| 0.055670965 | 0 | 0.007974537 | 0.00399548 | 0 |
| 0 | 0 | 0 | 0 | 0 |
| 0 | 0 | 0 | 0 | 0 |
| 0 | 0 | 0 | 0 | 0 |
| 0 | 0 | 0 | 0 | 0 |
| 0 | 0 | 0 | 0 | 0 |
| 0 | 0 | 0 | 0 | 0 |
| 0 | 0 | 0 | 0 | 0 |
| 0 | 0 | 0 | 0 | 0 |
| 0 | 0 | 0 | 0 | 0 |
| 0 | 0 | 0 | 0 | 0 |
| 0 | 0 | 0 | 0 | 0 |

| | | | | |
|---|---|---|---|---|
| 0.000458865 | 0.002680253 | 0 | 0.035548991 | 0 |
| 0 | 0 | 0 | 0 | 0 |
| 0 | 0 | 0 | 0 | 0 |
| 0 | 0 | 0 | 0 | 0 |
| 0 | 0 | 0 | 0 | 0 |
| 0 | 0 | 0 | 0 | 0 |
| 0 | 0 | 0 | 0 | 0 |
| 0.056538094 | 0 | 2.067401202 | 0 | 0 |
| 0.086430929 | 0.001352269 | 0.028704931 | 0.504269662 | 9.843866861 |
| 0.014374093 | 0 | 0.00295211 | 0 | 0 |
| 0 | 0 | 0 | 0 | 0 |
| 0 | 0 | 0 | 0 | 0 |
| 0 | 0 | 0 | 0 | 0 |
| 0 | 0 | 0 | 0 | 0.273904378 |
| 0 | 0 | 0 | 0 | 0 |
| 0 | 0 | 0 | 0 | 0 |
| 0 | 0 | 0 | 0 | 0 |
| 0 | 0 | 0 | 0 | 0 |
| 0 | 0 | 0 | 0 | 0 |
| 0 | 0 | 0 | 0 | 0 |
| 0 | 0 | 0 | 0 | 0 |
| 0 | 0 | 0 | 0 | 0 |
| 0 | 0 | 0 | 0 | 0 |
| 0 | 0 | 0 | 0 | 0 |
| 0 | 0 | 0 | 0 | 0 |
| 0 | 0 | 0 | 0 | 0 |
| 0 | 0 | 0 | 0 | 0 |
| 0 | 0 | 0 | 0 | 0 |
| 0 | 0.219728026 | 0 | 0 | 0 |
| 0 | 0 | 0 | 0 | 0 |
| 0 | 0 | 0 | 0 | 0 |
| 0 | 0 | 0 | 0 | 0 |
| 0 | 0 | 0 | 0 | 0 |
| 0 | 0 | 0 | 0 | 0 |
| 0 | 0 | 0 | 0 | 0 |
| 0 | 0 | 0 | 0 | 0 |
| 0 | 0 | 0 | 0 | 0 |
| 0 | 0 | 0 | 0 | 0 |
| 0 | 0 | 0 | 0 | 0 |
| 0.042144382 | 0 | 0.710141102 | 0 | 0 |
| 0 | 0 | 0 | 0 | 0 |
| 0.191561454 | 0 | 0.182570552 | 0 | 0 |
| 0 | 0 | 0 | 0 | 0 |
| 0 | 0 | 0 | 0 | 0 |
| 0 | 0 | 0 | 0 | 0 |
| 0 | 0 | 0 | 0 | 0 |
| 0 | 0 | 0 | 0 | 0 |
| 0 | 0 | 0 | 0 | 0 |

| | | | | |
|---|---|---|---|---|
| 0 | 0 | 0 | 0 | 0 |
| 0 | 0 | 0 | 0.014193524 | 0 |
| 0 | 0 | 0 | 0 | 0 |
| 0 | 0 | 0 | 0 | 0 |
| 0 | 0 | 0.126921563 | 0 | 0.007770264 |
| 0 | 0 | 0 | 0 | 0 |
| 0 | 0 | 0 | 0 | 0 |
| 0.033076882 | 0.052725116 | 0.033308366 | 0.051272758 | 0 |
| 0 | 0 | 0 | 0 | 0 |
| 0 | 0 | 0 | 0 | 0 |
| 0 | 0 | 0 | 0 | 0 |
| 0 | 0 | 0 | 0 | 0 |
| 0 | 0 | 0 | 0 | 0 |
| 0 | 0 | 0 | 0 | 0 |
| 0 | 0 | 0 | 0 | 0 |
| 0 | 0 | 0 | 0 | 0 |
| 0 | 0 | 0 | 0 | 0 |
| 0 | 0 | 0 | 0 | 0 |
| 0 | 0 | 0 | 0 | 0 |
| 0 | 0 | 0 | 0 | 0 |
| 0 | 0 | 0 | 0 | 0 |
| 0 | 0 | 0 | 0 | 0 |
| 0 | 0 | 0 | 0 | 0 |
| 0 | 0.005163587 | 0 | 0.008147054 | 0 |
| 0 | 0 | 0 | 0 | 0 |
| 0 | 0 | 0 | 0 | 0 |
| 0.425673906 | 0.007228601 | 0.019040903 | 0.157277888 | 0 |
| 0 | 0 | 0 | 0 | 0 |
| 0 | 0.000859554 | 0 | 0.001384449 | 0 |
| 0 | 0 | 0 | 0 | 0 |
| 0 | 0 | 0 | 0 | 0 |
| 0 | 0 | 0 | 0 | 0 |
| 0 | 0 | 0 | 0 | 0 |
| 0 | 0 | 0 | 0 | 0 |
| 0 | 0 | 0 | 0 | 0 |
| 0 | 0 | 0 | 0 | 0 |
| 0 | 0 | 0 | 0 | 0 |
| 0 | 0 | 0 | 0 | 0 |
| 0 | 0 | 0 | 0 | 0 |
| 0 | 0 | 0 | 0 | 0 |
| 0 | 0 | 0 | 0 | 0 |
| 0 | 0 | 0 | 0 | 0.000508666 |
| 0 | 0 | 0 | 0 | 0 |
| 0 | 0 | 0 | 0 | 0 |
| 0 | 0 | 0 | 0 | 0 |
| 0 | 0 | 0 | 0 | 0 |
| 0 | 0 | 0 | 0 | 0 |
| 0 | 0 | 0 | 0 | 0 |

| | | | | |
|---|---|---|---|---|
| 0 | 0 | 0.073789812 | 0 | 0 |
| 0 | 0 | 0 | 0 | 0 |
| 0 | 0 | 0 | 0 | 0 |
| 0.002593758 | 0 | 0 | 0 | 0 |
| 0 | 0 | 0 | 0 | 0 |
| 0 | 0 | 0 | 0 | 0 |
| 0 | 0 | 0 | 0 | 0 |
| 0 | 0 | 0 | 0 | 0 |
| 0 | 0 | 0 | 0 | 0 |
| 0 | 0 | 0 | 0 | 0 |
| 0 | 0 | 0 | 0 | 0 |
| 0.00046567 | 0 | 0 | 0 | 0 |
| 0 | 0 | 0 | 0 | 0 |
| 0 | 0 | 0 | 0 | 0 |
| 0 | 0 | 0 | 0 | 0 |
| 0 | 0 | 0 | 0 | 0 |
| 0 | 0 | 0 | 0 | 0 |
| 0 | 0 | 0 | 0 | 0 |
| 0 | 0 | 0.000834141 | 0 | 0 |
| 0 | 0 | 0 | 0 | 0 |
| 0.010899221 | 0.003987675 | 0 | 0 | 0 |
| 0 | 0 | 0 | 0 | 0 |
| 0 | 0 | 0.002014028 | 0 | 0 |
| 0 | 0 | 0 | 0 | 0 |
| 0 | 0 | 0 | 0 | 0 |
| 0 | 0 | 0 | 0 | 0 |
| 0 | 0 | 0 | 0 | 0 |
| 0 | 0 | 0 | 0 | 0 |
| 0 | 0 | 0 | 0 | 0 |
| 0 | 0 | 0 | 0 | 0 |
| 0 | 0 | 0 | 0 | 0 |
| 0 | 0 | 0.240119463 | 0 | 0 |
| 0.034309341 | 0.033386629 | 0.015089035 | 0.007025243 | 0 |
| 0 | 0 | 0.025620641 | 0 | 0 |
| 0 | 0 | 2.956340309 | 0 | 0 |
| 0 | 0.000697173 | 0 | 0 | 0 |
| 0 | 0 | 0 | 0 | 0 |
| 0 | 0 | 0 | 0 | 0 |
| 0.003313341 | 0.000460795 | 0.004251574 | 0 | 0 |
| 0 | 0 | 0 | 0 | 0 |
| 0 | 0 | 0 | 0 | 0 |
| 0 | 0 | 0 | 0 | 0 |
| 0 | 0 | 0 | 0 | 0.002015134 |
| 0 | 0 | 0 | 0 | 0 |
| 0 | 0 | 0 | 0 | 0 |
| 0 | 0 | 0.041803678 | 0 | 0 |
| 0 | 0 | 0 | 0 | 0 |
| 0 | 0 | 0 | 0 | 0 |

| | | | | |
|---|---|---|---|---|
| 0 | 0 | 0 | 0 | 0 |
| 0 | 0 | 0 | 0 | 0 |
| 0 | 0 | 0 | 0 | 0 |
| 0 | 0 | 0 | 0 | 0 |
| 0 | 0 | 0 | 0 | 0 |
| 0 | 0 | 0 | 0 | 0 |
| 0 | 0 | 0 | 0 | 0 |
| 0.665835842 | 0.993520786 | 0 | 0 | 0 |
| 0 | 0 | 0 | 0 | 0 |
| 0 | 0 | 0 | 0 | 0 |
| 0 | 0 | 0 | 0 | 0 |
| 0 | 0 | 0 | 0 | 0 |
| 0 | 0 | 0 | 0 | 0 |
| 0 | 0 | 0 | 0 | 0 |
| 0 | 0 | 0 | 0 | 0 |
| 0 | 0.100712516 | 0.05342527 | 0.004340332 | 0 |
| 0 | 0 | 0 | 0 | 0 |
| 0 | 0 | 0 | 0 | 0 |
| 0 | 0 | 0 | 0 | 0 |
| 0.001595336 | 0 | 0 | 0 | 0 |
| 0 | 0 | 0 | 0 | 0 |
| 0 | 0 | 0 | 0 | 0 |
| 0 | 0 | 0 | 0 | 0 |
| 0 | 0 | 0 | 0 | 0 |
| 0 | 0 | 0 | 0 | 0 |
| 0 | 0 | 0 | 0 | 0 |
| 0 | 0 | 0 | 0 | 0 |
| 0 | 0 | 0 | 0 | 0 |
| 0 | 0 | 0 | 0 | 0 |
| 0 | 0 | 0 | 0 | 0 |
| 0.007462052 | 0 | 8.881539349 | 0 | 0 |
| 0 | 0 | 0 | 0 | 0 |
| 0 | 0 | 0 | 0 | 0 |
| 0 | 0 | 0 | 0 | 0 |
| 0 | 0 | 0 | 0 | 0 |
| 0 | 0 | 0 | 0 | 0 |
| 0 | 0 | 0 | 0 | 0 |
| 0 | 0 | 0 | 0 | 0 |
| 0 | 0 | 0 | 0 | 0 |
| 0 | 0 | 0 | 0 | 0 |
| 0 | 0 | 0 | 0 | 0 |
| 0 | 0 | 0 | 0 | 0 |
| 0 | 0 | 0 | 0 | 0 |
| 0 | 0 | 0 | 0.001918384 | 0 |
| 0 | 0 | 0 | 0 | 0 |
| 0 | 0 | 0 | 0 | 0 |
| 2.554445228 | 0.193212897 | 0 | 0.000919276 | 0 |
| 0 | 0 | 0 | 0 | 0 |

| | | | | |
|---|---|---|---|---|
| 0 | 0 | 0 | 0 | 0 |
| 0 | 0 | 0 | 0 | 0 |
| 0 | 0 | 0 | 0 | 0 |
| 0 | 0 | 0 | 0 | 0 |
| 0 | 0 | 0 | 0 | 0 |
| 0 | 0 | 0 | 0 | 0 |
| 0 | 0 | 0 | 0 | 0 |
| 0 | 0 | 0 | 0 | 0 |
| 0 | 0 | 0 | 0 | 0 |
| 0 | 0 | 0 | 0 | 0 |
| 0 | 0 | 0 | 0 | 0 |
| 0.016185659 | 0.000805749 | 0.001249467 | 0 | 0.000688798 |
| 0.008092857 | 0 | 0 | 0 | 0 |
| 0 | 0 | 0 | 0 | 0 |
| 0 | 0 | 0 | 0 | 0 |
| 0.179216655 | 0 | 0 | 0 | 0 |
| 0 | 0.00185915 | 0 | 0 | 0 |
| 0 | 0.000843718 | 0 | 0 | 0 |
| 0.00111417 | 0.274291508 | 0.198303345 | 0 | 0 |
| 0.004546147 | 0 | 0.001729453 | 0 | 0 |
| 0 | 0 | 0 | 0 | 0 |
| 0 | 0 | 0 | 0 | 0 |
| 0 | 0 | 0 | 0 | 0 |
| 0 | 0 | 0 | 0 | 0 |
| 0 | 0 | 0 | 0 | 0 |
| 0 | 0 | 0 | 0 | 0 |
| 0.0040802 | 0.002918283 | 0 | 0.005791519 | 0 |
| 0 | 0 | 0 | 0 | 0 |
| 0 | 0 | 0 | 0 | 0 |
| 0 | 0 | 0 | 0 | 0 |
| 0 | 0 | 0.001122128 | 0.023405619 | 0 |
| 0 | 0 | 0 | 0 | 0 |
| 0 | 0 | 0 | 0 | 0 |
| 0 | 0 | 0 | 0 | 0 |
| 0 | 0 | 0 | 0 | 0 |
| 0 | 0 | 0 | 0 | 0 |
| 0 | 0 | 0 | 0 | 0 |
| 0.000580264 | 0.008570207 | 0.008514442 | 0 | 0.002716312 |
| 0 | 0 | 0 | 0 | 0 |
| 0 | 0 | 0 | 0 | 0 |
| 0 | 0 | 0 | 0 | 0 |
| 0 | 0 | 0 | 0 | 0 |
| 0 | 0 | 0 | 0 | 0 |
| 0.07468769 | 0.004954173 | 0 | 0 | 0 |
| 0 | 0 | 0 | 0 | 0 |
| 0 | 0 | 0 | 0 | 0 |
| 0 | 0 | 0 | 0 | 0 |
| 0 | 0 | 0 | 0 | 0 |

| | | | | |
|---|---|---|---|---|
| 0 | 0 | 0 | 0 | 0 |
| 0 | 0 | 0 | 0 | 0 |
| 0 | 0 | 0 | 0 | 0 |
| 0.001442821 | 0.002257389 | 0 | 0 | 0 |
| 0 | 0 | 0.000909067 | 0 | 0 |
| 0.001079417 | 0 | 0 | 0 | 0 |
| 0 | 0 | 0 | 0 | 0 |
| 0 | 0 | 0 | 0 | 0 |
| 0 | 0 | 0 | 0 | 0 |
| 0 | 0 | 0 | 0 | 0 |
| 0 | 0 | 0 | 0 | 0 |
| 0 | 0 | 0 | 0 | 0 |
| 0 | 0 | 0.004516094 | 0 | 0 |
| 0 | 0 | 0 | 0 | 0 |
| 0 | 0 | 0 | 0 | 0 |
| 0 | 0 | 0 | 0 | 0 |
| 0 | 0 | 0 | 0 | 0 |
| 0 | 0 | 0 | 0 | 0 |
| 0 | 0 | 0 | 0 | 0 |
| 0 | 0 | 0 | 0 | 0 |
| 0 | 0 | 0 | 0 | 0 |
| 0 | 0 | 0 | 0 | 0 |
| 0.004040206 | 0.021702685 | 0.003888197 | 0.010763045 | 0 |
| 0.002979739 | 0 | 0.019702299 | 0 | 0 |
| 0 | 0 | 0 | 0 | 0 |
| 0 | 0 | 0 | 0 | 0 |
| 0 | 0 | 0 | 0 | 0 |
| 0 | 0 | 0 | 0 | 0 |
| 0 | 0 | 0 | 0 | 0 |
| 0 | 0 | 0 | 0 | 0 |
| 0 | 0 | 0.096907216 | 0 | 0 |
| 0 | 0 | 0.000788831 | 0 | 0 |
| 0 | 0 | 0 | 0 | 0 |
| 0 | 0 | 0 | 0 | 0 |
| 0 | 0 | 0 | 0 | 0 |
| 0 | 0 | 0 | 0 | 0 |
| 0 | 0 | 0.062586912 | 0 | 0 |
| 0 | 0 | 0 | 0 | 0 |
| 0 | 0.00049693 | 0.006257149 | 0.000411627 | 0.006393287 |
| 0 | 0 | 0 | 0 | 0 |
| 0 | 0 | 0 | 0 | 0 |
| 0 | 0 | 0 | 0 | 0 |
| 0 | 0 | 0 | 0 | 0 |
| 0 | 0 | 0 | 0 | 0 |
| 0 | 0 | 0 | 0 | 0 |
| 0.012714927 | 0 | 0.58659465 | 0 | 0 |
| 0 | 0 | 0 | 0.606333345 | 0 |
| 0 | 0 | 0 | 0 | 0 |

| | | | | |
|---|---|---|---|---|
| 0 | 0 | 0 | 0 | 0 |
| 0 | 0 | 0 | 0 | 0 |
| 0 | 0 | 0 | 0 | 0 |
| 0 | 0 | 0 | 0 | 0 |
| 0 | 0 | 0 | 0 | 0 |
| 0 | 0 | 0 | 0 | 0 |
| 0 | 0 | 0 | 0 | 0 |
| 0 | 0 | 0 | 0 | 0 |
| 0 | 0 | 0 | 0 | 0 |
| 0 | 0 | 0 | 0 | 0 |
| 0 | 0 | 0 | 0 | 0 |
| 0 | 0.000601894 | 0 | 0 | 0 |
| 0 | 0 | 0 | 0 | 0 |
| 0 | 0 | 0 | 0 | 0 |
| 0 | 0 | 0 | 0 | 0 |
| 0 | 0 | 0 | 0 | 0 |
| 0 | 0 | 0 | 0 | 0 |
| 0 | 0 | 0 | 0 | 0 |
| 0 | 0 | 0 | 0 | 0 |
| 0 | 0 | 0 | 0 | 0.015516422 |
| 0 | 0 | 0 | 0 | 0 |
| 0 | 0 | 0 | 0 | 0 |
| 0 | 0 | 0 | 0 | 0 |
| 0 | 0 | 0 | 0 | 0 |
| 0 | 0 | 0 | 0 | 0 |
| 0 | 0 | 0 | 0 | 0 |
| 0 | 0 | 0 | 0 | 0 |
| 0 | 0 | 0 | 0 | 0 |
| 0 | 0 | 0 | 0 | 0 |
| 0 | 0 | 0 | 0 | 0 |
| 0 | 0 | 0.004188006 | 0 | 0 |
| 0 | 0 | 0 | 0 | 0 |
| 0.031018628 | 0 | 0.019682365 | 0 | 0 |
| 0.014107207 | 0.004795818 | 0.007572044 | 0.004815232 | 0 |
| 0 | 0 | 0 | 0 | 0 |
| 0 | 0 | 0 | 0 | 0 |
| 0 | 0 | 0 | 0 | 0 |
| 0 | 0 | 0 | 0 | 0 |
| 0 | 0 | 0 | 0 | 0 |
| 0 | 0 | 0 | 0 | 0 |
| 0 | 0 | 0 | 0 | 0 |
| 0.053000469 | 0.031852868 | 0.536638598 | 0.129138673 | 0 |
| 0.003245558 | 0 | 0.040614482 | 0 | 0 |
| 0 | 0 | 0 | 0 | 0 |
| 0 | 0 | 0 | 0 | 0 |
| 0 | 0 | 0.00939052 | 0 | 0 |
| 0 | 0 | 0 | 0 | 0 |
| 0 | 0 | 0 | 0 | 0 |

| | | | | |
|---|---|---|---|---|
| 0 | 0 | 0 | 0 | 0 |
| 0 | 0 | 0.007780399 | 0 | 0 |
| 0.003762698 | 0 | 0.017608635 | 0 | 0 |
| 0 | 0 | 0 | 0 | 0 |
| 0.002002889 | 0 | 0.001814148 | 0 | 0 |
| 0 | 0 | 0.00993308 | 0 | 0 |
| 0 | 0 | 0 | 0 | 0 |
| 0 | 0 | 0 | 0 | 0 |
| 0 | 0 | 0 | 0 | 0 |
| 0 | 0 | 0 | 0 | 0 |
| 0 | 0 | 0 | 0 | 0 |
| 0 | 0 | 0 | 0 | 0 |
| 0 | 0 | 0 | 0 | 0 |
| 0 | 0 | 0 | 0 | 0 |
| 0 | 0 | 0 | 0 | 0 |
| 0 | 0 | 0 | 0 | 0 |
| 0 | 0 | 0 | 0 | 0 |
| 0 | 0 | 0 | 0 | 0 |
| 0 | 0 | 0 | 0 | 0 |
| 0 | 0 | 0 | 0 | 0 |
| 0 | 0 | 0 | 0 | 0 |
| 0 | 0 | 0 | 0.423341872 | 0 |
| 0 | 0 | 0 | 0 | 0 |
| 0 | 0 | 0 | 0 | 0 |
| 0 | 0 | 0 | 0 | 0 |
| 0 | 0 | 0.002590095 | 0 | 0.136619424 |
| 0 | 0 | 0 | 0 | 0 |
| 0 | 0 | 0 | 0 | 0 |
| 0 | 0 | 0 | 0 | 0 |
| 0 | 0 | 0 | 0 | 0 |
| 0 | 0 | 0 | 0 | 0 |
| 0 | 0 | 0 | 0.002888817 | 0 |
| 0 | 0 | 0 | 0 | 0 |
| 0 | 0 | 0 | 0 | 0 |
| 0 | 0 | 0 | 0 | 0 |
| 0 | 0 | 0 | 0 | 0 |
| 0 | 0 | 0 | 0 | 0 |
| 0 | 0.0066937 | 0 | 0 | 0 |
| 2.564632512 | 0 | 0 | 0 | 0 |
| 0 | 0 | 0 | 0 | 0 |
| 0 | 0 | 0 | 0 | 0 |
| 0 | 0 | 0 | 0 | 0 |
| 0 | 0.002612437 | 0 | 0.029604515 | 0 |
| 0 | 0 | 0 | 0 | 0 |
| 0 | 0 | 0 | 0 | 0 |
| 0 | 0 | 0 | 0 | 0 |
| 0.003459073 | 0.002331302 | 0 | 0.002758728 | 0 |
| 0 | 0 | 0 | 0 | 0 |

| | | | | |
|---|---|---|---|---|
| 0 | 0 | 0 | 0 | 0 |
| 0.00604345 | 0.024163748 | 0.005702032 | 0.016534784 | 0 |
| 0 | 0 | 0 | 0 | 0 |
| 0 | 0 | 0 | 0 | 0 |
| 0 | 0 | 0 | 0 | 0 |
| 0 | 0 | 0 | 0 | 0 |
| 0 | 0 | 0 | 0 | 0 |
| 0 | 0 | 0 | 0 | 0 |
| 0 | 0 | 0 | 0 | 0 |
| 0 | 0 | 0 | 0 | 0 |
| 0 | 0 | 0 | 0 | 0 |
| 0 | 0 | 0 | 0 | 0 |
| 0 | 0 | 0 | 0 | 0 |
| 0 | 0 | 0 | 0 | 0 |
| 0.004814201 | 0 | 0 | 0 | 0 |
| 0 | 0 | 0 | 0 | 0 |
| 0 | 0 | 0 | 0 | 0 |
| 0 | 0 | 0 | 0 | 0 |
| 0 | 0 | 0 | 0 | 0 |
| 0 | 0 | 0 | 0 | 0 |
| 0 | 0 | 0 | 0 | 0 |
| 0 | 0 | 0 | 0 | 0 |
| 0 | 0 | 0 | 0 | 0 |
| 0 | 0 | 0 | 0 | 0 |
| 0 | 0 | 0 | 0 | 0 |
| 0 | 0 | 0 | 0 | 0 |
| 0 | 0 | 0 | 0 | 0 |
| 0 | 0 | 0 | 0 | 0 |
| 0 | 0 | 0 | 0 | 0 |
| 0 | 0 | 0 | 0 | 0 |
| 0 | 0 | 0 | 0 | 0 |
| 0 | 0 | 0 | 0 | 0 |
| 0 | 0 | 0 | 0 | 0 |
| 0 | 0 | 0 | 0 | 0 |
| 0 | 14.82272131 | 0 | 0 | 0 |
| 0 | 0 | 0 | 0 | 0 |
| 0 | 0 | 0 | 0 | 0 |
| 0 | 0 | 0 | 0 | 0 |
| 0 | 0 | 0.00195231 | 0 | 0 |
| 0 | 0 | 0 | 0 | 0 |
| 0.265597633 | 0.003455677 | 0.192555418 | 0 | 0 |
| 0 | 0 | 0 | 0 | 0 |
| 0 | 0 | 0 | 0 | 0 |
| 0 | 0 | 0 | 0 | 0 |
| 0 | 0 | 0 | 0 | 0 |
| 1.230273395 | 0.50379623 | 2.129041222 | 0.364148679 | 0.015389219 |
| 0 | 0 | 0 | 0 | 0 |
| 0 | 0 | 0 | 0 | 0 |

| | | | | |
|---|---|---|---|---|
| 0 | 0.001879434 | 0 | 0 | 0 |
| 0 | 0 | 0 | 0 | 0 |
| 0 | 0 | 0 | 0 | 0 |
| 0 | 0 | 0 | 0 | 0 |
| 0.012146842 | 0 | 0 | 0 | 0 |
| 0 | 0 | 0 | 0 | 0 |
| 0 | 0 | 0 | 0 | 0 |
| 0 | 0 | 0 | 0 | 0 |
| 0 | 0 | 0 | 0 | 0 |
| 0 | 0 | 0.005320321 | 0 | 0.00752534 |
| 0 | 0 | 0 | 0 | 0 |
| 0 | 0 | 0 | 0 | 0 |
| 0 | 0.000756418 | 0 | 0 | 0 |
| 0 | 0 | 0 | 0 | 0 |
| 0 | 0 | 0 | 0 | 0 |
| 0 | 0 | 0 | 0 | 0 |
| 0 | 0 | 0 | 0 | 0 |
| 0 | 0 | 0 | 0 | 0 |
| 0 | 0 | 0 | 0 | 0 |
| 0.063472289 | 0.196817855 | 0.082582446 | 0.055769224 | 0.001846537 |
| 0 | 0 | 0 | 0 | 0 |
| 0 | 0 | 0 | 0 | 0 |
| 0 | 0 | 0 | 0 | 0 |
| 0 | 0 | 0 | 0 | 0 |
| 0 | 0 | 0 | 0 | 0 |
| 0 | 0 | 0 | 0 | 0 |
| 0 | 0 | 0 | 0 | 0 |
| 0 | 0 | 0 | 0 | 0 |
| 0 | 0 | 0 | 0 | 0 |
| 0 | 0 | 0.006770889 | 0 | 0 |
| 0 | 0 | 0 | 0 | 0 |
| 0 | 0 | 0 | 0 | 0 |
| 0 | 0 | 0 | 0 | 0 |
| 0 | 0 | 0 | 0 | 0 |
| 0 | 0 | 0 | 0 | 0 |
| 0 | 0 | 0.010090957 | 0 | 0 |
| 0 | 0 | 0 | 0 | 0 |
| 0 | 0 | 0 | 0 | 0 |
| 0 | 0 | 0 | 0 | 0 |
| 0 | 0 | 0 | 0 | 0 |
| 0 | 0 | 0 | 0 | 0 |
| 0 | 0 | 0 | 0 | 0 |
| 0 | 0 | 0 | 0 | 0 |
| 0 | 0 | 0 | 0 | 0 |
| 0.006736215 | 0.000632355 | 0 | 0 | 0 |
| 0 | 0 | 0 | 0 | 0 |
| 0 | 0 | 0 | 0 | 0 |
| 0 | 0 | 0 | 0.006117364 | 0 |

| | | | | |
|---|---|---|---|---|
| 0 | 0 | 0 | 0 | 0 |
| 0 | 0 | 0 | 0 | 0 |
| 0 | 0 | 0 | 0 | 0 |
| 0 | 0 | 0 | 0 | 0 |
| 0 | 0 | 0 | 0 | 0 |
| 0.006286058 | 0.000865476 | 0 | 0.000543112 | 0 |
| 0 | 0 | 0 | 0 | 0 |
| 0 | 0 | 0 | 0 | 0 |
| 0 | 0 | 0 | 0 | 0 |
| 0 | 0 | 0 | 0 | 0 |
| 0 | 0 | 0 | 0 | 0 |
| 0 | 0 | 0 | 0 | 0 |
| 0 | 0 | 0 | 0 | 0 |
| 0 | 0 | 0 | 0 | 0 |
| 0 | 0 | 0 | 0 | 0 |
| 0 | 0 | 0 | 0 | 0 |
| 0 | 0 | 0 | 0 | 0 |
| 0 | 0 | 0 | 0 | 0 |
| 0 | 0 | 0 | 0 | 0 |
| 0 | 0 | 0 | 0 | 0 |
| 0 | 0 | 0 | 0 | 0 |
| 0 | 0 | 0 | 0 | 0 |
| 0 | 0 | 0 | 0 | 0 |
| 0 | 0 | 0 | 0 | 0 |
| 0 | 0 | 0 | 0 | 0 |
| 0 | 0 | 0.004007653 | 0 | 0 |
| 0 | 0 | 0 | 0 | 0 |
| 0 | 0 | 0 | 0 | 0 |
| 0 | 0 | 0 | 0 | 0 |
| 0 | 0 | 0 | 0 | 0 |
| 0 | 0 | 0 | 0 | 0 |
| 0 | 0 | 0 | 0 | 0 |
| 0 | 0 | 0 | 0 | 0 |
| 0 | 0 | 0 | 0 | 0 |
| 0 | 0 | 0 | 0 | 0 |
| 0 | 0 | 0 | 0 | 0 |
| 0 | 0 | 0 | 0 | 0 |
| 0 | 0 | 0 | 0 | 0 |
| 0 | 0 | 2.441529283 | 0 | 0 |
| 0 | 0 | 0 | 0 | 0 |
| 0 | 0 | 0 | 0 | 0 |
| 0 | 0 | 0 | 0 | 0 |
| 0 | 0 | 0 | 0 | 0 |
| 0 | 0 | 0 | 0 | 0 |
| 0 | 0 | 0 | 0 | 0 |
| 0 | 0 | 0 | 0 | 0 |
| 0 | 0 | 0 | 0 | 0 |
| 0 | 0 | 0 | 0 | 0 |

| | | | | |
|---|---|---|---|---|
| 0 | 0 | 0 | 0 | 0 |
| 0 | 0 | 0 | 0 | 0 |
| 0 | 0 | 0 | 0 | 0 |
| 0 | 0 | 0 | 0 | 0 |
| 0 | 0 | 0 | 0 | 0 |
| 0 | 0 | 0 | 0 | 0 |
| 0 | 0 | 0 | 0 | 0 |
| 0 | 0 | 0 | 0 | 0 |
| 0 | 0 | 0 | 0 | 0 |
| 0 | 0 | 0 | 0 | 0 |
| 0 | 0 | 0 | 0 | 0 |
| 0 | 0 | 0 | 0 | 0 |
| 0 | 0 | 0.513109226 | 0 | 0 |
| 0 | 0 | 0 | 0 | 0 |
| 0 | 0 | 0 | 0 | 0 |
| 0 | 0 | 0 | 0 | 0 |
| 0 | 0 | 0 | 0 | 0 |
| 0 | 0 | 0 | 0 | 0 |
| 0 | 0 | 0 | 0 | 0 |
| 0 | 0 | 0 | 0 | 0 |
| 0 | 0 | 0 | 0 | 0.003327525 |
| 0 | 0 | 0 | 0 | 0 |
| 0 | 0 | 0 | 0 | 0 |
| 0.156510875 | 0 | 3.727994654 | 0 | 0 |
| 0 | 0 | 0 | 0 | 0 |
| 0 | 0 | 0 | 0 | 0 |
| 0 | 0 | 0 | 0 | 0 |
| 0 | 0 | 0 | 0 | 0 |
| 0 | 0 | 0 | 0 | 0 |
| 0 | 0 | 0 | 0 | 0 |
| 0 | 0.028610581 | 0 | 0 | 0 |
| 0 | 0 | 0 | 0 | 0 |
| 0 | 0 | 0 | 0 | 0 |
| 0 | 0 | 0 | 0 | 0 |

| PHL018-0211_V2 | PHL018-0157_V1 | PHL018-0082_V2 | PHL018-0089_V3 | PHL018-0274_V1 |
|---|---|---|---|---|
| REF | REF | REF | CTRL | REF |
| 0.106243322 | 0.069456618 | 0.018862176 | 0.336464967 | 0.033148849 |
| 0.476661528 | 0.012683854 | 1.234803601 | 2.627953338 | 14.04387155 |
| 5.148650645 | 0 | 0 | 4.195966454 | 18.26192203 |
| 0 | 0 | 0 | 0 | 0 |
| 0 | 0 | 0 | 0 | 0 |
| 0 | 0 | 0 | 0 | 0 |
| 0 | 0 | 0 | 0 | 0 |
| 0 | 0 | 0 | 0 | 0 |
| 0 | 0 | 0 | 0 | 0 |
| 0 | 0 | 0 | 0 | 0 |
| 0 | 0 | 0 | 0 | 0 |
| 0 | 0 | 0 | 0 | 0 |
| 0 | 0 | 0 | 0 | 0 |
| 0.212805261 | 0.207136507 | 0.54399231 | 0.301522854 | 6.779858823 |
| 0 | 0 | 0 | 0 | 0 |
| 0 | 0 | 0 | 0 | 0 |
| 0 | 0 | 0 | 0 | 0 |
| 0 | 0 | 0 | 0 | 0 |
| 0 | 0 | 0 | 0 | 0 |
| 0 | 0 | 0 | 0 | 0 |
| 0 | 0 | 0 | 0 | 0 |
| 0 | 0 | 0 | 0 | 0 |
| 0 | 0 | 0 | 0 | 0 |
| 0 | 0 | 0 | 0 | 0 |
| 0 | 0.057388377 | 0.040995758 | 0 | 0 |
| 0 | 0 | 0 | 0 | 0 |
| 0 | 0 | 0 | 0 | 0 |
| 0 | 0 | 0 | 0 | 0 |
| 0 | 0 | 0 | 0 | 0 |
| 0 | 0 | 0 | 0 | 0 |
| 0 | 0 | 0 | 0 | 0 |
| 0 | 0 | 0 | 0 | 0 |
| 0 | 0 | 0 | 0 | 0 |
| 0 | 0 | 0 | 0 | 0 |
| 0 | 0 | 0 | 0 | 0 |
| 0 | 0.021044521 | 1.111109069 | 3.875008341 | 0 |
| 0 | 0 | 0 | 0 | 0 |
| 0 | 0 | 0 | 0 | 0 |
| 0 | 0 | 0 | 0 | 0 |
| 0 | 0 | 0 | 0 | 0 |
| 0 | 0 | 0 | 0 | 0 |
| 0.302379957 | 0 | 0 | 0.001742931 | 0 |
| 0.370993356 | 0.666070985 | 8.600510871 | 0.155726595 | 0.035420203 |
| 0.030649537 | 0 | 0.000813233 | 0.011146095 | 0.014948461 |
| 0 | 0 | 0.014853249 | 0 | 0 |
| 0 | 0 | 0 | 0 | 0 |

| | | | | |
|---|---|---|---|---|
| 0 | 0.077472603 | 0.02254292 | 0 | 0.004375403 |
| 0.000582331 | 0 | 0.00069702 | 0.014146207 | 0.012966643 |
| 0.009950845 | 0.00160459 | 0 | 0.027646486 | 0.002680321 |
| 0.590648252 | 0.090312099 | 0.404225136 | 0.718656865 | 0.642211694 |
| 0.029367009 | 0 | 0.014344603 | 0.00051093 | 0.001416687 |
| 0 | 0 | 0 | 0.033543278 | 0 |
| 0.002288607 | 0 | 0.000747093 | 0.00118149 | 0 |
| 0 | 0 | 0 | 0 | 0 |
| 0 | 0 | 0 | 0 | 0 |
| 0 | 0 | 0 | 0 | 0 |
| 0 | 0 | 0 | 0 | 0 |
| 0.937508305 | 0.088391283 | 0.241081523 | 2.653175109 | 0.153254339 |
| 0 | 0 | 0 | 0 | 0 |
| 0 | 0 | 0.094301544 | 0.054999478 | 0.006398626 |
| 0 | 0 | 0 | 0 | 0 |
| 0 | 0 | 0 | 0 | 0 |
| 0.004698134 | 0.928606827 | 0.357209129 | 0 | 0 |
| 0 | 0 | 0 | 0 | 0 |
| 0 | 0 | 0 | 0 | 0 |
| 0 | 0 | 0 | 0 | 0 |
| 0 | 0.000899497 | 0 | 0.011027425 | 0.865644037 |
| 0.017443943 | 0 | 0.007962508 | 0.040575223 | 0.022924322 |
| 0 | 0 | 0 | 0 | 0.016614102 |
| 0 | 0 | 0 | 0 | 0 |
| 0 | 0 | 0 | 0 | 0 |
| 0 | 0 | 0 | 0 | 0 |
| 0 | 0 | 0 | 0 | 0 |
| 0 | 0 | 0 | 0 | 0 |
| 0.06505637 | 0 | 0 | 0 | 0 |
| 0 | 0 | 0 | 0 | 0 |
| 0 | 0 | 0 | 0 | 0 |
| 0 | 0 | 0 | 0 | 0 |
| 0 | 0 | 0 | 0 | 0 |
| 0 | 0 | 0 | 0 | 0 |
| 0 | 0 | 0 | 0 | 0 |
| 0 | 0 | 0 | 0 | 0 |
| 0 | 0 | 0 | 0 | 0 |
| 0.000896297 | 52.95959617 | 17.13523936 | 0 | 0 |
| 0 | 0 | 0 | 0 | 0 |
| 0 | 0 | 0 | 0 | 0 |
| 0 | 0 | 0 | 0 | 0 |
| 0 | 0 | 0 | 0 | 0 |
| 0 | 0 | 0 | 0 | 0 |
| 0 | 0 | 0 | 0 | 0 |
| 0 | 0.014306319 | 0 | 0.470308911 | 0 |
| 0 | 0 | 0 | 0 | 0 |
| 0 | 0 | 0 | 0 | 0 |
| 0 | 0 | 0 | 0.353962904 | 0 |

| | | | | |
|---|---|---|---|---|
| 0 | 0 | 0 | 0 | 0 |
| 0 | 0 | 0 | 0 | 0 |
| 0 | 0 | 0 | 0 | 0 |
| 0 | 0 | 0 | 0 | 0 |
| 0.002187499 | 0 | 0 | 0 | 0 |
| 0 | 0 | 0 | 0 | 0 |
| 0 | 0 | 0 | 0 | 0 |
| 0 | 0 | 0 | 0 | 0.001275029 |
| 0.032435872 | 0 | 0 | 0 | 0 |
| 0 | 0 | 0 | 0 | 0 |
| 0 | 0 | 0 | 0 | 0 |
| 0.001493025 | 0 | 0 | 0 | 0 |
| 0 | 0 | 0 | 0 | 0 |
| 0 | 0 | 0 | 0 | 0 |
| 0 | 0 | 0 | 0 | 0 |
| 0 | 0 | 0 | 0.000728136 | 0 |
| 0 | 0 | 0 | 0 | 0 |
| 0 | 0 | 0 | 0 | 0 |
| 0.120323192 | 0.098048809 | 0 | 0.036190265 | 0 |
| 0 | 0 | 0 | 0 | 0 |
| 0 | 0 | 0 | 0 | 0 |
| 0 | 0 | 0 | 0 | 0 |
| 0 | 0 | 0 | 0 | 0 |
| 0 | 0 | 0 | 0 | 0 |
| 0 | 0 | 0 | 0 | 0 |
| 0 | 0 | 0 | 0 | 0 |
| 0 | 0 | 0 | 0.001585436 | 0 |
| 0 | 0 | 0 | 0 | 0 |
| 0 | 0 | 0 | 0 | 0 |
| 0 | 0 | 0 | 0 | 0 |
| 0.004098993 | 0.018601561 | 0.061368014 | 0.023902821 | 0.002839086 |
| 0.011236009 | 0 | 0.002241484 | 0.004857682 | 0.000496408 |
| 0 | 0 | 0 | 0 | 0 |
| 0 | 0 | 0 | 0 | 0 |
| 0 | 0 | 0 | 0 | 0 |
| 0 | 0 | 0 | 0 | 0 |
| 0 | 0 | 0 | 0 | 0 |
| 0 | 0 | 0 | 0 | 0 |
| 0.001951124 | 0 | 0.003025393 | 0 | 0 |
| 0 | 0 | 0 | 0 | 0 |
| 0 | 0 | 0 | 0 | 0 |
| 0 | 0 | 0 | 0 | 0 |
| 0.001443006 | 0 | 0 | 0 | 0.009371564 |
| 0 | 0 | 0 | 0 | 0 |
| 0 | 0 | 0 | 0 | 0 |
| 0 | 0 | 0 | 0 | 0 |
| 0 | 0 | 0 | 0 | 0 |
| 0 | 0 | 0 | 0 | 0 |

| | | | | |
|---|---|---|---|---|
| 0 | 0 | 0 | 0 | 0 |
| 0 | 0 | 0 | 0 | 0 |
| 0 | 0 | 0 | 0 | 0 |
| 0 | 0 | 0 | 0 | 0 |
| 0.018834929 | 0.010001497 | 0.325701576 | 0 | 0 |
| 0 | 0 | 0 | 0 | 0 |
| 0 | 0 | 0 | 0 | 0 |
| 0 | 0 | 0 | 0 | 0 |
| 0 | 0 | 0 | 0 | 0 |
| 0 | 0 | 0 | 0 | 0 |
| 0 | 0 | 0 | 0 | 0 |
| 0 | 0 | 0 | 0 | 0 |
| 0 | 0 | 0 | 0 | 0 |
| 0 | 0 | 0 | 0 | 0 |
| 0 | 0 | 0 | 0 | 0 |
| 14.51378804 | 0 | 0 | 0 | 0 |
| 0 | 0 | 0 | 0 | 0 |
| 0 | 0 | 0 | 0 | 0 |
| 0 | 0 | 0 | 0 | 0 |
| 0 | 0 | 0 | 0 | 0 |
| 0 | 0 | 0 | 0 | 0 |
| 0 | 0 | 0 | 0 | 0 |
| 0 | 0 | 0 | 0 | 0 |
| 0 | 0 | 0 | 0 | 0 |
| 0 | 0.003833079 | 0.004234481 | 0 | 0 |
| 0 | 0 | 0 | 0 | 0 |
| 0.001958878 | 0 | 0.018396655 | 0 | 0 |
| 0 | 0 | 0 | 0 | 0 |
| 0 | 0 | 0 | 0 | 0 |
| 0 | 0 | 0 | 0 | 0 |
| 0 | 0 | 0 | 0 | 0 |
| 0.000640194 | 0 | 0 | 0 | 0 |
| 0 | 0 | 0 | 0 | 0 |
| 0 | 0 | 0 | 0 | 0 |
| 0 | 0 | 0 | 0 | 0 |
| 0 | 0 | 0 | 0 | 0 |
| 0 | 0 | 0 | 0 | 0 |
| 0 | 0 | 0 | 0 | 0 |
| 0 | 0 | 0 | 0 | 0 |
| 0 | 0 | 0 | 0 | 0 |
| 0 | 0 | 0 | 0 | 0 |
| 0 | 0 | 0 | 0 | 0 |
| 0 | 0 | 0 | 0 | 0 |
| 0 | 0 | 0 | 0 | 0 |
| 0 | 0 | 0 | 0 | 0 |
| 0 | 0 | 0 | 0 | 0 |
| 0 | 0 | 0 | 0 | 0 |
| 0 | 0 | 0 | 0 | 0 |

| | | | | |
|---|---|---|---|---|
| 0 | 0 | 0 | 0 | 0 |
| 0 | 0 | 0 | 0 | 0 |
| 0 | 0 | 0 | 0 | 0 |
| 0 | 0 | 0 | 0 | 0 |
| 0 | 0 | 0 | 0 | 0 |
| 0 | 0 | 0 | 0 | 0 |
| 0 | 0 | 0 | 0 | 0 |
| 0 | 0 | 0 | 0 | 0 |
| 0 | 0 | 0 | 0.015254336 | 0 |
| 0 | 0 | 0 | 0 | 0 |
| 0 | 0 | 0 | 0 | 0 |
| 0 | 0 | 0 | 0 | 0 |
| 0.000765754 | 0.003499748 | 0.000962395 | 0.000579802 | 0 |
| 0 | 0 | 0 | 0 | 0 |
| 0 | 0 | 0 | 0 | 0 |
| 0 | 0 | 0 | 0 | 0 |
| 0 | 0 | 0 | 0 | 0 |
| 0 | 0 | 0 | 0 | 0 |
| 0 | 0 | 0 | 0 | 0 |
| 0 | 0 | 0 | 0 | 0 |
| 0.083042975 | 0.001635922 | 0.01902272 | 0.002439202 | 0.061143239 |
| 0 | 0 | 0 | 0 | 0 |
| 0 | 0 | 0 | 0 | 0 |
| 0.000895832 | 0 | 0 | 0 | 0 |
| 0 | 0 | 0 | 0 | 0 |
| 0 | 0 | 0 | 0 | 0 |
| 0.013341316 | 0 | 0.001646787 | 0.000631348 | 0.013328277 |
| 0 | 0 | 0 | 0 | 0 |
| 0 | 0.001146898 | 0 | 0 | 0 |
| 0 | 0 | 0 | 0 | 0 |
| 0 | 0 | 0 | 0 | 0 |
| 0 | 0 | 0 | 0 | 0 |
| 0 | 0 | 0 | 0 | 0 |
| 0.017150239 | 0.005748699 | 0.006755361 | 0 | 0 |
| 0.006893762 | 0.005322089 | 0.005017782 | 0.007230006 | 0.004667285 |
| 0 | 0 | 0 | 0 | 0 |
| 0 | 0 | 0 | 0 | 0 |
| 0 | 0 | 0 | 0 | 0 |
| 0.054358114 | 0 | 0 | 0 | 0 |
| 0 | 0 | 0 | 0 | 0 |
| 0 | 0 | 0 | 0 | 0 |
| 0 | 0 | 0 | 0 | 0 |
| 0 | 0 | 0 | 0 | 0 |
| 0.001672287 | 0 | 0 | 0 | 0 |
| 0 | 0 | 0 | 0 | 0 |
| 0 | 0.001773523 | 0 | 0.083276625 | 0 |
| 0 | 0 | 0.001267682 | 0 | 0 |
| 0.002257178 | 0.001232616 | 0 | 0.003644308 | 0.001425443 |

| | | | | |
|---|---|---|---|---|
| 0 | 0 | 0 | 0 | 0 |
| 0 | 0 | 0 | 0 | 0 |
| 0.000970617 | 0 | 0 | 0 | 0 |
| 0 | 0 | 0 | 0 | 0 |
| 0 | 0 | 0 | 0 | 0 |
| 0 | 0 | 0 | 0 | 0 |
| 0.115481298 | 0 | 0 | 0 | 0 |
| 0.003626468 | 0 | 0 | 0 | 0 |
| 0 | 0 | 0 | 0 | 0.003433135 |
| 0 | 0 | 0 | 0 | 0 |
| 0 | 0 | 0 | 0 | 0 |
| 0 | 0 | 0 | 0 | 0 |
| 0 | 0 | 0 | 0 | 0 |
| 0 | 0 | 0 | 0 | 0 |
| 0 | 0 | 0 | 0 | 0 |
| 0 | 0 | 0 | 0 | 0 |
| 0 | 0 | 0 | 0 | 0 |
| 0 | 0 | 0.06122524 | 0 | 0 |
| 0.001350315 | 0.002603553 | 0.038911323 | 0.000766808 | 0 |
| 0.064990736 | 0 | 0 | 0.07981765 | 0 |
| 0 | 0 | 0 | 0.011688234 | 0 |
| 0 | 0 | 0 | 0 | 0 |
| 0 | 0 | 0 | 0 | 0 |
| 0 | 0 | 0 | 0 | 0 |
| 0.013058872 | 0 | 0 | 0.001123604 | 0.025490362 |
| 0 | 0 | 0 | 0 | 0 |
| 0 | 0 | 0 | 0 | 0 |
| 0 | 0 | 0 | 0 | 0 |
| 0.03040039 | 0 | 0 | 0 | 0 |
| 0 | 0 | 0 | 0 | 0 |
| 0.079674653 | 0.003869819 | 0.128323767 | 0.01899075 | 0.029176169 |
| 0 | 0 | 0 | 0 | 0 |
| 0 | 0 | 0.025993981 | 0.003619589 | 0 |
| 0 | 0 | 0 | 0 | 0 |
| 0 | 0 | 0 | 0 | 0 |
| 0 | 0 | 0 | 0 | 0 |
| 0 | 0 | 0 | 0 | 0 |
| 0 | 0 | 0 | 0 | 0 |
| 0 | 0 | 0 | 0 | 0 |
| 0.000585176 | 0 | 0 | 0.000969225 | 0 |
| 0 | 0 | 0 | 0 | 0 |
| 0.007912497 | 0.002985131 | 0.010185615 | 0.003956362 | 0.001958939 |
| 0 | 0 | 0 | 0 | 0 |
| 0 | 0 | 0 | 0 | 0 |
| 0 | 0 | 0 | 0 | 0 |
| 0 | 0 | 0 | 0 | 0 |
| 0 | 0 | 0 | 0 | 0 |
| 0 | 0 | 0 | 0 | 0 |

| | | | | |
|---|---|---|---|---|
| 0 | 0 | 0 | 0 | 0 |
| 0 | 0 | 0 | 0 | 0 |
| 0.032430781 | 0 | 0.002356 | 0 | 0.012522465 |
| 0 | 0 | 0 | 0 | 0 |
| 0 | 0 | 0 | 0.000923839 | 0 |
| 0 | 0 | 0 | 0 | 0 |
| 0 | 0 | 0 | 0 | 0 |
| 0 | 0 | 0 | 0 | 0 |
| 0.001053746 | 0 | 0 | 0 | 0 |
| 0.002400308 | 0 | 0 | 0 | 0 |
| 0 | 0 | 0 | 0 | 0 |
| 0 | 0 | 0 | 0 | 0 |
| 0 | 0 | 0 | 0 | 0 |
| 0.004557926 | 0 | 0 | 0 | 0.002224219 |
| 0.003761971 | 0 | 0 | 0.000971056 | 0.003059669 |
| 0.198272023 | 0.012812288 | 0.001479148 | 0 | 0 |
| 0.004091591 | 0 | 0 | 0 | 0.003253804 |
| 0 | 0 | 0 | 0 | 0 |
| 0 | 0 | 0 | 0 | 0 |
| 0 | 0 | 0 | 0 | 0 |
| 0 | 0 | 0 | 0 | 0 |
| 0 | 0.001130404 | 0 | 0.002663444 | 0 |
| 0 | 0 | 0 | 0 | 0 |
| 0 | 0 | 0 | 0 | 0 |
| 0 | 0 | 0 | 0 | 0 |
| 0 | 0 | 0 | 0 | 0 |
| 0 | 0 | 0 | 0 | 0 |
| 0 | 0 | 0 | 0 | 0 |
| 0 | 0 | 0 | 0 | 0 |
| 0 | 0 | 0 | 0 | 0 |
| 0 | 0 | 0 | 0 | 0 |
| 0 | 0 | 0 | 0 | 0 |
| 0 | 0 | 0 | 0 | 0 |
| 0 | 0 | 0 | 0 | 0 |
| 0 | 0 | 0 | 0 | 0 |
| 0.284905021 | 0 | 0 | 0 | 0 |
| 0 | 0 | 0 | 0 | 0 |
| 0.415439637 | 0 | 0 | 0.010875612 | 0 |
| 0 | 0 | 0 | 0 | 0 |
| 0 | 0 | 0 | 0 | 0 |
| 0 | 0 | 0 | 0 | 0 |
| 0 | 0 | 0 | 0 | 0 |
| 0 | 0 | 0 | 0 | 0 |
| 0 | 0 | 0 | 0 | 0 |
| 0 | 0 | 0 | 0 | 0 |
| 0 | 0 | 0 | 0 | 0 |
| 0 | 0 | 0 | 0 | 0 |
| 0 | 0 | 0 | 0 | 0 |

| | | | | |
|---|---|---|---|---|
| 0 | 0 | 0 | 0.381109275 | 0 |
| 0 | 0 | 0 | 0 | 0 |
| 0.002763353 | 0 | 0 | 0.001046156 | 0 |
| 0 | 0 | 0 | 0 | 0 |
| 0.004633666 | 0 | 0 | 0.00298218 | 0.002432024 |
| 0 | 0 | 0 | 0 | 0 |
| 0 | 0 | 0 | 0 | 0 |
| 0.008053927 | 0 | 0.009329148 | 0.005901433 | 0.010227051 |
| 0 | 0 | 0 | 0 | 0 |
| 0.034812142 | 0.001529837 | 0.028847265 | 0.450122399 | 0.353930233 |
| 0 | 0 | 0 | 0 | 0 |
| 0.065400469 | 0 | 0.010708231 | 0.000583949 | 0.003189243 |
| 0.004836056 | 0 | 0 | 0.121072015 | 0.012840125 |
| 0.01634793 | 0 | 0 | 0 | 0.017522492 |
| 0 | 0 | 0 | 0.00045556 | 0 |
| 0 | 0 | 0 | 0 | 0 |
| 0.0005109 | 0 | 0 | 0 | 0 |
| 0 | 0 | 0 | 0 | 0 |
| 0 | 0 | 0 | 0 | 0 |
| 0 | 0 | 0 | 0 | 0 |
| 0 | 0 | 0 | 0 | 0 |
| 0 | 0 | 0 | 0 | 0 |
| 0 | 0 | 0 | 0 | 0 |
| 0.076483285 | 0.023698569 | 0 | 0 | 0 |
| 0 | 0 | 0 | 0 | 0 |
| 0 | 0 | 0.00489348 | 0 | 0.003467936 |
| 0 | 1.345919884 | 4.79229562 | 0 | 0 |
| 0 | 0 | 0 | 0 | 0 |
| 0 | 0 | 0 | 0 | 0 |
| 0 | 0 | 0.005034775 | 0 | 0 |
| 0 | 0 | 0 | 0 | 0 |
| 0 | 0 | 0 | 0 | 0 |
| 0 | 0 | 0 | 0 | 0 |
| 0 | 0 | 0 | 0.001349456 | 0 |
| 0 | 0 | 0 | 0 | 0 |
| 0 | 0 | 0 | 0 | 0 |
| 0 | 0 | 0 | 0 | 0 |
| 0 | 0 | 0 | 0 | 0 |
| 0 | 0 | 0 | 0 | 0 |
| 0 | 0 | 0 | 0 | 0.14065211 |
| 0 | 0 | 0 | 0 | 0 |
| 0 | 0 | 0 | 0 | 0 |
| 0 | 0 | 0 | 0 | 0 |
| 0 | 0 | 0 | 0.010806719 | 0.127820517 |
| 0 | 0 | 0 | 0 | 0 |
| 0 | 0 | 0.00167079 | 0.003774673 | 0 |
| 0.001581729 | 0.003227242 | 0.01785059 | 0 | 0 |
| 0 | 0 | 0 | 0 | 0 |

| | | | | |
|---|---|---|---|---|
| 0 | 0 | 0 | 0 | 0 |
| 0 | 0 | 0.001813571 | 0.00119503 | 0 |
| 0 | 0 | 0 | 0 | 0 |
| 0 | 0.00416795 | 0 | 0 | 0 |
| 0 | 0 | 0 | 0 | 0 |
| 0.005695099 | 0 | 0 | 0.027624561 | 0.330757138 |
| 0.014248432 | 0 | 0 | 0 | 0 |
| 0 | 0 | 0.000619917 | 0 | 0 |
| 0 | 0 | 0 | 0 | 0 |
| 0 | 0 | 0 | 0 | 0 |
| 0.041696306 | 0 | 0 | 0.002735108 | 0.001231138 |
| 0 | 0 | 0 | 0 | 0 |
| 0 | 0 | 0 | 0 | 0 |
| 0.012675828 | 0.001253352 | 0 | 0 | 0.253094315 |
| 0 | 0 | 0 | 0.005602578 | 0 |
| 0 | 0 | 0 | 0 | 0 |
| 0 | 0 | 0 | 0 | 0 |
| 0 | 0 | 0 | 0 | 0 |
| 0 | 0 | 0 | 0 | 0 |
| 0.032625251 | 0 | 0 | 0 | 0 |
| 0 | 0 | 0.001131751 | 0 | 0 |
| 0.6374165 | 0.045188169 | 0.080359503 | 0 | 0.002893066 |
| 0 | 0 | 0 | 0 | 0 |
| 0 | 0 | 0 | 0 | 0 |
| 0 | 0 | 0 | 0.00823037 | 0 |
| 0 | 0 | 0 | 0 | 0 |
| 0 | 0 | 0 | 0 | 0 |
| 0 | 0 | 0 | 0 | 0 |
| 0 | 0 | 0 | 0 | 0 |
| 0.569408887 | 0.062054907 | 0.076982526 | 0.073587505 | 0.821414211 |
| 0.000529203 | 0.022013319 | 0.016033625 | 0 | 0.000789083 |
| 0 | 0 | 0 | 0 | 0 |
| 0 | 0 | 0 | 0 | 0 |
| 0 | 0 | 0 | 0 | 0 |
| 0 | 0 | 0 | 0 | 0 |
| 0 | 0 | 0 | 0 | 0 |
| 1.205490862 | 0.06103154 | 0.51811129 | 0 | 0 |
| 0 | 0 | 0 | 0 | 0 |
| 0 | 0 | 0 | 0.005514895 | 0 |
| 0 | 0 | 0 | 0 | 0 |
| 0 | 0 | 0 | 0.037782606 | 0 |
| 1.359806332 | 0.57641121 | 0.010892579 | 0.118494495 | 0.661358042 |
| 0 | 0 | 0 | 0 | 0 |
| 0 | 0 | 0 | 0 | 0 |
| 0 | 0 | 0 | 0 | 0.001510115 |
| 0 | 0 | 0 | 0 | 0 |
| 0.001009615 | 0 | 0 | 0.000637037 | 0.003412274 |
| 0 | 0 | 0 | 0 | 0 |

| | | | | |
|---|---|---|---|---|
| 0 | 0 | 0.001677409 | 0 | 0 |
| 0.007561643 | 0 | 0 | 0 | 0 |
| 0 | 0.004040351 | 0.082535506 | 0.004016178 | 0 |
| 0 | 0 | 0 | 0 | 0 |
| 0 | 0 | 0 | 0 | 0 |
| 0 | 0 | 0 | 0 | 0 |
| 0 | 0 | 0 | 0 | 0 |
| 0 | 0 | 0 | 0 | 0 |
| 0 | 0 | 0.007643135 | 0.06101177 | 0 |
| 0 | 0 | 0 | 0 | 0.662903023 |
| 0 | 0 | 0 | 0 | 0 |
| 0 | 0 | 0 | 0 | 0 |
| 0 | 0 | 0 | 0 | 0 |
| 0.184631804 | 0 | 0 | 0 | 0 |
| 0 | 0 | 0 | 0 | 0 |
| 0.038016502 | 0.008909615 | 0.004875398 | 0.093217012 | 0 |
| 0.005206156 | 0.013244627 | 0.781954712 | 0.001693792 | 0.000776279 |
| 0 | 0 | 0 | 0 | 0 |
| 0 | 0 | 0 | 0 | 0 |
| 0 | 0 | 0 | 0 | 0 |
| 0.002255426 | 0 | 0 | 0 | 0.256567614 |
| 0.001509718 | 0 | 0.02558107 | 0 | 0.000675332 |
| 0 | 0 | 0 | 0 | 0 |
| 0 | 0 | 0.017089476 | 0 | 0 |
| 0 | 0 | 0.022522231 | 0.020579166 | 0 |
| 0 | 0 | 0 | 0 | 0 |
| 0 | 0 | 0 | 0.008014692 | 0 |
| 0 | 0 | 0 | 0 | 0 |
| 0 | 0 | 0 | 0 | 0 |
| 0 | 0 | 0 | 0 | 0 |
| 0.010838006 | 0.001233845 | 0.006486257 | 0.017244816 | 0.19148806 |
| 0.010537916 | 0 | 0 | 0.002438008 | 0.010894249 |
| 0.31795111 | 0.006368959 | 0.126901301 | 0.020047706 | 0.209583719 |
| 0.12174231 | 0 | 0 | 0 | 0 |
| 0 | 0 | 0.001842632 | 0 | 0.276675976 |
| 0 | 0 | 0 | 0 | 0 |
| 0 | 0 | 0 | 0 | 0.052178622 |
| 0.022227672 | 0 | 0.015142397 | 0.029542029 | 0.03760931 |
| 0 | 0.010453586 | 0.073275097 | 0 | 0 |
| 0 | 0 | 0 | 0.00060994 | 0 |
| 0 | 0 | 0 | 0 | 0 |
| 0 | 0 | 0 | 0 | 0 |
| 0 | 0 | 0 | 0.001785053 | 0 |
| 0.059437903 | 0 | 0 | 0 | 0.114205349 |
| 0 | 0 | 0 | 0 | 0.183559326 |
| 0 | 0 | 0 | 0 | 0 |
| 0 | 0 | 0 | 0 | 0 |
| 0 | 0 | 0 | 0 | 0 |

| | | | | |
|---|---|---|---|---|
| 0 | 0 | 0 | 0 | 0 |
| 0.515755618 | 0.010431058 | 0 | 0.288085847 | 0.019986508 |
| 0 | 0 | 0 | 0 | 0 |
| 0 | 0 | 0 | 0 | 0 |
| 0 | 0 | 0 | 0.000797814 | 0 |
| 0 | 0 | 0 | 0 | 0 |
| 0 | 0 | 0 | 0 | 0 |
| 0 | 0 | 0 | 0 | 0 |
| 0 | 0.002106347 | 0 | 0 | 0 |
| 0 | 0 | 0 | 0.001181913 | 0.001489621 |
| 0.005427295 | 0 | 0.007308205 | 0.001057555 | 0 |
| 0.019496971 | 0 | 0.001494126 | 0.003600589 | 1.060755789 |
| 0 | 0.013908992 | 0 | 0 | 0 |
| 0 | 0 | 0 | 0 | 0 |
| 0.000732605 | 0 | 0.004768084 | 0 | 0.19116503 |
| 0 | 0 | 0 | 0 | 0 |
| 0.029084414 | 0.003342233 | 0 | 0.007593685 | 0.034095549 |
| 0.000650958 | 0 | 0 | 0.015607949 | 0.061861492 |
| 0 | 0 | 0 | 0 | 0 |
| 0 | 0 | 0 | 0.005600914 | 0.057145066 |
| 0.010514917 | 0.021172497 | 0.012688955 | 0.019414367 | 0.001850585 |
| 0 | 0 | 0.007509577 | 0.002639116 | 0 |
| 0 | 0 | 0 | 0 | 0 |
| 0 | 0 | 0 | 0 | 0 |
| 0 | 0 | 0 | 0 | 0 |
| 0 | 0 | 0 | 0 | 0 |
| 0.036070403 | 0 | 0.002902049 | 0.07262597 | 0.137402397 |
| 0.035542721 | 0.014391606 | 0.024341462 | 0.028264208 | 0.312518323 |
| 0 | 0 | 0 | 0 | 0 |
| 0 | 0 | 0 | 0 | 0 |
| 0 | 0 | 0.001941233 | 0 | 0 |
| 0 | 0 | 0 | 0 | 0 |
| 0 | 0 | 0 | 0 | 0 |
| 0 | 0 | 0 | 0 | 0 |
| 0 | 2.142051127 | 0.021928507 | 0 | 0 |
| 0.430689772 | 0.011735614 | 0.070423699 | 0.821126917 | 0.153867234 |
| 0 | 0 | 0 | 0 | 0 |
| 0 | 0 | 0 | 0 | 0 |
| 0.075939329 | 0 | 0.009333001 | 0.003208625 | 0 |
| 0 | 0 | 0 | 0 | 0 |
| 0 | 0 | 0 | 0 | 0.182031929 |
| 0 | 0 | 0 | 0 | 0 |
| 0.019631307 | 0 | 0.020580961 | 0.000909518 | 0 |
| 0 | 0 | 0 | 0.003010707 | 0.004268854 |
| 0 | 0 | 0 | 0.00115365 | 0 |
| 0.049436894 | 0.221405314 | 0.021882653 | 0 | 0 |
| 0 | 0 | 0 | 0 | 0 |
| 0 | 0 | 0 | 0 | 0 |

| | | | | |
|---|---|---|---|---|
| 0 | 0 | 0 | 0 | 0 |
| 0 | 0 | 0 | 0.019783273 | 0 |
| 0 | 0 | 0 | 0 | 0 |
| 8.572818754 | 0.072732448 | 0.00932976 | 0.151328734 | 0 |
| 0 | 0 | 0 | 0 | 0 |
| 0 | 0 | 0 | 0 | 0 |
| 0 | 0 | 0 | 0 | 0.001330796 |
| 0 | 0 | 0 | 0 | 0 |
| 0.17888008 | 3.640612702 | 1.314264745 | 0 | 0 |
| 0.301095108 | 0.017552349 | 0 | 0 | 0 |
| 0 | 0 | 0 | 0 | 0 |
| 0.006893629 | 0.001422153 | 0.001787783 | 0.00724946 | 0 |
| 0 | 0 | 0 | 0 | 0 |
| 0 | 0 | 0 | 0 | 0 |
| 0.134504729 | 0.215303926 | 0.171586737 | 0.061497762 | 0.00814292 |
| 0 | 0 | 0 | 2.941254692 | 0 |
| 0.170502746 | 0.008360588 | 0.006069189 | 0 | 0 |
| 0 | 0 | 1.476734203 | 0 | 0 |
| 0 | 0 | 0 | 0 | 0 |
| 0 | 0 | 0 | 0.020587207 | 0 |
| 0 | 0 | 0 | 0.00947579 | 0 |
| 0 | 0 | 0.034090254 | 0 | 0 |
| 0.000423191 | 0 | 0 | 0 | 0 |
| 0.019604856 | 0 | 0 | 0.553542078 | 0 |
| 0.012908396 | 0 | 0.029485008 | 0 | 0 |
| 0 | 0.153586299 | 0.088375538 | 0 | 0 |
| 0 | 0 | 0.022060679 | 0.001615179 | 0 |
| 3.486733272 | 1.053960547 | 0.579727256 | 0 | 0 |
| 0 | 0 | 0 | 0 | 0 |
| 0 | 0 | 0 | 0 | 0 |
| 0 | 0.006806525 | 0.000534778 | 0.00267813 | 0 |
| 0.020065513 | 0 | 0.108078052 | 0 | 0 |
| 0 | 0 | 0 | 0 | 0 |
| 0 | 0 | 0 | 0 | 0 |
| 0 | 0 | 0.000493754 | 0.003123391 | 0 |
| 0 | 0.137335854 | 0 | 0 | 0 |
| 2.869634445 | 0.507303822 | 0.485763639 | 0.000926113 | 0 |
| 0 | 0 | 0 | 0 | 0 |
| 0 | 0 | 0.002004242 | 0.012980304 | 0.003804579 |
| 0 | 0 | 0 | 0 | 0 |
| 0.027628498 | 0.001784087 | 0 | 0.79087264 | 0.014550169 |
| 0.064329137 | 0.03372458 | 0 | 0 | 0 |
| 1.41354675 | 0.20646936 | 2.570670223 | 4.003065919 | 0.387158161 |
| 0 | 0 | 0.002528375 | 0.000605833 | 0 |
| 0 | 0 | 0 | 0 | 0 |
| 0 | 0 | 0 | 0 | 0 |
| 0 | 0 | 0 | 0 | 0 |
| 0.003709921 | 0.000630787 | 0 | 0.002173647 | 0 |

| | | | | |
|---|---|---|---|---|
| 0 | 0 | 0 | 0 | 0 |
| 0 | 0.129184728 | 0.179157465 | 0 | 0 |
| 0.003556378 | 0.009553958 | 0.001368255 | 0.038948896 | 2.621319468 |
| 0 | 0 | 0 | 0.003811694 | 0 |
| 0 | 0 | 0 | 0 | 3.489891997 |
| 0 | 0.136234471 | 0.001197621 | 0.002146508 | 0.002227932 |
| 0 | 0 | 0 | 0 | 0 |
| 0 | 0 | 0 | 0 | 0 |
| 0 | 0 | 0 | 0 | 0 |
| 0 | 0 | 0 | 0 | 0 |
| 0.095440687 | 0.008667866 | 0.276140045 | 0.01384856 | 0 |
| 1.946091087 | 0.104008714 | 0.646242574 | 1.281172777 | 6.343718607 |
| 0 | 0 | 0 | 0 | 0 |
| 0.00376842 | 0 | 0.062342895 | 0.009001526 | 0 |
| 0.017535132 | 0.000908861 | 0.006474311 | 0.188045935 | 1.08802175 |
| 0 | 1.419762014 | 0 | 0 | 0 |
| 0 | 0 | 0 | 0 | 0 |
| 0 | 0 | 0 | 0 | 0 |
| 0 | 0 | 0 | 0 | 0 |
| 0 | 0.000586853 | 0.004057514 | 0.001296315 | 0 |
| 0.509504684 | 0.010648622 | 0.031401595 | 0.150101324 | 0.003657254 |
| 0 | 0 | 0.032032957 | 0.012925221 | 3.364728413 |
| 1.270230139 | 0.135922399 | 0.538362515 | 7.20831564 | 2.529851665 |
| 0 | 0 | 0 | 0 | 0 |
| 0 | 0 | 0 | 0 | 0 |
| 0 | 0.472129002 | 0.104040638 | 0.006164139 | 0 |
| 0 | 0.048683415 | 0 | 0 | 0 |
| 0 | 0 | 0 | 0 | 0 |
| 0 | 0.032540894 | 0.016172543 | 0.012737887 | 0.000505643 |
| 0 | 0 | 0 | 0 | 0 |
| 0 | 0.007354442 | 0.001793852 | 0.007274072 | 0 |
| 0.484958509 | 0.009207526 | 0.105117557 | 0.212934303 | 0 |
| 0.011099834 | 0 | 0 | 0.001131362 | 0.035240311 |
| 0.025653894 | 0.032175908 | 0.032966449 | 0.008381008 | 0.000466014 |
| 0 | 0 | 0 | 0.002453485 | 0.001613344 |
| 0.087492552 | 0.096318217 | 0.120664179 | 0.181884079 | 0.002770795 |
| 0.00583389 | 0.010682214 | 0.007251425 | 0.001982083 | 0.008564961 |
| 0 | 0 | 0 | 0 | 0 |
| 1.130028922 | 0 | 0.674285756 | 0.009261418 | 0 |
| 0 | 0 | 0 | 0 | 0 |
| 0 | 0.014703523 | 0.019095781 | 0.007485229 | 0 |
| 0 | 0 | 0 | 0 | 0 |
| 0 | 0 | 0 | 0 | 0 |
| 0 | 0 | 0 | 0 | 0 |
| 0.026665618 | 0.042120043 | 0.118104643 | 0.019889411 | 0 |
| 0 | 0 | 0 | 0 | 0 |
| 0 | 0 | 0 | 0 | 0 |
| 0.017680661 | 9.134081731 | 0.651712407 | 1.950413768 | 0.024648054 |

| | | | | |
|---|---|---|---|---|
| 0 | 0 | 0 | 0 | 0 |
| 0 | 0 | 0 | 0 | 0 |
| 0 | 0 | 0 | 0 | 0 |
| 0.004364788 | 0.01708952 | 0.007701511 | 0.009259312 | 0.001436301 |
| 0 | 0 | 0 | 0 | 0 |
| 0.196070875 | 0.081637023 | 0.274702356 | 0 | 0.029493378 |
| 0 | 0 | 0.002662639 | 0 | 0 |
| 0.118552893 | 0 | 0.001616284 | 0.044567428 | 0.0023129 |
| 0 | 0 | 0 | 0 | 0 |
| 0 | 0 | 0 | 0 | 0 |
| 0.003855448 | 0.004668521 | 0.004213482 | 0.007615327 | 0.00079983 |
| 0 | 0 | 0 | 0 | 0 |
| 0 | 0.041190986 | 0.0388687 | 0 | 0 |
| 0 | 0 | 0 | 0.372302863 | 0 |
| 0 | 0 | 0 | 0 | 0 |
| 0 | 0 | 0 | 0 | 0 |
| 0 | 0.00169156 | 0.003917151 | 0 | 0 |
| 0 | 0.007872527 | 0.042216799 | 0.000528299 | 0 |
| 0 | 0 | 0 | 0 | 0 |
| 0 | 0 | 0 | 0 | 0 |
| 0 | 0.008056517 | 0 | 0 | 0 |
| 0 | 0 | 0 | 0 | 0 |
| 0.018019377 | 0.025391109 | 0.008521377 | 0 | 0 |
| 0 | 0 | 0 | 0 | 0 |
| 0 | 0 | 0 | 0 | 0 |
| 0 | 0 | 0 | 0 | 0 |
| 0 | 0 | 0 | 0 | 0 |
| 0 | 0 | 0 | 0 | 0 |
| 0 | 0 | 0 | 0 | 0 |
| 0.520378998 | 0.035993324 | 0 | 0 | 0 |
| 0 | 0.001979912 | 0.042074929 | 0 | 0 |
| 0 | 0 | 0 | 0 | 0 |
| 0 | 0 | 0 | 0 | 0 |
| 0 | 0 | 0 | 0 | 0 |
| 0 | 0.000761473 | 0.001914489 | 0.002270752 | 0 |
| 0 | 0 | 0 | 0 | 0 |
| 0.093392072 | 0.031374668 | 0.031143273 | 0 | 0 |
| 0 | 0 | 0.380627009 | 0.006043601 | 0 |
| 0 | 0 | 0 | 0 | 0 |
| 0 | 0 | 0 | 0.001192215 | 0 |
| 0 | 0 | 0.04697091 | 0 | 0.002064592 |
| 0 | 0 | 0 | 0 | 0 |
| 0.001759639 | 0 | 0 | 0 | 0 |
| 0 | 0.000688844 | 0.065193131 | 0.001565081 | 0 |
| 0 | 0.025889064 | 0.030664653 | 0 | 0.208730792 |
| 0 | 0.003121426 | 0 | 0 | 0 |
| 0 | 0 | 0.001089336 | 0 | 0 |
| 0 | 0 | 0 | 0 | 0 |

| | | | | |
|---|---|---|---|---|
| 0 | 0 | 0 | 0 | 0 |
| 0 | 0 | 0 | 0 | 0 |
| 0 | 0 | 0.006485916 | 0 | 0 |
| 0.285544647 | 0.385849293 | 0.005955415 | 0 | 0 |
| 0 | 0 | 0 | 0 | 0 |
| 0 | 0.047845025 | 0.467143164 | 0 | 0.625536509 |
| 0 | 0 | 0 | 0 | 0 |
| 0 | 0 | 0 | 0 | 0 |
| 0.003522673 | 0.006428732 | 0 | 0.016485045 | 0.095013384 |
| 0 | 0 | 0.000990351 | 0 | 0 |
| 0 | 0 | 0 | 0 | 0 |
| 0 | 0 | 0 | 0 | 0 |
| 0 | 0 | 0 | 0 | 0.000544757 |
| 0 | 0 | 0 | 0 | 0 |
| 2.183969277 | 0.006327195 | 0.462840648 | 4.10384465 | 8.109278518 |
| 0 | 0 | 0 | 0.000571033 | 0 |
| 0 | 0 | 0 | 0 | 0 |
| 0 | 0 | 0 | 0 | 0 |
| 0 | 0 | 0 | 0 | 0 |
| 0 | 0 | 0 | 0 | 0 |
| 0.041301698 | 0 | 0.083423118 | 0.069824206 | 0 |
| 0 | 0 | 0 | 0 | 0 |
| 0 | 0 | 0 | 0 | 0 |
| 0 | 0 | 0 | 0.000391424 | 0 |
| 0.011967406 | 0.704231008 | 0.271680475 | 0.008059325 | 0 |
| 4.201717614 | 0.135851511 | 2.674034668 | 6.626188065 | 0.034931844 |
| 0.004793564 | 0.277886554 | 0.057632535 | 0 | 0 |
| 0.00057144 | 0 | 0 | 0 | 0 |
| 0.010261725 | 0.251452448 | 0.265162579 | 0 | 0 |
| 0.555965045 | 0 | 0 | 0 | 0 |
| 0 | 0 | 0 | 0 | 0 |
| 0 | 0 | 0.057478578 | 0.002104155 | 0 |
| 0 | 0 | 0 | 0 | 0 |
| 0.001720752 | 0.000526634 | 0.019971226 | 0.004071538 | 0 |
| 0 | 0 | 0.046723269 | 0.001533396 | 0 |
| 0.00171787 | 0.004089184 | 0.000771074 | 0.000541963 | 0 |
| 0 | 0 | 0 | 0 | 0 |
| 1.28768539 | 0.089115738 | 1.032331846 | 3.91706329 | 0.964503167 |
| 0 | 0 | 0 | 0 | 0 |
| 0 | 0 | 0 | 0 | 0 |
| 0 | 0.258855275 | 0 | 0 | 0 |
| 0 | 0 | 0 | 0.001536475 | 0 |
| 3.770671731 | 0.030733687 | 0.301354458 | 7.365467215 | 0 |
| 0 | 0 | 0.116519217 | 0 | 0 |
| 0 | 0 | 0 | 0 | 0 |
| 0.001898711 | 0.000968497 | 0.000730496 | 0 | 0.002264901 |
| 0 | 0 | 0 | 0 | 0 |
| 1.064539389 | 0.024224034 | 0.125880908 | 0.098361358 | 0 |

| | | | | |
|---|---|---|---|---|
| 0 | 0.004717321 | 0.063351086 | 0.011894298 | 0.002114108 |
| 0 | 0 | 0 | 0 | 0 |
| 0 | 0 | 0 | 0 | 0 |
| 0 | 0 | 0 | 0 | 0 |
| 0.001924939 | 0 | 0.009072194 | 0.000455467 | 0 |
| 0.003861849 | 0 | 0 | 0 | 0 |
| 0 | 0 | 0 | 0 | 0 |
| 0 | 0 | 0 | 0 | 0 |
| 0 | 0 | 0 | 0 | 0 |
| 0 | 0 | 0 | 0 | 0 |
| 0 | 0 | 0 | 0 | 0 |
| 3.824503831 | 0.12206237 | 1.355622039 | 1.037047978 | 0 |
| 0.198309153 | 0.003744971 | 0 | 0.28089282 | 0.327609835 |
| 0 | 0 | 0 | 0 | 0 |
| 0.623333771 | 0.00331199 | 0.011990847 | 0.004038401 | 0.024785056 |
| 0 | 0 | 0 | 0 | 0 |
| 0 | 0 | 0 | 0.001343574 | 0 |
| 0 | 0.002918834 | 0 | 0 | 0 |
| 0 | 0 | 0.301576741 | 0 | 0 |
| 0 | 0 | 0 | 0.001706478 | 0 |
| 0.127217125 | 0.005191289 | 0 | 0.192648586 | 0 |
| 0 | 0 | 0 | 0 | 0 |
| 0.172972785 | 0.058035164 | 0.012348543 | 0.464496488 | 0.019426215 |
| 0 | 0 | 0 | 0 | 0 |
| 0 | 0 | 0 | 0 | 0 |
| 0 | 0 | 0 | 0 | 0 |
| 0 | 0 | 0 | 0 | 0 |
| 0 | 0 | 0 | 0 | 0 |
| 0 | 0 | 0 | 0 | 0 |
| 0 | 0 | 0 | 0 | 0 |
| 0 | 0 | 0 | 0 | 0 |
| 0.082330671 | 0 | 0 | 0.014581956 | 0.060878216 |
| 0 | 0 | 0 | 0 | 0 |
| 0.021046556 | 0.010477809 | 0.022456513 | 0.418881405 | 0.004016905 |
| 0.000623158 | 0.003337543 | 0.000978976 | 0.001032134 | 0.001115012 |
| 0.004016629 | 0.249544848 | 0.027944698 | 0.002090859 | 0 |
| 0 | 0.000493772 | 0.00099315 | 0 | 0 |
| 0 | 0.169254014 | 0 | 0 | 0 |
| 0 | 0.000766869 | 0.000803356 | 0 | 0 |
| 0 | 0 | 0 | 0 | 0 |
| 0.289358273 | 0.04236962 | 0 | 0 | 0 |
| 0 | 0 | 0.001886051 | 0 | 0 |
| 0 | 0 | 0 | 0 | 0 |
| 0.005073077 | 0 | 0 | 0.049814965 | 0 |
| 0.028238147 | 0.017754197 | 0.022732035 | 0.020625619 | 0.004027463 |
| 0 | 0 | 0 | 0 | 0 |
| 0 | 0.023882326 | 0.123682895 | 0 | 0 |
| 0.00043546 | 0 | 0.000651528 | 0.000686906 | 0 |

| | | | | |
|---|---|---|---|---|
| 0 | 0 | 0 | 0 | 0 |
| 0 | 0 | 0 | 0.006299627 | 0 |
| 0.004285562 | 0 | 0 | 0 | 0 |
| 0 | 0 | 0 | 0 | 0 |
| 0 | 0 | 0 | 0 | 0 |
| 0 | 0 | 0 | 0 | 0 |
| 0.157947465 | 0.275493234 | 2.493890329 | 0 | 0 |
| 0.001199204 | 0 | 0.018570264 | 0 | 0 |
| 0 | 0 | 0 | 0 | 0 |
| 0 | 0 | 0 | 0 | 0 |
| 0 | 0 | 0 | 0 | 0 |
| 0 | 0 | 0 | 0 | 0 |
| 0 | 0 | 0 | 0 | 0 |
| 0 | 0 | 0.152817166 | 0 | 0 |
| 0 | 0 | 0 | 0 | 0 |
| 0 | 0 | 0 | 0 | 0 |
| 0 | 0 | 0 | 0 | 0 |
| 0 | 0 | 0 | 0 | 0 |
| 1.721124616 | 0.946638248 | 0 | 0.00433407 | 0 |
| 0 | 0 | 0 | 0 | 0 |
| 0 | 0 | 0 | 0 | 0 |
| 0 | 0 | 0 | 0 | 0 |
| 0 | 0 | 0 | 0 | 0 |
| 0 | 0 | 0 | 0 | 0 |
| 0 | 0.002394815 | 0 | 0 | 0 |
| 0.001571604 | 0 | 0 | 0.022852693 | 0.00255642 |
| 0 | 0 | 0 | 0 | 0 |
| 0.743272622 | 0.009387293 | 0.006025903 | 0.359083068 | 0 |
| 0 | 0 | 0.26626676 | 0.049308997 | 0 |
| 0.182474211 | 0.040058842 | 0.138688419 | 0 | 0.972047951 |
| 0 | 0 | 0 | 0 | 0 |
| 0 | 0 | 0 | 0 | 0 |
| 0.00251028 | 0 | 0 | 0.001537327 | 0 |
| 0 | 0 | 0 | 0 | 0 |
| 0.001946388 | 0.001283025 | 0.00362899 | 0.003542634 | 0.002143176 |
| 0 | 0 | 0 | 0 | 0 |
| 0 | 0 | 0 | 0 | 0 |
| 0.01070312 | 0 | 0 | 0 | 0 |
| 0 | 0 | 0 | 0 | 0 |
| 0 | 0 | 0.000446266 | 0 | 0 |
| 7.584446151 | 0.138599058 | 1.157855261 | 2.393121341 | 9.800462729 |
| 0.112795272 | 0 | 0 | 1.507998139 | 0 |
| 0 | 2.498421486 | 0 | 0 | 0 |
| 0 | 0 | 0 | 0 | 0 |
| 0 | 0 | 0 | 0 | 0 |
| 0 | 0 | 0 | 0 | 0 |
| 0 | 0 | 0.001038419 | 0 | 0 |
| 0 | 0.005584669 | 0.006052123 | 0 | 0 |

| | | | | |
|---|---|---|---|---|
| 0 | 0 | 0 | 0 | 0 |
| 0 | 0 | 0.001369524 | 0.001312627 | 0.003639604 |
| 0.001619805 | 0 | 0 | 0.002299611 | 0 |
| 0 | 0 | 0 | 0 | 0 |
| 4.264723963 | 0.588713097 | 1.451586131 | 5.079777421 | 0 |
| 0 | 0 | 0 | 0 | 0 |
| 0 | 0 | 0 | 0 | 0 |
| 0 | 0 | 0 | 0 | 0 |
| 0.004569978 | 0 | 0.00087023 | 0.188870622 | 1.335582412 |
| 0 | 0 | 0 | 0 | 0 |
| 0 | 0 | 0 | 0 | 0 |
| 0 | 0 | 0 | 0 | 0 |
| 0.143653802 | 0.006062728 | 0.01475896 | 0.066705535 | 0 |
| 0.021371856 | 0.000591548 | 0.004164344 | 0.002508839 | 0.010869404 |
| 0 | 2.32185616 | 0 | 0 | 0 |
| 0.34816875 | 0.011164665 | 0.024042655 | 0.719141887 | 0.002270381 |
| 0.000573458 | 0 | 0 | 0.003527893 | 0 |
| 0 | 0 | 0 | 0 | 0 |
| 0 | 0 | 0 | 0 | 0 |
| 0 | 0 | 0 | 0 | 0 |
| 0 | 0 | 0 | 0 | 0 |
| 0 | 0 | 0 | 0 | 0 |
| 0 | 0 | 0 | 0 | 0 |
| 0 | 0 | 0 | 0 | 0 |
| 0 | 0 | 0 | 0.05745206 | 0 |
| 0 | 0 | 0 | 0 | 0 |
| 0.011593507 | 0.01673622 | 0.030775178 | 0.039997407 | 0 |
| 0 | 0 | 0.00050511 | 0 | 0 |
| 0 | 0 | 0 | 0 | 0 |
| 0 | 0 | 0 | 0 | 0 |
| 0 | 0 | 0 | 0 | 0 |
| 0 | 0 | 0 | 0.002112956 | 0 |
| 0 | 0 | 0 | 0 | 0 |
| 0 | 0 | 0.005030161 | 0.010327478 | 0 |
| 0.130800812 | 0.062022056 | 0.003914039 | 0 | 0 |
| 0 | 0 | 0.070701266 | 0 | 0.197016401 |
| 0 | 0 | 0 | 0 | 0 |
| 0 | 0 | 0 | 0.006835123 | 0 |
| 0 | 0 | 0 | 0 | 0 |
| 0 | 0 | 0 | 0 | 0 |
| 0 | 0.024051659 | 0.263350715 | 4.126093095 | 0 |
| 0.190593536 | 0 | 0 | 0 | 0 |
| 0 | 0 | 0 | 0 | 0 |
| 0 | 0 | 0 | 0 | 0 |
| 0 | 0 | 0 | 0 | 0 |
| 0 | 0 | 0 | 0 | 0 |
| 0 | 0 | 0 | 0 | 0 |
| 0 | 0 | 0 | 0 | 0 |

| | | | | |
|---|---|---|---|---|
| 0.566934887 | 0.419374298 | 0 | 0.049270201 | 0 |
| 0 | 0 | 0 | 0 | 0 |
| 0 | 0 | 0 | 0 | 0 |
| 0 | 0 | 0.010442488 | 0 | 0.004181752 |
| 0.031407653 | 0 | 0.000547475 | 0 | 0 |
| 0 | 0 | 0 | 0 | 0 |
| 0.139011496 | 0 | 0 | 0.009881152 | 0.020813005 |
| 0 | 0 | 0 | 0 | 0 |
| 0 | 0 | 0 | 0 | 0 |
| 0 | 0 | 0 | 0.001562141 | 0 |
| 0 | 0.023035913 | 0.00124552 | 0 | 0 |
| 0 | 0 | 0 | 0.006628907 | 0 |
| 0 | 0 | 0 | 0 | 0 |
| 0 | 0 | 0 | 0 | 0 |
| 0 | 0 | 0 | 0.024871138 | 0 |
| 0 | 0 | 0 | 0 | 0 |
| 0 | 0 | 0 | 0 | 0 |
| 0 | 0 | 0 | 0 | 0 |
| 0 | 0 | 0 | 0 | 0 |
| 0 | 0 | 0 | 0 | 0 |
| 0.006671798 | 0.00641739 | 0.029213307 | 0.410446474 | 0 |
| 0 | 0 | 0 | 0 | 0 |
| 0.012315011 | 0.019733605 | 0.046917645 | 0.005401381 | 0 |
| 0.004674052 | 0.005054393 | 0.005274899 | 0.002527876 | 0.001274401 |
| 0 | 0 | 0 | 0 | 0 |
| 0 | 0 | 0.132808635 | 0 | 0 |
| 0 | 0 | 0 | 0 | 0 |
| 0 | 0 | 0 | 0 | 0 |
| 0 | 0 | 0 | 0 | 0 |
| 0.000768806 | 0 | 0.034853338 | 0 | 1.537256354 |
| 0 | 0 | 0 | 0 | 0 |
| 0 | 0 | 0 | 0 | 0 |
| 0 | 0 | 0 | 0 | 0 |
| 0.216635682 | 0 | 0.039432919 | 4.18467943 | 0.950646641 |
| 0 | 0 | 0 | 0 | 0 |
| 0 | 0 | 0 | 0 | 0 |
| 0.00919042 | 0 | 0 | 0.001581514 | 0.012756841 |
| 0 | 0 | 0 | 0 | 0 |
| 0 | 0 | 0 | 0 | 0 |
| 0 | 0 | 0 | 0 | 0 |
| 0 | 0 | 0 | 0 | 0 |
| 0 | 0 | 0.000451529 | 0 | 0 |
| 0 | 0 | 0 | 0 | 0 |
| 0 | 0 | 0.035069792 | 0 | 0 |
| 0 | 0 | 0 | 0.035024729 | 0 |
| 0 | 0 | 0 | 0 | 0 |
| 0 | 0 | 0 | 0 | 0 |
| 0 | 0 | 0 | 0 | 0 |

| | | | | |
|---|---|---|---|---|
| 0 | 0 | 0.00528898 | 0.000952029 | 0.000514238 |
| 0 | 0 | 0 | 0 | 0 |
| 0 | 0 | 0 | 0.001616275 | 0 |
| 0 | 0 | 0 | 0 | 0 |
| 0 | 0 | 0 | 0 | 0 |
| 0 | 0 | 0 | 0 | 0 |
| 0 | 0 | 0 | 0 | 0 |
| 0.700547573 | 0.030344994 | 0.285834312 | 2.270776749 | 0.268578943 |
| 0.524869486 | 0.832747091 | 11.26641816 | 0.20984988 | 0.058654581 |
| 0 | 0 | 0 | 0.003418207 | 0.013860933 |
| 0 | 0.004210798 | 0 | 0.001918403 | 0 |
| 0 | 0 | 0 | 0 | 0 |
| 0 | 0 | 0 | 0.036552812 | 0 |
| 0 | 1.821707662 | 0 | 0 | 0 |
| 0 | 0 | 0 | 0 | 0 |
| 0 | 0 | 0 | 0 | 0 |
| 0 | 0 | 0 | 0 | 0 |
| 0 | 0 | 0 | 0 | 0 |
| 0 | 0 | 0 | 0 | 0 |
| 0 | 0 | 0 | 0 | 0 |
| 0 | 0 | 0 | 0 | 0 |
| 0 | 0 | 0 | 0 | 0 |
| 0 | 0 | 0 | 0 | 0 |
| 0 | 0 | 0 | 0 | 0 |
| 0.038482031 | 0.000484951 | 0 | 0 | 0 |
| 0 | 0 | 0 | 0 | 0 |
| 0 | 0 | 0 | 0 | 0 |
| 0 | 0 | 0 | 0 | 0 |
| 0 | 0 | 0 | 0 | 0 |
| 0 | 0 | 0 | 0 | 0 |
| 0 | 0 | 0 | 0 | 0 |
| 0 | 0 | 0 | 0 | 0 |
| 0 | 0 | 0 | 0 | 0 |
| 0 | 0 | 0 | 0 | 0 |
| 0 | 0.454318014 | 0.004997506 | 0 | 0 |
| 0 | 0 | 0.004389264 | 0 | 0 |
| 0 | 0 | 0 | 0 | 0 |
| 0.007573328 | 0 | 0 | 0.027693848 | 0 |
| 0 | 0.001952316 | 0.002147467 | 0 | 0 |
| 0 | 0 | 0 | 0.06381415 | 0 |
| 0 | 0 | 0 | 0 | 0 |
| 0.224664567 | 0.006235661 | 0.085164154 | 0.190643149 | 0 |
| 0 | 0 | 0 | 0 | 0 |
| 0 | 0 | 0 | 0 | 0 |
| 0 | 0 | 0 | 0 | 0 |
| 0 | 0 | 0 | 0 | 0 |
| 0 | 0 | 0 | 0 | 0 |
| 0 | 0 | 0 | 0 | 0 |

| | | | | |
|---|---|---|---|---|
| 0 | 0 | 0 | 0 | 0 |
| 0.011542778 | 0.001944945 | 0.001164277 | 0 | 0 |
| 0 | 0 | 0 | 0 | 0 |
| 0 | 0 | 0 | 0 | 0 |
| 0.134184665 | 2.858379203 | 0.700126645 | 0.231860507 | 0.008868405 |
| 0 | 0 | 0 | 0 | 0 |
| 0 | 0 | 0 | 0 | 0 |
| 0.004021486 | 0.009529155 | 0 | 0.008996374 | 0.027692242 |
| 0 | 0.092929375 | 0.023429762 | 0 | 0 |
| 0.050991998 | 0 | 0 | 0 | 0 |
| 0 | 0 | 0 | 0 | 0 |
| 0 | 0 | 0 | 0 | 0 |
| 0 | 0 | 0 | 0 | 0 |
| 0 | 0 | 0 | 0 | 0 |
| 0 | 0 | 0 | 0 | 0 |
| 0 | 0 | 0.004269596 | 0 | 0.005862484 |
| 0 | 0 | 0 | 0 | 0 |
| 0 | 0 | 0 | 0 | 0 |
| 0 | 0 | 0 | 0 | 0 |
| 0 | 0 | 0 | 0 | 0 |
| 0 | 0 | 0 | 0 | 0 |
| 0 | 0 | 0 | 0 | 0 |
| 0 | 0 | 0 | 0 | 0 |
| 0.003796653 | 0.000451874 | 0.002414207 | 0.006737557 | 0 |
| 0.070655597 | 0.057290861 | 0.100196381 | 0 | 0 |
| 0 | 0 | 0 | 0 | 0 |
| 0 | 0.989932838 | 0 | 0.003834295 | 0.016223539 |
| 0 | 0 | 0 | 0 | 0 |
| 0 | 0 | 0 | 0 | 0 |
| 0 | 0 | 0 | 0 | 0 |
| 0 | 0 | 0 | 0 | 0 |
| 0 | 0 | 0 | 0.00697583 | 0 |
| 0 | 0 | 0 | 0 | 0 |
| 0 | 0 | 0 | 0 | 0 |
| 0 | 0 | 0 | 0 | 0 |
| 0 | 0 | 0 | 0 | 0 |
| 0 | 0 | 0 | 0 | 0 |
| 0 | 0.059021544 | 0.005727857 | 0.024717279 | 0 |
| 0 | 0 | 0 | 0 | 0 |
| 0.000381897 | 0 | 0 | 0 | 0.004555506 |
| 0 | 0 | 0 | 0 | 0 |
| 0 | 0 | 1.125994997 | 0 | 0 |
| 0 | 0 | 0 | 0 | 0 |
| 0 | 0 | 0 | 0 | 0 |
| 0 | 0 | 0 | 0 | 0 |
| 0 | 0 | 0 | 0 | 0 |
| 0 | 0 | 0 | 0 | 0 |
| 0.00453965 | 0.001986927 | 0 | 0 | 0 |

| | | | | |
|---|---|---|---|---|
| 0.034288366 | 0.038089034 | 0.021742138 | 0.0181579 | 0 |
| 0 | 0 | 0 | 0 | 0 |
| 0 | 0 | 0 | 0 | 0 |
| 0 | 0 | 0.000850706 | 0 | 0 |
| 0 | 0 | 0 | 0 | 0 |
| 0.214704472 | 0 | 0.236753235 | 0 | 0 |
| 0 | 0 | 0 | 0 | 0 |
| 0 | 0 | 0 | 0 | 0 |
| 0 | 0 | 0 | 0 | 0 |
| 0 | 0 | 0 | 0.050960955 | 0 |
| 0 | 0 | 0 | 0 | 0 |
| 0 | 0 | 0 | 0.001104169 | 0 |
| 0 | 0 | 0 | 0 | 0 |
| 0 | 0 | 0 | 0 | 0 |
| 0 | 0 | 0.000639276 | 0 | 0 |
| 0 | 0 | 0 | 0 | 0 |
| 0 | 0 | 0 | 0 | 0 |
| 0 | 0 | 0 | 0 | 0 |
| 0 | 0 | 0 | 0 | 0 |
| 0 | 0 | 0 | 0 | 0 |
| 0.031979112 | 0 | 0 | 0.001341878 | 0 |
| 0 | 0 | 0 | 0 | 0 |
| 0 | 0.043616889 | 0.023218172 | 0.001318096 | 0 |
| 0.00528065 | 0 | 0 | 0.308491149 | 0 |
| 0 | 0 | 0 | 0 | 0 |
| 0 | 0 | 0.432010069 | 0 | 0 |
| 0 | 0 | 0 | 0 | 0 |
| 0 | 0 | 0 | 0 | 0 |
| 0 | 0.003443761 | 0 | 0 | 0 |
| 0 | 0 | 0 | 0 | 0 |
| 0 | 0 | 0 | 0 | 0 |
| 0 | 0 | 0 | 0 | 0 |
| 0.004657947 | 0.000399157 | 0 | 0.003879515 | 0.038671792 |
| 0.058551484 | 0 | 0 | 0 | 0 |
| 0.223158357 | 0.005236642 | 0 | 1.943474302 | 0.009144941 |
| 0.000598049 | 0.011637804 | 0.000671093 | 0 | 0 |
| 0 | 0 | 0 | 0 | 0 |
| 0 | 0 | 0 | 0 | 0 |
| 0.007782058 | 0 | 0 | 0.002455124 | 0.001444011 |
| 0 | 0 | 0 | 0 | 0 |
| 0 | 0 | 0 | 0 | 0 |
| 0 | 0 | 0 | 0 | 0 |
| 0.029320796 | 0.399273535 | 0.172451649 | 0 | 0 |
| 0 | 0 | 0 | 0 | 0 |
| 0 | 0 | 0 | 0 | 0 |
| 0.020395146 | 0.011009358 | 0 | 0.037086274 | 0 |
| 0 | 0 | 0 | 0 | 0 |
| 0 | 0 | 0 | 0 | 0 |

| | | | | |
|---|---|---|---|---|
| 0 | 0 | 0 | 0 | 0 |
| 0 | 0 | 0 | 0 | 0 |
| 0 | 0 | 0 | 0 | 0 |
| 0 | 0 | 0 | 0 | 0 |
| 0 | 0 | 0 | 0 | 0 |
| 0 | 0 | 0 | 0 | 0 |
| 0 | 0 | 0 | 0 | 0 |
| 0 | 0 | 0 | 0.163323127 | 0.00795979 |
| 0.034304774 | 0 | 0 | 0.066929541 | 0 |
| 0 | 0.004842919 | 0.205062077 | 0 | 0 |
| 0 | 0 | 0 | 0 | 0 |
| 0 | 0 | 0 | 0 | 0 |
| 0 | 0 | 0 | 0 | 0 |
| 0 | 0 | 0 | 0.004174099 | 0 |
| 0 | 0 | 0 | 0 | 0 |
| 0 | 0 | 0 | 0.000770145 | 0 |
| 0 | 0 | 0 | 0 | 0 |
| 0 | 0 | 0 | 0 | 0 |
| 0 | 0 | 0 | 0 | 0 |
| 2.48362846 | 0.021308818 | 0.000978529 | 1.816242737 | 0 |
| 0 | 0 | 0.083464971 | 0.004826168 | 0 |
| 0 | 0 | 0 | 0 | 0 |
| 0 | 0 | 0 | 0 | 0 |
| 0 | 0 | 0 | 0.004103394 | 0 |
| 0 | 0 | 0 | 0 | 0 |
| 0 | 0 | 0 | 0 | 0 |
| 0 | 0 | 0 | 0 | 0 |
| 0 | 0 | 0 | 0 | 0 |
| 0 | 0 | 0 | 0 | 0 |
| 0.08433525 | 0.0033726 | 0.010750619 | 0 | 0 |
| 0.3335921 | 0.007273259 | 0.032654224 | 0 | 0.033765637 |
| 0 | 0 | 0 | 0 | 0 |
| 0 | 0 | 0 | 0 | 0 |
| 0 | 0 | 0 | 0 | 0 |
| 0 | 0 | 0 | 0 | 0 |
| 0 | 0 | 0 | 0 | 0 |
| 0 | 0 | 0 | 0 | 0 |
| 0 | 0 | 0 | 0 | 0 |
| 0.012373363 | 0 | 0.001204448 | 0 | 0 |
| 0 | 0 | 0 | 0 | 0 |
| 0 | 0 | 0 | 0 | 0 |
| 0 | 0 | 0.073966912 | 0 | 0 |
| 0 | 0.000919481 | 0 | 0 | 0 |
| 0 | 0 | 0.003589179 | 0 | 0 |
| 0 | 0 | 0 | 0 | 0 |
| 0 | 0 | 0 | 0 | 0 |
| 0.014827284 | 0 | 0 | 0 | 0.007352987 |
| 0 | 0 | 0.002187811 | 0 | 0 |

| | | | | |
|---|---|---|---|---|
| 0 | 0 | 0 | 0 | 0 |
| 0 | 0 | 0 | 0 | 0 |
| 0 | 0 | 0 | 0 | 0 |
| 0 | 0 | 0 | 0 | 0 |
| 0.073556845 | 0.01110998 | 0.099663576 | 0.111789778 | 0 |
| 0 | 0 | 0.000664769 | 0 | 0 |
| 0 | 0 | 0.001171292 | 0 | 0 |
| 0 | 0 | 0 | 0 | 0 |
| 0 | 0 | 0 | 0 | 0 |
| 0 | 0 | 0 | 0 | 0 |
| 0.076957232 | 0.013654427 | 0 | 0 | 0 |
| 0.005011111 | 0.040597521 | 0.008841925 | 0.003597982 | 0.000824492 |
| 0.000596321 | 0 | 0 | 0.000846587 | 0 |
| 0 | 0 | 0 | 0 | 0 |
| 0.009086149 | 0 | 0 | 0 | 0 |
| 0 | 0 | 0 | 0 | 0 |
| 0.016346864 | 0 | 0.017776719 | 0.004528268 | 0.002695062 |
| 0 | 0 | 0 | 0 | 0 |
| 0 | 0 | 0.001174589 | 0.194300126 | 0 |
| 0.07875763 | 0 | 0 | 0.002425399 | 0.003260635 |
| 0 | 0 | 0 | 0 | 0 |
| 0 | 0 | 0 | 0 | 0 |
| 0 | 0 | 0 | 0 | 0 |
| 0 | 0 | 0 | 0 | 0 |
| 0 | 0 | 0 | 0 | 0 |
| 0 | 0 | 0 | 0 | 0 |
| 0.002874955 | 0 | 0.009586107 | 0.002850589 | 0.00081653 |
| 0 | 0 | 0 | 0 | 0 |
| 0 | 0 | 0 | 0 | 0 |
| 0 | 0 | 0 | 0 | 0 |
| 0.001396677 | 0 | 0.001811059 | 0.000881262 | 0.052387815 |
| 0 | 0 | 0 | 0 | 0 |
| 0 | 0 | 0 | 0 | 0 |
| 0 | 0 | 0 | 0 | 0 |
| 0 | 0 | 0 | 0 | 0 |
| 0 | 0 | 0 | 0 | 0 |
| 0 | 0 | 0 | 0 | 0 |
| 0.022460052 | 0.031610926 | 0.002055412 | 0.009596807 | 0.002080914 |
| 0 | 0.170722 | 0 | 0 | 0 |
| 0 | 0 | 0 | 0.006220161 | 0 |
| 0.006966174 | 0.017799895 | 0.014363744 | 0 | 0 |
| 0 | 0 | 0 | 0.001587431 | 0 |
| 0 | 0 | 0 | 0 | 0 |
| 0 | 0 | 0 | 0.004743616 | 0 |
| 0 | 0 | 0 | 0 | 0 |
| 0 | 0 | 0 | 0 | 0 |
| 0 | 0 | 0 | 0 | 0 |
| 0 | 0 | 0 | 0 | 0 |

| | | | | |
|---|---|---|---|---|
| 0 | 0 | 0 | 0 | 0 |
| 0 | 0.001578891 | 0 | 0.073240772 | 0 |
| 0 | 0 | 0 | 0 | 0 |
| 0.00084719 | 0 | 0.000456318 | 0.001603655 | 0 |
| 0 | 0 | 0.000846456 | 0 | 0 |
| 0.000811274 | 0 | 0 | 0 | 0 |
| 0 | 0 | 0.041625299 | 0 | 0 |
| 0 | 0 | 0 | 0 | 0 |
| 0 | 0 | 0 | 0 | 0 |
| 0 | 0 | 0 | 0 | 0 |
| 0 | 0.004699181 | 0.007918323 | 0.021467028 | 0 |
| 0 | 0 | 0 | 0 | 0 |
| 0.03495133 | 0.113474479 | 0.070407657 | 0.024099434 | 0.003295255 |
| 0 | 0.000484305 | 0.00578377 | 0 | 0 |
| 0 | 0 | 0 | 0 | 0 |
| 0 | 0 | 0 | 0 | 0 |
| 0 | 0 | 0 | 0 | 0 |
| 0 | 0 | 0 | 0 | 0 |
| 0 | 0 | 0 | 0 | 0 |
| 0 | 0 | 0 | 0 | 0 |
| 0 | 0 | 0 | 0 | 0 |
| 0 | 0 | 0 | 0 | 0 |
| 0 | 0 | 0 | 0.005688062 | 0 |
| 0.203423751 | 0.001732552 | 0.002010446 | 0.097678855 | 0 |
| 0 | 0 | 0 | 0 | 0 |
| 0 | 0 | 0.008166035 | 0 | 0 |
| 0.017527898 | 0 | 0 | 0 | 0.274512581 |
| 0 | 0 | 0 | 0 | 0 |
| 0 | 0 | 0 | 0 | 0 |
| 0 | 0 | 0 | 0 | 0 |
| 0.005995599 | 0.001888916 | 0.006173824 | 0 | 0.035969984 |
| 0 | 0 | 0 | 0 | 0 |
| 0 | 0 | 0 | 0 | 0 |
| 0 | 0 | 0 | 0 | 0 |
| 0 | 0 | 0 | 0 | 0 |
| 0 | 0 | 0 | 0 | 0 |
| 0 | 0 | 0 | 0.001063908 | 0 |
| 0 | 0 | 0 | 0 | 0 |
| 0.59350499 | 0 | 0 | 0 | 0 |
| 0 | 0 | 0 | 0 | 0 |
| 0 | 0 | 0 | 0 | 0 |
| 0 | 0 | 0 | 0 | 0 |
| 0 | 0 | 0 | 0 | 0 |
| 0 | 0 | 0 | 0 | 0 |
| 0 | 0 | 0 | 1.993879119 | 0 |
| 0.282227411 | 0.058681702 | 0.093330139 | 0.142130415 | 0.384573716 |
| 0 | 0 | 0 | 0 | 0 |
| 0 | 0 | 0 | 0 | 0 |

| | | | | |
|---|---|---|---|---|
| 0 | 0 | 0 | 0 | 0 |
| 0 | 0 | 0 | 0 | 0 |
| 0 | 0.048171865 | 0.043876373 | 0 | 0 |
| 0 | 0 | 0.001957067 | 0 | 0 |
| 0 | 0 | 0 | 0 | 0 |
| 0 | 0 | 0 | 0 | 0 |
| 0.001213248 | 0 | 0 | 0 | 0 |
| 0 | 0 | 0 | 0 | 0 |
| 0 | 0 | 0 | 0 | 0 |
| 0 | 0 | 0 | 0 | 0 |
| 0.001815579 | 0.000950789 | 0.001738521 | 0 | 0 |
| 0 | 0 | 0.000973356 | 0 | 0 |
| 0 | 0 | 0 | 0 | 0 |
| 0 | 0 | 0 | 0 | 0 |
| 0 | 0 | 0 | 0 | 0 |
| 0 | 0 | 0 | 0 | 0 |
| 0 | 0 | 0 | 0 | 0 |
| 0 | 0 | 0 | 0 | 0 |
| 0 | 0 | 0 | 0 | 0 |
| 0 | 0.000343884 | 0.00100869 | 0.000759616 | 0 |
| 0 | 0 | 0 | 0 | 0 |
| 0 | 0 | 0 | 0 | 0 |
| 0 | 0 | 0 | 0 | 0 |
| 0 | 0 | 0 | 0 | 0 |
| 0 | 0 | 0 | 0 | 0 |
| 0 | 0 | 0 | 0 | 0 |
| 0 | 0 | 0 | 0 | 0 |
| 0 | 0.003805663 | 0.005900369 | 0 | 0 |
| 0 | 0 | 0 | 0 | 0 |
| 0 | 0 | 0 | 0 | 0 |
| 0 | 0.062434703 | 0.004085254 | 0.007831062 | 0 |
| 0 | 0 | 0 | 0 | 0 |
| 0.030742607 | 0.001715135 | 0.016530017 | 0.273234452 | 0.23129919 |
| 0.009326509 | 0.001518991 | 0 | 0.008827127 | 0 |
| 0 | 0 | 0 | 0 | 0 |
| 0 | 0 | 0 | 0 | 0 |
| 0 | 0 | 0 | 0 | 0 |
| 0 | 0 | 0 | 0 | 0 |
| 0 | 0 | 0 | 0 | 0 |
| 0 | 0 | 0 | 0 | 0 |
| 0 | 0 | 0 | 0 | 0 |
| 0.158994991 | 0 | 0 | 0.008353169 | 0 |
| 0.196268429 | 0.012491602 | 0.095083229 | 0.046781589 | 0 |
| 0 | 0 | 0 | 0 | 0 |
| 0 | 0 | 0 | 0 | 0 |
| 0.03153927 | 0.340654916 | 0.109713316 | 0 | 0 |
| 0 | 0 | 0 | 0.000719516 | 0 |
| 0 | 0 | 0 | 0 | 0 |

| | | | | |
|---|---|---|---|---|
| 0 | 0 | 0 | 0 | 0 |
| 0.017653154 | 0 | 0 | 0 | 0 |
| 0.006873583 | 0.001262192 | 0 | 0.004646815 | 0.002576904 |
| 0 | 0 | 0 | 0 | 0 |
| 0 | 0.027882444 | 0.025619534 | 0.190410445 | 0 |
| 0 | 0.02190868 | 0 | 0.00332262 | 0.012562964 |
| 0 | 0 | 0 | 0 | 0 |
| 0 | 0 | 0 | 0 | 0 |
| 0 | 0 | 0 | 0 | 0 |
| 0 | 0 | 0 | 0 | 0 |
| 0 | 0 | 0 | 0 | 0 |
| 0 | 0 | 0 | 0 | 0 |
| 0 | 0 | 0 | 0 | 0 |
| 0 | 0 | 0 | 0 | 0 |
| 0 | 0 | 0 | 0 | 0 |
| 0 | 0 | 0 | 0 | 0 |
| 0 | 0 | 0 | 0 | 0 |
| 0 | 0 | 0 | 0 | 0 |
| 0 | 0 | 0 | 0 | 0 |
| 0 | 0 | 0 | 0 | 0 |
| 0 | 0 | 0 | 0 | 0 |
| 0 | 0 | 0 | 0 | 0 |
| 0 | 0 | 0 | 0 | 0 |
| 0 | 0 | 0 | 0 | 0 |
| 0.006150352 | 0.015372188 | 0.1085358 | 0 | 0 |
| 0.000814093 | 0 | 0 | 0 | 0.01184744 |
| 0 | 0 | 0 | 0 | 0 |
| 0.007739049 | 0.002030167 | 0 | 0 | 0 |
| 0 | 0 | 0 | 0 | 0 |
| 0 | 0 | 0 | 0 | 0 |
| 0 | 0 | 0 | 0 | 0 |
| 0 | 0 | 0 | 0 | 0 |
| 0 | 0 | 0 | 0 | 0 |
| 0 | 0 | 0 | 0 | 0 |
| 0 | 0 | 0 | 0 | 0 |
| 0 | 0 | 0 | 0 | 0 |
| 0 | 0 | 0 | 0 | 0 |
| 0 | 0 | 0 | 0.013652087 | 0 |
| 0 | 0 | 0 | 0 | 0.01816269 |
| 0 | 0 | 0 | 0 | 0 |
| 0 | 0 | 0 | 0 | 0 |
| 0.01854648 | 0 | 0.005391221 | 0.748110084 | 0 |
| 0 | 0 | 0.004547435 | 0.000618627 | 0 |
| 0 | 0 | 0 | 0 | 0 |
| 0 | 0 | 0 | 0 | 0 |
| 0 | 0 | 0 | 0 | 0 |
| 0.001249898 | 0 | 0 | 0 | 0.003280106 |
| 0 | 0 | 0 | 0 | 0 |

| | | | | |
|---|---|---|---|---|
| 0 | 0 | 0 | 0 | 0 |
| 0.007254852 | 0.002139893 | 0.031006364 | 0.00283612 | 0.005267678 |
| 0 | 0 | 0.000839378 | 0 | 0 |
| 0 | 0 | 0 | 0 | 0 |
| 0 | 0 | 0 | 0 | 0 |
| 0 | 0 | 0 | 0 | 0 |
| 0 | 0 | 0 | 0 | 0 |
| 0 | 0 | 0 | 0 | 0 |
| 0 | 0 | 0 | 0 | 0 |
| 0 | 0 | 0 | 0 | 0 |
| 0 | 0 | 0 | 0 | 0 |
| 0 | 0 | 0 | 0 | 0 |
| 0.080524725 | 0.281447393 | 0.129010596 | 0.02540436 | 0 |
| 0 | 0 | 0 | 0 | 0 |
| 0 | 0 | 0 | 0.001302816 | 0.000938286 |
| 0 | 0 | 0 | 0 | 0 |
| 0 | 0 | 0 | 0 | 0 |
| 0 | 0 | 0 | 0 | 0 |
| 0 | 0 | 0 | 0.067944715 | 0 |
| 0 | 0 | 0 | 0 | 0 |
| 0 | 0 | 0 | 0 | 0 |
| 0.036693123 | 0 | 0 | 0 | 0.000921471 |
| 0 | 0 | 0.000799311 | 0.001966333 | 0 |
| 0 | 0 | 0 | 0 | 0 |
| 0 | 0 | 0 | 0 | 0 |
| 0 | 0 | 0 | 0 | 0 |
| 0 | 0 | 0 | 0 | 0 |
| 0 | 0 | 0 | 0 | 0 |
| 0 | 0 | 0 | 0 | 0.006897766 |
| 0 | 0 | 0 | 0 | 0 |
| 0 | 0 | 0 | 0 | 0 |
| 0 | 0 | 0 | 0 | 0 |
| 0 | 0 | 0 | 0 | 0 |
| 0 | 0 | 0 | 0 | 0 |
| 0 | 2.055536959 | 17.11211895 | 0 | 0 |
| 0 | 0 | 0 | 0 | 0 |
| 0 | 0 | 0 | 0.001929321 | 0 |
| 0 | 0 | 0 | 0 | 0 |
| 0.010221335 | 0 | 0.009695178 | 0 | 0 |
| 0 | 0 | 0 | 0 | 0 |
| 0.418813112 | 0.005433335 | 0.073735349 | 0.045334222 | 0 |
| 0 | 0 | 0 | 0 | 0 |
| 0 | 0 | 0 | 0 | 0 |
| 0 | 0 | 0 | 0 | 0 |
| 0 | 0 | 0 | 0 | 0 |
| 0.076223087 | 0.010038106 | 0.008708791 | 0.313114024 | 2.474541273 |
| 0 | 0 | 0 | 0 | 0 |
| 0 | 0 | 0 | 0 | 0 |

| | | | | |
|---|---|---|---|---|
| 0 | 0 | 0 | 0 | 0 |
| 0 | 0 | 0 | 0 | 0 |
| 0 | 0 | 0 | 0 | 0 |
| 0 | 0 | 0.047263924 | 0 | 0 |
| 0.001501544 | 0 | 0.005122213 | 0 | 0 |
| 0 | 0 | 0 | 0 | 0 |
| 0 | 0 | 0 | 0 | 0 |
| 0 | 0 | 0 | 0 | 0 |
| 0 | 0 | 0 | 0 | 0 |
| 0.037176336 | 0 | 0 | 0 | 0 |
| 0 | 0.004374815 | 0.001283231 | 0 | 0 |
| 0 | 0 | 0 | 0 | 0 |
| 0.001703285 | 0 | 0 | 0 | 0 |
| 0 | 0 | 0 | 0 | 0 |
| 0 | 0 | 0 | 0 | 0 |
| 0 | 0 | 0 | 0 | 0 |
| 0 | 0 | 0 | 0 | 0 |
| 0 | 0 | 0 | 0 | 0 |
| 0 | 0 | 0 | 0 | 0 |
| 0.414722125 | 0.069769368 | 2.466526254 | 0.249029303 | 0.001004684 |
| 0 | 0 | 0 | 0 | 0 |
| 0 | 0 | 0 | 0 | 0 |
| 0 | 0 | 0 | 0 | 0 |
| 0 | 0 | 0 | 0 | 0 |
| 0 | 0 | 0 | 0 | 0 |
| 0 | 0 | 0.001813555 | 0 | 0 |
| 0 | 0 | 0 | 0 | 0 |
| 0 | 0 | 0 | 0 | 0 |
| 0.003322073 | 0 | 0 | 0 | 0 |
| 0 | 0 | 0 | 0 | 0 |
| 0 | 0 | 0 | 0 | 0 |
| 0 | 0 | 0 | 0 | 0 |
| 0 | 0 | 0 | 0 | 0 |
| 0 | 0 | 0 | 0 | 0 |
| 0 | 0 | 0 | 0 | 0 |
| 0 | 0 | 0 | 0 | 0 |
| 0 | 0 | 0 | 0 | 0 |
| 0 | 0 | 0 | 0 | 0 |
| 0 | 0 | 0 | 0 | 0 |
| 0 | 0 | 0 | 0 | 0 |
| 0 | 0 | 0 | 0 | 0 |
| 0 | 0 | 0 | 0 | 0 |
| 0 | 0 | 0 | 0.00675874 | 0 |
| 0 | 0 | 0 | 0 | 0 |
| 0.001423924 | 0.014800731 | 0.008805863 | 0.0038933 | 0 |
| 0 | 0 | 0 | 0 | 0 |
| 0 | 0 | 0 | 0 | 0 |
| 0 | 0 | 0 | 0 | 0 |

| | | | | |
|---|---|---|---|---|
| 0 | 0 | 0 | 0 | 0 |
| 0 | 0 | 0 | 0 | 0 |
| 0 | 0 | 0 | 0 | 0 |
| 0 | 0 | 0 | 0 | 0 |
| 0 | 0 | 0.00539559 | 0 | 0 |
| 0.078781183 | 0.101721232 | 0.028416295 | 0.012073152 | 0.001127138 |
| 0 | 0 | 0 | 0 | 0 |
| 0 | 0 | 0 | 0 | 0 |
| 0 | 0 | 0 | 0 | 0 |
| 0 | 0 | 0 | 0.035205885 | 0 |
| 0 | 0 | 0 | 0 | 0 |
| 0 | 0 | 0 | 0 | 0 |
| 0 | 0 | 0 | 0 | 0 |
| 0 | 0 | 0 | 0 | 0 |
| 0 | 0 | 0 | 0 | 0 |
| 0 | 0 | 0 | 0 | 0 |
| 0 | 0.028749408 | 0.000643074 | 0.009356304 | 0 |
| 0 | 0 | 0 | 0 | 0 |
| 0 | 0 | 0 | 0 | 0 |
| 0 | 0 | 0 | 0 | 0 |
| 0 | 0 | 0 | 0 | 0 |
| 0 | 0 | 0 | 0 | 0 |
| 0 | 0 | 0 | 0 | 0 |
| 0.007829435 | 0 | 0 | 0 | 0 |
| 0 | 0 | 0 | 0 | 0 |
| 0.002401725 | 0 | 0 | 0 | 0 |
| 0 | 0 | 0 | 0 | 0 |
| 0.006360475 | 0 | 0 | 0 | 0 |
| 0 | 0 | 0 | 0 | 0 |
| 0 | 0 | 0 | 0.000808371 | 0 |
| 0 | 0 | 0 | 0 | 0 |
| 0 | 0.00095343 | 0 | 0 | 0 |
| 0 | 0 | 0 | 0 | 0 |
| 0 | 0 | 0 | 0 | 0 |
| 0 | 0 | 0 | 0 | 0 |
| 0 | 0 | 0 | 0 | 0 |
| 0 | 0 | 0 | 0 | 0 |
| 0 | 0 | 0 | 0 | 0 |
| 0 | 0.008929642 | 0.001945739 | 1.830631038 | 0 |
| 0 | 0 | 0 | 0 | 0 |
| 0 | 0 | 0 | 0 | 0 |
| 0 | 0 | 0 | 0 | 0 |
| 0 | 0 | 0 | 0 | 0 |
| 0 | 0 | 0 | 0 | 0 |
| 0 | 0 | 0 | 0 | 0 |
| 0 | 0 | 0 | 0 | 0 |
| 0 | 0 | 0 | 0 | 0 |
| 0 | 0 | 0 | 0 | 0 |

| | | | | |
|---|---|---|---|---|
| 0 | 0 | 0 | 0 | 0 |
| 0 | 0 | 0.001305812 | 0.017396709 | 0.003785766 |
| 0 | 0 | 0 | 0 | 0 |
| 0 | 0 | 0 | 0 | 0 |
| 0 | 0 | 0 | 0 | 0 |
| 0 | 0.001294945 | 0 | 0 | 0 |
| 0 | 0 | 0 | 0 | 0 |
| 0 | 0 | 0 | 0 | 0 |
| 0 | 0 | 0 | 0 | 0 |
| 0 | 0 | 0.005963663 | 0 | 0 |
| 0 | 0 | 0.005906781 | 0 | 0 |
| 0 | 0 | 0 | 0 | 0 |
| 0.25953637 | 0.015756057 | 0.293553884 | 0.835416268 | 0.908844399 |
| 0 | 0 | 0 | 0 | 0 |
| 0 | 0 | 0 | 0 | 0 |
| 0 | 0 | 0 | 0 | 0 |
| 0 | 0 | 0 | 0 | 0 |
| 0 | 0 | 0 | 0 | 0 |
| 0 | 0 | 0 | 0 | 0 |
| 0 | 0 | 0 | 0 | 0 |
| 0.000702961 | 0 | 0.002524222 | 0 | 0 |
| 0 | 0 | 0 | 0 | 0 |
| 0 | 0.001004775 | 0 | 0 | 0 |
| 1.377217536 | 0.048758674 | 0.574519917 | 1.545568984 | 1.709343707 |
| 0 | 0 | 0 | 0 | 0 |
| 0 | 0 | 0 | 0 | 0 |
| 0 | 0 | 0 | 0 | 0 |
| 0 | 0 | 0 | 0 | 0 |
| 0 | 0 | 0 | 0 | 0 |
| 0 | 0 | 0 | 0 | 0 |
| 0 | 0 | 0 | 0 | 0 |
| 0 | 0 | 0 | 0 | 0 |
| 0 | 0.000924088 | 0.007467863 | 0 | 0 |
| 0 | 0 | 0 | 0 | 0 |

| PHL018-0032_V2 | PHL018-0275_V1 | PHL018-0205_V2 | PHL018-0277_V1 | PHL018-0156_V1 |
|---|---|---|---|---|
| EYCF | REF | REF | REF | CTRL |
| 0.021552745 | 0.200661966 | 0.172692331 | 0.100412369 | 0.743362449 |
| 0 | 4.727898433 | 13.01650347 | 27.48892397 | 0.0038554 |
| 0 | 5.772200161 | 0.005597916 | 0.604948313 | 18.84437467 |
| 0 | 0 | 0 | 0 | 0 |
| 0 | 0 | 0 | 0 | 0 |
| 0 | 0 | 0 | 0 | 0 |
| 0 | 0 | 0 | 0 | 0 |
| 0 | 0 | 0 | 0 | 0 |
| 0 | 0 | 0 | 0 | 0 |
| 0 | 0 | 0 | 0 | 0 |
| 0 | 0 | 0 | 0 | 0 |
| 0 | 0 | 0 | 0 | 0 |
| 0 | 0 | 0 | 0 | 0 |
| 15.36900281 | 6.288970202 | 15.64616104 | 1.766273378 | 0.011791812 |
| 0 | 0 | 0 | 0 | 0 |
| 0 | 0 | 0 | 0 | 0 |
| 0 | 0 | 0.008955587 | 0 | 0 |
| 0 | 0 | 0 | 0 | 0 |
| 0 | 0 | 0 | 0 | 0 |
| 0 | 0 | 0 | 0 | 0 |
| 0 | 0 | 0 | 0 | 0 |
| 0 | 0 | 0 | 0 | 0 |
| 0 | 0 | 0 | 0 | 0 |
| 0 | 0 | 0 | 0 | 0 |
| 0 | 0 | 0 | 0 | 0 |
| 0 | 0 | 0 | 0 | 0 |
| 0 | 0 | 0 | 0 | 0 |
| 0 | 0 | 0 | 0 | 0 |
| 0 | 0 | 0 | 0 | 0 |
| 0 | 0 | 0 | 0 | 0 |
| 0 | 0 | 0 | 0 | 0 |
| 0 | 0 | 0 | 0 | 0 |
| 0 | 0 | 0 | 0 | 0 |
| 0 | 0 | 0 | 0 | 0 |
| 0 | 2.75567513 | 0 | 0 | 0 |
| 0 | 0 | 0 | 0 | 0 |
| 0 | 0 | 0 | 0 | 0 |
| 0 | 0 | 0 | 0 | 0 |
| 0 | 0 | 0 | 0 | 0 |
| 0 | 0 | 0 | 0 | 0 |
| 0.00071704 | 0.002209498 | 0.006117866 | 0 | 0.054043369 |
| 0 | 0.002933955 | 0.163037763 | 0.032378741 | 2.33653098 |
| 0.096608916 | 0.010506781 | 0.000809015 | 0.021105916 | 0.077418754 |
| 0 | 0 | 0 | 0.002646369 | 0 |
| 0 | 0 | 0 | 0 | 0 |

| | | | | |
|---|---|---|---|---|
| 0 | 0 | 0 | 0 | 0 |
| 0.068526783 | 0 | 0.055125601 | 0.015042074 | 0.003338635 |
| 0 | 0.008280385 | 0 | 0.001219346 | 0.038647039 |
| 0.569534323 | 0.412229626 | 0 | 0.11545198 | 0.99440619 |
| 0 | 0 | 0 | 0.000656205 | 0 |
| 0 | 0 | 0 | 0 | 0 |
| 0 | 0 | 0.000668895 | 0 | 0.004920407 |
| 0 | 0 | 0 | 0 | 0 |
| 0 | 0 | 0 | 0 | 0 |
| 0 | 0 | 0 | 0 | 0 |
| 0 | 0 | 0 | 0 | 0.001438469 |
| 0 | 0.044570607 | 0 | 0 | 0 |
| 0 | 0 | 0 | 0 | 0 |
| 0.023980475 | 0 | 0.036087813 | 0.017659435 | 0 |
| 0 | 0 | 0.014459475 | 0 | 0 |
| 0 | 0 | 0 | 0 | 0 |
| 0.001590711 | 0.001715574 | 0.658685676 | 4.813288511 | 0.035572348 |
| 0 | 0 | 0 | 0 | 0 |
| 0 | 0 | 0 | 0 | 0 |
| 0 | 0 | 0 | 0 | 0 |
| 0.745281439 | 0.094266305 | 0.031215643 | 0.125600086 | 0 |
| 0.021873078 | 0.022171061 | 0.131967224 | 0.002920078 | 0 |
| 0 | 0 | 0 | 0 | 0.001960648 |
| 0 | 0 | 0 | 0 | 0 |
| 0 | 0 | 0 | 0 | 0 |
| 0 | 0 | 0 | 0 | 0 |
| 0 | 0 | 0 | 0 | 0 |
| 0 | 0 | 0 | 0 | 0 |
| 0 | 0 | 0 | 0 | 0.01646707 |
| 0 | 0 | 0 | 0 | 0 |
| 0.360463911 | 0.009044013 | 0.179691429 | 0.876569787 | 0 |
| 0 | 0 | 0 | 0 | 0 |
| 0 | 0 | 0 | 0 | 0 |
| 0 | 0 | 0 | 0 | 0 |
| 0 | 0 | 0 | 0 | 0 |
| 0 | 0 | 0 | 0 | 0 |
| 0 | 0 | 0 | 0 | 0 |
| 0.000697984 | 0.037208483 | 0 | 0 | 2.582181568 |
| 0 | 0 | 0 | 0 | 0 |
| 0 | 0 | 0 | 0 | 0 |
| 0 | 0 | 0 | 0 | 0 |
| 0 | 0 | 0 | 0 | 0 |
| 0 | 0 | 0 | 0 | 0 |
| 0 | 0 | 0 | 0 | 0 |
| 0 | 0 | 0 | 0 | 0 |
| 0 | 0 | 0 | 0 | 0 |
| 0 | 0 | 0 | 0 | 0 |
| 1.058614279 | 0 | 0 | 0 | 0.018303238 |

| | | | | |
|---|---|---|---|---|
| 0 | 0 | 0 | 0 | 0 |
| 0 | 0 | 0.067547486 | 0 | 0 |
| 0 | 0 | 0 | 0 | 0 |
| 0 | 0 | 0 | 0 | 0 |
| 0 | 0 | 0 | 0 | 0.192281549 |
| 0 | 0 | 0 | 0 | 0 |
| 0 | 0 | 0 | 0 | 0 |
| 0 | 0 | 0.092129443 | 0.008584635 | 0 |
| 0.004636413 | 0 | 0 | 0 | 0 |
| 0 | 0 | 0 | 0 | 0 |
| 0 | 0 | 0 | 0 | 0 |
| 0 | 0 | 0 | 0 | 0.00142664 |
| 0 | 0 | 0 | 0 | 0 |
| 0 | 0 | 0 | 0 | 0 |
| 0 | 0 | 0 | 0 | 0 |
| 0 | 0 | 0 | 0 | 0 |
| 0 | 0 | 0 | 0 | 0 |
| 0 | 0 | 0 | 0 | 0 |
| 0 | 0 | 0 | 0 | 0 |
| 0 | 0 | 0 | 0 | 0 |
| 0 | 0 | 0 | 0 | 0 |
| 0 | 0 | 0 | 0 | 0 |
| 0 | 0 | 0 | 0 | 0 |
| 0 | 0 | 0 | 0 | 0 |
| 0 | 0 | 0 | 0 | 0 |
| 0 | 0 | 0 | 0 | 0 |
| 0 | 0 | 0 | 0 | 0 |
| 0 | 0 | 0 | 0 | 0 |
| 0 | 0 | 0 | 0 | 0 |
| 0 | 0 | 0 | 0 | 0 |
| 0 | 0.001586461 | 0 | 0 | 0 |
| 0 | 0 | 0 | 0 | 0 |
| 0 | 0 | 0 | 0 | 0 |
| 0 | 0 | 0 | 0 | 0 |
| 0 | 0 | 0 | 0 | 0 |
| 0 | 0 | 0 | 0 | 0 |
| 0 | 0 | 0 | 0 | 0 |
| 0 | 0 | 0 | 0 | 0 |
| 0 | 0.333484008 | 0 | 0 | 0 |
| 0 | 0 | 0 | 0 | 0 |
| 0 | 0 | 0 | 0 | 0 |
| 0 | 0 | 0 | 0 | 0 |
| 0 | 0 | 0 | 0 | 0 |
| 0 | 0 | 0 | 0 | 0 |
| 0 | 0 | 0 | 0 | 0 |
| 0 | 0 | 0 | 0 | 0 |
| 0 | 0 | 0 | 0 | 0 |
| 0 | 0 | 0 | 0 | 0 |

| | | | | |
|---|---|---|---|---|
| 0 | 0 | 0.000834335 | 0 | 0 |
| 0 | 0 | 0 | 0 | 0 |
| 0 | 0 | 0 | 0 | 0 |
| 0 | 0 | 0 | 0 | 0 |
| 0 | 0 | 0 | 0 | 0 |
| 0 | 0 | 0 | 0 | 0 |
| 0 | 0 | 0 | 0 | 0 |
| 0 | 0 | 0 | 0 | 0 |
| 0 | 0 | 0 | 0 | 0 |
| 0 | 0 | 0 | 0 | 0 |
| 0 | 0 | 0 | 0 | 0 |
| 0 | 0 | 0 | 0 | 0 |
| 0 | 0 | 0 | 0 | 0 |
| 0 | 0 | 0 | 0 | 0 |
| 0 | 0 | 0 | 0 | 0 |
| 0 | 0 | 0 | 0 | 0 |
| 0 | 0 | 0 | 0 | 0 |
| 0 | 0 | 0 | 0 | 0 |
| 0 | 0 | 0 | 0 | 0 |
| 0 | 0 | 0 | 0 | 0 |
| 0 | 0 | 0 | 0 | 0 |
| 0 | 0 | 0 | 0 | 0 |
| 0 | 0 | 0 | 0 | 0 |
| 0 | 0 | 0 | 0 | 0 |
| 0 | 0 | 0 | 0 | 0 |
| 0 | 0 | 0 | 0 | 0 |
| 0 | 0 | 0 | 0 | 0 |
| 0 | 0 | 0 | 0 | 0 |
| 0 | 0 | 0 | 0 | 0 |
| 0 | 0 | 0 | 0 | 0 |
| 0 | 0 | 0 | 0.003591438 | 0 |
| 0.006231826 | 0 | 0 | 0 | 0 |
| 0 | 0 | 0 | 0 | 0 |
| 0 | 0 | 0 | 0 | 0 |
| 0 | 0 | 0 | 0 | 0 |
| 0 | 0 | 0 | 0 | 0 |
| 0 | 0 | 0 | 0 | 0 |
| 0 | 0 | 0 | 0 | 0 |
| 0 | 0 | 0 | 0 | 0 |
| 0 | 0 | 0 | 0 | 0 |
| 0 | 0 | 0 | 0 | 0 |
| 0 | 0 | 0 | 0 | 0 |
| 0 | 0 | 0 | 0 | 0 |
| 0 | 0 | 0 | 0 | 0 |
| 0 | 0 | 0 | 0 | 0 |
| 0 | 0 | 0 | 0 | 0 |
| 0 | 0 | 0 | 0 | 0 |
| 0 | 0 | 0 | 0 | 0 |

| | | | | |
|---|---|---|---|---|
| 0 | 0 | 0 | 0 | 0 |
| 0 | 0 | 0 | 0 | 0 |
| 0 | 0 | 0 | 0 | 0 |
| 0 | 0 | 0 | 0 | 0 |
| 0 | 0 | 0 | 0 | 0 |
| 0 | 0 | 0 | 0 | 0 |
| 0 | 0 | 0 | 0 | 0 |
| 0 | 0 | 0 | 0 | 0 |
| 0 | 0 | 0 | 0 | 0 |
| 0 | 0 | 0 | 0 | 0 |
| 0 | 0 | 0 | 0 | 0 |
| 0 | 0 | 0 | 0 | 0 |
| 0.001590199 | 0 | 0.000656504 | 0.001396236 | 0 |
| 0 | 0 | 0 | 0 | 0 |
| 0 | 0 | 0 | 0 | 0 |
| 0 | 0 | 0 | 0 | 0 |
| 0 | 0 | 0 | 0 | 0 |
| 0 | 0 | 0 | 0 | 0 |
| 0 | 0 | 0 | 0 | 0 |
| 0 | 0 | 0 | 0 | 0 |
| 0.27019775 | 0.122311899 | 0.037132031 | 0 | 0.100658992 |
| 0 | 0 | 0 | 0 | 0.00313708 |
| 0 | 0 | 0 | 0 | 0 |
| 0 | 0 | 0 | 0 | 0.002889002 |
| 0 | 0 | 0 | 0 | 0 |
| 0 | 0 | 0 | 0 | 0 |
| 0.004761829 | 0.003401507 | 0.001787175 | 0 | 0.002509786 |
| 0 | 0 | 0 | 0 | 0 |
| 0 | 0 | 0 | 0 | 0 |
| 0.016210117 | 0 | 0 | 0 | 0.005945759 |
| 0.088030356 | 0.005185815 | 0.054336061 | 0.005557009 | 0 |
| 0 | 0 | 0 | 0 | 0 |
| 0 | 0 | 0 | 0 | 0 |
| 0 | 0 | 0 | 0 | 0 |
| 0 | 0.018330833 | 0.02855282 | 0.008493062 | 0.017625325 |
| 0 | 0 | 0 | 0 | 0 |
| 0 | 0 | 0 | 0 | 0 |
| 0 | 0 | 0 | 0 | 0 |
| 0 | 0 | 0 | 0 | 0 |
| 0 | 0 | 0 | 0 | 0 |
| 0 | 0 | 0 | 0 | 0 |
| 0 | 0 | 0.002193513 | 0 | 0 |
| 0 | 0 | 0 | 0 | 0.000832707 |
| 0 | 0 | 0 | 0 | 0 |
| 0 | 0 | 0 | 0 | 0 |
| 0 | 0 | 0 | 0 | 0 |
| 0 | 0 | 0 | 0 | 0 |
| 0.007582491 | 0 | 0 | 0 | 0.020267729 |

| | | | | |
|---|---|---|---|---|
| 0 | 0 | 0 | 0 | 0 |
| 0 | 0 | 0 | 0 | 0 |
| 0.003149419 | 0 | 0 | 0 | 0 |
| 0 | 0 | 0 | 0 | 0 |
| 0 | 0 | 0 | 0 | 0 |
| 0 | 0 | 0 | 0 | 0 |
| 0 | 0 | 0 | 0 | 0.002641276 |
| 0 | 0 | 0 | 0 | 0.005744975 |
| 0 | 0 | 0.007150003 | 0 | 0 |
| 0 | 0 | 0 | 0 | 0 |
| 0 | 0 | 0 | 0 | 0 |
| 0 | 0 | 0 | 0 | 0 |
| 0 | 0 | 0 | 0 | 0 |
| 0 | 0 | 0 | 0 | 0 |
| 0 | 0 | 0 | 0 | 0.000675116 |
| 0 | 0 | 0 | 0 | 0 |
| 0 | 0 | 0 | 0 | 0 |
| 0 | 0 | 0 | 0 | 0 |
| 0 | 0 | 0 | 0 | 0.008854514 |
| 0 | 0 | 0 | 0 | 0 |
| 0.016804002 | 0 | 0.221784371 | 0.0090796 | 0.096139801 |
| 0 | 0 | 0 | 0 | 0 |
| 0 | 0 | 0 | 0 | 0 |
| 0 | 0 | 0 | 0 | 0 |
| 0.032871091 | 0 | 0 | 0 | 0.007231703 |
| 0 | 0 | 0 | 0 | 0 |
| 0 | 0 | 0 | 0 | 0 |
| 0 | 0 | 0 | 0 | 0 |
| 0 | 0 | 0 | 0 | 0.067795312 |
| 0 | 0 | 0 | 0 | 0 |
| 0.059688868 | 0.020559898 | 0.127919668 | 0.008456553 | 0.088010201 |
| 0 | 0 | 0 | 0 | 0 |
| 0 | 0 | 0 | 0 | 0 |
| 0.000737809 | 0 | 0 | 0 | 0 |
| 0 | 0 | 0 | 0 | 0 |
| 0 | 0 | 0 | 0 | 0 |
| 0 | 0 | 0 | 0 | 0 |
| 0 | 0 | 0 | 0 | 0 |
| 0 | 0 | 0 | 0 | 0 |
| 0 | 0 | 0 | 0 | 0 |
| 0 | 0 | 0 | 0 | 0 |
| 0 | 0.042093917 | 0 | 0.024499089 | 0.009950142 |
| 0 | 0 | 0 | 0 | 0 |
| 0 | 0 | 0 | 0 | 0 |
| 0 | 0 | 0 | 0 | 0 |
| 0 | 0 | 0 | 0 | 0 |
| 0 | 0 | 0 | 0 | 0 |
| 0 | 0 | 0 | 0 | 0 |

| | | | | |
|---|---|---|---|---|
| 0 | 0 | 0 | 0 | 0 |
| 0 | 0 | 0 | 0 | 0 |
| 0.107298169 | 0.007422309 | 0 | 0 | 0.085644635 |
| 0 | 0 | 0 | 0 | 0 |
| 0 | 0 | 0 | 0.252876761 | 0 |
| 0 | 0 | 0 | 0 | 0 |
| 0 | 0 | 0 | 0 | 0 |
| 0 | 0 | 0 | 0 | 0 |
| 0 | 0 | 0 | 0 | 0 |
| 0.002225263 | 0 | 0 | 0 | 0 |
| 0 | 0 | 0 | 0 | 0 |
| 0 | 0 | 0 | 0 | 0 |
| 0 | 0 | 0 | 0 | 0 |
| 0.038183462 | 0.000994302 | 0.013765585 | 0 | 0 |
| 0.009543411 | 0.001025833 | 0.008063134 | 0 | 0.007720438 |
| 0 | 0 | 0 | 0 | 0 |
| 0.023770765 | 0 | 0.001879204 | 0.016874708 | 0.005655052 |
| 0 | 0 | 0.00097182 | 0 | 0 |
| 0 | 0 | 0 | 0 | 0 |
| 0 | 0 | 0 | 0 | 0 |
| 0 | 0 | 0 | 0 | 0 |
| 0.00639181 | 0 | 0 | 0 | 0.017772608 |
| 0 | 0 | 0 | 0 | 0 |
| 0.006369919 | 0 | 0 | 0.002485758 | 0 |
| 0 | 0 | 0 | 0 | 0 |
| 0 | 0 | 0 | 0 | 0 |
| 0 | 0 | 0 | 0 | 0 |
| 0 | 0 | 0 | 0 | 0 |
| 0 | 0 | 0 | 0 | 0 |
| 0.020472868 | 0 | 0 | 0 | 0 |
| 0 | 0 | 0 | 0 | 0 |
| 0 | 0 | 0 | 0 | 0 |
| 0 | 0 | 0 | 0 | 0 |
| 0 | 0 | 0 | 0 | 0 |
| 0 | 0 | 0 | 0 | 0 |
| 0 | 0 | 0.001129429 | 0 | 0.023036103 |
| 0 | 0 | 0 | 0 | 0 |
| 0.001147235 | 0.000795399 | 0.001420888 | 0 | 0.014965477 |
| 0 | 0 | 0 | 0 | 0 |
| 0 | 0 | 0 | 0 | 0 |
| 0 | 0 | 0 | 0 | 0 |
| 0 | 0 | 0 | 0 | 0 |
| 0 | 0 | 0 | 0 | 0 |
| 0 | 0 | 0.005294894 | 0 | 0 |
| 0 | 0 | 0 | 0 | 0 |
| 0.001322892 | 0 | 0 | 0 | 0 |
| 0 | 0 | 0 | 0 | 0 |
| 0.014243294 | 0.002569141 | 0.002868419 | 0.002440192 | 0.031937952 |

| | | | | |
|---|---|---|---|---|
| 0 | 0 | 0 | 0 | 0 |
| 0 | 0 | 0 | 0.111117964 | 0 |
| 0 | 0 | 0 | 0.001959438 | 0 |
| 0 | 0 | 0 | 0 | 0 |
| 0.048352971 | 0.035136264 | 0.015890337 | 0 | 0 |
| 0 | 0 | 0 | 0 | 0 |
| 0 | 0 | 0 | 0 | 0 |
| 0.01011602 | 0.02587247 | 0.030069637 | 0 | 0 |
| 0 | 0 | 0 | 0 | 0 |
| 0 | 0.28691461 | 0.43189913 | 0.037433784 | 0.007484463 |
| 0 | 0.001161961 | 0 | 0 | 0 |
| 0 | 0 | 0 | 0 | 0.097497225 |
| 0.013768321 | 0.001310212 | 0.005349806 | 0.006162991 | 0.033875128 |
| 0 | 0 | 0 | 0 | 0 |
| 0 | 0 | 0 | 0 | 0 |
| 0 | 0 | 0 | 0 | 0 |
| 0 | 0 | 0 | 0 | 0 |
| 0 | 0 | 0 | 0 | 0 |
| 0 | 0.269679087 | 1.906747784 | 0 | 0 |
| 0 | 0 | 0 | 0.159587006 | 0 |
| 0 | 0 | 0 | 0.004201523 | 0 |
| 0.782173501 | 0 | 0 | 0 | 0 |
| 0 | 0 | 0 | 0 | 0.001211303 |
| 0 | 0 | 0 | 0 | 0 |
| 0 | 0 | 0 | 0 | 0 |
| 0 | 0 | 0 | 0.003036978 | 0.002148588 |
| 0 | 0 | 0 | 0 | 0 |
| 0.039926121 | 0.009754437 | 0.008477106 | 0.002670946 | 0 |
| 0 | 0 | 0 | 0 | 0 |
| 0 | 0 | 0 | 0 | 0 |
| 0.04858765 | 0.038365416 | 0 | 0 | 0.005659081 |
| 0 | 0 | 0 | 0 | 0.014654675 |
| 0 | 0 | 0 | 0 | 0 |
| 0 | 0 | 0.003055955 | 0 | 0 |
| 0 | 0 | 0 | 0 | 0 |
| 0.110724767 | 0 | 1.600810695 | 0 | 0 |
| 2.89315195 | 0 | 0.020749498 | 0.020343382 | 0 |
| 0 | 0 | 0 | 0 | 0.030503205 |
| 0 | 0 | 0 | 0 | 0 |
| 0 | 0 | 0 | 0.002322999 | 0.017256384 |
| 0 | 0 | 0 | 0 | 0 |
| 0 | 0 | 0 | 0 | 0 |
| 0.001081861 | 0 | 0 | 0 | 0 |
| 0 | 0 | 0 | 0 | 0 |
| 0 | 0 | 0 | 0 | 0 |
| 0 | 0 | 0 | 0 | 0 |
| 0.002346207 | 0.002168891 | 0.002663698 | 0.001373354 | 0.001214518 |
| 0 | 0 | 0.001978172 | 0 | 0 |

| | | | | |
|---|---|---|---|---|
| 0 | 0 | 0 | 0 | 0 |
| 0 | 0 | 0 | 0 | 0 |
| 0 | 0 | 0 | 0 | 0 |
| 0 | 0 | 0 | 0 | 0 |
| 0 | 0 | 0.005265574 | 0 | 0.020167116 |
| 0.137901189 | 0 | 0 | 0 | 0 |
| 0.019263635 | 0 | 0 | 0 | 0 |
| 0 | 0 | 0 | 0 | 0 |
| 0 | 0 | 0 | 0 | 0 |
| 0 | 0 | 0 | 0 | 0 |
| 0.025183523 | 0.025013899 | 0.010396867 | 0 | 0 |
| 0.001255463 | 0 | 0 | 0 | 0 |
| 0 | 0 | 0 | 0.002730188 | 0 |
| 0 | 0.019888211 | 0.001880892 | 0 | 0 |
| 0 | 0 | 0.741795831 | 0 | 0 |
| 0 | 0 | 0 | 0 | 0 |
| 0 | 0 | 0 | 0 | 0 |
| 1.747110155 | 0 | 0 | 0 | 0 |
| 0 | 0 | 0 | 0.003153802 | 0 |
| 0 | 0 | 0 | 0 | 0 |
| 0 | 0 | 0 | 0 | 0 |
| 0 | 0.472465904 | 0 | 0 | 0 |
| 0 | 0 | 0 | 0 | 0 |
| 0 | 0 | 0 | 0 | 0 |
| 0 | 0 | 0 | 0 | 0 |
| 0 | 0 | 0 | 0 | 0 |
| 0.044458598 | 0.002283256 | 0 | 0.005140517 | 0 |
| 0 | 0 | 0 | 0 | 0 |
| 0 | 0 | 0 | 0 | 0 |
| 0.582474574 | 0.412585863 | 0.621242689 | 0.618925356 | 0.070367207 |
| 0 | 0 | 0 | 0 | 0 |
| 0 | 0 | 0 | 0 | 0 |
| 0.003966349 | 0 | 0 | 0 | 0 |
| 0 | 0.001348948 | 0 | 0 | 0 |
| 0 | 0 | 0.111542437 | 0 | 0 |
| 0 | 0 | 0 | 0 | 0 |
| 0 | 0 | 0 | 0 | 0 |
| 0.004385731 | 0 | 0 | 0 | 0.002136735 |
| 0 | 0 | 0 | 0 | 0 |
| 0 | 0 | 3.406121929 | 0 | 0 |
| 0.126930076 | 0 | 0.055151301 | 0 | 0.045498402 |
| 0 | 0 | 0 | 0 | 0 |
| 0.004726604 | 0 | 0 | 0 | 0.002153126 |
| 0 | 0 | 0 | 0 | 0 |
| 0 | 0 | 0.000581435 | 0 | 0 |
| 0 | 0 | 0 | 0 | 0 |
| 0.013540619 | 0 | 0.000901639 | 0.001534066 | 0.003255943 |
| 0 | 0 | 0 | 0 | 0 |

| | | | | |
|---|---|---|---|---|
| 0 | 0 | 0 | 0 | 0 |
| 0 | 0.001273342 | 0 | 0 | 0 |
| 0.004773174 | 0 | 0.04183685 | 0.289714251 | 0.004561552 |
| 0 | 0 | 0.001006305 | 0 | 0 |
| 0 | 0 | 0 | 0 | 0 |
| 0 | 0 | 0.207307309 | 0 | 0 |
| 0 | 0 | 0 | 0 | 0 |
| 0 | 0 | 0 | 0 | 0 |
| 0.150195212 | 0 | 0.029048418 | 0.021385183 | 0 |
| 0 | 0 | 0 | 0 | 0 |
| 0 | 0 | 0 | 0 | 0 |
| 0.002272184 | 0 | 0 | 0 | 0 |
| 0 | 0 | 0 | 0 | 0 |
| 0 | 0 | 0 | 0 | 0 |
| 0 | 0 | 0 | 0 | 0 |
| 0.060010239 | 0 | 0 | 0.008698122 | 0 |
| 0.002322749 | 0.000585602 | 0.007497114 | 0 | 0.018538433 |
| 0 | 0 | 0.00826138 | 0 | 0.002974798 |
| 0 | 0 | 0 | 0 | 0 |
| 0 | 0 | 0 | 0 | 0 |
| 0 | 0 | 0 | 0 | 0 |
| 0.007348005 | 0.003849182 | 0.006269399 | 0 | 0.00142005 |
| 0 | 0 | 0 | 0 | 0 |
| 0 | 0.066366881 | 0 | 0 | 0 |
| 0 | 0 | 0 | 0 | 0 |
| 0 | 0 | 0 | 0 | 0 |
| 0.008578179 | 0 | 0.002656087 | 0 | 0.003108355 |
| 0 | 0 | 0 | 0 | 0 |
| 0 | 0 | 0 | 0 | 0 |
| 0 | 0 | 0.00662873 | 0 | 0 |
| 0 | 0 | 0 | 0 | 0.000844248 |
| 0.063522985 | 0.00112387 | 0.007779691 | 0.015300272 | 0.244182118 |
| 0 | 2.834363775 | 0.016248309 | 0.013212772 | 0 |
| 0 | 0 | 0 | 0 | 0.006182854 |
| 0 | 0 | 0 | 0 | 0.038981365 |
| 0 | 0 | 0 | 0 | 0 |
| 0.00313954 | 0 | 0 | 0.006432061 | 0.029903455 |
| 0.012276329 | 0 | 0.073151155 | 0.001437191 | 0.066260017 |
| 0 | 0 | 0 | 0.985259547 | 0 |
| 0 | 0 | 0 | 0 | 0 |
| 0 | 0 | 0 | 0 | 0 |
| 0 | 0 | 0 | 0 | 0 |
| 0 | 0 | 0 | 0 | 0 |
| 0 | 0.010335422 | 0 | 0 | 0.002381161 |
| 0 | 0 | 0.703508543 | 0.174172886 | 0 |
| 0 | 0 | 0 | 0 | 0 |
| 0 | 0 | 0 | 0 | 0 |
| 0 | 0 | 0 | 0 | 0 |

| | | | | |
|---|---|---|---|---|
| 0.14045662 | 0 | 0 | 0.007813219 | 0 |
| 0.516896926 | 0.020876161 | 1.298572913 | 3.30015563 | 0.002078233 |
| 0 | 0 | 0 | 0 | 0 |
| 0 | 0 | 0 | 0 | 0.000839105 |
| 0 | 0 | 0 | 0 | 0.002038844 |
| 0 | 0 | 0 | 0 | 0 |
| 0.001289636 | 0.004291815 | 0.001703739 | 0 | 0 |
| 0 | 0 | 0 | 0 | 0.017204589 |
| 0 | 0 | 0 | 0 | 0.000845534 |
| 0 | 0 | 0 | 0 | 0 |
| 0 | 0 | 0 | 0.001212727 | 0.00068638 |
| 0.059251172 | 0.005705545 | 0.020639385 | 0.046243615 | 0.177383286 |
| 0 | 0 | 0 | 0 | 0 |
| 0 | 0 | 0 | 0 | 0 |
| 0.000713138 | 0.028567159 | 0 | 0 | 0.018900837 |
| 0 | 0 | 0 | 0 | 0 |
| 0.073111112 | 0 | 0.007881741 | 0.068066649 | 0.002335901 |
| 0.67111731 | 0.001562058 | 0.01976557 | 0 | 0 |
| 0.001609649 | 0 | 0 | 0 | 0 |
| 0 | 0 | 0.18844401 | 0 | 0 |
| 0.005369448 | 0.010754572 | 0.030664968 | 0.007071772 | 0.003706004 |
| 0 | 0 | 0 | 0 | 0 |
| 0 | 0 | 0 | 0 | 0 |
| 0 | 0 | 0 | 0 | 0 |
| 0 | 0 | 0 | 0 | 0 |
| 0 | 0 | 0 | 0 | 0 |
| 0 | 0 | 0 | 0 | 0 |
| 0 | 0.005715261 | 0.006675538 | 0.124157313 | 0.027178654 |
| 0 | 0 | 0 | 0 | 0 |
| 0 | 0 | 0 | 0.002350537 | 0 |
| 0 | 0 | 0 | 0 | 0 |
| 0 | 0 | 0 | 0 | 0 |
| 0.013278999 | 0 | 0 | 0 | 0 |
| 0 | 0 | 0 | 0 | 0 |
| 0 | 0 | 0 | 0 | 0 |
| 1.20652508 | 1.955381388 | 18.20498555 | 1.642868352 | 0.326320927 |
| 0 | 0 | 0 | 0 | 0 |
| 0 | 0 | 0 | 0 | 0 |
| 0.018187022 | 0 | 0.004117512 | 0.00137365 | 0.051506645 |
| 0 | 0 | 0 | 0 | 0 |
| 0 | 0 | 0.17022418 | 0.035340863 | 0 |
| 0 | 0 | 0 | 0 | 0 |
| 0.018530577 | 0 | 0.00161832 | 0.002920315 | 0.009739957 |
| 0.011353855 | 0 | 0.005539621 | 0 | 0 |
| 0 | 0 | 3.079059673 | 0 | 0 |
| 0 | 0 | 0 | 0 | 0 |
| 0 | 0 | 0 | 0 | 0 |
| 0.031421921 | 0.016708561 | 0 | 0 | 0.001612166 |

| | | | | |
|---|---|---|---|---|
| 0 | 0 | 0.002877417 | 0 | 0 |
| 0 | 0 | 0 | 0 | 0 |
| 0 | 0 | 0 | 0 | 0 |
| 0 | 0 | 0 | 0.00121473 | 0 |
| 0 | 0 | 0 | 0 | 0 |
| 0 | 0 | 0 | 0 | 0 |
| 0 | 0 | 0 | 0 | 0 |
| 0 | 0.006178312 | 0 | 0.002390753 | 0 |
| 0 | 0 | 0 | 0 | 0 |
| 0 | 0 | 0 | 0 | 0 |
| 0 | 0 | 0 | 0 | 0 |
| 0 | 0 | 0 | 0 | 0 |
| 0 | 0 | 0 | 0 | 0 |
| 0 | 0 | 0 | 0 | 0 |
| 0 | 0 | 0 | 0.00576243 | 0.044436884 |
| 0 | 0 | 0 | 0 | 0 |
| 0 | 0 | 0 | 0 | 0 |
| 0 | 0 | 0 | 0 | 0 |
| 0 | 0 | 0 | 0 | 0 |
| 0 | 0 | 0 | 0 | 0 |
| 0 | 0 | 0 | 0 | 0 |
| 0 | 0 | 0 | 0 | 0 |
| 0 | 0 | 0 | 0 | 0 |
| 0.54177756 | 0.023668275 | 0.083517251 | 0.130256363 | 0.000392698 |
| 0.001861539 | 0 | 0 | 0 | 0 |
| 0 | 0 | 0 | 0.129369777 | 0 |
| 0 | 0 | 0 | 0 | 0 |
| 0 | 0 | 0 | 0 | 0 |
| 0 | 0 | 0 | 0 | 0 |
| 0 | 0 | 0 | 0 | 0 |
| 0 | 0 | 0 | 0 | 0 |
| 0 | 0 | 0 | 0 | 0 |
| 0 | 0 | 0 | 0 | 0 |
| 0 | 0 | 0 | 0 | 0 |
| 0 | 0 | 0 | 0 | 0 |
| 0 | 0 | 0 | 0 | 0 |
| 0 | 0 | 0 | 0 | 0 |
| 0 | 0 | 0 | 0 | 0.037181401 |
| 0 | 0 | 0.015210188 | 0 | 0 |
| 0 | 0 | 0 | 0 | 0 |
| 0.318345107 | 0.004408118 | 0.076698492 | 0.050912307 | 0.023064934 |
| 0 | 0 | 0 | 0 | 0 |
| 0.525819348 | 16.31964662 | 0.25543432 | 0.563073826 | 0 |
| 0 | 0 | 0 | 0 | 0 |
| 0 | 0 | 0 | 0 | 0 |
| 0 | 0 | 0 | 0 | 0 |
| 0 | 0 | 0 | 0 | 0 |
| 0.030725047 | 0 | 0.000757293 | 0 | 0.00512754 |

| | | | | |
|---|---|---|---|---|
| 0 | 0 | 0 | 0 | 0 |
| 0 | 0 | 0 | 0 | 0 |
| 2.008987433 | 0.04957837 | 0.419417927 | 0.530340882 | 1.327866126 |
| 0 | 0 | 0 | 0 | 0 |
| 0 | 0 | 0 | 0 | 0 |
| 0 | 0.000800326 | 0 | 0 | 0 |
| 0 | 0 | 0 | 0 | 0 |
| 0 | 0 | 0.020863509 | 0 | 0 |
| 0 | 0 | 0 | 0 | 0 |
| 0 | 0 | 0 | 0 | 0 |
| 0.010358701 | 0.001396476 | 0.08071947 | 0.050528937 | 0.255419362 |
| 0 | 0.086837601 | 0.295697473 | 2.004213145 | 0.071546279 |
| 0 | 0 | 0 | 0 | 0 |
| 0.001397442 | 0.002153049 | 0.002115393 | 0.040899667 | 0 |
| 3.701810108 | 0.001187679 | 0.530261057 | 1.246246224 | 0.390071241 |
| 0 | 0 | 0 | 0.261595749 | 0 |
| 0 | 0 | 0 | 0 | 0 |
| 0 | 0 | 0 | 0 | 0 |
| 0 | 0 | 0 | 0 | 0 |
| 0 | 0 | 0 | 0 | 0 |
| 0.884309813 | 0.003009735 | 0.105290586 | 0.05886746 | 0.051984203 |
| 0.016390629 | 1.120888019 | 0.644731889 | 0.002008102 | 6.869160191 |
| 0 | 9.744692725 | 4.113683155 | 0.879627561 | 0.277905093 |
| 0 | 0 | 0 | 0 | 0 |
| 0 | 0 | 0 | 0 | 0 |
| 0 | 0 | 0 | 0 | 0 |
| 0.187564912 | 0 | 0 | 0.483993221 | 0 |
| 0 | 0.006452593 | 0 | 0 | 0 |
| 0 | 0 | 0 | 0 | 0 |
| 0 | 0 | 0 | 0 | 0 |
| 0 | 0 | 0 | 0 | 0 |
| 0 | 0.076244793 | 0 | 0.109351585 | 0 |
| 1.937786899 | 0 | 0.467576269 | 0.821749959 | 0.004038552 |
| 0 | 0 | 0 | 0 | 0 |
| 0.05441775 | 0 | 0.02005034 | 0.000856276 | 0 |
| 0 | 0.012448334 | 0.006969961 | 0.016235297 | 0.019642853 |
| 0 | 0 | 0 | 0 | 0.050813678 |
| 0 | 0 | 0 | 0 | 0 |
| 0 | 0.031743157 | 0 | 0 | 0 |
| 0 | 0 | 0 | 0 | 0 |
| 0 | 0 | 0 | 0 | 0 |
| 0 | 0 | 0 | 0 | 0 |
| 0 | 0 | 0 | 0 | 0 |
| 0 | 0 | 0 | 0 | 0 |
| 0.012606073 | 0.036231359 | 0.02043887 | 0.001878284 | 0.187176593 |
| 0 | 0 | 0 | 0 | 0 |
| 0 | 0 | 0 | 0 | 0 |
| 0.061654912 | 0.051116865 | 0.236732342 | 0.043372425 | 0.812692639 |

| | | | | |
|---|---|---|---|---|
| 0 | 0 | 0 | 0 | 0 |
| 0 | 0 | 0 | 0 | 0 |
| 0.003751888 | 0 | 0 | 0 | 0 |
| 0 | 0 | 0 | 0 | 0 |
| 0 | 0 | 0 | 0 | 0 |
| 0 | 0 | 0 | 0 | 0 |
| 0 | 0 | 0 | 0 | 0 |
| 0 | 0 | 0.029832722 | 0.012836799 | 0 |
| 0 | 0 | 0 | 0 | 0 |
| 0 | 0 | 0 | 0 | 0 |
| 0.036333393 | 0 | 0.002514974 | 0 | 0.002162361 |
| 0 | 0 | 0 | 0 | 0 |
| 0 | 0 | 0 | 0 | 0 |
| 0 | 0 | 0 | 0 | 0 |
| 0 | 0 | 0 | 0 | 0 |
| 0 | 0 | 0 | 0 | 0 |
| 0 | 0 | 0 | 0 | 0 |
| 0 | 0 | 0 | 0.00657309 | 0 |
| 0 | 0 | 0 | 0 | 0 |
| 0 | 0 | 0 | 0 | 0 |
| 0 | 0 | 0 | 0 | 0 |
| 0 | 0 | 0 | 0 | 0 |
| 0 | 0 | 0 | 0 | 0 |
| 0 | 0 | 0 | 0 | 0 |
| 0 | 0 | 0 | 0 | 0 |
| 0 | 0 | 0 | 0 | 0 |
| 0.002915006 | 0 | 0 | 0 | 0 |
| 0 | 0 | 0 | 0 | 0 |
| 0 | 0 | 0 | 0 | 0 |
| 0 | 0.159982848 | 0 | 0 | 0 |
| 0 | 0 | 0 | 0 | 0 |
| 0 | 0 | 0 | 0 | 0 |
| 0 | 0 | 0 | 0 | 0 |
| 0 | 0 | 0 | 0 | 0 |
| 0 | 0 | 0 | 0 | 0 |
| 0 | 0 | 0 | 0 | 0 |
| 0 | 0 | 0 | 0 | 0 |
| 0.004235974 | 0.065547714 | 0.265969143 | 0.159552361 | 0 |
| 0 | 0 | 0 | 0 | 0 |
| 0 | 0 | 0 | 0 | 0 |
| 0 | 0 | 0 | 0.007687277 | 0 |
| 0 | 0 | 0 | 0 | 0 |
| 0 | 0 | 0 | 0 | 0 |
| 0 | 0 | 0 | 0.024497941 | 0 |
| 0 | 0 | 0 | 0 | 0 |
| 0 | 0 | 0 | 0 | 0 |
| 0 | 0 | 0 | 0 | 0 |
| 0 | 0 | 0 | 0 | 0 |

| | | | | |
|---|---|---|---|---|
| 0 | 0 | 0 | 0 | 0 |
| 0 | 0 | 0 | 0 | 0 |
| 0 | 0 | 0 | 0 | 0 |
| 0 | 0 | 0 | 0 | 0 |
| 0 | 0 | 0 | 0 | 0 |
| 0 | 0 | 0 | 0 | 0 |
| 0 | 0 | 0 | 0 | 0 |
| 0 | 0 | 0 | 0 | 0 |
| 2.417619697 | 0.027237778 | 0.426588608 | 0.257814579 | 0.960374402 |
| 0 | 0 | 0 | 0 | 0 |
| 0 | 0 | 0 | 0 | 0 |
| 0 | 0 | 0 | 0 | 0 |
| 0 | 0 | 0 | 0 | 0 |
| 0 | 0 | 0 | 0 | 0 |
| 0 | 0 | 0 | 2.364092422 | 0 |
| 1.38760873 | 0.012487164 | 0.048169884 | 0 | 0 |
| 0 | 0 | 0.002942865 | 0 | 0 |
| 0 | 0 | 0 | 0 | 0 |
| 0 | 0 | 0 | 0 | 0 |
| 0 | 0 | 0 | 0 | 0 |
| 0 | 0 | 0 | 0 | 0 |
| 0 | 0 | 0 | 0 | 0 |
| 0 | 0 | 0 | 0 | 0 |
| 0 | 0 | 0 | 0 | 0 |
| 0 | 0 | 0.052040033 | 0.017834271 | 0.116277171 |
| 0 | 2.569308155 | 0 | 0 | 0 |
| 0 | 0 | 0 | 0 | 0 |
| 0 | 0 | 0 | 0 | 0 |
| 0 | 0 | 0 | 0 | 0 |
| 0 | 0 | 0 | 0 | 0 |
| 0 | 0 | 0 | 0 | 0 |
| 0 | 0 | 0 | 0 | 0 |
| 0 | 0 | 0 | 0 | 0 |
| 0.000478579 | 0 | 0 | 0.003174882 | 0 |
| 0.002365642 | 0.001214921 | 0.005425785 | 0.069851939 | 0 |
| 0.004645061 | 0 | 0.001073904 | 0.001740153 | 0 |
| 0 | 0 | 0 | 0.002244738 | 0 |
| 0 | 0.247282543 | 0 | 15.95305314 | 0.000868839 |
| 0 | 0 | 0 | 0 | 26.5151286 |
| 0 | 0 | 0 | 0 | 0 |
| 0 | 0 | 0 | 0 | 0 |
| 0 | 0 | 0 | 0 | 0 |
| 0 | 0.263348594 | 0 | 0.010403534 | 0 |
| 0 | 0 | 0 | 0 | 0 |
| 0 | 0 | 0 | 0 | 0 |
| 0 | 0 | 0 | 0.02246186 | 0 |
| 0 | 0 | 0 | 0 | 0 |
| 0.007851988 | 0.007777038 | 0.42465568 | 0.967164661 | 0 |

| | | | | |
|---|---|---|---|---|
| 0 | 0 | 0.006733905 | 0.021788256 | 0 |
| 0 | 0 | 0 | 0 | 0 |
| 0 | 0 | 0 | 0 | 0 |
| 0 | 0 | 0 | 0 | 0 |
| 0 | 0 | 0.002578609 | 0 | 0.004828294 |
| 0 | 0 | 0 | 0 | 0 |
| 0 | 0 | 0 | 0 | 0 |
| 0 | 0 | 0 | 0 | 0 |
| 0 | 0 | 0 | 0 | 0 |
| 0 | 0 | 0 | 0 | 0 |
| 0 | 0 | 0 | 0 | 0 |
| 0 | 1.353009831 | 0 | 0 | 0 |
| 0 | 0.460487415 | 0 | 0 | 0 |
| 0 | 0 | 0 | 0 | 0 |
| 0.045266993 | 0.001780677 | 0.298613711 | 0 | 3.818804279 |
| 0 | 0 | 0 | 0 | 0 |
| 0 | 0 | 0 | 0 | 0 |
| 0 | 0 | 0 | 0 | 0 |
| 0 | 0.001365662 | 0.000871284 | 0 | 0 |
| 0 | 0 | 0 | 0 | 0 |
| 0 | 0 | 0 | 0 | 0 |
| 0 | 0 | 0 | 0 | 0 |
| 0 | 0.132602417 | 0.33905174 | 0.177058364 | 1.085908583 |
| 0 | 0 | 0 | 0 | 0 |
| 0 | 0 | 0 | 0 | 0 |
| 0 | 0 | 0 | 0 | 0 |
| 0 | 0 | 0 | 0 | 0 |
| 0 | 0 | 0 | 0 | 0 |
| 0 | 0 | 0 | 0 | 0 |
| 0 | 0 | 0 | 0 | 0 |
| 0 | 0 | 0 | 0 | 0 |
| 0.243084664 | 0 | 0.062174318 | 0.01902074 | 0.017597233 |
| 0 | 0 | 0 | 0 | 0 |
| 0 | 0 | 0.02732356 | 0 | 0 |
| 0.003235194 | 0 | 0.001836492 | 0 | 0 |
| 0 | 0.017590103 | 0 | 0 | 0.013601042 |
| 0 | 0 | 0 | 0 | 0 |
| 0 | 0 | 0 | 0 | 0 |
| 0 | 0 | 0.006329576 | 0 | 0 |
| 0 | 0 | 0 | 0 | 0 |
| 0 | 0 | 0 | 0 | 0 |
| 0 | 0 | 0 | 0 | 0 |
| 0 | 0 | 0 | 0 | 0 |
| 0 | 0 | 0 | 0 | 0 |
| 0.006174794 | 0.000736533 | 0 | 0.005829709 | 0.0006599 |
| 0 | 0 | 0 | 0 | 0 |
| 0 | 0 | 0 | 0 | 0 |
| 0.003202718 | 0 | 0 | 0 | 0 |

| | | | | |
|---|---|---|---|---|
| 0 | 0 | 0 | 0 | 0 |
| 0 | 0 | 0 | 0 | 0 |
| 0 | 0 | 0 | 0.002894102 | 0 |
| 0 | 0 | 0 | 0 | 0 |
| 0 | 0 | 0.090047738 | 0 | 0 |
| 0 | 0 | 0 | 0 | 0 |
| 0 | 0 | 0 | 0 | 0 |
| 0 | 0 | 0 | 0 | 0 |
| 0 | 0 | 0 | 0 | 0 |
| 0 | 0 | 0 | 0 | 0 |
| 0 | 0 | 0 | 0 | 0 |
| 0.012157338 | 0 | 0 | 0 | 0 |
| 0 | 0 | 0 | 0 | 0 |
| 0 | 0 | 0 | 0 | 0 |
| 0 | 0 | 0 | 0 | 0 |
| 0 | 0 | 0 | 0 | 0 |
| 0 | 0 | 0 | 0 | 0 |
| 0 | 0 | 0 | 0 | 0 |
| 0 | 0 | 0 | 0 | 4.856064882 |
| 0 | 0 | 0 | 0 | 0 |
| 0 | 0 | 0 | 0 | 0 |
| 0 | 0 | 0 | 0 | 0 |
| 0 | 0 | 0 | 0 | 0 |
| 0 | 0 | 0 | 0 | 0 |
| 0 | 0 | 0 | 0 | 0 |
| 0.062677192 | 0.002228474 | 0.039655973 | 0.001519627 | 0 |
| 0 | 0 | 0 | 0 | 0 |
| 0.238496849 | 0.012080674 | 0.000449598 | 0.06438371 | 0 |
| 0 | 0 | 0 | 0 | 0 |
| 0 | 0.096974799 | 0 | 3.058383267 | 0 |
| 0 | 0 | 0 | 0 | 0 |
| 0 | 0 | 0 | 0 | 0 |
| 0 | 0 | 0 | 0 | 0 |
| 0 | 0 | 0 | 0 | 0 |
| 0.005052448 | 0 | 0.001123162 | 0 | 0.00177043 |
| 0 | 0 | 0 | 0 | 0 |
| 0 | 0 | 0 | 0 | 0 |
| 0.001984517 | 0 | 0 | 0.00283956 | 0.009496708 |
| 0 | 0 | 0 | 0 | 0 |
| 0 | 0 | 0 | 0 | 0 |
| 0.704249876 | 9.314647806 | 16.55487754 | 12.32925128 | 10.3637158 |
| 13.87320987 | 8.613584707 | 0.834422 | 0.111298012 | 0 |
| 0 | 0 | 0 | 0 | 0 |
| 0 | 0 | 0 | 0 | 0 |
| 0 | 0 | 0 | 0 | 0 |
| 0 | 0 | 0 | 0 | 0 |
| 0 | 0 | 0 | 0 | 0 |
| 0.00140002 | 0.000970659 | 0.000866985 | 0.000819502 | 0.019422561 |

| | | | | |
|---|---|---|---|---|
| 0 | 0 | 0 | 0 | 0 |
| 0 | 0 | 0 | 0 | 0 |
| 0 | 0 | 0 | 0 | 0 |
| 0 | 0 | 0 | 0 | 0 |
| 0 | 7.11359227 | 0 | 0.099336338 | 0 |
| 0 | 0 | 0 | 0 | 0 |
| 0 | 0 | 0 | 0 | 0 |
| 0 | 0 | 0 | 0 | 0 |
| 6.994547282 | 0.060148187 | 0.977863201 | 0.010310619 | 0 |
| 0 | 0 | 0 | 0 | 0 |
| 0 | 0 | 0 | 0 | 0.036719415 |
| 0 | 0 | 0 | 0 | 0 |
| 0 | 0 | 0 | 0 | 0 |
| 0 | 0.024449582 | 0.021660615 | 0.035997892 | 0.007301907 |
| 0 | 0 | 0 | 0 | 0 |
| 0 | 0.267019327 | 0 | 0.493703964 | 0 |
| 0 | 0 | 0 | 0.007188598 | 0 |
| 0 | 0 | 0 | 0 | 0 |
| 0 | 0 | 0 | 0 | 0 |
| 0 | 0 | 0 | 0 | 0 |
| 0 | 0 | 0 | 0 | 0 |
| 0 | 0 | 0 | 0 | 0 |
| 0 | 0 | 0 | 0 | 0 |
| 0 | 0 | 0 | 0 | 0 |
| 0 | 0 | 0 | 0 | 0 |
| 0 | 0 | 0 | 0 | 0 |
| 0 | 0 | 0 | 0 | 0 |
| 0.004381714 | 0 | 0.000602988 | 0.000854946 | 0.000604854 |
| 0 | 0 | 0 | 0 | 0 |
| 0 | 0 | 0 | 0 | 0 |
| 0 | 0 | 0 | 0 | 0 |
| 0 | 0 | 0 | 0 | 0 |
| 0 | 0 | 0 | 0 | 0 |
| 0 | 0 | 0 | 0 | 0 |
| 0 | 0 | 0 | 0 | 0 |
| 0.13411025 | 0.015618871 | 0 | 0.001236244 | 0 |
| 0.001295214 | 0 | 0.002138886 | 0 | 0 |
| 0 | 0 | 0.000823335 | 0 | 0 |
| 0 | 0 | 0 | 0 | 0 |
| 0 | 0 | 0 | 0 | 0 |
| 0 | 2.829432691 | 0 | 0 | 0 |
| 0 | 0 | 0 | 0 | 0 |
| 0 | 0 | 0 | 0 | 0 |
| 0 | 0 | 0 | 0 | 0 |
| 0 | 0 | 0 | 0 | 0 |
| 0 | 0 | 0 | 0 | 0 |
| 0 | 0 | 0 | 0 | 0 |
| 0 | 0 | 0 | 0 | 0 |

| | | | | |
|---|---|---|---|---|
| 0 | 0 | 0.006106994 | 0 | 0 |
| 0 | 0 | 0 | 0 | 0 |
| 0 | 0 | 0 | 0 | 0 |
| 0 | 0 | 0.006659907 | 0.004116068 | 0 |
| 0 | 0 | 0 | 0 | 0 |
| 0 | 0 | 0 | 0 | 0 |
| 0.945738469 | 0 | 0 | 0.647139115 | 0 |
| 0 | 0 | 0 | 0 | 0 |
| 0 | 0 | 0 | 0 | 0 |
| 0 | 0 | 0 | 0 | 0 |
| 0 | 0 | 0 | 0 | 0 |
| 0 | 0 | 0 | 0 | 0 |
| 0 | 0 | 0 | 0 | 0 |
| 0 | 0 | 0 | 0 | 0 |
| 0 | 0 | 0 | 0 | 0 |
| 0 | 0 | 0 | 0 | 0 |
| 0 | 0 | 0 | 0 | 0.002371962 |
| 0 | 0 | 0 | 0 | 0 |
| 0 | 0 | 0 | 0 | 0 |
| 0 | 0 | 0 | 0 | 0 |
| 0 | 0.001633656 | 0 | 0.086272302 | 0.000439105 |
| 0 | 0 | 0 | 0 | 0 |
| 0 | 0 | 0 | 0 | 0 |
| 0 | 0 | 0.000572458 | 0.001623319 | 0 |
| 0 | 0 | 0 | 0 | 0 |
| 0 | 0 | 0 | 0 | 0 |
| 0 | 0 | 0 | 0 | 0 |
| 0 | 0 | 0 | 0 | 0 |
| 0 | 0 | 0 | 0 | 0 |
| 0.006236479 | 0.193709136 | 0.00679719 | 0.007593095 | 0.319629787 |
| 0 | 0 | 0 | 0 | 0 |
| 0 | 0 | 0 | 0 | 0 |
| 0 | 0 | 0 | 0 | 0 |
| 0 | 0 | 0 | 0 | 0 |
| 0 | 0 | 0 | 0 | 0 |
| 0 | 0 | 0 | 0 | 0 |
| 0.007807601 | 0 | 0 | 0 | 0 |
| 0 | 0 | 0 | 0 | 0 |
| 0 | 0 | 0 | 0 | 0 |
| 0 | 0 | 0 | 0 | 0 |
| 0 | 0 | 0 | 0 | 0 |
| 0 | 0 | 0 | 0 | 0 |
| 0 | 0 | 0 | 0 | 0 |
| 0 | 0 | 0 | 0 | 0 |
| 0 | 0 | 0 | 0 | 0 |
| 0 | 0 | 0 | 0 | 0 |
| 0 | 0 | 0 | 0 | 0 |
| 0 | 0 | 0 | 0 | 0 |

| | | | | |
|---|---|---|---|---|
| 0.004849175 | 0 | 0.000461989 | 0 | 0.001853673 |
| 0 | 0 | 0 | 0 | 0 |
| 0 | 0.022538313 | 0 | 0 | 0 |
| 0 | 0 | 0 | 0 | 0 |
| 0 | 0 | 0 | 0 | 0 |
| 0 | 0 | 0 | 0 | 0 |
| 0 | 0 | 0 | 0 | 0 |
| 0 | 4.382315706 | 0 | 6.492754469 | 0 |
| 0 | 0.005141885 | 0.17597248 | 0.043525841 | 2.820148122 |
| 0 | 0.00546364 | 0.02608317 | 0.001670155 | 0 |
| 0 | 0 | 0 | 0 | 0 |
| 0 | 0 | 0 | 0 | 0 |
| 0 | 0 | 0 | 0 | 0 |
| 0 | 0.033257903 | 0 | 0 | 1.681588664 |
| 0 | 0 | 0.013681858 | 0 | 0 |
| 0 | 0 | 0 | 0 | 0 |
| 0.003539245 | 0.091861078 | 0 | 0 | 0 |
| 0 | 0 | 0 | 0 | 0 |
| 0 | 0 | 0 | 0 | 0 |
| 0 | 0 | 0.002930352 | 0 | 0 |
| 0 | 0 | 0 | 0 | 0 |
| 0 | 0 | 0 | 0 | 0 |
| 0 | 0 | 0 | 0 | 0 |
| 0 | 0 | 0 | 0 | 0 |
| 0 | 0 | 0 | 0 | 0 |
| 0 | 0 | 0 | 0 | 0 |
| 0 | 0 | 0 | 0 | 0 |
| 0 | 0 | 0 | 0 | 0 |
| 0 | 0 | 0 | 0 | 0 |
| 0 | 0 | 0 | 0 | 0 |
| 0 | 0 | 0 | 0 | 0 |
| 0 | 0 | 0 | 0 | 0 |
| 0 | 0 | 0 | 0 | 0 |
| 0 | 0 | 0 | 0 | 0 |
| 0 | 0 | 0 | 0 | 0 |
| 0 | 0 | 0 | 0 | 0 |
| 0 | 0 | 0 | 0 | 0 |
| 0 | 0 | 0 | 0 | 0 |
| 0 | 0 | 0.018494486 | 0 | 0 |
| 0 | 0 | 0 | 0.012860545 | 0 |
| 0 | 0 | 0 | 0 | 0 |
| 0 | 0.838499103 | 0 | 0 | 0 |
| 0 | 0 | 0 | 0 | 0 |
| 0 | 0 | 0 | 0 | 0 |
| 0 | 0 | 0 | 0 | 0 |
| 0 | 0 | 0 | 0 | 0 |
| 0 | 0 | 0 | 0 | 0 |
| 0 | 0 | 0 | 0 | 0 |

| | | | | |
|---|---|---|---|---|
| 0 | 0 | 0 | 0 | 0 |
| 0 | 0 | 0 | 0 | 0 |
| 0 | 0 | 0 | 0 | 0 |
| 0 | 0 | 0 | 0 | 0 |
| 0 | 0.377254186 | 0 | 0 | 0.573474031 |
| 0 | 0 | 0 | 0 | 0 |
| 0 | 0 | 0 | 0.01099176 | 0 |
| 0.209254544 | 0.002631837 | 0.044272166 | 0.022219915 | 0.090783265 |
| 0 | 0 | 0 | 0 | 0 |
| 0 | 0 | 0 | 0 | 0 |
| 0 | 0 | 0 | 0 | 0 |
| 0 | 0 | 0 | 0 | 0 |
| 0 | 0 | 0 | 0 | 0 |
| 0 | 0 | 0 | 0 | 0 |
| 0 | 0 | 0.014341896 | 0 | 0 |
| 0 | 0 | 0 | 0 | 0.006646516 |
| 0 | 0 | 0 | 0 | 0 |
| 0 | 0 | 0 | 0 | 0 |
| 0 | 0 | 0 | 0 | 0 |
| 0 | 0 | 0 | 0 | 0 |
| 0 | 0 | 0 | 0 | 0 |
| 0 | 0 | 0 | 0 | 0 |
| 0 | 0 | 0 | 0 | 0 |
| 0.00533833 | 0 | 0 | 0 | 0.000850275 |
| 0 | 0 | 0 | 0 | 0 |
| 0 | 0 | 0 | 0 | 0 |
| 0.014647526 | 0 | 0.0234133 | 0.010772384 | 0.161133904 |
| 0 | 0 | 0 | 0 | 0 |
| 0.001395624 | 0 | 0 | 0 | 0 |
| 0 | 0 | 0 | 0 | 0 |
| 0 | 0 | 0 | 0 | 0 |
| 0 | 0 | 0 | 0 | 0 |
| 0 | 0 | 0 | 0 | 0 |
| 0 | 0 | 0 | 0 | 0 |
| 0 | 0 | 0 | 0 | 0 |
| 0 | 0 | 0 | 0 | 0 |
| 0 | 0 | 0 | 0 | 0 |
| 0 | 0 | 0 | 0 | 0 |
| 0 | 0 | 0 | 0 | 0 |
| 0 | 0 | 0 | 0 | 0 |
| 0 | 0 | 0 | 0 | 0 |
| 0 | 0 | 0 | 0 | 0 |
| 0 | 0 | 0 | 0 | 0 |
| 0 | 0 | 0 | 0 | 0 |
| 0 | 0 | 0 | 0 | 0 |
| 0 | 0 | 0 | 0 | 0 |
| 0 | 0 | 0 | 0 | 0 |
| 0 | 0 | 0 | 0 | 0 |

| | | | | |
|---|---|---|---|---|
| 0 | 0 | 0 | 0 | 0 |
| 0 | 0 | 0 | 0 | 0 |
| 0 | 0 | 0 | 0 | 0 |
| 0 | 0 | 0 | 0 | 0 |
| 0 | 0 | 0 | 0 | 0 |
| 0 | 0.011159116 | 0 | 0.011257717 | 0 |
| 0 | 0 | 0 | 0 | 0 |
| 0 | 0 | 0 | 0 | 0 |
| 0 | 0 | 0 | 0 | 0 |
| 0 | 0 | 0 | 0 | 0 |
| 0 | 0 | 0 | 0 | 0 |
| 0 | 0 | 0 | 0 | 0 |
| 0 | 0 | 0 | 0 | 0 |
| 0 | 0 | 0 | 0 | 0 |
| 0 | 0 | 0 | 0 | 0 |
| 0 | 0 | 0 | 0 | 0 |
| 0 | 0 | 0 | 0 | 0 |
| 0 | 0 | 0 | 0 | 0 |
| 0 | 0 | 0 | 0 | 0 |
| 0 | 0 | 0 | 0 | 0 |
| 0.205075467 | 0.000945049 | 0.066853474 | 0.00119682 | 0 |
| 0 | 0 | 0 | 0 | 0 |
| 0.005422635 | 0.008832105 | 0.004050986 | 0.003022993 | 0.190129871 |
| 0 | 0 | 0 | 0 | 0 |
| 0 | 0 | 0 | 0 | 0 |
| 0 | 0 | 0 | 0 | 0 |
| 0 | 0 | 0 | 0 | 0 |
| 0 | 0 | 0 | 0 | 0 |
| 0 | 0 | 0 | 0 | 0.003332574 |
| 0 | 0 | 0 | 0 | 0 |
| 0 | 0 | 0 | 0 | 0 |
| 0 | 0 | 0 | 0 | 0 |
| 0.063357531 | 0.005588669 | 0.032945562 | 0.026328492 | 0.355110241 |
| 0 | 0 | 0 | 0 | 0 |
| 0 | 0.482707062 | 0 | 0 | 0 |
| 0 | 0.021705814 | 0.029802197 | 0 | 0 |
| 0 | 0 | 0 | 0 | 0 |
| 0 | 0 | 0 | 0 | 0 |
| 0.119857607 | 0 | 0.035026938 | 0 | 0.6170056 |
| 0 | 0 | 0 | 0 | 0 |
| 0 | 0 | 0 | 0 | 0 |
| 0 | 0 | 0 | 0 | 0 |
| 0 | 0 | 0 | 0 | 0.002391114 |
| 0 | 0 | 0 | 0 | 0 |
| 0 | 0 | 0 | 0 | 0 |
| 0 | 0 | 0 | 0 | 0 |
| 0 | 0 | 0 | 0 | 0 |
| 0 | 0 | 0 | 0 | 0 |

| | | | | |
|---|---|---|---|---|
| 0 | 0 | 0 | 0 | 0 |
| 0 | 0 | 0 | 0 | 0 |
| 0 | 0 | 0 | 0 | 0 |
| 0 | 0 | 0 | 0 | 0 |
| 0 | 0 | 0 | 0 | 0 |
| 0 | 0 | 0 | 0 | 0 |
| 0 | 0 | 0 | 0 | 0 |
| 0 | 0 | 2.171589484 | 0 | 0 |
| 0 | 0 | 0 | 0 | 0 |
| 0 | 0.001984584 | 0 | 0 | 0 |
| 0 | 0 | 0 | 0 | 0 |
| 0 | 0 | 0 | 0 | 0 |
| 0 | 0 | 0 | 0 | 0 |
| 0 | 0.1628407 | 0 | 0 | 0 |
| 0 | 0 | 0 | 0 | 0 |
| 0.359926636 | 0.186474376 | 0.114235666 | 0 | 0.022043089 |
| 0 | 0 | 0 | 0 | 0 |
| 0 | 0 | 0 | 0 | 0 |
| 0 | 0 | 0 | 0 | 0 |
| 0 | 0 | 0 | 1.689375806 | 0 |
| 0 | 0.003670855 | 0 | 0 | 0 |
| 0 | 0 | 0 | 0 | 0 |
| 0 | 0 | 0 | 0 | 0 |
| 0 | 0 | 0 | 0 | 0 |
| 0 | 0 | 0 | 0 | 0 |
| 0 | 0 | 0 | 0 | 0 |
| 0 | 0 | 0 | 0 | 0 |
| 0 | 0 | 0 | 0 | 0 |
| 0 | 0 | 0 | 0 | 0 |
| 0 | 0.057878727 | 0 | 0 | 0 |
| 0 | 0 | 0 | 0.100893432 | 0 |
| 0 | 0 | 0 | 0 | 0 |
| 0 | 0 | 0 | 0 | 0 |
| 0 | 0 | 0 | 0 | 0 |
| 0 | 0 | 0 | 0 | 0 |
| 0 | 0 | 0 | 0 | 0 |
| 0 | 0 | 0 | 0 | 0 |
| 0 | 0 | 0 | 0 | 0 |
| 0 | 0 | 0 | 0 | 0 |
| 0 | 0 | 0 | 0 | 0 |
| 0 | 0 | 0 | 0 | 0 |
| 0 | 0 | 0 | 0 | 0 |
| 0 | 0 | 0 | 0 | 0 |
| 0 | 0 | 0.012854017 | 0 | 0 |
| 0 | 0 | 0 | 0 | 0 |
| 0 | 0 | 0 | 0 | 0 |
| 12.50063288 | 0.307826584 | 1.188704256 | 0.187829587 | 0.031340763 |
| 0 | 0 | 0 | 0 | 0 |

| | | | | |
|---|---|---|---|---|
| 0 | 0 | 0 | 0 | 0 |
| 0 | 0 | 0 | 0 | 0 |
| 0 | 0 | 0 | 0 | 0 |
| 0 | 0 | 0 | 0 | 0 |
| 0 | 0 | 0 | 0.057679828 | 0 |
| 0 | 0 | 0 | 0 | 0 |
| 0 | 0 | 0 | 0 | 0 |
| 0 | 0 | 0 | 0 | 0 |
| 0 | 0 | 0 | 0 | 0 |
| 0 | 0 | 0 | 0 | 0 |
| 0 | 0 | 0 | 0 | 0 |
| 0.003364106 | 0 | 0.000925901 | 0.00105023 | 0.016160514 |
| 0 | 0 | 0 | 0.011552571 | 0.002884645 |
| 0 | 0 | 0 | 0 | 0 |
| 0.008203788 | 0 | 0 | 0 | 0 |
| 0 | 0 | 0 | 0.084324565 | 0 |
| 0 | 0 | 0.001139404 | 0 | 0 |
| 0 | 0 | 0 | 0 | 0 |
| 0 | 0.266563919 | 0 | 0.151890892 | 0.325893597 |
| 0.149227613 | 0.000614932 | 0 | 0.006489634 | 0.001469202 |
| 0 | 0 | 0 | 0 | 0 |
| 0 | 0 | 0 | 0 | 0 |
| 0 | 0 | 0 | 0 | 0 |
| 0 | 0 | 0 | 0 | 0 |
| 0 | 0 | 0 | 0 | 0 |
| 0 | 0 | 0 | 0 | 0 |
| 0.015103341 | 0 | 0.001760562 | 0.000624053 | 0 |
| 0 | 0 | 0 | 0 | 0 |
| 0 | 0 | 0 | 0 | 0 |
| 0 | 0 | 0 | 0 | 0 |
| 0.002819836 | 0 | 0 | 0 | 0 |
| 0 | 0 | 0 | 0 | 0 |
| 0 | 0 | 0 | 0 | 0 |
| 0 | 0 | 0 | 0 | 0 |
| 0 | 0 | 0 | 0 | 0 |
| 0 | 0 | 0 | 0 | 0 |
| 0 | 0 | 0 | 0 | 0 |
| 0.012169827 | 0 | 0.013553765 | 0.005963958 | 0.085324715 |
| 0 | 0 | 0 | 0 | 0 |
| 0 | 0 | 0 | 0 | 0 |
| 0 | 0.000783573 | 0.003149463 | 0.000992325 | 0.001755115 |
| 0 | 0 | 0 | 0 | 0 |
| 0 | 0 | 0 | 0 | 0 |
| 0 | 0.006207423 | 0.005371155 | 0.04569293 | 0.045882958 |
| 0 | 0 | 0 | 0 | 0 |
| 0 | 0 | 0.020291674 | 0 | 0 |
| 0 | 0 | 0 | 0 | 0 |
| 0 | 0 | 0 | 0 | 0 |

| | | | | |
|---|---|---|---|---|
| 0 | 0 | 0 | 0 | 0 |
| 0 | 0.003813346 | 0 | 0 | 0 |
| 0 | 0 | 0 | 0 | 0 |
| 0.020452014 | 0 | 0.001271063 | 0.003346899 | 0 |
| 0 | 0.000754208 | 0 | 0 | 0.002196142 |
| 0 | 0 | 0 | 0 | 0.003052358 |
| 0 | 0 | 0 | 0 | 0 |
| 0 | 0 | 0 | 0 | 0.003846177 |
| 0 | 0 | 0 | 0 | 0 |
| 0 | 0 | 0 | 0 | 0 |
| 0 | 0 | 0 | 0 | 0 |
| 0 | 0 | 0 | 0 | 0 |
| 0 | 0.005764273 | 0 | 0.019709834 | 0 |
| 0 | 0 | 0 | 0 | 0 |
| 0 | 0 | 0 | 0 | 0 |
| 0 | 0 | 0 | 0 | 0 |
| 0 | 0 | 0 | 0 | 0 |
| 0 | 0 | 0 | 0 | 0 |
| 0 | 0 | 0 | 0 | 0 |
| 0 | 0 | 0 | 0 | 0 |
| 0 | 0 | 0 | 0 | 0 |
| 0 | 0 | 0 | 0 | 0 |
| 0 | 0 | 0.014236991 | 0 | 0 |
| 0 | 0.008508883 | 0 | 0.086205951 | 0 |
| 0 | 0 | 0 | 0 | 0 |
| 0 | 0 | 0 | 0 | 0 |
| 0 | 0 | 0 | 0 | 0 |
| 0 | 0 | 0 | 0 | 0 |
| 0 | 0 | 0 | 0 | 0 |
| 0 | 0 | 0 | 0 | 0 |
| 0 | 0.004654683 | 0 | 0.065645993 | 0 |
| 0 | 0 | 0.002192072 | 0 | 0 |
| 0 | 0 | 0 | 0 | 0 |
| 0 | 0 | 0 | 0 | 0 |
| 0 | 0 | 0 | 0 | 0 |
| 0.008374046 | 0.004202351 | 0 | 0 | 0.003566952 |
| 0 | 0 | 0 | 0 | 0 |
| 0 | 0 | 0 | 0 | 0 |
| 0.001742788 | 0.001969092 | 0 | 0 | 0.04619043 |
| 0 | 0 | 0 | 0 | 0 |
| 0 | 0 | 0 | 0 | 0 |
| 0 | 0 | 0 | 0 | 0 |
| 0 | 0 | 0 | 0 | 0 |
| 0 | 0 | 0 | 0 | 0 |
| 0 | 0 | 0 | 0 | 0 |
| 0 | 0.270266255 | 0.486738012 | 0 | 0 |
| 0 | 0 | 0 | 0 | 0 |
| 0 | 0 | 0 | 0 | 0 |

| | | | | |
|---|---|---|---|---|
| 0 | 0 | 0 | 0 | 0 |
| 0 | 0 | 0 | 0 | 0 |
| 0 | 0 | 0 | 0 | 0 |
| 0 | 0 | 0 | 0 | 0 |
| 0 | 0 | 0 | 0 | 0 |
| 0 | 0 | 0 | 0 | 0 |
| 0 | 0 | 0 | 0 | 0 |
| 0 | 0 | 0 | 0 | 0 |
| 0.004479284 | 0 | 0 | 0 | 0 |
| 0 | 0 | 0 | 0 | 0 |
| 0 | 0 | 0.000518851 | 0 | 0.003122734 |
| 0.002110909 | 0 | 0 | 0.001372913 | 0 |
| 0 | 0 | 0 | 0 | 0 |
| 0 | 0 | 0 | 0 | 0 |
| 0 | 0 | 0 | 0 | 0 |
| 0 | 0 | 0 | 0 | 0 |
| 0 | 0 | 0 | 0 | 0.001147571 |
| 0 | 0 | 0 | 0 | 0 |
| 0 | 0 | 0 | 0 | 0 |
| 0 | 0 | 0 | 0 | 0 |
| 0 | 0 | 0 | 0 | 0 |
| 0 | 0 | 0 | 0 | 0 |
| 0 | 0 | 0 | 0 | 0 |
| 0 | 0 | 0 | 0 | 0 |
| 0 | 0 | 0 | 0 | 0 |
| 0 | 0 | 0 | 0 | 0 |
| 0 | 0 | 0 | 0 | 0 |
| 0 | 0 | 0 | 0 | 0 |
| 0 | 0 | 0 | 0 | 0 |
| 0 | 0 | 0 | 0 | 0 |
| 0 | 0.012740125 | 0 | 0 | 0 |
| 0 | 0 | 0 | 0 | 0 |
| 0 | 0.19345624 | 0.037921907 | 0.02578896 | 0.001893355 |
| 0.014228578 | 0 | 0.012449762 | 0.021381158 | 0 |
| 0 | 0 | 0 | 0 | 0 |
| 0 | 0 | 0 | 0 | 0 |
| 0 | 0 | 0 | 0 | 0 |
| 0 | 0 | 0 | 0 | 0 |
| 0 | 0 | 0 | 0 | 0 |
| 0 | 0 | 0 | 0 | 0 |
| 0 | 0 | 0 | 0 | 0 |
| 0.344332529 | 0 | 0.011011062 | 0.007405705 | 0.000849625 |
| 0 | 0.056416406 | 0 | 0 | 0 |
| 0 | 0 | 0 | 0 | 0 |
| 0 | 0 | 0 | 0 | 0 |
| 0 | 0 | 0 | 0 | 0.02617592 |
| 0 | 0 | 0 | 0 | 0 |
| 0 | 0 | 0 | 0 | 0 |

| | | | | |
|---|---|---|---|---|
| 0 | 0 | 0 | 0 | 0 |
| 0 | 0.010892826 | 0 | 0 | 0 |
| 0.023450135 | 0 | 0.009681249 | 0.062067885 | 0.069245072 |
| 0 | 0 | 0 | 0 | 0 |
| 0 | 0 | 0 | 0 | 0 |
| 0.054478812 | 0 | 0.028625231 | 0.015770662 | 0.022642873 |
| 0 | 0 | 0 | 0 | 0 |
| 0 | 0 | 0 | 0 | 0 |
| 0 | 0 | 0 | 0 | 0 |
| 0 | 0 | 0 | 0 | 0 |
| 0 | 0 | 0 | 0 | 0 |
| 0 | 0 | 0 | 0 | 0 |
| 0 | 0 | 0 | 0 | 0 |
| 0 | 0 | 0 | 0 | 0 |
| 0 | 0 | 0 | 0 | 0 |
| 0 | 0 | 0 | 0 | 0 |
| 0 | 0 | 0 | 0 | 0 |
| 0 | 0 | 0 | 0 | 0 |
| 0 | 0 | 0 | 0 | 0 |
| 0 | 0 | 0 | 0 | 0 |
| 0 | 0 | 0 | 0 | 0 |
| 0 | 0 | 0 | 0.001680217 | 0 |
| 0 | 0 | 0 | 0 | 0 |
| 0 | 0 | 0 | 0 | 0 |
| 0 | 0 | 0 | 0 | 0 |
| 0.004649104 | 0 | 0 | 0 | 0.035880457 |
| 0 | 0 | 0 | 0 | 0 |
| 0 | 0 | 0 | 0 | 0 |
| 0 | 0 | 0 | 0 | 0 |
| 0 | 0 | 0 | 0 | 0 |
| 0 | 0 | 0 | 0 | 0 |
| 0 | 0 | 0 | 0 | 0 |
| 0 | 0 | 0 | 0 | 0 |
| 0 | 0 | 0 | 0 | 0 |
| 0 | 0 | 0 | 0 | 0 |
| 0 | 0 | 0 | 0 | 0 |
| 0 | 0 | 0 | 0 | 0 |
| 0 | 0 | 0.02570567 | 0 | 0 |
| 0 | 0 | 0 | 0 | 0 |
| 0 | 0 | 0 | 0 | 0 |
| 0 | 0 | 0 | 0 | 0 |
| 0 | 0.001483488 | 0 | 0 | 0 |
| 0.002120856 | 0 | 0.000875583 | 0 | 0.001405267 |
| 0 | 0 | 0 | 0 | 0 |
| 0 | 0 | 0 | 0 | 0 |
| 0 | 0 | 0 | 0 | 0 |
| 0 | 0 | 0 | 0 | 0.037621199 |
| 0 | 0 | 0 | 0 | 0 |

| | | | | |
|---|---|---|---|---|
| 0 | 0 | 0 | 0 | 0 |
| 0.133872349 | 0.01513821 | 0.021972148 | 0.006230639 | 0.018649334 |
| 0 | 0 | 0 | 0 | 0 |
| 0 | 0 | 0 | 0 | 0 |
| 0 | 0 | 0 | 0 | 0 |
| 0 | 0 | 0 | 0 | 0 |
| 0 | 0 | 0 | 0 | 0 |
| 0 | 0 | 0 | 0 | 0 |
| 0 | 0 | 0 | 0 | 0 |
| 0 | 0 | 0 | 0 | 0 |
| 0 | 0 | 0 | 0 | 0 |
| 0 | 0 | 0 | 0 | 0 |
| 0 | 0.001400213 | 0 | 0.023812143 | 0 |
| 0 | 0 | 0 | 0 | 0 |
| 0 | 0 | 0.006322144 | 0 | 0 |
| 0 | 0 | 0 | 0 | 0 |
| 0 | 0 | 0 | 0 | 0 |
| 0 | 0 | 0 | 0 | 0 |
| 0 | 0 | 0 | 0 | 0 |
| 0 | 0 | 0 | 0 | 0 |
| 0 | 0 | 0 | 0 | 0 |
| 0.179217135 | 0 | 0 | 0 | 0.001660814 |
| 0 | 0 | 0 | 0 | 0 |
| 0 | 0 | 0 | 0 | 0 |
| 0 | 0 | 0 | 0 | 0 |
| 0 | 0 | 0 | 0 | 0 |
| 0 | 0 | 0 | 0 | 0 |
| 0 | 0 | 0 | 0 | 0 |
| 0 | 0 | 0 | 0 | 0 |
| 0 | 0 | 0 | 0 | 0 |
| 0 | 0 | 0 | 0 | 0 |
| 0 | 0 | 0 | 0 | 0 |
| 0 | 0 | 0 | 0 | 0 |
| 0 | 0 | 0 | 0 | 0 |
| 0 | 0.010939752 | 0 | 0 | 0 |
| 0 | 0 | 0 | 0 | 0 |
| 0 | 0 | 0 | 0 | 0 |
| 0 | 0 | 0 | 0 | 0 |
| 0.029035691 | 0 | 0.02314772 | 0.003516426 | 0.001036577 |
| 0 | 0 | 0 | 0 | 0 |
| 14.24998448 | 0.013485694 | 0.142134638 | 0.023647059 | 0.244997393 |
| 0 | 0 | 0 | 0 | 0 |
| 0 | 0 | 0 | 0 | 0 |
| 0 | 0 | 0 | 0 | 0 |
| 0 | 0 | 0 | 0 | 0 |
| 4.775117284 | 0.013539869 | 2.849656974 | 0.906988172 | 1.276533412 |
| 0 | 0 | 0 | 0 | 0 |
| 0 | 0 | 0 | 0 | 0 |

| | | | | |
|---|---|---|---|---|
| 0 | 0 | 0 | 0 | 0 |
| 0 | 0 | 0 | 0 | 0 |
| 0 | 0 | 0 | 0 | 0 |
| 0 | 0 | 0 | 0 | 0 |
| 0 | 0 | 0 | 0 | 0 |
| 0 | 0 | 0 | 0 | 0 |
| 0 | 0 | 0 | 0 | 0 |
| 0.005832523 | 0 | 0 | 0 | 0 |
| 0 | 0 | 0 | 0 | 0 |
| 0 | 0 | 0 | 0 | 0.088253337 |
| 0 | 0 | 0 | 0 | 0 |
| 0 | 0 | 0 | 0 | 0 |
| 0.002210699 | 0 | 0 | 0 | 0 |
| 0 | 0 | 0 | 0 | 0 |
| 0 | 0 | 0 | 0 | 0 |
| 0 | 0 | 0 | 0 | 0 |
| 0 | 0 | 0 | 0 | 0 |
| 0 | 0 | 0 | 0 | 0 |
| 0 | 0 | 0 | 0 | 0 |
| 0.167252649 | 0.092565349 | 0.121851635 | 0.009726157 | 2.37992673 |
| 0 | 0 | 0 | 0 | 0 |
| 0 | 0 | 0 | 0 | 0 |
| 0 | 0 | 0 | 0 | 0 |
| 0 | 0 | 0 | 0 | 0 |
| 0 | 0 | 0 | 0 | 0 |
| 0 | 0 | 0 | 0 | 0 |
| 0 | 0 | 0 | 0 | 0 |
| 0 | 0 | 0 | 0 | 0 |
| 0 | 0 | 0 | 0 | 0 |
| 0 | 0 | 0.279048798 | 0 | 0 |
| 0 | 0 | 0 | 0 | 0 |
| 0 | 0 | 0 | 0 | 0 |
| 0 | 0 | 0 | 0 | 0 |
| 0 | 0 | 0 | 0 | 0 |
| 0 | 0 | 0 | 0 | 0 |
| 0 | 0 | 0 | 0 | 0 |
| 0 | 0 | 0 | 0 | 0 |
| 0 | 0 | 0 | 0 | 0 |
| 0 | 0 | 0 | 0 | 0 |
| 0 | 0 | 0 | 0 | 0 |
| 0 | 0 | 0 | 0 | 0 |
| 0 | 0 | 0 | 0 | 0 |
| 0 | 0 | 0 | 0 | 0 |
| 0 | 0 | 0 | 0 | 0 |
| 0.001026731 | 0.003796541 | 0.001525967 | 0.002163591 | 0.013095886 |
| 0 | 0 | 0 | 0 | 0 |
| 0 | 0 | 0 | 0 | 0 |
| 0.005961184 | 0 | 0 | 0 | 0 |

| | | | | |
|---|---|---|---|---|
| 0 | 0 | 0 | 0 | 0 |
| 0 | 0 | 0 | 0 | 0 |
| 0 | 0 | 0 | 0 | 0 |
| 0 | 0 | 0 | 0 | 0 |
| 0 | 0 | 0 | 0 | 0 |
| 0.001073092 | 0 | 0.011518515 | 0 | 0.01625199 |
| 0 | 0 | 0 | 0 | 0 |
| 0 | 0 | 0 | 0 | 0 |
| 0 | 0 | 0 | 0 | 0 |
| 0 | 0 | 0 | 0 | 0 |
| 0 | 0 | 0 | 0 | 0 |
| 0 | 0 | 0 | 0 | 0 |
| 0 | 0 | 0 | 0 | 0 |
| 0 | 0 | 0 | 0 | 0 |
| 0 | 0 | 0 | 0 | 0 |
| 0 | 0 | 0 | 0 | 0 |
| 0 | 0 | 0 | 0 | 0 |
| 0 | 0 | 0 | 0 | 0 |
| 0 | 0 | 0 | 0 | 0 |
| 0 | 0 | 0 | 0 | 0 |
| 0 | 0 | 0 | 0 | 0 |
| 0 | 0 | 0 | 0 | 0 |
| 0 | 0 | 0 | 0 | 0 |
| 0 | 0 | 0 | 0 | 0 |
| 0 | 0 | 0 | 0 | 0 |
| 0 | 0 | 0 | 0 | 0.002383204 |
| 0 | 0 | 0 | 0 | 0 |
| 0 | 0 | 0 | 0 | 0 |
| 0 | 0 | 0 | 0 | 0 |
| 0 | 0 | 0 | 0 | 0 |
| 0 | 0 | 0 | 0 | 0 |
| 0 | 0 | 0 | 0 | 0 |
| 0 | 0 | 0 | 0 | 0 |
| 0 | 0 | 0 | 0 | 0 |
| 0 | 0 | 0 | 0 | 0 |
| 0 | 0 | 0 | 0 | 0 |
| 0 | 0 | 0 | 0 | 0 |
| 0 | 0 | 0 | 0 | 0 |
| 0 | 0 | 0 | 0.332880992 | 0 |
| 0 | 0 | 0 | 0 | 0 |
| 0 | 0 | 0 | 0 | 0 |
| 0 | 0 | 0 | 0 | 0 |
| 0 | 0 | 0 | 0 | 0 |
| 0 | 0 | 0 | 0 | 0 |
| 0 | 0 | 0 | 0 | 0 |
| 0 | 0 | 0 | 0 | 0 |
| 0 | 0 | 0 | 0 | 0 |
| 0 | 0 | 0 | 0 | 0 |

| | | | | |
|---|---|---|---|---|
| 0 | 0 | 0 | 0 | 0 |
| 0 | 0 | 0 | 0 | 0 |
| 0 | 0 | 0 | 0 | 0 |
| 0 | 0 | 0 | 0 | 0 |
| 0 | 0 | 0 | 0 | 0 |
| 0 | 0 | 0 | 0 | 0 |
| 0 | 0 | 0.000906793 | 0 | 0 |
| 0 | 0 | 0 | 0 | 0 |
| 0 | 0 | 0 | 0 | 0 |
| 0 | 0 | 0 | 0 | 0 |
| 0 | 0 | 0.001484498 | 0.059460449 | 0 |
| 0 | 0 | 0 | 0 | 0 |
| 0 | 0 | 0 | 0 | 0 |
| 0 | 0 | 0 | 0 | 0 |
| 0 | 0 | 0 | 0 | 0 |
| 0 | 0 | 0 | 0 | 0 |
| 0 | 0 | 0 | 0 | 0 |
| 0 | 0 | 0 | 0 | 0 |
| 0 | 0 | 0 | 0 | 0 |
| 0 | 0 | 0 | 0 | 0 |
| 0 | 0.00147599 | 0 | 0.047264308 | 0.000566752 |
| 0 | 0 | 0 | 0 | 0 |
| 0 | 0 | 0 | 0 | 0 |
| 0 | 3.019742905 | 0 | 0.720472933 | 2.071576131 |
| 0 | 0 | 0 | 0 | 0 |
| 0 | 0 | 0 | 0 | 0 |
| 0 | 0 | 0 | 0 | 0 |
| 0 | 0 | 0 | 0 | 0 |
| 0 | 0 | 0 | 0 | 0 |
| 0 | 0 | 0 | 0 | 0 |
| 0 | 0 | 0 | 0 | 0 |
| 0 | 0 | 0 | 0 | 0 |
| 0 | 0 | 0 | 0 | 0 |
| 0 | 0 | 0 | 0 | 0 |

| PHL018-0083_V2 | PHL018-0141_V1 | PHL018-0206_V2 | PHL018-0212_V3 | PHL018-0230_V2 |
|---|---|---|---|---|
| REF | EYCF | REF | REF | REF |
| 0.51934345 | 0.004037858 | 0.018521206 | 0 | 0.018132131 |
| 2.563833555 | 0.494289013 | 0.058162279 | 6.227750313 | 0 |
| 2.069702561 | 4.209367824 | 0 | 0 | 0 |
| 0 | 0 | 0 | 0 | 0 |
| 0 | 0 | 0 | 0 | 0 |
| 0 | 0 | 0 | 0 | 0 |
| 0 | 0 | 0 | 0 | 0 |
| 0 | 0 | 0 | 0 | 0 |
| 0 | 0 | 0 | 0 | 0 |
| 0 | 0 | 0 | 0 | 0 |
| 0 | 0 | 0 | 0 | 0 |
| 0 | 0 | 0 | 0 | 0 |
| 0 | 0 | 0 | 0 | 0 |
| 0.144515516 | 3.852693154 | 0.074034488 | 18.27857648 | 15.0939946 |
| 0 | 0 | 0 | 0 | 0 |
| 0 | 0 | 0 | 0 | 0 |
| 0 | 0 | 0 | 0 | 0 |
| 0 | 0 | 0 | 0 | 0 |
| 0 | 0 | 0 | 0 | 0 |
| 0 | 0 | 0 | 0 | 0 |
| 0 | 0 | 0 | 0 | 0 |
| 0 | 0 | 0 | 0 | 0 |
| 0 | 0 | 0 | 0 | 0 |
| 0 | 0 | 0 | 0 | 0 |
| 0 | 0.004010668 | 0 | 0 | 0 |
| 0 | 0 | 0 | 0 | 0 |
| 0 | 0 | 0 | 0 | 0 |
| 0 | 0 | 0 | 0 | 0 |
| 0 | 0 | 0 | 0 | 0 |
| 0 | 0 | 0.018336189 | 0 | 0 |
| 0 | 0 | 0 | 0 | 0 |
| 0 | 0 | 0 | 0 | 0 |
| 0 | 0 | 0 | 0 | 0 |
| 0 | 0 | 0 | 0 | 0 |
| 0 | 0 | 0 | 0 | 0 |
| 0 | 0 | 1.389045641 | 1.363110745 | 0 |
| 0 | 0 | 0 | 0 | 0 |
| 0 | 0 | 0 | 0 | 0 |
| 0 | 0 | 0 | 0 | 0 |
| 0 | 0 | 0 | 0 | 0 |
| 0 | 0 | 0 | 0 | 0 |
| 0.036940196 | 0.07200809 | 0.048741318 | 0.002213234 | 0 |
| 0.066889443 | 0.027622229 | 21.40569025 | 0.002886435 | 0 |
| 0.045036226 | 0.123799245 | 0.121112966 | 0.001587755 | 0.124726844 |
| 0.010492337 | 0.003772994 | 0.017109642 | 0 | 0 |
| 0 | 0 | 0 | 0 | 0 |

| | | | | |
|---|---|---|---|---|
| 0.453889095 | 0.0051955 | 0 | 0 | 0 |
| 0.001769888 | 0.001471775 | 0 | 0.016913539 | 0.058990506 |
| 0.027386606 | 0 | 0 | 0.004581201 | 0 |
| 0.201098761 | 0.242623366 | 0.20384013 | 0.281540858 | 0.524038715 |
| 0.008297389 | 0.002455864 | 0.000487653 | 0.001459791 | 0 |
| 0 | 0 | 0 | 0 | 0 |
| 0.000813015 | 0.105016757 | 0.003570934 | 0.153780413 | 0 |
| 0 | 0 | 0 | 0 | 0 |
| 0 | 0 | 0 | 0 | 0 |
| 0 | 0 | 0 | 0 | 0 |
| 0 | 0 | 0 | 0 | 0 |
| 0.161993005 | 0 | 0.733481479 | 0 | 0 |
| 0 | 0.000780918 | 0 | 0 | 0 |
| 0.590795213 | 0.002905096 | 0.004441784 | 0.004029245 | 0.044431755 |
| 0 | 0.0104074 | 0 | 0 | 0 |
| 0 | 0 | 0 | 0 | 0 |
| 0.001064281 | 0.317278752 | 0.292835904 | 0.001841223 | 0.001457206 |
| 0 | 0 | 0 | 0 | 0 |
| 0 | 0 | 0 | 0 | 0 |
| 0 | 0 | 0 | 0 | 0 |
| 0.138024386 | 0.06617106 | 0.032712954 | 3.170545723 | 0.790299043 |
| 0.008857662 | 0.001280993 | 0.001869565 | 0.001332513 | 0.024677548 |
| 0.005615379 | 0.029633659 | 0.120423969 | 0 | 0 |
| 0 | 0 | 0 | 0 | 0 |
| 0 | 0 | 0 | 0 | 0 |
| 0 | 0.001484217 | 0 | 0 | 0 |
| 0 | 0 | 0 | 0 | 0 |
| 0 | 0 | 0 | 0 | 0 |
| 0.000985349 | 0.003482367 | 0 | 0.001704669 | 0 |
| 0 | 0 | 0 | 0 | 0 |
| 0.151205196 | 0.010523424 | 0 | 0.033720742 | 0.291334588 |
| 0 | 0 | 0 | 0 | 0 |
| 0 | 0 | 0 | 0 | 0 |
| 0 | 0 | 0 | 0 | 0 |
| 0 | 0 | 0 | 0 | 0 |
| 0 | 0 | 0.001862845 | 0 | 0 |
| 0 | 0 | 0 | 0 | 0 |
| 0 | 0 | 14.69432949 | 0 | 0 |
| 0 | 0 | 0 | 0 | 0 |
| 0 | 0 | 0 | 0 | 0 |
| 0 | 0 | 0 | 0 | 0 |
| 0 | 0 | 0 | 0 | 0 |
| 0 | 0 | 0 | 0 | 0 |
| 0 | 0 | 0 | 0 | 0 |
| 0 | 0.003225637 | 0 | 0 | 0 |
| 0 | 0 | 0 | 0 | 0 |
| 0 | 0 | 0 | 0 | 0 |
| 0 | 0 | 0 | 0 | 0.995374516 |

| | | | | |
|---|---|---|---|---|
| 0 | 0 | 0 | 0 | 0 |
| 0 | 0 | 0 | 0 | 0 |
| 0 | 0 | 0 | 0 | 0 |
| 0 | 0 | 0 | 0 | 0 |
| 0.00613707 | 0 | 0.144251533 | 0 | 0 |
| 0 | 0 | 0 | 0 | 0 |
| 0 | 0 | 0 | 0 | 0 |
| 0 | 0 | 0 | 0 | 0 |
| 0.003908568 | 0.003404996 | 0 | 0.006654553 | 0.005912228 |
| 0 | 0 | 0 | 0 | 0 |
| 0 | 0 | 0 | 0 | 0 |
| 0.00453777 | 0.021562546 | 0 | 0.011713316 | 0 |
| 0 | 0 | 0 | 0 | 0 |
| 0 | 0 | 0 | 0 | 0 |
| 0 | 0 | 0 | 0 | 0 |
| 0 | 0 | 0 | 0 | 0 |
| 0 | 0 | 0 | 0 | 0 |
| 0 | 0 | 0 | 0 | 0 |
| 0 | 0.83794837 | 0.051345485 | 0 | 0 |
| 0 | 0 | 0 | 0 | 0 |
| 0 | 0 | 0 | 0 | 0 |
| 0 | 0 | 0 | 0 | 0 |
| 0 | 0 | 0 | 0 | 0 |
| 0 | 0 | 0 | 0 | 0 |
| 0 | 0 | 0 | 0 | 0 |
| 0 | 0 | 0 | 0 | 0 |
| 0 | 0 | 0 | 0 | 0 |
| 0 | 0 | 0 | 0 | 0 |
| 0 | 0 | 0 | 0 | 0 |
| 0 | 0.0007169 | 0 | 0 | 0 |
| 0.002755714 | 0 | 0.00812221 | 0.000595929 | 0 |
| 0.013009428 | 0.003606057 | 0.002631456 | 0 | 0 |
| 0 | 0 | 0 | 0 | 0 |
| 0 | 0 | 0 | 0 | 0 |
| 0 | 0 | 0 | 0 | 0 |
| 0 | 0 | 0 | 0 | 0 |
| 0 | 0 | 0 | 0 | 0 |
| 0 | 0 | 0 | 0 | 0 |
| 0 | 0.305288429 | 0 | 0 | 0 |
| 0 | 0 | 0 | 0 | 0 |
| 0 | 0 | 0 | 0 | 0 |
| 0 | 0 | 0 | 0 | 0 |
| 0 | 0 | 0 | 0 | 0 |
| 0 | 0 | 0 | 0 | 0 |
| 0 | 0 | 0 | 0 | 0 |
| 0 | 0 | 0 | 0 | 0 |
| 0 | 0.001820732 | 0 | 0 | 0 |
| 0 | 0 | 0 | 0 | 0 |

| | | | | |
|---|---|---|---|---|
| 0 | 0 | 0 | 0 | 0 |
| 0 | 0 | 0 | 0 | 0 |
| 0 | 0 | 0 | 0 | 0 |
| 0 | 0 | 0 | 0 | 0 |
| 0 | 0.000915444 | 0.132078865 | 0 | 0 |
| 0 | 0 | 0 | 0 | 0 |
| 0 | 0 | 0 | 0 | 0 |
| 0 | 0 | 0 | 0 | 0 |
| 0 | 0 | 0 | 0 | 0 |
| 0 | 0 | 0 | 0 | 0 |
| 0 | 0 | 0 | 0 | 0 |
| 0 | 0 | 0 | 0 | 0 |
| 0 | 0 | 0 | 0 | 0 |
| 0 | 0 | 0 | 0 | 0 |
| 0 | 0 | 0 | 0 | 0 |
| 0 | 0 | 0 | 0 | 0 |
| 0 | 0 | 0 | 0 | 0 |
| 0 | 0 | 0 | 0 | 0 |
| 0 | 0 | 0 | 0 | 0 |
| 0 | 0 | 0 | 0 | 0 |
| 0 | 0 | 0 | 0 | 0 |
| 0 | 0 | 0 | 0 | 0 |
| 0 | 0 | 0 | 0 | 0 |
| 0 | 0 | 0 | 0 | 0 |
| 0 | 0 | 0.002203978 | 0 | 0 |
| 0 | 0 | 0 | 0 | 0 |
| 0 | 0 | 0 | 0 | 0 |
| 0 | 0 | 0 | 0 | 0 |
| 0 | 0 | 0 | 0 | 0 |
| 0 | 0 | 0 | 0 | 0 |
| 0 | 0 | 0 | 0 | 0 |
| 0.015427019 | 0.004507329 | 0 | 0 | 0.007535622 |
| 0 | 0 | 0 | 0 | 0 |
| 0 | 0 | 0 | 0 | 0 |
| 0 | 0 | 0 | 0 | 0 |
| 0 | 0 | 0 | 0 | 0 |
| 0 | 0 | 0 | 0 | 0 |
| 0 | 0 | 0 | 0 | 0 |
| 0 | 0 | 0 | 0 | 0 |
| 0 | 0 | 0 | 0 | 0 |
| 0 | 0 | 0 | 0 | 0 |
| 0 | 0 | 0 | 0 | 0 |
| 0 | 0 | 0 | 0 | 0 |
| 0 | 0 | 0 | 0 | 0 |
| 0 | 0 | 0 | 0 | 0 |
| 0 | 0 | 0 | 0 | 0 |
| 0 | 0 | 0 | 0 | 0 |
| 0.242301615 | 0 | 0.004608637 | 0 | 0 |

| | | | | |
|---|---|---|---|---|
| 0 | 0 | 0 | 0 | 0 |
| 0 | 0 | 0 | 0 | 0 |
| 0 | 0 | 0 | 0 | 0 |
| 0 | 0 | 0 | 0 | 0 |
| 0 | 0 | 0 | 0 | 0 |
| 0 | 0 | 0 | 0 | 0 |
| 0 | 0 | 0 | 0 | 0 |
| 0 | 0 | 0 | 0 | 0 |
| 0 | 0 | 0 | 0 | 0 |
| 0 | 0 | 0 | 0 | 0 |
| 0 | 0 | 0 | 0 | 0 |
| 0 | 0 | 0 | 0 | 0 |
| 0.010572887 | 0 | 0 | 0 | 0.00327766 |
| 0 | 0 | 0 | 0 | 0 |
| 0 | 0 | 0 | 0 | 0 |
| 0 | 0 | 0 | 0 | 0 |
| 0 | 0 | 0 | 0 | 0 |
| 0 | 0.002992551 | 0 | 0 | 0 |
| 0 | 0 | 0 | 0 | 0 |
| 0 | 0 | 0 | 0 | 0 |
| 0.018276777 | 0.049006787 | 0.032851744 | 0.064528807 | 0.303970426 |
| 0 | 0 | 0 | 0 | 0 |
| 0 | 0 | 0 | 0 | 0 |
| 0 | 0 | 0 | 0 | 0 |
| 0 | 0 | 0 | 0 | 0 |
| 0 | 0 | 0 | 0 | 0 |
| 0.00347558 | 0.003070802 | 0.002259695 | 0.003758006 | 0 |
| 0 | 0 | 0 | 0 | 0 |
| 0 | 0 | 0 | 0 | 0 |
| 0 | 0 | 0 | 0.001608253 | 0.040730448 |
| 0 | 0.00126044 | 0 | 0.104640878 | 0.071392052 |
| 0 | 0 | 0 | 0 | 0 |
| 0 | 0 | 0 | 0 | 0 |
| 0.037156119 | 0 | 0 | 0 | 0 |
| 0.102658174 | 0.001614503 | 0.001346463 | 0.002519153 | 0 |
| 0 | 0 | 0 | 0 | 0 |
| 0 | 0 | 0 | 0 | 0 |
| 0 | 0 | 0 | 0 | 0 |
| 0 | 0 | 0 | 0 | 0 |
| 0 | 0 | 0 | 0 | 0 |
| 0 | 0 | 0 | 0 | 0 |
| 0 | 0 | 0 | 0 | 0 |
| 0 | 0 | 0 | 0 | 0 |
| 0 | 0 | 0 | 0 | 0 |
| 0 | 0 | 0 | 0 | 0 |
| 0 | 0 | 0 | 0 | 0 |
| 0.006832007 | 0 | 0 | 0 | 0 |
| 0.033033061 | 0 | 0.002518755 | 0 | 0.007198697 |

| | | | | |
|---|---|---|---|---|
| 0 | 0 | 0 | 0 | 0 |
| 0 | 0 | 0 | 0 | 0 |
| 0 | 0 | 0 | 0 | 0.001731058 |
| 0 | 0 | 0 | 0 | 0 |
| 0 | 0 | 0 | 0 | 0 |
| 0 | 0 | 0 | 0 | 0 |
| 0 | 0 | 0 | 0 | 0 |
| 0 | 0.00062022 | 0 | 0 | 0 |
| 0 | 0 | 0 | 0 | 0 |
| 0 | 0 | 0 | 0 | 0 |
| 0 | 0.000533815 | 0 | 0 | 0 |
| 0 | 0 | 0 | 0 | 0 |
| 0 | 0 | 0 | 0 | 0 |
| 0 | 0 | 0 | 0 | 0 |
| 0 | 0 | 0 | 0 | 0 |
| 0 | 0 | 0 | 0 | 0 |
| 0 | 0 | 0 | 0 | 0 |
| 0 | 0 | 0.130207981 | 0 | 0 |
| 0 | 0 | 0.122100892 | 0 | 0 |
| 0 | 0 | 0 | 0 | 0 |
| 0.10896948 | 0.006904006 | 0 | 0.005685492 | 0.022451096 |
| 0 | 0 | 0 | 0 | 0 |
| 0 | 0 | 0 | 0 | 0 |
| 0 | 0 | 0 | 0 | 0 |
| 0.002061818 | 0.004714967 | 0.003932188 | 0.00653946 | 0.024842649 |
| 0 | 0 | 0 | 0 | 0 |
| 0 | 0 | 0 | 0 | 0 |
| 0 | 0 | 0 | 0 | 0 |
| 0 | 0 | 0 | 0 | 0 |
| 0 | 0 | 0 | 0 | 0 |
| 0.959213277 | 0.276987239 | 0.034266587 | 0.006820881 | 0.075645543 |
| 0 | 0 | 0 | 0 | 0 |
| 0 | 0 | 0 | 0 | 0 |
| 0 | 0 | 0.023279263 | 0 | 0.000811064 |
| 0 | 0 | 0 | 0 | 0 |
| 0 | 0 | 0 | 0 | 0 |
| 0 | 0 | 0 | 0 | 0 |
| 0 | 0 | 0 | 0 | 0 |
| 0 | 0 | 0 | 0 | 0 |
| 0 | 0 | 0 | 0 | 0 |
| 0 | 0 | 0 | 0 | 0 |
| 0.003111403 | 0.000808541 | 0.012811827 | 0.007849883 | 0 |
| 0 | 0 | 0 | 0 | 0 |
| 0 | 0 | 0 | 0 | 0 |
| 0 | 0 | 0 | 0 | 0 |
| 0 | 0 | 0 | 0 | 0 |
| 0 | 0 | 0 | 0 | 0 |
| 0 | 0 | 0 | 0 | 0 |

| | | | | |
|---|---|---|---|---|
| 0 | 0 | 0 | 0 | 0 |
| 0 | 0 | 0 | 0 | 0 |
| 0.043464025 | 0.116145283 | 0.108547328 | 0 | 0.137208863 |
| 0 | 0 | 0 | 0 | 0 |
| 0 | 0 | 0 | 0.000977602 | 0 |
| 0 | 0 | 0 | 0 | 0 |
| 0 | 0 | 0 | 0 | 0 |
| 0 | 0 | 0 | 0 | 0 |
| 0 | 0 | 0 | 0 | 0 |
| 0 | 0.000928545 | 0 | 0 | 0.002446202 |
| 0 | 0 | 0 | 0 | 0 |
| 0 | 0 | 0 | 0 | 0 |
| 0 | 0 | 0 | 0 | 0 |
| 0 | 0.004712573 | 0.00093576 | 0 | 0.028081587 |
| 0.002004628 | 0.007223565 | 0 | 0.010532563 | 0.006587339 |
| 0 | 0.001189812 | 0 | 0 | 0 |
| 0.00152273 | 0.002743535 | 0.001584037 | 0 | 0.014455387 |
| 0 | 0 | 0 | 0 | 0 |
| 0 | 0 | 0 | 0 | 0 |
| 0 | 0 | 0 | 0 | 0 |
| 0 | 0 | 0 | 0 | 0 |
| 0.039634019 | 0 | 0.002700985 | 0 | 0.00376416 |
| 0 | 0 | 0 | 0 | 0 |
| 0 | 0 | 0 | 0 | 0.012448656 |
| 0 | 0.002443772 | 0 | 0.011862932 | 0 |
| 0 | 0 | 0 | 0 | 0 |
| 0 | 0.564999352 | 0 | 0 | 0 |
| 0 | 0 | 0 | 0 | 0 |
| 0 | 0 | 0 | 0 | 0 |
| 0 | 0.001653446 | 0 | 0.06994443 | 0.0101638 |
| 0 | 0 | 0 | 0 | 0 |
| 0 | 0 | 0 | 0 | 0 |
| 0 | 0 | 0 | 0 | 0 |
| 0 | 0.005052119 | 0 | 0 | 0 |
| 0 | 0 | 0 | 0 | 0 |
| 0 | 0 | 0 | 0 | 0 |
| 0 | 0 | 0 | 0 | 0 |
| 0 | 0 | 0 | 0 | 0.001576426 |
| 0 | 0 | 0 | 0 | 0 |
| 0 | 0 | 0 | 0 | 0 |
| 0 | 0 | 0.010659896 | 0 | 0 |
| 0 | 0 | 0 | 0 | 0 |
| 0 | 0 | 0 | 0 | 0 |
| 0 | 0 | 0 | 0 | 0 |
| 0 | 0 | 0 | 0 | 0 |
| 0 | 0.021344353 | 0 | 0 | 0.004362714 |
| 0 | 0.002594532 | 0 | 0 | 0 |
| 0 | 0.011596806 | 0 | 0.004423177 | 0.010692905 |

| | | | | |
|---|---|---|---|---|
| 0.073779766 | 1.049874651 | 0 | 0 | 0 |
| 0.115415375 | 0 | 0 | 0 | 0 |
| 0 | 0 | 0 | 0 | 0 |
| 0 | 0 | 0 | 0 | 0 |
| 0 | 0.001003804 | 0 | 0.041767014 | 0.046013729 |
| 0 | 0.013749065 | 0 | 0 | 0.002156022 |
| 0 | 0 | 0 | 0 | 0 |
| 0.028426534 | 0.007598082 | 0.022764993 | 0.032395267 | 0.011120408 |
| 0 | 0 | 0 | 0 | 0 |
| 0.054762804 | 0.061781897 | 0 | 0.155688249 | 0 |
| 0.004415141 | 0 | 0 | 0 | 0 |
| 0.002678875 | 0.005569138 | 0 | 0.001544832 | 0.001173727 |
| 0.006299266 | 0.012842142 | 0.004509511 | 0.004218515 | 0.009348296 |
| 0.033947281 | 0 | 0 | 0 | 0 |
| 0.001462921 | 0 | 0 | 0 | 0 |
| 0 | 0.001775622 | 0 | 0.002462713 | 0 |
| 0 | 0 | 0 | 0 | 0 |
| 0.010850416 | 0 | 0 | 0 | 0 |
| 0.037745491 | 0 | 0 | 0.471297819 | 0 |
| 0.023941188 | 0 | 0 | 0 | 0 |
| 0 | 0.06539319 | 0 | 0 | 0 |
| 0 | 0 | 0 | 0 | 0.743726234 |
| 0 | 0 | 0 | 0 | 0 |
| 0 | 0 | 0 | 0 | 0 |
| 0 | 0 | 0 | 0 | 0 |
| 0.016567508 | 0.001377694 | 0.047271873 | 0 | 0 |
| 0 | 0 | 1.388251356 | 0 | 0 |
| 0 | 0 | 0 | 0.002310693 | 0.031036687 |
| 0 | 0.008060343 | 0 | 0 | 0 |
| 0 | 0 | 0 | 0 | 0 |
| 0 | 0 | 0 | 0 | 0.050351721 |
| 0 | 0 | 0 | 0 | 0 |
| 0 | 0 | 0 | 0 | 0 |
| 0 | 0 | 0 | 0.070685432 | 0 |
| 0 | 0 | 0 | 0 | 0 |
| 1.017620522 | 0 | 0 | 0 | 0.105681974 |
| 0.02924465 | 0.107315046 | 0 | 0.800525558 | 2.49448079 |
| 0 | 0 | 0 | 0.029129675 | 0 |
| 0 | 0 | 0 | 0 | 0 |
| 0 | 0 | 0 | 0 | 0 |
| 0 | 0 | 0 | 0 | 0 |
| 0 | 0 | 0 | 0.441765891 | 0 |
| 0 | 0 | 0 | 0 | 0 |
| 0.039470076 | 0.001427039 | 0 | 0.00945638 | 0 |
| 0 | 0 | 0 | 0.144152434 | 0 |
| 0 | 0 | 0 | 0 | 0 |
| 0.002648958 | 0 | 0 | 0 | 0.007737464 |
| 0 | 0 | 0 | 0 | 0 |

| | | | | |
|---|---|---|---|---|
| 0 | 0.002081976 | 0 | 0.00866284 | 0 |
| 0 | 0.015317606 | 0 | 0 | 0 |
| 0 | 0 | 0 | 0 | 0 |
| 0 | 0 | 0 | 0 | 0 |
| 0.002327305 | 0 | 0 | 0 | 0 |
| 0.00857732 | 0.042024446 | 0 | 0.022993612 | 0.191713065 |
| 0 | 0 | 0 | 0 | 0.019199812 |
| 0 | 0 | 0 | 0 | 0 |
| 0 | 0 | 0 | 0 | 0 |
| 0 | 0 | 0 | 0 | 0 |
| 0 | 0.036667753 | 0 | 0.044654598 | 0.017670586 |
| 0 | 0 | 0 | 0 | 0.001932159 |
| 0 | 0 | 0 | 0 | 0 |
| 0.007874506 | 0.000844923 | 0 | 0.00322265 | 0.000834714 |
| 0 | 0.150036775 | 0 | 0.081222136 | 0 |
| 0.061392766 | 0.002584912 | 0 | 0 | 0 |
| 0 | 0 | 0 | 0 | 0 |
| 0 | 0 | 0 | 0.104918774 | 1.357372704 |
| 0 | 0 | 0.000937488 | 0.009666407 | 0 |
| 0 | 0 | 0 | 0 | 0 |
| 0 | 0.026628324 | 0 | 0 | 0 |
| 0 | 0 | 0.032863132 | 0 | 0 |
| 0 | 0.001804031 | 0 | 0 | 0 |
| 0 | 0 | 0 | 0 | 0 |
| 0 | 0 | 0 | 0 | 0 |
| 0 | 0 | 0 | 0 | 0 |
| 0.055910451 | 0.00091612 | 0 | 0.087037492 | 0.04012393 |
| 0 | 0 | 0 | 0 | 0 |
| 0 | 0 | 0 | 0 | 0 |
| 0.954666223 | 0.275871509 | 0.119812653 | 0.592613398 | 0.611758001 |
| 0 | 0 | 0.002390241 | 0.001391289 | 0 |
| 0 | 0.000804773 | 0 | 0.002790467 | 0 |
| 0 | 0 | 0 | 0 | 0.004360156 |
| 0 | 0 | 0 | 0 | 0 |
| 0 | 0.001144563 | 0 | 0.001058307 | 0 |
| 0 | 0 | 0 | 0 | 0 |
| 0 | 0.007469157 | 0.273704025 | 0 | 0 |
| 0.004919296 | 0.000861202 | 0 | 0.002388901 | 0.007657164 |
| 0 | 0.763463712 | 0 | 0.673413991 | 0 |
| 0 | 0.065234363 | 0 | 0.097063776 | 0 |
| 0.005220726 | 0.038030378 | 0.000724123 | 0.015896214 | 0.142734966 |
| 0.000898009 | 1.513516639 | 0 | 0 | 0 |
| 0.005929465 | 0.000591687 | 0 | 0.000820645 | 0.005975279 |
| 0 | 0 | 0 | 0.007845233 | 0 |
| 0.018374464 | 0 | 0 | 0.006248946 | 0 |
| 0 | 0 | 0 | 0 | 0 |
| 0 | 0 | 0.000760019 | 0.007836526 | 0.007922675 |
| 0 | 0 | 0 | 0 | 0 |

| | | | | |
|---|---|---|---|---|
| 0.035967545 | 0.002023938 | 0.023443386 | 0 | 0 |
| 0 | 0 | 0 | 0 | 0 |
| 0.001473941 | 0.026352034 | 0.001277736 | 0 | 0 |
| 0 | 0 | 0 | 0 | 0 |
| 0 | 0.22952746 | 0 | 0 | 0 |
| 0 | 0.037647316 | 0 | 0 | 0 |
| 0 | 0 | 0 | 0 | 0 |
| 0 | 0 | 0 | 0 | 0 |
| 0 | 0 | 0.000941761 | 0.082747748 | 0.11775695 |
| 0 | 0 | 0 | 0 | 0 |
| 0 | 0 | 0 | 0 | 0 |
| 0 | 0 | 0 | 0 | 0.00424623 |
| 0.004323138 | 0 | 0 | 0 | 0 |
| 0 | 0.032347771 | 0 | 0.015237167 | 0 |
| 0 | 0 | 0 | 0 | 0 |
| 1.352687457 | 0 | 0.003314837 | 0.001543576 | 0.065340202 |
| 0.238194745 | 0.037711625 | 0.734389373 | 0.001466479 | 0.004410363 |
| 0.002317238 | 0 | 0 | 0.006236003 | 0 |
| 0 | 0 | 0 | 0 | 0 |
| 0 | 0.028906891 | 0 | 0 | 0 |
| 0 | 0.010660346 | 0 | 0 | 0 |
| 0 | 0 | 0.000681892 | 0.001984546 | 0.008077566 |
| 0 | 0 | 0 | 0 | 0 |
| 0 | 0 | 0 | 0 | 0 |
| 0.009704791 | 0.001482273 | 0 | 0 | 0 |
| 0 | 0.017178519 | 0 | 0 | 0 |
| 0 | 0.003132023 | 0 | 0.006515976 | 0.017091657 |
| 0 | 0.004672043 | 0 | 0.002159975 | 0 |
| 0 | 0 | 0 | 0 | 0 |
| 0 | 0 | 0 | 0 | 0 |
| 0.014321779 | 0.000680541 | 0.000567558 | 0 | 0 |
| 0.00244023 | 0.016994612 | 0.039769474 | 0.007739668 | 0.061143005 |
| 0.159735755 | 0.021735897 | 0.041491376 | 0.063978147 | 0 |
| 0 | 0.002768859 | 0 | 0 | 0 |
| 0 | 0 | 0.038319756 | 0 | 0 |
| 0 | 0 | 0 | 0 | 0 |
| 0.004988783 | 0.001091708 | 0.002003018 | 0 | 0.000862814 |
| 0.035318475 | 0.006830132 | 0.001139238 | 0.001894621 | 0.009221726 |
| 1.247326794 | 0 | 0.024860238 | 0 | 0 |
| 0.156134354 | 0 | 0 | 0 | 0 |
| 0 | 0 | 0 | 0 | 0 |
| 0 | 0 | 0 | 0 | 0 |
| 0.019346393 | 0 | 0 | 0.007083506 | 0 |
| 0.018594006 | 0.00199941 | 0.000333494 | 0 | 0 |
| 1.23212622 | 0.145289599 | 0 | 1.251029505 | 0 |
| 0 | 0 | 0.155853801 | 0 | 0 |
| 0 | 0 | 0 | 0 | 0 |
| 0 | 0 | 0 | 0 | 0 |

| | | | | |
|---|---|---|---|---|
| 0 | 0 | 0 | 0.004564778 | 0.112049928 |
| 0.038053039 | 1.299711547 | 0.383044079 | 0.180393584 | 0.399039863 |
| 0 | 0.003630749 | 0 | 0 | 0 |
| 0 | 0 | 0 | 0 | 0 |
| 0 | 0 | 0 | 0 | 0 |
| 0 | 0 | 0 | 0 | 0 |
| 0 | 0 | 0 | 0.091653813 | 0.00141768 |
| 0 | 0.001429739 | 0 | 0 | 0 |
| 0.007171793 | 0 | 0 | 0 | 0 |
| 0.016266117 | 0 | 0 | 0 | 0 |
| 0.039921273 | 0 | 0.001153571 | 0 | 0 |
| 0.218963309 | 0.038888795 | 0.05111873 | 0.030004753 | 0.058349245 |
| 0 | 0.140974253 | 0 | 0 | 0 |
| 0 | 0 | 0 | 0 | 0 |
| 0.021470939 | 0 | 0.001489026 | 0 | 0 |
| 0 | 0 | 0 | 0 | 0 |
| 0.002394983 | 0.016113702 | 0.005888782 | 0.007784491 | 0.054136531 |
| 0.045504725 | 0.20494754 | 0 | 0.11898229 | 0.605556022 |
| 0 | 0 | 0 | 0 | 0.001263904 |
| 0.028814269 | 0.010530589 | 0 | 0.00232841 | 0 |
| 0 | 0.046864497 | 0.019308453 | 0.012171142 | 0.006640385 |
| 0 | 0 | 0 | 0 | 0 |
| 0 | 0 | 0 | 0 | 0 |
| 0 | 0 | 0 | 0 | 0 |
| 0 | 0 | 0 | 0.001305306 | 0 |
| 0 | 0 | 0 | 0 | 0 |
| 0.019502776 | 0 | 0.014754897 | 0 | 0 |
| 0.074933707 | 0.111923759 | 0.050808587 | 0.106516621 | 0 |
| 0 | 0 | 0 | 0 | 0 |
| 0.021877254 | 0 | 0 | 0 | 0 |
| 0.001280317 | 0 | 0 | 0.003248622 | 0 |
| 0 | 0 | 0 | 0 | 0 |
| 0 | 0 | 0 | 0 | 0.019040126 |
| 0 | 0 | 0 | 0 | 0 |
| 0 | 0 | 0.035190162 | 0 | 0 |
| 0.821912064 | 0.17800509 | 0.010923875 | 0.010713901 | 1.344013323 |
| 0 | 0 | 0 | 0 | 0 |
| 0 | 0 | 0 | 0 | 0 |
| 0.002649529 | 0.015422766 | 0 | 0.022069795 | 0.008384059 |
| 0 | 0 | 0 | 0 | 0 |
| 1.116154066 | 0.145936068 | 0 | 1.056352319 | 0 |
| 0 | 0 | 0 | 0 | 0 |
| 0.080468208 | 0.001486988 | 0 | 0 | 0.021937377 |
| 0 | 0.001579225 | 0 | 0.000796479 | 0.009077196 |
| 0.008845838 | 0.184840036 | 0.000629195 | 0 | 0 |
| 0 | 0 | 0.020164218 | 0 | 0 |
| 0.003288963 | 0 | 0 | 0 | 0 |
| 0.000837205 | 0 | 0 | 0 | 0.040960964 |

| | | | | |
|---|---|---|---|---|
| 0.003497381 | 0.00498565 | 0 | 0 | 0 |
| 0 | 0 | 0 | 0 | 0 |
| 0 | 0 | 0 | 0 | 0 |
| 1.294377317 | 0.275713756 | 0.075394807 | 0.000960812 | 0 |
| 0 | 0 | 0 | 0 | 0 |
| 0 | 0.002252693 | 0 | 0 | 0 |
| 0.003875148 | 0 | 0 | 0 | 0 |
| 0 | 0.019366797 | 0.016409943 | 0 | 0 |
| 0 | 0.008416063 | 0.510658657 | 0 | 0 |
| 0.003164366 | 0 | 0.009294402 | 0 | 0 |
| 0 | 0 | 0 | 0 | 0 |
| 0 | 0.000829659 | 0.001798989 | 0 | 0 |
| 0 | 0 | 0 | 0 | 0 |
| 0 | 0 | 0 | 0 | 0 |
| 0.421865181 | 0 | 0.021126009 | 0 | 0 |
| 0 | 0 | 0 | 0 | 0 |
| 0 | 0 | 0.003574966 | 0 | 0 |
| 0 | 0.006079421 | 0 | 0 | 0 |
| 0 | 0 | 0 | 0 | 0 |
| 0 | 0 | 0 | 0 | 0 |
| 0 | 0 | 0 | 0 | 0 |
| 0 | 0 | 0 | 0 | 0 |
| 0 | 0 | 0 | 0 | 0 |
| 0.065030976 | 0.479442402 | 0.031899625 | 0.208728092 | 0.745329946 |
| 0 | 0 | 0 | 0 | 0.002864912 |
| 0 | 0.004963383 | 0.034984279 | 0 | 0 |
| 0 | 0 | 0 | 0 | 0 |
| 0 | 0 | 0.522673404 | 0.574024264 | 0 |
| 0 | 0 | 0 | 0 | 0 |
| 0 | 0 | 0 | 0.004149018 | 0 |
| 0 | 0.010808038 | 0.350592016 | 0 | 0 |
| 0 | 0 | 0 | 0 | 0 |
| 0.003248072 | 0.00135049 | 0 | 0 | 0 |
| 0 | 0.000512632 | 0 | 0 | 0 |
| 0 | 0.013851346 | 0 | 0 | 0 |
| 0 | 0 | 0.042775732 | 0 | 0 |
| 0 | 0 | 0.885247331 | 0 | 0 |
| 0.053565409 | 0 | 0 | 0 | 0 |
| 0 | 0 | 0 | 0 | 0 |
| 0 | 0 | 0 | 0 | 0 |
| 0.003828495 | 0.264401127 | 0.000929281 | 0.381946914 | 0.282016947 |
| 0 | 0 | 0.030900235 | 0 | 0 |
| 1.36335766 | 0.26218564 | 0.017518084 | 0.405751713 | 0.471375714 |
| 0 | 0 | 0 | 0 | 0 |
| 0 | 0 | 0 | 0 | 0 |
| 0 | 0 | 0 | 0 | 0 |
| 0 | 0 | 0 | 0 | 0 |
| 0.003451717 | 0 | 0 | 0 | 0.036800328 |

| | | | | |
|---|---|---|---|---|
| 0 | 0 | 0 | 0 | 0 |
| 0 | 0.004183914 | 0.130848737 | 0 | 0 |
| 0.112280677 | 0.014858257 | 0.003059626 | 0.508579532 | 1.685335292 |
| 0 | 0 | 0 | 0 | 0 |
| 3.448388987 | 0 | 0 | 0 | 0 |
| 0 | 0 | 0.038443579 | 0 | 0 |
| 0 | 0 | 0 | 0 | 0 |
| 0 | 0 | 0 | 0 | 0 |
| 0 | 0 | 0.027462252 | 0 | 0 |
| 0 | 0 | 0 | 0 | 0 |
| 0.325304617 | 0.001260706 | 0.002403208 | 0.002747716 | 0.011387185 |
| 0.339581328 | 2.855516894 | 0.835379018 | 2.040546858 | 0 |
| 0 | 0 | 0 | 0 | 0 |
| 0.026880446 | 0.058700417 | 0.243315904 | 0.005122136 | 0.00332841 |
| 0.051759766 | 0.065711123 | 0 | 0.782007013 | 4.329423707 |
| 0 | 0 | 0 | 0 | 0 |
| 0 | 0 | 0 | 0 | 0 |
| 0 | 0 | 0 | 0 | 0 |
| 0 | 0 | 0.017606275 | 0 | 0 |
| 0 | 0.00207698 | 0.001855886 | 0 | 0 |
| 0.038846724 | 0.383415751 | 0.006672179 | 0.723976477 | 0.543220538 |
| 0.104052443 | 0 | 0 | 1.864540044 | 0.018018005 |
| 0.063438872 | 2.047975398 | 0.023543449 | 13.56948251 | 0 |
| 0 | 0.000776802 | 0 | 0 | 0 |
| 0 | 0 | 0.004812002 | 0 | 0 |
| 0 | 0 | 0.001961104 | 0.001756156 | 0 |
| 0 | 0 | 0 | 0 | 0.185497552 |
| 0 | 0 | 0.002626023 | 0 | 0 |
| 0 | 0 | 0 | 0 | 0 |
| 0 | 0 | 0 | 0 | 0 |
| 0 | 0 | 0.002499365 | 0 | 0 |
| 0 | 0.037052442 | 0 | 0 | 0 |
| 0 | 0 | 0 | 1.090052279 | 3.067484281 |
| 0 | 0 | 0.014704385 | 0 | 0 |
| 0.191145837 | 0.000610407 | 0.012624864 | 0.003725078 | 0.073167821 |
| 0.502855895 | 0 | 0.147019571 | 0.001395826 | 0 |
| 0.021627944 | 0 | 0.001081019 | 0 | 0 |
| 0 | 0 | 0 | 0 | 0 |
| 0 | 0.054369766 | 1.23065288 | 0 | 0 |
| 0 | 0 | 0 | 0 | 0 |
| 0 | 0 | 0 | 0 | 0 |
| 0 | 0 | 0 | 0 | 0 |
| 0 | 0 | 0 | 0 | 0 |
| 0 | 0 | 0 | 0 | 0 |
| 0.397021586 | 0 | 0.012442828 | 0.001061188 | 0.014613565 |
| 0 | 0 | 0 | 0 | 0 |
| 0 | 0 | 0 | 0 | 0 |
| 7.496736238 | 0.013613402 | 0 | 0.054403495 | 0.052275999 |

| | | | | |
|---|---|---|---|---|
| 0 | 0 | 0 | 0 | 0 |
| 0 | 0 | 0 | 0 | 0 |
| 0 | 0 | 0 | 0 | 0.004992697 |
| 0 | 0 | 0.000679806 | 0 | 0 |
| 0 | 0 | 0 | 0 | 0 |
| 0 | 0.018142029 | 0.416917939 | 0 | 0 |
| 0 | 0 | 0.204968652 | 0 | 0 |
| 0.520815383 | 0.000750071 | 0.001000871 | 0 | 0 |
| 0 | 0 | 0 | 0 | 0 |
| 0 | 0 | 0 | 0 | 0 |
| 0.044760977 | 0 | 0.001514248 | 0.001007313 | 0.040419158 |
| 0 | 0 | 0 | 0 | 0 |
| 0.258091915 | 0 | 0 | 0 | 0 |
| 0 | 0 | 0 | 0 | 0 |
| 0 | 0 | 0 | 0 | 0 |
| 0 | 0 | 0 | 0 | 0 |
| 0 | 0.050774818 | 0.042908276 | 0 | 0 |
| 0 | 0.000453456 | 0.001134519 | 0 | 0 |
| 0 | 0.032446587 | 0 | 0 | 0 |
| 0.008524093 | 0 | 0.012959876 | 0.000756247 | 0 |
| 0 | 0 | 0 | 0 | 0 |
| 0 | 0 | 0 | 0 | 0 |
| 0 | 0.000522802 | 0.062784914 | 0 | 0 |
| 0 | 0 | 0 | 0 | 0 |
| 0 | 0 | 0 | 0 | 0 |
| 0 | 0 | 0 | 0 | 0 |
| 0 | 0 | 0 | 0 | 0.003204428 |
| 0 | 0 | 0 | 0 | 0 |
| 0 | 0 | 0 | 0 | 0 |
| 0 | 0.723057036 | 0.428201561 | 0 | 0 |
| 0.046432432 | 0 | 0 | 0 | 0 |
| 0 | 0 | 0 | 0 | 0 |
| 0 | 0.152874313 | 0 | 0 | 0 |
| 0 | 0 | 0 | 0 | 0 |
| 0 | 0 | 0 | 0 | 0 |
| 0 | 0 | 0 | 0 | 0 |
| 0 | 0 | 0.020060971 | 0 | 0 |
| 0.052862497 | 0 | 0 | 0.011298385 | 0.004049175 |
| 0 | 0 | 0 | 0 | 0 |
| 0 | 0 | 0 | 0 | 0 |
| 0.561186951 | 0 | 0 | 0 | 0 |
| 0 | 0 | 0 | 0 | 0 |
| 0 | 0 | 0 | 0 | 0 |
| 0.027821821 | 0.049853532 | 0.816848003 | 0 | 0 |
| 0 | 0 | 0.079616543 | 0 | 0 |
| 0 | 0 | 0 | 0 | 0 |
| 0 | 0 | 0.005968003 | 0 | 0 |
| 0 | 0 | 0 | 0 | 0 |

| | | | | |
|---|---|---|---|---|
| 0 | 0 | 0 | 0 | 0 |
| 0 | 0 | 0 | 0 | 0 |
| 0 | 0 | 0 | 0 | 0 |
| 0 | 0.078278472 | 0.094481362 | 0 | 0 |
| 0 | 0 | 0 | 0 | 0 |
| 0 | 0 | 0 | 0 | 0 |
| 0 | 0 | 0 | 0 | 0 |
| 0 | 0 | 0 | 0 | 0 |
| 0.018099095 | 0.064865764 | 0.001414294 | 0.393087063 | 2.301925939 |
| 0 | 0 | 0 | 0 | 0 |
| 0 | 0 | 0 | 0 | 0 |
| 0 | 0 | 0 | 0 | 0 |
| 0 | 0 | 0 | 0 | 0 |
| 0 | 0 | 0 | 0 | 0 |
| 0 | 0.035493857 | 0.73964791 | 14.19977552 | 0 |
| 0 | 0 | 0 | 0.019034333 | 1.836568081 |
| 0 | 0 | 0 | 0 | 0 |
| 0 | 0 | 0 | 0 | 0 |
| 0 | 0 | 0 | 0 | 0 |
| 0 | 0 | 0 | 0 | 0 |
| 0 | 0.001620888 | 0 | 0 | 0 |
| 0 | 0 | 0 | 0 | 0 |
| 0 | 0 | 0 | 0 | 0 |
| 0 | 0 | 0 | 0 | 0 |
| 0.079440356 | 0 | 0.593127756 | 0 | 0 |
| 15.30575317 | 0.040177933 | 0.926259428 | 0 | 0 |
| 0 | 0.020027118 | 0 | 0 | 0 |
| 0 | 0 | 0 | 0 | 0 |
| 0 | 0 | 0.052347081 | 0 | 0 |
| 0 | 0 | 0.201640039 | 0 | 0 |
| 0 | 0 | 0 | 0 | 0 |
| 0 | 0.001926468 | 0.028919436 | 0 | 0 |
| 0 | 0 | 0 | 0 | 0 |
| 0.010886736 | 0.00053253 | 0 | 0 | 0.002455114 |
| 0.452140646 | 0.012064825 | 0.589439774 | 0.001825462 | 0 |
| 0.038412705 | 0 | 0.000905226 | 0.00129038 | 0.003063753 |
| 0.665235022 | 0.002720322 | 0 | 0.023747525 | 0 |
| 0 | 6.052018772 | 0.252180558 | 0 | 0 |
| 0 | 1.503231526 | 0.44734079 | 0 | 0 |
| 0 | 0 | 0 | 0 | 0 |
| 0 | 0 | 0 | 0 | 0 |
| 0 | 0.003165133 | 0 | 0 | 0 |
| 0 | 0.082638644 | 0.273025129 | 0 | 0 |
| 0 | 0 | 0.461031258 | 0 | 0 |
| 0 | 0 | 0 | 0 | 0 |
| 0.007772881 | 0.003672527 | 0.000857588 | 0 | 0 |
| 0 | 0 | 0 | 0 | 0 |
| 0.10666379 | 1.377974604 | 0.430951147 | 0.439171797 | 0.006781962 |

| | | | | |
|---|---|---|---|---|
| 0.048688968 | 0 | 0 | 0 | 0 |
| 0 | 0.008454659 | 0 | 0 | 0 |
| 0 | 0 | 0 | 0 | 0 |
| 0 | 0 | 0 | 0 | 0 |
| 0.665284567 | 0.000695008 | 0 | 0 | 0 |
| 0 | 0 | 0.48169918 | 0 | 0 |
| 0 | 0.007054429 | 0.034607363 | 0 | 0 |
| 0 | 0 | 0 | 0 | 0 |
| 0 | 0 | 0 | 0 | 0 |
| 0 | 0 | 0 | 0 | 0 |
| 0 | 0 | 0.000554392 | 0 | 0 |
| 0 | 0 | 0.079007706 | 0.394405771 | 0 |
| 0.050852429 | 0.050071225 | 0.003333653 | 0 | 0 |
| 0 | 0 | 0 | 0 | 0 |
| 0.267744013 | 0.413342356 | 0.005362671 | 0.01393506 | 0.054525806 |
| 0 | 0 | 0 | 0 | 0 |
| 0 | 0 | 0 | 0 | 0 |
| 0 | 0 | 0 | 0 | 0 |
| 0.026051652 | 0 | 0 | 0 | 0 |
| 0 | 0 | 0.002605976 | 0 | 0 |
| 0 | 0.01531078 | 0.02201112 | 0 | 0 |
| 0 | 0 | 0 | 0 | 0 |
| 3.263051426 | 0.021578698 | 1.230168781 | 0 | 0 |
| 0 | 0.01217071 | 0 | 0 | 0 |
| 0 | 0 | 0 | 0 | 0 |
| 0 | 0 | 0 | 0 | 0 |
| 0 | 0 | 0 | 0 | 0 |
| 0 | 0 | 0 | 0 | 0 |
| 0 | 0 | 0.002113304 | 0 | 0 |
| 0 | 0.001311184 | 0 | 0 | 0 |
| 0 | 0 | 0 | 0 | 0 |
| 0.006584965 | 0.102215288 | 0.059149933 | 0.033272149 | 0.211577634 |
| 0 | 0 | 0 | 0 | 0 |
| 0.0773871 | 0.001172411 | 0.012276406 | 0.003252168 | 0 |
| 0.036526586 | 0 | 0 | 0 | 0.003556407 |
| 0.046040817 | 0 | 0.058961005 | 0 | 0 |
| 0 | 0 | 0 | 0 | 0 |
| 0 | 0 | 0 | 0 | 0 |
| 0 | 0 | 0.001940141 | 0 | 0 |
| 0 | 0 | 0 | 0 | 0 |
| 0 | 0 | 0 | 0 | 0 |
| 0 | 0 | 0.004609122 | 0 | 0 |
| 0 | 0 | 0 | 0 | 0 |
| 0 | 0.065802322 | 0 | 0 | 0 |
| 0.155123902 | 0 | 0.00318857 | 0 | 0.008758543 |
| 0 | 0 | 0 | 0 | 0 |
| 0 | 0 | 0 | 0 | 0 |
| 0.025713719 | 0 | 0 | 0 | 0.005177509 |

| | | | | |
|---|---|---|---|---|
| 0 | 0 | 0 | 0 | 0 |
| 0 | 0 | 0 | 0 | 0 |
| 0 | 0 | 0 | 0 | 0 |
| 0 | 0 | 0 | 0 | 0 |
| 0 | 0.01290238 | 0 | 0 | 0 |
| 0 | 0.076319029 | 0 | 0 | 0 |
| 0 | 0.431871065 | 0.274627027 | 0 | 0 |
| 0 | 0 | 0.067569908 | 0.081971303 | 0 |
| 0 | 0 | 0 | 0 | 0 |
| 0 | 0 | 0 | 0 | 0 |
| 0 | 0 | 0 | 0 | 0 |
| 0.022929427 | 0 | 0 | 0 | 0.010138512 |
| 0 | 0 | 0 | 0 | 0 |
| 0 | 0 | 0 | 0 | 0 |
| 0 | 0 | 0 | 0 | 0 |
| 0 | 0 | 0 | 0 | 0 |
| 0 | 0 | 0.002410767 | 0 | 0 |
| 0 | 0 | 0 | 0 | 0 |
| 0.119516468 | 0.697719815 | 0.328018489 | 0 | 0 |
| 0 | 0 | 0 | 0 | 0 |
| 0.072885661 | 0 | 0.000781651 | 0 | 0 |
| 0 | 0 | 0 | 0 | 0 |
| 0 | 0 | 0 | 0 | 0 |
| 0 | 0 | 0 | 0 | 0 |
| 0 | 0 | 0.002574975 | 0 | 0 |
| 0.211969205 | 0 | 0.004904377 | 0.011161209 | 0.075627165 |
| 0 | 0 | 0 | 0 | 0 |
| 0.003096652 | 0.283862938 | 0.066953076 | 0.017647436 | 0.135078748 |
| 0 | 0 | 0 | 0 | 0 |
| 0.501645678 | 0.524715736 | 0.244666979 | 0 | 0 |
| 0 | 0 | 0 | 0 | 0 |
| 0 | 0 | 0 | 0 | 0 |
| 0 | 0 | 0 | 0 | 0 |
| 0 | 0 | 0 | 0 | 0 |
| 0.029643418 | 0 | 0 | 0 | 0.004485996 |
| 0 | 0 | 0 | 0 | 0 |
| 0 | 0 | 0 | 0 | 0 |
| 0 | 0 | 0 | 0 | 0.001939159 |
| 0 | 0 | 0 | 0 | 0 |
| 0 | 0 | 0 | 0 | 0 |
| 13.78186869 | 3.31047805 | 1.577780509 | 12.84296971 | 0.780996524 |
| 0.328835182 | 0.6269086 | 0.002134455 | 0.033815799 | 12.78182065 |
| 0 | 0 | 0.301685175 | 0 | 0 |
| 0 | 0 | 0 | 0 | 0 |
| 0 | 0 | 0 | 0 | 0 |
| 0 | 0 | 0 | 0 | 0 |
| 0 | 0 | 0 | 0 | 0 |
| 0.024939558 | 0 | 0 | 0 | 0.001346646 |

| | | | | |
|---|---|---|---|---|
| 0.395109549 | 0 | 0 | 0 | 0 |
| 0.001806507 | 0 | 0.001002261 | 0.001041762 | 0 |
| 0 | 0 | 0.000438969 | 0 | 0 |
| 0 | 0 | 0 | 0 | 0 |
| 0 | 1.244288114 | 0.838081209 | 0 | 0 |
| 0 | 0 | 0 | 0 | 0 |
| 0 | 0 | 0 | 0 | 0 |
| 0 | 0 | 0 | 0 | 0 |
| 0.176776637 | 0.704255096 | 0 | 4.316105068 | 6.020212972 |
| 0 | 0 | 0 | 0 | 0 |
| 0 | 0 | 0 | 0 | 0 |
| 0 | 0 | 0 | 0 | 0 |
| 0 | 0 | 0.032137615 | 0 | 0 |
| 0.048123368 | 0.005024638 | 0.001047612 | 0.013937922 | 0 |
| 0 | 0 | 0 | 0 | 0 |
| 0.299271626 | 0.497285171 | 0.052807847 | 0 | 0 |
| 0.002614378 | 0.016615743 | 0 | 0.00775357 | 0 |
| 0 | 0 | 0 | 0 | 0 |
| 0 | 0 | 0 | 0 | 0 |
| 0 | 0 | 0 | 0 | 0 |
| 0 | 0 | 0 | 0 | 0 |
| 0 | 0 | 0 | 0 | 0 |
| 0 | 0 | 0 | 0 | 0 |
| 0 | 0 | 0 | 0 | 0 |
| 0 | 0 | 0 | 0 | 0 |
| 0 | 0 | 0 | 0 | 0 |
| 0 | 0 | 0.024324024 | 0 | 0 |
| 0.004947124 | 0 | 0 | 0 | 0.005619555 |
| 0 | 0 | 0 | 0 | 0 |
| 0 | 0 | 0 | 0 | 0 |
| 0 | 0 | 0 | 0 | 0 |
| 0 | 0.001399986 | 0 | 0 | 0 |
| 0 | 0 | 0 | 0 | 0 |
| 0 | 0.015333021 | 0 | 0 | 0 |
| 0 | 0 | 0.115951612 | 0 | 0 |
| 0.001483688 | 0 | 0 | 0 | 0.164838128 |
| 0 | 0.000900766 | 0 | 0 | 0.00118651 |
| 0 | 0 | 0 | 0 | 0 |
| 0 | 0 | 0 | 0 | 0 |
| 0 | 0 | 0 | 0 | 0 |
| 0 | 0 | 0.261760975 | 1.382018851 | 0 |
| 0 | 0 | 0.001428798 | 0 | 0 |
| 0 | 0 | 0 | 0 | 0 |
| 0 | 0.013240172 | 0.031881936 | 0 | 0 |
| 0 | 0 | 0 | 0 | 0 |
| 0 | 0 | 0 | 0 | 0 |
| 0 | 0 | 0 | 0 | 0 |
| 0 | 0 | 0 | 0 | 0 |

| | | | | |
|---|---|---|---|---|
| 0 | 0 | 0.050921155 | 0 | 0 |
| 0 | 0 | 0 | 0 | 0 |
| 0 | 0 | 0 | 0 | 0 |
| 0.00228316 | 0 | 0 | 0 | 0 |
| 0.036144172 | 0 | 0.0020659 | 0 | 0 |
| 0 | 0 | 0 | 0 | 0 |
| 0.004657181 | 0.213310696 | 0 | 0.452910649 | 0.73886734 |
| 0 | 0 | 0 | 0 | 0 |
| 0.002366797 | 0 | 0 | 0 | 0 |
| 0.020130899 | 0.037218343 | 0 | 0.002028744 | 0 |
| 0 | 0.001502827 | 0 | 0 | 0 |
| 0 | 0.024249396 | 0.005987966 | 0 | 0 |
| 0 | 0 | 0 | 0 | 0 |
| 0 | 0 | 0 | 0.002419693 | 0 |
| 0 | 0 | 0 | 0 | 0 |
| 0 | 0 | 0 | 0 | 0 |
| 0 | 0 | 0 | 0 | 0 |
| 0 | 0 | 0 | 0 | 0 |
| 0 | 0 | 0 | 0 | 0 |
| 0 | 0 | 0 | 0 | 0 |
| 0.093821231 | 0.034510951 | 0.326804613 | 0.020864331 | 0 |
| 0 | 0.001001607 | 0 | 0 | 0 |
| 0.008020537 | 0.003578804 | 0.010988939 | 0 | 0 |
| 0.054446307 | 0 | 0.000965084 | 0.00200624 | 0 |
| 0 | 0 | 0 | 0 | 0 |
| 0.016828182 | 0 | 0 | 0 | 0 |
| 0 | 0 | 0 | 0 | 0 |
| 0 | 0 | 0 | 0 | 0 |
| 0 | 0 | 0 | 0 | 0 |
| 0.673954682 | 0.006453762 | 0.214945052 | 1.463648437 | 0.004387635 |
| 0 | 0 | 0 | 0 | 0 |
| 0 | 0 | 0 | 0 | 0 |
| 0 | 0 | 0 | 0 | 0 |
| 0.587026798 | 0.014057492 | 0 | 0 | 0 |
| 0 | 0 | 0 | 0 | 0 |
| 0 | 0 | 0 | 0 | 0 |
| 0 | 0 | 0.001811356 | 0 | 0.009059615 |
| 0 | 0 | 0 | 0 | 0 |
| 0 | 0 | 0.001808232 | 0 | 0 |
| 0 | 0 | 0.081244639 | 0 | 0 |
| 0 | 0 | 0 | 0 | 0 |
| 0 | 0 | 0 | 0 | 0 |
| 0 | 0 | 0 | 0 | 0 |
| 0 | 0 | 0 | 0 | 0 |
| 0 | 0 | 0 | 0 | 0 |
| 0 | 0 | 0 | 0 | 0 |
| 0 | 0 | 0 | 0 | 0 |
| 0 | 0 | 0 | 0 | 0 |

| | | | | |
|---|---|---|---|---|
| 0.110059622 | 0.001089543 | 0.000908657 | 0.008635143 | 0.01025122 |
| 0 | 0 | 0.045796721 | 0 | 0 |
| 0 | 0 | 0 | 0 | 0 |
| 0 | 0 | 0 | 0 | 0 |
| 0 | 0 | 0 | 0 | 0 |
| 0 | 0 | 0 | 0 | 0 |
| 0 | 0 | 0 | 0 | 0 |
| 2.452122651 | 1.099376555 | 0.050247918 | 0.01041795 | 0 |
| 0.084908548 | 0.028096163 | 26.79682448 | 0.001355415 | 0 |
| 0.469613701 | 0 | 0.000425541 | 0.001887199 | 0 |
| 0 | 0 | 0 | 0 | 0 |
| 0 | 0 | 0 | 0 | 0 |
| 0 | 0 | 0 | 0 | 0 |
| 0 | 0 | 0 | 0 | 0 |
| 0 | 0 | 0 | 0 | 0 |
| 0 | 0 | 0 | 0 | 0 |
| 0 | 0 | 0 | 0 | 0.006883451 |
| 0 | 0 | 0 | 0 | 0 |
| 0 | 0 | 0 | 0 | 0 |
| 0 | 0.00070519 | 0 | 0 | 0 |
| 0 | 0 | 0 | 0 | 0 |
| 0 | 0 | 0 | 0 | 0 |
| 0 | 0 | 0 | 0 | 0 |
| 0 | 0 | 0 | 0 | 0 |
| 0.00796107 | 0.042908305 | 0.011962301 | 0 | 0 |
| 0 | 0 | 0 | 0 | 0 |
| 0 | 0 | 0 | 0 | 0 |
| 0 | 0 | 0 | 0 | 0 |
| 0 | 0 | 0 | 0 | 0 |
| 0 | 0 | 0 | 0 | 0 |
| 0 | 0.001918831 | 0 | 0 | 0 |
| 0 | 0 | 0 | 0 | 0 |
| 0 | 0 | 0 | 0 | 0 |
| 0 | 0 | 0 | 0 | 0 |
| 0 | 0 | 0.001414359 | 0 | 0 |
| 0 | 0 | 0 | 0 | 0 |
| 0 | 0.00081986 | 0 | 0 | 0 |
| 0 | 0 | 0 | 0 | 0 |
| 0 | 0 | 0 | 0 | 0 |
| 0 | 0.001864637 | 0 | 0 | 0 |
| 0 | 0 | 0 | 0 | 0 |
| 0 | 0.207599747 | 0.03743438 | 0 | 0 |
| 0 | 0 | 0 | 0 | 0 |
| 0 | 0 | 0 | 0 | 0 |
| 0 | 0 | 0 | 0 | 0 |
| 0 | 0.000968825 | 0 | 0 | 0 |
| 0 | 0 | 0 | 0 | 0 |
| 0 | 0 | 0 | 0 | 0 |

| | | | | |
|---|---|---|---|---|
| 0 | 0 | 0 | 0 | 0 |
| 0.001351478 | 0 | 0.079432789 | 0 | 0 |
| 0 | 0 | 0 | 0 | 0 |
| 0 | 0 | 0 | 0 | 0 |
| 2.796779166 | 0.018069795 | 0.201804884 | 0.078999917 | 0 |
| 0 | 0 | 0 | 0 | 0 |
| 0.010242112 | 0 | 0 | 0 | 0 |
| 0.021667256 | 0.018809695 | 0.000990752 | 0.056021088 | 0.186997822 |
| 0 | 0 | 0.00329743 | 0 | 0 |
| 0 | 0 | 0 | 0 | 0 |
| 0 | 0 | 0 | 0 | 0 |
| 0 | 0 | 0 | 0 | 0 |
| 0 | 0 | 0 | 0 | 0 |
| 0 | 0 | 0 | 0 | 0 |
| 0 | 0 | 0 | 0.039823506 | 0 |
| 0.200928238 | 0 | 0.019620039 | 0 | 0 |
| 0 | 0 | 0 | 0 | 0 |
| 0 | 0 | 0 | 0 | 0 |
| 0 | 0 | 0 | 0 | 0 |
| 0 | 0 | 0 | 0 | 0 |
| 0 | 0 | 0 | 0 | 0 |
| 0 | 0 | 0 | 0 | 0 |
| 0 | 0 | 0 | 0 | 0 |
| 0.01359979 | 0 | 0 | 0.001901241 | 0.004739826 |
| 0 | 0 | 0.026064327 | 0 | 0 |
| 0 | 0 | 0 | 0 | 0 |
| 0.003015406 | 0.000470157 | 0.03568122 | 0.036516909 | 0.013624629 |
| 0.001389117 | 0 | 0 | 0 | 0 |
| 0 | 0 | 0 | 0 | 0 |
| 0 | 0 | 0 | 0 | 0 |
| 0 | 0 | 0 | 0 | 0 |
| 0 | 0 | 0 | 0 | 0 |
| 0 | 0 | 0 | 0 | 0 |
| 0 | 0 | 0 | 0 | 0 |
| 0 | 0 | 0 | 0 | 0 |
| 0 | 0.013660252 | 0 | 0 | 0 |
| 0 | 0 | 0 | 0 | 0 |
| 0 | 0 | 0.050131319 | 0 | 0 |
| 0 | 0.148402266 | 0 | 0 | 0 |
| 0 | 0 | 0 | 0 | 0 |
| 0 | 0.000752017 | 0 | 0 | 0 |
| 0 | 0 | 0.102208398 | 0 | 0 |
| 0 | 0 | 0 | 0 | 0 |
| 0 | 0 | 0 | 0 | 0 |
| 0 | 0 | 0 | 0 | 0 |
| 0 | 0 | 0 | 0 | 0 |
| 0 | 0 | 0 | 0 | 0 |
| 0 | 0 | 0 | 0 | 0 |

| | | | | |
|---|---|---|---|---|
| 0 | 0.066321296 | 0.082474287 | 0 | 0 |
| 0 | 0 | 0 | 0 | 0 |
| 0 | 0 | 0 | 0 | 0 |
| 0 | 0 | 0 | 0 | 0 |
| 0 | 0 | 0 | 0 | 0 |
| 0 | 0 | 0 | 0 | 0 |
| 0 | 0 | 0 | 0 | 0 |
| 0 | 0 | 0 | 0 | 0 |
| 0 | 0 | 0 | 0 | 0 |
| 0 | 0 | 0 | 0 | 0 |
| 0 | 0 | 0 | 0 | 0 |
| 0 | 0 | 0 | 0 | 0 |
| 0 | 0 | 0 | 0 | 0 |
| 0 | 0 | 0 | 0 | 0 |
| 0 | 0 | 0 | 0 | 0 |
| 0 | 0 | 0 | 0 | 0 |
| 0 | 0 | 0 | 0 | 0 |
| 0.246593834 | 0 | 0 | 0 | 0 |
| 0 | 0 | 0.000312624 | 0 | 0 |
| 0 | 0 | 0 | 0 | 0 |
| 0.285427703 | 0 | 0 | 0 | 0.175764275 |
| 0 | 0 | 0 | 0 | 0 |
| 0.151083089 | 0 | 0.000718883 | 0 | 0.006812608 |
| 0 | 0 | 0.315162628 | 0 | 0 |
| 0 | 0 | 0 | 0 | 0 |
| 0 | 0 | 0 | 0 | 0 |
| 0 | 0 | 0 | 0 | 0 |
| 0 | 0 | 0 | 0 | 0 |
| 0 | 0.104282982 | 0 | 0 | 0 |
| 0 | 0 | 0 | 0 | 0 |
| 0 | 0 | 0 | 0 | 0 |
| 0 | 1.086664703 | 0 | 0 | 0 |
| 0.005339196 | 0 | 0 | 0.010916329 | 0.056888149 |
| 0 | 0 | 0 | 0 | 0 |
| 0.016064341 | 0.371511437 | 0.009635236 | 0 | 0 |
| 0.003115986 | 0 | 0 | 0.004492255 | 0 |
| 0 | 0 | 0 | 0 | 0 |
| 0 | 0 | 0.011171816 | 0 | 0 |
| 0.388570351 | 0.001311216 | 0.000624873 | 0.001818601 | 0.138913297 |
| 0 | 0 | 0 | 0 | 0 |
| 0 | 0 | 0 | 0 | 0 |
| 0 | 0 | 0 | 0 | 0 |
| 0 | 0.054100209 | 0.058453097 | 0 | 0 |
| 0 | 0 | 0 | 0 | 0 |
| 0 | 0 | 0 | 0 | 0 |
| 0.001882627 | 0.044878368 | 0.089942425 | 0 | 0 |
| 0 | 0 | 0 | 0 | 0 |
| 0 | 0 | 0 | 0 | 0 |

| | | | | |
|---|---|---|---|---|
| 0 | 0 | 0 | 0 | 0 |
| 0 | 0 | 0 | 0 | 0 |
| 0 | 0 | 0 | 0 | 0 |
| 0 | 0 | 0 | 0 | 0 |
| 0 | 0 | 0 | 0 | 0 |
| 0 | 0 | 0 | 0 | 0 |
| 0 | 0 | 0 | 0 | 0 |
| 0 | 0.00521073 | 0 | 2.811793499 | 0 |
| 0 | 0.001629724 | 0.005708458 | 0 | 0 |
| 0.017451764 | 0.008599855 | 0.022113994 | 0 | 0 |
| 0 | 0 | 0 | 0 | 0 |
| 0 | 0 | 0 | 0 | 0 |
| 0 | 0 | 0 | 0 | 0 |
| 0 | 0 | 0 | 0 | 0 |
| 0 | 0 | 0 | 0 | 0 |
| 0.014626813 | 0 | 0 | 0.007334672 | 0.619035593 |
| 0 | 0 | 0 | 0 | 0 |
| 0 | 0 | 0 | 0 | 0 |
| 0 | 0 | 0 | 0 | 0 |
| 3.254249428 | 7.244646263 | 0.017354676 | 0 | 0 |
| 0 | 0 | 0 | 0.002042812 | 0 |
| 0 | 0 | 0 | 0 | 0 |
| 0 | 0 | 0 | 0 | 0 |
| 0 | 0 | 0 | 0 | 0 |
| 0 | 0 | 0 | 0 | 0 |
| 0 | 0 | 0 | 0 | 0 |
| 0 | 0 | 0 | 0 | 0 |
| 0 | 0 | 0 | 0 | 0 |
| 0 | 0 | 0 | 0 | 0 |
| 0 | 0 | 0 | 0 | 0 |
| 0.208475332 | 0.254739722 | 0.790884117 | 0 | 0 |
| 0 | 0 | 0 | 0 | 0 |
| 0 | 0 | 0 | 0 | 0 |
| 0 | 0 | 0 | 0 | 0 |
| 0 | 0 | 0 | 0 | 0 |
| 0 | 0 | 0 | 0 | 0 |
| 0 | 0 | 0 | 0 | 0 |
| 0 | 0 | 0 | 0 | 0 |
| 0 | 0 | 0.003366662 | 0 | 0 |
| 0 | 0 | 0 | 0 | 0 |
| 0 | 0 | 0 | 0 | 0 |
| 0 | 0.05468234 | 0 | 0 | 0 |
| 0 | 0 | 0.000775414 | 0 | 0 |
| 0 | 0 | 0 | 0 | 0 |
| 0 | 0 | 0 | 0 | 0 |
| 0 | 0 | 0 | 0 | 0 |
| 0.001302031 | 0 | 0 | 0.000750845 | 12.3636112 |
| 0 | 0 | 0 | 0 | 0 |

| | | | | |
|---|---|---|---|---|
| 0 | 0 | 0 | 0 | 0 |
| 0 | 0 | 0 | 0 | 0 |
| 0 | 0 | 0 | 0 | 0 |
| 0 | 0 | 0.001618528 | 0 | 0 |
| 4.870382659 | 0 | 0.058576462 | 0 | 0 |
| 0 | 0 | 0 | 0 | 0 |
| 0 | 0 | 0 | 0 | 0 |
| 0 | 0 | 0 | 0 | 0 |
| 0 | 0 | 0 | 0 | 0 |
| 0 | 0 | 0 | 0 | 0 |
| 0 | 0 | 0 | 0 | 0 |
| 0.033536743 | 0 | 0.003590158 | 0 | 0 |
| 0.004077912 | 0 | 0 | 0 | 0 |
| 0 | 0 | 0 | 0 | 0 |
| 0 | 0 | 0.016296754 | 0 | 0.01296383 |
| 0.011727056 | 0 | 0.006312025 | 0 | 0 |
| 0 | 0.003886757 | 0.018368368 | 0 | 0 |
| 0 | 0 | 0 | 0 | 0 |
| 0.003919912 | 0 | 0.00390044 | 0 | 0 |
| 0.001335185 | 0.075869952 | 0.001234615 | 0.027718705 | 0.123825166 |
| 0 | 0 | 0 | 0 | 0 |
| 0 | 0 | 0 | 0 | 0 |
| 0 | 0 | 0 | 0 | 0 |
| 0 | 0 | 0 | 0 | 0 |
| 0 | 0 | 0 | 0 | 0 |
| 0 | 0 | 0 | 0 | 0 |
| 0.110204365 | 0 | 0.000989353 | 0.001439684 | 0.012110355 |
| 0 | 0.035621469 | 0 | 0 | 0 |
| 0 | 0 | 0 | 0 | 0 |
| 0 | 0.000424661 | 0 | 0 | 0 |
| 0.064684757 | 0 | 0 | 0 | 0.004206883 |
| 0 | 0.003812742 | 0 | 0 | 0 |
| 0 | 0 | 0 | 0 | 0 |
| 0 | 0 | 0 | 0 | 0 |
| 0 | 0 | 0 | 0 | 0 |
| 0 | 0 | 0 | 0 | 0 |
| 0 | 0 | 0 | 0 | 0 |
| 0.056096953 | 0 | 0.003250179 | 0 | 0.012600334 |
| 0 | 0.010937552 | 0.008055526 | 0 | 0 |
| 0 | 0 | 0 | 0 | 0 |
| 0 | 0 | 0 | 0 | 0 |
| 0 | 0 | 0 | 0 | 0 |
| 0 | 0 | 0 | 0 | 0 |
| 0.330843322 | 0.028369835 | 0 | 0.082338837 | 0 |
| 0 | 0 | 0 | 0 | 0 |
| 0 | 0 | 0.006006145 | 0 | 0 |
| 0 | 0 | 0 | 0 | 0 |
| 0 | 0 | 0 | 0 | 0 |

| | | | | |
|---|---|---|---|---|
| 0 | 0 | 0 | 0 | 0 |
| 0.21023552 | 0 | 0 | 0 | 0 |
| 0 | 0 | 0 | 0 | 0 |
| 0.02538091 | 0 | 0 | 0 | 0.016922411 |
| 0.003889281 | 0 | 0.014905833 | 0 | 0 |
| 0 | 0 | 0 | 0 | 0 |
| 0 | 0 | 0 | 0 | 0 |
| 0 | 0.001148285 | 0.000837941 | 0 | 0 |
| 0 | 0 | 0 | 0 | 0 |
| 0 | 0 | 0 | 0 | 0 |
| 0 | 0 | 0 | 0 | 0 |
| 0 | 0 | 0 | 0 | 0 |
| 0 | 0.002081541 | 0.065098617 | 0 | 0 |
| 0 | 0 | 0 | 0 | 0 |
| 0 | 0 | 0 | 0 | 0 |
| 0 | 0 | 0 | 0 | 0 |
| 0 | 0 | 0 | 0 | 0 |
| 0 | 0 | 0 | 0 | 0 |
| 0 | 0 | 0 | 0 | 0 |
| 0 | 0 | 0 | 0 | 0 |
| 0 | 0 | 0 | 0 | 0 |
| 0 | 0 | 0 | 0 | 0 |
| 0.09558662 | 0 | 0.020858504 | 0.000950381 | 0 |
| 0 | 0.003436516 | 0.002865985 | 0 | 0 |
| 0 | 0 | 0 | 0 | 0 |
| 0 | 0 | 0 | 0 | 0 |
| 0 | 0 | 0 | 0.005221042 | 0 |
| 0.003038133 | 0 | 0 | 0 | 0 |
| 0 | 0 | 0 | 0 | 0 |
| 0 | 0 | 0 | 0 | 0 |
| 0.015159895 | 0.018718625 | 0.014655183 | 0 | 0 |
| 0.004618246 | 0 | 0 | 0.000614588 | 0 |
| 0 | 0 | 0 | 0 | 0 |
| 0 | 0 | 0 | 0 | 0 |
| 0 | 0.027383162 | 0 | 0 | 0 |
| 0 | 0 | 0 | 0 | 0.017621917 |
| 0 | 0.003246885 | 0 | 0 | 0 |
| 0 | 0 | 0 | 0 | 0 |
| 0.000777353 | 0 | 0 | 0 | 0.003618779 |
| 0 | 0 | 0 | 0 | 0 |
| 0 | 0 | 0 | 0 | 0 |
| 0 | 0 | 0 | 0 | 0 |
| 0 | 0 | 5.489001984 | 0 | 0 |
| 0 | 0 | 0 | 0 | 0 |
| 0 | 0 | 0 | 0 | 0 |
| 0.095580912 | 0.190812792 | 0.221269814 | 0 | 0 |
| 0 | 0 | 0 | 0 | 0 |
| 0 | 0 | 0 | 0 | 0 |

| | | | | |
|---|---|---|---|---|
| 0 | 0 | 0 | 0 | 0 |
| 0 | 0 | 0 | 0 | 0 |
| 0 | 0.139424863 | 0.014838764 | 0 | 0 |
| 0 | 0 | 0 | 0 | 0 |
| 0 | 0 | 0 | 0 | 0 |
| 0 | 0 | 0 | 0 | 0 |
| 0 | 0.000492805 | 0 | 0 | 0 |
| 0 | 0 | 0 | 0 | 0 |
| 0 | 0.003738182 | 0 | 0 | 0.005120979 |
| 0 | 0 | 0 | 0 | 0 |
| 0.005833433 | 0 | 0 | 0 | 0 |
| 0.005884685 | 0 | 0 | 0 | 0.004383157 |
| 0 | 0 | 0 | 0 | 0 |
| 0 | 0 | 0 | 0 | 0 |
| 0 | 0 | 0 | 0 | 0 |
| 0 | 0 | 0 | 0 | 0 |
| 0 | 0 | 0 | 0 | 0 |
| 0 | 0 | 0 | 0 | 0 |
| 0 | 0 | 0 | 0 | 0 |
| 0.003345356 | 0 | 0.001015013 | 0 | 0 |
| 0 | 0 | 0 | 0 | 0 |
| 0 | 0 | 0 | 0 | 0 |
| 0 | 0 | 0 | 0 | 0 |
| 0 | 0 | 0 | 0 | 0 |
| 0 | 0.003308907 | 0 | 0 | 0 |
| 0 | 0 | 0 | 0 | 0 |
| 0 | 0 | 0 | 0 | 0 |
| 0 | 0 | 0.00657923 | 0 | 0 |
| 0 | 0 | 0 | 0 | 0 |
| 0 | 0 | 0 | 0 | 0 |
| 0.151783476 | 0 | 0.014076588 | 0.003936228 | 0 |
| 0 | 0 | 0 | 0 | 0 |
| 0.004796958 | 0.035380492 | 0.382573549 | 0.065909422 | 0 |
| 0 | 0.002126766 | 0.000739033 | 0 | 0.008170823 |
| 0 | 0 | 0 | 0 | 0 |
| 0 | 0 | 0 | 0 | 0 |
| 0 | 0 | 0 | 0 | 0 |
| 0 | 0 | 0 | 0 | 0 |
| 0 | 0 | 0 | 0 | 0 |
| 0 | 0 | 0 | 0 | 0 |
| 0 | 0 | 0 | 0 | 0 |
| 0.006691745 | 0.047227875 | 0 | 0.018206291 | 0.474184022 |
| 0 | 0 | 0.009712893 | 0.014706567 | 0 |
| 0 | 0 | 0 | 0 | 0 |
| 0 | 0 | 0 | 0 | 0 |
| 0 | 0 | 0.0282811 | 0 | 0 |
| 0 | 0 | 0 | 0 | 0 |
| 0 | 0 | 0 | 0 | 0 |

| | | | | |
|---|---|---|---|---|
| 0 | 0 | 0 | 0 | 0 |
| 0 | 0 | 0 | 0 | 0 |
| 0.001279039 | 0.01723036 | 0.00922504 | 0.001770206 | 0.012048613 |
| 0 | 0 | 0 | 0 | 0 |
| 0.834770538 | 0 | 0.009915433 | 0 | 0 |
| 0.029424756 | 0 | 0 | 0.018114948 | 0.061412266 |
| 0 | 0 | 0 | 0 | 0 |
| 0 | 0 | 0 | 0 | 0 |
| 0 | 0 | 0 | 0 | 0 |
| 0 | 0 | 0 | 0 | 0 |
| 0 | 0 | 0 | 0 | 0 |
| 0 | 0 | 0 | 0 | 0 |
| 0 | 0 | 0 | 0 | 0 |
| 0 | 0 | 0 | 0 | 0 |
| 0.003344504 | 0 | 0 | 0 | 0 |
| 0 | 0 | 0 | 0 | 0 |
| 0 | 0 | 0 | 0 | 0 |
| 0 | 0 | 0 | 0 | 0 |
| 0 | 0 | 0 | 0 | 0 |
| 0 | 0 | 0 | 0 | 0 |
| 0 | 0 | 0 | 0 | 0 |
| 0 | 0.000513327 | 0 | 0 | 0 |
| 0 | 0 | 0 | 0 | 0 |
| 0 | 0 | 0 | 0 | 0 |
| 0 | 0.008239645 | 0.011696507 | 0 | 0 |
| 0.000848326 | 0.001939953 | 0.044271117 | 0 | 0.006039918 |
| 0 | 0 | 0 | 0 | 0 |
| 0.00637752 | 0 | 0.028391897 | 0 | 0 |
| 0 | 0 | 0 | 0 | 0 |
| 0 | 0 | 0 | 0 | 0 |
| 0 | 0 | 0.003693117 | 0 | 0 |
| 0.000743932 | 0 | 0 | 0 | 0 |
| 0 | 0 | 0 | 0 | 0 |
| 0 | 0 | 0 | 0 | 0 |
| 0 | 0 | 0 | 0 | 0 |
| 0 | 0 | 0 | 0 | 0 |
| 0 | 0 | 0 | 0 | 0 |
| 0 | 0 | 0 | 0.278056055 | 0 |
| 0 | 0 | 0.003471967 | 0 | 0 |
| 0 | 0 | 0 | 0 | 0 |
| 0 | 0 | 0 | 0 | 0 |
| 0 | 0 | 0.01276474 | 0 | 0 |
| 0.096206819 | 0 | 0 | 0.004909715 | 0.00629486 |
| 0 | 0.000515423 | 0 | 0 | 0 |
| 0 | 0 | 0 | 0 | 0 |
| 0 | 0 | 0 | 0 | 0 |
| 0.016606323 | 0 | 0.001806527 | 0 | 0.001426652 |
| 0 | 0 | 0 | 0 | 0 |

| | | | | |
|---|---|---|---|---|
| 0 | 0 | 0 | 0 | 0 |
| 0.073339436 | 0.002904115 | 0.00455901 | 0.01492687 | 0.147839193 |
| 0 | 0 | 0 | 0 | 0 |
| 0 | 0 | 0 | 0 | 0 |
| 0 | 0 | 0 | 0 | 0 |
| 0 | 0 | 0 | 0 | 0 |
| 0 | 0 | 0 | 0 | 0 |
| 0 | 0 | 0 | 0 | 0 |
| 0 | 0 | 0 | 0 | 0 |
| 0 | 0 | 0 | 0 | 0 |
| 0 | 0 | 0 | 0 | 0 |
| 0 | 0 | 0 | 0 | 0 |
| 0 | 0.001444662 | 0.182831323 | 0 | 0 |
| 0 | 0 | 0 | 0 | 0 |
| 0 | 0.001277998 | 0.00763841 | 0.002954215 | 0 |
| 0 | 0 | 0 | 0 | 0 |
| 0 | 0 | 0 | 0 | 0 |
| 0 | 0 | 0 | 0 | 0 |
| 0 | 0 | 0 | 0 | 0 |
| 0.012282223 | 0 | 0 | 0 | 0 |
| 0 | 0 | 0 | 0 | 0 |
| 0.002263978 | 0.024892716 | 0 | 0.004061782 | 0.154302369 |
| 0 | 0.002089615 | 0 | 0 | 0 |
| 0 | 0 | 0 | 0 | 0 |
| 0 | 0 | 0 | 0 | 0 |
| 0 | 0 | 0 | 0 | 0 |
| 0 | 0 | 0 | 0 | 0 |
| 0 | 0 | 0 | 0 | 0 |
| 0 | 0 | 0 | 0 | 0 |
| 0 | 0 | 0.004504003 | 0 | 0 |
| 0 | 0 | 0 | 0 | 0 |
| 0 | 0 | 0 | 0 | 0 |
| 0 | 0 | 0 | 0 | 0 |
| 0 | 0 | 0 | 0 | 0 |
| 0 | 41.27833873 | 0 | 0 | 0 |
| 0 | 0 | 0 | 0 | 0 |
| 0 | 0 | 0 | 0 | 0 |
| 0 | 0 | 0 | 0 | 0 |
| 0.002512062 | 0.011071383 | 0.001219494 | 0.037374807 | 0.014308319 |
| 0 | 0 | 0 | 0 | 0 |
| 0.02838019 | 0.003933323 | 0.223529869 | 0 | 15.93177035 |
| 0 | 0 | 0 | 0 | 0 |
| 0 | 0 | 0 | 0 | 0 |
| 0 | 0 | 0 | 0 | 0 |
| 0 | 0 | 0 | 0 | 0 |
| 0.418287881 | 0.010293458 | 0.004828802 | 0.82447312 | 4.769582443 |
| 0 | 0 | 0 | 0 | 0 |
| 0 | 0 | 0 | 0 | 0 |

| | | | | |
|---|---|---|---|---|
| 0 | 0 | 0 | 0 | 0 |
| 0 | 0 | 0 | 0 | 0 |
| 0 | 0 | 0 | 0 | 0 |
| 0 | 0 | 0 | 0 | 0 |
| 0 | 0 | 0 | 0 | 0 |
| 0 | 0 | 0 | 0 | 0 |
| 0 | 0 | 0 | 0 | 0 |
| 0 | 0.017615807 | 0 | 0.000964436 | 0.003969096 |
| 0 | 0 | 0 | 0 | 0 |
| 0.006305274 | 0.040268071 | 0 | 0.004072404 | 0 |
| 0 | 0 | 0.017708908 | 0 | 0 |
| 0 | 0 | 0 | 0 | 0 |
| 0.01084666 | 0.000614979 | 0.00051288 | 0 | 0.003240257 |
| 0 | 0 | 0 | 0 | 0 |
| 0 | 0 | 0 | 0 | 0 |
| 0 | 0 | 0 | 0 | 0 |
| 0 | 0 | 0 | 0 | 0 |
| 0 | 0 | 0 | 0 | 0 |
| 0 | 0 | 0 | 0 | 0 |
| 5.252374069 | 0.001277207 | 0.070300907 | 0.142979929 | 0.209815173 |
| 0 | 0 | 0 | 0 | 0 |
| 0 | 0 | 0 | 0 | 0 |
| 0 | 0 | 0 | 0 | 0 |
| 0 | 0 | 0 | 0 | 0 |
| 0 | 0 | 0 | 0 | 0 |
| 0 | 0 | 0.002889461 | 0 | 0 |
| 0 | 0 | 0 | 0 | 0 |
| 0 | 0 | 0 | 0 | 0 |
| 0 | 0 | 0 | 0 | 0 |
| 0.002111031 | 0.003120812 | 0 | 0.002975799 | 0 |
| 0 | 0 | 0 | 0 | 0 |
| 0 | 0 | 0 | 0 | 0 |
| 0 | 0 | 0 | 0 | 0 |
| 0 | 0 | 0 | 0 | 0 |
| 0 | 0 | 0 | 0 | 0 |
| 0 | 0 | 0 | 0 | 0 |
| 0 | 0 | 0 | 0 | 0 |
| 0 | 0 | 0 | 0 | 0 |
| 0 | 0 | 0 | 0 | 0 |
| 0 | 0 | 0 | 0 | 0 |
| 0 | 0 | 0 | 0 | 0 |
| 0 | 0 | 0 | 0 | 0 |
| 0 | 0.200199719 | 0.000632434 | 0 | 0 |
| 0 | 0 | 0 | 0 | 0 |
| 0.0259665 | 0 | 0 | 0 | 0 |
| 0 | 0 | 0 | 0 | 0 |
| 0 | 0 | 0 | 0 | 0 |
| 0 | 0 | 0 | 0 | 0.00587515 |

| | | | | |
|---|---|---|---|---|
| 0 | 0 | 0 | 0 | 0 |
| 0 | 0 | 0 | 0 | 0 |
| 0 | 0 | 0 | 0 | 0 |
| 0 | 0 | 0 | 0 | 0 |
| 0 | 0 | 0 | 0 | 0 |
| 0.178465093 | 0.001151419 | 0.027740853 | 0 | 0.001179636 |
| 0 | 0 | 0 | 0 | 0 |
| 0 | 0 | 0 | 0 | 0 |
| 0 | 0 | 0 | 0 | 0 |
| 0 | 0 | 0 | 0 | 0 |
| 0 | 0 | 0 | 0 | 0 |
| 0 | 0 | 0 | 0 | 0 |
| 0 | 0 | 0 | 0 | 0 |
| 0 | 0 | 0 | 0 | 0 |
| 0 | 0 | 0 | 0 | 0 |
| 0 | 0 | 0 | 0 | 0 |
| 0 | 0 | 0.001164789 | 0 | 0 |
| 0 | 0 | 0 | 0 | 0 |
| 0 | 0 | 0 | 0 | 0 |
| 0 | 0 | 0 | 0 | 0 |
| 0 | 0 | 0 | 0 | 0 |
| 0 | 0 | 0 | 0 | 0 |
| 0 | 0 | 0 | 0 | 0 |
| 0 | 0 | 0 | 0 | 0 |
| 0 | 0 | 0 | 0 | 0 |
| 0 | 0.001400788 | 0 | 0 | 0 |
| 0 | 0 | 0 | 0 | 0 |
| 0 | 0.053168487 | 0 | 0 | 0 |
| 0 | 0 | 0 | 0 | 0 |
| 0 | 0 | 0 | 0.001283122 | 0 |
| 0 | 0 | 0 | 0 | 0 |
| 0 | 0 | 0 | 0 | 0 |
| 0 | 0 | 0 | 0 | 0 |
| 0 | 0 | 0 | 0 | 0 |
| 0 | 0 | 0 | 0 | 0 |
| 0 | 0 | 0 | 0 | 0 |
| 0 | 0 | 0 | 0 | 0 |
| 0 | 0 | 0 | 0 | 0 |
| 0.013464662 | 0 | 0.685277265 | 0 | 0 |
| 0 | 0 | 0 | 0 | 0 |
| 0 | 0 | 0 | 0 | 0 |
| 0 | 0 | 0 | 0 | 0 |
| 0 | 0 | 0 | 0 | 0 |
| 0 | 0 | 0 | 0 | 0 |
| 0 | 0 | 0 | 0 | 0 |
| 0 | 0 | 0 | 0 | 0 |
| 0 | 0 | 0 | 0 | 0 |
| 0 | 0 | 0 | 0 | 0 |

| | | | | |
|---|---|---|---|---|
| 0 | 0 | 0 | 0 | 0 |
| 0.107826371 | 0.001862043 | 0 | 0 | 0 |
| 0 | 0 | 0 | 0 | 0 |
| 0 | 0 | 0 | 0 | 0 |
| 0 | 0 | 0 | 0 | 0 |
| 0.007794643 | 0 | 0 | 0 | 0 |
| 0 | 0 | 0 | 0 | 0 |
| 0 | 0 | 0 | 0 | 0 |
| 0 | 0 | 0 | 0 | 0 |
| 0 | 0 | 0 | 0 | 0 |
| 0.005864125 | 0.001312875 | 0.068353771 | 0 | 0 |
| 0 | 0 | 0 | 0 | 0 |
| 0 | 0.224067517 | 0 | 0 | 0 |
| 0 | 0 | 0 | 0 | 0 |
| 0 | 0 | 0 | 0 | 0 |
| 0.051518412 | 0 | 0 | 0 | 0 |
| 0 | 0 | 0 | 0 | 0 |
| 0 | 0 | 0 | 0 | 0 |
| 0 | 0 | 0 | 0 | 0 |
| 0.009946913 | 0 | 0 | 0 | 0 |
| 0.012361294 | 0.001522845 | 0 | 0.006336366 | 0 |
| 0 | 0 | 0 | 0 | 0 |
| 0 | 0 | 0 | 0 | 0 |
| 0.561687929 | 0.682011595 | 0.201868979 | 0 | 0 |
| 0 | 0 | 0 | 0 | 0 |
| 0 | 0 | 0 | 0 | 0 |
| 0 | 0 | 0 | 0 | 0 |
| 0 | 0 | 0 | 0 | 0 |
| 0 | 0 | 0 | 0 | 0 |
| 0 | 0 | 0 | 0 | 0 |
| 0 | 0 | 0 | 0 | 0 |
| 0 | 0 | 0 | 0 | 0 |
| 0 | 0 | 0.008015652 | 0 | 0 |
| 0 | 0 | 0 | 0 | 0 |

| PHL018-0137_V1 | PHL018-0061_V3 | PHL018-0158_V1 | PHL018-0276_V1 | PHL018-0226_V3 |
|---|---|---|---|---|
| EYCF | REF | EYCF | REF | REF |
| 0.3133573 | 0.174421491 | 0.15367796 | 1.131112334 | 0.486408508 |
| 0.939212983 | 0.070767099 | 4.876948433 | 1.067603618 | 0.450378046 |
| 1.964150984 | 0 | 11.54583643 | 0 | 0.017005796 |
| 0 | 0 | 0 | 0 | 0 |
| 0 | 0 | 0 | 0 | 0 |
| 0 | 0 | 0 | 0 | 0 |
| 0 | 0 | 0 | 0 | 0 |
| 0 | 0 | 0 | 0 | 0 |
| 0 | 0 | 0 | 0 | 0 |
| 0 | 0 | 0 | 0 | 0 |
| 0 | 0 | 0 | 0 | 0 |
| 0 | 0 | 0 | 0 | 0 |
| 0 | 0 | 0 | 0 | 0 |
| 0.529787708 | 0.007314558 | 11.41655149 | 0.372120382 | 2.392543804 |
| 0 | 0 | 0 | 0 | 0 |
| 0 | 0 | 0 | 0 | 0 |
| 0 | 0 | 0.001128208 | 0 | 0 |
| 0 | 0 | 0 | 0 | 0 |
| 0 | 0 | 0 | 0 | 0 |
| 0 | 0 | 0 | 0 | 0 |
| 0 | 0 | 0 | 0 | 0 |
| 0 | 0 | 0 | 0 | 0 |
| 0 | 0 | 0 | 0 | 0 |
| 0 | 0 | 0 | 0 | 0 |
| 0 | 0.124318832 | 0 | 0 | 0 |
| 0 | 0 | 0 | 0 | 0 |
| 0 | 0 | 0 | 0 | 0 |
| 0 | 0 | 0 | 0 | 0 |
| 0 | 0 | 0 | 0 | 0 |
| 0 | 0 | 0 | 0 | 0 |
| 0 | 0 | 0 | 0 | 0 |
| 0 | 0 | 0 | 0 | 0 |
| 0 | 0 | 0 | 0 | 0 |
| 0 | 0 | 0 | 0 | 0 |
| 0 | 0 | 0 | 0 | 0 |
| 3.617731084 | 1.260985771 | 0 | 2.626549448 | 1.577093098 |
| 0 | 0 | 0 | 0 | 0 |
| 0 | 0 | 0 | 0 | 0 |
| 0 | 0 | 0 | 0 | 0 |
| 0 | 0 | 0 | 0 | 0 |
| 0 | 0 | 0.000915935 | 0 | 0 |
| 0.00211524 | 0.039812036 | 0 | 0.005945493 | 0 |
| 2.184392647 | 5.997170284 | 0.012185567 | 0.001166737 | 0.050643591 |
| 0.360913381 | 0.003338276 | 0 | 0.045047706 | 0.108019865 |
| 0.001111397 | 0.001961079 | 0 | 0 | 0 |
| 0 | 0 | 0 | 0 | 0 |

| | | | | |
|---|---|---|---|---|
| 0 | 0.006469531 | 0 | 0 | 0 |
| 0.016845941 | 0 | 0.117023416 | 0.005618655 | 0 |
| 0 | 0 | 0.008661845 | 0 | 0 |
| 0.626744066 | 0.271751159 | 0.173014918 | 0.635566099 | 2.44138721 |
| 0.000551173 | 0.002033524 | 0.001279062 | 0.065388251 | 0 |
| 0 | 0 | 0 | 0 | 0 |
| 0 | 0 | 0 | 0.01297109 | 0 |
| 0 | 0 | 0 | 0 | 0 |
| 0 | 0 | 0 | 0 | 0 |
| 0 | 0 | 0 | 0 | 0 |
| 0.012198595 | 0 | 0.033969919 | 0 | 0 |
| 0.15965817 | 0.258032586 | 0 | 0.104127074 | 0.162180185 |
| 0 | 0 | 0 | 0 | 0 |
| 0.006465616 | 0.014276119 | 0.125133224 | 0.215482921 | 0.118011395 |
| 0.003130897 | 0 | 0 | 0 | 0 |
| 0 | 0 | 0 | 0 | 0 |
| 0.009906461 | 0.057542226 | 0 | 0 | 0.003589439 |
| 0 | 0 | 0 | 0 | 0 |
| 0 | 0 | 0 | 0 | 0 |
| 0 | 0 | 0 | 0 | 0 |
| 0.214409428 | 0.000902549 | 0.073367489 | 0.044873723 | 0.221142957 |
| 0.109993882 | 0.002784333 | 0.045144921 | 0.053652537 | 0.046202225 |
| 0 | 0.117450214 | 0 | 0 | 0 |
| 0 | 0 | 0 | 0 | 0 |
| 0 | 0 | 0 | 0.000908818 | 0 |
| 0 | 0 | 0 | 0 | 0 |
| 0.00157694 | 0 | 0 | 0 | 0 |
| 0 | 0 | 0 | 0 | 0 |
| 0 | 0.000929209 | 0 | 0 | 0 |
| 0 | 0 | 0 | 0 | 0 |
| 0.028860639 | 0 | 0.032412345 | 0.008578329 | 0 |
| 0 | 0 | 0 | 0 | 0 |
| 0 | 0 | 0 | 0 | 0 |
| 0.058786875 | 0 | 0 | 0 | 0 |
| 0 | 0 | 0 | 0 | 0 |
| 0 | 0 | 0 | 0 | 0 |
| 0 | 0 | 0 | 0 | 0 |
| 0.000915123 | 0.299315975 | 0 | 0 | 0 |
| 0 | 0 | 0 | 0 | 0 |
| 0 | 0 | 0 | 0 | 0 |
| 0 | 0 | 0 | 0 | 0 |
| 0 | 0 | 0 | 0 | 0 |
| 0 | 0 | 0 | 0 | 0 |
| 0 | 0 | 0 | 0 | 0 |
| 0 | 0.367351379 | 0 | 0 | 0 |
| 0 | 0 | 0 | 0 | 0 |
| 0 | 0 | 0 | 0 | 0 |
| 0 | 0.001100769 | 0 | 0 | 0.000937333 |

| | | | | |
|---|---|---|---|---|
| 0 | 0 | 0 | 0 | 0 |
| 0 | 0 | 0 | 0 | 0 |
| 0 | 0 | 0 | 0 | 0 |
| 0 | 0 | 0 | 0 | 0 |
| 0.008160661 | 0 | 0 | 0.000936665 | 0 |
| 0 | 0 | 0 | 0 | 0 |
| 0 | 0 | 0 | 0 | 0 |
| 0 | 0 | 0 | 0 | 0 |
| 0 | 0.024923552 | 0 | 0.078584461 | 0.004125043 |
| 0 | 0 | 0 | 0 | 0 |
| 0 | 0 | 0 | 0 | 0 |
| 0 | 0.001086789 | 0.000655095 | 0 | 0 |
| 0 | 0 | 0 | 0 | 0 |
| 0 | 0 | 0 | 0 | 0 |
| 0 | 0 | 0 | 0 | 0 |
| 0 | 0 | 0 | 0 | 0 |
| 0 | 0 | 0 | 0 | 0 |
| 0 | 0 | 0 | 0 | 0 |
| 0 | 0.164906099 | 0 | 0.016600575 | 0 |
| 0 | 0 | 0 | 0 | 0 |
| 0 | 0 | 0 | 0 | 0 |
| 0 | 0 | 0 | 0 | 0 |
| 0 | 0 | 0 | 0 | 0 |
| 0 | 0 | 0 | 0 | 0 |
| 0 | 0 | 0 | 0 | 0 |
| 0 | 0 | 0 | 0 | 0 |
| 0 | 0 | 0 | 0 | 0 |
| 0 | 0 | 0 | 0 | 0 |
| 0 | 0 | 0 | 0 | 0 |
| 0 | 0 | 0 | 0 | 0 |
| 0.000506261 | 0.03497427 | 0.029936584 | 1.423629313 | 0.037342117 |
| 0 | 0.159373225 | 0 | 0 | 0.004004569 |
| 0 | 0 | 0 | 0 | 0 |
| 0 | 0 | 0 | 0 | 0 |
| 0 | 0 | 0 | 0 | 0 |
| 0 | 0 | 0 | 0 | 0 |
| 0 | 0 | 0 | 0 | 0 |
| 0 | 0 | 0 | 0 | 0 |
| 0 | 0 | 0 | 0 | 0 |
| 0 | 0 | 0 | 0 | 0 |
| 0 | 0 | 0 | 0 | 0 |
| 0 | 0 | 0 | 0 | 0 |
| 0 | 0 | 0 | 0.03034481 | 0.005634877 |
| 0 | 0 | 0 | 0 | 0 |
| 0 | 0 | 0 | 0 | 0 |
| 0 | 0 | 0 | 0 | 0 |
| 0 | 0 | 0 | 0.358369122 | 0 |
| 0 | 0 | 0 | 0 | 0 |

| | | | | |
|---|---|---|---|---|
| 0 | 0.000510038 | 0 | 0 | 0 |
| 0 | 0 | 0 | 0 | 0 |
| 0 | 0 | 0 | 0 | 0 |
| 0 | 0 | 0 | 0 | 0 |
| 0.006317723 | 0.065353893 | 0 | 0.273401156 | 0.003940158 |
| 0 | 0 | 0 | 0 | 0 |
| 0 | 0 | 0 | 0 | 0 |
| 0 | 0 | 0 | 0 | 0 |
| 0 | 0 | 0 | 0 | 0 |
| 0 | 0 | 0 | 0 | 0 |
| 0 | 0 | 0 | 0 | 0 |
| 0 | 0 | 0 | 0 | 0 |
| 0 | 0 | 0 | 0 | 0 |
| 0 | 0.001163019 | 0 | 0.001581126 | 0 |
| 0 | 0 | 0 | 0 | 0 |
| 0 | 0.002411144 | 0 | 0 | 0 |
| 0 | 0 | 0 | 0 | 0 |
| 0 | 0 | 0 | 0 | 0 |
| 0 | 0 | 0 | 0 | 0 |
| 0 | 0 | 0 | 0 | 0 |
| 0 | 0 | 0 | 0 | 0 |
| 0 | 0 | 0 | 0 | 0 |
| 0 | 0.001446651 | 0 | 0 | 0 |
| 0 | 0 | 0 | 0 | 0 |
| 0 | 0.020246055 | 0 | 0 | 0 |
| 0 | 0 | 0 | 0 | 0 |
| 0 | 0.034550361 | 0 | 0 | 0 |
| 0 | 0 | 0 | 0 | 0 |
| 0 | 0 | 0 | 0 | 0 |
| 0 | 0 | 0 | 0 | 0 |
| 0 | 0 | 0 | 0 | 0 |
| 0 | 0.0014417 | 0 | 0 | 0 |
| 0 | 0 | 0 | 0 | 0 |
| 0 | 0 | 0 | 0 | 0 |
| 0 | 0 | 0 | 0 | 0 |
| 0 | 0 | 0 | 0 | 0 |
| 0 | 0 | 0 | 0 | 0 |
| 0 | 0 | 0 | 0 | 0 |
| 0 | 0 | 0 | 0 | 0 |
| 0 | 0 | 0 | 0 | 0 |
| 0 | 0 | 0 | 0 | 0 |
| 0 | 0 | 0 | 0 | 0 |
| 0 | 0 | 0 | 0 | 0 |
| 0 | 0 | 0 | 0 | 0 |
| 0 | 0 | 0 | 0 | 0 |
| 0 | 0 | 0 | 0 | 0 |
| 0 | 0 | 0 | 0 | 0 |
| 0 | 0 | 0 | 0.036918671 | 0.001477752 |

| | | | | |
|---|---|---|---|---|
| 0 | 0.001005373 | 0 | 0 | 0 |
| 0 | 0 | 0 | 0 | 0 |
| 0 | 0 | 0 | 0 | 0 |
| 0 | 0 | 0 | 0 | 0 |
| 0 | 0 | 0 | 0 | 0 |
| 0 | 0 | 0 | 0 | 0 |
| 0 | 0 | 0 | 0 | 0 |
| 0 | 0 | 0 | 0 | 0 |
| 0 | 0 | 0 | 0 | 0 |
| 0 | 0 | 0 | 0 | 0 |
| 0 | 0 | 0 | 0 | 0 |
| 0 | 0 | 0 | 0 | 0 |
| 0 | 0.003260792 | 0.000806376 | 0 | 0 |
| 0 | 0 | 0 | 0 | 0 |
| 0 | 0.006804627 | 0 | 0 | 0 |
| 0 | 0 | 0 | 0 | 0 |
| 0 | 0 | 0 | 0 | 0 |
| 0 | 0 | 0 | 0 | 0 |
| 0 | 0 | 0 | 0 | 0 |
| 0 | 0 | 0 | 0 | 0 |
| 0.01279117 | 0.01360077 | 0.010931032 | 0.051326399 | 0.047803252 |
| 0 | 0 | 0 | 0 | 0 |
| 0 | 0.001234534 | 0 | 0 | 0.0011038 |
| 0 | 0 | 0 | 0.005319067 | 0 |
| 0 | 0 | 0 | 0 | 0 |
| 0 | 0 | 0 | 0 | 0 |
| 0.019580954 | 0 | 0.001975648 | 0.003898863 | 0.000837278 |
| 0 | 0 | 0 | 0 | 0 |
| 0 | 0 | 0 | 0 | 0 |
| 0.001366263 | 0 | 0 | 0 | 0.000895791 |
| 0.059405306 | 0 | 0 | 0.103455051 | 0 |
| 0 | 0 | 0 | 0 | 0 |
| 0 | 0 | 0 | 0 | 0 |
| 0.000670043 | 0.128405237 | 0 | 0.026496776 | 0.016693931 |
| 0 | 0.010832869 | 0.038259334 | 0.110973257 | 0.006314213 |
| 0 | 0 | 0 | 0 | 0 |
| 0 | 0 | 0 | 0 | 0 |
| 0 | 0 | 0 | 0 | 0 |
| 0 | 0 | 0 | 0 | 0 |
| 0 | 0 | 0 | 0 | 0 |
| 0 | 0 | 0 | 0 | 0 |
| 0 | 0.00426656 | 0 | 0 | 0 |
| 0 | 0.003647461 | 0 | 0 | 0 |
| 0 | 0 | 0 | 0 | 0 |
| 0 | 0 | 0 | 0 | 0 |
| 0 | 0.040239406 | 0 | 0 | 0 |
| 0 | 0 | 0 | 0.002470467 | 0.002278871 |
| 0.008811655 | 0.001413484 | 0.007165705 | 0.001478178 | 0.000999929 |

| | | | | |
|---|---|---|---|---|
| 0 | 0 | 0 | 0 | 0 |
| 0 | 0 | 0 | 0 | 0 |
| 0.001238755 | 0 | 0 | 0 | 0 |
| 0 | 0 | 0 | 0 | 0 |
| 0 | 0 | 0 | 0 | 0 |
| 0 | 0 | 0 | 0 | 0 |
| 0 | 0 | 0 | 0.001052084 | 0 |
| 0 | 0.001250405 | 0 | 0 | 0 |
| 0 | 0.010177283 | 0 | 0 | 0 |
| 0 | 0 | 0 | 0 | 0 |
| 0 | 0 | 0 | 0 | 0 |
| 0 | 0 | 0 | 0 | 0 |
| 0 | 0 | 0 | 0 | 0 |
| 0.001990286 | 0 | 0 | 0 | 0 |
| 0 | 0 | 0.033893825 | 0 | 0 |
| 0 | 0 | 0 | 0 | 0 |
| 0 | 0 | 0 | 0 | 0 |
| 0 | 0.039198328 | 0 | 0 | 0 |
| 0.008272057 | 0.028639494 | 0 | 0 | 0 |
| 0.002172362 | 0.127096536 | 0 | 0 | 0 |
| 0 | 0 | 0.261140698 | 0 | 0.002750113 |
| 0 | 0 | 0 | 0 | 0 |
| 0 | 0 | 0 | 0 | 0 |
| 0 | 0 | 0 | 0 | 0 |
| 0.013636183 | 0.003240577 | 0.002083579 | 0.001762227 | 0.003973586 |
| 0 | 0 | 0 | 0 | 0 |
| 0 | 0 | 0 | 0 | 0 |
| 0 | 0 | 0 | 0 | 0 |
| 0 | 0 | 0 | 0 | 0 |
| 0.040994692 | 0 | 0 | 0 | 0 |
| 0.181126665 | 0.050013581 | 1.053585327 | 1.016641312 | 0.028677862 |
| 0 | 0 | 0 | 0 | 0 |
| 0 | 0 | 0 | 0 | 0.004571625 |
| 0.014268222 | 0 | 0 | 0 | 0 |
| 0 | 0 | 0 | 0 | 0 |
| 0 | 0 | 0 | 0 | 0 |
| 0 | 0 | 0 | 0 | 0 |
| 0 | 0 | 0 | 0 | 0 |
| 0 | 0 | 0 | 0 | 0 |
| 0 | 0.002515801 | 0 | 0 | 0 |
| 0 | 0 | 0 | 0 | 0 |
| 0.017148159 | 0 | 0.012380452 | 0.01146824 | 0.042724149 |
| 0 | 0 | 0 | 0 | 0 |
| 0 | 0 | 0 | 0 | 0 |
| 0 | 0 | 0 | 0 | 0 |
| 0 | 0 | 0 | 0 | 0 |
| 0 | 0 | 0 | 0 | 0 |
| 0 | 0 | 0 | 0 | 0 |

| | | | | |
|---|---|---|---|---|
| 0 | 0 | 0 | 0 | 0 |
| 0 | 0 | 0 | 0 | 0 |
| 0.344756624 | 0 | 0 | 0.030470121 | 0.124000324 |
| 0 | 0 | 0 | 0 | 0 |
| 0 | 0 | 0.132340119 | 0 | 0 |
| 0 | 0 | 0 | 0 | 0 |
| 0 | 0 | 0 | 0 | 0 |
| 0 | 0 | 0 | 0 | 0 |
| 0 | 0 | 0 | 0 | 0 |
| 0 | 0 | 0 | 0 | 0 |
| 0 | 0 | 0 | 0 | 0 |
| 0 | 0 | 0 | 0 | 0 |
| 0 | 0 | 0 | 0 | 0 |
| 0.00978326 | 0.000848292 | 0 | 0 | 0.002080345 |
| 0.003710043 | 0.000840184 | 0.001575611 | 0.01427789 | 0.002146316 |
| 0 | 0.016641274 | 0 | 0 | 0 |
| 0.005719237 | 0.001116867 | 0.001538801 | 0.002169117 | 0 |
| 0 | 0 | 0 | 0.004543075 | 0 |
| 0 | 0 | 0 | 0 | 0 |
| 0 | 0 | 0 | 0 | 0 |
| 0 | 0 | 0 | 0 | 0 |
| 0.009427792 | 0.002160455 | 0.006250941 | 0 | 0 |
| 0 | 0 | 0 | 0 | 0 |
| 0.024706697 | 0 | 0 | 0 | 0 |
| 0 | 0.006466439 | 0 | 0.003139688 | 0 |
| 0 | 0 | 0 | 0 | 0 |
| 0 | 0.039349777 | 0 | 0 | 0 |
| 0 | 0 | 0 | 0 | 0 |
| 0 | 0 | 0 | 0 | 0 |
| 0 | 0 | 0 | 0.0184106 | 0 |
| 0 | 0 | 0 | 0 | 0 |
| 0 | 0 | 0 | 0 | 0 |
| 0 | 0.004173115 | 0 | 0 | 0 |
| 0 | 0 | 0 | 0 | 0 |
| 0 | 0 | 0 | 0 | 0 |
| 0 | 0.001726081 | 0 | 0 | 0 |
| 0 | 0 | 0 | 0 | 0 |
| 0.025664263 | 0.325364662 | 0 | 0 | 0 |
| 0 | 0 | 0 | 0 | 0 |
| 0 | 0 | 0 | 0 | 0 |
| 0.001221125 | 0 | 0 | 0 | 0 |
| 0 | 0 | 0 | 0 | 0 |
| 0 | 0 | 0 | 0 | 0 |
| 0 | 0 | 0 | 0 | 0 |
| 0 | 0 | 0 | 0 | 0 |
| 0 | 0.002503997 | 0 | 0 | 0 |
| 0 | 0 | 0 | 0 | 0 |
| 0.003416029 | 0 | 0 | 0 | 0 |

| | | | | |
|---|---|---|---|---|
| 0 | 1.443976461 | 0 | 0.74315596 | 0 |
| 0 | 0.00041781 | 0 | 0 | 0 |
| 0 | 0.000905163 | 0 | 0 | 0.001618617 |
| 0 | 0 | 0 | 0 | 0 |
| 0 | 0.000910682 | 0.001219868 | 0.004539598 | 0.001628484 |
| 0 | 0.005939796 | 0 | 0.001682324 | 0 |
| 0 | 0.002963352 | 0 | 0 | 0 |
| 0.097483397 | 0.001063767 | 0.004445776 | 0.019378975 | 0.076958907 |
| 0 | 0 | 0 | 0 | 0 |
| 0.022556342 | 0 | 0.166374843 | 0.014011843 | 0.045039451 |
| 0 | 0 | 0.001912174 | 0 | 0 |
| 0.002099815 | 0.005557741 | 0 | 0 | 0 |
| 0.017520627 | 0.005493242 | 0.021356066 | 0.006599686 | 0.002741313 |
| 0 | 0 | 0.237644032 | 0.025585589 | 0 |
| 0 | 0 | 0.001267167 | 0.008037979 | 0 |
| 0 | 0 | 0 | 0 | 0 |
| 0 | 0 | 0 | 0 | 0 |
| 0 | 0 | 0 | 0 | 0 |
| 0.005467073 | 0 | 0.106139454 | 0 | 0.16572987 |
| 0 | 0 | 0.253105098 | 0 | 0 |
| 0 | 0 | 0 | 0 | 0 |
| 0 | 0 | 0 | 0 | 0 |
| 0 | 0 | 0 | 0 | 0 |
| 0 | 0.342778188 | 0 | 0 | 0 |
| 0 | 0 | 0 | 0 | 0 |
| 0 | 0.000595184 | 0 | 0 | 0 |
| 2.206017576 | 0.015320718 | 0 | 0 | 0 |
| 0 | 0 | 0 | 0 | 0.020133089 |
| 0 | 0 | 0.048486714 | 0 | 0 |
| 0 | 0 | 0.004921654 | 0.001665033 | 0 |
| 0 | 0 | 0 | 0 | 0 |
| 0.018227075 | 0 | 0 | 0 | 0 |
| 0 | 0 | 0 | 0 | 0 |
| 0 | 0 | 0.057867836 | 0.018518905 | 0 |
| 0 | 0 | 0 | 0 | 0 |
| 0 | 0 | 0 | 0 | 0 |
| 0.716612669 | 0 | 0.080525979 | 0 | 0.045243922 |
| 0 | 0 | 0 | 0 | 0 |
| 0 | 0 | 0 | 0 | 0 |
| 0 | 0 | 0 | 0.006382662 | 0.025665827 |
| 0 | 0 | 0 | 0 | 0 |
| 0 | 0 | 0 | 0.010928318 | 0.027588216 |
| 0 | 0 | 0 | 0 | 0 |
| 0 | 0 | 0 | 0.011081994 | 0 |
| 0 | 0 | 0 | 0 | 0 |
| 0 | 0.080342388 | 0 | 0 | 0 |
| 0.002307072 | 0.000740158 | 0 | 0.008301525 | 0 |
| 0 | 0 | 0 | 0.025116892 | 0 |

| | | | | |
|---|---|---|---|---|
| 0 | 0 | 0 | 0 | 0 |
| 0.000966868 | 0 | 0 | 0.00112455 | 0 |
| 0 | 0 | 0 | 0 | 0 |
| 0 | 0 | 0 | 0 | 0 |
| 0 | 0 | 0 | 0 | 0 |
| 0 | 0 | 0.258629824 | 0.32771535 | 0.049367875 |
| 0 | 0 | 0 | 0 | 0 |
| 0 | 0 | 0 | 0 | 0 |
| 0 | 0.05884087 | 0 | 7.147674277 | 0 |
| 0 | 0 | 0 | 0 | 0 |
| 0.008693557 | 0 | 0.020649906 | 0.003676853 | 0.005181758 |
| 0 | 0 | 0 | 0 | 0 |
| 0 | 0 | 0 | 0 | 0 |
| 0 | 0.000479088 | 0 | 0.001736853 | 0.002447732 |
| 0.204736015 | 0 | 0 | 0.002196728 | 0 |
| 0 | 0 | 0 | 0 | 0 |
| 0 | 0 | 0 | 0 | 0 |
| 0 | 0 | 0 | 0 | 0 |
| 0 | 0 | 0 | 0 | 0 |
| 0 | 0.0157045 | 0 | 0 | 0 |
| 0 | 0 | 0 | 0 | 0 |
| 0 | 0.130359601 | 0 | 0.804626203 | 0 |
| 0 | 0 | 0 | 0.2305406 | 0 |
| 0 | 0 | 0 | 0 | 0 |
| 0 | 0 | 0 | 0 | 0 |
| 0 | 0 | 0 | 0 | 0 |
| 0.011873762 | 0 | 0.005566556 | 0 | 0 |
| 0 | 0 | 0 | 0 | 0 |
| 0.000560751 | 0 | 0.009831949 | 0 | 0 |
| 0.347668334 | 0.136234941 | 0.602399362 | 0.294047093 | 1.309211063 |
| 0 | 0.000975068 | 0 | 0 | 0 |
| 0 | 0 | 0 | 0 | 0 |
| 0 | 0 | 0 | 0 | 0 |
| 0 | 0 | 0 | 0 | 0.012465422 |
| 0 | 0 | 0 | 0.016665684 | 0.001768419 |
| 0 | 0 | 0 | 0 | 0 |
| 0 | 0.079056207 | 0 | 1.439074611 | 0 |
| 0.001522086 | 0.005045951 | 0 | 0 | 0 |
| 0.088707994 | 0 | 0 | 0 | 0 |
| 1.5670118 | 0 | 0 | 0 | 0 |
| 0.004092226 | 0 | 0.058456166 | 0.050332837 | 0 |
| 5.108887567 | 0.111218979 | 0 | 0.890174566 | 13.41265208 |
| 0 | 0.003429535 | 0 | 0 | 0 |
| 0 | 0 | 0 | 0 | 0 |
| 0 | 0 | 0.001190282 | 0 | 0 |
| 0 | 0 | 0.002293985 | 0 | 0 |
| 0 | 0 | 0 | 0.020419304 | 0.00190085 |
| 0 | 0 | 0 | 0 | 0 |

| | | | | |
|---|---|---|---|---|
| 0 | 0.001870182 | 0 | 0 | 0 |
| 0 | 0 | 0 | 0 | 0 |
| 0.026717173 | 0.006254835 | 0.002234246 | 0.001574713 | 0 |
| 0 | 0 | 0 | 0.07526857 | 0 |
| 0 | 0.000683632 | 0 | 0.066916662 | 0 |
| 0 | 0 | 0 | 0 | 0.028643022 |
| 0 | 0 | 0 | 0 | 0 |
| 0 | 0.000762216 | 0 | 0 | 0 |
| 0.008870269 | 0.007683584 | 0.001601016 | 0 | 0 |
| 0 | 0 | 0 | 0 | 0 |
| 0 | 0 | 0 | 0 | 0 |
| 0 | 0 | 0.011752482 | 0 | 0 |
| 0 | 0 | 0 | 0 | 0 |
| 0 | 0 | 0 | 0 | 0.006601033 |
| 0.002155843 | 0 | 0 | 0 | 0 |
| 0.001685979 | 0.036300278 | 0.033425411 | 0.03333593 | 0.136150025 |
| 0 | 0.029976462 | 0 | 0.001267872 | 0 |
| 0 | 0 | 0.072592548 | 0 | 0.003969634 |
| 0 | 0 | 0 | 0 | 0 |
| 0 | 0 | 0 | 0 | 0 |
| 0 | 0 | 0 | 0.055240961 | 0.009007511 |
| 0 | 0 | 0 | 0.003151425 | 0 |
| 0 | 0.001917239 | 0 | 0.008056418 | 0 |
| 0.209722635 | 0 | 0 | 0 | 0.199945391 |
| 0 | 0 | 0 | 0 | 0.099623746 |
| 0 | 0 | 0 | 0 | 0 |
| 0 | 0 | 0.003534307 | 0.002759268 | 0 |
| 0 | 0 | 0 | 0 | 0 |
| 0 | 0 | 0 | 0 | 0 |
| 0 | 0 | 0 | 0 | 0 |
| 0 | 0.005530945 | 0 | 0.055433152 | 0.08260669 |
| 0.033174357 | 0.001150599 | 0 | 0 | 0.024396144 |
| 0.005691256 | 0.011137848 | 0.041676134 | 0.032517873 | 0.077521378 |
| 0 | 0.020148277 | 0 | 0 | 0 |
| 0.640935957 | 0 | 0 | 0.003046851 | 0 |
| 0 | 0 | 0 | 0 | 0 |
| 0.003344437 | 0 | 0 | 0 | 0 |
| 0 | 0 | 0.016393067 | 0.082135466 | 0.021963358 |
| 0.009565439 | 0.17731895 | 0 | 0.126073587 | 0 |
| 0 | 0 | 0 | 0 | 0.000485332 |
| 0 | 0 | 0 | 0 | 0 |
| 0 | 0 | 0 | 0 | 0 |
| 0.003911483 | 0 | 0 | 0.001049859 | 0.066875979 |
| 0.172761259 | 0 | 0 | 0.58061374 | 0.103025107 |
| 0 | 0 | 0.210163302 | 0 | 0 |
| 0 | 0 | 0 | 0 | 0 |
| 0 | 0 | 0 | 0 | 0 |
| 0 | 0 | 0 | 0 | 0 |

| | | | | |
|---|---|---|---|---|
| 0 | 0 | 0 | 0 | 0 |
| 2.052565236 | 0.133337291 | 0.140811918 | 0.686107817 | 1.569999423 |
| 0 | 0 | 0 | 0 | 0 |
| 0 | 0.004154896 | 0 | 0 | 0 |
| 0 | 0 | 0 | 0 | 0 |
| 0 | 0 | 0.012492721 | 0 | 0 |
| 0.002029 | 0 | 0 | 0.008849621 | 0 |
| 0 | 0 | 0 | 0 | 0 |
| 0 | 0 | 0 | 0 | 0 |
| 0 | 0 | 0.023834801 | 0 | 0 |
| 0 | 0.000392154 | 0.009035064 | 0 | 0.008615369 |
| 0.112884307 | 0.013629564 | 0.003129764 | 0.011647051 | 0.013429712 |
| 0 | 1.289841925 | 0 | 0 | 0 |
| 0 | 0 | 0 | 0 | 0 |
| 0.001402486 | 0 | 0 | 0 | 0 |
| 0 | 0 | 0 | 0 | 0 |
| 0.055892006 | 0.002737611 | 0.007700827 | 0.002977427 | 0.016784251 |
| 0.048462673 | 0 | 0.006283664 | 0.020291833 | 0.0157965 |
| 0 | 0.002031184 | 0 | 0 | 0 |
| 0 | 0 | 0 | 0.050196023 | 0 |
| 0.034759251 | 0.010728036 | 0.001361399 | 0.025523348 | 0.187151365 |
| 0.001368744 | 0 | 0 | 0 | 0 |
| 0 | 0 | 0 | 0 | 0 |
| 0 | 0.000729448 | 0 | 0 | 0 |
| 0 | 0 | 0 | 0 | 0 |
| 0 | 0 | 0.004161471 | 0.007743215 | 0 |
| 0.025623265 | 0 | 0 | 0.119144992 | 0 |
| 0.066639408 | 0.022354545 | 0.060531429 | 0.301462565 | 0.100032033 |
| 0 | 0.000877598 | 0 | 0 | 0 |
| 0 | 0 | 0.002908967 | 0 | 0 |
| 0 | 0 | 0 | 0 | 0 |
| 0 | 0 | 0 | 0 | 0 |
| 0 | 0 | 0 | 0.004952279 | 0 |
| 0 | 0 | 0 | 0 | 0 |
| 0 | 0 | 0.019124113 | 0 | 0.030259342 |
| 0.457068851 | 0.234619928 | 0.440802787 | 0.131521666 | 0.309017953 |
| 0 | 0 | 0 | 0 | 0 |
| 0 | 0 | 0 | 0 | 0 |
| 0.00230757 | 0.011104764 | 0 | 0.004780696 | 0.001702079 |
| 0 | 0 | 0 | 0 | 0 |
| 0 | 0 | 0.214311239 | 0 | 0 |
| 0 | 0 | 0 | 0 | 0 |
| 0.019973555 | 0 | 0.01861267 | 0.101635363 | 0 |
| 0.023513058 | 0 | 0 | 0 | 0 |
| 0 | 0 | 0 | 0 | 0 |
| 0 | 1.781850069 | 0 | 0 | 0 |
| 0 | 0 | 0 | 0 | 0 |
| 0 | 0 | 0.001410068 | 0 | 0 |

| | | | | |
|---|---|---|---|---|
| 0 | 0 | 0 | 0.025835192 | 0 |
| 0 | 0.047683163 | 0 | 0 | 0 |
| 0 | 0.00051513 | 0 | 0 | 0 |
| 0 | 0.107889445 | 0 | 0.943780211 | 0.00702409 |
| 0 | 0 | 0 | 0 | 0 |
| 0 | 0 | 0 | 0 | 0 |
| 0 | 0 | 0 | 0 | 0 |
| 0 | 0.048376474 | 0 | 0 | 0 |
| 1.500234337 | 2.492221311 | 0 | 0 | 0.520349667 |
| 0.689030499 | 0.063075186 | 0 | 0 | 0.679258942 |
| 0 | 0 | 0 | 0 | 0 |
| 0 | 0.008154162 | 0 | 0.005798615 | 0.008844924 |
| 0 | 0 | 0 | 0 | 0 |
| 0 | 0.480448538 | 0 | 0 | 0 |
| 0.04001151 | 0.294722986 | 0.017638435 | 0.190838339 | 0.080763402 |
| 0 | 0 | 0 | 0 | 0 |
| 0 | 0.017824419 | 0 | 0 | 0 |
| 1.299272691 | 2.446933776 | 0 | 0 | 0 |
| 0 | 0 | 0 | 0 | 0 |
| 0 | 0.040666545 | 0 | 0 | 0 |
| 0 | 0.436912172 | 0 | 0 | 0 |
| 0 | 0.052062033 | 0 | 0 | 0 |
| 0 | 0.076934311 | 0 | 0 | 0 |
| 0.06325131 | 0.266343614 | 0.456815583 | 1.274447211 | 0 |
| 0 | 0.020436647 | 0.011327636 | 0.018362746 | 0 |
| 0 | 0.937868758 | 0 | 0 | 0 |
| 0 | 0.001257748 | 0 | 0 | 0 |
| 0.889563188 | 0.018739522 | 0.573702844 | 0 | 2.581901803 |
| 0 | 0 | 0 | 0 | 0 |
| 0 | 0 | 0 | 0 | 0 |
| 0 | 0.017927749 | 0 | 0.011274486 | 0 |
| 0 | 0 | 0 | 0.033586341 | 0 |
| 0 | 0.108133661 | 0 | 0.118741903 | 0.150234329 |
| 0 | 0 | 0 | 0 | 0 |
| 0 | 0.037045999 | 0 | 0 | 0 |
| 0.012409219 | 0.31869822 | 0 | 0 | 0 |
| 0.667912651 | 0.387962165 | 0 | 1.218819188 | 0.353103985 |
| 0 | 0 | 0 | 0 | 0 |
| 0 | 0.002873025 | 0 | 0.004616039 | 0 |
| 0 | 0 | 0 | 0 | 0 |
| 0.028508893 | 0.027679477 | 0.302548109 | 0.318057545 | 0.20567201 |
| 0 | 0.016619002 | 0 | 0 | 0 |
| 1.220622008 | 0.005197293 | 0.236006161 | 0.883607976 | 4.494668936 |
| 0 | 0.009120793 | 0 | 0 | 0 |
| 0 | 0 | 0 | 0 | 0 |
| 0 | 0 | 0 | 0 | 0 |
| 0 | 0 | 0 | 0 | 0 |
| 0.278000611 | 0.001591361 | 0.270447719 | 0.00983389 | 0.111314043 |

| | | | | |
|---|---|---|---|---|
| 0 | 0 | 0 | 0 | 0 |
| 0.168228068 | 0.124153436 | 0 | 0 | 0.061815808 |
| 0.0103745 | 0 | 0.51494264 | 0.019042326 | 0.005739225 |
| 0 | 0.015440555 | 0 | 0 | 0 |
| 0 | 0 | 0 | 0 | 0 |
| 0 | 0.047413682 | 0 | 0 | 0.004018791 |
| 0 | 0 | 0 | 0 | 0 |
| 0 | 0 | 0 | 0.025888381 | 0 |
| 0 | 0 | 0 | 0 | 0 |
| 0 | 0 | 0 | 0 | 0 |
| 0.036499544 | 0.059093812 | 0.027796086 | 0.203251621 | 0.138994236 |
| 1.105584196 | 0.53964971 | 1.498573841 | 1.579948015 | 2.543811426 |
| 0 | 0 | 0 | 0 | 0 |
| 0.006412618 | 0.023071205 | 0 | 0.003596024 | 0 |
| 0.009565343 | 0.001621235 | 0.957331052 | 0.267322218 | 0.284525881 |
| 0 | 3.593373411 | 0 | 0 | 0 |
| 0 | 0 | 0 | 0 | 0 |
| 0 | 0 | 0 | 0 | 0 |
| 0 | 0.332369648 | 0 | 0 | 0 |
| 0.002272432 | 0.091298818 | 0 | 0.00121986 | 0 |
| 0 | 0.067788973 | 0.021462787 | 0.174078391 | 0.037957939 |
| 0.907625215 | 0 | 2.838294096 | 0 | 0.0759369 |
| 0.654166614 | 0.037080378 | 31.94054853 | 1.618421018 | 3.243880016 |
| 0 | 0 | 0 | 0 | 0 |
| 0 | 0.146972944 | 0 | 0 | 0 |
| 0.007033295 | 0.059350978 | 0 | 0 | 2.916699215 |
| 0 | 0 | 0 | 0 | 0 |
| 0 | 0 | 0 | 0 | 0 |
| 0 | 0 | 0.005858696 | 0.12611917 | 0 |
| 0 | 0 | 0 | 0 | 0 |
| 0 | 0.021902171 | 0 | 0 | 0.057494257 |
| 0.578799124 | 0.042126621 | 0 | 0.364719188 | 0.686813889 |
| 0 | 0.0037757 | 0 | 0 | 0.007716254 |
| 0.003656344 | 0.018022058 | 0 | 0.047987163 | 0.018469979 |
| 0.000719221 | 0.000738373 | 0.004005689 | 0.010414618 | 0.016268742 |
| 0.167988245 | 0.049202252 | 0 | 0.002241176 | 0.014383191 |
| 0 | 0.012984646 | 0 | 0.006661371 | 0.002816345 |
| 0 | 0 | 0 | 0 | 0 |
| 0.234971121 | 0.24677793 | 0 | 0.197100637 | 0.09279926 |
| 0 | 0 | 0 | 0 | 0 |
| 0 | 0.015632753 | 0 | 0 | 0 |
| 0 | 0 | 0 | 0 | 0 |
| 0 | 0.233202707 | 0 | 0 | 0 |
| 0 | 0 | 0 | 0 | 0 |
| 0.079333149 | 0.086767396 | 0.068573289 | 0.011402831 | 0.11437347 |
| 0 | 0.001788383 | 0 | 0 | 0 |
| 0 | 0.731630306 | 0 | 0 | 0 |
| 0.003534281 | 0 | 0 | 0.014703547 | 0.105345872 |

| | | | | |
|---|---|---|---|---|
| 0 | 0 | 0 | 0 | 0 |
| 0 | 0 | 0 | 0 | 0 |
| 0 | 0 | 0 | 0 | 0 |
| 0 | 0.00049301 | 0 | 0.00368636 | 0 |
| 0 | 0 | 0 | 0 | 0 |
| 0 | 0.065531208 | 0 | 0 | 0.010678296 |
| 0 | 0.053165578 | 0 | 0.018161383 | 0 |
| 0.114361541 | 0.002154878 | 0.002734563 | 0.811641572 | 0 |
| 0 | 0 | 0 | 0 | 0 |
| 0 | 0 | 0 | 0 | 0 |
| 0.029309268 | 0.004118119 | 0.002647809 | 0.001306337 | 0.000631203 |
| 0 | 0 | 0 | 0 | 0 |
| 0 | 0 | 0 | 0 | 0.010316465 |
| 0 | 0.036781012 | 0 | 0 | 0 |
| 0 | 0 | 0 | 0 | 0 |
| 0 | 0 | 0 | 0 | 0 |
| 0.111219463 | 0.032261458 | 0 | 0 | 0 |
| 0 | 0.058508637 | 0 | 0 | 0.002452157 |
| 0 | 0.014429673 | 0 | 0.085825025 | 0 |
| 0 | 0 | 0 | 0.016392382 | 0 |
| 0 | 0.007715828 | 0 | 0 | 0.008558584 |
| 0 | 0.018379495 | 0 | 0 | 0 |
| 0 | 0.333065343 | 0 | 0.008239287 | 0 |
| 0 | 0 | 0 | 0 | 0 |
| 0 | 0 | 0 | 0 | 0 |
| 0 | 0 | 0 | 0 | 0 |
| 0 | 0 | 0 | 0 | 0 |
| 0 | 0 | 0 | 0 | 0 |
| 0 | 0 | 0 | 0 | 0 |
| 0.454590083 | 0.140113437 | 5.567710863 | 0.890115304 | 0.06411013 |
| 0 | 0.002486666 | 0.020506867 | 0 | 0 |
| 0 | 0 | 0 | 0 | 0 |
| 0 | 0.667432492 | 0 | 0 | 0 |
| 0 | 0 | 0 | 0 | 0 |
| 0 | 0 | 0.003509014 | 0.001187126 | 0 |
| 0 | 0 | 0 | 0 | 0 |
| 0 | 0.021528072 | 0 | 0 | 0.049269603 |
| 0.008692838 | 0 | 0.009339234 | 0.048182892 | 0.001068649 |
| 0 | 0 | 0 | 0 | 0 |
| 0 | 0 | 0 | 0 | 0 |
| 0 | 0.005560338 | 0 | 0 | 0 |
| 0 | 0 | 0 | 0 | 0 |
| 0 | 0 | 0 | 0 | 0 |
| 0 | 0.876925604 | 0 | 0.001687555 | 0 |
| 0 | 0.053438201 | 0 | 0 | 0 |
| 0 | 0.209292418 | 0 | 0 | 0 |
| 0 | 0.057524321 | 0 | 0 | 0 |
| 0 | 0 | 0 | 0 | 0 |

| | | | | |
|---|---|---|---|---|
| 0 | 0 | 0 | 0 | 0 |
| 0 | 0 | 0 | 0 | 0 |
| 0 | 0.071536036 | 0 | 0 | 0 |
| 0 | 0.301797492 | 0 | 0 | 0.117992191 |
| 0 | 0 | 0 | 0 | 0 |
| 0 | 0.000599624 | 0 | 0 | 1.068572939 |
| 0 | 0 | 0 | 0 | 0 |
| 0 | 0 | 0 | 0 | 0 |
| 0.173339154 | 0.001121834 | 0.524746034 | 0.010893812 | 0 |
| 0.002413647 | 0 | 0 | 0 | 0.00494534 |
| 0 | 0.007448499 | 0 | 0 | 0 |
| 0 | 0 | 0 | 0 | 0 |
| 0 | 0 | 0 | 0 | 0 |
| 0 | 0 | 0 | 0 | 0 |
| 0 | 0.578721631 | 0 | 3.548852352 | 0 |
| 0 | 0 | 0.000992725 | 0 | 0 |
| 0 | 0 | 0 | 0 | 0 |
| 0 | 0 | 0 | 0 | 0 |
| 0 | 0 | 0 | 0 | 0 |
| 0 | 0.256596414 | 0 | 0 | 0 |
| 0 | 0.252316575 | 0 | 0.109454646 | 0 |
| 0 | 0 | 0 | 0 | 0 |
| 0 | 0 | 0 | 0 | 0 |
| 0 | 0 | 0 | 0 | 0 |
| 0.085178896 | 0.188840444 | 0 | 0 | 0.004044135 |
| 9.646617057 | 0.06359323 | 0.104790417 | 4.177371943 | 1.081452319 |
| 0 | 0.04920828 | 0 | 0 | 0 |
| 0 | 0 | 0 | 0 | 0 |
| 0 | 0.085845694 | 0 | 0 | 0.06566356 |
| 0 | 0 | 0 | 0 | 0 |
| 0 | 0 | 0 | 0 | 0 |
| 0 | 0.066268864 | 0 | 0 | 0 |
| 0 | 0 | 0 | 0 | 0 |
| 0 | 0.007045613 | 0 | 0.00738914 | 0.007559396 |
| 0.019901791 | 0.005804473 | 0 | 0.001578237 | 0 |
| 0 | 0.007502735 | 0.002449687 | 0.003984365 | 0 |
| 0 | 0 | 0 | 0 | 0.937344321 |
| 0.318120143 | 0.023430057 | 0 | 2.420121549 | 1.617881069 |
| 1.61314461 | 0 | 0 | 0 | 0 |
| 0 | 0 | 0 | 0 | 0 |
| 0 | 0 | 0 | 0 | 0 |
| 0 | 0.020738645 | 0 | 0.002710981 | 0 |
| 9.976206038 | 0.051423305 | 0 | 3.30515376 | 0.294650024 |
| 0 | 0 | 0 | 0 | 0 |
| 0 | 0.1475741 | 0 | 0 | 0 |
| 0 | 0.001332731 | 0 | 0 | 0 |
| 0 | 0 | 0 | 0 | 0 |
| 0.002206005 | 1.170768213 | 0.015357877 | 0.676401406 | 0.305093245 |

| | | | | |
|---|---|---|---|---|
| 0 | 0.00224297 | 0.058110255 | 0.227802081 | 0 |
| 0 | 0.630434361 | 0 | 0 | 0 |
| 0 | 0 | 0 | 0 | 0 |
| 0 | 0 | 0 | 0 | 0 |
| 0 | 0.000525444 | 0 | 0.002857365 | 0.004832233 |
| 0 | 0.089940549 | 0 | 0 | 0.025787418 |
| 0 | 0.028339837 | 0 | 0 | 0 |
| 0 | 0 | 0 | 0 | 0 |
| 0 | 0 | 0 | 0 | 0 |
| 0 | 0.007383504 | 0 | 0 | 0 |
| 0 | 0 | 0 | 0 | 0.002156879 |
| 0.686175014 | 0.417374517 | 0 | 0.44204131 | 0.595891826 |
| 0 | 0.075074013 | 0 | 0.264239802 | 0 |
| 0 | 0 | 0 | 0 | 0 |
| 0.036935443 | 0.063654046 | 0 | 0.01053323 | 0 |
| 0 | 0 | 0 | 0 | 0 |
| 0 | 0 | 0 | 0.001580416 | 0.001781814 |
| 0.006481105 | 0 | 0 | 0 | 0 |
| 0 | 0.022902906 | 0 | 0.520088399 | 0 |
| 0.014266898 | 0.001476492 | 0 | 0.013047398 | 0 |
| 0 | 0.084224364 | 0 | 0 | 0 |
| 0 | 0 | 0 | 0 | 0 |
| 0.005044728 | 0.728434989 | 0.021644724 | 0.643541643 | 0.085137059 |
| 0 | 0 | 0 | 0 | 0 |
| 0 | 0 | 0 | 0 | 0 |
| 0 | 0 | 0 | 0 | 0 |
| 0 | 0.005049728 | 0 | 0 | 0 |
| 0 | 0 | 0 | 0 | 0 |
| 0 | 0.000410522 | 0 | 0 | 0 |
| 0 | 0 | 0 | 0 | 0 |
| 0 | 0 | 0 | 0 | 0 |
| 0.303242475 | 0.065646322 | 0.009823257 | 0.064053588 | 0.05771249 |
| 0 | 0 | 0 | 0 | 0 |
| 0 | 0.016841072 | 0 | 0.053422709 | 0.006943872 |
| 0 | 0.005613339 | 0.001845605 | 0.006070382 | 0.000782166 |
| 0.060233335 | 0.030260783 | 0 | 0.005813198 | 0.00075623 |
| 0 | 0 | 0 | 0 | 0 |
| 0.186322183 | 0 | 0 | 0 | 0 |
| 0 | 0 | 0 | 0 | 0 |
| 0 | 0 | 0 | 0 | 0 |
| 0.047601765 | 0.013478194 | 0 | 0 | 0.833176785 |
| 0 | 0 | 0 | 0 | 0 |
| 0 | 0 | 0 | 0 | 0 |
| 0 | 0.010281992 | 0 | 0 | 0 |
| 0.035059743 | 0.173054511 | 0.021413246 | 0.042884673 | 0.230383009 |
| 0 | 0.003921638 | 0 | 0 | 0 |
| 6.210665256 | 0.058157071 | 0 | 0 | 0 |
| 0 | 0.000713197 | 0 | 0.005655948 | 0 |

| | | | | |
|---|---|---|---|---|
| 0 | 0 | 0 | 0 | 0 |
| 0 | 0 | 0 | 0 | 0 |
| 0 | 0 | 0 | 0.056722982 | 0.028887696 |
| 0 | 0 | 0 | 0 | 0 |
| 0 | 0 | 0 | 0 | 0 |
| 0 | 0 | 0 | 0 | 0 |
| 0 | 0.002861573 | 0 | 0 | 0 |
| 0 | 0.018904027 | 0 | 0.00750975 | 0 |
| 0 | 0 | 0 | 0 | 0 |
| 0 | 0.003744616 | 0 | 0 | 0 |
| 0 | 0 | 0 | 0 | 0 |
| 0 | 0 | 0 | 0 | 0 |
| 0 | 0 | 0 | 0 | 0 |
| 0 | 0.317780576 | 0 | 0 | 0.467055757 |
| 0 | 0 | 0 | 0 | 0 |
| 0 | 0 | 0 | 0 | 0 |
| 0 | 0.160472677 | 0 | 0 | 0 |
| 0 | 0.269829632 | 0 | 0 | 0 |
| 0.653696455 | 0.002361084 | 0 | 3.920980833 | 0.052581105 |
| 0 | 0 | 0 | 0 | 0 |
| 0 | 0.004251526 | 0 | 0 | 0 |
| 0 | 0 | 0 | 0 | 0 |
| 0 | 0 | 0 | 0 | 0 |
| 0 | 0 | 0 | 0 | 0 |
| 0 | 0.00151042 | 0 | 0 | 0 |
| 0 | 0 | 0.008462939 | 0.000477179 | 0.001613964 |
| 0 | 0 | 0 | 0 | 0 |
| 0 | 0.193650708 | 0.003129335 | 0.024287313 | 0.064067625 |
| 0 | 0 | 0 | 0 | 0 |
| 0 | 0.208048174 | 0 | 2.326343292 | 0 |
| 0 | 0 | 0 | 0 | 0 |
| 0 | 0.001069071 | 0 | 0 | 0 |
| 0.00150765 | 0.003143961 | 0 | 0 | 0 |
| 0 | 0 | 0 | 0 | 0 |
| 0.003630588 | 0.00049043 | 0.001773727 | 0.000500054 | 0.002255112 |
| 0 | 6.649360942 | 0 | 0 | 0 |
| 0 | 0 | 0 | 0 | 0 |
| 0 | 0.001112988 | 0.000894516 | 0 | 0 |
| 0 | 0.328608164 | 0 | 0 | 0 |
| 0 | 0 | 0 | 0 | 0 |
| 5.887949656 | 0.984571565 | 9.602706499 | 3.581539344 | 29.85567201 |
| 0.140463734 | 0.769088301 | 0.065478974 | 0.507004672 | 0.016233712 |
| 0 | 0.870062569 | 0 | 0 | 0.041039028 |
| 0 | 0.030717843 | 0 | 0 | 0 |
| 0 | 0 | 0 | 0 | 0 |
| 0 | 0 | 0 | 0 | 0 |
| 0 | 0.000947256 | 0 | 0 | 0 |
| 0.006195004 | 0.013691596 | 0.038691586 | 0.001651218 | 0.006769595 |

| | | | | |
|---|---|---|---|---|
| 0 | 0 | 0 | 0 | 0 |
| 0 | 0.004997171 | 0 | 0.073340616 | 0.059186239 |
| 0 | 0.003979373 | 0 | 0 | 0 |
| 0 | 0 | 0 | 0 | 0 |
| 6.602933355 | 0.420024086 | 0 | 2.212011547 | 1.589991132 |
| 0 | 0 | 0 | 0 | 0 |
| 0 | 0 | 0 | 0 | 0 |
| 0 | 0 | 0 | 0 | 0 |
| 0.340580051 | 0 | 0.377518749 | 0.335019357 | 0.049886491 |
| 0 | 0 | 0 | 0 | 0 |
| 0.003399583 | 0 | 0.001856266 | 0 | 0 |
| 0 | 2.430553835 | 0 | 0 | 0 |
| 0.015478888 | 0.073716888 | 0 | 0.099618044 | 0.744071682 |
| 0.019875484 | 0.002306386 | 0.013084648 | 0.011251032 | 0.033063631 |
| 0 | 0.007862995 | 0 | 0 | 0.038785453 |
| 0.463611278 | 0.109325711 | 0 | 1.088377387 | 0.343327888 |
| 0 | 0.074432333 | 0 | 0.005426636 | 0.014215749 |
| 0 | 0 | 0 | 0 | 0 |
| 0 | 0 | 0 | 0 | 0 |
| 0 | 0.010837028 | 0 | 0 | 0 |
| 0 | 0 | 0 | 0 | 0 |
| 0 | 0.001466419 | 0 | 0 | 0 |
| 0 | 0 | 0 | 0 | 0 |
| 0 | 0 | 0 | 0 | 0 |
| 0 | 0.018391318 | 0 | 0 | 0 |
| 0 | 0 | 0 | 0 | 0 |
| 0 | 0.172115531 | 0 | 0.010270075 | 0 |
| 0.000897631 | 0 | 0.00592514 | 0.002662253 | 0 |
| 0 | 0 | 0 | 0 | 0 |
| 0 | 0 | 0 | 0 | 0 |
| 0 | 0 | 0 | 0 | 0 |
| 0 | 0 | 0 | 0 | 0 |
| 0 | 0.000496668 | 0.006187226 | 0 | 0 |
| 0 | 0.020648523 | 0 | 0.011163007 | 0 |
| 0.019587057 | 0.002713522 | 0 | 0 | 0.070924304 |
| 0 | 0 | 0.000642577 | 0 | 0 |
| 0 | 0 | 0 | 0 | 0 |
| 0 | 0 | 0 | 0.021896173 | 0 |
| 0 | 0 | 0 | 0 | 0 |
| 0 | 0 | 0 | 0 | 0 |
| 3.654215163 | 0.135945961 | 0 | 0.248717486 | 0.859998043 |
| 0 | 0 | 0 | 0 | 0 |
| 0 | 0 | 0 | 0 | 0 |
| 0 | 0.244890233 | 0 | 0 | 0 |
| 0 | 0 | 0 | 0 | 0 |
| 0 | 0 | 0 | 0 | 0 |
| 0 | 0.002024516 | 0 | 0 | 0 |
| 0 | 0 | 0 | 0 | 0 |

| | | | | |
|---|---|---|---|---|
| 0 | 0.237996122 | 0 | 0 | 0 |
| 0 | 0 | 0 | 0 | 0 |
| 0 | 0 | 0 | 0 | 0 |
| 0 | 0 | 0 | 0.001419205 | 0 |
| 0.084838325 | 0.015232062 | 0 | 0 | 0.082670946 |
| 0 | 0 | 0 | 0 | 0 |
| 0 | 0.04942282 | 0 | 0.280543603 | 0.077906748 |
| 0 | 0 | 0 | 0 | 0 |
| 0 | 0 | 0 | 0 | 0 |
| 0 | 0 | 0 | 0 | 0 |
| 0 | 0.047009264 | 0 | 0 | 0 |
| 0 | 0.06882616 | 0 | 0 | 0 |
| 0 | 0 | 0 | 0 | 0 |
| 0 | 0 | 0 | 0 | 0 |
| 0 | 0 | 0 | 0 | 0 |
| 0 | 0 | 0 | 0 | 0 |
| 0 | 0 | 2.414900929 | 0 | 0 |
| 0 | 0 | 0 | 0 | 0 |
| 0 | 0 | 0 | 0 | 0 |
| 0 | 0 | 0 | 0 | 0 |
| 0 | 0.838707969 | 0.025271103 | 0.854789016 | 0.085622016 |
| 0 | 0 | 0 | 0 | 0 |
| 0 | 0.024842938 | 0 | 0 | 0.004901119 |
| 0.006135717 | 0 | 0.001757858 | 0.00609564 | 0 |
| 0 | 0 | 0 | 0 | 0 |
| 0 | 0.007483131 | 0 | 0.150845787 | 0 |
| 0 | 0 | 0 | 0 | 0 |
| 0 | 0 | 0 | 0 | 0 |
| 0 | 0 | 0 | 0 | 0 |
| 0.002452982 | 0.002518301 | 0 | 0.004707493 | 0.002171203 |
| 0 | 0.138017065 | 0 | 0 | 0 |
| 0 | 0 | 0 | 0 | 0 |
| 0 | 0 | 0 | 0 | 0 |
| 0 | 0 | 0 | 0 | 0 |
| 0 | 0.133481686 | 0 | 0 | 0 |
| 0 | 0 | 0 | 0 | 0 |
| 0 | 0.005199803 | 0.001319721 | 0 | 0.009438133 |
| 0 | 0 | 0 | 0 | 0 |
| 0 | 0 | 0 | 0 | 0 |
| 0 | 0.000497638 | 0 | 0 | 0 |
| 0 | 0 | 0 | 0 | 0 |
| 0 | 0 | 0 | 0 | 0 |
| 0 | 0 | 0 | 0 | 0 |
| 0 | 0.025853951 | 0 | 0 | 0 |
| 0 | 0 | 0 | 0 | 0 |
| 0 | 0.631907363 | 0 | 0 | 0 |
| 0 | 0 | 0 | 0 | 0 |
| 0 | 0 | 0 | 0 | 0.012594406 |

| | | | | |
|---|---|---|---|---|
| 0.003667916 | 0.000588373 | 0.002458973 | 0.002239701 | 0.063668947 |
| 0.016175657 | 0.000334806 | 0 | 0 | 0 |
| 0 | 0 | 0 | 0 | 0 |
| 0 | 0 | 0 | 0 | 0 |
| 0 | 0 | 0 | 0 | 0 |
| 0 | 0 | 0 | 0 | 0 |
| 0 | 0.000633239 | 0 | 0 | 0 |
| 0.675265461 | 1.485062582 | 0.769678329 | 1.938783619 | 0.73336977 |
| 2.972517304 | 7.974026319 | 0.014548201 | 0.001255553 | 0.094275766 |
| 0 | 0 | 0.006407523 | 0.021152691 | 0.022862785 |
| 0 | 0.001961647 | 0 | 0 | 0 |
| 0 | 0.001463248 | 0 | 0 | 0 |
| 0 | 0.036749103 | 0 | 0 | 0 |
| 0 | 0 | 0 | 0 | 0 |
| 0 | 0 | 0 | 0 | 0 |
| 0 | 0 | 0 | 0 | 0 |
| 0 | 0 | 0.054117684 | 0.052543306 | 0 |
| 0 | 0.000557841 | 0 | 0 | 0 |
| 0 | 0 | 0 | 0 | 0 |
| 0 | 0.018553336 | 0 | 0.042908552 | 0 |
| 0 | 0 | 0 | 0 | 0 |
| 0 | 0.003149948 | 0 | 0 | 0 |
| 0 | 0 | 0 | 0 | 0 |
| 0 | 0 | 0 | 0 | 0 |
| 0.005055737 | 0.085548301 | 0 | 0 | 0 |
| 0 | 0 | 0 | 0 | 0 |
| 0 | 0 | 0 | 0 | 0 |
| 0 | 0 | 0 | 0 | 0 |
| 0 | 0 | 0 | 0 | 0 |
| 0 | 0 | 0 | 0 | 0 |
| 0.006782675 | 0 | 0 | 0 | 0 |
| 0 | 0 | 0 | 0 | 0 |
| 0 | 0 | 0 | 0 | 0 |
| 0 | 0.001309959 | 0 | 0 | 0 |
| 0 | 0 | 0.001144974 | 0 | 0.001310145 |
| 0 | 0.000500491 | 0 | 0 | 0 |
| 0 | 0 | 0 | 0 | 0 |
| 0 | 0 | 0 | 0 | 0.00496893 |
| 0 | 0 | 0 | 0 | 0 |
| 0.054376674 | 0 | 0.425628833 | 0.00127767 | 0 |
| 0 | 0 | 0 | 0 | 0 |
| 0.075554465 | 0.098897106 | 2.702270667 | 9.002098717 | 0 |
| 0 | 0 | 0 | 0 | 0 |
| 0 | 0 | 0 | 0 | 0 |
| 0 | 0 | 0 | 0 | 0 |
| 0 | 0 | 0 | 0.663328285 | 0 |
| 0 | 0 | 0 | 0 | 0 |
| 0 | 0 | 0 | 0 | 0 |

| | | | | |
|---|---|---|---|---|
| 0 | 0 | 0 | 0 | 0 |
| 0 | 0 | 0 | 0 | 0.00065115 |
| 0 | 0 | 0 | 0 | 0 |
| 0 | 0 | 0 | 0 | 0 |
| 0.521398391 | 0.782699031 | 0 | 0.530122087 | 0.048706378 |
| 0 | 4.334298299 | 0 | 0 | 0 |
| 0 | 0 | 0 | 0 | 0 |
| 0.270852874 | 0.003442884 | 0.046679253 | 0.02238549 | 0.002294377 |
| 0 | 0.000448382 | 0 | 0 | 0.004581701 |
| 0 | 0 | 0 | 0 | 0 |
| 0 | 0 | 0 | 0 | 0 |
| 0 | 3.669724181 | 0 | 0 | 0 |
| 0 | 0 | 0 | 0 | 0 |
| 0 | 0 | 0 | 0 | 0 |
| 0 | 0 | 0 | 0 | 0 |
| 0.018007642 | 0 | 0 | 0 | 0 |
| 0 | 0 | 0 | 0 | 0 |
| 0 | 0 | 0 | 0 | 0 |
| 0 | 0 | 0 | 0 | 0 |
| 0 | 0 | 0 | 0 | 0 |
| 0 | 0 | 0 | 0 | 0 |
| 0 | 0 | 0 | 0 | 0 |
| 0 | 0 | 0 | 0 | 0 |
| 0.028871079 | 0 | 0.001457625 | 0.002289512 | 0 |
| 0 | 0.027662072 | 0 | 0 | 0.265523356 |
| 0 | 0 | 0 | 0 | 0 |
| 0 | 0.004028439 | 0 | 0.086338014 | 0.009988283 |
| 0 | 0 | 0 | 0 | 0 |
| 0 | 0 | 0 | 0 | 0 |
| 0 | 0 | 0 | 0 | 0 |
| 0 | 0 | 0 | 0 | 0 |
| 0 | 0 | 0 | 0.297256232 | 0 |
| 0 | 0 | 0 | 0 | 0 |
| 0 | 0 | 0 | 0 | 0 |
| 0 | 0 | 0 | 0 | 0 |
| 0 | 0 | 0 | 0 | 0 |
| 0 | 0 | 0 | 0 | 0 |
| 0 | 0.051277967 | 0 | 0 | 0 |
| 0 | 0.956677911 | 0 | 0 | 0 |
| 0 | 0 | 0 | 0 | 0 |
| 0 | 0 | 0 | 0 | 0 |
| 0 | 0.209831518 | 0 | 0 | 0 |
| 0 | 0 | 0 | 0 | 0 |
| 0 | 0 | 0 | 0 | 0 |
| 0 | 0 | 0 | 0 | 0 |
| 0 | 0 | 0 | 0 | 0 |
| 0 | 0 | 0 | 0 | 0 |
| 0 | 0 | 0 | 0 | 0 |

| | | | | |
|---|---|---|---|---|
| 0.015281509 | 0.14095075 | 0.176596231 | 0.036961202 | 0.008197631 |
| 0 | 0 | 0 | 0 | 0 |
| 0 | 0 | 0 | 0 | 0 |
| 0 | 0.000332581 | 0 | 0.003315721 | 0 |
| 0 | 0 | 0 | 0 | 0 |
| 0 | 0.161879604 | 0 | 0 | 0.388405328 |
| 0 | 0 | 0 | 0 | 0 |
| 0 | 0 | 0 | 0 | 0 |
| 0 | 0 | 0 | 0 | 0 |
| 0 | 0 | 0 | 0 | 0 |
| 0 | 0 | 0 | 0 | 0 |
| 0 | 0 | 0 | 0.000649405 | 0 |
| 0 | 0 | 0 | 0 | 0 |
| 0 | 0 | 0 | 0 | 0 |
| 0 | 0 | 0 | 0 | 0 |
| 0 | 0 | 0 | 0 | 0 |
| 0 | 0 | 0 | 0 | 0 |
| 0 | 0.005525489 | 0 | 0 | 0 |
| 0 | 0 | 0 | 0 | 0 |
| 0 | 0 | 0 | 0 | 0 |
| 0 | 0 | 0.003732511 | 0 | 0.001384106 |
| 0 | 0 | 0 | 0 | 0 |
| 0.093694126 | 0.12007339 | 0.037449237 | 0.012846555 | 0.020227145 |
| 1.389257181 | 0.00656104 | 0 | 1.373234213 | 0 |
| 0 | 0 | 0 | 0 | 0 |
| 0 | 0 | 0 | 0 | 0 |
| 0 | 0 | 0 | 0 | 0 |
| 0 | 0 | 0 | 0 | 0 |
| 0 | 0 | 0 | 0 | 0 |
| 0 | 0 | 0 | 0 | 0 |
| 0 | 0 | 0 | 0 | 0 |
| 0 | 0.100256473 | 0 | 0.020043931 | 0 |
| 0.008322617 | 0.002136059 | 0.01373412 | 0.006430226 | 0 |
| 0 | 0 | 0 | 0 | 0 |
| 0 | 0.100447934 | 0 | 0.263284314 | 0 |
| 0 | 0 | 0 | 0.002663219 | 0 |
| 0 | 0 | 0 | 0 | 0 |
| 0 | 0.001365513 | 0 | 0 | 0 |
| 0 | 0 | 0 | 0.002887907 | 0.003907107 |
| 0 | 0 | 0 | 0 | 0 |
| 0 | 0 | 0 | 0 | 0 |
| 0 | 0 | 0 | 0 | 0 |
| 1.915945017 | 0.259647665 | 0 | 0 | 0.05228485 |
| 0 | 0 | 0 | 0 | 0 |
| 0 | 0 | 0 | 0 | 0 |
| 0 | 0.172144585 | 0 | 0.020739157 | 0.060269167 |
| 0 | 0 | 0 | 0 | 0 |
| 0 | 0 | 0 | 0 | 0 |

| | | | | |
|---|---|---|---|---|
| 0 | 0 | 0 | 0 | 0 |
| 0 | 0 | 0 | 0 | 0 |
| 0 | 0 | 0 | 0 | 0 |
| 0 | 0 | 0 | 0 | 0 |
| 0 | 0 | 0 | 0 | 0 |
| 0 | 0 | 0 | 0 | 0 |
| 0 | 0 | 0 | 0 | 0 |
| 0.030500059 | 0 | 0 | 0.045609632 | 0 |
| 0 | 0.060619931 | 0 | 0.00117254 | 0 |
| 0.021532437 | 0 | 0 | 0 | 0.005397963 |
| 0 | 0 | 0 | 0 | 0 |
| 0 | 0 | 0 | 0 | 0 |
| 0 | 0 | 0 | 0 | 0 |
| 0 | 0.001281516 | 0 | 0 | 0 |
| 0 | 0 | 0 | 0.048960549 | 0 |
| 0 | 0 | 0.087830247 | 0 | 0 |
| 0 | 0 | 0 | 0 | 0 |
| 0 | 0 | 0 | 0 | 0 |
| 0 | 0 | 0 | 0 | 0 |
| 0 | 0.728492175 | 0 | 0.109216968 | 0 |
| 0.005857092 | 0.076694308 | 0 | 0 | 0.005760306 |
| 0 | 0 | 0 | 0 | 0 |
| 0 | 0 | 0 | 0 | 0 |
| 0 | 0.000710074 | 0 | 0 | 0 |
| 0 | 0 | 0 | 0 | 0 |
| 0 | 0 | 0 | 0 | 0 |
| 0 | 0 | 0 | 0 | 0 |
| 0 | 0 | 0 | 0 | 0 |
| 0 | 0 | 0 | 0 | 0 |
| 0 | 0.096272561 | 0 | 0 | 0 |
| 0.021945797 | 0.131606569 | 0 | 0.688433986 | 0.193584295 |
| 0 | 0 | 0 | 0 | 0 |
| 0 | 0.274865253 | 0 | 0 | 0 |
| 0 | 0 | 0 | 0 | 0 |
| 0 | 0 | 0 | 0 | 0 |
| 0 | 0 | 0 | 0 | 0 |
| 0 | 0 | 0 | 0 | 0 |
| 0 | 0 | 0 | 0 | 0 |
| 0 | 0.001464946 | 0 | 0.013277297 | 0 |
| 0 | 0 | 0 | 0 | 0 |
| 0 | 0.001188626 | 0 | 0 | 0 |
| 0 | 0.00497754 | 0 | 0 | 0 |
| 0 | 0.011481251 | 0 | 0 | 0.001616129 |
| 0 | 0 | 0 | 0.001354688 | 0 |
| 0 | 0 | 0 | 0 | 0 |
| 0 | 0 | 0 | 0 | 0 |
| 0.009780628 | 0 | 1.031785655 | 0.029119322 | 0.039939797 |
| 0 | 0.01164183 | 0 | 0 | 0 |

| | | | | |
|---|---|---|---|---|
| 0 | 0 | 0 | 0 | 0 |
| 0 | 0 | 0 | 0 | 0 |
| 0 | 0 | 0 | 0 | 0 |
| 0 | 0.00048908 | 0 | 0 | 0 |
| 0 | 0.022830438 | 0 | 0.167660592 | 0.137942221 |
| 0 | 0.02405421 | 0 | 0 | 0 |
| 0 | 0 | 0 | 0 | 0 |
| 0 | 0 | 0 | 0 | 0 |
| 0 | 0 | 0 | 0 | 0 |
| 0 | 0 | 0 | 0 | 0 |
| 0 | 0.0051762 | 0 | 0 | 0 |
| 0.011026643 | 0.029857198 | 0.007733448 | 0.013658542 | 0.039039962 |
| 0 | 0 | 0.000588707 | 0 | 0 |
| 0 | 0 | 0 | 0 | 0 |
| 0 | 0.002480214 | 0 | 0.00894274 | 0 |
| 0 | 0.000440156 | 0 | 0 | 0 |
| 0 | 0.0367855 | 0 | 0.005030582 | 0 |
| 0 | 0 | 0 | 0 | 0 |
| 0.008182449 | 0 | 0.002411216 | 0.052002964 | 0 |
| 0 | 0.105485721 | 0 | 0.380202109 | 0.003645369 |
| 0 | 0 | 0 | 0 | 0 |
| 0 | 0 | 0 | 0 | 0 |
| 0 | 0 | 0 | 0 | 0 |
| 0 | 0 | 0 | 0 | 0 |
| 0 | 0 | 0 | 0 | 0 |
| 0 | 0 | 0 | 0 | 0 |
| 0.001572503 | 0.001905861 | 0.006127014 | 0.023623998 | 0.006873415 |
| 0 | 0 | 0 | 0 | 0 |
| 0 | 0 | 0 | 0 | 0 |
| 0 | 0 | 0 | 0 | 0 |
| 0 | 0 | 0 | 0.001036608 | 0 |
| 0 | 0 | 0 | 0 | 0 |
| 0 | 0 | 0 | 0 | 0 |
| 0 | 0 | 0 | 0 | 0 |
| 0 | 0 | 0 | 0 | 0 |
| 0 | 0 | 0 | 0 | 0 |
| 0 | 0 | 0 | 0 | 0 |
| 0 | 0.033749392 | 0.003587912 | 0.057899116 | 0.019432684 |
| 0 | 0.022552001 | 0 | 0 | 0 |
| 0 | 0.001699533 | 0 | 0.004428489 | 0 |
| 0 | 0.008556895 | 0 | 0 | 0.014208511 |
| 0 | 0 | 0 | 0 | 0 |
| 0 | 0 | 0 | 0 | 0 |
| 0 | 0 | 0.13918216 | 0.000899969 | 0 |
| 0 | 0 | 0 | 0 | 0 |
| 0 | 0 | 0 | 0 | 0 |
| 0 | 0 | 0 | 0 | 0 |
| 0 | 0.001154039 | 0 | 0 | 0 |

| | | | | |
|---|---|---|---|---|
| 0 | 0 | 0 | 0 | 0 |
| 0 | 0 | 0 | 0.364759144 | 0 |
| 0 | 0.182774775 | 0 | 0 | 0 |
| 0 | 0.000832516 | 0.002899428 | 0 | 0 |
| 0 | 0 | 0 | 0 | 0 |
| 0 | 0 | 0 | 0.002257966 | 0 |
| 0 | 0.002827627 | 0 | 0 | 0 |
| 0 | 0 | 0 | 0 | 0 |
| 0 | 0.004508802 | 0 | 0 | 0 |
| 0 | 0.041366885 | 0 | 0 | 0 |
| 0 | 0 | 0 | 0 | 0 |
| 0 | 0 | 0 | 0 | 0 |
| 0.002146031 | 0.226514113 | 0.008379244 | 0.00361031 | 0.026532873 |
| 0 | 0.002082636 | 0 | 0 | 0 |
| 0 | 0 | 0 | 0 | 0 |
| 0 | 0 | 0 | 0 | 0 |
| 0 | 0 | 0 | 0 | 0 |
| 0 | 0 | 0 | 0 | 0 |
| 0 | 0 | 0 | 0 | 0 |
| 0 | 0.944785182 | 0 | 0 | 0 |
| 0 | 0 | 0 | 0 | 0 |
| 0 | 0 | 0 | 0 | 0 |
| 0 | 0 | 0.011241714 | 0.077823861 | 0.010918025 |
| 0.001905472 | 0.03515067 | 0 | 0.01121964 | 0.022253568 |
| 0 | 0 | 0 | 0 | 0 |
| 0 | 0.016139793 | 0 | 0 | 0 |
| 0 | 0.003588397 | 0 | 0 | 0 |
| 0 | 0 | 0 | 0 | 0 |
| 0 | 0 | 0 | 0 | 0 |
| 0 | 0 | 0 | 0 | 0 |
| 0 | 0.005776227 | 0 | 0.051239414 | 0 |
| 0 | 0.003685103 | 0 | 0.015636959 | 0 |
| 0 | 0 | 0 | 0 | 0 |
| 0 | 0 | 0 | 0 | 0 |
| 0 | 0.213561247 | 0 | 0.166235831 | 0 |
| 0 | 0 | 0.501075517 | 0 | 0 |
| 0 | 0 | 0 | 0 | 0 |
| 0 | 0 | 0 | 0 | 0 |
| 0.003427437 | 0.00219919 | 0 | 0.0008305 | 0 |
| 0 | 0 | 0 | 0 | 0 |
| 0 | 0 | 0 | 0 | 0 |
| 0 | 0 | 0 | 0 | 0 |
| 0 | 5.536692202 | 0 | 0 | 0 |
| 0 | 0 | 0 | 0 | 0 |
| 0 | 0 | 0 | 0 | 0 |
| 1.627239395 | 0.075247034 | 0 | 0.090899423 | 0.016146846 |
| 0 | 0 | 0.007403496 | 0 | 0 |
| 0 | 0 | 0 | 0 | 0 |

| | | | | |
|---|---|---|---|---|
| 0 | 0 | 0 | 0 | 0 |
| 0 | 0 | 0 | 0 | 0 |
| 0 | 0.093169855 | 0 | 0 | 0.069966368 |
| 0 | 0.002434438 | 0 | 0 | 0 |
| 0 | 0 | 0 | 0 | 0 |
| 0 | 0 | 0 | 0 | 0 |
| 0 | 0 | 0 | 0.022117754 | 0 |
| 0 | 0 | 0 | 0 | 0 |
| 0 | 0 | 0 | 0 | 0 |
| 0 | 0 | 0 | 0 | 0 |
| 0.001081332 | 0.001189421 | 0.00111527 | 0 | 0.001063464 |
| 0.000922533 | 0 | 0 | 0 | 0 |
| 0 | 0 | 0 | 0 | 0 |
| 0 | 0 | 0 | 0 | 0 |
| 0 | 0 | 0 | 0 | 0 |
| 0 | 0 | 0 | 0 | 0 |
| 0 | 0.004225295 | 0 | 0 | 0 |
| 0 | 0 | 0 | 0 | 0 |
| 0 | 0 | 0 | 0 | 0 |
| 0 | 0 | 0 | 0 | 0 |
| 0 | 0 | 0 | 0 | 0 |
| 0 | 0 | 0 | 0 | 0 |
| 0 | 0 | 0 | 0 | 0 |
| 0 | 0 | 0 | 0 | 0 |
| 0 | 0 | 0 | 0 | 0 |
| 0 | 0 | 0 | 0 | 0 |
| 0 | 0 | 0 | 0 | 0 |
| 0 | 0.002909391 | 0 | 0.003559787 | 0.016276656 |
| 0 | 0 | 0 | 0 | 0 |
| 0 | 0 | 0 | 0 | 0 |
| 0.00281596 | 0.005194666 | 0 | 0.005680426 | 0.010039175 |
| 0 | 0 | 0 | 0 | 0 |
| 0.103605963 | 0.009047304 | 0.024448697 | 0.007308586 | 0.052655247 |
| 0.008770632 | 0.004421695 | 0.005599843 | 0.001275124 | 0.001026869 |
| 0 | 0 | 0 | 0 | 0 |
| 0 | 0 | 0 | 0 | 0 |
| 0 | 0 | 0 | 0 | 0 |
| 0 | 0 | 0 | 0 | 0 |
| 0 | 0 | 0 | 0 | 0 |
| 0 | 0 | 0 | 0 | 0 |
| 0 | 0 | 0 | 0 | 0 |
| 0.111630169 | 0 | 0.006762372 | 0.547889595 | 0 |
| 0.048541106 | 0.045470602 | 0 | 0.029836678 | 0.02940883 |
| 0 | 0 | 0 | 0 | 0 |
| 0 | 0 | 0 | 0 | 0 |
| 0.031964934 | 0.030575194 | 0 | 0.011101744 | 0.00349297 |
| 0 | 0 | 0 | 0.001523432 | 0.00229009 |
| 0 | 0 | 0 | 0 | 0 |

| | | | | |
|---|---|---|---|---|
| 0 | 0 | 0 | 0 | 0 |
| 0 | 0.002340521 | 0 | 0.025081171 | 0 |
| 0.022056424 | 0.011096725 | 0 | 0.004372755 | 0.045355904 |
| 0 | 0 | 0 | 0 | 0 |
| 0.030218863 | 0.008860201 | 0 | 0.620602417 | 0 |
| 0 | 0.002374853 | 0 | 0.048599138 | 0 |
| 0 | 0 | 0 | 0 | 0 |
| 0 | 0 | 0 | 0 | 0 |
| 0 | 0.000917026 | 0 | 0 | 0 |
| 0 | 0 | 0 | 0 | 0 |
| 0 | 0 | 0 | 0 | 0 |
| 0 | 0 | 0 | 0 | 0 |
| 0 | 0 | 0 | 0 | 0 |
| 0 | 0 | 0 | 0 | 0 |
| 0 | 0 | 0 | 0 | 0 |
| 0 | 0 | 0 | 0 | 0 |
| 0 | 0.163748884 | 0 | 0 | 0 |
| 0 | 0 | 0 | 0 | 0 |
| 0 | 0 | 0 | 0 | 0 |
| 0 | 0 | 0 | 0 | 0 |
| 0 | 0 | 0 | 0 | 0 |
| 0 | 0 | 0.009981096 | 0 | 0 |
| 0 | 0 | 0 | 0 | 0 |
| 0 | 0 | 0 | 0 | 0 |
| 0.006196907 | 0.022664343 | 0 | 0 | 0 |
| 0.014130269 | 0 | 0 | 0.001087591 | 0.005722201 |
| 0 | 0 | 0 | 0 | 0 |
| 0.000806286 | 0.028712772 | 0 | 0.097060076 | 0.647255303 |
| 0 | 0 | 0 | 0 | 0 |
| 0 | 0 | 0 | 0 | 0 |
| 0 | 0 | 0 | 0 | 0 |
| 0 | 0 | 0 | 0 | 0 |
| 0 | 0 | 0 | 0 | 0 |
| 0 | 0 | 0 | 0 | 0 |
| 0 | 0 | 0 | 0 | 0 |
| 0 | 0 | 0 | 0 | 0 |
| 0 | 0 | 0 | 0 | 0 |
| 0 | 0 | 0 | 0.118364817 | 0 |
| 0.022451054 | 0.012952895 | 0 | 2.548106473 | 0 |
| 0 | 0 | 0 | 0 | 0 |
| 0 | 0 | 0 | 0 | 0 |
| 0.003832291 | 0.003037541 | 0 | 0.002556381 | 0.000886815 |
| 0.00208548 | 0.000669067 | 0.001720747 | 0.002910709 | 0.053326494 |
| 0 | 0 | 0 | 0 | 0 |
| 0 | 0 | 0 | 0 | 0 |
| 0 | 0.00123158 | 0 | 0 | 0 |
| 0.003190377 | 0 | 0.021388301 | 0.006400922 | 0.007530379 |
| 0 | 0 | 0 | 0 | 0 |

| | | | | |
|---|---|---|---|---|
| 0 | 0 | 0 | 0 | 0 |
| 0.000805134 | 0.0094281 | 0.031970521 | 0.107105203 | 0.009106038 |
| 0 | 0 | 0 | 0 | 0 |
| 0 | 0 | 0 | 0 | 0 |
| 0 | 0 | 0 | 0 | 0 |
| 0 | 0 | 0 | 0 | 0 |
| 0 | 0 | 0 | 0 | 0 |
| 0 | 0 | 0 | 0 | 0 |
| 0 | 0 | 0 | 0 | 0 |
| 0 | 0 | 0 | 0 | 0 |
| 0 | 0 | 0 | 0 | 0 |
| 0 | 0 | 0 | 0 | 0 |
| 0.015745308 | 0.154136919 | 0 | 0.058651512 | 0.044990568 |
| 0 | 0 | 0 | 0 | 0 |
| 0.02233634 | 0 | 0 | 0.018608597 | 0.009626096 |
| 0 | 0 | 0 | 0 | 0 |
| 0 | 0 | 0 | 0 | 0 |
| 0 | 0 | 0 | 0 | 0 |
| 0 | 0 | 0 | 0.005328122 | 0 |
| 0 | 0 | 0 | 0 | 0 |
| 0 | 0 | 0 | 0 | 0 |
| 0 | 0.028466508 | 0.001016832 | 0.030745186 | 0.012119982 |
| 0 | 0.060032476 | 0.00722751 | 0.002147742 | 0.0078231 |
| 0 | 0 | 0 | 0 | 0 |
| 0 | 0 | 0 | 0 | 0 |
| 0 | 0 | 0 | 0 | 0 |
| 0 | 0 | 0 | 0 | 0 |
| 0 | 0 | 0 | 0 | 0 |
| 0 | 0 | 0 | 0 | 0 |
| 0 | 0 | 0 | 0 | 0 |
| 0 | 0 | 0 | 0 | 0 |
| 0 | 0 | 0 | 0 | 0 |
| 0 | 0 | 0 | 0 | 0 |
| 0 | 0 | 0 | 0 | 0 |
| 0 | 10.50898569 | 0 | 6.505542358 | 3.591133219 |
| 0 | 0 | 0 | 0 | 0 |
| 0 | 0 | 0 | 0 | 0 |
| 0 | 0 | 0 | 0 | 0 |
| 0 | 0.00773853 | 0 | 0.036714523 | 0.002420654 |
| 0 | 0 | 0 | 0 | 0 |
| 0.116303846 | 0.061456309 | 0.238770068 | 0.078159573 | 0.631813981 |
| 0 | 0 | 0 | 0 | 0 |
| 0 | 0 | 0 | 0 | 0 |
| 0 | 0 | 0 | 0 | 0 |
| 0 | 0 | 0 | 0 | 0 |
| 0.031079086 | 0.000972764 | 1.300162305 | 0.290282992 | 0.007206505 |
| 0 | 0 | 0 | 0 | 0 |
| 0 | 0 | 0 | 0 | 0 |

| | | | | |
|---|---|---|---|---|
| 0 | 0 | 0.002089018 | 0 | 0.113764164 |
| 0 | 0 | 0 | 0 | 0 |
| 0 | 0 | 0 | 0 | 0 |
| 0 | 0 | 0 | 0 | 0 |
| 0 | 0.000491845 | 0.001185899 | 0.002005993 | 0 |
| 0 | 0 | 0 | 0 | 0 |
| 0 | 0 | 0 | 0 | 0 |
| 0 | 0.006133288 | 0 | 0.000952938 | 0 |
| 0 | 0 | 0 | 0 | 0 |
| 0 | 0.037578922 | 0 | 0 | 0 |
| 0 | 0.034615588 | 0 | 0.080251927 | 0 |
| 0 | 0 | 0 | 0 | 0 |
| 0.010869124 | 0 | 0 | 0.005056686 | 0.010214428 |
| 0 | 0 | 0 | 0 | 0 |
| 0 | 0 | 0 | 0 | 0 |
| 0 | 0 | 0 | 0 | 0 |
| 0 | 0 | 0 | 0 | 0 |
| 0 | 0 | 0 | 0 | 0 |
| 0 | 0 | 0 | 0 | 0 |
| 2.11909898 | 0.006483323 | 0.072949668 | 0.025129507 | 0.21946608 |
| 0 | 0.001283652 | 0 | 0.002147846 | 0 |
| 0 | 0 | 0 | 0 | 0 |
| 0 | 0 | 0 | 0 | 0 |
| 0 | 0 | 0 | 0 | 0 |
| 0 | 0 | 0 | 0 | 0 |
| 0 | 0 | 0 | 0 | 0 |
| 0 | 0 | 0 | 0 | 0 |
| 0 | 0 | 0 | 0 | 0 |
| 0 | 0 | 0 | 0 | 0 |
| 0 | 0 | 0 | 0.006415248 | 0 |
| 0 | 0 | 0 | 0 | 0 |
| 0 | 0 | 0 | 0 | 0 |
| 0 | 0 | 0 | 0 | 0 |
| 0 | 0 | 0 | 0 | 0 |
| 0 | 0 | 0 | 0 | 0 |
| 0 | 0 | 0 | 0 | 0 |
| 0 | 0 | 0 | 0 | 0 |
| 0 | 0 | 0 | 0 | 0 |
| 0 | 0 | 0 | 0 | 0 |
| 0 | 0 | 0 | 0 | 0 |
| 0 | 0 | 0 | 0 | 0 |
| 0 | 0 | 0 | 0 | 0 |
| 0 | 0.002293274 | 0 | 0.200936218 | 0 |
| 0 | 0 | 0 | 0 | 0 |
| 0.008076841 | 0.006737181 | 0.006247752 | 0.011449001 | 0.009134882 |
| 0 | 0.039034634 | 0 | 0 | 0 |
| 0 | 0 | 0 | 0 | 0 |
| 0 | 0 | 0 | 0 | 0 |

| | | | | |
|---|---|---|---|---|
| 0 | 0 | 0 | 0 | 0 |
| 0 | 0 | 0 | 0 | 0 |
| 0 | 0 | 0 | 0 | 0 |
| 0 | 0 | 0 | 0 | 0 |
| 0 | 0 | 0 | 0 | 0 |
| 0 | 0.002031171 | 0.002176622 | 0.030638154 | 0.003409772 |
| 0 | 0 | 0 | 0 | 0 |
| 0 | 0 | 0 | 0 | 0 |
| 0 | 0 | 0 | 0 | 0 |
| 0 | 0 | 0 | 0 | 0 |
| 0 | 0 | 0 | 0 | 0 |
| 0 | 0 | 0 | 0 | 0 |
| 0.001370784 | 0 | 0 | 0 | 0 |
| 0 | 0 | 0 | 0 | 0 |
| 0 | 0 | 0 | 0 | 0 |
| 0 | 0 | 0 | 0 | 0 |
| 0 | 0.002463797 | 0 | 0 | 0 |
| 0 | 0 | 0 | 0 | 0 |
| 0 | 0 | 0 | 0 | 0 |
| 0 | 0 | 0 | 0 | 0 |
| 0 | 0 | 0 | 0 | 0 |
| 0 | 0 | 0 | 0 | 0 |
| 0 | 0 | 0 | 0 | 0 |
| 0 | 0 | 0 | 0 | 0 |
| 0 | 0 | 0 | 0 | 0 |
| 0 | 0 | 0 | 0 | 0 |
| 0 | 0 | 0 | 0 | 0 |
| 0 | 1.051002846 | 0 | 0 | 0 |
| 0 | 0 | 0 | 0 | 0 |
| 0 | 0 | 0 | 0 | 0 |
| 0 | 0 | 0 | 0 | 0 |
| 0 | 0 | 0 | 0.011147881 | 0 |
| 0 | 0 | 0 | 0 | 0 |
| 0 | 0 | 0 | 0 | 0 |
| 0 | 0 | 0 | 0 | 0 |
| 0 | 0 | 0 | 0 | 0 |
| 0 | 0 | 0 | 0 | 0 |
| 0 | 0 | 0 | 0 | 0 |
| 0 | 0.092295906 | 0 | 0 | 0 |
| 0 | 0 | 0 | 0 | 0 |
| 0 | 0 | 0 | 0 | 0 |
| 0 | 0 | 0 | 0 | 0 |
| 0 | 0 | 0 | 0 | 0 |
| 0 | 0 | 0 | 0 | 0 |
| 0 | 0 | 0 | 0 | 0 |
| 0 | 0 | 0 | 0 | 0 |
| 0 | 0 | 0 | 0 | 0 |
| 0 | 0 | 0 | 0 | 0 |

| | | | | |
|---|---|---|---|---|
| 0 | 0 | 0 | 0 | 0 |
| 0 | 0.083165626 | 0 | 0.089803213 | 0 |
| 0 | 0.005965989 | 0 | 0 | 0 |
| 0 | 0 | 0 | 0 | 0 |
| 0 | 0 | 0 | 0 | 0 |
| 0 | 0 | 0 | 0 | 0 |
| 0 | 0 | 0 | 0 | 0 |
| 0 | 0 | 0 | 0 | 0 |
| 0 | 0 | 0 | 0 | 0 |
| 0 | 0 | 0 | 0 | 0 |
| 0 | 0 | 0 | 0 | 0 |
| 0 | 0.00109689 | 0 | 0 | 0 |
| 0 | 0.02141008 | 0 | 0 | 0 |
| 0 | 0 | 0 | 0 | 0 |
| 0 | 0 | 0 | 0 | 0 |
| 0 | 0 | 0 | 0 | 0 |
| 0 | 0 | 0 | 0 | 0 |
| 0 | 0 | 0 | 0 | 0 |
| 0 | 0 | 0 | 0 | 0 |
| 0 | 0 | 0 | 0 | 0 |
| 0 | 0 | 0 | 0.014282506 | 0.001764666 |
| 0 | 0.004833167 | 0 | 0 | 0 |
| 0 | 0 | 0 | 0 | 0 |
| 2.276251665 | 0.013590858 | 0 | 2.639424129 | 3.573122571 |
| 0 | 0 | 0 | 0 | 0 |
| 0 | 0 | 0 | 0 | 0 |
| 0 | 0 | 0 | 0 | 0 |
| 0.001545616 | 0 | 0 | 0 | 0 |
| 0 | 0 | 0 | 0 | 0 |
| 0 | 0 | 0 | 0 | 0 |
| 0 | 0.003107527 | 0 | 0 | 0 |
| 0 | 0 | 0 | 0 | 0 |
| 0 | 0 | 0 | 0 | 0 |
| 0 | 0 | 0 | 0 | 0 |

| PHL018-0094_V3 | PHL018-0150_V1 | PHL018-0175_V1 | PHL018-0161_V1 | PHL018-0010_V3 |
|---|---|---|---|---|
| REF | EYCF | CTRL | EYCF | CTRL |
| 0.928534103 | 0.00211975 | 0.055776372 | 0.02271515 | 0.039432968 |
| 0.184211748 | 1.1855738 | 0 | 0.124673196 | 3.413078704 |
| 0 | 13.60301339 | 1.110202723 | 3.738246198 | 0 |
| 0 | 0 | 0 | 0 | 0 |
| 0 | 0 | 0 | 0 | 0 |
| 0 | 0 | 0 | 0 | 0 |
| 0 | 0 | 0 | 0 | 0 |
| 0 | 0 | 0 | 0 | 0 |
| 0 | 0 | 0 | 0 | 0 |
| 0 | 0 | 0 | 0 | 0 |
| 0 | 0 | 0 | 0 | 0 |
| 0 | 0 | 0 | 0 | 0 |
| 0 | 0 | 0 | 0 | 0 |
| 0.557074304 | 1.04992966 | 0 | 0.70799841 | 1.873894348 |
| 0 | 0 | 0 | 0 | 0 |
| 0 | 0 | 0 | 0 | 0 |
| 0 | 0 | 0 | 0 | 0 |
| 0 | 0 | 0 | 0 | 0 |
| 0 | 0 | 0 | 0 | 0 |
| 0 | 0 | 0 | 0 | 0 |
| 0 | 0 | 0 | 0 | 0 |
| 0 | 0 | 0 | 0 | 0 |
| 0 | 0 | 0 | 0 | 0 |
| 0 | 0 | 0 | 0 | 0 |
| 0 | 0 | 0 | 1.232820658 | 0 |
| 0 | 0 | 0 | 0 | 0 |
| 0 | 0 | 0 | 0 | 0 |
| 0 | 0 | 0 | 0 | 0 |
| 0 | 0 | 0 | 0 | 0 |
| 0 | 0 | 0 | 0 | 0 |
| 0 | 0 | 0 | 0 | 0 |
| 0 | 0 | 0 | 0 | 0 |
| 0 | 0 | 0 | 0 | 0 |
| 0 | 0 | 0 | 0 | 0 |
| 0 | 0 | 0 | 0 | 0 |
| 0 | 0.00436567 | 0 | 1.708043919 | 0.65217029 |
| 0 | 0 | 0 | 0 | 0 |
| 0 | 0 | 0 | 0 | 0 |
| 0 | 0 | 0 | 0 | 0 |
| 0 | 0 | 0 | 0 | 0 |
| 0 | 0 | 0 | 0 | 0 |
| 0.069403738 | 0.004031517 | 0 | 0.012709935 | 0.002661824 |
| 0.620727374 | 1.194860376 | 0.001398898 | 2.535073452 | 0.220154512 |
| 0.007330989 | 0.34690358 | 0 | 0.058715889 | 0.108245298 |
| 0.018373003 | 0 | 0 | 0.011847204 | 0.00597895 |
| 0 | 0 | 0 | 0 | 0 |

| | | | | |
|---|---|---|---|---|
| 0 | 0 | 0 | 0.007302624 | 0.001223503 |
| 0.000897624 | 0 | 0.01849994 | 0.003091651 | 0.005050345 |
| 0 | 0 | 0 | 0 | 0.001855914 |
| 0.761388723 | 0.588097336 | 0 | 0.502723764 | 0.899814182 |
| 0.03058034 | 0 | 0 | 0 | 0.00343331 |
| 0 | 0 | 0 | 0 | 0 |
| 0.002164743 | 0.194120231 | 0 | 0.000828438 | 0 |
| 0 | 0 | 0 | 0 | 0 |
| 0 | 0 | 0 | 0 | 0 |
| 0 | 0 | 0 | 0 | 0 |
| 0.001325987 | 0 | 0 | 0.007358014 | 0 |
| 0 | 0 | 0 | 0.152398649 | 0.061335449 |
| 0 | 0 | 0 | 0 | 0.001563155 |
| 0.172430141 | 0.004504649 | 0.015752227 | 0.021161069 | 0.163038082 |
| 0 | 0 | 0 | 0 | 0 |
| 0 | 0 | 0 | 0 | 0 |
| 0.001416884 | 0.006175397 | 0 | 0.919630957 | 0 |
| 0 | 0 | 0 | 0 | 0 |
| 0 | 0 | 0 | 0 | 0 |
| 0 | 0 | 0 | 0 | 0 |
| 0.037860895 | 0.1076751 | 0.021184867 | 0 | 0.260583614 |
| 0.005981583 | 0 | 0 | 0.011772645 | 0.019658481 |
| 0.383757236 | 0.175618439 | 0 | 0.037852597 | 0 |
| 0 | 0 | 0 | 0 | 0 |
| 0 | 0 | 0 | 0.001083495 | 0 |
| 0 | 0 | 0 | 0 | 0 |
| 0 | 0 | 0 | 0 | 0 |
| 0 | 0 | 0 | 0 | 0 |
| 0.004372671 | 0.06328572 | 0 | 0.001255051 | 0 |
| 0 | 0 | 0 | 0 | 0 |
| 0.032339756 | 0.021478764 | 0 | 0 | 0 |
| 0 | 0 | 0 | 0 | 0 |
| 0 | 0 | 0 | 0 | 0 |
| 0.001106106 | 0 | 0 | 0 | 0 |
| 0 | 0 | 0 | 0 | 0 |
| 0.000794678 | 0 | 0 | 0 | 0 |
| 0 | 0 | 0 | 0 | 0 |
| 0.000828948 | 0 | 0 | 0 | 0 |
| 0 | 0 | 0 | 0 | 0 |
| 0 | 0 | 0 | 0 | 0 |
| 0 | 0 | 0 | 0 | 0 |
| 0 | 0 | 0 | 0 | 0 |
| 0 | 0 | 0 | 0 | 0 |
| 0 | 0 | 0 | 0 | 0 |
| 0.516883353 | 0 | 0 | 0.801836061 | 0 |
| 0 | 0 | 0 | 0 | 0 |
| 0.035708611 | 0 | 0 | 0 | 0 |
| 0.000518 | 1.085550048 | 0 | 0 | 0 |

| | | | | |
|---|---|---|---|---|
| 0 | 0 | 0 | 0 | 0 |
| 0.001974253 | 0 | 0 | 0 | 0 |
| 0 | 0 | 0 | 0 | 0 |
| 0 | 0 | 0 | 0 | 0 |
| 0.203090842 | 0.024383079 | 0 | 0 | 0 |
| 0 | 0 | 0 | 0 | 0 |
| 0 | 0 | 0 | 0 | 0 |
| 0 | 0 | 0 | 0 | 0 |
| 0.011398146 | 0 | 0 | 0.031861714 | 0 |
| 0 | 0 | 0 | 0 | 0 |
| 0 | 0 | 0 | 0 | 0 |
| 0 | 0.006052878 | 0 | 0.00066055 | 0 |
| 0 | 0 | 0 | 0 | 0 |
| 0 | 0 | 0 | 0 | 0 |
| 0 | 0 | 0 | 0 | 0 |
| 0 | 0 | 0 | 0 | 0 |
| 0 | 0 | 0 | 0 | 0 |
| 0 | 0 | 0 | 0 | 0 |
| 0.256699478 | 0.002616435 | 0 | 0.560229573 | 0 |
| 0 | 0 | 0 | 0 | 0 |
| 0 | 0 | 0 | 0 | 0 |
| 0 | 0 | 0 | 0 | 0 |
| 0.001987491 | 0 | 0 | 0 | 0 |
| 0 | 0 | 0 | 0 | 0 |
| 0 | 0 | 0 | 0 | 0 |
| 0 | 0 | 0 | 0 | 0 |
| 0 | 0 | 0 | 0 | 0 |
| 0.004002763 | 0 | 0 | 0 | 0 |
| 0 | 0 | 0 | 0 | 0 |
| 0 | 0 | 0 | 0 | 0 |
| 0.12504152 | 0 | 0 | 0.031589894 | 0.001528991 |
| 0.028865893 | 0 | 0 | 0.14305671 | 0.001804551 |
| 0 | 0 | 0 | 0 | 0 |
| 0 | 0 | 0 | 0 | 0 |
| 0 | 0 | 0 | 0 | 0 |
| 0 | 0 | 0 | 0 | 0 |
| 0 | 0 | 0 | 0 | 0 |
| 0 | 0 | 0 | 0 | 0 |
| 0 | 0 | 0 | 0 | 0 |
| 0 | 0 | 0 | 0 | 0 |
| 0 | 0 | 0 | 0 | 0 |
| 0 | 0 | 0 | 0.008800147 | 0 |
| 0.000741431 | 0.002172892 | 0 | 0 | 0 |
| 0 | 0 | 0 | 0 | 0 |
| 0 | 0 | 0 | 0.003856989 | 0 |
| 0 | 0 | 0 | 0 | 0 |
| 0 | 0 | 0 | 0.000669319 | 0.008017985 |
| 0 | 0 | 0 | 0 | 0 |

| | | | | |
|---|---|---|---|---|
| 0 | 0 | 0 | 0 | 0 |
| 0 | 0 | 0 | 0 | 0 |
| 0 | 0 | 0 | 0 | 0 |
| 0 | 0 | 0 | 0 | 0 |
| 0 | 0 | 0 | 0.005128018 | 0.017626263 |
| 0 | 0 | 0 | 0 | 0 |
| 0 | 0 | 0 | 0 | 0 |
| 0 | 0 | 0 | 0 | 0 |
| 0 | 0 | 0 | 0 | 0 |
| 0 | 0 | 0 | 0 | 0 |
| 0 | 0 | 0 | 0 | 0 |
| 0 | 0 | 0 | 0 | 0 |
| 0 | 0 | 0 | 0 | 0 |
| 0 | 0 | 0 | 0 | 0 |
| 0 | 0 | 0 | 0 | 0 |
| 0 | 0 | 0 | 0.01107262 | 0 |
| 0 | 0 | 0 | 0 | 0 |
| 0 | 0 | 0 | 0 | 0 |
| 0 | 0 | 0 | 0 | 0 |
| 0 | 0 | 0 | 0 | 0 |
| 0 | 0 | 0 | 0 | 0 |
| 0 | 0 | 0 | 0 | 0 |
| 0 | 0 | 0 | 0.007268671 | 0 |
| 0 | 0 | 0 | 0 | 0 |
| 0 | 0 | 0 | 0 | 0 |
| 0 | 0 | 0 | 0 | 0 |
| 0 | 0 | 0 | 0.013399818 | 0 |
| 0 | 0 | 0 | 0 | 0 |
| 0 | 0 | 0 | 0 | 0 |
| 0 | 0 | 0 | 0 | 0 |
| 0 | 0 | 0 | 0 | 0 |
| 0 | 0 | 0 | 3.520357171 | 0.058066396 |
| 0 | 0 | 0 | 0 | 0 |
| 0 | 0 | 0 | 0 | 0 |
| 0 | 0 | 0 | 0 | 0 |
| 0 | 0 | 0 | 0 | 0 |
| 0 | 0 | 0 | 0 | 0 |
| 0 | 0 | 0 | 0 | 0 |
| 0 | 0 | 0 | 0 | 0 |
| 0 | 0 | 0 | 0 | 0 |
| 0 | 0 | 0 | 0 | 0 |
| 0 | 0 | 0 | 0 | 0 |
| 0 | 0 | 0 | 0 | 0 |
| 0 | 0 | 0 | 0 | 0 |
| 0 | 0 | 0 | 0 | 0 |
| 0 | 0 | 0 | 0 | 0 |
| 0 | 0 | 0 | 0 | 0 |
| 0 | 0 | 0 | 0 | 0 |

| | | | | |
|---|---|---|---|---|
| 0 | 0 | 0 | 3.99609894 | 0 |
| 0 | 0 | 0 | 0 | 0 |
| 0 | 0 | 0 | 0 | 0 |
| 0 | 0 | 0 | 0 | 0 |
| 0 | 0 | 0 | 0 | 0 |
| 0 | 0 | 0 | 0 | 0 |
| 0 | 0 | 0 | 0 | 0 |
| 0 | 0 | 0 | 0 | 0 |
| 0 | 0 | 0 | 0 | 0 |
| 0 | 0 | 0 | 0 | 0 |
| 0 | 0 | 0 | 0 | 0 |
| 0.011674393 | 0 | 0 | 0.01113498 | 0 |
| 0 | 0 | 0 | 0.001829455 | 0 |
| 0 | 0 | 0 | 0 | 0 |
| 0 | 0 | 0 | 0.00573505 | 0 |
| 0 | 0 | 0 | 0 | 0 |
| 0 | 0 | 0 | 0 | 0 |
| 0 | 0 | 0 | 0 | 0 |
| 0 | 0 | 0 | 0 | 0 |
| 0 | 0 | 0 | 0 | 0 |
| 0.015559233 | 0.062092272 | 0.016512458 | 0.02698506 | 0.100165867 |
| 0 | 0 | 0 | 0 | 0 |
| 0 | 0 | 0 | 0 | 0 |
| 0 | 0 | 0 | 0 | 0 |
| 0 | 0 | 0 | 0 | 0 |
| 0 | 0 | 0 | 0 | 0 |
| 0.009254125 | 0.124825251 | 0 | 0.023462522 | 0.001928405 |
| 0 | 0 | 0 | 0 | 0 |
| 0 | 0 | 0 | 0 | 0 |
| 0 | 0 | 0.054157581 | 0 | 0 |
| 0.000960916 | 0 | 0 | 0 | 0 |
| 0 | 0 | 0 | 0 | 0 |
| 0 | 0 | 0 | 0 | 0 |
| 0.01638757 | 0 | 0 | 0 | 0 |
| 0.04157165 | 0 | 0.008654797 | 0.021761927 | 0.010772438 |
| 0 | 0 | 0 | 0 | 0 |
| 0 | 0 | 0 | 0 | 0 |
| 0 | 0 | 0 | 0 | 0 |
| 0 | 0 | 0 | 0.000958778 | 0 |
| 0 | 0 | 0 | 0 | 0 |
| 0 | 0 | 0 | 0 | 0 |
| 0 | 0 | 0 | 0 | 0 |
| 0 | 0 | 0 | 0 | 0 |
| 0 | 0 | 0 | 0 | 0 |
| 0 | 0 | 0 | 0 | 0 |
| 0 | 0 | 0 | 0 | 0 |
| 0 | 0 | 0 | 0 | 0 |
| 0.014735792 | 0 | 0.000425355 | 0.003877034 | 0 |

| | | | | |
|---|---|---|---|---|
| 0 | 0 | 0 | 0 | 0 |
| 0 | 0 | 0 | 0 | 0 |
| 0 | 0.002529632 | 0.00058302 | 0 | 0 |
| 0 | 0 | 0 | 0 | 0 |
| 0 | 0 | 0 | 0 | 0 |
| 0 | 0 | 0 | 0 | 0 |
| 0 | 0 | 0 | 0 | 0.002458739 |
| 0 | 0 | 0 | 0 | 0 |
| 0 | 0 | 0 | 0 | 0 |
| 0 | 0 | 0 | 0 | 0 |
| 0 | 0 | 0 | 0 | 0 |
| 0 | 0 | 0 | 0 | 0 |
| 0 | 0 | 0 | 0 | 0 |
| 0 | 0 | 0 | 0 | 0 |
| 0.001210076 | 0 | 0 | 0.005557089 | 0 |
| 0 | 0 | 0 | 0 | 0 |
| 0 | 0 | 0 | 0 | 0 |
| 0 | 0 | 0 | 0.013852987 | 0 |
| 0.003278232 | 0.005771492 | 0 | 0.013620991 | 0.001171078 |
| 0 | 0 | 0 | 1.538105547 | 0 |
| 0.004835726 | 0 | 0.009731082 | 0 | 0 |
| 0 | 0 | 0 | 0 | 0 |
| 0 | 0 | 0 | 0 | 0 |
| 0 | 0 | 0 | 0 | 0 |
| 0.003202397 | 0.004537888 | 0.003486252 | 0.003151397 | 0.029743671 |
| 0 | 0 | 0 | 0 | 0 |
| 0 | 0 | 0 | 0 | 0 |
| 0 | 0 | 0 | 0 | 0 |
| 0 | 0.004921105 | 0 | 0 | 0 |
| 0 | 0 | 0 | 0 | 0 |
| 0.7290893 | 0.005343904 | 0.003284382 | 0.251545007 | 0.126384874 |
| 0 | 0 | 0 | 0 | 0 |
| 0 | 0 | 0 | 0 | 0 |
| 0.006352782 | 0 | 0 | 0 | 0 |
| 0 | 0 | 0 | 0 | 0 |
| 0 | 0 | 0 | 0 | 0 |
| 0 | 0 | 0 | 0 | 0 |
| 0 | 0.004057539 | 0 | 0 | 0 |
| 0 | 0 | 0 | 0 | 0 |
| 0 | 0 | 0 | 0 | 0 |
| 0 | 0 | 0 | 0 | 0 |
| 0.022954864 | 0.001711984 | 0 | 0.000990759 | 0.060422013 |
| 0.001448119 | 0 | 0 | 0 | 0 |
| 0 | 0 | 0 | 0 | 0 |
| 0 | 0 | 0 | 0 | 0 |
| 0 | 0 | 0 | 0 | 0 |
| 0 | 0 | 0 | 0 | 0 |
| 0 | 0 | 0 | 0 | 0 |

| | | | | |
|---|---|---|---|---|
| 0 | 0 | 0 | 0 | 0 |
| 0 | 0 | 0 | 0 | 0 |
| 0.00195047 | 0.401403414 | 0 | 0.072404313 | 0.12383032 |
| 0 | 0 | 0 | 0 | 0 |
| 0.000752298 | 0 | 0 | 0 | 0 |
| 0 | 0 | 0 | 0 | 0 |
| 0 | 0 | 0 | 0 | 0 |
| 0 | 0 | 0 | 0 | 0 |
| 0 | 0 | 0 | 0 | 0 |
| 0 | 0.001251141 | 0 | 0 | 0 |
| 0 | 0 | 0 | 0 | 0 |
| 0 | 0 | 0 | 0 | 0 |
| 0 | 0 | 0 | 0 | 0 |
| 0 | 0 | 0 | 0 | 0.033539923 |
| 0.005139863 | 0.011408854 | 0.001095613 | 0.007943665 | 0.033862015 |
| 0.055029016 | 0 | 0 | 0.03863583 | 0.001058504 |
| 0.00135148 | 0.003452957 | 0 | 0 | 0.014926165 |
| 0 | 0 | 0 | 0 | 0 |
| 0 | 0 | 0 | 0 | 0 |
| 0 | 0 | 0 | 0 | 0 |
| 0 | 0 | 0 | 0 | 0 |
| 0.017893342 | 0 | 0.000704316 | 0.002801333 | 0 |
| 0 | 0 | 0 | 0 | 0 |
| 0 | 0 | 0 | 0 | 0 |
| 0 | 0 | 0 | 0.001871573 | 0 |
| 0 | 0 | 0 | 0 | 0 |
| 0 | 0 | 0 | 0 | 0 |
| 0 | 0 | 0 | 0 | 0 |
| 0 | 0 | 0 | 0 | 0 |
| 0.002647113 | 0 | 0 | 0 | 0 |
| 0 | 0 | 0 | 0.003935628 | 0 |
| 0 | 0 | 0 | 0 | 0 |
| 0 | 0 | 0 | 0 | 0 |
| 0 | 0 | 0 | 0 | 0.001758742 |
| 0 | 0 | 0 | 0 | 0 |
| 0 | 0 | 0 | 0.004662721 | 0 |
| 0 | 0 | 0 | 0 | 0 |
| 0.022225664 | 0.001842932 | 0 | 0.010558755 | 0.042162223 |
| 0 | 0 | 0 | 0 | 0 |
| 0 | 0 | 0 | 0 | 0 |
| 0 | 0 | 0 | 0 | 0 |
| 0 | 0 | 0 | 0 | 0 |
| 0 | 0 | 0 | 0 | 0 |
| 0 | 0 | 0.012310869 | 0 | 0 |
| 0 | 0 | 0 | 0 | 0 |
| 0 | 0 | 0 | 0 | 0 |
| 0 | 0 | 0 | 0 | 0 |
| 0.012686839 | 0 | 0.01028963 | 0.001065775 | 0 |

| | | | | |
|---|---|---|---|---|
| 0.132392604 | 0 | 0 | 0.499088085 | 0 |
| 0 | 0 | 0 | 0 | 0 |
| 0.001916783 | 0 | 0 | 0 | 0.001065135 |
| 0 | 0 | 0 | 0 | 0 |
| 0.016499122 | 0.001352546 | 0.001039099 | 0.018942424 | 0.005893956 |
| 0.017120291 | 0 | 0 | 0.011632894 | 0 |
| 0 | 0 | 0 | 0 | 0 |
| 0.00810952 | 0.089648557 | 0 | 0.022069175 | 0.013143576 |
| 0 | 0 | 0 | 0 | 0 |
| 0 | 0.0591824 | 1.329699248 | 0.030921944 | 0.010837894 |
| 0 | 0 | 0 | 0 | 0 |
| 0 | 0.004073584 | 0 | 0.003002663 | 0.004756343 |
| 0.015149362 | 0.020166041 | 0 | 0.004555253 | 0.01398044 |
| 0.015886458 | 0 | 0 | 0 | 0.002397238 |
| 0.007419416 | 0 | 0 | 0.015332641 | 0 |
| 0 | 0.040770322 | 0 | 0 | 0 |
| 0 | 0 | 0 | 0 | 0 |
| 0 | 0 | 0 | 0 | 0 |
| 0 | 0 | 0.843969926 | 0 | 0 |
| 0 | 0 | 0 | 0 | 0 |
| 0 | 0 | 0 | 0 | 0 |
| 0 | 0 | 0 | 0 | 0 |
| 0 | 0 | 0 | 0 | 0 |
| 0 | 0 | 0 | 0.180832871 | 0 |
| 0 | 0 | 0 | 0 | 0 |
| 0.002100612 | 0.009660509 | 0 | 0 | 0.001575837 |
| 0 | 0 | 0 | 0.296691938 | 0 |
| 0.003302292 | 0 | 0 | 0 | 0 |
| 0 | 0 | 0 | 0 | 0 |
| 0 | 0 | 0 | 0 | 0 |
| 0.002479479 | 0 | 0.002186173 | 0 | 0 |
| 0 | 0 | 0 | 0 | 0 |
| 0 | 0 | 0.022926029 | 0 | 0 |
| 0.001098886 | 0 | 0 | 0 | 0 |
| 0 | 0 | 0 | 0 | 0 |
| 0 | 0 | 0 | 0 | 0 |
| 0 | 0 | 0.000989799 | 0 | 0.305171978 |
| 0 | 0 | 0 | 0 | 0 |
| 0 | 0 | 0 | 0 | 0 |
| 0.001767445 | 0 | 0.01836649 | 0 | 0.00552459 |
| 0 | 0 | 0 | 0 | 0 |
| 0.008141133 | 0 | 0 | 0 | 0 |
| 0 | 0 | 0 | 0 | 0 |
| 0 | 0.000763024 | 0 | 0 | 0 |
| 0 | 0 | 0 | 0 | 0 |
| 0 | 0 | 0 | 0.138512051 | 0 |
| 0.019069626 | 0 | 0.002895531 | 0 | 0.002612904 |
| 0 | 0 | 0 | 0 | 0 |

| | | | | |
|---|---|---|---|---|
| 0.002962823 | 0 | 0 | 0 | 0 |
| 0 | 0.012373022 | 0 | 0 | 0.006570231 |
| 0 | 0 | 0 | 0 | 0 |
| 0 | 0 | 0 | 0.00451889 | 0 |
| 0 | 0 | 0 | 0 | 0 |
| 0.016665629 | 0 | 0 | 0 | 0 |
| 0 | 0 | 0 | 0.002626605 | 0 |
| 0 | 0 | 0 | 0 | 0 |
| 0 | 0 | 0 | 0.096518332 | 0 |
| 0 | 0 | 0 | 0 | 0 |
| 0.005011306 | 0.018506061 | 0.002479772 | 0.002191777 | 0.018498537 |
| 0 | 0 | 0 | 0 | 0 |
| 0 | 0 | 0.000479686 | 0 | 0 |
| 0.00247994 | 0.001626381 | 0.001561843 | 0.005694372 | 0.001409394 |
| 0 | 0 | 0 | 0 | 0 |
| 0 | 0 | 0 | 0 | 0 |
| 0 | 0 | 0 | 0.026757026 | 0 |
| 0 | 0 | 0 | 0 | 0 |
| 0 | 0 | 0 | 0.006611763 | 0 |
| 0.241714434 | 0 | 0 | 0 | 0 |
| 0 | 0.048847002 | 0 | 0 | 0 |
| 0.362460113 | 0.955716335 | 0 | 0.144272072 | 0.249219172 |
| 0 | 0 | 0 | 0 | 0 |
| 0 | 0 | 0 | 0 | 0 |
| 0 | 0 | 0 | 0 | 0 |
| 0 | 0 | 0 | 0 | 0 |
| 0 | 0 | 0 | 0 | 0.00244505 |
| 0 | 0 | 0 | 0 | 0 |
| 0 | 0 | 0 | 0 | 0 |
| 0.038994408 | 0.232135563 | 0.840872544 | 0.107369637 | 1.721701644 |
| 0.003976681 | 0.001241292 | 0 | 0.000877995 | 0.003442183 |
| 0 | 0 | 0 | 0 | 0 |
| 0 | 0 | 0 | 0 | 0 |
| 0 | 0 | 0 | 0 | 0 |
| 0.000814403 | 0 | 0 | 0 | 0 |
| 0 | 0 | 0 | 0 | 0 |
| 0 | 0 | 0 | 0 | 0.044399717 |
| 0 | 0 | 0 | 0 | 0 |
| 0 | 0 | 0 | 0 | 0 |
| 0 | 0 | 0 | 0 | 0 |
| 0.081180518 | 0.002841257 | 0 | 0.096606931 | 0.012977164 |
| 0.321357177 | 1.307039059 | 11.00433241 | 0.640165119 | 3.372786895 |
| 0 | 0 | 0 | 0 | 0 |
| 0 | 0 | 0 | 0 | 0 |
| 0.00229985 | 0 | 0 | 0 | 0 |
| 0 | 0 | 0 | 0 | 0 |
| 0.001556252 | 0.000877089 | 0.001347655 | 0.001563373 | 0 |
| 0 | 0 | 0 | 0 | 0 |

| | | | | |
|---|---|---|---|---|
| 0.011040882 | 0.001082181 | 0 | 0.003582317 | 0.002400767 |
| 0.016870169 | 0 | 0 | 0 | 0 |
| 0.007194979 | 0.008257504 | 0 | 0.107385992 | 0 |
| 0 | 0 | 0 | 0 | 0 |
| 0.12063885 | 0 | 0 | 0.632501013 | 0 |
| 0.000811746 | 0.002378961 | 0.003233529 | 0 | 0 |
| 0 | 0 | 0 | 0 | 0 |
| 0 | 0.005336783 | 0 | 0 | 0.006054239 |
| 0.003213992 | 0 | 0 | 0.008071749 | 0 |
| 0 | 0 | 0 | 0 | 0 |
| 0 | 0 | 0 | 0 | 0 |
| 0 | 0 | 0.002663968 | 0 | 0 |
| 0 | 0 | 0 | 0 | 0 |
| 0 | 0.001174826 | 0 | 0 | 0 |
| 0 | 0 | 0 | 0 | 0 |
| 0.014423702 | 0 | 0 | 0.005454947 | 0 |
| 0.108336583 | 0.004918521 | 0 | 0.157634124 | 0.173079064 |
| 0 | 0 | 0.024854416 | 0 | 0 |
| 0 | 0 | 0.005652746 | 0 | 0 |
| 0 | 0 | 0 | 0 | 0 |
| 0.006874154 | 0.00448874 | 0 | 0 | 0 |
| 0.005454202 | 0 | 0 | 0 | 0.002454979 |
| 0 | 0 | 0 | 0 | 0 |
| 0 | 0 | 0 | 0 | 0 |
| 0 | 0 | 0 | 0 | 0 |
| 0 | 0 | 0 | 0 | 0 |
| 0.006924463 | 0 | 0 | 0.006305081 | 0 |
| 0.003475457 | 0 | 0 | 0 | 0 |
| 0 | 0 | 0 | 0.001350241 | 0 |
| 0.004086197 | 0 | 0 | 0 | 0 |
| 0 | 0 | 0 | 0.012300208 | 0.002724464 |
| 0.005685215 | 0.006591364 | 0 | 0.01771647 | 0.047388146 |
| 0.018816944 | 2.451075663 | 0.036785984 | 0.001775629 | 0.021915391 |
| 0.013298556 | 0 | 0 | 0 | 0 |
| 0.016083303 | 0 | 0 | 0.004843286 | 0 |
| 0 | 0 | 0 | 0 | 0 |
| 0 | 0.002311556 | 0 | 0 | 0 |
| 0.20757928 | 0 | 0 | 0.050844183 | 0 |
| 0.07662835 | 0.006104175 | 0 | 0.009170643 | 0.723471836 |
| 0 | 0 | 0 | 0.001539641 | 0.004657538 |
| 0.007633931 | 0 | 0 | 0 | 0 |
| 0 | 0 | 0 | 0 | 0 |
| 0 | 0 | 0 | 0.018774666 | 0 |
| 0 | 0 | 0 | 0 | 0 |
| 0 | 0 | 0.632413082 | 0 | 0.593832011 |
| 0 | 0 | 0 | 0 | 0.001490337 |
| 0 | 0 | 0 | 0 | 0 |
| 0 | 0 | 0 | 0 | 0 |

| | | | | |
|---|---|---|---|---|
| 0 | 0 | 0 | 0 | 0 |
| 0.91685132 | 0.714799111 | 0.023569301 | 1.116489468 | 0.823115957 |
| 0 | 0.002574813 | 0 | 0 | 0 |
| 0 | 0 | 0 | 0.001381386 | 0 |
| 0 | 0 | 0 | 0 | 0 |
| 0 | 0 | 0 | 0 | 0 |
| 0 | 0 | 0 | 0 | 0 |
| 0 | 0 | 0 | 0 | 0 |
| 0 | 0 | 0 | 0 | 0.015916905 |
| 0 | 0 | 0 | 0 | 0.001805031 |
| 0.011441465 | 0 | 0.002045497 | 0 | 0 |
| 0.012919228 | 0.029744308 | 0 | 0.004733743 | 0.039179387 |
| 0 | 0 | 0 | 0.083967739 | 0.082058888 |
| 0 | 0 | 0 | 0 | 0 |
| 0.001058681 | 0.004964238 | 0 | 0.000729275 | 0 |
| 0 | 0 | 0 | 0 | 0 |
| 0.006956631 | 0.009583918 | 0.005488697 | 0.001996704 | 0.026335227 |
| 0.00526789 | 0.004524091 | 0.013207479 | 0 | 0.069314444 |
| 0 | 0 | 0 | 0 | 0 |
| 0.009121847 | 0 | 0 | 0 | 0.007535473 |
| 0.053207478 | 0.151665834 | 0.000966381 | 0.04324122 | 0.197582588 |
| 0 | 0.010956712 | 0 | 0 | 0 |
| 0 | 0 | 0 | 0 | 0 |
| 0 | 0 | 0 | 0 | 0 |
| 0 | 0 | 0 | 0 | 0 |
| 0 | 0 | 0 | 0 | 0 |
| 0 | 0.001596346 | 0 | 0.020098576 | 0.040086877 |
| 0.006142319 | 0.091485817 | 0.183414342 | 0.06200822 | 0.048461798 |
| 0 | 0 | 0 | 0 | 0 |
| 0 | 0 | 0 | 0 | 0 |
| 0.00113633 | 0.019263987 | 0 | 0 | 0.001704903 |
| 0 | 0 | 0 | 0 | 0 |
| 0 | 0 | 0 | 0 | 0 |
| 0 | 0 | 0 | 0 | 0 |
| 0 | 0 | 0 | 0 | 0 |
| 0.040506635 | 0.049779708 | 0.21828582 | 0.075313623 | 1.401937147 |
| 0 | 0 | 0 | 0 | 0 |
| 0 | 0 | 0 | 0 | 0 |
| 0.012802918 | 0.001884893 | 0 | 0.006899466 | 0 |
| 0 | 0 | 0 | 0 | 0 |
| 0 | 0 | 0.614492232 | 0 | 0.55717771 |
| 0 | 0 | 0 | 0 | 0 |
| 0.090622222 | 0.001144913 | 0.001319377 | 0.025691676 | 0 |
| 0.001991984 | 0.002210784 | 0 | 0 | 0 |
| 0.003019621 | 0 | 0 | 0 | 0 |
| 0 | 0 | 0 | 0.470219405 | 0 |
| 0 | 0 | 0 | 0 | 0 |
| 0.001733785 | 0 | 0.001887465 | 0.001990536 | 0 |

| | | | | |
|---|---|---|---|---|
| 0.025941052 | 0 | 0 | 0 | 0 |
| 0 | 0 | 0 | 0 | 0 |
| 0 | 0 | 0 | 0 | 0 |
| 1.451768751 | 0.009334229 | 0 | 1.570834217 | 0.046453311 |
| 0 | 0 | 0 | 0 | 0 |
| 0 | 0 | 0 | 0 | 0 |
| 0 | 0 | 0 | 0 | 0 |
| 0 | 0 | 0 | 0 | 0.009924172 |
| 3.171043493 | 1.011957749 | 0 | 3.854451063 | 0.795895833 |
| 0.013381648 | 0 | 0 | 0.093697122 | 0 |
| 0 | 0 | 0 | 0 | 0 |
| 0.001593908 | 0.002714899 | 0 | 0.002439927 | 0.014392897 |
| 0 | 0 | 0 | 0 | 0 |
| 0 | 0 | 0 | 0.106477353 | 0 |
| 0.387720347 | 0.000922493 | 0 | 0.033221655 | 0.048239116 |
| 0 | 0 | 0 | 0 | 0 |
| 0.043654773 | 0 | 0 | 0.303562025 | 0 |
| 0 | 0 | 0 | 0 | 0.047721987 |
| 0 | 0 | 0 | 0 | 0 |
| 0 | 0 | 0 | 0.018702403 | 0 |
| 0 | 0 | 0 | 0 | 0 |
| 0 | 0 | 0 | 0.000755098 | 0 |
| 0 | 0 | 0 | 0.0018723 | 0 |
| 0.550568066 | 0 | 0 | 0.494883659 | 0 |
| 0.001326494 | 0 | 0 | 0.007805016 | 0 |
| 0.290677604 | 0 | 0 | 0.13242981 | 0.19699366 |
| 0 | 0 | 0 | 0.049151895 | 0 |
| 0 | 0 | 0 | 2.412286842 | 2.266843187 |
| 0 | 0 | 0 | 0 | 0 |
| 0 | 0 | 0 | 0 | 0 |
| 0 | 0 | 0 | 0.098834361 | 0.002152669 |
| 0.424687308 | 0 | 0 | 0 | 0 |
| 0.230491876 | 0 | 0 | 0.044530361 | 0.006225689 |
| 0 | 0 | 0 | 0 | 0 |
| 0 | 0 | 0 | 0 | 0 |
| 0 | 0 | 0 | 0 | 0 |
| 0 | 1.259795769 | 0 | 3.826753251 | 2.334183699 |
| 0 | 0 | 0 | 0.001269339 | 0 |
| 0 | 0 | 0 | 0.001481646 | 0 |
| 0 | 0 | 0 | 0 | 0 |
| 1.288326385 | 0.001072425 | 0.05320007 | 0.85610129 | 0.001062109 |
| 0 | 0 | 0 | 0.23615661 | 0 |
| 0.019769754 | 4.292515829 | 0 | 0.017315538 | 5.757027321 |
| 0 | 0 | 0 | 0 | 0 |
| 0 | 0 | 0 | 0 | 0 |
| 0 | 0 | 0 | 0 | 0 |
| 0 | 0 | 0 | 0 | 0 |
| 0.026550586 | 0 | 0.001273392 | 0 | 0 |

| | | | | |
|---|---|---|---|---|
| 0 | 0 | 0 | 0 | 0 |
| 0 | 0 | 0 | 0.073270893 | 0.017215056 |
| 0.017816214 | 0.017654602 | 0.78381874 | 0.030569452 | 0.00465095 |
| 0 | 0 | 0 | 0.95730746 | 0 |
| 0 | 0 | 0 | 0 | 0 |
| 0 | 0.003894132 | 0 | 0.033643183 | 0.002892505 |
| 0 | 0 | 0 | 0 | 0 |
| 0 | 0 | 0 | 0.00179443 | 0 |
| 0 | 0 | 0 | 0 | 0 |
| 0 | 0 | 0 | 0 | 0 |
| 0.420008928 | 0 | 0.019575521 | 0.06157239 | 0 |
| 0.780649827 | 3.311274902 | 1.904131349 | 0.578950222 | 1.623148517 |
| 0 | 0 | 0 | 0 | 0 |
| 0.206625901 | 0.008792395 | 0 | 0.100272592 | 0.030607052 |
| 0.082231982 | 0.123095775 | 0.854970481 | 0.096661767 | 0.029842809 |
| 17.83334156 | 0 | 0 | 0.318691351 | 0 |
| 0 | 0 | 0 | 0 | 0 |
| 0 | 0 | 0 | 0 | 0 |
| 0.035604274 | 0 | 0 | 0.004146921 | 0 |
| 0.045602436 | 0 | 0 | 0.001090741 | 0 |
| 0.495257742 | 0.030799748 | 0.006696804 | 0.246935427 | 0.004028761 |
| 0 | 0.181402145 | 25.72017402 | 0.002533718 | 0 |
| 0.733783127 | 3.167363083 | 8.025532484 | 1.117438667 | 0.603895104 |
| 0 | 0 | 0 | 0.022606849 | 0 |
| 0 | 0 | 0 | 0 | 0 |
| 0 | 2.495597544 | 0 | 0.430665894 | 4.299034636 |
| 0.012332624 | 0 | 0 | 0 | 0 |
| 0.000840184 | 0 | 0 | 0 | 0 |
| 0.078652735 | 0 | 0 | 0.075390794 | 0 |
| 0 | 0 | 0 | 0 | 0 |
| 0 | 0 | 0 | 0.00734463 | 0 |
| 0.004410955 | 0.145660932 | 0 | 0.090805622 | 0.294958243 |
| 0 | 0.00124615 | 0.025711967 | 0.003399809 | 0.078986449 |
| 0.296577848 | 0 | 0 | 0 | 0.009976137 |
| 0.021759911 | 0 | 0.002437212 | 0.000598378 | 0 |
| 0.025779266 | 0.16401579 | 0 | 0.024047428 | 0.03066498 |
| 0.015909879 | 0 | 0 | 0.002183969 | 0.001081093 |
| 0 | 0 | 0 | 0 | 0 |
| 1.142154132 | 0.004344908 | 0 | 1.685534656 | 0.131226234 |
| 0 | 0 | 0 | 0 | 0 |
| 0 | 0 | 0 | 0.048503353 | 0.001182572 |
| 0 | 0 | 0 | 0 | 0 |
| 0 | 0 | 0 | 0.001207588 | 0 |
| 0 | 0 | 0 | 0.013176093 | 0 |
| 0.27642593 | 0.001104575 | 0.020366271 | 0.030001627 | 0.008423414 |
| 0 | 0 | 0 | 0 | 0 |
| 0 | 0 | 0 | 0 | 0 |
| 0.238386237 | 0.158557917 | 0 | 0.31525023 | 1.296597359 |

| | | | | |
|---|---|---|---|---|
| 0 | 0 | 0 | 0 | 0 |
| 0 | 0 | 0 | 0 | 0 |
| 0 | 0 | 0 | 0 | 0 |
| 0 | 0 | 0 | 0.000998838 | 0 |
| 0.001411617 | 0 | 0 | 0 | 0 |
| 0 | 0 | 0 | 0.479546427 | 0.01882984 |
| 0 | 0 | 0 | 0 | 0 |
| 0.386830524 | 0.001155042 | 0.037491207 | 0.031801282 | 0 |
| 0 | 0 | 0 | 0 | 0 |
| 0 | 0 | 0 | 0 | 0 |
| 0.004650972 | 0 | 0.003624809 | 0.020246437 | 0 |
| 0 | 0 | 0 | 0 | 0 |
| 0 | 0 | 0 | 0 | 0 |
| 0 | 0 | 0 | 0 | 0 |
| 0 | 0 | 0 | 0 | 0 |
| 0 | 0 | 0 | 0 | 0 |
| 0.160703957 | 0 | 0 | 0.074628201 | 0 |
| 0.001451936 | 0 | 0 | 0.001852164 | 0.003025591 |
| 0.001188305 | 0 | 0 | 0.428188322 | 0 |
| 0.033753561 | 0 | 0 | 0.058128028 | 0 |
| 0 | 0 | 0 | 0.006417459 | 0 |
| 0 | 0 | 0 | 0.208526448 | 0 |
| 0.000557993 | 0 | 0 | 0.0098229 | 0.037673525 |
| 0 | 0 | 0 | 0 | 0 |
| 0 | 0 | 0 | 0 | 0 |
| 0 | 0 | 0 | 0 | 0 |
| 0 | 0 | 0 | 0 | 0 |
| 0 | 0 | 0 | 0 | 0 |
| 0 | 0 | 0 | 0 | 0 |
| 0.104261106 | 0 | 0 | 0.398704348 | 0.053848659 |
| 0 | 0 | 0 | 0.001401875 | 0.02156438 |
| 0 | 0 | 0 | 0 | 0 |
| 0 | 0 | 0 | 0 | 0 |
| 0 | 0 | 0 | 0 | 0 |
| 0 | 0 | 0 | 0 | 0.003467918 |
| 0 | 0 | 0 | 0 | 0 |
| 0.072857646 | 0.347641784 | 0 | 0 | 0 |
| 0.009350682 | 0.010650864 | 0.007614338 | 0 | 0 |
| 0 | 0 | 0 | 0 | 0 |
| 0.004368787 | 0 | 0 | 0 | 0 |
| 0 | 0 | 0 | 0 | 0.008275007 |
| 0 | 0 | 0 | 0 | 0 |
| 0 | 0 | 0 | 0 | 0 |
| 0.001593091 | 0 | 0 | 0.007316028 | 0 |
| 0 | 0 | 0 | 0.091103682 | 0.026084571 |
| 0 | 0 | 0 | 0.011944409 | 0 |
| 0.001714594 | 0 | 0 | 0 | 0 |
| 0 | 0 | 0 | 0 | 0 |

| | | | | |
|---|---|---|---|---|
| 0 | 0 | 0 | 0 | 0 |
| 0 | 0 | 0 | 0 | 0 |
| 0 | 0 | 0 | 0.21859675 | 0 |
| 0 | 0 | 0 | 1.026641557 | 0.068551491 |
| 0 | 0 | 0 | 0 | 0 |
| 0 | 0.288414864 | 0 | 0 | 1.036697332 |
| 0 | 0 | 0 | 0 | 0 |
| 0 | 0 | 0 | 0 | 0 |
| 0.068100732 | 0.007548678 | 1.181647531 | 0.022078977 | 0.026307665 |
| 0 | 0 | 0 | 0 | 0 |
| 0 | 0 | 0 | 0.006500593 | 0 |
| 0 | 0 | 0 | 0 | 0 |
| 0 | 0 | 0 | 0.003030696 | 0 |
| 0 | 0 | 0 | 0 | 0 |
| 0.073164593 | 0 | 0.024554657 | 0.565221985 | 0.634854943 |
| 0.006103154 | 0 | 0 | 0 | 0 |
| 0 | 0 | 0 | 0 | 0 |
| 0 | 0 | 0 | 0 | 0 |
| 0 | 0 | 0 | 0.003942079 | 0 |
| 0 | 0 | 0 | 0 | 0 |
| 1.189369884 | 0 | 0 | 0.008937817 | 0 |
| 0 | 0 | 0 | 0 | 0 |
| 0 | 0 | 0 | 0 | 0 |
| 0 | 0 | 0 | 0 | 0 |
| 0.023945538 | 1.138179272 | 0 | 0.299352503 | 0.015634861 |
| 0.668312939 | 2.047646623 | 0 | 5.846021053 | 0.987346038 |
| 2.38333174 | 0 | 0 | 0.113613674 | 0.038355853 |
| 0 | 0 | 0 | 0 | 0 |
| 1.018383498 | 0 | 0 | 0.022433116 | 0.014251013 |
| 0 | 0 | 0 | 0 | 0 |
| 0 | 0 | 0 | 0 | 0 |
| 0 | 0 | 0 | 0 | 0 |
| 0 | 0 | 0 | 0 | 0 |
| 0.035097204 | 0 | 0.000689069 | 0 | 0 |
| 0.006555526 | 0.025123514 | 0 | 0.020159767 | 0.007610916 |
| 0.0244938 | 0 | 0.001146525 | 0.009500341 | 0 |
| 0 | 0 | 0 | 0 | 0.128977371 |
| 0.413557875 | 0 | 0 | 1.976225 | 4.735886388 |
| 0 | 5.573928166 | 0 | 3.05025768 | 0 |
| 0 | 0 | 0.003832548 | 0 | 0 |
| 0 | 0 | 0 | 0 | 0 |
| 0 | 0 | 0 | 0.001723754 | 0 |
| 6.93962306 | 0.1784439 | 0 | 0.180714799 | 0.093312057 |
| 0 | 0 | 0 | 0 | 0 |
| 0 | 0 | 0 | 0 | 0 |
| 0.000940735 | 0 | 0 | 0.078123302 | 0.001176201 |
| 0 | 0 | 0 | 0 | 0 |
| 0.596234371 | 0.003603865 | 0.165428908 | 3.00882962 | 0 |

| | | | | |
|---|---|---|---|---|
| 0 | 0 | 0 | 0.081262035 | 0 |
| 0 | 0 | 0 | 1.035813501 | 0 |
| 0.004239699 | 0 | 0 | 0 | 0 |
| 0 | 0 | 0 | 0 | 0 |
| 0.040613021 | 0 | 0 | 0.00085164 | 0 |
| 0.004107415 | 0 | 0 | 0.037520322 | 0 |
| 0 | 0 | 0 | 0.210174138 | 0 |
| 0 | 0 | 0 | 0 | 0 |
| 0 | 0 | 0 | 0 | 0 |
| 0 | 0 | 0 | 0.005128794 | 0 |
| 0.001418999 | 0 | 0 | 0 | 0 |
| 0.416939573 | 6.200088383 | 0.191534877 | 0.820851667 | 0.1289879 |
| 0.04984888 | 0.322553759 | 0 | 0.008765084 | 0.195400533 |
| 0 | 0 | 0 | 0 | 0 |
| 0.003002582 | 0.155104737 | 0 | 0.114989557 | 0.160354832 |
| 0 | 0 | 0 | 0 | 0 |
| 0.004625043 | 0 | 0 | 0 | 0 |
| 0 | 0 | 0 | 0 | 0 |
| 0.001315876 | 0 | 0 | 0.020934539 | 0 |
| 0 | 0 | 0 | 0.0289565 | 0 |
| 0.065832379 | 0 | 0 | 0.005896412 | 0 |
| 0 | 0 | 0 | 0 | 0 |
| 1.271468029 | 0.012526893 | 0 | 0.502394345 | 0.202028299 |
| 0 | 0 | 0 | 0 | 0 |
| 0 | 0 | 0 | 0 | 0 |
| 0 | 0 | 0 | 0 | 0 |
| 0 | 0 | 0 | 0 | 0 |
| 0 | 0 | 0 | 0 | 0 |
| 0 | 0 | 0 | 0 | 0 |
| 0 | 0 | 0 | 0 | 0 |
| 0 | 0 | 0 | 0 | 0 |
| 0.069019633 | 0.050944922 | 0.002699218 | 0.021727242 | 0.187900873 |
| 0 | 0 | 0 | 0 | 0 |
| 0.042962237 | 0 | 0 | 0.041502713 | 0 |
| 0.019271128 | 0 | 0.000623853 | 0.009925201 | 0 |
| 0.119106236 | 0.007955814 | 0 | 0.03758469 | 0.040350229 |
| 0 | 0 | 0 | 0 | 0 |
| 0 | 0 | 0 | 0 | 0 |
| 0.002689868 | 0 | 0 | 0 | 0 |
| 0 | 0 | 0 | 0 | 0 |
| 0.021482898 | 0 | 0 | 0.45452655 | 0 |
| 0 | 0 | 0 | 0 | 0 |
| 0 | 0 | 0 | 0 | 0 |
| 0 | 0 | 0 | 0 | 0 |
| 0.097758693 | 0 | 0.000737469 | 0.018128777 | 0.000665486 |
| 0 | 0 | 0 | 0 | 0 |
| 0 | 0.195588877 | 0 | 0.231196532 | 0.103026226 |
| 0.022318469 | 0 | 0 | 0.001926582 | 0 |

| | | | | |
|---|---|---|---|---|
| 0 | 0 | 0 | 0 | 0 |
| 0 | 0 | 0 | 0 | 0 |
| 0.109364327 | 0 | 0 | 0 | 0 |
| 0 | 0 | 0 | 0 | 0 |
| 0 | 0 | 0 | 0 | 0 |
| 0 | 0 | 0 | 0 | 0 |
| 0 | 0 | 0 | 0.299617298 | 0 |
| 0.256824589 | 0 | 0 | 0.188413971 | 0 |
| 0 | 0 | 0 | 0 | 0 |
| 0.001875828 | 0 | 0 | 0.01096385 | 0.004690694 |
| 0 | 0 | 0 | 0 | 0 |
| 0 | 0 | 0 | 0 | 0 |
| 0 | 0 | 0 | 0 | 0 |
| 0 | 0 | 0 | 0 | 0 |
| 0 | 0 | 0 | 0 | 0 |
| 0 | 0 | 0 | 0 | 0 |
| 0 | 0 | 0 | 0 | 0 |
| 0 | 0 | 0 | 0 | 0 |
| 0.396851276 | 28.1442495 | 0 | 1.530513058 | 0 |
| 0 | 0 | 0 | 0 | 0 |
| 0 | 0 | 0 | 0 | 0 |
| 0 | 0 | 0 | 0 | 0 |
| 0 | 0 | 0 | 0 | 0 |
| 0 | 0 | 0 | 0 | 0 |
| 0 | 0 | 0.001342915 | 0 | 0 |
| 0.015195803 | 0.015490059 | 0.004233762 | 0 | 0.001858628 |
| 0 | 0 | 0 | 0 | 0 |
| 1.417201975 | 0.008601342 | 0 | 0.255215887 | 0.003234181 |
| 0 | 0 | 0 | 0 | 0 |
| 0 | 0.155491772 | 0 | 0.270696486 | 0.272785304 |
| 0 | 0 | 0 | 0 | 0 |
| 0 | 0 | 0 | 0 | 0 |
| 0 | 0 | 0 | 1.460711704 | 0 |
| 0 | 0 | 0 | 0 | 0 |
| 0.013674094 | 0 | 0 | 0.055443471 | 0.001731313 |
| 0 | 0 | 0 | 0 | 0 |
| 0 | 0 | 0 | 0 | 0.005698858 |
| 0.004517343 | 0 | 0.000680321 | 0 | 0 |
| 0 | 0 | 0 | 0 | 0 |
| 0 | 0 | 0 | 0 | 0 |
| 0.460525467 | 1.820333668 | 1.350907718 | 0.881956185 | 33.38806904 |
| 0.70418668 | 0.057562002 | 0 | 0.35488056 | 0.357714193 |
| 1.742640262 | 0 | 0 | 0 | 0 |
| 0 | 0 | 0 | 0.317734718 | 0 |
| 0 | 0 | 0 | 0 | 0 |
| 0 | 0 | 0 | 0 | 0 |
| 0 | 0 | 0 | 0 | 0 |
| 0.020887766 | 0 | 0 | 0.000894812 | 0 |

| | | | | |
|---|---|---|---|---|
| 0 | 0 | 0 | 0 | 0 |
| 0.109668637 | 0 | 0 | 0.023746017 | 0.004009317 |
| 0 | 0 | 0 | 0.002149923 | 0 |
| 0 | 0 | 0 | 0 | 0 |
| 4.571403477 | 2.051976689 | 0 | 1.235580173 | 0.910128832 |
| 0 | 0 | 0 | 0 | 0 |
| 0 | 0 | 0 | 0 | 0 |
| 0.00084998 | 0 | 0 | 0.00058551 | 0 |
| 0.008164992 | 0.021943862 | 0.629419749 | 0 | 0.09207834 |
| 0 | 0 | 0 | 0 | 0 |
| 0 | 0 | 0.002509826 | 0 | 0 |
| 0 | 0 | 0 | 0 | 0 |
| 0.009191732 | 0.019667867 | 0 | 0.083528934 | 0.159140772 |
| 0 | 0.007253893 | 0.002122985 | 0.002198936 | 0.108719604 |
| 0 | 0 | 0 | 0 | 0.245026708 |
| 0.086442698 | 0.043935059 | 0.005661844 | 0.298092049 | 0.183734752 |
| 0.046738663 | 0.001195642 | 0 | 0.223964077 | 0 |
| 0 | 0 | 0 | 0 | 0 |
| 0 | 0 | 0 | 0 | 0 |
| 0 | 0 | 0 | 0 | 0 |
| 0 | 0 | 0 | 0.006497105 | 0 |
| 0 | 0 | 0 | 0 | 0 |
| 0 | 0 | 0 | 0 | 0 |
| 0 | 0 | 0 | 0 | 0 |
| 0 | 0 | 0 | 0.032534018 | 0 |
| 0 | 0 | 0 | 0 | 0 |
| 0.019167653 | 0.138404687 | 0 | 0.098448473 | 0.017658648 |
| 0.001951448 | 0 | 0.001126587 | 0.001867027 | 0 |
| 0 | 0 | 0 | 0 | 0 |
| 0 | 0 | 0 | 0 | 0 |
| 0 | 0 | 0 | 0 | 0 |
| 0 | 0 | 0 | 0 | 0 |
| 0 | 0 | 0 | 0 | 1.2593345 |
| 0 | 0 | 0 | 0.023289994 | 0 |
| 0.962334438 | 0 | 0 | 0.733784579 | 0 |
| 0 | 0 | 0 | 0.433464312 | 0.075500591 |
| 0 | 0 | 0 | 0 | 0 |
| 0.065015174 | 0 | 0 | 0 | 0 |
| 0 | 0 | 0 | 0 | 0 |
| 0 | 0 | 0 | 0 | 0 |
| 0 | 0.012050027 | 0 | 0.316624842 | 0.704030352 |
| 0.204492698 | 0 | 0 | 0 | 0 |
| 0 | 0 | 0 | 0.018365539 | 0 |
| 0 | 0 | 0 | 0 | 0 |
| 0 | 0 | 0 | 0 | 0 |
| 0 | 0 | 0 | 0 | 0 |
| 0 | 0 | 0 | 0 | 0 |
| 0 | 0 | 0 | 0 | 0 |

| | | | | |
|---|---|---|---|---|
| 1.745823676 | 0 | 0 | 0.231405636 | 0.051425537 |
| 0 | 0 | 0 | 0 | 0 |
| 0 | 0 | 0 | 0.000626507 | 0 |
| 0.000921087 | 0 | 0 | 0 | 0 |
| 0.032960614 | 0 | 0 | 0.082765952 | 0 |
| 0 | 0 | 0 | 0 | 0 |
| 0.168478332 | 0.002683652 | 0 | 0.571361653 | 0.014883889 |
| 0 | 0 | 0 | 0 | 0 |
| 0 | 0 | 0 | 0 | 0 |
| 0.072335049 | 0 | 0 | 0 | 0 |
| 0 | 0 | 0 | 0.020256652 | 0 |
| 0 | 0 | 0 | 0.013778181 | 0 |
| 0 | 0 | 0 | 0 | 0 |
| 0 | 0 | 0 | 0 | 0 |
| 0 | 0 | 0 | 0 | 0 |
| 0 | 0 | 0 | 0 | 0 |
| 0 | 0 | 39.13982229 | 0 | 0 |
| 0 | 0 | 0 | 0 | 0 |
| 0 | 0 | 0 | 0 | 0 |
| 0 | 0 | 0 | 0 | 0 |
| 0.034787604 | 0.000851662 | 0.004689101 | 0.273429831 | 0.000984048 |
| 0 | 0 | 0 | 0 | 0 |
| 0.090283778 | 0 | 0 | 0.009966694 | 0.008031972 |
| 0.012505351 | 0 | 0.00160432 | 0.006026491 | 0.00154424 |
| 0 | 0 | 0 | 0 | 0 |
| 0 | 0 | 0 | 0.003136723 | 0 |
| 0 | 0 | 0 | 0 | 0 |
| 0 | 0 | 0 | 0 | 0 |
| 0 | 0 | 0 | 0.210347943 | 0 |
| 0 | 0.031657995 | 0 | 0.003826559 | 0.024447765 |
| 0 | 0 | 0 | 0 | 0 |
| 0 | 0 | 0 | 0 | 0 |
| 0 | 0 | 0 | 0 | 0 |
| 0 | 0.013826889 | 0 | 0 | 0 |
| 0 | 0 | 0 | 0 | 0 |
| 0 | 0 | 0 | 0 | 0 |
| 0.001545426 | 0.002612963 | 0.001204452 | 0.005766419 | 0.086226425 |
| 0 | 0 | 0 | 0 | 0 |
| 0 | 0 | 0 | 0 | 0 |
| 0 | 0 | 0 | 0 | 0 |
| 0 | 0 | 0 | 0 | 0 |
| 0 | 0 | 0 | 0 | 0 |
| 0 | 0 | 0 | 0 | 0 |
| 0 | 0 | 0 | 0.018835678 | 0 |
| 0 | 0 | 0 | 0 | 0 |
| 0 | 0 | 0 | 0 | 0 |
| 0 | 0 | 0 | 0 | 0 |
| 0 | 0 | 0 | 0 | 0 |

| | | | | |
|---|---|---|---|---|
| 0.001329008 | 0 | 0 | 0.017928339 | 0 |
| 0 | 0 | 0 | 0 | 0 |
| 0 | 0 | 0 | 0.005213182 | 0 |
| 0 | 0 | 0 | 0 | 0 |
| 0 | 0 | 0 | 0 | 0 |
| 0 | 0 | 0 | 0 | 0.033022973 |
| 0 | 0 | 0 | 0 | 0 |
| 0.201618238 | 0.019995837 | 0 | 0.303346145 | 1.440414436 |
| 0.901446043 | 1.588832719 | 0.00198711 | 3.478135158 | 0.286904328 |
| 0.066804156 | 0 | 0.006916833 | 0 | 0.027463451 |
| 0 | 0 | 0 | 0.485720225 | 0 |
| 0 | 0 | 0 | 0 | 0 |
| 0 | 0 | 0 | 0 | 0 |
| 0 | 0.52205799 | 0 | 0.000886529 | 0 |
| 0 | 0 | 0 | 0 | 0 |
| 0 | 0 | 0 | 0 | 0 |
| 0 | 0 | 0.051238811 | 0 | 0 |
| 0 | 0 | 0 | 0 | 0 |
| 0 | 0 | 0 | 0 | 0 |
| 0 | 0 | 0 | 0.029552783 | 0.001505676 |
| 0 | 0 | 0 | 0 | 0 |
| 0 | 0 | 0 | 0 | 0 |
| 0 | 0 | 0 | 0 | 0 |
| 0 | 0 | 0 | 0 | 0 |
| 0 | 0 | 0 | 0.022082922 | 0 |
| 0 | 0 | 0 | 0.070615542 | 0 |
| 0 | 0 | 0 | 0 | 0 |
| 0 | 0 | 0 | 0 | 0 |
| 0 | 0 | 0 | 0 | 0 |
| 0 | 0 | 0 | 0 | 0 |
| 0 | 0 | 0 | 0 | 0 |
| 0 | 0 | 0 | 0 | 0 |
| 0 | 0 | 0 | 0 | 0 |
| 0 | 0 | 0 | 0.001668215 | 0 |
| 0 | 0 | 0 | 0 | 0 |
| 0 | 0 | 0 | 0 | 0 |
| 0 | 0 | 0 | 0 | 0 |
| 0 | 0 | 0 | 0 | 0 |
| 0 | 0.000890639 | 0 | 0 | 0 |
| 0 | 0 | 0.052506056 | 0 | 0 |
| 0 | 0 | 0 | 0 | 0 |
| 0.138077133 | 0 | 0 | 0.281101126 | 0.190393582 |
| 0 | 0 | 0 | 0 | 0 |
| 0 | 0 | 0 | 0 | 0 |
| 0 | 0 | 0 | 0 | 0 |
| 0 | 0 | 0 | 0 | 0 |
| 0 | 0 | 0 | 0 | 0 |
| 0 | 0 | 0 | 0 | 0 |

| | | | | |
|---|---|---|---|---|
| 0 | 0 | 0 | 0 | 0 |
| 0.072868865 | 0.00067602 | 0 | 0.078495598 | 0.05005311 |
| 0 | 0 | 0 | 0 | 0 |
| 0 | 0 | 0 | 0 | 0 |
| 0.819239976 | 0.163325585 | 0 | 0.20625863 | 0.121353315 |
| 0 | 0 | 0 | 0 | 0 |
| 0 | 0 | 0 | 0 | 0 |
| 0.14306651 | 0 | 0.135126316 | 0.012858784 | 0.074773864 |
| 0.007806979 | 0 | 0 | 0.001211228 | 0 |
| 0 | 0 | 0 | 0 | 0 |
| 0 | 0 | 0 | 0 | 0 |
| 0 | 0 | 0 | 0 | 0 |
| 0 | 0 | 0 | 0 | 0 |
| 0 | 0 | 0 | 0 | 0 |
| 0 | 0 | 0 | 0 | 0 |
| 0.061398778 | 0 | 0.006983394 | 0.023988025 | 0 |
| 0 | 0 | 0 | 0 | 0 |
| 0 | 0 | 0 | 0 | 0 |
| 0 | 0 | 0 | 0 | 0 |
| 0 | 0 | 0 | 0 | 0 |
| 0 | 0 | 0 | 0 | 0 |
| 0 | 0 | 0 | 0 | 0 |
| 0 | 0 | 0 | 0 | 0 |
| 0.004754977 | 0 | 0.000633482 | 0.003149496 | 0 |
| 0.254486657 | 0.884181235 | 0 | 0 | 0.040078233 |
| 0 | 0 | 0 | 0 | 0 |
| 0.02575926 | 0 | 0.006141542 | 0 | 0.013384641 |
| 0 | 0 | 0 | 0 | 0 |
| 0 | 0 | 0 | 0 | 0 |
| 0 | 0 | 0 | 0 | 0 |
| 0 | 0 | 0 | 0 | 0 |
| 0 | 0 | 0 | 0.035254393 | 0 |
| 0 | 0 | 0 | 0 | 0 |
| 0 | 0 | 0 | 0 | 0 |
| 0 | 0 | 0 | 0 | 0 |
| 0 | 0 | 0 | 0 | 0 |
| 0 | 0 | 0 | 0 | 0 |
| 0.561975059 | 0 | 0 | 0.19970357 | 0 |
| 0 | 0 | 0 | 0 | 0 |
| 0 | 0 | 0 | 0 | 0 |
| 0 | 0 | 0 | 0 | 0 |
| 0 | 0 | 0 | 0 | 0.088043571 |
| 0 | 0 | 0 | 0 | 0 |
| 0 | 0 | 0 | 0.002584574 | 0.028146759 |
| 0 | 0 | 0 | 0 | 0 |
| 0 | 0 | 0 | 0 | 0 |
| 0 | 0 | 0 | 0 | 0 |
| 0 | 0 | 0 | 0.004470423 | 0.035772298 |

| | | | | |
|---|---|---|---|---|
| 0.025797375 | 0.016879615 | 0 | 0.030340007 | 0.036384588 |
| 0 | 0 | 0 | 0.025798168 | 0 |
| 0 | 0 | 0 | 0.002146676 | 0 |
| 0 | 0 | 0 | 0 | 0 |
| 0 | 0 | 0 | 0 | 0 |
| 2.336711903 | 0 | 0 | 0.12846727 | 0 |
| 0 | 0 | 0 | 0 | 0 |
| 0 | 0 | 0 | 0 | 0 |
| 0 | 0 | 0 | 0 | 0 |
| 0 | 0 | 0 | 0 | 0 |
| 0 | 0 | 0 | 0 | 0 |
| 0.005057689 | 0 | 0 | 0 | 0 |
| 0 | 0 | 0 | 0 | 0 |
| 0 | 0 | 0 | 0 | 0 |
| 0 | 0 | 0 | 0 | 0 |
| 0 | 0 | 0 | 0 | 0 |
| 0 | 0 | 0 | 0 | 0 |
| 0 | 0 | 0 | 0 | 0 |
| 0 | 0 | 0 | 0.003368476 | 0 |
| 0 | 0 | 0 | 0 | 0 |
| 0.050993381 | 0 | 0 | 0 | 0.022770358 |
| 0 | 0 | 0 | 0 | 0 |
| 0.13271191 | 0 | 0 | 0.001056255 | 0.001437865 |
| 0 | 0 | 0 | 0.026827026 | 0 |
| 0 | 0 | 0 | 0 | 0 |
| 0 | 0 | 0 | 0 | 0.10050443 |
| 0 | 0 | 0 | 0 | 0 |
| 0 | 0 | 0 | 0 | 0 |
| 0 | 0.000897732 | 0 | 0.156587866 | 0 |
| 0 | 0 | 0 | 0 | 0 |
| 0 | 0 | 0 | 0 | 0 |
| 0 | 0 | 0 | 0 | 0 |
| 0.008185092 | 0.003496201 | 0.010147003 | 0 | 0.021544877 |
| 0.172129677 | 0 | 0 | 0 | 0 |
| 0.046626558 | 0.313080976 | 0 | 0.068794403 | 1.238694295 |
| 0.003284097 | 0.019171289 | 0 | 0.001190661 | 0 |
| 0 | 0 | 0 | 0 | 0 |
| 0 | 0 | 0 | 0 | 0.021893236 |
| 0.05437963 | 0.027222512 | 0 | 0 | 0.060241872 |
| 0 | 0 | 0 | 0 | 0 |
| 0 | 0 | 0 | 0 | 0 |
| 0 | 0 | 0 | 0.013349892 | 0 |
| 0.309514607 | 0.015599432 | 0 | 0.59878024 | 0.104638201 |
| 0 | 0 | 0 | 0 | 0 |
| 0 | 0 | 0 | 0 | 0 |
| 0.047899196 | 0 | 0 | 0.112968956 | 0.003946131 |
| 0 | 0 | 0 | 0 | 0 |
| 0 | 0 | 0 | 0 | 0 |

| | | | | |
|---|---|---|---|---|
| 0 | 0 | 0 | 0 | 0 |
| 0 | 0 | 0 | 0 | 0 |
| 0 | 0 | 0 | 0 | 0 |
| 0 | 0 | 0 | 0 | 0 |
| 0 | 0 | 0 | 0.003798896 | 0 |
| 0 | 0 | 0 | 0 | 0 |
| 0 | 0 | 0 | 0 | 0 |
| 0 | 0 | 0 | 0 | 0 |
| 0.004870384 | 0 | 0 | 0.029156321 | 0 |
| 0.06253006 | 0 | 0 | 0.070692231 | 0 |
| 0 | 0 | 0 | 0 | 0 |
| 0.106700552 | 0 | 0 | 0 | 0 |
| 0 | 0 | 0 | 0 | 0 |
| 0 | 0 | 0 | 0.031156227 | 0 |
| 0 | 0 | 0 | 0 | 0 |
| 0 | 0 | 0.900776844 | 0 | 0 |
| 0 | 0 | 0 | 0 | 0 |
| 0 | 0 | 0 | 0 | 0 |
| 0 | 0 | 0 | 0 | 0 |
| 0.157519028 | 0 | 0 | 0.982533696 | 1.443023108 |
| 0 | 0 | 0 | 0.003835217 | 0.022603112 |
| 0 | 0 | 0 | 0 | 0 |
| 0 | 0 | 0 | 0 | 0 |
| 0 | 0 | 0 | 0 | 0 |
| 0.001779959 | 0 | 0 | 0 | 0 |
| 0 | 0 | 0 | 0 | 0 |
| 0 | 0 | 0 | 0 | 0 |
| 0 | 0 | 0 | 0.040443607 | 0 |
| 0 | 0 | 0 | 0 | 0 |
| 0.001754959 | 0 | 0 | 0.018357483 | 0 |
| 0.100313626 | 0.199943823 | 0 | 0.488062692 | 0 |
| 0 | 0 | 0 | 0 | 0 |
| 0 | 0 | 0 | 0.010008775 | 0 |
| 0 | 0 | 0 | 0 | 0 |
| 0 | 0 | 0 | 0 | 0 |
| 0 | 0 | 0 | 0 | 0 |
| 0 | 0 | 0 | 0 | 0 |
| 0 | 0 | 0 | 0 | 0 |
| 0 | 0 | 0 | 0 | 0 |
| 0 | 0 | 0 | 0 | 0 |
| 0 | 0 | 0 | 0 | 0 |
| 0 | 0 | 0 | 0 | 0 |
| 0 | 0 | 0 | 0.031521055 | 0 |
| 0 | 0 | 0 | 0.001615063 | 0 |
| 0 | 0 | 0 | 0 | 0 |
| 0 | 0 | 0 | 0 | 0 |
| 0.196644929 | 0 | 0.31729102 | 0 | 0 |
| 0 | 0 | 0 | 0 | 0 |

| | | | | |
|---|---|---|---|---|
| 0 | 0 | 0 | 0 | 0 |
| 0 | 0 | 0 | 0 | 0 |
| 0 | 0 | 0 | 0 | 0 |
| 0 | 0 | 0 | 0 | 0.006906192 |
| 0.027050723 | 0.000389248 | 0 | 0.085901174 | 0.042096982 |
| 0 | 0 | 0 | 0.021623082 | 0 |
| 0 | 0 | 0 | 0 | 0 |
| 0 | 0 | 0 | 0 | 0 |
| 0 | 0 | 0 | 0 | 0 |
| 0 | 0 | 0 | 0 | 0 |
| 0 | 0 | 0 | 0.022022667 | 0 |
| 0.027367931 | 0 | 0.000553567 | 0.001834789 | 0 |
| 0 | 0 | 0 | 0 | 0 |
| 0 | 0 | 0 | 0 | 0 |
| 0 | 0.0012355 | 0 | 0 | 0 |
| 0.290186927 | 0 | 0 | 0 | 0 |
| 0.010601415 | 0 | 0 | 0.062797085 | 0.004994617 |
| 0.001045897 | 0 | 0 | 0 | 0 |
| 0.037513447 | 0 | 0 | 0.002431297 | 0.009645384 |
| 1.509922374 | 0.001780988 | 0 | 0.223124343 | 0 |
| 0 | 0 | 0 | 0 | 0 |
| 0.019048489 | 0 | 0 | 0 | 0 |
| 0 | 0 | 0 | 0 | 0 |
| 0 | 0 | 0 | 0 | 0 |
| 0 | 0 | 0 | 0 | 0 |
| 0 | 0 | 0 | 0 | 0 |
| 0.024056957 | 0.000856312 | 0 | 0.008176818 | 0.00138519 |
| 0 | 0 | 0 | 0 | 0 |
| 0 | 0 | 0 | 0 | 0 |
| 0 | 0 | 0 | 0 | 0 |
| 0.004664575 | 0 | 0 | 0 | 0 |
| 0 | 0 | 0 | 0 | 0 |
| 0 | 0 | 0 | 0 | 0 |
| 0 | 0 | 0 | 0 | 0 |
| 0 | 0 | 0 | 0 | 0 |
| 0 | 0 | 0 | 0 | 0 |
| 0 | 0 | 0 | 0 | 0 |
| 0.06440949 | 0.001136614 | 0.00104785 | 0.003038946 | 0.000630381 |
| 0.508946274 | 0 | 0 | 0.059179902 | 0.29456808 |
| 0.005198481 | 0 | 0 | 0 | 0 |
| 0.14106048 | 0 | 0 | 0.005200882 | 0 |
| 0 | 0 | 0 | 0 | 0 |
| 0 | 0 | 0 | 0 | 0 |
| 0 | 0 | 0 | 0 | 0 |
| 0 | 0 | 0 | 0 | 0 |
| 0 | 0 | 0 | 0.003453193 | 0 |
| 0 | 0 | 0 | 0 | 0 |
| 0 | 0 | 0 | 0.006027062 | 0 |

| | | | | |
|---|---|---|---|---|
| 0 | 0 | 0 | 0 | 0 |
| 0.01182175 | 0 | 0 | 0.009170537 | 0.001398177 |
| 0 | 0 | 0 | 0 | 0 |
| 0.000783531 | 0 | 0.002171226 | 0.004497807 | 0 |
| 0 | 0.001474446 | 0 | 0 | 0 |
| 0 | 0.001057175 | 0 | 0 | 0 |
| 0.001330625 | 0 | 0 | 0.020514468 | 0.0083184 |
| 0.082573314 | 0 | 0 | 0.179753176 | 0 |
| 0 | 0 | 0 | 0.058462937 | 0 |
| 0 | 0 | 0 | 0 | 0 |
| 0.059726687 | 0 | 0 | 0.043112655 | 0 |
| 0 | 0 | 0 | 0 | 0 |
| 0.095669907 | 0 | 0 | 0.102823071 | 0.039582667 |
| 0 | 0 | 0 | 0.001350215 | 0 |
| 0 | 0 | 0 | 0 | 0 |
| 0 | 0 | 0 | 0 | 0 |
| 0 | 0 | 0 | 0 | 0 |
| 0 | 0 | 0 | 0 | 0 |
| 0 | 0 | 0 | 0 | 0 |
| 0 | 0 | 0 | 0 | 0 |
| 0 | 0 | 0 | 0 | 0 |
| 0 | 0 | 0 | 0 | 0 |
| 0.036567553 | 0 | 0 | 0 | 0.004343438 |
| 0.001941794 | 0 | 0 | 0.291796845 | 0.088750446 |
| 0 | 0 | 0 | 0 | 0 |
| 0 | 0 | 0 | 0.008049041 | 0 |
| 0 | 0 | 0 | 0 | 0 |
| 0 | 0 | 0 | 0 | 0 |
| 0 | 0 | 0 | 0 | 0 |
| 0 | 0 | 0 | 0 | 0 |
| 0 | 0.005331151 | 0 | 0.003510791 | 0.006372255 |
| 0 | 0 | 0 | 0 | 0 |
| 0 | 0 | 0 | 0 | 0 |
| 0 | 0 | 0 | 0 | 0 |
| 0 | 0 | 0 | 0 | 0 |
| 0 | 0 | 0.010039408 | 0 | 0 |
| 0.000866359 | 0 | 0 | 0 | 0 |
| 0 | 0 | 0 | 0 | 0 |
| 0 | 0 | 0 | 0.004158522 | 0.000862619 |
| 0 | 0 | 0 | 0 | 0 |
| 0 | 0 | 0 | 0 | 0 |
| 0 | 0 | 0 | 0 | 0 |
| 0 | 0 | 0 | 0 | 0 |
| 0 | 0 | 0 | 0 | 0 |
| 0 | 0 | 0 | 0 | 0 |
| 0.202746145 | 0.310775791 | 0 | 0.216218402 | 0.087090925 |
| 0 | 0 | 0 | 0 | 0 |
| 0 | 0 | 0 | 0 | 0 |

| | | | | |
|---|---|---|---|---|
| 0 | 0 | 0 | 0 | 0 |
| 0 | 0 | 0 | 0 | 0 |
| 0.293572383 | 0 | 0 | 0.157621995 | 0.134287835 |
| 0 | 0 | 0 | 0.013941609 | 0 |
| 0 | 0 | 0 | 0 | 0 |
| 0 | 0 | 0 | 0.00689739 | 0 |
| 0.001227279 | 0 | 0 | 0 | 0 |
| 0 | 0 | 0 | 0 | 0 |
| 0 | 0 | 0 | 0 | 0 |
| 0.001181084 | 0 | 0 | 0 | 0 |
| 0.003498238 | 0 | 0 | 0.000803256 | 0 |
| 0 | 0 | 0 | 0.000959411 | 0 |
| 0 | 0 | 0 | 0 | 0 |
| 0 | 0 | 0 | 0 | 0 |
| 0 | 0 | 0 | 0 | 0 |
| 0 | 0 | 0 | 0 | 0 |
| 0 | 0 | 0 | 0 | 0 |
| 0 | 0 | 0 | 0 | 0 |
| 0 | 0 | 0 | 0 | 0 |
| 0.001298994 | 0 | 0 | 0 | 0.008584695 |
| 0 | 0 | 0 | 0 | 0 |
| 0 | 0 | 0 | 0 | 0 |
| 0 | 0 | 0 | 0 | 0 |
| 0 | 0 | 0 | 0 | 0 |
| 0 | 0 | 0 | 0 | 0 |
| 0 | 0 | 0 | 0 | 0 |
| 0 | 0 | 0 | 0 | 0 |
| 0 | 0.005185239 | 0 | 0.004243987 | 0.009500417 |
| 0 | 0 | 0 | 0 | 0 |
| 0 | 0 | 0 | 0 | 0 |
| 0 | 0 | 0.005522217 | 0.001647297 | 0.012159017 |
| 0 | 0 | 0 | 0 | 0 |
| 0.010736256 | 0.265569966 | 0.018209673 | 0.019976871 | 0.018283782 |
| 0.005674805 | 0 | 0.0106146 | 0 | 0.001182534 |
| 0 | 0 | 0 | 0.002530474 | 0 |
| 0 | 0 | 0 | 0 | 0 |
| 0 | 0 | 0 | 0 | 0 |
| 0 | 0 | 0 | 0 | 0 |
| 0 | 0 | 0 | 0 | 0 |
| 0 | 0 | 0 | 0 | 0 |
| 0 | 0 | 0 | 0 | 0 |
| 0.671583749 | 0.051771034 | 0.041355814 | 0.217848391 | 0.000571211 |
| 0.032652924 | 0.285077468 | 0.008097275 | 0.056871593 | 0.033635003 |
| 0 | 0 | 0 | 0 | 0 |
| 0 | 0 | 0 | 0 | 0 |
| 0.102414514 | 0 | 0 | 0.072333709 | 0.009385777 |
| 0.008612951 | 0 | 0 | 0 | 0 |
| 0 | 0 | 0 | 0 | 0 |

| | | | | |
|---|---|---|---|---|
| 0 | 0 | 0 | 0 | 0 |
| 0.006219685 | 0.01647518 | 0 | 0.003347221 | 0 |
| 0.01157899 | 0.01330754 | 0 | 0.010687081 | 0.015328807 |
| 0 | 0 | 0 | 0 | 0 |
| 0 | 0 | 0 | 0.002497498 | 0 |
| 0 | 0 | 0.01894779 | 0 | 0 |
| 0 | 0 | 0 | 0 | 0 |
| 0 | 0 | 0 | 0 | 0 |
| 0 | 0 | 0 | 0 | 0 |
| 0 | 0 | 0 | 0 | 0 |
| 0 | 0 | 0 | 0 | 0 |
| 0 | 0 | 0 | 0 | 0 |
| 0 | 0 | 0 | 0 | 0 |
| 0 | 0 | 0 | 0 | 0 |
| 0 | 0 | 0 | 0 | 0 |
| 0 | 0 | 0 | 0 | 0 |
| 0.002103292 | 0 | 0 | 0.241037111 | 0 |
| 0 | 0 | 0 | 0 | 0 |
| 0 | 0 | 0 | 0 | 0 |
| 0 | 0 | 0 | 0 | 0 |
| 0 | 0 | 0 | 0 | 0 |
| 0.006757186 | 0 | 0 | 0 | 0 |
| 0 | 0 | 0 | 0 | 0 |
| 0 | 0 | 0 | 0 | 0 |
| 0.007671595 | 0 | 0 | 0.021052449 | 0 |
| 0 | 0.007638112 | 0 | 0.002161048 | 0.006118952 |
| 0 | 0 | 0 | 0 | 0 |
| 0.681060641 | 0 | 0 | 0.148627068 | 0 |
| 0 | 0 | 0 | 0 | 0 |
| 0 | 0 | 0 | 0 | 0 |
| 0 | 0 | 0 | 0 | 0 |
| 0 | 0 | 0 | 0 | 0 |
| 0 | 0 | 0 | 0 | 0 |
| 0 | 0 | 0 | 0 | 0 |
| 0 | 0 | 0 | 0 | 0 |
| 0 | 0 | 0 | 0 | 0 |
| 0.00344178 | 0 | 0 | 0 | 0 |
| 0.012366253 | 0 | 0 | 0 | 0 |
| 4.079343531 | 0 | 0 | 4.692465957 | 3.204684509 |
| 0 | 0 | 0 | 0 | 0 |
| 0 | 0 | 0 | 0 | 0 |
| 0.013273057 | 0 | 0 | 0.016176383 | 0.002808435 |
| 0.000755639 | 0 | 0 | 0.014097506 | 0 |
| 0 | 0 | 0 | 0.045263379 | 0 |
| 0 | 0 | 0 | 0 | 0 |
| 0 | 0 | 0 | 0 | 0 |
| 0.005201907 | 0 | 0 | 0.004645076 | 0 |
| 0 | 0 | 0 | 0 | 0 |

| | | | | |
|---|---|---|---|---|
| 0 | 0 | 0 | 0 | 0 |
| 0.077854553 | 0.00147973 | 0.004420925 | 0.018002355 | 0.034422876 |
| 0 | 0 | 0 | 0.040705754 | 0 |
| 0 | 0 | 0 | 0 | 0 |
| 0 | 0 | 0 | 0 | 0 |
| 0 | 0 | 0 | 0 | 0 |
| 0 | 0 | 0 | 0 | 0 |
| 0 | 0 | 0 | 0 | 0 |
| 0 | 0 | 0 | 0 | 0 |
| 0 | 0 | 0 | 0 | 0 |
| 0 | 0 | 0 | 0 | 0 |
| 0 | 0 | 0 | 0 | 0 |
| 0.006745833 | 0 | 0 | 0.031200487 | 0.02120627 |
| 0.007076191 | 0 | 0 | 0 | 0 |
| 0.024097721 | 0 | 0 | 0 | 0.009948377 |
| 0 | 0 | 0 | 0 | 0 |
| 0 | 0 | 0 | 0 | 0 |
| 0 | 0 | 0 | 0 | 0 |
| 0.009053769 | 0 | 0 | 0 | 0 |
| 0 | 0 | 0 | 0 | 0 |
| 0 | 0 | 0 | 0 | 0 |
| 0.495866821 | 0.000603979 | 0 | 0.061005385 | 0 |
| 0.044776961 | 0 | 0 | 0 | 0.001930504 |
| 0.001379758 | 0 | 0 | 0 | 0 |
| 0 | 0 | 0 | 0 | 0 |
| 0 | 0 | 0 | 0 | 0 |
| 0 | 0 | 0 | 0 | 0 |
| 0 | 0 | 0 | 0.023591005 | 0 |
| 0 | 0 | 0 | 0 | 0 |
| 0 | 0 | 0 | 0 | 0 |
| 0 | 0 | 0 | 0 | 0 |
| 0 | 0 | 0 | 0 | 0 |
| 0 | 0 | 0 | 0 | 0 |
| 0 | 0 | 0 | 0 | 0 |
| 8.638092145 | 0.025225438 | 0 | 0.002560019 | 0 |
| 0 | 0 | 0 | 0 | 0 |
| 0 | 0 | 0 | 0 | 0 |
| 0 | 0 | 0 | 0 | 0 |
| 0.006019785 | 0.001005243 | 0.000463369 | 0.233446168 | 0 |
| 0 | 0 | 0 | 0 | 0 |
| 0.039382 | 0.042903468 | 0.012048687 | 0.071837401 | 0.170462947 |
| 0 | 0 | 0 | 0 | 0 |
| 0 | 0 | 0 | 0 | 0 |
| 0 | 0 | 0 | 0 | 0 |
| 0 | 0 | 0 | 0 | 0 |
| 0.478362916 | 0.053455806 | 0.796300243 | 0.08461392 | 0.213769574 |
| 0 | 0 | 0 | 0 | 0 |
| 0 | 0 | 0 | 0 | 0 |

| | | | | |
|---|---|---|---|---|
| 0 | 0 | 0 | 0 | 0 |
| 0 | 0 | 0 | 0 | 0 |
| 0 | 0 | 0 | 0 | 0 |
| 0 | 0 | 0 | 0.001049252 | 0 |
| 0 | 0 | 0 | 0.000797183 | 0 |
| 0 | 0 | 0 | 0 | 0 |
| 0 | 0 | 0 | 0 | 0 |
| 0.016822442 | 0 | 0 | 0.013065099 | 0 |
| 0 | 0 | 0 | 0.001068077 | 0 |
| 0 | 0.004642613 | 0 | 0.001541973 | 0.004198132 |
| 0 | 0 | 0 | 0.056918073 | 0 |
| 0 | 0 | 0 | 0 | 0 |
| 0.007220114 | 0 | 0.000606288 | 0.001507149 | 0 |
| 0 | 0 | 0 | 0 | 0 |
| 0 | 0 | 0 | 0 | 0 |
| 0 | 0 | 0 | 0 | 0 |
| 0 | 0 | 0 | 0 | 0 |
| 0 | 0 | 0 | 0 | 0 |
| 0 | 0 | 0 | 0 | 0 |
| 1.760836674 | 0.013872846 | 0.076763675 | 0.077134451 | 0.126854643 |
| 0 | 0 | 0 | 0 | 0 |
| 0 | 0 | 0 | 0 | 0 |
| 0 | 0 | 0 | 0 | 0 |
| 0 | 0 | 0 | 0 | 0 |
| 0 | 0 | 0 | 0 | 0 |
| 0 | 0 | 0 | 0 | 0 |
| 0 | 0 | 0 | 0 | 0 |
| 0 | 0 | 0 | 0 | 0 |
| 0 | 0 | 0 | 0 | 0 |
| 0.101175443 | 0 | 0 | 0.001434052 | 0.009110054 |
| 0 | 0 | 0 | 0 | 0 |
| 0 | 0 | 0 | 0 | 0 |
| 0 | 0 | 0 | 0 | 0 |
| 0 | 0 | 0 | 0 | 0 |
| 0 | 0 | 0 | 0 | 0 |
| 0 | 0.001591222 | 0 | 0 | 0 |
| 0 | 0 | 0 | 0 | 0 |
| 0 | 0 | 0 | 0 | 0 |
| 0 | 0 | 0 | 0 | 0 |
| 0 | 0 | 0 | 0 | 0 |
| 0 | 0 | 0 | 0 | 0 |
| 0 | 0 | 0 | 0 | 0 |
| 0.003237507 | 0 | 0 | 0.047576819 | 0 |
| 0 | 0 | 0 | 0 | 0 |
| 0.021034272 | 0 | 0 | 0.002309921 | 0.001143441 |
| 0 | 0 | 0 | 0 | 0 |
| 0 | 0 | 0 | 0 | 0 |
| 0 | 0 | 0 | 0 | 0 |

| | | | | |
|---|---|---|---|---|
| 0 | 0 | 0 | 0 | 0 |
| 0 | 0 | 0 | 0 | 0 |
| 0 | 0 | 0 | 0.001717022 | 0 |
| 0 | 0 | 0 | 0 | 0 |
| 0 | 0 | 0 | 0 | 0 |
| 0.137775897 | 0.00627966 | 0.003216251 | 0.039662248 | 0.009560579 |
| 0 | 0 | 0 | 0 | 0 |
| 0 | 0 | 0 | 0 | 0 |
| 0 | 0 | 0 | 0 | 0 |
| 0 | 0 | 0 | 0 | 0 |
| 0 | 0 | 0 | 0 | 0 |
| 0 | 0 | 0 | 0 | 0 |
| 0 | 0 | 0 | 0 | 0 |
| 0 | 0 | 0 | 0 | 0 |
| 0 | 0 | 0 | 0 | 0 |
| 0 | 0 | 0 | 0 | 0 |
| 0.037598128 | 0 | 0 | 0.560966728 | 0 |
| 0 | 0 | 0 | 0 | 0 |
| 0 | 0 | 0 | 0 | 0 |
| 0 | 0 | 0 | 0 | 0 |
| 0 | 0 | 0 | 0 | 0 |
| 0 | 0 | 0 | 0 | 0 |
| 0 | 0 | 0 | 0 | 0 |
| 0 | 0 | 0 | 0 | 0 |
| 0 | 0 | 0 | 0 | 0 |
| 0 | 0.001733372 | 0 | 0 | 0.001068168 |
| 0 | 0 | 0 | 0 | 0 |
| 0 | 0 | 0 | 1.225203368 | 0 |
| 0 | 0 | 0 | 0 | 0 |
| 0.232781119 | 0 | 0 | 0 | 0 |
| 0 | 0 | 0 | 0 | 0 |
| 0 | 0 | 0 | 0 | 0 |
| 0 | 0 | 0 | 0 | 0 |
| 0 | 0 | 0 | 0 | 0 |
| 0 | 0 | 0 | 0 | 0 |
| 0 | 0 | 0 | 0 | 0 |
| 0 | 0 | 0 | 0 | 0 |
| 0 | 0 | 0 | 0 | 0 |
| 0.00231298 | 0 | 0 | 0.02367822 | 0 |
| 0 | 0 | 0 | 0 | 0 |
| 0 | 0 | 0 | 0.072454267 | 0 |
| 0 | 0 | 0 | 0 | 0 |
| 0 | 0 | 0 | 0 | 0 |
| 0 | 0 | 0 | 0 | 0 |
| 0 | 0 | 0 | 0 | 0 |
| 0 | 0 | 0 | 0 | 0 |
| 0 | 0 | 0 | 0 | 0 |
| 0 | 0 | 0 | 0 | 0 |

| | | | | |
|---|---|---|---|---|
| 0 | 0 | 0 | 0 | 0 |
| 0 | 0 | 0 | 0.042123399 | 0 |
| 0 | 0 | 0 | 0 | 0.004914259 |
| 0 | 0 | 0 | 0 | 0 |
| 0 | 0 | 0 | 0 | 0 |
| 0 | 0 | 0 | 0 | 0 |
| 0 | 0 | 0 | 0 | 0 |
| 0.213746038 | 0 | 0 | 0 | 0 |
| 0 | 0 | 0 | 0 | 0 |
| 0 | 0 | 0 | 0 | 0 |
| 0.010008907 | 0.000722039 | 0 | 0.004596438 | 0.00850961 |
| 0 | 0 | 0 | 0 | 0 |
| 0 | 0.049459084 | 0 | 0.127464407 | 0.398371321 |
| 0 | 0 | 0 | 0 | 0 |
| 0 | 0 | 0 | 0 | 0 |
| 0 | 0 | 0 | 0 | 0 |
| 0 | 0 | 0 | 0 | 0 |
| 0 | 0 | 0 | 0 | 0 |
| 0 | 0 | 0 | 0 | 0 |
| 0 | 0 | 0 | 0 | 0 |
| 0.103819241 | 0 | 0 | 0.005364874 | 0 |
| 0 | 0 | 0 | 0 | 0 |
| 0 | 0 | 0 | 0 | 0 |
| 1.563384443 | 2.837306096 | 0.127556639 | 0.297765369 | 3.366905109 |
| 0 | 0 | 0 | 0 | 0 |
| 0 | 0 | 0 | 0 | 0 |
| 0 | 0 | 0 | 0 | 0 |
| 0 | 0 | 0 | 0 | 0 |
| 0 | 0 | 0 | 0 | 0 |
| 0 | 0 | 0 | 0 | 0 |
| 0 | 0 | 0 | 0.007097141 | 0 |
| 0 | 0 | 0 | 0 | 0 |
| 0 | 0 | 0 | 0 | 0 |
| 0 | 0 | 0 | 0 | 0 |

| PHL018-0086_V1 | PHL018-0268_V1 | PHL018-0204_V2 | PHL018-0076_V1 | PHL018-0002_V3 |
|---|---|---|---|---|
| CTRL | REF | EYCF | EYCF | CTRL |
| 0.523380141 | 0.021442669 | 0.307916645 | 0.068825922 | 0.053298868 |
| 5.465116227 | 1.172713423 | 0.341879087 | 0.044204711 | 5.003612851 |
| 7.36354395 | 3.693152248 | 0 | 0 | 0 |
| 0 | 0 | 0 | 0 | 0 |
| 0 | 0 | 0 | 0 | 0 |
| 0 | 0 | 0 | 0.030875749 | 0 |
| 0 | 0 | 0 | 0 | 0 |
| 0 | 0 | 0 | 0 | 0 |
| 0 | 0 | 0 | 0 | 0 |
| 0 | 0 | 0 | 0 | 0 |
| 0 | 0 | 0 | 0 | 0 |
| 0 | 0 | 0 | 0 | 0 |
| 0 | 0 | 0 | 0 | 0 |
| 1.143447291 | 19.85292921 | 1.829659263 | 0.015149018 | 10.64277335 |
| 0 | 0 | 0 | 0 | 0 |
| 0 | 0 | 0 | 0 | 0 |
| 0 | 0 | 0 | 0 | 0 |
| 0 | 0 | 0 | 0 | 0 |
| 0 | 0 | 0 | 0 | 0 |
| 0 | 0 | 0 | 0 | 0 |
| 0 | 0 | 0 | 0 | 0 |
| 0 | 0 | 0 | 0 | 0 |
| 0 | 0 | 0 | 0 | 0 |
| 0 | 0 | 0 | 0 | 0 |
| 0.17700698 | 0 | 0 | 0.699224514 | 0.085525587 |
| 0 | 0 | 0 | 0 | 0 |
| 0 | 0 | 0 | 0 | 0 |
| 0 | 0 | 0 | 0 | 0 |
| 0 | 0 | 0 | 0 | 0 |
| 0 | 0 | 0 | 0 | 0 |
| 0 | 0 | 0 | 0 | 0 |
| 0 | 0 | 0 | 0 | 0 |
| 0 | 0 | 0 | 0 | 0 |
| 0 | 0 | 0 | 0 | 0 |
| 0 | 0 | 0 | 0 | 0 |
| 0 | 0 | 0 | 1.370023205 | 1.891066462 |
| 0 | 0 | 0 | 0 | 0 |
| 0 | 0 | 0 | 0 | 0 |
| 0 | 0 | 0 | 0 | 0 |
| 0 | 0 | 0 | 0 | 0 |
| 0 | 0 | 0 | 0 | 0 |
| 0.002542143 | 0.00339374 | 0.035526936 | 0.005455614 | 0.0040121 |
| 0.069924555 | 0.034549759 | 0.028080723 | 0.446239768 | 0.005232463 |
| 0.038732148 | 0.004637409 | 0 | 0.026837554 | 0.042351324 |
| 0.002754885 | 0 | 0 | 0.007468008 | 0 |
| 0 | 0 | 0 | 0 | 0 |

| | | | | |
|---|---|---|---|---|
| 0.004057262 | 0 | 0 | 0.006599114 | 0 |
| 0.018087265 | 0.022655839 | 0 | 0.001513315 | 0 |
| 0.021232688 | 0.005521205 | 0.006991696 | 0.003628662 | 0.005463616 |
| 0.774060412 | 0.561674142 | 0.148759294 | 0.325604746 | 1.123857421 |
| 0.010929803 | 0 | 0.000671904 | 0.00698633 | 0.018817966 |
| 0.102201267 | 0 | 0.06296315 | 0.023149703 | 0 |
| 0.022737405 | 0.076684436 | 0.002589552 | 0.000973218 | 1.027538029 |
| 0 | 0 | 0 | 0 | 0 |
| 0 | 0 | 0 | 0 | 0 |
| 0 | 0 | 0 | 0 | 0 |
| 0 | 0.002348605 | 0 | 0 | 0 |
| 0.037666731 | 0.000721575 | 0.076146367 | 0.372373268 | 0.19753455 |
| 0 | 0 | 0 | 0 | 0.00196342 |
| 0.01559818 | 0.010800733 | 0.026118327 | 0 | 0.031854093 |
| 0 | 0 | 0 | 0 | 0 |
| 0 | 0 | 0 | 0 | 0 |
| 0.065325274 | 0.008281692 | 0.109535048 | 0.567246368 | 0 |
| 0 | 0 | 0 | 0 | 0 |
| 0 | 0 | 0 | 0 | 0 |
| 0 | 0 | 0 | 0 | 0 |
| 0.366599177 | 0.308926076 | 0.009635164 | 0 | 0.15993844 |
| 0.046936487 | 0.084272941 | 0 | 0 | 0.000603887 |
| 0 | 0.020778528 | 0.954991981 | 0.003360939 | 0 |
| 0 | 0 | 0 | 0 | 0 |
| 0 | 0 | 0 | 0 | 0 |
| 0 | 0 | 0 | 0 | 0 |
| 0 | 0 | 0 | 0 | 0 |
| 0 | 0 | 0 | 0 | 0 |
| 0 | 0 | 0.009023067 | 0 | 0 |
| 0 | 0 | 0 | 0 | 0 |
| 0.038731985 | 0.366527271 | 0 | 0 | 0 |
| 0 | 0 | 0 | 0 | 0 |
| 0 | 0 | 0 | 0 | 0 |
| 0 | 0 | 0 | 0.000532799 | 0 |
| 0 | 0 | 0 | 0 | 0 |
| 0 | 0 | 0 | 0 | 0 |
| 0 | 0 | 0 | 0 | 0 |
| 4.126162128 | 0 | 0 | 0.003354081 | 0 |
| 0 | 0 | 0 | 0 | 0 |
| 0 | 0 | 0 | 0 | 0 |
| 0 | 0 | 0 | 0 | 0 |
| 0 | 0 | 0 | 0 | 0 |
| 0 | 0 | 0 | 0 | 0 |
| 0 | 0 | 0 | 0 | 0 |
| 0 | 0 | 0 | 0 | 0 |
| 0 | 0 | 0 | 0 | 0 |
| 0 | 0 | 0 | 0 | 0 |
| 0 | 0.001100986 | 0 | 0.005356254 | 0 |

| | | | | |
|---|---|---|---|---|
| 0 | 0 | 0 | 0 | 0 |
| 0 | 0 | 0 | 0 | 0 |
| 0 | 0 | 0 | 0 | 0 |
| 0 | 0 | 0 | 0 | 0 |
| 0.000774289 | 0 | 0 | 0 | 0.013289368 |
| 0 | 0 | 0 | 0 | 0 |
| 0 | 0 | 0 | 0 | 0 |
| 0.002107908 | 0 | 0 | 0 | 0 |
| 0.004109402 | 0.015448628 | 0.003556935 | 0 | 0 |
| 0 | 0 | 0 | 0 | 0 |
| 0 | 0 | 0 | 0 | 0 |
| 0.000954187 | 0 | 0.001032383 | 0 | 0 |
| 0 | 0 | 0 | 0 | 0 |
| 0 | 0 | 0 | 0 | 0 |
| 0 | 0 | 0 | 0 | 0 |
| 0 | 0 | 0 | 0 | 0 |
| 0 | 0 | 0 | 0 | 0 |
| 0 | 0 | 0 | 0 | 0 |
| 0 | 0 | 0.109333917 | 0.044428254 | 0.008845376 |
| 0 | 0 | 0 | 0 | 0 |
| 0 | 0 | 0 | 0 | 0 |
| 0 | 0 | 0 | 0 | 0 |
| 0 | 0 | 0 | 0.001827673 | 0 |
| 0 | 0 | 0 | 0 | 0 |
| 0 | 0 | 0 | 0 | 0 |
| 0 | 0 | 0 | 0 | 0 |
| 0 | 0 | 0 | 0 | 0 |
| 0 | 0 | 0 | 0 | 0 |
| 0 | 0 | 0 | 0 | 0 |
| 0 | 0 | 0 | 0 | 0 |
| 0.003194285 | 0 | 0.014948823 | 0.285545622 | 0 |
| 0.000478726 | 0 | 0.005898966 | 0 | 0.009444283 |
| 0 | 0 | 0 | 0 | 0 |
| 0 | 0 | 0 | 0 | 0 |
| 0 | 0 | 0 | 0 | 0 |
| 0 | 0 | 0 | 0 | 0 |
| 0 | 0 | 0 | 0 | 0 |
| 0 | 0 | 0 | 0 | 0 |
| 0 | 0 | 0 | 0.226647729 | 0 |
| 0 | 0 | 0 | 0 | 0 |
| 0 | 0 | 0 | 0 | 0 |
| 0 | 0 | 0 | 0.03747555 | 0 |
| 0 | 0.000590955 | 0 | 0.001249988 | 0.016592468 |
| 0 | 0 | 0 | 0 | 0 |
| 0 | 0 | 0 | 0.003464918 | 0 |
| 0 | 0 | 0 | 0 | 0 |
| 0 | 0 | 0 | 0 | 0 |
| 0 | 0 | 0 | 0 | 0 |

| | | | | |
|---|---|---|---|---|
| 0 | 0 | 0 | 0 | 0 |
| 0 | 0 | 0 | 0 | 0 |
| 0 | 0 | 0 | 0 | 0 |
| 0 | 0 | 0 | 0 | 0 |
| 0 | 0 | 0.158038793 | 0.065795588 | 0.034031542 |
| 0 | 0 | 0 | 0 | 0 |
| 0 | 0 | 0 | 0 | 0 |
| 0 | 0 | 0 | 0 | 0 |
| 0 | 0 | 0 | 0 | 0 |
| 0 | 0 | 0 | 0 | 0 |
| 0 | 0 | 0 | 0 | 0 |
| 0 | 0 | 0 | 0 | 0 |
| 0 | 0 | 0 | 0 | 0 |
| 0 | 0 | 0 | 0 | 0 |
| 0 | 0 | 0 | 0 | 0 |
| 0 | 0 | 0.006786487 | 0 | 0 |
| 0 | 0 | 0 | 0 | 0 |
| 0 | 0 | 0 | 0.00102436 | 0 |
| 0 | 0 | 0 | 0 | 0 |
| 0 | 0 | 0 | 0 | 0 |
| 0 | 0 | 0.018731527 | 0 | 0 |
| 0 | 0 | 0 | 0 | 0 |
| 0 | 0 | 0 | 0.003167679 | 0 |
| 0 | 0 | 0 | 0 | 0 |
| 0 | 0 | 0 | 0.012046753 | 0 |
| 0 | 0 | 0 | 0 | 0 |
| 0.000693366 | 0 | 0 | 0.090220531 | 0 |
| 0 | 0 | 0 | 0 | 0 |
| 0 | 0 | 0 | 0 | 0 |
| 0 | 0 | 0 | 0 | 0 |
| 0 | 0 | 0.017161009 | 0 | 0 |
| 0.002761734 | 0.000589903 | 0.002158042 | 0 | 0 |
| 0 | 0 | 0 | 0 | 0 |
| 0 | 0 | 0 | 0 | 0 |
| 0 | 0 | 0 | 0 | 0 |
| 0 | 0 | 0 | 0 | 0 |
| 0 | 0 | 0 | 0 | 0 |
| 0 | 0 | 0 | 0 | 0 |
| 0 | 0 | 0 | 0 | 0 |
| 0 | 0 | 0 | 0 | 0 |
| 0 | 0 | 0 | 0 | 0 |
| 0 | 0 | 0 | 0 | 0 |
| 0 | 0 | 0 | 0 | 0 |
| 0 | 0 | 0 | 0 | 0 |
| 0 | 0 | 0 | 0 | 0 |
| 0 | 0 | 0 | 0 | 0 |
| 0 | 0 | 0 | 0 | 0 |
| 0 | 0 | 0 | 0 | 0 |

| | | | | |
|---|---|---|---|---|
| 0 | 0 | 0.057514797 | 0 | 0 |
| 0 | 0 | 0 | 0 | 0 |
| 0 | 0 | 0 | 0 | 0 |
| 0 | 0 | 0 | 0 | 0 |
| 0 | 0 | 0 | 0 | 0 |
| 0 | 0 | 0 | 0 | 0 |
| 0 | 0 | 0 | 0 | 0 |
| 0 | 0 | 0 | 0 | 0 |
| 0 | 0 | 0 | 0 | 0 |
| 0 | 0 | 0 | 0 | 0 |
| 0 | 0 | 0 | 0 | 0 |
| 0 | 0 | 0 | 0 | 0 |
| 0 | 0 | 0 | 0.003104364 | 0.000625622 |
| 0 | 0 | 0 | 0 | 0 |
| 0 | 0 | 0 | 0 | 0 |
| 0 | 0 | 0 | 0 | 0 |
| 0 | 0 | 0 | 0 | 0 |
| 0 | 0 | 0 | 0 | 0 |
| 0 | 0 | 0 | 0 | 0 |
| 0 | 0 | 0 | 0 | 0 |
| 0.055671184 | 0.122079264 | 0.002376073 | 0.008260129 | 0.109177925 |
| 0 | 0 | 0 | 0 | 0 |
| 0 | 0 | 0 | 0 | 0 |
| 0 | 0 | 0 | 0 | 0 |
| 0 | 0 | 0 | 0 | 0 |
| 0 | 0 | 0 | 0 | 0 |
| 0.017841464 | 0.010039505 | 0.001902683 | 0.002210232 | 0.003179132 |
| 0 | 0 | 0 | 0 | 0 |
| 0 | 0 | 0 | 0 | 0 |
| 0 | 0 | 0 | 0 | 0 |
| 0 | 0.001633907 | 0 | 0 | 0 |
| 0 | 0 | 0 | 0 | 0 |
| 0 | 0 | 0 | 0 | 0 |
| 0 | 0 | 0.018514383 | 0.248447293 | 0 |
| 0.035365299 | 0.015565783 | 0.00908275 | 0.022514715 | 0.005581477 |
| 0 | 0 | 0 | 0 | 0 |
| 0 | 0 | 0 | 0 | 0 |
| 0 | 0 | 0 | 0 | 0 |
| 0 | 0 | 0 | 0 | 0 |
| 0 | 0 | 0 | 0 | 0 |
| 0 | 0 | 0 | 0 | 0 |
| 0 | 0 | 0 | 0 | 0 |
| 0 | 0 | 0 | 0 | 0 |
| 0 | 0 | 0 | 0 | 0 |
| 0 | 0 | 0 | 0 | 0 |
| 0 | 0 | 0 | 0 | 0 |
| 0 | 0 | 0 | 0 | 0 |
| 0.000458223 | 0.000815632 | 0 | 0.000517568 | 0 |

| | | | | |
|---|---|---|---|---|
| 0 | 0 | 0 | 0 | 0 |
| 0 | 0 | 0 | 0 | 0 |
| 0 | 0 | 0 | 0 | 0.003634567 |
| 0 | 0.00082942 | 0 | 0 | 0 |
| 0 | 0 | 0 | 0 | 0 |
| 0 | 0 | 0 | 0 | 0 |
| 0 | 0 | 0.00235243 | 0 | 0 |
| 0 | 0 | 0.002771553 | 0 | 0 |
| 0.035666857 | 0 | 0 | 0.007139304 | 0 |
| 0 | 0 | 0 | 0 | 0 |
| 0 | 0 | 0 | 0 | 0 |
| 0 | 0 | 0 | 0 | 0 |
| 0 | 0 | 0 | 0 | 0 |
| 0 | 0 | 0 | 0 | 0 |
| 0 | 0.001028785 | 0 | 0 | 0 |
| 0 | 0 | 0 | 0 | 0 |
| 0 | 0 | 0 | 0 | 0 |
| 0 | 0 | 0 | 0 | 0 |
| 0 | 0 | 0 | 0.002315996 | 0 |
| 0 | 0 | 0 | 0.149469879 | 0 |
| 0 | 0.122358596 | 0 | 0 | 0.001356122 |
| 0 | 0 | 0 | 0 | 0 |
| 0 | 0 | 0 | 0 | 0 |
| 0 | 0 | 0 | 0 | 0 |
| 0.004552297 | 0.006320372 | 0.00246268 | 0 | 0.002694224 |
| 0 | 0 | 0 | 0 | 0 |
| 0 | 0 | 0 | 0 | 0 |
| 0 | 0 | 0 | 0 | 0 |
| 0 | 0.00255184 | 0.011750855 | 0 | 0 |
| 0 | 0 | 0 | 0 | 0 |
| 0.008424228 | 0.337928168 | 0.00136719 | 0.133936673 | 0.064216858 |
| 0 | 0 | 0 | 0 | 0 |
| 0 | 0 | 0 | 0.00241362 | 0 |
| 0 | 0 | 0.00078615 | 0 | 0 |
| 0 | 0 | 0 | 0 | 0 |
| 0 | 0 | 0 | 0 | 0 |
| 0 | 0 | 0 | 0 | 0 |
| 0 | 0 | 0 | 0 | 0 |
| 0 | 0 | 0 | 0 | 0 |
| 0 | 0 | 0 | 0 | 0 |
| 0 | 0 | 0 | 0 | 0 |
| 0 | 0.000917097 | 0.122329085 | 0.004539233 | 0 |
| 0 | 0 | 0 | 0 | 0 |
| 0 | 0 | 0 | 0 | 0 |
| 0 | 0 | 0 | 0 | 0 |
| 0 | 0 | 0 | 0 | 0 |
| 0 | 0 | 0 | 0 | 0 |
| 0 | 0 | 0 | 0 | 0 |

| | | | | |
|---|---|---|---|---|
| 0 | 0 | 0 | 0 | 0 |
| 0 | 0 | 0 | 0 | 0 |
| 0.038817062 | 0.002533445 | 0 | 0.032590902 | 0.039820601 |
| 0 | 0 | 0 | 0 | 0 |
| 0 | 0 | 1.408277125 | 0 | 0 |
| 0 | 0 | 0 | 0 | 0 |
| 0 | 0 | 0 | 0 | 0 |
| 0 | 0 | 0 | 0 | 0 |
| 0 | 0 | 0 | 0 | 0 |
| 0 | 0 | 0 | 0.000801993 | 0.002334591 |
| 0 | 0 | 0 | 0 | 0 |
| 0 | 0 | 0 | 0 | 0 |
| 0 | 0 | 0 | 0 | 0 |
| 0.010486633 | 0 | 0 | 0 | 0 |
| 0.005507945 | 0.004832013 | 0.000709443 | 0.00213301 | 0.00186275 |
| 0.004633344 | 0 | 0 | 0.003996421 | 0 |
| 0.005827538 | 0.002154383 | 0 | 0 | 0.001857132 |
| 0 | 0 | 0 | 0 | 0 |
| 0 | 0 | 0 | 0 | 0 |
| 0 | 0 | 0 | 0 | 0 |
| 0 | 0 | 0 | 0 | 0 |
| 0.000809324 | 0 | 0 | 0 | 0 |
| 0 | 0 | 0 | 0 | 0 |
| 0 | 0 | 0 | 0 | 0 |
| 0 | 0 | 0 | 0 | 0 |
| 0 | 0 | 0 | 0 | 0 |
| 0 | 0 | 0 | 2.316385123 | 0 |
| 0 | 0 | 0 | 0 | 0 |
| 0 | 0 | 0 | 0 | 0 |
| 0 | 0 | 0 | 0 | 0 |
| 0 | 0 | 0 | 0 | 0 |
| 0 | 0 | 0 | 0 | 0 |
| 0 | 0 | 0 | 0 | 0 |
| 0 | 0.001744042 | 0 | 0 | 0 |
| 0 | 0 | 0 | 0 | 0 |
| 0 | 0 | 0.026234737 | 0 | 0 |
| 0 | 0 | 0 | 0 | 0 |
| 0.019065587 | 0 | 0.018336018 | 0.005340624 | 0.007823396 |
| 0 | 0 | 0 | 0 | 0 |
| 0 | 0 | 0 | 0 | 0 |
| 0 | 0 | 0 | 0 | 0 |
| 0 | 0 | 0 | 0 | 0 |
| 0 | 0 | 0 | 0 | 0 |
| 0 | 0 | 0 | 0 | 0 |
| 0 | 0 | 0 | 0 | 0 |
| 0 | 0 | 0 | 0 | 0 |
| 0 | 0 | 0 | 0 | 0 |
| 0 | 0 | 0 | 0 | 0 |

| | | | | |
|---|---|---|---|---|
| 1.281875404 | 0 | 0.017020861 | 0.408986273 | 7.330978627 |
| 0 | 0 | 0 | 0 | 0.034273695 |
| 0.003602739 | 0 | 0 | 0 | 0 |
| 0 | 0 | 0 | 0 | 0 |
| 0.002345396 | 0.145737793 | 0.004806065 | 0.002167485 | 0.006814286 |
| 0 | 0 | 0 | 0 | 0 |
| 0 | 0 | 0 | 0 | 0 |
| 0.045727375 | 0.018513201 | 0.003503114 | 0.008709529 | 0.047758608 |
| 0 | 0 | 0 | 0 | 0 |
| 0.057606017 | 0.162196634 | 0.00249974 | 0.002505237 | 0.012032981 |
| 0 | 0 | 0 | 0 | 0 |
| 0.003075643 | 0.002021395 | 0.021544681 | 0.000641348 | 0 |
| 0.13513364 | 0.027982727 | 0.005339598 | 0.002918913 | 0.00424846 |
| 0 | 0 | 0 | 0 | 0 |
| 0 | 0 | 0 | 0.001751187 | 0 |
| 0 | 0.001438586 | 0 | 0 | 0.000956647 |
| 0 | 0 | 0 | 0 | 0 |
| 0 | 0 | 0 | 0 | 0 |
| 0.242043782 | 0 | 0.00235221 | 0 | 0 |
| 0.038105562 | 0 | 0 | 0 | 0 |
| 0 | 0 | 0 | 0 | 0 |
| 0 | 0 | 0 | 0 | 0 |
| 0 | 0 | 0 | 0 | 0 |
| 0 | 0.002070453 | 0.014386769 | 0.334823681 | 0 |
| 0 | 0 | 0 | 0 | 0 |
| 0.003971485 | 0 | 0 | 0.001699895 | 0.000742256 |
| 0 | 0 | 0 | 0.003599081 | 0 |
| 0 | 0 | 0 | 0 | 0 |
| 0 | 0 | 0 | 0 | 0 |
| 0 | 0 | 0 | 0 | 0 |
| 0 | 0 | 0 | 0 | 0 |
| 0 | 0 | 0 | 0 | 0 |
| 0 | 0 | 0 | 0 | 0 |
| 0 | 0 | 0 | 0 | 0 |
| 0 | 0 | 0 | 0 | 0 |
| 0 | 0 | 0 | 0 | 0.011792099 |
| 0 | 0 | 0 | 0 | 0 |
| 0 | 0 | 0 | 0 | 0 |
| 0 | 0 | 0 | 0 | 0 |
| 0 | 0 | 0 | 0 | 0 |
| 0 | 0 | 0 | 0 | 0 |
| 0 | 0 | 0 | 0 | 0 |
| 0 | 0 | 0 | 0 | 0 |
| 0.017681827 | 0 | 0.011539249 | 0 | 0 |
| 0 | 0 | 0 | 0 | 0 |
| 0 | 0 | 0 | 0 | 0 |
| 0.001819578 | 0.000832842 | 0.007734154 | 0.000704651 | 0 |
| 0 | 0 | 0 | 0 | 0 |

| | | | | |
|---|---|---|---|---|
| 0 | 0 | 0 | 0 | 0 |
| 0 | 0.003412784 | 0 | 0 | 0 |
| 0 | 0 | 0 | 0 | 0 |
| 0 | 0 | 0 | 0 | 0 |
| 0 | 0 | 0 | 0 | 0 |
| 0.017402396 | 0.016399101 | 0 | 0 | 0 |
| 0.001821224 | 0 | 0 | 0 | 0.00107787 |
| 0 | 0.001131211 | 0 | 0 | 0 |
| 0.491220318 | 0 | 0 | 0 | 0 |
| 0 | 0 | 0 | 0 | 0 |
| 0.061263918 | 0.054270961 | 0.008349794 | 0.005471487 | 0.002810723 |
| 0 | 0 | 0 | 0 | 0 |
| 0 | 0 | 0 | 0 | 0 |
| 0.014581936 | 0.013656687 | 0 | 0.001064244 | 0.000796629 |
| 0.002042903 | 0 | 0 | 0 | 0 |
| 0 | 0 | 0 | 0 | 0 |
| 0 | 0 | 0 | 0 | 0 |
| 0 | 0 | 0 | 0 | 0 |
| 0 | 0 | 0 | 0 | 0 |
| 0 | 0 | 0 | 0.018780065 | 0.003821491 |
| 0 | 0.006453737 | 0 | 0 | 0 |
| 0 | 0 | 0 | 0.143529739 | 0 |
| 0 | 0 | 0 | 0 | 0 |
| 0 | 0 | 0 | 0 | 0 |
| 0 | 0 | 0 | 0 | 0 |
| 0 | 0 | 0 | 0 | 0 |
| 0 | 0 | 0.004166937 | 0 | 0 |
| 0 | 0 | 0 | 0 | 0 |
| 0 | 0 | 0 | 0 | 0 |
| 0.218344288 | 0.159089176 | 0.141718898 | 0.098277411 | 0.597468739 |
| 0.000760976 | 0 | 0.112385641 | 0.001237723 | 0.002341948 |
| 0 | 0 | 0 | 0 | 0 |
| 0.005859171 | 0 | 0 | 0 | 0 |
| 0.002802286 | 0 | 0 | 0 | 0 |
| 0 | 0 | 0 | 0 | 0 |
| 0 | 0 | 0 | 0 | 0 |
| 0 | 0 | 0 | 0.290791571 | 0 |
| 0 | 0 | 0 | 0 | 0 |
| 0 | 0 | 0 | 0 | 0.36979529 |
| 0 | 0 | 0 | 0 | 0 |
| 0.37771227 | 0.240697191 | 0.004988598 | 0 | 0.186432689 |
| 4.356586331 | 1.874607063 | 0.013586282 | 0.139529797 | 3.311940616 |
| 0 | 0 | 0 | 0 | 0 |
| 0 | 0 | 0 | 0 | 0 |
| 0 | 0 | 0 | 0 | 0 |
| 0 | 0 | 0 | 0 | 0 |
| 0 | 0 | 0 | 0 | 0.00229127 |
| 0 | 0 | 0 | 0 | 0 |

| | | | | |
|---|---|---|---|---|
| 0 | 0 | 0 | 0.001618604 | 0 |
| 0.011750894 | 0 | 0 | 0 | 0 |
| 0.014969874 | 0.014597493 | 0.227398776 | 0.02734786 | 0 |
| 0 | 0 | 0 | 0 | 0 |
| 0 | 0 | 0 | 0.099686473 | 0 |
| 0.019587778 | 0 | 0 | 0 | 0 |
| 0 | 0 | 0 | 0 | 0 |
| 0 | 0 | 0.006757881 | 0 | 0 |
| 0.019788528 | 0 | 0 | 0.003928418 | 0 |
| 0 | 0 | 0 | 0 | 0 |
| 0 | 0 | 0 | 0 | 0.010960626 |
| 0 | 0 | 0 | 0 | 0 |
| 0 | 0 | 0.001324012 | 0 | 0 |
| 0.012531984 | 0.00623052 | 0 | 0 | 0 |
| 0 | 0 | 0 | 0 | 0 |
| 0.09692217 | 0 | 0.219686658 | 0.03009597 | 0.032178824 |
| 0.017779915 | 0.016390364 | 0.001518709 | 0.007102896 | 0.008861334 |
| 0 | 0.007042634 | 0 | 0 | 0 |
| 0 | 0 | 0 | 0 | 0 |
| 0 | 0 | 0 | 0 | 0 |
| 0 | 0 | 0 | 0 | 0.004467121 |
| 0.001302554 | 0 | 0.00097868 | 0.001176999 | 0 |
| 0.025463919 | 0 | 0.011258699 | 0 | 0 |
| 0 | 1.843931601 | 0 | 0 | 0.538977173 |
| 0 | 0 | 0 | 0 | 0 |
| 0 | 0.469481647 | 0 | 0 | 0.004085636 |
| 0.028036484 | 0.023091471 | 0 | 0 | 0.007593436 |
| 0 | 0 | 0.00433824 | 0 | 0 |
| 0 | 0 | 0 | 0 | 0 |
| 0 | 0 | 0 | 0 | 0 |
| 0.006324223 | 0.006561252 | 0.00048875 | 0.004775789 | 0.043203961 |
| 0.033404327 | 0 | 0.002185998 | 0.002008238 | 0.014986905 |
| 0.072844708 | 0.228736156 | 0.00161884 | 0.017904337 | 0 |
| 0 | 0.001962882 | 0.00463991 | 0 | 0 |
| 0 | 0 | 0 | 0 | 0 |
| 0 | 0 | 0 | 0 | 0 |
| 0.003942123 | 0 | 0 | 0 | 0.001372411 |
| 0.000906742 | 0 | 0.014715748 | 0.001597712 | 0.040141017 |
| 0.087297434 | 0.075392203 | 0 | 0.165890976 | 0 |
| 0.001482708 | 0 | 0 | 0 | 0.001755047 |
| 0 | 0 | 0 | 0 | 0 |
| 0 | 0 | 0 | 0 | 0 |
| 0 | 0.004344698 | 0 | 0 | 0.009951634 |
| 0.346975255 | 0 | 0 | 0 | 0.006032409 |
| 0.0698111 | 0 | 0.018982947 | 0 | 0 |
| 0 | 0 | 0 | 0 | 0 |
| 0 | 0 | 0 | 0 | 0 |
| 0 | 0 | 0 | 0 | 0 |

| | | | | |
|---|---|---|---|---|
| 0 | 0 | 0 | 0 | 0 |
| 0.88391315 | 0.458151434 | 0.035425329 | 0.187396771 | 0.621851902 |
| 0 | 0.001517239 | 0 | 0 | 0 |
| 0 | 0 | 0.001079493 | 0 | 0 |
| 0 | 0 | 0 | 0 | 0 |
| 0 | 0 | 0 | 0 | 0 |
| 0 | 0.001220765 | 0 | 0 | 0 |
| 0 | 0 | 0 | 0 | 0 |
| 0 | 0 | 0 | 0 | 0 |
| 0 | 0 | 0 | 0 | 0 |
| 0.002570818 | 0.000588347 | 0.003973569 | 0 | 0.00130415 |
| 0.058533693 | 0 | 0.0017263 | 0.002039042 | 0.014893029 |
| 0 | 0 | 0 | 0.046966847 | 0 |
| 0 | 0 | 0 | 0 | 0 |
| 0.006110078 | 0 | 0 | 0 | 0 |
| 0 | 0 | 0 | 0 | 0 |
| 0.017690404 | 0.002669697 | 0.017683825 | 0.001042513 | 0.010469858 |
| 0.007738068 | 0.034656353 | 0.001350356 | 0 | 0.006500198 |
| 0 | 0 | 0 | 0 | 0 |
| 0 | 0.003462158 | 0.002192126 | 0 | 0.00402903 |
| 0.027959793 | 0.028166648 | 0.003396992 | 0.019754841 | 0.019481661 |
| 0 | 0 | 0 | 0 | 0 |
| 0 | 0 | 0 | 0 | 0 |
| 0 | 0 | 0 | 0 | 0 |
| 0 | 0 | 0 | 0 | 0.005915571 |
| 0 | 0 | 0 | 0 | 0 |
| 0.379908946 | 0.420372918 | 0 | 0.007693489 | 0.057919799 |
| 0.093155835 | 0.175795397 | 0.175584379 | 0.0344228 | 0.115754751 |
| 0 | 0 | 0 | 0 | 0 |
| 0 | 0 | 0 | 0 | 0 |
| 0.002261454 | 0.000724566 | 0 | 0 | 0 |
| 0 | 0 | 0 | 0 | 0 |
| 0 | 0 | 0.001153453 | 0 | 0 |
| 0 | 0 | 0 | 0 | 0 |
| 0 | 0 | 0 | 0.067213361 | 0 |
| 0.063492315 | 0.143999716 | 0.11481417 | 0.076388786 | 0.78067653 |
| 0 | 0 | 0 | 0 | 0 |
| 0 | 0 | 0 | 0 | 0 |
| 0.003379945 | 0.006941847 | 0.017112572 | 0.003347814 | 0.005231766 |
| 0 | 0 | 0 | 0 | 0 |
| 0.010767815 | 0 | 0.015592495 | 0 | 0 |
| 0 | 0 | 0 | 0 | 0 |
| 0.110705717 | 0.038961232 | 0.003702129 | 0 | 0.053836593 |
| 0 | 0 | 0 | 0 | 0.018228472 |
| 0 | 0 | 0 | 0 | 0 |
| 0.172818191 | 0 | 0 | 0.251108554 | 0 |
| 0 | 0 | 0 | 0 | 0.243054926 |
| 0.001478775 | 0 | 0 | 0 | 0 |

| | | | | |
|---|---|---|---|---|
| 0 | 0 | 0 | 0 | 0 |
| 0 | 0 | 0 | 0 | 0 |
| 0 | 0 | 0 | 0 | 0 |
| 0 | 0 | 2.563204181 | 0.05322666 | 0.034399341 |
| 0 | 0 | 0 | 0 | 0 |
| 0 | 0 | 0 | 0.000864742 | 0 |
| 0.019821298 | 0 | 0 | 0 | 0 |
| 0.004442794 | 0 | 0 | 0.000892121 | 0.012660149 |
| 0.586152042 | 0 | 0 | 0.444218228 | 0.427668067 |
| 0.308726279 | 0 | 0.038981619 | 0.11129736 | 0 |
| 0 | 0 | 0 | 0 | 0 |
| 0.012159713 | 0 | 0.059902049 | 0.000716584 | 0.016479133 |
| 0 | 0 | 0 | 0 | 0 |
| 1.692720035 | 0 | 0.010152102 | 0 | 0 |
| 0.002472201 | 0 | 0 | 0.367608013 | 0.02082827 |
| 0 | 0 | 0 | 0 | 0 |
| 0.184380909 | 0.197322822 | 0 | 0.024297011 | 0.362847467 |
| 0 | 0 | 0 | 0.803070796 | 0 |
| 0 | 0 | 0 | 0 | 0 |
| 0 | 0 | 0 | 0.242746237 | 0 |
| 0.106814493 | 0 | 0 | 0 | 0 |
| 0 | 0 | 0 | 0.982308482 | 0 |
| 0.017525799 | 0 | 0 | 0.013197045 | 0 |
| 0.002101202 | 0.318957273 | 0.030690857 | 0 | 0.326976542 |
| 0.061872817 | 0 | 0.009520828 | 0.005143607 | 0 |
| 0.787132548 | 0 | 0 | 0.081704935 | 0 |
| 0 | 0 | 0 | 0 | 0 |
| 0 | 0 | 0 | 2.446609678 | 4.910156345 |
| 0 | 0 | 0 | 0 | 0 |
| 0 | 0 | 0 | 0 | 0 |
| 0 | 0 | 0 | 0.001277176 | 0 |
| 0 | 0 | 0 | 0.002516067 | 0 |
| 0.04459327 | 0 | 0 | 0 | 0.006945264 |
| 0 | 0 | 0 | 0 | 0 |
| 0.039545189 | 0 | 0 | 0 | 0.001872349 |
| 0 | 0 | 0.213747727 | 1.664519312 | 0 |
| 0.332253858 | 0.004007284 | 2.93394447 | 0.176856067 | 0.114364636 |
| 0 | 0 | 0 | 0 | 0 |
| 0 | 0 | 0.00612002 | 0.059677082 | 0.021932059 |
| 0 | 0 | 0.020698898 | 0 | 0 |
| 0.088755971 | 0.278594718 | 0.009145672 | 0.252173637 | 0.008004447 |
| 0 | 0 | 0 | 0.011841436 | 0 |
| 0.026770526 | 7.752858913 | 0.02706268 | 0.005772629 | 0.239577796 |
| 0 | 0 | 0 | 0.037827024 | 0 |
| 0 | 0 | 0 | 0 | 0 |
| 0 | 0 | 0.024763625 | 0 | 0 |
| 0 | 0 | 0 | 0 | 0 |
| 0.002845196 | 0 | 0.294459916 | 0.032641785 | 0 |

| | | | | |
|---|---|---|---|---|
| 0 | 0 | 0 | 0 | 0 |
| 0 | 0 | 1.984499456 | 0.516455375 | 0 |
| 0.017728372 | 0.167699271 | 0.008958262 | 0 | 0.193704539 |
| 0 | 0 | 0 | 0.005280524 | 0 |
| 0 | 0 | 0 | 0 | 0 |
| 0 | 0.004917135 | 0.197595251 | 0.058036036 | 0.009264573 |
| 0 | 0.104714175 | 0 | 0 | 0 |
| 0 | 0 | 0 | 0 | 0 |
| 0 | 0 | 0 | 0 | 0 |
| 0 | 0 | 0 | 0 | 0 |
| 0.375666091 | 0.012461176 | 0.062602727 | 0.024759122 | 0.049583557 |
| 2.56833541 | 3.448462419 | 0.410324037 | 0.496310348 | 0.246888264 |
| 0 | 0 | 0 | 0 | 0 |
| 0.023120482 | 0.002866097 | 0.004094752 | 0.044348555 | 0.004642645 |
| 0.461514703 | 5.247583309 | 0.217957155 | 0 | 0.305973035 |
| 0 | 0.002301395 | 0 | 0.418728887 | 0 |
| 0 | 0 | 0 | 0 | 0 |
| 0 | 0 | 0 | 0 | 0 |
| 0 | 0 | 0 | 0.048716477 | 0 |
| 0.002363416 | 0 | 0 | 0 | 0.003170523 |
| 0.017955556 | 0.212652803 | 0.095545099 | 0.156453289 | 0.187297097 |
| 0 | 0.032654336 | 0.037772123 | 0 | 0 |
| 1.944583575 | 4.213118666 | 0 | 0.459741093 | 1.488740236 |
| 0 | 0 | 0 | 0 | 0 |
| 0 | 0 | 0 | 0 | 0 |
| 0 | 0 | 0 | 0.248147923 | 0.005002673 |
| 0 | 0.003686125 | 0 | 0 | 0 |
| 0 | 0 | 0 | 0.008272215 | 0 |
| 0 | 0 | 0 | 0 | 0.005916286 |
| 0 | 0 | 0 | 0 | 0 |
| 0.00265239 | 0 | 0.001434877 | 0.073099996 | 0 |
| 0.120778878 | 0.15275259 | 0 | 0.132215503 | 0.193304782 |
| 0.874182248 | 0.165324365 | 0 | 0 | 0 |
| 0.121491745 | 0 | 0 | 0.37086594 | 0.059934332 |
| 0.003371067 | 0 | 0.021509881 | 0 | 0.002148598 |
| 0.008550721 | 0.028347609 | 0.219882693 | 0.097064159 | 0.025092359 |
| 0.005506586 | 0 | 0 | 0.023907144 | 0 |
| 0 | 0 | 0 | 0 | 0 |
| 0.316441965 | 0 | 0.408045167 | 3.839981159 | 0 |
| 0 | 0 | 0 | 0 | 0 |
| 0.006745034 | 0 | 0 | 0.11619226 | 0 |
| 0 | 0 | 0 | 0 | 0 |
| 0 | 0 | 0 | 0 | 0 |
| 0 | 0 | 0 | 0 | 0 |
| 0.111934805 | 0.004122264 | 0.058246091 | 0.088249639 | 0.052420733 |
| 0 | 0.001844634 | 0 | 0.000744883 | 0 |
| 0 | 0 | 0 | 0 | 0 |
| 0.381301791 | 0.180844779 | 0.065178953 | 0 | 0.420301258 |

| | | | | |
|---|---|---|---|---|
| 0 | 0 | 0 | 0 | 0 |
| 0 | 0 | 0 | 0 | 0 |
| 0 | 0 | 0 | 0 | 0 |
| 0.00467485 | 0 | 0.002809976 | 0.005162951 | 0.003484062 |
| 0 | 0 | 0 | 0 | 0 |
| 0 | 0 | 0 | 0.072647269 | 0 |
| 0 | 0 | 0.031276616 | 0.301378263 | 0 |
| 0.244400786 | 0.002722488 | 0.051713729 | 0.000755817 | 0.001885864 |
| 0 | 0 | 0 | 0 | 0 |
| 0 | 0 | 0 | 0 | 0 |
| 0.00154268 | 0.005972451 | 0 | 0.003267137 | 0.000913017 |
| 0 | 0 | 0 | 0 | 0 |
| 0.064497698 | 0 | 0.000608929 | 0 | 0 |
| 0 | 0 | 0 | 0.321170844 | 10.93043635 |
| 0 | 0 | 0 | 0 | 0 |
| 0 | 0 | 0 | 0 | 0 |
| 0 | 0 | 0 | 0.004568192 | 0 |
| 0 | 0 | 0 | 0.004351707 | 0.004370379 |
| 0 | 0 | 0 | 0.505881541 | 0 |
| 0 | 0 | 0 | 0 | 0.002227724 |
| 0 | 0 | 0.002057421 | 0.047070357 | 0.007765476 |
| 0 | 0 | 0 | 0 | 0 |
| 0.139180518 | 0 | 0 | 0.165066089 | 0.867538678 |
| 0 | 0 | 0 | 0 | 0 |
| 0 | 0 | 0 | 0 | 0 |
| 0 | 0 | 0 | 0 | 0 |
| 0 | 0 | 0 | 0 | 0 |
| 0 | 0 | 0 | 0 | 0 |
| 0 | 0 | 0 | 0 | 0 |
| 1.702033397 | 0 | 0.024303168 | 2.139789697 | 0.025249205 |
| 0.00318946 | 0 | 0.00479283 | 0 | 0 |
| 0 | 0 | 0 | 0 | 0 |
| 0 | 0 | 0 | 0 | 0 |
| 0 | 0 | 0 | 0 | 0 |
| 0.002146662 | 0.00131007 | 0.036912466 | 0.003948759 | 0.004900411 |
| 0 | 0 | 0 | 0 | 0 |
| 0.049160836 | 0.110611225 | 0 | 0.07383931 | 0 |
| 0.001305906 | 0.011506268 | 0 | 0 | 0 |
| 0 | 0 | 0 | 0 | 0 |
| 0.004025229 | 0 | 0 | 0 | 0 |
| 0 | 0 | 0.269553638 | 0.010275804 | 0.209026635 |
| 0 | 0 | 0 | 0 | 0 |
| 0 | 0 | 0 | 0 | 0 |
| 0 | 0 | 0 | 0 | 0 |
| 0 | 1.599701017 | 0.059662215 | 0.029143009 | 0 |
| 0 | 0 | 0 | 0 | 0 |
| 0 | 0 | 0.009649331 | 0.063173923 | 0.001835926 |
| 0 | 0 | 0 | 0 | 0 |

| | | | | |
|---|---|---|---|---|
| 0 | 0 | 0 | 0 | 0 |
| 0 | 0 | 0 | 0 | 0 |
| 0 | 0 | 0 | 1.431726276 | 0 |
| 0.03101347 | 0 | 0.009076842 | 0.618876031 | 0.182011999 |
| 0 | 0 | 0 | 0 | 0 |
| 0.000701949 | 0.000449806 | 0 | 0.405119055 | 0 |
| 0 | 0 | 0 | 0 | 0.106270009 |
| 0 | 0 | 0 | 0 | 0 |
| 0.375071199 | 0.589078915 | 0.006495533 | 0 | 0.024516564 |
| 0.001269089 | 0 | 0 | 0.009460763 | 0 |
| 0 | 0 | 0 | 0.00087276 | 0 |
| 0 | 0 | 0 | 0 | 0 |
| 0 | 0 | 0.002368358 | 0 | 0 |
| 0 | 0 | 0 | 0 | 0 |
| 0.01375184 | 8.072599212 | 0.349652011 | 0.168741484 | 0.022788823 |
| 0.867059896 | 0.001297199 | 0 | 0 | 0 |
| 0.046835738 | 0 | 0 | 0 | 0 |
| 0 | 0 | 0 | 0 | 0 |
| 0 | 0 | 0 | 0 | 0 |
| 0 | 0 | 0 | 0 | 0 |
| 0 | 0 | 0.001241692 | 0.162747092 | 0 |
| 0 | 0 | 0 | 0 | 0 |
| 0 | 0 | 0 | 0 | 0 |
| 0 | 0 | 0 | 0 | 0 |
| 0.016149734 | 0 | 0.050844179 | 0.016865714 | 0 |
| 0.122023913 | 0 | 30.19561248 | 2.793242362 | 0.023925122 |
| 0.27729495 | 0 | 0 | 0.044178266 | 0 |
| 0.00131474 | 0 | 0 | 0 | 0.002139812 |
| 0 | 0 | 8.63794768 | 0.452560045 | 0 |
| 0 | 0 | 0 | 0.218500611 | 0 |
| 0 | 0 | 0 | 0 | 0 |
| 0.009207016 | 0 | 0 | 0.019065598 | 0 |
| 0.003076928 | 0 | 0 | 0 | 0 |
| 0 | 0 | 0 | 0.000958231 | 0.001171548 |
| 0.009784807 | 0.003234558 | 0.000840212 | 0.014683421 | 0.00275763 |
| 0.000823413 | 0.003517601 | 0.002821159 | 0 | 0.001364517 |
| 0 | 0 | 0.010113139 | 0 | 0 |
| 1.356729098 | 4.395988168 | 0.137147448 | 0.472964169 | 0.683664166 |
| 0 | 0 | 0 | 3.705535232 | 0 |
| 0 | 0 | 0 | 0 | 0 |
| 0 | 0 | 0 | 0 | 0 |
| 0.004855538 | 0 | 0 | 0.001771876 | 0.030726414 |
| 0.338462862 | 5.204632254 | 0.112611381 | 0.036907949 | 0.790606052 |
| 0 | 0 | 0.044061945 | 0 | 0 |
| 0 | 0 | 0 | 0 | 0 |
| 0.17848248 | 0 | 0.001828686 | 0.002008929 | 0 |
| 0 | 0 | 0 | 0 | 0.02392673 |
| 0.01938709 | 1.659608556 | 0.012773767 | 0.249973105 | 0.002942057 |

| | | | | |
|---|---|---|---|---|
| 0 | 0 | 0.003843587 | 0.041074465 | 0.01316337 |
| 0 | 0 | 0 | 0 | 0 |
| 0 | 0 | 0 | 0.09457034 | 0 |
| 0 | 0 | 0 | 0 | 0 |
| 0.007196796 | 0 | 0.001996558 | 0 | 0 |
| 0 | 0 | 0 | 0.092008699 | 0 |
| 0 | 0 | 0 | 0.067998348 | 0 |
| 0 | 0 | 0 | 0 | 0 |
| 0 | 0 | 0 | 0 | 0 |
| 0 | 0 | 0 | 0 | 0 |
| 0 | 0 | 0 | 0.001626769 | 0 |
| 0.397348769 | 0.005454575 | 2.082634075 | 0.358239927 | 0.755070956 |
| 1.244769744 | 3.955287681 | 0.026592301 | 0.004088472 | 0 |
| 0 | 0 | 0 | 0 | 0 |
| 0.033505791 | 0.030769658 | 0.521453555 | 0.010124175 | 0.021977209 |
| 0 | 0 | 0 | 0 | 0 |
| 0 | 0 | 0 | 0.000603671 | 0 |
| 0 | 0 | 0 | 0 | 0 |
| 0.010662161 | 0 | 0.000505962 | 0 | 0 |
| 0 | 0 | 0 | 0.029659458 | 0 |
| 0 | 0 | 0 | 0.052476394 | 0 |
| 0 | 0 | 0 | 0 | 0 |
| 0.47035723 | 0 | 1.09170167 | 0.784706591 | 0.034447074 |
| 0 | 0 | 0 | 0 | 0 |
| 0 | 0 | 0 | 0 | 0 |
| 0 | 0 | 0 | 0 | 0 |
| 0 | 0 | 0 | 0.067805509 | 0 |
| 0 | 0 | 0 | 0 | 0 |
| 0.011341619 | 0 | 0 | 0.000521104 | 0 |
| 0 | 0 | 0 | 0 | 0 |
| 0 | 0 | 0 | 0 | 0 |
| 0.082249042 | 0.006802537 | 0.010736654 | 0.00975931 | 0.072771349 |
| 1.190572774 | 0 | 0 | 0 | 0 |
| 0.022274488 | 0.000591031 | 0.016465764 | 0.039379709 | 0.061738525 |
| 0.00089608 | 0.003253827 | 0.002746957 | 0 | 0.000636401 |
| 0.811150422 | 0 | 2.172397781 | 0.650709984 | 0.002461196 |
| 0 | 0 | 0.001290911 | 0 | 0 |
| 0 | 0 | 0 | 0.490226428 | 0 |
| 0 | 0 | 0 | 0.002372091 | 0 |
| 0 | 0 | 0 | 0 | 0 |
| 0 | 0 | 0 | 0.136180684 | 1.602791366 |
| 0 | 0 | 0 | 0 | 0 |
| 0 | 0 | 0 | 0 | 0 |
| 0 | 0 | 0.00210473 | 0.071882983 | 0 |
| 0.039193152 | 0 | 0.006844659 | 0.001914338 | 0.035316445 |
| 0 | 0 | 0 | 0 | 0 |
| 0.30139089 | 0 | 0 | 0.197811707 | 0 |
| 0.004174517 | 0 | 0.00075277 | 0.000678983 | 0.006127184 |

| | | | | |
|---|---|---|---|---|
| 0 | 0 | 0 | 0 | 0 |
| 0.050727011 | 0 | 0 | 0 | 0 |
| 0 | 0 | 0 | 0 | 0 |
| 0.027485966 | 0 | 0 | 0 | 0 |
| 0 | 0 | 0 | 0 | 0 |
| 0 | 0 | 0 | 0 | 0 |
| 0 | 0 | 0 | 9.031725271 | 0 |
| 0.006380336 | 0 | 0 | 0.016945375 | 0 |
| 0 | 0 | 0 | 0 | 0 |
| 0 | 0 | 0 | 0.004599971 | 0 |
| 0.000853729 | 0 | 0 | 0 | 0 |
| 0 | 0 | 0 | 0 | 0.000659722 |
| 0 | 0 | 0 | 0 | 0 |
| 0 | 0 | 0 | 0 | 0 |
| 0 | 0 | 0 | 0 | 0 |
| 0 | 0 | 0 | 0 | 0 |
| 0 | 0 | 0 | 0.904163109 | 0 |
| 0 | 0 | 0 | 0 | 0 |
| 0.441915172 | 3.695894332 | 0.020757681 | 2.597815557 | 0.087527186 |
| 0 | 0 | 0 | 0.000702323 | 0 |
| 0 | 0 | 0 | 0.000562163 | 0.001963738 |
| 0 | 0 | 0 | 0 | 0 |
| 0 | 0 | 0 | 0 | 0 |
| 0 | 0 | 0 | 0 | 0 |
| 0 | 0 | 0.001391327 | 0 | 0 |
| 0.00065743 | 0.021941583 | 0.004742042 | 0 | 0.000583638 |
| 0 | 0 | 0 | 0 | 0 |
| 0.071105957 | 0.083328608 | 0.111251325 | 0.28586288 | 0.000761687 |
| 0 | 0.000727362 | 0 | 0.302087734 | 0 |
| 0.002844762 | 0 | 4.476712022 | 0.543874229 | 0.419491957 |
| 0 | 0 | 0 | 0 | 0 |
| 0 | 0 | 0 | 0 | 0 |
| 0 | 0 | 0 | 0.006101398 | 0 |
| 0 | 0 | 0 | 0 | 0 |
| 0 | 0.02593658 | 0 | 0 | 0.00468906 |
| 0 | 0 | 0.049474963 | 0 | 0 |
| 0 | 0 | 0 | 2.71168029 | 0 |
| 0.001758941 | 0 | 0.003348023 | 0.003576134 | 0 |
| 0 | 0 | 0 | 0 | 0 |
| 0 | 0 | 0 | 0 | 0 |
| 7.721647713 | 2.254295994 | 1.739908253 | 1.137244229 | 17.67168651 |
| 1.201286539 | 0.469672351 | 0.060495269 | 0.010277079 | 2.922950505 |
| 0 | 0 | 1.061737 | 0.012745192 | 0.180669122 |
| 0 | 0 | 0 | 0 | 0 |
| 0 | 0 | 0 | 0 | 0 |
| 0 | 0 | 0 | 0 | 0 |
| 0 | 0 | 0 | 0.002705441 | 0 |
| 0.003877757 | 0.00049697 | 0.001538365 | 0.003679171 | 0 |

| | | | | |
|---|---|---|---|---|
| 0 | 0 | 0 | 0 | 0 |
| 0.004786312 | 0 | 0 | 0.007460539 | 0.017185185 |
| 0.005590143 | 0 | 0.003192136 | 0.00050513 | 0 |
| 0 | 0 | 0 | 0 | 0 |
| 0.218216563 | 4.806368534 | 2.546094097 | 0.206037736 | 4.020028546 |
| 0 | 0 | 0.005752721 | 0.075282444 | 0 |
| 0 | 0 | 0 | 0 | 0 |
| 0 | 0 | 0 | 0 | 0 |
| 0.441125299 | 0.265419168 | 0.003590915 | 0.000647786 | 0.030359732 |
| 0 | 0 | 0 | 0 | 0 |
| 0 | 0 | 0 | 0 | 0 |
| 0 | 0 | 0 | 0 | 0 |
| 0 | 0 | 0.01551486 | 0.038347141 | 0.063123382 |
| 0.022489168 | 0.004274441 | 0.00704533 | 0.003099874 | 0.035192411 |
| 0.583417825 | 0 | 0 | 1.819591262 | 0 |
| 0 | 0 | 0.085028102 | 0.193535747 | 0.406337247 |
| 0.003958146 | 0.012505689 | 0.045799152 | 0.003017767 | 0.0179598 |
| 0 | 0 | 0 | 0 | 0 |
| 0 | 0 | 0 | 0 | 0 |
| 0 | 0 | 0 | 0 | 0 |
| 0 | 0 | 0 | 0 | 0 |
| 0 | 0 | 0 | 0 | 0 |
| 0 | 0 | 0 | 0 | 0 |
| 0 | 0 | 0 | 0 | 0 |
| 0 | 0 | 0 | 0 | 0 |
| 0 | 0 | 0 | 0 | 0 |
| 0.069552455 | 0 | 0.228730812 | 0.013023175 | 0 |
| 0 | 0.002592323 | 0 | 0 | 0.001340789 |
| 0.022972122 | 0 | 0 | 0 | 0 |
| 0 | 0 | 0 | 0 | 0 |
| 0 | 0 | 0 | 0 | 0 |
| 0 | 0 | 0 | 0 | 0.001439962 |
| 0 | 0 | 0 | 0.106625908 | 0 |
| 0 | 0 | 0 | 0.027705087 | 0.076098024 |
| 0 | 0 | 0 | 0.40612957 | 0 |
| 0 | 0.097960149 | 0 | 0 | 0 |
| 0 | 0 | 0.00603783 | 0.000778 | 0 |
| 0 | 0 | 0 | 0 | 0 |
| 0 | 0 | 0 | 0 | 0 |
| 0 | 0 | 0 | 0 | 0 |
| 0 | 0 | 0 | 0.169199387 | 1.985346392 |
| 0 | 0 | 0 | 0 | 0 |
| 0 | 0 | 0 | 0 | 0 |
| 0 | 0 | 0.064641967 | 0.185762194 | 0 |
| 0 | 0 | 0 | 0 | 0 |
| 0 | 0 | 0 | 0 | 0 |
| 0 | 0 | 0 | 0 | 0 |
| 0 | 0 | 0 | 0 | 0 |

| | | | | |
|---|---|---|---|---|
| 0.048723548 | 0 | 0 | 0.135443035 | 0.111990329 |
| 0 | 0 | 0 | 0 | 0 |
| 0 | 0 | 0 | 0 | 0 |
| 0 | 0 | 0 | 0 | 0.001518851 |
| 0.040866085 | 0 | 0.192927022 | 0.037441999 | 0 |
| 0 | 0 | 0 | 0.001875174 | 0 |
| 0.049850088 | 0.218756682 | 0.019432144 | 0.014606157 | 0.105159887 |
| 0 | 0 | 0 | 0.001293324 | 0 |
| 0 | 0 | 0 | 0 | 0 |
| 0 | 0 | 0 | 0 | 0.000612944 |
| 0.008578911 | 0 | 0 | 0.108978317 | 0 |
| 0.052515241 | 0 | 0.013880413 | 0.006142914 | 0.017030428 |
| 0 | 0 | 0 | 0 | 0 |
| 0 | 0 | 0 | 0 | 0 |
| 0 | 0 | 0 | 0 | 0 |
| 0 | 0 | 0 | 0 | 0 |
| 0 | 0 | 0 | 0 | 0 |
| 0 | 0 | 0.001140883 | 0 | 0 |
| 0 | 0 | 0 | 0 | 0 |
| 0 | 0 | 0 | 0 | 0 |
| 0.323762206 | 0.012044446 | 0.005790228 | 0.776181378 | 0 |
| 0 | 0 | 0 | 0 | 0 |
| 0.000863833 | 0 | 0.021807906 | 0.041682252 | 0.011042989 |
| 0.01075381 | 0 | 0.003878363 | 0.002082263 | 0.005818986 |
| 0 | 0 | 0 | 0.000622346 | 0 |
| 0.005981051 | 0 | 0 | 0 | 0 |
| 0 | 0 | 0 | 0 | 0 |
| 0 | 0 | 0 | 0 | 0 |
| 0 | 0 | 0 | 0 | 0 |
| 0.763470808 | 0 | 0 | 0 | 0.070139668 |
| 0 | 0 | 0 | 0.092398581 | 0 |
| 0 | 0 | 0 | 0 | 0 |
| 0 | 0 | 0 | 0 | 0 |
| 0.63197128 | 0 | 0 | 0 | 0 |
| 0 | 0 | 0 | 0.00288562 | 0 |
| 0 | 0 | 0 | 0 | 0 |
| 0 | 0.150276572 | 0 | 0.000651363 | 0 |
| 0 | 0 | 0 | 0 | 0 |
| 0 | 0 | 0 | 0 | 0 |
| 0 | 0 | 0 | 0.066011323 | 0 |
| 0 | 0 | 0 | 0 | 0 |
| 0 | 0 | 0 | 0 | 0 |
| 0 | 0 | 0 | 0 | 0 |
| 0 | 0 | 0 | 0 | 0 |
| 0 | 0 | 0 | 0 | 0 |
| 0 | 0 | 0 | 0 | 0 |
| 0 | 0 | 0 | 0 | 0 |
| 0 | 0 | 0 | 0.076985358 | 0 |

| | | | | |
|---|---|---|---|---|
| 0 | 0.019949776 | 0.000447135 | 0 | 0.01584926 |
| 0 | 0 | 0 | 2.585870324 | 0 |
| 0 | 0 | 0 | 0.007988153 | 0 |
| 0 | 0 | 0 | 0 | 0 |
| 0 | 0 | 0 | 0 | 0 |
| 0 | 0 | 0 | 0.077323887 | 0 |
| 0 | 0 | 0.000572893 | 0 | 0 |
| 1.680524433 | 0.394813796 | 0.516188382 | 0.057169385 | 1.031666361 |
| 0.092632206 | 0.048495396 | 0.029711435 | 0.606672413 | 0.005528399 |
| 0.059791175 | 0.004630085 | 0 | 0 | 0.060937833 |
| 0 | 0 | 0 | 0.050136318 | 0 |
| 0 | 0 | 0.000712819 | 0 | 0 |
| 0.040673766 | 0 | 0 | 0.047178313 | 0 |
| 0 | 0 | 0 | 1.701227142 | 0 |
| 0.008685708 | 0 | 0 | 0 | 0 |
| 0 | 0 | 0 | 0 | 0 |
| 0.207521055 | 0 | 0 | 0 | 0 |
| 0 | 0 | 0 | 0.092301761 | 0 |
| 0 | 0 | 0 | 0 | 0 |
| 0 | 0 | 0 | 0.330425336 | 0.005850971 |
| 0 | 0 | 0 | 0 | 0 |
| 0 | 0 | 0 | 0 | 0 |
| 0 | 0 | 0 | 0 | 0 |
| 0 | 0 | 0 | 0 | 0 |
| 0 | 0 | 0 | 0.067149679 | 0.012791724 |
| 0 | 0 | 0 | 0 | 0 |
| 0 | 0 | 0 | 0 | 0 |
| 0 | 0 | 0 | 0 | 0 |
| 0 | 0 | 0 | 0 | 0 |
| 0 | 0 | 0 | 0 | 0 |
| 0 | 0.003264684 | 0 | 0 | 0 |
| 0 | 0 | 0 | 0 | 0 |
| 0 | 0 | 0 | 0.01016335 | 0 |
| 0 | 0 | 0 | 0 | 0 |
| 0 | 0 | 0 | 0.003526313 | 0 |
| 0 | 0 | 0 | 0 | 0 |
| 0 | 0 | 0 | 0 | 0 |
| 0 | 0 | 0 | 0 | 0.05320248 |
| 0 | 0 | 0 | 0.002797448 | 0 |
| 0 | 0.001762489 | 0 | 0 | 0 |
| 0 | 0 | 0 | 0 | 0 |
| 0.463946781 | 0 | 0 | 0.127152416 | 0.32002596 |
| 0 | 0 | 0 | 0 | 0 |
| 0 | 0 | 0 | 0 | 0 |
| 0 | 0 | 0 | 0 | 0 |
| 0 | 0 | 0 | 0.000896553 | 0 |
| 0 | 0 | 0 | 0 | 0 |
| 0 | 0 | 0 | 0 | 0.001093287 |

| | | | | |
|---|---|---|---|---|
| 0 | 0 | 0 | 0 | 0 |
| 0 | 0 | 0.001210676 | 0 | 0.030905108 |
| 0 | 0 | 0 | 0 | 0 |
| 0 | 0 | 0 | 0 | 0 |
| 0.52128625 | 0 | 14.18478168 | 3.825420389 | 0.098518085 |
| 0.134970536 | 0 | 0 | 0 | 0 |
| 0 | 0 | 0 | 0 | 0.034262006 |
| 0.033224606 | 0.033013286 | 0.000947979 | 0 | 0.013440937 |
| 0.003779264 | 0 | 0 | 0.016932575 | 0.004224902 |
| 0 | 0 | 0 | 0.102133304 | 0 |
| 0 | 0 | 0 | 0 | 0 |
| 0 | 0 | 0 | 0 | 0 |
| 0 | 0 | 0 | 0 | 0 |
| 0 | 0 | 0 | 0 | 0 |
| 0 | 0 | 0 | 0 | 0 |
| 0 | 0 | 0.038971113 | 0.037944435 | 0.129956467 |
| 0 | 0 | 0 | 0 | 0 |
| 0 | 0 | 0 | 0 | 0 |
| 0 | 0 | 0 | 0.002241898 | 0 |
| 0 | 0 | 0 | 0 | 0 |
| 0 | 0 | 0 | 0 | 0 |
| 0 | 0 | 0 | 0 | 0 |
| 0 | 0 | 0 | 0 | 0 |
| 0 | 0.000583067 | 0 | 0.000739982 | 0.002154079 |
| 0 | 0 | 0.008905076 | 0.133760156 | 0 |
| 0 | 0 | 0 | 0 | 0 |
| 0.016644322 | 0 | 0.40053529 | 0 | 0.00078806 |
| 0 | 0 | 0 | 0 | 0 |
| 0 | 0 | 0 | 0 | 0 |
| 0 | 0 | 0 | 0 | 0 |
| 0 | 0 | 0 | 0 | 0 |
| 0 | 0 | 0 | 0 | 0.001136169 |
| 0 | 0 | 0 | 0 | 0 |
| 0 | 0 | 0 | 0 | 0 |
| 0 | 0 | 0 | 0 | 0 |
| 0 | 0 | 0 | 0 | 0 |
| 0 | 0 | 0 | 0 | 0 |
| 0.179744228 | 0 | 0.056944846 | 1.145488848 | 0.080011943 |
| 0 | 0 | 0 | 0.149293161 | 0 |
| 0 | 0 | 0 | 0 | 0 |
| 0 | 0 | 0 | 0 | 0 |
| 0 | 0 | 0 | 0.139411328 | 0 |
| 0 | 0 | 0 | 0 | 0 |
| 0 | 0 | 0 | 0 | 0 |
| 0 | 0 | 0 | 0 | 0 |
| 0 | 0 | 0 | 0 | 0 |
| 0 | 0 | 0 | 0 | 0 |
| 0 | 0 | 0 | 0 | 0 |

| | | | | |
|---|---|---|---|---|
| 0.013461193 | 0 | 1.693246054 | 0.083377527 | 0 |
| 0 | 0 | 0 | 0 | 0 |
| 0 | 0 | 0 | 0 | 0 |
| 0.001557342 | 0 | 0 | 0 | 0 |
| 0 | 0 | 0 | 0.038515738 | 0 |
| 0.058574063 | 0 | 0 | 0.028758091 | 0 |
| 0 | 0 | 0 | 0 | 0 |
| 0 | 0 | 0 | 0 | 0 |
| 0 | 0 | 0 | 0 | 0 |
| 0 | 0 | 0.001571152 | 0 | 0 |
| 0 | 0 | 0 | 0 | 0 |
| 0 | 0 | 0 | 0.001477981 | 0 |
| 0 | 0 | 0 | 0 | 0 |
| 0 | 0 | 0 | 0 | 0 |
| 0 | 0 | 0 | 0 | 0 |
| 0 | 0 | 0 | 0.019422514 | 0 |
| 0 | 0 | 0 | 0 | 0 |
| 0 | 0 | 0 | 0 | 0 |
| 0 | 0 | 0.002871619 | 0 | 0 |
| 0 | 0 | 0 | 0 | 0 |
| 0.030807619 | 0.005999839 | 0.016339371 | 0 | 0.014371995 |
| 0 | 0 | 0 | 0 | 0 |
| 0.00961251 | 0.004644188 | 0 | 0.003412334 | 0 |
| 0 | 0 | 0 | 0.169217734 | 0 |
| 0 | 0 | 0 | 0 | 0 |
| 0 | 0 | 0 | 0.271398222 | 0 |
| 0 | 0 | 0 | 0 | 0 |
| 0 | 0 | 0 | 0 | 0 |
| 0 | 0 | 0 | 0 | 0 |
| 0 | 0 | 0 | 0 | 0 |
| 0 | 0 | 0 | 0 | 0 |
| 0 | 0 | 0 | 0 | 0 |
| 0.34958062 | 0.039143366 | 0 | 0 | 0 |
| 0 | 0 | 0 | 0 | 0 |
| 1.464946034 | 3.756893055 | 0.440623216 | 0.002988413 | 0 |
| 0.00275192 | 0 | 0 | 0 | 0.001425105 |
| 0 | 0 | 0 | 0 | 0 |
| 0 | 0 | 0 | 0.298026424 | 0 |
| 0.003580909 | 0 | 0.054528114 | 0.001078579 | 0.001648359 |
| 0 | 0 | 0 | 0 | 0 |
| 0 | 0 | 0 | 0 | 0 |
| 0 | 0 | 0 | 0.007654774 | 0 |
| 0.050245867 | 0 | 0.088928009 | 0.203523331 | 0.040475944 |
| 0 | 0 | 0 | 0 | 0 |
| 0 | 0 | 0 | 0 | 0 |
| 0.029188988 | 0 | 0 | 0.03843631 | 0.01946189 |
| 0 | 0 | 0 | 0 | 0 |
| 0 | 0 | 0 | 0 | 0 |

| | | | | |
|---|---|---|---|---|
| 0 | 0 | 0 | 0 | 0 |
| 0 | 0 | 0 | 0 | 0 |
| 0 | 0 | 0 | 0 | 0 |
| 0 | 0 | 0 | 0 | 0 |
| 0 | 0 | 0 | 0 | 0 |
| 0 | 0 | 0 | 0 | 0 |
| 0 | 0 | 0 | 0.010383448 | 0 |
| 0.136209067 | 0.023831982 | 0.01673944 | 0 | 0.03697878 |
| 0.155776191 | 0 | 0.106275167 | 0.013724161 | 0.021921759 |
| 0 | 0 | 0 | 0.000644772 | 0.004054152 |
| 0 | 0 | 0 | 0 | 0 |
| 0 | 0 | 0 | 0 | 0 |
| 0.001264154 | 0 | 0 | 0 | 0 |
| 0 | 0 | 0 | 0.053570798 | 0 |
| 0 | 0 | 0 | 0 | 0.003276308 |
| 0 | 0 | 0 | 0 | 0 |
| 0 | 0 | 0 | 0 | 0 |
| 0 | 0 | 0 | 0 | 0 |
| 0 | 0 | 0 | 0 | 0 |
| 0 | 0.001339199 | 0.282645819 | 0.574997353 | 0 |
| 0 | 0 | 0 | 0.126418172 | 0.002083029 |
| 0 | 0 | 0 | 0 | 0 |
| 0 | 0 | 0 | 0 | 0 |
| 0 | 0 | 0 | 0 | 0 |
| 0 | 0 | 0 | 0 | 0 |
| 0 | 0 | 0 | 0 | 0 |
| 0 | 0 | 0 | 0 | 0 |
| 0 | 0 | 0 | 0.155037958 | 0 |
| 0 | 0 | 0 | 0 | 0 |
| 0 | 0 | 0 | 0.064171053 | 0 |
| 0.175748451 | 0.007475427 | 0.107560224 | 0.207378131 | 0.623907861 |
| 0 | 0 | 0 | 0 | 0 |
| 0 | 0 | 0.005040472 | 0.002612874 | 0 |
| 0 | 0 | 0 | 0.249920485 | 0 |
| 0 | 0 | 0 | 0 | 0 |
| 0 | 0 | 0 | 0 | 0 |
| 0 | 0 | 0 | 0 | 0 |
| 0 | 0 | 0 | 0 | 0 |
| 0 | 0 | 0 | 0.004648897 | 0 |
| 0 | 0 | 0 | 0 | 0 |
| 0 | 0 | 0 | 0 | 0 |
| 0 | 0.004356207 | 0.050610948 | 0 | 0 |
| 0 | 0 | 0 | 0.004238345 | 0 |
| 0 | 0 | 0 | 0.002981496 | 0 |
| 0 | 0 | 0 | 0 | 0 |
| 0 | 0 | 0.177811486 | 0 | 0 |
| 0.009199234 | 0 | 0 | 0 | 0 |
| 0 | 0 | 0 | 0 | 0 |

| | | | | |
|---|---|---|---|---|
| 0 | 0 | 0 | 0 | 0 |
| 0 | 0 | 0 | 0 | 0 |
| 0 | 0 | 0 | 0 | 0 |
| 0 | 0 | 0 | 0 | 0 |
| 0.098219694 | 0 | 0.16665825 | 0.017077665 | 0.006779029 |
| 0 | 0 | 0 | 0.114116497 | 0 |
| 0 | 0 | 0.006465769 | 0 | 0 |
| 0 | 0 | 0 | 0.002197363 | 0 |
| 0 | 0 | 0 | 0 | 0 |
| 0 | 0 | 0 | 0 | 0 |
| 0 | 0 | 0 | 0.008500604 | 0.051642068 |
| 0.00815002 | 0 | 0.016847241 | 0 | 0.006823479 |
| 0 | 0 | 0.004948059 | 0.033007947 | 0 |
| 0 | 0 | 0.450639718 | 0 | 0 |
| 0 | 0 | 0 | 0.012114151 | 0.005379277 |
| 0 | 0.00379709 | 0.19818926 | 0 | 0.034398959 |
| 0 | 0.001632815 | 0 | 0.001243343 | 0 |
| 0 | 0 | 0 | 0 | 0 |
| 0.135165251 | 0.111883652 | 0.002035664 | 0 | 0.017460145 |
| 0.074092102 | 0.04407766 | 0.010631828 | 0.129460771 | 0.003954691 |
| 0 | 0 | 0 | 0 | 0 |
| 0 | 0 | 0 | 0.003620131 | 0 |
| 0 | 0 | 0 | 0.077252708 | 0 |
| 0 | 0 | 0 | 0 | 0 |
| 0 | 0 | 0 | 0 | 0 |
| 0 | 0 | 0 | 0 | 0 |
| 0.004094717 | 0 | 0 | 0.000640388 | 0.00242341 |
| 0 | 0 | 0 | 0 | 0 |
| 0 | 0 | 0 | 0 | 0 |
| 0 | 0 | 0 | 0 | 0 |
| 0.001071133 | 0 | 0.001448641 | 0.001209856 | 0 |
| 0 | 0 | 0 | 0 | 0 |
| 0 | 0 | 0 | 0 | 0 |
| 0 | 0 | 0 | 0 | 0 |
| 0 | 0 | 0 | 0.022548604 | 0 |
| 0.005059259 | 0 | 0 | 0 | 0 |
| 0 | 0 | 0 | 0 | 0 |
| 0.030979853 | 0.002946957 | 0.001357031 | 0.061200681 | 0.030137796 |
| 0 | 0.00128894 | 0 | 0.005418652 | 0.035356745 |
| 0 | 0 | 0 | 0 | 0 |
| 0.001878211 | 0 | 0 | 0.001357733 | 0 |
| 0 | 0 | 0 | 0 | 0 |
| 0 | 0 | 0 | 0 | 0 |
| 0.002045875 | 0.006952214 | 0 | 0 | 0 |
| 0 | 0 | 0 | 0 | 0 |
| 0 | 0 | 0 | 0.008451419 | 0 |
| 0 | 0 | 0 | 0 | 0 |
| 0 | 0 | 0 | 0 | 0 |

| | | | | |
|---|---|---|---|---|
| 0 | 0 | 0 | 0 | 0 |
| 0 | 0 | 0.073527017 | 0.004955673 | 0.005456756 |
| 0 | 0 | 0 | 0 | 0 |
| 0.001364419 | 0.00166536 | 0 | 0.002113542 | 0 |
| 0.001265476 | 0 | 0 | 0 | 0 |
| 0 | 0 | 0 | 0.000632481 | 0 |
| 0 | 0 | 0 | 0.009486023 | 0 |
| 0 | 0 | 0.0015119 | 0.047729592 | 0 |
| 0 | 0 | 0 | 0.007511882 | 0 |
| 0 | 0 | 0 | 0.009616638 | 0 |
| 0.012885031 | 0 | 0 | 0.05828786 | 0 |
| 0 | 0 | 0 | 0 | 0 |
| 0.014231373 | 0.004131771 | 0.510887699 | 0.096259597 | 0.012920214 |
| 0 | 0 | 0 | 0 | 0 |
| 0 | 0 | 0 | 0 | 0 |
| 0 | 0 | 0 | 0 | 0 |
| 0 | 0 | 0 | 0 | 0 |
| 0 | 0 | 0 | 0 | 0 |
| 0 | 0 | 0 | 0 | 0 |
| 0 | 0 | 0 | 0 | 0 |
| 0 | 0 | 0 | 0 | 0 |
| 0 | 0 | 0 | 0 | 0 |
| 0.025653017 | 0 | 0.001640384 | 0 | 0.012275162 |
| 0.000858764 | 0.009171549 | 0 | 0.059508456 | 0.011943866 |
| 0 | 0 | 0 | 0 | 0 |
| 0.002061169 | 0 | 0 | 0.558854014 | 0 |
| 0 | 0 | 0 | 0.005418884 | 0 |
| 0 | 0 | 0 | 0 | 0 |
| 0 | 0 | 0 | 0 | 0 |
| 0 | 0 | 0 | 0 | 0 |
| 0 | 0 | 0.088158792 | 0.019384426 | 0.030254925 |
| 0 | 0 | 0 | 0.001807316 | 0 |
| 0 | 0 | 0 | 0 | 0 |
| 0 | 0 | 0 | 0 | 0 |
| 0 | 0 | 0 | 0.74514706 | 0 |
| 0 | 0 | 0 | 0 | 0.00527128 |
| 0 | 0 | 0 | 0 | 0 |
| 0 | 0 | 0 | 0 | 0 |
| 0 | 0.002932831 | 0.054316487 | 0 | 0 |
| 0 | 0 | 0 | 0 | 0 |
| 0 | 0 | 0 | 0 | 0 |
| 0 | 0 | 0 | 0 | 0 |
| 0 | 0 | 0 | 0 | 0 |
| 0 | 0 | 0 | 0 | 0 |
| 21.57592581 | 0 | 0 | 0 | 0 |
| 0.996198914 | 0.067896963 | 0.334935207 | 0.602060023 | 0.786475699 |
| 0 | 0 | 0 | 0 | 0 |
| 0 | 0 | 0 | 0 | 0 |

| | | | | |
|---|---|---|---|---|
| 0 | 0 | 0 | 0 | 0 |
| 0 | 0 | 0 | 0 | 0 |
| 0 | 0 | 0 | 0.058153796 | 0 |
| 0 | 0 | 0 | 0.002317653 | 0 |
| 0 | 0 | 0 | 0 | 0 |
| 0 | 0 | 0 | 0 | 0 |
| 0 | 0 | 0 | 0 | 0 |
| 0 | 0 | 0 | 0 | 0 |
| 0 | 0 | 0 | 0 | 0 |
| 0 | 0 | 0 | 0 | 0 |
| 0.004455667 | 0 | 0.001506503 | 0.000660545 | 0 |
| 0 | 0 | 0 | 0.00084531 | 0 |
| 0 | 0 | 0 | 0 | 0 |
| 0 | 0 | 0 | 0 | 0 |
| 0 | 0 | 0 | 0 | 0 |
| 0 | 0 | 0 | 0 | 0 |
| 0 | 0 | 0 | 0 | 0 |
| 0 | 0 | 0 | 0 | 0 |
| 0 | 0 | 0 | 0 | 0 |
| 0.032499388 | 0 | 0.019312861 | 0.000500569 | 0.054205986 |
| 0 | 0 | 0 | 0 | 0 |
| 0 | 0 | 0 | 0 | 0 |
| 0 | 0 | 0 | 0 | 0 |
| 0 | 0 | 0 | 0 | 0 |
| 0 | 0 | 0 | 0 | 0 |
| 0 | 0 | 0 | 0 | 0 |
| 0 | 0 | 0 | 0 | 0 |
| 0 | 0 | 0 | 0.011633247 | 0 |
| 0 | 0 | 0 | 0 | 0 |
| 0 | 0 | 0 | 0 | 0 |
| 0.000634553 | 0.000508274 | 0.072660408 | 0.001827672 | 0.007698833 |
| 0 | 0 | 0 | 0 | 0 |
| 0.069809981 | 0.07396646 | 0.002823276 | 0.007739473 | 0.002180275 |
| 0.008846693 | 0.001206155 | 0 | 0 | 0 |
| 0 | 0 | 0 | 0 | 0 |
| 0 | 0 | 0 | 0 | 0 |
| 0 | 0 | 0 | 0 | 0 |
| 0 | 0 | 0 | 0 | 0 |
| 0 | 0 | 0 | 0 | 0 |
| 0 | 0 | 0 | 0 | 0 |
| 0 | 0 | 0 | 0 | 0 |
| 0.001818429 | 0.010843226 | 0 | 0 | 0 |
| 0.023076655 | 0 | 0.112854439 | 0.021894864 | 0.038241414 |
| 0 | 0 | 0 | 0 | 0 |
| 0 | 0 | 0 | 0 | 0 |
| 0.032013475 | 0 | 0.559002585 | 0.259625521 | 0.00505248 |
| 0 | 0 | 0 | 0 | 0 |
| 0 | 0 | 0 | 0 | 0 |

| | | | | |
|---|---|---|---|---|
| 0 | 0 | 0 | 0 | 0 |
| 0 | 0 | 0 | 0.001835022 | 0 |
| 0.028917712 | 0.004825614 | 0.012017997 | 0.002296606 | 0.004813484 |
| 0 | 0 | 0 | 0 | 0 |
| 0.049966907 | 0 | 0.003415451 | 0.364789958 | 0.370881836 |
| 0.006145531 | 0.004500609 | 0 | 0 | 0.155462632 |
| 0 | 0 | 0 | 0 | 0 |
| 0 | 0 | 0 | 0 | 0 |
| 0 | 0 | 0 | 0.145048362 | 0 |
| 0 | 0 | 0 | 0 | 0 |
| 0 | 0 | 0 | 0 | 0 |
| 0 | 0 | 0 | 0 | 0 |
| 0 | 0 | 0 | 0 | 0 |
| 0 | 0 | 0 | 0 | 0 |
| 0 | 0 | 0 | 0 | 0 |
| 0 | 0 | 0 | 0 | 0 |
| 0 | 0 | 0 | 0.034299144 | 0 |
| 0 | 0 | 0 | 0 | 0 |
| 0 | 0 | 0 | 0 | 0 |
| 0 | 0 | 0 | 0 | 0 |
| 0 | 0 | 0 | 0 | 0 |
| 0 | 0 | 0 | 0 | 0 |
| 0 | 0 | 0 | 0.009002404 | 0 |
| 0 | 0 | 0 | 0 | 0 |
| 0 | 0 | 0 | 0.258167905 | 0 |
| 0.001498418 | 0 | 0 | 0.000761615 | 0.001330232 |
| 0 | 0 | 0 | 0 | 0 |
| 0.001453518 | 0 | 1.212566536 | 1.046993354 | 0 |
| 0 | 0 | 0 | 0 | 0 |
| 0 | 0 | 0 | 0 | 0 |
| 0 | 0 | 0 | 0.013944456 | 0 |
| 0 | 0 | 0 | 0 | 0 |
| 0 | 0 | 0 | 0 | 0 |
| 0 | 0 | 0 | 0 | 0 |
| 0 | 0 | 0 | 0 | 0 |
| 0 | 0 | 0 | 0 | 0 |
| 0 | 0 | 0 | 0 | 0 |
| 0.002983099 | 0 | 0 | 0 | 0 |
| 0 | 0.003450751 | 0.586014133 | 7.589759325 | 0 |
| 0 | 0 | 0 | 0 | 0 |
| 0 | 0 | 0 | 0 | 0 |
| 0.003454299 | 0 | 0.004396918 | 0.034839582 | 0 |
| 0.001315848 | 0.017666844 | 0.001016916 | 0 | 0.013127823 |
| 0 | 0 | 0 | 0 | 0 |
| 0 | 0 | 0 | 0 | 0 |
| 0 | 0 | 0 | 0.005862491 | 0 |
| 0.004025979 | 0 | 0.001037121 | 0.002923319 | 0.006127017 |
| 0 | 0 | 0 | 0 | 0 |

| | | | | |
|---|---|---|---|---|
| 0 | 0 | 0 | 0 | 0 |
| 0.034653168 | 0.006394288 | 0.013577311 | 0.014385892 | 0.007945923 |
| 0 | 0 | 0 | 0 | 0 |
| 0 | 0 | 0 | 0 | 0 |
| 0 | 0 | 0 | 0 | 0 |
| 0 | 0 | 0 | 0 | 0 |
| 0 | 0 | 0 | 0 | 0 |
| 0 | 0 | 0 | 0 | 0 |
| 0 | 0 | 0 | 0 | 0 |
| 0 | 0 | 0 | 0 | 0 |
| 0 | 0 | 0 | 0 | 0 |
| 0 | 0 | 0 | 0 | 0 |
| 0 | 0.000819313 | 0.27650026 | 0.022225853 | 0 |
| 0 | 0 | 0 | 0 | 0 |
| 0.00814378 | 0 | 0 | 0 | 0.004016501 |
| 0 | 0 | 0 | 0 | 0 |
| 0 | 0 | 0 | 0 | 0 |
| 0 | 0 | 0 | 0 | 0 |
| 0 | 0 | 0 | 0 | 0 |
| 0 | 0 | 0 | 0 | 0 |
| 0 | 0 | 0 | 0 | 0 |
| 0.026215125 | 0.042945452 | 0.017226399 | 0.00255953 | 0 |
| 0 | 0 | 0.000769598 | 0.003702192 | 0 |
| 0 | 0.000733154 | 0 | 0 | 0 |
| 0 | 0 | 0 | 0 | 0 |
| 0 | 0 | 0 | 0 | 0 |
| 0 | 0 | 0 | 0 | 0 |
| 0 | 0 | 0 | 0 | 0 |
| 0 | 0 | 0 | 0 | 0 |
| 0 | 0 | 0 | 0 | 0 |
| 0 | 0 | 0 | 0 | 0 |
| 0 | 0 | 0 | 0 | 0 |
| 0 | 0 | 0 | 0 | 0 |
| 0 | 0 | 0 | 0 | 0 |
| 0 | 0.006031924 | 0.028916944 | 6.69641708 | 0 |
| 0 | 0 | 0 | 0 | 0 |
| 0 | 0 | 0 | 0 | 0 |
| 0 | 0 | 0 | 0 | 0 |
| 0.011758353 | 0.008055977 | 0.468071922 | 0.027063533 | 0.044380314 |
| 0 | 0 | 0 | 0 | 0 |
| 0 | 0.019970158 | 0.022777959 | 0.057564431 | 0.382152364 |
| 0 | 0 | 0 | 0 | 0 |
| 0 | 0 | 0 | 0 | 0 |
| 0 | 0 | 0 | 0.017878258 | 0 |
| 0 | 0 | 0 | 0 | 0 |
| 1.391800329 | 0.374344921 | 0.166537338 | 0 | 0.00215669 |
| 0 | 0 | 0 | 0 | 0 |
| 0 | 0 | 0 | 0.000762514 | 0 |

| | | | | |
|---|---|---|---|---|
| 0 | 0 | 0 | 0 | 0 |
| 0 | 0 | 0 | 0 | 0 |
| 0 | 0 | 0 | 0 | 0 |
| 0.008366546 | 0 | 0 | 0 | 0 |
| 0.004491074 | 0 | 0.000778705 | 0 | 0 |
| 0 | 0 | 0 | 0 | 0 |
| 0 | 0 | 0 | 0 | 0 |
| 0 | 0 | 0 | 0 | 0 |
| 0 | 0 | 0.010154975 | 0 | 0 |
| 0.007796008 | 0.000713665 | 0.03514539 | 0 | 0.003427525 |
| 0 | 0 | 0 | 0.051740941 | 0.082584606 |
| 0 | 0 | 0 | 0 | 0 |
| 0.000653138 | 0 | 0 | 0.002213177 | 0 |
| 0 | 0 | 0 | 0 | 0 |
| 0 | 0 | 0 | 0 | 0 |
| 0 | 0 | 0 | 0 | 0 |
| 0 | 0 | 0 | 0 | 0 |
| 0 | 0 | 0 | 0 | 0 |
| 0 | 0 | 0 | 0 | 0 |
| 0.138746178 | 0.044288447 | 0.01939354 | 0.004859047 | 0.188314696 |
| 0 | 0 | 0 | 0 | 0 |
| 0 | 0 | 0 | 0 | 0 |
| 0 | 0 | 0 | 0 | 0 |
| 0 | 0 | 0 | 0 | 0 |
| 0 | 0 | 0 | 0 | 0 |
| 0 | 0 | 0 | 0 | 0 |
| 0 | 0 | 0 | 0 | 0 |
| 0 | 0 | 0 | 0 | 0 |
| 0 | 0 | 0 | 0 | 0 |
| 0 | 0 | 0.001307424 | 0 | 0.001716419 |
| 0 | 0 | 0 | 0 | 0 |
| 0 | 0 | 0 | 0.000980873 | 0 |
| 0 | 0 | 0 | 0 | 0 |
| 0 | 0 | 0 | 0 | 0 |
| 0 | 0 | 0 | 0 | 0 |
| 0.017809868 | 0.024646705 | 0 | 0 | 0 |
| 0 | 0 | 0 | 0 | 0 |
| 0 | 0 | 0 | 0 | 0 |
| 0 | 0 | 0 | 0 | 0 |
| 0 | 0 | 0 | 0 | 0 |
| 0 | 0 | 0 | 0 | 0 |
| 0 | 0 | 0 | 0 | 0 |
| 0 | 0 | 0 | 0.018776038 | 0 |
| 0 | 0 | 0 | 0 | 0 |
| 0.012194328 | 0 | 0.058747883 | 0.022078902 | 0.002154349 |
| 0 | 0 | 0 | 0.011721365 | 0 |
| 0 | 0 | 0 | 0 | 0 |
| 0 | 0 | 0 | 0 | 0 |

| | | | | |
|---|---|---|---|---|
| 0 | 0 | 0 | 0 | 0 |
| 0 | 0 | 0 | 0 | 0 |
| 0 | 0 | 0 | 0 | 0 |
| 0 | 0 | 0 | 0 | 0 |
| 0 | 0 | 0 | 0 | 0 |
| 0.00434796 | 0.004208256 | 0.004287752 | 0.022468118 | 0 |
| 0 | 0 | 0 | 0 | 0 |
| 0 | 0 | 0 | 0 | 0 |
| 0 | 0 | 0 | 0 | 0 |
| 0 | 0 | 0 | 0 | 0 |
| 0 | 0 | 0 | 0 | 0 |
| 0 | 0 | 0 | 0 | 0 |
| 0 | 0 | 0 | 0.002033587 | 0 |
| 0 | 0 | 0 | 0 | 0 |
| 0 | 0 | 0 | 0 | 0 |
| 0 | 0 | 0 | 0 | 0 |
| 0.032138707 | 0.001056121 | 0 | 0.08388322 | 0 |
| 0 | 0 | 0 | 0 | 0 |
| 0 | 0 | 0 | 0 | 0 |
| 0 | 0 | 0 | 0 | 0 |
| 0.003623511 | 0.002523839 | 0 | 0 | 0 |
| 0 | 0 | 0 | 0 | 0 |
| 0 | 0 | 0 | 0 | 0 |
| 0 | 0 | 0 | 0 | 0 |
| 0 | 0 | 0 | 0 | 0 |
| 0 | 0 | 0.001149731 | 0 | 0 |
| 0 | 0 | 0 | 0 | 0 |
| 0 | 0 | 0 | 0 | 0 |
| 0 | 0 | 0 | 0 | 0 |
| 0 | 0 | 0 | 0 | 0 |
| 0 | 0 | 0 | 0 | 0 |
| 0 | 0 | 0 | 0.012880907 | 0 |
| 0 | 0 | 0 | 0 | 0 |
| 0 | 0 | 0 | 0 | 0 |
| 0 | 0 | 0 | 0 | 0 |
| 0 | 0 | 0 | 0 | 0 |
| 0 | 0 | 0 | 0 | 0 |
| 0 | 0 | 0 | 0 | 0 |
| 0 | 0 | 0 | 0.396447039 | 0.467221047 |
| 0 | 0 | 0 | 0 | 0 |
| 0 | 0 | 0 | 0 | 0 |
| 0 | 0 | 0 | 0 | 0 |
| 0 | 0 | 0 | 0 | 0 |
| 0 | 0 | 0 | 0 | 0 |
| 0 | 0 | 0 | 0 | 0 |
| 0 | 0 | 0 | 0 | 0 |
| 0 | 0 | 0 | 0 | 0 |
| 0 | 0 | 0 | 0 | 0 |

| | | | | |
|---|---|---|---|---|
| 0 | 0 | 0 | 0 | 0 |
| 0.012626093 | 0 | 0.006172051 | 0.000515469 | 0 |
| 0 | 0 | 0 | 0 | 0 |
| 0 | 0 | 0 | 0 | 0 |
| 0 | 0 | 0 | 0 | 0 |
| 0 | 0 | 0.000564248 | 0 | 0 |
| 0.000973394 | 0 | 0 | 0 | 0 |
| 0 | 0 | 0 | 0 | 0 |
| 0 | 0 | 0 | 0 | 0 |
| 0 | 0 | 0 | 0 | 0 |
| 0.003585441 | 0.001276409 | 0 | 0.001484924 | 0.004244003 |
| 0 | 0 | 0 | 0 | 0 |
| 0 | 0 | 0.116936755 | 0.051902052 | 1.023218656 |
| 0 | 0 | 0 | 0 | 0 |
| 0 | 0 | 0 | 0 | 0 |
| 0 | 0 | 0 | 0 | 0 |
| 0 | 0 | 0 | 0 | 0 |
| 0 | 0 | 0 | 0 | 0 |
| 0 | 0 | 0 | 0 | 0 |
| 0 | 0 | 0 | 0 | 0 |
| 0 | 0.002375044 | 0 | 0 | 0 |
| 0 | 0 | 0 | 0 | 0 |
| 0 | 0 | 0 | 0.00603315 | 0 |
| 3.245165085 | 0.760522708 | 1.843405548 | 0.588832246 | 3.567263237 |
| 0 | 0 | 0 | 0 | 0 |
| 0 | 0 | 0 | 0 | 0 |
| 0 | 0 | 0 | 0 | 0 |
| 0 | 0 | 0 | 0 | 0 |
| 0 | 0 | 0 | 0 | 0 |
| 0 | 0 | 0 | 0 | 0 |
| 0 | 0 | 0 | 0 | 0 |
| 0 | 0 | 0 | 0 | 0 |
| 0.001063302 | 0 | 0 | 0 | 0.003524096 |
| 0 | 0 | 0 | 0 | 0 |

| PHL018-0151_V1 | PHL018-0269_V1 | PHL018-0085_V1 | PHL018-0144_V1 | PHL018-0273_V1 |
|---|---|---|---|---|
| EYCF | REF | CTRL | REF | REF |
| 0.165194004 | 0.206663601 | 0.101751177 | 3.985638511 | 0 |
| 7.57514321 | 5.419782615 | 4.15434979 | 1.209890565 | 2.641989512 |
| 38.69399097 | 0 | 0 | 0.975189476 | 0 |
| 0 | 0 | 0 | 0 | 0 |
| 0 | 0 | 0 | 0 | 0 |
| 0 | 0 | 0 | 0 | 0 |
| 0 | 0 | 0 | 0 | 0 |
| 0 | 0 | 0 | 0 | 0 |
| 0 | 0 | 0 | 0 | 0 |
| 0 | 0 | 0 | 0 | 0 |
| 0 | 0 | 0 | 0 | 0 |
| 0 | 0 | 0 | 0 | 0 |
| 0 | 0 | 0 | 0 | 0 |
| 0 | 2.911380824 | 1.103507482 | 0.199496284 | 3.355749509 |
| 0 | 0 | 0 | 0.565883843 | 0 |
| 0 | 0 | 0 | 0 | 0 |
| 0.015825494 | 0 | 0 | 0 | 0 |
| 0.003727266 | 0 | 0 | 0 | 0 |
| 0 | 0 | 0 | 0 | 0 |
| 0 | 0 | 0 | 0 | 0 |
| 0 | 0 | 0 | 0 | 0 |
| 0 | 0 | 0.002650407 | 0 | 0 |
| 0 | 0 | 0 | 0 | 0 |
| 0 | 0 | 0 | 0 | 0 |
| 0 | 0 | 0 | 0 | 0 |
| 0 | 0 | 0 | 0 | 0 |
| 0 | 0 | 0 | 0 | 0 |
| 0 | 0 | 0 | 0 | 0 |
| 0 | 0 | 0 | 0 | 0 |
| 0 | 0 | 0 | 0 | 0 |
| 0 | 0 | 0 | 0 | 0 |
| 0 | 0 | 0 | 0 | 0 |
| 0 | 0 | 0 | 0 | 0 |
| 0 | 0 | 0 | 0 | 0 |
| 0 | 0 | 0 | 0 | 0 |
| 0 | 0 | 1.128084102 | 3.848371336 | 0.200801896 |
| 0 | 0 | 0 | 0 | 0 |
| 0 | 0 | 0 | 0 | 0 |
| 0 | 0 | 0 | 0 | 0 |
| 0 | 0 | 0 | 0 | 0 |
| 0 | 0 | 0 | 0 | 0 |
| 0 | 0 | 0.004448709 | 0.007808757 | 0.055732899 |
| 0.002395759 | 1.3793559 | 0.056963874 | 3.629527274 | 0.081180894 |
| 0 | 0.115701149 | 0.012244804 | 0.014986993 | 0.003807838 |
| 0 | 0 | 0 | 0 | 0.026668788 |
| 0 | 0 | 0 | 0 | 0 |

| | | | | |
|---|---|---|---|---|
| 0 | 0 | 0.008033304 | 0.138404862 | 0.047948149 |
| 0.002252838 | 0.010009824 | 0.520506478 | 0.000561203 | 0 |
| 0.00846694 | 0.010008767 | 0.014678114 | 0.086926863 | 0 |
| 0.066313469 | 0.774534464 | 0.502445336 | 0.191295449 | 0.095080065 |
| 0.000797402 | 0.065386957 | 0.00428494 | 0.035168749 | 0 |
| 0 | 0 | 0 | 0 | 0 |
| 0 | 0 | 0.18453029 | 0 | 0.003498144 |
| 0 | 0 | 0 | 0 | 0 |
| 0 | 0 | 0 | 0 | 0 |
| 0 | 0.003004772 | 0 | 0 | 0 |
| 0 | 0 | 0 | 0 | 0 |
| 0.070642697 | 0.143509539 | 0 | 0.269844965 | 0.028037713 |
| 0 | 0 | 0 | 0 | 0 |
| 0.006760057 | 0.044494054 | 0.21237326 | 0.028575653 | 0.034894706 |
| 0 | 0 | 0.003880332 | 0 | 0 |
| 0 | 0 | 0 | 0 | 0 |
| 0.001077596 | 0.002204698 | 0.167423842 | 0.000787421 | 0 |
| 0 | 0 | 0 | 0 | 0 |
| 0 | 0 | 0 | 0 | 0 |
| 0 | 0 | 0 | 0 | 0 |
| 0 | 0.174093272 | 0.059771053 | 0.070254237 | 0 |
| 0.038681431 | 0.001595564 | 0.014539973 | 0.070378337 | 0.004793554 |
| 0 | 0 | 0 | 0.001597925 | 2.609210246 |
| 0 | 0 | 0 | 0 | 0 |
| 0 | 0 | 0 | 0 | 0 |
| 0.001626462 | 0 | 0 | 0 | 0 |
| 0 | 0 | 0 | 0 | 0 |
| 0 | 0 | 0 | 0 | 0 |
| 0 | 0 | 0 | 0 | 0 |
| 0 | 0 | 0 | 0 | 0 |
| 0.639427957 | 0 | 0.057230315 | 0.035514601 | 0 |
| 0 | 0 | 0 | 0 | 0 |
| 0 | 0 | 0 | 0 | 0 |
| 0 | 0 | 0 | 0 | 0 |
| 0 | 0 | 0 | 0 | 0 |
| 0 | 0 | 0 | 0 | 0 |
| 0 | 0 | 0 | 0 | 0 |
| 0 | 0 | 0.004021161 | 0 | 0 |
| 0 | 0 | 0 | 0 | 0 |
| 0 | 0 | 0 | 0 | 0 |
| 0 | 0 | 0 | 0 | 0 |
| 0 | 0 | 0 | 0 | 0 |
| 0 | 0 | 0 | 0 | 0 |
| 0 | 0 | 0 | 0 | 0 |
| 0 | 0 | 0 | 0 | 0 |
| 0 | 0 | 0 | 0 | 0 |
| 0 | 0 | 0 | 0 | 0 |
| 0 | 0 | 0 | 0 | 0 |

| | | | | |
|---|---|---|---|---|
| 0 | 0 | 0 | 0 | 0 |
| 0 | 0 | 0 | 0 | 0 |
| 0 | 0 | 0 | 0 | 0 |
| 0 | 0 | 0 | 0 | 0 |
| 0 | 0 | 0.001161423 | 0 | 0 |
| 0 | 0 | 0 | 0 | 0 |
| 0 | 0 | 0 | 0 | 0 |
| 0 | 0 | 0 | 0 | 0.000916923 |
| 0 | 0.001542237 | 0 | 0 | 0 |
| 0 | 0 | 0 | 0 | 0 |
| 0 | 0 | 0 | 0 | 0 |
| 0 | 0 | 0.003148787 | 0.000959235 | 0.004781532 |
| 0 | 0 | 0 | 0 | 0 |
| 0 | 0 | 0 | 0 | 0 |
| 0 | 0 | 0 | 0 | 0 |
| 0 | 0 | 0 | 0 | 0 |
| 0 | 0 | 0 | 0 | 0 |
| 0 | 0 | 0 | 0 | 0 |
| 0 | 0.057364826 | 0 | 0.000853675 | 0.178192344 |
| 0 | 0 | 0 | 0 | 0 |
| 0 | 0 | 0 | 0 | 0 |
| 0 | 0 | 0 | 0 | 0 |
| 0 | 0 | 0 | 0 | 0 |
| 0 | 0 | 0 | 0 | 0 |
| 0 | 0 | 0 | 0 | 0 |
| 0 | 0 | 0 | 0 | 0 |
| 0 | 0 | 0 | 0 | 0 |
| 0 | 0 | 0 | 0 | 0 |
| 0 | 0 | 0 | 0 | 0 |
| 0 | 0 | 0 | 0 | 0 |
| 0 | 0.013002843 | 0 | 0.059254029 | 0.001905584 |
| 0.001756291 | 0 | 0 | 0.000668415 | 0.028154319 |
| 0 | 0 | 0 | 0 | 0 |
| 0 | 0 | 0 | 0 | 0 |
| 0 | 0 | 0 | 0 | 0 |
| 0 | 0 | 0 | 0 | 0 |
| 0 | 0 | 0 | 0 | 0 |
| 0 | 0 | 0 | 0 | 0 |
| 0 | 0 | 0.096411504 | 0 | 0 |
| 0 | 0 | 0 | 0 | 0 |
| 0 | 0 | 0 | 0 | 0 |
| 0 | 0 | 0 | 0 | 0 |
| 0 | 0 | 0 | 0 | 0 |
| 0 | 0 | 0 | 0 | 0 |
| 0 | 0 | 0 | 0 | 0 |
| 0 | 0 | 0 | 0 | 0 |
| 0 | 0 | 0 | 0 | 0 |
| 0 | 0 | 0 | 0 | 0 |

| | | | | |
|---|---|---|---|---|
| 0 | 0 | 0 | 0 | 0 |
| 0 | 0 | 0.008656007 | 0 | 0 |
| 0 | 0 | 0 | 0 | 0 |
| 0 | 0 | 0 | 0 | 0 |
| 0 | 0 | 0 | 0.069348177 | 0.069310419 |
| 0 | 0.004140687 | 0 | 0 | 0 |
| 0 | 0 | 0 | 0 | 0 |
| 0 | 0 | 0 | 0 | 0 |
| 0 | 0 | 0 | 0 | 0 |
| 0 | 0 | 0 | 0 | 0 |
| 0 | 0 | 0 | 0 | 0 |
| 0 | 0 | 0 | 0 | 0 |
| 0 | 0 | 0 | 0 | 0 |
| 0 | 0 | 0 | 0 | 0 |
| 0 | 0 | 0 | 0 | 0 |
| 0 | 0 | 0 | 0 | 0 |
| 0 | 0 | 0 | 0 | 0 |
| 0 | 0 | 0 | 0 | 0 |
| 0 | 0 | 0 | 0 | 0 |
| 0 | 0 | 0 | 0 | 0 |
| 0 | 0 | 0 | 0 | 0 |
| 0 | 0 | 0 | 0 | 0 |
| 0.002050284 | 0 | 0 | 0.007661169 | 0.005304011 |
| 0 | 0 | 0 | 0 | 0 |
| 0 | 0 | 0 | 0 | 0 |
| 0 | 0 | 0 | 0 | 0 |
| 0 | 0 | 0 | 0 | 0.105140811 |
| 0 | 0 | 0 | 0 | 0 |
| 0 | 0 | 0 | 0 | 0 |
| 0 | 0 | 0 | 0 | 0 |
| 0 | 0.001090223 | 0 | 0 | 0.001091787 |
| 0 | 0 | 0 | 0 | 0 |
| 0 | 0 | 0 | 0 | 0 |
| 0 | 0 | 0 | 0 | 0 |
| 0 | 0 | 0 | 0 | 0 |
| 0 | 0 | 0 | 0 | 0 |
| 0 | 0 | 0 | 0 | 0 |
| 0 | 0 | 0 | 0 | 0 |
| 0 | 0 | 0 | 0 | 0 |
| 0 | 0 | 0 | 0 | 0 |
| 0 | 0 | 0 | 0 | 0 |
| 0 | 0 | 0 | 0 | 0 |
| 2.973335127 | 0 | 0 | 0 | 0 |
| 0 | 0 | 0 | 0 | 0 |
| 0 | 0 | 0 | 0 | 0 |
| 0 | 0 | 0 | 0 | 0 |
| 0 | 0 | 0 | 0 | 0 |
| 0.002484384 | 0 | 0 | 0 | 0 |

| | | | | |
|---|---|---|---|---|
| 0 | 0 | 0 | 0 | 0 |
| 0 | 0 | 0 | 0 | 0 |
| 0 | 0 | 0 | 0 | 0 |
| 0 | 0 | 0 | 0 | 0 |
| 0 | 0 | 0 | 0 | 0 |
| 0 | 0 | 0 | 0 | 0 |
| 0 | 0 | 0 | 0 | 0 |
| 0 | 0 | 0 | 0 | 0 |
| 0 | 0 | 0 | 0 | 0 |
| 0 | 0 | 0 | 0 | 0 |
| 0 | 0 | 0 | 0 | 0 |
| 0 | 0 | 0 | 0 | 0 |
| 0 | 0 | 0.000792805 | 0.005165788 | 0.004414297 |
| 0 | 0 | 0 | 0 | 0 |
| 0 | 0 | 0 | 0 | 0 |
| 0 | 0 | 0 | 0 | 0 |
| 0 | 0 | 0 | 0 | 0 |
| 0 | 0 | 0 | 0 | 0 |
| 0 | 0 | 0 | 0 | 0 |
| 0 | 0 | 0 | 0 | 0 |
| 0.013897956 | 0.048764322 | 0.06374127 | 0.005519304 | 0.001375613 |
| 0 | 0 | 0 | 0 | 0 |
| 0 | 0 | 0 | 0.000484285 | 0 |
| 0 | 0 | 0 | 0 | 0 |
| 0 | 0 | 0 | 0 | 0 |
| 0 | 0 | 0 | 0 | 0 |
| 0 | 0.006799806 | 0.001726577 | 0 | 0.001401968 |
| 0 | 0 | 0 | 0 | 0 |
| 0 | 0 | 0 | 0 | 0 |
| 0 | 0.004707359 | 0 | 0 | 0 |
| 0 | 0 | 0 | 0 | 0 |
| 0 | 0 | 0 | 0 | 0 |
| 0 | 0 | 0 | 0 | 0 |
| 0 | 0 | 0 | 0.757251004 | 0 |
| 0.0127778 | 0.061669803 | 0.004500996 | 0.12443346 | 0 |
| 0 | 0 | 0 | 0 | 0 |
| 0 | 0 | 0 | 0 | 0 |
| 0 | 0 | 0 | 0 | 0 |
| 0 | 0 | 0.009348571 | 0 | 0 |
| 0 | 0 | 0 | 0 | 0 |
| 0 | 0 | 0 | 0 | 0 |
| 0 | 0 | 0.008348119 | 0 | 0 |
| 0 | 0 | 0.002673303 | 0 | 0 |
| 0 | 0 | 0 | 0 | 0 |
| 0 | 0 | 0 | 0 | 0 |
| 0.015534865 | 0 | 0 | 0 | 0.193445337 |
| 0 | 0.001451565 | 0.002436691 | 0.001166479 | 0 |
| 0 | 0.011623813 | 0.000687328 | 0.012539849 | 0.013075609 |

| | | | | |
|---|---|---|---|---|
| 0 | 0 | 0 | 0 | 0 |
| 0.001161369 | 0 | 0 | 0 | 0 |
| 0 | 0 | 0.004187108 | 0 | 0 |
| 0 | 0 | 0 | 0 | 0 |
| 0 | 0 | 0 | 0 | 0 |
| 0 | 0 | 0 | 0 | 0 |
| 0 | 0 | 0.013045374 | 0 | 0 |
| 0 | 0 | 0 | 0 | 0.001146124 |
| 0 | 0 | 0.015350151 | 0 | 0 |
| 0 | 0 | 0 | 0 | 0 |
| 0 | 0 | 0 | 0 | 0 |
| 0 | 0 | 0 | 0 | 0 |
| 0 | 0 | 0 | 0 | 0 |
| 0 | 0 | 0 | 0 | 0 |
| 0 | 0 | 0 | 0 | 0 |
| 0 | 0 | 0 | 0 | 0 |
| 0 | 0 | 0 | 0 | 0 |
| 0 | 0 | 0 | 0 | 0 |
| 0 | 0.009392299 | 0 | 0.011972151 | 0 |
| 0 | 0 | 0 | 0 | 0.546362323 |
| 0.006304729 | 0.017676554 | 0.003437028 | 0.001151748 | 0.006698017 |
| 0 | 0 | 0 | 0 | 0 |
| 0 | 0 | 0 | 0 | 0 |
| 0 | 0 | 0 | 0 | 0 |
| 0 | 0.010440549 | 0.019802309 | 0.003432287 | 0 |
| 0 | 0 | 0 | 0 | 0 |
| 0 | 0 | 0 | 0 | 0 |
| 0 | 0 | 0 | 0 | 0 |
| 0 | 0 | 0 | 0.005641107 | 0 |
| 0 | 0 | 0 | 0 | 0 |
| 0.171990578 | 0.155443832 | 0.522634307 | 0.191959493 | 0.103953714 |
| 0 | 0 | 0 | 0 | 0 |
| 0 | 0 | 0 | 0 | 0 |
| 0 | 0 | 0 | 0 | 0 |
| 0 | 0 | 0 | 0 | 0 |
| 0 | 0 | 0 | 0 | 0 |
| 0 | 0 | 0 | 0 | 0 |
| 0 | 0 | 0 | 0 | 0 |
| 0 | 0 | 0 | 0 | 0 |
| 0 | 0 | 0 | 0.000422958 | 0.005095133 |
| 0 | 0 | 0 | 0 | 0 |
| 0 | 0.004834042 | 0.006440266 | 0.027192484 | 0.001792953 |
| 0 | 0 | 0 | 0 | 0 |
| 0 | 0 | 0 | 0 | 0 |
| 0 | 0 | 0 | 0 | 0 |
| 0.001799588 | 0 | 0 | 0 | 0 |
| 0 | 0 | 0 | 0 | 0 |
| 0 | 0 | 0 | 0 | 0 |

| | | | | |
|---|---|---|---|---|
| 0 | 0 | 0 | 0 | 0 |
| 0 | 0 | 0 | 0 | 0 |
| 0 | 0.117352069 | 0.007439868 | 0.012646162 | 0.00427756 |
| 0 | 0 | 0 | 0 | 0 |
| 0 | 0.082526626 | 0 | 0 | 0.045523113 |
| 0 | 0 | 0 | 0 | 0 |
| 0 | 0 | 0 | 0 | 0 |
| 0 | 0 | 0 | 0 | 0 |
| 0 | 0 | 0 | 0 | 0 |
| 0 | 0.000616835 | 0.006804447 | 0 | 0 |
| 0 | 0 | 0 | 0 | 0 |
| 0 | 0 | 0 | 0 | 0 |
| 0 | 0 | 0 | 0 | 0 |
| 0 | 0.000993833 | 0 | 0 | 0 |
| 0 | 0.007587582 | 0.013573026 | 0 | 0.000616092 |
| 0 | 0 | 0 | 0 | 0.112133248 |
| 0 | 0.009580004 | 0.007060221 | 0 | 0 |
| 0 | 0 | 0.004172749 | 0 | 0 |
| 0 | 0 | 0 | 0 | 0 |
| 0 | 0 | 0 | 0 | 0 |
| 0 | 0 | 0 | 0 | 0 |
| 0 | 0.006538739 | 0 | 0.0140686 | 0.011617624 |
| 0 | 0 | 0 | 0 | 0 |
| 0 | 0 | 0 | 0 | 0 |
| 0 | 0 | 0 | 0 | 0 |
| 0 | 0 | 0 | 0 | 0 |
| 0 | 0 | 0 | 0 | 0.009649079 |
| 0 | 0 | 0 | 0 | 0 |
| 0 | 0 | 0 | 0 | 0 |
| 0 | 0 | 0 | 0 | 0 |
| 0 | 0 | 0 | 0 | 0 |
| 0 | 0 | 0 | 0 | 0 |
| 0 | 0 | 0.001884295 | 0 | 0 |
| 0 | 0 | 0 | 0 | 0 |
| 0 | 0 | 0 | 0 | 0 |
| 0 | 0 | 0 | 0 | 0 |
| 0 | 0 | 0 | 0 | 0 |
| 0 | 0.133564043 | 0 | 0.016823896 | 0.007696253 |
| 0 | 0 | 0 | 0 | 0 |
| 0 | 0 | 0 | 0 | 0 |
| 0 | 0 | 0.005503355 | 0 | 0 |
| 0 | 0 | 0 | 0 | 0 |
| 0 | 0 | 0 | 0 | 0 |
| 0 | 0 | 0 | 0 | 0 |
| 0 | 0 | 0 | 0 | 0 |
| 0 | 0 | 0 | 0 | 0 |
| 0 | 0 | 0 | 0 | 0 |
| 0.001412025 | 0 | 0 | 0 | 0 |

| | | | | |
|---|---|---|---|---|
| 0.016941943 | 0 | 0 | 0.422580457 | 0.70398675 |
| 0 | 0 | 0 | 0 | 0 |
| 0 | 0 | 0 | 0.000887696 | 0 |
| 0 | 0 | 0 | 0 | 0 |
| 0 | 0.009113338 | 0 | 0.000714487 | 0.002671143 |
| 0 | 0 | 0 | 0 | 0 |
| 0 | 0 | 0 | 0 | 0 |
| 0.001918811 | 0.051097397 | 0.523170138 | 0 | 0.002184114 |
| 0 | 0 | 0 | 0 | 0 |
| 1.138907366 | 0.041427244 | 0.01062775 | 0.070452993 | 0.00160801 |
| 0 | 0 | 0 | 0.000699985 | 0 |
| 0 | 0 | 0.011356116 | 0.001783798 | 0.001234966 |
| 0.000658382 | 0.074459765 | 0.006460513 | 0.003157173 | 0.000936766 |
| 0.002499781 | 0 | 0 | 0 | 0 |
| 0 | 0 | 0 | 0 | 0 |
| 0 | 0 | 0 | 0 | 0 |
| 0 | 0 | 0 | 0 | 0 |
| 0.001500549 | 0 | 0 | 0 | 0 |
| 0.044133147 | 0 | 0 | 0 | 0 |
| 0.002557261 | 0 | 0.572275205 | 0 | 0 |
| 0 | 0 | 0 | 0 | 0 |
| 0 | 0 | 0 | 0 | 0 |
| 0 | 0 | 0.001944371 | 0 | 0 |
| 0 | 0 | 0 | 0 | 2.47958925 |
| 0 | 0 | 0 | 0 | 0 |
| 0 | 0.001743249 | 0.001881213 | 0.00437774 | 0.042334409 |
| 0 | 0 | 0 | 0.976434771 | 0 |
| 0 | 0 | 0.023507392 | 0 | 0 |
| 0.115808255 | 0 | 0 | 0 | 0 |
| 0 | 0 | 0 | 0.011170275 | 0 |
| 0 | 0 | 0 | 0 | 0 |
| 0 | 0 | 0 | 0 | 0 |
| 0.00316639 | 0 | 0 | 0 | 0 |
| 0.014541977 | 0 | 0.305074765 | 0 | 0 |
| 0 | 0 | 0 | 0 | 0 |
| 0.009525892 | 0 | 0.059137153 | 0.003196269 | 0 |
| 0.008910321 | 0 | 0.114802123 | 0 | 0 |
| 0 | 0 | 0 | 0 | 0 |
| 0 | 0 | 0 | 0 | 0 |
| 0 | 0.007104626 | 0 | 0 | 0 |
| 0 | 0 | 0 | 0 | 0 |
| 0.013734205 | 0 | 0 | 0 | 0 |
| 0 | 0 | 0 | 0 | 0 |
| 0 | 0 | 0 | 0 | 0 |
| 0 | 0 | 0 | 0 | 0 |
| 0 | 0 | 0 | 0 | 0 |
| 0 | 0.003522785 | 0 | 0.001742101 | 0 |
| 0.490788598 | 0 | 0 | 0 | 0 |

| | | | | |
|---|---|---|---|---|
| 0.018252077 | 0 | 0 | 0 | 0 |
| 0 | 0 | 0.171248696 | 0 | 0 |
| 0 | 0 | 0.003189218 | 0 | 0 |
| 0 | 0 | 0 | 0 | 0 |
| 0 | 0 | 0 | 0 | 0 |
| 0 | 0.026038744 | 0.010134243 | 0 | 0 |
| 0 | 0 | 0 | 0 | 0 |
| 0 | 0 | 0 | 0 | 0 |
| 0.870008651 | 0 | 0 | 0 | 0 |
| 0 | 0 | 0 | 0 | 0 |
| 0.001306729 | 0.041835201 | 0 | 0 | 0.005453812 |
| 0 | 0.252423172 | 0 | 0 | 0 |
| 0 | 0 | 0 | 0 | 0 |
| 0 | 0.03531416 | 0.002692025 | 0.025183564 | 0 |
| 0.009576623 | 0.420733473 | 0.008852498 | 0 | 0 |
| 0.00188843 | 0 | 0 | 0 | 0 |
| 0 | 0 | 0 | 0 | 0 |
| 0.002603032 | 0.008648493 | 0 | 0 | 0 |
| 0 | 0 | 0.015758857 | 0.007001053 | 0 |
| 0 | 0 | 0 | 0 | 0.004774851 |
| 0 | 0 | 0.005801093 | 0 | 0 |
| 1.16314383 | 0 | 0 | 0.153625359 | 0.300540024 |
| 0 | 0 | 0.018201546 | 0 | 0 |
| 0 | 0 | 0 | 0 | 0 |
| 0 | 0 | 0.332201066 | 0 | 0 |
| 0 | 0 | 0 | 0 | 0 |
| 0.248079755 | 0 | 0 | 0 | 0 |
| 0.065252613 | 0 | 0 | 0 | 0 |
| 0 | 0 | 0 | 0 | 0 |
| 0.112193076 | 1.51381537 | 0.482884979 | 4.37180958 | 0.161004863 |
| 0 | 0 | 0 | 0.002677507 | 0.012711102 |
| 0 | 0 | 0.003205129 | 0 | 0 |
| 0 | 0 | 0 | 0 | 0 |
| 0.00257017 | 0 | 0.001616688 | 0 | 0 |
| 0.000929079 | 0 | 0.024615311 | 0 | 0 |
| 0 | 0 | 0 | 0 | 0 |
| 0 | 0 | 0 | 0.948492048 | 0 |
| 0 | 0 | 0.002057918 | 0 | 0.007161487 |
| 0 | 0.354635314 | 0.336109594 | 0 | 0 |
| 0 | 2.194993926 | 0 | 0 | 0 |
| 0.009303432 | 0.017880686 | 0.029600713 | 0 | 0.003080658 |
| 0 | 5.916947933 | 2.475836603 | 0 | 0.200036957 |
| 0 | 0 | 0 | 0 | 0.005904347 |
| 0 | 0 | 0 | 0 | 0 |
| 0 | 0 | 0 | 0 | 0 |
| 0.119518169 | 0 | 0 | 0 | 0 |
| 0.004971079 | 0.000605389 | 0 | 0.000486491 | 0 |
| 0 | 0 | 0 | 0 | 0 |

| | | | | |
|---|---|---|---|---|
| 0 | 0 | 0 | 0.00900373 | 0.012217655 |
| 0 | 0 | 0 | 0 | 0 |
| 0 | 0.013231071 | 0.372453529 | 0.003544167 | 0 |
| 0 | 0 | 0 | 0 | 0 |
| 0 | 0.008176087 | 0 | 0 | 0.000668393 |
| 0 | 0.043576653 | 0 | 0 | 0 |
| 0 | 0 | 0 | 0 | 0 |
| 0 | 0 | 0 | 0 | 0.014252432 |
| 0 | 0 | 0 | 0 | 0 |
| 0.404933991 | 0 | 0 | 0 | 0 |
| 0 | 0 | 0 | 0 | 0 |
| 0 | 0 | 0 | 0 | 0 |
| 0 | 0 | 0 | 0 | 0 |
| 0.001189029 | 0.005856461 | 0 | 0 | 0.002030144 |
| 0 | 0 | 0 | 0 | 0 |
| 0.003407087 | 0.00246439 | 0 | 1.008016431 | 0.06592881 |
| 0.000686618 | 0.005463038 | 0.001122932 | 0.004233313 | 0.022665039 |
| 0.013347433 | 0.001185246 | 0.008526989 | 0 | 0 |
| 0 | 0 | 0.000869928 | 0 | 0 |
| 1.587126144 | 0 | 0 | 0 | 0 |
| 0.01427802 | 0.018687808 | 0 | 0.008891965 | 0 |
| 0 | 0.005657893 | 0.001953814 | 0.007820304 | 0 |
| 0 | 0 | 0 | 0 | 0.015081214 |
| 0 | 0.145378303 | 0.667201493 | 0 | 0 |
| 0.158342256 | 0 | 0 | 0 | 0 |
| 0 | 0 | 0 | 0 | 0 |
| 0.002179169 | 0 | 0.002851142 | 0 | 0.000992188 |
| 0 | 0 | 0 | 0 | 0 |
| 0 | 0 | 0 | 0 | 0 |
| 0 | 0 | 0 | 0 | 0 |
| 0.07291908 | 0.004144116 | 0.0509547 | 0.006660432 | 0.002075029 |
| 0 | 0.004493359 | 0.005253051 | 0.006093339 | 0 |
| 0.063987911 | 0 | 0.033087608 | 0.140314775 | 0.017941009 |
| 0 | 0 | 0 | 0 | 0 |
| 0 | 0 | 0 | 0 | 0 |
| 0.014004267 | 0 | 0 | 0 | 0 |
| 0 | 0.007252246 | 0.198959412 | 0.007576286 | 0 |
| 0.001663271 | 0.160884187 | 0.03400251 | 0.001975002 | 0.001703918 |
| 0 | 0.097466649 | 0 | 0 | 0.045005838 |
| 0.002719788 | 0 | 0.005782509 | 0 | 0 |
| 0 | 0 | 0 | 0 | 0 |
| 0.001952176 | 0.015310477 | 0 | 0 | 0 |
| 0 | 0.041561106 | 0.001220417 | 0 | 0 |
| 0 | 0.039846396 | 0 | 0 | 0 |
| 0.162612255 | 0 | 0.16081337 | 0 | 0 |
| 0 | 0 | 0 | 0 | 0 |
| 0 | 0 | 0 | 0 | 0 |
| 0.001689682 | 0 | 0 | 0 | 0 |

| | | | | |
|---|---|---|---|---|
| 0 | 0 | 0 | 0 | 0 |
| 0.009972239 | 0.425827396 | 0.016679784 | 0.004347309 | 0.277842549 |
| 0 | 0 | 0.001217684 | 0 | 0 |
| 0.001784417 | 0 | 0 | 0 | 0 |
| 0 | 0 | 0 | 0 | 0 |
| 0 | 0 | 0 | 0 | 0 |
| 0 | 0 | 0 | 0 | 0 |
| 0 | 0 | 0 | 0 | 0 |
| 0 | 0 | 0.003053794 | 0.00075802 | 0.002361578 |
| 0 | 0 | 0 | 0 | 0 |
| 0 | 0.00880585 | 0.00964048 | 0.000923007 | 0.000766824 |
| 0.000752597 | 0.465067727 | 0.352429752 | 0.044911626 | 0.003426619 |
| 0 | 0 | 0 | 0.215934284 | 1.003117282 |
| 0.086219908 | 0 | 0 | 0 | 0 |
| 0 | 0.018010787 | 0 | 0.002647589 | 0 |
| 0.065997703 | 0 | 0 | 0 | 0 |
| 0 | 0.045503612 | 0.018459381 | 0.001933044 | 0.00722678 |
| 0.035943457 | 0.023940254 | 0.587838105 | 0.01317413 | 0 |
| 0 | 0 | 0 | 0 | 0 |
| 0.011000926 | 0 | 0.08728304 | 0 | 0 |
| 0.000545614 | 0.005581471 | 0.004164186 | 0.024087593 | 0.157747401 |
| 0 | 0 | 0.002220712 | 0 | 0 |
| 0 | 0 | 0 | 0 | 0 |
| 0 | 0 | 0 | 0 | 0 |
| 0 | 0 | 0.003998063 | 0.001116459 | 0 |
| 0 | 0 | 0 | 0 | 0 |
| 0 | 0 | 0.016441039 | 0 | 0.020229334 |
| 0.016237398 | 0.054708805 | 0.063227358 | 0.003354687 | 0.054567225 |
| 0 | 0 | 0 | 0 | 0 |
| 0 | 0 | 0 | 0 | 0 |
| 0 | 0 | 0 | 0 | 0 |
| 0 | 0 | 0 | 0 | 0 |
| 0.009386707 | 0 | 0 | 0 | 0.00213691 |
| 0.000878417 | 0 | 0 | 0 | 0 |
| 0 | 0.002680517 | 0 | 0 | 0.049277199 |
| 0.087349773 | 0.697968019 | 1.54573824 | 0.046018498 | 0.027928964 |
| 0.000454552 | 0 | 0 | 0 | 0 |
| 0 | 0 | 0 | 0 | 0 |
| 0 | 0 | 0.006239841 | 0.003920572 | 0.003800028 |
| 0 | 0 | 0 | 0 | 0 |
| 0.041299242 | 0 | 0.047475836 | 0 | 0 |
| 0.003042108 | 0 | 0 | 0 | 0 |
| 0.000579377 | 0.120183538 | 0.000947544 | 0.018786719 | 0 |
| 0 | 0 | 0.032247957 | 0 | 0 |
| 0.083226597 | 0 | 0.369578703 | 0 | 0 |
| 0.003034251 | 0 | 0 | 0 | 0.442741548 |
| 0 | 0 | 0 | 0 | 0 |
| 0 | 0 | 0 | 0.002064721 | 0 |

| | | | | |
|---|---|---|---|---|
| 0 | 0 | 0.035079087 | 0 | 0 |
| 0 | 0 | 0 | 0 | 0.001630078 |
| 0 | 0 | 0 | 0 | 0 |
| 0 | 1.98938196 | 0 | 2.523355062 | 0.472951697 |
| 0 | 0 | 0 | 0 | 0 |
| 0 | 0 | 0 | 0 | 0 |
| 0 | 0 | 0 | 0 | 0 |
| 0.000603673 | 0 | 0 | 0.121306932 | 0 |
| 0 | 0.816465692 | 0 | 0.003827185 | 0.383416132 |
| 0 | 0.166748118 | 0 | 0 | 0.01029726 |
| 0 | 0 | 0 | 0 | 0 |
| 0 | 0 | 0 | 0 | 0.012326546 |
| 0 | 0 | 0 | 0 | 0 |
| 0 | 0 | 0 | 0 | 0 |
| 0 | 0.067766084 | 0.039482132 | 0.523492765 | 0.050859487 |
| 0 | 0 | 0 | 0 | 0 |
| 0 | 0.028080712 | 0 | 0 | 0.004158736 |
| 0 | 0 | 0 | 0 | 0 |
| 0.012374465 | 0 | 0 | 0 | 0 |
| 0 | 0 | 0 | 0 | 0.028916382 |
| 0 | 0 | 0 | 0 | 0.199558445 |
| 0 | 0 | 0 | 0.006579208 | 0.001366483 |
| 0 | 0 | 0 | 0 | 0.001694129 |
| 0 | 0.317618643 | 0.022062422 | 0 | 0 |
| 0 | 0.001204028 | 0 | 0 | 0.000689003 |
| 0 | 0 | 0 | 0 | 0 |
| 0 | 0 | 0 | 0 | 0.005123794 |
| 0 | 3.968708522 | 0 | 0 | 0.465944942 |
| 0 | 0 | 0 | 0 | 0 |
| 0 | 0 | 0 | 0 | 0 |
| 0 | 0 | 0 | 0 | 0.003398307 |
| 0 | 0 | 0 | 0 | 0.021401745 |
| 0.008849613 | 0 | 0.002738158 | 0.07045527 | 0 |
| 0 | 0 | 0 | 0 | 0 |
| 0 | 0 | 0 | 0 | 0 |
| 0 | 0 | 0 | 0 | 0 |
| 0 | 0.508700207 | 0 | 0 | 0.199972194 |
| 0.000840861 | 0 | 0 | 0 | 0 |
| 0 | 0 | 0.002476594 | 0 | 0 |
| 0 | 0 | 0 | 0.004226884 | 0 |
| 0.002635946 | 0.369402344 | 0 | 0.001982791 | 0 |
| 0 | 0 | 0 | 0 | 0 |
| 0 | 0.034673438 | 0.023728202 | 0.694551438 | 15.0920819 |
| 0 | 0 | 0 | 0.231330781 | 0.002152494 |
| 0 | 0 | 0 | 0 | 0 |
| 0 | 0 | 0 | 0 | 0 |
| 0 | 0 | 0 | 0 | 0 |
| 0.005405446 | 0 | 0 | 1.453279523 | 0 |

| | | | | |
|---|---|---|---|---|
| 0 | 0 | 0 | 0 | 0 |
| 0 | 0 | 0 | 0 | 0.027640412 |
| 0.06468229 | 0.010967088 | 0.863519588 | 0.023664973 | 0 |
| 0 | 0 | 0 | 0.001587768 | 0.007914597 |
| 0 | 0 | 0 | 0 | 0 |
| 0 | 0 | 0 | 0.037798979 | 0.043739844 |
| 0 | 0.020278767 | 0.005599859 | 0.259255137 | 0 |
| 0 | 0 | 0 | 0.265664217 | 0 |
| 0 | 0 | 0 | 0 | 0 |
| 0.211605986 | 0 | 0 | 0 | 0 |
| 0.01736781 | 0.008973113 | 0.013484834 | 0.151106366 | 0.013778502 |
| 1.299428299 | 0.309783495 | 2.166095646 | 0.235279978 | 1.15058456 |
| 0 | 0 | 0 | 0 | 0 |
| 0 | 0.003443253 | 0.002477172 | 0 | 0.416368986 |
| 0.063709389 | 0.085642172 | 0.261094205 | 0.253756069 | 0.005944107 |
| 0 | 0 | 0 | 0 | 1.326066411 |
| 0 | 0 | 0 | 0 | 0 |
| 0.001656921 | 0 | 0 | 0 | 0 |
| 0 | 0 | 0 | 0 | 0 |
| 0.032370273 | 0.09608928 | 0.017725459 | 0 | 0.008882498 |
| 0.000735191 | 0.048968698 | 0.001683318 | 0.001343048 | 0 |
| 2.399906815 | 0.057937479 | 0 | 0.003962435 | 0 |
| 11.68157136 | 2.717983035 | 0.092670469 | 0.727888817 | 0.312372854 |
| 0 | 0 | 0 | 0 | 0.025192623 |
| 0 | 0 | 0 | 0 | 0 |
| 0 | 0 | 0 | 0.007885932 | 3.31100282 |
| 0 | 0.023787346 | 0 | 0 | 0.008807933 |
| 0 | 0 | 0 | 0 | 0 |
| 0.006932235 | 0 | 0 | 0 | 0.093797685 |
| 0 | 0 | 0 | 0 | 0 |
| 0 | 0 | 0 | 0 | 0.074764181 |
| 0 | 0 | 0 | 0.076710362 | 0.050720734 |
| 0 | 0.010239578 | 1.361059725 | 0.494699537 | 0 |
| 0 | 0 | 0.001348231 | 0.159005844 | 0.004066227 |
| 0.001664836 | 0.007163744 | 0.009335177 | 0.006408504 | 0 |
| 0 | 0.510883841 | 0.003206491 | 0.034324157 | 0.239535422 |
| 0 | 0.002152701 | 0.003097426 | 0.041662072 | 0.002155788 |
| 0 | 0 | 0 | 0 | 0 |
| 0 | 0 | 0 | 0.014145486 | 0.263112208 |
| 0 | 0 | 0 | 0 | 1.729320293 |
| 0 | 0 | 0 | 0.013140934 | 0.002128881 |
| 0 | 0 | 0 | 0 | 0 |
| 0 | 0 | 0 | 0 | 0.239113092 |
| 0 | 0 | 0 | 0 | 0 |
| 0 | 0.049344592 | 0.002133039 | 0.25221581 | 0.01187665 |
| 0 | 0 | 0 | 0 | 1.467606828 |
| 0 | 0 | 0 | 0 | 0 |
| 0.006068375 | 0.022736277 | 0.004778481 | 0.064050777 | 0 |

| | | | | |
|---|---|---|---|---|
| 0 | 0 | 0 | 0 | 0 |
| 0 | 0 | 0 | 0 | 0 |
| 0 | 0 | 0 | 0 | 0 |
| 0 | 0 | 0 | 0.010878669 | 0.011568481 |
| 0 | 0 | 0 | 0 | 0 |
| 0 | 0 | 0 | 0 | 0.068646874 |
| 0 | 0.00118567 | 0 | 0 | 0.011280018 |
| 0 | 0.020263159 | 0 | 0.412693168 | 0.019959556 |
| 0 | 0 | 0 | 0 | 0 |
| 0 | 0 | 0 | 0 | 0 |
| 0 | 0.014876053 | 0.002892499 | 0.004038652 | 0.003422372 |
| 0 | 0 | 0 | 0 | 0 |
| 0 | 0 | 0 | 0.012164377 | 0.001233875 |
| 0 | 0 | 0 | 0 | 0 |
| 0 | 0 | 0 | 0 | 0 |
| 0 | 0 | 0 | 0 | 0 |
| 0 | 0 | 0 | 0.232005332 | 0.019164942 |
| 0 | 0.010710465 | 0 | 0.014927662 | 0.035529289 |
| 0 | 0 | 0 | 0 | 0 |
| 0 | 0 | 0 | 0.009338698 | 0 |
| 0 | 0 | 0 | 0 | 0.001637805 |
| 0 | 0 | 0 | 0 | 0 |
| 0 | 0 | 0 | 0.476623035 | 0.095450884 |
| 0 | 0 | 0 | 0 | 0 |
| 0 | 0 | 0 | 0 | 0 |
| 0 | 0 | 0 | 0 | 0 |
| 0 | 0 | 0 | 0 | 0 |
| 0 | 0 | 0 | 0 | 0 |
| 0 | 0 | 0 | 0 | 0 |
| 0.044464288 | 1.214366286 | 0 | 0 | 0 |
| 0 | 0 | 0 | 0 | 0 |
| 0 | 0 | 0 | 0 | 0 |
| 0 | 0 | 0 | 0 | 0 |
| 0 | 0 | 0 | 0 | 0 |
| 0 | 0 | 0 | 0 | 0 |
| 0 | 0 | 0 | 0 | 0 |
| 0 | 0.138632687 | 0 | 0 | 0.059828726 |
| 0 | 0.005956073 | 0.006611087 | 0.006564077 | 0.001192923 |
| 0 | 0 | 0 | 0 | 0 |
| 0 | 0 | 0.00313965 | 0 | 0.003529899 |
| 0 | 0 | 0 | 0 | 0 |
| 0 | 0 | 0 | 0 | 0 |
| 0 | 0 | 0 | 0.00154163 | 0 |
| 0 | 0.021648881 | 0 | 0 | 0.062391844 |
| 0 | 0 | 0.020018189 | 0.041149207 | 0.117652461 |
| 0 | 0 | 0 | 0 | 0 |
| 0 | 0 | 0 | 0 | 0.012630165 |
| 0 | 0 | 0 | 0 | 0 |

| | | | | |
|---|---|---|---|---|
| 0 | 0 | 0 | 0 | 0 |
| 0 | 0 | 0 | 0 | 0.001844145 |
| 0 | 0 | 0 | 0 | 0 |
| 0 | 0 | 0 | 0.122424983 | 0.046447244 |
| 0 | 0 | 0 | 0 | 0 |
| 0 | 0 | 0 | 0.160067931 | 0.8974129 |
| 0 | 0 | 0 | 0 | 0 |
| 0 | 0 | 0 | 0 | 0 |
| 0.026430054 | 0.221085221 | 0.247193502 | 0.015276867 | 0.000705103 |
| 0 | 0 | 0 | 0 | 0.003028353 |
| 0 | 0 | 0 | 0.003641145 | 0.003361136 |
| 0 | 0 | 0 | 0 | 0 |
| 0 | 0 | 0 | 0.008912237 | 0 |
| 0 | 0 | 0 | 0 | 0 |
| 0.349892829 | 0 | 0.015288669 | 1.961873543 | 1.406107491 |
| 0.362051632 | 0 | 0.108533311 | 0 | 0 |
| 0 | 0 | 0 | 0 | 0 |
| 0 | 0 | 0 | 0 | 0 |
| 0 | 0 | 0 | 0 | 0 |
| 0 | 0 | 0 | 0 | 0.022679682 |
| 0 | 0 | 0 | 0 | 0.040076974 |
| 0 | 0 | 0 | 0 | 0 |
| 0 | 0 | 0 | 0 | 0 |
| 0.013817779 | 0 | 0 | 0 | 0 |
| 0 | 0.045539637 | 0.016281958 | 0.117549815 | 0.003593116 |
| 0.002746418 | 9.555441426 | 0 | 10.68634014 | 0.05418659 |
| 0 | 0 | 0 | 0 | 0.003084061 |
| 0 | 0 | 0 | 0 | 0 |
| 0 | 0 | 0 | 0 | 0.217482309 |
| 0 | 0 | 0 | 0 | 0.947413136 |
| 0 | 0 | 0 | 0 | 0 |
| 0 | 0 | 0 | 0 | 0.088999491 |
| 0 | 0 | 0 | 0 | 0 |
| 0 | 0 | 0.001696705 | 0.108975125 | 0 |
| 0 | 0.001214348 | 0.001048362 | 0 | 0.127932561 |
| 0.000453126 | 0.009270683 | 0.022478976 | 0.004000888 | 0.001375404 |
| 0.610390811 | 0 | 0.223309552 | 0 | 0.011177608 |
| 0 | 1.054962305 | 0 | 0 | 2.83493298 |
| 0 | 0 | 0 | 0 | 0 |
| 0 | 0 | 0 | 0 | 0 |
| 0 | 0 | 0 | 0 | 0 |
| 0 | 0 | 0 | 0.003441892 | 0.003119438 |
| 0 | 0.197770199 | 0 | 0.339725799 | 0.15106594 |
| 0 | 0 | 0 | 0 | 0 |
| 0 | 0 | 0 | 0 | 0 |
| 0 | 0 | 0.002574243 | 0.253429947 | 0.00977267 |
| 0 | 0 | 0 | 0 | 0 |
| 0 | 0.006996033 | 0 | 0.054554411 | 0.76366112 |

| | | | | |
|---|---|---|---|---|
| 0 | 0 | 0.001946113 | 0.015993024 | 0 |
| 0 | 0 | 0 | 0 | 0.016144414 |
| 0 | 0 | 0 | 0 | 0.021344111 |
| 0 | 0 | 0 | 0 | 0 |
| 0 | 0 | 0 | 0.003091827 | 0 |
| 0 | 0 | 0 | 0 | 0.001113109 |
| 0 | 0 | 0 | 0.004553534 | 0 |
| 0 | 0 | 0 | 0 | 0 |
| 0 | 0 | 0 | 0 | 0 |
| 0 | 0 | 0 | 0 | 0 |
| 0 | 0 | 0 | 0 | 0 |
| 0.00562112 | 0 | 0 | 6.88671008 | 0.217187399 |
| 0 | 0.15583036 | 0 | 1.025950063 | 0.039421665 |
| 0 | 0 | 0 | 0 | 0 |
| 0 | 0 | 0.013444875 | 0.018772412 | 0.004678772 |
| 0 | 0 | 0 | 0 | 0 |
| 0 | 0.002631035 | 0 | 0.001865561 | 0 |
| 0 | 0 | 0.00149607 | 0 | 0 |
| 0.008578073 | 0.001170016 | 0 | 0 | 0 |
| 0 | 0 | 0 | 0 | 0.002598442 |
| 0 | 0 | 0 | 0 | 0.074209191 |
| 0 | 0 | 0 | 0 | 0.014122786 |
| 0 | 0.050247594 | 0.791469678 | 0 | 1.70099391 |
| 0 | 0 | 0 | 0.02125608 | 0 |
| 0 | 0 | 0 | 0 | 0 |
| 0 | 0 | 0 | 0 | 0 |
| 0 | 0 | 0 | 0 | 0 |
| 0 | 0 | 0 | 0 | 0 |
| 0 | 0 | 0 | 0 | 0.003612334 |
| 0 | 0 | 0 | 0 | 0.02035301 |
| 0 | 0 | 0.020687863 | 0 | 0 |
| 0.007365831 | 0.058894489 | 0.016200419 | 0.003711974 | 0.011564498 |
| 0 | 0 | 0 | 0 | 0.022669472 |
| 0.00342606 | 0 | 0.003735439 | 0.078581822 | 0.014587995 |
| 0 | 0.006352224 | 0.027150975 | 0.007056433 | 0.001683882 |
| 0 | 0.017340961 | 0.004505066 | 0.046450711 | 0.027013508 |
| 0 | 0 | 0 | 0 | 0 |
| 0 | 0 | 0 | 0 | 0.012881454 |
| 0 | 0 | 0 | 0 | 0.000644844 |
| 0 | 0 | 0 | 0 | 0 |
| 0 | 0 | 0 | 0 | 0.0797041 |
| 0 | 0 | 0 | 0 | 0.206540539 |
| 0 | 0 | 0 | 0 | 0 |
| 0.008028805 | 0 | 0.032170284 | 0.057445859 | 0 |
| 0.013277626 | 0.00202451 | 0.00688522 | 0.351557976 | 0.001290172 |
| 0 | 0 | 0 | 0 | 0 |
| 0 | 0 | 0 | 0.0125684 | 0.022485807 |
| 0.000612598 | 0.001218525 | 0.006512186 | 0.001678642 | 0 |

| | | | | |
|---|---|---|---|---|
| 6.05770437 | 0 | 0 | 0 | 0 |
| 0 | 0 | 0 | 0 | 0 |
| 0.003516831 | 0 | 0.008216585 | 0 | 0.000571869 |
| 0 | 0 | 0 | 0 | 0 |
| 0.003906249 | 0 | 0.01722113 | 0 | 0 |
| 0 | 0 | 0 | 0 | 0 |
| 0 | 0 | 0 | 0 | 0.42973965 |
| 0 | 0.357558168 | 0 | 0 | 0 |
| 0 | 0 | 0 | 0 | 0 |
| 0 | 0 | 0 | 0 | 0.015766532 |
| 0.711759532 | 0 | 0 | 0 | 0 |
| 0 | 0 | 0 | 0.002801496 | 0 |
| 0 | 0 | 0 | 0 | 0 |
| 0 | 0 | 0 | 0 | 0 |
| 0.004950134 | 0 | 0 | 0 | 0 |
| 0 | 0 | 0 | 0 | 0 |
| 0 | 0 | 0 | 0 | 0.075547927 |
| 0 | 0 | 0 | 0 | 0 |
| 11.7676156 | 0 | 0 | 0.089733293 | 0 |
| 0 | 0 | 0 | 0 | 0 |
| 0 | 0 | 0 | 0.002779657 | 0 |
| 0 | 0 | 0 | 0 | 0 |
| 0 | 0 | 0 | 0 | 0 |
| 0 | 0 | 0 | 0 | 0 |
| 0 | 0 | 0 | 0 | 0.004631625 |
| 0.000452231 | 0.137243528 | 0.003698007 | 0.015834256 | 0 |
| 0 | 0 | 0 | 0 | 0 |
| 0 | 0.203429816 | 0 | 0.004986502 | 0.002687176 |
| 0 | 0 | 0 | 0.003708566 | 1.684733409 |
| 0 | 0 | 0 | 0.522794 | 0.381394611 |
| 0 | 0 | 0 | 0 | 0 |
| 0 | 0 | 0 | 0 | 0 |
| 0 | 0 | 0 | 0 | 0.021280692 |
| 0 | 0 | 0 | 0 | 0 |
| 0.003001428 | 0.008977698 | 0.002583522 | 0.002452926 | 0 |
| 0 | 0 | 0 | 0 | 0 |
| 0 | 0 | 0 | 0 | 0 |
| 0 | 0.004686047 | 0 | 0.01637266 | 0 |
| 0 | 0 | 0 | 0 | 0 |
| 0 | 0 | 0 | 0 | 0 |
| 0.998327811 | 25.47054079 | 43.93988198 | 2.498172261 | 0.432272872 |
| 0.001312115 | 0.962993452 | 0 | 0.486345672 | 0.160704162 |
| 0 | 0 | 0 | 0 | 0 |
| 0 | 0 | 0 | 0 | 0 |
| 0 | 0 | 0 | 0 | 0 |
| 0 | 0 | 0 | 0 | 0 |
| 0 | 0 | 0 | 0 | 0.003473025 |
| 0 | 0 | 0.001163316 | 0.022090223 | 0 |

| | | | | |
|---|---|---|---|---|
| 0 | 0 | 0 | 0 | 0 |
| 0 | 0.00083161 | 0.001196567 | 0.017108042 | 0.001998727 |
| 0 | 0 | 0 | 0 | 0 |
| 0 | 0 | 0 | 0 | 0 |
| 0 | 0.834272694 | 0 | 2.837253725 | 0.62690489 |
| 0 | 0 | 0 | 0 | 0.004953053 |
| 0 | 0 | 0 | 0 | 0 |
| 0 | 0 | 0 | 0 | 0 |
| 0.042664948 | 0.497234091 | 0.681754528 | 0.012545195 | 0 |
| 0 | 0 | 0 | 0 | 0 |
| 0 | 0 | 0 | 0.004832131 | 0 |
| 0 | 0 | 0 | 0 | 0 |
| 0 | 0.173569391 | 0 | 0 | 0.02459998 |
| 0 | 0.081062805 | 0.054888309 | 0.188720642 | 0.002586587 |
| 0 | 0 | 0 | 0 | 0.576578771 |
| 0 | 0.023314451 | 0 | 0 | 0.048655027 |
| 0 | 0.001203507 | 0 | 0 | 0 |
| 0 | 0 | 0 | 0 | 0 |
| 0 | 0 | 0 | 0 | 0.174155322 |
| 0 | 0 | 0 | 0 | 0 |
| 0 | 0 | 0 | 0 | 0 |
| 0.006976113 | 0 | 0 | 0 | 0 |
| 0 | 0 | 0 | 0 | 0 |
| 0 | 0 | 0 | 0 | 0 |
| 0 | 0 | 0 | 0.00593656 | 0.014487829 |
| 0 | 0 | 0 | 0 | 0 |
| 0 | 0 | 0 | 0 | 0.071413694 |
| 0 | 0.003879959 | 0.000728178 | 0.001491188 | 0.004899138 |
| 0 | 0 | 0 | 0 | 0 |
| 0 | 0 | 0 | 0 | 0 |
| 0 | 0 | 0 | 0 | 0 |
| 0 | 0 | 0 | 0 | 0.001411133 |
| 0 | 0 | 0 | 0 | 0.003095681 |
| 0 | 0 | 0 | 0 | 0.043386998 |
| 0 | 0 | 0 | 1.8204013 | 0.053200307 |
| 0.096487345 | 0.003902876 | 0.309423664 | 0.00141136 | 0 |
| 0 | 0.412085332 | 0.000860984 | 0 | 0 |
| 0 | 0 | 0 | 0.011402192 | 0 |
| 0 | 0 | 0 | 0 | 0 |
| 0 | 0 | 0 | 0 | 0 |
| 0 | 0 | 1.258629413 | 4.043468725 | 0.020344212 |
| 0 | 0 | 0 | 0 | 0 |
| 0 | 0 | 0 | 0 | 0.06107059 |
| 0 | 0 | 0 | 0 | 0 |
| 0 | 0 | 0 | 0 | 0 |
| 0 | 0 | 0 | 0 | 0 |
| 0 | 0 | 0 | 0 | 0 |
| 0 | 0 | 0 | 0 | 0 |

| | | | | |
|---|---|---|---|---|
| 0 | 0 | 0 | 0.810863906 | 0.05660047 |
| 0 | 0 | 0 | 0 | 0 |
| 0 | 0 | 0 | 0 | 0 |
| 0 | 0 | 0 | 0 | 0 |
| 0 | 0.013896032 | 0 | 0 | 0.020690832 |
| 0 | 0 | 0 | 0 | 0 |
| 0 | 0.042191843 | 0 | 0.000689134 | 0 |
| 0 | 0 | 0 | 0 | 0 |
| 0 | 0 | 0 | 0 | 0 |
| 0.180793005 | 0 | 0.103824006 | 0 | 0 |
| 0 | 0 | 0 | 0.033427527 | 0.069566885 |
| 0.002243318 | 0 | 0 | 0 | 0.001351847 |
| 0 | 0 | 0 | 0 | 0 |
| 0 | 0 | 0 | 0 | 0 |
| 0 | 0 | 0 | 0 | 0 |
| 0 | 0 | 0 | 0 | 0 |
| 0 | 0 | 0 | 0.555951159 | 0 |
| 0 | 0 | 0.010439149 | 0 | 0 |
| 0 | 0 | 0 | 0 | 0 |
| 0 | 0 | 0 | 0 | 0 |
| 0 | 0.009144159 | 0 | 1.115753887 | 0.142755324 |
| 0 | 0 | 0 | 0 | 0.004600811 |
| 0 | 0 | 0 | 0.059630381 | 0.019659782 |
| 0.001972612 | 0.003843666 | 0.00345655 | 0.030501642 | 0.001283059 |
| 0 | 0 | 0 | 0 | 0 |
| 0.00249347 | 0 | 0 | 0 | 0 |
| 0 | 0 | 0 | 0 | 0 |
| 0 | 0 | 0 | 0 | 0 |
| 0 | 0 | 0 | 0 | 0 |
| 0 | 0.654381994 | 1.302730514 | 0.504375584 | 0.05655283 |
| 0 | 0 | 0 | 0 | 0.080602015 |
| 0 | 0 | 0 | 0 | 0 |
| 0 | 0 | 0 | 0 | 0 |
| 0.948134397 | 0 | 9.719914007 | 1.190940834 | 0 |
| 0 | 0 | 0 | 0 | 0.012138784 |
| 0 | 0 | 0 | 0 | 2.704538467 |
| 0 | 0.010019628 | 0 | 0.464909974 | 0 |
| 0 | 0 | 0 | 0 | 0 |
| 0 | 0 | 0 | 0 | 0 |
| 0 | 0 | 0 | 0 | 0 |
| 0 | 0 | 0 | 0 | 0 |
| 0 | 0 | 0 | 0 | 0 |
| 0 | 0 | 0 | 0 | 0 |
| 0 | 0 | 0 | 0 | 0.025660621 |
| 0 | 0 | 0 | 0 | 0.098382911 |
| 0 | 0 | 0 | 0 | 0 |
| 0 | 0 | 0 | 0 | 0 |
| 0 | 0 | 0 | 0 | 0 |

| | | | | |
|---|---|---|---|---|
| 0.001516143 | 0.03911892 | 0.003471412 | 0.044176586 | 0.001553194 |
| 0 | 0 | 0 | 0 | 0 |
| 0 | 0 | 0 | 0 | 0.029533073 |
| 0 | 0 | 0 | 0 | 0 |
| 0 | 0 | 0 | 0 | 0 |
| 0 | 0 | 0 | 0 | 0 |
| 0 | 0 | 0 | 0 | 0 |
| 0 | 1.147128005 | 0.667807527 | 0 | 0.234255406 |
| 0.002855778 | 1.779064615 | 0.061494762 | 4.589382895 | 0.091017618 |
| 0 | 0 | 0.040643173 | 0 | 0 |
| 0 | 0 | 0 | 0.001598228 | 0.00907323 |
| 0 | 0 | 0 | 0 | 0 |
| 0 | 0 | 0 | 0 | 0.006042757 |
| 0 | 0 | 0 | 0 | 0 |
| 0 | 0 | 0 | 0 | 0 |
| 0 | 0 | 0 | 0 | 0 |
| 0.021025033 | 0 | 0 | 0 | 0 |
| 0 | 0 | 0 | 0 | 0 |
| 0 | 0 | 0 | 0 | 0 |
| 0 | 0 | 0 | 0 | 0.004534938 |
| 0 | 0 | 0 | 0 | 0 |
| 0 | 0 | 0 | 0 | 0 |
| 0 | 0 | 0 | 0 | 0.005905647 |
| 0 | 0 | 0 | 0 | 0 |
| 0 | 0 | 0 | 0 | 0.045536026 |
| 0 | 0 | 0 | 0 | 0 |
| 0 | 0 | 0 | 0 | 0 |
| 0 | 0 | 0 | 0 | 0 |
| 0.001195955 | 0 | 0 | 0 | 0 |
| 0 | 0 | 0 | 0 | 0 |
| 0 | 0 | 0.034898586 | 0 | 0 |
| 0 | 0 | 0 | 0 | 0 |
| 0 | 0 | 0 | 0 | 0 |
| 0 | 0 | 0 | 0 | 0 |
| 0 | 0 | 0 | 0 | 0.002298218 |
| 0 | 0 | 0 | 0 | 0.722991426 |
| 0 | 0 | 0 | 0 | 0.005635967 |
| 0 | 0.059034935 | 0.042471368 | 0 | 0 |
| 0 | 0 | 0 | 0 | 0.002257285 |
| 0 | 0 | 1.661734393 | 0 | 0 |
| 0 | 0 | 0 | 0 | 0 |
| 0.878518166 | 0 | 0 | 0.479666162 | 0.573045478 |
| 0 | 0 | 0 | 0 | 0 |
| 0 | 0 | 0 | 0 | 0 |
| 0 | 0 | 0 | 0 | 0 |
| 0.00067408 | 0 | 0.040348768 | 0 | 0 |
| 0 | 0 | 0 | 0 | 0 |
| 0 | 0 | 0 | 0 | 0 |

| | | | | |
|---|---|---|---|---|
| 0 | 0 | 0 | 0 | 0 |
| 0 | 0 | 0 | 0.00074993 | 0 |
| 0 | 0 | 0 | 0 | 0 |
| 0 | 0 | 0 | 0 | 0 |
| 0.055291353 | 0.441661993 | 0.228725528 | 2.170062412 | 0.151133897 |
| 0 | 0 | 0 | 0 | 0.082593565 |
| 0 | 0 | 0 | 0 | 0 |
| 0.004243017 | 0.069929999 | 0.020502336 | 0.010393584 | 0.000878123 |
| 0 | 0 | 0 | 0.054526415 | 0.010521274 |
| 0 | 0 | 0 | 0 | 0 |
| 0 | 0 | 0 | 0 | 0 |
| 0 | 0 | 0 | 0 | 0 |
| 0 | 0 | 0 | 0 | 0 |
| 0 | 0 | 0 | 0 | 0 |
| 0 | 0 | 0 | 0 | 0 |
| 0.014385209 | 0 | 0.003829867 | 0.017875771 | 0 |
| 0 | 0 | 0 | 0 | 0 |
| 0 | 0 | 0 | 0 | 0 |
| 0 | 0 | 0 | 0 | 0 |
| 0 | 0 | 0 | 0 | 0 |
| 0 | 0 | 0 | 0 | 0 |
| 0 | 0 | 0 | 0 | 0 |
| 0 | 0 | 0 | 0 | 0 |
| 0 | 0.002276564 | 0.002456735 | 0.001067179 | 0 |
| 0 | 0 | 0 | 0.218518291 | 0 |
| 0.00106485 | 0 | 0 | 0 | 0 |
| 0.114797729 | 0.010237365 | 0 | 0.275108329 | 0 |
| 0 | 0 | 0 | 0 | 0 |
| 0 | 0 | 0 | 0.001243531 | 0 |
| 0 | 0 | 0 | 0 | 0 |
| 0 | 0 | 0 | 0 | 0 |
| 0 | 0 | 0.002399639 | 0 | 0 |
| 0 | 0 | 0 | 0 | 0 |
| 0 | 0 | 0 | 0 | 0 |
| 0 | 0 | 0 | 0 | 0 |
| 0 | 0 | 0 | 0 | 0 |
| 0 | 0 | 0 | 0 | 0 |
| 0 | 0 | 0 | 0.182468462 | 0.082330538 |
| 0 | 0 | 0 | 0 | 0.093422989 |
| 0 | 0 | 0 | 0 | 0 |
| 0 | 0 | 0 | 0 | 0 |
| 0 | 0 | 0 | 0 | 0 |
| 0 | 0 | 0 | 0 | 0 |
| 0 | 0 | 0 | 0 | 0.0722847 |
| 0 | 0 | 0 | 0 | 0 |
| 0 | 0 | 0 | 0 | 0 |
| 0 | 0 | 0 | 0 | 0 |
| 0 | 0 | 0 | 0 | 0.157051956 |

| | | | | |
|---|---|---|---|---|
| 0 | 0 | 0 | 0 | 0.096084829 |
| 0 | 0 | 0 | 0 | 0 |
| 0.000711022 | 0 | 0 | 0 | 0 |
| 0 | 0 | 0 | 0 | 0 |
| 0 | 0 | 0 | 0 | 0 |
| 0 | 0 | 0 | 0 | 0 |
| 0 | 0 | 0 | 0 | 0 |
| 0 | 0 | 0 | 0 | 0.001103306 |
| 0 | 0 | 0 | 0 | 0 |
| 0 | 0 | 0 | 0 | 0 |
| 0 | 0 | 0 | 0 | 0 |
| 0 | 0.001748857 | 0 | 0.00168646 | 0 |
| 0 | 0 | 0 | 0 | 0.002283391 |
| 0 | 0 | 0 | 0 | 0 |
| 0 | 0 | 0 | 0 | 0 |
| 0 | 0 | 0 | 0 | 0 |
| 0 | 0 | 0 | 0 | 0 |
| 0 | 0 | 5.333508417 | 0 | 0 |
| 0 | 0 | 0 | 0.002223464 | 0 |
| 0 | 0 | 0 | 0 | 0 |
| 0 | 0.180985929 | 0.022290013 | 0.032185157 | 0 |
| 0 | 0 | 0 | 0 | 0 |
| 0.002728854 | 0.00238593 | 0 | 0.196143404 | 0 |
| 0.002113345 | 0 | 0 | 0.001754845 | 0.005685832 |
| 0 | 0 | 0 | 0 | 0.001287674 |
| 0 | 0 | 0 | 0 | 0 |
| 0 | 0 | 0 | 0 | 0 |
| 0 | 0 | 0 | 0 | 0 |
| 0 | 0 | 0 | 0 | 0 |
| 0 | 0 | 0 | 0 | 0 |
| 0 | 0 | 0 | 0 | 0 |
| 0 | 0 | 0 | 0 | 0.004324482 |
| 0.008649588 | 0.017651864 | 0.067836447 | 1.643491284 | 0 |
| 0 | 0 | 0 | 0.156687975 | 0 |
| 0 | 0.161692561 | 0 | 0.981187727 | 0.119491814 |
| 0.016090342 | 0.003406744 | 0.029668858 | 0.000864525 | 0 |
| 0 | 0 | 0 | 0 | 0 |
| 0 | 0 | 0 | 0 | 0 |
| 0.000729844 | 0.023435235 | 0 | 0 | 0 |
| 0 | 0 | 0 | 0 | 0 |
| 0 | 0 | 0 | 0 | 0.071812375 |
| 0 | 0 | 0 | 0 | 0 |
| 0 | 0.059170674 | 0 | 0.047159861 | 0.038856079 |
| 0 | 0 | 0 | 0 | 0 |
| 0 | 0 | 0 | 0 | 0 |
| 0.004066517 | 0 | 0 | 0.011143074 | 0.03741589 |
| 0 | 0 | 0 | 0 | 0 |
| 0 | 0 | 0 | 0 | 0 |

| | | | | |
|---|---|---|---|---|
| 0 | 0 | 0 | 0 | 0 |
| 0 | 0 | 0 | 0 | 0 |
| 0 | 0 | 0 | 0 | 0 |
| 0 | 0 | 0 | 0 | 0 |
| 0 | 0 | 0 | 0 | 0 |
| 0 | 0 | 0 | 0 | 0 |
| 0 | 0 | 0 | 0 | 0 |
| 0.531634466 | 0.124130186 | 0 | 0 | 0 |
| 0 | 0 | 0 | 0.002320006 | 0 |
| 0 | 0.001785283 | 0 | 0.002550498 | 0 |
| 0 | 0 | 0 | 0 | 0.871227517 |
| 0 | 0 | 0 | 0 | 0 |
| 0 | 0 | 0 | 0 | 0 |
| 0 | 0 | 0 | 0 | 0.15342932 |
| 0 | 0 | 0 | 0 | 0.004494999 |
| 0.116417933 | 0.008977776 | 0.010951981 | 0 | 0 |
| 0 | 0 | 0 | 0 | 0 |
| 0 | 0 | 0 | 0 | 0 |
| 0 | 0 | 0 | 0 | 0 |
| 0 | 0.89004901 | 0 | 0 | 0 |
| 0 | 0 | 0 | 0.006552248 | 0.018167793 |
| 0 | 0 | 0 | 0 | 0 |
| 0 | 0 | 0 | 0 | 0 |
| 0 | 0 | 0 | 0 | 0.011194704 |
| 0 | 0.001055105 | 0 | 0 | 0 |
| 0 | 0 | 0 | 0 | 0 |
| 0 | 0 | 0 | 0 | 0 |
| 0 | 0 | 0 | 0 | 0 |
| 0 | 0 | 0 | 0 | 0 |
| 0 | 0 | 0 | 0 | 0.247537831 |
| 0 | 0.012214349 | 0 | 0.456100359 | 0.127560868 |
| 0 | 0 | 0 | 0 | 0 |
| 0 | 0 | 0 | 0 | 0 |
| 0 | 0 | 0 | 0 | 0 |
| 0 | 0 | 0 | 0 | 0 |
| 0 | 0 | 0 | 0 | 0 |
| 0.001829809 | 0 | 0 | 0 | 0 |
| 0 | 0 | 0 | 0 | 0 |
| 0 | 0 | 0 | 0.011924417 | 0 |
| 0 | 0 | 0 | 0 | 0 |
| 0 | 0 | 0 | 0.005395474 | 0.002241249 |
| 0 | 0 | 0 | 0 | 6.079173516 |
| 0 | 0 | 0 | 0 | 0 |
| 0 | 0 | 0 | 0 | 0 |
| 0.00514956 | 0 | 0 | 0 | 0 |
| 0 | 0 | 0 | 0 | 0 |
| 0.015468296 | 0.35203548 | 0 | 0 | 0 |
| 0 | 0 | 0 | 0 | 0 |

| | | | | |
|---|---|---|---|---|
| 0 | 0 | 0 | 0 | 0 |
| 0 | 0 | 0 | 0 | 0 |
| 0 | 0 | 0 | 0 | 0 |
| 0 | 0 | 0 | 0 | 0 |
| 0 | 0.713613892 | 0.079033193 | 0.170562916 | 0.215093681 |
| 0 | 0 | 0 | 0 | 0 |
| 0 | 0 | 0 | 0 | 0 |
| 0 | 0 | 0 | 0 | 0 |
| 0 | 0 | 0 | 0 | 0 |
| 0 | 0 | 0 | 0 | 0 |
| 0 | 0 | 0 | 0 | 0.002277367 |
| 0.005651788 | 0.001657811 | 0.00238535 | 0.091423476 | 0 |
| 0 | 0 | 0 | 0.009194949 | 0 |
| 0 | 0 | 0 | 0 | 0 |
| 0.003056621 | 0 | 0 | 0 | 0 |
| 0 | 0 | 0 | 0 | 0.426685861 |
| 0.003365339 | 0 | 0.006879814 | 0 | 0 |
| 0.001527256 | 0 | 0 | 0 | 0 |
| 0 | 0.091323342 | 0.016255804 | 0.016266308 | 0.004557001 |
| 0 | 0.148333492 | 0.003537521 | 0 | 0 |
| 0 | 0 | 0 | 0 | 0 |
| 0 | 0 | 0 | 0 | 0.058465145 |
| 0 | 0 | 0 | 0 | 0 |
| 0 | 0 | 0 | 0 | 0 |
| 0 | 0 | 0 | 0 | 0 |
| 0 | 0 | 0 | 0 | 0 |
| 0.013866637 | 0.01083589 | 0.005669556 | 0 | 0.000657662 |
| 0.009861568 | 0 | 0 | 0 | 0 |
| 0 | 0 | 0 | 0 | 0 |
| 0 | 0 | 0 | 0 | 0 |
| 0.00098241 | 0 | 0.006158958 | 0.003290224 | 0 |
| 0.004244469 | 0 | 0 | 0 | 0 |
| 0.00279849 | 0 | 0 | 0 | 0 |
| 0 | 0 | 0 | 0 | 0 |
| 0 | 0 | 0 | 0 | 0 |
| 0 | 0 | 0 | 0 | 0 |
| 0 | 0 | 0 | 0 | 0 |
| 0.008512624 | 0.068645455 | 0.007337266 | 0.391503635 | 0.002880696 |
| 0 | 0 | 0 | 0.220915411 | 2.284935653 |
| 0 | 0 | 0.003879602 | 0 | 0 |
| 0 | 0 | 0 | 0.005664447 | 0 |
| 0 | 0 | 0 | 0 | 0.006042923 |
| 0 | 0 | 0 | 0 | 0 |
| 0.005970405 | 0.003683919 | 0.006416549 | 0 | 0.000776674 |
| 0 | 0 | 0 | 0 | 0 |
| 0 | 0 | 0 | 0 | 0 |
| 0 | 0 | 0 | 0 | 0 |
| 0 | 0 | 0 | 0 | 0 |

| | | | | |
|---|---|---|---|---|
| 0 | 0 | 0 | 0 | 0 |
| 0 | 3.57705496 | 0 | 0 | 0.215910202 |
| 0 | 0 | 0 | 0.00484777 | 0 |
| 0 | 0.008127924 | 0 | 0 | 0.001424426 |
| 0 | 0.001130778 | 0 | 0 | 0 |
| 0.002353902 | 0 | 0 | 0 | 0 |
| 0 | 0 | 0 | 0 | 0.000789884 |
| 0 | 0 | 0 | 0 | 0.008912198 |
| 0 | 0 | 0 | 0 | 0.001157177 |
| 0 | 0 | 0 | 0.001698222 | 0.0096409 |
| 0 | 0 | 0 | 0 | 0.041364044 |
| 0 | 0 | 0 | 0 | 0 |
| 0 | 0.114078771 | 0 | 0.017478215 | 0.07486336 |
| 0 | 0 | 0 | 0 | 0 |
| 0 | 0 | 0 | 0 | 0 |
| 0 | 0 | 0 | 0 | 0 |
| 0 | 0.013025273 | 0 | 0 | 0 |
| 0 | 0 | 0 | 0 | 0 |
| 0 | 0 | 0 | 0 | 0 |
| 0 | 0 | 0 | 0 | 0 |
| 0 | 0 | 0 | 0 | 0 |
| 0 | 0 | 0 | 0 | 0 |
| 0.001168064 | 0 | 0.001637412 | 0 | 0.011016401 |
| 0 | 0.0235003 | 0 | 0 | 0.024206398 |
| 0 | 0 | 0 | 0 | 0 |
| 0 | 0 | 0 | 0 | 0.137385421 |
| 0 | 0 | 0 | 0 | 0 |
| 0 | 0 | 0 | 0 | 0 |
| 0 | 0 | 0 | 0 | 0 |
| 0 | 0 | 0 | 0 | 0 |
| 0 | 0 | 0 | 0.041296126 | 0.018424857 |
| 0 | 0 | 0 | 0 | 0 |
| 0 | 0 | 0 | 0 | 0 |
| 0 | 0 | 0 | 0 | 0 |
| 0 | 0 | 0 | 0 | 0.027698044 |
| 0.00466793 | 0.000663216 | 0 | 0 | 0 |
| 0 | 0 | 0.006465599 | 0 | 0 |
| 0.01821588 | 0 | 0.07350353 | 0 | 0 |
| 0 | 0.001610316 | 0 | 0.001294051 | 0 |
| 0 | 0 | 0 | 0 | 0 |
| 0 | 0 | 0 | 0 | 0 |
| 0 | 0 | 0 | 0 | 0 |
| 0 | 0 | 0 | 0 | 2.031312934 |
| 0 | 0 | 0 | 0 | 0 |
| 0 | 0 | 0 | 0 | 0 |
| 0.000415499 | 0.287612805 | 0 | 0.324107634 | 0.088283594 |
| 0.744749971 | 0 | 0 | 0 | 0 |
| 0 | 0 | 0 | 0 | 0 |

| | | | | |
|---|---|---|---|---|
| 0 | 0 | 0 | 0 | 0 |
| 0 | 0 | 0 | 0 | 0 |
| 0 | 0 | 0 | 0 | 0 |
| 0 | 0 | 0 | 0.780413306 | 0.063788581 |
| 0 | 0 | 0 | 0 | 0 |
| 0 | 0 | 0 | 0 | 0 |
| 0 | 0 | 0 | 0 | 0 |
| 0 | 0 | 0 | 0 | 0 |
| 0 | 0 | 0 | 0 | 0 |
| 0 | 0 | 0 | 0 | 0 |
| 0 | 0.000870932 | 0 | 0.002916173 | 0 |
| 0 | 0 | 0.002182774 | 0 | 0 |
| 0 | 0 | 0 | 0 | 0 |
| 0 | 0 | 0 | 0 | 0 |
| 0 | 0 | 0 | 0 | 0 |
| 0 | 0 | 0 | 0 | 0 |
| 0 | 0 | 0.00644724 | 0 | 0 |
| 0 | 0 | 0 | 0 | 0 |
| 0 | 0 | 0 | 0 | 0 |
| 0.001693604 | 0 | 0 | 0.001082857 | 0 |
| 0 | 0 | 0 | 0 | 0 |
| 0 | 0 | 0 | 0 | 0 |
| 0 | 0 | 0 | 0 | 0 |
| 0 | 0 | 0 | 0 | 0 |
| 0 | 0 | 0 | 0 | 0 |
| 0 | 0 | 0 | 0 | 0.043776097 |
| 0 | 0 | 0 | 0 | 0 |
| 0 | 0 | 0 | 0 | 0.008320267 |
| 0 | 0 | 0 | 0 | 0 |
| 0 | 0 | 0 | 0 | 0 |
| 0 | 0.005457479 | 0.006186836 | 0.006246208 | 0.006127765 |
| 0 | 0 | 0 | 0 | 0 |
| 0.019596823 | 0.011329231 | 0.057744663 | 0.014504966 | 0.016537473 |
| 0 | 0.007064082 | 0 | 0 | 0 |
| 0 | 0 | 0 | 0 | 0 |
| 0 | 0 | 0 | 0 | 0 |
| 0 | 0 | 0 | 0 | 0 |
| 0 | 0 | 0 | 0 | 0 |
| 0 | 0 | 0 | 0 | 0 |
| 0 | 0 | 0 | 0 | 0 |
| 0 | 0 | 0 | 0 | 0 |
| 0.052952852 | 0.342013314 | 2.220735494 | 0.127836552 | 0 |
| 0 | 0 | 0 | 0.321998699 | 0.020429658 |
| 0 | 0 | 0 | 0 | 0 |
| 0 | 0 | 0 | 0 | 0 |
| 0 | 1.895341747 | 0.005202142 | 0.132754294 | 0.027851204 |
| 0 | 0.006564202 | 0 | 0.006447221 | 0 |
| 0 | 0 | 0 | 0 | 0 |

| | | | | |
|---|---|---|---|---|
| 0 | 0 | 0 | 0 | 0 |
| 0.023237666 | 0 | 0 | 0.003240503 | 0.006461215 |
| 0 | 0.021196617 | 0.001355505 | 0 | 0.005424681 |
| 0 | 0 | 0 | 0 | 0 |
| 0 | 0 | 0.035445511 | 0.060900141 | 0 |
| 0 | 0 | 0.000790132 | 0.020152201 | 0 |
| 0 | 0 | 0 | 0 | 0.002155413 |
| 0 | 0 | 0 | 0 | 0 |
| 0 | 0 | 0 | 0 | 0 |
| 0 | 0 | 0 | 0 | 0 |
| 0 | 0 | 0 | 0 | 0 |
| 0 | 0 | 0 | 0 | 0 |
| 0 | 0 | 0 | 0 | 0 |
| 0 | 0 | 0 | 0 | 0 |
| 0 | 0 | 0 | 0 | 0 |
| 0 | 0 | 0 | 0 | 0 |
| 0 | 0 | 0 | 0 | 0 |
| 0 | 0 | 0 | 0 | 0 |
| 0 | 0 | 0 | 0 | 0 |
| 0 | 0 | 0 | 0 | 0 |
| 0 | 0 | 0 | 0 | 0 |
| 0.163173503 | 0 | 0 | 0 | 0 |
| 0 | 0 | 0 | 0 | 0 |
| 0 | 0 | 0.149311544 | 0 | 0 |
| 0 | 0 | 0 | 0.004991324 | 0.0044707 |
| 0 | 0.016011314 | 0.002528557 | 0.074062025 | 0 |
| 0 | 0 | 0 | 0 | 0 |
| 0 | 0 | 0 | 0.180824544 | 0.08042455 |
| 0 | 0 | 0 | 0 | 0 |
| 0 | 0 | 0 | 0 | 0 |
| 0 | 0 | 0 | 0 | 0 |
| 0.013558297 | 0.010273883 | 0 | 0 | 0 |
| 0 | 0 | 0 | 0 | 0 |
| 0 | 0 | 0 | 0 | 0 |
| 0 | 0 | 0 | 0 | 0 |
| 0 | 0 | 0 | 0 | 0 |
| 0 | 0 | 0 | 0 | 0 |
| 0 | 0 | 0.380621225 | 0 | 0 |
| 0 | 0 | 0 | 4.013038058 | 0.111136835 |
| 0 | 0 | 0 | 0 | 0 |
| 0 | 0 | 0 | 0 | 0 |
| 0 | 0 | 0.007010131 | 0 | 0.001697045 |
| 0.001206855 | 0.034293813 | 0.002255718 | 0.033070251 | 0.000784983 |
| 0 | 0 | 0 | 0 | 0 |
| 0 | 0 | 0 | 0 | 0 |
| 0 | 0 | 0 | 0.007790402 | 0 |
| 0.008440008 | 0.00119915 | 0.002156755 | 0.004336372 | 0 |
| 0 | 0 | 0 | 0 | 0 |

| | | | | |
|---|---|---|---|---|
| 0 | 0 | 0 | 0 | 0 |
| 0.01397778 | 0.028559895 | 0.005442856 | 0.093930926 | 0 |
| 0 | 0 | 0 | 0 | 0.11566189 |
| 0 | 0 | 0 | 0 | 0.011880901 |
| 0 | 0 | 0 | 0 | 0 |
| 0 | 0 | 0 | 0 | 0 |
| 0 | 0 | 0 | 0 | 0 |
| 0 | 0 | 0 | 0 | 0 |
| 0 | 0 | 0 | 0 | 0 |
| 0 | 0 | 0 | 0 | 0 |
| 0 | 0 | 0 | 0 | 0 |
| 0 | 0 | 0 | 0 | 0 |
| 0 | 1.106845841 | 0 | 0 | 1.106630988 |
| 0 | 0 | 0 | 0 | 0 |
| 0.00124487 | 0 | 0.006107779 | 0 | 0.001653157 |
| 0 | 0 | 0 | 0 | 0 |
| 0 | 0 | 0 | 0 | 0 |
| 0 | 0 | 0 | 0 | 0 |
| 0 | 0 | 0 | 0 | 0 |
| 0 | 0 | 0 | 0 | 0 |
| 0 | 0 | 0 | 0 | 0 |
| 0 | 0.032192508 | 0 | 0 | 0 |
| 0.003288033 | 0.004983055 | 0 | 0.007150692 | 0 |
| 0.029172209 | 0 | 0 | 0.001533577 | 0.001911118 |
| 0 | 0 | 0 | 0 | 0 |
| 0 | 0 | 0 | 0 | 0 |
| 0 | 0 | 0 | 0 | 0 |
| 0 | 0 | 0 | 0 | 0 |
| 0 | 0 | 0 | 0 | 0 |
| 0 | 0 | 0 | 0 | 0.029790184 |
| 0 | 0 | 0 | 0 | 0 |
| 0 | 0 | 0 | 0 | 0 |
| 0 | 0 | 0 | 0 | 0 |
| 0 | 0 | 0 | 0 | 0 |
| 0 | 0.354953684 | 0.001210254 | 0.053060258 | 14.76391442 |
| 0 | 0 | 0 | 0 | 0 |
| 0 | 0 | 0.002931224 | 0 | 0 |
| 0 | 0 | 0 | 0 | 0 |
| 0 | 0.06938442 | 0 | 0 | 0 |
| 0 | 0 | 0 | 0 | 0 |
| 0.029191359 | 0.73493475 | 1.158914639 | 0.197309155 | 0.425934713 |
| 0 | 0 | 0 | 0 | 0 |
| 0 | 0 | 0 | 0 | 0 |
| 0 | 0 | 0 | 0 | 0 |
| 0.274953715 | 0 | 0 | 0 | 0 |
| 0.5332917 | 1.30111385 | 3.958771057 | 0.194042186 | 0 |
| 0 | 0 | 0 | 0 | 0 |
| 0 | 0 | 0 | 0 | 0 |

| | | | | |
|---|---|---|---|---|
| 0 | 0 | 0 | 0 | 0 |
| 0 | 0 | 0 | 0 | 0 |
| 0 | 0 | 0 | 0 | 0 |
| 0 | 0 | 0 | 0 | 0.058673132 |
| 0 | 0 | 0 | 0 | 0 |
| 0 | 0 | 0 | 0 | 0 |
| 0 | 0 | 0 | 0 | 0 |
| 0 | 0 | 0 | 0 | 0 |
| 0 | 0 | 0 | 0 | 0.001771801 |
| 0 | 0 | 0.020380803 | 0.000559804 | 0.008603959 |
| 0 | 0 | 0 | 0 | 0 |
| 0 | 0 | 0 | 0 | 0 |
| 0 | 0 | 0 | 0 | 0 |
| 0 | 0 | 0 | 0 | 0 |
| 0 | 0 | 0 | 0 | 0 |
| 0 | 0 | 0 | 0 | 0 |
| 0 | 0 | 0 | 0 | 0 |
| 0 | 0 | 0 | 0 | 0 |
| 0 | 0 | 0 | 0 | 0 |
| 0.031813423 | 1.399542123 | 0.029648002 | 4.729862205 | 0.321660347 |
| 0 | 0 | 0 | 0 | 0 |
| 0 | 0 | 0 | 0 | 0 |
| 0 | 0 | 0 | 0 | 0 |
| 0 | 0 | 0 | 0 | 0 |
| 0 | 0 | 0 | 0 | 0 |
| 0 | 0 | 0 | 0 | 0 |
| 0 | 0 | 0 | 0 | 0 |
| 0 | 0 | 0 | 0 | 0 |
| 0 | 0 | 0 | 0 | 0 |
| 0 | 0 | 0 | 0 | 0 |
| 0 | 0 | 0 | 0 | 0 |
| 0 | 0.001131626 | 0 | 0 | 0 |
| 0 | 0 | 0 | 0 | 0 |
| 0 | 0 | 0 | 0 | 0 |
| 0 | 0 | 0 | 0 | 0 |
| 0 | 0 | 0 | 0.008195539 | 0 |
| 0 | 0 | 0 | 0 | 0 |
| 0 | 0 | 0 | 0 | 0.270726894 |
| 0 | 0 | 0 | 0 | 0 |
| 0 | 0 | 0 | 0 | 0.26684381 |
| 0 | 0 | 0 | 0 | 0 |
| 0 | 0 | 0 | 0 | 0 |
| 0 | 0 | 0 | 0 | 0.017488795 |
| 0 | 0 | 0 | 0 | 0 |
| 0.002670868 | 0.0060716 | 0.000819014 | 0.05595768 | 0 |
| 0 | 0 | 0 | 0 | 0.030777816 |
| 0 | 0 | 0 | 0 | 0 |
| 0 | 0 | 0 | 0 | 0 |

| | | | | |
|---|---|---|---|---|
| 0 | 0 | 0 | 0 | 0 |
| 0 | 0 | 0 | 0 | 0 |
| 0 | 0 | 0 | 0 | 0 |
| 0 | 0 | 0 | 0 | 0 |
| 0 | 0 | 0 | 0 | 0.004060287 |
| 0.002492383 | 0.004249393 | 0.001834278 | 0.052701995 | 0.070782917 |
| 0 | 0 | 0 | 0 | 0 |
| 0 | 0 | 0 | 0 | 0 |
| 0 | 0 | 0 | 0 | 0 |
| 0 | 0 | 0 | 0 | 0 |
| 0 | 0 | 0 | 0 | 0 |
| 0 | 0 | 0 | 0 | 0 |
| 0 | 0 | 0 | 0 | 0 |
| 0 | 0 | 0 | 0 | 0 |
| 0 | 0 | 0 | 0 | 0 |
| 0 | 0 | 0 | 0 | 0 |
| 0 | 0 | 0 | 0.038660047 | 0.342404345 |
| 0 | 0 | 0.006526034 | 0 | 0 |
| 0 | 0 | 0 | 0 | 0 |
| 0 | 0 | 0 | 0 | 0 |
| 0 | 0 | 0 | 0.004883307 | 0 |
| 0 | 0 | 0 | 0 | 0 |
| 0 | 0 | 0 | 0 | 0 |
| 0 | 0 | 0 | 0 | 0 |
| 0 | 0 | 0 | 0 | 0 |
| 0 | 0 | 0 | 0 | 0.002662518 |
| 0 | 0 | 0 | 0 | 0 |
| 0 | 0 | 0 | 0 | 0 |
| 0 | 0 | 0 | 0 | 0 |
| 0.091016518 | 0 | 0 | 0 | 0 |
| 0 | 0 | 0 | 0 | 0 |
| 0 | 0 | 0 | 0 | 0 |
| 0 | 0 | 0 | 0 | 0 |
| 0 | 0 | 0 | 0 | 0 |
| 0 | 0 | 0 | 0 | 0 |
| 0 | 0 | 0 | 0 | 0 |
| 0 | 0 | 0 | 0 | 0 |
| 0 | 0 | 0 | 0 | 0 |
| 0 | 0 | 0 | 0 | 0.098314566 |
| 0 | 0 | 0 | 0 | 0 |
| 0 | 0 | 0 | 0 | 0.001909498 |
| 0 | 0 | 0 | 0 | 0 |
| 0 | 0 | 0 | 0 | 0 |
| 0 | 0 | 0 | 0 | 0 |
| 0 | 0 | 0 | 0 | 0 |
| 0 | 0 | 0 | 0 | 0.001262616 |
| 0 | 0 | 0 | 0 | 0 |
| 0 | 0 | 0 | 0 | 0 |

| | | | | |
|---|---|---|---|---|
| 0 | 0 | 0 | 0 | 0 |
| 0 | 0.017127135 | 0.020536226 | 0 | 0.001429308 |
| 0 | 0 | 0 | 0 | 0 |
| 0 | 0 | 0 | 0 | 0 |
| 0 | 0 | 0 | 0 | 0 |
| 0 | 0 | 0 | 0 | 0 |
| 0 | 0 | 0 | 0 | 0 |
| 0 | 0 | 0 | 0 | 0 |
| 0 | 0 | 0 | 0 | 0 |
| 0 | 0 | 0 | 0 | 0 |
| 0 | 0 | 0 | 0.000667484 | 0.026409867 |
| 0 | 0 | 0 | 0 | 0 |
| 0 | 0 | 0 | 0.000614544 | 1.847959221 |
| 0 | 0 | 0 | 0 | 0 |
| 0 | 0 | 0 | 0 | 0 |
| 0 | 0 | 0.039082189 | 0 | 0 |
| 0 | 0 | 0 | 0 | 0 |
| 0 | 0 | 0 | 0 | 0 |
| 0 | 0 | 0 | 0 | 0 |
| 0 | 0 | 0 | 0 | 0 |
| 0 | 0.004004369 | 0 | 0 | 0 |
| 0 | 0 | 0 | 0 | 0 |
| 0 | 0 | 0 | 0 | 0 |
| 0 | 1.656513662 | 0 | 0.828196888 | 0.217501889 |
| 0 | 0 | 0 | 0 | 0 |
| 0 | 0 | 0 | 0 | 0 |
| 0 | 0 | 0 | 0 | 0 |
| 0 | 0 | 0 | 0 | 0 |
| 0 | 0 | 0 | 0 | 0 |
| 0 | 0 | 0 | 0 | 0 |
| 0.00382181 | 0 | 0 | 0 | 0 |
| 0 | 0 | 0 | 0 | 0 |
| 0 | 0 | 0 | 0 | 0.019759187 |
| 0 | 0 | 0 | 0 | 0 |

| PHL018-0102_V3 | PHL018-0143_V1 | PHL018-0166_V1 | PHL018-0256_V2 | PHL018-0240_V3 |
|---|---|---|---|---|
| CTRL | CTRL | EYCF | REF | REF |
| 0.057569734 | 0.5293295 | 0.293214991 | 0.000963367 | 0.617076941 |
| 2.842497497 | 0.963471755 | 0.049185022 | 12.58504782 | 0.205167656 |
| 0 | 2.54558277 | 15.29769874 | 1.176779077 | 0.989015942 |
| 0 | 0 | 0 | 0 | 0 |
| 0 | 0 | 0 | 0 | 0 |
| 0 | 0 | 0 | 0 | 0 |
| 0 | 0 | 3.565914821 | 0 | 0 |
| 0 | 0 | 0 | 0 | 0 |
| 0 | 0.00064326 | 0 | 0 | 0 |
| 0 | 0 | 0 | 0 | 0 |
| 0 | 0 | 0 | 0 | 0 |
| 0 | 0 | 0 | 0 | 0 |
| 0 | 0 | 0 | 0 | 0 |
| 15.43529351 | 0.653119038 | 0.241657572 | 6.741171026 | 0.024705993 |
| 0 | 0 | 0 | 0 | 0 |
| 0 | 0 | 0 | 0 | 0 |
| 0 | 0 | 0 | 0 | 0 |
| 0 | 0 | 0 | 0 | 0 |
| 0 | 0 | 0 | 0 | 0.029050628 |
| 0 | 0 | 0 | 0 | 0 |
| 0 | 0 | 0 | 0 | 0 |
| 0 | 0 | 0 | 0 | 0 |
| 0 | 0 | 0 | 0 | 0 |
| 0 | 0 | 0 | 0 | 0 |
| 0 | 0 | 0 | 0 | 0 |
| 0 | 0 | 0 | 0 | 0 |
| 0 | 0 | 0 | 0 | 0 |
| 0 | 0 | 0 | 0 | 0 |
| 0 | 0 | 0 | 0 | 0 |
| 0 | 0 | 0 | 0 | 0 |
| 0 | 0 | 0 | 0 | 0 |
| 0 | 0 | 0 | 0 | 0 |
| 0 | 0 | 0 | 0 | 0 |
| 0 | 0 | 0 | 0 | 0 |
| 0 | 0 | 0 | 0 | 0 |
| 1.578351574 | 0.016192114 | 0.967815358 | 0 | 0 |
| 0 | 0 | 0 | 0 | 0 |
| 0 | 0 | 0 | 0 | 0 |
| 0 | 0 | 0 | 0 | 0 |
| 0 | 0 | 0 | 0 | 0 |
| 0 | 0 | 0 | 0 | 0 |
| 0.004254503 | 0.001774612 | 0.006799357 | 0.067791827 | 0.003271382 |
| 0.004272718 | 0.87077454 | 1.078720727 | 0.011420028 | 0.057015157 |
| 0 | 0.053227344 | 0.002973135 | 0.048499141 | 0 |
| 0 | 0 | 0 | 0 | 0 |
| 0 | 0 | 0 | 0 | 0 |

| | | | | |
|---|---|---|---|---|
| 0 | 0.02175193 | 0 | 0 | 0 |
| 0 | 0.034917202 | 0.003105697 | 0.002758969 | 0.003232747 |
| 0.005671027 | 0.014435396 | 0 | 0 | 0.209915715 |
| 0.311859686 | 0.356686904 | 0.69100419 | 0.324352598 | 0.170563085 |
| 0.002347637 | 0.028438524 | 0 | 0.002301865 | 0.001198732 |
| 0 | 0 | 0 | 0 | 0 |
| 0.010461652 | 0.0023391 | 0.00128031 | 0.130115445 | 0.021020896 |
| 0 | 0 | 0.002326964 | 0 | 0 |
| 0 | 0 | 0 | 0 | 0 |
| 0 | 0 | 0 | 0 | 0 |
| 0 | 0 | 0 | 0 | 0 |
| 0.065050694 | 0.182880927 | 0 | 0.017530376 | 0 |
| 0 | 0 | 0.001330973 | 0 | 0 |
| 0.053863607 | 0.020580908 | 0.002017219 | 0.030073208 | 0.150214173 |
| 0 | 0 | 0 | 0 | 0 |
| 0 | 0 | 0 | 0 | 0 |
| 0.001943192 | 0.040681005 | 0.236818294 | 5.234981352 | 0 |
| 0 | 0 | 0 | 0 | 0 |
| 0 | 0 | 0 | 0 | 0 |
| 0 | 0 | 0 | 0 | 0 |
| 0.465180796 | 0 | 0.099602903 | 0.417591352 | 0.058037886 |
| 0 | 0.006766746 | 0.009097025 | 0.001891049 | 0.014443641 |
| 0 | 0.024766374 | 0 | 0 | 0 |
| 0 | 0 | 0 | 0 | 0 |
| 0 | 0 | 0 | 0 | 0 |
| 0 | 0 | 0 | 0 | 0 |
| 0 | 0 | 0.003475894 | 0 | 0 |
| 0 | 0 | 0 | 0 | 0 |
| 0 | 0 | 0.001629282 | 0.001881599 | 0 |
| 0 | 0 | 0 | 0 | 0 |
| 0 | 0 | 0.005978601 | 0.21975273 | 0.162360533 |
| 0 | 0 | 0 | 0 | 0 |
| 0 | 0 | 0 | 0 | 0 |
| 0 | 0 | 0 | 0 | 0 |
| 0 | 0 | 0 | 0 | 0 |
| 0 | 0 | 0 | 0 | 0 |
| 0 | 0 | 0 | 0 | 0 |
| 0 | 0.001151634 | 0 | 0.124846354 | 0 |
| 0 | 0 | 0 | 0 | 0 |
| 0 | 0 | 0 | 0 | 0 |
| 0 | 0 | 0 | 0 | 0 |
| 0 | 0 | 0 | 0 | 0 |
| 0 | 0 | 0 | 0 | 0 |
| 0 | 0 | 0 | 0 | 0 |
| 0 | 0 | 0 | 0 | 0 |
| 0 | 0 | 0 | 0 | 0 |
| 0 | 0 | 0 | 0 | 0 |
| 0 | 0 | 0 | 0 | 0.801772228 |

| | | | | |
|---|---|---|---|---|
| 0 | 0 | 0 | 0 | 0 |
| 0 | 0 | 0 | 0.01125363 | 0.023097283 |
| 0 | 0 | 0 | 0 | 0 |
| 0 | 0 | 0 | 0 | 0 |
| 0 | 0.085941699 | 0.210823467 | 0.018894232 | 0 |
| 0 | 0 | 0 | 0 | 0 |
| 0 | 0 | 0 | 0 | 0 |
| 0 | 0 | 0 | 0 | 0 |
| 0 | 0.000688483 | 0 | 0.003452601 | 0 |
| 0 | 0 | 0 | 0 | 0 |
| 0 | 0 | 0 | 0 | 0 |
| 0.007665201 | 0 | 0 | 0.000943153 | 0 |
| 0 | 0 | 0 | 0 | 0 |
| 0 | 0 | 0 | 0 | 0 |
| 0 | 0 | 0 | 0 | 0 |
| 0 | 0 | 0 | 0 | 0.006150017 |
| 0 | 0 | 0 | 0 | 0 |
| 0 | 0 | 0 | 0 | 0 |
| 0.008828045 | 1.069401199 | 0.006722955 | 0 | 0 |
| 0 | 0 | 0 | 0 | 0 |
| 0 | 0 | 0 | 0 | 0 |
| 0 | 0 | 0 | 0 | 0 |
| 0 | 0 | 0 | 0 | 0 |
| 0 | 0 | 0 | 0 | 0 |
| 0 | 0 | 0 | 0 | 0 |
| 0 | 0 | 0 | 0 | 0 |
| 0 | 0 | 0.001288532 | 0 | 0 |
| 0 | 0.001544701 | 0 | 0 | 0.002562799 |
| 0 | 0 | 0 | 0 | 0 |
| 0 | 0 | 0 | 0 | 0 |
| 0.015274062 | 0.320781007 | 0 | 0 | 0.086616157 |
| 0 | 0 | 0 | 0 | 0 |
| 0 | 0 | 0 | 0 | 0 |
| 0 | 0 | 0 | 0 | 0 |
| 0 | 0 | 0 | 0 | 0 |
| 0 | 0 | 0 | 0 | 0 |
| 0 | 0 | 0 | 0 | 0 |
| 0 | 0 | 0 | 0 | 0 |
| 0.079788502 | 0 | 1.078633263 | 0 | 0 |
| 0 | 0 | 0 | 0 | 0 |
| 0 | 0 | 0 | 0 | 0 |
| 0 | 0 | 0 | 0 | 0 |
| 0.017867321 | 0 | 0 | 0.003646223 | 0 |
| 0 | 0 | 0 | 0 | 0 |
| 0 | 0 | 0 | 0 | 0 |
| 0 | 0 | 0 | 0 | 0 |
| 0 | 0 | 0 | 0 | 0 |
| 0 | 0 | 0 | 0 | 0 |

| | | | | |
|---|---|---|---|---|
| 0 | 0.003751249 | 0 | 0.000663944 | 0 |
| 0 | 0 | 0 | 0 | 0 |
| 0 | 0 | 0 | 0 | 0 |
| 0 | 0 | 0 | 0 | 0 |
| 0.002789385 | 0.103453741 | 0 | 0 | 0 |
| 0 | 0 | 0 | 0 | 0 |
| 0 | 0 | 0 | 0 | 0 |
| 0 | 0 | 0 | 0 | 0 |
| 0 | 0 | 0 | 0 | 0 |
| 0 | 0 | 0 | 0 | 0 |
| 0 | 0 | 0 | 0 | 0 |
| 0 | 0 | 0 | 0 | 0 |
| 0 | 0 | 0 | 0 | 0 |
| 0 | 0 | 0 | 0 | 0 |
| 0 | 0 | 0 | 0 | 0 |
| 0 | 0 | 0 | 0 | 0 |
| 0 | 0 | 0 | 0 | 0 |
| 0 | 0 | 0 | 0 | 0 |
| 0 | 0 | 0 | 0 | 0 |
| 0 | 0 | 0 | 0 | 0 |
| 0 | 0 | 0 | 0 | 0 |
| 0 | 0 | 0 | 0 | 0 |
| 0 | 0 | 0 | 0 | 0 |
| 0 | 0 | 0 | 0 | 0 |
| 0 | 0 | 0 | 0 | 0 |
| 0 | 0 | 0 | 0 | 0 |
| 0 | 0 | 0 | 0 | 0 |
| 0 | 0 | 0 | 0 | 0 |
| 0 | 0 | 0 | 0 | 0 |
| 0 | 0 | 0 | 0 | 0 |
| 0 | 0 | 0 | 0 | 0 |
| 0.000652519 | 0.001156742 | 0.00118187 | 0 | 0 |
| 0 | 0 | 0 | 0 | 0 |
| 0 | 0 | 0 | 0 | 0 |
| 0 | 0 | 0 | 0 | 0 |
| 0 | 0 | 0 | 0 | 0 |
| 0 | 0 | 0 | 0 | 0 |
| 0 | 0 | 0.130551919 | 0 | 0 |
| 0 | 0 | 0 | 0 | 0 |
| 0 | 0 | 0 | 0 | 0 |
| 0 | 0 | 0 | 0 | 0 |
| 0 | 0 | 0 | 0 | 0 |
| 0 | 0 | 0 | 0 | 0 |
| 0 | 0 | 0 | 0 | 0 |
| 0 | 0 | 0 | 0 | 0 |
| 0 | 0 | 0 | 0 | 0 |
| 0 | 0 | 0 | 0 | 0 |
| 0 | 0 | 0 | 0 | 0 |

| | | | | |
|---|---|---|---|---|
| 0 | 0 | 0 | 0 | 0 |
| 0 | 0 | 0 | 0 | 0 |
| 0 | 0 | 0 | 0 | 0 |
| 0 | 0 | 0 | 0 | 0 |
| 0 | 0 | 0 | 0 | 0 |
| 0 | 0 | 0 | 0 | 0 |
| 0 | 0 | 0 | 0 | 0 |
| 0 | 0 | 0 | 0 | 0 |
| 0 | 0 | 0 | 0 | 0 |
| 0 | 0 | 0 | 0 | 0 |
| 0 | 0 | 0 | 0 | 0 |
| 0 | 0 | 0 | 0 | 0 |
| 0 | 0.003443654 | 0 | 0.000870717 | 0.002040477 |
| 0 | 0 | 0 | 0 | 0 |
| 0 | 0.142667092 | 0 | 0 | 0 |
| 0 | 0 | 0 | 0 | 0 |
| 0 | 0.001389693 | 0 | 0 | 0 |
| 0 | 0 | 0 | 0 | 0 |
| 0 | 0 | 0 | 0 | 0 |
| 0 | 0 | 0 | 0 | 0 |
| 0.028408472 | 0.037253091 | 0.069428515 | 0.043142817 | 0.097287648 |
| 0 | 0 | 0 | 0 | 0 |
| 0 | 0 | 0 | 0 | 0.895344244 |
| 0 | 0.004076587 | 0 | 0 | 0.001326163 |
| 0 | 0 | 0 | 0 | 0 |
| 0 | 0 | 0 | 0 | 0 |
| 0 | 0 | 0.002052463 | 0.007585014 | 0 |
| 0 | 0 | 0 | 0 | 0 |
| 0 | 0 | 0 | 0 | 0 |
| 0 | 0 | 0 | 0 | 0 |
| 0 | 0 | 0.002659745 | 0.002599084 | 0 |
| 0 | 0 | 0 | 0 | 0 |
| 0 | 0 | 0 | 0 | 0 |
| 0 | 0.066051756 | 0 | 0 | 0 |
| 0.009875049 | 0.009725463 | 0 | 0.001324105 | 0 |
| 0 | 0.00120046 | 0.004088459 | 0 | 0 |
| 0 | 0 | 0 | 0 | 0 |
| 0 | 0 | 0 | 0 | 0 |
| 0 | 0 | 0 | 0 | 0 |
| 0 | 0 | 0 | 0 | 0 |
| 0 | 0 | 0 | 0 | 0 |
| 0 | 0 | 0 | 0 | 0 |
| 0 | 0 | 0 | 0 | 0 |
| 0 | 0 | 0 | 0 | 0 |
| 0 | 0 | 0 | 0 | 0 |
| 0 | 0.026212074 | 0 | 0 | 0 |
| 0 | 0 | 0 | 0 | 0 |
| 0 | 0.003092126 | 0.009804712 | 0 | 0.000884503 |

| | | | | |
|---|---|---|---|---|
| 0 | 0 | 0 | 0 | 0 |
| 0 | 0 | 0 | 0 | 0 |
| 0 | 0 | 0 | 0 | 0 |
| 0 | 0 | 0 | 0 | 0 |
| 0 | 0 | 0 | 0 | 0 |
| 0 | 0 | 0 | 0 | 0 |
| 0 | 0 | 0 | 0 | 0 |
| 0 | 0 | 0 | 0.008138591 | 0 |
| 0 | 0 | 0 | 0 | 0 |
| 0 | 0 | 0 | 0 | 0 |
| 0 | 0 | 0 | 0 | 0 |
| 0 | 0 | 0 | 0 | 0 |
| 0 | 0 | 0 | 0 | 0 |
| 0 | 0.007983635 | 0 | 0 | 0 |
| 0 | 0 | 0 | 0 | 0 |
| 0 | 0 | 0 | 0 | 0 |
| 0 | 0 | 0 | 0 | 0 |
| 0 | 0 | 0.011295155 | 0 | 0 |
| 0 | 0.003903731 | 0.008974194 | 0 | 0 |
| 0 | 0 | 0 | 0 | 0 |
| 0.011910493 | 0 | 0.089723207 | 0.013872373 | 0.201247142 |
| 0 | 0 | 0 | 0 | 0 |
| 0 | 0 | 0 | 0 | 0 |
| 0 | 0 | 0 | 0 | 0 |
| 0.011293567 | 0 | 0 | 0.003374743 | 0 |
| 0 | 0 | 0 | 0 | 0 |
| 0 | 0 | 0 | 0 | 0 |
| 0 | 0 | 0 | 0 | 0 |
| 0 | 0 | 0 | 0 | 0 |
| 0 | 0 | 0.000835199 | 0 | 0 |
| 0.003582728 | 0.254401811 | 0.130324578 | 0.197137384 | 1.891828289 |
| 0 | 0 | 0 | 0 | 0 |
| 0 | 0 | 0 | 0 | 0.027296365 |
| 0 | 0 | 0 | 0 | 0 |
| 0 | 0 | 0 | 0 | 0 |
| 0 | 0 | 0 | 0 | 0 |
| 0 | 0 | 0.008117723 | 0 | 0 |
| 0 | 0.001480556 | 0.834263711 | 0 | 0 |
| 0 | 0 | 0 | 0 | 0 |
| 0 | 0 | 0 | 0 | 0 |
| 0 | 0 | 0 | 0 | 0 |
| 0.013999318 | 0.013187812 | 0.001837403 | 0 | 0 |
| 0 | 0 | 0 | 0 | 0 |
| 0 | 0 | 0 | 0 | 0 |
| 0 | 0 | 0 | 0 | 0 |
| 0 | 0 | 0 | 0 | 0 |
| 0 | 0 | 0 | 0 | 0 |
| 0 | 0 | 0 | 0 | 0 |

| | | | | |
|---|---|---|---|---|
| 0 | 0 | 0 | 0 | 0 |
| 0 | 0 | 0 | 0 | 0 |
| 0 | 0.062173277 | 0.001845729 | 0.04742737 | 0 |
| 0 | 0 | 0 | 0 | 0 |
| 0 | 0 | 0 | 0 | 0 |
| 0 | 0 | 0 | 0 | 0 |
| 0 | 0 | 0 | 0 | 0 |
| 0 | 0 | 0 | 0 | 0 |
| 0 | 0 | 0 | 0 | 0 |
| 0 | 0.00041305 | 0 | 0 | 0 |
| 0 | 0 | 0 | 0 | 0 |
| 0 | 0 | 0 | 0 | 0 |
| 0 | 0 | 0 | 0 | 0 |
| 0.001970891 | 0.000499124 | 0 | 0 | 0 |
| 0.001161938 | 0.013182761 | 0.005261378 | 0.000729141 | 0 |
| 0.004229626 | 0.098062105 | 0 | 0 | 0 |
| 0.000794354 | 0 | 0.000959179 | 0.000830791 | 0 |
| 0 | 0 | 0 | 0 | 0 |
| 0 | 0 | 0 | 0 | 0 |
| 0 | 0 | 0 | 0 | 0 |
| 0 | 0 | 0 | 0 | 0 |
| 0 | 0.003531066 | 0.012338574 | 0.000999956 | 0.001366951 |
| 0 | 0 | 0 | 0 | 0 |
| 0 | 0 | 0 | 0 | 0.020888126 |
| 0 | 0 | 0 | 0 | 0 |
| 0 | 0 | 0 | 0 | 0 |
| 0 | 1.187795638 | 0 | 0 | 0 |
| 0 | 0 | 0 | 0 | 0 |
| 0 | 0 | 0 | 0 | 0 |
| 0 | 0 | 0 | 0.002892892 | 0 |
| 0 | 0 | 0 | 0 | 0 |
| 0 | 0 | 0 | 0 | 0 |
| 0 | 0 | 0 | 0 | 0 |
| 0.002480356 | 0 | 0 | 0.008935336 | 0.001125778 |
| 0 | 0 | 0 | 0 | 0 |
| 0 | 0 | 0 | 0.003495224 | 0 |
| 0 | 0 | 0 | 0 | 0 |
| 0 | 0.011179774 | 0 | 0 | 0 |
| 0 | 0 | 0 | 0 | 0 |
| 0 | 0 | 0 | 0 | 0 |
| 0 | 0 | 0 | 0 | 0 |
| 0 | 0 | 0 | 0 | 0 |
| 0 | 0 | 0 | 0 | 0 |
| 0 | 0 | 0 | 0 | 0 |
| 0 | 0 | 0 | 0 | 0 |
| 0 | 0 | 0 | 0 | 0 |
| 0 | 0 | 0.000884408 | 0 | 0 |
| 0 | 0 | 0.004941311 | 0.003804363 | 0.010698381 |

| | | | | |
|---|---|---|---|---|
| 0 | 2.812643172 | 0 | 0 | 0 |
| 0.004160255 | 0 | 0 | 0 | 0 |
| 0 | 0 | 0 | 0 | 0 |
| 0 | 0 | 0 | 0 | 0 |
| 0.003274524 | 0 | 0 | 0.002107523 | 0.002675219 |
| 0 | 0 | 0 | 0 | 0 |
| 0 | 0 | 0 | 0 | 0 |
| 0 | 0 | 0 | 0.010339553 | 0.013845818 |
| 0 | 0 | 0 | 0 | 0 |
| 1.170007062 | 0.003225678 | 3.212366662 | 0.0026643 | 12.38946895 |
| 0 | 0 | 0 | 0 | 0 |
| 0 | 0 | 0.001012466 | 0.019292806 | 0 |
| 0.001484047 | 0.010523285 | 0.007487917 | 0.095788004 | 0.003464105 |
| 0 | 0 | 0 | 0.055564236 | 0.411679642 |
| 0 | 0 | 0 | 0 | 0 |
| 0 | 0 | 0 | 0 | 0 |
| 0 | 0 | 0 | 0 | 0 |
| 0 | 0 | 0.003500731 | 0 | 0.001353463 |
| 0 | 0 | 0 | 0.017191255 | 0 |
| 0 | 0 | 0 | 0 | 0 |
| 0 | 0 | 0 | 0 | 0 |
| 0 | 0 | 0 | 0 | 0 |
| 0 | 0 | 0 | 0 | 0 |
| 0 | 0 | 0 | 0 | 0 |
| 0 | 0 | 0 | 0 | 0 |
| 0 | 0.000583664 | 0 | 0 | 0 |
| 0 | 0 | 0 | 0 | 0 |
| 0 | 0 | 0 | 0 | 0 |
| 0.459129556 | 0 | 0.054264323 | 0.033052707 | 0.496345903 |
| 0 | 0.015613437 | 0 | 0 | 0 |
| 0 | 0.000626303 | 0.000719897 | 0 | 0.015153455 |
| 0 | 0 | 0 | 0 | 0 |
| 0 | 0 | 0 | 0 | 0.004613565 |
| 0.015824261 | 0 | 0 | 0.228999574 | 0 |
| 0 | 0 | 0 | 0 | 0 |
| 0 | 0 | 0 | 0 | 0 |
| 0.073620228 | 0 | 0 | 0.061773397 | 0.001829543 |
| 0.000964064 | 0 | 0 | 0 | 0 |
| 0 | 0 | 0 | 0 | 0 |
| 0.017400651 | 0 | 0.158524805 | 0.006247354 | 0.23212359 |
| 0 | 0 | 0 | 0 | 0 |
| 0 | 0 | 0 | 0.34273753 | 0.003080067 |
| 0 | 0 | 0 | 0 | 0 |
| 0 | 0 | 0 | 0 | 0.010401844 |
| 0 | 0 | 0 | 0 | 0 |
| 0 | 0 | 0 | 0 | 0 |
| 0.002149572 | 0.001814581 | 0 | 0.00128467 | 0 |
| 0 | 0 | 0 | 0 | 0.014573859 |

| | | | | |
|---|---|---|---|---|
| 0 | 0 | 0 | 0 | 0 |
| 0 | 0 | 0 | 0.51506051 | 0.003925256 |
| 0 | 0 | 0 | 0 | 0 |
| 0 | 0 | 0 | 0 | 0 |
| 0 | 0.000717409 | 0 | 0 | 0.011406537 |
| 0.167369944 | 0 | 0.119594348 | 0.01973075 | 0.004939951 |
| 0 | 0 | 0 | 0 | 0 |
| 0 | 0 | 0 | 0.004907554 | 0 |
| 0 | 0 | 0 | 0 | 0 |
| 0 | 0 | 0.001428663 | 0 | 0 |
| 0.053579628 | 0 | 0 | 0.056917481 | 0 |
| 0 | 0 | 0 | 0 | 0 |
| 0 | 0 | 0 | 0 | 0 |
| 0.001590136 | 0.054028695 | 0.146886255 | 0.003326151 | 0.000649555 |
| 0.005094952 | 0 | 0.018942082 | 0.004206834 | 0.070981105 |
| 0.007567429 | 0 | 0 | 0 | 0.014131282 |
| 0 | 0 | 0 | 0 | 0 |
| 0.024243531 | 0 | 0 | 0.111132954 | 0 |
| 0 | 0.004333728 | 0 | 0.005605279 | 0 |
| 0 | 0 | 0 | 0 | 0 |
| 0 | 0 | 0 | 0 | 0.000699868 |
| 0.223828776 | 0 | 0.10989982 | 0.401754697 | 0 |
| 0 | 0 | 0 | 0 | 0 |
| 0 | 0 | 0 | 0 | 0 |
| 0 | 0 | 0 | 0 | 0 |
| 0 | 0 | 0 | 0 | 0 |
| 0.092528242 | 0 | 0 | 0.013824642 | 0.092496633 |
| 0 | 0 | 0 | 0 | 0 |
| 0 | 0 | 0 | 0 | 0 |
| 2.439189261 | 0.87206046 | 0.071064552 | 5.64819808 | 0.002538962 |
| 0 | 0.00159366 | 0 | 0 | 0 |
| 0 | 0 | 0 | 0 | 0 |
| 0 | 0 | 0 | 0 | 0 |
| 0 | 0 | 0 | 0 | 0.003328753 |
| 0 | 0 | 0 | 0 | 0 |
| 0 | 0 | 0 | 0 | 0 |
| 0.038620901 | 0 | 2.502835066 | 0 | 0 |
| 0 | 0 | 0 | 0 | 0 |
| 0.115387625 | 0 | 0 | 0 | 0 |
| 0 | 0 | 0 | 0 | 0.403509356 |
| 0.002178775 | 0.054845915 | 0 | 0.016406748 | 0.006586056 |
| 8.77031396 | 0 | 0 | 0 | 0 |
| 0 | 0 | 0 | 0 | 0 |
| 0 | 0 | 0.002856489 | 0 | 0.472860767 |
| 0.001966217 | 0 | 0 | 0.001799357 | 0 |
| 0 | 0 | 0 | 0 | 0.054903447 |
| 0.001372068 | 0.004324111 | 0.004970298 | 0.000717502 | 0 |
| 0 | 0 | 0.013762854 | 0 | 0 |

| | | | | |
|---|---|---|---|---|
| 0 | 0 | 0 | 0 | 0 |
| 0 | 0 | 0 | 0 | 0 |
| 0.004613418 | 0.085418452 | 0.220738183 | 12.16230389 | 0 |
| 0 | 0 | 0 | 0 | 0 |
| 0 | 0.143912583 | 0 | 0 | 0 |
| 0 | 0 | 0 | 0.017964073 | 0 |
| 0 | 0 | 0 | 0 | 0 |
| 0 | 0 | 0 | 0 | 0 |
| 0 | 0 | 0 | 0 | 0.034918003 |
| 0 | 0 | 0 | 0.066594857 | 0 |
| 0 | 0 | 0 | 0 | 0 |
| 0 | 0 | 0 | 0 | 0.002138086 |
| 0 | 0 | 0 | 0 | 0 |
| 0.016846785 | 0 | 0 | 0 | 0 |
| 0 | 0 | 0 | 0 | 0 |
| 0.031716819 | 0.019566938 | 0 | 0.084285231 | 0 |
| 0 | 0.004311452 | 0.024828796 | 0 | 0.002709504 |
| 0 | 0 | 0.004865487 | 0 | 0.080542853 |
| 0 | 0 | 0 | 0 | 0 |
| 0 | 0 | 0 | 0 | 0 |
| 0.036781538 | 0 | 0 | 0.020691497 | 0 |
| 0 | 0.002727851 | 0 | 0 | 0 |
| 0.001446249 | 0 | 0 | 0.040839889 | 0 |
| 0 | 0.148692116 | 0 | 0 | 0 |
| 0.004752689 | 0 | 0 | 0.040197787 | 0.001857015 |
| 0 | 0 | 0.258759026 | 0 | 0.74241326 |
| 0 | 0.005971007 | 0 | 0.001467812 | 0 |
| 0 | 0 | 0 | 0.001857789 | 0 |
| 0 | 0 | 0 | 0 | 0 |
| 0 | 0 | 0 | 0 | 0 |
| 0.001707695 | 0.01198302 | 0.028030771 | 0.01786027 | 0.074117473 |
| 0 | 0.005453591 | 0.006340619 | 0.002662747 | 0 |
| 0.022422945 | 0.01271281 | 0.063115409 | 0 | 0.078722579 |
| 0 | 0 | 0 | 0 | 0 |
| 0 | 0 | 0.010385525 | 0.001944952 | 0 |
| 0 | 0 | 0 | 0 | 0 |
| 0.001369722 | 0 | 0.002232809 | 0 | 0.001119035 |
| 0.005141694 | 0.09557938 | 0 | 0.021734233 | 0.017502748 |
| 0.054270286 | 0 | 0 | 0 | 0 |
| 0.001050965 | 0 | 0 | 0.002198345 | 0.002862057 |
| 0 | 0 | 0 | 0 | 0 |
| 0.003520293 | 0 | 0 | 0 | 0 |
| 0 | 0 | 0 | 0.005361404 | 0 |
| 0.081445185 | 0.226799154 | 0 | 0.100048389 | 0.011067072 |
| 0.004909039 | 0 | 0 | 0 | 0.859334674 |
| 0 | 0 | 0 | 0 | 0 |
| 0 | 0 | 0 | 0 | 0 |
| 0 | 0 | 0 | 0 | 0 |

| | | | | |
|---|---|---|---|---|
| 0 | 0 | 0 | 0.022922693 | 0 |
| 0.059260861 | 0.020010235 | 0.011103703 | 0.55811761 | 0 |
| 0 | 0 | 0 | 0 | 0 |
| 0.007602448 | 0 | 0 | 0 | 0 |
| 0 | 0 | 0 | 0 | 0 |
| 0 | 0 | 0 | 0 | 0 |
| 0 | 0 | 0 | 0 | 0 |
| 0.000672689 | 0 | 0 | 0.001172574 | 0 |
| 0 | 0.005842722 | 0 | 0 | 0.025107318 |
| 0 | 0 | 0 | 0 | 0 |
| 0 | 0.003461072 | 0 | 0 | 0.001949524 |
| 0 | 0.102725432 | 0.453285126 | 0.011828211 | 0.079196489 |
| 0 | 0 | 0 | 0 | 0 |
| 0 | 0 | 0 | 0 | 0 |
| 0 | 0.016619989 | 0.023217415 | 0 | 0 |
| 0 | 0 | 0.00053462 | 0 | 0 |
| 0.037935661 | 0.004161167 | 0.002468647 | 0.02019425 | 0 |
| 0.405171442 | 0.001219754 | 0.210972488 | 0.034850431 | 1.078816169 |
| 0 | 0 | 0 | 0 | 0 |
| 0.413985489 | 0 | 0 | 0.001802393 | 0.065860161 |
| 0.00632499 | 0.161266558 | 0.002545798 | 0.011760207 | 0 |
| 0 | 0 | 0 | 0 | 0 |
| 0 | 0 | 0 | 0 | 0 |
| 0 | 0 | 0 | 0 | 0 |
| 0 | 0 | 0 | 0.017701077 | 0 |
| 0 | 0.000761644 | 0 | 0 | 0 |
| 0.097577053 | 0 | 0.009423113 | 0.061183369 | 0.118255362 |
| 0.062245658 | 0.174933584 | 0.518387668 | 0.012759767 | 0.141372559 |
| 0 | 0 | 0.002198268 | 0 | 0 |
| 0 | 0 | 0.016319189 | 0 | 0 |
| 0 | 0 | 0 | 0.01816185 | 0 |
| 0.000687341 | 0 | 0 | 0 | 0 |
| 0.003324895 | 0 | 0 | 0 | 0 |
| 0 | 0 | 0 | 0 | 0 |
| 0 | 0 | 0 | 0 | 0 |
| 0.533056136 | 0.134212657 | 0.016409582 | 0.822819087 | 0.021688736 |
| 0 | 0 | 0 | 0 | 0 |
| 0 | 0 | 0 | 0 | 0 |
| 0 | 0.007441384 | 0.003059757 | 0.007709682 | 0 |
| 0.007957586 | 0 | 0 | 0 | 0 |
| 0 | 0 | 0 | 0 | 0.470496894 |
| 0 | 0 | 0 | 0 | 0 |
| 0 | 0.038144478 | 0.004730837 | 0.024975885 | 0.020424402 |
| 0.003242273 | 0 | 0.001794388 | 0 | 0.001765914 |
| 0.003549663 | 0.001538193 | 0 | 0 | 0 |
| 0.017680221 | 0.869337168 | 0 | 0 | 0 |
| 0 | 0 | 0 | 0 | 0 |
| 0 | 0.001720498 | 0 | 0.001827094 | 0.005946799 |

| | | | | |
|---|---|---|---|---|
| 0 | 0 | 0 | 0.003271111 | 0 |
| 0 | 0.067796914 | 0 | 0 | 0 |
| 0 | 0.037684884 | 0 | 0 | 0 |
| 0.004128524 | 0.101307251 | 0.001771048 | 0.100220704 | 0 |
| 0 | 0 | 0 | 0 | 0 |
| 0 | 0 | 0 | 0 | 0 |
| 0 | 0 | 0 | 0 | 0 |
| 0 | 0.005973093 | 0 | 0.001626452 | 0 |
| 0.045052984 | 0.233361247 | 0 | 0.060126516 | 0 |
| 0 | 0.074821973 | 0 | 0 | 0 |
| 0 | 0 | 0 | 0 | 0 |
| 0 | 0.001722288 | 0 | 0.009144972 | 0 |
| 0 | 0 | 0 | 0 | 0 |
| 0 | 0.320572713 | 0 | 0 | 0 |
| 0 | 0 | 0.073435266 | 0.012397746 | 0.729423945 |
| 0 | 0 | 0 | 0 | 0 |
| 0 | 1.148212897 | 0 | 0 | 0 |
| 0 | 0 | 0 | 0 | 0 |
| 0 | 0 | 0 | 0 | 0 |
| 0 | 0.038011808 | 0 | 0 | 0 |
| 0 | 0 | 0 | 0 | 0 |
| 0 | 0.003426468 | 0 | 0 | 0 |
| 0 | 0 | 0 | 0 | 0 |
| 0.180543828 | 0.437302516 | 0.027125835 | 0.011422981 | 0.088013673 |
| 0 | 0.077745622 | 0 | 0.0038733 | 0 |
| 0.127560343 | 0.625568472 | 0 | 0 | 0 |
| 0 | 1.317960521 | 0 | 0 | 0 |
| 0.268194182 | 1.626263978 | 24.23397597 | 1.140783984 | 0 |
| 0 | 0 | 0 | 0 | 0 |
| 0 | 0 | 0 | 0 | 0 |
| 0.001214365 | 0.008371788 | 0 | 0 | 0 |
| 0 | 0 | 0 | 0 | 0 |
| 0.006469545 | 0.149913368 | 0.005021964 | 0 | 0 |
| 0 | 0 | 0 | 0 | 0 |
| 0 | 0 | 0 | 0 | 0 |
| 0 | 0 | 0 | 0 | 0 |
| 0.003546114 | 1.038289973 | 0.014852894 | 0 | 0 |
| 0 | 0 | 0 | 0 | 0 |
| 0 | 0.176600852 | 0 | 0 | 0 |
| 0 | 0 | 0 | 0 | 0 |
| 0.004992977 | 0.130171928 | 0.014831328 | 0.013786089 | 0.00913732 |
| 0 | 0 | 0 | 0 | 0 |
| 0.370881516 | 2.709603609 | 0.015622242 | 9.375101836 | 0.398914366 |
| 0 | 0 | 0 | 0 | 0 |
| 0 | 0 | 0 | 0 | 0 |
| 0 | 0 | 0 | 0 | 0 |
| 0 | 0 | 0 | 0 | 0 |
| 0.000960341 | 0.008086537 | 0.002391689 | 0 | 0.00372675 |

| | | | | |
|---|---|---|---|---|
| 0 | 0 | 0 | 0 | 0 |
| 0 | 0 | 0 | 0 | 0 |
| 0.002301488 | 0.008023854 | 0.023135471 | 0.006017642 | 1.217474171 |
| 0 | 0 | 0 | 0 | 0 |
| 0 | 0 | 0 | 0 | 0 |
| 0 | 0.037925297 | 0 | 0.000758475 | 0 |
| 0 | 0 | 0 | 0 | 0 |
| 0 | 0 | 0 | 0 | 0 |
| 0 | 0 | 0 | 0 | 0 |
| 0 | 0 | 0.002269001 | 0 | 0 |
| 0.005423167 | 0.536371744 | 0.00204639 | 0 | 0.023445352 |
| 2.052965327 | 0.421590192 | 2.347566098 | 1.044190726 | 0.311322437 |
| 0 | 0 | 0 | 0 | 0 |
| 0 | 0.000576424 | 0 | 0.003315745 | 0 |
| 0.014221296 | 0.088804813 | 0.095026941 | 0.573438512 | 0.412771132 |
| 0 | 1.582219663 | 0 | 0 | 0 |
| 0 | 0 | 0 | 0 | 0 |
| 0 | 0 | 0 | 0 | 0 |
| 0 | 0.019235995 | 0 | 0 | 0 |
| 0.001302952 | 0.042456074 | 0 | 0.00097337 | 0.037865252 |
| 0.024999771 | 0.261431086 | 0.080784799 | 0.018025242 | 0 |
| 0.679503941 | 0 | 2.543181338 | 0.011479294 | 5.037679698 |
| 0.305894254 | 0.299146503 | 2.85027284 | 0.081571663 | 53.07344812 |
| 0 | 0 | 0 | 0 | 0 |
| 0 | 0 | 0 | 0 | 0 |
| 0.003631179 | 4.217918658 | 0 | 0 | 0 |
| 0 | 0 | 0 | 0 | 0 |
| 0 | 0.001283967 | 0 | 0.000826374 | 0 |
| 0 | 0.101951261 | 0.003521469 | 0 | 0 |
| 0 | 0 | 0 | 0 | 0 |
| 0 | 0 | 0 | 0 | 0 |
| 0.255863888 | 0.13766897 | 0 | 0 | 0.101669307 |
| 0.763053593 | 0.007542377 | 0.05611528 | 0.011407716 | 3.473820511 |
| 0 | 0.202457941 | 0.000480812 | 0 | 0 |
| 0.004135605 | 0 | 0 | 0.007369038 | 0.024652038 |
| 0.00420894 | 0.125350354 | 0 | 0.011885409 | 0 |
| 0 | 0 | 0.003129747 | 0 | 0.032718337 |
| 0 | 0 | 0 | 0 | 0 |
| 0 | 0.945304801 | 0 | 0 | 0 |
| 0 | 0 | 0 | 0 | 0 |
| 0 | 0.019272226 | 0 | 0 | 0 |
| 0 | 0 | 0 | 0 | 0 |
| 0 | 0 | 0 | 0 | 0 |
| 0 | 0 | 0 | 0 | 0 |
| 0.006959737 | 0.115577875 | 0.00239075 | 0.003764989 | 0.136267057 |
| 0.301613339 | 0 | 0 | 0 | 0 |
| 0 | 0 | 0 | 0 | 0 |
| 0.024896538 | 0 | 0.223633151 | 0.015744232 | 0.011589051 |

| | | | | |
|---|---|---|---|---|
| 0 | 0 | 0 | 0 | 0 |
| 0 | 0 | 0 | 0 | 0 |
| 0 | 0 | 0 | 0 | 0 |
| 0.007363558 | 0.041578225 | 0 | 0 | 0 |
| 0 | 0 | 0 | 0 | 0 |
| 0 | 0 | 0 | 0 | 0 |
| 0 | 0.04657885 | 0 | 0 | 0 |
| 0 | 0.392715916 | 0.004943143 | 0 | 0 |
| 0 | 0 | 0 | 0 | 0 |
| 0 | 0 | 0 | 0 | 0 |
| 0 | 0.010499874 | 0.001650455 | 0 | 0.012469621 |
| 0 | 0 | 0 | 0 | 0 |
| 0 | 0 | 0 | 0 | 0 |
| 0.09855791 | 0 | 0 | 0 | 0 |
| 0 | 0 | 0 | 0 | 0 |
| 0 | 0 | 0 | 0 | 0 |
| 0 | 0 | 0 | 0 | 0 |
| 0 | 0.010982163 | 0 | 0 | 0 |
| 0 | 0.555874298 | 0 | 0 | 0.005543128 |
| 0 | 0 | 0 | 0 | 0 |
| 0 | 0 | 0 | 0 | 0 |
| 0 | 0 | 0 | 0 | 0 |
| 0 | 0.075323885 | 0 | 0 | 0 |
| 0 | 0 | 0.022020948 | 0 | 0 |
| 0 | 0 | 0 | 0 | 0 |
| 0 | 0 | 0 | 0 | 0 |
| 0 | 0 | 0 | 0 | 0 |
| 0 | 0 | 0 | 0 | 0 |
| 0 | 0 | 0 | 0 | 0 |
| 0.688181777 | 0.009611339 | 0.009750968 | 0 | 0 |
| 0 | 0 | 0 | 0 | 0 |
| 0 | 0 | 0 | 0 | 0 |
| 0 | 0 | 0 | 0 | 0 |
| 0 | 0 | 0 | 0 | 0 |
| 0 | 0.000321115 | 0 | 0.0011367 | 0 |
| 0 | 0 | 0 | 0 | 0 |
| 0 | 0.049838315 | 0 | 0 | 0 |
| 0.000771371 | 0.002962776 | 0.012748897 | 0 | 0 |
| 0 | 0 | 0 | 0 | 0 |
| 0 | 0 | 0 | 0 | 0 |
| 0.015922205 | 0 | 0 | 0 | 0 |
| 0 | 0 | 0 | 0 | 0 |
| 0 | 0 | 0 | 0 | 0 |
| 8.843409039 | 0.539586803 | 0.029510137 | 0 | 0 |
| 0 | 0 | 0 | 0.356347332 | 0 |
| 0 | 0 | 0 | 0 | 0 |
| 0 | 0 | 0 | 0 | 0 |
| 0 | 0 | 0 | 0 | 0 |

| | | | | |
|---|---|---|---|---|
| 0 | 0 | 0 | 0 | 0 |
| 0 | 0 | 0 | 0 | 0 |
| 0 | 0 | 0 | 0 | 0 |
| 0.03682981 | 0 | 0 | 0 | 0 |
| 0 | 0 | 0 | 0 | 0 |
| 0 | 0 | 0 | 0 | 0 |
| 0 | 0 | 0 | 0 | 0 |
| 0 | 0 | 0 | 0 | 0 |
| 0.021276979 | 0.002986042 | 0.296499982 | 0.006397723 | 1.262430204 |
| 0 | 0 | 0 | 0 | 0 |
| 0 | 0.002809359 | 0 | 0 | 0 |
| 0 | 0 | 0 | 0 | 0 |
| 0 | 0 | 0 | 0 | 0 |
| 0 | 0 | 0 | 0 | 0 |
| 0.037460987 | 3.628629594 | 0 | 4.340108557 | 0.002654504 |
| 0 | 0.001695787 | 0 | 0.288993797 | 0.223737475 |
| 0 | 0 | 0 | 0 | 0 |
| 0 | 0 | 0 | 0 | 0 |
| 0 | 0 | 0 | 0 | 0 |
| 0 | 0 | 0 | 0 | 0 |
| 0 | 0.05311577 | 0 | 0 | 0 |
| 0 | 0 | 0 | 0 | 0 |
| 1.115812237 | 0 | 0 | 0 | 0 |
| 0 | 0.04771403 | 0 | 0 | 0 |
| 0 | 1.09743616 | 1.149411313 | 0.006869319 | 0.086366387 |
| 0.154471597 | 2.069281228 | 0.026163152 | 0 | 0 |
| 0.177751767 | 1.785878893 | 0.000632104 | 0 | 0 |
| 0 | 0 | 0 | 0 | 0 |
| 0 | 0 | 0 | 0.018303236 | 0 |
| 0 | 0.578239395 | 0 | 0 | 0 |
| 0 | 0 | 0 | 0 | 0 |
| 0 | 0 | 0 | 0 | 0 |
| 0 | 0 | 0 | 0 | 0 |
| 0 | 0.055668773 | 0 | 0.002969866 | 0.03602678 |
| 0 | 0 | 0 | 0.004605553 | 0 |
| 0 | 0.005288217 | 0.016033088 | 0.001424311 | 0.075656695 |
| 0.811910864 | 0 | 0 | 0.026667269 | 0.008037196 |
| 6.690559526 | 1.387031703 | 0 | 0 | 0 |
| 0 | 0.016261368 | 12.58846065 | 0 | 0 |
| 0 | 0 | 0 | 0 | 0 |
| 0 | 0 | 0 | 0 | 0 |
| 0 | 0.021901651 | 0 | 0 | 0 |
| 2.89675351 | 0.508835826 | 0.055616425 | 0.131239348 | 0 |
| 0 | 0 | 0 | 0 | 0 |
| 0 | 0 | 0 | 0 | 0 |
| 0.001105865 | 0.001089113 | 0 | 0 | 0.000602313 |
| 0 | 0.004774355 | 0 | 0 | 0 |
| 0.006459843 | 0.670091081 | 0.257407548 | 0.000921294 | 0 |

| | | | | |
|---|---|---|---|---|
| 0 | 0.102991034 | 0 | 0.003206046 | 0 |
| 0 | 0 | 0 | 0 | 0 |
| 0 | 0.003624672 | 0 | 0 | 0 |
| 0 | 0 | 0 | 0 | 0 |
| 0.011117984 | 0.012495392 | 0 | 0 | 0 |
| 0 | 0 | 0 | 0 | 0 |
| 0 | 0 | 0 | 0 | 0 |
| 0 | 0 | 0 | 0 | 0 |
| 0 | 0 | 0 | 0 | 0 |
| 0 | 0 | 0 | 0 | 0 |
| 0 | 0 | 0 | 0 | 0 |
| 0.157878117 | 8.242374511 | 0.151173968 | 1.060133203 | 0 |
| 0.746477172 | 0.088438725 | 0 | 0 | 0 |
| 0 | 0 | 0 | 0 | 0 |
| 0.336323581 | 0.009385649 | 0.014155976 | 1.072654974 | 0.013388321 |
| 0 | 0 | 0 | 0 | 0 |
| 0 | 0 | 0 | 0 | 0 |
| 0 | 0 | 0 | 0 | 0 |
| 0 | 0 | 0 | 0.000693347 | 0.033218554 |
| 0 | 0.001882289 | 0 | 0 | 0 |
| 0 | 0 | 0 | 0 | 0 |
| 0 | 0 | 0 | 0 | 0 |
| 0.071982242 | 1.135624206 | 0.001167558 | 0.103599846 | 0 |
| 0 | 0 | 0 | 0 | 0 |
| 0 | 0 | 0 | 0 | 0 |
| 0 | 0 | 0 | 0 | 0 |
| 0 | 0 | 0 | 0 | 0 |
| 0 | 0 | 0 | 0 | 0 |
| 0 | 0 | 0 | 0 | 0 |
| 0 | 0 | 0 | 0 | 0 |
| 0 | 0 | 0 | 0 | 0 |
| 0.014067733 | 0.009279385 | 0.020739598 | 0.023951423 | 0.002138242 |
| 0 | 0 | 0 | 0 | 0 |
| 0.011114042 | 0.291960124 | 0 | 0.003418777 | 0.039658001 |
| 0 | 0.003002559 | 0.017256281 | 0 | 0.067112134 |
| 0 | 0.005483439 | 0.01285292 | 0 | 0 |
| 0 | 0 | 0 | 0 | 0 |
| 0 | 0 | 0 | 0 | 0 |
| 0 | 0 | 0 | 0 | 0 |
| 0 | 0 | 0 | 0 | 0 |
| 0 | 0.089003745 | 0 | 0 | 0 |
| 0 | 0 | 0 | 0 | 0 |
| 0 | 0 | 0 | 0 | 0 |
| 0 | 0 | 0 | 0 | 0 |
| 0.010428179 | 0.182645497 | 0 | 0.002399434 | 0.040382954 |
| 0 | 0 | 0 | 0 | 0 |
| 0 | 0.307243902 | 0 | 0 | 0 |
| 0.000591791 | 0 | 0 | 0 | 0.010958917 |

| | | | | |
|---|---|---|---|---|
| 0 | 0 | 0 | 0 | 0 |
| 0 | 0 | 0 | 0 | 0 |
| 0.004314139 | 0.005480938 | 0.015041858 | 0.003835223 | 0.003524564 |
| 0 | 0 | 0 | 0 | 0 |
| 0 | 0 | 0 | 0 | 0 |
| 0 | 0.746536142 | 0 | 0 | 0 |
| 0 | 0 | 0 | 0 | 0 |
| 0.003055726 | 0.002046418 | 0 | 0 | 0 |
| 0 | 0 | 0 | 0 | 0 |
| 0 | 0.004264419 | 0 | 0 | 0 |
| 0 | 0 | 0 | 0 | 0 |
| 0.002633719 | 0 | 0 | 0 | 0.00771024 |
| 0 | 0 | 0 | 0 | 0 |
| 0 | 0 | 0 | 0 | 0 |
| 0 | 0 | 0 | 0 | 0 |
| 0 | 0 | 0 | 0 | 0 |
| 0 | 0 | 0 | 0 | 0 |
| 0 | 0 | 0 | 0 | 0 |
| 0.100724102 | 0.45177243 | 6.187787765 | 0 | 0 |
| 0 | 0 | 0 | 0 | 0 |
| 0.000979945 | 0.003126928 | 0 | 0 | 0 |
| 0 | 0 | 0 | 0 | 0 |
| 0 | 0 | 0 | 0 | 0 |
| 0 | 0 | 0 | 0 | 0 |
| 0 | 0 | 0 | 0 | 0 |
| 0 | 0 | 0.034992119 | 0.001218427 | 0.007614178 |
| 0 | 0 | 0 | 0 | 0 |
| 0.019955117 | 0.379355956 | 0.013252638 | 0 | 0 |
| 0 | 0 | 0 | 0 | 0 |
| 1.451283224 | 0.613004824 | 0 | 0 | 0 |
| 0 | 0 | 0 | 0 | 0 |
| 0 | 0 | 0 | 0 | 0 |
| 0 | 0 | 0 | 0 | 0 |
| 0 | 0 | 0 | 0 | 0 |
| 0.000610418 | 0.002043983 | 0 | 0.001915254 | 0.003657128 |
| 0 | 0 | 0 | 0 | 0 |
| 0 | 0 | 0 | 0 | 0 |
| 0 | 0.010778028 | 0 | 0.004105031 | 0.002640762 |
| 0 | 0 | 0 | 0 | 0 |
| 0 | 0 | 0 | 0 | 0 |
| 16.24675228 | 2.47963803 | 1.164152999 | 24.82047631 | 0.421898307 |
| 0 | 0.558261672 | 0.070558816 | 0 | 0 |
| 0 | 0 | 0 | 0 | 0 |
| 0 | 0 | 0 | 0 | 0 |
| 0 | 0 | 0 | 0 | 0 |
| 0 | 0.009403871 | 0.016882892 | 0 | 0 |
| 0 | 0.004644608 | 0 | 0 | 0 |
| 0 | 0.010286503 | 0 | 0 | 0.00419171 |

| | | | | |
|---|---|---|---|---|
| 0 | 0 | 0 | 0 | 0 |
| 0.000753913 | 0.030850563 | 0 | 0 | 0.006775245 |
| 0 | 0 | 0 | 0 | 0 |
| 0 | 0 | 0 | 0 | 0 |
| 7.485393507 | 3.333669631 | 0.036347328 | 0.126466857 | 0 |
| 0 | 0 | 0 | 0 | 0 |
| 0 | 0 | 0 | 0 | 0 |
| 0 | 0 | 0 | 0 | 0 |
| 0.923686988 | 0.002557822 | 0.19668567 | 0.667657502 | 0.968661601 |
| 0 | 0 | 0 | 0 | 0 |
| 0 | 0 | 0.063638513 | 0 | 0 |
| 0 | 0 | 0 | 0 | 0 |
| 0.283498643 | 0.107894044 | 0.033668417 | 0 | 0 |
| 0.026567742 | 0.035788805 | 0.002242911 | 0.235713431 | 0 |
| 0 | 0 | 0 | 0 | 0 |
| 0.143001726 | 0.25459575 | 0.079978794 | 0 | 0 |
| 0.002532825 | 0.06988321 | 0 | 0.012429948 | 0 |
| 0 | 0 | 0 | 0 | 0 |
| 0 | 0 | 0 | 0 | 0 |
| 0 | 0 | 0 | 0 | 0 |
| 0 | 0 | 0 | 0 | 0 |
| 0 | 0 | 0 | 0 | 0 |
| 0 | 0 | 0 | 0 | 0 |
| 0 | 0 | 0 | 0 | 0 |
| 0 | 0.002576479 | 0 | 0 | 0 |
| 0 | 0 | 0 | 0 | 0 |
| 0.014061449 | 0.037040428 | 0.003682221 | 0.00088593 | 0 |
| 0 | 0 | 0.00086562 | 0 | 0.0095269 |
| 0 | 0 | 0 | 0 | 0 |
| 0 | 0 | 0 | 0 | 0 |
| 0 | 0 | 0 | 0 | 0 |
| 0 | 0 | 0 | 0 | 0 |
| 0 | 0.098385195 | 0 | 0 | 0 |
| 0 | 0.014446562 | 0 | 0 | 0 |
| 0.01493172 | 0 | 0 | 0 | 0 |
| 0 | 0 | 0.001201611 | 0 | 0.02258329 |
| 0 | 0.005209007 | 0.000818794 | 0 | 0 |
| 0 | 0.023321282 | 0 | 0 | 0 |
| 0 | 0 | 0 | 0 | 0 |
| 0 | 0 | 0 | 0 | 0 |
| 1.646699611 | 0.028967543 | 0.100318435 | 0 | 0 |
| 0 | 0 | 0 | 0 | 0 |
| 0 | 0 | 0 | 0 | 0 |
| 0 | 0 | 0 | 0 | 0 |
| 0 | 0 | 0 | 0 | 0 |
| 0 | 0 | 0 | 0 | 0 |
| 0 | 0 | 0 | 0 | 0 |
| 0 | 0 | 0 | 0 | 0 |

| | | | | |
|---|---|---|---|---|
| 0 | 0.168703515 | 0 | 0 | 0 |
| 0 | 0 | 0 | 0 | 0 |
| 0 | 0 | 0 | 0 | 0 |
| 0 | 0 | 0 | 0.00067946 | 0 |
| 0 | 0.007346186 | 0 | 0 | 0 |
| 0 | 0 | 0 | 0 | 0 |
| 0.006025128 | 0.4812169 | 0.004048359 | 0.043500671 | 0 |
| 0 | 0.000555083 | 0 | 0 | 0 |
| 0 | 0 | 0 | 0 | 0 |
| 0.017740533 | 0 | 0 | 0 | 0 |
| 0 | 0.024372792 | 0 | 0 | 0 |
| 0 | 0.16461712 | 0 | 0 | 0 |
| 0 | 0 | 0 | 0 | 0 |
| 0 | 0 | 0 | 0 | 0 |
| 0 | 0 | 0 | 0 | 0 |
| 0 | 0 | 0 | 0 | 0 |
| 0.011328289 | 0 | 0.246822849 | 0 | 0 |
| 0 | 0 | 0 | 0 | 0 |
| 0 | 0 | 0 | 0 | 0 |
| 0 | 0 | 0 | 0 | 0 |
| 0 | 0.331089259 | 0.002681227 | 0 | 0 |
| 0 | 0 | 0 | 0 | 0 |
| 0 | 0 | 0 | 0 | 0 |
| 0.002722305 | 0.009222875 | 0 | 0.001328683 | 0.004744679 |
| 0 | 0 | 0 | 0 | 0 |
| 0 | 0 | 0 | 0.000671806 | 0.010670528 |
| 0 | 0 | 0 | 0 | 0 |
| 0 | 0 | 0 | 0 | 0 |
| 0 | 0 | 0 | 0 | 0 |
| 0 | 0.526787781 | 1.830427292 | 0.10790755 | 0.030088982 |
| 0 | 0 | 0 | 0 | 0 |
| 0 | 0 | 0 | 0 | 0 |
| 0 | 0 | 0 | 0 | 0 |
| 0.029654032 | 4.513235388 | 0 | 0 | 0 |
| 0 | 0 | 0 | 0 | 0 |
| 0 | 0 | 0 | 0 | 0 |
| 0.033154741 | 0.008185494 | 0.063958727 | 0.016387756 | 0.618913692 |
| 0 | 0 | 0 | 0 | 0 |
| 0 | 0 | 0 | 0 | 0 |
| 0 | 0 | 0 | 0 | 0 |
| 0 | 0 | 0 | 0 | 0 |
| 0 | 0 | 0 | 0 | 0 |
| 0 | 0 | 0 | 0 | 0 |
| 0 | 0 | 0 | 0 | 0 |
| 0 | 0 | 0 | 0 | 0 |
| 0 | 0 | 0 | 0 | 0 |
| 0 | 0 | 0 | 0 | 0 |
| 0 | 0 | 0 | 0 | 0 |

| | | | | |
|---|---|---|---|---|
| 0 | 0.002769528 | 0.00601309 | 0.004493373 | 0.025208124 |
| 0 | 0 | 0 | 0 | 0 |
| 0 | 0.037987182 | 0 | 0 | 0 |
| 0 | 0 | 0 | 0 | 0 |
| 0 | 0 | 0 | 0 | 0 |
| 0 | 0 | 0 | 0 | 0 |
| 0 | 0 | 0 | 0 | 0 |
| 0.348896233 | 0.737342248 | 0.246418618 | 0.104436905 | 0 |
| 0.007969818 | 1.202179047 | 1.458517879 | 0.01122072 | 0.084895762 |
| 0.016645044 | 0.001008794 | 0 | 0.003347798 | 0.111230092 |
| 0 | 0 | 0 | 0 | 0 |
| 0 | 0 | 0 | 0 | 0 |
| 0 | 0 | 0 | 0 | 0 |
| 0 | 0 | 0.006987449 | 0 | 0 |
| 0 | 0.00628378 | 0 | 0 | 0 |
| 0 | 0 | 0 | 0 | 0 |
| 0.039339068 | 0.009096221 | 0 | 0.147878239 | 0 |
| 0 | 0 | 0 | 0 | 0 |
| 0 | 0 | 0 | 0 | 0 |
| 0 | 0 | 0 | 0 | 0.009830654 |
| 0 | 0 | 0 | 0 | 0 |
| 0 | 0 | 0 | 0 | 0 |
| 0 | 0 | 0 | 0 | 0 |
| 0 | 0 | 0 | 0 | 0 |
| 0 | 0.022450107 | 0 | 0 | 0 |
| 0 | 0 | 0 | 0 | 0 |
| 0 | 0 | 0 | 0 | 0 |
| 0 | 0 | 0 | 0 | 0 |
| 0 | 0 | 0 | 0 | 0 |
| 0 | 0 | 0 | 0 | 0 |
| 0 | 0 | 0 | 0 | 0 |
| 0 | 0 | 0 | 0 | 0 |
| 0 | 0 | 0 | 0 | 0 |
| 0 | 0 | 0 | 0 | 0 |
| 0 | 0 | 0 | 0 | 0 |
| 0 | 0.000858907 | 0 | 0 | 0 |
| 0 | 0 | 0 | 0 | 0 |
| 0 | 0.011472409 | 0 | 0 | 0 |
| 0.000696633 | 0 | 0 | 0 | 0 |
| 0 | 0 | 0.012005895 | 0 | 0 |
| 0 | 0 | 0 | 0 | 0 |
| 0 | 0.885920181 | 0 | 0 | 0 |
| 0 | 0 | 0 | 0 | 0 |
| 0 | 0 | 0 | 0 | 0 |
| 0 | 0 | 0 | 0 | 0 |
| 0 | 0 | 0 | 0 | 0 |
| 0 | 0 | 0 | 0 | 0 |
| 0 | 0.000429847 | 0 | 0 | 0 |

| | | | | |
|---|---|---|---|---|
| 0 | 0 | 0 | 0 | 0 |
| 0 | 0.002916223 | 0.381651211 | 0 | 0 |
| 0 | 0 | 0 | 0 | 0 |
| 0 | 0 | 0 | 0 | 0 |
| 0.011582149 | 0.194399358 | 0.263788215 | 0.068556799 | 0 |
| 0 | 0.005589315 | 0 | 0 | 0 |
| 0 | 0 | 0 | 0 | 0 |
| 0.005962045 | 0.008660806 | 0.04746933 | 0.008573844 | 0.094779007 |
| 0 | 0.035762516 | 0 | 0 | 0 |
| 0 | 0 | 0 | 0 | 0 |
| 0 | 0 | 0 | 0 | 0 |
| 0 | 0 | 0 | 0 | 0 |
| 0 | 0 | 0 | 0 | 0 |
| 0 | 0 | 0 | 0 | 0 |
| 0 | 0 | 0 | 0 | 0 |
| 0.004093579 | 0.864835513 | 0 | 0.002478677 | 0 |
| 0 | 0 | 0 | 0 | 0 |
| 0 | 0 | 0 | 0 | 0 |
| 0 | 0 | 0 | 0 | 0 |
| 0 | 0 | 0 | 0 | 0 |
| 0 | 0 | 0 | 0 | 0 |
| 0 | 0 | 0 | 0 | 0 |
| 0 | 0 | 0 | 0 | 0 |
| 0 | 0.001397413 | 0 | 0 | 0.004917893 |
| 0 | 0.14388438 | 0.635734329 | 0 | 0 |
| 0 | 0 | 0 | 0 | 0 |
| 0 | 0.227500488 | 0.031340554 | 0.005963799 | 0 |
| 0 | 0 | 0 | 0 | 0 |
| 0.004968498 | 0 | 0 | 0 | 0.008357099 |
| 0 | 0 | 0 | 0 | 0 |
| 0 | 0 | 0 | 0 | 0 |
| 0 | 0 | 0 | 0 | 0 |
| 0 | 0 | 0 | 0 | 0 |
| 0 | 0 | 0 | 0 | 0 |
| 0 | 0 | 0 | 0 | 0 |
| 0 | 0 | 0 | 0 | 0 |
| 0 | 0 | 0 | 0 | 0 |
| 0 | 0.374996204 | 0 | 0 | 0 |
| 0 | 1.512764038 | 0 | 0 | 0 |
| 0 | 0.001635638 | 0 | 0 | 0 |
| 0 | 0 | 0 | 0 | 0 |
| 0 | 0 | 0 | 0 | 0 |
| 0 | 0 | 0 | 0 | 0 |
| 0 | 0 | 0 | 0 | 0 |
| 0 | 0 | 0 | 0 | 0 |
| 0 | 0 | 0 | 0 | 0 |
| 0 | 0 | 0 | 0 | 0 |
| 0 | 0 | 0 | 0 | 0 |

| | | | | |
|---|---|---|---|---|
| 0 | 0.340584947 | 0 | 0 | 0 |
| 0 | 0 | 0 | 0 | 0 |
| 0.012455296 | 0 | 0 | 0 | 0 |
| 0 | 0.001956862 | 0 | 0 | 0 |
| 0 | 0 | 0 | 0 | 0 |
| 0.020137796 | 0.625238056 | 0 | 0 | 0.472562352 |
| 0 | 0 | 0 | 0 | 0 |
| 0 | 0 | 0 | 0 | 0 |
| 0 | 0 | 0 | 0 | 0 |
| 0 | 0 | 0 | 0 | 0 |
| 0 | 0 | 0 | 0 | 0 |
| 0 | 0 | 0 | 0 | 0 |
| 0 | 0 | 0 | 0 | 0 |
| 0 | 0 | 0 | 0 | 0 |
| 0 | 0 | 0 | 0 | 0 |
| 0 | 0 | 0 | 0 | 0 |
| 0 | 0 | 0 | 0 | 0 |
| 0 | 0 | 0 | 0.05343237 | 0 |
| 0 | 0 | 0 | 0 | 0 |
| 0 | 0 | 0 | 0 | 0 |
| 0.00363948 | 0 | 0 | 0.032690454 | 0.006121668 |
| 0 | 0 | 0 | 0 | 0 |
| 0 | 0.090269183 | 0.004407455 | 0.001696671 | 0.001546244 |
| 0 | 0.058052685 | 0 | 0 | 0 |
| 0 | 0 | 0 | 0 | 0 |
| 0 | 0 | 0 | 0 | 0 |
| 0 | 0 | 0 | 0 | 0 |
| 0 | 0 | 0 | 0 | 0 |
| 0 | 0 | 0.384301382 | 0 | 0 |
| 0 | 0 | 0 | 0 | 0 |
| 0 | 0 | 0 | 0 | 0 |
| 0 | 0.001285176 | 0 | 0 | 0 |
| 0.004557715 | 0.12523404 | 0.053887638 | 1.007113905 | 0.002896103 |
| 0 | 0 | 0 | 0 | 0 |
| 0.596579682 | 0.12446806 | 0 | 0 | 0 |
| 0 | 0.001200658 | 0 | 0 | 0.00083 |
| 0 | 0 | 0 | 0 | 0 |
| 0 | 0 | 0 | 0 | 0 |
| 0.015276202 | 0.003471877 | 0.015111481 | 0.00393278 | 0.006528182 |
| 0 | 0 | 0 | 0 | 0 |
| 0 | 0 | 0 | 0 | 0 |
| 0 | 0 | 0 | 0 | 0 |
| 0.006045755 | 0.021272642 | 0.004230809 | 0.007472722 | 0 |
| 0 | 0 | 0 | 0 | 0 |
| 0 | 0 | 0 | 0 | 0 |
| 0.010912223 | 0.187254437 | 0 | 0 | 0 |
| 0 | 0 | 0 | 0 | 0 |
| 0 | 0 | 0 | 0 | 0 |

| | | | | |
|---|---|---|---|---|
| 0 | 0 | 0 | 0 | 0 |
| 0 | 0 | 0 | 0 | 0 |
| 0 | 0 | 0 | 0 | 0 |
| 0 | 0 | 0 | 0 | 0 |
| 0 | 0 | 0 | 0 | 0 |
| 0 | 0 | 0 | 0 | 0 |
| 0 | 0 | 0 | 0 | 0 |
| 0 | 0 | 0 | 0.301735534 | 0 |
| 0 | 0 | 0 | 0 | 0 |
| 0 | 0.003984917 | 0.063922253 | 0 | 0 |
| 0 | 0 | 0 | 0 | 0 |
| 0 | 0 | 0 | 0 | 0.011729904 |
| 0 | 0 | 0 | 0 | 0 |
| 0 | 0.270878619 | 0 | 0 | 0 |
| 0 | 0 | 0 | 0 | 0.031463541 |
| 0.008846716 | 0 | 0.022232674 | 0.003469692 | 0.946633249 |
| 0 | 0 | 0 | 0 | 0 |
| 0 | 0 | 0 | 0 | 0 |
| 0 | 0 | 0 | 0 | 0 |
| 0.19109416 | 0.149684248 | 0 | 0 | 0 |
| 0.002540936 | 0 | 0 | 0 | 0 |
| 0 | 0 | 0 | 0 | 0 |
| 0 | 0 | 0 | 0 | 0 |
| 0 | 0 | 0 | 0 | 0 |
| 0 | 0 | 0 | 0 | 0 |
| 0 | 0 | 0 | 0 | 0 |
| 0 | 0 | 0 | 0 | 0 |
| 0 | 0 | 0 | 0 | 0 |
| 0 | 0 | 0 | 0 | 0 |
| 0.037821886 | 0.402830634 | 0 | 0 | 0 |
| 0.039906511 | 0.218497947 | 0 | 0 | 0 |
| 0 | 0 | 0 | 0 | 0 |
| 0 | 0 | 0 | 0 | 0 |
| 0 | 0 | 0 | 0 | 0 |
| 0 | 0 | 0 | 0 | 0 |
| 0 | 0 | 0 | 0 | 0 |
| 0 | 0 | 0 | 0 | 0 |
| 0 | 0 | 0 | 0 | 0 |
| 0.001012977 | 0 | 0.023668249 | 0 | 0 |
| 0 | 0 | 0 | 0 | 0 |
| 0 | 0 | 0 | 0 | 0 |
| 0 | 2.132677433 | 0 | 0 | 0 |
| 0 | 0 | 0 | 0 | 0 |
| 0 | 0.007259005 | 0 | 0 | 0 |
| 0 | 0 | 0 | 0 | 0 |
| 0 | 0 | 0 | 0 | 0 |
| 0 | 0 | 0.17264358 | 0.032203882 | 0.007398828 |
| 0 | 0 | 0 | 0 | 0 |

| | | | | |
|---|---|---|---|---|
| 0 | 0 | 0 | 0 | 0 |
| 0 | 0 | 0 | 0 | 0 |
| 0 | 0 | 0 | 0 | 0 |
| 0 | 0 | 0 | 0 | 0 |
| 0.005412597 | 0.119538882 | 0.000765901 | 0.118170699 | 0.005112917 |
| 0 | 0 | 0 | 0 | 0 |
| 0 | 0 | 0 | 0 | 0 |
| 0 | 0 | 0 | 0 | 0 |
| 0 | 0 | 0 | 0 | 0 |
| 0 | 0 | 0 | 0 | 0 |
| 0 | 0.038323911 | 0 | 0 | 0 |
| 0.003287639 | 0.040796763 | 0.001488678 | 0.001964824 | 0.016883016 |
| 0 | 0.011493005 | 0 | 0 | 0.004138008 |
| 0 | 0 | 0 | 0 | 0 |
| 0 | 0 | 0 | 0 | 0 |
| 0 | 0.248622498 | 0 | 0.128919038 | 0 |
| 0 | 0.042690548 | 0 | 0 | 0 |
| 0 | 0 | 0 | 0 | 0 |
| 0.055834114 | 0 | 0.032850863 | 0.082943519 | 0.139751448 |
| 0.002786077 | 0.043764784 | 0.026913392 | 0.09494371 | 0 |
| 0 | 0 | 0 | 0 | 0 |
| 0 | 0 | 0 | 0 | 0 |
| 0 | 0 | 0 | 0 | 0 |
| 0 | 0 | 0 | 0 | 0 |
| 0 | 0 | 0 | 0 | 0 |
| 0 | 0 | 0 | 0 | 0 |
| 0.003534967 | 0.001319275 | 0 | 0.000583755 | 0.023255972 |
| 0 | 0 | 0 | 0 | 0 |
| 0 | 0 | 0 | 0 | 0 |
| 0 | 0 | 0 | 0 | 0 |
| 0 | 0.003115557 | 0.000954971 | 0 | 0.002067597 |
| 0 | 0.007268756 | 0 | 0 | 0 |
| 0 | 0 | 0 | 0 | 0 |
| 0 | 0 | 0 | 0 | 0 |
| 0 | 0 | 0 | 0 | 0 |
| 0 | 0 | 0 | 0 | 0 |
| 0 | 0 | 0 | 0 | 0 |
| 0.006371359 | 0.23167319 | 0.006709348 | 0.026499482 | 0.036679005 |
| 0 | 1.904250342 | 0 | 0 | 0 |
| 0.003055492 | 0 | 0 | 0 | 0 |
| 0.002662606 | 0.022988597 | 0 | 0 | 0.030212434 |
| 0 | 0 | 0 | 0 | 0 |
| 0 | 0 | 0 | 0 | 0 |
| 0 | 0 | 0 | 0.007123722 | 0.00179506 |
| 0 | 0 | 0 | 0 | 0 |
| 0 | 0 | 0 | 0 | 0 |
| 0 | 0 | 0 | 0 | 0 |
| 0 | 0 | 0 | 0 | 0 |

| | | | | |
|---|---|---|---|---|
| 0 | 0 | 0 | 0 | 0 |
| 0 | 0.050935398 | 0 | 0 | 0 |
| 0 | 0 | 0 | 0 | 0 |
| 0 | 0.004082013 | 0.01897662 | 0 | 0.006772431 |
| 0 | 0 | 0.001160473 | 0.000893459 | 0 |
| 0 | 0.001628733 | 0 | 0 | 0 |
| 0 | 0.010167294 | 0 | 0 | 0 |
| 0 | 0 | 0 | 0.00376698 | 0.015154239 |
| 0 | 0 | 0 | 0 | 0 |
| 0 | 0 | 0 | 0 | 0 |
| 0 | 0 | 0 | 0 | 0 |
| 0 | 0 | 0 | 0 | 0 |
| 0.012241992 | 0.109473492 | 0 | 0.005633553 | 0 |
| 0 | 0 | 0 | 0 | 0 |
| 0 | 0 | 0 | 0 | 0 |
| 0 | 0 | 0 | 0 | 0 |
| 0 | 0 | 0 | 0 | 0 |
| 0 | 0 | 0 | 0 | 0 |
| 0 | 0 | 0 | 0 | 0 |
| 0 | 0 | 0 | 0 | 0 |
| 0 | 0 | 0 | 0 | 0 |
| 0 | 0 | 0 | 0 | 0 |
| 0 | 0 | 0 | 0.001798325 | 0.10816645 |
| 0.061884957 | 0.034470326 | 0 | 0 | 0 |
| 0 | 0 | 0 | 0 | 0 |
| 0 | 0.004205884 | 0 | 0 | 0 |
| 0 | 0.097559662 | 0 | 0 | 0 |
| 0 | 0 | 0 | 0 | 0 |
| 0 | 0 | 0 | 0 | 0 |
| 0 | 0 | 0 | 0 | 0 |
| 0.022047613 | 0.043899313 | 0 | 0 | 0 |
| 0 | 0 | 0 | 0 | 0 |
| 0 | 0 | 0 | 0 | 0 |
| 0 | 0 | 0 | 0 | 0 |
| 0 | 0.297401653 | 0 | 0 | 0 |
| 0 | 0 | 0 | 0 | 0.011666245 |
| 0 | 0 | 0 | 0 | 0.026417259 |
| 0 | 0 | 0 | 0.054353091 | 0 |
| 0 | 0 | 0 | 0.004877364 | 0 |
| 0 | 0 | 0 | 0 | 0 |
| 0 | 0 | 0 | 0 | 0 |
| 0 | 0 | 0 | 0 | 0 |
| 0 | 0 | 0 | 0 | 0 |
| 0 | 0 | 0 | 0 | 0 |
| 0 | 0 | 0 | 0 | 0 |
| 0.166977014 | 0.344813032 | 0.056545171 | 0 | 0.001603189 |
| 0 | 0 | 0 | 0 | 0.026539875 |
| 0 | 0 | 0 | 0 | 0 |

| | | | | |
|---|---|---|---|---|
| 0 | 0 | 0 | 0 | 0 |
| 0 | 0 | 0 | 0 | 0 |
| 0.194326809 | 0.082392079 | 0.371858483 | 0 | 0 |
| 0 | 0 | 0 | 0 | 0 |
| 0 | 0 | 0 | 0 | 0 |
| 0 | 0.005240737 | 0 | 0 | 0 |
| 0 | 0.003897195 | 0 | 0 | 0 |
| 0 | 0 | 0 | 0 | 0 |
| 0 | 0 | 0 | 0 | 0 |
| 0 | 0 | 0 | 0 | 0 |
| 0 | 0.001944 | 0 | 0 | 0.008063183 |
| 0 | 0.001306079 | 0.002446493 | 0 | 0 |
| 0 | 0 | 0 | 0 | 0 |
| 0 | 0 | 0 | 0 | 0 |
| 0 | 0 | 0 | 0 | 0 |
| 0 | 0 | 0 | 0 | 0 |
| 0 | 0 | 0 | 0 | 0 |
| 0 | 0 | 0 | 0 | 0 |
| 0 | 0 | 0 | 0 | 0 |
| 0.000872577 | 0 | 0.001580449 | 0 | 0 |
| 0 | 0 | 0 | 0 | 0 |
| 0 | 0 | 0 | 0 | 0 |
| 0 | 0 | 0 | 0 | 0 |
| 0 | 0 | 0 | 0 | 0 |
| 0 | 0 | 0 | 0 | 0 |
| 0 | 0 | 0 | 0 | 0 |
| 0 | 0 | 0 | 0 | 0 |
| 0 | 0.017118475 | 0.002623551 | 0 | 0 |
| 0 | 0 | 0 | 0 | 0 |
| 0 | 0 | 0 | 0 | 0 |
| 0.000937042 | 0.003322254 | 0 | 0 | 0 |
| 0 | 0 | 0 | 0 | 0 |
| 0.055923288 | 0.011186677 | 0.083554928 | 0.027082213 | 0.016355306 |
| 0 | 0.001182576 | 0 | 0 | 0 |
| 0 | 0 | 0 | 0 | 0 |
| 0 | 0 | 0 | 0 | 0 |
| 0 | 0 | 0 | 0 | 0 |
| 0 | 0 | 0 | 0 | 0 |
| 0 | 0 | 0 | 0 | 0 |
| 0 | 0 | 0 | 0 | 0 |
| 0 | 0 | 0 | 0 | 0 |
| 0.078767844 | 0.011741986 | 0.05901254 | 0.009361467 | 0.162049355 |
| 0.011122804 | 0.356079743 | 0.00533279 | 0.04744441 | 0 |
| 0 | 0 | 0 | 0 | 0 |
| 0 | 0 | 0 | 0 | 0 |
| 0.001890968 | 0.004842041 | 0 | 0 | 0 |
| 0 | 0 | 0 | 0 | 0 |
| 0 | 0 | 0 | 0 | 0 |

| | | | | |
|---|---|---|---|---|
| 0 | 0 | 0 | 0 | 0 |
| 0.242191543 | 0 | 0.007655992 | 0.009558521 | 0 |
| 0.001067567 | 0.002996478 | 0 | 0.017027178 | 0 |
| 0 | 0 | 0 | 0 | 0 |
| 0 | 0.156522714 | 0.054422652 | 0.052152776 | 0 |
| 0.007674929 | 0 | 0.206826692 | 0 | 0.05151782 |
| 0 | 0 | 0 | 0 | 0 |
| 0 | 0 | 0 | 0 | 0 |
| 0 | 0 | 0 | 0 | 0 |
| 0 | 0 | 0 | 0 | 0 |
| 0 | 0 | 0 | 0 | 0 |
| 0 | 0 | 0 | 0 | 0 |
| 0 | 0 | 0 | 0 | 0 |
| 0 | 0 | 0 | 0 | 0.84946866 |
| 0 | 0 | 0 | 0 | 0 |
| 0 | 0 | 0 | 0 | 0 |
| 0 | 0.006508185 | 0 | 0 | 0 |
| 0 | 0 | 0 | 0 | 0 |
| 0 | 0 | 0 | 0 | 0 |
| 0.100600512 | 0 | 0 | 0 | 0 |
| 0 | 0 | 0 | 0 | 0 |
| 0 | 0 | 0 | 0.002694373 | 0 |
| 0 | 0 | 0 | 0 | 0 |
| 0 | 0 | 0 | 0 | 0 |
| 0 | 0.000519899 | 0 | 0 | 0 |
| 0 | 0.002745781 | 0.031661247 | 0 | 0.00235006 |
| 0 | 0 | 0 | 0 | 0 |
| 0 | 0.62389392 | 0 | 0 | 0 |
| 0 | 0 | 0 | 0 | 0 |
| 0 | 0 | 0 | 0 | 0 |
| 0 | 0 | 0 | 0 | 0 |
| 0 | 0 | 0.003514562 | 0 | 0 |
| 0 | 0 | 0 | 0 | 0 |
| 0 | 0 | 0 | 0 | 0 |
| 0 | 0 | 0 | 0 | 0 |
| 0 | 0 | 0 | 0 | 0 |
| 0 | 0 | 0 | 0 | 0 |
| 0.007488724 | 0 | 0 | 0.001382158 | 0.009896983 |
| 0 | 0.091408286 | 0 | 0 | 1.471411383 |
| 0 | 0 | 0 | 0 | 0 |
| 0 | 0 | 0 | 0 | 0 |
| 0.032646094 | 0.010780234 | 0 | 0.011046447 | 0 |
| 0 | 0.003280584 | 0.006636659 | 0.002787069 | 0.014514101 |
| 0 | 0 | 0.592870591 | 0 | 0 |
| 0 | 0 | 0 | 0 | 0 |
| 0 | 0.000905807 | 0 | 0 | 0 |
| 0 | 0.018267893 | 0 | 0.004263671 | 0.05939499 |
| 0 | 0 | 0 | 0 | 0 |

| | | | | |
|---|---|---|---|---|
| 0 | 0 | 0 | 0 | 0 |
| 0.007501686 | 0.007725791 | 0.008346529 | 0.001344992 | 0.013833408 |
| 0 | 0 | 0 | 0 | 0 |
| 0 | 0 | 0 | 0 | 0 |
| 0 | 0 | 0 | 0 | 0 |
| 0 | 0 | 0 | 0 | 0 |
| 0 | 0 | 0 | 0 | 0 |
| 0 | 0 | 0 | 0 | 0 |
| 0 | 0 | 0 | 0 | 0 |
| 0 | 0 | 0 | 0 | 0 |
| 0 | 0 | 0 | 0 | 0 |
| 0 | 0 | 0 | 0 | 0 |
| 0 | 0.922719676 | 0 | 0 | 0 |
| 0 | 0 | 0 | 0 | 0 |
| 0.003741389 | 0.011527893 | 0 | 0.000838501 | 0.009169916 |
| 0 | 0 | 0 | 0 | 0 |
| 0 | 0 | 0 | 0 | 0 |
| 0 | 0 | 0 | 0 | 0 |
| 0 | 0 | 0 | 0 | 0 |
| 0 | 0 | 0 | 0 | 0 |
| 0 | 0 | 0 | 0 | 0 |
| 0.001574717 | 0.015818804 | 0 | 0.027723635 | 0 |
| 0 | 0.007031097 | 0 | 0.000632773 | 0.008073394 |
| 0 | 0 | 0 | 0 | 0 |
| 0 | 0 | 0 | 0 | 0 |
| 0 | 0 | 0 | 0 | 0 |
| 0 | 0 | 0 | 0 | 0 |
| 0 | 0 | 0 | 0 | 0 |
| 0 | 0 | 0 | 0 | 0 |
| 0 | 0 | 0 | 0 | 0 |
| 0 | 0 | 0 | 0 | 0 |
| 0 | 0 | 0 | 0 | 0 |
| 0 | 0 | 0 | 0 | 0 |
| 0 | 0 | 0 | 0 | 0 |
| 0.940302297 | 1.895017504 | 0 | 0 | 0 |
| 0 | 0 | 0 | 0 | 0 |
| 0 | 0 | 0 | 0 | 0 |
| 0 | 0 | 0 | 0 | 0 |
| 0 | 0.006659515 | 0.149533696 | 0.007401049 | 0 |
| 0 | 0 | 0 | 0 | 0 |
| 0.126664527 | 0.090400226 | 0.005320505 | 0.089463368 | 0.013247276 |
| 0 | 0 | 0 | 0 | 0 |
| 0 | 0 | 0 | 0 | 0 |
| 0 | 0 | 0 | 0 | 0 |
| 0 | 0 | 0 | 0 | 0 |
| 0.140717335 | 0.005087661 | 1.137913974 | 0.008160589 | 0.10177431 |
| 0 | 0 | 0 | 0 | 0 |
| 0 | 0 | 0 | 0 | 0 |

| | | | | |
|---|---|---|---|---|
| 0 | 0 | 0 | 0 | 0 |
| 0 | 0 | 0 | 0 | 0 |
| 0 | 0 | 0 | 0 | 0 |
| 0 | 0 | 0 | 0 | 0 |
| 0 | 0.007837793 | 0 | 0 | 0 |
| 0 | 0 | 0 | 0 | 0 |
| 0 | 0 | 0 | 0 | 0 |
| 0 | 0 | 0 | 0 | 0 |
| 0 | 0 | 0 | 0 | 0 |
| 0.001841974 | 0.000466476 | 0 | 0.084764461 | 0 |
| 0 | 0 | 0 | 0 | 0 |
| 0 | 0 | 0 | 0 | 0 |
| 0 | 0 | 0.002329225 | 0 | 0 |
| 0 | 0 | 0 | 0 | 0 |
| 0 | 0 | 0 | 0 | 0 |
| 0 | 0 | 0 | 0 | 0 |
| 0 | 0 | 0 | 0 | 0 |
| 0 | 0 | 0 | 0 | 0 |
| 0 | 0 | 0 | 0 | 0 |
| 0.051736579 | 0.146365531 | 0.061987924 | 0.026097168 | 0.138959852 |
| 0 | 0.008617982 | 0 | 0 | 0 |
| 0 | 0 | 0 | 0 | 0 |
| 0 | 0 | 0 | 0 | 0 |
| 0 | 0 | 0 | 0 | 0 |
| 0 | 0 | 0 | 0 | 0 |
| 0 | 0 | 0 | 0 | 0 |
| 0 | 0 | 0 | 0 | 0 |
| 0 | 0 | 0 | 0 | 0 |
| 0 | 0 | 0 | 0 | 0 |
| 0.011991393 | 0 | 0 | 0.042999206 | 0.016794384 |
| 0 | 0 | 0 | 0 | 0 |
| 0 | 0 | 0 | 0 | 0 |
| 0 | 0 | 0 | 0 | 0 |
| 0 | 0 | 0 | 0 | 0 |
| 0 | 0 | 0 | 0 | 0 |
| 0 | 0 | 0 | 0 | 0 |
| 0 | 0 | 0 | 0 | 0 |
| 0 | 0 | 0 | 0 | 0 |
| 0 | 0 | 0 | 0 | 0 |
| 0 | 0 | 0 | 0 | 0 |
| 0 | 0 | 0 | 0 | 0 |
| 0 | 0 | 0 | 0 | 0 |
| 0 | 0.040255082 | 0 | 0 | 0 |
| 0 | 0 | 0 | 0 | 0 |
| 0 | 0.01054544 | 0 | 0.000674626 | 0.011944955 |
| 0 | 0.018384951 | 0 | 0 | 0 |
| 0 | 0 | 0 | 0 | 0 |
| 0 | 0 | 0.001559369 | 0 | 0.001758424 |

| | | | | |
|---|---|---|---|---|
| 0 | 0 | 0 | 0 | 0 |
| 0 | 0 | 0 | 0 | 0 |
| 0 | 0 | 0 | 0 | 0 |
| 0 | 0 | 0 | 0 | 0 |
| 0 | 0 | 0 | 0 | 0 |
| 0 | 0.024755948 | 0.746019293 | 0.006211495 | 0.010491027 |
| 0 | 0 | 0 | 0 | 0 |
| 0 | 0 | 0 | 0 | 0 |
| 0 | 0 | 0 | 0 | 0 |
| 0 | 0 | 0 | 0 | 0 |
| 0 | 0 | 0 | 0 | 0 |
| 0 | 0 | 0 | 0 | 0 |
| 0 | 0 | 0 | 0 | 0 |
| 0 | 0 | 0 | 0 | 0 |
| 0 | 0 | 0 | 0 | 0 |
| 0 | 0 | 0 | 0 | 0 |
| 0 | 0.515551774 | 0 | 0 | 0 |
| 0 | 0 | 0 | 0 | 0.001130523 |
| 0 | 0 | 0 | 0 | 0 |
| 0 | 0 | 0 | 0 | 0 |
| 0 | 0 | 0 | 0 | 0 |
| 0 | 0 | 0 | 0 | 0 |
| 0 | 0 | 0 | 0 | 0 |
| 0 | 0 | 0 | 0 | 0 |
| 0 | 0 | 0 | 0 | 0 |
| 0 | 0 | 0 | 0.000787769 | 0 |
| 0 | 0 | 0 | 0 | 0 |
| 0 | 1.385984223 | 0 | 0 | 0 |
| 0 | 0 | 0 | 0 | 0 |
| 0 | 0 | 0.013402554 | 0 | 0 |
| 0 | 0 | 0 | 0 | 0 |
| 0 | 0 | 0 | 0 | 0 |
| 0 | 0 | 0 | 0 | 0 |
| 0 | 0 | 0 | 0 | 0 |
| 0 | 0 | 0 | 0 | 0 |
| 0 | 0 | 0 | 0 | 0 |
| 0 | 0 | 0 | 0 | 0 |
| 0 | 0 | 0 | 0 | 0 |
| 0.555352267 | 0.21770483 | 0 | 0 | 0 |
| 0 | 0 | 0 | 0 | 0 |
| 0 | 0 | 0 | 0 | 0 |
| 0 | 0 | 0 | 0 | 0 |
| 0 | 0 | 0 | 0 | 0 |
| 0 | 0 | 0 | 0 | 0 |
| 0 | 0 | 0 | 0 | 0 |
| 0 | 0.295073181 | 0 | 0 | 0 |
| 0 | 0 | 0 | 0 | 0 |
| 0 | 0 | 0 | 0 | 0 |

| | | | | |
|---|---|---|---|---|
| 0 | 0 | 0 | 0 | 0 |
| 0.00089855 | 0.144846762 | 0.003580482 | 0.001691579 | 0 |
| 0 | 0 | 0 | 0 | 0 |
| 0 | 0 | 0 | 0 | 0 |
| 0 | 0 | 0 | 0 | 0 |
| 0 | 0 | 0 | 0 | 0 |
| 0 | 0 | 0 | 0 | 0 |
| 0 | 0 | 0 | 0 | 0 |
| 0 | 0 | 0 | 0 | 0 |
| 0 | 0 | 0 | 0 | 0 |
| 0 | 0.000695254 | 0.00149175 | 0 | 0 |
| 0 | 0 | 0 | 0 | 0 |
| 0 | 0.396460002 | 0 | 0.396080062 | 0 |
| 0 | 0 | 0 | 0 | 0 |
| 0 | 0 | 0 | 0 | 0 |
| 0 | 0 | 0 | 0 | 0 |
| 0 | 0 | 0 | 0 | 0 |
| 0 | 0 | 0 | 0 | 0 |
| 0 | 0 | 0 | 0 | 0 |
| 0 | 0 | 0 | 0 | 0 |
| 0 | 0 | 0 | 0 | 0.105560192 |
| 0.000633226 | 0 | 0 | 0 | 0 |
| 0 | 0.00211858 | 0 | 0 | 0 |
| 2.096990238 | 2.346862915 | 1.460637094 | 1.334033714 | 0 |
| 0 | 0 | 0 | 0 | 0 |
| 0 | 0 | 0 | 0 | 0 |
| 0 | 0 | 0 | 0 | 0 |
| 0 | 0 | 0 | 0 | 0 |
| 0 | 0 | 0 | 0.005186062 | 0 |
| 0 | 0 | 0 | 0 | 0 |
| 0 | 0 | 0 | 0 | 0 |
| 0 | 0 | 0 | 0 | 0 |
| 0 | 0 | 0 | 0 | 0 |
| 0 | 0 | 0 | 0 | 0 |

| PHL018-0015_V3 | PHL018-0102_V1 | PHL018-0135_V1 | PHL018-0163_V1 | PHL018-0266_V1 |
|---|---|---|---|---|
| CTRL | CTRL | EYCF | CTRL | REF |
| 0.246652215 | 0.017150717 | 0.350476876 | 0.034211109 | 0.006328997 |
| 7.099133032 | 0.566558628 | 10.77767308 | 0.287082738 | 6.41160769 |
| 0 | 0 | 0 | 0.098871762 | 0 |
| 0 | 0 | 0 | 0 | 0 |
| 0 | 0 | 0 | 0 | 0 |
| 0 | 0 | 0 | 0 | 0 |
| 0 | 0 | 0 | 0 | 0 |
| 0 | 0 | 0 | 0 | 0 |
| 0 | 0 | 0 | 0 | 0 |
| 0 | 0 | 0 | 0 | 0 |
| 0 | 0 | 0 | 0 | 0 |
| 0 | 0 | 0 | 0 | 0 |
| 0 | 0 | 0 | 0 | 0 |
| 0 | 14.83681382 | 1.57948404 | 2.689750379 | 10.69940282 |
| 0 | 0 | 0 | 0 | 0 |
| 0 | 0 | 0 | 0 | 0 |
| 0 | 0 | 0 | 0 | 0 |
| 0 | 0 | 0 | 0 | 0 |
| 0 | 0 | 0 | 0 | 0 |
| 0 | 0 | 0 | 0 | 0 |
| 0 | 0 | 0 | 0 | 0 |
| 0 | 0 | 0 | 0 | 0 |
| 0 | 0 | 0 | 0 | 0 |
| 0 | 0 | 0 | 0 | 0 |
| 0 | 0 | 0 | 0 | 0 |
| 0 | 0 | 0 | 0 | 0 |
| 0 | 0 | 0 | 0 | 0 |
| 0 | 0 | 0 | 0 | 0 |
| 0 | 0 | 0 | 0 | 0 |
| 0 | 0 | 0 | 0 | 0 |
| 0 | 0 | 0 | 0 | 0 |
| 0 | 0 | 0 | 0 | 0 |
| 0 | 0 | 0 | 0 | 0 |
| 0 | 0 | 0 | 0 | 0 |
| 0 | 0 | 0 | 0 | 0 |
| 0 | 1.890203976 | 0 | 0.001660638 | 0.004901939 |
| 0 | 0 | 0 | 0 | 0 |
| 0 | 0 | 0 | 0 | 0 |
| 0 | 0 | 0 | 0 | 0 |
| 0 | 0 | 0 | 0 | 0 |
| 0 | 0.001266149 | 0 | 0 | 0 |
| 0 | 0.008377183 | 0.203029544 | 0.011548427 | 0.40899382 |
| 0.471889712 | 0.002185055 | 0.053974141 | 0.258343308 | 0.053195499 |
| 0 | 0.000858531 | 0.004454536 | 0.008313607 | 0.348771774 |
| 0 | 0 | 0 | 0.049282497 | 0.001251006 |
| 0 | 0 | 0 | 0 | 0 |

| | | | | |
|---|---|---|---|---|
| 0 | 0 | 0 | 0 | 0 |
| 1.610175448 | 0.007947119 | 0 | 0 | 0 |
| 0.022635661 | 0.008943809 | 0.223802879 | 0 | 0.001383397 |
| 0 | 1.504660271 | 0.266262466 | 0.474230319 | 1.718994873 |
| 0 | 0.033397802 | 0.011568071 | 0 | 0 |
| 0 | 0 | 0 | 0 | 0 |
| 0.005384022 | 2.103953163 | 0.16702829 | 0.00859214 | 8.480895808 |
| 0 | 0 | 0 | 0 | 0 |
| 0 | 0 | 0 | 0 | 0 |
| 0 | 0 | 0 | 0 | 0 |
| 0 | 0 | 0 | 0.004385841 | 0 |
| 0 | 0.758603992 | 0 | 0.5439356 | 0 |
| 0.000932845 | 0 | 0 | 0 | 0.002071422 |
| 0.011182015 | 0.097605511 | 0.33734343 | 0.006381325 | 0 |
| 0 | 0 | 0 | 0 | 0 |
| 0 | 0 | 0 | 0 | 0 |
| 0 | 0 | 0 | 0.058424947 | 0.012618085 |
| 0 | 0 | 0 | 0 | 0 |
| 0 | 0 | 0 | 0 | 0 |
| 0 | 0 | 0 | 0 | 0 |
| 2.004229005 | 0.004476518 | 0.130313791 | 0.002649073 | 0.026690183 |
| 0.004016801 | 0.00622046 | 0.060142985 | 0.020802216 | 0.020387374 |
| 0 | 0 | 0 | 0 | 0 |
| 0 | 0 | 0 | 0 | 0 |
| 0 | 0 | 0 | 0 | 0 |
| 0 | 0 | 0 | 0 | 0 |
| 0 | 0 | 0 | 0 | 0 |
| 0 | 0 | 0 | 0 | 0 |
| 0 | 0 | 0.003523982 | 0.012148979 | 0.365953906 |
| 0 | 0 | 0 | 0 | 0 |
| 0.276773269 | 0 | 0 | 0 | 0.212963674 |
| 0 | 0 | 0 | 0 | 0 |
| 0 | 0 | 0 | 0 | 0 |
| 0 | 0 | 0 | 0 | 0 |
| 0 | 0 | 0 | 0 | 0 |
| 0 | 0 | 0.07231628 | 0 | 0.003949964 |
| 0 | 0 | 0 | 0 | 0 |
| 0 | 0 | 0 | 3.53175655 | 0 |
| 0 | 0 | 0 | 0 | 0 |
| 0 | 0 | 0 | 0 | 0 |
| 0 | 0 | 0 | 0 | 0 |
| 0 | 0 | 0 | 0 | 0 |
| 0 | 0 | 0 | 0 | 0 |
| 0 | 0 | 0 | 0 | 0 |
| 0 | 0 | 0 | 0 | 0 |
| 0 | 0 | 0 | 0 | 0 |
| 0 | 0.122218324 | 0.003689481 | 0 | 0.007876794 |
| 0 | 0 | 0 | 0 | 0 |

| | | | | |
|---|---|---|---|---|
| 0 | 0 | 0 | 0 | 0 |
| 0 | 0 | 0 | 0 | 0 |
| 0 | 0 | 0 | 0 | 0 |
| 0 | 0 | 0 | 0 | 0 |
| 0 | 0 | 0.028480786 | 0 | 0.421820096 |
| 0 | 0 | 0 | 0 | 0 |
| 0 | 0 | 0 | 0 | 0 |
| 0 | 0 | 0 | 0 | 0 |
| 0 | 0 | 0 | 0 | 0 |
| 0 | 0 | 0 | 0 | 0 |
| 0 | 0 | 0 | 0 | 0 |
| 0 | 0 | 0 | 0 | 0.000714946 |
| 0 | 0 | 0 | 0 | 0 |
| 0 | 0 | 0 | 0 | 0 |
| 0 | 0 | 0 | 0 | 0 |
| 0 | 0 | 0 | 0 | 0 |
| 0 | 0 | 0 | 0 | 0 |
| 0 | 0 | 0 | 0 | 0 |
| 0 | 0.028542948 | 0 | 0 | 0.010604474 |
| 0 | 0 | 0 | 0 | 0 |
| 0 | 0 | 0 | 0 | 0 |
| 0 | 1.178205999 | 0 | 0 | 0 |
| 0 | 0 | 0 | 0 | 0 |
| 0 | 0 | 0 | 0 | 0 |
| 0 | 0 | 0 | 0 | 0 |
| 0 | 0 | 0 | 0 | 0 |
| 0 | 0 | 0 | 0 | 0 |
| 0 | 0 | 0 | 0 | 0 |
| 0 | 0 | 0 | 0 | 0 |
| 0 | 0 | 0 | 0 | 0 |
| 0 | 0.726157616 | 0 | 0.034179158 | 0.008167918 |
| 0.012204904 | 0.016248844 | 0.01249327 | 0.000848684 | 0 |
| 0 | 0 | 0 | 0 | 0 |
| 0 | 0 | 0 | 0 | 0 |
| 0 | 0 | 0 | 0 | 0 |
| 0 | 0 | 0 | 0 | 0 |
| 0 | 0 | 0 | 0 | 0 |
| 0 | 0 | 0 | 0 | 0 |
| 0 | 0.161779966 | 0 | 0 | 0 |
| 0 | 0 | 0 | 0 | 0 |
| 0 | 0 | 0 | 0 | 0 |
| 0 | 0 | 0 | 0 | 0 |
| 0 | 0.001641066 | 0.003485576 | 0 | 0 |
| 0 | 0 | 0 | 0 | 0 |
| 0 | 0 | 0 | 0 | 0 |
| 0 | 0 | 0 | 0 | 0 |
| 0 | 0 | 0 | 0.023396576 | 0 |
| 0 | 0 | 0 | 0 | 0 |

| | | | | |
|---|---|---|---|---|
| 0 | 0 | 0 | 0 | 0 |
| 0 | 0 | 0 | 0 | 0 |
| 0 | 0 | 0 | 0 | 0 |
| 0 | 0 | 0 | 0 | 0 |
| 0 | 0.183032284 | 0.019310684 | 0.216249645 | 0 |
| 0 | 0 | 0 | 0 | 0 |
| 0 | 0 | 0 | 0 | 0 |
| 0 | 0 | 0 | 0 | 0 |
| 0 | 0 | 0 | 0 | 0 |
| 0 | 0 | 0 | 0 | 0 |
| 0 | 0 | 0 | 0 | 0 |
| 0 | 0 | 0 | 0 | 0 |
| 0 | 0 | 0 | 0 | 0 |
| 0 | 0 | 0 | 0 | 0 |
| 0 | 0 | 0 | 0 | 0 |
| 0 | 0 | 0 | 0 | 0 |
| 0 | 0 | 0 | 0 | 0 |
| 0 | 0 | 0 | 0 | 0 |
| 0 | 0 | 0 | 0 | 0 |
| 0 | 0 | 0 | 0 | 0 |
| 0 | 0 | 0 | 0 | 0 |
| 0 | 0 | 0 | 0 | 0 |
| 0 | 0 | 0.075711749 | 0 | 0 |
| 0 | 0 | 0 | 0 | 0 |
| 0 | 0 | 0 | 0 | 0 |
| 0 | 0 | 0 | 0 | 0 |
| 0 | 0 | 0 | 0 | 0 |
| 0 | 0 | 0 | 0 | 0 |
| 0 | 0 | 0 | 0 | 0 |
| 0 | 0 | 0 | 0 | 0 |
| 0 | 0 | 0 | 0 | 0 |
| 0 | 0 | 0.003230845 | 0 | 0 |
| 0 | 0 | 0 | 0 | 0 |
| 0 | 0 | 0 | 0 | 0 |
| 0 | 0 | 0 | 0 | 0 |
| 0 | 0 | 0 | 0 | 0 |
| 0 | 0 | 0 | 0 | 0 |
| 0 | 0 | 0 | 0 | 0 |
| 0 | 0 | 0 | 0 | 0 |
| 0 | 0 | 0 | 0 | 0 |
| 0 | 0 | 0 | 0 | 0 |
| 0 | 0 | 0 | 0 | 0 |
| 0 | 0 | 0 | 0 | 0 |
| 0 | 0 | 0 | 0 | 0 |
| 0 | 0 | 0 | 0 | 0 |
| 0 | 0 | 0 | 0 | 0 |
| 0 | 0 | 0 | 0 | 0 |
| 0.248305645 | 0 | 0 | 0 | 0 |

| | | | | |
|---|---|---|---|---|
| 0 | 0 | 0 | 0 | 0 |
| 0 | 0 | 0 | 0 | 0 |
| 0 | 0 | 0 | 0 | 0 |
| 0 | 0 | 0 | 0 | 0 |
| 0 | 0 | 0 | 0 | 0 |
| 0 | 0 | 0 | 0 | 0 |
| 0 | 0 | 0 | 0 | 0 |
| 0 | 0 | 0 | 0 | 0 |
| 0 | 0 | 0 | 0 | 0 |
| 0 | 0 | 0 | 0 | 0 |
| 0 | 0 | 0 | 0 | 0 |
| 0 | 0 | 0 | 0 | 0 |
| 0.001387125 | 0 | 0.001189077 | 0.001171246 | 0 |
| 0 | 0 | 0 | 0 | 0 |
| 0 | 0 | 0 | 0 | 0 |
| 0 | 0 | 0 | 0 | 0 |
| 0 | 0 | 0 | 0 | 0 |
| 0 | 0 | 0 | 0 | 0 |
| 0 | 0 | 0 | 0 | 0 |
| 0 | 0 | 0 | 0 | 0 |
| 0.017043592 | 0.036148148 | 0.042687152 | 0.008759797 | 0.099140101 |
| 0 | 0 | 0 | 0 | 0 |
| 0 | 0 | 0 | 0 | 0 |
| 0.001545482 | 0.002716775 | 0 | 0 | 0 |
| 0 | 0 | 0 | 0 | 0 |
| 0 | 0 | 0 | 0 | 0 |
| 0.003668225 | 0.001137937 | 0.003107499 | 0.000765225 | 0 |
| 0 | 0 | 0 | 0 | 0 |
| 0 | 0 | 0 | 0 | 0 |
| 0.001385145 | 0 | 0 | 0 | 0 |
| 0.38975031 | 0 | 0 | 0 | 0.15809396 |
| 0 | 0 | 0 | 0 | 0 |
| 0 | 0 | 0 | 0 | 0 |
| 0 | 0.016319849 | 0 | 0 | 0 |
| 0.082447794 | 0.084544547 | 0.015912495 | 0.002849795 | 0 |
| 0 | 0 | 0 | 0 | 0 |
| 0 | 0 | 0 | 0 | 0 |
| 0 | 0 | 0 | 0 | 0 |
| 0 | 0 | 0 | 0 | 0 |
| 0 | 0 | 0 | 0 | 0 |
| 0 | 0 | 0 | 0 | 0 |
| 0 | 0 | 0 | 0 | 0 |
| 0 | 0 | 0 | 0 | 0 |
| 0 | 0 | 0 | 0 | 0 |
| 0 | 0 | 0 | 0 | 0.003084665 |
| 0 | 0 | 0 | 0.002984647 | 0 |
| 0 | 0 | 0 | 0 | 0 |
| 0 | 0 | 0.000824704 | 0 | 0 |

| | | | | |
|---|---|---|---|---|
| 0 | 0 | 0 | 0 | 0 |
| 0 | 0 | 0 | 0 | 0 |
| 0.00251175 | 0 | 0.004144783 | 0 | 0 |
| 0.002402138 | 0 | 0 | 0 | 0 |
| 0 | 0 | 0 | 0 | 0 |
| 0 | 0 | 0 | 0 | 0 |
| 0 | 0 | 0 | 0 | 0.00217214 |
| 0 | 0 | 0 | 0 | 0.030161318 |
| 0 | 0 | 0.045097416 | 0 | 0 |
| 0 | 0 | 0 | 0 | 0 |
| 0 | 0 | 0 | 0 | 0 |
| 0 | 0 | 0 | 0 | 0 |
| 0 | 0 | 0 | 0 | 0 |
| 0 | 0 | 0 | 0 | 0 |
| 0 | 0 | 0 | 0.001600982 | 0 |
| 0 | 0 | 0 | 0 | 0 |
| 0 | 0 | 0 | 0 | 0 |
| 0 | 0 | 0 | 0 | 0 |
| 0.002271312 | 0 | 0 | 0.001652282 | 0 |
| 0 | 0 | 0 | 0 | 0 |
| 0.05283348 | 0.006342685 | 0 | 0.002132623 | 0.012304184 |
| 0 | 0 | 0 | 0 | 0 |
| 0 | 0 | 0 | 0 | 0 |
| 0 | 0 | 0 | 0 | 0 |
| 0.004608214 | 0 | 0 | 0 | 0.012506674 |
| 0 | 0 | 0 | 0 | 0.00165495 |
| 0 | 0 | 0 | 0 | 0 |
| 0 | 0 | 0 | 0 | 0 |
| 0 | 0 | 0.005570933 | 0 | 0.040417268 |
| 0 | 0 | 0.006322597 | 0 | 0 |
| 0.135685269 | 0.179555292 | 0.238116235 | 0.165100312 | 0.009257663 |
| 0 | 0 | 0 | 0 | 0 |
| 0 | 0 | 0 | 0 | 0 |
| 0 | 0 | 1.136060226 | 0 | 0.010344072 |
| 0 | 0 | 0 | 0 | 0 |
| 0 | 0 | 0 | 0 | 0 |
| 0 | 0 | 0 | 0 | 0 |
| 0 | 0 | 0 | 0 | 0 |
| 0 | 0 | 0 | 0 | 0 |
| 0 | 0 | 0 | 0 | 0 |
| 0 | 0 | 0 | 0 | 0 |
| 0.049064772 | 0.00186762 | 0 | 0.011189041 | 0.003431508 |
| 0 | 0.004558552 | 0 | 0 | 0 |
| 0 | 0 | 0 | 0 | 0 |
| 0 | 0 | 0 | 0 | 0 |
| 0 | 0.001939738 | 0 | 0 | 0 |
| 0 | 0 | 0 | 0 | 0 |
| 0 | 0 | 0 | 0 | 0 |

| | | | | |
|---|---|---|---|---|
| 0 | 0 | 0 | 0 | 0 |
| 0 | 0 | 0 | 0 | 0 |
| 0 | 0 | 0.004948588 | 0.006308021 | 0.396411758 |
| 0 | 0 | 0 | 0 | 0 |
| 0 | 0 | 0 | 0 | 0 |
| 0 | 0 | 0 | 0 | 0 |
| 0 | 0 | 0 | 0 | 0 |
| 0 | 0 | 0 | 0 | 0 |
| 0 | 0 | 0 | 0 | 0 |
| 0 | 0 | 0 | 0 | 0 |
| 0 | 0 | 0 | 0 | 0 |
| 0 | 0 | 0 | 0 | 0 |
| 0 | 0 | 0 | 0 | 0 |
| 0 | 0 | 0.010938166 | 0 | 0.019048112 |
| 0.004203826 | 0 | 0.001593181 | 0.001046193 | 0.03463692 |
| 0 | 0.016031785 | 0 | 0 | 0 |
| 0.007562973 | 0 | 0 | 0 | 0.005877865 |
| 0 | 0 | 0.086601126 | 0 | 0.025403257 |
| 0 | 0 | 0 | 0 | 0 |
| 0 | 0 | 0 | 0 | 0 |
| 0 | 0 | 0 | 0 | 0 |
| 0 | 0 | 0 | 0 | 0 |
| 0 | 0 | 0 | 0 | 0 |
| 0 | 0 | 0.036411494 | 0 | 0 |
| 0 | 0 | 0 | 0 | 0 |
| 0 | 0 | 0 | 0 | 0 |
| 0 | 0 | 0 | 0 | 0 |
| 0 | 0 | 0 | 0 | 0 |
| 0 | 0 | 0 | 0 | 0 |
| 0 | 0 | 0 | 0 | 0 |
| 0 | 0 | 0 | 0 | 0 |
| 0 | 0 | 0 | 0 | 0 |
| 0 | 0 | 0 | 0 | 0 |
| 0 | 0.017758258 | 0 | 0 | 1.033915107 |
| 0 | 0 | 0 | 0 | 0 |
| 0 | 0 | 0 | 0 | 0.004542021 |
| 0 | 0 | 0 | 0 | 0 |
| 0 | 0 | 0 | 0.042587265 | 0.005079225 |
| 0 | 0 | 0 | 0 | 0 |
| 0 | 0 | 0 | 0 | 0 |
| 0 | 0 | 0.033676846 | 0 | 0 |
| 0 | 0 | 0 | 0 | 0 |
| 0 | 0 | 0 | 0 | 0 |
| 0 | 0 | 0 | 0 | 0 |
| 0 | 0 | 0 | 0 | 0 |
| 0 | 0 | 0 | 0 | 0 |
| 0 | 0 | 0 | 0 | 0 |
| 0.011082367 | 0 | 0.003740656 | 0 | 0 |

| | | | | |
|---|---|---|---|---|
| 0 | 0 | 5.120834244 | 0 | 0 |
| 0 | 0.007543104 | 0 | 0 | 0 |
| 0 | 0 | 0 | 0 | 0.003440455 |
| 0 | 0 | 0 | 0 | 0 |
| 0.026859697 | 0.00969618 | 0 | 0.005386385 | 0.003727693 |
| 0 | 0 | 0 | 0 | 0 |
| 0 | 0 | 0 | 0 | 0 |
| 0 | 0 | 0.00363086 | 0.021061097 | 0.059343141 |
| 0 | 0 | 0 | 0 | 0 |
| 24.730444 | 0.079789704 | 0.003742406 | 0.031947812 | 0.166956963 |
| 0.002506146 | 0 | 0 | 0 | 0 |
| 0 | 0.001169451 | 0.016287145 | 0.047185018 | 0.010931578 |
| 0.004036993 | 0.000709656 | 0.006056067 | 0.007635522 | 0.024203647 |
| 0 | 0 | 0 | 0 | 0 |
| 0 | 0 | 0 | 0 | 0 |
| 0 | 0.001065311 | 0.008363869 | 0 | 0.077377306 |
| 0 | 0 | 0 | 0 | 0 |
| 0 | 0 | 0 | 0 | 0 |
| 0.386027909 | 0 | 0 | 0 | 1.479451227 |
| 0 | 0 | 0 | 0 | 0.001450785 |
| 0 | 0 | 0 | 0 | 0 |
| 0 | 0 | 0 | 0 | 0 |
| 0 | 0 | 0 | 0 | 0 |
| 0 | 0 | 0 | 0 | 0 |
| 0 | 0 | 0 | 0 | 0 |
| 0 | 0 | 0.003103661 | 0.015007675 | 0 |
| 0 | 0 | 0 | 0.495389789 | 0 |
| 0.156652259 | 0 | 0 | 0 | 0 |
| 0 | 0.851787442 | 0 | 0 | 0 |
| 0 | 0 | 0 | 0 | 0 |
| 0.174324607 | 0 | 0 | 0 | 0.003641271 |
| 0 | 0 | 0.003696149 | 0 | 0 |
| 0 | 0 | 0 | 0 | 0 |
| 0 | 0.046482969 | 0 | 0 | 0.026627431 |
| 0 | 0 | 0 | 0 | 0 |
| 0 | 0 | 0 | 0 | 0 |
| 0.089015608 | 0.209419435 | 0 | 0 | 0 |
| 0 | 0.006454071 | 0 | 0 | 0 |
| 0 | 0 | 0 | 0 | 0 |
| 0 | 0.002608014 | 0 | 0 | 0 |
| 0 | 0 | 0 | 0 | 0 |
| 0 | 0 | 0 | 0 | 0 |
| 0 | 0 | 0 | 0 | 0 |
| 0 | 0 | 0.008183188 | 0 | 0 |
| 0 | 0 | 0 | 0 | 0 |
| 0 | 0 | 0 | 0 | 0 |
| 0.001461848 | 0.092511255 | 0.002807016 | 0 | 0 |
| 0 | 0 | 0 | 0 | 0 |

| | | | | |
|---|---|---|---|---|
| 0 | 0 | 0 | 0 | 0 |
| 0 | 0.015795358 | 0 | 0 | 0.579712692 |
| 0 | 0 | 0 | 0 | 0 |
| 0 | 0 | 0 | 0 | 0 |
| 0 | 0.003301917 | 0 | 0 | 0.00128337 |
| 0 | 0.031169523 | 0 | 0 | 0.00409071 |
| 0 | 0 | 0.017823115 | 0 | 0 |
| 0 | 0 | 0 | 0 | 0.065143296 |
| 0 | 0 | 0 | 0 | 0 |
| 0 | 0 | 0 | 0 | 0 |
| 0.060627613 | 0.002112744 | 0 | 0.004104391 | 0.008599469 |
| 0 | 0 | 0 | 0 | 0 |
| 0 | 0 | 0 | 0 | 0 |
| 0.006560467 | 0.001774235 | 0 | 0 | 0.009805245 |
| 0 | 0 | 0 | 0 | 0 |
| 0 | 0.01447466 | 0 | 0 | 0 |
| 0 | 0 | 0 | 0 | 0 |
| 0.152734341 | 0.004316543 | 0 | 0 | 0 |
| 0 | 0 | 0 | 0 | 0.001789059 |
| 0 | 0 | 0 | 0 | 0 |
| 0 | 0 | 0 | 0 | 0.021733198 |
| 0 | 0.524309114 | 0 | 0 | 0 |
| 0 | 0 | 0 | 0.026040797 | 0 |
| 0 | 0 | 0 | 0 | 0 |
| 0 | 0 | 0 | 0 | 0 |
| 0 | 0 | 0 | 0 | 0 |
| 0 | 0.004809351 | 0 | 0 | 0.005467619 |
| 0 | 0.00104045 | 0 | 0 | 0 |
| 0 | 0 | 0 | 0 | 0 |
| 0.003328709 | 0.811730032 | 0.944999571 | 0.528591689 | 0.178628838 |
| 0 | 0 | 0.003697903 | 0 | 0 |
| 0 | 0 | 0 | 0 | 0 |
| 0 | 0 | 0 | 0 | 0 |
| 0 | 0 | 0 | 0 | 0 |
| 0 | 0 | 0 | 0 | 0 |
| 0 | 0 | 0 | 0 | 0 |
| 0 | 0.663493942 | 0 | 0.554845703 | 0 |
| 0 | 0 | 0 | 0 | 0 |
| 0 | 0.177736681 | 0.040070217 | 0 | 0 |
| 0 | 0 | 0 | 0 | 0 |
| 0.05497137 | 0.001458614 | 0.038089456 | 0 | 0.028098267 |
| 0 | 5.173119859 | 0 | 2.315653227 | 0.004951987 |
| 0 | 0 | 0 | 0 | 0 |
| 0 | 0 | 0 | 0 | 0 |
| 0.027846256 | 0 | 0 | 0 | 0 |
| 0 | 0 | 0 | 0 | 0 |
| 0.005878501 | 0.00191365 | 0.00156775 | 0 | 0.062366524 |
| 0 | 0 | 0 | 0 | 0 |

| | | | | |
|---|---|---|---|---|
| 0 | 0 | 0.00225673 | 0.034613551 | 0 |
| 0 | 0.0140308 | 0 | 0 | 0 |
| 0.003660327 | 0 | 0 | 0.009951955 | 0.09347096 |
| 0.321970013 | 0 | 0 | 0 | 0 |
| 0 | 0 | 0 | 0 | 0 |
| 0 | 0 | 0 | 0 | 0.025721848 |
| 0 | 0 | 0 | 0 | 0 |
| 0 | 0 | 0 | 0 | 0 |
| 0.090153422 | 0 | 0.043979278 | 0 | 0.039938048 |
| 0 | 0 | 0 | 0 | 0 |
| 0 | 0 | 0 | 0 | 0 |
| 0 | 0 | 0 | 0 | 0 |
| 0.007928057 | 0 | 0 | 0 | 0 |
| 0.024302525 | 0 | 0 | 0 | 0.002968063 |
| 0 | 0 | 0 | 0 | 0 |
| 0 | 0.163589954 | 0.018234062 | 0.144358541 | 0.000843451 |
| 0.002105066 | 0 | 0.004547375 | 0.079381087 | 0.003973229 |
| 0.008951504 | 0 | 0 | 0 | 0 |
| 0 | 0 | 0 | 0 | 0 |
| 0 | 0 | 0 | 0 | 0 |
| 0 | 0.060393997 | 0 | 0.005958659 | 0 |
| 0.002685943 | 0.001287701 | 0 | 0.001154582 | 0 |
| 0 | 0 | 0 | 0 | 0 |
| 0 | 0 | 0 | 0 | 0.982451751 |
| 0 | 0.024727811 | 0 | 0 | 0 |
| 0 | 0 | 0 | 0 | 0 |
| 0.009086152 | 0 | 0.003207183 | 0 | 0.013055177 |
| 0 | 0 | 0 | 0.008996431 | 0.002628779 |
| 0 | 0 | 0 | 0 | 0 |
| 0 | 0 | 0 | 0 | 0 |
| 0 | 0.012325585 | 0.024878347 | 0.007207434 | 0.066339934 |
| 0.000908996 | 0 | 0.011272622 | 0.00644724 | 0.005718979 |
| 0 | 0.032710614 | 0.008309466 | 0.006138644 | 0.00576555 |
| 0 | 0 | 0 | 0 | 0.008721628 |
| 0 | 0 | 0.036122783 | 0.003837177 | 0.033418548 |
| 0 | 0 | 0 | 0 | 0 |
| 0 | 0 | 0 | 0 | 0.002895808 |
| 0.002039733 | 0.081214061 | 0.018848927 | 0.003857927 | 0.000679397 |
| 0 | 0 | 0 | 0.323134464 | 0 |
| 0 | 0.002198702 | 0 | 0 | 0.000740635 |
| 0 | 0 | 0 | 0 | 0 |
| 0 | 0.009258519 | 0 | 0 | 0 |
| 0 | 0 | 0 | 0 | 0 |
| 0 | 0.085369816 | 0.351605136 | 0.152838053 | 0.037478277 |
| 0.130867793 | 0 | 0 | 0 | 0 |
| 0 | 0 | 1.109797778 | 0 | 0 |
| 0 | 0 | 0 | 0 | 0 |
| 0 | 0 | 0 | 0 | 0 |

| | | | | |
|---|---|---|---|---|
| 0.025706096 | 0 | 0 | 0 | 0 |
| 0.475276469 | 0.052522868 | 0.010006774 | 0.615716109 | 2.135121265 |
| 0 | 0 | 0 | 0 | 0.104163991 |
| 0 | 0.061252443 | 0 | 0 | 0 |
| 0.003817403 | 0 | 0 | 0 | 0.00544931 |
| 0 | 0 | 0 | 0 | 0 |
| 0.007971036 | 0 | 0 | 0 | 0.011133884 |
| 0 | 0 | 0 | 0.001009466 | 0.001185141 |
| 0 | 0 | 0 | 0 | 0.057062381 |
| 0.01979337 | 0 | 0 | 0 | 0 |
| 0.005163485 | 0.003630719 | 0 | 0 | 0 |
| 0.011075267 | 0.001622418 | 0.192728015 | 0.001818371 | 0.080269133 |
| 0 | 0 | 0 | 1.086799279 | 0 |
| 0 | 0 | 0 | 0 | 0 |
| 0 | 0 | 0.041522528 | 0 | 0 |
| 0 | 0 | 0 | 0 | 0 |
| 0.038497229 | 0 | 0 | 0.007669931 | 0.072998186 |
| 0.120444694 | 0.499706706 | 0.163753012 | 0.006305866 | 0.056732351 |
| 0 | 0 | 0 | 0 | 0 |
| 0.008568515 | 0.038617533 | 0.455305024 | 0 | 0 |
| 0.020296249 | 0.008625551 | 0 | 0.102561584 | 0.031944281 |
| 0 | 0 | 0 | 0 | 0 |
| 0 | 0 | 0 | 0 | 0 |
| 0 | 0 | 0 | 0 | 0 |
| 0 | 0.00856377 | 0.001349198 | 0 | 0.029956661 |
| 0 | 0 | 0 | 0 | 0 |
| 0 | 0.027670011 | 0 | 0.015354167 | 0.023464634 |
| 0 | 0.048338714 | 0.098149724 | 0.021577384 | 0.073957577 |
| 0 | 0 | 0 | 0 | 0 |
| 0 | 0 | 0.00171582 | 0 | 0 |
| 0.004578465 | 0 | 0 | 0 | 0 |
| 0 | 0.000718985 | 0 | 0 | 0 |
| 0 | 0.034147351 | 0 | 0 | 0 |
| 0 | 0 | 0 | 0 | 0 |
| 0 | 0 | 0 | 0.074235963 | 0 |
| 0.04052118 | 0.368146754 | 1.446382915 | 0.09058469 | 0.399560147 |
| 0 | 0 | 0 | 0 | 0 |
| 0 | 0 | 0 | 0 | 0 |
| 0 | 0 | 0.011581437 | 0.012790524 | 0.011688462 |
| 0 | 0.011732804 | 0 | 0 | 0 |
| 0.139462287 | 0 | 0 | 0 | 0 |
| 0 | 0 | 0 | 0 | 0 |
| 0 | 0 | 0 | 0 | 0.004338736 |
| 0.0099468 | 0 | 0 | 0 | 0.003427338 |
| 0 | 0.003564564 | 0 | 0 | 0 |
| 0 | 0.496657116 | 0 | 0 | 0 |
| 0 | 0 | 0 | 0 | 0 |
| 0.042413265 | 0 | 0 | 0 | 0.001538896 |

| | | | | |
|---|---|---|---|---|
| 0.072956172 | 0 | 0 | 0 | 0 |
| 0 | 0 | 0 | 0 | 0 |
| 0 | 0 | 0 | 0 | 0 |
| 0 | 0.008364445 | 0.160637322 | 0.011005069 | 0 |
| 0 | 0 | 0 | 0 | 0 |
| 0 | 0 | 0 | 0 | 0 |
| 0 | 0 | 0 | 0 | 0 |
| 0 | 0 | 0 | 0.018596516 | 0 |
| 0 | 0.095039852 | 1.17140025 | 0.784628356 | 0 |
| 0 | 0 | 0 | 0 | 0 |
| 0 | 0 | 0 | 0 | 0 |
| 0 | 0 | 0.011656079 | 0.001405872 | 0 |
| 0 | 0 | 0 | 0 | 0 |
| 0 | 0 | 0 | 0 | 0 |
| 0 | 0 | 0 | 0.062066904 | 0.003087253 |
| 0 | 0 | 0 | 0 | 0 |
| 0 | 0 | 0 | 0.11904522 | 0 |
| 0 | 0 | 0 | 0.882713115 | 0 |
| 0 | 0 | 0 | 0 | 0 |
| 0 | 0 | 0 | 0 | 0 |
| 0 | 0 | 0 | 0 | 0 |
| 0 | 0 | 0 | 0 | 0 |
| 0 | 0 | 0 | 0 | 0 |
| 0.439424934 | 0.499507592 | 0.575577061 | 0 | 0.891794986 |
| 0 | 0.057089467 | 0.005567892 | 0 | 0.000824171 |
| 0 | 0.16356593 | 0 | 0 | 0.069437968 |
| 0 | 0 | 0 | 0 | 0.023780421 |
| 0 | 1.297654577 | 1.083636057 | 3.806175389 | 3.029855888 |
| 0 | 0 | 0 | 0 | 0 |
| 0 | 0 | 0 | 0 | 0 |
| 0 | 0 | 0 | 0.165148617 | 0.001283679 |
| 0 | 0 | 0 | 0 | 0 |
| 0 | 0.110598649 | 0 | 0 | 0 |
| 0 | 0 | 0 | 0 | 0 |
| 0 | 0 | 0 | 0 | 0 |
| 0 | 0 | 0 | 0.079960565 | 0.002176833 |
| 0 | 0.156535641 | 0.005824538 | 1.559768605 | 0 |
| 0 | 0 | 0.230923842 | 0 | 0 |
| 0 | 0 | 0 | 0 | 0 |
| 0 | 0 | 0 | 0 | 0 |
| 0 | 0.069025196 | 0.00273843 | 0 | 0.097959107 |
| 0 | 0 | 0 | 0 | 0 |
| 0 | 0.368709238 | 0.552989056 | 6.996923674 | 8.415165559 |
| 0 | 0 | 0 | 0.007832526 | 0 |
| 0 | 0 | 0 | 0 | 0 |
| 0 | 0 | 0 | 0 | 0 |
| 0 | 0 | 0 | 0 | 0 |
| 0 | 0 | 0.001645954 | 0.001351059 | 0.003806835 |

| | | | | |
|---|---|---|---|---|
| 0 | 0 | 0 | 0 | 0 |
| 0 | 0 | 0 | 0.287035028 | 0 |
| 0.850414486 | 0.454911242 | 0.051279557 | 0.002072229 | 0.070796047 |
| 0 | 0 | 0 | 0.001679985 | 0 |
| 0 | 0 | 0 | 0 | 0 |
| 0 | 0 | 0.010565123 | 0.018976906 | 0 |
| 0 | 0 | 0 | 0 | 0 |
| 0 | 0.004151341 | 0 | 0 | 0 |
| 0 | 0 | 0 | 0 | 0 |
| 0 | 0 | 0 | 0 | 0 |
| 0.035713149 | 0.148250406 | 0.210942828 | 0.037385063 | 0.000955452 |
| 0 | 1.57661679 | 2.200694455 | 0.446685872 | 0.890953865 |
| 0 | 0 | 0 | 0 | 0 |
| 0.006733431 | 0 | 0 | 0.564864782 | 0.008764887 |
| 1.194078347 | 0.034415913 | 0.802813353 | 0.44946101 | 0.022143245 |
| 0 | 0 | 0 | 0 | 0 |
| 0 | 0 | 0 | 0 | 0 |
| 0 | 0 | 0 | 0 | 0 |
| 0 | 0 | 0 | 0 | 0 |
| 0 | 0.055452138 | 0 | 0.002304422 | 0.002361124 |
| 0.001442548 | 0.002218851 | 0.321141874 | 0.004263155 | 0.216819261 |
| 0 | 1.956388998 | 0.012313161 | 0.028973675 | 0 |
| 0.695226785 | 24.46978044 | 0.017169412 | 0.641445841 | 8.697284751 |
| 0 | 0 | 0 | 0 | 0 |
| 0 | 0 | 0 | 0 | 0 |
| 0 | 0.003418521 | 0 | 1.793607808 | 0 |
| 0 | 0 | 0 | 0 | 0.076385017 |
| 0 | 0 | 0 | 0.002000877 | 0 |
| 0 | 0 | 0 | 0 | 0 |
| 0 | 0 | 0 | 0 | 0 |
| 0 | 0 | 0 | 0 | 0 |
| 0 | 0.591322431 | 0.21615036 | 0.114302758 | 0.303355378 |
| 0.751905858 | 1.361962429 | 1.391081108 | 0 | 0.008586152 |
| 0.044482521 | 0 | 0 | 0 | 0 |
| 0.024353989 | 0 | 0.020651818 | 0.001206736 | 0.0024287 |
| 0 | 0.001232761 | 0.019236837 | 0.225485546 | 0.001779672 |
| 0 | 0 | 0 | 0.0663515 | 0 |
| 0 | 0 | 0 | 0 | 0 |
| 0.061664838 | 0.05193285 | 0 | 2.450162226 | 0.008069982 |
| 0 | 0 | 0 | 0 | 0 |
| 0 | 0 | 0 | 0 | 0 |
| 0 | 0 | 0 | 0 | 0 |
| 0 | 0 | 0 | 0 | 0 |
| 0 | 0 | 0 | 0 | 0 |
| 0.006854793 | 0.047798011 | 0.080620052 | 0.080761618 | 0.001775825 |
| 0 | 0.709543915 | 0 | 0 | 0 |
| 0 | 0 | 0 | 0 | 0 |
| 0.691819058 | 0.005329681 | 0 | 4.485256585 | 0.050799086 |

| | | | | |
|---|---|---|---|---|
| 0 | 0 | 0 | 0 | 0 |
| 0 | 0 | 0 | 0 | 0 |
| 0 | 0 | 0.02669644 | 0 | 0 |
| 0 | 0.018315005 | 0 | 0.00368336 | 0 |
| 0 | 0 | 0 | 0 | 0 |
| 0 | 0 | 0 | 0.169365594 | 0 |
| 0 | 0 | 0 | 0 | 0 |
| 0 | 0 | 0 | 0 | 0.0039792 |
| 0 | 0 | 0 | 0 | 0 |
| 0 | 0 | 0 | 0 | 0 |
| 0.022556868 | 0 | 0.001301482 | 0.012563258 | 0 |
| 0 | 0 | 0 | 0 | 0 |
| 0 | 0 | 0 | 0 | 0 |
| 0 | 0.369923601 | 0 | 0 | 0 |
| 0 | 0 | 0 | 0 | 0 |
| 0 | 0 | 0 | 0 | 0 |
| 0 | 0 | 0.713579988 | 0 | 0 |
| 0 | 0 | 0.004117125 | 0 | 0 |
| 0 | 0 | 0 | 0 | 0 |
| 0 | 0 | 0 | 0 | 0 |
| 0 | 0 | 0 | 0 | 0 |
| 0 | 0 | 0 | 0 | 0 |
| 0 | 0 | 0 | 0.00098433 | 0 |
| 0 | 0 | 0 | 0 | 0 |
| 0 | 0 | 0 | 0 | 0 |
| 0 | 0 | 0 | 0 | 0 |
| 0 | 0 | 0 | 0 | 0 |
| 0 | 0 | 0 | 0 | 0 |
| 0 | 0 | 0 | 0 | 0 |
| 0 | 0.348334286 | 0.001570563 | 0.01186042 | 0.092991181 |
| 0 | 0 | 0.002460149 | 0 | 0.002655305 |
| 0 | 0 | 0 | 0 | 0 |
| 0 | 0 | 0 | 0.005575246 | 0 |
| 0 | 0 | 0 | 0 | 0 |
| 0 | 0 | 0 | 0.002446455 | 0 |
| 0 | 0 | 0 | 0 | 0 |
| 0 | 0 | 0 | 0 | 0 |
| 0.015973521 | 0.015492176 | 0.011457963 | 0 | 0.112525123 |
| 0 | 0 | 0 | 0 | 0 |
| 0 | 0 | 0 | 0 | 0 |
| 0 | 0 | 0 | 0 | 0 |
| 0 | 0 | 0 | 0 | 0 |
| 0 | 0 | 0 | 0 | 0 |
| 0 | 0.566841842 | 8.158259461 | 0.578149504 | 0 |
| 0 | 0 | 0 | 0.11509122 | 0 |
| 0 | 0 | 0 | 0 | 0 |
| 0 | 0 | 0.068881105 | 0.002887156 | 0 |
| 0 | 0 | 0 | 0 | 0 |

| | | | | |
|---|---|---|---|---|
| 0 | 0 | 0 | 0 | 0 |
| 0 | 0 | 0 | 0 | 0 |
| 0 | 0 | 0 | 0 | 0 |
| 0 | 0.768421898 | 0 | 0.423796692 | 0 |
| 0 | 0 | 0 | 0 | 0 |
| 0 | 0 | 0 | 0.407483157 | 0 |
| 0 | 0 | 0 | 0 | 0.001393978 |
| 0 | 0 | 0 | 0 | 0 |
| 0.706990668 | 0.263517668 | 0.779485936 | 0.017960162 | 0.101773767 |
| 0 | 0 | 0 | 0 | 0 |
| 0 | 0 | 0 | 0.000642104 | 0 |
| 0 | 0 | 0 | 0 | 0 |
| 0 | 0 | 0 | 0 | 0 |
| 0 | 0.001545869 | 0 | 0 | 0 |
| 0 | 0.393616063 | 8.822998229 | 0.982195781 | 7.243826794 |
| 0 | 0 | 0 | 0 | 0.753629496 |
| 0 | 0 | 0 | 0 | 0 |
| 0 | 0 | 0 | 0 | 0 |
| 0 | 0 | 0 | 0 | 0 |
| 0 | 0 | 0 | 0 | 0 |
| 0 | 0 | 0 | 0.012817578 | 0 |
| 0 | 0 | 0 | 0 | 0 |
| 0 | 0 | 0 | 0 | 0 |
| 0 | 0 | 0 | 0 | 0 |
| 0.015782324 | 0 | 0.007147394 | 0.194661888 | 0.00429801 |
| 0 | 0.33303446 | 0 | 3.448902022 | 1.817365019 |
| 0 | 0.61154238 | 0 | 0.002618542 | 0 |
| 0 | 0 | 0 | 0 | 0 |
| 0 | 0 | 0 | 0.041855083 | 0 |
| 0 | 0 | 0 | 0 | 0 |
| 0 | 0 | 0 | 0 | 0 |
| 0 | 0 | 0 | 0 | 0 |
| 0 | 0 | 0 | 0 | 0 |
| 0 | 0.039138695 | 0 | 0.001691967 | 0 |
| 0.002882406 | 0 | 0 | 0.433042376 | 0.003491184 |
| 0.00111137 | 0 | 0.004668199 | 0.001094809 | 0 |
| 0.049699136 | 0.155577306 | 0 | 0 | 0 |
| 0 | 3.205894367 | 1.858726845 | 3.449817993 | 0 |
| 0 | 0 | 0 | 0 | 0 |
| 0 | 0 | 0 | 0 | 0 |
| 0 | 0 | 0 | 0 | 0 |
| 0 | 0 | 0.002520845 | 0 | 0 |
| 0 | 0.942406894 | 0.977229214 | 0.995189318 | 0.50079438 |
| 0 | 0 | 0 | 0 | 0 |
| 0 | 0 | 0 | 0 | 0 |
| 0.005264418 | 0.163645576 | 0.050122038 | 0.000829755 | 0.000584493 |
| 0 | 0 | 0 | 0 | 0 |
| 0 | 0.007371546 | 0 | 0.815942183 | 0.290834667 |

| | | | | |
|---|---|---|---|---|
| 0 | 0 | 0 | 0 | 0 |
| 0 | 0 | 0 | 0 | 0 |
| 0 | 0 | 0 | 0 | 0 |
| 0 | 0 | 0 | 0 | 0 |
| 0 | 0.274920533 | 0.004483625 | 0 | 0 |
| 0 | 0 | 0 | 0 | 0 |
| 0 | 0 | 0 | 0 | 0 |
| 0 | 0 | 0 | 0 | 0 |
| 0 | 0 | 0 | 0 | 0 |
| 0 | 0 | 0 | 0 | 0 |
| 0 | 0 | 0 | 0 | 0 |
| 0 | 0.113267523 | 2.56826367 | 1.194931355 | 0.839550288 |
| 0 | 0 | 0 | 0 | 0.259773058 |
| 0 | 0 | 0 | 0 | 0 |
| 0 | 0.136714491 | 0.195313434 | 3.218326824 | 1.502034031 |
| 0 | 0 | 0 | 0 | 0 |
| 0 | 0.015997546 | 0 | 0 | 0 |
| 0 | 0 | 0 | 0 | 0 |
| 0 | 0 | 0 | 0.009202215 | 0 |
| 0 | 0 | 0 | 0 | 0 |
| 0 | 0 | 0 | 0 | 0 |
| 0 | 0 | 0 | 0 | 0 |
| 0 | 0.068688176 | 2.037299734 | 0.633608317 | 0 |
| 0 | 0 | 0 | 0 | 0 |
| 0 | 0 | 0 | 0 | 0 |
| 0 | 0 | 0 | 0 | 0 |
| 0 | 0 | 0 | 0 | 0 |
| 0 | 0 | 0 | 0 | 0 |
| 0 | 0 | 0 | 0 | 0 |
| 0 | 0 | 0 | 0 | 0 |
| 0 | 0 | 0 | 0 | 0 |
| 0.067903231 | 0.006844374 | 0.010093002 | 0.026879274 | 0.385600712 |
| 0 | 0 | 0 | 0 | 0 |
| 0.008403015 | 0 | 0.144732393 | 0 | 0 |
| 0.001814174 | 0.000885862 | 0.007015474 | 0.001429711 | 0 |
| 0.001818981 | 0.006395108 | 0 | 0.328066659 | 0.030581937 |
| 0 | 0 | 0 | 0.003172776 | 0 |
| 0 | 0 | 0 | 0.027114852 | 0 |
| 0 | 0.00325672 | 0 | 0 | 0 |
| 0 | 0 | 0 | 0 | 0 |
| 0 | 0 | 0 | 0 | 0 |
| 0 | 0 | 0 | 0 | 0 |
| 0 | 0 | 0 | 0 | 0 |
| 0 | 0 | 0 | 0 | 0 |
| 0.006751429 | 0.096865588 | 0.269050222 | 0.003286287 | 0.000661404 |
| 0 | 0 | 0 | 0 | 0 |
| 0 | 0 | 0 | 0.069915201 | 0 |
| 0 | 0.007923677 | 0 | 0 | 0 |

| | | | | |
|---|---|---|---|---|
| 0 | 0 | 0 | 0 | 0 |
| 0 | 0 | 0 | 0 | 0 |
| 0.002053726 | 0.030867275 | 0 | 0 | 0 |
| 0 | 0 | 0 | 0 | 0 |
| 0 | 0 | 0 | 0 | 0 |
| 0 | 0 | 0 | 0 | 0 |
| 0 | 0 | 0 | 0 | 0 |
| 0 | 0 | 0.426897249 | 0 | 0 |
| 0 | 0 | 0 | 0 | 0 |
| 0 | 0 | 0 | 0 | 0 |
| 0 | 0 | 0 | 0 | 0 |
| 0 | 0.001469319 | 0.001755445 | 0 | 0 |
| 0 | 0 | 0 | 0 | 0 |
| 0 | 0 | 0 | 0 | 0 |
| 0 | 0 | 0 | 0 | 0 |
| 0 | 0 | 0 | 0 | 0 |
| 0 | 0 | 0 | 0 | 0.001916915 |
| 0 | 0 | 0 | 0 | 0 |
| 0 | 0.349307952 | 0 | 2.2740104 | 0 |
| 0 | 0 | 0 | 0 | 0 |
| 0 | 0.002788167 | 0.010973088 | 0 | 0 |
| 0 | 0 | 0 | 0 | 0 |
| 0 | 0 | 0 | 0 | 0 |
| 0 | 0 | 0 | 0 | 0 |
| 0 | 0 | 0 | 0 | 0 |
| 0.035863324 | 0.002599732 | 0.002440418 | 0.009833815 | 0.001436732 |
| 0 | 0 | 0 | 0 | 0 |
| 0 | 0.003657892 | 0.006080285 | 0.038501398 | 0.001205379 |
| 0 | 0 | 0 | 0 | 0 |
| 0 | 1.035231012 | 1.550742615 | 0 | 0 |
| 0 | 0 | 0 | 0 | 0 |
| 0 | 0 | 0 | 0 | 0 |
| 0 | 0 | 0 | 0 | 0 |
| 0 | 0 | 0 | 0 | 0 |
| 0.003874489 | 0.001872996 | 0.000697475 | 0.00091602 | 0 |
| 0 | 0 | 0 | 0 | 0 |
| 0 | 0 | 0 | 0 | 0 |
| 0.005495513 | 0.001545674 | 0.001319049 | 0 | 0 |
| 0 | 0 | 0 | 0 | 0 |
| 0 | 0 | 0 | 0 | 0 |
| 0.036331371 | 1.957571329 | 16.78214155 | 4.612421789 | 10.96475355 |
| 0 | 0.026023134 | 1.724664965 | 0 | 0.550966746 |
| 0.624947182 | 0 | 0 | 0 | 0 |
| 0 | 0 | 0 | 0 | 0 |
| 0 | 0 | 0 | 0 | 0 |
| 0 | 0 | 0 | 0 | 0 |
| 0 | 0 | 0 | 0 | 0 |
| 0 | 0.00674703 | 0.003768733 | 0 | 0 |

| | | | | |
|---|---|---|---|---|
| 0 | 0 | 0 | 0 | 0.026458436 |
| 0.006998483 | 0.01514155 | 0.009045056 | 0 | 0 |
| 0 | 0 | 0 | 0 | 0 |
| 0 | 0 | 0 | 0 | 0 |
| 0 | 5.155230657 | 4.726048682 | 1.699337756 | 0.309855603 |
| 0 | 0 | 0 | 0 | 0 |
| 0 | 0 | 0 | 0 | 0 |
| 0 | 0 | 0 | 0 | 0 |
| 11.26080112 | 0.501926806 | 0.641897287 | 0.111838983 | 0.210481165 |
| 0 | 0 | 0 | 0 | 0 |
| 0 | 0 | 0 | 0 | 0 |
| 0 | 0 | 0 | 0 | 0 |
| 0 | 0.182987916 | 0.313467684 | 0.089455855 | 0 |
| 0 | 0.001507303 | 0.059427238 | 0.007855491 | 0.029036199 |
| 0.212334693 | 0 | 0 | 0 | 0 |
| 0 | 0.162031576 | 1.503499268 | 0.190889488 | 0.305843585 |
| 0 | 0 | 0 | 0 | 0.000823814 |
| 0 | 0 | 0 | 0 | 0 |
| 0 | 0 | 0 | 0 | 0 |
| 0 | 0 | 0 | 0 | 0 |
| 0 | 0 | 0 | 0 | 0 |
| 0 | 0 | 0 | 0 | 0 |
| 0 | 0 | 0 | 0 | 0 |
| 0 | 0 | 0 | 0 | 0 |
| 0 | 0 | 0 | 0 | 0 |
| 0 | 0 | 0 | 0 | 0 |
| 0 | 0.012475924 | 0 | 0.173894496 | 0 |
| 0 | 0 | 0 | 0.000860616 | 0 |
| 0 | 0 | 0 | 0 | 0 |
| 0 | 0 | 0 | 0 | 0 |
| 0 | 0 | 0 | 0 | 0 |
| 0 | 0 | 0 | 0 | 0 |
| 0 | 0 | 0 | 0 | 0 |
| 0 | 0 | 0.062280518 | 0.012404638 | 0 |
| 0.060927216 | 0.077748749 | 0 | 0.37238632 | 0 |
| 0 | 0.106222346 | 0.085405055 | 0 | 0 |
| 0 | 0 | 0 | 0 | 0 |
| 0 | 0 | 0 | 0 | 0 |
| 0 | 0 | 0 | 0 | 0 |
| 0 | 0 | 0 | 0 | 0 |
| 0 | 1.926721196 | 0 | 0.007925436 | 0.004867064 |
| 0 | 0 | 0 | 0.089310541 | 0 |
| 0 | 0 | 0 | 0 | 0 |
| 0 | 0 | 0 | 0.030282937 | 0 |
| 0 | 0 | 0 | 0 | 0 |
| 0 | 0 | 0 | 0 | 0 |
| 0 | 0 | 0 | 0 | 0 |
| 0 | 0 | 0 | 0 | 0 |

| | | | | |
|---|---|---|---|---|
| 0 | 0 | 0 | 0.205655691 | 0 |
| 0 | 0 | 0 | 0 | 0 |
| 0 | 0 | 0 | 0 | 0 |
| 0 | 0 | 0 | 0 | 0.004120454 |
| 0 | 0 | 0 | 0 | 0 |
| 0 | 0 | 0 | 0 | 0 |
| 0 | 0.030414748 | 0.003109092 | 0 | 0.002465432 |
| 0 | 0 | 0 | 0 | 0 |
| 0 | 0 | 0 | 0 | 0 |
| 0 | 0.083614492 | 0 | 0 | 0.003017747 |
| 0 | 0 | 0 | 0 | 0 |
| 0 | 0 | 0 | 0 | 0 |
| 0 | 0 | 0 | 0 | 0 |
| 0 | 0 | 0 | 0 | 0 |
| 0 | 0 | 0 | 0.008597514 | 0 |
| 0 | 0 | 0 | 0 | 0 |
| 0 | 0.075002503 | 0 | 0 | 0 |
| 0 | 0 | 0 | 0 | 0 |
| 0 | 0 | 0 | 0 | 0 |
| 0 | 0 | 0 | 0 | 0 |
| 0 | 0 | 0 | 0.789722074 | 0.022689881 |
| 0 | 0 | 0 | 0 | 0 |
| 0 | 0 | 0.013292812 | 0.020903619 | 0 |
| 0.006911684 | 0.001670615 | 0.004562146 | 0.022877186 | 0.00095923 |
| 0 | 0 | 0 | 0 | 0 |
| 0 | 0 | 0 | 0.001686871 | 0 |
| 0 | 0 | 0 | 0 | 0 |
| 0 | 0 | 0 | 0 | 0 |
| 0 | 0 | 0 | 0 | 0 |
| 0.349158415 | 0 | 1.325734648 | 0 | 0.015462241 |
| 0 | 0 | 0 | 0 | 0 |
| 0 | 0 | 0 | 0 | 0 |
| 0 | 0 | 0 | 0 | 0 |
| 0 | 2.405961398 | 0 | 0 | 0 |
| 0 | 0 | 0 | 0.150242843 | 0 |
| 0 | 0 | 0 | 0 | 0 |
| 0.00735106 | 0.000950169 | 0 | 0 | 0.009361915 |
| 0 | 0 | 0 | 0 | 0 |
| 0 | 0 | 0 | 0 | 0 |
| 0 | 0 | 0 | 0.1555338 | 0 |
| 0 | 0 | 0 | 0 | 0 |
| 0 | 0 | 0 | 0 | 0 |
| 0 | 0 | 0 | 0 | 0 |
| 0 | 0 | 0 | 0 | 0 |
| 0 | 0 | 0 | 0.048136096 | 0 |
| 0 | 0 | 0 | 0 | 0 |
| 0 | 0 | 0 | 0 | 0 |
| 0 | 0 | 0 | 0.005846643 | 0 |

| | | | | |
|---|---|---|---|---|
| 0.009110591 | 0 | 0.01606592 | 0 | 0 |
| 0 | 0 | 0 | 0.348110593 | 0 |
| 0 | 0 | 0 | 0 | 0 |
| 0 | 0 | 0 | 0 | 0 |
| 0 | 0 | 0 | 0 | 0 |
| 0 | 0 | 0 | 0 | 0 |
| 0 | 0 | 0 | 0 | 0 |
| 0 | 0.042788323 | 0.325568248 | 0.100651651 | 0.95085348 |
| 0.656652883 | 0.002565152 | 0.062694438 | 0.342236036 | 0.047308067 |
| 0.041650719 | 0.096967956 | 0.10777389 | 0 | 0.009925453 |
| 0 | 0 | 0 | 0 | 0 |
| 0 | 0 | 0 | 0 | 0 |
| 0 | 0 | 0 | 0 | 0 |
| 0 | 0 | 0 | 0 | 0 |
| 0 | 0 | 0.046470917 | 0 | 0 |
| 0 | 0 | 0 | 0 | 0 |
| 0 | 0.02290099 | 0 | 0 | 0 |
| 0 | 0 | 0 | 0 | 0 |
| 0 | 0 | 0 | 0 | 0 |
| 0 | 0 | 0 | 0.012347896 | 0 |
| 0 | 0 | 0 | 0 | 0 |
| 0 | 0 | 0 | 0 | 0 |
| 0 | 0 | 0 | 0 | 0 |
| 0 | 0 | 0 | 0 | 0 |
| 0 | 0 | 0 | 0 | 0 |
| 0 | 0 | 0 | 0 | 0 |
| 0 | 0 | 0 | 0 | 0 |
| 0 | 0 | 0 | 0 | 0 |
| 0 | 0 | 0 | 0 | 0 |
| 0 | 0 | 0 | 0 | 0 |
| 0 | 0 | 0 | 0 | 0 |
| 0 | 0 | 0 | 0 | 0 |
| 0 | 0 | 0 | 0 | 0 |
| 0 | 0 | 0 | 0 | 0 |
| 0 | 0 | 0 | 0.003193065 | 0 |
| 0 | 0 | 0 | 0 | 0 |
| 0 | 0 | 0 | 0 | 0 |
| 0 | 0 | 0.112254678 | 0 | 0 |
| 0 | 0.004663714 | 0 | 0 | 0 |
| 8.378741943 | 0 | 0 | 0 | 0 |
| 0 | 0 | 0 | 0 | 0 |
| 0 | 0 | 0 | 0.278889807 | 0 |
| 0 | 0 | 0 | 0 | 0 |
| 0 | 0 | 0 | 0 | 0 |
| 0 | 0 | 0 | 0 | 0 |
| 0 | 0.009881438 | 0 | 0 | 0 |
| 0 | 0 | 0 | 0 | 0 |
| 0 | 0 | 0 | 0.00081871 | 0 |

| | | | | |
|---|---|---|---|---|
| 0 | 0 | 0 | 0 | 0 |
| 0 | 0 | 0.098681887 | 0 | 0.605709939 |
| 0 | 0 | 0 | 0 | 0 |
| 0 | 0 | 0 | 0 | 0 |
| 0.062644194 | 0.091561374 | 0 | 1.537358547 | 0.401161545 |
| 0 | 0 | 0 | 0 | 0 |
| 0 | 0 | 0 | 0 | 0 |
| 0.038552265 | 0.018501719 | 0.116661287 | 0.030195817 | 0.094797855 |
| 0 | 0 | 0 | 0 | 0 |
| 0 | 0 | 0 | 0 | 0 |
| 0 | 0 | 0 | 0 | 0 |
| 0 | 0 | 0 | 0 | 0 |
| 0 | 0 | 0 | 0 | 0 |
| 0 | 0 | 0 | 0 | 0 |
| 0 | 0 | 0 | 0 | 0.001478877 |
| 0 | 0.389981665 | 0.096255888 | 0.025703632 | 0 |
| 0 | 0 | 0 | 0 | 0 |
| 0 | 0 | 0 | 0 | 0 |
| 0 | 0 | 0 | 0 | 0 |
| 0 | 0 | 0 | 0 | 0 |
| 0 | 0 | 0 | 0 | 0 |
| 0 | 0 | 0 | 0 | 0 |
| 0 | 0 | 0 | 0 | 0 |
| 0.002251546 | 0 | 0 | 0.008952627 | 0 |
| 0 | 0 | 0 | 0.115604905 | 0 |
| 0 | 0 | 0 | 0 | 0 |
| 0.011606929 | 0 | 0.015277673 | 0 | 0.010600463 |
| 0 | 0 | 0 | 0 | 0 |
| 0 | 0.000978305 | 0 | 0 | 0 |
| 0 | 0 | 0 | 0 | 0 |
| 0 | 0 | 0 | 0 | 0 |
| 0 | 0 | 0 | 0 | 0 |
| 0 | 0 | 0 | 0 | 0 |
| 0 | 0 | 0 | 0 | 0 |
| 0 | 0 | 0 | 0 | 0 |
| 0 | 0 | 0 | 0 | 0 |
| 0 | 0 | 0 | 0 | 0 |
| 0 | 0 | 0 | 0.076712044 | 0 |
| 0 | 0 | 0 | 0 | 0 |
| 0 | 0 | 0 | 0 | 0 |
| 0 | 0 | 0 | 0 | 0 |
| 0 | 0 | 0 | 0 | 0 |
| 0 | 0 | 0 | 0 | 0 |
| 0 | 0 | 0 | 0 | 0 |
| 0 | 0 | 0 | 0 | 0 |
| 0 | 0 | 0 | 0 | 0 |
| 0 | 0 | 0 | 0 | 0 |
| 0 | 0 | 0 | 0 | 0 |

| | | | | |
|---|---|---|---|---|
| 0 | 0.036054571 | 0 | 0.079500295 | 0.06665428 |
| 0 | 0 | 0 | 0 | 0 |
| 0 | 0.00183935 | 0 | 0 | 0 |
| 0 | 0.001077709 | 0 | 0 | 0 |
| 0 | 0 | 0 | 0 | 0 |
| 0 | 0.867516551 | 0 | 0 | 0 |
| 0 | 0 | 0 | 0 | 0 |
| 0 | 0 | 0 | 0 | 0 |
| 0 | 0 | 0 | 0 | 0 |
| 0 | 0 | 0 | 0 | 0 |
| 0 | 0 | 0 | 0 | 0 |
| 0 | 0.011609182 | 0 | 0 | 0 |
| 0 | 0 | 0 | 0 | 0 |
| 0 | 0 | 0 | 0 | 0 |
| 0 | 0 | 0 | 0 | 0 |
| 0 | 0 | 0 | 0 | 0 |
| 0 | 0 | 0 | 0 | 0 |
| 0.086805998 | 0 | 0 | 0 | 0 |
| 0 | 0 | 0 | 0 | 0 |
| 0 | 0 | 0 | 0 | 0 |
| 0.092131299 | 0.000716619 | 0.000733859 | 0 | 0.00362091 |
| 0 | 0 | 0 | 0 | 0 |
| 0 | 0.100006702 | 0.009885979 | 0.008520515 | 0 |
| 0 | 0 | 0 | 0.003063665 | 0 |
| 0 | 0 | 0 | 0 | 0 |
| 0 | 0 | 0 | 0.854312809 | 0 |
| 0 | 0 | 0 | 0 | 0 |
| 0 | 0 | 0 | 0 | 0 |
| 0 | 0.010381053 | 0 | 0 | 0 |
| 0 | 0 | 0 | 0 | 0 |
| 0 | 0 | 0 | 0 | 0 |
| 0 | 0 | 0 | 0 | 0 |
| 0.019768185 | 0 | 0.008968861 | 0 | 0.028907178 |
| 0 | 0 | 0 | 0 | 0 |
| 0 | 0 | 0 | 0 | 0.235110202 |
| 0.005029776 | 0 | 0 | 0 | 0 |
| 0 | 0 | 0 | 0 | 0 |
| 0 | 0 | 0 | 0 | 0 |
| 0.173189392 | 0 | 0.013963887 | 0 | 0.001490598 |
| 0 | 0.002227213 | 0 | 0 | 0 |
| 0 | 0 | 0 | 0 | 0 |
| 0 | 0 | 0 | 0.018864038 | 0 |
| 0 | 0.005059279 | 0.195307615 | 0.106705294 | 0 |
| 0 | 0 | 0 | 0 | 0 |
| 0 | 0 | 0 | 0 | 0 |
| 0 | 0.059173367 | 0 | 0.01080774 | 0 |
| 0 | 0 | 0 | 0 | 0 |
| 0 | 0 | 0 | 0 | 0 |

| | | | | |
|---|---|---|---|---|
| 0 | 0 | 0 | 0 | 0 |
| 0 | 0 | 0 | 0 | 0 |
| 0 | 0 | 0 | 0 | 0 |
| 0 | 0 | 0 | 0 | 0 |
| 0 | 0 | 0 | 0 | 0 |
| 0 | 0 | 0 | 0 | 0 |
| 0 | 0 | 0 | 0 | 0 |
| 0 | 0 | 0 | 0 | 0 |
| 0 | 0 | 0 | 0 | 0 |
| 0.003424318 | 0 | 0 | 0.054141287 | 0 |
| 0 | 0 | 0 | 0 | 0 |
| 0 | 0 | 0 | 0 | 0 |
| 0 | 0 | 0 | 0 | 0 |
| 0 | 0 | 0 | 0 | 0 |
| 0 | 0 | 0 | 0 | 0 |
| 0.038324083 | 0.002776204 | 0 | 0.001244604 | 0.003273086 |
| 0 | 0 | 0 | 0 | 0 |
| 0 | 0 | 0 | 0 | 0 |
| 0 | 0 | 0 | 0 | 0 |
| 0 | 0.012086495 | 0.393533397 | 0.013337877 | 0.013701656 |
| 0 | 0 | 0 | 0 | 0 |
| 0 | 0 | 0 | 0 | 0 |
| 0 | 0 | 0 | 0 | 0 |
| 0 | 0 | 0 | 0 | 0 |
| 0 | 0.005753242 | 0 | 0 | 0 |
| 0 | 0 | 0 | 0 | 0 |
| 0 | 0 | 0 | 0 | 0 |
| 0 | 0 | 0 | 0 | 0 |
| 0 | 0 | 0 | 0 | 0 |
| 0 | 0.003644609 | 0 | 0 | 0.002907688 |
| 0 | 0.030837751 | 0.015815482 | 0.306306119 | 0 |
| 0 | 0 | 0 | 0 | 0 |
| 0 | 0 | 0 | 0 | 0 |
| 0 | 0 | 0 | 0 | 0 |
| 0 | 0 | 0 | 0 | 0 |
| 0 | 0 | 0 | 0 | 0 |
| 0 | 0 | 0 | 0 | 0 |
| 0 | 0 | 0 | 0 | 0 |
| 0 | 0.007459685 | 0 | 0 | 0 |
| 0 | 0 | 0 | 0 | 0 |
| 0 | 0 | 0 | 0 | 0 |
| 0 | 0 | 0 | 0 | 0 |
| 0 | 0 | 0 | 0.003063513 | 0 |
| 0 | 0 | 0 | 0 | 0 |
| 0 | 0 | 0 | 0 | 0 |
| 0 | 0 | 0 | 0 | 0 |
| 29.13456464 | 0 | 0 | 0 | 0.15628316 |
| 0 | 0 | 0 | 0 | 0 |

| | | | | |
|---|---|---|---|---|
| 0 | 0 | 0 | 0 | 0 |
| 0 | 0 | 0 | 0 | 0 |
| 0 | 0 | 0 | 0 | 0 |
| 0 | 0 | 0 | 0 | 0 |
| 0 | 0.006652603 | 1.155154068 | 0.04949577 | 0.144468906 |
| 0 | 0 | 0 | 0 | 0 |
| 0 | 0 | 0 | 0 | 0 |
| 0 | 0 | 0 | 0 | 0 |
| 0 | 0 | 0 | 0 | 0 |
| 0 | 0 | 0 | 0 | 0 |
| 0 | 0 | 0.04784936 | 0 | 0 |
| 0.007154584 | 0.031638801 | 0.040516691 | 0.000792896 | 0.000992941 |
| 0.002893408 | 0 | 0.015741638 | 0 | 0 |
| 0 | 0 | 0 | 0 | 0 |
| 0 | 0.000898546 | 0 | 0 | 0.001135035 |
| 0 | 0 | 0 | 0 | 0.561161667 |
| 0 | 0 | 0 | 0.023783421 | 0 |
| 0 | 0.002057746 | 0 | 0 | 0 |
| 0.033690026 | 0.027677125 | 0.918352365 | 0 | 0.165409311 |
| 0 | 0 | 0 | 0 | 0.002945102 |
| 0 | 0 | 0 | 0 | 0 |
| 0 | 0 | 0 | 0 | 0 |
| 0 | 0 | 0 | 0 | 0 |
| 0 | 0 | 0 | 0 | 0 |
| 0 | 0 | 0 | 0 | 0 |
| 0 | 0 | 0 | 0 | 0 |
| 0.013462437 | 0.136543003 | 0.007653062 | 0.002303369 | 0.001770034 |
| 0 | 0 | 0 | 0 | 0 |
| 0 | 0 | 0 | 0 | 0 |
| 0 | 0 | 0 | 0 | 0 |
| 0 | 0.004941622 | 0.00722929 | 0 | 0 |
| 0 | 0 | 0 | 0 | 0 |
| 0 | 0 | 0 | 0 | 0 |
| 0 | 0 | 0 | 0 | 0 |
| 0 | 0 | 0 | 0 | 0 |
| 0 | 0 | 0 | 0 | 0 |
| 0 | 0 | 0 | 0 | 0 |
| 0.078013017 | 0.019963082 | 0.032506171 | 0.004502632 | 0.001879543 |
| 0 | 0 | 0 | 0.02667755 | 0 |
| 0 | 0.075134476 | 0 | 0 | 0 |
| 0 | 0.056075183 | 0.001183133 | 0 | 0 |
| 0 | 0 | 0 | 0.000641348 | 0 |
| 0 | 0 | 0 | 0 | 0 |
| 0 | 0 | 0.014059071 | 0.003214771 | 0.001161301 |
| 0 | 0 | 0 | 0 | 0 |
| 0 | 0 | 0 | 0 | 0 |
| 0 | 0 | 0 | 0 | 0 |
| 0 | 0 | 0 | 0 | 0 |

| | | | | |
|---|---|---|---|---|
| 0 | 0 | 0 | 0 | 0 |
| 0 | 0 | 0 | 0 | 0 |
| 0 | 0 | 0 | 0 | 0 |
| 0.000876938 | 0 | 0.002894173 | 0 | 0 |
| 0 | 0 | 0 | 0.001922942 | 0 |
| 0.002099403 | 0 | 0 | 0 | 0 |
| 0 | 0.017951538 | 0 | 0 | 0 |
| 0 | 0 | 0 | 0 | 0.000951838 |
| 0 | 0 | 0 | 0 | 0 |
| 0 | 0 | 0 | 0 | 0 |
| 0 | 0 | 0 | 0 | 0 |
| 0 | 0 | 0 | 0 | 0 |
| 0 | 0.000682966 | 0.030400404 | 0.0630121 | 0 |
| 0 | 0 | 0 | 0 | 0 |
| 0 | 0 | 0 | 0 | 0 |
| 0 | 0 | 0 | 0 | 0 |
| 0 | 0 | 0 | 0 | 0 |
| 0 | 0 | 0 | 0 | 0 |
| 0 | 0 | 0 | 0 | 0 |
| 0 | 0 | 0 | 0 | 0 |
| 0 | 0 | 0 | 0 | 0 |
| 0 | 0 | 0 | 0 | 0 |
| 0.041336172 | 0.005935436 | 0 | 0 | 0 |
| 0 | 0.030775231 | 0 | 0.081639496 | 0 |
| 0 | 0 | 0 | 0 | 0 |
| 0 | 0 | 0 | 0.105404812 | 0 |
| 0 | 0 | 0 | 0 | 0 |
| 0 | 0 | 0 | 0 | 0 |
| 0 | 0 | 0 | 0 | 0 |
| 0 | 0 | 0 | 0 | 0 |
| 0 | 0.022260493 | 0.029299382 | 0 | 0 |
| 0 | 0.010235455 | 0 | 0 | 0 |
| 0 | 0 | 0 | 0 | 0 |
| 0 | 0 | 0 | 0 | 0 |
| 0 | 0 | 0 | 0.001356762 | 0 |
| 0 | 0 | 0 | 0 | 0 |
| 0 | 0 | 0.011636803 | 0 | 0 |
| 0.043633782 | 0 | 0 | 0 | 0.015298524 |
| 0 | 0 | 0 | 0 | 0.001500324 |
| 0 | 0 | 0 | 0 | 0 |
| 0 | 0 | 0 | 0 | 0 |
| 0 | 0 | 0 | 0 | 0 |
| 0 | 0 | 0 | 0 | 0 |
| 0 | 0 | 0 | 0 | 0 |
| 0 | 0 | 0 | 0 | 0 |
| 0 | 0.04702507 | 0.084388276 | 0.777419094 | 0.011880335 |
| 0 | 0.019011571 | 0 | 0 | 0 |
| 0 | 0 | 0 | 0 | 0 |

| | | | | |
|---|---|---|---|---|
| 0 | 0 | 0 | 0 | 0 |
| 0 | 0 | 0 | 0 | 0 |
| 0 | 1.478881784 | 0 | 0 | 0 |
| 0 | 0 | 0 | 0 | 0 |
| 0 | 0 | 0 | 0 | 0 |
| 0 | 0 | 0 | 0 | 0 |
| 0 | 0.001552245 | 0 | 0 | 0 |
| 0 | 0 | 0 | 0 | 0 |
| 0 | 0 | 0.012004256 | 0 | 0.002181455 |
| 0 | 0 | 0 | 0 | 0 |
| 0 | 0 | 0.001315658 | 0 | 0 |
| 0.001870566 | 0 | 0 | 0 | 0 |
| 0 | 0 | 0 | 0 | 0 |
| 0 | 0 | 0 | 0 | 0 |
| 0 | 0 | 0 | 0 | 0 |
| 0 | 0 | 0 | 0 | 0 |
| 0 | 0 | 0 | 0 | 0 |
| 0 | 0 | 0 | 0 | 0 |
| 0 | 0 | 0 | 0 | 0 |
| 0 | 0.0010953 | 0 | 0.001964142 | 0.00368953 |
| 0 | 0 | 0 | 0 | 0 |
| 0 | 0 | 0 | 0 | 0 |
| 0 | 0 | 0 | 0 | 0 |
| 0 | 0 | 0 | 0 | 0 |
| 0 | 0 | 0 | 0 | 0 |
| 0 | 0 | 0 | 0 | 0 |
| 0 | 0 | 0 | 0 | 0 |
| 0 | 0 | 0 | 0.006792671 | 0 |
| 0 | 0 | 0 | 0 | 0 |
| 0 | 0 | 0 | 0 | 0 |
| 0 | 0.000470488 | 0.009850268 | 0.006116828 | 0.005150743 |
| 0 | 0 | 0 | 0 | 0 |
| 0 | 0.120728876 | 0.043510949 | 0.010530938 | 0.01012091 |
| 0.001905396 | 0 | 0 | 0 | 0.006816629 |
| 0 | 0 | 0 | 0 | 0 |
| 0 | 0 | 0 | 0 | 0 |
| 0 | 0 | 0 | 0 | 0 |
| 0 | 0 | 0 | 0 | 0 |
| 0 | 0 | 0 | 0 | 0 |
| 0 | 0 | 0 | 0 | 0 |
| 0 | 0 | 0 | 0 | 0 |
| 0 | 0.086439083 | 0 | 0 | 0.044659678 |
| 0 | 0.012045535 | 0.111640827 | 0.061610798 | 0.040344975 |
| 0 | 0 | 0 | 0 | 0 |
| 0 | 0 | 0 | 0 | 0 |
| 0.002700559 | 0.005802212 | 0 | 0.080518877 | 0 |
| 0 | 0.032334806 | 0 | 0 | 0 |
| 0 | 0 | 0 | 0 | 0 |

| | | | | |
|---|---|---|---|---|
| 0 | 0 | 0 | 0 | 0 |
| 0 | 0.311275948 | 0 | 0 | 0 |
| 0.007114945 | 0.000893374 | 0.015247764 | 0.012916436 | 0.033572948 |
| 0 | 0 | 0 | 0 | 0 |
| 0 | 0 | 0 | 0 | 0 |
| 0 | 0.001388675 | 0.202172738 | 0 | 0 |
| 0 | 0 | 0 | 0 | 0 |
| 0 | 0 | 0 | 0 | 0 |
| 0 | 0 | 0 | 0 | 0 |
| 0 | 0 | 0 | 0 | 0 |
| 0 | 0 | 0 | 0 | 0 |
| 0 | 0 | 0 | 0 | 0 |
| 0 | 0 | 0 | 0 | 0 |
| 0 | 0 | 0 | 0 | 0 |
| 0 | 0 | 0 | 0 | 0 |
| 0 | 0 | 0 | 0 | 0 |
| 0 | 0 | 0 | 0 | 0 |
| 0 | 0 | 0 | 0 | 0 |
| 0 | 0 | 0 | 0 | 0 |
| 0 | 0 | 0 | 0 | 0 |
| 0 | 0 | 0 | 0 | 0 |
| 0 | 0 | 0 | 0 | 0 |
| 0 | 0 | 0 | 0 | 0 |
| 0 | 0 | 0 | 0 | 0 |
| 0 | 0.000920332 | 0 | 0.0017329 | 0 |
| 0 | 0 | 0.024524319 | 0 | 0.008420427 |
| 0 | 0 | 0 | 0 | 0 |
| 0 | 0 | 0 | 0.008213525 | 0 |
| 0 | 0 | 0 | 0 | 0 |
| 0 | 0 | 0 | 0 | 0 |
| 0 | 0 | 0 | 0 | 0 |
| 0 | 0 | 0 | 0 | 0 |
| 0 | 0 | 0 | 0 | 0 |
| 0 | 0 | 0 | 0 | 0 |
| 0 | 0 | 0 | 0 | 0 |
| 0 | 0 | 0 | 0 | 0 |
| 0 | 0.123015872 | 0.00635655 | 0 | 0 |
| 0.015098704 | 0 | 0 | 0 | 0 |
| 0 | 0.479132323 | 0 | 0.180206267 | 0.001274181 |
| 0 | 0 | 0 | 0 | 0 |
| 0 | 0 | 0 | 0 | 0 |
| 0.001371266 | 0.006628943 | 0 | 0.02998847 | 0 |
| 0.004651466 | 0.000743339 | 0.009388399 | 0 | 0 |
| 0 | 0 | 0 | 0 | 0 |
| 0 | 0 | 0 | 0 | 0 |
| 0 | 0 | 0 | 0 | 0 |
| 0.00323447 | 0.006538689 | 0.006210774 | 0.0080294 | 0 |
| 0 | 0 | 0 | 0 | 0 |

| | | | | |
|---|---|---|---|---|
| 0 | 0 | 0 | 0 | 0 |
| 0.047343145 | 0.100801084 | 0.017632921 | 0.004583354 | 0.003171949 |
| 0 | 0 | 0 | 0 | 0 |
| 0 | 0 | 0 | 0 | 0 |
| 0 | 0 | 0 | 0 | 0 |
| 0 | 0 | 0 | 0 | 0 |
| 0 | 0 | 0 | 0 | 0 |
| 0 | 0 | 0 | 0 | 0 |
| 0 | 0 | 0 | 0 | 0 |
| 0 | 0 | 0 | 0 | 0 |
| 0 | 0 | 0 | 0 | 0 |
| 0 | 0 | 0 | 0.026140966 | 0 |
| 0 | 0 | 0 | 0.062220152 | 0.002874027 |
| 0 | 0 | 0 | 0 | 0 |
| 0.012213055 | 0.014760015 | 0 | 0.003910094 | 0 |
| 0 | 0 | 0 | 0 | 0 |
| 0 | 0 | 0 | 0 | 0 |
| 0 | 0 | 0 | 0 | 0 |
| 0 | 0 | 0 | 0 | 0 |
| 0 | 0 | 0 | 0 | 0 |
| 0 | 0 | 0 | 0 | 0 |
| 0 | 0 | 0.004798131 | 0 | 0.008322997 |
| 0 | 0.009282171 | 0 | 0.010668066 | 0 |
| 0 | 0 | 0 | 0 | 0 |
| 0 | 0 | 0 | 0 | 0 |
| 0 | 0 | 0 | 0 | 0 |
| 0 | 0 | 0 | 0 | 0 |
| 0 | 0 | 0 | 0 | 0 |
| 0 | 0 | 0 | 0 | 0 |
| 0 | 0 | 0 | 0 | 0 |
| 0 | 0 | 0 | 0 | 0 |
| 0 | 0 | 0 | 0 | 0 |
| 0 | 0 | 0 | 0 | 0 |
| 0 | 0 | 0 | 0 | 0 |
| 0 | 0.02552457 | 0.001361386 | 20.53643675 | 0 |
| 0 | 0 | 0 | 0 | 0 |
| 0 | 0 | 0 | 0 | 0 |
| 0 | 0 | 0 | 0 | 0 |
| 0 | 0 | 0 | 0 | 0 |
| 0 | 0 | 0 | 0 | 0 |
| 0 | 0.033431296 | 0.31657818 | 0.073263055 | 0.190284016 |
| 0 | 0 | 0 | 0 | 0 |
| 0 | 0 | 0 | 0 | 0 |
| 0 | 0 | 0 | 0 | 0 |
| 0 | 0 | 0 | 0 | 0 |
| 2.02859122 | 0.208044356 | 1.202975253 | 0.036035611 | 2.07907707 |
| 0 | 0 | 0 | 0 | 0 |
| 0 | 0 | 0 | 0 | 0 |

| | | | | |
|---|---|---|---|---|
| 0 | 0 | 0 | 0 | 0 |
| 0 | 0 | 0 | 0 | 0 |
| 0 | 0 | 0 | 0 | 0 |
| 0 | 0 | 0 | 0.019588162 | 0 |
| 0 | 0.004610627 | 0 | 0 | 0 |
| 0 | 0 | 0 | 0 | 0 |
| 0 | 0 | 0 | 0 | 0 |
| 0 | 0 | 0 | 0 | 0 |
| 0 | 0 | 0 | 0 | 0 |
| 0 | 0.003303047 | 0 | 0 | 0.544078567 |
| 0 | 0 | 0 | 0 | 0 |
| 0 | 0 | 0 | 0 | 0 |
| 0.002203868 | 0 | 0.000881631 | 0 | 0 |
| 0 | 0 | 0 | 0 | 0 |
| 0 | 0 | 0 | 0 | 0 |
| 0 | 0 | 0 | 0 | 0 |
| 0 | 0 | 0 | 0 | 0 |
| 0 | 0 | 0 | 0 | 0 |
| 0 | 0 | 0 | 0 | 0 |
| 0 | 0.224328378 | 0.206641379 | 0.207149709 | 0.124140626 |
| 0 | 0 | 0 | 0 | 0 |
| 0 | 0 | 0 | 0 | 0 |
| 0 | 0 | 0 | 0 | 0 |
| 0 | 0 | 0 | 0 | 0 |
| 0 | 0 | 0 | 0 | 0 |
| 0 | 0 | 0 | 0 | 0 |
| 0 | 0 | 0 | 0 | 0 |
| 0 | 0 | 0 | 0 | 0 |
| 0 | 0 | 0 | 0 | 0 |
| 0 | 0.005938949 | 0.007829473 | 0 | 0 |
| 0 | 0 | 0 | 0 | 0 |
| 0 | 0 | 0 | 0 | 0 |
| 0 | 0 | 0 | 0 | 0 |
| 0 | 0 | 0 | 0 | 0 |
| 0 | 0 | 0 | 0 | 0 |
| 0 | 0 | 0 | 0 | 0 |
| 0 | 0 | 0 | 0 | 0 |
| 0 | 0 | 0 | 0 | 0 |
| 0 | 0 | 0 | 0 | 0 |
| 0 | 0 | 0 | 0 | 0 |
| 0 | 0 | 0 | 0 | 0 |
| 0 | 0 | 0 | 0 | 0 |
| 0 | 0 | 0 | 0 | 0 |
| 0 | 0 | 0 | 0 | 0 |
| 0.004298945 | 0.005757744 | 0.006633291 | 0.008469766 | 0 |
| 0 | 0 | 0 | 0 | 0 |
| 0 | 0 | 0 | 0 | 0 |
| 0.021311982 | 0 | 0 | 0 | 0 |

| | | | | |
|---|---|---|---|---|
| 0 | 0 | 0 | 0 | 0 |
| 0 | 0 | 0 | 0 | 0 |
| 0 | 0 | 0 | 0 | 0 |
| 0 | 0 | 0 | 0 | 0 |
| 0 | 0 | 0 | 0 | 0 |
| 0.005043233 | 0.001746217 | 0.005318825 | 0.012284704 | 0.000848387 |
| 0 | 0 | 0 | 0 | 0 |
| 0 | 0 | 0 | 0 | 0 |
| 0 | 0 | 0 | 0 | 0 |
| 0 | 0 | 0 | 0 | 0 |
| 0 | 0 | 0 | 0 | 0 |
| 0 | 0 | 0 | 0 | 0 |
| 0 | 0 | 0 | 0 | 0 |
| 0 | 0 | 0 | 0 | 0 |
| 0 | 0 | 0 | 0 | 0 |
| 0 | 0 | 0 | 0 | 0 |
| 0 | 0 | 0 | 0 | 0 |
| 0 | 0 | 0 | 0 | 0 |
| 0 | 0 | 0 | 0 | 0 |
| 0 | 0 | 0 | 0 | 0 |
| 0 | 0 | 0 | 0 | 0 |
| 0 | 0 | 0 | 0 | 0 |
| 0 | 0 | 0 | 0 | 0 |
| 0 | 0 | 0 | 0 | 0 |
| 0 | 0 | 0 | 0 | 0 |
| 0 | 0 | 0.001147522 | 0 | 0.004246464 |
| 0 | 0 | 0 | 0 | 0 |
| 0 | 0 | 0 | 0 | 0 |
| 0 | 0 | 0 | 0 | 0 |
| 0 | 0 | 0 | 0 | 0 |
| 0 | 0 | 0 | 0 | 0 |
| 0 | 0 | 0 | 0 | 0 |
| 0 | 0 | 0 | 0 | 0 |
| 0 | 0 | 0 | 0 | 0 |
| 0 | 0 | 0 | 0 | 0 |
| 0 | 0 | 0 | 0 | 0 |
| 0 | 0 | 0 | 0 | 0 |
| 0 | 0 | 0 | 0 | 0 |
| 0 | 0.882828337 | 0 | 0 | 0 |
| 0 | 0 | 0 | 0 | 0 |
| 0 | 0 | 0 | 0 | 0 |
| 0 | 0 | 0 | 0 | 0 |
| 0 | 0 | 0 | 0 | 0 |
| 0 | 0 | 0 | 0 | 0 |
| 0 | 0 | 0 | 0 | 0 |
| 0 | 0 | 0 | 0 | 0 |
| 0 | 0 | 0 | 0 | 0 |
| 0 | 0 | 0 | 0 | 0 |

| | | | | |
|---|---|---|---|---|
| 0 | 0 | 0 | 0 | 0 |
| 0 | 0 | 0 | 0.006472337 | 0 |
| 0 | 0 | 0 | 0 | 0 |
| 0 | 0 | 0 | 0 | 0 |
| 0 | 0 | 0 | 0 | 0 |
| 0 | 0 | 0 | 0 | 0 |
| 0 | 0 | 0.002189876 | 0 | 0 |
| 0 | 0 | 0 | 0 | 0 |
| 0 | 0 | 0 | 0 | 0 |
| 0 | 0 | 0 | 0.00741478 | 0 |
| 0 | 0 | 0 | 0.101435305 | 0 |
| 0 | 0 | 0 | 0 | 0 |
| 0 | 0 | 0 | 0.544443442 | 0 |
| 0 | 0 | 0 | 0 | 0 |
| 0 | 0 | 0 | 0 | 0 |
| 0 | 0 | 0 | 0 | 0 |
| 0 | 0 | 0 | 0 | 0 |
| 0 | 0 | 0 | 0 | 0 |
| 0 | 0 | 0 | 0 | 0 |
| 0 | 0 | 0 | 0 | 0 |
| 0.221931909 | 0 | 0.001364466 | 0.001075203 | 0 |
| 0 | 0.009361634 | 0 | 0 | 0 |
| 0 | 0 | 0 | 0 | 0 |
| 0 | 0.519047516 | 2.260419361 | 1.963321267 | 5.629234437 |
| 0 | 0 | 0 | 0 | 0 |
| 0 | 0 | 0 | 0 | 0 |
| 0 | 0 | 0 | 0 | 0 |
| 0 | 0 | 0 | 0 | 0 |
| 0 | 0 | 0 | 0 | 0 |
| 0 | 0 | 0 | 0 | 0 |
| 0 | 0 | 0 | 0 | 0 |
| 0 | 0 | 0 | 0 | 0 |
| 0 | 0 | 0 | 0 | 0 |
| 0 | 0 | 0 | 0 | 0 |

| PHL018-0270_V1 | PHL018-0158_V3 | PHL018-0016_V3 | PHL018-0134_V1 | PHL018-0198_V3 |
|---|---|---|---|---|
| REF | EYCF | CTRL | CTRL | REF |
| 0.031968675 | 0.0392186 | 0 | 0 | 0.605402526 |
| 8.508601027 | 3.451739788 | 7.429773314 | 0.046895345 | 1.092752581 |
| 0 | 10.22001011 | 8.71572019 | 3.352857566 | 1.15427367 |
| 0 | 0 | 0 | 0 | 0 |
| 0 | 0 | 0 | 0 | 0 |
| 0 | 0 | 0 | 0 | 0 |
| 0 | 0 | 0 | 0 | 0 |
| 0 | 0 | 0 | 0 | 0 |
| 0 | 0 | 0 | 0 | 0 |
| 0 | 0 | 0 | 0 | 0 |
| 0 | 0 | 0 | 0 | 0 |
| 0 | 0 | 0 | 0 | 0 |
| 0 | 0 | 0 | 0 | 0 |
| 1.458354098 | 19.51349576 | 4.398047625 | 3.637967257 | 0.00941658 |
| 0 | 0 | 0 | 0 | 0 |
| 0 | 0 | 0 | 0 | 0 |
| 0 | 0 | 0 | 0 | 0 |
| 0 | 0 | 0 | 0 | 0 |
| 0 | 0 | 0 | 0 | 0 |
| 0 | 0 | 0 | 0 | 0 |
| 0 | 0 | 0 | 0 | 0 |
| 0 | 0 | 0 | 0 | 0 |
| 0 | 0 | 0 | 0 | 0 |
| 0 | 0 | 0 | 0 | 0 |
| 0 | 0 | 0 | 0 | 0 |
| 0 | 0 | 0 | 0 | 0 |
| 0 | 0 | 0 | 0 | 0 |
| 0 | 0 | 0 | 0 | 0 |
| 0 | 0 | 0 | 0 | 0 |
| 0 | 0 | 0 | 0 | 0 |
| 0 | 0 | 0 | 0 | 0 |
| 0 | 0 | 0 | 0 | 0 |
| 0 | 0 | 0 | 0 | 0 |
| 0 | 0 | 0 | 0 | 0 |
| 0 | 0 | 0 | 0 | 0 |
| 5.644847417 | 0 | 0.701486214 | 0.001621034 | 0 |
| 0 | 0 | 0 | 0 | 0 |
| 0 | 0 | 0 | 0 | 0 |
| 0 | 0 | 0 | 0 | 0 |
| 0 | 0 | 0 | 0 | 0 |
| 0 | 0 | 0 | 0 | 0 |
| 0.001198043 | 0 | 0 | 0.028345002 | 0.099834522 |
| 0.001136329 | 0.001225055 | 0.23983782 | 10.79444919 | 0.138957409 |
| 0.024310596 | 0 | 0.217858046 | 2.56493886 | 0.132343676 |
| 0 | 0 | 0.049315961 | 0.005998622 | 0.045219087 |
| 0 | 0 | 0 | 0 | 0 |

| | | | | |
|---|---|---|---|---|
| 0 | 0 | 0 | 0 | 0 |
| 0 | 0 | 0 | 0 | 0 |
| 0 | 0.002063734 | 0 | 0 | 0.010902458 |
| 0.578500007 | 0.386124027 | 0.427646667 | 0.020349677 | 0.178364693 |
| 0.001580396 | 0.00126207 | 0 | 0 | 0 |
| 0 | 0 | 0 | 0 | 0 |
| 0.00135354 | 0 | 0.001614029 | 0.004744291 | 0.001358943 |
| 0 | 0 | 0 | 0 | 0 |
| 0 | 0 | 0 | 0 | 0 |
| 0 | 0 | 0 | 0 | 0 |
| 0 | 0 | 0 | 0.000544886 | 0 |
| 0.0453733 | 0 | 0 | 0 | 0.339692204 |
| 0 | 0 | 0 | 0 | 0 |
| 0.078760705 | 0.040757012 | 0 | 0 | 0.130023828 |
| 0 | 0 | 0 | 0.000776948 | 0 |
| 0 | 0 | 0 | 0 | 0 |
| 0 | 0 | 0.111558596 | 7.270210697 | 0.01601038 |
| 0 | 0 | 0 | 0 | 0 |
| 0 | 0 | 0 | 0 | 0 |
| 0 | 0 | 0 | 0 | 0 |
| 0.216131028 | 0.121472748 | 0 | 0 | 0.02879539 |
| 0.060348838 | 0.047694085 | 0 | 0 | 0.017187213 |
| 0 | 0 | 0 | 0 | 0 |
| 0 | 0 | 0 | 0 | 0 |
| 0 | 0 | 0 | 0 | 0 |
| 0 | 0 | 0 | 0 | 0 |
| 0 | 0 | 0 | 0.001509398 | 0.001152929 |
| 0 | 0 | 0 | 0 | 0 |
| 0 | 0 | 0 | 0 | 0.003952794 |
| 0 | 0 | 0 | 0 | 0 |
| 0.007900697 | 0.150575385 | 0 | 0 | 0.025164337 |
| 0 | 0 | 0 | 0 | 0 |
| 0 | 0 | 0 | 0 | 0 |
| 0 | 0 | 0 | 0 | 0 |
| 0 | 0 | 0 | 0 | 0 |
| 0 | 0 | 0 | 0 | 0 |
| 0 | 0 | 0 | 0 | 0 |
| 0 | 0 | 0.003893793 | 9.831336854 | 0.00725933 |
| 0 | 0 | 0 | 0 | 0 |
| 0 | 0 | 0 | 0 | 0 |
| 0 | 0 | 0 | 0 | 0 |
| 0 | 0 | 0 | 0 | 0 |
| 0 | 0 | 0 | 0 | 0 |
| 0 | 0 | 0 | 0 | 0 |
| 0 | 0 | 0 | 0 | 0 |
| 0 | 0 | 0 | 0 | 0 |
| 0 | 0 | 0 | 0.000868265 | 0 |
| 0 | 0 | 0 | 0 | 0 |

| | | | | |
|---|---|---|---|---|
| 0 | 0 | 0 | 0 | 0 |
| 0 | 0 | 0 | 0 | 0 |
| 0 | 0 | 0 | 0 | 0 |
| 0 | 0 | 0 | 0 | 0 |
| 0 | 0 | 0 | 0 | 0 |
| 0 | 0 | 0 | 0 | 0 |
| 0 | 0 | 0 | 0 | 0 |
| 0 | 0 | 0 | 0 | 0 |
| 0 | 0 | 0 | 0 | 0 |
| 0 | 0 | 0 | 0 | 0 |
| 0 | 0 | 0 | 0 | 0 |
| 0 | 0 | 0.001286937 | 0 | 0.057861356 |
| 0 | 0 | 0 | 0 | 0 |
| 0 | 0 | 0 | 0 | 0 |
| 0 | 0 | 0 | 0 | 0 |
| 0 | 0 | 0 | 0 | 0 |
| 0 | 0 | 0 | 0 | 0 |
| 0 | 0 | 0 | 0 | 0 |
| 0 | 0 | 0 | 0 | 0.269812611 |
| 0 | 0 | 0 | 0 | 0 |
| 0 | 0 | 0 | 0 | 0 |
| 0 | 0 | 0 | 0 | 0 |
| 0 | 0 | 0 | 0 | 0 |
| 0 | 0 | 0 | 0 | 0 |
| 0 | 0 | 0 | 0 | 0 |
| 0 | 0 | 0 | 0 | 0 |
| 0 | 0 | 0 | 0 | 0 |
| 0 | 0 | 0 | 0 | 0 |
| 0 | 0 | 0 | 0 | 0 |
| 0 | 0 | 0 | 0 | 0 |
| 0.028387164 | 0 | 0 | 0 | 1.051408841 |
| 0 | 0 | 0 | 0 | 0 |
| 0 | 0 | 0 | 0 | 0 |
| 0 | 0 | 0 | 0 | 0 |
| 0 | 0 | 0 | 0 | 0 |
| 0 | 0 | 0 | 0 | 0 |
| 0 | 0 | 0 | 0 | 0 |
| 0 | 0 | 0 | 0 | 0 |
| 0 | 0 | 0 | 0 | 0 |
| 0 | 0 | 0 | 0 | 0 |
| 0 | 0 | 0 | 0 | 0 |
| 0 | 0 | 0 | 0 | 0 |
| 0.000782311 | 0 | 0 | 0 | 0.000628347 |
| 0 | 0 | 0 | 0 | 0 |
| 0 | 0 | 0 | 0 | 0 |
| 0 | 0 | 0 | 0 | 0 |
| 0 | 0 | 0 | 0 | 0 |
| 0 | 0 | 0 | 0 | 0 |

| | | | | |
|---|---|---|---|---|
| 0 | 0 | 0 | 0 | 0 |
| 0 | 0 | 0 | 0 | 0 |
| 0 | 0 | 0 | 0 | 0 |
| 0 | 0 | 0 | 0 | 0 |
| 0 | 0 | 0 | 0 | 0 |
| 0 | 0 | 0 | 0 | 0 |
| 0 | 0 | 0 | 0 | 0 |
| 0 | 0 | 0 | 0 | 0 |
| 0 | 0 | 0 | 0 | 0 |
| 0 | 0 | 0 | 0 | 0 |
| 0 | 0 | 0 | 0 | 0 |
| 0 | 0 | 0 | 0 | 0 |
| 0 | 0 | 0 | 0 | 0 |
| 0 | 0 | 0 | 0 | 0 |
| 0 | 0 | 0 | 0 | 0 |
| 0 | 0 | 0 | 0 | 0 |
| 0 | 0 | 0 | 0 | 0 |
| 0 | 0 | 0 | 0 | 0 |
| 0 | 0 | 0 | 0 | 0 |
| 0 | 0 | 0 | 0 | 0 |
| 0 | 0 | 0 | 0 | 0 |
| 0 | 0 | 0 | 0 | 0 |
| 0 | 0 | 0 | 0 | 0 |
| 0 | 0 | 0 | 0 | 0 |
| 0 | 0 | 0 | 0 | 0 |
| 0 | 0 | 0 | 0 | 0 |
| 0 | 0 | 0 | 0 | 0 |
| 0 | 0 | 0 | 0 | 0 |
| 0 | 0 | 0 | 0 | 0 |
| 0 | 0 | 0 | 0 | 0 |
| 0.008575649 | 0 | 0 | 0 | 0 |
| 0 | 0 | 0.00322818 | 0 | 0.030106963 |
| 0 | 0 | 0 | 0 | 0 |
| 0 | 0 | 0 | 0 | 0 |
| 0 | 0 | 0 | 0 | 0 |
| 0 | 0 | 0 | 0 | 0 |
| 0 | 0 | 0 | 0 | 0 |
| 0 | 0 | 0 | 0 | 0 |
| 0 | 0 | 0 | 0 | 0 |
| 0 | 0 | 0 | 0 | 0 |
| 0 | 0 | 0 | 0 | 0 |
| 0 | 0 | 0 | 0 | 0 |
| 0 | 0 | 0 | 0 | 0 |
| 0 | 0 | 0 | 0 | 0 |
| 0 | 0 | 0 | 0 | 0 |
| 0 | 0 | 0 | 0 | 0 |
| 0 | 0 | 0 | 0 | 0 |
| 0 | 0 | 0 | 0 | 0 |

| | | | | |
|---|---|---|---|---|
| 0.003327958 | 0 | 0 | 0 | 0 |
| 0 | 0 | 0 | 0 | 0 |
| 0 | 0 | 0 | 0 | 0 |
| 0 | 0 | 0 | 0 | 0 |
| 0 | 0 | 0 | 0 | 0 |
| 0 | 0 | 0 | 0 | 0 |
| 0 | 0 | 0 | 0 | 0 |
| 0 | 0 | 0 | 0 | 0 |
| 0 | 0 | 0 | 0 | 0 |
| 0 | 0 | 0 | 0 | 0 |
| 0 | 0 | 0 | 0 | 0 |
| 0 | 0 | 0 | 0 | 0 |
| 0.004732659 | 0.001432194 | 0 | 0 | 0.00820268 |
| 0 | 0 | 0 | 0 | 0 |
| 0 | 0 | 0 | 0 | 0.077938304 |
| 0 | 0 | 0 | 0 | 0 |
| 0 | 0 | 0 | 0 | 0 |
| 0 | 0 | 0 | 0 | 0 |
| 0 | 0 | 0 | 0 | 0 |
| 0 | 0 | 0 | 0 | 0 |
| 0.01071187 | 0.015732421 | 0.034654711 | 0.054958883 | 0.019451856 |
| 0 | 0 | 0 | 0 | 0 |
| 0 | 0 | 0 | 0 | 0 |
| 0.001456999 | 0 | 0 | 0 | 0 |
| 0 | 0 | 0 | 0 | 0 |
| 0 | 0 | 0 | 0 | 0 |
| 0.00244109 | 0.000974701 | 0.00879732 | 0 | 0.004574891 |
| 0 | 0 | 0 | 0 | 0 |
| 0 | 0 | 0 | 0 | 0 |
| 0 | 0 | 0 | 0 | 0 |
| 0 | 0 | 0 | 0 | 0 |
| 0 | 0 | 0 | 0 | 0 |
| 0 | 0 | 0 | 0 | 0 |
| 0 | 0 | 0 | 0 | 0 |
| 0.047272838 | 0.023086239 | 0 | 0 | 0.004867852 |
| 0 | 0 | 0 | 0 | 0 |
| 0 | 0 | 0 | 0 | 0 |
| 0 | 0 | 0 | 0 | 0 |
| 0 | 0 | 0 | 0 | 0 |
| 0 | 0 | 0 | 0 | 0 |
| 0 | 0 | 0 | 0 | 0 |
| 0 | 0 | 0 | 0 | 0 |
| 0 | 0 | 0 | 0 | 0 |
| 0 | 0 | 0 | 0 | 0 |
| 0 | 0 | 0 | 0 | 0 |
| 0 | 0 | 0 | 0 | 0 |
| 0.004593428 | 0 | 0 | 0 | 0.002108236 |
| 0.001511637 | 0.001086445 | 0 | 0 | 0.022721705 |

| | | | | |
|---|---|---|---|---|
| 0 | 0 | 0 | 0 | 0 |
| 0 | 0 | 0 | 0 | 0 |
| 0 | 0.001701892 | 0 | 0 | 0 |
| 0 | 0 | 0 | 0 | 0 |
| 0 | 0 | 0 | 0 | 0 |
| 0 | 0 | 0 | 0 | 0 |
| 0 | 0 | 0 | 0.005207749 | 0.024443221 |
| 0 | 0 | 0.001480686 | 0 | 0.002493349 |
| 0 | 0 | 0 | 0 | 0 |
| 0 | 0 | 0 | 0 | 0 |
| 0 | 0 | 0 | 0 | 0 |
| 0 | 0 | 0 | 0 | 0 |
| 0 | 0 | 0 | 0 | 0 |
| 0 | 0 | 0 | 0 | 0 |
| 0 | 0 | 0 | 0 | 0 |
| 0 | 0 | 0 | 0 | 0 |
| 0 | 0 | 0 | 0 | 0 |
| 0 | 0 | 0.002233614 | 0.121393453 | 0 |
| 0 | 0 | 0.000838026 | 0.121368986 | 0 |
| 0 | 0 | 0 | 0 | 0 |
| 0.013282302 | 0.030268653 | 0 | 0 | 0.002081612 |
| 0 | 0 | 0 | 0 | 0 |
| 0 | 0 | 0 | 0 | 0 |
| 0 | 0 | 0 | 0 | 0 |
| 0.007079734 | 0 | 0.016577471 | 0.010527649 | 0.003101671 |
| 0 | 0 | 0 | 0 | 0 |
| 0 | 0 | 0 | 0 | 0 |
| 0 | 0 | 0 | 0 | 0 |
| 0 | 0 | 0 | 0 | 0.007399925 |
| 0 | 0 | 0 | 0 | 0 |
| 0.041686158 | 0.998975725 | 0 | 0 | 0.1197701 |
| 0 | 0 | 0 | 0 | 0 |
| 0 | 0.067908346 | 0 | 0 | 0 |
| 0 | 0 | 0 | 0 | 0 |
| 0 | 0 | 0 | 0 | 0 |
| 0 | 0 | 0 | 0 | 0 |
| 0 | 0 | 0 | 0 | 0 |
| 0 | 0 | 0 | 0 | 0 |
| 0 | 0 | 0 | 0 | 0 |
| 0 | 0 | 0.008398289 | 0.001806971 | 0 |
| 0 | 0 | 0 | 0 | 0 |
| 0 | 0.005758965 | 0.00231633 | 0 | 0.002145276 |
| 0 | 0 | 0 | 0 | 0 |
| 0 | 0 | 0 | 0 | 0 |
| 0 | 0 | 0 | 0 | 0 |
| 0 | 0 | 0 | 0 | 0 |
| 0 | 0 | 0 | 0 | 0 |
| 0 | 0 | 0 | 0 | 0 |

| | | | | |
|---|---|---|---|---|
| 0 | 0 | 0 | 0 | 0 |
| 0 | 0 | 0 | 0 | 0 |
| 0.008232043 | 0 | 0.196326019 | 2.649948839 | 0.155012891 |
| 0 | 0 | 0 | 0 | 0 |
| 0.932688122 | 0 | 0 | 0 | 0 |
| 0 | 0 | 0 | 0 | 0 |
| 0 | 0 | 0 | 0 | 0 |
| 0 | 0 | 0 | 0 | 0 |
| 0 | 0 | 0 | 0 | 0 |
| 0 | 0 | 0 | 0 | 0 |
| 0 | 0 | 0 | 0 | 0 |
| 0 | 0 | 0 | 0 | 0 |
| 0 | 0 | 0 | 0 | 0 |
| 0 | 0 | 0 | 0 | 0 |
| 0 | 0 | 0.007428713 | 0.001624704 | 0.00335071 |
| 0 | 0 | 0 | 0 | 0 |
| 0.004753316 | 0.002733046 | 0.006348264 | 0.014994761 | 0.001527133 |
| 0 | 0 | 0 | 0 | 0 |
| 0 | 0 | 0 | 0 | 0 |
| 0 | 0 | 0 | 0 | 0 |
| 0 | 0 | 0 | 0 | 0 |
| 0.004290888 | 0.00123358 | 0 | 0 | 0.025273702 |
| 0 | 0 | 0 | 0 | 0 |
| 0 | 0 | 0 | 0 | 0.017809093 |
| 0 | 0 | 0 | 0 | 0 |
| 0 | 0 | 0 | 0 | 0 |
| 0 | 0 | 0 | 0 | 0 |
| 0 | 0 | 0 | 0 | 0 |
| 0 | 0 | 0 | 0 | 0 |
| 0 | 0 | 0 | 0 | 0 |
| 0 | 0 | 0 | 0 | 0 |
| 0 | 0 | 0 | 0 | 0 |
| 0 | 0 | 0 | 0 | 0 |
| 0 | 0 | 0 | 0 | 0 |
| 0 | 0 | 0 | 0 | 0 |
| 0 | 0 | 0 | 0 | 0.004130233 |
| 0 | 0 | 0 | 0 | 0 |
| 0.001437615 | 0 | 0.056571384 | 0.126604464 | 0.048208026 |
| 0 | 0 | 0 | 0 | 0 |
| 0 | 0 | 0 | 0 | 0 |
| 0 | 0 | 0 | 0 | 0 |
| 0 | 0 | 0 | 0 | 0 |
| 0 | 0 | 0 | 0 | 0 |
| 0 | 0 | 0 | 0 | 0.002151446 |
| 0 | 0 | 0 | 0 | 0 |
| 0 | 0 | 0 | 0 | 0 |
| 0 | 0 | 0 | 0 | 0 |
| 0 | 0 | 0 | 0 | 0 |

| | | | | |
|---|---|---|---|---|
| 0.001721938 | 0.096944742 | 0 | 0 | 0 |
| 0 | 0 | 0 | 0 | 0 |
| 0.001498124 | 0 | 0.00114332 | 0.002625536 | 0 |
| 0 | 0 | 0 | 0 | 0 |
| 0.001205805 | 0.001516616 | 0.002300581 | 0.00246544 | 0 |
| 0 | 0 | 0 | 0 | 0 |
| 0 | 0 | 0 | 0 | 0 |
| 0 | 0 | 0.006852636 | 0.01184865 | 0.006787796 |
| 0 | 0 | 0 | 0 | 0 |
| 0.021776593 | 0.157138971 | 0.014541802 | 0.012721521 | 0.009445043 |
| 0 | 0 | 0 | 0 | 0.014548846 |
| 0 | 0 | 0 | 0.00644836 | 0.071732803 |
| 0.002537244 | 0.002552998 | 0.014038491 | 0.005039532 | 0.02078656 |
| 0 | 0.035173063 | 0 | 0 | 0 |
| 0 | 0 | 0 | 0 | 0 |
| 0 | 0 | 0 | 0 | 0 |
| 0 | 0 | 0 | 0 | 0 |
| 0 | 0 | 0 | 0 | 0 |
| 0 | 0 | 0 | 0 | 0.003950051 |
| 0 | 0 | 0 | 0 | 0 |
| 0 | 0 | 0 | 0 | 0 |
| 0 | 0 | 0 | 0 | 0 |
| 0 | 0 | 0 | 0.001376509 | 0 |
| 0 | 0 | 0 | 0 | 0 |
| 0 | 0 | 0 | 0 | 0 |
| 0 | 0 | 0.039468565 | 0.117050116 | 0.0045099 |
| 0 | 0 | 0 | 0 | 0 |
| 0 | 0 | 0 | 0 | 0 |
| 0 | 0.097642397 | 0 | 0 | 0 |
| 0 | 0 | 0 | 0 | 0 |
| 0 | 0 | 0 | 0 | 0 |
| 0 | 0 | 0 | 0 | 0 |
| 0 | 0 | 0 | 0 | 0 |
| 0 | 0 | 0 | 0 | 0 |
| 0 | 0 | 0 | 0 | 0 |
| 0 | 0 | 0 | 0 | 0 |
| 0 | 0 | 0 | 0 | 0 |
| 0 | 0 | 0 | 0 | 0 |
| 0 | 0 | 0 | 0 | 0 |
| 0.001554079 | 0 | 0 | 0 | 0 |
| 0 | 0 | 0 | 0 | 0 |
| 0 | 0 | 0 | 0 | 0 |
| 0 | 0 | 0 | 0 | 0 |
| 0.296415479 | 0 | 0 | 0 | 0.032126326 |
| 0 | 0 | 0 | 0 | 0 |
| 0 | 0 | 0 | 0 | 0 |
| 0 | 0 | 0 | 0 | 0 |
| 0 | 0 | 0 | 0 | 0 |

| | | | | |
|---|---|---|---|---|
| 0 | 0 | 0 | 0 | 0 |
| 0 | 0 | 0 | 0.008637593 | 0 |
| 0 | 0 | 0 | 0 | 0 |
| 0 | 0 | 0 | 0.244923856 | 0 |
| 0.003269186 | 0 | 0 | 0 | 0 |
| 0.077863889 | 0.017267209 | 0 | 0 | 0.115082493 |
| 0.014483704 | 0 | 0 | 0 | 0 |
| 0 | 0 | 0 | 0 | 0 |
| 0.005280003 | 0 | 0 | 0 | 0 |
| 0 | 0 | 0 | 0 | 0 |
| 0.011750242 | 0.022681243 | 0.027542759 | 0 | 0.020223684 |
| 0 | 0 | 0 | 0 | 0 |
| 0 | 0 | 0 | 0 | 0 |
| 0.00253738 | 0.003419377 | 0.002723129 | 0.037242658 | 0.001019004 |
| 0.003851059 | 0 | 0 | 0 | 0.005155243 |
| 0 | 0 | 0 | 0 | 0 |
| 0 | 0 | 0 | 0 | 0 |
| 0 | 0 | 0 | 0 | 0 |
| 0 | 0 | 0 | 0 | 0.004609434 |
| 0 | 0 | 0 | 0 | 0 |
| 0 | 0 | 0 | 0 | 0 |
| 0 | 0 | 0.978029927 | 0 | 0 |
| 0 | 0 | 0 | 0 | 0 |
| 0 | 0 | 0 | 0 | 0 |
| 0 | 0 | 0 | 0 | 0 |
| 0 | 0 | 0 | 0 | 0 |
| 0.02338513 | 0 | 0 | 0 | 0.005524349 |
| 0 | 0 | 0 | 0 | 0 |
| 0 | 0.000856 | 0 | 0 | 0.001530573 |
| 0.843343947 | 1.372310548 | 1.029816631 | 1.29902322 | 0.208363624 |
| 0 | 0 | 0 | 0 | 0 |
| 0 | 0 | 0 | 0 | 0 |
| 0 | 0 | 0 | 0 | 0 |
| 0 | 0 | 0 | 0 | 0 |
| 0 | 0 | 0 | 0 | 0.002300631 |
| 0 | 0 | 0 | 0 | 0 |
| 0 | 0 | 1.736143575 | 0 | 0 |
| 0 | 0 | 0 | 0 | 0 |
| 0 | 0 | 0 | 0 | 0 |
| 0 | 0 | 0 | 0 | 0 |
| 0.020077774 | 0 | 0 | 0 | 0.002303763 |
| 4.830704411 | 0 | 0 | 0.033930749 | 0 |
| 0 | 0 | 0 | 0 | 0 |
| 0 | 0 | 0 | 0 | 0 |
| 0 | 0.001268427 | 0 | 0 | 0.004252524 |
| 0 | 0.00560219 | 0 | 0 | 0 |
| 0.001094705 | 0 | 0 | 0.000479631 | 0 |
| 0 | 0 | 0 | 0 | 0 |

| | | | | |
|---|---|---|---|---|
| 0 | 0 | 0.023837188 | 0.050696185 | 0.018442604 |
| 0 | 0 | 0 | 0 | 0 |
| 0 | 0 | 0.263351313 | 0.340817855 | 0.030672724 |
| 0 | 0 | 0 | 0 | 0 |
| 0 | 0 | 0 | 0 | 0 |
| 0 | 0 | 0 | 0 | 0 |
| 0 | 0 | 0 | 0 | 0 |
| 0 | 0 | 0 | 0 | 0 |
| 0 | 0.024170142 | 0 | 0 | 0 |
| 0 | 0 | 0 | 0 | 0 |
| 0 | 0 | 0 | 0 | 0 |
| 0 | 0.007571739 | 0 | 0 | 0 |
| 0 | 0 | 0 | 0 | 0 |
| 0.053764821 | 0.016905831 | 0 | 0 | 0.002698973 |
| 0 | 0 | 0 | 0 | 0 |
| 0.005013306 | 0.048900133 | 0 | 0 | 0.075164237 |
| 0 | 0.001331245 | 0.08405707 | 0.014994107 | 0.361088296 |
| 0 | 0.002772705 | 0 | 0 | 0.002582155 |
| 0.003006499 | 0 | 0 | 0 | 0 |
| 0.152093919 | 0 | 0 | 0 | 0 |
| 0.366493333 | 0 | 0 | 0 | 0 |
| 0 | 0 | 0 | 0 | 0.000739571 |
| 0 | 0 | 0 | 0 | 0 |
| 0.268432401 | 0 | 0.014768131 | 0.001194446 | 0 |
| 0 | 0 | 0 | 0 | 0 |
| 0.040782905 | 0.111233478 | 0 | 0 | 0 |
| 0 | 0 | 0 | 0 | 0 |
| 0 | 0 | 0 | 0 | 0 |
| 0 | 0 | 0 | 0 | 0 |
| 0 | 0 | 0 | 0 | 0 |
| 0.045983852 | 0 | 0 | 0 | 0.046372609 |
| 0 | 0 | 0.006903329 | 0.012014829 | 0.042215708 |
| 0.012692331 | 0.131099669 | 7.116377905 | 2.293456295 | 0.010485668 |
| 0 | 0 | 0 | 0 | 0.007930909 |
| 0.003709305 | 0 | 0 | 0.914004206 | 0.002085503 |
| 0 | 0 | 0 | 0 | 0 |
| 0.001967099 | 0 | 0 | 0.01321518 | 0 |
| 0.002563937 | 0.009213776 | 0 | 0 | 0.01565097 |
| 0 | 0 | 0.003270566 | 0 | 0 |
| 0 | 0 | 0 | 0 | 0 |
| 0 | 0 | 0 | 0 | 0 |
| 0 | 0 | 0 | 0 | 0 |
| 0 | 0.001102335 | 0 | 0 | 0 |
| 0.099473014 | 0.002876995 | 0 | 0 | 0.237598435 |
| 0 | 0.001970498 | 0 | 0 | 0 |
| 0 | 0 | 0 | 0 | 0 |
| 0 | 0 | 0 | 0 | 0 |
| 0 | 0 | 0 | 0 | 0 |

| | | | | |
|---|---|---|---|---|
| 0 | 0 | 0 | 0 | 0 |
| 0.139048856 | 0.06578788 | 2.684603366 | 2.448125543 | 0.846603215 |
| 0 | 0 | 0 | 0 | 0 |
| 0 | 0 | 0 | 0 | 0 |
| 0 | 0 | 0 | 0 | 0 |
| 0 | 0.001320725 | 0 | 0 | 0.002361524 |
| 0 | 0 | 0 | 0 | 0 |
| 0 | 0 | 0.001279987 | 0.000470301 | 0 |
| 0.000959456 | 0 | 0 | 0 | 0.004880648 |
| 0.029450035 | 0 | 0 | 0 | 0 |
| 0.023625361 | 0.002239126 | 0 | 0 | 0.001251148 |
| 0.012761443 | 0.003335242 | 0.05201579 | 0.712519277 | 0.015530166 |
| 0 | 0 | 0 | 0 | 0 |
| 0 | 0 | 0 | 0 | 0 |
| 0 | 0 | 0 | 0.007482728 | 0 |
| 0 | 0 | 0 | 0 | 0 |
| 0.065518004 | 0.004103204 | 0.059908139 | 0.043674244 | 0.010699413 |
| 0.033173583 | 0.008877544 | 0 | 0 | 0.027495177 |
| 0 | 0 | 0 | 0 | 0 |
| 0.00572904 | 0.010376303 | 0 | 0 | 0 |
| 0.03756759 | 0.002014971 | 0.065257225 | 0.030462852 | 0.009457538 |
| 0 | 0 | 0 | 0 | 0 |
| 0 | 0 | 0 | 0 | 0 |
| 0 | 0 | 0 | 0 | 0 |
| 0 | 0 | 0 | 0 | 0 |
| 0 | 0 | 0 | 0 | 0.009085819 |
| 0 | 0 | 0 | 0 | 0 |
| 0.068236535 | 0.156338015 | 0.225700346 | 0 | 0.01699261 |
| 0 | 0 | 0.001108502 | 0.845811333 | 0 |
| 0 | 0 | 0 | 0 | 0 |
| 0 | 0 | 0.018300503 | 0.001494241 | 0 |
| 0 | 0 | 0 | 0 | 0 |
| 0 | 0 | 0 | 0 | 0 |
| 0 | 0 | 0 | 0 | 0 |
| 0 | 0.003919167 | 0 | 0 | 0 |
| 0.97532942 | 0.312436936 | 0.063984788 | 0.068348962 | 0.184705553 |
| 0 | 0 | 0 | 0 | 0 |
| 0 | 0 | 0 | 0 | 0 |
| 0.013600715 | 0 | 0.00210398 | 0.001503167 | 0.020667042 |
| 0 | 0 | 0 | 0 | 0 |
| 0 | 0 | 0 | 0 | 0 |
| 0 | 0 | 0 | 0 | 0 |
| 0.002009501 | 0.015245076 | 0.000638995 | 0 | 0 |
| 0.008622824 | 0 | 0.003290332 | 0 | 0.006060073 |
| 0 | 0 | 0 | 0 | 0 |
| 0 | 0 | 0 | 0 | 0 |
| 0 | 0 | 0 | 0 | 0 |
| 0 | 0 | 0 | 0 | 0 |

| | | | | |
|---|---|---|---|---|
| 0 | 0 | 0 | 0 | 0 |
| 0 | 0 | 0 | 0 | 0 |
| 0 | 0 | 0 | 0 | 0 |
| 0.123522844 | 0 | 0 | 0 | 1.23044711 |
| 0 | 0 | 0 | 0 | 0 |
| 0 | 0 | 0 | 0 | 0 |
| 0 | 0 | 0 | 0 | 0 |
| 0.151216591 | 0 | 0 | 0 | 0.052319595 |
| 0.004212365 | 0 | 0.019824248 | 0.009351014 | 0 |
| 0.129457406 | 0 | 0 | 0 | 0.041069048 |
| 0 | 0 | 0 | 0 | 0 |
| 0.021178092 | 0 | 0 | 0 | 0 |
| 0 | 0 | 0 | 0 | 0 |
| 0 | 0 | 0 | 0 | 0 |
| 0.076278541 | 0.025564354 | 0 | 0 | 0 |
| 0 | 0 | 0 | 0 | 0 |
| 0 | 0 | 0.006140233 | 0 | 0 |
| 0 | 0 | 0.008452122 | 0 | 0 |
| 0 | 0 | 0 | 0 | 0 |
| 0 | 0 | 0 | 0 | 0 |
| 0 | 0 | 0 | 0 | 0 |
| 0 | 0 | 0 | 0 | 0 |
| 0 | 0 | 0 | 0 | 0 |
| 0.810016074 | 0.175791364 | 0 | 0 | 0.202497183 |
| 0 | 0.02900472 | 0 | 0 | 0 |
| 0 | 0 | 0 | 0 | 0.147525806 |
| 0 | 0 | 0 | 0 | 0 |
| 0 | 0.124668833 | 4.601981355 | 0.0079789 | 0.904217969 |
| 0 | 0 | 0 | 0 | 0 |
| 0 | 0 | 0 | 0 | 0 |
| 0.034395268 | 0 | 0 | 0 | 0 |
| 0 | 0 | 0 | 0 | 0 |
| 0 | 0 | 0 | 0 | 0.021321593 |
| 0 | 0 | 0 | 0 | 0 |
| 0 | 0 | 0 | 0 | 0 |
| 0 | 0 | 0 | 0 | 0 |
| 0.017240844 | 0 | 0 | 0 | 0 |
| 0 | 0 | 0 | 0 | 0 |
| 0 | 0.001118485 | 0 | 0 | 0 |
| 0 | 0 | 0 | 0 | 0 |
| 0.000956075 | 0.073525162 | 0 | 0 | 0.126705703 |
| 0 | 0 | 0 | 0 | 0 |
| 0.157129318 | 0.445546058 | 42.62628481 | 20.92127697 | 0.049054237 |
| 0 | 0 | 0 | 0 | 0 |
| 0 | 0 | 0 | 0 | 0 |
| 0 | 0 | 0 | 0 | 0 |
| 0 | 0 | 0 | 0 | 0 |
| 0.062637495 | 0.069799906 | 0 | 0 | 0.00553872 |

| | | | | |
|---|---|---|---|---|
| 0 | 0 | 0 | 0 | 0 |
| 0 | 0 | 0 | 0 | 0 |
| 0.025340138 | 0.248640026 | 0 | 0.001126337 | 0.00995529 |
| 0 | 0 | 0 | 0 | 0.031805302 |
| 0 | 0 | 0 | 0 | 0 |
| 0.006075386 | 0 | 0 | 0 | 0 |
| 0 | 0 | 0 | 0 | 0 |
| 0 | 0 | 0 | 0 | 0 |
| 0 | 0 | 0 | 0 | 0 |
| 0 | 0 | 0 | 0 | 0 |
| 0.004867733 | 0.013411074 | 0 | 0 | 0.097091673 |
| 1.704055691 | 1.963492037 | 3.358634312 | 0.125480767 | 0.373898453 |
| 0 | 0 | 0 | 0 | 0 |
| 0.003210445 | 0 | 0.489186593 | 0.072462041 | 0.50657945 |
| 0.125117714 | 0.411769217 | 0 | 0.002149755 | 0.093473966 |
| 0 | 0 | 0 | 0 | 0 |
| 0 | 0 | 0 | 0 | 0 |
| 0.002043319 | 0 | 0 | 0 | 0 |
| 0 | 0 | 0 | 0 | 0 |
| 0.000891051 | 0 | 0 | 0 | 0 |
| 0.57390266 | 0.021177668 | 0 | 0 | 0.002730776 |
| 0 | 23.69322507 | 1.720142731 | 1.312743416 | 0.022443646 |
| 1.053729774 | 2.813319806 | 0.904326986 | 0.145075713 | 0.456361306 |
| 0 | 0 | 0 | 0 | 0 |
| 0 | 0 | 0 | 0 | 0 |
| 0.012493897 | 0 | 0 | 0 | 0 |
| 0 | 0 | 0 | 0 | 0 |
| 0 | 0 | 0 | 0 | 0 |
| 0 | 0 | 0 | 0 | 0 |
| 0 | 0 | 0 | 0 | 0 |
| 0 | 0 | 0 | 0 | 0 |
| 0.36912282 | 0 | 0 | 0 | 0 |
| 0 | 0 | 0 | 0.005029751 | 0.036584723 |
| 0.002329766 | 0 | 0 | 0 | 0 |
| 0.001649796 | 0.003557229 | 0 | 0 | 0.035336153 |
| 0.023422763 | 0 | 0.080487831 | 0.030308991 | 0.066553566 |
| 0.086368329 | 0.001224009 | 0 | 0 | 0 |
| 0 | 0 | 0 | 0 | 0 |
| 0.031240779 | 0 | 0.202431796 | 0 | 0 |
| 0 | 0 | 0 | 0 | 0 |
| 0 | 0 | 0 | 0 | 0 |
| 0 | 0 | 0 | 0 | 0 |
| 0 | 0 | 0 | 0 | 0 |
| 0 | 0 | 0 | 0 | 0 |
| 0 | 0.005779969 | 0 | 0 | 0.076358166 |
| 0 | 0 | 0 | 0 | 0 |
| 0 | 0 | 0 | 0 | 0 |
| 0.092158327 | 0 | 0.022474606 | 0.267567577 | 3.707449369 |

| | | | | |
|---|---|---|---|---|
| 0 | 0 | 0 | 0 | 0 |
| 0 | 0 | 0 | 0 | 0 |
| 0 | 0 | 0 | 0 | 0 |
| 0.006119806 | 0 | 0 | 0 | 0 |
| 0 | 0 | 0 | 0 | 0 |
| 0 | 0 | 0 | 0 | 0 |
| 0 | 0 | 0 | 0 | 0.105260484 |
| 0 | 0.001618942 | 0 | 0 | 0 |
| 0 | 0 | 0 | 0 | 0 |
| 0 | 0 | 0 | 0 | 0 |
| 0 | 0 | 0 | 0 | 0.012481728 |
| 0 | 0 | 0 | 0 | 0 |
| 0.001670985 | 0 | 0 | 0 | 0 |
| 0.763312173 | 0 | 0 | 0 | 0 |
| 0 | 0 | 0 | 0 | 0 |
| 0 | 0 | 0 | 0 | 0 |
| 0 | 0 | 0 | 0 | 0.181265123 |
| 0.006354918 | 0 | 0 | 0 | 0.002187527 |
| 0 | 0 | 0 | 0 | 0 |
| 0.000818726 | 0 | 0 | 0 | 0 |
| 0.000604912 | 0 | 0 | 0 | 0 |
| 0 | 0 | 0 | 0 | 0 |
| 0.092631259 | 0 | 0 | 0 | 0.003993265 |
| 0 | 0 | 0 | 0 | 0 |
| 0 | 0 | 0 | 0 | 0 |
| 0 | 0 | 0 | 0 | 0 |
| 0 | 0 | 0 | 0 | 0.005029595 |
| 0 | 0 | 0 | 0 | 0 |
| 0 | 0 | 0 | 0 | 0 |
| 0 | 4.344553232 | 0.020923575 | 0 | 0 |
| 0.004079859 | 0.002160627 | 0 | 0 | 0 |
| 0 | 0 | 0 | 0 | 0 |
| 0 | 0 | 0 | 0 | 0 |
| 0 | 0 | 0 | 0 | 0 |
| 0 | 0.001713888 | 0 | 0 | 0 |
| 0 | 0 | 0 | 0 | 0 |
| 0.055056939 | 0 | 0 | 0 | 0 |
| 0.014308933 | 0.004146826 | 0 | 0 | 0.005004948 |
| 0 | 0 | 0 | 0 | 0 |
| 0.007284408 | 0 | 0 | 0 | 0 |
| 0.020787593 | 0 | 0 | 0 | 0 |
| 0 | 0 | 0 | 0 | 0 |
| 0 | 0 | 0 | 0 | 0 |
| 0.008068453 | 0 | 0 | 0 | 0 |
| 0 | 0 | 0 | 0 | 0 |
| 0 | 0 | 0 | 0 | 0 |
| 0.004385769 | 0 | 0 | 0 | 0 |
| 0 | 0 | 0 | 0 | 0 |

| | | | | |
|---|---|---|---|---|
| 0 | 0 | 0 | 0 | 0 |
| 0 | 0 | 0 | 0 | 0 |
| 0 | 0 | 0 | 0 | 0 |
| 0.005050488 | 0 | 0 | 0 | 0 |
| 0 | 0 | 0 | 0 | 0 |
| 0 | 0 | 0 | 0 | 0 |
| 0 | 0 | 0 | 0 | 0 |
| 0 | 0 | 0 | 0 | 0 |
| 0.13877745 | 0.127651776 | 0 | 0 | 0.095870267 |
| 0 | 0 | 0 | 0 | 0 |
| 0 | 0 | 0 | 0 | 0 |
| 0 | 0 | 0 | 0 | 0 |
| 0 | 0 | 0 | 0 | 0 |
| 0 | 0 | 0 | 0 | 0 |
| 0.017155242 | 0 | 0.016365417 | 0 | 1.13998101 |
| 0 | 0 | 0 | 0 | 0.005517111 |
| 0.000970927 | 0 | 0 | 0 | 0 |
| 0 | 0 | 0 | 0 | 0 |
| 0 | 0 | 0 | 0 | 0 |
| 0 | 0 | 0 | 0 | 0 |
| 0 | 0 | 0 | 0 | 0 |
| 0 | 0 | 0 | 0 | 0 |
| 0 | 0 | 0 | 0 | 0 |
| 0 | 0 | 0 | 0 | 0 |
| 0 | 0 | 0.015354225 | 0.748269957 | 0.038782802 |
| 1.946403008 | 0 | 0.125872721 | 0 | 13.28625192 |
| 0 | 0 | 0 | 0 | 0 |
| 0 | 0 | 0 | 0 | 0 |
| 0 | 0 | 0 | 0 | 0 |
| 0 | 0 | 0 | 0 | 0 |
| 0 | 0 | 0 | 0 | 0 |
| 0.014222332 | 0 | 0 | 0 | 0 |
| 0.001631332 | 0 | 0 | 0 | 0 |
| 0 | 0 | 0 | 0 | 0 |
| 0 | 0.001657124 | 0.299761088 | 0.052145282 | 0.229634229 |
| 0 | 0.000502022 | 0 | 0 | 0.096309487 |
| 0 | 0 | 0 | 0 | 0 |
| 0.350717824 | 0 | 0.181831973 | 0 | 0 |
| 0 | 0 | 0 | 8.23232094 | 0 |
| 0 | 0 | 0 | 0 | 0 |
| 0 | 0 | 0 | 0 | 0 |
| 0 | 0.001138595 | 0 | 0 | 0 |
| 0.006045197 | 0 | 0.339031687 | 0 | 0.088931656 |
| 0 | 0 | 0 | 0 | 0 |
| 0 | 0 | 0 | 0 | 0 |
| 0.01058777 | 0 | 0 | 0 | 0 |
| 0 | 0 | 0 | 0 | 0 |
| 0.002108448 | 0.003030772 | 0 | 0 | 0.143382346 |

| | | | | |
|---|---|---|---|---|
| 0 | 0.08060844 | 0 | 0 | 0 |
| 0 | 0 | 0 | 0 | 0 |
| 0 | 0 | 0 | 0 | 0 |
| 0 | 0 | 0 | 0 | 0 |
| 0 | 0 | 0 | 0 | 0.001466853 |
| 0 | 0 | 0 | 0.001908008 | 0 |
| 0 | 0 | 0 | 0 | 0 |
| 0 | 0 | 0 | 0 | 0 |
| 0 | 0 | 0 | 0 | 0 |
| 0 | 0 | 0 | 0 | 0 |
| 0 | 0 | 0 | 0 | 0 |
| 0.199775767 | 0 | 0.09207449 | 0 | 1.859580116 |
| 0.267251359 | 0 | 0 | 0.001476348 | 0.10605865 |
| 0 | 0 | 0 | 0 | 0 |
| 0 | 0 | 0.049219848 | 0.07896616 | 0.342675918 |
| 0 | 0 | 0 | 0 | 0 |
| 0 | 0 | 0 | 0 | 0 |
| 0 | 0 | 0 | 0 | 0 |
| 0.022479311 | 0 | 0 | 0 | 0 |
| 0 | 0 | 0 | 0 | 0.011776693 |
| 0 | 0 | 0 | 0 | 0 |
| 0 | 0 | 0 | 0 | 0 |
| 0.45310505 | 0 | 0 | 0 | 0.735917405 |
| 0 | 0 | 0 | 0 | 0 |
| 0 | 0 | 0 | 0 | 0 |
| 0 | 0 | 0 | 0 | 0 |
| 0 | 0 | 0 | 0 | 0 |
| 0 | 0 | 0 | 0 | 0 |
| 0 | 0 | 0 | 0 | 0 |
| 0 | 0 | 0 | 0 | 0 |
| 0 | 0 | 0 | 0 | 0 |
| 0.035237947 | 0.009849096 | 0.146041097 | 0.30420673 | 0.078619152 |
| 0 | 0 | 0 | 0 | 0 |
| 0.017213065 | 0.012371389 | 0 | 0 | 0.499286342 |
| 0.000760137 | 0 | 0 | 0 | 0.087917331 |
| 0.023517804 | 0 | 0.021500254 | 0 | 0.046961023 |
| 0 | 0 | 0 | 0 | 0 |
| 0 | 0 | 0 | 0 | 0 |
| 0.00698628 | 0 | 0 | 0.000850264 | 0 |
| 0 | 0 | 0 | 0 | 0 |
| 0 | 0 | 0 | 0 | 0 |
| 0 | 0 | 0 | 0 | 0 |
| 0 | 0 | 0 | 0 | 0 |
| 0 | 0 | 0 | 0 | 0 |
| 0 | 0.010943116 | 0 | 0 | 0 |
| 0 | 0 | 0 | 0 | 0 |
| 0 | 0 | 0.000601026 | 0 | 1.365628341 |
| 0 | 0 | 0 | 0 | 0 |

| | | | | |
|---|---|---|---|---|
| 0 | 0 | 0 | 0 | 0 |
| 0 | 0 | 0 | 0 | 0.059343067 |
| 0 | 0 | 0 | 0 | 0 |
| 0 | 0 | 0 | 0 | 0 |
| 0 | 0 | 0 | 0 | 0 |
| 0 | 0 | 0 | 0 | 0 |
| 0 | 0 | 0 | 0 | 0 |
| 0 | 0 | 0 | 0 | 0.062270578 |
| 0 | 0 | 0 | 0 | 0 |
| 0 | 0 | 0 | 0 | 0 |
| 0 | 0 | 0 | 0 | 0 |
| 0 | 0 | 0 | 0 | 0.003375521 |
| 0 | 0 | 0 | 0 | 0 |
| 0 | 0 | 0 | 0 | 0 |
| 0 | 0 | 0 | 0 | 0 |
| 0 | 0 | 0 | 0 | 0 |
| 3.947048182 | 0 | 0 | 0 | 0 |
| 0 | 0 | 0 | 0 | 0 |
| 0.001655067 | 0 | 0 | 1.755661228 | 1.977170925 |
| 0 | 0 | 0 | 0 | 0 |
| 0.008913082 | 0 | 0 | 0 | 0.014694646 |
| 0 | 0 | 0 | 0 | 0 |
| 0 | 0 | 0 | 0 | 0 |
| 0 | 0 | 0 | 0 | 0 |
| 0 | 0 | 0.001040632 | 0.001274518 | 0.002190419 |
| 0.001161858 | 0.007181442 | 0 | 0 | 0.007278935 |
| 0 | 0 | 0 | 0 | 0 |
| 0 | 0.002778987 | 0 | 0.005713404 | 0.020095108 |
| 0 | 0 | 0 | 0 | 0 |
| 0.149448212 | 0 | 0.060177276 | 0 | 0.639964512 |
| 0 | 0 | 0 | 0 | 0 |
| 0 | 0 | 0 | 0 | 0 |
| 0 | 0 | 0 | 0 | 0 |
| 0 | 0 | 0 | 0 | 0 |
| 0.000730533 | 0.001400132 | 0 | 0 | 0.022492447 |
| 0 | 0 | 0 | 0 | 0 |
| 0 | 0 | 0 | 0 | 0 |
| 0.028183995 | 0 | 0 | 0 | 0.007545735 |
| 0 | 0 | 0 | 0 | 0 |
| 0 | 0 | 0 | 0 | 0 |
| 24.7301989 | 7.180808161 | 0.752827416 | 0.845363661 | 3.455028382 |
| 2.192124917 | 0 | 0 | 0 | 0.82852684 |
| 0 | 0 | 0 | 0 | 0 |
| 0 | 0 | 0 | 0 | 0 |
| 0 | 0 | 0 | 0 | 0 |
| 0 | 0 | 0 | 0 | 0 |
| 0 | 0 | 0 | 0 | 0 |
| 0 | 0.012924341 | 0 | 0 | 0.043858515 |

| | | | | |
|---|---|---|---|---|
| 0 | 0 | 0 | 0 | 0.383490123 |
| 0.000902264 | 0 | 0 | 0 | 0.03913339 |
| 0 | 0 | 0 | 0 | 0 |
| 0 | 0 | 0 | 0 | 0 |
| 0.031406089 | 0 | 1.470005896 | 0 | 0.704161457 |
| 0 | 0 | 0 | 0 | 0 |
| 0 | 0 | 0 | 0 | 0 |
| 0 | 0 | 0 | 0 | 0 |
| 0.031307436 | 0.327806817 | 0 | 0 | 0.078875014 |
| 0 | 0 | 0 | 0 | 0 |
| 0 | 0 | 0 | 0 | 0 |
| 0 | 0 | 0 | 0 | 0 |
| 0.114190555 | 0 | 0 | 0 | 0 |
| 0.092961722 | 0.017623279 | 0.027761253 | 0.070519977 | 0.01644822 |
| 0 | 0 | 0.01273079 | 0 | 0 |
| 1.04121746 | 0 | 0 | 0 | 0.499577347 |
| 0.000699513 | 0 | 0 | 0 | 0 |
| 0 | 0 | 0 | 0 | 0 |
| 0 | 0 | 0 | 0 | 0 |
| 0 | 0 | 0 | 0 | 0 |
| 0 | 0 | 0 | 0 | 0 |
| 0 | 0 | 0 | 0 | 0 |
| 0 | 0 | 0 | 0 | 0 |
| 0 | 0 | 0 | 0 | 0 |
| 0 | 0 | 0 | 0 | 0 |
| 0 | 0 | 0 | 0 | 0 |
| 0 | 0 | 0 | 0 | 0.270329076 |
| 0 | 0.000986585 | 0 | 0 | 0.004042647 |
| 0 | 0 | 0 | 0 | 0 |
| 0 | 0 | 0 | 0 | 0 |
| 0 | 0 | 0 | 0 | 0 |
| 0 | 0 | 0 | 0 | 0 |
| 0 | 0 | 0 | 0 | 0 |
| 0 | 0 | 0 | 0 | 0 |
| 0 | 0 | 0 | 0.045899777 | 0 |
| 0.355430251 | 0.006467219 | 0 | 0 | 0.074611496 |
| 0 | 0 | 0 | 0 | 0 |
| 0 | 0 | 0 | 0 | 0.179648691 |
| 0 | 0 | 0 | 0 | 0 |
| 0 | 0 | 0 | 0 | 0 |
| 6.043459619 | 0 | 0.740557852 | 0.002082884 | 0 |
| 0 | 0 | 0 | 0 | 0 |
| 0 | 0 | 0 | 0 | 0 |
| 0 | 0 | 0 | 0 | 0 |
| 0 | 0 | 0 | 0 | 0 |
| 0 | 0 | 0 | 0 | 0 |
| 0 | 0 | 0 | 0 | 0 |
| 0 | 0 | 0 | 0 | 0 |

| | | | | |
|---|---|---|---|---|
| 0.014027747 | 0 | 0 | 0 | 0 |
| 0 | 0 | 0 | 0 | 0 |
| 0 | 0 | 0 | 0 | 0 |
| 0 | 0 | 0 | 0 | 0 |
| 0.015126228 | 0 | 0 | 0 | 0.005178386 |
| 0.150459801 | 0 | 0 | 0 | 0 |
| 0.142120984 | 0 | 0 | 0 | 0.000560478 |
| 0 | 0 | 0 | 0 | 0 |
| 0 | 0 | 0 | 0 | 0 |
| 0 | 0 | 0 | 0 | 0 |
| 0 | 0 | 0 | 0 | 0 |
| 0 | 0 | 0 | 0 | 0 |
| 0 | 0 | 0 | 0 | 0 |
| 0 | 0 | 0 | 0 | 0 |
| 0 | 0 | 0 | 0 | 0 |
| 0 | 0 | 0 | 0 | 0 |
| 0 | 13.86742331 | 0 | 0 | 0 |
| 0 | 0 | 0 | 0 | 0 |
| 0 | 0 | 0 | 0 | 0 |
| 0 | 0 | 0 | 0 | 0 |
| 0 | 0.002705755 | 0.060841805 | 0 | 0.060475371 |
| 0 | 0 | 0 | 0 | 0 |
| 0.002686867 | 0 | 0 | 0 | 0 |
| 0.004343982 | 0.000936633 | 0 | 0 | 0 |
| 0 | 0 | 0 | 0 | 0 |
| 0.01050609 | 0 | 0 | 0 | 0 |
| 0 | 0 | 0 | 0 | 0 |
| 0 | 0 | 0 | 0 | 0 |
| 0 | 0 | 0 | 0 | 0 |
| 0 | 0 | 0.152682671 | 2.477184957 | 0 |
| 0 | 0 | 0 | 0 | 0 |
| 0 | 0 | 0 | 0 | 0 |
| 0 | 0 | 0 | 0 | 0 |
| 0 | 0 | 0 | 0 | 8.837574187 |
| 0 | 0 | 0 | 0 | 0 |
| 0 | 0 | 0 | 0 | 0 |
| 0 | 0 | 0 | 0.003492826 | 0 |
| 0 | 0 | 0 | 0 | 0 |
| 0 | 0 | 0 | 0 | 0 |
| 0 | 0 | 0 | 0 | 0 |
| 0 | 0 | 0 | 0 | 0 |
| 0 | 0 | 0 | 0 | 0 |
| 0 | 0 | 0 | 0 | 0 |
| 0 | 0 | 0 | 0 | 0 |
| 0 | 0 | 0 | 0 | 0 |
| 0 | 0 | 0 | 0 | 0 |
| 0 | 0 | 0 | 0 | 0 |
| 0 | 0 | 0 | 0 | 0 |

| | | | | |
|---|---|---|---|---|
| 0 | 0.001679751 | 0 | 0 | 0 |
| 0 | 0 | 0 | 0 | 0 |
| 0 | 0 | 0 | 0 | 0 |
| 0 | 0 | 0 | 0 | 0 |
| 0 | 0 | 0 | 0 | 0 |
| 0 | 0 | 0 | 0 | 0 |
| 0 | 0 | 0 | 0 | 0 |
| 2.860938897 | 0 | 0.00801231 | 0 | 0.686166825 |
| 0.001834246 | 0 | 0.285916867 | 11.15832798 | 0.185040751 |
| 0.021197316 | 0.012848756 | 0 | 0 | 0.090461052 |
| 0.000899085 | 0 | 0 | 0 | 0 |
| 0 | 0 | 0 | 0 | 0 |
| 0 | 0 | 0 | 0 | 0 |
| 0 | 0 | 0 | 0 | 0 |
| 0 | 0 | 0 | 0 | 0 |
| 0 | 0 | 0 | 0 | 0 |
| 0 | 0 | 0 | 0 | 0 |
| 0 | 0 | 0 | 0 | 0 |
| 0 | 0 | 0 | 0 | 0 |
| 0 | 0 | 0 | 0 | 0 |
| 0 | 0 | 0 | 0 | 0 |
| 0 | 0 | 0 | 0 | 0 |
| 0 | 0 | 0 | 0 | 0 |
| 0 | 0 | 0 | 0 | 0 |
| 0 | 0 | 0 | 0 | 0 |
| 0 | 0 | 0 | 0 | 0 |
| 0 | 0 | 0 | 0 | 0 |
| 0 | 0 | 0 | 0 | 0 |
| 0 | 0 | 0 | 0 | 0 |
| 0 | 0 | 0 | 0 | 0 |
| 0 | 0 | 0 | 0 | 0 |
| 0 | 0 | 0 | 0 | 0 |
| 0 | 0 | 0 | 0 | 0 |
| 0 | 0 | 0 | 0 | 0 |
| 0 | 0 | 0 | 0 | 0 |
| 0 | 0 | 0 | 0 | 0 |
| 0 | 0 | 0 | 0 | 0 |
| 0 | 0 | 0 | 0 | 0 |
| 0 | 0 | 0.083053375 | 0 | 0 |
| 0.020009097 | 0 | 0 | 0 | 0 |
| 0 | 0 | 0 | 0 | 0 |
| 0 | 0 | 0 | 0 | 0 |
| 0 | 0.522298425 | 0 | 0 | 0.901531744 |
| 0 | 0 | 0 | 0 | 0 |
| 0 | 0 | 0 | 0 | 0 |
| 0 | 0 | 0 | 0 | 0 |
| 0 | 0 | 0 | 0 | 0 |
| 0 | 0 | 0 | 0 | 0.061310479 |
| 0 | 0 | 0 | 0 | 0 |

| | | | | |
|---|---|---|---|---|
| 0 | 0 | 0 | 0 | 0 |
| 0 | 0 | 0 | 0 | 0.002371926 |
| 0 | 0 | 0 | 0 | 0 |
| 0 | 0 | 0 | 0 | 0 |
| 0.918084867 | 0 | 0.146297629 | 0.000516861 | 1.576183433 |
| 0.018327715 | 0 | 0 | 0 | 0 |
| 0 | 0 | 0 | 0 | 0 |
| 0.014865052 | 0.027777941 | 0 | 0 | 0.074741367 |
| 0 | 0 | 0 | 0 | 0 |
| 0 | 0 | 0 | 0 | 0 |
| 0 | 0 | 0 | 0 | 0 |
| 0 | 0 | 0 | 0 | 0 |
| 0 | 0 | 0 | 0 | 0 |
| 0 | 0 | 0 | 0 | 0 |
| 0 | 0 | 0 | 0 | 0 |
| 0 | 0 | 0 | 0 | 0 |
| 0 | 0 | 0 | 0 | 0 |
| 0 | 0 | 0 | 0 | 0 |
| 0 | 0 | 0 | 0 | 0 |
| 0 | 0 | 0 | 0 | 0 |
| 0 | 0 | 0 | 0 | 0 |
| 0 | 0 | 0 | 0 | 0 |
| 0 | 0 | 0 | 0 | 0 |
| 0.001286448 | 0 | 0 | 0 | 0.00123992 |
| 0 | 0 | 0.270786691 | 0 | 0 |
| 0 | 0 | 0 | 0 | 0 |
| 0 | 0 | 0 | 0.007698337 | 0 |
| 0 | 0 | 0 | 0 | 0 |
| 0 | 0 | 0 | 0 | 0.003933118 |
| 0 | 0 | 0 | 0 | 0 |
| 0 | 0 | 0 | 0 | 0 |
| 0 | 0 | 0 | 0 | 0 |
| 0 | 0 | 0 | 0 | 0 |
| 0 | 0 | 0 | 0 | 0 |
| 0 | 0 | 0 | 0 | 0 |
| 0 | 0 | 0 | 0 | 0 |
| 0 | 0 | 0 | 0 | 0 |
| 0 | 0 | 0 | 0 | 0 |
| 0 | 0 | 0 | 0 | 0 |
| 0 | 0 | 0 | 0 | 0 |
| 0 | 0 | 0 | 0 | 0 |
| 0 | 0 | 0 | 0 | 0 |
| 0 | 0 | 0 | 0 | 0 |
| 0 | 0 | 0 | 0 | 0 |
| 0 | 0 | 0 | 0 | 0 |
| 0 | 0 | 0 | 0 | 0 |
| 0 | 0 | 0 | 0 | 0 |
| 0.006430681 | 0 | 0 | 0 | 0 |

| | | | | |
|---|---|---|---|---|
| 0.017039997 | 0.027356938 | 0 | 0 | 0.809987053 |
| 0 | 0 | 0 | 0 | 0 |
| 0 | 0 | 0 | 0 | 0 |
| 0 | 0 | 0 | 0 | 0 |
| 0 | 0 | 0 | 0 | 0 |
| 0 | 0 | 0 | 0 | 0.067750527 |
| 0 | 0 | 0 | 0 | 0 |
| 0 | 0 | 0 | 0 | 0 |
| 0 | 0 | 0 | 0 | 0 |
| 0 | 0 | 0 | 0 | 0 |
| 0 | 0 | 0 | 0 | 0 |
| 0 | 0 | 0 | 0 | 0 |
| 0 | 0 | 0 | 0 | 0 |
| 0 | 0 | 0 | 0 | 0 |
| 0 | 0 | 0 | 0 | 0 |
| 0 | 0 | 0 | 0 | 0 |
| 0 | 0 | 0 | 0 | 0 |
| 0 | 0 | 0 | 0 | 0 |
| 0 | 0 | 0 | 0 | 0 |
| 0 | 0 | 0 | 0 | 0 |
| 0 | 0.000552439 | 0 | 0 | 0.006379474 |
| 0 | 0 | 0 | 0 | 0 |
| 0 | 0.00674434 | 0 | 0 | 0.587627388 |
| 0 | 0 | 0 | 0 | 1.808298292 |
| 0 | 0 | 0 | 0 | 0 |
| 0 | 0 | 0.00307558 | 0 | 0 |
| 0 | 0 | 0 | 0 | 0 |
| 0 | 0 | 0 | 0 | 0 |
| 0 | 0 | 0 | 0 | 0 |
| 0 | 0 | 0 | 0 | 0 |
| 0 | 0 | 0 | 0 | 0 |
| 0 | 0 | 0 | 0 | 0.792602779 |
| 0.022727326 | 0.031580233 | 0.079785946 | 0.169944611 | 0.022878903 |
| 0 | 0 | 0 | 0 | 0 |
| 1.74833574 | 0 | 0 | 0.006175595 | 0.092310527 |
| 0.040366168 | 0 | 0 | 0 | 0.00253906 |
| 0 | 0 | 0 | 0 | 0 |
| 0 | 0 | 0 | 0 | 0 |
| 0.002250114 | 0 | 0 | 0 | 0.029142341 |
| 0 | 0 | 0 | 0 | 0 |
| 0 | 0 | 0 | 0 | 0 |
| 0 | 0 | 0 | 0 | 0 |
| 0 | 0 | 0.005333588 | 0.001729147 | 0.022717419 |
| 0 | 0 | 0 | 0 | 0 |
| 0 | 0 | 0 | 0 | 0 |
| 0.003134274 | 0 | 0 | 0 | 0.709914843 |
| 0 | 0 | 0 | 0 | 0 |
| 0 | 0 | 0 | 0 | 0 |

| | | | | |
|---|---|---|---|---|
| 0 | 0 | 0 | 0 | 0 |
| 0 | 0 | 0 | 0 | 0 |
| 0 | 0 | 0 | 0 | 0 |
| 0 | 0 | 0 | 0 | 0 |
| 0 | 0 | 0 | 0 | 0 |
| 0 | 0 | 0 | 0 | 0 |
| 0 | 0 | 0 | 0 | 0 |
| 0 | 0 | 0 | 0 | 0 |
| 0 | 0 | 0 | 0 | 0 |
| 0 | 0 | 0 | 0 | 0 |
| 0 | 0 | 0 | 0 | 0 |
| 0 | 0 | 0 | 0 | 0 |
| 0 | 0 | 0 | 0 | 0 |
| 0 | 0 | 0 | 0 | 0 |
| 0 | 0 | 0 | 0 | 0 |
| 0 | 0.067153628 | 0 | 0 | 0.000850383 |
| 0 | 0 | 0 | 0 | 0 |
| 0 | 0 | 0 | 0 | 0 |
| 0 | 0 | 0 | 0 | 0 |
| 10.30043612 | 0 | 0.006342079 | 0 | 0 |
| 0.008569888 | 0 | 0.001054882 | 0 | 0 |
| 0 | 0 | 0 | 0 | 0 |
| 0 | 0 | 0 | 0 | 0 |
| 0 | 0 | 0 | 0 | 0 |
| 0 | 0 | 0 | 0 | 0 |
| 0 | 0 | 0 | 0 | 0 |
| 0 | 0 | 0 | 0 | 0 |
| 0 | 0 | 0 | 0 | 0 |
| 0 | 0 | 0 | 0 | 0 |
| 2.913922318 | 0 | 0.001831892 | 0 | 0 |
| 0.709502399 | 0 | 0.105691651 | 0 | 2.182016569 |
| 0 | 0 | 0 | 0 | 0 |
| 0 | 0 | 0 | 0 | 0 |
| 0 | 0 | 0 | 0 | 0 |
| 0 | 0 | 0 | 0 | 0 |
| 0 | 0 | 0 | 0 | 0 |
| 0 | 0 | 0 | 0 | 0 |
| 0 | 0 | 0 | 0 | 0 |
| 0 | 0 | 0 | 0 | 0 |
| 0 | 0 | 0 | 0 | 0 |
| 0 | 0 | 0 | 0 | 0 |
| 0 | 0 | 0 | 0 | 0 |
| 0 | 0 | 0 | 0 | 0 |
| 0 | 0 | 0 | 0 | 0 |
| 0 | 0 | 0 | 0 | 0 |
| 0 | 0 | 0 | 0 | 0 |
| 0 | 0.907897153 | 0 | 0 | 0.097499503 |
| 0 | 0 | 0 | 0 | 0 |

| | | | | |
|---|---|---|---|---|
| 0 | 0 | 0 | 0 | 0 |
| 0 | 0 | 0 | 0 | 0 |
| 0 | 0 | 0 | 0 | 0 |
| 0 | 0 | 0 | 0 | 0 |
| 0.002429122 | 0 | 0 | 0 | 1.540683608 |
| 0 | 0 | 0 | 0 | 0 |
| 0 | 0 | 0 | 0 | 0 |
| 0 | 0 | 0 | 0 | 0 |
| 0 | 0 | 0 | 0 | 0 |
| 0 | 0 | 0 | 0 | 0 |
| 0 | 0 | 0 | 0 | 0 |
| 0 | 0.003029842 | 0 | 0 | 0.023024433 |
| 0 | 0 | 0 | 0 | 0.057645375 |
| 0 | 0 | 0 | 0 | 0 |
| 0 | 0 | 0 | 0 | 0 |
| 0.535881254 | 0 | 0 | 0 | 0 |
| 0.003458453 | 0 | 0 | 0 | 0 |
| 0 | 0 | 0 | 0 | 0 |
| 0.030431156 | 0 | 0 | 0 | 0.444572935 |
| 0 | 0 | 0 | 0 | 0.001115872 |
| 0 | 0 | 0 | 0 | 0 |
| 0 | 0 | 0 | 0 | 0 |
| 0 | 0 | 0 | 0 | 0 |
| 0 | 0 | 0 | 0 | 0 |
| 0 | 0 | 0 | 0 | 0 |
| 0 | 0 | 0 | 0 | 0 |
| 0.007125153 | 0.002560498 | 0 | 0 | 0.046319542 |
| 0 | 0 | 0 | 0 | 0 |
| 0 | 0 | 0 | 0 | 0 |
| 0 | 0 | 0 | 0 | 0 |
| 0 | 0 | 0 | 0 | 0.01682613 |
| 0 | 0 | 0 | 0 | 0 |
| 0 | 0 | 0 | 0 | 0 |
| 0 | 0 | 0 | 0 | 0 |
| 0 | 0 | 0 | 0 | 0 |
| 0 | 0 | 0 | 0 | 0 |
| 0 | 0 | 0 | 0 | 0 |
| 0.052488939 | 0.001019592 | 0 | 0 | 0.161086426 |
| 0 | 0 | 0 | 0 | 0 |
| 0 | 0 | 0 | 0 | 0 |
| 0 | 0 | 0 | 0 | 0.003412545 |
| 0 | 0 | 0 | 0 | 0 |
| 0 | 0 | 0 | 0 | 0 |
| 0 | 0.040444002 | 0 | 0 | 0.007603313 |
| 0 | 0 | 0 | 0 | 0 |
| 0 | 0 | 0 | 0 | 0 |
| 0 | 0 | 0 | 0 | 0 |
| 0 | 0 | 0 | 0 | 0 |

| | | | | |
|---|---|---|---|---|
| 0 | 0 | 0 | 0 | 0 |
| 0.043825994 | 0 | 0 | 0 | 0 |
| 0 | 0 | 0 | 0 | 0 |
| 0.001653463 | 0.000990315 | 0 | 0 | 0.00597623 |
| 0 | 0 | 0.008533908 | 0 | 0.001231746 |
| 0 | 0 | 0 | 0 | 0 |
| 0 | 0 | 0 | 0 | 0 |
| 0 | 0 | 0 | 0 | 0 |
| 0 | 0 | 0 | 0 | 0 |
| 0 | 0 | 0 | 0 | 0 |
| 0 | 0 | 0 | 0 | 0 |
| 0 | 0 | 0 | 0 | 0 |
| 0.006055708 | 0.006317948 | 0.005217849 | 0.001027058 | 0 |
| 0 | 0 | 0 | 0 | 0 |
| 0 | 0 | 0 | 0 | 0 |
| 0 | 0 | 0 | 0 | 0 |
| 0 | 0 | 0 | 0 | 0 |
| 0 | 0 | 0 | 0 | 0 |
| 0 | 0 | 0 | 0 | 0 |
| 0 | 0 | 0 | 0 | 0 |
| 0 | 0 | 0 | 0 | 0 |
| 0 | 0 | 0 | 0 | 0 |
| 0 | 0.013125977 | 0 | 0 | 0.017147917 |
| 0 | 0 | 0.005212072 | 0 | 0 |
| 0 | 0 | 0 | 0 | 0 |
| 0 | 0 | 0 | 0 | 0 |
| 0 | 0 | 0 | 0 | 0 |
| 0 | 0 | 0 | 0 | 0 |
| 0 | 0 | 0 | 0 | 0 |
| 0 | 0 | 0 | 0 | 0 |
| 0.004588874 | 0 | 0.001094401 | 0 | 0.025339563 |
| 0 | 0 | 0 | 0 | 0 |
| 0 | 0 | 0 | 0 | 0 |
| 0 | 0 | 0 | 0 | 0 |
| 0 | 0 | 0 | 0 | 0 |
| 0 | 0.422135708 | 0 | 0 | 0 |
| 0 | 0 | 0 | 0 | 0 |
| 0 | 0 | 0 | 0 | 0 |
| 0 | 0 | 0 | 0.000567024 | 0.00818581 |
| 0 | 0 | 0 | 0 | 0 |
| 0 | 0 | 0 | 0 | 0 |
| 0 | 0 | 0 | 0 | 0 |
| 0 | 0 | 0 | 0 | 0 |
| 0 | 0 | 0 | 0 | 0 |
| 0 | 0 | 0 | 0 | 25.62660707 |
| 0.046115511 | 0 | 0.006517387 | 0 | 0.474313738 |
| 0 | 0.013514518 | 0 | 0 | 0 |
| 0 | 0 | 0 | 0 | 0 |

| | | | | |
|---|---|---|---|---|
| 0 | 0 | 0 | 0 | 0 |
| 0 | 0 | 0 | 0 | 0 |
| 0 | 0 | 0 | 0 | 0 |
| 0 | 0 | 0 | 0 | 0 |
| 0 | 0 | 0 | 0 | 0 |
| 0 | 0 | 0 | 0 | 0 |
| 0 | 0 | 0 | 0 | 0 |
| 0 | 0 | 0 | 0 | 0 |
| 0 | 0 | 0 | 0 | 0.020377806 |
| 0 | 0 | 0 | 0 | 0 |
| 0 | 0 | 0 | 0 | 0.008538281 |
| 0.001469559 | 0 | 0 | 0 | 0.001180341 |
| 0 | 0 | 0 | 0 | 0 |
| 0 | 0 | 0 | 0 | 0 |
| 0 | 0 | 0 | 0 | 0 |
| 0 | 0 | 0 | 0 | 0 |
| 0 | 0 | 0 | 0 | 0 |
| 0 | 0 | 0 | 0 | 0 |
| 0 | 0 | 0 | 0 | 0 |
| 0 | 0 | 0.0019924 | 0 | 0.002725962 |
| 0 | 0 | 0 | 0 | 0 |
| 0 | 0 | 0 | 0 | 0 |
| 0 | 0 | 0 | 0 | 0 |
| 0 | 0 | 0 | 0 | 0 |
| 0 | 0 | 0 | 0 | 0 |
| 0 | 0 | 0 | 0 | 0 |
| 0 | 0 | 0 | 0 | 0 |
| 0 | 0 | 0 | 0 | 0 |
| 0 | 0 | 0 | 0 | 0 |
| 0 | 0 | 0 | 0 | 0 |
| 0.012111431 | 0 | 0.001925637 | 0 | 0.01585275 |
| 0 | 0 | 0 | 0 | 0 |
| 0.011979256 | 0.02620356 | 0.455121878 | 0.144981968 | 0.01401416 |
| 0.003459554 | 0.00267772 | 0.013201108 | 0.009483218 | 0.004702402 |
| 0 | 0 | 0 | 0 | 0 |
| 0 | 0 | 0 | 0 | 0 |
| 0 | 0 | 0 | 0 | 0 |
| 0 | 0 | 0 | 0 | 0 |
| 0 | 0 | 0 | 0 | 0 |
| 0 | 0 | 0 | 0 | 0 |
| 0 | 0 | 0 | 0 | 0 |
| 0.378140701 | 0.008161027 | 0 | 0 | 0.50797849 |
| 0.009657356 | 0 | 0.004730863 | 0 | 0.089726464 |
| 0 | 0 | 0 | 0 | 0 |
| 0 | 0 | 0 | 0 | 0 |
| 0 | 0 | 0 | 0.002203405 | 0.067859951 |
| 0 | 0 | 0 | 0 | 0 |
| 0 | 0 | 0 | 0 | 0 |

| | | | | |
|---|---|---|---|---|
| 0 | 0 | 0 | 0 | 0 |
| 0 | 0 | 0.02112909 | 0 | 0 |
| 0.002235866 | 0 | 0.038087976 | 0.022018044 | 0.02847678 |
| 0 | 0 | 0 | 0 | 0 |
| 0.002295297 | 0 | 0 | 0 | 0.605099972 |
| 0.001489486 | 0 | 0 | 0 | 0 |
| 0 | 0 | 0 | 0 | 0 |
| 0 | 0 | 0 | 0 | 0 |
| 0 | 0 | 0 | 0 | 0 |
| 0 | 0 | 0 | 0 | 0 |
| 0 | 0 | 0 | 0 | 0 |
| 0 | 0 | 0 | 0 | 0 |
| 0 | 0 | 0 | 0 | 0 |
| 0 | 0 | 0 | 0 | 0 |
| 0 | 0 | 0 | 0 | 0 |
| 0 | 0 | 0 | 0 | 0 |
| 0 | 0 | 0 | 0 | 0 |
| 0 | 0 | 0 | 0 | 0 |
| 0 | 0 | 0 | 0 | 0 |
| 0 | 0 | 0 | 0 | 0 |
| 0 | 0 | 0 | 0 | 0 |
| 0 | 0 | 0 | 0 | 0 |
| 0 | 0 | 0 | 0 | 0 |
| 0 | 0 | 0 | 0 | 0 |
| 0 | 0 | 0 | 0 | 0 |
| 0 | 0 | 0.00581025 | 0.570063587 | 0.005530066 |
| 0 | 0 | 0 | 0 | 0 |
| 0 | 0 | 0 | 0 | 1.09350356 |
| 0 | 0 | 0 | 0 | 0 |
| 0 | 0 | 0 | 0 | 0 |
| 0 | 0 | 0 | 0 | 0 |
| 0 | 0 | 0 | 0 | 0 |
| 0 | 0 | 0 | 0 | 0 |
| 0 | 0 | 0 | 0 | 0 |
| 0 | 0 | 0 | 0 | 0 |
| 0 | 0 | 0 | 0 | 0 |
| 0 | 0 | 0 | 0 | 0 |
| 0.066162978 | 0 | 0 | 0 | 0 |
| 1.705355584 | 0 | 0 | 0 | 0 |
| 0 | 0 | 0 | 0 | 0 |
| 0 | 0 | 0 | 0 | 0 |
| 0.000861839 | 0 | 0.003014586 | 0 | 0 |
| 0 | 0.001146074 | 0 | 0 | 0 |
| 0 | 0 | 0 | 0 | 0 |
| 0 | 0 | 0 | 0 | 0 |
| 0 | 0 | 0 | 0 | 0 |
| 0.002439431 | 0.003798751 | 0 | 0 | 0.033961808 |
| 0 | 0 | 0 | 0 | 0 |

| | | | | |
|---|---|---|---|---|
| 0 | 0 | 0 | 0 | 0 |
| 0.025650939 | 0.019726375 | 0 | 0 | 0.003708481 |
| 0 | 0 | 0 | 0 | 0 |
| 0 | 0 | 0 | 0 | 0 |
| 0 | 0 | 0 | 0 | 0 |
| 0 | 0 | 0 | 0 | 0 |
| 0 | 0 | 0 | 0 | 0 |
| 0 | 0 | 0 | 0 | 0 |
| 0 | 0 | 0 | 0 | 0 |
| 0 | 0 | 0 | 0 | 0 |
| 0 | 0 | 0 | 0 | 0 |
| 0 | 0 | 0 | 0 | 0 |
| 0.037147982 | 0 | 0.005432055 | 0 | 0.289876474 |
| 0 | 0 | 0 | 0 | 0 |
| 0 | 0 | 0 | 0 | 0.045468513 |
| 0 | 0 | 0 | 0 | 0 |
| 0 | 0 | 0 | 0 | 0 |
| 0 | 0 | 0 | 0 | 0 |
| 0 | 0 | 0 | 0 | 0 |
| 0 | 0 | 0 | 0 | 0 |
| 0 | 0 | 0 | 0 | 0 |
| 0 | 0 | 0 | 0 | 0.001009122 |
| 0 | 0 | 0 | 0 | 0 |
| 0 | 0 | 0 | 0 | 0.58855534 |
| 0 | 0 | 0 | 0 | 0 |
| 0 | 0 | 0 | 0 | 0 |
| 0 | 0 | 0 | 0 | 0 |
| 0 | 0 | 0 | 0 | 0 |
| 0 | 0 | 0 | 0 | 0 |
| 0 | 0 | 0 | 0 | 0 |
| 0 | 0 | 0 | 0 | 0 |
| 0 | 0 | 0 | 0 | 0 |
| 0 | 0 | 0 | 0 | 0 |
| 0 | 0 | 0 | 0 | 0 |
| 0 | 1.804730918 | 0 | 0 | 0 |
| 0 | 0 | 0 | 0 | 0 |
| 0 | 0 | 0 | 0 | 0 |
| 0 | 0 | 0 | 0 | 0 |
| 0 | 0 | 0 | 0 | 0.002519327 |
| 0 | 0 | 0 | 0 | 0 |
| 0.081841138 | 0.118652536 | 0.007780489 | 0.012322222 | 0.005873166 |
| 0 | 0 | 0 | 0 | 0 |
| 0 | 0 | 0 | 0 | 0 |
| 0 | 0 | 0 | 0 | 0 |
| 0 | 0 | 0 | 0 | 0 |
| 0.058604334 | 1.76858374 | 0 | 0.010157836 | 0.168368014 |
| 0 | 0 | 0 | 0 | 0 |
| 0 | 0 | 0 | 0 | 0 |

| | | | | |
|---|---|---|---|---|
| 0 | 0 | 0 | 0 | 0 |
| 0 | 0 | 0 | 0 | 0 |
| 0 | 0 | 0 | 0 | 0 |
| 0 | 0 | 0 | 0 | 0 |
| 0 | 0.002106261 | 0 | 0 | 0 |
| 0 | 0 | 0 | 0 | 0 |
| 0 | 0 | 0 | 0 | 0 |
| 0 | 0 | 0 | 0 | 0 |
| 0 | 0 | 0 | 0 | 0 |
| 0 | 0 | 0.029140693 | 0.00091985 | 0.031111666 |
| 0 | 0 | 0 | 0 | 0 |
| 0 | 0 | 0 | 0 | 0 |
| 0 | 0 | 0 | 0 | 0.009889117 |
| 0 | 0 | 0 | 0 | 0 |
| 0 | 0 | 0 | 0 | 0 |
| 0 | 0 | 0 | 0 | 0 |
| 0 | 0 | 0 | 0 | 0 |
| 0 | 0 | 0 | 0 | 0 |
| 0 | 0 | 0 | 0 | 0 |
| 0.005205414 | 0.095500069 | 0.746693165 | 0.085785833 | 1.089469005 |
| 0 | 0 | 0 | 0 | 0 |
| 0 | 0 | 0 | 0 | 0 |
| 0 | 0 | 0 | 0 | 0 |
| 0 | 0 | 0 | 0 | 0 |
| 0 | 0 | 0 | 0 | 0 |
| 0 | 0 | 0 | 0 | 0 |
| 0 | 0 | 0 | 0 | 0 |
| 0 | 0 | 0 | 0 | 0 |
| 0 | 0 | 0 | 0 | 0 |
| 0.004393162 | 0 | 0 | 0 | 0 |
| 0 | 0 | 0 | 0 | 0 |
| 0 | 0 | 0 | 0 | 0 |
| 0 | 0 | 0 | 0 | 0 |
| 0 | 0 | 0 | 0 | 0 |
| 0 | 0 | 0 | 0 | 0 |
| 0 | 0 | 0 | 0 | 0 |
| 0 | 0 | 0 | 0 | 0 |
| 0 | 0 | 0 | 0 | 0 |
| 0 | 0 | 0 | 0 | 0 |
| 0 | 0 | 0 | 0 | 0 |
| 0 | 0 | 0 | 0 | 0 |
| 0 | 0 | 0 | 0 | 0 |
| 0 | 0 | 0 | 0 | 0 |
| 0 | 0 | 0 | 0 | 0 |
| 0.004631794 | 0.002034369 | 0 | 0 | 0.010747325 |
| 0 | 0 | 0 | 0 | 0 |
| 0 | 0 | 0 | 0 | 0 |
| 0 | 0 | 0 | 0 | 0 |

| | | | | |
|---|---|---|---|---|
| 0 | 0 | 0 | 0 | 0 |
| 0 | 0 | 0 | 0 | 0 |
| 0 | 0 | 0 | 0 | 0 |
| 0 | 0 | 0 | 0 | 0 |
| 0 | 0 | 0 | 0 | 0 |
| 0.001152605 | 0 | 0 | 0 | 0 |
| 0 | 0 | 0 | 0 | 0 |
| 0 | 0 | 0 | 0 | 0 |
| 0 | 0 | 0 | 0 | 0 |
| 0 | 0 | 0 | 0 | 0 |
| 0 | 0 | 0 | 0 | 0 |
| 0 | 0 | 0 | 0 | 0 |
| 0 | 0 | 0 | 0 | 0 |
| 0 | 0 | 0 | 0 | 0 |
| 0 | 0 | 0 | 0 | 0 |
| 0 | 0 | 0 | 0 | 0 |
| 0 | 0 | 0 | 0 | 0 |
| 0 | 0 | 0 | 0 | 0 |
| 0 | 0 | 0 | 0 | 0 |
| 0 | 0 | 0 | 0 | 0 |
| 0 | 0 | 0 | 0 | 0 |
| 0 | 0 | 0 | 0 | 0 |
| 0 | 0 | 0 | 0 | 0 |
| 0 | 0 | 0 | 0 | 0 |
| 0 | 0 | 0 | 0 | 0 |
| 0 | 0 | 0 | 0 | 0.000724026 |
| 0 | 0 | 0 | 0 | 0 |
| 0.001349322 | 0 | 0 | 0 | 0 |
| 0 | 0 | 0 | 0 | 0 |
| 0 | 0 | 0 | 0 | 0 |
| 0 | 0 | 0 | 0 | 0 |
| 0 | 0 | 0 | 0 | 0 |
| 0 | 0 | 0 | 0 | 0 |
| 0 | 0 | 0 | 0 | 0 |
| 0 | 0 | 0 | 0 | 0 |
| 0 | 0 | 0 | 0 | 0 |
| 0 | 0 | 0 | 0 | 0 |
| 0 | 0 | 0 | 0 | 0 |
| 0 | 0 | 0 | 0 | 0 |
| 0 | 0 | 0 | 0 | 0 |
| 0 | 0 | 0 | 0 | 0 |
| 0 | 0 | 0 | 0 | 0 |
| 0 | 0 | 0 | 0 | 0 |
| 0 | 0 | 0 | 0 | 0 |
| 0 | 0 | 0 | 0 | 0 |
| 0 | 0 | 0 | 0 | 0 |
| 0 | 0 | 0 | 0 | 0 |
| 0 | 0 | 0 | 0 | 0 |

| | | | | |
|---|---|---|---|---|
| 0 | 0 | 0 | 0 | 0 |
| 0 | 0 | 0 | 0 | 0.221458752 |
| 0 | 0 | 0 | 0 | 0 |
| 0 | 0 | 0 | 0 | 0 |
| 0 | 0 | 0 | 0 | 0 |
| 0 | 0 | 0 | 0 | 0 |
| 0 | 0 | 0 | 0 | 0 |
| 0 | 0 | 0 | 0 | 0 |
| 0 | 0 | 0 | 0 | 0 |
| 0 | 0 | 0 | 0 | 0 |
| 0 | 0 | 0.017731193 | 0.002632285 | 0.02940542 |
| 0 | 0 | 0 | 0 | 0 |
| 0 | 0 | 0.486185215 | 0 | 0.003332089 |
| 0 | 0 | 0 | 0 | 0 |
| 0 | 0 | 0 | 0 | 0 |
| 0 | 0 | 0 | 0 | 0 |
| 0 | 0 | 0 | 0 | 0 |
| 0 | 0 | 0 | 0 | 0 |
| 0 | 0 | 0 | 0 | 0 |
| 0 | 0 | 0 | 0 | 0 |
| 0 | 0 | 0 | 0.006011124 | 0.000918299 |
| 0 | 0 | 0 | 0 | 0 |
| 0 | 0 | 0 | 0 | 0 |
| 1.472531205 | 0 | 1.324097606 | 0.003596387 | 0 |
| 0 | 0 | 0 | 0 | 0 |
| 0 | 0 | 0 | 0 | 0 |
| 0 | 0 | 0 | 0 | 0 |
| 0 | 0 | 0 | 0 | 0 |
| 0 | 0 | 0 | 0 | 0 |
| 0 | 0 | 0 | 0 | 0 |
| 0 | 0 | 0 | 0 | 0 |
| 0 | 0 | 0 | 0 | 0 |
| 0 | 0 | 0 | 0 | 0 |
| 0 | 0 | 0 | 0 | 0 |

| PHL018-0127_V1 EYCF | PHL018-0127_V3 EYCF | PHL018-0278_V3 REF | PHL018-0074_V3 CTRL | PHL018-0152_V1 REF |
|---|---|---|---|---|
| 0.034374296 | 0.214272184 | 0.375368 | 0.149055307 | 0.003027767 |
| 3.290534339 | 0.841672173 | 0.482583061 | 0.094734784 | 3.116839385 |
| 12.59953476 | 0.073999262 | 0.00102198 | 1.910622929 | 10.26026419 |
| 0 | 0 | 0 | 0.320983484 | 0 |
| 0 | 0 | 0 | 0 | 0 |
| 0 | 0 | 0 | 0 | 0 |
| 0 | 0 | 0 | 0 | 0 |
| 0 | 0 | 0 | 0 | 0 |
| 0.003627153 | 0 | 0 | 0 | 0 |
| 0 | 0 | 0 | 0 | 0 |
| 0 | 0 | 0 | 0 | 0 |
| 0 | 0 | 0 | 0 | 0 |
| 0 | 0 | 0 | 0 | 0 |
| 13.18365949 | 4.885227148 | 0.408131694 | 0 | 1.858431561 |
| 0 | 0 | 0 | 0 | 0 |
| 0 | 0 | 0 | 0 | 0 |
| 0 | 0 | 0 | 0 | 0 |
| 0 | 0 | 0 | 0 | 0 |
| 0 | 0 | 0 | 0 | 0 |
| 0 | 0 | 0 | 0 | 0 |
| 0 | 0 | 0 | 0 | 0 |
| 0 | 0 | 0 | 0 | 0 |
| 0 | 0 | 0 | 0 | 0 |
| 0 | 0 | 0 | 0 | 0 |
| 0 | 0 | 0.000715487 | 1.062578281 | 0 |
| 0 | 0 | 0 | 0 | 0 |
| 0 | 0 | 0 | 0 | 0 |
| 0 | 0 | 0 | 0 | 0 |
| 0 | 0 | 0 | 0 | 0 |
| 0 | 0 | 0 | 0 | 0 |
| 0 | 0 | 0 | 0 | 0 |
| 0 | 0 | 0 | 0 | 0 |
| 0 | 0 | 0 | 0 | 0 |
| 0 | 0 | 0 | 0 | 0 |
| 0 | 0 | 0 | 0 | 0 |
| 0 | 0 | 2.781576636 | 0.902729281 | 0.640203579 |
| 0 | 0 | 0 | 0 | 0 |
| 0 | 0 | 0 | 0 | 0 |
| 0 | 0 | 0 | 0 | 0 |
| 0 | 0 | 0 | 0 | 0 |
| 0 | 0 | 0 | 0 | 0 |
| 0.039692505 | 0 | 0.175053732 | 0.000952893 | 0.009871642 |
| 0.120694604 | 0.014817499 | 1.123615832 | 0.045371147 | 0.388661812 |
| 0.000717859 | 0 | 0.027570147 | 0.020937609 | 0.160283436 |
| 0 | 0 | 0.005452005 | 0 | 0.003661266 |
| 0 | 0 | 0 | 0 | 0 |

| | | | | |
|---|---|---|---|---|
| 0 | 0 | 0.008096364 | 0.008467917 | 0.002224256 |
| 0 | 0.020069056 | 0 | 0 | 0 |
| 0.003815481 | 0.009573354 | 0 | 0 | 0 |
| 0.122643648 | 0.160874382 | 0.309952893 | 0.206610693 | 0.496203392 |
| 0 | 0 | 0.033572233 | 0.010949929 | 0 |
| 0 | 0 | 0 | 0.014373612 | 0 |
| 0 | 0 | 0.001085479 | 0 | 2.421929949 |
| 0 | 0 | 0 | 0 | 0 |
| 0 | 0 | 0 | 0.001970331 | 0 |
| 0 | 0 | 0 | 0 | 0 |
| 0 | 0 | 0 | 0 | 0 |
| 0 | 0 | 0.165620109 | 0.027047594 | 0.010957512 |
| 0 | 0 | 0 | 0 | 0 |
| 0.068482788 | 0.137265632 | 0.009795067 | 0.016962146 | 0.000783075 |
| 0 | 0 | 0 | 0 | 0 |
| 0 | 0 | 0 | 0 | 0 |
| 0 | 0 | 0.009378285 | 0 | 0.369932314 |
| 0 | 0 | 0 | 0 | 0 |
| 0 | 0 | 0 | 0 | 0 |
| 0 | 0 | 0 | 0.056297746 | 0 |
| 0.375300992 | 0.270685769 | 0.002464374 | 0 | 0.128588622 |
| 0.049903614 | 0.005878509 | 0.004627613 | 0.012392019 | 0 |
| 0 | 0 | 0.501912449 | 0.012697939 | 0 |
| 0 | 0 | 0 | 0 | 0 |
| 0 | 0 | 0 | 0 | 0.001029645 |
| 0 | 0 | 0 | 0 | 0 |
| 0 | 0 | 0.006078086 | 0 | 0.001113193 |
| 0 | 0 | 0 | 0 | 0 |
| 0.005694751 | 0 | 0.011445441 | 0 | 0 |
| 0 | 0 | 0.003769489 | 0 | 0 |
| 0.047785607 | 0.066609999 | 0 | 0 | 0 |
| 0 | 0 | 0 | 0 | 0 |
| 0 | 0 | 0 | 0 | 0 |
| 0 | 0 | 0.002139329 | 0 | 0 |
| 0 | 0 | 0 | 0 | 0 |
| 0 | 0 | 0 | 0 | 0 |
| 0 | 0 | 0 | 0 | 0 |
| 0 | 0 | 0.003366881 | 0 | 0.008855627 |
| 0 | 0 | 0 | 0 | 0 |
| 0 | 0 | 0 | 0 | 0 |
| 0 | 0 | 0 | 0 | 0 |
| 0 | 0 | 0 | 0 | 0 |
| 0 | 0 | 0 | 0 | 0 |
| 0 | 0 | 0 | 0 | 0 |
| 0 | 0 | 0.000564978 | 0.093717153 | 0 |
| 0 | 0 | 0 | 0 | 0 |
| 0 | 0 | 0 | 0 | 0.007300016 |
| 0.030772038 | 0 | 0 | 0.020402881 | 0.001255893 |

| | | | | |
|---|---|---|---|---|
| 0 | 0 | 0 | 0 | 0 |
| 0.001722315 | 0 | 0.006155757 | 0 | 0 |
| 0 | 0 | 0 | 0 | 0 |
| 0 | 0 | 0 | 0 | 0 |
| 0 | 0 | 0.000877905 | 0.002960382 | 0.015564189 |
| 0 | 0 | 0 | 0 | 0 |
| 0 | 0 | 0 | 0 | 0 |
| 0 | 0 | 0 | 0 | 0 |
| 0 | 0 | 0.011629672 | 0.004140494 | 0 |
| 0 | 0 | 0 | 0 | 0 |
| 0 | 0 | 0 | 0 | 0 |
| 0 | 0 | 0.005712308 | 0 | 0 |
| 0 | 0 | 0 | 0 | 0 |
| 0 | 0 | 0 | 0 | 0 |
| 0 | 0 | 0 | 0 | 0 |
| 0 | 0 | 0 | 0 | 0 |
| 0 | 0 | 0 | 0 | 0 |
| 0 | 0 | 0 | 0 | 0 |
| 0.035799166 | 0 | 0.225838954 | 0.344381696 | 0 |
| 0 | 0 | 0 | 0 | 0 |
| 0 | 0 | 0 | 0 | 0 |
| 0 | 0 | 0 | 0 | 0 |
| 0 | 0 | 0 | 0.000808707 | 0 |
| 0 | 0 | 0 | 0 | 0 |
| 0 | 0 | 0 | 0 | 0 |
| 0 | 0 | 0 | 0 | 0 |
| 0 | 0 | 0 | 0 | 0 |
| 0.004572808 | 0 | 0 | 0.002985985 | 0 |
| 0 | 0 | 0 | 0 | 0 |
| 0 | 0 | 0 | 0 | 0 |
| 0.060143288 | 0.002804264 | 0.067054039 | 0.098524091 | 0 |
| 0 | 0 | 0.000434232 | 0 | 0.007581756 |
| 0 | 0 | 0 | 0 | 0 |
| 0 | 0 | 0 | 0 | 0 |
| 0 | 0 | 0 | 0 | 0 |
| 0 | 0 | 0 | 0 | 0 |
| 0 | 0 | 0 | 0 | 0 |
| 0 | 0 | 0 | 0 | 0 |
| 0 | 0 | 0 | 0 | 0 |
| 0 | 0 | 0 | 0 | 0 |
| 0 | 0 | 0 | 0 | 0 |
| 0 | 0 | 0 | 0.006003884 | 0 |
| 0 | 0.000637573 | 0 | 0 | 0 |
| 0 | 0 | 0 | 0 | 0 |
| 0 | 0 | 0 | 0.003361146 | 0 |
| 0 | 0 | 0 | 0 | 0 |
| 0 | 0 | 0 | 0 | 0 |
| 0 | 0 | 0 | 0 | 0 |

| | | | | |
|---|---|---|---|---|
| 0 | 0 | 0.014135287 | 0 | 0 |
| 0 | 0 | 0 | 0 | 0 |
| 0 | 0 | 0 | 0 | 0 |
| 0 | 0 | 0 | 0 | 0 |
| 0 | 0 | 0.041574446 | 0.003748479 | 0 |
| 0 | 0 | 0 | 0 | 0 |
| 0 | 0 | 0 | 0 | 0 |
| 0 | 0 | 0 | 0 | 0 |
| 0 | 0 | 0 | 0 | 0 |
| 0 | 0 | 0 | 0 | 0 |
| 0 | 0 | 0 | 0 | 0 |
| 0 | 0 | 0 | 0 | 0 |
| 0 | 0 | 0 | 0 | 0 |
| 0 | 0 | 0 | 0 | 0 |
| 0 | 0 | 0 | 0 | 0 |
| 0 | 0 | 0 | 0 | 0 |
| 0 | 0 | 0 | 0 | 0 |
| 0 | 0 | 0 | 0 | 0 |
| 0 | 0 | 0 | 0 | 0 |
| 0 | 0 | 0 | 0 | 0 |
| 0 | 0 | 0 | 0 | 0 |
| 0 | 0 | 0 | 0 | 0 |
| 0 | 0 | 0 | 0.021024487 | 0 |
| 0 | 0 | 0 | 0 | 0 |
| 0 | 0 | 0 | 0.127762204 | 0 |
| 0 | 0 | 0 | 0 | 0 |
| 0 | 0 | 0 | 0.050056836 | 0 |
| 0 | 0 | 0 | 0 | 0 |
| 0 | 0 | 0 | 0 | 0 |
| 0 | 0 | 0 | 0 | 0 |
| 0 | 0 | 0 | 0 | 0 |
| 0 | 0 | 0 | 0 | 0 |
| 0 | 0 | 0 | 0 | 0 |
| 0 | 0 | 0 | 0 | 0 |
| 0 | 0 | 0 | 0 | 0 |
| 0 | 0 | 0 | 0 | 0 |
| 0 | 0 | 0 | 0 | 0 |
| 0 | 0 | 0 | 0 | 0 |
| 0 | 0 | 0 | 0.014617545 | 0 |
| 0 | 0 | 0 | 0 | 0 |
| 0 | 0 | 0 | 0 | 0 |
| 0 | 0 | 0 | 0 | 0 |
| 0 | 0 | 0 | 0 | 0 |
| 0 | 0 | 0 | 0 | 0 |
| 0 | 0 | 0 | 0 | 0 |
| 0 | 0 | 0 | 0 | 0 |
| 0 | 0 | 0 | 0 | 0 |
| 0 | 0 | 0 | 0 | 0 |

| | | | | |
|---|---|---|---|---|
| 0 | 0 | 0.004697223 | 9.100086786 | 0 |
| 0 | 0 | 0 | 0 | 0 |
| 0 | 0 | 0 | 0 | 0 |
| 0 | 0 | 0 | 0 | 0 |
| 0 | 0 | 0 | 0 | 0 |
| 0 | 0 | 0 | 0 | 0 |
| 0 | 0 | 0 | 0 | 0 |
| 0 | 0 | 0 | 0 | 0 |
| 0 | 0 | 0 | 0 | 0 |
| 0 | 0 | 0 | 0 | 0 |
| 0 | 0 | 0 | 0 | 0 |
| 0 | 0 | 0 | 0.059488314 | 0 |
| 0 | 0.012992191 | 0.002077474 | 0.004596333 | 0 |
| 0 | 0 | 0 | 0 | 0 |
| 0 | 0 | 0 | 0 | 0 |
| 0 | 0 | 0 | 0 | 0 |
| 0 | 0 | 0 | 0 | 0 |
| 0 | 0 | 0 | 0 | 0 |
| 0 | 0 | 0 | 0 | 0 |
| 0 | 0 | 0 | 0 | 0 |
| 0.00907663 | 0.010627871 | 0.031971413 | 0.033487117 | 0.0078256 |
| 0 | 0 | 0 | 0 | 0 |
| 0 | 0 | 0.001573061 | 0 | 0 |
| 0 | 0 | 0 | 0 | 0 |
| 0 | 0 | 0 | 0 | 0 |
| 0 | 0 | 0 | 0 | 0 |
| 0.012422136 | 0.003978914 | 0.018619397 | 0.004314626 | 0.001752863 |
| 0 | 0 | 0 | 0 | 0 |
| 0 | 0 | 0 | 0 | 0 |
| 0 | 0 | 0 | 0 | 0 |
| 0 | 0 | 0 | 0 | 0 |
| 0 | 0 | 0 | 0 | 0 |
| 0 | 0 | 0 | 0 | 0 |
| 0 | 0 | 0.021912832 | 0.008512094 | 0 |
| 0.007677461 | 0.015311916 | 0.023135274 | 0 | 0.003329223 |
| 0 | 0 | 0 | 0.015298447 | 0 |
| 0 | 0 | 0 | 0 | 0 |
| 0 | 0 | 0 | 0 | 0 |
| 0 | 0 | 0 | 0.000747571 | 0 |
| 0 | 0 | 0 | 0 | 0 |
| 0 | 0 | 0 | 0 | 0 |
| 0 | 0 | 0 | 0 | 0 |
| 0 | 0 | 0 | 0 | 0 |
| 0 | 0 | 0 | 0 | 0 |
| 0 | 0 | 0 | 0 | 0 |
| 0 | 0 | 0 | 0.004589608 | 0 |
| 0 | 0.006684973 | 0 | 0 | 0 |
| 0.002525154 | 0.002287934 | 0.005541789 | 0 | 0 |

| | | | | |
|---|---|---|---|---|
| 0 | 0 | 0 | 0 | 0 |
| 0 | 0 | 0 | 0 | 0 |
| 0 | 0 | 0 | 0 | 0 |
| 0 | 0 | 0 | 0 | 0 |
| 0 | 0 | 0 | 0 | 0 |
| 0 | 0 | 0 | 0 | 0 |
| 0 | 0 | 0 | 0 | 0 |
| 0.000907489 | 0 | 0.009559708 | 0 | 0 |
| 0 | 0 | 0 | 0 | 0 |
| 0 | 0 | 0 | 0 | 0 |
| 0 | 0 | 0 | 0 | 0 |
| 0 | 0 | 0 | 0 | 0 |
| 0 | 0 | 0 | 0 | 0 |
| 0 | 0 | 0 | 0 | 0 |
| 0 | 0 | 0 | 0 | 0 |
| 0 | 0 | 0 | 0 | 0 |
| 0 | 0 | 0 | 0 | 0 |
| 0 | 0 | 0 | 0 | 0.002269739 |
| 0 | 0 | 0.002536183 | 0 | 0.003264391 |
| 0 | 0 | 0.085091452 | 0.83720567 | 0 |
| 0.193195998 | 0.034586895 | 0.002078405 | 0.041639235 | 0 |
| 0 | 0 | 0 | 0 | 0 |
| 0 | 0 | 0 | 0 | 0 |
| 0 | 0 | 0 | 0 | 0 |
| 0.003762987 | 0 | 0.003303352 | 0 | 0.004159404 |
| 0 | 0 | 0.001001727 | 0 | 0 |
| 0 | 0 | 0 | 0 | 0 |
| 0 | 0 | 0 | 0 | 0 |
| 0.000897768 | 0 | 0.171413757 | 0 | 0 |
| 0 | 0 | 0 | 0 | 0 |
| 1.561929971 | 0.952039712 | 0.131582334 | 0.091245654 | 0.024784865 |
| 0 | 0 | 0 | 0 | 0 |
| 0 | 0.12600666 | 0 | 0 | 0 |
| 0 | 0 | 0.000988607 | 0 | 0 |
| 0 | 0 | 0 | 0 | 0 |
| 0 | 0 | 0 | 0 | 0 |
| 0 | 0 | 0 | 0 | 0 |
| 0 | 0 | 0 | 0 | 0 |
| 0 | 0 | 0 | 0 | 0 |
| 0 | 0 | 0 | 0 | 0 |
| 0 | 0 | 0 | 0 | 0 |
| 0 | 0 | 0 | 0.014677622 | 0.009885932 |
| 0 | 0 | 0 | 0 | 0 |
| 0 | 0 | 0 | 0 | 0 |
| 0 | 0 | 0 | 0 | 0 |
| 0 | 0 | 0 | 0 | 0 |
| 0 | 0 | 0 | 0 | 0 |
| 0 | 0 | 0 | 0 | 0 |

| | | | | |
|---|---|---|---|---|
| 0 | 0 | 0 | 0 | 0 |
| 0 | 0 | 0 | 0 | 0 |
| 0 | 0 | 0.035991694 | 0.018430169 | 0.175955223 |
| 0 | 0 | 0 | 0 | 0 |
| 0 | 0 | 0 | 0 | 0 |
| 0 | 0 | 0 | 0 | 0 |
| 0 | 0 | 0 | 0 | 0 |
| 0 | 0 | 0 | 0 | 0 |
| 0 | 0 | 0 | 0 | 0 |
| 0 | 0 | 0.002146808 | 0 | 0 |
| 0 | 0 | 0 | 0 | 0 |
| 0 | 0 | 0 | 0 | 0 |
| 0 | 0 | 0 | 0 | 0 |
| 0.00164174 | 0 | 0 | 0.001500849 | 0 |
| 0 | 0 | 0.001784292 | 0 | 0.005032574 |
| 0 | 0 | 0.033239569 | 0.002841965 | 0 |
| 0 | 0.002582594 | 0.003862777 | 0 | 0.00122875 |
| 0 | 0 | 0 | 0 | 0 |
| 0 | 0 | 0 | 0 | 0 |
| 0 | 0 | 0 | 0 | 0 |
| 0 | 0 | 0 | 0 | 0 |
| 0.001951242 | 0.001398807 | 0.004037562 | 0 | 0 |
| 0 | 0 | 0 | 0 | 0 |
| 0.006481865 | 0 | 0 | 0 | 0 |
| 0 | 0 | 0 | 0 | 0 |
| 0 | 0 | 0 | 0 | 0 |
| 0 | 0 | 0 | 0 | 0 |
| 0 | 0 | 0 | 0 | 0 |
| 0 | 0 | 0 | 0 | 0 |
| 0.007257831 | 0 | 0 | 0 | 0 |
| 0 | 0 | 0 | 0 | 0 |
| 0 | 0 | 0 | 0 | 0 |
| 0 | 0 | 0 | 0 | 0 |
| 0 | 0 | 0 | 0 | 0 |
| 0 | 0 | 0 | 0 | 0 |
| 0 | 0.004190879 | 0.016128912 | 0 | 0 |
| 0 | 0 | 0 | 0 | 0 |
| 0.010506573 | 0 | 0.166248791 | 0.033159845 | 0.047150414 |
| 0 | 0 | 0 | 0 | 0 |
| 0 | 0 | 0 | 0 | 0 |
| 0 | 0 | 0 | 0 | 0 |
| 0 | 0 | 0 | 0 | 0 |
| 0 | 0 | 0 | 0 | 0 |
| 0 | 0.002183036 | 0 | 0 | 0 |
| 0 | 0 | 0 | 0 | 0 |
| 0 | 0 | 0 | 0 | 0 |
| 0 | 0 | 0 | 0 | 0 |
| 0.011877689 | 0 | 0 | 0 | 0 |

| | | | | |
|---|---|---|---|---|
| 0 | 0 | 0.000552367 | 4.98894506 | 0 |
| 0 | 0.017358067 | 0 | 0.000528011 | 0 |
| 0 | 0 | 0 | 0 | 0 |
| 0 | 0 | 0 | 0 | 0 |
| 0.009106178 | 0.00245679 | 0.009863431 | 0.013043316 | 0.008766709 |
| 0 | 0 | 0 | 0 | 0 |
| 0 | 0 | 0 | 0 | 0 |
| 0.001647008 | 0 | 0.071297597 | 0.001075475 | 0.006826929 |
| 0 | 0 | 0 | 0 | 0 |
| 0.053050216 | 0.542722968 | 0.005588439 | 0.020230409 | 0.004784931 |
| 0 | 0 | 0 | 0 | 0 |
| 0.005541054 | 0.005452141 | 0.408166544 | 0 | 0.006485066 |
| 0.002719644 | 0 | 0.004232297 | 0.000968669 | 0.012461853 |
| 0.040553391 | 0 | 0.001730553 | 0 | 0 |
| 0 | 0 | 0 | 0 | 0 |
| 0 | 0 | 0 | 0 | 0.002707637 |
| 0 | 0 | 0.003909402 | 0 | 0 |
| 0.001287993 | 0 | 0 | 0 | 0 |
| 0.079072149 | 0.476289986 | 0 | 0 | 0 |
| 0.045135114 | 0.02864658 | 0 | 0 | 0 |
| 0 | 0 | 0 | 0 | 0 |
| 0 | 0 | 0 | 0 | 0 |
| 0 | 0 | 0 | 0 | 0 |
| 0 | 0 | 0 | 0.509254808 | 0 |
| 0 | 0 | 0 | 0 | 0 |
| 0 | 0 | 0 | 0 | 0.04258972 |
| 0 | 0 | 0 | 0 | 0 |
| 0.016715434 | 0 | 0 | 0 | 0 |
| 2.177701488 | 0 | 0.062506952 | 0 | 0 |
| 0 | 0 | 0 | 0 | 0 |
| 0 | 0 | 0 | 0 | 0 |
| 0 | 0 | 0 | 0 | 0 |
| 0 | 0 | 0 | 0 | 0 |
| 0.08097211 | 0.057012377 | 0 | 0 | 0 |
| 0 | 0 | 0 | 0 | 0 |
| 0.002336152 | 0 | 0 | 0 | 0 |
| 0.019913192 | 0.010101976 | 0 | 0 | 0 |
| 0 | 0 | 0 | 0 | 0 |
| 0 | 0 | 0 | 0.006528243 | 0 |
| 0 | 0 | 0 | 0 | 0 |
| 0 | 0 | 0 | 0 | 0 |
| 0 | 0 | 0 | 0 | 0 |
| 0.005548405 | 0 | 0 | 0 | 0 |
| 0.063568109 | 0 | 0 | 0 | 0 |
| 0 | 0 | 0 | 0 | 0 |
| 0 | 0 | 0 | 0.293593605 | 0 |
| 0.005371739 | 0.004792225 | 0.00212202 | 0 | 0 |
| 0.00162526 | 0 | 0.001070056 | 0.002547058 | 0 |

| | | | | |
|---|---|---|---|---|
| 0 | 0 | 0 | 0 | 0 |
| 0 | 0 | 0 | 0 | 0.035832886 |
| 0 | 0 | 0 | 0 | 0 |
| 0 | 0 | 0 | 0.057129953 | 0 |
| 0 | 0 | 0 | 0 | 0 |
| 0.050488683 | 0.005661687 | 0.0103995 | 0 | 0 |
| 0 | 0 | 0 | 0 | 0 |
| 0 | 0 | 0 | 0 | 0 |
| 0 | 0 | 0.003613296 | 5.544344272 | 0 |
| 0 | 0 | 0 | 0 | 0 |
| 0.033041316 | 0.009302688 | 0.005169287 | 0.019866601 | 0 |
| 0 | 0.022053278 | 0 | 0 | 0 |
| 0.042399046 | 0 | 0 | 0 | 0 |
| 0.002472537 | 0.001292458 | 0.004680195 | 0.003430886 | 0.022342352 |
| 0 | 0 | 0 | 0 | 0 |
| 0.099912364 | 0 | 0 | 0 | 0 |
| 0 | 0 | 0 | 0 | 0 |
| 0.028573357 | 0 | 0 | 0 | 0 |
| 0 | 0 | 0 | 0 | 0 |
| 0 | 0 | 0.103715038 | 0 | 0 |
| 0 | 0 | 0 | 0 | 0.263698378 |
| 0 | 0 | 0.15529309 | 0.139187252 | 0.254690236 |
| 0 | 0 | 0 | 0.004021805 | 0 |
| 0 | 0 | 0 | 0 | 0 |
| 0 | 0 | 0 | 0 | 0 |
| 0 | 0 | 0 | 0 | 0 |
| 0.051942132 | 0.005044919 | 0 | 0 | 0 |
| 0 | 0 | 0 | 0 | 0 |
| 0 | 0 | 0 | 0 | 0 |
| 0.855769121 | 0.004167839 | 0.324814895 | 0.169505558 | 1.205411208 |
| 0 | 0 | 0.000690248 | 0.001916834 | 0 |
| 0 | 0 | 0.000581452 | 0 | 0 |
| 0 | 0 | 0 | 0 | 0 |
| 0.005345551 | 0 | 0 | 0 | 0 |
| 0.017305161 | 0 | 0 | 0 | 0 |
| 0 | 0 | 0 | 0 | 0 |
| 0 | 0 | 0.160695613 | 0 | 0 |
| 0 | 0 | 0 | 0 | 0 |
| 0 | 0 | 0 | 0 | 0.005249641 |
| 0 | 0 | 0 | 0 | 0 |
| 0.093503929 | 0.010200392 | 0.021914761 | 0.034344489 | 0 |
| 0 | 0 | 1.428534461 | 0.710474674 | 2.71464109 |
| 0 | 0 | 0 | 0 | 0 |
| 0 | 0 | 0.007265412 | 0.006204985 | 0 |
| 0.037261004 | 0 | 0 | 0 | 0 |
| 0.400833809 | 0 | 0 | 0 | 0 |
| 0.014134161 | 0.010036915 | 0 | 0 | 0 |
| 0 | 0 | 0.00065738 | 0 | 0 |

| | | | | |
|---|---|---|---|---|
| 0 | 0 | 0.002166375 | 0 | 0.001309336 |
| 0 | 0 | 0 | 0.003706803 | 0 |
| 0 | 0 | 0.002066297 | 0 | 0.925932783 |
| 0 | 0 | 0 | 0 | 0 |
| 0 | 0 | 0.166814458 | 0.043341249 | 0 |
| 0 | 0 | 0.027108748 | 0 | 0 |
| 0 | 0 | 0 | 0 | 0 |
| 0 | 0 | 0.002185261 | 0 | 0 |
| 0 | 0 | 0.003444817 | 0 | 0 |
| 0.004519943 | 0.008910712 | 0 | 0 | 0 |
| 0 | 0 | 0 | 0 | 0 |
| 0 | 0 | 0 | 0 | 0 |
| 0 | 0 | 0 | 0 | 0 |
| 0.026493098 | 0 | 0.000783946 | 0.130816484 | 0 |
| 0 | 0 | 0 | 0 | 0 |
| 0.025589089 | 0 | 0.242605542 | 0.012607962 | 0.000617123 |
| 0.013665731 | 0 | 0.029029296 | 0 | 0.005643137 |
| 0.282883144 | 0.001048028 | 0 | 0.000818246 | 0 |
| 0.006125667 | 0.001113754 | 0 | 0 | 0 |
| 0 | 0 | 0 | 0.000898166 | 0 |
| 0 | 0 | 0.010697419 | 0 | 0.011637749 |
| 0.006579883 | 0 | 0 | 0 | 0.001190135 |
| 0 | 0 | 0 | 0 | 0 |
| 0 | 0 | 0 | 0.062076577 | 0 |
| 0 | 0.029627303 | 0 | 0.001416212 | 0 |
| 0.014605551 | 0.128688955 | 0.004919892 | 0 | 0 |
| 0.006219365 | 0.00082131 | 0.005603369 | 0 | 0 |
| 0 | 0 | 0 | 0.30069937 | 0 |
| 0 | 0 | 0 | 0 | 0 |
| 0 | 0 | 0 | 0 | 0 |
| 0.058749212 | 0.003331222 | 0 | 0.005201693 | 0 |
| 0.00296908 | 0 | 0 | 0 | 0.02190643 |
| 0.11237189 | 0.046105075 | 0.001395935 | 0.025151375 | 6.581013519 |
| 0.012660368 | 0 | 0.053680381 | 0 | 0 |
| 0 | 0 | 0 | 0 | 0.02133484 |
| 0 | 0 | 0 | 0 | 0 |
| 0 | 0 | 0.001262022 | 0 | 0.001906884 |
| 0.129168064 | 0.026955531 | 0.018423241 | 0.001631433 | 0.001325575 |
| 0 | 0 | 0.048390038 | 0.014300922 | 0 |
| 0.001021355 | 0 | 0 | 0 | 0 |
| 0 | 0 | 0 | 0.043487316 | 0 |
| 0 | 0 | 0 | 0 | 0 |
| 0.044836511 | 0.000937487 | 0.012791967 | 0 | 0 |
| 0.153490261 | 0.019084708 | 0.09797472 | 0 | 0.00892493 |
| 1.091924714 | 11.7534128 | 0 | 0 | 0 |
| 0 | 0 | 0 | 0.000889198 | 0 |
| 0 | 0 | 0 | 0.017766406 | 0 |
| 0 | 0 | 0 | 0 | 0 |

| | | | | |
|---|---|---|---|---|
| 0 | 0 | 0 | 0 | 0 |
| 0.082727105 | 0.003416775 | 0.522701207 | 0.037346981 | 0.327159538 |
| 0 | 0 | 0 | 0 | 0 |
| 0 | 0 | 0 | 0 | 0 |
| 0 | 0 | 0 | 0 | 0 |
| 0 | 0 | 0 | 0 | 0 |
| 0 | 0 | 0 | 0 | 0 |
| 0 | 0.001093518 | 0.005681457 | 0 | 0 |
| 0 | 0 | 0.002667397 | 0 | 0 |
| 0.007916533 | 0.007862528 | 0 | 0 | 0 |
| 0.021250716 | 0.040624598 | 0.000999376 | 0 | 0 |
| 0 | 0.00315164 | 0.021336585 | 0.004429148 | 0.05223228 |
| 0 | 0 | 0 | 0.028907094 | 0 |
| 0 | 0 | 0 | 0 | 0 |
| 0.001451342 | 0 | 0 | 0 | 0.006160263 |
| 0 | 0 | 0 | 0 | 0 |
| 0.003708761 | 0.0057606 | 0.017964761 | 0 | 0.02793493 |
| 0.056398244 | 0.745504556 | 0 | 0 | 0.002280717 |
| 0 | 0 | 0 | 0 | 0 |
| 0 | 0.010832299 | 0 | 0 | 0 |
| 0.014479102 | 0 | 0.02626044 | 0.002854239 | 0.009602654 |
| 0 | 0 | 0 | 0 | 0.019614331 |
| 0 | 0 | 0 | 0 | 0 |
| 0 | 0 | 0 | 0 | 0 |
| 0 | 0 | 0 | 0 | 0.072451911 |
| 0 | 0 | 0.004288497 | 0 | 0 |
| 0 | 0 | 0 | 0.058106336 | 0 |
| 0.369629899 | 0.104181248 | 0.03880772 | 0.218990534 | 0.05873381 |
| 0 | 0 | 0 | 0 | 0 |
| 0 | 0 | 0 | 0 | 0 |
| 0 | 0 | 0 | 0 | 0 |
| 0 | 0 | 0 | 0 | 0 |
| 0 | 0 | 0 | 0 | 0 |
| 0 | 0 | 0 | 0 | 0 |
| 0.03491809 | 0.122318684 | 0 | 0 | 0.008390562 |
| 0.525818469 | 0.007809069 | 0.103211038 | 0.081827052 | 0.040413214 |
| 0 | 0 | 0 | 0 | 0 |
| 0 | 0 | 0 | 0 | 0 |
| 0.012894952 | 0.00209705 | 0.228284269 | 0.008186343 | 0.002850678 |
| 0 | 0 | 0 | 0 | 0 |
| 0.208584733 | 2.437065138 | 0 | 0 | 0 |
| 0 | 0 | 0 | 0 | 0 |
| 0 | 0.003821341 | 0 | 0.087232028 | 0 |
| 0.018905547 | 0 | 0 | 0.00109734 | 0 |
| 0 | 0 | 0 | 0 | 0 |
| 0 | 0 | 0 | 0.321851493 | 0 |
| 0 | 0 | 0 | 0 | 0 |
| 0 | 0.002839849 | 0 | 0 | 0 |

| | | | | |
|---|---|---|---|---|
| 0.006079055 | 0 | 0 | 0 | 0 |
| 0 | 0 | 0.000566514 | 0 | 0 |
| 0 | 0 | 0 | 0.003905994 | 0 |
| 0 | 0 | 0.457302228 | 1.616297112 | 0 |
| 0 | 0 | 0 | 0 | 0 |
| 0 | 0 | 0 | 0 | 0 |
| 0 | 0 | 0 | 0 | 0 |
| 0 | 0 | 0 | 0 | 0 |
| 0.108364531 | 0.016478514 | 11.27467401 | 5.022934156 | 0.288561737 |
| 0 | 0 | 0.23037789 | 0.077345538 | 0 |
| 0 | 0 | 0 | 0 | 0 |
| 0 | 0 | 0 | 0.009512201 | 0.004347489 |
| 0 | 0 | 0 | 0 | 0 |
| 0 | 0 | 0 | 0 | 0 |
| 0 | 0.235761749 | 0 | 0.066851513 | 0.004384795 |
| 0 | 0 | 0 | 0 | 0 |
| 0 | 0 | 0.003613294 | 0 | 0 |
| 0 | 0 | 0 | 0 | 0 |
| 0 | 0 | 0 | 0 | 0 |
| 0 | 0 | 0 | 0 | 0 |
| 0 | 0 | 0 | 0 | 0 |
| 0 | 0 | 0 | 0.001913466 | 0 |
| 0 | 0 | 0 | 0 | 0 |
| 0.104212855 | 0.039383194 | 0.105714428 | 0.132948873 | 0.012799034 |
| 0.005228142 | 0 | 0.372202008 | 0.023748937 | 0 |
| 0 | 0 | 0.753613127 | 0.113506351 | 0 |
| 0 | 0 | 0 | 0.110734346 | 0 |
| 0 | 0 | 0 | 0.479945054 | 0.003310031 |
| 0 | 0 | 0 | 0 | 0 |
| 0 | 0 | 0 | 0 | 0 |
| 0 | 0 | 0.032012396 | 0.006473237 | 0 |
| 0 | 0 | 0 | 0.069993172 | 0 |
| 0.112991236 | 0 | 0.086101139 | 0 | 0 |
| 0 | 0 | 0 | 0 | 0 |
| 0 | 0 | 0 | 0.105445348 | 0 |
| 0 | 0 | 0 | 0.076709456 | 0.036632357 |
| 0 | 0 | 5.75569668 | 3.147983481 | 0 |
| 0 | 0 | 0 | 0 | 0 |
| 0.117524614 | 0.03043911 | 0.142107551 | 0 | 0 |
| 0 | 0 | 0 | 0 | 0 |
| 0 | 0 | 0.07483287 | 0.025094456 | 0 |
| 0 | 0 | 0.293136828 | 0.297335731 | 0 |
| 22.2221818 | 14.66899392 | 0.015084507 | 0 | 9.724725352 |
| 0.004057911 | 0.003393877 | 0 | 0 | 0 |
| 0 | 0 | 0 | 0 | 0 |
| 0 | 0 | 0 | 0 | 0 |
| 0 | 0 | 0 | 0 | 0 |
| 0.06719645 | 0.368113043 | 0.302501182 | 0.006947417 | 0 |

| | | | | |
|---|---|---|---|---|
| 0 | 0 | 0 | 0 | 0 |
| 0 | 0 | 0.018137362 | 0 | 0.028927622 |
| 0.390786663 | 0.604292197 | 0.008305382 | 0 | 0.025988479 |
| 0 | 0 | 0 | 0 | 0 |
| 0 | 0 | 0 | 0 | 0 |
| 0 | 0 | 0 | 0.055087193 | 0 |
| 0 | 0 | 0 | 0 | 0 |
| 0 | 0 | 0 | 0 | 0 |
| 0 | 0 | 0.047958736 | 0 | 0 |
| 0 | 0 | 0 | 0 | 0 |
| 0.190524008 | 0.031957514 | 0.53628274 | 0.020820994 | 0 |
| 6.002283732 | 0.851745041 | 0.293830801 | 1.8796325 | 0.856258226 |
| 0 | 0 | 0 | 0 | 0 |
| 0.0011376 | 0 | 0.064974453 | 0.001857096 | 0.060734473 |
| 0.252132963 | 0.244426489 | 0.076729762 | 0.007756354 | 0.034632407 |
| 0 | 0 | 0 | 0 | 0 |
| 0 | 0 | 0 | 0 | 0 |
| 0 | 0 | 0 | 0 | 0 |
| 0 | 0 | 0 | 0.124101158 | 0 |
| 0.073803813 | 0.001997037 | 0 | 0.009922077 | 0 |
| 0.560782486 | 0.010898766 | 0.927614965 | 0.359837946 | 0 |
| 1.471525128 | 10.11888896 | 0.000612898 | 0 | 2.959421817 |
| 6.038143024 | 29.96678984 | 0.407379712 | 0.648147652 | 0.737984949 |
| 0 | 0 | 0 | 0 | 0 |
| 0 | 0 | 0 | 0.11576832 | 0 |
| 0 | 0 | 0.005576101 | 0 | 0 |
| 0.073966976 | 10.3010842 | 0 | 0 | 0 |
| 0 | 0 | 0 | 0.000752113 | 0.021236011 |
| 0.161737014 | 0.041016591 | 0 | 0 | 0 |
| 0 | 0 | 0 | 0.020367943 | 0 |
| 0 | 0 | 0 | 0.013203219 | 0 |
| 0 | 0 | 0.377533154 | 0.062632753 | 0.107035935 |
| 0 | 2.452988525 | 0.026906253 | 0 | 0 |
| 0 | 0 | 0 | 0.110777944 | 0.000684379 |
| 0.022149639 | 0.014342002 | 0.01423029 | 0.0080482 | 0 |
| 0.024787559 | 0.032431202 | 0.021490545 | 0.063461778 | 0.035320087 |
| 0 | 0.015069212 | 0 | 0.001548061 | 0 |
| 0 | 0 | 0 | 0 | 0 |
| 0 | 0 | 2.250487206 | 1.068000843 | 1.314993569 |
| 0 | 0 | 0 | 0 | 0 |
| 0 | 0 | 0 | 0.01905044 | 0 |
| 0 | 0 | 0 | 0 | 0 |
| 0 | 0 | 0 | 0.002667784 | 0 |
| 0 | 0 | 0 | 0 | 0 |
| 0.011474884 | 0.007490452 | 0.136173402 | 0.016630699 | 0 |
| 0 | 0 | 0.128608832 | 0.83368793 | 0 |
| 0 | 0 | 0 | 0 | 0 |
| 0.118837311 | 0.002541243 | 0.156482658 | 0.361321474 | 0.008508312 |

| | | | | |
|---|---|---|---|---|
| 0 | 0 | 0 | 0 | 0 |
| 0 | 0 | 0 | 0 | 0 |
| 0 | 0 | 0 | 0 | 0 |
| 0 | 0 | 0.004083303 | 0.000934568 | 0 |
| 0 | 0 | 0 | 0 | 0 |
| 0 | 0 | 0 | 0.014307008 | 0 |
| 0 | 0 | 0.187242632 | 0.267048338 | 0 |
| 0.194035146 | 0.218825518 | 0 | 0.015479487 | 0.001019004 |
| 0 | 0 | 0 | 0 | 0 |
| 0 | 0 | 0 | 0 | 0 |
| 0 | 0.005332632 | 0.008745612 | 0.001908245 | 0 |
| 0 | 0 | 0 | 0 | 0 |
| 0 | 0 | 0 | 0 | 0.00169697 |
| 0 | 0 | 0.036717379 | 0 | 0 |
| 0 | 0 | 0 | 0 | 0 |
| 0 | 0 | 0 | 0 | 0 |
| 0 | 0 | 0.039026598 | 0.010450708 | 0 |
| 0 | 0 | 0.002475378 | 0.012130902 | 0 |
| 0 | 0 | 0.19719519 | 0.395864688 | 0 |
| 0.046073153 | 0 | 0.055678203 | 0.255528662 | 0 |
| 0 | 0 | 0.029106768 | 0 | 0 |
| 0 | 0 | 0 | 0 | 0 |
| 0 | 0 | 0 | 0 | 0 |
| 0 | 0 | 0 | 0 | 0 |
| 0 | 0 | 0 | 0 | 0 |
| 0 | 0 | 0 | 0 | 0 |
| 0 | 0 | 0 | 0 | 0 |
| 0 | 0 | 0 | 0 | 0 |
| 0 | 0 | 0 | 0 | 0 |
| 0 | 0 | 0.012312729 | 1.433826247 | 0 |
| 0.011508308 | 0.000700007 | 0 | 0.034158081 | 0 |
| 0 | 0 | 0 | 0 | 0 |
| 0 | 0 | 0 | 0 | 0 |
| 0 | 0 | 0 | 0 | 0 |
| 0 | 0 | 0 | 0.00400027 | 0 |
| 0 | 0 | 0 | 0 | 0 |
| 0 | 0 | 0 | 0.069071617 | 0 |
| 0.006746741 | 0.001504722 | 0 | 0 | 0.000745749 |
| 0 | 0 | 0 | 0 | 0 |
| 0 | 0 | 0 | 0.001158767 | 0 |
| 0 | 0 | 0.208245298 | 0.040370345 | 0 |
| 0 | 0 | 0 | 0 | 0 |
| 0 | 0 | 0 | 0 | 0 |
| 0 | 0 | 0 | 0 | 0.002172631 |
| 0 | 0 | 0 | 0 | 0 |
| 0 | 0 | 0 | 0.032596202 | 0 |
| 0 | 0 | 0 | 0 | 0 |
| 0 | 0 | 0 | 0 | 0 |

| | | | | |
|---|---|---|---|---|
| 0 | 0 | 0 | 0 | 0 |
| 0 | 0 | 0 | 0 | 0 |
| 0 | 0 | 0 | 0.03857473 | 0 |
| 0 | 0 | 0.328587366 | 0.09041973 | 0 |
| 0 | 0 | 0 | 0 | 0 |
| 0 | 0 | 0 | 0 | 0 |
| 0 | 0 | 0 | 0 | 0 |
| 0 | 0 | 0 | 0 | 0 |
| 0.76082919 | 0.207526317 | 0.007351493 | 0 | 0.004731154 |
| 0 | 0 | 0 | 0 | 0.000993907 |
| 0 | 0 | 0 | 0 | 0 |
| 0 | 0 | 0 | 0 | 0 |
| 0 | 0 | 0 | 0 | 0 |
| 0 | 0 | 0 | 0 | 0 |
| 0 | 0 | 0.541798598 | 0.298658449 | 0 |
| 1.338207755 | 0 | 0.009443333 | 0.001873167 | 0 |
| 0 | 0 | 0 | 0 | 0 |
| 0 | 0 | 0 | 0 | 0 |
| 0 | 0 | 0 | 0 | 0 |
| 0 | 0 | 0 | 0 | 0 |
| 0 | 0 | 0 | 0.71361785 | 0 |
| 0 | 0 | 0 | 0 | 0 |
| 0 | 0 | 0 | 0 | 0 |
| 0.00196634 | 0 | 0.038694734 | 0 | 0 |
| 0.07878439 | 0 | 0.087412031 | 0.001905378 | 0.017900608 |
| 0 | 0 | 5.523716832 | 1.978181535 | 0.046968193 |
| 0 | 0 | 0.349951649 | 0.083447971 | 0 |
| 0 | 0 | 0 | 0 | 0 |
| 0 | 0 | 0.113164992 | 0 | 0.021571967 |
| 0 | 0 | 0 | 0 | 0 |
| 0 | 0 | 0 | 0 | 0 |
| 0 | 0 | 0 | 0.063040915 | 0 |
| 0 | 0 | 0 | 0 | 0 |
| 0.001363572 | 0.000814598 | 0.010131888 | 0.000508797 | 0 |
| 0.002567697 | 0 | 0.009931963 | 0 | 0.036399851 |
| 0.004764508 | 0 | 0 | 0 | 0 |
| 0.001404812 | 0.0309398 | 0 | 0 | 0 |
| 0 | 0 | 0.493027826 | 1.480830279 | 0.867200512 |
| 0 | 0 | 0.199220368 | 0.268686301 | 6.944108405 |
| 0 | 0 | 0 | 0 | 0 |
| 0 | 0 | 0 | 0 | 0 |
| 0 | 0 | 0 | 0.025536606 | 0 |
| 0.178271504 | 0.023596579 | 4.364409594 | 0.074906668 | 0.221451913 |
| 0 | 0 | 0 | 0 | 0 |
| 0 | 0 | 0 | 0 | 0 |
| 0.001934474 | 0 | 0.003396371 | 0 | 0 |
| 0 | 0 | 0 | 0 | 0 |
| 0 | 0 | 1.435447037 | 0.112694864 | 1.34261963 |

| | | | | |
|---|---|---|---|---|
| 0.065113926 | 0.016871526 | 0.078511628 | 0 | 0 |
| 0 | 0 | 0 | 0.458044529 | 0 |
| 0 | 0 | 0 | 0.002335558 | 0 |
| 0 | 0 | 0 | 0 | 0 |
| 0.002542292 | 0.001275764 | 0.006193136 | 0.006142312 | 0 |
| 0 | 0 | 0 | 0.004316319 | 0 |
| 0 | 0 | 0 | 0 | 0 |
| 0 | 0 | 0 | 0 | 0 |
| 0 | 0 | 0 | 0 | 0 |
| 0 | 0 | 0 | 0.016495803 | 0 |
| 0 | 0 | 0 | 0.00749451 | 0 |
| 0 | 0 | 0.587107239 | 0.385333905 | 0.115574063 |
| 0 | 0 | 0.435335861 | 0.003015107 | 0.133312107 |
| 0 | 0 | 0 | 0 | 0 |
| 0.024697353 | 0.032459261 | 1.833564613 | 0.002495854 | 0.039778732 |
| 0 | 0 | 0 | 0 | 0 |
| 0 | 0 | 0.005925084 | 0.000934891 | 0 |
| 0 | 0 | 0.029337754 | 0 | 0 |
| 0 | 0 | 0.09094304 | 0 | 0 |
| 0 | 0 | 0 | 0.010262606 | 0 |
| 0 | 0 | 0 | 0.043327993 | 0 |
| 0 | 0 | 0 | 0 | 0 |
| 0 | 0 | 0.292016151 | 0.009490364 | 0 |
| 0 | 0 | 0 | 0 | 0 |
| 0 | 0 | 0 | 0 | 0 |
| 0 | 0 | 0 | 0 | 0 |
| 0 | 0 | 0 | 0 | 0 |
| 0 | 0 | 0 | 0 | 0 |
| 0 | 0 | 0 | 0 | 0 |
| 0 | 0 | 0 | 0 | 0 |
| 0 | 0 | 0 | 0 | 0 |
| 0.003052216 | 0.001276377 | 0.075358103 | 0.010214423 | 0.02378498 |
| 0 | 0 | 0 | 0 | 0 |
| 0.045745037 | 0.105372619 | 0 | 0.026385983 | 0 |
| 0.00395046 | 0 | 0.000812795 | 0 | 0 |
| 0.003182889 | 0 | 0.05301825 | 0.181858744 | 0.039896327 |
| 0 | 0 | 0 | 0.003434746 | 0 |
| 0 | 0 | 0 | 0 | 0 |
| 0 | 0 | 0 | 0 | 0 |
| 0 | 0 | 0 | 0 | 0 |
| 0 | 0 | 0.017312611 | 0.06030292 | 0 |
| 0 | 0 | 0 | 0.116478326 | 0 |
| 0 | 0 | 0 | 0 | 0 |
| 0 | 0 | 0 | 0 | 0 |
| 0.009485773 | 0.01403624 | 0.309544702 | 0.003335278 | 0 |
| 0 | 0 | 0 | 0 | 0 |
| 0 | 0 | 0 | 0.098618865 | 0 |
| 0.005291087 | 0 | 0 | 0 | 0 |

| | | | | |
|---|---|---|---|---|
| 0 | 0 | 0 | 0 | 0 |
| 0 | 0 | 0 | 0 | 0 |
| 0.007798219 | 0.001683129 | 0.011096652 | 0 | 0 |
| 0 | 0 | 0 | 0 | 0 |
| 0 | 0 | 0 | 0 | 0 |
| 0.109720203 | 0 | 0 | 0.00042394 | 0 |
| 0 | 0 | 0 | 0 | 0.006243995 |
| 0 | 0 | 0.001094899 | 0.002171828 | 0 |
| 0 | 0 | 0 | 0 | 0 |
| 0 | 0 | 0 | 0.015723404 | 0 |
| 0 | 0 | 0 | 0 | 0 |
| 0 | 0 | 0 | 0 | 0 |
| 0 | 0 | 0 | 0 | 0 |
| 0 | 0 | 0 | 0 | 0 |
| 0 | 0 | 0 | 0 | 0 |
| 0 | 0 | 0 | 0 | 0 |
| 0 | 0 | 0 | 0 | 0 |
| 0 | 0 | 0 | 0 | 0 |
| 0 | 0 | 0.102643866 | 8.790034757 | 29.08142856 |
| 0 | 0 | 0 | 0 | 0 |
| 0 | 0 | 0 | 0.002387955 | 0 |
| 0 | 0 | 0 | 0 | 0 |
| 0 | 0 | 0 | 0 | 0 |
| 0 | 0 | 0 | 0 | 0 |
| 0 | 0 | 0.001224745 | 0.007288162 | 0 |
| 0.015171027 | 0.014961007 | 0.015206307 | 0 | 0 |
| 0 | 0 | 0 | 0 | 0 |
| 0.005762461 | 0 | 0.144315942 | 0.009117585 | 0 |
| 0 | 0 | 0 | 0 | 0 |
| 0 | 0 | 0.615347858 | 0.918612329 | 0.224492303 |
| 0 | 0 | 0 | 0.010080974 | 0 |
| 0 | 0 | 0 | 0 | 0 |
| 0 | 0 | 0.090753147 | 0.018643502 | 0 |
| 0 | 0 | 0 | 0 | 0 |
| 0.000711864 | 0.000595375 | 0.001406053 | 0 | 0 |
| 0 | 0 | 0 | 0 | 0 |
| 0 | 0 | 0 | 0 | 0 |
| 0 | 0 | 0.006559107 | 0.00105491 | 0 |
| 0 | 0 | 0 | 0 | 0 |
| 0 | 0 | 0 | 0 | 0 |
| 12.61833179 | 0.001431507 | 2.684101273 | 3.431494005 | 0.834165745 |
| 0.233162176 | 0 | 0.12055146 | 0.52303617 | 0 |
| 0 | 0 | 0 | 0 | 0.451060581 |
| 0 | 0 | 0.000426994 | 0.849097293 | 0 |
| 0 | 0 | 0 | 0 | 0 |
| 0 | 0 | 0.000349116 | 1.368844399 | 0 |
| 0 | 0 | 0 | 0.000997585 | 0 |
| 0.002136936 | 0 | 0.054167109 | 0.00683742 | 0.00042517 |

| | | | | |
|---|---|---|---|---|
| 0 | 0 | 0 | 0 | 0 |
| 0.001318809 | 0 | 0.031982054 | 0.000861165 | 0 |
| 0 | 0 | 0.004225479 | 0 | 0 |
| 0 | 0 | 0 | 0 | 0 |
| 1.815207438 | 0.194727004 | 2.876309071 | 0.514612722 | 1.368619446 |
| 0 | 0 | 0 | 0 | 0 |
| 0 | 0 | 0 | 0 | 0 |
| 0 | 0 | 0 | 0.006695764 | 0 |
| 0.080548225 | 0.653481775 | 0 | 0 | 0.005967926 |
| 0 | 0 | 0 | 0.022794681 | 0 |
| 0 | 0 | 0 | 0 | 0 |
| 0 | 0 | 0 | 0 | 0 |
| 0 | 0 | 0.053012028 | 0.11617143 | 0.012606349 |
| 0.044374041 | 0 | 0.010890962 | 0.012173215 | 0.048584319 |
| 0.058380379 | 0.016892771 | 3.308320814 | 0 | 0.334702485 |
| 0 | 0 | 0.159731934 | 0.332177805 | 0.03715292 |
| 0 | 0 | 0.094543568 | 0.005489565 | 0 |
| 0 | 0 | 0 | 0 | 0 |
| 0 | 0 | 0 | 0 | 0 |
| 0 | 0 | 0 | 0 | 0 |
| 0 | 0 | 0 | 0.001606252 | 0 |
| 0 | 0 | 0 | 0 | 0 |
| 0 | 0 | 0 | 0 | 0 |
| 0 | 0 | 0 | 0 | 0 |
| 0 | 0 | 0 | 0 | 0 |
| 0 | 0 | 0 | 0 | 0 |
| 0 | 0 | 0.005073056 | 0.0641184 | 0.000655151 |
| 0 | 0.000745819 | 0 | 0.000582297 | 0 |
| 0 | 0 | 0 | 0 | 0 |
| 0 | 0 | 0 | 0 | 0 |
| 0 | 0 | 0 | 0 | 0 |
| 0 | 0 | 0 | 0 | 0 |
| 0 | 0 | 0 | 0 | 0 |
| 0 | 0 | 0 | 0.07874199 | 0 |
| 0 | 0 | 0.700074911 | 0 | 0.049677286 |
| 0.007736719 | 0 | 0.013074037 | 0 | 0 |
| 0 | 0 | 0 | 0 | 0 |
| 0 | 0 | 0.039281445 | 0.002544267 | 0 |
| 0 | 0 | 0 | 0 | 0 |
| 0 | 0 | 0 | 0.005177688 | 0 |
| 0 | 0 | 2.915699979 | 0.132616048 | 0.556362896 |
| 0 | 0 | 0 | 0.111034692 | 0 |
| 0 | 0 | 0 | 0.052446387 | 0 |
| 0 | 0 | 0 | 0 | 0 |
| 0 | 0 | 0 | 0 | 0 |
| 0 | 0 | 0 | 0 | 0 |
| 0 | 0 | 0 | 0.503658469 | 0 |
| 0 | 0 | 0 | 0 | 0 |

| | | | | |
|---|---|---|---|---|
| 0 | 0 | 1.611108899 | 0.111732572 | 0 |
| 0 | 0 | 0 | 0.003498669 | 0 |
| 0 | 0 | 0 | 0 | 0 |
| 0 | 0 | 0 | 0 | 0 |
| 0 | 0 | 0.104680867 | 0.093408068 | 0 |
| 0 | 0 | 0 | 0 | 0 |
| 0.05077176 | 0 | 0.282941034 | 0.007696304 | 0 |
| 0 | 0 | 0 | 0.001001471 | 0 |
| 0 | 0 | 0 | 0 | 0 |
| 0.044469133 | 0.019888894 | 0 | 0.14068569 | 0 |
| 0 | 0 | 0 | 0.009153547 | 0 |
| 0 | 0 | 0 | 0 | 0 |
| 0 | 0 | 0 | 0 | 0 |
| 0 | 0 | 0 | 0 | 0 |
| 0 | 0 | 0 | 0.038538435 | 0 |
| 0 | 0 | 0 | 0.002384099 | 0 |
| 0 | 0 | 0 | 0 | 0 |
| 0 | 0 | 0 | 0 | 0 |
| 0 | 0 | 0 | 0 | 0 |
| 0 | 0 | 0 | 0 | 0 |
| 0.00410006 | 0 | 0.006677561 | 0.027618323 | 0 |
| 0 | 0 | 0 | 0 | 0 |
| 0.000714055 | 0 | 0 | 0.016785687 | 0 |
| 0.002328134 | 0.001062087 | 0.002508252 | 0.002902279 | 0 |
| 0 | 0 | 0 | 0 | 0 |
| 0 | 0 | 0.036825266 | 0 | 0 |
| 0 | 0 | 0 | 0 | 0 |
| 0 | 0 | 0 | 0 | 0 |
| 0 | 0 | 0 | 0 | 0 |
| 0 | 0.001783357 | 0.019654239 | 0 | 0.335758815 |
| 0 | 0 | 0 | 0.011966183 | 0 |
| 0 | 0 | 0 | 0 | 0 |
| 0 | 0 | 0 | 0.011740624 | 0 |
| 0 | 0 | 0.605526363 | 0.002361493 | 0 |
| 0 | 0 | 0 | 0.096645989 | 0 |
| 0 | 0 | 0 | 0 | 0 |
| 0.000794481 | 0.001771928 | 0.027548814 | 0 | 0 |
| 0 | 0 | 0 | 0 | 0 |
| 0 | 0 | 0 | 0 | 0 |
| 0 | 0 | 0 | 0 | 0 |
| 0 | 0 | 0 | 0 | 0 |
| 0 | 0 | 0 | 0 | 0 |
| 0 | 0 | 0 | 0 | 0 |
| 0 | 0 | 0 | 0 | 0 |
| 0 | 0 | 0 | 0 | 0 |
| 0 | 0 | 0 | 0 | 0 |
| 0 | 0 | 0 | 0 | 0 |
| 0 | 0 | 0 | 0 | 0 |

| | | | | |
|---|---|---|---|---|
| 0.145071213 | 0.003428529 | 0.010346044 | 0.001933258 | 0 |
| 0 | 0 | 0.000711025 | 1.477231735 | 0 |
| 0 | 0 | 0 | 0.003357863 | 0 |
| 0 | 0 | 0 | 0 | 0 |
| 0 | 0 | 0 | 0 | 0 |
| 0 | 0 | 0 | 0 | 0 |
| 0 | 0 | 0.002535914 | 0 | 0 |
| 0 | 0.01681933 | 0.23185958 | 0.436093426 | 0.062653838 |
| 0.161578333 | 0.020330459 | 1.525587488 | 0.051820587 | 0.494785296 |
| 0.099545122 | 0.472059363 | 0.001146942 | 0 | 0 |
| 0 | 0 | 0 | 1.32037799 | 0 |
| 0 | 0 | 0 | 0 | 0 |
| 0 | 0 | 0 | 0.02364318 | 0 |
| 0.173606728 | 0.044046293 | 0.000696957 | 0.777123906 | 1.24193877 |
| 0 | 0 | 0 | 0 | 0 |
| 0 | 0 | 0 | 0 | 0 |
| 0 | 0 | 0 | 0 | 0 |
| 0 | 0 | 0 | 0 | 0 |
| 0 | 0 | 0 | 0 | 0 |
| 0 | 0 | 0 | 0.042851267 | 0 |
| 0 | 0 | 0 | 0 | 0 |
| 0 | 0 | 0 | 0 | 0 |
| 0 | 0 | 0.483206853 | 0 | 0 |
| 0 | 0 | 0 | 0 | 0 |
| 0 | 0 | 0.002007711 | 0.05200636 | 0 |
| 0 | 0 | 0 | 0.055677055 | 0 |
| 0 | 0 | 0 | 0 | 0 |
| 0 | 0 | 0 | 0 | 0 |
| 0 | 0 | 0 | 0 | 0 |
| 0 | 0 | 0 | 0 | 0 |
| 0 | 0 | 0 | 0.010999883 | 0 |
| 0 | 0 | 0 | 0 | 0 |
| 0 | 0 | 0 | 0 | 0 |
| 0 | 0 | 0 | 0.002128463 | 0 |
| 0.00192999 | 0.005764885 | 0 | 0 | 0 |
| 0 | 0 | 0.101877831 | 0 | 0 |
| 0 | 0 | 0 | 0 | 0 |
| 0 | 0 | 0 | 0 | 0 |
| 0 | 0 | 0 | 0 | 0 |
| 0 | 0 | 0 | 0 | 0 |
| 0 | 0 | 0 | 0 | 0 |
| 0 | 0 | 0 | 0.257928489 | 0.024734283 |
| 0 | 0 | 0 | 0 | 0 |
| 0 | 0 | 0 | 0 | 0 |
| 0 | 0 | 0 | 0 | 0 |
| 0.019845814 | 0.00152433 | 0.012132984 | 0 | 0 |
| 0 | 0 | 0 | 0 | 0 |
| 0 | 0 | 0 | 0 | 0 |

| | | | | |
|---|---|---|---|---|
| 0 | 0 | 0 | 0 | 0 |
| 0 | 0 | 0.008525778 | 0 | 0 |
| 0 | 0 | 0 | 0 | 0 |
| 0 | 0 | 0 | 0 | 0 |
| 0.394289329 | 0 | 2.190106909 | 4.141410067 | 0.266641825 |
| 0 | 0 | 0 | 0 | 0 |
| 0 | 0.002918316 | 0 | 0.004882436 | 0 |
| 0.070687638 | 0.014780089 | 0.015259034 | 0 | 0.007877477 |
| 0 | 0 | 0.000761779 | 0 | 0 |
| 0 | 0 | 0 | 0 | 0 |
| 0 | 0 | 0 | 0 | 0 |
| 0 | 0 | 0 | 0 | 0 |
| 0 | 0 | 0 | 0.021524871 | 0 |
| 0 | 0 | 0 | 0 | 0 |
| 0 | 0 | 0 | 0 | 0 |
| 0 | 0 | 0.194540477 | 0.008859673 | 0 |
| 0 | 0 | 0 | 0 | 0 |
| 0 | 0 | 0 | 0 | 0 |
| 0 | 0 | 0 | 0 | 0 |
| 0 | 0 | 0 | 0 | 0 |
| 0 | 0 | 0 | 0 | 0 |
| 0 | 0 | 0 | 0 | 0 |
| 0 | 0 | 0 | 0 | 0 |
| 0.00225643 | 0.005032505 | 0.001155476 | 0 | 0 |
| 0 | 0 | 0.047730871 | 0 | 1.956195446 |
| 0 | 0 | 0 | 0 | 0 |
| 0.008942967 | 0 | 0.003623361 | 0 | 0 |
| 0.000965811 | 0 | 0 | 0.000630662 | 0 |
| 0 | 0 | 0 | 0 | 0 |
| 0 | 0 | 0 | 0 | 0 |
| 0 | 0 | 0 | 0 | 0 |
| 0 | 0 | 0 | 0 | 0 |
| 0 | 0 | 0 | 0 | 0 |
| 0 | 0 | 0 | 0 | 0 |
| 0 | 0 | 0 | 0 | 0 |
| 0.002562475 | 0 | 0 | 0 | 0 |
| 0 | 0 | 0 | 0 | 0 |
| 0 | 0 | 0 | 0.053285816 | 0 |
| 0 | 0 | 0 | 0 | 0 |
| 0 | 0 | 0 | 0 | 0 |
| 0 | 0 | 0 | 0 | 0 |
| 0 | 0 | 0 | 0 | 0 |
| 0 | 0 | 0 | 0 | 0 |
| 0 | 0 | 0 | 0 | 0 |
| 0 | 0 | 0 | 0 | 0 |
| 0 | 0 | 0 | 0 | 0 |
| 0 | 0 | 0 | 0 | 0 |
| 0 | 0 | 0 | 0 | 0 |

| | | | | |
|---|---|---|---|---|
| 0 | 0 | 0.049408156 | 0.029993025 | 0 |
| 0 | 0 | 0 | 0.059041671 | 0 |
| 0 | 0 | 0 | 0 | 0 |
| 0 | 0 | 0.007062987 | 0.000700502 | 0 |
| 0 | 0 | 0 | 0 | 0 |
| 0.013740939 | 0.126416184 | 0.431948385 | 0 | 0.000828464 |
| 0 | 0 | 0 | 0 | 0 |
| 0 | 0 | 0 | 0 | 0 |
| 0 | 0 | 0 | 0 | 0 |
| 0 | 0 | 0 | 0 | 0 |
| 0 | 0 | 0 | 0 | 0 |
| 0 | 0 | 0 | 0 | 0 |
| 0 | 0 | 0 | 0.000524701 | 0 |
| 0 | 0 | 0 | 0 | 0 |
| 0 | 0 | 0 | 0 | 0 |
| 0 | 0 | 0 | 0 | 0 |
| 0 | 0 | 0 | 0 | 0 |
| 0 | 0 | 0 | 0 | 0 |
| 0 | 0 | 0 | 0 | 0 |
| 0 | 0 | 0 | 0 | 0 |
| 0.020722312 | 0 | 0.056217253 | 0 | 0.006623229 |
| 0 | 0 | 0 | 0 | 0 |
| 0.007882762 | 0.001054853 | 0.519200991 | 0.181804148 | 0.00230028 |
| 0 | 0 | 0 | 1.291788114 | 0 |
| 0 | 0 | 0 | 0.486562567 | 0 |
| 0 | 0 | 0 | 0 | 0 |
| 0 | 0 | 0 | 0 | 0 |
| 0 | 0 | 0 | 0 | 0 |
| 0 | 0 | 0 | 0.000891193 | 0.004887768 |
| 0 | 0 | 0 | 0 | 0 |
| 0 | 0 | 0 | 0 | 0 |
| 0 | 0 | 0 | 0 | 0 |
| 0.001476435 | 0.019757311 | 0.002916211 | 0 | 0.184869781 |
| 0 | 0 | 0 | 0.0122229 | 0 |
| 0 | 0 | 0.391628343 | 0.003277031 | 0.160460243 |
| 0.018482523 | 0.112368162 | 0 | 0.001856746 | 0.009429041 |
| 0 | 0 | 0 | 0 | 0 |
| 0 | 0 | 0 | 0 | 0 |
| 0.005207452 | 0.005730668 | 0 | 0 | 0 |
| 0 | 0 | 0 | 0 | 0 |
| 0 | 0 | 0 | 0 | 0 |
| 0 | 0 | 0 | 0 | 0 |
| 0.009934801 | 0 | 0.934304652 | 0.527354559 | 0.032306561 |
| 0 | 0 | 0 | 0 | 0 |
| 0 | 0 | 0 | 0 | 0 |
| 0 | 0 | 0.059152208 | 0.009306911 | 0 |
| 0 | 0 | 0 | 0 | 0 |
| 0 | 0 | 0 | 0 | 0 |

| | | | | |
|---|---|---|---|---|
| 0 | 0 | 0 | 0 | 0 |
| 0 | 0 | 0 | 0 | 0 |
| 0 | 0 | 0 | 0 | 0 |
| 0 | 0 | 0 | 0 | 0 |
| 0 | 0 | 0 | 0.012370895 | 0 |
| 0 | 0 | 0 | 0 | 0 |
| 0 | 0 | 0 | 0 | 0 |
| 0 | 0 | 0 | 0 | 0 |
| 0 | 0 | 0.120574117 | 0.005294111 | 0 |
| 0 | 0 | 0.041077668 | 0.034578127 | 0.074237433 |
| 0 | 0 | 0 | 0 | 0 |
| 0 | 0 | 0 | 0 | 0 |
| 0 | 0 | 0 | 0 | 0 |
| 0 | 0 | 0 | 0.002355674 | 0 |
| 0 | 0 | 0 | 0.00636345 | 0 |
| 0.00128962 | 0.071186763 | 0 | 0 | 0 |
| 0 | 0 | 0 | 0 | 0 |
| 0 | 0 | 0 | 0 | 0 |
| 0 | 0 | 0 | 0 | 0 |
| 0 | 0 | 0.436616933 | 0.284412003 | 0.117006249 |
| 0 | 0 | 0 | 0 | 0.000714628 |
| 0 | 0 | 0 | 0 | 0 |
| 0 | 0 | 0 | 0 | 0 |
| 0 | 0 | 0 | 0 | 0 |
| 0 | 0 | 0 | 0 | 0 |
| 0 | 0 | 0 | 0 | 0 |
| 0 | 0 | 0 | 0 | 0 |
| 0 | 0 | 0 | 0 | 0 |
| 0 | 0 | 0 | 0 | 0 |
| 0 | 0 | 0.036255968 | 0.00349111 | 0 |
| 0 | 0 | 0.344913411 | 0.316621733 | 0.11247567 |
| 0 | 0 | 0 | 0 | 0 |
| 0 | 0 | 0 | 0.010058441 | 0 |
| 0 | 0 | 0 | 0 | 0 |
| 0 | 0 | 0 | 0 | 0 |
| 0 | 0 | 0 | 0 | 0 |
| 0 | 0 | 0 | 0.002485084 | 0 |
| 0 | 0 | 0 | 0 | 0 |
| 0 | 0 | 0 | 0 | 0 |
| 0 | 0 | 0 | 0 | 0 |
| 0 | 0 | 0 | 0 | 0 |
| 0 | 0 | 0 | 1.850951558 | 0 |
| 0 | 0 | 0 | 0.016730364 | 0 |
| 0.001928498 | 0 | 0.005622975 | 0 | 0 |
| 0 | 0 | 0 | 0 | 0 |
| 0 | 0 | 0 | 0 | 0 |
| 0.015314024 | 0.321305313 | 0 | 0 | 0 |
| 0 | 0 | 0 | 0 | 0 |

| | | | | |
|---|---|---|---|---|
| 0 | 0 | 0 | 0 | 0 |
| 0 | 0 | 0 | 0 | 0 |
| 0 | 0 | 0 | 0 | 0 |
| 0 | 0 | 0 | 0 | 0 |
| 0.069630588 | 0.302894655 | 0.131818027 | 0.006569014 | 0.013736127 |
| 0 | 0 | 0 | 0 | 0 |
| 0 | 0 | 0 | 0 | 0 |
| 0 | 0 | 0 | 0 | 0 |
| 0 | 0 | 0 | 0 | 0 |
| 0 | 0 | 0 | 0 | 0 |
| 0 | 0 | 0.007749818 | 0.049878655 | 0 |
| 0.003560156 | 0.001603309 | 0.069417643 | 0.010193076 | 0 |
| 0 | 0 | 0 | 0.010336873 | 0 |
| 0.002452401 | 0 | 0 | 0 | 0 |
| 0.018156818 | 0 | 0 | 0 | 0 |
| 0 | 0 | 0.315339346 | 0 | 0 |
| 0 | 0 | 0.017195871 | 0.031083663 | 0 |
| 0 | 0 | 0 | 0 | 0 |
| 0.065527968 | 0.054978445 | 0.032630289 | 0.003926832 | 0 |
| 0 | 0 | 0.127102588 | 0.033769101 | 0 |
| 0 | 0 | 0 | 0 | 0 |
| 0 | 0 | 0 | 0 | 0 |
| 0 | 0 | 0 | 0 | 0 |
| 0 | 0 | 0 | 0 | 0 |
| 0 | 0 | 0 | 0 | 0 |
| 0 | 0 | 0 | 0 | 0 |
| 0.040790524 | 0 | 0.01228523 | 0.002975263 | 0 |
| 0.007406576 | 0 | 0 | 0.046035358 | 0 |
| 0 | 0 | 0 | 0 | 0 |
| 0 | 0 | 0 | 0.001081959 | 0 |
| 0 | 0 | 0 | 0 | 0 |
| 0 | 0 | 0 | 0.036775129 | 0 |
| 0.002890001 | 0 | 0 | 0 | 0 |
| 0 | 0 | 0 | 0 | 0 |
| 0 | 0 | 0 | 0 | 0 |
| 0 | 0 | 0 | 0 | 0 |
| 0 | 0 | 0 | 0 | 0 |
| 0.005356671 | 0.003034912 | 0.165758667 | 0.004174839 | 0 |
| 0 | 0 | 0.116723248 | 0.181180253 | 0 |
| 0 | 0 | 0 | 0.010738989 | 0 |
| 0 | 0.004328315 | 0.018853656 | 0 | 0 |
| 0 | 0 | 0 | 0 | 0 |
| 0 | 0 | 0 | 0 | 0 |
| 0 | 0 | 0.001855724 | 0 | 0 |
| 0 | 0 | 0 | 0 | 0 |
| 0 | 0 | 0 | 0 | 0 |
| 0 | 0 | 0 | 0.261213548 | 0 |
| 0 | 0 | 0 | 0.003078899 | 0 |

| | | | | |
|---|---|---|---|---|
| 0 | 0 | 0 | 0 | 0 |
| 0 | 0 | 0.017302905 | 0.045619416 | 0 |
| 0 | 0 | 0 | 0 | 0 |
| 0.003490951 | 0.000673776 | 0.002298408 | 0 | 0 |
| 0 | 0 | 0 | 0 | 0.000660719 |
| 0.000964314 | 0 | 0 | 0 | 0 |
| 0 | 0 | 0 | 0 | 0 |
| 0.00567047 | 0.001229552 | 0.50096458 | 0.000959971 | 0 |
| 0.01810666 | 0 | 0 | 0.000569803 | 0 |
| 0 | 0 | 0 | 0.017425906 | 0 |
| 0 | 0 | 0 | 0.01956072 | 0 |
| 0 | 0 | 0 | 0 | 0 |
| 0 | 0 | 0.107781026 | 0.075449058 | 0.027394908 |
| 0 | 0 | 0 | 0 | 0 |
| 0 | 0 | 0 | 0 | 0 |
| 0 | 0 | 0 | 0 | 0 |
| 0 | 0 | 0 | 0 | 0 |
| 0 | 0 | 0 | 0 | 0 |
| 0 | 0 | 0 | 0 | 0 |
| 0 | 0 | 0 | 0 | 0 |
| 0 | 0 | 0 | 0 | 0 |
| 0 | 0 | 0 | 0 | 0 |
| 0.023811889 | 0.22892143 | 0.034160482 | 0 | 0 |
| 0 | 0 | 0.010710531 | 0.113179349 | 0.018243052 |
| 0 | 0 | 0.005329581 | 0 | 0 |
| 0 | 0 | 0 | 0.014263487 | 0 |
| 0 | 0 | 0 | 0.001250997 | 0 |
| 0 | 0 | 0 | 0 | 0 |
| 0 | 0 | 0 | 0 | 0 |
| 0 | 0 | 0 | 0 | 0 |
| 0 | 0 | 0.014720304 | 0.016606933 | 0.008526109 |
| 0 | 0 | 0 | 0.002257977 | 0 |
| 0 | 0 | 0 | 0 | 0 |
| 0 | 0 | 0 | 0 | 0 |
| 0 | 0 | 0.313002963 | 0.795287108 | 0 |
| 0 | 0 | 0 | 0 | 0 |
| 0 | 0 | 0 | 0 | 0 |
| 0 | 0 | 0 | 0 | 0 |
| 0 | 0.005932861 | 0.045925592 | 0 | 0 |
| 0 | 0 | 0 | 0 | 0 |
| 0 | 0 | 0 | 0 | 0 |
| 0 | 0 | 0 | 0 | 0 |
| 0 | 0 | 0 | 0 | 0.001619461 |
| 0 | 0 | 0 | 0 | 0 |
| 0 | 0 | 0 | 0 | 0 |
| 0.117543583 | 0.037061557 | 0.198747326 | 0.560782994 | 0.105412833 |
| 0 | 0 | 0 | 0 | 0 |
| 0 | 0 | 0 | 0.011439911 | 0 |

| | | | | |
|---|---|---|---|---|
| 0 | 0 | 0 | 0 | 0 |
| 0 | 0 | 0 | 0 | 0 |
| 0 | 0 | 0 | 0 | 0.199123073 |
| 0 | 0 | 0 | 0 | 0 |
| 0 | 0 | 0 | 0.000687716 | 0 |
| 0 | 0 | 0 | 0 | 0 |
| 0 | 0 | 0 | 0 | 0 |
| 0 | 0 | 0 | 0 | 0 |
| 0 | 0 | 0 | 0 | 0 |
| 0 | 0 | 0 | 0 | 0 |
| 0.000767315 | 0.007059271 | 0.007451592 | 0.001127355 | 0.002926113 |
| 0 | 0.001437207 | 0.000942818 | 0 | 0 |
| 0 | 0 | 0 | 0 | 0 |
| 0 | 0 | 0 | 0 | 0 |
| 0 | 0 | 0 | 0 | 0 |
| 0 | 0 | 0 | 0 | 0 |
| 0 | 0 | 0 | 0 | 0 |
| 0 | 0 | 0 | 0 | 0 |
| 0 | 0 | 0 | 0 | 0 |
| 0.007631935 | 0.000851073 | 0.017586771 | 0.000664474 | 0.001855905 |
| 0 | 0 | 0 | 0 | 0 |
| 0 | 0 | 0 | 0 | 0 |
| 0 | 0 | 0 | 0 | 0 |
| 0 | 0 | 0 | 0 | 0 |
| 0 | 0 | 0 | 0 | 0 |
| 0 | 0 | 0 | 0 | 0 |
| 0 | 0 | 0 | 0 | 0 |
| 0 | 0 | 0 | 0.004595956 | 0 |
| 0 | 0 | 0 | 0.001298349 | 0 |
| 0 | 0 | 0 | 0 | 0 |
| 0 | 0 | 0.001295045 | 0.006850225 | 0.001739361 |
| 0 | 0 | 0 | 0 | 0 |
| 0.190830177 | 0.029076582 | 0.004176886 | 0.012427825 | 0.394658722 |
| 0.003630463 | 0.006072753 | 0.000512199 | 0.000677327 | 0.000859911 |
| 0 | 0 | 0 | 0 | 0 |
| 0 | 0 | 0 | 0 | 0 |
| 0 | 0 | 0 | 0 | 0 |
| 0 | 0 | 0 | 0 | 0 |
| 0 | 0 | 0 | 0 | 0 |
| 0 | 0 | 0 | 0 | 0 |
| 0 | 0 | 0 | 0 | 0 |
| 0.000835076 | 0.033000559 | 0.118345698 | 0.089700834 | 0 |
| 0 | 0 | 0.049064204 | 0.034544725 | 0.005903797 |
| 0 | 0 | 0 | 0 | 0 |
| 0 | 0 | 0 | 0 | 0 |
| 0.011761222 | 0 | 0.113974262 | 0.131998755 | 0.007068872 |
| 0 | 0 | 0 | 0.000629397 | 0 |
| 0 | 0 | 0 | 0 | 0 |

| | | | | |
|---|---|---|---|---|
| 0 | 0 | 0 | 0 | 0 |
| 0 | 0 | 0.00192974 | 0.003827808 | 0.010072731 |
| 0 | 0 | 0.010451014 | 0.002235642 | 0.004541269 |
| 0 | 0 | 0 | 0 | 0 |
| 0.009443591 | 0.007690399 | 0.000490861 | 1.876576802 | 0 |
| 0 | 0 | 0 | 0.00063184 | 0 |
| 0 | 0 | 0 | 0 | 0 |
| 0 | 0 | 0 | 0 | 0 |
| 0 | 0 | 0 | 0 | 0 |
| 0 | 0 | 0 | 0 | 0 |
| 0 | 0 | 0 | 0 | 0 |
| 0 | 0 | 0 | 0 | 0 |
| 0 | 0 | 0 | 0 | 0 |
| 0 | 0 | 0 | 0 | 0 |
| 0 | 0 | 0 | 0 | 0 |
| 0 | 0 | 0 | 0 | 0 |
| 0 | 0 | 0 | 0.00479263 | 0 |
| 0 | 0 | 0 | 0 | 0 |
| 0 | 0 | 0 | 0 | 0 |
| 0 | 0 | 0 | 0 | 0 |
| 0 | 0 | 0 | 0 | 0 |
| 0 | 0 | 0 | 0.000653933 | 0 |
| 0 | 0 | 0 | 0 | 0 |
| 0 | 0 | 0 | 0 | 0 |
| 0 | 0 | 0.007937567 | 0.01306488 | 0.002211562 |
| 0 | 0 | 0.002208618 | 0 | 0.01146617 |
| 0 | 0 | 0 | 0 | 0 |
| 0 | 0 | 0.166450866 | 0.058842139 | 0 |
| 0 | 0 | 0 | 0 | 0 |
| 0 | 0 | 0 | 0 | 0 |
| 0 | 0 | 0 | 0 | 0 |
| 0.002262894 | 0.001324817 | 0 | 0 | 0 |
| 0 | 0 | 0 | 0 | 0 |
| 0 | 0 | 0 | 0 | 0 |
| 0 | 0 | 0 | 0 | 0 |
| 0 | 0 | 0 | 0 | 0 |
| 0 | 0 | 0 | 0 | 0 |
| 0 | 0 | 0.008793976 | 0 | 0 |
| 1.718471405 | 0 | 7.232943786 | 1.960493787 | 0 |
| 0 | 0 | 0 | 0 | 0 |
| 0 | 0 | 0 | 0.048261473 | 0 |
| 0 | 0 | 0.038704784 | 0.013344106 | 0.014852569 |
| 0.111359428 | 0.002599162 | 0.0098894 | 0.001352861 | 0.000687018 |
| 0 | 0 | 0 | 0.56566337 | 0 |
| 0 | 0 | 0 | 0 | 0 |
| 0 | 0 | 0 | 0 | 0 |
| 0.006735089 | 0.004970262 | 0.010172845 | 0 | 0 |
| 0 | 0 | 0 | 0 | 0 |

| | | | | |
|---|---|---|---|---|
| 0 | 0 | 0 | 0 | 0 |
| 0.034743636 | 0.016724149 | 0.025837086 | 0.001795386 | 0.001326173 |
| 0 | 0 | 0 | 0.14050211 | 0 |
| 0 | 0 | 0 | 0 | 0 |
| 0 | 0 | 0 | 0 | 0 |
| 0 | 0 | 0 | 0 | 0 |
| 0 | 0 | 0 | 0 | 0 |
| 0 | 0 | 0 | 0 | 0 |
| 0 | 0 | 0 | 0 | 0 |
| 0 | 0 | 0 | 0 | 0 |
| 0 | 0 | 0 | 0 | 0 |
| 0 | 0 | 0 | 0 | 0 |
| 0 | 0 | 0.186835565 | 0.332474428 | 0 |
| 0 | 0 | 0 | 0 | 0 |
| 0 | 0 | 0.029342281 | 0 | 0.001033461 |
| 0 | 0 | 0 | 0 | 0 |
| 0 | 0 | 0 | 0 | 0 |
| 0 | 0 | 0 | 0 | 0 |
| 0 | 0 | 0.004539883 | 0.001350788 | 0 |
| 0 | 0 | 0 | 0 | 0 |
| 0 | 0 | 0 | 0 | 0 |
| 0.005815331 | 0 | 0.166248668 | 0.002997897 | 0 |
| 0.028222773 | 0 | 0.001548467 | 0.002610791 | 0.000935878 |
| 0 | 0 | 0.00311337 | 0 | 0 |
| 0 | 0 | 0 | 0 | 0 |
| 0 | 0 | 0 | 0 | 0 |
| 0 | 0 | 0 | 0 | 0 |
| 0 | 0 | 0 | 0 | 0 |
| 0.043345502 | 0 | 0 | 0 | 0 |
| 0 | 0 | 0 | 0 | 0 |
| 0 | 0 | 0 | 0 | 0 |
| 0 | 0 | 0 | 0 | 0 |
| 0 | 0 | 0 | 0 | 0 |
| 0 | 0 | 0 | 0 | 0 |
| 0.072808376 | 0 | 0.007501486 | 0 | 0 |
| 0 | 0 | 0 | 0 | 0 |
| 0 | 0 | 0 | 0 | 0 |
| 0 | 0 | 0 | 0 | 0 |
| 0 | 0 | 0.073048541 | 0.003392932 | 0 |
| 0 | 0 | 0 | 0 | 0 |
| 0.098111271 | 0 | 0.195951732 | 0.076055299 | 0.039077249 |
| 0 | 0 | 0 | 0 | 0 |
| 0 | 0 | 0 | 0 | 0 |
| 0 | 0 | 0 | 0 | 0 |
| 0 | 0 | 0 | 0 | 0 |
| 0.039849169 | 0.799091331 | 0.033673368 | 0 | 0.005410514 |
| 0 | 0 | 0 | 0 | 0 |
| 0 | 0 | 0 | 0.005870698 | 0 |

| | | | | |
|---|---|---|---|---|
| 0.003074142 | 0 | 0 | 0.002007375 | 0 |
| 0.00127766 | 0 | 0 | 0.00350404 | 0 |
| 0 | 0 | 0 | 0.00206369 | 0 |
| 0 | 0 | 0 | 0.35849767 | 0 |
| 0.000713919 | 0 | 0.032902622 | 0.003107867 | 0 |
| 0 | 0 | 0 | 0 | 0 |
| 0 | 0 | 0 | 0 | 0 |
| 0 | 0 | 0 | 0 | 0 |
| 0 | 0 | 0 | 0 | 0 |
| 0.020867192 | 0.014115991 | 0.311142423 | 0 | 0 |
| 0 | 0 | 0 | 0.073754851 | 0 |
| 0 | 0 | 0 | 0 | 0 |
| 0 | 0 | 0.002962164 | 0.001175142 | 0 |
| 0 | 0 | 0 | 0 | 0 |
| 0 | 0 | 0 | 0 | 0 |
| 0 | 0 | 0 | 0 | 0 |
| 0 | 0 | 0 | 0 | 0 |
| 0 | 0 | 0 | 0 | 0 |
| 0 | 0 | 0 | 0 | 0 |
| 0.336379444 | 0.269277325 | 0.03902072 | 0.193676813 | 0.525145559 |
| 0 | 0 | 0 | 0 | 0 |
| 0 | 0 | 0 | 0 | 0 |
| 0 | 0 | 0 | 0 | 0 |
| 0 | 0 | 0 | 0 | 0 |
| 0 | 0 | 0 | 0 | 0 |
| 0 | 0 | 0 | 0 | 0 |
| 0 | 0 | 0 | 0 | 0 |
| 0 | 0 | 0 | 0 | 0 |
| 0 | 0 | 0 | 0 | 0 |
| 0.01084493 | 0 | 0.012777194 | 0 | 0 |
| 0 | 0 | 0 | 0 | 0 |
| 0 | 0 | 0 | 0.00732402 | 0 |
| 0 | 0 | 0 | 0 | 0 |
| 0 | 0 | 0 | 0 | 0 |
| 0 | 0 | 0 | 0 | 0 |
| 0 | 0 | 0 | 0 | 0 |
| 0 | 0 | 0 | 0 | 0 |
| 0 | 0 | 0 | 0 | 0 |
| 0 | 0 | 0 | 0 | 0 |
| 0 | 0 | 0 | 0 | 0 |
| 0 | 0 | 0 | 0 | 0 |
| 0 | 0 | 0 | 0 | 0 |
| 0 | 0 | 0 | 0.051152211 | 0 |
| 0 | 0 | 0 | 0 | 0 |
| 0.00250746 | 0 | 0.033513009 | 0.029635839 | 0 |
| 0 | 0 | 0 | 0.014852139 | 0 |
| 0 | 0 | 0 | 0 | 0 |
| 0 | 0 | 0 | 0 | 0 |

| | | | | |
|---|---|---|---|---|
| 0 | 0 | 0 | 0 | 0 |
| 0 | 0 | 0 | 0 | 0 |
| 0 | 0 | 0 | 0 | 0 |
| 0 | 0 | 0 | 0 | 0 |
| 0 | 0 | 0 | 0 | 0 |
| 0.015162521 | 0.011272298 | 0.100691282 | 0.003667012 | 0.003476115 |
| 0 | 0 | 0 | 0 | 0 |
| 0 | 0 | 0 | 0 | 0 |
| 0 | 0 | 0 | 0 | 0 |
| 0 | 0 | 0 | 0 | 0 |
| 0 | 0 | 0 | 0 | 0 |
| 0 | 0 | 0 | 0 | 0 |
| 0 | 0 | 0 | 0 | 0 |
| 0 | 0 | 0 | 0 | 0 |
| 0 | 0 | 0 | 0 | 0 |
| 0 | 0 | 0 | 0 | 0 |
| 0 | 0 | 0 | 0.029357195 | 0 |
| 0 | 0 | 0 | 0 | 0 |
| 0 | 0 | 0 | 0 | 0 |
| 0 | 0 | 0 | 0 | 0 |
| 0 | 0 | 0 | 0 | 0.001007628 |
| 0 | 0 | 0 | 0 | 0 |
| 0 | 0 | 0 | 0 | 0 |
| 0 | 0 | 0 | 0 | 0 |
| 0 | 0 | 0 | 0 | 0 |
| 0 | 0 | 0.004819402 | 0 | 0 |
| 0 | 0 | 0 | 0 | 0 |
| 0 | 0 | 0.000721398 | 2.364797058 | 0 |
| 0 | 0 | 0 | 0 | 0 |
| 0.002030447 | 0 | 0 | 0.03071566 | 0 |
| 0 | 0 | 0 | 0 | 0 |
| 0 | 0 | 0 | 0 | 0 |
| 0 | 0 | 0 | 0 | 0 |
| 0 | 0 | 0 | 0 | 0 |
| 0 | 0 | 0 | 0 | 0 |
| 0 | 0 | 0 | 0 | 0 |
| 0 | 0 | 0 | 0 | 0 |
| 0 | 0 | 0 | 0 | 0 |
| 0 | 0 | 6.836734688 | 0 | 0 |
| 0 | 0 | 0 | 0 | 0 |
| 0 | 0 | 0 | 0 | 0 |
| 0 | 0 | 0 | 0 | 0 |
| 0 | 0 | 0 | 0 | 0 |
| 0 | 0 | 0 | 0 | 0 |
| 0 | 0 | 0 | 0 | 0 |
| 0 | 0 | 0 | 0 | 0 |
| 0 | 0 | 0 | 0 | 0 |
| 0 | 0 | 0 | 0 | 0 |

| | | | | |
|---|---|---|---|---|
| 0 | 0 | 0 | 0 | 0 |
| 0 | 0 | 0 | 0.03968665 | 0 |
| 0 | 0 | 0 | 0 | 0 |
| 0 | 0 | 0 | 0 | 0 |
| 0 | 0 | 0 | 0 | 0 |
| 0 | 0 | 0 | 0 | 0 |
| 0 | 0 | 0 | 0 | 0 |
| 0 | 0 | 0 | 0 | 0 |
| 0 | 0 | 0 | 0.02415297 | 0 |
| 0 | 0 | 0 | 0.014864824 | 0 |
| 0 | 0 | 0.007949816 | 0 | 0.019837966 |
| 0 | 0 | 0 | 0 | 0 |
| 0 | 0 | 0.026615643 | 0.047383013 | 0 |
| 0 | 0 | 0 | 0 | 0 |
| 0 | 0 | 0 | 0 | 0 |
| 0 | 0 | 0 | 0 | 0 |
| 0 | 0 | 0 | 0.001840732 | 0 |
| 0 | 0 | 0 | 0 | 0 |
| 0 | 0 | 0 | 0 | 0 |
| 0 | 0 | 0 | 0 | 0 |
| 0.001392616 | 0.003960078 | 0 | 0 | 0.001293031 |
| 0 | 0 | 0 | 0 | 0 |
| 0 | 0 | 0 | 0.000728057 | 0 |
| 0.002076556 | 0.66594703 | 1.134611071 | 0.078796606 | 1.332353128 |
| 0 | 0 | 0 | 0 | 0 |
| 0 | 0 | 0 | 0 | 0 |
| 0 | 0 | 0 | 0 | 0 |
| 0 | 0 | 0 | 0 | 0 |
| 0 | 0 | 0 | 0 | 0 |
| 0 | 0 | 0 | 0 | 0 |
| 0.008474479 | 0 | 0.00539953 | 0.003748653 | 0 |
| 0 | 0 | 0 | 0 | 0 |
| 0 | 0 | 0 | 0 | 0 |
| 0 | 0 | 0 | 0 | 0 |

| PHL018-0248_V3 | PHL018-0085_V2 | PHL018-0184_V3 | PHL018-0145_V1 | PHL018-0170_V1 |
|---|---|---|---|---|
| REF | CTRL | EYCF | REF | EYCF |
| 0.098200513 | 0.080565164 | 11.88306949 | 1.474601925 | 0.965632743 |
| 0.002064868 | 0.18864585 | 1.45404579 | 1.032917352 | 4.381699701 |
| 2.165701327 | 6.30914357 | 3.840725729 | 1.862012885 | 0 |
| 0 | 0 | 0 | 0 | 0 |
| 0 | 0 | 0 | 0 | 0 |
| 0 | 0 | 0 | 0 | 0 |
| 0 | 0 | 0 | 0 | 0 |
| 0 | 0 | 0 | 0 | 0 |
| 0 | 0 | 0 | 0 | 0 |
| 0 | 0 | 0 | 0 | 0 |
| 0 | 0 | 0 | 0 | 0 |
| 0 | 0 | 0 | 0 | 0 |
| 0 | 0 | 0 | 0 | 0 |
| 0.262216916 | 1.699173316 | 2.328710575 | 0.057606802 | 0 |
| 0 | 0 | 0 | 0 | 0 |
| 0 | 0 | 0 | 0 | 0 |
| 0 | 0.001192961 | 0 | 0 | 0 |
| 0 | 0.003343539 | 0 | 0 | 0 |
| 0 | 0 | 0 | 0 | 0 |
| 0 | 0 | 0 | 0 | 0 |
| 0 | 0 | 0 | 0 | 0 |
| 0 | 0 | 0 | 0 | 0 |
| 0 | 0 | 0 | 0 | 0 |
| 0 | 0 | 0 | 0 | 0 |
| 0 | 0 | 0 | 0 | 0 |
| 0 | 0 | 0 | 0 | 0 |
| 0 | 0 | 0 | 0 | 0 |
| 0 | 0 | 0 | 0 | 0 |
| 0 | 0 | 0 | 0 | 0 |
| 0 | 0 | 0 | 0 | 0 |
| 0 | 0 | 0 | 0 | 0 |
| 0 | 0 | 0 | 0 | 0 |
| 0 | 0 | 0 | 0 | 0 |
| 0 | 0 | 0 | 0 | 0 |
| 0 | 0 | 0 | 0 | 0 |
| 2.244052544 | 1.914599741 | 0 | 0.822772435 | 8.566899634 |
| 0 | 0 | 0 | 0 | 0 |
| 0 | 0 | 0 | 0 | 0 |
| 0 | 0 | 0 | 0 | 0 |
| 0 | 0 | 0 | 0 | 0 |
| 0 | 0 | 0 | 0 | 0 |
| 0.001335897 | 0 | 0 | 0.002986643 | 0.103479693 |
| 0.662543363 | 0.036953071 | 0.035218747 | 0.166426701 | 0.063426626 |
| 0.000851877 | 0 | 0 | 0.048769471 | 0.380981815 |
| 0 | 0 | 0.003755887 | 0 | 0 |
| 0 | 0 | 0 | 0 | 0 |

| | | | | |
|---|---|---|---|---|
| 0 | 0 | 0 | 0 | 0 |
| 0 | 0.003025954 | 0.014441716 | 0 | 0.040131284 |
| 0 | 0.003216815 | 0 | 0 | 0.010422545 |
| 0.527664451 | 0.208532494 | 0.188324954 | 0.261208974 | 3.264755055 |
| 0 | 0 | 0 | 0.056470814 | 0 |
| 0 | 0 | 0 | 0 | 0 |
| 0.002012382 | 0.002316668 | 0 | 0 | 2.821018665 |
| 0 | 0 | 0 | 0 | 0 |
| 0 | 0 | 0 | 0 | 0 |
| 0 | 0 | 0 | 0 | 0 |
| 0 | 0 | 0 | 0 | 0 |
| 0.025563407 | 0.359848406 | 1.843316027 | 0.290917912 | 0.231174586 |
| 0 | 0 | 0 | 0 | 0 |
| 0.020056767 | 0.01078431 | 0.073904724 | 0.058802909 | 0 |
| 0 | 0 | 0 | 0 | 0 |
| 0 | 0 | 0 | 0 | 0 |
| 0.030788602 | 0.013874353 | 0 | 0.001932493 | 0.002707117 |
| 0 | 0 | 0 | 0 | 0 |
| 0 | 0 | 0 | 0 | 0 |
| 0 | 0 | 0 | 0 | 0 |
| 0.010406549 | 0.071003944 | 0.032935521 | 0.029791504 | 0.079501721 |
| 0.068990926 | 0.018600577 | 0.022156345 | 0.016582996 | 0.018024362 |
| 0.016973116 | 0 | 0 | 0 | 0 |
| 0 | 0 | 0 | 0 | 0 |
| 0 | 0 | 0 | 0 | 0 |
| 0 | 0 | 0 | 0 | 0 |
| 0 | 0 | 0 | 0 | 0 |
| 0 | 0 | 0 | 0 | 0 |
| 0 | 0 | 0 | 0.000766787 | 0.002506345 |
| 0 | 0 | 0 | 0 | 0 |
| 0 | 0.02984284 | 0.000541847 | 0 | 0.024863502 |
| 0 | 0 | 0.114571101 | 0 | 0 |
| 0 | 0 | 0 | 0 | 0 |
| 0 | 0 | 0 | 0 | 0 |
| 0 | 0 | 0 | 0 | 0 |
| 0 | 0 | 0 | 0 | 0 |
| 0 | 0 | 0 | 0 | 0 |
| 0.002022837 | 0 | 0 | 0 | 0 |
| 0 | 0 | 0 | 0 | 0 |
| 0 | 0 | 0 | 0 | 0 |
| 0 | 0 | 0 | 0 | 0 |
| 0 | 0 | 0 | 0 | 0 |
| 0 | 0 | 0 | 0 | 0 |
| 0 | 0 | 0 | 0 | 0 |
| 0 | 0 | 0 | 0 | 0 |
| 0 | 0 | 0 | 0 | 0 |
| 0 | 0 | 0 | 0 | 0 |
| 0 | 0 | 0 | 0 | 0.002969093 |

| | | | | |
|---|---|---|---|---|
| 0 | 0 | 0 | 0 | 0 |
| 0 | 0.001382929 | 0 | 0.003357121 | 0 |
| 0 | 0 | 0 | 0 | 0 |
| 0 | 0 | 0 | 0 | 0 |
| 0 | 0 | 0 | 0 | 0.12800465 |
| 0 | 0 | 0 | 0 | 0 |
| 0 | 0 | 0 | 0 | 0 |
| 0 | 0 | 0 | 0 | 0 |
| 0.002533805 | 0.04295847 | 0.000616283 | 0.008110942 | 0 |
| 0 | 0 | 0 | 0 | 0 |
| 0 | 0 | 0 | 0 | 0 |
| 0 | 0 | 0 | 0 | 0.039573684 |
| 0 | 0 | 0 | 0 | 0 |
| 0 | 0 | 0 | 0 | 0 |
| 0 | 0 | 0 | 0 | 0 |
| 0 | 0 | 0 | 0 | 0 |
| 0 | 0 | 0 | 0 | 0 |
| 0 | 0 | 0 | 0 | 0 |
| 0.022490769 | 0.022521516 | 0.283356002 | 0.699361992 | 0.003326213 |
| 0 | 0 | 0 | 0 | 0 |
| 0 | 0 | 0 | 0.001219526 | 0 |
| 0 | 0 | 0 | 0 | 0 |
| 0 | 0 | 0 | 0 | 0 |
| 0 | 0 | 0 | 0 | 0 |
| 0 | 0 | 0 | 0 | 0 |
| 0 | 0 | 0 | 0 | 0 |
| 0 | 0 | 0 | 0 | 0 |
| 0 | 0.001285105 | 0.005392573 | 0.010398825 | 0 |
| 0 | 0 | 0 | 0 | 0 |
| 0 | 0.001473939 | 0 | 0 | 0 |
| 0.627316268 | 0.178297391 | 0.022669051 | 0.024482647 | 0.14089027 |
| 0.066302901 | 0 | 0 | 0 | 0.000919193 |
| 0 | 0 | 0 | 0 | 0 |
| 0 | 0 | 0 | 0 | 0 |
| 0 | 0 | 0 | 0 | 0 |
| 0 | 0 | 0 | 0 | 0 |
| 0 | 0 | 0 | 0 | 0 |
| 0 | 0 | 0 | 0 | 0 |
| 0.10065043 | 0.327265357 | 0.201965322 | 0 | 0 |
| 0 | 0 | 0 | 0 | 0 |
| 0 | 0 | 0 | 0 | 0 |
| 0 | 0 | 0 | 0 | 0 |
| 0 | 0 | 0 | 0.000866778 | 0.065021403 |
| 0 | 0 | 0 | 0 | 0 |
| 0.001377813 | 0 | 0 | 0 | 0 |
| 0 | 0 | 0 | 0 | 0 |
| 0 | 0 | 0.187953851 | 0 | 0 |
| 0 | 0 | 0 | 0 | 0 |

| | | | | |
|---|---|---|---|---|
| 0.000376517 | 0 | 0.019979282 | 0 | 0 |
| 0 | 0 | 0 | 0 | 0 |
| 0 | 0 | 0 | 0 | 0 |
| 0 | 0 | 0 | 0 | 0 |
| 0.015960041 | 0 | 0 | 0.029535221 | 0.022721508 |
| 0 | 0 | 0 | 0 | 0 |
| 0 | 0 | 0 | 0 | 0 |
| 0 | 0 | 0 | 0 | 0 |
| 0 | 0 | 0 | 0 | 0 |
| 0 | 0 | 0 | 0 | 0 |
| 0 | 0 | 0 | 0 | 0 |
| 0 | 0 | 0 | 0 | 0 |
| 0 | 0 | 0 | 0 | 0 |
| 0 | 0 | 0 | 0 | 0 |
| 0 | 0 | 0 | 0 | 0 |
| 0.005273887 | 0 | 0 | 0 | 0 |
| 0 | 0 | 0 | 0 | 0 |
| 0 | 0 | 0 | 0 | 0 |
| 0 | 0 | 0 | 0 | 0 |
| 0 | 0 | 0 | 0 | 0 |
| 0 | 0 | 0 | 0 | 0 |
| 0 | 0 | 0 | 0 | 0 |
| 0 | 0 | 0 | 0 | 0 |
| 0 | 0 | 0 | 0 | 0 |
| 0 | 0 | 0 | 0 | 0 |
| 0 | 0 | 0 | 0 | 0 |
| 0 | 0 | 0 | 0 | 0 |
| 0 | 0 | 0 | 0 | 0 |
| 0 | 0 | 0 | 0 | 0 |
| 0 | 0 | 0 | 0 | 0 |
| 0 | 0 | 0 | 0 | 0 |
| 0.021027616 | 0 | 0 | 0 | 0 |
| 0 | 0 | 0 | 0 | 0 |
| 0 | 0 | 0 | 0 | 0 |
| 0 | 0 | 0 | 0 | 0 |
| 0 | 0 | 0 | 0 | 0 |
| 0 | 0 | 0 | 0 | 0 |
| 0 | 0 | 0 | 0 | 0 |
| 0 | 0 | 0 | 0 | 0 |
| 0 | 0 | 0 | 0 | 0 |
| 0 | 0 | 0 | 0 | 0 |
| 0 | 0 | 0 | 0 | 0 |
| 0 | 0 | 0 | 0 | 0 |
| 0 | 0 | 0 | 0 | 0 |
| 0 | 0 | 0 | 0 | 0 |
| 0 | 0 | 0 | 0 | 0 |
| 0 | 0 | 0 | 0 | 0 |
| 0 | 0 | 0 | 0 | 0 |

| | | | | |
|---|---|---|---|---|
| 0 | 0 | 0 | 0 | 0 |
| 0 | 0 | 0 | 0 | 0 |
| 0 | 0 | 0 | 0 | 0 |
| 0 | 0 | 0 | 0 | 0 |
| 0 | 0 | 0 | 0 | 0 |
| 0 | 0 | 0 | 0 | 0 |
| 0 | 0 | 0 | 0 | 0 |
| 0 | 0 | 0 | 0 | 0 |
| 0 | 0 | 0 | 0 | 0.006636135 |
| 0 | 0 | 0 | 0 | 0 |
| 0 | 0 | 0 | 0 | 0 |
| 0 | 0 | 0 | 0 | 0 |
| 0 | 0 | 0.005614598 | 0.006623558 | 0 |
| 0 | 0 | 0 | 0 | 0 |
| 0.001741397 | 0 | 0 | 0 | 0 |
| 0 | 0 | 0 | 0 | 0 |
| 0 | 0 | 0 | 0 | 0 |
| 0 | 0 | 0 | 0 | 0 |
| 0 | 0 | 0 | 0 | 0 |
| 0 | 0 | 0 | 0 | 0 |
| 0.018234034 | 0.067596792 | 0.008954134 | 0.017802664 | 0.074382437 |
| 0 | 0 | 0 | 0 | 0 |
| 0 | 0 | 0.007179395 | 0.002546859 | 0 |
| 0 | 0 | 0 | 0 | 0.010685101 |
| 0 | 0 | 0 | 0 | 0 |
| 0 | 0 | 0 | 0 | 0 |
| 0.001075349 | 0.000557077 | 0.002337614 | 0.002479269 | 0.000884053 |
| 0 | 0 | 0 | 0 | 0 |
| 0 | 0 | 0 | 0 | 0 |
| 0 | 0 | 0 | 0 | 0 |
| 0 | 0.003887179 | 0.301671773 | 0 | 0.298523553 |
| 0 | 0 | 0 | 0 | 0 |
| 0 | 0 | 0 | 0 | 0 |
| 0.00338537 | 0.009548288 | 0 | 0 | 0 |
| 0 | 0.020123874 | 0.377687897 | 0.14151847 | 0.032347093 |
| 0 | 0 | 0 | 0 | 0 |
| 0 | 0 | 0 | 0 | 0 |
| 0 | 0 | 0 | 0 | 0 |
| 0 | 0 | 0 | 0 | 0 |
| 0 | 0 | 0 | 0 | 0 |
| 0 | 0 | 0 | 0 | 0 |
| 0 | 0 | 0 | 0 | 0 |
| 0 | 0 | 0 | 0 | 0 |
| 0 | 0 | 0 | 0 | 0 |
| 0 | 0 | 0 | 0 | 0 |
| 0.554497692 | 0 | 0 | 0 | 0 |
| 0 | 0 | 0 | 0.009815244 | 0 |
| 0 | 0 | 0.001288487 | 0.004845101 | 0.00105579 |

| | | | | |
|---|---|---|---|---|
| 0 | 0 | 0 | 0 | 0 |
| 0 | 0 | 0 | 0 | 0 |
| 0 | 0 | 0 | 0 | 0 |
| 0 | 0 | 0 | 0 | 0 |
| 0 | 0 | 0 | 0 | 0 |
| 0 | 0 | 0 | 0 | 0 |
| 0 | 0 | 0 | 0 | 0 |
| 0 | 0 | 0 | 0 | 0 |
| 0 | 0 | 0 | 0 | 0 |
| 0 | 0 | 0 | 0 | 0 |
| 0 | 0 | 0 | 0 | 0 |
| 0 | 0 | 0 | 0 | 0 |
| 0 | 0 | 0 | 0 | 0 |
| 0 | 0 | 0 | 0 | 0 |
| 0 | 0 | 0 | 0 | 0 |
| 0 | 0 | 0 | 0 | 0 |
| 0 | 0 | 0 | 0 | 0 |
| 0 | 0 | 0 | 0 | 0 |
| 0.002394465 | 0 | 0 | 0.000912488 | 0 |
| 0.099280968 | 0 | 0 | 0 | 0 |
| 0.012362276 | 0.148599408 | 0.03779987 | 0.004576444 | 0 |
| 0 | 0 | 0 | 0 | 0 |
| 0 | 0 | 0 | 0 | 0 |
| 0 | 0 | 0 | 0 | 0 |
| 0 | 0.002203165 | 0.00341352 | 0 | 0.006817804 |
| 0 | 0 | 0 | 0 | 0 |
| 0 | 0 | 0 | 0 | 0 |
| 0 | 0 | 0 | 0 | 0 |
| 0 | 0 | 0 | 0 | 0 |
| 0 | 0 | 0 | 0 | 0 |
| 0.315315948 | 0.314748676 | 6.736310401 | 0.129455737 | 0.907622413 |
| 0 | 0 | 0 | 0 | 0 |
| 0 | 0 | 0.027883591 | 0 | 0 |
| 0 | 0 | 0 | 0 | 0.002762375 |
| 0 | 0 | 0 | 0 | 0 |
| 0 | 0 | 0 | 0 | 0 |
| 0 | 0 | 0 | 0 | 0 |
| 0 | 0 | 0 | 0 | 0 |
| 0 | 0 | 0 | 0 | 0 |
| 0 | 0 | 0 | 0 | 0 |
| 0 | 0 | 0 | 0 | 0 |
| 0.004211691 | 0.011802699 | 1.014947187 | 0.020177156 | 0.044715237 |
| 0.001413506 | 0 | 0 | 0 | 0 |
| 0 | 0 | 0 | 0 | 0 |
| 0 | 0 | 0 | 0 | 0 |
| 0 | 0 | 0 | 0 | 0 |
| 0 | 0 | 0 | 0 | 0 |
| 0 | 0 | 0 | 0 | 0 |

| | | | | |
|---|---|---|---|---|
| 0 | 0 | 0 | 0 | 0 |
| 0 | 0 | 0 | 0 | 0 |
| 0.001510992 | 0 | 0 | 0.049151334 | 0.387317259 |
| 0 | 0 | 0 | 0 | 0 |
| 0 | 0 | 0 | 0 | 0 |
| 0 | 0 | 0 | 0 | 0 |
| 0 | 0 | 0 | 0 | 0 |
| 0 | 0 | 0 | 0 | 0 |
| 0 | 0 | 0 | 0 | 0 |
| 0 | 0 | 0 | 0 | 0 |
| 0 | 0 | 0 | 0 | 0 |
| 0 | 0 | 0 | 0 | 0 |
| 0 | 0 | 0 | 0 | 0 |
| 0 | 0.006920718 | 0 | 0.002240045 | 0.005216841 |
| 0.001653959 | 0.075019521 | 0.001475039 | 0.00231108 | 0.011784347 |
| 0.036153457 | 0 | 0 | 0 | 0 |
| 0 | 0.001952544 | 0 | 0 | 0.002582155 |
| 0.000730934 | 0.02356077 | 0 | 0 | 0 |
| 0 | 0 | 0 | 0 | 0 |
| 0 | 0 | 0 | 0 | 0 |
| 0 | 0 | 0 | 0 | 0 |
| 0 | 0 | 0.002655046 | 0.00237709 | 0.00139857 |
| 0 | 0 | 0 | 0 | 0 |
| 0 | 0 | 0 | 0 | 0 |
| 0.000681944 | 0.002512186 | 0 | 0 | 0 |
| 0 | 0 | 0 | 0 | 0 |
| 0 | 0 | 0 | 0 | 0 |
| 0 | 0 | 0 | 0 | 0 |
| 0 | 0 | 0 | 0 | 0 |
| 0 | 0.001699735 | 0 | 0 | 0 |
| 0 | 0 | 0 | 0 | 0 |
| 0 | 0 | 0 | 0 | 0 |
| 0 | 0 | 0 | 0 | 0 |
| 0 | 0 | 0 | 0 | 0 |
| 0 | 0 | 0 | 0 | 0 |
| 0 | 0 | 0 | 0 | 0 |
| 0 | 0 | 0 | 0 | 0 |
| 0 | 0.00073817 | 0.006433311 | 0.00925836 | 0 |
| 0 | 0 | 0 | 0 | 0 |
| 0 | 0 | 0 | 0 | 0 |
| 0 | 0 | 0 | 0 | 0.004649471 |
| 0 | 0 | 0 | 0 | 0 |
| 0 | 0 | 0 | 0 | 0 |
| 0 | 0 | 0 | 0 | 0 |
| 0 | 0 | 0 | 0 | 0 |
| 0 | 0.001134925 | 0 | 0 | 0 |
| 0 | 0 | 0 | 0 | 0 |
| 0.023300149 | 0 | 0 | 0 | 0 |

| | | | | |
|---|---|---|---|---|
| 1.283747784 | 0 | 1.178869415 | 4.220895601 | 0 |
| 0 | 0 | 0 | 0 | 0 |
| 0 | 0 | 0 | 0 | 0 |
| 0 | 0 | 0 | 0 | 0 |
| 0.000896369 | 0.001651049 | 0.005995517 | 0.001502997 | 0 |
| 0 | 0 | 0 | 0 | 0 |
| 0 | 0 | 0 | 0 | 0 |
| 0.002512916 | 0.003471463 | 0 | 0.057936494 | 0.00378746 |
| 0 | 0 | 0 | 0 | 0 |
| 0.035829954 | 0 | 0.012127111 | 0.004343011 | 0.011001668 |
| 0 | 0 | 0 | 0 | 0 |
| 0 | 0 | 0 | 0 | 0.071138332 |
| 0.005281165 | 0.002953003 | 0.001009257 | 0.006536024 | 0.010957587 |
| 0.008975575 | 0.006595368 | 0 | 0 | 0 |
| 0 | 0 | 0 | 0.006722151 | 0 |
| 0 | 0 | 0 | 0 | 0 |
| 0 | 0 | 0 | 0 | 0 |
| 0 | 0.001809836 | 0 | 0 | 0 |
| 0 | 0 | 0.020920883 | 0.006130041 | 0 |
| 0 | 0 | 0 | 0 | 0 |
| 0 | 0 | 0 | 0 | 0 |
| 0 | 0 | 0 | 0 | 0 |
| 0 | 0 | 0 | 0 | 0 |
| 0.228410173 | 0 | 0 | 0 | 0 |
| 0 | 0 | 0 | 0 | 0 |
| 0 | 0 | 0 | 0 | 0.012281181 |
| 0.001370903 | 0 | 0 | 0 | 0 |
| 0 | 0 | 0 | 0 | 0 |
| 0.058331033 | 0.68980899 | 0.868576097 | 0 | 0 |
| 0 | 0 | 0 | 0 | 0 |
| 0 | 0.031697165 | 0 | 0 | 0 |
| 0 | 0 | 0 | 0 | 0 |
| 0 | 0.001028201 | 0 | 0 | 0 |
| 0 | 0.080438906 | 0 | 0.00160583 | 0 |
| 0 | 0 | 0 | 3.121967193 | 0 |
| 0 | 0 | 0 | 0 | 0 |
| 0 | 0.004816437 | 0.021543374 | 0 | 0 |
| 0 | 0 | 0 | 0 | 0 |
| 0 | 0 | 0 | 0 | 0 |
| 0 | 0.014257084 | 0 | 0 | 0.083071845 |
| 0 | 0.007056856 | 0 | 0 | 0 |
| 0 | 0.111915881 | 0.005982456 | 0.023937889 | 0 |
| 0 | 0 | 0 | 0 | 0 |
| 0 | 0 | 0.005524433 | 0.001780589 | 0.008924137 |
| 0 | 0 | 0 | 0 | 0 |
| 0 | 0 | 0 | 0 | 0 |
| 0 | 0.006038519 | 0.008040231 | 0 | 0.010780672 |
| 0 | 0.018498374 | 0 | 0 | 0 |

| | | | | |
|---|---|---|---|---|
| 0 | 0.120330272 | 0 | 0 | 0 |
| 0 | 0 | 0 | 0 | 0.959836361 |
| 0 | 0 | 0 | 0 | 0 |
| 0 | 0 | 0 | 0 | 0 |
| 0 | 0 | 0 | 0 | 0 |
| 0.026681791 | 0.030319918 | 0.034925556 | 0.063981611 | 0 |
| 0 | 0 | 0 | 0 | 0 |
| 0 | 0 | 0 | 0 | 0 |
| 37.5222999 | 0.001445929 | 0 | 0 | 0 |
| 0 | 0 | 0 | 0 | 0 |
| 0.014308544 | 0.003217804 | 0.011351077 | 0.005021587 | 0 |
| 0 | 0 | 0 | 0.004183441 | 0 |
| 0 | 0 | 0 | 0 | 0 |
| 0.002514975 | 0 | 0.002313004 | 0 | 0 |
| 0 | 0.120621308 | 0 | 0 | 0.171545621 |
| 0 | 0.003321596 | 0 | 0 | 0 |
| 0 | 0 | 0 | 0 | 0 |
| 0 | 0 | 0.003069445 | 0 | 0 |
| 0 | 0 | 0 | 0 | 0 |
| 1.040188654 | 0.049088111 | 0 | 0 | 0 |
| 0 | 0 | 0 | 0 | 0 |
| 0.170251711 | 0 | 0 | 0 | 0 |
| 0.025954181 | 0 | 0 | 0.031263644 | 0 |
| 0 | 0 | 0 | 0 | 0 |
| 0 | 0 | 0 | 0 | 0 |
| 0 | 0 | 0 | 0 | 0 |
| 0 | 0.072045204 | 0 | 0 | 0 |
| 0 | 0 | 0.046364069 | 0 | 0 |
| 0 | 0 | 0 | 0 | 0 |
| 0.048614758 | 0.554457646 | 0.068732232 | 0.144873331 | 0.823576362 |
| 0.000959744 | 0 | 0 | 0.006079434 | 0 |
| 0 | 0 | 0 | 0 | 0.002461664 |
| 0 | 0 | 0 | 0 | 0 |
| 0 | 0 | 0.016163705 | 0.008610561 | 0 |
| 0 | 0 | 0 | 0 | 0 |
| 0 | 0.030924554 | 0 | 0 | 0 |
| 1.076422995 | 0.006747559 | 0 | 0 | 0 |
| 0 | 0 | 0 | 0 | 0.02212786 |
| 0 | 0 | 0 | 0 | 0 |
| 0 | 0.004110995 | 0 | 0 | 0.830273045 |
| 0 | 0.003570324 | 0.02350996 | 0.010400538 | 0.018060121 |
| 0.51256875 | 0 | 0 | 0 | 8.898823894 |
| 0 | 0 | 0 | 0 | 0.021477132 |
| 0 | 0 | 0.015223978 | 0.009933229 | 0 |
| 0 | 0 | 0.003510061 | 0 | 0 |
| 0 | 0.152815796 | 0 | 0 | 0 |
| 0.017631877 | 0 | 0.000725747 | 0 | 0.005352111 |
| 0 | 0 | 0 | 0 | 0 |

| | | | | |
|---|---|---|---|---|
| 0 | 0.000693533 | 0.002910212 | 0 | 0.008804809 |
| 0 | 0 | 0 | 0 | 0 |
| 0.065213612 | 0.000944992 | 0 | 0.002676343 | 0 |
| 0 | 0 | 0 | 0 | 0 |
| 0.014803527 | 0.494060743 | 0 | 0.598924688 | 0 |
| 0 | 0.029319136 | 0 | 0 | 0 |
| 0 | 0 | 0 | 0 | 0 |
| 0 | 0 | 0 | 0 | 0 |
| 0.002240837 | 0 | 0 | 0 | 0 |
| 0 | 0.96470789 | 0.810297434 | 0 | 0 |
| 0 | 0 | 0 | 0 | 0 |
| 0 | 0 | 0 | 0 | 0 |
| 0 | 0 | 0 | 0 | 0 |
| 0.007872399 | 0 | 0.012353884 | 0 | 0 |
| 0 | 0 | 0 | 0 | 0 |
| 0.002011278 | 0 | 0.120758471 | 0.08748493 | 0 |
| 0 | 0.002717339 | 0.426980941 | 0.017810435 | 0.014446128 |
| 0.001593237 | 0.02024896 | 0.001705062 | 0 | 0 |
| 0.002031785 | 0 | 0.010328359 | 0.012491678 | 0 |
| 0 | 0 | 0 | 0 | 0.02961752 |
| 0 | 0 | 0 | 0 | 0.069020641 |
| 0 | 0.004202629 | 0.018923845 | 0.002040413 | 0 |
| 0 | 0 | 0 | 0 | 0 |
| 0 | 0.22437014 | 0 | 0.157526582 | 1.247651593 |
| 0 | 0 | 0 | 0 | 0 |
| 0 | 0 | 0 | 0.012165538 | 0 |
| 0 | 0 | 0.001558911 | 0 | 0 |
| 0 | 0 | 0 | 0.00953926 | 0 |
| 0 | 0 | 0 | 0 | 0 |
| 0 | 0 | 0.001905568 | 0 | 0 |
| 0.003291732 | 0 | 0 | 0.041608172 | 0 |
| 0.002642537 | 0 | 0.001010003 | 0 | 0.092484274 |
| 0.012400542 | 0.02532365 | 0.012020748 | 0.015971252 | 0.006205397 |
| 0 | 0 | 0 | 0 | 0 |
| 0 | 0 | 0 | 0 | 0.027202695 |
| 0 | 0.00200399 | 0 | 0 | 0 |
| 0 | 0.00269345 | 0 | 0 | 0.016028875 |
| 0.003557817 | 0.02755877 | 0.104707129 | 0.000639173 | 0.116160662 |
| 0.000377571 | 0 | 0.058211559 | 0 | 0.75071162 |
| 0 | 0.003444399 | 0 | 0 | 0.010761353 |
| 0 | 0 | 0 | 0 | 0 |
| 0 | 0 | 0.355715448 | 0 | 0 |
| 0.001330178 | 0 | 0 | 0 | 0 |
| 0 | 0 | 0 | 0.003492673 | 0 |
| 0 | 0.00938509 | 0 | 0 | 1.0487261 |
| 0 | 0 | 0 | 0 | 0.002049679 |
| 0 | 0.000780233 | 0.046088184 | 0 | 0 |
| 0 | 0 | 0.001208747 | 0 | 0 |

| | | | | |
|---|---|---|---|---|
| 0 | 0 | 0 | 0 | 0 |
| 0.447918501 | 0.002391868 | 0.236482076 | 0.990855592 | 0.009394532 |
| 0 | 0 | 0 | 0 | 0 |
| 0 | 0.001303745 | 0 | 0 | 0 |
| 0 | 0 | 0 | 0 | 0 |
| 0 | 0 | 0 | 0 | 0 |
| 0 | 0.005310694 | 0.305130317 | 0 | 0.00395053 |
| 0 | 0 | 0 | 0 | 0 |
| 0 | 0 | 0.005300683 | 0 | 0 |
| 0 | 0 | 0 | 0 | 0 |
| 0 | 0.002666129 | 0 | 0.003236071 | 0 |
| 0.004024606 | 0.212330797 | 0.002820114 | 0.008997743 | 0.121317502 |
| 0.001216996 | 0 | 0 | 0 | 0 |
| 0 | 0 | 0 | 0 | 0 |
| 0 | 0.007545655 | 0 | 0 | 0.010194563 |
| 0 | 0 | 0 | 0 | 0 |
| 0.001347292 | 0 | 0.007209278 | 0.00406636 | 0.004430477 |
| 0 | 0.249101286 | 0.020589004 | 0.002639349 | 0.024443328 |
| 0 | 0.002959152 | 0 | 0.001436694 | 0 |
| 0 | 0.050047838 | 0 | 0.014282126 | 0 |
| 0.015283381 | 0.010172722 | 0.004274886 | 0.021666124 | 0.035866176 |
| 0 | 0 | 0 | 0 | 0 |
| 0 | 0 | 0 | 0 | 0 |
| 0 | 0 | 0 | 0 | 0.489443267 |
| 0 | 0 | 0 | 0 | 3.893223612 |
| 0.005860939 | 0 | 0 | 0.00512734 | 0 |
| 0.092432734 | 0 | 0 | 0.020005834 | 0 |
| 0.033964681 | 0.03712835 | 0.003950933 | 0.003961788 | 0.0089838 |
| 0 | 0 | 0 | 0 | 0 |
| 0 | 0 | 0.00667205 | 0.002090744 | 0 |
| 0 | 0 | 0 | 0 | 0 |
| 0 | 0 | 0 | 0 | 0 |
| 0 | 0 | 0.149887626 | 0 | 0.00147382 |
| 0 | 0 | 0 | 0 | 0 |
| 0 | 0 | 0 | 0 | 0 |
| 0.014495763 | 0.336974819 | 0.05850878 | 0.150445441 | 0.098624083 |
| 0 | 0.005916455 | 0 | 0.004658091 | 0 |
| 0 | 0 | 0 | 0 | 0 |
| 0.008379847 | 0.003523229 | 0.001705871 | 0.003665481 | 0.023363156 |
| 0 | 0 | 0 | 0 | 0 |
| 0 | 0.011054221 | 0 | 0 | 0.492066573 |
| 0 | 0 | 0 | 0 | 0 |
| 0.007856377 | 0.043412705 | 0.004292713 | 0.079225197 | 0.050032857 |
| 0 | 0.017415034 | 0 | 0 | 0.001053944 |
| 0 | 0.64100866 | 0 | 0 | 0 |
| 0.061371895 | 0 | 0 | 0 | 0 |
| 0 | 0 | 0 | 0 | 0 |
| 0 | 0.012643668 | 0 | 0 | 0 |

| | | | | |
|---|---|---|---|---|
| 0 | 0 | 0.001033968 | 0 | 0 |
| 0 | 0 | 0 | 0 | 0 |
| 0.067131246 | 0 | 0 | 0 | 0 |
| 0.064180226 | 0.801159458 | 0 | 1.923887949 | 0.018859083 |
| 0 | 0 | 0 | 0 | 0 |
| 0 | 0 | 0 | 0 | 0 |
| 0 | 0 | 0 | 0 | 0 |
| 0 | 0 | 0 | 0 | 0 |
| 0.098394858 | 0.096321951 | 1.123942466 | 1.17752935 | 0 |
| 0.065308993 | 0 | 0.084900733 | 0 | 0 |
| 0 | 0 | 0 | 0 | 0 |
| 0.001111293 | 0 | 0 | 0 | 0.001015114 |
| 0 | 0 | 0 | 0 | 0 |
| 0 | 16.64793319 | 0 | 10.91124507 | 0 |
| 0.076216375 | 0.045324927 | 0 | 0 | 0 |
| 0 | 0 | 0 | 0 | 0 |
| 0.007044294 | 0 | 0 | 0 | 0 |
| 0.527174348 | 0 | 0 | 0 | 0 |
| 0 | 0 | 0 | 0 | 0 |
| 0 | 0 | 0 | 0 | 0 |
| 0 | 0 | 0 | 0 | 0 |
| 0.00183423 | 0 | 0 | 0 | 0 |
| 0 | 0 | 0 | 0 | 0 |
| 0.230357121 | 0 | 0 | 0.216414227 | 0.438144179 |
| 0.008208029 | 0 | 0 | 0 | 0 |
| 0.21553171 | 0.620015835 | 0 | 2.679001086 | 0 |
| 0.009628734 | 0 | 0 | 0 | 0 |
| 0.105424173 | 0.263925073 | 0 | 0 | 2.477955522 |
| 0 | 0 | 0 | 0 | 0 |
| 0 | 0 | 0 | 0 | 0 |
| 0.000480163 | 0 | 0 | 0.013284463 | 0 |
| 2.232832461 | 0 | 0 | 0 | 0 |
| 0.000730878 | 0.014640222 | 0 | 0.323074899 | 0.00465667 |
| 0 | 0 | 0 | 0 | 0 |
| 0 | 0 | 0 | 0 | 0 |
| 0.05262083 | 0 | 0 | 0 | 0 |
| 0.156557959 | 0 | 0 | 3.364145703 | 0 |
| 0 | 0 | 0 | 0 | 0.015420497 |
| 0.003599108 | 0 | 0 | 0 | 0 |
| 0.001325723 | 0 | 0.022936825 | 0 | 0 |
| 0.061240784 | 0.063655268 | 0.166541636 | 0.234172291 | 0.062714033 |
| 0.045266888 | 0 | 0 | 0 | 0 |
| 0.055277003 | 0.021004542 | 3.785120306 | 0.003097489 | 0 |
| 0.00526264 | 0 | 1.459767884 | 0 | 0 |
| 0 | 0 | 0 | 0 | 0 |
| 0.182739112 | 0 | 0 | 0 | 0 |
| 0 | 0 | 0 | 0 | 0 |
| 0.026485554 | 0 | 0.438120788 | 0.467498586 | 0.215164333 |

| | | | | |
|---|---|---|---|---|
| 0 | 0 | 0 | 0 | 0 |
| 0.009470019 | 0.162364374 | 0 | 0 | 0 |
| 0.000955515 | 0.005279976 | 0.017529944 | 0.024032568 | 1.394214145 |
| 0 | 0 | 0 | 0.006680072 | 0 |
| 0 | 0 | 0 | 0 | 0 |
| 0 | 0 | 0 | 0 | 0 |
| 0 | 0 | 0 | 0 | 0 |
| 0 | 0 | 0 | 0.085148033 | 0.005016882 |
| 0 | 0 | 0 | 0 | 0 |
| 0 | 0.715411598 | 0 | 0 | 0 |
| 0.074783645 | 0.167184008 | 10.43381044 | 1.047425245 | 0.022476633 |
| 1.069673721 | 3.39456943 | 0.557701107 | 0.610905185 | 1.767475704 |
| 0 | 0 | 0 | 0 | 0 |
| 0.001012485 | 0 | 0.067915328 | 0.017947088 | 0.002378206 |
| 0.187729154 | 0.001417148 | 0.185261133 | 0.031343932 | 0.098578573 |
| 0 | 0 | 0 | 0.027848298 | 0 |
| 0 | 0 | 0 | 0 | 0 |
| 0 | 0 | 0 | 0 | 0 |
| 0.019906973 | 0 | 0 | 0 | 0 |
| 0.00640308 | 0.003812725 | 0 | 0.409095478 | 0 |
| 0.260266813 | 0.099158094 | 0.050489602 | 0.071761125 | 0 |
| 0 | 0.014061723 | 1.234850421 | 0.02361696 | 0 |
| 1.658731045 | 0.313949565 | 0.414720965 | 0.479619969 | 0.483917115 |
| 0 | 0 | 0 | 0 | 0 |
| 0.01761498 | 0 | 0 | 0 | 0 |
| 0.001211437 | 0.003161126 | 0 | 0 | 0.015049632 |
| 0 | 0 | 0 | 0.899521747 | 0 |
| 0 | 0 | 0 | 0 | 0 |
| 0 | 0 | 0 | 0 | 0 |
| 0 | 0 | 0 | 0 | 0 |
| 0 | 0 | 0 | 0 | 0 |
| 0.244545301 | 0 | 0 | 0.071837777 | 0 |
| 0.008396218 | 0.034796818 | 0.044006994 | 0 | 0 |
| 0.160174127 | 0 | 0 | 0 | 0 |
| 0 | 0 | 0.243177281 | 0.094138218 | 0 |
| 0.004743042 | 0.004827986 | 0.072799736 | 0.107574176 | 0.020317359 |
| 0.002290857 | 0.002331884 | 0 | 0 | 0 |
| 0 | 0 | 0 | 0 | 0 |
| 0.041335258 | 0 | 0 | 0 | 0 |
| 0 | 0 | 0 | 0 | 0 |
| 0.057481657 | 0 | 0 | 0 | 0 |
| 0 | 0 | 0 | 0 | 0 |
| 0 | 0 | 0 | 0 | 0 |
| 0 | 0 | 0 | 0 | 0 |
| 0.002562111 | 0.002359616 | 2.131857979 | 0.719349949 | 0.118890687 |
| 0 | 0 | 0 | 0 | 0 |
| 0 | 0 | 0 | 0 | 0 |
| 0 | 0.444978018 | 0.552862552 | 0 | 0.049686968 |

| | | | | |
|---|---|---|---|---|
| 0 | 0 | 0 | 0 | 0 |
| 0 | 0 | 0 | 0 | 0 |
| 0 | 0 | 0 | 0 | 0 |
| 0.001698415 | 0 | 0 | 0.028478389 | 0 |
| 0 | 0 | 0 | 0 | 0 |
| 0.05592968 | 0 | 0 | 0 | 0 |
| 0 | 0 | 0 | 0 | 0 |
| 0.034159427 | 0 | 3.064634974 | 0 | 0 |
| 0 | 0 | 0 | 0 | 0 |
| 0 | 0 | 0 | 0 | 0 |
| 0.002972491 | 0.000746608 | 0.006326096 | 0.050294711 | 0.001110776 |
| 0 | 0.008720151 | 0 | 0 | 0 |
| 0.005915109 | 0 | 0 | 0 | 0 |
| 0 | 0 | 0 | 0 | 0 |
| 0 | 0 | 0 | 0 | 0 |
| 0 | 0 | 0 | 0 | 0 |
| 0.012963037 | 0 | 0 | 0 | 0 |
| 0 | 0 | 0.00616913 | 0.001886 | 0 |
| 0.13113999 | 0 | 0 | 0 | 0 |
| 0.006390539 | 0 | 0 | 0.020580401 | 0 |
| 0 | 0 | 0 | 0 | 0 |
| 0 | 0 | 0 | 0 | 0 |
| 0.000907759 | 0 | 0.026195059 | 0 | 0 |
| 0 | 0 | 0 | 0 | 0 |
| 0 | 0 | 0 | 0 | 0 |
| 0 | 0 | 0 | 0 | 0 |
| 0 | 0 | 0 | 0.197952968 | 0 |
| 0 | 0 | 0 | 0 | 0 |
| 0 | 0 | 0 | 0 | 0 |
| 0.191173633 | 0 | 0 | 0.00159479 | 0.062106543 |
| 0.050654352 | 0.002352146 | 0 | 0.033545878 | 0.001574748 |
| 0 | 0 | 0 | 0 | 0 |
| 0 | 0 | 0 | 0 | 0 |
| 0 | 0 | 0 | 0 | 0 |
| 0 | 0 | 0 | 0 | 0 |
| 0 | 0 | 0 | 0 | 0 |
| 0.061150253 | 0 | 0 | 0 | 0 |
| 0.000915006 | 0.001264033 | 0.403166381 | 0.0751781 | 0.014668545 |
| 0 | 0 | 0 | 0 | 0 |
| 0 | 0 | 0 | 0 | 0 |
| 0.026506241 | 0.097991228 | 0 | 0.507964743 | 0 |
| 0 | 0 | 0 | 0 | 0 |
| 0 | 0 | 0 | 0 | 0 |
| 0 | 0.003682577 | 0.000891512 | 0 | 0.023193611 |
| 0.089611975 | 0 | 0 | 0 | 0 |
| 0 | 0 | 0 | 0 | 0 |
| 0 | 0 | 0 | 0 | 0 |
| 0 | 0 | 0 | 0 | 0 |

| | | | | |
|---|---|---|---|---|
| 0 | 0 | 0 | 0 | 0 |
| 0 | 0 | 0 | 0 | 0 |
| 0.036926453 | 0 | 0 | 0 | 0 |
| 0.186810834 | 0.104359392 | 0 | 0 | 0 |
| 0 | 0 | 0 | 0 | 0 |
| 0 | 0 | 0 | 0 | 0 |
| 0 | 0 | 0 | 0 | 0 |
| 0 | 0 | 0 | 0 | 0 |
| 0.003628094 | 0.007191163 | 0.025955277 | 0.018514849 | 0.050835297 |
| 0 | 0 | 0 | 0 | 0 |
| 0.004173278 | 0 | 0 | 0 | 0 |
| 0 | 0 | 0 | 0 | 0 |
| 0.016840468 | 0 | 0.030037433 | 0 | 0 |
| 0 | 0 | 0 | 0 | 0 |
| 0.148386177 | 0 | 1.266981193 | 1.721320409 | 0 |
| 0 | 0 | 0 | 0.477838104 | 0 |
| 0 | 0 | 0 | 0 | 0 |
| 0 | 0 | 0 | 0 | 0 |
| 0 | 0 | 0 | 0 | 0 |
| 0 | 0 | 0 | 0 | 0 |
| 0.358836453 | 0 | 0 | 0 | 0 |
| 0 | 0 | 0 | 0 | 0 |
| 0 | 0 | 0 | 0 | 0 |
| 0 | 0 | 0 | 0 | 0 |
| 0.034317779 | 0.002050086 | 0.015385409 | 0.026127523 | 0.00274504 |
| 0.50966706 | 2.564874856 | 0.881310236 | 6.700400088 | 1.324704883 |
| 0.000965938 | 0 | 0 | 0 | 0 |
| 0 | 0 | 0 | 0 | 0 |
| 0 | 0 | 0 | 0 | 0 |
| 0 | 0 | 0 | 0 | 0 |
| 0 | 0 | 0 | 0 | 0 |
| 0 | 0 | 0 | 0 | 0 |
| 0 | 0 | 0 | 0 | 0 |
| 0.004953486 | 0 | 0.767210213 | 0 | 0 |
| 0 | 0 | 0.008297903 | 0.007663804 | 0.006709861 |
| 0.011077227 | 0.000637609 | 0.033804447 | 0.012963018 | 0.025043349 |
| 0 | 0 | 0 | 0 | 0 |
| 0.546317477 | 3.89504107 | 0.947304704 | 0 | 0 |
| 17.68864512 | 0.000603953 | 0 | 0 | 0 |
| 0 | 0 | 0 | 0 | 0 |
| 0 | 0 | 0 | 0 | 0 |
| 0.004710624 | 0 | 0.00455113 | 0.017333032 | 0 |
| 0.072217189 | 0 | 0.027843187 | 0 | 0.13377762 |
| 0 | 0 | 0 | 0 | 0 |
| 0 | 0 | 0 | 0 | 0 |
| 0 | 0.005738517 | 0 | 0.019612672 | 0.008088215 |
| 0 | 0 | 0 | 0 | 0 |
| 0.184079288 | 0.008901562 | 0.019336503 | 0.002920125 | 0 |

| | | | | |
|---|---|---|---|---|
| 0.002337584 | 0 | 0 | 0 | 0 |
| 0 | 0 | 0 | 0 | 0 |
| 0 | 0 | 0 | 0 | 0 |
| 0 | 0 | 0 | 0 | 0 |
| 0.001034371 | 0.003572326 | 0.207038251 | 0.01387516 | 0 |
| 0 | 0 | 0 | 0 | 0 |
| 0 | 0 | 0 | 0 | 0 |
| 0.000724726 | 0 | 0 | 0 | 0 |
| 0 | 0 | 0 | 0 | 0 |
| 0.010511862 | 0 | 0 | 0.035034184 | 0 |
| 0.020776236 | 0 | 0 | 0 | 0 |
| 0.11689852 | 0 | 0 | 0 | 0.027261917 |
| 0.005166325 | 0 | 0.038535014 | 0 | 0 |
| 0 | 0 | 0 | 0 | 0 |
| 0.002990626 | 0 | 0 | 0.002507285 | 0.925669141 |
| 0 | 0 | 0 | 0 | 0 |
| 0 | 0 | 0 | 0 | 0 |
| 0 | 0 | 0 | 0 | 0.003447121 |
| 0 | 0 | 0 | 0 | 0 |
| 0.003051902 | 0 | 0 | 0.004629951 | 0 |
| 0.008138152 | 0 | 0 | 0 | 0 |
| 0 | 0 | 0 | 0 | 0 |
| 0.004333014 | 0.24911634 | 0.124282123 | 0.172447245 | 0.42515954 |
| 0 | 0 | 0 | 0 | 0 |
| 0.001387492 | 0 | 0 | 0 | 0 |
| 0 | 0 | 0 | 0 | 0 |
| 0 | 0 | 0 | 0 | 0 |
| 0 | 0 | 0 | 0 | 0 |
| 0 | 0 | 0 | 0 | 0 |
| 0 | 0 | 0 | 0 | 0 |
| 0 | 0 | 0 | 0 | 0 |
| 0.005821134 | 0.006165222 | 0.012199839 | 0.028631262 | 0.155372423 |
| 0 | 0 | 0 | 0 | 0 |
| 0.024525297 | 0 | 0 | 0.367669722 | 0 |
| 0.009668953 | 0.001214287 | 0.033932042 | 0.009895963 | 0.025188784 |
| 0.009550684 | 0.018784357 | 0.040710857 | 0.035557014 | 0.026083581 |
| 0 | 0 | 0 | 0 | 0 |
| 0.006363965 | 0 | 0 | 0 | 0 |
| 0 | 0 | 0 | 0 | 0 |
| 0 | 0 | 0 | 0 | 0 |
| 0.064620024 | 0 | 0.010447754 | 0.059378562 | 0 |
| 0 | 0 | 0 | 0 | 0 |
| 0 | 0 | 0 | 0 | 0 |
| 0 | 0 | 0 | 0 | 0 |
| 0.016823178 | 0.033322556 | 1.645674113 | 0.57516243 | 0.038033772 |
| 0 | 0 | 0 | 0 | 0 |
| 0.864578042 | 0 | 0 | 0 | 0 |
| 0.002690952 | 0 | 0 | 0.002354132 | 0 |

| | | | | |
|---|---|---|---|---|
| 0 | 0 | 0 | 0.884932579 | 0 |
| 0 | 0 | 0 | 0 | 0 |
| 0 | 0.002120853 | 0.01848368 | 0.025527791 | 0 |
| 0 | 0 | 0 | 0 | 0 |
| 0 | 0.025451743 | 0 | 0 | 0 |
| 0 | 0.006081829 | 0 | 0.01199567 | 0 |
| 0.044643091 | 0 | 0 | 0 | 0 |
| 0 | 0.095640739 | 0 | 0.077390671 | 0 |
| 0 | 0 | 0 | 0 | 0 |
| 0 | 0 | 0 | 0 | 0 |
| 0 | 0.378332743 | 0.385699361 | 0 | 0 |
| 0 | 0 | 0 | 0 | 0.001712251 |
| 0 | 0 | 0 | 0 | 0 |
| 0.821616468 | 0 | 0 | 0 | 0 |
| 0 | 0 | 0 | 0 | 0 |
| 0 | 0 | 0 | 0 | 0 |
| 0 | 0 | 0 | 0 | 0 |
| 0 | 0 | 0 | 0 | 0 |
| 7.980121743 | 0 | 0 | 4.38384884 | 0.150147376 |
| 0 | 0 | 0 | 0 | 0 |
| 0 | 0 | 0 | 0 | 0 |
| 0 | 0 | 0 | 0 | 0 |
| 0 | 0 | 0 | 0 | 0 |
| 0 | 0 | 0 | 0 | 0 |
| 0 | 0 | 0.003073576 | 0 | 0 |
| 0 | 0.016226913 | 0.05777 | 0.028964361 | 0.003029562 |
| 0 | 0 | 0 | 0 | 0 |
| 0.076423219 | 0.288851149 | 0.005428353 | 0.029484362 | 0.002594671 |
| 0 | 0 | 0 | 0 | 0 |
| 0.279891733 | 0.148546583 | 0.574682624 | 0 | 0 |
| 0 | 0 | 0 | 0 | 0 |
| 0 | 0 | 0 | 0 | 0 |
| 0.002975528 | 0 | 0 | 0 | 0 |
| 0 | 0 | 0 | 0 | 0 |
| 0.004344495 | 0.013170385 | 0.127697746 | 0.001618819 | 0.016469241 |
| 0 | 0 | 0 | 0 | 0 |
| 0 | 0 | 0 | 0 | 0 |
| 0 | 0 | 0.10698019 | 0.015613527 | 0.004052768 |
| 0 | 0 | 0 | 0 | 0 |
| 0 | 0 | 0 | 0 | 0 |
| 0.283902146 | 16.77424102 | 1.900591953 | 6.987705847 | 15.27709749 |
| 0 | 0.229867205 | 0.289103298 | 0.498681693 | 0.26947023 |
| 0 | 0 | 0 | 0 | 0 |
| 0.656705556 | 0 | 0 | 0 | 0 |
| 0 | 0 | 0 | 0 | 0 |
| 0 | 0 | 0 | 0 | 0 |
| 0 | 0 | 0 | 0 | 0 |
| 0.001304169 | 0 | 0.782176441 | 0.101867548 | 0.005003446 |

| | | | | |
|---|---|---|---|---|
| 0 | 0.00089165 | 0 | 0 | 0 |
| 0.006819013 | 0.013280814 | 0 | 0.034301593 | 0 |
| 0 | 0 | 0 | 0.04575424 | 0 |
| 0 | 0 | 0 | 0 | 0 |
| 0.249335409 | 0.000562128 | 0.232977683 | 0 | 0.06439617 |
| 0 | 0 | 0 | 0 | 0 |
| 0 | 0 | 0 | 0 | 0 |
| 0 | 0 | 0 | 0 | 0 |
| 0.000781356 | 0.339086665 | 0.06077364 | 0.003368953 | 0.630796276 |
| 0.004472316 | 0 | 0 | 0 | 0 |
| 0 | 0 | 0.841205575 | 0 | 0 |
| 0 | 0 | 0 | 0 | 0 |
| 0.1176357 | 0 | 0 | 0 | 0 |
| 0.000667688 | 0.018816464 | 0.005716408 | 0.008060784 | 0.05972152 |
| 0.0033772 | 0 | 0 | 0 | 0 |
| 0.110785298 | 4.300070844 | 0.285994814 | 0.495729112 | 0.398542608 |
| 0.007973364 | 0.000478904 | 0.041119145 | 0.013950717 | 0 |
| 0 | 0 | 0 | 0 | 0 |
| 0 | 0 | 0 | 0 | 0 |
| 0 | 0 | 0 | 0 | 0 |
| 0 | 0 | 0 | 0 | 0 |
| 0 | 0 | 0 | 0 | 0 |
| 0 | 0 | 0 | 0 | 0 |
| 0 | 0 | 0 | 0 | 0 |
| 0.044893627 | 0 | 0 | 0 | 0 |
| 0 | 0 | 0 | 0 | 0 |
| 0 | 0.013325651 | 0.000403249 | 0.065708076 | 0.020816698 |
| 0 | 0.000939782 | 0.094796235 | 0.005323174 | 0.006338387 |
| 0 | 0 | 0 | 0 | 0 |
| 0 | 0 | 0 | 0 | 0 |
| 0 | 0 | 0 | 0 | 0 |
| 0 | 0 | 0 | 0 | 0 |
| 0.108405266 | 0 | 0 | 0 | 0 |
| 0.002020514 | 0 | 0 | 0 | 0.000781685 |
| 0.018696124 | 0.126419912 | 0 | 0 | 0 |
| 0.00183622 | 0 | 0 | 0.017813658 | 0 |
| 0 | 0 | 0 | 0 | 0 |
| 0 | 0 | 0 | 0.004984043 | 0 |
| 0 | 0 | 0 | 0 | 0 |
| 0 | 0 | 0 | 0 | 0 |
| 0.317415697 | 2.029141021 | 0 | 0.876456795 | 9.039482688 |
| 0.003930903 | 0 | 0 | 0 | 0 |
| 0.008922453 | 0 | 0 | 0 | 0 |
| 0 | 0 | 0 | 0 | 0 |
| 0 | 0 | 0 | 0 | 0 |
| 0 | 0 | 0 | 0 | 0 |
| 0 | 0 | 0 | 0 | 0 |
| 0 | 0 | 0 | 0 | 0 |

| | | | | |
|---|---|---|---|---|
| 0.009186495 | 0 | 0 | 0.837829252 | 0 |
| 0 | 0 | 0.017592365 | 0 | 0 |
| 0 | 0 | 0 | 0 | 0 |
| 0 | 0 | 0.010652646 | 0 | 0 |
| 0.060708008 | 0 | 0.004109878 | 0 | 0 |
| 0 | 0 | 0 | 0 | 0 |
| 0.010835852 | 0.160520426 | 0.026215253 | 0.049481937 | 0.111826798 |
| 0 | 0 | 0 | 0 | 0 |
| 0 | 0 | 0 | 0 | 0 |
| 0.219631143 | 0.011862357 | 0.066818703 | 0 | 0 |
| 0.043474641 | 0 | 0 | 0 | 0 |
| 0 | 0 | 0 | 0.001183243 | 0 |
| 0 | 0 | 0 | 0 | 0 |
| 0 | 0 | 0 | 0 | 0 |
| 0 | 0 | 0 | 0 | 0 |
| 0 | 0 | 0 | 0 | 0 |
| 0 | 0 | 0 | 0 | 0 |
| 0 | 0 | 0 | 0 | 0 |
| 0 | 0 | 0 | 0 | 0 |
| 0 | 0 | 0 | 0 | 0 |
| 0.087579101 | 0.339458149 | 0.021288032 | 0.099187709 | 0 |
| 0.010754179 | 0 | 0 | 0 | 0 |
| 0.04067345 | 0 | 0 | 0.059065818 | 0.026471712 |
| 0 | 0.005947997 | 0.074589192 | 0.007039022 | 0.003008734 |
| 0.000386059 | 0 | 0 | 0 | 0 |
| 0 | 0 | 0 | 0 | 0 |
| 0 | 0 | 0 | 0 | 0 |
| 0 | 0 | 0 | 0 | 0 |
| 0 | 0 | 0 | 0 | 0 |
| 0.005267283 | 0.564997148 | 0 | 0.003376934 | 1.508463614 |
| 0 | 0 | 0 | 0 | 0 |
| 0 | 0 | 0 | 0 | 0 |
| 0 | 0 | 0 | 0 | 0 |
| 0 | 0.000476408 | 0.030755507 | 0 | 0 |
| 0 | 0 | 0 | 0 | 0 |
| 0 | 0 | 0 | 0 | 0 |
| 0.010101473 | 0 | 0 | 0 | 0.001993079 |
| 0 | 0 | 0 | 0 | 0 |
| 0 | 0 | 0 | 0 | 0 |
| 0.004081809 | 0 | 0 | 0 | 0 |
| 0 | 0 | 0 | 0 | 0 |
| 0 | 0 | 0 | 0 | 0 |
| 0 | 0 | 0 | 0 | 0 |
| 0.010153341 | 0 | 0 | 0 | 0 |
| 0 | 0 | 0 | 0 | 0 |
| 0 | 0 | 0 | 0 | 0 |
| 0 | 0 | 0 | 0 | 0 |
| 0 | 0 | 0 | 0 | 0 |

| | | | | |
|---|---|---|---|---|
| 0.001969027 | 0 | 0.00898671 | 0.00310738 | 0 |
| 0.001428025 | 0 | 0 | 0 | 0 |
| 0 | 0 | 0 | 0 | 0 |
| 0 | 0 | 0 | 0 | 0 |
| 0 | 0 | 0 | 0 | 0 |
| 0 | 0 | 0 | 0 | 0 |
| 0 | 0 | 0.006312993 | 0.002388807 | 0 |
| 0.036510356 | 0.17579075 | 0.254705858 | 0.300711844 | 0.320686145 |
| 0.865209765 | 0.047970432 | 0.034292152 | 0.214609958 | 0.091770425 |
| 0.00354388 | 0.004720841 | 0.541463748 | 0.007427806 | 0.013110529 |
| 0 | 0 | 0 | 0 | 0 |
| 0 | 0 | 0 | 0 | 0 |
| 0 | 0 | 0 | 0 | 0 |
| 0 | 0 | 0 | 0 | 0 |
| 0 | 0 | 0 | 0 | 0 |
| 0 | 0 | 0 | 0 | 0 |
| 0 | 0 | 0.002940281 | 0.019844729 | 0 |
| 0 | 0 | 0 | 0 | 0 |
| 0 | 0 | 0 | 0 | 0 |
| 0 | 0 | 0 | 0.213816238 | 0.011734389 |
| 0 | 0 | 0 | 0 | 0 |
| 0 | 0 | 0 | 0 | 0 |
| 0 | 0 | 0 | 0.161795041 | 0 |
| 0 | 0 | 0 | 0 | 0 |
| 0.04324224 | 0 | 0 | 0 | 0 |
| 0 | 0 | 0 | 0 | 0 |
| 0 | 0 | 0 | 0 | 0 |
| 0.013706999 | 0 | 0 | 0 | 0 |
| 0 | 0.00072123 | 0 | 0 | 0 |
| 0 | 0 | 0 | 0 | 0 |
| 0 | 0 | 0 | 0 | 0 |
| 0 | 0 | 0 | 0 | 0 |
| 0 | 0 | 0 | 0 | 0 |
| 0 | 0 | 0 | 0 | 0 |
| 0 | 0 | 0 | 0 | 0 |
| 0 | 0 | 0 | 0 | 0.001012477 |
| 0.009273418 | 0 | 0 | 0 | 0 |
| 0 | 0 | 0 | 0 | 0 |
| 0 | 0 | 0 | 0 | 0 |
| 0 | 0 | 0.020572695 | 0 | 0 |
| 0 | 0 | 0 | 0 | 0 |
| 0.022427013 | 0 | 0 | 0 | 0.226348722 |
| 0 | 0 | 0 | 0 | 0 |
| 0 | 0 | 0 | 0 | 0 |
| 0 | 0 | 0 | 0 | 0 |
| 0 | 0 | 0 | 0 | 0 |
| 0 | 0 | 0 | 0.004359545 | 0 |
| 0 | 0 | 0 | 0 | 0 |

| | | | | |
|---|---|---|---|---|
| 0 | 0 | 0 | 0 | 0 |
| 0 | 0 | 0 | 0 | 0.03454823 |
| 0 | 0 | 0 | 0 | 0 |
| 0 | 0 | 0 | 0 | 0 |
| 0.073809678 | 0.015547022 | 1.501204829 | 1.321427433 | 0.177112038 |
| 0.002805026 | 0 | 0 | 0 | 0 |
| 0 | 0 | 0 | 0 | 0 |
| 0.005304167 | 0.014858401 | 0.004139081 | 0.002717558 | 0.024225532 |
| 0 | 0 | 0 | 0 | 0 |
| 0 | 0 | 0 | 0 | 0 |
| 0 | 0 | 0 | 0 | 0 |
| 0.644788524 | 0 | 0 | 0 | 0 |
| 0 | 0 | 0 | 0 | 0 |
| 0 | 0 | 0 | 0 | 0 |
| 0 | 0 | 0.008463798 | 0 | 0 |
| 0.04421365 | 0.00229487 | 0 | 0.210194226 | 0 |
| 0 | 0 | 0 | 0 | 0 |
| 0 | 0 | 0 | 0 | 0 |
| 0 | 0 | 0 | 0 | 0 |
| 0 | 0 | 0 | 0 | 0 |
| 0 | 0 | 0 | 0 | 0 |
| 0 | 0 | 0 | 0 | 0 |
| 0 | 0 | 0 | 0 | 0 |
| 0.000765053 | 0 | 0.005799483 | 0.021380208 | 0.001677216 |
| 0.188208834 | 0 | 0 | 0 | 0 |
| 0 | 0 | 0 | 0 | 0 |
| 0.326073259 | 0.096180471 | 0.007332394 | 1.890736815 | 0.217828654 |
| 0.002824365 | 0.001017838 | 0.005092425 | 0 | 0 |
| 0.000693373 | 0 | 0 | 0 | 0 |
| 0 | 0 | 0 | 0 | 0 |
| 0 | 0 | 0 | 0 | 0 |
| 0 | 0 | 0 | 0 | 0 |
| 0 | 0 | 0 | 0 | 0 |
| 0 | 0 | 0 | 0 | 0 |
| 0 | 0 | 0 | 0 | 0 |
| 0 | 0 | 0 | 0 | 0 |
| 0 | 0 | 0 | 0 | 0 |
| 0.002272438 | 0 | 0 | 0 | 0 |
| 0 | 0 | 0 | 0 | 0 |
| 0 | 0 | 0 | 0 | 0 |
| 0 | 0 | 0 | 0 | 0 |
| 0 | 0 | 0 | 0 | 0 |
| 0 | 0 | 0 | 0 | 0 |
| 0.105160944 | 0.098360808 | 0 | 0 | 0 |
| 0 | 0 | 0 | 0 | 0 |
| 0 | 0 | 0 | 0 | 0 |
| 0 | 0 | 0 | 0 | 0 |
| 0 | 0 | 0 | 0 | 0 |

| | | | | |
|---|---|---|---|---|
| 0.00471591 | 0 | 0 | 0 | 0 |
| 0 | 0 | 0 | 0 | 0 |
| 0 | 0 | 0 | 0 | 0.027329134 |
| 0 | 0 | 0 | 0 | 0 |
| 0 | 0 | 0 | 0 | 0 |
| 0 | 0 | 1.070671569 | 0 | 0 |
| 0 | 0 | 0 | 0 | 0 |
| 0 | 0 | 0 | 0 | 0 |
| 0 | 0 | 0 | 0.002381443 | 0 |
| 0 | 0 | 0 | 0 | 0 |
| 0 | 0 | 0 | 0 | 0 |
| 0 | 0 | 0 | 0 | 0 |
| 0 | 0 | 0 | 0 | 0 |
| 0 | 0 | 0 | 0 | 0 |
| 0 | 0 | 0 | 0 | 0 |
| 0 | 0 | 0 | 0 | 0 |
| 0 | 0 | 0 | 0 | 0 |
| 0 | 0 | 0 | 0 | 0 |
| 0.014226329 | 0 | 0 | 0 | 0 |
| 0 | 0 | 0.018831753 | 0 | 0 |
| 0 | 0.005262316 | 0.009342298 | 0 | 0.030272465 |
| 0 | 0 | 0 | 0 | 0 |
| 0.018601905 | 0.003766023 | 5.893314635 | 0.981171226 | 0.002636684 |
| 0.502690586 | 0 | 0 | 0.470543272 | 0 |
| 0 | 0 | 0 | 0 | 0 |
| 0.001351962 | 0 | 0 | 0 | 0 |
| 0 | 0 | 0 | 0 | 0 |
| 0 | 0 | 0 | 0 | 0 |
| 0.013049264 | 0.019944633 | 0 | 0 | 0 |
| 0 | 0 | 0 | 0 | 0 |
| 0 | 0 | 0 | 0 | 0 |
| 0 | 0 | 0 | 0 | 0 |
| 0 | 0.005601491 | 0.005826037 | 0 | 0.01185237 |
| 0 | 0 | 0 | 0 | 0 |
| 0.003895663 | 0 | 0.047081962 | 0 | 0 |
| 0 | 0.001997761 | 0.094147854 | 0.002020688 | 0.000990732 |
| 0 | 0 | 0 | 0 | 0 |
| 0 | 0 | 0 | 0 | 0 |
| 0.001533294 | 0.010205708 | 0.20418166 | 0.03809709 | 0.001833502 |
| 0 | 0 | 0 | 0 | 0 |
| 0 | 0 | 0 | 0 | 0 |
| 0 | 0 | 0 | 0 | 0 |
| 0.011898203 | 0.011483208 | 0.116867356 | 0.124348748 | 0 |
| 0.023825653 | 0 | 0 | 0 | 0 |
| 0 | 0 | 0 | 0 | 0 |
| 0.003624365 | 0.000715267 | 0.262508268 | 0 | 0 |
| 0 | 0 | 0 | 0 | 0 |
| 0 | 0 | 0 | 0 | 0 |

| | | | | |
|---|---|---|---|---|
| 0 | 0 | 0 | 0 | 0 |
| 0 | 0 | 0 | 0 | 0 |
| 0 | 0 | 0 | 0 | 0 |
| 0 | 0.000832855 | 0 | 0 | 0 |
| 0 | 0 | 0 | 0 | 0 |
| 0 | 0 | 0 | 0 | 0 |
| 0 | 0 | 0 | 0 | 0 |
| 0 | 0 | 0 | 0.021182942 | 0.228900754 |
| 0.009944532 | 0 | 0 | 1.160309261 | 0 |
| 0 | 0.001657614 | 0 | 0 | 0 |
| 0 | 0 | 0 | 0 | 0 |
| 0 | 0.001960637 | 0 | 0 | 0 |
| 0 | 0 | 0 | 0 | 0 |
| 0 | 0 | 0 | 0 | 0 |
| 0 | 0.021730942 | 0.034159334 | 0.007407071 | 0 |
| 0 | 0.001087273 | 0 | 0.005718704 | 0 |
| 0 | 0 | 0 | 0 | 0 |
| 0 | 0 | 0 | 0 | 0 |
| 0 | 0 | 0 | 0 | 0 |
| 0.118610225 | 3.13416026 | 0.412980107 | 0 | 1.396384579 |
| 0 | 0.004542317 | 0 | 0 | 0.017570534 |
| 0 | 0 | 0 | 0 | 0 |
| 0 | 0 | 0 | 0 | 0 |
| 0.026209271 | 0 | 0 | 0 | 0 |
| 0 | 0 | 0 | 0 | 0 |
| 0 | 1.805626317 | 0 | 0 | 0 |
| 0 | 0 | 0 | 0 | 0 |
| 0.026499689 | 0 | 0 | 0 | 0 |
| 0 | 0 | 0 | 0 | 0 |
| 0 | 0 | 0 | 0.308659795 | 0 |
| 0.257050617 | 0.117680031 | 0.487344796 | 0.475168804 | 0 |
| 0 | 0 | 0 | 0 | 0 |
| 0 | 0 | 0 | 0 | 0 |
| 0 | 0 | 0 | 0 | 0 |
| 0 | 0 | 0 | 0 | 0 |
| 0 | 0 | 0 | 0 | 0 |
| 0 | 0 | 0 | 0 | 0 |
| 0 | 0 | 0 | 0 | 0 |
| 0.020907865 | 0 | 0 | 0 | 0 |
| 0 | 0 | 0 | 0 | 0 |
| 0 | 0 | 0 | 0 | 0 |
| 0 | 0 | 0 | 0 | 0 |
| 0.002190972 | 0 | 0 | 0 | 0 |
| 0.000980799 | 0 | 0 | 0 | 0 |
| 0 | 0 | 0 | 0 | 0 |
| 0 | 0.020462799 | 0 | 0 | 0 |
| 0 | 0.06715301 | 0.233346572 | 0.242949108 | 0 |
| 0 | 0 | 0 | 0 | 0 |

| | | | | |
|---|---|---|---|---|
| 0 | 0 | 0 | 0 | 0 |
| 0 | 0 | 0 | 0 | 0 |
| 0 | 0 | 0 | 0 | 0 |
| 0 | 0 | 0 | 0 | 0 |
| 0.000902878 | 0.080518758 | 1.185264709 | 0.104628083 | 0 |
| 0 | 0 | 0 | 0 | 0 |
| 0 | 0 | 0 | 0 | 0 |
| 0 | 0 | 0 | 0 | 0 |
| 0 | 0 | 0 | 0 | 0 |
| 0 | 0 | 0 | 0 | 0 |
| 0.022035218 | 0 | 0 | 0 | 0 |
| 0.017549199 | 0.011159617 | 0.5403245 | 0.229334683 | 0.012137277 |
| 0 | 0 | 0 | 0 | 0.006124788 |
| 0 | 0.001148671 | 0 | 0 | 0 |
| 0 | 0 | 0 | 0 | 0.012390777 |
| 0 | 0 | 0 | 0 | 10.3748196 |
| 0.001071224 | 0 | 0 | 0.013291786 | 0 |
| 0 | 0.000805896 | 0 | 0 | 0 |
| 0.012444649 | 0.122235443 | 0 | 0.002829385 | 0.052888243 |
| 0.028504461 | 0.15846087 | 0.012342047 | 0.064881414 | 0.000905652 |
| 0 | 0 | 0 | 0 | 0 |
| 0 | 0 | 0 | 0 | 0 |
| 0 | 0 | 0 | 0 | 0 |
| 0 | 0 | 0 | 0 | 0 |
| 0 | 0 | 0 | 0 | 0 |
| 0 | 0 | 0 | 0 | 0 |
| 0 | 0.009298799 | 0.77055364 | 0.105835165 | 0.007075966 |
| 0 | 0 | 0 | 0 | 0 |
| 0 | 0 | 0 | 0 | 0 |
| 0 | 0 | 0 | 0 | 0 |
| 0.009756605 | 0.001382384 | 0.005019902 | 0.011116075 | 0 |
| 0 | 0.025010021 | 0.057112569 | 0.04644084 | 0 |
| 0 | 0 | 0 | 0 | 0 |
| 0 | 0 | 0 | 0 | 0 |
| 0 | 0 | 0 | 0 | 0 |
| 0 | 0 | 0 | 0 | 0 |
| 0 | 0 | 0 | 0 | 0 |
| 0.00404252 | 0.017967621 | 1.378053065 | 0.304730798 | 0.049995287 |
| 0.00317108 | 0 | 0 | 0 | 0 |
| 0 | 0.014635806 | 0 | 0.018933244 | 0 |
| 0 | 0 | 0.063963396 | 0.023684405 | 0 |
| 0 | 0 | 0 | 0 | 0 |
| 0 | 0 | 0 | 0 | 0 |
| 0 | 0.003780526 | 0 | 0 | 0 |
| 0 | 0 | 0 | 0 | 0 |
| 0 | 0 | 0 | 0 | 0 |
| 0 | 0 | 0 | 0 | 0 |
| 0 | 0 | 0 | 0 | 0 |

| | | | | |
|---|---|---|---|---|
| 0 | 0 | 0 | 0 | 0 |
| 0.00055691 | 0 | 0 | 0 | 0 |
| 0 | 0 | 0 | 0 | 0 |
| 0 | 0.000754668 | 0.007855971 | 0.035265797 | 0.000673661 |
| 0 | 0 | 0.001016691 | 0 | 0 |
| 0.009972133 | 0 | 0.115905489 | 0 | 0 |
| 0 | 0.018857964 | 0 | 0.004666742 | 0 |
| 0 | 0 | 0 | 0 | 0 |
| 0 | 0 | 0 | 0 | 0 |
| 0 | 0 | 0 | 0 | 0 |
| 0 | 0 | 0 | 0 | 0 |
| 0 | 0 | 0 | 0 | 0 |
| 0.001258534 | 0 | 0.095446253 | 0.031491599 | 0.014644278 |
| 0 | 0 | 0 | 0 | 0 |
| 0.005265841 | 0 | 0 | 0 | 0 |
| 0 | 0 | 0 | 0 | 0 |
| 0 | 0 | 0 | 0 | 0.071970965 |
| 0 | 0 | 0 | 0 | 0 |
| 0 | 0 | 0 | 0 | 0 |
| 0 | 0 | 0 | 0 | 0 |
| 0 | 0 | 0 | 0 | 0 |
| 0 | 0 | 0 | 0 | 0 |
| 0 | 0.006163592 | 0 | 0.036978464 | 0.001676795 |
| 0.012034164 | 0.005610793 | 0.006640638 | 0 | 0 |
| 0 | 0 | 0 | 0 | 0 |
| 0 | 0 | 0 | 0 | 0 |
| 0.001339724 | 0 | 0 | 0 | 0 |
| 0 | 0 | 0 | 0 | 0 |
| 0 | 0 | 0 | 0 | 0 |
| 0 | 0 | 0 | 0 | 0 |
| 0.006538261 | 0.008835914 | 0.012929544 | 0 | 0 |
| 0 | 0.008958646 | 0 | 0 | 0 |
| 0 | 0 | 0 | 0 | 0 |
| 0 | 0 | 0 | 0 | 0 |
| 0.013942154 | 0 | 0 | 0 | 0 |
| 0 | 0 | 0 | 0 | 0 |
| 0 | 0.002920559 | 0 | 0.043298319 | 0 |
| 0 | 0 | 0 | 0 | 0 |
| 0 | 0.000664515 | 0 | 0 | 0 |
| 0 | 0 | 0 | 0 | 0 |
| 0 | 0 | 0 | 0 | 0 |
| 0 | 0.001211398 | 0 | 0 | 0 |
| 0.023457857 | 0 | 0 | 0 | 0 |
| 0 | 0 | 0 | 0 | 0 |
| 0 | 0 | 0 | 0 | 0 |
| 0.005819346 | 0.059343353 | 0.009483228 | 0.229748545 | 0.029748582 |
| 0 | 0.287563813 | 0 | 0 | 0 |
| 0 | 0 | 0 | 0 | 0 |

| | | | | |
|---|---|---|---|---|
| 0 | 0 | 0 | 0 | 0 |
| 0 | 0 | 0 | 0 | 0 |
| 0.350760121 | 0 | 0 | 0 | 0 |
| 0 | 0 | 0 | 0 | 0 |
| 0 | 0.001294905 | 0 | 0 | 0 |
| 0 | 0 | 0 | 0.001633347 | 0 |
| 0 | 0 | 0 | 0 | 0 |
| 0 | 0 | 0 | 0 | 0 |
| 0.002447934 | 0 | 0.029025618 | 0.000932864 | 0 |
| 0 | 0 | 0 | 0 | 0 |
| 0 | 0 | 0.054814298 | 0.010469505 | 0 |
| 0 | 0.001006097 | 0.005455861 | 0.00659433 | 0 |
| 0 | 0 | 0 | 0 | 0 |
| 0 | 0 | 0 | 0 | 0 |
| 0 | 0 | 0 | 0 | 0 |
| 0 | 0 | 0 | 0 | 0 |
| 0 | 0 | 0 | 0 | 0 |
| 0 | 0 | 0 | 0 | 0 |
| 0 | 0 | 0 | 0 | 0 |
| 0 | 0 | 0 | 0 | 0 |
| 0 | 0 | 0 | 0 | 0 |
| 0 | 0 | 0 | 0 | 0 |
| 0 | 0 | 0 | 0 | 0 |
| 0 | 0 | 0 | 0 | 0 |
| 0 | 0.00340154 | 0 | 0.123110118 | 0 |
| 0 | 0 | 0 | 0 | 0 |
| 0 | 0 | 0 | 0 | 0 |
| 0.01016601 | 0 | 0 | 0 | 0 |
| 0 | 0 | 0 | 0 | 0 |
| 0 | 0 | 0 | 0 | 0 |
| 0.000778069 | 0.004145894 | 0.008475488 | 0.004286662 | 0.01516899 |
| 0 | 0 | 0 | 0 | 0 |
| 0.069175932 | 0.083974164 | 0.011912673 | 0.006492067 | 0.025252004 |
| 0.010287021 | 0 | 0.002117124 | 0 | 0.004120096 |
| 0 | 0 | 0 | 0 | 0 |
| 0 | 0 | 0 | 0 | 0 |
| 0 | 0 | 0 | 0 | 0 |
| 0 | 0 | 0 | 0 | 0 |
| 0 | 0 | 0 | 0 | 0 |
| 0 | 0 | 0 | 0 | 0 |
| 0 | 0 | 0 | 0 | 0 |
| 0.010617614 | 0.241136696 | 0.284213002 | 0.116789092 | 0.046611906 |
| 0.007890501 | 0 | 0 | 0 | 0.00191415 |
| 0 | 0 | 0 | 0 | 0 |
| 0 | 0 | 0 | 0 | 0 |
| 0.003177697 | 0.015235267 | 0.041008819 | 0.030715747 | 0.006146843 |
| 0 | 0 | 0 | 0 | 0 |
| 0 | 0 | 0 | 0 | 0 |

| | | | | |
|---|---|---|---|---|
| 0 | 0 | 0 | 0 | 0 |
| 0.005827097 | 0 | 0 | 0 | 0 |
| 0.006964959 | 0.003936162 | 0.00211756 | 0.013271114 | 0.004164324 |
| 0 | 0 | 0 | 0 | 0 |
| 0 | 0 | 0.61256596 | 7.094677318 | 0 |
| 0 | 0.065433262 | 0 | 0.495092139 | 0.040861407 |
| 0 | 0 | 0 | 0 | 0 |
| 0 | 0 | 0 | 0 | 0 |
| 0 | 0 | 0 | 0 | 0 |
| 0 | 0 | 0 | 0 | 0 |
| 0 | 0 | 0 | 0 | 0 |
| 0 | 0 | 0 | 0 | 0 |
| 0 | 0 | 0 | 0 | 0 |
| 0 | 0.039149946 | 0 | 0.038818412 | 0 |
| 0 | 0 | 0 | 0 | 0 |
| 0 | 0 | 0 | 0 | 0 |
| 0.085720199 | 0 | 0 | 0 | 0 |
| 0 | 0 | 0 | 0 | 0 |
| 0 | 0.016068317 | 0 | 0 | 0 |
| 0 | 0 | 0 | 0 | 0 |
| 0 | 0 | 0 | 0 | 0 |
| 0 | 0.007035971 | 0 | 0 | 0 |
| 0 | 0 | 0 | 0 | 0 |
| 0 | 0 | 0 | 0 | 0 |
| 0.001043655 | 0 | 0.004013885 | 0.002406198 | 0 |
| 0 | 0.030820411 | 0 | 0.002200526 | 0.009494373 |
| 0 | 0 | 0 | 0 | 0 |
| 0.001018433 | 0.014772573 | 0 | 0.001067293 | 0 |
| 0 | 0 | 0 | 0 | 0 |
| 0 | 0 | 0 | 0 | 0 |
| 0 | 0 | 0 | 0 | 0 |
| 0 | 0 | 0.001077688 | 0.001157838 | 0 |
| 0 | 0 | 0 | 0 | 0 |
| 0 | 0 | 0 | 0 | 0 |
| 0 | 0 | 0 | 0 | 0 |
| 0 | 0 | 0 | 0 | 0 |
| 0 | 0.019889978 | 0 | 0 | 0 |
| 0 | 0.004150706 | 0.031455889 | 0.010733148 | 0 |
| 0.263207904 | 0 | 0.317673087 | 0 | 0 |
| 0 | 0 | 0 | 0 | 0 |
| 0 | 0 | 0 | 0 | 0 |
| 0 | 0 | 0 | 0 | 0 |
| 0.000526841 | 0 | 0.00422864 | 0.003533548 | 0 |
| 0 | 0 | 0 | 0 | 0 |
| 0 | 0 | 0 | 0 | 0 |
| 0 | 0 | 0 | 0 | 0 |
| 0 | 0.001670095 | 0.016711562 | 0.009459866 | 0.020540253 |
| 0 | 0 | 0 | 0 | 0 |

| | | | | |
|---|---|---|---|---|
| 0 | 0 | 0 | 0 | 0 |
| 0 | 0.020371079 | 0.29470667 | 0.090377193 | 0.025918021 |
| 0.020800878 | 0 | 0 | 0 | 0 |
| 0 | 0 | 0 | 0 | 0 |
| 0 | 0 | 0 | 0 | 0 |
| 0 | 0 | 0 | 0 | 0 |
| 0 | 0 | 0 | 0 | 0 |
| 0 | 0 | 0 | 0 | 0 |
| 0 | 0 | 0 | 0 | 0 |
| 0 | 0 | 0 | 0 | 0 |
| 0 | 0 | 0 | 0 | 0 |
| 0 | 0 | 0 | 0 | 0 |
| 0 | 0 | 0 | 0.002929191 | 0.074459728 |
| 0 | 0 | 0 | 0 | 0 |
| 0.008559122 | 0.003065477 | 0 | 0 | 0.017982216 |
| 0 | 0 | 0 | 0 | 0 |
| 0 | 0 | 0 | 0 | 0 |
| 0 | 0 | 0 | 0 | 0 |
| 0.006487748 | 0 | 0 | 0.010290389 | 0 |
| 0 | 0 | 0 | 0 | 0 |
| 0 | 0 | 0 | 0 | 0 |
| 0.010273674 | 0.227725862 | 0.004164681 | 0.125022696 | 0 |
| 0.048921838 | 0.002974312 | 0.063524299 | 0.012033784 | 0 |
| 0 | 0.006423173 | 0.010724004 | 0 | 0 |
| 0 | 0 | 0 | 0 | 0 |
| 0 | 0 | 0 | 0 | 0 |
| 0 | 0 | 0 | 0 | 0 |
| 0 | 0 | 0 | 0 | 0 |
| 0 | 0 | 0 | 0 | 0 |
| 0 | 0 | 0 | 0 | 0 |
| 0 | 0 | 0 | 0 | 0 |
| 0 | 0 | 0 | 0 | 0 |
| 0 | 0 | 0 | 0 | 0 |
| 0 | 0 | 0 | 0 | 0 |
| 0.565328739 | 15.86740422 | 0 | 0 | 0 |
| 0 | 0 | 0 | 0 | 0 |
| 0 | 0 | 0 | 0 | 0 |
| 0 | 0 | 0 | 0 | 0 |
| 0.178919576 | 0.004080099 | 0.001871522 | 0.005473604 | 0.006389711 |
| 0 | 0 | 0 | 0 | 0 |
| 0.016864842 | 0.398759964 | 0.004288394 | 0.155881377 | 0.011687611 |
| 0 | 0 | 0 | 0 | 0 |
| 0 | 0 | 0 | 0 | 0 |
| 0 | 0 | 0 | 0 | 0 |
| 0 | 0 | 0 | 0 | 0 |
| 0.58773058 | 0.112429764 | 0.039919775 | 0.007492144 | 0.592460181 |
| 0 | 0 | 0 | 0 | 0 |
| 0 | 0 | 0 | 0 | 0 |

| | | | | |
|---|---|---|---|---|
| 0 | 0 | 0 | 0 | 0 |
| 0 | 0.001077189 | 0 | 0 | 0 |
| 0 | 0 | 0 | 0 | 0 |
| 0 | 0 | 0 | 0 | 0 |
| 0.000726173 | 0 | 0 | 0 | 0 |
| 0 | 0 | 0 | 0 | 0 |
| 0 | 0 | 0 | 0 | 0 |
| 0 | 0.001191379 | 0 | 0 | 0 |
| 0.002486392 | 0 | 0 | 0 | 0 |
| 0 | 0 | 0 | 0 | 0.014626811 |
| 0.023537349 | 0 | 0 | 0 | 0 |
| 0 | 0 | 0 | 0 | 0 |
| 0.014644236 | 0.002528781 | 0 | 0.016114155 | 0.008527722 |
| 0 | 0 | 0 | 0 | 0 |
| 0 | 0 | 0 | 0 | 0 |
| 0 | 0 | 0 | 0 | 0 |
| 0 | 0 | 0 | 0 | 0 |
| 0 | 0 | 0 | 0 | 0 |
| 0 | 0 | 0 | 0 | 0 |
| 0.005566776 | 0.146489167 | 0.302960412 | 0.280252489 | 0.089297351 |
| 0 | 0 | 0 | 0 | 0 |
| 0 | 0 | 0 | 0 | 0 |
| 0 | 0 | 0 | 0 | 0 |
| 0 | 0 | 0 | 0 | 0 |
| 0 | 0 | 0 | 0 | 0 |
| 0 | 0 | 0 | 0 | 0 |
| 0 | 0 | 0 | 0 | 0 |
| 0 | 0 | 0 | 0 | 0 |
| 0 | 0 | 0 | 0 | 0 |
| 0 | 0 | 0 | 0.077879952 | 0 |
| 0 | 0 | 0 | 0 | 0 |
| 0 | 0 | 0 | 0 | 0 |
| 0 | 0 | 0 | 0 | 0 |
| 0 | 0 | 0 | 0 | 0 |
| 0 | 0 | 0 | 0 | 0 |
| 0 | 0 | 0 | 0 | 0 |
| 0 | 0 | 0 | 0 | 0 |
| 0 | 0 | 0 | 0 | 0 |
| 0 | 0 | 0 | 0 | 0 |
| 0 | 0 | 0 | 0 | 0 |
| 0 | 0 | 0 | 0 | 0 |
| 0 | 0 | 0 | 0 | 0 |
| 0.004288739 | 0 | 0 | 0 | 0 |
| 0 | 0 | 0 | 0 | 0 |
| 0.002167923 | 0.001057014 | 0.164111867 | 0.038489211 | 0.013419417 |
| 0 | 0 | 0 | 0 | 0 |
| 0 | 0 | 0 | 0 | 0 |
| 0 | 0 | 0 | 0 | 0 |

| | | | | |
|---|---|---|---|---|
| 0 | 0 | 0 | 0 | 0 |
| 0 | 0 | 0 | 0 | 0 |
| 0 | 0 | 0 | 0 | 0 |
| 0 | 0 | 0 | 0 | 0 |
| 0 | 0 | 0 | 0 | 0 |
| 0.049124386 | 0 | 0.086856379 | 0.034799694 | 0.016592505 |
| 0 | 0 | 0 | 0 | 0 |
| 0 | 0 | 0 | 0 | 0 |
| 0 | 0 | 0 | 0 | 0 |
| 0 | 0 | 0 | 0 | 0 |
| 0 | 0.612678064 | 0 | 0 | 0 |
| 0 | 0 | 0 | 0 | 0 |
| 0 | 0 | 0 | 0 | 0 |
| 0 | 0 | 0 | 0 | 0 |
| 0 | 0 | 0 | 0 | 0 |
| 0 | 0 | 0 | 0 | 0 |
| 0.372422088 | 0 | 0 | 0 | 0 |
| 0 | 0.003077434 | 0 | 0 | 0.001927788 |
| 0 | 0 | 0 | 0 | 0 |
| 0 | 0 | 0 | 0 | 0 |
| 0.00055193 | 0 | 0 | 0 | 0 |
| 0 | 0 | 0 | 0 | 0 |
| 0 | 0 | 0 | 0 | 0 |
| 0 | 0 | 0 | 0 | 0 |
| 0 | 0 | 0 | 0 | 0 |
| 0 | 0 | 0 | 0 | 0 |
| 0 | 0 | 0 | 0 | 0 |
| 0.989792934 | 0 | 0 | 3.629509459 | 0 |
| 0 | 0 | 0 | 0.000717445 | 0 |
| 0 | 0.080600115 | 0 | 0 | 0 |
| 0 | 0 | 0 | 0 | 0 |
| 0 | 0 | 0 | 0 | 0 |
| 0 | 0 | 0 | 0 | 0 |
| 0 | 0 | 0 | 0 | 0 |
| 0 | 0 | 0 | 0 | 0 |
| 0 | 0 | 0 | 0 | 0 |
| 0 | 0 | 0 | 0 | 0 |
| 0 | 0.000782102 | 0 | 0 | 0 |
| 0 | 0.198086495 | 6.316523647 | 6.176414493 | 0.095012995 |
| 0 | 0 | 0 | 0 | 0 |
| 0 | 0 | 0 | 0 | 0 |
| 0 | 0 | 0 | 0 | 0 |
| 0 | 0 | 0 | 0 | 0 |
| 0 | 0 | 0 | 0 | 0 |
| 0 | 0 | 0 | 0 | 0 |
| 0 | 0 | 0 | 0 | 0 |
| 0 | 0 | 0 | 0 | 0 |
| 0 | 0 | 0 | 0 | 0 |

| | | | | |
|---|---|---|---|---|
| 0 | 0 | 0 | 0 | 0 |
| 0 | 0 | 0 | 0.073811554 | 0.012092345 |
| 0 | 0 | 0 | 0 | 0 |
| 0 | 0 | 0 | 0 | 0 |
| 0 | 0 | 0 | 0 | 0 |
| 0 | 0 | 0 | 0 | 0 |
| 0 | 0 | 0 | 0 | 0 |
| 0 | 0 | 0 | 0 | 0 |
| 0 | 0 | 0 | 0 | 0 |
| 0 | 0 | 0 | 0 | 0 |
| 0 | 0 | 0.009895273 | 0.003510304 | 0 |
| 0 | 0 | 0 | 0 | 0 |
| 0 | 0 | 0 | 0.288198924 | 0 |
| 0 | 0 | 0 | 0 | 0 |
| 0 | 0 | 0 | 0 | 0 |
| 0 | 0 | 0 | 0 | 0 |
| 0 | 0 | 0 | 0 | 0 |
| 0 | 0 | 0 | 0 | 0 |
| 0 | 0 | 0 | 0 | 0 |
| 0 | 0 | 0 | 0 | 0 |
| 0.002974689 | 0 | 0 | 0 | 0 |
| 0 | 0 | 0 | 0 | 0 |
| 0 | 0 | 0 | 0 | 0 |
| 0.168037105 | 0 | 0.674210102 | 1.144544653 | 3.442231122 |
| 0 | 0 | 0 | 0 | 0 |
| 0 | 0 | 0 | 0 | 0 |
| 0 | 0 | 0 | 0 | 0 |
| 0 | 0.525688667 | 0 | 0 | 0 |
| 0 | 0 | 0 | 0 | 0 |
| 0 | 0 | 0 | 0 | 0 |
| 0.003892874 | 0.002112709 | 0 | 0 | 0.002057375 |
| 0 | 0 | 0 | 0 | 0 |
| 0 | 0 | 0 | 0 | 0 |
| 0 | 0 | 0 | 0 | 0 |

| | | | | |
|---|---|---|---|---|
| PHL018-0072_V3 | PHL018-0183_V3 | PHL018-0181_V3 | PHL018-0159_V1 | PHL018-0208_V2 |
| CTRL | REF | CTRL | REF | REF |
| 0.048629695 | 0.003596262 | 0.003271117 | 1.677726284 | 4.160412591 |
| 4.191840473 | 7.579686492 | 15.14542131 | 3.756993573 | 0.392619168 |
| 8.220880587 | 0 | 0.753143595 | 1.075818031 | 0.627565852 |
| 0.000794962 | 0 | 0 | 0 | 0 |
| 0 | 0 | 0 | 0 | 0 |
| 0 | 0 | 0 | 0 | 0 |
| 0 | 0 | 0 | 0 | 0 |
| 0 | 0 | 0 | 0 | 0 |
| 0 | 0 | 0 | 0 | 0 |
| 0 | 0 | 0 | 0 | 0 |
| 0 | 0 | 0 | 0 | 0 |
| 0 | 0 | 0 | 0 | 0 |
| 0 | 0 | 0 | 0 | 0 |
| 7.110790373 | 1.050724285 | 1.119299307 | 0.251366161 | 0.007199564 |
| 0 | 0 | 0 | 0 | 0 |
| 0 | 0 | 0 | 0 | 0 |
| 0 | 0 | 0 | 0 | 0 |
| 0 | 0 | 0 | 0 | 0 |
| 0 | 0 | 0 | 0 | 0 |
| 0 | 0 | 0 | 0 | 0 |
| 0 | 0 | 0 | 0 | 0 |
| 0 | 0 | 0 | 0 | 0 |
| 0 | 0 | 0 | 0 | 0 |
| 0 | 0 | 0 | 0 | 0 |
| 0.010542154 | 0.011850121 | 0 | 0 | 0 |
| 0 | 0 | 0 | 0 | 0 |
| 0 | 0 | 0 | 0 | 0 |
| 0 | 0 | 0 | 0 | 0 |
| 0 | 0 | 0 | 0 | 0 |
| 0 | 0 | 0 | 0 | 0 |
| 0 | 0 | 0 | 0 | 0 |
| 0 | 0 | 0 | 0 | 0 |
| 0 | 0 | 0 | 0 | 0 |
| 0 | 0 | 0 | 0 | 0 |
| 0 | 0 | 0 | 0 | 0 |
| 0.008213678 | 1.004708175 | 7.661908373 | 0 | 0 |
| 0 | 0 | 0 | 0 | 0 |
| 0 | 0 | 0 | 0 | 0 |
| 0 | 0 | 0 | 0 | 0 |
| 0 | 0 | 0 | 0 | 0 |
| 0 | 0 | 0 | 0 | 0 |
| 0.000943755 | 0 | 0.003828481 | 0 | 0.005746853 |
| 0.082651274 | 0 | 0 | 0.000895141 | 0.120220862 |
| 0.001547524 | 0.109178091 | 0 | 0.01450805 | 0.046462647 |
| 0.005206646 | 0 | 0 | 0.006415339 | 0.006416504 |
| 0 | 0 | 0 | 0 | 0 |

| | | | | |
|---|---|---|---|---|
| 0 | 0 | 0 | 0.098327038 | 0 |
| 0 | 0 | 0.006629724 | 0.001492176 | 0.001682661 |
| 0 | 0 | 0.001430007 | 0 | 0.199092838 |
| 0.494368332 | 0.966137635 | 0.5262959 | 0.752823598 | 0.095483445 |
| 0 | 0 | 0.001763608 | 0.026282357 | 0 |
| 0 | 0 | 0 | 0 | 0 |
| 0 | 0.0072543 | 0.003707475 | 0 | 0 |
| 0 | 0 | 0 | 0 | 0 |
| 0 | 0 | 0 | 0 | 0 |
| 0 | 0 | 0 | 0 | 0 |
| 0 | 0.00260816 | 0.094529048 | 0 | 0.000497132 |
| 0.267129587 | 0.004451778 | 0.001090192 | 0.011475297 | 0.842724314 |
| 0 | 0 | 0 | 0 | 0 |
| 0.0020363 | 0.020781207 | 0.004204031 | 0.014699559 | 0.122964105 |
| 0 | 0 | 0 | 0 | 0 |
| 0 | 0 | 0 | 0 | 0 |
| 0 | 0.023224592 | 0.860852128 | 0.034283787 | 0 |
| 0 | 0 | 0 | 0 | 0 |
| 0 | 0 | 0 | 0 | 0 |
| 0 | 0 | 0 | 0 | 0 |
| 0.057290142 | 0.079721936 | 0.222585898 | 0.050633243 | 0.044904074 |
| 0.00277788 | 0.001120526 | 0.025464738 | 0.039584834 | 0.011276986 |
| 0.001416235 | 0 | 0 | 0 | 0 |
| 0 | 0 | 0 | 0 | 0 |
| 0 | 0 | 0 | 0 | 0 |
| 0 | 0 | 0 | 0 | 0 |
| 0 | 0 | 0 | 0 | 0 |
| 0 | 0 | 0 | 0 | 0 |
| 0 | 0 | 0 | 0 | 0.001147565 |
| 0 | 0 | 0 | 0 | 0 |
| 0 | 0.026917205 | 0 | 0 | 0.016408203 |
| 0 | 0 | 0 | 0 | 0 |
| 0 | 0 | 0 | 0 | 0 |
| 0 | 0 | 0 | 0 | 0 |
| 0 | 0 | 0 | 0 | 0 |
| 0 | 0 | 0 | 0 | 0 |
| 0 | 0 | 0 | 0 | 0 |
| 0.009186741 | 0 | 0.001597173 | 0 | 0 |
| 0 | 0 | 0 | 0 | 0 |
| 0 | 0 | 0 | 0 | 0 |
| 0 | 0 | 0 | 0 | 0 |
| 0 | 0 | 0 | 0 | 0 |
| 0 | 0 | 0 | 0 | 0 |
| 0 | 0 | 0 | 0 | 0 |
| 0.00138742 | 0 | 0 | 0 | 0 |
| 0 | 0 | 0.008864442 | 0 | 0 |
| 0 | 0 | 0 | 0 | 0 |
| 0 | 0 | 0 | 0.003253059 | 0 |

| | | | | |
|---|---|---|---|---|
| 0 | 0 | 0 | 0 | 0 |
| 0 | 0 | 0 | 0 | 0 |
| 0 | 0 | 0 | 0 | 0 |
| 0 | 0 | 0 | 0 | 0 |
| 0 | 0 | 0 | 0.021989963 | 0 |
| 0 | 0 | 0 | 0 | 0 |
| 0 | 0 | 0 | 0 | 0 |
| 0.001825946 | 0 | 0 | 0 | 0 |
| 0 | 0.007364917 | 0.085086892 | 0 | 0.026259309 |
| 0 | 0 | 0 | 0 | 0 |
| 0 | 0 | 0 | 0 | 0 |
| 0 | 0 | 0.00689765 | 0 | 0.005176959 |
| 0 | 0 | 0 | 0 | 0 |
| 0 | 0 | 0 | 0 | 0 |
| 0 | 0 | 0 | 0 | 0 |
| 0 | 0 | 0 | 0 | 0 |
| 0 | 0 | 0 | 0 | 0 |
| 0 | 0 | 0 | 0 | 0 |
| 0.014879965 | 0 | 0 | 0.022698276 | 0.068852819 |
| 0 | 0 | 0 | 0 | 0 |
| 0 | 0 | 0 | 0 | 0 |
| 0 | 0 | 0 | 0 | 0 |
| 0 | 0 | 0 | 0 | 0 |
| 0 | 0 | 0 | 0 | 0 |
| 0 | 0 | 0 | 0 | 0 |
| 0 | 0 | 0 | 0 | 0 |
| 0 | 0 | 0 | 0 | 0 |
| 0 | 0 | 0 | 0 | 0 |
| 0 | 0 | 0 | 0 | 0 |
| 0 | 0 | 0 | 0 | 0 |
| 0.00316229 | 0.028864714 | 0 | 0.001016452 | 0.263627722 |
| 0 | 0.004836644 | 0.004325785 | 0 | 0.001082224 |
| 0 | 0 | 0 | 0 | 0 |
| 0 | 0 | 0 | 0 | 0 |
| 0 | 0 | 0 | 0 | 0 |
| 0 | 0 | 0 | 0 | 0 |
| 0 | 0 | 0 | 0 | 0 |
| 0 | 0 | 0 | 0 | 0 |
| 0 | 0 | 0 | 0 | 0 |
| 0 | 0 | 0 | 0 | 0 |
| 0 | 0 | 0 | 0 | 0 |
| 0 | 0 | 0 | 0 | 0 |
| 0 | 0.001458364 | 0.002380916 | 0 | 0 |
| 0 | 0 | 0 | 0 | 0 |
| 0 | 0 | 0 | 0 | 0 |
| 0 | 0 | 0 | 0 | 0 |
| 0 | 0 | 0 | 0 | 0 |
| 0 | 0 | 0 | 0 | 0 |

| | | | | |
|---|---|---|---|---|
| 0 | 0 | 0 | 0 | 0.009583371 |
| 0 | 0 | 0 | 0 | 0 |
| 0 | 0 | 0 | 0 | 0 |
| 0 | 0 | 0 | 0 | 0 |
| 0.002887524 | 0 | 0 | 0.066825624 | 0 |
| 0 | 0 | 0 | 0 | 0 |
| 0 | 0 | 0 | 0 | 0 |
| 0 | 0 | 0 | 0 | 0 |
| 0 | 0 | 0 | 0 | 0 |
| 0 | 0 | 0 | 0 | 0 |
| 0 | 0 | 0 | 0 | 0 |
| 0 | 0 | 0 | 0 | 0 |
| 0 | 0 | 0 | 0 | 0 |
| 0 | 0 | 0 | 0 | 0 |
| 0 | 0 | 0 | 0 | 0 |
| 0.001956033 | 0 | 0 | 0 | 0 |
| 0 | 0 | 0 | 0 | 0 |
| 0 | 0 | 0 | 0 | 0 |
| 0 | 0 | 0 | 0 | 0 |
| 0 | 0 | 0 | 0 | 0 |
| 0 | 0 | 0 | 0 | 0 |
| 0 | 0 | 0 | 0 | 0 |
| 0 | 0 | 0 | 0 | 0 |
| 0 | 0 | 0 | 0 | 0 |
| 0 | 0 | 0 | 0 | 0 |
| 0 | 0 | 0 | 0 | 0 |
| 0 | 0 | 0 | 0 | 0 |
| 0 | 0 | 0 | 0 | 0 |
| 0 | 0 | 0 | 0 | 0 |
| 0 | 0 | 0 | 0 | 0 |
| 0 | 0 | 0 | 0 | 0 |
| 0 | 0 | 0 | 0 | 0 |
| 0 | 0 | 0 | 0 | 0 |
| 0 | 0 | 0 | 0 | 0 |
| 0 | 0 | 0 | 0 | 0 |
| 0 | 0 | 0 | 0 | 0 |
| 0 | 0 | 0 | 0 | 0 |
| 0 | 0 | 0 | 0 | 0 |
| 0 | 0 | 0 | 0 | 0 |
| 0 | 0 | 0 | 0 | 0 |
| 0 | 0 | 0 | 0 | 0 |
| 0 | 0 | 0 | 0 | 0 |
| 0 | 0 | 0 | 0 | 0 |
| 0 | 0 | 0 | 0 | 0 |
| 0 | 0 | 0 | 0 | 0 |
| 0 | 0 | 0 | 0 | 0 |
| 0 | 0 | 0 | 0 | 0 |
| 0 | 0 | 0 | 0 | 0 |

| | | | | |
|---|---|---|---|---|
| 0.117097685 | 0 | 0 | 0 | 0 |
| 0 | 0 | 0 | 0 | 0 |
| 0 | 0 | 0 | 0 | 0 |
| 0 | 0 | 0 | 0 | 0 |
| 0 | 0 | 0 | 0 | 0 |
| 0 | 0 | 0 | 0 | 0 |
| 0 | 0 | 0 | 0 | 0 |
| 0 | 0 | 0 | 0 | 0 |
| 0 | 0 | 0 | 0 | 0 |
| 0 | 0 | 0 | 0 | 0 |
| 0 | 0 | 0 | 0 | 0 |
| 0 | 0.002119665 | 0 | 0 | 0 |
| 0 | 0.000696514 | 0.000909699 | 0.001765963 | 0.003982796 |
| 0 | 0 | 0 | 0 | 0 |
| 0 | 0 | 0 | 0 | 0.1485286 |
| 0 | 0 | 0 | 0 | 0 |
| 0 | 0 | 0 | 0 | 0 |
| 0 | 0 | 0 | 0 | 0 |
| 0 | 0 | 0 | 0 | 0 |
| 0 | 0 | 0 | 0 | 0 |
| 0.006848431 | 0.079006993 | 0.113111069 | 0.022012838 | 0.006950414 |
| 0 | 0 | 0 | 0 | 0 |
| 0 | 0 | 0 | 0 | 0 |
| 0 | 0 | 0.001330287 | 0 | 0 |
| 0 | 0 | 0 | 0 | 0 |
| 0 | 0 | 0 | 0 | 0 |
| 0.015383693 | 0.077613328 | 0.016344483 | 0.004273253 | 0.001445628 |
| 0 | 0 | 0 | 0 | 0 |
| 0 | 0 | 0 | 0 | 0 |
| 0 | 0 | 0 | 0 | 0 |
| 0 | 0.00327614 | 0 | 0 | 0 |
| 0 | 0 | 0 | 0 | 0 |
| 0 | 0 | 0 | 0 | 0 |
| 0.002092668 | 0 | 0 | 0.010538089 | 0 |
| 0.004774237 | 0.046886938 | 0.008576988 | 0.203621421 | 0.098198721 |
| 0 | 0 | 0 | 0 | 0 |
| 0 | 0 | 0 | 0 | 0 |
| 0 | 0 | 0 | 0 | 0 |
| 0 | 0 | 0 | 0 | 0 |
| 0 | 0 | 0 | 0 | 0 |
| 0 | 0 | 0 | 0 | 0 |
| 0 | 0 | 0 | 0 | 0 |
| 0 | 0 | 0 | 0 | 0 |
| 0 | 0 | 0 | 0 | 0 |
| 0 | 0 | 0 | 0 | 0 |
| 0.001212158 | 0 | 0.0039514 | 0 | 0 |
| 0 | 0 | 0 | 0 | 0.01538887 |
| 0 | 0 | 0 | 0.003062028 | 0.023825109 |

| | | | | |
|---|---|---|---|---|
| 0 | 0 | 0 | 0 | 0 |
| 0 | 0 | 0 | 0 | 0 |
| 0 | 0 | 0.001801676 | 0 | 0 |
| 0 | 0 | 0 | 0 | 0 |
| 0 | 0 | 0 | 0 | 0 |
| 0 | 0 | 0 | 0 | 0 |
| 0 | 0 | 0 | 0 | 0.002949108 |
| 0.015976604 | 0 | 0 | 0 | 0 |
| 0 | 0 | 0 | 0 | 0 |
| 0 | 0 | 0 | 0 | 0 |
| 0 | 0 | 0 | 0 | 0 |
| 0 | 0 | 0 | 0 | 0 |
| 0 | 0 | 0 | 0 | 0 |
| 0 | 0 | 0 | 0 | 0 |
| 0 | 0 | 0.100410309 | 0 | 0.00145176 |
| 0 | 0 | 0 | 0 | 0 |
| 0 | 0 | 0 | 0 | 0 |
| 0 | 0 | 0 | 0 | 0 |
| 0 | 0 | 0 | 0 | 0.001404636 |
| 0.008987507 | 0.044570244 | 0 | 0.063639549 | 0 |
| 0.022797642 | 0.001509789 | 0.004140984 | 0 | 0 |
| 0 | 0 | 0 | 0 | 0 |
| 0 | 0 | 0 | 0 | 0 |
| 0 | 0 | 0 | 0 | 0 |
| 0 | 0.020396699 | 0.008814579 | 0.003042026 | 0 |
| 0 | 0 | 0 | 0 | 0 |
| 0 | 0 | 0 | 0 | 0 |
| 0 | 0 | 0 | 0 | 0 |
| 0 | 0 | 0 | 0 | 0 |
| 0 | 0 | 0 | 0 | 0 |
| 0.163878035 | 0.02664353 | 0.003914825 | 0.167568598 | 0.161366934 |
| 0 | 0 | 0 | 0 | 0 |
| 0.000933295 | 0 | 0 | 0 | 0 |
| 0 | 0 | 0 | 0 | 0 |
| 0 | 0 | 0 | 0 | 0 |
| 0 | 0 | 0 | 0 | 0 |
| 0 | 0 | 0 | 0 | 0 |
| 0 | 0 | 0 | 0 | 0 |
| 0 | 0 | 0 | 0 | 0 |
| 0.001999284 | 0 | 0 | 0 | 0 |
| 0 | 0 | 0 | 0 | 0 |
| 0 | 0.010410815 | 0.010197978 | 0.010520109 | 0.03093024 |
| 0 | 0 | 0 | 0 | 0 |
| 0 | 0 | 0 | 0 | 0 |
| 0 | 0 | 0 | 0 | 0 |
| 0 | 0 | 0 | 0 | 0 |
| 0 | 0 | 0 | 0 | 0 |
| 0 | 0 | 0 | 0 | 0 |

| | | | | |
|---|---|---|---|---|
| 0 | 0 | 0 | 0 | 0 |
| 0 | 0 | 0 | 0 | 0 |
| 0 | 0.102874827 | 0 | 0.013449905 | 0.057363247 |
| 0 | 0 | 0 | 0 | 0 |
| 0.16952462 | 0 | 0 | 0 | 0 |
| 0 | 0 | 0 | 0 | 0 |
| 0 | 0 | 0 | 0 | 0 |
| 0 | 0 | 0 | 0 | 0 |
| 0 | 0 | 0 | 0 | 0 |
| 0 | 0 | 0 | 0 | 0 |
| 0 | 0 | 0 | 0 | 0 |
| 0 | 0 | 0 | 0 | 0 |
| 0 | 0 | 0 | 0 | 0 |
| 0 | 0.002826678 | 0.009229632 | 0 | 0 |
| 0 | 0.008036069 | 0.007109996 | 0.00087634 | 0.000889389 |
| 0 | 0 | 0 | 0 | 0 |
| 0.00199702 | 0.00413514 | 0.002893287 | 0 | 0 |
| 0 | 0 | 0 | 0 | 0 |
| 0 | 0 | 0 | 0 | 0 |
| 0 | 0 | 0 | 0 | 0 |
| 0 | 0 | 0 | 0 | 0 |
| 0 | 0.001066529 | 0 | 0.002929454 | 0.020735285 |
| 0 | 0 | 0 | 0 | 0 |
| 0 | 0 | 0 | 0 | 0.003545341 |
| 0 | 0 | 0.002512733 | 0 | 0 |
| 0 | 0 | 0 | 0 | 0 |
| 0 | 0 | 0 | 0 | 0 |
| 0 | 0 | 0 | 0 | 0 |
| 0 | 0 | 0 | 0 | 0 |
| 0 | 0.002699802 | 0.019267853 | 0 | 0 |
| 0 | 0 | 0.013209723 | 0 | 0 |
| 0 | 0 | 0 | 0 | 0 |
| 0 | 0 | 0 | 0 | 0 |
| 0 | 0 | 0 | 0 | 0 |
| 0 | 0 | 0 | 0 | 0 |
| 0 | 0 | 0.007303413 | 0 | 0 |
| 0 | 0 | 0 | 0 | 0 |
| 0.003774924 | 0 | 0.001968883 | 0.161095066 | 0.041184733 |
| 0 | 0 | 0 | 0 | 0 |
| 0 | 0 | 0 | 0 | 0 |
| 0 | 0 | 0 | 0 | 0 |
| 0 | 0 | 0.004239797 | 0 | 0 |
| 0 | 0 | 0 | 0 | 0 |
| 0 | 0 | 0 | 0 | 0 |
| 0 | 0 | 0 | 0 | 0 |
| 0 | 0 | 0.010090422 | 0 | 0 |
| 0 | 0 | 0 | 0 | 0 |
| 0 | 0.002434583 | 0 | 0.003772218 | 0 |

| | | | | |
|---|---|---|---|---|
| 0.040392113 | 0.171734631 | 0 | 0 | 0 |
| 0 | 0 | 0 | 0 | 0 |
| 0 | 0 | 0 | 0 | 0 |
| 0 | 0 | 0 | 0 | 0 |
| 0 | 0.034279443 | 0.028349179 | 0.00831137 | 0.000482007 |
| 0 | 0 | 0.006731928 | 0 | 0 |
| 0 | 0 | 0 | 0 | 0 |
| 0 | 0.10397745 | 0.004243823 | 0.029957695 | 0.003152984 |
| 0 | 0 | 0.003567575 | 0 | 0 |
| 0.097437283 | 0.007713123 | 0.018689764 | 0.034995118 | 0.006267565 |
| 0 | 0 | 0 | 0 | 0.013851907 |
| 0 | 0 | 0.011605288 | 0 | 0.004813547 |
| 0.002131954 | 0.009459733 | 0.00926634 | 0.003797547 | 0.003921709 |
| 0 | 0 | 0 | 0 | 0 |
| 0 | 0 | 0 | 0 | 0.000834494 |
| 0 | 0 | 0 | 0 | 0 |
| 0 | 0 | 0 | 0 | 0 |
| 0 | 0 | 0 | 0 | 0 |
| 0.001721816 | 0.244094899 | 0 | 0 | 0.005897665 |
| 0.024497524 | 0.004899084 | 0 | 0.026136471 | 0 |
| 0 | 0 | 0 | 0 | 0 |
| 0 | 0 | 0 | 0 | 0 |
| 0 | 0.001464183 | 0 | 0 | 0 |
| 0.003543176 | 0 | 0 | 0 | 0 |
| 0 | 0 | 0 | 0 | 0 |
| 0.002793581 | 0.00743727 | 0 | 0.010941536 | 0.002205141 |
| 0.001245195 | 0 | 0 | 0 | 0 |
| 0 | 0 | 0 | 0 | 0 |
| 0.007945065 | 0 | 0 | 0 | 0 |
| 0 | 0.004251119 | 0 | 0 | 0 |
| 0 | 0 | 0 | 0 | 0 |
| 0 | 0 | 0 | 0 | 0 |
| 0 | 0 | 0 | 0 | 0 |
| 0 | 0 | 0 | 0 | 0 |
| 0.001420385 | 0 | 0 | 0 | 0 |
| 0 | 0 | 0 | 0 | 0 |
| 0 | 0.008743163 | 0.282319951 | 0 | 0.000714215 |
| 0 | 0 | 0 | 0 | 0 |
| 0 | 0 | 0 | 0 | 0 |
| 0 | 0 | 0 | 0 | 0 |
| 0 | 0 | 0.002125107 | 0 | 0 |
| 0 | 1.671526219 | 0 | 0 | 0 |
| 0 | 0 | 0 | 0 | 0 |
| 0 | 0.002219158 | 0.002825927 | 0 | 0.015988851 |
| 0 | 0 | 0 | 0 | 0 |
| 0.000908397 | 0 | 0 | 0 | 0 |
| 0 | 0.001370197 | 0.001677732 | 0.003474042 | 0.003917524 |
| 0.001401462 | 0 | 0 | 0 | 0 |

| | | | | |
|---|---|---|---|---|
| 0 | 0 | 0 | 0 | 0 |
| 0 | 0 | 0.00749993 | 0 | 0 |
| 0 | 0 | 0 | 0.008682592 | 0 |
| 0 | 0 | 0 | 0 | 0 |
| 0 | 0 | 0 | 0 | 0 |
| 0 | 0.057737009 | 0 | 0.03169465 | 0.059242067 |
| 0 | 0 | 0 | 0 | 0 |
| 0 | 0.002791614 | 0.004443631 | 0 | 0 |
| 0.059154602 | 0 | 0 | 0 | 0 |
| 0 | 0 | 0 | 0 | 0 |
| 0.091680659 | 0.090434324 | 0.08950507 | 0.005024811 | 0.001968279 |
| 0.001321933 | 0 | 0 | 0 | 0 |
| 0 | 0 | 0 | 0 | 0 |
| 0.004997035 | 0.006799548 | 0.013900256 | 0.004747189 | 0.00101429 |
| 0 | 0.010469445 | 0.001172047 | 0 | 0.002223604 |
| 0.001698628 | 0 | 0 | 0 | 0 |
| 0 | 0 | 0 | 0.001745163 | 0 |
| 0 | 0 | 0 | 0 | 0 |
| 0 | 0 | 0 | 0 | 0.02141118 |
| 0 | 0 | 0.001041884 | 0 | 0 |
| 0 | 0 | 0 | 0 | 0 |
| 1.934830562 | 0.213582324 | 0.235081375 | 0 | 0 |
| 0 | 0.003927576 | 0 | 0 | 0 |
| 0 | 0 | 0 | 0 | 0 |
| 0 | 0 | 0 | 0 | 0 |
| 0 | 0 | 0 | 0 | 0 |
| 0.002889658 | 0.019232636 | 0.002511926 | 0 | 0.010630998 |
| 0 | 0 | 0 | 0 | 0 |
| 0 | 0 | 0 | 0 | 0 |
| 1.106015661 | 0.26910127 | 0.802260259 | 0.869798019 | 0.152638234 |
| 0.001356036 | 0.003209011 | 0.000982314 | 0.000678019 | 0 |
| 0 | 0 | 0 | 0 | 0 |
| 0 | 0 | 0 | 0 | 0 |
| 0 | 0 | 0 | 0 | 0 |
| 0.019555356 | 0.00186888 | 0.001046099 | 0 | 0.00442731 |
| 0 | 0 | 0 | 0 | 0 |
| 0.62075629 | 0 | 0 | 0 | 0 |
| 0 | 0 | 0 | 0 | 0 |
| 0 | 0 | 0 | 0 | 0 |
| 0 | 0.001399236 | 0.011993013 | 0 | 0 |
| 0.001643243 | 0.001458253 | 0.012379816 | 0.006162167 | 0.001945665 |
| 0.077493651 | 1.211099409 | 4.220372287 | 6.951293944 | 0 |
| 0 | 0 | 0 | 0 | 0 |
| 0.082930919 | 0 | 0 | 0 | 0 |
| 0 | 0 | 0 | 0 | 0.013482413 |
| 0 | 0 | 0 | 0 | 0 |
| 0 | 0 | 0.006496753 | 0 | 0.001312788 |
| 0 | 0 | 0 | 0 | 0 |

| | | | | |
|---|---|---|---|---|
| 0.001773326 | 0 | 0 | 0.006117983 | 0.003959414 |
| 0 | 0 | 0 | 0.029743993 | 0 |
| 0 | 0.048031784 | 0.04494939 | 0.001449777 | 0 |
| 0 | 0.00142351 | 0 | 0 | 0 |
| 0 | 0 | 0 | 0 | 0 |
| 0.002099089 | 0 | 0.175692431 | 0 | 0 |
| 0 | 0 | 0 | 0 | 0 |
| 0 | 0 | 0 | 0 | 0 |
| 0 | 0 | 0.011869047 | 0 | 0 |
| 0.019487733 | 0 | 0 | 0 | 0 |
| 0 | 0 | 0 | 0 | 0 |
| 0 | 0 | 0 | 0 | 0 |
| 0 | 0 | 0 | 0 | 0 |
| 0.167487215 | 0.066628202 | 0.555040456 | 0.009866355 | 0.002605078 |
| 0 | 0 | 0 | 0 | 0 |
| 0.008525296 | 0.007788067 | 0.067133869 | 0 | 0.229284602 |
| 0.038631378 | 0.004192812 | 0.006281439 | 0.028765122 | 0.106337463 |
| 0.002363664 | 0 | 0 | 0 | 0.000685389 |
| 0 | 0 | 0 | 0 | 0 |
| 0 | 1.227861179 | 0.00354416 | 0 | 0 |
| 0 | 0.069315434 | 0 | 0 | 0.000710957 |
| 0.003868523 | 0 | 0.005044253 | 0 | 0.013087101 |
| 0 | 0.028716553 | 0 | 0 | 0 |
| 0 | 0 | 0 | 0.244130567 | 0 |
| 0 | 0 | 0 | 0 | 0 |
| 0.087135449 | 0 | 0.014639834 | 0 | 0 |
| 0 | 0 | 0 | 0 | 0 |
| 0 | 0 | 0 | 0 | 0 |
| 0 | 0 | 0 | 0 | 0 |
| 0 | 0 | 0 | 0 | 0 |
| 0.00643976 | 0.000952466 | 0.005831213 | 0.016099418 | 0.023010959 |
| 0.00280026 | 0.021300151 | 0.012866561 | 0 | 0.006292911 |
| 0.087604581 | 0.035829102 | 0.138068766 | 0.013997704 | 0 |
| 0 | 0 | 0 | 0 | 0.001938931 |
| 0 | 0 | 0 | 0 | 0 |
| 0 | 0 | 0 | 0 | 0 |
| 0 | 0.001527923 | 0.001496687 | 0 | 0 |
| 0.007271044 | 0.007647405 | 0.001872766 | 0.043828286 | 0.014621986 |
| 0.00060016 | 0.243041431 | 0 | 0 | 0 |
| 0 | 0 | 0 | 0 | 0 |
| 0 | 0 | 0 | 0 | 0 |
| 0 | 0 | 0 | 0 | 0 |
| 0.015303916 | 0 | 0 | 0.001812308 | 0 |
| 0.004835065 | 0.01175293 | 0.04367499 | 0.147417095 | 0.072539235 |
| 0.028796605 | 0 | 0.210986987 | 0 | 0 |
| 0 | 0 | 0 | 0 | 0 |
| 0 | 0 | 0 | 0 | 0 |
| 0 | 0 | 0 | 0 | 0.000555296 |

| | | | | |
|---|---|---|---|---|
| 0 | 0 | 0 | 0 | 0 |
| 0.257600726 | 1.916405967 | 0.32632174 | 0.526761024 | 0.146732305 |
| 0 | 0 | 0 | 0 | 0 |
| 0 | 0 | 0 | 0 | 0 |
| 0 | 0 | 0 | 0 | 0 |
| 0 | 0 | 0 | 0 | 0 |
| 0 | 0 | 0 | 0 | 0 |
| 0 | 0 | 0 | 0 | 0 |
| 0 | 0 | 0 | 0 | 0 |
| 0 | 0 | 0 | 0 | 0 |
| 0 | 0 | 0 | 0.002045146 | 0.004566318 |
| 0.002030867 | 0.15895754 | 0.020125502 | 0.007311129 | 0.00659555 |
| 0 | 0 | 0 | 0 | 0 |
| 0.00119655 | 0 | 0 | 0 | 0 |
| 0 | 0.001943557 | 0.001631848 | 0 | 0 |
| 0 | 0 | 0 | 0 | 0 |
| 0.004568664 | 0.058281155 | 0.044927082 | 0.019274072 | 0.002028555 |
| 0.004448068 | 0.013157737 | 0.019010895 | 0 | 0.021066759 |
| 0 | 0 | 0 | 0 | 0 |
| 0.000962781 | 0.076895524 | 0 | 0 | 0 |
| 0.006478237 | 0.030051287 | 0.046075183 | 0.012588406 | 0.001793102 |
| 0 | 0 | 0 | 0 | 0 |
| 0 | 0 | 0 | 0 | 0 |
| 0 | 0 | 0 | 0 | 0 |
| 0 | 0 | 0 | 0 | 0 |
| 0 | 0 | 0 | 0 | 0.015072977 |
| 0.002615868 | 0.024503492 | 0 | 0.087849658 | 0 |
| 0.202180667 | 0.129982246 | 0.065294824 | 0.048119676 | 0.0122289 |
| 0 | 0 | 0 | 0 | 0 |
| 0 | 0 | 0 | 0 | 0.002298844 |
| 0.004029846 | 0.033973719 | 0.002043458 | 0.001511194 | 0 |
| 0 | 0 | 0 | 0 | 0 |
| 0 | 0 | 0 | 0 | 0 |
| 0 | 0 | 0 | 0 | 0 |
| 0.001363672 | 0 | 0 | 0 | 0 |
| 0.851209285 | 0.609195314 | 0.128465078 | 0.735340734 | 0.096494926 |
| 0 | 0 | 0 | 0 | 0 |
| 0 | 0 | 0 | 0 | 0 |
| 0 | 0.008565579 | 0.036918048 | 0.002026961 | 0.004506123 |
| 0 | 0 | 0 | 0 | 0 |
| 0.005814524 | 0 | 0.202599531 | 0 | 0 |
| 0 | 0 | 0 | 0 | 0 |
| 0.003283215 | 0 | 0.021609147 | 0.076510728 | 0 |
| 0 | 0.005826989 | 0.000787291 | 0 | 0.000459584 |
| 0.001189855 | 0 | 0 | 0 | 0 |
| 0.001403632 | 0.924040089 | 0 | 0.192297849 | 0 |
| 0 | 0.005594482 | 0 | 0 | 0 |
| 0 | 0 | 0 | 0 | 0 |

| | | | | |
|---|---|---|---|---|
| 0 | 0.004361108 | 0 | 0 | 0 |
| 0 | 0 | 0 | 0 | 0 |
| 0 | 0 | 0 | 0 | 0 |
| 0.016388173 | 0.268080669 | 0 | 0.073951711 | 1.765232425 |
| 0 | 0 | 0 | 0 | 0 |
| 0 | 0 | 0 | 0 | 0 |
| 0 | 0 | 0 | 0 | 0 |
| 0 | 0 | 0 | 0.082101568 | 0.088779824 |
| 0.494054887 | 0.023296006 | 0.033588614 | 0 | 0 |
| 0.002685283 | 0 | 0 | 0.322417356 | 0.027624329 |
| 0 | 0 | 0 | 0 | 0 |
| 0.018841958 | 0 | 0.005232169 | 0.001766435 | 0 |
| 0 | 0 | 0 | 0 | 0 |
| 0.001967117 | 0 | 0 | 0 | 0 |
| 0.034120169 | 0.003257873 | 0.003754434 | 0.162772818 | 0 |
| 0 | 0 | 0 | 0 | 0 |
| 0 | 0 | 0 | 0.108379738 | 0 |
| 0.008177964 | 0 | 0 | 0 | 0 |
| 0 | 0 | 0 | 0 | 0 |
| 0 | 0 | 0 | 0 | 0 |
| 0 | 0 | 0 | 0 | 0 |
| 0 | 0 | 0 | 0 | 0 |
| 0 | 0 | 0 | 0 | 0 |
| 0.019553468 | 0.307724891 | 0.332716376 | 0.550721948 | 0.151684872 |
| 0 | 0 | 0 | 0 | 0 |
| 0.00606026 | 0.731186487 | 0 | 0 | 0.091488766 |
| 0.001457634 | 0 | 0 | 0 | 0 |
| 6.394922018 | 0.653718942 | 0 | 0 | 0.419124494 |
| 0 | 0 | 0 | 0 | 0 |
| 0 | 0 | 0 | 0 | 0 |
| 0 | 0 | 0 | 0 | 0 |
| 0 | 0 | 0 | 0 | 0 |
| 0.000580876 | 0.000515484 | 0 | 0.001452193 | 0.066813003 |
| 0 | 0 | 0 | 0 | 0 |
| 0.000939578 | 0 | 0 | 0 | 0 |
| 0.000862848 | 0 | 0 | 0 | 0 |
| 0.01894435 | 0.082574952 | 0.001937408 | 0.00835781 | 0 |
| 0 | 0 | 0 | 0 | 0 |
| 0 | 0 | 0 | 0 | 0 |
| 0 | 0 | 0 | 0 | 0 |
| 0.032636302 | 0.117631218 | 0.478364065 | 0.06571201 | 0.189051899 |
| 0.000654119 | 0 | 0 | 0 | 0 |
| 20.91489927 | 1.01478482 | 2.666652128 | 1.978194356 | 0.031481764 |
| 0 | 0 | 0 | 0 | 0 |
| 0 | 0 | 0 | 0 | 0 |
| 0 | 0 | 0 | 0 | 0 |
| 0 | 0 | 0 | 0 | 0 |
| 0.001810734 | 0 | 0 | 0.009053682 | 0.027871763 |

| | | | | |
|---|---|---|---|---|
| 0 | 0 | 0 | 0 | 0 |
| 0.017870995 | 0.009027514 | 0 | 0 | 0 |
| 0.010993358 | 0.001540384 | 0.012574108 | 0.009329899 | 0.008074191 |
| 0 | 0 | 0 | 0 | 0 |
| 0 | 0 | 0 | 0 | 0 |
| 0 | 0 | 0.001782972 | 0.005298646 | 0 |
| 0 | 0 | 0.010145559 | 0 | 0 |
| 0 | 0 | 0 | 0 | 0 |
| 0 | 0 | 0 | 0 | 0 |
| 0 | 0 | 0 | 0 | 0 |
| 0.009941407 | 0 | 0 | 0.499556259 | 0.340735082 |
| 1.190749202 | 4.980127609 | 3.550705635 | 0.825762897 | 0.292094864 |
| 0 | 0 | 0 | 0 | 0 |
| 0.018086319 | 0.01115355 | 0 | 0.100930963 | 0.082755974 |
| 0.708528924 | 0.005359405 | 0.414317246 | 0.041308754 | 0.055530716 |
| 0.006981688 | 0 | 0 | 0 | 0 |
| 0 | 0 | 0 | 0 | 0 |
| 0 | 0 | 0 | 0 | 0 |
| 0 | 0 | 0 | 0 | 0 |
| 0.02994872 | 0.009343549 | 0 | 0.027901474 | 0 |
| 0.010474971 | 0.241689208 | 1.105514507 | 0.037674226 | 0.002657739 |
| 1.186517254 | 0 | 0.108157091 | 0.011852643 | 0.038187719 |
| 2.464021874 | 0.233070433 | 1.083669597 | 0.75283392 | 0.26609172 |
| 0 | 0 | 0 | 0 | 0 |
| 0 | 0 | 0 | 0 | 0 |
| 0 | 0 | 0.00967143 | 0 | 0 |
| 0.035307741 | 0 | 0 | 0 | 0 |
| 0 | 0.001322085 | 0 | 0 | 0 |
| 0 | 0 | 0 | 0.032449737 | 0 |
| 0 | 0 | 0 | 0 | 0 |
| 0 | 0 | 0 | 0 | 0 |
| 0 | 0.135427938 | 0.132073144 | 0.097936658 | 0 |
| 0.025087636 | 0 | 0.483584514 | 0.357499196 | 0.039968201 |
| 0.00066737 | 0.001579306 | 0 | 0 | 0 |
| 0.006161173 | 0.05228365 | 0.001506317 | 0 | 0.055103863 |
| 0.12724065 | 0.002817041 | 0.010347936 | 0.171814741 | 0.373714185 |
| 0.001277679 | 0 | 0 | 0.005941214 | 0 |
| 0 | 0 | 0 | 0 | 0 |
| 0.056958617 | 0.026372052 | 0 | 0 | 0 |
| 0 | 0 | 0 | 0 | 0 |
| 0 | 0 | 0 | 0 | 0 |
| 0 | 0 | 0 | 0 | 0 |
| 0 | 0 | 0 | 0 | 0 |
| 0 | 0 | 0 | 0 | 0 |
| 0.003016705 | 0 | 0 | 0.041404322 | 0.499504595 |
| 0.006062356 | 0 | 0 | 0 | 0 |
| 0 | 0 | 0 | 0 | 0 |
| 0.29257325 | 0.006135688 | 0.384023374 | 0.086516623 | 9.896191341 |

| | | | | |
|---|---|---|---|---|
| 0 | 0 | 0 | 0 | 0 |
| 0 | 0 | 0 | 0 | 0 |
| 0 | 0 | 0 | 0 | 0 |
| 0 | 0.001369007 | 0 | 0 | 0 |
| 0 | 0 | 0 | 0 | 0 |
| 0 | 0 | 0 | 0 | 0 |
| 0.002533404 | 0 | 0 | 0 | 0.022425962 |
| 0.060803654 | 0 | 0.000822654 | 0 | 0 |
| 0 | 0 | 0 | 0 | 0 |
| 0 | 0 | 0 | 0 | 0 |
| 0 | 0.001270593 | 0 | 0.011382636 | 0.021360535 |
| 0 | 0 | 0 | 0 | 0 |
| 0 | 0 | 0 | 0 | 0 |
| 0 | 0 | 0 | 0.55309858 | 0 |
| 0 | 0 | 0 | 0 | 0 |
| 0 | 0 | 0 | 0 | 0 |
| 0 | 0 | 0.382855941 | 0.026674891 | 0.151733372 |
| 0.010488907 | 0 | 0.002279453 | 0.008581842 | 0.091693031 |
| 0.00727446 | 0.022705655 | 0 | 0 | 0 |
| 0.003654713 | 0.003243279 | 0 | 0 | 0 |
| 0 | 0 | 0 | 0 | 0 |
| 0 | 0 | 0 | 0 | 0 |
| 0 | 0 | 0 | 0 | 0.002649861 |
| 0 | 0 | 0 | 0 | 0 |
| 0 | 0 | 0 | 0 | 0 |
| 0 | 0 | 0 | 0 | 0 |
| 0 | 0 | 0 | 0 | 0.001251582 |
| 0 | 0 | 0 | 0 | 0 |
| 0 | 0 | 0 | 0 | 0 |
| 0.042043044 | 0 | 0.351454256 | 0.002159748 | 0 |
| 0 | 0 | 0 | 0.004905428 | 0 |
| 0 | 0 | 0 | 0 | 0 |
| 0 | 0 | 0 | 0 | 0 |
| 0 | 0 | 0 | 0 | 0 |
| 0 | 0 | 0 | 0.001878492 | 0.001155432 |
| 0 | 0 | 0 | 0 | 0 |
| 0 | 0 | 0.118185041 | 0.110273069 | 0 |
| 0.007272143 | 0 | 0 | 0.002545253 | 0.084136962 |
| 0 | 0 | 0 | 0 | 0 |
| 0 | 0 | 0 | 0 | 0 |
| 0.001137178 | 0.007064114 | 0 | 0 | 0 |
| 0.001050173 | 0 | 0 | 0 | 0 |
| 0 | 0 | 0 | 0 | 0 |
| 0.000706211 | 0 | 0 | 0.294313864 | 0 |
| 0.003578226 | 0.274794448 | 0 | 0 | 0 |
| 0 | 0 | 0 | 0 | 0 |
| 0 | 0 | 0 | 0.016756171 | 0 |
| 0 | 0 | 0 | 0 | 0 |

| | | | | |
|---|---|---|---|---|
| 0 | 0 | 0 | 0 | 0 |
| 0 | 0 | 0 | 0 | 0 |
| 0 | 0 | 0 | 0 | 0 |
| 0.000723365 | 0 | 0.008593707 | 0.038519234 | 0 |
| 0 | 0 | 0 | 0 | 0 |
| 0 | 0 | 0 | 0 | 0 |
| 0 | 0 | 0 | 0 | 0 |
| 0 | 0 | 0 | 0 | 0 |
| 0.023402167 | 0.043908725 | 0.141820255 | 0.064439608 | 0.059885584 |
| 0 | 0 | 0 | 0 | 0 |
| 0 | 0 | 0 | 0 | 0 |
| 0 | 0 | 0 | 0 | 0 |
| 0 | 0 | 0 | 0 | 0 |
| 0 | 0 | 0 | 0 | 0 |
| 0.008648956 | 4.410093531 | 0.045736794 | 0 | 0.269184357 |
| 0.008245349 | 1.054579781 | 0 | 0 | 0.002222785 |
| 0 | 0 | 0 | 0 | 0 |
| 0 | 0 | 0 | 0 | 0 |
| 0 | 0 | 0 | 0 | 0 |
| 0 | 0 | 0 | 0 | 0 |
| 0.006390826 | 0.001134274 | 0 | 0 | 0 |
| 0 | 0 | 0 | 0 | 0 |
| 0 | 0 | 0 | 0 | 0 |
| 0 | 0 | 0 | 0 | 0 |
| 0.082560846 | 0 | 0 | 0 | 0.01177054 |
| 0.03186793 | 9.756724168 | 0.104044309 | 0 | 3.184794705 |
| 0 | 0 | 0 | 0.000658024 | 0 |
| 0 | 0 | 0 | 0 | 0 |
| 0.002749782 | 0.005185473 | 0 | 0 | 0 |
| 0 | 0 | 0 | 6.386814923 | 0 |
| 0 | 0 | 0 | 0 | 0 |
| 0.001012755 | 0 | 0 | 0 | 0 |
| 0 | 0 | 0 | 0 | 0 |
| 0 | 0 | 0 | 0.043462942 | 0 |
| 0.013492327 | 0.031008069 | 0.00150367 | 0.00830298 | 0.050033015 |
| 0.001711848 | 0 | 0 | 0 | 0.177953244 |
| 0.011356585 | 0.421264762 | 0 | 0 | 0 |
| 0.032032115 | 4.208527458 | 0.001440255 | 1.440204108 | 0 |
| 0.001235421 | 0 | 0 | 0 | 0 |
| 0 | 0 | 0 | 0 | 0 |
| 0 | 0 | 0 | 0 | 0 |
| 0 | 0.000984406 | 0 | 0 | 0 |
| 0.40536225 | 0.065243351 | 0.012781905 | 6.959672954 | 0.061557175 |
| 0.003013285 | 0 | 0 | 0 | 0 |
| 0 | 0 | 0 | 0 | 0 |
| 0 | 0.001644788 | 0 | 0 | 0 |
| 0 | 0 | 0 | 0 | 0 |
| 0.00258366 | 0.040942884 | 0.401058527 | 0.173013138 | 0.057686954 |

| | | | | |
|---|---|---|---|---|
| 0 | 0 | 0 | 0 | 0 |
| 0.004197737 | 0 | 0 | 0 | 0 |
| 0 | 0 | 0 | 0 | 0 |
| 0 | 0 | 0 | 0 | 0 |
| 0.00411041 | 0 | 0 | 0.007604267 | 0.004032566 |
| 0 | 0 | 0 | 0 | 0 |
| 0 | 0 | 0 | 0 | 0 |
| 0 | 0 | 0 | 0 | 0 |
| 0 | 0 | 0 | 0 | 0 |
| 0 | 0 | 0 | 0.004950796 | 0 |
| 0 | 0 | 0 | 0 | 0 |
| 0.778647785 | 0.08570078 | 0.283931289 | 0.490321104 | 1.871567139 |
| 0 | 0.064483763 | 1.083619422 | 0.531293851 | 0.043271092 |
| 0 | 0 | 0 | 0 | 0 |
| 0.045635418 | 0.002249886 | 0.024518277 | 0 | 0.014795066 |
| 0 | 0 | 0 | 0 | 0 |
| 0 | 0 | 0 | 0 | 0 |
| 0 | 0 | 0 | 0 | 0 |
| 0 | 0 | 0 | 0.001666638 | 0 |
| 0.000924017 | 0 | 0 | 0 | 0 |
| 0.001379822 | 0 | 0 | 0.090205984 | 0 |
| 0 | 0 | 0 | 0 | 0 |
| 0.488604188 | 0.011984494 | 0.010565536 | 0 | 0.094707902 |
| 0 | 0 | 0 | 0 | 0 |
| 0 | 0 | 0 | 0 | 0 |
| 0 | 0 | 0 | 0 | 0 |
| 0 | 0 | 0 | 0 | 0 |
| 0.001659049 | 0 | 0 | 0 | 0 |
| 0 | 0 | 0 | 0 | 0 |
| 0 | 0 | 0 | 0 | 0 |
| 0 | 0 | 0 | 0 | 0 |
| 0.011925915 | 0.208382335 | 0.058269558 | 0.04102108 | 0.0321369 |
| 0 | 0 | 0 | 0 | 0 |
| 0 | 0.014038573 | 0.002321694 | 0.240219913 | 0.537843889 |
| 0.002395183 | 0 | 0 | 0 | 0.181368424 |
| 0.024358371 | 0.008829151 | 0.002982293 | 0.00514614 | 0.6131529 |
| 0 | 0 | 0.000821424 | 0 | 0 |
| 0 | 0 | 0 | 0 | 0 |
| 0.004586186 | 0 | 0 | 0 | 0 |
| 0 | 0 | 0 | 0 | 0 |
| 0 | 0 | 0 | 0.000510177 | 0 |
| 0.001538151 | 0 | 0 | 0 | 0 |
| 0 | 0 | 0 | 0 | 0 |
| 0 | 0 | 0 | 0 | 0 |
| 0.006816319 | 0 | 0.000911584 | 0.011010988 | 0 |
| 0 | 0 | 0 | 0 | 0 |
| 0.000661742 | 0 | 0 | 0.023160992 | 0.402509952 |
| 0 | 0.001760381 | 0 | 0 | 0 |

| | | | | |
|---|---|---|---|---|
| 0 | 0 | 0 | 0 | 0 |
| 0 | 0 | 0 | 0 | 0.022358052 |
| 0 | 0 | 0 | 0.000610078 | 0 |
| 0 | 0 | 0 | 0 | 0 |
| 0 | 0 | 0 | 0 | 0 |
| 0 | 0 | 0 | 0 | 0 |
| 0.187738368 | 0 | 0 | 0 | 0 |
| 0.000768214 | 0.046584998 | 0.034057517 | 0 | 0.000779654 |
| 0 | 0 | 0 | 0 | 0 |
| 0 | 0 | 0 | 0 | 0 |
| 0 | 0 | 0 | 0 | 0 |
| 0 | 0 | 0 | 0 | 0.004479874 |
| 0 | 0 | 0 | 0 | 0 |
| 0 | 0.344922001 | 0 | 0 | 0 |
| 0 | 0 | 0 | 0 | 0 |
| 0 | 0 | 0 | 0 | 0 |
| 0 | 21.33128977 | 0 | 0 | 0 |
| 0 | 0 | 0 | 0 | 0 |
| 0.29870926 | 0.076362044 | 0.068000826 | 0.679484621 | 1.141912632 |
| 0 | 0 | 0 | 0 | 0 |
| 0 | 0 | 0 | 0 | 0.077133781 |
| 0 | 0 | 0 | 0 | 0 |
| 0 | 0 | 0 | 0 | 0 |
| 0 | 0 | 0 | 0 | 0 |
| 0.004583028 | 0 | 0 | 0 | 0 |
| 0.006345735 | 0.003032269 | 0.000848651 | 0 | 0.056599727 |
| 0 | 0 | 0 | 0 | 0 |
| 0.005016748 | 0.338774632 | 0.398093923 | 0.008958489 | 0.014304563 |
| 0 | 0 | 0 | 0 | 0 |
| 0.385754608 | 0 | 0.071704425 | 0.322666845 | 0.559155361 |
| 0 | 0 | 0 | 0 | 0 |
| 0 | 0 | 0 | 0 | 0 |
| 0 | 0 | 0 | 0.065080605 | 0 |
| 0 | 0 | 0 | 0 | 0 |
| 0 | 0 | 0.000889333 | 0.000767301 | 0.023621375 |
| 0.100506969 | 0.012090002 | 0 | 0 | 0 |
| 0 | 0 | 0 | 0 | 0 |
| 0 | 0 | 0 | 0.007618295 | 0.042708621 |
| 0 | 0 | 0 | 0 | 0 |
| 0 | 0 | 0 | 0 | 0 |
| 18.65648414 | 15.2204945 | 26.66207783 | 8.002308815 | 2.52219333 |
| 0.012746561 | 0.037516812 | 0 | 2.673116008 | 1.513771388 |
| 0.007480447 | 0.036510799 | 0 | 0 | 0 |
| 0.004660318 | 0 | 0 | 0 | 0 |
| 0 | 0 | 0 | 0 | 0 |
| 0.013145655 | 0 | 0 | 0 | 0 |
| 0 | 0 | 0 | 0 | 0 |
| 0 | 0 | 0 | 0.004491537 | 0.129388927 |

| | | | | |
|---|---|---|---|---|
| 0 | 0 | 0 | 0 | 5.286131401 |
| 0 | 0 | 0 | 0.011016725 | 0.043520834 |
| 0 | 0 | 0 | 0 | 0 |
| 0 | 0 | 0 | 0 | 0 |
| 2.452661583 | 0.586803774 | 0.258072893 | 5.278693524 | 0.633346166 |
| 0 | 0 | 0 | 0 | 0 |
| 0 | 0 | 0 | 0 | 0 |
| 0 | 0 | 0 | 0 | 0 |
| 0.026968897 | 0.14548648 | 1.109456246 | 0.001419417 | 0.078510117 |
| 0 | 0 | 0 | 0 | 0 |
| 0 | 0 | 0 | 0 | 0 |
| 0 | 0 | 0 | 0 | 0 |
| 0.00069062 | 0 | 0 | 0.094442427 | 0 |
| 0.067074745 | 0.053998317 | 0.076512339 | 0.018254539 | 0.0094786 |
| 0.246637087 | 0 | 0 | 0 | 0 |
| 0.295642439 | 0 | 0 | 2.467966739 | 0.085912321 |
| 0.001469438 | 0.024124259 | 0.265264035 | 0.001285759 | 0 |
| 0 | 0 | 0 | 0 | 0 |
| 0 | 0 | 0 | 0 | 0 |
| 0 | 0 | 0 | 0 | 0 |
| 0 | 0 | 0 | 0 | 0 |
| 0 | 0 | 0 | 0 | 0 |
| 0 | 0 | 0 | 0 | 0 |
| 0 | 0 | 0 | 0 | 0 |
| 0 | 0 | 0 | 0 | 0 |
| 0 | 0 | 0 | 0 | 0 |
| 0.000851824 | 0.063876015 | 0.011662482 | 0.043602804 | 0.004214482 |
| 0.002162674 | 0.000639736 | 0 | 0 | 0.006706575 |
| 0 | 0 | 0 | 0 | 0 |
| 0 | 0 | 0 | 0 | 0 |
| 0 | 0 | 0 | 0 | 0 |
| 0 | 0 | 0 | 0 | 0 |
| 0 | 0 | 0 | 0 | 0 |
| 0.001511373 | 0 | 0.001532777 | 0 | 0 |
| 0.000893776 | 0.006345263 | 0 | 0 | 0 |
| 0.001945817 | 0 | 0 | 0 | 0.074054738 |
| 0 | 0.002773511 | 0.017535769 | 0 | 0 |
| 0 | 0 | 0.004563487 | 0.010433836 | 0.003463134 |
| 0 | 0 | 0 | 0 | 0 |
| 0 | 0 | 0 | 0 | 0 |
| 0.002723572 | 1.063765644 | 8.033394756 | 0 | 0 |
| 0.004278107 | 0 | 0 | 0 | 0 |
| 0 | 0 | 0 | 0 | 0 |
| 0 | 0 | 0 | 0 | 0 |
| 0 | 0 | 0 | 0 | 0 |
| 0 | 0 | 0 | 0 | 0 |
| 0.003318283 | 0 | 0 | 0 | 0 |
| 0 | 0 | 0 | 0 | 0 |

| | | | | |
|---|---|---|---|---|
| 0.001052411 | 0 | 0 | 0.423859123 | 0 |
| 0 | 0 | 0 | 0 | 0 |
| 0 | 0 | 0 | 0 | 0 |
| 0 | 0 | 0 | 0 | 0 |
| 0.01041805 | 0 | 0 | 0.051569401 | 0.00409711 |
| 0 | 0 | 0 | 0 | 0 |
| 0.037135237 | 0.02319837 | 1.574971656 | 0.022537693 | 0.042151311 |
| 0 | 0 | 0 | 0 | 0 |
| 0 | 0 | 0 | 0 | 0 |
| 0.009227583 | 0 | 0 | 0 | 0 |
| 0 | 0 | 0 | 0.003777406 | 0 |
| 0.001709235 | 0 | 0 | 0 | 0 |
| 0 | 0 | 0 | 0 | 0 |
| 0 | 0 | 0 | 0 | 0 |
| 0 | 0 | 0 | 0 | 0 |
| 0 | 0 | 0 | 0 | 0 |
| 0 | 0 | 0 | 0.000942337 | 0 |
| 0 | 0 | 0 | 0 | 0 |
| 0 | 0 | 0 | 0 | 0 |
| 0 | 0 | 0 | 0 | 0 |
| 0.002093377 | 0 | 0.017388569 | 0.033319624 | 0.082385345 |
| 0 | 0 | 0 | 0 | 0 |
| 0 | 0.015026372 | 0.001115089 | 0.000962079 | 0 |
| 0 | 0 | 0.003172956 | 0.023440449 | 0 |
| 0 | 0 | 0 | 0 | 0 |
| 0 | 0 | 0 | 0 | 0 |
| 0 | 0 | 0 | 0 | 0 |
| 0 | 0 | 0 | 0 | 0 |
| 0 | 0 | 0 | 0 | 0 |
| 0.002955001 | 1.639252966 | 0.120444793 | 0.001723753 | 0 |
| 0 | 0 | 0 | 0 | 0 |
| 0 | 0 | 0 | 0 | 0 |
| 0 | 0 | 0 | 0 | 0 |
| 0.035569948 | 0 | 0 | 0 | 4.084574523 |
| 0.001459133 | 0 | 0 | 0 | 0 |
| 0 | 0 | 0 | 0 | 0 |
| 0 | 0 | 0.330518269 | 0.295227518 | 0 |
| 0 | 0 | 0 | 0 | 0 |
| 0 | 0 | 0 | 0 | 0 |
| 0 | 0 | 0 | 0 | 0 |
| 0 | 0 | 0 | 0 | 0 |
| 0 | 0 | 0 | 0 | 0 |
| 0 | 0 | 0 | 0 | 0 |
| 0 | 0.003384141 | 0 | 0 | 0 |
| 0 | 0 | 0 | 0 | 0 |
| 0 | 0 | 0 | 0 | 0 |
| 0 | 0 | 0 | 0 | 0 |
| 0 | 0 | 0 | 0 | 0 |

| | | | | |
|---|---|---|---|---|
| 0.012273834 | 0 | 0.001920495 | 0.004234477 | 0 |
| 0.017460677 | 0.004338606 | 0 | 0 | 0 |
| 0 | 0 | 0 | 0 | 0 |
| 0 | 0 | 0 | 0 | 0 |
| 0 | 0 | 0 | 0 | 0 |
| 0 | 0 | 0 | 0 | 0 |
| 0 | 0 | 0 | 0 | 0 |
| 0.277001919 | 0.749980059 | 0.734647566 | 0.766166959 | 0.345205353 |
| 0.107887502 | 0 | 0.001004835 | 0.002600862 | 0.148599124 |
| 0.001341215 | 0.0099979 | 0.003031323 | 0.031384466 | 0.08616317 |
| 0.007869461 | 0 | 0 | 0 | 0 |
| 0 | 0 | 0 | 0 | 0 |
| 0 | 0 | 0 | 0.005575349 | 0 |
| 0.049634126 | 0 | 0 | 0 | 0 |
| 0 | 0 | 0 | 0 | 0 |
| 0 | 0 | 0 | 0 | 0 |
| 0 | 0 | 0 | 0 | 0 |
| 0 | 0 | 0 | 0 | 0 |
| 0 | 0 | 0 | 0 | 0 |
| 0.00689544 | 0 | 0 | 0.014847456 | 0 |
| 0 | 0 | 0 | 0 | 0 |
| 0 | 0 | 0 | 0 | 0 |
| 0.007531297 | 0 | 0 | 0 | 0 |
| 0 | 0 | 0 | 0 | 0 |
| 0.000580041 | 0 | 0 | 0.0084106 | 0 |
| 0.000611343 | 0 | 0 | 0 | 0 |
| 0 | 0 | 0 | 0 | 0 |
| 0 | 0 | 0 | 0 | 0 |
| 0 | 0 | 0 | 0 | 0 |
| 0 | 0 | 0 | 0 | 0 |
| 0 | 0.001118975 | 0 | 0 | 0 |
| 0 | 0 | 0 | 0 | 0 |
| 0 | 0 | 0 | 0 | 0 |
| 0 | 0 | 0 | 0 | 0 |
| 0 | 0 | 0 | 0 | 0 |
| 0 | 0 | 0 | 0 | 0 |
| 0 | 0 | 0 | 0 | 0 |
| 0 | 0 | 0.162558544 | 0 | 0 |
| 0.01225942 | 0 | 0 | 0 | 0 |
| 0 | 0 | 0 | 0 | 0 |
| 0 | 0 | 0 | 0 | 0 |
| 0.008848895 | 2.481235065 | 0.019560114 | 0 | 0.308036859 |
| 0 | 0 | 0 | 0 | 0 |
| 0 | 0 | 0 | 0 | 0 |
| 0 | 0 | 0 | 0.000716424 | 0 |
| 0 | 0.030024379 | 0 | 0 | 0 |
| 0 | 0 | 0 | 0 | 0.01356152 |
| 0 | 0 | 0 | 0 | 0 |

| | | | | |
|---|---|---|---|---|
| 0 | 0 | 0 | 0 | 0 |
| 0 | 0 | 0.085125791 | 0.000996992 | 0.000337279 |
| 0 | 0 | 0 | 0 | 0 |
| 0 | 0 | 0 | 0 | 0 |
| 0.29799186 | 0.054153513 | 0.068664129 | 0.324554373 | 23.14024715 |
| 0 | 0 | 0 | 0 | 0 |
| 0 | 0 | 0 | 0 | 0 |
| 0.025293392 | 0.003325327 | 0.010857816 | 0.02107785 | 0.042941865 |
| 0 | 0 | 0 | 0 | 0 |
| 0 | 0 | 0 | 0 | 0 |
| 0 | 0 | 0 | 0 | 0 |
| 0 | 0 | 0 | 0 | 0 |
| 0 | 0 | 0 | 0 | 0 |
| 0 | 0 | 0 | 0 | 0 |
| 0 | 0 | 0 | 0 | 0 |
| 0.001354122 | 0.001442016 | 0 | 0 | 0 |
| 0 | 0 | 0 | 0 | 0 |
| 0 | 0 | 0 | 0 | 0 |
| 0 | 0 | 0 | 0 | 0 |
| 0 | 0 | 0 | 0 | 0 |
| 0 | 0 | 0 | 0 | 0 |
| 0 | 0 | 0 | 0 | 0 |
| 0 | 0 | 0 | 0 | 0 |
| 0 | 0.000719449 | 0 | 0.006080381 | 0.002879762 |
| 0.217354905 | 0 | 0.134182676 | 0 | 0 |
| 0 | 0 | 0 | 0 | 0 |
| 0.023233427 | 0 | 0.001503986 | 0.721287448 | 0 |
| 0 | 0.000692872 | 0 | 0 | 0 |
| 0 | 0 | 0 | 0 | 0.007643419 |
| 0 | 0 | 0 | 0 | 0 |
| 0 | 0 | 0 | 0 | 0 |
| 0 | 0 | 0 | 0 | 0 |
| 0 | 0 | 0 | 0 | 0 |
| 0 | 0 | 0 | 0 | 0 |
| 0 | 0 | 0 | 0 | 0 |
| 0 | 0 | 0 | 0 | 0 |
| 0 | 0 | 0 | 0 | 0 |
| 0 | 0.001124727 | 0 | 0 | 0 |
| 0 | 0 | 0 | 20.26767338 | 0 |
| 0 | 0 | 0 | 0 | 0 |
| 0 | 0 | 0 | 0 | 0 |
| 0.213919864 | 0 | 0 | 0 | 0 |
| 0 | 0 | 0 | 0 | 0 |
| 0 | 0 | 0 | 0 | 0 |
| 0 | 0 | 0 | 0 | 0 |
| 0 | 0 | 0 | 0 | 0 |
| 0 | 0 | 0 | 0 | 0 |
| 0 | 0 | 0 | 0 | 0 |

| | | | | |
|---|---|---|---|---|
| 0 | 0.006188092 | 0.008486217 | 1.404906109 | 0.170907984 |
| 0 | 0 | 0 | 0 | 0 |
| 0 | 0 | 0 | 0 | 0 |
| 0 | 0 | 0 | 0 | 0 |
| 0 | 0 | 0 | 0 | 0 |
| 0 | 0 | 0 | 0 | 0.276854214 |
| 0 | 0 | 0 | 0 | 0 |
| 0 | 0 | 0 | 0 | 0 |
| 0 | 0 | 0 | 0 | 0 |
| 0 | 0.01753594 | 0 | 0 | 0 |
| 0 | 0 | 0 | 0 | 0 |
| 0 | 0 | 0 | 0 | 0 |
| 0 | 0 | 0 | 0 | 0 |
| 0 | 0 | 0 | 0 | 0 |
| 0 | 0 | 0 | 0 | 0 |
| 0 | 0.019103163 | 0 | 0 | 0 |
| 0 | 0 | 0 | 0 | 0 |
| 0.030298288 | 0 | 0 | 0 | 0 |
| 0 | 0 | 0 | 0 | 0 |
| 0 | 0 | 0 | 0 | 0 |
| 0 | 0.028657668 | 0 | 0 | 0.017206345 |
| 0 | 0 | 0 | 0 | 0 |
| 0.003738518 | 0 | 0 | 0.041548578 | 1.777922904 |
| 0.011820373 | 0 | 0 | 0 | 0.022730009 |
| 0.002747414 | 0 | 0 | 0 | 0 |
| 0 | 0 | 0 | 0 | 0 |
| 0 | 0 | 0 | 0 | 0 |
| 0 | 0 | 0 | 0 | 0 |
| 0.336288323 | 0 | 0 | 0 | 0 |
| 0 | 0 | 0 | 0 | 0 |
| 0 | 0 | 0 | 0 | 0 |
| 0 | 0 | 0 | 0 | 0.006763052 |
| 0 | 0.00197716 | 0.039011461 | 0.009548484 | 0.022611548 |
| 0 | 0 | 0 | 0 | 0 |
| 0 | 0.08463824 | 0.962969157 | 0.496449901 | 0.041752082 |
| 0 | 0.004533109 | 0.000666065 | 0.004214242 | 0.001296059 |
| 0 | 0 | 0 | 0 | 0 |
| 0 | 0 | 0 | 0 | 0 |
| 0 | 0.002621629 | 0.001027213 | 0 | 0.06790908 |
| 0 | 0 | 0 | 0 | 0 |
| 0 | 0 | 0 | 0 | 0 |
| 0 | 0 | 0 | 0 | 0 |
| 0.053196938 | 0.002452376 | 0.008708118 | 0 | 0.314819624 |
| 0 | 0 | 0 | 0 | 0 |
| 0 | 0 | 0 | 0 | 0 |
| 0 | 0 | 0.00166932 | 0 | 0.282318131 |
| 0 | 0 | 0 | 0 | 0 |
| 0 | 0 | 0 | 0 | 0 |

| | | | | |
|---|---|---|---|---|
| 0 | 0 | 0 | 0 | 0 |
| 0 | 0 | 0 | 0 | 0 |
| 0 | 0 | 0 | 0 | 0 |
| 0 | 0 | 0 | 0 | 0 |
| 0 | 0 | 0 | 0 | 0 |
| 0 | 0 | 0 | 0 | 0 |
| 0 | 0 | 0 | 0.338915047 | 0 |
| 0 | 0.171027632 | 0 | 0 | 0 |
| 0.000771079 | 0 | 0 | 0 | 0 |
| 0.006216365 | 0.00325978 | 0 | 0 | 0 |
| 0 | 0 | 0 | 0 | 0 |
| 0 | 0 | 0 | 0 | 0 |
| 0 | 0 | 0 | 0 | 0 |
| 0 | 0 | 0 | 0 | 0 |
| 0 | 0 | 0 | 0 | 0 |
| 0.007784275 | 0 | 0 | 0 | 0 |
| 0 | 0 | 0 | 0 | 0 |
| 0 | 0 | 0 | 0 | 0 |
| 0 | 0 | 0 | 0 | 0 |
| 0.002327586 | 0 | 1.690287761 | 0.807906033 | 0 |
| 0 | 0.002834411 | 0.006814959 | 0 | 0 |
| 0 | 0 | 0 | 0 | 0 |
| 0 | 0 | 0 | 0 | 0 |
| 0 | 0 | 0 | 0 | 0 |
| 0 | 0 | 0 | 0 | 0 |
| 0 | 0 | 0 | 0 | 0 |
| 0 | 0 | 0 | 0 | 0 |
| 0 | 0 | 0 | 0 | 0 |
| 0 | 0 | 0 | 0 | 0 |
| 0 | 0 | 0 | 0 | 0 |
| 0.128118515 | 0.172648602 | 0 | 0.549249409 | 1.416645344 |
| 0 | 0 | 0 | 0 | 0 |
| 0 | 0 | 0 | 0 | 0 |
| 0 | 0 | 0 | 0 | 0 |
| 0 | 0 | 0 | 0 | 0 |
| 0 | 0 | 0 | 0 | 0 |
| 0 | 0 | 0 | 0 | 0 |
| 0 | 0 | 0 | 0 | 0 |
| 0 | 0 | 0 | 0 | 0 |
| 0 | 0 | 0 | 0 | 0 |
| 0 | 0 | 0 | 0 | 0 |
| 0.01413305 | 0 | 0 | 0 | 0 |
| 0 | 0 | 0 | 0 | 0 |
| 0 | 0 | 0 | 0 | 0 |
| 0 | 0 | 0 | 0 | 0 |
| 0 | 0 | 0 | 0 | 0 |
| 0.006545723 | 0 | 0.009895783 | 0 | 0.100081209 |
| 0 | 0.001026267 | 0 | 0 | 0 |

| | | | | |
|---|---|---|---|---|
| 0 | 0 | 0 | 0 | 0 |
| 0 | 0 | 0 | 0 | 0 |
| 0 | 0 | 0 | 0 | 0 |
| 0 | 0 | 0 | 0 | 0 |
| 0.001488305 | 0.004528309 | 0.004435733 | 4.170871742 | 0.258505711 |
| 0 | 0 | 0 | 0 | 0 |
| 0 | 0 | 0 | 0 | 0 |
| 0 | 0 | 0 | 0 | 0 |
| 0 | 0 | 0 | 0 | 0 |
| 0 | 0 | 0 | 0 | 0 |
| 0 | 0 | 0 | 0 | 0 |
| 0.003247034 | 0 | 0 | 0.023910003 | 0.091372084 |
| 0.002546702 | 0 | 0 | 0 | 0.277330348 |
| 0 | 0 | 0 | 0 | 0 |
| 0.009897866 | 0.000598882 | 0 | 0 | 0 |
| 0.00367277 | 0 | 0.187569395 | 0.015322353 | 0 |
| 0 | 0.000805895 | 0 | 0.007662346 | 0 |
| 0 | 0 | 0 | 0 | 0 |
| 0 | 0.216998351 | 0.017098228 | 0.00989058 | 0.364198281 |
| 0.004377651 | 0.082358433 | 0.56802098 | 0.00153218 | 0.007849018 |
| 0 | 0 | 0 | 0 | 0 |
| 0 | 0 | 0 | 0 | 0 |
| 0 | 0 | 0 | 0 | 0 |
| 0 | 0 | 0 | 0 | 0 |
| 0 | 0 | 0 | 0 | 0 |
| 0 | 0 | 0 | 0 | 0 |
| 0.001637072 | 0.00498095 | 0 | 0 | 0.111673218 |
| 0 | 0 | 0 | 0 | 0 |
| 0 | 0 | 0 | 0 | 0 |
| 0 | 0 | 0 | 0 | 0 |
| 0 | 0 | 0.000691342 | 0 | 0.012376245 |
| 0 | 0.001270525 | 0 | 0 | 0 |
| 0 | 0 | 0 | 0 | 0 |
| 0 | 0 | 0 | 0 | 0 |
| 0 | 0 | 0 | 0 | 0 |
| 0 | 0 | 0 | 0 | 0 |
| 0 | 0 | 0 | 0 | 0 |
| 0.005401317 | 0 | 0.000647623 | 0.012991114 | 1.010532322 |
| 0.001792187 | 0.004572485 | 0 | 0.001008106 | 0 |
| 0 | 0 | 0 | 0 | 0 |
| 0 | 0 | 0 | 0.014503324 | 0.011794297 |
| 0 | 0 | 0 | 0 | 0 |
| 0 | 0 | 0 | 0 | 0 |
| 0 | 0.006127408 | 0 | 0 | 0.006587083 |
| 0 | 0 | 0 | 0 | 0 |
| 0 | 0 | 0 | 0 | 0 |
| 0.002779053 | 0 | 0 | 0 | 0 |
| 0 | 0 | 0 | 0 | 0 |

| | | | | |
|---|---|---|---|---|
| 0 | 0 | 0 | 0 | 0 |
| 0 | 0 | 0 | 0.041959747 | 0 |
| 0 | 0 | 0 | 0 | 0 |
| 0 | 0 | 0.002012886 | 0 | 0.009693986 |
| 0.002416116 | 0.006194122 | 0.001166825 | 0.003020148 | 0.001907185 |
| 0 | 0 | 0 | 0 | 0 |
| 0 | 0 | 0 | 0 | 0 |
| 0 | 0 | 0 | 0.00797964 | 0 |
| 0 | 0 | 0 | 0 | 0 |
| 0 | 0 | 0 | 0 | 0 |
| 0 | 0 | 0 | 0 | 0 |
| 0 | 0 | 0 | 0 | 0 |
| 0.082070838 | 0.019482457 | 0.003032063 | 0.027237289 | 0 |
| 0 | 0 | 0 | 0 | 0 |
| 0 | 0 | 0 | 0 | 0 |
| 0 | 0 | 0 | 0 | 0 |
| 0 | 0 | 0 | 0 | 0 |
| 0 | 0 | 0 | 0 | 0 |
| 0 | 0 | 0 | 0 | 0 |
| 0 | 0 | 0 | 0 | 0 |
| 0 | 0 | 0 | 0 | 0 |
| 0 | 0 | 0 | 0 | 0 |
| 0.002161368 | 0.002877074 | 0.001409128 | 0 | 0.028104902 |
| 0.000956433 | 0.00565841 | 0.032424971 | 0.017215814 | 0 |
| 0 | 0 | 0 | 0 | 0 |
| 0 | 0 | 0 | 0 | 0 |
| 0 | 0 | 0 | 0.081890233 | 0 |
| 0 | 0 | 0 | 0 | 0 |
| 0 | 0 | 0 | 0 | 0 |
| 0 | 0 | 0 | 0 | 0 |
| 0.025002871 | 0 | 0.002094895 | 0.009263122 | 0.020177868 |
| 0 | 0 | 0 | 0.001048277 | 0 |
| 0 | 0 | 0 | 0 | 0 |
| 0 | 0 | 0 | 0 | 0 |
| 0.01186996 | 0 | 0 | 0 | 0 |
| 0 | 0 | 0 | 0 | 0 |
| 0 | 0 | 0 | 0 | 0 |
| 0 | 0 | 0 | 0 | 0 |
| 0.001019478 | 0 | 0.017281155 | 0 | 0.001810653 |
| 0 | 0 | 0 | 0 | 0 |
| 0 | 0 | 0 | 0 | 0 |
| 0 | 0 | 0 | 0 | 0 |
| 0 | 0 | 0 | 0 | 0 |
| 0 | 0 | 0 | 0 | 0 |
| 0.005606974 | 0 | 0 | 0 | 9.958470576 |
| 0.446467813 | 0.09512132 | 0.058089223 | 0.460315313 | 0.404755083 |
| 0 | 0 | 0 | 0 | 0 |
| 0 | 0 | 0 | 0 | 0 |

| | | | | |
|---|---|---|---|---|
| 0 | 0 | 0 | 0 | 0 |
| 0 | 0 | 0 | 0 | 0 |
| 0 | 0.191256818 | 0 | 0 | 0 |
| 0 | 0 | 0 | 0 | 0 |
| 0 | 0 | 0 | 0 | 0 |
| 0 | 0 | 0 | 0 | 0 |
| 0 | 0 | 0 | 0 | 0 |
| 0 | 0 | 0 | 0 | 0 |
| 0 | 0 | 0 | 0 | 0.020343366 |
| 0 | 0 | 0 | 0 | 0 |
| 0 | 0 | 0 | 0 | 0.034939431 |
| 0 | 0 | 0 | 0 | 0.001253206 |
| 0 | 0 | 0 | 0 | 0 |
| 0 | 0 | 0 | 0 | 0 |
| 0 | 0 | 0 | 0 | 0 |
| 0 | 0 | 0 | 0 | 0 |
| 0 | 0 | 0 | 0 | 0 |
| 0 | 0 | 0 | 0 | 0 |
| 0 | 0 | 0 | 0 | 0 |
| 0 | 0 | 0 | 0.001850913 | 0.024350587 |
| 0 | 0 | 0 | 0 | 0 |
| 0 | 0 | 0 | 0 | 0 |
| 0 | 0 | 0 | 0 | 0 |
| 0 | 0 | 0 | 0 | 0 |
| 0 | 0 | 0 | 0 | 0 |
| 0 | 0 | 0 | 0 | 0 |
| 0 | 0 | 0 | 0 | 0 |
| 0 | 0 | 0 | 0 | 0 |
| 0 | 0 | 0 | 0 | 0 |
| 0 | 0 | 0 | 0 | 0 |
| 0.023792972 | 0 | 0.013105926 | 0.003886975 | 0.117867422 |
| 0 | 0 | 0 | 0 | 0 |
| 0.006291084 | 0.082286465 | 0.056113846 | 0.012103511 | 0.012908332 |
| 0.004472212 | 0.002232421 | 0.007046295 | 0.002725257 | 0 |
| 0 | 0 | 0 | 0 | 0 |
| 0 | 0 | 0 | 0 | 0 |
| 0 | 0 | 0 | 0 | 0 |
| 0 | 0 | 0 | 0 | 0 |
| 0 | 0 | 0 | 0 | 0 |
| 0 | 0 | 0 | 0 | 0 |
| 0 | 0 | 0 | 0 | 0 |
| 0.020252416 | 0.000599083 | 0.038144274 | 0.669849388 | 0.946283411 |
| 0.036461438 | 0.007541798 | 0.013107059 | 0.031663914 | 0.105865965 |
| 0 | 0 | 0 | 0 | 0 |
| 0 | 0 | 0 | 0 | 0 |
| 0.027731217 | 0.003164058 | 0 | 0.012776182 | 0.899005273 |
| 0 | 0 | 0 | 0 | 0 |
| 0 | 0 | 0 | 0 | 0 |

| | | | | |
|---|---|---|---|---|
| 0 | 0 | 0 | 0 | 0 |
| 0.036762169 | 0.009685139 | 0 | 0 | 0 |
| 0.003019366 | 0.023221964 | 0.013415026 | 0.00427744 | 0.00374529 |
| 0 | 0 | 0 | 0 | 0 |
| 0.020358023 | 0.01236108 | 0.005821333 | 0 | 3.430435736 |
| 0.001042967 | 0 | 0.007253057 | 0.061795876 | 0 |
| 0 | 0 | 0 | 0 | 0 |
| 0 | 0 | 0 | 0 | 0 |
| 0 | 0 | 0 | 0 | 0 |
| 0 | 0 | 0 | 0 | 0 |
| 0 | 0 | 0 | 0 | 0 |
| 0 | 0 | 0 | 0 | 0 |
| 0 | 0 | 0 | 0 | 0 |
| 0 | 0 | 0 | 0 | 0 |
| 0 | 0 | 0 | 0 | 0 |
| 0 | 0 | 0 | 0 | 0 |
| 0 | 0 | 0 | 0 | 0 |
| 0 | 0 | 0 | 0 | 0 |
| 0 | 0 | 0 | 0 | 0 |
| 0 | 0 | 0 | 0 | 0 |
| 0 | 0 | 0 | 0 | 0 |
| 0 | 0 | 0 | 0 | 0 |
| 0 | 0 | 0 | 0 | 0 |
| 0 | 0 | 0 | 0 | 0 |
| 0.00718866 | 0 | 0 | 0 | 0 |
| 0 | 0.006911168 | 0 | 0.000834419 | 0.001552546 |
| 0 | 0 | 0 | 0 | 0 |
| 0.017807118 | 0 | 0 | 0.023675398 | 0.032037241 |
| 0 | 0 | 0 | 0 | 0 |
| 0 | 0 | 0 | 0 | 0 |
| 0 | 0 | 0 | 0 | 0 |
| 0 | 0 | 0 | 0 | 0 |
| 0 | 0 | 0 | 0 | 0 |
| 0 | 0 | 0 | 0 | 0 |
| 0 | 0 | 0 | 0 | 0 |
| 0 | 0 | 0 | 0 | 0 |
| 0 | 0 | 0 | 0 | 0 |
| 0 | 0.020884157 | 0 | 0 | 0 |
| 7.747073057 | 0.174349731 | 3.092356304 | 0 | 0 |
| 0 | 0 | 0 | 0 | 0 |
| 0.001011612 | 0 | 0 | 0 | 0 |
| 0 | 0 | 0 | 0 | 0 |
| 0.011165725 | 0.000743155 | 0 | 0.002931006 | 0 |
| 0.002167687 | 0 | 0 | 0 | 0 |
| 0 | 0 | 0 | 0 | 0 |
| 0 | 0 | 0 | 0 | 0 |
| 0 | 0.009474012 | 0.001856065 | 0.020817948 | 0.030120847 |
| 0 | 0 | 0 | 0 | 0 |

| | | | | |
|---|---|---|---|---|
| 0 | 0 | 0 | 0 | 0 |
| 0.014279229 | 0.002151803 | 0.018970305 | 0.204691619 | 0.077107663 |
| 0 | 0 | 0 | 0 | 0 |
| 0 | 0 | 0 | 0 | 0 |
| 0 | 0 | 0 | 0 | 0 |
| 0 | 0 | 0 | 0 | 0 |
| 0 | 0 | 0 | 0 | 0 |
| 0 | 0 | 0 | 0 | 0 |
| 0 | 0 | 0 | 0 | 0 |
| 0 | 0 | 0 | 0 | 0 |
| 0 | 0 | 0 | 0 | 0 |
| 0 | 0 | 0 | 0 | 0 |
| 0.000854401 | 0 | 0 | 0.085012949 | 0.613779096 |
| 0 | 0 | 0 | 0 | 0 |
| 0 | 0.001490541 | 0.003212151 | 0 | 0.062048856 |
| 0 | 0 | 0 | 0 | 0 |
| 0 | 0 | 0 | 0 | 0 |
| 0 | 0 | 0 | 0 | 0 |
| 0 | 0 | 0 | 0 | 0 |
| 0 | 0 | 0 | 0 | 0 |
| 0 | 0 | 0 | 0 | 0 |
| 0.003628958 | 0.021664668 | 0.069687666 | 0.001484575 | 0.032309936 |
| 0.046644955 | 0 | 0 | 0 | 0 |
| 0 | 0 | 0 | 0 | 0.497634341 |
| 0 | 0 | 0 | 0 | 0 |
| 0 | 0 | 0 | 0 | 0 |
| 0 | 0 | 0 | 0 | 0 |
| 0 | 0 | 0 | 0 | 0 |
| 0 | 0 | 0 | 0 | 0 |
| 0 | 0 | 0 | 0 | 0 |
| 0 | 0 | 0 | 0 | 0 |
| 0 | 0 | 0 | 0 | 0 |
| 0 | 0 | 0 | 0 | 0 |
| 0 | 0 | 0 | 0 | 0 |
| 0.005391641 | 0 | 0 | 0 | 0 |
| 0 | 0 | 0 | 0 | 0 |
| 0 | 0 | 0 | 0 | 0 |
| 0 | 0 | 0 | 0 | 0 |
| 0.001317801 | 0.022219516 | 0.171258258 | 0.008153904 | 0.001337425 |
| 0 | 0 | 0 | 0 | 0 |
| 0.150947246 | 0.33685151 | 0.00873059 | 0.053171298 | 0.004197125 |
| 0 | 0 | 0 | 0 | 0 |
| 0 | 0 | 0 | 0 | 0 |
| 0 | 0 | 0 | 0 | 0 |
| 0 | 0 | 0 | 0 | 0 |
| 0.030776934 | 0.033614996 | 1.452350104 | 0.435781566 | 0.174711421 |
| 0 | 0 | 0 | 0 | 0 |
| 0 | 0 | 0 | 0 | 0 |

| | | | | |
|---|---|---|---|---|
| 0 | 0 | 0 | 0 | 0 |
| 0 | 0 | 0 | 0 | 0 |
| 0 | 0 | 0 | 0 | 0 |
| 0.004051343 | 0 | 0 | 0 | 0 |
| 0 | 0 | 0 | 0 | 0 |
| 0 | 0 | 0 | 0 | 0 |
| 0 | 0 | 0 | 0 | 0 |
| 0 | 0 | 0.009215338 | 0 | 0.000556498 |
| 0 | 0 | 0 | 0.009622745 | 0 |
| 0.508059689 | 0 | 0.001437653 | 0 | 0.010406502 |
| 0 | 0 | 0 | 0 | 0 |
| 0 | 0 | 0 | 0 | 0 |
| 0 | 0.00258212 | 0 | 0.004364526 | 0.016405612 |
| 0 | 0 | 0 | 0 | 0 |
| 0 | 0 | 0 | 0 | 0 |
| 0 | 0 | 0 | 0 | 0 |
| 0 | 0 | 0 | 0 | 0 |
| 0 | 0 | 0 | 0 | 0 |
| 0 | 0 | 0 | 0 | 0 |
| 0.228192049 | 0.063840813 | 0.31292844 | 1.441021385 | 3.123254336 |
| 0 | 0 | 0 | 0 | 0 |
| 0 | 0 | 0 | 0 | 0 |
| 0 | 0 | 0 | 0 | 0 |
| 0 | 0 | 0 | 0 | 0 |
| 0 | 0 | 0 | 0 | 0 |
| 0 | 0 | 0 | 0 | 0 |
| 0 | 0 | 0 | 0 | 0 |
| 0 | 0 | 0 | 0 | 0 |
| 0 | 0 | 0 | 0 | 0 |
| 0.000922854 | 0.03739928 | 0.001871846 | 0 | 0 |
| 0 | 0 | 0 | 0 | 0 |
| 0 | 0 | 0 | 0 | 0.001363297 |
| 0 | 0 | 0 | 0 | 0 |
| 0 | 0 | 0 | 0 | 0 |
| 0 | 0 | 0 | 0 | 0 |
| 0 | 0 | 0 | 0.005308074 | 0 |
| 0 | 0 | 0 | 0 | 0 |
| 0 | 0 | 0 | 0 | 0 |
| 0 | 0 | 0 | 0 | 0 |
| 0 | 0 | 0 | 0 | 0 |
| 0 | 0 | 0 | 0 | 0 |
| 0 | 0 | 0 | 0 | 0 |
| 0 | 0 | 0 | 0 | 0 |
| 0 | 0 | 0 | 0 | 0 |
| 0 | 0 | 0 | 0.012364995 | 0.156075312 |
| 0 | 0 | 0 | 0 | 0 |
| 0 | 0 | 0 | 0 | 0 |
| 0 | 0 | 0 | 0 | 0 |

| | | | | |
|---|---|---|---|---|
| 0 | 0 | 0 | 0 | 0 |
| 0 | 0 | 0 | 0 | 0 |
| 0 | 0 | 0 | 0 | 0 |
| 0 | 0 | 0 | 0 | 0 |
| 0 | 0 | 0 | 0 | 0 |
| 0.009079613 | 0.001432438 | 0 | 0.003026541 | 0 |
| 0 | 0 | 0 | 0 | 0 |
| 0 | 0 | 0 | 0 | 0 |
| 0 | 0 | 0 | 0 | 0 |
| 0 | 0 | 0 | 0 | 0 |
| 0 | 0 | 0 | 0 | 0 |
| 0 | 0 | 0 | 0 | 0 |
| 0 | 0 | 0 | 0 | 0 |
| 0 | 0 | 0 | 0 | 0 |
| 0 | 0 | 0 | 0 | 0 |
| 0 | 0 | 0 | 0 | 0 |
| 0 | 0 | 0 | 0 | 0 |
| 0 | 0 | 0 | 0 | 0 |
| 0 | 0 | 0 | 0 | 0 |
| 0 | 0 | 0 | 0 | 0 |
| 0 | 0 | 0 | 0 | 0 |
| 0 | 0 | 0 | 0 | 0 |
| 0 | 0 | 0 | 0 | 0 |
| 0 | 0 | 0 | 0 | 0 |
| 0 | 0 | 0 | 0 | 0 |
| 0.004418412 | 0 | 0 | 0 | 0 |
| 0 | 0 | 0 | 0 | 0 |
| 0.027163624 | 0 | 0 | 0.247130196 | 0 |
| 0 | 0 | 0 | 0 | 0 |
| 0 | 0 | 0 | 0 | 0 |
| 0 | 0 | 0 | 0 | 0 |
| 0 | 0 | 0 | 0 | 0 |
| 0 | 0 | 0 | 0 | 0 |
| 0 | 0 | 0 | 0 | 0 |
| 0 | 0 | 0 | 0 | 0 |
| 0 | 0 | 0 | 0 | 0 |
| 0 | 0 | 0 | 0 | 0 |
| 0 | 0.052601066 | 0 | 0 | 0 |
| 0.001708892 | 0.478712032 | 0 | 0 | 0 |
| 0 | 0 | 0 | 0 | 0 |
| 0 | 0 | 0 | 0 | 0 |
| 0 | 0 | 0 | 0 | 0 |
| 0 | 0 | 0 | 0 | 0 |
| 0 | 0 | 0 | 0 | 0 |
| 0 | 0 | 0 | 0 | 0 |
| 0 | 0 | 0 | 0 | 0 |
| 0 | 0 | 0 | 0 | 0 |
| 0 | 0 | 0 | 0 | 0 |

| | | | | |
|---|---|---|---|---|
| 0 | 0 | 0 | 0 | 0 |
| 0.003388453 | 0.003006994 | 0 | 0.055062418 | 0.142370935 |
| 0 | 0 | 0 | 0 | 0 |
| 0 | 0 | 0 | 0 | 0 |
| 0 | 0 | 0 | 0 | 0 |
| 0 | 0 | 0 | 0 | 0 |
| 0 | 0 | 0 | 0 | 0 |
| 0 | 0 | 0 | 0 | 0 |
| 0 | 0 | 0 | 0 | 0 |
| 0 | 0 | 0 | 0 | 0 |
| 0.00443691 | 0 | 0 | 0.003105841 | 0.003602386 |
| 0 | 0 | 0 | 0 | 0 |
| 0.006318151 | 0 | 0 | 0.10245219 | 0.00552779 |
| 0 | 0 | 0 | 0 | 0 |
| 0 | 0 | 0 | 0 | 0 |
| 0 | 0 | 0.003764938 | 0 | 0 |
| 0 | 0 | 0 | 0 | 0 |
| 0 | 0 | 0 | 0 | 0 |
| 0 | 0 | 0 | 0 | 0 |
| 0 | 0 | 0 | 0 | 0 |
| 0.003902771 | 0.001065665 | 0.010438788 | 0 | 0.001066393 |
| 0 | 0 | 0 | 0 | 0 |
| 0 | 0 | 0 | 0 | 0 |
| 2.133069376 | 2.025356041 | 2.014432324 | 2.274269223 | 0 |
| 0 | 0 | 0 | 0 | 0 |
| 0 | 0 | 0 | 0 | 0 |
| 0 | 0 | 0 | 0 | 0 |
| 0 | 0 | 0 | 0 | 0 |
| 0 | 0 | 0 | 0 | 0 |
| 0 | 0 | 0 | 0 | 0 |
| 0.003535908 | 0 | 0 | 0 | 0 |
| 0 | 0 | 0 | 0 | 0 |
| 0 | 0.001961711 | 0 | 0 | 0 |
| 0 | 0 | 0 | 0 | 0 |

| PHL018-0040_V1 | PHL018-0104_V1 | PHL018-0149_V1 | PHL018-0133_V3 | PHL018-0139_V3 |
|---|---|---|---|---|
| EYCF | EYCF | REF | EYCF | CTRL |
| 0.00695835 | 0.003188374 | 0.080113894 | 1.356264265 | 0.173271741 |
| 2.466343983 | 0.35027721 | 3.944679278 | 0 | 0.038786103 |
| 0 | 1.391397722 | 1.157219056 | 0 | 0.026872162 |
| 0 | 0 | 0 | 0 | 0 |
| 0 | 0 | 0 | 0 | 0 |
| 0 | 0 | 0 | 0 | 0 |
| 0 | 0 | 0 | 0 | 0 |
| 0 | 0 | 0 | 0 | 0 |
| 0 | 0 | 0 | 0 | 0 |
| 0 | 0 | 0 | 0 | 0 |
| 0 | 0 | 0 | 0 | 0 |
| 0 | 0 | 0 | 0 | 0 |
| 0 | 0 | 0 | 0 | 0 |
| 1.649171112 | 1.027886337 | 2.931243905 | 0 | 0.158735565 |
| 0 | 0 | 0 | 0 | 0 |
| 0 | 0 | 0 | 0 | 0 |
| 0 | 0 | 0 | 0.002401719 | 0 |
| 0 | 0 | 0 | 0 | 0 |
| 0 | 0 | 0 | 0 | 0 |
| 0 | 0 | 0 | 0 | 0 |
| 0 | 0 | 0 | 0 | 0 |
| 0 | 0 | 0 | 0 | 0 |
| 0 | 0 | 0 | 0 | 0 |
| 0 | 0 | 0 | 0 | 0 |
| 0 | 0 | 0 | 0 | 0.41394477 |
| 0 | 0 | 0 | 0 | 0 |
| 0 | 0 | 0 | 0 | 0 |
| 0 | 0 | 0 | 0 | 0 |
| 0 | 0 | 0 | 0 | 0 |
| 0 | 0 | 0 | 0 | 0 |
| 0 | 0 | 0 | 0 | 0 |
| 0 | 0 | 0 | 0 | 0 |
| 0 | 0 | 0 | 0 | 0 |
| 0 | 0 | 0 | 0 | 0 |
| 0 | 0 | 0 | 0 | 0 |
| 0 | 2.021321569 | 0 | 0 | 0.472827086 |
| 0 | 0 | 0 | 0 | 0 |
| 0 | 0 | 0 | 0 | 0 |
| 0 | 0 | 0 | 0 | 0 |
| 0 | 0 | 0 | 0 | 0 |
| 0 | 0 | 0 | 0 | 0 |
| 0.065959776 | 0.123357332 | 0.00201311 | 0.001238461 | 0.028323955 |
| 11.13843515 | 2.043607355 | 0.054895545 | 0.279178731 | 7.901017036 |
| 0.619323042 | 0.019318434 | 0.011759812 | 0 | 0.002721343 |
| 0.016886921 | 0.022940085 | 0 | 0 | 0 |
| 0 | 0 | 0 | 0 | 0 |

| | | | | |
|---|---|---|---|---|
| 0 | 0 | 0 | 0 | 0.001017114 |
| 0.038527245 | 0 | 0 | 0.014142089 | 0 |
| 0 | 0.005435956 | 0.001315883 | 0 | 0 |
| 0.147169161 | 1.015669935 | 0.171070748 | 0 | 0.299639836 |
| 0 | 0 | 0 | 0.005929773 | 0.002205483 |
| 0 | 0 | 0 | 0 | 0.015969841 |
| 0 | 0.042280228 | 0.009097584 | 0 | 0 |
| 0.003649824 | 0 | 0 | 0 | 0 |
| 0 | 0 | 0 | 0 | 0 |
| 0 | 0 | 0 | 0.005429508 | 0 |
| 0.00566951 | 0 | 0 | 0.006642252 | 0 |
| 0.039862163 | 0 | 0 | 0.558075807 | 0.09844927 |
| 0 | 0 | 0 | 0 | 0 |
| 0.006118795 | 0 | 0.038481601 | 0.038245501 | 0 |
| 0 | 0 | 0 | 0 | 0 |
| 0 | 0 | 0 | 0 | 0 |
| 0.083416181 | 0.059959405 | 0 | 0.004121177 | 0.982287822 |
| 0 | 0 | 0 | 0 | 0 |
| 0 | 0 | 0 | 0 | 0 |
| 0 | 0 | 0 | 0 | 0 |
| 0.09914045 | 0 | 0.017857385 | 0.188303214 | 0 |
| 0 | 0 | 0.007474154 | 0.008450531 | 0.006039565 |
| 0 | 0 | 0 | 0 | 0.002789326 |
| 0 | 0 | 0 | 0 | 0 |
| 0 | 0 | 0 | 0 | 0 |
| 0 | 0 | 0 | 0 | 0 |
| 0 | 0 | 0 | 0.004154801 | 0 |
| 0 | 0 | 0 | 0 | 0 |
| 0.002588703 | 0.007116992 | 0 | 0.000635922 | 0.001363474 |
| 0 | 0 | 0 | 0 | 0 |
| 0.238844113 | 0 | 0 | 0.031917948 | 0 |
| 0 | 0 | 0 | 0.009347364 | 0 |
| 0 | 0 | 0 | 0 | 0 |
| 0 | 0 | 0 | 0.003064039 | 0 |
| 0 | 0 | 0 | 0 | 0 |
| 0 | 0 | 0 | 0 | 0 |
| 0 | 0 | 0 | 0 | 0 |
| 0 | 0.008095216 | 0 | 0 | 31.18973584 |
| 0 | 0 | 0 | 0 | 0 |
| 0 | 0 | 0 | 0 | 0 |
| 0 | 0 | 0 | 0 | 0 |
| 0 | 0 | 0 | 0 | 0 |
| 0 | 0 | 0 | 0 | 0 |
| 0 | 0 | 0 | 0 | 0 |
| 0 | 0 | 0 | 0 | 0.224298774 |
| 0 | 0 | 0 | 0 | 0 |
| 0 | 0 | 0 | 0 | 0 |
| 0 | 0 | 0 | 0 | 0.010588617 |

| | | | | |
|---|---|---|---|---|
| 0 | 0 | 0 | 0 | 0 |
| 0 | 0 | 0 | 0.012180748 | 0 |
| 0 | 0 | 0 | 0 | 0 |
| 0 | 0 | 0 | 0 | 0 |
| 0.012668291 | 0.008865385 | 0 | 0 | 0.0034373 |
| 0 | 0 | 0 | 0 | 0 |
| 0 | 0 | 0 | 0 | 0 |
| 0 | 0 | 0 | 0 | 0.008073249 |
| 0.172120784 | 0 | 0 | 0.008648577 | 0 |
| 0 | 0 | 0 | 0 | 0 |
| 0 | 0 | 0 | 0 | 0 |
| 0.003406173 | 0.077412545 | 0 | 0 | 0 |
| 0 | 0 | 0 | 0 | 0 |
| 0 | 0 | 0 | 0 | 0 |
| 0 | 0 | 0 | 0 | 0 |
| 0 | 0 | 0 | 0.01828098 | 0 |
| 0 | 0 | 0 | 0 | 0 |
| 0 | 0 | 0 | 0 | 0 |
| 0 | 0.060004056 | 0 | 0.062551066 | 0.218025703 |
| 0 | 0 | 0 | 0 | 0.008579203 |
| 0 | 0 | 0 | 0 | 0 |
| 0 | 0 | 0 | 0 | 0 |
| 0 | 0 | 0 | 0 | 0 |
| 0 | 0 | 0 | 0 | 0 |
| 0 | 0 | 0 | 0 | 0 |
| 0 | 0 | 0 | 0 | 0 |
| 0 | 0 | 0 | 0 | 0 |
| 0 | 0 | 0 | 0.003449635 | 0 |
| 0 | 0 | 0 | 0 | 0 |
| 0 | 0 | 0 | 0 | 0 |
| 0 | 0 | 0 | 0.141240619 | 0.031141224 |
| 0 | 0 | 0.001137302 | 0 | 0.000833415 |
| 0 | 0 | 0 | 0 | 0 |
| 0 | 0 | 0 | 0 | 0 |
| 0 | 0 | 0 | 0 | 0 |
| 0 | 0 | 0 | 0 | 0 |
| 0 | 0 | 0 | 0 | 0 |
| 0 | 0 | 0 | 0 | 0 |
| 0 | 0 | 0 | 0 | 0 |
| 0 | 0 | 0 | 0 | 0 |
| 0 | 0 | 0 | 0 | 0 |
| 0 | 0 | 0 | 0 | 0 |
| 0 | 0 | 0 | 0 | 0.001926591 |
| 0 | 0 | 0 | 0 | 0 |
| 0 | 0 | 0 | 0 | 0 |
| 0 | 0 | 0 | 0 | 0 |
| 0 | 0 | 0 | 0 | 0 |
| 0 | 0 | 0 | 0 | 0 |

| | | | | |
|---|---|---|---|---|
| 0 | 0 | 0 | 0 | 0 |
| 0 | 0 | 0 | 0 | 0 |
| 0 | 0 | 0 | 0 | 0 |
| 0 | 0 | 0 | 0 | 0 |
| 0 | 0.054523754 | 0 | 0.001082631 | 0.023067503 |
| 0 | 0 | 0 | 0 | 0 |
| 0 | 0 | 0 | 0 | 0 |
| 0 | 0 | 0 | 0 | 0 |
| 0 | 0 | 0 | 0 | 0 |
| 0 | 0 | 0 | 0 | 0 |
| 0 | 0 | 0 | 0 | 0 |
| 0 | 0 | 0 | 0 | 0 |
| 0 | 0 | 0 | 0 | 0 |
| 0 | 0 | 0 | 0.003449051 | 0 |
| 0 | 0 | 0 | 0 | 0 |
| 0 | 0 | 0 | 0 | 1.815971919 |
| 0 | 0 | 0 | 0 | 0 |
| 0 | 0 | 0 | 0 | 0 |
| 0 | 0 | 0 | 0 | 0 |
| 0 | 0 | 0 | 0 | 0 |
| 0 | 0 | 0.001921024 | 0 | 0 |
| 0 | 0 | 0 | 0 | 0 |
| 0 | 0 | 0 | 0 | 0.003608662 |
| 0 | 0 | 0 | 0 | 0 |
| 0 | 0 | 0 | 0 | 0.043584809 |
| 0 | 0 | 0 | 0 | 0 |
| 0 | 0 | 0 | 0 | 0.045084512 |
| 0 | 0 | 0 | 0 | 0 |
| 0 | 0 | 0 | 0 | 0 |
| 0 | 0 | 0 | 0 | 0 |
| 0 | 0 | 0 | 0.040962242 | 0 |
| 0 | 0 | 0 | 0.006637501 | 0 |
| 0 | 0 | 0 | 0 | 0 |
| 0 | 0 | 0 | 0 | 0 |
| 0 | 0 | 0 | 0 | 0 |
| 0 | 0 | 0 | 0 | 0 |
| 0 | 0 | 0 | 0 | 0 |
| 0 | 0 | 0 | 0 | 0 |
| 0 | 0 | 0 | 0 | 0 |
| 0 | 0 | 0 | 0 | 0 |
| 0 | 0 | 0 | 0 | 0 |
| 0 | 0 | 0 | 0 | 0 |
| 0 | 0 | 0 | 0 | 0 |
| 0 | 0 | 0 | 0 | 0 |
| 0 | 0 | 0 | 0 | 0 |
| 0 | 0 | 0 | 0 | 0 |
| 0 | 0 | 0 | 0 | 0 |
| 0 | 0 | 0 | 0 | 0 |

| | | | | |
|---|---|---|---|---|
| 0 | 0 | 0 | 0 | 0 |
| 0 | 0 | 0 | 0 | 0 |
| 0 | 0 | 0 | 0 | 0 |
| 0 | 0 | 0 | 0 | 0 |
| 0 | 0 | 0 | 0 | 0 |
| 0 | 0 | 0 | 0 | 0 |
| 0 | 0 | 0 | 0 | 0 |
| 0 | 0 | 0 | 0 | 0 |
| 0 | 0 | 0 | 0 | 0 |
| 0 | 0 | 0 | 0 | 0 |
| 0 | 0 | 0 | 0 | 0 |
| 0 | 0 | 0 | 0 | 0 |
| 0 | 0 | 0.001255648 | 0 | 0 |
| 0 | 0 | 0 | 0 | 0 |
| 0 | 0 | 0 | 0 | 0 |
| 0 | 0 | 0 | 0 | 0 |
| 0 | 0 | 0 | 0 | 0 |
| 0 | 0 | 0 | 0 | 0 |
| 0 | 0 | 0 | 0 | 0 |
| 0 | 0 | 0 | 0 | 0 |
| 0.050303164 | 0.046338127 | 0.023086312 | 0.014764336 | 0.00137635 |
| 0 | 0 | 0 | 0 | 0 |
| 0 | 0 | 0 | 0 | 0 |
| 0 | 0 | 0 | 0.007028715 | 0.001076442 |
| 0 | 0 | 0 | 0 | 0 |
| 0 | 0 | 0 | 0 | 0 |
| 0.017120675 | 0 | 0.004329721 | 0 | 0.004208158 |
| 0 | 0 | 0 | 0 | 0 |
| 0 | 0 | 0.004572075 | 0 | 0 |
| 0 | 0 | 0.01438452 | 0 | 0 |
| 0.002275521 | 0 | 0 | 0 | 0 |
| 0 | 0 | 0 | 0.003710381 | 0 |
| 0 | 0 | 0 | 0 | 0 |
| 0 | 0 | 0 | 0.037465253 | 0.033645592 |
| 0.002975447 | 0.042771008 | 0.025714232 | 0.038008183 | 0.014776193 |
| 0 | 0 | 0 | 0 | 0.001397092 |
| 0 | 0 | 0 | 0 | 0 |
| 0 | 0 | 0 | 0 | 0 |
| 0 | 0 | 0 | 0 | 0 |
| 0 | 0 | 0 | 0 | 0 |
| 0 | 0 | 0 | 0 | 0 |
| 0 | 0.021707809 | 0 | 0 | 0 |
| 0 | 0 | 0 | 0 | 0 |
| 0 | 0 | 0 | 0 | 0 |
| 0 | 0 | 0 | 0 | 0 |
| 0 | 0 | 0 | 0 | 0.344466194 |
| 0 | 0 | 0.002480941 | 0 | 0 |
| 0 | 0 | 0.004172942 | 0.002976446 | 0 |

| | | | | |
|---|---|---|---|---|
| 0 | 0 | 0 | 0 | 0 |
| 0 | 0 | 0 | 0 | 0 |
| 0 | 0 | 0 | 0 | 0 |
| 0 | 0 | 0 | 0.000567518 | 0 |
| 0 | 0 | 0 | 0 | 0 |
| 0 | 0 | 0 | 0 | 0 |
| 0.056917269 | 0.056190468 | 0 | 0 | 0 |
| 0.002612649 | 0.00287313 | 0 | 0 | 0 |
| 0 | 0 | 0.001966382 | 0 | 0 |
| 0 | 0 | 0 | 0 | 0 |
| 0 | 0 | 0 | 0 | 0 |
| 0 | 0 | 0 | 0 | 0 |
| 0 | 0 | 0 | 0 | 0 |
| 0 | 0 | 0 | 0 | 0 |
| 0.001432775 | 0 | 0 | 0.001407861 | 0 |
| 0 | 0 | 0 | 0 | 0 |
| 0 | 0 | 0 | 0 | 0 |
| 0.002463237 | 0 | 0 | 0 | 0 |
| 0.059147428 | 0.008638712 | 0 | 0.002118918 | 0.039752562 |
| 0 | 0 | 0 | 0 | 0.416313351 |
| 0 | 0 | 0 | 0.022325857 | 0 |
| 0 | 0 | 0 | 0 | 0 |
| 0 | 0 | 0 | 0 | 0 |
| 0 | 0 | 0 | 0 | 0 |
| 0.015347595 | 0 | 0 | 0 | 0 |
| 0 | 0.027457121 | 0 | 0 | 0 |
| 0 | 0 | 0 | 0 | 0 |
| 0 | 0 | 0 | 0 | 0 |
| 0.009907881 | 0.039082387 | 0 | 0 | 0 |
| 0 | 0 | 0 | 0 | 0 |
| 0.062817234 | 0.002204684 | 0.802934005 | 1.455952218 | 0.031502019 |
| 0 | 0 | 0 | 0 | 0 |
| 0 | 0 | 0 | 0.023678209 | 0.041796231 |
| 0 | 0 | 0 | 0 | 0.029599743 |
| 0 | 0 | 0 | 0 | 0 |
| 0 | 0 | 0 | 0 | 0 |
| 0 | 0 | 0 | 0 | 0 |
| 0 | 0 | 0 | 0 | 0 |
| 0 | 0 | 0 | 0 | 0 |
| 0.008010087 | 0.00165163 | 0 | 0 | 0 |
| 0 | 0 | 0 | 0 | 0 |
| 0.001702971 | 0 | 0.009588114 | 0 | 0.005022961 |
| 0 | 0 | 0 | 0.006598843 | 0 |
| 0 | 0 | 0 | 0 | 0 |
| 0 | 0 | 0 | 0 | 0 |
| 0 | 0 | 0 | 0 | 0 |
| 0 | 0 | 0 | 0 | 0 |
| 0 | 0 | 0 | 0 | 0 |

| | | | | |
|---|---|---|---|---|
| 0 | 0 | 0 | 0 | 0 |
| 0 | 0 | 0 | 0 | 0 |
| 0.562388894 | 0.032451461 | 0.007172441 | 0 | 0.002703066 |
| 0 | 0 | 0 | 0 | 0 |
| 0 | 0 | 0 | 6.519594761 | 0 |
| 0 | 0 | 0 | 0 | 0 |
| 0 | 0 | 0 | 0 | 0 |
| 0 | 0 | 0 | 0 | 0 |
| 0 | 0 | 0 | 0 | 0 |
| 0 | 0.007742572 | 0 | 0 | 0 |
| 0 | 0 | 0 | 0 | 0 |
| 0 | 0 | 0 | 0 | 0 |
| 0 | 0 | 0 | 0 | 0 |
| 0 | 0.00389834 | 0 | 0.000928872 | 0 |
| 0.024967347 | 0.00514811 | 0.001401979 | 0 | 0.003493058 |
| 0 | 0 | 0 | 0 | 0.005719663 |
| 0.023558544 | 0.009531941 | 0.001331192 | 0 | 0 |
| 0 | 0 | 0 | 0 | 0 |
| 0 | 0 | 0 | 0 | 0 |
| 0 | 0 | 0 | 0 | 0 |
| 0 | 0 | 0 | 0 | 0 |
| 0 | 0 | 0.005287419 | 0.002562819 | 0 |
| 0 | 0 | 0 | 0 | 0 |
| 0 | 0 | 0 | 0 | 0 |
| 0 | 0 | 0 | 0 | 0 |
| 0 | 0 | 0 | 0 | 0 |
| 0 | 0 | 0 | 0 | 0.119935533 |
| 0 | 0 | 0 | 0 | 0 |
| 0 | 0 | 0 | 0 | 0 |
| 0.001741268 | 0 | 0 | 0 | 0 |
| 0.003689879 | 0 | 0 | 0 | 0 |
| 0 | 0 | 0 | 0 | 0 |
| 0 | 0 | 0 | 0 | 0 |
| 0 | 0 | 0 | 0 | 0 |
| 0 | 0 | 0 | 0 | 0 |
| 0.133041462 | 0.009254275 | 0 | 0 | 0 |
| 0 | 0 | 0 | 0 | 0 |
| 1.025417448 | 0.284407669 | 0 | 0.001238429 | 0 |
| 0 | 0 | 0 | 0 | 0 |
| 0 | 0 | 0 | 0 | 0 |
| 0 | 0 | 0 | 0 | 0 |
| 0 | 0 | 0 | 0 | 0 |
| 0 | 0 | 0 | 0 | 0 |
| 0 | 0 | 0 | 0.001845988 | 0 |
| 0 | 0 | 0 | 0 | 0 |
| 0.031004192 | 0 | 0 | 0 | 0 |
| 0.014208157 | 0 | 0 | 0 | 0 |
| 0 | 0 | 0.002560235 | 0.002100068 | 0 |

| | | | | |
|---|---|---|---|---|
| 0 | 0 | 0 | 0 | 0.020354898 |
| 0 | 0 | 0 | 0 | 0 |
| 0.002942001 | 0.006239542 | 0 | 0 | 0 |
| 0 | 0 | 0 | 0 | 0 |
| 0.092180882 | 0 | 0.007091537 | 0 | 0.001113573 |
| 0 | 0 | 0 | 0.001016254 | 0 |
| 0 | 0 | 0 | 0 | 0 |
| 0.148178664 | 0 | 0.035501249 | 0 | 0.000936551 |
| 0 | 0 | 0 | 0 | 0 |
| 0.053761298 | 0.011085242 | 0.025248416 | 0.463731999 | 0 |
| 0 | 0 | 0 | 0.012754604 | 0 |
| 0.203161454 | 0.040245952 | 0 | 0 | 0.008155146 |
| 0.062994494 | 0.008219072 | 0.009805847 | 0 | 0.008810318 |
| 0 | 0 | 0 | 0.003186736 | 0 |
| 0 | 0 | 0 | 0 | 0 |
| 0 | 0 | 0 | 0 | 0 |
| 0.003496687 | 0 | 0 | 0 | 0 |
| 0 | 0 | 0 | 0.003643637 | 0 |
| 0 | 0 | 0 | 0.036716658 | 0 |
| 0 | 0 | 0 | 0 | 0 |
| 0 | 0 | 0 | 0 | 0 |
| 0.033794154 | 0 | 0 | 0 | 0 |
| 0 | 0.046645615 | 0 | 0 | 0 |
| 0 | 0 | 0 | 0 | 0.374778113 |
| 0 | 0 | 0 | 0 | 0 |
| 0.003316272 | 0.068379456 | 0 | 0 | 0 |
| 0 | 0 | 0 | 0 | 2.148818182 |
| 0.005012869 | 0 | 0 | 0.001970279 | 0 |
| 0 | 0 | 0 | 0.481475746 | 0 |
| 0 | 0 | 0 | 0 | 0 |
| 0 | 0 | 0 | 0 | 0 |
| 0 | 0 | 0 | 0 | 0 |
| 0 | 0 | 0 | 0 | 0 |
| 0 | 0 | 0 | 0 | 0 |
| 0 | 0 | 0 | 0 | 0 |
| 0 | 0 | 0 | 0 | 0 |
| 0.403658534 | 0 | 0 | 0.003001344 | 0 |
| 0 | 0 | 0 | 0.028889631 | 0 |
| 0 | 0 | 0 | 0 | 0 |
| 0 | 0 | 0 | 0 | 0 |
| 0 | 0 | 0 | 0 | 0 |
| 0 | 0 | 0 | 0 | 0 |
| 0 | 0 | 0 | 0.003550291 | 0 |
| 0 | 0 | 0.009801504 | 0 | 0 |
| 0 | 0 | 0 | 0 | 0 |
| 0 | 0 | 0 | 0 | 0.00958458 |
| 0 | 0 | 0.004013977 | 0 | 0.001357588 |
| 0 | 0 | 0 | 0.056322312 | 0 |

| | | | | |
|---|---|---|---|---|
| 0 | 0 | 0 | 0 | 0 |
| 0 | 0 | 0.004140831 | 0 | 0 |
| 0 | 0 | 0 | 0 | 0 |
| 0 | 0 | 0 | 0 | 0.007305462 |
| 0 | 0 | 0 | 0 | 0 |
| 0 | 0 | 0 | 0 | 0 |
| 0 | 0 | 0 | 0 | 0 |
| 0 | 0 | 0.002201769 | 0 | 0 |
| 0 | 0 | 0 | 0 | 0 |
| 0 | 0 | 0 | 0 | 0 |
| 0.318811925 | 0 | 0.046540153 | 0 | 0 |
| 0 | 0 | 0 | 0.028623089 | 0.02464117 |
| 0.070598084 | 0 | 0 | 0 | 0 |
| 0.01690621 | 0.032290947 | 0.001598865 | 0 | 0 |
| 0.005401909 | 0 | 0.007819194 | 0 | 0 |
| 0 | 0 | 0 | 0.069992428 | 0 |
| 0 | 0 | 0 | 0 | 0 |
| 0 | 0 | 0 | 0 | 0 |
| 0 | 0 | 0.002268995 | 0 | 0 |
| 0 | 0 | 0 | 0 | 0 |
| 0 | 0 | 0 | 0 | 0 |
| 0 | 0.379189695 | 0.149435758 | 0 | 0.057225954 |
| 0 | 0 | 0 | 0 | 0 |
| 0 | 0 | 0 | 0 | 0 |
| 0 | 0 | 0 | 0 | 0 |
| 0 | 0 | 0 | 0 | 0 |
| 0.005788676 | 0 | 0 | 0.106176286 | 0 |
| 0 | 0 | 0 | 0 | 0 |
| 0 | 0 | 0 | 0 | 0 |
| 1.160140343 | 0.134465241 | 0.340212844 | 0.168848305 | 0.060795614 |
| 0 | 0 | 0 | 0 | 0 |
| 0.021187957 | 0 | 0 | 0 | 0 |
| 0 | 0 | 0 | 0 | 0 |
| 0 | 0 | 0 | 0 | 0 |
| 0 | 0 | 0 | 0.001579194 | 0 |
| 0 | 0 | 0 | 0 | 0 |
| 0 | 0.057405217 | 0 | 0 | 0.476315585 |
| 0.011336797 | 0 | 0 | 0 | 0.001433065 |
| 0 | 0 | 0 | 0 | 0 |
| 0 | 0 | 0.037206715 | 0 | 0 |
| 0 | 0 | 0 | 0.024618639 | 0 |
| 0 | 0.102221771 | 1.863022765 | 2.65034277 | 0.250843016 |
| 0.001661638 | 0 | 0 | 0 | 0.001312781 |
| 0 | 0 | 0 | 3.215620406 | 0 |
| 0 | 0 | 0 | 0.002635215 | 0 |
| 0 | 0 | 0 | 0 | 0 |
| 0 | 0 | 0.002759201 | 0.012825211 | 0 |
| 0 | 0 | 0 | 0 | 0 |

| | | | | |
|---|---|---|---|---|
| 0.022735339 | 0.342215564 | 0 | 0 | 0 |
| 0 | 0 | 0 | 0.008326846 | 0 |
| 0.006453884 | 0.119235231 | 0.003092496 | 0.013000394 | 0.753278015 |
| 0 | 0 | 0 | 0 | 0 |
| 0 | 0 | 0 | 0 | 0.004179695 |
| 0.032438389 | 0 | 0 | 0.101919088 | 0 |
| 0 | 0 | 0 | 0 | 0 |
| 0 | 0.002627084 | 0 | 0 | 0 |
| 0 | 0 | 0 | 0.001168535 | 0 |
| 0 | 0 | 0.032568566 | 0.689105233 | 0 |
| 0 | 0 | 0 | 0 | 0 |
| 0 | 0 | 0 | 0 | 0 |
| 0 | 0 | 0 | 0 | 0 |
| 0.005142022 | 0 | 0 | 0 | 0.005416619 |
| 0 | 0 | 0 | 0 | 0 |
| 0 | 0 | 0.129168287 | 0.174064113 | 0.0195776 |
| 0.129907718 | 0.527151569 | 0.001556191 | 0 | 0.00254141 |
| 0 | 0 | 0.004051525 | 0.039879827 | 0 |
| 0 | 0 | 0 | 0.008099448 | 0 |
| 0 | 0 | 0 | 0 | 0 |
| 0 | 0 | 0.008965685 | 0 | 0 |
| 0 | 0 | 0 | 0 | 0 |
| 0 | 0 | 0 | 0 | 0 |
| 1.485232922 | 0 | 0 | 0 | 0 |
| 0 | 0 | 0 | 0 | 0.000913536 |
| 0 | 0 | 0 | 0 | 0 |
| 0 | 0 | 0 | 0 | 0 |
| 0 | 0 | 0 | 0 | 0 |
| 0 | 0 | 0 | 0 | 0 |
| 0 | 0 | 0 | 0 | 0 |
| 0 | 0 | 0.053658429 | 0.21021118 | 0.005095985 |
| 0.008013701 | 0.006168866 | 0 | 0 | 0 |
| 0.070705841 | 6.398861887 | 1.017313674 | 0.434601865 | 0.014467672 |
| 0.002186943 | 0.013227396 | 0 | 0 | 0.005375374 |
| 0.006763991 | 0 | 0 | 0 | 0.00602902 |
| 0 | 0 | 0 | 0 | 0 |
| 0.030811784 | 0.033377988 | 0 | 0 | 0 |
| 0 | 0 | 0.015078931 | 0.244546885 | 0 |
| 0 | 0 | 0.80492099 | 0 | 0.037994039 |
| 0 | 0 | 0.005107549 | 0 | 0.000464625 |
| 0 | 0 | 0 | 0 | 0 |
| 0 | 0 | 0 | 0 | 0 |
| 0 | 0 | 0.114363336 | 0 | 0 |
| 0 | 0 | 0.027914 | 0.227451175 | 0 |
| 0 | 0 | 0 | 0.038868588 | 0 |
| 0 | 0 | 0 | 0 | 0 |
| 0 | 0 | 0 | 0.018456873 | 0 |
| 0 | 0 | 0 | 0 | 0 |

| | | | | |
|---|---|---|---|---|
| 0.006932052 | 0 | 0 | 0 | 0 |
| 1.576041034 | 0.012125738 | 0.394795082 | 0.007704651 | 0.035103853 |
| 0 | 0 | 0 | 0 | 0 |
| 0 | 0 | 0 | 0 | 0 |
| 0 | 0 | 0 | 0 | 0 |
| 0 | 0 | 0 | 0 | 0 |
| 0 | 0 | 0 | 0 | 0 |
| 0 | 0 | 0.000676381 | 0 | 0 |
| 0.003588895 | 0 | 0.003493109 | 0 | 0 |
| 0 | 0 | 0 | 0 | 0 |
| 0 | 0 | 0.010906168 | 0.002147024 | 0 |
| 0.248980688 | 0.297963216 | 0.004873514 | 0 | 0.003428456 |
| 0 | 0 | 0 | 0 | 0.009615641 |
| 0 | 0 | 0 | 0.100230815 | 0 |
| 0.01002815 | 0.015163438 | 0 | 0 | 0 |
| 0 | 0 | 0 | 0 | 0 |
| 0.13995126 | 0 | 0.005481759 | 0.003372366 | 0.001405961 |
| 0 | 0 | 0 | 0.157357501 | 0 |
| 0 | 0 | 0 | 0 | 0 |
| 0 | 0 | 0 | 0.02842713 | 0 |
| 0.199380936 | 0.131685255 | 0.024025576 | 0 | 0.015120388 |
| 0.002936066 | 0 | 0 | 0 | 0 |
| 0 | 0 | 0 | 0 | 0 |
| 0 | 0 | 0 | 0 | 0 |
| 0 | 0 | 0 | 0 | 0 |
| 0 | 0 | 0 | 0 | 0 |
| 0 | 0 | 0.042314008 | 0.10755867 | 0.004088926 |
| 0.078578994 | 0.044125946 | 0.004756639 | 0.076391153 | 0.01607074 |
| 0 | 0 | 0 | 0 | 0 |
| 0 | 0 | 0 | 0 | 0 |
| 0.053369808 | 0 | 0 | 0 | 0 |
| 0 | 0 | 0 | 0.028061331 | 0 |
| 0 | 0 | 0 | 0 | 0 |
| 0 | 0 | 0 | 0 | 0 |
| 0.003278112 | 0.018024699 | 0 | 0 | 0.001918429 |
| 0.408166915 | 0 | 0.248289053 | 5.764413908 | 0.067438044 |
| 0 | 0 | 0.008830487 | 0 | 0 |
| 0 | 0 | 0 | 0 | 0 |
| 0.142310267 | 0.033170896 | 0 | 0.004053206 | 0.008418859 |
| 0 | 0 | 0 | 0 | 0 |
| 0 | 0 | 0 | 0.039104892 | 0 |
| 0 | 0 | 0 | 0 | 0 |
| 0.007203452 | 0 | 0.008629163 | 0.002000358 | 0 |
| 0 | 0.008479515 | 0 | 0.001634184 | 0 |
| 0 | 0 | 0 | 0.046647118 | 0 |
| 0 | 0 | 0 | 0 | 0.091327389 |
| 0 | 0 | 0 | 0 | 0 |
| 0 | 0 | 0 | 0 | 0 |

| | | | | |
|---|---|---|---|---|
| 0 | 0 | 0 | 0 | 0.00115226 |
| 0 | 0 | 0 | 0 | 0 |
| 0 | 0 | 0 | 0 | 0 |
| 0 | 0 | 1.395236049 | 0.001971356 | 0.575416135 |
| 0 | 0 | 0 | 0 | 0 |
| 0 | 0 | 0 | 0 | 0 |
| 0 | 0 | 0 | 0 | 0 |
| 0 | 0 | 0 | 0 | 0 |
| 0 | 7.285999934 | 0.708760974 | 0 | 0.239633917 |
| 0 | 0 | 0.058581173 | 0 | 0.038807144 |
| 0 | 0 | 0 | 0 | 0 |
| 0 | 0 | 0.042075479 | 0 | 0.001656693 |
| 0 | 0 | 0 | 0 | 0 |
| 0 | 0 | 0 | 0 | 0 |
| 0 | 0.021407875 | 0.084469987 | 0 | 0.20825597 |
| 0 | 0 | 0 | 0 | 0 |
| 0 | 0.003723293 | 0.244251795 | 0 | 0.002972119 |
| 0 | 0 | 0 | 0 | 2.950994998 |
| 0 | 0 | 0 | 0 | 0 |
| 0 | 0 | 0 | 0 | 0.020055059 |
| 0 | 0 | 0 | 0 | 0 |
| 0 | 0 | 0 | 0 | 0.17295279 |
| 0 | 0 | 0 | 0 | 0.004576601 |
| 0 | 0 | 0.391855856 | 0.397038564 | 0 |
| 0 | 0 | 0 | 0 | 0.006376694 |
| 0 | 0 | 0 | 0 | 0.25417074 |
| 0 | 0 | 0 | 0 | 0.014969501 |
| 7.209815068 | 0.692892881 | 11.03509459 | 0 | 1.144044081 |
| 0 | 0 | 0 | 0 | 0 |
| 0 | 0 | 0 | 0 | 0 |
| 0 | 0.031385638 | 0 | 0 | 0.045096458 |
| 0 | 0 | 0 | 0 | 0 |
| 0 | 0 | 0 | 0.050690719 | 0.002042959 |
| 0 | 0 | 0 | 0 | 0 |
| 0 | 0 | 0 | 0 | 0.242221589 |
| 0 | 0.046570144 | 0.047278599 | 0 | 0.164478732 |
| 0 | 0.01166088 | 1.422443781 | 0 | 0.405841823 |
| 0 | 0 | 0 | 0 | 0 |
| 0 | 0 | 0 | 0 | 0 |
| 0 | 0 | 0 | 0 | 0.011726407 |
| 0.058337572 | 0.013273205 | 0.013253818 | 0.914863896 | 0.00829968 |
| 0 | 0 | 0 | 0 | 0.05107231 |
| 1.846507565 | 7.637654725 | 0.739089654 | 0 | 0.001694721 |
| 0 | 0 | 0 | 0 | 0 |
| 0 | 0 | 0 | 0 | 0 |
| 0 | 0 | 0 | 0 | 0 |
| 0 | 0 | 0 | 0 | 0 |
| 0.003224297 | 0 | 0 | 0.008118586 | 0.013585931 |

| | | | | |
|---|---|---|---|---|
| 0 | 0 | 0 | 0 | 0 |
| 0.009913812 | 0.084189347 | 0 | 0 | 0.006962157 |
| 0.075725885 | 0 | 0.000925649 | 0.084469477 | 0 |
| 0 | 0 | 0 | 0 | 0.023976493 |
| 0 | 0 | 0 | 0 | 0 |
| 0 | 0.006024621 | 0 | 0 | 0.009618308 |
| 0 | 0 | 0 | 0 | 0 |
| 0 | 0 | 0 | 0 | 0 |
| 0 | 0 | 0 | 0 | 0 |
| 0 | 0 | 0 | 0 | 0 |
| 0 | 0.004380108 | 0.052938939 | 0.056283331 | 0.003196728 |
| 3.829028841 | 0.601460146 | 0.06749598 | 3.687008622 | 0.448107902 |
| 0 | 0 | 0 | 0 | 0 |
| 0.313594508 | 0.111426648 | 0 | 0 | 0.005821942 |
| 0.032261684 | 0 | 0.050241009 | 0.463146891 | 0.002208995 |
| 0 | 0 | 0 | 0 | 2.880482305 |
| 0 | 0 | 0 | 0 | 0 |
| 0 | 0 | 0 | 0 | 0 |
| 0 | 0 | 0 | 0 | 0.040939821 |
| 0 | 0 | 0 | 1.011688317 | 0.003949894 |
| 1.79698239 | 0.298414125 | 0.022470961 | 0.1572978 | 0.001842039 |
| 0.19229426 | 0.573610673 | 0.803828393 | 0 | 0 |
| 1.314711709 | 0.065387412 | 0.792856021 | 33.15138101 | 0.112299423 |
| 0 | 0 | 0 | 0 | 0.001938933 |
| 0 | 0 | 0 | 0 | 0.085181022 |
| 0 | 2.976980835 | 0 | 0 | 0.427617101 |
| 0 | 0 | 0 | 0.12928477 | 0.002203164 |
| 0 | 0 | 0.018074146 | 0 | 0 |
| 0 | 0 | 0 | 0 | 0 |
| 0 | 0 | 0 | 0 | 0 |
| 0 | 0 | 0 | 0 | 0.031399325 |
| 0 | 0 | 0 | 0 | 0.062446834 |
| 0.066619123 | 0.085471216 | 0.044802474 | 0.602735774 | 0.058480633 |
| 0 | 0 | 0 | 0 | 0.045847828 |
| 0.001028525 | 0 | 0 | 0.016549227 | 0 |
| 0.032855287 | 0.632291716 | 0.195943683 | 0.007810635 | 0.013583434 |
| 0 | 0 | 0.002861668 | 0 | 0.005572106 |
| 0 | 0 | 0 | 0 | 0 |
| 0 | 1.876027321 | 0.085427991 | 0 | 1.522470722 |
| 0 | 0 | 0 | 0 | 0 |
| 0 | 0 | 0 | 0 | 0.147463118 |
| 0 | 0 | 0 | 0 | 0 |
| 0 | 0 | 0 | 0 | 0 |
| 0 | 0 | 0 | 0 | 0 |
| 0 | 0 | 0.159263508 | 0.099166186 | 0.011458626 |
| 0 | 0 | 0 | 0 | 0.041166855 |
| 0 | 0 | 0 | 0 | 0.060963769 |
| 0.270205926 | 12.77271073 | 0 | 0 | 0 |

| | | | | |
|---|---|---|---|---|
| 0 | 0 | 0 | 0 | 0 |
| 0 | 0 | 0 | 0 | 0 |
| 0 | 0 | 0 | 0 | 0 |
| 0 | 0 | 0 | 0 | 0.001085127 |
| 0 | 0 | 0 | 0 | 0 |
| 0 | 0 | 0 | 0 | 0.133693595 |
| 0 | 0 | 0 | 0 | 0.059796357 |
| 0 | 0.010423978 | 0.073239775 | 0.061317815 | 0.016309045 |
| 0 | 0 | 0 | 0 | 0 |
| 0 | 0 | 0 | 0 | 0 |
| 0 | 0 | 0.001374346 | 0.002630427 | 0.003424207 |
| 0 | 0 | 0 | 0 | 0 |
| 0.00200906 | 0 | 0 | 0 | 0.123100978 |
| 0 | 0 | 0 | 0 | 0.034891813 |
| 0 | 0 | 0 | 0 | 0 |
| 0 | 0 | 0 | 0 | 0 |
| 0 | 0 | 0 | 0 | 0.034155859 |
| 0 | 0.013128785 | 0.010106317 | 0 | 0.001676809 |
| 0 | 0 | 0 | 0.090522907 | 0.001411558 |
| 0 | 0 | 0 | 0 | 0 |
| 0 | 0 | 0 | 0 | 0 |
| 0 | 0 | 0 | 0 | 0 |
| 0 | 0 | 0.004616775 | 0 | 0.006959665 |
| 0 | 0 | 0.008364243 | 0 | 0 |
| 0 | 0 | 0 | 0 | 0 |
| 0 | 0 | 0 | 0 | 0 |
| 0 | 0 | 0 | 0 | 0 |
| 0 | 0 | 0 | 0 | 0 |
| 0 | 0 | 0 | 0 | 0 |
| 0.002307459 | 0 | 0.0175063 | 0 | 0.497682495 |
| 0 | 0 | 0 | 0 | 0.010311847 |
| 0 | 0 | 0 | 0 | 0 |
| 0 | 0 | 0 | 0 | 0.080766251 |
| 0 | 0 | 0 | 0 | 0 |
| 0 | 0 | 0 | 0 | 0 |
| 0 | 0 | 0 | 0 | 0 |
| 0 | 0 | 0 | 0 | 0.010962462 |
| 0 | 0 | 0.039555859 | 0.007475359 | 0 |
| 0 | 0 | 0 | 0 | 0 |
| 0 | 0 | 0 | 0 | 0 |
| 0 | 0 | 0 | 0 | 0.026076668 |
| 0 | 0 | 0 | 0 | 1.428394504 |
| 0 | 0 | 0 | 0 | 0 |
| 0 | 0 | 0.012616174 | 0 | 0 |
| 0 | 0.046689802 | 0.040291638 | 0.18764324 | 0.021684444 |
| 0 | 0 | 0 | 0 | 0.057775296 |
| 0 | 0 | 0 | 0 | 0.003564264 |
| 0 | 0 | 0 | 0 | 0 |

| | | | | |
|---|---|---|---|---|
| 0 | 0 | 0 | 0 | 0 |
| 0 | 0 | 0 | 0 | 0 |
| 0 | 0 | 0 | 0 | 0.221183292 |
| 0 | 0 | 0 | 0 | 0.837177156 |
| 0 | 0 | 0 | 0 | 0 |
| 0 | 0 | 0 | 0 | 0.341384546 |
| 0 | 0 | 0 | 0 | 0 |
| 0 | 0 | 0 | 0 | 0 |
| 0.059827981 | 0 | 0.012568845 | 0.175924135 | 0.010582212 |
| 0 | 0.003558538 | 0 | 0 | 0 |
| 0 | 0 | 0 | 0 | 0 |
| 0 | 0 | 0 | 0 | 0 |
| 0 | 0 | 0 | 0 | 0 |
| 0 | 0 | 0 | 0 | 0 |
| 0 | 0 | 0 | 0.158266111 | 0.028052734 |
| 0 | 0 | 0 | 0.36264307 | 0 |
| 0 | 0 | 0 | 0 | 0 |
| 0 | 0 | 0 | 0 | 0 |
| 0 | 0 | 0 | 0 | 0 |
| 0 | 0 | 0 | 0 | 0.237213818 |
| 0 | 0 | 0 | 0 | 0.040997535 |
| 0 | 0 | 0 | 0 | 0 |
| 0 | 0 | 0 | 0 | 0 |
| 0 | 0 | 0 | 0 | 0 |
| 3.071512007 | 0.219120149 | 0.005346176 | 0.009544421 | 0.41315342 |
| 0 | 0.032652944 | 41.6123054 | 0 | 0.889605801 |
| 0 | 0 | 0 | 0 | 0.253645763 |
| 0 | 0 | 0 | 0 | 0 |
| 0 | 0.010600915 | 0 | 0 | 0 |
| 0 | 0 | 0 | 0 | 0 |
| 0 | 0 | 0 | 0 | 0 |
| 0 | 0 | 0 | 0 | 0.013000894 |
| 0 | 0 | 0 | 0 | 0 |
| 0 | 0 | 0 | 0 | 0 |
| 0.113657705 | 0.02667456 | 0.005811404 | 0 | 0.001216741 |
| 0.002286161 | 0 | 0.002934256 | 0.006578761 | 0 |
| 0 | 0 | 0 | 0.211290006 | 0 |
| 0 | 0 | 4.244755692 | 0.608417002 | 0.062350304 |
| 0 | 2.298377868 | 0.000988222 | 0 | 0 |
| 0 | 0 | 0 | 0 | 0 |
| 0 | 0 | 0 | 0 | 0 |
| 0 | 0 | 0 | 0 | 0 |
| 0 | 0.281960472 | 0.611614822 | 0 | 0.055005267 |
| 0 | 0 | 0 | 0 | 3.89858154 |
| 0 | 0 | 0 | 0 | 0.061196858 |
| 0 | 0 | 0.001667903 | 0 | 0.003259303 |
| 0 | 0 | 0 | 0 | 0.000952522 |
| 1.567987211 | 1.409916465 | 0.004723858 | 0.001453054 | 0 |

| | | | | |
|---|---|---|---|---|
| 0 | 0 | 0.02047503 | 0 | 0.006001637 |
| 0 | 0 | 0 | 0 | 0.919856652 |
| 0 | 0 | 0 | 0 | 0.001864374 |
| 0 | 0 | 0 | 0 | 0 |
| 0 | 0 | 0.001095982 | 0.106620727 | 0 |
| 0 | 0.005231942 | 0 | 0 | 0.00482606 |
| 0 | 0 | 0 | 0 | 0 |
| 0 | 0 | 0 | 0 | 0 |
| 0 | 0 | 0 | 0 | 0 |
| 0 | 0 | 0 | 0 | 0 |
| 0 | 0 | 0 | 0 | 0 |
| 0.158345122 | 0.307979933 | 2.520907663 | 0 | 0.202188623 |
| 0 | 0 | 0.010085519 | 0 | 0.006768292 |
| 0 | 0 | 0 | 0 | 0 |
| 0.500686403 | 1.557564591 | 0.013689033 | 0.152418098 | 0.022737653 |
| 0 | 0 | 0 | 0 | 0 |
| 0 | 0 | 0 | 0 | 0 |
| 0 | 0 | 0 | 0 | 0 |
| 0 | 0 | 0 | 0 | 0.005666222 |
| 0 | 0 | 0 | 0 | 0.600345355 |
| 0 | 0 | 0 | 0.087818888 | 0.01747035 |
| 0 | 0 | 0 | 0 | 0 |
| 0 | 0 | 0.181267548 | 0.136636276 | 0.128051429 |
| 0 | 0 | 0 | 0 | 0 |
| 0 | 0 | 0 | 0 | 0 |
| 0 | 0 | 0 | 0 | 0 |
| 0 | 0.42254469 | 0 | 0 | 0 |
| 0 | 0 | 0 | 0.046082371 | 0 |
| 0 | 0 | 0 | 0 | 0.00140555 |
| 0 | 0 | 0 | 0 | 0 |
| 0 | 0 | 0 | 0 | 0 |
| 0.753510549 | 0.007247541 | 0.035855823 | 0.002698278 | 0.002892675 |
| 0 | 0 | 0 | 0 | 0 |
| 0 | 0.135777987 | 0.019556341 | 0 | 0.094656049 |
| 0.001421666 | 0 | 0 | 0.006984722 | 0 |
| 0.001374525 | 0.064997296 | 0.012074858 | 0 | 0.016651175 |
| 0 | 0 | 0 | 0 | 0.001329361 |
| 0 | 0 | 0 | 0 | 0 |
| 0 | 1.660291146 | 0 | 0 | 0 |
| 0 | 0 | 0 | 0 | 0 |
| 0 | 0 | 0 | 0 | 0.087243045 |
| 0 | 0 | 0 | 0 | 0.046343267 |
| 0 | 0 | 0 | 0 | 0 |
| 0 | 0 | 0 | 0 | 0 |
| 0.001050363 | 0.017903798 | 0.026632978 | 0.033371159 | 0.004057001 |
| 0 | 0 | 0 | 0 | 0 |
| 0 | 0 | 0 | 0 | 0.07788916 |
| 0 | 0 | 0 | 0.004392809 | 0 |

| | | | | |
|---|---|---|---|---|
| 0 | 0 | 0 | 0 | 0 |
| 0 | 0 | 0 | 0 | 0 |
| 0 | 0 | 0 | 0.006938407 | 0 |
| 0 | 0 | 0 | 0 | 0 |
| 0 | 0 | 0 | 0.00252594 | 0 |
| 0 | 0 | 0 | 0 | 0 |
| 0 | 0.021040658 | 0 | 0 | 0.373494537 |
| 0 | 0 | 0.013109333 | 0.006888712 | 0.013689281 |
| 0 | 0 | 0 | 0 | 0 |
| 0 | 0 | 0 | 0 | 0 |
| 0 | 0 | 0 | 0 | 0 |
| 0 | 0 | 0 | 0 | 0 |
| 0 | 0 | 0 | 0 | 0 |
| 0 | 0 | 0 | 0 | 0.100128468 |
| 0 | 0 | 0 | 0 | 0 |
| 0 | 0 | 0 | 0 | 0 |
| 0 | 0 | 0 | 0 | 0 |
| 0 | 0 | 0 | 0 | 0 |
| 0 | 0 | 0 | 13.23726148 | 1.765282088 |
| 0 | 0 | 0 | 0 | 0 |
| 0 | 0.003256328 | 0.008670858 | 0 | 0 |
| 0 | 0 | 0 | 0 | 0 |
| 0 | 0 | 0 | 0 | 0 |
| 0 | 0 | 0 | 0 | 0 |
| 0.001912687 | 0.003786088 | 0 | 0 | 0 |
| 0.011734176 | 0 | 0.01151861 | 0.010729424 | 0 |
| 0 | 0 | 0 | 0 | 0 |
| 0.002233277 | 0.219455282 | 0 | 0.1548649 | 0 |
| 0 | 0 | 0 | 0 | 0 |
| 0 | 0.018612392 | 0.952380729 | 0.792436418 | 0.356732637 |
| 0 | 0 | 0 | 0 | 0 |
| 0 | 0 | 0 | 0 | 0 |
| 0 | 0 | 0 | 0 | 0.007452236 |
| 0 | 0 | 0 | 0 | 0 |
| 0.005123621 | 0 | 0.006956049 | 0.021648458 | 0 |
| 0 | 0 | 0 | 0 | 0.017357875 |
| 0 | 0 | 0 | 0 | 0 |
| 0 | 0.006393445 | 0 | 0.001713813 | 0 |
| 0 | 0 | 0 | 0 | 0 |
| 0 | 0 | 0 | 0 | 0 |
| 23.78927831 | 0.084767616 | 5.145668875 | 4.908710057 | 0.849622359 |
| 0 | 0 | 0.572169059 | 0.158069416 | 0.001942634 |
| 0 | 0 | 0 | 0 | 0 |
| 0 | 0 | 0 | 0 | 0.244381953 |
| 0 | 0 | 0 | 0 | 0.020941836 |
| 0 | 0 | 0 | 0 | 0 |
| 0 | 0 | 0 | 0 | 0 |
| 0 | 0 | 0.013081507 | 0.00181357 | 0.146627163 |

| | | | | |
|---|---|---|---|---|
| 0 | 0 | 0 | 0 | 0 |
| 0 | 0 | 0 | 0.004974416 | 0.003832947 |
| 0 | 0 | 0 | 0 | 0 |
| 0 | 0 | 0 | 0 | 0 |
| 0 | 1.37880241 | 0.406832338 | 0 | 0.553840523 |
| 0 | 0 | 0 | 0 | 0.025974751 |
| 0 | 0 | 0 | 0 | 0 |
| 0 | 0 | 0 | 0 | 0.005830839 |
| 0.317516207 | 0 | 0.005487769 | 0.249440646 | 0 |
| 0 | 0 | 0 | 0 | 0 |
| 0 | 0 | 0 | 0 | 0 |
| 0 | 0 | 0 | 0 | 0 |
| 0 | 0 | 0 | 0 | 0.040320262 |
| 0.096003245 | 0.00623473 | 0.017658132 | 0.004271019 | 0.005175949 |
| 0.80891764 | 4.696833034 | 0 | 0 | 0 |
| 0 | 0 | 0.385474766 | 0.236800387 | 0.189496973 |
| 0 | 0.038305861 | 0 | 0.002249681 | 0.00223949 |
| 0 | 0 | 0 | 0 | 0 |
| 0 | 0 | 0 | 0 | 0 |
| 0 | 0 | 0 | 0 | 0 |
| 0 | 0 | 0 | 0 | 0.001721556 |
| 0 | 0 | 0 | 0 | 0 |
| 0 | 0 | 0 | 0 | 0 |
| 0 | 0 | 0 | 0 | 0 |
| 0 | 0 | 0 | 0 | 0 |
| 0 | 0 | 0 | 0 | 0 |
| 0 | 0 | 0 | 0.003074013 | 0.000449384 |
| 0 | 0 | 0.003459876 | 0 | 0 |
| 0 | 0 | 0 | 0 | 0 |
| 0 | 0 | 0 | 0 | 0 |
| 0 | 0 | 0 | 0 | 0 |
| 0 | 0 | 0 | 0 | 0 |
| 0 | 0 | 0 | 0 | 0 |
| 0 | 0 | 0 | 0 | 0.011694194 |
| 0.187002197 | 5.461878124 | 0 | 0 | 0.042750719 |
| 0 | 0 | 0.005336478 | 0.077515035 | 0 |
| 0 | 0 | 0 | 0.001677805 | 0 |
| 0 | 0 | 0 | 0 | 0 |
| 0 | 0 | 0 | 0 | 0 |
| 0 | 0 | 0 | 0 | 0 |
| 0 | 0.224997175 | 0 | 0 | 0.056275924 |
| 0 | 0 | 0 | 0 | 0 |
| 0 | 0 | 0 | 0 | 0.017250263 |
| 0 | 0 | 0 | 0 | 0.121641687 |
| 0 | 0 | 0 | 0.010916284 | 0 |
| 0 | 0 | 0 | 0 | 0 |
| 0 | 0 | 0 | 0 | 0 |
| 0 | 0 | 0 | 0 | 0 |

| | | | | |
|---|---|---|---|---|
| 0 | 0 | 0 | 0 | 0.086796914 |
| 0 | 0 | 0 | 0 | 0 |
| 0 | 0 | 0 | 0 | 0 |
| 0 | 0 | 0.000870969 | 0 | 0 |
| 0 | 0 | 0.006041766 | 0 | 0.003664058 |
| 0 | 0 | 0 | 0 | 0 |
| 0.160527422 | 0.003229244 | 0.000586278 | 0.009137144 | 0.000859248 |
| 0 | 0 | 0 | 0 | 0 |
| 0 | 0 | 0 | 0 | 0 |
| 0 | 0 | 0 | 0.354863189 | 0 |
| 0 | 0 | 0 | 0 | 0.013962517 |
| 0 | 0 | 0 | 0.034906329 | 0.002855429 |
| 0 | 0 | 0 | 0 | 0 |
| 0 | 0 | 0 | 0 | 0 |
| 0 | 0 | 0 | 0 | 0 |
| 0 | 0 | 0 | 0 | 0 |
| 0 | 4.537524154 | 0 | 0 | 0 |
| 0 | 0 | 0 | 0 | 0 |
| 0 | 0 | 0 | 0 | 0 |
| 0 | 0 | 0 | 0 | 0 |
| 0.249748304 | 0.207010945 | 0.02083831 | 0.000915692 | 0 |
| 0 | 0 | 0 | 0 | 0 |
| 0 | 0 | 0.013339267 | 0 | 0.02021171 |
| 0 | 0 | 0.005109532 | 0.00134716 | 0.00401171 |
| 0 | 0 | 0 | 0 | 0.000575529 |
| 0 | 0 | 0 | 0 | 0.001893166 |
| 0 | 0 | 0 | 0 | 0 |
| 0 | 0 | 0 | 0 | 0 |
| 0 | 0 | 0.024197646 | 0 | 0 |
| 1.357117145 | 1.661434104 | 0.003939544 | 0 | 0 |
| 0 | 0 | 0 | 0 | 0.088649354 |
| 0 | 0 | 0 | 0 | 0 |
| 0 | 0 | 0 | 0 | 0 |
| 0 | 0 | 0 | 1.637863676 | 0 |
| 0 | 0 | 0 | 0 | 0.488206579 |
| 0 | 0 | 0 | 0 | 0.329542073 |
| 0 | 0.1100464 | 0.04909178 | 0.01779293 | 0.054814998 |
| 0 | 0 | 0 | 0 | 0 |
| 0 | 0 | 0 | 0 | 0.017947208 |
| 0 | 0 | 0 | 0 | 0 |
| 0 | 0 | 0 | 0 | 0 |
| 0 | 0 | 0 | 0 | 0 |
| 0 | 0 | 0 | 0 | 0 |
| 0 | 0 | 0 | 0 | 0 |
| 0 | 0 | 0 | 0 | 0.086273734 |
| 0 | 0 | 0 | 0 | 0 |
| 0 | 0 | 0 | 0 | 0 |
| 0 | 0 | 0 | 0 | 0 |

| | | | | |
|---|---|---|---|---|
| 0 | 0 | 0.092288495 | 0.003382381 | 0.001208688 |
| 0 | 0.423630098 | 0 | 0 | 0.007205405 |
| 0 | 0 | 0.024035313 | 0 | 0 |
| 0 | 0 | 0 | 0 | 0 |
| 0 | 0 | 0 | 0 | 0 |
| 0 | 0 | 0 | 0 | 0 |
| 0 | 0 | 0 | 0 | 0.001327403 |
| 0 | 0.006107799 | 0.222180613 | 1.015421787 | 0.118608856 |
| 14.23025144 | 2.354650826 | 0.077978182 | 0.35798875 | 10.56587376 |
| 0.003224129 | 0 | 0.110717837 | 0 | 0.002264204 |
| 0.003363074 | 0 | 0 | 0 | 0.048933148 |
| 0 | 0 | 0 | 0 | 0.000660645 |
| 0 | 0 | 0 | 0 | 0.00947751 |
| 0 | 0.931292888 | 0 | 1.008069558 | 0.035314228 |
| 0 | 0 | 0 | 0.001230963 | 0 |
| 0 | 0 | 0 | 0 | 0 |
| 0 | 0 | 0 | 0.031269986 | 0 |
| 0 | 0 | 0 | 0 | 0.148484583 |
| 0 | 0 | 0 | 0 | 0 |
| 0 | 0 | 0 | 0 | 0 |
| 0 | 0 | 0 | 0 | 0 |
| 0 | 0 | 0 | 0 | 0 |
| 0 | 0 | 0 | 0.123918747 | 0 |
| 0 | 0 | 0 | 0 | 0 |
| 0 | 0 | 0 | 0 | 0.01128811 |
| 0 | 0 | 0 | 0 | 0 |
| 0 | 0 | 0 | 0 | 0 |
| 0 | 0 | 0 | 0 | 0.023508267 |
| 0 | 0 | 0 | 0.001996515 | 0 |
| 0 | 0 | 0 | 0 | 0 |
| 0.003704704 | 0 | 0 | 0 | 0 |
| 0 | 0 | 0 | 0 | 0 |
| 0 | 0 | 0 | 0 | 0.002776014 |
| 0 | 0 | 0 | 0 | 0 |
| 0 | 0 | 0 | 0 | 0 |
| 0 | 0 | 0 | 0 | 0 |
| 0 | 0 | 0 | 0 | 0 |
| 0 | 0 | 0 | 0 | 0 |
| 0 | 0.044368696 | 0 | 0 | 0 |
| 0.14956664 | 0 | 0 | 0 | 0 |
| 0 | 0 | 0 | 0 | 0 |
| 0 | 0 | 1.126459676 | 0 | 0.048727873 |
| 0 | 0 | 0 | 0 | 0.001643122 |
| 0 | 0 | 0 | 0 | 0 |
| 0 | 0 | 0 | 0 | 0 |
| 0 | 0 | 0 | 0 | 0 |
| 0 | 0 | 0 | 0 | 0 |
| 0 | 0 | 0 | 0.00464969 | 0 |

| | | | | |
|---|---|---|---|---|
| 0 | 0 | 0 | 0 | 0 |
| 0 | 0 | 0 | 0 | 0.000623368 |
| 0 | 0 | 0 | 0 | 0 |
| 0 | 0 | 0 | 0 | 0 |
| 0.121623688 | 1.043383025 | 0.622922592 | 0 | 0.14990761 |
| 0 | 0 | 0 | 0 | 0 |
| 0 | 0 | 0 | 0 | 0 |
| 0.028774823 | 0 | 0.025477788 | 0.016186098 | 0.011421715 |
| 0 | 0 | 0.001995186 | 0 | 0 |
| 0 | 0 | 0 | 0 | 0 |
| 0 | 0 | 0 | 0 | 0 |
| 0 | 0 | 0 | 0 | 0 |
| 0 | 0 | 0 | 0 | 0 |
| 0 | 0 | 0 | 0 | 0 |
| 0 | 0 | 0 | 0 | 0 |
| 0 | 0 | 0 | 0.028076164 | 0.000571498 |
| 0 | 0 | 0 | 0 | 0 |
| 0 | 0 | 0 | 0 | 0 |
| 0 | 0 | 0 | 0 | 0 |
| 0 | 0 | 0 | 0 | 0 |
| 0 | 0 | 0 | 0 | 0 |
| 0 | 0 | 0 | 0 | 0.013002499 |
| 0 | 0 | 0 | 0 | 0 |
| 0 | 0 | 0 | 0.001063879 | 0 |
| 0 | 0.059323394 | 0 | 0 | 0.098430551 |
| 0 | 0 | 0 | 0 | 0 |
| 0.038620046 | 0.844508771 | 0.032424161 | 1.010661867 | 0.258698708 |
| 0 | 0 | 0 | 0 | 0 |
| 0 | 0 | 0 | 0 | 0 |
| 0 | 0 | 0 | 0 | 0 |
| 0 | 0 | 0 | 0 | 0 |
| 0 | 0 | 0 | 0 | 0 |
| 0 | 0 | 0 | 0 | 0 |
| 0 | 0 | 0 | 0 | 0 |
| 0 | 0 | 0 | 0 | 0 |
| 0 | 0 | 0 | 0 | 0 |
| 0 | 0 | 0 | 0 | 0 |
| 0 | 0 | 0 | 0 | 0.527055367 |
| 0 | 0 | 0 | 0 | 0 |
| 0 | 0 | 0 | 0 | 0 |
| 0 | 0 | 0 | 0 | 0 |
| 0 | 3.220044606 | 0 | 0 | 0.001022231 |
| 0 | 0 | 0 | 0 | 0 |
| 0 | 0 | 0.287617773 | 0 | 0 |
| 0 | 0 | 0 | 0 | 0 |
| 0 | 0 | 0 | 0 | 0.060027346 |
| 0 | 0 | 0 | 0 | 0 |
| 0 | 0 | 0 | 0 | 0.027098512 |

| | | | | |
|---|---|---|---|---|
| 0 | 0 | 0 | 0 | 0.044144245 |
| 0 | 0 | 0 | 0 | 0 |
| 0 | 0 | 0 | 0 | 0 |
| 0 | 0 | 0 | 0 | 0 |
| 0 | 0 | 0 | 0 | 0 |
| 0 | 0 | 0 | 0 | 0 |
| 0 | 0 | 0 | 0 | 0 |
| 0 | 0 | 0 | 0 | 0 |
| 0 | 0 | 0 | 0 | 0 |
| 0 | 0 | 0 | 0 | 0 |
| 0 | 0 | 0 | 0 | 0.058731747 |
| 0 | 0 | 0 | 0 | 0 |
| 0 | 0 | 0 | 0 | 0 |
| 0 | 0 | 0 | 0 | 0 |
| 0 | 0 | 0 | 0 | 0.001711379 |
| 0 | 0 | 0 | 0 | 0 |
| 0 | 0 | 0 | 0 | 0 |
| 0 | 0 | 0 | 0.963087871 | 0 |
| 0 | 0 | 0 | 0 | 0.000554466 |
| 0 | 0 | 0 | 0 | 0 |
| 0 | 0 | 0.061134497 | 0.001236003 | 0 |
| 0 | 0 | 0 | 0 | 0 |
| 0 | 0 | 0.062527944 | 0.002675969 | 0.122639427 |
| 0 | 0 | 0 | 0 | 0.199329632 |
| 0 | 0 | 0 | 0 | 0 |
| 0 | 0 | 0 | 0 | 0.001282576 |
| 0 | 0 | 0 | 0 | 0 |
| 0 | 0 | 0 | 0 | 0 |
| 0 | 0.771941291 | 0 | 0.001737405 | 0 |
| 0 | 0 | 0 | 0 | 0 |
| 0 | 0 | 0 | 0 | 0 |
| 0 | 0 | 0 | 0 | 0.007211336 |
| 0.04803222 | 0 | 0.025714232 | 0.003967887 | 0 |
| 0 | 0 | 0 | 0 | 0 |
| 0 | 0 | 0.013208643 | 0 | 0.003804961 |
| 0.143601398 | 0 | 0.219829925 | 0 | 0 |
| 0 | 0 | 0 | 0 | 0 |
| 0 | 0 | 0 | 0 | 0 |
| 0 | 0 | 0.006616642 | 0.000581506 | 0 |
| 0 | 0 | 0 | 0 | 0 |
| 0 | 0 | 0 | 0 | 0 |
| 0 | 0 | 0 | 0 | 0.006186868 |
| 0.001383976 | 1.01210252 | 0.077368366 | 0 | 0.030858552 |
| 0 | 0 | 0 | 0 | 0 |
| 0 | 0 | 0 | 0 | 0 |
| 0 | 0 | 0 | 0 | 0.026436731 |
| 0 | 0 | 0 | 0 | 0 |
| 0 | 0 | 0 | 0 | 0 |

| | | | | |
|---|---|---|---|---|
| 0 | 0 | 0 | 0 | 0 |
| 0 | 0 | 0 | 0 | 0 |
| 0 | 0 | 0 | 0 | 0 |
| 0 | 0 | 0 | 0 | 0 |
| 0 | 0 | 0 | 0 | 0 |
| 0 | 0 | 0 | 0 | 0 |
| 0 | 0 | 0 | 0 | 0 |
| 0.252424693 | 0 | 0 | 0 | 0 |
| 0 | 0 | 0 | 0 | 0.021333681 |
| 0 | 0.084033908 | 0 | 0 | 0.079303554 |
| 0 | 0 | 0 | 0 | 0.005474411 |
| 0 | 0 | 0 | 0 | 0 |
| 0 | 0 | 0 | 0 | 0 |
| 0 | 0 | 0.149688744 | 0 | 0 |
| 0 | 0 | 0 | 0 | 0 |
| 0 | 0 | 0.013787684 | 0.20448374 | 0 |
| 0 | 0 | 0 | 0 | 0 |
| 0 | 0 | 0 | 0 | 0 |
| 0 | 0 | 0 | 0 | 0 |
| 0 | 0 | 0.569354467 | 0 | 0.054683733 |
| 0 | 0 | 0 | 0 | 0 |
| 0 | 0 | 0 | 0 | 0 |
| 0 | 0 | 0 | 0 | 0 |
| 0 | 0 | 0 | 0 | 0.023182859 |
| 0 | 0 | 0 | 0 | 0 |
| 0 | 0 | 0 | 0 | 0 |
| 0 | 0 | 0 | 0 | 0 |
| 0 | 0 | 0 | 0 | 0.126802003 |
| 0 | 0 | 0 | 0 | 0 |
| 0 | 0 | 0 | 0 | 0.007296351 |
| 0 | 0.003484447 | 0.180384351 | 0 | 0.097112685 |
| 0 | 0 | 0 | 0 | 0 |
| 0 | 0 | 0 | 0 | 0.001208158 |
| 0 | 0 | 0 | 0 | 0 |
| 0 | 0 | 0 | 0 | 0 |
| 0 | 0 | 0 | 0 | 0 |
| 0 | 0 | 0 | 0 | 0 |
| 0 | 0 | 0 | 0 | 0.003207182 |
| 0 | 0 | 0 | 0 | 0.003761777 |
| 0 | 0 | 0 | 0 | 0 |
| 0 | 0 | 0 | 0 | 0 |
| 0 | 0 | 0 | 0 | 0.169001572 |
| 0 | 0 | 0 | 0 | 0.011861666 |
| 0 | 0 | 0.000950143 | 0.002143262 | 0 |
| 0 | 0 | 0 | 0 | 0 |
| 0 | 0 | 0 | 0 | 0 |
| 0.203341044 | 0 | 0 | 0.027636592 | 0 |
| 0 | 0 | 0 | 0 | 0 |

| | | | | |
|---|---|---|---|---|
| 0 | 0 | 0 | 0 | 0 |
| 0 | 0 | 0 | 0 | 0 |
| 0 | 0 | 0 | 0 | 0 |
| 0 | 0.279823154 | 0 | 0 | 0 |
| 0 | 0 | 0 | 0.004045617 | 0.077768377 |
| 0 | 0 | 0 | 0 | 0 |
| 0 | 0 | 0 | 0 | 0 |
| 0 | 0 | 0 | 0 | 0 |
| 0 | 0 | 0 | 0 | 0 |
| 0 | 0 | 0 | 0 | 0 |
| 0 | 0 | 0 | 0 | 0.009304234 |
| 0 | 0.035765458 | 0.008972578 | 0.035249954 | 0.012458086 |
| 0 | 0 | 0 | 0 | 0.000716546 |
| 0 | 0 | 0 | 0 | 0 |
| 0 | 0 | 0.011516178 | 0 | 0.001265857 |
| 0 | 0 | 0 | 0.063860542 | 0.022120768 |
| 0 | 0 | 0.005629744 | 0.007001295 | 0.004790873 |
| 0 | 0 | 0 | 0 | 0 |
| 0 | 0 | 0.117823774 | 0 | 0 |
| 0.417817065 | 0.08229374 | 0 | 0.19953111 | 0.012522344 |
| 0 | 0 | 0 | 0 | 0 |
| 0 | 0 | 0 | 0 | 0.001439914 |
| 0 | 0 | 0 | 0 | 0 |
| 0 | 0 | 0 | 0 | 0 |
| 0 | 0 | 0 | 0 | 0 |
| 0 | 0 | 0 | 0 | 0 |
| 0 | 0 | 0.000748289 | 0.088539831 | 0 |
| 0 | 0 | 0 | 0 | 0 |
| 0 | 0 | 0 | 0 | 0 |
| 0 | 0 | 0 | 0 | 0 |
| 0 | 0 | 0 | 0.006957681 | 0 |
| 0 | 0 | 0.006642317 | 0.001503021 | 0 |
| 0 | 0 | 0 | 0 | 0 |
| 0 | 0 | 0 | 0 | 0 |
| 0 | 0 | 0 | 0 | 0 |
| 0 | 0 | 0 | 0 | 0 |
| 0 | 0 | 0 | 0 | 0 |
| 0 | 0.03405546 | 0.005363443 | 0.006232538 | 0.011135931 |
| 0 | 0 | 0 | 0 | 0.113141962 |
| 0 | 0 | 0 | 0 | 0 |
| 0 | 0 | 0 | 0.017080224 | 0.004839273 |
| 0 | 0 | 0 | 0 | 0 |
| 0 | 0 | 0 | 0 | 0 |
| 0 | 0 | 0.003313878 | 0 | 0 |
| 0 | 0 | 0 | 0 | 0 |
| 0 | 0 | 0 | 0 | 0 |
| 0 | 0 | 0 | 0 | 0 |
| 0 | 0 | 0 | 0 | 0.005644595 |

| | | | | |
|---|---|---|---|---|
| 0 | 0 | 0 | 0 | 0 |
| 0 | 0 | 0 | 0 | 0.005645568 |
| 0 | 0 | 0 | 0 | 0 |
| 0.005411759 | 0 | 0.003472959 | 0.003988239 | 0 |
| 0.001792614 | 0.004139809 | 0 | 0 | 0 |
| 0 | 0 | 0 | 0.005001282 | 0.000974836 |
| 0 | 0 | 0 | 0 | 0.005729732 |
| 0 | 0 | 0 | 0 | 0 |
| 0 | 0 | 0 | 0 | 0 |
| 0 | 0 | 0 | 0 | 0.014351471 |
| 0 | 0 | 0 | 0 | 0 |
| 0 | 0 | 0 | 0 | 0 |
| 0.037813726 | 0 | 0.26505798 | 0.001480862 | 0.062347427 |
| 0 | 0 | 0 | 0 | 0.00855667 |
| 0 | 0 | 0 | 0 | 0 |
| 0 | 0 | 0 | 0 | 0.013935829 |
| 0 | 0 | 0 | 0 | 0 |
| 0 | 0 | 0 | 0 | 0 |
| 0 | 0 | 0 | 0 | 0 |
| 0 | 0 | 0 | 0 | 0 |
| 0 | 0 | 0 | 0 | 0 |
| 0 | 0 | 0 | 0 | 0 |
| 0 | 0 | 0.052515126 | 0.024817596 | 0 |
| 0 | 0 | 0.019636475 | 0.000627549 | 0.03072273 |
| 0 | 0 | 0 | 0 | 0 |
| 0 | 0 | 0 | 0 | 0.016064497 |
| 0 | 0 | 0 | 0 | 0.004176031 |
| 0 | 0 | 0 | 0 | 0 |
| 0 | 0 | 0 | 0 | 0 |
| 0 | 0 | 0 | 0 | 0 |
| 0 | 0 | 0.080240848 | 0.02767151 | 0.006992487 |
| 0 | 0 | 0 | 0 | 0.01474727 |
| 0 | 0.010536559 | 0 | 0 | 0 |
| 0 | 0 | 0 | 0 | 0 |
| 0 | 0 | 0 | 0 | 0.077453994 |
| 0 | 0 | 0 | 0 | 0 |
| 0 | 0 | 0 | 0 | 0 |
| 0 | 0 | 0 | 0 | 0 |
| 0.214096671 | 0.030880719 | 0 | 0 | 0 |
| 0 | 0 | 0 | 0 | 0 |
| 0 | 0 | 0 | 0 | 0 |
| 0 | 0 | 0 | 0 | 0 |
| 0 | 0 | 0 | 0 | 0 |
| 0 | 0 | 0 | 0 | 0 |
| 0 | 0 | 0 | 0 | 0 |
| 0.074269771 | 0.102257741 | 0.382962504 | 0 | 0.082337052 |
| 0 | 0 | 0 | 0.031345866 | 0 |
| 0 | 0 | 0 | 0 | 0 |

| | | | | |
|---|---|---|---|---|
| 0 | 0 | 0 | 0 | 0 |
| 0 | 0 | 0 | 0 | 0 |
| 0.27307977 | 0.004053622 | 0 | 0 | 0.051125698 |
| 0 | 0 | 0 | 0 | 0.018813434 |
| 0 | 0 | 0 | 0.010614039 | 0 |
| 0 | 0 | 0 | 0 | 0 |
| 0 | 0 | 0 | 0 | 0 |
| 0 | 0 | 0 | 0 | 0 |
| 0 | 0 | 0 | 0.001702041 | 0.002156421 |
| 0 | 0 | 0 | 0 | 0 |
| 0 | 0 | 0.001984736 | 0.001763674 | 0.002181621 |
| 0 | 0 | 0 | 0 | 0 |
| 0 | 0 | 0 | 0 | 0 |
| 0 | 0 | 0 | 0 | 0 |
| 0 | 0 | 0 | 0 | 0 |
| 0 | 0 | 0 | 0 | 0 |
| 0 | 0.012052193 | 0 | 0 | 0 |
| 0 | 0 | 0 | 0 | 0 |
| 0 | 0 | 0 | 0 | 0 |
| 0 | 0.326198941 | 0.000877367 | 0 | 0.000771521 |
| 0 | 0 | 0 | 0 | 0 |
| 0 | 0 | 0 | 0 | 0 |
| 0 | 0 | 0 | 0 | 0 |
| 0 | 0 | 0 | 0 | 0 |
| 0 | 0 | 0 | 0 | 0 |
| 0 | 0 | 0 | 0 | 0 |
| 0 | 0 | 0 | 0 | 0 |
| 0 | 0.011364634 | 0 | 0 | 0.003415274 |
| 0 | 0 | 0 | 0 | 0 |
| 0 | 0 | 0 | 0 | 0 |
| 0 | 0.456685206 | 0.001507498 | 0.000772841 | 0.000497112 |
| 0 | 0 | 0 | 0 | 0 |
| 0.124198988 | 0.362744697 | 0.069795559 | 0.079863927 | 0.008273171 |
| 0.025383579 | 0 | 0.003353769 | 0.004401561 | 0.001179667 |
| 0 | 0 | 0 | 0 | 0.066245135 |
| 0 | 0 | 0 | 0 | 0 |
| 0 | 0 | 0 | 0 | 0 |
| 0 | 0 | 0 | 0 | 0 |
| 0 | 0 | 0 | 0 | 0 |
| 0 | 0 | 0 | 0 | 0 |
| 0 | 0 | 0 | 0 | 0 |
| 0.018031302 | 0 | 0.086580261 | 0.056106213 | 0 |
| 0 | 0.023553406 | 0.149556746 | 0 | 0.019090762 |
| 0 | 0 | 0 | 0 | 0 |
| 0 | 0 | 0 | 0 | 0 |
| 0.007936037 | 0.49570829 | 0.01172498 | 0 | 0.00808118 |
| 0 | 0 | 0 | 0 | 0 |
| 0 | 0 | 0 | 0 | 0 |

| | | | | |
|---|---|---|---|---|
| 0 | 0 | 0 | 0 | 0 |
| 0 | 0.006327004 | 0.006662368 | 0 | 0 |
| 0.043011641 | 0.00812967 | 0.006440565 | 0.001100617 | 0.001179911 |
| 0 | 0 | 0 | 0 | 0 |
| 0 | 0 | 0.003214051 | 0 | 0.102500698 |
| 0 | 0 | 0 | 0.024464691 | 0 |
| 0 | 0 | 0 | 0 | 0 |
| 0 | 0 | 0 | 0 | 0 |
| 0 | 0 | 0 | 0 | 0 |
| 0 | 0 | 0 | 0 | 0 |
| 0 | 0 | 0 | 0 | 0 |
| 0 | 0 | 0 | 0 | 0 |
| 0 | 0 | 0 | 0 | 0 |
| 0 | 0 | 0.006992389 | 0 | 0 |
| 0 | 0 | 0 | 0 | 0 |
| 0 | 0 | 0 | 0 | 0 |
| 0 | 0 | 0 | 0 | 0.381580998 |
| 0 | 0 | 0 | 0 | 0 |
| 0 | 0 | 0 | 0 | 0 |
| 0 | 0 | 0 | 0 | 0 |
| 0 | 0 | 0 | 0 | 0 |
| 0 | 0 | 0 | 0.010800895 | 0 |
| 0 | 0 | 0 | 0 | 0 |
| 0 | 0 | 0 | 0 | 0 |
| 0 | 0.018881792 | 0 | 0 | 0.011279984 |
| 0.018201196 | 0.571064563 | 0 | 0 | 0 |
| 0 | 0 | 0 | 0 | 0 |
| 0 | 0 | 0.013380763 | 0.828138427 | 0.013664323 |
| 0 | 0 | 0 | 0 | 0 |
| 0 | 0 | 0 | 0 | 0 |
| 0 | 0 | 0 | 0 | 0 |
| 0 | 0 | 0 | 0 | 0 |
| 0 | 0 | 0 | 0 | 0 |
| 0 | 0 | 0 | 0 | 0 |
| 0 | 0 | 0 | 0 | 0 |
| 0 | 0 | 0 | 0 | 0 |
| 0 | 0 | 0 | 0 | 0 |
| 0 | 0 | 0 | 0 | 0 |
| 0 | 0 | 2.425406846 | 0 | 1.250693426 |
| 0 | 0 | 0 | 0 | 0 |
| 0 | 0 | 0 | 0 | 0 |
| 0 | 0.023265124 | 0 | 0 | 0.019526487 |
| 0 | 0 | 0.081277029 | 0.005128354 | 0 |
| 0 | 0 | 0 | 0 | 0 |
| 0 | 0 | 0 | 0 | 0 |
| 0 | 0 | 0 | 0 | 0.002258947 |
| 0 | 0 | 0.017762564 | 0.022135174 | 0.001802271 |
| 0 | 0 | 0 | 0 | 0 |

| | | | | |
|---|---|---|---|---|
| 0 | 0 | 0 | 0 | 0 |
| 0 | 0.021366744 | 0.034481675 | 0.035355086 | 0.005685335 |
| 0 | 0 | 0 | 0 | 0.057300191 |
| 0 | 0 | 0 | 0 | 0 |
| 0 | 0 | 0 | 0 | 0 |
| 0 | 0 | 0 | 0 | 0 |
| 0 | 0 | 0 | 0 | 0 |
| 0 | 0 | 0 | 0 | 0 |
| 0 | 0 | 0 | 0 | 0 |
| 0 | 0 | 0 | 0 | 0 |
| 0 | 0 | 0 | 0 | 0 |
| 0 | 0 | 0 | 0 | 0 |
| 0 | 0 | 0 | 0 | 0.013221786 |
| 0 | 0 | 0 | 0 | 0 |
| 0 | 0 | 0.007792563 | 0 | 0 |
| 0 | 0 | 0 | 0 | 0 |
| 0 | 0 | 0 | 0 | 0 |
| 0 | 0 | 0 | 0 | 0 |
| 0 | 0 | 0 | 0.002438331 | 0.0033982 |
| 0 | 0 | 0 | 0 | 0 |
| 0 | 0 | 0 | 0 | 0 |
| 0.038330987 | 0.01090153 | 0 | 0.068402085 | 0 |
| 0 | 0 | 0 | 0.180471911 | 0.009985753 |
| 0 | 0.032188436 | 0 | 0 | 0 |
| 0 | 0 | 0 | 0 | 0 |
| 0 | 0 | 0 | 0 | 0 |
| 0 | 0 | 0 | 0 | 0 |
| 0 | 0 | 0 | 0 | 0 |
| 0 | 0 | 0 | 0 | 0 |
| 0 | 0 | 0 | 0 | 0 |
| 0 | 0 | 0 | 0 | 0 |
| 0 | 0 | 0 | 0 | 0 |
| 0 | 0 | 0 | 0 | 0 |
| 0 | 0 | 0 | 0 | 0 |
| 0 | 0.021115696 | 0 | 0 | 1.632044052 |
| 0 | 0 | 0 | 0 | 0 |
| 0 | 0 | 0 | 0 | 0 |
| 0 | 0 | 0 | 0 | 0 |
| 0.036078236 | 0.119751464 | 0 | 0.002810129 | 0 |
| 0 | 0 | 0 | 0 | 0 |
| 1.375223191 | 0 | 0.028590987 | 0.053300344 | 0.021817225 |
| 0 | 0 | 0 | 0 | 0 |
| 0 | 0 | 0 | 0 | 0 |
| 0 | 0 | 0 | 0 | 0 |
| 0 | 0 | 0 | 0 | 0 |
| 0.228547247 | 0 | 0.031382059 | 0.238109492 | 0.002141078 |
| 0 | 0 | 0 | 0 | 0 |
| 0 | 0 | 0 | 0 | 0 |

| | | | | |
|---|---|---|---|---|
| 0 | 0 | 0 | 0 | 0 |
| 0 | 0 | 0 | 0 | 0 |
| 0 | 0 | 0 | 0 | 0 |
| 0 | 0 | 0 | 0 | 0 |
| 0 | 0 | 0 | 0 | 0.001082564 |
| 0 | 0 | 0 | 0 | 0 |
| 0 | 0 | 0 | 0 | 0 |
| 0.002440981 | 0 | 0 | 0 | 0 |
| 0 | 0 | 0 | 0 | 0 |
| 0.149749276 | 0.106385718 | 0 | 0 | 0.003257298 |
| 0 | 0 | 0 | 0 | 0.02306549 |
| 0 | 0 | 0 | 0 | 0 |
| 0.00215881 | 0 | 0 | 0.002969778 | 0 |
| 0 | 0 | 0 | 0 | 0 |
| 0 | 0 | 0 | 0 | 0 |
| 0 | 0 | 0 | 0 | 0 |
| 0 | 0 | 0 | 0 | 0.06756391 |
| 0 | 0 | 0 | 0 | 0 |
| 0 | 0 | 0 | 0 | 0 |
| 0.034202578 | 2.526170146 | 0.336522551 | 0.376860724 | 0.080357005 |
| 0 | 0 | 0 | 0 | 0 |
| 0 | 0 | 0 | 0 | 0 |
| 0 | 0 | 0 | 0 | 0 |
| 0 | 0 | 0 | 0 | 0 |
| 0 | 0 | 0 | 0 | 0 |
| 0 | 0 | 0 | 0 | 0.055225571 |
| 0 | 0 | 0 | 0 | 0 |
| 0 | 0 | 0 | 0 | 0 |
| 0 | 0 | 0 | 0 | 0 |
| 0 | 0 | 0 | 0.043799015 | 0 |
| 0 | 0 | 0 | 0 | 0 |
| 0 | 0 | 0 | 0 | 0 |
| 0 | 0 | 0 | 0 | 0 |
| 0 | 0 | 0 | 0 | 0 |
| 0 | 0 | 0 | 0 | 0 |
| 0 | 0 | 0 | 0 | 0 |
| 0 | 0 | 0 | 0 | 0 |
| 0 | 0 | 0 | 0 | 0.058044016 |
| 0 | 0 | 0 | 0 | 0.002558114 |
| 0 | 0 | 0 | 0 | 0.001286684 |
| 0 | 0 | 0 | 0 | 0 |
| 0 | 0 | 0 | 0 | 0 |
| 0 | 0 | 0 | 0 | 0.005384053 |
| 0 | 0 | 0 | 0 | 0 |
| 0 | 0 | 0.007134373 | 0.015960185 | 0.002661561 |
| 0 | 0 | 0 | 0 | 0.006774751 |
| 0 | 0 | 0 | 0 | 0 |
| 0 | 0 | 0 | 0 | 0 |

| | | | | |
|---|---|---|---|---|
| 0 | 0 | 0 | 0 | 0 |
| 0 | 0 | 0 | 0 | 0 |
| 0 | 0 | 0 | 0 | 0.061401195 |
| 0 | 0 | 0 | 0 | 0 |
| 0 | 0 | 0 | 0 | 0.00180554 |
| 0.000808384 | 0.043115509 | 0.003066535 | 0.03733335 | 0.047687002 |
| 0 | 0 | 0 | 0 | 0.017004881 |
| 0 | 0 | 0 | 0 | 0 |
| 0 | 0 | 0 | 0 | 0 |
| 0 | 0 | 0 | 0 | 0 |
| 0 | 0 | 0 | 0 | 0 |
| 0 | 0 | 0 | 0 | 0 |
| 0 | 0 | 0 | 0 | 0 |
| 0 | 0 | 0 | 0 | 0 |
| 0 | 0 | 0 | 0 | 0 |
| 0 | 0 | 0 | 0 | 0 |
| 0 | 0 | 0 | 0 | 0.125328662 |
| 0 | 0 | 0 | 0 | 0 |
| 0 | 0 | 0 | 0 | 0 |
| 0 | 0 | 0 | 0 | 0.181559913 |
| 0 | 0 | 0 | 0 | 0.002797537 |
| 0 | 0 | 0 | 0 | 0 |
| 0 | 0 | 0 | 0 | 0 |
| 0 | 0 | 0 | 0 | 0 |
| 0 | 0 | 0 | 0 | 0 |
| 0.004214826 | 0.002781026 | 0 | 0 | 0 |
| 0 | 0 | 0 | 0 | 0 |
| 0 | 0 | 0 | 0 | 0.342099262 |
| 0 | 0 | 0 | 0 | 0 |
| 0 | 0 | 0 | 0 | 0 |
| 0 | 0 | 0 | 0 | 0 |
| 0 | 0 | 0 | 0 | 0 |
| 0 | 0 | 0 | 0 | 0.003443461 |
| 0 | 0 | 0 | 0 | 0 |
| 0 | 0 | 0 | 0 | 0.13762007 |
| 0 | 0 | 0 | 0 | 0 |
| 0 | 0 | 0 | 0 | 0 |
| 0 | 0 | 0 | 0 | 0 |
| 0 | 0 | 0 | 0 | 0.033056217 |
| 0 | 0 | 0 | 0 | 0 |
| 0 | 0 | 0 | 0 | 0 |
| 0 | 0 | 0 | 0 | 0 |
| 0 | 0 | 0 | 0 | 0 |
| 0 | 0 | 0 | 0 | 0 |
| 0 | 0 | 0 | 0 | 0 |
| 0 | 0 | 0 | 0 | 0 |
| 0 | 0 | 0 | 0 | 0 |
| 0 | 0 | 0 | 0 | 0 |

| | | | | |
|---|---|---|---|---|
| 0 | 0 | 0 | 0 | 0 |
| 0 | 0 | 0 | 0.056619571 | 0 |
| 0 | 0 | 0 | 0 | 0 |
| 0 | 0 | 0 | 0 | 0 |
| 0 | 0 | 0 | 0 | 0 |
| 0 | 0 | 0 | 0 | 0.002396848 |
| 0 | 0 | 0 | 0 | 0 |
| 0 | 0 | 0 | 0 | 0 |
| 0 | 0 | 0 | 0 | 0 |
| 0 | 0 | 0 | 0 | 0.001941699 |
| 0.025281969 | 0.04887172 | 0 | 0 | 0.003120948 |
| 0 | 0 | 0 | 0 | 0 |
| 0 | 0 | 0 | 0.241728916 | 0.011493681 |
| 0 | 0 | 0 | 0 | 0 |
| 0 | 0 | 0 | 0 | 0 |
| 0 | 0 | 0 | 0 | 0 |
| 0 | 0 | 0 | 0 | 0 |
| 0 | 0 | 0 | 0 | 0 |
| 0 | 0 | 0 | 0 | 0 |
| 0 | 0 | 0 | 0 | 0.001038793 |
| 0.002806527 | 0.969550918 | 0 | 0 | 0.006123978 |
| 0 | 0 | 0 | 0 | 0 |
| 0 | 0 | 0 | 0 | 0 |
| 5.100223228 | 1.515952811 | 1.126563334 | 0 | 0.197151189 |
| 0 | 0 | 0 | 0 | 0 |
| 0 | 0 | 0 | 0 | 0 |
| 0 | 0 | 0 | 0 | 0 |
| 0 | 0 | 0 | 0 | 0 |
| 0 | 0 | 0 | 0 | 0 |
| 0 | 0 | 0 | 0 | 0 |
| 0 | 0 | 0 | 0.091641272 | 0 |
| 0 | 0 | 0 | 0 | 0 |
| 0 | 0 | 0 | 0 | 0 |
| 0 | 0 | 0 | 0 | 0 |

| | | | | |
|---|---|---|---|---|
| PHL018-0178_V3 | PHL018-0246_V2 | PHL018-0142_V1 | PHL018-0138_V1 | PHL018-0077_V1 |
| EYCF | REF | EYCF | EYCF | EYCF |
| 0.00109187 | 0.279240109 | 0.132665257 | 0.00282879 | 0.00868637 |
| 0.044679999 | 3.781383177 | 2.318780851 | 0.48279117 | 7.944584752 |
| 0 | 1.281595373 | 0 | 0 | 9.587124263 |
| 0 | 0 | 0 | 0 | 0 |
| 0 | 0 | 0 | 0 | 0 |
| 0 | 0 | 0 | 0 | 0 |
| 0 | 0 | 0 | 0 | 0 |
| 0 | 0 | 0 | 0 | 0 |
| 0 | 0 | 0 | 0 | 0 |
| 0 | 0 | 0 | 0 | 0 |
| 0 | 0 | 0 | 0 | 0 |
| 0 | 0 | 0 | 0 | 0 |
| 0 | 0 | 0 | 0 | 0 |
| 0.573343421 | 0.11107774 | 0.28991583 | 0.33878093 | 3.762032613 |
| 0 | 0 | 0 | 0 | 0 |
| 0 | 0 | 0 | 0 | 0 |
| 0 | 0 | 0 | 0 | 0 |
| 0 | 0 | 0 | 0 | 0 |
| 0 | 0 | 0 | 0 | 0 |
| 0 | 0 | 0 | 0 | 0 |
| 0 | 0 | 0 | 0 | 0 |
| 0 | 0 | 0 | 0 | 0 |
| 0 | 0 | 0 | 0 | 0 |
| 0 | 0 | 0 | 0 | 0 |
| 0 | 0 | 0 | 0 | 0 |
| 0 | 0 | 0 | 0 | 0 |
| 0 | 0 | 0 | 0 | 0 |
| 0 | 0 | 0 | 0 | 0 |
| 0 | 0 | 0 | 0 | 0 |
| 0 | 0 | 0 | 0 | 0 |
| 0 | 0 | 0 | 0 | 0 |
| 0 | 0 | 0 | 0 | 0 |
| 0 | 0 | 0 | 0 | 0 |
| 0 | 0 | 0 | 0 | 0 |
| 0 | 0 | 0 | 0 | 0 |
| 1.778994723 | 0 | 0 | 0.106377572 | 2.860125528 |
| 0 | 0 | 0 | 0 | 0 |
| 0 | 0 | 0 | 0 | 0 |
| 0 | 0 | 0 | 0 | 0 |
| 0 | 0 | 0 | 0 | 0 |
| 0 | 0 | 0 | 0 | 0 |
| 0.504082024 | 0.153394244 | 0.02172565 | 0.004867644 | 0.007670213 |
| 10.04650691 | 0.116582604 | 1.373544634 | 0.408717244 | 0.21013858 |
| 0.006810246 | 0.073611453 | 0.015267714 | 0 | 0.03894108 |
| 0.006172504 | 0.016058462 | 0 | 0.002120095 | 0.016740458 |
| 0 | 0 | 0 | 0 | 0 |

| | | | | |
|---|---|---|---|---|
| 0 | 0.220303864 | 0 | 0.002747689 | 0.00565 |
| 0 | 0.001746093 | 0.004312402 | 0 | 0 |
| 0 | 0.015303557 | 0 | 0.002009538 | 0.024682762 |
| 0.205865961 | 0.317661527 | 0.972840779 | 0.186581128 | 0.797316349 |
| 0 | 0.003690563 | 0 | 0 | 0 |
| 0.009104003 | 0 | 0 | 0 | 0 |
| 0.008043898 | 0.011977789 | 0.623837495 | 0 | 0.014998409 |
| 0 | 0 | 0 | 0 | 0 |
| 0 | 0 | 0 | 0 | 0 |
| 0 | 0 | 0 | 0 | 0 |
| 0 | 0 | 0.001025115 | 0 | 0 |
| 0.619337074 | 0.097209848 | 1.00076018 | 0.006128029 | 1.490569281 |
| 0 | 0 | 0 | 0 | 0 |
| 0 | 0.432926769 | 0.09576964 | 0 | 0.036282025 |
| 0 | 0 | 0.001096277 | 0 | 0 |
| 0 | 0 | 0 | 0 | 0 |
| 0.84291887 | 0.002204942 | 5.4162225 | 0 | 0.223170204 |
| 0 | 0 | 0 | 0 | 0 |
| 0 | 0 | 0 | 0 | 0 |
| 0 | 0 | 0 | 0 | 0 |
| 0 | 0.107356801 | 0.081658801 | 0 | 0.010593651 |
| 0 | 0.003723393 | 0.035107228 | 0 | 0.008525491 |
| 0.002350534 | 0 | 0 | 0.000692012 | 0 |
| 0 | 0 | 0 | 0 | 0 |
| 0 | 0 | 0 | 0 | 0 |
| 0 | 0 | 0 | 0 | 0 |
| 0 | 0 | 0.006355501 | 0 | 0.003110475 |
| 0 | 0 | 0 | 0 | 0 |
| 0.016329506 | 0.002381648 | 0.004636108 | 0 | 0 |
| 0 | 0 | 0 | 0 | 0 |
| 0 | 0.09764007 | 0.114079817 | 0 | 0 |
| 0 | 0 | 0 | 0 | 0 |
| 0 | 0 | 0 | 0 | 0 |
| 0 | 0 | 0 | 0 | 0 |
| 0 | 0 | 0 | 0 | 0 |
| 0 | 0.00123667 | 0 | 0 | 0 |
| 0 | 0 | 0 | 0 | 0 |
| 38.98156135 | 0 | 0.157101483 | 0 | 0.0063177 |
| 0 | 0 | 0 | 0 | 0 |
| 0 | 0 | 0 | 0 | 0 |
| 0 | 0 | 0 | 0 | 0 |
| 0 | 0 | 0 | 0 | 0 |
| 0 | 0 | 0 | 0 | 0 |
| 0 | 0 | 0 | 0 | 0 |
| 0 | 0 | 0 | 0 | 0 |
| 0 | 0 | 0 | 0 | 0 |
| 0 | 0 | 0.000840087 | 0 | 0 |
| 0 | 0 | 0.001258601 | 0 | 0 |

| | | | | |
|---|---|---|---|---|
| 0 | 0 | 0 | 0 | 0 |
| 0 | 0 | 0 | 0 | 0 |
| 0 | 0 | 0 | 0 | 0 |
| 0 | 0 | 0 | 0 | 0 |
| 0 | 0.023310059 | 0 | 0 | 0.008625984 |
| 0 | 0 | 0.00069599 | 0 | 0 |
| 0 | 0 | 0 | 0 | 0 |
| 0 | 0 | 0 | 0 | 0 |
| 0.000552443 | 0.021850776 | 0 | 0 | 0 |
| 0 | 0 | 0 | 0 | 0 |
| 0 | 0 | 0 | 0 | 0 |
| 0.014324062 | 0.008058172 | 0 | 0 | 0 |
| 0 | 0 | 0 | 0 | 0 |
| 0 | 0 | 0 | 0 | 0 |
| 0 | 0 | 0 | 0 | 0 |
| 0 | 0 | 0 | 0 | 0 |
| 0 | 0 | 0 | 0 | 0 |
| 0 | 0 | 0 | 0 | 0 |
| 0.014650394 | 0 | 0.056437073 | 0 | 0 |
| 0 | 0 | 0 | 0 | 0 |
| 0 | 0 | 0 | 0 | 0 |
| 0 | 0 | 0 | 0 | 0 |
| 0 | 0 | 0 | 0 | 0 |
| 0 | 0 | 0 | 0 | 0 |
| 0 | 0 | 0 | 0 | 0 |
| 0 | 0 | 0 | 0 | 0 |
| 0 | 0 | 0 | 0 | 0 |
| 0 | 0 | 0 | 0 | 0 |
| 0 | 0 | 0 | 0 | 0 |
| 0 | 0 | 0 | 0 | 0 |
| 0.00102243 | 0.003568248 | 0.409231537 | 0.002575306 | 0 |
| 0 | 0.032442824 | 0 | 0 | 0.005777696 |
| 0 | 0 | 0 | 0 | 0 |
| 0 | 0 | 0 | 0 | 0 |
| 0 | 0 | 0 | 0 | 0 |
| 0 | 0 | 0 | 0 | 0 |
| 0 | 0 | 0 | 0 | 0 |
| 0 | 0 | 0 | 0 | 0 |
| 0 | 0 | 0 | 0.022208543 | 0 |
| 0 | 0 | 0 | 0 | 0 |
| 0 | 0 | 0 | 0 | 0 |
| 0 | 0 | 0 | 0 | 0 |
| 0 | 0.001442261 | 0.007983826 | 0 | 0 |
| 0 | 0 | 0 | 0 | 0 |
| 0 | 0 | 0 | 0 | 0 |
| 0 | 0 | 0 | 0 | 0 |
| 0 | 0 | 0 | 0 | 0 |
| 0 | 0 | 0 | 0 | 0 |

| | | | | |
|---|---|---|---|---|
| 0 | 0 | 0 | 0 | 0 |
| 0 | 0 | 0 | 0 | 0 |
| 0 | 0 | 0 | 0 | 0 |
| 0 | 0 | 0 | 0 | 0 |
| 0.00124479 | 0 | 0 | 0.137901152 | 0 |
| 0 | 0 | 0 | 0 | 0 |
| 0 | 0 | 0 | 0 | 0 |
| 0 | 0 | 0 | 0 | 0 |
| 0 | 0 | 0 | 0 | 0 |
| 0 | 0 | 0 | 0 | 0 |
| 0 | 0 | 0 | 0 | 0 |
| 0 | 0 | 0 | 0 | 0 |
| 0 | 0 | 0 | 0 | 0 |
| 0 | 0 | 0 | 0 | 0 |
| 0 | 0 | 0 | 0 | 0 |
| 0.348043885 | 0 | 0 | 0 | 0 |
| 0 | 0 | 0 | 0 | 0 |
| 0 | 0 | 0 | 0 | 0 |
| 0 | 0 | 0 | 0 | 0 |
| 0 | 0 | 0 | 0 | 0 |
| 0 | 0 | 0 | 0 | 0 |
| 0 | 0 | 0 | 0 | 0 |
| 0 | 0 | 0 | 0 | 0 |
| 0 | 0 | 0 | 0 | 0 |
| 0 | 0 | 0 | 0.008372464 | 0 |
| 0 | 0 | 0 | 0 | 0 |
| 0 | 0 | 0 | 0 | 0 |
| 0 | 0 | 0 | 0 | 0 |
| 0 | 0 | 0 | 0 | 0 |
| 0 | 0 | 0 | 0 | 0 |
| 0 | 0 | 0 | 0 | 0 |
| 0.004744018 | 0.008926097 | 0 | 1.135997744 | 0 |
| 0 | 0 | 0 | 0 | 0 |
| 0 | 0 | 0 | 0 | 0 |
| 0 | 0 | 0 | 0 | 0 |
| 0 | 0 | 0 | 0 | 0 |
| 0 | 0 | 0 | 0 | 0 |
| 0 | 0 | 0 | 0 | 0 |
| 0 | 0 | 0 | 0 | 0 |
| 0 | 0 | 0 | 0 | 0 |
| 0 | 0 | 0 | 0 | 0 |
| 0 | 0 | 0 | 0 | 0 |
| 0 | 0 | 0 | 0 | 0 |
| 0 | 0 | 0 | 0 | 0 |
| 0 | 0 | 0 | 0 | 0 |
| 0 | 0 | 0 | 0 | 0 |
| 0 | 0 | 0 | 0 | 0 |
| 0 | 0.160623454 | 0 | 0 | 0 |

| | | | | |
|---|---|---|---|---|
| 0 | 0 | 0 | 0 | 0 |
| 0 | 0 | 0 | 0 | 0 |
| 0 | 0 | 0 | 0 | 0 |
| 0 | 0 | 0 | 0 | 0 |
| 0 | 0 | 0 | 0 | 0 |
| 0 | 0 | 0 | 0 | 0 |
| 0 | 0 | 0 | 0 | 0 |
| 0 | 0 | 0 | 0 | 0 |
| 0 | 0 | 0 | 0 | 0 |
| 0 | 0 | 0 | 0 | 0 |
| 0 | 0 | 0 | 0 | 0 |
| 0 | 0 | 0 | 0 | 0 |
| 0 | 0.006612696 | 0.000586626 | 0 | 0 |
| 0 | 0 | 0 | 0 | 0 |
| 0 | 0 | 0.017314694 | 0 | 0 |
| 0 | 0 | 0 | 0 | 0 |
| 0 | 0 | 0 | 0 | 0 |
| 0.001898947 | 0 | 0 | 0 | 0 |
| 0 | 0 | 0 | 0 | 0 |
| 0 | 0 | 0 | 0 | 0 |
| 0.066426942 | 0.019061402 | 0.021059514 | 0.001593493 | 0.066974888 |
| 0 | 0 | 0 | 0 | 0 |
| 0 | 0 | 0 | 0 | 0 |
| 0 | 0.003223331 | 0.000686276 | 0 | 0 |
| 0 | 0 | 0 | 0 | 0 |
| 0 | 0 | 0 | 0 | 0 |
| 0.002579032 | 0.009300764 | 0.003449409 | 0 | 0 |
| 0 | 0 | 0 | 0 | 0 |
| 0 | 0 | 0 | 0 | 0 |
| 0 | 0 | 0 | 0 | 0 |
| 0 | 0 | 0 | 0 | 0 |
| 0 | 0 | 0 | 0 | 0 |
| 0 | 0 | 0 | 0 | 0 |
| 0.001353201 | 0.043133322 | 0.009116439 | 0 | 0 |
| 0 | 0.049944657 | 0.002141007 | 0 | 0.002984756 |
| 0 | 0 | 0 | 0 | 0 |
| 0 | 0 | 0 | 0 | 0 |
| 0 | 0 | 0 | 0 | 0 |
| 0.006749362 | 0 | 0 | 0 | 0 |
| 0 | 0 | 0 | 0 | 0 |
| 0 | 0 | 0 | 0 | 0 |
| 0 | 0 | 0 | 0 | 0 |
| 0 | 0 | 0 | 0 | 0 |
| 0 | 0 | 0 | 0 | 0 |
| 0 | 0 | 0 | 0 | 0 |
| 0 | 0 | 0 | 0 | 0 |
| 0 | 0.00508104 | 0.001545427 | 0 | 0 |
| 0 | 0.017915392 | 0.000712012 | 0 | 0 |

| | | | | |
|---|---|---|---|---|
| 0 | 0 | 0 | 0 | 0 |
| 0 | 0 | 0 | 0 | 0 |
| 0 | 0 | 0 | 0 | 0 |
| 0 | 0 | 0 | 0 | 0 |
| 0 | 0 | 0 | 0 | 0 |
| 0 | 0 | 0 | 0 | 0 |
| 0 | 0 | 0.000434374 | 0 | 0 |
| 0.003443698 | 0 | 0 | 0 | 0 |
| 0 | 0 | 0 | 0 | 0.018023135 |
| 0 | 0 | 0 | 0 | 0 |
| 0.000635131 | 0 | 0 | 0 | 0 |
| 0 | 0 | 0 | 0 | 0 |
| 0 | 0 | 0 | 0 | 0 |
| 0 | 0 | 0.000896007 | 0 | 0 |
| 0 | 0 | 0.001443351 | 0 | 0 |
| 0 | 0 | 0 | 0 | 0 |
| 0 | 0 | 0 | 0 | 0 |
| 0.066558578 | 0 | 0.009374247 | 0 | 0 |
| 0.04750775 | 0.001943449 | 0.010654778 | 0.002629955 | 0 |
| 0.031109542 | 0 | 0 | 0 | 0 |
| 0 | 0.060202491 | 0 | 0 | 0 |
| 0 | 0 | 0 | 0 | 0 |
| 0 | 0 | 0 | 0 | 0 |
| 0 | 0 | 0 | 0 | 0 |
| 0.002039946 | 0.002135803 | 0.004547317 | 0 | 0.005071495 |
| 0 | 0 | 0 | 0 | 0 |
| 0 | 0 | 0 | 0 | 0 |
| 0 | 0 | 0 | 0 | 0 |
| 0.086143673 | 0 | 0.000723262 | 0 | 0 |
| 0 | 0 | 0 | 0 | 0 |
| 0.013023785 | 0.610250384 | 0.437805035 | 0 | 0.051383021 |
| 0 | 0 | 0 | 0 | 0 |
| 0 | 0 | 0 | 0 | 0 |
| 0 | 0 | 0 | 0 | 0 |
| 0 | 0 | 0 | 0 | 0 |
| 0 | 0 | 0 | 0 | 0 |
| 0 | 0 | 0 | 0 | 0 |
| 0 | 0 | 0 | 0 | 0 |
| 0 | 0 | 0 | 0 | 0 |
| 0 | 0 | 0 | 0.001017611 | 0 |
| 0 | 0 | 0 | 0 | 0 |
| 0.001539198 | 0.003223051 | 0.005604101 | 0.003323097 | 0.003587428 |
| 0 | 0 | 0 | 0 | 0 |
| 0 | 0 | 0 | 0 | 0 |
| 0 | 0 | 0 | 0 | 0 |
| 0 | 0 | 0 | 0 | 0 |
| 0 | 0 | 0 | 0 | 0 |
| 0 | 0 | 0 | 0 | 0 |

| | | | | |
|---|---|---|---|---|
| 0 | 0 | 0 | 0 | 0 |
| 0 | 0 | 0 | 0 | 0 |
| 0.003623843 | 0.070149223 | 0.016515106 | 0 | 0.040241146 |
| 0 | 0 | 0 | 0 | 0 |
| 0.115731268 | 0 | 0 | 0 | 0 |
| 0 | 0 | 0 | 0 | 0 |
| 0 | 0 | 0 | 0 | 0 |
| 0 | 0 | 0 | 0 | 0 |
| 0 | 0 | 0 | 0 | 0 |
| 0 | 0 | 0 | 0 | 0 |
| 0 | 0 | 0 | 0 | 0 |
| 0 | 0 | 0 | 0 | 0 |
| 0 | 0 | 0 | 0 | 0 |
| 0 | 0 | 0 | 0 | 0.002323255 |
| 0.003085231 | 0.003384026 | 0.00890787 | 0 | 0.004656889 |
| 0 | 0 | 0 | 0.006367344 | 0 |
| 0.002762056 | 0.005257901 | 0.002238909 | 0 | 0.00156062 |
| 0 | 0 | 0 | 0 | 0 |
| 0 | 0 | 0 | 0 | 0 |
| 0 | 0 | 0 | 68.72389171 | 0 |
| 0 | 0 | 0 | 0 | 0 |
| 0 | 0.018985536 | 0.001347397 | 0 | 0 |
| 0 | 0 | 0 | 0 | 0 |
| 0 | 0 | 0.028198433 | 0 | 0 |
| 0 | 0 | 0 | 0 | 0 |
| 0 | 0 | 0 | 0 | 0 |
| 0 | 0 | 0 | 0 | 0 |
| 0 | 0 | 0 | 0 | 0 |
| 0 | 0 | 0 | 0 | 0 |
| 0 | 0 | 0 | 0 | 0 |
| 0 | 0 | 0 | 0 | 0 |
| 0 | 0 | 0 | 0 | 0 |
| 0 | 0 | 0 | 0 | 0 |
| 0.000784042 | 0 | 0.023613875 | 0 | 0 |
| 0 | 0 | 0 | 0 | 0 |
| 0 | 0.003160076 | 0 | 0 | 0 |
| 0 | 0 | 0 | 0 | 0 |
| 0.017371878 | 0.013516904 | 0 | 0 | 0.014868036 |
| 0 | 0 | 0 | 0 | 0 |
| 0 | 0 | 0 | 0 | 0 |
| 0 | 0 | 0 | 0 | 0.007152926 |
| 0 | 0 | 0 | 0 | 0 |
| 0 | 0 | 0 | 0 | 0 |
| 0 | 0 | 0 | 0 | 0 |
| 0 | 0 | 0 | 0 | 0 |
| 0.00394067 | 0.003667416 | 0 | 0 | 0 |
| 0 | 0 | 0 | 0 | 0 |
| 0 | 0 | 0.011277662 | 0 | 0 |

| | | | | |
|---|---|---|---|---|
| 1.153399363 | 0.105077524 | 3.026498841 | 0 | 0 |
| 0 | 0.047119021 | 0 | 0 | 0 |
| 0.007359929 | 0 | 0.001975811 | 0 | 0 |
| 0 | 0 | 0 | 0 | 0 |
| 0.012420952 | 0 | 0.000709949 | 0.001031407 | 0 |
| 0.004284392 | 0 | 0 | 0 | 0 |
| 0 | 0 | 0 | 0 | 0 |
| 0.004687496 | 0.036457662 | 0.013334999 | 0.003252924 | 0.018312722 |
| 0 | 0 | 0 | 0 | 0 |
| 0.002760859 | 0.059257146 | 0.041849451 | 0.008940957 | 0.421691796 |
| 0 | 0 | 0.004451449 | 0 | 0 |
| 0.081634044 | 0.006290003 | 0.00173308 | 0.577664145 | 0 |
| 0.001005232 | 0.003087241 | 0.104690183 | 0 | 0.010496226 |
| 0 | 0.040493518 | 0 | 0 | 0.002277294 |
| 0 | 0.001154603 | 0.003318641 | 0 | 0 |
| 0.009054125 | 0 | 0 | 0 | 0 |
| 0.541220764 | 0 | 0 | 0 | 0 |
| 0 | 0.011331146 | 0 | 0 | 0 |
| 0 | 0.052133234 | 0 | 0 | 0 |
| 0 | 0.014898336 | 0 | 0 | 0 |
| 0 | 0 | 0 | 0 | 0 |
| 0 | 0 | 0 | 0 | 0 |
| 0 | 0 | 0 | 0 | 0 |
| 0 | 0 | 0 | 0 | 0 |
| 0 | 0 | 0 | 0 | 0 |
| 0 | 0.065052176 | 0.000556792 | 0.0027806 | 0.035803038 |
| 2.463706559 | 0 | 0 | 3.682058065 | 0 |
| 0 | 0 | 0 | 0 | 0 |
| 0 | 0 | 0 | 0 | 0 |
| 0 | 0 | 0.005585468 | 0 | 0 |
| 0 | 0 | 0 | 0 | 0 |
| 0 | 0 | 0 | 0 | 0 |
| 0 | 0 | 0 | 0 | 0 |
| 0 | 0 | 0 | 0 | 0 |
| 0 | 0 | 0 | 0 | 0 |
| 0 | 0.754801274 | 0 | 0 | 0 |
| 0 | 0.020010746 | 0.059488752 | 0 | 0 |
| 0 | 0 | 0 | 0 | 0 |
| 0 | 0 | 0 | 0 | 0 |
| 0 | 0 | 0 | 0 | 0 |
| 0 | 0 | 0 | 0 | 0 |
| 0 | 0 | 0 | 0 | 0 |
| 0 | 0 | 0 | 0 | 0 |
| 0 | 0.04661444 | 0.001009285 | 0 | 0 |
| 0 | 0 | 0 | 0 | 0 |
| 0 | 0 | 0 | 0 | 0 |
| 0 | 0 | 0 | 0 | 0 |
| 0 | 0 | 0 | 0 | 0 |

| | | | | |
|---|---|---|---|---|
| 0 | 0 | 0 | 0 | 0 |
| 0.00097633 | 0 | 0.062292635 | 0 | 0 |
| 0 | 0 | 0 | 0 | 0 |
| 0 | 0 | 0 | 0 | 0 |
| 0 | 0.003214421 | 0 | 0 | 0 |
| 0 | 0.003522023 | 0 | 0 | 0.003991476 |
| 0 | 0 | 0 | 0 | 0 |
| 0 | 0 | 0.019749965 | 0 | 0 |
| 0 | 0 | 0.36591897 | 0 | 0 |
| 0 | 0 | 0 | 0 | 0 |
| 0.011438828 | 0 | 0.002846367 | 0 | 0 |
| 0 | 0 | 0.00874411 | 0 | 0 |
| 0 | 0 | 0 | 0 | 0 |
| 0.001759251 | 0.005262626 | 0 | 0 | 0.006872898 |
| 0 | 0 | 0 | 0 | 0 |
| 0 | 0.031842595 | 0 | 0 | 0 |
| 0 | 0 | 0.008660965 | 0 | 0 |
| 0 | 0 | 0 | 0 | 0 |
| 0 | 0 | 0.029416423 | 0 | 0 |
| 0 | 0 | 0 | 0 | 0 |
| 0 | 0 | 0.132475564 | 0.0064309 | 0.017503356 |
| 0.239535361 | 0 | 0 | 0.054905214 | 0.161415653 |
| 0 | 0 | 0 | 0 | 0 |
| 0 | 0 | 0 | 0 | 0 |
| 0 | 0 | 0 | 0 | 0 |
| 0 | 0 | 0 | 0 | 0 |
| 0 | 0.059723631 | 0.005021486 | 0 | 0 |
| 0 | 0 | 0 | 0 | 0 |
| 0 | 0 | 0 | 0 | 0 |
| 0.12083023 | 1.165828453 | 0.569351487 | 0.037380759 | 6.213967713 |
| 0 | 0 | 0.001722991 | 0 | 0 |
| 0.051227131 | 0 | 0 | 0 | 0 |
| 0 | 0 | 0 | 0 | 0 |
| 0 | 0 | 0 | 0 | 0 |
| 0 | 0 | 0.050054177 | 0 | 0 |
| 0 | 0 | 0 | 0 | 0 |
| 0 | 0 | 0 | 0.729663221 | 1.107345441 |
| 0.059942301 | 0.014304047 | 0 | 0 | 0 |
| 0 | 0 | 0 | 0 | 0 |
| 0 | 0 | 0 | 0 | 0 |
| 0.003925661 | 0.004038021 | 0.019036899 | 0 | 0 |
| 0.290711499 | 0 | 6.547535721 | 0.328206684 | 4.385009765 |
| 0.052095075 | 0.015724118 | 0 | 0 | 0 |
| 0 | 0 | 0 | 0 | 0 |
| 0 | 0.009110196 | 0.001385459 | 0 | 0 |
| 0 | 0 | 0 | 0 | 0 |
| 0 | 0.001816368 | 0.001546884 | 0 | 0 |
| 0 | 0 | 0 | 0 | 0 |

| | | | | |
|---|---|---|---|---|
| 0.004281021 | 0.094872964 | 0.000954298 | 0.002021827 | 0.0116408 |
| 0 | 0 | 0 | 0 | 0 |
| 0.00291661 | 0.007634154 | 11.4179698 | 0.001180669 | 0.535664879 |
| 0 | 0 | 0 | 0 | 0 |
| 0 | 0 | 0 | 0 | 0 |
| 0 | 0 | 0 | 0 | 0 |
| 0 | 0 | 0 | 0 | 0 |
| 0.007197234 | 0 | 0 | 0 | 0 |
| 0 | 0 | 0 | 0 | 0 |
| 0 | 0 | 0 | 0 | 0 |
| 0 | 0 | 0 | 0 | 0 |
| 0 | 0 | 0 | 0 | 0 |
| 0 | 0.002755856 | 0 | 0 | 0 |
| 0 | 0 | 0.046763667 | 0 | 0 |
| 0 | 0 | 0 | 0 | 0 |
| 0 | 1.070015452 | 0.007758784 | 0 | 0.002821681 |
| 0.041724118 | 0.420895561 | 0 | 0.095739897 | 0.48164099 |
| 0 | 0.002133679 | 0 | 0 | 0 |
| 0 | 0 | 0.019182017 | 0 | 0 |
| 0 | 0 | 0 | 0 | 0 |
| 0 | 0 | 0.184720616 | 0 | 0 |
| 0 | 0 | 0 | 0 | 0.002720837 |
| 0 | 0 | 0 | 0 | 0 |
| 0.130999659 | 0 | 0.599991604 | 0.031230738 | 0 |
| 0 | 0.006018141 | 0.178335935 | 0 | 0 |
| 0 | 0 | 0 | 0 | 0 |
| 0 | 0 | 0.001265799 | 0 | 0 |
| 0 | 0 | 0 | 0 | 0 |
| 0 | 0 | 0 | 0 | 0 |
| 0 | 0 | 0 | 0 | 0 |
| 0 | 0.016672493 | 0.100475223 | 0 | 0.00603901 |
| 0 | 0.006740784 | 0.007714057 | 0 | 0.013129993 |
| 0 | 0.238674493 | 0.24186681 | 0.037222661 | 1.570906303 |
| 0.005765161 | 0.003449181 | 0 | 0.002222654 | 0 |
| 0 | 0.002872144 | 0.015200275 | 0 | 0.008768503 |
| 0 | 0 | 0 | 0 | 0 |
| 0 | 0.009791615 | 0.008493315 | 0 | 0 |
| 0.001625293 | 0.024390547 | 0.019201899 | 0 | 0.007323642 |
| 0.010413664 | 0.669349684 | 0 | 0.024600839 | 0 |
| 0 | 0.079303293 | 0 | 0 | 0 |
| 0 | 0 | 0 | 0 | 0 |
| 0 | 0 | 0 | 0 | 0 |
| 0 | 0.011027635 | 0 | 0 | 0 |
| 0 | 0.011512454 | 0.001414099 | 0 | 0.064069885 |
| 0 | 1.131202057 | 0 | 0 | 0 |
| 0 | 0 | 0 | 0 | 0.001376443 |
| 0 | 0 | 0 | 0 | 0 |
| 0 | 0 | 0 | 0 | 0 |

| | | | | |
|---|---|---|---|---|
| 0 | 0 | 0 | 0 | 0 |
| 0.141185089 | 0.163470879 | 0.010202714 | 0 | 0.384378615 |
| 0 | 0 | 0 | 0 | 0 |
| 0 | 0 | 0.001096301 | 0 | 0 |
| 0 | 0 | 0 | 0 | 0 |
| 0 | 0 | 0 | 0 | 0 |
| 0 | 0 | 0 | 0 | 0 |
| 0.001275824 | 0 | 0 | 0 | 0.002907499 |
| 0.005575202 | 0 | 0 | 0 | 0 |
| 0 | 0.01516482 | 0 | 0 | 0 |
| 0 | 0.041066604 | 0.014552034 | 0 | 0.002045732 |
| 0.015014685 | 0.267778596 | 0.656463055 | 0 | 0.061324415 |
| 0 | 0 | 0.390955386 | 0.166780069 | 0 |
| 0 | 0 | 0 | 0 | 0 |
| 0 | 0.057003854 | 0.042232675 | 0 | 0 |
| 0 | 0 | 0 | 0 | 0 |
| 0.020464518 | 0.003307897 | 0.011012394 | 0 | 0.017405186 |
| 0 | 0.04489294 | 0.080454354 | 0 | 0 |
| 0 | 0 | 0 | 0 | 0 |
| 0 | 0.02382788 | 0.005504929 | 0 | 0 |
| 0.014661783 | 0 | 0 | 0.029496226 | 0.054675835 |
| 0 | 0 | 0 | 0 | 0 |
| 0 | 0 | 0 | 0 | 0 |
| 0 | 0 | 0 | 0 | 0 |
| 0 | 0.005210528 | 0.001774989 | 0 | 0.006495548 |
| 0 | 0 | 0 | 0 | 0 |
| 0 | 0.009183001 | 0 | 0 | 0 |
| 0.004486105 | 0.101519129 | 0.779940628 | 0.012998836 | 0.041383031 |
| 0 | 0 | 0 | 0 | 0 |
| 0 | 0.008746846 | 0 | 0 | 0 |
| 0 | 0.009902737 | 0 | 0 | 0 |
| 0 | 0 | 0 | 0 | 0 |
| 0 | 0 | 0 | 0 | 0 |
| 0 | 0 | 0 | 0 | 0 |
| 0.04670618 | 0 | 0 | 0.041978682 | 0.01202081 |
| 0.0943064 | 1.023080641 | 0.032067735 | 0.026313432 | 0.136102817 |
| 0.000555217 | 0 | 0 | 0 | 0 |
| 0 | 0 | 0 | 0 | 0 |
| 0.020971371 | 0.019517174 | 0.003116536 | 0.002515375 | 0 |
| 0 | 0 | 0 | 0 | 0 |
| 0 | 1.080298747 | 0 | 0 | 0 |
| 0 | 0 | 0 | 0 | 0 |
| 0.019284418 | 0.083232187 | 0.08171587 | 0 | 0.152845349 |
| 0.001195699 | 0 | 0 | 0 | 0 |
| 0 | 0.009398211 | 0 | 0 | 0 |
| 0.239668302 | 0 | 0 | 0.033149622 | 0.015576022 |
| 0 | 0.001397736 | 0 | 0 | 0 |
| 0 | 0.001927215 | 0.0011489 | 0 | 0 |

| | | | | |
|---|---|---|---|---|
| 0 | 0.003795397 | 0 | 0 | 0 |
| 0 | 0 | 0 | 0 | 0 |
| 0 | 0 | 0 | 0 | 0 |
| 0.068823135 | 0.783855231 | 0.000979904 | 0.006895538 | 0 |
| 0 | 0 | 0 | 0 | 0 |
| 0 | 0 | 0 | 0 | 0 |
| 0 | 0.002230112 | 0.005412844 | 0 | 0 |
| 0 | 0 | 0.004383144 | 0 | 0 |
| 0.086782805 | 0 | 0 | 0.324280881 | 0.346306618 |
| 0 | 0.005398917 | 0.008538979 | 0 | 0 |
| 0 | 0 | 0 | 0 | 0 |
| 0.017768253 | 0 | 0.001525636 | 0 | 0.079266946 |
| 0 | 0 | 0 | 0 | 0 |
| 0.018055542 | 0 | 0 | 0 | 0 |
| 0.002117903 | 0.234250957 | 0.018593852 | 0.02444534 | 0.015795914 |
| 0 | 0 | 0 | 0 | 0 |
| 0 | 0 | 0 | 0.009176059 | 0 |
| 0.034464414 | 0 | 0 | 0.031232971 | 0.167396734 |
| 0 | 0 | 0 | 0 | 0 |
| 0 | 0 | 0 | 0 | 0 |
| 0.067635344 | 0 | 0 | 0 | 0 |
| 0 | 0 | 0 | 0 | 0 |
| 0 | 0 | 0 | 0 | 0 |
| 0.020086978 | 0.034175164 | 1.003555811 | 0.00355748 | 0.224721437 |
| 0 | 0 | 0.038126896 | 0.00438973 | 0 |
| 0 | 0 | 0 | 0.064159621 | 0 |
| 0 | 0 | 0 | 0.007834651 | 0 |
| 0.575216719 | 0 | 0.596527441 | 1.65632817 | 1.530938979 |
| 0 | 0 | 0 | 0 | 0 |
| 0 | 0 | 0 | 0 | 0 |
| 0 | 0 | 0 | 0.001035937 | 0.008588841 |
| 0 | 0 | 0 | 0.00175688 | 0 |
| 0 | 0.002243083 | 0 | 0 | 0 |
| 0 | 0 | 0 | 0 | 0 |
| 0 | 0 | 0 | 0 | 0 |
| 0.000976415 | 0 | 0 | 0 | 0 |
| 0.548134403 | 0 | 0 | 0.144296265 | 0.083863289 |
| 0 | 0.092622572 | 0 | 0 | 0 |
| 0 | 0 | 0 | 0 | 0 |
| 0 | 0 | 0 | 0 | 0 |
| 0.113257629 | 0.003965874 | 0.043682085 | 0 | 0.033901285 |
| 0.186040135 | 0 | 0.184803929 | 0 | 0 |
| 0.175626671 | 1.127222817 | 4.843810714 | 1.478927171 | 20.60013541 |
| 0 | 0 | 0 | 0 | 0 |
| 0 | 0 | 0 | 0 | 0 |
| 0 | 0 | 0 | 0 | 0 |
| 0 | 0 | 0 | 0 | 0 |
| 0 | 0.001589145 | 0.000947361 | 0.003195023 | 0 |

| | | | | |
|---|---|---|---|---|
| 0 | 0 | 0 | 0 | 0 |
| 0 | 0 | 0 | 0.036944222 | 0 |
| 0 | 0.175188131 | 0.124676385 | 0 | 0.003255548 |
| 0 | 0 | 0 | 0 | 0 |
| 0 | 7.863052753 | 0 | 0 | 0 |
| 0 | 0 | 0 | 0 | 0 |
| 0 | 0 | 0 | 0 | 0 |
| 0 | 0 | 0 | 0 | 0 |
| 0 | 0 | 0 | 0 | 0 |
| 0 | 0 | 0 | 0 | 0 |
| 0.0012857 | 0.122646101 | 0.156929495 | 0.046263343 | 0.027169151 |
| 0.380211973 | 0.507950771 | 3.247848075 | 0.519635089 | 0.971299865 |
| 0 | 0 | 0 | 0 | 0 |
| 0.072847963 | 0.028087138 | 0 | 0.038944843 | 0.449029912 |
| 0.002915911 | 0.061567419 | 0.1252326 | 0 | 0.002944998 |
| 0 | 0 | 0 | 0.269836129 | 0 |
| 0 | 0 | 0 | 0 | 0 |
| 0 | 0 | 0 | 0 | 0 |
| 0 | 0 | 0 | 0 | 0 |
| 0 | 0 | 0.091390382 | 0 | 0 |
| 0.015266088 | 0.465337647 | 0.131530026 | 0 | 0 |
| 0 | 0.084802374 | 0.023623609 | 0 | 2.590685122 |
| 0.386773577 | 0.073411655 | 0.327725824 | 0.152357479 | 4.024614214 |
| 0 | 0 | 0 | 0 | 0 |
| 0 | 0 | 0 | 0.000570057 | 0 |
| 0.004519534 | 0 | 0 | 0.580343765 | 0.700643785 |
| 0 | 0 | 3.172497528 | 0 | 0 |
| 0 | 0 | 0 | 0.000808843 | 0 |
| 0 | 0 | 0 | 0 | 0 |
| 0 | 0 | 0 | 0 | 0 |
| 0.001386667 | 0 | 0 | 0 | 0 |
| 0.056801645 | 0 | 0.318600133 | 0.11348182 | 0 |
| 0.001760597 | 0 | 0 | 0 | 0 |
| 0.000671296 | 0 | 0.01227056 | 0.001086985 | 0 |
| 0.000581008 | 0.138897371 | 0.002503949 | 0 | 0 |
| 0 | 0.362171451 | 0.006287395 | 0.010556114 | 0.306616376 |
| 0 | 0.012379402 | 0.001375141 | 0.000832413 | 0.001198168 |
| 0 | 0 | 0 | 0 | 0 |
| 0.007006023 | 0 | 0 | 0.087982081 | 0.179982127 |
| 0 | 0 | 0 | 0 | 0 |
| 0 | 0 | 0 | 0 | 0.001092197 |
| 0 | 0 | 0 | 0 | 0 |
| 0 | 0 | 0 | 0 | 0 |
| 0 | 0 | 0 | 0 | 0 |
| 0.000910336 | 0.164253059 | 0.615948716 | 0.147896039 | 0.026875252 |
| 0.03307003 | 0 | 0 | 0 | 0 |
| 0 | 0 | 0 | 0 | 0 |
| 0 | 13.73141952 | 0 | 0.042970832 | 0.014673817 |

| | | | | |
|---|---|---|---|---|
| 0 | 0 | 0 | 0 | 0 |
| 0 | 0 | 0 | 0 | 0 |
| 0 | 0 | 0 | 0 | 0 |
| 0 | 0 | 0 | 0 | 0.001687786 |
| 0 | 0 | 0 | 0 | 0 |
| 0 | 0 | 0 | 0.018492959 | 0 |
| 0.142917569 | 0 | 0 | 0 | 0 |
| 0.001249405 | 0.295011072 | 0.012307464 | 0.003468137 | 0.005546674 |
| 0 | 0 | 0 | 0 | 0 |
| 0 | 0 | 0 | 0 | 0 |
| 0 | 0.022316548 | 0.002054657 | 0.002565223 | 0 |
| 0 | 0 | 0 | 0 | 0 |
| 0 | 0.148133433 | 0.001686574 | 0.033894985 | 0 |
| 0 | 0 | 0 | 0 | 0 |
| 0 | 0 | 0 | 0 | 0 |
| 0 | 0 | 0 | 0 | 0 |
| 0.071819216 | 0 | 0 | 0.002081291 | 0 |
| 0 | 0 | 0 | 0.001358948 | 0.001564846 |
| 0 | 0 | 0 | 0 | 0 |
| 0 | 0.006339463 | 0 | 0 | 0 |
| 0 | 0 | 0 | 0 | 0 |
| 0 | 0 | 0 | 0 | 0 |
| 0.001658741 | 0 | 0 | 0 | 0.002319629 |
| 0 | 0 | 0 | 0 | 0 |
| 0 | 0 | 0 | 0 | 0 |
| 0 | 0 | 0 | 0 | 0 |
| 0 | 0 | 0 | 0 | 0 |
| 0 | 0 | 0 | 0 | 0 |
| 0 | 0 | 0 | 0 | 0 |
| 0.008472559 | 0 | 0.103827258 | 0 | 0.052118832 |
| 0 | 0.02327734 | 0 | 0 | 0 |
| 0 | 0 | 0 | 0 | 0 |
| 0 | 0 | 0 | 0.005679476 | 0 |
| 0 | 0 | 0 | 0 | 0 |
| 0 | 0 | 0 | 0 | 0 |
| 0 | 0 | 0 | 0 | 0 |
| 0.012840306 | 0 | 0 | 0.022544963 | 0.08653607 |
| 0 | 0.02195481 | 0.025002478 | 0 | 0.005228355 |
| 0 | 0 | 0 | 0 | 0 |
| 0 | 0 | 0 | 0 | 0 |
| 0 | 0.415094719 | 0 | 0.001111315 | 0.031565769 |
| 0 | 0 | 0 | 0 | 0 |
| 0 | 0 | 0 | 0 | 0 |
| 0.004439786 | 0.045616428 | 0.002111336 | 0.081667672 | 0 |
| 0.02740987 | 0 | 0 | 0.029364515 | 0.144288672 |
| 0 | 0 | 0 | 0 | 0 |
| 0 | 0 | 0 | 0 | 0 |
| 0 | 0 | 0 | 0 | 0 |

| | | | | |
|---|---|---|---|---|
| 0 | 0 | 0 | 0 | 0 |
| 0 | 0 | 0 | 0 | 0 |
| 0 | 0 | 0 | 0 | 0.014204653 |
| 0 | 0 | 0 | 0 | 0.026003405 |
| 0 | 0 | 0 | 0 | 0 |
| 0 | 0 | 0 | 0 | 0 |
| 0 | 0 | 0 | 0 | 0 |
| 0 | 0 | 0 | 0 | 0 |
| 0 | 0.021829299 | 0.072113596 | 0 | 0.010972919 |
| 0 | 0 | 0 | 0 | 0 |
| 0 | 0 | 0 | 0 | 0 |
| 0 | 0 | 0 | 0 | 0 |
| 0 | 0 | 0 | 0 | 0 |
| 0 | 0 | 0.00520663 | 0 | 0 |
| 0.007612357 | 0 | 0 | 0.237523767 | 0.006308343 |
| 0 | 0 | 0.000808856 | 0 | 0 |
| 0 | 0 | 0 | 0 | 0 |
| 0 | 0 | 0 | 0 | 0 |
| 0 | 0 | 0 | 0 | 0 |
| 0 | 0 | 0 | 0 | 0 |
| 0 | 0 | 0.007336873 | 0 | 0 |
| 0 | 0 | 0 | 0 | 0 |
| 0 | 0 | 0 | 0 | 0 |
| 0 | 0 | 0 | 0 | 0 |
| 0.287400358 | 0.197498263 | 0.996133003 | 0.181536972 | 0.011060434 |
| 10.37896265 | 7.008931185 | 0.188610257 | 0.008751462 | 0.004864963 |
| 0 | 0 | 0 | 0.192678977 | 0 |
| 0 | 0 | 0.008208134 | 0 | 0 |
| 0.002852395 | 0 | 0 | 0.05150547 | 0 |
| 0 | 0 | 0 | 0 | 0 |
| 0 | 0 | 0 | 0 | 0 |
| 0 | 0 | 0 | 0 | 0 |
| 0 | 0 | 0 | 0 | 0 |
| 0 | 0.00796051 | 0 | 0.003419838 | 0 |
| 0.020879541 | 0.710107608 | 0 | 0.004226109 | 0.021087821 |
| 0 | 0.032193432 | 0.011076502 | 0 | 0.001605319 |
| 0 | 0.489835949 | 0 | 0 | 0 |
| 0.154783667 | 0 | 0.196525358 | 0.306514252 | 0 |
| 0 | 0 | 0 | 0 | 0 |
| 0 | 0 | 0 | 0 | 0 |
| 0 | 0 | 0 | 0 | 0.317245441 |
| 0.00271979 | 0 | 0.009078628 | 0 | 0 |
| 0.039329261 | 0 | 0.095613061 | 0.082189556 | 0.159199716 |
| 0 | 0 | 0 | 0.597743722 | 0 |
| 0 | 0 | 0 | 0 | 0 |
| 0 | 0.007319817 | 0.003324703 | 0 | 0.001520838 |
| 0 | 0 | 0 | 0 | 0 |
| 0.165955742 | 1.000543417 | 0.045187054 | 0 | 0 |

| | | | | |
|---|---|---|---|---|
| 0 | 0.024058595 | 0 | 0.000896581 | 0 |
| 0 | 0 | 0 | 0 | 0 |
| 0 | 0 | 0 | 0 | 0 |
| 0 | 0 | 0 | 0 | 0 |
| 0 | 0.264061495 | 0.316435499 | 0 | 0 |
| 0 | 0 | 0 | 0 | 0 |
| 0 | 0 | 0 | 0 | 0 |
| 0 | 0 | 0 | 0 | 0 |
| 0 | 0 | 0 | 0 | 0 |
| 0 | 0 | 0 | 0 | 0 |
| 0 | 0 | 0 | 0.001195309 | 0.001376414 |
| 0.190911944 | 0 | 0.429224977 | 0.221528596 | 3.063013591 |
| 0.002753446 | 0.115662675 | 0 | 0.001351053 | 0 |
| 0 | 0 | 0 | 0 | 0 |
| 4.033804039 | 0.834391 | 0.017480695 | 0.01367863 | 0.018128649 |
| 0 | 0 | 0 | 0 | 0 |
| 0 | 0 | 0.031340085 | 0 | 0 |
| 0 | 0 | 0 | 0 | 0 |
| 0 | 0.014919351 | 0.001370239 | 0.002262126 | 0.007033144 |
| 0 | 0 | 0 | 0 | 0 |
| 0.001561432 | 0 | 0 | 0.004307517 | 0 |
| 0 | 0 | 0 | 0 | 0 |
| 0.008558085 | 1.275481849 | 0.05196249 | 0.001319766 | 0 |
| 0 | 0 | 0 | 0 | 0 |
| 0 | 0 | 0 | 0 | 0 |
| 0 | 0 | 0 | 0 | 0 |
| 0 | 0 | 0 | 0 | 0 |
| 0 | 0 | 0 | 0 | 0 |
| 0 | 0 | 0 | 0 | 0 |
| 0 | 0 | 0 | 0 | 0 |
| 0 | 0 | 0 | 0 | 0 |
| 0.045605764 | 0.010394294 | 0.039097016 | 0 | 0.053219265 |
| 0.096610377 | 0 | 0 | 0 | 0 |
| 0 | 0.077459421 | 0 | 0.00200775 | 0.021963527 |
| 0 | 0.030269826 | 0.008115571 | 0 | 0.00074871 |
| 0.004658768 | 0.039744097 | 0 | 0.008102426 | 0.031207408 |
| 0 | 0 | 0 | 0 | 0 |
| 0 | 0 | 0 | 0 | 0 |
| 0 | 0 | 0 | 0 | 0.03670003 |
| 0 | 0 | 0 | 0 | 0 |
| 0 | 0 | 0 | 0 | 0 |
| 0 | 0 | 0 | 0 | 0 |
| 0 | 0 | 0 | 0 | 0 |
| 0 | 0 | 0.004469321 | 0 | 0 |
| 0 | 0.075099232 | 0.003879757 | 0.010675144 | 0.006637987 |
| 0 | 0 | 0 | 0 | 0 |
| 0.084702038 | 0 | 0 | 0.041316394 | 0 |
| 0 | 0.013318207 | 0 | 0 | 0 |

| | | | | |
|---|---|---|---|---|
| 0 | 0 | 0 | 0 | 0 |
| 0 | 0 | 0 | 0 | 0 |
| 0 | 0 | 0.018968842 | 0 | 0 |
| 0 | 0 | 0 | 0 | 0 |
| 0 | 0 | 0 | 0 | 0 |
| 0 | 0 | 0 | 0 | 0 |
| 0.097723904 | 0 | 0 | 0.033057384 | 0.365265772 |
| 0 | 0 | 0 | 0 | 0 |
| 0 | 0 | 0 | 0 | 0 |
| 0.005702996 | 0 | 0.001921038 | 0 | 0 |
| 0 | 0 | 0 | 0 | 0 |
| 0 | 0.007844767 | 0 | 0 | 0 |
| 0 | 0 | 0 | 0 | 0 |
| 0 | 0 | 0 | 0.048287483 | 0.051106314 |
| 0 | 0 | 0 | 0 | 0 |
| 0 | 0 | 0 | 0 | 0 |
| 0 | 0 | 0 | 0 | 0 |
| 0 | 0 | 0 | 0 | 0 |
| 0 | 0.088486716 | 0 | 0.135198487 | 0.001358489 |
| 0 | 0 | 0 | 0 | 0 |
| 0 | 0.038918064 | 0 | 0 | 0 |
| 0 | 0 | 0 | 0 | 0 |
| 0 | 0 | 0 | 0 | 0 |
| 0 | 0 | 0 | 0 | 0 |
| 0 | 0 | 0 | 0 | 0 |
| 0 | 0.071713818 | 0.000985066 | 0 | 0 |
| 0 | 0 | 0 | 0 | 0 |
| 0.004866038 | 0.049563197 | 0.198514617 | 0 | 0 |
| 0 | 0 | 0 | 0 | 0 |
| 0.006206147 | 0.6881788 | 0.271901011 | 0.018468814 | 0.066511754 |
| 0 | 0 | 0 | 0 | 0 |
| 0 | 0 | 0 | 0 | 0 |
| 0 | 0 | 0 | 0 | 0 |
| 0 | 0 | 0 | 0 | 0 |
| 0 | 0.027205434 | 0.002408671 | 0 | 0.001439102 |
| 0 | 0 | 0 | 0 | 0 |
| 0 | 0 | 0 | 0 | 0 |
| 0 | 0 | 0.001431643 | 0.002836197 | 0 |
| 0 | 0 | 0 | 0 | 0 |
| 0 | 0 | 0 | 0.000414949 | 0 |
| 1.338116459 | 17.23438815 | 1.097166534 | 0.321520866 | 4.765360823 |
| 0.011058585 | 0.22999847 | 0.030581645 | 0.002768146 | 0.115947351 |
| 0.004702784 | 0 | 0 | 0 | 0.291821291 |
| 0 | 0 | 0 | 0 | 0 |
| 0 | 0 | 0 | 0 | 0 |
| 0 | 0 | 0 | 0 | 0 |
| 0 | 0 | 0 | 0 | 0 |
| 0 | 0.011401129 | 0.014254535 | 0.00712805 | 0.00583203 |

| | | | | |
|---|---|---|---|---|
| 0 | 0.160199788 | 0 | 0 | 0 |
| 0 | 0.001497064 | 0.005843537 | 0 | 0.001555225 |
| 0 | 0 | 0 | 0 | 0 |
| 0 | 0 | 0 | 0 | 0 |
| 0.077421875 | 0 | 1.622192077 | 0.32218944 | 2.525060482 |
| 0 | 0 | 0 | 0 | 0 |
| 0 | 0 | 0 | 0 | 0 |
| 0 | 0 | 0 | 0 | 0 |
| 0 | 0.171161121 | 0.096965795 | 0.000963287 | 0 |
| 0 | 0 | 0 | 0 | 0 |
| 0 | 0 | 0 | 0 | 0 |
| 0 | 0 | 0 | 0 | 0 |
| 0.005383794 | 0 | 0 | 0.008623888 | 0.00259057 |
| 0.007045849 | 0.066466929 | 0.003426798 | 0 | 0.320076706 |
| 0 | 0 | 0 | 0 | 0 |
| 0.015242855 | 0.366142576 | 0.206109594 | 0.004539174 | 0.04726005 |
| 0.007575171 | 0.002063384 | 0 | 0 | 0 |
| 0 | 0 | 0 | 0 | 0 |
| 0 | 0 | 0 | 0 | 0 |
| 0 | 0 | 0 | 0 | 0 |
| 0 | 0 | 0 | 0 | 0 |
| 0 | 0 | 0 | 0 | 0 |
| 0 | 0 | 0 | 0 | 0 |
| 0 | 0 | 0 | 0 | 0 |
| 0 | 0 | 0 | 0 | 0 |
| 0 | 0 | 0 | 0 | 0 |
| 0.012531219 | 0 | 0.008595014 | 0.044223938 | 0.110036739 |
| 0 | 0.00430217 | 0 | 0 | 0 |
| 0 | 0 | 0 | 0 | 0 |
| 0 | 0 | 0 | 0 | 0 |
| 0 | 0 | 0 | 0 | 0 |
| 0 | 0 | 0.00144022 | 0 | 0 |
| 0 | 0 | 0 | 0 | 0 |
| 0.010831883 | 0 | 0 | 0.006564439 | 0 |
| 0 | 0 | 0 | 0 | 0 |
| 0 | 0.001756488 | 0.28683666 | 0 | 0.09775328 |
| 0 | 0.00239379 | 0 | 0 | 0 |
| 0 | 0 | 0 | 0 | 0 |
| 0 | 0 | 0 | 0 | 0 |
| 0 | 0 | 0 | 0 | 0 |
| 1.802481885 | 0 | 0 | 0.037429137 | 0.6161724 |
| 0.00917277 | 0 | 0 | 0 | 0 |
| 0.026748636 | 0 | 0 | 0 | 0 |
| 0 | 0 | 0 | 0 | 0.044128102 |
| 0 | 0 | 0 | 0 | 0 |
| 0 | 0 | 0 | 0 | 0 |
| 0 | 0 | 0 | 0 | 0 |
| 0 | 0 | 0 | 0 | 0 |

| | | | | |
|---|---|---|---|---|
| 0.007939519 | 0 | 0 | 0.009641958 | 0 |
| 0 | 0 | 0 | 0 | 0 |
| 0.001520801 | 0 | 0 | 0 | 0 |
| 0 | 0 | 0 | 0 | 0 |
| 0.001768388 | 0.04333847 | 0.074401194 | 0.004878445 | 0 |
| 0 | 0 | 0 | 0 | 0 |
| 0.021011456 | 0.020326367 | 0.005368497 | 0 | 0 |
| 0 | 0 | 0 | 0 | 0 |
| 0 | 0.002905749 | 0 | 0 | 0 |
| 0 | 0.009987971 | 0 | 0 | 0 |
| 0 | 0 | 0 | 0.001930191 | 0.00250047 |
| 0.001128284 | 0 | 0 | 0 | 0 |
| 0 | 0 | 0 | 0 | 0 |
| 0 | 0 | 0 | 0 | 0 |
| 0 | 0 | 0 | 0 | 0 |
| 0 | 0 | 0 | 0 | 0 |
| 0 | 0 | 0 | 0 | 0 |
| 0 | 0 | 0.003159947 | 0 | 0 |
| 0 | 0 | 0 | 0 | 0 |
| 0 | 0 | 0 | 0 | 0 |
| 1.424149936 | 0.569286877 | 0.0025034 | 0 | 0 |
| 0 | 0 | 0 | 0 | 0 |
| 0 | 0.011347978 | 0.026461899 | 0.005850114 | 0 |
| 0 | 0.019460711 | 0.017903408 | 0 | 0.005348349 |
| 0 | 0 | 0 | 0 | 0 |
| 0 | 0.009353774 | 0 | 0.001315108 | 0.002776334 |
| 0 | 0 | 0 | 0 | 0 |
| 0 | 0 | 0 | 0 | 0 |
| 0.012081919 | 0 | 0 | 0 | 0.027414517 |
| 0 | 2.922222865 | 0.7016041 | 0.024064945 | 0.012623945 |
| 0 | 0 | 0 | 0 | 0 |
| 0 | 0 | 0 | 0 | 0 |
| 0 | 0 | 0 | 0 | 0 |
| 0 | 0.437979678 | 0 | 0 | 0 |
| 0 | 0 | 0 | 0 | 0 |
| 0 | 0 | 0 | 0 | 0 |
| 0.000646042 | 0 | 0 | 0 | 0 |
| 0 | 0 | 0 | 0 | 0 |
| 0 | 0 | 0 | 0.003427413 | 0 |
| 0 | 0 | 0 | 0 | 0 |
| 0 | 0 | 0 | 0 | 0 |
| 0 | 0 | 0 | 0 | 0 |
| 0 | 0 | 0 | 0 | 0 |
| 0 | 0 | 0.009650069 | 0 | 0 |
| 0.067664924 | 0 | 0 | 0 | 0 |
| 0 | 0 | 0 | 0 | 0 |
| 0 | 0 | 0 | 0 | 0 |
| 0 | 0 | 0 | 0 | 0 |

| | | | | |
|---|---|---|---|---|
| 0 | 0.062304201 | 0 | 0 | 0 |
| 0 | 0 | 0 | 0 | 0 |
| 0 | 0 | 0 | 0 | 0 |
| 0 | 0 | 0 | 0 | 0 |
| 0 | 0 | 0 | 0 | 0 |
| 0 | 0 | 0 | 0 | 0 |
| 0 | 0 | 0 | 0 | 0 |
| 0.000950744 | 1.644168587 | 0.435283941 | 0.100168713 | 0.487146012 |
| 12.486641 | 0.155418223 | 1.818132134 | 0.581140677 | 0.28798343 |
| 0 | 0.217525009 | 0.147239342 | 0 | 0.382104593 |
| 0.003799563 | 0 | 0 | 0.000769048 | 0 |
| 0 | 0 | 0 | 0 | 0 |
| 0 | 0 | 0 | 0 | 0 |
| 0 | 0 | 0 | 0 | 0 |
| 0 | 0 | 0 | 0 | 0 |
| 0 | 0 | 0 | 0 | 0 |
| 0 | 0 | 0 | 0 | 0 |
| 0 | 0 | 0 | 0 | 0.281112462 |
| 0 | 0 | 0 | 0 | 0 |
| 0 | 0 | 0 | 0 | 0.00750929 |
| 0 | 0 | 0 | 0 | 0 |
| 0 | 0 | 0 | 0 | 0 |
| 0 | 0 | 0 | 0 | 0 |
| 0 | 0 | 0 | 0 | 0 |
| 0.003646582 | 0.008144929 | 0 | 0.001259663 | 0 |
| 0 | 0 | 0 | 0 | 0 |
| 0 | 0 | 0 | 0 | 0 |
| 0 | 0 | 0 | 0 | 0 |
| 0 | 0 | 0 | 0 | 0 |
| 0 | 0 | 0 | 0 | 0 |
| 0 | 0 | 0 | 0 | 0 |
| 0 | 0 | 0 | 0 | 0 |
| 0 | 0 | 0 | 0 | 0 |
| 0 | 0 | 0 | 0 | 0 |
| 0.004035587 | 0 | 0 | 0.01210104 | 0 |
| 0 | 0 | 0 | 0 | 0 |
| 0 | 0 | 0 | 0 | 0 |
| 0 | 0 | 0 | 0 | 0 |
| 0 | 0 | 0 | 0 | 0.064325705 |
| 0 | 0 | 0 | 0 | 0 |
| 0 | 0 | 0 | 0 | 0 |
| 0.179099863 | 0 | 0 | 0.168011234 | 0.283325037 |
| 0 | 0 | 0 | 0 | 0 |
| 0 | 0 | 0 | 0 | 0 |
| 0 | 0 | 0 | 0 | 0.00134368 |
| 0 | 0 | 0 | 0 | 0 |
| 0 | 0 | 0 | 0 | 0 |
| 0 | 0 | 0 | 0 | 0 |

| | | | | |
|---|---|---|---|---|
| 0 | 0 | 0 | 0 | 0 |
| 0 | 0.003266605 | 0.000496778 | 0 | 0 |
| 0 | 0 | 0 | 0 | 0 |
| 0 | 0 | 0 | 0 | 0 |
| 0.004440088 | 2.234652307 | 0 | 0.354179666 | 0.15018133 |
| 0 | 0 | 0 | 0 | 0 |
| 0 | 0.006790166 | 0 | 0 | 0 |
| 0 | 0.013812154 | 0.15417836 | 0 | 0.00673513 |
| 0 | 0 | 0 | 0 | 0 |
| 0.006662417 | 0 | 0 | 0 | 0 |
| 0 | 0 | 0 | 0 | 0 |
| 0 | 0 | 0 | 0 | 0 |
| 0 | 0 | 0 | 0 | 0 |
| 0 | 0 | 0 | 0 | 0 |
| 0 | 0 | 0 | 0 | 0 |
| 0 | 0.107242165 | 0 | 0.001543878 | 0.003809562 |
| 0 | 0 | 0 | 0 | 0 |
| 0 | 0 | 0 | 0 | 0 |
| 0 | 0 | 0 | 0 | 0 |
| 0 | 0 | 0 | 0 | 0 |
| 0 | 0 | 0 | 0 | 0 |
| 0 | 0 | 0 | 0 | 0 |
| 0 | 0 | 0 | 0 | 0 |
| 0 | 0.009961068 | 0.00084832 | 0.000880309 | 0.00152053 |
| 0 | 0 | 0 | 0.17857598 | 0.199770249 |
| 0 | 0 | 0 | 0 | 0 |
| 0 | 0.001822105 | 0.006428763 | 0 | 0.004172093 |
| 0 | 0.001096353 | 0 | 0 | 0 |
| 0 | 0 | 0 | 0 | 0 |
| 0 | 0 | 0 | 0 | 0 |
| 0 | 0 | 0 | 0 | 0 |
| 0 | 0 | 0 | 0 | 0 |
| 0 | 0 | 0 | 0 | 0 |
| 0 | 0 | 0 | 0 | 0 |
| 0 | 0 | 0 | 0 | 0 |
| 0 | 0 | 0 | 0 | 0 |
| 0 | 0 | 0 | 0 | 0 |
| 0 | 0 | 0.109581432 | 0 | 0 |
| 0 | 0 | 0 | 0 | 0 |
| 0 | 0 | 0 | 0 | 0 |
| 0 | 0 | 0 | 0 | 0 |
| 0 | 0 | 0 | 0.080478413 | 0 |
| 0 | 0 | 0 | 0 | 0 |
| 0 | 0 | 0 | 0 | 0 |
| 0 | 0 | 0 | 0 | 0 |
| 0 | 0 | 0 | 0 | 0 |
| 0 | 0 | 0 | 0 | 0 |
| 0 | 0 | 0 | 0 | 0 |

| | | | | |
|---|---|---|---|---|
| 0.021393061 | 0 | 0 | 0.003596543 | 0.247180123 |
| 0 | 0 | 0 | 0 | 0 |
| 0 | 0 | 0 | 0 | 0 |
| 0 | 0 | 0.001037092 | 0 | 0 |
| 0 | 0 | 0 | 0 | 0 |
| 0 | 0 | 0 | 0 | 0.044950883 |
| 0 | 0 | 0 | 0 | 0 |
| 0 | 0 | 0 | 0 | 0 |
| 0 | 0 | 0 | 0 | 0 |
| 0 | 0 | 0 | 0 | 0 |
| 0 | 0 | 0 | 0 | 0 |
| 0 | 0 | 0.022343316 | 0 | 0 |
| 0 | 0 | 0 | 0 | 0 |
| 0 | 0 | 0 | 0 | 0 |
| 0 | 0 | 0 | 0 | 0 |
| 0 | 0 | 0 | 0 | 0 |
| 0 | 0 | 0 | 0 | 0 |
| 0 | 0.162317809 | 0 | 0 | 0 |
| 0 | 0 | 0 | 0 | 0 |
| 0 | 0 | 0 | 0 | 0 |
| 0 | 0.206607515 | 0.00060341 | 0 | 0 |
| 0 | 0 | 0 | 0 | 0 |
| 0 | 0.068006511 | 0.017527449 | 0.038749139 | 0.017848057 |
| 0 | 0 | 0.000557984 | 0 | 0 |
| 0 | 0 | 0 | 0 | 0 |
| 0 | 0 | 0 | 0.039456556 | 0 |
| 0 | 0 | 0 | 0 | 0 |
| 0 | 0 | 0 | 0 | 0 |
| 0 | 0 | 0 | 0 | 0 |
| 0 | 0 | 0 | 0 | 0 |
| 0 | 0 | 0 | 0 | 0 |
| 0.052850743 | 0 | 0.267575797 | 0 | 0 |
| 0 | 0.008044777 | 0.007993091 | 0 | 1.312397041 |
| 0 | 0 | 0 | 0 | 0 |
| 0.00236261 | 0.046774096 | 0 | 0 | 0 |
| 0.000963416 | 0.002958815 | 0.002634371 | 0.002287994 | 0.006227367 |
| 0 | 0 | 0 | 0 | 0 |
| 0 | 0 | 0 | 0 | 0 |
| 0 | 0.168627904 | 0 | 0 | 0 |
| 0 | 0 | 0 | 0 | 0 |
| 0 | 0 | 0 | 0 | 0 |
| 0.014351197 | 0 | 0 | 0 | 0 |
| 0.005733202 | 0 | 0 | 0.032069926 | 0.04859071 |
| 0 | 0 | 0 | 0 | 0 |
| 0 | 0 | 0 | 0 | 0 |
| 0.007036697 | 0 | 0 | 0 | 0 |
| 0 | 0 | 0 | 0 | 0 |
| 0 | 0 | 0 | 0 | 0 |

| | | | | |
|---|---|---|---|---|
| 0 | 0 | 0 | 0 | 0 |
| 0 | 0 | 0 | 0 | 0 |
| 0 | 0 | 0 | 0 | 0 |
| 0 | 0 | 0 | 0 | 0 |
| 0 | 0 | 0 | 0 | 0 |
| 0 | 0 | 0 | 0 | 0 |
| 0 | 0 | 2.410768304 | 0 | 0 |
| 0 | 0 | 0 | 0 | 0 |
| 0 | 0 | 0 | 0 | 0 |
| 0 | 0.006844343 | 0 | 0 | 0.36566972 |
| 0 | 0 | 0 | 0 | 0 |
| 0 | 0 | 0 | 0 | 0 |
| 0 | 0 | 0 | 0 | 0 |
| 0.000916722 | 0 | 0 | 0 | 0 |
| 0 | 0 | 0.087144932 | 0 | 0 |
| 0 | 0.004099003 | 0.001122062 | 0 | 0 |
| 0 | 0 | 0 | 0 | 0 |
| 0 | 0 | 0 | 0 | 0 |
| 0 | 0 | 0 | 0 | 0 |
| 0.343992437 | 4.844006855 | 1.710589362 | 0.002464202 | 0 |
| 0.003723889 | 0 | 0 | 0.141642341 | 0.002722922 |
| 0 | 0 | 0 | 0 | 0 |
| 0 | 0 | 0 | 0 | 0 |
| 0 | 0 | 0 | 0 | 0 |
| 0 | 0 | 0.007750981 | 0 | 0 |
| 0 | 0 | 0 | 0 | 0 |
| 0 | 0 | 0 | 0 | 0 |
| 0 | 0 | 0 | 0 | 0 |
| 0 | 0 | 0 | 0 | 0 |
| 0.074776378 | 0 | 0 | 0.001173261 | 0 |
| 0.009205159 | 0.213457462 | 0.121496023 | 0.070846232 | 0.080097085 |
| 0 | 0 | 0 | 0 | 0 |
| 0 | 0 | 0 | 0 | 0 |
| 0 | 0 | 0 | 0 | 0 |
| 0 | 0 | 0 | 0 | 0 |
| 0 | 0 | 0 | 0 | 0 |
| 0 | 0 | 0 | 0 | 0 |
| 0 | 0 | 0 | 0 | 0 |
| 0 | 0 | 0 | 0 | 0.010030283 |
| 0 | 0 | 0 | 0 | 0 |
| 0 | 0 | 0 | 0 | 0 |
| 0 | 0 | 0 | 0 | 0 |
| 0 | 0 | 0 | 0 | 0 |
| 0 | 0 | 0 | 0 | 0 |
| 0 | 0 | 0 | 0 | 0 |
| 0 | 0 | 0 | 0 | 0 |
| 0 | 0.00419612 | 0.008040524 | 0 | 0 |
| 0 | 0 | 0 | 0.001318514 | 0 |

| | | | | |
|---|---|---|---|---|
| 0 | 0 | 0 | 0 | 0 |
| 0 | 0 | 0 | 0 | 0 |
| 0 | 0 | 0 | 0 | 0 |
| 0 | 0 | 0 | 0 | 0 |
| 0.001122795 | 1.036280168 | 0.002955762 | 0.001212046 | 0 |
| 0 | 0 | 0 | 0 | 0 |
| 0 | 0 | 0 | 0 | 0 |
| 0 | 0 | 0 | 0 | 0 |
| 0 | 0 | 0 | 0 | 0 |
| 0 | 0 | 0 | 0 | 0 |
| 0 | 0 | 0 | 0 | 0 |
| 0 | 0.015854567 | 0.013502319 | 0.006009818 | 0.001937708 |
| 0 | 0.00321847 | 0 | 0 | 0.001671754 |
| 0 | 0 | 0 | 0 | 0 |
| 0 | 0 | 0 | 0 | 0 |
| 0.030478604 | 0.015955401 | 3.522635971 | 0 | 0 |
| 0 | 0 | 0.009570385 | 0.001479124 | 0.012561329 |
| 0 | 0 | 0 | 0 | 0 |
| 0 | 0.004237118 | 0.114368841 | 0 | 0.003772911 |
| 0.24681081 | 0.009220643 | 0 | 0 | 0 |
| 0 | 0 | 0 | 0 | 0 |
| 0.001351286 | 0 | 0 | 0 | 0 |
| 0 | 0 | 0 | 0 | 0 |
| 0 | 0 | 0 | 0 | 0 |
| 0 | 0 | 0 | 0 | 0 |
| 0 | 0 | 0 | 0 | 0 |
| 0 | 0.06182066 | 0.014158514 | 0 | 0.002193138 |
| 0 | 0 | 0 | 0 | 0 |
| 0 | 0 | 0 | 0 | 0 |
| 0 | 0 | 0 | 0 | 0 |
| 0 | 0.053046308 | 0.001426615 | 0 | 0 |
| 0 | 0 | 0 | 0 | 0 |
| 0 | 0 | 0 | 0 | 0 |
| 0 | 0 | 0 | 0 | 0 |
| 0 | 0 | 0 | 0 | 0 |
| 0 | 0 | 0 | 0 | 0 |
| 0 | 0 | 0 | 0 | 0 |
| 0 | 0.033345765 | 0.012695778 | 0.013651218 | 0.017291512 |
| 0.252663917 | 0 | 0 | 0.006567847 | 0.001050411 |
| 0 | 0 | 0 | 0 | 0 |
| 0 | 0 | 0 | 0 | 0.002789899 |
| 0 | 0 | 0 | 0 | 0 |
| 0 | 0 | 0 | 0 | 0 |
| 0 | 0.185864582 | 0.000990854 | 0 | 0 |
| 0 | 0 | 0 | 0 | 0 |
| 0 | 0 | 0.000442915 | 0 | 0 |
| 0 | 0 | 0 | 0 | 0 |
| 0 | 0 | 0 | 0 | 0 |

| | | | | |
|---|---|---|---|---|
| 0 | 0 | 0 | 0 | 0 |
| 0.001780867 | 0.658061959 | 0.035992809 | 0 | 0 |
| 0 | 0 | 0 | 0 | 0 |
| 0 | 0.01066908 | 0 | 0 | 0.000814303 |
| 0 | 0.014701739 | 0 | 0.006121538 | 0 |
| 0 | 0.001459539 | 0.000466123 | 0 | 0.000974727 |
| 0 | 0 | 0 | 0 | 0.001580439 |
| 0 | 0 | 0 | 0 | 0 |
| 0 | 0 | 0 | 0 | 0 |
| 0 | 0 | 0 | 0 | 0 |
| 0 | 0 | 0 | 0.003895579 | 0 |
| 0 | 0 | 0 | 0 | 0 |
| 0.078013202 | 0 | 0.005888747 | 0.042775592 | 0.028091686 |
| 0 | 0 | 0 | 0 | 0 |
| 0 | 0 | 0 | 0 | 0 |
| 0 | 0 | 0 | 0 | 0 |
| 0 | 0 | 0 | 0 | 0 |
| 0 | 0 | 0 | 0 | 0 |
| 0 | 0 | 0 | 0 | 0 |
| 0 | 0 | 0 | 0 | 0 |
| 0 | 0 | 0 | 0 | 0 |
| 0 | 0 | 0 | 0 | 0 |
| 0 | 0.050361886 | 0.011025393 | 0 | 0 |
| 0.001443089 | 0 | 0 | 0.017135802 | 0.049330253 |
| 0 | 0 | 0 | 0 | 0 |
| 0 | 0 | 0 | 0 | 0 |
| 0 | 0 | 0 | 0 | 0 |
| 0 | 0.001864979 | 0 | 0 | 0 |
| 0 | 0 | 0 | 0 | 0 |
| 0 | 0 | 0 | 0 | 0 |
| 0 | 0.021890261 | 0.01148272 | 0.001226617 | 0.001977452 |
| 0 | 0.004170635 | 0 | 0 | 0.003713712 |
| 0 | 0 | 0 | 0 | 0 |
| 0 | 0 | 0 | 0 | 0 |
| 0 | 0 | 0 | 0 | 0 |
| 0 | 0 | 0 | 0 | 0 |
| 0 | 0 | 0 | 0 | 0 |
| 0 | 0 | 0 | 0 | 0 |
| 0 | 0.002684156 | 0.00091437 | 0 | 0 |
| 0 | 0 | 0 | 0 | 0 |
| 0 | 0 | 0 | 0 | 0 |
| 0 | 0.000978633 | 0 | 0 | 0 |
| 0.163483416 | 0 | 0 | 0 | 0 |
| 0 | 0 | 0 | 0 | 0 |
| 0 | 0 | 0 | 0 | 0.07361268 |
| 0.069529548 | 0.150907149 | 0.017698873 | 0.002921905 | 0.354335671 |
| 0 | 0 | 0 | 0 | 0 |
| 0 | 0 | 0 | 0 | 0 |

| | | | | |
|---|---|---|---|---|
| 0 | 0 | 0 | 0 | 0 |
| 0 | 0 | 0 | 0 | 0 |
| 0 | 0 | 0 | 0.237515906 | 0.100945784 |
| 0.002043308 | 0 | 0 | 0 | 0 |
| 0 | 0 | 0 | 0 | 0 |
| 0 | 0 | 0 | 0.001681286 | 0 |
| 0 | 0 | 0.000580901 | 0 | 0 |
| 0 | 0 | 0 | 0 | 0 |
| 0 | 0 | 0 | 0 | 0 |
| 0 | 0 | 0 | 0 | 0 |
| 0 | 0.000871028 | 0.003708994 | 0.003199358 | 0 |
| 0 | 0 | 0 | 0 | 0 |
| 0 | 0 | 0 | 0 | 0 |
| 0 | 0 | 0 | 0 | 0 |
| 0 | 0 | 0 | 0 | 0 |
| 0 | 0 | 0 | 0 | 0 |
| 0 | 0 | 0.000817809 | 0 | 0 |
| 0 | 0 | 0 | 0 | 0 |
| 0 | 0 | 0 | 0 | 0 |
| 0 | 0.001443916 | 0.000860783 | 0 | 0.010285796 |
| 0 | 0 | 0 | 0 | 0 |
| 0 | 0 | 0 | 0 | 0 |
| 0 | 0 | 0 | 0 | 0 |
| 0 | 0 | 0 | 0 | 0 |
| 0 | 0 | 0 | 0 | 0 |
| 0 | 0 | 0 | 0 | 0 |
| 0 | 0 | 0 | 0 | 0 |
| 0 | 0 | 0 | 0 | 0.012236695 |
| 0 | 0 | 0 | 0 | 0 |
| 0 | 0 | 0 | 0 | 0 |
| 0 | 0.112634966 | 0.000422573 | 0 | 0.007069251 |
| 0 | 0 | 0 | 0 | 0 |
| 0.018984281 | 0.009794147 | 0.876177948 | 0.01470174 | 0.118504728 |
| 0.001686946 | 0 | 0.00275765 | 0 | 0.002883309 |
| 0 | 0 | 0 | 0 | 0 |
| 0 | 0 | 0 | 0 | 0 |
| 0 | 0 | 0 | 0 | 0 |
| 0 | 0 | 0 | 0 | 0 |
| 0 | 0 | 0 | 0 | 0 |
| 0 | 0 | 0 | 0 | 0 |
| 0 | 0 | 0 | 0 | 0 |
| 0 | 0.004028778 | 0.184570412 | 0 | 0 |
| 0.015097801 | 0 | 0.019424653 | 0.017190957 | 0.141397234 |
| 0 | 0 | 0 | 0 | 0 |
| 0 | 0 | 0 | 0 | 0 |
| 0 | 0 | 0 | 0.024842129 | 0.007058632 |
| 0 | 0 | 0.051949042 | 0 | 0 |
| 0 | 0 | 0 | 0 | 0 |

| | | | | |
|---|---|---|---|---|
| 0 | 0 | 0 | 0 | 0 |
| 0.00411674 | 0 | 0 | 0 | 0.003770642 |
| 0.002024752 | 0.002473212 | 0 | 0 | 0.012584308 |
| 0 | 0 | 0 | 0 | 0 |
| 0 | 0.47139885 | 0.055017586 | 0 | 0 |
| 0 | 0.019222009 | 0.150371556 | 0 | 0 |
| 0 | 0 | 0 | 0 | 0 |
| 0 | 0 | 0 | 0 | 0 |
| 0 | 0 | 0 | 0 | 0 |
| 0 | 0 | 0 | 0 | 0 |
| 0 | 0 | 0 | 0 | 0 |
| 0 | 0 | 0 | 0 | 0 |
| 0 | 0 | 0 | 0 | 0 |
| 0 | 0 | 0 | 0 | 0 |
| 0 | 0.003804175 | 0 | 0 | 0 |
| 0 | 0 | 0 | 0 | 0 |
| 0.035169989 | 0 | 0 | 0 | 0 |
| 0 | 0 | 0 | 0 | 0 |
| 0 | 0 | 0 | 0 | 0 |
| 0 | 0 | 0 | 0 | 0 |
| 0 | 0 | 0 | 0 | 0 |
| 0 | 0 | 0 | 0 | 0 |
| 0 | 0 | 0 | 0 | 0 |
| 0 | 0 | 0 | 0 | 0 |
| 0.002294436 | 0 | 0 | 0.002927151 | 0.009334115 |
| 0 | 0.001464611 | 0.007359163 | 0 | 0.007824916 |
| 0 | 0 | 0 | 0 | 0 |
| 0 | 0.012502373 | 0.083485842 | 0 | 0 |
| 0 | 0 | 0 | 0 | 0 |
| 0 | 0 | 0 | 0 | 0 |
| 0 | 0 | 0 | 0 | 0 |
| 0 | 0 | 0 | 0 | 0 |
| 0 | 0 | 0 | 0 | 0 |
| 0 | 0 | 0 | 0 | 0 |
| 0 | 0 | 0 | 0 | 0 |
| 0 | 0 | 0 | 0 | 0 |
| 0 | 0 | 0 | 0 | 0 |
| 0 | 0 | 0 | 0 | 0 |
| 0.064583116 | 0 | 0 | 0 | 0 |
| 0 | 0 | 0 | 0 | 0 |
| 0 | 0 | 0 | 0 | 0 |
| 0 | 0 | 0 | 0 | 0.002829609 |
| 0 | 0.049976511 | 0 | 0 | 0 |
| 0 | 0 | 0 | 0 | 0 |
| 0 | 0 | 0 | 0 | 0 |
| 0 | 0 | 0 | 0 | 0 |
| 0 | 0.006296235 | 0 | 0 | 0 |
| 0 | 0 | 0 | 0 | 0 |

| | | | | |
|---|---|---|---|---|
| 0 | 0 | 0 | 0 | 0 |
| 0 | 0.024685288 | 0.002416423 | 0 | 0.004295106 |
| 0.065552244 | 0 | 0 | 0 | 0 |
| 0 | 0 | 0 | 0 | 0 |
| 0 | 0 | 0 | 0 | 0 |
| 0.002480169 | 0 | 0 | 0 | 0 |
| 0 | 0 | 0 | 0 | 0 |
| 0 | 0 | 0 | 0 | 0 |
| 0 | 0 | 0 | 0 | 0 |
| 0 | 0 | 0 | 0 | 0 |
| 0 | 0 | 0 | 0 | 0 |
| 0 | 0 | 0 | 0 | 0 |
| 0.121179198 | 0.00209956 | 0 | 0.018090979 | 0 |
| 0 | 0 | 0 | 0 | 0 |
| 0 | 0 | 0 | 0 | 0.001260082 |
| 0 | 0 | 0 | 0 | 0 |
| 0 | 0 | 0 | 0 | 0 |
| 0 | 0 | 0 | 0 | 0 |
| 0 | 0 | 0 | 0 | 0 |
| 0 | 0.003887568 | 0 | 0 | 0 |
| 0 | 0 | 0 | 0 | 0 |
| 0 | 0.009380869 | 0.008876761 | 0 | 0 |
| 0 | 0 | 0 | 0 | 0.004992318 |
| 0 | 0 | 0 | 0 | 0 |
| 0 | 0 | 0 | 0 | 0 |
| 0 | 0 | 0 | 0 | 0 |
| 0 | 0 | 0 | 0 | 0 |
| 0 | 0 | 0 | 0 | 0 |
| 0 | 0 | 0 | 0 | 0 |
| 0 | 0 | 0 | 0 | 0 |
| 0 | 0 | 0 | 0 | 0 |
| 0 | 0 | 0 | 0 | 0 |
| 0 | 0 | 0 | 0 | 0 |
| 0 | 0 | 0 | 0 | 0 |
| 0 | 0 | 22.82829403 | 10.84758399 | 0.456735833 |
| 0 | 0 | 0 | 0 | 0 |
| 0 | 0 | 0 | 0 | 0 |
| 0 | 0 | 0 | 0 | 0 |
| 0.22020762 | 0.019776747 | 0 | 0 | 0 |
| 0 | 0 | 0 | 0 | 0 |
| 0.038775931 | 0.031420691 | 0.011922322 | 0.007457499 | 0.121331755 |
| 0 | 0 | 0 | 0 | 0 |
| 0 | 0 | 0 | 0 | 0 |
| 0 | 0 | 0 | 0 | 0 |
| 0 | 0 | 0 | 0 | 0 |
| 0.353125226 | 0.470162468 | 0.005915107 | 0 | 0.011100642 |
| 0 | 0 | 0 | 0 | 0 |
| 0 | 0 | 0 | 0 | 0 |

| | | | | |
|---|---|---|---|---|
| 0 | 0 | 0 | 0 | 0 |
| 0 | 0 | 0 | 0 | 0 |
| 0 | 0 | 0 | 0 | 0 |
| 0 | 0 | 0 | 0 | 0 |
| 0 | 0 | 0.00529137 | 0 | 0 |
| 0 | 0 | 0 | 0 | 0 |
| 0 | 0 | 0 | 0 | 0 |
| 0.003309351 | 0 | 0 | 0 | 0 |
| 0 | 0 | 0 | 0 | 0.011172455 |
| 0.04965755 | 0.021945937 | 0 | 0 | 0 |
| 0.011053171 | 0 | 0 | 0.011363604 | 0.011449672 |
| 0 | 0 | 0 | 0 | 0 |
| 0 | 0.007150102 | 0.004639454 | 0 | 0 |
| 0 | 0 | 0 | 0 | 0 |
| 0 | 0 | 0 | 0 | 0 |
| 0 | 0 | 0 | 0 | 0 |
| 0 | 0 | 0 | 0 | 0 |
| 0 | 0 | 0 | 0 | 0 |
| 0 | 0 | 0 | 0 | 0 |
| 0.204062809 | 5.375539165 | 0.029938452 | 0.037729442 | 0.981176796 |
| 0 | 0 | 0 | 0 | 0 |
| 0 | 0 | 0 | 0 | 0 |
| 0 | 0 | 0 | 0 | 0 |
| 0 | 0 | 0 | 0 | 0 |
| 0 | 0 | 0 | 0 | 0 |
| 0 | 0 | 0 | 0.007494606 | 0 |
| 0 | 0 | 0 | 0 | 0 |
| 0 | 0 | 0 | 0 | 0 |
| 0 | 0 | 0 | 0 | 0 |
| 0.012067676 | 0 | 0 | 0 | 0 |
| 0 | 0 | 0 | 0 | 0 |
| 0 | 0 | 0 | 0 | 0 |
| 0 | 0 | 0 | 0 | 0 |
| 0 | 0 | 0 | 0 | 0 |
| 0 | 0 | 0 | 0 | 0 |
| 0 | 0 | 0 | 0 | 0 |
| 0 | 0 | 0 | 0 | 0 |
| 0 | 0 | 0 | 0 | 0 |
| 0 | 0 | 0 | 0 | 0 |
| 0 | 0 | 0 | 0 | 0 |
| 0 | 0 | 0 | 0 | 0 |
| 0 | 0 | 0 | 0 | 0 |
| 0 | 0 | 0 | 0 | 0 |
| 0 | 0 | 0 | 0 | 0 |
| 0 | 0.015085799 | 0.018180578 | 0 | 0.001774175 |
| 0 | 0 | 0 | 0 | 0 |
| 0 | 0 | 0 | 0 | 0 |
| 0 | 0 | 0 | 0 | 0 |

| | | | | |
|---|---|---|---|---|
| 0 | 0 | 0 | 0 | 0 |
| 0 | 0 | 0 | 0 | 0 |
| 0 | 0 | 0 | 0 | 0 |
| 0 | 0 | 0 | 0 | 0 |
| 0 | 0 | 0 | 0 | 0 |
| 0.002435474 | 0.0618355 | 0.001900151 | 0.068520113 | 0.0102175 |
| 0 | 0 | 0 | 0 | 0 |
| 0 | 0 | 0.001616708 | 0 | 0 |
| 0 | 0 | 0 | 0 | 0 |
| 0 | 0 | 0 | 0 | 0 |
| 0 | 0 | 0 | 0 | 0 |
| 0 | 0 | 0 | 0 | 0 |
| 0 | 0 | 0 | 0 | 0 |
| 0 | 0 | 0 | 0 | 0 |
| 0 | 0 | 0 | 0 | 0 |
| 0 | 0 | 0 | 0 | 0 |
| 0 | 0 | 0 | 0 | 0 |
| 0 | 0 | 0 | 0 | 0 |
| 0 | 0 | 0 | 0 | 0 |
| 0 | 0 | 0 | 0 | 0 |
| 0 | 0 | 0 | 0 | 0 |
| 0 | 0 | 0 | 0 | 0 |
| 0 | 0 | 0 | 0 | 0 |
| 0.050656631 | 0 | 0 | 0 | 0 |
| 0 | 0 | 0 | 0 | 0 |
| 0.003333304 | 0 | 0 | 0 | 0 |
| 0 | 0 | 0 | 0 | 0 |
| 0 | 0 | 0 | 0 | 0 |
| 0 | 0 | 0 | 0 | 0 |
| 0 | 0 | 0 | 0 | 0 |
| 0 | 0 | 0 | 0 | 0 |
| 0 | 0 | 0 | 0 | 0 |
| 0 | 0 | 0 | 0 | 0 |
| 0 | 0 | 0 | 0 | 0 |
| 0 | 0 | 0 | 0 | 0 |
| 0 | 0 | 0 | 0 | 0 |
| 0 | 0 | 0 | 0 | 0 |
| 0 | 0 | 0 | 0 | 0 |
| 0.051138612 | 0.020996717 | 1.024868753 | 0 | 0 |
| 0 | 0 | 0 | 0 | 0 |
| 0 | 0 | 0 | 0 | 0 |
| 0 | 0 | 0 | 0 | 0 |
| 0 | 0 | 0 | 0 | 0 |
| 0 | 0 | 0 | 0 | 0 |
| 0 | 0 | 0.010802936 | 0 | 0 |
| 0 | 0 | 0 | 0 | 0 |
| 0 | 0 | 0 | 0 | 0 |
| 0 | 0 | 0 | 0 | 0 |

| | | | | |
|---|---|---|---|---|
| 0 | 0 | 0 | 0 | 0 |
| 0.001363355 | 0.083504017 | 0.093300476 | 0 | 0.029021966 |
| 0 | 0 | 0 | 0 | 0 |
| 0 | 0 | 0 | 0 | 0 |
| 0 | 0 | 0 | 0 | 0 |
| 0 | 0.006198484 | 0 | 0 | 0 |
| 0 | 0 | 0 | 0 | 0 |
| 0 | 0 | 0 | 0 | 0 |
| 0 | 0 | 0 | 0 | 0 |
| 0 | 0 | 0 | 0.015986032 | 0 |
| 0.007587121 | 0.011214553 | 0 | 0.003372443 | 0.004992959 |
| 0 | 0 | 0 | 0 | 0 |
| 0 | 0 | 0 | 0.011887594 | 0.720803067 |
| 0 | 0 | 0 | 0 | 0 |
| 0 | 0 | 0 | 0 | 0 |
| 0 | 0.030104499 | 0 | 0 | 0 |
| 0 | 0 | 0 | 0 | 0 |
| 0 | 0 | 0 | 0 | 0 |
| 0 | 0 | 0 | 0 | 0 |
| 0 | 0.007517268 | 0 | 0 | 0 |
| 0 | 0.007904234 | 0 | 0 | 0.000844592 |
| 0 | 0 | 0.002022734 | 0 | 0 |
| 0 | 0 | 0 | 0 | 0 |
| 0.466234762 | 0.277629426 | 1.069552774 | 0.188153574 | 3.053315213 |
| 0 | 0 | 0 | 0 | 0 |
| 0 | 0 | 0 | 0 | 0 |
| 0 | 0 | 0 | 0 | 0 |
| 0 | 0 | 0 | 0 | 0 |
| 0 | 0 | 0 | 0 | 0 |
| 0 | 0 | 0 | 0 | 0 |
| 0 | 0 | 0 | 0 | 0 |
| 0 | 0 | 0 | 0 | 0 |
| 0 | 0 | 0 | 0 | 0 |
| 0 | 0 | 0 | 0 | 0 |

| PHL018-0143_V3 | PHL018-0039_V1 | PHL018-0081_V3 | PHL018-0148_V1 | PHL018-0130_V1 |
|---|---|---|---|---|
| CTRL | EYCF | EYCF | CTRL | REF |
| 1.48089402 | 0.167997591 | 0.110158262 | 0.263958817 | 0.624833541 |
| 0.037177866 | 0.252524424 | 0.089939032 | 0.842613501 | 0.477032966 |
| 0 | 0.010416728 | 1.32117561 | 0.317445597 | 0 |
| 0 | 0 | 0 | 0 | 0 |
| 0 | 0 | 0 | 0 | 0 |
| 0 | 0 | 0 | 0 | 0 |
| 0 | 0 | 0 | 0 | 5.275427602 |
| 0 | 0 | 0 | 0 | 0 |
| 0 | 0 | 0 | 0 | 0 |
| 0 | 0 | 0 | 0 | 0 |
| 0 | 0 | 0 | 0 | 0 |
| 0 | 0 | 0 | 0 | 0 |
| 0 | 0 | 0 | 0 | 0 |
| 0.115313665 | 0.188534834 | 15.34463577 | 2.794791162 | 0.078916672 |
| 0 | 0 | 0 | 0 | 0 |
| 0 | 0 | 0 | 0 | 0 |
| 0 | 0 | 0 | 0 | 0.000944911 |
| 0 | 0 | 0 | 0 | 0 |
| 0 | 0 | 0 | 0 | 0 |
| 0 | 0 | 0 | 0 | 0 |
| 0 | 0 | 0 | 0 | 0 |
| 0 | 0 | 0 | 0 | 0 |
| 0 | 0 | 0 | 0 | 0 |
| 0 | 0 | 0 | 0 | 0 |
| 0 | 0.029455561 | 0.012202666 | 0 | 0.735743728 |
| 0 | 0 | 0 | 0 | 0 |
| 0 | 0 | 0 | 0 | 0.005172034 |
| 0 | 0 | 0 | 0 | 0 |
| 0 | 0 | 0 | 0 | 0 |
| 0 | 0 | 0 | 0 | 0 |
| 0 | 0 | 0 | 0 | 0 |
| 0 | 0 | 0 | 0 | 0 |
| 0 | 0 | 0 | 0 | 0 |
| 0 | 0 | 0 | 0 | 0 |
| 0 | 0 | 0 | 0 | 0 |
| 0 | 0.647208387 | 0.935658525 | 1.485788599 | 0 |
| 0 | 0 | 0 | 0 | 0 |
| 0 | 0 | 0 | 0 | 0 |
| 0 | 0 | 0 | 0 | 0 |
| 0 | 0 | 0 | 0 | 0 |
| 0 | 0 | 0 | 0 | 0 |
| 0.054106459 | 0.044999638 | 0 | 0.090687264 | 0 |
| 3.047901023 | 2.566605593 | 0 | 1.514930773 | 0.024802049 |
| 0.012703255 | 0.012219706 | 0.00603185 | 0.025669242 | 0.005060128 |
| 0 | 0 | 0.001186023 | 0.000546994 | 0.001920092 |
| 0 | 0 | 0 | 0 | 0 |

| | | | | |
|---|---|---|---|---|
| 0.001695678 | 0.001317452 | 0 | 0.001535984 | 0 |
| 0 | 0 | 0.001409967 | 0.000975415 | 0.001369585 |
| 0.380892397 | 0.001498819 | 0 | 0.00134418 | 0 |
| 0.093302605 | 0.692502857 | 0.23729601 | 1.047322621 | 0.435588399 |
| 0.037633785 | 0.002520644 | 0.000588182 | 0.018355912 | 0.006094258 |
| 0.025104973 | 0.012672198 | 0 | 0.535982193 | 0 |
| 0 | 0 | 0 | 0.011151866 | 0.001100981 |
| 0 | 0 | 0 | 0 | 0 |
| 0 | 0 | 0 | 0.008117629 | 0.010838282 |
| 0 | 0 | 0 | 0 | 0 |
| 0 | 0 | 0 | 0 | 0 |
| 0.192170573 | 0.101924503 | 0.010398693 | 0.0072553 | 0 |
| 0 | 0 | 0 | 0 | 0 |
| 0.016715507 | 0.068877862 | 0.056550953 | 0.130905624 | 0.010250342 |
| 0 | 0.00445459 | 0 | 0 | 0 |
| 0 | 0 | 0 | 0 | 0 |
| 0.812670861 | 0.043873896 | 0 | 1.601775874 | 0.010088711 |
| 0 | 0 | 0 | 0 | 0 |
| 0 | 0 | 0 | 0 | 0 |
| 0 | 0 | 0 | 0 | 0 |
| 0.043947584 | 0 | 0.036311634 | 0.001582386 | 0 |
| 0.009673983 | 0.027380358 | 0.003489843 | 0.036276 | 0.038331873 |
| 0.00907899 | 1.228152548 | 0 | 0.053046847 | 0 |
| 0 | 0 | 0 | 0 | 0 |
| 0 | 0 | 0 | 0 | 0 |
| 0 | 0 | 0 | 0 | 0 |
| 0 | 0 | 0 | 0 | 0 |
| 0 | 0 | 0 | 0 | 0 |
| 0.007577034 | 0 | 0 | 0.005701945 | 0 |
| 0 | 0 | 0 | 0 | 0 |
| 0 | 0 | 0 | 0.000701446 | 0 |
| 0 | 0 | 0 | 0 | 0 |
| 0 | 0 | 0 | 0 | 0 |
| 0 | 0 | 0 | 0 | 0 |
| 0 | 0 | 0 | 0 | 0 |
| 0 | 0 | 0 | 0 | 0 |
| 0 | 0 | 0 | 0 | 0 |
| 35.05522619 | 0 | 0.001302093 | 0.039334395 | 0.002055301 |
| 0 | 0 | 0 | 0 | 0 |
| 0 | 0 | 0 | 0 | 0 |
| 0 | 0 | 0 | 0 | 0 |
| 0 | 0 | 0 | 0 | 0 |
| 0 | 0 | 0 | 0 | 0 |
| 0 | 0 | 0 | 0 | 0 |
| 0 | 0 | 0 | 0 | 0 |
| 0 | 0 | 0 | 0 | 0 |
| 0 | 0 | 0 | 0 | 0 |
| 0 | 0 | 0 | 0.230160137 | 0 |

| | | | | |
|---|---|---|---|---|
| 0 | 0 | 0 | 0 | 0 |
| 0 | 0 | 0 | 0 | 0 |
| 0 | 0 | 0 | 0 | 0 |
| 0 | 0 | 0 | 0 | 0 |
| 0 | 0.167091603 | 0 | 0 | 0 |
| 0 | 0 | 0 | 0 | 0 |
| 0 | 0 | 0 | 0 | 0 |
| 0 | 0 | 0 | 0 | 0 |
| 0.011258961 | 0.006227121 | 0 | 0 | 0.012098207 |
| 0 | 0 | 0 | 0 | 0 |
| 0 | 0 | 0 | 0 | 0 |
| 0.011963683 | 0.021688692 | 0 | 0.001111485 | 0 |
| 0 | 0 | 0 | 0 | 0 |
| 0 | 0 | 0 | 0 | 0 |
| 0 | 0 | 0 | 0 | 0 |
| 0 | 0 | 0 | 0 | 0 |
| 0 | 0 | 0 | 0 | 0 |
| 0 | 0 | 0 | 0 | 0 |
| 0.275839232 | 0.070773711 | 0 | 0 | 2.750670165 |
| 0.04162478 | 0 | 0 | 0 | 0 |
| 0 | 0 | 0 | 0 | 0 |
| 0 | 0 | 0 | 0 | 0 |
| 0 | 0 | 0 | 0 | 0.007443391 |
| 0 | 0 | 0 | 0 | 0 |
| 0 | 0 | 0 | 0 | 0 |
| 0 | 0 | 0 | 0 | 0 |
| 0 | 0.001303611 | 0 | 0.000876834 | 0.000554025 |
| 0 | 0 | 0 | 0 | 0 |
| 0 | 0 | 0 | 0 | 0 |
| 0 | 0 | 0 | 0 | 0 |
| 0.069399232 | 0.006791395 | 0.13542367 | 0.187704358 | 0.37096186 |
| 0.002593592 | 0.036919236 | 0.004282289 | 0.001974991 | 0 |
| 0 | 0 | 0 | 0 | 0 |
| 0 | 0 | 0.003514583 | 0 | 0 |
| 0 | 0 | 0 | 0 | 0 |
| 0 | 0 | 0 | 0 | 0 |
| 0 | 0 | 0 | 0 | 0 |
| 0 | 0 | 0 | 0 | 0 |
| 0 | 0 | 0 | 0 | 0 |
| 0 | 0 | 0 | 0 | 0 |
| 0 | 0 | 0 | 0 | 0 |
| 0 | 0 | 0 | 0 | 0 |
| 0 | 0.003244131 | 0.003785025 | 0 | 0.005797749 |
| 0 | 0 | 0 | 0 | 0 |
| 0 | 0 | 0 | 0 | 0 |
| 0 | 0 | 0 | 0 | 0 |
| 0 | 0 | 0 | 0 | 0 |
| 0 | 0 | 0 | 0 | 0 |

| | | | | |
|---|---|---|---|---|
| 0 | 0 | 0 | 0 | 0.00411987 |
| 0 | 0 | 0 | 0 | 0 |
| 0 | 0 | 0 | 0 | 0 |
| 0 | 0 | 0 | 0 | 0 |
| 0.002741163 | 0.447245379 | 0 | 0.025987328 | 0.012352289 |
| 0 | 0 | 0 | 0 | 0 |
| 0 | 0 | 0 | 0 | 0 |
| 0 | 0 | 0 | 0 | 0 |
| 0 | 0 | 0 | 0 | 0 |
| 0 | 0 | 0 | 0 | 0 |
| 0 | 0 | 0 | 0 | 0 |
| 0 | 0 | 0 | 0 | 0 |
| 0 | 0 | 0 | 0 | 0 |
| 0 | 0 | 0 | 0 | 0 |
| 0 | 0 | 0 | 0 | 0 |
| 0 | 0 | 0 | 0 | 1.63080599 |
| 0 | 0 | 0 | 0 | 0 |
| 0 | 0 | 0 | 0 | 0 |
| 0 | 0 | 0 | 0 | 0 |
| 0 | 0 | 0 | 0 | 0 |
| 0 | 0 | 0 | 0 | 0 |
| 0 | 0 | 0 | 0 | 0 |
| 0.015335329 | 0 | 0 | 0.012575962 | 0.009192534 |
| 0 | 0 | 0 | 0 | 0 |
| 0 | 0 | 0.025992546 | 0 | 0 |
| 0 | 0 | 0 | 0 | 0 |
| 0 | 0.003048855 | 0.007114383 | 0 | 0.008771178 |
| 0 | 0 | 0 | 0 | 0 |
| 0 | 0 | 0 | 0 | 0 |
| 0 | 0 | 0 | 0 | 0 |
| 0 | 0 | 0 | 0 | 0 |
| 0.000641238 | 0.000996417 | 0.000871912 | 1.606358924 | 0 |
| 0 | 0 | 0 | 0 | 0 |
| 0 | 0 | 0 | 0 | 0 |
| 0 | 0 | 0 | 0 | 0 |
| 0 | 0 | 0 | 0 | 0 |
| 0 | 0 | 0 | 0 | 0 |
| 0 | 0.014057394 | 0 | 0 | 0 |
| 0 | 0 | 0 | 0 | 0 |
| 0 | 0 | 0 | 0 | 0 |
| 0 | 0 | 0 | 0 | 0 |
| 0 | 0 | 0 | 0 | 0 |
| 0 | 0 | 0 | 0 | 0 |
| 0 | 0 | 0 | 0 | 0 |
| 0 | 0 | 0 | 0 | 0 |
| 0 | 0 | 0 | 0 | 0 |
| 0.110100372 | 0 | 0 | 0 | 0 |
| 0 | 0 | 0 | 0 | 0 |

| | | | | |
|---|---|---|---|---|
| 0 | 0 | 0.010404071 | 0 | 0 |
| 0 | 0 | 0 | 0 | 0 |
| 0 | 0 | 0 | 0 | 0 |
| 0 | 0 | 0 | 0 | 0 |
| 0 | 0 | 0 | 0 | 0 |
| 0 | 0 | 0 | 0 | 0 |
| 0 | 0 | 0 | 0 | 0 |
| 0 | 0 | 0 | 0 | 0 |
| 0 | 0 | 0 | 0 | 0 |
| 0 | 0 | 0 | 0 | 0 |
| 0 | 0 | 0 | 0 | 0 |
| 0 | 0 | 0 | 0 | 0 |
| 0 | 0 | 0.001668669 | 0.00128265 | 0.009590197 |
| 0 | 0 | 0 | 0 | 0 |
| 0 | 0.001034649 | 0 | 0 | 0 |
| 0 | 0 | 0 | 0 | 0 |
| 0 | 0 | 0 | 0 | 0 |
| 0 | 0 | 0 | 0 | 0 |
| 0 | 0 | 0 | 0 | 0 |
| 0 | 0 | 0 | 0 | 0 |
| 0.024093064 | 0.029415644 | 0.158080549 | 0.054919872 | 0.00631386 |
| 0 | 0 | 0 | 0 | 0 |
| 0 | 0 | 0 | 0 | 0.001994407 |
| 0 | 0 | 0 | 0 | 0 |
| 0 | 0 | 0 | 0 | 0 |
| 0 | 0 | 0 | 0 | 0 |
| 0.00111359 | 0.000778681 | 0.025438291 | 0.003910712 | 0.001617899 |
| 0 | 0 | 0 | 0 | 0 |
| 0 | 0 | 0 | 0 | 0 |
| 0 | 0 | 0.003564005 | 0 | 0 |
| 0 | 0.00103495 | 0 | 0 | 0 |
| 0 | 0 | 0 | 0 | 0 |
| 0 | 0 | 0 | 0 | 0 |
| 0.007478951 | 0 | 0 | 0.04470263 | 0.037197283 |
| 0.019906326 | 0.063507962 | 0 | 0.006241711 | 0.013967656 |
| 0 | 0 | 0 | 0 | 0 |
| 0 | 0 | 0 | 0 | 0 |
| 0 | 0 | 0 | 0 | 0 |
| 0 | 0 | 0 | 0 | 0 |
| 0 | 0 | 0 | 0 | 0 |
| 0 | 0 | 0 | 0 | 0 |
| 0 | 0 | 0 | 0 | 0 |
| 0 | 0 | 0 | 0 | 0 |
| 0 | 0 | 0 | 0 | 0 |
| 0 | 0 | 0 | 0 | 0.004418831 |
| 0 | 0.109060576 | 0 | 0.36340153 | 2.948406631 |
| 0.001616494 | 0 | 0 | 0 | 0 |
| 0.001241256 | 0 | 0 | 0 | 0.026113901 |

| | | | | |
|---|---|---|---|---|
| 0 | 0 | 0 | 0 | 0 |
| 0 | 0 | 0 | 0 | 0 |
| 0 | 0 | 0 | 0 | 0 |
| 0 | 0 | 0 | 0.0005654 | 0 |
| 0 | 0 | 0 | 0 | 0 |
| 0 | 0 | 0 | 0 | 0 |
| 0.00126208 | 0 | 0 | 0 | 0 |
| 0.001019616 | 0 | 0 | 0 | 0 |
| 0 | 0 | 0 | 0 | 0.009576478 |
| 0 | 0 | 0 | 0 | 0 |
| 0 | 0 | 0 | 0 | 0 |
| 0 | 0 | 0 | 0 | 0 |
| 0 | 0 | 0 | 0 | 0 |
| 0 | 0 | 0 | 0 | 0 |
| 0 | 0 | 0 | 0 | 0 |
| 0 | 0 | 0 | 0 | 0 |
| 0 | 0 | 0 | 0 | 0 |
| 0 | 0 | 0 | 0 | 0 |
| 0.013525172 | 0.019545529 | 0 | 0.009160288 | 0 |
| 0 | 0.085980487 | 0 | 0.120005812 | 0.712261597 |
| 0 | 0 | 0.002893656 | 0 | 0.010189081 |
| 0 | 0 | 0 | 0 | 0 |
| 0 | 0 | 0 | 0 | 0 |
| 0 | 0 | 0 | 0 | 0 |
| 0 | 0.008622819 | 0 | 0.008285541 | 0 |
| 0 | 0 | 0 | 0 | 0 |
| 0 | 0 | 0 | 0 | 0 |
| 0 | 0 | 0 | 0 | 0 |
| 0.00806956 | 0 | 0 | 0.013758201 | 0 |
| 0 | 0 | 0 | 0 | 0 |
| 0.116968536 | 0.141560603 | 0.39655084 | 0.470286507 | 0.082670644 |
| 0 | 0 | 0 | 0 | 0 |
| 0 | 0 | 0.005952674 | 0 | 0 |
| 0 | 0 | 0 | 0.000793487 | 0 |
| 0 | 0 | 0 | 0 | 0 |
| 0 | 0 | 0 | 0 | 0 |
| 0 | 0 | 0 | 0 | 0 |
| 0 | 0 | 0 | 0 | 0 |
| 0 | 0 | 0 | 0 | 0 |
| 0 | 0 | 0 | 0 | 0 |
| 0 | 0 | 0 | 0 | 0 |
| 0.004386387 | 0.165907265 | 0.030634872 | 0.025631892 | 0.002106724 |
| 0 | 0 | 0 | 0 | 0 |
| 0 | 0 | 0 | 0 | 0 |
| 0 | 0 | 0 | 0 | 0 |
| 0 | 0 | 0 | 0 | 0 |
| 0 | 0 | 0 | 0 | 0 |
| 0 | 0 | 0 | 0 | 0 |

| | | | | |
|---|---|---|---|---|
| 0 | 0 | 0 | 0 | 0 |
| 0 | 0 | 0 | 0 | 0 |
| 0.009513526 | 0.010503725 | 0.007829591 | 0.023864043 | 0 |
| 0 | 0 | 0 | 0 | 0 |
| 0 | 0.001266031 | 0 | 0 | 0 |
| 0 | 0 | 0 | 0 | 0 |
| 0 | 0 | 0 | 0 | 0 |
| 0 | 0 | 0 | 0 | 0 |
| 0 | 0 | 0 | 0 | 0 |
| 0 | 0 | 0 | 0 | 0 |
| 0 | 0 | 0 | 0 | 0 |
| 0 | 0 | 0 | 0 | 0 |
| 0 | 0 | 0 | 0 | 0 |
| 0 | 0.003224587 | 0.001128667 | 0 | 0.010597968 |
| 0.003197182 | 0.002661474 | 0 | 0.013605217 | 0.002563845 |
| 0.022738537 | 0.123951492 | 0 | 0 | 0.011302671 |
| 0 | 0 | 0 | 0 | 0.000515518 |
| 0 | 0 | 0 | 0.0006329 | 0 |
| 0 | 0 | 0 | 0 | 0 |
| 0 | 0 | 0 | 0 | 0 |
| 0 | 0 | 0 | 0 | 0 |
| 0.000704679 | 0 | 0 | 0.00044191 | 0.026370743 |
| 0 | 0 | 0 | 0 | 0 |
| 0 | 0 | 0 | 0 | 0 |
| 0 | 0.023263867 | 0 | 0 | 0.000994916 |
| 0 | 0 | 0 | 0 | 0 |
| 0 | 0 | 0 | 0 | 0.910967219 |
| 0 | 0 | 0 | 0 | 0 |
| 0 | 0 | 0 | 0 | 0 |
| 0 | 0.044745827 | 0 | 0 | 0 |
| 0.001800022 | 0 | 0 | 0 | 0 |
| 0 | 0 | 0 | 0 | 0 |
| 0 | 0 | 0 | 0 | 0 |
| 0 | 0 | 0 | 0 | 0 |
| 0 | 0 | 0 | 0 | 0 |
| 0.013605792 | 0.024058107 | 0 | 0.00117687 | 0 |
| 0 | 0 | 0 | 0 | 0 |
| 0.001475593 | 0.220463973 | 0.007223082 | 0.000555214 | 0.016566054 |
| 0 | 0 | 0 | 0 | 0 |
| 0 | 0 | 0 | 0 | 0 |
| 0 | 0 | 0 | 0 | 0 |
| 0 | 0 | 0 | 0 | 0 |
| 0 | 0 | 0 | 0 | 0 |
| 0 | 0 | 0 | 0 | 0 |
| 0 | 0 | 0 | 0 | 0 |
| 0 | 0.004759187 | 0 | 0 | 0 |
| 0 | 0 | 0 | 0 | 0 |
| 0 | 0 | 0 | 0 | 0 |

| | | | | |
|---|---|---|---|---|
| 0 | 0 | 0 | 0.139358811 | 0.907769639 |
| 0 | 0 | 0 | 0.10910514 | 0 |
| 0 | 0 | 0 | 0.001080021 | 0 |
| 0 | 0 | 0 | 0 | 0 |
| 0 | 0.001153916 | 0 | 0.002017981 | 0.013567907 |
| 0 | 0.017404474 | 0 | 0 | 0 |
| 0 | 0 | 0 | 0 | 0 |
| 0.005204558 | 0 | 0 | 0.010444228 | 0 |
| 0 | 0 | 0 | 0 | 0 |
| 0.00178815 | 0 | 0.624384866 | 0.013232093 | 0.004487362 |
| 0 | 0 | 0 | 0 | 0 |
| 0.082673595 | 0.10435202 | 0 | 0.034620946 | 0 |
| 0.007083618 | 0 | 0.003399467 | 0.034100385 | 0.00426522 |
| 0 | 0 | 0 | 0 | 0.019015385 |
| 0.001285648 | 0 | 0 | 0 | 0 |
| 0 | 0 | 0 | 0 | 0 |
| 0 | 0.001590359 | 0.006803575 | 0 | 0 |
| 0 | 0 | 0 | 0 | 0 |
| 0 | 0 | 0 | 0 | 0 |
| 0 | 0 | 0 | 0 | 0 |
| 0 | 0 | 0 | 0 | 0 |
| 0 | 0 | 0 | 0 | 0 |
| 0 | 0.018289733 | 0 | 0.000674084 | 0 |
| 0 | 0 | 0 | 0.998286918 | 0 |
| 0 | 0 | 0 | 0 | 0 |
| 0 | 0 | 0.001319838 | 0.003652253 | 0.000801273 |
| 3.221883818 | 0 | 0 | 0.722395876 | 0 |
| 0 | 0 | 0 | 0 | 0 |
| 0 | 0 | 0 | 0 | 0 |
| 0 | 0 | 0.001697433 | 0 | 0 |
| 0 | 0 | 0 | 0 | 0 |
| 0 | 0 | 0 | 0 | 0 |
| 0 | 0 | 0 | 0 | 0 |
| 0 | 0 | 0.004746789 | 0 | 0 |
| 0 | 0 | 0 | 0 | 0 |
| 0 | 0 | 0 | 0 | 0 |
| 0 | 0 | 0 | 0 | 0 |
| 0 | 0 | 0 | 0 | 0 |
| 0 | 0 | 0 | 0 | 0 |
| 0 | 0 | 0 | 0 | 0 |
| 0 | 0 | 0 | 0 | 0 |
| 0 | 0 | 0 | 0 | 0 |
| 0 | 0 | 0 | 0 | 0 |
| 0 | 0 | 0 | 0 | 0.001936602 |
| 0 | 0 | 0 | 0 | 0 |
| 0 | 0 | 0.007242341 | 0 | 0 |
| 0 | 0.016881252 | 0.024209503 | 0.011733153 | 0.000797157 |
| 0 | 0 | 0 | 0 | 0.004884016 |

| | | | | |
|---|---|---|---|---|
| 0 | 0 | 0 | 0 | 0 |
| 0 | 0 | 0 | 0.003172408 | 0 |
| 0 | 0 | 0 | 0 | 0 |
| 0 | 0 | 0 | 0 | 0 |
| 0 | 0 | 0 | 0 | 0 |
| 0.006892803 | 0 | 0.007433255 | 0 | 0.005336792 |
| 0 | 0 | 0.03270212 | 0 | 0.00418886 |
| 0 | 0 | 0 | 0 | 0 |
| 0 | 0 | 1.852677872 | 0 | 0 |
| 0 | 0 | 0 | 0 | 0 |
| 0 | 0 | 0 | 0.004840535 | 0.002366681 |
| 0 | 0 | 0 | 0 | 0 |
| 0 | 0 | 0 | 0 | 0 |
| 0.01145941 | 0 | 0.081095811 | 0 | 0 |
| 0 | 0 | 0 | 0 | 0 |
| 0 | 0 | 0.002889084 | 0 | 0 |
| 0 | 0 | 0 | 0 | 0.454707313 |
| 0 | 0 | 0 | 0 | 0 |
| 0 | 0 | 0 | 0 | 0 |
| 0 | 0 | 0 | 0.04583367 | 0.339515454 |
| 0 | 0 | 0 | 0 | 0 |
| 0.286212147 | 0.040493632 | 0.8772382 | 0 | 0 |
| 0 | 0.005760588 | 0 | 0 | 0 |
| 0 | 0 | 0 | 0 | 0 |
| 0 | 0 | 0 | 0 | 0 |
| 0 | 0 | 0 | 0 | 0 |
| 0 | 0 | 0 | 0 | 0 |
| 0 | 0 | 0 | 0 | 0 |
| 0 | 0 | 0 | 0 | 0 |
| 1.139530174 | 0.102327476 | 0.260059415 | 0.228626941 | 0.261182893 |
| 0 | 0.001441417 | 0.014895449 | 0 | 0.036872205 |
| 0 | 0 | 0 | 0.000777819 | 0 |
| 0 | 0 | 0 | 0 | 0 |
| 0 | 0 | 0 | 0 | 0.001983177 |
| 0.001411211 | 0 | 0.001279246 | 0.005604888 | 0 |
| 0 | 0 | 0 | 0 | 0 |
| 0 | 0 | 0.674296273 | 0 | 0 |
| 0.001592753 | 0.002784343 | 0 | 0.001331772 | 0 |
| 0 | 0 | 0 | 0 | 0 |
| 0 | 0 | 0 | 0.000966275 | 0 |
| 0.037683527 | 0.002994353 | 0 | 0.07317756 | 0.011736009 |
| 1.105235023 | 0.030259427 | 2.113517795 | 1.495100323 | 1.429015542 |
| 0.000729533 | 0.002834043 | 0 | 0.001982482 | 0 |
| 0 | 0 | 0 | 0 | 0 |
| 0 | 0 | 0 | 0 | 0 |
| 0 | 0 | 0 | 0 | 0 |
| 0 | 0 | 0 | 0 | 0 |
| 0 | 0 | 0 | 0 | 0 |

| | | | | |
|---|---|---|---|---|
| 0 | 0 | 0 | 0.00121716 | 0.001831094 |
| 0 | 0 | 0 | 0.003333126 | 0.02281529 |
| 0.029091008 | 0.003082116 | 0.002054856 | 3.680627299 | 0.001247503 |
| 0 | 0 | 0 | 0 | 0 |
| 0.142150546 | 0 | 0 | 0.147291169 | 0 |
| 0 | 0 | 0 | 0.017347853 | 0 |
| 0 | 0 | 0 | 0 | 0 |
| 0 | 0 | 0 | 0 | 0 |
| 0 | 0 | 0.011043518 | 0 | 0 |
| 0 | 0 | 0 | 0 | 0 |
| 0 | 0 | 0 | 0 | 0.000519459 |
| 0 | 0 | 0 | 0 | 0 |
| 0 | 0 | 0 | 0 | 0 |
| 0 | 0 | 0 | 0 | 0.016896752 |
| 0 | 0 | 0 | 0 | 0 |
| 0.05488795 | 0.046604953 | 0.017058951 | 0.117522064 | 0.071200844 |
| 0 | 0.244356597 | 0.022157892 | 0.025207402 | 0.011192108 |
| 0 | 0 | 0 | 0 | 0 |
| 0.0029176 | 0 | 0 | 0 | 0.039572832 |
| 0 | 0 | 0 | 0 | 0 |
| 0.002190647 | 0 | 0 | 0.036404983 | 0.017721969 |
| 0 | 0 | 0 | 0.002212702 | 0 |
| 0.039510674 | 0 | 0 | 0 | 0 |
| 0.002878159 | 0 | 0 | 0.075054469 | 0 |
| 0 | 0 | 0.031477213 | 0.056158976 | 0 |
| 0 | 0 | 0.008172953 | 0 | 0 |
| 0.001930845 | 0 | 0 | 0.006054243 | 0.002732398 |
| 0 | 0 | 0 | 0.001642022 | 0 |
| 0 | 0 | 0 | 0 | 0 |
| 0 | 0 | 0 | 0 | 0 |
| 0.016781736 | 0.005622865 | 0.0034228 | 0 | 0.011221099 |
| 0 | 0 | 0 | 0.001176746 | 0.001239206 |
| 0.00803991 | 0.045114245 | 0.070046569 | 0.007282736 | 0.044245938 |
| 0.016216112 | 0.00298399 | 0 | 0.00133806 | 0 |
| 0 | 0.006389429 | 0 | 0 | 0 |
| 0 | 0 | 0 | 0 | 0 |
| 0 | 0.001568707 | 0 | 0.004051751 | 0 |
| 0.007158143 | 0 | 0.012023312 | 0.022444654 | 0.036844342 |
| 0.036128189 | 0.026246998 | 0 | 0.011475262 | 0 |
| 0 | 0 | 0 | 0.001187405 | 0 |
| 0 | 0 | 0 | 0 | 0 |
| 0 | 0 | 0 | 0 | 0 |
| 0.001574263 | 0 | 0 | 0 | 0.006653664 |
| 0.057192908 | 0 | 0.019665227 | 0 | 0.016171588 |
| 0 | 0 | 0 | 0 | 0 |
| 0 | 0 | 0 | 0 | 0 |
| 0 | 0 | 0 | 0.001907267 | 0 |
| 0 | 0 | 0 | 0 | 0 |

| | | | | |
|---|---|---|---|---|
| 0 | 0 | 0 | 0 | 0.001443002 |
| 0.006311333 | 0.045469506 | 0.004680939 | 0.372221474 | 0.445593851 |
| 0 | 0 | 0 | 0 | 0 |
| 0 | 0 | 0 | 0 | 0 |
| 0 | 0 | 0 | 0 | 0 |
| 0 | 0 | 0 | 0 | 0 |
| 0 | 0 | 0 | 0 | 0 |
| 0 | 0 | 0 | 0.011607563 | 0 |
| 0 | 0 | 0.001071254 | 0 | 0 |
| 0 | 0 | 0 | 0 | 0 |
| 0.001279094 | 0.002484472 | 0.002029096 | 0.003609586 | 0.001407844 |
| 0.006668362 | 0.01221228 | 0.022452081 | 0.017324539 | 0.037956132 |
| 0.038312323 | 0.081544991 | 0 | 1.56295291 | 1.282773329 |
| 0 | 0 | 0 | 0 | 0 |
| 0.0080718 | 0 | 0.006319219 | 0 | 0 |
| 0 | 0 | 0 | 0 | 0 |
| 0.00089293 | 0.007804801 | 0.001214146 | 0.020718672 | 0.002063902 |
| 0 | 0.001013171 | 0.008668713 | 0 | 0.008037692 |
| 0 | 0 | 0 | 0 | 0 |
| 0 | 0 | 0 | 0 | 0.000745608 |
| 0.033150118 | 0.004560941 | 0.010642775 | 0.015735901 | 0.009121732 |
| 0 | 0 | 0 | 0 | 0 |
| 0 | 0 | 0 | 0 | 0 |
| 0 | 0 | 0 | 0 | 0 |
| 0 | 0 | 0 | 0.009459915 | 0 |
| 0 | 0 | 0 | 0 | 0 |
| 0 | 0.007944488 | 0.034604542 | 0.046881356 | 0.025210095 |
| 0.033031576 | 0.099803669 | 0.586212123 | 0.022248773 | 0.126197294 |
| 0 | 0 | 0 | 0 | 0 |
| 0 | 0 | 0 | 0 | 0 |
| 0 | 0 | 0 | 0 | 0 |
| 0 | 0 | 0 | 0 | 0 |
| 0 | 0 | 0 | 0 | 0 |
| 0 | 0 | 0 | 0 | 0 |
| 0 | 0.332232847 | 0.082337927 | 0.232525373 | 0 |
| 0.440606122 | 0.123064279 | 0.485929266 | 0.22592833 | 0.131266412 |
| 0.003260393 | 0 | 0 | 0 | 0 |
| 0 | 0 | 0 | 0 | 0 |
| 0.025656239 | 0.052061744 | 0 | 0.011924966 | 0.015946579 |
| 0 | 0 | 0 | 0 | 0 |
| 0 | 0 | 0 | 0 | 0 |
| 0 | 0 | 0 | 0 | 0 |
| 0 | 0 | 0 | 0 | 0 |
| 0 | 0 | 0 | 0 | 0 |
| 0.001627872 | 0 | 0 | 0 | 0 |
| 1.135199812 | 0.049662596 | 0 | 0.00057346 | 0.036354698 |
| 0 | 0 | 0 | 0 | 0 |
| 0.001144507 | 0 | 0 | 0 | 0 |

| | | | | |
|---|---|---|---|---|
| 0 | 0 | 0 | 0 | 0 |
| 0 | 0 | 0 | 0 | 0 |
| 0.021170596 | 0 | 0 | 0 | 0 |
| 0.038863249 | 0.261549183 | 0 | 0.070837941 | 0.692534049 |
| 0 | 0 | 0 | 0 | 0 |
| 0 | 0 | 0 | 0.014631074 | 0.754486991 |
| 0 | 0 | 0 | 0 | 0.065377707 |
| 0.004011617 | 0.019591412 | 0 | 0.012458794 | 0 |
| 0.620990798 | 16.06095634 | 2.940440536 | 0 | 0 |
| 0.249246419 | 0.00333626 | 0 | 0.00646282 | 0.078897558 |
| 0 | 0 | 0 | 0 | 0 |
| 0.002659655 | 0.003814915 | 0.00139093 | 0.024248579 | 0 |
| 0 | 0 | 0 | 0 | 0 |
| 0.011021377 | 0 | 0.059595968 | 0 | 0 |
| 1.221802716 | 0.117069136 | 0.595248612 | 0.061088698 | 0.178499632 |
| 0 | 0 | 0 | 0 | 0 |
| 0.130590661 | 0.001283247 | 0.002994407 | 0.006490794 | 0.005380995 |
| 0 | 0.820641479 | 0.024281415 | 0.143507802 | 0 |
| 0 | 0 | 0 | 0 | 0 |
| 0.03818501 | 0 | 0 | 0 | 0 |
| 0.295454524 | 0 | 0 | 0 | 0 |
| 0 | 0.001328201 | 0.00387413 | 0 | 0.022328166 |
| 0 | 0 | 0 | 0 | 0.001492954 |
| 0 | 0.367300127 | 0 | 0 | 0.458688047 |
| 0.256482028 | 0.009152572 | 0 | 0 | 0.005565863 |
| 0.324146683 | 0.012193468 | 0 | 0 | 0.755484133 |
| 0.01686548 | 0.00318736 | 0.509784047 | 0.00814675 | 0.088199896 |
| 0.033645673 | 0.712666878 | 0.904688512 | 5.385114909 | 0 |
| 0 | 0 | 0 | 0 | 0 |
| 0 | 0 | 0.001475057 | 0 | 0 |
| 0.000596686 | 0.055167625 | 0.005408885 | 0.011350326 | 0.006961515 |
| 0 | 0 | 0.003057704 | 0 | 0 |
| 0.261422698 | 0.008644303 | 0 | 0.041578384 | 0 |
| 0 | 0 | 0 | 0 | 0 |
| 0 | 0 | 0.026717667 | 0 | 0 |
| 0.54875757 | 0 | 0.106856822 | 0.131560615 | 0 |
| 0.292288498 | 3.790690178 | 0 | 1.134471726 | 0.248977302 |
| 0 | 0 | 0 | 0 | 0 |
| 0.130767882 | 0 | 0 | 0 | 0 |
| 0.003514542 | 0 | 0.085222565 | 0.001928498 | 0 |
| 0.04337488 | 0.513048265 | 0 | 0.115941645 | 0.478532223 |
| 0 | 0.031388011 | 0.159233293 | 0.034741589 | 0.194297874 |
| 0.001412674 | 0.960376863 | 6.464008236 | 2.687967011 | 0.001710357 |
| 0.162766972 | 0.003785872 | 0 | 0.017369483 | 0 |
| 0 | 0 | 0 | 0 | 0 |
| 0 | 0 | 0 | 0 | 0.907049456 |
| 0 | 0 | 0 | 0 | 0 |
| 0.056624343 | 0.001924744 | 0 | 0.005770309 | 0.996249328 |

| | | | | |
|---|---|---|---|---|
| 0 | 0 | 0 | 0 | 0 |
| 0 | 0.015280465 | 0.00555304 | 0 | 0 |
| 0.048400402 | 0.07037678 | 0.004920492 | 0.002836663 | 0.003883399 |
| 0 | 0 | 0 | 0.003066297 | 0.097840299 |
| 0 | 0 | 0 | 0 | 0 |
| 0.006948555 | 0.010589685 | 0.002422609 | 0.005474804 | 0.124132602 |
| 0 | 0 | 0 | 0.015449353 | 0 |
| 0 | 0 | 0 | 0 | 0.015262528 |
| 0 | 0 | 0 | 0 | 0 |
| 0 | 0 | 0 | 0 | 0 |
| 0.19585598 | 0.080225582 | 0.040158109 | 0.329477325 | 1.758438294 |
| 0.221617291 | 1.952875204 | 7.098023932 | 0.659145848 | 0.732563938 |
| 0 | 0 | 0 | 0 | 0 |
| 0 | 0.014244254 | 0.0068432 | 0.008867091 | 0.033236047 |
| 0.128989586 | 0.005282362 | 0.014894135 | 0.056256161 | 0.295213138 |
| 1.5373909 | 10.9715612 | 0 | 0 | 0 |
| 0 | 0 | 0 | 0 | 0 |
| 0 | 0 | 0 | 0 | 0 |
| 0.359245544 | 0.114393894 | 0 | 0 | 0 |
| 0.032925261 | 0.002771297 | 0.007710309 | 0.056781186 | 0.00579831 |
| 0.122093465 | 0.150098934 | 0 | 0.028945616 | 0.596488878 |
| 0 | 0.008684961 | 0.141062109 | 0 | 0 |
| 0.055550602 | 0.105304696 | 21.81687858 | 0.93837373 | 0.523810568 |
| 0 | 0 | 0 | 0 | 0 |
| 0.008026213 | 0.026573634 | 0 | 0 | 0 |
| 0.876266389 | 0 | 0.286575655 | 0.002657359 | 0 |
| 0 | 0 | 0 | 0 | 0 |
| 0 | 0.002488519 | 0 | 0.003165055 | 0 |
| 0.492090142 | 0 | 0 | 0.023874528 | 0 |
| 0 | 0 | 0 | 0 | 0 |
| 0.060763895 | 0.002631658 | 0.012281741 | 0.013517169 | 0.001491249 |
| 0.176764 | 0.096039881 | 1.000589344 | 0.20671463 | 0.041077365 |
| 0.008210171 | 0 | 0 | 0.001716222 | 0.009488416 |
| 0.795133179 | 0.020065583 | 0.107596566 | 0.046133517 | 0 |
| 0 | 0 | 0.000614011 | 0.004530912 | 0.088171639 |
| 0.010754063 | 0.083874781 | 0.003936866 | 0.031774436 | 0.079077197 |
| 0.048345447 | 0.027705762 | 0.019286433 | 0.003006727 | 0.007783872 |
| 0 | 0 | 0 | 0 | 0 |
| 0.360834191 | 1.129858136 | 0.021718375 | 0 | 0 |
| 0 | 0 | 0.003791113 | 0 | 0 |
| 0 | 0 | 0.001980919 | 0.021926392 | 0.025735977 |
| 0 | 0 | 0 | 0 | 0 |
| 0 | 0 | 0 | 0 | 0 |
| 0 | 0 | 0 | 0 | 0 |
| 0.126883036 | 0.060193325 | 0.006413641 | 0.030910791 | 0.06338977 |
| 0.003828034 | 0 | 0 | 0 | 0 |
| 0 | 0 | 0 | 0 | 0 |
| 0.00426735 | 0.947571846 | 0.068468942 | 0 | 0 |

| | | | | |
|---|---|---|---|---|
| 0 | 0 | 0 | 0 | 0 |
| 0 | 0 | 0 | 0 | 0 |
| 0 | 0 | 0 | 0 | 0 |
| 0.00944734 | 0.005153678 | 0 | 0.000504213 | 0.004646043 |
| 0 | 0 | 0 | 0 | 0 |
| 0 | 0.110275185 | 0.01255233 | 0.323412149 | 0.343861216 |
| 1.175585383 | 0 | 0.280078327 | 0.190032015 | 0 |
| 0.185887865 | 0.01702923 | 0 | 0.023198372 | 0.014657831 |
| 0 | 0 | 0 | 0 | 0 |
| 0 | 0 | 0 | 0 | 0 |
| 0.000671607 | 0.002348114 | 0 | 0.0014039 | 0.029420511 |
| 0 | 0 | 0 | 0 | 0 |
| 0 | 0.34540078 | 0.00213222 | 0.005285664 | 0 |
| 0 | 0 | 0 | 0.064811162 | 0 |
| 0 | 0 | 0 | 0 | 0 |
| 0 | 0 | 0 | 0 | 0 |
| 0 | 0.0267778 | 0.114329318 | 0.460044569 | 0.114353561 |
| 0 | 0.002823529 | 0.001267037 | 0.002688044 | 0 |
| 0.051968129 | 0.015998239 | 0 | 0.003935353 | 0.013857295 |
| 0.002184923 | 0 | 0 | 0.006218524 | 0.281700674 |
| 0 | 0 | 0 | 0 | 0 |
| 0.007805816 | 0 | 0 | 0 | 0 |
| 0.115383214 | 0.016276635 | 0 | 0.110355705 | 0 |
| 0 | 0 | 0 | 0 | 0 |
| 0 | 0 | 0 | 0 | 0 |
| 0 | 0 | 0 | 0 | 0 |
| 0 | 0 | 0 | 0 | 0 |
| 0 | 0 | 0 | 0 | 0 |
| 0 | 0 | 0 | 0 | 0 |
| 0.095679445 | 0.035944564 | 0 | 0.064095584 | 1.160841701 |
| 0 | 0 | 0 | 0 | 0.051816625 |
| 0 | 0 | 0 | 0 | 0 |
| 0.000584161 | 0.053555792 | 0 | 0 | 0 |
| 0 | 0 | 0 | 0 | 0 |
| 0 | 0 | 0 | 0.002120995 | 0 |
| 0 | 0 | 0 | 0 | 0 |
| 0.067332741 | 0.140874141 | 0.259186404 | 0.075502178 | 0 |
| 0 | 0 | 0.033756255 | 0.000356528 | 0 |
| 0 | 0.006536056 | 0 | 0 | 0 |
| 0 | 0.002614102 | 0 | 0.004806011 | 0.020120993 |
| 0.060276317 | 0.007667109 | 0 | 0 | 0 |
| 0 | 0 | 0 | 0 | 0 |
| 0.003204562 | 0 | 0 | 0 | 0 |
| 0.210904784 | 0 | 0 | 0.010040691 | 0.002309186 |
| 0.039594111 | 0.020814875 | 0.433917625 | 0 | 0 |
| 0 | 0 | 0 | 0 | 0 |
| 0.030971212 | 0.004197036 | 0.001958723 | 0 | 0 |
| 0 | 0 | 0 | 0 | 0 |

| | | | | |
|---|---|---|---|---|
| 0 | 0 | 0 | 0 | 0 |
| 0 | 0 | 0 | 0 | 0 |
| 1.593399245 | 0.063892834 | 0 | 2.402644503 | 0 |
| 0.937062354 | 0.18673537 | 0 | 1.103405295 | 0.066974654 |
| 0 | 0 | 0.014289332 | 0 | 0 |
| 0.448856301 | 0 | 0.156237711 | 2.12194537 | 0 |
| 0 | 0 | 0 | 0 | 0 |
| 0 | 0 | 0 | 0 | 0 |
| 0.008102297 | 0.002132198 | 0.015281581 | 0.003278076 | 0.009838407 |
| 0 | 0.004660562 | 0.001717143 | 0.001055928 | 0 |
| 0.009904836 | 0.000871193 | 0.007115129 | 0.000703178 | 0 |
| 0 | 0 | 0 | 0 | 0 |
| 0 | 0 | 0 | 0 | 0 |
| 0 | 0 | 0 | 0 | 0 |
| 0.662828662 | 0.144907393 | 0.184646585 | 0.033215536 | 0.447899511 |
| 0 | 0 | 0 | 0 | 0.003591821 |
| 0 | 0 | 0 | 0 | 0 |
| 0 | 0 | 0 | 0 | 0 |
| 0 | 0 | 0 | 0 | 0 |
| 0 | 0 | 0 | 0 | 0 |
| 0.071146805 | 0 | 0 | 0.30429627 | 0.047248923 |
| 0 | 0 | 0.009670228 | 0 | 0 |
| 0 | 0 | 0 | 0 | 0 |
| 0 | 0 | 0 | 0 | 0 |
| 0.088518848 | 0.097788739 | 0 | 0.055510903 | 0 |
| 3.757368614 | 0.636411641 | 0 | 1.358538116 | 4.405010466 |
| 0.237782771 | 0.053824666 | 0 | 0 | 0.667530384 |
| 0 | 0 | 0 | 0.000957172 | 0 |
| 0.114205152 | 0.708389803 | 0 | 0.417749399 | 0 |
| 0 | 0.003017307 | 0 | 0 | 0 |
| 0 | 0 | 0 | 0 | 0 |
| 0.003562914 | 0.02675924 | 0 | 0 | 0 |
| 0 | 0 | 0 | 0.001586738 | 0 |
| 0.047603045 | 0.004017343 | 0 | 0.006382208 | 0 |
| 0 | 0.000945616 | 0.007355193 | 0.006445206 | 0.013396009 |
| 0 | 0 | 0.002599615 | 0.000719365 | 0.010605677 |
| 0 | 0 | 0.624003111 | 0 | 0 |
| 0.342388552 | 0.154478256 | 0.089530138 | 2.641830156 | 0.649821164 |
| 0 | 0 | 0 | 0 | 0 |
| 0 | 0 | 0 | 0 | 0 |
| 0 | 0 | 0 | 0 | 0 |
| 0 | 0 | 0 | 0.000407883 | 0.003293086 |
| 0.127494294 | 0.671675281 | 0.166253248 | 0.246247849 | 0.136991905 |
| 0 | 0 | 0.138270828 | 0 | 0 |
| 0 | 0 | 0.114893159 | 0 | 0 |
| 0 | 0 | 0.008373549 | 0.002385279 | 0.00717682 |
| 0 | 0 | 0 | 0 | 0 |
| 0.138505493 | 0.089452057 | 0 | 0.052838582 | 0.607882893 |

| | | | | |
|---|---|---|---|---|
| 0.079614517 | 0 | 0.021065724 | 0 | 0 |
| 0 | 0 | 0 | 0 | 0 |
| 0 | 0 | 0 | 0 | 0 |
| 0 | 0 | 0 | 0 | 0 |
| 0.004070389 | 0 | 0 | 0.009135531 | 0.033247094 |
| 0 | 0.011540539 | 0 | 0 | 0 |
| 0.00562805 | 0 | 0 | 0 | 4.284830907 |
| 0 | 0 | 0 | 0 | 0.009289431 |
| 0 | 0 | 0 | 0 | 2.114888365 |
| 0 | 0 | 0 | 0 | 0.014626188 |
| 0 | 0.001337286 | 0 | 0 | 0 |
| 0.897688971 | 0.369228499 | 0.277070817 | 3.372979329 | 3.013873217 |
| 0.070555765 | 0.629098453 | 0.247601959 | 0.017730976 | 0.001541735 |
| 0 | 0 | 0 | 0 | 0 |
| 0.184827587 | 0.944477123 | 0.015833582 | 0.375532178 | 0.001472563 |
| 0 | 0 | 0 | 0 | 0 |
| 0.003102329 | 0.005222417 | 0 | 0.000972748 | 0.003642242 |
| 0 | 0 | 0 | 0 | 0 |
| 0 | 0 | 0 | 0.014707688 | 0.001577518 |
| 0 | 0 | 0 | 0.002114062 | 0.052953522 |
| 0.002876606 | 0.064953862 | 0 | 0.028750346 | 0 |
| 0 | 0 | 0 | 0 | 0 |
| 0.340175148 | 2.409937223 | 0.062304604 | 0.298736768 | 0.802519971 |
| 0 | 0 | 0 | 0 | 0 |
| 0 | 0 | 0 | 0 | 0 |
| 0 | 0 | 0 | 0 | 0 |
| 0 | 0 | 0.015863717 | 0 | 0 |
| 0 | 0 | 0 | 0 | 0 |
| 0.004240181 | 0.001820588 | 0 | 0 | 0.001179028 |
| 0 | 0 | 0 | 0 | 0 |
| 0 | 0 | 0 | 0 | 0 |
| 0.003536509 | 0.003746839 | 0.026666458 | 0.016129268 | 0.057962654 |
| 0 | 0 | 0 | 7.127516796 | 0 |
| 0.540634424 | 0 | 0.031885689 | 0.02064838 | 0.003076033 |
| 0.000832232 | 0.001697327 | 0.001697419 | 0.000391425 | 0.009205827 |
| 0.12605967 | 0.24412501 | 0.070751133 | 0.059037501 | 0 |
| 0 | 0 | 0 | 0 | 0 |
| 0 | 0 | 0 | 0 | 0 |
| 0 | 0 | 0 | 0 | 0 |
| 0 | 0 | 0 | 0 | 0 |
| 0.093596567 | 0 | 0 | 0.001222816 | 0.02528618 |
| 0 | 0 | 0 | 0 | 0 |
| 0 | 0 | 0 | 0 | 0 |
| 0.010840218 | 0 | 0 | 0 | 0.024608447 |
| 0.183232487 | 0.058282439 | 0.027311427 | 0.012467496 | 0.517319821 |
| 0 | 0 | 0 | 0 | 0 |
| 0.073979792 | 0.041400536 | 0.015944764 | 0.304206216 | 0.391074574 |
| 0.003101658 | 0.002485133 | 0.009225615 | 0.002309777 | 0.003072477 |

| | | | | |
|---|---|---|---|---|
| 0 | 0 | 0 | 0.719194817 | 0 |
| 0 | 0.016575401 | 0 | 0 | 0 |
| 0 | 0 | 0 | 0 | 0.01175908 |
| 0 | 0 | 0 | 0 | 0 |
| 0 | 0 | 0 | 0 | 0 |
| 0 | 0 | 0 | 0 | 0.002529055 |
| 0 | 0.979529358 | 0 | 0 | 0 |
| 0.00060058 | 0.006066048 | 0.001088837 | 0.01694829 | 0.97171831 |
| 0 | 0 | 0 | 0 | 0 |
| 0.001181987 | 0 | 0 | 0.002717858 | 0 |
| 0 | 0 | 0 | 0 | 0 |
| 0 | 0 | 0 | 0 | 0 |
| 0 | 0 | 0 | 0 | 0 |
| 0 | 0 | 0.153636282 | 0 | 0 |
| 0 | 0 | 0 | 0 | 0 |
| 0 | 0 | 0 | 0 | 0 |
| 0 | 0 | 0 | 0 | 0 |
| 0 | 0.001649692 | 0 | 0 | 0 |
| 0 | 0 | 0.509409817 | 0.345875619 | 0.633034795 |
| 0 | 0 | 0 | 0.000565857 | 0 |
| 0 | 0 | 0 | 0.001690943 | 0 |
| 0 | 0 | 0 | 0 | 0 |
| 0 | 0 | 0.256540302 | 0 | 0 |
| 0 | 0 | 0 | 0 | 0 |
| 0 | 0 | 0 | 0 | 0 |
| 0 | 0 | 0 | 0.004666638 | 0.001134078 |
| 0 | 0 | 0 | 0 | 0 |
| 0.098050713 | 0.170426117 | 0 | 0.084326969 | 1.091289675 |
| 0 | 0 | 0 | 0 | 0 |
| 0.357200365 | 1.470279711 | 1.608353683 | 5.677257583 | 0 |
| 0 | 0 | 0 | 0 | 0 |
| 0 | 0 | 0 | 0 | 0 |
| 0 | 0.002298279 | 0 | 0.004946777 | 0.020576921 |
| 0 | 0 | 0 | 0 | 0 |
| 0.008598073 | 0.001631224 | 0 | 0 | 0.010431895 |
| 0.235323196 | 0 | 0 | 0 | 0 |
| 0 | 0 | 0 | 0 | 0 |
| 0.015655449 | 0 | 0 | 0.001280565 | 0 |
| 0 | 0 | 0 | 0 | 0 |
| 0 | 0 | 0 | 0 | 0 |
| 6.333360235 | 1.687134332 | 13.40443474 | 2.218044386 | 3.083616056 |
| 0.238664027 | 0.001161353 | 0 | 0.801979858 | 1.062485408 |
| 0.242306947 | 2.52651285 | 0.099398394 | 0.24204854 | 0 |
| 0 | 0 | 0 | 0.265150028 | 0.210290137 |
| 0 | 0 | 0 | 0 | 0 |
| 0 | 0 | 0 | 0 | 0 |
| 0.015705864 | 0 | 0 | 0 | 0 |
| 0.007202916 | 0.004407077 | 0.001469105 | 0.000903402 | 0.00570812 |

| | | | | |
|---|---|---|---|---|
| 0 | 0 | 0 | 0 | 0 |
| 0.01166885 | 0.001942732 | 0.013096167 | 0.07573169 | 0.006972136 |
| 0.010816316 | 0 | 0 | 0 | 0 |
| 0 | 0 | 0 | 0 | 0 |
| 1.362076737 | 6.546793677 | 1.973070201 | 3.482318744 | 0.287517546 |
| 0 | 0 | 0 | 0 | 0 |
| 0 | 0 | 0 | 0 | 0 |
| 0 | 0 | 0 | 0 | 0 |
| 0 | 0.001293245 | 0.080221503 | 0.003015523 | 0.007083995 |
| 0 | 0 | 0 | 0 | 0 |
| 0 | 0 | 0 | 0 | 0 |
| 0 | 0 | 0 | 0 | 0 |
| 0.139658774 | 0.019506187 | 0 | 0.121778463 | 0.101262718 |
| 0.013275484 | 0.005415036 | 0.041517535 | 0.004440089 | 0.001461176 |
| 0.082536217 | 2.112092314 | 0.178485546 | 0 | 0 |
| 0.1102086 | 0.015869783 | 0 | 0.287153701 | 1.100110595 |
| 0.058396684 | 0.160212252 | 0 | 0.00600345 | 0.192444932 |
| 0 | 0 | 0 | 0 | 0 |
| 0 | 0 | 0 | 0 | 0 |
| 0 | 0 | 0 | 0 | 0 |
| 0 | 0 | 0 | 0 | 0 |
| 0 | 0 | 0 | 0 | 0 |
| 0 | 0 | 0 | 0 | 0 |
| 0 | 0 | 0 | 0 | 0 |
| 0 | 0 | 0 | 0 | 0 |
| 0 | 0 | 0 | 0 | 0 |
| 0.001997835 | 0.017462399 | 0.015846373 | 0.027980521 | 0.705197716 |
| 0 | 0 | 0 | 0 | 0.003597812 |
| 0 | 0 | 0 | 0 | 0 |
| 0 | 0 | 0 | 0 | 0 |
| 0 | 0 | 0 | 0 | 0 |
| 0 | 0 | 0.001280229 | 0 | 0.000310891 |
| 0.014579918 | 0 | 0 | 5.326160575 | 0.038647722 |
| 0.004726287 | 0.020655433 | 0 | 0 | 0.025619967 |
| 0.049377787 | 0.541257405 | 0.090259721 | 0 | 0 |
| 0.48765791 | 0 | 0 | 0 | 0.008897901 |
| 0 | 0.001812072 | 0 | 0.011282934 | 0 |
| 0 | 0.03419903 | 0 | 0.000514751 | 0.021005353 |
| 0 | 0 | 0 | 0 | 0 |
| 0 | 0 | 0 | 0 | 0 |
| 0.002129253 | 0.695433844 | 0.936239739 | 1.629532814 | 0 |
| 0 | 0.859369139 | 0 | 0 | 0 |
| 0 | 0 | 0 | 0.076774772 | 0 |
| 0 | 0 | 0 | 0 | 0 |
| 0 | 0 | 0 | 0 | 0 |
| 0 | 0 | 0 | 0 | 0 |
| 0 | 0 | 0 | 0 | 0 |
| 0 | 0 | 0 | 0 | 0 |

| | | | | |
|---|---|---|---|---|
| 0.044429119 | 1.08039967 | 0.007271779 | 0.152079367 | 0.093184042 |
| 0 | 0 | 0 | 0 | 0 |
| 0 | 0 | 0 | 0 | 0 |
| 0 | 0 | 0 | 0 | 0 |
| 0.037058364 | 0.005695201 | 0 | 0.002936873 | 0.111070411 |
| 0.069433785 | 0 | 0 | 0 | 0 |
| 0.077354585 | 0.547804489 | 0 | 0.022637861 | 0.333626235 |
| 0.003507891 | 0 | 0 | 0 | 0 |
| 0 | 0 | 0 | 0 | 0.000979338 |
| 0 | 0 | 0.002724754 | 0 | 0.002911388 |
| 0.047018359 | 0.005668572 | 0 | 0 | 0.00152959 |
| 0.009520836 | 0 | 0 | 0 | 0.468474523 |
| 0 | 0 | 0 | 0 | 0 |
| 0 | 0 | 0 | 0 | 0 |
| 0 | 0 | 0 | 0.002000745 | 0 |
| 0 | 0 | 0 | 0 | 0 |
| 0 | 0 | 0 | 0 | 0 |
| 0 | 0 | 0 | 0 | 0.000848857 |
| 0 | 0 | 0 | 0 | 0 |
| 0 | 0 | 0 | 0 | 0 |
| 0.111287102 | 0.09769611 | 0 | 0 | 0.187696368 |
| 0 | 0 | 0 | 0 | 0 |
| 0.044526702 | 0.031789917 | 0.008999841 | 0.008175672 | 0.204246866 |
| 0.013019518 | 0.011639733 | 0 | 0.002907957 | 0.038985537 |
| 0 | 0 | 0 | 0 | 0 |
| 0 | 0 | 0 | 0.004750249 | 0.001111642 |
| 0 | 0 | 0 | 0 | 0 |
| 0 | 0 | 0 | 0 | 0 |
| 0 | 0 | 0 | 0 | 0 |
| 0.103958169 | 0.044273947 | 0.184285379 | 0.339003751 | 0.00186466 |
| 0 | 0 | 0.050210671 | 0 | 0 |
| 0 | 0 | 0 | 0 | 0 |
| 0 | 0 | 0 | 0 | 0 |
| 0 | 0 | 0 | 0 | 2.083596989 |
| 0 | 0 | 0 | 0 | 0 |
| 0 | 0 | 0 | 0 | 1.036689633 |
| 0.007587491 | 0 | 0 | 0.000699732 | 0.012821593 |
| 0 | 0 | 0 | 0 | 0 |
| 0 | 0 | 0 | 0 | 0 |
| 0 | 0 | 0 | 0 | 0 |
| 0 | 0 | 0 | 0 | 0 |
| 0 | 0 | 0 | 0 | 0 |
| 0 | 0 | 0 | 0 | 0 |
| 0 | 0 | 0 | 0 | 0 |
| 0 | 0 | 0 | 0 | 0 |
| 0 | 0 | 0 | 0 | 0 |
| 0 | 0 | 0 | 0 | 0 |
| 0 | 0 | 0 | 0 | 0 |

| | | | | |
|---|---|---|---|---|
| 0.00403012 | 0 | 0.001565681 | 0.001925578 | 0.010772613 |
| 0 | 0.276809937 | 0 | 0 | 0 |
| 0.018017253 | 0 | 0 | 0 | 0 |
| 0 | 0 | 0 | 0 | 0 |
| 0 | 0 | 0 | 0 | 0 |
| 0 | 0 | 0 | 0 | 0 |
| 0 | 0 | 0 | 0 | 0 |
| 0.651442905 | 0.072108438 | 0.072342021 | 0.339201995 | 1.346583869 |
| 4.199479402 | 3.172200896 | 0 | 2.040743605 | 0.037001382 |
| 0 | 0 | 0.03279201 | 0.001841143 | 0 |
| 0 | 0 | 0.036807764 | 0 | 0.013488851 |
| 0 | 0 | 0 | 0 | 0 |
| 0 | 0 | 0 | 0.053642897 | 0 |
| 0 | 0 | 0 | 0 | 0.013990932 |
| 0.002200045 | 0 | 0 | 0 | 0.003632244 |
| 0 | 0 | 0 | 0 | 0 |
| 0 | 0 | 0 | 0 | 0 |
| 0 | 0 | 0 | 0 | 0 |
| 0 | 0 | 0 | 0 | 0 |
| 0 | 0 | 0 | 0 | 0 |
| 0 | 0 | 0 | 0 | 0 |
| 0 | 0 | 0 | 0 | 0 |
| 0 | 0 | 0 | 0 | 0 |
| 0 | 0 | 0 | 0 | 0 |
| 0.009220522 | 0.001585445 | 0 | 0.033745681 | 0.088742344 |
| 0 | 0 | 0.00974805 | 0 | 0 |
| 0 | 0 | 0 | 0 | 0 |
| 0 | 0 | 0 | 0 | 0 |
| 0 | 0 | 0 | 0 | 0 |
| 0 | 0 | 0 | 0 | 0 |
| 0 | 0 | 0 | 0 | 0 |
| 0 | 0 | 0 | 0 | 0 |
| 0 | 0 | 0 | 0 | 0 |
| 0 | 0 | 0 | 0.003923486 | 0.079006939 |
| 0 | 0.008393799 | 0.002843216 | 0.00845054 | 0 |
| 0.006733893 | 0 | 0 | 0.04030931 | 0 |
| 0 | 0 | 0 | 0 | 0 |
| 0 | 0 | 0 | 0 | 0 |
| 0 | 0 | 0 | 0 | 0 |
| 0 | 0.001339676 | 0 | 0 | 0 |
| 0 | 0 | 0 | 0 | 0 |
| 0.206747888 | 0.213766825 | 0.407707162 | 1.273726058 | 0.620412717 |
| 0 | 0 | 0 | 0 | 0 |
| 0 | 0 | 0 | 0 | 0 |
| 0 | 0 | 0 | 0 | 0 |
| 0 | 0.042957239 | 0.002552382 | 0.039702342 | 0 |
| 0 | 0 | 0 | 0 | 0 |
| 0 | 0 | 0.004860053 | 0 | 0 |

| | | | | |
|---|---|---|---|---|
| 0 | 0 | 0 | 0 | 0 |
| 0 | 0 | 0.037447031 | 0.00032586 | 0 |
| 0 | 0 | 0 | 0 | 0 |
| 0 | 0 | 0 | 0 | 0 |
| 0.446558817 | 1.997894712 | 0.421813275 | 2.865254093 | 0 |
| 0 | 0 | 0.001203724 | 0 | 0 |
| 0 | 0 | 0 | 0.00698049 | 0 |
| 0 | 0 | 0 | 0.008266978 | 0.015960598 |
| 0.010724946 | 0.00965838 | 0 | 0.005197134 | 0.000804852 |
| 0 | 0 | 0 | 0 | 0 |
| 0 | 0 | 0 | 0 | 0 |
| 0 | 0.13156408 | 0.045628368 | 0 | 0 |
| 0 | 0 | 0.159795824 | 0 | 0 |
| 0 | 0 | 0 | 0 | 0 |
| 0 | 0 | 0 | 0 | 0 |
| 0.604268647 | 0.006415528 | 0 | 0.05696078 | 0 |
| 0 | 0 | 0 | 0 | 0 |
| 0 | 0 | 0 | 0 | 0 |
| 0 | 0 | 0 | 0 | 0 |
| 0 | 0 | 0 | 0 | 0 |
| 0 | 0 | 0 | 0 | 0 |
| 0.148126654 | 0 | 0 | 0 | 0 |
| 0 | 0 | 0 | 0 | 0 |
| 0.001901423 | 0.000984871 | 0 | 0.001589864 | 0.027625166 |
| 0.036654226 | 0 | 0.614436863 | 0 | 0.03277154 |
| 0 | 0 | 0 | 0.000483005 | 0 |
| 0.743547814 | 0.004278686 | 0 | 0.008118788 | 0.022586586 |
| 0 | 0 | 0 | 0.003572641 | 0 |
| 0 | 0 | 0 | 0 | 0 |
| 0 | 0 | 0 | 0 | 0 |
| 0 | 0 | 0 | 0 | 0 |
| 0 | 0 | 0 | 0 | 0 |
| 0 | 0 | 0 | 0 | 0 |
| 0 | 0 | 0 | 0 | 0 |
| 0 | 0 | 0 | 0 | 0 |
| 0 | 0 | 0 | 0 | 0 |
| 0 | 0 | 0 | 0 | 0 |
| 0.078474671 | 0.027944904 | 0.135805768 | 0.014912763 | 1.097339128 |
| 0.089147897 | 0 | 0 | 0 | 0 |
| 0 | 0 | 0 | 0 | 0 |
| 0 | 0 | 0 | 0 | 0 |
| 0 | 0 | 0 | 0 | 0 |
| 0 | 0 | 0 | 0 | 0 |
| 0 | 0 | 0 | 0 | 0 |
| 0 | 0 | 0 | 0 | 0 |
| 0 | 0 | 0 | 0 | 0 |
| 0 | 0 | 0 | 0 | 0 |
| 0 | 0.004330557 | 0 | 0 | 0 |

| | | | | |
|---|---|---|---|---|
| 0.067053153 | 0.280602574 | 0 | 0.070310558 | 0.137524772 |
| 0 | 0 | 0 | 0 | 0 |
| 0 | 0 | 0 | 0.001354551 | 0.074175362 |
| 0.005604715 | 0 | 0 | 0 | 0 |
| 0 | 0 | 0 | 0 | 0 |
| 0 | 0.23872964 | 0 | 0 | 0 |
| 0 | 0 | 0 | 0 | 0 |
| 0 | 0 | 0 | 0 | 0 |
| 0 | 0 | 0 | 0 | 0 |
| 0 | 0 | 0 | 0 | 0 |
| 0 | 0 | 0 | 0 | 0 |
| 0.002142322 | 0.006128282 | 0 | 0.000977067 | 0 |
| 0 | 0 | 0 | 0 | 0 |
| 0 | 0 | 0 | 0 | 0 |
| 0 | 0.001773379 | 0.043708787 | 0 | 0 |
| 0 | 0 | 0 | 0 | 0 |
| 0 | 0 | 0 | 0 | 0 |
| 0.003861979 | 0 | 0 | 0 | 0 |
| 0 | 0 | 0 | 0 | 0 |
| 0 | 0 | 0 | 0 | 0 |
| 0 | 0 | 0.005435305 | 0 | 0 |
| 0 | 0 | 0 | 0 | 0 |
| 0.073333825 | 0.051402741 | 0.001445136 | 0.002166113 | 0.015441213 |
| 0 | 0.014454035 | 0 | 0.047733455 | 5.79549126 |
| 0 | 0 | 0 | 0 | 0 |
| 0 | 0 | 0 | 0 | 0 |
| 0 | 0 | 0 | 0 | 0 |
| 0 | 0 | 0 | 0 | 0 |
| 0 | 0 | 0 | 0 | 0 |
| 0 | 0 | 0 | 0 | 0 |
| 0 | 0 | 0 | 0 | 0 |
| 0.003740288 | 0 | 0 | 0 | 0 |
| 0.017915693 | 0.007829749 | 0.015902022 | 0 | 0.01347468 |
| 0 | 0 | 0 | 0 | 0 |
| 0.075898561 | 0.61113532 | 0.212456312 | 0.052900639 | 0.043949489 |
| 0 | 0 | 0 | 0 | 0.006856939 |
| 0 | 0 | 0 | 0 | 0 |
| 0 | 0 | 0 | 0 | 0 |
| 0 | 0 | 0.000942113 | 0.015062754 | 0 |
| 0 | 0 | 0 | 0 | 0 |
| 0 | 0 | 0 | 0 | 0 |
| 0 | 0 | 0 | 0 | 0 |
| 0.052390925 | 1.453105489 | 0.156061806 | 0 | 0 |
| 0 | 0 | 0 | 0 | 0 |
| 0 | 0 | 0 | 0 | 0 |
| 0.081713661 | 0.021995574 | 0.00758222 | 0.089978417 | 0.019829018 |
| 0 | 0 | 0 | 0 | 0 |
| 0 | 0 | 0 | 0 | 0 |

| | | | | |
|---|---|---|---|---|
| 0 | 0 | 0 | 0 | 0 |
| 0 | 0 | 0 | 0 | 0 |
| 0 | 0 | 0 | 0 | 0 |
| 0 | 0 | 0 | 0 | 0 |
| 0 | 0 | 0 | 0 | 0 |
| 0 | 0 | 0 | 0 | 0 |
| 0 | 0 | 0 | 0 | 0 |
| 0 | 0 | 0 | 0 | 0.008211906 |
| 0.225856513 | 0 | 0 | 0.009702856 | 0 |
| 0.00066271 | 0.081095312 | 0 | 0.001385303 | 0 |
| 0 | 0 | 0 | 0 | 0 |
| 0 | 0 | 0 | 0 | 0 |
| 0 | 0 | 0 | 0 | 0 |
| 0.119318286 | 0 | 0 | 0 | 0 |
| 0 | 0 | 0 | 0 | 0 |
| 0 | 0 | 0 | 0 | 0 |
| 0 | 0 | 0 | 0 | 0 |
| 0 | 0 | 0 | 0 | 0 |
| 0 | 0 | 0 | 0 | 0 |
| 0.004185256 | 0.126393106 | 0 | 0.479276378 | 0.362674406 |
| 0 | 0.000793655 | 0.003395264 | 0.001992958 | 0 |
| 0 | 0 | 0 | 0 | 0 |
| 0 | 0 | 0 | 0 | 0 |
| 0 | 0.003373979 | 0 | 0.025417339 | 0.175893913 |
| 0 | 0.004108066 | 0 | 0 | 0 |
| 0 | 0 | 0 | 0 | 0 |
| 0 | 0 | 0 | 0 | 0 |
| 0 | 0 | 0 | 0 | 0 |
| 0 | 0 | 0 | 0 | 0 |
| 0 | 0.032290425 | 0 | 0.198050649 | 0.041950612 |
| 0.563164893 | 0.013175896 | 0 | 0.140911654 | 1.291305876 |
| 0 | 0 | 0 | 0 | 0 |
| 0 | 0 | 0 | 0 | 0.004729425 |
| 0 | 0.00119579 | 0 | 0 | 0 |
| 0 | 0 | 0 | 0 | 0 |
| 0 | 0 | 0 | 0 | 0 |
| 0 | 0 | 0 | 0 | 0 |
| 0 | 0 | 0 | 0 | 0 |
| 0 | 0 | 0 | 0 | 0 |
| 0 | 0 | 0 | 0 | 0 |
| 0.006369288 | 0 | 0 | 0.001041974 | 0.001645921 |
| 0 | 0 | 0.159435474 | 0 | 0 |
| 0 | 0 | 0 | 0 | 0.006056529 |
| 0.005803873 | 0 | 0 | 0 | 0 |
| 0 | 0 | 0 | 0 | 0 |
| 0 | 0 | 0 | 0 | 0 |
| 0 | 0 | 0 | 0 | 0 |
| 0 | 0.001475125 | 0 | 0 | 0.001313542 |

| | | | | |
|---|---|---|---|---|
| 0 | 0 | 0 | 0 | 0 |
| 0 | 0 | 0 | 0 | 0 |
| 0 | 0 | 0 | 0 | 0 |
| 0 | 0 | 0.002440235 | 0 | 0 |
| 0.025764036 | 0.01259153 | 0.13899868 | 0.055037495 | 1.428114324 |
| 0 | 0 | 0 | 0 | 0 |
| 0 | 0 | 0 | 0 | 0 |
| 0 | 0 | 0 | 0 | 0 |
| 0 | 0 | 0 | 0 | 0 |
| 0 | 0 | 0 | 0 | 0 |
| 0 | 0.009838072 | 0 | 0 | 0.019929967 |
| 0.051923635 | 0.013447313 | 0.004706816 | 0.003183815 | 0.09662184 |
| 0 | 0 | 0 | 0.00549366 | 0 |
| 0 | 0 | 0 | 0 | 0 |
| 0 | 0 | 0 | 0.021726317 | 0.026944418 |
| 0 | 0.031999007 | 0 | 0 | 0.826153419 |
| 0.007277124 | 0 | 0.014480399 | 0 | 0 |
| 0 | 0 | 0 | 0 | 0 |
| 0 | 0 | 0.004514435 | 0.012821087 | 0.069008267 |
| 0.037418136 | 0.182674519 | 0 | 0.028902287 | 0.114663828 |
| 0 | 0 | 0 | 0 | 0 |
| 0 | 0 | 0 | 0 | 0.017174173 |
| 0 | 0 | 0 | 0 | 0 |
| 0 | 0 | 0 | 0 | 0 |
| 0 | 0 | 0 | 0 | 0 |
| 0 | 0 | 0 | 0 | 0 |
| 0 | 0 | 0.004226302 | 0.004012997 | 0.014972125 |
| 0 | 0 | 0 | 0 | 0.001373905 |
| 0 | 0 | 0 | 0 | 0 |
| 0 | 0 | 0 | 0 | 0 |
| 0 | 0 | 0.000845424 | 0 | 0.004516659 |
| 0 | 0 | 0 | 0 | 0.015079057 |
| 0 | 0 | 0 | 0 | 0 |
| 0 | 0 | 0 | 0 | 0 |
| 0 | 0 | 0 | 0 | 0.045713736 |
| 0 | 0 | 0 | 0.003541635 | 0 |
| 0 | 0 | 0 | 0 | 0 |
| 0.500605946 | 0.009842401 | 0.004157792 | 0.014610082 | 0.01009678 |
| 0.014886777 | 0.118792135 | 0 | 0.136190243 | 0 |
| 0 | 0 | 0.008479719 | 0.023754763 | 0 |
| 0.027474052 | 0.011520008 | 0 | 0 | 0 |
| 0 | 0.001305251 | 0 | 0 | 0 |
| 0 | 0 | 0 | 0 | 0 |
| 0 | 0 | 0 | 0 | 0 |
| 0 | 0 | 0 | 0 | 0 |
| 0 | 0 | 0 | 0 | 0 |
| 0 | 0 | 0 | 0 | 0 |
| 0 | 0 | 0 | 0 | 0 |

| | | | | |
|---|---|---|---|---|
| 0 | 0 | 0 | 0 | 0 |
| 0.000738195 | 0.017421244 | 0 | 0.066083016 | 0.051065632 |
| 0.883290918 | 0 | 0 | 0 | 0.003614944 |
| 0.000678857 | 0.001054873 | 0 | 0.000709529 | 0.006724706 |
| 0 | 0 | 0 | 0 | 0 |
| 0 | 0 | 0 | 0 | 0 |
| 0.006807389 | 0.474217092 | 0 | 0.027954867 | 0 |
| 0 | 0 | 0 | 0 | 0 |
| 0.003676606 | 0 | 0 | 0 | 0 |
| 0 | 0 | 0 | 0 | 0 |
| 0 | 0.007844923 | 0 | 0.034356854 | 0 |
| 0 | 0 | 0 | 0 | 0 |
| 0.036732694 | 0.705882182 | 0 | 0.127449007 | 0.122985242 |
| 0 | 0 | 0 | 0 | 0 |
| 0 | 0 | 0 | 0 | 0 |
| 0 | 0 | 0 | 0 | 0 |
| 0 | 0 | 0 | 0 | 0 |
| 0 | 0 | 0 | 0 | 0 |
| 0 | 0 | 0 | 0 | 0 |
| 0 | 0 | 0 | 0 | 0 |
| 0 | 0 | 0 | 0 | 0 |
| 0 | 0 | 0 | 0 | 0 |
| 0 | 0 | 0 | 0 | 0 |
| 0.010966661 | 0 | 0 | 0.002969736 | 0.015801432 |
| 0 | 0 | 0 | 0 | 1.287629821 |
| 0.012884748 | 0.004719364 | 0 | 0.001269735 | 0 |
| 0.000605396 | 0 | 0 | 0 | 0 |
| 0 | 0.00181513 | 0 | 0 | 0 |
| 0 | 0 | 0 | 0 | 0 |
| 0 | 0 | 0 | 0 | 0 |
| 0.011539747 | 0.048579878 | 0.051556029 | 0.137496752 | 0 |
| 0 | 0 | 0 | 0.010849701 | 0 |
| 0 | 0 | 0 | 0 | 0 |
| 0 | 0 | 0 | 0 | 0 |
| 0.006910475 | 0.008743924 | 0 | 0 | 0.232745631 |
| 0 | 0 | 0 | 0 | 0 |
| 0 | 0 | 0 | 0 | 0.005287528 |
| 0 | 0 | 0 | 0 | 0 |
| 0.033673859 | 0.058750239 | 0.001354658 | 0.001999257 | 0 |
| 0 | 0 | 0 | 0 | 0 |
| 0 | 0 | 0 | 0 | 0 |
| 0 | 0 | 0 | 0 | 0 |
| 0 | 0 | 0 | 0 | 0 |
| 0 | 0 | 0 | 0 | 0 |
| 0.401893971 | 6.462348878 | 0.16312482 | 0 | 0 |
| 0.365171259 | 0.155071514 | 0.145647116 | 0.724494801 | 0.434268033 |
| 0 | 0 | 0 | 0 | 0 |
| 0 | 0 | 0 | 0 | 0 |

| | | | | |
|---|---|---|---|---|
| 0.041749597 | 0 | 0.001910471 | 0 | 0 |
| 0 | 0 | 0 | 0 | 0 |
| 0.073725638 | 0 | 0.310766068 | 0 | 0 |
| 0.001323407 | 0 | 0 | 0 | 0.266561123 |
| 0 | 0 | 0 | 0 | 0 |
| 0 | 0 | 0 | 0 | 0.0017054 |
| 0.013367501 | 0 | 0 | 0 | 0.012572963 |
| 0 | 0 | 0 | 0 | 0 |
| 0 | 0 | 0 | 0 | 0.057710467 |
| 0 | 0 | 0 | 0 | 0 |
| 0.002263073 | 0 | 0 | 0.000304112 | 0.002989042 |
| 0 | 0 | 0 | 0.000605388 | 0.004781409 |
| 0 | 0 | 0 | 0 | 0 |
| 0 | 0 | 0 | 0 | 0 |
| 0 | 0 | 0 | 0 | 0 |
| 0 | 0 | 0 | 0 | 0 |
| 0 | 0 | 0 | 0 | 0 |
| 0 | 0 | 0 | 0 | 0 |
| 0 | 0 | 0 | 0 | 0 |
| 0 | 0.00099934 | 0.006704269 | 0.002823138 | 0.009343674 |
| 0 | 0 | 0 | 0 | 0 |
| 0 | 0 | 0 | 0 | 0 |
| 0 | 0 | 0 | 0 | 0 |
| 0 | 0 | 0 | 0 | 0 |
| 0 | 0 | 0 | 0 | 0 |
| 0 | 0 | 0 | 0 | 0.015661915 |
| 0 | 0 | 0 | 0 | 0 |
| 0.005693761 | 0 | 0 | 0 | 0 |
| 0 | 0 | 0 | 0 | 0 |
| 0 | 0 | 0 | 0 | 0 |
| 0.003867542 | 0.003648779 | 0.010768062 | 0.00438876 | 0.005351455 |
| 0 | 0 | 0 | 0 | 0 |
| 0.009408897 | 0.026416474 | 0.07676148 | 0.006034497 | 0.007484545 |
| 0 | 0 | 0.000891385 | 0.000959249 | 0.001154474 |
| 0 | 0 | 0 | 0 | 0 |
| 0 | 0 | 0 | 0 | 0 |
| 0 | 0 | 0 | 0 | 0 |
| 0 | 0 | 0 | 0 | 0 |
| 0 | 0 | 0 | 0 | 0 |
| 0 | 0 | 0 | 0 | 0 |
| 0 | 0 | 0 | 0 | 0 |
| 0 | 0 | 0.002152874 | 0 | 0.009178112 |
| 0.066654729 | 0.057199073 | 0 | 0.152280055 | 0.141113711 |
| 0 | 0 | 0 | 0 | 0 |
| 0 | 0 | 0 | 0 | 0 |
| 0.010530239 | 0.187691927 | 0.025688723 | 0.061568861 | 0 |
| 0 | 0.01135903 | 0 | 0.000509354 | 0.003218339 |
| 0 | 0 | 0 | 0 | 0 |

| | | | | |
|---|---|---|---|---|
| 0 | 0 | 0 | 0 | 0 |
| 0.013921027 | 0 | 0.025340258 | 0 | 0 |
| 0.000786834 | 0 | 0 | 0.008881739 | 0.012470896 |
| 0 | 0 | 0 | 0 | 0 |
| 0.234127397 | 0.003172756 | 0 | 0.028629217 | 0.103308133 |
| 0 | 0 | 0 | 0 | 0.070539743 |
| 0 | 0 | 0 | 0 | 0 |
| 0 | 0 | 0 | 0 | 0 |
| 0 | 0 | 0 | 0 | 0 |
| 0 | 0 | 0 | 0 | 0 |
| 0 | 0 | 0 | 0 | 0 |
| 0 | 0 | 0 | 0 | 0 |
| 0 | 0 | 0 | 0 | 0 |
| 0 | 0 | 0 | 0.014377677 | 0 |
| 0 | 0 | 0 | 0 | 0 |
| 0 | 0 | 0 | 0 | 0 |
| 0.267051069 | 0 | 0 | 0 | 2.587766665 |
| 0 | 0 | 0 | 0 | 0 |
| 0 | 0 | 0 | 0 | 0 |
| 0 | 0 | 0 | 0 | 0 |
| 0 | 0 | 0 | 0 | 0 |
| 0 | 0 | 0 | 0 | 0.006130296 |
| 0 | 0 | 0 | 0 | 0 |
| 0 | 0 | 0 | 0 | 0 |
| 0 | 0.036526994 | 0.001763468 | 0 | 0 |
| 0 | 0 | 0 | 0 | 0 |
| 0 | 0 | 0 | 0 | 0 |
| 0.029319254 | 0 | 0 | 0.168387196 | 0.040674709 |
| 0 | 0 | 0 | 0 | 0 |
| 0 | 0 | 0 | 0 | 0 |
| 0 | 0.006628291 | 0 | 0 | 0 |
| 0 | 0 | 0 | 0 | 0 |
| 0 | 0 | 0 | 0 | 0 |
| 0 | 0 | 0 | 0 | 0 |
| 0 | 0 | 0 | 0 | 0 |
| 0 | 0 | 0 | 0 | 0 |
| 0 | 0 | 0 | 0 | 0 |
| 0 | 0 | 0 | 0 | 0 |
| 0 | 0 | 0 | 1.738730981 | 0 |
| 0 | 0 | 0 | 0 | 0 |
| 0 | 0 | 0 | 0 | 0 |
| 0.004246111 | 0.011546517 | 0 | 0.158139017 | 0 |
| 0.001964074 | 0.001017322 | 0.00178041 | 0.002052812 | 0.007494147 |
| 0 | 0 | 0 | 0 | 0 |
| 0 | 0 | 0 | 0 | 0 |
| 0 | 0 | 0 | 0.001731904 | 0 |
| 0.020030994 | 0.001556303 | 0 | 0.007536962 | 0.005952762 |
| 0 | 0 | 0 | 0.248235906 | 0 |

| | | | | |
|---|---|---|---|---|
| 0 | 0 | 0 | 0 | 0 |
| 0.044232003 | 0.059403989 | 0.014606321 | 0.024039821 | 0.002642861 |
| 0 | 0 | 0 | 0.161003031 | 0 |
| 0 | 0 | 0 | 0 | 0 |
| 0 | 0 | 0 | 0 | 0 |
| 0 | 0 | 0 | 0 | 0 |
| 0 | 0 | 0 | 0 | 0 |
| 0 | 0 | 0 | 0 | 0 |
| 0 | 0 | 0 | 0 | 0 |
| 0 | 0 | 0 | 0 | 0 |
| 0 | 0 | 0 | 0 | 0 |
| 0 | 0 | 0 | 0 | 0 |
| 0.052991433 | 0.134932007 | 0 | 0.199807082 | 0.002940778 |
| 0 | 0 | 0 | 0 | 0 |
| 0.014706838 | 0.036727845 | 0.002142574 | 0.023386334 | 0 |
| 0 | 0 | 0 | 0 | 0 |
| 0 | 0 | 0 | 0 | 0 |
| 0 | 0 | 0 | 0 | 0 |
| 0 | 0 | 0 | 0 | 0 |
| 0 | 0 | 0 | 0 | 0 |
| 0 | 0 | 0 | 0 | 0 |
| 0.029144466 | 0.098289777 | 0 | 0.001940892 | 0.161298794 |
| 0.024377008 | 0.0032336 | 0.005928589 | 0 | 0.023951342 |
| 0 | 0.008049505 | 0 | 0 | 0.018245245 |
| 0 | 0 | 0 | 0 | 0 |
| 0 | 0 | 0.867891963 | 0 | 0 |
| 0 | 0 | 0 | 0 | 0 |
| 0 | 0 | 0 | 0 | 0 |
| 0 | 0 | 0 | 0 | 0 |
| 0 | 0 | 0 | 0 | 0 |
| 0 | 0 | 0 | 0 | 0 |
| 0 | 0 | 0 | 0 | 0 |
| 0 | 0 | 0 | 0 | 0 |
| 0 | 0 | 0 | 0 | 0 |
| 0.00234173 | 0 | 0.00732349 | 0 | 0.686476251 |
| 0 | 0 | 0 | 0 | 0 |
| 0 | 0 | 0 | 0 | 0 |
| 0 | 0 | 0 | 0 | 0 |
| 0.161747658 | 0.160888899 | 0 | 0 | 0.046264805 |
| 0 | 0 | 0 | 0 | 0 |
| 0.462796893 | 0.026644701 | 0 | 0.028964448 | 0.03873735 |
| 0 | 0 | 0 | 0 | 0 |
| 0 | 0 | 0 | 0 | 0 |
| 0 | 0 | 0 | 0 | 0 |
| 0 | 0 | 0 | 0 | 0 |
| 0.00079322 | 0.410757472 | 0.848831989 | 0.021389697 | 0.009254474 |
| 0 | 0 | 0 | 0 | 0 |
| 0 | 0 | 0 | 0 | 0 |

| | | | | |
|---|---|---|---|---|
| 0 | 0 | 0 | 0 | 0 |
| 0 | 0 | 0 | 0 | 0 |
| 0 | 0 | 0 | 0 | 0 |
| 0 | 0 | 0.028711078 | 0 | 0 |
| 0.023863361 | 0 | 0 | 0 | 0.002913472 |
| 0 | 0 | 0 | 0 | 0 |
| 0 | 0 | 0 | 0 | 0 |
| 0 | 0.014321638 | 0 | 0 | 0 |
| 0 | 0 | 0 | 0 | 0 |
| 0.035685438 | 0.031643476 | 0.001758067 | 0.009405513 | 0 |
| 0.077997983 | 0 | 0 | 0 | 0 |
| 0 | 0 | 0 | 0 | 0 |
| 0 | 0.013549774 | 0 | 0.003962542 | 0.013019352 |
| 0 | 0 | 0 | 0 | 0 |
| 0 | 0 | 0 | 0 | 0 |
| 0 | 0 | 0 | 0 | 0 |
| 0 | 0 | 0 | 0 | 0 |
| 0 | 0 | 0 | 0 | 0 |
| 0 | 0 | 0 | 0 | 0 |
| 0.026545918 | 1.297264003 | 0.199747929 | 0.089020011 | 1.417913223 |
| 0 | 0 | 0 | 0 | 2.218494592 |
| 0 | 0 | 0 | 0 | 0 |
| 0 | 0 | 0 | 0 | 0 |
| 0 | 0 | 0 | 0 | 0 |
| 0 | 0 | 0 | 0 | 0 |
| 0 | 0 | 0 | 0 | 0 |
| 0 | 0 | 0 | 0 | 0 |
| 0 | 0 | 0 | 0 | 0 |
| 0 | 0 | 0 | 0 | 0 |
| 0 | 0 | 0 | 0.004826058 | 0 |
| 0 | 0 | 0 | 0 | 0 |
| 0 | 0 | 0 | 0 | 0.407238401 |
| 0 | 0 | 0 | 0 | 0 |
| 0 | 0 | 0 | 0 | 0 |
| 0 | 0 | 0.005058099 | 0 | 0 |
| 0 | 0.003733238 | 0 | 0 | 0 |
| 0 | 0 | 0 | 0 | 0 |
| 0 | 0 | 0 | 0 | 0 |
| 0 | 0 | 0 | 0 | 0 |
| 0.434261668 | 0 | 0 | 0 | 0 |
| 0 | 0 | 0 | 0 | 0 |
| 0 | 0 | 0 | 0 | 0 |
| 0 | 0 | 0 | 0.132514336 | 3.785951207 |
| 0 | 0 | 0 | 0 | 0 |
| 0.02451031 | 0.008126203 | 0 | 0.016165651 | 0.009349085 |
| 0 | 0.001861413 | 0 | 0 | 0 |
| 0 | 0 | 0 | 0 | 0 |
| 0 | 0 | 0 | 0 | 0 |

| | | | | |
|---|---|---|---|---|
| 0 | 0 | 0 | 0 | 0 |
| 0 | 0 | 0 | 0 | 0 |
| 0 | 0 | 0 | 0 | 0 |
| 0 | 0 | 0 | 0 | 0 |
| 0 | 0 | 0 | 0 | 0 |
| 0.022714634 | 0.437525269 | 0.004718667 | 0.018794873 | 0.128338641 |
| 0 | 0 | 0 | 0 | 0 |
| 0 | 0 | 0 | 0 | 0 |
| 0 | 0 | 0 | 0 | 0 |
| 0 | 0 | 0 | 0 | 0 |
| 0 | 0 | 0 | 0 | 0 |
| 0 | 0 | 0 | 0 | 0 |
| 0 | 0 | 0 | 0 | 0 |
| 0 | 0 | 0 | 0 | 0 |
| 0 | 0 | 0 | 0 | 0 |
| 0 | 0 | 0 | 0 | 0 |
| 0 | 0 | 0.002601686 | 0.156826576 | 0.250316505 |
| 0 | 0 | 0 | 0 | 0 |
| 0 | 0 | 0 | 0 | 0 |
| 0 | 0 | 0 | 0 | 0 |
| 0 | 0 | 0 | 0 | 0.000362355 |
| 0 | 0 | 0 | 0 | 0 |
| 0 | 0 | 0 | 0 | 0 |
| 0 | 0 | 0 | 0 | 0 |
| 0 | 0 | 0 | 0 | 0 |
| 0 | 0.002012832 | 0 | 0.00077364 | 0 |
| 0 | 0 | 0 | 0 | 0 |
| 0 | 0 | 0.002762004 | 0 | 1.343279169 |
| 0 | 0 | 0 | 0 | 0 |
| 0 | 0 | 0 | 0 | 0 |
| 0 | 0 | 0 | 0 | 0 |
| 0 | 0 | 0 | 0 | 0 |
| 0 | 0 | 0 | 0 | 0 |
| 0 | 0 | 0 | 0 | 0 |
| 0 | 0 | 0 | 0 | 0 |
| 0 | 0 | 0 | 0 | 0 |
| 0 | 0 | 0 | 0 | 0 |
| 0 | 0 | 0 | 0 | 0 |
| 0.098863205 | 0 | 0.145327002 | 0.081267564 | 0.009558034 |
| 0 | 0 | 0 | 0 | 0 |
| 0 | 0 | 0 | 0 | 0 |
| 0 | 0 | 0 | 0 | 0 |
| 0 | 0 | 0 | 0 | 0 |
| 0 | 0 | 0 | 0 | 0 |
| 0 | 0 | 0 | 0 | 0 |
| 0 | 0 | 0 | 0 | 0 |
| 0 | 0 | 0 | 0 | 0 |
| 0 | 0 | 0 | 0 | 0 |

| | | | | |
|---|---|---|---|---|
| 0 | 0 | 0 | 0 | 0 |
| 0.05103833 | 0.044456503 | 0 | 0.140097917 | 0.199316914 |
| 0 | 0 | 0.023813556 | 0 | 0 |
| 0 | 0 | 0 | 0 | 0 |
| 0 | 0 | 0 | 0 | 0 |
| 0 | 0.006209203 | 0 | 0 | 0 |
| 0 | 0 | 0 | 0 | 0 |
| 0 | 0 | 0 | 0 | 0 |
| 0 | 0 | 0 | 0 | 0 |
| 0.005035484 | 0 | 0 | 0 | 0 |
| 0 | 0 | 0.000943306 | 0.000725087 | 0.00534502 |
| 0 | 0 | 0 | 0 | 0 |
| 0.147161478 | 0.018659121 | 0 | 0.000961314 | 0.214728557 |
| 0 | 0 | 0 | 0 | 0 |
| 0 | 0 | 0 | 0 | 0 |
| 0 | 0 | 0 | 0 | 0 |
| 0 | 0 | 0 | 0 | 0 |
| 0 | 0 | 0 | 0 | 0 |
| 0 | 0 | 0 | 0 | 0 |
| 0 | 0.003363834 | 0 | 0 | 0 |
| 0 | 0.003555858 | 0 | 0 | 0 |
| 0 | 0 | 0 | 0 | 0 |
| 0 | 0 | 0.007665181 | 0 | 0 |
| 1.964107972 | 2.19701759 | 1.505055583 | 3.940076865 | 0.905982992 |
| 0 | 0 | 0 | 0 | 0 |
| 0 | 0 | 0 | 0 | 0 |
| 0 | 0 | 0 | 0 | 0 |
| 0 | 0 | 0 | 0 | 0 |
| 0 | 0 | 0 | 0 | 0 |
| 0 | 0 | 0 | 0 | 0 |
| 0 | 0 | 0 | 0 | 0.010192634 |
| 0 | 0 | 0 | 0 | 0 |
| 0 | 0.010645819 | 0 | 0 | 0.007662946 |
| 0 | 0 | 0 | 0 | 0 |

| PHL018-0079_V3 EYCF | PHL018-0265_V1 REF | PHL018-0078_V1 CTRL | PHL018-0162_V1 REF | PHL018-0079_V1 EYCF |
|---|---|---|---|---|
| 0.078400268 | 0.006427041 | 0.108510316 | 0.00774885 | 2.889890047 |
| 5.617828135 | 1.296259346 | 0.436115345 | 1.27840634 | 2.193464201 |
| 5.292127513 | 5.366177912 | 0.312306339 | 5.253699423 | 3.805820988 |
| 0 | 0 | 0 | 0 | 0 |
| 0 | 0 | 0 | 0 | 0 |
| 0 | 0 | 0 | 0 | 0 |
| 0 | 0 | 0 | 0 | 0 |
| 0 | 0 | 0 | 0 | 0 |
| 0 | 0 | 0 | 0 | 0 |
| 0 | 0 | 0 | 0 | 0 |
| 0 | 0 | 0 | 0 | 0 |
| 0 | 0 | 0 | 0 | 0 |
| 0 | 0 | 0 | 0 | 0 |
| 5.886192547 | 3.750015746 | 1.045731642 | 3.766706135 | 7.22813575 |
| 0 | 0 | 0 | 0 | 0 |
| 0 | 0 | 0 | 0 | 0 |
| 0 | 0 | 0 | 0 | 0 |
| 0 | 0 | 0 | 0 | 0 |
| 0 | 0 | 0 | 0 | 0 |
| 0 | 0 | 0 | 0 | 0 |
| 0 | 0 | 0 | 0 | 0 |
| 0 | 0 | 0 | 0 | 0 |
| 0 | 0 | 0 | 0 | 0 |
| 0 | 0 | 0 | 0 | 0 |
| 0 | 0 | 0 | 0 | 0 |
| 0 | 0 | 0 | 0 | 0 |
| 0 | 0 | 0 | 0 | 0 |
| 0 | 0 | 0 | 0 | 0 |
| 0 | 0 | 0 | 0 | 0 |
| 0 | 0 | 0 | 0 | 0 |
| 0 | 0 | 0 | 0 | 0 |
| 0 | 0 | 0 | 0 | 0 |
| 0 | 0 | 0 | 0 | 0 |
| 0 | 0 | 0 | 0 | 0 |
| 0 | 0 | 0 | 0 | 0 |
| 0.263903048 | 11.01605213 | 1.697606576 | 11.23227104 | 0.001454787 |
| 0 | 0 | 0 | 0 | 0 |
| 0 | 0 | 0 | 0 | 0 |
| 0 | 0 | 0 | 0 | 0 |
| 0 | 0 | 0 | 0 | 0 |
| 0 | 0 | 0 | 0 | 0 |
| 0.187323373 | 0 | 0.780251064 | 0 | 0.008118193 |
| 0.899198602 | 0 | 7.311436181 | 0 | 0.329298199 |
| 0.000520029 | 0.029716957 | 0.004547127 | 0.031501693 | 0.055796348 |
| 0 | 0 | 0.002415603 | 0.001866708 | 0.030221448 |
| 0 | 0 | 0 | 0 | 0 |

| | | | | |
|---|---|---|---|---|
| 0 | 0 | 0.004472391 | 0 | 0.06880804 |
| 0 | 0.060889959 | 0 | 0.04641781 | 0.001092401 |
| 0.0022112 | 0.003290588 | 0.032309302 | 0.001681988 | 0 |
| 0 | 0 | 0.763456223 | 0 | 0.12459942 |
| 0 | 0 | 0.018825177 | 0 | 0 |
| 0 | 0 | 0 | 0 | 0 |
| 0.242450392 | 0.010024242 | 0.044521294 | 0.00911797 | 0.00156117 |
| 0 | 0 | 0 | 0 | 0 |
| 0 | 0 | 0 | 0 | 0 |
| 0 | 0 | 0 | 0 | 0 |
| 0 | 0 | 0.005757204 | 0.000728491 | 0 |
| 0.003034346 | 0 | 0.822500803 | 0 | 0 |
| 0 | 0 | 0 | 0 | 0 |
| 0.009408812 | 0.072554195 | 0.036608417 | 0.072255909 | 0 |
| 0.004459459 | 0.000838273 | 0 | 0 | 0 |
| 0 | 0 | 0 | 0 | 0 |
| 0 | 0 | 0.132103196 | 0 | 0 |
| 0 | 0 | 0 | 0 | 0 |
| 0 | 0 | 0 | 0 | 0 |
| 0 | 0 | 0 | 0 | 0 |
| 0.00596533 | 0.004680749 | 0.05315906 | 0.00510015 | 0 |
| 0.001188062 | 0 | 0.053426049 | 0 | 0 |
| 0 | 0 | 0.31643837 | 0 | 0 |
| 0 | 0 | 0 | 0 | 0 |
| 0 | 0 | 0 | 0 | 0 |
| 0 | 0 | 0 | 0 | 0 |
| 0 | 0.049037043 | 0 | 0.050113868 | 0 |
| 0 | 0 | 0 | 0 | 0 |
| 0 | 0 | 0.120507201 | 0 | 0.000709534 |
| 0 | 0 | 0 | 0 | 0 |
| 0 | 0 | 0 | 0 | 0 |
| 0 | 0 | 0 | 0 | 0 |
| 0 | 0 | 0 | 0 | 0 |
| 0 | 0 | 0.001083275 | 0 | 0.012820201 |
| 0 | 0 | 0 | 0 | 0 |
| 0 | 0 | 0 | 0 | 0 |
| 0 | 0 | 0 | 0 | 0 |
| 0 | 0.000918813 | 6.694987509 | 0 | 0 |
| 0 | 0 | 0 | 0 | 0 |
| 0 | 0 | 0 | 0 | 0 |
| 0 | 0 | 0 | 0 | 0 |
| 0 | 0 | 0 | 0 | 0 |
| 0 | 0 | 0 | 0 | 0 |
| 0 | 0 | 0 | 0 | 0 |
| 0 | 0 | 0 | 0 | 0 |
| 0 | 0 | 0 | 0 | 0 |
| 0.000839343 | 0 | 0 | 0 | 0.004114279 |
| 0 | 0 | 0 | 0 | 0 |

| | | | | |
|---|---|---|---|---|
| 0 | 0 | 0 | 0 | 0 |
| 0 | 0 | 0 | 0 | 0 |
| 0 | 0 | 0 | 0 | 0 |
| 0 | 0 | 0 | 0 | 0 |
| 0 | 0 | 0 | 0 | 0.005681835 |
| 0 | 0 | 0 | 0 | 0 |
| 0 | 0 | 0 | 0 | 0 |
| 0 | 0 | 0 | 0 | 0 |
| 0 | 0 | 0 | 0 | 0 |
| 0 | 0 | 0 | 0 | 0 |
| 0 | 0 | 0 | 0 | 0 |
| 0.083040401 | 0 | 0.001577725 | 0 | 0.051658799 |
| 0 | 0 | 0 | 0 | 0 |
| 0 | 0 | 0 | 0 | 0 |
| 0 | 0 | 0 | 0 | 0 |
| 0 | 0 | 0 | 0 | 0 |
| 0 | 0 | 0 | 0 | 0 |
| 0 | 0 | 0 | 0 | 0 |
| 0 | 0 | 0.685435619 | 0 | 0.012185857 |
| 0 | 0 | 0 | 0 | 0 |
| 0 | 0 | 0 | 0 | 0 |
| 0 | 0 | 0 | 0 | 0 |
| 0 | 0 | 0 | 0 | 0 |
| 0 | 0 | 0 | 0 | 0 |
| 0 | 0 | 0 | 0 | 0 |
| 0 | 0 | 0 | 0 | 0 |
| 0 | 0 | 0 | 0 | 0 |
| 0 | 0 | 0 | 0 | 0 |
| 0 | 0 | 0 | 0 | 0 |
| 0 | 0 | 0 | 0 | 0 |
| 0.003947001 | 0 | 0.066230521 | 0 | 0 |
| 0 | 0 | 0.002638541 | 0 | 0 |
| 0 | 0 | 0 | 0 | 0 |
| 0 | 0 | 0 | 0 | 0 |
| 0 | 0 | 0 | 0 | 0 |
| 0 | 0 | 0 | 0 | 0 |
| 0 | 0 | 0 | 0 | 0 |
| 0 | 0 | 0 | 0 | 0 |
| 0 | 0 | 0 | 0 | 0 |
| 0 | 0 | 0 | 0 | 0 |
| 0 | 0 | 0 | 0 | 0 |
| 0 | 0 | 0 | 0 | 0 |
| 0.00165671 | 0 | 0 | 0 | 0 |
| 0 | 0 | 0 | 0 | 0 |
| 0 | 0 | 0 | 0 | 0 |
| 0 | 0 | 0 | 0 | 0 |
| 0 | 0 | 0 | 0 | 0 |
| 0 | 0 | 0 | 0 | 0 |

| | | | | |
|---|---|---|---|---|
| 0 | 0 | 0 | 0 | 0.005841897 |
| 0 | 0 | 0 | 0 | 0 |
| 0 | 0 | 0 | 0 | 0 |
| 0 | 0 | 0 | 0 | 0 |
| 0 | 0 | 0.021434481 | 0 | 0 |
| 0 | 0 | 0 | 0 | 0 |
| 0 | 0 | 0 | 0 | 0 |
| 0 | 0 | 0 | 0 | 0 |
| 0 | 0 | 0 | 0 | 0 |
| 0 | 0 | 0 | 0 | 0 |
| 0 | 0 | 0 | 0 | 0 |
| 0 | 0 | 0 | 0 | 0 |
| 0 | 0 | 0 | 0 | 0 |
| 0 | 0 | 0 | 0 | 0 |
| 0 | 0 | 0 | 0 | 0 |
| 0 | 0 | 0 | 0 | 0 |
| 0 | 0 | 0 | 0 | 0 |
| 0 | 0 | 0 | 0 | 0 |
| 0 | 0 | 0 | 0 | 0 |
| 0 | 0 | 0 | 0 | 0 |
| 0 | 0 | 0 | 0 | 0 |
| 0 | 0 | 0 | 0 | 0 |
| 0 | 0 | 0 | 0 | 0 |
| 0 | 0 | 0 | 0 | 0 |
| 0 | 0 | 0 | 0 | 0 |
| 0 | 0 | 0 | 0 | 0 |
| 0 | 0 | 0 | 0 | 0 |
| 0 | 0 | 0 | 0 | 0 |
| 0 | 0 | 0 | 0 | 0 |
| 0 | 0 | 0 | 0 | 0 |
| 0 | 0 | 0 | 0 | 0 |
| 0 | 0 | 0.541125602 | 0 | 0.0016513 |
| 0 | 0 | 0 | 0 | 0 |
| 0 | 0 | 0 | 0 | 0 |
| 0 | 0 | 0 | 0 | 0 |
| 0 | 0 | 0 | 0 | 0 |
| 0 | 0 | 0 | 0 | 0 |
| 0 | 0 | 0 | 0 | 0 |
| 0 | 0 | 0 | 0 | 0 |
| 0 | 0 | 0 | 0 | 0 |
| 0 | 0 | 0 | 0 | 0 |
| 0 | 0 | 0 | 0 | 0 |
| 0 | 0 | 0 | 0 | 0 |
| 0 | 0 | 0 | 0 | 0 |
| 0 | 0 | 0 | 0 | 0 |
| 0 | 0 | 0 | 0 | 0 |
| 0 | 0 | 0 | 0 | 0 |
| 0 | 0 | 0.01648469 | 0 | 0 |

| | | | | |
|---|---|---|---|---|
| 0 | 0 | 0 | 0 | 0 |
| 0 | 0 | 0 | 0 | 0 |
| 0 | 0 | 0 | 0 | 0 |
| 0 | 0 | 0 | 0 | 0 |
| 0 | 0 | 0 | 0 | 0 |
| 0 | 0 | 0 | 0 | 0 |
| 0 | 0 | 0 | 0 | 0 |
| 0 | 0 | 0 | 0 | 0 |
| 0 | 0 | 0 | 0 | 0 |
| 0 | 0 | 0 | 0 | 0 |
| 0 | 0 | 0 | 0 | 0 |
| 0 | 0 | 0 | 0 | 0 |
| 0 | 0 | 0.005583448 | 0 | 0 |
| 0 | 0 | 0 | 0 | 0 |
| 0 | 0 | 0 | 0 | 0 |
| 0 | 0 | 0 | 0 | 0 |
| 0 | 0 | 0 | 0 | 0 |
| 0 | 0 | 0 | 0 | 0.002073094 |
| 0 | 0 | 0 | 0 | 0 |
| 0 | 0 | 0 | 0 | 0 |
| 0.036492724 | 0.034638744 | 0.026326217 | 0.027554223 | 0.019338335 |
| 0 | 0 | 0 | 0 | 0 |
| 0 | 0 | 0 | 0 | 0 |
| 0 | 0 | 0 | 0 | 0 |
| 0 | 0 | 0 | 0 | 0 |
| 0 | 0 | 0 | 0 | 0 |
| 0 | 0.007693012 | 0 | 0.007308503 | 0.001251354 |
| 0 | 0 | 0 | 0 | 0 |
| 0 | 0 | 0 | 0 | 0 |
| 0 | 0.048286404 | 0 | 0.048275407 | 0 |
| 0 | 0.395189917 | 0 | 0.399901618 | 0.004989552 |
| 0 | 0 | 0 | 0 | 0 |
| 0 | 0 | 0 | 0 | 0 |
| 0 | 0 | 0 | 0 | 0.004103607 |
| 0.073585354 | 0.006684945 | 0.033667817 | 0.007100296 | 0.000873788 |
| 0 | 0 | 0 | 0 | 0 |
| 0 | 0 | 0 | 0 | 0 |
| 0 | 0 | 0 | 0 | 0 |
| 0 | 0 | 0 | 0 | 0 |
| 0 | 0 | 0 | 0 | 0 |
| 0 | 0 | 0 | 0 | 0 |
| 0 | 0 | 0 | 0 | 0 |
| 0 | 0 | 0 | 0 | 0 |
| 0 | 0 | 0 | 0 | 0 |
| 0 | 0 | 0 | 0 | 0 |
| 0.001832998 | 0 | 0 | 0 | 0 |
| 0 | 0 | 0 | 0 | 0 |
| 0.020731663 | 0.002449991 | 0.001683691 | 0.002023945 | 0 |

| | | | | |
|---|---|---|---|---|
| 0 | 0 | 0 | 0 | 0 |
| 0 | 0 | 0 | 0 | 0 |
| 0 | 0 | 0 | 0 | 0 |
| 0 | 0 | 0 | 0 | 0 |
| 0 | 0 | 0 | 0 | 0 |
| 0 | 0 | 0 | 0 | 0 |
| 0 | 0 | 0 | 0 | 0 |
| 0.002410468 | 0 | 0 | 0 | 0.00375951 |
| 0 | 0.00630305 | 0.001451547 | 0.005857839 | 0 |
| 0 | 0 | 0 | 0 | 0 |
| 0 | 0 | 0 | 0 | 0 |
| 0 | 0 | 0 | 0 | 0 |
| 0 | 0 | 0 | 0 | 0 |
| 0 | 0 | 0 | 0 | 0 |
| 0 | 0 | 0 | 0 | 0 |
| 0 | 0 | 0 | 0 | 0 |
| 0 | 0 | 0 | 0 | 0 |
| 0 | 0 | 0.055621854 | 0 | 0 |
| 0.00728637 | 0 | 0.031249522 | 0 | 0.001393187 |
| 0 | 0 | 0 | 0 | 0 |
| 0.104279803 | 0 | 0.002525821 | 0 | 0 |
| 0 | 0 | 0 | 0 | 0 |
| 0 | 0 | 0 | 0 | 0 |
| 0 | 0 | 0 | 0 | 0 |
| 0 | 0.00365098 | 0 | 0.004901139 | 0.011506285 |
| 0 | 0 | 0 | 0 | 0.032235988 |
| 0 | 0 | 0 | 0 | 0 |
| 0 | 0 | 0 | 0 | 0 |
| 0 | 0 | 0.035616874 | 0 | 0 |
| 0 | 0 | 0 | 0 | 0 |
| 0.12359105 | 0.041588789 | 0.358446334 | 0.039209337 | 0 |
| 0 | 0 | 0 | 0 | 0.001579367 |
| 0.001881744 | 0 | 0 | 0 | 0.00375789 |
| 0 | 0 | 0 | 0 | 0.148227314 |
| 0 | 0 | 0 | 0 | 0 |
| 0 | 0 | 0 | 0 | 0 |
| 0 | 0 | 0 | 0 | 0 |
| 0 | 0 | 0 | 0 | 0 |
| 0 | 0 | 0 | 0 | 0 |
| 0 | 0 | 0 | 0 | 0.037871899 |
| 0 | 0 | 0 | 0 | 0 |
| 0 | 0.015151237 | 0.036206344 | 0.015645601 | 0 |
| 0 | 0 | 0 | 0 | 0 |
| 0 | 0 | 0 | 0 | 0 |
| 0 | 0 | 0 | 0 | 0 |
| 0 | 0 | 0 | 0 | 0 |
| 0 | 0 | 0 | 0 | 0 |
| 0 | 0 | 0 | 0 | 0 |

| | | | | |
|---|---|---|---|---|
| 0 | 0 | 0 | 0 | 0 |
| 0 | 0 | 0 | 0 | 0 |
| 0.000860894 | 0.032476801 | 0.006240023 | 0.031790196 | 0.056441431 |
| 0 | 0 | 0 | 0 | 0 |
| 0 | 0 | 0 | 0 | 0 |
| 0 | 0 | 0 | 0 | 0 |
| 0 | 0 | 0 | 0 | 0 |
| 0 | 0 | 0 | 0 | 0 |
| 0 | 0 | 0 | 0 | 0 |
| 0 | 0 | 0 | 0 | 0 |
| 0 | 0 | 0 | 0 | 0 |
| 0 | 0 | 0 | 0 | 0 |
| 0 | 0 | 0 | 0 | 0 |
| 0.001189304 | 0.003185745 | 0 | 0.004934538 | 0.00097162 |
| 0 | 0.001752944 | 0.005963085 | 0.001629126 | 0.00753828 |
| 0 | 0 | 0 | 0 | 0 |
| 0 | 0.003794912 | 0 | 0.00389811 | 0.002923981 |
| 0 | 0 | 0 | 0 | 0.000425284 |
| 0 | 0 | 0 | 0 | 0.001457793 |
| 0 | 0 | 0 | 0.000945479 | 0 |
| 0 | 0 | 0 | 0 | 0 |
| 0.022414278 | 0.001081807 | 0.003345493 | 0.000837829 | 0 |
| 0 | 0 | 0 | 0 | 0 |
| 0.002191268 | 0 | 0 | 0 | 0 |
| 0 | 0 | 0 | 0 | 0 |
| 0 | 0 | 0 | 0 | 0 |
| 0 | 0 | 0 | 0 | 0 |
| 0 | 0 | 0 | 0 | 0 |
| 0 | 0 | 0 | 0 | 0 |
| 0 | 0 | 0 | 0 | 0 |
| 0 | 0 | 0 | 0 | 0 |
| 0 | 0 | 0 | 0 | 0 |
| 0 | 0 | 0 | 0 | 0 |
| 0.002095426 | 0 | 0 | 0 | 0 |
| 0 | 0 | 0 | 0 | 0 |
| 0 | 0 | 0 | 0 | 0.015486696 |
| 0 | 0 | 0 | 0 | 0 |
| 0.001268523 | 0.001359179 | 0.053241371 | 0.002736877 | 0.301781862 |
| 0 | 0 | 0 | 0 | 0 |
| 0 | 0 | 0 | 0 | 0 |
| 0 | 0 | 0 | 0 | 0.003589751 |
| 0 | 0 | 0 | 0 | 0 |
| 0 | 0 | 0 | 0 | 0 |
| 0 | 0.009004333 | 0 | 0.010198889 | 0 |
| 0 | 0 | 0 | 0 | 0 |
| 0 | 0 | 0 | 0 | 0 |
| 0 | 0 | 0 | 0 | 0 |
| 0 | 0 | 0 | 0.0073951 | 0 |

| | | | | |
|---|---|---|---|---|
| 0 | 0 | 0 | 0 | 0 |
| 0 | 0 | 0 | 0 | 0 |
| 0 | 0.002266217 | 0.00146006 | 0.002808193 | 0.007775696 |
| 0 | 0 | 0 | 0 | 0 |
| 0 | 0.031160453 | 0.005875844 | 0.032667682 | 0 |
| 0 | 0 | 0.012455425 | 0 | 0 |
| 0 | 0 | 0 | 0 | 0 |
| 0.012527767 | 0 | 0 | 0.00173263 | 0.019007381 |
| 0 | 0 | 0 | 0 | 0 |
| 0.257023051 | 2.570812512 | 0.005942525 | 2.486937184 | 0.370979209 |
| 0 | 0.007942242 | 0 | 0.008534568 | 0 |
| 0.002361197 | 0 | 0.002281956 | 0 | 0.007330254 |
| 0 | 0.001599208 | 0.057863017 | 0.001188999 | 0.022094709 |
| 0 | 0 | 0 | 0 | 0 |
| 0 | 0.008223744 | 0 | 0.007337151 | 0 |
| 0 | 0 | 0 | 0 | 0.003734126 |
| 0 | 0 | 0 | 0 | 0 |
| 0 | 0 | 0 | 0 | 0 |
| 0 | 0 | 0 | 0 | 0.141808262 |
| 0 | 0 | 0 | 0 | 0 |
| 0 | 0 | 0 | 0 | 0 |
| 0 | 0 | 0 | 0 | 0 |
| 0.000739253 | 0 | 0 | 0 | 0.001207888 |
| 0 | 0 | 0 | 0 | 0 |
| 0 | 0 | 0 | 0 | 0 |
| 0.000834447 | 0.000931335 | 0 | 0.001731101 | 0.026245988 |
| 0 | 0 | 1.091473478 | 0 | 0 |
| 0 | 0 | 0.005224365 | 0 | 0 |
| 0.008970714 | 0.074949213 | 0 | 0.076213104 | 0 |
| 0 | 0 | 0 | 0 | 0 |
| 0 | 0.016389712 | 0 | 0.014674768 | 0 |
| 0 | 0.005767456 | 0 | 0.008246271 | 0 |
| 0 | 0 | 0 | 0 | 0 |
| 0.002455433 | 0.012667352 | 0 | 0.010867018 | 0 |
| 0 | 0 | 0 | 0 | 0 |
| 0 | 0.019455116 | 0 | 0.022645034 | 0 |
| 0 | 0 | 0 | 0 | 0 |
| 0.001357415 | 0 | 0 | 0 | 0 |
| 0 | 0 | 0 | 0 | 0 |
| 0 | 0 | 0 | 0 | 0 |
| 0 | 0 | 0 | 0 | 0 |
| 0 | 0 | 0 | 0 | 0 |
| 0 | 0 | 0 | 0 | 0 |
| 0 | 0 | 0 | 0 | 0 |
| 0 | 0 | 0 | 0 | 0 |
| 0 | 0 | 0 | 0 | 0 |
| 0.021791724 | 0.002084739 | 0 | 0.001291657 | 0 |
| 0 | 0 | 0.001950608 | 0 | 0 |

| | | | | |
|---|---|---|---|---|
| 0 | 0 | 0 | 0 | 0 |
| 0.039719849 | 0 | 0.001200841 | 0 | 0 |
| 0 | 0 | 0 | 0 | 0 |
| 0 | 0 | 0 | 0 | 0 |
| 0 | 0 | 0 | 0 | 0 |
| 0 | 0 | 0.020311375 | 0 | 0 |
| 0 | 0 | 0 | 0 | 0 |
| 0 | 0 | 0 | 0 | 0 |
| 0 | 0 | 0 | 0 | 0 |
| 0 | 0 | 0 | 0 | 0 |
| 0 | 0.006982893 | 0.003926296 | 0.011209416 | 0 |
| 1.231604696 | 0 | 0 | 0 | 0 |
| 0 | 0 | 0 | 0 | 0 |
| 0.003582295 | 0.004797881 | 0.001236456 | 0.003715822 | 0 |
| 0 | 0.57021242 | 0 | 0.581068421 | 0 |
| 0 | 0 | 0 | 0 | 0 |
| 0 | 0 | 0 | 0 | 0 |
| 0 | 0 | 0 | 0 | 0 |
| 0 | 0 | 0 | 0 | 0 |
| 0 | 0 | 0 | 0 | 0 |
| 0 | 0 | 0 | 0 | 0 |
| 1.150249181 | 0 | 0.178587753 | 0 | 0 |
| 0.041005587 | 0 | 0 | 0 | 0 |
| 0 | 0 | 0 | 0 | 0 |
| 0 | 0 | 0 | 0 | 0 |
| 0 | 0 | 0 | 0 | 0 |
| 0 | 0 | 0 | 0 | 0 |
| 0 | 0 | 0 | 0 | 0 |
| 0 | 0 | 0 | 0 | 0 |
| 12.23359957 | 0.190175615 | 0.112820967 | 0.197167519 | 1.047246276 |
| 0 | 0 | 0.010485461 | 0 | 0 |
| 0 | 0 | 0.012365858 | 0 | 0 |
| 0 | 0 | 0 | 0 | 0 |
| 0 | 0 | 0 | 0 | 0 |
| 0 | 0 | 0 | 0 | 0 |
| 0 | 0 | 0 | 0 | 0 |
| 2.751006371 | 0 | 2.083385283 | 0 | 0 |
| 0 | 0 | 0.013863055 | 0 | 0 |
| 0.521568194 | 0 | 0.016163321 | 0 | 0 |
| 0 | 0 | 0 | 0 | 0 |
| 0.002208777 | 0.021036694 | 0.030495033 | 0.020467226 | 0 |
| 0 | 0 | 0 | 0 | 0.014484775 |
| 0 | 0 | 0 | 0 | 0 |
| 0 | 0 | 0 | 0 | 0 |
| 0.001661077 | 0 | 0 | 0 | 0 |
| 0 | 0 | 0 | 0 | 0 |
| 0 | 0.00137997 | 0.002400504 | 0.00224437 | 0 |
| 0 | 0 | 0 | 0 | 0 |

| | | | | |
|---|---|---|---|---|
| 0 | 0 | 0.002632729 | 0 | 0.034662686 |
| 0 | 0 | 0 | 0 | 0 |
| 0 | 0 | 0.016591236 | 0 | 0.003714764 |
| 0 | 0 | 0 | 0 | 0 |
| 0 | 0 | 0.009703954 | 0 | 0 |
| 0 | 0 | 0.001947727 | 0 | 0 |
| 0 | 0 | 0 | 0 | 0 |
| 0 | 0 | 0 | 0 | 0 |
| 0 | 0 | 0.023961514 | 0 | 0 |
| 0 | 0 | 0 | 0 | 0 |
| 0 | 0 | 0 | 0 | 0 |
| 0 | 0.000897318 | 0 | 0.000667149 | 0 |
| 0 | 0 | 0 | 0 | 0 |
| 0 | 0.203518179 | 0 | 0.193079577 | 0 |
| 0 | 0 | 0 | 0 | 0 |
| 0 | 0 | 0.026290957 | 0 | 0 |
| 0.008031822 | 0.007004736 | 0.103668986 | 0.005734966 | 0.29756615 |
| 0.006808152 | 0 | 0 | 0 | 0 |
| 0 | 0 | 0 | 0 | 0 |
| 0 | 0 | 0 | 0 | 0 |
| 0.008709956 | 0 | 0 | 0 | 0 |
| 0 | 0.003095287 | 0.001196524 | 0.003236236 | 0.000708023 |
| 0 | 0 | 0 | 0 | 0 |
| 0 | 0 | 0 | 0 | 0 |
| 0 | 0 | 0.042177912 | 0 | 0 |
| 0.351125303 | 0 | 0.016985773 | 0 | 0 |
| 0 | 0.002752462 | 0.004583385 | 0.003345134 | 0 |
| 0 | 0 | 0 | 0 | 0 |
| 0 | 0 | 0 | 0 | 0 |
| 0 | 0 | 0 | 0 | 0 |
| 0.055182342 | 0 | 0.018424155 | 0 | 0 |
| 0 | 0 | 0 | 0.002231023 | 0.012300192 |
| 2.989169209 | 0.005942776 | 0.013076698 | 0 | 0.029278615 |
| 0 | 0 | 0.02431159 | 0 | 0 |
| 0 | 0 | 0 | 0 | 0.995561402 |
| 0 | 0 | 0 | 0 | 0 |
| 0 | 0.001033208 | 0 | 0 | 0.000945353 |
| 0.025881491 | 0 | 0.089456819 | 0 | 0 |
| 0 | 0 | 0.141448439 | 0 | 0.00395432 |
| 0 | 0 | 0 | 0 | 0 |
| 0 | 0 | 0 | 0 | 0 |
| 0 | 0.005310849 | 0 | 0.003878067 | 0 |
| 0 | 0 | 0.005231719 | 0 | 0 |
| 0 | 0.01211052 | 0.005266665 | 0.011489584 | 0 |
| 0 | 0 | 0 | 0 | 0 |
| 0.000986466 | 0 | 0 | 0 | 0.142162205 |
| 0 | 0 | 0 | 0 | 0 |
| 0 | 0 | 0 | 0 | 0 |

| | | | | |
|---|---|---|---|---|
| 0 | 0 | 0 | 0 | 0 |
| 0 | 0.884563195 | 0.189313751 | 0.899645453 | 0.368105268 |
| 0 | 0 | 0 | 0 | 0 |
| 0 | 0 | 0.000618645 | 0 | 0 |
| 0 | 0 | 0 | 0 | 0 |
| 0 | 0 | 0 | 0 | 0 |
| 0 | 0.005704111 | 0 | 0.005301205 | 0.002423147 |
| 0.000568293 | 0 | 0 | 0 | 0.013463984 |
| 0 | 0 | 0 | 0 | 0 |
| 0 | 0 | 0 | 0 | 0 |
| 0.001099599 | 0 | 0.002024184 | 0 | 0 |
| 0 | 0.004387337 | 0.00226131 | 0.006342686 | 0.045495155 |
| 0 | 0 | 0 | 0 | 0 |
| 0 | 0 | 0 | 0 | 0 |
| 0.002313028 | 0 | 0 | 0 | 0.001202513 |
| 0 | 0 | 0 | 0 | 0 |
| 0 | 0.018505893 | 0 | 0.019109715 | 0.019597594 |
| 0 | 0.050938094 | 0.001719719 | 0.052094806 | 0.004477508 |
| 0 | 0 | 0 | 0 | 0 |
| 0 | 0 | 0 | 0 | 0 |
| 0 | 0.065381209 | 0.070767088 | 0.058628139 | 0.036054754 |
| 0 | 0 | 0 | 0 | 0 |
| 0 | 0 | 0 | 0 | 0 |
| 0 | 0 | 0 | 0 | 0 |
| 0.001662578 | 0.011282162 | 0 | 0.009657472 | 0 |
| 0 | 0 | 0 | 0 | 0 |
| 0 | 0 | 0.259714972 | 0 | 0 |
| 0.019141181 | 1.380045221 | 0.026136475 | 1.329345217 | 0.063753573 |
| 0 | 0 | 0 | 0 | 0 |
| 0.000761167 | 0 | 0.002101781 | 0 | 0 |
| 0.001805583 | 0 | 0 | 0 | 0.002950195 |
| 0 | 0 | 0 | 0 | 0 |
| 0.003830317 | 0.002736034 | 0 | 0.002542777 | 0 |
| 0 | 0 | 0 | 0 | 0 |
| 0 | 0 | 0.001518408 | 0 | 0 |
| 0.46207812 | 8.014234807 | 0.04079397 | 8.030407287 | 2.294673996 |
| 0 | 0 | 0 | 0 | 0 |
| 0 | 0 | 0 | 0 | 0 |
| 0.000778444 | 0.000695063 | 0.001791237 | 0.00258387 | 0.007631531 |
| 0 | 0 | 0 | 0 | 0 |
| 0 | 0 | 0 | 0 | 0 |
| 0 | 0 | 0 | 0 | 0 |
| 0 | 0 | 0 | 0 | 0.003862924 |
| 0 | 0.00652189 | 0.006722991 | 0.007955353 | 0 |
| 0 | 0 | 0 | 0 | 0 |
| 0 | 0 | 0.337109505 | 0 | 0 |
| 0 | 0 | 0 | 0 | 0 |
| 0 | 0.005271065 | 0 | 0.004082289 | 0 |

| | | | | |
|---|---|---|---|---|
| 0 | 0 | 0 | 0 | 0 |
| 0 | 0 | 0 | 0 | 0 |
| 0 | 0 | 0 | 0 | 0 |
| 0 | 0 | 3.366708856 | 0 | 0.000899817 |
| 0 | 0 | 0 | 0 | 0 |
| 0 | 0 | 0 | 0 | 0 |
| 0 | 0.002160243 | 0 | 0.002007656 | 0 |
| 0 | 0 | 0.00136038 | 0 | 0 |
| 0.107245207 | 0 | 1.499269879 | 0 | 0 |
| 0 | 0 | 0.160371793 | 0 | 0 |
| 0 | 0 | 0 | 0 | 0 |
| 0 | 0 | 0.002428236 | 0 | 0 |
| 0 | 0 | 0 | 0 | 0 |
| 0 | 0 | 0 | 0 | 0 |
| 0.316541016 | 0 | 0.172325656 | 0 | 0 |
| 0 | 0 | 0 | 0 | 0 |
| 0 | 0 | 0 | 0 | 0 |
| 0 | 0 | 0 | 0 | 0 |
| 0 | 0 | 0 | 0 | 0 |
| 0 | 0 | 0 | 0 | 0 |
| 0 | 0 | 0 | 0 | 0 |
| 0 | 0 | 0 | 0 | 0 |
| 0 | 0 | 0 | 0 | 0 |
| 0.001677635 | 0 | 0.647761753 | 0 | 0 |
| 0 | 0 | 0 | 0 | 0 |
| 0 | 0 | 0 | 0 | 0 |
| 0 | 0 | 0 | 0 | 0 |
| 0.326602184 | 0 | 3.768065644 | 0 | 0 |
| 0 | 0 | 0 | 0 | 0 |
| 0 | 0 | 0 | 0 | 0 |
| 0 | 0 | 0.001652462 | 0 | 0.036458601 |
| 0 | 0 | 0 | 0 | 0 |
| 0.000650658 | 0 | 0.11498466 | 0 | 0 |
| 0 | 0 | 0 | 0 | 0 |
| 0 | 0 | 0 | 0 | 0 |
| 0 | 0 | 0 | 0 | 0 |
| 0 | 0 | 0.850221455 | 0 | 0.171780577 |
| 0 | 0 | 0 | 0 | 0 |
| 0 | 0 | 0 | 0 | 0 |
| 0.002517789 | 0 | 0.007995107 | 0 | 0 |
| 0.001855968 | 0.044592967 | 0.279302084 | 0.044383402 | 0 |
| 0 | 0 | 0 | 0 | 0 |
| 5.123088149 | 0.092169987 | 0.89283365 | 0.096912955 | 0.160397479 |
| 0 | 0 | 0.00547901 | 0 | 0 |
| 0 | 0 | 0 | 0 | 0 |
| 0 | 0 | 0 | 0 | 0 |
| 0 | 0 | 0 | 0 | 0 |
| 0.013183694 | 0.013763667 | 0 | 0.01464288 | 0.024523855 |

| | | | | |
|---|---|---|---|---|
| 0 | 0 | 0 | 0 | 0 |
| 0.000923901 | 0 | 0.19975357 | 0 | 0 |
| 0.004277497 | 0.253117028 | 0.012616591 | 0.263473351 | 0.020173164 |
| 0 | 0 | 0 | 0 | 0 |
| 0 | 0 | 0 | 0 | 0 |
| 0 | 0 | 0.045676482 | 0 | 0 |
| 0 | 0 | 0 | 0 | 0 |
| 0 | 0 | 0 | 0 | 0 |
| 0 | 0 | 0 | 0 | 0 |
| 0 | 0 | 0 | 0 | 0 |
| 0.136491657 | 0.005113499 | 0.044804966 | 0.003643438 | 0 |
| 2.034188227 | 0.209732121 | 1.114584793 | 0.197761295 | 1.003022675 |
| 0 | 0 | 0 | 0 | 0 |
| 0 | 0.017107958 | 0.047786459 | 0.016583402 | 0.357500124 |
| 0.007955397 | 0.516509611 | 0.051554996 | 0.511417353 | 0.050535203 |
| 0 | 0 | 0 | 0 | 0 |
| 0 | 0 | 0 | 0 | 0 |
| 0 | 0 | 0 | 0 | 0 |
| 0 | 0 | 0 | 0 | 0 |
| 0 | 0 | 0 | 0 | 0 |
| 0 | 0.062573627 | 0.009719641 | 0.062800779 | 0 |
| 29.53436194 | 2.065455036 | 0 | 2.148718847 | 8.714886251 |
| 5.890512081 | 33.52396224 | 1.041979563 | 33.1166144 | 6.118084835 |
| 0 | 0 | 0 | 0 | 0 |
| 0 | 0 | 0 | 0 | 0 |
| 0 | 0 | 1.180851606 | 0.016228172 | 1.018754991 |
| 0.113098272 | 0 | 0 | 0 | 0 |
| 0.00183565 | 0 | 0.000691187 | 0 | 0 |
| 0 | 0 | 0.098098371 | 0 | 0 |
| 0 | 0 | 0 | 0 | 0 |
| 0 | 0 | 0 | 0 | 0 |
| 0 | 0 | 0 | 0 | 0 |
| 0 | 0.002100679 | 0.006767039 | 0.001789607 | 0 |
| 0.014053786 | 0 | 0.00949512 | 0 | 0 |
| 0 | 0.000924316 | 0.002143838 | 0.000966406 | 0 |
| 0 | 0 | 0.122974744 | 0 | 0.011183945 |
| 0.012537036 | 0 | 0.013278139 | 0 | 0 |
| 0 | 0 | 0 | 0 | 0 |
| 0.015603539 | 0 | 2.761200592 | 0 | 0 |
| 0 | 0 | 0 | 0 | 0 |
| 0 | 0 | 0 | 0 | 0 |
| 0 | 0 | 0 | 0 | 0 |
| 0 | 0 | 0 | 0 | 0 |
| 0 | 0 | 0 | 0 | 0 |
| 0.023113087 | 0 | 0.022673329 | 0.000672971 | 0.019710759 |
| 0 | 0 | 0 | 0 | 0 |
| 0 | 0 | 0 | 0 | 0 |
| 0.001222838 | 0.06048886 | 0 | 0.056013329 | 8.533188063 |

| | | | | |
|---|---|---|---|---|
| 0 | 0 | 0 | 0 | 0 |
| 0 | 0 | 0 | 0 | 0 |
| 0 | 0 | 0 | 0 | 1.2869029 |
| 0 | 0 | 0.001431436 | 0 | 0 |
| 0 | 0 | 0 | 0 | 0 |
| 0 | 0 | 0 | 0 | 0 |
| 0 | 0 | 0.193543004 | 0 | 0.042811964 |
| 0.019239871 | 0 | 0.010317912 | 0 | 0 |
| 0 | 0 | 0 | 0 | 0 |
| 0 | 0 | 0 | 0 | 0 |
| 0.00173208 | 0.001374714 | 0.0156767 | 0.000958209 | 0 |
| 0 | 0 | 0 | 0 | 0 |
| 0 | 0 | 0.027452353 | 0 | 0 |
| 0 | 0 | 0 | 0 | 0 |
| 0 | 0 | 0 | 0 | 0 |
| 0 | 0 | 0 | 0 | 0 |
| 0 | 0 | 0 | 0 | 0 |
| 0 | 0 | 0.009511378 | 0 | 0.013088838 |
| 0 | 0 | 0 | 0 | 0 |
| 0 | 0 | 0.071015274 | 0 | 0 |
| 0 | 0 | 0.00235817 | 0 | 0 |
| 0 | 0 | 0 | 0 | 0 |
| 0 | 0 | 0.007140611 | 0 | 0 |
| 0 | 0 | 0 | 0 | 0 |
| 0 | 0 | 0 | 0 | 0 |
| 0 | 0 | 0 | 0 | 0 |
| 0 | 0 | 0 | 0 | 0 |
| 0 | 0 | 0 | 0 | 0 |
| 0 | 0 | 0 | 0 | 0 |
| 0 | 0 | 0 | 0 | 0 |
| 0 | 0 | 0 | 0 | 0 |
| 0 | 0 | 0 | 0 | 0 |
| 0 | 0 | 0 | 0 | 0 |
| 0 | 0 | 0 | 0 | 0 |
| 0 | 0 | 0.002957879 | 0 | 0 |
| 0 | 0 | 0 | 0 | 0 |
| 0 | 0 | 0.208353163 | 0 | 0 |
| 0.001792066 | 0.010328008 | 0.004048658 | 0.011491159 | 0 |
| 0 | 0 | 0 | 0 | 0 |
| 0 | 0 | 0.005768205 | 0 | 0 |
| 0.003821368 | 0 | 0.011184894 | 0 | 0 |
| 0 | 0 | 0 | 0 | 0 |
| 0 | 0 | 0 | 0 | 0 |
| 0 | 0 | 0.001747434 | 0 | 0 |
| 0.170929319 | 0 | 0.309800815 | 0 | 0 |
| 0 | 0 | 0 | 0 | 0 |
| 0 | 0 | 0 | 0 | 0 |
| 0 | 0 | 0 | 0 | 0 |

| | | | | |
|---|---|---|---|---|
| 0 | 0 | 0 | 0 | 0 |
| 0 | 0 | 0 | 0 | 0 |
| 0 | 0 | 0 | 0 | 0 |
| 0 | 0 | 0.006488312 | 0 | 0 |
| 0 | 0 | 0 | 0 | 0 |
| 0 | 0 | 0 | 0 | 0 |
| 0 | 0 | 0 | 0 | 0 |
| 0 | 0 | 0 | 0 | 0 |
| 0.036848696 | 0.956959322 | 0.055527544 | 0.968792823 | 0 |
| 0 | 0 | 0 | 0 | 0 |
| 0 | 0 | 0.015526674 | 0 | 0 |
| 0 | 0 | 0 | 0 | 0 |
| 0 | 0 | 0 | 0 | 0 |
| 0 | 0 | 0 | 0 | 0 |
| 0 | 0 | 2.975601683 | 0 | 0.204595704 |
| 0.14771627 | 0 | 0 | 0 | 0 |
| 0 | 0 | 0 | 0 | 0 |
| 0 | 0 | 0 | 0 | 0 |
| 0 | 0 | 0 | 0 | 0 |
| 0 | 0 | 0 | 0 | 0 |
| 0 | 0 | 0 | 0 | 0 |
| 0 | 0 | 0 | 0 | 0 |
| 0 | 0 | 0 | 0 | 0 |
| 0 | 0 | 0 | 0 | 0 |
| 0.035670471 | 0 | 0.251345702 | 0 | 0.029357378 |
| 0 | 0 | 6.107235532 | 0 | 0.021566754 |
| 0 | 0 | 0 | 0 | 0 |
| 0.002132224 | 0 | 0 | 0 | 0 |
| 0.005313203 | 0 | 0.286087042 | 0 | 0 |
| 0 | 0 | 0 | 0 | 0 |
| 0 | 0 | 0 | 0 | 0 |
| 0 | 0 | 0 | 0 | 0 |
| 0 | 0 | 0 | 0 | 0 |
| 0 | 0 | 0.003506858 | 0 | 0 |
| 0.001162552 | 0.003944509 | 0.008988303 | 0.00347295 | 0.223954394 |
| 0.001643567 | 0.00073376 | 0.001134579 | 0.000545545 | 0 |
| 0 | 0 | 0 | 0 | 0 |
| 0 | 0 | 2.450299576 | 0 | 0.139337064 |
| 0.000622725 | 0 | 0 | 0 | 0 |
| 0 | 0 | 0 | 0 | 0 |
| 0 | 0 | 0 | 0 | 0 |
| 0 | 0 | 0 | 0 | 0 |
| 0 | 0.180485399 | 0.145429578 | 0.186560765 | 0 |
| 0 | 0 | 0 | 0 | 0 |
| 0 | 0 | 0 | 0 | 0 |
| 0.004048391 | 0 | 0 | 0 | 0 |
| 0 | 0 | 0 | 0 | 0 |
| 0.050108216 | 0.005758744 | 0.078085354 | 0.007822123 | 0 |

| | | | | |
|---|---|---|---|---|
| 0 | 0 | 0 | 0 | 0 |
| 0 | 0 | 0 | 0 | 0 |
| 0 | 0 | 0 | 0 | 0 |
| 0 | 0 | 0 | 0 | 0 |
| 0.00184168 | 0.008057625 | 0.018815815 | 0.009780873 | 0 |
| 0.014542785 | 0 | 0.59891796 | 0 | 0 |
| 0 | 0 | 0 | 0 | 0 |
| 0 | 0 | 0 | 0 | 0 |
| 0 | 0 | 0 | 0 | 0 |
| 0 | 0 | 0 | 0 | 0 |
| 0 | 0 | 0 | 0 | 0 |
| 0.046384536 | 0 | 0.504577808 | 0 | 0.920712095 |
| 0 | 0 | 0.007388969 | 0 | 0 |
| 0 | 0 | 0 | 0 | 0 |
| 0.015761281 | 0.001521411 | 0.940994144 | 0.001413947 | 0.382290156 |
| 0 | 0 | 0 | 0 | 0 |
| 0 | 0 | 0 | 0 | 0 |
| 0 | 0 | 0 | 0 | 0 |
| 0 | 0 | 0 | 0 | 0 |
| 0 | 0 | 0 | 0 | 0 |
| 0 | 0 | 0 | 0 | 0 |
| 0 | 0 | 0 | 0 | 0 |
| 0 | 0.022043197 | 0.14811652 | 0.01837732 | 0.001334703 |
| 0 | 0 | 0 | 0 | 0 |
| 0 | 0 | 0 | 0 | 0 |
| 0 | 0 | 0 | 0 | 0 |
| 0 | 0 | 0 | 0 | 0 |
| 0 | 0 | 0 | 0 | 0 |
| 0 | 0 | 0 | 0 | 0 |
| 0 | 0 | 0 | 0 | 0 |
| 0 | 0 | 0 | 0 | 0 |
| 0 | 0.029120095 | 0.009921206 | 0.024999075 | 0.018289518 |
| 0 | 0 | 0 | 0 | 0 |
| 0.000552308 | 0 | 0.036029677 | 0 | 0 |
| 0 | 0.000638812 | 0.001975529 | 0.002077916 | 0 |
| 0.002305739 | 0 | 0.081494303 | 0 | 0.273702657 |
| 0 | 0 | 0 | 0 | 0 |
| 0 | 0 | 0 | 0 | 0 |
| 0 | 0 | 0 | 0 | 0 |
| 0 | 0 | 0 | 0 | 0 |
| 0 | 0 | 0.080370808 | 0 | 0.06673073 |
| 0 | 0 | 0 | 0 | 0 |
| 0 | 0 | 0 | 0 | 0 |
| 0 | 0 | 0 | 0 | 0 |
| 0.004933496 | 0 | 0.002919142 | 0 | 0 |
| 0 | 0 | 0 | 0 | 0 |
| 0 | 0 | 0.102542172 | 0 | 0 |
| 0.001833152 | 0 | 0 | 0 | 0 |

| | | | | |
|---|---|---|---|---|
| 0 | 0 | 0 | 0 | 0 |
| 0 | 0 | 0 | 0 | 0 |
| 0 | 0 | 0 | 0 | 0 |
| 0 | 0 | 0 | 0 | 0 |
| 0 | 0 | 0 | 0 | 0 |
| 0 | 0 | 0 | 0 | 0 |
| 0 | 0 | 0.27805679 | 0 | 0 |
| 0 | 0 | 0.006890592 | 0 | 0 |
| 0 | 0 | 0 | 0 | 0 |
| 0 | 0 | 0 | 0 | 0 |
| 0 | 0 | 0 | 0 | 0 |
| 0.000927078 | 0 | 0 | 0 | 0 |
| 0 | 0 | 0 | 0 | 0 |
| 0 | 0 | 0 | 0 | 0 |
| 0 | 0 | 0 | 0 | 0 |
| 0 | 0 | 0 | 0 | 0 |
| 0 | 0 | 0.005438829 | 0 | 0 |
| 0 | 0 | 0 | 0 | 0 |
| 0 | 0 | 4.893352823 | 0 | 9.43419495 |
| 0 | 0 | 0 | 0 | 0 |
| 0 | 0 | 0.001371572 | 0 | 0 |
| 0 | 0 | 0 | 0 | 0 |
| 0 | 0 | 0 | 0 | 0 |
| 0 | 0 | 0 | 0 | 0 |
| 0 | 0 | 0 | 0.000638993 | 0.004403663 |
| 0 | 0.002782782 | 0 | 0.002586223 | 0.009916623 |
| 0 | 0 | 0 | 0 | 0 |
| 0 | 0.004014007 | 0.002438334 | 0.003597249 | 0 |
| 0 | 0 | 0 | 0 | 0 |
| 0 | 0 | 1.00965502 | 0 | 0.073612653 |
| 0 | 0 | 0 | 0 | 0 |
| 0 | 0 | 0 | 0 | 0 |
| 0 | 0 | 0 | 0 | 0 |
| 0 | 0 | 0 | 0 | 0 |
| 0 | 0.001534834 | 0.011391551 | 0.001426422 | 0 |
| 0 | 0 | 0 | 0 | 0 |
| 0 | 0 | 0 | 0 | 0 |
| 0.002535663 | 0 | 0 | 0 | 0 |
| 0 | 0 | 0 | 0 | 0 |
| 0 | 0 | 0 | 0 | 0 |
| 10.59888572 | 7.455026012 | 2.00433131 | 7.639707837 | 21.48933888 |
| 0 | 0 | 0.055194678 | 0 | 0.130525503 |
| 0 | 0 | 0.391475036 | 0 | 0 |
| 0 | 0 | 0 | 0 | 0 |
| 0 | 0 | 0 | 0 | 0 |
| 0 | 0 | 0 | 0 | 0 |
| 0 | 0 | 0 | 0 | 0 |
| 0 | 0 | 0.001923534 | 0.001156129 | 0 |

| | | | | |
|---|---|---|---|---|
| 0 | 0 | 0 | 0 | 0 |
| 0 | 0 | 0.009233063 | 0 | 0 |
| 0 | 0 | 0 | 0 | 0 |
| 0 | 0 | 0 | 0 | 0 |
| 0 | 1.807688876 | 1.959520835 | 1.750224777 | 0 |
| 0 | 0 | 0 | 0 | 0 |
| 0 | 0 | 0 | 0 | 0 |
| 0 | 0 | 0 | 0 | 0 |
| 0.00397483 | 0.194631372 | 0.019975475 | 0.192230198 | 0.00493589 |
| 0 | 0 | 0 | 0 | 0 |
| 0 | 0.003051885 | 0 | 0.003134877 | 0 |
| 0 | 0 | 0 | 0 | 0 |
| 0 | 0 | 0.146748009 | 0 | 0.03412757 |
| 0.18107891 | 0.006963244 | 0.008403459 | 0.006787079 | 0.075832264 |
| 0.064540003 | 0 | 0 | 0 | 0 |
| 0 | 0 | 0.480336778 | 0 | 0.003060364 |
| 0 | 0.006907404 | 0 | 0.009151209 | 0 |
| 0 | 0 | 0 | 0 | 0 |
| 0 | 0 | 0 | 0 | 0 |
| 0 | 0 | 0 | 0 | 0 |
| 0 | 0 | 0 | 0 | 0 |
| 0 | 0 | 0 | 0 | 0 |
| 0 | 0 | 0 | 0 | 0 |
| 0 | 0 | 0 | 0 | 0 |
| 0 | 0 | 0 | 0 | 0 |
| 0 | 0 | 0 | 0 | 0 |
| 0 | 0 | 0.093860108 | 0 | 0.002221607 |
| 0 | 0 | 0 | 0 | 0 |
| 0 | 0 | 0 | 0 | 0 |
| 0 | 0 | 0 | 0 | 0 |
| 0 | 0 | 0 | 0 | 0 |
| 0 | 0 | 0 | 0 | 0 |
| 0 | 0 | 0 | 0 | 0 |
| 0 | 0 | 0.045110241 | 0 | 0 |
| 0.064473827 | 0 | 0.148449598 | 0 | 0 |
| 0 | 0.000667238 | 0 | 0.000775136 | 0 |
| 0 | 0 | 0 | 0 | 0 |
| 0 | 0 | 0 | 0 | 0 |
| 0 | 0 | 0 | 0 | 0 |
| 0 | 0 | 0 | 0 | 0 |
| 0.031346544 | 11.61602398 | 0.283991897 | 11.74791474 | 0.00261698 |
| 0 | 0 | 0.004805331 | 0 | 0 |
| 0 | 0 | 0 | 0 | 0 |
| 0 | 0 | 0 | 0 | 0 |
| 0 | 0 | 0 | 0 | 0 |
| 0 | 0 | 0 | 0 | 0 |
| 0 | 0 | 0 | 0 | 0 |
| 0 | 0 | 0 | 0 | 0 |

| | | | | |
|---|---|---|---|---|
| 0 | 0 | 0.067136025 | 0 | 0.005345038 |
| 0 | 0 | 0 | 0 | 0 |
| 0 | 0 | 0.004115133 | 0 | 0 |
| 0 | 0 | 0 | 0 | 0 |
| 0 | 0 | 0.03987565 | 0 | 0 |
| 0 | 0 | 0 | 0 | 0 |
| 0 | 0.014074433 | 0.001360164 | 0.011309006 | 0 |
| 0 | 0 | 0 | 0 | 0 |
| 0 | 0 | 0 | 0 | 0 |
| 0 | 0.037838825 | 0 | 0.033450689 | 0 |
| 0 | 0 | 0 | 0 | 0 |
| 0 | 0 | 0 | 0 | 0 |
| 0 | 0 | 0 | 0 | 0 |
| 0 | 0 | 0 | 0 | 0 |
| 0 | 0 | 0 | 0 | 0 |
| 0 | 0 | 0 | 0 | 0 |
| 0 | 0 | 0 | 0 | 0.392996833 |
| 0 | 0 | 0 | 0 | 0 |
| 0 | 0 | 0 | 0 | 0 |
| 0 | 0 | 0 | 0 | 0 |
| 0 | 0.000837478 | 0.116114021 | 0.001037764 | 0.085183276 |
| 0 | 0 | 0 | 0 | 0 |
| 0.012587 | 0 | 0.108790939 | 0 | 0 |
| 0 | 0.002053487 | 0.011642432 | 0.001145064 | 0 |
| 0 | 0 | 0 | 0 | 0 |
| 0 | 0 | 0 | 0 | 0 |
| 0 | 0 | 0 | 0 | 0 |
| 0 | 0 | 0 | 0 | 0 |
| 0 | 0 | 0 | 0 | 0 |
| 0.003971988 | 0 | 0.003351238 | 0.002014242 | 0.507658122 |
| 0 | 0 | 0 | 0 | 0 |
| 0 | 0 | 0 | 0 | 0 |
| 0 | 0 | 0 | 0 | 0 |
| 0 | 0 | 0 | 0 | 0 |
| 0 | 0 | 0 | 0 | 0 |
| 0 | 0 | 0 | 0 | 0 |
| 0 | 0.002912037 | 0.00476761 | 0.002865544 | 0 |
| 0 | 0 | 0 | 0 | 0 |
| 0 | 0 | 0 | 0 | 0 |
| 0 | 0 | 0 | 0 | 0 |
| 0 | 0 | 0 | 0 | 0 |
| 0 | 0 | 0 | 0 | 0 |
| 0 | 0 | 0 | 0 | 0 |
| 0 | 0 | 0 | 0 | 0 |
| 0 | 0 | 0 | 0 | 0 |
| 0 | 0 | 0 | 0 | 0 |
| 0 | 0 | 0 | 0 | 0 |
| 0 | 0 | 0 | 0 | 0 |

| | | | | |
|---|---|---|---|---|
| 0 | 0 | 0.015488756 | 0 | 0 |
| 0 | 0 | 0.443276353 | 0 | 0 |
| 0 | 0 | 0 | 0 | 0 |
| 0 | 0 | 0 | 0 | 0 |
| 0 | 0 | 0 | 0 | 0 |
| 0 | 0 | 0.031615087 | 0 | 0 |
| 0 | 0 | 0 | 0 | 0 |
| 0.000677587 | 0 | 0.36671489 | 0 | 0.345286144 |
| 1.163767996 | 0 | 9.533493892 | 0 | 0.398158727 |
| 0 | 0 | 0.068945445 | 0 | 0 |
| 0 | 0 | 0 | 0 | 0 |
| 0 | 0 | 0 | 0 | 0 |
| 0 | 0 | 0 | 0 | 0 |
| 0.004409397 | 0 | 0 | 0 | 0 |
| 0 | 0 | 0 | 0 | 0 |
| 0 | 0 | 0 | 0 | 0 |
| 0 | 0 | 0 | 0 | 0 |
| 0 | 0 | 0 | 0 | 0 |
| 0 | 0 | 0 | 0 | 0 |
| 0 | 0 | 0.026831259 | 0 | 0 |
| 0 | 0 | 0 | 0 | 0 |
| 0 | 0 | 0 | 0 | 0 |
| 0 | 0 | 0 | 0 | 0 |
| 0 | 0 | 0 | 0 | 0 |
| 0 | 0 | 0 | 0 | 0 |
| 0 | 0 | 0 | 0 | 0 |
| 0 | 0 | 0 | 0 | 0 |
| 0 | 0 | 0 | 0 | 0 |
| 0 | 0 | 0 | 0 | 0 |
| 0 | 0 | 0 | 0 | 0 |
| 0 | 0.009685373 | 0.008658017 | 0.008157387 | 0 |
| 0 | 0 | 0 | 0 | 0 |
| 0 | 0 | 0 | 0 | 0 |
| 0 | 0 | 0 | 0 | 0 |
| 0 | 0 | 0 | 0 | 0 |
| 0 | 0 | 0 | 0 | 0 |
| 0 | 0 | 0 | 0 | 0 |
| 0 | 0 | 0 | 0 | 0 |
| 0 | 0 | 0.001354216 | 0 | 0 |
| 0 | 0.083677049 | 0 | 0.075033138 | 0 |
| 0 | 0 | 0 | 0 | 0 |
| 0.075613828 | 0 | 0.119408751 | 0 | 0 |
| 0 | 0 | 0 | 0 | 0 |
| 0 | 0 | 0 | 0 | 0 |
| 0 | 0 | 0 | 0 | 0 |
| 0.001320304 | 0 | 0.089319697 | 0 | 0 |
| 0 | 0 | 0 | 0 | 0 |
| 0 | 0 | 0 | 0 | 0 |

| | | | | |
|---|---|---|---|---|
| 0 | 0 | 0 | 0 | 0 |
| 0.115100767 | 0 | 0 | 0 | 0.000608236 |
| 0 | 0 | 0 | 0 | 0 |
| 0 | 0 | 0 | 0 | 0 |
| 0.038722422 | 0 | 1.621947449 | 0 | 11.62155561 |
| 0 | 0 | 0 | 0 | 0 |
| 0 | 0 | 0 | 0 | 0 |
| 0.00377759 | 0.06202505 | 0.005794947 | 0.066699806 | 0 |
| 0 | 0 | 0.003471624 | 0 | 0 |
| 0 | 0 | 0 | 0 | 0 |
| 0 | 0 | 0 | 0 | 0 |
| 0 | 0 | 0 | 0 | 0 |
| 0 | 0 | 0 | 0 | 0 |
| 0 | 0 | 0 | 0 | 0 |
| 0 | 0 | 0 | 0 | 0 |
| 0 | 0 | 0.009800533 | 0 | 0 |
| 0 | 0 | 0 | 0 | 0 |
| 0 | 0 | 0 | 0 | 0 |
| 0 | 0 | 0 | 0 | 0 |
| 0 | 0 | 0 | 0 | 0 |
| 0 | 0 | 0 | 0 | 0 |
| 0 | 0 | 0 | 0 | 0 |
| 0 | 0 | 0 | 0 | 0 |
| 0.001634596 | 0 | 0.004764297 | 0 | 0 |
| 0.724580857 | 0 | 0.562879803 | 0 | 0 |
| 0 | 0 | 0.000685613 | 0 | 0 |
| 0.000830569 | 0 | 0.002293416 | 0 | 0 |
| 0.00069965 | 0 | 0 | 0 | 0 |
| 0 | 0 | 0 | 0 | 0 |
| 0 | 0 | 0 | 0 | 0 |
| 0 | 0 | 0 | 0 | 0.011067946 |
| 0 | 0 | 0 | 0 | 0 |
| 0 | 0 | 0 | 0 | 0 |
| 0 | 0 | 0 | 0 | 0 |
| 0 | 0 | 0 | 0 | 0 |
| 0 | 0 | 0 | 0 | 0 |
| 0 | 0 | 0 | 0 | 0 |
| 0 | 0 | 0 | 0 | 0 |
| 0 | 0 | 0 | 0 | 0 |
| 0 | 0 | 0 | 0 | 0 |
| 0 | 0 | 0 | 0 | 0 |
| 0 | 0 | 0 | 0 | 0 |
| 0 | 0 | 0 | 0 | 0 |
| 0 | 0 | 0 | 0 | 0 |
| 0 | 0 | 0 | 0 | 0 |
| 0 | 0 | 0 | 0 | 0 |
| 0 | 0 | 0 | 0 | 0 |
| 0 | 0 | 0 | 0 | 0 |

| | | | | |
|---|---|---|---|---|
| 0 | 0 | 0.02264598 | 0 | 0.017739555 |
| 0 | 0 | 0 | 0 | 0 |
| 0 | 0 | 0.000640917 | 0 | 0 |
| 0 | 0 | 0 | 0 | 0 |
| 0 | 0 | 0 | 0 | 0 |
| 0 | 0 | 0.119939099 | 0 | 0.137064629 |
| 0 | 0 | 0 | 0 | 0 |
| 0 | 0 | 0 | 0 | 0 |
| 0 | 0 | 0 | 0 | 0 |
| 0 | 0 | 0 | 0 | 0 |
| 0 | 0 | 0 | 0 | 0 |
| 0 | 0 | 0 | 0 | 0 |
| 0 | 0 | 0 | 0 | 0 |
| 0 | 0 | 0 | 0 | 0 |
| 0 | 0 | 0 | 0 | 0 |
| 0 | 0 | 0 | 0 | 0 |
| 0 | 0 | 0 | 0 | 0 |
| 0 | 0 | 0 | 0 | 0 |
| 0 | 0 | 0 | 0 | 0 |
| 0 | 0 | 0 | 0 | 0 |
| 0 | 0.00403725 | 0.03296096 | 0.003301832 | 0.001034308 |
| 0 | 0 | 0 | 0 | 0 |
| 0 | 0.010401454 | 0.006307166 | 0.007960858 | 0 |
| 0 | 0 | 0 | 0 | 0 |
| 0 | 0 | 0 | 0 | 0 |
| 0 | 0 | 0 | 0 | 0 |
| 0 | 0 | 0 | 0 | 0 |
| 0 | 0 | 0 | 0 | 0 |
| 0 | 0 | 0 | 0 | 0 |
| 0 | 0 | 0 | 0 | 0 |
| 0 | 0 | 0 | 0 | 0 |
| 0 | 0 | 0 | 0 | 0 |
| 0 | 2.676269807 | 0 | 2.778700681 | 0.009611671 |
| 0 | 0 | 0 | 0 | 0 |
| 0 | 0 | 0.010038567 | 0 | 0 |
| 0.003604745 | 0 | 0 | 0 | 0 |
| 0 | 0 | 0 | 0 | 0 |
| 0 | 0 | 0 | 0 | 0 |
| 0 | 0 | 0 | 0 | 0 |
| 0 | 0 | 0 | 0 | 0 |
| 0 | 0 | 0 | 0 | 0 |
| 0 | 0 | 0.065480649 | 0 | 0 |
| 0.01392956 | 0 | 0.248407678 | 0 | 0.157043427 |
| 0 | 0 | 0 | 0 | 0 |
| 0 | 0 | 0 | 0 | 0 |
| 0 | 0 | 0.013491323 | 0 | 0 |
| 0 | 0 | 0 | 0 | 0 |
| 0 | 0 | 0 | 0 | 0 |

| | | | | |
|---|---|---|---|---|
| 0 | 0 | 0 | 0 | 0 |
| 0 | 0 | 0 | 0 | 0 |
| 0 | 0 | 0 | 0 | 0 |
| 0 | 0 | 0 | 0 | 0 |
| 0 | 0 | 0 | 0 | 0 |
| 0 | 0 | 0 | 0 | 0 |
| 0 | 0 | 0 | 0 | 0 |
| 0 | 0 | 0.007871331 | 0 | 0 |
| 0 | 0 | 0.006200813 | 0 | 0 |
| 0 | 0 | 0.216828691 | 0 | 0 |
| 0 | 0 | 0 | 0 | 0 |
| 0 | 0.001052941 | 0 | 0.001118362 | 0 |
| 0 | 0 | 0 | 0 | 0 |
| 0 | 0 | 0 | 0 | 0 |
| 0 | 0 | 0 | 0 | 0 |
| 0 | 0 | 0 | 0 | 0 |
| 0 | 0 | 0 | 0 | 0 |
| 0 | 0 | 0 | 0 | 0 |
| 0 | 0 | 0 | 0 | 0 |
| 0 | 0 | 0.644036988 | 0 | 0 |
| 0 | 0.009060653 | 0 | 0.008096789 | 0 |
| 0 | 0 | 0 | 0 | 0 |
| 0 | 0 | 0 | 0 | 0 |
| 0 | 0 | 0 | 0 | 0 |
| 0 | 0 | 0 | 0 | 0 |
| 0 | 0 | 0 | 0 | 0 |
| 0 | 0 | 0 | 0 | 0 |
| 0 | 0 | 0 | 0 | 0 |
| 0 | 0 | 0 | 0 | 0 |
| 0 | 0 | 0.029142123 | 0 | 0 |
| 0.01913461 | 0 | 0.13775 | 0 | 0 |
| 0 | 0 | 0 | 0 | 0 |
| 0 | 0 | 0 | 0 | 0 |
| 0 | 0 | 0 | 0 | 0 |
| 0 | 0 | 0 | 0 | 0 |
| 0 | 0 | 0 | 0 | 0 |
| 0 | 0 | 0 | 0 | 0 |
| 0 | 0 | 0 | 0 | 0 |
| 0.030636638 | 0 | 0.021030773 | 0 | 0 |
| 0 | 0 | 0 | 0 | 0 |
| 0 | 0 | 0.008216976 | 0 | 0 |
| 0 | 0 | 0 | 0 | 0 |
| 0 | 0 | 0 | 0 | 0 |
| 0 | 0 | 0 | 0 | 0 |
| 0 | 0 | 0 | 0 | 0 |
| 0 | 0 | 0 | 0 | 0 |
| 0 | 0.004098813 | 0 | 0.005872665 | 0.003906549 |
| 0 | 0 | 0.001287681 | 0 | 0 |

| | | | | |
|---|---|---|---|---|
| 0 | 0 | 0 | 0 | 0 |
| 0 | 0 | 0 | 0 | 0 |
| 0 | 0 | 0 | 0 | 0 |
| 0 | 0 | 0 | 0 | 0 |
| 0 | 0 | 0.008285915 | 0 | 0 |
| 0 | 0 | 0 | 0 | 0 |
| 0 | 0 | 0 | 0 | 0 |
| 0 | 0 | 0 | 0 | 0 |
| 0 | 0 | 0 | 0 | 0 |
| 0 | 0 | 0 | 0 | 0 |
| 0 | 0 | 0.007764933 | 0 | 0.004001897 |
| 0.001785488 | 0 | 0 | 0 | 0 |
| 0.011467617 | 0 | 0.009215951 | 0 | 0 |
| 0 | 0 | 0 | 0 | 0 |
| 0 | 0 | 0 | 0 | 0 |
| 0 | 0 | 0 | 0 | 0 |
| 0 | 0 | 0.020223492 | 0 | 0 |
| 0 | 0 | 0 | 0 | 0 |
| 0.002553743 | 0 | 0.010162522 | 0 | 0 |
| 0 | 0.026970417 | 0.093154904 | 0.029622725 | 0 |
| 0 | 0 | 0 | 0 | 0 |
| 0 | 0 | 0 | 0 | 0 |
| 0 | 0 | 0 | 0 | 0 |
| 0 | 0 | 0 | 0 | 0 |
| 0 | 0 | 0 | 0 | 0 |
| 0 | 0 | 0 | 0 | 0 |
| 0.075130796 | 0 | 0.015841274 | 0 | 0 |
| 0 | 0 | 0 | 0 | 0 |
| 0 | 0 | 0 | 0 | 0 |
| 0 | 0 | 0 | 0 | 0 |
| 0 | 0 | 0.048213091 | 0 | 0 |
| 0 | 0 | 0 | 0 | 0 |
| 0 | 0 | 0 | 0 | 0 |
| 0 | 0 | 0 | 0 | 0 |
| 0 | 0 | 0 | 0 | 0 |
| 0 | 0 | 0 | 0 | 0 |
| 0 | 0 | 0 | 0 | 0 |
| 0.001251763 | 0.005253112 | 0.007258525 | 0.004051073 | 0 |
| 0 | 0 | 0 | 0 | 0 |
| 0 | 0 | 0 | 0 | 0 |
| 0 | 0 | 0.003450642 | 0 | 0.008575813 |
| 0 | 0 | 0 | 0 | 0 |
| 0 | 0 | 0 | 0 | 0 |
| 0.155735607 | 0 | 0 | 0 | 0 |
| 0 | 0 | 0 | 0 | 0 |
| 0 | 0 | 0 | 0 | 0 |
| 0 | 0 | 0 | 0 | 0 |
| 0 | 0 | 0 | 0 | 0 |

| | | | | |
|---|---|---|---|---|
| 0 | 0 | 0 | 0 | 0 |
| 0 | 0 | 0.024970373 | 0 | 0 |
| 0 | 0 | 0 | 0 | 0 |
| 0.001167187 | 0 | 0.015577375 | 0.000968554 | 0 |
| 0 | 0.023198231 | 0 | 0.022907119 | 0.072521023 |
| 0 | 0 | 0 | 0 | 0 |
| 0 | 0 | 0.002606321 | 0 | 0 |
| 0.00851985 | 0.000407532 | 0 | 0.000504995 | 0 |
| 0 | 0 | 0 | 0 | 0 |
| 0 | 0 | 0 | 0 | 0 |
| 0 | 0 | 0 | 0 | 0 |
| 0 | 0 | 0 | 0 | 0 |
| 0 | 0 | 0.334501476 | 0 | 0.057904926 |
| 0 | 0 | 0 | 0 | 0 |
| 0 | 0 | 0 | 0 | 0 |
| 0 | 0 | 0 | 0 | 0 |
| 0 | 0 | 0 | 0 | 0 |
| 0 | 0 | 0 | 0 | 0 |
| 0 | 0 | 0 | 0 | 0.001612714 |
| 0 | 0 | 0 | 0 | 0 |
| 0 | 0 | 0 | 0 | 0 |
| 0 | 0 | 0 | 0 | 0 |
| 0 | 0 | 0.075708214 | 0 | 0 |
| 0 | 0 | 0.021890841 | 0 | 0 |
| 0 | 0 | 0 | 0 | 0 |
| 0 | 0 | 0 | 0 | 0 |
| 0 | 0 | 0 | 0 | 0 |
| 0 | 0 | 0 | 0 | 0 |
| 0 | 0 | 0 | 0 | 0 |
| 0 | 0 | 0 | 0 | 0 |
| 0 | 0 | 0.023199963 | 0 | 0.003804198 |
| 0 | 0 | 0 | 0 | 0 |
| 0 | 0 | 0 | 0 | 0 |
| 0 | 0 | 0 | 0 | 0 |
| 0 | 0 | 0.001406048 | 0 | 0 |
| 0 | 0.043085877 | 0 | 0.043028167 | 0 |
| 0 | 0 | 0 | 0 | 0 |
| 0 | 0 | 0 | 0 | 0 |
| 0.001541631 | 0 | 0.001655439 | 0 | 0.033025795 |
| 0 | 0 | 0 | 0 | 0 |
| 0 | 0 | 0 | 0 | 0 |
| 0 | 0 | 0 | 0 | 0 |
| 0 | 0 | 0 | 0 | 0 |
| 0 | 0 | 0 | 0 | 0 |
| 0 | 0 | 0 | 0 | 0 |
| 0.00150709 | 0 | 0.906575243 | 0 | 0.22494739 |
| 0 | 0.009929857 | 0 | 0.00640039 | 0 |
| 0 | 0 | 0 | 0 | 0 |

| | | | | |
|---|---|---|---|---|
| 0 | 0 | 0 | 0 | 0 |
| 0 | 0 | 0 | 0 | 0 |
| 0 | 0 | 0 | 0 | 0 |
| 0 | 0 | 0.471534154 | 0 | 0 |
| 0 | 0 | 0 | 0 | 0 |
| 0 | 0 | 0 | 0 | 0 |
| 0 | 0 | 0 | 0 | 0 |
| 0 | 0 | 0 | 0 | 0 |
| 0 | 0.000990603 | 0.00743979 | 0.001315189 | 0 |
| 0 | 0 | 0 | 0 | 0 |
| 0 | 0 | 0 | 0 | 0.001021758 |
| 0 | 0 | 0.003150887 | 0 | 0 |
| 0 | 0 | 0 | 0 | 0 |
| 0 | 0 | 0 | 0 | 0 |
| 0 | 0 | 0 | 0 | 0 |
| 0 | 0 | 0 | 0 | 0 |
| 0 | 0 | 0 | 0 | 0 |
| 0 | 0 | 0 | 0 | 0 |
| 0 | 0 | 0 | 0 | 0 |
| 0.001842901 | 0 | 0 | 0 | 0 |
| 0 | 0 | 0 | 0 | 0 |
| 0 | 0 | 0 | 0 | 0 |
| 0 | 0 | 0 | 0 | 0 |
| 0 | 0 | 0 | 0 | 0 |
| 0 | 0 | 0 | 0 | 0 |
| 0 | 0 | 0 | 0 | 0 |
| 0 | 0 | 0 | 0 | 0 |
| 0.000815793 | 0 | 0.020836689 | 0 | 0 |
| 0 | 0 | 0 | 0 | 0 |
| 0 | 0 | 0 | 0 | 0 |
| 0.003483134 | 0 | 0.003497395 | 0 | 0.084979898 |
| 0 | 0 | 0 | 0 | 0 |
| 0.213243526 | 0.03332044 | 0.042638735 | 0.035695808 | 0.05331501 |
| 0 | 0.005535202 | 0 | 0.007794283 | 0.000767354 |
| 0 | 0 | 0 | 0 | 0 |
| 0 | 0 | 0 | 0 | 0 |
| 0 | 0 | 0 | 0 | 0 |
| 0 | 0 | 0 | 0 | 0 |
| 0 | 0 | 0 | 0 | 0 |
| 0 | 0 | 0 | 0 | 0 |
| 0 | 0 | 0 | 0 | 0 |
| 0 | 0.033894179 | 0.012110435 | 0.029617614 | 0 |
| 0.002026726 | 0 | 0.031542872 | 0 | 0.050576054 |
| 0 | 0 | 0 | 0 | 0 |
| 0 | 0 | 0 | 0 | 0 |
| 0.005591269 | 0 | 0.107337418 | 0 | 0 |
| 0 | 0 | 0 | 0 | 0 |
| 0 | 0 | 0 | 0 | 0 |

| | | | | |
|---|---|---|---|---|
| 0 | 0 | 0 | 0 | 0 |
| 0.036481543 | 0 | 0.006129378 | 0 | 0 |
| 0 | 0.009864757 | 0.002801645 | 0.005987244 | 0.004973482 |
| 0 | 0 | 0 | 0 | 0 |
| 0.005400867 | 0 | 0.024109663 | 0 | 0 |
| 0 | 0 | 0 | 0 | 0 |
| 0 | 0 | 0 | 0 | 0 |
| 0 | 0 | 0 | 0 | 0 |
| 0 | 0 | 0 | 0 | 0 |
| 0 | 0 | 0 | 0 | 0 |
| 0 | 0 | 0 | 0 | 0 |
| 0 | 0 | 0 | 0 | 0 |
| 0 | 0 | 0 | 0 | 0 |
| 0 | 0 | 0 | 0 | 0 |
| 0 | 0 | 0 | 0 | 0 |
| 0 | 0 | 0 | 0 | 0 |
| 0 | 0 | 0 | 0 | 0 |
| 0 | 0 | 0 | 0 | 0 |
| 0 | 0 | 0 | 0 | 0 |
| 0 | 0 | 0 | 0 | 0 |
| 0 | 0 | 0 | 0 | 0 |
| 0 | 0 | 0 | 0.000451504 | 0 |
| 0 | 0 | 0 | 0 | 0 |
| 0 | 0 | 0 | 0 | 0 |
| 0 | 0 | 1.490042245 | 0 | 0 |
| 0 | 0.001001453 | 0 | 0.000775597 | 0.003207046 |
| 0 | 0 | 0 | 0 | 0 |
| 0 | 0 | 0 | 0 | 0 |
| 0 | 0 | 0 | 0 | 0 |
| 0 | 0 | 0 | 0 | 0 |
| 0 | 0 | 0 | 0 | 0 |
| 0 | 0 | 0 | 0 | 0 |
| 0 | 0 | 0 | 0 | 0 |
| 0 | 0 | 0 | 0 | 0 |
| 0 | 0 | 0 | 0 | 0 |
| 0 | 0 | 0 | 0 | 0 |
| 0 | 0 | 0 | 0 | 0 |
| 0 | 0 | 0 | 0 | 0 |
| 0 | 0 | 2.948439392 | 0 | 0 |
| 0 | 0 | 0 | 0 | 0 |
| 0 | 0 | 0 | 0 | 0 |
| 0 | 0 | 0.02883799 | 0 | 0 |
| 0 | 0 | 0.011137631 | 0 | 0 |
| 0 | 0 | 0 | 0 | 0 |
| 0 | 0 | 0 | 0 | 0 |
| 0 | 0 | 0 | 0 | 0 |
| 0 | 0.003587632 | 0.066568614 | 0.002381588 | 0 |
| 0 | 0 | 0 | 0 | 0 |

| | | | | |
|---|---|---|---|---|
| 0 | 0 | 0 | 0 | 0 |
| 0.025168882 | 0.018107728 | 0.026999147 | 0.013973834 | 0 |
| 0 | 0 | 0 | 0 | 0 |
| 0 | 0 | 0 | 0 | 0 |
| 0 | 0 | 0 | 0 | 0 |
| 0 | 0 | 0 | 0 | 0 |
| 0 | 0 | 0 | 0 | 0 |
| 0 | 0 | 0 | 0 | 0 |
| 0 | 0 | 0 | 0 | 0 |
| 0 | 0 | 0 | 0 | 0 |
| 0 | 0 | 0 | 0 | 0 |
| 0 | 0 | 0 | 0 | 0 |
| 0 | 0 | 0.371290643 | 0 | 0.014855504 |
| 0 | 0 | 0 | 0 | 0 |
| 0.00135461 | 0 | 0.032417057 | 0 | 0 |
| 0 | 0 | 0 | 0 | 0 |
| 0 | 0 | 0 | 0 | 0 |
| 0 | 0 | 0 | 0 | 0 |
| 0 | 0 | 0 | 0 | 0 |
| 0 | 0 | 0 | 0 | 0 |
| 0 | 0 | 0 | 0 | 0 |
| 0 | 0 | 0 | 0 | 0 |
| 0.000681502 | 0 | 0.075977811 | 0 | 0 |
| 0 | 0 | 0 | 0 | 0 |
| 0 | 0 | 0 | 0 | 0 |
| 0 | 0 | 0 | 0 | 0 |
| 0 | 0 | 0 | 0 | 0 |
| 0 | 0 | 0 | 0 | 0 |
| 0 | 0 | 0 | 0 | 0 |
| 0 | 0 | 0 | 0 | 0 |
| 0 | 0 | 0 | 0 | 0 |
| 0 | 0 | 0 | 0 | 0 |
| 0 | 0 | 0 | 0 | 0 |
| 0 | 0 | 0 | 0 | 0 |
| 0.181784336 | 0 | 0 | 0 | 0 |
| 0 | 0 | 0 | 0 | 0 |
| 0 | 0 | 0 | 0 | 0 |
| 0 | 0 | 0 | 0 | 0 |
| 0 | 0.002570102 | 1.15307888 | 0.002204829 | 0 |
| 0 | 0 | 0 | 0 | 0 |
| 0.198132205 | 0.071082997 | 0.029328204 | 0.059637111 | 0.171598496 |
| 0 | 0 | 0 | 0 | 0 |
| 0 | 0 | 0 | 0 | 0 |
| 0 | 0 | 0 | 0 | 0 |
| 0 | 0 | 0 | 0 | 0 |
| 0.00818289 | 0.955920855 | 0.057742979 | 0.927821697 | 0.023397962 |
| 0 | 0 | 0 | 0 | 0 |
| 0 | 0 | 0 | 0 | 0 |

| | | | | |
|---|---|---|---|---|
| 0 | 0 | 0 | 0 | 0 |
| 0 | 0 | 0 | 0 | 0 |
| 0 | 0 | 0 | 0 | 0 |
| 0 | 0 | 0 | 0 | 0 |
| 0 | 0 | 0 | 0 | 0 |
| 0 | 0 | 0 | 0 | 0 |
| 0 | 0 | 0 | 0 | 0 |
| 0 | 0 | 0.026796457 | 0 | 0 |
| 0 | 0 | 0 | 0 | 0 |
| 0.028454682 | 0 | 0.017187351 | 0 | 0.105153952 |
| 0 | 0 | 0 | 0 | 0 |
| 0 | 0 | 0 | 0 | 0 |
| 0 | 0 | 0.003599826 | 0 | 0 |
| 0 | 0 | 0 | 0 | 0 |
| 0 | 0 | 0 | 0 | 0 |
| 0 | 0 | 0 | 0 | 0 |
| 0 | 0 | 0 | 0 | 0 |
| 0 | 0 | 0 | 0 | 0 |
| 0 | 0 | 0 | 0 | 0 |
| 0.358556241 | 0.097392045 | 0.153529758 | 0.094043476 | 0.473835179 |
| 0 | 0 | 0 | 0 | 0 |
| 0 | 0 | 0 | 0 | 0 |
| 0 | 0 | 0 | 0 | 0 |
| 0 | 0 | 0 | 0 | 0 |
| 0 | 0 | 0 | 0 | 0 |
| 0 | 0 | 0 | 0 | 0 |
| 0 | 0 | 0 | 0 | 0 |
| 0 | 0 | 0 | 0 | 0 |
| 0 | 0 | 0 | 0 | 0 |
| 0 | 0 | 0.016555297 | 0 | 0 |
| 0 | 0 | 0 | 0 | 0 |
| 0 | 0 | 0 | 0 | 0 |
| 0 | 0 | 0 | 0 | 0 |
| 0 | 0 | 0 | 0 | 0 |
| 0 | 0 | 0 | 0 | 0 |
| 0 | 0 | 0 | 0 | 0 |
| 0 | 0 | 0 | 0 | 0 |
| 0 | 0 | 0 | 0 | 0 |
| 0 | 0 | 0 | 0 | 0 |
| 0 | 0 | 0 | 0 | 0 |
| 0 | 0 | 0 | 0 | 0 |
| 0 | 0 | 0 | 0 | 0 |
| 0 | 0 | 0.015536346 | 0 | 0 |
| 0 | 0 | 0 | 0 | 0 |
| 0.000908223 | 0 | 0.006018816 | 0 | 0.0761278 |
| 0 | 0 | 0 | 0 | 0 |
| 0 | 0 | 0 | 0 | 0 |
| 0 | 0.024515699 | 0 | 0.02489648 | 0 |

| | | | | |
|---|---|---|---|---|
| 0 | 0 | 0 | 0 | 0 |
| 0 | 0 | 0 | 0 | 0 |
| 0 | 0 | 0 | 0 | 0 |
| 0 | 0 | 0 | 0 | 0 |
| 0 | 0 | 0 | 0 | 0 |
| 0 | 0.001574038 | 0.113829756 | 0.001687912 | 0 |
| 0 | 0 | 0 | 0 | 0 |
| 0 | 0 | 0 | 0 | 0 |
| 0 | 0 | 0 | 0 | 0 |
| 0 | 0 | 0 | 0 | 0 |
| 0 | 0 | 0 | 0 | 0 |
| 0 | 0 | 0 | 0 | 0 |
| 0 | 0 | 0 | 0 | 0 |
| 0 | 0 | 0 | 0 | 0 |
| 0 | 0 | 0 | 0 | 0 |
| 0 | 0 | 0 | 0 | 0 |
| 0 | 0 | 0 | 0 | 0 |
| 0 | 0 | 0 | 0 | 0 |
| 0 | 0 | 0 | 0 | 0 |
| 0 | 0 | 0 | 0 | 0 |
| 0 | 0 | 0 | 0 | 0.103276285 |
| 0 | 0 | 0 | 0 | 0 |
| 0 | 0 | 0 | 0 | 0 |
| 0 | 0 | 0 | 0 | 0 |
| 0 | 0 | 0 | 0 | 0 |
| 0 | 0 | 0 | 0 | 0.001386282 |
| 0 | 0 | 0 | 0 | 0 |
| 0 | 0 | 0 | 0 | 0 |
| 0 | 0 | 0 | 0 | 0 |
| 0 | 0 | 0 | 0 | 0 |
| 0 | 0 | 0 | 0 | 0 |
| 0 | 0 | 0 | 0 | 0 |
| 0 | 0 | 0 | 0 | 0 |
| 0 | 0 | 0 | 0 | 0 |
| 0 | 0 | 0 | 0 | 0 |
| 0 | 0 | 0 | 0 | 0 |
| 0 | 0 | 0 | 0 | 0 |
| 0 | 0 | 0 | 0 | 0 |
| 0 | 0 | 0 | 0 | 0 |
| 0 | 0 | 0 | 0 | 0 |
| 0 | 0 | 0 | 0 | 0 |
| 0 | 0 | 0 | 0 | 0 |
| 0 | 0 | 0 | 0 | 0 |
| 0 | 0 | 0 | 0 | 0 |
| 0 | 0 | 0 | 0 | 0 |
| 0 | 0 | 0 | 0 | 0 |
| 0 | 0 | 0 | 0 | 0 |
| 0 | 0 | 0 | 0 | 0 |

| | | | | |
|---|---|---|---|---|
| 0 | 0 | 0 | 0 | 0 |
| 0 | 0 | 0.001048039 | 0 | 0 |
| 0 | 0 | 0.010778148 | 0 | 0 |
| 0 | 0 | 0 | 0 | 0 |
| 0 | 0 | 0 | 0 | 0 |
| 0 | 0 | 0 | 0 | 0 |
| 0 | 0 | 0 | 0 | 0 |
| 0 | 0 | 0 | 0 | 0 |
| 0 | 0 | 0 | 0 | 0 |
| 0 | 0 | 0 | 0 | 0 |
| 0 | 0 | 0.037052761 | 0 | 0.009419783 |
| 0 | 0 | 0 | 0 | 0 |
| 0 | 0 | 0.000606472 | 0 | 0 |
| 0 | 0 | 0 | 0 | 0 |
| 0 | 0 | 0 | 0 | 0 |
| 0 | 0 | 0 | 0 | 0 |
| 0 | 0 | 0 | 0 | 0 |
| 0 | 0 | 0 | 0 | 0 |
| 0 | 0 | 0 | 0 | 0 |
| 0 | 0 | 0 | 0 | 0 |
| 0.001210601 | 0.001801552 | 0.005571307 | 0.003348601 | 0 |
| 0 | 0 | 0 | 0 | 0 |
| 0 | 0 | 0 | 0 | 0 |
| 0.03877732 | 0 | 1.886490975 | 0 | 0.047929202 |
| 0 | 0 | 0 | 0 | 0 |
| 0 | 0 | 0 | 0 | 0 |
| 0 | 0 | 0 | 0 | 0 |
| 0 | 0 | 0 | 0 | 0 |
| 0 | 0 | 0 | 0 | 0 |
| 0 | 0 | 0 | 0 | 0 |
| 0 | 0 | 0 | 0 | 0 |
| 0 | 0 | 0 | 0 | 0 |
| 0 | 0 | 0.009083748 | 0 | 0 |
| 0 | 0 | 0 | 0 | 0 |

| PHL018-0272_V1 | PHL018-0271_V1 | PHL018-0068_V3 | PHL018-0037_V1 | PHL018-0119_V1 |
|---|---|---|---|---|
| REF | REF | REF | CTRL | REF |
| 0.045792286 | 0.061397006 | 0.014635213 | 0.070213023 | 0.047281111 |
| 1.530897398 | 0.020733613 | 3.134098157 | 3.305878151 | 0.30345787 |
| 0 | 0 | 10.82093323 | 6.434329335 | 0 |
| 0 | 0 | 0 | 0 | 0 |
| 0 | 0 | 0 | 0 | 0 |
| 0 | 0 | 0 | 0 | 0 |
| 0 | 0 | 0 | 0 | 0 |
| 0 | 0 | 0 | 0 | 0 |
| 0 | 0 | 0 | 0 | 0 |
| 0 | 0 | 0 | 0 | 0 |
| 0 | 0 | 0 | 0 | 0 |
| 0 | 0 | 0 | 0 | 0 |
| 0 | 0 | 0 | 0 | 0 |
| 3.807223926 | 0.002341445 | 17.54614555 | 5.054631236 | 0.014726251 |
| 0 | 0 | 0 | 0 | 0 |
| 0 | 0 | 0 | 0 | 0 |
| 0 | 0 | 0 | 0.007711122 | 0 |
| 0 | 0 | 0 | 0.001452917 | 0 |
| 0 | 0 | 0 | 0 | 0 |
| 0 | 0 | 0 | 0 | 0 |
| 0 | 0 | 0 | 0 | 0 |
| 0 | 0 | 0 | 0.001754779 | 0 |
| 0 | 0 | 0 | 0 | 0 |
| 0 | 0 | 0 | 0 | 0 |
| 0 | 0 | 0 | 0 | 0.639021177 |
| 0 | 0 | 0 | 0 | 0 |
| 0.005125313 | 0 | 0 | 0 | 0 |
| 0 | 0 | 0 | 0 | 0 |
| 0 | 0 | 0 | 0 | 0 |
| 0 | 0 | 0 | 0 | 0 |
| 0 | 0 | 0 | 0 | 0 |
| 0 | 0 | 0 | 0 | 0 |
| 0 | 0 | 0 | 0 | 0 |
| 0 | 0 | 0 | 0 | 0 |
| 0 | 0 | 0 | 0 | 0 |
| 0 | 0.374855118 | 0 | 0 | 1.84953568 |
| 0 | 0 | 0 | 0 | 0 |
| 0 | 0 | 0 | 0 | 0 |
| 0 | 0 | 0 | 0 | 0 |
| 0 | 0 | 0 | 0 | 0 |
| 0 | 0 | 0 | 0 | 0 |
| 0.00217559 | 0.010146429 | 0 | 1.902179713 | 0.01544202 |
| 0.004900861 | 0.953894517 | 0.007088643 | 0.203120028 | 0.80202586 |
| 0.017168238 | 0.000563987 | 0.012412002 | 0.03617115 | 0.100629441 |
| 0 | 0.002236382 | 0 | 0 | 0.015422835 |
| 0 | 0 | 0 | 0 | 0 |

| | | | | |
|---|---|---|---|---|
| 0 | 0.003425381 | 0 | 0.00174066 | 0.03528552 |
| 0.051310848 | 0 | 0.031510877 | 0.013172599 | 0.003648284 |
| 0 | 0 | 0 | 0.003135454 | 0.032717767 |
| 2.872951447 | 0.109768799 | 0 | 0.319710535 | 0.212516272 |
| 0 | 0.029037864 | 0.002022321 | 0.015985706 | 0 |
| 0 | 0 | 0 | 0 | 0 |
| 0 | 0.002137236 | 0 | 1.822621829 | 0 |
| 0 | 0 | 0 | 0 | 0 |
| 0 | 0 | 0 | 0 | 0 |
| 0 | 0 | 0 | 0 | 0 |
| 0 | 0 | 0 | 0 | 0 |
| 0.220300286 | 0.07500374 | 0 | 0.193997384 | 0.071963152 |
| 0 | 0 | 0 | 0 | 0 |
| 0.018483325 | 0.00320039 | 0.02620398 | 0.02711724 | 0.014864114 |
| 0 | 0 | 0 | 0 | 0 |
| 0 | 0 | 0 | 0 | 0 |
| 0 | 0 | 0 | 0.996370366 | 0.529509976 |
| 0 | 0 | 0 | 0 | 0 |
| 0 | 0 | 0 | 0 | 0 |
| 0 | 0 | 0 | 0 | 0 |
| 0.123697491 | 0 | 0.14434605 | 0.313419495 | 0.004325059 |
| 0.019866034 | 0.002761048 | 0.008768481 | 0.021279882 | 0.011541245 |
| 0 | 0.004645257 | 0 | 0 | 0 |
| 0 | 0 | 0 | 0 | 0 |
| 0 | 0 | 0 | 0 | 0 |
| 0 | 0 | 0 | 0 | 0 |
| 0.00208534 | 0 | 0 | 0 | 0 |
| 0 | 0 | 0 | 0 | 0 |
| 0 | 0.000529827 | 0 | 0.021972949 | 0.002187347 |
| 0 | 0 | 0 | 0 | 0 |
| 0 | 0 | 0.024054748 | 0 | 0 |
| 0 | 0 | 0 | 0 | 0 |
| 0 | 0 | 0 | 0 | 0 |
| 0 | 0 | 0 | 0 | 0 |
| 0 | 0 | 0 | 0 | 0 |
| 0 | 0 | 0 | 0 | 0 |
| 0 | 0 | 0 | 0 | 0 |
| 0.001323607 | 0 | 0 | 0.035019842 | 0 |
| 0 | 0 | 0 | 0 | 0 |
| 0 | 0 | 0 | 0 | 0 |
| 0 | 0 | 0 | 0 | 0 |
| 0 | 0 | 0 | 0 | 0 |
| 0 | 0 | 0 | 0 | 0 |
| 0 | 0 | 0 | 0 | 0 |
| 0 | 0 | 0 | 0 | 0 |
| 0 | 0 | 0 | 0 | 0.002157591 |
| 0 | 0 | 0.155195047 | 0 | 0 |
| 0.004631798 | 0 | 0 | 0.006757007 | 0 |

| | | | | |
|---|---|---|---|---|
| 0 | 0 | 0 | 0 | 0 |
| 0 | 0 | 0 | 0.016920341 | 0 |
| 0 | 0 | 0 | 0 | 0 |
| 0 | 0 | 0 | 0 | 0 |
| 0 | 0 | 0 | 0 | 0.00072983 |
| 0 | 0 | 0 | 0 | 0 |
| 0 | 0 | 0 | 0 | 0 |
| 0 | 0 | 0 | 0 | 0.00286993 |
| 0.001055059 | 0.003602839 | 0 | 0 | 0.007643596 |
| 0 | 0 | 0 | 0 | 0 |
| 0 | 0 | 0 | 0 | 0 |
| 0 | 0 | 0 | 0.070786619 | 0.003837433 |
| 0 | 0 | 0 | 0 | 0 |
| 0 | 0 | 0 | 0 | 0 |
| 0 | 0 | 0 | 0 | 0 |
| 0 | 0 | 0 | 0 | 0 |
| 0 | 0 | 0 | 0 | 0 |
| 0 | 0 | 0 | 0 | 0 |
| 0 | 0.848044868 | 0.002918987 | 0 | 0.251225926 |
| 0 | 0 | 0 | 0 | 0.005161181 |
| 0 | 0 | 0 | 0 | 0 |
| 0 | 0 | 0 | 0 | 0 |
| 0 | 0 | 0 | 0 | 0 |
| 0 | 0 | 0 | 0 | 0 |
| 0 | 0 | 0 | 0 | 0 |
| 0 | 0 | 0 | 0 | 0 |
| 0 | 0 | 0 | 0 | 0 |
| 0 | 0 | 0 | 0.001582233 | 0 |
| 0 | 0 | 0 | 0 | 0 |
| 0 | 0 | 0 | 0 | 0 |
| 0 | 0.006914884 | 0 | 0.01549883 | 0.148727536 |
| 0 | 0.020726617 | 0 | 0 | 0 |
| 0 | 0 | 0 | 0 | 0 |
| 0 | 0 | 0 | 0 | 0 |
| 0 | 0 | 0 | 0 | 0 |
| 0 | 0 | 0 | 0 | 0 |
| 0 | 0 | 0 | 0 | 0 |
| 0 | 0 | 0 | 0 | 0 |
| 0 | 0 | 0 | 0.154291178 | 0 |
| 0 | 0 | 0 | 0 | 0 |
| 0 | 0 | 0 | 0 | 0 |
| 0 | 0.000464379 | 0 | 0 | 0 |
| 0.358237911 | 0 | 0 | 0 | 0 |
| 0 | 0 | 0 | 0 | 0 |
| 0 | 0 | 0 | 0 | 0 |
| 0 | 0 | 0 | 0 | 0 |
| 0 | 0 | 0 | 0.004320861 | 0 |
| 0 | 0 | 0 | 0 | 0 |

| | | | | |
|---|---|---|---|---|
| 0 | 0 | 0 | 0 | 0 |
| 0 | 0 | 0 | 0 | 0 |
| 0 | 0 | 0 | 0 | 0 |
| 0 | 0 | 0 | 0 | 0 |
| 0 | 0.09707869 | 0 | 0 | 0 |
| 0 | 0 | 0 | 0 | 0 |
| 0 | 0 | 0 | 0 | 0 |
| 0 | 0 | 0 | 0 | 0 |
| 0 | 0 | 0 | 0 | 0 |
| 0 | 0 | 0 | 0 | 0 |
| 0 | 0 | 0 | 0 | 0 |
| 0 | 0 | 0 | 0 | 0 |
| 0 | 0 | 0 | 0 | 0 |
| 0 | 0 | 0 | 0 | 0 |
| 0 | 0 | 0 | 0 | 0 |
| 0 | 0 | 0 | 0 | 0 |
| 0 | 0 | 0 | 0 | 0 |
| 0 | 0 | 0 | 0 | 0 |
| 0 | 0 | 0 | 0 | 0 |
| 0 | 0 | 0 | 0 | 0 |
| 0 | 0 | 0 | 0 | 0 |
| 0 | 0 | 0 | 0 | 0 |
| 0 | 0 | 0 | 0 | 0 |
| 0 | 0 | 0 | 0 | 0 |
| 0 | 0.007594822 | 0 | 0 | 0 |
| 0 | 0 | 0 | 0 | 0 |
| 0 | 0 | 0 | 0 | 0 |
| 0 | 0 | 0 | 0 | 0 |
| 0 | 0 | 0 | 0 | 0 |
| 0 | 0 | 0 | 0 | 0 |
| 0 | 0 | 0.010027651 | 0 | 0 |
| 0.000709055 | 0 | 0 | 0.00148106 | 0.7316608 |
| 0 | 0 | 0 | 0 | 0 |
| 0 | 0 | 0 | 0 | 0 |
| 0 | 0 | 0 | 0 | 0 |
| 0 | 0 | 0 | 0 | 0 |
| 0 | 0 | 0 | 0 | 0 |
| 0 | 0 | 0 | 0 | 0 |
| 0 | 0 | 0 | 0 | 0 |
| 0 | 0 | 0 | 0 | 0 |
| 0 | 0 | 0 | 0 | 0 |
| 0 | 0 | 0 | 0 | 0 |
| 0 | 0 | 0 | 0 | 0 |
| 0 | 0 | 0 | 0 | 0 |
| 0 | 0 | 0 | 0 | 0 |
| 0 | 0 | 0 | 0 | 0 |
| 0 | 0 | 0 | 0 | 0 |
| 0 | 0 | 0 | 0 | 0 |

| | | | | |
|---|---|---|---|---|
| 0 | 0 | 0 | 0 | 0 |
| 0 | 0 | 0 | 0 | 0 |
| 0 | 0 | 0 | 0 | 0 |
| 0 | 0 | 0 | 0 | 0 |
| 0 | 0 | 0 | 0 | 0 |
| 0 | 0 | 0 | 0 | 0 |
| 0 | 0 | 0 | 0 | 0 |
| 0 | 0 | 0 | 0 | 0 |
| 0 | 0 | 0 | 0 | 0 |
| 0 | 0 | 0 | 0 | 0 |
| 0 | 0 | 0 | 0 | 0 |
| 0 | 0 | 0 | 0 | 0 |
| 0.002713985 | 0.000572085 | 0.000796849 | 0.00062988 | 0.005314062 |
| 0 | 0 | 0 | 0 | 0 |
| 0.001963364 | 0 | 0 | 0.065445683 | 0.097789984 |
| 0 | 0 | 0 | 0 | 0 |
| 0 | 0 | 0 | 0 | 0 |
| 0 | 0 | 0 | 0 | 0 |
| 0 | 0 | 0 | 0 | 0 |
| 0 | 0 | 0 | 0 | 0 |
| 0.076963254 | 0.003743814 | 0.024434622 | 0.006771911 | 0.014076027 |
| 0 | 0 | 0 | 0 | 0 |
| 0 | 0 | 0 | 0 | 0 |
| 0.003086815 | 0 | 0 | 0.019343031 | 0 |
| 0 | 0 | 0 | 0 | 0 |
| 0 | 0 | 0 | 0 | 0 |
| 0.005910539 | 0.000467209 | 0.002429537 | 0.010116708 | 0.009403068 |
| 0 | 0 | 0 | 0 | 0 |
| 0 | 0 | 0 | 0 | 0 |
| 0.005269663 | 0 | 0 | 0.00201798 | 0 |
| 0.442130752 | 0 | 0.236648003 | 0 | 0 |
| 0 | 0 | 0 | 0 | 0 |
| 0 | 0 | 0 | 0 | 0 |
| 0 | 0 | 0 | 0.001259565 | 0.00809637 |
| 0.016508702 | 0.003131904 | 0 | 0.008046043 | 0.01104421 |
| 0 | 0 | 0 | 0 | 0.000720411 |
| 0 | 0 | 0 | 0 | 0 |
| 0 | 0 | 0 | 0 | 0 |
| 0 | 0 | 0 | 0.007984447 | 0 |
| 0 | 0 | 0 | 0 | 0 |
| 0 | 0 | 0 | 0 | 0 |
| 0 | 0 | 0 | 0 | 0 |
| 0 | 0 | 0 | 0 | 0 |
| 0 | 0 | 0 | 0 | 0 |
| 0 | 0 | 0 | 0.131731866 | 0 |
| 0 | 0 | 0 | 0.857634654 | 0.850456202 |
| 0.002085362 | 0 | 0 | 0 | 0 |
| 0.006862652 | 0.00148792 | 0.0004145 | 0.008191183 | 0.015164922 |

| | | | | |
|---|---|---|---|---|
| 0 | 0 | 0 | 0 | 0 |
| 0 | 0 | 0 | 0 | 0 |
| 0 | 0 | 0 | 0 | 0 |
| 0 | 0 | 0 | 0 | 0 |
| 0 | 0 | 0 | 0 | 0 |
| 0 | 0 | 0 | 0 | 0 |
| 0 | 0 | 0 | 0.014186388 | 0.022680091 |
| 0 | 0.000712971 | 0 | 0.020802456 | 0.002483528 |
| 0 | 0 | 0 | 0 | 0 |
| 0 | 0 | 0 | 0 | 0 |
| 0 | 0 | 0 | 0 | 0 |
| 0 | 0 | 0 | 0 | 0 |
| 0 | 0 | 0 | 0 | 0 |
| 0 | 0 | 0 | 0 | 0 |
| 0 | 0 | 0 | 0 | 0 |
| 0 | 0 | 0 | 0 | 0 |
| 0 | 0 | 0 | 0 | 0 |
| 0 | 0 | 0 | 0 | 0.004162666 |
| 0 | 0.004035212 | 0 | 0.001110717 | 0.122014442 |
| 0 | 0 | 0 | 0 | 0 |
| 0.026179063 | 0 | 0 | 0.019865853 | 0 |
| 0 | 0 | 0 | 0 | 0 |
| 0 | 0 | 0 | 0 | 0 |
| 0 | 0 | 0 | 0 | 0 |
| 0 | 0.002217299 | 0.002882549 | 0 | 0.008581817 |
| 0 | 0.001255121 | 0 | 0 | 0 |
| 0 | 0 | 0 | 0 | 0 |
| 0 | 0 | 0 | 0 | 0 |
| 0 | 0 | 0 | 0.136291543 | 0.019655406 |
| 0 | 0 | 0 | 0 | 0 |
| 0.998376186 | 0.023114758 | 0.072241456 | 0.006023651 | 0.093592233 |
| 0 | 0 | 0 | 0 | 0 |
| 0 | 0 | 0 | 0.006179198 | 0 |
| 0 | 0 | 0 | 0 | 0 |
| 0 | 0 | 0 | 0 | 0 |
| 0 | 0 | 0 | 0 | 0 |
| 0 | 0 | 0 | 0 | 0 |
| 0 | 0 | 0 | 0 | 0 |
| 0 | 0 | 0 | 0 | 0 |
| 0 | 0 | 0 | 0 | 0.001269036 |
| 0 | 0 | 0 | 0 | 0 |
| 0 | 0.007807412 | 0 | 0 | 0.007122753 |
| 0.026452207 | 0.002807446 | 0 | 0 | 0 |
| 0 | 0 | 0 | 0 | 0 |
| 0 | 0 | 0 | 0 | 0 |
| 0 | 0 | 0 | 0 | 0 |
| 0 | 0 | 0 | 0 | 0 |
| 0 | 0 | 0 | 0 | 0 |

| | | | | |
|---|---|---|---|---|
| 0 | 0 | 0 | 0 | 0 |
| 0 | 0 | 0 | 0 | 0 |
| 0.015502669 | 0 | 0.011509334 | 0.041055319 | 0.114101056 |
| 0 | 0 | 0 | 0 | 0 |
| 0 | 0 | 1.297778166 | 0 | 0 |
| 0 | 0 | 0 | 0 | 0 |
| 0 | 0 | 0 | 0 | 0 |
| 0 | 0 | 0 | 0 | 0 |
| 0 | 0 | 0 | 0 | 0 |
| 0 | 0 | 0 | 0 | 0.001487258 |
| 0 | 0 | 0 | 0 | 0 |
| 0 | 0 | 0 | 0 | 0 |
| 0 | 0 | 0 | 0 | 0 |
| 0.003059509 | 0 | 0 | 0.022367239 | 0.002995303 |
| 0.001515134 | 0 | 0.005160332 | 0.0116042 | 0.003708346 |
| 0 | 0.039322596 | 0 | 0 | 0.015881097 |
| 0 | 0.000727804 | 0.003649491 | 0.00160266 | 0.003098574 |
| 0 | 0 | 0.002359143 | 0 | 0 |
| 0 | 0 | 0 | 0 | 0 |
| 0 | 0 | 0 | 0 | 0 |
| 0 | 0 | 0 | 0 | 0 |
| 0.007272581 | 0.001478248 | 0.000549075 | 0.006148665 | 0.017291336 |
| 0 | 0 | 0 | 0 | 0 |
| 0.017313888 | 0 | 0 | 0 | 0 |
| 0 | 0 | 0 | 0 | 0.003261841 |
| 0 | 0 | 0 | 0 | 0 |
| 0 | 1.76898077 | 11.34383182 | 7.604577293 | 0 |
| 0 | 0 | 0 | 0 | 0 |
| 0 | 0 | 0 | 0 | 0 |
| 0.003757091 | 0 | 0 | 0 | 0 |
| 0 | 0 | 0 | 0 | 0.007015037 |
| 0 | 0 | 0 | 0 | 0 |
| 0 | 0 | 0 | 0 | 0 |
| 0 | 0 | 0 | 0 | 0 |
| 0 | 0 | 0 | 0 | 0 |
| 0 | 0 | 0 | 0.009391417 | 0.027426429 |
| 0 | 0 | 0 | 0 | 0 |
| 0.001631651 | 0.216474596 | 0 | 0.002726527 | 0.199676224 |
| 0 | 0 | 0 | 0 | 0 |
| 0 | 0 | 0 | 0 | 0 |
| 0 | 0 | 0 | 0 | 0 |
| 0 | 0 | 0 | 0 | 0 |
| 0 | 0 | 0 | 0 | 0 |
| 0 | 0 | 0.008569085 | 0.001693387 | 0 |
| 0 | 0 | 0 | 0 | 0 |
| 0 | 0 | 0 | 0 | 0.008841573 |
| 0 | 0 | 0 | 0 | 0.001385227 |
| 0.037550429 | 0 | 0 | 0.001926463 | 0 |

| | | | | |
|---|---|---|---|---|
| 0 | 1.521120895 | 0 | 0.307163137 | 0 |
| 0 | 0 | 0 | 0.042492483 | 1.375945649 |
| 0 | 0 | 0 | 0 | 0.001598058 |
| 0 | 0 | 0 | 0 | 0 |
| 0.263309721 | 0 | 0.003664588 | 0.003049181 | 0 |
| 0 | 0 | 0 | 0 | 0.003058609 |
| 0 | 0 | 0 | 0 | 0 |
| 0.054096882 | 0 | 0.007299559 | 0.092160243 | 0.014273348 |
| 0 | 0 | 0 | 0 | 0 |
| 0.251903359 | 0 | 0.019784843 | 0.074616558 | 0 |
| 0 | 0 | 0 | 0.001991528 | 0 |
| 0 | 0.151726767 | 0 | 0.936142123 | 0.144803467 |
| 0.007832776 | 0.001311153 | 0.003733742 | 0.024701817 | 0.005412989 |
| 0 | 0 | 0.005424044 | 0 | 0 |
| 0.00189549 | 0 | 0.000667838 | 0.00412423 | 0 |
| 0 | 0 | 0 | 0 | 0 |
| 0 | 0 | 0 | 0 | 0.000738639 |
| 0 | 0 | 0 | 0 | 0 |
| 0.106796651 | 0 | 0.004982061 | 0 | 0 |
| 0 | 0 | 0.120853822 | 0.072478572 | 0 |
| 0 | 0 | 0 | 0 | 0 |
| 0 | 0 | 0.003121919 | 0 | 0 |
| 0 | 0 | 0 | 0.071832872 | 0 |
| 0 | 0 | 0 | 0 | 0 |
| 0 | 0 | 0 | 0 | 0 |
| 0 | 0 | 0 | 0.004857495 | 0.001576188 |
| 0 | 0 | 0 | 0 | 3.267259889 |
| 0 | 0 | 0 | 0.015137006 | 0 |
| 0 | 0 | 0 | 0 | 0 |
| 0.188560454 | 0 | 0.000778161 | 0 | 0 |
| 0 | 0 | 0.044230883 | 0 | 0 |
| 0 | 0 | 0 | 0 | 0 |
| 0 | 0 | 0.012491742 | 0 | 0 |
| 0 | 0 | 0.012116863 | 0.026876856 | 0 |
| 0 | 0 | 0 | 0 | 0 |
| 0 | 0 | 0 | 0 | 0 |
| 0 | 0 | 0 | 0.051887878 | 0 |
| 0 | 0 | 0 | 0 | 0 |
| 0 | 0 | 0 | 0 | 0 |
| 0 | 0 | 0 | 0.034386435 | 0 |
| 0 | 0 | 0 | 0.578904218 | 0 |
| 0 | 0 | 0 | 0.063849427 | 0 |
| 0 | 0 | 0 | 0 | 0 |
| 0 | 0 | 0 | 0.011740109 | 0.006772441 |
| 0 | 0 | 0 | 0 | 0 |
| 0 | 0.004345174 | 0 | 0 | 0 |
| 0.00200213 | 0.001477112 | 0 | 0.003949675 | 0.002940172 |
| 0.005148957 | 0 | 0 | 0 | 0 |

| | | | | |
|---|---|---|---|---|
| 0 | 0 | 0 | 0 | 0 |
| 0 | 0 | 0 | 0.097108837 | 0 |
| 0 | 0 | 0 | 0 | 0 |
| 0 | 0 | 0 | 0 | 0 |
| 0 | 0 | 0 | 0 | 0 |
| 0.103025437 | 0.001994407 | 0.054633642 | 0.394528651 | 0.018268626 |
| 0 | 0 | 0 | 0 | 0 |
| 0 | 0 | 0 | 0.005443553 | 0 |
| 0 | 0 | 0 | 0 | 0 |
| 0 | 0 | 0 | 0 | 0 |
| 0.032006829 | 0 | 0.013317563 | 0.023134027 | 0.00328273 |
| 0 | 0 | 0.004630268 | 0 | 0 |
| 0 | 0 | 0 | 0 | 0 |
| 0.00835157 | 0 | 0 | 0.001002563 | 0 |
| 4.224269711 | 0 | 0 | 0.019882499 | 0 |
| 0 | 0 | 0 | 0 | 0 |
| 0.003218423 | 0 | 0 | 0 | 0 |
| 0 | 0 | 0 | 0 | 0 |
| 0 | 0 | 0 | 0 | 0 |
| 0 | 0.017144731 | 0 | 0 | 0 |
| 0 | 0 | 0 | 0.001512308 | 0 |
| 0 | 0.035003019 | 0 | 0.477659335 | 0 |
| 0 | 0 | 0 | 0.007230518 | 0 |
| 0 | 0 | 0 | 0 | 0 |
| 0 | 0 | 0 | 0 | 0 |
| 0 | 0 | 0 | 0 | 0 |
| 0 | 0 | 0.075470863 | 0.005000399 | 0 |
| 0 | 0 | 0 | 0 | 0 |
| 0 | 0 | 0 | 0 | 0 |
| 0.219083361 | 0.101704754 | 0.974463701 | 0.932868339 | 0.268210554 |
| 0.006252001 | 0.028045888 | 0 | 0 | 0.039593926 |
| 0 | 0.001387685 | 0 | 0 | 0 |
| 0 | 0 | 0 | 0 | 0 |
| 0 | 0 | 0 | 0.001156006 | 0 |
| 0.011703449 | 0 | 0.006597564 | 0.014848637 | 0 |
| 0 | 0 | 0 | 0 | 0 |
| 0 | 0.104984946 | 0 | 0 | 0 |
| 0 | 0.002227477 | 0 | 0 | 0 |
| 0 | 0 | 0 | 0 | 0.00361041 |
| 16.70338134 | 0 | 0 | 0 | 0.00400331 |
| 0.062266603 | 0.000898307 | 0 | 0.008242156 | 0.028509767 |
| 1.846013639 | 0.086529203 | 0 | 0.008506322 | 0.714656716 |
| 0 | 0.001870471 | 0 | 0 | 0 |
| 0 | 0 | 0 | 0.032415351 | 0 |
| 0 | 0 | 0 | 0 | 0 |
| 0 | 0 | 0 | 0 | 0 |
| 0.023606694 | 0 | 0 | 0.002076176 | 0 |
| 0 | 0 | 0 | 0 | 0 |

| | | | | |
|---|---|---|---|---|
| 0 | 0.004071563 | 0 | 0 | 0.009004882 |
| 0 | 0 | 0 | 0 | 0 |
| 0.005848663 | 0 | 0 | 2.244067484 | 0.023721695 |
| 0.027456393 | 0 | 0 | 0 | 0 |
| 0 | 0.026506479 | 0 | 0 | 0 |
| 0 | 0 | 0 | 0 | 0 |
| 0 | 0 | 0 | 0 | 0 |
| 0 | 0 | 0 | 0.045219722 | 0.008074117 |
| 0 | 0 | 0.287852083 | 0 | 0 |
| 0 | 0 | 0 | 0 | 0 |
| 0 | 0 | 0 | 0 | 0 |
| 0 | 0 | 0.002186096 | 0 | 0 |
| 0 | 0 | 0 | 0.004200061 | 0 |
| 0.005824779 | 0 | 0.014854343 | 0.021243409 | 0.002172397 |
| 0 | 0 | 0 | 0 | 0 |
| 0.087571116 | 0.067577275 | 0.040171218 | 0.041050629 | 0.017613544 |
| 0.001922049 | 0.022637834 | 0 | 0.001003684 | 0.050806262 |
| 0.07326734 | 0 | 0 | 0 | 0 |
| 0 | 0 | 0.000524619 | 0.001382306 | 0 |
| 0 | 0 | 0 | 0 | 0 |
| 0.288257039 | 0 | 0 | 0.009697777 | 0.008003793 |
| 0.002786846 | 0 | 0 | 0.000776148 | 0 |
| 0 | 0 | 0 | 0.005033376 | 0 |
| 0 | 0 | 0 | 0 | 0.263022309 |
| 0 | 0.002697904 | 0 | 0 | 0 |
| 0 | 0 | 0 | 0.055090699 | 0 |
| 0.007320168 | 0 | 0.002149263 | 0.001486549 | 0.00238885 |
| 0 | 0 | 0 | 0 | 0 |
| 0 | 0 | 0 | 0 | 0 |
| 0 | 0 | 0 | 0 | 0 |
| 0.498011617 | 0.001026785 | 0 | 0.052811266 | 0.012944087 |
| 0.003804013 | 0.001968186 | 0 | 0 | 0.030470597 |
| 0.037207149 | 0.008745235 | 0.007424679 | 0.036680845 | 0.003868284 |
| 0 | 0.002983994 | 0 | 0.056181224 | 0 |
| 0 | 0 | 0 | 0 | 0 |
| 0 | 0 | 0 | 0 | 0 |
| 0 | 0 | 0 | 0 | 0 |
| 0.015830364 | 0 | 0.009842671 | 0.056731129 | 0.019600609 |
| 0 | 0.018810374 | 0.5850678 | 0 | 0.057734879 |
| 0.001903374 | 0.010351343 | 0 | 0.02080631 | 0.015839157 |
| 0 | 0 | 0 | 0 | 0 |
| 0.015027977 | 0 | 0 | 0 | 0 |
| 0.004874117 | 0 | 0 | 0.04581431 | 0.012611269 |
| 0 | 0 | 0 | 0.095150858 | 0.042699604 |
| 0 | 0 | 0.055632033 | 0.003714111 | 0 |
| 0 | 0 | 0 | 0 | 0 |
| 0 | 0 | 0 | 0 | 0 |
| 0 | 0 | 0 | 0 | 0 |

| | | | | |
|---|---|---|---|---|
| 0.154288467 | 0 | 0 | 0 | 0 |
| 0.368608262 | 0.172115691 | 0.138365979 | 0.29043977 | 2.206231181 |
| 0 | 0 | 0 | 0 | 0 |
| 0 | 0 | 0 | 0.00909604 | 0.015987483 |
| 0 | 0 | 0.001535064 | 0 | 0 |
| 0 | 0 | 0 | 0 | 0 |
| 0.426703523 | 0 | 0.013855314 | 0 | 0 |
| 0 | 0 | 0 | 0.005259117 | 0 |
| 0 | 0 | 0 | 0 | 0.022459209 |
| 0.002305167 | 0 | 0.001949231 | 0.005456985 | 0 |
| 0.007307576 | 0 | 0.000830542 | 0.000492384 | 0.010385163 |
| 0.010884842 | 0.001776334 | 0.008164961 | 0.008067626 | 0.029906722 |
| 0 | 0.035250045 | 0 | 0 | 0.606192326 |
| 0 | 0 | 0 | 0 | 0 |
| 0 | 0 | 0 | 0.002259798 | 0 |
| 0 | 0 | 0 | 0 | 0 |
| 0 | 0.005307272 | 0.008175151 | 0.001718659 | 0.007733166 |
| 0.133941574 | 0 | 0.091447929 | 0.039935906 | 0 |
| 0 | 0 | 0 | 0 | 0 |
| 0.109030433 | 0 | 0.020966803 | 0.015066806 | 0 |
| 0.018328023 | 0.007243857 | 0.003408132 | 0.001417896 | 0.069530677 |
| 0 | 0 | 0 | 0.001764344 | 0 |
| 0 | 0 | 0 | 0 | 0 |
| 0 | 0 | 0 | 0 | 0 |
| 0 | 0 | 0 | 0.010124905 | 0 |
| 0 | 0 | 0 | 0 | 0 |
| 0 | 0.00047667 | 0 | 0.118960443 | 0.003689799 |
| 0.293552519 | 0.007356966 | 0.020208853 | 0.125835717 | 0.007946327 |
| 0 | 0 | 0 | 0 | 0.012395137 |
| 0.003263531 | 0 | 0 | 0 | 0.003514548 |
| 0 | 0 | 0 | 0 | 0 |
| 0 | 0 | 0 | 0 | 0 |
| 0.001642261 | 0 | 0 | 0 | 0 |
| 0 | 0 | 0 | 0 | 0 |
| 0 | 0.741747508 | 0 | 0 | 0 |
| 0.053574152 | 0.012451227 | 2.436105711 | 1.172607212 | 0.195005934 |
| 0 | 0 | 0 | 0 | 0 |
| 0 | 0 | 0 | 0 | 0 |
| 0.087111424 | 0.04622245 | 0 | 0.354617803 | 0.070906079 |
| 0 | 0 | 0 | 0 | 0 |
| 0 | 0 | 0.043093772 | 0.001166577 | 0 |
| 0 | 0 | 0 | 0 | 0 |
| 0.030612392 | 0 | 0.028428446 | 0.123368175 | 0.024611106 |
| 0.031121552 | 0.001361542 | 0.003310207 | 0 | 0 |
| 0 | 0 | 0.065051459 | 0.008056878 | 0 |
| 0 | 0.158579496 | 0 | 0 | 0 |
| 0 | 0 | 0 | 0 | 0 |
| 0 | 0 | 0.014491459 | 0.000881151 | 0 |

| | | | | |
|---|---|---|---|---|
| 0 | 0 | 0 | 0 | 0 |
| 0 | 0 | 0 | 0 | 0 |
| 0 | 0 | 0 | 0 | 0 |
| 0 | 0.048377956 | 0.002994887 | 0.188730262 | 2.045556997 |
| 0 | 0 | 0 | 0 | 0 |
| 0 | 0 | 0 | 0 | 0 |
| 0 | 0.000810322 | 0 | 0 | 0 |
| 0 | 0.023776883 | 0 | 0 | 0.001033998 |
| 0.078789407 | 4.026262969 | 0 | 0 | 2.347280115 |
| 0.003587527 | 0.020951744 | 0 | 0 | 0 |
| 0 | 0 | 0 | 0 | 0 |
| 0 | 0.005436261 | 0 | 0.013073503 | 0.001993307 |
| 0 | 0 | 0 | 0 | 0 |
| 0 | 0 | 0 | 0 | 0 |
| 0 | 0.027624419 | 0 | 0.020146803 | 0.207072342 |
| 0 | 0 | 0 | 0 | 0 |
| 0 | 0.058824138 | 0 | 0 | 0.048872095 |
| 0 | 6.397466698 | 0 | 0 | 0 |
| 0 | 0 | 0 | 0 | 0 |
| 0 | 0 | 0 | 0 | 0 |
| 0 | 0.289839948 | 0 | 0 | 0 |
| 0 | 0 | 0 | 0 | 0 |
| 0 | 0 | 0 | 0 | 0 |
| 0 | 0.053332507 | 0.259763731 | 0.503192682 | 0.248581464 |
| 0.105267254 | 0.000669701 | 0 | 0 | 0.005806094 |
| 0 | 0.002760789 | 0 | 0 | 0 |
| 0 | 0.011633881 | 0 | 0.028074944 | 0.016283981 |
| 0 | 0.068031843 | 0 | 0 | 0 |
| 0 | 0 | 0 | 0 | 0 |
| 0 | 0 | 0 | 0 | 0 |
| 0 | 0 | 0 | 0 | 0.00925854 |
| 0 | 0.497418344 | 0 | 0 | 0 |
| 0.016236157 | 0 | 0 | 0.015267017 | 0.117495107 |
| 0 | 0 | 0 | 0 | 0 |
| 0 | 0 | 0 | 0 | 0 |
| 0 | 0.194218841 | 0 | 0 | 0 |
| 0 | 0.175904106 | 0 | 0.112683524 | 0 |
| 0 | 0 | 0 | 0.005080498 | 0.002995751 |
| 0 | 0.002531724 | 0 | 0 | 0.007608452 |
| 0 | 0.005939095 | 0 | 0 | 0.270132434 |
| 0.193150824 | 0.14300411 | 0.075087525 | 0.018888087 | 0.375645291 |
| 0 | 0.049465985 | 0 | 0 | 0 |
| 6.344376714 | 0 | 0.482121336 | 4.666035761 | 0.01503808 |
| 0 | 0.005619038 | 0 | 0.029353922 | 0.041089606 |
| 0 | 0 | 0 | 0 | 0 |
| 0 | 0 | 0 | 0 | 0 |
| 0 | 0 | 0 | 0 | 0 |
| 0.063395546 | 0.000659913 | 0.003309061 | 0.004541129 | 0.050571612 |

| | | | | |
|---|---|---|---|---|
| 0 | 0 | 0 | 0 | 0 |
| 0.02044009 | 1.125295101 | 0 | 0 | 2.054632662 |
| 3.756346737 | 0 | 0.489209564 | 0.176042809 | 0.035502004 |
| 0 | 0 | 0 | 0 | 0 |
| 0 | 0 | 0 | 0 | 0 |
| 0 | 0.040116273 | 0 | 0 | 0.204834973 |
| 0 | 0 | 0 | 0 | 0 |
| 0 | 0 | 0 | 0 | 0 |
| 0 | 0 | 0 | 0.006222024 | 0.317040547 |
| 0 | 0 | 0 | 0.020309986 | 0 |
| 0.043952322 | 0.036489751 | 0.027857004 | 0.083258609 | 0.574053337 |
| 2.441077424 | 0.169181287 | 3.336838056 | 0.811376152 | 0.399351156 |
| 0 | 0 | 0 | 0 | 0 |
| 0.004505013 | 0.015417331 | 0 | 0.023432696 | 0.171230135 |
| 1.939620183 | 0 | 0.337572949 | 0.392145132 | 0.011653556 |
| 0 | 2.411016823 | 0 | 0 | 12.2398114 |
| 0 | 0 | 0 | 0 | 0 |
| 0 | 0 | 0 | 0 | 0 |
| 0 | 0 | 0 | 0 | 0 |
| 0 | 0.00166278 | 0 | 0.003943181 | 0.007524714 |
| 2.524158817 | 0.012493801 | 0.001305183 | 0.00186279 | 0.350580294 |
| 0 | 0 | 0.51404082 | 1.225241793 | 0 |
| 6.393315005 | 0 | 10.13780903 | 0.58982184 | 0.00380611 |
| 0 | 0 | 0 | 0 | 0 |
| 0 | 0 | 0 | 0.006179347 | 0 |
| 0 | 0.115559889 | 0 | 0 | 0.001448622 |
| 0 | 0 | 0.62615832 | 0.070267171 | 0 |
| 0 | 0 | 0 | 0.002540658 | 0 |
| 0 | 0 | 0 | 0 | 0 |
| 0 | 0 | 0 | 0.046678416 | 0 |
| 0 | 0.011052981 | 0 | 0.001580468 | 0 |
| 1.275919865 | 0.012655166 | 0 | 0.111604505 | 0.003233661 |
| 0.019165663 | 0 | 0.001599309 | 0.009657047 | 0.043452228 |
| 0 | 0.00522922 | 0 | 0 | 0.040478255 |
| 0.047269466 | 0 | 0.003870402 | 0.002201848 | 0.113575398 |
| 0.043449358 | 0.105132845 | 0 | 0.01098627 | 0.149105315 |
| 0 | 0.001257238 | 0 | 0.000615222 | 0.008001872 |
| 0 | 0 | 0 | 0 | 0 |
| 0.032647536 | 3.136925986 | 0 | 0 | 0.617241328 |
| 0 | 0 | 0 | 0 | 0 |
| 0 | 0 | 0 | 0 | 0 |
| 0 | 0 | 0 | 0 | 0 |
| 0 | 0 | 0 | 0 | 0 |
| 0 | 0 | 0 | 0 | 0 |
| 0.011735304 | 0.049968774 | 0.009188224 | 0.009804986 | 0 |
| 0 | 0 | 0 | 0 | 0 |
| 0 | 0 | 0 | 0 | 0 |
| 0 | 0.004774336 | 0 | 0.087830085 | 0.002155831 |

| | | | | |
|---|---|---|---|---|
| 0 | 0 | 0 | 0 | 0 |
| 0 | 0 | 0 | 0 | 0 |
| 0 | 0 | 0 | 0 | 0 |
| 0 | 0.003513876 | 0 | 0 | 0.004787227 |
| 0 | 0 | 0 | 0 | 0 |
| 0 | 0.070137597 | 0 | 0.14528904 | 0 |
| 0 | 0 | 0 | 0 | 0 |
| 0 | 0.065185394 | 0 | 0.001424023 | 0.508590643 |
| 0 | 0 | 0 | 0 | 0 |
| 0 | 0 | 0 | 0 | 0 |
| 0.009159166 | 0.002035035 | 0 | 0.00758365 | 0.019145663 |
| 0 | 0 | 0 | 0 | 0 |
| 0 | 0.129388407 | 0 | 0.009960118 | 0 |
| 0 | 0.002938783 | 0 | 0 | 0 |
| 0 | 0 | 0 | 0 | 0 |
| 0 | 0 | 0 | 0 | 0 |
| 0 | 0.00454059 | 0 | 0 | 0 |
| 0 | 0.015507712 | 0 | 0 | 0.008070039 |
| 0 | 0 | 0 | 0.001811776 | 0 |
| 0 | 0.006698884 | 0 | 0.00271734 | 0 |
| 0 | 0.007872789 | 0 | 0 | 0 |
| 0 | 0 | 0 | 0 | 0 |
| 0 | 0.081283126 | 0.001171943 | 0 | 0.033029832 |
| 0 | 0 | 0 | 0 | 0 |
| 0 | 0 | 0 | 0 | 0 |
| 0 | 0 | 0 | 0 | 0 |
| 0 | 0 | 0 | 0 | 0 |
| 0 | 0 | 0 | 0 | 0 |
| 0 | 0 | 0 | 0 | 0 |
| 0 | 0.082646374 | 0 | 0.000866626 | 0 |
| 0 | 0.016398044 | 0 | 0 | 0 |
| 0 | 0 | 0 | 0 | 0 |
| 0 | 0 | 0 | 0 | 0 |
| 0 | 0 | 0 | 0 | 0 |
| 0 | 0 | 0 | 0.003015074 | 0.010598778 |
| 0 | 0 | 0 | 0 | 0 |
| 0 | 0.020178287 | 0 | 0 | 0 |
| 0.017183198 | 0 | 0 | 0.003939357 | 0.006359763 |
| 0 | 0 | 0 | 0 | 0 |
| 0 | 0 | 0 | 0.002302556 | 0.001416466 |
| 0 | 0 | 0 | 0 | 0.105674214 |
| 0 | 0.004401396 | 0 | 0 | 0 |
| 0 | 0 | 0 | 0 | 0 |
| 0.08648709 | 0.001544251 | 0 | 0 | 0 |
| 0.707405172 | 0.021517093 | 0 | 0 | 0 |
| 0 | 0 | 0 | 0 | 0 |
| 0 | 0.025434073 | 0 | 0.002356734 | 0.00584791 |
| 0 | 0 | 0 | 0 | 0 |

| | | | | |
|---|---|---|---|---|
| 0 | 0 | 0 | 0 | 0 |
| 0 | 0 | 0 | 0 | 0 |
| 0 | 0.000460029 | 0 | 0 | 0.121310889 |
| 0 | 0.515390332 | 0 | 0 | 1.550302779 |
| 0 | 0 | 0 | 0 | 0 |
| 0 | 0 | 0 | 0 | 0 |
| 0 | 0 | 0 | 0 | 0 |
| 0 | 0 | 0 | 0 | 0 |
| 4.335669435 | 0 | 0.161698965 | 0.077470993 | 0.003678235 |
| 0 | 0 | 0 | 0 | 0 |
| 0 | 0 | 0 | 0 | 0 |
| 0 | 0 | 0 | 0 | 0 |
| 0 | 0 | 0 | 0 | 0.003961501 |
| 0 | 0 | 0 | 0 | 0 |
| 0.022637927 | 0.124638042 | 7.637956796 | 4.654382365 | 0.032329309 |
| 0 | 0 | 0.075026669 | 0.04668162 | 0 |
| 0 | 0.010815267 | 0 | 0 | 0 |
| 0 | 0 | 0 | 0 | 0 |
| 0 | 0 | 0 | 0 | 0 |
| 0 | 0 | 0 | 0 | 0 |
| 0 | 0.009502746 | 0 | 0.066161569 | 0 |
| 0 | 0 | 0 | 0 | 0 |
| 0 | 0 | 0 | 0 | 0 |
| 0 | 0 | 0 | 0 | 0 |
| 0 | 0.047282532 | 0 | 0.015617774 | 0.095826455 |
| 0 | 1.781820006 | 1.011484077 | 0 | 12.58527353 |
| 0 | 0.099442541 | 0 | 0 | 1.038066422 |
| 0 | 0 | 0 | 0 | 0 |
| 0 | 0.127397599 | 0 | 0 | 0 |
| 0 | 0 | 0 | 0 | 0 |
| 0 | 0 | 0 | 0 | 0 |
| 0 | 0.022976069 | 0 | 0 | 0 |
| 0 | 0 | 0 | 0.001833436 | 0 |
| 0.056812201 | 0.000459126 | 0 | 0.001769281 | 0.164194413 |
| 0.002392551 | 0.002080358 | 0 | 0.011452632 | 0.033963962 |
| 0.006553569 | 0.019250983 | 0.000446907 | 0.024139675 | 0.018213272 |
| 0 | 0 | 0.347280336 | 0.049215356 | 0 |
| 0.027213315 | 0.057212384 | 0 | 0 | 1.037644754 |
| 0 | 0.011481191 | 0 | 0 | 0 |
| 0.003230519 | 0 | 0 | 0 | 0 |
| 0 | 0 | 0 | 0 | 0 |
| 0 | 0.004244872 | 0 | 0.001168427 | 0 |
| 0 | 0.043844685 | 0 | 0 | 0.414906951 |
| 0 | 0 | 0 | 0 | 0 |
| 0 | 0 | 0 | 0 | 0 |
| 0 | 0.00063326 | 0.003669365 | 0.013386914 | 0.005098006 |
| 0 | 0 | 0.01759773 | 0 | 0 |
| 0.090935019 | 0.018159465 | 0.205275357 | 0 | 0.07028424 |

| | | | | |
|---|---|---|---|---|
| 0 | 0.000752307 | 0 | 0.00811743 | 0.234801434 |
| 0 | 0 | 0 | 0 | 0 |
| 0 | 0 | 0 | 0 | 0 |
| 0 | 0 | 0 | 0 | 0 |
| 0.000710664 | 0.000599207 | 0.000500776 | 0.002309098 | 0.011363918 |
| 0 | 0.018609144 | 0 | 0.367057236 | 0 |
| 0 | 0 | 0 | 0 | 0 |
| 0 | 0 | 0 | 0 | 0 |
| 0 | 0 | 0 | 0 | 0 |
| 0 | 0 | 0 | 0 | 0 |
| 0 | 0.001031622 | 0 | 0 | 0 |
| 1.195436822 | 0.112219821 | 0 | 1.409947664 | 2.086090028 |
| 0 | 0.078176398 | 0 | 0 | 0.159219529 |
| 0 | 0 | 0 | 0 | 0 |
| 0.001369804 | 0.020616252 | 0 | 5.008278451 | 0.19498987 |
| 0 | 0 | 0 | 0 | 0 |
| 0.001524635 | 0 | 0 | 0 | 0.017351935 |
| 0 | 0 | 0 | 0 | 0 |
| 0.022571817 | 0.000607397 | 0 | 0.003678178 | 0 |
| 0 | 0 | 0 | 0 | 0 |
| 0 | 0 | 0 | 0.001384179 | 0 |
| 0 | 0 | 0 | 0 | 0 |
| 0 | 2.140228016 | 0.243670178 | 0 | 1.181351723 |
| 0 | 0 | 0 | 0 | 0 |
| 0 | 0 | 0 | 0 | 0 |
| 0 | 0 | 0 | 0 | 0 |
| 0 | 0 | 0 | 0 | 0 |
| 0 | 0 | 0 | 0 | 0 |
| 0 | 0 | 0 | 0 | 0 |
| 0 | 0 | 0 | 0 | 0 |
| 0 | 0 | 0 | 0 | 0 |
| 0.019908141 | 0.012589398 | 0.053358304 | 0.017821595 | 0.128427544 |
| 0 | 0 | 0 | 0 | 0 |
| 0.029126881 | 0.03728743 | 0 | 0 | 0 |
| 0.005291428 | 0.021240864 | 0 | 0.019542328 | 0.012838346 |
| 0 | 1.291269922 | 0 | 0.00309743 | 0.691042784 |
| 0 | 0.006844571 | 0 | 0 | 0 |
| 0 | 0.039509439 | 0 | 0 | 0 |
| 0 | 0 | 0 | 0.002576373 | 0 |
| 0 | 0 | 0 | 0 | 0 |
| 0 | 0 | 0 | 0 | 0 |
| 0 | 0 | 0 | 0 | 0 |
| 0 | 0 | 0 | 0 | 0 |
| 0 | 0 | 0 | 0 | 0 |
| 0.026516205 | 0.038265818 | 0.003194002 | 0.006627446 | 0.117151688 |
| 0 | 0 | 0 | 0 | 0 |
| 0 | 0 | 0 | 0 | 0.464468066 |
| 0.021435543 | 0.001084423 | 0 | 0.002089459 | 0.028120848 |

| | | | | |
|---|---|---|---|---|
| 0 | 0.013435824 | 0 | 0 | 0 |
| 0 | 0 | 0 | 0 | 0 |
| 0 | 0.002223395 | 0.002973058 | 0.016320089 | 0.001835821 |
| 0 | 0 | 0 | 0 | 0 |
| 0 | 0 | 0.00217758 | 0.002648149 | 0 |
| 0 | 0 | 0 | 0 | 0 |
| 0.139588173 | 28.8944699 | 0 | 0 | 0 |
| 0 | 0 | 0 | 0.031442109 | 0.025572975 |
| 0 | 0 | 0 | 0 | 0 |
| 0 | 0 | 0 | 0 | 0 |
| 0 | 0 | 0 | 0 | 0 |
| 0 | 0.00045245 | 0.004705578 | 0 | 0 |
| 0 | 0 | 0 | 0 | 0 |
| 0 | 0 | 0 | 0 | 0 |
| 0 | 0 | 0 | 0 | 0 |
| 0 | 0 | 0 | 0 | 0 |
| 0 | 0 | 0 | 0 | 0 |
| 0 | 0 | 0 | 0 | 0 |
| 0 | 0.044347716 | 0 | 0 | 0.031876562 |
| 0 | 0 | 0 | 0 | 0 |
| 0 | 0.021413627 | 0 | 0 | 0 |
| 0 | 0 | 0 | 0 | 0 |
| 0 | 0 | 0 | 0 | 0 |
| 0 | 0 | 0 | 0 | 0 |
| 0 | 0 | 0 | 0 | 0.006787794 |
| 0.007384576 | 0 | 0.006095666 | 0.003525658 | 0.012806733 |
| 0 | 0 | 0 | 0 | 0 |
| 0.135473963 | 0.020633995 | 0 | 0 | 0.150422936 |
| 0 | 0 | 0 | 0 | 0 |
| 0 | 0.126041513 | 0 | 0 | 0.101611649 |
| 0 | 0 | 0 | 0 | 0 |
| 0 | 0 | 0 | 0 | 0 |
| 0 | 0 | 0 | 0 | 0 |
| 0 | 0 | 0 | 0 | 0 |
| 0.004643148 | 0.000838916 | 0.000934812 | 0 | 0.032036425 |
| 0 | 0.021499782 | 0 | 0 | 0 |
| 0 | 0 | 0 | 0 | 0 |
| 0.001505318 | 0.004918276 | 0 | 0 | 0.100213507 |
| 0 | 0.009122414 | 0 | 0 | 0 |
| 0 | 0 | 0 | 0 | 0 |
| 0.617658817 | 0.377768772 | 18.07616919 | 24.10015117 | 3.918150693 |
| 0.175202022 | 0.017420324 | 1.289578483 | 0.719002196 | 0.007012041 |
| 0.057552811 | 3.954642136 | 0 | 0 | 0 |
| 0 | 0 | 0 | 0.081626076 | 0 |
| 0 | 0 | 0 | 0 | 0 |
| 0 | 0 | 0 | 0 | 0 |
| 0 | 0.001260274 | 0 | 0 | 0 |
| 0.005177049 | 0.001259166 | 0 | 0 | 0.100003546 |

| | | | | |
|---|---|---|---|---|
| 0 | 0 | 0 | 0 | 0 |
| 0.03645592 | 0.004662564 | 0.00346369 | 0.0017112 | 0.002406126 |
| 0 | 0.00151267 | 0 | 0 | 0 |
| 0 | 0 | 0 | 0 | 0 |
| 0 | 0.510103735 | 0 | 0 | 0.905405025 |
| 0 | 0 | 0 | 0 | 0 |
| 0 | 0 | 0 | 0 | 0 |
| 0 | 0 | 0 | 0 | 0 |
| 5.685692021 | 0 | 0.324386314 | 0.053111372 | 0 |
| 0 | 0 | 0 | 0.040972709 | 0 |
| 0 | 0 | 0 | 0 | 0 |
| 0 | 0 | 0 | 0 | 0 |
| 0 | 0.002013545 | 0 | 0.000969921 | 0.076763081 |
| 0.000978633 | 0 | 0 | 0.088239047 | 0.010778574 |
| 0 | 5.434875073 | 0 | 0 | 0 |
| 0 | 0.067034057 | 0 | 0 | 0.718592458 |
| 0.001905423 | 0.198011906 | 0 | 0.014003735 | 0.074410199 |
| 0 | 0 | 0 | 0 | 0 |
| 0 | 0 | 0 | 0 | 0 |
| 0 | 0 | 0 | 0 | 0 |
| 0 | 0 | 0 | 0 | 0 |
| 0 | 0 | 0 | 0 | 0 |
| 0 | 0 | 0 | 0 | 0 |
| 0 | 0 | 0 | 0 | 0 |
| 0 | 0 | 0 | 0 | 0 |
| 0 | 0 | 0 | 0 | 0 |
| 0 | 0.043423225 | 0.02023687 | 0 | 0.083086231 |
| 0.008932355 | 0 | 0 | 0 | 0.002237069 |
| 0 | 0 | 0 | 0 | 0 |
| 0 | 0 | 0 | 0 | 0 |
| 0 | 0 | 0 | 0 | 0 |
| 0 | 0 | 0 | 0 | 0 |
| 0 | 0 | 0 | 0.003585769 | 0 |
| 0 | 0.00358028 | 0 | 0 | 0 |
| 0 | 1.13501346 | 0 | 0 | 0 |
| 0 | 0.0041029 | 0 | 0 | 0 |
| 0 | 0 | 0 | 0 | 0.005771057 |
| 0 | 0 | 0 | 0 | 0 |
| 0 | 0 | 0 | 0 | 0 |
| 0 | 0 | 0 | 0 | 0 |
| 0 | 0.034244426 | 0 | 0 | 2.020648589 |
| 0 | 0.950743813 | 0 | 0.001084197 | 0 |
| 0 | 0 | 0 | 0 | 0 |
| 0 | 0.158043238 | 0 | 0.001770189 | 0 |
| 0 | 0 | 0 | 0 | 0 |
| 0 | 0 | 0 | 0 | 0 |
| 0 | 0 | 0 | 0 | 0 |
| 0 | 0 | 0 | 0 | 0 |

| | | | | |
|---|---|---|---|---|
| 0 | 0.175328469 | 0.016187344 | 0.128746797 | 0 |
| 0 | 0 | 0 | 0 | 0 |
| 0 | 0 | 0 | 0 | 0 |
| 0 | 0 | 0 | 0 | 0 |
| 0.024991848 | 0.002705224 | 0 | 0 | 0 |
| 0 | 0 | 0 | 0 | 0 |
| 0.015628716 | 0.082938738 | 0 | 0.016175424 | 0.136052525 |
| 0 | 0 | 0 | 0 | 0 |
| 0 | 0 | 0 | 0.000518965 | 0 |
| 0 | 0 | 0 | 0.013885079 | 0 |
| 0 | 0.015872023 | 0 | 0.000534964 | 0 |
| 0 | 0 | 0 | 0.000900182 | 0 |
| 0 | 0 | 0 | 0 | 0 |
| 0 | 0 | 0 | 0 | 0 |
| 0 | 0 | 0 | 0 | 0 |
| 0 | 0 | 0 | 0 | 0 |
| 0 | 0 | 0 | 0 | 0 |
| 0.00182741 | 0 | 0 | 0.014419992 | 0 |
| 0 | 0 | 0 | 0 | 0 |
| 0 | 0 | 0 | 0 | 0 |
| 1.672123831 | 0.056837598 | 0 | 0 | 0.080119364 |
| 0 | 0 | 0 | 0 | 0 |
| 0 | 0.031696461 | 0 | 0 | 0.047116927 |
| 0.012424335 | 0.014715968 | 0 | 0.009199809 | 0.008881362 |
| 0 | 0 | 0 | 0 | 0 |
| 0.008841286 | 0 | 0 | 0.001781952 | 0 |
| 0 | 0 | 0 | 0 | 0 |
| 0 | 0 | 0 | 0 | 0 |
| 0 | 0 | 0 | 0 | 0 |
| 0.004825175 | 0 | 0.000800026 | 0.078455978 | 0.003056652 |
| 0 | 0 | 0 | 0 | 0 |
| 0 | 0 | 0 | 0 | 0 |
| 0 | 0 | 0 | 0 | 0 |
| 0 | 0.066725419 | 0 | 0 | 0.67424266 |
| 0 | 0.080696675 | 0 | 0 | 0 |
| 0 | 0 | 0 | 0 | 0 |
| 0 | 0 | 0.003129911 | 0 | 0.044934703 |
| 0 | 0 | 0 | 0 | 0 |
| 0 | 0 | 0 | 0 | 0 |
| 0 | 1.441067358 | 0 | 0 | 0 |
| 0 | 0 | 0 | 0 | 0 |
| 0 | 0 | 0 | 0 | 0 |
| 0 | 0 | 0 | 0 | 0 |
| 0 | 0 | 0 | 0 | 0 |
| 0 | 0.077978335 | 0 | 0 | 0 |
| 0 | 0 | 0 | 0 | 0 |
| 0 | 0 | 0 | 0 | 0 |
| 0 | 0 | 0 | 0 | 0 |

| | | | | |
|---|---|---|---|---|
| 0.021008468 | 0 | 0 | 0.037972021 | 0.012257456 |
| 0 | 0.680635212 | 0 | 0 | 0 |
| 0 | 0 | 0 | 0 | 0.021232915 |
| 0 | 0 | 0 | 0 | 0 |
| 0 | 0 | 0 | 0 | 0 |
| 0 | 0 | 0 | 0 | 1.71993659 |
| 0 | 0 | 0 | 0 | 0.001277689 |
| 0.912734387 | 0.059110629 | 0.753407339 | 0.099368045 | 0.654239795 |
| 0.006328717 | 1.162299383 | 0.008449759 | 0.265313586 | 1.064065283 |
| 0.108755247 | 0 | 0.004248449 | 0.037295871 | 0.08740318 |
| 0 | 0 | 0 | 0 | 0 |
| 0 | 0 | 0 | 0 | 0 |
| 0 | 0 | 0 | 0 | 0 |
| 0 | 0 | 0 | 0 | 0 |
| 0 | 0 | 0 | 0 | 0 |
| 0 | 0 | 0 | 0 | 0 |
| 0 | 0 | 0.111758791 | 0 | 0 |
| 0 | 0 | 0 | 0 | 0 |
| 0 | 0 | 0 | 0 | 0 |
| 0 | 0.006566248 | 0 | 0 | 0 |
| 0 | 0 | 0 | 0 | 0 |
| 0 | 0 | 0 | 0 | 0 |
| 0 | 0 | 0 | 0 | 0 |
| 0 | 0 | 0 | 0 | 0 |
| 0 | 0.002325322 | 0 | 0 | 0.01472712 |
| 0 | 0 | 0 | 0 | 0 |
| 0 | 0 | 0 | 0 | 0 |
| 0 | 0 | 0 | 0 | 0 |
| 0 | 0 | 0 | 0 | 0 |
| 0 | 0 | 0 | 0 | 0 |
| 0 | 0 | 0 | 0 | 0.029453473 |
| 0 | 0 | 0 | 0 | 0 |
| 0 | 0 | 0 | 0 | 0 |
| 0 | 0 | 0 | 0 | 0 |
| 0 | 0.035091544 | 0 | 0 | 0 |
| 0 | 0 | 0 | 0 | 0 |
| 0 | 0 | 0 | 0 | 0 |
| 0 | 0 | 0 | 0 | 0 |
| 0 | 0 | 0 | 0.008784377 | 0 |
| 0 | 0 | 0.119126566 | 0 | 0 |
| 0 | 0 | 0 | 0 | 0 |
| 0 | 0.072975672 | 0 | 0.251087608 | 0.038943094 |
| 0 | 0 | 0 | 0 | 0 |
| 0 | 0 | 0 | 0 | 0 |
| 0 | 0 | 0 | 0 | 0 |
| 0 | 0 | 0.003723042 | 0.016028447 | 0 |
| 0 | 0 | 0 | 0 | 0 |
| 0 | 0 | 0 | 0 | 0 |

| | | | | |
|---|---|---|---|---|
| 0 | 0 | 0 | 0 | 0 |
| 0 | 0 | 0 | 0 | 0.001875066 |
| 0 | 0 | 0 | 0 | 0 |
| 0 | 0 | 0 | 0 | 0 |
| 0.071497286 | 2.440488629 | 0 | 0.398059111 | 2.941966266 |
| 0 | 0 | 0 | 0 | 0 |
| 0 | 0 | 0 | 0.001616961 | 0.115727066 |
| 0.31772215 | 0.001707046 | 0.015407628 | 0.036274358 | 0.007664047 |
| 0 | 0.002727076 | 0 | 0 | 0.021637444 |
| 0 | 0 | 0 | 0 | 0 |
| 0 | 0 | 0 | 0 | 0 |
| 0 | 0.267676949 | 0 | 0 | 0 |
| 0 | 0 | 0 | 0 | 0 |
| 0 | 0 | 0 | 0 | 0 |
| 0.047127414 | 0 | 0 | 0.004895477 | 0.000744168 |
| 0 | 0.043082794 | 0 | 0.012877606 | 0.200096738 |
| 0 | 0 | 0 | 0 | 0 |
| 0 | 0 | 0 | 0 | 0 |
| 0 | 0 | 0 | 0 | 0 |
| 0 | 0 | 0 | 0 | 0 |
| 0 | 0 | 0 | 0 | 0 |
| 0 | 0.005263116 | 0 | 0 | 0 |
| 0 | 0 | 0 | 0 | 0 |
| 0.00327058 | 0.001034116 | 0 | 0.001951863 | 0.002515814 |
| 0 | 0.003680292 | 0 | 0 | 0 |
| 0 | 0 | 0 | 0 | 0 |
| 0.008760285 | 0 | 0 | 0.054002318 | 0 |
| 0.001349897 | 0 | 0 | 0 | 0 |
| 0 | 0 | 0.002461703 | 0 | 0 |
| 0 | 0 | 0 | 0 | 0 |
| 0 | 0 | 0 | 0 | 0 |
| 0 | 0 | 0 | 0 | 0 |
| 0 | 0 | 0 | 0 | 0 |
| 0 | 0 | 0 | 0 | 0 |
| 0 | 0 | 0 | 0 | 0 |
| 0 | 0 | 0 | 0 | 0 |
| 0 | 0 | 0 | 0 | 0 |
| 0 | 0.091733316 | 0 | 0.05416199 | 0.188570115 |
| 0 | 0 | 0 | 0 | 0 |
| 0 | 0 | 0 | 0 | 0 |
| 0 | 0 | 0 | 0 | 0 |
| 0 | 1.700917218 | 0 | 0 | 0 |
| 0 | 0 | 0 | 0 | 0 |
| 0 | 0 | 0 | 0 | 0 |
| 0 | 0 | 0 | 0 | 0 |
| 0 | 0 | 0 | 0 | 0 |
| 0 | 0 | 0 | 0 | 0 |
| 0 | 0 | 0 | 0 | 0 |

| | | | | |
|---|---|---|---|---|
| 0 | 0.424652874 | 0.01599969 | 0.566324408 | 0.584250322 |
| 0 | 0 | 0 | 0 | 0 |
| 0 | 0 | 0 | 0 | 0 |
| 0.002598937 | 0 | 0 | 0 | 0 |
| 0 | 0.000470775 | 0 | 0 | 0 |
| 0 | 0.025173376 | 0 | 0 | 0.458779513 |
| 0 | 0 | 0 | 0 | 0 |
| 0 | 0 | 0 | 0 | 0 |
| 0 | 0 | 0 | 0 | 0 |
| 0 | 0 | 0 | 0 | 0 |
| 0 | 0 | 0 | 0 | 0 |
| 0 | 0 | 0 | 0 | 0.00843338 |
| 0 | 0 | 0 | 0 | 0 |
| 0 | 0 | 0 | 0 | 0 |
| 0 | 0 | 0 | 0 | 0 |
| 0 | 0 | 0 | 0 | 0 |
| 0 | 0 | 0 | 0 | 0 |
| 0 | 0 | 0 | 0 | 0 |
| 0 | 0.000323187 | 0 | 0 | 0.015844207 |
| 0 | 0 | 0 | 0 | 0 |
| 0.005815908 | 0 | 0 | 0.006479013 | 0 |
| 0 | 0 | 0 | 0 | 0 |
| 0.022622809 | 0.006688559 | 0 | 0.001431941 | 0.220330033 |
| 0 | 0 | 0 | 0 | 0.003422386 |
| 0 | 0.004839526 | 0 | 0 | 0 |
| 0 | 0 | 0 | 0 | 0 |
| 0 | 0 | 0 | 0 | 0 |
| 0 | 0 | 0 | 0 | 0 |
| 0 | 0 | 0 | 0 | 0 |
| 0 | 0 | 0 | 0 | 0 |
| 0 | 0 | 0 | 0 | 0 |
| 0 | 0 | 0 | 0 | 0 |
| 0.175267383 | 0 | 0.131453181 | 0.168200615 | 0.009966726 |
| 0 | 0 | 0 | 0 | 0 |
| 0 | 0.088402039 | 0 | 0 | 0.138787809 |
| 0 | 0 | 0 | 0.002613391 | 0.001296954 |
| 0 | 0 | 0 | 0 | 0 |
| 0 | 0 | 0 | 0 | 0 |
| 0.001787669 | 0 | 0 | 0.026493951 | 0 |
| 0 | 0 | 0 | 0 | 0 |
| 0 | 0 | 0 | 0 | 0 |
| 0 | 0 | 0 | 0 | 0 |
| 0.009257134 | 3.805272142 | 0 | 0 | 0.332110296 |
| 0 | 0 | 0 | 0 | 0 |
| 0 | 0 | 0 | 0 | 0 |
| 0 | 0.005398921 | 0 | 0.001651207 | 0 |
| 0 | 0 | 0 | 0 | 0 |
| 0 | 0 | 0 | 0 | 0 |

| | | | | |
|---|---|---|---|---|
| 0 | 0 | 0 | 0 | 0 |
| 0 | 0 | 0 | 0 | 0 |
| 0 | 0 | 0 | 0 | 0 |
| 0 | 0 | 0 | 0 | 0 |
| 0 | 0 | 0 | 0 | 0 |
| 0 | 0 | 0 | 0 | 0 |
| 0 | 0 | 0 | 0 | 0 |
| 0.112520397 | 0 | 0 | 0.835450637 | 0 |
| 0 | 0.034564971 | 0 | 0 | 0 |
| 0.004641052 | 0.019308434 | 0 | 0.014286093 | 0.004065377 |
| 0 | 0 | 0 | 0 | 0 |
| 0 | 0 | 0 | 0 | 0 |
| 0 | 0 | 0 | 0 | 0 |
| 0 | 0 | 0 | 0 | 0.152616307 |
| 0 | 0.014355542 | 0 | 0.005909998 | 0.013528033 |
| 0 | 0 | 0.020322171 | 0.003681317 | 0 |
| 0 | 0 | 0 | 0 | 0 |
| 0 | 0 | 0 | 0 | 0 |
| 0 | 0 | 0 | 0 | 0 |
| 12.60072524 | 0.033973496 | 0 | 0 | 0 |
| 0 | 0.001428581 | 0 | 0 | 0.004669074 |
| 0 | 0 | 0 | 0 | 0 |
| 0 | 0 | 0 | 0 | 0 |
| 0 | 0 | 0 | 0.055053939 | 0.044817251 |
| 0.00097444 | 0 | 0 | 0 | 0.003179969 |
| 0 | 0 | 0 | 0 | 0 |
| 0 | 0 | 0 | 0 | 0 |
| 0 | 0 | 0 | 0 | 0 |
| 0 | 0 | 0 | 0 | 0 |
| 0 | 0.011656072 | 0 | 0 | 0.008291141 |
| 0 | 0.004570395 | 0 | 0 | 0.24325067 |
| 0 | 0 | 0 | 0 | 0 |
| 0 | 0 | 0 | 0 | 0 |
| 0 | 0 | 0 | 0 | 0 |
| 0 | 0 | 0 | 0 | 0 |
| 0 | 0 | 0 | 0 | 0 |
| 0 | 0 | 0 | 0 | 0 |
| 0 | 0 | 0 | 0 | 0 |
| 0 | 0.002505899 | 0 | 0 | 0 |
| 0 | 0 | 0 | 0 | 0 |
| 0 | 0 | 0 | 0 | 0 |
| 0 | 0.381731347 | 0 | 0 | 0 |
| 0 | 0 | 0 | 0 | 0 |
| 0 | 0 | 0 | 0 | 0.004021125 |
| 0 | 0 | 0 | 0 | 0 |
| 0 | 0 | 0 | 0 | 0 |
| 0.02558657 | 0 | 0.079747356 | 0.016101931 | 0 |
| 0 | 0.015425616 | 0 | 0 | 0 |

| | | | | |
|---|---|---|---|---|
| 0 | 0 | 0 | 0 | 0 |
| 0 | 0 | 0 | 0 | 0 |
| 0 | 0 | 0 | 0 | 0 |
| 0 | 0 | 0 | 0 | 0 |
| 0 | 0.02766266 | 0 | 0.073583761 | 0.120740895 |
| 0 | 0 | 0 | 0.004822205 | 0 |
| 0 | 0 | 0 | 0 | 0 |
| 0 | 0 | 0 | 0 | 0 |
| 0 | 0 | 0 | 0 | 0 |
| 0 | 0 | 0 | 0 | 0 |
| 0 | 0 | 0 | 0 | 0.007767757 |
| 0.005103559 | 0.011295758 | 0.000899069 | 0.003731073 | 0.052712631 |
| 0 | 0.001809859 | 0 | 0 | 0.150227348 |
| 0 | 0 | 0 | 0.025810157 | 0 |
| 0 | 0 | 0 | 0.006363662 | 0 |
| 0.010715599 | 0.385117997 | 0 | 0 | 0.278975622 |
| 0 | 0 | 0 | 0.009018863 | 0.024017932 |
| 0 | 0 | 0 | 0 | 0 |
| 0.278046732 | 0 | 0.034175152 | 0.437769947 | 0.006999191 |
| 0.009334704 | 0.096682018 | 0 | 0.000702636 | 0.274411326 |
| 0 | 0 | 0 | 0 | 0 |
| 0 | 0 | 0 | 0 | 0 |
| 0 | 0 | 0 | 0 | 0 |
| 0 | 0 | 0 | 0 | 0 |
| 0 | 0 | 0 | 0 | 0 |
| 0 | 0 | 0 | 0 | 0 |
| 0.031740844 | 0.001534173 | 0.001709542 | 0.004645197 | 0.0342416 |
| 0 | 0 | 0 | 0.003915301 | 0 |
| 0 | 0 | 0 | 0 | 0 |
| 0 | 0 | 0 | 0 | 0 |
| 0.002750057 | 0 | 0 | 0 | 0.007403951 |
| 0 | 0 | 0 | 0.000689386 | 0 |
| 0 | 0 | 0 | 0 | 0 |
| 0 | 0 | 0 | 0 | 0 |
| 0 | 0 | 0 | 0 | 0 |
| 0 | 0 | 0 | 0 | 0 |
| 0 | 0 | 0 | 0 | 0 |
| 0.012558717 | 0.117396249 | 0.00034037 | 0.022532954 | 0.46327556 |
| 0 | 1.553239922 | 0 | 0 | 0 |
| 0 | 0 | 0 | 0.00248957 | 0 |
| 0 | 0.001321414 | 0 | 0 | 0.026909566 |
| 0 | 0 | 0 | 0 | 0 |
| 0 | 0 | 0 | 0 | 0 |
| 0.003581343 | 0 | 1.833333084 | 0.008644292 | 0 |
| 0 | 0 | 0 | 0 | 0 |
| 0 | 0 | 0 | 0 | 0 |
| 0 | 0 | 0 | 0 | 0 |
| 0 | 0 | 0 | 0 | 0 |

| | | | | |
|---|---|---|---|---|
| 0 | 0 | 0 | 0 | 0 |
| 0 | 0.002580929 | 0.158896293 | 0 | 0 |
| 0 | 0 | 0 | 0 | 0 |
| 0.005254569 | 0.002057005 | 0 | 0.001219515 | 0.020332232 |
| 0 | 0 | 0 | 0.001777413 | 0.001136013 |
| 0.001497561 | 0 | 0 | 0 | 0 |
| 0.008984235 | 0.019040367 | 0 | 0 | 0 |
| 0 | 0 | 0 | 0.003133804 | 0.085638553 |
| 0 | 0.005398849 | 0 | 0 | 0 |
| 0 | 0 | 0 | 0 | 0 |
| 0 | 0 | 0 | 0 | 0 |
| 0 | 0 | 0 | 0.155620987 | 0 |
| 0.025541143 | 0.130221983 | 0 | 0.014325415 | 0.304576619 |
| 0 | 0 | 0 | 0 | 0 |
| 0 | 0 | 0 | 0 | 0 |
| 0 | 0 | 0 | 0 | 0 |
| 0 | 0 | 0 | 0 | 0 |
| 0 | 0 | 0 | 0 | 0 |
| 0 | 0 | 0 | 0 | 0 |
| 0 | 0 | 0 | 0 | 0 |
| 0 | 0 | 0 | 0 | 0 |
| 0 | 0 | 0 | 0 | 0 |
| 0 | 0 | 0.002633221 | 0.015285754 | 0.071339711 |
| 0 | 0.001568552 | 0 | 0 | 0.008904004 |
| 0 | 0 | 0 | 0 | 0 |
| 0 | 0 | 0 | 0 | 0 |
| 0 | 0.002116628 | 0 | 0 | 0.004806078 |
| 0 | 0 | 0 | 0 | 0 |
| 0 | 0 | 0 | 0 | 0 |
| 0 | 0 | 0 | 0 | 0 |
| 0 | 0.0029642 | 0 | 0 | 0.003824205 |
| 0 | 0 | 0 | 0 | 0 |
| 0 | 0 | 0 | 0 | 0 |
| 0 | 0 | 0 | 0 | 0 |
| 0 | 0.018224203 | 0 | 0 | 0.02693147 |
| 0 | 0 | 0.08689803 | 0 | 0 |
| 0 | 0 | 0.019105816 | 0 | 0 |
| 0 | 0 | 0 | 0 | 0 |
| 0.002423591 | 0.001393288 | 0 | 0.028379834 | 0.047238921 |
| 0 | 0 | 0 | 0 | 0 |
| 0 | 0 | 0 | 0 | 0 |
| 0 | 0 | 0 | 0.010766656 | 0 |
| 0 | 1.103590332 | 0 | 0 | 0 |
| 0 | 0 | 0 | 0 | 0 |
| 0 | 0 | 0 | 0 | 0 |
| 0.171945764 | 0.065951454 | 0 | 0.498986397 | 0.268732532 |
| 0 | 0 | 0.06324301 | 0 | 0 |
| 0 | 0 | 0 | 0 | 0 |

| | | | | |
|---|---|---|---|---|
| 0 | 0 | 0 | 0 | 0 |
| 0 | 0 | 0 | 0 | 0 |
| 0 | 0.000377217 | 0 | 0.370331287 | 0 |
| 0 | 0 | 0 | 0 | 0 |
| 0 | 0 | 0 | 0.002562267 | 0 |
| 0 | 0.003385781 | 0 | 0 | 0 |
| 0.00604687 | 0 | 0 | 0 | 0 |
| 0 | 0 | 0 | 0 | 0 |
| 0 | 0 | 0.002873049 | 0 | 0 |
| 0 | 0 | 0 | 0.001050446 | 0 |
| 0.022164233 | 0.000565165 | 0 | 0.000746713 | 0.001224949 |
| 0.006404762 | 0 | 0 | 0.00074323 | 0.003657707 |
| 0 | 0 | 0 | 0 | 0 |
| 0 | 0 | 0 | 0 | 0 |
| 0 | 0 | 0 | 0 | 0 |
| 0 | 0 | 0 | 0 | 0 |
| 0 | 0 | 0 | 0 | 0 |
| 0 | 0 | 0 | 0 | 0 |
| 0 | 0 | 0 | 0 | 0 |
| 0 | 0.000599605 | 0 | 0 | 0.001392423 |
| 0 | 0 | 0 | 0 | 0 |
| 0 | 0 | 0 | 0 | 0 |
| 0 | 0 | 0 | 0 | 0 |
| 0 | 0 | 0 | 0 | 0 |
| 0 | 0 | 0 | 0 | 0 |
| 0 | 0 | 0 | 0 | 0 |
| 0 | 0 | 0 | 0 | 0 |
| 0 | 0.001990692 | 0 | 0 | 0 |
| 0 | 0 | 0 | 0 | 0 |
| 0 | 0 | 0 | 0 | 0 |
| 0 | 0.027816615 | 0 | 0.007798487 | 0.028310905 |
| 0 | 0 | 0 | 0 | 0 |
| 0.060059306 | 0.002392439 | 0.074645623 | 0.00674997 | 0.005324313 |
| 0.001691412 | 0.002444813 | 0.001532405 | 0 | 0.007569906 |
| 0 | 0 | 0 | 0 | 0 |
| 0 | 0 | 0 | 0 | 0 |
| 0 | 0 | 0 | 0 | 0 |
| 0 | 0 | 0 | 0 | 0 |
| 0 | 0 | 0 | 0 | 0 |
| 0 | 0 | 0 | 0 | 0 |
| 0 | 0 | 0 | 0 | 0 |
| 0.027039466 | 0.000984119 | 0.00041123 | 0.030609986 | 0.191779377 |
| 0.057351725 | 0.00974134 | 0 | 0.057422455 | 0.08723848 |
| 0 | 0 | 0 | 0 | 0 |
| 0 | 0 | 0 | 0 | 0 |
| 0.012671312 | 1.225557786 | 0 | 0.017645053 | 0.171999171 |
| 0.002245317 | 0 | 0 | 0.00062533 | 0.015167564 |
| 0 | 0 | 0 | 0 | 0 |

| | | | | |
|---|---|---|---|---|
| 0 | 0 | 0 | 0 | 0 |
| 0 | 0 | 0 | 0.009334806 | 0 |
| 0.006670372 | 0.002750982 | 0.001634903 | 0.00242312 | 0.024417985 |
| 0 | 0 | 0 | 0 | 0 |
| 0 | 0.011421939 | 0 | 0 | 1.630458628 |
| 0.00450806 | 0 | 0.012547768 | 0.193976677 | 0 |
| 0 | 0 | 0 | 0 | 0 |
| 0 | 0 | 0 | 0 | 0 |
| 0 | 0 | 0 | 0 | 0 |
| 0 | 0 | 0 | 0 | 0 |
| 0 | 0 | 0 | 0 | 0.000600526 |
| 0 | 0 | 0 | 0 | 0 |
| 0 | 0 | 0 | 0 | 0 |
| 0 | 0 | 0 | 0.01476604 | 0 |
| 0 | 0 | 0 | 0 | 0 |
| 0 | 0 | 0 | 0 | 0 |
| 0 | 0 | 0 | 0 | 0.476089882 |
| 0 | 0 | 0 | 0 | 0 |
| 0 | 0 | 0 | 0 | 0 |
| 0 | 0 | 0 | 0 | 0 |
| 0 | 0 | 0 | 0 | 0 |
| 0 | 0 | 0 | 0.000812133 | 0 |
| 0 | 0 | 0 | 0 | 0 |
| 0 | 0 | 0 | 0 | 0 |
| 0.005019296 | 0.830247123 | 0 | 0 | 0.003743973 |
| 0 | 0 | 0 | 0 | 0.003295554 |
| 0 | 0 | 0 | 0 | 0 |
| 0 | 0.050147793 | 0.00049306 | 0 | 0.004110175 |
| 0 | 0 | 0 | 0 | 0 |
| 0 | 0 | 0 | 0 | 0 |
| 0 | 0.028793338 | 0 | 0 | 0 |
| 0 | 0 | 0 | 0 | 0 |
| 0 | 0 | 0 | 0 | 0 |
| 0 | 0 | 0 | 0 | 0 |
| 0 | 0 | 0 | 0 | 0 |
| 0 | 0 | 0 | 0 | 0 |
| 0.058436516 | 0 | 0 | 0 | 0 |
| 0 | 0 | 0 | 0 | 0 |
| 0 | 0.407797262 | 0 | 0 | 1.597570016 |
| 0 | 0 | 0 | 0 | 0 |
| 0 | 0 | 0 | 0 | 0 |
| 0 | 0.042392272 | 0 | 0 | 0.00612886 |
| 0.013995999 | 0 | 0 | 0.029066545 | 0.008504867 |
| 0 | 0 | 0 | 0 | 0 |
| 0 | 0 | 0 | 0 | 0 |
| 0 | 0 | 0 | 0 | 0 |
| 0.019565352 | 0.004202024 | 0 | 0.00719683 | 0.011926551 |
| 0 | 0 | 0 | 0 | 0 |

| | | | | |
|---|---|---|---|---|
| 0 | 0 | 0 | 0 | 0 |
| 0.016303314 | 0.006038596 | 0.027407793 | 0.005837834 | 0.013909029 |
| 0 | 0 | 0 | 0 | 0 |
| 0 | 0 | 0 | 0 | 0 |
| 0 | 0 | 0 | 0 | 0 |
| 0 | 0 | 0 | 0 | 0 |
| 0 | 0 | 0 | 0 | 0 |
| 0 | 0 | 0 | 0 | 0 |
| 0 | 0 | 0 | 0 | 0 |
| 0 | 0 | 0 | 0 | 0 |
| 0 | 0 | 0 | 0 | 0 |
| 0 | 0 | 0 | 0 | 0 |
| 0 | 0.648608675 | 0 | 0.00514259 | 0.006266885 |
| 0 | 0 | 0 | 0 | 0 |
| 0.011615872 | 0 | 0.001227786 | 0.003437258 | 0.010803502 |
| 0 | 0 | 0 | 0 | 0 |
| 0 | 0 | 0 | 0 | 0 |
| 0 | 0 | 0 | 0 | 0 |
| 0 | 0 | 0 | 0.004697203 | 0 |
| 0 | 0 | 0 | 0 | 0 |
| 0 | 0 | 0 | 0 | 0 |
| 0.057038514 | 0.01875624 | 0 | 0.001985682 | 0.061425686 |
| 0 | 0 | 0 | 0.002593917 | 0.014374732 |
| 0 | 0 | 0 | 0 | 0.003450991 |
| 0 | 0 | 0 | 0 | 0 |
| 0 | 0 | 0 | 0 | 0 |
| 0 | 0 | 0 | 0 | 0 |
| 0 | 0 | 0 | 0 | 0 |
| 0 | 0 | 0 | 0 | 0 |
| 0 | 0 | 0 | 0 | 0 |
| 0 | 0 | 0 | 0 | 0 |
| 0 | 0 | 0 | 0 | 0 |
| 0 | 0 | 0 | 0 | 0 |
| 0 | 0 | 0 | 0 | 0 |
| 0 | 0.752414562 | 0 | 5.433905465 | 9.904187665 |
| 0 | 0.001617383 | 0 | 0 | 0 |
| 0 | 0 | 0 | 0 | 0 |
| 0 | 0 | 0 | 0 | 0 |
| 0.017087957 | 0.009545287 | 0 | 0 | 0.25484389 |
| 0 | 0 | 0 | 0 | 0 |
| 0 | 0.181345698 | 0.084572828 | 0.137970544 | 0.070250385 |
| 0 | 0 | 0 | 0 | 0 |
| 0 | 0 | 0 | 0 | 0 |
| 0 | 0 | 0 | 0 | 0 |
| 0 | 0 | 0 | 0 | 0 |
| 3.791162883 | 0 | 0.241045224 | 0.493850074 | 0 |
| 0 | 0 | 0 | 0 | 0 |
| 0 | 0 | 0 | 0 | 0 |

| | | | | |
|---|---|---|---|---|
| 0 | 0 | 0 | 0 | 0 |
| 0 | 0 | 0 | 0 | 0 |
| 0 | 0 | 0 | 0.034067392 | 0 |
| 0 | 0.028053336 | 0 | 0 | 0 |
| 0.008426142 | 0 | 0 | 0 | 0 |
| 0 | 0 | 0 | 0 | 0 |
| 0 | 0 | 0 | 0 | 0 |
| 0 | 0 | 0 | 0.001320154 | 0.001856275 |
| 0 | 0 | 0 | 0 | 0 |
| 0 | 0.009583466 | 0 | 0.418879994 | 0.032752728 |
| 0 | 0.002860509 | 0 | 0 | 0 |
| 0 | 0 | 0 | 0 | 0 |
| 0.004471691 | 0 | 0.000590817 | 0.002140503 | 0.003557004 |
| 0 | 0 | 0 | 0 | 0 |
| 0 | 0 | 0 | 0 | 0 |
| 0 | 0 | 0 | 0 | 0 |
| 0 | 0.393747782 | 0 | 0 | 0 |
| 0 | 0 | 0 | 0 | 0 |
| 0 | 0 | 0 | 0 | 0 |
| 0.193284623 | 0.120340541 | 0.025066405 | 0.388314215 | 0.234282844 |
| 0 | 0 | 0 | 0 | 0 |
| 0 | 0 | 0 | 0 | 0 |
| 0 | 0 | 0 | 0 | 0 |
| 0 | 0 | 0 | 0 | 0 |
| 0 | 0 | 0 | 0 | 0 |
| 0 | 0 | 0 | 0 | 0 |
| 0 | 0 | 0 | 0 | 0 |
| 0 | 0 | 0 | 0 | 0 |
| 0 | 0 | 0 | 0 | 0 |
| 0 | 0 | 0.009744143 | 0.012590439 | 0 |
| 0 | 0 | 0 | 0 | 0 |
| 0 | 0.002643604 | 0 | 0 | 0 |
| 0 | 0 | 0 | 0 | 0 |
| 0 | 0 | 0 | 0 | 0 |
| 0 | 0 | 0 | 0 | 0 |
| 0 | 0 | 0 | 0 | 0 |
| 0 | 0 | 0 | 0 | 0 |
| 0 | 0 | 0 | 0 | 0 |
| 0 | 0 | 0 | 0 | 0 |
| 0 | 0 | 0 | 0 | 0 |
| 0 | 0 | 0 | 0 | 0 |
| 0 | 0 | 0 | 0 | 0 |
| 0 | 0.000653803 | 0 | 0 | 0 |
| 0 | 0 | 0 | 0 | 0 |
| 0.004672845 | 0.008126215 | 0 | 0.004229569 | 0.017841653 |
| 0 | 0 | 0 | 0 | 0 |
| 0 | 0 | 0 | 0 | 0 |
| 0 | 0 | 0 | 0 | 0 |

| | | | | |
|---|---|---|---|---|
| 0 | 0 | 0 | 0 | 0 |
| 0 | 0 | 0 | 0 | 0 |
| 0 | 0 | 0 | 0 | 0 |
| 0 | 0 | 0 | 0 | 0 |
| 0 | 0 | 0 | 0 | 0 |
| 0.009767689 | 0.083497585 | 0 | 0.009836944 | 0.029029653 |
| 0 | 0.006872175 | 0 | 0 | 0 |
| 0 | 0 | 0 | 0 | 0.000610219 |
| 0 | 0 | 0 | 0 | 0 |
| 0 | 0 | 0 | 0 | 0 |
| 0 | 0 | 0 | 0 | 0 |
| 0 | 0 | 0 | 0 | 0 |
| 0 | 0 | 0 | 0 | 0 |
| 0 | 0.042448183 | 0 | 0 | 0 |
| 0 | 0 | 0 | 0 | 0 |
| 0 | 0 | 0 | 0 | 0 |
| 0 | 0 | 0 | 0.01060635 | 0 |
| 0 | 0 | 0 | 0 | 0 |
| 0 | 0 | 0 | 0 | 0 |
| 0 | 0 | 0 | 0 | 0 |
| 0 | 0 | 0 | 0 | 0 |
| 0 | 0 | 0 | 0 | 0 |
| 0 | 0 | 0 | 0 | 0 |
| 0 | 0 | 0 | 0 | 0 |
| 0 | 0 | 0 | 0 | 0 |
| 0 | 0 | 0 | 0.011017588 | 0.001602609 |
| 0 | 0 | 0 | 0 | 0 |
| 0 | 0.043257149 | 0 | 0.074355299 | 0 |
| 0 | 0 | 0 | 0 | 0 |
| 0 | 0 | 0 | 0 | 0 |
| 0 | 0 | 0 | 0 | 0 |
| 0 | 0 | 0 | 0 | 0 |
| 0 | 0 | 0 | 0 | 0 |
| 0 | 0 | 0 | 0 | 0 |
| 0 | 0 | 0 | 0 | 0 |
| 0 | 0 | 0 | 0 | 0 |
| 0 | 0 | 0 | 0 | 0 |
| 0 | 0 | 0 | 0 | 0 |
| 0 | 0.359976647 | 0 | 0.022285737 | 0 |
| 0 | 0 | 0 | 0 | 0 |
| 0 | 0 | 0 | 0 | 0 |
| 0 | 0 | 0 | 0 | 0 |
| 0 | 0 | 0 | 0 | 0 |
| 0 | 0 | 0 | 0 | 0 |
| 0 | 0 | 0 | 0 | 0 |
| 0 | 0 | 0 | 0 | 0 |
| 0 | 0 | 0 | 0 | 0 |
| 0 | 0 | 0 | 0 | 0 |

| | | | | |
|---|---|---|---|---|
| 0 | 0 | 0 | 0 | 0 |
| 0 | 0 | 0 | 0.017675589 | 0.000573548 |
| 0.00406439 | 0 | 0 | 0 | 0 |
| 0 | 0 | 0 | 0 | 0 |
| 0 | 0 | 0 | 0 | 0 |
| 0 | 0 | 0 | 0 | 0 |
| 0 | 0 | 0 | 0 | 0 |
| 0 | 0 | 0 | 0 | 0 |
| 0 | 0 | 0 | 0 | 0 |
| 0 | 0 | 0 | 0 | 0 |
| 0.001789932 | 0.058050647 | 0 | 0 | 0.017273351 |
| 0 | 0 | 0 | 0 | 0 |
| 0.886136386 | 0 | 0 | 0 | 0 |
| 0 | 0 | 0 | 0 | 0 |
| 0 | 0 | 0 | 0 | 0 |
| 0 | 0 | 0 | 0 | 0 |
| 0 | 0 | 0 | 0 | 0 |
| 0 | 0 | 0 | 0 | 0 |
| 0 | 0 | 0 | 0 | 0 |
| 0 | 0.001614643 | 0 | 0 | 0 |
| 0 | 0 | 0 | 0.014997821 | 0.005843804 |
| 0 | 0 | 0 | 0 | 0 |
| 0 | 0 | 0 | 0 | 0 |
| 1.269731752 | 0.664952219 | 0 | 1.445247442 | 0.28646119 |
| 0 | 0 | 0 | 0 | 0 |
| 0 | 0 | 0 | 0 | 0 |
| 0 | 0 | 0 | 0 | 0 |
| 0 | 0 | 0 | 0 | 0 |
| 0 | 0 | 0 | 0 | 0 |
| 0 | 0 | 0 | 0 | 0 |
| 0 | 0 | 0 | 0 | 0 |
| 0 | 0 | 0 | 0 | 0 |
| 0 | 0.003970608 | 0 | 0 | 0 |
| 0 | 0 | 0 | 0 | 0 |

| PHL018-0174_V1 | PHL018-0140_V1 | PHL018-0134_V2 | PHL018-0267_V1 | PHL018-0210_V2 |
|---|---|---|---|---|
| EYCF | CTRL | CTRL | REF | EYCF |
| 0.012819555 | 0.013777427 | 0.008012274 | 0.475673068 | 0.816571045 |
| 0.499603939 | 2.155360381 | 2.023275738 | 0.442201997 | 1.654220667 |
| 0 | 0.282885936 | 9.602466953 | 0 | 0 |
| 0 | 0 | 0 | 0 | 0 |
| 0 | 0 | 0 | 0 | 0 |
| 0 | 0 | 0 | 0 | 0 |
| 0 | 0 | 0 | 0 | 0 |
| 0 | 0 | 0 | 0 | 0 |
| 0 | 0 | 0 | 0 | 0 |
| 0 | 0 | 0 | 0 | 0 |
| 0 | 0 | 0 | 0 | 0 |
| 0 | 0 | 0 | 0 | 0 |
| 0 | 0 | 0 | 0 | 0 |
| 17.3897103 | 0.758487108 | 5.538153265 | 0.136325349 | 0.065452417 |
| 0 | 0 | 0 | 0 | 0 |
| 0 | 0 | 0 | 0 | 0 |
| 0 | 0 | 0 | 0 | 0 |
| 0 | 0 | 0 | 0 | 0 |
| 0 | 0 | 0 | 0 | 0 |
| 0 | 0 | 0 | 0 | 0 |
| 0 | 0 | 0 | 0 | 0 |
| 0 | 0 | 0 | 0 | 0 |
| 0 | 0 | 0 | 0 | 0 |
| 0 | 0 | 0 | 0 | 0 |
| 0 | 0 | 0 | 0 | 0 |
| 0 | 0 | 0 | 0 | 0 |
| 0 | 0 | 0 | 0 | 0 |
| 0 | 0 | 0 | 0 | 0 |
| 0 | 0 | 0 | 0 | 0 |
| 0 | 0 | 0 | 0 | 0 |
| 0 | 0 | 0 | 0 | 0 |
| 0 | 0 | 0 | 0 | 0 |
| 0 | 0 | 0 | 0 | 0 |
| 0 | 0 | 0 | 0 | 0 |
| 0 | 0 | 0 | 0 | 0 |
| 0 | 5.80010161 | 3.149305142 | 0.863040787 | 0.572597929 |
| 0 | 0 | 0 | 0 | 0 |
| 0 | 0 | 0 | 0 | 0 |
| 0 | 0 | 0 | 0 | 0 |
| 0 | 0 | 0 | 0 | 0 |
| 0 | 0 | 0 | 0 | 0 |
| 0.040867693 | 1.405716802 | 0.000757188 | 0.02133645 | 0.161829185 |
| 1.887616778 | 0.038985535 | 0.010413662 | 3.791247094 | 0.230631754 |
| 0.111180425 | 0.118428663 | 0.010553601 | 0.025943361 | 0.054925377 |
| 0.002562101 | 0.002776679 | 0 | 0.05450952 | 0.002281439 |
| 0 | 0 | 0 | 0 | 0 |

| | | | | |
|---|---|---|---|---|
| 0.009487253 | 0.000899659 | 0 | 0.4585033 | 0 |
| 0.039813944 | 0.003713591 | 0.126303857 | 0.001524752 | 0.052307074 |
| 0.007015651 | 0.003326408 | 0 | 0.260969003 | 0.259976712 |
| 0 | 1.197955265 | 0.518557057 | 0.252858931 | 0.44774787 |
| 0.003811856 | 0 | 0 | 0 | 0.007111847 |
| 0 | 0 | 0 | 0 | 0 |
| 0 | 0.438172495 | 0 | 0 | 0.004983533 |
| 0 | 0 | 0 | 0 | 0 |
| 0 | 0 | 0 | 0 | 0 |
| 0 | 0 | 0 | 0 | 0 |
| 0 | 0 | 0 | 0 | 0 |
| 0 | 0.060628641 | 0 | 0.880877948 | 0 |
| 0 | 0 | 0 | 0 | 0 |
| 0.063879309 | 0 | 0 | 0.02516416 | 0.089226237 |
| 0 | 0 | 0 | 0 | 0 |
| 0 | 0 | 0 | 0 | 0 |
| 0.050482549 | 0.042009852 | 0.162518361 | 0.007220386 | 0 |
| 0 | 0 | 0 | 0 | 0 |
| 0 | 0 | 0 | 0 | 0 |
| 0 | 0 | 0 | 0 | 0 |
| 0.910598464 | 0.078568835 | 0.182887679 | 0.003864937 | 0.445730708 |
| 0 | 0.000942728 | 0 | 0.00217728 | 0.010180272 |
| 0 | 0 | 0 | 2.04645723 | 0 |
| 0 | 0 | 0 | 0 | 0 |
| 0 | 0 | 0 | 0 | 0 |
| 0 | 0 | 0 | 0 | 0 |
| 0 | 0 | 0.034292997 | 0 | 0 |
| 0 | 0 | 0 | 0 | 0 |
| 0.001271797 | 0.022009901 | 0 | 0.739608671 | 0.106169934 |
| 0 | 0 | 0 | 0 | 0 |
| 0.010983122 | 0 | 0 | 0 | 0 |
| 0 | 0 | 0 | 0 | 0 |
| 0 | 0 | 0 | 0 | 0 |
| 0 | 0 | 0 | 0.004428813 | 0 |
| 0 | 0 | 0 | 0 | 0 |
| 0 | 0 | 0 | 0 | 0.001029069 |
| 0 | 0 | 0 | 0 | 0 |
| 3.782567855 | 2.196290456 | 0 | 0 | 0 |
| 0 | 0 | 0 | 0 | 0 |
| 0 | 0 | 0 | 0 | 0 |
| 0 | 0 | 0 | 0 | 0 |
| 0 | 0 | 0 | 0 | 0 |
| 0 | 0 | 0 | 0 | 0 |
| 0 | 0 | 0 | 0 | 0 |
| 0 | 0 | 0 | 0 | 0 |
| 0 | 0 | 0 | 0 | 0 |
| 0 | 0 | 0 | 0 | 0 |
| 0 | 0 | 0 | 0 | 0 |

| | | | | |
|---|---|---|---|---|
| 0 | 0 | 0 | 0 | 0 |
| 0 | 0 | 0 | 0 | 0 |
| 0 | 0 | 0 | 0 | 0 |
| 0 | 0 | 0 | 0 | 0 |
| 0.001131594 | 0 | 0 | 0 | 0.02090843 |
| 0 | 0 | 0 | 0 | 0 |
| 0 | 0 | 0 | 0 | 0 |
| 0 | 0 | 0 | 0 | 0 |
| 0.002882752 | 0.019363439 | 0 | 0 | 0.022460939 |
| 0 | 0 | 0 | 0 | 0 |
| 0 | 0 | 0 | 0 | 0 |
| 0.009761553 | 0.101559281 | 0 | 0 | 0 |
| 0 | 0 | 0 | 0 | 0 |
| 0 | 0 | 0 | 0 | 0 |
| 0 | 0 | 0 | 0 | 0 |
| 0 | 0 | 0 | 0 | 0 |
| 0 | 0 | 0 | 0 | 0 |
| 0 | 0 | 0 | 0 | 0 |
| 0 | 0 | 0.014568903 | 0.380813304 | 0 |
| 0 | 0 | 0 | 0 | 0 |
| 0 | 0 | 0 | 0 | 0 |
| 0 | 0 | 0 | 0 | 0 |
| 0 | 0 | 0 | 0 | 0 |
| 0 | 0 | 0 | 0 | 0 |
| 0 | 0 | 0 | 0 | 0 |
| 0 | 0 | 0 | 0 | 0 |
| 0 | 0 | 0 | 0 | 0 |
| 0 | 0 | 0 | 0 | 0 |
| 0 | 0 | 0 | 0 | 0 |
| 0 | 0 | 0 | 0 | 0 |
| 0 | 0 | 0.200616089 | 0.047388035 | 0 |
| 0.153687601 | 0.003096128 | 0.001283316 | 0.477937104 | 0 |
| 0 | 0 | 0 | 0 | 0 |
| 0 | 0 | 0 | 0 | 0 |
| 0 | 0 | 0 | 0 | 0 |
| 0 | 0 | 0 | 0 | 0 |
| 0 | 0 | 0 | 0 | 0 |
| 0 | 0 | 0 | 0 | 0 |
| 0 | 0 | 0 | 0 | 0 |
| 0 | 0 | 0 | 0 | 0 |
| 0 | 0 | 0 | 0 | 0 |
| 0 | 0 | 0 | 0 | 0 |
| 0.048789953 | 0.003067401 | 0 | 0.001574294 | 0 |
| 0 | 0 | 0 | 0 | 0 |
| 0 | 0 | 0 | 0 | 0 |
| 0 | 0 | 0 | 0 | 0 |
| 0 | 0 | 0 | 0 | 0 |
| 0 | 0 | 0 | 0 | 0 |

| | | | | |
|---|---|---|---|---|
| 0 | 0 | 0 | 0 | 0 |
| 0 | 0 | 0 | 0 | 0 |
| 0 | 0 | 0 | 0 | 0 |
| 0 | 0 | 0 | 0 | 0 |
| 0 | 0 | 0 | 0.300997156 | 0 |
| 0 | 0 | 0 | 0 | 0 |
| 0 | 0 | 0 | 0 | 0 |
| 0 | 0 | 0 | 0 | 0 |
| 0 | 0 | 0 | 0 | 0 |
| 0 | 0 | 0 | 0 | 0 |
| 0 | 0 | 0 | 0 | 0 |
| 0 | 0 | 0 | 0 | 0 |
| 0 | 0 | 0 | 0 | 0 |
| 0 | 0 | 0 | 0 | 0 |
| 0 | 0 | 0 | 0 | 0 |
| 0.002475078 | 0 | 0 | 0.126562955 | 0 |
| 0 | 0 | 0 | 0 | 0 |
| 0 | 0 | 0 | 0 | 0 |
| 0 | 0 | 0 | 0 | 0 |
| 0 | 0 | 0 | 0 | 0 |
| 0 | 0 | 0 | 0.00467202 | 0 |
| 0 | 0 | 0 | 0 | 0 |
| 0 | 0 | 0 | 0 | 0 |
| 0 | 0 | 0 | 0 | 0 |
| 0 | 0 | 0 | 0.005110654 | 0 |
| 0 | 0 | 0 | 0 | 0 |
| 0 | 0 | 0 | 0 | 0 |
| 0 | 0 | 0 | 0 | 0 |
| 0 | 0 | 0 | 0 | 0 |
| 0 | 0 | 0 | 0 | 0 |
| 0 | 0 | 0 | 0 | 0 |
| 0 | 0 | 0 | 0 | 0.002635623 |
| 0 | 0 | 0 | 0 | 0 |
| 0 | 0 | 0 | 0 | 0 |
| 0 | 0 | 0 | 0 | 0 |
| 0 | 0 | 0 | 0 | 0 |
| 0 | 0 | 0 | 0 | 0 |
| 0 | 0 | 0 | 0 | 0 |
| 0 | 0 | 0 | 0 | 0 |
| 0 | 0 | 0 | 0 | 0 |
| 0 | 0 | 0 | 0 | 0 |
| 0 | 0 | 0 | 0 | 0 |
| 0 | 0 | 0 | 0 | 0 |
| 0 | 0 | 0 | 0 | 0 |
| 0 | 0 | 0 | 0 | 0 |
| 0 | 0 | 0 | 0 | 0 |
| 0 | 0 | 0 | 0 | 0 |
| 0 | 0 | 0 | 0 | 0 |

| | | | | |
|---|---|---|---|---|
| 0 | 0 | 0 | 0 | 0 |
| 0 | 0 | 0 | 0 | 0 |
| 0 | 0 | 0 | 0 | 0 |
| 0 | 0 | 0 | 0 | 0 |
| 0 | 0 | 0 | 0 | 0 |
| 0 | 0 | 0 | 0 | 0 |
| 0 | 0 | 0 | 0 | 0 |
| 0 | 0 | 0 | 0 | 0 |
| 0 | 0 | 0 | 0 | 0 |
| 0 | 0 | 0 | 0 | 0 |
| 0 | 0 | 0 | 0 | 0 |
| 0 | 0 | 0 | 0 | 0 |
| 0.001287406 | 0 | 0 | 0.028120369 | 0.005044065 |
| 0 | 0 | 0 | 0 | 0 |
| 0 | 0 | 0 | 0 | 0 |
| 0 | 0 | 0 | 0 | 0 |
| 0 | 0 | 0 | 0 | 0 |
| 0.002167607 | 0 | 0 | 0 | 0 |
| 0 | 0 | 0 | 0 | 0 |
| 0 | 0 | 0 | 0 | 0 |
| 0.135281237 | 0.148372055 | 0.001177413 | 0.003374011 | 0.036867378 |
| 0 | 0 | 0 | 0 | 0 |
| 0 | 0 | 0 | 0.001356889 | 0.001504597 |
| 0.010040649 | 0 | 0.000920854 | 0.00234561 | 0 |
| 0 | 0 | 0 | 0 | 0 |
| 0 | 0 | 0 | 0 | 0 |
| 0.071214552 | 0.01355947 | 0 | 0.000982472 | 0.004244207 |
| 0 | 0 | 0 | 0 | 0 |
| 0 | 0 | 0 | 0 | 0 |
| 0 | 0 | 0.028060862 | 0 | 0 |
| 0 | 0 | 0.000683524 | 0 | 0 |
| 0 | 0 | 0 | 0 | 0 |
| 0 | 0 | 0 | 0 | 0 |
| 0 | 0 | 0 | 0.002921147 | 1.308110938 |
| 0.034143448 | 0.005643805 | 0.004979569 | 0.032014955 | 0.035702893 |
| 0 | 0 | 0 | 0 | 0 |
| 0 | 0 | 0 | 0 | 0 |
| 0 | 0 | 0 | 0 | 0 |
| 0 | 0 | 0 | 0 | 0 |
| 0 | 0 | 0 | 0 | 0 |
| 0 | 0 | 0 | 0 | 0 |
| 0 | 0 | 0 | 0 | 0 |
| 0 | 0 | 0 | 0 | 0 |
| 0 | 0 | 0 | 0 | 0 |
| 0 | 0 | 0 | 0 | 0 |
| 0 | 0 | 0 | 0 | 0 |
| 0 | 0 | 0 | 0.008121206 | 0 |
| 0.003571603 | 0 | 0.004776929 | 0.048497581 | 0.032598647 |

| | | | | |
|---|---|---|---|---|
| 0 | 0 | 0 | 0 | 0 |
| 0 | 0 | 0 | 0 | 0 |
| 0 | 0.008510882 | 0 | 0 | 0 |
| 0 | 0 | 0 | 0 | 0 |
| 0 | 0 | 0 | 0 | 0 |
| 0 | 0 | 0 | 0 | 0 |
| 0 | 0.012655625 | 0 | 0 | 0 |
| 0.001283561 | 0.099199998 | 0 | 0.000749636 | 0 |
| 0.003299098 | 0 | 0 | 0 | 0.179199847 |
| 0 | 0 | 0 | 0 | 0 |
| 0 | 0 | 0 | 0 | 0 |
| 0 | 0 | 0 | 0 | 0 |
| 0 | 0 | 0 | 0 | 0 |
| 0 | 0 | 0 | 0 | 0 |
| 0 | 0 | 0 | 0 | 0 |
| 0 | 0 | 0 | 0 | 0 |
| 0 | 0 | 0 | 0 | 0 |
| 0 | 0 | 0 | 0 | 0 |
| 0.009080741 | 0 | 0 | 0.014982135 | 0 |
| 0 | 0 | 0 | 0 | 0 |
| 0.109169394 | 0 | 0.020065358 | 0.001173461 | 0.102578017 |
| 0 | 0 | 0 | 0 | 0 |
| 0 | 0 | 0 | 0 | 0 |
| 0 | 0 | 0 | 0 | 0 |
| 0.006653009 | 0.018926785 | 0 | 0.001942773 | 0 |
| 0.001291197 | 0 | 0 | 0 | 0 |
| 0 | 0 | 0 | 0 | 0 |
| 0 | 0 | 0 | 0 | 0 |
| 0 | 0.091815702 | 0 | 0.021877394 | 0.104873577 |
| 0 | 0 | 0 | 0 | 0 |
| 0.106126757 | 0.015177439 | 0 | 0.054646857 | 0.504079635 |
| 0 | 0 | 0 | 0 | 0 |
| 0 | 0 | 0 | 0 | 0 |
| 0 | 0 | 0 | 0 | 0 |
| 0 | 0 | 0 | 0 | 0 |
| 0 | 0 | 0 | 0 | 0 |
| 0 | 0 | 0 | 0 | 0 |
| 0 | 0 | 0 | 0 | 0 |
| 0 | 0 | 0 | 0 | 0 |
| 0 | 0 | 0 | 0 | 0 |
| 0 | 0 | 0 | 0 | 0 |
| 0.010039776 | 0.013328754 | 0.002301934 | 0.319121753 | 0 |
| 0 | 0 | 0 | 0 | 0 |
| 0 | 0 | 0 | 0 | 0 |
| 0 | 0 | 0 | 0 | 0 |
| 0 | 0 | 0 | 0 | 0 |
| 0 | 0 | 0 | 0 | 0 |
| 0 | 0 | 0 | 0 | 0 |

| | | | | |
|---|---|---|---|---|
| 0 | 0 | 0 | 0 | 0 |
| 0 | 0 | 0 | 0 | 0 |
| 0.102113382 | 0.115959646 | 0.0132961 | 0.014909279 | 0.060898969 |
| 0 | 0 | 0 | 0 | 0 |
| 0 | 0 | 0 | 0.000798684 | 0 |
| 0 | 0 | 0 | 0 | 0 |
| 0 | 0 | 0 | 0 | 0 |
| 0 | 0 | 0 | 0 | 0 |
| 0 | 0.001259972 | 0 | 0 | 0 |
| 0 | 0 | 0 | 0 | 0 |
| 0 | 0 | 0 | 0 | 0 |
| 0 | 0 | 0 | 0 | 0 |
| 0 | 0 | 0 | 0 | 0 |
| 0.058168433 | 0.005615098 | 0 | 0 | 0.001550792 |
| 0.006037239 | 0.008178579 | 0 | 0.002854315 | 0.002559951 |
| 0 | 0.015761247 | 0 | 0 | 0 |
| 0.012775086 | 0.007765631 | 0 | 0.00095654 | 0.00087505 |
| 0 | 0 | 0.000466081 | 0 | 0 |
| 0 | 0 | 0 | 0 | 0 |
| 0 | 0 | 0 | 0 | 0 |
| 0 | 0 | 0 | 0 | 0 |
| 0 | 0 | 0.003796702 | 0.049391216 | 0.032913365 |
| 0 | 0 | 0 | 0 | 0 |
| 0 | 0 | 0.015975621 | 0 | 0 |
| 0 | 0 | 0 | 0 | 0 |
| 0 | 0 | 0 | 0 | 0 |
| 0 | 0 | 0 | 0 | 0 |
| 0 | 0 | 0 | 0 | 0 |
| 0 | 0 | 0 | 0 | 0 |
| 0.003849581 | 0.001216829 | 0 | 0 | 0 |
| 0 | 0 | 0 | 0 | 0 |
| 0 | 0 | 0 | 0 | 0 |
| 0 | 0 | 0 | 0 | 0 |
| 0 | 0 | 0 | 0 | 0 |
| 0 | 0 | 0 | 0 | 0 |
| 0.010040481 | 0.015681988 | 0 | 0 | 0 |
| 0 | 0 | 0 | 0 | 0 |
| 0.28569508 | 0.03734391 | 0 | 0.058800318 | 0.008270447 |
| 0 | 0 | 0 | 0 | 0 |
| 0 | 0 | 0 | 0 | 0 |
| 0 | 0 | 0 | 0 | 0 |
| 0 | 0 | 0 | 0 | 0 |
| 0 | 0 | 0 | 0 | 0 |
| 0 | 0 | 0 | 0 | 0 |
| 0 | 0 | 0 | 0 | 0 |
| 0 | 0 | 0 | 0 | 0 |
| 0 | 0 | 0 | 0 | 0 |
| 0 | 0.002133625 | 0.008529228 | 0.001839681 | 0 |

| | | | | |
|---|---|---|---|---|
| 0 | 0 | 0 | 0.117988867 | 13.66464007 |
| 0.038028159 | 0 | 0 | 0 | 0 |
| 0 | 0.010279618 | 0 | 0 | 0 |
| 0 | 0 | 0 | 0 | 0 |
| 0.045495023 | 0.003250433 | 0 | 0.000909945 | 0 |
| 0 | 0 | 0 | 0 | 0 |
| 0 | 0 | 0 | 0 | 0 |
| 0.164669697 | 0.087396149 | 0 | 0.008163122 | 0.005834131 |
| 0 | 0 | 0 | 0 | 0 |
| 0.163875912 | 0.017076986 | 0.009634281 | 0.003681084 | 0.037683753 |
| 0 | 0 | 0 | 0 | 0 |
| 0.012101749 | 0.419033208 | 0 | 0 | 0 |
| 0.009704155 | 0.008704855 | 0 | 0.001531757 | 0.006072142 |
| 0 | 0 | 0 | 0 | 0 |
| 0 | 0 | 0.00263881 | 0.000630151 | 0 |
| 0 | 0 | 0 | 0 | 0 |
| 0 | 0 | 0 | 0 | 0 |
| 0 | 0 | 0 | 0 | 0 |
| 0 | 0 | 0 | 0 | 0 |
| 0 | 0 | 0.012664831 | 0 | 0 |
| 0 | 0 | 0 | 0 | 0 |
| 0 | 0 | 0 | 0 | 0 |
| 0.0008119 | 0 | 0 | 0 | 0 |
| 0 | 0 | 0 | 0 | 0 |
| 0 | 0 | 0 | 0 | 0 |
| 0.011302866 | 0.001738103 | 0 | 0.005887552 | 0.009520669 |
| 0 | 0 | 0 | 0 | 0 |
| 0.612487931 | 0.018216035 | 0 | 0 | 0 |
| 0 | 0 | 0 | 0 | 0 |
| 0 | 0 | 0.011722399 | 0 | 0 |
| 0 | 0 | 0 | 0 | 0 |
| 0 | 0 | 0 | 0 | 0 |
| 0 | 0 | 0 | 0 | 0 |
| 0.045544763 | 0 | 0 | 0 | 0 |
| 0 | 0 | 0 | 0 | 0 |
| 0 | 0 | 0 | 0 | 0 |
| 0.299892452 | 0 | 0 | 0 | 0.021585307 |
| 0 | 0 | 0 | 0 | 0 |
| 0 | 0 | 0 | 0 | 0 |
| 0 | 0 | 0 | 0 | 0 |
| 0 | 0 | 0 | 0 | 0 |
| 0 | 0 | 0 | 0 | 0 |
| 0 | 0 | 0 | 0 | 0 |
| 0 | 0 | 0 | 0.002012275 | 0.037036044 |
| 0 | 0 | 0 | 0 | 0 |
| 0 | 0 | 0 | 0 | 0 |
| 0.002659245 | 0 | 0.001393635 | 0 | 0.002367941 |
| 0 | 0 | 0 | 0 | 0 |

| | | | | |
|---|---|---|---|---|
| 0 | 0 | 0 | 0 | 0 |
| 0 | 0.028181916 | 0.001038323 | 0 | 0 |
| 0 | 0 | 0 | 0 | 0 |
| 0 | 0 | 0 | 0 | 0 |
| 0.001126451 | 0 | 0 | 0 | 0 |
| 0 | 0 | 0.237276714 | 0.014504063 | 0 |
| 0 | 0 | 0 | 0 | 0.178052444 |
| 0 | 0 | 0.00189291 | 0 | 0 |
| 0 | 0 | 0 | 0 | 0 |
| 0 | 0 | 0 | 0 | 0 |
| 0 | 0.011847114 | 0 | 0.001418746 | 0 |
| 0.007806008 | 0 | 0 | 0 | 0.00492356 |
| 0.184608024 | 0 | 0 | 0 | 0 |
| 0.024589551 | 0.018654263 | 0.00521195 | 0 | 0 |
| 0.117766309 | 0 | 0.029410218 | 0 | 0.007975704 |
| 0 | 0 | 0 | 0 | 0 |
| 0 | 0 | 0 | 0 | 0 |
| 0 | 0 | 0 | 0 | 0 |
| 0 | 0 | 0 | 0.02547512 | 0 |
| 0 | 0 | 0 | 0 | 0 |
| 0 | 0 | 0 | 0 | 0 |
| 0 | 0.371547243 | 0.414073687 | 0.075121628 | 0 |
| 0 | 0 | 0 | 0 | 0 |
| 0 | 0 | 0 | 0 | 0 |
| 0 | 0 | 0 | 0 | 0 |
| 0 | 0 | 0 | 0 | 0 |
| 0 | 0 | 0 | 0 | 0 |
| 0 | 0 | 0 | 0 | 0 |
| 0 | 0 | 0.002860412 | 0 | 0 |
| 0.238845816 | 0.3919198 | 0.066112153 | 0.562461667 | 0.358606224 |
| 0 | 0.007804185 | 0.000543984 | 0.018576261 | 0.017825568 |
| 0.001561406 | 0.098611482 | 0 | 0 | 0 |
| 0 | 0 | 0 | 0 | 0 |
| 0 | 0 | 0 | 0.000919942 | 0 |
| 0 | 0 | 0 | 0 | 0 |
| 0 | 0 | 0 | 0 | 0 |
| 0 | 0 | 0 | 0 | 0 |
| 0.002005064 | 0.1524262 | 0 | 0 | 0 |
| 0 | 0 | 0.157869852 | 0 | 0.176644826 |
| 0.840672675 | 0 | 0.168408459 | 0.001359419 | 0.03005383 |
| 0.097033375 | 0 | 0 | 0.003778016 | 0.012240562 |
| 0 | 2.95905632 | 0.885983095 | 3.934304561 | 0 |
| 0 | 0.046157026 | 0 | 0 | 0 |
| 0 | 0 | 0 | 0 | 0 |
| 0 | 0 | 0 | 0.019710822 | 0 |
| 0 | 0 | 0 | 0 | 0 |
| 0.060823195 | 0.003219212 | 0 | 0.001321766 | 0 |
| 0 | 0 | 0 | 0 | 0 |

| | | | | |
|---|---|---|---|---|
| 0 | 0.000992992 | 0 | 0.017939218 | 0.002797319 |
| 0 | 0 | 0 | 0 | 0 |
| 0 | 0.007667155 | 0.009886994 | 0 | 0 |
| 0 | 0 | 0 | 0 | 0 |
| 0 | 0 | 0 | 0.050137985 | 0 |
| 0 | 0 | 0 | 0.00534314 | 0 |
| 0 | 0 | 0 | 0 | 0 |
| 0 | 0.208862178 | 0 | 0 | 0 |
| 0 | 0 | 0 | 0 | 0 |
| 0 | 0 | 0 | 0.010001046 | 0 |
| 0 | 0 | 0 | 0 | 0 |
| 0 | 0 | 0 | 0 | 0 |
| 0 | 0 | 0 | 0 | 0.002675433 |
| 0.005052419 | 0.001497224 | 0 | 0 | 0 |
| 0 | 0 | 0 | 0 | 0 |
| 0.001974189 | 0.005616264 | 0.001357934 | 0.258556306 | 0.096466341 |
| 0.007658639 | 0.060175411 | 0.00050171 | 0.172045471 | 0.070875987 |
| 0.004320154 | 0 | 0.001015929 | 0 | 0.002663244 |
| 0.002542756 | 0 | 0 | 0 | 0.008302113 |
| 0 | 0.019025171 | 0.34346828 | 0 | 0 |
| 0 | 0 | 0 | 0 | 0.085947406 |
| 0.004441812 | 0 | 0.002327828 | 0 | 0.007062926 |
| 0 | 0 | 0 | 0 | 0 |
| 0 | 0 | 0.011261094 | 0 | 0.393072945 |
| 0 | 0 | 0 | 0 | 0 |
| 0.414005513 | 0 | 0 | 0.001683488 | 0.004491865 |
| 0.008680986 | 0 | 0 | 0 | 0.001236806 |
| 0 | 0 | 0 | 0 | 0 |
| 0 | 0 | 0 | 0 | 0 |
| 0.256710684 | 0 | 0 | 0 | 0 |
| 0 | 0 | 0 | 0.008019808 | 0 |
| 0.044882123 | 0.009706874 | 0 | 0 | 0.016477031 |
| 0 | 0.008175842 | 15.93934999 | 0.081660109 | 0 |
| 0.012355796 | 0.022924133 | 0 | 0.004392431 | 0.007653771 |
| 0.004984593 | 0 | 0 | 0 | 0 |
| 0 | 0 | 0 | 0 | 0 |
| 0 | 0.004499189 | 0 | 0 | 0 |
| 0.012986654 | 0.00150796 | 0 | 0.063153195 | 0.033040134 |
| 0 | 0.348948047 | 0 | 0 | 0 |
| 0 | 0 | 0 | 0 | 0 |
| 0 | 0 | 0 | 0 | 0 |
| 0 | 0 | 0 | 0 | 0 |
| 0.003170861 | 0 | 0 | 0.010648268 | 0 |
| 0 | 0 | 0 | 0.059931939 | 0.0178701 |
| 0 | 0 | 0 | 0 | 0.035330523 |
| 0 | 0 | 0 | 0 | 0 |
| 0 | 0 | 0 | 0 | 0 |
| 0 | 0 | 0 | 0 | 0 |

| | | | | |
|---|---|---|---|---|
| 0 | 0 | 0 | 0.003442472 | 0 |
| 5.082714259 | 0.356505622 | 0.003974552 | 0.439340644 | 0.314184871 |
| 0 | 0 | 0 | 0 | 0 |
| 0 | 0 | 0 | 0 | 0.007790472 |
| 0 | 0 | 0 | 0 | 0 |
| 0 | 0 | 0.000929123 | 0 | 0 |
| 0 | 0 | 0 | 0 | 0 |
| 0.012482785 | 0 | 0 | 0 | 0 |
| 0 | 0.002194487 | 0 | 0.002316929 | 0.003826955 |
| 0 | 0 | 0 | 0 | 0 |
| 0.010197987 | 0 | 0 | 0.003918365 | 0.00095588 |
| 0.402540068 | 0.412805077 | 0.030551124 | 0.007470739 | 0.042358363 |
| 0 | 0 | 0 | 0 | 0 |
| 0 | 0 | 0 | 0 | 0 |
| 0.010777159 | 0.036791228 | 0.000941334 | 0 | 0.001370941 |
| 0 | 0 | 0 | 0 | 0 |
| 0.026696911 | 0.030112954 | 0 | 0.005087836 | 0.008758264 |
| 0.035385885 | 0.014183378 | 0 | 0 | 0 |
| 0 | 0 | 0 | 0 | 0 |
| 0 | 0 | 0 | 0 | 0.011601414 |
| 0.149827965 | 0.079696242 | 0.011340177 | 0.016079032 | 0.009806149 |
| 0 | 0 | 0 | 0 | 0 |
| 0 | 0 | 0 | 0 | 0 |
| 0 | 0 | 0 | 0 | 0 |
| 0 | 0.58687133 | 0 | 0 | 0 |
| 0 | 0 | 0 | 0 | 0.001893161 |
| 0 | 0 | 0 | 0.012195463 | 0 |
| 0.068687886 | 0.022782815 | 0.026742448 | 0.023565711 | 0 |
| 0 | 0 | 0 | 0 | 0 |
| 0.007430823 | 0 | 0 | 0.000650972 | 0.001985047 |
| 0.090888312 | 0 | 0 | 0 | 0.00559167 |
| 0 | 0 | 0 | 0 | 0 |
| 0 | 0 | 0 | 0 | 0 |
| 0 | 0 | 0 | 0 | 0 |
| 0 | 0.019599104 | 0 | 0 | 0 |
| 0.016681636 | 0.119136051 | 0.00796831 | 0.229863264 | 0.265984195 |
| 0 | 0 | 0 | 0 | 0 |
| 0 | 0 | 0 | 0 | 0 |
| 0.02507828 | 0.241776365 | 0 | 0 | 0.012857306 |
| 0 | 0 | 0 | 0 | 0 |
| 0 | 0 | 0 | 0 | 0.031422838 |
| 0 | 0 | 0 | 0 | 0 |
| 0.085396732 | 0.005252769 | 0 | 0.027093777 | 0.000616557 |
| 0 | 0 | 0 | 0 | 0 |
| 0.007026084 | 0 | 0 | 0 | 0 |
| 0 | 0.338843798 | 0 | 0.015059908 | 0 |
| 0 | 0 | 0 | 0 | 0 |
| 0 | 0 | 0 | 0.001051822 | 0 |

| | | | | |
|---|---|---|---|---|
| 0 | 0 | 0 | 0 | 0 |
| 0 | 0 | 0 | 0 | 0 |
| 0 | 0 | 0.01474296 | 0 | 0 |
| 0.074729533 | 0.033138155 | 0 | 2.388183832 | 1.7047551 |
| 0 | 0 | 0 | 0 | 0 |
| 0 | 0 | 0 | 0 | 0 |
| 0.001042017 | 0 | 0 | 0 | 0 |
| 0 | 0 | 0 | 0 | 1.096383278 |
| 0 | 2.00344822 | 0.020233497 | 1.32289123 | 0 |
| 0 | 0 | 0 | 0.135963785 | 0 |
| 0 | 0 | 0 | 0 | 0 |
| 0 | 0.008303824 | 0 | 0.033091645 | 0 |
| 0 | 0 | 0 | 0 | 0 |
| 0 | 0 | 0 | 0.014887035 | 0 |
| 0.085437459 | 0.00886773 | 0.007406869 | 0.153913927 | 0.480315324 |
| 0 | 0 | 0 | 0 | 0 |
| 0 | 0.112779971 | 0 | 0.157699422 | 0 |
| 0 | 0 | 0 | 0 | 0 |
| 0 | 0 | 0 | 0 | 0 |
| 0 | 0 | 0 | 0 | 0 |
| 0 | 0 | 0 | 0 | 0 |
| 0 | 0 | 0 | 0 | 0 |
| 0 | 0 | 0 | 0 | 0 |
| 0.194485726 | 0 | 0.07710497 | 0.263637197 | 0.107189936 |
| 0.004099256 | 0 | 0 | 0 | 0 |
| 0 | 0.010369036 | 0 | 0 | 0 |
| 0 | 0 | 0 | 0 | 0 |
| 0 | 0.823380055 | 0.528511024 | 0.177898584 | 0 |
| 0 | 0 | 0 | 0 | 0 |
| 0 | 0 | 0 | 0.058701215 | 0 |
| 0 | 0.002374333 | 0.005511175 | 0.001169844 | 0 |
| 0 | 0 | 0 | 0 | 0 |
| 0 | 0 | 0 | 0 | 0 |
| 0 | 0 | 0 | 0 | 0 |
| 0 | 0 | 0 | 0 | 0 |
| 0.004670522 | 0 | 0.019989456 | 0 | 0 |
| 0 | 0.333582398 | 0 | 0.361573728 | 0 |
| 0 | 0 | 0 | 0 | 0.162022288 |
| 0 | 0 | 0 | 0.063103215 | 0 |
| 0 | 0 | 0 | 0 | 0 |
| 0.007165125 | 0.359174446 | 0.14607967 | 0.003463147 | 0.070622544 |
| 0 | 0 | 0 | 0 | 0 |
| 0.280093013 | 4.863000098 | 33.74367206 | 0.00415447 | 0.695710292 |
| 0 | 0 | 0 | 0 | 0.028173382 |
| 0 | 0 | 0 | 0 | 0 |
| 0 | 0 | 0 | 0 | 0 |
| 0 | 0 | 0 | 0 | 0 |
| 0 | 0 | 0 | 0.362536666 | 0.007140819 |

| | | | | |
|---|---|---|---|---|
| 0 | 0 | 0 | 0 | 0 |
| 0 | 0 | 0 | 0 | 0 |
| 3.551287847 | 0.313731832 | 0.005570606 | 0.000665134 | 0.106989205 |
| 0 | 0 | 0 | 0 | 0 |
| 0 | 0 | 0 | 0.090055831 | 0 |
| 0.020410379 | 0.002764971 | 0 | 0.169372214 | 0.053724763 |
| 0 | 0 | 0 | 0 | 0 |
| 0 | 0 | 0 | 0 | 0.009310289 |
| 0 | 0 | 0 | 0 | 0 |
| 0 | 0 | 0 | 0 | 0.021291315 |
| 0.018449812 | 0.007952557 | 0.022048307 | 0.926994929 | 0.16901698 |
| 5.721483987 | 1.638974888 | 1.399591902 | 0.429172559 | 0.050712734 |
| 0 | 0 | 0 | 0 | 0 |
| 0.06637263 | 0.032900439 | 0 | 0.500401556 | 0.058564629 |
| 1.39081564 | 0.026152258 | 0.190279009 | 0.023465746 | 0.167033376 |
| 0 | 0.003257888 | 0 | 0 | 0 |
| 0 | 0 | 0 | 0 | 0 |
| 0.003168263 | 0 | 0 | 0 | 0 |
| 0 | 0 | 0 | 0 | 0 |
| 0 | 0 | 0 | 0.004975906 | 0 |
| 0.299432675 | 0.41058947 | 0 | 0.001505203 | 0.311696187 |
| 0.19404553 | 0.031370641 | 1.268390538 | 0 | 0.005495835 |
| 9.708874211 | 0.721900576 | 0.289014894 | 0.077547295 | 0.553732886 |
| 0 | 0 | 0 | 0 | 0 |
| 0 | 0 | 0 | 0 | 0 |
| 0 | 0.134184367 | 0.001716609 | 0 | 0 |
| 0.653499072 | 0 | 0.004112115 | 0 | 1.073493431 |
| 0 | 0 | 0 | 0.001998053 | 0 |
| 0.009977179 | 0 | 0 | 0.008948539 | 0 |
| 0 | 0 | 0 | 0 | 0 |
| 0 | 0 | 0 | 0 | 0 |
| 0 | 0.116414233 | 0 | 0 | 0 |
| 1.897714714 | 0 | 0.249612653 | 0 | 0.054708624 |
| 0.001532539 | 0.001868501 | 0.003881941 | 0 | 0 |
| 0.24728176 | 0 | 0 | 0.056771831 | 0.008099074 |
| 0.007810345 | 0.019750426 | 0.002706048 | 0.288740636 | 0.797943978 |
| 0.005281281 | 0 | 0 | 0.007637606 | 0.027544674 |
| 0 | 0 | 0 | 0 | 0 |
| 0.19619849 | 0.008089195 | 0.010058682 | 7.700714283 | 0 |
| 0 | 0 | 0 | 0 | 0 |
| 0 | 0 | 0 | 0 | 0 |
| 0 | 0 | 0 | 0 | 0 |
| 0 | 0 | 0 | 0 | 0 |
| 0 | 0 | 0 | 0 | 0 |
| 0.0095006 | 0 | 0.002541362 | 1.097413002 | 1.517488784 |
| 0 | 0 | 0 | 0 | 0 |
| 0 | 0 | 0 | 0 | 0 |
| 0.140030779 | 0.04245162 | 0.002846608 | 4.073410481 | 0.026787895 |

| | | | | |
|---|---|---|---|---|
| 0 | 0 | 0 | 0 | 0 |
| 0 | 0 | 0 | 0 | 0 |
| 0 | 0 | 0 | 0 | 0 |
| 0 | 0 | 0 | 0.0017734 | 0 |
| 0 | 0 | 0 | 0 | 0 |
| 0 | 0 | 0 | 0 | 0 |
| 0 | 0 | 0 | 0.0059864 | 0 |
| 0.023750071 | 0 | 0.001138917 | 0.903500339 | 0.496779999 |
| 0 | 0 | 0 | 0 | 0 |
| 0 | 0 | 0 | 0 | 0 |
| 0.103428321 | 0 | 0.001206171 | 0.038679 | 0.036889545 |
| 0 | 0 | 0 | 0 | 0 |
| 0 | 0 | 0 | 0 | 0 |
| 0 | 0 | 0 | 0 | 0 |
| 0 | 0 | 0 | 0 | 0 |
| 0 | 0 | 0 | 0 | 0 |
| 0 | 0 | 0 | 0.085444088 | 0 |
| 0.009149774 | 0.000667428 | 0 | 0.222929358 | 0.104245868 |
| 0 | 0 | 0 | 0 | 0 |
| 0 | 0 | 0 | 0.107380713 | 0 |
| 0 | 0 | 0 | 0 | 0 |
| 0 | 0 | 0 | 0 | 0 |
| 0 | 0.030779875 | 0 | 0.204732631 | 0 |
| 0 | 0 | 0 | 0 | 0 |
| 0 | 0 | 0 | 0 | 0 |
| 0 | 0 | 0 | 0 | 0 |
| 0 | 0 | 0 | 0 | 0 |
| 0 | 0 | 0 | 0 | 0 |
| 0 | 0 | 0 | 0 | 0 |
| 0.002550657 | 0.059931044 | 0 | 0.047337953 | 0 |
| 0 | 0 | 0 | 0 | 0 |
| 0 | 0 | 0 | 0 | 0 |
| 0 | 0 | 0 | 0 | 0 |
| 0 | 0 | 0 | 0 | 0 |
| 0 | 0 | 0 | 0.006282007 | 0.192957743 |
| 0 | 0 | 0 | 0 | 0 |
| 0 | 0.013587908 | 0 | 0 | 0 |
| 0.004294196 | 0 | 0 | 0 | 0.001487031 |
| 0 | 0 | 0 | 0 | 0 |
| 0 | 0 | 0 | 0 | 0.003981111 |
| 0.001119172 | 0 | 0 | 0 | 0 |
| 0 | 0 | 0 | 0 | 0 |
| 0 | 0 | 0 | 0 | 0 |
| 0.321334999 | 0.060635666 | 0 | 0 | 0 |
| 0 | 0.142825196 | 0.547230262 | 0 | 0 |
| 0 | 0 | 0 | 0 | 0 |
| 0 | 0 | 0 | 0 | 0 |
| 0 | 0 | 0 | 0 | 0 |

| | | | | |
|---|---|---|---|---|
| 0 | 0 | 0 | 0 | 0 |
| 0 | 0 | 0 | 0 | 0 |
| 0 | 0 | 0 | 0 | 0 |
| 0 | 0 | 0.002611647 | 2.2201067 | 0 |
| 0 | 0 | 0 | 0 | 0 |
| 0 | 0 | 0 | 0 | 0 |
| 0 | 0 | 0 | 0 | 0 |
| 0 | 0 | 0 | 0 | 0 |
| 0.236896536 | 0 | 0.067392032 | 0.01690997 | 0.046290925 |
| 0 | 0 | 0 | 0 | 0 |
| 0 | 0 | 0 | 0 | 0 |
| 0 | 0 | 0 | 0 | 0 |
| 0 | 0 | 0 | 0 | 0 |
| 0 | 0 | 0 | 0 | 0 |
| 0 | 0 | 0 | 0.351163768 | 0 |
| 0 | 0 | 0 | 0.005924084 | 0 |
| 0 | 0 | 0 | 0 | 0 |
| 0 | 0 | 0 | 0 | 0 |
| 0 | 0 | 0 | 0 | 0 |
| 0 | 0 | 0 | 0 | 0 |
| 0 | 0 | 0 | 0.012195433 | 0 |
| 0 | 0 | 0 | 0 | 0 |
| 0 | 0 | 0 | 0 | 0 |
| 0 | 0 | 0 | 0.036005461 | 0 |
| 0.101760391 | 0.003669108 | 0.000709711 | 0.158859177 | 0.04478978 |
| 1.021860738 | 3.303701986 | 0.020513934 | 0.09903997 | 0 |
| 0 | 0.112177595 | 0 | 0 | 0 |
| 0 | 0 | 0 | 0 | 0 |
| 0 | 0 | 0 | 0 | 0 |
| 0 | 0 | 0 | 0 | 0 |
| 0 | 0 | 0 | 0 | 0 |
| 0 | 0 | 0 | 0 | 0 |
| 0 | 0 | 0 | 0 | 0 |
| 0 | 0 | 0 | 0.022326579 | 0.110586857 |
| 0.006809582 | 0.012914808 | 0 | 0.171209382 | 0.045477257 |
| 0 | 0 | 0.002354469 | 0.070421792 | 0.052292303 |
| 0 | 0 | 0 | 0 | 0 |
| 0.755510672 | 1.50838372 | 0.851251123 | 0.103326272 | 2.345426183 |
| 0 | 0 | 0 | 0 | 0 |
| 0 | 0 | 0 | 0 | 0 |
| 0 | 0 | 0 | 0 | 0 |
| 0 | 0 | 0 | 0 | 0.026490525 |
| 0.01874673 | 6.370470544 | 0.306459565 | 1.047226181 | 0.01640028 |
| 0 | 0 | 0.119369074 | 0 | 0 |
| 0 | 0 | 0 | 0 | 0 |
| 0 | 0 | 0 | 0.065916817 | 0 |
| 0 | 0 | 0 | 0 | 0 |
| 1.489687879 | 0.020667801 | 0 | 0 | 1.025038813 |

| | | | | |
|---|---|---|---|---|
| 0.001625255 | 0 | 0.001821799 | 0.044770371 | 0 |
| 0 | 0 | 0 | 0 | 0 |
| 0 | 0 | 0 | 0 | 0 |
| 0 | 0 | 0 | 0 | 0 |
| 0 | 0 | 0 | 0.010867888 | 0.00600364 |
| 0.016621172 | 0 | 0.007463062 | 0 | 0 |
| 0 | 0 | 0 | 0 | 0 |
| 0 | 0 | 0 | 0 | 0 |
| 0 | 0 | 0 | 0 | 1.953589591 |
| 0 | 0 | 0 | 0 | 0 |
| 0 | 0 | 0 | 0.02398337 | 0 |
| 0 | 0.008965605 | 0 | 1.002486708 | 0 |
| 0 | 1.581727815 | 0 | 0.045770806 | 4.42529301 |
| 0 | 0 | 0 | 0 | 0 |
| 0.293492728 | 3.710358648 | 0.018879064 | 0.023133716 | 4.652783722 |
| 0 | 0 | 0 | 0 | 0 |
| 0 | 0 | 0 | 0 | 0 |
| 0 | 0 | 0 | 0 | 0 |
| 0 | 0 | 0 | 0.012932332 | 0 |
| 0 | 0 | 0 | 0.010799178 | 0 |
| 0 | 0.018886883 | 0 | 0 | 0 |
| 0 | 0 | 0 | 0 | 0 |
| 0 | 0.010839088 | 0.006013483 | 5.087415426 | 2.731288188 |
| 0 | 0 | 0 | 0 | 0 |
| 0 | 0 | 0 | 0 | 0 |
| 0 | 0 | 0 | 0 | 0 |
| 0 | 0 | 0 | 0 | 0 |
| 0 | 0 | 0 | 0 | 0 |
| 0 | 0 | 0 | 0 | 0 |
| 0 | 0 | 0 | 0 | 0 |
| 0 | 0 | 0 | 0 | 0 |
| 0.821595271 | 0.163112907 | 0.001484741 | 0.031910207 | 0.019461137 |
| 0 | 0 | 0 | 0 | 0 |
| 0.016377755 | 0.003067798 | 0 | 0 | 0.064456233 |
| 0.002095339 | 0 | 0.002882533 | 0.059963134 | 0.052709062 |
| 0.006077578 | 0.064996745 | 0 | 0.679133524 | 0 |
| 0 | 0 | 0 | 0 | 0.00558974 |
| 0.012272233 | 0 | 0 | 0 | 0 |
| 0 | 0 | 0 | 0.000702947 | 0 |
| 0 | 0 | 0 | 0 | 0 |
| 0 | 0 | 0 | 0 | 0 |
| 0 | 0 | 0 | 0 | 0 |
| 0 | 0 | 0 | 0 | 0 |
| 0 | 0 | 0 | 0.000622647 | 0.129299689 |
| 0.007740446 | 0.005138088 | 0.001735301 | 0.756755149 | 0.464096868 |
| 0 | 0 | 0 | 0 | 0 |
| 0 | 0 | 0 | 0 | 0 |
| 0 | 0 | 0 | 0.019098201 | 0 |

| | | | | |
|---|---|---|---|---|
| 0 | 0 | 0 | 0.026868586 | 0 |
| 0 | 0 | 0 | 0 | 0 |
| 0 | 0 | 0 | 0.007324908 | 0 |
| 0 | 0 | 0 | 0 | 0 |
| 0.004600649 | 0 | 0 | 0 | 0 |
| 0 | 0 | 0 | 0 | 0 |
| 3.443573328 | 0 | 0 | 0.718302694 | 0 |
| 0 | 0.020552529 | 0 | 0 | 0 |
| 0 | 0 | 0 | 0 | 0 |
| 0 | 0 | 0 | 0 | 0 |
| 0 | 0 | 0 | 0 | 0 |
| 0 | 0 | 0 | 0 | 0 |
| 0 | 0 | 0 | 0 | 0 |
| 0 | 0 | 0 | 0 | 0 |
| 0 | 0 | 0 | 0 | 0 |
| 0 | 0 | 0 | 0 | 0 |
| 0 | 0 | 0 | 0 | 0 |
| 0 | 0 | 0 | 0 | 0 |
| 0 | 0 | 0 | 0.192008949 | 0 |
| 0 | 0 | 0 | 0 | 0 |
| 0 | 0 | 0 | 0.006230526 | 0 |
| 0 | 0 | 0 | 0 | 0 |
| 0 | 0 | 0 | 0 | 0 |
| 0 | 0 | 0 | 0 | 0 |
| 0 | 0 | 0 | 0.001317118 | 0 |
| 0.004083435 | 0 | 0.001468636 | 0.002244557 | 0.023313947 |
| 0 | 0 | 0 | 0 | 0 |
| 0 | 0.033888496 | 0 | 0.002746219 | 1.468417414 |
| 0 | 0 | 0 | 0 | 0 |
| 0 | 0.5608715 | 0 | 0.314375645 | 1.387696036 |
| 0 | 0 | 0 | 0 | 0 |
| 0 | 0 | 0 | 0 | 0 |
| 0 | 0 | 0 | 0 | 0 |
| 0 | 0 | 0 | 0 | 0 |
| 0.006292923 | 0 | 0.000615616 | 0.005145344 | 0.026224718 |
| 0 | 0 | 0 | 0 | 0 |
| 0 | 0 | 0 | 0 | 0 |
| 0 | 0 | 0 | 0.013345048 | 0.00584974 |
| 0 | 0 | 0 | 0 | 0 |
| 0 | 0 | 0 | 0 | 0 |
| 0.140325179 | 9.172580241 | 0.447382467 | 3.916365803 | 10.2394827 |
| 1.886517804 | 0 | 0 | 0.407718421 | 11.85435201 |
| 0 | 0.154518355 | 0 | 0 | 0 |
| 0 | 0 | 0 | 0 | 0 |
| 0 | 0 | 0 | 0 | 0 |
| 0 | 0 | 0 | 0 | 0 |
| 0 | 0 | 0 | 0 | 0 |
| 0 | 0.00107482 | 0.003049217 | 0.077581772 | 0.06560305 |

| | | | | |
|---|---|---|---|---|
| 0 | 0 | 0 | 0 | 0 |
| 0 | 0 | 0 | 0.001906467 | 0 |
| 0 | 0 | 0 | 0 | 0 |
| 0 | 0 | 0 | 0 | 0 |
| 0 | 7.782273842 | 3.956886537 | 1.506278062 | 0.008124511 |
| 0 | 0 | 0 | 0 | 0 |
| 0 | 0 | 0 | 0 | 0 |
| 0 | 0 | 0 | 0 | 0 |
| 1.159460203 | 0.240211007 | 0 | 0.001631705 | 0.116927904 |
| 0 | 0 | 0 | 0 | 0 |
| 0 | 0 | 0 | 0 | 0 |
| 0 | 0 | 0 | 0 | 0 |
| 0.461959057 | 0 | 0 | 0.013364158 | 0 |
| 0.007520429 | 0.027205305 | 0 | 0.014640481 | 0.02852261 |
| 0 | 1.151447327 | 0 | 0 | 0 |
| 0 | 0.083662601 | 0.120602103 | 0.605232207 | 0 |
| 0.079539351 | 0.5005526 | 0 | 0 | 0.064602177 |
| 0 | 0 | 0 | 0 | 0 |
| 0 | 0 | 0 | 0 | 0 |
| 0 | 0 | 0 | 0 | 0 |
| 0 | 0 | 0 | 0 | 0 |
| 0 | 0 | 0 | 0 | 0 |
| 0 | 0 | 0 | 0 | 0 |
| 0 | 0 | 0 | 0 | 0 |
| 0 | 0 | 0 | 0 | 0 |
| 0 | 0 | 0 | 0 | 0 |
| 0 | 0.160784524 | 0 | 0.142966743 | 0 |
| 0.007331257 | 0 | 0 | 0.008010852 | 0.000842344 |
| 0 | 0 | 0 | 0 | 0 |
| 0 | 0 | 0 | 0 | 0 |
| 0 | 0 | 0 | 0 | 0 |
| 0 | 0 | 0 | 0 | 0 |
| 0 | 0 | 0 | 0.005061919 | 0 |
| 0 | 0 | 0 | 0 | 0 |
| 0 | 0.297784779 | 0.00161345 | 0.508416552 | 0 |
| 0.0692138 | 0 | 0.002174468 | 0 | 0 |
| 0 | 0 | 0 | 0 | 0 |
| 0.013949811 | 0 | 0 | 0 | 0 |
| 0 | 0 | 0 | 0 | 0 |
| 0 | 0 | 0 | 0 | 0 |
| 0 | 4.337295455 | 0.593885587 | 0.891331759 | 0.602075221 |
| 0.626901289 | 0 | 0 | 0 | 0 |
| 0 | 0 | 0 | 0 | 0 |
| 0 | 0 | 0 | 0 | 0 |
| 0 | 0 | 0 | 0 | 0 |
| 0 | 0 | 0 | 0 | 0 |
| 0 | 0 | 0 | 0 | 0 |
| 0 | 0 | 0 | 0 | 0 |

| | | | | |
|---|---|---|---|---|
| 0 | 0 | 0 | 0.24120614 | 0 |
| 0 | 0 | 0 | 0 | 0 |
| 0 | 0 | 0 | 0 | 0 |
| 0 | 0 | 0 | 0 | 0 |
| 0 | 0 | 0 | 0.021257731 | 0 |
| 0 | 0 | 0 | 0 | 0 |
| 0.048569336 | 0.975870228 | 0 | 0.004914879 | 0.333572934 |
| 0.001162096 | 0 | 0 | 0 | 0 |
| 0 | 0 | 0 | 0 | 0 |
| 0 | 0 | 0 | 0.002209932 | 0 |
| 0 | 0 | 0 | 0 | 0 |
| 0 | 0 | 0 | 0 | 0 |
| 0 | 0 | 0 | 0 | 0 |
| 0 | 0 | 0 | 0 | 0 |
| 0 | 0 | 0 | 0 | 0 |
| 0 | 0 | 0 | 0 | 0 |
| 0 | 0 | 0 | 0 | 0 |
| 0.096624878 | 0 | 0 | 0.000810024 | 0 |
| 0 | 0 | 0 | 0 | 0 |
| 0 | 0 | 0 | 0 | 0 |
| 0.000915658 | 0.013024526 | 0 | 0 | 0.833494684 |
| 0 | 0 | 0 | 0 | 0 |
| 0 | 0.004070369 | 0 | 0.246557116 | 0 |
| 0.004265848 | 0.001064533 | 0 | 0.089165055 | 0.059977121 |
| 0 | 0 | 0 | 0 | 0 |
| 0 | 0 | 0 | 0.003867453 | 0 |
| 0 | 0 | 0 | 0 | 0 |
| 0 | 0 | 0 | 0 | 0 |
| 0 | 0 | 0 | 0 | 0 |
| 0.023911954 | 2.564757532 | 0.339227601 | 0.001321038 | 0.2526332 |
| 0 | 0 | 0 | 0 | 0 |
| 0 | 0 | 0 | 0 | 0 |
| 0 | 0 | 0 | 0 | 0 |
| 0 | 0 | 0 | 0 | 0 |
| 0 | 0 | 0 | 0 | 0 |
| 0 | 0 | 0 | 0 | 0 |
| 0.062366589 | 0 | 0 | 0 | 0 |
| 0 | 0 | 0 | 0 | 0 |
| 0 | 0 | 0 | 0 | 0 |
| 0 | 0 | 0 | 0 | 0 |
| 0 | 0 | 0 | 0 | 0 |
| 0 | 0 | 0 | 0 | 0 |
| 0 | 0 | 0 | 0 | 0 |
| 0 | 0 | 0 | 0 | 0 |
| 0 | 0 | 0 | 0 | 0 |
| 0 | 0 | 0 | 0 | 0 |
| 0 | 0 | 0 | 0 | 0 |
| 0 | 0 | 0 | 0 | 0 |

| | | | | |
|---|---|---|---|---|
| 0.011837895 | 0 | 0.00502221 | 0.011146524 | 0.037001507 |
| 0 | 0 | 0 | 0 | 0 |
| 0 | 0 | 0 | 0 | 0 |
| 0 | 0 | 0 | 0 | 0 |
| 0 | 0 | 0 | 0 | 0 |
| 0 | 0 | 0 | 0 | 0 |
| 0 | 0 | 0 | 0 | 0 |
| 0 | 2.68866689 | 0.08159679 | 0.25656995 | 3.343533131 |
| 2.193737436 | 0.029487139 | 0.016461794 | 4.793943389 | 0.295799847 |
| 0.120645961 | 0.022030115 | 0.047777765 | 0.009096673 | 0.044899604 |
| 0 | 0 | 0 | 0 | 0 |
| 0 | 0 | 0 | 0 | 0 |
| 0 | 0 | 0 | 0 | 0 |
| 0 | 0 | 0 | 0.002623332 | 0 |
| 0 | 0 | 0 | 0 | 0.06165437 |
| 0 | 0 | 0 | 0 | 0 |
| 0 | 0 | 0 | 0 | 0 |
| 0 | 0 | 0 | 0 | 0 |
| 0 | 0 | 0 | 0 | 0 |
| 0 | 0 | 0 | 0 | 0 |
| 0 | 0 | 0 | 0 | 0 |
| 0 | 0 | 0 | 0 | 0 |
| 0 | 0 | 0 | 0 | 0 |
| 0 | 0 | 0 | 0 | 0 |
| 0 | 0 | 0 | 0 | 0 |
| 0 | 0 | 0 | 0 | 0 |
| 0 | 0 | 0 | 0 | 0 |
| 0 | 0 | 0 | 0 | 0 |
| 0 | 0 | 0 | 0 | 0 |
| 0 | 0 | 0 | 0 | 0 |
| 0.022337266 | 0 | 0 | 0 | 0 |
| 0 | 0 | 0 | 0 | 0 |
| 0 | 0 | 0 | 0 | 0 |
| 0 | 0 | 0 | 0 | 0 |
| 0 | 0 | 0 | 0 | 0 |
| 0 | 0 | 0 | 0 | 0 |
| 0 | 0 | 0 | 0 | 0 |
| 0 | 0 | 0 | 0 | 0 |
| 0 | 0 | 0 | 0.000838866 | 0 |
| 0 | 0.019440241 | 0.426020288 | 0 | 0 |
| 0 | 0 | 0 | 0 | 0 |
| 0 | 0.042471653 | 0 | 0.204754809 | 0 |
| 0 | 0 | 0 | 0 | 0 |
| 0 | 0 | 0 | 0 | 0 |
| 0 | 0 | 0 | 0 | 0 |
| 0 | 0 | 0 | 0 | 0 |
| 0 | 0 | 0 | 0 | 0 |
| 0 | 0 | 0 | 0 | 0 |

| | | | | |
|---|---|---|---|---|
| 0 | 0 | 0 | 0 | 0 |
| 0.02485716 | 0 | 0 | 0 | 0.000776639 |
| 0 | 0 | 0 | 0 | 0 |
| 0 | 0 | 0 | 0 | 0 |
| 0.065849315 | 0.301522219 | 0.005964651 | 3.324647216 | 0 |
| 0 | 0 | 0 | 0 | 0 |
| 0.004230262 | 0 | 0 | 0 | 0 |
| 0.022741649 | 0 | 0.004133809 | 0.001435867 | 0.028186427 |
| 0 | 0.000872933 | 0 | 0.011648487 | 0 |
| 0 | 0.001873165 | 0 | 0 | 0 |
| 0 | 0 | 0 | 0 | 0 |
| 0 | 0 | 0 | 0 | 0 |
| 0 | 0 | 0 | 0 | 0 |
| 0 | 0 | 0 | 0 | 0 |
| 0 | 0 | 0 | 0.002274299 | 0 |
| 0 | 0 | 0 | 1.862526729 | 0.017325719 |
| 0 | 0 | 0 | 0 | 0 |
| 0 | 0 | 0 | 0 | 0 |
| 0 | 0 | 0 | 0 | 0 |
| 0 | 0 | 0 | 0 | 0 |
| 0 | 0 | 0 | 0 | 0 |
| 0 | 0 | 0 | 0 | 0 |
| 0 | 0 | 0 | 0 | 0 |
| 0.012234151 | 0 | 0.001300899 | 0.015688135 | 0.008288894 |
| 0 | 0.004712225 | 0 | 0.137645652 | 0 |
| 0 | 0 | 0 | 0 | 0 |
| 0 | 0 | 0.099944845 | 0.001846847 | 0.206856666 |
| 0 | 0 | 0 | 0 | 0 |
| 0 | 0 | 0 | 0 | 0 |
| 0 | 0 | 0 | 0 | 0 |
| 0 | 0 | 0 | 0 | 0 |
| 0 | 0 | 0 | 0 | 0 |
| 0 | 0 | 0 | 0 | 0 |
| 0 | 0 | 0 | 0 | 0 |
| 0 | 0 | 0 | 0 | 0 |
| 0 | 0 | 0 | 0 | 0 |
| 0 | 0 | 0 | 0 | 0 |
| 0 | 0 | 0 | 0 | 0 |
| 0 | 0 | 0 | 0 | 0 |
| 0 | 0 | 0 | 0 | 0 |
| 0 | 0 | 0 | 0 | 0 |
| 0 | 0 | 0 | 2.603647875 | 0 |
| 0 | 0 | 0 | 0 | 0 |
| 0 | 0 | 0 | 0 | 0 |
| 0 | 0 | 0 | 0 | 0 |
| 0 | 0 | 0 | 0 | 0 |
| 0 | 0 | 0 | 0 | 0 |
| 0 | 0 | 0 | 0.144506377 | 0 |

| | | | | |
|---|---|---|---|---|
| 0 | 0.081997743 | 0 | 0.901265807 | 6.340687764 |
| 0 | 0 | 0 | 0 | 0 |
| 0 | 0 | 0 | 0 | 0 |
| 0 | 0 | 0 | 0 | 0 |
| 0 | 0 | 0 | 0 | 0 |
| 0 | 0 | 0 | 0.062222913 | 0 |
| 0 | 0 | 0 | 0 | 0 |
| 0 | 0 | 0 | 0 | 0 |
| 0 | 0 | 0 | 0 | 0 |
| 0 | 0 | 0 | 0 | 0 |
| 0 | 0 | 0 | 0 | 0 |
| 0 | 0 | 0 | 0 | 0 |
| 0 | 0 | 0 | 0 | 0 |
| 0 | 0 | 0 | 0 | 0 |
| 0 | 0 | 0 | 0 | 0 |
| 0 | 0 | 0 | 0 | 0 |
| 0 | 0 | 0 | 0 | 0 |
| 0 | 0 | 0 | 0 | 0 |
| 0 | 0 | 0 | 0 | 0 |
| 0 | 0 | 0 | 0 | 0 |
| 0.064093217 | 0.013059831 | 0 | 0.001856145 | 0.008018408 |
| 0 | 0 | 0 | 0 | 0 |
| 0.001003451 | 0 | 0.005726254 | 0.206287491 | 0.072673721 |
| 0 | 0 | 0 | 1.280870706 | 0 |
| 0 | 0 | 0 | 0 | 0 |
| 0 | 0 | 0 | 0 | 0 |
| 0 | 0 | 0 | 0 | 0 |
| 0 | 0 | 0 | 0 | 0 |
| 0 | 0 | 0 | 0 | 0 |
| 0 | 0 | 0 | 0 | 0 |
| 0 | 0 | 0 | 0 | 0 |
| 0 | 0 | 0 | 0 | 0 |
| 0.051371794 | 0 | 0.005745657 | 0 | 0.037655395 |
| 0 | 0 | 0 | 0 | 0 |
| 0 | 1.613614195 | 0 | 0.420067181 | 4.246499056 |
| 0.004021825 | 0.117990202 | 0 | 0.000587215 | 0.003805088 |
| 0 | 0 | 0 | 0 | 0 |
| 0 | 0 | 0 | 0 | 0 |
| 0.054368817 | 0 | 0 | 0.013753944 | 0.003624506 |
| 0 | 0 | 0 | 0 | 0 |
| 0 | 0 | 0 | 0 | 0 |
| 0 | 0 | 0.013046922 | 0 | 0 |
| 0.041815684 | 0.34204794 | 0.013303062 | 0.158045132 | 0 |
| 0 | 0 | 0 | 0 | 0 |
| 0 | 0 | 0 | 0 | 0 |
| 0 | 0.005120554 | 0 | 0.322776014 | 0 |
| 0 | 0 | 0 | 0 | 0 |
| 0 | 0 | 0 | 0 | 0 |

| | | | | |
|---|---|---|---|---|
| 0 | 0 | 0 | 0 | 0 |
| 0 | 0 | 0 | 0 | 0 |
| 0 | 0 | 0 | 0 | 0 |
| 0 | 0 | 0 | 0 | 0 |
| 0 | 0 | 0 | 0 | 0 |
| 0 | 0 | 0 | 0 | 0 |
| 0 | 0 | 0 | 0 | 0 |
| 0 | 0 | 0 | 0 | 0.000696952 |
| 0 | 0 | 0 | 0.00192054 | 0 |
| 0 | 0 | 0 | 0.005846803 | 0 |
| 0 | 0 | 0 | 0 | 0 |
| 0 | 0 | 0 | 0 | 0 |
| 0 | 0 | 0 | 0 | 0 |
| 0 | 0 | 0 | 0 | 0 |
| 0 | 0 | 0 | 0.001706255 | 0 |
| 0 | 0 | 0.000501865 | 0 | 0 |
| 0 | 0 | 0 | 0 | 0 |
| 0 | 0 | 0 | 0 | 0 |
| 0 | 0 | 0 | 0 | 0 |
| 15.79893741 | 3.031603604 | 0 | 0 | 0 |
| 0 | 0.011652342 | 0.001048325 | 0 | 0 |
| 0 | 0 | 0 | 0 | 0 |
| 0 | 0 | 0 | 0 | 0 |
| 0 | 0 | 0 | 0 | 0 |
| 0 | 0 | 0 | 0 | 0 |
| 0 | 0 | 0 | 0 | 0 |
| 0 | 0 | 0 | 0 | 0 |
| 0 | 0 | 0 | 0 | 0 |
| 0 | 0 | 0 | 0 | 0 |
| 0 | 0 | 0 | 0.079826387 | 0 |
| 0.029799009 | 0.485023026 | 0 | 1.085768549 | 0 |
| 0 | 0 | 0 | 0 | 0 |
| 0 | 0 | 0 | 0 | 0 |
| 0 | 0 | 0 | 0 | 0 |
| 0 | 0 | 0 | 0 | 0 |
| 0 | 0 | 0 | 0 | 0 |
| 0 | 0 | 0 | 0 | 0 |
| 0 | 0 | 0 | 0 | 0 |
| 0 | 0 | 0 | 0 | 0 |
| 0 | 0 | 0 | 0 | 0 |
| 0 | 0 | 0 | 0.001221597 | 0 |
| 0 | 0 | 0 | 0 | 0 |
| 0 | 0 | 0 | 0 | 0 |
| 0 | 0 | 0 | 0.001536152 | 0 |
| 0 | 0 | 0 | 0 | 0 |
| 0 | 0 | 0 | 0 | 0 |
| 0.020726676 | 0 | 0 | 0 | 0.11073718 |
| 0 | 0 | 0 | 0 | 0 |

| | | | | |
|---|---|---|---|---|
| 0 | 0 | 0 | 0 | 0 |
| 0 | 0 | 0 | 0 | 0 |
| 0 | 0 | 0 | 0 | 0 |
| 0 | 0 | 0 | 0 | 0 |
| 0.009206926 | 0.084331221 | 0 | 0.265800125 | 0.83623311 |
| 0 | 0 | 0 | 0 | 0 |
| 0 | 0 | 0 | 0 | 0 |
| 0 | 0 | 0 | 0 | 0 |
| 0 | 0 | 0 | 0 | 0 |
| 0 | 0 | 0 | 0 | 0 |
| 0 | 0 | 0 | 0 | 0 |
| 0.003776641 | 0 | 0.004085335 | 0.149985251 | 0.095196877 |
| 0 | 0 | 0 | 0 | 0.015622729 |
| 0 | 0 | 0 | 0 | 0 |
| 0.04782014 | 0.001679518 | 0 | 0 | 0 |
| 0 | 0.079050709 | 0 | 1.049317328 | 0.294708104 |
| 0 | 0 | 0 | 0.039800588 | 0 |
| 0 | 0 | 0 | 0 | 0 |
| 0 | 0 | 0 | 0.017729205 | 0.029186031 |
| 0.009765556 | 0.758816339 | 0 | 0.002012951 | 0.341182261 |
| 0 | 0 | 0 | 0 | 0 |
| 0 | 0 | 0 | 0 | 0 |
| 0 | 0 | 0 | 0 | 0 |
| 0 | 0 | 0 | 0 | 0 |
| 0 | 0 | 0 | 0 | 0 |
| 0 | 0 | 0 | 0 | 0 |
| 0.017032161 | 0 | 0 | 0.030648332 | 0.003894074 |
| 0 | 0 | 0 | 0 | 0 |
| 0 | 0 | 0 | 0 | 0 |
| 0 | 0 | 0 | 0 | 0 |
| 0.002087227 | 0 | 0 | 0.003199874 | 0.026484824 |
| 0 | 0 | 0 | 0 | 0 |
| 0 | 0 | 0 | 0 | 0 |
| 0 | 0 | 0 | 0 | 0 |
| 0 | 0 | 0 | 0 | 0 |
| 0 | 0 | 0 | 0 | 0 |
| 0 | 0 | 0 | 0 | 0 |
| 0.007698732 | 0.004345576 | 0.00336224 | 0.460975534 | 0.104299758 |
| 0 | 0.009617382 | 0 | 3.792108781 | 0 |
| 0 | 0 | 0 | 0.002547237 | 0 |
| 0 | 0 | 0 | 0.002671866 | 0.005540285 |
| 0 | 0 | 0 | 0 | 0 |
| 0 | 0 | 0 | 0 | 0 |
| 0.001630892 | 0 | 0 | 0 | 0 |
| 0 | 0 | 0 | 0 | 0 |
| 0 | 0 | 0 | 0 | 0 |
| 0 | 0 | 0 | 0 | 0 |
| 0 | 0 | 0 | 0 | 0 |

| | | | | |
|---|---|---|---|---|
| 0 | 0 | 0 | 0 | 0 |
| 0 | 0 | 0 | 0.234595631 | 0 |
| 0 | 0 | 0 | 0 | 0 |
| 0.006551864 | 0 | 0 | 0.02279248 | 0.002282928 |
| 0.015588183 | 0.002755965 | 0.000807702 | 0 | 0 |
| 0.006478693 | 0 | 0 | 0.00139401 | 0.000910894 |
| 0 | 0 | 0 | 0 | 0 |
| 0 | 0 | 0 | 0.141564623 | 0 |
| 0 | 0 | 0 | 0 | 0 |
| 0 | 0 | 0 | 0 | 0 |
| 0 | 0 | 0 | 0 | 0 |
| 0 | 0 | 0 | 0 | 0 |
| 0.001009641 | 0.009574246 | 0.012099312 | 0.791322547 | 0.036321266 |
| 0 | 0 | 0 | 0 | 0 |
| 0 | 0 | 0 | 0 | 0 |
| 0 | 0 | 0 | 0 | 0 |
| 0.210528737 | 0 | 0 | 0 | 0 |
| 0 | 0 | 0 | 0 | 0 |
| 0 | 0 | 0 | 0 | 0 |
| 0 | 0 | 0 | 0 | 0 |
| 0 | 0 | 0 | 0 | 0 |
| 0 | 0 | 0 | 0 | 0 |
| 0.06833449 | 0.001512849 | 0.005364859 | 0.008540897 | 0 |
| 0.004706443 | 0.010413748 | 0 | 0.022172815 | 0.053363883 |
| 0 | 0 | 0 | 0 | 0 |
| 0 | 0 | 0 | 0 | 0 |
| 0 | 0 | 0 | 0 | 0 |
| 0 | 0 | 0 | 0 | 0 |
| 0 | 0 | 0 | 0 | 0 |
| 0 | 0 | 0 | 0 | 0 |
| 0 | 0.019117293 | 0 | 0.008830476 | 0.041710905 |
| 0 | 0 | 0 | 0 | 0 |
| 0 | 0 | 0 | 0 | 0 |
| 0 | 0 | 0 | 0 | 0 |
| 0 | 0 | 0 | 0.018871087 | 0 |
| 0 | 0 | 0 | 0 | 0 |
| 0 | 0 | 0 | 0.000919778 | 0 |
| 0 | 0 | 0 | 0 | 0 |
| 0.028845879 | 0.030684083 | 0 | 0 | 0 |
| 0 | 0 | 0 | 0 | 0 |
| 0 | 0 | 0 | 0 | 0 |
| 0 | 0 | 0 | 0 | 0 |
| 0 | 0 | 0 | 0 | 0 |
| 0 | 0 | 0 | 0 | 0 |
| 0 | 0.305430319 | 0 | 0 | 0 |
| 0.047890217 | 0.173491862 | 0.071650484 | 0.242314212 | 0.704316124 |
| 0 | 0 | 0 | 0 | 0 |
| 0 | 0 | 0 | 0 | 0 |

| | | | | |
|---|---|---|---|---|
| 0 | 0 | 0 | 0 | 0 |
| 0 | 0 | 0 | 0 | 0 |
| 0 | 0 | 0 | 0.013881542 | 0 |
| 0 | 0 | 0 | 0 | 0 |
| 0 | 0 | 0 | 0 | 0 |
| 0 | 0 | 0 | 0 | 0 |
| 0 | 0 | 0 | 0 | 0 |
| 0 | 0 | 0 | 0 | 0 |
| 0 | 0 | 0 | 0 | 0.020873076 |
| 0.001145066 | 0 | 0 | 0 | 0 |
| 0 | 0 | 0 | 0.009151134 | 0.029898316 |
| 0 | 0 | 0 | 0.005677994 | 0.003516955 |
| 0 | 0 | 0 | 0 | 0 |
| 0 | 0 | 0 | 0 | 0 |
| 0 | 0 | 0 | 0 | 0 |
| 0 | 0 | 0 | 0 | 0 |
| 0 | 0 | 0 | 0 | 0.001065436 |
| 0 | 0 | 0 | 0 | 0 |
| 0 | 0 | 0 | 0 | 0 |
| 0.002698671 | 0 | 0 | 0.049331954 | 0 |
| 0 | 0 | 0 | 0 | 0 |
| 0 | 0 | 0 | 0 | 0 |
| 0 | 0 | 0 | 0 | 0 |
| 0 | 0 | 0 | 0 | 0 |
| 0 | 0 | 0 | 0 | 0 |
| 0 | 0 | 0 | 0 | 0 |
| 0 | 0 | 0 | 0 | 0 |
| 0 | 1.563307313 | 0 | 0 | 0 |
| 0 | 0 | 0 | 0 | 0 |
| 0 | 0 | 0 | 0 | 0 |
| 0.018547504 | 0 | 0 | 0.089501625 | 0.025186504 |
| 0 | 0 | 0 | 0 | 0 |
| 0.079412309 | 0.025271936 | 1.034612491 | 0.005188164 | 0.203030728 |
| 0.024482774 | 0.011216996 | 0 | 0.001767247 | 0 |
| 0 | 0 | 0 | 0 | 0 |
| 0 | 0 | 0 | 0 | 0 |
| 0 | 0 | 0 | 0 | 0 |
| 0 | 0 | 0 | 0 | 0 |
| 0 | 0 | 0 | 0 | 0 |
| 0 | 0 | 0 | 0 | 0 |
| 0 | 0 | 0 | 0 | 0 |
| 0.106745494 | 0 | 0.02884162 | 0.694044229 | 0 |
| 0 | 0.008443097 | 0 | 0.077368787 | 0 |
| 0 | 0 | 0 | 0 | 0 |
| 0 | 0 | 0 | 0 | 0 |
| 0.024562899 | 0.037711755 | 0 | 0.109298518 | 0 |
| 0.001278106 | 0 | 0 | 0 | 0 |
| 0 | 0 | 0 | 0 | 0 |

| | | | | |
|---|---|---|---|---|
| 0 | 0 | 0 | 0 | 0 |
| 0 | 0.010185455 | 0.010196447 | 0.001650796 | 0 |
| 0.034998329 | 0.019725113 | 0 | 0.010220009 | 0.03322251 |
| 0 | 0 | 0 | 0 | 0 |
| 0 | 0 | 0.002417792 | 0.881607944 | 0 |
| 0 | 0 | 0 | 0 | 0 |
| 0 | 0 | 0 | 0 | 0 |
| 0 | 0 | 0 | 0 | 0 |
| 0 | 0 | 0 | 0 | 0 |
| 0 | 0 | 0 | 0 | 0 |
| 0 | 0 | 0 | 0 | 0 |
| 0 | 0 | 0 | 0 | 0 |
| 0 | 0 | 0 | 0 | 0 |
| 0 | 0 | 0 | 0 | 0 |
| 0 | 0 | 0 | 0 | 0 |
| 0 | 0 | 0 | 0 | 0 |
| 0 | 0 | 0 | 0 | 0 |
| 0 | 0 | 0 | 0 | 0 |
| 0 | 0 | 0 | 0 | 0 |
| 0 | 0 | 0 | 0 | 0 |
| 0 | 0 | 0 | 0 | 0 |
| 0 | 0 | 0 | 0 | 0 |
| 0 | 0 | 0 | 0 | 0 |
| 0 | 0 | 0 | 0.002443932 | 0 |
| 0.117823155 | 0.004902684 | 0 | 0.028764398 | 0 |
| 0.002189881 | 0.007268186 | 0.031632234 | 0 | 0.001949993 |
| 0 | 0 | 0 | 0 | 0 |
| 0 | 0 | 0 | 0.034582525 | 0 |
| 0 | 0 | 0 | 0 | 0 |
| 0 | 0 | 0 | 0 | 0 |
| 0 | 0 | 0 | 0 | 0 |
| 0 | 0 | 0 | 0 | 0 |
| 0 | 0 | 0 | 0 | 0 |
| 0 | 0 | 0 | 0 | 0 |
| 0 | 0 | 0 | 0 | 0 |
| 0 | 0 | 0 | 0 | 0 |
| 0 | 0 | 0 | 0 | 0 |
| 0 | 0 | 0 | 0 | 0 |
| 0.337503714 | 0 | 0 | 0 | 0 |
| 0 | 0 | 0 | 0 | 0 |
| 0 | 0 | 0 | 0 | 0 |
| 0.001484802 | 0 | 0 | 0.090358754 | 0 |
| 0.008516415 | 0 | 0.003191437 | 0.010428995 | 0.032535638 |
| 0 | 0 | 0 | 0 | 0 |
| 0 | 0 | 0 | 0 | 0 |
| 0 | 0 | 0 | 0 | 0.010694599 |
| 0 | 0 | 0 | 0.093271416 | 0 |
| 0 | 0 | 0 | 0 | 0 |

| | | | | |
|---|---|---|---|---|
| 0 | 0 | 0 | 0 | 0 |
| 0.037121464 | 0.006788883 | 0.006160532 | 0.022609132 | 0.023138526 |
| 0 | 0 | 0 | 0 | 0 |
| 0 | 0 | 0 | 0 | 0 |
| 0 | 0 | 0 | 0 | 0 |
| 0 | 0 | 0 | 0 | 0 |
| 0 | 0 | 0 | 0 | 0 |
| 0 | 0 | 0 | 0 | 0 |
| 0 | 0 | 0 | 0 | 0 |
| 0 | 0 | 0 | 0 | 0 |
| 0 | 0 | 0 | 0 | 0 |
| 0 | 0 | 0 | 0 | 0 |
| 0 | 0.033490105 | 0 | 0.004419836 | 0 |
| 0 | 0 | 0 | 0 | 0 |
| 0 | 0.002508063 | 0.014553956 | 0.009461088 | 0.007065369 |
| 0 | 0 | 0 | 0 | 0 |
| 0 | 0 | 0 | 0 | 0 |
| 0 | 0 | 0 | 0 | 0 |
| 0 | 0 | 0 | 0.010893613 | 0 |
| 0 | 0 | 0 | 0 | 0 |
| 0 | 0 | 0 | 0 | 0 |
| 0.365591263 | 0.32174456 | 0 | 0.000948116 | 0.185900666 |
| 0 | 0 | 0 | 0.001019969 | 0 |
| 0 | 0 | 0 | 0.000976555 | 0 |
| 0 | 0 | 0 | 0 | 0 |
| 0 | 0 | 0 | 0 | 0 |
| 0 | 0 | 0 | 0 | 0 |
| 0 | 0 | 0 | 0 | 0 |
| 0 | 0 | 0 | 0 | 0 |
| 0 | 0 | 0 | 0 | 0 |
| 0 | 0 | 0 | 0 | 0 |
| 0 | 0 | 0 | 0 | 0 |
| 0 | 0 | 0 | 0 | 0 |
| 0 | 0 | 0 | 0 | 0 |
| 0 | 0.001397733 | 11.82725036 | 0 | 0 |
| 0 | 0 | 0 | 0 | 0 |
| 0 | 0 | 0 | 0 | 0 |
| 0 | 0 | 0 | 0 | 0 |
| 0.019129787 | 0.003689579 | 0 | 0.000568085 | 0.009238908 |
| 0 | 0 | 0 | 0 | 0 |
| 0.002035029 | 0.038871302 | 0.019374768 | 0.623630525 | 0.088534214 |
| 0 | 0 | 0 | 0 | 0 |
| 0 | 0 | 0 | 0 | 0 |
| 0 | 0 | 0 | 0 | 0 |
| 0 | 0 | 0 | 0 | 0 |
| 2.290359748 | 0.486714206 | 0.368763452 | 0.013218891 | 0.385604415 |
| 0 | 0 | 0 | 0 | 0 |
| 0 | 0 | 0 | 0 | 0 |

| | | | | |
|---|---|---|---|---|
| 0 | 0 | 0 | 0 | 0 |
| 0 | 0 | 0 | 0 | 0 |
| 0 | 0 | 0 | 0 | 0 |
| 0 | 0 | 0 | 0 | 0 |
| 0.002019549 | 0 | 0 | 0 | 0 |
| 0 | 0 | 0 | 0 | 0 |
| 0 | 0 | 0 | 0 | 0 |
| 0 | 0.002729283 | 0 | 0 | 0 |
| 0 | 0 | 0 | 0 | 0 |
| 0.028321155 | 1.405178378 | 0 | 0.001330833 | 0.001159483 |
| 0 | 0 | 0 | 0 | 0 |
| 0 | 0 | 0 | 0 | 0 |
| 0 | 0 | 0 | 0.001114952 | 0 |
| 0 | 0 | 0 | 0 | 0 |
| 0 | 0 | 0 | 0 | 0 |
| 0 | 0 | 0 | 0 | 0 |
| 0 | 0 | 0 | 0 | 0 |
| 0 | 0 | 0 | 0 | 0 |
| 0 | 0 | 0 | 0 | 0 |
| 1.180383714 | 0.110107489 | 0.26544539 | 6.747564512 | 0.563114587 |
| 0 | 0 | 0 | 0 | 0 |
| 0 | 0 | 0 | 0 | 0 |
| 0 | 0 | 0 | 0 | 0 |
| 0 | 0 | 0 | 0 | 0 |
| 0 | 0 | 0 | 0 | 0 |
| 0 | 0 | 0.001384418 | 0 | 0 |
| 0 | 0 | 0 | 0 | 0 |
| 0 | 0 | 0 | 0 | 0 |
| 0 | 0 | 0 | 0 | 0 |
| 0.018770311 | 0 | 0 | 0 | 0 |
| 0 | 0 | 0 | 0 | 0 |
| 0 | 0 | 0 | 0 | 0 |
| 0 | 0 | 0 | 0 | 0 |
| 0 | 0 | 0 | 0 | 0.005248953 |
| 0 | 0 | 0 | 0 | 0 |
| 0 | 0 | 0 | 0 | 0.02709446 |
| 0 | 0 | 0 | 0 | 0 |
| 0 | 0 | 0 | 0 | 0 |
| 0 | 0 | 0 | 0 | 0 |
| 0 | 0 | 0 | 0 | 0 |
| 0 | 0 | 0 | 0 | 0 |
| 0 | 0 | 0 | 0 | 0 |
| 0 | 0 | 0 | 0.069155 | 0 |
| 0 | 0 | 0 | 0 | 0 |
| 0.00079798 | 0 | 0 | 0.041167113 | 0.036475718 |
| 0 | 0 | 0 | 0.03489305 | 0 |
| 0 | 0 | 0 | 0 | 0 |
| 0 | 0 | 0.028815705 | 0 | 0 |

| | | | | |
|---|---|---|---|---|
| 0 | 0 | 0 | 0 | 0 |
| 0 | 0 | 0 | 0 | 0 |
| 0 | 0 | 0 | 0 | 0 |
| 0 | 0 | 0 | 0 | 0 |
| 0 | 0 | 0 | 0 | 0 |
| 0 | 0 | 0 | 0.277639412 | 0.002298682 |
| 0 | 0 | 0 | 0 | 0 |
| 0 | 0 | 0 | 0 | 0 |
| 0 | 0 | 0 | 0 | 0 |
| 0 | 0 | 0 | 0 | 0 |
| 0 | 0 | 0 | 0 | 0 |
| 0 | 0 | 0 | 0 | 0 |
| 0 | 0 | 0 | 0 | 0 |
| 0 | 0 | 0 | 0 | 0 |
| 0 | 0 | 0 | 0 | 0 |
| 0 | 0 | 0 | 0 | 0 |
| 0 | 0 | 0 | 0 | 0 |
| 0 | 0 | 0 | 0 | 0 |
| 0 | 0 | 0 | 0 | 0 |
| 0 | 0 | 0 | 0 | 0 |
| 0 | 0 | 0 | 0 | 0 |
| 0 | 0 | 0 | 0 | 0 |
| 0 | 0 | 0 | 0 | 0 |
| 0 | 0 | 0 | 0 | 0 |
| 0 | 0 | 0 | 0 | 0 |
| 0 | 0.035639305 | 0 | 0 | 0 |
| 0 | 0 | 0 | 0 | 0 |
| 0 | 0 | 0 | 0 | 0 |
| 0 | 0 | 0 | 0 | 0 |
| 0 | 0 | 0 | 0 | 0 |
| 0 | 0 | 0 | 0 | 0 |
| 0 | 0 | 0 | 0 | 0.000999378 |
| 0 | 0 | 0 | 0 | 0 |
| 0 | 0 | 0 | 0 | 0 |
| 0 | 0 | 0 | 0 | 0 |
| 0 | 0 | 0 | 0 | 0 |
| 0 | 0 | 0 | 0 | 0 |
| 0 | 0 | 0 | 0 | 0 |
| 0 | 1.059778757 | 0 | 0 | 0 |
| 0 | 0 | 0 | 0 | 0 |
| 0 | 0 | 0 | 0 | 0 |
| 0 | 0 | 0 | 0 | 0 |
| 0 | 0 | 0 | 0 | 0 |
| 0 | 0 | 0 | 0 | 0 |
| 0 | 0 | 0 | 0 | 0 |
| 0 | 0 | 0 | 0 | 0 |
| 0 | 0 | 0 | 0 | 0 |
| 0 | 0 | 0 | 0 | 0 |

| | | | | |
|---|---|---|---|---|
| 0 | 0 | 0 | 0 | 0 |
| 0 | 0.01707658 | 0 | 0.021293948 | 0 |
| 0 | 0 | 0 | 0 | 0 |
| 0 | 0 | 0 | 0 | 0 |
| 0 | 0 | 0 | 0 | 0 |
| 0 | 0 | 0 | 0 | 0 |
| 0 | 0 | 0 | 0 | 0.011189565 |
| 0 | 0 | 0 | 0 | 0 |
| 0 | 0 | 0 | 0 | 0 |
| 0 | 0 | 0 | 0 | 0 |
| 0.002328882 | 0.001656328 | 0 | 0.075997459 | 0.002332989 |
| 0 | 0 | 0 | 0 | 0 |
| 0 | 0.001016641 | 0 | 0 | 0 |
| 0 | 0 | 0 | 0 | 0 |
| 0 | 0 | 0 | 0 | 0 |
| 0 | 0 | 0 | 0 | 0 |
| 0 | 0 | 0 | 0 | 0 |
| 0 | 0 | 0 | 0 | 0 |
| 0 | 0 | 0 | 0 | 0 |
| 0 | 0 | 0 | 0 | 0 |
| 0.010045621 | 0.015129659 | 0 | 0.260733182 | 0 |
| 0 | 0 | 0 | 0 | 0 |
| 0 | 0 | 0 | 0 | 0 |
| 2.204050121 | 4.095173467 | 0 | 0.711727652 | 1.508409137 |
| 0 | 0 | 0 | 0 | 0 |
| 0 | 0 | 0 | 0 | 0 |
| 0 | 0 | 0 | 0 | 0 |
| 0 | 0 | 0 | 0 | 0 |
| 0 | 0 | 0 | 0 | 0 |
| 0 | 0 | 0 | 0 | 0 |
| 0 | 0 | 0 | 0 | 0 |
| 0 | 0 | 0 | 0 | 0 |
| 0 | 0 | 0 | 0 | 0 |
| 0 | 0 | 0 | 0 | 0 |

| PHL018-0146_V1 | PHL018-0252_V2 | PHL018-0194_V2 | PHL018-0166_V2 | PHL018-0278_V1 |
|---|---|---|---|---|
| CTRL | REF | EYCF | EYCF | REF |
| 0.390403839 | 0.1093524 | 0.601552275 | 0.003835067 | 0.005950162 |
| 1.086014236 | 3.792488242 | 0.616599732 | 0.16517942 | 0.031185181 |
| 10.6345844 | 0 | 2.367618133 | 0.071427197 | 0.001719626 |
| 0 | 0 | 0 | 0 | 0 |
| 0 | 0 | 0 | 0 | 0 |
| 0 | 0 | 0 | 0 | 0 |
| 0 | 0 | 0 | 0 | 0 |
| 0 | 0 | 0 | 0 | 0 |
| 0 | 0 | 0 | 0 | 0 |
| 0 | 0 | 0 | 0 | 0 |
| 0 | 0 | 0 | 0 | 0 |
| 0 | 0 | 0 | 0 | 0 |
| 0 | 0 | 0 | 0 | 0 |
| 4.057511194 | 2.76173118 | 15.22143826 | 1.091430367 | 0 |
| 0 | 0 | 0 | 0 | 0 |
| 0 | 0 | 0 | 0 | 0 |
| 0 | 0 | 0 | 0 | 0 |
| 0 | 0 | 0 | 0 | 0 |
| 0 | 0 | 0 | 0 | 0 |
| 0 | 0 | 0 | 0 | 0 |
| 0 | 0 | 0 | 0 | 0 |
| 0 | 0 | 0 | 0 | 0 |
| 0 | 0 | 0 | 0 | 0 |
| 0 | 0 | 0 | 0 | 0 |
| 0 | 0 | 0.032529404 | 0 | 0 |
| 0 | 0 | 0 | 0 | 0 |
| 0 | 0 | 0 | 0 | 0 |
| 0 | 0 | 0 | 0 | 0 |
| 0 | 0 | 0 | 0 | 0 |
| 0 | 0 | 0 | 0 | 0 |
| 0 | 0 | 0 | 0 | 0 |
| 0 | 0 | 0 | 0 | 0 |
| 0 | 0 | 0 | 0 | 0 |
| 0 | 0 | 0 | 0 | 0 |
| 0 | 0 | 0 | 0 | 0 |
| 0.031543881 | 0.330977468 | 4.291687192 | 16.96061846 | 0 |
| 0 | 0 | 0 | 0 | 0 |
| 0 | 0 | 0 | 0 | 0 |
| 0 | 0 | 0 | 0 | 0 |
| 0 | 0 | 0 | 0 | 0 |
| 0 | 0 | 0 | 0 | 0 |
| 0.014365222 | 0.01223785 | 0.015485739 | 0.002778609 | 0.007113236 |
| 0.02360086 | 0.00232149 | 0.160182728 | 0 | 0 |
| 0.007361061 | 0.010702418 | 0.014732594 | 0.022325521 | 0.028099488 |
| 0 | 0 | 0.001036261 | 0 | 0 |
| 0 | 0 | 0 | 0 | 0 |

| | | | | |
|---|---|---|---|---|
| 0 | 0.004375082 | 0 | 0 | 0 |
| 0.003244701 | 0 | 0.003079819 | 0 | 0 |
| 0 | 0 | 0.025083073 | 0.000726503 | 0.001127179 |
| 0.33457385 | 0.384445817 | 0.274689855 | 0.685377886 | 0.853713175 |
| 0 | 0 | 0.00899345 | 0 | 0 |
| 0 | 0.060471732 | 0.026691076 | 0 | 0 |
| 0.428058787 | 0.106308338 | 0.016835455 | 0 | 16.33012485 |
| 0 | 0.002577351 | 0 | 0 | 0 |
| 0 | 0 | 0 | 0 | 0 |
| 0 | 0 | 0 | 0 | 0 |
| 0 | 0 | 0 | 0 | 0.012754127 |
| 0 | 0.515413967 | 0.178684061 | 0 | 0.018561428 |
| 0.003012846 | 0 | 0 | 0 | 0 |
| 0.208387601 | 0.063592268 | 0.001773092 | 0.00134894 | 0.012557396 |
| 0.010251729 | 0 | 0 | 0 | 0 |
| 0 | 0 | 0 | 0 | 0 |
| 0 | 0 | 1.524710177 | 0 | 0 |
| 0 | 0 | 0 | 0 | 0 |
| 0 | 0 | 0 | 0 | 0 |
| 0 | 0 | 0 | 0 | 0 |
| 0.092993471 | 0.108120454 | 0.044716869 | 0.175801061 | 0.001382219 |
| 0.092665833 | 0.123127349 | 0.00258007 | 0 | 0 |
| 0 | 0.146182101 | 0 | 0 | 0 |
| 0 | 0 | 0 | 0 | 0 |
| 0 | 0 | 0 | 0 | 0 |
| 0.001192965 | 0 | 0 | 0 | 0 |
| 0.014261074 | 0 | 0 | 0 | 0 |
| 0 | 0 | 0 | 0 | 0 |
| 0.002634364 | 0 | 0.003300656 | 0 | 0 |
| 0 | 0 | 0 | 0 | 0 |
| 0.013533412 | 0.004452666 | 0.049433803 | 0 | 0.050291621 |
| 0 | 0 | 0 | 0 | 0 |
| 0 | 0 | 0 | 0 | 0 |
| 0 | 0 | 0 | 0 | 0 |
| 0 | 0 | 0 | 0 | 0 |
| 0 | 0 | 0 | 0 | 0 |
| 0 | 0 | 0 | 0 | 0 |
| 0 | 0 | 0.029721764 | 0.183653612 | 0 |
| 0 | 0 | 0 | 0 | 0 |
| 0 | 0 | 0 | 0 | 0 |
| 0 | 0 | 0 | 0 | 0 |
| 0 | 0 | 0 | 0 | 0 |
| 0 | 0 | 0 | 0 | 0 |
| 0 | 0 | 0 | 0 | 0 |
| 0 | 0 | 0 | 0 | 0 |
| 0 | 0 | 0 | 0.001125876 | 0 |
| 0 | 0.0040486 | 0.00913456 | 0 | 0 |
| 0 | 0 | 0 | 0 | 0.008915932 |

| | | | | |
|---|---|---|---|---|
| 0 | 0 | 0 | 0 | 0 |
| 0 | 0 | 0 | 0 | 0 |
| 0 | 0 | 0 | 0 | 0 |
| 0 | 0 | 0 | 0 | 0 |
| 0 | 0.044977548 | 0.004271699 | 0 | 0 |
| 0 | 0 | 0 | 0 | 0 |
| 0 | 0 | 0 | 0 | 0 |
| 0 | 0 | 0 | 0 | 0 |
| 0 | 0 | 0.007028095 | 0.001077997 | 0.017310654 |
| 0 | 0 | 0 | 0 | 0 |
| 0 | 0 | 0 | 0 | 0 |
| 0.003004086 | 0 | 0.001974068 | 0 | 0 |
| 0 | 0 | 0 | 0 | 0 |
| 0 | 0 | 0 | 0 | 0 |
| 0 | 0 | 0 | 0 | 0 |
| 0 | 0 | 0 | 0 | 0 |
| 0 | 0 | 0 | 0 | 0 |
| 0 | 0 | 0 | 0 | 0 |
| 0 | 0.114244631 | 0 | 0 | 0 |
| 0 | 0 | 0 | 0 | 0 |
| 0 | 0 | 0 | 0 | 0 |
| 0 | 0 | 0 | 0 | 0 |
| 0 | 0 | 0 | 0 | 0 |
| 0 | 0 | 0 | 0 | 0 |
| 0 | 0 | 0 | 0 | 0 |
| 0 | 0 | 0 | 0 | 0 |
| 0 | 0 | 0.000664453 | 0 | 0 |
| 0.001875619 | 0 | 0 | 0 | 0 |
| 0 | 0 | 0 | 0 | 0 |
| 0 | 0 | 0 | 0 | 0 |
| 0.075516848 | 0 | 0.007762355 | 0 | 0 |
| 0 | 0 | 0 | 0 | 0 |
| 0 | 0 | 0 | 0 | 0 |
| 0 | 0 | 0 | 0 | 0 |
| 0 | 0 | 0 | 0 | 0 |
| 0 | 0 | 0 | 0 | 0 |
| 0 | 0 | 0 | 0 | 0 |
| 0 | 0 | 0 | 0 | 0 |
| 0 | 0 | 0 | 0.010972958 | 0 |
| 0 | 0 | 0 | 0 | 0 |
| 0 | 0 | 0 | 0 | 0 |
| 0 | 0 | 0 | 0 | 0 |
| 0 | 0 | 0.001907932 | 0 | 0.001407538 |
| 0 | 0 | 0 | 0 | 0 |
| 0 | 0 | 0 | 0 | 0 |
| 0 | 0 | 0 | 0 | 0 |
| 0 | 0 | 0 | 0 | 0 |
| 0 | 0 | 0 | 0 | 0 |

| | | | | |
|---|---|---|---|---|
| 0 | 0 | 0 | 0 | 0 |
| 0 | 0 | 0 | 0 | 0 |
| 0 | 0 | 0 | 0 | 0 |
| 0 | 0 | 0 | 0 | 0 |
| 0 | 0.059730613 | 0 | 0 | 0 |
| 0 | 0 | 0 | 0 | 0 |
| 0 | 0 | 0 | 0 | 0 |
| 0 | 0 | 0 | 0 | 0 |
| 0 | 0 | 0 | 0 | 0 |
| 0 | 0 | 0 | 0 | 0 |
| 0 | 0 | 0 | 0 | 0 |
| 0 | 0 | 0 | 0 | 0 |
| 0 | 0 | 0 | 0 | 0 |
| 0 | 0 | 0 | 0 | 0 |
| 0 | 0 | 0 | 0 | 0 |
| 1.993758764 | 0 | 0 | 0 | 0 |
| 0 | 0 | 0 | 0 | 0 |
| 0 | 0 | 0 | 0 | 0 |
| 0 | 0 | 0 | 0 | 0 |
| 0 | 0 | 0 | 0 | 0 |
| 0 | 0 | 0 | 0 | 0 |
| 0 | 0 | 0 | 0 | 0 |
| 0 | 0 | 0 | 0 | 0 |
| 0 | 0 | 0 | 0 | 0 |
| 0 | 0 | 0 | 0 | 0 |
| 0 | 0 | 0 | 0 | 0 |
| 0 | 0 | 0 | 0 | 0 |
| 0 | 0 | 0 | 0 | 0 |
| 0 | 0 | 0 | 0 | 0 |
| 0 | 0 | 0 | 0 | 0 |
| 0 | 0 | 0 | 0 | 0 |
| 0 | 0 | 0 | 0 | 0 |
| 0 | 0.000945419 | 0.000761814 | 0.00096596 | 0 |
| 0 | 0 | 0 | 0 | 0 |
| 0 | 0 | 0 | 0 | 0 |
| 0 | 0 | 0 | 0 | 0 |
| 0 | 0 | 0 | 0 | 0 |
| 0 | 0 | 0 | 0 | 0 |
| 0 | 0 | 0 | 0 | 0 |
| 0 | 0 | 0 | 0 | 0 |
| 0 | 0 | 0 | 0 | 0 |
| 0 | 0 | 0 | 0 | 0 |
| 0 | 0 | 0 | 0 | 0 |
| 0 | 0 | 0 | 0 | 0 |
| 0 | 0 | 0 | 0 | 0 |
| 0 | 0 | 0 | 0 | 0 |
| 0 | 0 | 0 | 0 | 0 |
| 0 | 0 | 0 | 0 | 0 |

| | | | | |
|---|---|---|---|---|
| 0 | 0 | 0 | 0 | 0 |
| 0 | 0 | 0 | 0 | 0 |
| 0 | 0 | 0 | 0 | 0 |
| 0 | 0 | 0 | 0 | 0 |
| 0 | 0 | 0 | 0 | 0 |
| 0 | 0 | 0 | 0 | 0 |
| 0 | 0 | 0 | 0 | 0 |
| 0 | 0 | 0 | 0 | 0 |
| 0 | 0 | 0 | 0 | 0 |
| 0 | 0 | 0 | 0 | 0 |
| 0 | 0 | 0 | 0 | 0 |
| 0 | 0 | 0 | 0 | 0 |
| 0 | 0 | 0.002672933 | 0 | 0 |
| 0 | 0 | 0 | 0 | 0 |
| 0 | 0 | 0 | 0 | 0 |
| 0 | 0 | 0 | 0 | 0 |
| 0 | 0 | 0 | 0 | 0 |
| 0 | 0 | 0 | 0 | 0 |
| 0 | 0 | 0 | 0 | 0 |
| 0 | 0 | 0 | 0 | 0 |
| 0.03909088 | 0.079078658 | 0.032712492 | 0.118818821 | 0.081951552 |
| 0 | 0 | 0 | 0 | 0 |
| 0 | 0 | 0 | 0 | 0 |
| 0 | 0 | 0.003079605 | 0 | 0 |
| 0 | 0 | 0 | 0 | 0 |
| 0 | 0 | 0 | 0 | 0 |
| 0.001161511 | 0.000985103 | 0.008467104 | 0.005284156 | 0.001171206 |
| 0 | 0 | 0 | 0 | 0 |
| 0 | 0 | 0 | 0 | 0 |
| 0 | 0 | 0 | 0 | 0 |
| 0.126666473 | 0 | 0 | 0 | 0.030354836 |
| 0 | 0 | 0 | 0 | 0 |
| 0 | 0 | 0 | 0 | 0 |
| 0 | 0.056856428 | 0 | 0.003432694 | 0 |
| 0.026991831 | 0.006053269 | 0.042125555 | 0 | 0.030423009 |
| 0 | 0 | 0 | 0 | 0 |
| 0 | 0 | 0 | 0 | 0 |
| 0 | 0 | 0 | 0 | 0 |
| 0 | 0 | 0 | 0 | 0 |
| 0 | 0 | 0 | 0 | 0 |
| 0 | 0 | 0 | 0 | 0 |
| 0 | 0 | 0 | 0 | 0 |
| 0 | 0 | 0 | 0 | 0 |
| 0 | 0 | 0 | 0 | 0 |
| 0 | 0 | 0 | 0 | 0 |
| 0 | 0 | 0.076979437 | 0 | 0 |
| 0.050300679 | 0 | 0 | 0 | 0 |
| 0.001664577 | 0 | 0.000631996 | 0 | 0 |

| | | | | |
|---|---|---|---|---|
| 0 | 0 | 0 | 0 | 0 |
| 0 | 0 | 0 | 0 | 0 |
| 0.004056148 | 0 | 0 | 0 | 0 |
| 0 | 0 | 0 | 0 | 0 |
| 0 | 0 | 0 | 0 | 0 |
| 0 | 0 | 0 | 0 | 0 |
| 0 | 0 | 0 | 0 | 0 |
| 0.00079762 | 0 | 0.017564445 | 0 | 0 |
| 0 | 0.002897889 | 0.003009691 | 0 | 0 |
| 0 | 0 | 0 | 0 | 0 |
| 0 | 0 | 0 | 0 | 0 |
| 0 | 0 | 0 | 0 | 0 |
| 0 | 0 | 0 | 0 | 0 |
| 0 | 0 | 0 | 0 | 0 |
| 0 | 0 | 0 | 0 | 0.001837773 |
| 0 | 0 | 0 | 0 | 0 |
| 0 | 0 | 0 | 0 | 0 |
| 0 | 0 | 0 | 0 | 0 |
| 0 | 0 | 0.00203533 | 0 | 0 |
| 0 | 0 | 0 | 0 | 0 |
| 0.009988584 | 0 | 0 | 0.004508124 | 0.025179907 |
| 0 | 0 | 0 | 0 | 0 |
| 0 | 0 | 0 | 0 | 0 |
| 0 | 0 | 0 | 0 | 0 |
| 0 | 0.005259531 | 0 | 0.038213723 | 0 |
| 0 | 0 | 0 | 0 | 0 |
| 0 | 0 | 0 | 0 | 0 |
| 0 | 0 | 0 | 0 | 0 |
| 0 | 0 | 0 | 0 | 0.012597998 |
| 0 | 0 | 0 | 0 | 0 |
| 0.159949107 | 0.089110992 | 0.319986349 | 0.002651857 | 0.692246886 |
| 0 | 0 | 0 | 0 | 0 |
| 0 | 0 | 0.047242586 | 0 | 0 |
| 0 | 0.004197441 | 0.004209057 | 0 | 0 |
| 0 | 0 | 0 | 0 | 0 |
| 0 | 0 | 0 | 0 | 0 |
| 0 | 0 | 0 | 0 | 0 |
| 0 | 0 | 0 | 0 | 0 |
| 0 | 0 | 0 | 0 | 0 |
| 0 | 0 | 0 | 0 | 0 |
| 0 | 0 | 0 | 0 | 0 |
| 0 | 0.002866116 | 0.021673797 | 0.002027346 | 0.007077276 |
| 0 | 0 | 0 | 0 | 0 |
| 0 | 0 | 0 | 0 | 0 |
| 0 | 0 | 0 | 0 | 0 |
| 0 | 0 | 0 | 0 | 0 |
| 0 | 0 | 0 | 0 | 0 |
| 0 | 0 | 0 | 0 | 0 |

| | | | | |
|---|---|---|---|---|
| 0 | 0 | 0 | 0 | 0 |
| 0 | 0 | 0 | 0 | 0 |
| 0.005483718 | 0.009689294 | 0.014722879 | 0.018385367 | 0.026660052 |
| 0 | 0 | 0 | 0 | 0 |
| 0 | 0 | 0 | 0 | 0 |
| 0 | 0 | 0 | 0 | 0 |
| 0 | 0 | 0 | 0 | 0 |
| 0 | 0 | 0 | 0 | 0 |
| 0 | 0 | 0 | 0 | 0 |
| 0.004537712 | 0 | 0 | 0 | 0 |
| 0 | 0 | 0 | 0 | 0 |
| 0 | 0 | 0 | 0 | 0 |
| 0 | 0 | 0 | 0 | 0 |
| 0.003174547 | 0.004589323 | 0.003944591 | 0.005939444 | 0 |
| 0.000714593 | 0.023232361 | 0.003527056 | 0.007224344 | 0.002702091 |
| 0 | 0 | 0 | 0 | 0 |
| 0 | 0.002014112 | 0 | 0.005585655 | 0 |
| 0 | 0 | 0 | 0 | 0.00159218 |
| 0 | 0 | 0 | 0 | 0 |
| 0 | 0 | 0 | 0 | 0 |
| 0 | 0 | 0 | 0 | 0 |
| 0.001960008 | 0 | 0.000697653 | 0 | 0 |
| 0 | 0 | 0 | 0 | 0 |
| 0 | 0 | 0 | 0 | 0 |
| 0 | 0 | 0.015661047 | 0 | 0 |
| 0 | 0 | 0 | 0 | 0 |
| 0 | 0 | 0 | 0 | 0 |
| 0 | 0 | 0 | 0 | 0 |
| 0 | 0 | 0 | 0 | 0 |
| 0 | 0 | 0.029467525 | 0 | 0 |
| 0 | 0 | 0 | 0 | 0 |
| 0 | 0 | 0 | 0 | 0 |
| 0 | 0 | 0 | 0 | 0 |
| 0 | 0 | 0.000804392 | 0 | 0 |
| 0 | 0 | 0 | 0 | 0 |
| 0 | 0 | 0.011426398 | 0 | 0 |
| 0 | 0 | 0 | 0 | 0 |
| 0.02277856 | 0.006526688 | 0.00210367 | 0.00500137 | 0 |
| 0 | 0 | 0 | 0 | 0 |
| 0 | 0 | 0 | 0 | 0 |
| 0 | 0 | 0 | 0 | 0 |
| 0 | 0 | 0 | 0 | 0 |
| 0 | 0 | 0 | 0 | 0 |
| 0 | 0 | 0 | 0 | 0 |
| 0 | 0 | 0 | 0 | 0 |
| 0 | 0 | 0.007007778 | 0 | 0 |
| 0 | 0 | 0.001710226 | 0 | 0 |
| 0.006711215 | 0 | 0 | 0 | 0 |

| | | | | |
|---|---|---|---|---|
| 0.639566012 | 0 | 0 | 0 | 0 |
| 0.002369037 | 0 | 0.329472527 | 0 | 0.008659444 |
| 0 | 0 | 0.001315327 | 0.001111867 | 0 |
| 0 | 0 | 0 | 0 | 0 |
| 0.003614575 | 0.002463429 | 0.003823 | 0.037474611 | 0.002277967 |
| 0 | 0 | 0.013426534 | 0 | 0 |
| 0 | 0 | 0 | 0 | 0 |
| 0.064418479 | 0.024171262 | 0.0457557 | 0.210899676 | 0 |
| 0 | 0 | 0 | 0 | 0 |
| 0.037674998 | 0.013445566 | 0.547454522 | 0.043637393 | 2.550185367 |
| 0 | 0 | 0 | 0 | 0 |
| 0.006016134 | 0.003340867 | 0.065098783 | 0 | 0 |
| 0.002607686 | 0.005298711 | 0.028588269 | 0.022596882 | 0.000821704 |
| 0 | 0.007347604 | 0 | 0 | 0 |
| 0 | 0 | 0 | 0 | 0 |
| 0 | 0 | 0 | 0 | 0.002055858 |
| 0 | 0 | 0.000540407 | 0 | 0 |
| 0 | 0 | 0 | 0 | 0 |
| 0.01895426 | 0 | 0.573115997 | 0 | 0.03229655 |
| 0 | 0 | 0 | 0 | 0 |
| 0 | 0 | 0 | 0 | 0 |
| 0 | 0 | 0 | 0 | 0 |
| 0 | 0 | 0 | 0 | 0 |
| 0 | 0 | 0 | 0 | 0 |
| 0 | 0 | 0 | 0 | 0 |
| 0 | 0 | 0.002450505 | 0 | 0.000957076 |
| 0 | 0 | 0 | 0 | 0.057591212 |
| 0 | 0 | 0 | 0 | 0 |
| 0 | 0 | 0 | 0 | 0 |
| 0.002604158 | 0 | 0.001038167 | 0 | 0 |
| 0 | 0 | 0 | 0 | 0.016431914 |
| 0 | 0 | 0 | 0 | 0 |
| 0 | 0 | 0 | 0 | 0 |
| 0 | 0 | 0.009802951 | 0 | 0 |
| 0 | 0 | 0 | 0 | 0 |
| 0 | 0 | 0.000584693 | 0 | 0 |
| 0 | 0 | 0.0336616 | 0 | 0.038696328 |
| 0.189398539 | 0 | 0 | 0 | 0 |
| 0 | 0 | 0 | 0 | 0 |
| 0 | 0 | 0 | 0 | 0 |
| 0 | 0 | 0 | 0 | 0 |
| 0 | 0 | 0.006771607 | 0 | 0 |
| 0 | 0 | 0 | 0 | 0 |
| 0 | 0 | 0 | 0 | 0 |
| 0 | 0 | 0 | 0 | 0 |
| 0 | 0 | 0 | 0 | 0 |
| 0 | 0 | 0.002330363 | 0 | 0 |
| 0 | 0 | 0 | 0 | 0 |

| | | | | |
|---|---|---|---|---|
| 0 | 0 | 0 | 0 | 0 |
| 0.035176538 | 0 | 0.00340568 | 0.00304822 | 0.004433771 |
| 0 | 0 | 0 | 0 | 0 |
| 0 | 0 | 0.007138247 | 0 | 0 |
| 0 | 0 | 0 | 0 | 0 |
| 0 | 0.001051294 | 0.002823758 | 0 | 0.001562377 |
| 0 | 0 | 0 | 0 | 0 |
| 0.002885779 | 0 | 0.001095654 | 0 | 0.002020742 |
| 0 | 0 | 0 | 0 | 0 |
| 0 | 0 | 0 | 0 | 0 |
| 0 | 0 | 0.025218147 | 0.047341002 | 0 |
| 0.003503315 | 0 | 0 | 0 | 0 |
| 0 | 0 | 0 | 0 | 0 |
| 0.017657159 | 0 | 0 | 0.011475596 | 0.1496267 |
| 0 | 0.004370895 | 0.106913687 | 0.037513238 | 0 |
| 0 | 0 | 0 | 0 | 0 |
| 0 | 0 | 0 | 0 | 0 |
| 0 | 0 | 0 | 0 | 0 |
| 0.109868703 | 0 | 0 | 0 | 0 |
| 0 | 0 | 0.041096006 | 0 | 0 |
| 0 | 0.002792666 | 0 | 0 | 0 |
| 1.164322974 | 0.269273428 | 0 | 1.678688436 | 0 |
| 0 | 0.03935343 | 0 | 0 | 0 |
| 0 | 0 | 0 | 0 | 0 |
| 0 | 0.007491153 | 0 | 0 | 0 |
| 0 | 0 | 0 | 0 | 0 |
| 0 | 0 | 0.066258509 | 0 | 0.090955525 |
| 0 | 0 | 0 | 0 | 0 |
| 0 | 0 | 0 | 0 | 0 |
| 0.174004635 | 0.239770651 | 1.63567521 | 0.865193959 | 0.912919068 |
| 0.001105752 | 0.000976888 | 0.001049562 | 0 | 0 |
| 0 | 0 | 0 | 0 | 0 |
| 0 | 0 | 0 | 0 | 0 |
| 0 | 0 | 0 | 0 | 0 |
| 0 | 0 | 0.001257428 | 0 | 0 |
| 0 | 0 | 0 | 0 | 0 |
| 0 | 0 | 0.506265868 | 0 | 0 |
| 0 | 0 | 0 | 0 | 0.079919149 |
| 0 | 0 | 0.984276617 | 0 | 0 |
| 0 | 0 | 0.453799944 | 0.337375156 | 0 |
| 0.023672419 | 0.013258461 | 0 | 0.001693316 | 0.019985514 |
| 3.743034277 | 3.441823785 | 1.059394998 | 2.599952856 | 3.966885664 |
| 0 | 0 | 0 | 0 | 0.067456526 |
| 0 | 0 | 0 | 0 | 0 |
| 0.001007688 | 0 | 0 | 0 | 0.004445434 |
| 0 | 0 | 0 | 0 | 0 |
| 0.000937581 | 0 | 0.000934434 | 0.005077876 | 0.002658959 |
| 0 | 0 | 0 | 0 | 0 |

| | | | | |
|---|---|---|---|---|
| 0 | 0 | 0 | 0 | 0 |
| 0 | 0 | 0 | 0 | 0 |
| 0.007487202 | 0 | 3.429186026 | 0 | 0.231457921 |
| 0 | 0 | 0 | 0 | 0 |
| 0 | 0.006164169 | 0 | 0 | 0 |
| 0.088761715 | 0.052623945 | 0.182567518 | 0.19732336 | 0 |
| 0 | 0 | 0 | 0 | 0 |
| 0 | 0 | 0 | 0 | 0 |
| 0 | 0 | 0 | 0 | 0 |
| 0.108114658 | 0 | 0 | 0 | 0 |
| 0 | 0 | 0 | 0 | 0 |
| 0 | 0 | 0 | 0 | 0 |
| 0.003556326 | 0 | 0 | 0 | 0 |
| 0 | 0 | 0 | 0.001700403 | 0 |
| 0 | 0 | 0 | 0 | 0 |
| 0 | 0.707514458 | 0.009431997 | 0.002436199 | 0.002319421 |
| 0.001359762 | 0.042045359 | 0.005549859 | 0.003191231 | 0 |
| 0 | 0 | 0 | 0 | 0 |
| 0 | 0 | 0 | 0 | 0 |
| 0 | 0.176549469 | 0.0277124 | 0.001309513 | 0 |
| 0.054552378 | 0.003330744 | 0.002277247 | 0 | 0.064079707 |
| 0.001840129 | 0 | 0.002095946 | 0 | 0.002319362 |
| 0.075306288 | 0 | 0 | 0 | 0 |
| 0 | 0 | 0.392656081 | 0 | 0 |
| 0.103360776 | 0 | 0 | 0 | 0 |
| 0.034929317 | 0 | 0.00289038 | 0.016168786 | 0 |
| 0.002589344 | 0.001525055 | 0 | 0 | 0 |
| 0 | 0 | 0 | 0 | 0 |
| 0 | 0 | 0 | 0 | 0 |
| 0 | 0 | 0 | 0 | 0 |
| 0.062357634 | 0.012989759 | 0.013706899 | 0.000947999 | 0.004136721 |
| 0 | 0.00449571 | 0 | 0.017313542 | 0.002878094 |
| 2.967361988 | 0 | 0.026709392 | 0.089489091 | 2.929808741 |
| 0 | 0.001415633 | 0.007858226 | 0 | 0 |
| 0 | 0.002020803 | 0 | 0 | 0 |
| 0 | 0 | 0 | 0 | 0 |
| 0.001123173 | 0.001190735 | 0 | 0.003041515 | 0.001415686 |
| 0.005489836 | 0.023047448 | 0.054151313 | 0 | 0.001660698 |
| 0.152036551 | 0.548226048 | 0.021367893 | 0 | 0.002056138 |
| 0 | 0 | 0 | 0 | 0.002036686 |
| 0.006271564 | 0 | 0 | 0 | 0 |
| 0 | 0 | 0 | 0 | 0 |
| 0 | 0.001740779 | 0 | 0 | 0 |
| 0.095480792 | 0.123577111 | 0 | 0.002785193 | 0.055744333 |
| 0 | 0 | 0 | 0 | 1.44940774 |
| 0 | 0.001776416 | 0 | 0 | 0 |
| 0 | 0 | 0 | 0 | 0 |
| 0 | 0 | 0 | 0 | 0 |

| | | | | |
|---|---|---|---|---|
| 0 | 0 | 0 | 0 | 0 |
| 0.01226818 | 0.444746562 | 0.229884658 | 3.183459867 | 0.035075141 |
| 0 | 0 | 0 | 0 | 0 |
| 0.00126855 | 0 | 0 | 0 | 0 |
| 0 | 0 | 0 | 0 | 0 |
| 0 | 0 | 0 | 0 | 0 |
| 0.13536551 | 0 | 0 | 0 | 0.027913182 |
| 0.012640953 | 0 | 0.000392681 | 0 | 0 |
| 0 | 0 | 0 | 0 | 0 |
| 0 | 0.017058458 | 0 | 0 | 0.002192552 |
| 0.006670682 | 0.002121582 | 0.001013073 | 0 | 0.001121061 |
| 0.008942553 | 0.068118929 | 0.000943126 | 0.135969079 | 0.006679408 |
| 0.174356161 | 0.016722929 | 0 | 0 | 0 |
| 0 | 0 | 0 | 0 | 0 |
| 0.003316635 | 0.007032278 | 0 | 0 | 0.014792187 |
| 0.001008489 | 0 | 0 | 0 | 0 |
| 0.002794067 | 0.04023569 | 0.00159125 | 0.092812514 | 0 |
| 0.029318935 | 0.004165701 | 0.027197896 | 0.006548014 | 0.076195031 |
| 0 | 0 | 0 | 0 | 0 |
| 0 | 0 | 0.005700668 | 0 | 0.028944087 |
| 0.733553314 | 0.346709613 | 0.04170826 | 0.123542746 | 0 |
| 0 | 0 | 0 | 0 | 0 |
| 0 | 0 | 0 | 0 | 0 |
| 0 | 0 | 0 | 0 | 0 |
| 0.002017195 | 0.146703473 | 0 | 0 | 20.27779122 |
| 0 | 0 | 0 | 0 | 0 |
| 0.021567559 | 0.039950759 | 0.047107146 | 0 | 0.299029397 |
| 0.040577991 | 0.003787824 | 0.256385675 | 0.038977654 | 1.088860612 |
| 0 | 0 | 0 | 0 | 0 |
| 0 | 0 | 0 | 0 | 0 |
| 0 | 0 | 0.012008423 | 0 | 0.001035464 |
| 0 | 0 | 0 | 0 | 0 |
| 0 | 0 | 0 | 0 | 0 |
| 0 | 0 | 0 | 0 | 0 |
| 0 | 0.023105773 | 0 | 0 | 0 |
| 0.064356594 | 0.119055219 | 0.663667407 | 0.123513345 | 0.382176603 |
| 0 | 0 | 0 | 0 | 0 |
| 0 | 0 | 0 | 0 | 0 |
| 0.004092751 | 0.003003881 | 0.034783673 | 0.0146637 | 0 |
| 0 | 0 | 0 | 0 | 0 |
| 0 | 0 | 0 | 0 | 0.898412935 |
| 0 | 0 | 0 | 0 | 0 |
| 0 | 0 | 0.00261379 | 0 | 0.001687236 |
| 0.002769445 | 0.003914712 | 0 | 0 | 0 |
| 0 | 0 | 0 | 0 | 0.013146461 |
| 0 | 0 | 0 | 0 | 0 |
| 0 | 0 | 0 | 0 | 0 |
| 0 | 0 | 0 | 0 | 0.003385471 |

| | | | | |
|---|---|---|---|---|
| 0 | 0 | 0 | 0 | 0 |
| 0 | 0 | 0 | 0 | 0 |
| 0 | 0 | 0 | 0 | 0 |
| 0 | 0.208966747 | 0 | 0 | 0.00280728 |
| 0 | 0 | 0 | 0 | 0 |
| 0 | 0 | 0 | 0 | 0 |
| 0 | 0 | 0 | 0 | 0 |
| 0 | 0 | 0 | 0 | 0 |
| 0.237150264 | 1.200980377 | 0 | 0.149020459 | 0 |
| 0 | 0.037352976 | 0 | 0 | 0.030860926 |
| 0 | 0 | 0 | 0 | 0 |
| 0 | 0.004072121 | 0.003159766 | 0.000924577 | 0 |
| 0 | 0 | 0 | 0 | 0 |
| 0 | 0 | 0 | 0 | 0 |
| 0.06092794 | 0.015868199 | 0.320214562 | 0.001525926 | 0.004661008 |
| 0 | 0 | 0 | 0 | 0 |
| 0 | 0 | 0 | 0 | 0 |
| 0 | 0 | 0.402567887 | 0 | 0 |
| 0 | 0 | 0 | 0 | 0.229946312 |
| 0 | 0 | 0 | 0 | 0 |
| 0 | 0 | 0 | 0 | 0 |
| 0 | 0 | 0 | 0 | 0 |
| 0 | 0 | 0 | 0 | 0 |
| 0.037825713 | 0.460352526 | 0.32197291 | 0.031418217 | 0.049814799 |
| 0 | 0 | 0 | 0 | 0 |
| 0 | 5.357282811 | 0 | 0 | 0 |
| 0 | 0 | 0.001895373 | 0 | 0 |
| 0 | 1.840692395 | 9.482589425 | 0 | 0 |
| 0 | 0 | 0 | 0 | 0 |
| 0 | 0 | 0 | 0 | 0.010553715 |
| 0 | 0.001539532 | 0.003544421 | 0 | 0 |
| 0 | 0 | 0 | 0 | 0 |
| 0.036472104 | 0.005523713 | 0.010970096 | 0 | 0.000796029 |
| 0 | 0 | 0 | 0 | 0 |
| 0 | 0 | 0 | 0 | 0 |
| 0 | 0 | 0 | 0 | 0 |
| 0.857124484 | 4.537151163 | 0 | 0 | 0 |
| 0 | 0 | 0 | 0 | 0 |
| 0.002887846 | 0 | 0.002656756 | 0 | 0 |
| 0 | 0 | 0 | 0 | 0 |
| 0.003275397 | 0.164939928 | 0.066687078 | 0.135484085 | 0.002064211 |
| 0 | 0 | 0 | 0 | 0 |
| 22.06317493 | 1.303570414 | 8.139373063 | 1.234002784 | 0 |
| 0 | 0.006427979 | 0.00416402 | 0 | 0 |
| 0 | 0 | 0 | 0 | 0 |
| 0 | 0 | 0 | 0 | 0 |
| 0 | 0 | 0 | 0 | 0 |
| 0.001722614 | 0 | 0.003223439 | 0 | 0 |

| | | | | |
|---|---|---|---|---|
| 0 | 0 | 0 | 0 | 0 |
| 0 | 0 | 0 | 0 | 0 |
| 0.065816879 | 0.003251208 | 0.003493077 | 0 | 0.061252032 |
| 0 | 0 | 0 | 0 | 0 |
| 0 | 0 | 0 | 0 | 5.335903256 |
| 0 | 0.000788054 | 0.000529175 | 0 | 0 |
| 0 | 0 | 0.169918701 | 0 | 0 |
| 0.008286356 | 0 | 0 | 0 | 0 |
| 0 | 0 | 0 | 0 | 0 |
| 0 | 0 | 0 | 0 | 0 |
| 0.65014249 | 0.020871486 | 0.084288583 | 0 | 0.053424126 |
| 0.484758337 | 0.151178198 | 2.861850999 | 2.046745265 | 6.942988176 |
| 0 | 0 | 0.000866797 | 0 | 0 |
| 0 | 0 | 0.02235043 | 0.004061429 | 0 |
| 0.057519444 | 0.057011542 | 0.008189482 | 0.086415177 | 0.412154568 |
| 0 | 0 | 0 | 0 | 0 |
| 0 | 0 | 0 | 0 | 0 |
| 0 | 0 | 0 | 0 | 0 |
| 0 | 0 | 0 | 0 | 0 |
| 0.524862082 | 0.012945025 | 0.006954027 | 0 | 0 |
| 0.001747145 | 0.027989406 | 0.167494619 | 0.626622379 | 0.020553508 |
| 1.242435965 | 0 | 4.650529441 | 0.185893566 | 5.912108837 |
| 3.767637659 | 0.616985278 | 6.238095375 | 1.913415925 | 0.013404412 |
| 0 | 0 | 0 | 0 | 0 |
| 0 | 0 | 0 | 0 | 0 |
| 13.72543459 | 9.988666412 | 0.443546564 | 0.030740789 | 0 |
| 2.274021988 | 0 | 0 | 0.001257499 | 0 |
| 0 | 0 | 0 | 0 | 0 |
| 0 | 0 | 0 | 0 | 0 |
| 0 | 0 | 0.092743915 | 0 | 0 |
| 0 | 0.001361982 | 0 | 0 | 0 |
| 0 | 0.258412592 | 0.070557337 | 0.013896164 | 0 |
| 0 | 0.091290266 | 0.005960765 | 0.039422322 | 0 |
| 0.063670558 | 0.010577001 | 0.085848819 | 0 | 0 |
| 0 | 0.002829547 | 0.011623711 | 0.007652712 | 0.010290186 |
| 0.106344254 | 0.012806293 | 0.030097814 | 0.001635567 | 0.001631318 |
| 0.128356778 | 0 | 0.013291007 | 0 | 0 |
| 0 | 0 | 0 | 0 | 0 |
| 0 | 0.022478924 | 0.03680185 | 0 | 0 |
| 0 | 0 | 0 | 0 | 0 |
| 0 | 0 | 0 | 0 | 0 |
| 0 | 0 | 0 | 0 | 0 |
| 0 | 0 | 0 | 0 | 0 |
| 0 | 0 | 0 | 0 | 0 |
| 0.002213919 | 0.002086303 | 0.020173601 | 0 | 0 |
| 0.003422369 | 0 | 0 | 0 | 0 |
| 0 | 0 | 0 | 0 | 0 |
| 0.051631427 | 0.020133216 | 0.077229431 | 0.030534559 | 0.047125462 |

| | | | | |
|---|---|---|---|---|
| 0 | 0 | 0 | 0 | 0 |
| 0 | 0 | 0 | 0 | 0 |
| 0 | 0 | 0 | 0 | 0 |
| 0 | 0 | 0.001671627 | 0 | 0 |
| 0 | 0 | 0 | 0 | 0 |
| 0 | 0.028268082 | 0.209117324 | 0 | 0 |
| 0 | 0 | 0 | 0 | 0 |
| 0.030481754 | 0 | 0.519471937 | 0 | 0 |
| 0 | 0 | 0 | 0 | 0 |
| 0 | 0 | 0 | 0 | 0 |
| 0.001167513 | 0 | 0.003989464 | 0 | 0.002354516 |
| 0 | 0 | 0 | 0 | 0 |
| 0 | 0.001950731 | 0 | 0 | 0.003092345 |
| 0 | 0 | 0 | 0 | 0 |
| 0 | 0 | 0 | 0 | 0 |
| 0 | 0 | 0 | 0 | 0 |
| 0 | 0 | 0 | 0 | 0 |
| 0 | 0 | 0 | 0 | 0 |
| 0 | 0 | 0 | 0 | 0 |
| 0 | 0 | 0 | 0 | 0 |
| 0 | 0 | 0 | 0 | 0 |
| 0 | 0 | 0 | 0 | 0 |
| 0 | 0.00475187 | 0 | 0 | 0 |
| 0 | 0 | 0 | 0 | 0 |
| 0 | 0 | 0 | 0 | 0 |
| 0 | 0 | 0 | 0 | 0 |
| 0 | 0 | 0 | 0 | 0 |
| 0 | 0 | 0 | 0 | 0 |
| 0 | 0 | 0 | 0 | 0 |
| 0 | 0.012695976 | 0.000936111 | 0 | 0 |
| 0 | 0 | 0 | 0 | 0.001854444 |
| 0 | 0 | 0 | 0 | 0 |
| 0.269513372 | 0 | 0 | 0 | 0 |
| 0 | 0 | 0 | 0 | 0 |
| 0 | 0.00118103 | 0 | 0 | 0 |
| 0 | 0 | 0 | 0 | 0 |
| 0 | 0.1464277 | 0.072946661 | 0.026263104 | 0 |
| 0.00197664 | 0.015926098 | 0.029493787 | 0 | 0.006477703 |
| 0 | 0 | 0 | 0 | 0 |
| 0 | 0.002273616 | 0.002442759 | 0 | 0 |
| 0.256302556 | 0 | 0.128011585 | 0 | 0 |
| 0 | 0 | 0 | 0 | 0 |
| 0 | 0 | 0 | 0 | 0 |
| 0 | 0 | 0 | 0 | 0 |
| 0 | 0.063769611 | 0.105071333 | 0 | 0 |
| 0 | 0 | 0 | 0 | 0 |
| 0 | 0 | 0 | 0 | 0 |
| 0 | 0 | 0 | 0 | 0 |

| | | | | |
|---|---|---|---|---|
| 0 | 0 | 0 | 0 | 0 |
| 0 | 0 | 0 | 0 | 0 |
| 0 | 0 | 0 | 0 | 0 |
| 0 | 0 | 0.004031131 | 0 | 0 |
| 0 | 0 | 0 | 0 | 0 |
| 0 | 0 | 0.122180639 | 0 | 0 |
| 0 | 0 | 0 | 0 | 0 |
| 0 | 0 | 0 | 0 | 0 |
| 0.092142723 | 0.002312079 | 0.043316196 | 0.012697434 | 0.134694732 |
| 0 | 0.001629169 | 0 | 0 | 0 |
| 0 | 0 | 0 | 0 | 0 |
| 0 | 0 | 0 | 0 | 0 |
| 0 | 0 | 0 | 0 | 0 |
| 0 | 0 | 0 | 0 | 0 |
| 0 | 6.236310898 | 0.545802745 | 0 | 0 |
| 0.116610688 | 0.019601818 | 0 | 0 | 0.01615457 |
| 0.002328395 | 0 | 0 | 0 | 0 |
| 0 | 0 | 0 | 0 | 0 |
| 0 | 0 | 0 | 0 | 0 |
| 0 | 0 | 0 | 0 | 0 |
| 0 | 0 | 0 | 0 | 0 |
| 0 | 0 | 0 | 0 | 0 |
| 0 | 0 | 0 | 0 | 0 |
| 0 | 0 | 0 | 0 | 0 |
| 0 | 0 | 0.008398481 | 0 | 0 |
| 1.514442453 | 8.101377122 | 0.534152402 | 0 | 0.026659988 |
| 0 | 0 | 0 | 0 | 0 |
| 0 | 0 | 0 | 0 | 0 |
| 0 | 0.03298268 | 0 | 0 | 0 |
| 0 | 0 | 0.007140383 | 0 | 0 |
| 0 | 0 | 0 | 0 | 0 |
| 0 | 0 | 0.03668488 | 0 | 0 |
| 0 | 0 | 0 | 0 | 0 |
| 0 | 0.002359643 | 0 | 0 | 0 |
| 0 | 0.001794437 | 0.002891897 | 0 | 0.001066719 |
| 0.010170064 | 0 | 0.002839195 | 0 | 0.001759429 |
| 1.26683537 | 0 | 0.107705738 | 0 | 0 |
| 0.127447221 | 0.774634698 | 0 | 0.001707321 | 0 |
| 0 | 0 | 0 | 0 | 0 |
| 0 | 0 | 0 | 0 | 0 |
| 0 | 0 | 0 | 0 | 0 |
| 0 | 0 | 0 | 0 | 0 |
| 3.21933208 | 0.441717497 | 0.107082786 | 0.035715551 | 0 |
| 0 | 0 | 0 | 0 | 0 |
| 0 | 0 | 0 | 0 | 0 |
| 0.001700269 | 0 | 0 | 0 | 0 |
| 0 | 0 | 0 | 0 | 0 |
| 0.010533961 | 0.007657785 | 0.009255911 | 5.095814132 | 0.00113806 |

| | | | | |
|---|---|---|---|---|
| 0.003590942 | 0.000713802 | 0.001405992 | 0 | 0 |
| 0 | 0 | 0 | 0 | 0 |
| 0 | 0 | 0 | 0 | 0 |
| 0 | 0 | 0 | 0 | 0 |
| 0.010949047 | 0 | 0 | 0 | 0.12392334 |
| 0 | 0 | 0 | 0 | 0 |
| 0 | 0 | 0 | 0 | 0 |
| 0 | 0 | 0 | 0 | 0 |
| 0 | 0 | 0 | 0 | 0 |
| 0 | 0 | 0 | 0 | 0 |
| 0 | 0 | 0 | 0 | 0 |
| 1.22492442 | 3.193190972 | 0.282471104 | 0.040193176 | 0 |
| 0.130138731 | 0.1646424 | 0 | 0.911433629 | 0.080140039 |
| 0 | 0 | 0 | 0 | 0 |
| 0.411145964 | 0.030136086 | 0.664779628 | 0.052344449 | 0.094459153 |
| 0 | 0 | 0 | 0 | 0 |
| 0 | 0 | 0 | 0 | 0 |
| 0 | 0.000731645 | 0 | 0 | 0 |
| 0 | 0.002401288 | 0 | 0 | 0 |
| 0 | 0 | 0 | 0 | 0 |
| 0 | 0 | 0.014097235 | 0 | 0 |
| 0 | 0 | 0 | 0 | 0 |
| 0.047572796 | 0.866491265 | 0.036626004 | 0.152920647 | 0.021653082 |
| 0 | 0 | 0 | 0 | 0 |
| 0 | 0 | 0 | 0 | 0 |
| 0 | 0 | 0 | 0 | 0 |
| 0 | 0.077974159 | 0 | 0 | 0 |
| 0 | 0 | 0 | 0 | 0 |
| 0 | 0 | 0 | 0 | 0 |
| 0 | 0 | 0 | 0 | 0 |
| 0 | 0 | 0 | 0 | 0 |
| 0.004024031 | 0.044082886 | 0.006111274 | 0.4627566 | 0 |
| 0 | 0 | 0 | 0 | 0 |
| 0.018763139 | 0 | 0.04093045 | 0 | 0.001266948 |
| 0.009982522 | 0 | 0.003460527 | 0.000940255 | 0.0019147 |
| 0.109383786 | 0 | 0.011311919 | 0.013636153 | 0 |
| 0 | 0 | 0 | 0 | 0 |
| 0 | 0 | 0 | 0 | 0 |
| 0 | 0 | 0 | 0 | 0 |
| 0 | 0 | 0 | 0 | 0 |
| 0 | 0.089187417 | 0 | 0 | 0 |
| 0 | 0 | 0 | 0 | 0 |
| 0 | 0 | 0 | 0 | 0 |
| 0 | 0 | 0 | 0 | 0 |
| 0.001496445 | 0.028782921 | 0.010470395 | 0 | 0.002155624 |
| 0 | 0 | 0 | 0 | 0 |
| 0 | 0.071126401 | 0 | 0 | 0 |
| 0 | 0 | 0.002878814 | 0 | 0 |

| | | | | |
|---|---|---|---|---|
| 0 | 0 | 0 | 0 | 0 |
| 0 | 0 | 0 | 0 | 0 |
| 0.086007848 | 0.023674355 | 0.013095545 | 0 | 0 |
| 0 | 0 | 0.004898422 | 0 | 0 |
| 0 | 0 | 0.001532397 | 0 | 0 |
| 0 | 0 | 0.001083266 | 0 | 0 |
| 0 | 0 | 0 | 0 | 0 |
| 0 | 0.08965423 | 0.26423678 | 0 | 0 |
| 0 | 0 | 0 | 0 | 0 |
| 0 | 0 | 0.001404243 | 0 | 0 |
| 0 | 0 | 0 | 0 | 0 |
| 0 | 0 | 0 | 0 | 0 |
| 0 | 0 | 0 | 0 | 0 |
| 0 | 0 | 0.255109767 | 0 | 0 |
| 0 | 0 | 0 | 0 | 0 |
| 0 | 0 | 0 | 0 | 0 |
| 0 | 0 | 2.172347157 | 0 | 0 |
| 0 | 0 | 0 | 0 | 0 |
| 0 | 0 | 0 | 0 | 3.436984704 |
| 0 | 0 | 0 | 0 | 0 |
| 0 | 0 | 0.001601718 | 0 | 0.001519244 |
| 0 | 0 | 0 | 0 | 0 |
| 0 | 0 | 0 | 0 | 0 |
| 0 | 0 | 0 | 0 | 0 |
| 0 | 0 | 0 | 0 | 0 |
| 0.015324475 | 0 | 0.022895389 | 0 | 0.002508506 |
| 0 | 0 | 0 | 0 | 0 |
| 0 | 0.04564045 | 0.122145766 | 0.021522498 | 0.017678363 |
| 0 | 0 | 0 | 0 | 0 |
| 0.533022183 | 1.350821651 | 0.010738239 | 0.114294178 | 0.002970719 |
| 0 | 0 | 0 | 0 | 0 |
| 0 | 0 | 0 | 0 | 0 |
| 0 | 0 | 0 | 0 | 0 |
| 0 | 0 | 0 | 0 | 0 |
| 0 | 0.001105526 | 0.002850648 | 0 | 0 |
| 0 | 0 | 0 | 0 | 0 |
| 0 | 0 | 0 | 0 | 0 |
| 0.002129888 | 0 | 0 | 0 | 0 |
| 0 | 0 | 0 | 0 | 0 |
| 0 | 0 | 0 | 0 | 0 |
| 2.340583982 | 5.446677891 | 6.303630631 | 34.24234165 | 13.76367509 |
| 0.297092854 | 0.134433489 | 0.662780681 | 0 | 0.708974426 |
| 0 | 0 | 0 | 0 | 0 |
| 0 | 0 | 0 | 0 | 0 |
| 0 | 0 | 0 | 0 | 0 |
| 0 | 0 | 0 | 0 | 0 |
| 0 | 0 | 0 | 0 | 0 |
| 0.01183278 | 0 | 0.000427866 | 0 | 0 |

| | | | | |
|---|---|---|---|---|
| 0 | 0 | 0 | 0 | 0.390013873 |
| 0.003091045 | 0.001433678 | 0.001870407 | 0 | 0 |
| 0 | 0 | 0 | 0 | 0 |
| 0 | 0 | 0 | 0 | 0 |
| 1.951273767 | 3.543856689 | 0.984314336 | 0.374958533 | 0 |
| 0 | 0 | 0 | 0 | 0 |
| 0 | 0 | 0 | 0 | 0 |
| 0 | 0 | 0 | 0 | 0 |
| 0.279905896 | 0.10532226 | 0.080199056 | 0.335368943 | 0.253599938 |
| 0 | 0 | 0 | 0 | 0 |
| 0 | 0 | 0 | 0 | 0 |
| 0 | 0 | 0 | 0 | 0 |
| 0.028345318 | 0.017910809 | 0.009194002 | 0 | 0 |
| 0.005077146 | 0.011499082 | 0.076755871 | 0.101240712 | 0.048577333 |
| 0 | 0 | 0 | 0 | 0 |
| 0 | 0.757695808 | 0 | 0 | 0 |
| 0.000998519 | 0.000635149 | 0.023315345 | 0.017954259 | 0.002517135 |
| 0 | 0 | 0 | 0 | 0 |
| 0 | 0 | 0 | 0 | 0 |
| 0 | 0 | 0 | 0 | 0 |
| 0 | 0 | 0 | 0 | 0 |
| 0 | 0 | 0 | 0 | 0 |
| 0 | 0 | 0 | 0 | 0 |
| 0 | 0 | 0 | 0 | 0 |
| 0 | 0 | 0 | 0 | 0 |
| 0 | 0 | 0 | 0 | 0 |
| 0.014933954 | 0.007732037 | 0.003164667 | 0.000752384 | 0 |
| 0.002547289 | 0 | 0.002120265 | 0 | 0 |
| 0 | 0 | 0 | 0 | 0 |
| 0 | 0 | 0 | 0 | 0 |
| 0 | 0 | 0 | 0 | 0 |
| 0 | 0 | 0 | 0 | 0 |
| 0 | 0 | 0 | 0 | 0 |
| 0 | 0 | 0.008656458 | 0 | 0 |
| 0 | 0 | 0 | 0 | 0 |
| 0.132600809 | 0 | 0.018976207 | 0 | 0.046569128 |
| 0 | 0 | 0 | 0.001254774 | 0.014016959 |
| 0 | 0.005219019 | 0 | 0 | 0 |
| 0 | 0 | 0 | 0 | 0 |
| 0 | 0 | 0 | 0 | 0 |
| 0.060518988 | 0.447938226 | 4.680288795 | 17.71353512 | 0 |
| 0 | 0 | 0 | 0 | 0 |
| 0 | 0 | 0 | 0 | 0 |
| 0 | 0 | 0 | 0 | 0 |
| 0 | 0 | 0 | 0 | 0 |
| 0 | 0 | 0 | 0 | 0 |
| 0 | 0 | 0 | 0 | 0 |
| 0 | 0 | 0 | 0 | 0 |

| | | | | |
|---|---|---|---|---|
| 0 | 0.012964477 | 0 | 0 | 0 |
| 0 | 0 | 0 | 0 | 0 |
| 0 | 0 | 0 | 0 | 0 |
| 0 | 0 | 0 | 0 | 0 |
| 0 | 0.025217328 | 0 | 0 | 0.196752508 |
| 0 | 5.023087303 | 0 | 0 | 0 |
| 0 | 0.005913645 | 0.030633347 | 0.012947423 | 0.002008811 |
| 0 | 0 | 0 | 0 | 0 |
| 0.005765251 | 0 | 0 | 0 | 0.001063422 |
| 0 | 0 | 0 | 0 | 0.164917338 |
| 0 | 0 | 0 | 0 | 0 |
| 0 | 0.048576048 | 0 | 0 | 0 |
| 0 | 0 | 0 | 0 | 0 |
| 0 | 0 | 0 | 0 | 0 |
| 0 | 0 | 0 | 0 | 0 |
| 0 | 0 | 0 | 0 | 0 |
| 0 | 0 | 0 | 0 | 0.005165486 |
| 0 | 0 | 0 | 0 | 0 |
| 0 | 0 | 0 | 0 | 0 |
| 0 | 0 | 0 | 0 | 0 |
| 0 | 0.027346296 | 0.006913103 | 0.020544448 | 0 |
| 0 | 0 | 0 | 0 | 0 |
| 0 | 0.039699659 | 0.00226371 | 0 | 0 |
| 0 | 0.001774928 | 0.001059429 | 0.00221649 | 0.001172358 |
| 0 | 0 | 0 | 0 | 0 |
| 0 | 0.001628679 | 0 | 0 | 0 |
| 0 | 0 | 0 | 0 | 0 |
| 0 | 0 | 0 | 0 | 0 |
| 0 | 0 | 0 | 0 | 0 |
| 0.442829952 | 0.225366983 | 0 | 0.002030031 | 0.399889529 |
| 0 | 0 | 0 | 0 | 0 |
| 0 | 0 | 0 | 0 | 0 |
| 0 | 0 | 0 | 0 | 0 |
| 0 | 0 | 0 | 0 | 3.299790675 |
| 0 | 0 | 0 | 0 | 0 |
| 0 | 0 | 0 | 0 | 0 |
| 0 | 0.093030811 | 0.00676066 | 0 | 0 |
| 0 | 0 | 0 | 0 | 0 |
| 0 | 0 | 0 | 0 | 0 |
| 0 | 0 | 0 | 0 | 0 |
| 0 | 0 | 0 | 0 | 0 |
| 0 | 0 | 0 | 0 | 0 |
| 0 | 0 | 0 | 0 | 0 |
| 0 | 0 | 0 | 0 | 0 |
| 0 | 0 | 0 | 0 | 0 |
| 0 | 0 | 0 | 0 | 0 |
| 0 | 0 | 0 | 0 | 0 |
| 0 | 0 | 0 | 0 | 0 |

| | | | | |
|---|---|---|---|---|
| 0.016413847 | 0.010186056 | 0.000797988 | 0.003469123 | 0 |
| 0 | 0 | 0 | 0 | 0 |
| 0 | 0 | 0 | 0 | 0 |
| 0 | 0 | 0 | 0 | 0 |
| 0 | 0 | 0 | 0 | 0 |
| 0 | 0 | 0 | 0 | 0 |
| 0 | 0 | 0 | 0 | 0 |
| 0.655018338 | 3.022804695 | 0.764690708 | 0.428553749 | 0.030309408 |
| 0.035504047 | 0.001998568 | 0.214545968 | 0 | 0 |
| 0.305789022 | 0 | 0.256989693 | 0.001421573 | 0.009098061 |
| 0 | 0 | 0 | 0 | 0 |
| 0 | 0 | 0 | 0 | 0 |
| 0 | 0 | 0 | 0 | 0 |
| 0 | 0 | 0 | 0 | 0 |
| 0 | 0.003040939 | 0 | 0 | 0 |
| 0 | 0 | 0 | 0 | 0 |
| 0 | 0 | 0 | 0 | 0.020992511 |
| 0 | 0 | 0 | 0 | 0 |
| 0 | 0 | 0 | 0 | 0 |
| 0 | 0 | 0.001652755 | 0 | 0 |
| 0 | 0 | 0 | 0 | 0 |
| 0 | 0 | 0 | 0 | 0 |
| 0 | 0.006009831 | 0 | 0 | 0 |
| 0 | 0 | 0 | 0 | 0 |
| 0 | 0 | 0 | 0 | 0.008346282 |
| 0 | 0 | 0 | 0 | 0 |
| 0 | 0 | 0 | 0 | 0 |
| 0 | 0 | 0 | 0 | 0 |
| 0 | 0 | 0 | 0 | 0 |
| 0 | 0 | 0 | 0 | 0 |
| 0 | 0 | 0.033845836 | 0 | 0 |
| 0 | 0 | 0 | 0 | 0 |
| 0 | 0 | 0 | 0 | 0 |
| 0 | 0 | 0 | 0 | 0 |
| 0 | 0.008221103 | 0 | 0 | 0 |
| 0 | 0 | 0 | 0 | 0 |
| 0 | 0 | 0 | 0 | 0 |
| 0 | 0 | 0 | 0 | 0 |
| 0 | 0 | 0.000542211 | 0 | 0 |
| 0 | 0 | 0 | 0 | 0 |
| 0 | 0 | 0 | 0 | 0 |
| 0 | 0.601921043 | 0.038980475 | 0.004019601 | 0 |
| 0 | 0 | 0 | 0 | 0 |
| 0 | 0 | 0 | 0 | 0 |
| 0 | 0 | 0 | 0 | 0 |
| 0 | 0 | 0 | 0 | 0 |
| 0 | 0 | 0 | 0 | 0 |
| 0 | 0 | 0 | 0 | 0 |

| | | | | |
|---|---|---|---|---|
| 0 | 0 | 0 | 0 | 0 |
| 0 | 0 | 0.089615825 | 0.023482764 | 0.013890402 |
| 0 | 0 | 0 | 0 | 0 |
| 0 | 0 | 0 | 0 | 0 |
| 0.410963443 | 0.03213403 | 0.241528339 | 0.152396153 | 0.045527298 |
| 0 | 0 | 0 | 0 | 0 |
| 0 | 0 | 0.016218493 | 0 | 0 |
| 0.02546293 | 0 | 0.016821632 | 0.026477854 | 0.014121514 |
| 0 | 0 | 0 | 0 | 0 |
| 0 | 0 | 0 | 0 | 0 |
| 0 | 0 | 0 | 0 | 0 |
| 0 | 0 | 0 | 0 | 0 |
| 0 | 0 | 0 | 0 | 0 |
| 0 | 0 | 0 | 0 | 0 |
| 0 | 0 | 0.003675052 | 0 | 0 |
| 0.115277482 | 0.017559152 | 0.004779245 | 0 | 0 |
| 0 | 0 | 0 | 0 | 0 |
| 0 | 0 | 0 | 0 | 0 |
| 0 | 0 | 0 | 0 | 0 |
| 0 | 0 | 0 | 0 | 0 |
| 0 | 0 | 0 | 0 | 0 |
| 0 | 0 | 0 | 0 | 0 |
| 0 | 0 | 0 | 0 | 0 |
| 0 | 0 | 0.001380475 | 0 | 0.00083325 |
| 0.373296801 | 0.100184992 | 0 | 0.261213493 | 0 |
| 0 | 0 | 0 | 0 | 0 |
| 0 | 0 | 0.001606947 | 0.001309864 | 0 |
| 0 | 0 | 0 | 0 | 0 |
| 0 | 0 | 0 | 0 | 0 |
| 0 | 0 | 0 | 0 | 0 |
| 0 | 0 | 0 | 0 | 0 |
| 0 | 0 | 0 | 0 | 0 |
| 0 | 0 | 0 | 0 | 0 |
| 0 | 0 | 0 | 0 | 0 |
| 0 | 0 | 0 | 0 | 0 |
| 0 | 0 | 0 | 0 | 0 |
| 0 | 0 | 0 | 0 | 0 |
| 0 | 0 | 0.002118881 | 0 | 0 |
| 0 | 0 | 0 | 0 | 0 |
| 0 | 0 | 0 | 0 | 0 |
| 0 | 0 | 0 | 0 | 0 |
| 0 | 0 | 0 | 0 | 0 |
| 0 | 0 | 0 | 0 | 0 |
| 0 | 0 | 0 | 0 | 0 |
| 0 | 0 | 0 | 0 | 0 |
| 0 | 0 | 0 | 0 | 0 |
| 0 | 0 | 0 | 0 | 0 |
| 0 | 0 | 0 | 0 | 0 |

| | | | | |
|---|---|---|---|---|
| 0.102159672 | 1.34125175 | 0.022991753 | 0.01847721 | 0 |
| 0 | 0 | 0 | 0 | 0 |
| 0 | 0 | 0 | 0 | 0 |
| 0 | 0 | 0 | 0 | 0 |
| 0 | 0 | 0 | 0 | 0 |
| 0.00636805 | 0.051680861 | 0 | 0 | 0 |
| 0 | 0 | 0 | 0 | 0 |
| 0 | 0 | 0 | 0 | 0 |
| 0 | 0 | 0 | 0 | 0 |
| 0 | 0 | 0 | 0 | 0 |
| 0.299575291 | 0.133106267 | 0 | 0 | 0 |
| 0 | 0 | 0 | 0 | 0 |
| 0 | 0 | 0 | 0 | 0 |
| 0 | 0 | 0 | 0 | 0 |
| 0 | 0 | 0 | 0 | 0 |
| 0 | 0 | 0 | 0 | 0 |
| 0 | 0 | 0 | 0 | 0 |
| 0 | 0 | 0 | 0 | 0 |
| 0 | 0 | 0 | 0 | 0 |
| 0 | 0 | 0 | 0 | 0 |
| 0.052007165 | 0.006281262 | 0.02886879 | 0 | 0.003319069 |
| 0 | 0 | 0 | 0 | 0 |
| 0.038521922 | 0.001469033 | 0.000946992 | 0 | 0 |
| 0 | 0 | 0 | 0 | 0 |
| 0 | 0 | 0 | 0 | 0 |
| 0 | 0 | 0 | 0 | 0 |
| 0 | 0 | 0 | 0 | 0 |
| 0 | 0 | 0 | 0 | 0 |
| 0 | 0 | 0 | 0 | 0 |
| 0 | 0 | 0 | 0 | 0 |
| 0 | 0 | 0 | 0 | 0 |
| 0 | 0 | 0 | 0 | 0 |
| 0.0049312 | 0.087772406 | 0.226147714 | 0.007590422 | 0 |
| 0 | 0 | 0 | 0 | 0 |
| 0.148811016 | 0.139057304 | 0 | 0.746855649 | 0.026042775 |
| 0.036446801 | 0 | 0 | 0 | 0 |
| 0 | 0 | 0 | 0 | 0 |
| 0 | 0 | 0.053134579 | 0 | 0 |
| 0.004817885 | 0.001021538 | 0.005624864 | 0 | 0.005161733 |
| 0 | 0 | 0 | 0 | 0 |
| 0 | 0 | 0 | 0 | 0 |
| 0 | 0 | 0 | 0 | 0 |
| 0.08365823 | 0.248154156 | 0 | 0.039053986 | 0 |
| 0 | 0 | 0 | 0 | 0 |
| 0 | 0 | 0 | 0 | 0 |
| 0 | 0.34933221 | 0.003312399 | 0 | 0 |
| 0 | 0 | 0 | 0 | 0 |
| 0 | 0 | 0 | 0 | 0 |

| | | | | |
|---|---|---|---|---|
| 0 | 0 | 0 | 0 | 0 |
| 0 | 0 | 0 | 0 | 0 |
| 0 | 0 | 0 | 0 | 0 |
| 0 | 0 | 0 | 0 | 0 |
| 0 | 0 | 0 | 0 | 0 |
| 0 | 0 | 0 | 0 | 0 |
| 0 | 0 | 0 | 0 | 0 |
| 0 | 0.104042616 | 0 | 0 | 0 |
| 0 | 0 | 0 | 0 | 0 |
| 0 | 0 | 0.006298587 | 0 | 0.001452079 |
| 0 | 0 | 0 | 0 | 0 |
| 0 | 0 | 0 | 0 | 0 |
| 0 | 0 | 0 | 0 | 0 |
| 0 | 0 | 0 | 0 | 0 |
| 0 | 0 | 0 | 0 | 0 |
| 0 | 0 | 0 | 0 | 0.001714423 |
| 0 | 0 | 0 | 0 | 0 |
| 0 | 0 | 0 | 0 | 0 |
| 0 | 0 | 0 | 0 | 0 |
| 0 | 0.600224392 | 0 | 0 | 0 |
| 0 | 0.028615316 | 0.008360242 | 0.016157312 | 0 |
| 0 | 0 | 0 | 0 | 0 |
| 0 | 0 | 0 | 0 | 0 |
| 0 | 0 | 0.004643259 | 0 | 0 |
| 0 | 0 | 0 | 0 | 0 |
| 0 | 0 | 0 | 0 | 0 |
| 0 | 0 | 0 | 0 | 0 |
| 0 | 0 | 0 | 0 | 0 |
| 0 | 0 | 0 | 0 | 0 |
| 0.079649545 | 0.918385561 | 0 | 0 | 0 |
| 0 | 0.647345238 | 0.035152945 | 0 | 0.65282412 |
| 0 | 0 | 0 | 0 | 0 |
| 0 | 0 | 0 | 0 | 0 |
| 0 | 0 | 0 | 0 | 0 |
| 0 | 0 | 0 | 0 | 0 |
| 0 | 0 | 0 | 0 | 0 |
| 0 | 0 | 0 | 0 | 0 |
| 0 | 0 | 0 | 0 | 0 |
| 0 | 0 | 0.00697763 | 0 | 0 |
| 0 | 0 | 0 | 0 | 0 |
| 0 | 0 | 0 | 0 | 0 |
| 0 | 0 | 0.056123135 | 0 | 0 |
| 0 | 0 | 0 | 0 | 0 |
| 0.003147887 | 0 | 0 | 0 | 0 |
| 0 | 0 | 0 | 0 | 0 |
| 0 | 0 | 0 | 0 | 0 |
| 0.136688306 | 0 | 0.158730664 | 0.005027467 | 0.002047549 |
| 0 | 0 | 0 | 0 | 0 |

| | | | | |
|---|---|---|---|---|
| 0 | 0 | 0 | 0 | 0 |
| 0 | 0 | 0 | 0 | 0 |
| 0 | 0 | 0 | 0 | 0 |
| 0 | 0 | 0 | 0 | 0 |
| 0.00069349 | 0.013968893 | 0.045616695 | 0.002629125 | 0 |
| 0 | 0 | 0 | 0 | 0 |
| 0 | 0 | 0 | 0 | 0 |
| 0 | 0 | 0 | 0 | 0 |
| 0 | 0 | 0 | 0 | 0 |
| 0 | 0 | 0 | 0 | 0 |
| 0 | 0 | 0 | 0 | 0 |
| 0.001684917 | 0.005869169 | 0.003564147 | 0 | 0 |
| 0 | 0 | 0.087281572 | 0 | 0 |
| 0 | 0 | 0 | 0 | 0 |
| 0 | 0 | 0.011386847 | 0 | 0 |
| 0 | 2.29336642 | 0.084001687 | 0.104214061 | 0.315200347 |
| 0 | 0 | 0.013600826 | 0 | 0 |
| 0 | 0 | 0 | 0 | 0 |
| 0.035358682 | 0.003478045 | 0.003736789 | 0.065149555 | 0 |
| 0 | 0 | 0.597013099 | 0.06341244 | 0.018897177 |
| 0 | 0 | 0 | 0 | 0 |
| 0 | 0 | 0 | 0 | 0 |
| 0 | 0 | 0 | 0 | 0 |
| 0 | 0 | 0 | 0 | 0 |
| 0 | 0 | 0 | 0 | 0 |
| 0 | 0 | 0 | 0 | 0 |
| 0.036805564 | 0.006873903 | 0.026500115 | 0 | 0.004086253 |
| 0 | 0 | 0 | 0 | 0 |
| 0 | 0 | 0 | 0 | 0 |
| 0 | 0 | 0 | 0 | 0 |
| 0 | 0.000916698 | 0 | 0 | 0 |
| 0 | 0 | 0 | 0 | 0 |
| 0 | 0 | 0 | 0 | 0 |
| 0 | 0 | 0 | 0 | 0 |
| 0 | 0 | 0 | 0 | 0 |
| 0 | 0 | 0 | 0 | 0 |
| 0 | 0 | 0 | 0 | 0 |
| 0.030982623 | 0.002254158 | 0.008044012 | 0.000822548 | 0.004020013 |
| 0 | 0 | 0 | 0 | 0 |
| 0 | 0 | 0 | 0 | 0 |
| 0 | 0.002089635 | 0 | 0 | 0 |
| 0 | 0 | 0 | 0 | 0 |
| 0 | 0 | 0 | 0 | 0 |
| 0 | 0 | 0 | 0.002683415 | 0.006812766 |
| 0 | 0 | 0 | 0 | 0 |
| 0 | 0 | 0 | 0 | 0 |
| 0 | 0 | 0 | 0 | 0 |
| 0 | 0 | 0 | 0 | 0 |

| | | | | |
|---|---|---|---|---|
| 0 | 0 | 0 | 0 | 0 |
| 0 | 0 | 0 | 0 | 0 |
| 0 | 0 | 0 | 0 | 0 |
| 0.008496845 | 0 | 0 | 0 | 0 |
| 0 | 0 | 0.002493408 | 0 | 0 |
| 0.005932957 | 0 | 0 | 0 | 0 |
| 0 | 0 | 0 | 0 | 0 |
| 0.0260273 | 0 | 0 | 0 | 0 |
| 0 | 0 | 0.013540616 | 0 | 0 |
| 0 | 0 | 0 | 0 | 0 |
| 0 | 0 | 0 | 0 | 0 |
| 0 | 0 | 0 | 0 | 0 |
| 0.001338459 | 0.024299876 | 0.0022868 | 0 | 0 |
| 0 | 0 | 0 | 0 | 0 |
| 0 | 0 | 0 | 0 | 0 |
| 0 | 0 | 0 | 0 | 0 |
| 0 | 0 | 0 | 0 | 0 |
| 0 | 0 | 0 | 0 | 0 |
| 0 | 0 | 0 | 0 | 0 |
| 0 | 0 | 0 | 0 | 0 |
| 0 | 0 | 0 | 0 | 0 |
| 0 | 0 | 0 | 0 | 0 |
| 0.007490381 | 0 | 0.03362494 | 0 | 0.002776804 |
| 0.004159486 | 0.013780277 | 0 | 0.039141513 | 0 |
| 0 | 0 | 0 | 0 | 0 |
| 0 | 0.001221224 | 0 | 0 | 0 |
| 0 | 0 | 0 | 0 | 0 |
| 0 | 0 | 0 | 0 | 0 |
| 0 | 0 | 0 | 0 | 0 |
| 0 | 0 | 0 | 0 | 0 |
| 0.013510169 | 0.041145609 | 0 | 0.003991099 | 0 |
| 0 | 0 | 0 | 0 | 0 |
| 0 | 0 | 0 | 0 | 0 |
| 0 | 0 | 0 | 0 | 0 |
| 0 | 0 | 0 | 0 | 0 |
| 0 | 0 | 0 | 0 | 0 |
| 0 | 0 | 0 | 0 | 0 |
| 0 | 0 | 0 | 0 | 0 |
| 0 | 0.00330494 | 0.021186483 | 0 | 0 |
| 0 | 0 | 0 | 0 | 0 |
| 0 | 0 | 0 | 0 | 0 |
| 0 | 0 | 0 | 0 | 0 |
| 0 | 0 | 0 | 0 | 0 |
| 0 | 0 | 0 | 0 | 0 |
| 0 | 0 | 0 | 0 | 0 |
| 0.600676413 | 0.236113489 | 0.032803299 | 0.001584522 | 0.003687613 |
| 0 | 0 | 0 | 0 | 0 |
| 0 | 0 | 0 | 0 | 0 |

| | | | | |
|---|---|---|---|---|
| 0 | 0 | 0 | 0 | 0 |
| 0 | 0 | 0 | 0 | 0 |
| 0 | 0 | 0.110233608 | 0 | 0 |
| 0 | 0 | 0 | 0 | 0 |
| 0 | 0 | 0 | 0 | 0 |
| 0 | 0 | 0 | 0 | 0 |
| 0 | 0 | 0 | 0 | 0 |
| 0 | 0 | 0 | 0 | 0 |
| 0 | 0 | 0 | 0 | 0 |
| 0 | 0 | 0 | 0 | 0 |
| 0 | 0.001072477 | 0 | 0 | 0 |
| 0.001762086 | 0 | 0.000430083 | 0 | 0 |
| 0 | 0 | 0 | 0 | 0 |
| 0 | 0 | 0 | 0 | 0 |
| 0 | 0 | 0 | 0 | 0 |
| 0 | 0 | 0 | 0 | 0 |
| 0 | 0 | 0 | 0 | 0 |
| 0 | 0 | 0 | 0 | 0 |
| 0 | 0 | 0 | 0 | 0 |
| 0 | 0 | 0.004202269 | 0 | 0 |
| 0 | 0 | 0 | 0 | 0 |
| 0 | 0 | 0 | 0 | 0 |
| 0 | 0 | 0 | 0 | 0 |
| 0 | 0 | 0 | 0 | 0 |
| 0 | 0 | 0 | 0 | 0 |
| 0 | 0 | 0 | 0 | 0 |
| 0 | 0 | 0 | 0 | 0 |
| 0.01954848 | 0.016264694 | 0.01789746 | 0 | 0 |
| 0 | 0 | 0 | 0 | 0 |
| 0 | 0 | 0 | 0 | 0 |
| 0 | 0 | 0.002406789 | 0.002704956 | 0.003631826 |
| 0 | 0 | 0 | 0 | 0 |
| 0.173971673 | 0.002601027 | 0.04153684 | 0.038655126 | 0.139439499 |
| 0 | 0.001449801 | 0 | 0.047895382 | 0 |
| 0 | 0 | 0 | 0 | 0 |
| 0 | 0 | 0 | 0 | 0 |
| 0 | 0 | 0 | 0 | 0 |
| 0 | 0 | 0 | 0 | 0 |
| 0 | 0 | 0 | 0 | 0 |
| 0 | 0 | 0 | 0 | 0 |
| 0 | 0 | 0 | 0 | 0 |
| 0.084040015 | 0.099794434 | 0.328030119 | 0.011925458 | 0.091587463 |
| 0.064381479 | 0.175849312 | 0.018078341 | 0 | 0 |
| 0 | 0 | 0 | 0 | 0 |
| 0 | 0 | 0 | 0 | 0 |
| 0.022612847 | 0.004109668 | 0.005335277 | 0 | 0 |
| 0 | 0 | 0 | 0 | 0 |
| 0 | 0 | 0 | 0 | 0 |

| | | | | |
|---|---|---|---|---|
| 0 | 0 | 0 | 0 | 0 |
| 0.019906568 | 0.00645534 | 0 | 0.053779468 | 0 |
| 0 | 0.007540519 | 0.002336967 | 0.094230147 | 0 |
| 0 | 0 | 0 | 0 | 0 |
| 1.398594513 | 0.120183261 | 0.243445025 | 0.001892786 | 0.034689582 |
| 0 | 0 | 0.000847611 | 0 | 0.001607935 |
| 0 | 0 | 0 | 0 | 0 |
| 0 | 0 | 0 | 0 | 0 |
| 0 | 0 | 0 | 0 | 0 |
| 0 | 0 | 0 | 0 | 0 |
| 0 | 0 | 0 | 0 | 0 |
| 0 | 0 | 0 | 0 | 0 |
| 0 | 0 | 0 | 0 | 0 |
| 0 | 0.004387848 | 0 | 0 | 0 |
| 0 | 0 | 0 | 0 | 1.203939648 |
| 0 | 0 | 0 | 0 | 0 |
| 0 | 0.030775475 | 0 | 0 | 0 |
| 0 | 0 | 0 | 0 | 0 |
| 0 | 0 | 0 | 0 | 0 |
| 0 | 0 | 0 | 0 | 0 |
| 0 | 0 | 0 | 0 | 0 |
| 0 | 0 | 0 | 0 | 0 |
| 0 | 0 | 0 | 0 | 0 |
| 0 | 0 | 0 | 0 | 0 |
| 0.001127278 | 0 | 0 | 0 | 0 |
| 0 | 0 | 0 | 0 | 0 |
| 0 | 0 | 0 | 0 | 0 |
| 0.008360257 | 0.873486378 | 0.002631219 | 0 | 0 |
| 0 | 0 | 0 | 0 | 0 |
| 0 | 0 | 0 | 0 | 0 |
| 0 | 0 | 0 | 0 | 0 |
| 0 | 0 | 0.001019444 | 0 | 0 |
| 0 | 0 | 0 | 0 | 0 |
| 0 | 0 | 0 | 0 | 0 |
| 0 | 0 | 0 | 0 | 0 |
| 0 | 0 | 0 | 0 | 0 |
| 0 | 0.012090471 | 0 | 0 | 0 |
| 0 | 0 | 0 | 0 | 0 |
| 0 | 3.308860631 | 1.101312372 | 0.002676587 | 0 |
| 0 | 0 | 0 | 0 | 0 |
| 0 | 0 | 0 | 0 | 0 |
| 0 | 0 | 0.00070063 | 0 | 0 |
| 0.011836318 | 0.009411225 | 0.000648164 | 0.001479339 | 0 |
| 0 | 0 | 0 | 0 | 0 |
| 0 | 0 | 0 | 0 | 0 |
| 0 | 0 | 0 | 0 | 0 |
| 0.009053625 | 0 | 0.003767945 | 0 | 0.00351123 |
| 0 | 0 | 0 | 0 | 0 |

| | | | | |
|---|---|---|---|---|
| 0 | 0 | 0 | 0 | 0 |
| 0.020211708 | 0.002561983 | 0.02177039 | 0.000713904 | 0.024644806 |
| 0 | 0 | 0 | 0 | 0 |
| 0 | 0 | 0 | 0 | 0 |
| 0 | 0 | 0 | 0 | 0 |
| 0 | 0 | 0 | 0 | 0 |
| 0 | 0 | 0 | 0 | 0 |
| 0 | 0 | 0 | 0 | 0 |
| 0 | 0 | 0 | 0 | 0 |
| 0 | 0 | 0 | 0 | 0 |
| 0 | 0 | 0 | 0 | 0 |
| 0 | 0 | 0 | 0 | 0 |
| 0 | 0.05835032 | 0 | 0 | 0 |
| 0 | 0 | 0 | 0 | 0 |
| 0.004108851 | 0.017424034 | 0.012012175 | 0 | 0 |
| 0 | 0 | 0 | 0 | 0 |
| 0 | 0 | 0 | 0 | 0 |
| 0 | 0 | 0 | 0 | 0 |
| 0 | 0 | 0.00465963 | 0 | 0 |
| 0 | 0 | 0 | 0 | 0 |
| 0 | 0 | 0 | 0 | 0 |
| 0 | 0 | 0.047647234 | 0.032927379 | 0.032212123 |
| 0.024392436 | 0 | 0.044736141 | 0 | 0 |
| 0 | 0 | 0 | 0 | 0 |
| 0 | 0 | 0 | 0 | 0 |
| 0 | 0 | 0 | 0 | 0 |
| 0 | 0 | 0 | 0 | 0 |
| 0 | 0 | 0 | 0 | 0 |
| 0 | 0 | 0 | 0 | 0 |
| 0 | 0 | 0 | 0 | 0 |
| 0 | 0 | 0 | 0 | 0 |
| 0 | 0 | 0 | 0 | 0 |
| 0 | 0 | 0 | 0 | 0 |
| 0 | 0 | 0 | 0 | 0 |
| 0 | 0 | 0 | 0 | 0 |
| 0 | 0 | 0 | 0 | 0 |
| 0 | 0 | 0 | 0 | 0 |
| 0 | 0 | 0 | 0 | 0 |
| 0 | 0 | 0.052171387 | 0.006692791 | 0 |
| 0 | 0 | 0 | 0 | 0 |
| 0.021437819 | 0.033963352 | 0.118112405 | 0.010697385 | 0.018519985 |
| 0 | 0 | 0 | 0 | 0 |
| 0 | 0 | 0 | 0 | 0 |
| 0 | 0 | 0 | 0 | 0 |
| 0 | 0 | 0 | 0 | 0 |
| 0.447598678 | 0.178055698 | 0.708979357 | 0.773999818 | 0.013556734 |
| 0 | 0 | 0 | 0 | 0 |
| 0 | 0 | 0 | 0 | 0 |

| | | | | |
|---|---|---|---|---|
| 0 | 0 | 0 | 0 | 0 |
| 0 | 0 | 0 | 0 | 0 |
| 0 | 0 | 0.004867577 | 0 | 0 |
| 0 | 0 | 0 | 0 | 0 |
| 0 | 0 | 0 | 0 | 0 |
| 0 | 0 | 0 | 0 | 0 |
| 0 | 0 | 0 | 0 | 0 |
| 0 | 0 | 0.01086478 | 0 | 0.002254294 |
| 0.002241903 | 0 | 0.004575154 | 0 | 0 |
| 0.011058435 | 0.001429711 | 0.218122233 | 0 | 0.003739579 |
| 0 | 0 | 0.017496079 | 0 | 0 |
| 0 | 0 | 0 | 0 | 0 |
| 0.002636266 | 0 | 0.003603315 | 0 | 0 |
| 0 | 0 | 0 | 0 | 0 |
| 0 | 0 | 0 | 0 | 0 |
| 0 | 0 | 0 | 0 | 0 |
| 0 | 0 | 0 | 0 | 0 |
| 0 | 0 | 0 | 0 | 0 |
| 0 | 0 | 0 | 0 | 0 |
| 0.141960985 | 0.034826511 | 0.795119409 | 0.087687138 | 0.0526446 |
| 0 | 0 | 0 | 0 | 0 |
| 0 | 0 | 0 | 0 | 0 |
| 0 | 0 | 0 | 0 | 0 |
| 0 | 0 | 0 | 0 | 0 |
| 0 | 0 | 0 | 0 | 0 |
| 0 | 0 | 0 | 0 | 0 |
| 0 | 0 | 0 | 0 | 0 |
| 0 | 0 | 0 | 0 | 0 |
| 0 | 0 | 0 | 0 | 0 |
| 0 | 0.047601347 | 0.007285676 | 0 | 0 |
| 0 | 0 | 0 | 0 | 0 |
| 0 | 0 | 0 | 0 | 0 |
| 0 | 0 | 0 | 0 | 0 |
| 0 | 0 | 0 | 0 | 0 |
| 0 | 0 | 0 | 0 | 0 |
| 0 | 0 | 0 | 0 | 0 |
| 0 | 0 | 0 | 0 | 0 |
| 0 | 0 | 0 | 0 | 0 |
| 0 | 0 | 0 | 0 | 0 |
| 0 | 0 | 0 | 0 | 0 |
| 0 | 0 | 0 | 0 | 0 |
| 0 | 0 | 0 | 0 | 0 |
| 0 | 0.001447458 | 0 | 0 | 0 |
| 0 | 0 | 0 | 0 | 0 |
| 0.006391263 | 0 | 0.006652216 | 0 | 0 |
| 0 | 0 | 0 | 0 | 0 |
| 0 | 0 | 0 | 0 | 0 |
| 0 | 0 | 0 | 0 | 0 |

| | | | | |
|---|---|---|---|---|
| 0 | 0 | 0 | 0 | 0 |
| 0 | 0 | 0 | 0 | 0 |
| 0 | 0 | 0 | 0 | 0 |
| 0 | 0 | 0 | 0 | 0 |
| 0 | 0 | 0 | 0 | 0 |
| 0.003784155 | 0.002441954 | 0.008714166 | 0 | 0.003318038 |
| 0 | 0 | 0 | 0 | 0 |
| 0 | 0 | 0 | 0 | 0 |
| 0 | 0 | 0 | 0 | 0 |
| 0 | 0 | 0 | 0 | 0 |
| 0 | 0 | 0 | 0 | 0 |
| 0 | 0 | 0 | 0 | 0 |
| 0 | 0 | 0 | 0 | 0 |
| 0 | 0 | 0 | 0 | 0 |
| 0 | 0 | 0 | 0 | 0 |
| 0 | 0 | 0 | 0 | 0 |
| 0 | 0 | 0 | 0 | 0 |
| 0 | 0 | 0 | 0 | 0 |
| 0 | 0 | 0 | 0 | 0 |
| 0 | 0 | 0 | 0 | 0 |
| 0 | 0.000808976 | 0 | 0 | 0.351540142 |
| 0 | 0 | 0 | 0 | 0 |
| 0 | 0 | 0 | 0 | 0 |
| 0 | 0.017697356 | 0 | 0 | 0 |
| 0 | 0 | 0 | 0 | 0 |
| 0 | 0 | 0 | 0 | 0 |
| 0 | 0 | 0 | 0 | 0 |
| 0 | 0 | 0 | 0 | 0 |
| 0 | 0 | 0 | 0 | 0 |
| 0 | 0 | 0 | 0 | 0 |
| 0 | 0 | 0 | 0 | 0 |
| 0 | 0 | 0 | 0 | 0 |
| 0 | 0 | 0 | 0 | 0 |
| 0 | 0 | 0 | 0 | 0 |
| 0 | 0 | 0 | 0 | 0 |
| 0 | 0 | 0 | 0 | 0 |
| 0 | 0 | 0 | 0 | 0 |
| 0 | 0 | 0 | 0 | 0 |
| 0 | 0.425216634 | 0.06692708 | 0.058363423 | 0 |
| 0 | 0 | 0 | 0 | 0 |
| 0 | 0 | 0 | 0 | 0 |
| 0 | 0 | 0 | 0 | 0 |
| 0 | 0 | 0 | 0 | 0 |
| 0 | 0 | 0 | 0 | 0 |
| 0 | 0 | 0 | 0 | 0 |
| 0 | 0 | 0 | 0 | 0 |
| 0 | 0 | 0 | 0 | 0 |
| 0 | 0 | 0 | 0 | 0 |

| | | | | |
|---|---|---|---|---|
| 0 | 0 | 0 | 0 | 0 |
| 0 | 0 | 0.003461882 | 0 | 0.005804392 |
| 0 | 0 | 0 | 0 | 0 |
| 0 | 0 | 0 | 0 | 0 |
| 0 | 0 | 0 | 0 | 0 |
| 0 | 0 | 0 | 0 | 0 |
| 0 | 0 | 0 | 0 | 0 |
| 0 | 0 | 0 | 0 | 0 |
| 0 | 0 | 0 | 0 | 0 |
| 0 | 0 | 0 | 0 | 0 |
| 0 | 0 | 0.000824193 | 0.000783791 | 0 |
| 0 | 0 | 0 | 0 | 0 |
| 0 | 0 | 0.150449592 | 0 | 0 |
| 0 | 0 | 0 | 0 | 0 |
| 0 | 0 | 0 | 0 | 0 |
| 0 | 0 | 0 | 0.002295293 | 0.010090009 |
| 0 | 0 | 0 | 0 | 0 |
| 0 | 0 | 0 | 0 | 0 |
| 0 | 0 | 0 | 0 | 0 |
| 0 | 0 | 0 | 0 | 0 |
| 0.002692828 | 0 | 0.027883531 | 0 | 0 |
| 0 | 0 | 0 | 0 | 0 |
| 0 | 0 | 0 | 0 | 0 |
| 0.957594985 | 2.852956236 | 0.877404316 | 0.889419869 | 0.172024832 |
| 0 | 0 | 0 | 0 | 0 |
| 0 | 0 | 0 | 0 | 0 |
| 0 | 0 | 0 | 0 | 0 |
| 0 | 0 | 0 | 0 | 0 |
| 0 | 0 | 0 | 0 | 0 |
| 0 | 0 | 0 | 0 | 0 |
| 0.001681915 | 0 | 0 | 0 | 0 |
| 0 | 0 | 0 | 0 | 0 |
| 0 | 0 | 0 | 0 | 0 |
| 0 | 0 | 0 | 0 | 0 |

| PHL018-0120_V1 | PHL018-0261_V2 | PHL018-0153_V1 | PHL018-0080_V1 | PHL018-0223_V2 |
|---|---|---|---|---|
| CTRL | REF | CTRL | CTRL | REF |
| 0 | 0.725999297 | 0.021515501 | 0.004829959 | 0.159678345 |
| 0.356169925 | 0.093400196 | 2.517649229 | 1.660022506 | 0.186880208 |
| 0 | 4.442096392 | 3.262702826 | 0 | 0 |
| 0 | 0 | 0 | 0 | 0 |
| 0 | 0 | 0 | 0 | 0 |
| 0 | 0 | 0 | 0 | 0 |
| 0 | 0 | 0 | 0 | 0 |
| 0 | 0 | 0 | 0 | 0 |
| 0 | 0 | 0 | 0 | 0 |
| 0 | 0 | 0 | 0 | 0 |
| 0 | 0 | 0 | 0 | 0 |
| 0 | 0 | 0 | 0 | 0 |
| 0 | 0 | 0 | 0 | 0 |
| 0.293163902 | 0.566135215 | 3.27744841 | 0.734887195 | 0.779886205 |
| 0 | 0 | 0 | 0 | 0 |
| 0 | 0 | 0 | 0 | 0 |
| 0 | 0 | 0 | 0 | 0 |
| 0 | 0 | 0 | 0 | 0 |
| 0 | 0 | 0 | 0 | 0 |
| 0 | 0 | 0 | 0 | 0 |
| 0 | 0 | 0 | 0 | 0 |
| 0 | 0 | 0 | 0 | 0 |
| 0 | 0 | 0 | 0 | 0 |
| 0 | 0 | 0 | 0 | 0 |
| 0.016823515 | 0 | 0 | 0.019917393 | 0 |
| 0 | 0 | 0 | 0 | 0 |
| 0 | 0 | 0 | 0 | 0 |
| 0 | 0 | 0 | 0 | 0 |
| 0 | 0 | 0 | 0 | 0 |
| 0.598350017 | 0 | 0 | 0 | 0 |
| 0 | 0 | 0 | 0 | 0 |
| 0 | 0 | 0 | 0 | 0 |
| 0 | 0 | 0 | 0 | 0 |
| 0 | 0 | 0 | 0 | 0 |
| 0 | 0 | 0 | 0 | 0 |
| 1.484124945 | 0.479991502 | 0 | 4.250540515 | 0.195441457 |
| 0 | 0 | 0 | 0 | 0 |
| 0 | 0 | 0 | 0 | 0 |
| 0 | 0 | 0 | 0 | 0 |
| 0 | 0 | 0 | 0 | 0 |
| 0 | 0 | 0 | 0 | 0 |
| 0 | 0 | 0.186052327 | 0.001249798 | 0.049948936 |
| 3.026621654 | 3.046916394 | 1.899122771 | 0.032006297 | 0.332714589 |
| 0.046304793 | 0.006630512 | 0.012307322 | 0.139356371 | 0.004680219 |
| 0 | 0 | 0.001535737 | 0.039006397 | 0 |
| 0 | 0 | 0 | 0 | 0 |

| | | | | |
|---|---|---|---|---|
| 0 | 0.064568278 | 0 | 0.425871902 | 0.001093283 |
| 0 | 0.043954381 | 0.163401354 | 0 | 0 |
| 0 | 0 | 0.000628986 | 0 | 0.000995031 |
| 0.246323156 | 0.31676293 | 0.598743442 | 0.218208542 | 0.627362702 |
| 0 | 0.007556665 | 0 | 0.003077515 | 0.012550482 |
| 0 | 0 | 0 | 0 | 0 |
| 0 | 0 | 4.593752976 | 0 | 1.401768937 |
| 0 | 0 | 0 | 0 | 0 |
| 0 | 0 | 0 | 0 | 0 |
| 0 | 0 | 0 | 0 | 0 |
| 0 | 0 | 0 | 0 | 0.004740573 |
| 0.045031072 | 0 | 0 | 2.06692584 | 0.02412284 |
| 0 | 0.001236774 | 0 | 0 | 0 |
| 0.022848714 | 0 | 0.067619911 | 0.003235964 | 0.053347493 |
| 0 | 0 | 0 | 0 | 0.214826663 |
| 0 | 0 | 0 | 0 | 0 |
| 0.657786514 | 0.00476558 | 0 | 0 | 0 |
| 0 | 0 | 0 | 0 | 0 |
| 0 | 0 | 0 | 0 | 0 |
| 0 | 0 | 0 | 0 | 0 |
| 0 | 0.305858956 | 0.323576638 | 0.132502434 | 0.240617598 |
| 0 | 0 | 0.003360183 | 0.024881375 | 0.016726068 |
| 0 | 0 | 0 | 0 | 0.10947754 |
| 0 | 0 | 0 | 0 | 0 |
| 0 | 0 | 0 | 0 | 0 |
| 0 | 0 | 0 | 0 | 0 |
| 0 | 0.116760514 | 0 | 0 | 0 |
| 0 | 0 | 0 | 0 | 0 |
| 0 | 0 | 0.025198962 | 0.04979929 | 0.014655807 |
| 0 | 0 | 0 | 0 | 0 |
| 0 | 0 | 1.362676307 | 0 | 0.007788691 |
| 0 | 0 | 0 | 0 | 0 |
| 0 | 0 | 0 | 0 | 0 |
| 0 | 0 | 0 | 0 | 0 |
| 0 | 0 | 0 | 0 | 0 |
| 0 | 0 | 0 | 0 | 0 |
| 0 | 0 | 0 | 0 | 0 |
| 32.53532019 | 0.239735914 | 0 | 0.000973267 | 0 |
| 0 | 0 | 0 | 0 | 0 |
| 0 | 0 | 0 | 0 | 0 |
| 0 | 0 | 0 | 0 | 0 |
| 0.038577404 | 0 | 0 | 0 | 0 |
| 0 | 0 | 0 | 0 | 0 |
| 0 | 0 | 0 | 0 | 0 |
| 0 | 0 | 0 | 0 | 0 |
| 0 | 0 | 0 | 0 | 0 |
| 0 | 0 | 0.072629179 | 0 | 0 |
| 0 | 0 | 0.808916227 | 0 | 0 |

| | | | | |
|---|---|---|---|---|
| 0 | 0 | 0 | 0 | 0 |
| 0 | 0 | 0 | 0 | 0 |
| 0 | 0 | 0 | 0 | 0 |
| 0 | 0 | 0 | 0 | 0 |
| 0 | 0 | 0.092453804 | 0 | 0 |
| 0 | 0 | 0 | 0 | 0 |
| 0 | 0 | 0 | 0 | 0 |
| 0 | 0 | 0 | 0 | 0 |
| 0 | 0 | 0.008287695 | 0 | 0.050937297 |
| 0 | 0 | 0 | 0 | 0 |
| 0 | 0 | 0 | 0 | 0 |
| 0 | 0 | 0.005201 | 0.001351033 | 0 |
| 0 | 0 | 0 | 0 | 0 |
| 0 | 0 | 0 | 0 | 0 |
| 0 | 0 | 0 | 0 | 0 |
| 0 | 0 | 0 | 0 | 0 |
| 0 | 0 | 0 | 0 | 0 |
| 0 | 0 | 0 | 0 | 0 |
| 0.058861748 | 0.102014148 | 2.442421342 | 0.077151167 | 0.883257597 |
| 0 | 0 | 0 | 0 | 0 |
| 0 | 0 | 0 | 0 | 0 |
| 0 | 0 | 0 | 0 | 0 |
| 0 | 0 | 0 | 0 | 0 |
| 0 | 0 | 0 | 0 | 0 |
| 0 | 0 | 0 | 0 | 0 |
| 0 | 0 | 0 | 0 | 0 |
| 0 | 0 | 0 | 0 | 0.010709782 |
| 0 | 0 | 0 | 0 | 0.002385066 |
| 0 | 0 | 0 | 0 | 0 |
| 0 | 0 | 0 | 0 | 0 |
| 0 | 0 | 0.005492799 | 0 | 0.217234968 |
| 0 | 0.020750223 | 0.0034792 | 0.003765712 | 0.000859994 |
| 0 | 0 | 0 | 0 | 0 |
| 0 | 0 | 0 | 0 | 0 |
| 0 | 0 | 0 | 0 | 0 |
| 0 | 0 | 0 | 0 | 0 |
| 0 | 0 | 0 | 0 | 0 |
| 0 | 0 | 0 | 0 | 0 |
| 0.625587577 | 0.182810588 | 0 | 0 | 0 |
| 0 | 0 | 0 | 0 | 0 |
| 0 | 0 | 0 | 0 | 0 |
| 0 | 0 | 0 | 0 | 0 |
| 0 | 0.007261204 | 0 | 0.001741027 | 0.000745513 |
| 0 | 0 | 0 | 0 | 0 |
| 0 | 0 | 0 | 0 | 0 |
| 0 | 0 | 0 | 0 | 0 |
| 0 | 0.000461372 | 0 | 0 | 0 |
| 0 | 0 | 0 | 0 | 0 |

| | | | | |
|---|---|---|---|---|
| 0 | 0 | 0 | 0 | 0 |
| 0 | 0 | 0 | 0 | 0 |
| 0 | 0 | 0 | 0 | 0 |
| 0 | 0 | 0 | 0 | 0 |
| 0 | 0 | 0 | 0.862835092 | 0 |
| 0 | 0 | 0 | 0 | 0 |
| 0 | 0 | 0 | 0 | 0 |
| 0 | 0 | 0 | 0 | 0 |
| 0 | 0 | 0 | 0 | 0 |
| 0 | 0 | 0 | 0 | 0 |
| 0 | 0 | 0 | 0 | 0 |
| 0 | 0 | 0 | 0 | 0 |
| 0 | 0 | 0 | 0 | 0 |
| 0 | 0 | 0 | 0 | 0 |
| 0 | 0 | 0 | 0 | 0 |
| 0.882939243 | 0 | 0 | 0 | 0 |
| 0 | 0 | 0 | 0 | 0 |
| 0 | 0 | 0 | 0 | 0 |
| 0 | 0 | 0 | 0 | 0 |
| 0 | 0 | 0 | 0 | 0 |
| 0 | 0 | 0 | 0 | 0 |
| 0 | 0 | 0 | 0 | 0 |
| 0 | 0 | 0 | 0 | 0 |
| 0 | 0 | 0 | 0 | 0 |
| 0 | 0.018750579 | 0 | 0 | 0 |
| 0 | 0 | 0 | 0 | 0 |
| 0 | 0 | 0 | 0 | 0 |
| 0 | 0 | 0 | 0 | 0 |
| 0 | 0 | 0 | 0 | 0 |
| 0 | 0 | 0 | 0 | 0 |
| 0 | 0 | 0 | 0 | 0 |
| 2.115568059 | 0 | 0 | 0 | 0 |
| 0 | 0 | 0 | 0 | 0 |
| 0 | 0 | 0 | 0 | 0 |
| 0 | 0 | 0 | 0 | 0 |
| 0 | 0 | 0 | 0 | 0 |
| 0 | 0 | 0 | 0 | 0 |
| 0.088402951 | 0 | 0 | 0 | 0 |
| 0 | 0 | 0 | 0 | 0 |
| 0 | 0 | 0 | 0 | 0 |
| 0 | 0 | 0 | 0 | 0 |
| 0 | 0 | 0 | 0 | 0 |
| 0 | 0 | 0 | 0 | 0 |
| 0 | 0 | 0 | 0 | 0 |
| 0 | 0 | 0 | 0 | 0 |
| 0 | 0 | 0 | 0 | 0 |
| 0 | 0 | 0 | 0 | 0 |
| 0 | 0 | 0 | 0 | 0 |

| | | | | |
|---|---|---|---|---|
| 0 | 0 | 0 | 0 | 0 |
| 0 | 0 | 0 | 0 | 0 |
| 0 | 0 | 0 | 0 | 0 |
| 0 | 0 | 0 | 0 | 0 |
| 0 | 0 | 0 | 0 | 0 |
| 0 | 0 | 0 | 0 | 0 |
| 0 | 0 | 0 | 0 | 0 |
| 0 | 0 | 0 | 0 | 0 |
| 0 | 0 | 0 | 0 | 0 |
| 0 | 0 | 0 | 0 | 0 |
| 0 | 0 | 0 | 0 | 0 |
| 0 | 0 | 0 | 0 | 0 |
| 0 | 0 | 0 | 0.003326053 | 0 |
| 0 | 0 | 0 | 0 | 0 |
| 0 | 0 | 0 | 0 | 0 |
| 0 | 0 | 0 | 0 | 0 |
| 0 | 0 | 0 | 0 | 0 |
| 0 | 0 | 0 | 0 | 0 |
| 0 | 0 | 0 | 0 | 0 |
| 0 | 0 | 0 | 0 | 0 |
| 0 | 0.013164941 | 0.015037722 | 0.005441554 | 0.018196887 |
| 0 | 0 | 0 | 0 | 0 |
| 0 | 0 | 0 | 0 | 0 |
| 0 | 0.006556841 | 0.002574545 | 0 | 0.010644901 |
| 0 | 0 | 0 | 0 | 0 |
| 0 | 0 | 0 | 0 | 0 |
| 0 | 0.001945346 | 0.005097716 | 0 | 0.016542342 |
| 0 | 0 | 0 | 0 | 0 |
| 0 | 0 | 0 | 0 | 0 |
| 0 | 0.009182188 | 0 | 0 | 0.009678813 |
| 0 | 0 | 0.008599559 | 0 | 0.003349512 |
| 0 | 0 | 0 | 0 | 0 |
| 0 | 0 | 0 | 0 | 0 |
| 0 | 0 | 0 | 0.07173665 | 0.004611066 |
| 0 | 0.043851844 | 0.020006792 | 0.028324523 | 0.006930652 |
| 0 | 0 | 0 | 0 | 0 |
| 0 | 0 | 0 | 0 | 0 |
| 0 | 0 | 0 | 0 | 0 |
| 0 | 0 | 0 | 0 | 0 |
| 0 | 0 | 0 | 0 | 0 |
| 0 | 0 | 0 | 0 | 0 |
| 0 | 0 | 0 | 0 | 0 |
| 0 | 0 | 0 | 0 | 0 |
| 0 | 0 | 0 | 0 | 0 |
| 0 | 0.002455661 | 0 | 0 | 0 |
| 0.027202527 | 0 | 0 | 0.055621563 | 0 |
| 0 | 0 | 0 | 0.001095283 | 0.001563345 |
| 0 | 0.005010934 | 0 | 0.001802216 | 0 |

| | | | | |
|---|---|---|---|---|
| 0 | 0 | 0 | 0 | 0 |
| 0 | 0 | 0 | 0 | 0 |
| 0 | 0 | 0.000760765 | 0 | 0 |
| 0 | 0.00046324 | 0 | 0 | 0 |
| 0 | 0 | 0 | 0 | 0 |
| 0 | 0 | 0 | 0 | 0 |
| 0 | 0 | 0.362054533 | 0 | 0.003515281 |
| 0 | 0 | 0.045927302 | 0.002072575 | 0 |
| 0 | 0 | 0 | 0 | 0 |
| 0 | 0 | 0 | 0 | 0 |
| 0 | 0 | 0 | 0 | 0.001018459 |
| 0 | 0 | 0 | 0 | 0 |
| 0 | 0 | 0 | 0 | 0 |
| 0 | 0.023006774 | 0 | 0 | 0 |
| 0 | 0 | 0 | 0 | 0.005515876 |
| 0 | 0 | 0 | 0 | 0 |
| 0 | 0 | 0 | 0 | 0 |
| 0 | 0 | 0 | 0 | 0 |
| 0 | 0.012971858 | 0.008255282 | 0 | 0.001465013 |
| 0 | 0 | 0 | 0 | 0.418270264 |
| 0 | 0.3589144 | 0.087895713 | 0 | 0 |
| 0 | 0 | 0 | 0 | 0 |
| 0 | 0 | 0 | 0 | 0 |
| 0 | 0 | 0 | 0 | 0 |
| 0 | 0 | 0.006203318 | 0.002148532 | 0.003066691 |
| 0 | 0 | 0 | 0 | 0 |
| 0 | 0 | 0 | 0 | 0 |
| 0 | 0 | 0 | 0 | 0 |
| 0 | 0 | 0.004143942 | 0.004357047 | 0.052678587 |
| 0 | 0 | 0.00195028 | 0 | 0 |
| 0 | 0.231046534 | 0.103774836 | 0.040157132 | 0.304636644 |
| 0 | 0 | 0 | 0 | 0 |
| 0 | 0 | 0 | 0 | 0 |
| 0.034868224 | 0 | 0.003861501 | 0 | 0 |
| 0 | 0 | 0 | 0 | 0 |
| 0 | 0 | 0 | 0 | 0 |
| 0 | 0 | 0 | 0 | 0 |
| 0 | 0 | 0 | 0 | 0 |
| 0 | 0 | 0 | 0 | 0 |
| 0 | 0 | 0 | 0.006155709 | 0 |
| 0 | 0 | 0 | 0 | 0 |
| 0 | 0 | 0.00152119 | 0.001215847 | 0.026378547 |
| 0 | 0 | 0 | 0 | 0 |
| 0 | 0 | 0 | 0 | 0 |
| 0 | 0 | 0 | 0 | 0 |
| 0 | 0 | 0 | 0 | 0 |
| 0 | 0 | 0 | 0 | 0 |
| 0 | 0 | 0 | 0 | 0 |

| | | | | |
|---|---|---|---|---|
| 0 | 0 | 0 | 0 | 0 |
| 0 | 0 | 0 | 0 | 0 |
| 0.103486067 | 0.002829914 | 0.00955052 | 0.142746086 | 0.001743295 |
| 0 | 0 | 0 | 0 | 0 |
| 0 | 0 | 0.115652337 | 0 | 0 |
| 0 | 0 | 0 | 0 | 0 |
| 0 | 0 | 0 | 0 | 0 |
| 0 | 0 | 0 | 0 | 0 |
| 0 | 0 | 0 | 0 | 0 |
| 0 | 0 | 0 | 0 | 0 |
| 0 | 0 | 0 | 0 | 0 |
| 0 | 0 | 0 | 0 | 0 |
| 0 | 0 | 0 | 0 | 0 |
| 0 | 0 | 0.006982571 | 0 | 0.003532202 |
| 0 | 0 | 0.005897226 | 0.00116052 | 0.003445438 |
| 0.067377097 | 0 | 0 | 0 | 0.004540057 |
| 0 | 0.002941269 | 0.003970531 | 0.003173541 | 0.00422775 |
| 0 | 0 | 0 | 0 | 0 |
| 0 | 0 | 0 | 0 | 0 |
| 0 | 0 | 0 | 0 | 0 |
| 0 | 0 | 0 | 0 | 0 |
| 0 | 0.005068878 | 0 | 0.000716201 | 0 |
| 0 | 0 | 0 | 0 | 0 |
| 0 | 0.008232105 | 0 | 0 | 0 |
| 0 | 0 | 0.003978793 | 0 | 0.003933937 |
| 0 | 0 | 0 | 0 | 0 |
| 8.529466058 | 0 | 0 | 0 | 0 |
| 0 | 0 | 0 | 0 | 0 |
| 0 | 0 | 0 | 0 | 0 |
| 0 | 0 | 0.00837522 | 0 | 0.001971619 |
| 0 | 0 | 0.001056413 | 0 | 0 |
| 0 | 0 | 0 | 0 | 0 |
| 0 | 0 | 0 | 0 | 0 |
| 0 | 0 | 0 | 0 | 0 |
| 0 | 0 | 0 | 0 | 0 |
| 0 | 0 | 0.071865772 | 0.004291526 | 0.006533845 |
| 0 | 0 | 0 | 0 | 0 |
| 0 | 0.001213052 | 0.018532594 | 0.048290977 | 0.006507468 |
| 0 | 0 | 0 | 0 | 0 |
| 0 | 0 | 0 | 0 | 0 |
| 0 | 0 | 0.005249562 | 0 | 0 |
| 0 | 0 | 0 | 0 | 0 |
| 0 | 0 | 0 | 0 | 0 |
| 0 | 0 | 0.000860575 | 0 | 0 |
| 0 | 0 | 0 | 0 | 0 |
| 0 | 0 | 0 | 0 | 0 |
| 0 | 0 | 0 | 0 | 0.001485023 |
| 0 | 0 | 0 | 0.001089922 | 0 |

| | | | | |
|---|---|---|---|---|
| 0.63857761 | 0 | 0 | 0.05532681 | 0.832778015 |
| 0 | 0.117632425 | 0 | 0 | 0 |
| 0 | 0.000758465 | 0 | 0.001250274 | 0 |
| 0 | 0 | 0 | 0 | 0 |
| 0 | 0 | 0.000871642 | 0 | 0.008618155 |
| 0 | 0 | 0.001895056 | 0 | 0 |
| 0 | 0 | 0 | 0 | 0 |
| 0.079679034 | 0 | 0.019956042 | 0 | 0.023757807 |
| 0 | 0 | 0.001412271 | 0 | 0 |
| 0 | 0.194958415 | 0.063176598 | 0.006542594 | 0.011621275 |
| 0 | 0 | 0 | 0 | 0.002251529 |
| 0 | 0 | 0.021277961 | 0.030986089 | 0.024544422 |
| 0 | 0.002890231 | 0.010393239 | 0.005293705 | 0.004594006 |
| 0 | 0 | 0 | 0 | 0 |
| 0 | 0 | 0 | 0 | 0.002238078 |
| 0 | 0 | 0.249264523 | 0 | 0 |
| 0 | 0 | 0 | 0 | 0 |
| 0 | 0.001672955 | 0 | 0 | 0 |
| 0 | 0.072620143 | 0 | 0 | 0.043351081 |
| 0 | 0 | 0.001741409 | 0 | 0.053844639 |
| 0 | 0 | 0 | 0 | 0 |
| 0 | 0.002444599 | 0 | 0 | 0 |
| 0 | 0 | 0 | 0 | 0.003492942 |
| 0.031805525 | 0 | 0 | 0 | 0 |
| 0 | 0 | 0 | 0 | 0 |
| 0 | 0.000374043 | 0.000996921 | 0.01677102 | 0 |
| 0.991077204 | 0 | 0 | 0 | 0 |
| 0 | 0 | 2.566282273 | 0 | 0 |
| 0 | 0 | 0 | 0 | 0 |
| 0 | 0 | 0 | 0 | 0 |
| 0 | 0 | 0.001681037 | 0 | 0 |
| 0 | 0.00564233 | 0 | 0 | 0 |
| 0 | 0.021068017 | 0 | 0 | 0 |
| 0 | 0 | 0 | 0 | 0.040514294 |
| 0 | 0 | 0 | 0 | 0 |
| 0 | 0 | 0 | 0 | 0.010280926 |
| 0 | 0 | 0.005004784 | 0 | 0 |
| 0 | 0 | 0 | 0 | 0 |
| 0 | 0 | 0 | 0 | 0 |
| 0 | 0 | 0 | 0 | 0.008885875 |
| 0 | 0 | 0 | 0 | 0 |
| 0 | 0 | 0 | 0 | 0.096908407 |
| 0 | 0 | 0 | 0 | 0 |
| 0 | 0 | 0 | 0 | 0 |
| 0 | 0 | 0 | 0 | 0 |
| 0 | 0 | 0 | 0 | 0 |
| 0 | 0 | 0.000708429 | 0.001226829 | 0 |
| 0 | 0 | 0 | 0 | 0 |

| | | | | |
|---|---|---|---|---|
| 0 | 0 | 0 | 0 | 0 |
| 0 | 0 | 0.130831532 | 0 | 0.018395637 |
| 0 | 0 | 0 | 0 | 0 |
| 0 | 0 | 0 | 0 | 0 |
| 0 | 0 | 0 | 0 | 0 |
| 0 | 0 | 0 | 0 | 0.039997018 |
| 0 | 0 | 0 | 0 | 0 |
| 0 | 0 | 0 | 0 | 0 |
| 0 | 0 | 0 | 0 | 0 |
| 0 | 0 | 0 | 0 | 0 |
| 0 | 0 | 0.003882932 | 0 | 0.030393321 |
| 0 | 0 | 0 | 0 | 0 |
| 0 | 0 | 0 | 0 | 0 |
| 0 | 0.198045012 | 0 | 0 | 0.005138309 |
| 0 | 0 | 0 | 0.002678283 | 0.014373832 |
| 0 | 0 | 0.000467621 | 0 | 0 |
| 0 | 0 | 0 | 0 | 0 |
| 0 | 0 | 0 | 0 | 0.006471112 |
| 0 | 0 | 0.003579076 | 0 | 0 |
| 0 | 0.036589266 | 0 | 0 | 0 |
| 0 | 0 | 1.664682116 | 0 | 0 |
| 0.101429559 | 0.044630981 | 0 | 0.051175413 | 0.265866213 |
| 0 | 0.026412521 | 0 | 0 | 0 |
| 0 | 0 | 0 | 0 | 0 |
| 0 | 0 | 0 | 0 | 0 |
| 0 | 0 | 0 | 0 | 0 |
| 0 | 0 | 0.101758113 | 0 | 0.000983168 |
| 0 | 0 | 0 | 0 | 0 |
| 0 | 0 | 0 | 0 | 0 |
| 0.227932536 | 0.030009969 | 0.187302479 | 0.143945753 | 0.397850205 |
| 0 | 0 | 0 | 0.14779231 | 0.00328088 |
| 0 | 0 | 0 | 0 | 0 |
| 0 | 0 | 0 | 0 | 0 |
| 0 | 0 | 0 | 0 | 0 |
| 0 | 0 | 0 | 0.002629523 | 0.001637773 |
| 0 | 0 | 0 | 0 | 0 |
| 0 | 0 | 0 | 0.382050716 | 0.576489934 |
| 0 | 0 | 0 | 0.040200088 | 0 |
| 0 | 0 | 0 | 0.023727298 | 0.053864642 |
| 0 | 0 | 0 | 0 | 0 |
| 0 | 0.032892939 | 0.016335709 | 0.000870447 | 0 |
| 0.046516048 | 0 | 0 | 1.15624536 | 1.307250703 |
| 0 | 0 | 0 | 0.036825882 | 0 |
| 0 | 0 | 0 | 0 | 0 |
| 0 | 0 | 0 | 0 | 0.000840913 |
| 0 | 0 | 0 | 0 | 0 |
| 0 | 0.005311308 | 0.010155411 | 0 | 0.032078783 |
| 0 | 0 | 0 | 0 | 0.027096595 |

| | | | | |
|---|---|---|---|---|
| 0 | 0.005983414 | 0.003254564 | 0.012399474 | 0 |
| 0 | 0 | 0 | 0 | 0.011925529 |
| 0 | 0 | 0.071618682 | 0 | 0 |
| 0 | 0 | 0 | 0 | 0 |
| 0 | 0.182162125 | 0 | 0 | 0 |
| 0 | 0 | 0 | 0 | 0.0171405 |
| 0 | 0 | 0 | 0 | 0 |
| 0 | 0 | 0.18001432 | 0 | 0 |
| 0 | 0 | 0.009805603 | 0 | 0.03608715 |
| 0 | 0 | 0 | 0 | 0 |
| 0 | 0 | 0 | 0 | 0 |
| 0 | 0 | 0 | 0 | 0 |
| 0 | 0 | 0 | 0 | 0 |
| 0 | 0 | 0 | 0 | 0 |
| 0 | 0 | 0 | 0 | 0 |
| 0.027009821 | 0.014906418 | 0 | 0.100612932 | 0.022067466 |
| 0.024947975 | 0.005693464 | 0.08212123 | 0.094735896 | 0.000756479 |
| 0 | 0.738527484 | 0.003253503 | 0 | 0 |
| 0 | 0 | 0 | 0 | 0 |
| 0 | 0.034838312 | 0 | 0 | 0 |
| 0 | 0 | 0 | 0.01475033 | 0.045444384 |
| 0 | 0 | 0 | 0.004610715 | 0.000877476 |
| 0 | 0 | 0 | 0 | 0 |
| 0 | 0 | 0.368518137 | 0.005606667 | 0 |
| 0 | 0 | 0 | 0.005159665 | 0 |
| 0 | 0.016617948 | 0 | 0 | 0 |
| 0 | 0.001700682 | 0.003723342 | 0 | 0 |
| 0 | 0 | 0 | 0 | 0 |
| 0 | 0 | 0 | 0 | 0 |
| 0 | 0 | 0 | 0 | 0 |
| 0 | 0.019077118 | 0.003693379 | 0.001279209 | 0.172605598 |
| 0 | 0.003053058 | 0.006974742 | 0.010171435 | 0.01742175 |
| 0 | 0.025244337 | 0.014155597 | 0.037527751 | 0.004492546 |
| 0 | 0 | 0.003756732 | 0 | 0 |
| 0.027978089 | 0 | 0.01573059 | 0 | 0 |
| 0 | 0 | 0 | 0 | 0 |
| 0 | 0.008990729 | 0.000947973 | 0 | 0.002499428 |
| 0 | 0 | 0.01247954 | 0.032096388 | 0.014660016 |
| 0.039507393 | 0 | 0 | 0.040374893 | 0 |
| 0 | 0 | 0.004950115 | 0.050209839 | 0 |
| 0 | 0 | 0 | 0 | 0 |
| 0 | 0 | 0 | 0 | 0.042155707 |
| 0 | 0.001617708 | 0.000923917 | 0.003200006 | 0.002192403 |
| 0 | 0.340890029 | 0.030961623 | 0.003674761 | 0.62331379 |
| 0 | 0 | 0.503585548 | 0 | 0 |
| 0 | 0 | 0 | 0 | 0 |
| 0 | 0 | 0 | 0 | 0 |
| 0 | 0 | 0 | 0 | 0 |

| | | | | |
|---|---|---|---|---|
| 0 | 0 | 0 | 0 | 0 |
| 0.191050365 | 0.008106879 | 1.801346816 | 0.729798044 | 1.282467677 |
| 0 | 0 | 0 | 0 | 0 |
| 0 | 0 | 0 | 0 | 0 |
| 0 | 0 | 0.001321399 | 0 | 0 |
| 0 | 0.002325841 | 0 | 0 | 0 |
| 0 | 0 | 0 | 0 | 0 |
| 0 | 0 | 0 | 0 | 0 |
| 0 | 0 | 0.002928377 | 0 | 0 |
| 0 | 0.001499555 | 0 | 0 | 0 |
| 0 | 0 | 0.003878543 | 0 | 0.018555563 |
| 0.109381187 | 0.725700273 | 0.024413325 | 0.019041246 | 0.016951943 |
| 0.083350627 | 0 | 0 | 0 | 0 |
| 0 | 0 | 0 | 0 | 0 |
| 0.028085229 | 0.017091808 | 0 | 0 | 0 |
| 0 | 0 | 0 | 0 | 0 |
| 0 | 0.00034409 | 0.034456342 | 0.012024776 | 0.047669131 |
| 0 | 0 | 2.16247299 | 0 | 0.058854324 |
| 0 | 0 | 0 | 0 | 0 |
| 0 | 0 | 0 | 0.003251234 | 0.039963982 |
| 0.100444848 | 0.003193589 | 0.038370431 | 0.03930749 | 0.16457545 |
| 0 | 0 | 0.016307558 | 0 | 0 |
| 0 | 0 | 0 | 0 | 0 |
| 0 | 0 | 0 | 0 | 0 |
| 0 | 0 | 0.05788633 | 0 | 0.004040028 |
| 0 | 0 | 0 | 0 | 0.001960004 |
| 0 | 0.040745511 | 0 | 0.011085273 | 0.271619349 |
| 0.016856533 | 0.092275033 | 0.095062509 | 0.015418093 | 0.077521101 |
| 0 | 0 | 0 | 0 | 0 |
| 0 | 0 | 0 | 0 | 0 |
| 0 | 0 | 0.000616327 | 0 | 0.001218758 |
| 0 | 0 | 0 | 0 | 0.002743647 |
| 0 | 0 | 0 | 0 | 0 |
| 0 | 0 | 0 | 0 | 0 |
| 0 | 0 | 0 | 0.118104972 | 0 |
| 0.351853685 | 0.033404207 | 0.077527421 | 0.219696356 | 0.663207977 |
| 0 | 0 | 0 | 0 | 0 |
| 0 | 0 | 0 | 0 | 0 |
| 0 | 0 | 0.05208074 | 0.018713169 | 0.017164529 |
| 0 | 0 | 0 | 0 | 0 |
| 0 | 0 | 0.474873988 | 0 | 0 |
| 0 | 0 | 0 | 0 | 0 |
| 0 | 0 | 0.08274502 | 0 | 0.004042734 |
| 0 | 0 | 0.000623319 | 0 | 0.014585579 |
| 0 | 0 | 0 | 0 | 0.016193299 |
| 0.092398459 | 0 | 0 | 0.305959433 | 2.167944574 |
| 0 | 0 | 0 | 0 | 0 |
| 0 | 0 | 0.001007547 | 0 | 0 |

| | | | | |
|---|---|---|---|---|
| 0 | 0 | 0.013528859 | 0 | 0 |
| 0 | 0 | 0 | 0 | 0 |
| 0 | 0.000604301 | 0 | 0 | 0 |
| 0.04903647 | 0 | 0.453912267 | 0.130432447 | 0.053528272 |
| 0 | 0 | 0 | 0 | 0 |
| 0 | 0 | 0 | 0 | 0 |
| 0 | 0 | 0 | 0 | 0 |
| 0 | 0.002846366 | 0 | 0.080379245 | 0 |
| 0.518951018 | 0 | 0 | 0.970270002 | 0.764576258 |
| 0.043826545 | 0 | 0 | 0 | 0 |
| 0 | 0 | 0 | 0 | 0 |
| 0 | 0 | 0 | 0.057181837 | 0.004986123 |
| 0 | 0 | 0 | 0 | 0 |
| 0 | 0 | 0 | 0 | 0 |
| 0 | 0 | 0.007629376 | 0.003774928 | 0.187310517 |
| 0 | 0 | 0 | 0 | 0 |
| 0.171944316 | 0 | 0 | 0 | 0 |
| 0.334141976 | 0 | 0 | 0.918453111 | 0 |
| 0 | 0 | 0 | 0 | 0 |
| 0 | 0 | 0 | 0 | 0 |
| 0 | 0 | 0 | 0 | 0 |
| 0 | 0 | 0 | 0 | 0 |
| 0 | 0 | 0 | 0 | 0 |
| 0.115770404 | 0.208889687 | 0.076926358 | 0.147101747 | 0.353684246 |
| 0 | 0 | 0 | 0 | 0 |
| 0.104267208 | 0 | 0 | 0 | 5.026283876 |
| 0 | 0 | 0 | 0.001853107 | 0 |
| 0.888543193 | 0 | 3.939050914 | 0 | 0 |
| 0 | 0 | 0 | 0 | 0 |
| 0 | 0 | 0 | 0 | 0 |
| 0.013702416 | 0 | 0 | 0 | 0.009694724 |
| 0 | 0 | 0 | 0 | 1.469088368 |
| 0 | 0.003188801 | 0 | 0 | 0 |
| 0 | 0 | 0 | 0 | 0 |
| 0 | 0 | 0 | 0 | 0 |
| 0 | 0 | 0.006037381 | 0.036507831 | 0.08748473 |
| 0.705228417 | 0 | 0 | 0.089430164 | 2.454370916 |
| 0 | 0 | 0 | 0 | 0 |
| 0 | 0 | 0 | 0 | 0 |
| 0 | 0 | 0 | 0 | 0.001019695 |
| 0 | 0.163161298 | 2.81343149 | 0 | 0.53504559 |
| 0 | 0 | 0 | 0 | 0.197431332 |
| 0.059475058 | 0.006653437 | 3.735430933 | 1.425242788 | 0.012569277 |
| 0 | 0 | 0 | 0 | 0 |
| 0 | 0 | 0 | 0 | 0 |
| 0 | 0 | 0 | 0 | 0 |
| 0 | 0 | 0 | 0 | 0 |
| 0 | 0.144982548 | 0.003738621 | 0 | 0 |

| | | | | |
|---|---|---|---|---|
| 0 | 0 | 0 | 0 | 0 |
| 0.034552011 | 0 | 0 | 0.219113026 | 0.069347429 |
| 0.015581452 | 0.142816932 | 0.051767305 | 0 | 0.956216262 |
| 0 | 0 | 0 | 0 | 0 |
| 0 | 0 | 0 | 0 | 0 |
| 0 | 0 | 0 | 0.109373047 | 0 |
| 0 | 0.111576713 | 0 | 0 | 0 |
| 0 | 0.001020465 | 0 | 0 | 0 |
| 0 | 0 | 0 | 0 | 0 |
| 0 | 0 | 0 | 0 | 0 |
| 0 | 0.434924388 | 0.004431012 | 0.000677069 | 0.047418502 |
| 0.109840267 | 2.607462662 | 1.942080981 | 0.993530763 | 1.488595311 |
| 0 | 0 | 0 | 0 | 0 |
| 0.099062795 | 0 | 0.045852176 | 0.360975933 | 0 |
| 0 | 0.025423062 | 0.00443353 | 0.005374467 | 0.075397013 |
| 0.579711706 | 0 | 0 | 0 | 0.019795234 |
| 0 | 0 | 0 | 0 | 0 |
| 0 | 0 | 0 | 0 | 0 |
| 0 | 0 | 0 | 0 | 0.046811306 |
| 0 | 0 | 0.005474981 | 0.001487271 | 0 |
| 0 | 0.00334695 | 0.45734854 | 0 | 0.42357267 |
| 0 | 1.80100829 | 0.495024323 | 0.000597952 | 0 |
| 0.040369481 | 0.162760528 | 2.935007267 | 0.057888214 | 0.379469073 |
| 0 | 0 | 0 | 0 | 0 |
| 0 | 0 | 0 | 0 | 0 |
| 2.99101405 | 0 | 0 | 0 | 7.194977355 |
| 0 | 0 | 0 | 0 | 0 |
| 0 | 0 | 0 | 0 | 0 |
| 0 | 0 | 0 | 0 | 0 |
| 0 | 0 | 0 | 0 | 0 |
| 0 | 0 | 0 | 0 | 0 |
| 0.072630632 | 0 | 0.040217605 | 0.129115264 | 0.208813009 |
| 0 | 0 | 2.974593976 | 0.002317892 | 0.004234786 |
| 0 | 0.027075027 | 0 | 0 | 0 |
| 0 | 0 | 0.002650201 | 0.001070886 | 0.004716581 |
| 0 | 0.011050837 | 0.104988344 | 0.262107308 | 0.001920087 |
| 0 | 0 | 0.000820718 | 0 | 0.01182421 |
| 0 | 0 | 0 | 0 | 0 |
| 1.00035472 | 0 | 0 | 0.445688763 | 0.045218658 |
| 0.104771867 | 0 | 0 | 0 | 0 |
| 0.025091472 | 0 | 0 | 0 | 0 |
| 0 | 0 | 0 | 0 | 0 |
| 0 | 0 | 0 | 0 | 0 |
| 0 | 0 | 0 | 0 | 0 |
| 0 | 0.527597014 | 0.00498287 | 0.007670342 | 0.277537069 |
| 0 | 0 | 0 | 0 | 0.296173551 |
| 0 | 0 | 0 | 0 | 0 |
| 0 | 0.230364642 | 0.037066 | 0.46754182 | 0.046883183 |

| | | | | |
|---|---|---|---|---|
| 0 | 0 | 0 | 0 | 0 |
| 0 | 0 | 0 | 0 | 0 |
| 0 | 0 | 0 | 0 | 0 |
| 0 | 0 | 0 | 0.002247232 | 0 |
| 0 | 0 | 0 | 0 | 0 |
| 0 | 0 | 0 | 0 | 0 |
| 0.030321664 | 0 | 0 | 0 | 0 |
| 0 | 0.962121082 | 0.01074672 | 0.001127924 | 0.107972888 |
| 0 | 0 | 0 | 0 | 0 |
| 0 | 0 | 0 | 0 | 0 |
| 0 | 0.005406101 | 0 | 0.001137646 | 0 |
| 0 | 0 | 0 | 0 | 0 |
| 0 | 0.009869776 | 0 | 0.000597659 | 0 |
| 0.491337041 | 0 | 0 | 0 | 0.248750874 |
| 0 | 0 | 0 | 0 | 0 |
| 0 | 0 | 0 | 0 | 0 |
| 0 | 0 | 0 | 0.147899859 | 0 |
| 0 | 0 | 0.007875067 | 0 | 0 |
| 0 | 0 | 0 | 0 | 0 |
| 0 | 0 | 0 | 0 | 0.002340646 |
| 0 | 0 | 0 | 0.003870403 | 0 |
| 0 | 0 | 0 | 0 | 0 |
| 0.083881995 | 0 | 0 | 0 | 0 |
| 0 | 0.352384589 | 0 | 0 | 0 |
| 0 | 0 | 0 | 0 | 0 |
| 0 | 0 | 0 | 0 | 0 |
| 0 | 0 | 0 | 0 | 0 |
| 0 | 0 | 0 | 0 | 0 |
| 0 | 0 | 0 | 0 | 0 |
| 0.537668432 | 0 | 0 | 0.503379877 | 1.238164028 |
| 0 | 0 | 0.003311398 | 0 | 0 |
| 0 | 0 | 0 | 0 | 0 |
| 0 | 0 | 0 | 0 | 0 |
| 0 | 0 | 0 | 0 | 0 |
| 0 | 0 | 0 | 0.001266441 | 0 |
| 0 | 0 | 0 | 0 | 0 |
| 0.011044594 | 0 | 0 | 0 | 0 |
| 0 | 0 | 0.020130934 | 0.001733467 | 0 |
| 0 | 0 | 0 | 0 | 0 |
| 0 | 0 | 0 | 0.004939411 | 0 |
| 0 | 0 | 0 | 0 | 0 |
| 0 | 0 | 0 | 0 | 0 |
| 0 | 0 | 0 | 0 | 0 |
| 0.185954191 | 0 | 0 | 0 | 0 |
| 0.044474059 | 0.018463833 | 0 | 0 | 0.210274508 |
| 0 | 0 | 0 | 0 | 0 |
| 0 | 0 | 0 | 0.01354269 | 0 |
| 0 | 0 | 0 | 0 | 0 |

| | | | | |
|---|---|---|---|---|
| 0 | 0 | 0 | 0 | 0 |
| 0 | 0 | 0 | 0 | 0 |
| 0.060431301 | 0 | 0 | 0 | 0 |
| 0.194799897 | 0 | 0 | 0.539894829 | 0.231676745 |
| 0 | 0 | 0 | 0 | 0 |
| 0 | 0 | 0 | 0 | 0 |
| 0 | 0.000856076 | 0 | 0 | 0 |
| 0 | 0 | 0 | 0 | 0 |
| 0 | 0.122336895 | 0.056448191 | 0.003187654 | 0.00788645 |
| 0 | 0 | 0 | 0.00213917 | 0 |
| 0 | 0 | 0 | 0 | 0 |
| 0.043909522 | 0 | 0 | 0 | 0 |
| 0 | 0 | 0 | 0 | 0.016987594 |
| 0 | 0 | 0 | 0 | 0 |
| 0.465827215 | 0 | 0 | 0.11587287 | 1.027981204 |
| 0 | 1.126907366 | 0.180999941 | 0 | 0 |
| 0 | 0.113549852 | 0 | 0 | 0 |
| 0 | 0 | 0 | 0 | 0 |
| 0 | 0 | 0 | 0 | 0 |
| 0 | 0 | 0 | 0 | 0 |
| 0.119324668 | 0 | 0 | 0 | 0 |
| 0 | 0 | 0 | 0 | 0 |
| 0 | 0 | 0 | 0 | 0 |
| 0 | 0 | 0.000648306 | 0 | 0 |
| 0.091756766 | 0.175132774 | 0.058624936 | 0.004060973 | 0.009630942 |
| 4.570377211 | 0 | 0 | 39.09724076 | 0.368481881 |
| 0.231020863 | 0 | 0 | 0 | 0.215989782 |
| 0 | 0 | 0 | 0 | 0 |
| 0 | 0 | 0 | 0.514815452 | 0 |
| 0 | 0 | 0 | 0 | 0 |
| 0 | 0 | 0 | 0 | 0 |
| 0 | 0 | 0 | 0 | 0 |
| 0 | 0 | 0 | 0 | 0 |
| 0 | 0.005735064 | 0 | 0 | 0.010287083 |
| 0.018633028 | 0.013618503 | 0.015397073 | 0.117926507 | 0.001255545 |
| 0 | 0.031040973 | 0.000448819 | 0.000777247 | 0.002218796 |
| 0 | 0 | 0 | 0 | 0 |
| 0.059490659 | 0 | 0 | 0 | 4.87050585 |
| 0.016634739 | 0 | 0 | 1.725946485 | 0 |
| 0 | 0 | 0 | 0 | 0 |
| 0 | 0 | 0 | 0 | 0.146160561 |
| 0 | 0 | 0 | 0 | 0.002767746 |
| 0.149990038 | 0.479619198 | 3.998995536 | 0.00557614 | 0.454803649 |
| 0.198358859 | 0 | 0 | 0 | 0 |
| 0 | 0 | 0 | 0 | 0 |
| 0 | 0 | 0.0005315 | 0 | 0 |
| 0 | 0 | 0 | 0 | 0 |
| 0.185539026 | 0 | 4.248278045 | 0.913832629 | 0.065301345 |

| | | | | |
|---|---|---|---|---|
| 0 | 0.016804545 | 0 | 0 | 0.051941637 |
| 0.026598517 | 0 | 0 | 0 | 0 |
| 0 | 0 | 0 | 0 | 0 |
| 0 | 0 | 0 | 0 | 0 |
| 0 | 0.007044577 | 0 | 0 | 0 |
| 0.085275818 | 0 | 0 | 0.273415386 | 0 |
| 0.027415335 | 0 | 0 | 0 | 0 |
| 0 | 0 | 0 | 0 | 0 |
| 0 | 0 | 0 | 0 | 0 |
| 0 | 0 | 0 | 0 | 0 |
| 0 | 0 | 0 | 0 | 0 |
| 0.049925558 | 0 | 0 | 0.168130951 | 0 |
| 0.339539215 | 0 | 0 | 0.113627729 | 0.21072856 |
| 0 | 0 | 0 | 0 | 0 |
| 0 | 0 | 0.007561136 | 0.090147657 | 2.4239145 |
| 0 | 0 | 0 | 0 | 0 |
| 0 | 0 | 0 | 0 | 0 |
| 0 | 0 | 0 | 0 | 0 |
| 0 | 0.01193863 | 0 | 0 | 0 |
| 0 | 0.003340439 | 0 | 0 | 0 |
| 0.028900801 | 0 | 0 | 0 | 0 |
| 0 | 0 | 0 | 0 | 0 |
| 0.329736813 | 0.03580252 | 2.75004418 | 2.631980308 | 4.428932351 |
| 0 | 0 | 0 | 0 | 0 |
| 0 | 0 | 0 | 0 | 0 |
| 0 | 0 | 0 | 0 | 0 |
| 0 | 0 | 0 | 0 | 0 |
| 0 | 0 | 0 | 0 | 0 |
| 0 | 0 | 0 | 0 | 0 |
| 0 | 0 | 0 | 0 | 0 |
| 0 | 0 | 0 | 0 | 0 |
| 0 | 0.015857911 | 0.112456394 | 0.086263985 | 0.092532772 |
| 0 | 0 | 0 | 0 | 0 |
| 0 | 0.064203957 | 0 | 0.002611879 | 0 |
| 0 | 0.025121468 | 0 | 0 | 0.001207303 |
| 0.172459777 | 0 | 0.014796599 | 0.11040196 | 0.008093072 |
| 0 | 0 | 0 | 0 | 0 |
| 0 | 0 | 0 | 0.010919053 | 0 |
| 0 | 0 | 0 | 0 | 0 |
| 0 | 0 | 0 | 0 | 0 |
| 0.455927198 | 0 | 0 | 0 | 0 |
| 0 | 0 | 0 | 0 | 0 |
| 0 | 0 | 0 | 0 | 0 |
| 0 | 0 | 0 | 0 | 0 |
| 0 | 0.085404043 | 0.001202876 | 0 | 0.033538669 |
| 0 | 0 | 0 | 0 | 0 |
| 0.047521325 | 0 | 0 | 0.147048712 | 0 |
| 0 | 0 | 0 | 0.000985116 | 0.002474733 |

| | | | | |
|---|---|---|---|---|
| 0.250904725 | 0 | 0 | 0 | 0 |
| 0 | 0 | 0 | 0 | 0 |
| 0 | 0 | 0 | 0 | 0.006642264 |
| 0 | 0 | 0 | 0 | 0 |
| 0 | 0 | 0 | 0 | 0 |
| 0 | 0 | 0 | 0 | 0 |
| 0 | 0 | 0 | 1.205464365 | 0 |
| 0 | 0.084447217 | 0 | 0.215674543 | 0.118257613 |
| 0 | 0 | 0 | 0 | 0 |
| 0 | 0 | 0 | 0.014415758 | 0 |
| 0 | 0 | 0 | 0 | 0 |
| 0 | 0 | 0 | 0 | 0.000750927 |
| 0 | 0 | 0 | 0 | 0 |
| 0 | 0 | 0 | 0 | 0 |
| 0 | 0 | 0 | 0 | 0 |
| 0 | 0 | 0 | 0 | 0 |
| 0 | 0 | 0 | 0 | 0 |
| 0.016253291 | 0 | 0 | 0 | 0 |
| 0.234068213 | 0 | 0 | 0 | 0.180591347 |
| 0 | 0 | 0 | 0 | 0 |
| 0 | 0 | 0 | 0 | 0 |
| 0 | 0 | 0 | 0 | 0 |
| 0 | 0 | 0 | 0 | 0 |
| 0 | 0 | 0 | 0 | 0 |
| 0 | 0 | 0.000657123 | 0 | 0 |
| 0 | 0.010293876 | 0.03213917 | 0 | 0.00487171 |
| 0 | 0 | 0 | 0 | 0 |
| 0.025733223 | 0 | 0.52656284 | 0.019361325 | 0.625748754 |
| 0 | 0 | 0 | 0 | 0 |
| 0.54685491 | 0.006698136 | 0 | 2.549588193 | 0.681228618 |
| 0 | 0 | 0 | 0 | 0 |
| 0 | 0 | 0 | 0 | 0 |
| 0 | 0 | 0 | 0 | 0.027006207 |
| 0 | 0 | 0 | 0 | 0 |
| 0 | 1.109452754 | 0 | 0.002845143 | 0.003248796 |
| 0 | 0 | 0 | 0 | 0 |
| 0.544125284 | 0 | 0 | 0.342970429 | 0 |
| 0 | 0.010025534 | 0 | 0 | 0.001579899 |
| 0.173446516 | 0 | 0 | 0 | 0 |
| 0 | 0 | 0 | 0 | 0 |
| 3.061359599 | 0.684944658 | 1.520383877 | 9.672331962 | 16.9274778 |
| 0 | 0.084224796 | 2.287699835 | 0.033760094 | 0.061679654 |
| 0.255164983 | 0 | 0 | 3.338001621 | 0 |
| 0 | 0 | 0 | 0 | 0 |
| 0.069717556 | 0 | 0 | 0 | 0 |
| 0 | 0 | 0 | 0 | 0.013828444 |
| 0 | 0 | 0 | 0 | 0 |
| 0 | 0.039876612 | 0 | 0 | 0.010867095 |

| | | | | |
|---|---|---|---|---|
| 0 | 0 | 0.041424302 | 0 | 0 |
| 0 | 0 | 0 | 0 | 0 |
| 0 | 0 | 0 | 0 | 0 |
| 0 | 0 | 0 | 0 | 0 |
| 1.594722663 | 0.291416511 | 2.617966725 | 0 | 1.982540088 |
| 0.21807797 | 0 | 0 | 0 | 0 |
| 0 | 0 | 0 | 0 | 0 |
| 0 | 0 | 0 | 0 | 0 |
| 0 | 0.040575814 | 2.49790399 | 0.006954917 | 0.089719075 |
| 0 | 0 | 0 | 0 | 0 |
| 0 | 0.015374981 | 0 | 0 | 0 |
| 0 | 0 | 0 | 0 | 0 |
| 0.095057376 | 0 | 0 | 0 | 0.349015497 |
| 0 | 0.004433598 | 0.0033762 | 0.007420899 | 0.021569504 |
| 0.679980561 | 0 | 0 | 0.253078936 | 0 |
| 0.058372234 | 0 | 0.035163856 | 0.084884438 | 0.817982633 |
| 0 | 0 | 1.922173432 | 0 | 0.003110843 |
| 0 | 0 | 0 | 0 | 0 |
| 0 | 0 | 0 | 0 | 0 |
| 0 | 0 | 0 | 0 | 0 |
| 0 | 0 | 0 | 0 | 0 |
| 0 | 0 | 0 | 0 | 0 |
| 0 | 0 | 0 | 0 | 0 |
| 0 | 0 | 0 | 0 | 0 |
| 0 | 0 | 0 | 0 | 0 |
| 0 | 0 | 0 | 0 | 0 |
| 0 | 0 | 0 | 0 | 0.011592883 |
| 0 | 0.03146641 | 0 | 0 | 0 |
| 0 | 0 | 0 | 0 | 0 |
| 0 | 0 | 0 | 0 | 0 |
| 0 | 0 | 0 | 0 | 0 |
| 0 | 0 | 0 | 0 | 0 |
| 0.581278251 | 0 | 0 | 0 | 0 |
| 0.022611551 | 0 | 0 | 0 | 0 |
| 0.123019818 | 0 | 0 | 0.135641892 | 0.279497585 |
| 0 | 0.00792763 | 0.015177322 | 0 | 0.002017642 |
| 0 | 0 | 0 | 0.001806042 | 0.333314931 |
| 0 | 0 | 0.034685108 | 0 | 0.088116714 |
| 0 | 0 | 0 | 0 | 0 |
| 0 | 0 | 0 | 0 | 0 |
| 0.37283665 | 0.042255999 | 0 | 0.422804607 | 0.344717421 |
| 0 | 0 | 0 | 0.003578161 | 0 |
| 0 | 0 | 0 | 0 | 0 |
| 0 | 0 | 0 | 0.050709672 | 0 |
| 0 | 0 | 0 | 0 | 0 |
| 0 | 0 | 0 | 0 | 0 |
| 0 | 0 | 0 | 0 | 0 |
| 0 | 0 | 0 | 0 | 0 |

| | | | | |
|---|---|---|---|---|
| 0.037788132 | 0 | 0 | 0.020905348 | 0.188583112 |
| 0 | 0 | 0 | 0 | 0 |
| 0 | 0 | 0 | 0 | 0 |
| 0 | 0 | 0 | 0 | 0.001728827 |
| 0.065462769 | 0 | 0 | 0 | 0 |
| 0 | 0 | 0 | 0 | 0 |
| 0 | 0 | 1.621568338 | 0 | 0.703557697 |
| 0 | 0 | 0 | 0 | 0 |
| 0 | 0.001662418 | 0 | 0 | 0 |
| 0 | 0 | 0.00117606 | 0 | 0 |
| 0 | 0 | 0 | 0 | 0 |
| 0 | 0 | 0 | 0 | 0.000775393 |
| 0 | 0 | 0 | 0 | 0 |
| 0 | 0 | 0 | 0 | 0 |
| 0 | 0 | 0 | 0 | 0 |
| 0 | 0 | 0 | 0 | 0 |
| 0 | 55.27849526 | 0 | 0 | 0 |
| 0 | 0 | 0.001724283 | 0 | 0 |
| 0 | 0 | 0 | 0 | 0 |
| 0 | 0 | 0 | 0 | 0 |
| 0.701542806 | 0 | 0.066273631 | 1.460406945 | 1.480834913 |
| 0 | 0 | 0 | 0 | 0 |
| 0 | 0 | 0 | 0.002446202 | 0.005819281 |
| 0 | 0.009988291 | 0.001988756 | 0.000543797 | 0.003311721 |
| 0 | 0 | 0 | 0 | 0 |
| 0 | 0.002918957 | 0 | 0 | 0.000976772 |
| 0 | 0 | 0 | 0 | 0 |
| 0 | 0 | 0 | 0 | 0 |
| 0 | 0 | 0 | 0.002827786 | 0 |
| 0.436201141 | 1.274934673 | 0.03193708 | 0.276275904 | 0.002085275 |
| 0 | 0 | 0 | 0 | 0 |
| 0 | 0 | 0 | 0 | 0 |
| 0 | 0 | 0 | 0 | 0 |
| 0 | 0 | 0.0058127 | 0 | 0 |
| 0 | 0 | 0 | 0 | 0.048762526 |
| 0 | 0 | 0 | 0 | 0 |
| 0 | 0 | 0.005762722 | 0.004536205 | 0.002330899 |
| 0 | 0 | 0 | 0 | 0 |
| 0 | 0 | 0 | 0 | 0 |
| 0 | 0 | 0 | 0 | 0 |
| 0 | 0 | 0 | 0 | 0 |
| 0 | 0 | 0 | 0 | 0 |
| 0 | 0 | 0 | 0 | 0 |
| 0 | 0 | 0 | 0 | 0 |
| 0 | 0 | 0 | 0 | 0 |
| 0 | 0 | 0 | 0 | 0 |
| 0 | 0 | 0 | 0 | 0 |
| 0 | 0 | 0 | 0 | 0 |

| | | | | |
|---|---|---|---|---|
| 0 | 0.097025606 | 0.001689451 | 0.000975241 | 0.00734978 |
| 0.062694721 | 0 | 0 | 0.447843704 | 0 |
| 0.052373666 | 0 | 0 | 0 | 0 |
| 0 | 0 | 0 | 0 | 0 |
| 0 | 0 | 0 | 0 | 0 |
| 0 | 0 | 0 | 0 | 0 |
| 0 | 0 | 0 | 0 | 0 |
| 0.325804806 | 0.186692574 | 0 | 0.299404383 | 1.421269375 |
| 3.3688523 | 3.699375258 | 2.452782131 | 0.040412797 | 0.489081233 |
| 0 | 0 | 0.001107682 | 0 | 0.015819471 |
| 0 | 0 | 0 | 0 | 0 |
| 0 | 0 | 0 | 0 | 0 |
| 0 | 0 | 0 | 0 | 0 |
| 0 | 0 | 0.010339568 | 0 | 0 |
| 0 | 0 | 0 | 0 | 0 |
| 0 | 0 | 0 | 0 | 0 |
| 0 | 0 | 0.028278454 | 0 | 0.144024686 |
| 0.396946221 | 0 | 0 | 0 | 0 |
| 0 | 0 | 0 | 0 | 0 |
| 0 | 0 | 0 | 0.000883695 | 0 |
| 0 | 0 | 0 | 0 | 0 |
| 0 | 0 | 0 | 0 | 0 |
| 0 | 0 | 0 | 0 | 0 |
| 0 | 0 | 0 | 0 | 0 |
| 0.02776944 | 0 | 0 | 0 | 0 |
| 0 | 0 | 0 | 0 | 0 |
| 0 | 0 | 0 | 0 | 0 |
| 0 | 0 | 0 | 0 | 0 |
| 0 | 0 | 0 | 0 | 0 |
| 0 | 0 | 0 | 0 | 0 |
| 0 | 0 | 0 | 0 | 0 |
| 0 | 0 | 0 | 0 | 0 |
| 0 | 0 | 0 | 0 | 0 |
| 0.014017068 | 0 | 0 | 0 | 0 |
| 0 | 0 | 0 | 0.007084002 | 0 |
| 0 | 0 | 0 | 0 | 0 |
| 0 | 0 | 0 | 0 | 0 |
| 0 | 0 | 0 | 0 | 0 |
| 0 | 0 | 0 | 0 | 0 |
| 0 | 0.000689059 | 0.814516053 | 0 | 0 |
| 0 | 0 | 0 | 0 | 0 |
| 0.313191502 | 0 | 0 | 0.062877778 | 0.987975161 |
| 0 | 0 | 0 | 0 | 0 |
| 0 | 0 | 0 | 0 | 0 |
| 0 | 0 | 0 | 0 | 0 |
| 0 | 0 | 0 | 0 | 0.001188263 |
| 0 | 0 | 0 | 0 | 0 |
| 0 | 0 | 0 | 0 | 0 |

| | | | | |
|---|---|---|---|---|
| 0 | 0 | 0 | 0 | 0 |
| 0.035798176 | 0.044051908 | 0 | 0 | 0 |
| 0 | 0 | 0 | 0 | 0 |
| 0 | 0 | 0 | 0 | 0 |
| 0.155713858 | 0 | 0.168135048 | 0.441307688 | 1.058143306 |
| 0.015247063 | 0 | 0 | 0 | 0 |
| 0 | 0.006474576 | 0 | 0 | 0 |
| 0 | 0.128064526 | 0.029227819 | 0.005458542 | 0.006799602 |
| 0 | 0 | 0 | 0.006936556 | 0.002828813 |
| 0 | 0 | 0 | 0 | 0 |
| 0 | 0 | 0 | 0 | 0 |
| 0.096325899 | 0 | 0 | 0.325300196 | 0 |
| 0 | 0 | 0 | 0 | 0 |
| 0 | 0 | 0 | 0 | 0 |
| 0 | 0 | 0.019163319 | 0 | 0 |
| 0 | 0 | 0.000621301 | 0 | 0 |
| 0 | 0 | 0 | 0 | 0 |
| 0 | 0 | 0 | 0 | 0 |
| 0 | 0 | 0 | 0 | 0 |
| 0 | 0 | 0 | 0 | 0 |
| 0 | 0 | 0 | 0 | 0 |
| 0.110621508 | 0 | 0 | 0 | 0 |
| 0 | 0 | 0 | 0 | 0 |
| 0 | 0 | 0 | 0.001288341 | 0 |
| 0.099281597 | 0 | 0 | 0 | 0 |
| 0 | 0 | 0 | 0 | 0 |
| 0 | 4.996022071 | 0.012814688 | 0.005695283 | 0 |
| 0 | 0 | 0 | 0 | 0 |
| 0 | 0 | 0 | 0 | 0.003999868 |
| 0 | 0 | 0 | 0 | 0 |
| 0 | 0 | 0 | 0 | 0 |
| 0 | 0 | 0 | 0 | 0 |
| 0 | 0 | 0 | 0 | 0 |
| 0 | 0 | 0 | 0 | 0 |
| 0 | 0 | 0 | 0 | 0 |
| 0 | 0 | 0 | 0 | 0 |
| 0 | 0 | 0 | 0 | 0 |
| 0 | 0 | 0 | 0 | 0 |
| 0 | 0 | 0 | 0 | 0 |
| 0 | 0 | 0 | 0 | 0 |
| 0 | 0 | 0 | 0 | 0 |
| 0 | 0 | 0 | 0.050345617 | 0 |
| 0 | 0 | 0 | 0 | 0 |
| 0 | 0 | 0 | 0.020904776 | 0 |
| 0 | 0 | 0 | 0 | 0 |
| 0 | 0 | 0 | 0 | 0 |
| 0 | 0 | 0 | 0 | 0 |
| 0.031438133 | 0 | 0 | 0 | 0 |

| | | | | |
|---|---|---|---|---|
| 0 | 0 | 0.309276265 | 0.125771837 | 0 |
| 0 | 0 | 0 | 0 | 0 |
| 0 | 0 | 0 | 0 | 0 |
| 0 | 0 | 0 | 0 | 0 |
| 0 | 0 | 0 | 0 | 0 |
| 0 | 0 | 0 | 0 | 0 |
| 0 | 0 | 0 | 0 | 0 |
| 0 | 0 | 0 | 0 | 0.006882081 |
| 0 | 0 | 0 | 0 | 0 |
| 0 | 0 | 0 | 0 | 0 |
| 0 | 0 | 0 | 0 | 0 |
| 0 | 0 | 0 | 0 | 0 |
| 0 | 0 | 0 | 0.001720473 | 0 |
| 0 | 0 | 0 | 0 | 0 |
| 0 | 0 | 0 | 0 | 0 |
| 0 | 0 | 0 | 0 | 0 |
| 0 | 0 | 0 | 0 | 0 |
| 0 | 0 | 0 | 0 | 0 |
| 0 | 0 | 0 | 0 | 0 |
| 0 | 0 | 0 | 0 | 0 |
| 0 | 0 | 0.170886909 | 0.003421215 | 0.059575615 |
| 0 | 0 | 0 | 0 | 0 |
| 0 | 0.377607012 | 0.001403439 | 0.001080186 | 0.007708982 |
| 0 | 0.554518337 | 0 | 0 | 0 |
| 0 | 0 | 0 | 0 | 0 |
| 0 | 0 | 0 | 0 | 0 |
| 0 | 0 | 0 | 0 | 0 |
| 0 | 0 | 0 | 0 | 0 |
| 0 | 0 | 0 | 0 | 0 |
| 0 | 0 | 0 | 0 | 0 |
| 0 | 0 | 0 | 0 | 0 |
| 0.036811177 | 0 | 0 | 0 | 0 |
| 0 | 0.012529098 | 0.133037862 | 0.122402402 | 0 |
| 0 | 0 | 0 | 0 | 0 |
| 0.45490494 | 0 | 0 | 0.803065502 | 0.769763271 |
| 0 | 0.000615556 | 0.004687478 | 0 | 0.118183111 |
| 0 | 0 | 0 | 0 | 0 |
| 0 | 0 | 0 | 0 | 0 |
| 0 | 0 | 0.151946215 | 0 | 0 |
| 0 | 0 | 0 | 0 | 0 |
| 0 | 0 | 0 | 0 | 0 |
| 0.02937828 | 0 | 0 | 0 | 0 |
| 0.043411407 | 0 | 0 | 0.111710037 | 0.091516403 |
| 0 | 0 | 0 | 0 | 0 |
| 0 | 0 | 0 | 0 | 0 |
| 0.162530591 | 0 | 0 | 0.004577543 | 0.131670086 |
| 0 | 0 | 0 | 0 | 0 |
| 0 | 0 | 0 | 0 | 0 |

| | | | | |
|---|---|---|---|---|
| 0 | 0 | 0 | 0 | 0 |
| 0 | 0 | 0 | 0 | 0 |
| 0 | 0 | 0 | 0 | 0 |
| 0 | 0 | 0 | 0 | 0 |
| 0 | 0 | 0 | 0 | 0 |
| 0 | 0 | 0 | 0 | 0 |
| 0 | 0 | 0 | 0 | 0 |
| 0 | 0 | 0 | 0 | 0.027900306 |
| 0 | 0 | 0 | 0.000816902 | 0 |
| 0.055801571 | 0 | 0 | 0.004714819 | 0 |
| 0 | 0 | 0 | 0 | 0 |
| 0 | 0 | 0 | 0 | 0 |
| 0 | 0 | 0 | 0 | 0 |
| 0.231246259 | 0 | 0 | 0 | 0 |
| 0 | 0 | 0 | 0 | 0 |
| 0 | 0.028364485 | 0.008673877 | 0 | 0.000756714 |
| 0 | 0 | 0 | 0 | 0 |
| 0 | 0 | 0 | 0 | 0 |
| 0 | 0 | 0 | 0 | 0 |
| 0 | 0 | 0 | 0.007582659 | 0 |
| 0 | 0 | 0 | 0 | 0.018704565 |
| 0 | 0 | 0 | 0 | 0 |
| 0 | 0 | 0 | 0 | 0 |
| 0 | 0 | 0 | 0.003236649 | 0 |
| 0 | 0 | 0 | 0 | 0 |
| 0 | 0 | 0 | 0 | 0 |
| 0 | 0 | 0 | 0 | 0 |
| 0 | 0 | 0 | 0 | 0 |
| 0 | 0 | 0 | 0 | 0 |
| 0.263819792 | 0 | 0 | 0 | 0 |
| 0.109529853 | 0 | 0 | 0.052964238 | 1.408841604 |
| 0 | 0 | 0 | 0 | 0 |
| 0 | 0 | 0 | 0 | 0 |
| 0 | 0 | 0 | 0 | 0 |
| 0 | 0 | 0 | 0 | 0 |
| 0 | 0 | 0 | 0 | 0 |
| 0 | 0 | 0 | 0 | 0 |
| 0 | 0 | 0 | 0 | 0 |
| 0 | 0 | 0 | 0.002023484 | 0 |
| 0 | 0 | 0 | 0 | 0 |
| 0 | 0 | 0 | 0.001970174 | 0 |
| 0.264088755 | 0 | 0 | 0 | 0 |
| 0 | 0 | 0 | 0 | 0.005986936 |
| 0 | 0 | 0 | 0 | 0 |
| 0 | 0 | 0 | 0 | 0 |
| 0 | 0 | 0 | 0 | 0 |
| 0 | 0.407082817 | 0.22603256 | 0 | 0.034342474 |
| 0 | 0 | 0 | 0.002205325 | 0 |

| | | | | |
|---|---|---|---|---|
| 0 | 0 | 0 | 0 | 0 |
| 0 | 0 | 0 | 0 | 0 |
| 0 | 0 | 0 | 0 | 0 |
| 0 | 0 | 0 | 0 | 0 |
| 0 | 0.124603462 | 0.472641375 | 0.226882848 | 0.121916092 |
| 0 | 0 | 0 | 0 | 0 |
| 0 | 0 | 0 | 0 | 0 |
| 0 | 0 | 0 | 0 | 0 |
| 0 | 0 | 0 | 0 | 0 |
| 0 | 0 | 0 | 0 | 0 |
| 0.029078353 | 0 | 0 | 0 | 0 |
| 0 | 0.190305011 | 0 | 0.001172725 | 0.00669552 |
| 0 | 0.075903722 | 0 | 0 | 0 |
| 0 | 0 | 0 | 0 | 0 |
| 0 | 0 | 0.004954209 | 0 | 0.001836883 |
| 0.020605522 | 0 | 0 | 0.162146417 | 0.48089231 |
| 0 | 0.004833307 | 0 | 0.008298072 | 0.010093314 |
| 0 | 0 | 0 | 0 | 0 |
| 0 | 0.00368106 | 0.048713106 | 0.011899116 | 0.196914385 |
| 0 | 0 | 0.176083965 | 0 | 0.977336466 |
| 0 | 0 | 0 | 0 | 0 |
| 0.045938889 | 0 | 0 | 0 | 0 |
| 0 | 0 | 0 | 0 | 0 |
| 0 | 0 | 0 | 0 | 0 |
| 0 | 0 | 0 | 0 | 0 |
| 0 | 0 | 0 | 0 | 0 |
| 0 | 0.003945483 | 0.001073035 | 0 | 0.012731257 |
| 0 | 0 | 0 | 0 | 0 |
| 0 | 0 | 0 | 0 | 0 |
| 0 | 0 | 0 | 0 | 0 |
| 0 | 0 | 0.000729805 | 0 | 0 |
| 0 | 0 | 0 | 0 | 0.019744488 |
| 0 | 0 | 0 | 0 | 0 |
| 0 | 0 | 0 | 0 | 0 |
| 0 | 0 | 0 | 0 | 0 |
| 0 | 0 | 0 | 0 | 0 |
| 0 | 0 | 0 | 0 | 0 |
| 0 | 0.147533165 | 0.010169342 | 0 | 0.034980196 |
| 0 | 0 | 0 | 0.064614776 | 0 |
| 0 | 0 | 0 | 0 | 0.028682028 |
| 0 | 0.083277639 | 0 | 0 | 0 |
| 0 | 0 | 0 | 0 | 0 |
| 0 | 0 | 0 | 0 | 0 |
| 0 | 0 | 0.00088705 | 0 | 0.042098361 |
| 0 | 0 | 0 | 0 | 0 |
| 0 | 0 | 0 | 0 | 0 |
| 0 | 0 | 0 | 0 | 0 |
| 0 | 0 | 0 | 0 | 0 |

| | | | | |
|---|---|---|---|---|
| 0 | 0 | 0 | 0 | 0 |
| 0.038142153 | 0 | 0 | 0.058333099 | 0 |
| 0 | 0 | 0 | 0 | 0 |
| 0 | 0 | 0 | 0.0016099 | 0.001050459 |
| 0 | 0.004636869 | 0.000862219 | 0 | 0 |
| 0 | 0.004870962 | 0.005086977 | 0 | 0 |
| 0 | 0 | 0 | 0 | 0 |
| 0 | 0 | 0 | 0 | 0 |
| 0 | 0 | 0 | 0 | 0 |
| 0 | 0 | 0 | 0 | 0 |
| 0.01717571 | 0 | 0 | 0.034587412 | 0 |
| 0 | 0 | 0 | 0 | 0 |
| 0.02946855 | 0.074421371 | 0.103553729 | 0.281874472 | 0.010983255 |
| 0 | 0 | 0 | 0 | 0 |
| 0 | 0 | 0 | 0 | 0 |
| 0 | 0 | 0 | 0 | 0 |
| 0 | 0 | 0 | 0 | 0 |
| 0 | 0 | 0 | 0 | 0 |
| 0 | 0 | 0 | 0 | 0 |
| 0 | 0 | 0 | 0 | 0 |
| 0 | 0 | 0 | 0 | 0 |
| 0 | 0 | 0 | 0 | 0 |
| 0 | 0.044277171 | 0.001363564 | 0.043577927 | 0 |
| 0.028618261 | 0 | 0 | 0 | 0.029504189 |
| 0 | 0 | 0 | 0 | 0 |
| 0 | 0 | 0 | 0 | 0 |
| 0.023170741 | 0 | 0 | 0 | 0 |
| 0 | 0 | 0 | 0 | 0 |
| 0 | 0 | 0 | 0 | 0 |
| 0 | 0 | 0 | 0 | 0 |
| 0 | 0 | 0 | 0.074200249 | 0.022685067 |
| 0 | 0 | 0 | 0 | 0 |
| 0 | 0 | 0 | 0 | 0 |
| 0 | 0 | 0 | 0 | 0 |
| 0.045341024 | 0 | 0 | 0 | 1.864515127 |
| 0 | 0.000885446 | 0 | 0 | 0 |
| 0 | 0 | 0 | 0 | 0.000871129 |
| 0 | 0 | 0 | 0 | 0 |
| 0 | 0 | 0.164651083 | 0.008505482 | 0.006243552 |
| 0 | 0 | 0 | 0 | 0 |
| 0 | 0 | 0 | 0 | 0 |
| 0 | 0 | 0 | 0 | 0 |
| 0 | 0 | 0 | 0 | 0 |
| 0 | 0 | 0 | 0 | 0 |
| 0 | 0 | 0 | 0 | 0 |
| 0.688422269 | 0 | 0.08261875 | 0.325706902 | 0.988386 |
| 0 | 0 | 0 | 0 | 0 |
| 0 | 0 | 0 | 0 | 0 |

| | | | | |
|---|---|---|---|---|
| 0 | 0 | 0 | 0 | 0 |
| 0 | 0 | 0 | 0 | 0 |
| 0.024776355 | 0 | 0 | 0.075479356 | 0.101630813 |
| 0 | 0 | 0 | 0.052187761 | 0 |
| 0 | 0 | 0 | 0 | 0 |
| 0 | 0 | 0 | 0 | 0 |
| 0 | 0 | 0 | 0 | 0 |
| 0 | 0 | 0 | 0 | 0 |
| 0 | 0.004736258 | 0 | 0.052090799 | 0 |
| 0 | 0 | 0 | 0 | 0 |
| 0 | 0.005315487 | 0 | 0 | 0.001875997 |
| 0 | 0 | 0 | 0 | 0 |
| 0 | 0 | 0 | 0 | 0 |
| 0 | 0 | 0 | 0 | 0 |
| 0 | 0 | 0 | 0 | 0 |
| 0 | 0 | 0 | 0 | 0 |
| 0 | 0 | 0 | 0 | 0 |
| 0 | 0 | 0 | 0 | 0 |
| 0 | 0 | 0 | 0 | 0 |
| 0 | 0 | 0 | 0.000871513 | 0 |
| 0 | 0 | 0 | 0 | 0 |
| 0 | 0 | 0 | 0 | 0 |
| 0 | 0 | 0 | 0 | 0 |
| 0 | 0 | 0 | 0 | 0 |
| 0 | 0 | 0 | 0 | 0 |
| 0 | 0 | 0 | 0 | 0 |
| 0 | 0 | 0 | 0 | 0 |
| 0 | 0 | 0 | 0 | 0.074338457 |
| 0 | 0 | 0 | 0 | 0 |
| 0 | 0 | 0 | 0 | 0 |
| 0 | 0 | 0.000432346 | 0.057651407 | 0.001709887 |
| 0 | 0 | 0 | 0 | 0 |
| 0 | 0.052627675 | 0.017444423 | 0.016734789 | 0.044918618 |
| 0 | 0 | 0.001410715 | 0.000888371 | 0.023331381 |
| 0 | 0 | 0 | 0 | 0 |
| 0 | 0 | 0 | 0 | 0 |
| 0 | 0 | 0 | 0 | 0 |
| 0 | 0 | 0 | 0 | 0 |
| 0 | 0 | 0 | 0 | 0 |
| 0 | 0 | 0 | 0 | 0 |
| 0 | 0 | 0 | 0 | 0 |
| 0 | 0.570625488 | 0.095813453 | 0 | 0.302778942 |
| 0 | 0 | 0 | 0.017426253 | 0.045020561 |
| 0 | 0 | 0 | 0 | 0 |
| 0 | 0 | 0 | 0 | 0 |
| 0 | 0 | 0 | 0.044068757 | 0.02444162 |
| 0 | 0 | 0 | 0 | 0 |
| 0 | 0 | 0 | 0 | 0 |

| | | | | |
|---|---|---|---|---|
| 0 | 0 | 0 | 0 | 0 |
| 0 | 0.185160029 | 0 | 0.000912816 | 0.012768428 |
| 0 | 0.004177496 | 0.015546722 | 0.014394577 | 0.017958723 |
| 0 | 0 | 0 | 0 | 0 |
| 0 | 0.567685546 | 0 | 0.003618285 | 0.377922355 |
| 0 | 0.406685899 | 0.001914145 | 0 | 0 |
| 0 | 0 | 0 | 0 | 0 |
| 0 | 0 | 0 | 0 | 0 |
| 0 | 0 | 0 | 0 | 0 |
| 0 | 0 | 0 | 0 | 0 |
| 0 | 0 | 0 | 0 | 0 |
| 0 | 0 | 0 | 0 | 0 |
| 0 | 0 | 0 | 0 | 0 |
| 0 | 0 | 0 | 0 | 0 |
| 0 | 0 | 0 | 0 | 0 |
| 0 | 0 | 0 | 0 | 0 |
| 0 | 0 | 0 | 0 | 0.03178714 |
| 0 | 0 | 0 | 0 | 0 |
| 0 | 0 | 0 | 0 | 0 |
| 0 | 0 | 0 | 0 | 0 |
| 0 | 0 | 0 | 0 | 0 |
| 0 | 0 | 0 | 0 | 0 |
| 0 | 0 | 0 | 0 | 0 |
| 0 | 0 | 0 | 0 | 0 |
| 0 | 0 | 0 | 0.048551061 | 0 |
| 0 | 0.102226733 | 0 | 0.00663003 | 0 |
| 0 | 0 | 0 | 0 | 0 |
| 0.101720596 | 0 | 0 | 0.029155499 | 0.712838666 |
| 0 | 0 | 0 | 0 | 0 |
| 0 | 0 | 0 | 0 | 0 |
| 0 | 0 | 0 | 0 | 0 |
| 0 | 0 | 0 | 0 | 0 |
| 0 | 0 | 0 | 0 | 0 |
| 0 | 0 | 0 | 0 | 0 |
| 0 | 0 | 0 | 0 | 0 |
| 0 | 0 | 0 | 0 | 0 |
| 0 | 0 | 0 | 0 | 0 |
| 0 | 0 | 0 | 0 | 0 |
| 0 | 0 | 0.513516661 | 1.068068114 | 3.86454735 |
| 0 | 0 | 0 | 0 | 0 |
| 0 | 0 | 0 | 0 | 0 |
| 0 | 0 | 0 | 0.004315536 | 0 |
| 0 | 0.074900455 | 0.003330002 | 0.001996188 | 0.002026129 |
| 0 | 0 | 0 | 0 | 0 |
| 0 | 0 | 0 | 0 | 0 |
| 0 | 0 | 0 | 0.001020686 | 0 |
| 0 | 0.040996027 | 0 | 0.003393085 | 0.001162343 |
| 0 | 0 | 0 | 0 | 0 |

| | | | | |
|---|---|---|---|---|
| 0 | 0 | 0 | 0 | 0 |
| 0 | 0.024782517 | 0.013721325 | 0.011773984 | 0.012955523 |
| 0 | 0 | 0 | 0 | 0 |
| 0 | 0 | 0 | 0 | 0 |
| 0 | 0 | 0 | 0 | 0 |
| 0 | 0 | 0 | 0 | 0 |
| 0 | 0 | 0 | 0 | 0 |
| 0 | 0 | 0 | 0 | 0 |
| 0 | 0 | 0 | 0 | 0 |
| 0 | 0 | 0 | 0 | 0 |
| 0 | 0 | 0 | 0 | 0 |
| 0 | 0 | 0 | 0 | 0 |
| 0 | 0 | 0.005357576 | 2.392375304 | 0 |
| 0 | 0 | 0 | 0 | 0 |
| 0.036185452 | 0 | 0 | 0.006672902 | 0.009143538 |
| 0 | 0 | 0 | 0 | 0 |
| 0 | 0 | 0 | 0 | 0 |
| 0 | 0 | 0 | 0 | 0 |
| 0 | 0 | 0 | 0 | 0 |
| 0 | 0 | 0 | 0 | 0 |
| 0 | 0 | 0 | 0 | 0 |
| 0 | 0 | 0.066299084 | 0 | 0.130205304 |
| 0 | 0 | 0 | 0.025178436 | 0.027600585 |
| 0 | 0.001091932 | 0 | 0 | 0 |
| 0 | 0 | 0 | 0 | 0 |
| 0 | 0 | 0 | 0 | 0 |
| 0 | 0 | 0 | 0 | 0 |
| 0 | 0 | 0 | 0 | 0 |
| 0 | 0 | 0 | 0 | 0 |
| 0 | 0 | 0 | 0 | 0 |
| 0 | 0 | 0 | 0 | 0 |
| 0 | 0 | 0 | 0 | 0 |
| 0 | 0 | 0 | 0 | 0 |
| 0 | 0 | 0 | 0 | 0 |
| 7.345799564 | 0 | 4.546987787 | 0 | 0 |
| 0 | 0 | 0 | 0 | 0 |
| 0 | 0 | 0 | 0 | 0 |
| 0 | 0 | 0 | 0 | 0 |
| 0 | 0 | 0.189099954 | 0 | 0.021521504 |
| 0 | 0 | 0 | 0 | 0 |
| 0.074245859 | 0.024917509 | 0.017890497 | 0.114304974 | 0.271496302 |
| 0 | 0 | 0 | 0 | 0 |
| 0 | 0 | 0 | 0 | 0 |
| 0 | 0 | 0 | 0 | 0 |
| 0 | 0 | 0 | 0 | 0 |
| 0 | 0.566501497 | 8.998885848 | 0.230301445 | 0.028844433 |
| 0 | 0 | 0 | 0 | 0 |
| 0 | 0 | 0 | 0 | 0 |

| | | | | |
|---|---|---|---|---|
| 0 | 0 | 0 | 0 | 0 |
| 0 | 0 | 0 | 0 | 0 |
| 0 | 0 | 0 | 0 | 0 |
| 0 | 0 | 0 | 0 | 0 |
| 0 | 0 | 0 | 0 | 0 |
| 0 | 0 | 0 | 0 | 0 |
| 0 | 0 | 0 | 0 | 0 |
| 0 | 0 | 0.003019045 | 0 | 0.00132667 |
| 0 | 0 | 0 | 0 | 0.006444082 |
| 0 | 0 | 0.625266149 | 0.042313726 | 0.002100737 |
| 0.025051216 | 0 | 0 | 0.027163571 | 0 |
| 0 | 0 | 0 | 0 | 0 |
| 0 | 0.007402165 | 0 | 0 | 0 |
| 0 | 0 | 0 | 0 | 0 |
| 0 | 0 | 0 | 0 | 0 |
| 0 | 0 | 0 | 0 | 0 |
| 0 | 0 | 0 | 0 | 0 |
| 0 | 0 | 0 | 0 | 0 |
| 0 | 0 | 0 | 0 | 0 |
| 0.030996915 | 0.869212977 | 0.309609548 | 0.291977975 | 0.193200933 |
| 0 | 0 | 0 | 0 | 0.025042998 |
| 0 | 0 | 0 | 0 | 0 |
| 0 | 0 | 0 | 0 | 0 |
| 0 | 0 | 0 | 0 | 0 |
| 0 | 0 | 0 | 0 | 0 |
| 0 | 0 | 0 | 0 | 0 |
| 0 | 0 | 0 | 0 | 0 |
| 0 | 0 | 0 | 0 | 0 |
| 0 | 0 | 0 | 0 | 0 |
| 0 | 0 | 0 | 0.006843866 | 0 |
| 0 | 0 | 0 | 0 | 0 |
| 0 | 0 | 0 | 0 | 0 |
| 0 | 0 | 0 | 0 | 0 |
| 0 | 0 | 0 | 0 | 0 |
| 0 | 0 | 0 | 0 | 0 |
| 0 | 0 | 0 | 0 | 0 |
| 0 | 0 | 0 | 0 | 0 |
| 0 | 0 | 0 | 0 | 0 |
| 0 | 0 | 0 | 0 | 0 |
| 0 | 0 | 0 | 0 | 0 |
| 0 | 0 | 0 | 0 | 0 |
| 0 | 0 | 0 | 0 | 0 |
| 0 | 0 | 0 | 0 | 0.086374731 |
| 0 | 0 | 0 | 0 | 0 |
| 0 | 0.019541412 | 0 | 0 | 0.012260907 |
| 0.015719351 | 0 | 0 | 0 | 0 |
| 0 | 0 | 0 | 0 | 0 |
| 0 | 0 | 0 | 0 | 0 |

| | | | | |
|---|---|---|---|---|
| 0 | 0 | 0 | 0 | 0 |
| 0 | 0 | 0 | 0 | 0 |
| 0 | 0 | 0 | 0 | 0 |
| 0 | 0 | 0 | 0 | 0 |
| 0 | 0 | 0 | 0 | 0 |
| 0.010867158 | 0.114032847 | 0.000925761 | 0 | 0.005125816 |
| 0 | 0 | 0 | 0 | 0 |
| 0 | 0 | 0 | 0 | 0 |
| 0 | 0 | 0 | 0 | 0 |
| 0 | 0 | 0 | 0 | 0 |
| 0 | 0 | 0 | 0 | 0 |
| 0 | 0 | 0 | 0 | 0 |
| 0 | 0 | 0 | 0 | 0 |
| 0 | 0 | 0 | 0 | 0 |
| 0 | 0 | 0 | 0 | 0 |
| 0 | 0 | 0 | 0 | 0 |
| 0 | 0 | 0 | 0 | 0 |
| 0 | 0 | 0 | 0 | 0 |
| 0 | 0 | 0 | 0 | 0 |
| 0 | 0 | 0 | 0 | 0 |
| 0 | 0 | 0 | 0 | 0 |
| 0 | 0 | 0 | 0 | 0 |
| 0 | 0 | 0 | 0 | 0 |
| 0 | 0 | 0 | 0 | 0 |
| 0 | 0 | 0 | 0 | 0 |
| 0 | 0 | 0.02678885 | 0 | 0 |
| 0 | 0 | 0 | 0 | 0 |
| 0 | 0 | 0 | 0 | 0 |
| 0 | 0 | 0 | 0 | 0 |
| 0 | 0 | 0 | 0 | 0 |
| 0 | 0 | 0 | 0 | 0 |
| 0 | 0 | 0 | 0 | 0.005690653 |
| 0 | 0 | 0 | 0 | 0 |
| 0 | 0 | 0 | 0 | 0 |
| 0 | 0 | 0 | 0 | 0 |
| 0 | 0 | 0 | 0 | 0 |
| 0 | 0 | 0 | 0 | 0 |
| 0 | 0 | 0 | 0 | 0 |
| 0 | 0 | 0 | 0.29668655 | 0 |
| 0 | 0 | 0 | 0 | 0 |
| 0 | 0 | 0 | 0 | 0 |
| 0 | 0 | 0 | 0 | 0 |
| 0 | 0 | 0 | 0 | 0 |
| 0 | 0 | 0 | 0 | 0 |
| 0 | 0 | 0 | 0 | 0 |
| 0 | 0 | 0 | 0 | 0 |
| 0 | 0 | 0 | 0 | 0 |
| 0 | 0 | 0 | 0 | 0 |

| | | | | |
|---|---|---|---|---|
| 0 | 0 | 0 | 0 | 0 |
| 0 | 0.015153317 | 0 | 0 | 0 |
| 0 | 0 | 0 | 0 | 0 |
| 0 | 0 | 0 | 0 | 0 |
| 0 | 0 | 0 | 0 | 0 |
| 0 | 0 | 0 | 0 | 0 |
| 0 | 0 | 0 | 0 | 0 |
| 0 | 0 | 0 | 0 | 0 |
| 0 | 0 | 0 | 0 | 0 |
| 0 | 0 | 0 | 0 | 0 |
| 0 | 0 | 0.004614371 | 0.029143584 | 0 |
| 0 | 0 | 0 | 0 | 0 |
| 0.035202572 | 0 | 0 | 0 | 0 |
| 0 | 0 | 0 | 0 | 0 |
| 0 | 0 | 0 | 0 | 0 |
| 0 | 0 | 0 | 0 | 0 |
| 0 | 0 | 0 | 0 | 0 |
| 0 | 0 | 0 | 0 | 0 |
| 0 | 0 | 0 | 0 | 0 |
| 0 | 0 | 0 | 0 | 0 |
| 0 | 0 | 0.009779851 | 0 | 0.00653719 |
| 0 | 0 | 0 | 0 | 0 |
| 0 | 0 | 0 | 0 | 0 |
| 1.727907304 | 1.067994315 | 0 | 1.150070412 | 0.79403385 |
| 0 | 0 | 0 | 0 | 0 |
| 0.140356703 | 0 | 0 | 0 | 0 |
| 0 | 0 | 0 | 0 | 0 |
| 0 | 0 | 0 | 0 | 0 |
| 0 | 0 | 0 | 0 | 0 |
| 0 | 0 | 0 | 0 | 0 |
| 0 | 0 | 0 | 0 | 0 |
| 0 | 0 | 0 | 0 | 0 |
| 0 | 0 | 0 | 0 | 0 |
| 0 | 0 | 0 | 0 | 0 |

| PHL018-0133_V2 | PHL018-0139_V1 | PHL018-0075_V1 | PHL018-0125_V2 | PHL018-0118_V1 |
|---|---|---|---|---|
| EYCF | CTRL | EYCF | CTRL | EYCF |
| 0.111359035 | 0.036080025 | 0.014124833 | 0.155152184 | 2.119932465 |
| 0 | 0.074165359 | 0.599082153 | 0.869747461 | 0 |
| 0 | 0.015320817 | 0 | 2.227285065 | 1.02711978 |
| 0 | 0 | 0 | 0 | 0 |
| 0 | 0 | 0 | 0 | 0 |
| 0 | 0 | 0 | 0 | 0 |
| 0 | 0 | 0 | 0 | 0 |
| 0 | 0 | 0 | 0 | 0 |
| 0 | 0 | 0 | 0 | 0 |
| 0 | 0 | 0 | 0 | 0 |
| 0 | 0 | 0 | 0 | 0 |
| 0 | 0 | 0 | 0 | 0 |
| 0 | 0 | 0 | 0 | 0 |
| 0 | 0.525747545 | 3.385878309 | 2.420212783 | 0.398518011 |
| 0 | 0 | 0 | 0 | 0 |
| 0 | 0 | 0 | 0 | 0 |
| 0 | 0 | 0 | 0 | 0 |
| 0 | 0 | 0 | 0 | 0 |
| 0 | 0 | 0 | 0 | 0 |
| 0 | 0 | 0 | 0 | 0 |
| 0 | 0 | 0 | 0 | 0 |
| 0 | 0 | 0 | 0 | 0 |
| 0 | 0 | 0 | 0 | 0 |
| 0 | 0 | 0 | 0 | 0 |
| 0 | 0 | 0 | 0.010292542 | 0 |
| 0 | 0 | 0 | 0 | 0 |
| 0 | 0 | 0 | 0 | 0 |
| 0 | 0 | 0 | 0 | 0 |
| 0 | 0 | 0 | 0 | 0 |
| 0 | 0 | 0 | 0 | 0 |
| 0 | 0 | 0 | 0 | 0 |
| 0 | 0 | 0 | 0 | 0 |
| 0 | 0 | 0 | 0 | 0 |
| 0 | 0 | 0 | 0 | 0 |
| 0 | 0 | 0 | 0 | 0 |
| 0 | 1.506575102 | 2.72383896 | 1.260589724 | 0.005132498 |
| 0 | 0 | 0 | 0 | 0 |
| 0 | 0 | 0 | 0 | 0 |
| 0 | 0 | 0 | 0 | 0 |
| 0 | 0 | 0 | 0 | 0 |
| 0 | 0 | 0 | 0 | 0 |
| 0 | 0.002321729 | 0.002074423 | 0.020857354 | 0.059358492 |
| 0.231948448 | 0.14583005 | 0.392332497 | 0.174232941 | 1.00635019 |
| 0 | 0.001269019 | 0.272633305 | 0 | 0.066985948 |
| 0 | 0 | 0 | 0 | 0.016009228 |
| 0 | 0 | 0 | 0 | 0 |

| | | | | |
|---|---|---|---|---|
| 0 | 0.004940641 | 0 | 0.002310131 | 0 |
| 0.036379503 | 0 | 0 | 0 | 0 |
| 0 | 0 | 0 | 0.007008411 | 0.626112234 |
| 0 | 0.340212878 | 1.481478885 | 1.270831917 | 0.372963476 |
| 0.008653032 | 0 | 0 | 0.003388597 | 0 |
| 0 | 0.035893553 | 0.279190978 | 0 | 0 |
| 0 | 0 | 0.002109301 | 0 | 0.001019966 |
| 0 | 0 | 0 | 0 | 0 |
| 0 | 0 | 0 | 0 | 0 |
| 0 | 0 | 0 | 0 | 0 |
| 0 | 0 | 0.001076688 | 0 | 0.000702864 |
| 0.110616416 | 0.257929433 | 0.007608334 | 0.009016293 | 3.00559157 |
| 0 | 0 | 0 | 0 | 0.002650817 |
| 0.043541781 | 0 | 0.000671384 | 0 | 0.016983371 |
| 0 | 0 | 0 | 0 | 0 |
| 0 | 0 | 0 | 0 | 0 |
| 0 | 0.002575308 | 0.012655456 | 0 | 0.001752439 |
| 0 | 0 | 0 | 0 | 0 |
| 0 | 0 | 0 | 0 | 0 |
| 0 | 0 | 0 | 0 | 0 |
| 0.174334209 | 0 | 0.003724594 | 0.007949597 | 0.013121948 |
| 0.00710872 | 0 | 0.004496182 | 0.029855588 | 0.004710676 |
| 0 | 0.224374429 | 0.009338876 | 0 | 0 |
| 0 | 0 | 0 | 0 | 0 |
| 0 | 0 | 0 | 0 | 0 |
| 0 | 0 | 0 | 0 | 0.001889309 |
| 0.010664487 | 0 | 0 | 0 | 0 |
| 0 | 0 | 0 | 0 | 0 |
| 0 | 0 | 0.000639102 | 0 | 0.041488877 |
| 0 | 0 | 0 | 0 | 0 |
| 0.01259879 | 0 | 0 | 0 | 0 |
| 0 | 0 | 0 | 0 | 0 |
| 0 | 0 | 0 | 0 | 0 |
| 0.00185383 | 0 | 0 | 0 | 0 |
| 0 | 0 | 0 | 0 | 0 |
| 0 | 0 | 0 | 0 | 0 |
| 0 | 0 | 0 | 0 | 0 |
| 0 | 0 | 0 | 0 | 0 |
| 0 | 0 | 0 | 0 | 0 |
| 0 | 0 | 0 | 0 | 0 |
| 0 | 0 | 0 | 0 | 0 |
| 0 | 0 | 0 | 0 | 0 |
| 0 | 0 | 0 | 0 | 0 |
| 0 | 0 | 0 | 0 | 0 |
| 0 | 0 | 0 | 0 | 0 |
| 0 | 0 | 0 | 0 | 0 |
| 0 | 0 | 0 | 0 | 0 |
| 0 | 0.055444463 | 0 | 0 | 0 |

| | | | | |
|---|---|---|---|---|
| 0 | 0 | 0 | 0 | 0 |
| 0.0111024 | 0 | 0 | 0 | 0.001331895 |
| 0 | 0 | 0 | 0 | 0 |
| 0 | 0 | 0 | 0 | 0 |
| 0 | 0.017914602 | 0.083970292 | 0 | 2.192017897 |
| 0 | 0 | 0 | 0 | 0 |
| 0 | 0 | 0 | 0 | 0 |
| 0 | 0 | 0 | 0 | 0 |
| 0.001585647 | 0 | 0 | 0 | 0 |
| 0 | 0 | 0 | 0 | 0 |
| 0 | 0 | 0 | 0 | 0 |
| 0 | 0.000929552 | 0 | 0.004527472 | 0.044729546 |
| 0 | 0 | 0 | 0 | 0 |
| 0 | 0 | 0 | 0 | 0 |
| 0 | 0 | 0 | 0 | 0 |
| 0 | 0 | 0 | 0.001259781 | 0 |
| 0 | 0 | 0 | 0 | 0 |
| 0 | 0 | 0 | 0 | 0 |
| 0.021298143 | 1.38999992 | 0.070513959 | 0.098917811 | 1.370996303 |
| 0 | 0 | 0 | 0.019298253 | 0 |
| 0 | 0 | 0 | 0 | 0 |
| 0 | 0 | 0 | 0 | 0 |
| 0 | 0 | 0 | 0 | 0 |
| 0.018393852 | 0 | 0 | 0 | 0 |
| 0 | 0 | 0 | 0 | 0 |
| 0 | 0 | 0 | 0 | 0 |
| 0 | 0 | 0 | 0 | 0 |
| 0 | 0 | 0 | 0 | 0.002121738 |
| 0 | 0 | 0 | 0 | 0 |
| 0 | 0 | 0 | 0 | 0 |
| 0.008314763 | 0 | 0.052578445 | 0.016960734 | 0 |
| 0 | 0.000777278 | 0 | 0 | 0 |
| 0 | 0 | 0 | 0 | 0 |
| 0 | 0 | 0 | 0 | 0 |
| 0 | 0 | 0 | 0 | 0 |
| 0 | 0 | 0 | 0 | 0 |
| 0 | 0 | 0 | 0 | 0 |
| 0 | 0 | 0 | 0 | 0 |
| 0 | 0 | 0 | 0 | 0 |
| 0 | 0 | 0 | 0 | 0 |
| 0 | 0 | 0 | 0 | 0 |
| 0 | 0 | 0 | 0 | 0 |
| 0 | 0 | 0 | 0.001312738 | 0 |
| 0 | 0 | 0 | 0 | 0 |
| 0 | 0 | 0 | 0 | 0 |
| 0 | 0 | 0 | 0 | 0 |
| 0 | 0.001883783 | 0 | 0 | 0 |
| 0 | 0 | 0 | 0 | 0 |

| | | | | |
|---|---|---|---|---|
| 0 | 0 | 0 | 0 | 0.004770891 |
| 0 | 0 | 0 | 0 | 0 |
| 0 | 0 | 0 | 0 | 0 |
| 0 | 0 | 0 | 0 | 0 |
| 0.002233031 | 0.059534841 | 0.010472434 | 0 | 0 |
| 0 | 0 | 0 | 0 | 0 |
| 0 | 0 | 0 | 0 | 0 |
| 0 | 0 | 0 | 0 | 0 |
| 0 | 0 | 0 | 0 | 0 |
| 0 | 0 | 0 | 0 | 0 |
| 0 | 0 | 0 | 0 | 0 |
| 0 | 0 | 0 | 0 | 0 |
| 0 | 0 | 0 | 0 | 0 |
| 0 | 0 | 0 | 0 | 0 |
| 0 | 0 | 0 | 0 | 0 |
| 0 | 0 | 0 | 0 | 0 |
| 0 | 0 | 0 | 0 | 0 |
| 0 | 0 | 0 | 0 | 0 |
| 0 | 0 | 0 | 0 | 0 |
| 0 | 0 | 0 | 0 | 0 |
| 0 | 0 | 0 | 0 | 0 |
| 0 | 0 | 0 | 0 | 0 |
| 0 | 0.005196867 | 0 | 0 | 0 |
| 0 | 0 | 0 | 0 | 0 |
| 0 | 0.0790651 | 0.200264456 | 0 | 0 |
| 0 | 0 | 0 | 0 | 0 |
| 0 | 0.060580798 | 0.011372508 | 0 | 0 |
| 0 | 0 | 0 | 0 | 0 |
| 0 | 0 | 0 | 0 | 0 |
| 0 | 0 | 0 | 0 | 0 |
| 0.013002553 | 0 | 0 | 0 | 0 |
| 0.000986702 | 0 | 0 | 0.060496842 | 0 |
| 0 | 0 | 0 | 0 | 0 |
| 0 | 0 | 0 | 0 | 0 |
| 0 | 0 | 0 | 0 | 0 |
| 0 | 0 | 0 | 0 | 0 |
| 0 | 0 | 0 | 0 | 0 |
| 0 | 0 | 0 | 0 | 0 |
| 0 | 0 | 0 | 0 | 0 |
| 0 | 0 | 0 | 0 | 0 |
| 0 | 0 | 0 | 0 | 0 |
| 0 | 0 | 0 | 0 | 0 |
| 0 | 0 | 0 | 0 | 0 |
| 0 | 0 | 0 | 0 | 0 |
| 0 | 0 | 0 | 0 | 0 |
| 0 | 0 | 0 | 0 | 0 |
| 0 | 0 | 0 | 0 | 0 |
| 0 | 0 | 0 | 0 | 0 |

| | | | | |
|---|---|---|---|---|
| 0 | 0 | 0 | 0 | 0 |
| 0 | 0 | 0 | 0 | 0 |
| 0 | 0 | 0 | 0 | 0 |
| 0 | 0 | 0 | 0 | 0 |
| 0 | 0 | 0 | 0 | 0 |
| 0 | 0 | 0 | 0 | 0 |
| 0 | 0 | 0 | 0 | 0 |
| 0 | 0 | 0 | 0 | 0 |
| 0 | 0 | 0 | 0 | 0 |
| 0 | 0 | 0 | 0 | 0 |
| 0 | 0 | 0 | 0 | 0 |
| 0 | 0 | 0 | 0 | 0 |
| 0.001321847 | 0 | 0 | 0 | 0.006381831 |
| 0 | 0 | 0 | 0 | 0 |
| 0 | 0 | 0 | 0 | 0 |
| 0 | 0 | 0 | 0 | 0 |
| 0 | 0 | 0 | 0 | 0 |
| 0 | 0 | 0 | 0 | 0 |
| 0 | 0 | 0 | 0 | 0 |
| 0 | 0 | 0 | 0 | 0 |
| 0.010592301 | 0.005214797 | 0.082577623 | 0.025789927 | 0.033691737 |
| 0 | 0 | 0 | 0 | 0 |
| 0 | 0 | 0 | 0 | 0 |
| 0.001104564 | 0 | 0 | 0 | 0 |
| 0 | 0 | 0 | 0 | 0 |
| 0 | 0 | 0 | 0 | 0 |
| 0 | 0.001401681 | 0.041704141 | 0.001365404 | 0.008993084 |
| 0 | 0 | 0 | 0 | 0 |
| 0 | 0 | 0 | 0 | 0 |
| 0 | 0 | 0 | 0 | 0.001530703 |
| 0 | 0 | 0 | 0 | 0 |
| 0 | 0 | 0 | 0 | 0 |
| 0 | 0 | 0 | 0 | 0 |
| 0 | 0.023289323 | 0.001379938 | 0.010746274 | 0.087937726 |
| 0.006662185 | 0.059769437 | 0.000629641 | 0.003813705 | 0.008905666 |
| 0 | 0.00186141 | 0 | 0 | 0.002239252 |
| 0 | 0 | 0 | 0 | 0 |
| 0 | 0 | 0 | 0 | 0 |
| 0 | 0 | 0 | 0 | 0 |
| 0 | 0 | 0 | 0 | 0 |
| 0 | 0 | 0 | 0 | 0 |
| 0 | 0 | 0 | 0 | 0 |
| 0 | 0 | 0 | 0 | 0 |
| 0 | 0 | 0 | 0 | 0 |
| 0 | 0 | 0 | 0.004395144 | 0 |
| 0 | 0 | 0.003397094 | 0 | 0 |
| 0 | 0 | 0 | 0 | 0.006675555 |
| 0.005729926 | 0 | 0 | 0.001630649 | 0.009763721 |

| | | | | |
|---|---|---|---|---|
| 0 | 0 | 0 | 0 | 0 |
| 0 | 0 | 0 | 0 | 0 |
| 0 | 0 | 0 | 0 | 0 |
| 0.000832398 | 0 | 0 | 0 | 0 |
| 0 | 0 | 0 | 0 | 0 |
| 0 | 0 | 0 | 0 | 0 |
| 0 | 0.001059058 | 0 | 0 | 0.097753014 |
| 0 | 0 | 0 | 0 | 0.058481363 |
| 0 | 0 | 0 | 0.001963684 | 0 |
| 0 | 0 | 0 | 0 | 0.000627192 |
| 0 | 0 | 0 | 0 | 0 |
| 0 | 0 | 0 | 0 | 0 |
| 0 | 0 | 0 | 0 | 0 |
| 0 | 0 | 0 | 0 | 0 |
| 0 | 0 | 0 | 0 | 0 |
| 0 | 0 | 0 | 0 | 0 |
| 0 | 0 | 0 | 0 | 0 |
| 0 | 0 | 0 | 0 | 0 |
| 0.001831436 | 0.000567473 | 0.001521081 | 0 | 0.007943711 |
| 0 | 0.622552166 | 0.040613984 | 0 | 0.662821534 |
| 0.006631089 | 0 | 0 | 0.025006105 | 0.00488241 |
| 0 | 0 | 0 | 0 | 0 |
| 0 | 0 | 0 | 0 | 0 |
| 0 | 0 | 0 | 0 | 0 |
| 0 | 0 | 0.017384966 | 0.002159996 | 0.012124924 |
| 0 | 0 | 0 | 0 | 0 |
| 0 | 0 | 0 | 0 | 0 |
| 0 | 0 | 0 | 0 | 0 |
| 0 | 0 | 0.001701613 | 0 | 0.02429908 |
| 0 | 0 | 0 | 0 | 0 |
| 1.173270899 | 0.030775257 | 0.076387058 | 0.094732914 | 0.227967148 |
| 0 | 0 | 0 | 0 | 0 |
| 0.047603588 | 0 | 0 | 0 | 0 |
| 0 | 0.029198621 | 0 | 0 | 0 |
| 0 | 0 | 0 | 0 | 0 |
| 0 | 0 | 0 | 0 | 0 |
| 0 | 0 | 0 | 0 | 0 |
| 0 | 0 | 0 | 0 | 0 |
| 0 | 0 | 0 | 0 | 0 |
| 0 | 0 | 0 | 0 | 0 |
| 0 | 0 | 0 | 0 | 0 |
| 0 | 0.003555296 | 0.011772081 | 0.003463282 | 0.018297213 |
| 0.001853379 | 0 | 0 | 0.000683723 | 0 |
| 0 | 0 | 0 | 0 | 0 |
| 0 | 0 | 0 | 0 | 0 |
| 0 | 0 | 0 | 0 | 0 |
| 0 | 0 | 0 | 0 | 0 |
| 0 | 0 | 0 | 0 | 0 |

| | | | | |
|---|---|---|---|---|
| 0 | 0 | 0 | 0 | 0 |
| 0 | 0 | 0 | 0 | 0 |
| 0 | 0.001313017 | 0.338502859 | 0 | 0.067317223 |
| 0 | 0 | 0 | 0 | 0 |
| 1.942712597 | 0 | 0 | 0 | 0 |
| 0 | 0 | 0 | 0 | 0 |
| 0 | 0 | 0 | 0 | 0 |
| 0 | 0 | 0 | 0 | 0 |
| 0 | 0 | 0 | 0 | 0 |
| 0 | 0 | 0 | 0 | 0 |
| 0 | 0 | 0 | 0 | 0 |
| 0 | 0.001837645 | 0 | 0 | 0 |
| 0 | 0 | 0 | 0 | 0 |
| 0 | 0 | 0.024038053 | 0.008481391 | 0.011172349 |
| 0 | 0.001916337 | 0.005778718 | 0.007933648 | 0.002724478 |
| 0 | 0.3141653 | 0.007630486 | 0 | 0.070679065 |
| 0.001201177 | 0 | 0.005267465 | 0.000797619 | 0.00859652 |
| 0 | 0 | 0 | 0.00969338 | 0 |
| 0 | 0 | 0 | 0 | 0 |
| 0 | 0 | 0 | 0 | 0 |
| 0 | 0 | 0 | 0 | 0 |
| 0.007373377 | 0 | 0 | 0.002160067 | 0.008191327 |
| 0 | 0 | 0 | 0 | 0 |
| 0 | 0 | 0 | 0 | 0 |
| 0 | 0 | 0 | 0 | 0 |
| 0 | 0 | 0 | 0 | 0 |
| 0 | 0 | 0 | 1.722560957 | 0 |
| 0 | 0 | 0 | 0 | 0 |
| 0 | 0 | 0 | 0 | 0 |
| 0 | 0 | 0 | 0 | 0 |
| 0 | 0 | 0 | 0 | 0 |
| 0 | 0 | 0 | 0 | 0 |
| 0 | 0 | 0 | 0 | 0 |
| 0 | 0 | 0 | 0 | 0 |
| 0 | 0 | 0 | 0 | 0 |
| 0 | 0 | 0 | 0 | 0.401275295 |
| 0 | 0 | 0 | 0 | 0 |
| 0 | 0.067173652 | 0.002738169 | 0 | 0.662451054 |
| 0 | 0 | 0 | 0 | 0 |
| 0 | 0 | 0 | 0 | 0 |
| 0 | 0 | 0 | 0 | 0 |
| 0 | 0 | 0 | 0 | 0 |
| 0 | 0 | 0 | 0 | 0 |
| 0.002707575 | 0 | 0 | 0 | 0 |
| 0 | 0 | 0 | 0 | 0 |
| 0 | 0 | 0 | 0 | 0 |
| 0 | 0 | 0 | 0 | 0 |
| 0.00429034 | 0 | 0 | 0 | 0.006888913 |

| | | | | |
|---|---|---|---|---|
| 0 | 0 | 0 | 0.094148117 | 0 |
| 0 | 0 | 0 | 0 | 0 |
| 0 | 0 | 0.001452649 | 0 | 0.002257836 |
| 0 | 0 | 0 | 0 | 0 |
| 0.001142665 | 0 | 0.007098738 | 0.004805511 | 0.003861721 |
| 0 | 0 | 0 | 0 | 0 |
| 0 | 0 | 0 | 0 | 0 |
| 0 | 0 | 0.094236549 | 0.003545253 | 0.003184148 |
| 0 | 0 | 0 | 0 | 0 |
| 0.357915765 | 0.020256775 | 0.011764393 | 0.020097931 | 0.00886132 |
| 0 | 0 | 0 | 0 | 0 |
| 0.001775072 | 0.004609596 | 0.001158354 | 0.02638049 | 7.675427318 |
| 0 | 0 | 0.004393263 | 0.002341659 | 0.052961062 |
| 0.055504913 | 0 | 0.090475478 | 0.000969921 | 0 |
| 0 | 0 | 0 | 0 | 0.00078656 |
| 0 | 0 | 0 | 0 | 0 |
| 0 | 0 | 0 | 0 | 0.016280075 |
| 0.001252559 | 0 | 0 | 0 | 0 |
| 0.848505096 | 0 | 0 | 0 | 0 |
| 0.017432829 | 0 | 0 | 0 | 0.036389013 |
| 0 | 0 | 0 | 0 | 0 |
| 0 | 0 | 0 | 0 | 0 |
| 0 | 0 | 0 | 0 | 0 |
| 0 | 0 | 0 | 0 | 0 |
| 0 | 0 | 0 | 0 | 0 |
| 0 | 0 | 0 | 0 | 0.003785792 |
| 0 | 5.279426179 | 0 | 0 | 0 |
| 0.010566117 | 0 | 0 | 0 | 0 |
| 0.413120893 | 0 | 0 | 0 | 0 |
| 0.001152542 | 0 | 0 | 0 | 0 |
| 0 | 0 | 0 | 0 | 0 |
| 0 | 0 | 0 | 0 | 0 |
| 0 | 0 | 0 | 0 | 0 |
| 0 | 0 | 0 | 0 | 0 |
| 0 | 0 | 0 | 0 | 0 |
| 0 | 0 | 0 | 0 | 0 |
| 0 | 0 | 0 | 0 | 0 |
| 0.027989793 | 0 | 0 | 0 | 0 |
| 0 | 0 | 0 | 0 | 0 |
| 0 | 0 | 0 | 0 | 0 |
| 0 | 0 | 0 | 0 | 0 |
| 0 | 0 | 0 | 0 | 0 |
| 0 | 0 | 0 | 0 | 0 |
| 0 | 0 | 0 | 0 | 0 |
| 0 | 0 | 0 | 0 | 0 |
| 0 | 0 | 0 | 0.000544227 | 0 |
| 0.001950276 | 0.002215752 | 0 | 0 | 0 |
| 0 | 0 | 0 | 0 | 0 |

| | | | | |
|---|---|---|---|---|
| 0 | 0 | 0 | 0 | 0 |
| 0 | 0 | 0 | 0 | 0 |
| 0 | 0 | 0 | 0 | 0 |
| 0 | 0 | 0.003287323 | 0.005973342 | 0.005961024 |
| 0 | 0 | 0 | 0 | 0 |
| 0 | 0 | 0.220727323 | 0.039343007 | 0 |
| 0 | 0 | 0 | 0.011341882 | 0 |
| 0 | 0 | 0 | 0 | 0 |
| 0 | 0 | 0 | 4.834302338 | 0 |
| 0 | 0 | 0 | 0 | 0 |
| 0.002290619 | 0.002313265 | 0.007905742 | 0.008731909 | 0.002276858 |
| 0 | 0 | 0 | 0.003431911 | 0 |
| 0 | 0 | 0 | 0 | 0 |
| 0 | 0 | 0.009665697 | 0.002395007 | 0.01195034 |
| 0 | 0 | 0 | 0 | 0 |
| 0.034819515 | 0 | 0 | 0 | 0.011211799 |
| 0 | 0 | 0 | 0.140892043 | 0 |
| 0 | 0 | 0 | 0 | 0 |
| 0 | 0 | 0 | 0 | 0.011956142 |
| 0 | 0 | 0 | 0.044998707 | 0 |
| 0 | 0 | 0 | 0 | 0 |
| 0 | 0.165241123 | 0 | 0 | 0 |
| 0 | 0 | 0 | 0.141667899 | 0 |
| 0 | 0 | 0 | 0 | 0 |
| 0 | 0 | 0 | 0 | 0 |
| 0 | 0 | 0 | 0 | 0 |
| 0.055010417 | 0 | 0 | 0 | 0 |
| 0 | 0 | 0 | 0 | 0 |
| 0 | 0 | 0 | 0 | 0 |
| 0.503652837 | 0.241858298 | 0.661060741 | 0.565983265 | 1.201645293 |
| 0 | 0.032618484 | 0.000894192 | 0 | 0 |
| 0 | 0 | 0 | 0 | 0.013886618 |
| 0 | 0 | 0 | 0 | 0 |
| 0 | 0 | 0 | 0 | 0 |
| 0.003257239 | 0 | 0 | 0 | 0 |
| 0 | 0 | 0 | 0 | 0 |
| 0 | 0.401319243 | 0.931422968 | 0 | 0 |
| 0 | 0 | 0 | 0 | 0.273332214 |
| 0 | 0 | 0.001172105 | 0.258985275 | 0.000765152 |
| 0 | 0 | 0 | 0 | 0 |
| 0.003953543 | 0 | 0.253738359 | 0.001312638 | 0 |
| 0 | 0.132696034 | 6.056267957 | 4.356704624 | 0.513727609 |
| 0 | 0 | 0 | 0 | 0.232536872 |
| 0.013593259 | 0 | 0 | 0 | 0 |
| 0 | 0 | 0 | 0 | 0 |
| 0 | 0 | 0 | 0 | 0 |
| 0.003734574 | 0 | 0 | 0 | 0 |
| 0 | 0.016106286 | 0 | 0 | 0 |

| | | | | |
|---|---|---|---|---|
| 0 | 0.001745022 | 0 | 0 | 0.006106875 |
| 0.000654283 | 0 | 0 | 0.004163614 | 0 |
| 0 | 0 | 0.006054699 | 0.003474283 | 0.006934238 |
| 0 | 0 | 0 | 0 | 0 |
| 0 | 0 | 0.000783661 | 0.074996134 | 0 |
| 0 | 0 | 0 | 0.006467547 | 0 |
| 0 | 0 | 0 | 0 | 0 |
| 0 | 0 | 0 | 0 | 0.003137094 |
| 0.002356656 | 0 | 0 | 0.00142263 | 0.002129544 |
| 0.181600724 | 0 | 0 | 0 | 0.070406296 |
| 0 | 0 | 0 | 0 | 0 |
| 0 | 0 | 0 | 0 | 0 |
| 0 | 0 | 0 | 0 | 0 |
| 0 | 0 | 0.024543048 | 0.00845801 | 0 |
| 0 | 0 | 0 | 0 | 0 |
| 0.057914394 | 0.053871966 | 0.005621715 | 0.010215129 | 0.009894238 |
| 0.001203601 | 0.029855729 | 0.003848611 | 0 | 0.121232226 |
| 0.009017693 | 0 | 0 | 0 | 0.005087403 |
| 0.045377866 | 0 | 0 | 0 | 0 |
| 0 | 0 | 0.160088763 | 0 | 0 |
| 0 | 0 | 0.114560534 | 0 | 0 |
| 0 | 0.010045661 | 0 | 0 | 0 |
| 0 | 0 | 0 | 0 | 0 |
| 0 | 0 | 0.422410676 | 0.623210144 | 0 |
| 0 | 0 | 0 | 0 | 0 |
| 0 | 0 | 0 | 0 | 0 |
| 0 | 0 | 0.019776031 | 0 | 0 |
| 0 | 0 | 0 | 0.006064274 | 0 |
| 0 | 0 | 0 | 0 | 0 |
| 0 | 0 | 0 | 0 | 0 |
| 0.126854462 | 0.008141251 | 0.064228005 | 0.11017034 | 0.029068626 |
| 0.001732438 | 0 | 0.010551634 | 0.005432408 | 0.01664629 |
| 0.289764768 | 0.021863919 | 0.016828024 | 0.005172387 | 0.038803994 |
| 0 | 0 | 0 | 0 | 0.011356953 |
| 0 | 0.017252137 | 0 | 0 | 0.010621622 |
| 0 | 0 | 0 | 0 | 0 |
| 0 | 0 | 0 | 0 | 0.002964633 |
| 0.266681643 | 0 | 0.003906745 | 0.001075588 | 0.007148647 |
| 0 | 0.151910487 | 0.076644548 | 0.039631506 | 0 |
| 0 | 0 | 0 | 0 | 0.011689498 |
| 0 | 0 | 0 | 0 | 0 |
| 0 | 0 | 0 | 0 | 0.00340149 |
| 0 | 0 | 0 | 0 | 0 |
| 0.053713538 | 0 | 0 | 0.05860613 | 0 |
| 0.076607087 | 0 | 0 | 0 | 0 |
| 0 | 0 | 0 | 0 | 0.029274936 |
| 0.00496788 | 0 | 0 | 0 | 0 |
| 0 | 0 | 0 | 0 | 0 |

| | | | | |
|---|---|---|---|---|
| 0 | 0 | 0 | 0 | 0 |
| 0.017745603 | 0.02708217 | 1.047748898 | 0.332403916 | 0.961983476 |
| 0 | 0 | 0 | 0 | 0 |
| 0 | 0 | 0 | 0.001420222 | 0 |
| 0 | 0 | 0 | 0 | 0 |
| 0 | 0 | 0 | 0 | 0 |
| 0 | 0 | 0 | 0 | 0 |
| 0.000610321 | 0.006009456 | 0 | 0 | 0 |
| 0 | 0 | 0 | 0.005366639 | 0.005543017 |
| 0 | 0 | 0 | 0 | 0 |
| 0.001968197 | 0 | 0.000719254 | 0 | 0.002152011 |
| 0 | 0 | 0.025712366 | 0.005840191 | 0.536481011 |
| 0 | 0.018612732 | 0.018368748 | 0 | 0 |
| 0.003454591 | 0 | 0 | 0 | 0.013449199 |
| 0 | 0 | 0.002475761 | 0 | 0.008080899 |
| 0 | 0 | 0 | 0 | 0 |
| 0.011129329 | 0 | 0.024477775 | 0.00570233 | 0.02990964 |
| 0.146614422 | 0 | 0 | 0.002072675 | 0 |
| 0 | 0 | 0 | 0 | 0 |
| 0.024843313 | 0 | 0 | 0.002307235 | 0 |
| 0 | 0.038393903 | 0.030873839 | 0.017406401 | 0.150207929 |
| 0.002115773 | 0 | 0 | 0 | 0 |
| 0 | 0 | 0 | 0 | 0 |
| 0 | 0 | 0 | 0 | 0 |
| 0 | 0 | 0 | 0 | 0 |
| 0 | 0 | 0 | 0 | 0 |
| 0.048251089 | 0 | 0.145087008 | 0.09356673 | 0.018558907 |
| 0.042242995 | 0.023868275 | 0.054690555 | 0.023500552 | 0.050072707 |
| 0 | 0 | 0 | 0 | 0 |
| 0 | 0 | 0 | 0 | 0.004062741 |
| 0 | 0 | 0.001107227 | 0 | 0.007709856 |
| 0 | 0 | 0 | 0 | 0 |
| 0 | 0 | 0 | 0 | 0 |
| 0 | 0 | 0 | 0 | 0 |
| 0 | 0.001789208 | 0 | 0 | 0 |
| 19.88061453 | 0.224999326 | 0.222843772 | 0.216214334 | 0.030781841 |
| 0 | 0 | 0 | 0 | 0 |
| 0 | 0 | 0 | 0 | 0 |
| 0.008081456 | 0.007915104 | 0.008401541 | 0.007093435 | 3.241416468 |
| 0 | 0 | 0 | 0 | 0 |
| 0.069946975 | 0 | 0 | 0 | 0 |
| 0 | 0 | 0 | 0 | 0 |
| 0 | 0 | 0 | 0.005994658 | 0.041390012 |
| 0.003595372 | 0 | 0.005076372 | 0.001266066 | 0.002923996 |
| 0 | 0 | 0 | 0 | 0 |
| 0 | 0.033187858 | 0.280758476 | 0 | 0 |
| 0 | 0 | 0 | 0 | 0.000714144 |
| 0 | 0 | 0 | 0.005262422 | 0 |

| | | | | |
|---|---|---|---|---|
| 0 | 0 | 0 | 0 | 0 |
| 0 | 0 | 0 | 0 | 0 |
| 0 | 0.001057441 | 0.001417208 | 0 | 0 |
| 0.001577158 | 0.088696292 | 0.041425378 | 0.049527711 | 0 |
| 0 | 0 | 0 | 0 | 0 |
| 0 | 0 | 0 | 0 | 0 |
| 0 | 0 | 0 | 0 | 0 |
| 0 | 0 | 0.001772408 | 0.002732643 | 0 |
| 0.001277373 | 8.358481846 | 0.060684018 | 0 | 3.706462658 |
| 0 | 0.027224958 | 0.213130036 | 0.067275902 | 0.012785252 |
| 0 | 0 | 0 | 0 | 0 |
| 0 | 0.004291949 | 0.001380521 | 0.003344696 | 0 |
| 0 | 0 | 0 | 0 | 0 |
| 0 | 0 | 10.9040436 | 0.065734871 | 0 |
| 0 | 0.43209068 | 0.036739492 | 0 | 0 |
| 0 | 0 | 0 | 0 | 0 |
| 0 | 0 | 0 | 0.045453097 | 0 |
| 0 | 0 | 0 | 0.03626738 | 0 |
| 0 | 0 | 0 | 0 | 0 |
| 0 | 0 | 0 | 0 | 0 |
| 0 | 0 | 0 | 0 | 0 |
| 0 | 0.030682649 | 0 | 0 | 0 |
| 0 | 0.010868436 | 0 | 0 | 0 |
| 0.119813244 | 0 | 0.320151947 | 0.283809497 | 0.089096241 |
| 0 | 0 | 0 | 0 | 0 |
| 0 | 0.181804866 | 1.169397121 | 0 | 0 |
| 0 | 0 | 0 | 0.042150258 | 0 |
| 0 | 1.124332678 | 0 | 0 | 0 |
| 0 | 0 | 0 | 0 | 0 |
| 0 | 0 | 0 | 0 | 0 |
| 0 | 0.025869536 | 0.004026304 | 0.002641937 | 0 |
| 0 | 0.003931274 | 0 | 0.054187851 | 0 |
| 0 | 0 | 0.009703658 | 0.021963117 | 0.380768196 |
| 0 | 0 | 0 | 0 | 0 |
| 0 | 0 | 0 | 0 | 0 |
| 0 | 0.224120792 | 0.041677486 | 0 | 0 |
| 0 | 2.078945773 | 0.225227078 | 0.001112715 | 0 |
| 0 | 0 | 0 | 0.002827553 | 0 |
| 0 | 0 | 0 | 0 | 0 |
| 0 | 0 | 0 | 0 | 0 |
| 0.490695685 | 0.01935158 | 0.165710877 | 0.031885837 | 0.082852291 |
| 0 | 0.017343553 | 1.851726511 | 0 | 0.20289177 |
| 0 | 0.02938869 | 0.3600472 | 3.506339454 | 0.016205135 |
| 0 | 0 | 0 | 0.000873482 | 0 |
| 0 | 0 | 0 | 0.063167329 | 0 |
| 0 | 0 | 0 | 0 | 0 |
| 0 | 0 | 0 | 0 | 0 |
| 0 | 0.014106178 | 0.017910403 | 0.000723216 | 0.01364059 |

| | | | | |
|---|---|---|---|---|
| 0 | 0 | 0 | 0 | 0 |
| 0 | 0.404697782 | 0 | 0 | 0 |
| 0.212569298 | 0.007591502 | 0.034529055 | 0 | 0.001452894 |
| 0 | 0.012309076 | 0 | 0 | 0 |
| 0 | 0 | 0 | 24.44953163 | 0 |
| 0 | 0.117363249 | 0.001803359 | 0.001638427 | 0.172007085 |
| 0 | 0 | 0 | 0 | 0 |
| 0 | 0 | 0 | 0 | 0 |
| 0 | 0 | 0 | 0 | 0 |
| 0 | 0 | 0 | 0 | 0 |
| 0.015786161 | 0.002562144 | 0.008428527 | 0.089623114 | 0.137146267 |
| 2.36365635 | 0.974362401 | 0.514052244 | 0.921450739 | 0.83674019 |
| 0 | 0 | 0 | 0 | 0 |
| 0 | 0.016339641 | 0 | 0.00073462 | 0.187601497 |
| 0.204526468 | 0.002773349 | 0.030903424 | 0.416042201 | 0.051474374 |
| 0 | 0.22638434 | 0 | 0 | 0.222482313 |
| 0 | 0 | 0 | 0 | 0 |
| 0 | 0 | 0 | 0 | 0 |
| 0 | 0.009170398 | 0 | 0.014008665 | 0 |
| 0.02651392 | 0 | 0 | 0.002990414 | 0 |
| 0.248642105 | 0.035921045 | 0.293719846 | 0.049444343 | 0.120926787 |
| 0 | 0 | 0 | 0 | 0.02735556 |
| 23.53646089 | 1.609750205 | 1.083901921 | 1.524620451 | 0.150355138 |
| 0 | 0 | 0 | 0 | 0 |
| 0 | 0 | 0 | 0 | 0 |
| 0 | 0.981594606 | 0.001693041 | 0.00159517 | 2.623873091 |
| 0 | 0 | 0 | 1.330371644 | 0.001429998 |
| 0 | 0.002036141 | 0 | 0.004561922 | 0 |
| 0 | 0 | 0 | 0 | 0 |
| 0 | 0 | 0 | 0 | 0 |
| 0 | 0.016364775 | 0.034803336 | 0.003356051 | 0 |
| 0 | 0.128276658 | 0 | 0.376127035 | 0 |
| 0.497332872 | 0.263617274 | 0 | 0.020739315 | 0.003348655 |
| 0 | 0.030829247 | 0 | 0.066104539 | 0 |
| 0.017649122 | 0 | 0 | 0 | 0 |
| 0.027494719 | 0.070500978 | 0.00732638 | 0.004226238 | 0.116909718 |
| 0 | 0.013830144 | 0.002022052 | 0 | 0 |
| 0 | 0 | 0 | 0 | 0 |
| 0 | 0.94659669 | 0.04286672 | 0.013847544 | 1.072921017 |
| 0 | 0 | 0 | 0 | 0 |
| 0 | 0.147654085 | 0.021196952 | 0 | 0 |
| 0 | 0 | 0 | 0 | 0 |
| 0 | 0 | 0 | 0 | 0 |
| 0 | 0 | 0 | 0 | 0 |
| 0.181595423 | 0.029685549 | 0.004376379 | 0.099041633 | 0.230067475 |
| 0 | 0.012619524 | 0 | 0 | 0 |
| 0 | 0 | 0 | 0 | 0 |
| 0 | 0.182326905 | 0.011758015 | 0.003197528 | 0.920555495 |

| | | | | |
|---|---|---|---|---|
| 0 | 0 | 0 | 0 | 0 |
| 0 | 0 | 0 | 0 | 0 |
| 0 | 0 | 0 | 0 | 0 |
| 0 | 0.013072118 | 0.00542541 | 0 | 0.018077503 |
| 0 | 0 | 0 | 0 | 0 |
| 0 | 0.663477248 | 0 | 0.013060351 | 0.825829933 |
| 0 | 0.018466426 | 0.328359095 | 0 | 0 |
| 0 | 0.001746104 | 0.001872135 | 0.015875191 | 0 |
| 0 | 0 | 0 | 0 | 0 |
| 0 | 0 | 0 | 0 | 0 |
| 0.005373838 | 0.018550828 | 0 | 0.01463957 | 0.025680551 |
| 0 | 0 | 0 | 0 | 0 |
| 0 | 0.154202854 | 0.004794657 | 0.005407631 | 0.088862027 |
| 0 | 0.001202275 | 0 | 0.5311207 | 0 |
| 0 | 0 | 0 | 0 | 0 |
| 0 | 0 | 0 | 0 | 0 |
| 0 | 0.008208301 | 0 | 0 | 0 |
| 0 | 0.048675217 | 0 | 0.006283977 | 0.027193418 |
| 0 | 0 | 0.155981762 | 0 | 0.110139552 |
| 0 | 0 | 0 | 0 | 0 |
| 0 | 0 | 0 | 0.000845882 | 0.000759724 |
| 0 | 0 | 0 | 0.003828373 | 0 |
| 0 | 0.0252423 | 0.002537274 | 0.003073626 | 0 |
| 0 | 0 | 0 | 0 | 2.131910674 |
| 0 | 0 | 0 | 0 | 0 |
| 0 | 0 | 0 | 0 | 0 |
| 0 | 0 | 0 | 0 | 0 |
| 0 | 0 | 0 | 0 | 0 |
| 0 | 0 | 0 | 0 | 0 |
| 0.001247092 | 2.589999729 | 2.43476143 | 0.227269599 | 0.986308574 |
| 0 | 0.029961476 | 0 | 0.001621447 | 0.015857415 |
| 0 | 0 | 0 | 0 | 0 |
| 0 | 0.346197375 | 0.018887752 | 0 | 0 |
| 0 | 0 | 0 | 0 | 0 |
| 0 | 0 | 0.001801754 | 0.00163697 | 0 |
| 0 | 0 | 0 | 0 | 0 |
| 0 | 0.013957179 | 0 | 0.539251686 | 0 |
| 0.005073938 | 0 | 0.000479537 | 0.004066339 | 0 |
| 0 | 0 | 0 | 0 | 0 |
| 0 | 0 | 0.00110364 | 0.002100902 | 0 |
| 0 | 0 | 0 | 0.02797837 | 0 |
| 0 | 0 | 0 | 0 | 0 |
| 0 | 0.00179271 | 0 | 0 | 0 |
| 0 | 0.000772155 | 0.009003279 | 0 | 0 |
| 0.354227018 | 0.036791114 | 0 | 0 | 0 |
| 0 | 0 | 0 | 0 | 0 |
| 0 | 0.016809791 | 0.034021127 | 0 | 0 |
| 0 | 0 | 0 | 0 | 0 |

| | | | | |
|---|---|---|---|---|
| 0 | 0 | 0 | 0 | 0 |
| 0 | 0 | 0 | 0 | 0 |
| 0 | 0.20541078 | 0 | 0 | 0 |
| 0 | 0.939996991 | 0.025280914 | 0.026002385 | 0 |
| 0 | 0 | 0 | 0 | 0 |
| 0 | 0 | 0 | 0 | 0 |
| 0 | 0 | 0 | 0 | 0 |
| 0 | 0 | 0 | 0 | 0 |
| 0.208245112 | 0.003838107 | 0.006613611 | 0.004272884 | 0.008874601 |
| 0 | 0 | 0 | 0.001505409 | 0 |
| 0 | 0 | 0.01891575 | 0 | 0 |
| 0 | 0 | 0 | 0 | 0 |
| 0 | 0 | 0 | 0 | 0 |
| 0 | 0 | 0 | 0 | 0 |
| 2.781625265 | 0.248037823 | 0.494916086 | 0.278465706 | 0.007584081 |
| 0.372330937 | 0 | 0 | 0 | 0.003697244 |
| 0 | 0 | 0 | 0 | 0 |
| 0 | 0 | 0 | 0 | 0 |
| 0 | 0 | 0 | 0 | 0 |
| 0 | 0 | 0 | 0 | 0 |
| 0 | 0 | 0.043659232 | 0.002246223 | 0 |
| 0 | 0 | 0 | 0 | 0 |
| 0 | 0 | 0 | 0 | 0 |
| 0 | 0 | 0.116933492 | 0.154820169 | 0 |
| 0.028944163 | 0.006447863 | 0.017369567 | 0.008165284 | 0.45947832 |
| 0.000802385 | 3.826014487 | 0.506296006 | 0.971191207 | 10.54938425 |
| 0 | 0.016787563 | 0.051490751 | 0 | 2.268788038 |
| 0 | 0 | 0 | 0 | 0 |
| 0 | 0 | 0 | 0 | 0 |
| 0 | 0 | 0 | 0 | 0 |
| 0 | 0 | 0 | 0 | 0 |
| 0 | 0 | 0 | 0.002629244 | 0 |
| 0 | 0 | 0 | 0 | 0 |
| 0 | 0 | 0 | 0.00201267 | 0.042932114 |
| 0 | 0.00425544 | 0.000912518 | 0 | 0.01042463 |
| 0.002824185 | 0 | 0 | 0.001172092 | 0.18808367 |
| 0.398008811 | 0 | 0 | 0 | 0 |
| 0 | 0.079254997 | 0.820135021 | 1.278274652 | 0.236152737 |
| 0 | 0.007091594 | 8.55734852 | 0 | 4.44192947 |
| 0 | 0 | 0 | 0 | 0 |
| 0 | 0 | 0 | 0 | 0 |
| 0 | 0 | 0 | 0 | 0 |
| 0 | 0.017264501 | 0.381230774 | 0.373726286 | 0 |
| 0 | 1.171249963 | 0 | 0 | 0 |
| 0 | 0 | 0 | 0 | 0 |
| 0 | 0.002849779 | 0.001374962 | 0.003701366 | 0 |
| 0 | 0 | 0 | 0 | 0 |
| 0 | 0.004540039 | 0.512572202 | 0.016510813 | 0.002912857 |

| | | | | |
|---|---|---|---|---|
| 0 | 0 | 0.000725975 | 0 | 0 |
| 0 | 0 | 0 | 0 | 0 |
| 0 | 0 | 0 | 0 | 0 |
| 0 | 0 | 0 | 0 | 0 |
| 0.033030594 | 0 | 0 | 0.001751165 | 0.002948997 |
| 0 | 0.183501796 | 0 | 0 | 0 |
| 0 | 0 | 0 | 0 | 0 |
| 0 | 0 | 0 | 0 | 0 |
| 0 | 0 | 0 | 0 | 0 |
| 0 | 0 | 0.001269578 | 0 | 0.00868248 |
| 0 | 0.000859718 | 0 | 0 | 0 |
| 0 | 1.108696603 | 0.451604943 | 7.141184654 | 2.682367287 |
| 0 | 0.017957675 | 0.669070694 | 0.643527292 | 0.020472116 |
| 0 | 0 | 0 | 0 | 0 |
| 0.024704114 | 0.016892109 | 0.036779986 | 0.014936012 | 0.801016173 |
| 0 | 0 | 0 | 0 | 0 |
| 0 | 0 | 0 | 0 | 0 |
| 0 | 0 | 0 | 0 | 0 |
| 0 | 0 | 0 | 0.00088755 | 0 |
| 0 | 0.007324664 | 0.012592436 | 0 | 0.008618118 |
| 0 | 0.069964353 | 0.554114889 | 0 | 0 |
| 0 | 0 | 0 | 0 | 0 |
| 0.07642064 | 0.306317481 | 0.205177836 | 0.167641966 | 0.065885013 |
| 0 | 0 | 0 | 0 | 0 |
| 0 | 0 | 0 | 0 | 0 |
| 0 | 0 | 0 | 0 | 0 |
| 0 | 0 | 0 | 0 | 0 |
| 0 | 0 | 0 | 0 | 0 |
| 0 | 0.026451591 | 0 | 0 | 0 |
| 0 | 0 | 0 | 0 | 0 |
| 0 | 0 | 0 | 0 | 0 |
| 0.004155543 | 0.001348915 | 0.091386693 | 0.038763129 | 0.132178503 |
| 0 | 0 | 0 | 0 | 0 |
| 0 | 0.021733109 | 0.009618755 | 0.016083121 | 0 |
| 0.00653102 | 0 | 0 | 0.001062941 | 0.195708226 |
| 0 | 0.541003595 | 0.003845901 | 0.003562676 | 0.170819896 |
| 0 | 0.001743495 | 0 | 0 | 0 |
| 0 | 0 | 0 | 0 | 0 |
| 0 | 0 | 0 | 0 | 0 |
| 0 | 0 | 0 | 0 | 0 |
| 0 | 0.062103349 | 0.053378349 | 0 | 0 |
| 0 | 0 | 0.093651927 | 0 | 0 |
| 0 | 0 | 0 | 0 | 0 |
| 0 | 0 | 0.011715015 | 0.018518104 | 0 |
| 0.102371669 | 0.011179203 | 0.0029389 | 0.098427737 | 0.218748633 |
| 0 | 0 | 0 | 0 | 0 |
| 0 | 0 | 0 | 0 | 0 |
| 0.001789746 | 0 | 0.005722857 | 0.000594224 | 0 |

| | | | | |
|---|---|---|---|---|
| 0 | 0 | 0 | 0 | 0 |
| 0 | 0.046620152 | 0 | 0.004995495 | 0 |
| 0.004892695 | 0 | 0 | 0 | 0.00739222 |
| 0 | 0 | 0 | 0 | 0 |
| 0 | 0 | 0 | 0 | 0 |
| 0 | 0 | 0 | 0 | 0 |
| 0 | 0.768044376 | 0 | 0 | 0 |
| 0.063765567 | 0.009449409 | 0.006078872 | 0.003272836 | 0.004409217 |
| 0 | 0 | 0 | 0 | 0 |
| 0 | 0 | 0 | 0.004428319 | 0 |
| 0 | 0 | 0 | 0 | 0 |
| 0 | 0 | 0 | 0.0030853 | 0 |
| 0 | 0 | 0 | 0 | 0 |
| 0 | 0.089022298 | 0 | 0.050324495 | 0 |
| 0 | 0 | 0 | 0 | 0 |
| 0 | 0 | 0 | 0 | 0 |
| 0 | 0 | 2.822210379 | 0 | 0 |
| 0 | 0 | 0 | 0.058215735 | 0 |
| 11.86192567 | 7.269896648 | 0.584998232 | 0.032169768 | 2.503305228 |
| 0 | 0 | 0 | 0 | 0 |
| 0 | 0.002222253 | 0 | 0 | 0 |
| 0 | 0 | 0 | 0 | 0 |
| 0 | 0 | 0 | 0 | 0 |
| 0 | 0 | 0 | 0 | 0 |
| 0 | 0 | 0 | 0 | 0 |
| 0.026424442 | 0 | 0 | 0 | 0.016109628 |
| 0 | 0 | 0 | 0 | 0 |
| 0.247090021 | 0.140264263 | 0.8913029 | 0.044844997 | 0.00325646 |
| 0 | 0 | 0 | 0 | 0 |
| 0.941689515 | 0.459819843 | 0.13209786 | 0.023976651 | 0.72451582 |
| 0 | 0 | 0 | 0 | 0 |
| 0 | 0 | 0 | 0 | 0 |
| 0 | 0.021926447 | 0.043414146 | 0 | 0.017011725 |
| 0 | 0 | 0 | 0 | 0 |
| 0.01476863 | 0 | 0 | 0 | 0.00954194 |
| 0 | 0.016652425 | 0 | 0 | 0 |
| 0 | 0 | 0 | 0 | 0 |
| 0.001047379 | 0.001189951 | 0 | 0 | 0.022279266 |
| 0 | 0 | 8.929360944 | 0 | 0 |
| 0 | 0 | 0 | 0 | 0 |
| 16.53981884 | 3.059222165 | 3.783449456 | 6.73736986 | 1.070982822 |
| 0.448051535 | 0.025086221 | 3.981302645 | 0.164949554 | 0.310014367 |
| 0 | 0 | 0.140418242 | 0 | 0 |
| 0 | 0.043184212 | 0 | 0 | 0 |
| 0 | 0 | 1.426746192 | 0 | 0 |
| 0 | 0 | 0 | 0 | 0 |
| 0 | 0 | 0 | 0 | 0 |
| 0.005153801 | 0.189071034 | 0 | 0.023183211 | 0.005783851 |

| | | | | |
|---|---|---|---|---|
| 0 | 0 | 0 | 0 | 0 |
| 0.001881039 | 0 | 0 | 0.012301429 | 0.014617956 |
| 0 | 0 | 0 | 0.002210365 | 0 |
| 0 | 0 | 0 | 0 | 0 |
| 0 | 0.046851651 | 4.370584385 | 1.402411897 | 0 |
| 0 | 0 | 0 | 0 | 0 |
| 0 | 0 | 0 | 0 | 0 |
| 0 | 0 | 0 | 0 | 0 |
| 0.202852681 | 0 | 0 | 0.002834606 | 0 |
| 0 | 0 | 0 | 0 | 0.006800315 |
| 0 | 0 | 0 | 0 | 0 |
| 0 | 0 | 0 | 0 | 0 |
| 0 | 0.045684032 | 0.128120869 | 0.045421152 | 0.055989537 |
| 0.016137779 | 0.011372024 | 0.037511643 | 0.019668582 | 0.059493211 |
| 0 | 0 | 0.498725377 | 0 | 0 |
| 0 | 0.113314718 | 0.257127433 | 0.445052995 | 0.208275164 |
| 0.001060614 | 0.004217453 | 0.162787031 | 0.001369434 | 0.001581363 |
| 0 | 0 | 0 | 0 | 0 |
| 0 | 0 | 0 | 0 | 0 |
| 0 | 0 | 0 | 0 | 0 |
| 0 | 0 | 0 | 0 | 0 |
| 0 | 0 | 0 | 0 | 0 |
| 0 | 0 | 0 | 0 | 0 |
| 0 | 0 | 0 | 0 | 0 |
| 0 | 0 | 0 | 0 | 0 |
| 0 | 0 | 0 | 0 | 0 |
| 0.00353528 | 0.082600402 | 0.003370238 | 0.00663434 | 0.1031295 |
| 0.001560979 | 0 | 0 | 0 | 0.02206722 |
| 0 | 0 | 0 | 0 | 0 |
| 0 | 0 | 0 | 0 | 0 |
| 0 | 0 | 0 | 0 | 0 |
| 0 | 0 | 0 | 0 | 0 |
| 0 | 0 | 0.02544953 | 0 | 0 |
| 0 | 0.020449685 | 0 | 0.000804866 | 0 |
| 0 | 2.375410157 | 0 | 0 | 0 |
| 0 | 0 | 0.114746032 | 0.032863122 | 0 |
| 0 | 0.000931959 | 0.001873551 | 0.065137497 | 0.00305764 |
| 0 | 0.019156995 | 0 | 0 | 0 |
| 0 | 0 | 0 | 0 | 0 |
| 0 | 0 | 0 | 0 | 0 |
| 0 | 0.173927353 | 0.239761505 | 1.352688338 | 0.004885038 |
| 0 | 0 | 0 | 0 | 0 |
| 0 | 0.047489627 | 0.001558695 | 0 | 0 |
| 0 | 0.001929391 | 0 | 0 | 0 |
| 0 | 0 | 0 | 0 | 0 |
| 0 | 0 | 0 | 0 | 0 |
| 0 | 0 | 0 | 0 | 0 |
| 0 | 0 | 0 | 0 | 0 |

| | | | | |
|---|---|---|---|---|
| 0 | 1.187072098 | 0.017175922 | 0 | 0 |
| 0 | 0 | 0 | 0 | 0 |
| 0 | 0 | 0 | 0 | 0 |
| 0 | 0 | 0 | 0 | 0 |
| 0 | 0.002135784 | 0.316870746 | 0.010610594 | 0.001868596 |
| 0 | 0 | 0 | 0 | 0 |
| 0 | 0.025042842 | 1.043943438 | 0.026736607 | 0.006134791 |
| 0 | 0 | 0 | 0 | 0 |
| 0 | 0 | 0 | 0 | 0 |
| 0.173722893 | 0 | 0 | 0 | 0 |
| 0 | 0.021865353 | 0.00195363 | 0.002603268 | 0 |
| 0 | 0 | 0 | 0.006826764 | 0 |
| 0 | 0 | 0 | 0 | 0 |
| 0 | 0 | 0 | 0 | 0 |
| 0 | 0 | 0 | 0.001342311 | 0 |
| 0 | 0 | 0 | 0 | 0 |
| 0 | 0 | 0 | 0 | 0.027043013 |
| 0 | 0 | 0 | 0 | 0 |
| 0 | 0.082623823 | 0 | 0 | 0 |
| 0 | 0 | 0 | 0 | 0 |
| 0.018635196 | 0.023079231 | 0.007515544 | 0.003344418 | 0 |
| 0 | 0 | 0 | 0 | 0 |
| 0 | 0.069215922 | 0.003891045 | 0.010246894 | 0.064974498 |
| 0.001317285 | 0.017772078 | 0.000902598 | 0.000728932 | 0.012602696 |
| 0 | 0.002907463 | 0 | 0 | 0 |
| 0 | 0 | 0 | 0 | 0 |
| 0 | 0 | 0 | 0 | 0 |
| 0 | 0 | 0 | 0 | 0 |
| 0 | 0 | 0 | 0 | 0 |
| 0 | 0 | 0 | 0.004196413 | 0.077499727 |
| 0 | 0 | 0 | 0.013891837 | 0 |
| 0 | 0 | 0 | 0 | 0 |
| 0 | 0 | 0 | 0 | 0 |
| 0 | 0 | 0 | 0.00155691 | 0 |
| 0 | 0 | 0 | 0 | 0 |
| 0 | 0 | 0 | 0 | 0 |
| 0.012774047 | 0.323262629 | 0 | 0.028502566 | 0.004915087 |
| 0 | 0 | 0 | 0 | 0 |
| 0 | 0.004529701 | 0 | 0 | 0 |
| 0 | 0 | 0 | 0 | 0 |
| 0 | 0 | 0 | 0.091043372 | 0 |
| 0 | 0 | 0 | 0 | 0 |
| 0 | 0 | 0 | 0 | 0 |
| 0 | 0 | 0 | 0 | 0 |
| 0 | 0.020159939 | 0 | 0 | 0 |
| 0 | 0 | 0 | 0 | 0 |
| 0 | 0 | 0 | 0 | 0 |
| 0 | 0 | 0 | 0 | 0 |

| | | | | |
|---|---|---|---|---|
| 0.020907313 | 0 | 0 | 0.001176534 | 0.023599572 |
| 0 | 0.826644322 | 0 | 0 | 0 |
| 0 | 0 | 0.019621517 | 0 | 0 |
| 0 | 0 | 0 | 0 | 0 |
| 0 | 0 | 0 | 0 | 0 |
| 0 | 0 | 0 | 0.02375301 | 0 |
| 0 | 0 | 0 | 0.001055208 | 0 |
| 0.916899229 | 0.23645151 | 0.341551548 | 1.096895594 | 0.375767944 |
| 0.323165669 | 0.199376261 | 0.52467758 | 0.242847264 | 1.206111385 |
| 0 | 0 | 0 | 0.025284451 | 0.05311615 |
| 0 | 0.197726427 | 0 | 0 | 0 |
| 0 | 0.000770182 | 0 | 0 | 0 |
| 0 | 0 | 0 | 0.01614443 | 0 |
| 2.980476418 | 0.690991338 | 0 | 0 | 4.554983992 |
| 0 | 0 | 0 | 0 | 0 |
| 0 | 0 | 0 | 0 | 0 |
| 0.057934469 | 0 | 0 | 0.093313341 | 0 |
| 0 | 0.376171396 | 0 | 0 | 0 |
| 0 | 0 | 0 | 0 | 0 |
| 0 | 0 | 0 | 0.002132184 | 0 |
| 0 | 0 | 0 | 0 | 0 |
| 0 | 0 | 0 | 0 | 0 |
| 0.243781606 | 0 | 0 | 0 | 0 |
| 0 | 0 | 0 | 0 | 0 |
| 0 | 0.008403188 | 0.133615815 | 0.040310747 | 0 |
| 0 | 0 | 0 | 0 | 0 |
| 0 | 0 | 0 | 0 | 0 |
| 0 | 0.096764267 | 0 | 0 | 0 |
| 0 | 0 | 0 | 0 | 0 |
| 0 | 0 | 0 | 0 | 0 |
| 0 | 0 | 0 | 0.059225641 | 0 |
| 0 | 0 | 0 | 0 | 0 |
| 0 | 0 | 0 | 0 | 0 |
| 0 | 0.00080031 | 0 | 0.004209826 | 0 |
| 0 | 0 | 0 | 0 | 0 |
| 0 | 0 | 0 | 0 | 0 |
| 0 | 0 | 0 | 0 | 0 |
| 0 | 0 | 0.005428873 | 0 | 0 |
| 0 | 0 | 0 | 0 | 0 |
| 0 | 0 | 0 | 0 | 0 |
| 0 | 0 | 0 | 0 | 0 |
| 0 | 0.160884305 | 0.645862304 | 0.065838766 | 0 |
| 0 | 0 | 0 | 0 | 0 |
| 0 | 0 | 0 | 0 | 0 |
| 0 | 0 | 0 | 0 | 0 |
| 0 | 0 | 0 | 0.000697452 | 0 |
| 0 | 0 | 0 | 0 | 0 |
| 0 | 0 | 0 | 0 | 0 |

| | | | | |
|---|---|---|---|---|
| 0 | 0 | 0 | 0 | 0 |
| 0 | 0.03143081 | 0 | 0.003185621 | 0.043870912 |
| 0 | 0 | 0 | 0 | 0 |
| 0 | 0 | 0 | 0 | 0 |
| 0.002459393 | 10.91758853 | 0.105399458 | 0.086604504 | 8.307258505 |
| 0 | 0 | 0 | 0 | 0 |
| 0 | 0 | 0 | 0 | 0 |
| 0.014650128 | 0.007938067 | 0.013590184 | 0.007233745 | 0.01657841 |
| 0 | 0.009459904 | 0 | 0 | 0 |
| 0 | 0 | 0 | 0 | 0 |
| 0 | 0 | 0 | 0 | 0 |
| 0 | 0 | 0 | 0 | 0 |
| 0 | 0 | 0 | 0 | 0 |
| 0 | 0 | 0 | 0 | 0 |
| 0 | 0 | 0 | 0 | 0 |
| 0.013487851 | 0 | 0.065898174 | 0.002379707 | 0.078497917 |
| 0 | 0 | 0 | 0 | 0 |
| 0 | 0 | 0 | 0 | 0 |
| 0 | 0 | 0 | 0 | 0 |
| 0 | 0 | 0 | 0 | 0 |
| 0 | 0 | 0 | 0 | 0 |
| 0 | 0 | 0 | 0.209800814 | 0 |
| 0 | 0 | 0 | 0 | 0 |
| 0.002535696 | 0 | 0 | 0.000647608 | 0.002520463 |
| 0 | 0.061318622 | 0.282532271 | 0 | 0 |
| 0 | 0 | 0 | 0 | 0 |
| 0.343417272 | 0.985442642 | 0.001303871 | 0.234357707 | 0.590321103 |
| 0 | 0 | 0 | 0 | 0 |
| 0 | 0 | 0 | 0.002641191 | 0 |
| 0 | 0 | 0 | 0 | 0 |
| 0 | 0 | 0 | 0 | 0 |
| 0 | 0 | 0 | 0 | 0 |
| 0 | 0 | 0 | 0 | 0 |
| 0 | 0 | 0 | 0 | 0 |
| 0 | 0 | 0 | 0 | 0 |
| 0 | 0 | 0 | 0 | 0 |
| 0 | 0 | 0 | 0 | 0 |
| 0 | 0.442332146 | 0 | 0 | 0 |
| 0 | 0 | 0.067299437 | 0 | 0 |
| 0 | 0 | 0 | 0 | 0 |
| 0 | 0 | 0 | 0 | 0 |
| 0 | 0.017875802 | 0 | 0 | 0 |
| 0 | 0 | 0 | 0 | 0 |
| 0 | 0 | 0 | 0 | 0 |
| 0 | 0 | 0 | 0 | 0 |
| 0 | 0 | 0 | 0 | 0 |
| 0 | 0 | 0 | 0 | 0 |
| 0 | 0.001641118 | 0 | 0 | 0 |

| | | | | |
|---|---|---|---|---|
| 0 | 0.113982081 | 0.021764671 | 0.018381575 | 0 |
| 0 | 0 | 0 | 0 | 0 |
| 0 | 0 | 0 | 0 | 0 |
| 0 | 0 | 0 | 0 | 0 |
| 0 | 0 | 0 | 0 | 0 |
| 0 | 0 | 0 | 0 | 0 |
| 0 | 0 | 0 | 0 | 0 |
| 0 | 0 | 0 | 0 | 0 |
| 0 | 0 | 0 | 0 | 0 |
| 0 | 0 | 0 | 0 | 0 |
| 0 | 0.05756091 | 0 | 0 | 0 |
| 0 | 0 | 0 | 0.010745842 | 0 |
| 0 | 0 | 0 | 0 | 0 |
| 0 | 0 | 0 | 0 | 0 |
| 0 | 0 | 0 | 0 | 0 |
| 0 | 0 | 0 | 0 | 0 |
| 0 | 0 | 0 | 0 | 0 |
| 1.090347631 | 0 | 0 | 0 | 0 |
| 0 | 0 | 0 | 0 | 0 |
| 0 | 0 | 0 | 0 | 0 |
| 0.00213662 | 0 | 0 | 0 | 0 |
| 0 | 0 | 0 | 0 | 0 |
| 0.002453086 | 0.153006599 | 0.004034026 | 0.330943341 | 0.004389029 |
| 0 | 0.851971413 | 0 | 0.032550823 | 1.253771541 |
| 0 | 0 | 0 | 0 | 0 |
| 0 | 0.353942351 | 0 | 0 | 0 |
| 0 | 0 | 0 | 0 | 0 |
| 0 | 0 | 0 | 0 | 0 |
| 0.003185391 | 0 | 0 | 0 | 0.005488176 |
| 0 | 0 | 0 | 0 | 0 |
| 0 | 0 | 0 | 0 | 0 |
| 0 | 0 | 0 | 0 | 0 |
| 0.007351377 | 0 | 0.00419761 | 0 | 0.246161745 |
| 0 | 0 | 0 | 0 | 0.018479086 |
| 0 | 0.011206318 | 0.790748634 | 0.876259739 | 0.123780566 |
| 0 | 0 | 0.003873667 | 0.108539996 | 0.001282691 |
| 0 | 0 | 0 | 0 | 0 |
| 0 | 0 | 0 | 0 | 0 |
| 0.011301132 | 0 | 0 | 0.003067797 | 0 |
| 0 | 0 | 0 | 0 | 0 |
| 0 | 0 | 0 | 0 | 0 |
| 0 | 0.001509628 | 0 | 0 | 0 |
| 0 | 0.895408867 | 0.005922412 | 0 | 0.392563746 |
| 0 | 0 | 0 | 0 | 0 |
| 0 | 0 | 0 | 0 | 0 |
| 0 | 0.058265572 | 0.001447207 | 0.0127102 | 0.002165026 |
| 0 | 0 | 0 | 0 | 0 |
| 0 | 0 | 0 | 0 | 0 |

| | | | | |
|---|---|---|---|---|
| 0 | 0 | 0 | 0 | 0 |
| 0 | 0 | 0 | 0 | 0 |
| 0 | 0 | 0 | 0 | 0 |
| 0 | 0 | 0 | 0 | 0 |
| 0 | 0.001415105 | 0 | 0 | 0 |
| 0 | 0 | 0 | 0 | 0 |
| 0 | 0 | 0 | 0 | 0 |
| 0 | 0 | 0.026745552 | 0.00804688 | 0 |
| 0 | 0.023184704 | 0.023050277 | 0.076787865 | 0 |
| 0 | 0.008341561 | 0.000745304 | 0 | 0 |
| 0 | 0 | 0 | 0 | 0 |
| 0 | 0 | 0 | 0 | 0 |
| 0 | 0 | 0 | 0 | 0 |
| 0 | 0 | 0.00256412 | 0 | 0 |
| 0 | 0 | 0 | 0.077158186 | 0 |
| 0.393900409 | 0 | 0.010999444 | 0 | 0.007978279 |
| 0 | 0 | 0 | 0 | 0 |
| 0 | 0 | 0 | 0 | 0 |
| 0 | 0 | 0 | 0 | 0 |
| 0 | 0.195259596 | 1.978110939 | 0.925000491 | 0.414306393 |
| 0 | 0.005238329 | 0 | 0 | 0 |
| 0 | 0 | 0 | 0 | 0 |
| 0 | 0 | 0 | 0 | 0 |
| 0 | 0 | 0.000732574 | 0 | 0 |
| 0 | 0 | 0 | 0.000900428 | 0 |
| 0 | 0 | 0.016127476 | 0 | 0 |
| 0 | 0 | 0 | 0 | 0 |
| 0 | 0.380798292 | 0 | 0 | 0 |
| 0 | 0 | 0 | 0 | 0 |
| 0 | 0.938506999 | 0.007980056 | 0.184132474 | 0 |
| 0 | 0.096908419 | 0.278631229 | 0.497812969 | 0.152548366 |
| 0 | 0 | 0 | 0 | 0 |
| 0 | 0 | 0 | 0 | 0 |
| 0 | 0 | 0 | 0 | 0 |
| 0 | 0 | 0 | 0 | 0 |
| 0 | 0 | 0 | 0 | 0 |
| 0 | 0 | 0 | 0 | 0 |
| 0 | 0 | 0 | 0 | 0 |
| 0 | 0.005011988 | 0 | 0 | 0 |
| 0 | 0 | 0 | 0 | 0 |
| 0 | 0.004938039 | 0 | 0 | 0 |
| 0 | 0 | 0 | 0 | 0 |
| 0 | 0.023448093 | 0 | 0 | 0 |
| 0 | 0 | 0 | 0 | 0 |
| 0 | 0 | 0 | 0 | 0 |
| 0 | 0 | 0 | 0 | 0 |
| 0.21959045 | 0 | 0 | 0 | 0 |
| 0 | 0 | 0 | 0 | 0 |

| | | | | |
|---|---|---|---|---|
| 0 | 0 | 0 | 0 | 0 |
| 0 | 0 | 0 | 0 | 0 |
| 0 | 0 | 0 | 0 | 0 |
| 0 | 0 | 0 | 0 | 0 |
| 0.001227693 | 0.052130987 | 0.077952117 | 0.217053972 | 0.098235564 |
| 0 | 0 | 0 | 0 | 0 |
| 0 | 0 | 0 | 0 | 0 |
| 0 | 0 | 0 | 0 | 0 |
| 0 | 0 | 0 | 0 | 0 |
| 0 | 0 | 0 | 0 | 0 |
| 0 | 0.009297344 | 0 | 0 | 0 |
| 0.024501799 | 0.038487726 | 0.000583949 | 0.03584106 | 0.059509947 |
| 0 | 0 | 0 | 0 | 0.01242442 |
| 0 | 0 | 0 | 0 | 0 |
| 0 | 0.002767013 | 0 | 0.00323448 | 0 |
| 0 | 0 | 0 | 5.980638132 | 0.006258156 |
| 0 | 0.074283334 | 0.020360374 | 0.008342351 | 0 |
| 0 | 0 | 0 | 0 | 0.283186035 |
| 0 | 0 | 0 | 0.021247024 | 0.024535141 |
| 0.011796248 | 0.008854878 | 0.038874122 | 0.002564399 | 0.004187636 |
| 0 | 0 | 0 | 0 | 0 |
| 0 | 0.014688235 | 0 | 0 | 0 |
| 0 | 0 | 0 | 0 | 0 |
| 0 | 0 | 0 | 0 | 0 |
| 0 | 0 | 0 | 0 | 0 |
| 0 | 0 | 0 | 0 | 0 |
| 0.028358622 | 0 | 0.002004807 | 0.007472621 | 0 |
| 0 | 0 | 0 | 0 | 0 |
| 0 | 0 | 0 | 0 | 0 |
| 0 | 0 | 0 | 0 | 0 |
| 0.001722106 | 0 | 0 | 0 | 0.002852935 |
| 0 | 0 | 0 | 0 | 0.000821734 |
| 0 | 0 | 0 | 0 | 0 |
| 0 | 0 | 0 | 0 | 0 |
| 0 | 0 | 0 | 0 | 0 |
| 0 | 0 | 0 | 0 | 0 |
| 0 | 0 | 0 | 0 | 0 |
| 0.001344336 | 0.019549815 | 0.002087901 | 0.007290224 | 0.180127781 |
| 0 | 0.157803813 | 0 | 0 | 0 |
| 0 | 0 | 0 | 0 | 0 |
| 0 | 0 | 0 | 0.00311914 | 0 |
| 0 | 0 | 0 | 0 | 0 |
| 0 | 0 | 0 | 0 | 0 |
| 0 | 0 | 0 | 0 | 0 |
| 0 | 0 | 0 | 0 | 0 |
| 0 | 0 | 0 | 0 | 0 |
| 0 | 0 | 0 | 0 | 0 |
| 0 | 0 | 0 | 0 | 0 |

| | | | | |
|---|---|---|---|---|
| 0 | 0 | 0 | 0 | 0 |
| 0 | 0.00193577 | 0.010118019 | 0.046010369 | 0.027944467 |
| 0 | 0 | 0 | 0 | 0 |
| 0.004596185 | 0 | 0 | 0 | 0.013498059 |
| 0 | 0 | 0 | 0 | 0.001733432 |
| 0 | 0 | 0 | 0 | 0 |
| 0 | 0.036393054 | 0 | 0.001570622 | 0 |
| 0 | 0 | 0 | 0 | 0 |
| 0 | 0 | 0 | 0 | 0 |
| 0 | 0 | 0 | 0 | 0 |
| 0 | 0 | 0 | 0 | 0 |
| 0 | 0 | 0 | 0 | 0 |
| 0 | 0.094052787 | 0.003382425 | 0.004916921 | 0 |
| 0 | 0.001662565 | 0 | 0 | 0 |
| 0 | 0 | 0 | 0 | 0 |
| 0 | 0 | 0 | 0 | 0 |
| 0 | 0 | 0 | 0 | 0 |
| 0 | 0 | 0 | 0 | 0 |
| 0 | 0 | 0 | 0 | 0 |
| 0 | 0 | 0 | 0 | 0 |
| 0 | 0 | 0 | 0 | 0 |
| 0 | 0 | 0 | 0 | 0 |
| 0.036270838 | 0 | 0 | 0.000647445 | 0 |
| 0 | 0.015424686 | 0.387662317 | 0.021392213 | 0 |
| 0 | 0 | 0 | 0 | 0 |
| 0 | 0 | 0.002173586 | 0 | 0 |
| 0 | 0.008890174 | 0 | 0 | 0 |
| 0 | 0 | 0 | 0 | 0 |
| 0 | 0 | 0 | 0 | 0 |
| 0 | 0 | 0 | 0 | 0 |
| 0.030222653 | 0.009634016 | 0.009932101 | 0.001203164 | 0.017505959 |
| 0 | 0 | 0 | 0 | 0.120500149 |
| 0 | 0 | 0 | 0 | 0 |
| 0 | 0 | 0 | 0 | 0 |
| 0 | 0 | 0.08410119 | 0.136780509 | 0.099112365 |
| 0 | 0 | 0 | 0 | 0 |
| 0.004158046 | 0 | 0 | 0 | 0 |
| 0 | 0 | 0 | 0 | 0 |
| 0 | 0 | 0 | 0 | 0.910252228 |
| 0 | 0 | 0 | 0 | 0 |
| 0 | 0 | 0 | 0 | 0 |
| 0 | 0 | 0 | 0 | 0 |
| 0 | 0 | 0 | 0 | 0 |
| 0 | 0 | 0 | 0 | 0 |
| 0.001491004 | 10.0977139 | 0 | 0.924539805 | 0 |
| 0 | 0.220479929 | 0.5074877 | 0.038333262 | 0.106664898 |
| 0 | 0 | 0 | 0 | 0 |
| 0 | 0 | 0 | 0 | 0 |

| | | | | |
|---|---|---|---|---|
| 0 | 0 | 0 | 0 | 0 |
| 0 | 0 | 0 | 0 | 0 |
| 0 | 0.056773247 | 0 | 0.047403354 | 0 |
| 0 | 0.018601195 | 0 | 0 | 0 |
| 0 | 0 | 0 | 0 | 0 |
| 0 | 0 | 0 | 0 | 0 |
| 0 | 0 | 0 | 0 | 0 |
| 0 | 0 | 0 | 0 | 0 |
| 0 | 0 | 0 | 0 | 0.005075685 |
| 0 | 0 | 0 | 0 | 0 |
| 0.001790892 | 0.002373783 | 0 | 0.003303354 | 0.001186755 |
| 0.000668452 | 0 | 0 | 0 | 0.003765137 |
| 0 | 0 | 0 | 0 | 0 |
| 0 | 0 | 0 | 0 | 0 |
| 0 | 0 | 0 | 0 | 0 |
| 0 | 0 | 0 | 0 | 0 |
| 0 | 0 | 0 | 0 | 0 |
| 0 | 0 | 0 | 0 | 0 |
| 0 | 0 | 0 | 0 | 0 |
| 0 | 0 | 0.000723271 | 0 | 0.010820159 |
| 0 | 0 | 0 | 0 | 0 |
| 0 | 0 | 0 | 0 | 0 |
| 0 | 0 | 0 | 0 | 0 |
| 0 | 0 | 0 | 0 | 0 |
| 0 | 0 | 0 | 0 | 0 |
| 0 | 0 | 0 | 0 | 0 |
| 0 | 0 | 0 | 0 | 0 |
| 0 | 0.003234997 | 0 | 0 | 0.004354303 |
| 0 | 0 | 0 | 0 | 0 |
| 0 | 0 | 0 | 0 | 0 |
| 0 | 0.004056744 | 0.000932047 | 0.006962611 | 0.167828482 |
| 0 | 0 | 0 | 0 | 0 |
| 0.044223349 | 0.015700976 | 0.017856329 | 0.047413331 | 0.01628789 |
| 0.008271652 | 0 | 0.000737262 | 0.004688834 | 0.004211248 |
| 0 | 0 | 0 | 0 | 0 |
| 0 | 0 | 0 | 0 | 0 |
| 0 | 0 | 0 | 0 | 0 |
| 0 | 0 | 0 | 0 | 0 |
| 0 | 0 | 0 | 0 | 0 |
| 0 | 0 | 0 | 0 | 0 |
| 0 | 0 | 0 | 0 | 0 |
| 0.325490292 | 0 | 0 | 0.024985742 | 0.514200947 |
| 0 | 0.056202178 | 0.026574116 | 0.327828766 | 0.111323713 |
| 0 | 0 | 0 | 0 | 0 |
| 0 | 0 | 0 | 0 | 0 |
| 0 | 0.588978823 | 0.003134812 | 0 | 0.15888094 |
| 0 | 0 | 0 | 0 | 0 |
| 0 | 0 | 0 | 0 | 0 |

| | | | | |
|---|---|---|---|---|
| 0 | 0 | 0 | 0 | 0 |
| 0 | 0 | 0 | 0 | 0 |
| 0 | 0 | 0.011503658 | 0.00401983 | 0.138879512 |
| 0 | 0 | 0 | 0 | 0 |
| 0 | 0.084789153 | 0 | 0 | 0 |
| 0.015432295 | 0 | 0 | 0.008956168 | 0.00299309 |
| 0 | 0 | 0 | 0 | 0 |
| 0 | 0 | 0 | 0 | 0 |
| 0 | 0 | 0 | 0 | 0 |
| 0 | 0 | 0 | 0 | 0 |
| 0 | 0 | 0 | 0 | 0 |
| 0 | 0 | 0 | 0 | 0 |
| 0 | 0 | 0 | 0 | 0 |
| 0 | 0 | 0 | 0 | 0 |
| 0 | 0 | 0 | 0 | 0 |
| 0 | 0 | 0 | 0 | 0 |
| 0 | 0 | 0 | 0 | 0 |
| 0 | 0 | 0 | 0 | 0 |
| 0 | 0 | 0 | 0 | 0 |
| 0 | 0 | 0 | 0 | 0 |
| 0 | 0 | 0 | 0 | 0 |
| 0.001558235 | 0 | 0 | 0 | 0 |
| 0 | 0 | 0 | 0 | 0 |
| 0 | 0 | 0 | 0.15404069 | 0 |
| 0 | 0.047386196 | 0 | 0 | 0.008331304 |
| 0 | 0 | 0.006052512 | 0 | 0 |
| 0 | 0 | 0 | 0 | 0 |
| 0.004479025 | 0.069029587 | 0.064404708 | 0.00086209 | 0.005032829 |
| 0 | 0 | 0 | 0 | 0 |
| 0 | 0 | 0 | 0 | 0 |
| 0 | 0.011454139 | 0 | 0 | 0 |
| 0 | 0 | 0 | 0 | 0.012599526 |
| 0 | 0 | 0 | 0 | 0 |
| 0 | 0 | 0 | 0 | 0 |
| 0 | 0 | 0 | 0 | 0 |
| 0 | 0 | 0 | 0 | 0 |
| 0 | 0 | 0 | 0 | 0 |
| 0 | 0 | 0.010588984 | 0 | 0 |
| 0 | 0 | 0.590651271 | 0 | 0.793367951 |
| 0 | 0 | 0 | 0 | 0 |
| 0 | 0 | 0 | 0 | 0 |
| 0 | 0.010392275 | 0 | 0 | 0.014071255 |
| 0.014305113 | 0 | 0.000552214 | 0.000891929 | 0.019426197 |
| 0 | 0 | 0 | 0 | 0 |
| 0 | 0 | 0 | 0 | 0 |
| 0 | 0 | 0 | 0 | 0.000921614 |
| 0.007089191 | 0.012956734 | 0.015206033 | 0 | 0.021752555 |
| 0 | 0 | 0 | 0 | 0 |

| | | | | |
|---|---|---|---|---|
| 0 | 0 | 0 | 0 | 0 |
| 0.009334179 | 0.034023626 | 0.001776594 | 0.002152147 | 0.043491111 |
| 0 | 0.09954617 | 0.002106538 | 0 | 0 |
| 0 | 0 | 0 | 0 | 0 |
| 0 | 0 | 0 | 0 | 0 |
| 0 | 0 | 0 | 0 | 0 |
| 0 | 0 | 0 | 0 | 0 |
| 0 | 0 | 0 | 0 | 0 |
| 0 | 0 | 0 | 0 | 0 |
| 0 | 0 | 0 | 0 | 0 |
| 0 | 0 | 0 | 0 | 0 |
| 0 | 0 | 0 | 0 | 0 |
| 0 | 0.024522264 | 0 | 0 | 0 |
| 0 | 0 | 0 | 0 | 0 |
| 0 | 0 | 0.001993631 | 0.001073361 | 0 |
| 0 | 0 | 0 | 0 | 0 |
| 0 | 0 | 0 | 0.117577613 | 0 |
| 0 | 0 | 0 | 0 | 0 |
| 0 | 0 | 0 | 0 | 0.009598232 |
| 0 | 0 | 0 | 0 | 0.0024076 |
| 0 | 0 | 0 | 0 | 0 |
| 0.02690752 | 0.003516929 | 0.100940759 | 0.00184472 | 0.003550337 |
| 0.032752607 | 0 | 0 | 0 | 0.016550737 |
| 0 | 0 | 0 | 0 | 0 |
| 0 | 0 | 0 | 0 | 0 |
| 0 | 0 | 0 | 0 | 0 |
| 0 | 0 | 0 | 0 | 0 |
| 0 | 0 | 0.00232812 | 0 | 0 |
| 0 | 0 | 0 | 0 | 0 |
| 0 | 0 | 0 | 0.003742078 | 0 |
| 0 | 0 | 0 | 0 | 0 |
| 0 | 0 | 0 | 0 | 0 |
| 0 | 0 | 0 | 0 | 0 |
| 0 | 0 | 0 | 0 | 0 |
| 0 | 0 | 5.373688775 | 0.00526398 | 0.003223509 |
| 0 | 0 | 0 | 0 | 0 |
| 0 | 0 | 0 | 0 | 0 |
| 0 | 0 | 0 | 0 | 0 |
| 0 | 0.004592719 | 0.020855496 | 0.002368512 | 0.120308639 |
| 0 | 0 | 0 | 0 | 0 |
| 0.163319879 | 0.06976342 | 0.264522982 | 0.218975425 | 0.049379799 |
| 0 | 0 | 0 | 0 | 0 |
| 0 | 0 | 0 | 0 | 0 |
| 0 | 0 | 0 | 0 | 0 |
| 0 | 0 | 0 | 0 | 0 |
| 0.493595399 | 0 | 0.014496283 | 0.007834749 | 0.075948186 |
| 0 | 0 | 0 | 0 | 0 |
| 0 | 0 | 0 | 0 | 0 |

| | | | | |
|---|---|---|---|---|
| 0 | 0 | 0 | 0 | 0 |
| 0 | 0 | 0 | 0 | 0 |
| 0 | 0 | 0 | 0 | 0 |
| 0 | 0 | 0.005164937 | 0 | 0 |
| 0 | 0 | 0 | 0 | 0 |
| 0 | 0 | 0 | 0 | 0 |
| 0 | 0 | 0 | 0 | 0 |
| 0 | 0 | 0 | 0 | 0 |
| 0 | 0 | 0 | 0 | 0 |
| 0 | 0.001356202 | 0 | 0.001056882 | 1.11700938 |
| 0 | 0.011442473 | 0 | 0 | 0 |
| 0 | 0 | 0 | 0 | 0 |
| 0 | 0 | 0 | 0 | 0 |
| 0 | 0 | 0 | 0 | 0 |
| 0 | 0 | 0 | 0 | 0 |
| 0 | 0 | 0 | 0 | 0 |
| 0.000540226 | 5.954110247 | 0 | 0 | 0 |
| 0 | 0 | 0 | 0 | 0 |
| 0 | 0 | 0 | 0 | 0 |
| 0.536069761 | 0.093680416 | 0.016128931 | 0.076314754 | 0.843970577 |
| 0 | 0 | 0 | 0 | 0 |
| 0 | 0 | 0 | 0 | 0 |
| 0 | 0 | 0 | 0 | 0 |
| 0 | 0 | 0 | 0 | 0 |
| 0 | 0 | 0 | 0 | 0 |
| 0 | 0.020045716 | 0 | 0 | 0 |
| 0 | 0 | 0 | 0 | 0 |
| 0 | 0 | 0 | 0 | 0 |
| 0 | 0 | 0 | 0 | 0 |
| 0.11301481 | 0 | 0 | 0.002703008 | 0 |
| 0 | 0 | 0 | 0 | 0 |
| 0 | 0 | 0 | 0 | 0 |
| 0 | 0 | 0 | 0 | 0 |
| 0 | 0 | 0 | 0 | 0 |
| 0 | 0 | 0 | 0 | 0 |
| 0 | 0 | 0.002947557 | 0 | 0 |
| 0 | 0 | 0 | 0 | 0 |
| 0 | 0 | 0 | 0 | 0 |
| 0 | 0 | 0 | 0 | 0 |
| 0 | 0.101001319 | 0 | 0 | 0 |
| 0 | 0 | 0 | 0 | 0 |
| 0 | 0 | 0 | 0 | 0 |
| 0 | 0 | 0.064353618 | 0.014903599 | 0 |
| 0 | 0 | 0 | 0 | 0 |
| 0.006047419 | 0.043218331 | 0 | 0.00669279 | 0.064958547 |
| 0 | 0.012445369 | 0 | 0 | 0 |
| 0 | 0 | 0 | 0 | 0 |
| 0 | 0 | 0 | 0 | 0 |

| | | | | |
|---|---|---|---|---|
| 0 | 0 | 0 | 0 | 0 |
| 0 | 0 | 0 | 0 | 0 |
| 0 | 0 | 0 | 0 | 0 |
| 0 | 0 | 0 | 0 | 0 |
| 0 | 0.002631129 | 0 | 0 | 0 |
| 0.0026214 | 0.010920179 | 0.011575331 | 0.005963479 | 0.037058163 |
| 0 | 0 | 0 | 0 | 0 |
| 0 | 0 | 0 | 0 | 0 |
| 0 | 0 | 0 | 0 | 0 |
| 0 | 0 | 0 | 0 | 0 |
| 0 | 0 | 0 | 0 | 0 |
| 0 | 0 | 0 | 0 | 0 |
| 0 | 0 | 0 | 0 | 0 |
| 0 | 0 | 0 | 0 | 0 |
| 0 | 0 | 0 | 0 | 0 |
| 0 | 0 | 0 | 0 | 0 |
| 0 | 0.024485215 | 0 | 0 | 0 |
| 0 | 0 | 0 | 0 | 0 |
| 0 | 0 | 0 | 0 | 0 |
| 0 | 0 | 0 | 0 | 0 |
| 0 | 0.00978413 | 0 | 0 | 0 |
| 0 | 0 | 0 | 0 | 0 |
| 0 | 0 | 0 | 0 | 0 |
| 0 | 0 | 0 | 0 | 0 |
| 0 | 0 | 0 | 0 | 0 |
| 0 | 0 | 0 | 0 | 0.00973634 |
| 0 | 0 | 0 | 0 | 0 |
| 0 | 0 | 0 | 0.006415225 | 1.231838135 |
| 0 | 0 | 0 | 0 | 0 |
| 0 | 0 | 0 | 0 | 0.040562957 |
| 0 | 0 | 0 | 0 | 0 |
| 0 | 0 | 0 | 0 | 0 |
| 0 | 0.003512597 | 0 | 0 | 0 |
| 0 | 0 | 0 | 0 | 0 |
| 0 | 0.299930254 | 0 | 0 | 0 |
| 0 | 0 | 0 | 0 | 0 |
| 0 | 0 | 0 | 0 | 0 |
| 0 | 0 | 0 | 0 | 0 |
| 0 | 0 | 0.792001928 | 0.335354302 | 0 |
| 0 | 0 | 0 | 0 | 0 |
| 0 | 0 | 0 | 0 | 0 |
| 0 | 0 | 0 | 0 | 0 |
| 0 | 0 | 0 | 0 | 0 |
| 0 | 0 | 0 | 0 | 0 |
| 0 | 0 | 0 | 0 | 0 |
| 0 | 0 | 0 | 0 | 0 |
| 0 | 0 | 0 | 0 | 0 |
| 0 | 0 | 0 | 0 | 0 |

| | | | | |
|---|---|---|---|---|
| 0 | 0 | 0 | 0 | 0 |
| 0.029348687 | 0 | 0.014002236 | 0.008661528 | 0.21830661 |
| 0 | 0 | 0 | 0 | 0 |
| 0 | 0 | 0 | 0 | 0 |
| 0 | 0 | 0 | 0 | 0 |
| 0 | 0 | 0 | 0 | 0.002889178 |
| 0 | 0 | 0 | 0 | 0 |
| 0 | 0 | 0 | 0 | 0 |
| 0 | 0 | 0 | 0 | 0 |
| 0 | 0 | 0.00202252 | 0 | 0 |
| 0 | 0.01091523 | 0 | 0 | 0.014006268 |
| 0 | 0 | 0 | 0 | 0 |
| 0.63184113 | 0.023404224 | 0.336463697 | 0.098155398 | 0 |
| 0 | 0 | 0 | 0 | 0 |
| 0 | 0 | 0 | 0 | 0 |
| 0 | 0 | 0 | 0 | 0 |
| 0 | 0 | 0 | 0 | 0 |
| 0 | 0 | 0 | 0 | 0 |
| 0 | 0 | 0 | 0 | 0 |
| 0 | 0 | 0 | 0 | 0.000529764 |
| 0 | 0.051698742 | 0 | 0.001438878 | 0.005600053 |
| 0 | 0 | 0 | 0 | 0 |
| 0 | 0 | 0 | 0 | 0 |
| 0 | 0.124402867 | 0 | 5.666607157 | 0.217144707 |
| 0 | 0 | 0 | 0 | 0 |
| 0 | 0 | 0 | 0 | 0 |
| 0 | 0 | 0 | 0 | 0 |
| 0 | 0 | 0 | 0 | 0 |
| 0 | 0 | 0 | 0 | 0 |
| 0 | 0 | 0 | 0 | 0 |
| 0.01424953 | 0 | 0 | 0 | 0.003171028 |
| 0 | 0 | 0 | 0 | 0 |
| 0 | 0.0020717 | 0 | 0 | 0 |
| 0 | 0 | 0 | 0 | 0 |

| | | | | |
|---|---|---|---|---|
| PHL018-0155_V1 | PHL018-0106_V2 | PHL018-0123_V1 | PHL018-0184_V2 | PHL018-0255_V2 |
| CTRL | CTRL | CTRL | EYCF | REF |
| 0.354193486 | 0.138221071 | 2.128274044 | 0.356204201 | 0.057831529 |
| 0.085668346 | 4.968048728 | 1.025918681 | 0.025358089 | 0.193722631 |
| 0 | 4.911996149 | 0.923054226 | 0.192410138 | 0.557353809 |
| 0 | 0 | 0 | 0 | 0 |
| 0 | 0 | 0 | 0 | 0 |
| 0 | 0 | 0 | 0 | 0 |
| 0 | 0 | 0 | 0 | 0 |
| 0 | 0 | 0 | 0 | 0 |
| 0 | 0 | 0 | 0 | 0 |
| 0 | 0 | 0 | 0 | 0 |
| 0 | 0 | 0 | 0 | 0 |
| 0 | 0 | 0 | 0 | 0 |
| 0 | 0 | 0 | 0 | 0 |
| 1.776698733 | 5.354791049 | 1.442670879 | 0.194711844 | 0.124942731 |
| 0 | 0 | 0 | 0 | 0 |
| 0 | 0 | 0 | 0 | 0 |
| 0 | 0.002311244 | 0 | 0 | 0 |
| 0 | 0 | 0 | 0 | 0 |
| 0 | 0 | 0 | 0 | 0 |
| 0 | 0 | 0 | 0 | 0 |
| 0.005449374 | 0 | 0 | 0 | 0 |
| 0 | 0.000690319 | 0 | 0 | 0 |
| 0 | 0 | 0 | 0 | 0 |
| 0 | 0 | 0 | 0 | 0 |
| 0 | 0 | 0 | 0 | 0.074926815 |
| 0 | 0 | 0 | 0 | 0 |
| 0 | 0 | 0 | 0 | 0 |
| 0 | 0 | 0 | 0 | 0 |
| 0 | 0 | 0 | 0 | 0 |
| 0 | 0 | 0 | 0 | 0 |
| 0 | 0 | 0 | 0 | 0 |
| 0 | 0 | 0 | 0 | 0 |
| 0 | 0 | 0 | 0 | 0 |
| 0 | 0 | 0 | 0 | 0 |
| 0 | 0 | 0 | 0 | 0 |
| 11.16035747 | 0 | 0 | 0 | 1.919918735 |
| 0 | 0 | 0 | 0 | 0 |
| 0 | 0 | 0 | 0 | 0 |
| 0 | 0 | 0 | 0 | 0 |
| 0 | 0 | 0 | 0 | 0 |
| 0 | 0 | 0 | 0 | 0 |
| 0.366965388 | 0.002234638 | 0.1590211 | 0.248005774 | 0.016636961 |
| 0.594346231 | 0.001177515 | 0.857539149 | 2.628939293 | 0.055605558 |
| 0.003527395 | 0 | 0.001682291 | 0.006686028 | 0.00866047 |
| 0.006881402 | 0.00205473 | 0.003335399 | 0.079961767 | 0.003434141 |
| 0 | 0 | 0 | 0 | 0 |

| | | | | |
|---|---|---|---|---|
| 0.050020857 | 0.002092337 | 0 | 0 | 0.020230554 |
| 0.0109373 | 0.010207007 | 0.8656724 | 0 | 0 |
| 0 | 0.030512106 | 0 | 0 | 0.027158383 |
| 0.449923726 | 0 | 0.418158327 | 0.007804155 | 0.201648849 |
| 0 | 0 | 0.00270674 | 0 | 0.001083783 |
| 0 | 0 | 0 | 0 | 0.006696251 |
| 0 | 0.002805196 | 0.24341142 | 0 | 0 |
| 0 | 0 | 0.080012886 | 0 | 0 |
| 0 | 0 | 0 | 0 | 0 |
| 0 | 0 | 0 | 0 | 0 |
| 0.001838983 | 0 | 0 | 0.322677653 | 0 |
| 2.584477733 | 0.157979921 | 0.356651388 | 0 | 0.08590689 |
| 0 | 0 | 0 | 0.001408523 | 0 |
| 0.075151388 | 0.18944806 | 0.141949219 | 0 | 0.002991406 |
| 0 | 0 | 0 | 0 | 0 |
| 0 | 0 | 0 | 0 | 0 |
| 0.002526489 | 0.002203294 | 0.073969752 | 0.119987373 | 0 |
| 0 | 0 | 0 | 0 | 0 |
| 0 | 0 | 0 | 0 | 0 |
| 0 | 0 | 0 | 0 | 0.082244814 |
| 0.31613283 | 0.19488104 | 0.165785728 | 0 | 0 |
| 0.031693084 | 0.016942065 | 0.004735586 | 0 | 0 |
| 0 | 0 | 0 | 0 | 0.198777388 |
| 0 | 0 | 0 | 0 | 0 |
| 0 | 0 | 0 | 0 | 0 |
| 0 | 0 | 0 | 0 | 0 |
| 0.009460664 | 0 | 0 | 0 | 0 |
| 0 | 0 | 0 | 0 | 0 |
| 0.212599354 | 0.002039887 | 0.00368759 | 0.014778966 | 0.005966336 |
| 0 | 0 | 0 | 0 | 0 |
| 0.017955881 | 0.076563068 | 0.697572345 | 0 | 0 |
| 0 | 0 | 0 | 0 | 0 |
| 0 | 0 | 0 | 0 | 0 |
| 0 | 0 | 0 | 0 | 0.000588014 |
| 0 | 0 | 0 | 0 | 0 |
| 0 | 0 | 0 | 0 | 0 |
| 0 | 0 | 0 | 0 | 0 |
| 0.001231769 | 0 | 0 | 0.00770473 | 0 |
| 0 | 0 | 0 | 0 | 0 |
| 0 | 0 | 0 | 0 | 0 |
| 0 | 0 | 0 | 0 | 0 |
| 0 | 0 | 0 | 0 | 0 |
| 0 | 0 | 0 | 0 | 0 |
| 0 | 0 | 0 | 0 | 0 |
| 0.138218006 | 0 | 0 | 0 | 0.006599805 |
| 0 | 0 | 0 | 0 | 0 |
| 0 | 0 | 0 | 0 | 0 |
| 0 | 0 | 0 | 0 | 0 |

| | | | | |
|---|---|---|---|---|
| 0 | 0 | 0 | 0 | 0 |
| 0 | 0.111567245 | 0 | 0 | 0 |
| 0 | 0 | 0 | 0 | 0 |
| 0 | 0 | 0 | 0 | 0 |
| 0 | 0 | 0 | 0 | 0 |
| 0 | 0 | 0 | 0 | 0 |
| 0 | 0 | 0 | 0 | 0 |
| 0 | 0 | 0.007018249 | 0 | 0 |
| 0 | 0.001926569 | 0 | 0 | 0.043439938 |
| 0 | 0 | 0 | 0 | 0 |
| 0 | 0 | 0 | 0 | 0 |
| 0.057451628 | 0 | 0.030036624 | 0.006914104 | 0.002616802 |
| 0 | 0 | 0 | 0 | 0 |
| 0 | 0 | 0 | 0 | 0 |
| 0 | 0 | 0 | 0 | 0 |
| 0 | 0 | 0 | 0 | 0 |
| 0 | 0 | 0 | 0 | 0 |
| 0 | 0 | 0 | 0 | 0 |
| 0.105301934 | 0.077034892 | 0 | 0 | 0.996739801 |
| 0 | 0 | 0 | 0 | 0 |
| 0 | 0 | 0 | 0 | 0.0147512 |
| 0 | 0 | 0 | 0 | 0 |
| 0 | 0 | 0 | 0 | 0 |
| 0 | 0 | 0 | 0 | 0 |
| 0 | 0 | 0 | 0 | 0 |
| 0 | 0 | 0 | 0 | 0 |
| 0 | 0 | 0 | 0 | 0.00180162 |
| 0 | 0 | 0 | 0 | 0 |
| 0 | 0 | 0 | 0 | 0 |
| 0 | 0 | 0 | 0 | 0 |
| 0 | 0 | 0.013996271 | 0 | 0.051195631 |
| 0 | 0.001122181 | 0.001545611 | 0 | 0.001193526 |
| 0 | 0 | 0 | 0 | 0 |
| 0 | 0 | 0 | 0 | 0 |
| 0 | 0 | 0 | 0 | 0 |
| 0 | 0 | 0 | 0 | 0 |
| 0 | 0 | 0 | 0 | 0 |
| 0 | 0 | 0 | 0 | 0 |
| 0 | 0 | 0 | 0 | 0 |
| 0 | 0 | 0 | 0 | 0 |
| 0 | 0 | 0 | 0 | 0 |
| 0 | 0 | 0 | 0 | 0.001426181 |
| 0 | 0 | 0 | 0 | 0 |
| 0 | 0 | 0 | 0 | 0 |
| 0 | 0 | 0 | 0 | 0 |
| 0 | 0 | 0 | 0 | 0 |
| 0 | 0 | 0 | 0 | 0 |
| 0 | 0 | 0 | 0 | 0 |

| | | | | |
|---|---|---|---|---|
| 0 | 0 | 0 | 0 | 0 |
| 0 | 0 | 0 | 0 | 0 |
| 0 | 0 | 0 | 0 | 0 |
| 0 | 0 | 0 | 0 | 0 |
| 0.003982244 | 0 | 0 | 0 | 0.026663289 |
| 0 | 0 | 0 | 0 | 0 |
| 0 | 0 | 0 | 0 | 0 |
| 0 | 0 | 0 | 0 | 0 |
| 0 | 0 | 0 | 0 | 0 |
| 0 | 0 | 0 | 0 | 0 |
| 0 | 0 | 0 | 0 | 0 |
| 0 | 0 | 0 | 0 | 0 |
| 0 | 0 | 0 | 0 | 0 |
| 0 | 0 | 0 | 0 | 0 |
| 0 | 0 | 0 | 0 | 0 |
| 0 | 0 | 0 | 0 | 0 |
| 0 | 0 | 0 | 0 | 0 |
| 0 | 0 | 0 | 0 | 0 |
| 0 | 0 | 0 | 0 | 0 |
| 0 | 0 | 0 | 0 | 0 |
| 0 | 0 | 0 | 0 | 0 |
| 0 | 0 | 0 | 0 | 0 |
| 0 | 0 | 0 | 0 | 0.003546621 |
| 0 | 0 | 0 | 0 | 0 |
| 0 | 0 | 0 | 0 | 0.018659921 |
| 0 | 0 | 0 | 0 | 0 |
| 0 | 0 | 0 | 0 | 0.018366926 |
| 0 | 0 | 0 | 0 | 0 |
| 0 | 0 | 0 | 0 | 0 |
| 0 | 0 | 0 | 0 | 0 |
| 0 | 0 | 0 | 0 | 0 |
| 0 | 0 | 0 | 0.227424799 | 0 |
| 0 | 0 | 0 | 0 | 0 |
| 0 | 0 | 0 | 0 | 0 |
| 0 | 0 | 0 | 0 | 0 |
| 0 | 0 | 0 | 0 | 0 |
| 0 | 0 | 0 | 0 | 0 |
| 0 | 0 | 0 | 0 | 0 |
| 0 | 0.002271337 | 0 | 0 | 0 |
| 0 | 0 | 0 | 0 | 0 |
| 0 | 0 | 0 | 0 | 0 |
| 0 | 0 | 0 | 0 | 0 |
| 0 | 0 | 0 | 0 | 0 |
| 0 | 0 | 0 | 0 | 0 |
| 0 | 0 | 0 | 0 | 0 |
| 0 | 0 | 0 | 0 | 0 |
| 0 | 0 | 0 | 0 | 0 |
| 0 | 0 | 0 | 0 | 0 |

| | | | | |
|---|---|---|---|---|
| 0 | 0 | 0 | 0 | 0.279341783 |
| 0 | 0 | 0 | 0 | 0 |
| 0 | 0 | 0 | 0 | 0 |
| 0 | 0 | 0 | 0 | 0 |
| 0 | 0 | 0 | 0 | 0 |
| 0 | 0 | 0 | 0 | 0 |
| 0 | 0 | 0 | 0 | 0 |
| 0 | 0 | 0 | 0 | 0 |
| 0 | 0 | 0 | 0 | 0 |
| 0 | 0 | 0 | 0 | 0 |
| 0 | 0 | 0 | 0 | 0 |
| 0 | 0 | 0 | 0 | 0.010025399 |
| 0.05935338 | 0.000825969 | 0.002346362 | 0 | 0 |
| 0 | 0 | 0 | 0 | 0 |
| 0 | 0 | 0 | 0 | 0.00691123 |
| 0 | 0 | 0 | 0 | 0 |
| 0 | 0 | 0 | 0 | 0 |
| 0 | 0 | 0 | 0 | 0 |
| 0 | 0 | 0 | 0 | 0 |
| 0 | 0 | 0 | 0 | 0 |
| 0.020463763 | 0.071040745 | 0.004785963 | 0.011934831 | 0.002463824 |
| 0 | 0 | 0 | 0 | 0 |
| 0 | 0 | 0 | 0 | 0 |
| 0.007386781 | 0 | 0 | 0 | 0 |
| 0 | 0 | 0 | 0 | 0 |
| 0 | 0 | 0 | 0 | 0 |
| 0 | 0.001349102 | 0.008361697 | 0.004126898 | 0.001195728 |
| 0 | 0 | 0 | 0 | 0 |
| 0 | 0 | 0 | 0 | 0 |
| 0.008091647 | 0 | 0.000994006 | 0 | 0 |
| 0 | 0.13739633 | 0.009955924 | 0 | 0 |
| 0 | 0 | 0 | 0 | 0 |
| 0 | 0 | 0 | 0 | 0 |
| 0.015392223 | 0.007550555 | 0 | 0 | 0.004266253 |
| 0.135708691 | 0.00728513 | 0.05527355 | 0 | 0.006679565 |
| 0 | 0 | 0 | 0 | 0 |
| 0 | 0 | 0 | 0 | 0 |
| 0 | 0 | 0 | 0 | 0 |
| 0.010423761 | 0 | 0 | 0.010114112 | 0 |
| 0 | 0 | 0 | 0 | 0 |
| 0 | 0 | 0 | 0.001067609 | 0 |
| 0.001278605 | 0 | 0 | 0.032717819 | 0 |
| 0 | 0 | 0 | 0.051454546 | 0 |
| 0 | 0 | 0 | 0 | 0 |
| 0 | 0 | 0 | 0 | 0 |
| 0 | 0 | 0 | 0 | 0.057237058 |
| 0.005267532 | 0 | 0.001685818 | 0 | 0 |
| 0.078097629 | 0.0008951 | 0.003143757 | 0 | 0 |

| | | | | |
|---|---|---|---|---|
| 0 | 0 | 0 | 0 | 0 |
| 0 | 0 | 0 | 0 | 0 |
| 0.00100043 | 0 | 0.001689823 | 0.012641816 | 0.00139188 |
| 0 | 0 | 0 | 0 | 0 |
| 0 | 0 | 0 | 0 | 0 |
| 0 | 0 | 0 | 0 | 0 |
| 0 | 0 | 0.010266402 | 0.000738504 | 0 |
| 0.001311528 | 0 | 0.038812024 | 0.00273454 | 0.004652997 |
| 0 | 0 | 0 | 0 | 0 |
| 0 | 0 | 0 | 0 | 0 |
| 0 | 0 | 0.00526243 | 0 | 0.024970934 |
| 0 | 0 | 0 | 0 | 0 |
| 0 | 0 | 0 | 0 | 0 |
| 0 | 0 | 0 | 0 | 0 |
| 0 | 0 | 0 | 0.315192919 | 0 |
| 0 | 0 | 0 | 0 | 0 |
| 0 | 0 | 0 | 0 | 0 |
| 0 | 0 | 0.010777225 | 0 | 0 |
| 0.002041291 | 0 | 0.009967679 | 0.024094445 | 0 |
| 0 | 0 | 0 | 0 | 0.186617246 |
| 0.02354142 | 0.020679139 | 0 | 0 | 0 |
| 0 | 0 | 0 | 0 | 0 |
| 0 | 0 | 0 | 0 | 0 |
| 0 | 0 | 0 | 0 | 0 |
| 0.002175349 | 0.00320131 | 0.017637009 | 0.008761976 | 0.002837366 |
| 0 | 0 | 0 | 0.00500147 | 0 |
| 0 | 0 | 0 | 0 | 0 |
| 0 | 0 | 0 | 0 | 0 |
| 0.071880369 | 0 | 0.007100685 | 0.035414178 | 0.002436966 |
| 0 | 0 | 0 | 0 | 0 |
| 0 | 0.383886043 | 0.230506376 | 0.005722739 | 0.008821129 |
| 0 | 0 | 0 | 0 | 0 |
| 0 | 0 | 0 | 0.382828809 | 0 |
| 0 | 0 | 0 | 0 | 0 |
| 0 | 0 | 0 | 0 | 0 |
| 0 | 0 | 0 | 0 | 0 |
| 0 | 0 | 0 | 0 | 0 |
| 0 | 0 | 0 | 0 | 0 |
| 0 | 0 | 0 | 0 | 0 |
| 0 | 0 | 0 | 0.015433796 | 0 |
| 0 | 0 | 0 | 0 | 0 |
| 0.013951594 | 0 | 0.156156773 | 0.02741689 | 0.001284525 |
| 0 | 0 | 0 | 0 | 0 |
| 0 | 0 | 0 | 0 | 0 |
| 0 | 0 | 0 | 0 | 0 |
| 0 | 0 | 0 | 0 | 0 |
| 0 | 0 | 0 | 0 | 0 |
| 0 | 0 | 0 | 0 | 0 |

| | | | | |
|---|---|---|---|---|
| 0 | 0 | 0 | 0 | 0 |
| 0 | 0 | 0 | 0 | 0 |
| 0 | 0 | 0.002349833 | 0 | 0.005376441 |
| 0 | 0 | 0 | 0 | 0 |
| 0.106644979 | 0 | 0 | 0 | 0 |
| 0 | 0 | 0 | 0 | 0 |
| 0 | 0 | 0 | 0 | 0 |
| 0 | 0 | 0 | 0 | 0 |
| 0 | 0 | 0 | 0.001372452 | 0 |
| 0 | 0 | 0 | 0 | 0.002704489 |
| 0 | 0 | 0 | 0 | 0 |
| 0 | 0 | 0 | 0 | 0 |
| 0 | 0 | 0 | 0 | 0 |
| 0.017652796 | 0.056426186 | 0 | 0 | 0.001485485 |
| 0.00399502 | 0.022363988 | 0.002619803 | 0.008017826 | 0 |
| 0 | 0 | 0 | 0 | 0.117352978 |
| 0.001338814 | 0.006304765 | 0.004884596 | 0.013195858 | 0 |
| 0 | 0.101074729 | 0 | 0 | 0 |
| 0 | 0 | 0 | 0 | 0 |
| 0 | 0 | 0 | 0 | 0 |
| 0 | 0 | 0 | 0 | 0 |
| 0.086291665 | 0.001422851 | 0.003919463 | 0 | 0 |
| 0 | 0 | 0 | 0 | 0 |
| 0.037845795 | 0.014704958 | 0 | 0 | 0 |
| 0 | 0 | 0 | 0 | 0.012536948 |
| 0 | 0 | 0 | 0 | 0 |
| 0 | 0 | 0 | 0 | 0.153988997 |
| 0 | 0 | 0 | 0 | 0 |
| 0 | 0 | 0 | 0 | 0 |
| 0 | 0 | 0 | 0 | 0.001824182 |
| 0 | 0 | 0 | 0.193803538 | 0.001546233 |
| 0.00539957 | 0 | 0 | 0 | 0 |
| 0 | 0 | 0 | 0.063528391 | 0 |
| 0 | 0 | 0 | 0 | 0 |
| 0 | 0 | 0 | 0 | 0 |
| 0.002896728 | 0.0033156 | 0 | 0.016014327 | 0.009067872 |
| 0 | 0 | 0 | 0 | 0 |
| 0.003947941 | 0.001191777 | 0.013234324 | 0.918125752 | 0.050701835 |
| 0 | 0 | 0 | 0 | 0 |
| 0 | 0 | 0 | 0 | 0 |
| 0 | 0 | 0 | 0 | 0 |
| 0 | 0 | 0 | 0 | 0 |
| 0 | 0 | 0 | 0 | 0 |
| 0 | 0 | 0.009557616 | 0 | 0 |
| 0 | 0 | 0 | 0 | 0 |
| 0 | 0 | 0 | 0.001770036 | 0.004384872 |
| 0 | 0 | 0 | 0.001871877 | 0.002404453 |
| 0.001103526 | 0 | 0 | 0 | 0 |

| | | | | |
|---|---|---|---|---|
| 0 | 6.100806107 | 0 | 0 | 0.925869192 |
| 0 | 0 | 0 | 0 | 0 |
| 0 | 0 | 0.001539499 | 0 | 0 |
| 0 | 0 | 0 | 0 | 0 |
| 0.005094387 | 0.000999607 | 0.032268436 | 0.004828098 | 0 |
| 0 | 0 | 0 | 0.002361796 | 0.02383669 |
| 0 | 0 | 0 | 0 | 0 |
| 0.001785226 | 0 | 0.003256655 | 0 | 0.001862815 |
| 0 | 0 | 0 | 0 | 0 |
| 0.119236621 | 0 | 0 | 0.032087555 | 0.008064182 |
| 0 | 0 | 0.001011632 | 0 | 0 |
| 0.239394631 | 0 | 0.050413822 | 0.130450007 | 0.065767634 |
| 0.005574171 | 0.000631009 | 0.00695285 | 0.014222931 | 0.001789671 |
| 0 | 0.011180636 | 0.050488018 | 0 | 0 |
| 0 | 0 | 0 | 0 | 0 |
| 0 | 0 | 0.011742054 | 0 | 0 |
| 0 | 0 | 0 | 0 | 0.012699057 |
| 0 | 0 | 0 | 0 | 0 |
| 0 | 0.108038987 | 0.86931631 | 0 | 0 |
| 0 | 0 | 0.07342254 | 0 | 0 |
| 0.025017352 | 0 | 0 | 0 | 0 |
| 0 | 0 | 0 | 0 | 0 |
| 0 | 0 | 0.020626523 | 0.000838642 | 0 |
| 0 | 0 | 0 | 0 | 0.049819909 |
| 0 | 0 | 0 | 0 | 0 |
| 0 | 0 | 0 | 0.008993019 | 0 |
| 0 | 0.106142073 | 0 | 0.015190093 | 0 |
| 0 | 0.059548044 | 0 | 0 | 0 |
| 0 | 0 | 0 | 0 | 0 |
| 0.030316708 | 0 | 0 | 0 | 0 |
| 0 | 0.002891743 | 0 | 0 | 0 |
| 0 | 0 | 0 | 0 | 0 |
| 0 | 0 | 0 | 0 | 0 |
| 0 | 0 | 0.077116336 | 0 | 0 |
| 0 | 0 | 0 | 0 | 0 |
| 0 | 0 | 0 | 0 | 0 |
| 0 | 0 | 0.12077766 | 0 | 0 |
| 0 | 0 | 0 | 0 | 0 |
| 0 | 0 | 0 | 0 | 0 |
| 0 | 0 | 0 | 0 | 0 |
| 0 | 0 | 0 | 0 | 0 |
| 0 | 0 | 0.04844582 | 0 | 0 |
| 0 | 0 | 0 | 0 | 0 |
| 0.044459903 | 0.006118507 | 0 | 0 | 0 |
| 0 | 0 | 0 | 0 | 0 |
| 0 | 0 | 0 | 0 | 0.014107274 |
| 0.001242142 | 0 | 0.029268473 | 0 | 0 |
| 0 | 0 | 0.000856974 | 0 | 0 |

| | | | | |
|---|---|---|---|---|
| 0 | 0 | 0 | 0 | 0 |
| 0 | 0 | 0.001055147 | 0 | 0 |
| 0 | 0 | 0 | 0 | 0 |
| 0 | 0 | 0 | 0 | 0 |
| 0 | 0.009937184 | 0.071855463 | 0 | 0 |
| 0 | 0.047511773 | 0.004461768 | 0 | 0 |
| 0.196358191 | 0 | 0 | 0 | 0 |
| 0 | 0 | 0 | 0 | 0 |
| 0 | 0 | 0 | 0 | 0 |
| 0 | 0 | 0 | 0 | 0 |
| 0 | 0.011410767 | 0.048586558 | 0.013173577 | 0.002960055 |
| 0 | 0 | 0 | 0 | 0 |
| 0 | 0.003230186 | 0.053666351 | 0 | 0 |
| 0 | 0.001314673 | 0.002172882 | 0.002539945 | 0 |
| 0 | 0.367936886 | 0 | 0 | 0 |
| 0 | 0 | 0 | 0 | 0 |
| 0 | 0 | 0 | 0 | 0 |
| 0 | 0 | 0 | 0 | 0 |
| 0.088997493 | 0 | 0 | 0 | 0 |
| 0 | 0 | 0 | 0 | 0.000838442 |
| 0 | 0 | 0.006438277 | 0 | 0 |
| 0 | 0 | 0 | 0 | 0.05691167 |
| 0 | 0 | 0.004897158 | 0 | 0 |
| 0 | 0 | 0 | 0 | 0 |
| 0 | 0 | 0 | 0 | 0 |
| 0 | 0 | 0 | 0 | 0 |
| 0 | 0 | 0.055365522 | 0 | 0 |
| 0 | 0 | 0 | 0 | 0 |
| 0 | 0 | 0 | 0 | 0 |
| 0.076996012 | 0.106372353 | 0.249623612 | 0.003723028 | 0.065585601 |
| 0 | 0.000713521 | 0.014557013 | 0 | 0.006829959 |
| 0.005360632 | 0 | 0 | 0.016692827 | 0 |
| 0 | 0 | 0.001702518 | 0 | 0 |
| 0 | 0.001768533 | 0 | 0 | 0 |
| 0 | 0.093319315 | 0 | 0 | 0 |
| 0 | 0 | 0 | 0 | 0 |
| 0 | 0 | 0 | 0 | 0.117463534 |
| 0.009287672 | 0 | 0 | 0.030807685 | 0 |
| 0 | 0 | 0 | 0 | 0 |
| 0 | 2.176588138 | 0 | 0 | 0 |
| 0.010355413 | 0.069603799 | 0.020542975 | 0 | 0 |
| 0.757775777 | 5.44151163 | 0 | 0 | 0.097083889 |
| 0.003503339 | 0 | 0 | 0.009249172 | 0 |
| 0 | 0 | 0.778243232 | 0 | 0 |
| 0 | 0 | 0.005289614 | 0 | 0 |
| 0 | 0 | 0 | 0 | 0 |
| 0.013643748 | 0.001134382 | 0.000937449 | 0.001753298 | 0 |
| 0.025108377 | 0 | 0 | 0 | 0 |

| | | | | |
|---|---|---|---|---|
| 0.001141295 | 0 | 0 | 0.05867886 | 0.002381794 |
| 0.192705257 | 0 | 0 | 0 | 0 |
| 0 | 0.006865604 | 0.180061504 | 0.263812814 | 0 |
| 0 | 0 | 0 | 0 | 0 |
| 0.477650456 | 0 | 0 | 0 | 0 |
| 0 | 0 | 0 | 0 | 0 |
| 0 | 0 | 0 | 0 | 0.002750191 |
| 0.00223853 | 0 | 0.002268657 | 0 | 0.002002131 |
| 0 | 0.167740235 | 0 | 0 | 0.000747505 |
| 0 | 0 | 0 | 0 | 0 |
| 0 | 0 | 0 | 0 | 0 |
| 0 | 0.004484763 | 0 | 0 | 0 |
| 0 | 0 | 0.184547292 | 0 | 0 |
| 0 | 0.001519459 | 0.025375111 | 0 | 0.003232123 |
| 0 | 0 | 0 | 0 | 0 |
| 0.460124045 | 0 | 0.001226611 | 0 | 0.01431314 |
| 0.112463728 | 0.005045217 | 0.012689324 | 0.155321872 | 0.002333033 |
| 0.003531427 | 0 | 0 | 0 | 0 |
| 0 | 0.004758169 | 0.007489779 | 0 | 0 |
| 0 | 0 | 0 | 0 | 0 |
| 0 | 0 | 0 | 0 | 0 |
| 0 | 0 | 0.039425775 | 0 | 0 |
| 0 | 0 | 0 | 0 | 0 |
| 0.561000918 | 0 | 0 | 0 | 0 |
| 0 | 0 | 0 | 0 | 0.001308267 |
| 0 | 0 | 0.004178987 | 0 | 0 |
| 0 | 0.005847992 | 0 | 0 | 0 |
| 0 | 0 | 0 | 0 | 0.020617924 |
| 0 | 0 | 0 | 0 | 0 |
| 0 | 0 | 0.001722999 | 0 | 0 |
| 0 | 0.086406267 | 0.002843982 | 0 | 0 |
| 0.001930949 | 0.001262952 | 0 | 0.00457504 | 0.005708799 |
| 0.0088862 | 0 | 0 | 0.010454535 | 0.002238167 |
| 0.115491379 | 0 | 0.024922282 | 0.116113355 | 0.002749296 |
| 0 | 0 | 0 | 0 | 0.002616397 |
| 0 | 0 | 0 | 0 | 0 |
| 0.000831076 | 0 | 0 | 0.001837815 | 0 |
| 0 | 0.013603154 | 0.018882371 | 0 | 0 |
| 0 | 0 | 0 | 0 | 0.023175031 |
| 0 | 0 | 0.057444716 | 0 | 0 |
| 0 | 0.003642231 | 0 | 0 | 0 |
| 0 | 0 | 0 | 0 | 0 |
| 0.442900926 | 0 | 0 | 0 | 0 |
| 0 | 0 | 0.075756836 | 0 | 0 |
| 0 | 0 | 0 | 0 | 0 |
| 0 | 0 | 0 | 0 | 0 |
| 0 | 0 | 0 | 0 | 0 |
| 0 | 0 | 0 | 0 | 0 |

| | | | | |
|---|---|---|---|---|
| 0 | 0 | 0.063479959 | 0 | 0 |
| 0.920460532 | 0.191731988 | 0.323415415 | 10.64554434 | 0.332373862 |
| 0 | 0 | 0 | 0 | 0 |
| 0 | 0 | 0 | 0 | 0 |
| 0 | 0.001136544 | 0 | 0 | 0 |
| 0 | 0 | 0 | 0 | 0 |
| 0 | 0.110123826 | 0 | 0 | 0 |
| 0 | 0 | 0 | 0 | 0.000709819 |
| 0.013511961 | 0 | 0.021910098 | 0 | 0.001409921 |
| 0 | 0 | 0.033481061 | 0 | 0 |
| 0.025885931 | 0.003658796 | 0.022677121 | 0 | 0 |
| 0.015357779 | 0.00256465 | 0.018876061 | 0.047071528 | 0.009887919 |
| 0 | 0 | 0 | 0 | 0.151561868 |
| 0 | 0 | 0 | 0 | 0 |
| 0 | 0 | 0.001530539 | 0 | 0 |
| 0 | 0 | 0 | 0 | 0 |
| 0.020444587 | 0.006310356 | 0.048190855 | 0.038098733 | 0.005033661 |
| 0 | 0.657092508 | 0.091873188 | 0 | 0 |
| 0 | 0 | 0 | 0 | 0 |
| 0.001742726 | 0.018237501 | 0 | 0 | 0 |
| 0.054248625 | 0.006042743 | 0.007442535 | 0.09631517 | 0.021258277 |
| 0 | 0 | 0.001194973 | 0 | 0 |
| 0 | 0 | 0 | 0 | 0 |
| 0 | 0 | 0 | 0 | 0 |
| 0 | 0 | 0.001210147 | 0 | 0 |
| 0 | 0 | 0 | 0 | 0 |
| 0 | 0.073179695 | 0.006872209 | 0 | 0 |
| 0.021653328 | 0.000741063 | 0.171220074 | 0.003102085 | 0.009720851 |
| 0 | 0 | 0.003980311 | 0 | 0.002817475 |
| 0 | 0.001489825 | 0 | 0 | 0 |
| 0 | 0 | 0 | 0.029188734 | 0.00112762 |
| 0 | 0 | 0 | 0 | 0 |
| 0 | 0 | 0 | 0 | 0 |
| 0 | 0 | 0 | 0 | 0 |
| 0 | 0 | 0.379353315 | 0 | 0.166444583 |
| 0.233091745 | 0.432282475 | 0.6547036 | 0 | 0.051134471 |
| 0 | 0 | 0 | 0 | 0 |
| 0 | 0 | 0 | 0 | 0 |
| 0.073612756 | 0.010970205 | 0.050679996 | 0.17691447 | 0.024631717 |
| 0 | 0 | 0 | 0 | 0 |
| 0 | 0 | 0 | 0 | 0 |
| 0 | 0 | 0 | 0 | 0 |
| 0.018300467 | 0.001480771 | 0 | 0 | 0.012008718 |
| 0 | 0.002680606 | 0 | 0 | 0 |
| 0 | 0 | 0 | 0 | 0 |
| 0.000752803 | 0 | 0 | 0 | 0.314993763 |
| 0 | 0 | 0 | 0 | 0 |
| 0 | 0 | 0 | 0 | 0 |

| | | | | |
|---|---|---|---|---|
| 0 | 0.024048281 | 0 | 0 | 0 |
| 0 | 0 | 0 | 0 | 0.011289085 |
| 0 | 0 | 0 | 0 | 0 |
| 0.538569427 | 0 | 0 | 0 | 2.347644248 |
| 0 | 0 | 0 | 0 | 0 |
| 0 | 0 | 0 | 0 | 0 |
| 0 | 0 | 0 | 0 | 0 |
| 0 | 0 | 0 | 0 | 0.021537558 |
| 0 | 0 | 0 | 0.011244331 | 4.138931125 |
| 0 | 0 | 0 | 0 | 0.068444608 |
| 0 | 0 | 0 | 0 | 0 |
| 0.001684235 | 0 | 0 | 0 | 0.053821409 |
| 0 | 0 | 0 | 0 | 0 |
| 0 | 0 | 0 | 0 | 0 |
| 0.237140555 | 0.044144893 | 0.563591996 | 0 | 0.072330661 |
| 0 | 0 | 0 | 0 | 0 |
| 0 | 0 | 0 | 0 | 0.186096919 |
| 0 | 0 | 0 | 0 | 0.303480415 |
| 0 | 0 | 0 | 0 | 0 |
| 0 | 0 | 0 | 0 | 0.211873873 |
| 0 | 0 | 0 | 0 | 0 |
| 0 | 0 | 0 | 0 | 0.00101978 |
| 0 | 0 | 0 | 0 | 0.008091496 |
| 0 | 0 | 0.304767799 | 0 | 0 |
| 0 | 0 | 0.016180726 | 0 | 0 |
| 0 | 0 | 0 | 0 | 0.35399076 |
| 0 | 0 | 0 | 0 | 0 |
| 1.32200692 | 0 | 0 | 0 | 1.106701515 |
| 0 | 0 | 0 | 0 | 0 |
| 0 | 0 | 0 | 0 | 0 |
| 0 | 0.002811193 | 0 | 0 | 1.147061165 |
| 0 | 0 | 0 | 0 | 0 |
| 0 | 0.000916939 | 0.014050041 | 0 | 0 |
| 0 | 0 | 0 | 0 | 0 |
| 0 | 0 | 0 | 0 | 0.037267496 |
| 0 | 0 | 0.076504831 | 0 | 0.603176806 |
| 0 | 0 | 0 | 0 | 0.679847128 |
| 0 | 0 | 0 | 0 | 0 |
| 0 | 0 | 0.005552766 | 0 | 0 |
| 0 | 0.001478413 | 0.001221755 | 0 | 0 |
| 0.661234277 | 0.491202688 | 0.015351256 | 0.005550842 | 0.191776079 |
| 0 | 0 | 0 | 0 | 0.094994434 |
| 0.062508789 | 0.13952981 | 0.034867089 | 2.116157538 | 0.243457985 |
| 0 | 0 | 0 | 0 | 0 |
| 0 | 0 | 0 | 0 | 0 |
| 0 | 0 | 0 | 0 | 0 |
| 0 | 0 | 0 | 0 | 0 |
| 0.002185069 | 0.000714581 | 0.095960647 | 0.004371801 | 0 |

| | | | | |
|---|---|---|---|---|
| 0 | 0 | 0 | 0 | 0 |
| 0 | 0 | 0 | 0 | 1.653893885 |
| 0.026764782 | 0.106404331 | 0.562784679 | 0.544150667 | 0.00315708 |
| 0 | 0 | 0 | 0 | 0.013650609 |
| 0 | 0 | 0 | 0 | 0 |
| 0.009717095 | 0 | 0.005574267 | 0 | 0.119185931 |
| 0 | 0 | 0 | 0 | 0 |
| 0 | 0 | 0 | 0 | 0 |
| 0 | 0 | 0 | 0 | 0 |
| 0 | 1.741603755 | 0 | 0 | 0 |
| 0.23399085 | 0.030264967 | 0.060442913 | 0 | 0.004053444 |
| 1.166101564 | 0.711055651 | 0.22156068 | 0.378036105 | 0.221913967 |
| 0 | 0 | 0 | 0 | 0 |
| 0.084341897 | 0 | 0.014995967 | 0.933022127 | 0.0298505 |
| 0.07015713 | 0.347583222 | 0.130779193 | 0.001473461 | 0.00466411 |
| 0 | 0 | 0 | 0 | 3.013909293 |
| 0 | 0 | 0 | 0 | 0 |
| 0 | 0 | 0 | 0 | 0 |
| 0 | 0 | 0 | 0 | 0.097289245 |
| 0.596875334 | 0 | 0.003433728 | 0 | 0.016498446 |
| 0.297051018 | 0.360580094 | 0.058229956 | 2.67057411 | 0.3735123 |
| 0.07365884 | 0.329254548 | 0.169343412 | 3.578217492 | 0 |
| 1.40557289 | 0.786574078 | 0 | 0.958359192 | 0.156292828 |
| 0 | 0 | 0 | 0 | 0.001542628 |
| 0 | 0 | 0 | 0 | 0.013319071 |
| 0.0206551 | 0 | 0 | 0 | 0.151109062 |
| 0 | 0 | 0 | 0.000651288 | 0 |
| 0 | 0 | 0 | 0 | 0.001667487 |
| 0 | 0 | 0 | 0 | 0 |
| 0 | 0 | 0 | 0 | 0 |
| 0 | 0 | 0 | 0 | 0.018295278 |
| 0.391999745 | 0 | 0 | 0 | 0.023583751 |
| 0.052334158 | 0.046969731 | 2.884057424 | 0 | 0.00122441 |
| 0 | 0 | 0.032646963 | 0 | 0.020541557 |
| 0 | 0.001823565 | 0.125268344 | 0 | 0 |
| 0.124921731 | 0.02296678 | 0.204462747 | 0 | 0.130350995 |
| 0.006372833 | 0.002823621 | 0.020625858 | 0 | 0.002788628 |
| 0 | 0 | 0 | 0 | 0 |
| 0 | 0 | 0 | 0 | 4.077700871 |
| 0 | 0 | 0 | 0 | 0 |
| 0 | 0 | 0.001329407 | 0 | 0.038520921 |
| 0 | 0 | 0 | 0 | 0 |
| 0 | 0 | 0 | 0 | 0 |
| 0 | 0 | 0 | 0 | 0 |
| 0.012134502 | 0.069287113 | 0.012051858 | 0.000690013 | 0.029122332 |
| 0 | 0 | 0 | 0 | 0.001565862 |
| 0 | 0 | 0 | 0 | 0 |
| 1.679509668 | 0.107705145 | 26.92466162 | 56.46125036 | 0.641495053 |

| | | | | |
|---|---|---|---|---|
| 0 | 0 | 0 | 0 | 0 |
| 0 | 0 | 0 | 0 | 0 |
| 0 | 0 | 0 | 0 | 0 |
| 0 | 0 | 0 | 0 | 0.004964169 |
| 0 | 0 | 0 | 0 | 0 |
| 0 | 0 | 0 | 0 | 0.2180724 |
| 0 | 0 | 0 | 0 | 0.002835558 |
| 0 | 0.005415283 | 0.023919015 | 0.000541154 | 0.033762992 |
| 0 | 0 | 0 | 0 | 0 |
| 0 | 0 | 0 | 0 | 0 |
| 0.146975519 | 0.043846363 | 0.008871844 | 0 | 0 |
| 0 | 0 | 0 | 0 | 0 |
| 0 | 0 | 0.219343587 | 0 | 0 |
| 0 | 0 | 0 | 0 | 0.068101254 |
| 0 | 0 | 0 | 0 | 0 |
| 0 | 0 | 0 | 0 | 0 |
| 0 | 0 | 0 | 0 | 0.078663375 |
| 0.004602649 | 0 | 0.031680369 | 0 | 0.035019591 |
| 0 | 0 | 0 | 0 | 0 |
| 0 | 0 | 0 | 0 | 0.006135906 |
| 0 | 0 | 0 | 0 | 0.000888918 |
| 0 | 0 | 0 | 0 | 0 |
| 0 | 0 | 0 | 0 | 0.073367293 |
| 0 | 0 | 0 | 0 | 0 |
| 0 | 0 | 0 | 0 | 0 |
| 0 | 0 | 0 | 0 | 0 |
| 0 | 0 | 0 | 0 | 0 |
| 0 | 0 | 0 | 0 | 0 |
| 0 | 0 | 0 | 0 | 0 |
| 0 | 0 | 0 | 0 | 0.011119739 |
| 0.031026473 | 0 | 0.042477039 | 0 | 0 |
| 0 | 0 | 0 | 0 | 0 |
| 0 | 0 | 0 | 0 | 0 |
| 0 | 0 | 0 | 0 | 0 |
| 0.001648605 | 0.002695708 | 0 | 0 | 0.043579807 |
| 0 | 0 | 0 | 0 | 0 |
| 0 | 0 | 0 | 0 | 0.045961616 |
| 0.019696163 | 0.005739698 | 0.005731446 | 0 | 0 |
| 0 | 0 | 0 | 0 | 0 |
| 0 | 0 | 0 | 0 | 0 |
| 0 | 0 | 0 | 0 | 0.006873161 |
| 0 | 0 | 0 | 0 | 0 |
| 0 | 0 | 0 | 0 | 0 |
| 0 | 0 | 0 | 0 | 0.007311569 |
| 0 | 0 | 0 | 0 | 0.041590324 |
| 0 | 0 | 0 | 0 | 0.007005574 |
| 0.000926468 | 0 | 0 | 0 | 0.007927215 |
| 0 | 0 | 0 | 0 | 0.094335003 |

| | | | | |
|---|---|---|---|---|
| 0 | 0 | 0 | 0 | 0 |
| 0 | 0 | 0 | 0 | 0 |
| 0 | 0 | 0 | 0 | 0.485304402 |
| 0 | 0 | 0 | 0 | 0.474448808 |
| 0 | 0 | 0 | 0 | 0 |
| 0 | 0 | 0 | 0 | 0.175004983 |
| 0 | 0 | 0 | 0 | 0 |
| 0 | 0 | 0 | 0 | 0 |
| 0.03039174 | 0.157528443 | 0.006269183 | 0 | 0 |
| 0 | 0 | 0.001756016 | 0 | 0.005876005 |
| 0 | 0 | 0 | 0 | 0.007023385 |
| 0 | 0 | 0 | 0 | 0.00205312 |
| 0 | 0.00815855 | 0.009063801 | 0 | 0 |
| 0 | 0 | 0 | 0 | 0 |
| 1.400485504 | 0.002085838 | 0.757265375 | 0 | 0.374111071 |
| 0 | 1.023151623 | 0.026890203 | 0 | 0 |
| 0 | 0 | 0 | 0 | 0 |
| 0 | 0 | 0 | 0 | 0 |
| 0 | 0 | 0 | 0 | 0 |
| 0 | 0 | 0 | 0 | 0 |
| 0 | 0 | 0 | 0 | 0.011587939 |
| 0 | 0 | 0 | 0 | 0 |
| 0 | 0 | 0 | 0 | 0 |
| 0 | 0 | 0 | 0 | 0 |
| 0.096782171 | 0 | 0.118919699 | 0.213721204 | 0.119139971 |
| 2.38934037 | 0.96369019 | 0 | 0 | 1.48315691 |
| 0 | 0 | 0 | 0 | 0.77283858 |
| 0 | 0 | 0 | 0 | 0 |
| 0 | 0 | 1.964777442 | 0 | 6.897224668 |
| 0 | 0 | 0 | 0 | 0 |
| 0 | 0 | 0 | 0 | 0 |
| 0 | 0 | 0 | 0 | 0.004038253 |
| 0 | 0 | 0 | 0 | 0 |
| 0.018242786 | 0 | 0.012154349 | 0 | 0.000528768 |
| 0.006122997 | 0 | 0.006769531 | 0.295686244 | 0.002613727 |
| 0.105844567 | 0.012353039 | 0.014754482 | 0 | 0.001231725 |
| 0 | 0 | 0.004732473 | 0 | 0 |
| 0 | 0 | 0 | 0 | 0.230182513 |
| 1.373790515 | 0 | 0 | 0 | 0.029297376 |
| 0 | 0 | 0 | 0 | 0 |
| 0 | 0 | 0 | 0 | 0 |
| 0 | 0.021012667 | 0 | 0 | 0.012803876 |
| 0 | 0.185950878 | 0.216608095 | 0.015032726 | 4.917956013 |
| 0 | 0 | 0 | 0 | 0.00484822 |
| 0 | 0 | 0 | 0 | 0 |
| 0 | 0 | 0.000566676 | 0 | 0.000583453 |
| 0 | 0 | 0 | 0 | 0 |
| 0.083437781 | 5.146966509 | 0 | 0.002251285 | 0.619053622 |

| | | | | |
|---|---|---|---|---|
| 0 | 0.004561924 | 0.003141631 | 0 | 0 |
| 0 | 0 | 0 | 0 | 0.079181746 |
| 0 | 0 | 0 | 0 | 0 |
| 0 | 0 | 0 | 0 | 0 |
| 0.003086321 | 0 | 0.000670255 | 0 | 0 |
| 0 | 0 | 2.037887597 | 0 | 0 |
| 0 | 0 | 0 | 0 | 0.034427771 |
| 0 | 0 | 0 | 0 | 0 |
| 0 | 0 | 0 | 0 | 0 |
| 0 | 0 | 0 | 0 | 0.124447516 |
| 0 | 0 | 0 | 0 | 0.000704061 |
| 7.318242481 | 2.434115503 | 0 | 0.004625991 | 0.168502476 |
| 0 | 0 | 0 | 0 | 0.005013524 |
| 0 | 0 | 0 | 0 | 0 |
| 2.68617557 | 0.004752474 | 0.472841607 | 1.265392798 | 0.03272202 |
| 0 | 0 | 0 | 0 | 0 |
| 0 | 0.000696007 | 0 | 0 | 0 |
| 0 | 0 | 0 | 0 | 0 |
| 0.038435915 | 0 | 0.000905888 | 0 | 0 |
| 0 | 0 | 0 | 0 | 0.018357457 |
| 0 | 0 | 0 | 0 | 0.057528579 |
| 0 | 0 | 0 | 0 | 0 |
| 0.554086909 | 0 | 0 | 0 | 0.370934618 |
| 0 | 0 | 0 | 0 | 0 |
| 0 | 0 | 0 | 0 | 0 |
| 0 | 0 | 0 | 0 | 0 |
| 0 | 0 | 0 | 0 | 0 |
| 0 | 0 | 0 | 0 | 0 |
| 0 | 0 | 0 | 0 | 0.012700292 |
| 0 | 0 | 0 | 0 | 0 |
| 0 | 0 | 0 | 0 | 0 |
| 0.066167207 | 0.008114473 | 0.058675502 | 0.199726821 | 0.038318821 |
| 0 | 0 | 0 | 0 | 0 |
| 0 | 0.014756018 | 0.021775572 | 0 | 0.008795627 |
| 0.105765044 | 0.009872351 | 0.016490536 | 0 | 0.003351065 |
| 0.019871995 | 0 | 0.090347774 | 0.003662024 | 0.620053785 |
| 0 | 0 | 0 | 0 | 0.014278253 |
| 0 | 0 | 0 | 0 | 0.011963074 |
| 0 | 0 | 0 | 0 | 0.004363196 |
| 0 | 0 | 0 | 0 | 0 |
| 0 | 0 | 0 | 0 | 0.078420614 |
| 0 | 0 | 0 | 0 | 0.036584066 |
| 0 | 0 | 0 | 0 | 0 |
| 0 | 0 | 0 | 0 | 0 |
| 0.080778242 | 0.001448442 | 0.015817335 | 0.003997696 | 0.006602269 |
| 0 | 0 | 0 | 0 | 0 |
| 0 | 0 | 0 | 0 | 0.05406867 |
| 0 | 0 | 0.001617337 | 0 | 0.000832609 |

| | | | | |
|---|---|---|---|---|
| 0 | 0 | 0 | 0 | 0 |
| 0 | 0 | 0 | 0 | 0.015748968 |
| 0 | 0 | 0 | 0 | 0 |
| 0 | 0 | 0 | 0 | 0 |
| 0 | 0 | 0 | 0 | 0 |
| 0 | 0 | 0 | 0 | 0 |
| 0 | 0 | 0 | 0 | 1.085391182 |
| 0.002472074 | 0.034156587 | 0 | 0 | 0.012682609 |
| 0 | 0 | 0 | 0 | 0 |
| 0 | 0 | 0 | 0 | 0 |
| 0 | 0 | 0 | 0 | 0 |
| 0 | 0 | 0.001349592 | 0.000631031 | 0 |
| 0 | 0 | 0 | 0 | 0 |
| 0 | 0 | 0 | 0 | 0.185371674 |
| 0 | 0 | 0 | 0 | 0 |
| 0 | 0 | 0 | 0 | 0.001752656 |
| 0 | 0 | 0 | 0 | 0 |
| 0 | 0 | 0 | 0 | 0.001519941 |
| 1.81477802 | 17.02362995 | 0 | 0 | 1.277107596 |
| 0 | 0 | 0 | 0 | 0 |
| 0 | 0.004472232 | 0 | 0 | 0.001861264 |
| 0 | 0 | 0 | 0 | 0 |
| 0 | 0 | 0 | 0 | 0 |
| 0 | 0 | 0 | 0 | 0 |
| 0.00138262 | 0 | 0 | 0.009172443 | 0.002164066 |
| 0.039269708 | 0.01772242 | 0.201577976 | 0 | 0 |
| 0 | 0 | 0 | 0 | 0 |
| 1.702192864 | 0.023191816 | 0 | 0.002914246 | 3.108151342 |
| 0 | 0 | 0 | 0 | 0.163852907 |
| 1.345796893 | 0.464857902 | 0 | 0 | 0.366376317 |
| 0 | 0 | 0 | 0 | 0 |
| 0 | 0 | 0 | 0 | 0 |
| 0 | 0 | 0 | 0 | 0.181682939 |
| 0 | 0 | 0 | 0 | 0 |
| 0.125102763 | 0.013929 | 0.002710895 | 0 | 0 |
| 0 | 0 | 0 | 0 | 0.145652734 |
| 0 | 0 | 0 | 0 | 0 |
| 0.007004358 | 0 | 0 | 0 | 0.001705381 |
| 0 | 0 | 0 | 0 | 0 |
| 0 | 0 | 0 | 0 | 0 |
| 1.874098381 | 14.45128873 | 7.86115577 | 0 | 0.388720434 |
| 1.139781439 | 0.214456361 | 0.156233212 | 0 | 0.001070012 |
| 0 | 0 | 1.792471287 | 0 | 0 |
| 0 | 0 | 0.013678646 | 0 | 0.024450685 |
| 0 | 0 | 0 | 0 | 0 |
| 0 | 0.127062228 | 0 | 0 | 0 |
| 0 | 0 | 0 | 0 | 0.004146964 |
| 0.016121234 | 0.019452272 | 0.001314568 | 0 | 0.012761429 |

| | | | | |
|---|---|---|---|---|
| 0 | 0 | 0 | 0 | 0 |
| 0 | 0.008040706 | 0 | 0 | 0 |
| 0.00934952 | 0 | 0 | 0 | 0 |
| 0 | 0 | 0 | 0 | 0 |
| 0 | 1.741656511 | 4.059673103 | 0.078245296 | 3.553466952 |
| 0 | 0 | 0 | 0 | 0 |
| 0 | 0 | 0 | 0 | 0 |
| 0 | 0 | 0 | 0 | 0 |
| 0.145211815 | 1.968748944 | 0.442077072 | 0 | 0 |
| 0 | 0 | 0 | 0 | 0 |
| 0 | 0 | 0 | 0 | 0 |
| 0 | 0 | 0 | 0 | 0 |
| 0.001296389 | 0 | 0.034159737 | 0 | 0.040775189 |
| 0.001593778 | 0.013402584 | 0.017767506 | 0 | 0 |
| 0 | 0 | 3.832562516 | 0 | 6.297064065 |
| 0 | 0 | 0 | 0 | 0.091628733 |
| 0.003940479 | 0.000966488 | 0.004792214 | 0 | 0.866959827 |
| 0 | 0 | 0 | 0 | 0 |
| 0 | 0 | 0 | 0 | 0 |
| 0 | 0 | 0 | 0 | 0.003724506 |
| 0 | 0 | 0 | 0 | 0.007533247 |
| 0 | 0 | 0 | 0 | 0 |
| 0 | 0 | 0 | 0 | 0 |
| 0 | 0 | 0 | 0 | 0 |
| 0 | 0 | 0 | 0 | 0.002392441 |
| 0 | 0 | 0 | 0 | 0 |
| 0 | 0 | 0 | 0.001623653 | 0.036825844 |
| 0.051422038 | 0.006448431 | 0.024881544 | 0.000916056 | 0 |
| 0 | 0 | 0 | 0 | 0 |
| 0 | 0 | 0 | 0 | 0.000992549 |
| 0 | 0 | 0 | 0 | 0 |
| 0 | 0 | 0 | 0 | 0 |
| 0 | 0 | 0 | 0 | 0.219826862 |
| 0 | 0 | 0 | 0 | 0.018396497 |
| 0 | 0 | 0 | 0 | 2.792296405 |
| 0.202829152 | 0.000877588 | 0.000906544 | 0 | 0 |
| 0 | 0 | 0 | 0.015813652 | 0 |
| 0.003378668 | 0 | 0 | 0 | 0.25670112 |
| 0 | 0 | 0 | 0 | 0 |
| 0 | 0 | 0 | 0 | 0 |
| 11.40292023 | 0 | 0 | 0 | 0.200909111 |
| 0 | 0 | 0.024966749 | 0 | 0 |
| 0 | 0 | 0 | 0 | 0.038692942 |
| 0 | 0 | 0 | 0 | 0 |
| 0 | 0 | 0 | 0 | 0 |
| 0 | 0 | 0 | 0 | 0 |
| 0 | 0 | 0 | 0 | 0 |
| 0 | 0 | 0 | 0 | 0 |

| | | | | |
|---|---|---|---|---|
| 0 | 0 | 0 | 0 | 0.964725164 |
| 0 | 0 | 0 | 0 | 0 |
| 0 | 0 | 0 | 0 | 0 |
| 0.001094945 | 0 | 0 | 0 | 0 |
| 0 | 0 | 0 | 0 | 0.020114522 |
| 0 | 0 | 0 | 0 | 0.642076705 |
| 0.351029133 | 0.002892414 | 0.008764363 | 0.002607798 | 0.687451518 |
| 0 | 0 | 0 | 0 | 0 |
| 0 | 0 | 0 | 0 | 0.001013308 |
| 0 | 0 | 0 | 0 | 0.001721357 |
| 0 | 0 | 0 | 0 | 0.014175987 |
| 0 | 0.008600201 | 0 | 0 | 0.000896762 |
| 0 | 0 | 0 | 0 | 0 |
| 0 | 0 | 0 | 0 | 0 |
| 0 | 0 | 0 | 0 | 0 |
| 0 | 0 | 0 | 0 | 0 |
| 0 | 0 | 0 | 0 | 0 |
| 0 | 0.010196093 | 0.023363042 | 0 | 0 |
| 0 | 0 | 0 | 0 | 0 |
| 0 | 0 | 0 | 0 | 0 |
| 0.181695139 | 0.049383729 | 0 | 0 | 0.666207412 |
| 0 | 0 | 0 | 0 | 0.04489594 |
| 0.160574957 | 0 | 0.006065997 | 0 | 0.058579206 |
| 0.003487035 | 0.0012604 | 0.029759708 | 0 | 0.007468677 |
| 0 | 0 | 0 | 0 | 0 |
| 0.011042621 | 0 | 0 | 0 | 0 |
| 0 | 0 | 0 | 0 | 0 |
| 0 | 0 | 0 | 0 | 0 |
| 0 | 0 | 0 | 0 | 0 |
| 0.00211312 | 0 | 0.052468216 | 0.013517992 | 0 |
| 0 | 0 | 0 | 0 | 0 |
| 0 | 0 | 0 | 0 | 0 |
| 0 | 0 | 0 | 0 | 0 |
| 0.279883745 | 0 | 0 | 0 | 0.455046568 |
| 0 | 0 | 0 | 0 | 0.261510013 |
| 0 | 0 | 0 | 0 | 0 |
| 0.044320764 | 0.033344113 | 0.002792783 | 0.034604391 | 0.000958487 |
| 0 | 0 | 0 | 0 | 0 |
| 0 | 0 | 0 | 0 | 0.660878194 |
| 0 | 0 | 0 | 0 | 0 |
| 0 | 0 | 0 | 0 | 0 |
| 0 | 0 | 0 | 0 | 0 |
| 0 | 0 | 0 | 0 | 0 |
| 0 | 0 | 0 | 0 | 0.009832256 |
| 0 | 0 | 0 | 0 | 0.045810354 |
| 0 | 0 | 0 | 0 | 0 |
| 0 | 0 | 0 | 0 | 0 |
| 0 | 0 | 0 | 0 | 0 |

| | | | | |
|---|---|---|---|---|
| 0.498446634 | 0.011431122 | 0.011207876 | 0 | 0 |
| 0 | 0 | 0.42669855 | 0 | 0.463564256 |
| 0 | 0 | 0 | 0 | 0 |
| 0 | 0 | 0 | 0 | 0 |
| 0 | 0 | 0 | 0 | 0 |
| 0 | 0 | 0 | 0 | 0 |
| 0.002429047 | 0 | 0 | 0 | 0 |
| 4.106096991 | 0 | 0 | 0 | 0.043586074 |
| 0.792616161 | 0.002737047 | 1.099841918 | 3.212147032 | 0.094124253 |
| 0.204144948 | 0.101623995 | 1.09810453 | 0 | 0 |
| 0 | 0 | 0 | 0 | 0.043864027 |
| 0 | 0 | 0 | 0 | 0 |
| 0 | 0 | 0 | 0 | 0.020671009 |
| 0.317374771 | 0 | 0 | 0 | 0 |
| 0 | 0 | 0 | 0 | 0 |
| 0 | 0 | 0 | 0 | 0 |
| 0 | 0.004242295 | 0 | 0 | 0 |
| 0 | 0 | 0 | 0 | 0 |
| 0 | 0 | 0 | 0 | 0 |
| 0 | 0 | 0 | 0 | 0.03622409 |
| 0 | 0 | 0 | 0 | 0 |
| 0 | 0 | 0 | 0 | 0 |
| 0 | 0 | 0 | 0 | 0 |
| 0 | 0 | 0 | 0 | 0 |
| 0.001088816 | 0 | 0 | 0 | 0.017853582 |
| 0 | 0 | 0 | 0 | 0 |
| 0 | 0 | 0 | 0 | 0 |
| 0 | 0 | 0 | 0 | 0 |
| 0 | 0 | 0 | 0 | 0 |
| 0 | 0 | 0 | 0 | 0 |
| 0 | 0 | 0 | 0 | 0 |
| 0 | 0 | 0 | 0 | 0 |
| 0 | 0 | 0 | 0 | 0.028396475 |
| 0 | 0 | 0 | 0 | 0.005898685 |
| 0 | 0 | 0.01986847 | 0 | 0.010727275 |
| 0 | 0 | 0 | 0 | 0 |
| 0 | 0 | 0 | 0 | 0.044285268 |
| 0 | 0 | 0 | 0 | 0 |
| 0 | 0 | 0 | 0 | 0.012741466 |
| 0 | 0.001353947 | 3.217827873 | 0 | 0 |
| 0 | 0 | 0 | 0 | 0 |
| 0 | 0 | 0.07379536 | 0 | 0.258289649 |
| 0 | 0 | 0 | 0 | 0 |
| 0 | 0 | 0 | 0 | 0 |
| 0 | 0 | 0 | 0 | 0 |
| 0 | 0 | 0 | 0 | 0 |
| 0 | 0 | 0 | 0 | 0 |
| 0 | 0 | 0 | 0 | 0 |

| | | | | |
|---|---|---|---|---|
| 0 | 0 | 0 | 0 | 0 |
| 0 | 0 | 0 | 0 | 0 |
| 0 | 0 | 0 | 0 | 0 |
| 0 | 0 | 0 | 0 | 0 |
| 0.385263538 | 0 | 4.463893284 | 0.067309665 | 6.067818515 |
| 0 | 0 | 0 | 0 | 0.002851686 |
| 0 | 0 | 0 | 0 | 0 |
| 0.039942779 | 0.024892588 | 0.004328088 | 0 | 0.000786391 |
| 0 | 0 | 0 | 0 | 0.018320929 |
| 0 | 0 | 0 | 0 | 0 |
| 0 | 0 | 0 | 0 | 0 |
| 0 | 0 | 0 | 0 | 0 |
| 0 | 0 | 0 | 0 | 0 |
| 0 | 0 | 0 | 0 | 0 |
| 0 | 0 | 0.027421389 | 0 | 0 |
| 0 | 0.033986889 | 0 | 0 | 0.015914066 |
| 0 | 0 | 0 | 0 | 0 |
| 0 | 0 | 0 | 0 | 0 |
| 0 | 0 | 0 | 0 | 0.061031055 |
| 0 | 0 | 0 | 0 | 0 |
| 0 | 0 | 0 | 0 | 0 |
| 0 | 0 | 0 | 0 | 0.003879327 |
| 0 | 0 | 0 | 0 | 0 |
| 0.005869899 | 0.009384845 | 0.006389559 | 0 | 0 |
| 0 | 0 | 0 | 0 | 0.024825704 |
| 0 | 0 | 0 | 0 | 0 |
| 8.16836131 | 0.004681912 | 0.250686442 | 0 | 0 |
| 0 | 0 | 0 | 0 | 0 |
| 0 | 0 | 0.001198117 | 0 | 0 |
| 0 | 0 | 0 | 0 | 0 |
| 0 | 0 | 0 | 0 | 0 |
| 0 | 0 | 0 | 0 | 0 |
| 0 | 0 | 0 | 0 | 0 |
| 0 | 0 | 0 | 0 | 0 |
| 0 | 0 | 0 | 0 | 0 |
| 0 | 0 | 0 | 0 | 0 |
| 0 | 0 | 0 | 0 | 0 |
| 0 | 0 | 0 | 0 | 0.041280322 |
| 0 | 0 | 0 | 0 | 0 |
| 0 | 0 | 0 | 0 | 0 |
| 0 | 0 | 0 | 0 | 0 |
| 0 | 0 | 4.722392348 | 0 | 1.162359585 |
| 0 | 0 | 0 | 0 | 0 |
| 0 | 0 | 0 | 0 | 0 |
| 0 | 0 | 0 | 0 | 0 |
| 0 | 0 | 0 | 0 | 0 |
| 0 | 0 | 0 | 0 | 0 |
| 0 | 0 | 0 | 0 | 0.008819898 |

| | | | | |
|---|---|---|---|---|
| 0.104528643 | 0 | 0 | 0 | 0.103022904 |
| 0 | 0 | 0 | 0 | 0.002838908 |
| 0 | 0 | 0 | 0 | 0 |
| 0 | 0 | 0 | 0 | 0 |
| 0 | 0 | 0 | 0 | 0 |
| 0 | 0 | 0 | 0 | 0.008590159 |
| 0 | 0 | 0 | 0 | 0 |
| 0 | 0 | 0 | 0 | 0 |
| 0 | 0 | 0 | 0 | 0 |
| 0 | 0 | 0 | 0 | 0 |
| 0 | 0 | 0 | 0 | 0 |
| 0 | 0 | 0 | 0 | 0 |
| 0 | 0 | 0 | 0 | 0 |
| 0 | 0 | 0 | 0 | 0 |
| 0 | 0 | 0 | 0 | 0 |
| 0 | 0 | 0 | 0 | 0.006231199 |
| 0 | 0 | 0 | 0 | 0 |
| 0 | 0 | 0 | 0 | 0 |
| 0 | 0.005443816 | 0.001928036 | 0 | 0 |
| 0 | 0 | 0 | 0 | 0 |
| 0 | 0.033134448 | 0.294007478 | 0 | 0 |
| 0 | 0 | 0 | 0 | 0 |
| 0.155027586 | 0.01314406 | 0.033805839 | 0 | 0.07674579 |
| 0 | 0 | 0 | 0 | 0.149361632 |
| 0 | 0 | 0 | 0 | 0.019731848 |
| 0 | 0 | 0 | 0 | 0 |
| 0 | 0 | 0 | 0 | 0 |
| 0 | 0 | 0 | 0 | 0 |
| 0.001420155 | 0 | 0 | 0 | 0 |
| 0 | 0 | 0 | 0 | 0 |
| 0 | 0 | 0 | 0 | 0 |
| 0 | 0 | 0 | 0 | 0.004876777 |
| 0.050954773 | 6.369464667 | 0.574274019 | 0 | 0 |
| 0 | 0 | 0 | 0 | 0 |
| 0 | 0 | 0 | 0 | 0.014578607 |
| 0 | 0.055436317 | 0 | 0.002531522 | 0 |
| 0 | 0 | 0 | 0 | 0 |
| 0 | 0 | 0 | 0 | 0 |
| 0.337241941 | 0.007694504 | 0.116335505 | 0 | 0 |
| 0 | 0 | 0 | 0 | 0 |
| 0 | 0 | 0 | 0 | 0 |
| 0 | 0 | 0 | 0 | 0 |
| 0 | 0 | 0 | 0.005004063 | 0.526885551 |
| 0 | 0 | 0 | 0 | 0 |
| 0 | 0 | 0 | 0 | 0 |
| 0 | 0 | 0 | 0 | 0.05573039 |
| 0 | 0 | 0 | 0 | 0 |
| 0 | 0 | 0 | 0 | 0 |

| | | | | |
|---|---|---|---|---|
| 0 | 0 | 0 | 0 | 0 |
| 0 | 0 | 0 | 0 | 0 |
| 0 | 0 | 0 | 0 | 0 |
| 0 | 0 | 0 | 0 | 0 |
| 0 | 0 | 0 | 0 | 0 |
| 0 | 0 | 0 | 0 | 0 |
| 0 | 0 | 0 | 0 | 0 |
| 0.027511204 | 0 | 0 | 0 | 0 |
| 0.264878132 | 0.009737461 | 0 | 0 | 0.007767403 |
| 0 | 0 | 0.464670166 | 0 | 0.023719705 |
| 0 | 0 | 0 | 0 | 0 |
| 0 | 0 | 0 | 0 | 0 |
| 0 | 0 | 0 | 0 | 0 |
| 0 | 0 | 0 | 0 | 0 |
| 0 | 0 | 0 | 0 | 0 |
| 0.001118277 | 0.001755403 | 0.006573296 | 0 | 0 |
| 0 | 0 | 0 | 0 | 0 |
| 0 | 0 | 0 | 0 | 0 |
| 0 | 0 | 0 | 0 | 0 |
| 0 | 0 | 0 | 0 | 0.054904449 |
| 0.016351448 | 0 | 0 | 0 | 0 |
| 0 | 0 | 0 | 0 | 0 |
| 0 | 0 | 0 | 0 | 0 |
| 0 | 0 | 0 | 0 | 0.004973779 |
| 0 | 0 | 0 | 0 | 0 |
| 0 | 0 | 0 | 0 | 0 |
| 0 | 0 | 0 | 0 | 0 |
| 0 | 0 | 0 | 0 | 0 |
| 0 | 0 | 0 | 0 | 0 |
| 0 | 0 | 0 | 0 | 0.047407692 |
| 0.027630676 | 0 | 0.089165993 | 0 | 0.053300417 |
| 0 | 0 | 0 | 0 | 0 |
| 0 | 0 | 0 | 0 | 0 |
| 0 | 0 | 0 | 0 | 0 |
| 0 | 0 | 0 | 0 | 0 |
| 0 | 0 | 0 | 0 | 0 |
| 0 | 0 | 0 | 0 | 0 |
| 0 | 0 | 0 | 0 | 0 |
| 0 | 0 | 0 | 0 | 0 |
| 0 | 0 | 0 | 0 | 0 |
| 0 | 0 | 0 | 0 | 0 |
| 0 | 0 | 0 | 0 | 0.342361316 |
| 0 | 0 | 0 | 0 | 0.003692824 |
| 0 | 0 | 0 | 0 | 0 |
| 0 | 0 | 0 | 0 | 0 |
| 0 | 0 | 0 | 0 | 0 |
| 0 | 0.919611113 | 0 | 0 | 0 |
| 0 | 0 | 0.001508601 | 0 | 0.000970789 |

| | | | | |
|---|---|---|---|---|
| 0 | 0 | 0 | 0 | 0 |
| 0 | 0 | 0 | 0 | 0 |
| 0 | 0 | 0 | 0 | 0 |
| 0 | 0 | 0 | 0 | 0 |
| 0.441298624 | 0.00688025 | 0 | 0 | 0.066751469 |
| 0 | 0 | 0 | 0 | 0.000884926 |
| 0 | 0 | 0 | 0 | 0 |
| 0 | 0 | 0 | 0 | 0 |
| 0 | 0 | 0 | 0 | 0 |
| 0 | 0 | 0 | 0 | 0 |
| 0 | 0 | 0 | 0 | 0.020847881 |
| 0.260744752 | 0.001630868 | 0.010589413 | 0.000675179 | 0.005451453 |
| 0 | 0.001608032 | 0 | 0 | 0.001924049 |
| 0 | 0 | 0 | 0 | 0 |
| 0 | 0.002663218 | 0 | 0 | 0 |
| 0 | 0 | 0 | 0 | 0.142979269 |
| 0 | 0 | 0.003887155 | 0 | 0 |
| 0 | 0 | 0 | 0 | 0 |
| 0.632051483 | 0.741823607 | 0.045558886 | 0 | 0 |
| 0.136409652 | 0.011286845 | 0.002379441 | 0 | 0.678617565 |
| 0 | 0 | 0 | 0 | 0 |
| 0 | 0 | 0 | 0 | 0.02727971 |
| 0 | 0 | 0 | 0 | 0 |
| 0 | 0 | 0 | 0 | 0 |
| 0 | 0 | 0 | 0 | 0 |
| 0 | 0 | 0 | 0 | 0 |
| 0 | 0.006275891 | 0.023643763 | 0 | 0.007067532 |
| 0 | 0 | 0 | 0 | 0 |
| 0 | 0 | 0 | 0 | 0 |
| 0 | 0 | 0 | 0 | 0 |
| 0.065687285 | 0.001255421 | 0.001080704 | 0 | 0 |
| 0 | 0 | 0 | 0 | 0 |
| 0 | 0 | 0 | 0 | 0 |
| 0 | 0 | 0 | 0 | 0 |
| 0 | 0 | 0 | 0 | 0 |
| 0 | 0 | 0 | 0 | 0 |
| 0 | 0 | 0 | 0 | 0 |
| 0.117922189 | 0.003381086 | 0.104931241 | 0 | 0.011100831 |
| 0 | 0 | 0 | 0 | 0.16774657 |
| 0 | 0 | 0 | 0 | 0 |
| 0.141059804 | 0 | 0 | 0 | 0 |
| 0 | 0 | 0 | 0 | 0.005411662 |
| 0 | 0 | 0 | 0 | 0 |
| 0.01399798 | 0.141038155 | 0 | 0 | 0 |
| 0 | 0 | 0 | 0 | 0 |
| 0 | 0.004092533 | 0 | 0 | 0 |
| 0 | 0 | 0 | 0 | 0 |
| 0 | 0 | 0 | 0 | 0 |

| | | | | |
|---|---|---|---|---|
| 0 | 0 | 0 | 0 | 0 |
| 0.050135552 | 0 | 0 | 0 | 0.049738417 |
| 0 | 0 | 0 | 0 | 0 |
| 0 | 0.004112141 | 0.001651931 | 0 | 0 |
| 0 | 0 | 0.002188771 | 0.034386488 | 0 |
| 0.032890215 | 0.001093836 | 0 | 0.005547383 | 0 |
| 0 | 0 | 0 | 0 | 0.006602128 |
| 0 | 0 | 0 | 0 | 0.002470373 |
| 0 | 0 | 0 | 0 | 0 |
| 0 | 0 | 0 | 0 | 0.004211602 |
| 0 | 0 | 0.007800094 | 0 | 0.016463574 |
| 0 | 0 | 0 | 0 | 0 |
| 0.244622371 | 0.000809703 | 0.281036813 | 0.002346515 | 0.150189929 |
| 0 | 0 | 0 | 0 | 0 |
| 0 | 0 | 0 | 0 | 0 |
| 0 | 0 | 0 | 0 | 0.483673035 |
| 0 | 0 | 0 | 0 | 0 |
| 0 | 0 | 0 | 0 | 0 |
| 0 | 0.016034572 | 0 | 0 | 0 |
| 0 | 0 | 0 | 0 | 0 |
| 0 | 0 | 0 | 0 | 0 |
| 0 | 0 | 0 | 0 | 0 |
| 0 | 0.015139921 | 0.163663759 | 0 | 0 |
| 0 | 0 | 0 | 0 | 0.003479142 |
| 0 | 0 | 0 | 0 | 0 |
| 0 | 0 | 0 | 0 | 0.011463386 |
| 0 | 0 | 0 | 0 | 0.00065005 |
| 0 | 0 | 0 | 0 | 0 |
| 0 | 0 | 0 | 0 | 0 |
| 0 | 0 | 0 | 0 | 0 |
| 0.032231649 | 0.018545264 | 0 | 0 | 0.008092025 |
| 0 | 0 | 0.003608626 | 0 | 0 |
| 0 | 0 | 0 | 0 | 0 |
| 0 | 0 | 0 | 0 | 0 |
| 0 | 0 | 0 | 0 | 0.099642525 |
| 0 | 0.146394559 | 0 | 0 | 0 |
| 0 | 0.079822352 | 0 | 0 | 0 |
| 0 | 0 | 0 | 0 | 0 |
| 0.001640313 | 0.006437161 | 0.001870188 | 0.035172203 | 0.016046284 |
| 0 | 0 | 0 | 0 | 0 |
| 0 | 0 | 0 | 0 | 0 |
| 0 | 0 | 0 | 0 | 0 |
| 0 | 0 | 0 | 0 | 1.443509287 |
| 0 | 0 | 0 | 0 | 0 |
| 0 | 0 | 0 | 0 | 2.830261573 |
| 0.365130821 | 0.070972464 | 0.083552634 | 0.024448835 | 0.413942076 |
| 0 | 0 | 0 | 0 | 0 |
| 0 | 0 | 0 | 0 | 0 |

| | | | | |
|---|---|---|---|---|
| 0 | 0 | 0 | 0 | 0 |
| 0 | 0 | 0 | 0 | 0 |
| 0 | 0 | 0 | 0 | 0.122413965 |
| 0 | 0 | 0 | 0 | 0.019894295 |
| 0 | 0 | 0 | 0 | 0 |
| 0.006145188 | 0 | 0 | 0 | 0 |
| 0 | 0 | 0 | 0 | 0 |
| 0 | 0 | 0 | 0 | 0 |
| 0 | 0 | 0 | 0 | 0.008116404 |
| 0.018486231 | 0.001147886 | 0 | 0.001330628 | 0 |
| 0.005156593 | 0 | 0 | 0 | 0 |
| 0.004470276 | 0 | 0.001510147 | 0 | 0 |
| 0 | 0 | 0 | 0 | 0 |
| 0 | 0 | 0 | 0 | 0 |
| 0 | 0 | 0 | 0 | 0 |
| 0 | 0 | 0 | 0 | 0 |
| 0.001222576 | 0 | 0 | 0.206011581 | 0 |
| 0 | 0 | 0 | 0 | 0 |
| 0 | 0 | 0 | 0 | 0 |
| 0.004853149 | 0 | 0 | 0 | 0.002532037 |
| 0 | 0 | 0 | 0 | 0 |
| 0 | 0 | 0 | 0 | 0 |
| 0 | 0 | 0 | 0 | 0 |
| 0 | 0 | 0 | 0 | 0 |
| 0 | 0 | 0 | 0 | 0 |
| 0 | 0 | 0 | 0 | 0 |
| 0 | 0 | 0 | 0 | 0 |
| 0 | 0 | 0 | 0 | 0.008406379 |
| 0 | 0 | 0 | 0 | 0 |
| 0 | 0 | 0 | 0 | 0 |
| 0.053254061 | 0.005577957 | 0.014020879 | 0.00251454 | 0.03045394 |
| 0 | 0 | 0 | 0 | 0 |
| 0.023105002 | 2.663168247 | 0.002007082 | 0.00792473 | 0.002525723 |
| 0.00652108 | 0.073022558 | 0 | 0.012360433 | 0.006100581 |
| 0 | 0 | 0 | 0 | 0 |
| 0 | 0 | 0 | 0 | 0 |
| 0 | 0 | 0 | 0 | 0 |
| 0 | 0 | 0 | 0 | 0 |
| 0 | 0 | 0 | 0 | 0 |
| 0 | 0 | 0 | 0 | 0 |
| 0 | 0 | 0 | 0 | 0 |
| 1.409508913 | 0.164819627 | 0.004036287 | 0.006519603 | 0.059314303 |
| 0.307222033 | 0.114679195 | 0 | 0 | 0.02784601 |
| 0 | 0 | 0 | 0 | 0.001772251 |
| 0 | 0 | 0 | 0 | 0 |
| 0.018803643 | 0 | 0.095606552 | 0 | 0.35384085 |
| 0 | 0.001025003 | 0 | 0 | 0 |
| 0 | 0 | 0 | 0 | 0 |

| | | | | |
|---|---|---|---|---|
| 0 | 0 | 0 | 0 | 0 |
| 0 | 0 | 0 | 0 | 0.001205471 |
| 0.00782691 | 0.004236623 | 0.003008784 | 0.044250952 | 0.006477338 |
| 0 | 0 | 0 | 0 | 0 |
| 0.249976446 | 0 | 0 | 0 | 0.008544783 |
| 1.027840802 | 0.211764264 | 0 | 0 | 0 |
| 0 | 0 | 0 | 0 | 0 |
| 0 | 0 | 0 | 0 | 0 |
| 0 | 0 | 0 | 0 | 0 |
| 0 | 0 | 0 | 0 | 0 |
| 0 | 0 | 0 | 0 | 0 |
| 0 | 0 | 0 | 0 | 0 |
| 0 | 0 | 0 | 0 | 0 |
| 0 | 0 | 0 | 0 | 0 |
| 0 | 0 | 0 | 0 | 0 |
| 0 | 0 | 0 | 0 | 0 |
| 0 | 0 | 0 | 0 | 0.030121651 |
| 0 | 0 | 0 | 0 | 0 |
| 0 | 0 | 0 | 0 | 0 |
| 0 | 0 | 0 | 0 | 0 |
| 0 | 0 | 0 | 0 | 0 |
| 0 | 0 | 0 | 0 | 0 |
| 0.005188014 | 0 | 0 | 0 | 0 |
| 0 | 0 | 0 | 0 | 0 |
| 0 | 0 | 0 | 0 | 0.046419453 |
| 0 | 0 | 0 | 0 | 0 |
| 0 | 0 | 0 | 0 | 0 |
| 0.333612763 | 0 | 0 | 0 | 0.060925265 |
| 0 | 0 | 0 | 0 | 0 |
| 0 | 0 | 0 | 0 | 0 |
| 0 | 0 | 0.003084314 | 0 | 0 |
| 0.00156979 | 0 | 0.001789782 | 0 | 0 |
| 0 | 0 | 0 | 0 | 0 |
| 0 | 0 | 0 | 0 | 0 |
| 0 | 0 | 0 | 0 | 0 |
| 0 | 0 | 0 | 0 | 0 |
| 0 | 0 | 0.001382164 | 0 | 0 |
| 0 | 0.963896974 | 0 | 0 | 0 |
| 0.581838845 | 0 | 0 | 0 | 1.150021077 |
| 0 | 0 | 0 | 0 | 0 |
| 0 | 0 | 0 | 0 | 0 |
| 0 | 0 | 0 | 0 | 0.056484681 |
| 0.380641159 | 0.008151833 | 0.008875986 | 0 | 0 |
| 0 | 0 | 0 | 0 | 0 |
| 0 | 0 | 0 | 0 | 0 |
| 0 | 0 | 0 | 0 | 0 |
| 0.013741746 | 0.014830033 | 0.005222508 | 0 | 0 |
| 0 | 0 | 0 | 0 | 0 |

| | | | | |
|---|---|---|---|---|
| 0 | 0 | 0 | 0 | 0 |
| 0.073909856 | 0.006592 | 0.022844796 | 0 | 0.004297124 |
| 0 | 0 | 0 | 0 | 0.085545291 |
| 0 | 0 | 0 | 0 | 0 |
| 0 | 0 | 0 | 0 | 0 |
| 0 | 0 | 0 | 0 | 0 |
| 0 | 0 | 0 | 0 | 0 |
| 0 | 0 | 0 | 0 | 0 |
| 0 | 0 | 0 | 0 | 0 |
| 0 | 0 | 0 | 0 | 0 |
| 0 | 0 | 0 | 0 | 0 |
| 0 | 0 | 0 | 0 | 0.017393104 |
| 0 | 0 | 0 | 0 | 0.492974427 |
| 0 | 0 | 0 | 0 | 0 |
| 0 | 0 | 0 | 0 | 0.003665909 |
| 0 | 0 | 0 | 0 | 0 |
| 0 | 0 | 0 | 0 | 0 |
| 0 | 0 | 0 | 0 | 0 |
| 0 | 0 | 0 | 0 | 0 |
| 0 | 0 | 0 | 0 | 0 |
| 0 | 0 | 0 | 0 | 0 |
| 0.193214497 | 0.003385004 | 0.014793706 | 0.008300541 | 0.290509391 |
| 0 | 0 | 0 | 0 | 0 |
| 1.448836656 | 0.19417336 | 0 | 0.028498394 | 0 |
| 0 | 0 | 0 | 0 | 0 |
| 0 | 0 | 0 | 0 | 0 |
| 0 | 0 | 0 | 0 | 0 |
| 0 | 0 | 0 | 0 | 0 |
| 0 | 0 | 0 | 0 | 0 |
| 0 | 0 | 0 | 0 | 0.048977097 |
| 0 | 0 | 0 | 0 | 0 |
| 0 | 0 | 0 | 0 | 0 |
| 0 | 0 | 0 | 0 | 0 |
| 0 | 0 | 0 | 0 | 0 |
| 0.000722918 | 0.00094566 | 0.075951073 | 0 | 2.132760982 |
| 0 | 0 | 0 | 0 | 0 |
| 0 | 0 | 0 | 0 | 0 |
| 0 | 0 | 0 | 0 | 0 |
| 0.002915424 | 0.001560155 | 0.059361827 | 3.647704158 | 0.580494115 |
| 0 | 0 | 0 | 0 | 0 |
| 0 | 0.40730882 | 0.128127095 | 0 | 0.033723967 |
| 0 | 0 | 0 | 0 | 0 |
| 0 | 0 | 0 | 0 | 0 |
| 0 | 0 | 0 | 0 | 0 |
| 0 | 0 | 0 | 0 | 0 |
| 0.618719092 | 0.38278925 | 3.325770955 | 0.722529094 | 0.005678183 |
| 0 | 0 | 0 | 0 | 0 |
| 0 | 0 | 0 | 0 | 0 |

| | | | | |
|---|---|---|---|---|
| 0 | 0 | 0 | 0 | 0 |
| 0 | 0 | 0 | 0 | 0 |
| 0 | 0 | 0 | 0 | 0 |
| 0 | 0 | 0 | 0 | 0 |
| 0 | 0 | 0.001045662 | 0 | 0 |
| 0 | 0 | 0 | 0 | 0 |
| 0 | 0 | 0 | 0 | 0 |
| 0 | 0 | 0 | 0 | 0.023935524 |
| 0 | 0.007233289 | 0 | 0 | 0 |
| 0.029004749 | 0 | 0.565788425 | 0.041106866 | 0.073303194 |
| 0.056969714 | 0 | 0.018487038 | 0 | 0.014934632 |
| 0 | 0 | 0 | 0 | 0 |
| 0.021327297 | 0.00076551 | 0.003426662 | 0 | 0 |
| 0 | 0 | 0 | 0 | 0 |
| 0 | 0 | 0 | 0 | 0 |
| 0 | 0 | 0 | 0 | 0 |
| 0 | 0 | 0 | 0 | 0 |
| 0 | 0 | 0 | 0 | 0 |
| 0 | 0 | 0 | 0 | 0 |
| 1.148922371 | 0.151488536 | 1.351606425 | 3.316404114 | 0.048553308 |
| 0 | 0 | 0 | 0 | 0 |
| 0 | 0 | 0 | 0 | 0 |
| 0 | 0 | 0 | 0 | 0 |
| 0 | 0 | 0 | 0 | 0 |
| 0 | 0 | 0 | 0 | 0 |
| 0 | 0 | 0 | 0 | 0 |
| 0 | 0 | 0 | 0 | 0 |
| 0 | 0 | 0 | 0 | 0 |
| 0 | 0 | 0 | 0 | 0 |
| 0 | 0 | 0.143209805 | 0 | 0 |
| 0 | 0 | 0 | 0 | 0 |
| 0 | 0 | 0 | 0 | 0 |
| 0 | 0 | 0 | 0 | 0 |
| 0 | 0 | 0 | 0 | 0 |
| 0 | 0 | 0 | 0 | 0 |
| 0 | 0 | 0 | 0 | 0 |
| 0 | 0 | 0 | 0 | 0 |
| 0 | 0 | 0 | 0 | 0 |
| 0 | 0 | 0 | 0 | 0 |
| 0 | 0 | 0 | 0 | 0 |
| 0 | 0 | 0 | 0 | 0 |
| 0 | 0 | 0 | 0 | 0 |
| 0 | 0 | 0 | 0 | 0.001003966 |
| 0 | 0 | 0 | 0 | 0 |
| 0.002826603 | 0.002559825 | 0.049360036 | 0 | 0.0319902 |
| 0 | 0 | 0 | 0 | 0.01225008 |
| 0 | 0 | 0 | 0 | 0 |
| 0 | 0 | 0 | 0 | 0 |

| | | | | |
|---|---|---|---|---|
| 0 | 0 | 0 | 0 | 0 |
| 0 | 0 | 0 | 0 | 0 |
| 0 | 0 | 0 | 0 | 0 |
| 0 | 0 | 0 | 0 | 0 |
| 0 | 0 | 0 | 0 | 0.001885405 |
| 0.014284222 | 0.001114755 | 0.047871041 | 0.002769049 | 0.010840024 |
| 0 | 0 | 0 | 0 | 0 |
| 0 | 0 | 0 | 0 | 0 |
| 0 | 0 | 0 | 0 | 0 |
| 0 | 0 | 0 | 0 | 0 |
| 0 | 0 | 0 | 0 | 0 |
| 0 | 0 | 0 | 0 | 0 |
| 0 | 0 | 0 | 0 | 0 |
| 0 | 0 | 0 | 0 | 0.044393431 |
| 0 | 0 | 0 | 0 | 0 |
| 0 | 0 | 0 | 0 | 0 |
| 0 | 0 | 0 | 0 | 0.302834982 |
| 0 | 0 | 0 | 0 | 0 |
| 0 | 0 | 0 | 0 | 0 |
| 0 | 0 | 0 | 0 | 0 |
| 0 | 0 | 0 | 0 | 0 |
| 0 | 0 | 0 | 0 | 0 |
| 0 | 0 | 0 | 0 | 0 |
| 0 | 0 | 0 | 0 | 0 |
| 0 | 0 | 0 | 0 | 0 |
| 0 | 0 | 0.006690161 | 0.003609381 | 0.003973973 |
| 0 | 0 | 0 | 0 | 0 |
| 0 | 0 | 0 | 0 | 0 |
| 0 | 0 | 0 | 0 | 0 |
| 0 | 0 | 0 | 0 | 0 |
| 0 | 0 | 0 | 0 | 0 |
| 0 | 0 | 0 | 0 | 0 |
| 0 | 0 | 0 | 0 | 0 |
| 0 | 0 | 0 | 0 | 0 |
| 0 | 0 | 0 | 0 | 0 |
| 0 | 0 | 0 | 0 | 0 |
| 0 | 0 | 0 | 0 | 0 |
| 0 | 0 | 0 | 0 | 0 |
| 0 | 0 | 0 | 0 | 0.293361625 |
| 0 | 0 | 0 | 0 | 0 |
| 0 | 0 | 0 | 0 | 0 |
| 0 | 0 | 0 | 0 | 0 |
| 0 | 0 | 0 | 0 | 0 |
| 0 | 0 | 0 | 0 | 0 |
| 0 | 0 | 0 | 0 | 0 |
| 0 | 0 | 0 | 0 | 0 |
| 0 | 0 | 0 | 0 | 0 |
| 0 | 0 | 0 | 0 | 0 |

| | | | | |
|---|---|---|---|---|
| 0 | 0 | 0 | 0 | 0 |
| 0.002907698 | 0.404370864 | 0 | 0 | 0 |
| 0 | 0 | 0 | 0 | 0 |
| 0 | 0 | 0 | 0 | 0 |
| 0 | 0 | 0 | 0 | 0 |
| 0 | 0 | 0 | 0 | 0 |
| 0 | 0 | 0 | 0 | 0 |
| 0 | 0 | 0 | 0 | 0 |
| 0 | 0 | 0 | 0 | 0 |
| 0 | 0 | 0 | 0 | 0.008239084 |
| 0.0019037 | 0 | 0.003858642 | 0.070137945 | 0.002483047 |
| 0 | 0 | 0 | 0 | 0 |
| 0.780308291 | 0 | 0 | 0 | 0.082665797 |
| 0 | 0 | 0 | 0 | 0 |
| 0 | 0 | 0 | 0 | 0 |
| 0 | 0 | 0 | 0 | 0 |
| 0 | 0 | 0 | 0 | 0 |
| 0 | 0 | 0 | 0 | 0 |
| 0 | 0 | 0 | 0 | 0 |
| 0 | 0 | 0.002006767 | 0 | 0 |
| 0.283783183 | 0 | 2.058009875 | 0.04188746 | 0 |
| 0 | 0 | 0 | 0 | 0 |
| 0 | 0 | 0 | 0 | 0.001008846 |
| 1.642800073 | 0 | 1.369682719 | 0.078121402 | 0.311482637 |
| 0 | 0 | 0 | 0 | 0 |
| 0 | 0 | 0 | 0 | 0 |
| 0 | 0 | 0 | 0 | 0 |
| 0 | 0 | 0 | 0 | 0 |
| 0 | 0 | 0 | 0 | 0 |
| 0 | 0 | 0 | 0 | 0 |
| 0 | 0 | 0.002162157 | 0 | 0 |
| 0 | 0 | 0 | 0 | 0 |
| 0 | 0 | 0 | 0 | 0.020677405 |
| 0 | 0 | 0 | 0 | 0 |

| PHL018-0147_V1 | PHL018-0088_V1 | PHL018-0155_V2 | PHL018-0169_V1 | PHL018-0147_V2 |
|---|---|---|---|---|
| CTRL | EYCF | CTRL | REF | CTRL |
| 0.007659387 | 2.209938279 | 0.066239226 | 0.295575935 | 0.166711827 |
| 0.039261073 | 0.788685914 | 0.899947781 | 0.270644513 | 1.433634212 |
| 3.189270655 | 0.637499873 | 0 | 28.50227506 | 8.540530731 |
| 0 | 0 | 0 | 0 | 0 |
| 0 | 0 | 0 | 0 | 0 |
| 0 | 0 | 0 | 0 | 0 |
| 0 | 0 | 0 | 0 | 0 |
| 0 | 0 | 0 | 0 | 0 |
| 0 | 0 | 0 | 0 | 0 |
| 0 | 0 | 0 | 0 | 0 |
| 0 | 0 | 0 | 0 | 0 |
| 0 | 0 | 0 | 0 | 0 |
| 0 | 0 | 0 | 0 | 0 |
| 0.035619445 | 0.558362526 | 7.816006341 | 3.690519693 | 0.731288139 |
| 0 | 0 | 0 | 0 | 0 |
| 0 | 0 | 0 | 0 | 0 |
| 0 | 0 | 0 | 0 | 0 |
| 0 | 0 | 0 | 0 | 0 |
| 0 | 0 | 0 | 0 | 0 |
| 0 | 0 | 0 | 0 | 0 |
| 0 | 0 | 0 | 0 | 0 |
| 0 | 0 | 0 | 0 | 0 |
| 0 | 0 | 0 | 0 | 0 |
| 0 | 0 | 0 | 0 | 0 |
| 0.363790222 | 0 | 0 | 0 | 0 |
| 0 | 0 | 0 | 0 | 0 |
| 0 | 0 | 0 | 0 | 0 |
| 0 | 0 | 0 | 0 | 0 |
| 0 | 0 | 0 | 0 | 0 |
| 0 | 0 | 0 | 0 | 0 |
| 0 | 0 | 0 | 0 | 0 |
| 0 | 0 | 0 | 0 | 0 |
| 0 | 0 | 0 | 0 | 0 |
| 0 | 0 | 0 | 0 | 0 |
| 0 | 0 | 0 | 0 | 0 |
| 1.167225363 | 0 | 12.77824807 | 0 | 2.80358828 |
| 0 | 0 | 0 | 0 | 0 |
| 0 | 0 | 0 | 0 | 0 |
| 0 | 0 | 0 | 0 | 0 |
| 0 | 0 | 0 | 0 | 0 |
| 0 | 0 | 0 | 0 | 0 |
| 0 | 0.001235717 | 0.069090791 | 0.126274586 | 0.777707569 |
| 11.78685988 | 0.01406475 | 0.287705634 | 1.213927474 | 0.144636506 |
| 0.001949933 | 0.001418388 | 0.003471843 | 0.089699196 | 0.002542866 |
| 0.001124666 | 0 | 0.004742671 | 0.007407131 | 0.002444422 |
| 0 | 0 | 0 | 0 | 0 |

| | | | | |
|---|---|---|---|---|
| 0.030245039 | 0.014767872 | 0.000512217 | 0 | 0 |
| 0 | 0 | 0 | 0.081283768 | 0.018525598 |
| 0 | 0.325247812 | 0 | 0.003360416 | 0.002703111 |
| 0.237917488 | 0.33825361 | 0.313676962 | 0.998137669 | 0.401337911 |
| 0 | 0.00565099 | 0 | 0 | 0 |
| 0 | 0 | 0 | 0 | 0 |
| 0 | 0.001675324 | 0 | 0.03049308 | 2.262467507 |
| 0.007513313 | 0 | 0 | 0 | 0 |
| 0 | 0 | 0 | 0 | 0 |
| 0 | 0 | 0 | 0 | 0 |
| 0 | 0 | 0 | 0 | 0 |
| 0.636207291 | 0.147198699 | 0.011017537 | 0 | 0.82430608 |
| 0 | 0 | 0.000872549 | 0 | 0 |
| 0 | 0.043593258 | 0 | 0.332837332 | 0.08574165 |
| 0 | 0 | 0 | 0.017121229 | 0.003156146 |
| 0 | 0 | 0 | 0 | 0 |
| 0.287551734 | 0.418058049 | 0 | 0.002940323 | 0.437488279 |
| 0 | 0 | 0 | 0 | 0 |
| 0 | 0 | 0 | 0 | 0 |
| 0 | 0 | 0 | 0 | 0 |
| 0.01240406 | 0.025567994 | 0.154730803 | 0.140311756 | 0.14606894 |
| 0.017182923 | 0.005951865 | 0 | 0.084537403 | 0.032574412 |
| 0 | 0.141302538 | 0 | 0 | 0 |
| 0 | 0 | 0 | 0 | 0 |
| 0 | 0 | 0 | 0.051703151 | 0.004124617 |
| 0 | 0 | 0 | 0 | 0 |
| 0 | 0 | 0.000640884 | 0 | 0 |
| 0 | 0 | 0 | 0 | 0 |
| 0 | 0.022081053 | 0.001373287 | 7.450813363 | 1.628131368 |
| 0 | 0 | 0 | 0 | 0 |
| 0 | 0 | 0 | 0.000657599 | 0 |
| 0 | 0 | 0 | 0 | 0 |
| 0 | 0 | 0 | 0 | 0 |
| 0 | 0.003118396 | 0 | 0 | 0 |
| 0 | 0 | 0 | 0 | 0 |
| 0 | 0 | 0 | 0.000899519 | 0 |
| 0 | 0 | 0 | 0 | 0 |
| 13.76178191 | 0.014193948 | 0 | 0.038705334 | 0.01610187 |
| 0 | 0 | 0 | 0 | 0 |
| 0 | 0 | 0 | 0 | 0 |
| 0 | 0 | 0 | 0 | 0 |
| 0 | 0 | 0 | 0 | 0 |
| 0 | 0 | 0 | 0 | 0 |
| 0 | 0 | 0 | 0 | 0 |
| 0 | 0 | 0 | 0 | 0 |
| 0 | 0 | 0 | 0 | 0 |
| 0 | 0 | 0 | 0 | 0 |
| 0 | 0 | 0 | 0 | 0 |

| | | | | |
|---|---|---|---|---|
| 0 | 0 | 0 | 0 | 0 |
| 0 | 0 | 0 | 0 | 0 |
| 0 | 0 | 0 | 0 | 0 |
| 0 | 0 | 0 | 0 | 0 |
| 0.001521228 | 0 | 0 | 0.005064506 | 0 |
| 0 | 0 | 0 | 0 | 0 |
| 0 | 0 | 0 | 0 | 0 |
| 0 | 0 | 0 | 0 | 0 |
| 0 | 0.004602355 | 0 | 0 | 0.002206006 |
| 0 | 0 | 0 | 0 | 0 |
| 0 | 0 | 0 | 0 | 0 |
| 0 | 0 | 0 | 0.006729628 | 0.037252791 |
| 0 | 0 | 0 | 0 | 0 |
| 0 | 0 | 0 | 0 | 0 |
| 0 | 0 | 0 | 0 | 0 |
| 0 | 0 | 0 | 0.018725361 | 0 |
| 0 | 0.002354138 | 0 | 0 | 0 |
| 0 | 0 | 0 | 0 | 0 |
| 0 | 0.509602848 | 0.239606797 | 0 | 0 |
| 0 | 0.156443075 | 0 | 0 | 0 |
| 0 | 0.335200093 | 0 | 0 | 0 |
| 0 | 0 | 0 | 0 | 0 |
| 0 | 0.003146208 | 0 | 0 | 0 |
| 0 | 0 | 0 | 0 | 0 |
| 0 | 0 | 0 | 0 | 0 |
| 0 | 0 | 0 | 0 | 0 |
| 0 | 0 | 0 | 0 | 0 |
| 0 | 0 | 0 | 0 | 0 |
| 0 | 0 | 0 | 0 | 0 |
| 0 | 0 | 0 | 0 | 0 |
| 0.093922601 | 0.009227584 | 0 | 0 | 0.084438637 |
| 0.01218226 | 0.002233971 | 0 | 0.014340325 | 0.002141579 |
| 0 | 0 | 0 | 0 | 0 |
| 0 | 0 | 0 | 0 | 0 |
| 0 | 0 | 0 | 0 | 0 |
| 0 | 0 | 0 | 0 | 0 |
| 0 | 0 | 0 | 0 | 0 |
| 0 | 0 | 0 | 0 | 0 |
| 0.063912868 | 0 | 0 | 0 | 0 |
| 0 | 0 | 0 | 0 | 0 |
| 0 | 0 | 0 | 0 | 0 |
| 0 | 0 | 0 | 0 | 0 |
| 0 | 0 | 0 | 0.00419624 | 0.001350178 |
| 0 | 0 | 0 | 0 | 0 |
| 0 | 0 | 0 | 0 | 0 |
| 0 | 0 | 0 | 0 | 0 |
| 0 | 0 | 0 | 0 | 0 |
| 0 | 0 | 0 | 0 | 0 |

| | | | | |
|---|---|---|---|---|
| 0 | 0 | 0 | 0 | 0 |
| 0 | 0 | 0 | 0 | 0 |
| 0 | 0 | 0 | 0 | 0 |
| 0 | 0 | 0 | 0 | 0 |
| 0.132073197 | 0.066110234 | 0 | 0.023067316 | 0 |
| 0 | 0 | 0 | 0 | 0 |
| 0 | 0 | 0 | 0 | 0 |
| 0 | 0 | 0 | 0 | 0 |
| 0 | 0 | 0 | 0 | 0 |
| 0 | 0 | 0 | 0 | 0 |
| 0 | 0 | 0 | 0 | 0 |
| 0 | 0 | 0 | 0 | 0 |
| 0 | 0 | 0 | 0 | 0 |
| 0 | 0 | 0 | 0 | 0 |
| 0 | 0 | 0 | 0 | 0 |
| 0 | 0 | 0 | 0 | 0 |
| 0 | 0 | 0 | 0 | 0 |
| 0 | 0 | 0 | 0 | 0 |
| 0 | 0 | 0 | 0 | 0 |
| 0 | 0 | 0 | 0 | 0 |
| 0 | 0 | 0 | 0 | 0 |
| 0 | 0 | 0 | 0 | 0 |
| 0 | 0 | 0 | 0 | 0 |
| 0 | 0 | 0 | 0 | 0 |
| 0 | 0 | 0 | 0 | 0.047027047 |
| 0 | 0 | 0 | 0 | 0 |
| 0 | 0.016177935 | 0 | 0 | 0 |
| 0 | 0 | 0 | 0 | 0 |
| 0 | 0 | 0 | 0 | 0 |
| 0 | 0 | 0 | 0 | 0 |
| 0 | 0 | 0 | 0 | 0 |
| 0 | 0.005369833 | 0 | 0.024294864 | 0 |
| 0 | 0 | 0 | 0 | 0 |
| 0 | 0 | 0 | 0 | 0 |
| 0 | 0 | 0 | 0 | 0 |
| 0 | 0 | 0 | 0 | 0 |
| 0 | 0 | 0 | 0 | 0 |
| 0 | 0 | 0 | 0 | 0 |
| 0 | 0 | 0 | 0 | 0 |
| 0 | 0 | 0 | 0 | 0 |
| 0 | 0 | 0 | 0 | 0 |
| 0 | 0 | 0 | 0 | 0 |
| 0 | 0 | 0 | 0 | 0 |
| 0 | 0 | 0 | 0 | 0 |
| 0 | 0 | 0 | 0 | 0 |
| 0 | 0 | 0 | 0 | 0 |
| 0 | 0 | 0 | 0 | 0 |
| 0 | 0 | 0 | 0 | 0 |

| | | | | |
|---|---|---|---|---|
| 0 | 0 | 0 | 0 | 0 |
| 0 | 0 | 0 | 0 | 0 |
| 0 | 0 | 0 | 0 | 0 |
| 0 | 0 | 0 | 0 | 0 |
| 0 | 0 | 0 | 0 | 0 |
| 0 | 0 | 0 | 0 | 0 |
| 0 | 0 | 0 | 0 | 0 |
| 0 | 0 | 0 | 0 | 0 |
| 0 | 0 | 0 | 0 | 0 |
| 0 | 0 | 0 | 0 | 0 |
| 0 | 0 | 0 | 0 | 0 |
| 0 | 0 | 0 | 0 | 0 |
| 0.000791172 | 0.004110723 | 0 | 0.002605357 | 0.001504636 |
| 0 | 0 | 0 | 0 | 0 |
| 0 | 0 | 0 | 0 | 0 |
| 0 | 0 | 0 | 0 | 0 |
| 0 | 0 | 0 | 0 | 0 |
| 0 | 0 | 0 | 0 | 0.001809541 |
| 0 | 0 | 0 | 0 | 0 |
| 0 | 0 | 0 | 0 | 0 |
| 0.039941149 | 0.006148852 | 0.049212097 | 0.034849189 | 0.002813308 |
| 0 | 0 | 0 | 0 | 0 |
| 0 | 0.004383629 | 0 | 0 | 0.00105793 |
| 0 | 0 | 0 | 0.002735321 | 0 |
| 0 | 0 | 0 | 0 | 0 |
| 0 | 0 | 0 | 0 | 0 |
| 0 | 0.00268571 | 0.000605492 | 0.000872918 | 0.003744924 |
| 0 | 0 | 0 | 0 | 0 |
| 0 | 0 | 0 | 0 | 0 |
| 0 | 0 | 0 | 0 | 0 |
| 0 | 0 | 0 | 0.253866594 | 0 |
| 0 | 0 | 0 | 0 | 0 |
| 0 | 0 | 0 | 0 | 0 |
| 0 | 0.197988897 | 0 | 0 | 0.318074001 |
| 0 | 0.04925949 | 0.009921694 | 0.001462886 | 0 |
| 0 | 0 | 0 | 0 | 0 |
| 0 | 0 | 0 | 0 | 0 |
| 0 | 0 | 0 | 0 | 0 |
| 0 | 0 | 0 | 0 | 0 |
| 0 | 0 | 0 | 0 | 0 |
| 0 | 0 | 0 | 0 | 0 |
| 0 | 0 | 0 | 0 | 0.050142351 |
| 0 | 0 | 0 | 0 | 0 |
| 0 | 0 | 0 | 0 | 0 |
| 0 | 0 | 0 | 0 | 0 |
| 0 | 0.072612294 | 0 | 0 | 0 |
| 0 | 0 | 0 | 0.018215045 | 0 |
| 0.010117209 | 0.002494675 | 0.000482077 | 0.003822472 | 0.000931754 |

| | | | | |
|---|---|---|---|---|
| 0 | 0 | 0 | 0 | 0 |
| 0 | 0 | 0 | 0 | 0 |
| 0 | 0 | 0 | 0.00158768 | 0 |
| 0 | 0 | 0 | 0 | 0 |
| 0 | 0 | 0 | 0 | 0 |
| 0 | 0 | 0 | 0 | 0 |
| 0 | 0 | 0 | 0 | 0.001856875 |
| 0 | 0 | 0 | 0.004495806 | 0.02062696 |
| 0 | 0 | 0 | 0 | 0 |
| 0 | 0 | 0 | 0 | 0 |
| 0 | 0 | 0 | 0 | 0 |
| 0 | 0 | 0 | 0 | 0 |
| 0 | 0 | 0 | 0 | 0 |
| 0 | 0 | 0 | 0 | 0 |
| 0 | 0 | 0 | 0 | 0 |
| 0 | 0 | 0 | 0 | 0 |
| 0 | 0 | 0 | 0 | 0 |
| 0 | 0 | 0 | 0 | 0 |
| 0.052317636 | 0 | 0.001470808 | 0.008481663 | 0 |
| 0 | 0.885084551 | 0 | 0 | 0 |
| 0 | 0.001871217 | 0.008196237 | 0.041704389 | 0.013418763 |
| 0 | 0 | 0 | 0 | 0 |
| 0 | 0 | 0 | 0 | 0 |
| 0 | 0 | 0 | 0 | 0 |
| 0 | 0.002124325 | 0 | 0.007249776 | 0.006664802 |
| 0 | 0 | 0 | 0 | 0 |
| 0 | 0 | 0 | 0 | 0 |
| 0 | 0 | 0 | 0 | 0 |
| 0 | 0 | 0 | 0.283661229 | 0.147877385 |
| 0 | 0 | 0 | 0 | 0 |
| 0.05466521 | 0.054249971 | 0 | 0.94276386 | 0.398166937 |
| 0 | 0 | 0 | 0 | 0 |
| 0 | 0 | 0 | 0 | 0 |
| 0.001713052 | 0 | 0 | 0.004959242 | 0 |
| 0 | 0 | 0 | 0 | 0 |
| 0 | 0 | 0 | 0 | 0 |
| 0 | 0 | 0 | 0 | 0 |
| 0 | 0 | 0 | 0 | 0 |
| 0 | 0 | 0 | 0 | 0 |
| 0 | 0 | 0 | 0 | 0 |
| 0 | 0 | 0 | 0 | 0 |
| 0.0077124 | 0.100179031 | 0.002890921 | 0.032820997 | 0.025353466 |
| 0 | 0 | 0 | 0 | 0 |
| 0 | 0 | 0 | 0 | 0 |
| 0 | 0 | 0 | 0 | 0 |
| 0 | 0 | 0 | 0 | 0 |
| 0 | 0 | 0 | 0 | 0 |
| 0 | 0 | 0 | 0 | 0 |

| | | | | |
|---|---|---|---|---|
| 0 | 0 | 0 | 0 | 0 |
| 0 | 0 | 0 | 0 | 0 |
| 0.001452628 | 0.000754748 | 0 | 0.0841415 | 0 |
| 0 | 0 | 0 | 0 | 0 |
| 0 | 0 | 0 | 0 | 0 |
| 0 | 0 | 0 | 0 | 0 |
| 0 | 0 | 0 | 0 | 0 |
| 0 | 0 | 0 | 0 | 0 |
| 0 | 0 | 0 | 0 | 0 |
| 0.000885708 | 0 | 0 | 0.002019232 | 0 |
| 0 | 0 | 0 | 0 | 0 |
| 0 | 0 | 0 | 0 | 0 |
| 0 | 0 | 0 | 0 | 0 |
| 0.001337847 | 0 | 0 | 0.024671228 | 0 |
| 0.006625305 | 0.002983357 | 0.007450302 | 0.006489266 | 0.002639974 |
| 0 | 0 | 0 | 0 | 0 |
| 0.003774471 | 0 | 0.002122238 | 0.003569487 | 0.002734558 |
| 0 | 0 | 0 | 0.000791117 | 0 |
| 0 | 0 | 0 | 0 | 0 |
| 0 | 0 | 0 | 0 | 0 |
| 0 | 0 | 0 | 0 | 0 |
| 0.009767507 | 0.003304613 | 0 | 0.003912705 | 0 |
| 0 | 0 | 0 | 0 | 0 |
| 0 | 0 | 0.002309913 | 0 | 0 |
| 0 | 0 | 0 | 0 | 0 |
| 0 | 0 | 0 | 0 | 0 |
| 0 | 0.167337761 | 0 | 0 | 0 |
| 0 | 0 | 0 | 0 | 0 |
| 0 | 0 | 0 | 0 | 0 |
| 0 | 0 | 0 | 0 | 0.002856596 |
| 0 | 0 | 0 | 0 | 0.001513336 |
| 0 | 0 | 0 | 0 | 0 |
| 0 | 0 | 0 | 0 | 0.002043506 |
| 0 | 0 | 0 | 0 | 0.01280791 |
| 0 | 0 | 0 | 0 | 0 |
| 0 | 0 | 0.001700661 | 0 | 0.005423582 |
| 0 | 0 | 0 | 0 | 0 |
| 0.004566274 | 0.016014485 | 0 | 0.021109491 | 0.007443473 |
| 0 | 0 | 0 | 0 | 0 |
| 0 | 0 | 0 | 0 | 0 |
| 0 | 0 | 0 | 0 | 0 |
| 0 | 0 | 0 | 0 | 0 |
| 0 | 0 | 0 | 0 | 0 |
| 0 | 0 | 0 | 0 | 0 |
| 0 | 0 | 0 | 0 | 0 |
| 0 | 0 | 0 | 0 | 0 |
| 0 | 0 | 0 | 0 | 0 |
| 0 | 0 | 0.001619693 | 0 | 0.028174727 |

| | | | | |
|---|---|---|---|---|
| 0.18185632 | 0.437864434 | 0.004484681 | 0 | 0 |
| 0 | 0 | 0 | 0 | 0 |
| 0 | 0 | 0 | 0.001205369 | 0 |
| 0 | 0 | 0 | 0 | 0 |
| 0 | 0.004726149 | 0.002916129 | 0.006063586 | 0.004682432 |
| 0 | 0 | 0 | 0 | 0 |
| 0 | 0 | 0 | 0 | 0 |
| 0 | 0.002789364 | 0.056597483 | 0.008839428 | 0.001823183 |
| 0 | 0 | 0 | 0 | 0 |
| 0.010029454 | 0.002156293 | 0.163017415 | 0.111784838 | 0.002255033 |
| 0 | 0 | 0 | 0 | 0 |
| 0 | 0.001656053 | 0.001244521 | 0.017224179 | 0.956289677 |
| 0 | 0.010468123 | 0.003209646 | 0.005511861 | 0.012699071 |
| 0 | 0 | 0 | 0.004340568 | 0 |
| 0 | 0 | 0 | 0 | 0 |
| 0 | 0 | 0 | 0 | 0.003286795 |
| 0 | 0.001371308 | 0 | 0 | 0 |
| 0 | 0 | 0 | 0 | 0 |
| 0 | 0 | 0 | 0 | 0 |
| 0 | 0 | 0 | 0 | 0 |
| 0 | 0 | 0 | 0 | 0 |
| 0 | 0 | 0 | 0 | 0 |
| 0 | 0 | 0 | 0 | 0 |
| 0 | 0.245650903 | 0 | 0 | 0 |
| 0 | 0 | 0 | 0 | 0 |
| 0.003285341 | 0 | 0 | 0.000951098 | 0 |
| 1.076012108 | 0 | 0 | 0 | 0 |
| 0 | 0 | 0 | 0 | 0 |
| 0 | 0 | 0 | 0 | 0 |
| 0.00241443 | 0.004265214 | 0 | 0.019571206 | 0 |
| 0 | 0 | 0 | 0 | 0 |
| 0 | 0 | 0 | 0 | 0 |
| 0 | 0 | 0 | 0 | 0 |
| 0 | 0 | 0 | 0 | 0 |
| 0 | 0 | 0 | 0 | 0 |
| 0 | 0 | 0 | 0 | 0 |
| 0 | 0 | 0 | 0.005121294 | 0 |
| 0 | 0 | 0 | 0.005569826 | 0 |
| 0 | 0 | 0 | 0 | 0 |
| 0 | 0 | 0 | 0.00125039 | 0 |
| 0 | 0 | 0 | 0 | 0 |
| 0 | 0 | 0 | 0 | 0 |
| 0 | 0 | 0 | 0 | 0 |
| 0 | 0 | 0.1215351 | 0.004022751 | 0.001438174 |
| 0 | 0 | 0 | 0 | 0 |
| 0 | 0 | 0 | 0 | 0 |
| 0 | 0.001819509 | 0 | 0 | 0.003805655 |
| 0 | 0 | 0 | 0 | 0 |

| | | | | |
|---|---|---|---|---|
| 0 | 0 | 0 | 0 | 0 |
| 0 | 0 | 0 | 0.001652279 | 0.108808116 |
| 0 | 0 | 0 | 0 | 0 |
| 0 | 0 | 0 | 0 | 0 |
| 0 | 0 | 0 | 0 | 0 |
| 0 | 0.002627321 | 0.000646176 | 0 | 0 |
| 0 | 0 | 0 | 0 | 0 |
| 0 | 0 | 0 | 0 | 0 |
| 0 | 0.095269389 | 0 | 0 | 0 |
| 0 | 0 | 0 | 0 | 0 |
| 0 | 0.01329709 | 0 | 0 | 0.01158832 |
| 0 | 0 | 0 | 0 | 0 |
| 0 | 0 | 0 | 0 | 0 |
| 0.02921547 | 0 | 0.002360161 | 0.00204154 | 0.003284419 |
| 0 | 0 | 0 | 0 | 0 |
| 0 | 0 | 0 | 0 | 0 |
| 0 | 0 | 0 | 0.006238625 | 0 |
| 0 | 0 | 0 | 0 | 0 |
| 0 | 0 | 0 | 0.004074197 | 0 |
| 0 | 0 | 0 | 0 | 0 |
| 0 | 0 | 0 | 0 | 0 |
| 0.142091353 | 0.258616238 | 0 | 0 | 0.248487785 |
| 0 | 0 | 0 | 0 | 0 |
| 0 | 0 | 0 | 0 | 0 |
| 0 | 0 | 0 | 0 | 0 |
| 0 | 0 | 0 | 0 | 0 |
| 0 | 0 | 0 | 0 | 0 |
| 0 | 0 | 0 | 0 | 0 |
| 0 | 0 | 0 | 0 | 0 |
| 3.199599343 | 0.158819709 | 0.644505138 | 0.128302124 | 1.134161774 |
| 0 | 0.001420434 | 0 | 0 | 0 |
| 0 | 0 | 0 | 0 | 0.013295475 |
| 0 | 0 | 0 | 0 | 0 |
| 0 | 0 | 0 | 0 | 0 |
| 0 | 0 | 0 | 0 | 0 |
| 0 | 0 | 0 | 0 | 0 |
| 0 | 0 | 0 | 0.38719653 | 0 |
| 0 | 0 | 0 | 0 | 0.012553866 |
| 0 | 0 | 0 | 0 | 0 |
| 0 | 0 | 0 | 0 | 0 |
| 0 | 0.002366759 | 0.005626896 | 0.028322454 | 0.210386657 |
| 0 | 0.115469105 | 0.201751598 | 0 | 4.255509653 |
| 0 | 0 | 0 | 0 | 0.003322267 |
| 0 | 0 | 0 | 0 | 0 |
| 0 | 0 | 0 | 0 | 0 |
| 0 | 0 | 0 | 0 | 0 |
| 0.000869273 | 0 | 0 | 0.000660588 | 0.001417 |
| 0 | 0.002536494 | 0 | 0 | 0 |

| | | | | |
|---|---|---|---|---|
| 0.020646344 | 0 | 0.001256346 | 0.002173478 | 0.003496681 |
| 0 | 0 | 0 | 0 | 0 |
| 0.001826768 | 0.89484989 | 0 | 0.005182679 | 0.960443047 |
| 0 | 0 | 0 | 0 | 0 |
| 0 | 0.000746912 | 0 | 0 | 0 |
| 0 | 0 | 0 | 0 | 0 |
| 0 | 0 | 0 | 0 | 0 |
| 0 | 0 | 0 | 0 | 0.002612714 |
| 0.001122022 | 0 | 0 | 0 | 0 |
| 0 | 0 | 0 | 0 | 0 |
| 0 | 0 | 0 | 0 | 0 |
| 0 | 0 | 0 | 0 | 0 |
| 0 | 0 | 0 | 0 | 0 |
| 0 | 0 | 0 | 0 | 0.001845293 |
| 0 | 0 | 0 | 0 | 0 |
| 0.030709963 | 1.779990809 | 0.004796393 | 0.001536623 | 0.021630969 |
| 0.064925845 | 0.007205271 | 0.27566009 | 0.185221462 | 0.003196757 |
| 0 | 0 | 0 | 0.00336262 | 0 |
| 0 | 0 | 0 | 0 | 0 |
| 0 | 0 | 0 | 0.115274406 | 0 |
| 0 | 0 | 0 | 0 | 0 |
| 0 | 0 | 0.001141969 | 0 | 0 |
| 0 | 0 | 0 | 0 | 0 |
| 0 | 0 | 0.220613227 | 0.552515329 | 0 |
| 0 | 0.022038619 | 0 | 0 | 0 |
| 0 | 0.004026771 | 0 | 0.157896669 | 0.057753096 |
| 0 | 0 | 0 | 0.001081104 | 0.014783837 |
| 0 | 0.279436777 | 0 | 0 | 0 |
| 0 | 0 | 0 | 0 | 0 |
| 0 | 0 | 0 | 0 | 0 |
| 0.004260019 | 0 | 0 | 0.120037783 | 0.012124875 |
| 0.001814619 | 0.000942828 | 0.003684378 | 0.003983734 | 0.017748034 |
| 1.365796009 | 0.160688299 | 0.026984815 | 0.037930537 | 5.099913088 |
| 0 | 0 | 0.033644528 | 0.057285136 | 0.501386524 |
| 0 | 0 | 0 | 0.026263191 | 0 |
| 0 | 0 | 0 | 0 | 0 |
| 0.003644704 | 0 | 0 | 0 | 0 |
| 0.000610785 | 0.015655817 | 0 | 0.345330657 | 0.010177638 |
| 0 | 0.227732576 | 0.916577321 | 0 | 0 |
| 0 | 0 | 0 | 0 | 0.001989866 |
| 0 | 0 | 0 | 0 | 0 |
| 0 | 0 | 0 | 0 | 0 |
| 0 | 0 | 0 | 0.045967608 | 0.001985301 |
| 0.019389565 | 0.087530894 | 0 | 0.019807345 | 0.094993376 |
| 0 | 0 | 0 | 0 | 0 |
| 0 | 0 | 0 | 0.00179899 | 0 |
| 0 | 0 | 0 | 0 | 0 |
| 0 | 0 | 0 | 0 | 0 |

| | | | | |
|---|---|---|---|---|
| 0 | 0 | 0 | 0 | 0.004920671 |
| 0.003329085 | 0.202375562 | 0.747426131 | 0.518624462 | 0.286411005 |
| 0 | 0 | 0 | 0 | 0 |
| 0 | 0 | 0 | 0 | 0 |
| 0 | 0 | 0 | 0 | 0 |
| 0 | 0 | 0 | 0 | 0 |
| 0 | 0 | 0 | 0.005201032 | 0 |
| 0 | 0 | 0 | 0.004750071 | 0 |
| 0 | 0 | 0 | 0.012866071 | 0 |
| 0 | 0 | 0 | 0 | 0 |
| 0 | 0 | 0.001545515 | 0.160215566 | 0 |
| 0.081067938 | 0.002871871 | 0.012949245 | 0.011201099 | 0.008342719 |
| 0 | 0.040728977 | 0 | 0 | 0 |
| 0 | 0 | 0 | 0 | 0 |
| 0.008278847 | 0 | 0 | 0 | 0 |
| 0 | 0 | 0 | 0 | 0 |
| 0.002374628 | 0.003589221 | 0.019015929 | 0.02362325 | 0.012199126 |
| 0 | 0 | 0.015231357 | 0.005678934 | 0.010658947 |
| 0 | 0 | 0 | 0 | 0 |
| 0 | 0 | 0.007673623 | 0 | 0.002966299 |
| 0.012466827 | 0.006014763 | 0.062168571 | 0.00789494 | 0.027821986 |
| 0 | 0 | 0 | 0 | 0.00963339 |
| 0 | 0 | 0 | 0 | 0 |
| 0 | 0 | 0 | 0 | 0 |
| 0 | 0 | 0 | 0 | 0.041461781 |
| 0 | 0 | 0.001147859 | 0 | 0 |
| 0 | 0.026030897 | 0 | 0.031393452 | 0.023879731 |
| 0.006388584 | 0.042044985 | 0.05166346 | 0.135217489 | 0.362304916 |
| 0 | 0 | 0 | 0 | 0 |
| 0 | 0 | 0 | 0 | 0 |
| 0 | 0 | 0 | 0 | 0 |
| 0 | 0 | 0 | 0 | 0 |
| 0 | 0 | 0 | 0 | 0 |
| 0 | 0 | 0 | 0 | 0 |
| 0.015876847 | 0 | 0 | 0 | 0 |
| 0.117333447 | 0.177688506 | 0.10882757 | 0.02475124 | 0.327696331 |
| 0 | 0 | 0 | 0 | 0 |
| 0 | 0 | 0 | 0 | 0 |
| 0 | 0.004549754 | 0.005744145 | 0.006802368 | 0.319109511 |
| 0 | 0 | 0 | 0 | 0 |
| 0 | 0 | 0 | 0 | 0 |
| 0 | 0 | 0 | 0 | 0 |
| 0.000709194 | 0 | 0.000830734 | 0.001976108 | 0.029094066 |
| 0 | 0 | 0 | 0.00069378 | 0 |
| 0 | 0 | 0 | 0 | 0 |
| 0 | 0 | 2.598609132 | 0 | 0 |
| 0 | 0 | 0 | 0 | 0 |
| 0 | 0 | 0 | 0.003364325 | 0 |

| | | | | |
|---|---|---|---|---|
| 0 | 0 | 0 | 0 | 0 |
| 0 | 0 | 0 | 0 | 0 |
| 0 | 0.017728598 | 0 | 0 | 0 |
| 0.086109106 | 0.151064681 | 3.72832575 | 0 | 0 |
| 0 | 0 | 0 | 0 | 0 |
| 0 | 0 | 0 | 0 | 0 |
| 0 | 0 | 0 | 0.009732726 | 0 |
| 0.005911469 | 0.333059765 | 0 | 0 | 0 |
| 2.148105863 | 2.510036426 | 0 | 0.097157314 | 0.17545069 |
| 0.005905831 | 0.257371721 | 0 | 0 | 0.110310319 |
| 0 | 0 | 0 | 0 | 0 |
| 0 | 0.000822365 | 0.000741607 | 0 | 0.010750277 |
| 0 | 0 | 0 | 0 | 0 |
| 0 | 0 | 0 | 0 | 0 |
| 0 | 1.117513397 | 0.001070958 | 0.151694587 | 0.017742312 |
| 0 | 0 | 0 | 0 | 0 |
| 0.001703698 | 0.580689292 | 0 | 0 | 0.393898825 |
| 0 | 0 | 0 | 0 | 0 |
| 0 | 0 | 0 | 0 | 0 |
| 0 | 0 | 0 | 0 | 0 |
| 0 | 0 | 0 | 0 | 0 |
| 0 | 0.001336134 | 0 | 0 | 0 |
| 0 | 0 | 0 | 0 | 0 |
| 0.028307544 | 0.098379231 | 0.032864064 | 0.638820233 | 0.368810704 |
| 0 | 0 | 0 | 0.398647558 | 0 |
| 0 | 0.31680783 | 0 | 0 | 0 |
| 0 | 0.001832228 | 0 | 0 | 0 |
| 0.567766759 | 1.502313875 | 2.585320734 | 3.031096953 | 0 |
| 0 | 0 | 0 | 0 | 0 |
| 0 | 0 | 0 | 0 | 0 |
| 0.015002523 | 0.009593746 | 0 | 0 | 0 |
| 0 | 0 | 0 | 0 | 0 |
| 0 | 0.162307256 | 0 | 0.028626384 | 0.000636325 |
| 0 | 0 | 0 | 0 | 0 |
| 0 | 0 | 0 | 0 | 0 |
| 0.0389768 | 0 | 0 | 0 | 0.027647462 |
| 0.874714749 | 0.952950179 | 0 | 0 | 0 |
| 0 | 0 | 0 | 0 | 0 |
| 0 | 0 | 0 | 0 | 0 |
| 0 | 0 | 0 | 0 | 0 |
| 0 | 0.031359252 | 0.00071144 | 0.045962342 | 0.086216423 |
| 0 | 0.034773017 | 0 | 0 | 0 |
| 0.098360282 | 0.318935722 | 0.145514785 | 8.926272149 | 4.20810072 |
| 0 | 0.010652305 | 0 | 0 | 0 |
| 0 | 0 | 0 | 0 | 0 |
| 0 | 0 | 0 | 0 | 0 |
| 0 | 0 | 0 | 0 | 0 |
| 0 | 0.01375126 | 0 | 0.516456007 | 0.00198358 |

| | | | | |
|---|---|---|---|---|
| 0 | 0 | 0 | 0 | 0 |
| 0.073997853 | 0 | 0 | 0.054758986 | 0 |
| 0.011154515 | 0 | 0.041401741 | 0.395355952 | 0.396459608 |
| 0 | 0.001965415 | 0 | 0 | 0 |
| 0 | 0 | 0 | 0 | 0 |
| 0.042384819 | 0.09363844 | 0.000807294 | 0.001222042 | 0.001872396 |
| 0 | 1.534448317 | 0 | 0 | 0 |
| 0 | 0 | 0 | 0.04741394 | 0 |
| 0.022887634 | 0 | 0 | 0 | 0 |
| 0 | 0 | 0 | 0 | 0 |
| 0 | 0.622133492 | 0.006238239 | 0.211273835 | 0.022869782 |
| 0.086534742 | 1.970882114 | 2.466890061 | 1.252914525 | 7.635466463 |
| 0 | 0 | 0 | 0 | 0 |
| 0 | 0.006743218 | 0.030187936 | 0.05706254 | 0.033245145 |
| 0.002922128 | 0.102292848 | 0.074448377 | 0.760560909 | 0.0390992 |
| 0 | 2.768520481 | 0 | 0 | 0 |
| 0 | 0 | 0 | 0.009921203 | 0 |
| 0 | 0 | 0 | 0 | 0 |
| 0 | 0.162332722 | 0 | 0 | 0 |
| 0 | 0.012315558 | 0 | 0 | 0.011726118 |
| 0.001079904 | 0.037780049 | 0 | 0.025531413 | 0.396078539 |
| 0.058601014 | 0 | 0 | 0.247115816 | 0.159518113 |
| 0.209439071 | 0.316211222 | 4.184028084 | 1.599860539 | 0.519498988 |
| 0 | 0 | 0 | 0 | 0 |
| 0 | 0 | 0 | 0 | 0 |
| 0.196479756 | 0 | 0.020817251 | 0 | 0.004218852 |
| 0 | 0 | 0.044582147 | 0 | 0.029710373 |
| 0 | 0 | 0 | 0 | 0 |
| 0 | 0.232104035 | 0 | 0 | 0 |
| 0 | 0 | 0 | 0 | 0 |
| 0 | 0.006394965 | 0 | 0 | 0 |
| 0 | 0.058974581 | 0.09187634 | 0 | 0 |
| 0.01587916 | 0.000916711 | 0.027074038 | 0.016089422 | 0.004793441 |
| 0.013622725 | 0.55225963 | 0 | 0 | 0 |
| 0.013100553 | 0 | 0 | 0.000589957 | 0.023253421 |
| 0.148794702 | 0.142773801 | 0.033921874 | 0.107197123 | 0.028906038 |
| 0 | 0.048783061 | 0 | 0.005872455 | 0 |
| 0 | 0 | 0 | 0 | 0 |
| 2.378935848 | 5.151332276 | 0 | 0.470230708 | 0.127165044 |
| 0 | 0 | 0 | 0 | 0 |
| 0 | 0.030926636 | 0 | 0 | 0 |
| 0 | 0 | 0 | 0 | 0 |
| 0 | 0 | 0 | 0 | 0 |
| 0 | 0 | 0 | 0 | 0 |
| 0.001596484 | 0.227991639 | 0 | 0.040440598 | 0.016853803 |
| 0 | 0.01318899 | 0 | 0 | 0 |
| 0 | 0 | 0 | 0 | 0 |
| 3.019098314 | 0.006289444 | 2.29398951 | 5.044101442 | 0.113311478 |

| | | | | |
|---|---|---|---|---|
| 0 | 0 | 0 | 0 | 0 |
| 0.010839298 | 0.00551688 | 0 | 0 | 0 |
| 0 | 0 | 0 | 0 | 0 |
| 0 | 0.016159363 | 0 | 0 | 0 |
| 0 | 0 | 0 | 0 | 0 |
| 0 | 0 | 0 | 0 | 0 |
| 0 | 0.128262839 | 0 | 0 | 0 |
| 0 | 0.945888596 | 0 | 0.353952883 | 0.032072781 |
| 0 | 0 | 0 | 0 | 0 |
| 0 | 0 | 0 | 0 | 0 |
| 0.004329811 | 0.005399176 | 0.000811495 | 0.026542221 | 0.002352672 |
| 0 | 0 | 0 | 0 | 0 |
| 0.185952879 | 0 | 0 | 0 | 0 |
| 0 | 0 | 0 | 0 | 0 |
| 0 | 0 | 0 | 0 | 0 |
| 0 | 0 | 0 | 0 | 0 |
| 0 | 0 | 0.010712806 | 0 | 0 |
| 0.023068602 | 0.004869248 | 0.003715529 | 0 | 0.011555419 |
| 0 | 0 | 0 | 0 | 0 |
| 0 | 0 | 0 | 0 | 0 |
| 0.006404573 | 0.164718609 | 0 | 0 | 0 |
| 0 | 0 | 0 | 0 | 0 |
| 0 | 0.027637635 | 0 | 0 | 0.015354828 |
| 0 | 0 | 0 | 0 | 0 |
| 0 | 0 | 0 | 0 | 0 |
| 0 | 0 | 0 | 0 | 0 |
| 0 | 0 | 0 | 0 | 0 |
| 0 | 0 | 0 | 0 | 0 |
| 0 | 0 | 0 | 0 | 0 |
| 0.00653125 | 1.192464051 | 1.522970758 | 0 | 0 |
| 0 | 0 | 0 | 0 | 0 |
| 0 | 0 | 0 | 0 | 0 |
| 0 | 0 | 0 | 0 | 0 |
| 0 | 0 | 0 | 0 | 0 |
| 0 | 0 | 0.002097098 | 0 | 0 |
| 0 | 0 | 0 | 0 | 0 |
| 0 | 0.050102564 | 0 | 0 | 0.032853324 |
| 0.01740999 | 0 | 0.005152083 | 0.007056209 | 0.001194947 |
| 0 | 0 | 0 | 0 | 0 |
| 0 | 0 | 0 | 0.00091577 | 0.019643824 |
| 0 | 0.823702711 | 0 | 0 | 0 |
| 0 | 0 | 0 | 0 | 0 |
| 0 | 0 | 0 | 0 | 0 |
| 0 | 0 | 0.000833881 | 0 | 0 |
| 0.048069551 | 0 | 0.06513153 | 0 | 0.198326193 |
| 0 | 0 | 0 | 0 | 0 |
| 0 | 0.008504527 | 0 | 0 | 0 |
| 0 | 0 | 0 | 0 | 0 |

| | | | | |
|---|---|---|---|---|
| 0 | 0 | 0 | 0 | 0 |
| 0 | 0 | 0 | 0 | 0 |
| 0 | 0 | 0 | 0 | 0 |
| 0 | 0.238491562 | 0 | 0 | 0 |
| 0 | 0 | 0 | 0 | 0 |
| 0 | 0 | 0 | 0 | 0 |
| 0 | 0 | 0 | 0 | 0 |
| 0 | 0 | 0 | 0 | 0 |
| 0.004296756 | 0.001838515 | 0.022027309 | 0.135219279 | 0.004669422 |
| 0 | 0 | 0 | 0 | 0 |
| 0 | 0.009765564 | 0 | 0 | 0 |
| 0 | 0 | 0 | 0 | 0 |
| 0 | 0 | 0 | 0 | 0 |
| 0 | 0 | 0 | 0 | 0 |
| 0.590489119 | 2.598244088 | 0.007318972 | 1.523099714 | 0.019146485 |
| 0 | 0 | 0 | 0.001184287 | 0 |
| 0 | 0 | 0 | 0 | 0 |
| 0 | 0 | 0 | 0 | 0 |
| 0 | 0 | 0 | 0 | 0 |
| 0 | 0 | 0 | 0 | 0 |
| 0 | 0.04880162 | 0 | 0 | 0 |
| 0 | 0 | 0 | 0 | 0 |
| 0 | 0 | 0 | 0 | 0 |
| 0 | 0.146713057 | 0 | 0 | 0 |
| 1.707273125 | 0 | 0.057656151 | 0.118055864 | 0.002907057 |
| 2.072262435 | 7.506431472 | 8.053050704 | 0 | 0 |
| 0 | 0.017461566 | 0 | 0 | 0 |
| 0 | 0 | 0 | 0 | 0 |
| 0.337820055 | 0 | 0 | 0.032649511 | 0 |
| 0 | 0 | 0 | 0 | 0 |
| 0 | 0 | 0 | 0 | 0 |
| 0 | 0 | 0 | 0 | 0 |
| 0 | 0 | 0 | 0 | 0 |
| 0 | 0.033815293 | 0 | 0.002895126 | 0.00086253 |
| 0.007846523 | 0.002446109 | 0.005147093 | 0.008215436 | 0.006821653 |
| 0.130898357 | 0.001152735 | 0 | 0.059009837 | 0.001607361 |
| 0 | 0 | 0 | 0 | 0 |
| 0.024634478 | 0.266618633 | 0.918900838 | 0 | 1.939087737 |
| 0.131461009 | 0.418314655 | 0 | 0 | 0 |
| 0 | 0 | 0 | 0 | 0 |
| 0.034614169 | 0 | 0 | 0 | 0 |
| 0.001048304 | 0.002178684 | 0 | 0 | 0 |
| 0.00429502 | 0.278028583 | 0.028174092 | 1.003239035 | 1.164687172 |
| 0 | 0 | 0 | 0 | 0 |
| 0 | 0 | 0 | 0 | 0 |
| 0 | 0.003094203 | 0 | 0.006744024 | 0.008946288 |
| 0 | 0 | 0 | 0 | 0 |
| 0.001395215 | 0.149332918 | 0 | 1.913852692 | 0.839381801 |

| | | | | |
|---|---|---|---|---|
| 0 | 0 | 0 | 0 | 0 |
| 0 | 0 | 0 | 0 | 0 |
| 0 | 0 | 0 | 0 | 0 |
| 0 | 0 | 0 | 0 | 0 |
| 0 | 0.009257066 | 0 | 0.004618099 | 0 |
| 0 | 0 | 0 | 0 | 0 |
| 0 | 0 | 0 | 0 | 0 |
| 0 | 0 | 0 | 0 | 0 |
| 0 | 0 | 0 | 0 | 0 |
| 0 | 0 | 0 | 0 | 0 |
| 0 | 0 | 0 | 0 | 0 |
| 0.370405853 | 0.114995245 | 3.366302381 | 0 | 1.264506617 |
| 0 | 0.254671724 | 0.305824965 | 0.076504738 | 0 |
| 0 | 0 | 0 | 0 | 0 |
| 0.002395925 | 0.013195517 | 0.612722088 | 0.657165045 | 3.195035039 |
| 0 | 0 | 0 | 0 | 0 |
| 0 | 0 | 0 | 0.004728589 | 0 |
| 0 | 0 | 0 | 0 | 0 |
| 0.009870094 | 0 | 0 | 0.014469225 | 0 |
| 0 | 0 | 0 | 0 | 0 |
| 0 | 0.000722675 | 0 | 0 | 0 |
| 0 | 0 | 0 | 0 | 0 |
| 0.05390838 | 2.202343796 | 0.004401158 | 0.00268973 | 0 |
| 0 | 0 | 0 | 0 | 0 |
| 0 | 0 | 0 | 0 | 0 |
| 0 | 0 | 0 | 0 | 0 |
| 0 | 0 | 0 | 0 | 0 |
| 0 | 0 | 0 | 0 | 0 |
| 0 | 0.033639119 | 0 | 0 | 0 |
| 0 | 0 | 0 | 0 | 0 |
| 0 | 0 | 0 | 0 | 0 |
| 0.001865429 | 0.010338432 | 0.047489957 | 0.040322768 | 0.025678132 |
| 0 | 0 | 0 | 0 | 0 |
| 0 | 0.417227275 | 0 | 0 | 0.023968764 |
| 0.137621393 | 0 | 0 | 0.048316016 | 0.000655954 |
| 0.024899742 | 0.107271398 | 0.02479194 | 0.008672623 | 0.032978574 |
| 0 | 0.017074419 | 0 | 0 | 0 |
| 0.019431862 | 0 | 0 | 0 | 0.031989458 |
| 0 | 0 | 0 | 0 | 0.010047928 |
| 0 | 0 | 0 | 0 | 0 |
| 0 | 0.004453368 | 0 | 0 | 0 |
| 0 | 0.12929854 | 0 | 0 | 0 |
| 0 | 0 | 0 | 0 | 0 |
| 0 | 0.014127285 | 0 | 0 | 0 |
| 0.001387421 | 1.047111816 | 0.001857364 | 0.167891793 | 0.112650818 |
| 0 | 0 | 0 | 0 | 0 |
| 0.001500872 | 0.272588228 | 0 | 0 | 0 |
| 0 | 0.001363624 | 0 | 0 | 0 |

| | | | | |
|---|---|---|---|---|
| 0 | 0 | 0 | 0 | 0 |
| 0.006232244 | 0 | 0 | 0 | 0 |
| 0 | 0 | 0 | 0 | 0 |
| 0 | 0 | 0 | 0 | 0 |
| 0 | 0 | 0 | 0 | 0 |
| 0 | 0 | 0 | 0 | 0 |
| 0.002707805 | 0 | 0 | 0 | 0 |
| 0 | 0.001005871 | 0 | 0 | 0 |
| 0 | 0 | 0 | 0 | 0 |
| 0 | 0.027912787 | 0 | 0 | 0 |
| 0 | 0 | 0 | 0 | 0 |
| 0 | 0 | 0 | 0 | 0 |
| 0 | 0 | 0 | 0 | 0 |
| 0.027028134 | 0 | 0 | 0 | 0 |
| 0 | 0 | 0 | 0 | 0 |
| 0 | 0 | 0 | 0 | 0 |
| 0 | 0 | 0 | 0 | 4.182481079 |
| 0 | 0 | 0 | 0 | 0 |
| 4.582320196 | 0.007283682 | 7.347180559 | 0 | 0 |
| 0 | 0 | 0 | 0 | 0 |
| 0 | 0 | 0 | 0 | 0 |
| 0 | 0 | 0 | 0 | 0 |
| 0 | 0 | 0 | 0 | 0 |
| 0 | 0 | 0 | 0 | 0 |
| 0 | 0 | 0 | 0 | 0.01035479 |
| 0.006827222 | 0 | 0.007608196 | 0.011591695 | 0 |
| 0 | 0 | 0 | 0 | 0 |
| 0 | 0.059681693 | 0 | 0.05270383 | 0.077723857 |
| 0 | 0 | 0 | 0 | 0 |
| 0.112789299 | 0.649778846 | 2.424055182 | 0 | 0.275284033 |
| 0 | 0.003440279 | 0 | 0 | 0 |
| 0 | 0 | 0 | 0 | 0 |
| 0 | 0.02417699 | 0 | 0 | 0 |
| 0 | 0 | 0 | 0 | 0 |
| 0.00154692 | 0.001004674 | 0 | 0.013910708 | 0 |
| 0 | 0.375009533 | 0 | 0 | 0 |
| 0 | 0 | 0 | 0 | 0 |
| 0 | 0.00319203 | 0 | 0.00444636 | 0 |
| 0 | 0 | 0 | 0 | 0 |
| 0 | 0 | 0 | 0 | 0 |
| 2.363733028 | 3.770845923 | 6.357439351 | 1.487532405 | 17.15577976 |
| 0.009315443 | 1.632268546 | 1.187214338 | 0 | 0.164243247 |
| 0 | 0 | 0 | 0 | 0 |
| 0 | 0 | 0 | 0 | 0 |
| 0 | 1.254640046 | 0 | 0 | 0 |
| 0 | 0.321947115 | 0 | 0 | 0 |
| 0 | 0 | 0 | 0 | 0 |
| 0.002437933 | 0.128116239 | 0.00065274 | 0.005999092 | 0.003595586 |

| | | | | |
|---|---|---|---|---|
| 0 | 0 | 0 | 0 | 0 |
| 0 | 0.002419655 | 0 | 0.005989074 | 0.038540772 |
| 0 | 0.019129984 | 0 | 0 | 0 |
| 0 | 0 | 0 | 0 | 0 |
| 0.023797681 | 0.975113703 | 0.014816316 | 6.751417507 | 0.89098862 |
| 0 | 0 | 0 | 0 | 0 |
| 0 | 0 | 0 | 0 | 0 |
| 0 | 0 | 0 | 0 | 0 |
| 0 | 0 | 0.009888509 | 0.172158547 | 0.204275917 |
| 0 | 0 | 0 | 0 | 0 |
| 0 | 0 | 0 | 0 | 0 |
| 0 | 0 | 0 | 0 | 0 |
| 0.015489658 | 0.059862805 | 0.050069922 | 0 | 0.051444993 |
| 0.168391107 | 0.00956071 | 0.022055852 | 0.011707148 | 0.055805635 |
| 5.756445929 | 1.431310911 | 0 | 0 | 0.064686392 |
| 0.003687155 | 0.446278341 | 0.90922153 | 0.00284646 | 0.002480492 |
| 0 | 0.003463247 | 0 | 0 | 0.024145561 |
| 0 | 0 | 0 | 0 | 0 |
| 0 | 0 | 0 | 0 | 0 |
| 0 | 0 | 0 | 0 | 0 |
| 0 | 0 | 0 | 0 | 0 |
| 0 | 0 | 0 | 0 | 0 |
| 0 | 0 | 0 | 0 | 0 |
| 0 | 0 | 0 | 0 | 0 |
| 0 | 0 | 0 | 0 | 0 |
| 0 | 0 | 0 | 0 | 0 |
| 0.001448993 | 0.034296901 | 0.003922689 | 0 | 0 |
| 0.001816696 | 0 | 0 | 0.022273096 | 0.000987131 |
| 0 | 0 | 0 | 0 | 0 |
| 0 | 0 | 0 | 0 | 0 |
| 0 | 0 | 0 | 0 | 0 |
| 0 | 0 | 0 | 0 | 0 |
| 0 | 0 | 0 | 0 | 0 |
| 0 | 0.007915738 | 0.0008923 | 0 | 0.001448687 |
| 0.66129787 | 0 | 0 | 0 | 0 |
| 0 | 0.006769813 | 0 | 0.001064684 | 0.025807062 |
| 0 | 0 | 0 | 0 | 0 |
| 0 | 0.001443495 | 0 | 0 | 0 |
| 0 | 0 | 0 | 0 | 0 |
| 0 | 0 | 0 | 0 | 0 |
| 0.104498155 | 0 | 13.02649002 | 0 | 2.849945444 |
| 0 | 0 | 0 | 0 | 0 |
| 0 | 0 | 0 | 0 | 0 |
| 0.002223478 | 0 | 0 | 0.015770443 | 0 |
| 0 | 0 | 0 | 0 | 0 |
| 0 | 0 | 0 | 0 | 0 |
| 0 | 2.068456533 | 0 | 0 | 0 |
| 0 | 0 | 0 | 0 | 0 |

| | | | | |
|---|---|---|---|---|
| 0 | 0.242870283 | 0 | 0 | 0 |
| 0 | 0 | 0 | 0 | 0 |
| 0 | 0.002533931 | 0 | 0 | 0 |
| 0 | 0 | 0 | 0 | 0 |
| 0 | 0.016573813 | 0 | 0 | 0.010913216 |
| 0 | 0 | 0 | 0 | 0 |
| 0 | 0.092320613 | 0 | 0.001310048 | 0.493780404 |
| 0 | 0.001113182 | 0 | 0 | 0 |
| 0 | 0 | 0 | 0 | 0 |
| 0 | 0 | 0 | 0 | 0 |
| 0.000671952 | 0.026999268 | 0 | 0 | 0.007058902 |
| 0 | 0.061936857 | 0 | 0 | 0 |
| 0 | 0 | 0 | 0 | 0 |
| 0 | 0 | 0 | 0 | 0 |
| 0 | 0 | 0 | 0 | 0 |
| 0 | 0 | 0 | 0 | 0 |
| 0 | 0 | 0 | 0 | 0 |
| 0 | 0 | 0 | 0 | 0 |
| 0 | 0 | 0 | 0 | 0 |
| 0 | 0 | 0 | 0 | 0 |
| 0.004044516 | 0.09081809 | 0.034605469 | 0.079466943 | 0.106060605 |
| 0 | 0 | 0 | 0 | 0 |
| 0 | 0.106420191 | 0.001454084 | 0 | 0 |
| 0 | 0.004301361 | 0 | 0.0066407 | 0.004685757 |
| 0.012783955 | 0 | 0 | 0 | 0 |
| 0.00427303 | 0 | 0 | 0.004948126 | 0 |
| 0 | 0 | 0 | 0 | 0 |
| 0 | 0 | 0 | 0 | 0 |
| 0.005381192 | 0 | 0 | 0 | 0 |
| 0.214219845 | 0.02734211 | 0.005117505 | 0.044517944 | 0.002158049 |
| 0 | 0 | 0 | 0 | 0 |
| 0 | 0 | 0 | 0 | 0 |
| 0 | 0 | 0 | 0 | 0 |
| 0.027259998 | 0.158670392 | 0 | 0 | 0 |
| 0 | 0 | 0 | 0 | 0 |
| 0 | 0 | 0 | 0 | 0 |
| 0.014459037 | 0 | 0.023863454 | 0.019461105 | 0.00797381 |
| 0 | 0 | 0 | 0 | 0 |
| 0 | 0 | 0 | 0 | 0 |
| 0 | 0 | 0 | 0 | 0 |
| 0 | 0 | 0 | 0 | 0 |
| 0 | 0 | 0 | 0 | 0 |
| 0 | 0 | 0 | 0 | 0 |
| 0 | 0 | 0 | 0 | 0 |
| 0 | 0.186771448 | 0 | 0 | 0 |
| 0 | 0 | 0 | 0 | 0 |
| 0 | 0 | 0 | 0 | 0 |
| 0.033322927 | 0 | 0 | 0 | 0 |

| | | | | |
|---|---|---|---|---|
| 0 | 0.001735656 | 0.00069565 | 0.167170116 | 0.008470629 |
| 0.137110589 | 0 | 0 | 0 | 0 |
| 0 | 0 | 0 | 0 | 0 |
| 0 | 0 | 0 | 0 | 0 |
| 0 | 0 | 0 | 0 | 0 |
| 0 | 0 | 0 | 0 | 0 |
| 0 | 0.006078435 | 0 | 0 | 0 |
| 0.048210245 | 0.433056791 | 0.66731155 | 0.390209695 | 0.82004257 |
| 15.20113023 | 0.016648947 | 0.345594855 | 1.555206133 | 0.16556591 |
| 0 | 0 | 0 | 0.208975811 | 0.04253455 |
| 0 | 0 | 0 | 0 | 0 |
| 0 | 0.118610778 | 0 | 0 | 0 |
| 0 | 0.007482646 | 0 | 0 | 0 |
| 3.55856683 | 0 | 0.215229202 | 0 | 0 |
| 0 | 0 | 0 | 0 | 0 |
| 0 | 0 | 0 | 0 | 0 |
| 0 | 0.00084453 | 0 | 0 | 0 |
| 0.145338924 | 0 | 0 | 0 | 0 |
| 0 | 0 | 0 | 0 | 0 |
| 0 | 0.257053758 | 0 | 0 | 0 |
| 0 | 0 | 0 | 0 | 0 |
| 0 | 0 | 0 | 0 | 0 |
| 0 | 0 | 0 | 0 | 0 |
| 0 | 0 | 0 | 0 | 0 |
| 0 | 0.018531392 | 0.003835445 | 0 | 0 |
| 0 | 0 | 0 | 0 | 0 |
| 0 | 0 | 0 | 0 | 0 |
| 0 | 0 | 0 | 0 | 0 |
| 0 | 0 | 0 | 0 | 0 |
| 0 | 0 | 0 | 0 | 0 |
| 0 | 0 | 0 | 0 | 0 |
| 0 | 0 | 0 | 0 | 0 |
| 0 | 0 | 0 | 0 | 0 |
| 0 | 0.011807528 | 0 | 0 | 0 |
| 0 | 0 | 0 | 0 | 0 |
| 0 | 0 | 0 | 0 | 0 |
| 0 | 0 | 0 | 0 | 0 |
| 0 | 0.011087778 | 0 | 0 | 0 |
| 0 | 0 | 0 | 0 | 0.013429652 |
| 0 | 0 | 0 | 0.364187818 | 0 |
| 0 | 0 | 0 | 0 | 0 |
| 0.097864384 | 0.08123726 | 0.07379699 | 0 | 0.361224071 |
| 0 | 0 | 0 | 0 | 0 |
| 0 | 0 | 0 | 0 | 0 |
| 0 | 0.009755802 | 0 | 0 | 0 |
| 0 | 0 | 0 | 0 | 0.002690039 |
| 0 | 0 | 0 | 0 | 0 |
| 0 | 0 | 0 | 0 | 0 |

| | | | | |
|---|---|---|---|---|
| 0 | 0 | 0 | 0 | 0 |
| 0 | 0 | 0 | 0.000509151 | 0 |
| 0 | 0 | 0 | 0 | 0 |
| 0 | 0 | 0 | 0 | 0 |
| 0.090864614 | 5.425356866 | 0.525819395 | 0.1736949 | 0.380220959 |
| 0 | 0 | 0 | 0 | 0 |
| 0 | 0 | 0 | 0 | 0 |
| 0.00377725 | 0.00171724 | 0.019910631 | 0.247337097 | 0.007440056 |
| 0 | 0.010532548 | 0 | 0 | 0 |
| 0 | 0.064176475 | 0 | 0 | 0 |
| 0 | 0 | 0 | 0 | 0 |
| 0.202112735 | 0 | 0 | 0 | 0 |
| 0 | 0 | 0 | 0 | 0 |
| 0 | 0 | 0 | 0 | 0 |
| 0 | 0 | 0 | 0.000505174 | 0 |
| 0 | 0.427017846 | 0 | 0.0041492 | 0.023808255 |
| 0 | 0 | 0 | 0 | 0 |
| 0 | 0 | 0 | 0 | 0 |
| 0 | 0 | 0 | 0 | 0 |
| 0 | 0 | 0 | 0 | 0 |
| 0 | 0 | 0 | 0 | 0 |
| 0 | 0 | 0 | 0 | 0 |
| 0 | 0 | 0 | 0 | 0 |
| 0 | 0 | 0 | 0.030223679 | 0 |
| 1.215349311 | 0 | 0 | 0 | 0.109635228 |
| 0 | 0.002708941 | 0 | 0 | 0 |
| 0.129307954 | 0.371265237 | 0.062513539 | 0 | 0.032287816 |
| 0 | 0 | 0 | 0 | 0 |
| 0 | 0 | 0 | 0 | 0 |
| 0 | 0 | 0 | 0 | 0 |
| 0 | 0 | 0 | 0 | 0 |
| 0 | 0 | 0 | 0 | 0 |
| 0 | 0 | 0 | 0 | 0 |
| 0 | 0 | 0 | 0 | 0 |
| 0 | 0 | 0 | 0 | 0 |
| 0 | 0 | 0 | 0 | 0 |
| 0 | 0 | 0 | 0 | 0 |
| 0 | 0.372589549 | 0 | 0 | 0.002499098 |
| 0 | 0 | 0 | 0 | 0 |
| 0 | 0.000512525 | 0 | 0 | 0 |
| 0 | 0 | 0 | 0 | 0 |
| 0.421459293 | 0 | 0 | 0.2611668 | 0.132771303 |
| 0 | 0 | 0 | 0 | 0 |
| 0 | 0 | 0 | 0 | 0 |
| 0 | 0 | 0 | 0 | 0 |
| 0 | 0 | 0 | 0 | 0 |
| 0 | 0 | 0 | 0 | 0 |
| 0 | 0 | 0 | 0 | 0 |

| | | | | |
|---|---|---|---|---|
| 0 | 0.035425593 | 0.057800553 | 0.110571365 | 0.02730842 |
| 0 | 0 | 0 | 0 | 0 |
| 0 | 0 | 0 | 0 | 0 |
| 0 | 0 | 0 | 0.014526599 | 0 |
| 0 | 0 | 0 | 0 | 0 |
| 0 | 0 | 0 | 0.211648972 | 0 |
| 0 | 0 | 0 | 0 | 0 |
| 0 | 0 | 0 | 0 | 0 |
| 0 | 0 | 0 | 0 | 0 |
| 0 | 0 | 0 | 0 | 0 |
| 0 | 0 | 0 | 0 | 0 |
| 0 | 0 | 0 | 0 | 0 |
| 0 | 0 | 0 | 0 | 0 |
| 0 | 0 | 0 | 0 | 0 |
| 0 | 0.047784964 | 0 | 0 | 0 |
| 0 | 0 | 0 | 0 | 0 |
| 0 | 0 | 0 | 0 | 0 |
| 0 | 0 | 0 | 0 | 0 |
| 0 | 0 | 0 | 0 | 0 |
| 0 | 0 | 0 | 0 | 0 |
| 0 | 0 | 0 | 0.028448166 | 0.006190742 |
| 0 | 0 | 0 | 0 | 0 |
| 0.035715387 | 1.074157218 | 0.00505646 | 0.133037597 | 0.015916142 |
| 0 | 0.085775977 | 0 | 0 | 0 |
| 0 | 0 | 0 | 0 | 0 |
| 0.310559247 | 0 | 0 | 0 | 0 |
| 0 | 0 | 0 | 0 | 0 |
| 0 | 0 | 0 | 0 | 0 |
| 0.024467634 | 0 | 0.001459097 | 0 | 0 |
| 0 | 0 | 0 | 0 | 0 |
| 0 | 0 | 0 | 0 | 0 |
| 0 | 0.030337373 | 0 | 0 | 0 |
| 0.659561763 | 0.001750337 | 0.138678226 | 0.02267474 | 0.00601446 |
| 0 | 0 | 0 | 0 | 0 |
| 0.001291444 | 0.234626582 | 0.343902698 | 0.088981057 | 0 |
| 0 | 0 | 0 | 0.034434548 | 0.000629524 |
| 0 | 0 | 0 | 0 | 0 |
| 0.11701013 | 0 | 0 | 0 | 0 |
| 0.003796844 | 0.073803711 | 0.041440572 | 0.028740245 | 0 |
| 0 | 0 | 0 | 0 | 0 |
| 0 | 0 | 0 | 0 | 0 |
| 0 | 0.084687259 | 0 | 0 | 0 |
| 0.379981662 | 0.26323803 | 0 | 0.009790712 | 0.016460766 |
| 0 | 0 | 0 | 0 | 0 |
| 0 | 0 | 0 | 0 | 0 |
| 0.001244418 | 0.73018962 | 0.003109723 | 0 | 0 |
| 0 | 0 | 0 | 0 | 0 |
| 0 | 0 | 0 | 0 | 0 |

| | | | | |
|---|---|---|---|---|
| 0 | 0 | 0 | 0 | 0 |
| 0.066654886 | 0 | 0 | 0 | 0 |
| 0 | 0 | 0 | 0 | 0 |
| 0 | 0 | 0 | 0 | 0 |
| 0 | 0 | 0 | 0 | 0 |
| 0 | 0 | 0 | 0 | 0 |
| 0 | 0 | 0 | 0 | 0 |
| 0 | 0 | 0 | 0.073714696 | 0 |
| 0 | 0.503397609 | 0 | 0 | 0 |
| 0.133941486 | 0 | 0 | 0 | 0.002785808 |
| 0 | 0 | 0 | 0 | 0 |
| 0 | 0 | 0 | 0 | 0 |
| 0 | 0 | 0 | 0 | 0 |
| 0 | 0 | 0 | 0 | 0 |
| 0 | 0 | 0 | 0 | 0 |
| 0 | 0 | 0 | 0.001916679 | 0 |
| 0 | 0 | 0 | 0 | 0 |
| 0 | 0 | 0 | 0 | 0 |
| 0 | 0 | 0 | 0 | 0 |
| 0.043464677 | 0.206265149 | 0.085584198 | 0 | 0.092556572 |
| 0 | 0 | 0.02201118 | 0 | 0.006202526 |
| 0 | 0 | 0 | 0 | 0 |
| 0 | 0 | 0 | 0 | 0 |
| 0 | 0.00203648 | 0 | 0 | 0 |
| 0 | 0 | 0 | 0 | 0 |
| 0 | 0 | 0 | 0 | 0 |
| 0 | 0 | 0 | 0 | 0 |
| 2.185767565 | 0 | 0 | 0 | 0 |
| 0 | 0 | 0 | 0 | 0 |
| 0.038557094 | 0.075153007 | 0 | 0 | 0 |
| 0.006491517 | 0.194736092 | 0.01776938 | 0.441209983 | 0 |
| 0 | 0 | 0 | 0 | 0 |
| 0 | 0 | 0 | 0 | 0 |
| 0 | 0 | 0 | 0 | 0 |
| 0 | 0 | 0 | 0 | 0 |
| 0 | 0 | 0 | 0 | 0 |
| 0 | 0 | 0 | 0 | 0 |
| 0 | 0 | 0 | 0 | 0 |
| 0 | 0 | 0 | 0.005267181 | 0 |
| 0 | 0 | 0 | 0 | 0 |
| 0 | 0 | 0 | 0 | 0 |
| 0 | 1.14022733 | 0 | 0 | 0 |
| 0 | 0.002496003 | 0 | 0 | 0 |
| 0 | 0 | 0 | 0.003639621 | 0 |
| 0 | 0 | 0 | 0 | 0 |
| 0 | 0 | 0 | 0 | 0 |
| 0 | 0 | 0 | 0.02354508 | 0 |
| 0 | 0 | 0 | 0 | 0 |

| | | | | |
|---|---|---|---|---|
| 0 | 0 | 0 | 0 | 0 |
| 0 | 0 | 0 | 0 | 0 |
| 0 | 0 | 0 | 0 | 0 |
| 0 | 0 | 0 | 0.000976931 | 0 |
| 0.018806733 | 0.037081538 | 0.549501731 | 0.057981632 | 0.079829995 |
| 0 | 0 | 0 | 0 | 0 |
| 0 | 0 | 0 | 0 | 0 |
| 0 | 0 | 0 | 0 | 0 |
| 0 | 0 | 0 | 0 | 0 |
| 0 | 0 | 0 | 0 | 0 |
| 0 | 0.017178081 | 0 | 0 | 0 |
| 0.005802314 | 0.24581648 | 0.001673033 | 0.033465901 | 0.025222238 |
| 0 | 0.042615611 | 0 | 0 | 0 |
| 0 | 0.833904963 | 0 | 0 | 0 |
| 0 | 0 | 0 | 0 | 0.010534645 |
| 0 | 0.132908327 | 0 | 0 | 0 |
| 0 | 0.038347491 | 0 | 0 | 0 |
| 0 | 0 | 0 | 0 | 0 |
| 0.012842671 | 0 | 0.017260564 | 0.143996086 | 0.150032801 |
| 0 | 0.008941811 | 0 | 0.003129858 | 0.008392166 |
| 0 | 0 | 0 | 0 | 0 |
| 0 | 0 | 0 | 0 | 0 |
| 0 | 0 | 0 | 0 | 0 |
| 0 | 0 | 0 | 0 | 0 |
| 0 | 0 | 0 | 0 | 0 |
| 0 | 0 | 0 | 0 | 0 |
| 0 | 0.008084134 | 0 | 0 | 0.014026485 |
| 0 | 0 | 0 | 0 | 0 |
| 0 | 0 | 0 | 0 | 0 |
| 0 | 0 | 0 | 0 | 0 |
| 0 | 0.002707484 | 0 | 0.003249213 | 0 |
| 0 | 0 | 0 | 0 | 0 |
| 0 | 0 | 0 | 0 | 0 |
| 0 | 0 | 0 | 0 | 0 |
| 0 | 0 | 0 | 0 | 0 |
| 0 | 0 | 0 | 0 | 0 |
| 0 | 0 | 0 | 0 | 0 |
| 0.000563243 | 0.127154715 | 0.002639078 | 0.053217834 | 0.003213491 |
| 0 | 0.252203158 | 0 | 0 | 0 |
| 0 | 0 | 0 | 0 | 0 |
| 0 | 0.009787196 | 0 | 0.011394864 | 0 |
| 0 | 0 | 0 | 0 | 0 |
| 0 | 0 | 0 | 0 | 0 |
| 0 | 0 | 0 | 0.03046596 | 0 |
| 0 | 0 | 0 | 0 | 0 |
| 0 | 0 | 0 | 0 | 0 |
| 0 | 0 | 0 | 0 | 0 |
| 0 | 0 | 0 | 0 | 0 |

| | | | | |
|---|---|---|---|---|
| 0 | 0 | 0 | 0 | 0 |
| 0 | 0.185638413 | 1.264677861 | 0 | 0 |
| 0 | 0 | 0 | 0 | 0 |
| 0 | 0.006594439 | 0 | 0 | 0 |
| 0.002435512 | 0 | 0.021682064 | 0 | 0 |
| 0 | 0 | 0 | 0 | 0 |
| 0 | 0.002868698 | 0 | 0 | 0 |
| 0 | 0 | 0 | 0.040577587 | 0 |
| 0 | 0 | 0 | 0 | 0 |
| 0 | 0.0210214 | 0 | 0 | 0 |
| 0 | 0.026869639 | 0 | 0 | 0 |
| 0 | 0 | 0 | 0 | 0 |
| 0.286503218 | 0.159095177 | 0.004506206 | 0.097132375 | 0.035231513 |
| 0 | 0.001365249 | 0 | 0 | 0 |
| 0 | 0 | 0 | 0 | 0 |
| 0 | 0 | 0 | 0 | 0 |
| 0 | 0 | 0 | 0 | 0 |
| 0 | 0 | 0 | 0 | 0 |
| 0 | 0 | 0 | 0 | 0 |
| 0 | 0 | 0 | 0 | 0 |
| 0 | 0 | 0 | 0 | 0 |
| 0 | 0 | 0 | 0 | 0 |
| 0 | 0 | 0 | 0.006208784 | 0.002441673 |
| 0 | 0.005510195 | 0 | 0 | 0.051862642 |
| 0 | 0 | 0 | 0 | 0 |
| 0 | 0.054011136 | 0 | 0 | 0 |
| 0 | 0.015817418 | 0.004937581 | 0 | 0 |
| 0 | 0 | 0 | 0 | 0 |
| 0 | 0 | 0 | 0 | 0 |
| 0 | 0 | 0 | 0 | 0 |
| 0.006604554 | 0.037392101 | 0.057196183 | 0 | 0.019057181 |
| 0 | 0.005307282 | 0 | 0 | 0 |
| 0 | 0 | 0 | 0 | 0 |
| 0 | 0 | 0 | 0 | 0 |
| 0 | 0.174997154 | 0 | 0 | 0 |
| 0 | 0 | 0 | 0 | 0 |
| 0 | 0 | 0 | 0 | 0 |
| 0 | 0 | 0 | 0 | 0 |
| 0 | 0.007408503 | 0.003972472 | 0 | 0.001884588 |
| 0 | 0 | 0 | 0 | 0 |
| 0 | 0 | 0 | 0 | 0 |
| 0 | 0 | 0 | 0 | 0 |
| 0 | 0 | 0 | 0 | 0 |
| 0 | 0 | 0 | 0 | 0 |
| 2.910177237 | 0 | 0 | 0 | 0 |
| 0.067473882 | 0.170884084 | 0.180951871 | 0.159409254 | 0.060061009 |
| 0 | 0 | 0 | 0 | 0 |
| 0 | 0 | 0 | 0 | 0 |

| | | | | |
|---|---|---|---|---|
| 0 | 0 | 0 | 0 | 0 |
| 0 | 0 | 0 | 0 | 0 |
| 0.031648372 | 0 | 0 | 0 | 0 |
| 0 | 0 | 0 | 0 | 0 |
| 0 | 0 | 0 | 0 | 0 |
| 0 | 0 | 0 | 0 | 0 |
| 0 | 0 | 0 | 0.012502737 | 0 |
| 0 | 0 | 0 | 0 | 0 |
| 0 | 0.000926329 | 0 | 0 | 0 |
| 0 | 0 | 0 | 0 | 0 |
| 0 | 0.020142562 | 0 | 0.002217465 | 0 |
| 0 | 0.001455141 | 0 | 0.002128296 | 0 |
| 0 | 0 | 0 | 0 | 0 |
| 0 | 0 | 0 | 0 | 0 |
| 0 | 0 | 0 | 0 | 0 |
| 0 | 0 | 0 | 0 | 0 |
| 0 | 0 | 0 | 0 | 0.008490273 |
| 0 | 0 | 0 | 0 | 0 |
| 0 | 0 | 0 | 0 | 0 |
| 0.023633085 | 0 | 0.000777074 | 0 | 0 |
| 0 | 0 | 0 | 0 | 0 |
| 0 | 0 | 0 | 0 | 0 |
| 0 | 0 | 0 | 0 | 0 |
| 0 | 0 | 0 | 0 | 0 |
| 0 | 0 | 0 | 0 | 0 |
| 0 | 0 | 0 | 0 | 0 |
| 0 | 0 | 0 | 0 | 0 |
| 0.010094528 | 0 | 0 | 0 | 0 |
| 0 | 0 | 0 | 0 | 0 |
| 0 | 0 | 0 | 0 | 0 |
| 0.068746146 | 0 | 0.00150207 | 0.00433097 | 0.00135482 |
| 0 | 0 | 0 | 0 | 0 |
| 0.096571336 | 0.078863251 | 0.065886586 | 0.00460964 | 0.331021728 |
| 0 | 0 | 0.006732895 | 0.00321174 | 0.00275575 |
| 0 | 0 | 0 | 0 | 0 |
| 0 | 0 | 0 | 0 | 0 |
| 0 | 0 | 0 | 0 | 0 |
| 0 | 0 | 0 | 0 | 0 |
| 0 | 0 | 0 | 0 | 0 |
| 0 | 0 | 0 | 0 | 0 |
| 0 | 0 | 0 | 0 | 0 |
| 0 | 0.001237495 | 0.001275395 | 0.174618575 | 0.131634721 |
| 0.02259136 | 0.015937671 | 0.171693904 | 0 | 0.049101501 |
| 0 | 0 | 0 | 0 | 0 |
| 0 | 0 | 0 | 0 | 0 |
| 0.042230258 | 0.149390546 | 0.015156018 | 0.011228436 | 0.006509641 |
| 0 | 0 | 0 | 0 | 0 |
| 0 | 0 | 0 | 0 | 0 |

| | | | | |
|---|---|---|---|---|
| 0 | 0 | 0 | 0 | 0 |
| 0.006948241 | 0.006768983 | 0 | 0 | 0.008258769 |
| 0 | 0.002108503 | 0.031136158 | 0.012335625 | 0.008820219 |
| 0 | 0 | 0 | 0 | 0 |
| 0.00891063 | 0.238010216 | 0.021444746 | 0 | 0 |
| 0 | 0 | 0.003879274 | 0 | 0 |
| 0 | 0 | 0 | 0 | 0 |
| 0 | 0 | 0 | 0 | 0 |
| 0 | 0 | 0 | 0 | 0 |
| 0 | 0 | 0 | 0 | 0 |
| 0 | 0.007804276 | 0 | 0 | 0 |
| 0 | 0 | 0 | 0 | 0 |
| 0 | 0 | 0 | 0 | 0 |
| 0 | 0 | 0 | 0 | 0 |
| 0 | 0 | 0 | 0 | 0 |
| 0 | 0 | 0 | 0 | 0 |
| 0 | 2.199481988 | 0 | 0 | 0 |
| 0 | 0 | 0 | 0 | 0 |
| 0 | 0 | 0 | 0 | 0 |
| 0 | 0 | 0 | 0 | 0 |
| 0 | 0 | 0 | 0 | 0 |
| 0 | 0 | 0 | 0 | 0 |
| 0 | 0 | 0 | 0 | 0 |
| 0 | 0 | 0 | 0 | 0 |
| 0.006688955 | 0.007602451 | 0 | 0 | 0 |
| 0.037639956 | 0 | 0 | 0.003195952 | 0 |
| 0 | 0 | 0 | 0 | 0 |
| 0.002651715 | 1.271355906 | 0.011277742 | 0 | 0 |
| 0 | 0 | 0 | 0 | 0 |
| 0 | 0 | 0 | 0 | 0 |
| 0.004400013 | 0 | 0 | 0 | 0 |
| 0 | 0 | 0 | 0 | 0 |
| 0 | 0 | 0 | 0 | 0 |
| 0 | 0 | 0 | 0 | 0 |
| 0 | 0 | 0 | 0 | 0 |
| 0 | 0 | 0 | 0 | 0 |
| 0 | 0 | 0 | 0 | 0 |
| 0 | 0 | 0 | 0 | 0 |
| 0.499247655 | 4.228502138 | 0 | 0.621925326 | 7.141097839 |
| 0 | 0 | 0 | 0 | 0 |
| 0 | 0 | 0 | 0 | 0 |
| 0.021443316 | 0.050728862 | 0 | 0.005547384 | 0.002479057 |
| 0 | 0.002192996 | 0.000593292 | 0.139418757 | 0.00665091 |
| 0 | 0 | 0 | 0 | 0 |
| 0 | 0 | 0 | 0 | 0 |
| 0 | 0.003784447 | 0 | 0 | 0.003430044 |
| 0.003228469 | 0.004361312 | 0 | 0.004579705 | 0.001403392 |
| 0 | 0 | 0 | 0 | 0 |

| | | | | |
|---|---|---|---|---|
| 0 | 0 | 0 | 0 | 0 |
| 0.009573274 | 0.037887234 | 0.008589381 | 0.007842583 | 0.003541645 |
| 0 | 0 | 0 | 0 | 0 |
| 0 | 0 | 0 | 0 | 0 |
| 0 | 0 | 0 | 0 | 0 |
| 0 | 0 | 0 | 0 | 0 |
| 0 | 0 | 0 | 0 | 0 |
| 0 | 0 | 0 | 0 | 0 |
| 0 | 0 | 0 | 0 | 0 |
| 0 | 0 | 0 | 0 | 0 |
| 0 | 0 | 0 | 0 | 0 |
| 0 | 0 | 0 | 0 | 0 |
| 0.349886655 | 0 | 0 | 0 | 0 |
| 0 | 0 | 0 | 0 | 0 |
| 0.006603131 | 0 | 0.008329738 | 0.022902322 | 0.005243878 |
| 0 | 0 | 0 | 0 | 0 |
| 0 | 0.001490702 | 0 | 0 | 0 |
| 0 | 0 | 0 | 0 | 0 |
| 0 | 0 | 0 | 0 | 0 |
| 0 | 0 | 0 | 0 | 0 |
| 0 | 0 | 0 | 0 | 0 |
| 0 | 0.00336933 | 0 | 0 | 0.023310083 |
| 0.086244837 | 0.050187826 | 0 | 0.019419297 | 0.004373834 |
| 0.001798276 | 0.029364874 | 0 | 0 | 0 |
| 0 | 0 | 0 | 0 | 0 |
| 0 | 0 | 0 | 0 | 0 |
| 0 | 0 | 0 | 0 | 0 |
| 0 | 0 | 0 | 0 | 0 |
| 0 | 0 | 0 | 0 | 0 |
| 0 | 0 | 0 | 0 | 0 |
| 0 | 0 | 0 | 0 | 0 |
| 0 | 0 | 0 | 0 | 0 |
| 0 | 0 | 0 | 0 | 0 |
| 0 | 0 | 0 | 0 | 0 |
| 0.000679365 | 1.781374923 | 0.790487862 | 0.080308806 | 0.2104121 |
| 0 | 0 | 0 | 0 | 0 |
| 0 | 0 | 0 | 0 | 0 |
| 0 | 0 | 0 | 0 | 0 |
| 0 | 0 | 0 | 0.00757108 | 0.028150065 |
| 0 | 0 | 0 | 0 | 0 |
| 0.039081936 | 0.036434678 | 0.009417536 | 0.007919887 | 0.023541393 |
| 0 | 0 | 0 | 0 | 0 |
| 0 | 0 | 0 | 0 | 0 |
| 0 | 0 | 0 | 0 | 0 |
| 0 | 0 | 0 | 0 | 0 |
| 0.06366739 | 0.035338376 | 0.500065478 | 0.716091736 | 0.247303407 |
| 0 | 0 | 0 | 0 | 0 |
| 0 | 0 | 0 | 0 | 0 |

| | | | | |
|---|---|---|---|---|
| 0 | 0 | 0 | 0 | 0 |
| 0 | 0.003029372 | 0 | 0 | 0 |
| 0 | 0 | 0 | 0 | 0 |
| 0 | 0 | 0 | 0 | 0 |
| 0 | 0 | 0 | 0.039101729 | 0 |
| 0 | 0 | 0 | 0 | 0 |
| 0 | 0 | 0 | 0 | 0 |
| 0 | 0 | 0 | 0 | 0 |
| 0 | 0.254512783 | 0 | 0 | 0 |
| 0 | 0.002078856 | 0 | 0.076013623 | 0.46714911 |
| 0 | 0 | 0 | 0 | 0 |
| 0 | 0 | 0 | 0 | 0 |
| 0 | 0 | 0.016720382 | 0.021546106 | 0.000796857 |
| 0 | 0 | 0 | 0 | 0 |
| 0 | 0 | 0 | 0 | 0 |
| 0 | 0 | 0 | 0 | 0 |
| 0 | 0 | 0 | 0 | 0 |
| 0 | 0 | 0 | 0 | 0 |
| 0 | 0 | 0 | 0 | 0 |
| 0.581296229 | 0.324406992 | 0.995484501 | 0.130936408 | 0.621074244 |
| 0 | 0.055829203 | 0 | 0 | 0 |
| 0 | 0 | 0 | 0 | 0 |
| 0 | 0 | 0 | 0 | 0 |
| 0 | 0 | 0 | 0 | 0 |
| 0 | 0 | 0 | 0 | 0 |
| 0 | 0 | 0 | 0 | 0 |
| 0 | 0 | 0 | 0 | 0 |
| 0 | 0 | 0 | 0 | 0 |
| 0 | 0 | 0 | 0 | 0 |
| 0 | 0 | 0 | 0 | 0 |
| 0 | 0 | 0 | 0 | 0 |
| 0 | 0.252220081 | 0 | 0 | 0 |
| 0 | 0 | 0 | 0 | 0 |
| 0 | 0 | 0 | 0 | 0 |
| 0 | 0 | 0 | 0 | 0 |
| 0 | 0 | 0.003430722 | 0 | 0 |
| 0 | 0 | 0 | 0 | 0 |
| 0 | 0 | 0 | 0 | 0 |
| 0 | 0 | 0 | 0 | 0 |
| 0 | 0 | 0 | 0 | 0 |
| 0 | 0 | 0 | 0 | 0 |
| 0 | 0 | 0 | 0 | 0 |
| 0 | 0.007140819 | 0 | 0 | 0 |
| 0 | 0 | 0 | 0 | 0 |
| 0.001430323 | 0.043740184 | 0 | 0.019875602 | 0.004885187 |
| 0 | 0 | 0 | 0 | 0 |
| 0 | 0 | 0 | 0 | 0 |
| 0 | 0 | 0 | 0 | 0 |

| | | | | |
|---|---|---|---|---|
| 0 | 0 | 0 | 0 | 0 |
| 0 | 0 | 0 | 0 | 0 |
| 0 | 0 | 0 | 0 | 0 |
| 0 | 0 | 0 | 0 | 0 |
| 0 | 0 | 0 | 0 | 0 |
| 0.040423548 | 0.175474265 | 0.000571788 | 0.033694419 | 0.008288596 |
| 0 | 0 | 0 | 0 | 0 |
| 0 | 0 | 0 | 0 | 0 |
| 0 | 0 | 0 | 0 | 0 |
| 0 | 0 | 0 | 0 | 0 |
| 0 | 0 | 0 | 0 | 0 |
| 0 | 0 | 0 | 0 | 0 |
| 0 | 0 | 0 | 0 | 0 |
| 0 | 0 | 0 | 0 | 0 |
| 0 | 0 | 0 | 0 | 0 |
| 0 | 0 | 0 | 0 | 0 |
| 0 | 0.000576827 | 0 | 0 | 0 |
| 0 | 0 | 0 | 0 | 0 |
| 0 | 0 | 0 | 0 | 0 |
| 0 | 0 | 0 | 0 | 0 |
| 0 | 0 | 0 | 0 | 0 |
| 0 | 0 | 0 | 0 | 0 |
| 0 | 0 | 0 | 0 | 0 |
| 0.002269964 | 0 | 0 | 0 | 0 |
| 0 | 0 | 0 | 0 | 0 |
| 0 | 0 | 0 | 0.019582577 | 0.001815066 |
| 0 | 0 | 0 | 0 | 0 |
| 0 | 0 | 0 | 0 | 0 |
| 0 | 0 | 0 | 0 | 0 |
| 0 | 0 | 0 | 0 | 0 |
| 0 | 0 | 0 | 0 | 0 |
| 0 | 0 | 0 | 0 | 0 |
| 0 | 0 | 0 | 0 | 0 |
| 0 | 0 | 0 | 0 | 0 |
| 0 | 0 | 0 | 0 | 0 |
| 0 | 0 | 0 | 0 | 0 |
| 0 | 0 | 0 | 0 | 0 |
| 0 | 0 | 0 | 0 | 0 |
| 0 | 1.46425787 | 0 | 3.08939331 | 0 |
| 0 | 0 | 0 | 0 | 0 |
| 0 | 0 | 0 | 0 | 0 |
| 0 | 0 | 0 | 0 | 0 |
| 0 | 0 | 0 | 0 | 0 |
| 0 | 0 | 0 | 0 | 0 |
| 0 | 0 | 0 | 0 | 0 |
| 0 | 0 | 0 | 0 | 0 |
| 0 | 0 | 0 | 0 | 0 |
| 0 | 0 | 0 | 0 | 0 |

| | | | | |
|---|---|---|---|---|
| 0 | 0 | 0 | 0 | 0 |
| 0 | 0.472598502 | 0.003840979 | 0 | 0 |
| 0 | 0 | 0 | 0 | 0 |
| 0 | 0 | 0 | 0 | 0 |
| 0 | 0 | 0 | 0 | 0 |
| 0 | 0 | 0 | 0 | 0 |
| 0 | 0 | 0 | 0 | 0 |
| 0 | 0 | 0 | 0 | 0 |
| 0 | 0 | 0 | 0 | 0 |
| 0 | 0 | 0 | 0 | 0 |
| 0 | 0.001626666 | 0.001257364 | 0.009743265 | 0.001944176 |
| 0 | 0 | 0 | 0 | 0 |
| 0 | 0 | 1.202248206 | 0 | 0 |
| 0 | 0 | 0 | 0 | 0 |
| 0 | 0 | 0 | 0 | 0 |
| 0 | 0 | 0 | 0 | 0 |
| 0 | 0 | 0 | 0 | 0 |
| 0 | 0 | 0 | 0 | 0 |
| 0 | 0 | 0 | 0 | 0 |
| 0.001488661 | 0 | 0 | 0 | 0 |
| 0.020880998 | 0 | 0.001488841 | 0.001149864 | 0 |
| 0 | 0 | 0 | 0 | 0 |
| 0 | 0 | 0 | 0 | 0 |
| 0.534351923 | 1.242031947 | 0.02079084 | 1.58637111 | 0.267872659 |
| 0 | 0 | 0 | 0 | 0 |
| 0 | 0 | 0 | 0 | 0 |
| 0 | 0 | 0 | 0 | 0 |
| 0 | 0 | 0 | 0 | 0 |
| 0 | 0 | 0 | 0 | 0 |
| 0 | 0 | 0 | 0 | 0 |
| 0 | 0 | 0 | 0 | 0 |
| 0 | 0 | 0 | 0 | 0 |
| 0 | 0 | 0 | 0 | 0 |
| 0 | 0 | 0 | 0 | 0 |

| PHL018-0124_V1 | PHL018-0192_V2 | PHL018-0167_V1 | PHL018-0131_V1 | PHL018-0168_V2 |
|---|---|---|---|---|
| EYCF | CTRL | REF | EYCF | EYCF |
| 0.629523457 | 0.074533642 | 0.224836515 | 0.022240562 | 0 |
| 0.167274372 | 3.861342518 | 1.077172838 | 0.324462162 | 0 |
| 0.146167781 | 2.67177314 | 1.609287355 | 0 | 0 |
| 0 | 0 | 0 | 0 | 0 |
| 0 | 0 | 0 | 0 | 0 |
| 0 | 0 | 0 | 0 | 0 |
| 0 | 0 | 0 | 0 | 0 |
| 0 | 0 | 0 | 0 | 0 |
| 0 | 0 | 0 | 0 | 0 |
| 0 | 0 | 0 | 0 | 0 |
| 0 | 0 | 0 | 0 | 0 |
| 0 | 0 | 0 | 0 | 0 |
| 0 | 0 | 0 | 0 | 0 |
| 1.102572289 | 2.846322523 | 0.716161306 | 0.007206343 | 0 |
| 0 | 0 | 0 | 0 | 0 |
| 0 | 0 | 0 | 0 | 0 |
| 0 | 0 | 0 | 0 | 0 |
| 0 | 0 | 0 | 0 | 0 |
| 0 | 0 | 0 | 0 | 0 |
| 0 | 0 | 0 | 0 | 0 |
| 0 | 0 | 0 | 0 | 0 |
| 0 | 0 | 0 | 0 | 0 |
| 0 | 0 | 0 | 0 | 0 |
| 0 | 0 | 0 | 0 | 0 |
| 0 | 0 | 0.020747869 | 0 | 0 |
| 0 | 0 | 0 | 0 | 0 |
| 0 | 0 | 0 | 0 | 0 |
| 0 | 0 | 0 | 0 | 0 |
| 0 | 0 | 0 | 0 | 0 |
| 0 | 0 | 0 | 0 | 0 |
| 0 | 0 | 0 | 0 | 0 |
| 0 | 0 | 0 | 0 | 0 |
| 0 | 0 | 0 | 0 | 0 |
| 0 | 0 | 0 | 0 | 0 |
| 0 | 0 | 0 | 0 | 0 |
| 0.840354101 | 0.003519483 | 0.022075094 | 0.253357209 | 0.001873775 |
| 0 | 0 | 0 | 0 | 0 |
| 0 | 0 | 0 | 0 | 0 |
| 0 | 0 | 0 | 0 | 0 |
| 0.003881134 | 0 | 0 | 0 | 0 |
| 0 | 0 | 0 | 0 | 0 |
| 0.048595561 | 0 | 0.001031884 | 0 | 0.026418805 |
| 0.442796805 | 0.013414732 | 0.220018384 | 0.466379731 | 36.67793073 |
| 0.342166448 | 0.048491252 | 0.019119977 | 0.082305948 | 0.011172071 |
| 0.000860671 | 0.001140016 | 0.001463876 | 0 | 0 |
| 0 | 0 | 0 | 0 | 0 |

| | | | | |
|---|---|---|---|---|
| 0.000836587 | 0 | 0.003320126 | 0 | 0 |
| 0 | 0 | 0 | 0.003533407 | 0 |
| 0.003489767 | 0.005322793 | 0.002338259 | 0 | 0 |
| 0.491905776 | 1.727038439 | 0.272028626 | 0.186019129 | 0 |
| 0 | 0.007915125 | 0 | 0 | 0 |
| 0 | 0 | 0 | 0 | 0 |
| 0.014805274 | 0.017794764 | 0.000932652 | 0 | 0 |
| 0 | 0 | 0 | 0 | 0 |
| 0 | 0 | 0 | 0 | 0 |
| 0 | 0 | 0 | 0 | 0 |
| 0.002267197 | 0 | 0 | 0 | 0 |
| 0.023944383 | 0.590492512 | 0.044263683 | 0.094671742 | 0 |
| 0.001068828 | 0 | 0 | 0 | 0 |
| 0.016935461 | 0.189600255 | 0 | 0.015702813 | 0 |
| 0 | 0 | 0 | 0 | 0 |
| 0 | 0 | 0 | 0 | 0 |
| 0.892127834 | 0.009983282 | 0.031361672 | 1.236426486 | 0.653022548 |
| 0 | 0 | 0 | 0 | 0 |
| 0 | 0 | 0 | 0 | 0 |
| 0 | 0 | 0 | 0 | 0 |
| 0.219259597 | 0.099487472 | 0 | 0.018252341 | 0 |
| 0.038718098 | 0.030190215 | 0.006626797 | 0.000566518 | 0 |
| 0.513420227 | 0 | 0.002415636 | 0 | 0 |
| 0 | 0 | 0 | 0 | 0 |
| 0 | 0 | 0 | 0 | 0 |
| 0 | 0 | 0 | 0 | 0 |
| 0 | 0 | 0 | 0 | 0 |
| 0 | 0 | 0 | 0 | 0 |
| 0.004112061 | 0 | 0 | 0 | 0 |
| 0 | 0 | 0 | 0 | 0 |
| 0.000662217 | 0 | 0 | 0 | 0 |
| 0 | 0 | 0 | 0 | 0 |
| 0 | 0 | 0 | 0 | 0 |
| 0 | 0 | 0 | 0 | 0 |
| 0 | 0 | 0 | 0 | 0 |
| 0 | 0 | 0 | 0 | 0 |
| 0 | 0 | 0 | 0 | 0 |
| 0.014350683 | 0 | 0.040781119 | 66.85434675 | 0 |
| 0 | 0 | 0 | 0 | 0 |
| 0 | 0 | 0 | 0 | 0 |
| 0 | 0 | 0 | 0 | 0 |
| 0 | 0 | 0 | 0 | 0 |
| 0 | 0 | 0 | 0 | 0 |
| 0 | 0 | 0 | 0 | 0 |
| 0.035185181 | 0 | 0.051425591 | 0 | 0 |
| 0 | 0 | 0 | 0 | 0 |
| 0 | 0 | 0 | 0 | 0 |
| 0 | 0.338909863 | 0.007699491 | 0 | 0 |

| | | | | |
|---|---|---|---|---|
| 0 | 0 | 0 | 0 | 0 |
| 0 | 0 | 0 | 0 | 0 |
| 0 | 0 | 0 | 0 | 0 |
| 0 | 0 | 0 | 0 | 0 |
| 0 | 0.07049095 | 0.005845843 | 0 | 0 |
| 0 | 0 | 0 | 0 | 0 |
| 0 | 0 | 0 | 0 | 0 |
| 0.000603667 | 0.001066128 | 0 | 0 | 0 |
| 0.000847338 | 0 | 0 | 0 | 0 |
| 0 | 0 | 0 | 0 | 0 |
| 0 | 0 | 0 | 0 | 0 |
| 0.206421898 | 0 | 0 | 0 | 0 |
| 0 | 0 | 0 | 0 | 0 |
| 0 | 0 | 0 | 0 | 0 |
| 0 | 0 | 0 | 0 | 0 |
| 0 | 0 | 0 | 0 | 0 |
| 0 | 0 | 0 | 0 | 0 |
| 0 | 0 | 0 | 0 | 0 |
| 0.575922904 | 2.565116345 | 1.481790684 | 0 | 0 |
| 0 | 0.017734907 | 0 | 0 | 0 |
| 0 | 0 | 0 | 0 | 0 |
| 0 | 0 | 0 | 0 | 0 |
| 0 | 0 | 0 | 0 | 0 |
| 0 | 0 | 0 | 0 | 0 |
| 0 | 0 | 0 | 0 | 0 |
| 0 | 0 | 0 | 0 | 0 |
| 0 | 0 | 0 | 0 | 0 |
| 0 | 0 | 0 | 0 | 0 |
| 0 | 0 | 0 | 0 | 0 |
| 0 | 0 | 0 | 0 | 0 |
| 0.032017477 | 0.197102029 | 0.020449169 | 0.000886757 | 0 |
| 0.041540832 | 0.001210639 | 0 | 0.010631833 | 0 |
| 0 | 0 | 0 | 0 | 0 |
| 0 | 0 | 0 | 0 | 0 |
| 0 | 0 | 0 | 0 | 0 |
| 0 | 0 | 0 | 0 | 0 |
| 0 | 0 | 0 | 0 | 0 |
| 0 | 0 | 0 | 0 | 0 |
| 0.26032996 | 0 | 0 | 0.00458825 | 0 |
| 0 | 0 | 0 | 0 | 0 |
| 0 | 0 | 0 | 0 | 0 |
| 0 | 0.005207868 | 0 | 0 | 0 |
| 0 | 0 | 0 | 0 | 0.001210613 |
| 0 | 0 | 0 | 0 | 0 |
| 0 | 0 | 0 | 0 | 0 |
| 0 | 0 | 0 | 0 | 0 |
| 0 | 0 | 0 | 0 | 0 |
| 0 | 0 | 0 | 0 | 0 |

| | | | | |
|---|---|---|---|---|
| 0 | 0 | 0 | 0 | 0 |
| 0 | 0 | 0 | 0 | 0 |
| 0 | 0 | 0 | 0 | 0 |
| 0 | 0 | 0 | 0 | 0 |
| 0.01551273 | 0.04425643 | 0.018672373 | 0.026951714 | 0 |
| 0 | 0 | 0 | 0 | 0 |
| 0 | 0 | 0 | 0 | 0 |
| 0 | 0 | 0 | 0 | 0 |
| 0 | 0 | 0 | 0 | 0 |
| 0 | 0 | 0 | 0 | 0 |
| 0 | 0 | 0 | 0 | 0 |
| 0 | 0 | 0 | 0 | 0 |
| 0 | 0 | 0 | 0 | 0 |
| 0 | 0 | 0 | 0 | 0 |
| 0 | 0 | 0 | 0 | 0 |
| 0 | 0.003997291 | 0.689452671 | 0 | 0 |
| 0 | 0 | 0 | 0 | 0 |
| 0 | 0 | 0 | 0 | 0 |
| 0 | 0 | 0 | 0 | 0 |
| 0 | 0 | 0 | 0.149154952 | 0 |
| 0 | 0 | 0 | 0 | 0 |
| 0 | 0 | 0 | 0 | 0 |
| 0.003666553 | 0 | 0 | 0 | 0 |
| 0 | 0 | 0 | 0 | 0 |
| 0.03375911 | 0 | 0 | 0 | 0 |
| 0 | 0 | 0 | 0 | 0 |
| 0 | 0 | 0.007936753 | 0 | 0 |
| 0 | 0 | 0 | 0 | 0 |
| 0 | 0 | 0 | 0 | 0 |
| 0 | 0 | 0 | 0 | 0 |
| 0 | 0 | 0 | 0 | 0 |
| 0 | 0.00167618 | 0 | 0 | 0 |
| 0 | 0 | 0 | 0 | 0 |
| 0 | 0 | 0 | 0 | 0 |
| 0 | 0 | 0 | 0 | 0 |
| 0 | 0 | 0 | 0 | 0 |
| 0 | 0 | 0 | 0 | 0 |
| 0 | 0 | 0 | 0 | 0 |
| 0 | 0 | 0 | 0 | 0 |
| 0 | 0 | 0 | 0 | 0 |
| 0 | 0 | 0 | 0 | 0 |
| 0 | 0 | 0 | 0 | 0 |
| 0 | 0 | 0 | 0 | 0 |
| 0 | 0 | 0 | 0 | 0 |
| 0 | 0 | 0 | 0 | 0 |
| 0 | 0 | 0 | 0 | 0 |
| 0 | 0 | 0 | 0 | 0 |
| 0 | 0 | 0 | 0 | 0 |

| | | | | |
|---|---|---|---|---|
| 0 | 0 | 0 | 0 | 0 |
| 0 | 0 | 0 | 0 | 0 |
| 0 | 0 | 0 | 0 | 0 |
| 0 | 0 | 0 | 0 | 0 |
| 0 | 0 | 0 | 0 | 0 |
| 0 | 0 | 0 | 0 | 0 |
| 0 | 0 | 0 | 0 | 0 |
| 0 | 0 | 0 | 0 | 0 |
| 0 | 0 | 0 | 0 | 0 |
| 0 | 0 | 0 | 0 | 0 |
| 0 | 0 | 0 | 0 | 0 |
| 0 | 0 | 0 | 0 | 0 |
| 0 | 0.004544496 | 0 | 0.000586908 | 0 |
| 0 | 0 | 0 | 0 | 0 |
| 0 | 0 | 0.009126008 | 0 | 0 |
| 0 | 0 | 0 | 0 | 0 |
| 0 | 0 | 0 | 0 | 0 |
| 0 | 0 | 0 | 0 | 0 |
| 0 | 0 | 0 | 0 | 0 |
| 0 | 0 | 0 | 0 | 0 |
| 0.05547106 | 0.041485917 | 0.018933807 | 0.023319268 | 0.004756717 |
| 0 | 0 | 0 | 0 | 0 |
| 0.04606496 | 0 | 0 | 0 | 0 |
| 0 | 0 | 0 | 0 | 0 |
| 0 | 0 | 0 | 0 | 0 |
| 0 | 0 | 0 | 0 | 0 |
| 0.001153751 | 0.029400029 | 0.000560675 | 0.011663332 | 0.005792236 |
| 0 | 0 | 0 | 0 | 0 |
| 0 | 0 | 0 | 0 | 0 |
| 0 | 0 | 0 | 0 | 0 |
| 0 | 0 | 0 | 0 | 0 |
| 0 | 0 | 0 | 0 | 0 |
| 0 | 0 | 0 | 0 | 0 |
| 0.009685848 | 0.630480275 | 0.008825466 | 0 | 0 |
| 0.028174191 | 0.002276514 | 0.042595699 | 0.001071018 | 0 |
| 0 | 0 | 0 | 0 | 0 |
| 0 | 0 | 0 | 0 | 0 |
| 0 | 0 | 0 | 0 | 0 |
| 0 | 0 | 0 | 0 | 0 |
| 0 | 0 | 0 | 0 | 0 |
| 0 | 0 | 0 | 0 | 0 |
| 0 | 0 | 0 | 0 | 0 |
| 0 | 0 | 0 | 0 | 0 |
| 0 | 0 | 0 | 0 | 0 |
| 0 | 0 | 0 | 0 | 0 |
| 0.251593006 | 0 | 0.038170113 | 0 | 0 |
| 0 | 0.001760614 | 0 | 0 | 0 |
| 0.004068029 | 0.007416252 | 0.000743991 | 0.000763237 | 0 |

| | | | | |
|---|---|---|---|---|
| 0 | 0 | 0 | 0 | 0 |
| 0 | 0 | 0 | 0 | 0 |
| 0 | 0 | 0 | 0 | 0.001832179 |
| 0.000834031 | 0 | 0.001323994 | 0 | 0 |
| 0 | 0 | 0 | 0 | 0 |
| 0 | 0 | 0 | 0 | 0 |
| 0 | 0 | 0 | 0 | 0 |
| 0.004716021 | 0 | 0 | 0 | 0 |
| 0 | 0 | 0.020818385 | 0 | 0 |
| 0 | 0 | 0 | 0 | 0 |
| 0 | 0 | 0 | 0 | 0 |
| 0 | 0 | 0 | 0 | 0 |
| 0 | 0 | 0 | 0 | 0 |
| 0 | 0 | 0 | 0 | 0 |
| 0.000620703 | 0 | 0 | 0 | 0 |
| 0 | 0 | 0 | 0 | 0 |
| 0 | 0 | 0 | 0 | 0 |
| 0 | 0 | 0 | 0 | 0.291312953 |
| 0.002535678 | 0 | 0.001210615 | 0.001681781 | 0.197183766 |
| 0 | 0.291434733 | 0.009017508 | 0 | 0 |
| 0 | 0.002086056 | 0.002232225 | 0 | 0 |
| 0 | 0 | 0 | 0 | 0 |
| 0 | 0 | 0 | 0 | 0 |
| 0 | 0 | 0 | 0 | 0 |
| 0.004302193 | 0.002762927 | 0.002660872 | 0.009857259 | 0.00239035 |
| 0 | 0 | 0.000645518 | 0 | 0 |
| 0 | 0 | 0 | 0 | 0 |
| 0 | 0 | 0 | 0 | 0 |
| 0.004292266 | 0 | 0 | 0 | 0 |
| 0 | 0 | 0 | 0 | 0 |
| 0.235946955 | 0.564721987 | 0.021994147 | 0.036342276 | 0 |
| 0 | 0 | 0 | 0 | 0 |
| 0 | 0 | 0 | 0 | 0.341013116 |
| 0 | 0 | 0 | 0 | 0 |
| 0 | 0 | 0 | 0 | 0 |
| 0 | 0 | 0 | 0 | 0 |
| 0 | 0 | 0 | 0 | 0 |
| 0.001214777 | 0 | 0 | 0 | 0 |
| 0 | 0 | 0 | 0 | 0 |
| 0 | 0 | 0.002787124 | 0 | 0 |
| 0 | 0 | 0 | 0 | 0 |
| 0 | 0 | 0.075288948 | 0.023313977 | 0 |
| 0 | 0 | 0 | 0 | 0 |
| 0 | 0 | 0 | 0 | 0 |
| 0 | 0 | 0 | 0 | 0 |
| 0 | 0 | 0 | 0 | 0 |
| 0 | 0 | 0 | 0 | 0 |
| 0 | 0 | 0 | 0 | 0 |

| | | | | |
|---|---|---|---|---|
| 0 | 0 | 0 | 0 | 0 |
| 0 | 0 | 0 | 0 | 0 |
| 0.293290309 | 0.055626012 | 0.023949541 | 0.09249316 | 0.007077193 |
| 0 | 0 | 0 | 0 | 0 |
| 0 | 0 | 0 | 0 | 0 |
| 0 | 0 | 0 | 0 | 0 |
| 0 | 0 | 0 | 0 | 0 |
| 0 | 0 | 0 | 0 | 0 |
| 0 | 0 | 0 | 0 | 0 |
| 0 | 0 | 0 | 0 | 0 |
| 0 | 0 | 0 | 0 | 0 |
| 0 | 0 | 0 | 0 | 0 |
| 0 | 0 | 0 | 0 | 0 |
| 0.014742887 | 0 | 0.004643608 | 0 | 0 |
| 0.002197058 | 0.001193907 | 0.00210798 | 0.000819132 | 0.002324047 |
| 0 | 0 | 0.017439457 | 0 | 0 |
| 0.003273613 | 0.002040525 | 0.003711956 | 0.00205332 | 0.007649904 |
| 0.002539397 | 0.006859101 | 0 | 0 | 0 |
| 0 | 0 | 0 | 0 | 0 |
| 0.002550169 | 0 | 0 | 0 | 0 |
| 0 | 0 | 0 | 0 | 0 |
| 0.005910276 | 0.00644704 | 0.000591324 | 0 | 0 |
| 0 | 0 | 0 | 0 | 0 |
| 0 | 0 | 0 | 0 | 0 |
| 0 | 0 | 0 | 0 | 0 |
| 0 | 0 | 0 | 0 | 0 |
| 8.84382874 | 0 | 0 | 0 | 0 |
| 0 | 0 | 0 | 0 | 0 |
| 0 | 0 | 0 | 0 | 0 |
| 0 | 0 | 0 | 0 | 0 |
| 0 | 0 | 0 | 0 | 0 |
| 0 | 0 | 0 | 0 | 0 |
| 0 | 0 | 0 | 0 | 0 |
| 0 | 0 | 0 | 0 | 0 |
| 0 | 0 | 0 | 0 | 0 |
| 0.045136522 | 0 | 0 | 0 | 0 |
| 0 | 0 | 0 | 0 | 0 |
| 0.058749997 | 0.011185767 | 0.026993402 | 0 | 0.252946693 |
| 0 | 0 | 0 | 0 | 0 |
| 0 | 0 | 0 | 0 | 0 |
| 0 | 0 | 0 | 0 | 0 |
| 0 | 0 | 0 | 0 | 0 |
| 0 | 0 | 0 | 0 | 0 |
| 0 | 0 | 0 | 0 | 0 |
| 0 | 0 | 0 | 0 | 0 |
| 0 | 0 | 0 | 0 | 0 |
| 0 | 0 | 0 | 0 | 0 |
| 0 | 0 | 0 | 0.001282169 | 0 |

| | | | | |
|---|---|---|---|---|
| 0.091384047 | 2.6374076 | 0.000988746 | 0 | 0 |
| 0 | 0 | 0 | 0 | 0 |
| 0.003459427 | 0 | 0 | 0 | 0 |
| 0 | 0 | 0 | 0 | 0 |
| 0 | 0 | 0 | 0.001065435 | 0.001679367 |
| 0 | 0 | 0 | 0 | 0 |
| 0 | 0 | 0 | 0 | 0 |
| 0.014892907 | 0.051697381 | 0.000873471 | 0.144117141 | 0.032563705 |
| 0 | 0 | 0 | 0 | 0 |
| 0 | 0 | 0.008702949 | 0.028990581 | 0 |
| 0 | 0 | 0 | 0 | 0 |
| 0.141877868 | 0 | 0.000691442 | 0 | 0.006211457 |
| 0.027444489 | 0.005990596 | 0.005944148 | 0 | 0.00447603 |
| 0.002575798 | 0.003308426 | 0 | 0 | 0 |
| 0 | 0 | 0 | 0 | 0 |
| 0 | 0 | 0 | 0 | 0 |
| 0 | 0 | 0 | 0 | 0 |
| 0 | 0 | 0 | 0 | 0 |
| 0 | 0 | 0 | 0 | 0 |
| 0 | 0 | 0 | 0 | 0 |
| 0 | 0 | 0 | 0 | 0 |
| 0 | 0 | 0 | 0 | 0 |
| 0 | 0 | 0 | 0 | 0 |
| 0 | 0 | 0 | 0 | 0 |
| 0 | 0 | 0 | 0 | 0 |
| 0.001257083 | 0.003805918 | 0.0456131 | 0.002437135 | 0 |
| 0 | 0 | 0 | 8.539364324 | 0 |
| 0 | 0.003068275 | 0 | 0.004841786 | 0 |
| 0 | 0 | 0 | 0 | 0 |
| 0 | 0.005547399 | 0.001047548 | 0 | 0 |
| 0 | 0 | 0 | 0 | 0 |
| 0 | 0 | 0 | 0 | 0 |
| 0 | 0 | 0 | 0 | 0 |
| 0 | 0 | 0 | 0 | 0 |
| 0 | 0 | 0 | 0 | 0 |
| 0 | 0 | 0 | 0 | 0 |
| 0 | 0 | 0 | 0.002368065 | 0 |
| 0 | 0 | 0 | 0 | 0 |
| 0 | 0 | 0 | 0 | 0 |
| 0 | 0.001000713 | 0 | 0 | 0 |
| 0 | 0 | 0 | 0 | 0 |
| 0 | 0 | 0 | 0 | 0 |
| 0 | 0 | 0 | 0 | 0 |
| 0 | 0 | 0 | 0 | 0 |
| 0 | 0 | 0 | 0 | 0 |
| 0 | 0 | 0.000595935 | 0 | 0 |
| 0.00111663 | 0.00709944 | 0 | 0.000865934 | 0 |
| 0 | 0 | 0 | 0 | 0 |

| | | | | |
|---|---|---|---|---|
| 0 | 0 | 0 | 0 | 0 |
| 0 | 0.003967061 | 0 | 0 | 0 |
| 0 | 0 | 0 | 0 | 0 |
| 0 | 0 | 0 | 0 | 0 |
| 0 | 0 | 0 | 0 | 0 |
| 0.005100993 | 0 | 0.008975214 | 0 | 0 |
| 0 | 0 | 0 | 0 | 0 |
| 0 | 0 | 0 | 0 | 0 |
| 0 | 0 | 0.545967668 | 0 | 0 |
| 0 | 0 | 0 | 0 | 0 |
| 0.010404516 | 0.019816484 | 0 | 0.004152949 | 0.007169416 |
| 0 | 0 | 0 | 0 | 0 |
| 0 | 0 | 0 | 0.00076509 | 0 |
| 0.060712628 | 0.001021179 | 0.008741864 | 0.002241998 | 0 |
| 0.016771438 | 0 | 0 | 0 | 0 |
| 0 | 0 | 0 | 0 | 0 |
| 0 | 0 | 0 | 0 | 0 |
| 0 | 0 | 0 | 0 | 0 |
| 0 | 0.036229607 | 0 | 0 | 0 |
| 0 | 0.018165918 | 0 | 0 | 0 |
| 0 | 0.001100279 | 0 | 0 | 0 |
| 0.138156066 | 0.305956748 | 0.204343939 | 0.175747686 | 0 |
| 0.056333084 | 0 | 0 | 0 | 0 |
| 0 | 0 | 0 | 0 | 0 |
| 0 | 0 | 0 | 0 | 0 |
| 0 | 0 | 0 | 0 | 0 |
| 0 | 0 | 0 | 0 | 0 |
| 0 | 0 | 0 | 0 | 0 |
| 0 | 0 | 0 | 0 | 0 |
| 0.604580549 | 1.246364409 | 0.526324409 | 0.354372705 | 0 |
| 0.001961366 | 0.043414771 | 0.002223997 | 0 | 0 |
| 0 | 0 | 0 | 0 | 0 |
| 0 | 0 | 0 | 0 | 0 |
| 0 | 0 | 0 | 0 | 0 |
| 0 | 0.020288776 | 0 | 0 | 0 |
| 0 | 0 | 0 | 0 | 0 |
| 0 | 0 | 0 | 0 | 0 |
| 0.007072268 | 0 | 0 | 0 | 0 |
| 0 | 0 | 0 | 0 | 0 |
| 0 | 0 | 0 | 0 | 0 |
| 0 | 0.013152486 | 0.004491727 | 0 | 0 |
| 2.2116139 | 1.069316016 | 5.254863304 | 1.204752896 | 0 |
| 0.007558431 | 0 | 0 | 0 | 0 |
| 0 | 0 | 0 | 0 | 0 |
| 0 | 0 | 0 | 0 | 0 |
| 0 | 0 | 0 | 0 | 0 |
| 0 | 0 | 0 | 0 | 0.00228695 |
| 0.003268949 | 0 | 0 | 0 | 0 |

| | | | | |
|---|---|---|---|---|
| 0.002257141 | 0.003623917 | 0.107959174 | 0.000795631 | 0 |
| 0 | 0 | 0 | 0 | 0 |
| 1.971692808 | 0.0291334 | 0.004438442 | 0.033065293 | 0.032894395 |
| 0 | 0 | 0 | 0 | 0 |
| 0.549092244 | 0 | 0.103223728 | 0 | 0 |
| 0 | 0.078745439 | 0 | 0 | 0 |
| 0 | 0 | 0 | 0 | 0 |
| 0 | 0 | 0 | 0 | 0 |
| 0.001373836 | 0 | 0 | 0 | 0 |
| 0.017314863 | 0 | 0 | 0 | 0 |
| 0 | 0 | 0 | 0 | 0 |
| 0 | 0 | 0 | 0 | 0 |
| 0 | 0 | 0 | 0 | 0 |
| 0 | 0 | 0 | 0.000719787 | 0 |
| 0 | 0 | 0 | 0 | 0 |
| 0.040619625 | 0.370986964 | 0.045894182 | 0.003234382 | 0 |
| 0.011255655 | 0.01533482 | 0.138020678 | 0.001402816 | 0.006633469 |
| 0 | 0 | 0 | 0 | 0 |
| 0 | 0.013786416 | 0 | 0 | 0 |
| 0 | 0 | 0 | 0 | 0 |
| 0.010942967 | 0.051178739 | 0.001608477 | 0.001964386 | 0 |
| 0.001865139 | 0 | 0.00148042 | 0.002350391 | 0 |
| 0.000631073 | 0 | 0 | 0 | 0 |
| 0.00141998 | 0 | 0 | 0 | 0 |
| 0 | 0.051753983 | 0 | 0 | 0 |
| 0 | 0.006733641 | 0 | 0 | 0 |
| 0 | 0 | 0 | 0 | 0 |
| 0 | 0 | 0 | 0 | 0 |
| 0 | 0 | 0 | 0 | 0 |
| 0.001746664 | 0 | 0 | 0 | 0 |
| 0.005278186 | 0.008499232 | 0.008977419 | 0 | 0 |
| 0.004166009 | 0 | 0.005511154 | 0.002019184 | 0.001768161 |
| 0.090130051 | 0.070053691 | 0.004747615 | 0.013671365 | 0 |
| 0.002842265 | 0 | 0 | 0 | 0 |
| 0 | 0.00238852 | 0.00460059 | 0 | 0 |
| 0 | 0 | 0 | 0 | 0 |
| 0.016535828 | 0.002462964 | 0.023042181 | 0.002896841 | 0 |
| 0.11919054 | 0.159320303 | 0 | 0.000906186 | 0 |
| 0.083450392 | 0 | 0.346869272 | 0.309100927 | 0 |
| 0 | 0 | 0 | 0.003581014 | 0 |
| 0 | 0 | 0 | 0 | 0 |
| 0 | 0 | 0 | 0 | 0 |
| 0.000932022 | 0.004938099 | 0 | 0 | 0 |
| 0.001459497 | 0 | 0 | 0 | 0 |
| 0 | 0 | 0 | 0 | 0 |
| 0 | 0 | 0 | 0 | 0 |
| 0 | 0 | 0 | 0 | 0 |
| 0 | 0 | 0 | 0 | 0 |

| | | | | |
|---|---|---|---|---|
| 0 | 0 | 0 | 0.033365386 | 0 |
| 0.042673018 | 0.151103509 | 0.011314379 | 0.011113136 | 3.55396831 |
| 0 | 0 | 0 | 0 | 0 |
| 0 | 0 | 0.004373883 | 0 | 0 |
| 0 | 0 | 0 | 0 | 0 |
| 0 | 0 | 0 | 0 | 0 |
| 0 | 0 | 0 | 0 | 0 |
| 0 | 0 | 0 | 0 | 0 |
| 0.001166079 | 0.009610519 | 0 | 0 | 0 |
| 0 | 0 | 0 | 0.0008973 | 0 |
| 0 | 0.008358791 | 0.000536669 | 0.003823276 | 0 |
| 0.164026575 | 0.044407342 | 0.150284243 | 0.014806682 | 0.127824182 |
| 0.002238386 | 0 | 0.093233274 | 0 | 0 |
| 0 | 0 | 0.003139884 | 0 | 0 |
| 0.00506843 | 0.007352845 | 0.011288952 | 0.000526408 | 0 |
| 0 | 0 | 0 | 0 | 0 |
| 0.004460467 | 0.02013158 | 0.004495762 | 0.014252536 | 0.012116077 |
| 0.005790302 | 0.006817454 | 0 | 0 | 0 |
| 0 | 0 | 0 | 0 | 0 |
| 0.000974794 | 0.000983755 | 0 | 0 | 0 |
| 0.034073087 | 0.09267112 | 0.151258155 | 0.110812715 | 0.017700871 |
| 0 | 0 | 0 | 0 | 0 |
| 0 | 0 | 0 | 0 | 0 |
| 0 | 0 | 0 | 0 | 0 |
| 0 | 0 | 0 | 0 | 0 |
| 0 | 0 | 0 | 0 | 0 |
| 0.02942786 | 0.095034832 | 0.013728677 | 0 | 0 |
| 0.019193771 | 0.124079023 | 0.0137564 | 0.017377017 | 0 |
| 0 | 0 | 0 | 0 | 0 |
| 0 | 0.003471689 | 0 | 0 | 0 |
| 0.000971459 | 0.006176454 | 0 | 0.005461829 | 0 |
| 0 | 0 | 0 | 0 | 0 |
| 0 | 0 | 0 | 0 | 0 |
| 0 | 0 | 0 | 0 | 0 |
| 0 | 0.121502672 | 0 | 0.089633496 | 0 |
| 0.066194446 | 0.254377016 | 0.02501934 | 0.162792334 | 0 |
| 0 | 0 | 0 | 0 | 0 |
| 0 | 0 | 0 | 0 | 0 |
| 0.057853912 | 0.005917475 | 0 | 0 | 0.008873819 |
| 0 | 0 | 0 | 0 | 0 |
| 0 | 0 | 0 | 0 | 0 |
| 0 | 0 | 0 | 0 | 0 |
| 0.012346961 | 0 | 0.007230895 | 0.02091228 | 0 |
| 0 | 0.014575224 | 0.005941526 | 0 | 0 |
| 0.054039443 | 0 | 0 | 0 | 0 |
| 0 | 0 | 0.495249431 | 0.260432734 | 0 |
| 0 | 0 | 0 | 0 | 0 |
| 0 | 0 | 0 | 0 | 0 |

| | | | | |
|---|---|---|---|---|
| 0 | 0 | 0 | 0 | 0 |
| 0 | 0 | 0 | 0 | 0 |
| 0 | 0 | 0.009446519 | 0 | 0 |
| 0 | 0.053305521 | 1.017951207 | 0.005820976 | 0 |
| 0 | 0 | 0 | 0 | 0 |
| 0 | 0.001694074 | 0 | 0 | 0 |
| 0 | 0 | 0 | 0 | 0 |
| 0.000848223 | 0 | 0.067326165 | 0.005344526 | 0 |
| 2.553956263 | 0.389387886 | 2.081297006 | 0.218687903 | 0 |
| 1.440887163 | 0 | 0 | 0.000547634 | 0 |
| 0 | 0 | 0 | 0 | 0 |
| 0.000454216 | 0.002406554 | 0 | 0 | 0 |
| 0 | 0 | 0 | 0 | 0 |
| 0 | 0 | 0.051619418 | 0 | 0 |
| 0.423171697 | 0.075684639 | 1.493198616 | 0.017561233 | 0 |
| 0 | 0 | 0 | 0 | 0 |
| 0.340451121 | 0.010073876 | 1.006397426 | 0 | 0 |
| 7.011654589 | 0 | 0 | 0.013601242 | 0 |
| 0 | 0 | 0 | 0 | 0 |
| 0 | 0 | 0 | 0 | 0 |
| 0 | 0 | 0 | 0 | 0 |
| 0 | 0 | 0.001115738 | 0 | 0 |
| 0 | 0 | 0 | 0 | 0 |
| 0.164035985 | 0.89397372 | 0.298174341 | 0.035115217 | 0 |
| 0 | 0 | 0 | 0.01209215 | 0 |
| 1.493501602 | 0 | 0.182385456 | 0 | 0 |
| 0.00944526 | 0 | 0.00860625 | 0 | 0 |
| 1.579656684 | 1.329493951 | 1.144865677 | 0 | 0 |
| 0 | 0.061091214 | 0 | 0 | 0 |
| 0 | 0 | 0 | 0 | 0 |
| 0.0017663 | 0.127897075 | 0.007343643 | 0.001712183 | 0 |
| 0 | 0 | 0.449425271 | 0 | 0 |
| 0.089394901 | 0.033435573 | 0.044331338 | 0 | 0 |
| 0 | 0 | 0 | 0 | 0 |
| 0.017802934 | 0 | 0.011865514 | 0 | 0 |
| 0 | 0.213358251 | 0 | 0.002782501 | 0 |
| 2.244951026 | 0 | 0.589052454 | 0.162493893 | 0 |
| 0 | 0 | 0 | 0 | 0 |
| 0.006462127 | 0.013917904 | 0 | 0 | 0 |
| 0.002709296 | 0 | 0.093022456 | 0.002101029 | 0 |
| 0.047059784 | 0.184692655 | 0.004446766 | 0.000422388 | 0.005104309 |
| 0 | 0 | 0 | 0 | 0 |
| 0.620471143 | 0.152938988 | 0.01876933 | 0.044252366 | 0 |
| 0.012020204 | 0.013854541 | 0 | 0 | 0 |
| 0 | 0 | 0 | 0 | 0 |
| 0 | 0 | 0 | 0 | 0 |
| 0 | 0 | 0 | 0 | 0 |
| 0.03753957 | 0.036078536 | 0.001385876 | 0 | 0 |

| | | | | |
|---|---|---|---|---|
| 0 | 0 | 0 | 0 | 0 |
| 0.098622136 | 0 | 0 | 0.00107936 | 0 |
| 0.000669505 | 0.005616424 | 0 | 0.017685046 | 0 |
| 0.017911657 | 0.020130443 | 0 | 0 | 0 |
| 0 | 0 | 0 | 0.554579693 | 0 |
| 0.024656443 | 0.038422443 | 0.084472009 | 0.016999105 | 0 |
| 0 | 0 | 0 | 0 | 0 |
| 0 | 0 | 0 | 0 | 0 |
| 0 | 0 | 0 | 0 | 0 |
| 0 | 0.704971099 | 0 | 0 | 0 |
| 0.043548483 | 0.006385017 | 0.405845356 | 0.005256863 | 0 |
| 1.717501999 | 1.814224748 | 0.314907953 | 0.33226108 | 0 |
| 0 | 0 | 0 | 0 | 0 |
| 0.01578467 | 0.022552267 | 0.001206625 | 0.00653303 | 0.006774695 |
| 0.004472423 | 0.48058588 | 0.038034669 | 0.003522506 | 0 |
| 0.005554059 | 0 | 2.312231584 | 0.036871478 | 0 |
| 0 | 0 | 0 | 0 | 0 |
| 0 | 0 | 0 | 0 | 0 |
| 0 | 0 | 0 | 0 | 0 |
| 0.025584529 | 0 | 0.015963368 | 0 | 0 |
| 0.077132163 | 0.120410647 | 0 | 0 | 0.411432388 |
| 0 | 0.008458364 | 0.332749144 | 0 | 0.001108752 |
| 0.049175908 | 0.425513928 | 0.211831519 | 0.105679366 | 0.002385317 |
| 0 | 0 | 0 | 0 | 0 |
| 0 | 0 | 0 | 0 | 0 |
| 0.699642921 | 2.015369171 | 2.85721081 | 1.992381984 | 0 |
| 0 | 0.003782258 | 0 | 0 | 0 |
| 0.001580218 | 0 | 0 | 0 | 0 |
| 0 | 0.068995365 | 0 | 0 | 0 |
| 0 | 0 | 0 | 0 | 0 |
| 0.021268695 | 0 | 0.005167846 | 0 | 0 |
| 0 | 0.230109565 | 0.043938635 | 0.04827687 | 0 |
| 0.022784651 | 0 | 0 | 0.001799645 | 0 |
| 0 | 0.089102693 | 0 | 0.002869498 | 0 |
| 0 | 0.012000595 | 0 | 0 | 0 |
| 0.091675851 | 0.451124768 | 0.116619977 | 0.016467842 | 0 |
| 0.01700188 | 0.015143955 | 0.059176513 | 0.001003189 | 0 |
| 0 | 0 | 0 | 0 | 0 |
| 1.179892751 | 0 | 2.897527684 | 0 | 0 |
| 0 | 0 | 0 | 0 | 0 |
| 0 | 0 | 0.005042801 | 0 | 0 |
| 0 | 0 | 0 | 0 | 0 |
| 0 | 0 | 0 | 0 | 0 |
| 0 | 0 | 0.037189932 | 0 | 0 |
| 0.005934158 | 0.032981938 | 0.005937136 | 0.010151192 | 0 |
| 0.007419559 | 0 | 0 | 0 | 0 |
| 0 | 0 | 0 | 0 | 0 |
| 0.176850047 | 0.01784758 | 0 | 0 | 0 |

| | | | | |
|---|---|---|---|---|
| 0 | 0 | 0 | 0 | 0 |
| 0 | 0 | 0.00211148 | 0 | 0 |
| 0 | 0 | 0 | 0 | 0 |
| 0.002677584 | 0.017601821 | 0.006746929 | 0 | 0 |
| 0 | 0 | 0 | 0 | 0 |
| 0 | 0 | 0 | 0 | 0 |
| 0 | 0 | 0 | 3.538932496 | 0 |
| 0.162751775 | 0.066237915 | 0.021263757 | 0 | 0 |
| 0 | 0 | 0 | 0 | 0 |
| 0 | 0 | 0 | 0 | 0 |
| 0.007455299 | 0.013751897 | 0.003193574 | 0 | 0 |
| 0 | 0 | 0 | 0 | 0 |
| 0 | 0.002049508 | 0 | 0 | 0 |
| 0.58019804 | 0 | 0.014747971 | 0.004385352 | 0 |
| 0 | 0 | 0 | 0 | 0 |
| 0 | 0 | 0 | 0 | 0 |
| 0 | 0 | 0 | 0 | 0 |
| 0.000551677 | 0.00292293 | 0.000625548 | 0.012968275 | 0 |
| 0.066758711 | 0.105342626 | 0 | 0 | 0 |
| 0 | 0 | 0 | 0 | 0 |
| 0.026589094 | 0.004111588 | 0 | 0.000475105 | 0 |
| 0 | 0 | 0 | 0 | 0 |
| 0 | 0 | 0.00306515 | 0 | 0 |
| 0.004653623 | 0 | 0 | 0 | 0 |
| 0 | 0 | 0 | 0 | 0 |
| 0 | 0 | 0 | 0 | 0 |
| 0 | 0 | 0 | 0 | 0 |
| 0 | 0 | 0 | 0 | 0 |
| 0 | 0 | 0 | 0 | 0 |
| 1.360664884 | 0.001765436 | 0.175312203 | 0 | 0 |
| 0 | 0 | 0.001775503 | 0.000885418 | 0 |
| 0 | 0 | 0 | 0 | 0 |
| 0 | 0 | 0.004084943 | 0.00139687 | 0 |
| 0 | 0 | 0 | 0 | 0 |
| 0 | 0.008142965 | 0 | 0 | 0 |
| 0 | 0 | 0 | 0 | 0 |
| 0.035592391 | 0.17139629 | 0.053407465 | 0.073949028 | 0 |
| 0 | 0.017090304 | 0.000477074 | 0 | 0 |
| 0 | 0 | 0 | 0 | 0 |
| 0 | 0 | 0 | 0 | 0 |
| 0.084082723 | 0.045316121 | 0 | 0 | 0 |
| 0 | 0 | 0 | 0 | 0 |
| 0 | 0 | 0 | 0 | 0 |
| 0.41280662 | 0.683832574 | 0 | 0 | 0 |
| 0 | 0 | 0 | 0.044457983 | 0 |
| 0 | 0 | 0 | 0 | 0 |
| 0.00426421 | 0.024710991 | 0.007403896 | 0 | 0 |
| 0 | 0 | 0 | 0 | 0 |

| | | | | |
|---|---|---|---|---|
| 0 | 0 | 0 | 0 | 0 |
| 0 | 0 | 0 | 0 | 0 |
| 0 | 0 | 0 | 0 | 0 |
| 0.87203342 | 0 | 0.714195215 | 0 | 0 |
| 0 | 0 | 0 | 0 | 0 |
| 0 | 0 | 0 | 0 | 0 |
| 0 | 0 | 0 | 0 | 0 |
| 0 | 0 | 0 | 0 | 0 |
| 0.015667164 | 0.007515195 | 0.001973892 | 0.002437442 | 0 |
| 0 | 0 | 0.001412948 | 0 | 0 |
| 0 | 0 | 0 | 0 | 0 |
| 0 | 0 | 0 | 0 | 0 |
| 0 | 0 | 0 | 0.001531339 | 0 |
| 0.002149513 | 0 | 0 | 0 | 0 |
| 1.561781431 | 0.681214243 | 0.557624574 | 0.126250674 | 0 |
| 0.003279662 | 0 | 0 | 0 | 0 |
| 0 | 0 | 0 | 0 | 0 |
| 0 | 0 | 0 | 0 | 0 |
| 0 | 0 | 0 | 0 | 0 |
| 0 | 0 | 0 | 0 | 0 |
| 0.069808179 | 0 | 0.000503108 | 0 | 0 |
| 0 | 0 | 0.002131375 | 0 | 0 |
| 0 | 0 | 0 | 0 | 0 |
| 0 | 0 | 0 | 0 | 0 |
| 0.074833846 | 0.001205135 | 0.308209218 | 0.005512236 | 1.351240502 |
| 0.260911552 | 13.68709324 | 4.894743242 | 0.519551172 | 0 |
| 0 | 0 | 0.058900621 | 0 | 0 |
| 0.001411941 | 0 | 0 | 0 | 0 |
| 0 | 0 | 0.247590263 | 0 | 0 |
| 0 | 0.001691912 | 0 | 0 | 0 |
| 0 | 0 | 0 | 0 | 0 |
| 0 | 0.169192738 | 0 | 0 | 0 |
| 0 | 0 | 0 | 0 | 0 |
| 0.000728865 | 0.000858159 | 0.018870863 | 0 | 0 |
| 0.016612985 | 0.02509806 | 0.025419275 | 0.012223504 | 0.009174768 |
| 0.008489181 | 0.0054973 | 0.010748921 | 0.000822908 | 0 |
| 0 | 0.040742895 | 0 | 0 | 0 |
| 1.185278282 | 2.246232919 | 0.056882694 | 0.975356095 | 0 |
| 2.94089296 | 0 | 1.239818647 | 0 | 0 |
| 0 | 0 | 0 | 0 | 0 |
| 0 | 0 | 0 | 0 | 0 |
| 0 | 0.038537344 | 0 | 0 | 0 |
| 2.419695306 | 0.120194529 | 0.192047866 | 0.005247754 | 0 |
| 0 | 0 | 0 | 0 | 0 |
| 0 | 0 | 0.358694564 | 0 | 0 |
| 0.003753135 | 0 | 0.013678998 | 0 | 0 |
| 0 | 0 | 0 | 0 | 0 |
| 0.524033706 | 0.230429502 | 0.001937092 | 0.022562995 | 0.163139414 |

| | | | | |
|---|---|---|---|---|
| 0.00636956 | 0.00421845 | 0 | 0 | 0 |
| 0.008000214 | 0 | 0 | 0 | 0 |
| 0 | 0 | 0 | 0 | 0 |
| 0 | 0 | 0 | 0 | 0 |
| 0 | 0 | 0.006831253 | 0 | 0 |
| 0.001679397 | 0 | 0 | 0.025011118 | 0 |
| 0 | 0 | 0 | 0 | 0 |
| 0.001142545 | 0 | 0 | 0 | 0 |
| 0 | 0 | 0 | 0 | 0 |
| 0.003500838 | 0 | 0 | 0 | 0 |
| 0 | 0 | 0.000687777 | 0 | 0 |
| 1.751658536 | 3.227389426 | 1.109862419 | 0.10123285 | 0 |
| 0.074674463 | 0.014578175 | 0.002176697 | 0.357838761 | 0 |
| 0 | 0 | 0 | 0 | 0 |
| 0.134580745 | 0.003238166 | 0.003326461 | 0.002666022 | 0.004762555 |
| 0 | 0 | 0 | 0 | 0 |
| 0.007015149 | 0 | 0 | 0 | 0 |
| 0 | 0 | 0 | 0 | 0 |
| 0 | 0.000851472 | 0 | 0 | 0 |
| 0.132045696 | 0.00786789 | 0 | 0 | 0 |
| 0.22711864 | 0 | 0 | 0 | 0 |
| 0 | 0 | 0 | 0 | 0 |
| 1.03151531 | 0.126950959 | 2.613566271 | 0.024993984 | 0 |
| 0 | 0 | 0.014302288 | 0 | 0 |
| 0 | 0 | 0 | 0 | 0 |
| 0 | 0 | 0 | 0 | 0 |
| 0 | 0 | 0 | 0 | 0 |
| 0 | 0 | 0 | 0 | 0 |
| 0 | 0 | 0 | 0 | 0 |
| 0 | 0 | 0 | 0 | 0 |
| 0 | 0 | 0 | 0 | 0 |
| 0.035450904 | 0.08992222 | 0.011330939 | 0.017297681 | 0.020357915 |
| 0 | 0 | 0 | 0 | 0 |
| 0.00273386 | 0.047232748 | 0 | 0.000806552 | 0 |
| 0.006466849 | 0.004350865 | 0.008904082 | 0 | 0 |
| 0.036125062 | 0.034003307 | 0.088901657 | 0.006157051 | 0 |
| 0 | 0 | 0 | 0 | 0 |
| 0 | 0 | 0 | 0.007707011 | 0 |
| 0 | 0.006248128 | 0.00100289 | 0 | 0 |
| 0 | 0 | 0.002950848 | 0 | 0 |
| 0.047751419 | 0 | 0.086275691 | 0 | 0 |
| 0 | 0 | 0 | 0 | 0 |
| 0 | 0 | 0 | 0 | 0 |
| 0 | 0 | 0 | 0 | 0 |
| 0.00970742 | 0.020090805 | 0.128647399 | 0.01867296 | 0 |
| 0 | 0 | 0 | 0 | 0 |
| 1.865276632 | 0 | 0 | 0 | 0 |
| 0 | 0 | 0 | 0 | 0 |

| | | | | |
|---|---|---|---|---|
| 0 | 0 | 0 | 0 | 0 |
| 0 | 0 | 0 | 0 | 0 |
| 0.0230604 | 0 | 0 | 0 | 0 |
| 0 | 0 | 0 | 0 | 0 |
| 0.002863663 | 0 | 0 | 0 | 0 |
| 0 | 0 | 0 | 0 | 0 |
| 0.866350878 | 0 | 0 | 0.070639523 | 0 |
| 0.109484568 | 0.320782856 | 0.012431277 | 0 | 0 |
| 0 | 0 | 0 | 0 | 0 |
| 0 | 0 | 0.006116426 | 0 | 0 |
| 0 | 0 | 0 | 0 | 0 |
| 0 | 0.003100834 | 0 | 0 | 0 |
| 0 | 0 | 0 | 0 | 0 |
| 0.490356639 | 0 | 0 | 0 | 0 |
| 0 | 0 | 0 | 0 | 0 |
| 0 | 0 | 0 | 0 | 0 |
| 0 | 0 | 3.384937481 | 0 | 0 |
| 0 | 0 | 0 | 0 | 0 |
| 0.336757506 | 0 | 1.313381655 | 0.028922973 | 0 |
| 0 | 0 | 0 | 0 | 0 |
| 0 | 0 | 0 | 0.000828999 | 0 |
| 0 | 0 | 0 | 0 | 0 |
| 0 | 0 | 0 | 0 | 0 |
| 0 | 0 | 0 | 0 | 0 |
| 0 | 0 | 0 | 0 | 0 |
| 0.006636724 | 0.028180361 | 0 | 0.002600729 | 0 |
| 0 | 0 | 0 | 0 | 0 |
| 0.017414897 | 0.01415763 | 0.004074734 | 0 | 0.017739194 |
| 0 | 0 | 0 | 0 | 0 |
| 0.990746359 | 0.147213007 | 0.331100913 | 0.390540712 | 0 |
| 0 | 0 | 0 | 0 | 0 |
| 0 | 0 | 0 | 0 | 0 |
| 0 | 0.004897163 | 0 | 0 | 0 |
| 0 | 0 | 0 | 0 | 0 |
| 0.004587259 | 0.000784016 | 0.002516856 | 0 | 0 |
| 0.272750828 | 0.083260347 | 0 | 0 | 0 |
| 0 | 0 | 0 | 0 | 0 |
| 0.002014914 | 0 | 0.000761572 | 0 | 0 |
| 0 | 0 | 0 | 0 | 0 |
| 0 | 0.023775294 | 0 | 0 | 0 |
| 1.422653483 | 19.09173081 | 0.550684022 | 3.398293926 | 0 |
| 0.269051155 | 0.545498795 | 0.24375552 | 0 | 0 |
| 0 | 0 | 0 | 0 | 0 |
| 0 | 0 | 0 | 0 | 0 |
| 0 | 0 | 0 | 0 | 0 |
| 0 | 0 | 0 | 0 | 0 |
| 0.020388002 | 0 | 0.001512392 | 0 | 0 |
| 0.18230281 | 0.001412117 | 0.019039405 | 0 | 0 |

| | | | | |
|---|---|---|---|---|
| 0 | 0 | 0 | 0 | 0 |
| 0.009320846 | 0.005083677 | 0.006683292 | 0 | 0 |
| 0 | 0 | 0 | 0 | 0 |
| 0 | 0 | 0.064266447 | 0 | 0 |
| 2.854606752 | 1.257011778 | 1.434762984 | 0.003627457 | 0 |
| 0 | 0 | 0 | 0 | 0 |
| 0 | 0 | 0 | 0 | 0 |
| 0 | 0 | 0.004778946 | 0 | 0 |
| 0.002874253 | 0.015953714 | 0 | 0.005307026 | 0 |
| 0 | 0 | 0 | 0 | 0 |
| 0 | 0 | 0 | 0 | 0 |
| 0 | 0 | 0 | 0 | 0 |
| 0.163155417 | 0.227459505 | 0.005738846 | 0 | 0 |
| 0.018502758 | 0.07721141 | 0 | 0.013174122 | 0 |
| 0 | 0 | 2.033762539 | 0.214591465 | 0 |
| 0.824328407 | 0.145227656 | 0.031537815 | 0 | 0 |
| 0.007509709 | 0.001751691 | 0 | 0 | 0.00064949 |
| 0 | 0 | 0 | 0 | 0 |
| 0 | 0 | 0 | 0 | 0 |
| 0.175776246 | 0 | 0 | 0 | 0 |
| 0 | 0 | 0 | 0 | 0 |
| 0 | 0 | 0 | 0 | 0 |
| 0 | 0 | 0 | 0 | 0 |
| 0 | 0 | 0 | 0 | 0 |
| 0 | 0 | 0 | 0 | 0 |
| 0 | 0 | 0 | 0 | 0 |
| 0.032280381 | 0 | 0.025007251 | 0 | 0 |
| 0 | 0.00122766 | 0 | 0 | 0 |
| 0 | 0 | 0 | 0 | 0 |
| 0 | 0 | 0 | 0 | 0 |
| 0 | 0 | 0 | 0 | 0 |
| 0 | 0 | 0 | 0 | 0 |
| 0.429494562 | 0 | 0 | 0.033411418 | 0 |
| 0 | 0 | 0 | 0 | 0 |
| 0 | 0 | 0.395194935 | 0.197996999 | 0 |
| 0 | 0.001420148 | 0.014771068 | 0 | 0 |
| 0.12394378 | 0 | 0 | 0 | 3.237003347 |
| 0.010174862 | 0 | 0 | 0 | 0 |
| 0 | 0 | 0 | 0 | 0 |
| 0 | 0 | 0 | 0 | 0 |
| 0.1258627 | 0.016001704 | 0.023227639 | 0.008210134 | 0 |
| 0.064874602 | 0 | 0 | 0 | 0 |
| 0 | 0 | 0 | 0 | 0 |
| 0.22715787 | 0 | 0 | 0 | 0 |
| 0 | 0 | 0 | 0 | 0 |
| 0 | 0 | 0 | 0 | 0 |
| 0.656927766 | 0 | 15.0472489 | 0 | 0 |
| 0 | 0 | 0 | 0 | 0 |

| | | | | |
|---|---|---|---|---|
| 0.257556915 | 0 | 0.306715578 | 0.427028609 | 0 |
| 0 | 0 | 0 | 0 | 0 |
| 0 | 0 | 0 | 0 | 0 |
| 0 | 0 | 0 | 0 | 0 |
| 0.207344343 | 0 | 0.000854318 | 0.004089944 | 0 |
| 0 | 0 | 0 | 0 | 0 |
| 0.028552314 | 0.05491928 | 0 | 0 | 0.001727763 |
| 0 | 0 | 0 | 0 | 0 |
| 0.000698383 | 0 | 0 | 0 | 0 |
| 0 | 0 | 0 | 0 | 0 |
| 0 | 0 | 0.005442067 | 0 | 0 |
| 0.068728036 | 0 | 0 | 0 | 0 |
| 0 | 0 | 0 | 0 | 0 |
| 0 | 0 | 0 | 0 | 0 |
| 0 | 0 | 0 | 0 | 0 |
| 0 | 0 | 0 | 0 | 0 |
| 0 | 0 | 0.154412575 | 0 | 0 |
| 0 | 0.014159765 | 0 | 0 | 0 |
| 0 | 0 | 0 | 0 | 0 |
| 0 | 0 | 0 | 0 | 0 |
| 0.435203452 | 0.020439157 | 0.007782122 | 0 | 0.020355967 |
| 0 | 0 | 0 | 0 | 0 |
| 0.084606028 | 0.012845019 | 0.015989159 | 0.006618657 | 0 |
| 0.001583843 | 0.007925416 | 0.016462629 | 0.001663259 | 0 |
| 0.022248782 | 0 | 0 | 0 | 0 |
| 0 | 0.000825018 | 0 | 0 | 0 |
| 0 | 0 | 0 | 0 | 0 |
| 0 | 0 | 0 | 0 | 0 |
| 0 | 0 | 0 | 0 | 0 |
| 1.776317345 | 0.768934453 | 1.077411826 | 0.053953014 | 0.260350954 |
| 0 | 0 | 0.008591802 | 0 | 0 |
| 0 | 0 | 0 | 0 | 0 |
| 0 | 0 | 0 | 0 | 0 |
| 0.005356256 | 0 | 0.43409324 | 0 | 0 |
| 0 | 0 | 0 | 0 | 0 |
| 0 | 0 | 0.033901462 | 0 | 0 |
| 0.02097403 | 0 | 0.000561791 | 0.002401348 | 0 |
| 0 | 0 | 0 | 0 | 0 |
| 0 | 0 | 0 | 0 | 0 |
| 0 | 0 | 0 | 0 | 0 |
| 0 | 0 | 0 | 0 | 0 |
| 0 | 0 | 0 | 0 | 0 |
| 0 | 0 | 0 | 0 | 0 |
| 0 | 0 | 0 | 0 | 0 |
| 0 | 0 | 0.036332357 | 0 | 0 |
| 0 | 0 | 0 | 0 | 0 |
| 0 | 0 | 0 | 0 | 0 |
| 0 | 0 | 0 | 0.056763642 | 0 |

| | | | | |
|---|---|---|---|---|
| 0.002272362 | 0.010032976 | 0.008051988 | 0 | 0 |
| 0 | 0 | 0 | 0.012534412 | 0 |
| 0 | 0 | 0 | 0 | 0 |
| 0 | 0 | 0 | 0 | 0 |
| 0 | 0 | 0 | 0 | 0 |
| 0.031181778 | 0 | 0 | 0 | 0 |
| 0.001637704 | 0 | 0 | 0 | 0 |
| 0.71133505 | 0.372660319 | 0.080691499 | 0.353958484 | 0 |
| 0.584734505 | 0.025197219 | 0.280328678 | 0.608741441 | 51.70103527 |
| 0 | 0.006852167 | 0 | 0 | 0 |
| 0 | 0 | 0.002065037 | 0 | 0 |
| 0 | 0 | 0 | 0 | 0 |
| 0 | 0 | 0.013411146 | 0 | 0 |
| 0.009737119 | 0 | 0.011040939 | 0 | 0 |
| 0 | 0 | 0.005076901 | 0 | 0 |
| 0 | 0 | 0 | 0 | 0 |
| 0 | 0 | 0 | 0 | 0 |
| 0 | 0 | 0 | 0 | 0 |
| 0 | 0 | 0 | 0 | 0 |
| 0 | 0 | 0.169562303 | 0 | 0 |
| 0 | 0 | 0 | 0 | 0 |
| 0 | 0 | 0 | 0 | 0 |
| 0 | 0 | 0.490844221 | 0 | 0 |
| 0 | 0 | 0 | 0 | 0 |
| 0.013423501 | 0 | 0.004185757 | 0 | 0 |
| 0 | 0 | 0 | 0 | 0 |
| 0 | 0 | 0 | 0 | 0 |
| 0 | 0 | 0 | 0 | 0 |
| 0 | 0 | 0 | 0 | 0 |
| 0 | 0 | 0 | 0 | 0 |
| 0 | 0 | 0 | 0 | 0 |
| 0 | 0 | 0 | 0 | 0 |
| 0 | 0 | 0 | 0 | 0 |
| 0 | 0 | 0 | 0 | 0 |
| 0 | 0.011235354 | 0 | 0.002833397 | 0 |
| 0 | 0 | 0 | 0 | 0 |
| 0 | 0 | 0 | 0 | 0 |
| 0 | 0 | 0 | 0 | 0 |
| 0 | 0 | 0 | 0 | 0 |
| 0 | 0 | 0 | 0.016767556 | 0 |
| 0 | 0 | 0 | 0 | 0 |
| 0.191473179 | 1.13615016 | 0.089896574 | 0 | 0 |
| 0 | 0 | 0 | 0 | 0 |
| 0 | 0 | 0.000418502 | 0 | 0 |
| 0 | 0 | 0 | 0 | 0 |
| 0 | 0 | 0 | 0 | 0 |
| 0 | 0 | 0 | 0 | 0 |
| 0 | 0 | 0 | 0 | 0 |

| | | | | |
|---|---|---|---|---|
| 0 | 0 | 0 | 0 | 0 |
| 0.007690898 | 0.00430122 | 0.005232435 | 0 | 0 |
| 0 | 0 | 0 | 0 | 0 |
| 0 | 0 | 0 | 0 | 0 |
| 1.302720731 | 0.93434059 | 0.906737908 | 0.076438987 | 0 |
| 0 | 0.839425145 | 0 | 0 | 0 |
| 0 | 0 | 0 | 0 | 0 |
| 0.012646415 | 0.001914401 | 0.004916502 | 0.002101533 | 0 |
| 0.005591947 | 0 | 0 | 0 | 0 |
| 0 | 0 | 0 | 0 | 0 |
| 0 | 0 | 0 | 0 | 0 |
| 0 | 0 | 0 | 0 | 0 |
| 0 | 0 | 0.028090562 | 0 | 0 |
| 0 | 0 | 0 | 0 | 0 |
| 0 | 0.000673835 | 0 | 0 | 0 |
| 0.077717115 | 0.070011233 | 0 | 0 | 0 |
| 0 | 0 | 0 | 0 | 0 |
| 0 | 0 | 0 | 0 | 0 |
| 0 | 0 | 0 | 0 | 0 |
| 0 | 0 | 0 | 0 | 0 |
| 0 | 0 | 0 | 0 | 0 |
| 0 | 0 | 0.014905776 | 0 | 0 |
| 0 | 0 | 0 | 0 | 0 |
| 0.004065074 | 0.007455403 | 0.003013836 | 0 | 0 |
| 0.360286597 | 0 | 0 | 0 | 0 |
| 0 | 0 | 0 | 0 | 0 |
| 0.153580834 | 0.003030583 | 0.020592639 | 0 | 0.001747942 |
| 0 | 0 | 0 | 0 | 0 |
| 0 | 0.001876912 | 0 | 0 | 0 |
| 0 | 0 | 0 | 0 | 0 |
| 0 | 0 | 0 | 0 | 0 |
| 0 | 0 | 0 | 0 | 0 |
| 0 | 0 | 0 | 0 | 0 |
| 0 | 0 | 0 | 0 | 0 |
| 0 | 0 | 0 | 0 | 0 |
| 0 | 0 | 0 | 0 | 0 |
| 0 | 0 | 0 | 0 | 0 |
| 0 | 0.085989122 | 0.186412223 | 0 | 0 |
| 0 | 0 | 0 | 0 | 0 |
| 0 | 0 | 0 | 0 | 0 |
| 0 | 0 | 0 | 0 | 0 |
| 0 | 0 | 0 | 0 | 0 |
| 0 | 0 | 0 | 0 | 0 |
| 0 | 0 | 0 | 0 | 0 |
| 0 | 0 | 0 | 0 | 0 |
| 0 | 0 | 0 | 0 | 0 |
| 0 | 0 | 0 | 0 | 0 |
| 0 | 0 | 0 | 0 | 0 |

| | | | | |
|---|---|---|---|---|
| 0.027971456 | 0.062937549 | 0.290179068 | 0.108718483 | 0 |
| 0 | 0 | 0 | 0 | 0 |
| 0 | 0 | 0 | 0 | 0 |
| 0 | 0 | 0 | 0 | 0 |
| 0 | 0 | 0.002824769 | 0 | 0 |
| 0 | 0 | 1.615757649 | 0 | 0 |
| 0 | 0 | 0 | 0 | 0 |
| 0 | 0 | 0 | 0 | 0 |
| 0 | 0 | 0 | 0 | 0 |
| 0 | 0 | 0 | 0 | 0 |
| 0 | 0 | 0 | 0 | 0 |
| 0.005909283 | 0 | 0 | 0 | 0 |
| 0 | 0 | 0.015057214 | 0 | 0 |
| 0 | 0 | 0 | 0 | 0 |
| 0 | 0 | 0 | 0 | 0 |
| 0 | 0 | 0 | 0 | 0 |
| 0 | 0 | 0 | 0 | 0 |
| 0 | 0 | 0 | 0 | 0 |
| 0 | 0 | 0 | 0 | 0 |
| 0 | 0 | 0 | 0 | 0 |
| 0.008563248 | 0.025022428 | 0 | 0.106100338 | 0 |
| 0 | 0 | 0 | 0 | 0 |
| 0.338207012 | 0.004167231 | 0.23477837 | 0 | 0 |
| 0.702594441 | 0.790147613 | 0.012039888 | 0 | 0 |
| 0 | 0 | 0 | 0 | 0 |
| 0 | 0 | 0 | 0.008180879 | 0 |
| 0 | 0 | 0 | 0 | 0 |
| 0 | 0 | 0 | 0 | 0 |
| 0.367166483 | 0.00541125 | 0 | 0 | 0 |
| 0 | 0 | 0 | 0 | 0 |
| 0 | 0 | 0 | 0 | 0 |
| 0 | 0.188207466 | 0 | 0 | 0 |
| 0.070343406 | 0 | 0 | 0.004462576 | 0 |
| 0.00149018 | 0 | 0.00915264 | 0 | 0 |
| 0.076099192 | 0.01192708 | 0.016249233 | 0.392788501 | 0 |
| 0 | 0 | 0 | 0 | 0 |
| 0 | 0 | 0 | 0 | 0 |
| 0 | 0 | 0 | 0 | 0 |
| 0.009058646 | 0.007848253 | 0 | 0.000662724 | 0 |
| 0 | 0 | 0 | 0 | 0 |
| 0 | 0 | 0 | 0 | 0 |
| 0 | 0 | 0 | 0 | 0 |
| 0.418294802 | 0.050826235 | 0.183558208 | 0.029834554 | 0 |
| 0 | 0 | 0 | 0 | 0 |
| 0 | 0 | 0 | 0 | 0 |
| 0.007935944 | 0.001401557 | 0.501401073 | 0.005231101 | 0 |
| 0 | 0 | 0 | 0 | 0 |
| 0 | 0 | 0 | 0 | 0 |

| | | | | |
|---|---|---|---|---|
| 0 | 0 | 0 | 0 | 0 |
| 0 | 0 | 0 | 0 | 0 |
| 0 | 0 | 0 | 0 | 0 |
| 0 | 0 | 0 | 0 | 0 |
| 0 | 0 | 0 | 0 | 0 |
| 0.001118539 | 0 | 0 | 0 | 0 |
| 0 | 0 | 0 | 0 | 0 |
| 0 | 0 | 0 | 0 | 0 |
| 0.002527983 | 0 | 0.007924988 | 0 | 0 |
| 0.038254015 | 0 | 0.040225037 | 0 | 0 |
| 0 | 0 | 0 | 0 | 0 |
| 0 | 0 | 0 | 0 | 0 |
| 0 | 0 | 0 | 0 | 0 |
| 0 | 0 | 0 | 0 | 0 |
| 0 | 0 | 0 | 0 | 0 |
| 0 | 0 | 0.000911912 | 0 | 0 |
| 0 | 0 | 0 | 0 | 0 |
| 0 | 0 | 0 | 0 | 0 |
| 0 | 0 | 0 | 0 | 0 |
| 0.054000461 | 0.013318439 | 0.38250435 | 0 | 0 |
| 0.004031788 | 0 | 0 | 0 | 0 |
| 0 | 0 | 0 | 0 | 0 |
| 0 | 0 | 0 | 0 | 0 |
| 0.012854928 | 0 | 0.002186434 | 0 | 0 |
| 0.003695567 | 0 | 0 | 0 | 0 |
| 0 | 0 | 0 | 0 | 0 |
| 0 | 0 | 0 | 0 | 0 |
| 0.023617173 | 0 | 0 | 0 | 0 |
| 0 | 0 | 0 | 0 | 0 |
| 0.212213508 | 0 | 0.000756102 | 0.34791587 | 0 |
| 0.775361242 | 0.207562104 | 0.54432012 | 0.158532776 | 0 |
| 0 | 0 | 0 | 0 | 0 |
| 0 | 0 | 0 | 0 | 0 |
| 0 | 0 | 0.003228774 | 0 | 0 |
| 0 | 0 | 0 | 0 | 0 |
| 0 | 0 | 0 | 0 | 0 |
| 0 | 0 | 0 | 0 | 0 |
| 0 | 0 | 0 | 0 | 0 |
| 0 | 0 | 0 | 0 | 0 |
| 0 | 0 | 0 | 0 | 0 |
| 0 | 0 | 0 | 0 | 0 |
| 0 | 0 | 0.058657615 | 0 | 0 |
| 0 | 0 | 0 | 0 | 0 |
| 0 | 0 | 0 | 0 | 0 |
| 0 | 0 | 0 | 0 | 0 |
| 0 | 0 | 0 | 0 | 0 |
| 0 | 0 | 0 | 0 | 0 |
| 0 | 0.003781281 | 0 | 0 | 0 |

| | | | | |
|---|---|---|---|---|
| 0 | 0 | 0 | 0 | 0 |
| 0 | 0 | 0 | 0 | 0 |
| 0 | 0 | 0 | 0 | 0 |
| 0 | 0.007297346 | 0 | 0 | 0 |
| 0.028538365 | 1.513992681 | 0.734649137 | 0.012997301 | 0 |
| 0 | 0 | 0 | 0 | 0 |
| 0 | 0 | 0 | 0 | 0 |
| 0 | 0 | 0 | 0 | 0 |
| 0 | 0 | 0 | 0 | 0 |
| 0 | 0 | 0 | 0 | 0 |
| 0.049977663 | 0 | 0 | 0 | 0 |
| 0.022884755 | 0.004825848 | 0.066034577 | 0.003145435 | 0 |
| 0.001178733 | 0.000780655 | 0 | 0 | 0 |
| 0 | 0 | 0 | 0 | 0 |
| 0.012233877 | 0 | 0.001918471 | 0 | 0 |
| 0.165610359 | 0.593173968 | 0 | 0 | 0 |
| 0 | 0 | 0.009383199 | 0 | 0 |
| 0 | 0 | 0 | 0 | 0 |
| 0.017716418 | 0.031060359 | 0 | 0.001253548 | 0 |
| 0.001688485 | 0 | 0 | 0 | 0.008255653 |
| 0 | 0 | 0.001146598 | 0 | 0 |
| 0 | 0 | 0 | 0 | 0 |
| 0 | 0 | 0 | 0 | 0 |
| 0 | 0 | 0 | 0 | 0 |
| 0 | 0 | 0 | 0 | 0 |
| 0 | 0 | 0 | 0 | 0 |
| 0.017860397 | 0.003823397 | 0.002761628 | 0 | 0 |
| 0 | 0 | 0 | 0 | 0 |
| 0 | 0 | 0 | 0 | 0 |
| 0 | 0 | 0 | 0 | 0 |
| 0 | 0.001354382 | 0 | 0 | 0 |
| 0 | 0 | 0 | 0 | 0 |
| 0 | 0 | 0 | 0 | 0 |
| 0 | 0 | 0 | 0 | 0 |
| 0.018255061 | 0 | 0 | 0 | 0 |
| 0 | 0 | 0 | 0 | 0 |
| 0 | 0 | 0 | 0 | 0 |
| 0.050322929 | 0.008183323 | 0.053517855 | 0.004178252 | 0 |
| 0 | 0 | 0 | 0 | 0 |
| 0 | 0 | 0 | 0 | 0 |
| 0.00408793 | 0 | 0.087217054 | 0 | 0 |
| 0 | 0 | 0 | 0 | 0 |
| 0 | 0 | 0 | 0 | 0 |
| 0 | 0.000846616 | 0 | 0 | 0 |
| 0 | 0 | 0 | 0 | 0 |
| 0 | 0 | 0 | 0 | 0 |
| 0 | 0 | 0 | 0 | 0 |
| 0 | 0 | 0 | 0 | 0 |

| | | | | |
|---|---|---|---|---|
| 0 | 0 | 0 | 0 | 0 |
| 0.121005144 | 0 | 0.004336144 | 0 | 0 |
| 0 | 0 | 0 | 0 | 0 |
| 0.002009544 | 0.01064709 | 0.001329198 | 0 | 0 |
| 0 | 0.006309031 | 0.002817852 | 0.034779249 | 0 |
| 0 | 0 | 0 | 0 | 0 |
| 0 | 0 | 0 | 0 | 0 |
| 0 | 0 | 0 | 0 | 0 |
| 0 | 0 | 0 | 0 | 0 |
| 0 | 0 | 0 | 0 | 0 |
| 0 | 0 | 0 | 0 | 0 |
| 0 | 0 | 0 | 0 | 0 |
| 0.165952338 | 0.205453946 | 0.173905881 | 0.051781462 | 0 |
| 0 | 0 | 0 | 0 | 0 |
| 0 | 0 | 0 | 0 | 0 |
| 0 | 0 | 0 | 0 | 0 |
| 0 | 0 | 0 | 0 | 0 |
| 0 | 0 | 0 | 0 | 0 |
| 0 | 0 | 0 | 0 | 0 |
| 0 | 0 | 0 | 0 | 0 |
| 0 | 0 | 0 | 0 | 0 |
| 0 | 0 | 0 | 0 | 0 |
| 0.002188336 | 0.012422549 | 0 | 0 | 0 |
| 0.04795722 | 0 | 0.116077804 | 0.009118773 | 0 |
| 0.004559548 | 0 | 0 | 0 | 0 |
| 0 | 0 | 0.003861467 | 0 | 0 |
| 0 | 0 | 0 | 0 | 0 |
| 0 | 0 | 0 | 0 | 0 |
| 0 | 0 | 0 | 0 | 0 |
| 0 | 0 | 0 | 0 | 0 |
| 0.024051281 | 0.004001425 | 0.009288221 | 0.032268448 | 0 |
| 0.002156406 | 0 | 0 | 0 | 0 |
| 0 | 0 | 0 | 0 | 0 |
| 0 | 0 | 0 | 0 | 0 |
| 0.00277621 | 0 | 0.019550429 | 0 | 0 |
| 0 | 0 | 0 | 0 | 0 |
| 0 | 0 | 0 | 0 | 0 |
| 0 | 0 | 0 | 0 | 0 |
| 0.090391077 | 0 | 0 | 0 | 0.000901217 |
| 0 | 0 | 0 | 0 | 0 |
| 0 | 0 | 0 | 0 | 0 |
| 0 | 0 | 0 | 0 | 0 |
| 0 | 0 | 0 | 0 | 0 |
| 0 | 0 | 0 | 0 | 0 |
| 0 | 0 | 0 | 0 | 0 |
| 0.260902531 | 0.43488521 | 0.213456402 | 0.254795206 | 0 |
| 0 | 0 | 0 | 0 | 0 |
| 0 | 0 | 0 | 0 | 0 |

| | | | | |
|---|---|---|---|---|
| 0 | 0 | 0 | 0 | 0 |
| 0 | 0 | 0 | 0 | 0 |
| 0.031804713 | 0 | 0 | 0 | 0 |
| 0 | 0 | 0 | 0 | 0 |
| 0 | 0 | 0 | 0 | 0 |
| 0 | 0 | 0 | 0 | 0 |
| 0 | 0 | 0 | 0.000726475 | 0 |
| 0 | 0 | 0 | 0 | 0 |
| 0.001637242 | 0 | 0 | 0 | 0 |
| 0.001346293 | 0 | 0 | 0 | 0 |
| 0.008613147 | 0 | 0.003391134 | 0 | 0 |
| 0 | 0 | 0 | 0 | 0 |
| 0 | 0 | 0 | 0 | 0 |
| 0 | 0 | 0 | 0 | 0 |
| 0 | 0 | 0 | 0 | 0 |
| 0 | 0 | 0 | 0 | 0 |
| 0 | 0 | 0 | 0 | 0 |
| 0 | 0 | 0 | 0 | 0 |
| 0 | 0 | 0 | 0 | 0 |
| 0 | 0.002521645 | 0 | 0 | 0 |
| 0 | 0 | 0 | 0 | 0 |
| 0 | 0 | 0 | 0 | 0 |
| 0 | 0 | 0 | 0 | 0 |
| 0 | 0 | 0 | 0 | 0 |
| 0 | 0 | 0 | 0 | 0 |
| 0 | 0.833316096 | 0 | 0 | 0 |
| 0 | 0 | 0 | 0 | 0 |
| 0.003686941 | 0.010852435 | 0.011546503 | 0.004254735 | 0 |
| 0 | 0 | 0 | 0 | 0 |
| 0 | 0 | 0 | 0 | 0 |
| 0.00081776 | 0.002166354 | 0 | 0.00105694 | 0 |
| 0 | 0 | 0 | 0 | 0 |
| 0.045259629 | 0.076578653 | 0.009151466 | 0.005215662 | 0 |
| 0 | 0 | 0 | 0 | 0 |
| 0 | 0 | 0 | 0 | 0 |
| 0 | 0 | 0 | 0 | 0 |
| 0 | 0 | 0 | 0 | 0 |
| 0 | 0 | 0 | 0 | 0 |
| 0 | 0 | 0 | 0 | 0 |
| 0 | 0 | 0 | 0 | 0 |
| 0 | 0 | 0 | 0 | 0 |
| 0.334200836 | 0.170837294 | 0.000590496 | 0 | 0 |
| 0.071374321 | 0.158827151 | 0.054134175 | 0.009686249 | 0 |
| 0 | 0 | 0 | 0 | 0 |
| 0 | 0 | 0 | 0 | 0 |
| 0.077928647 | 0.023477855 | 0.05275818 | 0.013997303 | 0 |
| 0.01532767 | 0 | 0 | 0.000582669 | 0 |
| 0 | 0 | 0 | 0 | 0 |

| | | | | |
|---|---|---|---|---|
| 0 | 0 | 0 | 0 | 0 |
| 0.00398795 | 0.00733653 | 0.007348161 | 0.002416102 | 0 |
| 0.00174688 | 0.012683366 | 0.011884746 | 0 | 0.048933781 |
| 0 | 0 | 0 | 0 | 0 |
| 0.027586038 | 0 | 0.000527189 | 0.000600918 | 0 |
| 0 | 0 | 0.000684217 | 0 | 0 |
| 0 | 0 | 0 | 0 | 0 |
| 0 | 0 | 0 | 0 | 0 |
| 0 | 0 | 0 | 0 | 0 |
| 0 | 0 | 0 | 0 | 0 |
| 0 | 0 | 0 | 0 | 0 |
| 0 | 0 | 0 | 0 | 0 |
| 0 | 0 | 0 | 0 | 0 |
| 0 | 0 | 0 | 0 | 0 |
| 0 | 0 | 0 | 0 | 0 |
| 0 | 0 | 0 | 0 | 0 |
| 0 | 0 | 0 | 0 | 0 |
| 0 | 0 | 0 | 0 | 0 |
| 0 | 0 | 0 | 0 | 0 |
| 0 | 0 | 0 | 0 | 0 |
| 0 | 0 | 0 | 0 | 0 |
| 0 | 0 | 0 | 0 | 0 |
| 0 | 0 | 0 | 0 | 0 |
| 0 | 0 | 0 | 0 | 0 |
| 0.021755086 | 0.001130041 | 0.003809051 | 0.002946192 | 0 |
| 0.006275875 | 0.002841986 | 0.478611924 | 0.004679684 | 0 |
| 0 | 0 | 0 | 0 | 0 |
| 0.062283057 | 1.03298241 | 0.36479642 | 0 | 0 |
| 0 | 0 | 0 | 0 | 0 |
| 0 | 0 | 0 | 0 | 0 |
| 0 | 0 | 0 | 0 | 0 |
| 0 | 0 | 0 | 0.004514186 | 0 |
| 0 | 0 | 0 | 0 | 0 |
| 0 | 0 | 0 | 0 | 0 |
| 0 | 0 | 0 | 0 | 0 |
| 0 | 0 | 0 | 0 | 0 |
| 0 | 0 | 0 | 0.002139194 | 0 |
| 0.00945075 | 0 | 0 | 0 | 0 |
| 4.377964313 | 4.999072534 | 0.676414433 | 0.107494267 | 0 |
| 0 | 0 | 0 | 0 | 0 |
| 0 | 0 | 0 | 0 | 0 |
| 0.012220127 | 0 | 0.031473886 | 0 | 0 |
| 0.000807503 | 0.007130607 | 0.006958785 | 0 | 0 |
| 0 | 0 | 0 | 0 | 0 |
| 0 | 0 | 0 | 0 | 0 |
| 0 | 0.012469335 | 0 | 0 | 0 |
| 0.035824337 | 0 | 0.001961028 | 0 | 0 |
| 0 | 0 | 0 | 0 | 0 |

| | | | | |
|---|---|---|---|---|
| 0 | 0 | 0 | 0 | 0 |
| 0.024939985 | 0.00440462 | 0.031637692 | 0.002115387 | 0 |
| 0 | 0 | 0 | 0 | 0 |
| 0 | 0 | 0 | 0 | 0 |
| 0 | 0 | 0 | 0 | 0 |
| 0 | 0 | 0 | 0 | 0 |
| 0 | 0 | 0 | 0 | 0 |
| 0 | 0 | 0 | 0 | 0 |
| 0 | 0 | 0 | 0 | 0 |
| 0 | 0 | 0 | 0 | 0 |
| 0 | 0 | 0 | 0 | 0 |
| 0 | 0 | 0 | 0 | 0 |
| 0.046466153 | 0 | 0.26512187 | 0.306352762 | 0 |
| 0 | 0 | 0 | 0 | 0 |
| 0.010689389 | 0.013043255 | 0 | 0.007347513 | 0 |
| 0 | 0 | 0 | 0 | 0 |
| 0 | 0 | 0 | 0 | 0 |
| 0 | 0 | 0 | 0 | 0 |
| 0 | 0 | 0 | 0 | 0 |
| 0 | 0 | 0 | 0 | 0 |
| 0 | 0 | 0 | 0 | 0 |
| 0 | 0.001011277 | 0 | 0 | 0 |
| 0 | 0 | 0.032596112 | 0 | 0 |
| 0.000589784 | 0 | 0 | 0 | 0 |
| 0 | 0 | 0 | 0 | 0 |
| 0 | 0 | 0 | 0 | 0 |
| 0 | 0 | 0 | 0 | 0 |
| 0 | 0 | 0 | 0 | 0 |
| 0 | 0 | 0 | 0 | 0 |
| 0 | 0 | 0 | 0 | 0 |
| 0 | 0 | 0 | 0 | 0 |
| 0 | 0 | 0 | 0 | 0 |
| 0 | 0 | 0 | 0 | 0 |
| 0 | 0 | 0 | 0 | 0 |
| 4.144789701 | 0 | 11.91560825 | 0 | 0 |
| 0 | 0 | 0 | 0 | 0 |
| 0 | 0 | 0 | 0 | 0 |
| 0 | 0 | 0 | 0 | 0 |
| 0.015057898 | 0 | 0 | 0 | 0.4237449 |
| 0 | 0 | 0 | 0 | 0 |
| 0.350865422 | 0.025051972 | 0.609076092 | 0.016401078 | 0 |
| 0 | 0 | 0 | 0 | 0 |
| 0 | 0 | 0 | 0 | 0 |
| 0 | 0 | 0 | 0 | 0 |
| 0 | 0 | 0 | 0 | 0 |
| 0.136579876 | 0.007257095 | 0 | 0.031107186 | 0 |
| 0 | 0 | 0 | 0 | 0 |
| 0 | 0 | 0 | 0 | 0 |

| | | | | |
|---|---|---|---|---|
| 0 | 0 | 0 | 0 | 0 |
| 0 | 0 | 0 | 0 | 0 |
| 0 | 0 | 0 | 0 | 0 |
| 0 | 0 | 0 | 0 | 0 |
| 0.000742016 | 0 | 0 | 0.001150851 | 0 |
| 0 | 0 | 0 | 0 | 0 |
| 0 | 0 | 0 | 0 | 0 |
| 0 | 0 | 0 | 0 | 0 |
| 0 | 0.02013294 | 0 | 0 | 0 |
| 0.143143769 | 0 | 0.00130196 | 0 | 0 |
| 0 | 0 | 0.012129074 | 0 | 0 |
| 0 | 0 | 0 | 0 | 0 |
| 0.00692073 | 0.00363375 | 0.009119987 | 0 | 0 |
| 0 | 0 | 0 | 0 | 0 |
| 0 | 0 | 0 | 0 | 0 |
| 0 | 0 | 0 | 0 | 0 |
| 0 | 0 | 0 | 0 | 0 |
| 0 | 0 | 0 | 0 | 0 |
| 0 | 0 | 0 | 0 | 0 |
| 0.14084616 | 1.929697608 | 0.167068075 | 0.096184811 | 0 |
| 0.004171063 | 0 | 0 | 0 | 0 |
| 0 | 0 | 0 | 0 | 0 |
| 0 | 0 | 0 | 0 | 0 |
| 0 | 0 | 0 | 0 | 0 |
| 0 | 0 | 0 | 0 | 0 |
| 0 | 0 | 0 | 0 | 0 |
| 0 | 0 | 0 | 0 | 0 |
| 0 | 0 | 0 | 0 | 0 |
| 0 | 0 | 0 | 0 | 0 |
| 0.072435811 | 0.014144371 | 0 | 0 | 0 |
| 0 | 0 | 0 | 0 | 0 |
| 0 | 0 | 0 | 0 | 0 |
| 0 | 0 | 0 | 0 | 0 |
| 0 | 0 | 0 | 0 | 0 |
| 0 | 0 | 0 | 0 | 0 |
| 0 | 0 | 0 | 0 | 0 |
| 0 | 0 | 0 | 0 | 0 |
| 0 | 0 | 0 | 0 | 0 |
| 0 | 0 | 0 | 0 | 0 |
| 0 | 0 | 0 | 0 | 0 |
| 0 | 0 | 0 | 0 | 0 |
| 0 | 0 | 0 | 0 | 0 |
| 0.083171638 | 0.024929477 | 0.002667628 | 0 | 0 |
| 0 | 0 | 0 | 0 | 0 |
| 0.015949604 | 0.006627858 | 0.006914963 | 0.001515777 | 0 |
| 0 | 0 | 0 | 0 | 0 |
| 0 | 0 | 0 | 0 | 0 |
| 0 | 0 | 0 | 0 | 0 |

| | | | | |
|---|---|---|---|---|
| 0 | 0 | 0 | 0 | 0 |
| 0 | 0 | 0 | 0 | 0 |
| 0 | 0 | 0 | 0 | 0 |
| 0 | 0 | 0 | 0 | 0 |
| 0 | 0 | 0 | 0 | 0 |
| 0.244910631 | 0.001443153 | 0.047784265 | 0.000565793 | 0 |
| 0 | 0 | 0 | 0 | 0 |
| 0.000973356 | 0 | 0 | 0 | 0 |
| 0 | 0 | 0 | 0 | 0 |
| 0 | 0 | 0 | 0 | 0 |
| 0 | 0 | 0 | 0 | 0 |
| 0 | 0 | 0 | 0 | 0 |
| 0 | 0 | 0 | 0 | 0 |
| 0 | 0 | 0 | 0 | 0 |
| 0 | 0 | 0 | 0 | 0 |
| 0 | 0 | 0 | 0 | 0 |
| 0.02152306 | 0 | 0.005137904 | 0 | 0 |
| 0 | 0 | 0 | 0 | 0 |
| 0 | 0 | 0 | 0 | 0 |
| 0 | 0 | 0 | 0 | 0 |
| 0 | 0 | 0 | 0 | 0 |
| 0 | 0 | 0 | 0 | 0 |
| 0 | 0 | 0 | 0 | 0 |
| 0 | 0 | 0 | 0 | 0 |
| 0 | 0 | 0 | 0 | 0 |
| 0.001278155 | 0 | 0 | 0 | 0 |
| 0 | 0 | 0 | 0 | 0 |
| 0.526409213 | 0 | 0 | 0 | 0 |
| 0 | 0 | 0 | 0 | 0 |
| 0 | 0 | 0 | 0 | 0 |
| 0 | 0 | 0 | 0 | 0 |
| 0 | 0 | 0 | 0 | 0 |
| 0 | 0 | 0 | 0 | 0 |
| 0 | 0 | 0 | 0 | 0 |
| 0 | 0 | 0 | 0 | 0 |
| 0 | 0 | 0 | 0 | 0 |
| 0 | 0 | 0 | 0 | 0 |
| 0 | 0 | 0 | 0 | 0 |
| 0.831491597 | 0.225831515 | 1.567272781 | 0 | 0 |
| 0 | 0 | 0 | 0 | 0 |
| 0 | 0 | 0 | 0 | 0 |
| 0 | 0 | 0 | 0 | 0 |
| 0 | 0 | 0 | 0 | 0 |
| 0 | 0 | 0 | 0 | 0 |
| 0 | 0 | 0 | 0 | 0 |
| 0 | 0 | 0 | 0 | 0 |
| 0 | 0 | 0 | 0 | 0 |
| 0 | 0 | 0 | 0 | 0 |

| | | | | |
|---|---|---|---|---|
| 0 | 0 | 0 | 0 | 0 |
| 0.15644139 | 0.005539633 | 0.146120107 | 0 | 0 |
| 0 | 0 | 0 | 0 | 0 |
| 0 | 0 | 0 | 0 | 0 |
| 0 | 0 | 0 | 0 | 0 |
| 0 | 0 | 0 | 0 | 0 |
| 0 | 0 | 0 | 0 | 0 |
| 0 | 0 | 0 | 0 | 0 |
| 0 | 0 | 0 | 0 | 0 |
| 0 | 0 | 0 | 0 | 0 |
| 0 | 0.001208952 | 0.006791729 | 0.000995345 | 0 |
| 0 | 0 | 0 | 0 | 0 |
| 0 | 0 | 0 | 0 | 0 |
| 0 | 0 | 0 | 0 | 0 |
| 0 | 0 | 0 | 0 | 0 |
| 0 | 0 | 0 | 0 | 0 |
| 0 | 0 | 0 | 0 | 0 |
| 0 | 0 | 0 | 0 | 0 |
| 0 | 0 | 0 | 0 | 0 |
| 0 | 0 | 0 | 0 | 0 |
| 0.010247762 | 0 | 0 | 0.021551256 | 0 |
| 0 | 0 | 0 | 0 | 0 |
| 0 | 0 | 0 | 0 | 0 |
| 2.029216585 | 3.122048289 | 1.904636652 | 0.655402257 | 0 |
| 0 | 0 | 0 | 0 | 0 |
| 0 | 0 | 0 | 0 | 0 |
| 0 | 0 | 0 | 0 | 0 |
| 0 | 0 | 0 | 0 | 0 |
| 0 | 0 | 0 | 0 | 0 |
| 0 | 0 | 0 | 0 | 0 |
| 0 | 0 | 0 | 0 | 0 |
| 0 | 0 | 0 | 0 | 0 |
| 0 | 0 | 0 | 0 | 0 |
| 0 | 0 | 0 | 0 | 0 |

| | | | | |
|---|---|---|---|---|
| PHL018-0127_V2 | PHL018-0148_V2 | PHL018-0128_V1 | PHL018-0180_V2 | PHL018-0133_V1 |
| EYCF | CTRL | CTRL | CTRL | EYCF |
| 0.228471627 | 0.294674953 | 0.274863778 | 0.07440259 | 0.380970915 |
| 0.574673011 | 0.15825107 | 15.21650424 | 9.935850756 | 0 |
| 3.884511436 | 0.262847186 | 2.18024043 | 0 | 0 |
| 0 | 0 | 0 | 0 | 0 |
| 0 | 0 | 0 | 0 | 0 |
| 0 | 0 | 0 | 0 | 0 |
| 0 | 0 | 0 | 0 | 0 |
| 0 | 0 | 0 | 0 | 0 |
| 0 | 0 | 0 | 0 | 0 |
| 0 | 0 | 0 | 0 | 0 |
| 0 | 0 | 0 | 0 | 0 |
| 0 | 0 | 0 | 0 | 0 |
| 0 | 0 | 0 | 0 | 0 |
| 11.67347159 | 2.864398134 | 9.091555178 | 0 | 0 |
| 0 | 0 | 0 | 0 | 0 |
| 0 | 0 | 0 | 0 | 0 |
| 0 | 0 | 0 | 0 | 0.001847585 |
| 0 | 0 | 0 | 0 | 0 |
| 0 | 0 | 0 | 0 | 0 |
| 0 | 0 | 0 | 0 | 0 |
| 0 | 0 | 0 | 0 | 0 |
| 0 | 0 | 0 | 0 | 0.003678895 |
| 0 | 0 | 0 | 0 | 0 |
| 0 | 0 | 0 | 0 | 0 |
| 0 | 0 | 0 | 0 | 0 |
| 0 | 0 | 0 | 0 | 0 |
| 0 | 0 | 0 | 0 | 0 |
| 0 | 0 | 0 | 0 | 0 |
| 0 | 0 | 0 | 0 | 0 |
| 0 | 0 | 0 | 0 | 0 |
| 0 | 0 | 0 | 0 | 0 |
| 0 | 0 | 0 | 0 | 0 |
| 0 | 0 | 0 | 0 | 0 |
| 0 | 0 | 0 | 0 | 0 |
| 0 | 0 | 0 | 0 | 0 |
| 0 | 1.448315188 | 0 | 0 | 0 |
| 0 | 0 | 0 | 0 | 0 |
| 0 | 0 | 0 | 0 | 0 |
| 0 | 0 | 0 | 0 | 0 |
| 0 | 0 | 0 | 0 | 0 |
| 0 | 0 | 0 | 0 | 0 |
| 0 | 0.003234847 | 0 | 0.027447894 | 0.001786347 |
| 0.092280502 | 1.675500135 | 0.10262952 | 0.001876324 | 0.005459502 |
| 0 | 0.002210144 | 0.01201534 | 0.007906762 | 0.000650925 |
| 0 | 0 | 0.009817654 | 0 | 0 |
| 0 | 0 | 0 | 0 | 0 |

| | | | | |
|---|---|---|---|---|
| 0 | 0 | 0.048837298 | 0 | 0 |
| 0.025421529 | 0 | 0.016734754 | 0.177501628 | 0.0056488 |
| 0.061625147 | 0.001644595 | 0.014688395 | 0 | 0 |
| 0.280611052 | 0.529815544 | 0.514378473 | 0 | 0 |
| 0 | 0.013433909 | 0 | 0 | 0.050620156 |
| 0 | 0.369993794 | 0 | 0 | 0 |
| 0.004954141 | 0.010659557 | 0 | 0 | 0 |
| 0 | 0 | 0 | 0 | 0 |
| 0 | 0.010894647 | 0 | 0 | 0 |
| 0 | 0 | 0 | 0 | 0 |
| 0 | 0 | 0 | 0 | 0 |
| 0 | 0.258351728 | 0 | 0 | 0 |
| 0 | 0.002374578 | 0 | 0 | 0 |
| 0.051224254 | 0.055619432 | 0.822927098 | 0 | 0.11739632 |
| 0 | 0 | 0 | 0 | 0.003745782 |
| 0 | 0 | 0 | 0 | 0 |
| 0 | 1.892753197 | 0.005418568 | 0.002194289 | 0 |
| 0 | 0 | 0 | 0 | 0 |
| 0 | 0 | 0 | 0 | 0 |
| 0 | 0 | 0 | 0 | 0 |
| 0.45247478 | 0 | 0.13114711 | 0.068500437 | 0.067880579 |
| 0.005060116 | 0.000865596 | 0.037842246 | 0 | 0.028998628 |
| 0 | 0.053154971 | 0 | 0 | 0 |
| 0 | 0 | 0 | 0 | 0 |
| 0 | 0 | 0 | 0 | 0 |
| 0 | 0 | 0 | 0 | 0 |
| 0 | 0 | 0.041439068 | 0 | 0 |
| 0 | 0 | 0 | 0 | 0 |
| 0 | 0 | 0 | 0 | 0 |
| 0 | 0 | 0 | 0 | 0 |
| 0.250037852 | 0 | 0 | 1.170968506 | 0.321545315 |
| 0 | 0 | 0 | 0 | 0 |
| 0 | 0 | 0 | 0 | 0 |
| 0 | 0 | 0 | 0 | 0.021655783 |
| 0 | 0 | 0 | 0 | 0 |
| 0 | 0 | 0 | 0 | 0 |
| 0 | 0 | 0 | 0 | 0 |
| 0 | 0.05615501 | 0.224921072 | 0 | 0 |
| 0 | 0 | 0 | 0 | 0 |
| 0 | 0 | 0 | 0 | 0 |
| 0 | 0 | 0 | 0 | 0 |
| 0 | 0 | 0 | 0 | 0 |
| 0 | 0 | 0 | 0 | 0 |
| 0 | 0 | 0 | 0 | 0 |
| 0 | 0 | 0 | 0 | 0 |
| 0 | 0 | 0 | 0 | 0 |
| 0 | 0 | 0.233444729 | 0 | 0 |
| 0 | 0 | 0 | 0 | 0 |

| | | | | |
|---|---|---|---|---|
| 0 | 0 | 0 | 0 | 0 |
| 0 | 0 | 0 | 0 | 0.00920303 |
| 0 | 0 | 0 | 0 | 0 |
| 0 | 0 | 0 | 0 | 0 |
| 0 | 0 | 0 | 0 | 0 |
| 0 | 0 | 0 | 0 | 0 |
| 0 | 0 | 0 | 0 | 0 |
| 0 | 0 | 0 | 0 | 0 |
| 0 | 0 | 0 | 0 | 0.002156111 |
| 0 | 0 | 0 | 0 | 0 |
| 0 | 0 | 0 | 0 | 0 |
| 0 | 0 | 0 | 0 | 0 |
| 0 | 0 | 0 | 0 | 0 |
| 0 | 0 | 0 | 0 | 0 |
| 0 | 0 | 0 | 0 | 0 |
| 0 | 0 | 0 | 0 | 0 |
| 0 | 0 | 0 | 0 | 0 |
| 0 | 0 | 0 | 0 | 0 |
| 0 | 0.079962458 | 0 | 0 | 0.586052966 |
| 0 | 0 | 0 | 0 | 0 |
| 0 | 0 | 0 | 0 | 0 |
| 0 | 0 | 0 | 0 | 0 |
| 0 | 0 | 0 | 0 | 0 |
| 0 | 0 | 0 | 0 | 0.034261236 |
| 0 | 0 | 0 | 0 | 0 |
| 0 | 0 | 0 | 0 | 0 |
| 0 | 0 | 0 | 0 | 0 |
| 0.003148942 | 0 | 0 | 0 | 0 |
| 0 | 0 | 0 | 0 | 0 |
| 0 | 0 | 0 | 0 | 0 |
| 0.337087668 | 0.04161469 | 0 | 0 | 0.095627205 |
| 0 | 0.006294793 | 0 | 0 | 0 |
| 0 | 0 | 0 | 0 | 0 |
| 0 | 0 | 0 | 0 | 0 |
| 0 | 0 | 0 | 0 | 0 |
| 0 | 0 | 0 | 0 | 0 |
| 0 | 0 | 0 | 0 | 0 |
| 0 | 0 | 0 | 0 | 0 |
| 0 | 0 | 0 | 0 | 0 |
| 0 | 0 | 0 | 0 | 0 |
| 0 | 0 | 0 | 0 | 0 |
| 0 | 0 | 0 | 0 | 0 |
| 0 | 0.00258173 | 0 | 0 | 0 |
| 0 | 0 | 0 | 0 | 0 |
| 0 | 0 | 0 | 0 | 0 |
| 0 | 0 | 0 | 0 | 0 |
| 0 | 0 | 0 | 0 | 0.052359206 |
| 0 | 0 | 0 | 0 | 0 |

| | | | | |
|---|---|---|---|---|
| 0 | 0 | 0 | 0 | 0 |
| 0 | 0 | 0 | 0 | 0 |
| 0 | 0 | 0 | 0 | 0 |
| 0 | 0 | 0 | 0 | 0 |
| 0 | 0.095262249 | 0 | 0 | 0 |
| 0 | 0 | 0 | 0 | 0 |
| 0 | 0 | 0 | 0 | 0 |
| 0 | 0 | 0 | 0 | 0 |
| 0 | 0 | 0 | 0 | 0 |
| 0 | 0 | 0 | 0 | 0 |
| 0 | 0 | 0 | 0 | 0 |
| 0 | 0 | 0 | 0 | 0 |
| 0 | 0 | 0 | 0 | 0 |
| 0 | 0 | 0 | 0 | 0 |
| 0 | 0 | 0 | 0 | 0 |
| 0 | 0 | 0 | 0 | 0 |
| 0 | 0 | 0 | 0 | 0 |
| 0 | 0 | 0 | 0 | 0 |
| 0 | 0 | 0 | 0 | 0 |
| 0 | 0 | 0 | 0 | 0 |
| 0 | 0 | 0 | 0 | 0 |
| 0 | 0 | 0 | 0 | 0 |
| 0 | 0 | 0 | 0 | 0 |
| 0 | 0 | 0 | 0 | 0 |
| 0 | 0 | 0 | 0 | 0 |
| 0 | 0 | 0 | 0 | 0 |
| 0 | 0 | 0 | 0 | 0 |
| 0 | 0 | 0 | 0 | 0 |
| 0 | 0 | 0 | 0 | 0 |
| 0 | 0 | 0 | 0 | 0 |
| 0 | 0 | 0 | 0 | 0 |
| 0 | 0.102382512 | 0 | 0 | 0 |
| 0 | 0 | 0 | 0 | 0 |
| 0 | 0 | 0 | 0 | 0 |
| 0 | 0 | 0 | 0 | 0 |
| 0 | 0 | 0 | 0 | 0 |
| 0 | 0 | 0 | 0 | 0 |
| 0 | 0 | 0 | 0 | 0 |
| 0 | 0 | 0 | 0 | 0 |
| 0 | 0 | 0 | 0 | 0 |
| 0 | 0 | 0 | 0 | 0 |
| 0 | 0 | 0 | 0 | 0 |
| 0 | 0 | 0 | 0 | 0 |
| 0 | 0 | 0 | 0 | 0 |
| 0 | 0 | 0 | 0 | 0 |
| 0 | 0 | 0 | 0 | 0 |
| 0 | 0 | 0 | 0 | 0 |
| 0 | 0 | 0 | 0 | 0 |

| | | | | |
|---|---|---|---|---|
| 0 | 0 | 0 | 0 | 0 |
| 0 | 0 | 0 | 0 | 0 |
| 0 | 0 | 0 | 0 | 0 |
| 0 | 0 | 0 | 0 | 0 |
| 0 | 0 | 0 | 0 | 0 |
| 0 | 0 | 0 | 0 | 0 |
| 0 | 0 | 0 | 0 | 0 |
| 0 | 0 | 0 | 0 | 0 |
| 0 | 0 | 0 | 0 | 0 |
| 0 | 0 | 0 | 0 | 0 |
| 0 | 0 | 0 | 0 | 0 |
| 0 | 0 | 0 | 0 | 0 |
| 0 | 0.000896751 | 0.000609393 | 0 | 0.000660271 |
| 0 | 0 | 0 | 0 | 0 |
| 0 | 0 | 0 | 0 | 0 |
| 0 | 0 | 0 | 0 | 0 |
| 0 | 0 | 0 | 0 | 0 |
| 0 | 0 | 0 | 0 | 0 |
| 0 | 0 | 0 | 0 | 0 |
| 0 | 0 | 0 | 0 | 0 |
| 0.011506378 | 0.019701325 | 0.02202875 | 0.010381853 | 0.032098272 |
| 0 | 0 | 0 | 0 | 0 |
| 0 | 0 | 0 | 0 | 0.00081243 |
| 0 | 0 | 0.001980306 | 0 | 0.001609233 |
| 0 | 0 | 0 | 0 | 0 |
| 0 | 0 | 0 | 0 | 0 |
| 0.001489121 | 0.000976476 | 0.009953554 | 0.002687176 | 0 |
| 0 | 0 | 0 | 0 | 0 |
| 0 | 0 | 0 | 0 | 0 |
| 0 | 0 | 0 | 0 | 0 |
| 0 | 0 | 0 | 0.020982824 | 0.118612725 |
| 0 | 0 | 0 | 0 | 0 |
| 0 | 0 | 0 | 0 | 0 |
| 0 | 0.015114327 | 0 | 0 | 0 |
| 0.004159257 | 0.009545864 | 0.027430547 | 0 | 0.151415139 |
| 0 | 0 | 0 | 0 | 0 |
| 0 | 0 | 0 | 0 | 0 |
| 0 | 0 | 0 | 0 | 0 |
| 0 | 0 | 0 | 0 | 0 |
| 0 | 0 | 0 | 0 | 0 |
| 0 | 0 | 0 | 0 | 0 |
| 0 | 0 | 0 | 0 | 0 |
| 0 | 0 | 0 | 0 | 0 |
| 0 | 0 | 0 | 0 | 0 |
| 0 | 0 | 0 | 0 | 0 |
| 0 | 0.306419119 | 0 | 0 | 0 |
| 0.003002246 | 0 | 0 | 0 | 0 |
| 0 | 0 | 0.021484914 | 0.001426307 | 0.002146602 |

| | | | | |
|---|---|---|---|---|
| 0 | 0 | 0 | 0 | 0 |
| 0 | 0 | 0 | 0 | 0 |
| 0 | 0.002841651 | 0 | 0 | 0 |
| 0 | 0 | 0 | 0 | 0 |
| 0 | 0 | 0 | 0 | 0 |
| 0 | 0 | 0 | 0 | 0 |
| 0 | 0 | 0 | 0 | 0 |
| 0 | 0 | 0 | 0 | 0 |
| 0 | 0 | 0 | 0 | 0 |
| 0 | 0 | 0 | 0 | 0 |
| 0 | 0 | 0 | 0 | 0 |
| 0 | 0 | 0 | 0 | 0 |
| 0 | 0 | 0 | 0 | 0 |
| 0 | 0 | 3.986069817 | 0.006543864 | 0 |
| 0 | 0 | 0 | 0 | 0 |
| 0 | 0 | 0 | 0 | 0 |
| 0 | 0 | 0 | 0 | 0 |
| 0 | 0 | 0 | 0 | 0 |
| 0 | 0.010080865 | 0 | 0 | 0 |
| 0 | 0.366725996 | 0 | 0 | 0 |
| 0.057508053 | 0 | 0.082162624 | 0 | 0.142979773 |
| 0 | 0 | 0 | 0 | 0 |
| 0 | 0 | 0 | 0 | 0 |
| 0 | 0 | 0 | 0 | 0 |
| 0.001766782 | 0.004634196 | 0.007872995 | 0.018597989 | 0 |
| 0 | 0 | 0 | 0 | 0 |
| 0 | 0 | 0 | 0 | 0 |
| 0 | 0 | 0 | 0 | 0 |
| 0 | 0 | 0 | 0 | 0 |
| 0 | 0 | 0 | 0 | 0 |
| 0.352235087 | 0.022725834 | 3.320835331 | 0 | 2.055460038 |
| 0 | 0 | 0 | 0 | 0 |
| 0.027644569 | 0 | 0 | 0 | 0 |
| 0 | 0 | 0 | 0 | 0 |
| 0 | 0 | 0 | 0 | 0 |
| 0 | 0 | 0 | 0 | 0 |
| 0 | 0 | 0 | 0 | 0 |
| 0 | 0 | 0 | 0 | 0 |
| 0 | 0 | 0 | 0 | 0 |
| 0.001088595 | 0 | 0 | 0 | 0 |
| 0 | 0 | 0 | 0 | 0 |
| 0 | 0.017046097 | 0 | 0.004209817 | 0 |
| 0 | 0 | 0.001329125 | 0 | 0.016471077 |
| 0 | 0 | 0 | 0 | 0 |
| 0 | 0 | 0 | 0 | 0 |
| 0 | 0 | 0 | 0 | 0 |
| 0 | 0 | 0 | 0 | 0 |
| 0 | 0 | 0 | 0 | 0 |

| | | | | |
|---|---|---|---|---|
| 0 | 0 | 0 | 0 | 0 |
| 0 | 0 | 0 | 0 | 0 |
| 0 | 0 | 0.010442814 | 0.00100688 | 0 |
| 0 | 0 | 0 | 0 | 0 |
| 0 | 0 | 0 | 0 | 0 |
| 0 | 0 | 0 | 0 | 0 |
| 0 | 0 | 0 | 0 | 0 |
| 0 | 0 | 0 | 0 | 0 |
| 0 | 0 | 0 | 0 | 0 |
| 0 | 0.001003903 | 0 | 0 | 0 |
| 0 | 0 | 0 | 0 | 0 |
| 0 | 0 | 0 | 0 | 0 |
| 0 | 0 | 0 | 0 | 0 |
| 0 | 0 | 0.007968921 | 0.022812065 | 0 |
| 0 | 0 | 0.004762873 | 0.002755372 | 0.001658741 |
| 0 | 0 | 0 | 0 | 0 |
| 0 | 0 | 0.002842644 | 0.003401124 | 0.001259991 |
| 0 | 0 | 0.015636073 | 0 | 0 |
| 0 | 0 | 0 | 0 | 0 |
| 0 | 0 | 0 | 0 | 0 |
| 0 | 0 | 0 | 0 | 0 |
| 0 | 0 | 0.015629862 | 0.002125554 | 0.003412238 |
| 0 | 0 | 0 | 0 | 0 |
| 0 | 0 | 0.005786233 | 0 | 0.002350999 |
| 0 | 0 | 0 | 0 | 0.001519812 |
| 0 | 0 | 0 | 0 | 0 |
| 0 | 0 | 0 | 0 | 0 |
| 0 | 0 | 0 | 0 | 0 |
| 0 | 0 | 0 | 0 | 0 |
| 0 | 0 | 0 | 0 | 0 |
| 0 | 0 | 0 | 0 | 0 |
| 0 | 0 | 0 | 0 | 0 |
| 0 | 0 | 0 | 0 | 0 |
| 0 | 0 | 0 | 0 | 0 |
| 0 | 0 | 0 | 0 | 0 |
| 0 | 0 | 0.0069892 | 0 | 0 |
| 0 | 0 | 0 | 0 | 0 |
| 0.003617535 | 0 | 0.003810223 | 0.003560718 | 0 |
| 0 | 0 | 0 | 0 | 0 |
| 0 | 0 | 0 | 0 | 0 |
| 0 | 0 | 0 | 0 | 0 |
| 0 | 0 | 0 | 0 | 0 |
| 0 | 0 | 0 | 0 | 0 |
| 0 | 0 | 0 | 0.055655681 | 0.007987917 |
| 0 | 0 | 0 | 0 | 0 |
| 0 | 0 | 0 | 0 | 0.015746086 |
| 0 | 0 | 0 | 0 | 0 |
| 0 | 0 | 0.004615136 | 0.114292436 | 0.003173367 |

| | | | | |
|---|---|---|---|---|
| 0 | 0 | 0 | 0 | 0 |
| 0.1688197 | 0.045060766 | 0 | 0 | 0 |
| 0.001096668 | 0 | 0 | 0 | 0.00138991 |
| 0 | 0 | 0 | 0 | 0 |
| 0.004965091 | 0.002713172 | 0.027779223 | 0.004230969 | 0.00199769 |
| 0 | 0 | 0 | 0 | 0.005537605 |
| 0 | 0 | 0 | 0 | 0 |
| 0 | 0 | 0.023087545 | 0.004884051 | 0 |
| 0 | 0 | 0 | 0 | 0 |
| 0.822559011 | 0.010191859 | 0.206357606 | 0.090254179 | 0.038386848 |
| 0 | 0 | 0 | 0 | 0.003653334 |
| 0.001224288 | 0 | 0 | 0.233630912 | 0.030146442 |
| 0.015787325 | 0.036309432 | 0.006621198 | 0.003770582 | 0.012106136 |
| 0 | 0 | 0 | 0 | 0.123340079 |
| 0 | 0 | 0 | 0 | 0 |
| 0 | 0 | 0 | 0 | 0 |
| 0 | 0 | 0 | 0 | 0 |
| 0 | 0 | 0 | 0 | 0 |
| 0.829138961 | 0 | 0 | 1.606517705 | 0.725957138 |
| 0.025249625 | 0 | 0.120284308 | 0 | 0.054206135 |
| 0 | 0 | 0.002750657 | 0 | 0 |
| 0 | 0 | 0 | 0 | 0 |
| 0 | 0 | 0 | 0 | 0 |
| 0 | 0.239478514 | 0 | 0 | 0 |
| 0 | 0 | 0 | 0 | 0 |
| 0 | 0 | 0.011086011 | 0 | 0 |
| 0 | 0.446724836 | 0 | 0 | 0.009951414 |
| 0.001635033 | 0 | 0 | 0.344616217 | 0.057548811 |
| 0.006921644 | 0 | 0 | 0 | 0.864411898 |
| 0.003060451 | 0 | 0 | 0 | 0.012089754 |
| 0.008124755 | 0 | 0 | 0 | 0 |
| 0 | 0 | 0.192007763 | 0.006682974 | 0 |
| 0 | 0 | 0 | 0.003099838 | 0 |
| 0.084169391 | 0 | 0.020802146 | 0 | 0 |
| 0 | 0 | 0 | 0 | 0 |
| 0 | 0 | 0 | 0 | 0.621398713 |
| 0.006130858 | 0 | 0 | 0.004425349 | 0.187817078 |
| 0 | 0 | 0 | 0 | 0.053521345 |
| 0 | 0 | 0 | 0 | 0 |
| 0 | 0 | 0 | 0.263540416 | 0 |
| 0 | 0 | 0 | 0 | 0 |
| 0 | 0 | 0 | 0 | 0 |
| 0 | 0 | 0 | 0 | 0 |
| 0.001089279 | 0 | 0.005048116 | 0 | 0 |
| 0 | 0 | 0 | 0 | 0 |
| 0 | 0 | 0 | 0 | 0 |
| 0.001008847 | 0.001653852 | 0.016183953 | 0 | 0.020457691 |
| 0.003357576 | 0 | 0 | 0 | 0 |

| | | | | |
|---|---|---|---|---|
| 0.001907042 | 0 | 0 | 0 | 0 |
| 0 | 0.049903967 | 0 | 0 | 0.031572577 |
| 0 | 0 | 0 | 0 | 0 |
| 0 | 0 | 0 | 0 | 0 |
| 0 | 0 | 0.025179116 | 0 | 0.014202344 |
| 0.015626915 | 0 | 0 | 0 | 0.014194706 |
| 0 | 0 | 0.007874315 | 0 | 0 |
| 0 | 0 | 0 | 0 | 0 |
| 0 | 0 | 0 | 0 | 0 |
| 0 | 0 | 0 | 0 | 0 |
| 0.016588608 | 0.003021613 | 0.064064734 | 0.008038045 | 0.001557355 |
| 0 | 0 | 0 | 0 | 0 |
| 0 | 0 | 0 | 0 | 0.001721452 |
| 0.001451119 | 0.001427334 | 0.002586544 | 0.00157116 | 0 |
| 0 | 0 | 0.002617118 | 0 | 0 |
| 0.008582982 | 0 | 0 | 0 | 0 |
| 0 | 0 | 0 | 0 | 0 |
| 0 | 0 | 0 | 0 | 0 |
| 0 | 0 | 0.025878001 | 0 | 0 |
| 0 | 0.02601872 | 0 | 0 | 0 |
| 0 | 0 | 0 | 0 | 0 |
| 0 | 0 | 0 | 0 | 0 |
| 0 | 0 | 0 | 0 | 0 |
| 0 | 0 | 0 | 0 | 0.000740954 |
| 0.057518369 | 0 | 0 | 0 | 0 |
| 0 | 0 | 0 | 0 | 0 |
| 0.084334078 | 0.001547608 | 0 | 0.005110661 | 4.493477616 |
| 0 | 0 | 0 | 0 | 0 |
| 0 | 0 | 0 | 0 | 0 |
| 0.201643034 | 0.624304421 | 0.115635637 | 0.038383194 | 0.079179261 |
| 0 | 0.002904998 | 0 | 0 | 0 |
| 0 | 0 | 0 | 0 | 0 |
| 0 | 0 | 0 | 0 | 0 |
| 0.005019642 | 0 | 0 | 0 | 0 |
| 0.001834688 | 0 | 0.052323789 | 0 | 0.010629814 |
| 0 | 0 | 0 | 0 | 0 |
| 0 | 0 | 0 | 0 | 0 |
| 0 | 0 | 0 | 0 | 0 |
| 0 | 0 | 0 | 0 | 0 |
| 0 | 0.002026682 | 0 | 0 | 0 |
| 0.048434889 | 0.03614146 | 0.006591905 | 0 | 0.011577386 |
| 0 | 4.731909758 | 0 | 0 | 0 |
| 0 | 0 | 0 | 0 | 0 |
| 0 | 0 | 0 | 0 | 0 |
| 0.013995705 | 0 | 0 | 0 | 0 |
| 0 | 0 | 0 | 0 | 0 |
| 0.013522848 | 0 | 0.002455015 | 0.001581644 | 0.005078154 |
| 0 | 0 | 0 | 0 | 0 |

| | | | | |
|---|---|---|---|---|
| 0 | 0.001823495 | 0.00495667 | 0 | 0 |
| 0 | 0.003107091 | 0 | 0 | 0 |
| 0 | 4.426405696 | 0.006003415 | 0.003418772 | 0 |
| 0 | 0 | 0 | 0 | 0 |
| 0 | 0 | 0 | 0 | 0 |
| 0 | 0 | 0 | 0 | 0.003405575 |
| 0 | 0 | 0 | 0.004159981 | 0 |
| 0 | 0 | 0 | 0 | 0 |
| 0 | 0 | 0.000691381 | 0 | 0.001872763 |
| 0.02698206 | 0 | 0 | 0 | 0.386649898 |
| 0 | 0 | 0 | 0 | 0 |
| 0 | 0 | 0 | 0 | 0 |
| 0 | 0 | 0 | 0 | 0.002201357 |
| 0.044724238 | 0.021720647 | 0.070252059 | 0 | 0 |
| 0 | 0 | 0 | 0 | 0 |
| 0.936911157 | 0.317999755 | 0 | 0.000591289 | 0.191109704 |
| 0 | 0.007382809 | 0.015536625 | 0.009175352 | 0.006137319 |
| 0.063835062 | 0 | 0.004719098 | 0 | 0.291659708 |
| 0 | 0 | 0 | 0 | 0.017750284 |
| 0 | 0 | 0 | 0 | 0 |
| 0 | 0.00750358 | 0 | 0 | 0 |
| 0.003089359 | 0 | 0 | 0.001520419 | 0 |
| 0 | 0 | 0 | 0 | 0 |
| 0 | 0.151426556 | 0 | 0 | 0 |
| 0 | 0 | 0 | 0 | 0 |
| 0.007654926 | 0 | 0.308991299 | 0 | 0.006005878 |
| 0.003381157 | 0.003023398 | 0 | 0 | 0.000890443 |
| 0 | 0.003252672 | 0 | 0 | 0 |
| 0 | 0 | 0 | 0 | 0 |
| 0 | 0 | 0 | 0 | 0 |
| 0.017298243 | 0 | 0 | 0 | 0.098360301 |
| 0.009409703 | 0 | 0.003105984 | 0.002830029 | 0 |
| 0.097225104 | 0.022520737 | 0 | 0 | 0.318900469 |
| 0 | 0 | 0 | 0 | 0 |
| 0 | 0 | 0 | 0 | 0 |
| 0 | 0 | 0 | 0 | 0 |
| 0 | 0 | 0 | 0.005413505 | 0.001086315 |
| 0.024633955 | 0.002769165 | 0.015263519 | 0 | 0.184182159 |
| 0 | 0.12719915 | 0 | 0 | 0 |
| 0 | 0.002452748 | 0.034190373 | 0 | 0 |
| 0 | 0 | 0 | 0 | 0 |
| 0 | 0 | 0 | 0 | 0 |
| 0.002456003 | 0 | 0.027204267 | 0 | 0.003303297 |
| 0.073256525 | 0 | 0 | 0 | 0 |
| 0.480600086 | 0 | 0 | 0.79644484 | 0.221919069 |
| 0 | 0 | 0 | 0 | 0 |
| 0 | 0 | 0 | 0 | 0 |
| 0 | 0 | 0 | 0 | 0 |

| | | | | |
|---|---|---|---|---|
| 0 | 0 | 0 | 0 | 0 |
| 0.067453461 | 0.055971183 | 0.041596962 | 0.014133642 | 0.007640278 |
| 0 | 0 | 0 | 0 | 0 |
| 0 | 0 | 0 | 0 | 0 |
| 0 | 0 | 0 | 0 | 0 |
| 0 | 0 | 0 | 0 | 0 |
| 0 | 0 | 0 | 0 | 0.002998653 |
| 0.000982206 | 0 | 0.000656524 | 0 | 0.000711338 |
| 0 | 0 | 0 | 0 | 0 |
| 0.005110789 | 0 | 0.03850926 | 0.049885972 | 0 |
| 0.019004852 | 0 | 0 | 0 | 0.003612994 |
| 0.006015506 | 0.001740267 | 0.265219989 | 0.203694913 | 0.002050155 |
| 0 | 0.415738047 | 0 | 0 | 0 |
| 0 | 0 | 0 | 0 | 0 |
| 0 | 0 | 0 | 0 | 0 |
| 0 | 0 | 0 | 0 | 0 |
| 0.003980165 | 0.012478832 | 0.042344927 | 0.114693333 | 0.018195997 |
| 0.150483151 | 0 | 2.13633029 | 0.512803349 | 0.61367613 |
| 0 | 0 | 0 | 0 | 0 |
| 0 | 0 | 0 | 0 | 0.129691229 |
| 0.001026138 | 0.005803587 | 0.008916565 | 0.001157317 | 0 |
| 0 | 0.002511872 | 0 | 0 | 0 |
| 0 | 0 | 0.001434203 | 0 | 0 |
| 0 | 0 | 0 | 0 | 0 |
| 0 | 0.004663574 | 0 | 0 | 0 |
| 0 | 0 | 0.0020966 | 0 | 0 |
| 0 | 0.013200296 | 0 | 0 | 0.041994657 |
| 0.341641004 | 0.048542283 | 0.002915997 | 0 | 0.045022251 |
| 0 | 0 | 0.008528513 | 0 | 0 |
| 0 | 0 | 0 | 0 | 0 |
| 0 | 0 | 0.001825173 | 0 | 0 |
| 0 | 0 | 0 | 0 | 0 |
| 0 | 0 | 0 | 0 | 0 |
| 0 | 0 | 0 | 0 | 0 |
| 0.065103176 | 0.035289968 | 0 | 0 | 0 |
| 0.095983288 | 0.302747682 | 0.013160405 | 0.346827491 | 3.016818413 |
| 0 | 0 | 0 | 0 | 0.000464343 |
| 0 | 0 | 0 | 0 | 0.004230092 |
| 0 | 0.000992525 | 0.006594883 | 0.092562168 | 0.016564556 |
| 0 | 0 | 0 | 0 | 0 |
| 0.080204432 | 0 | 0 | 0.80897575 | 0.175480999 |
| 0 | 0 | 0 | 0 | 0.000423768 |
| 0.001634456 | 0 | 0.052986243 | 0.00073736 | 0.051047643 |
| 0.001183527 | 0 | 0.00175798 | 0.400625592 | 0.007571402 |
| 0 | 0 | 0 | 0 | 0.001126083 |
| 0 | 0 | 0 | 0 | 0 |
| 0 | 0 | 0 | 0 | 0 |
| 0.00095654 | 0 | 0 | 0 | 0 |

| | | | | |
|---|---|---|---|---|
| 0 | 0 | 0 | 0 | 0.204848781 |
| 0 | 0 | 0 | 0 | 0 |
| 0 | 0 | 0 | 0 | 0 |
| 0 | 0.023873384 | 0 | 0 | 0.293825125 |
| 0 | 0 | 0 | 0 | 0 |
| 0 | 0 | 0 | 0 | 0 |
| 0 | 0 | 0 | 0 | 0 |
| 0 | 0.007119101 | 0 | 0 | 0 |
| 0.713845853 | 0 | 0 | 0 | 0 |
| 0 | 0.007949054 | 0 | 0 | 0 |
| 0 | 0 | 0 | 0 | 0 |
| 0 | 0.001569734 | 0 | 0 | 0 |
| 0 | 0 | 0 | 0 | 0 |
| 0 | 0 | 0 | 0 | 0 |
| 0 | 0.078831212 | 0 | 0 | 0 |
| 0 | 0 | 0 | 0 | 0 |
| 0 | 0.011344918 | 0 | 0 | 0 |
| 0 | 0.237055874 | 0 | 0 | 0 |
| 0 | 0 | 0 | 0 | 0 |
| 0 | 0 | 0 | 0 | 0 |
| 0 | 0 | 0 | 0 | 0 |
| 0 | 0 | 0 | 0 | 0 |
| 0 | 0.001858448 | 0 | 0 | 0 |
| 0.139539925 | 0 | 0.308480777 | 0 | 0.5839069 |
| 0.024546953 | 0.003569204 | 0 | 0 | 0 |
| 0 | 0 | 0 | 0 | 0 |
| 0 | 0.00674492 | 0 | 0 | 0 |
| 0 | 1.746512611 | 0 | 0 | 0 |
| 0 | 0 | 0 | 0 | 0 |
| 0 | 0 | 0 | 0 | 0 |
| 0 | 0.017876576 | 0 | 0.000799915 | 0 |
| 0 | 0 | 0 | 0 | 0 |
| 0.268039155 | 0.036336339 | 0 | 0 | 0 |
| 0 | 0 | 0 | 0 | 0 |
| 0 | 0 | 0 | 0 | 0 |
| 0 | 0.111277039 | 0 | 0 | 0 |
| 0 | 0.937011594 | 0 | 0.177598657 | 0 |
| 0 | 0 | 0 | 0 | 0 |
| 0.551796657 | 0 | 0 | 0 | 0 |
| 0 | 0 | 0 | 0 | 0 |
| 0 | 0.031408339 | 0.279320347 | 0.031376374 | 0.368111147 |
| 0 | 0.056612522 | 0 | 0 | 0 |
| 23.70565009 | 17.82563968 | 0.226114051 | 0 | 0 |
| 0.012765197 | 0.005059868 | 0 | 0 | 0 |
| 0 | 0 | 0 | 0 | 0 |
| 0 | 0 | 0 | 0 | 0 |
| 0 | 0 | 0 | 0 | 0 |
| 0.410673187 | 0.002327451 | 0.016870766 | 0 | 0.00437942 |

| | | | | |
|---|---|---|---|---|
| 0 | 0 | 0 | 0 | 0 |
| 0 | 0 | 0 | 0 | 0 |
| 0.176927646 | 0.009916109 | 0.53459208 | 0.856169577 | 0.036323284 |
| 0 | 0 | 0 | 0 | 0 |
| 0 | 0 | 0 | 0 | 0 |
| 0 | 0.002929323 | 0 | 0 | 0 |
| 0 | 0.026503056 | 0 | 0 | 0 |
| 0 | 0 | 0 | 0 | 0 |
| 0 | 0 | 0 | 0 | 0 |
| 0 | 0 | 0 | 0 | 0 |
| 1.35232727 | 0.063283092 | 0.014775882 | 0 | 0.093189809 |
| 5.662318493 | 1.557523036 | 3.509477406 | 0 | 2.492225093 |
| 0 | 0 | 0 | 0 | 0 |
| 0 | 0.009719287 | 0.036177666 | 0.005542096 | 0.001740708 |
| 0.967878276 | 0.049888113 | 0.245143227 | 1.340786935 | 0.559214579 |
| 0 | 0 | 0 | 0 | 0 |
| 0 | 0 | 0 | 0 | 0 |
| 0 | 0 | 0.008331641 | 0 | 0 |
| 0 | 0 | 0 | 0 | 0 |
| 0.002649863 | 0 | 0 | 0 | 0.396957812 |
| 0.023228972 | 0.029172303 | 0.008872381 | 0.18731919 | 0.20653236 |
| 2.113907194 | 3.493447718 | 5.244836242 | 18.68784958 | 0 |
| 28.5905969 | 0.539132472 | 2.928872521 | 0 | 2.196471528 |
| 0 | 0 | 0 | 0 | 0 |
| 0 | 0 | 0 | 0 | 0 |
| 0 | 0.003911292 | 0 | 0 | 0 |
| 0.662325268 | 0 | 0 | 0.169430724 | 0 |
| 0 | 0.01255346 | 0 | 0 | 0 |
| 0.156134385 | 0.015978032 | 0 | 0 | 0 |
| 0 | 0 | 0 | 0 | 0 |
| 0 | 0.007650308 | 0 | 0 | 0 |
| 0 | 0.167171276 | 0 | 0 | 0 |
| 0 | 0 | 0.015628207 | 1.806218014 | 0.19693836 |
| 0 | 0.035646866 | 0 | 0 | 0 |
| 0.02012827 | 0 | 0.00074745 | 0.00227014 | 0.002429566 |
| 0.004609169 | 0.072991315 | 0.976630114 | 0 | 0.033547651 |
| 0 | 0.00262765 | 0 | 0 | 0 |
| 0 | 0 | 0 | 0 | 0 |
| 0 | 0 | 0 | 0 | 0 |
| 0 | 0 | 0 | 0 | 0 |
| 0 | 0.008982186 | 0 | 0 | 0 |
| 0 | 0 | 0 | 0 | 0 |
| 0 | 0 | 0 | 0 | 0 |
| 0 | 0 | 0 | 0 | 0 |
| 0.061235123 | 0.005945594 | 0.051295087 | 0.008773697 | 0.282076118 |
| 0 | 0 | 0 | 0 | 0 |
| 0 | 0 | 0 | 0 | 0 |
| 0.016168158 | 0 | 0.317007994 | 0 | 0 |

| | | | | |
|---|---|---|---|---|
| 0 | 0 | 0 | 0 | 0 |
| 0 | 0 | 0 | 0 | 0 |
| 0 | 0 | 0 | 0 | 0 |
| 0 | 0.002423542 | 0 | 0 | 0 |
| 0 | 0 | 0 | 0 | 0 |
| 0 | 0.715496275 | 0 | 0 | 0 |
| 0 | 0.254235971 | 0 | 0 | 0 |
| 0.1830296 | 0 | 0 | 0.035520409 | 0 |
| 0 | 0 | 0 | 0 | 0 |
| 0 | 0 | 0 | 0 | 0 |
| 0.001995754 | 0 | 0.005558326 | 0.005402124 | 0.004697468 |
| 0 | 0 | 0 | 0 | 0 |
| 0 | 0.003222745 | 0 | 0 | 0 |
| 0 | 0.586920384 | 0 | 0 | 0 |
| 0 | 0 | 0 | 0 | 0 |
| 0 | 0 | 0 | 0 | 0 |
| 0 | 0.115521463 | 0 | 0 | 0 |
| 0 | 0.005311112 | 0.008328923 | 0 | 0 |
| 0 | 0.045354434 | 0 | 0.008872854 | 0 |
| 0.003933104 | 0 | 0 | 0 | 0 |
| 0 | 0 | 0 | 0 | 0 |
| 0 | 0 | 0 | 0 | 0 |
| 0 | 0.009656023 | 0 | 0 | 0 |
| 0 | 0 | 0 | 0 | 0 |
| 0 | 0 | 0 | 0 | 0 |
| 0 | 0 | 0 | 0 | 0 |
| 0 | 0 | 0 | 0 | 0 |
| 0 | 0 | 0 | 0 | 0 |
| 0 | 0 | 0 | 0 | 0 |
| 0 | 0 | 0 | 0 | 0 |
| 0.006680484 | 0 | 0.002451568 | 0 | 0 |
| 0 | 0 | 0 | 0 | 0 |
| 0 | 0.010458122 | 0 | 0 | 0 |
| 0 | 0 | 0 | 0 | 0 |
| 0 | 0 | 0 | 0 | 0 |
| 0 | 0 | 0 | 0 | 0 |
| 0 | 0.053946276 | 0 | 0 | 0 |
| 0.00063354 | 0.00498525 | 0.00959865 | 0.040394764 | 0 |
| 0 | 0 | 0 | 0 | 0 |
| 0 | 0.001229294 | 0 | 0 | 0 |
| 0 | 0 | 0.101713795 | 0 | 0 |
| 0 | 0 | 0 | 0 | 0 |
| 0 | 0 | 0 | 0 | 0 |
| 0 | 0.000605159 | 0 | 0 | 0 |
| 0 | 0 | 0 | 0 | 0.044978956 |
| 0 | 0 | 0 | 0 | 0 |
| 0 | 0 | 0 | 0 | 0 |
| 0 | 0 | 0 | 0 | 0 |

| | | | | |
|---|---|---|---|---|
| 0 | 0 | 0 | 0 | 0 |
| 0 | 0 | 0 | 0 | 0 |
| 0 | 0.19277454 | 0 | 0 | 0 |
| 0 | 0.007851532 | 0 | 0 | 0 |
| 0 | 0 | 0 | 0 | 0 |
| 0 | 2.361500743 | 0 | 0 | 0 |
| 0 | 0 | 0 | 0 | 0 |
| 0 | 0 | 0 | 0 | 0 |
| 0.380375927 | 0 | 0.468785932 | 0.010774321 | 0.120864263 |
| 0 | 0 | 0 | 0 | 0 |
| 0 | 0 | 0 | 0 | 0 |
| 0 | 0 | 0 | 0 | 0 |
| 0 | 0 | 0 | 0 | 0 |
| 0 | 0 | 0 | 0 | 0 |
| 0 | 0.07040797 | 0 | 0 | 0 |
| 0.069363238 | 0 | 0 | 0 | 1.726021234 |
| 0 | 0 | 0 | 0 | 0 |
| 0 | 0 | 0 | 0 | 0 |
| 0 | 0 | 0 | 0 | 0 |
| 0 | 0 | 0 | 0 | 0 |
| 0 | 0.308647308 | 0 | 0 | 0 |
| 0 | 0 | 0 | 0 | 0 |
| 0 | 0 | 0 | 0 | 0 |
| 0 | 0 | 0 | 0 | 0 |
| 0.003425048 | 0.049859862 | 0.006715468 | 0 | 0 |
| 0 | 13.50883921 | 0 | 0 | 0.002571777 |
| 0 | 0.004009711 | 0 | 0.016091803 | 0 |
| 0 | 0 | 0 | 0 | 0 |
| 0 | 0.025330357 | 0 | 0 | 0 |
| 0 | 1.091744385 | 0 | 0 | 0 |
| 0 | 0 | 0 | 0 | 0 |
| 0 | 0 | 0 | 0 | 0 |
| 0 | 0.000652559 | 0 | 0 | 0 |
| 0.023413702 | 0.000539764 | 0 | 0 | 0 |
| 0.000904181 | 0.001778722 | 0.11630783 | 0 | 0 |
| 0.002769634 | 0.000838227 | 0.002278491 | 0.00076891 | 0.009874894 |
| 0.003737627 | 0 | 0 | 0 | 0 |
| 0 | 0.418117838 | 0 | 0 | 0 |
| 0 | 0 | 0 | 0 | 0 |
| 0 | 0 | 0 | 0 | 0 |
| 0 | 0 | 0.77942963 | 0 | 0 |
| 0 | 0 | 0 | 0 | 0 |
| 0.083555725 | 0.121704307 | 0 | 0 | 0 |
| 0 | 0 | 0 | 0 | 0 |
| 0 | 0 | 0 | 0 | 0 |
| 0.024018598 | 0.001191172 | 0 | 0 | 0 |
| 0 | 0 | 0 | 0 | 0 |
| 0 | 0.447852607 | 0 | 0.067018909 | 0.012808117 |

| | | | | |
|---|---|---|---|---|
| 0.349839804 | 0 | 0 | 0 | 0 |
| 0 | 0 | 0 | 0 | 0 |
| 0 | 0 | 0 | 0 | 0 |
| 0 | 0 | 0 | 0 | 0 |
| 0.018859615 | 0.001643715 | 0 | 0 | 0.162174293 |
| 0 | 0 | 0 | 0 | 0 |
| 0 | 0 | 0 | 0 | 0 |
| 0 | 0 | 0 | 0 | 0 |
| 0 | 0 | 0 | 0 | 0 |
| 0 | 0 | 0 | 0 | 0 |
| 0 | 0 | 0 | 0 | 0 |
| 0 | 1.543727607 | 0 | 0 | 0 |
| 0 | 0.03980501 | 0 | 0 | 0 |
| 0 | 0 | 0 | 0 | 0 |
| 0 | 0.010047911 | 0 | 0.539567809 | 0.646843293 |
| 0 | 0 | 0 | 0 | 0 |
| 0.000960304 | 0 | 0 | 0 | 0 |
| 0 | 0 | 0 | 0 | 0 |
| 0 | 0.005712622 | 0.003666379 | 0 | 0 |
| 0 | 0 | 0 | 0 | 0 |
| 0 | 0.046704077 | 0 | 0 | 0 |
| 0 | 0 | 0 | 0 | 0 |
| 0 | 0.017358588 | 0.158330418 | 0.003821547 | 1.598478168 |
| 0 | 0 | 0 | 0 | 0 |
| 0 | 0 | 0 | 0 | 0 |
| 0 | 0 | 0 | 0 | 0 |
| 0 | 0 | 0 | 0 | 0 |
| 0 | 0 | 0 | 0 | 0 |
| 0 | 0 | 0 | 0 | 0 |
| 0 | 0 | 0 | 0 | 0 |
| 0 | 0 | 0 | 0 | 0 |
| 0 | 0.006343088 | 0.051087245 | 0.028122958 | 0.001556791 |
| 0 | 0.330524144 | 0 | 0 | 0 |
| 0.101483639 | 0.013379789 | 0 | 0 | 0 |
| 0.004868859 | 0.000912201 | 0.002272938 | 0.001255149 | 0.007723945 |
| 0 | 0.037336029 | 0.008523906 | 0 | 0 |
| 0 | 0 | 0 | 0 | 0 |
| 0 | 0 | 0 | 0 | 0 |
| 0 | 0 | 0 | 0 | 0 |
| 0 | 0 | 0 | 0 | 0 |
| 0 | 0 | 0 | 0 | 0 |
| 0 | 0.005052542 | 0 | 0 | 0 |
| 0 | 0 | 0 | 0 | 0 |
| 0 | 0.00092827 | 0 | 0 | 0 |
| 0.197562473 | 0.003594438 | 0.010991807 | 0 | 0.109501426 |
| 0 | 0 | 0 | 0 | 0 |
| 0 | 0.414326256 | 0 | 0 | 0 |
| 0.002160217 | 0.002549772 | 0 | 0 | 0 |

| | | | | |
|---|---|---|---|---|
| 0 | 0.11149855 | 1.659821034 | 0 | 0 |
| 0 | 0 | 0 | 0 | 0 |
| 0.005669245 | 0 | 0.01999975 | 0 | 0 |
| 0 | 0 | 0 | 0 | 0 |
| 0 | 0 | 0 | 0 | 0.063325722 |
| 0.156273454 | 0 | 0 | 0 | 0 |
| 0 | 0 | 0 | 0 | 0 |
| 0 | 0.033792193 | 0 | 0 | 0 |
| 0 | 0 | 0 | 0 | 0 |
| 0 | 0.000863709 | 0 | 0 | 0 |
| 0 | 0 | 0 | 0 | 0 |
| 0 | 0 | 0 | 0 | 0 |
| 0 | 0 | 0 | 0 | 0 |
| 0 | 0 | 0 | 0 | 0 |
| 0 | 0 | 0 | 0 | 0 |
| 0 | 0 | 0 | 0 | 0 |
| 0 | 0 | 0 | 0 | 0 |
| 0 | 0 | 0 | 0 | 0 |
| 0 | 0 | 0.109101457 | 0 | 11.64835696 |
| 0.002513783 | 0 | 0 | 0 | 0 |
| 0 | 0.000844431 | 0 | 0 | 0 |
| 0 | 0 | 0 | 0 | 0 |
| 0 | 0 | 0 | 0 | 0 |
| 0 | 0 | 0 | 0 | 0 |
| 0 | 0 | 0 | 0.001350927 | 0 |
| 0.004677815 | 0 | 0.118815996 | 0.000575543 | 0.062366139 |
| 0 | 0 | 0 | 0 | 0 |
| 0 | 0 | 0 | 0 | 0.130378023 |
| 0 | 0 | 0 | 0 | 0 |
| 0 | 1.725561843 | 0 | 0 | 3.022621086 |
| 0 | 0 | 0 | 0 | 0 |
| 0 | 0 | 0 | 0 | 0 |
| 0 | 0.038691802 | 0 | 0 | 0 |
| 0 | 0 | 0 | 0 | 0 |
| 0.000891284 | 0 | 0.004170252 | 0.019099231 | 0.008714114 |
| 0 | 0 | 0 | 0 | 0 |
| 0 | 0 | 0 | 0 | 0 |
| 0 | 0.00099477 | 0.008788027 | 0 | 0.002929768 |
| 0 | 0 | 0 | 0 | 0 |
| 0 | 0 | 0 | 0 | 0 |
| 2.277455306 | 4.717161495 | 0 | 1.138790761 | 2.473502693 |
| 0 | 0.021299144 | 0.430837735 | 0.467905274 | 5.648751539 |
| 0 | 0.028631601 | 0 | 0 | 0 |
| 0 | 0.209057991 | 0 | 0 | 0 |
| 0 | 0 | 0 | 0 | 0 |
| 0 | 0 | 0 | 0 | 0 |
| 0 | 0 | 0 | 0 | 0 |
| 0.001404655 | 0 | 0 | 0 | 0.007411658 |

| | | | | |
|---|---|---|---|---|
| 0 | 0 | 0 | 0 | 0 |
| 0 | 0.032888845 | 0 | 0 | 0.006128698 |
| 0 | 0 | 0 | 0 | 0 |
| 0 | 0 | 0 | 0 | 0 |
| 0.804842095 | 1.006328883 | 0 | 0 | 0 |
| 0 | 0 | 0 | 0 | 0 |
| 0 | 0 | 0 | 0 | 0 |
| 0 | 0 | 0 | 0 | 0 |
| 0.163022152 | 0.000608155 | 1.965170799 | 23.36646685 | 1.358863479 |
| 0 | 0 | 0 | 0 | 0 |
| 0 | 0 | 0 | 0.003155999 | 0 |
| 0 | 0 | 0 | 0 | 0 |
| 0 | 0.037677837 | 0 | 0 | 0 |
| 0.030573431 | 0.012125934 | 0 | 0 | 0.003392732 |
| 0.017927254 | 0 | 0 | 0 | 0 |
| 0 | 0.46866638 | 0.017490769 | 0 | 0 |
| 0 | 0.000629587 | 0 | 0 | 0.000927121 |
| 0 | 0 | 0 | 0 | 0 |
| 0 | 0 | 0 | 0 | 0 |
| 0 | 0 | 0 | 0 | 0 |
| 0 | 0 | 0 | 0 | 0 |
| 0 | 0 | 0 | 0 | 0 |
| 0 | 0 | 0 | 0 | 0 |
| 0 | 0 | 0 | 0 | 0 |
| 0 | 0 | 0 | 0 | 0 |
| 0 | 0 | 0 | 0 | 0 |
| 0 | 0.00656942 | 0 | 0 | 0 |
| 0.002930815 | 0.000823651 | 0.003731448 | 0 | 0.011522518 |
| 0 | 0 | 0 | 0 | 0 |
| 0 | 0 | 0 | 0 | 0 |
| 0 | 0 | 0 | 0 | 0 |
| 0.002273264 | 0 | 0 | 0 | 0 |
| 0 | 0 | 0 | 0 | 0 |
| 0 | 0 | 0 | 0 | 0 |
| 0 | 0 | 0 | 0 | 0 |
| 0 | 0 | 0 | 0.008958512 | 0 |
| 0 | 0 | 0 | 0 | 0 |
| 0 | 0 | 0 | 0 | 0 |
| 0 | 0 | 0 | 0 | 0 |
| 0 | 0 | 0 | 0 | 0 |
| 0 | 1.526922845 | 0 | 0 | 0 |
| 0 | 0 | 0 | 0 | 0 |
| 0 | 0.009368023 | 0 | 0 | 0 |
| 0 | 0 | 0 | 0 | 0 |
| 0 | 0 | 0 | 0 | 0 |
| 0 | 0 | 0 | 0 | 0 |
| 0 | 0 | 0 | 0 | 0 |
| 0 | 0 | 0 | 0 | 0 |

| | | | | |
|---|---|---|---|---|
| 0 | 0.041032826 | 0 | 0 | 0 |
| 0 | 0 | 0 | 0 | 0 |
| 0 | 0 | 0 | 0 | 0 |
| 0 | 0 | 0 | 0 | 0 |
| 0 | 0.004240475 | 0 | 0 | 0 |
| 0 | 0 | 0 | 0 | 0 |
| 0 | 0.002302872 | 0 | 0.009025858 | 0.351757616 |
| 0 | 0 | 0 | 0 | 0 |
| 0 | 0 | 0 | 0 | 0 |
| 0.109195853 | 0 | 0 | 0 | 0.836267588 |
| 0 | 0.001523242 | 0 | 0 | 0 |
| 0 | 0 | 0 | 0 | 0 |
| 0 | 0 | 0 | 0 | 0 |
| 0 | 0 | 0 | 0 | 0 |
| 0 | 0 | 0 | 0 | 0 |
| 0 | 0 | 0 | 0 | 0 |
| 0 | 0 | 0.689379116 | 9.805156567 | 0 |
| 0 | 0 | 0 | 0 | 0 |
| 0 | 0 | 0 | 0 | 0 |
| 0 | 0 | 0 | 0 | 0 |
| 0 | 0 | 0 | 0 | 0 |
| 0 | 0 | 0 | 0 | 0 |
| 0.0064817 | 0.01692795 | 0 | 0 | 0 |
| 0.006359829 | 0.000586461 | 0.003719647 | 0 | 0.004893818 |
| 0 | 0 | 0 | 0 | 0 |
| 0 | 0.001153154 | 0.000835876 | 0 | 0 |
| 0 | 0 | 0 | 0 | 0 |
| 0 | 0 | 0 | 0 | 0 |
| 0 | 0 | 0 | 0 | 0 |
| 0 | 0 | 1.24990169 | 0.635664008 | 0 |
| 0 | 0 | 0 | 0 | 0 |
| 0 | 0 | 0 | 0 | 0 |
| 0 | 0 | 0 | 0 | 0 |
| 0 | 0 | 6.557418489 | 0 | 0 |
| 0 | 0 | 0 | 0 | 0 |
| 0 | 0 | 0 | 0 | 0 |
| 0 | 0 | 0 | 0.00740445 | 0 |
| 0 | 0 | 0 | 0 | 0 |
| 0 | 0 | 0 | 0 | 0 |
| 0 | 0 | 0 | 0 | 0 |
| 0 | 0 | 0 | 0 | 0 |
| 0 | 0 | 0 | 0 | 0 |
| 0 | 0 | 0 | 0 | 0 |
| 0 | 0.029876778 | 0 | 0 | 0 |
| 0 | 0.008797007 | 0 | 0 | 0 |
| 0 | 0 | 0 | 0 | 0 |
| 0 | 0 | 0 | 0 | 0 |
| 0 | 0 | 0 | 0 | 0 |

| | | | | |
|---|---|---|---|---|
| 0.018605498 | 0.000841404 | 0.294467588 | 0 | 0.113062436 |
| 0 | 0 | 0 | 0 | 0 |
| 0 | 0.264230274 | 0 | 0 | 0 |
| 0 | 0 | 0 | 0 | 0 |
| 0 | 0 | 0 | 0 | 0 |
| 0 | 0 | 0 | 0 | 0 |
| 0 | 0 | 0 | 0 | 0 |
| 0 | 1.324407858 | 0.37612938 | 0 | 2.624101334 |
| 0.116480812 | 2.34459122 | 0.118469643 | 0.001211494 | 0.006077687 |
| 0.039960998 | 0.012182567 | 0.138499704 | 0 | 0 |
| 0 | 0 | 0 | 0 | 0 |
| 0 | 0 | 0 | 0 | 0 |
| 0 | 0 | 0 | 0 | 0 |
| 0.034045797 | 0 | 0 | 0 | 5.431878004 |
| 0 | 0 | 0 | 0 | 0 |
| 0 | 0 | 0 | 0 | 0 |
| 0 | 0 | 0 | 0.026194735 | 0.257282676 |
| 0 | 0 | 0 | 0 | 0 |
| 0 | 0 | 0 | 0 | 0 |
| 0 | 0 | 0 | 0 | 0 |
| 0 | 0 | 0 | 0 | 0 |
| 0 | 0 | 0 | 0 | 0 |
| 0 | 0 | 0 | 0 | 0 |
| 0 | 0 | 0 | 0 | 0 |
| 0 | 0.016402383 | 0 | 0 | 0 |
| 0 | 0 | 0 | 0 | 0 |
| 0 | 0 | 0 | 0 | 0 |
| 0 | 0 | 0 | 0 | 0 |
| 0 | 0 | 0 | 0 | 0 |
| 0 | 0 | 0 | 0 | 0 |
| 0 | 0 | 0 | 0 | 0 |
| 0 | 0 | 0 | 0 | 0 |
| 0 | 0 | 0 | 0 | 0 |
| 0 | 0 | 0 | 0 | 0 |
| 0.001553419 | 0 | 0 | 0 | 0 |
| 0 | 0.031760454 | 0 | 0 | 0 |
| 0 | 0 | 0 | 0 | 0 |
| 0 | 0 | 0 | 0 | 0 |
| 0 | 0 | 0 | 0 | 0 |
| 0 | 0 | 0 | 0 | 0 |
| 0 | 0 | 0 | 0 | 0 |
| 0 | 0.150137343 | 0 | 0 | 0 |
| 0 | 0 | 0 | 0 | 0 |
| 0 | 0 | 0 | 0 | 0 |
| 0 | 0 | 0 | 0 | 0 |
| 0.012170349 | 0.035164433 | 0 | 0 | 0 |
| 0 | 0 | 0 | 0 | 0 |
| 0 | 0 | 0 | 0 | 0 |

| | | | | |
|---|---|---|---|---|
| 0 | 0 | 0 | 0 | 0 |
| 0 | 0.001328957 | 0.007568851 | 1.744995227 | 0 |
| 0 | 0 | 0 | 0 | 0 |
| 0 | 0 | 0 | 0 | 0 |
| 0.096340602 | 2.763745224 | 0.303039097 | 0 | 0 |
| 0 | 0 | 0 | 0 | 0 |
| 0.015446668 | 0.001841635 | 0 | 0 | 0 |
| 0.035637331 | 0 | 0.030791055 | 0.013009062 | 0.013791305 |
| 0 | 0.017633254 | 0 | 0 | 0 |
| 0 | 0 | 0 | 0 | 0 |
| 0 | 0 | 0 | 0 | 0 |
| 0 | 0 | 0 | 0 | 0 |
| 0 | 0 | 0 | 0 | 0 |
| 0 | 0 | 0 | 0 | 0 |
| 0 | 0 | 0 | 0 | 0 |
| 0 | 0.068925295 | 0 | 0 | 0 |
| 0 | 0 | 0 | 0 | 0 |
| 0 | 0 | 0 | 0 | 0 |
| 0 | 0 | 0 | 0 | 0 |
| 0 | 0 | 0 | 0 | 0 |
| 0 | 0 | 0 | 0 | 0 |
| 0 | 0 | 0 | 0 | 0 |
| 0 | 0 | 0 | 0 | 0 |
| 0.002589717 | 0 | 0.009022254 | 0.002124202 | 0.001705034 |
| 0 | 0.21592196 | 0 | 0 | 0 |
| 0 | 0 | 0 | 0 | 0 |
| 0 | 0.019273533 | 0 | 0.010685532 | 0.205691082 |
| 0.001133658 | 0.002007139 | 0 | 0 | 0 |
| 0 | 0 | 0 | 0 | 0 |
| 0 | 0 | 0 | 0 | 0 |
| 0 | 0 | 0 | 0 | 0 |
| 0 | 0 | 0 | 0 | 0 |
| 0 | 0 | 0 | 0 | 0 |
| 0 | 0 | 0 | 0 | 0 |
| 0 | 0 | 0 | 0 | 0 |
| 0.002406245 | 0 | 0 | 0 | 0 |
| 0 | 0 | 0 | 0 | 0 |
| 0 | 0.122506609 | 0 | 0 | 0 |
| 0 | 0 | 0 | 0 | 0 |
| 0 | 0 | 0 | 0 | 0 |
| 0 | 0 | 0 | 0 | 0 |
| 0 | 0 | 0 | 0 | 0 |
| 0 | 0 | 0 | 0 | 0 |
| 0 | 0 | 0 | 0 | 0 |
| 0 | 0 | 0 | 0 | 0 |
| 0 | 0 | 0 | 0 | 0 |
| 0 | 0 | 0 | 0 | 0 |
| 0 | 0 | 0 | 0 | 0 |

| | | | | |
|---|---|---|---|---|
| 0 | 0.132651804 | 0 | 0 | 0 |
| 0 | 0 | 0 | 0 | 0 |
| 0 | 0 | 0 | 0 | 0 |
| 0 | 0 | 0 | 0 | 0 |
| 0 | 0 | 0 | 0 | 0 |
| 0.037771117 | 0 | 0.100360351 | 0 | 0 |
| 0 | 0 | 0 | 0 | 0 |
| 0 | 0 | 0 | 0 | 0 |
| 0 | 0 | 0 | 0 | 0 |
| 0 | 0 | 0 | 0 | 0 |
| 0 | 0 | 0 | 0 | 0 |
| 0 | 0 | 0 | 0 | 0 |
| 0 | 0 | 0 | 0 | 0 |
| 0 | 0 | 0 | 0 | 0 |
| 0 | 0 | 0 | 0 | 0 |
| 0 | 0 | 0 | 0 | 0 |
| 0 | 0 | 0 | 0 | 0 |
| 0 | 0 | 0 | 0 | 4.253164072 |
| 0.000858402 | 0 | 0 | 0 | 0 |
| 0 | 0 | 0 | 0 | 0 |
| 0.007736674 | 0 | 0 | 0.001057661 | 0.003395813 |
| 0 | 0 | 0 | 0 | 0 |
| 0.011251288 | 0 | 0 | 0 | 0.003645365 |
| 0 | 0.001865865 | 0 | 0 | 0.064962214 |
| 0 | 0 | 0 | 0 | 0 |
| 0 | 0 | 0 | 0 | 0 |
| 0 | 0 | 0 | 0 | 0 |
| 0 | 0 | 0 | 0 | 0 |
| 0 | 0 | 0 | 0 | 0.004826409 |
| 0 | 0 | 0 | 0 | 0 |
| 0 | 0 | 0 | 0 | 0 |
| 0 | 0 | 0 | 0 | 0 |
| 0.013032339 | 0 | 3.384336426 | 0.065798575 | 0.01927832 |
| 0 | 0 | 0 | 0 | 0 |
| 0 | 0.15516093 | 0 | 0 | 0 |
| 0.05384709 | 0 | 0.001586441 | 0.014454917 | 0.002256048 |
| 0 | 0 | 0 | 0 | 0 |
| 0 | 0 | 0 | 0 | 0 |
| 0 | 0 | 0.012615416 | 0.060143389 | 0.230379493 |
| 0 | 0 | 0 | 0 | 0 |
| 0 | 0 | 0 | 0 | 0 |
| 0 | 0 | 0 | 0 | 0 |
| 0.071623358 | 0.00799216 | 0.004559462 | 0 | 0 |
| 0 | 0 | 0 | 0 | 0 |
| 0 | 0 | 0 | 0 | 0 |
| 0 | 0.005641921 | 0 | 0 | 0 |
| 0 | 0 | 0 | 0 | 0 |
| 0 | 0 | 0 | 0 | 0 |

| | | | | |
|---|---|---|---|---|
| 0 | 0 | 0 | 0 | 0 |
| 0 | 0 | 0 | 0 | 0 |
| 0 | 0 | 0 | 0 | 0 |
| 0 | 0 | 0 | 0 | 0 |
| 0 | 0 | 0 | 0 | 0 |
| 0 | 0 | 0 | 0 | 0 |
| 0 | 0 | 0 | 0 | 0 |
| 0 | 0 | 0.123083764 | 0 | 0 |
| 0 | 0 | 0 | 0 | 0 |
| 0 | 0.001452779 | 0 | 0 | 0 |
| 0 | 0 | 0 | 0 | 0 |
| 0 | 0 | 0 | 0 | 0 |
| 0 | 0 | 0 | 0 | 0 |
| 0 | 1.567944747 | 0 | 0 | 0 |
| 0 | 0 | 0 | 0 | 0.026077284 |
| 0.003875181 | 0 | 0.347524008 | 0.035838682 | 0.07910828 |
| 0 | 0 | 0 | 0 | 0 |
| 0 | 0 | 0 | 0 | 0 |
| 0 | 0 | 0 | 0 | 0 |
| 0 | 0.059038049 | 0 | 0 | 0 |
| 0 | 0.001244067 | 0 | 0 | 0 |
| 0 | 0 | 0 | 0 | 0 |
| 0 | 0 | 0 | 0 | 0 |
| 0 | 0.018404905 | 0 | 0 | 0 |
| 0 | 0 | 0 | 0 | 0 |
| 0 | 0 | 0 | 0 | 0 |
| 0 | 0 | 0 | 0 | 0 |
| 0 | 0 | 0 | 0 | 0 |
| 0 | 0 | 0 | 0 | 0 |
| 0 | 0.004197404 | 0 | 0 | 0 |
| 0 | 0.100298194 | 0 | 0 | 0 |
| 0 | 0 | 0 | 0 | 0 |
| 0 | 0 | 0 | 0 | 0 |
| 0 | 0 | 0 | 0 | 0 |
| 0 | 0 | 0 | 0 | 0 |
| 0 | 0 | 0 | 0 | 0 |
| 0 | 0 | 0 | 0 | 0 |
| 0 | 0 | 0 | 0 | 0 |
| 0 | 0 | 0 | 0 | 0 |
| 0 | 0 | 0 | 0 | 0 |
| 0 | 0 | 0 | 0 | 0 |
| 0 | 0 | 0 | 0 | 0 |
| 0 | 0 | 0 | 0 | 0 |
| 0.011900334 | 0 | 0 | 0 | 0 |
| 0 | 0 | 0 | 0 | 0 |
| 0 | 0 | 0 | 0 | 0 |
| 0.057273477 | 0 | 2.35838691 | 11.8228665 | 0.04417231 |
| 0 | 0.000594587 | 0 | 0 | 0 |

| | | | | |
|---|---|---|---|---|
| 0 | 0 | 0 | 0 | 0 |
| 0 | 0 | 0 | 0 | 0 |
| 0 | 0 | 0 | 0 | 0 |
| 0 | 0 | 0 | 0 | 0 |
| 0.149663862 | 0.262902445 | 0 | 0.009191883 | 0.233683032 |
| 0 | 0 | 0 | 0 | 0 |
| 0 | 0 | 0 | 0 | 0 |
| 0 | 0 | 0 | 0 | 0 |
| 0 | 0 | 0 | 0 | 0 |
| 0 | 0 | 0 | 0 | 0 |
| 0 | 0 | 0 | 0 | 0 |
| 0.046289071 | 0.003541251 | 0.002062697 | 0 | 0.029985084 |
| 0.009096489 | 0.003928124 | 0 | 0 | 0 |
| 0 | 0 | 0 | 0 | 0 |
| 0.00117585 | 0.004626309 | 0 | 0 | 0 |
| 0 | 0 | 0 | 0 | 0 |
| 0 | 0 | 0 | 0 | 0 |
| 0 | 0 | 0 | 0 | 0 |
| 0.070684871 | 0.001340728 | 0.130157167 | 0 | 0 |
| 0 | 0.001000333 | 0 | 0.001147012 | 0.214700809 |
| 0 | 0 | 0 | 0 | 0 |
| 0 | 0 | 0 | 0 | 0 |
| 0 | 0 | 0 | 0 | 0 |
| 0 | 0 | 0 | 0 | 0 |
| 0 | 0 | 0 | 0 | 0 |
| 0 | 0 | 0 | 0 | 0 |
| 0.044212126 | 0.001202418 | 0.035408152 | 0 | 0.027297749 |
| 0.011591682 | 0 | 0 | 0 | 0 |
| 0 | 0 | 0 | 0 | 0 |
| 0 | 0 | 0 | 0 | 0 |
| 0.001385717 | 0.002271674 | 0.015025646 | 0 | 0 |
| 0 | 0 | 0 | 0 | 0 |
| 0.001233547 | 0 | 0 | 0 | 0 |
| 0 | 0 | 0 | 0 | 0 |
| 0 | 0 | 0 | 0 | 0 |
| 0 | 0 | 0 | 0 | 0 |
| 0 | 0 | 0 | 0 | 0 |
| 0.00843756 | 0.011810491 | 0.060591893 | 0.023278065 | 0.00857847 |
| 0 | 0.061429622 | 0 | 0 | 0 |
| 0.002059101 | 0.004557041 | 0 | 0 | 0 |
| 0.001943863 | 0 | 0.001876785 | 0 | 0.004106065 |
| 0 | 0 | 0 | 0 | 0 |
| 0 | 0 | 0 | 0 | 0 |
| 0.001684285 | 0 | 0.004503227 | 0 | 0 |
| 0 | 0 | 0 | 0 | 0 |
| 0 | 0 | 0 | 0 | 0.006075698 |
| 0 | 0 | 0 | 0 | 0 |
| 0 | 0 | 0 | 0 | 0 |

| | | | | |
|---|---|---|---|---|
| 0 | 0 | 0 | 0 | 0 |
| 0 | 0.043490636 | 0 | 0 | 0 |
| 0 | 0 | 0 | 0 | 0 |
| 0 | 0 | 0.003146268 | 0 | 0.005478673 |
| 0 | 0 | 0.043146387 | 0 | 0 |
| 0 | 0 | 0.002959083 | 0 | 0 |
| 0 | 0.001925551 | 0 | 0 | 0 |
| 0.003549834 | 0 | 0 | 0 | 0 |
| 0 | 0 | 0 | 0 | 0 |
| 0 | 0 | 0 | 0 | 0 |
| 0 | 0.025413882 | 0 | 0 | 0 |
| 0 | 0 | 0 | 0 | 0 |
| 0 | 0.032174698 | 0 | 0 | 0 |
| 0 | 0 | 0 | 0 | 0 |
| 0 | 0 | 0 | 0 | 0 |
| 0 | 0 | 0 | 0 | 0 |
| 0 | 0 | 0 | 0 | 0 |
| 0 | 0 | 0 | 0 | 0 |
| 0 | 0 | 0 | 0 | 0 |
| 0 | 0 | 0 | 0 | 0 |
| 0 | 0 | 0 | 0 | 0 |
| 0 | 0 | 0 | 0 | 0 |
| 0.06119611 | 0 | 0 | 0.013591477 | 0.043637903 |
| 0 | 0 | 0 | 0 | 0 |
| 0 | 0 | 0 | 0 | 0 |
| 0 | 0.010087743 | 0 | 0 | 0 |
| 0 | 0 | 0 | 0 | 0 |
| 0 | 0 | 0 | 0 | 0 |
| 0 | 0 | 0 | 0 | 0 |
| 0 | 0 | 0 | 0 | 0 |
| 0 | 0.039752737 | 0 | 0 | 0.102443831 |
| 0 | 0.007385833 | 0 | 0 | 0 |
| 0 | 0 | 0 | 0 | 0 |
| 0 | 0 | 0 | 0 | 0 |
| 0 | 0 | 0 | 0 | 0 |
| 0 | 0 | 0 | 0 | 0 |
| 0 | 0 | 0.005218298 | 0 | 0.001009639 |
| 0 | 0 | 0 | 0 | 0 |
| 0.004218744 | 0 | 0.015633047 | 0 | 0 |
| 0 | 0 | 0 | 0 | 0 |
| 0 | 0 | 0 | 0 | 0 |
| 0 | 0 | 0 | 0 | 0 |
| 0 | 0 | 0 | 0 | 0 |
| 0 | 0 | 0 | 0 | 0 |
| 0 | 0 | 0 | 0 | 0 |
| 0.031061887 | 0.230202603 | 0.184205822 | 0 | 0 |
| 0 | 0 | 0 | 0 | 0 |
| 0 | 0 | 0 | 0 | 0 |

| | | | | |
|---|---|---|---|---|
| 0 | 0 | 0 | 0 | 0 |
| 0 | 0 | 0 | 0 | 0 |
| 0 | 0 | 0 | 0 | 0 |
| 0 | 0 | 0 | 0 | 0 |
| 0 | 0 | 0 | 0 | 0 |
| 0 | 0 | 0 | 0 | 0 |
| 0.001805592 | 0 | 0 | 0 | 0 |
| 0 | 0 | 0 | 0 | 0 |
| 0 | 0 | 0 | 0 | 0 |
| 0 | 0 | 0 | 0 | 0 |
| 0.001080799 | 0 | 0 | 0 | 0.003913709 |
| 0 | 0 | 0 | 0 | 0.000779091 |
| 0 | 0 | 0 | 0 | 0 |
| 0 | 0 | 0 | 0 | 0 |
| 0 | 0 | 0 | 0 | 0 |
| 0 | 0 | 0 | 0 | 0 |
| 0 | 0 | 0 | 0 | 0 |
| 0 | 0 | 0 | 0 | 0 |
| 0 | 0 | 0 | 0 | 0 |
| 0.00501664 | 0 | 0.003832244 | 0 | 0 |
| 0 | 0 | 0 | 0 | 0 |
| 0 | 0 | 0 | 0 | 0 |
| 0 | 0 | 0 | 0 | 0 |
| 0 | 0 | 0 | 0 | 0 |
| 0 | 0 | 0 | 0 | 0 |
| 0 | 0 | 0 | 0 | 0 |
| 0 | 0 | 0 | 0 | 0 |
| 0 | 0 | 0 | 0 | 0 |
| 0 | 0 | 0 | 0 | 0 |
| 0 | 0 | 0 | 0 | 0 |
| 0 | 0.007065288 | 0.017558901 | 0 | 0.001932217 |
| 0 | 0 | 0 | 0 | 0 |
| 0.186582626 | 0.024610557 | 14.61201483 | 0 | 0.049357018 |
| 0.008766308 | 0.011736349 | 0.003255311 | 0.030320025 | 0.008817748 |
| 0 | 0 | 0 | 0 | 0 |
| 0 | 0 | 0 | 0 | 0 |
| 0 | 0 | 0 | 0 | 0 |
| 0 | 0 | 0 | 0 | 0 |
| 0 | 0 | 0 | 0 | 0 |
| 0 | 0 | 0 | 0 | 0 |
| 0 | 0 | 0 | 0 | 0 |
| 0.08664997 | 0 | 0 | 0 | 1.420627074 |
| 0 | 0.079637271 | 0 | 0 | 0 |
| 0 | 0 | 0 | 0 | 0 |
| 0 | 0 | 0 | 0 | 0 |
| 0.01967241 | 0.059747272 | 0.009842823 | 0 | 0 |
| 0 | 0 | 0 | 0 | 0 |
| 0 | 0 | 0 | 0 | 0 |

| | | | | |
|---|---|---|---|---|
| 0 | 0 | 0 | 0 | 0 |
| 0 | 0 | 0 | 0 | 0 |
| 0.003214976 | 0 | 0 | 0.001318534 | 0.000635009 |
| 0 | 0 | 0 | 0 | 0 |
| 0.003967188 | 0.019281278 | 0 | 0 | 0 |
| 0 | 0 | 0.046765055 | 0 | 0.135886397 |
| 0 | 0 | 0 | 0 | 0 |
| 0 | 0 | 0 | 0 | 0 |
| 0 | 0 | 0 | 0 | 0 |
| 0 | 0 | 0 | 0 | 0 |
| 0 | 0 | 0 | 0 | 0 |
| 0 | 0 | 0 | 0 | 0 |
| 0 | 0 | 0 | 0 | 0 |
| 0 | 0.110185329 | 0 | 0 | 0 |
| 0 | 0 | 0 | 0 | 0 |
| 0 | 0 | 0 | 0 | 0 |
| 0 | 0 | 0 | 0 | 0 |
| 0 | 0 | 0 | 0 | 0 |
| 0 | 0 | 0 | 0 | 0 |
| 0 | 0 | 0 | 0 | 0 |
| 0 | 0 | 0 | 0 | 0 |
| 0 | 0 | 0.0008381 | 0.001060608 | 0 |
| 0 | 0 | 0 | 0 | 0 |
| 0 | 0 | 0 | 0 | 0 |
| 0 | 0 | 0 | 0 | 0 |
| 0 | 0 | 0 | 0 | 0 |
| 0 | 0 | 0 | 0 | 0 |
| 0 | 0.052250765 | 0 | 0 | 0.008851937 |
| 0 | 0 | 0 | 0 | 0 |
| 0 | 0 | 0 | 0 | 0 |
| 0 | 0 | 0 | 0 | 0 |
| 0 | 0 | 0 | 0 | 0 |
| 0 | 0 | 0 | 0 | 0 |
| 0 | 0 | 0 | 0 | 0 |
| 0 | 0 | 0 | 0 | 0 |
| 0 | 0 | 0 | 0 | 0 |
| 0 | 0 | 0 | 0 | 0 |
| 0 | 0 | 0 | 0 | 0 |
| 1.447159131 | 3.594519994 | 0 | 0.928579011 | 0 |
| 0 | 0 | 0 | 0 | 0 |
| 0 | 0 | 0 | 0 | 0 |
| 0 | 0.123851251 | 0 | 0 | 0 |
| 0.015563911 | 0 | 0.233205084 | 0 | 0.08982193 |
| 0 | 0 | 0 | 0 | 0 |
| 0 | 0 | 0 | 0 | 0 |
| 0 | 0 | 0 | 0 | 0 |
| 0.017113249 | 0 | 0.004973394 | 0 | 0.025057113 |
| 0 | 0 | 0 | 0 | 0 |

| | | | | |
|---|---|---|---|---|
| 0 | 0 | 0 | 0 | 0 |
| 0.026522786 | 0.006002563 | 0.026147953 | 0.045531859 | 0.096042308 |
| 0 | 0.018614587 | 0 | 0 | 0 |
| 0 | 0 | 0 | 0 | 0 |
| 0 | 0 | 0 | 0 | 0 |
| 0 | 0 | 0 | 0 | 0 |
| 0 | 0 | 0 | 0 | 0 |
| 0 | 0 | 0 | 0 | 0 |
| 0 | 0 | 0 | 0 | 0 |
| 0 | 0 | 0 | 0 | 0 |
| 0 | 0 | 0 | 0 | 0 |
| 0 | 0 | 0 | 0 | 0 |
| 0 | 0.011958339 | 0 | 0 | 0 |
| 0 | 0 | 0 | 0 | 0 |
| 0 | 0.027634308 | 0 | 0 | 0 |
| 0 | 0 | 0 | 0 | 0 |
| 0 | 0 | 0 | 0 | 0 |
| 0 | 0 | 0 | 0 | 0 |
| 0 | 0 | 0 | 0 | 0 |
| 0 | 0 | 0 | 0 | 0 |
| 0 | 0 | 0 | 0 | 0 |
| 0 | 0 | 0 | 0 | 0 |
| 0.02760633 | 0 | 0 | 0 | 0 |
| 0 | 0 | 0 | 0 | 0 |
| 0 | 0 | 0 | 0 | 0 |
| 0 | 0 | 0 | 0 | 0 |
| 0 | 0 | 0 | 0 | 0 |
| 0 | 0 | 0 | 0 | 0 |
| 0.006047273 | 0 | 0 | 0 | 0 |
| 0 | 0 | 0 | 0 | 0 |
| 0 | 0 | 0 | 0 | 0 |
| 0 | 0 | 0 | 0 | 0 |
| 0 | 0 | 0 | 0 | 0 |
| 0 | 0 | 0 | 0 | 0 |
| 0 | 0 | 0 | 0 | 26.92641185 |
| 0 | 0 | 0 | 0 | 0 |
| 0 | 0 | 0 | 0 | 0 |
| 0 | 0 | 0 | 0 | 0 |
| 0 | 0.001411544 | 0 | 0 | 0.002910069 |
| 0 | 0 | 0 | 0 | 0 |
| 0.012867261 | 0.055434856 | 0 | 0.008823399 | 0.022365097 |
| 0 | 0 | 0 | 0 | 0 |
| 0 | 0 | 0 | 0 | 0 |
| 0 | 0 | 0 | 0 | 0 |
| 0 | 0 | 0 | 0 | 0 |
| 0.881865788 | 0.002028694 | 3.738270596 | 5.62532562 | 0.309838823 |
| 0 | 0 | 0 | 0 | 0 |
| 0 | 0 | 0 | 0 | 0 |

| | | | | |
|---|---|---|---|---|
| 0 | 0 | 0 | 0 | 0 |
| 0.001439715 | 0 | 0 | 0 | 0 |
| 0 | 0 | 0 | 0 | 0 |
| 0 | 0 | 0 | 0 | 0 |
| 0.002681571 | 0 | 0 | 0 | 0 |
| 0 | 0 | 0 | 0 | 0 |
| 0 | 0 | 0 | 0 | 0 |
| 0 | 0 | 0 | 0 | 0.005304763 |
| 0 | 0 | 0 | 0 | 0 |
| 0 | 0 | 0 | 0.003899984 | 0 |
| 0 | 0.041251763 | 0 | 0 | 0 |
| 0 | 0 | 0 | 0 | 0 |
| 0.001126613 | 0 | 0.001004065 | 0 | 0.001835822 |
| 0 | 0 | 0 | 0 | 0 |
| 0 | 0 | 0 | 0 | 0 |
| 0 | 0 | 0 | 0 | 0 |
| 0 | 0 | 0 | 0 | 0 |
| 0 | 0 | 0 | 0 | 0 |
| 0 | 0 | 0 | 0 | 0 |
| 0.270997214 | 0.240171126 | 1.849488057 | 0.035059608 | 0.279415949 |
| 0 | 0 | 0 | 0 | 0 |
| 0 | 0 | 0 | 0 | 0 |
| 0 | 0 | 0 | 0 | 0 |
| 0 | 0 | 0 | 0 | 0 |
| 0 | 0 | 0 | 0 | 0 |
| 0 | 0 | 0 | 0 | 0 |
| 0 | 0 | 0 | 0 | 0 |
| 0 | 0 | 0 | 0 | 0 |
| 0 | 0 | 0 | 0 | 0 |
| 0.011523698 | 0 | 0.10079125 | 0 | 0.132367455 |
| 0 | 0 | 0 | 0 | 0 |
| 0 | 0 | 0 | 0 | 0 |
| 0 | 0 | 0 | 0 | 0 |
| 0 | 0 | 0 | 0 | 0 |
| 0 | 0 | 0 | 0 | 0 |
| 0 | 0 | 0 | 0 | 0 |
| 0 | 0 | 0 | 0 | 0 |
| 0 | 0 | 0 | 0 | 0 |
| 0 | 0 | 0 | 0 | 0 |
| 0 | 0 | 0 | 0 | 0 |
| 0 | 0 | 0 | 0 | 0 |
| 0 | 0 | 0 | 0 | 0 |
| 0 | 0 | 0 | 0 | 0 |
| 0 | 0 | 0 | 0 | 0 |
| 0 | 0.018759529 | 0.008184003 | 0 | 0.01057254 |
| 0 | 0.008003095 | 0 | 0 | 0 |
| 0 | 0 | 0 | 0 | 0 |
| 0 | 0 | 0 | 0 | 0 |

| | | | | |
|---|---|---|---|---|
| 0 | 0 | 0 | 0 | 0 |
| 0 | 0 | 0 | 0 | 0 |
| 0 | 0 | 0 | 0 | 0 |
| 0 | 0 | 0 | 0 | 0 |
| 0 | 0 | 0 | 0 | 0 |
| 0.005449145 | 0.003457955 | 0.023498756 | 0 | 0.003309889 |
| 0 | 0 | 0 | 0 | 0 |
| 0 | 0 | 0 | 0 | 0 |
| 0 | 0 | 0 | 0 | 0 |
| 0 | 0 | 0 | 0 | 0 |
| 0 | 0 | 0 | 0 | 0 |
| 0 | 0 | 0 | 0 | 0 |
| 0 | 0 | 0 | 0 | 0 |
| 0 | 0 | 0 | 0 | 0 |
| 0 | 0 | 0 | 0 | 0 |
| 0 | 0 | 0 | 0 | 0 |
| 0 | 0 | 0 | 0 | 0 |
| 0 | 0 | 0 | 0 | 0 |
| 0 | 0 | 0 | 0 | 0 |
| 0 | 0 | 0 | 0 | 0 |
| 0 | 0 | 0 | 0 | 0 |
| 0 | 0 | 0 | 0 | 0 |
| 0 | 0 | 0 | 0 | 0 |
| 0 | 0 | 0 | 0 | 0 |
| 0 | 0 | 0 | 0 | 0 |
| 0 | 0 | 0 | 0 | 0 |
| 0 | 0 | 0 | 0 | 0 |
| 0 | 0 | 0 | 0 | 0 |
| 0 | 0 | 0 | 0 | 0 |
| 0 | 0 | 0 | 0 | 0 |
| 0 | 0 | 0 | 0 | 0 |
| 0 | 0 | 0 | 0 | 0 |
| 0 | 0 | 0 | 0 | 0 |
| 0 | 0 | 0 | 0 | 0 |
| 0 | 0 | 0 | 0 | 0 |
| 0 | 0 | 0 | 0 | 0 |
| 0 | 0 | 0 | 0 | 0 |
| 0 | 0 | 0 | 0 | 0 |
| 0 | 0.14106707 | 0 | 0 | 0 |
| 0 | 0 | 0 | 0 | 0 |
| 0 | 0 | 0 | 0 | 0 |
| 0 | 0 | 0 | 0 | 0 |
| 0 | 0 | 0 | 0 | 0 |
| 0 | 0 | 0 | 0 | 0 |
| 0 | 0 | 0 | 0 | 0 |
| 0 | 0 | 0 | 0 | 0 |
| 0 | 0 | 0 | 0 | 0 |
| 0 | 0 | 0 | 0 | 0 |

| | | | | |
|---|---|---|---|---|
| 0 | 0 | 0 | 0 | 0 |
| 0 | 0 | 0 | 0.000887827 | 0.64564478 |
| 0 | 0 | 0 | 0 | 0 |
| 0 | 0 | 0 | 0 | 0 |
| 0 | 0 | 0 | 0 | 0 |
| 0 | 0 | 0 | 0 | 0 |
| 0 | 0 | 0 | 0 | 0 |
| 0 | 0 | 0 | 0 | 0 |
| 0 | 0 | 0 | 0 | 0 |
| 0 | 0 | 0 | 0 | 0 |
| 0 | 0.001267342 | 0.006889845 | 0 | 0 |
| 0 | 0 | 0 | 0 | 0 |
| 0 | 0.439660454 | 0 | 0 | 0 |
| 0 | 0 | 0 | 0 | 0 |
| 0 | 0 | 0 | 0 | 0 |
| 0 | 0 | 0 | 0 | 0 |
| 0 | 0 | 0 | 0 | 0 |
| 0 | 0 | 0 | 0 | 0 |
| 0 | 0 | 0 | 0 | 0 |
| 0 | 0 | 0 | 0 | 0 |
| 0.002353878 | 0 | 0 | 0 | 0 |
| 0 | 0 | 0 | 0 | 0 |
| 0.005496681 | 0 | 0 | 0 | 0 |
| 1.677825252 | 1.106041559 | 1.308017493 | 2.25122687 | 0 |
| 0 | 0 | 0 | 0 | 0 |
| 0 | 0 | 0 | 0 | 0 |
| 0 | 0 | 0 | 0 | 0 |
| 0 | 0 | 0 | 0 | 0 |
| 0 | 0 | 0 | 0 | 0 |
| 0 | 0 | 0 | 0 | 0 |
| 0.005134067 | 0 | 0 | 0 | 0 |
| 0 | 0 | 0 | 0 | 0 |
| 0 | 0 | 0 | 0 | 0 |
| 0 | 0 | 0 | 0 | 0 |

| PHL018-0190_V2 | PHL018-0200_V2 | PHL018-0173_V1 | PHL018-0137_V2 | PHL018-0132_V1 |
|---|---|---|---|---|
| REF | EYCF | CTRL | EYCF | CTRL |
| 0.047476927 | 0.011494893 | 0 | 0.01204127 | 0.189485568 |
| 0.248244549 | 4.189778647 | 2.818977438 | 0.060910789 | 17.84904035 |
| 0 | 0.008518153 | 8.7311056 | 0.121252319 | 0 |
| 0 | 0 | 0 | 0 | 0 |
| 0 | 0 | 0 | 0 | 0 |
| 0 | 0 | 0 | 0 | 0 |
| 0 | 0 | 0 | 0 | 0 |
| 0 | 0 | 0 | 0 | 0 |
| 0 | 0 | 0 | 0 | 0 |
| 0 | 0 | 0 | 0 | 0 |
| 0 | 0 | 0 | 0 | 0 |
| 0 | 0 | 0 | 0 | 0 |
| 0 | 0 | 0 | 0 | 0 |
| 2.140117535 | 1.376648861 | 18.32391111 | 0.204226069 | 3.556697139 |
| 0 | 0 | 0 | 0 | 0 |
| 0 | 0 | 0 | 0 | 0 |
| 0 | 0 | 0 | 0 | 0 |
| 0 | 0 | 0 | 0 | 0 |
| 0 | 0 | 0 | 0 | 0 |
| 0 | 0 | 0 | 0 | 0 |
| 0 | 0 | 0 | 0 | 0 |
| 0 | 0 | 0 | 0.027001524 | 0 |
| 0 | 0 | 0 | 0 | 0 |
| 0 | 0 | 0 | 0 | 0 |
| 0 | 0 | 0 | 0 | 0 |
| 0 | 0 | 0 | 0 | 0 |
| 0 | 0 | 0 | 0 | 0 |
| 0 | 0 | 0 | 0 | 0 |
| 0 | 0 | 0 | 0 | 0 |
| 0 | 0 | 0 | 0 | 0 |
| 0 | 0 | 0 | 0 | 0 |
| 0 | 0 | 0 | 0 | 0 |
| 0 | 0 | 0 | 0 | 0 |
| 0 | 0 | 0 | 0 | 0 |
| 0 | 0 | 0 | 0 | 0 |
| 0 | 10.0211724 | 0 | 0.199017865 | 0 |
| 0 | 0 | 0 | 0 | 0 |
| 0 | 0 | 0 | 0 | 0 |
| 0 | 0 | 0 | 0 | 0 |
| 0 | 0 | 0 | 0 | 0 |
| 0 | 0 | 0 | 0 | 0 |
| 0 | 0.029549656 | 1.095950806 | 0.011604863 | 0.756974222 |
| 2.360853238 | 0.00159506 | 0.162608303 | 9.337509797 | 0.00457999 |
| 0.005911407 | 0.007090843 | 0.007329769 | 0.09352596 | 0.050767806 |
| 0 | 0 | 0 | 0 | 0.001678981 |
| 0 | 0 | 0 | 0 | 0 |

| | | | | |
|---|---|---|---|---|
| 0.049884431 | 0 | 0 | 0 | 0 |
| 0 | 0 | 0.021618578 | 0 | 0.103892102 |
| 0.000589118 | 0 | 0 | 0 | 0.001485332 |
| 0.656847999 | 0.801809563 | 0.705013806 | 0.095210345 | 0.419431948 |
| 0 | 0 | 0 | 0.006948553 | 0 |
| 0 | 0 | 0 | 0 | 0 |
| 0 | 0.239937534 | 0.169034058 | 0 | 0.006739098 |
| 0.005338434 | 0 | 0 | 0 | 0 |
| 0 | 0 | 0 | 0 | 0 |
| 0 | 0 | 0 | 0 | 0 |
| 0 | 0 | 0 | 0 | 0 |
| 0 | 0.010918339 | 0 | 0.101584219 | 0.056392013 |
| 0 | 0.001410815 | 0 | 0 | 0 |
| 0 | 0.013995669 | 0.033402336 | 0.017780584 | 0.112154716 |
| 0 | 0 | 0 | 0 | 0 |
| 0 | 0 | 0 | 0 | 0 |
| 0.001999404 | 0.00906033 | 0 | 0 | 0 |
| 0 | 0 | 0 | 0 | 0 |
| 0 | 0 | 0 | 0 | 0 |
| 0 | 0 | 0 | 0 | 0 |
| 0 | 0.490505554 | 0.305973459 | 0.000633324 | 0.035213903 |
| 0.002532233 | 0.003857093 | 0.005065909 | 0.097717397 | 0.00205215 |
| 0 | 0 | 0 | 0 | 0 |
| 0 | 0 | 0 | 0 | 0 |
| 0 | 0 | 0 | 0 | 0 |
| 0 | 0 | 0 | 0 | 0 |
| 0 | 0 | 0 | 0 | 0 |
| 0 | 0 | 0 | 0 | 0 |
| 0 | 0.002467169 | 0.007020826 | 0 | 0.026836337 |
| 0 | 0 | 0 | 0 | 0 |
| 0 | 0 | 0 | 0.012577236 | 0.514669241 |
| 0 | 0 | 0 | 0 | 0 |
| 0 | 0 | 0 | 0 | 0 |
| 0 | 0 | 0 | 0 | 0 |
| 0 | 0 | 0 | 0 | 0 |
| 0 | 0 | 0 | 0 | 0.125109928 |
| 0 | 0 | 0 | 0 | 0 |
| 0 | 0 | 0 | 0 | 0 |
| 0 | 0 | 0 | 0 | 0 |
| 0 | 0 | 0 | 0 | 0 |
| 0 | 0 | 0 | 0 | 0 |
| 0 | 0 | 0 | 0 | 0 |
| 0 | 0 | 0 | 0 | 0 |
| 0 | 0 | 0 | 0 | 0 |
| 0 | 0 | 0 | 0.007078243 | 0 |
| 0 | 0 | 0 | 0 | 0 |
| 0 | 0 | 0 | 0.00514615 | 0 |
| 0 | 0 | 0 | 0.029437628 | 0 |

| | | | | |
|---|---|---|---|---|
| 0 | 0 | 0 | 0 | 0 |
| 0 | 0 | 0.005098425 | 0 | 0 |
| 0 | 0 | 0 | 0 | 0 |
| 0 | 0 | 0 | 0 | 0 |
| 0 | 0 | 0 | 0 | 0.499878822 |
| 0 | 0 | 0 | 0 | 0 |
| 0 | 0 | 0 | 0 | 0 |
| 0 | 0 | 0 | 0 | 0 |
| 0.000874144 | 0 | 0.001428174 | 0.007471824 | 0 |
| 0 | 0 | 0 | 0 | 0 |
| 0 | 0 | 0 | 0 | 0 |
| 0 | 0.006276102 | 0 | 0 | 0.001023502 |
| 0 | 0 | 0 | 0 | 0 |
| 0 | 0 | 0 | 0 | 0 |
| 0 | 0 | 0 | 0 | 0 |
| 0 | 0 | 0 | 0 | 0.000949306 |
| 0 | 0 | 0 | 0 | 0 |
| 0 | 0 | 0 | 0 | 0 |
| 0 | 0 | 2.072607694 | 0.180331848 | 0 |
| 0 | 0 | 0 | 0 | 0 |
| 0 | 0 | 0 | 0 | 0 |
| 0 | 0 | 0 | 0 | 0 |
| 0 | 0 | 0 | 0 | 0 |
| 0 | 0 | 0.033643019 | 0 | 0 |
| 0 | 0 | 0 | 0 | 0 |
| 0 | 0 | 0 | 0 | 0 |
| 0 | 0 | 0 | 0 | 0 |
| 0 | 0 | 0 | 0 | 0 |
| 0 | 0 | 0 | 0 | 0 |
| 0 | 0 | 0 | 0 | 0 |
| 0 | 0 | 0 | 0.129419796 | 0.000611844 |
| 0.00678891 | 0.00180965 | 0 | 0.013391274 | 0 |
| 0 | 0 | 0 | 0 | 0 |
| 0 | 0 | 0 | 0 | 0 |
| 0 | 0 | 0 | 0 | 0 |
| 0 | 0 | 0 | 0 | 0 |
| 0 | 0 | 0 | 0 | 0 |
| 0 | 0 | 0 | 0 | 0 |
| 0 | 0 | 0 | 0 | 0 |
| 0 | 0 | 0 | 0 | 0 |
| 0 | 0 | 0 | 0 | 0 |
| 0 | 0 | 0 | 0 | 0 |
| 0.000588518 | 0 | 0 | 0 | 0.001731122 |
| 0 | 0 | 0 | 0 | 0 |
| 0 | 0 | 0 | 0 | 0 |
| 0 | 0 | 0 | 0 | 0 |
| 0 | 0 | 0 | 0 | 0 |
| 0 | 0 | 0 | 0 | 0 |

| | | | | |
|---|---|---|---|---|
| 0 | 0 | 0 | 0 | 0 |
| 0 | 0 | 0 | 0 | 0 |
| 0 | 0 | 0 | 0 | 0 |
| 0 | 0 | 0 | 0 | 0 |
| 0.002363591 | 0.001260076 | 0 | 0.000647533 | 0 |
| 0 | 0 | 0 | 0 | 0 |
| 0 | 0 | 0 | 0 | 0 |
| 0 | 0 | 0 | 0 | 0 |
| 0 | 0 | 0 | 0 | 0 |
| 0 | 0 | 0 | 0 | 0 |
| 0 | 0 | 0 | 0 | 0 |
| 0 | 0 | 0 | 0 | 0 |
| 0 | 0 | 0 | 0 | 0 |
| 0 | 0 | 0 | 0 | 0 |
| 0 | 0 | 0 | 0 | 0 |
| 0 | 0.416336829 | 0 | 0 | 0 |
| 0 | 0 | 0 | 0 | 0 |
| 0 | 0 | 0 | 0 | 0 |
| 0 | 0 | 0 | 0 | 0 |
| 0 | 0 | 0.01074834 | 0 | 0 |
| 0 | 0 | 0 | 0 | 0 |
| 0 | 0 | 0 | 0 | 0 |
| 0.000648436 | 0 | 0 | 0 | 0 |
| 0 | 0 | 0 | 0 | 0 |
| 0 | 0 | 0 | 0 | 0 |
| 0 | 0 | 0 | 0 | 0 |
| 0 | 0 | 0 | 0 | 0 |
| 0 | 0 | 0 | 0 | 0 |
| 0 | 0 | 0 | 0 | 0 |
| 0 | 0 | 0 | 0 | 0 |
| 0 | 0 | 0 | 0 | 0.006730568 |
| 0 | 0 | 0 | 0.002575107 | 0 |
| 0 | 0 | 0 | 0 | 0 |
| 0 | 0 | 0 | 0 | 0 |
| 0 | 0 | 0 | 0 | 0 |
| 0 | 0 | 0 | 0 | 0 |
| 0 | 0 | 0 | 0 | 0 |
| 0 | 0 | 0 | 0 | 0 |
| 0 | 0 | 0 | 0 | 0 |
| 0 | 0 | 0 | 0 | 0 |
| 0 | 0 | 0 | 0 | 0 |
| 0 | 0 | 0 | 0 | 0 |
| 0 | 0 | 0 | 0 | 0 |
| 0 | 0 | 0 | 0 | 0 |
| 0 | 0 | 0 | 0 | 0 |
| 0 | 0 | 0 | 0 | 0 |
| 0 | 0 | 0 | 0 | 0 |
| 0 | 0 | 0 | 0 | 0 |

| | | | | |
|---|---|---|---|---|
| 0 | 0 | 0 | 0 | 0 |
| 0 | 0 | 0 | 0 | 0 |
| 0 | 0 | 0 | 0 | 0 |
| 0 | 0 | 0 | 0 | 0 |
| 0 | 0 | 0 | 0 | 0 |
| 0 | 0 | 0 | 0 | 0 |
| 0 | 0 | 0 | 0 | 0 |
| 0 | 0 | 0 | 0 | 0 |
| 0 | 0 | 0 | 0 | 0 |
| 0 | 0 | 0 | 0 | 0 |
| 0 | 0 | 0 | 0 | 0 |
| 0 | 0 | 0 | 0 | 0 |
| 0.002623374 | 0 | 0.001968091 | 0 | 0 |
| 0 | 0 | 0 | 0 | 0 |
| 0 | 0 | 0 | 0 | 0 |
| 0 | 0 | 0 | 0 | 0 |
| 0 | 0.003668574 | 0 | 0 | 0 |
| 0 | 0 | 0 | 0 | 0 |
| 0 | 0 | 0 | 0 | 0 |
| 0 | 0 | 0 | 0 | 0 |
| 0.003503627 | 0.038104174 | 0.026576743 | 0.16509567 | 0.013692118 |
| 0 | 0 | 0 | 0 | 0 |
| 0 | 0 | 0 | 0 | 0 |
| 0 | 0 | 0.011512051 | 0.005605261 | 0.023950403 |
| 0 | 0 | 0 | 0 | 0 |
| 0 | 0 | 0 | 0 | 0 |
| 0.001224258 | 0.009137459 | 0.005476714 | 0.002906795 | 0.002057798 |
| 0 | 0 | 0 | 0 | 0 |
| 0 | 0 | 0 | 0 | 0 |
| 0 | 0 | 0.007897523 | 0 | 0 |
| 0 | 0 | 0.041063833 | 0 | 0 |
| 0 | 0 | 0 | 0 | 0 |
| 0 | 0 | 0 | 0 | 0 |
| 0 | 0 | 0 | 0.122248675 | 0 |
| 0 | 0.026737125 | 0 | 0 | 0.058913175 |
| 0 | 0 | 0 | 0 | 0 |
| 0 | 0 | 0 | 0 | 0 |
| 0 | 0 | 0 | 0 | 0 |
| 0 | 0 | 0 | 0 | 0 |
| 0 | 0 | 0 | 0 | 0 |
| 0 | 0 | 0 | 0 | 0 |
| 0 | 0 | 0 | 0 | 0 |
| 0 | 0 | 0 | 0 | 0 |
| 0 | 0 | 0 | 0 | 0 |
| 0 | 0 | 0 | 0 | 0 |
| 0 | 0 | 0 | 0.006897655 | 0 |
| 0 | 0 | 0 | 0 | 0 |
| 0.019007093 | 0 | 0.0011375 | 0.004094558 | 0 |

| | | | | |
|---|---|---|---|---|
| 0 | 0 | 0 | 0 | 0 |
| 0 | 0 | 0 | 0 | 0 |
| 0 | 0 | 0 | 0 | 0 |
| 0 | 0.018935329 | 0 | 0 | 0 |
| 0 | 0 | 0 | 0 | 0 |
| 0 | 0 | 0 | 0 | 0 |
| 0 | 0 | 0 | 0 | 0 |
| 0 | 0 | 0.003270356 | 0 | 0.001471988 |
| 0 | 0 | 0 | 0 | 0.002219598 |
| 0 | 0 | 0 | 0 | 0 |
| 0 | 0 | 0 | 0 | 0 |
| 0 | 0 | 0 | 0 | 0 |
| 0 | 0 | 0 | 0 | 0 |
| 0 | 0 | 0 | 0 | 0 |
| 0 | 0 | 0 | 0 | 0 |
| 0 | 0 | 0 | 0 | 0 |
| 0 | 0 | 0 | 0 | 0 |
| 0 | 0 | 0 | 0 | 0 |
| 0.010573724 | 0 | 0 | 0.046710766 | 0 |
| 0 | 0 | 0 | 0.379678128 | 0 |
| 0 | 0 | 0.01877087 | 0 | 0.001536141 |
| 0 | 0 | 0 | 0 | 0 |
| 0 | 0 | 0 | 0 | 0 |
| 0 | 0 | 0 | 0 | 0 |
| 0.008715198 | 0.008259984 | 0.007910488 | 0 | 0.003662242 |
| 0 | 0 | 0 | 0 | 0 |
| 0 | 0 | 0 | 0 | 0 |
| 0 | 0 | 0 | 0 | 0 |
| 0 | 0 | 0.026961048 | 0 | 0.051550225 |
| 0 | 0 | 0 | 0.016681188 | 0 |
| 0 | 0.059804395 | 0.249694578 | 2.149704297 | 0.019879616 |
| 0 | 0 | 0 | 0 | 0 |
| 0 | 0.074124495 | 0 | 0 | 0 |
| 0 | 0 | 0 | 0.000762191 | 0 |
| 0 | 0 | 0 | 0 | 0 |
| 0 | 0 | 0 | 0 | 0 |
| 0 | 0 | 0 | 0 | 0 |
| 0 | 0 | 0 | 0 | 0 |
| 0 | 0 | 0 | 0 | 0 |
| 0 | 0.000763403 | 0 | 0 | 0 |
| 0 | 0 | 0 | 0 | 0 |
| 0 | 0 | 0 | 0.001201023 | 0.014507155 |
| 0 | 0 | 0.013771872 | 0 | 0 |
| 0 | 0 | 0 | 0 | 0 |
| 0 | 0 | 0 | 0 | 0 |
| 0 | 0 | 0 | 0 | 0 |
| 0 | 0 | 0 | 0 | 0 |
| 0 | 0 | 0 | 0 | 0 |

| | | | | |
|---|---|---|---|---|
| 0 | 0 | 0 | 0 | 0 |
| 0 | 0 | 0 | 0 | 0 |
| 0.006422178 | 0.004891125 | 0.001873669 | 0.094255109 | 0.05580494 |
| 0 | 0 | 0 | 0 | 0 |
| 0 | 0 | 0 | 0.013741928 | 0.184933802 |
| 0 | 0 | 0 | 0 | 0 |
| 0 | 0 | 0 | 0 | 0 |
| 0 | 0 | 0 | 0 | 0 |
| 0 | 0 | 0 | 0 | 0 |
| 0 | 0 | 0 | 0 | 0 |
| 0 | 0 | 0 | 0 | 0 |
| 0 | 0 | 0 | 0 | 0 |
| 0 | 0 | 0 | 0 | 0 |
| 0.001013952 | 0 | 0 | 0.014444756 | 0 |
| 0.001882991 | 0 | 0.00244161 | 0.006419676 | 0.001318763 |
| 0 | 0 | 0 | 0 | 0 |
| 0.003337444 | 0.003812691 | 0.001112798 | 0 | 0.004507837 |
| 0 | 0 | 0 | 0 | 0 |
| 0 | 0 | 0 | 0 | 0 |
| 0 | 0 | 0 | 0 | 0 |
| 0 | 0 | 0 | 0 | 0 |
| 0.014203008 | 0 | 0.001255674 | 0.003301519 | 0 |
| 0 | 0 | 0 | 0 | 0 |
| 0 | 0 | 0 | 0 | 0 |
| 0 | 0 | 0 | 0 | 0 |
| 0 | 0 | 0 | 0 | 0 |
| 0 | 0 | 0 | 0 | 0 |
| 0 | 0 | 0 | 0 | 0 |
| 0 | 0 | 0 | 0 | 0 |
| 0 | 0 | 0 | 0 | 0 |
| 0 | 0 | 0 | 0 | 0 |
| 0 | 0 | 0 | 0 | 0 |
| 0 | 0 | 0 | 0 | 0 |
| 0 | 0 | 0 | 0 | 0 |
| 0 | 0 | 0 | 0 | 0 |
| 0 | 0 | 0 | 0 | 0 |
| 0 | 0 | 0 | 0 | 0 |
| 0.016763117 | 0.163455906 | 0.001893149 | 0 | 0 |
| 0 | 0 | 0 | 0 | 0 |
| 0 | 0 | 0 | 0 | 0 |
| 0 | 0 | 0 | 0 | 0 |
| 0 | 0 | 0 | 0 | 0 |
| 0 | 0 | 0 | 0 | 0 |
| 0 | 0 | 0 | 0 | 0 |
| 0 | 0 | 0 | 0 | 0 |
| 0 | 0 | 0 | 0 | 0 |
| 0 | 0 | 0 | 0 | 0 |
| 0 | 0 | 0 | 0.010407454 | 0 |

| | | | | |
|---|---|---|---|---|
| 0 | 0 | 6.364801218 | 0.945412662 | 0 |
| 0 | 0 | 0 | 0 | 0 |
| 0 | 0 | 0.002893485 | 0 | 0 |
| 0 | 0 | 0 | 0 | 0 |
| 0.003174869 | 0.016684041 | 0.004499012 | 0.002485118 | 0 |
| 0 | 0 | 0 | 0 | 0 |
| 0 | 0 | 0 | 0 | 0 |
| 0 | 0 | 0.01446752 | 0.457027309 | 0 |
| 0 | 0 | 0 | 0 | 0 |
| 0.024245555 | 0.008034941 | 0.040147656 | 0.044880711 | 0.010325984 |
| 0 | 0 | 0 | 0 | 0 |
| 0.001509795 | 0 | 0 | 0 | 0.00158609 |
| 0.005535294 | 0.008547644 | 0.006236935 | 0.00292833 | 0.012752917 |
| 0 | 0 | 0 | 0 | 0.24082586 |
| 0 | 0 | 0 | 0.003226844 | 0 |
| 0 | 0 | 0 | 0 | 0.010836365 |
| 0 | 0 | 0 | 0 | 0 |
| 0 | 0 | 0.005511896 | 0 | 0 |
| 0 | 0 | 0.218034256 | 0 | 0 |
| 0 | 0 | 0.00144198 | 0.095325846 | 0.018692223 |
| 0 | 0 | 0 | 0 | 0 |
| 0 | 0 | 0 | 0 | 0 |
| 0 | 0 | 0 | 0 | 0 |
| 0 | 0 | 0 | 0 | 0 |
| 0 | 0 | 0 | 0 | 0 |
| 0 | 0 | 0 | 0 | 0.00756708 |
| 0 | 0 | 0 | 9.641363244 | 0.001124302 |
| 0 | 0 | 0.299935631 | 0 | 0.008133958 |
| 0 | 0 | 0.311853697 | 0 | 0 |
| 0.00274484 | 0 | 0 | 0 | 0 |
| 0 | 0 | 0.006681547 | 0 | 0 |
| 0 | 0 | 0 | 0 | 0 |
| 0 | 0 | 0 | 0 | 0 |
| 0 | 0 | 0 | 0 | 0.015760854 |
| 0 | 0 | 0 | 0 | 0 |
| 0 | 0 | 0 | 0 | 0 |
| 0 | 0 | 0 | 0 | 0.143285377 |
| 0 | 0 | 0 | 0 | 0 |
| 0 | 0 | 0.001154757 | 0 | 0 |
| 0 | 0 | 0.013097709 | 0 | 0 |
| 0 | 0 | 0.103043941 | 0 | 0 |
| 0 | 0 | 0 | 0 | 0 |
| 0 | 0 | 0 | 0 | 0 |
| 0 | 0 | 0.004180348 | 0.000981367 | 0.121474547 |
| 0 | 0 | 0 | 0 | 0 |
| 0 | 0 | 0 | 0.060425435 | 0 |
| 0 | 0 | 0 | 0 | 0 |
| 0 | 0 | 0.011128338 | 0 | 0.088577877 |

| | | | | |
|---|---|---|---|---|
| 0 | 0 | 0.012197829 | 0 | 0 |
| 0 | 0.006918236 | 0 | 0 | 0 |
| 0 | 0 | 0 | 0 | 0 |
| 0 | 0 | 0 | 0 | 0 |
| 0 | 0 | 0 | 0 | 0 |
| 0 | 0 | 0.021599997 | 0.031498329 | 0.005764672 |
| 0 | 0 | 0 | 0.003449374 | 0 |
| 0 | 0 | 0 | 0 | 0 |
| 0 | 0 | 0 | 0 | 0 |
| 0 | 0 | 0 | 0 | 0 |
| 0.005303695 | 0.007001435 | 0.003831537 | 0.020480486 | 0 |
| 0 | 0 | 0 | 0 | 0 |
| 0 | 0 | 0 | 0 | 0.191846384 |
| 0.048675024 | 0 | 0.004176743 | 0.005229441 | 0.13054395 |
| 0 | 0 | 0 | 0 | 0 |
| 0 | 0 | 0.053668015 | 0 | 0 |
| 0 | 0 | 0 | 0 | 0 |
| 0 | 0 | 0.012193937 | 0 | 0 |
| 0 | 0 | 0.003260034 | 0.003061265 | 0 |
| 0 | 0 | 0 | 0 | 0 |
| 0 | 0 | 0 | 0 | 0 |
| 0.04963706 | 1.06051258 | 0 | 0 | 0 |
| 0 | 0 | 0 | 0 | 0 |
| 0 | 0 | 0 | 0 | 0 |
| 0 | 0 | 0 | 0 | 0 |
| 0 | 0 | 0 | 0 | 0 |
| 0 | 0 | 0 | 0 | 0.003913665 |
| 0 | 0 | 0 | 0 | 0 |
| 0 | 0 | 0 | 0 | 0 |
| 0.152837301 | 1.525569031 | 0.52120588 | 0.054040125 | 0.140205825 |
| 0.00453245 | 0.007076415 | 0 | 0.011707609 | 0.002244702 |
| 0 | 0 | 0 | 0 | 0 |
| 0 | 0 | 0 | 0 | 0 |
| 0 | 0 | 0 | 0 | 0 |
| 0 | 0 | 0 | 0 | 0 |
| 0 | 0 | 0 | 0 | 0 |
| 0.575508703 | 0 | 0 | 0 | 0 |
| 0 | 0.00414895 | 0.046829352 | 0 | 0 |
| 0 | 0 | 0 | 0.001953167 | 0.008345576 |
| 0 | 0 | 0 | 0 | 0 |
| 0 | 0.111268287 | 0 | 0.532667155 | 0.01780446 |
| 0.725257991 | 9.193955835 | 0 | 3.408140779 | 1.350059806 |
| 0 | 0.012352295 | 0.05043849 | 0 | 0 |
| 0 | 0 | 0 | 0 | 0 |
| 0 | 0 | 0 | 0 | 0 |
| 0 | 0 | 0 | 0 | 0 |
| 0.001235289 | 0 | 0.002642899 | 0.167857088 | 0.004671765 |
| 0 | 0 | 0 | 0 | 0 |

| | | | | |
|---|---|---|---|---|
| 0 | 0 | 0 | 0 | 0.024978086 |
| 0 | 0 | 0 | 0 | 0 |
| 0.002422883 | 0 | 0.004443218 | 0 | 0 |
| 0 | 0 | 0 | 0 | 0 |
| 0 | 0 | 0 | 0 | 0.000858429 |
| 0 | 0 | 0.001754511 | 0 | 0 |
| 0 | 0 | 0 | 0 | 0 |
| 0 | 0 | 0.048509178 | 0 | 0.025841879 |
| 0 | 0 | 0 | 0.222720228 | 0 |
| 0 | 0 | 0 | 0 | 0 |
| 0 | 0 | 0 | 0 | 0 |
| 0 | 0 | 0 | 0 | 0 |
| 0 | 0 | 0 | 0 | 1.021876693 |
| 0.001608659 | 0.105118163 | 0 | 0 | 0 |
| 0 | 0 | 0 | 0 | 0 |
| 0 | 0.003638258 | 0 | 0.092891657 | 0.0013584 |
| 0.003184933 | 0 | 0.001393805 | 0 | 0.030614777 |
| 0 | 0 | 0.001411182 | 0 | 0 |
| 0 | 0 | 0 | 0 | 0 |
| 0 | 0 | 0 | 0 | 0 |
| 0 | 0.044670395 | 0 | 0 | 0 |
| 0 | 0 | 0.002425112 | 0 | 0.00174647 |
| 0 | 0 | 0 | 0 | 0 |
| 0 | 0 | 0 | 2.702693914 | 0 |
| 0 | 0 | 0 | 0.194288319 | 0 |
| 0 | 0 | 0.059366984 | 0 | 0.001322281 |
| 0 | 0.008083351 | 0 | 0.031569657 | 0 |
| 0 | 0 | 0 | 0 | 0.000604817 |
| 0 | 0 | 0 | 0 | 0 |
| 0 | 0 | 0 | 0 | 0 |
| 0 | 0.00942596 | 0.018390674 | 0.118147758 | 0 |
| 0.002005641 | 0.002443991 | 0 | 0 | 0.007585174 |
| 0.025370953 | 0.274005449 | 0.021967135 | 0.016741377 | 0.237297924 |
| 0 | 0.015840454 | 0.006387459 | 0 | 1.034013184 |
| 0.062784946 | 0 | 0 | 0.007033154 | 0.159705501 |
| 0 | 0 | 0 | 0 | 0 |
| 0.002219718 | 0.001840805 | 0 | 0 | 0.023318878 |
| 0 | 0 | 0.052671322 | 0.001479569 | 0 |
| 0 | 0 | 0 | 0.002355273 | 0 |
| 0 | 0 | 0 | 0 | 0.001391615 |
| 0.001836213 | 0 | 0 | 0 | 0 |
| 0 | 0 | 0.00396283 | 0 | 0 |
| 0 | 0 | 0 | 0.001264396 | 0 |
| 0 | 0 | 0 | 0.806180205 | 0.001138863 |
| 0 | 0 | 0 | 0 | 2.255757454 |
| 0 | 0 | 0 | 0 | 0.001325281 |
| 0 | 0 | 0 | 0 | 0 |
| 0 | 0 | 0.001829323 | 0 | 0 |

| | | | | |
|---|---|---|---|---|
| 0 | 0 | 0 | 0 | 0 |
| 0.078321516 | 0.06809662 | 0.473261564 | 0.184905214 | 0.019548248 |
| 0 | 0 | 0 | 0 | 0 |
| 0 | 0 | 0 | 0 | 0.002407965 |
| 0 | 0 | 0 | 0 | 0 |
| 0 | 0 | 0 | 0 | 0 |
| 0 | 0 | 0.033198928 | 0 | 0 |
| 0 | 0 | 0 | 0 | 0 |
| 0 | 0 | 0.002526953 | 0 | 0.002123113 |
| 0 | 0.047244444 | 0 | 0 | 0.02552144 |
| 0 | 0.003540144 | 0 | 0 | 0 |
| 0.213531711 | 0.153851756 | 0.015277395 | 0.003506778 | 0.433852473 |
| 0 | 0 | 0 | 0 | 0 |
| 0 | 0 | 0 | 0 | 0 |
| 0.004929985 | 0 | 0 | 0 | 0.032204664 |
| 0 | 0 | 0 | 0 | 0 |
| 0.014725044 | 0.067598996 | 0.011694742 | 0.002913512 | 0.03764159 |
| 0 | 0.007926063 | 0.044924525 | 0 | 0.013722078 |
| 0 | 0 | 0 | 0 | 0 |
| 0 | 0 | 0 | 0 | 0.010431688 |
| 0.282403185 | 0.041377219 | 0.00861445 | 0.001617843 | 0 |
| 0 | 0 | 0 | 0 | 0 |
| 0 | 0 | 0 | 0 | 0 |
| 0 | 0 | 0 | 0 | 0 |
| 0 | 0.009823683 | 0.001654158 | 0 | 0 |
| 0 | 0 | 0 | 0 | 0 |
| 0 | 0.181788546 | 0.762830917 | 0.205314664 | 0.006298402 |
| 0.059627088 | 0.013862591 | 0.134199164 | 0.257683585 | 0.012716438 |
| 0 | 0 | 0 | 0 | 0 |
| 0 | 0 | 0 | 0 | 0 |
| 0 | 0 | 0 | 0 | 0 |
| 0 | 0 | 0 | 0 | 0 |
| 0 | 0 | 0.00476365 | 0 | 0 |
| 0 | 0 | 0 | 0 | 0 |
| 0 | 0 | 0 | 0 | 0 |
| 0.022004085 | 1.097841473 | 0.764164056 | 0.012472281 | 0.021521813 |
| 0 | 0 | 0.000820197 | 0 | 0 |
| 0 | 0 | 0 | 0.005262217 | 0 |
| 0.003594875 | 0.005012379 | 0.007745014 | 0.010909185 | 0.003486033 |
| 0 | 0 | 0 | 0 | 0 |
| 0 | 0 | 0 | 0 | 0.882001548 |
| 0 | 0 | 0 | 0 | 0 |
| 0 | 0 | 0.001764165 | 0.005522005 | 0.080675597 |
| 0 | 0 | 0 | 0 | 0 |
| 0 | 0 | 0 | 0 | 0 |
| 0 | 0 | 0 | 0.024787919 | 0 |
| 0 | 0 | 0 | 0 | 0 |
| 0 | 0 | 0.00244729 | 0.016948303 | 0 |

| | | | | |
|---|---|---|---|---|
| 0 | 0 | 0 | 0 | 0 |
| 0 | 0 | 0.041861463 | 0 | 0 |
| 0 | 0 | 0 | 0 | 0 |
| 0 | 0 | 0 | 0.066457676 | 0.003452655 |
| 0 | 0 | 0 | 0 | 0 |
| 0 | 0 | 0 | 0 | 0 |
| 0 | 0 | 0 | 0 | 0 |
| 0 | 0.001679436 | 0 | 0 | 0 |
| 1.072350739 | 0.026805053 | 0 | 0.467181579 | 0 |
| 0 | 0 | 0 | 0.080089903 | 0.291610418 |
| 0 | 0 | 0 | 0 | 0 |
| 0.002624076 | 0.003797139 | 0 | 0.006572758 | 0.004016875 |
| 0 | 0 | 0 | 0 | 0 |
| 0 | 0 | 0 | 0 | 0 |
| 0 | 0.030674419 | 0 | 0.011695192 | 0.08345334 |
| 0 | 0 | 0 | 0 | 0 |
| 0 | 0 | 0 | 0.045766058 | 0 |
| 0 | 0 | 0 | 0.002213356 | 0 |
| 0 | 0 | 0 | 0 | 0 |
| 0 | 0 | 0 | 0 | 0 |
| 0 | 0 | 0 | 0 | 0 |
| 0 | 0 | 0 | 0 | 0 |
| 0 | 0 | 0 | 0 | 0 |
| 0 | 0 | 0 | 0.019755384 | 0 |
| 0 | 0 | 0.048945321 | 0 | 0.001327349 |
| 0 | 0 | 0 | 0 | 0.132921253 |
| 0 | 0 | 0 | 0.907705389 | 0 |
| 5.43229953 | 0 | 0 | 3.608426473 | 0.158348618 |
| 0 | 0 | 0 | 0 | 0 |
| 0 | 0 | 0 | 0 | 0 |
| 0 | 0 | 0.001913832 | 0.001397778 | 0.004134798 |
| 0 | 0 | 0 | 0 | 0 |
| 0 | 0 | 0.000809204 | 0 | 0 |
| 0 | 0 | 0 | 0 | 0 |
| 0 | 0 | 0 | 0 | 0 |
| 0.034917074 | 0 | 0 | 0.000507925 | 0 |
| 0 | 0.014254596 | 0 | 0.017711688 | 0.273933112 |
| 0 | 0 | 0.004551762 | 0 | 0 |
| 0 | 0 | 0.001366221 | 0 | 0.001229873 |
| 0 | 0 | 0 | 0 | 0 |
| 0.164705431 | 0 | 0.082466105 | 0.061871608 | 0.001133375 |
| 0 | 0 | 0 | 0 | 0 |
| 4.478435545 | 0.04760822 | 0.246691225 | 0.037111294 | 0.0881035 |
| 0 | 0 | 0 | 0 | 0 |
| 0 | 0 | 0 | 0 | 0 |
| 0 | 0 | 0 | 0.100339852 | 0 |
| 0 | 0 | 0 | 0 | 0 |
| 0 | 0 | 0 | 0.021318183 | 0.002724892 |

| | | | | |
|---|---|---|---|---|
| 0 | 0 | 0 | 0 | 0 |
| 0.133708808 | 0 | 0 | 0.016831939 | 0 |
| 0.01388278 | 0.226674909 | 0.103735978 | 0.016121642 | 0.052503553 |
| 0 | 0 | 0 | 0 | 0 |
| 0 | 0 | 0 | 0 | 0 |
| 0.184774816 | 0.001740407 | 0 | 0.001430987 | 0.002057725 |
| 0 | 0 | 0 | 0 | 0 |
| 0 | 0 | 0 | 0 | 0 |
| 0 | 0 | 0 | 0 | 0 |
| 0 | 0 | 0 | 0 | 0 |
| 0.004068784 | 0 | 2.112504137 | 0.00869458 | 0.015900742 |
| 0.806660882 | 1.510516584 | 2.879304177 | 1.057049201 | 1.378972537 |
| 0 | 0 | 0 | 0 | 0 |
| 0.005049878 | 0 | 0.001537357 | 0 | 0.00138393 |
| 0.001903238 | 0.246468427 | 0.946380566 | 2.925794049 | 0.517375753 |
| 0 | 0 | 0 | 0.531083804 | 0 |
| 0 | 0 | 0 | 0 | 0 |
| 0 | 0 | 0 | 0 | 0 |
| 0 | 0 | 0 | 0 | 0 |
| 0 | 0 | 0.033637374 | 0.051052528 | 0.000845034 |
| 0.116630225 | 0.337432915 | 0.208538756 | 0.234876515 | 0.074804077 |
| 6.100046328 | 0.018390497 | 1.77612259 | 0 | 0 |
| 1.663030588 | 0.3530577 | 1.310752047 | 0.96892412 | 1.30750993 |
| 0 | 0 | 0 | 0 | 0 |
| 0 | 0 | 0 | 0 | 0 |
| 5.26360478 | 0.016557265 | 0 | 0 | 0 |
| 0 | 0 | 0.001903985 | 0 | 0.016454098 |
| 0 | 0 | 0 | 0 | 0 |
| 0 | 0 | 0 | 0 | 0 |
| 0 | 0 | 0 | 1.288111727 | 0 |
| 0 | 0 | 0 | 0 | 0 |
| 0.047970974 | 0.074834734 | 0 | 0.072149759 | 0.194613019 |
| 0 | 0 | 0.000975319 | 0 | 0 |
| 0.00175264 | 0 | 0.002975025 | 0.002968235 | 0 |
| 0 | 0.005146248 | 0 | 0 | 0 |
| 0.563856487 | 0.004646415 | 0 | 0.003114413 | 0.026512437 |
| 0 | 0 | 0 | 0 | 0.004153079 |
| 0 | 0 | 0 | 0 | 0 |
| 9.33511831 | 0.013238505 | 0 | 0.010933477 | 0 |
| 0 | 0 | 0 | 0 | 0 |
| 0 | 0 | 0 | 0.004022183 | 0 |
| 0 | 0 | 0 | 0 | 0 |
| 0 | 0 | 0 | 0 | 0 |
| 0 | 0 | 0 | 0 | 0 |
| 0.003457066 | 0 | 0.251645253 | 0 | 0.001361911 |
| 0 | 0 | 6.538812212 | 0 | 0 |
| 0 | 0 | 0 | 0 | 0 |
| 12.94263327 | 0.011393883 | 0.011558128 | 0.005997939 | 0.106455828 |

| | | | | |
|---|---|---|---|---|
| 0 | 0 | 0 | 0 | 0 |
| 0 | 0 | 0 | 0 | 0 |
| 0 | 0 | 0 | 0 | 0 |
| 0 | 0 | 0 | 0 | 0.000928601 |
| 0 | 0 | 0 | 0 | 0 |
| 0 | 0 | 0 | 0 | 0 |
| 0 | 0.077602978 | 0 | 0 | 0 |
| 0.001694538 | 0 | 0 | 0.005570856 | 0 |
| 0 | 0 | 0 | 0 | 0 |
| 0 | 0 | 0 | 0 | 0 |
| 0.012305842 | 0 | 0.01675439 | 0.00089902 | 0 |
| 0 | 0 | 0 | 0 | 0 |
| 0.019753666 | 0 | 0 | 0 | 0 |
| 0 | 0 | 0 | 0.027617746 | 7.216855144 |
| 0 | 0 | 0 | 0 | 0 |
| 0 | 0 | 0 | 0 | 0 |
| 0 | 0 | 0 | 0.028751272 | 0 |
| 0.033123342 | 0.003094823 | 0 | 0 | 0.001291441 |
| 0 | 0 | 0 | 0.072649614 | 0 |
| 0 | 0 | 0 | 0 | 0 |
| 0 | 0 | 0 | 0 | 0.000573672 |
| 0 | 0 | 0 | 0.030092986 | 0 |
| 0 | 0 | 0 | 0 | 0 |
| 0 | 0 | 0 | 0 | 0 |
| 0 | 0 | 0 | 0 | 0 |
| 0 | 0 | 0 | 0 | 0 |
| 0 | 0 | 0 | 0 | 0 |
| 0 | 0 | 0 | 0 | 0 |
| 0 | 0 | 0 | 0 | 0 |
| 0.0198001 | 2.364942798 | 0 | 0.007006597 | 1.960462348 |
| 0 | 0 | 0.00640946 | 0 | 0.00162912 |
| 0 | 0 | 0 | 0 | 0 |
| 0 | 0 | 0 | 0 | 0 |
| 0 | 0 | 0 | 0 | 0 |
| 0.002120085 | 0 | 0 | 0 | 0 |
| 0 | 0 | 0 | 0 | 0 |
| 0.04538391 | 0 | 0 | 0 | 0 |
| 0 | 0 | 0.010333244 | 0 | 0.036113617 |
| 0 | 0 | 0 | 0 | 0 |
| 0 | 0 | 0 | 0 | 0 |
| 0 | 0 | 0 | 0.006604873 | 0 |
| 0 | 0 | 0 | 0 | 0 |
| 0 | 0 | 0 | 0 | 0 |
| 0 | 0 | 0 | 0 | 0 |
| 0.095613881 | 0.229749106 | 0 | 0 | 0 |
| 0 | 0 | 0 | 0 | 0 |
| 0 | 0 | 0 | 0 | 0 |
| 0 | 0 | 0 | 0 | 0 |

| | | | | |
|---|---|---|---|---|
| 0 | 0 | 0 | 0 | 0 |
| 0 | 0 | 0 | 0 | 0 |
| 0 | 0 | 0 | 0 | 0 |
| 0.038339296 | 0 | 0 | 0.000567756 | 0 |
| 0 | 0 | 0 | 0 | 0 |
| 0 | 0.015602362 | 0 | 0 | 0.232470468 |
| 0 | 0 | 0 | 0 | 0 |
| 0 | 0 | 0 | 0 | 0 |
| 0 | 0.013276105 | 0.254566793 | 0.001836793 | 0.008150189 |
| 0 | 0 | 0 | 0 | 0 |
| 0 | 0 | 0 | 0.008443056 | 0 |
| 0 | 0 | 0 | 0 | 0 |
| 0 | 0 | 0 | 0 | 0 |
| 0 | 0 | 0 | 0 | 0 |
| 0 | 0 | 0 | 0.018102418 | 0 |
| 0 | 0 | 0.237121931 | 0.006876099 | 0 |
| 0 | 0 | 0 | 0 | 0 |
| 0 | 0 | 0 | 0 | 0 |
| 0 | 0 | 0 | 0 | 0 |
| 0 | 0 | 0 | 0 | 0 |
| 0 | 0 | 0 | 0 | 0 |
| 0 | 0 | 0 | 0 | 0 |
| 0 | 0 | 0 | 0 | 0 |
| 0 | 0 | 0 | 0 | 0 |
| 0.136850366 | 0 | 0.012158553 | 0.265682012 | 0 |
| 0 | 1.126000691 | 0 | 0.022685804 | 11.57482955 |
| 0 | 0 | 0 | 0.004361304 | 0 |
| 0 | 0 | 0 | 0.001340823 | 0 |
| 0.002954231 | 0 | 0 | 0 | 0 |
| 0 | 0 | 0 | 0 | 0 |
| 0 | 0 | 0 | 0 | 0 |
| 0 | 0 | 0 | 0 | 0 |
| 0 | 0 | 0 | 0 | 0 |
| 0 | 0 | 0.007370941 | 0 | 0 |
| 0.000991145 | 0 | 0 | 0 | 0 |
| 0.001926703 | 0 | 0.00408811 | 0 | 0 |
| 0 | 0 | 0 | 0 | 0 |
| 0.337017206 | 0.116827937 | 0 | 0.245345567 | 0 |
| 0 | 0 | 0 | 0.08314825 | 0 |
| 0 | 0 | 0 | 0 | 0 |
| 0 | 0 | 0 | 0 | 0 |
| 0 | 0 | 0.041491746 | 0.004135624 | 0 |
| 1.471058389 | 1.414609903 | 0.321501619 | 0.0149507 | 0.512650926 |
| 0 | 0 | 0 | 0 | 0 |
| 0 | 0 | 0 | 0 | 0 |
| 0.016261845 | 0 | 0 | 0 | 0 |
| 0 | 0 | 0 | 0 | 0 |
| 0.874231368 | 0.438587493 | 0.832347387 | 0.582867007 | 0.473895733 |

| | | | | |
|---|---|---|---|---|
| 0 | 0 | 0.004831108 | 0 | 0 |
| 0 | 0 | 0 | 0 | 0 |
| 0 | 0 | 0 | 0 | 0 |
| 0 | 0 | 0 | 0 | 0 |
| 0 | 0 | 0.036418025 | 0 | 0.000989691 |
| 0 | 0.006247187 | 0 | 0 | 0 |
| 0 | 0 | 0 | 0 | 0 |
| 0 | 0 | 0 | 0.022905467 | 0 |
| 0 | 0 | 0 | 0 | 0 |
| 0 | 0 | 0 | 0 | 0 |
| 0 | 0 | 0 | 0 | 0 |
| 0 | 0 | 0 | 0.608765246 | 1.208882743 |
| 0.121199328 | 0 | 0 | 0 | 0.100361202 |
| 0 | 0 | 0 | 0 | 0 |
| 0.008852359 | 0.249521241 | 0.366872074 | 0 | 0.171400558 |
| 0 | 0 | 0 | 0 | 0 |
| 0 | 0 | 0.004053917 | 0 | 0 |
| 0 | 0 | 0 | 0.001162487 | 0 |
| 0 | 0 | 0.003018272 | 0 | 0 |
| 0 | 0 | 0 | 0 | 0 |
| 0 | 0 | 0 | 0.056315855 | 0 |
| 0 | 0 | 0 | 0 | 0 |
| 0.04584815 | 0.023308073 | 0 | 0.010811674 | 0.071211041 |
| 0 | 0 | 0 | 0.008555725 | 0 |
| 0 | 0 | 0 | 0 | 0 |
| 0 | 0 | 0 | 0 | 0 |
| 0 | 0 | 0 | 0 | 0 |
| 0 | 0 | 0 | 0 | 0 |
| 0 | 0 | 0 | 0 | 0 |
| 0 | 0 | 0 | 0 | 0 |
| 0 | 0 | 0 | 0 | 0 |
| 0.055079524 | 0.087620804 | 0.0725273 | 0.020334731 | 0.024506626 |
| 0 | 0 | 0 | 0 | 0 |
| 0.002354377 | 0 | 0.000858611 | 0 | 0.00983157 |
| 0.001524899 | 0 | 0.004671332 | 0 | 0 |
| 0.019903514 | 0 | 0 | 0.003769833 | 0.001548837 |
| 0 | 0 | 0 | 0.001854177 | 0 |
| 0.006561705 | 0 | 0 | 0.009587485 | 0 |
| 0 | 0 | 0 | 0 | 0 |
| 0 | 0 | 0 | 0 | 0 |
| 0 | 0 | 0 | 0.051076794 | 0 |
| 0 | 0 | 0 | 0 | 0 |
| 0 | 0 | 0 | 0 | 0 |
| 0 | 0 | 0 | 0 | 0 |
| 0.010703018 | 0.00260273 | 0 | 0.001440385 | 0.02248775 |
| 0 | 0 | 0 | 0 | 0 |
| 0 | 3.704432461 | 0 | 0.949962602 | 0 |
| 0 | 0 | 0.00186532 | 0.0157643 | 0 |

| | | | | |
|---|---|---|---|---|
| 0 | 0 | 0 | 0 | 0 |
| 0 | 0 | 0 | 0 | 0.001796622 |
| 0 | 0 | 0.001586447 | 0.009789582 | 0 |
| 0 | 0 | 0 | 0 | 0 |
| 0 | 0 | 0 | 0 | 0 |
| 0 | 0 | 0 | 0 | 0 |
| 0.016246912 | 0.005698384 | 0 | 0 | 0 |
| 0 | 0 | 1.263453513 | 0.003014787 | 0.062657967 |
| 0 | 0 | 0 | 0 | 0 |
| 0 | 0 | 0 | 0 | 0 |
| 0 | 0 | 0.412735824 | 0 | 0 |
| 0 | 0 | 0.003689534 | 0 | 0 |
| 0 | 0 | 0 | 0 | 0 |
| 0.091181384 | 0 | 0 | 0 | 0 |
| 0 | 0 | 0 | 0 | 0 |
| 0 | 0 | 0 | 0 | 0 |
| 0 | 0 | 0 | 0 | 0 |
| 0 | 0 | 0 | 0 | 0 |
| 2.360665526 | 0.016151847 | 0.253306914 | 25.03991069 | 0 |
| 0 | 0 | 0 | 0 | 0 |
| 0.001764513 | 0 | 0 | 0 | 0 |
| 0 | 0 | 0 | 0 | 0 |
| 0 | 0 | 0 | 0 | 0 |
| 0 | 0 | 0 | 0 | 0 |
| 0 | 0 | 0 | 0 | 0 |
| 0 | 0 | 0.043452376 | 0 | 0.00661113 |
| 0 | 0 | 0 | 0 | 0 |
| 0.134315935 | 0 | 0.263174483 | 0.089626221 | 0 |
| 0 | 0 | 0 | 0 | 0 |
| 0.037422635 | 0.03192117 | 0 | 0.247627116 | 17.15697525 |
| 0 | 0 | 0 | 0 | 0 |
| 0 | 0 | 0 | 0 | 0 |
| 0 | 0 | 0 | 0.359147504 | 0 |
| 0 | 0 | 0 | 0 | 0 |
| 0.000549567 | 0 | 0.005985874 | 0.001405227 | 0 |
| 0 | 0 | 0 | 0 | 0 |
| 0 | 0 | 0 | 0 | 0 |
| 0.001870791 | 0 | 0 | 0 | 0 |
| 0 | 0 | 0 | 0 | 0 |
| 0 | 0 | 0 | 0 | 0 |
| 0.544591467 | 32.08931285 | 9.814376282 | 0.495013473 | 0.189183456 |
| 0 | 0.082903332 | 4.031881364 | 0.065529746 | 0.059463344 |
| 0 | 0 | 0 | 0 | 0 |
| 0 | 0 | 0 | 0 | 0 |
| 0 | 0 | 0 | 0 | 0 |
| 0 | 0 | 0 | 0 | 0 |
| 0 | 0 | 0 | 0 | 0 |
| 0.003134501 | 0 | 0.008086015 | 0 | 0 |

| | | | | |
|---|---|---|---|---|
| 0 | 0 | 0 | 0 | 0 |
| 0 | 0 | 0.003366459 | 0.023058175 | 0 |
| 0 | 0 | 0 | 0.014334055 | 0 |
| 0 | 0 | 0 | 0 | 0 |
| 5.930182396 | 5.562314908 | 3.119755992 | 0.007276469 | 4.73665462 |
| 0 | 0 | 0 | 0 | 0 |
| 0 | 0 | 0 | 0 | 0 |
| 0 | 0 | 0 | 0 | 0 |
| 0 | 0.072626262 | 0.102822227 | 0 | 0.710652015 |
| 0 | 0 | 0 | 0 | 0 |
| 0 | 0 | 0 | 0 | 0 |
| 0 | 0 | 0 | 0 | 0 |
| 0 | 0.045708897 | 0.28015914 | 0.01463549 | 0.057160582 |
| 0.001013528 | 0.120818047 | 0.032171768 | 0.001332803 | 0 |
| 15.18740012 | 0 | 0 | 0 | 0.239117688 |
| 0 | 0.5161081 | 0.366493014 | 0.238059605 | 0.256245026 |
| 0.04016893 | 0 | 0.054860537 | 0.005766673 | 0.002432419 |
| 0 | 0 | 0 | 0 | 0 |
| 0 | 0 | 0 | 0 | 0 |
| 0 | 0 | 0 | 0 | 0 |
| 0 | 0 | 0 | 0 | 0 |
| 0 | 0 | 0 | 0 | 0 |
| 0 | 0 | 0 | 0 | 0 |
| 0 | 0 | 0 | 0 | 0 |
| 0 | 0 | 0 | 0 | 0 |
| 0 | 0 | 0 | 0 | 0 |
| 0 | 0 | 0 | 0.0106973 | 0.014228779 |
| 0.001204763 | 0 | 0 | 0.000754419 | 0 |
| 0 | 0 | 0 | 0 | 0 |
| 0 | 0 | 0 | 0 | 0 |
| 0 | 0 | 0 | 0 | 0 |
| 0 | 0 | 0 | 0 | 0 |
| 0 | 0 | 0 | 0 | 1.108090109 |
| 0 | 0 | 0 | 0.001581665 | 0 |
| 0.472432549 | 0.001820148 | 0 | 0.036010755 | 0 |
| 0 | 0 | 0 | 0 | 0 |
| 0 | 0 | 0 | 0.016056198 | 0 |
| 0 | 0 | 0.011189274 | 0 | 0 |
| 0 | 0 | 0 | 0 | 0 |
| 0 | 0 | 0 | 0 | 0 |
| 0.036332056 | 10.50519289 | 0 | 0.210828841 | 0 |
| 0 | 0 | 0 | 0 | 0 |
| 0 | 0 | 0 | 0 | 0 |
| 0.002738403 | 0 | 0 | 0 | 0 |
| 0 | 0 | 0 | 0 | 0 |
| 0 | 0 | 0 | 0 | 0 |
| 0 | 0 | 0 | 0 | 0 |
| 0 | 0 | 0 | 0 | 0 |

| | | | | |
|---|---|---|---|---|
| 0 | 0 | 0 | 0.015900853 | 0 |
| 0 | 0 | 0 | 0 | 0 |
| 0 | 0 | 0 | 0 | 0 |
| 0 | 0 | 0 | 0 | 0 |
| 0 | 0 | 0 | 0.007768089 | 0.04468131 |
| 0 | 0 | 0 | 0 | 0 |
| 0.205430805 | 0 | 0.519499414 | 0.046212963 | 0.005955966 |
| 0 | 0 | 0.01243569 | 0 | 0 |
| 0 | 0 | 0 | 0 | 0 |
| 0 | 0 | 0.357360757 | 0.004426009 | 0.000694309 |
| 0 | 0 | 0 | 0.005580825 | 0 |
| 0 | 0 | 0 | 0.005030802 | 0 |
| 0 | 0 | 0 | 0 | 0 |
| 0 | 0 | 0 | 0 | 0 |
| 0 | 0 | 0 | 0 | 0 |
| 0 | 0 | 0 | 0 | 0 |
| 0 | 0 | 0.274100971 | 0 | 0 |
| 0 | 0 | 0 | 0 | 0 |
| 0.050095414 | 0 | 0 | 0 | 0 |
| 0 | 0 | 0 | 0 | 0 |
| 0.161596361 | 0 | 0.228632334 | 0.015882849 | 0.011760855 |
| 0 | 0 | 0 | 0 | 0 |
| 0.001653775 | 0 | 0 | 0.001208188 | 0 |
| 0 | 0.005400757 | 0 | 0.002327722 | 0 |
| 0 | 0 | 0 | 0 | 0 |
| 0 | 0 | 0.001799692 | 0 | 0 |
| 0 | 0 | 0 | 0 | 0 |
| 0 | 0 | 0 | 0 | 0 |
| 0 | 0 | 0 | 0 | 0 |
| 0.607192313 | 0.326714871 | 0.001921049 | 0 | 0.487967468 |
| 0 | 0 | 0 | 0 | 0 |
| 0 | 0 | 0 | 0 | 0 |
| 0 | 0 | 0 | 0 | 0 |
| 0.00383891 | 0 | 0 | 1.062612537 | 0 |
| 0 | 0 | 0 | 0 | 0 |
| 0 | 0 | 0 | 0 | 0 |
| 0 | 0 | 0 | 0.001344269 | 0 |
| 0 | 0 | 0 | 0 | 0 |
| 0 | 0 | 0 | 0 | 0 |
| 0 | 0 | 0 | 0 | 0 |
| 0 | 0 | 0 | 0 | 0 |
| 0 | 0 | 0 | 0 | 0 |
| 0 | 0 | 0 | 0 | 0 |
| 0 | 0 | 0 | 0 | 0 |
| 0 | 0 | 0 | 0.002120416 | 0 |
| 0 | 0 | 0 | 0 | 0 |
| 0 | 0 | 0 | 0 | 0 |
| 0.096266725 | 0 | 0 | 0 | 0 |

| | | | | |
|---|---|---|---|---|
| 0 | 0 | 0.013952257 | 0.001541361 | 0 |
| 0 | 0 | 0 | 0.019003662 | 0 |
| 0 | 0 | 0 | 0 | 0.22494524 |
| 0 | 0 | 0 | 0 | 0 |
| 0 | 0 | 0 | 0 | 0 |
| 0.003993297 | 0 | 0 | 0 | 0 |
| 0 | 0 | 0 | 0 | 0 |
| 0.171680297 | 3.207936605 | 0 | 0.030465616 | 0 |
| 2.782092549 | 0 | 0.205153002 | 12.00021226 | 0.003131133 |
| 0 | 0.006760077 | 0 | 0 | 0.121342238 |
| 0 | 0 | 0 | 0 | 0 |
| 0 | 0 | 0 | 0 | 0 |
| 0 | 0 | 0 | 0 | 0 |
| 0.011441264 | 0 | 0 | 0 | 0 |
| 0 | 0 | 0 | 0 | 0.001905681 |
| 0 | 0 | 0 | 0 | 0 |
| 0 | 0 | 0.292319769 | 0 | 0 |
| 0 | 0 | 0 | 0 | 0 |
| 0 | 0 | 0 | 0 | 0 |
| 0 | 0 | 0 | 0 | 0 |
| 0 | 0 | 0 | 0 | 0 |
| 0 | 0 | 0 | 0 | 0 |
| 0 | 0 | 0 | 0 | 0 |
| 0 | 0 | 0 | 0 | 0 |
| 0 | 0 | 0 | 0.000607018 | 0.046262487 |
| 0 | 0 | 0 | 0 | 0 |
| 0 | 0 | 0 | 0 | 0 |
| 0 | 0 | 0 | 0 | 0 |
| 0 | 0 | 0.00231213 | 0 | 0 |
| 0 | 0 | 0 | 0 | 0 |
| 0 | 0 | 0 | 0 | 0 |
| 0 | 0 | 0 | 0 | 0 |
| 0 | 0 | 0 | 0 | 0 |
| 0 | 0 | 0 | 0 | 0 |
| 0 | 0 | 0 | 0 | 0 |
| 0 | 0 | 0 | 0 | 0 |
| 0 | 0 | 0 | 0 | 0 |
| 0 | 0 | 0 | 0 | 0 |
| 0.001254373 | 0 | 0 | 0 | 0 |
| 0.000702089 | 0 | 0 | 0 | 0.296280774 |
| 0 | 0 | 0 | 0 | 0 |
| 0 | 0.127623823 | 0 | 0.039687627 | 0 |
| 0 | 0 | 0 | 0 | 0 |
| 0 | 0 | 0 | 0 | 0 |
| 0 | 0 | 0 | 0 | 0 |
| 0 | 0 | 0 | 0 | 0 |
| 0 | 0 | 0 | 0 | 0 |
| 0 | 0 | 0 | 0 | 0 |

| | | | | |
|---|---|---|---|---|
| 0 | 0 | 0 | 0 | 0 |
| 0 | 0 | 0 | 0 | 0 |
| 0 | 0 | 0 | 0 | 0 |
| 0 | 0 | 0 | 0 | 0 |
| 0.867039209 | 0.002602144 | 0 | 0.017018902 | 0.156066393 |
| 0 | 0 | 0 | 0.009109888 | 0.015591961 |
| 0 | 0 | 0 | 0 | 0 |
| 0 | 0.016215871 | 0.017487278 | 0 | 0.003383368 |
| 0 | 0 | 0 | 0 | 0 |
| 0 | 0 | 0 | 0 | 0 |
| 0 | 0 | 0 | 0 | 0 |
| 0.031578536 | 0 | 0 | 0 | 0 |
| 0 | 0 | 0 | 0 | 0 |
| 0 | 0 | 0 | 0 | 0 |
| 0 | 0 | 0 | 0.000517605 | 0 |
| 0 | 0 | 0 | 0.00212565 | 0.000733592 |
| 0 | 0 | 0 | 0 | 0 |
| 0 | 0 | 0 | 0 | 0 |
| 0 | 0 | 0 | 0 | 0 |
| 0 | 0 | 0 | 0 | 0 |
| 0 | 0 | 0 | 0 | 0 |
| 0 | 0 | 0 | 1.394637816 | 0 |
| 0 | 0 | 0 | 0 | 0 |
| 0 | 0 | 0.001581143 | 0.001696844 | 0 |
| 0.121062934 | 0 | 0 | 0 | 0 |
| 0 | 0 | 0 | 0 | 0 |
| 0 | 0 | 0.072719924 | 0 | 0.259320833 |
| 0 | 0 | 0.0006526 | 0 | 0 |
| 0 | 0 | 0 | 0 | 0 |
| 0 | 0 | 0 | 0 | 0 |
| 0 | 0 | 0 | 0 | 0 |
| 0 | 0 | 0 | 0 | 0 |
| 0 | 0 | 0 | 0 | 0 |
| 0 | 0 | 0 | 0 | 0 |
| 0 | 0 | 0 | 0 | 0 |
| 0 | 0 | 0 | 0 | 0 |
| 0 | 0 | 0 | 0 | 0 |
| 0 | 0 | 0 | 0.018701563 | 0 |
| 0 | 0 | 0 | 0 | 0 |
| 0 | 0 | 0 | 0 | 0 |
| 0 | 0 | 0 | 0 | 0 |
| 1.088337847 | 0 | 0 | 0 | 0 |
| 0 | 0 | 0 | 0 | 0 |
| 0 | 0 | 0 | 0 | 0 |
| 0 | 0 | 0 | 0 | 0 |
| 0 | 0 | 0 | 0 | 0 |
| 0 | 0 | 0 | 0 | 0 |
| 0 | 0 | 0 | 0 | 0 |

| | | | | |
|---|---|---|---|---|
| 0 | 0 | 0 | 0.000729741 | 0.008394798 |
| 0 | 0 | 0 | 0 | 0 |
| 0 | 0 | 0 | 0 | 0 |
| 0 | 0 | 0.00135309 | 0 | 0 |
| 0 | 0 | 0 | 0 | 0 |
| 0.011958279 | 0 | 0 | 0 | 0 |
| 0 | 0 | 0 | 0 | 0 |
| 0 | 0 | 0 | 0 | 0 |
| 0 | 0 | 0 | 0 | 0 |
| 0 | 0 | 0 | 0 | 0 |
| 0 | 0 | 0 | 2.169531036 | 0 |
| 0 | 0 | 0.002915124 | 0 | 0 |
| 0 | 0 | 0 | 0 | 0 |
| 0 | 0 | 0 | 0 | 0 |
| 0 | 0 | 0 | 0 | 0 |
| 0 | 0 | 0 | 0 | 0 |
| 0 | 0 | 0 | 0 | 0 |
| 0 | 0 | 0 | 0 | 0 |
| 0 | 0 | 0 | 0 | 0 |
| 0 | 0 | 0 | 0 | 0 |
| 0 | 0 | 0.087948943 | 0 | 0.036690306 |
| 0 | 0 | 0 | 0 | 0 |
| 0.01679619 | 0 | 0.003692968 | 0.006135343 | 0 |
| 0 | 0 | 0 | 0.169194004 | 0.07218161 |
| 0 | 0 | 0 | 0.005271187 | 0 |
| 3.716220076 | 0.003182793 | 0 | 0 | 0 |
| 0 | 0 | 0 | 0 | 0 |
| 0 | 0 | 0 | 0 | 0 |
| 0.283217236 | 0 | 0 | 0 | 0 |
| 0 | 0 | 0 | 0 | 0 |
| 0 | 0 | 0 | 0 | 0 |
| 0 | 0 | 0 | 0 | 0.51652324 |
| 0.012082128 | 0.029653902 | 0.334405429 | 0 | 0.066672471 |
| 0 | 0 | 0 | 0 | 0.000581403 |
| 0.140700191 | 0 | 0 | 0 | 2.946435995 |
| 0 | 0.018529683 | 0.027752605 | 0.000801861 | 0.002536726 |
| 0 | 0 | 0 | 0 | 0 |
| 0.386792807 | 0 | 0 | 0 | 0 |
| 0 | 0 | 0.063706704 | 0 | 0.005068034 |
| 0 | 0 | 0 | 0 | 0 |
| 0 | 0 | 0 | 0 | 0 |
| 0 | 0 | 0 | 0 | 0 |
| 0.156116974 | 0 | 0 | 0.716855337 | 0 |
| 0 | 0 | 0 | 0 | 0 |
| 0 | 0 | 0 | 0 | 0 |
| 0 | 0 | 0 | 0.003229808 | 0 |
| 0 | 0 | 0 | 0 | 0 |
| 0 | 0 | 0 | 0 | 0 |

| | | | | |
|---|---|---|---|---|
| 0 | 0 | 0 | 0 | 0 |
| 0 | 0 | 0 | 0 | 0 |
| 0 | 0 | 0 | 0 | 0 |
| 0 | 0 | 0 | 0 | 0 |
| 0 | 0 | 0 | 0 | 0 |
| 0 | 0 | 0 | 0 | 0 |
| 0 | 0 | 0 | 0 | 0 |
| 0 | 0 | 0 | 0 | 0 |
| 0 | 0 | 0 | 0.258019536 | 0.003713161 |
| 0.010321406 | 0 | 0 | 0 | 0 |
| 0 | 0 | 0 | 0 | 0 |
| 0 | 0 | 0 | 0 | 0 |
| 0 | 0 | 0 | 0 | 0 |
| 0 | 0 | 0 | 0 | 1.469552497 |
| 0 | 0 | 0 | 0 | 0 |
| 0 | 0.054563546 | 0.007900679 | 0.001963843 | 0.006777504 |
| 0 | 0 | 0 | 0 | 0 |
| 0 | 0 | 0 | 0 | 0 |
| 0 | 0 | 0 | 0 | 0 |
| 0 | 0 | 0 | 0.010230526 | 0 |
| 0.012062072 | 0.017739378 | 0 | 0 | 0 |
| 0 | 0 | 0 | 0 | 0 |
| 0 | 0 | 0 | 0 | 0 |
| 0 | 0 | 0 | 0 | 0 |
| 0 | 0 | 0 | 0 | 0 |
| 0 | 0 | 0 | 0 | 0 |
| 0 | 0 | 0 | 0 | 0 |
| 0 | 0 | 0 | 0 | 0 |
| 0 | 0 | 0 | 0 | 0 |
| 0 | 0 | 0 | 0 | 0 |
| 0.738310377 | 0.071266955 | 0 | 0.352041213 | 0.143018665 |
| 0 | 0 | 0 | 0 | 0 |
| 0 | 0 | 0 | 0 | 0 |
| 0 | 0 | 0 | 0 | 0 |
| 0 | 0 | 0 | 0 | 0 |
| 0 | 0 | 0 | 0 | 0.006732413 |
| 0 | 0 | 0 | 0 | 0 |
| 0 | 0 | 0 | 0 | 0 |
| 0.007660762 | 0 | 0 | 0 | 0 |
| 0 | 0 | 0 | 0 | 0 |
| 0 | 0 | 0 | 0 | 0 |
| 0 | 0 | 0 | 0 | 0 |
| 0 | 0 | 0 | 0 | 0 |
| 0 | 0 | 0 | 0 | 0 |
| 0 | 0 | 0 | 0 | 0 |
| 0 | 0 | 0 | 0 | 0 |
| 0 | 0.535525031 | 2.505811568 | 0 | 0 |
| 0 | 0 | 0 | 0 | 0 |

| | | | | |
|---|---|---|---|---|
| 0 | 0 | 0 | 0 | 0 |
| 0 | 0 | 0 | 0 | 0 |
| 0 | 0 | 0 | 0 | 0 |
| 0 | 0 | 0 | 0 | 0 |
| 0 | 0 | 0 | 0.015352735 | 0.007486938 |
| 0 | 0 | 0 | 0 | 0 |
| 0 | 0 | 0 | 0 | 0 |
| 0.001825695 | 0 | 0 | 0 | 0 |
| 0 | 0 | 0 | 0 | 0 |
| 0 | 0 | 0 | 0 | 0 |
| 0 | 0 | 0 | 0.003813777 | 0 |
| 0.002325636 | 0 | 0 | 0 | 0.011460629 |
| 0 | 0 | 0 | 0 | 0 |
| 0 | 0 | 0 | 0 | 0 |
| 0 | 0 | 0.038545006 | 0 | 0 |
| 0 | 0 | 0.354761081 | 0.009368741 | 0.58489323 |
| 0 | 0 | 0 | 0.001247354 | 0 |
| 0 | 0 | 0 | 0 | 0 |
| 0 | 0.081932949 | 0 | 0.096312918 | 0 |
| 0.004389591 | 0.004903226 | 0.035126729 | 0.068031569 | 0.00632422 |
| 0 | 0 | 0 | 0 | 0 |
| 0 | 0 | 0 | 0 | 0 |
| 0 | 0 | 0 | 0 | 0 |
| 0 | 0 | 0 | 0 | 0 |
| 0 | 0 | 0 | 0 | 0 |
| 0 | 0 | 0 | 0 | 0 |
| 0 | 0 | 0 | 0.010829939 | 0.004645564 |
| 0 | 0 | 0 | 0 | 0 |
| 0 | 0 | 0 | 0 | 0 |
| 0 | 0 | 0 | 0 | 0 |
| 0.000949372 | 0 | 0.002880582 | 0 | 0 |
| 0 | 0 | 0 | 0 | 0 |
| 0 | 0 | 0 | 0 | 0 |
| 0 | 0 | 0 | 0 | 0 |
| 0 | 0 | 0 | 0 | 0 |
| 0 | 0 | 0 | 0 | 0 |
| 0 | 0 | 0 | 0 | 0 |
| 0.002934805 | 0.001280128 | 0.039697872 | 0.001754232 | 0.003531562 |
| 0 | 0.002052976 | 0 | 0.050991232 | 0 |
| 0 | 0 | 0 | 0.021333784 | 0 |
| 0.000665882 | 0 | 0.000932501 | 0 | 0 |
| 0 | 0 | 0 | 0 | 0 |
| 0 | 0 | 0 | 0 | 0 |
| 0 | 0.09554616 | 0 | 0.001950978 | 0 |
| 0 | 0 | 0 | 0 | 0 |
| 0 | 0 | 0 | 0 | 0 |
| 0 | 0 | 0 | 0 | 0 |
| 0 | 0 | 0 | 0 | 0 |

| | | | | |
|---|---|---|---|---|
| 0 | 0 | 0 | 0 | 0 |
| 0 | 0 | 0 | 0 | 0 |
| 0 | 0 | 0 | 0 | 0 |
| 0.003109675 | 0 | 0.001451595 | 0.001590271 | 0 |
| 0 | 0 | 0.002019497 | 0 | 0.004363085 |
| 0 | 0 | 0.003475139 | 0 | 0.004692483 |
| 0.014283796 | 0 | 0 | 0 | 0 |
| 0 | 0 | 0.001135273 | 0 | 0 |
| 0 | 0 | 0 | 0.001291906 | 0 |
| 0 | 0 | 0 | 0 | 0 |
| 0 | 0 | 0 | 0 | 0 |
| 0 | 0 | 0 | 0 | 0 |
| 0.059810597 | 0 | 0 | 0.001932477 | 0.002223085 |
| 0 | 0 | 0 | 0 | 0 |
| 0 | 0 | 0 | 0 | 0 |
| 0 | 0 | 0 | 0 | 0 |
| 0 | 0 | 0 | 0 | 0 |
| 0 | 0 | 0 | 0 | 0 |
| 0 | 0 | 0 | 0 | 0 |
| 0 | 0 | 0 | 0 | 0 |
| 0 | 0 | 0 | 0 | 0 |
| 0 | 0 | 0 | 0 | 0 |
| 0 | 0.004332786 | 0 | 0 | 0.002683349 |
| 0.00981876 | 0 | 0 | 0.001251145 | 0 |
| 0 | 0 | 0 | 0 | 0 |
| 0 | 0 | 0 | 0 | 0 |
| 0 | 0 | 0 | 0 | 0 |
| 0 | 0 | 0 | 0 | 0 |
| 0 | 0 | 0 | 0 | 0 |
| 0 | 0 | 0 | 0 | 0 |
| 0.001726063 | 0 | 0 | 0.006383807 | 0.438180657 |
| 0 | 0 | 0 | 0 | 0 |
| 0 | 0 | 0 | 0 | 0 |
| 0 | 0 | 0 | 0 | 0 |
| 0 | 0 | 0 | 1.619884066 | 0 |
| 0 | 0 | 0 | 0 | 0 |
| 0 | 0 | 0 | 0 | 0 |
| 0 | 0 | 0 | 0 | 0 |
| 0.000730184 | 0 | 0 | 0 | 0 |
| 0 | 0 | 0 | 0 | 0 |
| 0 | 0 | 0 | 0 | 0 |
| 0 | 0 | 0 | 0 | 0 |
| 0 | 0 | 0 | 0 | 0 |
| 0 | 0 | 0 | 0 | 0 |
| 0 | 0 | 0 | 0 | 1.030107698 |
| 0.333748477 | 0.11542187 | 0.172812501 | 0.005104122 | 0.019842246 |
| 0 | 0 | 0.424520192 | 0 | 0 |
| 0 | 0 | 0 | 0 | 0 |

| | | | | |
|---|---|---|---|---|
| 0 | 0 | 0 | 0 | 0 |
| 0 | 0 | 0 | 0 | 0 |
| 0.13360477 | 0 | 0 | 0 | 0 |
| 0 | 0 | 0 | 0 | 0 |
| 0 | 0 | 0.013283918 | 0 | 0 |
| 0 | 0 | 0 | 0 | 0 |
| 0 | 0 | 0.050237868 | 0 | 0 |
| 0 | 0 | 0 | 0 | 0 |
| 0 | 0 | 0 | 0.009069526 | 0 |
| 0 | 0 | 0 | 0 | 0 |
| 0 | 0 | 0.001728253 | 0 | 0 |
| 0.000884418 | 0 | 0 | 0 | 0 |
| 0 | 0 | 0 | 0 | 0 |
| 0 | 0 | 0 | 0 | 0 |
| 0 | 0 | 0 | 0 | 0 |
| 0 | 0 | 0 | 0 | 0 |
| 0 | 0 | 0 | 0 | 0 |
| 0 | 0 | 0 | 0 | 0 |
| 0 | 0 | 0 | 0 | 0 |
| 0.000589193 | 0 | 0 | 0 | 0 |
| 0 | 0 | 0 | 0 | 0 |
| 0 | 0 | 0 | 0 | 0 |
| 0 | 0 | 0 | 0 | 0 |
| 0 | 0 | 0 | 0 | 0 |
| 0 | 0 | 0 | 0 | 0 |
| 0 | 0 | 0 | 0 | 0 |
| 0 | 0 | 0 | 0 | 0 |
| 0.026516383 | 0 | 0 | 0 | 0 |
| 0 | 0 | 0 | 0 | 0 |
| 0 | 0 | 0 | 0 | 0 |
| 0.041000433 | 0 | 0 | 0.007026112 | 0.001701622 |
| 0 | 0 | 0 | 0 | 0 |
| 0.025859875 | 0.017964097 | 0.069273255 | 0.033061497 | 0.008396966 |
| 0.004204134 | 0.010886341 | 0.003037184 | 0 | 0 |
| 0 | 0 | 0 | 0 | 0 |
| 0 | 0 | 0 | 0 | 0 |
| 0 | 0 | 0 | 0 | 0 |
| 0 | 0 | 0 | 0 | 0 |
| 0 | 0 | 0 | 0 | 0 |
| 0 | 0 | 0 | 0 | 0 |
| 0 | 0 | 0 | 0 | 0 |
| 0 | 0 | 0 | 0 | 0.154213352 |
| 0 | 0 | 0.007332591 | 0.029048257 | 0.050991201 |
| 0 | 0 | 0 | 0 | 0 |
| 0 | 0 | 0 | 0 | 0 |
| 0.056748677 | 0 | 0 | 0.005907839 | 0 |
| 0 | 0 | 0.003473553 | 0 | 0 |
| 0 | 0 | 0 | 0 | 0 |

| | | | | |
|---|---|---|---|---|
| 0 | 0 | 0 | 0 | 0 |
| 0 | 0.025881085 | 0 | 0 | 0 |
| 0 | 0.002049616 | 0.008692812 | 0.012902489 | 0.333508152 |
| 0 | 0 | 0 | 0 | 0 |
| 0.000959283 | 0 | 0 | 0.007989319 | 0 |
| 0 | 0 | 0.03269093 | 0 | 0.02895754 |
| 0 | 0 | 0 | 0 | 0 |
| 0 | 0 | 0 | 0 | 0 |
| 0 | 0 | 0 | 0 | 0 |
| 0 | 0 | 0 | 0 | 0 |
| 0 | 0 | 0 | 0 | 0 |
| 0 | 0 | 0 | 0 | 0 |
| 0 | 0 | 0 | 0 | 0 |
| 0 | 0 | 0 | 0 | 0 |
| 0 | 0 | 0 | 0 | 0 |
| 0 | 0 | 0 | 0 | 0 |
| 0 | 0 | 0 | 0 | 0 |
| 0 | 0 | 0 | 0 | 0 |
| 0 | 0 | 0 | 0 | 0 |
| 0 | 0 | 0 | 0 | 0 |
| 0 | 0 | 0 | 0 | 0 |
| 0 | 0 | 0 | 0 | 0 |
| 0 | 0 | 0 | 0 | 0 |
| 0 | 0 | 0 | 0 | 0 |
| 0.004356644 | 0 | 0 | 0.000868038 | 0 |
| 0 | 0 | 0 | 0.002183064 | 0.10748602 |
| 0 | 0 | 0 | 0 | 0 |
| 0 | 0 | 0 | 0.045741206 | 0 |
| 0 | 0 | 0 | 0 | 0 |
| 0 | 0 | 0 | 0 | 0 |
| 0.0229265 | 0 | 0 | 0 | 0 |
| 0 | 0 | 0.002242594 | 0 | 0 |
| 0 | 0 | 0 | 0 | 0 |
| 0 | 0 | 0 | 0 | 0 |
| 0 | 0 | 0 | 0 | 0 |
| 0 | 0 | 0 | 0 | 0 |
| 0 | 0 | 0 | 0 | 0 |
| 0 | 0 | 0 | 0 | 0 |
| 0 | 0.604006588 | 0 | 0.044794347 | 0 |
| 0 | 0 | 0 | 0 | 0 |
| 0 | 0 | 0 | 0 | 0 |
| 0.103519159 | 0 | 0 | 0 | 0 |
| 0 | 0 | 0.011666009 | 0 | 0 |
| 0 | 0 | 0 | 0 | 0 |
| 0 | 0 | 0 | 0 | 0 |
| 0 | 0 | 0 | 0.003024723 | 0 |
| 0.002752708 | 0 | 0 | 0.005362743 | 0 |
| 0 | 0 | 0 | 0 | 0 |

| | | | | |
|---|---|---|---|---|
| 0 | 0 | 0 | 0 | 0 |
| 0.00713979 | 0.017077185 | 0.132638475 | 0 | 0.032353882 |
| 0 | 0 | 0 | 0 | 0 |
| 0 | 0 | 0 | 0 | 0 |
| 0 | 0 | 0 | 0 | 0 |
| 0 | 0 | 0 | 0 | 0 |
| 0 | 0 | 0 | 0 | 0 |
| 0 | 0 | 0 | 0 | 0 |
| 0 | 0 | 0 | 0 | 0 |
| 0 | 0 | 0 | 0 | 0 |
| 0 | 0 | 0 | 0 | 0 |
| 0 | 0 | 0 | 0 | 0 |
| 0 | 0.009352292 | 0 | 0.037330251 | 0 |
| 0 | 0 | 0 | 0 | 0 |
| 0 | 0 | 0 | 0.002109293 | 0 |
| 0 | 0 | 0 | 0 | 0 |
| 0 | 0 | 0 | 0 | 0 |
| 0 | 0 | 0 | 0 | 0 |
| 0 | 0 | 0 | 0.021875856 | 0 |
| 0 | 0 | 0 | 0 | 0 |
| 0 | 0 | 0 | 0 | 0 |
| 0.00708868 | 0 | 0.019853957 | 0.045313901 | 0.005361763 |
| 0 | 0 | 0.008051852 | 0.006168119 | 0 |
| 0 | 0 | 0 | 0 | 0 |
| 0 | 0 | 0 | 0 | 0 |
| 0 | 0 | 0 | 0 | 0 |
| 0 | 0 | 0.003157967 | 0 | 0 |
| 0 | 0 | 0 | 0 | 0 |
| 0 | 0 | 0 | 0 | 0 |
| 0 | 0 | 0 | 0 | 0 |
| 0 | 0 | 0 | 0 | 0 |
| 0 | 0 | 0 | 0 | 0 |
| 0 | 0 | 0 | 0 | 0 |
| 0 | 0 | 0 | 0 | 0 |
| 0 | 0 | 6.496974631 | 2.134002704 | 0.305072763 |
| 0 | 0 | 0 | 0 | 0 |
| 0 | 0 | 0 | 0 | 0 |
| 0 | 0 | 0 | 0 | 0 |
| 0.023360384 | 0 | 0.053972352 | 0.003361534 | 0 |
| 0 | 0 | 0 | 0 | 0 |
| 0.27546735 | 0.208667754 | 0.148389819 | 0.006955522 | 0.02000375 |
| 0 | 0 | 0 | 0 | 0 |
| 0 | 0 | 0 | 0 | 0 |
| 0 | 0 | 0 | 0 | 0 |
| 0 | 0 | 0 | 0 | 0 |
| 0.389515608 | 0.262413992 | 0.719161471 | 0 | 2.284491185 |
| 0 | 0 | 0 | 0 | 0 |
| 0 | 0 | 0 | 0 | 0 |

| | | | | |
|---|---|---|---|---|
| 0 | 0 | 0.002014257 | 0.01796873 | 0 |
| 0 | 0 | 0 | 0 | 0 |
| 0 | 0 | 0 | 0.100777832 | 0 |
| 0 | 0 | 0 | 0 | 0 |
| 0 | 0 | 0.003859171 | 0 | 0 |
| 0 | 0 | 0 | 0 | 0 |
| 0 | 0 | 0 | 0 | 0 |
| 0 | 0 | 0 | 0 | 0 |
| 0 | 0 | 0 | 0 | 0 |
| 0 | 0.031322363 | 0.095658807 | 0 | 0.008959642 |
| 0.013492037 | 0 | 0 | 0 | 0 |
| 0 | 0 | 0 | 0 | 0 |
| 0 | 0 | 0 | 0 | 0 |
| 0 | 0 | 0 | 0 | 0 |
| 0 | 0 | 0 | 0 | 0 |
| 0 | 0 | 0 | 0 | 0 |
| 0 | 0 | 0 | 0 | 0 |
| 0 | 0 | 0 | 0 | 0 |
| 0 | 0 | 0 | 0 | 0 |
| 0.868513763 | 0.027688994 | 0.873331063 | 0.009260093 | 0.045728384 |
| 0 | 0 | 0 | 0 | 0 |
| 0 | 0 | 0 | 0 | 0 |
| 0 | 0 | 0 | 0 | 0 |
| 0 | 0 | 0 | 0 | 0 |
| 0 | 0 | 0 | 0 | 0 |
| 0 | 0 | 0 | 0 | 0 |
| 0 | 0 | 0 | 0 | 0 |
| 0 | 0 | 0 | 0 | 0 |
| 0 | 0 | 0 | 0 | 0 |
| 0 | 0 | 0 | 0 | 0 |
| 0 | 0 | 0 | 0 | 0 |
| 0 | 0 | 0 | 0 | 0 |
| 0 | 0 | 0 | 0 | 0 |
| 0 | 0 | 0 | 0 | 0 |
| 0 | 0 | 0 | 0 | 0 |
| 0 | 0 | 0 | 0 | 0.01345326 |
| 0 | 0 | 0 | 0 | 0 |
| 0 | 0 | 0 | 0 | 0 |
| 0 | 0 | 0 | 0 | 0 |
| 0 | 0 | 0 | 0 | 0 |
| 0 | 0 | 0 | 0 | 0 |
| 0 | 0 | 0 | 0 | 0 |
| 0 | 0 | 0.125956141 | 0.004881249 | 0 |
| 0 | 0 | 0 | 0 | 0 |
| 0.005033043 | 0 | 0 | 0 | 0 |
| 0 | 0 | 0 | 0 | 0 |
| 0 | 0 | 0 | 0 | 0 |
| 0 | 0 | 0 | 0 | 0 |

| | | | | |
|---|---|---|---|---|
| 0 | 0 | 0 | 0 | 0 |
| 0 | 0 | 0 | 0 | 0 |
| 0 | 0 | 0 | 0 | 0 |
| 0 | 0 | 0 | 0 | 0 |
| 0 | 0 | 0 | 0 | 0 |
| 0.00072257 | 0.00169495 | 0 | 0.006334597 | 0.00291488 |
| 0 | 0 | 0 | 0 | 0 |
| 0 | 0 | 0 | 0 | 0 |
| 0 | 0 | 0 | 0 | 0 |
| 0 | 0 | 0 | 0 | 0 |
| 0 | 0 | 0 | 0 | 0 |
| 0 | 0 | 0 | 0 | 0 |
| 0 | 0 | 0 | 0 | 0 |
| 0 | 0 | 0 | 0 | 0 |
| 0 | 0 | 0 | 0 | 0 |
| 0 | 0 | 0 | 0 | 0 |
| 0 | 0 | 0 | 0.016904407 | 0 |
| 0 | 0 | 0 | 0 | 0 |
| 0 | 0 | 0 | 0 | 0 |
| 0 | 0 | 0 | 0 | 0 |
| 0 | 0 | 0 | 0 | 0 |
| 0 | 0 | 0 | 0 | 0 |
| 0 | 0 | 0 | 0 | 0 |
| 0 | 0 | 0 | 0 | 0 |
| 0 | 0 | 0 | 0 | 0 |
| 0 | 0 | 0.00923276 | 0 | 0.002849601 |
| 0 | 0 | 0 | 0 | 0 |
| 0 | 0 | 0 | 0.01965169 | 0 |
| 0 | 0 | 0 | 0 | 0 |
| 0 | 0 | 0.013414881 | 0 | 0 |
| 0 | 0 | 0 | 0 | 0 |
| 0 | 0 | 0 | 0 | 0 |
| 0 | 0 | 0 | 0 | 0 |
| 0 | 0 | 0 | 0 | 0 |
| 0 | 0 | 0 | 0 | 0 |
| 0 | 0 | 0 | 0 | 0 |
| 0 | 0 | 0 | 0 | 0 |
| 0 | 0 | 0 | 0 | 0 |
| 0 | 0 | 0.449697619 | 0.018107249 | 0 |
| 0 | 0 | 0 | 0 | 0 |
| 0 | 0 | 0 | 0 | 0 |
| 0 | 0 | 0 | 0 | 0 |
| 0 | 0 | 0 | 0 | 0 |
| 0 | 0 | 0 | 0 | 0 |
| 0 | 0 | 0 | 0 | 0 |
| 0 | 0 | 0 | 0 | 0 |
| 0 | 0 | 0 | 0 | 0 |
| 0 | 0 | 0 | 0 | 0 |

| | | | | |
|---|---|---|---|---|
| 0 | 0 | 0 | 0 | 0 |
| 0 | 0 | 0 | 0 | 0 |
| 0 | 0 | 0 | 0 | 0 |
| 0 | 0 | 0 | 0 | 0 |
| 0 | 0 | 0 | 0 | 0 |
| 0 | 0 | 0 | 0 | 0 |
| 0 | 0 | 0 | 0 | 0 |
| 0 | 0 | 0 | 0 | 0 |
| 0 | 0 | 0 | 0 | 0 |
| 0 | 0 | 0 | 0 | 0 |
| 0.001483004 | 0 | 0 | 0 | 0 |
| 0 | 0 | 0 | 0 | 0 |
| 0 | 0 | 0 | 0.012996001 | 0.813220491 |
| 0 | 0 | 0 | 0 | 0 |
| 0 | 0 | 0 | 0 | 0 |
| 0 | 0 | 0 | 0 | 0 |
| 0 | 0 | 0 | 0 | 0 |
| 0 | 0 | 0 | 0 | 0 |
| 0 | 0 | 0 | 0 | 0 |
| 0 | 0 | 0 | 0 | 0 |
| 0 | 0 | 0 | 0 | 0 |
| 0 | 0 | 0 | 0 | 0 |
| 0 | 0 | 0 | 0 | 0 |
| 1.231198716 | 0 | 0 | 0.012687822 | 0 |
| 0 | 0 | 0 | 0 | 0 |
| 0 | 0 | 0 | 0 | 0 |
| 0 | 0 | 0 | 0 | 0 |
| 0 | 0 | 0 | 0.007036371 | 0 |
| 0 | 0 | 0 | 0 | 0 |
| 0 | 0 | 0 | 0 | 0 |
| 0 | 0 | 0.034726754 | 0 | 0 |
| 0 | 0 | 0 | 0 | 0 |
| 0 | 0 | 0 | 0 | 0 |
| 0 | 0 | 0 | 0 | 0 |

| PHL018-0177_V2 | PHL018-0201_V2 | PHL018-0160_V2 | PHL018-0125_V1 | PHL018-0193_V2 |
|---|---|---|---|---|
| CTRL | CTRL | REF | CTRL | REF |
| 0.168453904 | 0.002789226 | 1.016825235 | 0.011358644 | 0.008652047 |
| 7.752379016 | 12.12015316 | 0.03971986 | 10.9579214 | 3.348424782 |
| 4.553742792 | 3.465960497 | 5.858563905 | 0.614249955 | 19.73411848 |
| 0 | 0 | 0 | 0 | 0 |
| 0 | 0 | 0 | 0 | 0 |
| 0 | 0 | 0 | 0 | 0 |
| 0.165043297 | 0 | 0 | 0 | 0 |
| 0 | 0 | 0 | 0 | 0 |
| 0 | 0 | 0 | 0 | 0 |
| 0 | 0 | 0 | 0 | 0 |
| 0 | 0 | 0 | 0 | 0 |
| 0 | 0 | 0 | 0 | 0 |
| 0 | 0 | 0 | 0 | 0 |
| 0.880268841 | 3.864577162 | 4.073507494 | 0.143372098 | 0 |
| 0.008550338 | 0 | 0 | 0 | 0 |
| 0 | 0 | 0 | 0 | 0 |
| 0 | 0 | 0 | 0 | 0 |
| 0.006332239 | 0 | 0 | 0 | 0 |
| 0 | 0 | 0 | 0 | 0 |
| 0 | 0 | 0 | 0 | 0 |
| 0 | 0 | 0 | 0 | 0 |
| 0 | 0 | 0 | 0 | 0 |
| 0 | 0 | 0 | 0 | 0 |
| 0 | 0 | 0 | 0 | 0 |
| 0 | 0 | 0.319775828 | 0 | 0 |
| 0 | 0 | 0 | 0 | 0 |
| 0 | 0 | 0 | 0 | 0 |
| 0 | 0 | 0 | 0 | 0 |
| 0 | 0 | 0 | 0 | 0 |
| 0 | 0 | 0 | 0 | 0 |
| 0 | 0 | 0 | 0 | 0 |
| 0 | 0 | 0 | 0 | 0 |
| 0 | 0 | 0 | 0 | 0 |
| 0 | 0 | 0 | 0 | 0 |
| 0 | 0 | 0 | 0 | 0 |
| 0 | 0.344412855 | 1.262346719 | 0 | 0 |
| 0 | 0 | 0 | 0 | 0 |
| 0 | 0 | 0 | 0 | 0 |
| 0 | 0 | 0 | 0 | 0 |
| 0 | 0 | 0 | 0 | 0 |
| 0 | 0 | 0 | 0 | 0 |
| 0 | 0.010912693 | 0.002899876 | 0.006005415 | 0.009830368 |
| 0.051298296 | 0.141170096 | 0.262355103 | 0.200628002 | 3.749255206 |
| 0.211302986 | 0.029823508 | 0.001097325 | 0.130751429 | 0.012615151 |
| 0 | 0.012542656 | 0 | 0.002760963 | 0.001263205 |
| 0 | 0 | 0 | 0 | 0 |

| | | | | |
|---|---|---|---|---|
| 0 | 0 | 0 | 0 | 0 |
| 0.011241132 | 0 | 0 | 0.013011921 | 0.036266612 |
| 0.020138235 | 0 | 0.139005039 | 0.003780091 | 0 |
| 0.439634766 | 0.730500044 | 0.876835698 | 1.851000909 | 0.562336897 |
| 0.053146019 | 0 | 0.016216932 | 0.017213301 | 0 |
| 0 | 0 | 0 | 0 | 0 |
| 0.00172124 | 0.004794641 | 0 | 0.334720326 | 0 |
| 0 | 0 | 0 | 0 | 0 |
| 0 | 0 | 0 | 0 | 0 |
| 0 | 0 | 0 | 0 | 0 |
| 0.001054324 | 0.001573338 | 0 | 0.002424327 | 0 |
| 0 | 0.170393467 | 0.098888372 | 0.14976602 | 0 |
| 0 | 0 | 0 | 0.000979633 | 0 |
| 0.079098115 | 0.012508737 | 0.013717138 | 0.002969457 | 0.080577111 |
| 0 | 0 | 0 | 0 | 0 |
| 0 | 0 | 0 | 0 | 0 |
| 0.001126599 | 0.313037869 | 0.002226877 | 0.166903257 | 0.474325829 |
| 0 | 0 | 0 | 0 | 0 |
| 0 | 0 | 0 | 0 | 0 |
| 0 | 0 | 0 | 0 | 0 |
| 0.133948856 | 0.001036697 | 0.003432977 | 0.017115282 | 0.262561464 |
| 0.024256119 | 0 | 0.223119178 | 0.076866165 | 0.00428879 |
| 0 | 0 | 0 | 0 | 0 |
| 0 | 0 | 0 | 0 | 0 |
| 0 | 0 | 0 | 0 | 0 |
| 0 | 0 | 0 | 0 | 0 |
| 0 | 0 | 0 | 0 | 0.070874163 |
| 0 | 0 | 0 | 0 | 0 |
| 0 | 0 | 0 | 0 | 0 |
| 0 | 0 | 0 | 0 | 0 |
| 0.25406173 | 0 | 0 | 0 | 0.106718394 |
| 0 | 0 | 0 | 0 | 0.005839963 |
| 0 | 0 | 0 | 0 | 0 |
| 0 | 0 | 0 | 0 | 0.016654612 |
| 0 | 0 | 0 | 0 | 0 |
| 0 | 0.005940386 | 0 | 0 | 0 |
| 0 | 0 | 0 | 0 | 0 |
| 0 | 0.03157297 | 0 | 0 | 0 |
| 0 | 0 | 0 | 0 | 0 |
| 0 | 0 | 0 | 0 | 0 |
| 0 | 0 | 0 | 0 | 0 |
| 0 | 0 | 0 | 0 | 0 |
| 0 | 0 | 0 | 0 | 0 |
| 0 | 0 | 0 | 0 | 0 |
| 0.188695449 | 0 | 0.008854184 | 0 | 0 |
| 0 | 0 | 0 | 0 | 0 |
| 0 | 0 | 0 | 0 | 0.573720264 |
| 0 | 0 | 0 | 0 | 0.38962925 |

| | | | | |
|---|---|---|---|---|
| 0 | 0 | 0 | 0 | 0 |
| 0 | 0 | 0 | 0 | 0 |
| 0 | 0 | 0 | 0 | 0 |
| 0 | 0 | 0 | 0 | 0 |
| 0 | 0.563107621 | 0.195877262 | 0 | 0 |
| 0 | 0 | 0 | 0 | 0 |
| 0 | 0 | 0 | 0 | 0 |
| 0 | 0 | 0.081199312 | 0 | 0 |
| 0.024430543 | 0 | 0 | 0.002847627 | 0 |
| 0 | 0 | 0 | 0 | 0 |
| 0 | 0 | 0 | 0 | 0 |
| 0 | 0 | 0 | 0 | 0 |
| 0 | 0 | 0 | 0 | 0 |
| 0 | 0 | 0 | 0 | 0 |
| 0 | 0 | 0 | 0 | 0 |
| 0 | 0 | 0 | 0 | 0 |
| 0 | 0 | 0 | 0 | 0 |
| 0 | 0 | 0 | 0 | 0 |
| 0.519498286 | 0 | 0.156300144 | 0 | 0 |
| 0 | 0 | 0 | 0.005472589 | 0 |
| 0 | 0 | 0 | 0 | 0 |
| 0 | 0 | 0 | 0 | 0 |
| 0 | 0 | 0 | 0 | 0 |
| 0 | 0 | 0 | 0 | 0 |
| 0 | 0 | 0 | 0 | 0 |
| 0 | 0 | 0 | 0 | 0 |
| 0 | 0 | 0 | 0 | 0 |
| 0.019891841 | 0 | 0.001398007 | 0 | 0 |
| 0 | 0 | 0 | 0 | 0 |
| 0 | 0 | 0 | 0 | 0 |
| 0.094440222 | 0 | 0.286858236 | 0.004072444 | 0 |
| 0.002868997 | 0.032423883 | 0.008065374 | 0 | 0.000804876 |
| 0 | 0 | 0 | 0 | 0 |
| 0 | 0 | 0 | 0 | 0 |
| 0 | 0 | 0 | 0 | 0 |
| 0 | 0 | 0 | 0 | 0 |
| 0 | 0 | 0 | 0 | 0 |
| 0 | 0 | 0 | 0 | 0 |
| 0 | 0 | 0.009023971 | 0 | 0 |
| 0 | 0 | 0 | 0 | 0 |
| 0 | 0 | 0 | 0 | 0 |
| 0 | 0 | 0 | 0 | 0 |
| 0 | 0.020585892 | 0 | 0 | 0.022327434 |
| 0 | 0 | 0 | 0 | 0 |
| 0.001414172 | 0 | 0 | 0 | 0 |
| 0 | 0 | 0 | 0 | 0 |
| 0 | 0 | 0 | 0 | 0 |
| 0 | 0 | 0 | 0 | 0 |

| | | | | |
|---|---|---|---|---|
| 0 | 0 | 0.004149456 | 0 | 0 |
| 0 | 0 | 0 | 0 | 0 |
| 0 | 0 | 0 | 0 | 0 |
| 0 | 0 | 0 | 0 | 0 |
| 0 | 0 | 0.025739984 | 0 | 0 |
| 0 | 0 | 0 | 0 | 0 |
| 0 | 0 | 0 | 0 | 0 |
| 0 | 0 | 0 | 0 | 0 |
| 0 | 0 | 0 | 0 | 0 |
| 0 | 0 | 0 | 0 | 0 |
| 0 | 0 | 0 | 0 | 0 |
| 0 | 0 | 0 | 0 | 0 |
| 0 | 0 | 0 | 0 | 0 |
| 0 | 0 | 0 | 0 | 0 |
| 0 | 0 | 0 | 0 | 0 |
| 0 | 0 | 1.357271929 | 0 | 0 |
| 0 | 0 | 0 | 0 | 0 |
| 0 | 0 | 0 | 0 | 0 |
| 0 | 0 | 0 | 0 | 0 |
| 0 | 0 | 0 | 0 | 0 |
| 0 | 0 | 0 | 0 | 0 |
| 0 | 0 | 0 | 0 | 0 |
| 0 | 0 | 0 | 0 | 0 |
| 0 | 0 | 0 | 0 | 0 |
| 0 | 0 | 0.005319688 | 0 | 0 |
| 0 | 0 | 0 | 0 | 0 |
| 0 | 0 | 0 | 0 | 0 |
| 0 | 0 | 0 | 0 | 0 |
| 0 | 0 | 0 | 0 | 0 |
| 0 | 0 | 0 | 0 | 0 |
| 0 | 0 | 0 | 0 | 0 |
| 0.004192924 | 0.281366583 | 0 | 0 | 0 |
| 0 | 0 | 0 | 0 | 0 |
| 0 | 0 | 0 | 0 | 0 |
| 0 | 0 | 0 | 0 | 0 |
| 0 | 0 | 0 | 0 | 0 |
| 0 | 0 | 0 | 0 | 0 |
| 0 | 0 | 0 | 0 | 0 |
| 0 | 0 | 0 | 0 | 0 |
| 0 | 0 | 0 | 0 | 0 |
| 0 | 0 | 0 | 0 | 0 |
| 0 | 0 | 0 | 0 | 0 |
| 0 | 0 | 0 | 0 | 0 |
| 0 | 0 | 0 | 0 | 0 |
| 0 | 0 | 0 | 0 | 0 |
| 0 | 0 | 0 | 0 | 0 |
| 0 | 0 | 0 | 0 | 0 |
| 0 | 0 | 0 | 0 | 0 |

| | | | | |
|---|---|---|---|---|
| 0 | 0 | 0 | 0 | 0 |
| 0 | 0 | 0 | 0 | 0 |
| 0 | 0 | 0 | 0 | 0 |
| 0 | 0 | 0 | 0 | 0 |
| 0 | 0 | 0 | 0 | 0 |
| 0 | 0 | 0 | 0 | 0 |
| 0 | 0 | 0 | 0 | 0 |
| 0 | 0 | 0 | 0 | 0 |
| 0 | 0 | 0 | 0 | 0 |
| 0 | 0 | 0 | 0 | 0 |
| 0 | 0 | 0 | 0 | 0 |
| 0 | 0 | 0 | 0 | 0 |
| 0.001689355 | 0 | 0.001669621 | 0 | 0.00088863 |
| 0 | 0 | 0 | 0 | 0 |
| 0 | 0 | 0.076496844 | 0 | 0.013500002 |
| 0 | 0 | 0 | 0 | 0 |
| 0 | 0 | 0 | 0 | 0 |
| 0 | 0 | 0 | 0 | 0 |
| 0 | 0 | 0 | 0 | 0 |
| 0 | 0 | 0 | 0 | 0 |
| 0.035140394 | 0.120197292 | 0.038502071 | 0.038390812 | 0.031236678 |
| 0 | 0 | 0 | 0 | 0 |
| 0 | 0 | 0.001369588 | 0 | 0 |
| 0.001646938 | 0 | 0.001446844 | 0 | 0 |
| 0 | 0 | 0 | 0 | 0 |
| 0 | 0 | 0 | 0 | 0 |
| 0.001609602 | 0.031843169 | 0 | 0.00392774 | 0.001935266 |
| 0 | 0 | 0 | 0 | 0 |
| 0.026942649 | 0 | 0 | 0 | 0.33485483 |
| 0 | 0 | 0 | 0 | 0.038097425 |
| 0.031173137 | 0 | 0 | 0 | 0.003858264 |
| 0 | 0 | 0 | 0 | 0 |
| 0 | 0 | 0 | 0 | 0 |
| 0.010375864 | 0 | 0 | 0 | 0 |
| 0.024919423 | 0 | 0.01105877 | 0.006075999 | 0.022486402 |
| 0 | 0 | 0 | 0 | 0 |
| 0 | 0 | 0 | 0 | 0 |
| 0 | 0 | 0 | 0 | 0 |
| 0 | 0 | 0 | 0 | 0 |
| 0 | 0 | 0 | 0 | 0 |
| 0 | 0 | 0 | 0 | 0 |
| 0 | 0.002450354 | 0 | 0 | 0 |
| 0 | 0.008925649 | 0 | 0 | 0 |
| 0 | 0 | 0 | 0 | 0 |
| 0 | 0 | 0 | 0 | 0 |
| 0.265144223 | 0 | 0 | 0 | 0 |
| 0 | 0 | 0 | 0 | 0 |
| 0.001464598 | 0 | 0.001125825 | 0 | 0.005855076 |

| | | | | |
|---|---|---|---|---|
| 0 | 0 | 0 | 0 | 0 |
| 0 | 0 | 0 | 0 | 0 |
| 0.002676639 | 0 | 0 | 0 | 0 |
| 0 | 0 | 0 | 0 | 0 |
| 0 | 0 | 0 | 0 | 0 |
| 0 | 0 | 0 | 0 | 0 |
| 0 | 0 | 0 | 0 | 0 |
| 0 | 0 | 0 | 0 | 0 |
| 0 | 0 | 0 | 0 | 0 |
| 0 | 0 | 0 | 0 | 0 |
| 0 | 0 | 0 | 0 | 0 |
| 0 | 0 | 0 | 0 | 0 |
| 0 | 0.002055322 | 0 | 0 | 0 |
| 0 | 0.006738403 | 0 | 0 | 0 |
| 0 | 0.001378373 | 0 | 0.003034156 | 0 |
| 0.000726828 | 0 | 0 | 0 | 0 |
| 0 | 0 | 0 | 0 | 0 |
| 0 | 0 | 0 | 0 | 0 |
| 0 | 0 | 0.001799218 | 0 | 0.023348195 |
| 0 | 0 | 0.16944424 | 0 | 0 |
| 0.21971447 | 0 | 0.004584234 | 0 | 0.148858937 |
| 0 | 0 | 0 | 0 | 0 |
| 0 | 0 | 0 | 0 | 0 |
| 0 | 0 | 0 | 0 | 0 |
| 0 | 0.10422246 | 0.001917379 | 0.01075409 | 0.00137767 |
| 0 | 0 | 0 | 0 | 0 |
| 0 | 0 | 0 | 0 | 0 |
| 0 | 0 | 0 | 0 | 0 |
| 0 | 0 | 0 | 0 | 0.004601555 |
| 0 | 0 | 0 | 0 | 0.075863328 |
| 0.362087296 | 0.130909364 | 0.107510311 | 0.022554379 | 0.209733851 |
| 0 | 0 | 0 | 0 | 0 |
| 0 | 0 | 0 | 0 | 0.002747357 |
| 0 | 0.023081383 | 0 | 0 | 0 |
| 0 | 0 | 0 | 0 | 0 |
| 0 | 0 | 0 | 0 | 0 |
| 0 | 0 | 0 | 0 | 0 |
| 0 | 0 | 0 | 0 | 0 |
| 0 | 0 | 0 | 0 | 0 |
| 0 | 0 | 0 | 0 | 0 |
| 0 | 0 | 0 | 0 | 0 |
| 0.011939259 | 0.012287323 | 0.039965584 | 0 | 0 |
| 0.006390461 | 0 | 0 | 0 | 0 |
| 0 | 0 | 0 | 0 | 0 |
| 0 | 0 | 0 | 0 | 0 |
| 0 | 0 | 0 | 0 | 0 |
| 0 | 0 | 0 | 0 | 0 |
| 0 | 0 | 0 | 0 | 0 |

| | | | | |
|---|---|---|---|---|
| 0 | 0 | 0 | 0 | 0 |
| 0 | 0 | 0 | 0 | 0 |
| 0.222290793 | 0 | 0.00113537 | 0.118190134 | 0.008919228 |
| 0 | 0 | 0 | 0 | 0 |
| 0 | 0 | 0 | 0 | 0 |
| 0 | 0 | 0 | 0 | 0 |
| 0 | 0 | 0 | 0 | 0 |
| 0 | 0 | 0 | 0 | 0 |
| 0 | 0 | 0 | 0 | 0 |
| 0.000945607 | 0 | 0 | 0 | 0 |
| 0 | 0 | 0 | 0 | 0 |
| 0 | 0 | 0 | 0 | 0 |
| 0 | 0 | 0 | 0 | 0 |
| 0.003427974 | 0.067524183 | 0.004517241 | 0 | 0 |
| 0 | 0.017733028 | 0.007663915 | 0 | 0.004762512 |
| 0.005048643 | 0 | 0.023285121 | 0 | 0 |
| 0.003223783 | 0.015073717 | 0 | 0.001764955 | 0.001130512 |
| 0.004793091 | 0 | 0.000915382 | 0 | 0 |
| 0 | 0 | 0 | 0 | 0 |
| 0 | 0 | 0 | 0 | 0 |
| 0 | 0 | 0 | 0 | 0 |
| 0.002182617 | 0 | 0.001917441 | 0 | 0.007551917 |
| 0 | 0 | 0 | 0 | 0 |
| 0 | 0 | 0 | 0 | 0 |
| 0.001166566 | 0 | 0 | 0 | 0 |
| 0 | 0 | 0 | 0 | 0 |
| 0 | 0 | 0 | 0 | 0 |
| 0 | 0 | 0 | 0 | 0 |
| 0 | 0 | 0 | 0 | 0 |
| 0.024205016 | 0 | 0 | 0 | 0 |
| 0 | 0 | 0.001959154 | 0 | 0 |
| 0 | 0 | 0 | 0 | 0 |
| 0 | 0.005991716 | 0 | 0 | 0 |
| 0 | 0 | 0.001473869 | 0 | 0 |
| 0 | 0 | 0 | 0 | 0 |
| 0 | 0.00231307 | 0.002127676 | 0 | 0 |
| 0 | 0 | 0 | 0 | 0 |
| 0.000914078 | 0.090573057 | 0.025964384 | 0.003603158 | 0.003077254 |
| 0 | 0 | 0 | 0 | 0 |
| 0 | 0 | 0 | 0 | 0 |
| 0 | 0 | 0 | 0 | 0 |
| 0 | 0 | 0 | 0 | 0 |
| 0 | 0 | 0 | 0 | 0 |
| 0.006055612 | 0 | 0 | 0 | 0 |
| 0 | 0 | 0 | 0 | 0 |
| 0.00140538 | 0 | 0 | 0 | 0 |
| 0 | 0 | 0 | 0 | 0 |
| 0.001476236 | 0 | 0 | 0 | 0 |

| | | | | |
|---|---|---|---|---|
| 0.949606935 | 0 | 0 | 0 | 0 |
| 0 | 0 | 0.306745517 | 0 | 0 |
| 0 | 0 | 0 | 0.001001283 | 0 |
| 0 | 0 | 0 | 0 | 0 |
| 0 | 0.368248987 | 0 | 0.003357959 | 0.001935795 |
| 0 | 0 | 0 | 0 | 0 |
| 0 | 0 | 0 | 0 | 0 |
| 0.003940491 | 0.002993599 | 0 | 0.01788629 | 0.009044808 |
| 0 | 0 | 0 | 0 | 0 |
| 0.134031148 | 0.078902366 | 0.144994367 | 0.016495177 | 1.055327283 |
| 0.002336833 | 0 | 0 | 0 | 0.004214454 |
| 0 | 0.009478975 | 0 | 0 | 0 |
| 0.017853339 | 0.02259758 | 0.001511736 | 0.00621789 | 0 |
| 0 | 0 | 0 | 0.167405204 | 0 |
| 0.003760841 | 0 | 0.001943482 | 0 | 0 |
| 0 | 0 | 0 | 0 | 0 |
| 0 | 0 | 0.001031432 | 0 | 0 |
| 0.001120561 | 0 | 0 | 0 | 0 |
| 0.529499175 | 0 | 0 | 0 | 0 |
| 0 | 0 | 0 | 0 | 0 |
| 0 | 0 | 0 | 0.018786254 | 0 |
| 0 | 0 | 0 | 0 | 0 |
| 0 | 0.001059899 | 0 | 0 | 0 |
| 0 | 0 | 0.379531111 | 0 | 0 |
| 0 | 0 | 0 | 0 | 0 |
| 0.000751612 | 0 | 0 | 0.004937902 | 0.012862413 |
| 0 | 0.002399709 | 0 | 0 | 0 |
| 0 | 0 | 0 | 0.010350246 | 0 |
| 0.018212491 | 0 | 0.001802791 | 0 | 0.134283227 |
| 0 | 0 | 0.003623256 | 0 | 0.01453549 |
| 0 | 0 | 0 | 0 | 0.007014166 |
| 0 | 0 | 0 | 0 | 0 |
| 0.041061445 | 0 | 0 | 0 | 0 |
| 1.993462968 | 0 | 0 | 0 | 0 |
| 0 | 0 | 0 | 0 | 0 |
| 0.013719139 | 0 | 0 | 0 | 0 |
| 0.01420048 | 0 | 0 | 0 | 0 |
| 0 | 0 | 0 | 0 | 0 |
| 0 | 0 | 0 | 0 | 0 |
| 0 | 0 | 0 | 0.005539636 | 0 |
| 0.030725748 | 0 | 0 | 2.59603764 | 0 |
| 0.006178976 | 0 | 0 | 0 | 0 |
| 0.00114932 | 0 | 0 | 0 | 0 |
| 0 | 0 | 0 | 0.007425876 | 0 |
| 0 | 0 | 0 | 0 | 0 |
| 0 | 0 | 0.008695764 | 0 | 0 |
| 0.003427191 | 0.002603644 | 0.001094839 | 0 | 0 |
| 0 | 0 | 0 | 0 | 0 |

| | | | | |
|---|---|---|---|---|
| 0 | 0 | 0 | 0 | 0 |
| 0 | 0.001662717 | 0 | 0.011437716 | 0 |
| 0 | 0.01622581 | 0 | 0 | 0 |
| 0 | 0.011608903 | 0 | 0 | 1.078050355 |
| 0 | 0 | 0 | 0 | 0.156024635 |
| 0.264288755 | 0.048044615 | 0 | 0.004514088 | 0 |
| 0.01828189 | 0 | 0 | 0 | 0 |
| 0 | 0 | 0 | 0 | 0 |
| 0 | 0 | 0 | 0 | 0 |
| 0 | 0 | 0 | 0 | 0 |
| 0 | 0.012571802 | 0 | 0.006357492 | 0.001437241 |
| 0 | 0 | 0.056710433 | 0 | 0 |
| 0 | 0 | 0 | 0 | 0 |
| 0 | 0.01669229 | 0.015590611 | 0.054409456 | 0.002036749 |
| 0.03890578 | 0 | 0 | 0 | 0 |
| 0 | 0 | 0 | 0 | 0 |
| 0 | 0 | 0 | 0 | 0 |
| 0.098650884 | 0 | 0 | 0 | 0 |
| 0 | 0 | 0 | 0 | 0.035889242 |
| 0.073362271 | 0 | 0 | 0 | 0 |
| 0 | 0 | 0 | 0 | 0 |
| 0.13802024 | 0.039922478 | 0.089469751 | 0 | 0 |
| 0.07144586 | 0 | 0.010983977 | 0 | 0 |
| 0 | 0 | 0 | 0 | 0 |
| 0 | 0 | 0 | 0 | 0 |
| 0 | 0 | 0 | 0 | 0 |
| 0.434406499 | 0 | 0 | 0 | 0 |
| 0 | 0 | 0.001329843 | 0 | 0 |
| 0 | 0 | 0 | 0 | 0.002889297 |
| 0.133218127 | 0.310077263 | 0.039818873 | 0.830154833 | 0.057055228 |
| 0 | 0.018293249 | 0 | 0 | 0 |
| 0 | 0.006420788 | 0 | 0 | 0 |
| 0 | 0 | 0 | 0 | 0 |
| 0 | 0 | 0 | 0 | 0 |
| 0.017969569 | 0 | 0.001919963 | 0.002552554 | 0 |
| 0 | 0 | 0 | 0 | 0 |
| 0 | 0.180925061 | 3.092571262 | 0 | 0 |
| 0 | 0.004253473 | 0.007223175 | 0 | 0 |
| 0 | 0 | 0.066296549 | 0 | 0 |
| 0.251669089 | 0 | 0 | 0 | 0 |
| 0.046421942 | 0.020056516 | 0.000970998 | 0.015103803 | 0 |
| 0 | 1.657309002 | 1.045043642 | 7.946826027 | 0 |
| 0 | 0.000899182 | 0.005072954 | 0 | 0 |
| 0 | 0 | 0 | 0 | 0 |
| 0 | 0 | 0 | 0 | 0 |
| 0 | 0 | 0 | 0 | 0 |
| 0.008352541 | 0.001107934 | 0 | 0 | 0.004686491 |
| 0 | 0 | 0 | 0 | 0 |

| | | | | |
|---|---|---|---|---|
| 0 | 0.00820203 | 0 | 0.001128424 | 0 |
| 0 | 0 | 0 | 0 | 0 |
| 0.002730431 | 0.6826609 | 0.004797398 | 0.387466291 | 0.023636656 |
| 0 | 0 | 0 | 0 | 0 |
| 0.423019919 | 0 | 0 | 0 | 0 |
| 0.038000206 | 0 | 0 | 0 | 0 |
| 0 | 0 | 0 | 0 | 0 |
| 0 | 0 | 0 | 0 | 0 |
| 0 | 0 | 0.002946617 | 0 | 0.053635603 |
| 0.349138685 | 0 | 0 | 0 | 0.015365991 |
| 0 | 0 | 0 | 0 | 0 |
| 0 | 0 | 0 | 0 | 0 |
| 0 | 0 | 0 | 0.004162845 | 0 |
| 0 | 0.010821081 | 0 | 0 | 0.014385634 |
| 0 | 0 | 0 | 0 | 0 |
| 0.050192148 | 0 | 0.126948278 | 0.004254838 | 0 |
| 0.006056793 | 0.135508754 | 0.001379499 | 0.011200606 | 0.024166252 |
| 0.478864949 | 0 | 0 | 0 | 0.039453915 |
| 0 | 0 | 0.001832038 | 0 | 0 |
| 0.262070332 | 0 | 0 | 0 | 0 |
| 0.113368377 | 0 | 0 | 0.158618701 | 0 |
| 0.000780618 | 0.003727663 | 0 | 0 | 0.000656991 |
| 0 | 0 | 0 | 0 | 0 |
| 0 | 0 | 0 | 0.662778761 | 0 |
| 0.0287226 | 0 | 0.089512554 | 0.007713299 | 0 |
| 0.004432643 | 0 | 0 | 0 | 0 |
| 0.002278266 | 0 | 0 | 0 | 0 |
| 0 | 0 | 0 | 0 | 0 |
| 0 | 0 | 0 | 0 | 0 |
| 0 | 0 | 0 | 0 | 0 |
| 0.017542724 | 0 | 0.031393552 | 0.011951991 | 0 |
| 0.022279375 | 0.188109887 | 0.003120262 | 0 | 0 |
| 0.022442715 | 0.078909425 | 0.013774226 | 0.054129783 | 0 |
| 0 | 0.004733787 | 0 | 0 | 0 |
| 0 | 0.126889677 | 0.001381294 | 0 | 0 |
| 0.023884854 | 0 | 0 | 0 | 0 |
| 0 | 0.003318119 | 0.0083019 | 0.045285036 | 0 |
| 0.176933872 | 0 | 0.004010055 | 0.017136289 | 0 |
| 0 | 0 | 0.057876732 | 3.066240732 | 0.179524006 |
| 0.004442921 | 0 | 0 | 0 | 0 |
| 0 | 0 | 0 | 0 | 0 |
| 0 | 0 | 0 | 0 | 0 |
| 0.010077069 | 0 | 0 | 0 | 0.05225481 |
| 0.090778257 | 0 | 0 | 0 | 0 |
| 0 | 0 | 0 | 0.094238749 | 0 |
| 0 | 0.020932216 | 0 | 0 | 0 |
| 0.004347737 | 0 | 0 | 0 | 0 |
| 0.00088326 | 0 | 0 | 0 | 0 |

| | | | | |
|---|---|---|---|---|
| 0.004189613 | 0 | 0 | 0 | 0 |
| 2.094326358 | 9.366276408 | 0 | 0.003891728 | 0.026174187 |
| 0 | 0 | 0 | 0 | 0 |
| 0.00161443 | 0 | 0 | 0 | 0 |
| 0 | 0 | 0 | 0 | 0 |
| 0 | 0 | 0 | 0 | 0.003408984 |
| 0.02356486 | 0 | 0 | 0 | 0 |
| 0 | 0 | 0 | 0 | 0 |
| 0 | 0 | 0 | 0 | 0 |
| 0 | 0 | 0 | 0 | 0 |
| 0.000660294 | 0 | 0.008507724 | 0 | 0 |
| 0.035406971 | 0.052054052 | 0.343597471 | 0.110707514 | 0.063737844 |
| 0 | 0 | 0.098578742 | 0 | 0 |
| 0 | 0 | 0 | 0 | 0 |
| 0 | 0.001205923 | 0.002662241 | 0.013604557 | 0 |
| 0 | 0 | 0 | 0 | 0 |
| 0.008066606 | 0.103179149 | 0.001012365 | 0.022106735 | 0.002133712 |
| 0.196267115 | 0 | 0.000788514 | 0.002751824 | 0.003273463 |
| 0 | 0 | 0 | 0 | 0 |
| 0.288112556 | 0 | 0 | 0 | 0 |
| 0.010933156 | 0.10271507 | 0.011901647 | 0 | 0 |
| 0 | 0.001059219 | 0 | 0.011269471 | 0 |
| 0 | 0 | 0 | 0 | 0 |
| 0 | 0 | 0 | 0 | 0 |
| 0 | 0 | 0 | 0.015216759 | 0 |
| 0 | 0 | 0 | 0 | 0 |
| 0.158119979 | 0.002800688 | 0.011541423 | 0 | 0.298340139 |
| 0.017935728 | 0.358273745 | 0.063248469 | 0.134097592 | 0 |
| 0 | 0 | 0 | 0 | 0 |
| 0.001828284 | 0 | 0 | 0 | 0.002308103 |
| 0 | 0 | 0 | 0 | 0 |
| 0 | 0 | 0 | 0 | 0 |
| 0.032814106 | 0 | 0 | 0 | 0 |
| 0 | 0 | 0 | 0 | 0 |
| 0 | 0 | 0.002514092 | 0 | 0 |
| 0.39034157 | 0.542556757 | 0.0492028 | 1.046564112 | 0.166959979 |
| 0 | 0 | 0.00556648 | 0 | 0 |
| 0 | 0 | 0 | 0 | 0 |
| 0.003427929 | 0.002232175 | 0.004380298 | 0.002047331 | 0 |
| 0 | 0 | 0 | 0 | 0 |
| 0 | 0 | 0 | 0.051388443 | 0 |
| 0 | 0 | 0 | 0 | 0 |
| 0.049404375 | 0.00542343 | 0.056206442 | 0.02089215 | 0 |
| 0.052085108 | 0.002181758 | 0.014610147 | 0.002401307 | 0.009843942 |
| 0.051620939 | 0 | 0 | 0 | 0 |
| 0 | 0 | 0.046612942 | 0 | 0 |
| 0 | 0 | 0 | 0 | 0 |
| 0 | 0 | 0 | 0 | 0.004226617 |

| | | | | |
|---|---|---|---|---|
| 0 | 0 | 0 | 0 | 0 |
| 0 | 0 | 0 | 0 | 0 |
| 0 | 0 | 0 | 0 | 0 |
| 0.296079855 | 0 | 2.721932131 | 0.006372862 | 0 |
| 0 | 0 | 0 | 0 | 0 |
| 0 | 0 | 0 | 0 | 0 |
| 0 | 0 | 0.032883555 | 0 | 0 |
| 0 | 0 | 0.001212852 | 0 | 0 |
| 0 | 0.225100783 | 4.27649564 | 0.044420275 | 0 |
| 0 | 0.052926269 | 0.078413994 | 0.121682379 | 0.403306128 |
| 0 | 0 | 0 | 0 | 0 |
| 0 | 0.011851759 | 0.008350339 | 0.001387701 | 0 |
| 0 | 0 | 0 | 0 | 0 |
| 0 | 0 | 0 | 0 | 0 |
| 0 | 0.001248524 | 1.490845723 | 0.00297735 | 0 |
| 0 | 0 | 0 | 0 | 0 |
| 0 | 0.026600299 | 0 | 0 | 0 |
| 0 | 0 | 0 | 0 | 0 |
| 0 | 0 | 0 | 0 | 0 |
| 0 | 0 | 0.019222693 | 0 | 0 |
| 0 | 0 | 0 | 0 | 0 |
| 0 | 0 | 0.001378254 | 0 | 0 |
| 0 | 0 | 0.005638783 | 0 | 0 |
| 0.711390342 | 0 | 0 | 0.341286234 | 0 |
| 0.003361944 | 0 | 0 | 0 | 0 |
| 0 | 0 | 0.101880325 | 0 | 0 |
| 0.007529776 | 0 | 0 | 0 | 0 |
| 0 | 4.091689711 | 0.686119159 | 6.252698043 | 0 |
| 0 | 0 | 0 | 0 | 0 |
| 0 | 0 | 0 | 0 | 0 |
| 0.006981817 | 0 | 0.01046314 | 0.002158532 | 0 |
| 0.375148961 | 0 | 0 | 0 | 0 |
| 0.127215634 | 0 | 0.68648426 | 0.002464202 | 0 |
| 0 | 0 | 0 | 0 | 0 |
| 0 | 0 | 0.030147381 | 0 | 0 |
| 0 | 0 | 1.900810834 | 0 | 0 |
| 0 | 0 | 7.618569468 | 0 | 0 |
| 0 | 0 | 0 | 0 | 0 |
| 0 | 0 | 0.215579043 | 0 | 0 |
| 0 | 0 | 0.020985735 | 0 | 0 |
| 0.645387547 | 0.009192479 | 0.002670219 | 0.055912562 | 0.053379666 |
| 0 | 0.002521204 | 0.091064414 | 0 | 0 |
| 7.281815333 | 0.114047633 | 1.851054641 | 0.812553382 | 0 |
| 0 | 0 | 0 | 0 | 0 |
| 0 | 0 | 0 | 0 | 0 |
| 0 | 0 | 0 | 0 | 0 |
| 0 | 0 | 0 | 0 | 0 |
| 0.001217943 | 0 | 0.662525362 | 0 | 0.015785857 |

| | | | | |
|---|---|---|---|---|
| 0 | 0 | 0 | 0 | 0 |
| 0 | 0.006093329 | 0.420517024 | 0.002915868 | 0 |
| 0.023117215 | 0.139661254 | 0.010256869 | 0.074249691 | 0.292627675 |
| 0 | 0 | 0.001330462 | 0 | 0 |
| 0 | 0 | 0 | 0 | 0 |
| 0 | 0.001537202 | 0.040561431 | 0 | 0 |
| 0.089258243 | 0 | 0 | 0 | 0 |
| 0 | 0 | 0 | 0 | 0 |
| 0 | 0 | 0 | 0 | 0 |
| 0 | 0 | 0 | 0 | 0.37608286 |
| 0.006190093 | 0.004379164 | 0.360722691 | 0.010844632 | 0.053448254 |
| 0.098148205 | 8.346654985 | 0.836930647 | 3.616069178 | 0 |
| 0 | 0 | 0 | 0 | 0 |
| 0.004701163 | 0.127311277 | 0 | 0.03738768 | 0.010203946 |
| 0.088522756 | 0.062151014 | 0.002055535 | 1.040425573 | 0.396311414 |
| 0 | 0 | 0 | 0 | 0 |
| 0 | 0 | 0 | 0 | 0.013063927 |
| 0 | 0 | 0 | 0 | 0 |
| 0 | 0 | 0 | 0 | 0 |
| 0.303107643 | 0 | 0.435175702 | 0 | 0 |
| 0.138544399 | 0.001146995 | 0.060602795 | 0.056808743 | 0.008733088 |
| 0.059122028 | 0.144787536 | 1.842768251 | 0 | 14.09655914 |
| 2.027672174 | 2.162829403 | 0.586439259 | 0.529895004 | 21.01655031 |
| 0 | 0 | 0 | 0 | 0 |
| 0 | 0 | 0.151552498 | 0 | 0 |
| 0 | 0.476382392 | 0.268634523 | 0 | 0 |
| 0 | 0 | 3.09964003 | 0 | 0 |
| 0 | 0 | 0 | 0 | 0 |
| 0 | 0 | 0.381118295 | 0.009215636 | 0 |
| 0 | 0 | 0 | 0 | 0 |
| 0 | 0 | 0.008751074 | 0 | 0 |
| 0 | 0.095507492 | 0.025180396 | 0.606873709 | 0 |
| 0.130679608 | 0 | 0 | 0.008043909 | 0 |
| 0.033935995 | 0.003644061 | 0.274153031 | 0.002359271 | 0 |
| 0.009479748 | 0 | 0.014062051 | 0 | 0.063304222 |
| 0.00619202 | 0.005862732 | 0.137306131 | 0.003927727 | 0 |
| 0 | 0 | 0.053996561 | 0 | 0 |
| 0 | 0 | 0 | 0 | 0 |
| 0 | 0.094556702 | 0.490588306 | 0 | 0 |
| 0 | 0 | 0 | 0 | 0 |
| 0 | 0 | 0.014535023 | 0 | 0 |
| 0 | 0 | 0 | 0 | 0 |
| 0 | 0 | 0 | 0 | 0 |
| 0 | 0 | 0 | 0 | 0 |
| 0.022888339 | 0 | 0.012406775 | 0 | 0 |
| 0 | 0 | 0.141196515 | 0 | 0 |
| 0 | 0 | 0 | 0 | 0 |
| 0.65734324 | 0 | 0.055735303 | 0.028944919 | 0 |

| | | | | |
|---|---|---|---|---|
| 0 | 0 | 0 | 0 | 0 |
| 0 | 0 | 0 | 0 | 0 |
| 0 | 0 | 0 | 0 | 0.001589017 |
| 0 | 0 | 0.006198695 | 0 | 0 |
| 0 | 0 | 0 | 0 | 0 |
| 0 | 0 | 0 | 0 | 0 |
| 0.083383762 | 0 | 0.041516741 | 0 | 0 |
| 0 | 0 | 0.128338228 | 0 | 0 |
| 0 | 0 | 0 | 0 | 0 |
| 0 | 0 | 0 | 0 | 0 |
| 0.055471515 | 0 | 0.022335515 | 0 | 0.033069534 |
| 0 | 0 | 0 | 0 | 0 |
| 0 | 0.005315135 | 0.185075505 | 0 | 0 |
| 0 | 0 | 0 | 0 | 0 |
| 0 | 0 | 0 | 0 | 0 |
| 0 | 0 | 0 | 0 | 0 |
| 0 | 0 | 0 | 0 | 0 |
| 0 | 0.002067338 | 0 | 0.000758458 | 0 |
| 0.327795649 | 0 | 0.000889346 | 0 | 0 |
| 0 | 0 | 0 | 0 | 0 |
| 0 | 0 | 0 | 0 | 0 |
| 0 | 0 | 0 | 0 | 0 |
| 0 | 0 | 0.005067005 | 0 | 0 |
| 0 | 0 | 0.030038463 | 0 | 0 |
| 0 | 0 | 0 | 0 | 0 |
| 0 | 0 | 0 | 0 | 0 |
| 0 | 0 | 0 | 0 | 0 |
| 0 | 0 | 0 | 0 | 0 |
| 0 | 0 | 0 | 0 | 0 |
| 2.926311647 | 0.19923112 | 1.181660791 | 0 | 0 |
| 0 | 0 | 0.01391344 | 0.000717581 | 0 |
| 0 | 0 | 0 | 0 | 0 |
| 0 | 0 | 0.017661238 | 0.002113224 | 0 |
| 0 | 0 | 0 | 0 | 0 |
| 0 | 0 | 0 | 0.001207418 | 0 |
| 0 | 0 | 0 | 0 | 0 |
| 0 | 0.091289744 | 0.203715822 | 0 | 0 |
| 0.008413238 | 0 | 0.000859427 | 0.008226662 | 0.000494011 |
| 0 | 0 | 0 | 0 | 0 |
| 0 | 0 | 0 | 0.001775009 | 0 |
| 0 | 0 | 0 | 0 | 0 |
| 0 | 0 | 0 | 0 | 0 |
| 0 | 0 | 0 | 0 | 0 |
| 0 | 0.032437014 | 0 | 0 | 0 |
| 0 | 0.671816988 | 0.037278272 | 0 | 0 |
| 0 | 0 | 0 | 0 | 0 |
| 0 | 0 | 0 | 0 | 0 |
| 0 | 0 | 0 | 0 | 0 |

| | | | | |
|---|---|---|---|---|
| 0 | 0 | 0 | 0 | 0 |
| 0 | 0 | 0 | 0 | 0 |
| 0 | 0 | 0 | 0 | 0 |
| 0 | 0 | 0.362981379 | 0 | 0 |
| 0 | 0 | 0 | 0 | 0 |
| 0 | 0.194456185 | 0.286488233 | 0 | 0 |
| 0 | 0 | 0 | 0 | 0 |
| 0 | 0 | 0 | 0 | 0 |
| 0.144905834 | 0.005154297 | 0.059027641 | 0.026237489 | 0.481468435 |
| 0 | 0 | 0 | 0 | 0 |
| 0 | 0 | 0.001186531 | 0 | 0 |
| 0 | 0 | 0 | 0 | 0 |
| 0 | 0 | 0 | 0 | 0 |
| 0 | 0 | 0 | 0 | 0 |
| 0 | 0.305349653 | 0.000681427 | 0 | 0 |
| 0.018509838 | 0 | 0.003836864 | 0 | 0.423942667 |
| 0 | 0 | 0 | 0 | 0 |
| 0 | 0 | 0 | 0 | 0 |
| 0 | 0 | 0 | 0 | 0 |
| 0 | 0 | 0 | 0 | 0 |
| 0.013618036 | 0 | 0.007431883 | 0.001626675 | 0 |
| 0 | 0 | 0 | 0 | 0 |
| 0 | 0 | 0 | 0 | 0 |
| 0.074131494 | 0 | 0 | 0.007242944 | 0 |
| 0.010154496 | 0.008334297 | 0.028992538 | 0.01417641 | 0.126859319 |
| 3.581361899 | 0 | 1.415206457 | 0.8987814 | 0 |
| 0.227320425 | 0 | 1.432547394 | 0 | 0 |
| 0 | 0 | 0 | 0 | 0 |
| 0 | 0 | 2.132624751 | 0 | 0 |
| 0 | 0 | 0 | 0 | 0 |
| 0 | 0 | 0 | 0 | 0 |
| 0 | 0 | 0 | 0.009129686 | 0 |
| 0 | 0 | 0 | 0 | 0 |
| 0.004575785 | 0 | 0.016674944 | 0 | 0 |
| 0 | 0.038336419 | 0 | 0.012107897 | 0.176496512 |
| 0.027042157 | 0 | 0.005549006 | 0.00259358 | 0.011795042 |
| 2.653993747 | 0 | 0 | 0 | 0 |
| 1.335305077 | 14.68092894 | 0.97589639 | 3.382240755 | 0 |
| 0 | 0.006249792 | 0 | 0 | 0 |
| 0 | 0 | 0 | 0 | 0 |
| 0 | 0 | 0 | 0 | 0 |
| 0.007162867 | 0 | 0 | 0 | 0 |
| 0.047473223 | 0.027049125 | 0 | 0.046428677 | 0 |
| 0 | 0 | 0 | 0 | 0 |
| 0 | 0 | 0 | 0 | 0 |
| 0.016830032 | 0 | 0.001807089 | 0 | 0 |
| 0 | 0 | 0 | 0 | 0 |
| 0.045878753 | 0 | 0.005496101 | 0 | 0 |

| | | | | |
|---|---|---|---|---|
| 0 | 0 | 0.131735838 | 0 | 0 |
| 0 | 0 | 0 | 0 | 0 |
| 0 | 0 | 0 | 0 | 0 |
| 0 | 0 | 0 | 0 | 0 |
| 0 | 0 | 0.0068008 | 0 | 0 |
| 0 | 0 | 0.014220836 | 0 | 0 |
| 0 | 0 | 0 | 0 | 0 |
| 0 | 0 | 0 | 0 | 0 |
| 0 | 0 | 0 | 0 | 0 |
| 0.030858185 | 0 | 0 | 0 | 0 |
| 0 | 0 | 0.000743401 | 0 | 0 |
| 0.436674328 | 0 | 0.130861304 | 0.41745672 | 0 |
| 0.007230902 | 0 | 0 | 0 | 0 |
| 0 | 0 | 0 | 0 | 0 |
| 0 | 0.073900324 | 0.017078557 | 0.011763596 | 0.024111903 |
| 0 | 0 | 0 | 0 | 0 |
| 0 | 0 | 0 | 0 | 0 |
| 0 | 0 | 0 | 0 | 0 |
| 0.028922308 | 0 | 0.003348395 | 0 | 0 |
| 0 | 0 | 0.062681396 | 0 | 0 |
| 0 | 0.00066479 | 0 | 0 | 0 |
| 0 | 0 | 0 | 0 | 0 |
| 0.289950466 | 0.152513284 | 0.666493489 | 0.160126398 | 0 |
| 0 | 0 | 0 | 0 | 0 |
| 0 | 0 | 0 | 0 | 0 |
| 0 | 0 | 0 | 0 | 0 |
| 0 | 0 | 0 | 0 | 0 |
| 0 | 0 | 0 | 0 | 0 |
| 0 | 0 | 0 | 0 | 0 |
| 0 | 0 | 0 | 0 | 0 |
| 0 | 0 | 0 | 0 | 0 |
| 0.159658609 | 0.17316758 | 0.016912965 | 0.020498594 | 0 |
| 0 | 0 | 0 | 0 | 0 |
| 0.033331186 | 0.009898345 | 0.537266944 | 0.006754534 | 0 |
| 0.028569412 | 0 | 0.003774193 | 0.004798191 | 0.008316256 |
| 0.006922826 | 0.009917508 | 0.063493379 | 0 | 0 |
| 0 | 0.001138174 | 0 | 0 | 0 |
| 0 | 0.001226086 | 0 | 0 | 0 |
| 0 | 0.001178452 | 0 | 0 | 0 |
| 0.001210198 | 0 | 0 | 0 | 0 |
| 0 | 0 | 0.13762882 | 0 | 0 |
| 0 | 0 | 0 | 0 | 0 |
| 0 | 0 | 0 | 0 | 0 |
| 0 | 0 | 0 | 0 | 0 |
| 0.029624984 | 0 | 0.480546181 | 0.00750711 | 0.001424756 |
| 0 | 0 | 0 | 0 | 0 |
| 0 | 0 | 0.99054663 | 0.410092486 | 0 |
| 0.008005695 | 0 | 0 | 0.002366794 | 0 |

| | | | | |
|---|---|---|---|---|
| 0 | 0 | 0 | 0 | 0 |
| 0 | 0 | 0 | 0 | 0 |
| 0.094326342 | 0 | 0.002114915 | 0 | 0 |
| 0 | 0 | 0 | 0 | 0 |
| 0.004217054 | 0 | 0 | 0 | 0 |
| 0 | 0 | 0 | 0 | 0 |
| 0 | 0 | 0 | 0 | 0 |
| 0.011574513 | 0 | 0.025420684 | 0 | 0 |
| 0 | 0 | 0 | 0 | 0 |
| 0 | 0 | 0 | 0 | 0 |
| 0 | 0 | 0 | 0 | 0 |
| 0 | 0 | 0.001369375 | 0 | 0.002623785 |
| 0 | 0 | 0 | 0 | 0 |
| 0 | 0 | 0.132469942 | 0 | 0 |
| 0 | 0 | 0 | 0 | 0 |
| 0 | 0 | 0 | 0 | 0 |
| 0 | 0 | 0 | 0 | 0 |
| 0 | 0 | 0 | 0 | 0 |
| 0.021280654 | 0.573152785 | 0.586329895 | 0 | 0 |
| 0 | 0 | 0 | 0 | 0 |
| 0 | 0 | 0 | 0 | 0 |
| 0 | 0 | 0 | 0 | 0 |
| 0 | 0 | 0 | 0 | 0 |
| 0 | 0 | 0 | 0 | 0 |
| 0 | 0.00496817 | 0 | 0 | 0 |
| 0.007682928 | 0 | 0.003288215 | 0.001035383 | 0.14839031 |
| 0 | 0 | 0 | 0 | 0 |
| 0.276313925 | 0 | 0.033879038 | 0.039270573 | 0.006653663 |
| 0 | 0 | 0 | 0 | 0 |
| 0.74260126 | 0.190031983 | 0.186349044 | 0.802428048 | 0 |
| 0 | 0 | 0 | 0 | 0 |
| 0 | 0 | 0 | 0 | 0 |
| 0 | 0 | 0.004113934 | 0 | 0 |
| 0 | 0 | 0 | 0 | 0 |
| 0.021057071 | 0 | 0.003445846 | 0 | 0.010598588 |
| 0 | 0 | 0 | 0 | 0 |
| 0 | 0 | 0 | 0 | 0 |
| 0.001639757 | 0 | 0.023254366 | 0 | 0 |
| 0 | 0 | 0 | 0 | 0 |
| 0 | 0 | 0.107666688 | 0 | 0 |
| 9.024114863 | 10.04961903 | 0.584381005 | 23.81286657 | 7.331766021 |
| 3.339776374 | 0 | 0 | 0.993828251 | 0 |
| 0 | 0.074483392 | 0 | 0 | 0 |
| 0 | 0 | 0.31657683 | 0 | 0 |
| 0 | 0 | 0 | 0 | 0 |
| 0 | 0 | 0 | 0 | 0 |
| 0 | 0 | 0 | 0 | 0 |
| 0 | 0 | 0.137031354 | 0 | 0 |

| | | | | |
|---|---|---|---|---|
| 0 | 0 | 0 | 0 | 0 |
| 0.005545627 | 0 | 0.010079718 | 0 | 0 |
| 0 | 0 | 0.000588627 | 0 | 0 |
| 0 | 0 | 0 | 0 | 0 |
| 7.502392258 | 0.350888497 | 0 | 1.922301923 | 0 |
| 0 | 0 | 0 | 0 | 0 |
| 0 | 0 | 0 | 0 | 0 |
| 0 | 0 | 0 | 0 | 0 |
| 0.550498981 | 0.000683866 | 0.000838738 | 0.187167164 | 0.189633149 |
| 0.051003761 | 0 | 0 | 0 | 0 |
| 0 | 0.003868835 | 0 | 0 | 0 |
| 0 | 0 | 0 | 0 | 0 |
| 0 | 0.018189609 | 0.088963658 | 0 | 0 |
| 0.023528891 | 0.038724915 | 0.000802729 | 0.033917329 | 0.018264518 |
| 0 | 0.094545357 | 0 | 0.033526734 | 0 |
| 3.411828349 | 0.352012862 | 0.129931192 | 0.160589992 | 0 |
| 0.040721212 | 0.009203563 | 0.003473185 | 0.001038945 | 0 |
| 0 | 0 | 0 | 0 | 0 |
| 0 | 0 | 0 | 0 | 0 |
| 0 | 0 | 0 | 0 | 0 |
| 0 | 0 | 0 | 0 | 0 |
| 0 | 0 | 0 | 0 | 0 |
| 0 | 0 | 0 | 0 | 0 |
| 0 | 0 | 0 | 0 | 0 |
| 0 | 0 | 0 | 0 | 0 |
| 0 | 0 | 0 | 0 | 0 |
| 0 | 0 | 0.005134132 | 0.001355108 | 0 |
| 0.000969778 | 0 | 0.011756978 | 0 | 0.0021221 |
| 0 | 0 | 0 | 0 | 0 |
| 0 | 0 | 0 | 0 | 0 |
| 0 | 0 | 0 | 0 | 0 |
| 0 | 0 | 0 | 0 | 0 |
| 0 | 0 | 0 | 0 | 0 |
| 0 | 0 | 0 | 0 | 0 |
| 0 | 0.012057245 | 0 | 0 | 0 |
| 0 | 0 | 1.07245918 | 0 | 0.082708859 |
| 0 | 0.002463996 | 0 | 0 | 0 |
| 0 | 0 | 0 | 0 | 0 |
| 0 | 0 | 0 | 0 | 0 |
| 0 | 0 | 0 | 0 | 0 |
| 0 | 0.371680866 | 0.143394783 | 0 | 0 |
| 0 | 0 | 0.908305657 | 0.00119399 | 0 |
| 0 | 0 | 0 | 0 | 0 |
| 0 | 0 | 0 | 0 | 0 |
| 0 | 0 | 0 | 0 | 0 |
| 0 | 0 | 0 | 0 | 0 |
| 0 | 0 | 0 | 0 | 0 |
| 0 | 0 | 0 | 0 | 0 |

| | | | | |
|---|---|---|---|---|
| 0 | 0 | 0.134138198 | 0 | 0 |
| 0 | 0 | 0.027063276 | 0 | 0 |
| 0 | 0 | 0 | 0 | 0 |
| 0 | 0 | 0 | 0 | 0 |
| 0 | 0.041409674 | 0.044323671 | 0.015192234 | 0.059979115 |
| 0 | 0 | 0 | 0 | 0 |
| 0.124430122 | 0.000706245 | 0.005543584 | 0.03549733 | 0 |
| 0 | 0 | 0 | 0 | 0 |
| 0 | 0 | 0.004524263 | 0 | 0 |
| 0 | 0 | 0 | 0 | 0.233674819 |
| 0 | 0 | 0.162810631 | 0 | 0 |
| 0.049148495 | 0 | 0.003938975 | 0 | 0 |
| 0 | 0 | 0 | 0 | 0 |
| 0 | 0 | 0 | 0 | 0 |
| 0 | 0 | 0 | 0 | 0 |
| 0 | 0 | 0 | 0 | 0 |
| 0.027381803 | 0 | 7.656044238 | 0 | 1.923286986 |
| 0.015166593 | 0 | 0 | 0 | 0 |
| 0 | 0 | 0 | 0 | 0 |
| 0 | 0 | 0 | 0 | 0 |
| 0.016708926 | 0 | 0.000989588 | 0.010607327 | 0 |
| 0 | 0 | 0 | 0 | 0 |
| 0 | 0 | 0.035292214 | 0 | 0 |
| 0.007549549 | 0 | 0.004367626 | 0.002177502 | 0 |
| 0 | 0 | 0 | 0 | 0 |
| 0.013686014 | 0 | 0.001908451 | 0 | 0 |
| 0 | 0 | 0 | 0 | 0 |
| 0 | 0 | 0 | 0 | 0 |
| 0 | 0 | 0 | 0 | 0 |
| 0.523137524 | 0 | 0.212328387 | 0.398706364 | 1.540782816 |
| 0 | 0 | 0 | 0 | 0 |
| 0 | 0 | 0 | 0 | 0 |
| 0 | 0 | 0 | 0 | 0 |
| 0 | 0 | 0 | 0.073380595 | 0 |
| 0 | 0 | 0 | 0 | 0 |
| 0 | 0 | 0.272739636 | 0 | 0 |
| 0.154368681 | 0 | 0 | 0 | 0 |
| 0 | 0 | 0 | 0 | 0 |
| 0 | 0 | 0 | 0 | 0 |
| 0 | 0 | 0 | 0 | 0 |
| 0 | 0 | 0 | 0 | 0 |
| 0 | 0 | 0 | 0 | 0 |
| 0 | 0 | 0 | 0 | 0 |
| 0 | 0 | 0 | 0 | 0 |
| 0 | 0 | 0 | 0 | 0 |
| 0 | 0 | 0 | 0 | 0 |
| 0 | 0 | 0 | 0 | 0 |
| 0 | 0 | 0 | 0 | 0 |

| | | | | |
|---|---|---|---|---|
| 0.020606162 | 0.00591346 | 0.002262828 | 0.002603411 | 0.065535513 |
| 0 | 0.151199774 | 0 | 0 | 0 |
| 0 | 0 | 0 | 0 | 0 |
| 0 | 0 | 0 | 0 | 0 |
| 0 | 0 | 0 | 0 | 0 |
| 0 | 2.185670917 | 0 | 0 | 0 |
| 0 | 0 | 0.009233026 | 0 | 0 |
| 6.46352632 | 0.361392765 | 0.212860411 | 2.259851163 | 0 |
| 0.069042996 | 0.170418692 | 0.363381484 | 0.244369385 | 4.292179847 |
| 0.003031168 | 0.096928416 | 0.004184554 | 0.023896825 | 0.017675596 |
| 0 | 0 | 0 | 0 | 0 |
| 0 | 0 | 0 | 0 | 0 |
| 0 | 0 | 0 | 0 | 0 |
| 0.047890351 | 0 | 0.056635301 | 0 | 0 |
| 0 | 0 | 0 | 0 | 0 |
| 0 | 0 | 0 | 0 | 0 |
| 0.413592466 | 0 | 0 | 0 | 0.046980135 |
| 0 | 0 | 0 | 0 | 0 |
| 0 | 0 | 0 | 0 | 0 |
| 0 | 0 | 0 | 0 | 0 |
| 0 | 0 | 0 | 0 | 0 |
| 0 | 0 | 0 | 0 | 0 |
| 0 | 0 | 0 | 0 | 0 |
| 0 | 0 | 0 | 0 | 0 |
| 0 | 0 | 0.008363058 | 0 | 0 |
| 0 | 0 | 0 | 0 | 0 |
| 0 | 0 | 0 | 0 | 0 |
| 0 | 0 | 0 | 0 | 0 |
| 0 | 0 | 0 | 0 | 0 |
| 0 | 0 | 0 | 0 | 0 |
| 0 | 0 | 0 | 0.010163316 | 0 |
| 0 | 0 | 0 | 0 | 0 |
| 0 | 0 | 0 | 0 | 0 |
| 0 | 0 | 0 | 0 | 0 |
| 0 | 0 | 0 | 0 | 0 |
| 0 | 0 | 0 | 0 | 0 |
| 0 | 0 | 0 | 0 | 0 |
| 0 | 0 | 0 | 0 | 0 |
| 0 | 0 | 0 | 0 | 0 |
| 0.000593406 | 0 | 0 | 0.052500194 | 0 |
| 0 | 0 | 0 | 0 | 0 |
| 0 | 0 | 0 | 0.346698536 | 0 |
| 0 | 0 | 0 | 0 | 0 |
| 0 | 0 | 0 | 0 | 0 |
| 0 | 0 | 0 | 0 | 0 |
| 0 | 0 | 0 | 0 | 0 |
| 0 | 0 | 0 | 0 | 0 |
| 0 | 0 | 0 | 0 | 0 |

| | | | | |
|---|---|---|---|---|
| 0 | 0 | 0 | 0 | 0 |
| 0 | 0.023270002 | 0 | 0 | 0 |
| 0 | 0 | 0 | 0 | 0 |
| 0 | 0 | 0 | 0 | 0 |
| 0.361785712 | 0.007462095 | 3.194561424 | 0.00394224 | 0 |
| 0 | 0 | 0 | 0 | 0 |
| 0 | 0 | 0.024002013 | 0 | 0 |
| 0.118460476 | 0 | 0.008635445 | 0.047026528 | 0.074032329 |
| 0 | 0 | 0.003537301 | 0 | 0 |
| 0 | 0 | 0 | 0 | 0 |
| 0 | 0 | 0 | 0 | 0 |
| 0 | 0 | 0 | 0 | 0 |
| 0 | 0 | 0 | 0 | 0 |
| 0 | 0 | 0 | 0 | 0 |
| 0.098828387 | 0 | 0 | 0 | 0.101843017 |
| 0 | 0 | 0.25560172 | 0 | 0 |
| 0 | 0 | 0 | 0 | 0 |
| 0 | 0 | 0 | 0 | 0 |
| 0 | 0 | 0 | 0 | 0 |
| 0 | 0 | 0 | 0 | 0 |
| 0 | 0 | 0 | 0 | 0 |
| 0 | 0 | 0 | 0 | 0 |
| 0 | 0 | 0 | 0 | 0 |
| 0.001963107 | 0 | 0.008623 | 0 | 0.011565443 |
| 0 | 0.319489594 | 0.34217355 | 0 | 0 |
| 0 | 0 | 0 | 0 | 0 |
| 0 | 0 | 1.776963821 | 0 | 0 |
| 0.003361042 | 0 | 0 | 0 | 0 |
| 0 | 0 | 0.001563017 | 0 | 0.001497406 |
| 0 | 0 | 0 | 0 | 0 |
| 0 | 0 | 0 | 0 | 0 |
| 0 | 0 | 0 | 0 | 0 |
| 0 | 0 | 0 | 0 | 0 |
| 0 | 0 | 0 | 0 | 0 |
| 0 | 0 | 0 | 0 | 0 |
| 0.001783494 | 0 | 0 | 0 | 0 |
| 0 | 0 | 0 | 0 | 0 |
| 0 | 0 | 0.170213469 | 0 | 0 |
| 0 | 0 | 0 | 0 | 0 |
| 0 | 0 | 0 | 0 | 0 |
| 0 | 0 | 0 | 0 | 0 |
| 0 | 0 | 0 | 0 | 0 |
| 0 | 0 | 0 | 0 | 0 |
| 0 | 0 | 0 | 0 | 0 |
| 0 | 0 | 0 | 0 | 0 |
| 0 | 0 | 0 | 0 | 0 |
| 0 | 0 | 0 | 0 | 0 |
| 0 | 0 | 0 | 0 | 0 |

| | | | | |
|---|---|---|---|---|
| 0 | 0.062040547 | 0.434128437 | 0.060395281 | 0 |
| 0 | 0 | 0 | 0 | 0 |
| 0 | 0 | 0 | 0 | 0 |
| 0 | 0 | 0 | 0 | 0 |
| 0 | 0 | 0 | 0 | 0 |
| 0.531657751 | 0.002335372 | 0 | 0.008282332 | 0.051587583 |
| 0 | 0 | 0 | 0 | 0 |
| 0 | 0 | 0 | 0 | 0 |
| 0 | 0 | 0 | 0 | 0 |
| 0 | 0 | 0 | 0 | 0 |
| 0 | 0 | 0 | 0 | 0 |
| 0 | 0 | 0 | 0 | 0 |
| 0 | 0 | 0 | 0 | 0 |
| 0 | 0 | 0 | 0 | 0 |
| 0 | 0 | 0 | 0 | 0 |
| 0 | 0 | 0 | 0 | 0 |
| 0 | 0 | 0 | 0 | 0 |
| 0 | 0 | 0 | 0 | 0 |
| 0 | 0 | 0.00125762 | 0 | 0 |
| 0 | 0 | 0 | 0 | 0 |
| 0.100351543 | 0.0746814 | 0 | 0.000856214 | 0 |
| 0 | 0 | 0 | 0 | 0 |
| 0.002468897 | 0 | 1.198218449 | 0.005046234 | 0 |
| 0 | 0 | 0.003529113 | 0 | 0 |
| 0 | 0 | 0 | 0 | 0 |
| 0 | 0 | 0 | 0 | 0 |
| 0 | 0 | 0 | 0 | 0 |
| 0 | 0 | 0 | 0 | 0 |
| 0 | 0 | 0.039221252 | 0 | 0 |
| 0 | 0 | 0 | 0 | 0 |
| 0 | 0 | 0 | 0 | 0 |
| 0 | 0 | 0 | 0 | 0 |
| 0.005908729 | 0.017174819 | 0 | 0.218060864 | 0.05643222 |
| 0 | 0 | 0 | 0 | 0 |
| 0.279203315 | 0 | 0 | 0 | 0 |
| 0.002886135 | 0 | 0.000905531 | 0 | 0 |
| 0 | 0 | 0 | 0 | 0 |
| 0 | 0 | 0 | 0 | 0 |
| 0.019552734 | 0 | 0.008379115 | 0 | 0 |
| 0 | 0 | 0 | 0 | 0 |
| 0 | 0 | 0 | 0 | 0 |
| 0 | 0 | 0 | 0 | 0 |
| 0 | 0.035615671 | 0.630481726 | 0.006227982 | 0 |
| 0 | 0 | 0 | 0 | 0 |
| 0 | 0 | 0 | 0 | 0 |
| 0 | 0 | 0.091622096 | 0.022402913 | 0 |
| 0 | 0 | 0 | 0 | 0 |
| 0 | 0 | 0 | 0 | 0 |

| | | | | |
|---|---|---|---|---|
| 0 | 0 | 0 | 0 | 0 |
| 0 | 0 | 0 | 0 | 0 |
| 0 | 0 | 0 | 0 | 0 |
| 0 | 0 | 0 | 0 | 0 |
| 0 | 0 | 0 | 0 | 0 |
| 0 | 0 | 0 | 0 | 0 |
| 0 | 0 | 0 | 0 | 0 |
| 0.000855883 | 0 | 0 | 0 | 0 |
| 0 | 0 | 0.020230141 | 0 | 0 |
| 0 | 0.005717742 | 0.054698389 | 0.003895738 | 0 |
| 0 | 0 | 0 | 0 | 0 |
| 0 | 0 | 0.019906955 | 0 | 0 |
| 0 | 0 | 0 | 0 | 0 |
| 0 | 0 | 0 | 0 | 0 |
| 0.007925388 | 0 | 0 | 0 | 0 |
| 0.013463716 | 0 | 0 | 0 | 0.369971702 |
| 0 | 0 | 0 | 0 | 0 |
| 0 | 0 | 0 | 0 | 0 |
| 0 | 0 | 0 | 0 | 0 |
| 0 | 0 | 0 | 0 | 0 |
| 0 | 0.001678734 | 0.067326321 | 0 | 0 |
| 0 | 0 | 0 | 0 | 0 |
| 0 | 0 | 0 | 0 | 0 |
| 0.005977982 | 0 | 0 | 0 | 0 |
| 0 | 0 | 0 | 0 | 0 |
| 0 | 0 | 0 | 0 | 0 |
| 0 | 0 | 0 | 0 | 0 |
| 0 | 0 | 0 | 0 | 0 |
| 0 | 0 | 0 | 0 | 0 |
| 0 | 0.008884617 | 0.156503521 | 0.00091675 | 0 |
| 0.591465391 | 0.033966037 | 0.141477569 | 0.991155683 | 0 |
| 0 | 0 | 0 | 0 | 0 |
| 0 | 0 | 0.002029853 | 0 | 0 |
| 0 | 0 | 0 | 0 | 0 |
| 0 | 0 | 0 | 0 | 0 |
| 0 | 0 | 0 | 0 | 0 |
| 0 | 0 | 0 | 0 | 0 |
| 0 | 0 | 0 | 0 | 0 |
| 0 | 0 | 0.02997602 | 0 | 0 |
| 0 | 0 | 0 | 0 | 0 |
| 0 | 0 | 0 | 0 | 0 |
| 0 | 0 | 0.322938637 | 0 | 0 |
| 0 | 0 | 0 | 0 | 0 |
| 0 | 0 | 0.008422627 | 0 | 0 |
| 0 | 0 | 0 | 0 | 0 |
| 0 | 0 | 0 | 0 | 0 |
| 0.369376577 | 0.037844283 | 0 | 0.00120732 | 3.960018787 |
| 0 | 0 | 0.003608114 | 0 | 0 |

| | | | | |
|---|---|---|---|---|
| 0 | 0 | 0 | 0 | 0 |
| 0 | 0 | 0 | 0 | 0 |
| 0 | 0 | 0 | 0 | 0 |
| 0 | 0 | 0 | 0 | 0 |
| 0.086492383 | 0 | 0.119554248 | 0.154009315 | 0 |
| 0 | 0 | 0 | 0 | 0 |
| 0 | 0 | 0 | 0 | 0 |
| 0 | 0 | 0 | 0 | 0 |
| 0 | 0 | 0 | 0 | 0 |
| 0 | 0 | 0 | 0 | 0 |
| 0 | 0 | 0.013207657 | 0 | 0 |
| 0.002620839 | 0 | 0.206380713 | 0.001878356 | 0 |
| 0.002672237 | 0 | 0.005598064 | 0 | 0 |
| 0 | 0 | 0 | 0 | 0 |
| 0.00272354 | 0 | 0 | 0 | 0 |
| 0 | 0 | 0 | 1.263495168 | 0 |
| 0.027120817 | 0 | 0 | 0 | 0 |
| 0.007235081 | 0 | 0 | 0 | 0 |
| 0.117086354 | 0.004092164 | 0 | 0.27568952 | 0 |
| 0.42400984 | 0.000843651 | 0 | 0.004952248 | 0 |
| 0 | 0 | 0 | 0 | 0 |
| 0 | 0 | 0 | 0 | 0 |
| 0 | 0 | 0 | 0 | 0 |
| 0 | 0 | 0 | 0 | 0 |
| 0 | 0 | 0 | 0 | 0 |
| 0 | 0 | 0 | 0 | 0 |
| 0.02906994 | 0 | 0.004311628 | 0.002728318 | 0 |
| 0 | 0 | 0 | 0 | 0 |
| 0 | 0 | 0 | 0 | 0 |
| 0 | 0 | 0 | 0 | 0 |
| 0.017760008 | 0 | 0 | 0 | 0.003061501 |
| 0.062633249 | 0 | 0.031471094 | 0 | 0 |
| 0 | 0 | 0 | 0 | 0 |
| 0 | 0 | 0 | 0 | 0 |
| 0 | 0 | 0 | 0 | 0 |
| 0 | 0 | 0 | 0 | 0 |
| 0 | 0 | 0 | 0 | 0 |
| 0.05381933 | 0.000717882 | 0.308512379 | 0.004148141 | 0.00582014 |
| 0 | 0.00172693 | 0.102300383 | 0 | 0 |
| 0.010969511 | 0 | 0 | 0 | 0 |
| 0.018459977 | 0 | 0.030456677 | 0 | 0.0010105 |
| 0 | 0 | 0.001523744 | 0 | 0 |
| 0 | 0 | 0 | 0 | 0 |
| 0.002675103 | 0 | 0 | 0.007322818 | 0 |
| 0 | 0 | 0 | 0 | 0 |
| 0 | 0 | 0 | 0 | 0 |
| 0 | 0 | 0 | 0 | 0 |
| 0 | 0 | 0 | 0 | 0 |

| | | | | |
|---|---|---|---|---|
| 0 | 0 | 0 | 0 | 0 |
| 0 | 0 | 0 | 0 | 0 |
| 0 | 0 | 0 | 0 | 0 |
| 0.00420528 | 0 | 0.001847181 | 0 | 0.003539283 |
| 0 | 0.14744892 | 0 | 0 | 0.342122835 |
| 0.003076181 | 0.002670835 | 0.001474059 | 0.490545607 | 0 |
| 0 | 0 | 0 | 0.002383158 | 0 |
| 0 | 0 | 0 | 0 | 0 |
| 0 | 0 | 0 | 0 | 0 |
| 0 | 0 | 0 | 0 | 0 |
| 0 | 0 | 0.184688875 | 0 | 0 |
| 0 | 0 | 0 | 0 | 0 |
| 0.010598939 | 0.03341235 | 0.061541389 | 0.006528037 | 0 |
| 0 | 0 | 0 | 0 | 0 |
| 0 | 0 | 0 | 0 | 0 |
| 0 | 0 | 0 | 0 | 0 |
| 0 | 0 | 0 | 0 | 0 |
| 0 | 0 | 0 | 0 | 0 |
| 0 | 0 | 0 | 0 | 0 |
| 0 | 0 | 0 | 0 | 0 |
| 0 | 0 | 0 | 0 | 0 |
| 0 | 0 | 0 | 0 | 0 |
| 0 | 0 | 0 | 0.009742044 | 0 |
| 0.029850036 | 0.34498842 | 0 | 0.044293207 | 0 |
| 0 | 0 | 0 | 0 | 0 |
| 0 | 0 | 0 | 0 | 0 |
| 0 | 0 | 0 | 0 | 0 |
| 0 | 0 | 0 | 0 | 0 |
| 0 | 0 | 0 | 0 | 0 |
| 0 | 0 | 0 | 0 | 0 |
| 0.01993789 | 0.006966497 | 0 | 0.056228561 | 0 |
| 0 | 0 | 0 | 0 | 0 |
| 0 | 0 | 0 | 0 | 0 |
| 0 | 0 | 0 | 0 | 0 |
| 0 | 0 | 0 | 0 | 0 |
| 0 | 0 | 0 | 0 | 0 |
| 0 | 0 | 0 | 0 | 0 |
| 1.575376938 | 0 | 1.104875572 | 0 | 0 |
| 0 | 0.007122086 | 0.009759095 | 0 | 0 |
| 0 | 0 | 0 | 0 | 0 |
| 0 | 0 | 0 | 0 | 0 |
| 0 | 0 | 0 | 0 | 0 |
| 0 | 0 | 0 | 0 | 0 |
| 0 | 0 | 0 | 0 | 0 |
| 0 | 0 | 0 | 0 | 0 |
| 0 | 0.082109599 | 0.141834473 | 1.061398061 | 0 |
| 0 | 0 | 0 | 0 | 0 |
| 0 | 0 | 0 | 0 | 0 |

| | | | | |
|---|---|---|---|---|
| 0 | 0 | 0 | 0 | 0 |
| 0 | 0 | 0 | 0 | 0 |
| 0 | 0.445042414 | 0.137490243 | 0 | 0 |
| 0 | 0 | 0 | 0 | 0 |
| 0.041003354 | 0 | 0 | 0 | 0 |
| 0 | 0 | 0 | 0 | 0 |
| 0.00125465 | 0 | 0 | 0 | 0 |
| 0 | 0 | 0 | 0 | 0 |
| 0 | 0 | 0 | 0 | 0 |
| 0 | 0 | 0 | 0 | 0 |
| 0 | 0 | 0.029909577 | 0 | 0 |
| 0 | 0 | 0.001094488 | 0 | 0.001048544 |
| 0 | 0 | 0 | 0 | 0 |
| 0 | 0 | 0 | 0 | 0 |
| 0 | 0 | 0 | 0 | 0 |
| 0 | 0 | 0 | 0 | 0 |
| 0 | 0.002635849 | 0 | 0 | 0 |
| 0 | 0 | 0 | 0 | 0 |
| 0 | 0 | 0 | 0 | 0 |
| 0 | 0 | 0.008166372 | 0.00087244 | 0 |
| 0 | 0 | 0 | 0 | 0 |
| 0 | 0 | 0 | 0 | 0 |
| 0 | 0 | 0 | 0 | 0 |
| 0 | 0 | 0 | 0 | 0 |
| 0 | 0 | 0 | 0 | 0 |
| 0 | 0 | 0 | 0 | 0 |
| 0 | 0 | 0 | 0 | 0 |
| 0 | 0.001754455 | 0.004518736 | 0 | 0 |
| 0 | 0 | 0 | 0 | 0 |
| 0 | 0 | 0 | 0 | 0 |
| 0.010457878 | 0 | 0.001002251 | 0.003747581 | 0 |
| 0 | 0 | 0 | 0 | 0 |
| 0.013688091 | 0.097255782 | 0.006788342 | 0.073682415 | 0 |
| 0.004512175 | 0.051173777 | 0 | 0 | 0 |
| 0 | 0 | 0 | 0 | 0 |
| 0 | 0 | 0 | 0 | 0 |
| 0 | 0 | 0 | 0 | 0 |
| 0 | 0 | 0 | 0 | 0 |
| 0 | 0 | 0 | 0 | 0 |
| 0 | 0 | 0 | 0 | 0 |
| 0 | 0 | 0 | 0 | 0 |
| 0.444993812 | 0 | 0.104513811 | 0.063004384 | 0.372837907 |
| 0.01947235 | 0 | 0.060456164 | 0.022096742 | 0 |
| 0 | 0 | 0 | 0 | 0 |
| 0 | 0 | 0 | 0 | 0 |
| 0.01406945 | 0.017559824 | 0.097757072 | 0 | 0 |
| 0 | 0 | 0 | 0 | 0 |
| 0 | 0 | 0 | 0 | 0 |

| | | | | |
|---|---|---|---|---|
| 0 | 0 | 0 | 0 | 0 |
| 0.448333493 | 0 | 0.002851131 | 0 | 0 |
| 0.036826948 | 0.104739312 | 0 | 0 | 0 |
| 0 | 0 | 0 | 0 | 0 |
| 0.223128609 | 0.00103246 | 5.350464504 | 0 | 0 |
| 0 | 0 | 0 | 0.104252035 | 0 |
| 0 | 0 | 0 | 0 | 0 |
| 0 | 0 | 0 | 0 | 0 |
| 0 | 0 | 0 | 0 | 0 |
| 0 | 0 | 0 | 0 | 0 |
| 0 | 0 | 0 | 0 | 0 |
| 0 | 0 | 0 | 0 | 0 |
| 0 | 0 | 0 | 0 | 0 |
| 0.129506102 | 0 | 0 | 0 | 0 |
| 0 | 0 | 0 | 0 | 0 |
| 0 | 0 | 0 | 0 | 0 |
| 0 | 0 | 0 | 0 | 0 |
| 0 | 0 | 0 | 0 | 0 |
| 0 | 0 | 0 | 0 | 0 |
| 0 | 0 | 0 | 0 | 0 |
| 0 | 0 | 0 | 0 | 0 |
| 0.001306898 | 0 | 0 | 0 | 0 |
| 0 | 0 | 0 | 0 | 0 |
| 0 | 0 | 0 | 0 | 0 |
| 0 | 0 | 0.005489473 | 0 | 0 |
| 0.012347487 | 0 | 0.021694672 | 0.047787011 | 0 |
| 0 | 0 | 0 | 0 | 0 |
| 0 | 0 | 0.002487088 | 0.130480932 | 0.008797613 |
| 0 | 0 | 0 | 0 | 0 |
| 0 | 0 | 0 | 0 | 0 |
| 0 | 0 | 0 | 0 | 0 |
| 0 | 0 | 0 | 0 | 0 |
| 0 | 0 | 0 | 0 | 0 |
| 0 | 0 | 0 | 0 | 0 |
| 0.000638057 | 0 | 0 | 0 | 0 |
| 0 | 0 | 0 | 0 | 0 |
| 0 | 0 | 0 | 0 | 0 |
| 0.046481896 | 0 | 0 | 0 | 0 |
| 0 | 0.029686697 | 0.044228846 | 3.112604832 | 0 |
| 0 | 0 | 0 | 0 | 0 |
| 0 | 0 | 0 | 0 | 0 |
| 0 | 0 | 0.006418738 | 0 | 0 |
| 0.015771701 | 0.005648559 | 0.002177297 | 0.001480231 | 0.045131299 |
| 0 | 0 | 0 | 0 | 0 |
| 0 | 0 | 0 | 0 | 0 |
| 0 | 0 | 0 | 0 | 0 |
| 0.006548925 | 0 | 0.013928961 | 0 | 0 |
| 0 | 0 | 0 | 0 | 0 |

| | | | | |
|---|---|---|---|---|
| 0 | 0 | 0 | 0 | 0 |
| 0.021746153 | 0.022845622 | 0.017766827 | 0.007429075 | 0.034957163 |
| 0 | 0 | 0 | 0 | 0 |
| 0 | 0 | 0 | 0 | 0 |
| 0 | 0 | 0 | 0 | 0 |
| 0 | 0 | 0 | 0 | 0 |
| 0 | 0 | 0 | 0 | 0 |
| 0 | 0 | 0 | 0 | 0 |
| 0 | 0 | 0 | 0 | 0 |
| 0 | 0 | 0 | 0 | 0 |
| 0 | 0 | 0 | 0 | 0 |
| 0 | 0 | 0 | 0 | 0 |
| 0 | 0.003842021 | 0.013126583 | 0 | 0 |
| 0 | 0 | 0 | 0 | 0 |
| 0.005422835 | 0 | 0 | 0 | 0 |
| 0 | 0 | 0 | 0 | 0 |
| 0 | 0 | 0 | 0 | 0 |
| 0 | 0 | 0 | 0 | 0 |
| 0.003899389 | 0 | 0.005006697 | 0 | 0 |
| 0 | 0 | 0 | 0 | 0 |
| 0 | 0 | 0 | 0 | 0 |
| 0.263619775 | 0 | 0 | 0.008047289 | 0.000672333 |
| 0.022507802 | 0 | 0 | 0 | 0 |
| 0.005485396 | 0 | 0 | 0 | 0 |
| 0 | 0 | 0 | 0 | 0 |
| 0 | 0 | 0 | 0 | 0 |
| 0 | 0 | 0 | 0 | 0 |
| 0 | 0 | 0 | 0 | 0 |
| 0 | 0 | 0 | 0 | 0 |
| 0 | 0 | 0 | 0 | 0 |
| 0 | 0 | 0 | 0 | 0 |
| 0 | 0 | 0 | 0 | 0 |
| 0 | 0 | 0 | 0 | 0 |
| 0 | 0 | 0 | 0 | 0 |
| 0.005318935 | 0 | 0 | 0.019378083 | 0 |
| 0 | 0 | 0 | 0 | 0 |
| 0 | 0 | 0 | 0 | 0 |
| 0 | 0 | 0 | 0 | 0 |
| 0.027123371 | 0.003174544 | 0.094844907 | 0 | 0.000895208 |
| 0 | 0 | 0 | 0 | 0 |
| 0.269900862 | 0.003131039 | 0.065980089 | 0.02098996 | 0.183009256 |
| 0 | 0 | 0 | 0 | 0 |
| 0 | 0 | 0 | 0 | 0 |
| 0 | 0 | 0 | 0 | 0 |
| 0 | 0 | 0 | 0 | 0 |
| 0.373988359 | 0.522732818 | 0.032375448 | 2.826100322 | 4.139658094 |
| 0 | 0 | 0 | 0 | 0 |
| 0 | 0 | 0 | 0 | 0 |

| | | | | |
|---|---|---|---|---|
| 0.001701967 | 0 | 0 | 0 | 0 |
| 0 | 0 | 0 | 0 | 0 |
| 0 | 0 | 0 | 0 | 0 |
| 0 | 0 | 0.003830326 | 0 | 0 |
| 0.000621114 | 0 | 0 | 0 | 0 |
| 0 | 0 | 0 | 0 | 0 |
| 0 | 0 | 0 | 0 | 0 |
| 0.002360459 | 0 | 0 | 0 | 0 |
| 0 | 0 | 0.020794919 | 0 | 0 |
| 0 | 0.002231069 | 0.00093817 | 0 | 0.005168033 |
| 0 | 0.014454025 | 0.033393407 | 0 | 0 |
| 0 | 0 | 0 | 0 | 0 |
| 0.002609495 | 0 | 0.003438683 | 0 | 0 |
| 0 | 0 | 0 | 0 | 0 |
| 0 | 0 | 0 | 0 | 0 |
| 0 | 0 | 0 | 0 | 0 |
| 0 | 0 | 0 | 0 | 0 |
| 0 | 0 | 0 | 0 | 0 |
| 0 | 0 | 0 | 0 | 0 |
| 0.085937476 | 0.249829402 | 0.511948211 | 0.045472214 | 0 |
| 0 | 0 | 0 | 0 | 0 |
| 0 | 0 | 0 | 0 | 0 |
| 0 | 0 | 0 | 0 | 0 |
| 0 | 0 | 0 | 0 | 0 |
| 0 | 0 | 0 | 0 | 0 |
| 0 | 0 | 0 | 0 | 0 |
| 0 | 0 | 0 | 0 | 0 |
| 0 | 0 | 0 | 0 | 0 |
| 0 | 0 | 0 | 0 | 0 |
| 0 | 0 | 0 | 0.000978736 | 0 |
| 0 | 0 | 0 | 0 | 0 |
| 0 | 0 | 0 | 0 | 0 |
| 0 | 0 | 0 | 0 | 0 |
| 0 | 0 | 0 | 0 | 0 |
| 0 | 0 | 0 | 0 | 0 |
| 0 | 0 | 0 | 0 | 0 |
| 0 | 0 | 0 | 0 | 0 |
| 0 | 0 | 0 | 0 | 0 |
| 0 | 0 | 0 | 0 | 0 |
| 0 | 0 | 0 | 0 | 0 |
| 0 | 0 | 0 | 0 | 0 |
| 0 | 0 | 0 | 0 | 0 |
| 0.364509623 | 0 | 0.013399192 | 0 | 0 |
| 0 | 0 | 0 | 0 | 0 |
| 0.003708556 | 0 | 0.032963164 | 0 | 0 |
| 0 | 0 | 0 | 0 | 0 |
| 0 | 0 | 0 | 0 | 0 |
| 0 | 0 | 0 | 0 | 0.010843668 |

| | | | | |
|---|---|---|---|---|
| 0 | 0 | 0 | 0 | 0 |
| 0 | 0 | 0 | 0 | 0 |
| 0 | 0 | 0 | 0 | 0 |
| 0 | 0 | 0 | 0 | 0 |
| 0 | 0 | 0 | 0 | 0 |
| 0.004071443 | 0.000972112 | 0.240789193 | 0.018616867 | 0 |
| 0 | 0 | 0 | 0 | 0 |
| 0 | 0 | 0 | 0 | 0 |
| 0 | 0 | 0 | 0 | 0 |
| 0 | 0 | 0 | 0 | 0 |
| 0 | 0 | 0 | 0 | 0 |
| 0 | 0 | 0 | 0 | 0 |
| 0 | 0 | 0 | 0 | 2.071190925 |
| 0 | 0 | 0 | 0 | 0 |
| 0 | 0 | 0 | 0 | 0 |
| 0 | 0 | 0 | 0 | 0 |
| 0 | 0 | 0.312206774 | 0 | 0 |
| 0 | 0 | 0 | 0 | 0 |
| 0 | 0 | 0 | 0 | 0 |
| 0 | 0 | 0 | 0 | 0 |
| 0 | 0 | 0 | 0 | 0 |
| 0 | 0 | 0 | 0 | 0 |
| 0 | 0 | 0 | 0 | 0 |
| 0 | 0 | 0 | 0 | 0 |
| 0 | 0 | 0 | 0 | 0 |
| 0 | 0 | 0 | 0 | 0 |
| 0 | 0 | 0 | 0 | 0 |
| 4.244124326 | 0 | 0.066158404 | 0 | 0 |
| 0 | 0 | 0 | 0 | 0 |
| 0.020903582 | 0 | 0 | 0 | 0 |
| 0 | 0 | 0 | 0 | 0 |
| 0 | 0 | 0 | 0 | 0 |
| 0 | 0 | 0 | 0 | 0 |
| 0 | 0 | 0 | 0 | 0 |
| 0 | 0 | 0 | 0 | 0 |
| 0 | 0 | 0 | 0 | 0 |
| 0 | 0 | 0 | 0 | 0 |
| 0 | 0 | 0 | 0 | 0 |
| 0 | 0 | 0 | 0 | 0 |
| 0 | 0 | 0 | 0 | 0 |
| 0 | 0 | 0 | 0 | 0 |
| 0 | 0 | 0 | 0 | 0 |
| 0 | 0 | 0 | 0 | 0 |
| 0 | 0 | 0 | 0 | 0 |
| 0 | 0 | 0 | 0 | 0 |
| 0 | 0 | 0 | 0 | 0 |
| 0 | 0 | 0 | 0 | 0 |
| 0 | 0 | 0 | 0 | 0 |

| | | | | |
|---|---|---|---|---|
| 0 | 0 | 0 | 0 | 0 |
| 0.057981689 | 0 | 0 | 0 | 0.104963962 |
| 0 | 0 | 0 | 0 | 0 |
| 0 | 0 | 0 | 0 | 0 |
| 0 | 0 | 0 | 0 | 0 |
| 0 | 0 | 0 | 0 | 0 |
| 0 | 0 | 0 | 0 | 0 |
| 0 | 0 | 0 | 0 | 0 |
| 0 | 0 | 0 | 0 | 0 |
| 0 | 0 | 0 | 0 | 0 |
| 0 | 0.007410614 | 0 | 0 | 0 |
| 0 | 0 | 0 | 0 | 0 |
| 0 | 1.900955133 | 0 | 0 | 0 |
| 0 | 0 | 0 | 0 | 0 |
| 0 | 0 | 0 | 0 | 0 |
| 0 | 0 | 0 | 0 | 0 |
| 0 | 0 | 0 | 0 | 0 |
| 0 | 0 | 0 | 0 | 0 |
| 0 | 0 | 0 | 0 | 0 |
| 0 | 0 | 0.00191983 | 0 | 0 |
| 0 | 0.002024973 | 0 | 0.00159196 | 0 |
| 0 | 0 | 0 | 0 | 0 |
| 0 | 0 | 0 | 0 | 0 |
| 0.942852338 | 9.74547795 | 0.1826953 | 5.46370215 | 0 |
| 0 | 0 | 0 | 0 | 0 |
| 0 | 0 | 0 | 0 | 0 |
| 0 | 0 | 0 | 0 | 0 |
| 0 | 0 | 0 | 0.063628692 | 0 |
| 0 | 0 | 0 | 0 | 0 |
| 0 | 0 | 0 | 0 | 0 |
| 0 | 0 | 0.001880444 | 0 | 0 |
| 0 | 0 | 0 | 0 | 0 |
| 0 | 0 | 0 | 0 | 0 |
| 0 | 0 | 0 | 0 | 0 |

| PHL018-0202_V2 | PHL018-0126_V1 | PHL018-0203_V2 | PHL018-0154_V1 | PHL018-0163_V2 |
|---|---|---|---|---|
| REF | CTRL | EYCF | EYCF | CTRL |
| 0.011720626 | 0.12686034 | 0.097132386 | 0.085841252 | 0.003333425 |
| 2.285410316 | 0.147745692 | 0 | 0.203071366 | 1.749229396 |
| 0 | 13.74066911 | 0 | 10.8193739 | 0 |
| 0 | 0 | 0 | 0 | 0 |
| 0 | 0 | 0 | 0 | 0 |
| 0 | 0 | 0 | 0 | 0 |
| 0 | 0 | 0 | 0 | 0 |
| 0 | 0 | 0 | 0 | 0 |
| 0 | 0 | 0 | 0 | 0 |
| 0 | 0 | 0 | 0 | 0 |
| 0 | 0 | 0 | 0 | 0 |
| 0 | 0 | 0 | 0 | 0 |
| 0 | 0 | 0 | 0 | 0 |
| 0.024590541 | 0.968465272 | 0 | 2.327929296 | 1.673716416 |
| 0 | 0 | 0 | 0 | 0 |
| 0 | 0 | 0 | 0 | 0 |
| 0 | 0 | 0.055946965 | 0 | 0 |
| 0 | 0 | 0 | 0 | 0 |
| 0 | 0 | 0 | 0 | 0 |
| 0 | 0 | 0 | 0 | 0 |
| 0 | 0 | 0 | 0 | 0 |
| 0 | 0 | 0 | 0 | 0 |
| 0 | 0 | 0 | 0 | 0 |
| 0 | 0 | 0 | 0 | 0 |
| 0 | 0 | 0 | 0 | 0 |
| 0 | 0 | 0 | 0 | 0 |
| 0 | 0 | 0 | 0 | 0 |
| 0 | 0 | 0 | 0 | 0 |
| 0 | 0 | 0 | 0 | 0 |
| 0 | 0 | 0 | 0 | 0 |
| 0 | 0 | 0 | 0 | 0 |
| 0 | 0 | 0 | 0 | 0 |
| 0 | 0 | 0 | 0 | 0 |
| 0 | 0 | 0 | 0 | 0 |
| 0 | 0 | 0 | 0 | 0 |
| 5.887963999 | 3.426688683 | 0 | 0 | 1.879773226 |
| 0 | 0 | 0 | 0 | 0 |
| 0 | 0 | 0 | 0 | 0 |
| 0 | 0 | 0 | 0 | 0 |
| 0 | 0 | 0 | 0 | 0 |
| 0 | 0 | 0 | 0 | 0 |
| 0.001140704 | 0.030075441 | 0.006556679 | 0.002359094 | 0 |
| 0 | 0.011015064 | 0.030265461 | 0.006712716 | 0.145053313 |
| 0.003740942 | 0.000976559 | 0.001254321 | 0.002109997 | 0.01166861 |
| 0.00247233 | 0 | 0 | 0 | 0.001835489 |
| 0 | 0 | 0 | 0 | 0 |

| | | | | |
|---|---|---|---|---|
| 0 | 0 | 0 | 0 | 0 |
| 0.014628971 | 0.276839714 | 0.009675672 | 0 | 0 |
| 0.000828477 | 0.078636051 | 0.186480663 | 0.000934568 | 0.003608418 |
| 0.341806624 | 1.452100749 | 0.269471827 | 0.908043773 | 0.420264962 |
| 0.003789759 | 0 | 0 | 0.005029485 | 0 |
| 0 | 0 | 0 | 0 | 0 |
| 0 | 0.007064941 | 0 | 0.02859125 | 0 |
| 0.00225223 | 0 | 0 | 0 | 0 |
| 0 | 0 | 0 | 0 | 0 |
| 0 | 0 | 0 | 0 | 0 |
| 0.00059206 | 0 | 0.001361246 | 0 | 0 |
| 0.649996325 | 0 | 0 | 0.616783929 | 0.391184022 |
| 0 | 0 | 0 | 0 | 0 |
| 0.101988011 | 0.06264376 | 0 | 0.012667451 | 0 |
| 0 | 0 | 0.001213118 | 0.011903998 | 0 |
| 0 | 0 | 0 | 0 | 0 |
| 0 | 0 | 0 | 0.314010532 | 0.034730539 |
| 0 | 0 | 0 | 0 | 0 |
| 0 | 0 | 0 | 0 | 0 |
| 0 | 0 | 0 | 0 | 0 |
| 0.29665174 | 0.172616617 | 0.143137598 | 0.350225836 | 0.373901654 |
| 0.006562556 | 0.02294799 | 0.015088425 | 0.551604549 | 0.083090465 |
| 0 | 0 | 0 | 0 | 0 |
| 0 | 0 | 0 | 0 | 0 |
| 0 | 0 | 0 | 0 | 0 |
| 0 | 0 | 0 | 0 | 0 |
| 0 | 0 | 0 | 0.0006167 | 0 |
| 0 | 0 | 0 | 0 | 0 |
| 0.000780968 | 0 | 0 | 0 | 0 |
| 0 | 0 | 0 | 0.000647245 | 0 |
| 0 | 0.296863077 | 0 | 0 | 0 |
| 0 | 0 | 0 | 0 | 0 |
| 0 | 0 | 0 | 0 | 0 |
| 0 | 0 | 0 | 0 | 0 |
| 0 | 0 | 0 | 0 | 0 |
| 0 | 0 | 0 | 0 | 0 |
| 0 | 0 | 0 | 0 | 0 |
| 0 | 0 | 0 | 0.009394361 | 2.025697901 |
| 0 | 0 | 0 | 0 | 0 |
| 0 | 0 | 0 | 0 | 0 |
| 0 | 0 | 0 | 0 | 0 |
| 0 | 0 | 0 | 0 | 0 |
| 0 | 0 | 0 | 0 | 0 |
| 0 | 0 | 0 | 0 | 0 |
| 0 | 0 | 0 | 0 | 0 |
| 0 | 0 | 0 | 0 | 0 |
| 0.069436937 | 0 | 0 | 0 | 0 |
| 0 | 0 | 0 | 1.593624986 | 0 |

| | | | | |
|---|---|---|---|---|
| 0 | 0 | 0 | 0 | 0 |
| 0 | 0 | 0.178354021 | 0.001084797 | 0 |
| 0 | 0 | 0 | 0 | 0 |
| 0 | 0 | 0 | 0 | 0 |
| 0 | 0 | 0 | 0 | 0 |
| 0 | 0 | 0 | 0 | 0 |
| 0 | 0 | 0 | 0 | 0 |
| 0 | 0 | 0 | 0 | 0 |
| 0 | 0.051986792 | 0 | 0.00249611 | 0 |
| 0 | 0 | 0 | 0 | 0 |
| 0 | 0 | 0 | 0 | 0 |
| 0 | 0 | 0 | 0 | 0 |
| 0 | 0 | 0 | 0 | 0 |
| 0 | 0 | 0 | 0 | 0 |
| 0 | 0 | 0 | 0 | 0 |
| 0 | 0 | 0 | 0 | 0 |
| 0 | 0 | 0 | 0 | 0 |
| 0 | 0 | 0 | 0 | 0 |
| 0.183662581 | 0.021724206 | 0 | 0 | 0.003941612 |
| 0 | 0 | 0 | 0 | 0 |
| 0 | 0 | 0 | 0 | 0 |
| 0 | 0 | 0 | 0 | 0 |
| 0 | 0 | 0 | 0 | 0 |
| 0 | 0 | 0 | 0 | 0 |
| 0 | 0 | 0 | 0 | 0 |
| 0 | 0 | 0 | 0 | 0 |
| 0 | 0 | 0 | 0 | 0 |
| 0 | 0 | 0 | 0.002240138 | 0 |
| 0 | 0 | 0 | 0 | 0 |
| 0 | 0 | 0 | 0 | 0 |
| 0.112755433 | 0 | 0.155985682 | 0 | 0 |
| 0 | 0 | 0 | 0 | 0.006432349 |
| 0 | 0 | 0 | 0 | 0 |
| 0 | 0 | 0 | 0 | 0 |
| 0 | 0 | 0 | 0 | 0 |
| 0 | 0 | 0 | 0 | 0 |
| 0 | 0 | 0 | 0 | 0 |
| 0 | 0 | 0 | 0 | 0 |
| 0 | 0 | 0 | 0 | 0.018443888 |
| 0 | 0 | 0 | 0 | 0 |
| 0 | 0 | 0 | 0 | 0 |
| 0 | 0 | 0 | 0 | 0 |
| 0 | 0 | 0 | 0.017365253 | 0 |
| 0 | 0 | 0 | 0 | 0 |
| 0 | 0 | 0 | 0 | 0 |
| 0 | 0 | 0 | 0 | 0 |
| 0 | 0 | 0 | 0 | 0 |
| 0 | 0 | 0 | 0 | 0 |

| | | | | |
|---|---|---|---|---|
| 0 | 0 | 0 | 0 | 0 |
| 0 | 0 | 0 | 0 | 0 |
| 0 | 0 | 0 | 0 | 0 |
| 0 | 0 | 0 | 0 | 0 |
| 0 | 0 | 0 | 0 | 0.015608306 |
| 0 | 0 | 0 | 0 | 0 |
| 0 | 0 | 0 | 0 | 0 |
| 0 | 0 | 0 | 0 | 0 |
| 0 | 0 | 0 | 0 | 0 |
| 0 | 0 | 0 | 0 | 0 |
| 0 | 0 | 0 | 0 | 0 |
| 0 | 0 | 0 | 0 | 0 |
| 0 | 0 | 0 | 0 | 0 |
| 0 | 0 | 0 | 0 | 0 |
| 0 | 0 | 0 | 0 | 0 |
| 0 | 0 | 0 | 0 | 0 |
| 0 | 0 | 0 | 0 | 0 |
| 0 | 0 | 0 | 0 | 0 |
| 0 | 0 | 0 | 0 | 0 |
| 0 | 0 | 0 | 0 | 0 |
| 0 | 0 | 0.001411781 | 0 | 0 |
| 0 | 0 | 0 | 0 | 0 |
| 0 | 0 | 0 | 0 | 0 |
| 0 | 0 | 0 | 0 | 0 |
| 0 | 0 | 0 | 0 | 0.005028257 |
| 0 | 0 | 0 | 0 | 0 |
| 0 | 0 | 0 | 0 | 0 |
| 0 | 0 | 0 | 0 | 0 |
| 0 | 0 | 0 | 0 | 0 |
| 0 | 0 | 0 | 0 | 0 |
| 0 | 0 | 0.008199793 | 0 | 0 |
| 0 | 0 | 0 | 0.025908314 | 0 |
| 0 | 0 | 0 | 0 | 0 |
| 0 | 0 | 0 | 0 | 0 |
| 0 | 0 | 0 | 0 | 0 |
| 0 | 0 | 0 | 0 | 0 |
| 0 | 0 | 0 | 0 | 0 |
| 0 | 0 | 0 | 0 | 0 |
| 0 | 0 | 0 | 0 | 0 |
| 0 | 0 | 0 | 0 | 0 |
| 0 | 0 | 0 | 0 | 0 |
| 0 | 0 | 0 | 0 | 0 |
| 0 | 0 | 0 | 0 | 0 |
| 0 | 0 | 0 | 0 | 0 |
| 0 | 0 | 0 | 0 | 0 |
| 0 | 0 | 0 | 0 | 0 |
| 0 | 0 | 0 | 0 | 0 |
| 0 | 0 | 0 | 0 | 0 |

| | | | | |
|---|---|---|---|---|
| 0 | 0 | 0 | 0 | 0 |
| 0 | 0 | 0 | 0 | 0 |
| 0 | 0 | 0 | 0 | 0 |
| 0 | 0 | 0 | 0 | 0 |
| 0 | 0 | 0 | 0 | 0 |
| 0 | 0 | 0 | 0 | 0 |
| 0 | 0 | 0 | 0 | 0 |
| 0 | 0 | 0 | 0 | 0 |
| 0 | 0 | 0 | 0 | 0 |
| 0 | 0 | 0 | 0 | 0 |
| 0 | 0 | 0 | 0 | 0 |
| 0 | 0 | 0 | 0 | 0 |
| 0 | 0 | 0.000727046 | 0 | 0 |
| 0 | 0 | 0 | 0 | 0 |
| 0 | 0 | 0 | 0 | 0 |
| 0 | 0 | 0 | 0 | 0 |
| 0 | 0 | 0 | 0 | 0 |
| 0 | 0 | 0 | 0 | 0 |
| 0 | 0 | 0 | 0 | 0 |
| 0 | 0 | 0 | 0 | 0 |
| 0.015963958 | 0.022688812 | 0.103994209 | 0.082704525 | 0.050297161 |
| 0 | 0 | 0 | 0 | 0 |
| 0 | 0.001218859 | 0 | 0 | 0 |
| 0.000924844 | 0 | 0 | 0.051468303 | 0 |
| 0 | 0 | 0 | 0 | 0 |
| 0 | 0 | 0 | 0 | 0 |
| 0.003443345 | 0 | 0 | 0.000971072 | 0.02601119 |
| 0 | 0 | 0 | 0 | 0 |
| 0 | 0 | 0 | 0 | 0 |
| 0 | 0.004904621 | 0.008046595 | 0 | 0 |
| 0 | 0.508582944 | 0.002564815 | 0 | 0 |
| 0 | 0 | 0 | 0 | 0 |
| 0 | 0 | 0 | 0 | 0 |
| 0 | 0 | 0 | 0 | 0 |
| 0.047126211 | 0.01566638 | 0.002653497 | 0.012585058 | 0 |
| 0 | 0 | 0 | 0 | 0 |
| 0 | 0 | 0 | 0 | 0 |
| 0 | 0 | 0 | 0 | 0 |
| 0 | 0 | 0 | 0 | 0 |
| 0 | 0 | 0 | 0 | 0 |
| 0 | 0 | 0 | 0 | 0 |
| 0 | 0 | 0 | 0 | 0 |
| 0 | 0 | 0 | 0 | 0 |
| 0 | 0 | 0 | 0 | 0 |
| 0 | 0 | 0.011042959 | 0 | 0 |
| 0 | 0 | 0 | 0.01714715 | 0 |
| 0.000833064 | 0 | 0.002394193 | 0 | 0 |
| 0 | 0.001502885 | 0.004254646 | 0.000618514 | 0 |

| | | | | |
|---|---|---|---|---|
| 0 | 0 | 0 | 0 | 0 |
| 0 | 0 | 0 | 0 | 0 |
| 0 | 0 | 0 | 0.003673704 | 0.004773585 |
| 0 | 0 | 0 | 0.00196553 | 0 |
| 0 | 0 | 0 | 0 | 0 |
| 0 | 0 | 0 | 0 | 0 |
| 0 | 0.000916859 | 0 | 0 | 0 |
| 0 | 0 | 0 | 0 | 0 |
| 0 | 0 | 0 | 0.003047055 | 0 |
| 0 | 0 | 0 | 0 | 0 |
| 0 | 0 | 0 | 0 | 0 |
| 0 | 0 | 0 | 0 | 0 |
| 0 | 0 | 0 | 0 | 0 |
| 0 | 0 | 0 | 0 | 0 |
| 0 | 0 | 0 | 0 | 0 |
| 0 | 0 | 0 | 0 | 0 |
| 0 | 0 | 0 | 0 | 0 |
| 0 | 0 | 0 | 0 | 0 |
| 0 | 0 | 0 | 0 | 0.00094871 |
| 0 | 0 | 0 | 0.071362497 | 0 |
| 0.001850723 | 0.15170107 | 0.067372807 | 0.244263184 | 0 |
| 0 | 0 | 0 | 0 | 0 |
| 0 | 0 | 0 | 0 | 0 |
| 0 | 0 | 0 | 0 | 0 |
| 0.006128087 | 0.007038745 | 0 | 0 | 0.001668175 |
| 0 | 0 | 0 | 0 | 0 |
| 0 | 0 | 0 | 0 | 0 |
| 0 | 0 | 0 | 0 | 0 |
| 0.003508873 | 0.001221305 | 0 | 0 | 0 |
| 0 | 0 | 0 | 0 | 0 |
| 0.686012399 | 2.28704281 | 0.654786518 | 0.651734942 | 0.770317321 |
| 0 | 0 | 0 | 0 | 0 |
| 0 | 0 | 0.050359154 | 0 | 0 |
| 0 | 0 | 0 | 0 | 0 |
| 0 | 0 | 0 | 0 | 0 |
| 0 | 0 | 0 | 0 | 0 |
| 0 | 0 | 0 | 0 | 0 |
| 0 | 0 | 0 | 0 | 0 |
| 0 | 0 | 0 | 0 | 0 |
| 0 | 0 | 0 | 0 | 0 |
| 0 | 0 | 0 | 0 | 0 |
| 0.019882422 | 0.007242187 | 0.005492653 | 0.011475042 | 0.006922774 |
| 0 | 0 | 0.005500514 | 0.00102115 | 0 |
| 0 | 0 | 0 | 0 | 0 |
| 0 | 0 | 0 | 0 | 0 |
| 0 | 0 | 0.016396561 | 0 | 0 |
| 0 | 0 | 0 | 0 | 0 |
| 0 | 0 | 0 | 0 | 0 |

| | | | | |
|---|---|---|---|---|
| 0 | 0 | 0 | 0 | 0 |
| 0 | 0 | 0 | 0 | 0 |
| 0 | 0.001212501 | 0.00133489 | 0 | 0.008429268 |
| 0 | 0 | 0 | 0 | 0 |
| 0 | 0 | 0.96383387 | 0 | 0 |
| 0 | 0 | 0 | 0 | 0 |
| 0 | 0 | 0 | 0 | 0 |
| 0 | 0 | 0 | 0 | 0 |
| 0 | 0 | 0 | 0 | 0 |
| 0 | 0 | 0 | 0 | 0.000722751 |
| 0 | 0 | 0 | 0 | 0 |
| 0 | 0 | 0 | 0 | 0 |
| 0 | 0 | 0 | 0 | 0 |
| 0 | 0 | 0.003196471 | 0 | 0 |
| 0.003354195 | 0 | 0.001420606 | 0 | 0.006007051 |
| 0 | 0 | 0.003911004 | 0 | 0 |
| 0.003218361 | 0.001575265 | 0 | 0.001134529 | 0 |
| 0 | 0 | 0 | 0 | 0 |
| 0 | 0 | 0 | 0 | 0 |
| 0 | 0 | 0 | 0 | 0 |
| 0 | 0 | 0 | 0 | 0 |
| 0 | 0.003981642 | 0.005009766 | 0.001843481 | 0 |
| 0 | 0 | 0 | 0 | 0 |
| 0 | 0 | 0 | 0 | 0 |
| 0 | 0.007752401 | 0 | 0 | 0 |
| 0 | 0 | 0 | 0 | 0 |
| 0 | 0 | 0 | 0 | 0 |
| 0 | 0 | 0 | 0 | 0 |
| 0 | 0 | 0 | 0 | 0 |
| 0 | 0.010284788 | 0.007246673 | 0 | 0 |
| 0 | 0.00174354 | 0 | 0 | 0 |
| 0 | 0 | 0 | 0 | 0 |
| 0 | 0 | 0 | 0 | 0 |
| 0.004815276 | 0 | 0 | 0 | 0.001709745 |
| 0 | 0 | 0 | 0 | 0 |
| 0 | 0.015905527 | 0 | 0.001227363 | 0 |
| 0 | 0 | 0 | 0 | 0 |
| 0.000684405 | 0 | 0 | 0.001286746 | 0 |
| 0 | 0 | 0 | 0 | 0 |
| 0 | 0 | 0 | 0 | 0 |
| 0 | 0 | 0 | 0 | 0 |
| 0 | 0 | 0 | 0 | 0 |
| 0 | 0 | 0 | 0 | 0 |
| 0 | 0 | 0 | 0 | 0 |
| 0 | 0 | 0 | 0 | 0 |
| 0 | 0 | 0 | 0 | 0 |
| 0 | 0 | 0 | 0 | 0 |
| 0.000828986 | 0.002596846 | 0.002541306 | 0.008728003 | 0.01617265 |

| | | | | |
|---|---|---|---|---|
| 0 | 0 | 0 | 0 | 0 |
| 0 | 0 | 0 | 0 | 0 |
| 0 | 0 | 0 | 0 | 0 |
| 0 | 0 | 0 | 0 | 0 |
| 0.01396849 | 0.002397648 | 0.001539804 | 0.009929222 | 0.017710147 |
| 0 | 0.003909583 | 0 | 0 | 0 |
| 0 | 0 | 0 | 0 | 0 |
| 0.006437232 | 0 | 0 | 0.006353865 | 0.11682197 |
| 0 | 0 | 0 | 0 | 0 |
| 0.010781865 | 0.006928164 | 0.202764023 | 0.045843463 | 0 |
| 0 | 0 | 0 | 0 | 0 |
| 0.001061612 | 0.001330223 | 0 | 0.007185346 | 0 |
| 0.016588563 | 0.002270298 | 0 | 0.021438004 | 0 |
| 0 | 0 | 0.027837594 | 0.011588725 | 0 |
| 0 | 0 | 0 | 0.00560555 | 0 |
| 0 | 0 | 0 | 0 | 0 |
| 0 | 0 | 0 | 0 | 0 |
| 0 | 0 | 0.058352854 | 0 | 0 |
| 0.110040074 | 0.266882861 | 0 | 0 | 0 |
| 0 | 0 | 0 | 0 | 0 |
| 0 | 0 | 0 | 0 | 0 |
| 0 | 0 | 0 | 0 | 0 |
| 0 | 0 | 0 | 0 | 0 |
| 0 | 0 | 0 | 0 | 0 |
| 0 | 0 | 0 | 0 | 0 |
| 0 | 0 | 0 | 0 | 0.00204258 |
| 0 | 0 | 0 | 0 | 0.00170709 |
| 0 | 0.001065905 | 0 | 0 | 0 |
| 0 | 0 | 0 | 0.032281838 | 0 |
| 0.000965016 | 0 | 0 | 0 | 0 |
| 0 | 0 | 0.09070916 | 0 | 0 |
| 0 | 0 | 0 | 0 | 0 |
| 0 | 0 | 0.366856491 | 0 | 0 |
| 0 | 0.710869302 | 0 | 0.001660471 | 0 |
| 0 | 0 | 0 | 0 | 0 |
| 0 | 0.002681481 | 0 | 0 | 0 |
| 0 | 0.987877802 | 0 | 0.067646152 | 0 |
| 0 | 0 | 0 | 0 | 0 |
| 0 | 0 | 0.000767857 | 0 | 0 |
| 0 | 0 | 0 | 0.007566973 | 0 |
| 0.287041747 | 0 | 0 | 0 | 0 |
| 0 | 0.001725307 | 0 | 0 | 0 |
| 0 | 0 | 0 | 0 | 0 |
| 0 | 0.180843591 | 0.000868664 | 0.028981482 | 0 |
| 0 | 0 | 0 | 0 | 0 |
| 0 | 0 | 0 | 0 | 0 |
| 0 | 0 | 0 | 0.000789456 | 0 |
| 0 | 0 | 0.177692416 | 0 | 0 |

| | | | | |
|---|---|---|---|---|
| 0 | 0 | 0 | 0 | 0 |
| 0.028416921 | 0 | 0.00089911 | 0.002352726 | 0.004258125 |
| 0 | 0 | 0 | 0 | 0 |
| 0 | 0 | 0 | 0 | 0 |
| 0 | 0 | 0 | 0 | 0 |
| 0.073494216 | 0 | 0.048791714 | 0.168091134 | 0 |
| 0 | 0 | 0 | 0 | 0 |
| 0 | 0 | 0 | 0 | 0 |
| 0 | 0 | 0 | 0 | 0 |
| 0 | 0 | 0 | 0 | 0 |
| 0.003409637 | 0 | 0.000979916 | 0 | 0.002610461 |
| 0 | 0 | 0 | 0.00450603 | 0 |
| 0 | 0 | 0 | 0 | 0 |
| 0.000805313 | 0.001576681 | 0.002082995 | 0.003406645 | 0.007398698 |
| 0.01914856 | 0.011486287 | 0 | 0.006664032 | 0 |
| 0 | 0 | 0 | 0 | 0 |
| 0 | 0 | 0 | 0 | 0 |
| 0 | 0.135024125 | 0 | 0 | 0 |
| 0 | 0 | 0 | 0 | 0 |
| 0 | 0 | 0.04732459 | 0 | 0 |
| 0 | 0 | 0 | 0 | 0 |
| 0.112673636 | 0 | 0 | 0 | 0.505309803 |
| 0 | 0 | 0 | 0 | 0 |
| 0 | 0 | 0.007342995 | 0 | 0 |
| 0 | 0 | 0 | 0 | 0 |
| 0 | 0 | 0 | 0 | 0 |
| 0 | 0.009915329 | 0 | 0.00911114 | 0 |
| 0 | 0 | 0 | 0 | 0 |
| 0 | 0 | 0 | 0 | 0 |
| 0.440296905 | 0.948006569 | 0.79112713 | 0.02663162 | 0.593258481 |
| 0.011199992 | 0 | 0 | 0 | 0 |
| 0 | 0 | 0 | 0 | 0 |
| 0 | 0 | 0 | 0 | 0 |
| 0 | 0 | 0 | 0 | 0 |
| 0 | 0.038729679 | 0 | 0 | 0 |
| 0 | 0 | 0 | 0 | 0 |
| 0 | 0 | 0 | 0 | 0.359102471 |
| 0 | 0 | 0 | 0 | 0 |
| 0 | 0 | 0 | 0 | 0 |
| 0 | 0 | 0 | 0 | 0 |
| 0.002979248 | 0 | 0.011606598 | 0.18204116 | 0 |
| 0.061210006 | 0 | 7.255069857 | 0.011561581 | 4.337664039 |
| 0 | 0 | 0 | 0 | 0 |
| 0 | 0 | 2.001630477 | 0 | 0 |
| 0.004854413 | 0.021347897 | 0 | 0 | 0.016264098 |
| 0 | 0 | 0.041624697 | 0 | 0 |
| 0 | 0.001360458 | 0.00379438 | 0.046443555 | 0 |
| 0 | 0 | 0 | 0 | 0 |

| | | | | |
|---|---|---|---|---|
| 0.000857358 | 0 | 0 | 0 | 0.002625618 |
| 0 | 0 | 0 | 0 | 0 |
| 0.006133137 | 0 | 0 | 0.673732435 | 0 |
| 0 | 0 | 0 | 0 | 0 |
| 0.010629532 | 0 | 0 | 0.00048611 | 0 |
| 0 | 0 | 0 | 0 | 0 |
| 0 | 0 | 0 | 0 | 0 |
| 0 | 0 | 0 | 0 | 0 |
| 0 | 0.019386496 | 0 | 0.096522933 | 0 |
| 0 | 0 | 0 | 0 | 0 |
| 0 | 0 | 0 | 0 | 0 |
| 0 | 0 | 0 | 0 | 0 |
| 0 | 0 | 0 | 0 | 0 |
| 0 | 0.003948273 | 0.049486734 | 0 | 0.002639256 |
| 0 | 0 | 0 | 0 | 0 |
| 0.004091458 | 0.009731214 | 0.000522609 | 0 | 0.022481612 |
| 0.033760214 | 0.001578444 | 0 | 0 | 0.010744684 |
| 0 | 0.007031744 | 0.002815099 | 0.00989853 | 0 |
| 0 | 0 | 0 | 0 | 0 |
| 0 | 0 | 0 | 0 | 0 |
| 0.095678901 | 0 | 0.018250717 | 0.000716357 | 0 |
| 0.000779305 | 0 | 0 | 0.002417526 | 0 |
| 0 | 0 | 0 | 0 | 0 |
| 0 | 0 | 0 | 0.804394046 | 0 |
| 0 | 0 | 0 | 0 | 0 |
| 0.008629115 | 0.527454723 | 0 | 0 | 0 |
| 0 | 0 | 0.015687925 | 0.015394137 | 0 |
| 0 | 0 | 0 | 0.007763213 | 0 |
| 0 | 0 | 0 | 0 | 0 |
| 0 | 0 | 0 | 0 | 0 |
| 0 | 0 | 0.097868386 | 0.076828486 | 0 |
| 0.0246595 | 0 | 0 | 0.002454468 | 0 |
| 0.074350731 | 0.055162384 | 0 | 0.048557365 | 0.003133065 |
| 0 | 0.006200241 | 0.010239132 | 0.002977003 | 0 |
| 0 | 0 | 0 | 0 | 0 |
| 0 | 0 | 0 | 0 | 0 |
| 0 | 0 | 0.000717706 | 0 | 0 |
| 0.009602156 | 0.028549839 | 0.000561279 | 0.013402024 | 0.004873333 |
| 0.019948648 | 0 | 0 | 0.076238109 | 0.103506309 |
| 0.003193523 | 0 | 0 | 0 | 0 |
| 0 | 0 | 0 | 0 | 0 |
| 0 | 0 | 0.035573659 | 0 | 0 |
| 0 | 0 | 0 | 0 | 0 |
| 0.156646726 | 0 | 0 | 1.390720375 | 0 |
| 0 | 0 | 0 | 0.178367158 | 0.243977012 |
| 0 | 0 | 0 | 0 | 0 |
| 0 | 0 | 0 | 0 | 0 |
| 0 | 0 | 0.00095032 | 0 | 0 |

| | | | | |
|---|---|---|---|---|
| 0 | 0.044597611 | 0 | 0.00898267 | 0 |
| 0.589674091 | 0.100035713 | 0 | 0.060289479 | 0.041956105 |
| 0 | 0 | 0 | 0 | 0 |
| 0 | 0 | 0 | 0 | 0 |
| 0 | 0 | 0.008003398 | 0 | 0 |
| 0 | 0 | 0 | 0 | 0 |
| 0 | 0.008354853 | 0 | 0 | 0 |
| 0 | 0 | 0 | 0 | 0.054020315 |
| 0 | 0 | 0 | 0 | 0 |
| 0 | 0 | 0 | 0.002908626 | 0 |
| 0.00082398 | 0 | 0.00056834 | 0.000557697 | 0 |
| 0.048848168 | 0.0026913 | 0.145043826 | 0 | 0 |
| 0 | 0 | 0 | 0 | 0 |
| 0 | 0 | 0.000831295 | 0 | 0 |
| 0 | 0 | 0 | 0 | 0.002573585 |
| 0 | 0 | 0.016669072 | 0 | 0 |
| 0.036066107 | 0.014325102 | 0 | 0 | 0.008925308 |
| 0.011424681 | 0.155785034 | 0.081892085 | 0.187503153 | 0 |
| 0 | 0 | 0 | 0 | 0 |
| 0.004945727 | 0 | 0 | 0.090249477 | 0 |
| 0.039862627 | 0.020626165 | 0.002045779 | 0 | 0.083080417 |
| 0 | 0 | 0 | 0 | 0 |
| 0 | 0 | 0 | 0 | 0 |
| 0 | 0 | 0 | 0 | 0 |
| 0 | 0 | 0 | 0 | 0 |
| 0.007833246 | 0 | 0 | 0.051913514 | 0 |
| 0 | 0 | 0.245949107 | 0.147619624 | 0 |
| 0.073387581 | 0.023700074 | 0 | 0.411792978 | 0.046339357 |
| 0 | 0 | 0 | 0 | 0 |
| 0 | 0 | 0 | 0 | 0 |
| 0.001217705 | 0 | 0 | 0 | 0 |
| 0 | 0 | 0 | 0 | 0 |
| 0 | 0 | 0 | 0 | 0 |
| 0 | 0 | 0 | 0 | 0 |
| 0.070050699 | 0 | 0 | 0.057638945 | 0.089063693 |
| 0.264413888 | 0.375538882 | 0.096783402 | 0.041888324 | 0.142039281 |
| 0 | 0 | 0 | 0.000668971 | 0 |
| 0 | 0 | 0 | 0.002056798 | 0 |
| 0.003266611 | 0.008405549 | 0 | 0.001316043 | 0 |
| 0 | 0 | 0 | 0 | 0 |
| 0 | 0 | 0 | 0.181020614 | 0.174971721 |
| 0 | 0 | 0 | 0 | 0 |
| 0.000992095 | 0.290578589 | 0 | 0.055797057 | 0 |
| 0 | 0 | 0 | 0 | 0 |
| 0 | 0.678302328 | 0 | 0 | 0 |
| 0 | 0 | 0 | 0 | 0 |
| 0 | 0 | 0 | 0 | 0 |
| 0 | 0 | 0.045005747 | 0.00124754 | 0 |

| | | | | |
|---|---|---|---|---|
| 0 | 0 | 0 | 0 | 0 |
| 0 | 0 | 0 | 0 | 0 |
| 0 | 0 | 0 | 0 | 0 |
| 0.008253403 | 0 | 0.02428924 | 0 | 0.001123362 |
| 0 | 0 | 0 | 0 | 0 |
| 0 | 0 | 0 | 0 | 0 |
| 0 | 0 | 0 | 0.000577446 | 0 |
| 0 | 0 | 0 | 0 | 0.003617884 |
| 0.087880202 | 0 | 0 | 0 | 0.005095069 |
| 0 | 0 | 0 | 0 | 0 |
| 0 | 0 | 0 | 0 | 0 |
| 0.022299565 | 0 | 0 | 0 | 0.011624056 |
| 0 | 0 | 0 | 0 | 0 |
| 0 | 0 | 0 | 0 | 0 |
| 0.011615231 | 0 | 0 | 0.030327446 | 0 |
| 0 | 0 | 0 | 0 | 0 |
| 0 | 0 | 0 | 0 | 0 |
| 0 | 0 | 0 | 0 | 0.026443713 |
| 0 | 0 | 0 | 0 | 0 |
| 0 | 0 | 0 | 0 | 0 |
| 0 | 0 | 0 | 0 | 0 |
| 0 | 0 | 0 | 0 | 0 |
| 0 | 0 | 0 | 0 | 0 |
| 0.0175998 | 0.032291812 | 0.343229198 | 0.071756457 | 0 |
| 0 | 0 | 0 | 0 | 0 |
| 0 | 0 | 0.042804999 | 0 | 0 |
| 0 | 0 | 0 | 0.460488839 | 0 |
| 1.857263119 | 0 | 0 | 0 | 1.23164748 |
| 0 | 0 | 0 | 0 | 0 |
| 0 | 0 | 0 | 0 | 0 |
| 0.020910221 | 0.000963271 | 0 | 0 | 0.006278091 |
| 0 | 0 | 0 | 0 | 0 |
| 0.005499765 | 0 | 0.002286842 | 0.056232404 | 0 |
| 0 | 0 | 0 | 0 | 0 |
| 0 | 0 | 0 | 0 | 0 |
| 0.026072837 | 0 | 0 | 0 | 0.009936489 |
| 0.009260133 | 0 | 0 | 0 | 0.182641514 |
| 0 | 0 | 0 | 0 | 0 |
| 0 | 0 | 0 | 0.012443244 | 0 |
| 0.002377235 | 0 | 0 | 0.001532374 | 0 |
| 0.526011421 | 0.091727068 | 0.185053643 | 1.564978343 | 0.000826015 |
| 0 | 0 | 0 | 0 | 0 |
| 0.15634327 | 3.594475671 | 0 | 0.051122979 | 4.106564183 |
| 0 | 0.007659409 | 0 | 0 | 0.001259242 |
| 0 | 0 | 0 | 0 | 0 |
| 0 | 0 | 0 | 0 | 0 |
| 0 | 0 | 0 | 0 | 0 |
| 0 | 0 | 0.168990858 | 0.038884797 | 0 |

| | | | | |
|---|---|---|---|---|
| 0 | 0 | 0 | 0 | 0 |
| 0.03838231 | 0 | 0 | 0 | 0.008367727 |
| 0.018008094 | 0.00520299 | 0.023515667 | 0.629541231 | 0.00428341 |
| 0 | 0 | 0 | 0 | 0 |
| 0 | 8.703450951 | 0 | 0 | 0 |
| 0.000964103 | 0 | 0 | 0 | 0.000749846 |
| 0 | 0 | 0 | 0 | 0 |
| 0 | 0 | 0 | 0.005825602 | 0 |
| 0 | 0 | 0 | 0 | 0 |
| 0 | 0 | 0 | 0 | 0 |
| 0 | 0.051352999 | 0.130438546 | 0.057123705 | 0.001869133 |
| 2.762477067 | 2.359703811 | 0 | 12.01549773 | 3.923956996 |
| 0 | 0 | 0 | 0 | 0 |
| 0.020008091 | 0.001160675 | 0 | 0 | 0.030258668 |
| 0.018686985 | 0.368377993 | 0.769669423 | 0.382403852 | 0 |
| 0 | 0 | 0 | 0 | 0 |
| 0 | 0 | 0.000408997 | 0.001605349 | 0 |
| 0 | 0 | 0 | 0 | 0 |
| 0 | 0 | 0 | 0 | 0 |
| 0 | 0 | 0 | 0.007815898 | 0 |
| 0.184303264 | 0.30557123 | 0.026628745 | 0.030999026 | 0 |
| 0.033503404 | 0 | 55.04367985 | 0.153911102 | 0.009491637 |
| 0.592765151 | 0.174959567 | 10.80014559 | 3.925269914 | 0.141311339 |
| 0 | 0 | 0 | 0 | 0 |
| 0 | 0 | 0 | 0 | 0 |
| 0.013619982 | 0.009181153 | 0 | 0 | 1.289447866 |
| 0 | 0 | 0 | 0.415409616 | 0 |
| 0 | 0 | 0 | 0 | 0 |
| 0 | 0 | 0 | 0 | 0 |
| 0 | 0 | 0 | 0 | 0 |
| 0 | 0 | 0 | 0 | 0 |
| 0.113281034 | 0 | 0 | 1.080063894 | 0 |
| 0.032262359 | 0.037001446 | 0.164566707 | 0.214583959 | 0 |
| 0.002419923 | 0.030743316 | 0 | 0.048074969 | 0 |
| 0 | 0.006014229 | 0 | 0.002100138 | 0 |
| 0.004796073 | 0.002253595 | 0.000551349 | 0.003426486 | 0.004569519 |
| 0 | 0 | 0 | 0.03205414 | 0 |
| 0 | 0 | 0 | 0 | 0 |
| 0.094849122 | 0 | 0 | 0.00253243 | 0.007638943 |
| 0 | 0 | 0 | 0 | 0 |
| 0 | 0 | 0 | 0 | 0 |
| 0 | 0 | 0 | 0 | 0 |
| 0 | 0 | 0 | 0 | 0 |
| 0 | 0 | 0 | 0 | 0 |
| 0.001823126 | 0.003141074 | 0.073144715 | 0.008432012 | 0 |
| 0 | 3.172917188 | 0 | 0 | 0 |
| 0 | 0 | 0 | 0 | 0 |
| 0.728810819 | 0.399566595 | 0 | 0.124536543 | 0.01676243 |

| | | | | |
|---|---|---|---|---|
| 0 | 0 | 0 | 0 | 0 |
| 0 | 0 | 0 | 0 | 0 |
| 0 | 0 | 0 | 0 | 0 |
| 0 | 0 | 0 | 0 | 0 |
| 0 | 0 | 0 | 0 | 0 |
| 0 | 0 | 0 | 0 | 0 |
| 0 | 0 | 0 | 0 | 0 |
| 0 | 0.018139871 | 0.071829468 | 0.050322901 | 0.001946108 |
| 0 | 0 | 0 | 0 | 0 |
| 0 | 0 | 0 | 0 | 0 |
| 0 | 0 | 0.018103864 | 0.002928269 | 0 |
| 0 | 0 | 0 | 0 | 0 |
| 0.001969823 | 0 | 0 | 0 | 0 |
| 0 | 0 | 0 | 0 | 1.565574259 |
| 0 | 0 | 0 | 0 | 0 |
| 0 | 0 | 0 | 0 | 0 |
| 0 | 0 | 0 | 0 | 0 |
| 0 | 0 | 0.01192435 | 0.000975087 | 0 |
| 0 | 0 | 0.024746614 | 0 | 0 |
| 0 | 0 | 0 | 0 | 0 |
| 0.000511965 | 0 | 0 | 0 | 0 |
| 0 | 0 | 0 | 0 | 0 |
| 0.027904416 | 0 | 0.036279241 | 0 | 0 |
| 0 | 0 | 0 | 0 | 0 |
| 0 | 0 | 0 | 0 | 0 |
| 0 | 0 | 0 | 0 | 0 |
| 0 | 0 | 0 | 0 | 0 |
| 0 | 0 | 0 | 0 | 0 |
| 0 | 0 | 0 | 0 | 0 |
| 0.030803375 | 0 | 0 | 0.002159477 | 0 |
| 0 | 0 | 0 | 0.001230047 | 0 |
| 0 | 0 | 0 | 0 | 0 |
| 0 | 0 | 0 | 0 | 0 |
| 0 | 0 | 0 | 0 | 0 |
| 0.000412819 | 0 | 0 | 0 | 0 |
| 0 | 0 | 0 | 0 | 0 |
| 0.071011214 | 0 | 0 | 0.380532032 | 0 |
| 0.003369401 | 0.004130154 | 0.004715458 | 0.005783939 | 0.06360661 |
| 0 | 0 | 0 | 0 | 0 |
| 0 | 0 | 0 | 0 | 0 |
| 0.000962145 | 0.017222698 | 0 | 0.097216649 | 0 |
| 0 | 0 | 0 | 0 | 0 |
| 0 | 0 | 0 | 0 | 0 |
| 0 | 0 | 0 | 0 | 0.276897999 |
| 0.129748605 | 0 | 0 | 0 | 0.316294144 |
| 0 | 0 | 0 | 0 | 0 |
| 0 | 0 | 0 | 0 | 0 |
| 0 | 0 | 0 | 0 | 0 |

| | | | | |
|---|---|---|---|---|
| 0 | 0 | 0 | 0 | 0 |
| 0 | 0 | 0 | 0 | 0 |
| 0 | 0 | 0 | 0 | 0 |
| 0 | 0 | 0 | 0 | 0 |
| 0 | 0 | 0 | 0 | 0 |
| 0 | 0 | 0 | 0 | 0 |
| 0 | 0 | 0 | 0 | 0 |
| 0 | 0 | 0 | 0 | 0 |
| 0 | 0.047784561 | 0.114274097 | 0.443294315 | 0.035199654 |
| 0 | 0.00280323 | 0 | 0 | 0 |
| 0.009823712 | 0 | 0 | 0.003096358 | 0 |
| 0 | 0 | 0 | 0 | 0 |
| 0 | 0 | 0 | 0.003722914 | 0 |
| 0 | 0 | 0 | 0 | 0 |
| 1.476465095 | 0 | 0 | 0.526243137 | 0.555313086 |
| 0 | 3.681913387 | 0.472951427 | 0 | 0 |
| 0 | 0 | 0.015445197 | 0 | 0 |
| 0 | 0 | 0 | 0 | 0 |
| 0 | 0 | 0 | 0 | 0 |
| 0 | 0 | 0 | 0 | 0 |
| 0 | 0 | 0 | 0 | 0 |
| 0 | 0 | 0 | 0 | 0 |
| 0 | 0 | 0 | 0 | 0 |
| 0 | 0 | 0 | 0 | 0 |
| 0 | 0 | 0 | 0 | 0.173336499 |
| 0.21744948 | 1.413556155 | 0.00102977 | 0 | 1.417964962 |
| 0 | 0 | 0.21638786 | 0 | 0 |
| 0 | 0 | 0 | 0.006821306 | 0 |
| 0.01765676 | 0 | 0 | 0.014059637 | 0.002827345 |
| 0 | 0 | 0.460317909 | 0 | 0 |
| 0 | 0 | 0 | 0 | 0 |
| 0.060440113 | 0 | 0 | 0 | 0 |
| 0 | 0 | 0 | 0 | 0 |
| 0 | 0.00278246 | 0.006272511 | 0.011451248 | 0 |
| 0.000627231 | 0 | 0 | 0 | 0.00369944 |
| 0.001625712 | 0.001157417 | 0.004077586 | 0.00366779 | 0 |
| 0 | 0 | 0 | 0 | 0 |
| 0.296049944 | 0 | 1.3970211 | 0 | 2.549672317 |
| 6.351854207 | 0 | 0 | 0.00800116 | 0 |
| 0 | 0 | 0 | 0 | 0 |
| 0 | 0 | 0 | 0 | 0 |
| 0 | 0 | 0 | 0 | 0 |
| 0.054933231 | 0 | 0 | 0 | 4.792365498 |
| 0 | 0 | 0 | 0 | 0 |
| 0 | 0 | 0 | 0 | 0 |
| 0 | 0 | 0.001448625 | 0 | 0 |
| 0 | 0 | 0 | 0 | 0 |
| 0.470432866 | 0.024106771 | 0.007949205 | 0.028433499 | 0 |

| | | | | |
|---|---|---|---|---|
| 0 | 0 | 0 | 0.006004359 | 0 |
| 0 | 0 | 0 | 0 | 0 |
| 0 | 0 | 0 | 0 | 0 |
| 0 | 0 | 0 | 0 | 0 |
| 0 | 0.011412984 | 0.002094167 | 0 | 0 |
| 0 | 0 | 0 | 0 | 0 |
| 0 | 0 | 0 | 0 | 0 |
| 0 | 0 | 0 | 0 | 0 |
| 0 | 0 | 0 | 0 | 0 |
| 0 | 0 | 0 | 0 | 0 |
| 0 | 0 | 0 | 0 | 0 |
| 0.052557236 | 0 | 0 | 0 | 0.284684731 |
| 0.000601562 | 0.11966104 | 0.319263754 | 0 | 0.319596832 |
| 0 | 0 | 0 | 0 | 0 |
| 0.02183374 | 0.067495478 | 0.001541212 | 0.010802495 | 0.069341234 |
| 0 | 0 | 0 | 0 | 0 |
| 0 | 0 | 0 | 0.001202351 | 0 |
| 0 | 0 | 0 | 0 | 0 |
| 0 | 0.014198291 | 0.001350866 | 0 | 0 |
| 0 | 0 | 0 | 0 | 0 |
| 0 | 0 | 0 | 0 | 0 |
| 0 | 0 | 0 | 0 | 0 |
| 0.059416045 | 0 | 0.027584214 | 1.35945867 | 0.416350025 |
| 0 | 0 | 0 | 0 | 0 |
| 0 | 0 | 0 | 0 | 0 |
| 0 | 0 | 0 | 0 | 0 |
| 0 | 0 | 0 | 0 | 0 |
| 0 | 0 | 0 | 0 | 0 |
| 0 | 0 | 0 | 0 | 0 |
| 0 | 0 | 0 | 0 | 0 |
| 0 | 0 | 0 | 0 | 0 |
| 0.239085108 | 0.007785317 | 0 | 0.008130294 | 0.004397516 |
| 0 | 0 | 0 | 0 | 0 |
| 0 | 0 | 0.019839952 | 0.048180813 | 0 |
| 0 | 0.001511472 | 0.003882756 | 0.004717197 | 0 |
| 0.077128798 | 0.00259796 | 0 | 0.005145494 | 0.005079638 |
| 0 | 0 | 0 | 0 | 0.0029148 |
| 0 | 0 | 0 | 0 | 0.011513103 |
| 0 | 0 | 0 | 0 | 0 |
| 0 | 0 | 0 | 0 | 0 |
| 0 | 0 | 0 | 0 | 0 |
| 0 | 0 | 0 | 0 | 0 |
| 0 | 0 | 0 | 0 | 0 |
| 0 | 0 | 0 | 0 | 0 |
| 0.000792192 | 0.005955804 | 0.01366037 | 0.041822204 | 0 |
| 0 | 0 | 0 | 0 | 0 |
| 0 | 0 | 0 | 0 | 0 |
| 0.004945192 | 0 | 0.000689081 | 0.0057475 | 0 |

| | | | | |
|---|---|---|---|---|
| 1.931811173 | 0 | 0 | 0 | 0 |
| 0 | 0 | 0 | 0 | 0 |
| 0 | 0 | 0 | 0.003327279 | 0 |
| 0 | 0 | 0 | 0 | 0 |
| 0 | 0.03826238 | 0 | 0 | 0 |
| 0 | 0 | 0.162058465 | 0 | 0 |
| 0 | 0 | 0 | 0 | 0 |
| 0.008821039 | 0 | 0.028998358 | 0 | 0 |
| 0 | 0 | 0 | 0 | 0 |
| 0 | 0 | 0 | 0 | 0 |
| 0 | 0 | 0 | 0 | 0 |
| 0 | 0.005484009 | 0 | 0 | 0 |
| 0 | 0 | 0 | 0 | 0 |
| 0.190366297 | 0 | 0 | 0 | 0 |
| 0 | 0 | 0 | 0 | 0 |
| 0 | 0 | 0 | 0 | 0 |
| 0 | 0 | 0 | 0 | 0 |
| 0 | 0 | 0 | 0 | 0 |
| 0 | 0 | 0 | 32.60650602 | 0 |
| 0 | 0 | 0 | 0 | 0 |
| 0 | 0 | 0 | 0 | 0 |
| 0 | 0 | 0 | 0 | 0 |
| 0 | 0 | 0 | 0 | 0 |
| 0 | 0 | 0 | 0 | 0 |
| 0.009347813 | 0 | 0 | 0 | 0 |
| 0.000885001 | 0.000693079 | 0 | 0.001497496 | 0 |
| 0 | 0 | 0 | 0 | 0 |
| 0.023677215 | 0.011080305 | 0.023899575 | 0.004560113 | 0.001375534 |
| 0 | 0 | 0 | 0 | 0 |
| 0.263226627 | 0 | 0 | 0.633733015 | 0 |
| 0 | 0 | 0 | 0 | 0 |
| 0 | 0 | 0 | 0 | 0 |
| 0 | 0 | 0 | 0.001203771 | 0 |
| 0 | 0 | 0 | 0 | 0 |
| 0 | 0.002663113 | 0.003553848 | 0.002266742 | 0 |
| 0 | 0 | 0 | 0 | 0 |
| 0 | 0 | 0 | 0 | 0.024115689 |
| 0 | 0 | 0 | 0.000593559 | 0 |
| 0 | 0 | 0 | 0 | 0 |
| 0 | 0 | 0 | 0 | 0 |
| 4.541792367 | 25.99279511 | 4.570976587 | 1.723790158 | 3.251209404 |
| 0 | 0.914565563 | 0.046344274 | 0.086463065 | 0 |
| 0.062989235 | 0 | 0 | 0 | 0.056198829 |
| 0 | 0 | 0 | 2.003793305 | 0 |
| 0 | 0 | 0 | 0 | 0 |
| 0 | 0 | 0 | 0 | 0 |
| 0 | 0 | 0 | 0 | 0 |
| 0 | 0 | 0.002400357 | 0 | 0 |

| | | | | |
|---|---|---|---|---|
| 0 | 0 | 0 | 0 | 0 |
| 0.006299927 | 0 | 0.004773333 | 0.004845458 | 0 |
| 0 | 0 | 0 | 0.004338737 | 0 |
| 0 | 0 | 0 | 0 | 0 |
| 2.013337042 | 0 | 0 | 0.000734907 | 3.734703938 |
| 0 | 0 | 0 | 0 | 0 |
| 0 | 0 | 0 | 0 | 0 |
| 0 | 0 | 0 | 0 | 0 |
| 0.224461426 | 1.451061928 | 0.103050601 | 0.786064842 | 0.038529547 |
| 0 | 0 | 0 | 0 | 0 |
| 0 | 0 | 0.007438445 | 0 | 0 |
| 0 | 0 | 0 | 0 | 0 |
| 0 | 0.575709653 | 0.027028967 | 0 | 0 |
| 0.007183626 | 0.098852958 | 0.013960266 | 0 | 0.00907916 |
| 0.004686083 | 0.086100431 | 0 | 0 | 0 |
| 0.002526356 | 0.177404456 | 0.510343075 | 0 | 0 |
| 0.00118406 | 0.018777491 | 0.00238206 | 0.000834804 | 0 |
| 0 | 0 | 0 | 0 | 0 |
| 0 | 0 | 0 | 0 | 0 |
| 0 | 0 | 0 | 0 | 0 |
| 0 | 0 | 0 | 0 | 0 |
| 0 | 0 | 0 | 0 | 0 |
| 0 | 0 | 0 | 0 | 0 |
| 0 | 0 | 0 | 0 | 0 |
| 0 | 0 | 0 | 0 | 0 |
| 0 | 0 | 0 | 0 | 0 |
| 0.13114386 | 0 | 0.017753989 | 0 | 0.001226192 |
| 0 | 0.00227458 | 0 | 0.005242194 | 0 |
| 0 | 0 | 0 | 0 | 0 |
| 0 | 0 | 0 | 0 | 0 |
| 0 | 0 | 0 | 0 | 0 |
| 0 | 0 | 0 | 0.001778905 | 0 |
| 0 | 0 | 0 | 0 | 0 |
| 0 | 0 | 0 | 0 | 0 |
| 0.018905161 | 0 | 0 | 0.009342857 | 0.007596953 |
| 0 | 0 | 0 | 0 | 0 |
| 0 | 0 | 0.016186188 | 0 | 0.004440481 |
| 0 | 0 | 0 | 0 | 0 |
| 0 | 0 | 0 | 0 | 0 |
| 0 | 0 | 0 | 0 | 0 |
| 6.262925487 | 3.641174413 | 0 | 0 | 0.220951142 |
| 0.037556997 | 0 | 0 | 0 | 0.014076089 |
| 0 | 0 | 0 | 0 | 0 |
| 0 | 0 | 0 | 0.144159281 | 0 |
| 0 | 0 | 0 | 0 | 0 |
| 0 | 0 | 0 | 0 | 0 |
| 0 | 0 | 0 | 0 | 0 |
| 0 | 0 | 0 | 0 | 0 |

| | | | | |
|---|---|---|---|---|
| 0 | 0 | 0.002193469 | 0 | 0 |
| 0 | 0 | 0 | 0 | 0 |
| 0 | 0 | 0 | 0 | 0 |
| 0 | 0 | 0 | 0 | 0 |
| 0 | 0 | 0 | 0 | 0 |
| 0 | 0 | 0 | 0 | 0 |
| 0.024804795 | 0.0016188 | 0.176692456 | 0.003331096 | 0 |
| 0 | 0 | 0 | 0 | 0 |
| 0 | 0 | 0 | 0 | 0 |
| 0 | 0 | 4.34171839 | 0 | 0 |
| 0 | 0 | 0 | 0 | 0 |
| 0 | 0 | 0 | 0 | 0 |
| 0 | 0 | 0 | 0 | 0 |
| 0 | 0 | 0 | 0 | 0 |
| 0 | 0 | 0 | 0 | 0 |
| 0 | 0 | 0 | 0 | 0 |
| 0 | 0 | 0 | 0 | 0 |
| 0 | 0.004001931 | 0 | 0 | 0 |
| 0 | 0 | 0 | 0 | 0 |
| 0 | 0 | 0 | 0 | 0 |
| 0.040624364 | 0 | 0.003878295 | 0.008087042 | 0.140242406 |
| 0 | 0 | 0 | 0 | 0 |
| 0.001705515 | 0 | 0 | 0 | 0 |
| 0 | 0.002159416 | 0.003011355 | 0.002799437 | 0.001125914 |
| 0 | 0 | 0 | 0 | 0 |
| 0 | 0.003311912 | 0 | 0 | 0 |
| 0 | 0 | 0 | 0 | 0 |
| 0 | 0 | 0 | 0 | 0 |
| 0 | 0 | 0 | 0.757152309 | 0 |
| 0.005357506 | 0 | 0.097401963 | 0 | 0 |
| 0 | 0 | 0 | 0 | 0 |
| 0 | 0 | 0 | 0 | 0 |
| 0 | 0 | 0 | 0 | 0 |
| 0 | 0 | 0 | 0 | 0 |
| 0 | 0 | 0 | 0 | 0 |
| 0 | 0 | 0 | 0 | 0 |
| 0.04295502 | 0.002161591 | 0.200496436 | 0.195768738 | 0 |
| 0 | 0 | 0 | 0 | 0 |
| 0 | 0 | 0 | 0 | 0 |
| 0.011920067 | 0 | 0 | 0 | 0.000853909 |
| 0 | 0 | 0 | 0 | 0 |
| 0 | 0 | 0 | 0.351653099 | 0 |
| 0 | 0 | 0 | 0 | 0 |
| 0 | 0 | 0 | 0 | 0 |
| 0 | 0 | 0 | 0 | 0 |
| 0 | 0 | 0 | 0 | 0 |
| 0 | 0 | 0 | 0 | 0 |
| 0 | 0 | 0 | 0 | 0.002014222 |

| | | | | |
|---|---|---|---|---|
| 0.000593409 | 0 | 0.001875977 | 0.015730865 | 0 |
| 0 | 0 | 0 | 0 | 0.004213014 |
| 0 | 0 | 0 | 0 | 0 |
| 0 | 0 | 0 | 0 | 0 |
| 0 | 0 | 0 | 0 | 0 |
| 0 | 0 | 0 | 0 | 0 |
| 0 | 0 | 0 | 0 | 0 |
| 2.996368778 | 0.332106164 | 0.01453409 | 1.852845523 | 0.006634458 |
| 0 | 0.019330433 | 0.037209391 | 0.00709237 | 0.204112387 |
| 0.013941535 | 0.043672643 | 0.004845364 | 0.026881919 | 0 |
| 0 | 0 | 0 | 0 | 0 |
| 0 | 0 | 0 | 0 | 0 |
| 0 | 0 | 0 | 0 | 0 |
| 0 | 0 | 0.004161741 | 0.003889338 | 0 |
| 0 | 0 | 0 | 0 | 0 |
| 0 | 0 | 0 | 0 | 0 |
| 0 | 0 | 0 | 0 | 0 |
| 0 | 0 | 0 | 0 | 0 |
| 0 | 0 | 0 | 0 | 0 |
| 0.005780328 | 0 | 0 | 0 | 0 |
| 0 | 0 | 0 | 0 | 0 |
| 0 | 0 | 0 | 0 | 0 |
| 0 | 0 | 0 | 0 | 0 |
| 0 | 0 | 0 | 0 | 0 |
| 0 | 0 | 0 | 0 | 0 |
| 0 | 0 | 0 | 0 | 0 |
| 0 | 0 | 0 | 0 | 0 |
| 0 | 0 | 0 | 0 | 0 |
| 0 | 0 | 0 | 0 | 0 |
| 0 | 0 | 0 | 0 | 0 |
| 0 | 0 | 0 | 0 | 0 |
| 0 | 0 | 0 | 0 | 0 |
| 0 | 0 | 0 | 0 | 0 |
| 0 | 0 | 0 | 0 | 0 |
| 0.003412423 | 0 | 0 | 0.003849405 | 0.005133519 |
| 0 | 0 | 0 | 0 | 0 |
| 0 | 0 | 0 | 0 | 0 |
| 0 | 0 | 0 | 0 | 0 |
| 0 | 0 | 0 | 0 | 0 |
| 0.000592408 | 1.001177497 | 0 | 0 | 0 |
| 0 | 0 | 0 | 0 | 0 |
| 1.404477317 | 0 | 0 | 0 | 0 |
| 0 | 0 | 0 | 0 | 0 |
| 0 | 0 | 0 | 0 | 0 |
| 0 | 0 | 0 | 0 | 0 |
| 0 | 0 | 0 | 0 | 0 |
| 0 | 0 | 0 | 0 | 0 |
| 0 | 0 | 0 | 0 | 0 |

| | | | | |
|---|---|---|---|---|
| 0 | 0 | 0 | 0 | 0 |
| 0 | 0 | 0 | 0 | 0.000728969 |
| 0 | 0 | 0 | 0 | 0 |
| 0 | 0 | 0 | 0 | 0 |
| 0.084241401 | 0.104383521 | 0 | 0.135152419 | 0.036691225 |
| 0 | 0 | 0 | 0 | 0 |
| 0 | 0 | 0 | 0 | 0 |
| 0 | 0.027488917 | 0.009979403 | 0.052155807 | 0.004109722 |
| 0 | 0 | 0 | 0 | 0 |
| 0 | 0 | 0 | 0 | 0 |
| 0 | 0 | 0 | 0 | 0 |
| 0 | 0 | 0 | 0 | 0 |
| 0 | 0 | 0 | 0 | 0 |
| 0 | 0 | 0 | 0 | 0 |
| 0 | 0.125262786 | 0 | 0 | 0 |
| 0 | 0.021277391 | 0 | 0.00092315 | 0 |
| 0 | 0 | 0 | 0 | 0 |
| 0 | 0 | 0 | 0 | 0 |
| 0 | 0 | 0 | 0 | 0 |
| 0 | 0 | 0 | 0 | 0 |
| 0 | 0 | 0 | 0 | 0 |
| 0 | 0 | 0 | 0 | 0 |
| 0 | 0 | 0 | 0 | 0 |
| 0 | 0.002302199 | 0.003191695 | 0 | 0 |
| 0.260098463 | 0 | 0 | 0 | 0.148924579 |
| 0.00089309 | 0 | 0 | 0 | 0 |
| 0.001045604 | 0.022927891 | 0.023525055 | 0.020388501 | 0 |
| 0.000786419 | 0 | 0 | 0.00106455 | 0 |
| 0 | 0.003825248 | 0 | 0 | 0 |
| 0 | 0 | 0 | 0 | 0 |
| 0 | 0 | 0 | 0 | 0 |
| 0 | 0 | 0 | 0 | 0 |
| 0 | 0 | 0 | 0 | 0 |
| 0 | 0 | 0 | 0 | 0 |
| 0 | 0 | 0 | 0 | 0 |
| 0 | 0 | 0.000767561 | 0 | 0 |
| 0 | 0 | 0 | 0 | 0 |
| 0 | 0 | 0 | 0 | 0 |
| 0 | 0 | 0 | 0 | 0 |
| 0 | 0 | 0 | 0 | 0 |
| 0 | 0 | 0 | 0 | 0 |
| 0.050939542 | 0 | 0 | 0 | 0 |
| 0 | 0 | 0 | 0 | 0 |
| 0 | 0 | 0 | 0 | 0 |
| 0 | 0 | 0 | 0 | 0 |
| 0 | 0 | 0 | 0 | 0 |
| 0 | 0 | 0 | 0 | 0 |
| 0 | 0 | 0 | 0 | 0 |

| | | | | |
|---|---|---|---|---|
| 0.009411589 | 0 | 0 | 0 | 0.005162244 |
| 0 | 0 | 0 | 0 | 0 |
| 0 | 0 | 0 | 0 | 0 |
| 0 | 0 | 0 | 0 | 0 |
| 0 | 0 | 0 | 0 | 0 |
| 0 | 0 | 0 | 0 | 0.000886038 |
| 0 | 0 | 0 | 0 | 0 |
| 0 | 0 | 0 | 0 | 0 |
| 0 | 0 | 0 | 0 | 0 |
| 0 | 0 | 0 | 0 | 0 |
| 0 | 0 | 0 | 0 | 0 |
| 0 | 0 | 0 | 0.00118882 | 0 |
| 0 | 0 | 0 | 0 | 0 |
| 0 | 0 | 0 | 0 | 0 |
| 0 | 0 | 0 | 0 | 0 |
| 0 | 0 | 0 | 0 | 0 |
| 0 | 0 | 0 | 0 | 0 |
| 0 | 0 | 0 | 0 | 0 |
| 0 | 0.000932677 | 0 | 0.004030367 | 0 |
| 0 | 0 | 0 | 0 | 0 |
| 0.004228493 | 0.002292576 | 0 | 0.007154967 | 0.005755357 |
| 0 | 0 | 0 | 0 | 0 |
| 0.001026977 | 0.001286825 | 0.009444772 | 0.006950925 | 0 |
| 0 | 0 | 0 | 0 | 0 |
| 0 | 0 | 0 | 0 | 0 |
| 0 | 0 | 0 | 0 | 0 |
| 0 | 0 | 0 | 0 | 0 |
| 0 | 0 | 0 | 0 | 0 |
| 0 | 0 | 0 | 0 | 0 |
| 0 | 0 | 0 | 0 | 0 |
| 0 | 0 | 0 | 0 | 0 |
| 0 | 0 | 0 | 0 | 0 |
| 0.005770556 | 0.510663739 | 0.022112475 | 0.482137908 | 0 |
| 0 | 0 | 0 | 0 | 0 |
| 0.003441157 | 0.041501527 | 0.293725639 | 0 | 0.377742194 |
| 0 | 0.011846328 | 0 | 0.001915324 | 0 |
| 0 | 0 | 0 | 0 | 0 |
| 0 | 0 | 0 | 0 | 0 |
| 0.052310811 | 0.005872351 | 0 | 0.001611182 | 0 |
| 0 | 0 | 0 | 0 | 0 |
| 0 | 0 | 0 | 0 | 0 |
| 0 | 0 | 0 | 0 | 0 |
| 0.007932923 | 0 | 0 | 0 | 0.001420713 |
| 0 | 0 | 0 | 0 | 0 |
| 0 | 0 | 0 | 0 | 0 |
| 0.002652681 | 0.028304819 | 0.01715333 | 0 | 0 |
| 0 | 0 | 0 | 0 | 0 |
| 0 | 0 | 0 | 0 | 0 |

| | | | | |
|---|---|---|---|---|
| 0 | 0 | 0 | 0 | 0 |
| 0 | 0 | 0 | 0 | 0 |
| 0 | 0 | 0 | 0 | 0 |
| 0 | 0 | 0 | 0 | 0 |
| 0 | 0 | 0 | 0 | 0 |
| 0 | 0 | 0 | 0 | 0 |
| 0 | 0 | 0 | 0 | 0 |
| 0 | 0.865710145 | 0 | 0.298555695 | 0 |
| 0 | 0 | 0 | 0.118626325 | 0 |
| 0.001024587 | 0 | 0 | 0 | 0.056479456 |
| 0 | 0 | 0 | 0 | 0 |
| 0 | 0 | 0 | 0 | 0 |
| 0 | 0 | 0 | 0 | 0 |
| 0 | 0 | 0 | 0 | 0 |
| 0 | 0 | 0 | 0.015878637 | 0 |
| 0 | 0.099736006 | 0.528446349 | 0.003221986 | 0 |
| 0 | 0 | 0 | 0 | 0 |
| 0 | 0 | 0 | 0 | 0 |
| 0 | 0 | 0 | 0 | 0 |
| 0 | 0 | 0 | 0 | 0.003446265 |
| 0.008247469 | 0.007146034 | 0 | 0 | 0 |
| 0 | 0 | 0 | 0 | 0 |
| 0 | 0 | 0 | 0 | 0 |
| 0 | 0 | 0 | 0 | 0 |
| 0 | 0 | 0 | 0 | 0 |
| 0 | 0 | 0 | 0 | 0 |
| 0 | 0 | 0 | 0 | 0 |
| 0 | 0 | 0 | 0 | 0 |
| 0 | 0 | 0 | 0 | 0 |
| 2.177707048 | 0 | 0 | 0 | 0 |
| 0.030861783 | 0 | 0 | 0.551937837 | 1.297900111 |
| 0 | 0 | 0 | 0 | 0 |
| 0 | 0 | 0 | 0 | 0 |
| 0 | 0 | 0 | 0 | 0 |
| 0 | 0 | 0 | 0 | 0 |
| 0 | 0 | 0 | 0 | 0 |
| 0 | 0 | 0 | 0 | 0 |
| 0 | 0 | 0 | 0 | 0 |
| 0 | 0 | 0 | 0 | 0 |
| 0 | 0 | 0 | 0 | 0 |
| 0 | 0 | 0 | 0 | 0 |
| 0 | 0 | 0 | 0 | 0 |
| 0 | 0 | 0 | 0 | 0 |
| 0 | 0 | 0 | 0 | 0 |
| 0 | 0 | 0 | 0 | 0 |
| 0 | 0 | 0 | 0 | 0 |
| 0.008255716 | 0.129307469 | 0 | 0.156573339 | 0 |
| 0 | 0 | 0 | 0 | 0 |

| | | | | |
|---|---|---|---|---|
| 0 | 0 | 0 | 0 | 0 |
| 0 | 0 | 0 | 0 | 0 |
| 0 | 0 | 0 | 0 | 0 |
| 0.002158038 | 0 | 0 | 0.003303813 | 0 |
| 0.780334479 | 0.005031372 | 0 | 0 | 0.036551942 |
| 0 | 0 | 0 | 0 | 0 |
| 0 | 0 | 0 | 0 | 0.142394268 |
| 0 | 0 | 0 | 0 | 0 |
| 0 | 0 | 0 | 0 | 0 |
| 0 | 0 | 0 | 0 | 0 |
| 0 | 0 | 0 | 0 | 0 |
| 0 | 0.002514718 | 0.005332025 | 0.00925692 | 0 |
| 0 | 0 | 0 | 0 | 0 |
| 0 | 0 | 0 | 0 | 0 |
| 0 | 0 | 0 | 0.005367493 | 0 |
| 0 | 0 | 0 | 0 | 0 |
| 0.009329968 | 0 | 0 | 0 | 0 |
| 0 | 0 | 0 | 0 | 0 |
| 0 | 0.205860722 | 0 | 0.104683755 | 0.052031322 |
| 0.001587363 | 0.243652617 | 0.092254155 | 0.006764619 | 0 |
| 0 | 0 | 0 | 0 | 0 |
| 0 | 0 | 0 | 0 | 0 |
| 0 | 0 | 0 | 0 | 0 |
| 0 | 0 | 0 | 0 | 0 |
| 0 | 0 | 0 | 0 | 0 |
| 0 | 0 | 0 | 0 | 0 |
| 0.007490822 | 0.01173271 | 0.013485673 | 0.003826446 | 0 |
| 0 | 0 | 0 | 0 | 0 |
| 0 | 0 | 0 | 0 | 0 |
| 0 | 0 | 0 | 0 | 0 |
| 0 | 0 | 0 | 0.008674956 | 0 |
| 0 | 0 | 0 | 0.005856227 | 0 |
| 0 | 0 | 0 | 0 | 0 |
| 0 | 0 | 0 | 0 | 0 |
| 0 | 0 | 0 | 0 | 0 |
| 0 | 0 | 0 | 0 | 0 |
| 0 | 0 | 0 | 0 | 0 |
| 0.002251207 | 0.013575163 | 0.002458555 | 0.026029756 | 0 |
| 0 | 0 | 0 | 0 | 0 |
| 0 | 0 | 0 | 0 | 0 |
| 0 | 0 | 0 | 0 | 0 |
| 0 | 0 | 0 | 0 | 0 |
| 0 | 0 | 0 | 0 | 0 |
| 0 | 0.003921476 | 0 | 0.004895461 | 0 |
| 0 | 0 | 0 | 0 | 0 |
| 0 | 0 | 0 | 0 | 0 |
| 0 | 0 | 0 | 0 | 0 |
| 0 | 0 | 0 | 0 | 0 |

| | | | | |
|---|---|---|---|---|
| 0 | 0 | 0 | 0 | 0 |
| 0 | 0 | 0.28241034 | 0 | 0 |
| 0 | 0 | 0 | 0 | 0 |
| 0 | 0 | 0.00844584 | 0 | 0 |
| 0 | 0 | 0 | 0 | 0.017665896 |
| 0 | 0 | 0 | 0.004251602 | 0 |
| 0.010015351 | 0 | 0 | 0 | 0 |
| 0 | 0 | 0.0210743 | 0 | 0 |
| 0 | 0 | 0 | 0 | 0 |
| 0 | 0 | 0 | 0 | 0 |
| 0 | 0 | 0 | 0 | 0 |
| 0 | 0 | 0 | 0 | 0 |
| 0.025047477 | 0.006797505 | 0 | 0 | 0.003037888 |
| 0 | 0 | 0 | 0 | 0 |
| 0 | 0 | 0 | 0 | 0 |
| 0 | 0 | 0 | 0 | 0 |
| 0 | 0 | 0 | 0 | 0 |
| 0 | 0 | 0 | 0 | 0 |
| 0 | 0 | 0 | 0 | 0 |
| 0 | 0 | 0 | 0 | 0 |
| 0 | 0 | 0 | 0 | 0 |
| 0 | 0 | 0 | 0 | 0 |
| 0.011429321 | 0.029921071 | 0 | 0.008472489 | 0 |
| 0.005972811 | 0 | 0.021041737 | 0 | 0.051137363 |
| 0 | 0 | 0 | 0 | 0 |
| 0 | 0 | 0 | 0 | 0.002324027 |
| 0 | 0 | 0 | 0 | 0 |
| 0 | 0 | 0 | 0 | 0 |
| 0 | 0 | 0 | 0 | 0 |
| 0 | 0 | 0 | 0 | 0 |
| 0.006189769 | 0 | 0 | 0.050314412 | 0 |
| 0 | 0 | 0 | 0 | 0 |
| 0 | 0 | 0 | 0 | 0 |
| 0 | 0 | 0 | 0 | 0 |
| 0 | 0 | 0 | 0 | 0 |
| 0 | 0.000894246 | 0.022315564 | 0 | 0 |
| 0 | 0.085430421 | 0 | 0.164075291 | 0 |
| 0 | 0 | 0 | 0 | 0 |
| 0.002926544 | 0.020988898 | 0 | 0.00173753 | 0 |
| 0 | 0 | 0 | 0 | 0 |
| 0 | 0 | 0 | 0 | 0 |
| 0 | 0 | 0 | 0 | 0 |
| 0 | 0 | 0 | 0 | 0 |
| 0 | 0 | 0 | 0 | 0 |
| 0 | 0 | 0 | 0 | 0 |
| 0.032118184 | 0 | 0 | 0.007032201 | 0.223743324 |
| 0 | 0 | 1.687701816 | 0 | 0 |
| 0 | 0 | 0 | 0 | 0 |

| | | | | |
|---|---|---|---|---|
| 0 | 0 | 0 | 0 | 0 |
| 0 | 0 | 0 | 0 | 0 |
| 0.294551306 | 0 | 0 | 0 | 0.032731262 |
| 0 | 0 | 0 | 0 | 0 |
| 0 | 0 | 0.039039357 | 0 | 0 |
| 0 | 0 | 0 | 0 | 0 |
| 0 | 0 | 0 | 0 | 0 |
| 0 | 0 | 0 | 0 | 0 |
| 0 | 0 | 0 | 0 | 0 |
| 0 | 0 | 0 | 0 | 0 |
| 0 | 0 | 0 | 0 | 0 |
| 0 | 0 | 0.001072352 | 0.000631362 | 0 |
| 0 | 0 | 0 | 0 | 0 |
| 0 | 0 | 0 | 0 | 0 |
| 0 | 0 | 0 | 0 | 0 |
| 0 | 0 | 0 | 0 | 0 |
| 0 | 0 | 0 | 0 | 0 |
| 0 | 0 | 0 | 0 | 0 |
| 0 | 0 | 0 | 0 | 0 |
| 0.001160015 | 0 | 0.00076202 | 0 | 0 |
| 0 | 0 | 0 | 0 | 0 |
| 0 | 0 | 0 | 0 | 0 |
| 0 | 0 | 0 | 0 | 0 |
| 0 | 0 | 0 | 0 | 0 |
| 0 | 0 | 0 | 0 | 0 |
| 0 | 0 | 0 | 0 | 0 |
| 0 | 0 | 0 | 0 | 0 |
| 0 | 0 | 0 | 0 | 0.008486896 |
| 0 | 0 | 0 | 0 | 0 |
| 0 | 0 | 0 | 0 | 0 |
| 0.00056947 | 0.00156091 | 0 | 0.00353317 | 0.002131525 |
| 0 | 0 | 0 | 0 | 0 |
| 0.026283215 | 0.075601967 | 0 | 0.18852255 | 0.083922198 |
| 0.078548723 | 0.002381209 | 0.00058257 | 0 | 0 |
| 0 | 0 | 0 | 0 | 0 |
| 0 | 0 | 0 | 0 | 0 |
| 0 | 0 | 0 | 0 | 0 |
| 0 | 0 | 0 | 0 | 0 |
| 0 | 0 | 0 | 0 | 0 |
| 0 | 0 | 0 | 0 | 0 |
| 0 | 0 | 0 | 0 | 0 |
| 0 | 1.292505871 | 0.00938016 | 0.233027208 | 0 |
| 0.004638995 | 0 | 0 | 0 | 0.012402671 |
| 0 | 0 | 0 | 0 | 0 |
| 0 | 0 | 0 | 0 | 0 |
| 0.006464254 | 0.009374833 | 0 | 0.004861362 | 0 |
| 0 | 0 | 0 | 0.000531208 | 0 |
| 0 | 0 | 0 | 0 | 0 |

| | | | | |
|---|---|---|---|---|
| 0 | 0 | 0 | 0 | 0 |
| 0.002603551 | 0 | 0 | 0 | 0.328852227 |
| 0.013584129 | 0.005716081 | 0 | 0 | 0.000827875 |
| 0 | 0 | 0 | 0 | 0 |
| 0 | 0 | 0.003349802 | 0.183345495 | 0 |
| 0 | 0.211516436 | 0.042932254 | 0 | 0 |
| 0 | 0 | 0 | 0 | 0 |
| 0 | 0 | 0 | 0 | 0 |
| 0 | 0 | 0 | 0 | 0 |
| 0 | 0 | 0 | 0 | 0 |
| 0 | 0 | 0 | 0 | 0 |
| 0 | 0 | 0 | 0 | 0 |
| 0 | 0 | 0 | 0 | 0 |
| 0 | 0 | 0 | 0 | 0 |
| 0 | 0 | 0 | 0 | 0 |
| 0 | 0 | 0 | 0 | 0 |
| 0 | 0 | 0 | 0 | 0 |
| 0 | 0 | 0 | 0 | 0 |
| 0 | 0 | 0 | 0 | 0 |
| 0 | 0 | 0 | 0 | 0 |
| 0 | 0 | 0 | 0 | 0 |
| 0 | 0 | 0 | 0 | 0 |
| 0 | 0 | 0 | 0 | 0 |
| 0 | 0 | 0 | 0 | 0 |
| 0 | 0 | 0 | 0 | 0 |
| 0 | 0 | 0 | 0 | 0 |
| 0 | 0 | 0 | 0 | 0 |
| 0 | 0 | 0 | 0 | 0 |
| 0 | 0 | 0 | 0 | 0 |
| 0 | 0 | 0 | 0 | 0 |
| 0 | 0 | 0 | 0 | 0 |
| 0 | 0 | 0 | 0 | 0 |
| 0 | 0 | 0 | 0 | 0 |
| 0 | 0 | 0 | 0 | 0 |
| 0 | 0 | 0 | 0 | 0 |
| 0 | 0 | 0 | 0 | 0 |
| 0 | 0 | 0 | 0 | 0 |
| 0 | 0.156456601 | 0 | 0 | 0 |
| 0.035864288 | 4.052376483 | 0 | 3.664881962 | 0 |
| 0 | 0 | 0 | 0 | 0 |
| 0 | 0 | 0 | 0 | 0 |
| 0.000729414 | 0 | 0 | 0 | 0 |
| 0 | 0 | 0.002715061 | 0.008944155 | 0 |
| 0 | 0 | 0 | 0 | 0 |
| 0 | 0 | 0 | 0 | 0 |
| 0 | 0 | 0 | 0 | 0 |
| 0.002064603 | 0 | 0.010680461 | 0.006404718 | 0.002810109 |
| 0 | 0 | 0 | 0 | 0 |

| | | | | |
|---|---|---|---|---|
| 0 | 0 | 0 | 0 | 0 |
| 0.032238722 | 0.004335412 | 0.006551211 | 0.010285643 | 0.003767456 |
| 0 | 0 | 0 | 0 | 0 |
| 0 | 0 | 0 | 0 | 0 |
| 0 | 0 | 0 | 0 | 0 |
| 0 | 0 | 0 | 0 | 0 |
| 0 | 0 | 0 | 0 | 0 |
| 0 | 0 | 0 | 0 | 0 |
| 0 | 0 | 0 | 0 | 0 |
| 0 | 0 | 0 | 0 | 0 |
| 0 | 0 | 0 | 0 | 0 |
| 0 | 0 | 0 | 0 | 0 |
| 0.016695359 | 0 | 0 | 0 | 0 |
| 0 | 0 | 0.001874672 | 0 | 0 |
| 0.006496457 | 0.004133699 | 0 | 0.006641335 | 0 |
| 0 | 0 | 0 | 0 | 0 |
| 0 | 0 | 0 | 0 | 0 |
| 0 | 0 | 0 | 0 | 0 |
| 0 | 0 | 0 | 0 | 0 |
| 0 | 0 | 0 | 0 | 0 |
| 0 | 0 | 0 | 0 | 0 |
| 0.012959443 | 0.54024152 | 0.034379944 | 0.002473981 | 0 |
| 0 | 0 | 0 | 0.010715022 | 0 |
| 0 | 0 | 0 | 0 | 0 |
| 0 | 0 | 0 | 0 | 0 |
| 0 | 0 | 0 | 0 | 0 |
| 0 | 0 | 0.001166607 | 0 | 0 |
| 0 | 0 | 0 | 0 | 0 |
| 0 | 0 | 0 | 0 | 0 |
| 0 | 0 | 0 | 0 | 0 |
| 0 | 0 | 0 | 0 | 0 |
| 0 | 0 | 0 | 0 | 0 |
| 0 | 0 | 0 | 0 | 0 |
| 0 | 0 | 0 | 0 | 0 |
| 39.29144509 | 0 | 0 | 0 | 45.9241861 |
| 0 | 0 | 0 | 0 | 0 |
| 0 | 0 | 0 | 0 | 0 |
| 0 | 0 | 0 | 0 | 0 |
| 0.234541103 | 0.108523001 | 0 | 0 | 0 |
| 0 | 0 | 0 | 0 | 0 |
| 0.058376195 | 0.565656514 | 0.04822773 | 0.045109377 | 0.095006089 |
| 0 | 0 | 0 | 0 | 0 |
| 0 | 0 | 0 | 0 | 0 |
| 0 | 0 | 0 | 0 | 0 |
| 0 | 0 | 0 | 0 | 0 |
| 0.128972484 | 1.382673968 | 0.140510834 | 0.599245481 | 0.039504148 |
| 0 | 0 | 0 | 0 | 0 |
| 0 | 0 | 0 | 0 | 0 |

| | | | | |
|---|---|---|---|---|
| 0 | 0 | 0 | 0.002875029 | 0 |
| 0 | 0 | 0 | 0 | 0 |
| 0 | 0 | 0 | 0 | 0 |
| 0 | 0 | 0 | 0 | 0 |
| 0 | 0 | 0 | 0 | 0 |
| 0 | 0 | 0 | 0 | 0 |
| 0 | 0 | 0 | 0 | 0 |
| 0 | 0.006920468 | 0 | 0 | 0 |
| 0.000553851 | 0 | 0 | 0 | 0 |
| 0 | 0 | 0 | 0 | 0.001904548 |
| 0.007800355 | 0 | 0 | 0 | 0 |
| 0 | 0 | 0 | 0 | 0 |
| 0 | 0 | 0 | 0 | 0 |
| 0 | 0 | 0 | 0 | 0 |
| 0 | 0 | 0 | 0 | 0 |
| 0 | 0 | 0 | 0 | 0 |
| 0 | 0 | 0 | 0 | 0 |
| 0 | 0 | 0 | 0 | 0 |
| 0 | 0 | 0 | 0 | 0 |
| 0.243814243 | 0.028600211 | 0.000599754 | 0.07278062 | 0.035504955 |
| 0 | 0 | 0 | 0 | 0 |
| 0 | 0 | 0 | 0 | 0 |
| 0 | 0 | 0 | 0 | 0 |
| 0 | 0 | 0 | 0 | 0 |
| 0 | 0 | 0 | 0 | 0 |
| 0 | 0 | 0 | 0 | 0 |
| 0 | 0 | 0 | 0 | 0 |
| 0 | 0 | 0 | 0 | 0 |
| 0 | 0 | 0 | 0 | 0 |
| 0.001301348 | 0 | 0 | 0 | 0 |
| 0 | 0 | 0 | 0 | 0 |
| 0 | 0 | 0 | 0 | 0 |
| 0 | 0 | 0 | 0 | 0 |
| 0 | 0 | 0 | 0 | 0 |
| 0 | 0 | 0 | 0 | 0 |
| 0 | 0 | 0 | 0 | 0 |
| 0 | 0 | 0 | 0 | 0 |
| 0 | 0 | 0 | 0 | 0 |
| 0 | 0 | 0 | 0 | 0 |
| 0 | 0 | 0 | 0 | 0 |
| 0 | 0 | 0 | 0 | 0 |
| 0 | 0 | 0 | 0 | 0 |
| 0 | 0 | 0 | 0.007501115 | 0 |
| 0 | 0 | 0 | 0 | 0 |
| 0 | 0 | 0.003943178 | 0.001289778 | 0 |
| 0 | 0 | 0 | 0 | 0 |
| 0 | 0 | 0 | 0 | 0 |
| 0 | 0 | 0 | 0 | 0 |

| | | | | |
|---|---|---|---|---|
| 0 | 0 | 0 | 0 | 0 |
| 0 | 0 | 0 | 0 | 0 |
| 0 | 0 | 0 | 0 | 0 |
| 0 | 0 | 0 | 0 | 0 |
| 0 | 0 | 0 | 0 | 0 |
| 0.002926509 | 0.007639544 | 0.000560712 | 0.007015191 | 0 |
| 0 | 0 | 0 | 0 | 0 |
| 0 | 0 | 0 | 0 | 0 |
| 0 | 0 | 0 | 0 | 0 |
| 0 | 0 | 0 | 0 | 0 |
| 0 | 0 | 0 | 0 | 0 |
| 0 | 0 | 0 | 0 | 0 |
| 0 | 0 | 0 | 0 | 0 |
| 0 | 0 | 0 | 0 | 0 |
| 0 | 0 | 0 | 0 | 0 |
| 0 | 0 | 0 | 0 | 0 |
| 0 | 0 | 0 | 0 | 0 |
| 0 | 0 | 0 | 0 | 0 |
| 0 | 0 | 0 | 0 | 0 |
| 0 | 0 | 0 | 0 | 0 |
| 0 | 0 | 0 | 0 | 0 |
| 0 | 0 | 0 | 0 | 0 |
| 0 | 0 | 0 | 0 | 0 |
| 0 | 0 | 0 | 0 | 0 |
| 0 | 0 | 0 | 0 | 0 |
| 0 | 0 | 0 | 0 | 0 |
| 0 | 0 | 0 | 0 | 0 |
| 0 | 0 | 0 | 0 | 0 |
| 0 | 0 | 0 | 0 | 0 |
| 0 | 0 | 0.000760247 | 0 | 0 |
| 0 | 0 | 0 | 0 | 0 |
| 0 | 0 | 0 | 0 | 0 |
| 0 | 0 | 0 | 0 | 0 |
| 0 | 0 | 0 | 0 | 0 |
| 0 | 0 | 0 | 0 | 0 |
| 0 | 0 | 0 | 0 | 0 |
| 0 | 0 | 0 | 0 | 0 |
| 0 | 0 | 0 | 0 | 0 |
| 2.628195248 | 0 | 0 | 0 | 0.340876196 |
| 0 | 0 | 0 | 0 | 0 |
| 0 | 0 | 0 | 0 | 0 |
| 0 | 0 | 0 | 0 | 0 |
| 0 | 0 | 0 | 0 | 0 |
| 0 | 0 | 0 | 0 | 0 |
| 0 | 0 | 0 | 0 | 0 |
| 0 | 0 | 0 | 0 | 0 |
| 0 | 0 | 0 | 0 | 0 |
| 0 | 0 | 0 | 0 | 0 |

| | | | | |
|---|---|---|---|---|
| 0 | 0 | 0 | 0 | 0 |
| 0.001365193 | 0 | 0 | 0.020482188 | 0 |
| 0 | 0 | 0 | 0.013198057 | 0 |
| 0 | 0 | 0 | 0 | 0 |
| 0 | 0 | 0 | 0 | 0 |
| 0 | 0 | 0 | 0 | 0 |
| 0 | 0 | 0 | 0 | 0 |
| 0 | 0 | 0 | 0 | 0 |
| 0 | 0 | 0 | 0 | 0 |
| 0 | 0 | 0 | 0 | 0 |
| 0.001072567 | 0 | 0 | 0 | 0.003892959 |
| 0 | 0 | 0 | 0 | 0 |
| 0.337988235 | 0 | 0 | 0 | 0.231180967 |
| 0 | 0 | 0 | 0 | 0 |
| 0 | 0 | 0 | 0 | 0 |
| 0 | 0 | 0 | 0 | 0 |
| 0 | 0 | 0 | 0 | 0 |
| 0 | 0 | 0 | 0 | 0 |
| 0 | 0 | 0 | 0 | 0 |
| 0 | 0 | 0 | 0 | 0 |
| 0 | 0.001420863 | 0 | 0 | 0.00098778 |
| 0 | 0 | 0 | 0 | 0 |
| 0 | 0 | 0 | 0 | 0 |
| 1.921560409 | 0 | 0 | 2.386001871 | 2.019096673 |
| 0 | 0 | 0 | 0 | 0 |
| 0 | 0 | 0 | 0 | 0 |
| 0 | 0 | 0 | 0 | 0 |
| 0 | 0 | 0 | 0 | 0 |
| 0 | 0 | 0 | 0 | 0 |
| 0 | 0 | 0 | 0 | 0 |
| 0 | 0 | 0.002047125 | 0 | 0 |
| 0 | 0 | 0 | 0 | 0 |
| 0 | 0.002690302 | 0 | 0 | 0 |
| 0 | 0 | 0 | 0 | 0 |

| PHL018-0160_V1 | PHL018-0198_V2 | PHL018-0168_V1 | PHL018-0117_V2 | PHL018-0251_V2 |
|---|---|---|---|---|
| REF | REF | EYCF | EYCF | REF |
| 0.099238341 | 0.081734462 | 0.041334785 | 2.142053036 | 0.128880589 |
| 0.15675094 | 0 | 1.037641789 | 0.358479844 | 21.02548503 |
| 12.1966446 | 0.011332142 | 0 | 0 | 0 |
| 0 | 0 | 0 | 0 | 0 |
| 0 | 0 | 0 | 0 | 0 |
| 0 | 0 | 0 | 0 | 0 |
| 0 | 0 | 0 | 0 | 0 |
| 0 | 0 | 0 | 0 | 0 |
| 0 | 0 | 0 | 0 | 0 |
| 0 | 0 | 0 | 0 | 0 |
| 0 | 0 | 0 | 0 | 0 |
| 0 | 0 | 0 | 0 | 0 |
| 0 | 0 | 0 | 0 | 0 |
| 3.523778205 | 0 | 3.32757156 | 1.262194166 | 0 |
| 0 | 0 | 0 | 0 | 0 |
| 0 | 0 | 0 | 0 | 0 |
| 0 | 0 | 0 | 0 | 0 |
| 0 | 0 | 0 | 0 | 0 |
| 0 | 0 | 0 | 0 | 0 |
| 0 | 0 | 0 | 0 | 0 |
| 0 | 0 | 0 | 0 | 0 |
| 0 | 0 | 0 | 0 | 0 |
| 0 | 0 | 0 | 0 | 0 |
| 0 | 0 | 0 | 0 | 0 |
| 0 | 0 | 0 | 0 | 0 |
| 0 | 0 | 0 | 0 | 0 |
| 0 | 0 | 0 | 0 | 0 |
| 0 | 0 | 0 | 0 | 0 |
| 0 | 0 | 0 | 0 | 0 |
| 0 | 0 | 0 | 0 | 0 |
| 0 | 0 | 0 | 0 | 0 |
| 0 | 0 | 0 | 0 | 0 |
| 0 | 0 | 0 | 0 | 0 |
| 0 | 0 | 0 | 0 | 0 |
| 0 | 0 | 0 | 0 | 0 |
| 1.871311773 | 0 | 3.00447437 | 0.38755067 | 0 |
| 0 | 0 | 0 | 0 | 0 |
| 0 | 0 | 0 | 0 | 0 |
| 0 | 0 | 0 | 0 | 0 |
| 0 | 0 | 0 | 0 | 0 |
| 0 | 0 | 0 | 0 | 0 |
| 0.007243742 | 0 | 0.440437632 | 0 | 0.005423975 |
| 0.29142799 | 0.005226581 | 2.251796094 | 0.014651346 | 0.554100899 |
| 0 | 0.017167914 | 0.008675666 | 0.001191969 | 0.054410173 |
| 0 | 0 | 0.005900801 | 0.001933577 | 0 |
| 0 | 0 | 0 | 0 | 0 |

| | | | | |
|---|---|---|---|---|
| 0 | 0 | 0.001529513 | 0 | 0 |
| 0.006150744 | 0.059477821 | 0.003577643 | 0 | 0.118716921 |
| 0.009119105 | 0.005763093 | 0 | 0.144130757 | 0.000834216 |
| 1.322084461 | 0 | 0.15356058 | 0.131345171 | 1.020463534 |
| 0.018875193 | 0 | 0.02259161 | 0 | 0 |
| 0 | 0 | 0 | 0 | 0 |
| 0 | 0 | 0.007443744 | 0.051945264 | 0 |
| 0 | 0 | 0.009776207 | 0 | 0 |
| 0 | 0 | 0 | 0 | 0 |
| 0 | 0 | 0 | 0 | 0 |
| 0 | 0.001830453 | 0 | 0.002452409 | 0 |
| 0.024706746 | 0 | 0.237350925 | 0 | 0.08751078 |
| 0.003544903 | 0 | 0 | 0 | 0 |
| 0.019374087 | 0 | 0.063971707 | 0 | 0.0436284 |
| 0 | 0 | 0.001773124 | 0 | 0 |
| 0 | 0 | 0 | 0 | 0 |
| 0.000892767 | 0.006356777 | 0.533055897 | 0.285435404 | 0 |
| 0 | 0 | 0 | 0 | 0 |
| 0 | 0 | 0 | 0 | 0 |
| 0 | 0 | 0 | 0 | 0 |
| 0 | 0.186382219 | 0.063330819 | 0 | 0.252877829 |
| 0.075109731 | 0.04051951 | 0.008975316 | 0 | 0.013574611 |
| 0 | 0.103596158 | 0.281332882 | 0 | 0 |
| 0 | 0 | 0 | 0 | 0 |
| 0 | 0 | 0 | 0 | 0 |
| 0 | 0 | 0 | 0 | 0 |
| 0.037416408 | 0.001267643 | 0 | 0.002264488 | 0 |
| 0 | 0 | 0 | 0 | 0 |
| 0.002479666 | 0 | 0.005535976 | 0 | 0 |
| 0 | 0 | 0 | 0 | 0 |
| 0 | 0.259037967 | 0.153639992 | 0 | 1.2369074 |
| 0 | 0 | 0 | 0 | 0 |
| 0 | 0 | 0 | 0 | 0 |
| 0 | 0 | 0 | 0 | 0 |
| 0 | 0 | 0 | 0 | 0 |
| 0 | 0 | 0 | 0 | 0 |
| 0 | 0 | 0 | 0 | 0 |
| 0 | 0.001287358 | 0.001943485 | 0.005130114 | 0 |
| 0 | 0 | 0 | 0 | 0 |
| 0 | 0 | 0 | 0 | 0 |
| 0 | 0 | 0 | 0 | 0 |
| 0 | 0 | 0 | 0 | 0 |
| 0 | 0 | 0 | 0 | 0 |
| 0 | 0 | 0 | 0 | 0 |
| 0 | 0 | 0 | 0 | 0 |
| 0 | 0 | 0 | 0 | 0 |
| 0 | 0 | 0 | 0 | 0 |
| 0 | 0.029139146 | 0.012468464 | 0 | 0 |

| | | | | |
|---|---|---|---|---|
| 0 | 0 | 0 | 0 | 0 |
| 0 | 0 | 0 | 0 | 0 |
| 0 | 0 | 0 | 0 | 0 |
| 0 | 0 | 0 | 0 | 0 |
| 0.21474738 | 0 | 0 | 0.001328438 | 0 |
| 0 | 0 | 0 | 0 | 0 |
| 0 | 0 | 0 | 0 | 0 |
| 0.004171116 | 0 | 0 | 0 | 0 |
| 0 | 0 | 0.037644805 | 0 | 0.00321834 |
| 0 | 0 | 0 | 0 | 0 |
| 0 | 0 | 0 | 0 | 0 |
| 0 | 0 | 0.156114533 | 0 | 0 |
| 0 | 0 | 0 | 0 | 0 |
| 0 | 0 | 0 | 0 | 0 |
| 0 | 0 | 0 | 0 | 0 |
| 0 | 0 | 0 | 0 | 0 |
| 0 | 0 | 0 | 0 | 0 |
| 0 | 0 | 0 | 0 | 0 |
| 0 | 0.002827338 | 0.198157923 | 0 | 0 |
| 0 | 0 | 0 | 0 | 0 |
| 0 | 0 | 0 | 0 | 0 |
| 0 | 0 | 0 | 0 | 0 |
| 0 | 0 | 0 | 0 | 0 |
| 0 | 0 | 0 | 0 | 0 |
| 0 | 0 | 0 | 0 | 0 |
| 0 | 0 | 0 | 0 | 0 |
| 0 | 0 | 0.001362099 | 0 | 0 |
| 0 | 0 | 0.002294002 | 0 | 0 |
| 0 | 0 | 0 | 0 | 0 |
| 0 | 0 | 0 | 0 | 0 |
| 0 | 0 | 0.019066292 | 0 | 0 |
| 0.001970385 | 0 | 0 | 0.000410672 | 0.210773282 |
| 0 | 0 | 0 | 0 | 0 |
| 0 | 0 | 0 | 0 | 0 |
| 0 | 0 | 0 | 0 | 0 |
| 0 | 0 | 0 | 0 | 0 |
| 0 | 0 | 0 | 0 | 0 |
| 0 | 0 | 0 | 0 | 0 |
| 0.016861339 | 0 | 0.827297434 | 0.002100489 | 0 |
| 0 | 0 | 0 | 0 | 0 |
| 0 | 0 | 0 | 0 | 0 |
| 0 | 0 | 0 | 0 | 0 |
| 0 | 0 | 0 | 0 | 0 |
| 0 | 0 | 0 | 0 | 0 |
| 0 | 0 | 0 | 0 | 0 |
| 0 | 0 | 0 | 0 | 0 |
| 0 | 0 | 0 | 0 | 0 |
| 0 | 0 | 0 | 0 | 0 |

| | | | | |
|---|---|---|---|---|
| 0 | 0 | 0 | 0 | 0 |
| 0 | 0 | 0 | 0 | 0 |
| 0 | 0 | 0 | 0 | 0 |
| 0 | 0 | 0 | 0 | 0 |
| 0.004089608 | 0 | 0 | 0 | 0 |
| 0 | 0 | 0 | 0 | 0 |
| 0 | 0 | 0 | 0 | 0 |
| 0 | 0 | 0 | 0 | 0 |
| 0 | 0 | 0 | 0 | 0 |
| 0 | 0 | 0 | 0 | 0 |
| 0 | 0 | 0 | 0 | 0 |
| 0 | 0 | 0 | 0 | 0 |
| 0 | 0 | 0 | 0 | 0 |
| 0 | 0 | 0 | 0 | 0 |
| 0 | 0 | 0 | 0 | 0 |
| 0 | 0 | 3.499195795 | 0 | 0 |
| 0 | 0 | 0 | 0 | 0 |
| 0 | 0 | 0 | 0 | 0 |
| 0 | 0 | 0 | 0 | 0 |
| 0 | 0 | 0 | 0 | 0 |
| 0 | 0 | 0 | 0 | 0 |
| 0 | 0 | 0 | 0 | 0 |
| 0 | 0 | 0 | 0 | 0 |
| 0 | 0 | 0 | 0 | 0 |
| 0.004265383 | 0 | 0 | 0 | 0 |
| 0 | 0 | 0 | 0 | 0 |
| 0 | 0 | 0 | 0 | 0 |
| 0 | 0 | 0 | 0 | 0 |
| 0 | 0 | 0 | 0 | 0 |
| 0 | 0 | 0 | 0 | 0 |
| 0 | 0 | 0 | 0 | 0 |
| 0 | 0 | 0.000867602 | 0 | 0 |
| 0 | 0 | 0 | 0 | 0 |
| 0 | 0 | 0 | 0 | 0 |
| 0 | 0 | 0 | 0 | 0 |
| 0 | 0 | 0 | 0 | 0 |
| 0 | 0 | 0 | 0 | 0 |
| 0 | 0 | 0.01354222 | 0 | 0 |
| 0 | 0 | 0 | 0 | 0 |
| 0 | 0 | 0 | 0 | 0 |
| 0 | 0 | 0 | 0 | 0 |
| 0 | 0 | 0 | 0 | 0 |
| 0 | 0 | 0 | 0 | 0 |
| 0 | 0 | 0 | 0 | 0 |
| 0 | 0 | 0 | 0 | 0 |
| 0 | 0 | 0 | 0 | 0 |
| 0 | 0 | 0 | 0 | 0 |
| 0 | 0 | 0.002978355 | 0 | 0.042835987 |

| | | | | |
|---|---|---|---|---|
| 0 | 0 | 0 | 0 | 0 |
| 0 | 0 | 0 | 0 | 0 |
| 0 | 0 | 0 | 0 | 0 |
| 0 | 0 | 0 | 0 | 0 |
| 0 | 0 | 0 | 0 | 0 |
| 0 | 0 | 0 | 0 | 0 |
| 0 | 0 | 0 | 0 | 0 |
| 0 | 0 | 0 | 0 | 0 |
| 0 | 0 | 0 | 0 | 0 |
| 0 | 0 | 0 | 0 | 0 |
| 0 | 0 | 0 | 0 | 0 |
| 0 | 0 | 0 | 0 | 0 |
| 0.00267744 | 0.002932953 | 0 | 0.002418166 | 0.004245491 |
| 0 | 0 | 0 | 0 | 0 |
| 0 | 0 | 0 | 0 | 0 |
| 0 | 0 | 0 | 0 | 0 |
| 0 | 0 | 0 | 0 | 0 |
| 0 | 0 | 0.002595963 | 0 | 0 |
| 0 | 0 | 0 | 0 | 0 |
| 0 | 0 | 0 | 0 | 0 |
| 0.043491092 | 0.06409329 | 0.02049028 | 0.001412934 | 0.150326804 |
| 0 | 0 | 0 | 0 | 0 |
| 0 | 0 | 0 | 0.00161169 | 0 |
| 0.009135747 | 0 | 0 | 0 | 0.013451406 |
| 0 | 0 | 0 | 0 | 0 |
| 0 | 0 | 0 | 0 | 0 |
| 0 | 0.000998033 | 0.005424117 | 0.000822859 | 0.005778666 |
| 0 | 0 | 0 | 0 | 0 |
| 0 | 0 | 0 | 0 | 0 |
| 0 | 0.002349116 | 0 | 0 | 0 |
| 0 | 0.002188712 | 0 | 0 | 0 |
| 0 | 0 | 0 | 0 | 0 |
| 0 | 0 | 0 | 0 | 0 |
| 0 | 0 | 0.003225463 | 0 | 0 |
| 0.011400481 | 0 | 0.038178196 | 0.070236828 | 0.00484211 |
| 0 | 0 | 0 | 0 | 0 |
| 0 | 0 | 0 | 0 | 0 |
| 0 | 0 | 0 | 0 | 0 |
| 0 | 0 | 0 | 0 | 0 |
| 0 | 0 | 0 | 0 | 0 |
| 0 | 0 | 0 | 0 | 0 |
| 0 | 0 | 0 | 0 | 0 |
| 0 | 0 | 0 | 0 | 0 |
| 0 | 0 | 0 | 0 | 0 |
| 0 | 0 | 0 | 0 | 0 |
| 0 | 0 | 0 | 0 | 0 |
| 0.002865499 | 0 | 0 | 0 | 0 |
| 0 | 0.024791767 | 0 | 0.003668782 | 0.009431691 |

| | | | | |
|---|---|---|---|---|
| 0 | 0 | 0 | 0 | 0 |
| 0 | 0 | 0 | 0 | 0 |
| 0 | 0 | 0 | 0 | 0 |
| 0.000922057 | 0 | 0 | 0 | 0 |
| 0 | 0 | 0 | 0 | 0 |
| 0 | 0 | 0 | 0 | 0 |
| 0.001239088 | 0 | 0.001434391 | 0 | 0 |
| 0.002502604 | 0 | 0.00372479 | 0 | 0 |
| 0 | 0 | 0 | 0 | 0 |
| 0 | 0 | 0 | 0 | 0 |
| 0 | 0 | 0.002137251 | 0 | 0 |
| 0 | 0 | 0 | 0 | 0 |
| 0 | 0 | 0 | 0 | 0 |
| 0 | 0 | 0 | 0.107149527 | 0 |
| 0 | 0.0015034 | 0 | 0 | 0 |
| 0 | 0 | 0 | 0 | 0 |
| 0 | 0 | 0 | 0 | 0 |
| 0 | 0 | 0 | 0 | 0 |
| 0.00147542 | 0 | 0.012882991 | 0 | 0.003041356 |
| 0 | 0 | 0 | 0 | 0 |
| 0.007835019 | 0.00309932 | 0 | 0.020442654 | 0.063153429 |
| 0 | 0 | 0 | 0 | 0 |
| 0 | 0 | 0 | 0 | 0 |
| 0 | 0 | 0 | 0 | 0 |
| 0 | 0.005210153 | 0 | 0.003124127 | 0 |
| 0 | 0 | 0 | 0 | 0 |
| 0 | 0 | 0 | 0 | 0 |
| 0 | 0 | 0 | 0 | 0 |
| 0 | 0 | 0.105633589 | 0 | 0 |
| 0 | 0 | 0 | 0 | 0 |
| 0.373350405 | 0.356389352 | 0.154025306 | 0.208995322 | 0.566121626 |
| 0 | 0 | 0 | 0 | 0 |
| 0 | 0 | 0.029221362 | 0.017791881 | 0 |
| 0 | 0 | 0 | 0 | 0 |
| 0 | 0 | 0 | 0 | 0 |
| 0 | 0 | 0 | 0 | 0 |
| 0 | 0 | 0 | 0 | 0 |
| 0 | 0 | 0 | 0.005256058 | 0 |
| 0 | 0 | 0 | 0 | 0 |
| 0 | 0 | 0 | 0 | 0 |
| 0 | 0 | 0 | 0 | 0 |
| 0.012234312 | 0.003037754 | 0.005665063 | 0.028434263 | 0.009053044 |
| 0.000684336 | 0 | 0 | 0 | 0 |
| 0 | 0 | 0 | 0 | 0 |
| 0 | 0 | 0 | 0 | 0 |
| 0 | 0 | 0 | 0 | 0 |
| 0 | 0 | 0 | 0 | 0 |
| 0 | 0 | 0 | 0 | 0 |

| | | | | |
|---|---|---|---|---|
| 0 | 0 | 0 | 0 | 0 |
| 0 | 0 | 0 | 0 | 0 |
| 0 | 0.021091391 | 0.001625924 | 0.001109966 | 0.054239624 |
| 0 | 0 | 0 | 0 | 0 |
| 0 | 0 | 0 | 0 | 0 |
| 0 | 0 | 0 | 0 | 0 |
| 0 | 0 | 0 | 0 | 0 |
| 0 | 0 | 0 | 0 | 0 |
| 0 | 0 | 0 | 0 | 0 |
| 0.000936676 | 0.001026065 | 0.000557647 | 0 | 0 |
| 0 | 0 | 0 | 0 | 0 |
| 0 | 0 | 0 | 0 | 0 |
| 0 | 0 | 0 | 0 | 0 |
| 0.003621972 | 0 | 0.00808623 | 0 | 0.002512647 |
| 0.007847343 | 0.007163526 | 0.00315167 | 0 | 0.014517028 |
| 0.006401232 | 0 | 0 | 0 | 0 |
| 0.001490224 | 0.001865645 | 0 | 0 | 0 |
| 0 | 0 | 0 | 0 | 0 |
| 0 | 0 | 0 | 0 | 0 |
| 0 | 0 | 0 | 0 | 0 |
| 0 | 0 | 0 | 0 | 0 |
| 0 | 0.018315075 | 0 | 0.004165639 | 0.013103301 |
| 0 | 0 | 0 | 0 | 0 |
| 0 | 1.98388826 | 0 | 0 | 0.008503161 |
| 0 | 0 | 0.001528782 | 0 | 0 |
| 0 | 0 | 0 | 0 | 0 |
| 0 | 0 | 0 | 0 | 0 |
| 0 | 0 | 0 | 0 | 0 |
| 0 | 0 | 0 | 0 | 0 |
| 0 | 0 | 0.007723278 | 0 | 0 |
| 0 | 0 | 0 | 0 | 0 |
| 0 | 0 | 0 | 0 | 0 |
| 0 | 0 | 0 | 0 | 0 |
| 0 | 0 | 0 | 0 | 0 |
| 0 | 0 | 0 | 0 | 0 |
| 0.014586243 | 0 | 0.001904357 | 0 | 0 |
| 0 | 0 | 0 | 0 | 0 |
| 0.03501053 | 0.003438424 | 0.30642182 | 0 | 0 |
| 0 | 0 | 0 | 0 | 0 |
| 0 | 0 | 0 | 0 | 0 |
| 0 | 0 | 0 | 0 | 0 |
| 0 | 0 | 0 | 0 | 0 |
| 0 | 0 | 0 | 0 | 0 |
| 0 | 0 | 0 | 0 | 0 |
| 0 | 0 | 0 | 0 | 0 |
| 0 | 0 | 0.004788541 | 0 | 0 |
| 0 | 0 | 0.001817866 | 0 | 0 |
| 0 | 0.017299916 | 0 | 0 | 0.025041874 |

| | | | | |
|---|---|---|---|---|
| 0 | 0 | 0.119376067 | 0 | 0 |
| 0.029174779 | 0 | 0 | 0 | 0 |
| 0.000805166 | 0 | 0 | 0 | 0 |
| 0 | 0 | 0 | 0 | 0 |
| 0 | 0.010204896 | 0.012659726 | 0 | 0.00417462 |
| 0 | 0 | 0.011795904 | 0 | 0 |
| 0 | 0 | 0 | 0 | 0 |
| 0 | 0.001554827 | 0.09633213 | 0 | 0.016654692 |
| 0 | 0 | 0.000976753 | 0 | 0 |
| 0.443575144 | 0.108976379 | 0 | 0.269811295 | 0.009743065 |
| 0 | 0 | 0 | 0 | 0.093123607 |
| 0.021572753 | 0 | 0.04704736 | 0 | 0 |
| 0.014318237 | 0.023713706 | 0 | 0.004926365 | 0.01405471 |
| 0 | 0 | 0.001541218 | 0 | 0.119863076 |
| 0 | 0 | 0 | 0 | 0.000833939 |
| 0 | 0 | 0 | 0 | 0 |
| 0.000744312 | 0 | 0 | 0 | 0 |
| 0 | 0 | 0 | 0 | 0 |
| 0 | 0.076908589 | 0 | 0 | 0.026194418 |
| 0 | 0 | 0 | 0 | 0 |
| 0 | 0 | 0 | 0 | 0 |
| 0 | 0 | 0 | 0 | 0 |
| 0 | 0 | 0 | 0 | 0 |
| 0 | 0 | 0 | 0 | 0 |
| 0 | 0 | 0 | 0 | 0 |
| 0 | 0 | 0.001575976 | 0.023669084 | 0 |
| 0 | 0 | 0 | 0 | 0 |
| 0 | 0 | 0 | 0.001084186 | 0.111162527 |
| 0 | 0 | 0 | 0 | 0 |
| 0.003268311 | 0 | 0 | 0 | 0 |
| 0 | 0.002100346 | 0 | 0 | 0 |
| 0 | 0 | 0 | 0 | 0 |
| 0 | 0.043380896 | 0 | 0 | 0 |
| 0 | 0 | 0 | 0 | 0 |
| 0 | 0 | 0 | 0 | 0 |
| 0 | 0 | 0 | 0 | 0 |
| 0 | 0.113901471 | 0.001339898 | 0 | 1.143411879 |
| 0 | 0 | 0 | 0 | 0 |
| 0 | 0 | 0 | 0 | 0 |
| 0 | 0 | 0 | 0 | 0.607237812 |
| 0 | 0 | 0 | 0 | 0 |
| 0 | 0 | 0 | 0 | 0 |
| 0 | 0 | 0 | 0 | 0 |
| 0 | 0 | 0 | 0 | 0.321593402 |
| 0 | 0 | 0 | 0 | 0 |
| 0 | 0 | 0 | 0 | 0 |
| 0 | 0 | 0 | 0 | 0 |
| 0 | 0 | 0 | 0 | 0 |

| | | | | |
|---|---|---|---|---|
| 0 | 0 | 0 | 0 | 0 |
| 0 | 0 | 0 | 0.022428391 | 0 |
| 0 | 0 | 0 | 0 | 0 |
| 1.638429358 | 0 | 0 | 0 | 0 |
| 0 | 0 | 0 | 0 | 0 |
| 0.027807882 | 0 | 0 | 0 | 0 |
| 0 | 0 | 0 | 0 | 0 |
| 0 | 0 | 0 | 0 | 0 |
| 0 | 0 | 7.703597133 | 0 | 0 |
| 0 | 0 | 0 | 0 | 0 |
| 0 | 0.01852992 | 0.016113051 | 0.000814804 | 0.027537584 |
| 0.041008485 | 0 | 0 | 0 | 0 |
| 0 | 0 | 0 | 0 | 0.075406178 |
| 0 | 0.004201473 | 0.00190285 | 0.038874189 | 0.002702974 |
| 0 | 0 | 0 | 0 | 0 |
| 0 | 0 | 0 | 0 | 0 |
| 0 | 0 | 0 | 0 | 0 |
| 0 | 0 | 0.003497807 | 0 | 0 |
| 0 | 0.002484345 | 0 | 0 | 0.009709538 |
| 0 | 0 | 0 | 0 | 0.034441929 |
| 0 | 0 | 0 | 0 | 0 |
| 0.290779031 | 0 | 0.006215093 | 0 | 0 |
| 0.001011016 | 0 | 0 | 0 | 0 |
| 0 | 0 | 0 | 0 | 0 |
| 0 | 0 | 0 | 0 | 0 |
| 0 | 0 | 0 | 0 | 0 |
| 0 | 0 | 0.004584874 | 0 | 0.011356615 |
| 0 | 0 | 0 | 0 | 0 |
| 0 | 0 | 0 | 0 | 0 |
| 0.077770494 | 0.358218199 | 0.263422518 | 5.533403595 | 1.841831842 |
| 0 | 0 | 0.002008117 | 0.031856553 | 0 |
| 0 | 0 | 0.018728432 | 0 | 0 |
| 0 | 0 | 0 | 0 | 0 |
| 0 | 0 | 0 | 0 | 0 |
| 0 | 0.021922678 | 0 | 0 | 0 |
| 0 | 0 | 0 | 0 | 0 |
| 0.909855267 | 0 | 0.225869508 | 0 | 0 |
| 0 | 0 | 0.016162593 | 0 | 0 |
| 0.008185818 | 0 | 0 | 0 | 0 |
| 0 | 0 | 0 | 0 | 0 |
| 0.001051052 | 0 | 0 | 0 | 0.03555422 |
| 0.616493199 | 0 | 0 | 0 | 1.422360825 |
| 0 | 0 | 0.001184478 | 0 | 0 |
| 0 | 0 | 0 | 0 | 0 |
| 0 | 0 | 0 | 0 | 0 |
| 0 | 0 | 0 | 0 | 0 |
| 0.002574027 | 0.000604215 | 0 | 0.000830274 | 0.040232072 |
| 0 | 0 | 0 | 0 | 0 |

| | | | | |
|---|---|---|---|---|
| 0 | 0 | 0.005627294 | 0.001229303 | 0 |
| 0 | 0 | 0 | 0 | 0 |
| 0.004018328 | 0.013205417 | 0.02637661 | 0.287824071 | 0.002940774 |
| 0 | 0 | 0 | 0 | 0 |
| 0 | 0 | 0 | 0 | 0 |
| 0 | 0.676964946 | 0 | 0 | 0 |
| 0 | 0 | 0 | 0.000436751 | 0 |
| 0 | 0 | 0.018921166 | 0 | 0 |
| 0 | 0 | 0 | 0 | 0 |
| 0 | 0 | 0 | 0 | 0 |
| 0 | 0 | 0 | 0 | 0 |
| 0 | 0 | 0 | 0 | 0 |
| 0 | 0 | 0 | 0 | 0.129160241 |
| 0 | 0.002697741 | 0.001221809 | 0 | 0.000976255 |
| 0 | 0 | 0 | 0 | 0 |
| 0.00465105 | 0.038826722 | 0.043921841 | 0 | 0.003306014 |
| 0.004444141 | 0.045372135 | 0.125411078 | 0.011238601 | 0.003946246 |
| 0 | 0.009463584 | 0 | 0.006632167 | 0.20924721 |
| 0 | 0 | 0 | 0 | 0.003493864 |
| 0 | 0 | 1.655110266 | 0 | 0 |
| 0 | 0 | 0.049127785 | 0 | 0.081705741 |
| 0 | 0.000903508 | 0 | 0 | 0.00228872 |
| 0 | 0 | 0.855872259 | 0 | 0 |
| 0 | 0 | 0.038357666 | 0.005671316 | 0 |
| 0.027412965 | 0 | 0.005110369 | 0 | 0.025026722 |
| 0 | 0 | 0 | 0 | 0.003713203 |
| 0 | 0.007416346 | 0 | 0 | 0.001789211 |
| 0 | 0 | 0 | 0 | 0 |
| 0 | 0 | 0 | 0 | 0 |
| 0 | 0 | 0 | 0 | 0 |
| 0.015789513 | 0 | 0.002213822 | 0.002945106 | 0.058237532 |
| 0.003582208 | 0.001681743 | 0 | 0 | 0.018663321 |
| 0.032158468 | 0.068586715 | 0.007010321 | 0.029924487 | 0.018542614 |
| 0 | 0 | 0.018707226 | 0 | 0 |
| 0.004734719 | 0 | 0 | 0 | 0 |
| 0 | 0 | 0 | 0 | 0 |
| 0.003744311 | 0 | 0.000874181 | 0.010543025 | 0 |
| 0.000516746 | 0.085286316 | 0.102376611 | 0 | 0.01693384 |
| 0 | 0 | 0.41467024 | 0 | 0 |
| 0 | 0 | 0 | 0 | 0 |
| 0 | 0 | 0 | 0 | 0 |
| 0 | 0 | 0 | 0 | 0 |
| 0 | 0 | 0 | 0 | 0 |
| 0 | 0.007806534 | 0.004269386 | 0 | 0.242631676 |
| 0 | 3.459431785 | 0 | 0 | 0.015437455 |
| 0 | 0 | 0 | 0 | 0 |
| 0 | 0 | 0 | 0 | 0 |
| 0 | 0 | 0 | 0 | 0 |

| | | | | |
|---|---|---|---|---|
| 0 | 0 | 0.020779339 | 0 | 0 |
| 0.004694209 | 0.186918495 | 0.028645472 | 0 | 2.857889059 |
| 0 | 0 | 0 | 0 | 0 |
| 0 | 0 | 0 | 0 | 0 |
| 0 | 0.009332745 | 0 | 0 | 0 |
| 0 | 0 | 0 | 0 | 0 |
| 0 | 0 | 0 | 0 | 0 |
| 0 | 0 | 0.000536652 | 0 | 0 |
| 0 | 0 | 0.001492342 | 0 | 0 |
| 0 | 0 | 0 | 0 | 0.027044825 |
| 0.021627525 | 0 | 0.002249816 | 0 | 0.003318751 |
| 0.408400674 | 0.059479272 | 0.017013603 | 0.038480804 | 0.032543877 |
| 0 | 0 | 0 | 0 | 0 |
| 0 | 0 | 0 | 0 | 0 |
| 0.00160096 | 0 | 0 | 0.011928911 | 0 |
| 0 | 0 | 0 | 0 | 0 |
| 0.001095829 | 0.021207184 | 0.002718325 | 0.00148457 | 0.001158404 |
| 0 | 0.196864428 | 0.001176254 | 0 | 0.195489853 |
| 0 | 0 | 0 | 0 | 0 |
| 0 | 0.161393974 | 0.001985874 | 0 | 0.066888087 |
| 0.011300765 | 0.00577697 | 0.023360658 | 0.00187118 | 0.02926786 |
| 0 | 0 | 0 | 0 | 0 |
| 0 | 0 | 0 | 0 | 0 |
| 0 | 0 | 0 | 0 | 0 |
| 0 | 0 | 0.016328109 | 0 | 0 |
| 0 | 0 | 0 | 0 | 0 |
| 0 | 0 | 0 | 0 | 0.021519238 |
| 0.097689834 | 0.11008261 | 0.011520624 | 0.075393167 | 0.063801941 |
| 0 | 0 | 0 | 0 | 0 |
| 0 | 0 | 0 | 0 | 0.001148658 |
| 0 | 0 | 0 | 0 | 0.002384163 |
| 0 | 0 | 0 | 0 | 0 |
| 0 | 0 | 0 | 0 | 0 |
| 0 | 0 | 0 | 0 | 0 |
| 0 | 0 | 0.119088313 | 0.048684096 | 0 |
| 0.035235161 | 0.365193861 | 0.030255469 | 0.10097718 | 0.654869719 |
| 0 | 0 | 0 | 0 | 0 |
| 0 | 0 | 0 | 0 | 0 |
| 0.020002906 | 0 | 0.023768308 | 0 | 0.024669305 |
| 0 | 0 | 0 | 0 | 0 |
| 0 | 3.444235987 | 0 | 0 | 0.011794375 |
| 0 | 0 | 0 | 0 | 0 |
| 0.029850174 | 0.000657263 | 0 | 0 | 0.008800428 |
| 0.021723453 | 0 | 0 | 0 | 0.008726287 |
| 0 | 0 | 0.01548063 | 0 | 0 |
| 0 | 0 | 0.356480027 | 0 | 0 |
| 0 | 0 | 0 | 0 | 0 |
| 0 | 0.01051385 | 0 | 0 | 0 |

| | | | | |
|---|---|---|---|---|
| 0 | 0 | 0 | 0 | 0 |
| 0 | 0 | 0 | 0 | 0 |
| 0 | 0 | 0 | 0 | 0 |
| 0.100629246 | 0 | 0.016121414 | 0 | 0 |
| 0 | 0 | 0 | 0 | 0 |
| 0 | 0 | 0 | 0 | 0 |
| 0.008532958 | 0 | 0 | 0 | 0 |
| 0.009690046 | 0 | 0.461669775 | 0 | 0 |
| 1.962832195 | 0 | 0.24878619 | 0.540653831 | 0.028120237 |
| 0.083535976 | 0 | 0.020295956 | 0 | 0 |
| 0 | 0 | 0 | 0 | 0 |
| 0 | 0 | 0.001660865 | 0 | 0 |
| 0 | 0 | 0 | 0 | 0 |
| 0 | 0 | 0 | 0 | 0 |
| 0.292551606 | 0 | 0.065512306 | 0.019211604 | 0.082351718 |
| 0 | 0 | 0 | 0 | 0 |
| 0 | 0 | 0.905323095 | 0 | 0 |
| 0 | 0 | 0 | 0 | 0 |
| 0 | 0 | 0 | 0 | 0 |
| 0 | 0 | 0 | 0 | 0 |
| 0 | 0 | 0.000875372 | 0 | 0 |
| 0 | 0 | 0 | 0 | 0 |
| 0 | 0 | 0 | 0 | 0 |
| 0 | 0 | 0.310905039 | 0 | 0.34093221 |
| 0 | 0 | 0 | 0 | 0 |
| 0 | 0.041223401 | 0 | 0 | 0 |
| 0 | 0 | 0 | 0 | 0 |
| 1.346771987 | 0 | 1.340195805 | 0.00766567 | 0 |
| 0 | 0 | 0 | 0 | 0 |
| 0 | 0 | 0 | 0 | 0 |
| 0 | 0 | 0 | 0 | 0 |
| 0 | 0 | 0.01658211 | 0 | 0 |
| 0.036039434 | 0 | 0 | 0 | 0 |
| 0 | 0 | 0 | 0 | 0 |
| 0 | 0 | 0 | 0 | 0 |
| 0.054775645 | 0 | 0.098570724 | 0 | 0.033691939 |
| 1.871750427 | 0 | 0.138974216 | 0 | 0 |
| 0 | 0 | 0 | 0 | 0 |
| 0.011151845 | 0 | 0.000691584 | 0 | 0 |
| 0.002276382 | 0 | 0.004992973 | 0 | 0.008738917 |
| 0.013167214 | 0.02409828 | 0.148814541 | 0 | 0.055761242 |
| 0 | 0 | 0 | 0 | 0 |
| 2.561983744 | 0 | 15.92031632 | 3.737541649 | 0 |
| 0 | 0 | 0 | 0 | 0 |
| 0 | 0 | 0 | 0 | 0 |
| 0 | 0 | 0 | 0 | 0 |
| 0 | 0 | 0 | 0 | 0 |
| 0.150563795 | 0 | 0 | 0.015864768 | 0.001224318 |

| | | | | |
|---|---|---|---|---|
| 0 | 0 | 0 | 0 | 0 |
| 0.020926895 | 0 | 0.036992369 | 0 | 0 |
| 0.012212741 | 0.060810129 | 0.002386882 | 0.039106726 | 0.350627042 |
| 0 | 0 | 0 | 0 | 0 |
| 0 | 8.271472001 | 0 | 0 | 0 |
| 0.067928161 | 0 | 0.001880294 | 0 | 0 |
| 0 | 0 | 0 | 0 | 0 |
| 0 | 0 | 0 | 0 | 0 |
| 0 | 0 | 0 | 0 | 0 |
| 0 | 0 | 0 | 0 | 0 |
| 0.067039171 | 0.003980318 | 0.019649306 | 0.088277684 | 0.055022957 |
| 1.470425765 | 2.671368391 | 0.611403927 | 2.795971453 | 3.845384203 |
| 0 | 0 | 0 | 0 | 0 |
| 0 | 0.012457591 | 0.104280681 | 0.009031443 | 0.003109059 |
| 0.008189244 | 0.609334708 | 0 | 0.005582058 | 0.552977943 |
| 0 | 0 | 0.203456553 | 0 | 0 |
| 0 | 0 | 0 | 0 | 0 |
| 0 | 0 | 0 | 0 | 0 |
| 0 | 0 | 0 | 0 | 0 |
| 0.00404068 | 0 | 0.009949105 | 0 | 0 |
| 0.270741204 | 0 | 0.173302359 | 0 | 1.047901869 |
| 13.36839925 | 0.008084974 | 4.774844635 | 52.14974788 | 1.217378389 |
| 0.984628946 | 8.574870595 | 0.00085626 | 0.225242937 | 0.76718478 |
| 0 | 0 | 0 | 0 | 0 |
| 0 | 0 | 0 | 0 | 0 |
| 0.185388099 | 0 | 0.363011997 | 0.000494397 | 0 |
| 1.933990394 | 0 | 0 | 0 | 0 |
| 0 | 0 | 0.006461029 | 0 | 0 |
| 0.0163265 | 0 | 0.003216996 | 0 | 0 |
| 0 | 0 | 0 | 0 | 0 |
| 0 | 0 | 0.033330081 | 0 | 0 |
| 0.154251565 | 0 | 0.098137899 | 0 | 0 |
| 0 | 1.456515128 | 0.001110845 | 0.000505558 | 0.204441973 |
| 0.013232766 | 0 | 0.0295075 | 0 | 0.010378481 |
| 0 | 0 | 0.015640991 | 0 | 0.025385602 |
| 0.090014998 | 0.064315873 | 0.014438436 | 0.047525863 | 0.021195254 |
| 0.010787434 | 0 | 0.002270496 | 0.001033329 | 0.107555002 |
| 0 | 0 | 0 | 0 | 0 |
| 0.054251858 | 0 | 0.257760491 | 0 | 0.025116686 |
| 0 | 0 | 0 | 0 | 0 |
| 0 | 0 | 0 | 0 | 0 |
| 0 | 0 | 0 | 0 | 0 |
| 0 | 0 | 0 | 0 | 0 |
| 0 | 0 | 0 | 0 | 0 |
| 0.003859093 | 0.035087211 | 0.019911643 | 0.305320377 | 0.033349559 |
| 0.041460572 | 0 | 0 | 0 | 0 |
| 0 | 0 | 0 | 0 | 0 |
| 0.036775458 | 0 | 0.006697573 | 5.726515474 | 0.006274198 |

| | | | | |
|---|---|---|---|---|
| 0 | 0 | 0 | 0 | 0 |
| 0 | 0 | 0 | 0 | 0 |
| 0 | 0 | 0 | 0 | 0 |
| 0 | 0 | 0.010606633 | 0 | 0 |
| 0 | 0 | 0 | 0 | 0 |
| 0 | 0 | 0 | 0 | 0 |
| 0.036189316 | 0 | 0.134344723 | 0 | 0 |
| 0.016192085 | 0 | 0 | 0 | 0.032873644 |
| 0 | 0 | 0 | 0 | 0 |
| 0 | 0 | 0 | 0 | 0 |
| 0.018682193 | 0.250596826 | 0 | 0.001323378 | 0.054600147 |
| 0 | 0 | 0 | 0 | 0 |
| 0.041054671 | 0 | 0.104545763 | 0 | 0.035600732 |
| 0 | 0 | 0 | 0 | 0 |
| 0 | 0 | 0 | 0 | 0 |
| 0 | 0 | 0 | 0 | 0 |
| 0 | 0 | 0 | 0 | 0 |
| 0 | 0 | 0.000907757 | 0.030020843 | 0 |
| 0 | 0 | 0 | 0 | 0 |
| 0 | 0 | 0.001091529 | 0 | 0 |
| 0 | 0 | 0 | 0 | 0.002147965 |
| 0 | 0 | 0 | 0 | 0 |
| 0 | 0 | 0.015873117 | 0 | 0 |
| 0.021608049 | 0 | 0 | 0 | 0 |
| 0 | 0 | 0 | 0 | 0 |
| 0 | 0 | 0 | 0 | 0 |
| 0 | 0 | 0 | 0 | 0 |
| 0 | 0 | 0 | 0 | 0 |
| 0 | 0 | 0 | 0 | 0 |
| 0.185109964 | 0 | 0.036003807 | 0 | 0 |
| 0.031880999 | 0 | 0 | 0 | 0 |
| 0 | 0 | 0 | 0 | 0 |
| 0 | 0 | 0 | 0 | 0 |
| 0 | 0 | 0 | 0 | 0 |
| 0 | 0 | 0 | 0 | 0 |
| 0 | 0 | 0 | 0 | 0 |
| 0.043414921 | 0 | 0 | 0 | 0 |
| 0 | 0.035157616 | 0.002769202 | 0 | 0.053841359 |
| 0 | 0 | 0 | 0 | 0 |
| 0 | 0 | 0 | 0 | 0 |
| 0 | 0 | 0.005340737 | 0 | 0 |
| 0 | 0 | 0 | 0 | 0 |
| 0 | 0 | 0 | 0 | 0 |
| 0 | 7.059850331 | 1.375686898 | 0 | 0 |
| 0.037147471 | 0 | 0.067429907 | 0.038161411 | 0 |
| 0 | 0 | 0 | 0 | 0 |
| 0 | 0 | 0 | 0 | 0 |
| 0 | 0 | 0 | 0 | 0 |

| | | | | |
|---|---|---|---|---|
| 0 | 0 | 0 | 0 | 0 |
| 0 | 0 | 0 | 0 | 0 |
| 0 | 0 | 0 | 0 | 0 |
| 0.127853693 | 0 | 0.037186344 | 0 | 0 |
| 0 | 0 | 0 | 0 | 0 |
| 0.397102025 | 0 | 0 | 0 | 0 |
| 0 | 0 | 0 | 0 | 0 |
| 0 | 0 | 0 | 0 | 0 |
| 0.017320702 | 2.778351796 | 0.002546128 | 0.077058359 | 0.112910153 |
| 0 | 0 | 0 | 0 | 0 |
| 0 | 0 | 0 | 0 | 0 |
| 0 | 0 | 0 | 0 | 0 |
| 0 | 0 | 0 | 0 | 0 |
| 0 | 0 | 0 | 0 | 0 |
| 0 | 0 | 4.032080985 | 0 | 0 |
| 0 | 0 | 0 | 0.164330041 | 0 |
| 0 | 0 | 0 | 0 | 0 |
| 0 | 0 | 0 | 0 | 0 |
| 0 | 0 | 0 | 0 | 0 |
| 0 | 0 | 0 | 0 | 0 |
| 0.034336713 | 0 | 0 | 0 | 0 |
| 0 | 0 | 0 | 0 | 0 |
| 0 | 0 | 0 | 0 | 0 |
| 0 | 0 | 0 | 0 | 0 |
| 0.009555664 | 0 | 0.079096088 | 0.007948986 | 0.018341861 |
| 0.098598697 | 0.00207708 | 6.053711478 | 0 | 0 |
| 0.3943226 | 0 | 0 | 0 | 0 |
| 0 | 0 | 0 | 0 | 0 |
| 0.04726822 | 0 | 0.016579172 | 0 | 0 |
| 0 | 0 | 6.280144401 | 0 | 0 |
| 0 | 0 | 0 | 0 | 0 |
| 0 | 0 | 0 | 0 | 0 |
| 0 | 0 | 0.000483311 | 0 | 0 |
| 0 | 0 | 0 | 0 | 0.000638852 |
| 0 | 0.022058275 | 0.04149779 | 0.001998532 | 0.001403502 |
| 0.022055086 | 0.027244085 | 0.000776029 | 0 | 0.021330272 |
| 0 | 0 | 0.337314124 | 0 | 0 |
| 0.850445254 | 0 | 1.329128142 | 0 | 0 |
| 2.311731379 | 0 | 0 | 0 | 0 |
| 0 | 0 | 0 | 0 | 0 |
| 0 | 0 | 0 | 0 | 0 |
| 0 | 0 | 0.001936052 | 0 | 0 |
| 0 | 0 | 0.109227157 | 0 | 0 |
| 0 | 0 | 0 | 0 | 0 |
| 0 | 0 | 0 | 0 | 0 |
| 0 | 0 | 0 | 0 | 0 |
| 0 | 0 | 0 | 0 | 0 |
| 0.041786216 | 0.005948029 | 0.081752993 | 0 | 0 |

| | | | | |
|---|---|---|---|---|
| 0.00492927 | 0 | 0 | 0 | 0 |
| 0 | 0 | 0 | 0 | 0 |
| 0 | 0 | 0 | 0 | 0 |
| 0 | 0 | 0 | 0 | 0 |
| 0 | 0 | 0 | 0.002691111 | 0 |
| 0 | 0 | 0 | 0 | 0 |
| 0 | 0 | 0 | 0 | 0 |
| 0 | 0 | 0 | 0 | 0 |
| 0 | 0 | 0 | 0 | 0 |
| 0 | 0 | 0 | 0 | 0 |
| 0 | 0 | 0 | 0 | 0 |
| 0.015361598 | 0 | 0.100600184 | 0 | 0 |
| 0 | 0.128793172 | 0.006317385 | 0 | 0 |
| 0 | 0 | 0 | 0 | 0 |
| 0.282974566 | 0 | 5.110755536 | 0.000732299 | 0.007071197 |
| 0 | 0 | 0 | 0 | 0 |
| 0 | 0 | 0 | 0 | 0.001565149 |
| 0 | 0 | 0 | 0 | 0 |
| 0 | 0 | 0 | 0 | 0 |
| 0 | 0 | 0 | 0 | 0 |
| 0 | 0 | 0 | 0 | 0 |
| 0 | 0 | 0 | 0 | 0 |
| 0.001036555 | 0 | 0.024512949 | 0 | 0.10957446 |
| 0 | 0 | 0 | 0 | 0 |
| 0 | 0 | 0 | 0 | 0 |
| 0 | 0 | 0 | 0 | 0 |
| 0 | 0 | 0 | 0 | 0 |
| 0 | 0 | 0 | 0 | 0 |
| 0 | 0 | 0 | 0 | 0 |
| 0 | 0 | 0 | 0 | 0 |
| 0 | 0 | 0 | 0 | 0 |
| 0.028144914 | 0.040627558 | 0.01618178 | 0.00237565 | 0.20270325 |
| 0 | 0 | 0 | 0 | 0 |
| 0.070829205 | 0 | 0.002738176 | 0 | 0 |
| 0.026554807 | 0.02274908 | 0.00118232 | 0 | 0.020513532 |
| 0.007680339 | 0.054085431 | 0.007838506 | 0.005152904 | 0.013918121 |
| 0 | 0 | 0 | 0 | 0 |
| 0 | 0 | 0.010498169 | 0 | 0 |
| 0 | 0 | 0.003104712 | 0 | 0.000930277 |
| 0 | 0 | 0 | 0 | 0 |
| 0.141404956 | 0 | 0.191250885 | 0 | 0 |
| 0 | 0 | 0.002277805 | 0 | 0 |
| 0 | 0 | 0 | 0 | 0 |
| 0 | 0 | 0 | 0 | 0 |
| 0.177077301 | 0 | 0.00133109 | 0.374078124 | 0.061687227 |
| 0 | 0 | 0 | 0 | 0 |
| 0.612885923 | 0 | 0 | 0.039756744 | 0 |
| 0 | 0 | 0.003147432 | 0 | 0 |

| | | | | |
|---|---|---|---|---|
| 0 | 0 | 0 | 0 | 0 |
| 0 | 0.035850326 | 0 | 0 | 0 |
| 0.001907732 | 0 | 0.041644575 | 0 | 0 |
| 0 | 0 | 0 | 0 | 0 |
| 0 | 0 | 0.000872595 | 0 | 0 |
| 0 | 0 | 0 | 0 | 0 |
| 0 | 0 | 0 | 0 | 0 |
| 0 | 0 | 0.036566599 | 0 | 0 |
| 0 | 0 | 0 | 0 | 0 |
| 0 | 0 | 0 | 0 | 0 |
| 0 | 0 | 0 | 0 | 0 |
| 0.007940744 | 0.005605729 | 0 | 0 | 0.001958642 |
| 0 | 0 | 0 | 0 | 0 |
| 0.137758328 | 0 | 0.125068461 | 0 | 0 |
| 0 | 0 | 0 | 0 | 0 |
| 0 | 0 | 0 | 0 | 0 |
| 0 | 0 | 0 | 0 | 0 |
| 0 | 0 | 0 | 0 | 0 |
| 0.234795126 | 6.023239038 | 0 | 0 | 0 |
| 0 | 0 | 0 | 0 | 0 |
| 0 | 0.009666424 | 0 | 0 | 0 |
| 0 | 0 | 0 | 0 | 0 |
| 0 | 0 | 0 | 0 | 0 |
| 0 | 0 | 0 | 0 | 0 |
| 0 | 0 | 0.000545374 | 0 | 0 |
| 0 | 0.004446208 | 0.000464698 | 0.121958904 | 0.030446994 |
| 0 | 0 | 0 | 0 | 0 |
| 0.04920184 | 0 | 0.279124346 | 0 | 0.020594568 |
| 0 | 0 | 0 | 0 | 0 |
| 0 | 0 | 2.298371907 | 0 | 0 |
| 0 | 0 | 0 | 0 | 0 |
| 0 | 0 | 0 | 0 | 0 |
| 0 | 0 | 0 | 0 | 0 |
| 0 | 0 | 0 | 0 | 0 |
| 0.144616863 | 0.005913792 | 0 | 0.001625271 | 0.003112836 |
| 0 | 0 | 0 | 0 | 0 |
| 0 | 0 | 0 | 0 | 0 |
| 0.003341358 | 0.009150576 | 0 | 0.002850137 | 0.00323781 |
| 0 | 0 | 0 | 0 | 0 |
| 0.001520885 | 0 | 0 | 0 | 0 |
| 0.427168493 | 6.367707382 | 1.262031821 | 4.231196162 | 32.21914872 |
| 0 | 0.12056873 | 0 | 0 | 0.462383383 |
| 0 | 0 | 0 | 0 | 0 |
| 0.08897794 | 0 | 0 | 0 | 0 |
| 0 | 0 | 0 | 0 | 0 |
| 0 | 0 | 0 | 0 | 0 |
| 0 | 0 | 0 | 0 | 0 |
| 0.015322009 | 0 | 0.001315659 | 0.007185261 | 0.00584024 |

| | | | | |
|---|---|---|---|---|
| 0 | 0 | 0 | 0 | 0 |
| 0 | 0 | 0 | 0 | 0.002883441 |
| 0 | 0 | 0 | 0 | 0 |
| 0 | 0 | 0 | 0 | 0 |
| 0 | 0 | 1.18688424 | 0 | 0 |
| 0 | 0 | 0 | 0 | 0 |
| 0 | 0 | 0 | 0 | 0 |
| 0 | 0 | 0 | 0 | 0 |
| 0 | 1.546989928 | 0.031980027 | 0.009429664 | 2.041105658 |
| 0 | 0 | 0 | 0 | 0 |
| 0 | 0.002438084 | 0 | 0.066799017 | 0 |
| 0 | 0 | 0 | 0 | 0 |
| 0.051094011 | 0 | 0 | 0 | 0 |
| 0 | 0.014277464 | 0.002514662 | 0.293470171 | 0.022963147 |
| 0 | 0 | 0.469024009 | 0 | 0 |
| 0.091058667 | 0.277441006 | 0.062494979 | 0 | 0 |
| 0 | 0 | 0.371638245 | 0 | 0 |
| 0 | 0 | 0 | 0 | 0 |
| 0 | 0 | 0 | 0 | 0 |
| 0 | 0 | 0 | 0 | 0 |
| 0 | 0 | 0 | 0 | 0 |
| 0 | 0 | 0 | 0 | 0 |
| 0 | 0 | 0 | 0 | 0 |
| 0 | 0 | 0 | 0 | 0 |
| 0 | 0 | 0 | 0 | 0 |
| 0 | 0 | 0 | 0 | 0 |
| 0.000681055 | 0 | 0.062171131 | 0 | 0 |
| 0.065475767 | 0 | 0 | 0 | 0 |
| 0 | 0 | 0 | 0 | 0 |
| 0 | 0 | 0 | 0 | 0 |
| 0 | 0 | 0 | 0 | 0 |
| 0 | 0 | 0 | 0 | 0 |
| 0 | 0 | 0 | 0 | 0 |
| 0 | 0 | 0.024140057 | 0 | 0 |
| 0 | 0 | 0.004159785 | 0.168491834 | 0 |
| 0 | 0.014217877 | 0 | 0 | 0.015505916 |
| 0 | 0 | 0.041834535 | 0 | 0 |
| 0 | 0 | 0 | 0 | 0 |
| 0 | 0 | 0 | 0 | 0 |
| 0 | 0 | 0 | 0 | 0 |
| 0.194456631 | 0 | 0.575604232 | 0.381932757 | 0 |
| 0.027363653 | 0 | 0.014004416 | 0 | 0 |
| 0 | 0 | 0 | 0 | 0 |
| 0 | 0 | 0.119086018 | 0 | 0 |
| 0 | 0 | 0 | 0 | 0 |
| 0 | 0 | 0 | 0 | 0 |
| 0 | 0 | 0 | 0 | 0 |
| 0 | 0 | 0 | 0 | 0 |

| | | | | |
|---|---|---|---|---|
| 0.163405699 | 0 | 0.009851865 | 0 | 0 |
| 0 | 0 | 0 | 0 | 0 |
| 0 | 0 | 0 | 0 | 0 |
| 0 | 0.000762906 | 0 | 0 | 0.001380397 |
| 0.016992178 | 0 | 0.005785426 | 0 | 0 |
| 0 | 0 | 0 | 0 | 0 |
| 0.042816939 | 0 | 0.300805136 | 0 | 0.007185737 |
| 0 | 0 | 0.000599522 | 0 | 0 |
| 0 | 0 | 0 | 0 | 0 |
| 0 | 0 | 0.012200781 | 0 | 0 |
| 0.010611908 | 0 | 0 | 0 | 0 |
| 0 | 0 | 0.023594021 | 0 | 0 |
| 0 | 0 | 0 | 0 | 0 |
| 0 | 0 | 0 | 0 | 0 |
| 0 | 0 | 0 | 0 | 0 |
| 0 | 0 | 0 | 0 | 0 |
| 30.14653404 | 0 | 0 | 11.15331074 | 0 |
| 0 | 0 | 0.000447205 | 0 | 0 |
| 0 | 0 | 0 | 0 | 0 |
| 0 | 0 | 0 | 0 | 0 |
| 0 | 0.075456017 | 0.015352504 | 0 | 0 |
| 0 | 0 | 0 | 0 | 0 |
| 0 | 0 | 0.015956811 | 0 | 0 |
| 0.001313254 | 0 | 0.002750921 | 0 | 0.001156869 |
| 0 | 0 | 0 | 0 | 0 |
| 0 | 0 | 0 | 0 | 0 |
| 0 | 0 | 0 | 0 | 0 |
| 0 | 0 | 0 | 0 | 0 |
| 0 | 0 | 0 | 0 | 0 |
| 0.440388576 | 0.001380303 | 0.014169841 | 0.046090378 | 3.692318178 |
| 0 | 0 | 0 | 0 | 0 |
| 0 | 0 | 0 | 0 | 0 |
| 0 | 0 | 0 | 0 | 0 |
| 0 | 0 | 0 | 0 | 0 |
| 0 | 0 | 0 | 0 | 0 |
| 0 | 0 | 0 | 0 | 0 |
| 0 | 0.124802511 | 0.002173967 | 0 | 0.006079682 |
| 0 | 0 | 0 | 0 | 0 |
| 0 | 0 | 0 | 0 | 0 |
| 0 | 0 | 0 | 0 | 0 |
| 0 | 0 | 0 | 0 | 0 |
| 0 | 0 | 0 | 0 | 0 |
| 0 | 0 | 0 | 0 | 0 |
| 0 | 0 | 0 | 0 | 0 |
| 0 | 0 | 0 | 0 | 0 |
| 0 | 0 | 0 | 0 | 0 |
| 0 | 0 | 0 | 0 | 0 |
| 0 | 0 | 0.028146403 | 0 | 0 |

| | | | | |
|---|---|---|---|---|
| 0.037996888 | 0 | 0 | 0 | 0 |
| 0 | 0 | 0 | 0 | 0 |
| 0 | 0 | 0 | 0 | 0 |
| 0 | 0 | 0 | 0 | 0 |
| 0 | 0 | 0 | 0 | 0 |
| 0 | 0 | 0.012384707 | 0 | 0 |
| 0 | 0 | 0 | 0.000763104 | 0 |
| 0.037240843 | 0.166358074 | 0.125412853 | 0 | 0 |
| 0.381017599 | 0.003509654 | 2.933040429 | 0.021813633 | 0.774155687 |
| 0.032427439 | 0 | 0.199014088 | 0 | 0.204586503 |
| 0 | 0 | 0 | 0 | 0 |
| 0 | 0 | 0 | 0 | 0 |
| 0 | 0 | 0 | 0 | 0 |
| 0.098707655 | 0 | 0 | 0 | 0 |
| 0 | 0 | 0 | 0 | 0 |
| 0 | 0 | 0 | 0 | 0 |
| 0 | 0 | 0 | 0 | 0 |
| 0 | 0 | 0 | 0 | 0 |
| 0 | 0 | 0 | 0 | 0 |
| 0 | 0 | 0 | 0 | 0 |
| 0 | 0 | 0 | 0 | 0 |
| 0 | 0 | 0 | 0 | 0 |
| 0 | 0 | 0 | 0 | 0 |
| 0 | 0 | 0 | 0 | 0 |
| 0.005194084 | 0 | 0.019388099 | 0 | 0 |
| 0 | 0 | 0 | 0 | 0 |
| 0 | 0 | 0 | 0 | 0 |
| 0 | 0 | 0 | 0 | 0 |
| 0 | 0 | 0 | 0 | 0 |
| 0 | 0 | 0 | 0 | 0 |
| 0 | 0 | 0 | 0 | 0 |
| 0 | 0 | 0 | 0 | 0 |
| 0 | 0 | 0.01019773 | 0 | 0 |
| 0 | 0 | 0 | 0 | 0 |
| 0 | 0 | 0.006601376 | 0.002746846 | 0 |
| 0 | 0 | 0 | 0 | 0 |
| 0 | 0 | 0 | 0 | 0 |
| 0 | 0 | 0 | 0 | 0 |
| 0 | 0 | 0.004075531 | 0 | 0 |
| 0 | 0.165180963 | 0.000466594 | 0 | 0.534872196 |
| 0 | 0 | 0 | 0 | 0 |
| 0 | 0 | 0.388629724 | 0 | 0 |
| 0 | 0 | 0 | 0 | 0 |
| 0 | 0 | 0 | 0 | 0 |
| 0 | 0 | 0 | 0 | 0 |
| 0 | 0 | 0.011636743 | 0 | 0 |
| 0 | 0 | 0 | 0 | 0 |
| 0 | 0 | 0 | 0 | 0 |

| | | | | |
|---|---|---|---|---|
| 0 | 0 | 0 | 0 | 0 |
| 0 | 0.001086637 | 0.002530999 | 0 | 0 |
| 0 | 0 | 0 | 0 | 0 |
| 0 | 0 | 0 | 0 | 0 |
| 0.473334238 | 0.409552068 | 0.09121961 | 0.182344361 | 0.201694978 |
| 0 | 0 | 0 | 0 | 0 |
| 0.00171831 | 0 | 0 | 0 | 0 |
| 0.00359515 | 0.365162603 | 0.005350899 | 0.072877296 | 0.019952317 |
| 0 | 0 | 0.012466239 | 0 | 0 |
| 0 | 0 | 0 | 0 | 0 |
| 0 | 0 | 0 | 0 | 0 |
| 0 | 0 | 0.082021176 | 0 | 0 |
| 0 | 0 | 0 | 0 | 0 |
| 0 | 0 | 0 | 0 | 0 |
| 0 | 0 | 0 | 0 | 0 |
| 0.007621892 | 0 | 0.000687525 | 0 | 0.031587571 |
| 0 | 0 | 0 | 0 | 0 |
| 0 | 0 | 0 | 0 | 0 |
| 0 | 0 | 0 | 0 | 0 |
| 0 | 0 | 0 | 0 | 0 |
| 0 | 0 | 0 | 0 | 0 |
| 0 | 0 | 0 | 0 | 0 |
| 0 | 0 | 0 | 0 | 0 |
| 0.022989096 | 0.329272368 | 0 | 0 | 0.035630498 |
| 0.091965437 | 0 | 0.263227352 | 0.195426717 | 0 |
| 0 | 0 | 0 | 0 | 0 |
| 0.541462559 | 0 | 0.001725516 | 0 | 0.014789999 |
| 0 | 0 | 0 | 0 | 0 |
| 0.003054781 | 0.006435202 | 0 | 0 | 0.001117806 |
| 0 | 0 | 0 | 0 | 0 |
| 0 | 0 | 0 | 0 | 0 |
| 0 | 0 | 0 | 0 | 0 |
| 0 | 0 | 0 | 0 | 0 |
| 0 | 0 | 0 | 0 | 0 |
| 0 | 0 | 0 | 0 | 0 |
| 0 | 0 | 0 | 0 | 0 |
| 0 | 0 | 0 | 0 | 0 |
| 0 | 0 | 0.008687215 | 0 | 0 |
| 0 | 0 | 0 | 0 | 0 |
| 0 | 0 | 0 | 0 | 0 |
| 0 | 0 | 0 | 0 | 0 |
| 0 | 0 | 0 | 1.063002872 | 0 |
| 0 | 0 | 0 | 0 | 0 |
| 0 | 0 | 0.007677066 | 0 | 0 |
| 0 | 0 | 0 | 0 | 0 |
| 0 | 0 | 0 | 0 | 0 |
| 0 | 0 | 0 | 0 | 0 |
| 0 | 0 | 0 | 0 | 0 |

| | | | | |
|---|---|---|---|---|
| 0.285746053 | 0 | 0.015194364 | 0 | 0 |
| 0 | 0 | 0 | 0 | 0 |
| 0 | 0 | 0 | 0 | 0 |
| 0 | 0 | 0 | 0 | 0 |
| 0 | 0 | 0 | 0 | 0 |
| 0 | 0.133586724 | 0.002563624 | 0.055692152 | 1.817064725 |
| 0 | 0 | 0 | 0 | 0 |
| 0 | 0 | 0 | 0 | 0 |
| 0 | 0 | 0 | 0 | 0 |
| 0 | 0 | 0 | 0 | 0 |
| 0 | 0 | 0 | 0 | 0 |
| 0 | 0 | 0 | 0 | 0.001515952 |
| 0 | 0 | 0 | 0 | 0 |
| 0 | 0 | 0.006614368 | 0 | 0 |
| 0 | 0 | 0 | 0 | 0 |
| 0 | 0 | 0 | 0 | 0 |
| 0 | 0 | 0 | 0 | 0 |
| 0 | 0 | 0 | 0 | 0 |
| 0 | 0 | 0 | 0 | 0 |
| 0 | 0 | 0.010293614 | 0 | 0 |
| 0 | 0.060148819 | 0 | 0 | 0.130190002 |
| 0 | 0 | 0 | 0 | 0 |
| 0.150647701 | 0.001190652 | 0.00539247 | 0.005890022 | 0.068594712 |
| 0 | 0 | 0 | 0 | 0 |
| 0 | 0 | 0 | 0 | 0 |
| 0 | 0 | 0.034386128 | 0 | 0 |
| 0 | 0 | 0 | 0 | 0 |
| 0 | 0 | 0 | 0 | 0 |
| 0.003010981 | 0 | 0 | 0 | 0 |
| 0 | 0 | 0 | 0 | 0 |
| 0 | 0 | 0 | 0 | 0 |
| 0 | 0 | 0.083283827 | 0 | 0 |
| 0.00346086 | 0.148077365 | 0.038784199 | 1.20174845 | 0.325712628 |
| 0 | 0 | 0 | 0 | 0 |
| 0 | 0.045481364 | 0.018068897 | 0 | 0 |
| 0 | 0.008231913 | 0 | 0 | 0 |
| 0 | 0 | 0 | 0 | 0 |
| 0 | 0 | 0 | 0 | 0 |
| 0 | 0 | 0 | 0.216395474 | 0.101571185 |
| 0 | 0 | 0 | 0 | 0 |
| 0 | 0 | 0 | 0 | 0 |
| 0 | 0 | 0 | 0 | 0 |
| 0.254752034 | 0.006534879 | 0.058316097 | 0.04330265 | 0.004554493 |
| 0 | 0 | 0 | 0 | 0 |
| 0 | 0 | 0 | 0 | 0 |
| 0.004562229 | 0 | 0.003830406 | 0 | 0 |
| 0 | 0 | 0 | 0 | 0 |
| 0 | 0 | 0 | 0 | 0 |

| | | | | |
|---|---|---|---|---|
| 0 | 0 | 0 | 0 | 0 |
| 0 | 0 | 0 | 0 | 0 |
| 0 | 0 | 0 | 0 | 0 |
| 0 | 0 | 0 | 0 | 0 |
| 0 | 0 | 0 | 0 | 0 |
| 0 | 0 | 0 | 0 | 0 |
| 0 | 0 | 0 | 0 | 0 |
| 0 | 0 | 0.011272403 | 0.023736753 | 0 |
| 0 | 0.001260619 | 0 | 0 | 0 |
| 0.057834403 | 0 | 0.006993899 | 0.044888527 | 0 |
| 0 | 0 | 0 | 0 | 0 |
| 0 | 0 | 0 | 0 | 0 |
| 0 | 0 | 0 | 0 | 0 |
| 0 | 0 | 0 | 0 | 0 |
| 0 | 0 | 0 | 0 | 0.016211653 |
| 0 | 0.064475695 | 0 | 0.006424051 | 0 |
| 0 | 0 | 0 | 0 | 0 |
| 0 | 0 | 0 | 0 | 0 |
| 0 | 0 | 0 | 0 | 0 |
| 0 | 0.111713134 | 0.024965104 | 0 | 0 |
| 0 | 0 | 0.001289965 | 0 | 0 |
| 0 | 0 | 0 | 0 | 0 |
| 0 | 0 | 0 | 0 | 0 |
| 0 | 0 | 0 | 0 | 0 |
| 0 | 0 | 0 | 0 | 0 |
| 0 | 0 | 0 | 0 | 0 |
| 0 | 0 | 0 | 0 | 0 |
| 0 | 0 | 0 | 0 | 0 |
| 0 | 0 | 0 | 0 | 0 |
| 0.091227324 | 0 | 0 | 0 | 0 |
| 0.242373058 | 18.59488304 | 0.130500186 | 0 | 0 |
| 0 | 0 | 0 | 0 | 0 |
| 0 | 0 | 0 | 0 | 0 |
| 0 | 0 | 0 | 0 | 0 |
| 0 | 0 | 0 | 0 | 0 |
| 0 | 0 | 0 | 0 | 0 |
| 0 | 0 | 0 | 0 | 0 |
| 0 | 0 | 0 | 0 | 0 |
| 0 | 0 | 0.005495253 | 0 | 0 |
| 0 | 0 | 0 | 0 | 0 |
| 0 | 0 | 0 | 0 | 0 |
| 0 | 0 | 0.19275653 | 0 | 0 |
| 0 | 0 | 0 | 0 | 0 |
| 0 | 0 | 0 | 0 | 0 |
| 0 | 0 | 0 | 0 | 0 |
| 0 | 0 | 0 | 0 | 0 |
| 0 | 0.652455575 | 0 | 0.043896495 | 0.863618773 |
| 0 | 0 | 0 | 0 | 0 |

| | | | | |
|---|---|---|---|---|
| 0 | 0 | 0 | 0 | 0 |
| 0 | 0 | 0 | 0 | 0 |
| 0 | 0 | 0 | 0 | 0 |
| 0 | 0 | 0 | 0 | 0.001552134 |
| 0 | 0 | 0.227505682 | 0 | 0 |
| 0 | 0 | 0 | 0 | 0 |
| 0 | 0 | 0 | 0 | 0 |
| 0 | 0 | 0 | 0 | 0 |
| 0 | 0 | 0 | 0 | 0 |
| 0 | 0 | 0 | 0 | 0 |
| 0 | 0 | 0.019873645 | 0 | 0 |
| 0.062306094 | 0 | 0.00093671 | 0.125163884 | 0.018561649 |
| 0 | 0 | 0 | 0 | 0.000774872 |
| 0 | 0 | 0 | 0 | 0 |
| 0 | 0 | 0.015561674 | 0 | 0 |
| 0 | 0 | 0 | 0 | 0 |
| 0 | 0 | 0.004466734 | 0 | 0 |
| 0 | 0 | 0 | 0 | 0 |
| 0 | 0.699571135 | 0.00156042 | 0.002711541 | 0.044205201 |
| 0.010080129 | 0 | 0.286352031 | 0 | 0.006807826 |
| 0 | 0 | 0 | 0 | 0 |
| 0 | 0 | 0 | 0 | 0 |
| 0 | 0 | 0 | 0 | 0 |
| 0 | 0 | 0 | 0 | 0 |
| 0 | 0 | 0 | 0 | 0 |
| 0 | 0 | 0 | 0 | 0 |
| 0.000747932 | 0.000655447 | 0.010241412 | 0 | 0.005692615 |
| 0 | 0 | 0 | 0 | 0 |
| 0 | 0 | 0 | 0 | 0 |
| 0 | 0 | 0 | 0 | 0 |
| 0 | 0.007801331 | 0 | 0 | 0.006721748 |
| 0 | 0 | 0 | 0 | 0 |
| 0 | 0 | 0 | 0 | 0 |
| 0 | 0 | 0 | 0 | 0 |
| 0 | 0 | 0 | 0 | 0 |
| 0 | 0 | 0 | 0 | 0 |
| 0 | 0 | 0 | 0 | 0 |
| 0.019656586 | 0.302367867 | 0.00449186 | 0.038626466 | 0.112395624 |
| 0 | 0 | 0.089145985 | 0 | 0 |
| 0 | 0 | 0 | 0 | 0 |
| 0.009990201 | 0.007816847 | 0 | 0.003115009 | 0.078450651 |
| 0 | 0 | 0 | 0 | 0 |
| 0 | 0 | 0 | 0 | 0 |
| 0 | 0.03618722 | 0.089571021 | 0 | 0 |
| 0 | 0 | 0 | 0 | 0 |
| 0 | 0 | 0 | 0 | 0 |
| 0 | 0 | 0 | 0 | 0 |
| 0 | 0 | 0 | 0 | 0 |

| | | | | |
|---|---|---|---|---|
| 0 | 0 | 0 | 0 | 0 |
| 0 | 0 | 0.062773697 | 0 | 0 |
| 0 | 0 | 0 | 0 | 0 |
| 0.012959533 | 0 | 0 | 0 | 0.042566467 |
| 0 | 0.009781074 | 0.01073388 | 0.006203319 | 0 |
| 0.002437703 | 0 | 0 | 0 | 0.004216747 |
| 0 | 0 | 0.009982961 | 0 | 0 |
| 0 | 0 | 0 | 0 | 0 |
| 0 | 0 | 0 | 0 | 0 |
| 0 | 0 | 0 | 0 | 0 |
| 0 | 0 | 0.005161009 | 0 | 0 |
| 0 | 0 | 0 | 0 | 0 |
| 0 | 0 | 0.078651447 | 0 | 0 |
| 0 | 0 | 0 | 0 | 0 |
| 0 | 0 | 0 | 0 | 0 |
| 0 | 0 | 0 | 0 | 0 |
| 0 | 0 | 0 | 0 | 0 |
| 0 | 0 | 0 | 0 | 0 |
| 0 | 0 | 0 | 0 | 0 |
| 0 | 0 | 0 | 0 | 0 |
| 0 | 0 | 0 | 0 | 0 |
| 0 | 0 | 0 | 0 | 0 |
| 0 | 0.002839473 | 0.056241047 | 0 | 0.005206226 |
| 0.001223509 | 0 | 0.012342526 | 0 | 0 |
| 0 | 0 | 0 | 0 | 0 |
| 0 | 0 | 0 | 0 | 0 |
| 0 | 0 | 0 | 0 | 0 |
| 0 | 0 | 0 | 0 | 0 |
| 0 | 0 | 0 | 0 | 0 |
| 0 | 0 | 0 | 0 | 0 |
| 0 | 0 | 0.045693097 | 0 | 0 |
| 0 | 0 | 0 | 0 | 0 |
| 0 | 0 | 0 | 0 | 0 |
| 0 | 0 | 0 | 0 | 0 |
| 0 | 0 | 0 | 0 | 0 |
| 0 | 0 | 0 | 0 | 0.033854751 |
| 0 | 0.123557842 | 0 | 0 | 0.005842706 |
| 0 | 0 | 0 | 0 | 0 |
| 0.022170685 | 0 | 0.001779309 | 0 | 0 |
| 0 | 0 | 0 | 0 | 0 |
| 0 | 0 | 0 | 0 | 0 |
| 0 | 0 | 0 | 0 | 0 |
| 0 | 0 | 0 | 0 | 0 |
| 0 | 0 | 0 | 0 | 0 |
| 0 | 14.47415637 | 0 | 0 | 0 |
| 0.284565997 | 0.830998724 | 0.255033834 | 0 | 0 |
| 0 | 0 | 0 | 0 | 0 |
| 0 | 0 | 0 | 0 | 0 |

| | | | | |
|---|---|---|---|---|
| 0 | 0 | 0 | 0 | 0 |
| 0 | 0 | 0 | 0 | 0 |
| 0.164626136 | 0 | 0.003795426 | 0 | 0 |
| 0 | 0 | 0 | 0 | 0 |
| 0 | 0 | 0 | 0 | 0 |
| 0 | 0 | 0 | 0 | 0 |
| 0 | 0 | 0 | 0 | 0 |
| 0 | 0 | 0 | 0 | 0 |
| 0 | 0 | 0 | 0 | 0 |
| 0 | 0 | 0 | 0 | 0 |
| 0 | 0 | 0.000524908 | 0.003463928 | 0 |
| 0.002961805 | 0 | 0 | 0 | 0.007827328 |
| 0 | 0 | 0 | 0 | 0 |
| 0 | 0 | 0 | 0 | 0 |
| 0 | 0 | 0 | 0 | 0 |
| 0 | 0 | 0 | 0 | 0 |
| 0 | 0 | 0 | 0 | 0 |
| 0 | 0 | 0 | 0 | 0 |
| 0 | 0 | 0 | 0 | 0 |
| 0.010172607 | 0.001729148 | 0.000522088 | 0.001584056 | 0.001390537 |
| 0 | 0 | 0 | 0 | 0 |
| 0 | 0 | 0 | 0 | 0 |
| 0 | 0 | 0 | 0 | 0 |
| 0 | 0 | 0 | 0 | 0 |
| 0 | 0 | 0 | 0 | 0 |
| 0 | 0 | 0 | 0 | 0 |
| 0 | 0 | 0 | 0 | 0 |
| 0.005434756 | 0 | 0.012133359 | 0 | 0 |
| 0 | 0 | 0 | 0 | 0 |
| 0 | 0 | 0 | 0 | 0 |
| 0.010999492 | 0.00132046 | 0 | 0 | 0.00860123 |
| 0 | 0 | 0 | 0 | 0 |
| 0.018195056 | 0.130512658 | 0.011804496 | 0.300062289 | 0.269646397 |
| 0 | 0.004308569 | 0 | 0 | 0.008221123 |
| 0 | 0 | 0 | 0 | 0 |
| 0 | 0 | 0 | 0 | 0 |
| 0 | 0 | 0 | 0 | 0 |
| 0 | 0 | 0 | 0 | 0 |
| 0 | 0 | 0 | 0 | 0 |
| 0 | 0 | 0 | 0 | 0 |
| 0 | 0 | 0 | 0 | 0 |
| 0.1226299 | 0.001734343 | 0.002427863 | 0.713406832 | 1.292900129 |
| 0.020691244 | 0 | 0.010961339 | 0 | 0 |
| 0 | 0 | 0 | 0 | 0 |
| 0 | 0 | 0 | 0 | 0 |
| 0.016470496 | 0.007494498 | 0.01558847 | 0.010756168 | 0.004821508 |
| 0 | 0 | 0 | 0 | 0.003161117 |
| 0 | 0 | 0 | 0 | 0 |

| | | | | |
|---|---|---|---|---|
| 0 | 0 | 0 | 0 | 0 |
| 0.005327355 | 0 | 0 | 0 | 0 |
| 0.001072916 | 0.011753066 | 0.002129189 | 0 | 0.026539858 |
| 0 | 0 | 0 | 0 | 0 |
| 0.181271746 | 0 | 0.013600438 | 0 | 0.080416367 |
| 0 | 0 | 0 | 0 | 0 |
| 0 | 0 | 0 | 0 | 0 |
| 0 | 0 | 0 | 0 | 0 |
| 0 | 0 | 0 | 0 | 0 |
| 0 | 0 | 0 | 0 | 0 |
| 0 | 0 | 0 | 0 | 0 |
| 0 | 0 | 0 | 0 | 0 |
| 0 | 0 | 0 | 0 | 0 |
| 0 | 0 | 0 | 0 | 0 |
| 0 | 0 | 0 | 0 | 0 |
| 0 | 0 | 0 | 0 | 0 |
| 0 | 0 | 0 | 0 | 0 |
| 0 | 0 | 0 | 0 | 0 |
| 0 | 0 | 0 | 0 | 0 |
| 0 | 0 | 0 | 0 | 0 |
| 0 | 0 | 0 | 0 | 0 |
| 0 | 0 | 0 | 0 | 0 |
| 0 | 0 | 0.002166788 | 0 | 0 |
| 0 | 0 | 0 | 0 | 0 |
| 0.002652701 | 0 | 0 | 0.002555544 | 0 |
| 0.027219254 | 0 | 0 | 0.021691268 | 0.003385122 |
| 0 | 0 | 0 | 0 | 0 |
| 0 | 0 | 0 | 0 | 0 |
| 0 | 0 | 0 | 0 | 0 |
| 0 | 0 | 0 | 0 | 0 |
| 0 | 0 | 0 | 0 | 0 |
| 0 | 0 | 0.003096018 | 0 | 0.001484276 |
| 0 | 0 | 0 | 0 | 0 |
| 0 | 0 | 0 | 0 | 0 |
| 0 | 0 | 0 | 0 | 0 |
| 0 | 0 | 0 | 0 | 0 |
| 0 | 0 | 0 | 0 | 0 |
| 0 | 0.004073748 | 0 | 0 | 0 |
| 0 | 0 | 0 | 0 | 0.002561187 |
| 0 | 0 | 0 | 0 | 0 |
| 0 | 0 | 0 | 0 | 0 |
| 0 | 0 | 0.003255519 | 0 | 0 |
| 0.028745858 | 0 | 0 | 0 | 0 |
| 0 | 0 | 0 | 0 | 0 |
| 0 | 0 | 0 | 0 | 0 |
| 0 | 0 | 0 | 0 | 0 |
| 0.021031778 | 0.004488094 | 0.01626129 | 0 | 0.003031738 |
| 0 | 0 | 0 | 0 | 0 |

| | | | | |
|---|---|---|---|---|
| 0 | 0 | 0 | 0 | 0 |
| 0.008443984 | 0.111752819 | 0.025819435 | 0.058831444 | 0.004098745 |
| 0 | 0 | 0 | 0 | 0 |
| 0 | 0 | 0 | 0 | 0 |
| 0 | 0 | 0 | 0 | 0 |
| 0 | 0 | 0 | 0 | 0 |
| 0 | 0 | 0 | 0 | 0 |
| 0 | 0 | 0 | 0 | 0 |
| 0 | 0 | 0 | 0 | 0 |
| 0 | 0 | 0 | 0 | 0 |
| 0 | 0 | 0 | 0 | 0 |
| 0 | 0 | 0 | 0 | 0 |
| 0 | 0 | 0.092363861 | 0 | 0 |
| 0 | 0 | 0 | 0 | 0 |
| 0 | 0.024478459 | 0.018761451 | 0 | 0.006473321 |
| 0 | 0 | 0 | 0 | 0 |
| 0 | 0 | 0 | 0 | 0 |
| 0 | 0 | 0 | 0 | 0 |
| 0 | 0 | 0.000707558 | 0 | 0 |
| 0 | 0 | 0 | 0.001550865 | 0 |
| 0 | 0 | 0 | 0 | 0 |
| 0.082717566 | 0 | 0.270515811 | 0 | 0.1164392 |
| 0 | 0 | 0.048749525 | 0.08054496 | 0.001028437 |
| 0 | 0 | 0 | 0 | 0 |
| 0 | 0 | 0 | 0 | 0 |
| 0 | 0 | 0 | 0 | 0 |
| 0 | 0 | 0 | 0 | 0 |
| 0 | 0 | 0 | 0 | 0 |
| 0 | 0 | 0 | 0 | 0 |
| 0 | 0 | 0 | 0 | 0 |
| 0 | 0 | 0 | 0 | 0 |
| 0 | 0 | 0 | 0 | 0 |
| 0 | 0 | 0 | 0 | 0 |
| 0 | 0 | 0 | 0 | 0 |
| 0.09234597 | 0 | 0 | 0 | 0 |
| 0 | 0 | 0 | 0 | 0 |
| 0 | 0 | 0 | 0 | 0 |
| 0 | 0 | 0 | 0 | 0 |
| 0.00063217 | 0 | 0.160162044 | 0 | 0.01002402 |
| 0 | 0 | 0 | 0 | 0 |
| 0.147374158 | 0.073268188 | 0.015373015 | 0.020989313 | 0.421231017 |
| 0 | 0 | 0 | 0 | 0 |
| 0 | 0 | 0 | 0 | 0 |
| 0 | 0 | 0 | 0 | 0 |
| 0 | 0 | 0 | 0 | 0 |
| 0.392845924 | 1.773951261 | 0.001717176 | 0.287203763 | 5.820985758 |
| 0 | 0 | 0 | 0 | 0 |
| 0 | 0 | 0 | 0 | 0 |

| | | | | |
|---|---|---|---|---|
| 0 | 0 | 0 | 0 | 0 |
| 0 | 0 | 0 | 0 | 0 |
| 0 | 0 | 0 | 0 | 0 |
| 0 | 0 | 0 | 0 | 0 |
| 0 | 0 | 0 | 0 | 0 |
| 0 | 0 | 0 | 0 | 0 |
| 0 | 0 | 0 | 0 | 0 |
| 0 | 0 | 0.007888105 | 0 | 0 |
| 0.000586181 | 0 | 0 | 0 | 0 |
| 0.009943604 | 0 | 0.06717604 | 0 | 0 |
| 0 | 0 | 0.046714561 | 0 | 0 |
| 0 | 0 | 0 | 0 | 0 |
| 0.002274668 | 0.004530444 | 0 | 0.000747053 | 0.009836823 |
| 0 | 0 | 0 | 0 | 0 |
| 0 | 0 | 0 | 0 | 0 |
| 0 | 0 | 0 | 0 | 0 |
| 0 | 0 | 0 | 0 | 0 |
| 0 | 0 | 0 | 0 | 0 |
| 0 | 0 | 0 | 0 | 0 |
| 2.395487635 | 0.044759805 | 0.604682501 | 0.289577154 | 0.103314508 |
| 0 | 0 | 0 | 0 | 0 |
| 0 | 0 | 0 | 0 | 0 |
| 0 | 0 | 0 | 0 | 0 |
| 0 | 0 | 0 | 0 | 0 |
| 0 | 0 | 0 | 0 | 0 |
| 0 | 0 | 0 | 0 | 0 |
| 0 | 0 | 0 | 0 | 0 |
| 0 | 0 | 0 | 0 | 0 |
| 0 | 0 | 0 | 0 | 0 |
| 0 | 0 | 0.003904653 | 0 | 0 |
| 0 | 0 | 0 | 0 | 0 |
| 0 | 0 | 0 | 0 | 0 |
| 0 | 0 | 0 | 0 | 0 |
| 0 | 0 | 0 | 0 | 0 |
| 0 | 0 | 0 | 0 | 0 |
| 0 | 0 | 0 | 0 | 0 |
| 0 | 0 | 0 | 0 | 0 |
| 0 | 0 | 0 | 0 | 0 |
| 0 | 0 | 0 | 0 | 0 |
| 0 | 0 | 0 | 0 | 0 |
| 0 | 0 | 0 | 0 | 0 |
| 0 | 0 | 0 | 0 | 0 |
| 0 | 0 | 0.007742203 | 0 | 0 |
| 0 | 0 | 0 | 0 | 0 |
| 0.004494674 | 0.002083066 | 0.001200719 | 0.020921654 | 0.002192922 |
| 0 | 0 | 0.046678286 | 0 | 0 |
| 0 | 0 | 0 | 0 | 0 |
| 0 | 0.009288685 | 0 | 0.009064985 | 0 |

| | | | | |
|---|---|---|---|---|
| 0 | 0 | 0 | 0 | 0 |
| 0 | 0 | 0 | 0 | 0 |
| 0 | 0 | 0 | 0 | 0 |
| 0 | 0 | 0 | 0 | 0 |
| 0 | 0 | 0 | 0 | 0 |
| 0.095888402 | 0 | 0.000896383 | 0.034151617 | 0.050954777 |
| 0 | 0 | 0 | 0 | 0 |
| 0 | 0 | 0 | 0 | 0 |
| 0 | 0 | 0 | 0 | 0 |
| 0 | 0 | 0 | 0 | 0 |
| 0 | 0 | 0 | 0 | 0 |
| 0 | 0 | 0 | 0 | 0 |
| 0 | 0 | 0 | 0 | 0 |
| 0 | 0 | 0.000818616 | 0 | 0 |
| 0 | 0 | 0 | 0 | 0 |
| 0 | 0 | 0 | 0 | 0 |
| 0.013306213 | 0 | 0 | 0 | 0 |
| 0 | 0 | 0 | 0 | 0 |
| 0 | 0 | 0 | 0 | 0 |
| 0 | 0 | 0 | 0 | 0 |
| 0.004040241 | 0 | 0 | 0 | 0 |
| 0 | 0 | 0 | 0 | 0 |
| 0 | 0 | 0 | 0 | 0 |
| 0 | 0 | 0 | 0 | 0 |
| 0 | 0 | 0 | 0 | 0 |
| 0 | 0 | 0.004406581 | 0 | 0 |
| 0 | 0 | 0 | 0 | 0 |
| 0.003474883 | 0 | 0 | 0 | 0.024914601 |
| 0 | 0 | 0 | 0 | 0 |
| 0 | 0 | 0 | 0 | 0 |
| 0 | 0 | 0 | 0 | 0 |
| 0 | 0 | 0 | 0 | 0 |
| 0 | 0 | 0 | 0 | 0 |
| 0 | 0 | 0 | 0 | 0 |
| 0 | 0 | 0 | 0 | 0 |
| 0 | 0 | 0 | 0 | 0 |
| 0 | 0 | 0 | 0 | 0 |
| 0 | 0 | 0 | 0 | 0 |
| 0 | 0 | 0.629409014 | 0 | 0 |
| 0 | 0 | 0 | 0 | 0 |
| 0 | 0 | 0 | 0 | 0 |
| 0 | 0 | 0 | 0 | 0 |
| 0 | 0 | 0 | 0 | 0 |
| 0 | 0 | 0 | 0 | 0 |
| 0 | 0 | 0 | 0 | 0 |
| 0 | 0 | 0 | 0 | 0 |
| 0 | 0 | 0 | 0 | 0 |
| 0 | 0 | 0 | 0 | 0 |

| | | | | |
|---|---|---|---|---|
| 0 | 0 | 0 | 0 | 0 |
| 0 | 0.007439024 | 0 | 0 | 0 |
| 0 | 0 | 0 | 0 | 0 |
| 0 | 0 | 0 | 0 | 0 |
| 0 | 0 | 0 | 0 | 0 |
| 0 | 0 | 0 | 0 | 0 |
| 0 | 0 | 0 | 0 | 0 |
| 0 | 0 | 0 | 0 | 0 |
| 0 | 0 | 0 | 0 | 0 |
| 0 | 0 | 0 | 0 | 0 |
| 0 | 0 | 0.01163917 | 0.00666412 | 0 |
| 0 | 0 | 0 | 0 | 0 |
| 0 | 0 | 0.315950524 | 0 | 0 |
| 0 | 0 | 0 | 0 | 0 |
| 0 | 0 | 0 | 0 | 0 |
| 0 | 0 | 0 | 0 | 0 |
| 0 | 0 | 0 | 0 | 0 |
| 0 | 0 | 0 | 0 | 0 |
| 0 | 0 | 0 | 0 | 0 |
| 0 | 0 | 0.001405903 | 0 | 0 |
| 0 | 0 | 0 | 0 | 0.01583302 |
| 0 | 0 | 0 | 0 | 0 |
| 0 | 0 | 0 | 0 | 0 |
| 1.675498703 | 0 | 2.054098537 | 0 | 1.499519638 |
| 0 | 0 | 0 | 0 | 0 |
| 0 | 0 | 0 | 0 | 0 |
| 0 | 0 | 0 | 0 | 0 |
| 0 | 0 | 0 | 0 | 0.006294692 |
| 0 | 0 | 0 | 0 | 0 |
| 0 | 0 | 0 | 0 | 0 |
| 0 | 0 | 0 | 0 | 0 |
| 0 | 0 | 0 | 0 | 0 |
| 0 | 0 | 0 | 0 | 0 |
| 0 | 0 | 0 | 0 | 0 |

PHL018-0039_V3
EYCF
0.024202686
0.100456421
0.093629756
0
0
0
0
0
0
0
0
0
0
0.685755926
0
0
0
0
0
0
0
0
0
0
0.478130148
0
0
0
0
0
0
0
0
0
0
0.539190903
0
0
0
0
0
0.017190309
1.126209818
0.086601178
0
0

| |
|---|
| 0 |
| 0.001091553 |
| 0 |
| 0.791321135 |
| 0 |
| 0 |
| 0.004913868 |
| 0 |
| 0 |
| 0 |
| 0 |
| 0.020972186 |
| 0.000810843 |
| 0.038878314 |
| 0 |
| 0 |
| 0.816930477 |
| 0 |
| 0 |
| 0 |
| 0.153173623 |
| 0.010474395 |
| 0.127366009 |
| 0 |
| 0 |
| 0 |
| 0 |
| 0 |
| 0.005317373 |
| 0 |
| 0 |
| 0 |
| 0 |
| 0 |
| 0 |
| 0 |
| 0 |
| 0.241123192 |
| 0 |
| 0 |
| 0 |
| 0 |
| 0 |
| 0 |
| 0 |
| 0 |
| 0 |
| 0.00050393 |

| |
|---|
| 0 |
| 0 |
| 0 |
| 0 |
| 2.069421524 |
| 0 |
| 0 |
| 0 |
| 0.003053367 |
| 0 |
| 0 |
| 0.000932868 |
| 0 |
| 0 |
| 0 |
| 0 |
| 0 |
| 0 |
| 1.935549483 |
| 0 |
| 0 |
| 0 |
| 0 |
| 0 |
| 0 |
| 0 |
| 0 |
| 0 |
| 0 |
| 0 |
| 0.048033452 |
| 0.848851163 |
| 0 |
| 0 |
| 0 |
| 0 |
| 0 |
| 0 |
| 0.379427207 |
| 0 |
| 0 |
| 0 |
| 0.000721292 |
| 0 |
| 0 |
| 0 |
| 0 |
| 0 |

| |
|---|
| 0 |
| 0 |
| 0 |
| 0 |
| 0.059747235 |
| 0 |
| 0 |
| 0 |
| 0 |
| 0 |
| 0 |
| 0 |
| 0 |
| 0 |
| 0 |
| 0 |
| 0 |
| 0 |
| 0 |
| 0 |
| 0 |
| 0 |
| 0.00278155 |
| 0 |
| 0 |
| 0 |
| 0.035757912 |
| 0 |
| 0 |
| 0 |
| 0 |
| 0 |
| 0 |
| 0 |
| 0 |
| 0 |
| 0 |
| 0 |
| 0 |
| 0 |
| 0 |
| 0 |
| 0 |
| 0 |
| 0 |
| 0 |
| 0 |
| 0 |

| |
|---|
| 0 |
| 0 |
| 0 |
| 0 |
| 0 |
| 0 |
| 0 |
| 0 |
| 0 |
| 0 |
| 0 |
| 0 |
| 0.004650596 |
| 0 |
| 0 |
| 0 |
| 0 |
| 0 |
| 0 |
| 0 |
| 0.034137878 |
| 0 |
| 0 |
| 0 |
| 0 |
| 0 |
| 0 |
| 0 |
| 0 |
| 0 |
| 0 |
| 0 |
| 0 |
| 0.026373919 |
| 0.013410972 |
| 0 |
| 0 |
| 0 |
| 0 |
| 0 |
| 0 |
| 0 |
| 0 |
| 0 |
| 0 |
| 0.235023999 |
| 0.001134415 |
| 0.00194127 |

| |
|---|
| 0 |
| 0 |
| 0 |
| 0 |
| 0 |
| 0 |
| 0 |
| 0 |
| 0 |
| 0 |
| 0 |
| 0 |
| 0 |
| 0 |
| 0 |
| 0 |
| 0 |
| 0 |
| 0.004100383 |
| 0 |
| 0 |
| 0 |
| 0 |
| 0 |
| 0 |
| 0 |
| 0 |
| 0 |
| 0.004247262 |
| 0 |
| 0.272117877 |
| 0 |
| 0 |
| 0 |
| 0 |
| 0 |
| 0 |
| 0 |
| 0 |
| 0.001316261 |
| 0 |
| 0.001343239 |
| 0 |
| 0 |
| 0 |
| 0 |
| 0 |
| 0 |

| |
|---|
| 0 |
| 0 |
| 0.094874515 |
| 0 |
| 0 |
| 0 |
| 0 |
| 0 |
| 0 |
| 0 |
| 0 |
| 0 |
| 0 |
| 0 |
| 0.001153904 |
| 0.00370622 |
| 0.000657385 |
| 0 |
| 0 |
| 0 |
| 0 |
| 0.004351829 |
| 0 |
| 0 |
| 0 |
| 0 |
| 0.067944417 |
| 0 |
| 0 |
| 0 |
| 0 |
| 0 |
| 0 |
| 0 |
| 0 |
| 0 |
| 0 |
| 0.088476071 |
| 0 |
| 0 |
| 0 |
| 0 |
| 0 |
| 0 |
| 0 |
| 0 |
| 0 |
| 0 |

| |
|---|
| 0.128796499 |
| 0 |
| 0.001035955 |
| 0 |
| 0 |
| 0 |
| 0 |
| 0 |
| 0 |
| 0.003915223 |
| 0 |
| 0.003700833 |
| 0.003333566 |
| 0 |
| 0 |
| 0 |
| 0 |
| 0 |
| 0 |
| 0 |
| 0 |
| 0 |
| 0 |
| 0 |
| 0 |
| 0 |
| 0 |
| 0 |
| 0 |
| 0 |
| 0 |
| 0 |
| 0 |
| 0 |
| 0 |
| 0 |
| 0 |
| 0 |
| 0 |
| 0 |
| 0.012645986 |
| 0 |
| 0 |
| 0 |
| 0 |
| 0 |
| 0.00355785 |
| 0 |

| |
|---|
| 0 |
| 0 |
| 0 |
| 0.426671803 |
| 0 |
| 0.011208953 |
| 0 |
| 0 |
| 0 |
| 0 |
| 0 |
| 0 |
| 0 |
| 0 |
| 0 |
| 0 |
| 0 |
| 0 |
| 0 |
| 0 |
| 0 |
| 0.144567115 |
| 0.034341322 |
| 0 |
| 0 |
| 0 |
| 0 |
| 0 |
| 0 |
| 0.658079706 |
| 0.001190358 |
| 0.000835613 |
| 0 |
| 0 |
| 0 |
| 0 |
| 0.011515564 |
| 0.000894204 |
| 0 |
| 0 |
| 0 |
| 1.52840519 |
| 0 |
| 0 |
| 0 |
| 0 |
| 0.001135485 |
| 0 |

| |
|---|
| 0 |
| 0 |
| 0.035952215 |
| 0 |
| 0.094672017 |
| 0 |
| 0 |
| 0 |
| 0 |
| 0 |
| 0 |
| 0 |
| 0 |
| 0 |
| 0 |
| 0.036482989 |
| 0.023603842 |
| 0 |
| 0 |
| 0 |
| 0.002997816 |
| 0 |
| 0 |
| 0 |
| 0 |
| 0 |
| 0 |
| 0 |
| 0 |
| 0 |
| 0.010245993 |
| 0.00526742 |
| 0.066954201 |
| 0.005031186 |
| 0.141843968 |
| 0 |
| 0 |
| 0 |
| 0 |
| 0 |
| 0 |
| 0 |
| 0 |
| 0.002352833 |
| 0 |
| 0 |
| 0 |
| 0 |

| |
|---|
| 0 |
| 0.467475144 |
| 0 |
| 0 |
| 0 |
| 0 |
| 0 |
| 0 |
| 0.001990395 |
| 0 |
| 0.00897636 |
| 0.003743752 |
| 0.263885215 |
| 0 |
| 0 |
| 0 |
| 0.003947809 |
| 0.002928458 |
| 0 |
| 0 |
| 0.011438097 |
| 0 |
| 0 |
| 0 |
| 0 |
| 0 |
| 0.102183822 |
| 0.073148099 |
| 0 |
| 0.00173982 |
| 0 |
| 0 |
| 0 |
| 0 |
| 0.004668536 |
| 0.027200862 |
| 0 |
| 0 |
| 0.004829552 |
| 0 |
| 0 |
| 0 |
| 0 |
| 0.00059627 |
| 0 |
| 0.848019685 |
| 0 |
| 0 |

| |
|---|
| 0 |
| 0 |
| 0 |
| 0.002877179 |
| 0 |
| 0 |
| 0 |
| 0 |
| 1.627898936 |
| 0 |
| 0 |
| 0.003101227 |
| 0 |
| 0 |
| 0.127814865 |
| 0 |
| 0.030970845 |
| 0.39712348 |
| 0 |
| 0 |
| 0 |
| 0.021434508 |
| 0 |
| 0.204604006 |
| 0.000967847 |
| 0 |
| 0.003646698 |
| 2.50095058 |
| 0 |
| 0 |
| 0 |
| 0 |
| 0 |
| 0 |
| 0 |
| 0 |
| 0 |
| 0 |
| 0 |
| 0 |
| 0.324778845 |
| 0 |
| 0.095767247 |
| 0.002447696 |
| 0 |
| 0 |
| 0 |
| 0 |

| |
|---|
| 0 |
| 0.045614563 |
| 0.015427636 |
| 0 |
| 0 |
| 0.033459076 |
| 0 |
| 0 |
| 0 |
| 0 |
| 0.02169229 |
| 3.14553086 |
| 0 |
| 0.024420307 |
| 0.011451058 |
| 0 |
| 0 |
| 0 |
| 0 |
| 0.030192005 |
| 0.158929932 |
| 0.006110577 |
| 0.092081256 |
| 0 |
| 0.026114763 |
| 0 |
| 0 |
| 0.000490417 |
| 0 |
| 0 |
| 0.014002903 |
| 0.09484906 |
| 0.015172459 |
| 0.00629769 |
| 0.001774635 |
| 0.241734063 |
| 0.00572003 |
| 0 |
| 7.393949483 |
| 0 |
| 0 |
| 0 |
| 0 |
| 0 |
| 0.054220447 |
| 0 |
| 0 |
| 0.110684837 |

| |
|---|
| 0 |
| 0 |
| 0 |
| 0.002369839 |
| 0 |
| 0.039020005 |
| 0 |
| 0.000934578 |
| 0 |
| 0 |
| 0.018475615 |
| 0 |
| 0.059755285 |
| 0 |
| 0 |
| 0 |
| 0.054823757 |
| 0.006748599 |
| 0.002146909 |
| 0 |
| 0 |
| 0 |
| 0 |
| 0 |
| 0 |
| 0 |
| 0 |
| 0 |
| 0 |
| 0.090812241 |
| 0.013660689 |
| 0 |
| 0.054277354 |
| 0 |
| 0 |
| 0 |
| 0.024706274 |
| 0 |
| 0.001805828 |
| 0.001259295 |
| 0.014674126 |
| 0 |
| 0 |
| 0 |
| 0.223074533 |
| 0 |
| 0.012889256 |
| 0 |

| |
|---|
| 0 |
| 0 |
| 0.318749246 |
| 0.318445281 |
| 0 |
| 0 |
| 0 |
| 0 |
| 0.002861337 |
| 0 |
| 0.001101663 |
| 0 |
| 0.001412442 |
| 0 |
| 0.000632687 |
| 0.000508918 |
| 0 |
| 0 |
| 0 |
| 0 |
| 0.803128435 |
| 0 |
| 0 |
| 0 |
| 0.459949185 |
| 0 |
| 0.228547737 |
| 0 |
| 0.110205911 |
| 0 |
| 0 |
| 0.01400204 |
| 0 |
| 0 |
| 0.008882894 |
| 0.00241505 |
| 0 |
| 2.879539054 |
| 0.001016699 |
| 0 |
| 0 |
| 0.000730316 |
| 0.010340653 |
| 1.75735649 |
| 0 |
| 0.001220243 |
| 0 |
| 0.4816245 |

| |
|---|
| 0 |
| 0 |
| 0 |
| 0 |
| 0 |
| 0.001737325 |
| 0 |
| 0 |
| 0 |
| 0 |
| 0 |
| 0.947543386 |
| 0.017257181 |
| 0 |
| 0.092638198 |
| 0 |
| 0 |
| 0 |
| 0 |
| 0 |
| 0.131570732 |
| 0 |
| 0.056369667 |
| 0 |
| 0 |
| 0 |
| 0 |
| 0 |
| 0 |
| 0 |
| 0 |
| 0.017327714 |
| 0 |
| 0.004328314 |
| 0.003329027 |
| 0.121517757 |
| 0 |
| 0.043393699 |
| 0 |
| 0 |
| 0 |
| 0 |
| 0 |
| 0 |
| 0.014843714 |
| 0 |
| 1.802360866 |
| 0 |

| |
|---|
| 0 |
| 0.077291253 |
| 0 |
| 0 |
| 0 |
| 0 |
| 0.674596029 |
| 0 |
| 0 |
| 0 |
| 0 |
| 0 |
| 0 |
| 0 |
| 0 |
| 0 |
| 0 |
| 0 |
| 0.404383537 |
| 0 |
| 0 |
| 0 |
| 0 |
| 0 |
| 0 |
| 0 |
| 0 |
| 0.353248533 |
| 0 |
| 0.44572452 |
| 0 |
| 0 |
| 0 |
| 0 |
| 0.012123955 |
| 0 |
| 0 |
| 0 |
| 0 |
| 0 |
| 0.388327215 |
| 0.075247423 |
| 0.586060047 |
| 0 |
| 0 |
| 0 |
| 0 |
| 0.002426317 |

| |
|---|
| 0 |
| 0.007642981 |
| 0 |
| 0 |
| 0.144214636 |
| 0 |
| 0 |
| 0 |
| 0.003737982 |
| 0 |
| 0 |
| 0 |
| 0.080175438 |
| 0.031862099 |
| 0.606314213 |
| 0.277206725 |
| 0.169944894 |
| 0 |
| 0 |
| 0 |
| 0 |
| 0 |
| 0 |
| 0 |
| 0 |
| 0 |
| 0.054258601 |
| 0.001898442 |
| 0 |
| 0 |
| 0 |
| 0 |
| 0.043485245 |
| 0 |
| 0.138869919 |
| 0 |
| 0 |
| 0.028686797 |
| 0 |
| 0 |
| 0.563708303 |
| 0.145571752 |
| 0 |
| 0.01703921 |
| 0 |
| 0 |
| 0.477642918 |
| 0 |

| |
|---|
| 0 |
| 0 |
| 0 |
| 0 |
| 0 |
| 0 |
| 0.147656606 |
| 0 |
| 0 |
| 0 |
| 0.008777344 |
| 0 |
| 0 |
| 0 |
| 0 |
| 0 |
| 0 |
| 0 |
| 0 |
| 0 |
| 0.227098217 |
| 0 |
| 0.00202688 |
| 0.015469933 |
| 0 |
| 0 |
| 0 |
| 0 |
| 0 |
| 0.001945473 |
| 0 |
| 0 |
| 0 |
| 0 |
| 0 |
| 0 |
| 0.015786204 |
| 0 |
| 0 |
| 0 |
| 0 |
| 0 |
| 0 |
| 0 |
| 0 |
| 0 |
| 0 |
| 0 |

| |
|---|
| 0.004040339 |
| 0.348084374 |
| 0 |
| 0 |
| 0 |
| 0 |
| 0 |
| 0.702260643 |
| 1.492126728 |
| 0 |
| 0 |
| 0 |
| 0 |
| 0.006573047 |
| 0 |
| 0 |
| 0 |
| 0 |
| 0 |
| 0 |
| 0 |
| 0 |
| 0 |
| 0 |
| 0.018202911 |
| 0 |
| 0 |
| 0 |
| 0 |
| 0 |
| 0 |
| 0 |
| 0 |
| 0 |
| 0 |
| 0 |
| 0 |
| 0 |
| 0 |
| 0 |
| 0 |
| 0.299446832 |
| 0 |
| 0 |
| 0 |
| 0.017532291 |
| 0 |
| 0 |

| |
|---|
| 0 |
| 0.020129137 |
| 0 |
| 0 |
| 1.320494773 |
| 0.004286674 |
| 0 |
| 0.004522852 |
| 0 |
| 0 |
| 0 |
| 0.167558963 |
| 0 |
| 0 |
| 0.001302516 |
| 0 |
| 0 |
| 0 |
| 0 |
| 0 |
| 0 |
| 0 |
| 0 |
| 0.006049149 |
| 0 |
| 0 |
| 0 |
| 0 |
| 0 |
| 0 |
| 0 |
| 0 |
| 0 |
| 0.122857264 |
| 0 |
| 0 |
| 0 |
| 0.049063758 |
| 0 |
| 0 |
| 0 |
| 0 |
| 0 |
| 0 |
| 0 |
| 0 |
| 0 |
| 0.035409912 |

| |
|---|
| 0 |
| 0 |
| 0 |
| 0 |
| 0.00085045 |
| 0 |
| 0 |
| 0 |
| 0 |
| 0 |
| 0 |
| 0 |
| 0 |
| 0 |
| 0.01041169 |
| 0 |
| 0 |
| 0 |
| 0 |
| 0 |
| 0 |
| 0 |
| 0.026179436 |
| 0 |
| 0 |
| 0 |
| 0 |
| 0 |
| 0.217526864 |
| 0 |
| 0 |
| 0.020473756 |
| 0.11126916 |
| 0 |
| 0.016682008 |
| 0 |
| 0 |
| 0 |
| 0 |
| 0 |
| 0 |
| 0 |
| 0.125082395 |
| 0 |
| 0 |
| 0.06086656 |
| 0 |
| 0 |

0
0
0
0
0
0
0
0
0
0.049111743
0
0
0
0
0
0
0
0
0
0
0
0
0
0.012677745
0
0
0
0
0
0
0.269261957
0
0
1.066051569
0
0
0
0
0
0
0
0
0
0
0
0
0.001311582
0

| |
|---|
| 0 |
| 0 |
| 0 |
| 0 |
| 0.201429074 |
| 0 |
| 0 |
| 0 |
| 0 |
| 0 |
| 0 |
| 0.003303776 |
| 0 |
| 0 |
| 0.016142954 |
| 0 |
| 0 |
| 0 |
| 0.003531725 |
| 0.062445454 |
| 0 |
| 0 |
| 0 |
| 0 |
| 0 |
| 0 |
| 0.000461911 |
| 0 |
| 0 |
| 0 |
| 0.000872668 |
| 0 |
| 0 |
| 0 |
| 0 |
| 0 |
| 0 |
| 0.009932403 |
| 0 |
| 0 |
| 0 |
| 0.001729146 |
| 0 |
| 0 |
| 0 |
| 0 |
| 0 |
| 0 |

| |
|---|
| 0 |
| 0.035745239 |
| 0 |
| 0.015054404 |
| 0 |
| 0.001140521 |
| 0.003143742 |
| 0 |
| 0 |
| 0 |
| 0.043302711 |
| 0 |
| 0.274621061 |
| 0 |
| 0 |
| 0 |
| 0 |
| 0 |
| 0 |
| 0 |
| 0 |
| 0 |
| 0.004446786 |
| 0.004827572 |
| 0 |
| 0.002325136 |
| 0 |
| 0 |
| 0 |
| 0 |
| 0.015271092 |
| 0 |
| 0 |
| 0 |
| 0.055867377 |
| 0 |
| 0 |
| 0 |
| 0 |
| 0 |
| 0 |
| 0 |
| 0 |
| 0 |
| 33.64944089 |
| 0.453531746 |
| 0 |
| 0 |

| |
|---|
| 0 |
| 0 |
| 0.077078211 |
| 0.000668689 |
| 0 |
| 0 |
| 0 |
| 0 |
| 0 |
| 0 |
| 0.000952901 |
| 0.001016203 |
| 0 |
| 0 |
| 0 |
| 0 |
| 0 |
| 0 |
| 0 |
| 0 |
| 0 |
| 0 |
| 0 |
| 0 |
| 0 |
| 0 |
| 0 |
| 0 |
| 0 |
| 0 |
| 0.002171316 |
| 0 |
| 0.052763407 |
| 0.001472177 |
| 0 |
| 0 |
| 0 |
| 0 |
| 0 |
| 0 |
| 0 |
| 0 |
| 0.048190452 |
| 0 |
| 0 |
| 0.051120436 |
| 0.000513 |
| 0 |

| |
|---|
| 0 |
| 0.126876523 |
| 0.003533954 |
| 0 |
| 0.004408893 |
| 0 |
| 0 |
| 0 |
| 0 |
| 0 |
| 0.00093431 |
| 0 |
| 0 |
| 0 |
| 0 |
| 0 |
| 0.262094661 |
| 0 |
| 0 |
| 0 |
| 0 |
| 0 |
| 0 |
| 0 |
| 0.022389648 |
| 0.004394821 |
| 0 |
| 1.0098288 |
| 0 |
| 0 |
| 0 |
| 0 |
| 0 |
| 0 |
| 0 |
| 0 |
| 0 |
| 0 |
| 2.802416724 |
| 0 |
| 0 |
| 0.08422942 |
| 0.003675569 |
| 0 |
| 0 |
| 0.001480014 |
| 0.006747481 |
| 0 |

| |
|---|
| 0 |
| 0.012416366 |
| 0 |
| 0 |
| 0 |
| 0 |
| 0 |
| 0 |
| 0 |
| 0 |
| 0 |
| 0 |
| 0.504757664 |
| 0 |
| 0 |
| 0 |
| 0 |
| 0 |
| 0 |
| 0 |
| 0 |
| 0.025629423 |
| 0.000667597 |
| 0 |
| 0 |
| 0 |
| 0 |
| 0 |
| 0 |
| 0 |
| 0 |
| 0 |
| 0 |
| 0 |
| 0 |
| 0 |
| 0 |
| 0 |
| 0.07895123 |
| 0 |
| 0.001555829 |
| 0 |
| 0 |
| 0 |
| 0 |
| 0.072366057 |
| 0 |
| 0 |

| |
|---|
| 0 |
| 0 |
| 0 |
| 0 |
| 0 |
| 0 |
| 0 |
| 0.001203346 |
| 0 |
| 0.008710661 |
| 0 |
| 0 |
| 0 |
| 0 |
| 0 |
| 0 |
| 0 |
| 0 |
| 0 |
| 0.243064515 |
| 0 |
| 0 |
| 0 |
| 0 |
| 0 |
| 0.031808891 |
| 0 |
| 0 |
| 0 |
| 0 |
| 0 |
| 0 |
| 0 |
| 0 |
| 0 |
| 0 |
| 0 |
| 0 |
| 0 |
| 0 |
| 0 |
| 0 |
| 0 |
| 0 |
| 0.005871971 |
| 0 |
| 0 |
| 0 |

| |
|---|
| 0 |
| 0 |
| 0 |
| 0 |
| 0 |
| 0.098698653 |
| 0 |
| 0 |
| 0 |
| 0 |
| 0 |
| 0 |
| 0 |
| 0.001188872 |
| 0 |
| 0 |
| 0.034159897 |
| 0 |
| 0 |
| 0 |
| 0 |
| 0 |
| 0 |
| 0 |
| 0 |
| 0 |
| 0 |
| 0 |
| 0 |
| 0 |
| 0 |
| 0 |
| 0 |
| 0 |
| 0 |
| 0 |
| 0 |
| 0 |
| 0.069754755 |
| 0 |
| 0 |
| 0 |
| 0 |
| 0 |
| 0 |
| 0 |
| 0 |
| 0 |

| |
|---|
| 0 |
| 0 |
| 0 |
| 0 |
| 0 |
| 0.001631546 |
| 0 |
| 0 |
| 0 |
| 0 |
| 0.01187919 |
| 0 |
| 0.047190596 |
| 0 |
| 0 |
| 0 |
| 0 |
| 0 |
| 0 |
| 0 |
| 0.000592951 |
| 0 |
| 0 |
| 1.470652977 |
| 0 |
| 0 |
| 0 |
| 0 |
| 0 |
| 0 |
| 0 |
| 0 |
| 0.001455364 |
| 0 |
