## Supplemental table S3. Demographics and baseline characteristics of subjects. for "A YOUNG CHILD FORMULA SUPPLEMENTED WITH L. REUTERI AND GALACTO-OLIGOSACCHARIDES MODULATES THE COMPOSITION AND FUNCTION OF THE GUT MICROBIOME SUPPORTING BONE AND MUSCLE DEVELOPMENT IN TODDLERS"

**Demographic and baseline charac**

| | CM<br>(n=91) | EYCF<br>(n=91) |
|---|---|---|
| | N (%) or<br>mean (SD)<br>range | N (%) or<br>mean (SD)<br>range |
| Sex, n (%) | | |
| Male | 43 (47%) | 43 (47%) |
| Female | 48 (53%) | 48 (53%) |
| Age at enrollment in months, mean (SD); range | 29.7 (3.8) | 29.5 (3.4) |
| | (24, 36) | (25, 36) |
| Consumed Growing Up Milk in the past day | 9 (9.9%) | 9 (9.9%) |
| Consumed fortified cow milk in the past day | 80 (87.9%) | 81 (89.0%) |
| Consumed cow milk in the past day | 6 (6.6%) | 4 (4.4%) |
| Weight (kg) | 12.17 (1.7) | 12.13 (1.7) |
| Height (cm) | 86.95 (4.16) | 86.94 (3.96) |

cteristics of subjects.

| REF (n=91) |
| --- |
| N (%) or mean (SD) range |
| 42 (47%) |
| 49 (53%) |
| 29.7 (3.7) |
| (25, 36) |
| 0 (0.0%) |
| 91 (100.0%) |
| 0 (0.0%) |
| 11.93 (1.66) |
| 86.22 (4.1) |
