## Supplemental table S4. Descriptive statistics of subjects at all visits. for "A YOUNG CHILD FORMULA SUPPLEMENTED WITH L. REUTERI AND GALACTO-OLIGOSACCHARIDES MODULATES THE COMPOSITION AND FUNCTION OF THE GUT MICROBIOME SUPPORTING BONE AND MUSCLE DEVELOPMENT IN TODDLERS"

**Descriptive statistics of subjects a**

| | Visit | Arm | n | min | Q1 |
|---|---|---|---|---|---|
| Height (cm) | V1 | REF | 91 | 76,1 | 83,85 |
| | V1 | CM | 91 | 77,9 | 83,5 |
| | V1 | EYCF | 91 | 79,4 | 84,4 |
| | V3 | REF | 87 | 80 | 87,35 |
| | V3 | CM | 69 | 81,9 | 88,1 |
| | V3 | EYCF | 69 | 84 | 88,9 |
| Weight (kg) | V1 | REF | 91 | 9,32 | 10,62 |
| | V1 | CM | 91 | 9,13 | 10,94 |
| | V1 | EYCF | 91 | 9,31 | 11 |
| | V3 | REF | 87 | 10,14 | 11,57 |
| | V3 | CM | 69 | 10,58 | 12,29 |
| | V3 | EYCF | 69 | 10,7 | 12,15 |
| Head circ (cm) | V1 | REF | 91 | 45 | 46,8 |
| | V1 | CM | 91 | 43,6 | 46,8 |
| | V1 | EYCF | 91 | 40,2 | 46,8 |
| | V3 | REF | 87 | 46 | 47,7 |
| | V3 | CM | 69 | 44,9 | 47,6 |
| | V3 | EYCF | 69 | 45,2 | 47,6 |
| BMI (kg/m2) | V1 | REF | 91 | 13,89 | 15,01 |
| | V1 | CM | 91 | 13,1 | 15,11 |
| | V1 | EYCF | 91 | 13,46 | 15,13 |
| | V3 | REF | 87 | 13,85 | 14,83 |
| | V3 | CM | 69 | 13,17 | 15,35 |
| | V3 | EYCF | 69 | 13,66 | 15,23 |
| Tibia SOS | V1 | REF | 91 | 3103 | 3306 |
| | V1 | CM | 91 | 3128 | 3321 |
| | V1 | EYCF | 91 | 3114 | 3308 |
| | V2 | REF | 84 | 3068 | 3310 |
| | V2 | CM | 45 | 2988 | 3304 |
| | V2 | EYCF | 43 | 3213 | 3366 |
| | V3 | REF | 87 | 3133 | 3310 |
| | V3 | CM | 69 | 3032 | 3341 |
| | V3 | EYCF | 69 | 2889 | 3374 |
| Radius SOS | V1 | REF | 91 | 2916 | 3193 |
| | V1 | CM | 91 | 2861 | 3142 |
| | V1 | EYCF | 91 | 2890 | 3154 |
| | V2 | REF | 87 | 2877 | 3248 |
| | V2 | CM | 71 | 2964 | 3244 |
| | V2 | EYCF | 70 | 3090 | 3253 |
| | V3 | REF | 87 | 2950 | 3284 |
| | V3 | CM | 69 | 3015 | 3276 |
| | V3 | EYCF | 69 | 3130 | 3293 |
| | V1 | REF | 91 | 13 | 23,6 |
| | V1 | CM | 91 | 15 | 22,7 |
| | V1 | EYCF | 91 | 13,4 | 23 |

| | | | | | |
|---|---|---|---|---|---|
| Tibia length | V2 | REF | 87 | 16 | 24,55 |
| | V2 | CM | 72 | 23 | 24,87 |
| | V2 | EYCF | 70 | 17 | 24,9 |
| | V3 | REF | 87 | 23,3 | 25,35 |
| | V3 | CM | 69 | 23,3 | 25,8 |
| | V3 | EYCF | 69 | 23,6 | 25,6 |
| Radius length | V1 | REF | 91 | 11,5 | 13,15 |
| | V1 | CM | 91 | 11 | 13 |
| | V1 | EYCF | 91 | 11 | 13 |
| | V2 | REF | 87 | 12,1 | 13,5 |
| | V2 | CM | 72 | 11,5 | 13,6 |
| | V2 | EYCF | 70 | 11,2 | 13,9 |
| | V3 | REF | 87 | 12,8 | 14 |
| | V3 | CM | 69 | 11,9 | 14,1 |
| | V3 | EYCF | 69 | 11,5 | 14,3 |
| Handgrip (right hand) | V1 | REF | 91 | 1,07 | 1,58 |
| | V1 | CM | 91 | 1,1 | 1,63 |
| | V1 | EYCF | 91 | 1,07 | 1,6 |
| | V3 | REF | 87 | 1,1 | 2,1 |
| | V3 | CM | 69 | 1,1 | 1,97 |
| | V3 | EYCF | 69 | 1,27 | 2,2 |
| Bone turnover | V1 | REF | 91 | -4026 | -1506 |
| | V1 | CM | 91 | -2846 | -870,6 |
| | V1 | EYCF | 91 | -3928 | -1121 |
| | V3 | REF | 87 | -3621 | -1160 |
| | V3 | CM | 68 | -3275 | -1387 |
| | V3 | EYCF | 68 | -3327 | -1002 |

**t all visits**

| Median | Q3 | max | mean | sd |
|---|---|---|---|---|
| 86,7 | 88,85 | 97,1 | 86,22 | 4,1 |
| 86,5 | 90,25 | 98,3 | 86,95 | 4,16 |
| 87 | 89,5 | 97,5 | 86,94 | 3,96 |
| 90,2 | 93 | 98,6 | 90,19 | 4,07 |
| 91,4 | 93,7 | 104,1 | 91,49 | 4,34 |
| 92 | 94,1 | 101,5 | 91,63 | 4,1 |
| 11,72 | 12,93 | 18,86 | 11,93 | 1,66 |
| 11,81 | 12,88 | 18 | 12,17 | 1,7 |
| 11,69 | 13,04 | 18,74 | 12,13 | 1,7 |
| 12,52 | 13,98 | 20,5 | 12,94 | 1,97 |
| 12,97 | 14,43 | 20,26 | 13,73 | 2,13 |
| 13,16 | 14,58 | 20,52 | 13,78 | 2,25 |
| 47,4 | 48,2 | 50,2 | 47,55 | 1,14 |
| 47,7 | 48,5 | 50,8 | 47,68 | 1,35 |
| 47,7 | 48,5 | 51,2 | 47,53 | 1,62 |
| 48,2 | 49,1 | 51,2 | 48,42 | 1,07 |
| 48,5 | 49,4 | 51,6 | 48,54 | 1,3 |
| 48,5 | 49,5 | 51,4 | 48,54 | 1,36 |
| 15,76 | 16,55 | 22,53 | 15,99 | 1,37 |
| 15,69 | 16,66 | 22,54 | 16,05 | 1,49 |
| 15,75 | 16,66 | 22 | 16 | 1,35 |
| 15,52 | 16,2 | 23,25 | 15,84 | 1,53 |
| 15,92 | 16,91 | 23,37 | 16,36 | 1,86 |
| 16,18 | 16,89 | 21,45 | 16,33 | 1,67 |
| 3372 | 3469 | 3709 | 3384 | 125,5 |
| 3414 | 3486 | 3721 | 3408 | 131 |
| 3396 | 3493 | 3724 | 3399 | 140,9 |
| 3412 | 3495 | 3897 | 3406 | 144,6 |
| 3391 | 3427 | 3712 | 3370 | 135,8 |
| 3421 | 3456 | 3651 | 3417 | 87,36 |
| 3397 | 3482 | 3699 | 3399 | 127,9 |
| 3400 | 3469 | 3760 | 3406 | 131,3 |
| 3466 | 3535 | 3719 | 3458 | 133,9 |
| 3284 | 3372 | 3668 | 3287 | 144,2 |
| 3298 | 3410 | 3719 | 3285 | 169,1 |
| 3272 | 3343 | 3521 | 3249 | 140,7 |
| 3361 | 3448 | 3721 | 3345 | 149,6 |
| 3328 | 3424 | 3683 | 3333 | 141,9 |
| 3336 | 3445 | 3726 | 3354 | 131,1 |
| 3356 | 3472 | 3693 | 3357 | 159,7 |
| 3384 | 3478 | 3683 | 3377 | 134,1 |
| 3372 | 3469 | 3616 | 3375 | 123,5 |
| 24,4 | 25,75 | 28 | 24,21 | 2,37 |
| 24,1 | 25,4 | 30 | 23,29 | 3,53 |
| 24,4 | 25,6 | 29 | 23,52 | 3,57 |

| | | | | |
|---|---|---|---|---|
| 25,4 | 26,55 | 28,7 | 25,46 | 1,7 |
| 25,55 | 26,63 | 30 | 25,71 | 1,46 |
| 25,8 | 26,6 | 30 | 25,72 | 1,8 |
| 26,2 | 27,35 | 29,4 | 26,33 | 1,36 |
| 26,6 | 27,5 | 31,5 | 26,72 | 1,47 |
| 26,7 | 27,6 | 30,7 | 26,77 | 1,47 |
| 13,7 | 14,2 | 16 | 13,71 | 0,91 |
| 13,9 | 14,15 | 16 | 13,7 | 1,04 |
| 13,7 | 14,05 | 18 | 13,65 | 1,26 |
| 14,2 | 14,6 | 16,4 | 14,14 | 0,85 |
| 14,2 | 15 | 17 | 14,29 | 1,11 |
| 14,3 | 14,9 | 19 | 14,35 | 1,22 |
| 14,5 | 15 | 16,7 | 14,6 | 0,87 |
| 14,8 | 15,5 | 17,4 | 14,78 | 1,11 |
| 14,9 | 15,7 | 19,1 | 14,95 | 1,17 |
| 1,9 | 2,15 | 4,2 | 1,98 | 0,61 |
| 1,9 | 2,25 | 3,5 | 1,97 | 0,49 |
| 1,8 | 2,1 | 3,9 | 1,92 | 0,52 |
| 2,5 | 3,1 | 5,47 | 2,59 | 0,79 |
| 2,27 | 3,03 | 5,03 | 2,52 | 0,77 |
| 2,67 | 3,43 | 6,4 | 2,82 | 0,89 |
| -755,2 | -346,1 | -61,54 | -1011 | 790,8 |
| -424,3 | -256,5 | -39,68 | -691,4 | 628,2 |
| -531,2 | -299,2 | -39,99 | -779,9 | 725,6 |
| -715,1 | -359,4 | -59,59 | -926 | 785,5 |
| -782,7 | -418,1 | -107,8 | -1038 | 766,2 |
| -568,1 | -382,4 | -120,7 | -791,6 | 607,7 |
