## Supplemental table S5. Clinical outcomes comparisons between the experimental and control blends versus the habitual diet arm. for "A YOUNG CHILD FORMULA SUPPLEMENTED WITH L. REUTERI AND GALACTO-OLIGOSACCHARIDES MODULATES THE COMPOSITION AND FUNCTION OF THE GUT MICROBIOME SUPPORTING BONE AND MUSCLE DEVELOPMENT IN TODDLERS"

**Clinical outcomes comparisons between of the experimental and control blends versus the habitual**

| | Model | Treatment | Visit | Estimate | 95% CI | p-value |
|---|---|---|---|---|---|---|
| Height | Model 1 | EYCF - REF | V3 | 0,52 | [0.24; 0.8] | < 0.001 |
| | Model 2 | EYCF - REF | V3 | 0,51 | [0.23; 0.79] | < 0.001 |
| | Model 3 | EYCF - REF | V3 | 0,52 | [0.23; 0.81] | < 0.001 |
| | Model 1 | CM - REF | V3 | 0,72 | [0.42; 1.02] | < 0.001 |
| | Model 2 | CM - REF | V3 | 0,72 | [0.42; 1.02] | < 0.001 |
| | Model 3 | CM - REF | V3 | 0,74 | [0.43; 1.05] | < 0.001 |
| Weight | Model 1 | EYCF - REF | V3 | 0,54 | [0.31; 0.77] | < 0.001 |
| | Model 2 | EYCF - REF | V3 | 0,52 | [0.28; 0.76] | < 0.001 |
| | Model 3 | EYCF - REF | V3 | 0,54 | [0.3; 0.78] | < 0.001 |
| | Model 1 | CM - REF | V3 | 0,73 | [0.48; 0.98] | < 0.001 |
| | Model 2 | CM - REF | V3 | 0,73 | [0.48; 0.98] | < 0.001 |
| | Model 3 | CM - REF | V3 | 0,75 | [0.49; 1.01] | < 0.001 |
| Head circumference | Model 1 | EYCF - REF | V3 | 0,03 | [-0.11; 0.17] | 0,671 |
| | Model 2 | EYCF - REF | V3 | 0,03 | [-0.11; 0.17] | 0,676 |
| | Model 3 | EYCF - REF | V3 | 0,04 | [-0.1; 0.18] | 0,582 |
| | Model 1 | CM - REF | V3 | 0,03 | [-0.11; 0.17] | 0,665 |
| | Model 2 | CM - REF | V3 | 0,03 | [-0.11; 0.17] | 0,665 |
| | Model 3 | CM - REF | V3 | 0,03 | [-0.11; 0.17] | 0,675 |
| BMI | Model 1 | EYCF - REF | V3 | 0,51 | [0.26; 0.76] | < 0.001 |
| | Model 2 | EYCF - REF | V3 | 0,48 | [0.22; 0.74] | < 0.001 |
| | Model 3 | EYCF - REF | V3 | 0,52 | [0.26; 0.78] | < 0.001 |
| | Model 1 | CM - REF | V3 | 0,63 | [0.36; 0.9] | < 0.001 |
| | Model 2 | CM - REF | V3 | 0,63 | [0.36; 0.9] | < 0.001 |
| | Model 3 | CM - REF | V3 | 0,64 | [0.36; 0.92] | < 0.001 |
| Tibia length | Model 1 | EYCF - REF | V2 | 0,24 | [-0.14; 0.62] | 0,211 |
| | Model 1 | EYCF - REF | V3 | 0,42 | [0.04; 0.8] | 0,03 |
| | Model 2 | EYCF - REF | V2 | 0,26 | [-0.12; 0.64] | 0,181 |
| | Model 2 | EYCF - REF | V3 | 0,44 | [0.06; 0.82] | 0,025 |
| | Model 3 | EYCF - REF | V2 | 0,23 | [-0.15; 0.61] | 0,23 |
| | Model 3 | EYCF - REF | V3 | 0,44 | [0.06; 0.82] | 0,023 |
| | Model 1 | CM - REF | V2 | 0,3 | [-0.07; 0.67] | 0,109 |
| | Model 1 | CM - REF | V3 | 0,42 | [0.05; 0.79] | 0,026 |
| | Model 2 | CM - REF | V2 | 0,3 | [-0.07; 0.67] | 0,111 |
| | Model 2 | CM - REF | V3 | 0,42 | [0.05; 0.79] | 0,027 |
| | Model 3 | CM - REF | V2 | 0,23 | [-0.15; 0.61] | 0,23 |
| | Model 3 | CM - REF | V3 | 0,44 | [0.06; 0.82] | 0,023 |
| Tibia SoS | Model 1 | EYCF - REF | V2 | 10,14 | [-34.51; 54.79] | 0,655 |
| | Model 1 | EYCF - REF | V3 | 53,38 | [13.82; 92.94] | 0,008 |
| | Model 2 | EYCF - REF | V2 | 4,11 | [-40.54; 48.76] | 0,856 |
| | Model 2 | EYCF - REF | V3 | 47,35 | [7.78; 86.92] | 0,019 |
| | Model 3 | EYCF - REF | V2 | 9,76 | [-34.54; 54.06] | 0,663 |
| | Model 3 | EYCF - REF | V3 | 55,01 | [15.65; 94.37] | 0,007 |
| | Model 1 | CM - REF | V2 | -42,03 | [-84.1; 0.04] | 0,05 |

| | | | | | | |
|---|---|---|---|---|---|---|
| | Model 1 | CM - REF | V3 | -5,83 | [-43.85; 32.19] | 0,763 |
| | Model 2 | CM - REF | V2 | -41,81 | [-83.33; -0.29] | 0,048 |
| | Model 2 | CM - REF | V3 | -6,89 | [-44.34; 30.56] | 0,717 |
| | Model 3 | CM - REF | V2 | -40,62 | [-82.72; 1.48] | 0,058 |
| | Model 3 | CM - REF | V3 | -5,06 | [-43.29; 33.17] | 0,794 |
| Radius length | Model 1 | EYCF - REF | V2 | 0,13 | [-0.02; 0.28] | 0,096 |
| | Model 1 | EYCF - REF | V3 | 0,25 | [0.1; 0.4] | 0,002 |
| | Model 2 | EYCF - REF | V2 | 0,13 | [-0.03; 0.29] | 0,101 |
| | Model 2 | EYCF - REF | V3 | 0,25 | [0.09; 0.41] | 0,002 |
| | Model 3 | EYCF - REF | V2 | 0,13 | [-0.02; 0.28] | 0,097 |
| | Model 3 | EYCF - REF | V3 | 0,25 | [0.1; 0.4] | 0,002 |
| | Model 1 | CM - REF | V2 | 0,11 | [-0.05; 0.27] | 0,173 |
| | Model 1 | CM - REF | V3 | 0,16 | [0; 0.32] | 0,049 |
| | Model 2 | CM - REF | V2 | 0,11 | [-0.05; 0.27] | 0,173 |
| | Model 2 | CM - REF | V3 | 0,16 | [0; 0.32] | 0,049 |
| | Model 3 | CM - REF | V2 | 0,11 | [-0.05; 0.27] | 0,175 |
| | Model 3 | CM - REF | V3 | 0,16 | [0; 0.32] | 0,05 |
| Radius SoS | Model 1 | EYCF - REF | V2 | 15,01 | [-23.49; 53.51] | 0,444 |
| | Model 1 | EYCF - REF | V3 | 24,55 | [-14.1; 63.2] | 0,212 |
| | Model 2 | EYCF - REF | V2 | 16,31 | [-22.64; 55.26] | 0,41 |
| | Model 2 | EYCF - REF | V3 | 25,87 | [-13.23; 64.97] | 0,194 |
| | Model 3 | EYCF - REF | V2 | 14,11 | [-24.64; 52.86] | 0,472 |
| | Model 3 | EYCF - REF | V3 | 26,23 | [-12.66; 65.12] | 0,184 |
| | Model 1 | CM - REF | V2 | -20,14 | [-57.54; 17.26] | 0,29 |
| | Model 1 | CM - REF | V3 | 14,14 | [-23.51; 51.79] | 0,46 |
| | Model 2 | CM - REF | V2 | -20 | [-57.46; 17.46] | 0,294 |
| | Model 2 | CM - REF | V3 | 14,27 | [-23.44; 51.98] | 0,457 |
| | Model 3 | CM - REF | V2 | -19,4 | [-57.03; 18.23] | 0,309 |
| | Model 3 | CM - REF | V3 | 13,88 | [-24; 51.76] | 0,469 |
| Handgrip (right hand) | Model 1 | (EYCF/REF-1)% | V3 | 7% | [-2%; 16%] | 0,12 |
| | Model 2 | (EYCF/REF-1)% | V3 | 8% | [-1%; 17%] | 0,079 |
| | Model 3 | (EYCF/REF-1)% | V3 | 8% | [-1%; 17%] | 0,079 |
| | Model 1 | (CM/REF-1)% | V3 | -3% | [-12%; 6%] | 0,52 |
| | Model 2 | (CM/REF-1)% | V3 | -3% | [-12%; 6%] | 0,521 |
| | Model 3 | (CM/REF-1)% | V3 | -3% | [-12%; 6%] | 0,532 |
| Bone turnover index | Model 1 | (EYCF- REF) | V3 | 121,7 | [-112.15; 355.47] | 0,305 |
| | Model 2 | (EYCF- REF) | V3 | 119,8 | [-117.24; 356.84] | 0,32 |
| | Model 3 | (EYCF- REF) | V3 | 115,9 | [-124.48; 356.26] | 0,342 |
| | Model 1 | (CM- REF) | V3 | -194,5 | [-448.56; 59.52] | 0,132 |
| | Model 2 | (CM- REF) | V3 | -194,8 | [-449.72; 60.04] | 0,133 |
| | Model 3 | (CM- REF) | V3 | -186,6 | [-448.62; 75.5] | 0,162 |
