## Supplemental table S6. Comparisons of the blood vitamin D level between Experimental group and Habitual diet group. for "A YOUNG CHILD FORMULA SUPPLEMENTED WITH L. REUTERI AND GALACTO-OLIGOSACCHARIDES MODULATES THE COMPOSITION AND FUNCTION OF THE GUT MICROBIOME SUPPORTING BONE AND MUSCLE DEVELOPMENT IN TODDLERS"

**Comparisons of the blood vitamin level between Control milk group and Habitual diet group**

| | Model | Treatment | Visit | Estimate | 95% CI | p-value |
|---|---|---|---|---|---|---|
| Vitamin D | Model 1 | (EYCF/REF-1)% | V3 | 10% | [5%; 15%] | < 0.001 |
| | Model 2 | (EYCF/REF-1)% | V3 | 10% | [5%; 15%] | < 0.001 |
| | Model 3 | (EYCF/REF-1)% | V3 | 10% | [5%; 15%] | < 0.001 |
