## Supplemental table S7. Correlation of the vitamin B3/B6 ratio and mineral excretion (adjusted to intake) and the clinical outcome. for "A YOUNG CHILD FORMULA SUPPLEMENTED WITH L. REUTERI AND GALACTO-OLIGOSACCHARIDES MODULATES THE COMPOSITION AND FUNCTION OF THE GUT MICROBIOME SUPPORTING BONE AND MUSCLE DEVELOPMENT IN TODDLERS"

**Correlation of ratio B3/B6 and mineral excretion (adjusted to intake) in the o**

| | Ratio B3/B6 | Calcium |
|---|---|---|
| *(p-value)* | n=222 | n=137 |
| **TIBIA SOS V3** | -0.11 (0.11) | -0.05 (0.58) |
| **RADIUS SOS V3** | -0.07 (0.29) | -0.14 (0.1) |
| **TIBIA LENGTH V3** | -0.13 (0.05) | -0.02 (0.81) |
| **RADIUS LENGTH V3** | -0.11 (0.09) | -0.06 (0.46) |
| **HANDGRIP V3** | -0.05 (0.45) | -0.16 (0.06) |

**verall population and the clinical outcomes**

| Magnesium | Phosphorus |
|---|---|
| n=137 | n=137 |
| -0.07 (0.42) | -0.05 (0.53) |
| -0.03 (0.69) | -0.13 (0.15) |
| -0.1 (0.25) | -0.09 (0.28) |
| -0.11 (0.2) | -0.13 (0.13) |
| -0.16 (0.06) | -0.18 (0.03) |
