## Supplemental table S8. PERMANOVA analysis on microbiome beta diversity. for "A YOUNG CHILD FORMULA SUPPLEMENTED WITH L. REUTERI AND GALACTO-OLIGOSACCHARIDES MODULATES THE COMPOSITION AND FUNCTION OF THE GUT MICROBIOME SUPPORTING BONE AND MUSCLE DEVELOPMENT IN TODDLERS"

| Comparisons | Baseline | 3 months | 6 months |
|---|---|---|---|
| All | 0,088911 | 0,000999 | 0,000999 |
| CM vs. REF | 0,334665 | 0,038961 | 0,120879 |
| CM vs. EYCF | 0,05994 | 0,012987 | 0,000999 |
| REF vs. EYCF | 0,120879 | 0,001998 | 0,000999 |
