## Supplemental table S9. Comparison of microbiome taxonomic composition between EYCF and CM. for "A YOUNG CHILD FORMULA SUPPLEMENTED WITH L. REUTERI AND GALACTO-OLIGOSACCHARIDES MODULATES THE COMPOSITION AND FUNCTION OF THE GUT MICROBIOME SUPPORTING BONE AND MUSCLE DEVELOPMENT IN TODDLERS"

| Visit | Category | Item |
|---|---|---|
| baseline | MGS | Hgn3C.0001 - Phocaeicola vulgatus |
| baseline | MGS | Hgn3C.0002 - Bacteroides uniformis |
| baseline | MGS | Hgn3C.0003 - Lachnospiraceae sp. |
| baseline | MGS | Hgn3C.0004 - Blautia wexlerae |
| baseline | MGS | Hgn3C.0005 - Fusicatenibacter saccharivorans |
| baseline | MGS | Hgn3C.0006 - Bacteroides ovatus |
| baseline | MGS | Hgn3C.0007 - Parabacteroides distasonis |
| baseline | MGS | Hgn3C.0008 - Alistipes putredinis |
| baseline | MGS | Hgn3C.0009 - Faecalibacterium longum CLA-AA-H243 |
| baseline | MGS | Hgn3C.0010 - Anaerostipes hadrus |
| baseline | MGS | Hgn3C.0011 - Roseburia faecis |
| baseline | MGS | Hgn3C.0012 - Lachnospira sp. |
| baseline | MGS | Hgn3C.0013 - Ruminococcus bromii |
| baseline | MGS | Hgn3C.0014 - Lacrimispora saccharolytica |
| baseline | MGS | Hgn3C.0015 - Faecalibacterium sp. Marseille-Q4896 |
| baseline | MGS | Hgn3C.0016 - Parabacteroides merdae |
| baseline | MGS | Hgn3C.0017 - Subdoligranulum sp. APC924/74 |
| baseline | MGS | Hgn3C.0018 - Faecalibacterium prausnitzii |
| baseline | MGS | Hgn3C.0019 - Phocaeicola dorei |
| baseline | MGS | Hgn3C.0020 - Collinsella aerofaciens |
| baseline | MGS | Hgn3C.0021 - Bifidobacterium longum subsp. longum |
| baseline | MGS | Hgn3C.0022 - Alistipes shahii |
| baseline | MGS | Hgn3C.0023 - Bacteroides xylanisolvens |
| baseline | MGS | Hgn3C.0024 - Bacteroides caccae |
| baseline | MGS | Hgn3C.0025 - Ruminococcus bicirculans |
| baseline | MGS | Hgn3C.0026 - Dorea longicatena |
| baseline | MGS | Hgn3C.0027 - Barnesiella intestinihominis |
| baseline | MGS | Hgn3C.0028 - Oscillibacter sp. ER4 |
| baseline | MGS | Hgn3C.0029 - Anaerobutyricum hallii |
| baseline | MGS | Hgn3C.0030 - Faecalibacterium prausnitzii |
| baseline | MGS | Hgn3C.0031 - Blautia massiliensis |
| baseline | MGS | Hgn3C.0032 - Alistipes onderdonkii subsp. vulgaris |
| baseline | MGS | Hgn3C.0033 - Gemmiger formicilis |
| baseline | MGS | Hgn3C.0034 - Roseburia inulinivorans |
| baseline | MGS | Hgn3C.0035 - Odoribacter splanchnicus |
| baseline | MGS | Hgn3C.0036 - Bacteroides thetaiotaomicron |
| baseline | MGS | Hgn3C.0037 - Roseburia intestinalis |
| baseline | MGS | Hgn3C.0038 - Bifidobacterium adolescentis |
| baseline | MGS | Hgn3C.0039 - Coprococcus comes |
| baseline | MGS | Hgn3C.0040 - Eubacteriales sp. |
| baseline | MGS | Hgn3C.0041 - Ruminococcus sp. |
| baseline | MGS | Hgn3C.0042 - Blautia faecis |
| baseline | MGS | Hgn3C.0043 - Lachnoclostridium sp. 210928-DFI.6.3 |
| baseline | MGS | Hgn3C.0044 - Lachnospira pectinoschiza |
| baseline | MGS | Hgn3C.0045 - Faecalibacterium sp. |
| baseline | MGS | Hgn3C.0046 - Clostridium sp. AF37-5 |
| baseline | MGS | Hgn3C.0047 - Dysosmobacter sp. BX15 |

| | | |
|---|---|---|
| baseline | MGS | Hgn3C.0048 - Akkermansia muciniphila |
| baseline | MGS | Hgn3C.0050 - Escherichia coli |
| baseline | MGS | Hgn3C.0051 - Alistipes communis |
| baseline | MGS | Hgn3C.0052 - Phocaeicola massiliensis |
| baseline | MGS | Hgn3C.0053 - Eubacteriales sp. |
| baseline | MGS | Hgn3C.0054 - Prevotella copri |
| baseline | MGS | Hgn3C.0055 - Alistipes finegoldii |
| baseline | MGS | Hgn3C.0056 - Bacteroides fragilis |
| baseline | MGS | Hgn3C.0057 - Oscillibacter sp. MSJ-31 |
| baseline | MGS | Hgn3C.0058 - Lachnospiraceae sp. |
| baseline | MGS | Hgn3C.0059 - Bacteroides cellulosilyticus |
| baseline | MGS | Hgn3C.0060 - Eubacteriales sp. |
| baseline | MGS | Hgn3C.0061 - Faecalibacillus sp. TM498 |
| baseline | MGS | Hgn3C.0062 - Waltera intestinalis |
| baseline | MGS | Hgn3C.0063 - Eubacteriales sp. |
| baseline | MGS | Hgn3C.0064 - Blautia sp. DFI.9.9 |
| baseline | MGS | Hgn3C.0065 - Blautia obeum |
| baseline | MGS | Hgn3C.0067 - Roseburia hominis |
| baseline | MGS | Hgn3C.0068 - [Ruminococcus] lactaris |
| baseline | MGS | Hgn3C.0069 - Oscillibacter sp. KLE 1728 |
| baseline | MGS | Hgn3C.0070 - Faecalibacterium sp. OF04-11AC |
| baseline | MGS | Hgn3C.0071 - Faecalicatena fissicatena |
| baseline | MGS | Hgn3C.0072 - Clostridium fessum |
| baseline | MGS | Hgn3C.0073 - Blautia sp. |
| baseline | MGS | Hgn3C.0074 - Clostridium sp. D43t1_170807_D5 |
| baseline | MGS | Hgn3C.0075 - Agathobaculum butyriciproducens |
| baseline | MGS | Hgn3C.0076 - Lachnospira pectinoschiza |
| baseline | MGS | Hgn3C.0077 - Sutterella wadsworthensis |
| baseline | MGS | Hgn3C.0078 - Clostridium sp. MCC328 |
| baseline | MGS | Hgn3C.0079 - [Ruminococcus] torques |
| baseline | MGS | Hgn3C.0080 - Phascolarctobacterium faecium |
| baseline | MGS | Hgn3C.0082 - Roseburia sp. CLA-AA-H204 |
| baseline | MGS | Hgn3C.0083 - Eubacteriales sp. |
| baseline | MGS | Hgn3C.0084 - Flavonifractor plautii |
| baseline | MGS | Hgn3C.0085 - Faecalibacterium sp. |
| baseline | MGS | Hgn3C.0086 - Dialister invisus |
| baseline | MGS | Hgn3C.0087 - Eubacteriales sp. |
| baseline | MGS | Hgn3C.0088 - [Ruminococcus] gnavus |
| baseline | MGS | Hgn3C.0089 - Paraprevotella clara |
| baseline | MGS | Hgn3C.0092 - Lachnospiraceae sp. |
| baseline | MGS | Hgn3C.0093 - Bilophila wadsworthia |
| baseline | MGS | Hgn3C.0094 - Eubacteriales sp. |
| baseline | MGS | Hgn3C.0095 - Eubacteriales sp. |
| baseline | MGS | Hgn3C.0096 - Eubacteriales sp. |
| baseline | MGS | Hgn3C.0097 - Eubacterium ventriosum |
| baseline | MGS | Hgn3C.0098 - Coprococcus catus |
| baseline | MGS | Hgn3C.0099 - Dorea longicatena |
| baseline | MGS | Hgn3C.0100 - Bifidobacterium bifidum |

| | | |
|---|---|---|
| baseline | MGS | Hgn3C.0101 - Bifidobacterium pseudocatenulatum |
| baseline | MGS | Hgn3C.0102 - Eubacteriales sp. |
| baseline | MGS | Hgn3C.0103 - Holdemanella porci |
| baseline | MGS | Hgn3C.0104 - Clostridium sp. AM32-2 |
| baseline | MGS | Hgn3C.0105 - Vescimonas coprocola |
| baseline | MGS | Hgn3C.0106 - Faecalibacterium sp. CLA-AA-H233 |
| baseline | MGS | Hgn3C.0111 - Eubacteriales sp. |
| baseline | MGS | Hgn3C.0113 - Clostridium sp. MCC345 |
| baseline | MGS | Hgn3C.0114 - Eubacteriales sp. |
| baseline | MGS | Hgn3C.0115 - Eubacteriales sp. |
| baseline | MGS | Hgn3C.0120 - Flavonifractor plautii |
| baseline | MGS | Hgn3C.0121 - Oscillospiraceae sp. |
| baseline | MGS | Hgn3C.0122 - Bacteroides finegoldii |
| baseline | MGS | Hgn3C.0123 - Eubacteriales sp. |
| baseline | MGS | Hgn3C.0124 - Ruthenibacterium lactatiformans |
| baseline | MGS | Hgn3C.0125 - Sutterella sp. KLE1602 |
| baseline | MGS | Hgn3C.0126 - Ruminococcus sp. BSD2780120874_150323_B10 |
| baseline | MGS | Hgn3C.0127 - Faecalibacterium sp. |
| baseline | MGS | Hgn3C.0128 - Eubacteriales sp. |
| baseline | MGS | Hgn3C.0129 - Haemophilus parainfluenzae |
| baseline | MGS | Hgn3C.0131 - Streptococcus thermophilus |
| baseline | MGS | Hgn3C.0132 - Eubacteriales sp. |
| baseline | MGS | Hgn3C.0136 - Alistipes ihumii |
| baseline | MGS | Hgn3C.0137 - Clostridium sp. OF03-18AA |
| baseline | MGS | Hgn3C.0138 - Anaerobutyricum soehngenii |
| baseline | MGS | Hgn3C.0139 - Ruminococcus callidus |
| baseline | MGS | Hgn3C.0140 - Veillonella parvula |
| baseline | MGS | Hgn3C.0141 - Adlercreutzia equolifaciens subsp. celatus |
| baseline | MGS | Hgn3C.0142 - Eubacterium ramulus |
| baseline | MGS | Hgn3C.0143 - Streptococcus salivarius |
| baseline | MGS | Hgn3C.0144 - Eubacteriales sp. |
| baseline | MGS | Hgn3C.0146 - Phascolarctobacterium succinatutens |
| baseline | MGS | Hgn3C.0147 - Oscillospiraceae sp. |
| baseline | MGS | Hgn3C.0153 - Lachnospiraceae sp. |
| baseline | MGS | Hgn3C.0154 - Prevotella copri |
| baseline | MGS | Hgn3C.0155 - Blautia sp. |
| baseline | MGS | Hgn3C.0157 - Eubacteriales sp. |
| baseline | MGS | Hgn3C.0161 - Blautia sp. |
| baseline | MGS | Hgn3C.0162 - Prevotella sp. |
| baseline | MGS | Hgn3C.0168 - Evtepia gabavorous |
| baseline | MGS | Hgn3C.0169 - Bacteroides intestinalis |
| baseline | MGS | Hgn3C.0170 - Oscillospiraceae sp. |
| baseline | MGS | Hgn3C.0171 - Catenibacterium mitsuokai |
| baseline | MGS | Hgn3C.0174 - Lachnospiraceae sp. |
| baseline | MGS | Hgn3C.0175 - Oscillospiraceae sp. |
| baseline | MGS | Hgn3C.0176 - Alistipes indistinctus |
| baseline | MGS | Hgn3C.0179 - Bacteroidales sp. |
| baseline | MGS | Hgn3C.0181 - Alistipes onderdonkii |

| | | |
|---|---|---|
| baseline | MGS | Hgn3C.0182 - Butyricimonas faecihominis |
| baseline | MGS | Hgn3C.0183 - Intestinibacter bartlettii |
| baseline | MGS | Hgn3C.0184 - Eubacteriales sp. |
| baseline | MGS | Hgn3C.0185 - Bifidobacterium catenulatum subsp. kashiwanohense |
| baseline | MGS | Hgn3C.0186 - Butyricimonas virosa |
| baseline | MGS | Hgn3C.0191 - Prevotella sp. |
| baseline | MGS | Hgn3C.0192 - Erysipelatoclostridium ramosum |
| baseline | MGS | Hgn3C.0193 - Eubacteriales sp. |
| baseline | MGS | Hgn3C.0195 - Eubacteriales sp. |
| baseline | MGS | Hgn3C.0201 - Veillonella dispar |
| baseline | MGS | Hgn3C.0202 - Eggerthella lenta |
| baseline | MGS | Hgn3C.0203 - Faecalibacterium sp. Marseille-Q3530 |
| baseline | MGS | Hgn3C.0206 - Gemmiger formicilis |
| baseline | MGS | Hgn3C.0207 - Eubacteriales sp. |
| baseline | MGS | Hgn3C.0208 - Romboutsia timonensis |
| baseline | MGS | Hgn3C.0209 - Bifidobacterium breve |
| baseline | MGS | Hgn3C.0212 - Holdemanella biformis |
| baseline | MGS | Hgn3C.0213 - Akkermansia sp. GGCC_0220 |
| baseline | MGS | Hgn3C.0214 - Duodenibacillus massiliensis |
| baseline | MGS | Hgn3C.0215 - Alistipes sp. cv1 |
| baseline | MGS | Hgn3C.0217 - Eubacteriales sp. |
| baseline | MGS | Hgn3C.0218 - Eubacteriales sp. |
| baseline | MGS | Hgn3C.0219 - Tyzzerella nexilis |
| baseline | MGS | Hgn3C.0220 - Clostridium sp. MCC334 |
| baseline | MGS | Hgn3C.0221 - Clostridium sp. |
| baseline | MGS | Hgn3C.0222 - Parabacteroides merdae |
| baseline | MGS | Hgn3C.0224 - Veillonellales sp. |
| baseline | MGS | Hgn3C.0226 - Eubacteriales sp. |
| baseline | MGS | Hgn3C.0232 - Megamonas funiformis |
| baseline | MGS | Hgn3C.0236 - Oscillospiraceae sp. |
| baseline | MGS | Hgn3C.0238 - Oscillospiraceae sp. |
| baseline | MGS | Hgn3C.0239 - Blautia sp. BIOML-A1 |
| baseline | MGS | Hgn3C.0242 - Enterocloster bolteae |
| baseline | MGS | Hgn3C.0244 - Desulfovibrio piger |
| baseline | MGS | Hgn3C.0245 - Blautia stercoris |
| baseline | MGS | Hgn3C.0248 - Coprococcus catus |
| baseline | MGS | Hgn3C.0249 - Clostridium sp. AM33-3 |
| baseline | MGS | Hgn3C.0250 - Eubacteriales sp. |
| baseline | MGS | Hgn3C.0251 - Alistipes senegalensis |
| baseline | MGS | Hgn3C.0253 - Waltera sp. |
| baseline | MGS | Hgn3C.0260 - Hydrogenoanaerobacterium saccharovorans |
| baseline | MGS | Hgn3C.0262 - Oscillospiraceae sp. |
| baseline | MGS | Hgn3C.0264 - Clostridium sp. AF27-2AA |
| baseline | MGS | Hgn3C.0266 - Dorea sp. AF36-15AT |
| baseline | MGS | Hgn3C.0267 - Oscillospiraceae sp. |
| baseline | MGS | Hgn3C.0269 - Flintibacter sp. NSJ-23 |
| baseline | MGS | Hgn3C.0272 - Blautia faecicola |
| baseline | MGS | Hgn3C.0273 - Slackia isoflavoniconvertens |

| | | |
|---|---|---|
| baseline | MGS | Hgn3C.0275 - Lachnospiraceae sp. |
| baseline | MGS | Hgn3C.0281 - Eubacteriales sp. |
| baseline | MGS | Hgn3C.0284 - Erysipelotrichaceae sp. |
| baseline | MGS | Hgn3C.0285 - Colidextribacter sp. |
| baseline | MGS | Hgn3C.0286 - Clostridia sp. |
| baseline | MGS | Hgn3C.0290 - Sellimonas intestinalis |
| baseline | MGS | Hgn3C.0295 - Eubacteriales sp. |
| baseline | MGS | Hgn3C.0299 - Escherichia coli |
| baseline | MGS | Hgn3C.0300 - Clostridia sp. |
| baseline | MGS | Hgn3C.0301 - Prevotella hominis |
| baseline | MGS | Hgn3C.0304 - Eubacteriales sp. |
| baseline | MGS | Hgn3C.0306 - Bacteroides nordii |
| baseline | MGS | Hgn3C.0307 - Veillonella atypica |
| baseline | MGS | Hgn3C.0308 - [Clostridium] symbiosum |
| baseline | MGS | Hgn3C.0309 - Eubacteriales sp. |
| baseline | MGS | Hgn3C.0310 - Ligilactobacillus ruminis |
| baseline | MGS | Hgn3C.0313 - Ruminococcus bromii |
| baseline | MGS | Hgn3C.0314 - Ruminococcus sp. |
| baseline | MGS | Hgn3C.0317 - Intestinimonas butyriciproducens |
| baseline | MGS | Hgn3C.0319 - Clostridiaceae sp. |
| baseline | MGS | Hgn3C.0323 - Bacteroides sp. |
| baseline | MGS | Hgn3C.0324 - Prevotella stercorea |
| baseline | MGS | Hgn3C.0330 - Faecalibacillus intestinalis |
| baseline | MGS | Hgn3C.0332 - Klebsiella pneumoniae subsp. pneumoniae |
| baseline | MGS | Hgn3C.0334 - Bacteroides cellulosilyticus |
| baseline | MGS | Hgn3C.0336 - Parabacteroides goldsteinii |
| baseline | MGS | Hgn3C.0337 - Eubacteriales sp. |
| baseline | MGS | Hgn3C.0338 - Eubacteriales sp. |
| baseline | MGS | Hgn3C.0345 - Oscillospiraceae sp. |
| baseline | MGS | Hgn3C.0348 - Streptococcus sp. |
| baseline | MGS | Hgn3C.0350 - [Clostridium] symbiosum |
| baseline | MGS | Hgn3C.0351 - Prevotella sp. |
| baseline | MGS | Hgn3C.0354 - Sutterella seckii |
| baseline | MGS | Hgn3C.0356 - Blautia sp. M29 |
| baseline | MGS | Hgn3C.0357 - Eubacteriales sp. |
| baseline | MGS | Hgn3C.0361 - Dorea sp. AF24-7LB |
| baseline | MGS | Hgn3C.0362 - Dysosmobacter sp. NSJ-60 |
| baseline | MGS | Hgn3C.0365 - Eubacteriales sp. |
| baseline | MGS | Hgn3C.0367 - Lachnospiraceae sp. |
| baseline | MGS | Hgn3C.0369 - Prevotella sp. |
| baseline | MGS | Hgn3C.0371 - Lachnospiraceae sp. |
| baseline | MGS | Hgn3C.0373 - Enterocloster clostridioformis |
| baseline | MGS | Hgn3C.0375 - Ellagibacter isourolithinifaciens |
| baseline | MGS | Hgn3C.0381 - Clostridium sp. AM30-24 |
| baseline | MGS | Hgn3C.0385 - Veillonella rogosae |
| baseline | MGS | Hgn3C.0386 - Clostridium sp. AT4 |
| baseline | MGS | Hgn3C.0388 - Clostridium sp. |
| baseline | MGS | Hgn3C.0392 - Clostridia sp. |

| | | |
|---|---|---|
| baseline | MGS | Hgn3C.0394 - Hungatella hathewayi |
| baseline | MGS | Hgn3C.0397 - Parabacteroides sp. |
| baseline | MGS | Hgn3C.0400 - Prevotellamassilia timonensis |
| baseline | MGS | Hgn3C.0401 - Eubacteriales sp. |
| baseline | MGS | Hgn3C.0405 - Bacteroides ovatus |
| baseline | MGS | Hgn3C.0407 - Eubacteriales sp. |
| baseline | MGS | Hgn3C.0408 - Eubacteriales sp. |
| baseline | MGS | Hgn3C.0413 - Blautia caecimuris |
| baseline | MGS | Hgn3C.0415 - Clostridium sp. C5-48 |
| baseline | MGS | Hgn3C.0417 - Eubacteriales sp. |
| baseline | MGS | Hgn3C.0418 - Blautia sp. |
| baseline | MGS | Hgn3C.0419 - Prevotella sp. 885 |
| baseline | MGS | Hgn3C.0420 - Clostridium sp. 1001270J_160509_D11 |
| baseline | MGS | Hgn3C.0430 - Enterocloster aldenensis |
| baseline | MGS | Hgn3C.0435 - Eubacteriales sp. |
| baseline | MGS | Hgn3C.0436 - Eubacteriales sp. |
| baseline | MGS | Hgn3C.0437 - Prevotella sp. |
| baseline | MGS | Hgn3C.0438 - Coprobacillus cateniformis |
| baseline | MGS | Hgn3C.0439 - Enterococcus faecalis |
| baseline | MGS | Hgn3C.0442 - Holdemanella sp. |
| baseline | MGS | Hgn3C.0443 - Prevotella stercorea |
| baseline | MGS | Hgn3C.0445 - Aeromonadales sp. |
| baseline | MGS | Hgn3C.0452 - Oscillospiraceae sp. |
| baseline | MGS | Hgn3C.0453 - Blautia sp. Marseille-P3087 |
| baseline | MGS | Hgn3C.0455 - Peptostreptococcaceae sp. |
| baseline | MGS | Hgn3C.0458 - Eubacteriales sp. |
| baseline | MGS | Hgn3C.0459 - Eubacteriales sp. |
| baseline | MGS | Hgn3C.0464 - Bifidobacterium longum subsp. infantis |
| baseline | MGS | Hgn3C.0465 - Eisenbergiella massiliensis |
| baseline | MGS | Hgn3C.0468 - Clostridium sp. NSJ-42 |
| baseline | MGS | Hgn3C.0470 - Eubacteriales sp. |
| baseline | MGS | Hgn3C.0472 - Parabacteroides goldsteinii |
| baseline | MGS | Hgn3C.0476 - Streptococcus sp. |
| baseline | MGS | Hgn3C.0482 - Lachnospiraceae sp. |
| baseline | MGS | Hgn3C.0493 - Eisenbergiella tayi |
| baseline | MGS | Hgn3C.0494 - Prevotellaceae sp. |
| baseline | MGS | Hgn3C.0500 - Anaerotignum sp. |
| baseline | MGS | Hgn3C.0501 - Oscillospiraceae sp. |
| baseline | MGS | Hgn3C.0504 - Megasphaera sp. BL7 |
| baseline | MGS | Hgn3C.0507 - [Clostridium] spiroforme |
| baseline | MGS | Hgn3C.0514 - Blautia glucerasea |
| baseline | MGS | Hgn3C.0523 - Phascolarctobacterium succinatutens |
| baseline | MGS | Hgn3C.0527 - Sutterella sp. |
| baseline | MGS | Hgn3C.0528 - Coprococcus sp. AM27-12LB |
| baseline | MGS | Hgn3C.0533 - Flintibacter sp. KGMB00164 |
| baseline | MGS | Hgn3C.0537 - Bifidobacterium dentium |
| baseline | MGS | Hgn3C.0542 - [Clostridium] innocuum |
| baseline | MGS | Hgn3C.0543 - Oscillospiraceae sp. |

| | | |
|---|---|---|
| baseline | MGS | Hgn3C.0549 - Longicatena caecimuris |
| baseline | MGS | Hgn3C.0557 - Enterocloster bolteae |
| baseline | MGS | Hgn3C.0558 - Olsenella sp. |
| baseline | MGS | Hgn3C.0560 - Oscillospiraceae sp. |
| baseline | MGS | Hgn3C.0573 - Prevotella sp. Marseille-P4119 |
| baseline | MGS | Hgn3C.0582 - Bacteroides sp. |
| baseline | MGS | Hgn3C.0584 - Bacteroides fragilis |
| baseline | MGS | Hgn3C.0598 - Eubacteriales sp. |
| baseline | MGS | Hgn3C.0604 - Megasphaera elsdenii |
| baseline | MGS | Hgn3C.0607 - Eubacteriales sp. |
| baseline | MGS | Hgn3C.0608 - Lachnospiraceae sp. |
| baseline | MGS | Hgn3C.0610 - Eubacteriales sp. |
| baseline | MGS | Hgn3C.0613 - Clostridiaceae sp. |
| baseline | MGS | Hgn3C.0616 - Lachnospiraceae sp. |
| baseline | MGS | Hgn3C.0617 - Prevotella sp. |
| baseline | MGS | Hgn3C.0629 - Collinsella intestinalis |
| baseline | MGS | Hgn3C.0630 - Allisonella histaminiformans |
| baseline | MGS | Hgn3C.0635 - Eubacteriales sp. |
| baseline | MGS | Hgn3C.0636 - Anaerostipes caccae |
| baseline | MGS | Hgn3C.0640 - Eubacterium sp. AF22-8LB |
| baseline | MGS | Hgn3C.0643 - Eubacteriales sp. |
| baseline | MGS | Hgn3C.0644 - Bacteroides cellulosilyticus |
| baseline | MGS | Hgn3C.0649 - Lachnospiraceae sp. |
| baseline | MGS | Hgn3C.0657 - Parolsenella catena |
| baseline | MGS | Hgn3C.0664 - Blautia hansenii |
| baseline | MGS | Hgn3C.0675 - Eubacteriales sp. |
| baseline | MGS | Hgn3C.0676 - Bacteroides sp. |
| baseline | MGS | Hgn3C.0677 - Bacteroides sp. |
| baseline | MGS | Hgn3C.0679 - Holdemania filiformis |
| baseline | MGS | Hgn3C.0680 - Eubacteriales sp. |
| baseline | MGS | Hgn3C.0682 - [Clostridium] scindens |
| baseline | MGS | Hgn3C.0683 - Clostridium disporicum |
| baseline | MGS | Hgn3C.0693 - Hungatella hathewayi |
| baseline | MGS | Hgn3C.0694 - Anaeromassilibacillus sp. An250 |
| baseline | MGS | Hgn3C.0701 - Bacteroides sp. |
| baseline | MGS | Hgn3C.0704 - [Clostridium] scindens |
| baseline | MGS | Hgn3C.0711 - Enterocloster asparagiformis |
| baseline | MGS | Hgn3C.0712 - Megasphaera micronuciformis |
| baseline | MGS | Hgn3C.0713 - Veillonellales sp. |
| baseline | MGS | Hgn3C.0714 - Intestinimonas sp. MSJ-38 |
| baseline | MGS | Hgn3C.0715 - Desulfovibrio fairfieldensis |
| baseline | MGS | Hgn3C.0718 - Eubacteriales sp. |
| baseline | MGS | Hgn3C.0729 - Eubacteriales sp. |
| baseline | MGS | Hgn3C.0731 - Eubacteriales sp. |
| baseline | MGS | Hgn3C.0732 - Eubacterium ramulus |
| baseline | MGS | Hgn3C.0736 - Coprococcus sp. |
| baseline | MGS | Hgn3C.0737 - Alistipes timonensis |
| baseline | MGS | Hgn3C.0740 - Anaerostipes sp. NSJ-7 |

| | | |
|---|---|---|
| baseline | MGS | Hgn3C.0741 - Blautia sp. OF01-4LB |
| baseline | MGS | Hgn3C.0743 - Enterobacter sp. |
| baseline | MGS | Hgn3C.0749 - Eubacteriales sp. |
| baseline | MGS | Hgn3C.0754 - Collinsella bouchesdurhonensis |
| baseline | MGS | Hgn3C.0761 - Bacteroides sp. |
| baseline | MGS | Hgn3C.0762 - Enterocloster lavalensis |
| baseline | MGS | Hgn3C.0767 - Citrobacter freundii |
| baseline | MGS | Hgn3C.0777 - Holdemanella sp. |
| baseline | MGS | Hgn3C.0781 - Collinsella sp. |
| baseline | MGS | Hgn3C.0782 - Streptococcus sp. |
| baseline | MGS | Hgn3C.0790 - Oscillospiraceae sp. |
| baseline | MGS | Hgn3C.0793 - Prevotellaceae sp. |
| baseline | MGS | Hgn3C.0794 - Bacteroides sp. |
| baseline | MGS | Hgn3C.0797 - Senegalimassilia sp. |
| baseline | MGS | Hgn3C.0798 - Streptococcus vestibularis |
| baseline | MGS | Hgn3C.0814 - Massilimicrobiota timonensis |
| baseline | MGS | Hgn3C.0817 - Prevotella sp. |
| baseline | MGS | Hgn3C.0818 - Rothia sp. |
| baseline | MGS | Hgn3C.0823 - Anaerotignum lactatifermentans |
| baseline | MGS | Hgn3C.0829 - Dorea phocaeensis |
| baseline | MGS | Hgn3C.0832 - Veillonellales sp. |
| baseline | MGS | Hgn3C.0833 - Eisenbergiella tayi |
| baseline | MGS | Hgn3C.0837 - Anaerotruncus colihominis |
| baseline | MGS | Hgn3C.0851 - Oscillospiraceae sp. |
| baseline | MGS | Hgn3C.0855 - Eubacteriales sp. |
| baseline | MGS | Hgn3C.0858 - Turicibacter sanguinis |
| baseline | MGS | Hgn3C.0859 - Hydrogeniiclostidium mannosilyticum |
| baseline | MGS | Hgn3C.0873 - Clostridium perfringens |
| baseline | MGS | Hgn3C.0875 - Lachnospiraceae sp. |
| baseline | MGS | Hgn3C.0886 - Bacteroides sp. |
| baseline | MGS | Hgn3C.0889 - Clostridium paraputrificum |
| baseline | MGS | Hgn3C.0890 - Enterobacter roggenkampii |
| baseline | MGS | Hgn3C.0891 - Eubacteriales sp. |
| baseline | MGS | Hgn3C.0893 - Phocea massiliensis |
| baseline | MGS | Hgn3C.0908 - Eubacteriales sp. |
| baseline | MGS | Hgn3C.0913 - Erysipelotrichaceae sp. |
| baseline | MGS | Hgn3C.0921 - Bacteroides sp. |
| baseline | MGS | Hgn3C.0922 - Eubacteriales sp. |
| baseline | MGS | Hgn3C.0940 - Eubacteriales sp. |
| baseline | MGS | Hgn3C.0948 - Enterobacter sp. |
| baseline | MGS | Hgn3C.0978 - Veillonella sp. |
| baseline | MGS | Hgn3C.0987 - Fusobacterium mortiferum |
| baseline | MGS | Hgn3C.0996 - Oscillospiraceae sp. |
| baseline | MGS | Hgn3C.0998 - Eubacteriales sp. |
| baseline | MGS | Hgn3C.1012 - Erysipelotrichaceae sp. |
| baseline | MGS | Hgn3C.1035 - Klebsiella sp. |
| baseline | MGS | Hgn3C.1036 - Escherichia coli |
| baseline | MGS | Hgn3C.1037 - Rothia sp. |

| | | |
|---|---|---|
| baseline | MGS | Hgn3C.1038 - Veillonella sp. |
| baseline | MGS | Hgn3C.1040 - Lachnospiraceae sp. |
| baseline | MGS | Hgn3C.1041 - Bacteroides stercoris |
| baseline | MGS | Hgn3C.1042 - Dorea formicigenerans |
| baseline | MGS | Hgn3C.1043 - Clostridium phoceensis |
| baseline | MGS | Hgn3C.1044 - Eubacteriales sp. |
| baseline | MGS | Hgn3C.1045 - Lactococcus lactis subsp. lactis |
| baseline | MGS | Hgn3C.1049 - Enterococcus faecium |
| baseline | MGS | Hgn3C.1050 - Staphylococcus epidermidis |
| baseline | MGS | Hgn3C.1051 - Holdemania massiliensis |
| baseline | MGS | Hgn3C.1052 - Enterococcus gallinarum |
| baseline | MGS | Hgn3C.1054 - Streptococcus lutetiensis |
| baseline | MGS | Hgn3C.1058 - Mediterraneibacter glycyrrhizinilyticus |
| baseline | MGS | Hgn3C.1059 - Faecalicatena sp. |
| baseline | MGS | Hgn3C.1063 - Enterococcus avium |
| baseline | MGS | Hgn3C.1065 - Klebsiella variicola subsp. variicola |
| baseline | MGS | Hgn3C.1067 - Lachnospiraceae sp. |
| baseline | MGS | Hgn3C.1073 - Streptococcus gallolyticus subsp. gallolyticus |
| baseline | MGS | Hgn3C.1081 - Lacticaseibacillus rhamnosus |
| baseline | MGS | Hgn3C.1082 - Lacticaseibacillus paracasei subsp. paracasei |
| baseline | MGS | Hgn3C.1084 - Massilimicrobiota timonensis |
| baseline | MGS | Hgn3C.1087 - Ligilactobacillus salivarius |
| baseline | MGS | Hgn3C.1090 - Turicibacter sanguinis |
| baseline | MGS | Hgn3C.1093 - Enterobacter kobei |
| baseline | MGS | Hgn3C.1107 - Eubacteriales sp. |
| baseline | MGS | Hgn3C.1110 - Porphyromonas sp. |
| baseline | MGS | Hgn3C.1117 - Atopobiaceae sp. |
| baseline | MGS | Hgn3C.1120 - Actinomycetaceae sp. |
| baseline | MGS | Hgn3C.1146 - Bacteria sp. |
| baseline | MGS | Hgn3C.1148 - Eggerthellales sp. |
| baseline | MGS | Hgn3C.1154 - Lactococcus garvieae |
| baseline | MGS | Hgn3C.1180 - Enterocloster citroniae |
| baseline | MGS | Hgn3C.1188 - Actinomyces oris |
| baseline | MGS | Hgn3C.1190 - Fenollaria massiliensis |
| baseline | MGS | Hgn3C.1193 - Atopobium sp. |
| baseline | MGS | Hgn3C.1196 - Blautia marasmi |
| baseline | MGS | Hgn3C.1197 - Blautia hansenii |
| baseline | MGS | Hgn3C.1210 - Dielma sp. |
| baseline | MGS | Hgn3C.1211 - Lachnoclostridium pacaense |
| baseline | MGS | Hgn3C.1214 - Corynebacterium tuberculostearicum |
| baseline | MGS | Hgn3C.1218 - Enterobacter bugandensis |
| baseline | MGS | Hgn3C.1219 - Enterobacter sp. NFIX58 |
| baseline | MGS | Hgn3C.1225 - Enterococcus faecium |
| baseline | MGS | Hgn3C.1229 - Escherichia sp. 93.0750 |
| baseline | MGS | Hgn3C.1230 - Eubacterium limosum |
| baseline | MGS | Hgn3C.1234 - Granulicatella adiacens |
| baseline | MGS | Hgn3C.1238 - Klebsiella quasivariicola |
| baseline | MGS | Hgn3C.1242 - Limosilactobacillus vaginalis |

| | | |
|---|---|---|
| baseline | MGS | Hgn3C.1248 - Neisseria sp. HMSC061E12 |
| baseline | MGS | Hgn3C.1258 - Rothia sp. HMSC072E10 |
| baseline | MGS | Hgn3C.1264 - Staphylococcus hominis subsp. hominis |
| baseline | MGS | Hgn3C.1268 - Streptococcus sp. |
| baseline | MGS | Hgn3C.1269 - Streptococcus constellatus subsp. constellatus |
| baseline | MGS | Hgn3C.1270 - Streptococcus sp. |
| baseline | MGS | Hgn3C.1271 - Streptococcus agalactiae |
| baseline | MGS | Hgn3C.1275 - Lachnospiraceae sp. |
| baseline | MGS | Hgn3C.1277 - Lachnospiraceae sp. |
| baseline | MGS | Hgn3C.1278 - Clostridium sp. NSJ-6 |
| baseline | MGS | Hgn3C.1282 - Terrisporobacter othiniensis |
| baseline | MGS | Hgn3C.1286 - Corynebacterium variabile |
| baseline | MGS | Hgn3C.1288 - Enterobacter hormaechei |
| baseline | MGS | Hgn3C.1292 - Limosilactobacillus portuensis |
| baseline | MGS | Hgn3C.1295 - Butyricicoccus pullicaecorum |
| baseline | MGS | Hgn3C.1299 - Erysipelotrichaceae sp. |
| baseline | MGS | Hgn3C.1300 - Rothia sp. |
| baseline | MGS | Hgn3C.1302 - Schaalia sp. |
| baseline | MGS | Hgn3C.1303 - Clostridium paraputrificum |
| baseline | MGS | Hgn3C.1305 - Isoptericola variabilis |
| baseline | MGS | Hgn3C.1307 - Bifidobacterium longum subsp. spp |
| baseline | MGS | Hgn3C.1308 - Eisenbergiella sp. |
| baseline | MGS | Hgn3C.1309 - Enterobacter cloacae subsp. cloacae |
| baseline | MGS | Hgn3C.1310 - Erysipelotrichaceae sp. |
| baseline | MGS | Hgn3C.1311 - Lachnospiraceae sp. |
| baseline | MGS | Hgn3C.1312 - Candida tropicalis |
| baseline | MGS | Hgn3C.1315 - Eubacteriales sp. |
| baseline | MGS | Hgn3C.1316 - Eubacteriales sp. |
| baseline | MGS | Hgn3C.1319 - Atlantibacter hermannii |
| baseline | MGS | Hgn3C.1321 - Veillonellaceae sp. |
| baseline | MGS | Hgn3C.1322 - Enterococcus hirae |
| baseline | MGS | Hgn3C.1323 - Lachnospiraceae sp. |
| baseline | MGS | Hgn3C.1324 - Eubacterium sp. c-25 |
| baseline | MGS | Hgn3C.1325 - Luxibacter massiliensis |
| baseline | MGS | Hgn3C.1326 - Eggerthella guodeyinii |
| baseline | MGS | Hgn3C.1329 - Eubacteriales sp. |
| baseline | MGS | Hgn3C.1330 - Lachnospiraceae sp. |
| baseline | MGS | Hgn3C.1331 - Lachnospiraceae sp. |
| baseline | MGS | Hgn3C.1334 - Eubacteriales sp. |
| baseline | MGS | Hgn3C.1338 - Erysipelatoclostridium sp. |
| baseline | MGS | Hgn3C.1340 - Clostridiaceae sp. |
| baseline | MGS | Hgn3C.1343 - Enterococcus italicus |
| baseline | MGS | Hgn3C.1345 - Limosilactobacillus mucosae |
| baseline | MGS | Hgn3C.1346 - Erysipelotrichales sp. |
| baseline | MGS | Hgn3C.1347 - [Clostridium] hylemonae |
| baseline | MGS | Hgn3C.1348 - Eubacteriales sp. |
| baseline | MGS | Hgn3C.1351 - Actinomyces sp. ICM47 |
| baseline | MGS | Hgn3C.1352 - Akkermansia sp. |

| | | |
|---|---|---|
| baseline | MGS | Hgn3C.1354 - Actinomyces sp. |
| baseline | MGS | Hgn3C.1355 - Collinsella stercoris |
| baseline | MGS | Hgn3C.1358 - Lachnospiraceae sp. |
| baseline | MGS | Hgn3C.1359 - Extibacter muris |
| baseline | MGS | Hgn3C.1360 - Bifidobacterium crudilactis |
| baseline | MGS | Hgn3C.1361 - Senegalimassilia anaerobia |
| baseline | MGS | Hgn3C.1362 - Christensenella intestinihominis |
| baseline | MGS | Hgn3C.1366 - Lachnospiraceae sp. |
| baseline | MGS | Hgn3C.1369 - Lachnospiraceae sp. |
| baseline | MGS | Hgn3C.1370 - Kluyvera genomosp. 3 |
| baseline | MGS | Hgn3C.1372 - Eubacteriales sp. |
| baseline | MGS | Hgn3C.1373 - Eubacteriales sp. |
| baseline | MGS | Hgn3C.1375 - Atopobiaceae sp. |
| baseline | MGS | Hgn3C.1376 - Kluyvera sp. CRP |
| baseline | MGS | Hgn3C.1377 - Collinsella tanakaei |
| baseline | MGS | Hgn3C.1378 - Collinsella sp. AF08-23 |
| baseline | MGS | Hgn3C.1379 - Gordonibacter urolithinfaciens |
| baseline | MGS | Hgn3C.1380 - Kluyvera georgiana |
| baseline | MGS | Hgn3C.1382 - Oscillospiraceae sp. |
| baseline | MGS | Hgn3C.1384 - Anaerococcus sp. |
| baseline | MGS | Hgn3C.1386 - Veillonella sp. |
| baseline | MGS | Hgn3C.1387 - Alistipes sp. |
| baseline | MGS | Hgn3C.1388 - Streptococcus infantarius subsp. infantarius |
| baseline | MGS | Hgn3C.1390 - Erysipelatoclostridium sp. |
| baseline | MGS | Hgn3C.1392 - Enterobacter sp. JBIWA005 |
| baseline | MGS | Hgn3C.1393 - Anaerofustis stercorihominis |
| baseline | MGS | Hgn3C.1394 - Eubacteriales sp. |
| baseline | MGS | Hgn3C.1398 - Actinomyces sp. |
| baseline | MGS | Hgn3C.1400 - Eubacterium sp. |
| baseline | MGS | Hgn3C.1401 - Eubacteriales sp. |
| baseline | MGS | Hgn3C.1403 - Eubacteriaceae sp. |
| baseline | MGS | Hgn3C.1404 - Clostridium sp. |
| baseline | MGS | Hgn3C.1406 - Slackia piriformis |
| baseline | MGS | Hgn3C.1407 - Christensenella minuta |
| baseline | MGS | Hgn3C.1408 - Eubacterium callanderi |
| baseline | MGS | Hgn3C.1409 - Eggerthella timonensis |
| baseline | MGS | Hgn3C.1412 - Christensenella hongkongensis |
| baseline | MGS | Hgn3C.1413 - Streptococcus sp. |
| baseline | MGS | Hgn3C.1414 - Collinsella sp. |
| baseline | MGS | Hgn3C.1415 - Klebsiella quasipneumoniae subsp. quasipneumoniae |
| baseline | MGS | Hgn3C.1416 - Limosilactobacillus fermentum |
| baseline | MGS | Hgn3C.1418 - Carnobacteriaceae sp. |
| baseline | MGS | Hgn3C.1419 - Agathobaculum sp. NSJ-28 |
| baseline | MGS | Hgn3C.1420 - Eubacteriales sp. |
| baseline | MGS | Hgn3C.1423 - Lachnospiraceae sp. |
| baseline | MGS | Hgn3C.1424 - Mogibacterium sp. NSJ-24 |
| baseline | MGS | Hgn3C.1425 - Eubacteriales sp. |
| baseline | MGS | Hgn3C.1426 - Limosilactobacillus oris |

| | | |
|---|---|---|
| baseline | MGS | Hgn3C.1429 - Lachnospiraceae sp. |
| baseline | MGS | Hgn3C.1431 - Bacteroidales sp. |
| baseline | MGS | Hgn3C.1434 - Blautia sp. |
| baseline | MGS | Hgn3C.1435 - Acinetobacter baumannii |
| baseline | MGS | Hgn3C.1436 - Bacteria sp. |
| baseline | MGS | Hgn3C.1437 - Candidatus Stoquefichus sp. SB1 |
| baseline | MGS | Hgn3C.1438 - Dielma fastidiosa |
| baseline | MGS | Hgn3C.1439 - Streptococcus sp. IMAU 99161 |
| baseline | MGS | Hgn3C.1441 - Lancefieldella sp. |
| baseline | MGS | Hgn3C.1443 - Streptococcus sp. |
| baseline | MGS | Hgn3C.1444 - Ruminococcus sp. 1001175B_160314_E1 |
| baseline | MGS | Hgn3C.1446 - Eubacteriales sp. |
| baseline | MGS | Hgn3C.1447 - Bacteria sp. |
| baseline | MGS | Hgn3C.1448 - Leuconostoc falkenbergense |
| baseline | MGS | Hgn3C.1450 - Lactococcus sp. NH2-7C |
| baseline | MGS | Hgn3C.1452 - Raoultibacter timonensis |
| baseline | MGS | Hgn3C.1453 - Collinsella sp. |
| baseline | MGS | Hgn3C.1455 - Enterocloster sp. |
| baseline | MGS | Hgn3C.1456 - Eubacteriales sp. |
| baseline | MGS | Hgn3C.1460 - Blautia sp. |
| baseline | MGS | Hgn3C.1463 - Enterobacter sp. |
| baseline | MGS | Hgn3C.1465 - Clostridia sp. |
| baseline | MGS | Hgn3C.1467 - Eubacteriales sp. |
| baseline | MGS | Hgn3C.1468 - Staphylococcus aureus |
| baseline | MGS | Hgn3C.1469 - Lachnospiraceae sp. |
| baseline | MGS | Hgn3C.1471 - Eggerthella sp. |
| baseline | MGS | Hgn3C.1472 - Amedibacillus dolichus |
| baseline | genus | Acinetobacter |
| baseline | genus | Actinomyces |
| baseline | genus | Adlercreutzia |
| baseline | genus | Agathobaculum |
| baseline | genus | Akkermansia |
| baseline | genus | Alistipes |
| baseline | genus | Allisonella |
| baseline | genus | Amedibacillus |
| baseline | genus | Anaerobutyricum |
| baseline | genus | Anaerococcus |
| baseline | genus | Anaerofustis |
| baseline | genus | Anaeromassilibacillus |
| baseline | genus | Anaerostipes |
| baseline | genus | Anaerotignum |
| baseline | genus | Anaerotruncus |
| baseline | genus | Atlantibacter |
| baseline | genus | Atopobium |
| baseline | genus | Bacteroides |
| baseline | genus | Barnesiella |
| baseline | genus | Bifidobacterium |
| baseline | genus | Bilophila |

| | | |
|---|---|---|
| baseline | genus | Blautia |
| baseline | genus | Butyricicoccus |
| baseline | genus | Butyricimonas |
| baseline | genus | Candida |
| baseline | genus | Candidatus Stoquefichus |
| baseline | genus | Catenibacterium |
| baseline | genus | Christensenella |
| baseline | genus | Citrobacter |
| baseline | genus | Clostridium |
| baseline | genus | Colidextribacter |
| baseline | genus | Collinsella |
| baseline | genus | Coprobacillus |
| baseline | genus | Coprobacter |
| baseline | genus | Coprococcus |
| baseline | genus | Corynebacterium |
| baseline | genus | Desulfovibrio |
| baseline | genus | Dialister |
| baseline | genus | Dielma |
| baseline | genus | Dorea |
| baseline | genus | Duodenibacillus |
| baseline | genus | Dysosmobacter |
| baseline | genus | Eggerthella |
| baseline | genus | Eisenbergiella |
| baseline | genus | Ellagibacter |
| baseline | genus | Enterobacter |
| baseline | genus | Enterocloster |
| baseline | genus | Enterococcus |
| baseline | genus | Erysipelatoclostridium |
| baseline | genus | Escherichia |
| baseline | genus | Eubacterium |
| baseline | genus | Evtepia |
| baseline | genus | Extibacter |
| baseline | genus | Faecalibacillus |
| baseline | genus | Faecalibacterium |
| baseline | genus | Faecalicatena |
| baseline | genus | Fenollaria |
| baseline | genus | Flavonifractor |
| baseline | genus | Flintibacter |
| baseline | genus | Fusicatenibacter |
| baseline | genus | Fusobacterium |
| baseline | genus | Gemmiger |
| baseline | genus | Gordonibacter |
| baseline | genus | Granulicatella |
| baseline | genus | Haemophilus |
| baseline | genus | Holdemanella |
| baseline | genus | Holdemania |
| baseline | genus | Hungatella |
| baseline | genus | Hydrogeniiclostidium |

| | | |
|---|---|---|
| baseline | genus | Hydrogenoanaerobacterium |
| baseline | genus | Intestinibacter |
| baseline | genus | Intestinimonas |
| baseline | genus | Isoptericola |
| baseline | genus | Klebsiella |
| baseline | genus | Kluyvera |
| baseline | genus | Lachnoclostridium |
| baseline | genus | Lachnospira |
| baseline | genus | Lacrimispora |
| baseline | genus | Lacticaseibacillus |
| baseline | genus | Lactobacillus |
| baseline | genus | Lactococcus |
| baseline | genus | Lancefieldella |
| baseline | genus | Leuconostoc |
| baseline | genus | Ligilactobacillus |
| baseline | genus | Limosilactobacillus |
| baseline | genus | Longicatena |
| baseline | genus | Luxibacter |
| baseline | genus | Massilimicrobiota |
| baseline | genus | Mediterraneibacter |
| baseline | genus | Megamonas |
| baseline | genus | Megasphaera |
| baseline | genus | Mogibacterium |
| baseline | genus | Neisseria |
| baseline | genus | Odoribacter |
| baseline | genus | Olsenella |
| baseline | genus | Oscillibacter |
| baseline | genus | Parabacteroides |
| baseline | genus | Paraprevotella |
| baseline | genus | Parasutterella |
| baseline | genus | Parolsenella |
| baseline | genus | Phascolarctobacterium |
| baseline | genus | Phocaeicola |
| baseline | genus | Phocea |
| baseline | genus | Porphyromonas |
| baseline | genus | Prevotella |
| baseline | genus | Prevotellamassilia |
| baseline | genus | Raoultibacter |
| baseline | genus | Romboutsia |
| baseline | genus | Roseburia |
| baseline | genus | Rothia |
| baseline | genus | Ruminococcus |
| baseline | genus | Ruthenibacterium |
| baseline | genus | Schaalia |
| baseline | genus | Sellimonas |
| baseline | genus | Senegalimassilia |
| baseline | genus | Slackia |
| baseline | genus | Staphylococcus |

| | | |
|---|---|---|
| baseline | genus | Streptococcus |
| baseline | genus | Subdoligranulum |
| baseline | genus | Sutterella |
| baseline | genus | Terrisporobacter |
| baseline | genus | Turicibacter |
| baseline | genus | Tyzzerella |
| baseline | genus | Veillonella |
| baseline | genus | Vescimonas |
| baseline | genus | Waltera |
| baseline | family | Acidaminococcaceae |
| baseline | family | Actinomycetaceae |
| baseline | family | Akkermansiaceae |
| baseline | family | Atopobiaceae |
| baseline | family | Bacteroidaceae |
| baseline | family | Barnesiellaceae |
| baseline | family | Bifidobacteriaceae |
| baseline | family | Carnobacteriaceae |
| baseline | family | Christensenellaceae |
| baseline | family | Clostridiaceae |
| baseline | family | Coprobacillaceae |
| baseline | family | Coriobacteriaceae |
| baseline | family | Corynebacteriaceae |
| baseline | family | Debaryomycetaceae |
| baseline | family | Desulfovibrionaceae |
| baseline | family | Eggerthellaceae |
| baseline | family | Enterobacteriaceae |
| baseline | family | Enterococcaceae |
| baseline | family | Erysipelotrichaceae |
| baseline | family | Eubacteriaceae |
| baseline | family | Eubacteriales Family XIII. Incertae Sedis |
| baseline | family | Fusobacteriaceae |
| baseline | family | Lachnospiraceae |
| baseline | family | Lactobacillaceae |
| baseline | family | Micrococcaceae |
| baseline | family | Moraxellaceae |
| baseline | family | Neisseriaceae |
| baseline | family | Odoribacteraceae |
| baseline | family | Oscillospiraceae |
| baseline | family | Pasteurellaceae |
| baseline | family | Peptoniphilaceae |
| baseline | family | Peptostreptococcaceae |
| baseline | family | Porphyromonadaceae |
| baseline | family | Prevotellaceae |
| baseline | family | Promicromonosporaceae |
| baseline | family | Rikenellaceae |
| baseline | family | Selenomonadaceae |
| baseline | family | Staphylococcaceae |
| baseline | family | Streptococcaceae |

| | | |
|---|---|---|
| baseline | family | Sutterellaceae |
| baseline | family | Tannerellaceae |
| baseline | family | Turicibacteraceae |
| baseline | family | Veillonellaceae |
| baseline | phylum | Actinobacteria |
| baseline | phylum | Ascomycota |
| baseline | phylum | Bacteroidetes |
| baseline | phylum | Firmicutes |
| baseline | phylum | Fusobacteria |
| baseline | phylum | Proteobacteria |
| baseline | phylum | Verrucomicrobia |
| 3 months | MGS | Hgn3C.0001 - Phocaeicola vulgatus |
| 3 months | MGS | Hgn3C.0002 - Bacteroides uniformis |
| 3 months | MGS | Hgn3C.0003 - Lachnospiraceae sp. |
| 3 months | MGS | Hgn3C.0004 - Blautia wexlerae |
| 3 months | MGS | Hgn3C.0005 - Fusicatenibacter saccharivorans |
| 3 months | MGS | Hgn3C.0006 - Bacteroides ovatus |
| 3 months | MGS | Hgn3C.0007 - Parabacteroides distasonis |
| 3 months | MGS | Hgn3C.0008 - Alistipes putredinis |
| 3 months | MGS | Hgn3C.0009 - Faecalibacterium longum CLA-AA-H243 |
| 3 months | MGS | Hgn3C.0010 - Anaerostipes hadrus |
| 3 months | MGS | Hgn3C.0011 - Roseburia faecis |
| 3 months | MGS | Hgn3C.0012 - Lachnospira sp. |
| 3 months | MGS | Hgn3C.0013 - Ruminococcus bromii |
| 3 months | MGS | Hgn3C.0014 - Lacrimispora saccharolytica |
| 3 months | MGS | Hgn3C.0015 - Faecalibacterium sp. Marseille-Q4896 |
| 3 months | MGS | Hgn3C.0016 - Parabacteroides merdae |
| 3 months | MGS | Hgn3C.0017 - Subdoligranulum sp. APC924/74 |
| 3 months | MGS | Hgn3C.0018 - Faecalibacterium prausnitzii |
| 3 months | MGS | Hgn3C.0019 - Phocaeicola dorei |
| 3 months | MGS | Hgn3C.0020 - Collinsella aerofaciens |
| 3 months | MGS | Hgn3C.0021 - Bifidobacterium longum subsp. longum |
| 3 months | MGS | Hgn3C.0022 - Alistipes shahii |
| 3 months | MGS | Hgn3C.0023 - Bacteroides xylanisolvens |
| 3 months | MGS | Hgn3C.0024 - Bacteroides caccae |
| 3 months | MGS | Hgn3C.0025 - Ruminococcus bicirculans |
| 3 months | MGS | Hgn3C.0026 - Dorea longicatena |
| 3 months | MGS | Hgn3C.0027 - Barnesiella intestinihominis |
| 3 months | MGS | Hgn3C.0028 - Oscillibacter sp. ER4 |
| 3 months | MGS | Hgn3C.0029 - Anaerobutyricum hallii |
| 3 months | MGS | Hgn3C.0030 - Faecalibacterium prausnitzii |
| 3 months | MGS | Hgn3C.0031 - Blautia massiliensis |
| 3 months | MGS | Hgn3C.0032 - Alistipes onderdonkii subsp. vulgaris |
| 3 months | MGS | Hgn3C.0033 - Gemmiger formicilis |
| 3 months | MGS | Hgn3C.0034 - Roseburia inulinivorans |
| 3 months | MGS | Hgn3C.0035 - Odoribacter splanchnicus |
| 3 months | MGS | Hgn3C.0036 - Bacteroides thetaiotaomicron |
| 3 months | MGS | Hgn3C.0037 - Roseburia intestinalis |

| | | |
|---|---|---|
| 3 months | MGS | Hgn3C.0038 - Bifidobacterium adolescentis |
| 3 months | MGS | Hgn3C.0039 - Coprococcus comes |
| 3 months | MGS | Hgn3C.0040 - Eubacteriales sp. |
| 3 months | MGS | Hgn3C.0041 - Ruminococcus sp. |
| 3 months | MGS | Hgn3C.0042 - Blautia faecis |
| 3 months | MGS | Hgn3C.0043 - Lachnoclostridium sp. 210928-DFI.6.3 |
| 3 months | MGS | Hgn3C.0045 - Faecalibacterium sp. |
| 3 months | MGS | Hgn3C.0046 - Clostridium sp. AF37-5 |
| 3 months | MGS | Hgn3C.0047 - Dysosmobacter sp. BX15 |
| 3 months | MGS | Hgn3C.0048 - Akkermansia muciniphila |
| 3 months | MGS | Hgn3C.0049 - Eubacteriales sp. |
| 3 months | MGS | Hgn3C.0050 - Escherichia coli |
| 3 months | MGS | Hgn3C.0051 - Alistipes communis |
| 3 months | MGS | Hgn3C.0053 - Eubacteriales sp. |
| 3 months | MGS | Hgn3C.0054 - Prevotella copri |
| 3 months | MGS | Hgn3C.0055 - Alistipes finegoldii |
| 3 months | MGS | Hgn3C.0056 - Bacteroides fragilis |
| 3 months | MGS | Hgn3C.0057 - Oscillibacter sp. MSJ-31 |
| 3 months | MGS | Hgn3C.0058 - Lachnospiraceae sp. |
| 3 months | MGS | Hgn3C.0059 - Bacteroides cellulosilyticus |
| 3 months | MGS | Hgn3C.0060 - Eubacteriales sp. |
| 3 months | MGS | Hgn3C.0061 - Faecalibacillus sp. TM498 |
| 3 months | MGS | Hgn3C.0062 - Waltera intestinalis |
| 3 months | MGS | Hgn3C.0063 - Eubacteriales sp. |
| 3 months | MGS | Hgn3C.0064 - Blautia sp. DFI.9.9 |
| 3 months | MGS | Hgn3C.0065 - Blautia obeum |
| 3 months | MGS | Hgn3C.0067 - Roseburia hominis |
| 3 months | MGS | Hgn3C.0068 - [Ruminococcus] lactaris |
| 3 months | MGS | Hgn3C.0069 - Oscillibacter sp. KLE 1728 |
| 3 months | MGS | Hgn3C.0070 - Faecalibacterium sp. OF04-11AC |
| 3 months | MGS | Hgn3C.0071 - Faecalicatena fissicatena |
| 3 months | MGS | Hgn3C.0072 - Clostridium fessum |
| 3 months | MGS | Hgn3C.0073 - Blautia sp. |
| 3 months | MGS | Hgn3C.0075 - Agathobaculum butyriciproducens |
| 3 months | MGS | Hgn3C.0077 - Sutterella wadsworthensis |
| 3 months | MGS | Hgn3C.0078 - Clostridium sp. MCC328 |
| 3 months | MGS | Hgn3C.0079 - [Ruminococcus] torques |
| 3 months | MGS | Hgn3C.0080 - Phascolarctobacterium faecium |
| 3 months | MGS | Hgn3C.0082 - Roseburia sp. CLA-AA-H204 |
| 3 months | MGS | Hgn3C.0083 - Eubacteriales sp. |
| 3 months | MGS | Hgn3C.0084 - Flavonifractor plautii |
| 3 months | MGS | Hgn3C.0085 - Faecalibacterium sp. |
| 3 months | MGS | Hgn3C.0086 - Dialister invisus |
| 3 months | MGS | Hgn3C.0087 - Eubacteriales sp. |
| 3 months | MGS | Hgn3C.0088 - [Ruminococcus] gnavus |
| 3 months | MGS | Hgn3C.0089 - Paraprevotella clara |
| 3 months | MGS | Hgn3C.0092 - Lachnospiraceae sp. |
| 3 months | MGS | Hgn3C.0093 - Bilophila wadsworthia |

| | | |
|---|---|---|
| 3 months | MGS | Hgn3C.0094 - Eubacteriales sp. |
| 3 months | MGS | Hgn3C.0095 - Eubacteriales sp. |
| 3 months | MGS | Hgn3C.0096 - Eubacteriales sp. |
| 3 months | MGS | Hgn3C.0097 - Eubacterium ventriosum |
| 3 months | MGS | Hgn3C.0098 - Coprococcus catus |
| 3 months | MGS | Hgn3C.0099 - Dorea longicatena |
| 3 months | MGS | Hgn3C.0100 - Bifidobacterium bifidum |
| 3 months | MGS | Hgn3C.0101 - Bifidobacterium pseudocatenulatum |
| 3 months | MGS | Hgn3C.0102 - Eubacteriales sp. |
| 3 months | MGS | Hgn3C.0103 - Holdemanella porci |
| 3 months | MGS | Hgn3C.0104 - Clostridium sp. AM32-2 |
| 3 months | MGS | Hgn3C.0105 - Vescimonas coprocola |
| 3 months | MGS | Hgn3C.0106 - Faecalibacterium sp. CLA-AA-H233 |
| 3 months | MGS | Hgn3C.0111 - Eubacteriales sp. |
| 3 months | MGS | Hgn3C.0113 - Clostridium sp. MCC345 |
| 3 months | MGS | Hgn3C.0114 - Eubacteriales sp. |
| 3 months | MGS | Hgn3C.0120 - Flavonifractor plautii |
| 3 months | MGS | Hgn3C.0121 - Oscillospiraceae sp. |
| 3 months | MGS | Hgn3C.0122 - Bacteroides finegoldii |
| 3 months | MGS | Hgn3C.0123 - Eubacteriales sp. |
| 3 months | MGS | Hgn3C.0124 - Ruthenibacterium lactatiformans |
| 3 months | MGS | Hgn3C.0125 - Sutterella sp. KLE1602 |
| 3 months | MGS | Hgn3C.0126 - Ruminococcus sp. BSD2780120874_150323_B10 |
| 3 months | MGS | Hgn3C.0127 - Faecalibacterium sp. |
| 3 months | MGS | Hgn3C.0128 - Eubacteriales sp. |
| 3 months | MGS | Hgn3C.0129 - Haemophilus parainfluenzae |
| 3 months | MGS | Hgn3C.0131 - Streptococcus thermophilus |
| 3 months | MGS | Hgn3C.0135 - Firmicutes sp. |
| 3 months | MGS | Hgn3C.0136 - Alistipes ihumii |
| 3 months | MGS | Hgn3C.0137 - Clostridium sp. OF03-18AA |
| 3 months | MGS | Hgn3C.0138 - Anaerobutyricum soehngenii |
| 3 months | MGS | Hgn3C.0139 - Ruminococcus callidus |
| 3 months | MGS | Hgn3C.0140 - Veillonella parvula |
| 3 months | MGS | Hgn3C.0141 - Adlercreutzia equolifaciens subsp. celatus |
| 3 months | MGS | Hgn3C.0142 - Eubacterium ramulus |
| 3 months | MGS | Hgn3C.0143 - Streptococcus salivarius |
| 3 months | MGS | Hgn3C.0144 - Eubacteriales sp. |
| 3 months | MGS | Hgn3C.0146 - Phascolarctobacterium succinatutens |
| 3 months | MGS | Hgn3C.0147 - Oscillospiraceae sp. |
| 3 months | MGS | Hgn3C.0153 - Lachnospiraceae sp. |
| 3 months | MGS | Hgn3C.0154 - Prevotella copri |
| 3 months | MGS | Hgn3C.0155 - Blautia sp. |
| 3 months | MGS | Hgn3C.0157 - Eubacteriales sp. |
| 3 months | MGS | Hgn3C.0161 - Blautia sp. |
| 3 months | MGS | Hgn3C.0162 - Prevotella sp. |
| 3 months | MGS | Hgn3C.0168 - Evtepia gabavorous |
| 3 months | MGS | Hgn3C.0169 - Bacteroides intestinalis |
| 3 months | MGS | Hgn3C.0170 - Oscillospiraceae sp. |

| | | |
|---|---|---|
| 3 months | MGS | Hgn3C.0171 - Catenibacterium mitsuokai |
| 3 months | MGS | Hgn3C.0174 - Lachnospiraceae sp. |
| 3 months | MGS | Hgn3C.0175 - Oscillospiraceae sp. |
| 3 months | MGS | Hgn3C.0176 - Alistipes indistinctus |
| 3 months | MGS | Hgn3C.0179 - Bacteroidales sp. |
| 3 months | MGS | Hgn3C.0181 - Alistipes onderdonkii |
| 3 months | MGS | Hgn3C.0182 - Butyricimonas faecihominis |
| 3 months | MGS | Hgn3C.0183 - Intestinibacter bartlettii |
| 3 months | MGS | Hgn3C.0185 - Bifidobacterium catenulatum subsp. kashiwanohense |
| 3 months | MGS | Hgn3C.0186 - Butyricimonas virosa |
| 3 months | MGS | Hgn3C.0191 - Prevotella sp. |
| 3 months | MGS | Hgn3C.0192 - Erysipelatoclostridium ramosum |
| 3 months | MGS | Hgn3C.0195 - Eubacteriales sp. |
| 3 months | MGS | Hgn3C.0201 - Veillonella dispar |
| 3 months | MGS | Hgn3C.0202 - Eggerthella lenta |
| 3 months | MGS | Hgn3C.0203 - Faecalibacterium sp. Marseille-Q3530 |
| 3 months | MGS | Hgn3C.0207 - Eubacteriales sp. |
| 3 months | MGS | Hgn3C.0208 - Romboutsia timonensis |
| 3 months | MGS | Hgn3C.0209 - Bifidobacterium breve |
| 3 months | MGS | Hgn3C.0212 - Holdemanella biformis |
| 3 months | MGS | Hgn3C.0213 - Akkermansia sp. GGCC_0220 |
| 3 months | MGS | Hgn3C.0214 - Duodenibacillus massiliensis |
| 3 months | MGS | Hgn3C.0215 - Alistipes sp. cv1 |
| 3 months | MGS | Hgn3C.0217 - Eubacteriales sp. |
| 3 months | MGS | Hgn3C.0218 - Eubacteriales sp. |
| 3 months | MGS | Hgn3C.0219 - Tyzzerella nexilis |
| 3 months | MGS | Hgn3C.0220 - Clostridium sp. MCC334 |
| 3 months | MGS | Hgn3C.0221 - Clostridium sp. |
| 3 months | MGS | Hgn3C.0222 - Parabacteroides merdae |
| 3 months | MGS | Hgn3C.0224 - Veillonellales sp. |
| 3 months | MGS | Hgn3C.0226 - Eubacteriales sp. |
| 3 months | MGS | Hgn3C.0232 - Megamonas funiformis |
| 3 months | MGS | Hgn3C.0236 - Oscillospiraceae sp. |
| 3 months | MGS | Hgn3C.0238 - Oscillospiraceae sp. |
| 3 months | MGS | Hgn3C.0239 - Blautia sp. BIOML-A1 |
| 3 months | MGS | Hgn3C.0242 - Enterocloster bolteae |
| 3 months | MGS | Hgn3C.0244 - Desulfovibrio piger |
| 3 months | MGS | Hgn3C.0245 - Blautia stercoris |
| 3 months | MGS | Hgn3C.0248 - Coprococcus catus |
| 3 months | MGS | Hgn3C.0249 - Clostridium sp. AM33-3 |
| 3 months | MGS | Hgn3C.0250 - Eubacteriales sp. |
| 3 months | MGS | Hgn3C.0251 - Alistipes senegalensis |
| 3 months | MGS | Hgn3C.0253 - Waltera sp. |
| 3 months | MGS | Hgn3C.0260 - Hydrogenoanaerobacterium saccharovorans |
| 3 months | MGS | Hgn3C.0264 - Clostridium sp. AF27-2AA |
| 3 months | MGS | Hgn3C.0266 - Dorea sp. AF36-15AT |
| 3 months | MGS | Hgn3C.0267 - Oscillospiraceae sp. |
| 3 months | MGS | Hgn3C.0269 - Flintibacter sp. NSJ-23 |

| | | |
|---|---|---|
| 3 months | MGS | Hgn3C.0272 - Blautia faecicola |
| 3 months | MGS | Hgn3C.0273 - Slackia isoflavoniconvertens |
| 3 months | MGS | Hgn3C.0275 - Lachnospiraceae sp. |
| 3 months | MGS | Hgn3C.0281 - Eubacteriales sp. |
| 3 months | MGS | Hgn3C.0284 - Erysipelotrichaceae sp. |
| 3 months | MGS | Hgn3C.0290 - Sellimonas intestinalis |
| 3 months | MGS | Hgn3C.0295 - Eubacteriales sp. |
| 3 months | MGS | Hgn3C.0299 - Escherichia coli |
| 3 months | MGS | Hgn3C.0301 - Prevotella hominis |
| 3 months | MGS | Hgn3C.0304 - Eubacteriales sp. |
| 3 months | MGS | Hgn3C.0306 - Bacteroides nordii |
| 3 months | MGS | Hgn3C.0307 - Veillonella atypica |
| 3 months | MGS | Hgn3C.0308 - [Clostridium] symbiosum |
| 3 months | MGS | Hgn3C.0309 - Eubacteriales sp. |
| 3 months | MGS | Hgn3C.0310 - Ligilactobacillus ruminis |
| 3 months | MGS | Hgn3C.0313 - Ruminococcus bromii |
| 3 months | MGS | Hgn3C.0314 - Ruminococcus sp. |
| 3 months | MGS | Hgn3C.0317 - Intestinimonas butyriciproducens |
| 3 months | MGS | Hgn3C.0319 - Clostridiaceae sp. |
| 3 months | MGS | Hgn3C.0323 - Bacteroides sp. |
| 3 months | MGS | Hgn3C.0324 - Prevotella stercorea |
| 3 months | MGS | Hgn3C.0330 - Faecalibacillus intestinalis |
| 3 months | MGS | Hgn3C.0332 - Klebsiella pneumoniae subsp. pneumoniae |
| 3 months | MGS | Hgn3C.0334 - Bacteroides cellulosilyticus |
| 3 months | MGS | Hgn3C.0336 - Parabacteroides goldsteinii |
| 3 months | MGS | Hgn3C.0338 - Eubacteriales sp. |
| 3 months | MGS | Hgn3C.0348 - Streptococcus sp. |
| 3 months | MGS | Hgn3C.0350 - [Clostridium] symbiosum |
| 3 months | MGS | Hgn3C.0351 - Prevotella sp. |
| 3 months | MGS | Hgn3C.0354 - Sutterella seckii |
| 3 months | MGS | Hgn3C.0356 - Blautia sp. M29 |
| 3 months | MGS | Hgn3C.0361 - Dorea sp. AF24-7LB |
| 3 months | MGS | Hgn3C.0362 - Dysosmobacter sp. NSJ-60 |
| 3 months | MGS | Hgn3C.0365 - Eubacteriales sp. |
| 3 months | MGS | Hgn3C.0367 - Lachnospiraceae sp. |
| 3 months | MGS | Hgn3C.0369 - Prevotella sp. |
| 3 months | MGS | Hgn3C.0371 - Lachnospiraceae sp. |
| 3 months | MGS | Hgn3C.0373 - Enterocloster clostridioformis |
| 3 months | MGS | Hgn3C.0375 - Ellagibacter isourolithinifaciens |
| 3 months | MGS | Hgn3C.0381 - Clostridium sp. AM30-24 |
| 3 months | MGS | Hgn3C.0385 - Veillonella rogosae |
| 3 months | MGS | Hgn3C.0386 - Clostridium sp. AT4 |
| 3 months | MGS | Hgn3C.0388 - Clostridium sp. |
| 3 months | MGS | Hgn3C.0392 - Clostridia sp. |
| 3 months | MGS | Hgn3C.0394 - Hungatella hathewayi |
| 3 months | MGS | Hgn3C.0397 - Parabacteroides sp. |
| 3 months | MGS | Hgn3C.0400 - Prevotellamassilia timonensis |
| 3 months | MGS | Hgn3C.0405 - Bacteroides ovatus |

| | | |
|---|---|---|
| 3 months | MGS | Hgn3C.0407 - Eubacteriales sp. |
| 3 months | MGS | Hgn3C.0408 - Eubacteriales sp. |
| 3 months | MGS | Hgn3C.0413 - Blautia caecimuris |
| 3 months | MGS | Hgn3C.0415 - Clostridium sp. C5-48 |
| 3 months | MGS | Hgn3C.0417 - Eubacteriales sp. |
| 3 months | MGS | Hgn3C.0418 - Blautia sp. |
| 3 months | MGS | Hgn3C.0419 - Prevotella sp. 885 |
| 3 months | MGS | Hgn3C.0420 - Clostridium sp. 1001270J_160509_D11 |
| 3 months | MGS | Hgn3C.0430 - Enterocloster aldenensis |
| 3 months | MGS | Hgn3C.0436 - Eubacteriales sp. |
| 3 months | MGS | Hgn3C.0437 - Prevotella sp. |
| 3 months | MGS | Hgn3C.0438 - Coprobacillus cateniformis |
| 3 months | MGS | Hgn3C.0439 - Enterococcus faecalis |
| 3 months | MGS | Hgn3C.0442 - Holdemanella sp. |
| 3 months | MGS | Hgn3C.0453 - Blautia sp. Marseille-P3087 |
| 3 months | MGS | Hgn3C.0455 - Peptostreptococcaceae sp. |
| 3 months | MGS | Hgn3C.0458 - Eubacteriales sp. |
| 3 months | MGS | Hgn3C.0459 - Eubacteriales sp. |
| 3 months | MGS | Hgn3C.0464 - Bifidobacterium longum subsp. infantis |
| 3 months | MGS | Hgn3C.0468 - Clostridium sp. NSJ-42 |
| 3 months | MGS | Hgn3C.0470 - Eubacteriales sp. |
| 3 months | MGS | Hgn3C.0472 - Parabacteroides goldsteinii |
| 3 months | MGS | Hgn3C.0476 - Streptococcus sp. |
| 3 months | MGS | Hgn3C.0482 - Lachnospiraceae sp. |
| 3 months | MGS | Hgn3C.0493 - Eisenbergiella tayi |
| 3 months | MGS | Hgn3C.0494 - Prevotellaceae sp. |
| 3 months | MGS | Hgn3C.0499 - Bacteroides sp. |
| 3 months | MGS | Hgn3C.0500 - Anaerotignum sp. |
| 3 months | MGS | Hgn3C.0504 - Megasphaera sp. BL7 |
| 3 months | MGS | Hgn3C.0507 - [Clostridium] spiroforme |
| 3 months | MGS | Hgn3C.0514 - Blautia glucerasea |
| 3 months | MGS | Hgn3C.0523 - Phascolarctobacterium succinatutens |
| 3 months | MGS | Hgn3C.0527 - Sutterella sp. |
| 3 months | MGS | Hgn3C.0528 - Coprococcus sp. AM27-12LB |
| 3 months | MGS | Hgn3C.0533 - Flintibacter sp. KGMB00164 |
| 3 months | MGS | Hgn3C.0537 - Bifidobacterium dentium |
| 3 months | MGS | Hgn3C.0542 - [Clostridium] innocuum |
| 3 months | MGS | Hgn3C.0543 - Oscillospiraceae sp. |
| 3 months | MGS | Hgn3C.0549 - Longicatena caecimuris |
| 3 months | MGS | Hgn3C.0557 - Enterocloster bolteae |
| 3 months | MGS | Hgn3C.0558 - Olsenella sp. |
| 3 months | MGS | Hgn3C.0560 - Oscillospiraceae sp. |
| 3 months | MGS | Hgn3C.0573 - Prevotella sp. Marseille-P4119 |
| 3 months | MGS | Hgn3C.0584 - Bacteroides fragilis |
| 3 months | MGS | Hgn3C.0598 - Eubacteriales sp. |
| 3 months | MGS | Hgn3C.0604 - Megasphaera elsdenii |
| 3 months | MGS | Hgn3C.0608 - Lachnospiraceae sp. |
| 3 months | MGS | Hgn3C.0610 - Eubacteriales sp. |

| | | |
|---|---|---|
| 3 months | MGS | Hgn3C.0613 - Clostridiaceae sp. |
| 3 months | MGS | Hgn3C.0617 - Prevotella sp. |
| 3 months | MGS | Hgn3C.0629 - Collinsella intestinalis |
| 3 months | MGS | Hgn3C.0630 - Allisonella histaminiformans |
| 3 months | MGS | Hgn3C.0634 - Blautia schinkii |
| 3 months | MGS | Hgn3C.0636 - Anaerostipes caccae |
| 3 months | MGS | Hgn3C.0640 - Eubacterium sp. AF22-8LB |
| 3 months | MGS | Hgn3C.0649 - Lachnospiraceae sp. |
| 3 months | MGS | Hgn3C.0657 - Parolsenella catena |
| 3 months | MGS | Hgn3C.0664 - Blautia hansenii |
| 3 months | MGS | Hgn3C.0675 - Eubacteriales sp. |
| 3 months | MGS | Hgn3C.0676 - Bacteroides sp. |
| 3 months | MGS | Hgn3C.0677 - Bacteroides sp. |
| 3 months | MGS | Hgn3C.0679 - Holdemania filiformis |
| 3 months | MGS | Hgn3C.0680 - Eubacteriales sp. |
| 3 months | MGS | Hgn3C.0682 - [Clostridium] scindens |
| 3 months | MGS | Hgn3C.0683 - Clostridium disporicum |
| 3 months | MGS | Hgn3C.0693 - Hungatella hathewayi |
| 3 months | MGS | Hgn3C.0701 - Bacteroides sp. |
| 3 months | MGS | Hgn3C.0704 - [Clostridium] scindens |
| 3 months | MGS | Hgn3C.0711 - Enterocloster asparagiformis |
| 3 months | MGS | Hgn3C.0712 - Megasphaera micronuciformis |
| 3 months | MGS | Hgn3C.0713 - Veillonellales sp. |
| 3 months | MGS | Hgn3C.0714 - Intestinimonas sp. MSJ-38 |
| 3 months | MGS | Hgn3C.0715 - Desulfovibrio fairfieldensis |
| 3 months | MGS | Hgn3C.0718 - Eubacteriales sp. |
| 3 months | MGS | Hgn3C.0729 - Eubacteriales sp. |
| 3 months | MGS | Hgn3C.0731 - Eubacteriales sp. |
| 3 months | MGS | Hgn3C.0732 - Eubacterium ramulus |
| 3 months | MGS | Hgn3C.0736 - Coprococcus sp. |
| 3 months | MGS | Hgn3C.0737 - Alistipes timonensis |
| 3 months | MGS | Hgn3C.0740 - Anaerostipes sp. NSJ-7 |
| 3 months | MGS | Hgn3C.0741 - Blautia sp. OF01-4LB |
| 3 months | MGS | Hgn3C.0743 - Enterobacter sp. |
| 3 months | MGS | Hgn3C.0754 - Collinsella bouchesdurhonensis |
| 3 months | MGS | Hgn3C.0761 - Bacteroides sp. |
| 3 months | MGS | Hgn3C.0762 - Enterocloster lavalensis |
| 3 months | MGS | Hgn3C.0777 - Holdemanella sp. |
| 3 months | MGS | Hgn3C.0781 - Collinsella sp. |
| 3 months | MGS | Hgn3C.0782 - Streptococcus sp. |
| 3 months | MGS | Hgn3C.0790 - Oscillospiraceae sp. |
| 3 months | MGS | Hgn3C.0793 - Prevotellaceae sp. |
| 3 months | MGS | Hgn3C.0797 - Senegalimassilia sp. |
| 3 months | MGS | Hgn3C.0798 - Streptococcus vestibularis |
| 3 months | MGS | Hgn3C.0807 - Enorma massiliensis |
| 3 months | MGS | Hgn3C.0817 - Prevotella sp. |
| 3 months | MGS | Hgn3C.0818 - Rothia sp. |
| 3 months | MGS | Hgn3C.0829 - Dorea phocaeensis |

| | | |
|---|---|---|
| 3 months | MGS | Hgn3C.0832 - Veillonellales sp. |
| 3 months | MGS | Hgn3C.0833 - Eisenbergiella tayi |
| 3 months | MGS | Hgn3C.0837 - Anaerotruncus colihominis |
| 3 months | MGS | Hgn3C.0851 - Oscillospiraceae sp. |
| 3 months | MGS | Hgn3C.0855 - Eubacteriales sp. |
| 3 months | MGS | Hgn3C.0858 - Turicibacter sanguinis |
| 3 months | MGS | Hgn3C.0859 - Hydrogeniiclostidium mannosilyticum |
| 3 months | MGS | Hgn3C.0873 - Clostridium perfringens |
| 3 months | MGS | Hgn3C.0875 - Lachnospiraceae sp. |
| 3 months | MGS | Hgn3C.0886 - Bacteroides sp. |
| 3 months | MGS | Hgn3C.0889 - Clostridium paraputrificum |
| 3 months | MGS | Hgn3C.0890 - Enterobacter roggenkampii |
| 3 months | MGS | Hgn3C.0891 - Eubacteriales sp. |
| 3 months | MGS | Hgn3C.0893 - Phocea massiliensis |
| 3 months | MGS | Hgn3C.0908 - Eubacteriales sp. |
| 3 months | MGS | Hgn3C.0913 - Erysipelotrichaceae sp. |
| 3 months | MGS | Hgn3C.0921 - Bacteroides sp. |
| 3 months | MGS | Hgn3C.0948 - Enterobacter sp. |
| 3 months | MGS | Hgn3C.0978 - Veillonella sp. |
| 3 months | MGS | Hgn3C.0987 - Fusobacterium mortiferum |
| 3 months | MGS | Hgn3C.0996 - Oscillospiraceae sp. |
| 3 months | MGS | Hgn3C.1035 - Klebsiella sp. |
| 3 months | MGS | Hgn3C.1036 - Escherichia coli |
| 3 months | MGS | Hgn3C.1037 - Rothia sp. |
| 3 months | MGS | Hgn3C.1038 - Veillonella sp. |
| 3 months | MGS | Hgn3C.1040 - Lachnospiraceae sp. |
| 3 months | MGS | Hgn3C.1041 - Bacteroides stercoris |
| 3 months | MGS | Hgn3C.1042 - Dorea formicigenerans |
| 3 months | MGS | Hgn3C.1043 - Clostridium phoceensis |
| 3 months | MGS | Hgn3C.1045 - Lactococcus lactis subsp. lactis |
| 3 months | MGS | Hgn3C.1049 - Enterococcus faecium |
| 3 months | MGS | Hgn3C.1050 - Staphylococcus epidermidis |
| 3 months | MGS | Hgn3C.1051 - Holdemania massiliensis |
| 3 months | MGS | Hgn3C.1054 - Streptococcus lutetiensis |
| 3 months | MGS | Hgn3C.1058 - Mediterraneibacter glycyrrhizinilyticus |
| 3 months | MGS | Hgn3C.1059 - Faecalicatena sp. |
| 3 months | MGS | Hgn3C.1063 - Enterococcus avium |
| 3 months | MGS | Hgn3C.1065 - Klebsiella variicola subsp. variicola |
| 3 months | MGS | Hgn3C.1067 - Lachnospiraceae sp. |
| 3 months | MGS | Hgn3C.1073 - Streptococcus gallolyticus subsp. gallolyticus |
| 3 months | MGS | Hgn3C.1081 - Lacticaseibacillus rhamnosus |
| 3 months | MGS | Hgn3C.1082 - Lacticaseibacillus paracasei subsp. paracasei |
| 3 months | MGS | Hgn3C.1084 - Massilimicrobiota timonensis |
| 3 months | MGS | Hgn3C.1087 - Ligilactobacillus salivarius |
| 3 months | MGS | Hgn3C.1090 - Turicibacter sanguinis |
| 3 months | MGS | Hgn3C.1093 - Enterobacter kobei |
| 3 months | MGS | Hgn3C.1110 - Porphyromonas sp. |
| 3 months | MGS | Hgn3C.1117 - Atopobiaceae sp. |

| | | |
|---|---|---|
| 3 months | MGS | Hgn3C.1120 - Actinomycetaceae sp. |
| 3 months | MGS | Hgn3C.1148 - Eggerthellales sp. |
| 3 months | MGS | Hgn3C.1153 - Eubacterium sp. OM08-24 |
| 3 months | MGS | Hgn3C.1154 - Lactococcus garvieae |
| 3 months | MGS | Hgn3C.1180 - Enterocloster citroniae |
| 3 months | MGS | Hgn3C.1188 - Actinomyces oris |
| 3 months | MGS | Hgn3C.1190 - Fenollaria massiliensis |
| 3 months | MGS | Hgn3C.1193 - Atopobium sp. |
| 3 months | MGS | Hgn3C.1196 - Blautia marasmi |
| 3 months | MGS | Hgn3C.1197 - Blautia hansenii |
| 3 months | MGS | Hgn3C.1210 - Dielma sp. |
| 3 months | MGS | Hgn3C.1211 - Lachnoclostridium pacaense |
| 3 months | MGS | Hgn3C.1214 - Corynebacterium tuberculostearicum |
| 3 months | MGS | Hgn3C.1219 - Enterobacter sp. NFIX58 |
| 3 months | MGS | Hgn3C.1229 - Escherichia sp. 93.0750 |
| 3 months | MGS | Hgn3C.1230 - Eubacterium limosum |
| 3 months | MGS | Hgn3C.1234 - Granulicatella adiacens |
| 3 months | MGS | Hgn3C.1238 - Klebsiella quasivariicola |
| 3 months | MGS | Hgn3C.1241 - Limosilactobacillus reuteri subsp. reuteri |
| 3 months | MGS | Hgn3C.1242 - Limosilactobacillus vaginalis |
| 3 months | MGS | Hgn3C.1248 - Neisseria sp. HMSC061E12 |
| 3 months | MGS | Hgn3C.1258 - Rothia sp. HMSC072E10 |
| 3 months | MGS | Hgn3C.1264 - Staphylococcus hominis subsp. hominis |
| 3 months | MGS | Hgn3C.1268 - Streptococcus sp. |
| 3 months | MGS | Hgn3C.1269 - Streptococcus constellatus subsp. constellatus |
| 3 months | MGS | Hgn3C.1270 - Streptococcus sp. |
| 3 months | MGS | Hgn3C.1275 - Lachnospiraceae sp. |
| 3 months | MGS | Hgn3C.1277 - Lachnospiraceae sp. |
| 3 months | MGS | Hgn3C.1282 - Terrisporobacter othiniensis |
| 3 months | MGS | Hgn3C.1286 - Corynebacterium variabile |
| 3 months | MGS | Hgn3C.1288 - Enterobacter hormaechei |
| 3 months | MGS | Hgn3C.1299 - Erysipelotrichaceae sp. |
| 3 months | MGS | Hgn3C.1300 - Rothia sp. |
| 3 months | MGS | Hgn3C.1302 - Schaalia sp. |
| 3 months | MGS | Hgn3C.1303 - Clostridium paraputrificum |
| 3 months | MGS | Hgn3C.1305 - Isoptericola variabilis |
| 3 months | MGS | Hgn3C.1307 - Bifidobacterium longum subsp. spp |
| 3 months | MGS | Hgn3C.1309 - Enterobacter cloacae subsp. cloacae |
| 3 months | MGS | Hgn3C.1310 - Erysipelotrichaceae sp. |
| 3 months | MGS | Hgn3C.1311 - Lachnospiraceae sp. |
| 3 months | MGS | Hgn3C.1312 - Candida tropicalis |
| 3 months | MGS | Hgn3C.1315 - Eubacteriales sp. |
| 3 months | MGS | Hgn3C.1316 - Eubacteriales sp. |
| 3 months | MGS | Hgn3C.1321 - Veillonellaceae sp. |
| 3 months | MGS | Hgn3C.1322 - Enterococcus hirae |
| 3 months | MGS | Hgn3C.1323 - Lachnospiraceae sp. |
| 3 months | MGS | Hgn3C.1324 - Eubacterium sp. c-25 |
| 3 months | MGS | Hgn3C.1325 - Luxibacter massiliensis |

| | | |
|---|---|---|
| 3 months | MGS | Hgn3C.1326 - Eggerthella guodeyinii |
| 3 months | MGS | Hgn3C.1329 - Eubacteriales sp. |
| 3 months | MGS | Hgn3C.1330 - Lachnospiraceae sp. |
| 3 months | MGS | Hgn3C.1331 - Lachnospiraceae sp. |
| 3 months | MGS | Hgn3C.1334 - Eubacteriales sp. |
| 3 months | MGS | Hgn3C.1336 - Eubacteriales sp. |
| 3 months | MGS | Hgn3C.1338 - Erysipelatoclostridium sp. |
| 3 months | MGS | Hgn3C.1340 - Clostridiaceae sp. |
| 3 months | MGS | Hgn3C.1343 - Enterococcus italicus |
| 3 months | MGS | Hgn3C.1347 - [Clostridium] hylemonae |
| 3 months | MGS | Hgn3C.1349 - Corynebacterium falsenii |
| 3 months | MGS | Hgn3C.1351 - Actinomyces sp. ICM47 |
| 3 months | MGS | Hgn3C.1352 - Akkermansia sp. |
| 3 months | MGS | Hgn3C.1354 - Actinomyces sp. |
| 3 months | MGS | Hgn3C.1355 - Collinsella stercoris |
| 3 months | MGS | Hgn3C.1356 - Eubacteriales sp. |
| 3 months | MGS | Hgn3C.1358 - Lachnospiraceae sp. |
| 3 months | MGS | Hgn3C.1359 - Extibacter muris |
| 3 months | MGS | Hgn3C.1360 - Bifidobacterium crudilactis |
| 3 months | MGS | Hgn3C.1361 - Senegalimassilia anaerobia |
| 3 months | MGS | Hgn3C.1362 - Christensenella intestinihominis |
| 3 months | MGS | Hgn3C.1366 - Lachnospiraceae sp. |
| 3 months | MGS | Hgn3C.1369 - Lachnospiraceae sp. |
| 3 months | MGS | Hgn3C.1372 - Eubacteriales sp. |
| 3 months | MGS | Hgn3C.1373 - Eubacteriales sp. |
| 3 months | MGS | Hgn3C.1375 - Atopobiaceae sp. |
| 3 months | MGS | Hgn3C.1376 - Kluyvera sp. CRP |
| 3 months | MGS | Hgn3C.1377 - Collinsella tanakaei |
| 3 months | MGS | Hgn3C.1378 - Collinsella sp. AF08-23 |
| 3 months | MGS | Hgn3C.1379 - Gordonibacter urolithinfaciens |
| 3 months | MGS | Hgn3C.1380 - Kluyvera georgiana |
| 3 months | MGS | Hgn3C.1382 - Oscillospiraceae sp. |
| 3 months | MGS | Hgn3C.1384 - Anaerococcus sp. |
| 3 months | MGS | Hgn3C.1386 - Veillonella sp. |
| 3 months | MGS | Hgn3C.1387 - Alistipes sp. |
| 3 months | MGS | Hgn3C.1388 - Streptococcus infantarius subsp. infantarius |
| 3 months | MGS | Hgn3C.1390 - Erysipelatoclostridium sp. |
| 3 months | MGS | Hgn3C.1393 - Anaerofustis stercorihominis |
| 3 months | MGS | Hgn3C.1394 - Eubacteriales sp. |
| 3 months | MGS | Hgn3C.1398 - Actinomyces sp. |
| 3 months | MGS | Hgn3C.1400 - Eubacterium sp. |
| 3 months | MGS | Hgn3C.1401 - Eubacteriales sp. |
| 3 months | MGS | Hgn3C.1402 - Collinsella phocaeensis |
| 3 months | MGS | Hgn3C.1403 - Eubacteriaceae sp. |
| 3 months | MGS | Hgn3C.1404 - Clostridium sp. |
| 3 months | MGS | Hgn3C.1406 - Slackia piriformis |
| 3 months | MGS | Hgn3C.1407 - Christensenella minuta |
| 3 months | MGS | Hgn3C.1408 - Eubacterium callanderi |

| | | |
|---|---|---|
| 3 months | MGS | Hgn3C.1409 - Eggerthella timonensis |
| 3 months | MGS | Hgn3C.1412 - Christensenella hongkongensis |
| 3 months | MGS | Hgn3C.1413 - Streptococcus sp. |
| 3 months | MGS | Hgn3C.1414 - Collinsella sp. |
| 3 months | MGS | Hgn3C.1415 - Klebsiella quasipneumoniae subsp. quasipneumoniae |
| 3 months | MGS | Hgn3C.1416 - Limosilactobacillus fermentum |
| 3 months | MGS | Hgn3C.1418 - Carnobacteriaceae sp. |
| 3 months | MGS | Hgn3C.1419 - Agathobaculum sp. NSJ-28 |
| 3 months | MGS | Hgn3C.1420 - Eubacteriales sp. |
| 3 months | MGS | Hgn3C.1423 - Lachnospiraceae sp. |
| 3 months | MGS | Hgn3C.1424 - Mogibacterium sp. NSJ-24 |
| 3 months | MGS | Hgn3C.1425 - Eubacteriales sp. |
| 3 months | MGS | Hgn3C.1426 - Limosilactobacillus oris |
| 3 months | MGS | Hgn3C.1429 - Lachnospiraceae sp. |
| 3 months | MGS | Hgn3C.1431 - Bacteroidales sp. |
| 3 months | MGS | Hgn3C.1434 - Blautia sp. |
| 3 months | MGS | Hgn3C.1435 - Acinetobacter baumannii |
| 3 months | MGS | Hgn3C.1436 - Bacteria sp. |
| 3 months | MGS | Hgn3C.1437 - Candidatus Stoquefichus sp. SB1 |
| 3 months | MGS | Hgn3C.1438 - Dielma fastidiosa |
| 3 months | MGS | Hgn3C.1439 - Streptococcus sp. IMAU 99161 |
| 3 months | MGS | Hgn3C.1441 - Lancefieldella sp. |
| 3 months | MGS | Hgn3C.1443 - Streptococcus sp. |
| 3 months | MGS | Hgn3C.1444 - Ruminococcus sp. 1001175B_160314_E1 |
| 3 months | MGS | Hgn3C.1446 - Eubacteriales sp. |
| 3 months | MGS | Hgn3C.1447 - Bacteria sp. |
| 3 months | MGS | Hgn3C.1448 - Leuconostoc falkenbergense |
| 3 months | MGS | Hgn3C.1450 - Lactococcus sp. NH2-7C |
| 3 months | MGS | Hgn3C.1452 - Raoultibacter timonensis |
| 3 months | MGS | Hgn3C.1453 - Collinsella sp. |
| 3 months | MGS | Hgn3C.1455 - Enterocloster sp. |
| 3 months | MGS | Hgn3C.1456 - Eubacteriales sp. |
| 3 months | MGS | Hgn3C.1460 - Blautia sp. |
| 3 months | MGS | Hgn3C.1463 - Enterobacter sp. |
| 3 months | MGS | Hgn3C.1465 - Clostridia sp. |
| 3 months | MGS | Hgn3C.1467 - Eubacteriales sp. |
| 3 months | MGS | Hgn3C.1468 - Staphylococcus aureus |
| 3 months | MGS | Hgn3C.1469 - Lachnospiraceae sp. |
| 3 months | MGS | Hgn3C.1471 - Eggerthella sp. |
| 3 months | MGS | Hgn3C.1472 - Amedibacillus dolichus |
| 3 months | genus | Acinetobacter |
| 3 months | genus | Actinomyces |
| 3 months | genus | Adlercreutzia |
| 3 months | genus | Agathobaculum |
| 3 months | genus | Akkermansia |
| 3 months | genus | Alistipes |
| 3 months | genus | Allisonella |
| 3 months | genus | Amedibacillus |

| | | |
|---|---|---|
| 3 months | genus | Anaerobutyricum |
| 3 months | genus | Anaerococcus |
| 3 months | genus | Anaerofustis |
| 3 months | genus | Anaerostipes |
| 3 months | genus | Anaerotignum |
| 3 months | genus | Anaerotruncus |
| 3 months | genus | Atopobium |
| 3 months | genus | Bacteroides |
| 3 months | genus | Barnesiella |
| 3 months | genus | Bifidobacterium |
| 3 months | genus | Bilophila |
| 3 months | genus | Blautia |
| 3 months | genus | Butyricicoccus |
| 3 months | genus | Butyricimonas |
| 3 months | genus | Candida |
| 3 months | genus | Candidatus Stoquefichus |
| 3 months | genus | Catenibacterium |
| 3 months | genus | Christensenella |
| 3 months | genus | Citrobacter |
| 3 months | genus | Clostridium |
| 3 months | genus | Collinsella |
| 3 months | genus | Coprobacillus |
| 3 months | genus | Coprococcus |
| 3 months | genus | Corynebacterium |
| 3 months | genus | Desulfovibrio |
| 3 months | genus | Dialister |
| 3 months | genus | Dielma |
| 3 months | genus | Dorea |
| 3 months | genus | Duodenibacillus |
| 3 months | genus | Dysosmobacter |
| 3 months | genus | Eggerthella |
| 3 months | genus | Eisenbergiella |
| 3 months | genus | Ellagibacter |
| 3 months | genus | Enorma |
| 3 months | genus | Enterobacter |
| 3 months | genus | Enterocloster |
| 3 months | genus | Enterococcus |
| 3 months | genus | Erysipelatoclostridium |
| 3 months | genus | Escherichia |
| 3 months | genus | Eubacterium |
| 3 months | genus | Evtepia |
| 3 months | genus | Extibacter |
| 3 months | genus | Faecalibacillus |
| 3 months | genus | Faecalibacterium |
| 3 months | genus | Faecalicatena |
| 3 months | genus | Fenollaria |
| 3 months | genus | Flavonifractor |
| 3 months | genus | Flintibacter |

| | | |
|---|---|---|
| 3 months | genus | Fusicatenibacter |
| 3 months | genus | Fusobacterium |
| 3 months | genus | Gemmiger |
| 3 months | genus | Gordonibacter |
| 3 months | genus | Granulicatella |
| 3 months | genus | Haemophilus |
| 3 months | genus | Holdemanella |
| 3 months | genus | Holdemania |
| 3 months | genus | Hungatella |
| 3 months | genus | Hydrogeniiclostidium |
| 3 months | genus | Hydrogenoanaerobacterium |
| 3 months | genus | Intestinibacter |
| 3 months | genus | Intestinimonas |
| 3 months | genus | Isoptericola |
| 3 months | genus | Klebsiella |
| 3 months | genus | Kluyvera |
| 3 months | genus | Lachnoclostridium |
| 3 months | genus | Lachnospira |
| 3 months | genus | Lacrimispora |
| 3 months | genus | Lacticaseibacillus |
| 3 months | genus | Lactobacillus |
| 3 months | genus | Lactococcus |
| 3 months | genus | Lancefieldella |
| 3 months | genus | Leuconostoc |
| 3 months | genus | Ligilactobacillus |
| 3 months | genus | Limosilactobacillus |
| 3 months | genus | Longicatena |
| 3 months | genus | Luxibacter |
| 3 months | genus | Massilimicrobiota |
| 3 months | genus | Mediterraneibacter |
| 3 months | genus | Megamonas |
| 3 months | genus | Megasphaera |
| 3 months | genus | Mogibacterium |
| 3 months | genus | Neisseria |
| 3 months | genus | Odoribacter |
| 3 months | genus | Olsenella |
| 3 months | genus | Oscillibacter |
| 3 months | genus | Parabacteroides |
| 3 months | genus | Paraprevotella |
| 3 months | genus | Parolsenella |
| 3 months | genus | Phascolarctobacterium |
| 3 months | genus | Phocaeicola |
| 3 months | genus | Phocea |
| 3 months | genus | Porphyromonas |
| 3 months | genus | Prevotella |
| 3 months | genus | Prevotellamassilia |
| 3 months | genus | Raoultibacter |
| 3 months | genus | Romboutsia |

| | | |
|---|---|---|
| 3 months | genus | Roseburia |
| 3 months | genus | Rothia |
| 3 months | genus | Ruminococcus |
| 3 months | genus | Ruthenibacterium |
| 3 months | genus | Schaalia |
| 3 months | genus | Sellimonas |
| 3 months | genus | Senegalimassilia |
| 3 months | genus | Slackia |
| 3 months | genus | Staphylococcus |
| 3 months | genus | Streptococcus |
| 3 months | genus | Subdoligranulum |
| 3 months | genus | Sutterella |
| 3 months | genus | Terrisporobacter |
| 3 months | genus | Turicibacter |
| 3 months | genus | Tyzzerella |
| 3 months | genus | Veillonella |
| 3 months | genus | Vescimonas |
| 3 months | genus | Waltera |
| 3 months | family | Acidaminococcaceae |
| 3 months | family | Actinomycetaceae |
| 3 months | family | Akkermansiaceae |
| 3 months | family | Atopobiaceae |
| 3 months | family | Bacteroidaceae |
| 3 months | family | Barnesiellaceae |
| 3 months | family | Bifidobacteriaceae |
| 3 months | family | Carnobacteriaceae |
| 3 months | family | Christensenellaceae |
| 3 months | family | Clostridiaceae |
| 3 months | family | Coprobacillaceae |
| 3 months | family | Coriobacteriaceae |
| 3 months | family | Corynebacteriaceae |
| 3 months | family | Debaryomycetaceae |
| 3 months | family | Desulfovibrionaceae |
| 3 months | family | Eggerthellaceae |
| 3 months | family | Enterobacteriaceae |
| 3 months | family | Enterococcaceae |
| 3 months | family | Erysipelotrichaceae |
| 3 months | family | Eubacteriaceae |
| 3 months | family | Eubacteriales Family XIII. Incertae Sedis |
| 3 months | family | Fusobacteriaceae |
| 3 months | family | Lachnospiraceae |
| 3 months | family | Lactobacillaceae |
| 3 months | family | Micrococcaceae |
| 3 months | family | Moraxellaceae |
| 3 months | family | Neisseriaceae |
| 3 months | family | Odoribacteraceae |
| 3 months | family | Oscillospiraceae |
| 3 months | family | Pasteurellaceae |

| | | |
|---|---|---|
| 3 months | family | Peptoniphilaceae |
| 3 months | family | Peptostreptococcaceae |
| 3 months | family | Porphyromonadaceae |
| 3 months | family | Prevotellaceae |
| 3 months | family | Promicromonosporaceae |
| 3 months | family | Rikenellaceae |
| 3 months | family | Selenomonadaceae |
| 3 months | family | Staphylococcaceae |
| 3 months | family | Streptococcaceae |
| 3 months | family | Sutterellaceae |
| 3 months | family | Tannerellaceae |
| 3 months | family | Turicibacteraceae |
| 3 months | family | Veillonellaceae |
| 3 months | phylum | Actinobacteria |
| 3 months | phylum | Ascomycota |
| 3 months | phylum | Bacteroidetes |
| 3 months | phylum | Firmicutes |
| 3 months | phylum | Fusobacteria |
| 3 months | phylum | Proteobacteria |
| 3 months | phylum | Verrucomicrobia |
| 6 months | MGS | Hgn3C.0001 - Phocaeicola vulgatus |
| 6 months | MGS | Hgn3C.0002 - Bacteroides uniformis |
| 6 months | MGS | Hgn3C.0003 - Lachnospiraceae sp. |
| 6 months | MGS | Hgn3C.0004 - Blautia wexlerae |
| 6 months | MGS | Hgn3C.0005 - Fusicatenibacter saccharivorans |
| 6 months | MGS | Hgn3C.0006 - Bacteroides ovatus |
| 6 months | MGS | Hgn3C.0007 - Parabacteroides distasonis |
| 6 months | MGS | Hgn3C.0008 - Alistipes putredinis |
| 6 months | MGS | Hgn3C.0009 - Faecalibacterium longum CLA-AA-H243 |
| 6 months | MGS | Hgn3C.0010 - Anaerostipes hadrus |
| 6 months | MGS | Hgn3C.0011 - Roseburia faecis |
| 6 months | MGS | Hgn3C.0012 - Lachnospira sp. |
| 6 months | MGS | Hgn3C.0013 - Ruminococcus bromii |
| 6 months | MGS | Hgn3C.0014 - Lacrimispora saccharolytica |
| 6 months | MGS | Hgn3C.0015 - Faecalibacterium sp. Marseille-Q4896 |
| 6 months | MGS | Hgn3C.0016 - Parabacteroides merdae |
| 6 months | MGS | Hgn3C.0017 - Subdoligranulum sp. APC924/74 |
| 6 months | MGS | Hgn3C.0018 - Faecalibacterium prausnitzii |
| 6 months | MGS | Hgn3C.0019 - Phocaeicola dorei |
| 6 months | MGS | Hgn3C.0020 - Collinsella aerofaciens |
| 6 months | MGS | Hgn3C.0021 - Bifidobacterium longum subsp. longum |
| 6 months | MGS | Hgn3C.0022 - Alistipes shahii |
| 6 months | MGS | Hgn3C.0023 - Bacteroides xylanisolvens |
| 6 months | MGS | Hgn3C.0024 - Bacteroides caccae |
| 6 months | MGS | Hgn3C.0025 - Ruminococcus bicirculans |
| 6 months | MGS | Hgn3C.0026 - Dorea longicatena |
| 6 months | MGS | Hgn3C.0027 - Barnesiella intestinihominis |
| 6 months | MGS | Hgn3C.0028 - Oscillibacter sp. ER4 |

| | | |
|---|---|---|
| 6 months | MGS | Hgn3C.0029 - Anaerobutyricum hallii |
| 6 months | MGS | Hgn3C.0030 - Faecalibacterium prausnitzii |
| 6 months | MGS | Hgn3C.0031 - Blautia massiliensis |
| 6 months | MGS | Hgn3C.0032 - Alistipes onderdonkii subsp. vulgaris |
| 6 months | MGS | Hgn3C.0033 - Gemmiger formicilis |
| 6 months | MGS | Hgn3C.0034 - Roseburia inulinivorans |
| 6 months | MGS | Hgn3C.0035 - Odoribacter splanchnicus |
| 6 months | MGS | Hgn3C.0036 - Bacteroides thetaiotaomicron |
| 6 months | MGS | Hgn3C.0037 - Roseburia intestinalis |
| 6 months | MGS | Hgn3C.0038 - Bifidobacterium adolescentis |
| 6 months | MGS | Hgn3C.0039 - Coprococcus comes |
| 6 months | MGS | Hgn3C.0040 - Eubacteriales sp. |
| 6 months | MGS | Hgn3C.0041 - Ruminococcus sp. |
| 6 months | MGS | Hgn3C.0042 - Blautia faecis |
| 6 months | MGS | Hgn3C.0043 - Lachnoclostridium sp. 210928-DFI.6.3 |
| 6 months | MGS | Hgn3C.0044 - Lachnospira pectinoschiza |
| 6 months | MGS | Hgn3C.0045 - Faecalibacterium sp. |
| 6 months | MGS | Hgn3C.0046 - Clostridium sp. AF37-5 |
| 6 months | MGS | Hgn3C.0047 - Dysosmobacter sp. BX15 |
| 6 months | MGS | Hgn3C.0048 - Akkermansia muciniphila |
| 6 months | MGS | Hgn3C.0049 - Eubacteriales sp. |
| 6 months | MGS | Hgn3C.0050 - Escherichia coli |
| 6 months | MGS | Hgn3C.0051 - Alistipes communis |
| 6 months | MGS | Hgn3C.0052 - Phocaeicola massiliensis |
| 6 months | MGS | Hgn3C.0053 - Eubacteriales sp. |
| 6 months | MGS | Hgn3C.0054 - Prevotella copri |
| 6 months | MGS | Hgn3C.0055 - Alistipes finegoldii |
| 6 months | MGS | Hgn3C.0056 - Bacteroides fragilis |
| 6 months | MGS | Hgn3C.0057 - Oscillibacter sp. MSJ-31 |
| 6 months | MGS | Hgn3C.0058 - Lachnospiraceae sp. |
| 6 months | MGS | Hgn3C.0059 - Bacteroides cellulosilyticus |
| 6 months | MGS | Hgn3C.0060 - Eubacteriales sp. |
| 6 months | MGS | Hgn3C.0061 - Faecalibacillus sp. TM498 |
| 6 months | MGS | Hgn3C.0062 - Waltera intestinalis |
| 6 months | MGS | Hgn3C.0063 - Eubacteriales sp. |
| 6 months | MGS | Hgn3C.0064 - Blautia sp. DFI.9.9 |
| 6 months | MGS | Hgn3C.0065 - Blautia obeum |
| 6 months | MGS | Hgn3C.0066 - Oscillospiraceae sp. |
| 6 months | MGS | Hgn3C.0067 - Roseburia hominis |
| 6 months | MGS | Hgn3C.0068 - [Ruminococcus] lactaris |
| 6 months | MGS | Hgn3C.0069 - Oscillibacter sp. KLE 1728 |
| 6 months | MGS | Hgn3C.0070 - Faecalibacterium sp. OF04-11AC |
| 6 months | MGS | Hgn3C.0071 - Faecalicatena fissicatena |
| 6 months | MGS | Hgn3C.0072 - Clostridium fessum |
| 6 months | MGS | Hgn3C.0073 - Blautia sp. |
| 6 months | MGS | Hgn3C.0074 - Clostridium sp. D43t1_170807_D5 |
| 6 months | MGS | Hgn3C.0075 - Agathobaculum butyriciproducens |
| 6 months | MGS | Hgn3C.0077 - Sutterella wadsworthensis |

| | | |
|---|---|---|
| 6 months | MGS | Hgn3C.0078 - Clostridium sp. MCC328 |
| 6 months | MGS | Hgn3C.0079 - [Ruminococcus] torques |
| 6 months | MGS | Hgn3C.0080 - Phascolarctobacterium faecium |
| 6 months | MGS | Hgn3C.0081 - Coprococcus eutactus |
| 6 months | MGS | Hgn3C.0082 - Roseburia sp. CLA-AA-H204 |
| 6 months | MGS | Hgn3C.0083 - Eubacteriales sp. |
| 6 months | MGS | Hgn3C.0084 - Flavonifractor plautii |
| 6 months | MGS | Hgn3C.0085 - Faecalibacterium sp. |
| 6 months | MGS | Hgn3C.0087 - Eubacteriales sp. |
| 6 months | MGS | Hgn3C.0088 - [Ruminococcus] gnavus |
| 6 months | MGS | Hgn3C.0089 - Paraprevotella clara |
| 6 months | MGS | Hgn3C.0092 - Lachnospiraceae sp. |
| 6 months | MGS | Hgn3C.0093 - Bilophila wadsworthia |
| 6 months | MGS | Hgn3C.0094 - Eubacteriales sp. |
| 6 months | MGS | Hgn3C.0095 - Eubacteriales sp. |
| 6 months | MGS | Hgn3C.0096 - Eubacteriales sp. |
| 6 months | MGS | Hgn3C.0097 - Eubacterium ventriosum |
| 6 months | MGS | Hgn3C.0098 - Coprococcus catus |
| 6 months | MGS | Hgn3C.0099 - Dorea longicatena |
| 6 months | MGS | Hgn3C.0100 - Bifidobacterium bifidum |
| 6 months | MGS | Hgn3C.0101 - Bifidobacterium pseudocatenulatum |
| 6 months | MGS | Hgn3C.0102 - Eubacteriales sp. |
| 6 months | MGS | Hgn3C.0103 - Holdemanella porci |
| 6 months | MGS | Hgn3C.0104 - Clostridium sp. AM32-2 |
| 6 months | MGS | Hgn3C.0105 - Vescimonas coprocola |
| 6 months | MGS | Hgn3C.0106 - Faecalibacterium sp. CLA-AA-H233 |
| 6 months | MGS | Hgn3C.0108 - Eubacteriales sp. |
| 6 months | MGS | Hgn3C.0111 - Eubacteriales sp. |
| 6 months | MGS | Hgn3C.0113 - Clostridium sp. MCC345 |
| 6 months | MGS | Hgn3C.0114 - Eubacteriales sp. |
| 6 months | MGS | Hgn3C.0115 - Eubacteriales sp. |
| 6 months | MGS | Hgn3C.0120 - Flavonifractor plautii |
| 6 months | MGS | Hgn3C.0121 - Oscillospiraceae sp. |
| 6 months | MGS | Hgn3C.0123 - Eubacteriales sp. |
| 6 months | MGS | Hgn3C.0124 - Ruthenibacterium lactatiformans |
| 6 months | MGS | Hgn3C.0125 - Sutterella sp. KLE1602 |
| 6 months | MGS | Hgn3C.0126 - Ruminococcus sp. BSD2780120874_150323_B10 |
| 6 months | MGS | Hgn3C.0127 - Faecalibacterium sp. |
| 6 months | MGS | Hgn3C.0129 - Haemophilus parainfluenzae |
| 6 months | MGS | Hgn3C.0130 - Phocaeicola plebeius |
| 6 months | MGS | Hgn3C.0131 - Streptococcus thermophilus |
| 6 months | MGS | Hgn3C.0132 - Eubacteriales sp. |
| 6 months | MGS | Hgn3C.0134 - Eubacteriales sp. |
| 6 months | MGS | Hgn3C.0135 - Firmicutes sp. |
| 6 months | MGS | Hgn3C.0136 - Alistipes ihumii |
| 6 months | MGS | Hgn3C.0137 - Clostridium sp. OF03-18AA |
| 6 months | MGS | Hgn3C.0138 - Anaerobutyricum soehngenii |
| 6 months | MGS | Hgn3C.0139 - Ruminococcus callidus |

| | | |
|---|---|---|
| 6 months | MGS | Hgn3C.0140 - Veillonella parvula |
| 6 months | MGS | Hgn3C.0141 - Adlercreutzia equolifaciens subsp. celatus |
| 6 months | MGS | Hgn3C.0142 - Eubacterium ramulus |
| 6 months | MGS | Hgn3C.0143 - Streptococcus salivarius |
| 6 months | MGS | Hgn3C.0144 - Eubacteriales sp. |
| 6 months | MGS | Hgn3C.0146 - Phascolarctobacterium succinatutens |
| 6 months | MGS | Hgn3C.0147 - Oscillospiraceae sp. |
| 6 months | MGS | Hgn3C.0149 - Eggerthellales sp. |
| 6 months | MGS | Hgn3C.0153 - Lachnospiraceae sp. |
| 6 months | MGS | Hgn3C.0154 - Prevotella copri |
| 6 months | MGS | Hgn3C.0155 - Blautia sp. |
| 6 months | MGS | Hgn3C.0157 - Eubacteriales sp. |
| 6 months | MGS | Hgn3C.0159 - Firmicutes sp. |
| 6 months | MGS | Hgn3C.0161 - Blautia sp. |
| 6 months | MGS | Hgn3C.0162 - Prevotella sp. |
| 6 months | MGS | Hgn3C.0168 - Evtepia gabavorous |
| 6 months | MGS | Hgn3C.0169 - Bacteroides intestinalis |
| 6 months | MGS | Hgn3C.0170 - Oscillospiraceae sp. |
| 6 months | MGS | Hgn3C.0171 - Catenibacterium mitsuokai |
| 6 months | MGS | Hgn3C.0174 - Lachnospiraceae sp. |
| 6 months | MGS | Hgn3C.0175 - Oscillospiraceae sp. |
| 6 months | MGS | Hgn3C.0176 - Alistipes indistinctus |
| 6 months | MGS | Hgn3C.0179 - Bacteroidales sp. |
| 6 months | MGS | Hgn3C.0181 - Alistipes onderdonkii |
| 6 months | MGS | Hgn3C.0182 - Butyricimonas faecihominis |
| 6 months | MGS | Hgn3C.0183 - Intestinibacter bartlettii |
| 6 months | MGS | Hgn3C.0184 - Eubacteriales sp. |
| 6 months | MGS | Hgn3C.0185 - Bifidobacterium catenulatum subsp. kashiwanohense |
| 6 months | MGS | Hgn3C.0186 - Butyricimonas virosa |
| 6 months | MGS | Hgn3C.0191 - Prevotella sp. |
| 6 months | MGS | Hgn3C.0192 - Erysipelatoclostridium ramosum |
| 6 months | MGS | Hgn3C.0193 - Eubacteriales sp. |
| 6 months | MGS | Hgn3C.0195 - Eubacteriales sp. |
| 6 months | MGS | Hgn3C.0201 - Veillonella dispar |
| 6 months | MGS | Hgn3C.0202 - Eggerthella lenta |
| 6 months | MGS | Hgn3C.0203 - Faecalibacterium sp. Marseille-Q3530 |
| 6 months | MGS | Hgn3C.0207 - Eubacteriales sp. |
| 6 months | MGS | Hgn3C.0208 - Romboutsia timonensis |
| 6 months | MGS | Hgn3C.0209 - Bifidobacterium breve |
| 6 months | MGS | Hgn3C.0211 - Eubacteriales sp. |
| 6 months | MGS | Hgn3C.0212 - Holdemanella biformis |
| 6 months | MGS | Hgn3C.0213 - Akkermansia sp. GGCC_0220 |
| 6 months | MGS | Hgn3C.0214 - Duodenibacillus massiliensis |
| 6 months | MGS | Hgn3C.0215 - Alistipes sp. cv1 |
| 6 months | MGS | Hgn3C.0217 - Eubacteriales sp. |
| 6 months | MGS | Hgn3C.0218 - Eubacteriales sp. |
| 6 months | MGS | Hgn3C.0219 - Tyzzerella nexilis |
| 6 months | MGS | Hgn3C.0220 - Clostridium sp. MCC334 |

| | | |
|---|---|---|
| 6 months | MGS | Hgn3C.0221 - Clostridium sp. |
| 6 months | MGS | Hgn3C.0222 - Parabacteroides merdae |
| 6 months | MGS | Hgn3C.0224 - Veillonellales sp. |
| 6 months | MGS | Hgn3C.0226 - Eubacteriales sp. |
| 6 months | MGS | Hgn3C.0230 - Gemmiger sp. |
| 6 months | MGS | Hgn3C.0232 - Megamonas funiformis |
| 6 months | MGS | Hgn3C.0236 - Oscillospiraceae sp. |
| 6 months | MGS | Hgn3C.0238 - Oscillospiraceae sp. |
| 6 months | MGS | Hgn3C.0239 - Blautia sp. BIOML-A1 |
| 6 months | MGS | Hgn3C.0242 - Enterocloster bolteae |
| 6 months | MGS | Hgn3C.0244 - Desulfovibrio piger |
| 6 months | MGS | Hgn3C.0245 - Blautia stercoris |
| 6 months | MGS | Hgn3C.0248 - Coprococcus catus |
| 6 months | MGS | Hgn3C.0249 - Clostridium sp. AM33-3 |
| 6 months | MGS | Hgn3C.0250 - Eubacteriales sp. |
| 6 months | MGS | Hgn3C.0251 - Alistipes senegalensis |
| 6 months | MGS | Hgn3C.0253 - Waltera sp. |
| 6 months | MGS | Hgn3C.0260 - Hydrogenoanaerobacterium saccharovorans |
| 6 months | MGS | Hgn3C.0262 - Oscillospiraceae sp. |
| 6 months | MGS | Hgn3C.0264 - Clostridium sp. AF27-2AA |
| 6 months | MGS | Hgn3C.0266 - Dorea sp. AF36-15AT |
| 6 months | MGS | Hgn3C.0267 - Oscillospiraceae sp. |
| 6 months | MGS | Hgn3C.0269 - Flintibacter sp. NSJ-23 |
| 6 months | MGS | Hgn3C.0272 - Blautia faecicola |
| 6 months | MGS | Hgn3C.0273 - Slackia isoflavoniconvertens |
| 6 months | MGS | Hgn3C.0275 - Lachnospiraceae sp. |
| 6 months | MGS | Hgn3C.0281 - Eubacteriales sp. |
| 6 months | MGS | Hgn3C.0284 - Erysipelotrichaceae sp. |
| 6 months | MGS | Hgn3C.0286 - Clostridia sp. |
| 6 months | MGS | Hgn3C.0288 - Butyricimonas paravirosa |
| 6 months | MGS | Hgn3C.0290 - Sellimonas intestinalis |
| 6 months | MGS | Hgn3C.0295 - Eubacteriales sp. |
| 6 months | MGS | Hgn3C.0299 - Escherichia coli |
| 6 months | MGS | Hgn3C.0300 - Clostridia sp. |
| 6 months | MGS | Hgn3C.0301 - Prevotella hominis |
| 6 months | MGS | Hgn3C.0304 - Eubacteriales sp. |
| 6 months | MGS | Hgn3C.0306 - Bacteroides nordii |
| 6 months | MGS | Hgn3C.0307 - Veillonella atypica |
| 6 months | MGS | Hgn3C.0308 - [Clostridium] symbiosum |
| 6 months | MGS | Hgn3C.0309 - Eubacteriales sp. |
| 6 months | MGS | Hgn3C.0310 - Ligilactobacillus ruminis |
| 6 months | MGS | Hgn3C.0313 - Ruminococcus bromii |
| 6 months | MGS | Hgn3C.0314 - Ruminococcus sp. |
| 6 months | MGS | Hgn3C.0315 - Bacteroidia sp. |
| 6 months | MGS | Hgn3C.0317 - Intestinimonas butyriciproducens |
| 6 months | MGS | Hgn3C.0319 - Clostridiaceae sp. |
| 6 months | MGS | Hgn3C.0323 - Bacteroides sp. |
| 6 months | MGS | Hgn3C.0324 - Prevotella stercorea |

| | | |
|---|---|---|
| 6 months | MGS | Hgn3C.0330 - Faecalibacillus intestinalis |
| 6 months | MGS | Hgn3C.0332 - Klebsiella pneumoniae subsp. pneumoniae |
| 6 months | MGS | Hgn3C.0334 - Bacteroides cellulosilyticus |
| 6 months | MGS | Hgn3C.0336 - Parabacteroides goldsteinii |
| 6 months | MGS | Hgn3C.0337 - Eubacteriales sp. |
| 6 months | MGS | Hgn3C.0338 - Eubacteriales sp. |
| 6 months | MGS | Hgn3C.0345 - Oscillospiraceae sp. |
| 6 months | MGS | Hgn3C.0348 - Streptococcus sp. |
| 6 months | MGS | Hgn3C.0350 - [Clostridium] symbiosum |
| 6 months | MGS | Hgn3C.0351 - Prevotella sp. |
| 6 months | MGS | Hgn3C.0353 - Prevotella sp. P4-67 |
| 6 months | MGS | Hgn3C.0354 - Sutterella seckii |
| 6 months | MGS | Hgn3C.0356 - Blautia sp. M29 |
| 6 months | MGS | Hgn3C.0359 - Eubacterium sp. BX4 |
| 6 months | MGS | Hgn3C.0361 - Dorea sp. AF24-7LB |
| 6 months | MGS | Hgn3C.0362 - Dysosmobacter sp. NSJ-60 |
| 6 months | MGS | Hgn3C.0365 - Eubacteriales sp. |
| 6 months | MGS | Hgn3C.0367 - Lachnospiraceae sp. |
| 6 months | MGS | Hgn3C.0369 - Prevotella sp. |
| 6 months | MGS | Hgn3C.0371 - Lachnospiraceae sp. |
| 6 months | MGS | Hgn3C.0373 - Enterocloster clostridioformis |
| 6 months | MGS | Hgn3C.0375 - Ellagibacter isourolithinifaciens |
| 6 months | MGS | Hgn3C.0385 - Veillonella rogosae |
| 6 months | MGS | Hgn3C.0386 - Clostridium sp. AT4 |
| 6 months | MGS | Hgn3C.0388 - Clostridium sp. |
| 6 months | MGS | Hgn3C.0392 - Clostridia sp. |
| 6 months | MGS | Hgn3C.0394 - Hungatella hathewayi |
| 6 months | MGS | Hgn3C.0397 - Parabacteroides sp. |
| 6 months | MGS | Hgn3C.0400 - Prevotellamassilia timonensis |
| 6 months | MGS | Hgn3C.0405 - Bacteroides ovatus |
| 6 months | MGS | Hgn3C.0407 - Eubacteriales sp. |
| 6 months | MGS | Hgn3C.0413 - Blautia caecimuris |
| 6 months | MGS | Hgn3C.0417 - Eubacteriales sp. |
| 6 months | MGS | Hgn3C.0418 - Blautia sp. |
| 6 months | MGS | Hgn3C.0419 - Prevotella sp. 885 |
| 6 months | MGS | Hgn3C.0420 - Clostridium sp. 1001270J_160509_D11 |
| 6 months | MGS | Hgn3C.0430 - Enterocloster aldenensis |
| 6 months | MGS | Hgn3C.0435 - Eubacteriales sp. |
| 6 months | MGS | Hgn3C.0436 - Eubacteriales sp. |
| 6 months | MGS | Hgn3C.0437 - Prevotella sp. |
| 6 months | MGS | Hgn3C.0438 - Coprobacillus cateniformis |
| 6 months | MGS | Hgn3C.0439 - Enterococcus faecalis |
| 6 months | MGS | Hgn3C.0442 - Holdemanella sp. |
| 6 months | MGS | Hgn3C.0443 - Prevotella stercorea |
| 6 months | MGS | Hgn3C.0445 - Aeromonadales sp. |
| 6 months | MGS | Hgn3C.0453 - Blautia sp. Marseille-P3087 |
| 6 months | MGS | Hgn3C.0455 - Peptostreptococcaceae sp. |
| 6 months | MGS | Hgn3C.0458 - Eubacteriales sp. |

| | | |
|---|---|---|
| 6 months | MGS | Hgn3C.0459 - Eubacteriales sp. |
| 6 months | MGS | Hgn3C.0468 - Clostridium sp. NSJ-42 |
| 6 months | MGS | Hgn3C.0470 - Eubacteriales sp. |
| 6 months | MGS | Hgn3C.0472 - Parabacteroides goldsteinii |
| 6 months | MGS | Hgn3C.0476 - Streptococcus sp. |
| 6 months | MGS | Hgn3C.0482 - Lachnospiraceae sp. |
| 6 months | MGS | Hgn3C.0489 - Eubacteriales sp. |
| 6 months | MGS | Hgn3C.0493 - Eisenbergiella tayi |
| 6 months | MGS | Hgn3C.0494 - Prevotellaceae sp. |
| 6 months | MGS | Hgn3C.0495 - Ruminococcus sp. |
| 6 months | MGS | Hgn3C.0500 - Anaerotignum sp. |
| 6 months | MGS | Hgn3C.0501 - Oscillospiraceae sp. |
| 6 months | MGS | Hgn3C.0503 - Eubacteriales sp. |
| 6 months | MGS | Hgn3C.0504 - Megasphaera sp. BL7 |
| 6 months | MGS | Hgn3C.0507 - [Clostridium] spiroforme |
| 6 months | MGS | Hgn3C.0514 - Blautia glucerasea |
| 6 months | MGS | Hgn3C.0523 - Phascolarctobacterium succinatutens |
| 6 months | MGS | Hgn3C.0526 - Anaerostipes sp. |
| 6 months | MGS | Hgn3C.0527 - Sutterella sp. |
| 6 months | MGS | Hgn3C.0528 - Coprococcus sp. AM27-12LB |
| 6 months | MGS | Hgn3C.0533 - Flintibacter sp. KGMB00164 |
| 6 months | MGS | Hgn3C.0537 - Bifidobacterium dentium |
| 6 months | MGS | Hgn3C.0542 - [Clostridium] innocuum |
| 6 months | MGS | Hgn3C.0543 - Oscillospiraceae sp. |
| 6 months | MGS | Hgn3C.0549 - Longicatena caecimuris |
| 6 months | MGS | Hgn3C.0557 - Enterocloster bolteae |
| 6 months | MGS | Hgn3C.0558 - Olsenella sp. |
| 6 months | MGS | Hgn3C.0560 - Oscillospiraceae sp. |
| 6 months | MGS | Hgn3C.0573 - Prevotella sp. Marseille-P4119 |
| 6 months | MGS | Hgn3C.0584 - Bacteroides fragilis |
| 6 months | MGS | Hgn3C.0596 - Butyricimonas sp. |
| 6 months | MGS | Hgn3C.0598 - Eubacteriales sp. |
| 6 months | MGS | Hgn3C.0607 - Eubacteriales sp. |
| 6 months | MGS | Hgn3C.0608 - Lachnospiraceae sp. |
| 6 months | MGS | Hgn3C.0610 - Eubacteriales sp. |
| 6 months | MGS | Hgn3C.0613 - Clostridiaceae sp. |
| 6 months | MGS | Hgn3C.0616 - Lachnospiraceae sp. |
| 6 months | MGS | Hgn3C.0617 - Prevotella sp. |
| 6 months | MGS | Hgn3C.0629 - Collinsella intestinalis |
| 6 months | MGS | Hgn3C.0630 - Allisonella histaminiformans |
| 6 months | MGS | Hgn3C.0634 - Blautia schinkii |
| 6 months | MGS | Hgn3C.0635 - Eubacteriales sp. |
| 6 months | MGS | Hgn3C.0636 - Anaerostipes caccae |
| 6 months | MGS | Hgn3C.0640 - Eubacterium sp. AF22-8LB |
| 6 months | MGS | Hgn3C.0649 - Lachnospiraceae sp. |
| 6 months | MGS | Hgn3C.0657 - Parolsenella catena |
| 6 months | MGS | Hgn3C.0660 - Eubacteriales sp. |
| 6 months | MGS | Hgn3C.0661 - Alistipes sp. |

| | | |
|---|---|---|
| 6 months | MGS | Hgn3C.0664 - Blautia hansenii |
| 6 months | MGS | Hgn3C.0675 - Eubacteriales sp. |
| 6 months | MGS | Hgn3C.0676 - Bacteroides sp. |
| 6 months | MGS | Hgn3C.0677 - Bacteroides sp. |
| 6 months | MGS | Hgn3C.0679 - Holdemania filiformis |
| 6 months | MGS | Hgn3C.0680 - Eubacteriales sp. |
| 6 months | MGS | Hgn3C.0682 - [Clostridium] scindens |
| 6 months | MGS | Hgn3C.0683 - Clostridium disporicum |
| 6 months | MGS | Hgn3C.0693 - Hungatella hathewayi |
| 6 months | MGS | Hgn3C.0694 - Anaeromassilibacillus sp. An250 |
| 6 months | MGS | Hgn3C.0700 - Eubacteriales sp. |
| 6 months | MGS | Hgn3C.0701 - Bacteroides sp. |
| 6 months | MGS | Hgn3C.0704 - [Clostridium] scindens |
| 6 months | MGS | Hgn3C.0711 - Enterocloster asparagiformis |
| 6 months | MGS | Hgn3C.0712 - Megasphaera micronuciformis |
| 6 months | MGS | Hgn3C.0713 - Veillonellales sp. |
| 6 months | MGS | Hgn3C.0714 - Intestinimonas sp. MSJ-38 |
| 6 months | MGS | Hgn3C.0715 - Desulfovibrio fairfieldensis |
| 6 months | MGS | Hgn3C.0716 - Eubacteriales sp. |
| 6 months | MGS | Hgn3C.0718 - Eubacteriales sp. |
| 6 months | MGS | Hgn3C.0729 - Eubacteriales sp. |
| 6 months | MGS | Hgn3C.0731 - Eubacteriales sp. |
| 6 months | MGS | Hgn3C.0736 - Coprococcus sp. |
| 6 months | MGS | Hgn3C.0737 - Alistipes timonensis |
| 6 months | MGS | Hgn3C.0740 - Anaerostipes sp. NSJ-7 |
| 6 months | MGS | Hgn3C.0741 - Blautia sp. OF01-4LB |
| 6 months | MGS | Hgn3C.0743 - Enterobacter sp. |
| 6 months | MGS | Hgn3C.0749 - Eubacteriales sp. |
| 6 months | MGS | Hgn3C.0754 - Collinsella bouchesdurhonensis |
| 6 months | MGS | Hgn3C.0755 - Oscillibacter sp. |
| 6 months | MGS | Hgn3C.0761 - Bacteroides sp. |
| 6 months | MGS | Hgn3C.0762 - Enterocloster lavalensis |
| 6 months | MGS | Hgn3C.0777 - Holdemanella sp. |
| 6 months | MGS | Hgn3C.0781 - Collinsella sp. |
| 6 months | MGS | Hgn3C.0782 - Streptococcus sp. |
| 6 months | MGS | Hgn3C.0790 - Oscillospiraceae sp. |
| 6 months | MGS | Hgn3C.0793 - Prevotellaceae sp. |
| 6 months | MGS | Hgn3C.0794 - Bacteroides sp. |
| 6 months | MGS | Hgn3C.0797 - Senegalimassilia sp. |
| 6 months | MGS | Hgn3C.0798 - Streptococcus vestibularis |
| 6 months | MGS | Hgn3C.0814 - Massilimicrobiota timonensis |
| 6 months | MGS | Hgn3C.0817 - Prevotella sp. |
| 6 months | MGS | Hgn3C.0818 - Rothia sp. |
| 6 months | MGS | Hgn3C.0823 - Anaerotignum lactatifermentans |
| 6 months | MGS | Hgn3C.0829 - Dorea phocaeensis |
| 6 months | MGS | Hgn3C.0832 - Veillonellales sp. |
| 6 months | MGS | Hgn3C.0833 - Eisenbergiella tayi |
| 6 months | MGS | Hgn3C.0837 - Anaerotruncus colihominis |

| | | |
|---|---|---|
| 6 months | MGS | Hgn3C.0851 - Oscillospiraceae sp. |
| 6 months | MGS | Hgn3C.0855 - Eubacteriales sp. |
| 6 months | MGS | Hgn3C.0858 - Turicibacter sanguinis |
| 6 months | MGS | Hgn3C.0859 - Hydrogeniiclostidium mannosilyticum |
| 6 months | MGS | Hgn3C.0873 - Clostridium perfringens |
| 6 months | MGS | Hgn3C.0875 - Lachnospiraceae sp. |
| 6 months | MGS | Hgn3C.0886 - Bacteroides sp. |
| 6 months | MGS | Hgn3C.0889 - Clostridium paraputrificum |
| 6 months | MGS | Hgn3C.0890 - Enterobacter roggenkampii |
| 6 months | MGS | Hgn3C.0891 - Eubacteriales sp. |
| 6 months | MGS | Hgn3C.0893 - Phocea massiliensis |
| 6 months | MGS | Hgn3C.0908 - Eubacteriales sp. |
| 6 months | MGS | Hgn3C.0913 - Erysipelotrichaceae sp. |
| 6 months | MGS | Hgn3C.0921 - Bacteroides sp. |
| 6 months | MGS | Hgn3C.0922 - Eubacteriales sp. |
| 6 months | MGS | Hgn3C.0948 - Enterobacter sp. |
| 6 months | MGS | Hgn3C.0978 - Veillonella sp. |
| 6 months | MGS | Hgn3C.0987 - Fusobacterium mortiferum |
| 6 months | MGS | Hgn3C.0998 - Eubacteriales sp. |
| 6 months | MGS | Hgn3C.1035 - Klebsiella sp. |
| 6 months | MGS | Hgn3C.1036 - Escherichia coli |
| 6 months | MGS | Hgn3C.1037 - Rothia sp. |
| 6 months | MGS | Hgn3C.1038 - Veillonella sp. |
| 6 months | MGS | Hgn3C.1040 - Lachnospiraceae sp. |
| 6 months | MGS | Hgn3C.1041 - Bacteroides stercoris |
| 6 months | MGS | Hgn3C.1042 - Dorea formicigenerans |
| 6 months | MGS | Hgn3C.1043 - Clostridium phoceensis |
| 6 months | MGS | Hgn3C.1044 - Eubacteriales sp. |
| 6 months | MGS | Hgn3C.1045 - Lactococcus lactis subsp. lactis |
| 6 months | MGS | Hgn3C.1049 - Enterococcus faecium |
| 6 months | MGS | Hgn3C.1050 - Staphylococcus epidermidis |
| 6 months | MGS | Hgn3C.1051 - Holdemania massiliensis |
| 6 months | MGS | Hgn3C.1052 - Enterococcus gallinarum |
| 6 months | MGS | Hgn3C.1054 - Streptococcus lutetiensis |
| 6 months | MGS | Hgn3C.1058 - Mediterraneibacter glycyrrhizinilyticus |
| 6 months | MGS | Hgn3C.1059 - Faecalicatena sp. |
| 6 months | MGS | Hgn3C.1063 - Enterococcus avium |
| 6 months | MGS | Hgn3C.1065 - Klebsiella variicola subsp. variicola |
| 6 months | MGS | Hgn3C.1067 - Lachnospiraceae sp. |
| 6 months | MGS | Hgn3C.1073 - Streptococcus gallolyticus subsp. gallolyticus |
| 6 months | MGS | Hgn3C.1081 - Lacticaseibacillus rhamnosus |
| 6 months | MGS | Hgn3C.1082 - Lacticaseibacillus paracasei subsp. paracasei |
| 6 months | MGS | Hgn3C.1087 - Ligilactobacillus salivarius |
| 6 months | MGS | Hgn3C.1090 - Turicibacter sanguinis |
| 6 months | MGS | Hgn3C.1093 - Enterobacter kobei |
| 6 months | MGS | Hgn3C.1107 - Eubacteriales sp. |
| 6 months | MGS | Hgn3C.1110 - Porphyromonas sp. |
| 6 months | MGS | Hgn3C.1117 - Atopobiaceae sp. |

| | | |
|---|---|---|
| 6 months | MGS | Hgn3C.1120 - Actinomycetaceae sp. |
| 6 months | MGS | Hgn3C.1148 - Eggerthellales sp. |
| 6 months | MGS | Hgn3C.1154 - Lactococcus garvieae |
| 6 months | MGS | Hgn3C.1180 - Enterocloster citroniae |
| 6 months | MGS | Hgn3C.1188 - Actinomyces oris |
| 6 months | MGS | Hgn3C.1190 - Fenollaria massiliensis |
| 6 months | MGS | Hgn3C.1193 - Atopobium sp. |
| 6 months | MGS | Hgn3C.1196 - Blautia marasmi |
| 6 months | MGS | Hgn3C.1197 - Blautia hansenii |
| 6 months | MGS | Hgn3C.1203 - Citrobacter sp. CRE-46 |
| 6 months | MGS | Hgn3C.1210 - Dielma sp. |
| 6 months | MGS | Hgn3C.1211 - Lachnoclostridium pacaense |
| 6 months | MGS | Hgn3C.1214 - Corynebacterium tuberculostearicum |
| 6 months | MGS | Hgn3C.1218 - Enterobacter bugandensis |
| 6 months | MGS | Hgn3C.1219 - Enterobacter sp. NFIX58 |
| 6 months | MGS | Hgn3C.1225 - Enterococcus faecium |
| 6 months | MGS | Hgn3C.1229 - Escherichia sp. 93.0750 |
| 6 months | MGS | Hgn3C.1230 - Eubacterium limosum |
| 6 months | MGS | Hgn3C.1234 - Granulicatella adiacens |
| 6 months | MGS | Hgn3C.1238 - Klebsiella quasivariicola |
| 6 months | MGS | Hgn3C.1241 - Limosilactobacillus reuteri subsp. reuteri |
| 6 months | MGS | Hgn3C.1242 - Limosilactobacillus vaginalis |
| 6 months | MGS | Hgn3C.1248 - Neisseria sp. HMSC061E12 |
| 6 months | MGS | Hgn3C.1258 - Rothia sp. HMSC072E10 |
| 6 months | MGS | Hgn3C.1264 - Staphylococcus hominis subsp. hominis |
| 6 months | MGS | Hgn3C.1268 - Streptococcus sp. |
| 6 months | MGS | Hgn3C.1269 - Streptococcus constellatus subsp. constellatus |
| 6 months | MGS | Hgn3C.1270 - Streptococcus sp. |
| 6 months | MGS | Hgn3C.1275 - Lachnospiraceae sp. |
| 6 months | MGS | Hgn3C.1282 - Terrisporobacter othiniensis |
| 6 months | MGS | Hgn3C.1283 - Clostridium baratii |
| 6 months | MGS | Hgn3C.1286 - Corynebacterium variabile |
| 6 months | MGS | Hgn3C.1288 - Enterobacter hormaechei |
| 6 months | MGS | Hgn3C.1299 - Erysipelotrichaceae sp. |
| 6 months | MGS | Hgn3C.1300 - Rothia sp. |
| 6 months | MGS | Hgn3C.1302 - Schaalia sp. |
| 6 months | MGS | Hgn3C.1303 - Clostridium paraputrificum |
| 6 months | MGS | Hgn3C.1305 - Isoptericola variabilis |
| 6 months | MGS | Hgn3C.1307 - Bifidobacterium longum subsp. spp |
| 6 months | MGS | Hgn3C.1309 - Enterobacter cloacae subsp. cloacae |
| 6 months | MGS | Hgn3C.1310 - Erysipelotrichaceae sp. |
| 6 months | MGS | Hgn3C.1311 - Lachnospiraceae sp. |
| 6 months | MGS | Hgn3C.1312 - Candida tropicalis |
| 6 months | MGS | Hgn3C.1315 - Eubacteriales sp. |
| 6 months | MGS | Hgn3C.1316 - Eubacteriales sp. |
| 6 months | MGS | Hgn3C.1321 - Veillonellaceae sp. |
| 6 months | MGS | Hgn3C.1322 - Enterococcus hirae |
| 6 months | MGS | Hgn3C.1323 - Lachnospiraceae sp. |

| | | |
|---|---|---|
| 6 months | MGS | Hgn3C.1324 - Eubacterium sp. c-25 |
| 6 months | MGS | Hgn3C.1325 - Luxibacter massiliensis |
| 6 months | MGS | Hgn3C.1326 - Eggerthella guodeyinii |
| 6 months | MGS | Hgn3C.1329 - Eubacteriales sp. |
| 6 months | MGS | Hgn3C.1330 - Lachnospiraceae sp. |
| 6 months | MGS | Hgn3C.1331 - Lachnospiraceae sp. |
| 6 months | MGS | Hgn3C.1334 - Eubacteriales sp. |
| 6 months | MGS | Hgn3C.1336 - Eubacteriales sp. |
| 6 months | MGS | Hgn3C.1338 - Erysipelatoclostridium sp. |
| 6 months | MGS | Hgn3C.1340 - Clostridiaceae sp. |
| 6 months | MGS | Hgn3C.1343 - Enterococcus italicus |
| 6 months | MGS | Hgn3C.1347 - [Clostridium] hylemonae |
| 6 months | MGS | Hgn3C.1349 - Corynebacterium falsenii |
| 6 months | MGS | Hgn3C.1351 - Actinomyces sp. ICM47 |
| 6 months | MGS | Hgn3C.1352 - Akkermansia sp. |
| 6 months | MGS | Hgn3C.1354 - Actinomyces sp. |
| 6 months | MGS | Hgn3C.1355 - Collinsella stercoris |
| 6 months | MGS | Hgn3C.1358 - Lachnospiraceae sp. |
| 6 months | MGS | Hgn3C.1361 - Senegalimassilia anaerobia |
| 6 months | MGS | Hgn3C.1362 - Christensenella intestinihominis |
| 6 months | MGS | Hgn3C.1366 - Lachnospiraceae sp. |
| 6 months | MGS | Hgn3C.1369 - Lachnospiraceae sp. |
| 6 months | MGS | Hgn3C.1370 - Kluyvera genomosp. 3 |
| 6 months | MGS | Hgn3C.1372 - Eubacteriales sp. |
| 6 months | MGS | Hgn3C.1373 - Eubacteriales sp. |
| 6 months | MGS | Hgn3C.1375 - Atopobiaceae sp. |
| 6 months | MGS | Hgn3C.1376 - Kluyvera sp. CRP |
| 6 months | MGS | Hgn3C.1377 - Collinsella tanakaei |
| 6 months | MGS | Hgn3C.1378 - Collinsella sp. AF08-23 |
| 6 months | MGS | Hgn3C.1379 - Gordonibacter urolithinfaciens |
| 6 months | MGS | Hgn3C.1380 - Kluyvera georgiana |
| 6 months | MGS | Hgn3C.1381 - Eubacteriales sp. |
| 6 months | MGS | Hgn3C.1382 - Oscillospiraceae sp. |
| 6 months | MGS | Hgn3C.1384 - Anaerococcus sp. |
| 6 months | MGS | Hgn3C.1386 - Veillonella sp. |
| 6 months | MGS | Hgn3C.1388 - Streptococcus infantarius subsp. infantarius |
| 6 months | MGS | Hgn3C.1390 - Erysipelatoclostridium sp. |
| 6 months | MGS | Hgn3C.1392 - Enterobacter sp. JBIWA005 |
| 6 months | MGS | Hgn3C.1393 - Anaerofustis stercorihominis |
| 6 months | MGS | Hgn3C.1398 - Actinomyces sp. |
| 6 months | MGS | Hgn3C.1400 - Eubacterium sp. |
| 6 months | MGS | Hgn3C.1401 - Eubacteriales sp. |
| 6 months | MGS | Hgn3C.1402 - Collinsella phocaeensis |
| 6 months | MGS | Hgn3C.1403 - Eubacteriaceae sp. |
| 6 months | MGS | Hgn3C.1404 - Clostridium sp. |
| 6 months | MGS | Hgn3C.1406 - Slackia piriformis |
| 6 months | MGS | Hgn3C.1407 - Christensenella minuta |
| 6 months | MGS | Hgn3C.1408 - Eubacterium callanderi |

| | | |
|---|---|---|
| 6 months | MGS | Hgn3C.1409 - Eggerthella timonensis |
| 6 months | MGS | Hgn3C.1412 - Christensenella hongkongensis |
| 6 months | MGS | Hgn3C.1413 - Streptococcus sp. |
| 6 months | MGS | Hgn3C.1414 - Collinsella sp. |
| 6 months | MGS | Hgn3C.1415 - Klebsiella quasipneumoniae subsp. quasipneumoniae |
| 6 months | MGS | Hgn3C.1416 - Limosilactobacillus fermentum |
| 6 months | MGS | Hgn3C.1418 - Carnobacteriaceae sp. |
| 6 months | MGS | Hgn3C.1419 - Agathobaculum sp. NSJ-28 |
| 6 months | MGS | Hgn3C.1420 - Eubacteriales sp. |
| 6 months | MGS | Hgn3C.1423 - Lachnospiraceae sp. |
| 6 months | MGS | Hgn3C.1424 - Mogibacterium sp. NSJ-24 |
| 6 months | MGS | Hgn3C.1425 - Eubacteriales sp. |
| 6 months | MGS | Hgn3C.1426 - Limosilactobacillus oris |
| 6 months | MGS | Hgn3C.1434 - Blautia sp. |
| 6 months | MGS | Hgn3C.1435 - Acinetobacter baumannii |
| 6 months | MGS | Hgn3C.1436 - Bacteria sp. |
| 6 months | MGS | Hgn3C.1437 - Candidatus Stoquefichus sp. SB1 |
| 6 months | MGS | Hgn3C.1438 - Dielma fastidiosa |
| 6 months | MGS | Hgn3C.1439 - Streptococcus sp. IMAU 99161 |
| 6 months | MGS | Hgn3C.1441 - Lancefieldella sp. |
| 6 months | MGS | Hgn3C.1443 - Streptococcus sp. |
| 6 months | MGS | Hgn3C.1444 - Ruminococcus sp. 1001175B_160314_E1 |
| 6 months | MGS | Hgn3C.1446 - Eubacteriales sp. |
| 6 months | MGS | Hgn3C.1447 - Bacteria sp. |
| 6 months | MGS | Hgn3C.1450 - Lactococcus sp. NH2-7C |
| 6 months | MGS | Hgn3C.1452 - Raoultibacter timonensis |
| 6 months | MGS | Hgn3C.1453 - Collinsella sp. |
| 6 months | MGS | Hgn3C.1455 - Enterocloster sp. |
| 6 months | MGS | Hgn3C.1456 - Eubacteriales sp. |
| 6 months | MGS | Hgn3C.1460 - Blautia sp. |
| 6 months | MGS | Hgn3C.1463 - Enterobacter sp. |
| 6 months | MGS | Hgn3C.1465 - Clostridia sp. |
| 6 months | MGS | Hgn3C.1467 - Eubacteriales sp. |
| 6 months | MGS | Hgn3C.1468 - Staphylococcus aureus |
| 6 months | MGS | Hgn3C.1469 - Lachnospiraceae sp. |
| 6 months | MGS | Hgn3C.1471 - Eggerthella sp. |
| 6 months | MGS | Hgn3C.1472 - Amedibacillus dolichus |
| 6 months | genus | Acinetobacter |
| 6 months | genus | Actinomyces |
| 6 months | genus | Adlercreutzia |
| 6 months | genus | Agathobaculum |
| 6 months | genus | Akkermansia |
| 6 months | genus | Alistipes |
| 6 months | genus | Allisonella |
| 6 months | genus | Amedibacillus |
| 6 months | genus | Anaerobutyricum |
| 6 months | genus | Anaerococcus |
| 6 months | genus | Anaerofustis |

| | | |
|---|---|---|
| 6 months | genus | Anaeromassilibacillus |
| 6 months | genus | Anaerostipes |
| 6 months | genus | Anaerotignum |
| 6 months | genus | Anaerotruncus |
| 6 months | genus | Atopobium |
| 6 months | genus | Bacteroides |
| 6 months | genus | Barnesiella |
| 6 months | genus | Bifidobacterium |
| 6 months | genus | Bilophila |
| 6 months | genus | Blautia |
| 6 months | genus | Butyricicoccus |
| 6 months | genus | Butyricimonas |
| 6 months | genus | Candida |
| 6 months | genus | Candidatus Stoquefichus |
| 6 months | genus | Catenibacterium |
| 6 months | genus | Christensenella |
| 6 months | genus | Citrobacter |
| 6 months | genus | Clostridium |
| 6 months | genus | Collinsella |
| 6 months | genus | Coprobacillus |
| 6 months | genus | Coprococcus |
| 6 months | genus | Corynebacterium |
| 6 months | genus | Desulfovibrio |
| 6 months | genus | Dielma |
| 6 months | genus | Dorea |
| 6 months | genus | Duodenibacillus |
| 6 months | genus | Dysosmobacter |
| 6 months | genus | Eggerthella |
| 6 months | genus | Eisenbergiella |
| 6 months | genus | Ellagibacter |
| 6 months | genus | Enterobacter |
| 6 months | genus | Enterocloster |
| 6 months | genus | Enterococcus |
| 6 months | genus | Erysipelatoclostridium |
| 6 months | genus | Escherichia |
| 6 months | genus | Eubacterium |
| 6 months | genus | Evtepia |
| 6 months | genus | Faecalibacillus |
| 6 months | genus | Faecalibacterium |
| 6 months | genus | Faecalicatena |
| 6 months | genus | Fenollaria |
| 6 months | genus | Flavonifractor |
| 6 months | genus | Flintibacter |
| 6 months | genus | Fusicatenibacter |
| 6 months | genus | Fusobacterium |
| 6 months | genus | Gemmiger |
| 6 months | genus | Gordonibacter |
| 6 months | genus | Granulicatella |

| | | |
|---|---|---|
| 6 months | genus | Haemophilus |
| 6 months | genus | Holdemanella |
| 6 months | genus | Holdemania |
| 6 months | genus | Hungatella |
| 6 months | genus | Hydrogeniiclostidium |
| 6 months | genus | Hydrogenoanaerobacterium |
| 6 months | genus | Intestinibacter |
| 6 months | genus | Intestinimonas |
| 6 months | genus | Isoptericola |
| 6 months | genus | Klebsiella |
| 6 months | genus | Kluyvera |
| 6 months | genus | Lachnoclostridium |
| 6 months | genus | Lachnospira |
| 6 months | genus | Lacrimispora |
| 6 months | genus | Lacticaseibacillus |
| 6 months | genus | Lactobacillus |
| 6 months | genus | Lactococcus |
| 6 months | genus | Lancefieldella |
| 6 months | genus | Ligilactobacillus |
| 6 months | genus | Limosilactobacillus |
| 6 months | genus | Longicatena |
| 6 months | genus | Luxibacter |
| 6 months | genus | Massilimicrobiota |
| 6 months | genus | Mediterraneibacter |
| 6 months | genus | Megamonas |
| 6 months | genus | Megasphaera |
| 6 months | genus | Mogibacterium |
| 6 months | genus | Neisseria |
| 6 months | genus | Odoribacter |
| 6 months | genus | Olsenella |
| 6 months | genus | Oscillibacter |
| 6 months | genus | Parabacteroides |
| 6 months | genus | Paraprevotella |
| 6 months | genus | Parolsenella |
| 6 months | genus | Phascolarctobacterium |
| 6 months | genus | Phocaeicola |
| 6 months | genus | Phocea |
| 6 months | genus | Porphyromonas |
| 6 months | genus | Prevotella |
| 6 months | genus | Prevotellamassilia |
| 6 months | genus | Raoultibacter |
| 6 months | genus | Romboutsia |
| 6 months | genus | Roseburia |
| 6 months | genus | Rothia |
| 6 months | genus | Ruminococcus |
| 6 months | genus | Ruthenibacterium |
| 6 months | genus | Schaalia |
| 6 months | genus | Sellimonas |

| | | |
|---|---|---|
| 6 months | genus | Senegalimassilia |
| 6 months | genus | Slackia |
| 6 months | genus | Staphylococcus |
| 6 months | genus | Streptococcus |
| 6 months | genus | Subdoligranulum |
| 6 months | genus | Sutterella |
| 6 months | genus | Terrisporobacter |
| 6 months | genus | Turicibacter |
| 6 months | genus | Tyzzerella |
| 6 months | genus | Veillonella |
| 6 months | genus | Vescimonas |
| 6 months | genus | Waltera |
| 6 months | family | Acidaminococcaceae |
| 6 months | family | Actinomycetaceae |
| 6 months | family | Akkermansiaceae |
| 6 months | family | Atopobiaceae |
| 6 months | family | Bacteroidaceae |
| 6 months | family | Barnesiellaceae |
| 6 months | family | Bifidobacteriaceae |
| 6 months | family | Carnobacteriaceae |
| 6 months | family | Christensenellaceae |
| 6 months | family | Clostridiaceae |
| 6 months | family | Coprobacillaceae |
| 6 months | family | Coriobacteriaceae |
| 6 months | family | Corynebacteriaceae |
| 6 months | family | Debaryomycetaceae |
| 6 months | family | Desulfovibrionaceae |
| 6 months | family | Eggerthellaceae |
| 6 months | family | Enterobacteriaceae |
| 6 months | family | Enterococcaceae |
| 6 months | family | Erysipelotrichaceae |
| 6 months | family | Eubacteriaceae |
| 6 months | family | Eubacteriales Family XIII. Incertae Sedis |
| 6 months | family | Fusobacteriaceae |
| 6 months | family | Lachnospiraceae |
| 6 months | family | Lactobacillaceae |
| 6 months | family | Micrococcaceae |
| 6 months | family | Moraxellaceae |
| 6 months | family | Neisseriaceae |
| 6 months | family | Odoribacteraceae |
| 6 months | family | Oscillospiraceae |
| 6 months | family | Pasteurellaceae |
| 6 months | family | Peptoniphilaceae |
| 6 months | family | Peptostreptococcaceae |
| 6 months | family | Porphyromonadaceae |
| 6 months | family | Prevotellaceae |
| 6 months | family | Promicromonosporaceae |
| 6 months | family | Rikenellaceae |

| | | |
|---|---|---|
| 6 months | family | Selenomonadaceae |
| 6 months | family | Staphylococcaceae |
| 6 months | family | Streptococcaceae |
| 6 months | family | Sutterellaceae |
| 6 months | family | Tannerellaceae |
| 6 months | family | Turicibacteraceae |
| 6 months | family | Veillonellaceae |
| 6 months | phylum | Actinobacteria |
| 6 months | phylum | Ascomycota |
| 6 months | phylum | Bacteroidetes |
| 6 months | phylum | Firmicutes |
| 6 months | phylum | Fusobacteria |
| 6 months | phylum | Proteobacteria |
| 6 months | phylum | Verrucomicrobia |

| Feature | Contrast |
|---|---|
| Intervention | Experimental - |
| Intervention | Experimental - |
| Intervention | Experimental - |
| Intervention | Experimental - |
| Intervention | Experimental - |
| Intervention | Experimental - |
| Intervention | Experimental - |
| Intervention | Experimental - |
| Intervention | Experimental - |
| Intervention | Experimental - |
| Intervention | Experimental - |
| Intervention | Experimental - |
| Intervention | Experimental - |
| Intervention | Experimental - |
| Intervention | Experimental - |
| Intervention | Experimental - |
| Intervention | Experimental - |
| Intervention | Experimental - |
| Intervention | Experimental - |
| Intervention | Experimental - |
| Intervention | Experimental - |
| Intervention | Experimental - |
| Intervention | Experimental - |
| Intervention | Experimental - |
| Intervention | Experimental - |
| Intervention | Experimental - |
| Intervention | Experimental - |
| Intervention | Experimental - |
| Intervention | Experimental - |
| Intervention | Experimental - |
| Intervention | Experimental - |
| Intervention | Experimental - |
| Intervention | Experimental - |
| Intervention | Experimental - |
| Intervention | Experimental - |
| Intervention | Experimental - |
| Intervention | Experimental - |
| Intervention | Experimental - |
| Intervention | Experimental - |
| Intervention | Experimental - |
| Intervention | Experimental - |
| Intervention | Experimental - |
| Intervention | Experimental - |
| Intervention | Experimental - |
| Intervention | Experimental - |
| Intervention | Experimental - |
| Intervention | Experimental - |

| | |
|---|---|
| Intervention | Experimental - |
| Intervention | Experimental - |
| Intervention | Experimental - |
| Intervention | Experimental - |
| Intervention | Experimental - |
| Intervention | Experimental - |
| Intervention | Experimental - |
| Intervention | Experimental - |
| Intervention | Experimental - |
| Intervention | Experimental - |
| Intervention | Experimental - |
| Intervention | Experimental - |
| Intervention | Experimental - |
| Intervention | Experimental - |
| Intervention | Experimental - |
| Intervention | Experimental - |
| Intervention | Experimental - |
| Intervention | Experimental - |
| Intervention | Experimental - |
| Intervention | Experimental - |
| Intervention | Experimental - |
| Intervention | Experimental - |
| Intervention | Experimental - |
| Intervention | Experimental - |
| Intervention | Experimental - |
| Intervention | Experimental - |
| Intervention | Experimental - |
| Intervention | Experimental - |
| Intervention | Experimental - |
| Intervention | Experimental - |
| Intervention | Experimental - |
| Intervention | Experimental - |
| Intervention | Experimental - |
| Intervention | Experimental - |
| Intervention | Experimental - |
| Intervention | Experimental - |
| Intervention | Experimental - |
| Intervention | Experimental - |
| Intervention | Experimental - |
| Intervention | Experimental - |
| Intervention | Experimental - |
| Intervention | Experimental - |
| Intervention | Experimental - |
| Intervention | Experimental - |
| Intervention | Experimental - |
| Intervention | Experimental - |
| Intervention | Experimental - |
| Intervention | Experimental - |

| | |
|---|---|
| Intervention | Experimental - |
| Intervention | Experimental - |
| Intervention | Experimental - |
| Intervention | Experimental - |
| Intervention | Experimental - |
| Intervention | Experimental - |
| Intervention | Experimental - |
| Intervention | Experimental - |
| Intervention | Experimental - |
| Intervention | Experimental - |
| Intervention | Experimental - |
| Intervention | Experimental - |
| Intervention | Experimental - |
| Intervention | Experimental - |
| Intervention | Experimental - |
| Intervention | Experimental - |
| Intervention | Experimental - |
| Intervention | Experimental - |
| Intervention | Experimental - |
| Intervention | Experimental - |
| Intervention | Experimental - |
| Intervention | Experimental - |
| Intervention | Experimental - |
| Intervention | Experimental - |
| Intervention | Experimental - |
| Intervention | Experimental - |
| Intervention | Experimental - |
| Intervention | Experimental - |
| Intervention | Experimental - |
| Intervention | Experimental - |
| Intervention | Experimental - |
| Intervention | Experimental - |
| Intervention | Experimental - |
| Intervention | Experimental - |
| Intervention | Experimental - |
| Intervention | Experimental - |
| Intervention | Experimental - |
| Intervention | Experimental - |
| Intervention | Experimental - |
| Intervention | Experimental - |
| Intervention | Experimental - |
| Intervention | Experimental - |
| Intervention | Experimental - |
| Intervention | Experimental - |
| Intervention | Experimental - |
| Intervention | Experimental - |
| Intervention | Experimental - |
| Intervention | Experimental - |

| | |
|---|---|
| Intervention | Experimental - |
| Intervention | Experimental - |
| Intervention | Experimental - |
| Intervention | Experimental - |
| Intervention | Experimental - |
| Intervention | Experimental - |
| Intervention | Experimental - |
| Intervention | Experimental - |
| Intervention | Experimental - |
| Intervention | Experimental - |
| Intervention | Experimental - |
| Intervention | Experimental - |
| Intervention | Experimental - |
| Intervention | Experimental - |
| Intervention | Experimental - |
| Intervention | Experimental - |
| Intervention | Experimental - |
| Intervention | Experimental - |
| Intervention | Experimental - |
| Intervention | Experimental - |
| Intervention | Experimental - |
| Intervention | Experimental - |
| Intervention | Experimental - |
| Intervention | Experimental - |
| Intervention | Experimental - |
| Intervention | Experimental - |
| Intervention | Experimental - |
| Intervention | Experimental - |
| Intervention | Experimental - |
| Intervention | Experimental - |
| Intervention | Experimental - |
| Intervention | Experimental - |
| Intervention | Experimental - |
| Intervention | Experimental - |
| Intervention | Experimental - |
| Intervention | Experimental - |
| Intervention | Experimental - |
| Intervention | Experimental - |
| Intervention | Experimental - |
| Intervention | Experimental - |
| Intervention | Experimental - |
| Intervention | Experimental - |
| Intervention | Experimental - |
| Intervention | Experimental - |
| Intervention | Experimental - |
| Intervention | Experimental - |
| Intervention | Experimental - |
| Intervention | Experimental - |

| | |
|---|---|
| Intervention | Experimental - |
| Intervention | Experimental - |
| Intervention | Experimental - |
| Intervention | Experimental - |
| Intervention | Experimental - |
| Intervention | Experimental - |
| Intervention | Experimental - |
| Intervention | Experimental - |
| Intervention | Experimental - |
| Intervention | Experimental - |
| Intervention | Experimental - |
| Intervention | Experimental - |
| Intervention | Experimental - |
| Intervention | Experimental - |
| Intervention | Experimental - |
| Intervention | Experimental - |
| Intervention | Experimental - |
| Intervention | Experimental - |
| Intervention | Experimental - |
| Intervention | Experimental - |
| Intervention | Experimental - |
| Intervention | Experimental - |
| Intervention | Experimental - |
| Intervention | Experimental - |
| Intervention | Experimental - |
| Intervention | Experimental - |
| Intervention | Experimental - |
| Intervention | Experimental - |
| Intervention | Experimental - |
| Intervention | Experimental - |
| Intervention | Experimental - |
| Intervention | Experimental - |
| Intervention | Experimental - |
| Intervention | Experimental - |
| Intervention | Experimental - |
| Intervention | Experimental - |
| Intervention | Experimental - |
| Intervention | Experimental - |
| Intervention | Experimental - |
| Intervention | Experimental - |
| Intervention | Experimental - |
| Intervention | Experimental - |
| Intervention | Experimental - |
| Intervention | Experimental - |
| Intervention | Experimental - |
| Intervention | Experimental - |
| Intervention | Experimental - |
| Intervention | Experimental - |

| | |
|---|---|
| Intervention | Experimental - |
| Intervention | Experimental - |
| Intervention | Experimental - |
| Intervention | Experimental - |
| Intervention | Experimental - |
| Intervention | Experimental - |
| Intervention | Experimental - |
| Intervention | Experimental - |
| Intervention | Experimental - |
| Intervention | Experimental - |
| Intervention | Experimental - |
| Intervention | Experimental - |
| Intervention | Experimental - |
| Intervention | Experimental - |
| Intervention | Experimental - |
| Intervention | Experimental - |
| Intervention | Experimental - |
| Intervention | Experimental - |
| Intervention | Experimental - |
| Intervention | Experimental - |
| Intervention | Experimental - |
| Intervention | Experimental - |
| Intervention | Experimental - |
| Intervention | Experimental - |
| Intervention | Experimental - |
| Intervention | Experimental - |
| Intervention | Experimental - |
| Intervention | Experimental - |
| Intervention | Experimental - |
| Intervention | Experimental - |
| Intervention | Experimental - |
| Intervention | Experimental - |
| Intervention | Experimental - |
| Intervention | Experimental - |
| Intervention | Experimental - |
| Intervention | Experimental - |
| Intervention | Experimental - |
| Intervention | Experimental - |
| Intervention | Experimental - |
| Intervention | Experimental - |
| Intervention | Experimental - |
| Intervention | Experimental - |
| Intervention | Experimental - |
| Intervention | Experimental - |
| Intervention | Experimental - |
| Intervention | Experimental - |
| Intervention | Experimental - |
| Intervention | Experimental - |

| | |
|---|---|
| Intervention | Experimental - |
| Intervention | Experimental - |
| Intervention | Experimental - |
| Intervention | Experimental - |
| Intervention | Experimental - |
| Intervention | Experimental - |
| Intervention | Experimental - |
| Intervention | Experimental - |
| Intervention | Experimental - |
| Intervention | Experimental - |
| Intervention | Experimental - |
| Intervention | Experimental - |
| Intervention | Experimental - |
| Intervention | Experimental - |
| Intervention | Experimental - |
| Intervention | Experimental - |
| Intervention | Experimental - |
| Intervention | Experimental - |
| Intervention | Experimental - |
| Intervention | Experimental - |
| Intervention | Experimental - |
| Intervention | Experimental - |
| Intervention | Experimental - |
| Intervention | Experimental - |
| Intervention | Experimental - |
| Intervention | Experimental - |
| Intervention | Experimental - |
| Intervention | Experimental - |
| Intervention | Experimental - |
| Intervention | Experimental - |
| Intervention | Experimental - |
| Intervention | Experimental - |
| Intervention | Experimental - |
| Intervention | Experimental - |
| Intervention | Experimental - |
| Intervention | Experimental - |
| Intervention | Experimental - |
| Intervention | Experimental - |
| Intervention | Experimental - |
| Intervention | Experimental - |
| Intervention | Experimental - |
| Intervention | Experimental - |
| Intervention | Experimental - |
| Intervention | Experimental - |
| Intervention | Experimental - |
| Intervention | Experimental - |
| Intervention | Experimental - |
| Intervention | Experimental - |

| | |
|---|---|
| Intervention | Experimental - |
| Intervention | Experimental - |
| Intervention | Experimental - |
| Intervention | Experimental - |
| Intervention | Experimental - |
| Intervention | Experimental - |
| Intervention | Experimental - |
| Intervention | Experimental - |
| Intervention | Experimental - |
| Intervention | Experimental - |
| Intervention | Experimental - |
| Intervention | Experimental - |
| Intervention | Experimental - |
| Intervention | Experimental - |
| Intervention | Experimental - |
| Intervention | Experimental - |
| Intervention | Experimental - |
| Intervention | Experimental - |
| Intervention | Experimental - |
| Intervention | Experimental - |
| Intervention | Experimental - |
| Intervention | Experimental - |
| Intervention | Experimental - |
| Intervention | Experimental - |
| Intervention | Experimental - |
| Intervention | Experimental - |
| Intervention | Experimental - |
| Intervention | Experimental - |
| Intervention | Experimental - |
| Intervention | Experimental - |
| Intervention | Experimental - |
| Intervention | Experimental - |
| Intervention | Experimental - |
| Intervention | Experimental - |
| Intervention | Experimental - |
| Intervention | Experimental - |
| Intervention | Experimental - |
| Intervention | Experimental - |
| Intervention | Experimental - |
| Intervention | Experimental - |
| Intervention | Experimental - |
| Intervention | Experimental - |
| Intervention | Experimental - |
| Intervention | Experimental - |
| Intervention | Experimental - |
| Intervention | Experimental - |
| Intervention | Experimental - |
| Intervention | Experimental - |

| | |
|---|---|
| Intervention | Experimental - |
| Intervention | Experimental - |
| Intervention | Experimental - |
| Intervention | Experimental - |
| Intervention | Experimental - |
| Intervention | Experimental - |
| Intervention | Experimental - |
| Intervention | Experimental - |
| Intervention | Experimental - |
| Intervention | Experimental - |
| Intervention | Experimental - |
| Intervention | Experimental - |
| Intervention | Experimental - |
| Intervention | Experimental - |
| Intervention | Experimental - |
| Intervention | Experimental - |
| Intervention | Experimental - |
| Intervention | Experimental - |
| Intervention | Experimental - |
| Intervention | Experimental - |
| Intervention | Experimental - |
| Intervention | Experimental - |
| Intervention | Experimental - |
| Intervention | Experimental - |
| Intervention | Experimental - |
| Intervention | Experimental - |
| Intervention | Experimental - |
| Intervention | Experimental - |
| Intervention | Experimental - |
| Intervention | Experimental - |
| Intervention | Experimental - |
| Intervention | Experimental - |
| Intervention | Experimental - |
| Intervention | Experimental - |
| Intervention | Experimental - |
| Intervention | Experimental - |
| Intervention | Experimental - |
| Intervention | Experimental - |
| Intervention | Experimental - |
| Intervention | Experimental - |
| Intervention | Experimental - |
| Intervention | Experimental - |
| Intervention | Experimental - |
| Intervention | Experimental - |
| Intervention | Experimental - |
| Intervention | Experimental - |
| Intervention | Experimental - |
| Intervention | Experimental - |

| | |
|---|---|
| Intervention | Experimental - |
| Intervention | Experimental - |
| Intervention | Experimental - |
| Intervention | Experimental - |
| Intervention | Experimental - |
| Intervention | Experimental - |
| Intervention | Experimental - |
| Intervention | Experimental - |
| Intervention | Experimental - |
| Intervention | Experimental - |
| Intervention | Experimental - |
| Intervention | Experimental - |
| Intervention | Experimental - |
| Intervention | Experimental - |
| Intervention | Experimental - |
| Intervention | Experimental - |
| Intervention | Experimental - |
| Intervention | Experimental - |
| Intervention | Experimental - |
| Intervention | Experimental - |
| Intervention | Experimental - |
| Intervention | Experimental - |
| Intervention | Experimental - |
| Intervention | Experimental - |
| Intervention | Experimental - |
| Intervention | Experimental - |
| Intervention | Experimental - |
| Intervention | Experimental - |
| Intervention | Experimental - |
| Intervention | Experimental - |
| Intervention | Experimental - |
| Intervention | Experimental - |
| Intervention | Experimental - |
| Intervention | Experimental - |
| Intervention | Experimental - |
| Intervention | Experimental - |
| Intervention | Experimental - |
| Intervention | Experimental - |
| Intervention | Experimental - |
| Intervention | Experimental - |
| Intervention | Experimental - |
| Intervention | Experimental - |
| Intervention | Experimental - |
| Intervention | Experimental - |
| Intervention | Experimental - |
| Intervention | Experimental - |
| Intervention | Experimental - |
| Intervention | Experimental - |

| | |
|---|---|
| Intervention | Experimental - |
| Intervention | Experimental - |
| Intervention | Experimental - |
| Intervention | Experimental - |
| Intervention | Experimental - |
| Intervention | Experimental - |
| Intervention | Experimental - |
| Intervention | Experimental - |
| Intervention | Experimental - |
| Intervention | Experimental - |
| Intervention | Experimental - |
| Intervention | Experimental - |
| Intervention | Experimental - |
| Intervention | Experimental - |
| Intervention | Experimental - |
| Intervention | Experimental - |
| Intervention | Experimental - |
| Intervention | Experimental - |
| Intervention | Experimental - |
| Intervention | Experimental - |
| Intervention | Experimental - |
| Intervention | Experimental - |
| Intervention | Experimental - |
| Intervention | Experimental - |
| Intervention | Experimental - |
| Intervention | Experimental - |
| Intervention | Experimental - |
| Intervention | Experimental - |
| Intervention | Experimental - |
| Intervention | Experimental - |
| Intervention | Experimental - |
| Intervention | Experimental - |
| Intervention | Experimental - |
| Intervention | Experimental - |
| Intervention | Experimental - |
| Intervention | Experimental - |
| Intervention | Experimental - |
| Intervention | Experimental - |
| Intervention | Experimental - |
| Intervention | Experimental - |
| Intervention | Experimental - |
| Intervention | Experimental - |
| Intervention | Experimental - |
| Intervention | Experimental - |
| Intervention | Experimental - |
| Intervention | Experimental - |
| Intervention | Experimental - |
| Intervention | Experimental - |

| | |
|---|---|
| Intervention | Experimental - |
| Intervention | Experimental - |
| Intervention | Experimental - |
| Intervention | Experimental - |
| Intervention | Experimental - |
| Intervention | Experimental - |
| Intervention | Experimental - |
| Intervention | Experimental - |
| Intervention | Experimental - |
| Intervention | Experimental - |
| Intervention | Experimental - |
| Intervention | Experimental - |
| Intervention | Experimental - |
| Intervention | Experimental - |
| Intervention | Experimental - |
| Intervention | Experimental - |
| Intervention | Experimental - |
| Intervention | Experimental - |
| Intervention | Experimental - |
| Intervention | Experimental - |
| Intervention | Experimental - |
| Intervention | Experimental - |
| Intervention | Experimental - |
| Intervention | Experimental - |
| Intervention | Experimental - |
| Intervention | Experimental - |
| Intervention | Experimental - |
| Intervention | Experimental - |
| Intervention | Experimental - |
| Intervention | Experimental - |
| Intervention | Experimental - |
| Intervention | Experimental - |
| Intervention | Experimental - |
| Intervention | Experimental - |
| Intervention | Experimental - |
| Intervention | Experimental - |
| Intervention | Experimental - |
| Intervention | Experimental - |
| Intervention | Experimental - |
| Intervention | Experimental - |
| Intervention | Experimental - |
| Intervention | Experimental - |
| Intervention | Experimental - |
| Intervention | Experimental - |
| Intervention | Experimental - |
| Intervention | Experimental - |
| Intervention | Experimental - |
| Intervention | Experimental - |

| | |
|---|---|
| Intervention | Experimental - |
| Intervention | Experimental - |
| Intervention | Experimental - |
| Intervention | Experimental - |
| Intervention | Experimental - |
| Intervention | Experimental - |
| Intervention | Experimental - |
| Intervention | Experimental - |
| Intervention | Experimental - |
| Intervention | Experimental - |
| Intervention | Experimental - |
| Intervention | Experimental - |
| Intervention | Experimental - |
| Intervention | Experimental - |
| Intervention | Experimental - |
| Intervention | Experimental - |
| Intervention | Experimental - |
| Intervention | Experimental - |
| Intervention | Experimental - |
| Intervention | Experimental - |
| Intervention | Experimental - |
| Intervention | Experimental - |
| Intervention | Experimental - |
| Intervention | Experimental - |
| Intervention | Experimental - |
| Intervention | Experimental - |
| Intervention | Experimental - |
| Intervention | Experimental - |
| Intervention | Experimental - |
| Intervention | Experimental - |
| Intervention | Experimental - |
| Intervention | Experimental - |
| Intervention | Experimental - |
| Intervention | Experimental - |
| Intervention | Experimental - |
| Intervention | Experimental - |
| Intervention | Experimental - |
| Intervention | Experimental - |
| Intervention | Experimental - |
| Intervention | Experimental - |
| Intervention | Experimental - |
| Intervention | Experimental - |
| Intervention | Experimental - |
| Intervention | Experimental - |
| Intervention | Experimental - |
| Intervention | Experimental - |
| Intervention | Experimental - |
| Intervention | Experimental - |

| | |
|---|---|
| Intervention | Experimental - |
| Intervention | Experimental - |
| Intervention | Experimental - |
| Intervention | Experimental - |
| Intervention | Experimental - |
| Intervention | Experimental - |
| Intervention | Experimental - |
| Intervention | Experimental - |
| Intervention | Experimental - |
| Intervention | Experimental - |
| Intervention | Experimental - |
| Intervention | Experimental - |
| Intervention | Experimental - |
| Intervention | Experimental - |
| Intervention | Experimental - |
| Intervention | Experimental - |
| Intervention | Experimental - |
| Intervention | Experimental - |
| Intervention | Experimental - |
| Intervention | Experimental - |
| Intervention | Experimental - |
| Intervention | Experimental - |
| Intervention | Experimental - |
| Intervention | Experimental - |
| Intervention | Experimental - |
| Intervention | Experimental - |
| Intervention | Experimental - |
| Intervention | Experimental - |
| Intervention | Experimental - |
| Intervention | Experimental - |
| Intervention | Experimental - |
| Intervention | Experimental - |
| Intervention | Experimental - |
| Intervention | Experimental - |
| Intervention | Experimental - |
| Intervention | Experimental - |
| Intervention | Experimental - |
| Intervention | Experimental - |
| Intervention | Experimental - |
| Intervention | Experimental - |
| Intervention | Experimental - |
| Intervention | Experimental - |
| Intervention | Experimental - |
| Intervention | Experimental - |
| Intervention | Experimental - |
| Intervention | Experimental - |
| Intervention | Experimental - |
| Intervention | Experimental - |

| | |
|---|---|
| Intervention | Experimental - |
| Intervention | Experimental - |
| Intervention | Experimental - |
| Intervention | Experimental - |
| Intervention | Experimental - |
| Intervention | Experimental - |
| Intervention | Experimental - |
| Intervention | Experimental - |
| Intervention | Experimental - |
| Intervention | Experimental - |
| Intervention | Experimental - |
| Intervention | Experimental - |
| Intervention | Experimental - |
| Intervention | Experimental - |
| Intervention | Experimental - |
| Intervention | Experimental - |
| Intervention | Experimental - |
| Intervention | Experimental - |
| Intervention | Experimental - |
| Intervention | Experimental - |
| Intervention | Experimental - |
| Intervention | Experimental - |
| Intervention | Experimental - |
| Intervention | Experimental - |
| Intervention | Experimental - |
| Intervention | Experimental - |
| Intervention | Experimental - |
| Intervention | Experimental - |
| Intervention | Experimental - |
| Intervention | Experimental - |
| Intervention | Experimental - |
| Intervention | Experimental - |
| Intervention | Experimental - |
| Intervention | Experimental - |
| Intervention | Experimental - |
| Intervention | Experimental - |
| Intervention | Experimental - |
| Intervention | Experimental - |
| Intervention | Experimental - |
| Intervention | Experimental - |
| Intervention | Experimental - |
| Intervention | Experimental - |
| Intervention | Experimental - |
| Intervention | Experimental - |
| Intervention | Experimental - |
| Intervention | Experimental - |
| Intervention | Experimental - |
| Intervention | Experimental - |

| | |
|---|---|
| Intervention | Experimental - |
| Intervention | Experimental - |
| Intervention | Experimental - |
| Intervention | Experimental - |
| Intervention | Experimental - |
| Intervention | Experimental - |
| Intervention | Experimental - |
| Intervention | Experimental - |
| Intervention | Experimental - |
| Intervention | Experimental - |
| Intervention | Experimental - |
| Intervention | Experimental - |
| Intervention | Experimental - |
| Intervention | Experimental - |
| Intervention | Experimental - |
| Intervention | Experimental - |
| Intervention | Experimental - |
| Intervention | Experimental - |
| Intervention | Experimental - |
| Intervention | Experimental - |
| Intervention | Experimental - |
| Intervention | Experimental - |
| Intervention | Experimental - |
| Intervention | Experimental - |
| Intervention | Experimental - |
| Intervention | Experimental - |
| Intervention | Experimental - |
| Intervention | Experimental - |
| Intervention | Experimental - |
| Intervention | Experimental - |
| Intervention | Experimental - |
| Intervention | Experimental - |
| Intervention | Experimental - |
| Intervention | Experimental - |
| Intervention | Experimental - |
| Intervention | Experimental - |
| Intervention | Experimental - |
| Intervention | Experimental - |
| Intervention | Experimental - |
| Intervention | Experimental - |
| Intervention | Experimental - |
| Intervention | Experimental - |
| Intervention | Experimental - |
| Intervention | Experimental - |
| Intervention | Experimental - |
| Intervention | Experimental - |
| Intervention | Experimental - |
| Intervention | Experimental - |

| | |
|---|---|
| Intervention | Experimental - |
| Intervention | Experimental - |
| Intervention | Experimental - |
| Intervention | Experimental - |
| Intervention | Experimental - |
| Intervention | Experimental - |
| Intervention | Experimental - |
| Intervention | Experimental - |
| Intervention | Experimental - |
| Intervention | Experimental - |
| Intervention | Experimental - |
| Intervention | Experimental - |
| Intervention | Experimental - |
| Intervention | Experimental - |
| Intervention | Experimental - |
| Intervention | Experimental - |
| Intervention | Experimental - |
| Intervention | Experimental - |
| Intervention | Experimental - |
| Intervention | Experimental - |
| Intervention | Experimental - |
| Intervention | Experimental - |
| Intervention | Experimental - |
| Intervention | Experimental - |
| Intervention | Experimental - |
| Intervention | Experimental - |
| Intervention | Experimental - |
| Intervention | Experimental - |
| Intervention | Experimental - |
| Intervention | Experimental - |
| Intervention | Experimental - |
| Intervention | Experimental - |
| Intervention | Experimental - |
| Intervention | Experimental - |
| Intervention | Experimental - |
| Intervention | Experimental - |
| Intervention | Experimental - |
| Intervention | Experimental - |
| Intervention | Experimental - |
| Intervention | Experimental - |
| Intervention | Experimental - |
| Intervention | Experimental - |
| Intervention | Experimental - |
| Intervention | Experimental - |
| Intervention | Experimental - |
| Intervention | Experimental - |
| Intervention | Experimental - |
| Intervention | Experimental - |

| | |
|---|---|
| Intervention | Experimental - |
| Intervention | Experimental - |
| Intervention | Experimental - |
| Intervention | Experimental - |
| Intervention | Experimental - |
| Intervention | Experimental - |
| Intervention | Experimental - |
| Intervention | Experimental - |
| Intervention | Experimental - |
| Intervention | Experimental - |
| Intervention | Experimental - |
| Intervention | Experimental - |
| Intervention | Experimental - |
| Intervention | Experimental - |
| Intervention | Experimental - |
| Intervention | Experimental - |
| Intervention | Experimental - |
| Intervention | Experimental - |
| Intervention | Experimental - |
| Intervention | Experimental - |
| Intervention | Experimental - |
| Intervention | Experimental - |
| Intervention | Experimental - |
| Intervention | Experimental - |
| Intervention | Experimental - |
| Intervention | Experimental - |
| Intervention | Experimental - |
| Intervention | Experimental - |
| Intervention | Experimental - |
| Intervention | Experimental - |
| Intervention | Experimental - |
| Intervention | Experimental - |
| Intervention | Experimental - |
| Intervention | Experimental - |
| Intervention | Experimental - |
| Intervention | Experimental - |
| Intervention | Experimental - |
| Intervention | Experimental - |
| Intervention | Experimental - |
| Intervention | Experimental - |
| Intervention | Experimental - |
| Intervention | Experimental - |
| Intervention | Experimental - |
| Intervention | Experimental - |
| Intervention | Experimental - |
| Intervention | Experimental - |
| Intervention | Experimental - |
| Intervention | Experimental - |

| | |
|---|---|
| Intervention | Experimental - |
| Intervention | Experimental - |
| Intervention | Experimental - |
| Intervention | Experimental - |
| Intervention | Experimental - |
| Intervention | Experimental - |
| Intervention | Experimental - |
| Intervention | Experimental - |
| Intervention | Experimental - |
| Intervention | Experimental - |
| Intervention | Experimental - |
| Intervention | Experimental - |
| Intervention | Experimental - |
| Intervention | Experimental - |
| Intervention | Experimental - |
| Intervention | Experimental - |
| Intervention | Experimental - |
| Intervention | Experimental - |
| Intervention | Experimental - |
| Intervention | Experimental - |
| Intervention | Experimental - |
| Intervention | Experimental - |
| Intervention | Experimental - |
| Intervention | Experimental - |
| Intervention | Experimental - |
| Intervention | Experimental - |
| Intervention | Experimental - |
| Intervention | Experimental - |
| Intervention | Experimental - |
| Intervention | Experimental - |
| Intervention | Experimental - |
| Intervention | Experimental - |
| Intervention | Experimental - |
| Intervention | Experimental - |
| Intervention | Experimental - |
| Intervention | Experimental - |
| Intervention | Experimental - |
| Intervention | Experimental - |
| Intervention | Experimental - |
| Intervention | Experimental - |
| Intervention | Experimental - |
| Intervention | Experimental - |
| Intervention | Experimental - |
| Intervention | Experimental - |
| Intervention | Experimental - |
| Intervention | Experimental - |
| Intervention | Experimental - |
| Intervention | Experimental - |

| | |
|---|---|
| Intervention | Experimental - |
| Intervention | Experimental - |
| Intervention | Experimental - |
| Intervention | Experimental - |
| Intervention | Experimental - |
| Intervention | Experimental - |
| Intervention | Experimental - |
| Intervention | Experimental - |
| Intervention | Experimental - |
| Intervention | Experimental - |
| Intervention | Experimental - |
| Intervention | Experimental - |
| Intervention | Experimental - |
| Intervention | Experimental - |
| Intervention | Experimental - |
| Intervention | Experimental - |
| Intervention | Experimental - |
| Intervention | Experimental - |
| Intervention | Experimental - |
| Intervention | Experimental - |
| Intervention | Experimental - |
| Intervention | Experimental - |
| Intervention | Experimental - |
| Intervention | Experimental - |
| Intervention | Experimental - |
| Intervention | Experimental - |
| Intervention | Experimental - |
| Intervention | Experimental - |
| Intervention | Experimental - |
| Intervention | Experimental - |
| Intervention | Experimental - |
| Intervention | Experimental - |
| Intervention | Experimental - |
| Intervention | Experimental - |
| Intervention | Experimental - |
| Intervention | Experimental - |
| Intervention | Experimental - |
| Intervention | Experimental - |
| Intervention | Experimental - |
| Intervention | Experimental - |
| Intervention | Experimental - |
| Intervention | Experimental - |
| Intervention | Experimental - |
| Intervention | Experimental - |
| Intervention | Experimental - |
| Intervention | Experimental - |
| Intervention | Experimental - |
| Intervention | Experimental - |

| | |
|---|---|
| Intervention | Experimental - |
| Intervention | Experimental - |
| Intervention | Experimental - |
| Intervention | Experimental - |
| Intervention | Experimental - |
| Intervention | Experimental - |
| Intervention | Experimental - |
| Intervention | Experimental - |
| Intervention | Experimental - |
| Intervention | Experimental - |
| Intervention | Experimental - |
| Intervention | Experimental - |
| Intervention | Experimental - |
| Intervention | Experimental - |
| Intervention | Experimental - |
| Intervention | Experimental - |
| Intervention | Experimental - |
| Intervention | Experimental - |
| Intervention | Experimental - |
| Intervention | Experimental - |
| Intervention | Experimental - |
| Intervention | Experimental - |
| Intervention | Experimental - |
| Intervention | Experimental - |
| Intervention | Experimental - |
| Intervention | Experimental - |
| Intervention | Experimental - |
| Intervention | Experimental - |
| Intervention | Experimental - |
| Intervention | Experimental - |
| Intervention | Experimental - |
| Intervention | Experimental - |
| Intervention | Experimental - |
| Intervention | Experimental - |
| Intervention | Experimental - |
| Intervention | Experimental - |
| Intervention | Experimental - |
| Intervention | Experimental - |
| Intervention | Experimental - |
| Intervention | Experimental - |
| Intervention | Experimental - |
| Intervention | Experimental - |
| Intervention | Experimental - |
| Intervention | Experimental - |
| Intervention | Experimental - |
| Intervention | Experimental - |
| Intervention | Experimental - |
| Intervention | Experimental - |

| | |
|---|---|
| Intervention | Experimental - |
| Intervention | Experimental - |
| Intervention | Experimental - |
| Intervention | Experimental - |
| Intervention | Experimental - |
| Intervention | Experimental - |
| Intervention | Experimental - |
| Intervention | Experimental - |
| Intervention | Experimental - |
| Intervention | Experimental - |
| Intervention | Experimental - |
| Intervention | Experimental - |
| Intervention | Experimental - |
| Intervention | Experimental - |
| Intervention | Experimental - |
| Intervention | Experimental - |
| Intervention | Experimental - |
| Intervention | Experimental - |
| Intervention | Experimental - |
| Intervention | Experimental - |
| Intervention | Experimental - |
| Intervention | Experimental - |
| Intervention | Experimental - |
| Intervention | Experimental - |
| Intervention | Experimental - |
| Intervention | Experimental - |
| Intervention | Experimental - |
| Intervention | Experimental - |
| Intervention | Experimental - |
| Intervention | Experimental - |
| Intervention | Experimental - |
| Intervention | Experimental - |
| Intervention | Experimental - |
| Intervention | Experimental - |
| Intervention | Experimental - |
| Intervention | Experimental - |
| Intervention | Experimental - |
| Intervention | Experimental - |
| Intervention | Experimental - |
| Intervention | Experimental - |
| Intervention | Experimental - |
| Intervention | Experimental - |
| Intervention | Experimental - |
| Intervention | Experimental - |
| Intervention | Experimental - |
| Intervention | Experimental - |
| Intervention | Experimental - |
| Intervention | Experimental - |

| | |
|---|---|
| Intervention | Experimental - |
| Intervention | Experimental - |
| Intervention | Experimental - |
| Intervention | Experimental - |
| Intervention | Experimental - |
| Intervention | Experimental - |
| Intervention | Experimental - |
| Intervention | Experimental - |
| Intervention | Experimental - |
| Intervention | Experimental - |
| Intervention | Experimental - |
| Intervention | Experimental - |
| Intervention | Experimental - |
| Intervention | Experimental - |
| Intervention | Experimental - |
| Intervention | Experimental - |
| Intervention | Experimental - |
| Intervention | Experimental - |
| Intervention | Experimental - |
| Intervention | Experimental - |
| Intervention | Experimental - |
| Intervention | Experimental - |
| Intervention | Experimental - |
| Intervention | Experimental - |
| Intervention | Experimental - |
| Intervention | Experimental - |
| Intervention | Experimental - |
| Intervention | Experimental - |
| Intervention | Experimental - |
| Intervention | Experimental - |
| Intervention | Experimental - |
| Intervention | Experimental - |
| Intervention | Experimental - |
| Intervention | Experimental - |
| Intervention | Experimental - |
| Intervention | Experimental - |
| Intervention | Experimental - |
| Intervention | Experimental - |
| Intervention | Experimental - |
| Intervention | Experimental - |
| Intervention | Experimental - |
| Intervention | Experimental - |
| Intervention | Experimental - |
| Intervention | Experimental - |
| Intervention | Experimental - |
| Intervention | Experimental - |
| Intervention | Experimental - |
| Intervention | Experimental - |

| | |
|---|---|
| Intervention | Experimental - |
| Intervention | Experimental - |
| Intervention | Experimental - |
| Intervention | Experimental - |
| Intervention | Experimental - |
| Intervention | Experimental - |
| Intervention | Experimental - |
| Intervention | Experimental - |
| Intervention | Experimental - |
| Intervention | Experimental - |
| Intervention | Experimental - |
| Intervention | Experimental - |
| Intervention | Experimental - |
| Intervention | Experimental - |
| Intervention | Experimental - |
| Intervention | Experimental - |
| Intervention | Experimental - |
| Intervention | Experimental - |
| Intervention | Experimental - |
| Intervention | Experimental - |
| Intervention | Experimental - |
| Intervention | Experimental - |
| Intervention | Experimental - |
| Intervention | Experimental - |
| Intervention | Experimental - |
| Intervention | Experimental - |
| Intervention | Experimental - |
| Intervention | Experimental - |
| Intervention | Experimental - |
| Intervention | Experimental - |
| Intervention | Experimental - |
| Intervention | Experimental - |
| Intervention | Experimental - |
| Intervention | Experimental - |
| Intervention | Experimental - |
| Intervention | Experimental - |
| Intervention | Experimental - |
| Intervention | Experimental - |
| Intervention | Experimental - |
| Intervention | Experimental - |
| Intervention | Experimental - |
| Intervention | Experimental - |
| Intervention | Experimental - |
| Intervention | Experimental - |
| Intervention | Experimental - |
| Intervention | Experimental - |
| Intervention | Experimental - |
| Intervention | Experimental - |

| | |
|---|---|
| Intervention | Experimental - |
| Intervention | Experimental - |
| Intervention | Experimental - |
| Intervention | Experimental - |
| Intervention | Experimental - |
| Intervention | Experimental - |
| Intervention | Experimental - |
| Intervention | Experimental - |
| Intervention | Experimental - |
| Intervention | Experimental - |
| Intervention | Experimental - |
| Intervention | Experimental - |
| Intervention | Experimental - |
| Intervention | Experimental - |
| Intervention | Experimental - |
| Intervention | Experimental - |
| Intervention | Experimental - |
| Intervention | Experimental - |
| Intervention | Experimental - |
| Intervention | Experimental - |
| Intervention | Experimental - |
| Intervention | Experimental - |
| Intervention | Experimental - |
| Intervention | Experimental - |
| Intervention | Experimental - |
| Intervention | Experimental - |
| Intervention | Experimental - |
| Intervention | Experimental - |
| Intervention | Experimental - |
| Intervention | Experimental - |
| Intervention | Experimental - |
| Intervention | Experimental - |
| Intervention | Experimental - |
| Intervention | Experimental - |
| Intervention | Experimental - |
| Intervention | Experimental - |
| Intervention | Experimental - |
| Intervention | Experimental - |
| Intervention | Experimental - |
| Intervention | Experimental - |
| Intervention | Experimental - |
| Intervention | Experimental - |
| Intervention | Experimental - |
| Intervention | Experimental - |
| Intervention | Experimental - |
| Intervention | Experimental - |
| Intervention | Experimental - |
| Intervention | Experimental - |

| | |
|---|---|
| Intervention | Experimental - |
| Intervention | Experimental - |
| Intervention | Experimental - |
| Intervention | Experimental - |
| Intervention | Experimental - |
| Intervention | Experimental - |
| Intervention | Experimental - |
| Intervention | Experimental - |
| Intervention | Experimental - |
| Intervention | Experimental - |
| Intervention | Experimental - |
| Intervention | Experimental - |
| Intervention | Experimental - |
| Intervention | Experimental - |
| Intervention | Experimental - |
| Intervention | Experimental - |
| Intervention | Experimental - |
| Intervention | Experimental - |
| Intervention | Experimental - |
| Intervention | Experimental - |
| Intervention | Experimental - |
| Intervention | Experimental - |
| Intervention | Experimental - |
| Intervention | Experimental - |
| Intervention | Experimental - |
| Intervention | Experimental - |
| Intervention | Experimental - |
| Intervention | Experimental - |
| Intervention | Experimental - |
| Intervention | Experimental - |
| Intervention | Experimental - |
| Intervention | Experimental - |
| Intervention | Experimental - |
| Intervention | Experimental - |
| Intervention | Experimental - |
| Intervention | Experimental - |
| Intervention | Experimental - |
| Intervention | Experimental - |
| Intervention | Experimental - |
| Intervention | Experimental - |
| Intervention | Experimental - |
| Intervention | Experimental - |
| Intervention | Experimental - |
| Intervention | Experimental - |
| Intervention | Experimental - |
| Intervention | Experimental - |
| Intervention | Experimental - |
| Intervention | Experimental - |

| | |
|---|---|
| Intervention | Experimental - |
| Intervention | Experimental - |
| Intervention | Experimental - |
| Intervention | Experimental - |
| Intervention | Experimental - |
| Intervention | Experimental - |
| Intervention | Experimental - |
| Intervention | Experimental - |
| Intervention | Experimental - |
| Intervention | Experimental - |
| Intervention | Experimental - |
| Intervention | Experimental - |
| Intervention | Experimental - |
| Intervention | Experimental - |
| Intervention | Experimental - |
| Intervention | Experimental - |
| Intervention | Experimental - |
| Intervention | Experimental - |
| Intervention | Experimental - |
| Intervention | Experimental - |
| Intervention | Experimental - |
| Intervention | Experimental - |
| Intervention | Experimental - |
| Intervention | Experimental - |
| Intervention | Experimental - |
| Intervention | Experimental - |
| Intervention | Experimental - |
| Intervention | Experimental - |
| Intervention | Experimental - |
| Intervention | Experimental - |
| Intervention | Experimental - |
| Intervention | Experimental - |
| Intervention | Experimental - |
| Intervention | Experimental - |
| Intervention | Experimental - |
| Intervention | Experimental - |
| Intervention | Experimental - |
| Intervention | Experimental - |
| Intervention | Experimental - |
| Intervention | Experimental - |
| Intervention | Experimental - |
| Intervention | Experimental - |
| Intervention | Experimental - |
| Intervention | Experimental - |
| Intervention | Experimental - |
| Intervention | Experimental - |
| Intervention | Experimental - |
| Intervention | Experimental - |

| | |
|---|---|
| Intervention | Experimental - |
| Intervention | Experimental - |
| Intervention | Experimental - |
| Intervention | Experimental - |
| Intervention | Experimental - |
| Intervention | Experimental - |
| Intervention | Experimental - |
| Intervention | Experimental - |
| Intervention | Experimental - |
| Intervention | Experimental - |
| Intervention | Experimental - |
| Intervention | Experimental - |
| Intervention | Experimental - |
| Intervention | Experimental - |
| Intervention | Experimental - |
| Intervention | Experimental - |
| Intervention | Experimental - |
| Intervention | Experimental - |
| Intervention | Experimental - |
| Intervention | Experimental - |
| Intervention | Experimental - |
| Intervention | Experimental - |
| Intervention | Experimental - |
| Intervention | Experimental - |
| Intervention | Experimental - |
| Intervention | Experimental - |
| Intervention | Experimental - |
| Intervention | Experimental - |
| Intervention | Experimental - |
| Intervention | Experimental - |
| Intervention | Experimental - |
| Intervention | Experimental - |
| Intervention | Experimental - |
| Intervention | Experimental - |
| Intervention | Experimental - |
| Intervention | Experimental - |
| Intervention | Experimental - |
| Intervention | Experimental - |
| Intervention | Experimental - |
| Intervention | Experimental - |
| Intervention | Experimental - |
| Intervention | Experimental - |
| Intervention | Experimental - |
| Intervention | Experimental - |
| Intervention | Experimental - |
| Intervention | Experimental - |
| Intervention | Experimental - |
| Intervention | Experimental - |

| | |
|---|---|
| Intervention | Experimental - |
| Intervention | Experimental - |
| Intervention | Experimental - |
| Intervention | Experimental - |
| Intervention | Experimental - |
| Intervention | Experimental - |
| Intervention | Experimental - |
| Intervention | Experimental - |
| Intervention | Experimental - |
| Intervention | Experimental - |
| Intervention | Experimental - |
| Intervention | Experimental - |
| Intervention | Experimental - |
| Intervention | Experimental - |
| Intervention | Experimental - |
| Intervention | Experimental - |
| Intervention | Experimental - |
| Intervention | Experimental - |
| Intervention | Experimental - |
| Intervention | Experimental - |
| Intervention | Experimental - |
| Intervention | Experimental - |
| Intervention | Experimental - |
| Intervention | Experimental - |
| Intervention | Experimental - |
| Intervention | Experimental - |
| Intervention | Experimental - |
| Intervention | Experimental - |
| Intervention | Experimental - |
| Intervention | Experimental - |
| Intervention | Experimental - |
| Intervention | Experimental - |
| Intervention | Experimental - |
| Intervention | Experimental - |
| Intervention | Experimental - |
| Intervention | Experimental - |
| Intervention | Experimental - |
| Intervention | Experimental - |
| Intervention | Experimental - |
| Intervention | Experimental - |
| Intervention | Experimental - |
| Intervention | Experimental - |
| Intervention | Experimental - |
| Intervention | Experimental - |
| Intervention | Experimental - |
| Intervention | Experimental - |
| Intervention | Experimental - |
| Intervention | Experimental - |

| | |
|---|---|
| Intervention | Experimental - |
| Intervention | Experimental - |
| Intervention | Experimental - |
| Intervention | Experimental - |
| Intervention | Experimental - |
| Intervention | Experimental - |
| Intervention | Experimental - |
| Intervention | Experimental - |
| Intervention | Experimental - |
| Intervention | Experimental - |
| Intervention | Experimental - |
| Intervention | Experimental - |
| Intervention | Experimental - |
| Intervention | Experimental - |
| Intervention | Experimental - |
| Intervention | Experimental - |
| Intervention | Experimental - |
| Intervention | Experimental - |
| Intervention | Experimental - |
| Intervention | Experimental - |
| Intervention | Experimental - |
| Intervention | Experimental - |
| Intervention | Experimental - |
| Intervention | Experimental - |
| Intervention | Experimental - |
| Intervention | Experimental - |
| Intervention | Experimental - |
| Intervention | Experimental - |
| Intervention | Experimental - |
| Intervention | Experimental - |
| Intervention | Experimental - |
| Intervention | Experimental - |
| Intervention | Experimental - |
| Intervention | Experimental - |
| Intervention | Experimental - |
| Intervention | Experimental - |
| Intervention | Experimental - |
| Intervention | Experimental - |
| Intervention | Experimental - |
| Intervention | Experimental - |
| Intervention | Experimental - |
| Intervention | Experimental - |
| Intervention | Experimental - |
| Intervention | Experimental - |
| Intervention | Experimental - |
| Intervention | Experimental - |
| Intervention | Experimental - |
| Intervention | Experimental - |

| | |
|---|---|
| Intervention | Experimental - |
| Intervention | Experimental - |
| Intervention | Experimental - |
| Intervention | Experimental - |
| Intervention | Experimental - |
| Intervention | Experimental - |
| Intervention | Experimental - |
| Intervention | Experimental - |
| Intervention | Experimental - |
| Intervention | Experimental - |
| Intervention | Experimental - |
| Intervention | Experimental - |
| Intervention | Experimental - |
| Intervention | Experimental - |
| Intervention | Experimental - |
| Intervention | Experimental - |
| Intervention | Experimental - |
| Intervention | Experimental - |
| Intervention | Experimental - |
| Intervention | Experimental - |
| Intervention | Experimental - |
| Intervention | Experimental - |
| Intervention | Experimental - |
| Intervention | Experimental - |
| Intervention | Experimental - |
| Intervention | Experimental - |
| Intervention | Experimental - |
| Intervention | Experimental - |
| Intervention | Experimental - |
| Intervention | Experimental - |
| Intervention | Experimental - |
| Intervention | Experimental - |
| Intervention | Experimental - |
| Intervention | Experimental - |
| Intervention | Experimental - |
| Intervention | Experimental - |
| Intervention | Experimental - |
| Intervention | Experimental - |
| Intervention | Experimental - |
| Intervention | Experimental - |
| Intervention | Experimental - |
| Intervention | Experimental - |
| Intervention | Experimental - |
| Intervention | Experimental - |
| Intervention | Experimental - |
| Intervention | Experimental - |
| Intervention | Experimental - |
| Intervention | Experimental - |

| | |
|---|---|
| Intervention | Experimental - |
| Intervention | Experimental - |
| Intervention | Experimental - |
| Intervention | Experimental - |
| Intervention | Experimental - |
| Intervention | Experimental - |
| Intervention | Experimental - |
| Intervention | Experimental - |
| Intervention | Experimental - |
| Intervention | Experimental - |
| Intervention | Experimental - |
| Intervention | Experimental - |
| Intervention | Experimental - |
| Intervention | Experimental - |
| Intervention | Experimental - |
| Intervention | Experimental - |
| Intervention | Experimental - |
| Intervention | Experimental - |
| Intervention | Experimental - |
| Intervention | Experimental - |
| Intervention | Experimental - |
| Intervention | Experimental - |
| Intervention | Experimental - |
| Intervention | Experimental - |
| Intervention | Experimental - |
| Intervention | Experimental - |
| Intervention | Experimental - |
| Intervention | Experimental - |
| Intervention | Experimental - |
| Intervention | Experimental - |
| Intervention | Experimental - |
| Intervention | Experimental - |
| Intervention | Experimental - |
| Intervention | Experimental - |
| Intervention | Experimental - |
| Intervention | Experimental - |
| Intervention | Experimental - |
| Intervention | Experimental - |
| Intervention | Experimental - |
| Intervention | Experimental - |
| Intervention | Experimental - |
| Intervention | Experimental - |
| Intervention | Experimental - |
| Intervention | Experimental - |
| Intervention | Experimental - |
| Intervention | Experimental - |
| Intervention | Experimental - |
| Intervention | Experimental - |

| | |
|---|---|
| Intervention | Experimental - |
| Intervention | Experimental - |
| Intervention | Experimental - |
| Intervention | Experimental - |
| Intervention | Experimental - |
| Intervention | Experimental - |
| Intervention | Experimental - |
| Intervention | Experimental - |
| Intervention | Experimental - |
| Intervention | Experimental - |
| Intervention | Experimental - |
| Intervention | Experimental - |
| Intervention | Experimental - |
| Intervention | Experimental - |
| Intervention | Experimental - |
| Intervention | Experimental - |
| Intervention | Experimental - |
| Intervention | Experimental - |
| Intervention | Experimental - |
| Intervention | Experimental - |
| Intervention | Experimental - |
| Intervention | Experimental - |
| Intervention | Experimental - |
| Intervention | Experimental - |
| Intervention | Experimental - |
| Intervention | Experimental - |
| Intervention | Experimental - |
| Intervention | Experimental - |
| Intervention | Experimental - |
| Intervention | Experimental - |
| Intervention | Experimental - |
| Intervention | Experimental - |
| Intervention | Experimental - |
| Intervention | Experimental - |
| Intervention | Experimental - |
| Intervention | Experimental - |
| Intervention | Experimental - |
| Intervention | Experimental - |
| Intervention | Experimental - |
| Intervention | Experimental - |
| Intervention | Experimental - |
| Intervention | Experimental - |
| Intervention | Experimental - |
| Intervention | Experimental - |
| Intervention | Experimental - |
| Intervention | Experimental - |
| Intervention | Experimental - |
| Intervention | Experimental - |

| | |
|---|---|
| Intervention | Experimental - |
| Intervention | Experimental - |
| Intervention | Experimental - |
| Intervention | Experimental - |
| Intervention | Experimental - |
| Intervention | Experimental - |
| Intervention | Experimental - |
| Intervention | Experimental - |
| Intervention | Experimental - |
| Intervention | Experimental - |
| Intervention | Experimental - |
| Intervention | Experimental - |
| Intervention | Experimental - |
| Intervention | Experimental - |
| Intervention | Experimental - |
| Intervention | Experimental - |
| Intervention | Experimental - |
| Intervention | Experimental - |
| Intervention | Experimental - |
| Intervention | Experimental - |
| Intervention | Experimental - |
| Intervention | Experimental - |
| Intervention | Experimental - |
| Intervention | Experimental - |
| Intervention | Experimental - |
| Intervention | Experimental - |
| Intervention | Experimental - |
| Intervention | Experimental - |
| Intervention | Experimental - |
| Intervention | Experimental - |
| Intervention | Experimental - |
| Intervention | Experimental - |
| Intervention | Experimental - |
| Intervention | Experimental - |
| Intervention | Experimental - |
| Intervention | Experimental - |
| Intervention | Experimental - |
| Intervention | Experimental - |
| Intervention | Experimental - |
| Intervention | Experimental - |
| Intervention | Experimental - |
| Intervention | Experimental - |
| Intervention | Experimental - |
| Intervention | Experimental - |
| Intervention | Experimental - |
| Intervention | Experimental - |
| Intervention | Experimental - |
| Intervention | Experimental - |

| | |
|---|---|
| Intervention | Experimental - |
| Intervention | Experimental - |
| Intervention | Experimental - |
| Intervention | Experimental - |
| Intervention | Experimental - |
| Intervention | Experimental - |
| Intervention | Experimental - |
| Intervention | Experimental - |
| Intervention | Experimental - |
| Intervention | Experimental - |
| Intervention | Experimental - |
| Intervention | Experimental - |
| Intervention | Experimental - |
| Intervention | Experimental - |
| Intervention | Experimental - |
| Intervention | Experimental - |
| Intervention | Experimental - |
| Intervention | Experimental - |
| Intervention | Experimental - |
| Intervention | Experimental - |
| Intervention | Experimental - |
| Intervention | Experimental - |
| Intervention | Experimental - |
| Intervention | Experimental - |
| Intervention | Experimental - |
| Intervention | Experimental - |
| Intervention | Experimental - |
| Intervention | Experimental - |
| Intervention | Experimental - |
| Intervention | Experimental - |
| Intervention | Experimental - |
| Intervention | Experimental - |
| Intervention | Experimental - |
| Intervention | Experimental - |
| Intervention | Experimental - |
| Intervention | Experimental - |
| Intervention | Experimental - |
| Intervention | Experimental - |
| Intervention | Experimental - |
| Intervention | Experimental - |
| Intervention | Experimental - |
| Intervention | Experimental - |
| Intervention | Experimental - |
| Intervention | Experimental - |
| Intervention | Experimental - |
| Intervention | Experimental - |
| Intervention | Experimental - |
| Intervention | Experimental - |

| | |
|---|---|
| Intervention | Experimental - |
| Intervention | Experimental - |
| Intervention | Experimental - |
| Intervention | Experimental - |
| Intervention | Experimental - |
| Intervention | Experimental - |
| Intervention | Experimental - |
| Intervention | Experimental - |
| Intervention | Experimental - |
| Intervention | Experimental - |
| Intervention | Experimental - |
| Intervention | Experimental - |
| Intervention | Experimental - |
| Intervention | Experimental - |
| Intervention | Experimental - |
| Intervention | Experimental - |
| Intervention | Experimental - |
| Intervention | Experimental - |
| Intervention | Experimental - |
| Intervention | Experimental - |
| Intervention | Experimental - |
| Intervention | Experimental - |
| Intervention | Experimental - |
| Intervention | Experimental - |
| Intervention | Experimental - |
| Intervention | Experimental - |
| Intervention | Experimental - |
| Intervention | Experimental - |
| Intervention | Experimental - |
| Intervention | Experimental - |
| Intervention | Experimental - |
| Intervention | Experimental - |
| Intervention | Experimental - |
| Intervention | Experimental - |
| Intervention | Experimental - |
| Intervention | Experimental - |
| Intervention | Experimental - |
| Intervention | Experimental - |
| Intervention | Experimental - |
| Intervention | Experimental - |
| Intervention | Experimental - |
| Intervention | Experimental - |
| Intervention | Experimental - |
| Intervention | Experimental - |
| Intervention | Experimental - |
| Intervention | Experimental - |
| Intervention | Experimental - |
| Intervention | Experimental - |

| | |
|---|---|
| Intervention | Experimental - |
| Intervention | Experimental - |
| Intervention | Experimental - |
| Intervention | Experimental - |
| Intervention | Experimental - |
| Intervention | Experimental - |
| Intervention | Experimental - |
| Intervention | Experimental - |
| Intervention | Experimental - |
| Intervention | Experimental - |
| Intervention | Experimental - |
| Intervention | Experimental - |
| Intervention | Experimental - |
| Intervention | Experimental - |
| Intervention | Experimental - |
| Intervention | Experimental - |
| Intervention | Experimental - |
| Intervention | Experimental - |
| Intervention | Experimental - |
| Intervention | Experimental - |
| Intervention | Experimental - |
| Intervention | Experimental - |
| Intervention | Experimental - |
| Intervention | Experimental - |
| Intervention | Experimental - |
| Intervention | Experimental - |
| Intervention | Experimental - |
| Intervention | Experimental - |
| Intervention | Experimental - |
| Intervention | Experimental - |
| Intervention | Experimental - |
| Intervention | Experimental - |
| Intervention | Experimental - |
| Intervention | Experimental - |
| Intervention | Experimental - |
| Intervention | Experimental - |
| Intervention | Experimental - |
| Intervention | Experimental - |
| Intervention | Experimental - |
| Intervention | Experimental - |
| Intervention | Experimental - |
| Intervention | Experimental - |
| Intervention | Experimental - |
| Intervention | Experimental - |
| Intervention | Experimental - |
| Intervention | Experimental - |
| Intervention | Experimental - |
| Intervention | Experimental - |

| | |
|---|---|
| Intervention | Experimental - |
| Intervention | Experimental - |
| Intervention | Experimental - |
| Intervention | Experimental - |
| Intervention | Experimental - |
| Intervention | Experimental - |
| Intervention | Experimental - |
| Intervention | Experimental - |
| Intervention | Experimental - |
| Intervention | Experimental - |
| Intervention | Experimental - |
| Intervention | Experimental - |
| Intervention | Experimental - |
| Intervention | Experimental - |
| Intervention | Experimental - |
| Intervention | Experimental - |
| Intervention | Experimental - |
| Intervention | Experimental - |
| Intervention | Experimental - |
| Intervention | Experimental - |
| Intervention | Experimental - |
| Intervention | Experimental - |
| Intervention | Experimental - |
| Intervention | Experimental - |
| Intervention | Experimental - |
| Intervention | Experimental - |
| Intervention | Experimental - |
| Intervention | Experimental - |
| Intervention | Experimental - |
| Intervention | Experimental - |
| Intervention | Experimental - |
| Intervention | Experimental - |
| Intervention | Experimental - |
| Intervention | Experimental - |
| Intervention | Experimental - |
| Intervention | Experimental - |
| Intervention | Experimental - |
| Intervention | Experimental - |
| Intervention | Experimental - |
| Intervention | Experimental - |
| Intervention | Experimental - |
| Intervention | Experimental - |
| Intervention | Experimental - |
| Intervention | Experimental - |
| Intervention | Experimental - |
| Intervention | Experimental - |
| Intervention | Experimental - |
| Intervention | Experimental - |

| | |
|---|---|
| Intervention | Experimental - |
| Intervention | Experimental - |
| Intervention | Experimental - |
| Intervention | Experimental - |
| Intervention | Experimental - |
| Intervention | Experimental - |
| Intervention | Experimental - |
| Intervention | Experimental - |
| Intervention | Experimental - |
| Intervention | Experimental - |
| Intervention | Experimental - |
| Intervention | Experimental - |
| Intervention | Experimental - |
| Intervention | Experimental - |
| Intervention | Experimental - |
| Intervention | Experimental - |
| Intervention | Experimental - |
| Intervention | Experimental - |
| Intervention | Experimental - |
| Intervention | Experimental - |
| Intervention | Experimental - |
| Intervention | Experimental - |
| Intervention | Experimental - |
| Intervention | Experimental - |
| Intervention | Experimental - |
| Intervention | Experimental - |
| Intervention | Experimental - |
| Intervention | Experimental - |
| Intervention | Experimental - |
| Intervention | Experimental - |
| Intervention | Experimental - |
| Intervention | Experimental - |
| Intervention | Experimental - |
| Intervention | Experimental - |
| Intervention | Experimental - |
| Intervention | Experimental - |
| Intervention | Experimental - |
| Intervention | Experimental - |
| Intervention | Experimental - |
| Intervention | Experimental - |
| Intervention | Experimental - |
| Intervention | Experimental - |
| Intervention | Experimental - |
| Intervention | Experimental - |
| Intervention | Experimental - |
| Intervention | Experimental - |
| Intervention | Experimental - |
| Intervention | Experimental - |

| | |
|---|---|
| Intervention | Experimental - |
| Intervention | Experimental - |
| Intervention | Experimental - |
| Intervention | Experimental - |
| Intervention | Experimental - |
| Intervention | Experimental - |
| Intervention | Experimental - |
| Intervention | Experimental - |
| Intervention | Experimental - |
| Intervention | Experimental - |
| Intervention | Experimental - |
| Intervention | Experimental - |
| Intervention | Experimental - |
| Intervention | Experimental - |
| Intervention | Experimental - |
| Intervention | Experimental - |
| Intervention | Experimental - |
| Intervention | Experimental - |
| Intervention | Experimental - |
| Intervention | Experimental - |
| Intervention | Experimental - |
| Intervention | Experimental - |
| Intervention | Experimental - |
| Intervention | Experimental - |
| Intervention | Experimental - |
| Intervention | Experimental - |
| Intervention | Experimental - |
| Intervention | Experimental - |
| Intervention | Experimental - |
| Intervention | Experimental - |
| Intervention | Experimental - |
| Intervention | Experimental - |
| Intervention | Experimental - |
| Intervention | Experimental - |
| Intervention | Experimental - |
| Intervention | Experimental - |
| Intervention | Experimental - |
| Intervention | Experimental - |
| Intervention | Experimental - |
| Intervention | Experimental - |
| Intervention | Experimental - |
| Intervention | Experimental - |
| Intervention | Experimental - |
| Intervention | Experimental - |
| Intervention | Experimental - |
| Intervention | Experimental - |
| Intervention | Experimental - |
| Intervention | Experimental - |

| | |
|---|---|
| Intervention | Experimental - |
| Intervention | Experimental - |
| Intervention | Experimental - |
| Intervention | Experimental - |
| Intervention | Experimental - |
| Intervention | Experimental - |
| Intervention | Experimental - |
| Intervention | Experimental - |
| Intervention | Experimental - |
| Intervention | Experimental - |
| Intervention | Experimental - |
| Intervention | Experimental - |
| Intervention | Experimental - |
| Intervention | Experimental - |
| Intervention | Experimental - |
| Intervention | Experimental - |
| Intervention | Experimental - |
| Intervention | Experimental - |
| Intervention | Experimental - |
| Intervention | Experimental - |
| Intervention | Experimental - |
| Intervention | Experimental - |
| Intervention | Experimental - |
| Intervention | Experimental - |
| Intervention | Experimental - |
| Intervention | Experimental - |
| Intervention | Experimental - |
| Intervention | Experimental - |
| Intervention | Experimental - |
| Intervention | Experimental - |
| Intervention | Experimental - |
| Intervention | Experimental - |
| Intervention | Experimental - |
| Intervention | Experimental - |
| Intervention | Experimental - |
| Intervention | Experimental - |
| Intervention | Experimental - |
| Intervention | Experimental - |
| Intervention | Experimental - |
| Intervention | Experimental - |
| Intervention | Experimental - |
| Intervention | Experimental - |
| Intervention | Experimental - |
| Intervention | Experimental - |
| Intervention | Experimental - |
| Intervention | Experimental - |
| Intervention | Experimental - |
| Intervention | Experimental - |

| | |
|---|---|
| Intervention | Experimental - |
| Intervention | Experimental - |
| Intervention | Experimental - |
| Intervention | Experimental - |
| Intervention | Experimental - |
| Intervention | Experimental - |
| Intervention | Experimental - |
| Intervention | Experimental - |
| Intervention | Experimental - |
| Intervention | Experimental - |
| Intervention | Experimental - |
| Intervention | Experimental - |
| Intervention | Experimental - |
| Intervention | Experimental - |
| Intervention | Experimental - |
| Intervention | Experimental - |
| Intervention | Experimental - |
| Intervention | Experimental - |
| Intervention | Experimental - |
| Intervention | Experimental - |
| Intervention | Experimental - |
| Intervention | Experimental - |
| Intervention | Experimental - |
| Intervention | Experimental - |
| Intervention | Experimental - |
| Intervention | Experimental - |
| Intervention | Experimental - |
| Intervention | Experimental - |
| Intervention | Experimental - |
| Intervention | Experimental - |
| Intervention | Experimental - |
| Intervention | Experimental - |
| Intervention | Experimental - |
| Intervention | Experimental - |
| Intervention | Experimental - |
| Intervention | Experimental - |
| Intervention | Experimental - |
| Intervention | Experimental - |
| Intervention | Experimental - |
| Intervention | Experimental - |
| Intervention | Experimental - |
| Intervention | Experimental - |
| Intervention | Experimental - |
| Intervention | Experimental - |
| Intervention | Experimental - |
| Intervention | Experimental - |
| Intervention | Experimental - |
| Intervention | Experimental - |

| | |
|---|---|
| Intervention | Experimental - |
| Intervention | Experimental - |
| Intervention | Experimental - |
| Intervention | Experimental - |
| Intervention | Experimental - |
| Intervention | Experimental - |
| Intervention | Experimental - |
| Intervention | Experimental - |
| Intervention | Experimental - |
| Intervention | Experimental - |
| Intervention | Experimental - |
| Intervention | Experimental - |
| Intervention | Experimental - |
| Intervention | Experimental - |
| Intervention | Experimental - |
| Intervention | Experimental - |
| Intervention | Experimental - |
| Intervention | Experimental - |
| Intervention | Experimental - |
| Intervention | Experimental - |
| Intervention | Experimental - |
| Intervention | Experimental - |
| Intervention | Experimental - |
| Intervention | Experimental - |
| Intervention | Experimental - |
| Intervention | Experimental - |
| Intervention | Experimental - |
| Intervention | Experimental - |
| Intervention | Experimental - |
| Intervention | Experimental - |
| Intervention | Experimental - |
| Intervention | Experimental - |
| Intervention | Experimental - |
| Intervention | Experimental - |
| Intervention | Experimental - |
| Intervention | Experimental - |
| Intervention | Experimental - |
| Intervention | Experimental - |
| Intervention | Experimental - |
| Intervention | Experimental - |
| Intervention | Experimental - |
| Intervention | Experimental - |
| Intervention | Experimental - |
| Intervention | Experimental - |
| Intervention | Experimental - |
| Intervention | Experimental - |
| Intervention | Experimental - |
| Intervention | Experimental - |

| | |
|---|---|
| Intervention | Experimental - |
| Intervention | Experimental - |
| Intervention | Experimental - |
| Intervention | Experimental - |
| Intervention | Experimental - |
| Intervention | Experimental - |
| Intervention | Experimental - |
| Intervention | Experimental - |
| Intervention | Experimental - |
| Intervention | Experimental - |
| Intervention | Experimental - |
| Intervention | Experimental - |
| Intervention | Experimental - |
| Intervention | Experimental - |
| Intervention | Experimental - |
| Intervention | Experimental - |
| Intervention | Experimental - |
| Intervention | Experimental - |
| Intervention | Experimental - |
| Intervention | Experimental - |
| Intervention | Experimental - |
| Intervention | Experimental - |
| Intervention | Experimental - |
| Intervention | Experimental - |
| Intervention | Experimental - |
| Intervention | Experimental - |
| Intervention | Experimental - |
| Intervention | Experimental - |
| Intervention | Experimental - |
| Intervention | Experimental - |
| Intervention | Experimental - |
| Intervention | Experimental - |
| Intervention | Experimental - |
| Intervention | Experimental - |
| Intervention | Experimental - |
| Intervention | Experimental - |
| Intervention | Experimental - |
| Intervention | Experimental - |
| Intervention | Experimental - |
| Intervention | Experimental - |
| Intervention | Experimental - |
| Intervention | Experimental - |
| Intervention | Experimental - |
| Intervention | Experimental - |
| Intervention | Experimental - |
| Intervention | Experimental - |
| Intervention | Experimental - |
| Intervention | Experimental - |

| | |
|---|---|
| Intervention | Experimental - |
| Intervention | Experimental - |
| Intervention | Experimental - |
| Intervention | Experimental - |
| Intervention | Experimental - |
| Intervention | Experimental - |
| Intervention | Experimental - |
| Intervention | Experimental - |
| Intervention | Experimental - |
| Intervention | Experimental - |
| Intervention | Experimental - |
| Intervention | Experimental - |
| Intervention | Experimental - |
| Intervention | Experimental - |

| Alternative group | Reference group | FDR | P-value | Effect size |
|---|---|---|---|---|
| Experimental | Control | 0,970062739 | 0,650759425 | -0,21189443 |
| Experimental | Control | 0,970062739 | 0,534073748 | -0,306524892 |
| Experimental | Control | 0,970062739 | 0,276437863 | -0,99281447 |
| Experimental | Control | 0,970062739 | 0,575902513 | -0,180460269 |
| Experimental | Control | 0,99640621 | 0,988121404 | -0,006772247 |
| Experimental | Control | 0,970062739 | 0,34355193 | -0,472971223 |
| Experimental | Control | 0,87952483 | 0,111131296 | -0,697111087 |
| Experimental | Control | 0,78357626 | 0,044978235 | -1,347641877 |
| Experimental | Control | 0,78357626 | 0,030410529 | -0,96636063 |
| Experimental | Control | 0,988586169 | 0,961916922 | -0,026610055 |
| Experimental | Control | 0,970062739 | 0,253816224 | -0,78605949 |
| Experimental | Control | 0,870685429 | 0,07464048 | -0,748855096 |
| Experimental | Control | 0,970062739 | 0,481102054 | -0,528283637 |
| Experimental | Control | 0,970062739 | 0,222440006 | -0,657904262 |
| Experimental | Control | 0,694796407 | 0,012122597 | -1,826717914 |
| Experimental | Control | 0,970062739 | 0,555539844 | -0,336364912 |
| Experimental | Control | 0,970062739 | 0,400557698 | 0,386744088 |
| Experimental | Control | 0,970062739 | 0,497580853 | -0,553270509 |
| Experimental | Control | 0,98331311 | 0,938203422 | -0,042008688 |
| Experimental | Control | 0,970062739 | 0,435144181 | 0,264269499 |
| Experimental | Control | 0,970062739 | 0,435100331 | 0,409757933 |
| Experimental | Control | 0,970062739 | 0,18944268 | 0,709529109 |
| Experimental | Control | 0,98331311 | 0,923229872 | 0,044301136 |
| Experimental | Control | 0,970062739 | 0,727676874 | -0,220911337 |
| Experimental | Control | 0,970062739 | 0,500369741 | 0,210869963 |
| Experimental | Control | 0,899730559 | 0,122821576 | 1,02439221 |
| Experimental | Control | 0,970062739 | 0,331056629 | -0,478116209 |
| Experimental | Control | 0,87952483 | 0,09316543 | -1,146680364 |
| Experimental | Control | 0,78357626 | 0,030568796 | -1,719346343 |
| Experimental | Control | 0,970062739 | 0,69751051 | -0,233777301 |
| Experimental | Control | 0,970062739 | 0,699190003 | 0,221093584 |
| Experimental | Control | 0,78357626 | 0,044022783 | -0,872886816 |
| Experimental | Control | 0,87952483 | 0,10486208 | -1,58367612 |
| Experimental | Control | 0,870685429 | 0,07672779 | -1,207287048 |
| Experimental | Control | 0,970062739 | 0,187646107 | -0,723762982 |
| Experimental | Control | 0,970062739 | 0,701589708 | -0,180049764 |
| Experimental | Control | 0,012359826 | 2,23102E-05 | -2,106345056 |
| Experimental | Control | 0,970062739 | 0,653878028 | -0,403758709 |
| Experimental | Control | 0,98331311 | 0,914038379 | 0,052748297 |
| Experimental | Control | 0,98331311 | 0,878825237 | 0,104004172 |
| Experimental | Control | 0,970062739 | 0,332035898 | -0,728177334 |
| Experimental | Control | 0,970062739 | 0,528293441 | -0,386212096 |
| Experimental | Control | 0,970062739 | 0,542802211 | -0,320499787 |
| Experimental | Control | 0,970062739 | 0,425111436 | -0,308916984 |
| Experimental | Control | 0,970062739 | 0,190676924 | -0,845080399 |
| Experimental | Control | 0,970062739 | 0,256847058 | -0,3653499 |
| Experimental | Control | 0,970062739 | 0,721549482 | -0,16694102 |

| | | | | |
|---|---|---|---|---|
| Experimental | Control | 0,868668495 | 0,059680137 | -0,867010148 |
| Experimental | Control | 0,78357626 | 0,042116188 | -1,179658157 |
| Experimental | Control | 0,899730559 | 0,124969413 | -0,687439475 |
| Experimental | Control | 0,96190808 | 0,177024859 | 0,355522695 |
| Experimental | Control | 0,98331311 | 0,937623388 | 0,039467392 |
| Experimental | Control | 0,899730559 | 0,133224922 | -1,079930265 |
| Experimental | Control | 0,694796407 | 0,004635114 | -1,494725287 |
| Experimental | Control | 0,78357626 | 0,047891032 | -1,394480252 |
| Experimental | Control | 0,885468273 | 0,113480591 | -0,71193324 |
| Experimental | Control | 0,991002471 | 0,967747901 | -0,020859062 |
| Experimental | Control | 0,970062739 | 0,64514097 | 0,123245791 |
| Experimental | Control | 0,98331311 | 0,905345751 | -0,070308762 |
| Experimental | Control | 0,970062739 | 0,610233203 | 0,336204672 |
| Experimental | Control | 0,870685429 | 0,077764411 | -0,859386195 |
| Experimental | Control | 0,899730559 | 0,149413739 | -0,66408938 |
| Experimental | Control | 0,973116013 | 0,790270458 | -0,172442958 |
| Experimental | Control | 0,970062739 | 0,203825972 | -0,774664506 |
| Experimental | Control | 0,87952483 | 0,107139099 | -1,051742395 |
| Experimental | Control | 0,970062739 | 0,233685495 | -0,779545538 |
| Experimental | Control | 0,970062739 | 0,626244742 | 0,207052713 |
| Experimental | Control | 0,98331311 | 0,916072869 | -0,056338555 |
| Experimental | Control | 0,973116013 | 0,764301455 | 0,137289429 |
| Experimental | Control | 0,87952483 | 0,109774845 | -0,979394965 |
| Experimental | Control | 0,970062739 | 0,341663554 | 0,343466593 |
| Experimental | Control | 0,970062739 | 0,582953175 | 0,189718322 |
| Experimental | Control | 0,970062739 | 0,552662958 | -0,382062012 |
| Experimental | Control | 0,868668495 | 0,061151753 | 0,395298829 |
| Experimental | Control | 0,981816738 | 0,850482586 | -0,056843318 |
| Experimental | Control | 0,694796407 | 0,01486486 | -1,007305099 |
| Experimental | Control | 0,970062739 | 0,723601049 | 0,276067665 |
| Experimental | Control | 0,973116013 | 0,772444835 | 0,162985945 |
| Experimental | Control | 0,970062739 | 0,242804369 | -0,596977076 |
| Experimental | Control | 0,970062739 | 0,665009309 | -0,336984912 |
| Experimental | Control | 0,970062739 | 0,505456658 | -0,248006812 |
| Experimental | Control | 0,927436375 | 0,165349054 | -0,874115275 |
| Experimental | Control | 0,87952483 | 0,110796941 | -0,548704728 |
| Experimental | Control | 0,870685429 | 0,068747236 | -1,864977817 |
| Experimental | Control | 0,970062739 | 0,504232443 | -0,44448198 |
| Experimental | Control | 0,970062739 | 0,486984094 | 0,264306925 |
| Experimental | Control | 0,973116013 | 0,769108995 | -0,101914543 |
| Experimental | Control | 0,970062739 | 0,285910984 | -0,553721381 |
| Experimental | Control | 0,927436375 | 0,160379658 | -0,560713755 |
| Experimental | Control | 0,988586169 | 0,958437185 | -0,025596259 |
| Experimental | Control | 0,899730559 | 0,146305762 | -1,004226586 |
| Experimental | Control | 0,970062739 | 0,697958191 | 0,198170134 |
| Experimental | Control | 0,970062739 | 0,216521768 | -0,829240459 |
| Experimental | Control | 0,899730559 | 0,142622986 | -1,070175769 |
| Experimental | Control | 0,999707234 | 0,99817574 | -0,002522927 |

| | | | | |
|---|---|---|---|---|
| Experimental | Control | 0,970062739 | 0,629112373 | -0,365372005 |
| Experimental | Control | 0,970062739 | 0,302358093 | -0,482599553 |
| Experimental | Control | 0,970062739 | 0,326100559 | -0,999950187 |
| Experimental | Control | 0,694796407 | 0,005463171 | -1,211942473 |
| Experimental | Control | 0,87952483 | 0,102710494 | -1,250611368 |
| Experimental | Control | 0,981816738 | 0,829285624 | 0,119814711 |
| Experimental | Control | 0,98331311 | 0,891473711 | -0,08113974 |
| Experimental | Control | 0,970062739 | 0,332565734 | -0,508061903 |
| Experimental | Control | 0,970062739 | 0,419076159 | 0,346216361 |
| Experimental | Control | 0,970062739 | 0,400357668 | -0,263175865 |
| Experimental | Control | 0,981816738 | 0,859526936 | -0,074162372 |
| Experimental | Control | 0,970062739 | 0,229927096 | -0,416866041 |
| Experimental | Control | 0,981816738 | 0,834354347 | 0,05882421 |
| Experimental | Control | 0,970062739 | 0,527302198 | 0,243376092 |
| Experimental | Control | 0,970062739 | 0,437526315 | 0,339059207 |
| Experimental | Control | 0,986356047 | 0,951265488 | -0,026684967 |
| Experimental | Control | 0,970062739 | 0,356527044 | -0,49231504 |
| Experimental | Control | 0,970062739 | 0,412574491 | -0,307017947 |
| Experimental | Control | 0,970062739 | 0,598154161 | 0,276979874 |
| Experimental | Control | 0,694796407 | 0,009426922 | -1,399437904 |
| Experimental | Control | 0,970062739 | 0,635821852 | 0,197782343 |
| Experimental | Control | 0,970062739 | 0,267486882 | 0,393610093 |
| Experimental | Control | 0,970062739 | 0,230175971 | -0,430798163 |
| Experimental | Control | 0,970062739 | 0,676439189 | -0,160080124 |
| Experimental | Control | 0,98331311 | 0,871925533 | -0,124198165 |
| Experimental | Control | 0,972536115 | 0,756351388 | -0,140402215 |
| Experimental | Control | 0,970062739 | 0,41320437 | -0,394581111 |
| Experimental | Control | 0,970062739 | 0,254994776 | 0,328148376 |
| Experimental | Control | 0,981816738 | 0,842945414 | -0,147208788 |
| Experimental | Control | 0,99640621 | 0,983955894 | -0,010267509 |
| Experimental | Control | 0,970062739 | 0,449834829 | 0,386745037 |
| Experimental | Control | 0,970062739 | 0,712580932 | 0,26724192 |
| Experimental | Control | 0,981816738 | 0,829844364 | 0,142541396 |
| Experimental | Control | 0,99640621 | 0,988022128 | -0,007344202 |
| Experimental | Control | 0,870685429 | 0,080153352 | -1,259856595 |
| Experimental | Control | 0,970062739 | 0,651544126 | 0,264117133 |
| Experimental | Control | 0,970062739 | 0,660383043 | -0,201193301 |
| Experimental | Control | 0,970062739 | 0,341053772 | -0,530555826 |
| Experimental | Control | 0,970062739 | 0,554363074 | -0,277852125 |
| Experimental | Control | 0,735000532 | 0,018574021 | 1,570420003 |
| Experimental | Control | 0,970062739 | 0,63720742 | -0,206342924 |
| Experimental | Control | 0,970062739 | 0,539748039 | -0,208439752 |
| Experimental | Control | 0,970062739 | 0,341617363 | -0,841030219 |
| Experimental | Control | 0,870685429 | 0,07650319 | -0,856263566 |
| Experimental | Control | 0,970062739 | 0,494107315 | -0,188752225 |
| Experimental | Control | 0,970062739 | 0,386676726 | -0,436640043 |
| Experimental | Control | 0,970062739 | 0,737917996 | -0,168733761 |
| Experimental | Control | 0,78357626 | 0,044020493 | -0,756782651 |

| | | | | |
|---|---|---|---|---|
| Experimental | Control | 0,970062739 | 0,655758741 | 0,154835618 |
| Experimental | Control | 0,970062739 | 0,436261443 | 0,454385194 |
| Experimental | Control | 0,970062739 | 0,568500283 | 0,237836162 |
| Experimental | Control | 0,973959896 | 0,79639681 | -0,18188184 |
| Experimental | Control | 0,973116013 | 0,777626737 | 0,097135012 |
| Experimental | Control | 0,970062739 | 0,715837793 | -0,183791297 |
| Experimental | Control | 0,981816738 | 0,820459992 | -0,14649311 |
| Experimental | Control | 0,970062739 | 0,72126241 | -0,072151035 |
| Experimental | Control | 0,87952483 | 0,105434381 | -0,806937604 |
| Experimental | Control | 0,970062739 | 0,491446827 | -0,374441306 |
| Experimental | Control | 0,970062739 | 0,315405799 | 0,526050906 |
| Experimental | Control | 0,970062739 | 0,374502775 | 0,604915906 |
| Experimental | Control | 0,78357626 | 0,042619729 | 0,702382571 |
| Experimental | Control | 0,970062739 | 0,553576255 | 0,327898374 |
| Experimental | Control | 0,970062739 | 0,592894029 | 0,327260551 |
| Experimental | Control | 0,899730559 | 0,14680433 | 1,443481912 |
| Experimental | Control | 0,973116013 | 0,790437195 | -0,216613414 |
| Experimental | Control | 0,98331311 | 0,868483423 | 0,112149004 |
| Experimental | Control | 0,970062739 | 0,364292095 | -0,138817412 |
| Experimental | Control | 0,970062739 | 0,276539835 | -0,409995632 |
| Experimental | Control | 0,970062739 | 0,410381043 | 0,206245949 |
| Experimental | Control | 0,98331311 | 0,871769253 | -0,079174738 |
| Experimental | Control | 0,970062739 | 0,258914605 | -0,792255883 |
| Experimental | Control | 0,970062739 | 0,435854797 | 0,431265877 |
| Experimental | Control | 0,870685429 | 0,075362443 | -0,849313126 |
| Experimental | Control | 0,970062739 | 0,444726523 | -0,323539678 |
| Experimental | Control | 0,970062739 | 0,514417255 | -0,409490027 |
| Experimental | Control | 0,970062739 | 0,34387338 | -0,276584076 |
| Experimental | Control | 0,78357626 | 0,040520613 | -1,41185282 |
| Experimental | Control | 0,98331311 | 0,907791096 | 0,040267991 |
| Experimental | Control | 0,972536115 | 0,751739086 | -0,077624066 |
| Experimental | Control | 0,927436375 | 0,164435758 | 0,737371524 |
| Experimental | Control | 0,970062739 | 0,298464306 | -0,456839161 |
| Experimental | Control | 0,970062739 | 0,363913179 | 0,459617007 |
| Experimental | Control | 0,98331311 | 0,934357992 | 0,041327997 |
| Experimental | Control | 0,98331311 | 0,939628161 | -0,033602758 |
| Experimental | Control | 0,970062739 | 0,683320785 | -0,174948495 |
| Experimental | Control | 0,970062739 | 0,40028721 | -0,285615969 |
| Experimental | Control | 0,981816738 | 0,856246029 | -0,055281321 |
| Experimental | Control | 0,970062739 | 0,366835579 | -0,385503137 |
| Experimental | Control | 0,970062739 | 0,449807398 | -0,262455673 |
| Experimental | Control | 0,970062739 | 0,328641132 | -0,284366833 |
| Experimental | Control | 0,694796407 | 0,002727909 | -0,940298606 |
| Experimental | Control | 0,98331311 | 0,885562413 | -0,069451549 |
| Experimental | Control | 0,970062739 | 0,564589213 | 0,302965 |
| Experimental | Control | 0,970062739 | 0,321754823 | -0,20806914 |
| Experimental | Control | 0,98331311 | 0,925366727 | -0,053340081 |
| Experimental | Control | 0,972536115 | 0,754601096 | 0,100342022 |

| | | | | |
|---|---|---|---|---|
| Experimental | Control | 0,981816738 | 0,850982853 | 0,043529249 |
| Experimental | Control | 0,970062739 | 0,673995303 | -0,121430102 |
| Experimental | Control | 0,87952483 | 0,085033409 | 1,024183533 |
| Experimental | Control | 0,970062739 | 0,70337637 | 0,066068875 |
| Experimental | Control | 0,970062739 | 0,240208389 | 0,214543341 |
| Experimental | Control | 0,970062739 | 0,436177848 | -0,601790286 |
| Experimental | Control | 0,986356047 | 0,952527951 | -0,023988885 |
| Experimental | Control | 0,870685429 | 0,066088219 | -1,068958325 |
| Experimental | Control | 0,973116013 | 0,780023747 | -0,038867107 |
| Experimental | Control | 0,868668495 | 0,057155333 | -0,938651603 |
| Experimental | Control | 0,970062739 | 0,492988572 | 0,231896197 |
| Experimental | Control | 0,970062739 | 0,188566573 | -0,595894716 |
| Experimental | Control | 0,981816738 | 0,859532704 | 0,086450283 |
| Experimental | Control | 0,970062739 | 0,520138615 | -0,315794832 |
| Experimental | Control | 0,970062739 | 0,671384997 | -0,204835468 |
| Experimental | Control | 0,981816738 | 0,856634255 | 0,141756756 |
| Experimental | Control | 0,970062739 | 0,379800212 | -0,257455104 |
| Experimental | Control | 0,970062739 | 0,250814477 | -0,827321026 |
| Experimental | Control | 0,970062739 | 0,396945041 | -0,319774346 |
| Experimental | Control | 0,970062739 | 0,311962399 | -0,746860298 |
| Experimental | Control | 0,99640621 | 0,989108206 | 0,006247958 |
| Experimental | Control | 0,970062739 | 0,719907828 | -0,155387972 |
| Experimental | Control | 0,970062739 | 0,739849459 | 0,244170613 |
| Experimental | Control | 0,970062739 | 0,706922443 | 0,213562901 |
| Experimental | Control | 0,997096582 | 0,993496955 | -0,002294959 |
| Experimental | Control | 0,970062739 | 0,29556379 | -0,35270128 |
| Experimental | Control | 0,98331311 | 0,927147988 | -0,021196247 |
| Experimental | Control | 0,970062739 | 0,575251302 | 0,258777405 |
| Experimental | Control | 0,970062739 | 0,393361804 | 0,116152096 |
| Experimental | Control | 0,970062739 | 0,732877364 | 0,101893001 |
| Experimental | Control | 0,970062739 | 0,539639455 | -0,320323503 |
| Experimental | Control | 0,78357626 | 0,041941598 | -1,035736386 |
| Experimental | Control | 0,970062739 | 0,411961029 | 0,185059495 |
| Experimental | Control | 0,972536115 | 0,758511599 | -0,139855653 |
| Experimental | Control | 0,98331311 | 0,912305191 | -0,031772257 |
| Experimental | Control | 0,970062739 | 0,686715364 | 0,265197474 |
| Experimental | Control | 0,98331311 | 0,886459339 | -0,036116886 |
| Experimental | Control | 0,78357626 | 0,03230963 | 0,942641345 |
| Experimental | Control | 0,970062739 | 0,416978425 | 0,264604472 |
| Experimental | Control | 0,970062739 | 0,520940117 | 0,349808904 |
| Experimental | Control | 0,970062739 | 0,233903982 | 0,238125767 |
| Experimental | Control | 0,981816738 | 0,822179402 | 0,054099412 |
| Experimental | Control | 0,970062739 | 0,301127033 | 0,372768694 |
| Experimental | Control | 0,981816738 | 0,814527675 | 0,041280242 |
| Experimental | Control | 0,970062739 | 0,382273808 | -0,209242752 |
| Experimental | Control | 0,970062739 | 0,333017594 | -0,590808949 |
| Experimental | Control | 0,927436375 | 0,165669147 | -0,991068234 |
| Experimental | Control | 0,970062739 | 0,656465424 | 0,119913674 |

| | | | | |
|---|---|---|---|---|
| Experimental | Control | 0,970062739 | 0,485656002 | -0,286499863 |
| Experimental | Control | 0,899730559 | 0,124424835 | -0,614929226 |
| Experimental | Control | 0,99640621 | 0,989211941 | -0,007554744 |
| Experimental | Control | 0,993132522 | 0,975205942 | 0,008685328 |
| Experimental | Control | 0,970062739 | 0,33831839 | -0,431001131 |
| Experimental | Control | 0,970062739 | 0,23287107 | -0,488661671 |
| Experimental | Control | 0,970062739 | 0,557050182 | -0,112398044 |
| Experimental | Control | 0,970062739 | 0,452205903 | 0,612466323 |
| Experimental | Control | 0,78357626 | 0,035307865 | 0,745090156 |
| Experimental | Control | 0,970062739 | 0,713140057 | -0,205617799 |
| Experimental | Control | 0,970062739 | 0,333009193 | -0,376705623 |
| Experimental | Control | 0,970062739 | 0,543072737 | -0,190756959 |
| Experimental | Control | 0,981816738 | 0,851359086 | 0,092812734 |
| Experimental | Control | 0,981816738 | 0,836297828 | -0,075928185 |
| Experimental | Control | 0,970062739 | 0,255202334 | -0,392680075 |
| Experimental | Control | 0,970062739 | 0,73562113 | 0,127607777 |
| Experimental | Control | 0,868668495 | 0,05934936 | -0,952442575 |
| Experimental | Control | 0,98331311 | 0,930695808 | 0,04772251 |
| Experimental | Control | 0,970062739 | 0,523252587 | 0,169609961 |
| Experimental | Control | 0,970062739 | 0,21967439 | -1,118758265 |
| Experimental | Control | 0,972536115 | 0,761331715 | 0,118069119 |
| Experimental | Control | 0,970062739 | 0,601170404 | 0,192545652 |
| Experimental | Control | 0,98331311 | 0,93763168 | -0,015050488 |
| Experimental | Control | 0,970062739 | 0,287626988 | 0,407649117 |
| Experimental | Control | 0,970062739 | 0,3842793 | 0,533221913 |
| Experimental | Control | 0,970062739 | 0,507061189 | 0,325523513 |
| Experimental | Control | 0,970062739 | 0,468844472 | 0,206018595 |
| Experimental | Control | 0,98331311 | 0,933510972 | -0,050040564 |
| Experimental | Control | 0,87952483 | 0,101313729 | -0,454962571 |
| Experimental | Control | 0,970062739 | 0,417492476 | -0,546110201 |
| Experimental | Control | 0,970062739 | 0,398052614 | -0,302204145 |
| Experimental | Control | 0,970062739 | 0,369554774 | -0,245841925 |
| Experimental | Control | 0,970062739 | 0,44161322 | 0,60639475 |
| Experimental | Control | 0,970062739 | 0,655359706 | -0,251854556 |
| Experimental | Control | 0,972536115 | 0,756261318 | 0,171740815 |
| Experimental | Control | 0,927436375 | 0,165733215 | -0,419779205 |
| Experimental | Control | 0,973116013 | 0,789587281 | 0,188508081 |
| Experimental | Control | 0,970062739 | 0,600465947 | 0,136797714 |
| Experimental | Control | 0,970062739 | 0,532094128 | -0,408503998 |
| Experimental | Control | 0,981816738 | 0,838160263 | -0,114647507 |
| Experimental | Control | 0,970062739 | 0,466491741 | 0,254601561 |
| Experimental | Control | 0,970062739 | 0,309182017 | 0,333823483 |
| Experimental | Control | 0,970062739 | 0,298099435 | 0,226558381 |
| Experimental | Control | 0,981816738 | 0,849047548 | 0,121992807 |
| Experimental | Control | 0,970062739 | 0,349599321 | -0,326728293 |
| Experimental | Control | 0,970062739 | 0,588069376 | -0,27514946 |
| Experimental | Control | 0,970062739 | 0,211154586 | 0,544900792 |
| Experimental | Control | 0,970062739 | 0,67396112 | 0,129695901 |

| | | | | |
|---|---|---|---|---|
| Experimental | Control | 0,970062739 | 0,338007157 | -0,313903697 |
| Experimental | Control | 0,970062739 | 0,45554879 | -0,339665016 |
| Experimental | Control | 0,98331311 | 0,941788555 | -0,045462864 |
| Experimental | Control | 0,981816738 | 0,833585721 | -0,034816488 |
| Experimental | Control | 0,87952483 | 0,108179387 | -0,582093847 |
| Experimental | Control | 0,870685429 | 0,063843747 | -0,310180804 |
| Experimental | Control | 0,970062739 | 0,257156655 | 0,668946027 |
| Experimental | Control | 0,970062739 | 0,430417435 | -0,538789328 |
| Experimental | Control | 0,970062739 | 0,654029975 | -0,112382741 |
| Experimental | Control | 0,970062739 | 0,555618908 | 0,188377295 |
| Experimental | Control | 0,981816738 | 0,857563769 | -0,11207261 |
| Experimental | Control | 0,970062739 | 0,598044611 | -0,296888656 |
| Experimental | Control | 0,970062739 | 0,426424346 | -0,478807054 |
| Experimental | Control | 0,970062739 | 0,611906228 | 0,145948901 |
| Experimental | Control | 0,970062739 | 0,196586356 | -0,826791299 |
| Experimental | Control | 0,98331311 | 0,928826155 | 0,054360779 |
| Experimental | Control | 0,970062739 | 0,63629629 | -0,236064499 |
| Experimental | Control | 0,899730559 | 0,124872605 | 0,356504384 |
| Experimental | Control | 0,87952483 | 0,110133268 | 1,026218232 |
| Experimental | Control | 0,970062739 | 0,394051994 | -0,325193703 |
| Experimental | Control | 0,970062739 | 0,244059405 | -0,239481409 |
| Experimental | Control | 0,890654702 | 0,115752958 | 0,215572695 |
| Experimental | Control | 0,970062739 | 0,402321577 | -0,438749888 |
| Experimental | Control | 0,899730559 | 0,140927985 | 0,287149863 |
| Experimental | Control | 0,942205427 | 0,170073182 | 1,179047638 |
| Experimental | Control | 0,970062739 | 0,421473436 | 0,258944107 |
| Experimental | Control | 0,970062739 | 0,381707996 | -0,374196204 |
| Experimental | Control | 0,970062739 | 0,678516461 | -0,174290303 |
| Experimental | Control | 0,870685429 | 0,067784555 | -0,796222022 |
| Experimental | Control | 0,970062739 | 0,533291318 | -0,269454596 |
| Experimental | Control | 0,970062739 | 0,514432342 | -0,397729622 |
| Experimental | Control | 0,970062739 | 0,490819515 | -0,392614815 |
| Experimental | Control | 0,870685429 | 0,072501533 | -0,654172364 |
| Experimental | Control | 0,970062739 | 0,73998404 | 0,093633265 |
| Experimental | Control | 0,98331311 | 0,87998176 | -0,083060314 |
| Experimental | Control | 0,98331311 | 0,919405913 | 0,044197035 |
| Experimental | Control | 0,98331311 | 0,94372116 | -0,022921671 |
| Experimental | Control | 0,855140552 | 0,054025125 | 0,817579736 |
| Experimental | Control | 0,970062739 | 0,245150688 | -0,365733594 |
| Experimental | Control | 0,970062739 | 0,33362099 | -0,266115942 |
| Experimental | Control | 0,78357626 | 0,048089518 | -0,404766263 |
| Experimental | Control | 0,970062739 | 0,408610482 | 0,241775131 |
| Experimental | Control | 0,970062739 | 0,730158671 | 0,169998691 |
| Experimental | Control | 0,970062739 | 0,705409266 | 0,058927783 |
| Experimental | Control | 0,981816738 | 0,812532015 | 0,049282941 |
| Experimental | Control | 0,999707234 | 0,999707234 | 0,000153082 |
| Experimental | Control | 0,970062739 | 0,242493762 | 0,266315299 |
| Experimental | Control | 0,981816738 | 0,829381631 | 0,053752174 |

| | | | | |
|---|---|---|---|---|
| Experimental | Control | 0,970062739 | 0,486407184 | -0,211511573 |
| Experimental | Control | 0,981816738 | 0,85485549 | -0,095975419 |
| Experimental | Control | 0,970062739 | 0,47850376 | -0,380107984 |
| Experimental | Control | 0,970062739 | 0,508929559 | 0,269112605 |
| Experimental | Control | 0,970062739 | 0,642420724 | 0,157212381 |
| Experimental | Control | 0,98331311 | 0,896463501 | -0,031576581 |
| Experimental | Control | 0,970062739 | 0,631465782 | -0,072129451 |
| Experimental | Control | 0,970062739 | 0,552968044 | -0,141553326 |
| Experimental | Control | 0,973116013 | 0,776691011 | 0,124991589 |
| Experimental | Control | 0,970062739 | 0,625677089 | -0,174033671 |
| Experimental | Control | 0,78357626 | 0,047254501 | -0,851837031 |
| Experimental | Control | 0,78357626 | 0,024760057 | -0,89421457 |
| Experimental | Control | 0,970062739 | 0,303371218 | 0,301736847 |
| Experimental | Control | 0,970062739 | 0,337398752 | -0,382339081 |
| Experimental | Control | 0,985213025 | 0,947867405 | -0,026038716 |
| Experimental | Control | 0,970062739 | 0,717487136 | 0,091739021 |
| Experimental | Control | 0,970062739 | 0,28573923 | -0,429556358 |
| Experimental | Control | 0,973959896 | 0,795361342 | -0,130829273 |
| Experimental | Control | 0,694796407 | 0,008893436 | 0,581402325 |
| Experimental | Control | 0,970062739 | 0,44778176 | 0,384093909 |
| Experimental | Control | 0,694796407 | 0,012590433 | -0,904098187 |
| Experimental | Control | 0,98331311 | 0,942311623 | -0,035896498 |
| Experimental | Control | 0,970062739 | 0,596548259 | -0,199509639 |
| Experimental | Control | 0,970062739 | 0,679150762 | -0,183431768 |
| Experimental | Control | 0,973116013 | 0,787259007 | 0,054926728 |
| Experimental | Control | 0,973116013 | 0,777984336 | -0,139409212 |
| Experimental | Control | 0,970062739 | 0,349357128 | 0,329801056 |
| Experimental | Control | 0,981816738 | 0,811884724 | 0,13501512 |
| Experimental | Control | 0,970062739 | 0,685287659 | -0,17900167 |
| Experimental | Control | 0,970062739 | 0,362222342 | -0,222153552 |
| Experimental | Control | 0,970062739 | 0,721710496 | 0,076077164 |
| Experimental | Control | 0,970062739 | 0,288320329 | -0,611843366 |
| Experimental | Control | 0,98331311 | 0,944264575 | 0,019483667 |
| Experimental | Control | 0,970062739 | 0,64871399 | 0,12670613 |
| Experimental | Control | 0,981816738 | 0,846558723 | -0,079745656 |
| Experimental | Control | 0,970062739 | 0,33451972 | 0,166782607 |
| Experimental | Control | 0,970062739 | 0,710954323 | 0,150217049 |
| Experimental | Control | 0,970062739 | 0,693525105 | 0,080810367 |
| Experimental | Control | 0,970062739 | 0,722747449 | 0,093615925 |
| Experimental | Control | 0,981816738 | 0,81861247 | 0,059857089 |
| Experimental | Control | 0,970062739 | 0,459934079 | -0,281464209 |
| Experimental | Control | 0,970062739 | 0,194835746 | -0,618610401 |
| Experimental | Control | 0,970062739 | 0,303010467 | 0,245217797 |
| Experimental | Control | 0,981816738 | 0,842070732 | 0,042596103 |
| Experimental | Control | 0,981816738 | 0,820377318 | 0,043203821 |
| Experimental | Control | 0,981816738 | 0,845751663 | -0,109985684 |
| Experimental | Control | 0,78357626 | 0,03968095 | -1,163859747 |
| Experimental | Control | 0,98331311 | 0,920919496 | -0,048530005 |

| | | | | |
|---|---|---|---|---|
| Experimental | Control | 0,970062739 | 0,420260501 | -0,321127914 |
| Experimental | Control | 0,970062739 | 0,566013634 | 0,328121888 |
| Experimental | Control | 0,87952483 | 0,09449919 | 0,851697502 |
| Experimental | Control | 0,970062739 | 0,593050527 | -0,267230568 |
| Experimental | Control | 0,970062739 | 0,431371615 | 0,38641063 |
| Experimental | Control | 0,970062739 | 0,59244661 | -0,146989384 |
| Experimental | Control | 0,970062739 | 0,4277195 | -0,563076191 |
| Experimental | Control | 0,970062739 | 0,329326049 | 0,273507945 |
| Experimental | Control | 0,972536115 | 0,761721807 | 0,046279533 |
| Experimental | Control | 0,87952483 | 0,094026069 | 0,706351249 |
| Experimental | Control | 0,78357626 | 0,045546627 | 0,416222615 |
| Experimental | Control | 0,970062739 | 0,292634172 | -0,883403175 |
| Experimental | Control | 0,98331311 | 0,873045722 | 0,102288475 |
| Experimental | Control | 0,970062739 | 0,21632939 | 0,494143599 |
| Experimental | Control | 0,970062739 | 0,418338181 | -0,228953275 |
| Experimental | Control | 0,970062739 | 0,538036884 | 0,269653549 |
| Experimental | Control | 0,96190808 | 0,178838506 | 0,865710648 |
| Experimental | Control | 0,694796407 | 0,015049742 | -1,791430705 |
| Experimental | Control | 0,970062739 | 0,725854999 | -0,129776951 |
| Experimental | Control | 0,970062739 | 0,466068507 | 0,374781522 |
| Experimental | Control | 0,981816738 | 0,82396388 | -0,069502117 |
| Experimental | Control | 0,970062739 | 0,430835859 | 0,516034812 |
| Experimental | Control | 0,972536115 | 0,748764858 | -0,121413135 |
| Experimental | Control | 0,973116013 | 0,784420637 | -0,113335019 |
| Experimental | Control | 0,970062739 | 0,73761458 | 0,051562926 |
| Experimental | Control | 0,98331311 | 0,937505802 | 0,034572789 |
| Experimental | Control | 0,970062739 | 0,469741424 | 0,257561758 |
| Experimental | Control | 0,970062739 | 0,632648579 | -0,124602573 |
| Experimental | Control | 0,981816738 | 0,852484511 | -0,023259788 |
| Experimental | Control | 0,970062739 | 0,56987082 | -0,155194947 |
| Experimental | Control | 0,970062739 | 0,571590594 | 0,301744773 |
| Experimental | Control | 0,970062739 | 0,287128539 | -0,30126831 |
| Experimental | Control | 0,970062739 | 0,217312519 | 0,295838896 |
| Experimental | Control | 0,899730559 | 0,125118253 | -0,390369457 |
| Experimental | Control | 0,694796407 | 0,011916826 | 1,029180014 |
| Experimental | Control | 0,970062739 | 0,578025401 | 0,217151156 |
| Experimental | Control | 0,78357626 | 0,038151972 | 1,225313303 |
| Experimental | Control | 0,970062739 | 0,203089521 | -0,295179759 |
| Experimental | Control | 0,98331311 | 0,887183336 | -0,057344025 |
| Experimental | Control | 0,98331311 | 0,916750508 | 0,026092472 |
| Experimental | Control | 0,899730559 | 0,137019573 | -0,400525224 |
| Experimental | Control | 0,899730559 | 0,148625118 | -0,54949257 |
| Experimental | Control | 0,899730559 | 0,136299905 | 0,363009254 |
| Experimental | Control | 0,87952483 | 0,08892748 | -0,725752885 |
| Experimental | Control | 0,87952483 | 0,0884033 | 1,040680333 |
| Experimental | Control | 0,98331311 | 0,89945553 | 0,043438554 |
| Experimental | Control | 0,98331311 | 0,942054464 | 0,033688526 |
| Experimental | Control | 0,970062739 | 0,311085016 | 0,325000827 |

| | | | | |
|---|---|---|---|---|
| Experimental | Control | 0,98331311 | 0,891447281 | -0,028598727 |
| Experimental | Control | 0,98331311 | 0,930436999 | -0,049622519 |
| Experimental | Control | 0,970062739 | 0,429887158 | -0,157074633 |
| Experimental | Control | 0,970062739 | 0,562234382 | -0,218504398 |
| Experimental | Control | 0,970062739 | 0,738444879 | -0,09694129 |
| Experimental | Control | 0,988586169 | 0,962577675 | -0,018072918 |
| Experimental | Control | 0,992708935 | 0,971206214 | -0,011672181 |
| Experimental | Control | 0,970062739 | 0,653055181 | -0,180782373 |
| Experimental | Control | 0,78357626 | 0,032641491 | 0,601258382 |
| Experimental | Control | 0,970062739 | 0,480521953 | 0,135624695 |
| Experimental | Control | 0,970062739 | 0,419272786 | 0,199537736 |
| Experimental | Control | 0,87952483 | 0,099414643 | 0,303417569 |
| Experimental | Control | 0,970062739 | 0,622365193 | -0,211817311 |
| Experimental | Control | 0,970062739 | 0,546900995 | 0,119822839 |
| Experimental | Control | 0,987814822 | 0,955719756 | -0,016879399 |
| Experimental | Control | 0,970062739 | 0,197255063 | 0,494413847 |
| Experimental | Control | 0,970062739 | 0,199094591 | -0,387703047 |
| Experimental | Control | 0,98331311 | 0,921413101 | 0,040854451 |
| Experimental | Control | 0,970062739 | 0,285996343 | 0,206147023 |
| Experimental | Control | 0,970062739 | 0,244823189 | 0,595801261 |
| Experimental | Control | 0,899730559 | 0,147142593 | 0,77896506 |
| Experimental | Control | 0,970062739 | 0,317223473 | 0,154816752 |
| Experimental | Control | 0,970062739 | 0,685590024 | -0,229894067 |
| Experimental | Control | 0,970062739 | 0,577317996 | -0,23437144 |
| Experimental | Control | 0,970062739 | 0,30754081 | 0,231243513 |
| Experimental | Control | 0,899730559 | 0,130062469 | 0,415985825 |
| Experimental | Control | 0,970062739 | 0,312138827 | 0,655495143 |
| Experimental | Control | 0,87952483 | 0,103048087 | 0,682134933 |
| Experimental | Control | 0,981816738 | 0,823422754 | 0,069154338 |
| Experimental | Control | 0,970062739 | 0,438906392 | 0,393780225 |
| Experimental | Control | 0,970062739 | 0,568181092 | -0,32159584 |
| Experimental | Control | 0,973116013 | 0,777234635 | -0,150621082 |
| Experimental | Control | 0,970062739 | 0,451553747 | 0,312280388 |
| Experimental | Control | 0,970062739 | 0,577057209 | -0,175625247 |
| Experimental | Control | 0,993132522 | 0,974583465 | 0,011147511 |
| Experimental | Control | 0,96190808 | 0,177718137 | -0,602110917 |
| Experimental | Control | 0,970062739 | 0,453012347 | 0,291566368 |
| Experimental | Control | 0,970062739 | 0,359524817 | 0,612246007 |
| Experimental | Control | 0,899730559 | 0,128439897 | 0,583209017 |
| Experimental | Control | 0,970062739 | 0,594496552 | -0,248394132 |
| Experimental | Control | 0,970062739 | 0,669993382 | 0,146000216 |
| Experimental | Control | 0,970062739 | 0,63442013 | -0,215075285 |
| Experimental | Control | 0,99640621 | 0,986257652 | -0,005605439 |
| Experimental | Control | 0,970062739 | 0,594891155 | 0,118326315 |
| Experimental | Control | 0,970062739 | 0,542422378 | -0,331098928 |
| Experimental | Control | 0,98331311 | 0,923643761 | -0,021744813 |
| Experimental | Control | 0,870685429 | 0,078847319 | 0,788718278 |
| Experimental | Control | 0,970062739 | 0,622925408 | -0,192700266 |

| | | | | |
|---|---|---|---|---|
| Experimental | Control | 0,970062739 | 0,474193929 | -0,297274395 |
| Experimental | Control | 0,970062739 | 0,305662237 | 0,479498226 |
| Experimental | Control | 0,970062739 | 0,261673906 | 0,430849005 |
| Experimental | Control | 0,98331311 | 0,89931009 | 0,041435144 |
| Experimental | Control | 0,983153927 | 0,862477994 | 0,042268262 |
| Experimental | Control | 0,970062739 | 0,471607735 | -0,344997122 |
| Experimental | Control | 0,970062739 | 0,381418448 | 0,225738382 |
| Experimental | Control | 0,899730559 | 0,140349349 | 0,800701553 |
| Experimental | Control | 0,904627753 | 0,151859893 | -0,484131976 |
| Experimental | Control | 0,970062739 | 0,660172653 | 0,103829177 |
| Experimental | Control | 0,98331311 | 0,892363347 | 0,033502741 |
| Experimental | Control | 0,970062739 | 0,553569912 | 0,221117235 |
| Experimental | Control | 0,99681224 | 0,99141434 | -0,005206995 |
| Experimental | Control | 0,970062739 | 0,60829948 | -0,220395648 |
| Experimental | Control | 0,970062739 | 0,467959419 | 0,435919589 |
| Experimental | Control | 0,970062739 | 0,535265978 | 0,264718526 |
| Experimental | Control | 0,899730559 | 0,148507133 | -0,717191566 |
| Experimental | Control | 0,972536115 | 0,744798348 | -0,10683126 |
| Experimental | Control | 0,970062739 | 0,390979369 | 0,275992113 |
| Experimental | Control | 0,970062739 | 0,505738693 | 0,291637183 |
| Experimental | Control | 0,970062739 | 0,340674893 | -0,383973856 |
| Experimental | Control | 0,98331311 | 0,930711245 | 0,024865673 |
| Experimental | Control | 0,973116013 | 0,774826486 | -0,213828612 |
| Experimental | Control | 0,970062739 | 0,284469053 | 0,515529506 |
| Experimental | Control | 0,970062739 | 0,311694913 | -0,279370906 |
| Experimental | Control | 0,970062739 | 0,397921301 | 0,401329141 |
| Experimental | Control | 0,973116013 | 0,785654074 | -0,109477747 |
| Experimental | Control | 0,87952483 | 0,084977792 | 0,824260679 |
| Experimental | Control | 0,970062739 | 0,196945714 | 0,632656563 |
| Experimental | Control | 0,899730559 | 0,139462618 | 0,644158771 |
| Experimental | Control | 0,970062739 | 0,289469502 | 0,58389163 |
| Experimental | Control | 0,970062739 | 0,717682983 | -0,117936981 |
| Experimental | Control | 0,970062739 | 0,624920268 | -0,290240405 |
| Experimental | Control | 0,970062739 | 0,707443487 | -0,156042328 |
| Experimental | Control | 0,970062739 | 0,587950258 | 0,253481993 |
| Experimental | Control | 0,970062739 | 0,582275735 | -0,183028707 |
| Experimental | Control | 0,98331311 | 0,869985979 | 0,083866982 |
| Experimental | Control | 0,972536115 | 0,755261655 | 0,120505322 |
| Experimental | Control | 0,970062739 | 0,637628373 | 0,268766915 |
| Experimental | Control | 0,970062739 | 0,634410657 | -0,210585494 |
| Experimental | Control | 0,970062739 | 0,715072235 | 0,14968039 |
| Experimental | Control | 0,970062739 | 0,388592499 | -0,335955463 |
| Experimental | Control | 0,970062739 | 0,655420373 | 0,202437345 |
| Experimental | Control | 0,78357626 | 0,046712087 | -0,623369571 |
| Experimental | Control | 0,970062739 | 0,525485075 | -0,263460194 |
| Experimental | Control | 0,970062739 | 0,725375626 | 0,236501109 |
| Experimental | Control | 0,970062739 | 0,670334205 | 0,241520928 |
| Experimental | Control | 0,694796407 | 0,014184723 | 0,828971555 |

| | | | | |
|---|---|---|---|---|
| Experimental | Control | 0,970062739 | 0,740679672 | 0,083711835 |
| Experimental | Control | 0,970062739 | 0,631280219 | 0,101496797 |
| Experimental | Control | 0,735000532 | 0,018262501 | 1,200223098 |
| Experimental | Control | 0,970062739 | 0,679482379 | 0,108848986 |
| Experimental | Control | 0,988586169 | 0,963603847 | -0,014141703 |
| Experimental | Control | 0,970062739 | 0,30071299 | 0,303600754 |
| Experimental | Control | 0,970062739 | 0,327449997 | -0,379666203 |
| Experimental | Control | 0,970062739 | 0,509686026 | 0,328270173 |
| Experimental | Control | 0,970062739 | 0,36835808 | 0,435343173 |
| Experimental | Control | 0,970062739 | 0,665765394 | 0,158081297 |
| Experimental | Control | 0,970062739 | 0,284177693 | 0,748898592 |
| Experimental | Control | 0,970062739 | 0,667710818 | 0,213944568 |
| Experimental | Control | 0,972536115 | 0,761878473 | -0,129060741 |
| Experimental | Control | 0,970062739 | 0,614412715 | -0,100898625 |
| Experimental | Control | 0,970062739 | 0,568430971 | -0,223516811 |
| Experimental | Control | 0,927436375 | 0,163369338 | -0,728596343 |
| Experimental | Control | 0,970062739 | 0,496653748 | 0,271616212 |
| Experimental | Control | 0,970062739 | 0,512395264 | 0,16960232 |
| Experimental | Control | 0,970062739 | 0,387353509 | 0,276592974 |
| Experimental | Control | 0,973116013 | 0,781468048 | 0,107445089 |
| Experimental | Control | 0,970062739 | 0,398966914 | -0,317793027 |
| Experimental | Control | 0,970062739 | 0,62245394 | 0,272134378 |
| Experimental | Control | 0,973959896 | 0,795832408 | -0,142398942 |
| Experimental | Control | 0,981816738 | 0,852515284 | -0,078282034 |
| Experimental | Control | 0,970062739 | 0,447509896 | 0,373325555 |
| Experimental | Control | 0,970062739 | 0,52772037 | -0,189173635 |
| Experimental | Control | 0,98331311 | 0,934186691 | 0,024487068 |
| Experimental | Control | 0,995080182 | 0,909544426 | 0,032914098 |
| Experimental | Control | 0,714386358 | 0,141743325 | 0,360948896 |
| Experimental | Control | 0,795738716 | 0,326827618 | 0,282590331 |
| Experimental | Control | 0,994641719 | 0,8405716 | -0,073874006 |
| Experimental | Control | 0,77418217 | 0,198133343 | -1,102436784 |
| Experimental | Control | 0,159397495 | 0,002530119 | -1,745831312 |
| Experimental | Control | 0,884317616 | 0,572569687 | -0,283225157 |
| Experimental | Control | 0,995080182 | 0,912257255 | -0,033141388 |
| Experimental | Control | 0,491272929 | 0,066282856 | -1,506424363 |
| Experimental | Control | 0,880511036 | 0,538090077 | 0,268036215 |
| Experimental | Control | 0,829721397 | 0,455319928 | 0,3492079 |
| Experimental | Control | 0,994995072 | 0,860749705 | 0,050667518 |
| Experimental | Control | 0,795738716 | 0,371965721 | 0,364389449 |
| Experimental | Control | 0,905060951 | 0,617740014 | 0,333851842 |
| Experimental | Control | 0,795738716 | 0,401399771 | -0,304586244 |
| Experimental | Control | 0,995080182 | 0,992388111 | 0,002976695 |
| Experimental | Control | 0,275596245 | 0,015743348 | 0,983909396 |
| Experimental | Control | 0,795738716 | 0,245152096 | -0,466583568 |
| Experimental | Control | 0,795738716 | 0,353569769 | -0,469880315 |
| Experimental | Control | 0,795738716 | 0,393518703 | 0,343211494 |
| Experimental | Control | 0,275596245 | 0,014487276 | -1,156594731 |

| | | | | |
|---|---|---|---|---|
| Experimental | Control | 0,934383027 | 0,702293087 | 0,102391991 |
| Experimental | Control | 0,995080182 | 0,890590445 | 0,05290861 |
| Experimental | Control | 0,994995072 | 0,860502336 | -0,081559063 |
| Experimental | Control | 0,77418217 | 0,186185304 | 0,369199852 |
| Experimental | Control | 0,795738716 | 0,400938772 | 0,246228189 |
| Experimental | Control | 0,795738716 | 0,328736626 | -0,868582136 |
| Experimental | Control | 0,995080182 | 0,964922073 | -0,02339825 |
| Experimental | Control | 0,930626776 | 0,686891192 | -0,088065449 |
| Experimental | Control | 0,491272929 | 0,064825599 | -0,549432348 |
| Experimental | Control | 0,995080182 | 0,874526358 | 0,030215514 |
| Experimental | Control | 0,795738716 | 0,358059258 | 0,268776883 |
| Experimental | Control | 0,995080182 | 0,981282885 | -0,012712082 |
| Experimental | Control | 0,795738716 | 0,361768694 | -0,180530538 |
| Experimental | Control | 0,930626776 | 0,681086844 | 0,274693846 |
| Experimental | Control | 0,96948655 | 0,76846744 | 0,100881985 |
| Experimental | Control | 0,930626776 | 0,676580535 | -0,22391642 |
| Experimental | Control | 0,415491504 | 0,042868171 | -0,771220716 |
| Experimental | Control | 0,795738716 | 0,26480878 | -0,428799561 |
| Experimental | Control | 0,930626776 | 0,668338921 | -0,155168012 |
| Experimental | Control | 0,795738716 | 0,3230444 | -0,176977307 |
| Experimental | Control | 0,829721397 | 0,49946856 | -0,30251442 |
| Experimental | Control | 0,829721397 | 0,484646362 | 0,361888816 |
| Experimental | Control | 0,995080182 | 0,877251878 | 0,085539477 |
| Experimental | Control | 0,795738716 | 0,373444604 | 0,336546241 |
| Experimental | Control | 0,795738716 | 0,376331793 | -0,502587101 |
| Experimental | Control | 0,795738716 | 0,221559666 | -0,616936145 |
| Experimental | Control | 0,980766794 | 0,810799084 | -0,170425505 |
| Experimental | Control | 0,829721397 | 0,476842145 | 0,36992483 |
| Experimental | Control | 0,415491504 | 0,037959652 | -1,186988966 |
| Experimental | Control | 0,795738716 | 0,358854228 | 0,447948045 |
| Experimental | Control | 0,275596245 | 0,01919754 | 1,525153693 |
| Experimental | Control | 0,995080182 | 0,983046833 | -0,006948524 |
| Experimental | Control | 0,920122287 | 0,649927647 | 0,349395188 |
| Experimental | Control | 0,159397495 | 0,002335977 | -1,280301771 |
| Experimental | Control | 0,733100473 | 0,162911216 | 0,575139755 |
| Experimental | Control | 0,632698353 | 0,095406895 | -0,431927016 |
| Experimental | Control | 0,795738716 | 0,404184745 | -0,318451183 |
| Experimental | Control | 0,795738716 | 0,21817927 | -0,433472821 |
| Experimental | Control | 0,995080182 | 0,929016968 | -0,039583536 |
| Experimental | Control | 0,720110796 | 0,154309456 | -0,675510248 |
| Experimental | Control | 0,415491504 | 0,042748919 | -1,95645076 |
| Experimental | Control | 0,713226318 | 0,120005831 | -0,752279226 |
| Experimental | Control | 0,995080182 | 0,995080182 | -0,002216143 |
| Experimental | Control | 0,275596245 | 0,008750763 | -1,434710524 |
| Experimental | Control | 0,795738716 | 0,385660497 | -0,919613674 |
| Experimental | Control | 0,77418217 | 0,188668471 | 0,645605004 |
| Experimental | Control | 0,77418217 | 0,202761997 | -0,556369691 |
| Experimental | Control | 0,795738716 | 0,392599428 | 0,297993664 |

| | | | | |
|---|---|---|---|---|
| Experimental | Control | 0,795738716 | 0,396977831 | -0,305548029 |
| Experimental | Control | 0,829721397 | 0,495054933 | 0,389898268 |
| Experimental | Control | 0,795738716 | 0,256595318 | -0,489203033 |
| Experimental | Control | 0,795738716 | 0,270261209 | 0,564037505 |
| Experimental | Control | 0,96948655 | 0,774514402 | 0,162663818 |
| Experimental | Control | 0,884317616 | 0,575092164 | -0,240621873 |
| Experimental | Control | 0,713226318 | 0,122418237 | -0,53248115 |
| Experimental | Control | 0,795738716 | 0,350996749 | -0,501883724 |
| Experimental | Control | 0,491272929 | 0,058723176 | -0,963836936 |
| Experimental | Control | 0,895182615 | 0,596782103 | 0,302218943 |
| Experimental | Control | 0,96948655 | 0,792008663 | 0,089542377 |
| Experimental | Control | 0,795738716 | 0,351627937 | -0,711470391 |
| Experimental | Control | 0,807413917 | 0,422931099 | 0,388613005 |
| Experimental | Control | 0,829721397 | 0,448736153 | -0,161521673 |
| Experimental | Control | 0,995080182 | 0,973238428 | -0,02965955 |
| Experimental | Control | 0,77418217 | 0,194749535 | 0,735121419 |
| Experimental | Control | 0,795738716 | 0,293651185 | -0,358734747 |
| Experimental | Control | 0,829721397 | 0,481400345 | -0,21712037 |
| Experimental | Control | 0,980766794 | 0,817305662 | -0,096656137 |
| Experimental | Control | 0,96948655 | 0,773585523 | -0,145602481 |
| Experimental | Control | 0,415491504 | 0,033539064 | -1,469154677 |
| Experimental | Control | 0,895182615 | 0,603893034 | 0,392060502 |
| Experimental | Control | 0,906259401 | 0,625750539 | 0,324001608 |
| Experimental | Control | 0,96948655 | 0,792516783 | -0,061401722 |
| Experimental | Control | 0,714386358 | 0,132268612 | -0,805301949 |
| Experimental | Control | 0,994641719 | 0,844656063 | -0,137510942 |
| Experimental | Control | 0,275596245 | 0,019685446 | -1,427276337 |
| Experimental | Control | 0,720110796 | 0,153617199 | -0,532919546 |
| Experimental | Control | 0,884317616 | 0,57550829 | 0,215096359 |
| Experimental | Control | 0,713226318 | 0,124531579 | 0,474707484 |
| Experimental | Control | 0,795738716 | 0,248133767 | 0,239596775 |
| Experimental | Control | 0,950058396 | 0,723854016 | 0,249997247 |
| Experimental | Control | 0,934383027 | 0,704495139 | -0,161069879 |
| Experimental | Control | 0,96948655 | 0,785576437 | 0,07373105 |
| Experimental | Control | 0,995080182 | 0,990689153 | -0,005076574 |
| Experimental | Control | 0,275596245 | 0,019633043 | -1,509638103 |
| Experimental | Control | 0,995080182 | 0,937470703 | -0,044308156 |
| Experimental | Control | 0,714386358 | 0,141243982 | -0,759059884 |
| Experimental | Control | 0,915171283 | 0,639167245 | 0,281715915 |
| Experimental | Control | 0,275596245 | 0,012066572 | -1,530435843 |
| Experimental | Control | 0,96948655 | 0,769339228 | -0,149868628 |
| Experimental | Control | 0,829721397 | 0,469335403 | -0,35518597 |
| Experimental | Control | 0,829721397 | 0,500466874 | 0,278183643 |
| Experimental | Control | 0,995080182 | 0,994312481 | -0,002924171 |
| Experimental | Control | 0,795738716 | 0,352526242 | -0,724946295 |
| Experimental | Control | 0,795738716 | 0,372087123 | -0,452641595 |
| Experimental | Control | 0,884317616 | 0,555796061 | -0,381492891 |
| Experimental | Control | 0,895182615 | 0,597395994 | -0,224494498 |

| | | | | |
|---|---|---|---|---|
| Experimental | Control | 0,807413917 | 0,418164788 | -0,315826569 |
| Experimental | Control | 0,829721397 | 0,453214657 | 0,35503001 |
| Experimental | Control | 0,995080182 | 0,924695831 | -0,038336247 |
| Experimental | Control | 0,882163331 | 0,54610111 | 0,150662636 |
| Experimental | Control | 0,995080182 | 0,918662057 | 0,056604391 |
| Experimental | Control | 0,795738716 | 0,234419903 | -0,825543626 |
| Experimental | Control | 0,795738716 | 0,32438001 | -0,495894212 |
| Experimental | Control | 0,564787144 | 0,080683878 | -1,333680512 |
| Experimental | Control | 0,491272929 | 0,058271863 | -1,061305723 |
| Experimental | Control | 0,882414063 | 0,714006675 | 0,261815686 |
| Experimental | Control | 0,767190285 | 0,413546284 | 0,19030758 |
| Experimental | Control | 0,435518578 | 0,140823427 | -1,252853989 |
| Experimental | Control | 0,982189463 | 0,982189463 | 0,013190388 |
| Experimental | Control | 0,435518578 | 0,13742448 | -0,567836218 |
| Experimental | Control | 0,592615022 | 0,23428966 | -0,611289948 |
| Experimental | Control | 0,882414063 | 0,633187101 | 0,188004478 |
| Experimental | Control | 0,725868593 | 0,320732634 | -0,398049213 |
| Experimental | Control | 0,882414063 | 0,710267517 | -0,201637888 |
| Experimental | Control | 0,107641242 | 0,017522993 | -0,688126979 |
| Experimental | Control | 0,435518578 | 0,11370904 | -1,418696069 |
| Experimental | Control | 0,904982648 | 0,797562197 | 0,073476795 |
| Experimental | Control | 0,945044287 | 0,91992781 | -0,034156897 |
| Experimental | Control | 0,767190285 | 0,463882498 | 0,206095732 |
| Experimental | Control | 0,103128262 | 0,011991658 | -1,217938595 |
| Experimental | Control | 0,767190285 | 0,406400449 | 0,265728522 |
| Experimental | Control | 0,182301359 | 0,033916532 | -0,948438375 |
| Experimental | Control | 0,882414063 | 0,644435894 | -0,320846441 |
| Experimental | Control | 0,767190285 | 0,45640675 | -0,225175193 |
| Experimental | Control | 0,767190285 | 0,463068724 | 0,316949611 |
| Experimental | Control | 0,945044287 | 0,923066513 | 0,063595458 |
| Experimental | Control | 0,309455441 | 0,07078204 | -0,839177232 |
| Experimental | Control | 0,839436686 | 0,566131718 | -0,121801766 |
| Experimental | Control | 0,904982648 | 0,820798215 | 0,191069508 |
| Experimental | Control | 0,839436686 | 0,543935653 | -0,303241455 |
| Experimental | Control | 0,882414063 | 0,67342171 | -0,119891593 |
| Experimental | Control | 0,738724983 | 0,343593015 | -0,216594566 |
| Experimental | Control | 0,532423493 | 0,198111067 | -0,699198177 |
| Experimental | Control | 0,070709936 | 0,006577668 | -0,557078509 |
| Experimental | Control | 0,062463066 | 0,002905259 | -1,604769396 |
| Experimental | Control | 0,904982648 | 0,781653967 | 0,119194762 |
| Experimental | Control | 0,839436686 | 0,563568623 | 0,299065987 |
| Experimental | Control | 0,882414063 | 0,718244005 | -0,154175407 |
| Experimental | Control | 0,070709936 | 0,00653399 | -1,693885769 |
| Experimental | Control | 0,767190285 | 0,404840074 | 0,416387108 |
| Experimental | Control | 0,052489085 | 0,001220676 | -1,915132044 |
| Experimental | Control | 0,107641242 | 0,01607936 | -1,521883672 |
| Experimental | Control | 0,597791958 | 0,250238494 | -0,494515841 |
| Experimental | Control | 0,435518578 | 0,151608457 | -0,518925138 |

| | | | | |
|---|---|---|---|---|
| Experimental | Control | 0,904982648 | 0,8074438 | 0,088385198 |
| Experimental | Control | 0,309455441 | 0,071966382 | -0,680053677 |
| Experimental | Control | 0,915182732 | 0,851332774 | -0,10395336 |
| Experimental | Control | 0,435518578 | 0,151925085 | -0,703863159 |
| Experimental | Control | 0,064287302 | 0,0183678 | 0,755843292 |
| Experimental | Control | 0,026609416 | 0,003801345 | 0,773823932 |
| Experimental | Control | 0,247388387 | 0,106023595 | -0,619396985 |
| Experimental | Control | 0,289345287 | 0,165340164 | 0,242697719 |
| Experimental | Control | 0,534121185 | 0,534121185 | -0,262898224 |
| Experimental | Control | 0,452121961 | 0,356059881 | -0,361807175 |
| Experimental | Control | 0,452121961 | 0,38753311 | -0,650989951 |
| Experimental | Control | 0,624192061 | 0,016348642 | 1,058685604 |
| Experimental | Control | 0,704891642 | 0,072012506 | 0,875288911 |
| Experimental | Control | 0,790420277 | 0,217700715 | -1,127208754 |
| Experimental | Control | 0,772193668 | 0,160537896 | -0,651079864 |
| Experimental | Control | 0,858120076 | 0,3123778 | -0,826346418 |
| Experimental | Control | 0,915172456 | 0,449900366 | -0,376356802 |
| Experimental | Control | 0,983491561 | 0,957148622 | -0,021734766 |
| Experimental | Control | 0,885436121 | 0,406929077 | 0,502835618 |
| Experimental | Control | 0,983491561 | 0,932354773 | 0,040212878 |
| Experimental | Control | 0,637944107 | 0,03539273 | -1,169489815 |
| Experimental | Control | 0,870817871 | 0,362081185 | -0,603240996 |
| Experimental | Control | 0,772193668 | 0,154733844 | -0,654305596 |
| Experimental | Control | 0,983491561 | 0,846685214 | -0,133686513 |
| Experimental | Control | 0,983491561 | 0,889993121 | -0,076831204 |
| Experimental | Control | 0,772193668 | 0,122610713 | -1,163195578 |
| Experimental | Control | 0,942203073 | 0,615893087 | 0,29952553 |
| Experimental | Control | 0,772193668 | 0,186268349 | -0,638142521 |
| Experimental | Control | 0,814348357 | 0,25530163 | -0,839788111 |
| Experimental | Control | 0,621747057 | 0,008074685 | 1,35767187 |
| Experimental | Control | 0,885436121 | 0,401941417 | -0,339891053 |
| Experimental | Control | 0,783663238 | 0,197222228 | 0,745324408 |
| Experimental | Control | 0,790420277 | 0,233910526 | 0,569899645 |
| Experimental | Control | 0,858120076 | 0,304693374 | 0,499434205 |
| Experimental | Control | 0,885436121 | 0,401812809 | 0,49717511 |
| Experimental | Control | 0,772193668 | 0,171773355 | -0,523869293 |
| Experimental | Control | 0,692638815 | 0,066678393 | -1,252403406 |
| Experimental | Control | 0,983491561 | 0,811311758 | -0,103621343 |
| Experimental | Control | 0,885436121 | 0,394977169 | -0,506266484 |
| Experimental | Control | 0,624192061 | 0,022370334 | -1,893469818 |
| Experimental | Control | 0,858120076 | 0,304348228 | -0,486980784 |
| Experimental | Control | 0,98145184 | 0,722336944 | -0,267904374 |
| Experimental | Control | 0,783663238 | 0,2005763 | 0,448072623 |
| Experimental | Control | 0,908003536 | 0,428025954 | -0,520620495 |
| Experimental | Control | 0,790420277 | 0,230393432 | -0,805597592 |
| Experimental | Control | 0,983491561 | 0,828668761 | 0,095568752 |
| Experimental | Control | 0,772193668 | 0,142749167 | 0,676178771 |
| Experimental | Control | 0,983491561 | 0,967412026 | -0,020038111 |

| | | | | |
|---|---|---|---|---|
| Experimental | Control | 0,942203073 | 0,621965094 | -0,437346032 |
| Experimental | Control | 0,772193668 | 0,149271601 | -1,036512848 |
| Experimental | Control | 0,772193668 | 0,151098587 | -1,15728352 |
| Experimental | Control | 0,818172512 | 0,270045151 | -0,971891143 |
| Experimental | Control | 0,621747057 | 0,006483081 | -1,682559409 |
| Experimental | Control | 0,919104202 | 0,463444218 | 0,317493314 |
| Experimental | Control | 0,754607597 | 0,095579528 | -0,777487051 |
| Experimental | Control | 0,98145184 | 0,725510866 | -0,101987283 |
| Experimental | Control | 0,840958856 | 0,27962138 | 0,391655602 |
| Experimental | Control | 0,989996372 | 0,978326474 | -0,02023123 |
| Experimental | Control | 0,92316567 | 0,531442631 | -0,172018156 |
| Experimental | Control | 0,815638754 | 0,261803018 | 0,790616212 |
| Experimental | Control | 0,772193668 | 0,14334935 | 0,487674256 |
| Experimental | Control | 0,92316567 | 0,530965087 | 0,458629541 |
| Experimental | Control | 0,933825177 | 0,558700894 | 0,353753719 |
| Experimental | Control | 0,885436121 | 0,408441945 | -0,390925824 |
| Experimental | Control | 0,790420277 | 0,229321097 | -0,815378179 |
| Experimental | Control | 0,919104202 | 0,475340443 | 0,261041853 |
| Experimental | Control | 0,816993398 | 0,268050879 | -0,732965478 |
| Experimental | Control | 0,98145184 | 0,745989921 | -0,058445157 |
| Experimental | Control | 0,917990807 | 0,459897163 | -0,322082726 |
| Experimental | Control | 0,98145184 | 0,720261294 | -0,242673951 |
| Experimental | Control | 0,983491561 | 0,946200026 | -0,031235978 |
| Experimental | Control | 0,790420277 | 0,228714408 | 0,508747014 |
| Experimental | Control | 0,68521937 | 0,049809659 | -1,334790534 |
| Experimental | Control | 0,624192061 | 0,020054334 | -1,369505097 |
| Experimental | Control | 0,885436121 | 0,397885331 | -0,488081538 |
| Experimental | Control | 0,983491561 | 0,817610499 | -0,171423368 |
| Experimental | Control | 0,814348357 | 0,255983766 | 0,46499518 |
| Experimental | Control | 0,885436121 | 0,410536197 | -0,306479628 |
| Experimental | Control | 0,948299071 | 0,643125122 | -0,245115833 |
| Experimental | Control | 0,692638815 | 0,054544813 | -1,00767304 |
| Experimental | Control | 0,948260112 | 0,633415399 | -0,214777283 |
| Experimental | Control | 0,983491561 | 0,878062814 | -0,082548054 |
| Experimental | Control | 0,692638815 | 0,058009445 | 0,570723943 |
| Experimental | Control | 0,983491561 | 0,830927383 | -0,082056355 |
| Experimental | Control | 0,878603114 | 0,376297601 | -0,595082485 |
| Experimental | Control | 0,790420277 | 0,233439987 | -0,748654127 |
| Experimental | Control | 0,98145184 | 0,736278543 | 0,188838343 |
| Experimental | Control | 0,870817871 | 0,346063501 | -0,744684588 |
| Experimental | Control | 0,983491561 | 0,949965945 | -0,023629083 |
| Experimental | Control | 0,964525247 | 0,691685464 | -0,220647994 |
| Experimental | Control | 0,772193668 | 0,159720134 | -0,475429585 |
| Experimental | Control | 0,772193668 | 0,166306056 | -1,445042588 |
| Experimental | Control | 0,919104202 | 0,47734995 | -0,43686723 |
| Experimental | Control | 0,772193668 | 0,147616407 | 0,292637769 |
| Experimental | Control | 0,885436121 | 0,403799038 | -0,193367027 |
| Experimental | Control | 0,98145184 | 0,752311032 | 0,129695415 |

| | | | | |
|---|---|---|---|---|
| Experimental | Control | 0,983491561 | 0,792766776 | 0,063798178 |
| Experimental | Control | 0,999642631 | 0,999642631 | 0,00011395 |
| Experimental | Control | 0,783663238 | 0,195063602 | -0,829316339 |
| Experimental | Control | 0,919104202 | 0,468960085 | -0,344997541 |
| Experimental | Control | 0,858120076 | 0,323892619 | -0,734225895 |
| Experimental | Control | 0,772193668 | 0,145593503 | -1,142650772 |
| Experimental | Control | 0,959244846 | 0,669120087 | -0,395311504 |
| Experimental | Control | 0,983491561 | 0,792150172 | 0,144948955 |
| Experimental | Control | 0,983491561 | 0,968033933 | 0,012824579 |
| Experimental | Control | 0,624192061 | 0,028027581 | -1,955635743 |
| Experimental | Control | 0,942203073 | 0,578110478 | -0,191514411 |
| Experimental | Control | 0,942203073 | 0,618752693 | -0,359965088 |
| Experimental | Control | 0,915172456 | 0,445641024 | -0,372219535 |
| Experimental | Control | 0,98145184 | 0,719913042 | 0,205708117 |
| Experimental | Control | 0,983491561 | 0,919888557 | 0,050464368 |
| Experimental | Control | 0,772193668 | 0,178977137 | 0,802711034 |
| Experimental | Control | 0,983491561 | 0,887177613 | -0,055399524 |
| Experimental | Control | 0,98145184 | 0,767422067 | 0,109992355 |
| Experimental | Control | 0,637944107 | 0,036346521 | 0,435333794 |
| Experimental | Control | 0,772193668 | 0,11931435 | -0,627902458 |
| Experimental | Control | 0,983491561 | 0,929321832 | 0,043932601 |
| Experimental | Control | 0,920859684 | 0,495639423 | -0,312749309 |
| Experimental | Control | 0,959244846 | 0,676824843 | 0,207924576 |
| Experimental | Control | 0,858120076 | 0,322602496 | -0,250601393 |
| Experimental | Control | 0,933825177 | 0,559481713 | 0,373577311 |
| Experimental | Control | 0,736049836 | 0,079805917 | 0,923004518 |
| Experimental | Control | 0,983491561 | 0,894580057 | -0,054010213 |
| Experimental | Control | 0,983491561 | 0,840300106 | -0,072213566 |
| Experimental | Control | 0,920859684 | 0,499326666 | 0,234916098 |
| Experimental | Control | 0,878603114 | 0,375365515 | 0,312149444 |
| Experimental | Control | 0,942203073 | 0,604653141 | -0,452200819 |
| Experimental | Control | 0,983491561 | 0,794658375 | 0,105700885 |
| Experimental | Control | 0,797872703 | 0,23876264 | 0,605607343 |
| Experimental | Control | 0,893662802 | 0,416106256 | 0,243606192 |
| Experimental | Control | 0,754607597 | 0,090963582 | -1,014690087 |
| Experimental | Control | 0,983491561 | 0,819024535 | -0,12832342 |
| Experimental | Control | 0,925818599 | 0,544807915 | 0,176942106 |
| Experimental | Control | 0,624192061 | 0,028205142 | 1,064831287 |
| Experimental | Control | 0,692638815 | 0,064180158 | -1,04282722 |
| Experimental | Control | 0,922188811 | 0,502775289 | -0,212127696 |
| Experimental | Control | 0,983491561 | 0,873313074 | 0,138914336 |
| Experimental | Control | 0,772193668 | 0,178134855 | -0,609450842 |
| Experimental | Control | 0,983491561 | 0,88125794 | -0,047973224 |
| Experimental | Control | 0,624192061 | 0,016823282 | -1,035278091 |
| Experimental | Control | 0,772193668 | 0,161069644 | 0,867839532 |
| Experimental | Control | 0,959244846 | 0,669174406 | 0,284917592 |
| Experimental | Control | 0,624192061 | 0,013261292 | 0,995424437 |
| Experimental | Control | 0,772193668 | 0,164104376 | 0,316614727 |

| | | | | |
|---|---|---|---|---|
| Experimental | Control | 0,771535076 | 0,105375239 | -1,267988834 |
| Experimental | Control | 0,692638815 | 0,055393973 | -0,838029448 |
| Experimental | Control | 0,942203073 | 0,609045563 | -0,172613382 |
| Experimental | Control | 0,98145184 | 0,751246018 | 0,162029503 |
| Experimental | Control | 0,983491561 | 0,940178522 | -0,044152572 |
| Experimental | Control | 0,797872703 | 0,24296713 | 0,346218609 |
| Experimental | Control | 0,621747057 | 0,002974348 | 0,928170466 |
| Experimental | Control | 0,624192061 | 0,01665875 | -1,341998486 |
| Experimental | Control | 0,885436121 | 0,407388989 | 0,606266479 |
| Experimental | Control | 0,983491561 | 0,956474312 | 0,019526798 |
| Experimental | Control | 0,870817871 | 0,345026141 | 0,557033714 |
| Experimental | Control | 0,983491561 | 0,837907713 | 0,13722634 |
| Experimental | Control | 0,858120076 | 0,322704443 | 0,393929142 |
| Experimental | Control | 0,844224726 | 0,286936891 | 0,576855178 |
| Experimental | Control | 0,983491561 | 0,788738184 | 0,162450398 |
| Experimental | Control | 0,922188811 | 0,505257604 | -0,393583229 |
| Experimental | Control | 0,983491561 | 0,873080876 | 0,082571853 |
| Experimental | Control | 0,789234341 | 0,215527649 | -0,724731109 |
| Experimental | Control | 0,624192061 | 0,027690139 | 2,150789641 |
| Experimental | Control | 0,772193668 | 0,15974216 | -1,163150227 |
| Experimental | Control | 0,843858323 | 0,283496607 | 0,693049548 |
| Experimental | Control | 0,98145184 | 0,753661426 | -0,056307084 |
| Experimental | Control | 0,990728512 | 0,981201648 | 0,00950379 |
| Experimental | Control | 0,983491561 | 0,781245555 | -0,061793604 |
| Experimental | Control | 0,983491561 | 0,873890212 | 0,088446238 |
| Experimental | Control | 0,98145184 | 0,74319096 | -0,251246602 |
| Experimental | Control | 0,950766093 | 0,650032614 | 0,248086487 |
| Experimental | Control | 0,942203073 | 0,60394764 | -0,254660189 |
| Experimental | Control | 0,959244846 | 0,672358564 | 0,15190845 |
| Experimental | Control | 0,933692218 | 0,553978487 | -0,351379846 |
| Experimental | Control | 0,983491561 | 0,821104872 | -0,060476888 |
| Experimental | Control | 0,915172456 | 0,451781956 | -0,552762718 |
| Experimental | Control | 0,98145184 | 0,731718657 | -0,083317534 |
| Experimental | Control | 0,637944107 | 0,034371645 | 0,933114743 |
| Experimental | Control | 0,92316567 | 0,535669522 | 0,272059989 |
| Experimental | Control | 0,925818599 | 0,545669116 | 0,250706945 |
| Experimental | Control | 0,736049836 | 0,080979943 | 0,636506663 |
| Experimental | Control | 0,772193668 | 0,189200981 | -0,444555234 |
| Experimental | Control | 0,990728512 | 0,985306837 | 0,005475975 |
| Experimental | Control | 0,942203073 | 0,58851632 | 0,219231796 |
| Experimental | Control | 0,942203073 | 0,616634943 | 0,102077822 |
| Experimental | Control | 0,68521937 | 0,047750541 | 0,455595519 |
| Experimental | Control | 0,98145184 | 0,762881437 | -0,132587177 |
| Experimental | Control | 0,676638733 | 0,042397727 | 0,662685889 |
| Experimental | Control | 0,772193668 | 0,141928249 | -0,375397769 |
| Experimental | Control | 0,772193668 | 0,189634987 | -0,629612632 |
| Experimental | Control | 0,816993398 | 0,267555772 | -0,52942343 |
| Experimental | Control | 0,92316567 | 0,536244405 | 0,134784489 |

| | | | | |
|---|---|---|---|---|
| Experimental | Control | 0,626448651 | 0,031999342 | -1,271023014 |
| Experimental | Control | 0,983491561 | 0,814865144 | 0,105762148 |
| Experimental | Control | 0,624192061 | 0,010653513 | 0,67682121 |
| Experimental | Control | 0,983491561 | 0,827425286 | -0,144638547 |
| Experimental | Control | 0,915172456 | 0,442212543 | 0,476057881 |
| Experimental | Control | 0,983491561 | 0,908145909 | 0,079998175 |
| Experimental | Control | 0,983491561 | 0,919558308 | -0,036051065 |
| Experimental | Control | 0,783663238 | 0,200210503 | 0,829444152 |
| Experimental | Control | 0,870817871 | 0,365391141 | -0,462591039 |
| Experimental | Control | 0,919104202 | 0,477848424 | 0,236739635 |
| Experimental | Control | 0,885436121 | 0,39878601 | -0,325538149 |
| Experimental | Control | 0,772193668 | 0,18428159 | 0,640267278 |
| Experimental | Control | 0,769746662 | 0,101322252 | 0,572441233 |
| Experimental | Control | 0,990728512 | 0,987358296 | 0,010811463 |
| Experimental | Control | 0,920859684 | 0,498440416 | 0,401988385 |
| Experimental | Control | 0,983491561 | 0,951755796 | 0,018387654 |
| Experimental | Control | 0,866291532 | 0,33792363 | -0,523057122 |
| Experimental | Control | 0,983491561 | 0,845151096 | -0,068213344 |
| Experimental | Control | 0,919104202 | 0,484953509 | -0,49366141 |
| Experimental | Control | 0,754607597 | 0,09570564 | 0,65685969 |
| Experimental | Control | 0,98145184 | 0,736606048 | 0,121814214 |
| Experimental | Control | 0,933825177 | 0,563230509 | -0,37644608 |
| Experimental | Control | 0,98145184 | 0,718616132 | 0,22997942 |
| Experimental | Control | 0,919104202 | 0,486561202 | 0,110801825 |
| Experimental | Control | 0,942203073 | 0,597270774 | -0,158609137 |
| Experimental | Control | 0,790420277 | 0,227349326 | -0,491147343 |
| Experimental | Control | 0,772193668 | 0,177803912 | -0,562334334 |
| Experimental | Control | 0,692638815 | 0,063315241 | 0,710780857 |
| Experimental | Control | 0,919104202 | 0,489849092 | 0,362800951 |
| Experimental | Control | 0,92316567 | 0,534872517 | -0,148641633 |
| Experimental | Control | 0,951184868 | 0,65218766 | -0,109799235 |
| Experimental | Control | 0,790420277 | 0,234486173 | 0,828798408 |
| Experimental | Control | 0,815638754 | 0,258059594 | 0,251068889 |
| Experimental | Control | 0,624192061 | 0,023349422 | 0,735046966 |
| Experimental | Control | 0,983491561 | 0,861020604 | -0,037406523 |
| Experimental | Control | 0,98145184 | 0,724524495 | 0,155451881 |
| Experimental | Control | 0,692638815 | 0,065804624 | 0,326089775 |
| Experimental | Control | 0,983491561 | 0,854552289 | 0,044235236 |
| Experimental | Control | 0,692638815 | 0,056882621 | 0,749701555 |
| Experimental | Control | 0,870817871 | 0,366132702 | -0,2050025 |
| Experimental | Control | 0,772193668 | 0,1341956 | 0,340185571 |
| Experimental | Control | 0,919104202 | 0,468863468 | 0,345922051 |
| Experimental | Control | 0,704891642 | 0,070247487 | -1,159940538 |
| Experimental | Control | 0,858120076 | 0,311787904 | -0,181256939 |
| Experimental | Control | 0,915172456 | 0,451093022 | 0,343374869 |
| Experimental | Control | 0,942203073 | 0,581991683 | 0,154298219 |
| Experimental | Control | 0,885436121 | 0,395383933 | 0,263919493 |
| Experimental | Control | 0,983491561 | 0,9573986 | 0,022501918 |

| | | | | |
|---|---|---|---|---|
| Experimental | Control | 0,92316567 | 0,515832079 | -0,25403641 |
| Experimental | Control | 0,626448651 | 0,031594205 | 0,402970914 |
| Experimental | Control | 0,942203073 | 0,604114036 | -0,410063944 |
| Experimental | Control | 0,754607597 | 0,093779051 | 0,58855212 |
| Experimental | Control | 0,790420277 | 0,23286364 | -0,600117049 |
| Experimental | Control | 0,937006646 | 0,568831147 | 0,214740413 |
| Experimental | Control | 0,772193668 | 0,130553048 | 0,898752679 |
| Experimental | Control | 0,964525247 | 0,688330326 | -0,182611377 |
| Experimental | Control | 0,98145184 | 0,721050075 | 0,12031103 |
| Experimental | Control | 0,983491561 | 0,943910394 | -0,021619049 |
| Experimental | Control | 0,858120076 | 0,299030654 | 0,538127426 |
| Experimental | Control | 0,866291532 | 0,339848962 | 0,365641311 |
| Experimental | Control | 0,92316567 | 0,525571759 | 0,244226602 |
| Experimental | Control | 0,624192061 | 0,020469953 | -1,758645288 |
| Experimental | Control | 0,870817871 | 0,349691838 | -0,397333929 |
| Experimental | Control | 0,917990807 | 0,457192737 | -0,431319046 |
| Experimental | Control | 0,92316567 | 0,526060411 | 0,311658646 |
| Experimental | Control | 0,983491561 | 0,935645558 | 0,022861956 |
| Experimental | Control | 0,959244846 | 0,681545245 | 0,244545225 |
| Experimental | Control | 0,772193668 | 0,115642282 | -0,927165136 |
| Experimental | Control | 0,772193668 | 0,135199828 | 0,472598331 |
| Experimental | Control | 0,98145184 | 0,759472812 | -0,069035188 |
| Experimental | Control | 0,908003536 | 0,432526352 | -0,613647562 |
| Experimental | Control | 0,772193668 | 0,186881078 | -0,756517679 |
| Experimental | Control | 0,866291532 | 0,337485642 | 0,450524664 |
| Experimental | Control | 0,983491561 | 0,788875886 | 0,075456389 |
| Experimental | Control | 0,772193668 | 0,124756756 | 0,232869849 |
| Experimental | Control | 0,942203073 | 0,609040401 | -0,327138644 |
| Experimental | Control | 0,624192061 | 0,020059391 | 1,151767008 |
| Experimental | Control | 0,942203073 | 0,618072722 | -0,228342567 |
| Experimental | Control | 0,797872703 | 0,241302767 | -0,375568126 |
| Experimental | Control | 0,621747057 | 0,005987037 | 0,700034009 |
| Experimental | Control | 0,983491561 | 0,780223066 | -0,074375026 |
| Experimental | Control | 0,92316567 | 0,521242974 | -0,371522194 |
| Experimental | Control | 0,983491561 | 0,947288149 | -0,017485935 |
| Experimental | Control | 0,937006646 | 0,568084145 | 0,293424364 |
| Experimental | Control | 0,92316567 | 0,538862597 | -0,308441406 |
| Experimental | Control | 0,942203073 | 0,585685871 | 0,134069696 |
| Experimental | Control | 0,989996372 | 0,977775524 | -0,009175624 |
| Experimental | Control | 0,772193668 | 0,189140119 | 0,502638039 |
| Experimental | Control | 0,858120076 | 0,316768906 | 0,402739091 |
| Experimental | Control | 0,870817871 | 0,367429992 | 0,128768103 |
| Experimental | Control | 0,983491561 | 0,952669674 | -0,033770677 |
| Experimental | Control | 0,983491561 | 0,875292158 | -0,075052127 |
| Experimental | Control | 0,92316567 | 0,535406146 | -0,327646046 |
| Experimental | Control | 0,983491561 | 0,942647934 | 0,019226494 |
| Experimental | Control | 0,866291532 | 0,338949486 | 0,593316662 |
| Experimental | Control | 0,68521937 | 0,048898102 | -0,928857998 |

| | | | | |
|---|---|---|---|---|
| Experimental | Control | 0,92316567 | 0,538039936 | -0,290797007 |
| Experimental | Control | 0,959244846 | 0,67552195 | 0,267129431 |
| Experimental | Control | 0,983491561 | 0,837881229 | -0,120687485 |
| Experimental | Control | 0,983491561 | 0,953114283 | 0,023973229 |
| Experimental | Control | 0,941268639 | 0,575116987 | 0,18734564 |
| Experimental | Control | 0,990728512 | 0,988782091 | -0,008661516 |
| Experimental | Control | 0,983491561 | 0,951876423 | -0,021018883 |
| Experimental | Control | 0,626448651 | 0,030955739 | -1,033314615 |
| Experimental | Control | 0,948299071 | 0,641874252 | 0,071516285 |
| Experimental | Control | 0,858120076 | 0,315684179 | 0,860837183 |
| Experimental | Control | 0,983491561 | 0,916769685 | 0,024938663 |
| Experimental | Control | 0,908003536 | 0,433486953 | 0,316402338 |
| Experimental | Control | 0,624192061 | 0,02580597 | 0,713426735 |
| Experimental | Control | 0,919104202 | 0,483116305 | -0,208668745 |
| Experimental | Control | 0,754607597 | 0,092841861 | 0,633857029 |
| Experimental | Control | 0,871454353 | 0,36981167 | -0,554710698 |
| Experimental | Control | 0,772193668 | 0,134326283 | -0,834060144 |
| Experimental | Control | 0,919104202 | 0,477899201 | 0,181019651 |
| Experimental | Control | 0,983491561 | 0,866022618 | -0,051909435 |
| Experimental | Control | 0,866291532 | 0,334150078 | 0,514770001 |
| Experimental | Control | 0,919104202 | 0,489263361 | 0,242695498 |
| Experimental | Control | 0,858120076 | 0,314437287 | 0,352972166 |
| Experimental | Control | 0,942203073 | 0,62183084 | 0,073755296 |
| Experimental | Control | 0,885436121 | 0,407686088 | -0,231735724 |
| Experimental | Control | 0,919104202 | 0,479179186 | 0,194903117 |
| Experimental | Control | 0,947375571 | 0,629102049 | -0,093155659 |
| Experimental | Control | 0,983491561 | 0,833973846 | -0,102021646 |
| Experimental | Control | 0,983491561 | 0,868079876 | 0,035856675 |
| Experimental | Control | 0,983491561 | 0,788419119 | -0,074487604 |
| Experimental | Control | 0,772193668 | 0,131140577 | -0,621669277 |
| Experimental | Control | 0,92316567 | 0,525739397 | -0,127992601 |
| Experimental | Control | 0,948299071 | 0,638585026 | -0,131796136 |
| Experimental | Control | 0,959244846 | 0,669225259 | -0,132659914 |
| Experimental | Control | 0,920859684 | 0,496348792 | 0,270940399 |
| Experimental | Control | 0,98145184 | 0,767072906 | 0,089931991 |
| Experimental | Control | 0,813614216 | 0,252104492 | 0,379141354 |
| Experimental | Control | 0,772193668 | 0,135092081 | 0,341739539 |
| Experimental | Control | 0,915172456 | 0,453091275 | 0,216860924 |
| Experimental | Control | 0,98145184 | 0,736655817 | -0,137836944 |
| Experimental | Control | 0,885436121 | 0,404722112 | -0,370664316 |
| Experimental | Control | 0,858120076 | 0,327063447 | -0,527963145 |
| Experimental | Control | 0,915172456 | 0,443929679 | 0,426896543 |
| Experimental | Control | 0,919104202 | 0,486642784 | -0,221382058 |
| Experimental | Control | 0,983491561 | 0,967444709 | 0,016187073 |
| Experimental | Control | 0,942203073 | 0,581032361 | 0,175053316 |
| Experimental | Control | 0,933825177 | 0,561276759 | 0,216995969 |
| Experimental | Control | 0,983491561 | 0,804586581 | -0,111594519 |
| Experimental | Control | 0,98145184 | 0,767048279 | 0,10403229 |

| | | | | |
|---|---|---|---|---|
| Experimental | Control | 0,983491561 | 0,87828715 | 0,054788217 |
| Experimental | Control | 0,790420277 | 0,230327743 | 0,503124539 |
| Experimental | Control | 0,754607597 | 0,097499462 | 0,666244418 |
| Experimental | Control | 0,870817871 | 0,358635231 | 0,295714432 |
| Experimental | Control | 0,813614216 | 0,252370308 | 0,183163649 |
| Experimental | Control | 0,858120076 | 0,324534173 | -0,425763518 |
| Experimental | Control | 0,707023515 | 0,073619344 | 0,80424213 |
| Experimental | Control | 0,98145184 | 0,748944771 | 0,159711263 |
| Experimental | Control | 0,783663238 | 0,204768587 | -0,47287906 |
| Experimental | Control | 0,621747057 | 0,00804813 | 0,566205778 |
| Experimental | Control | 0,816993398 | 0,265640687 | -0,260007169 |
| Experimental | Control | 0,983491561 | 0,956056598 | 0,026833081 |
| Experimental | Control | 0,942203073 | 0,611261837 | -0,128608954 |
| Experimental | Control | 0,942203073 | 0,590518708 | -0,144760213 |
| Experimental | Control | 0,948260112 | 0,631947743 | 0,162953477 |
| Experimental | Control | 0,983491561 | 0,870427036 | -0,023959726 |
| Experimental | Control | 0,621747057 | 0,008550549 | 0,96685568 |
| Experimental | Control | 0,983491561 | 0,816977097 | 0,048046572 |
| Experimental | Control | 0,772193668 | 0,155224466 | 0,557934245 |
| Experimental | Control | 0,772193668 | 0,170466052 | -0,590533949 |
| Experimental | Control | 0,858120076 | 0,316019991 | 0,152784485 |
| Experimental | Control | 0,858120076 | 0,323275145 | 0,535234424 |
| Experimental | Control | 0,772193668 | 0,179161288 | 0,914644 |
| Experimental | Control | 0,983491561 | 0,8709438 | 0,087084365 |
| Experimental | Control | 0,870817871 | 0,354293474 | 0,281707716 |
| Experimental | Control | 0,983491561 | 0,964436938 | -0,025617492 |
| Experimental | Control | 0,789234341 | 0,215107327 | 0,669550866 |
| Experimental | Control | 0,771535076 | 0,109136592 | -0,836409714 |
| Experimental | Control | 0,919104202 | 0,491151951 | -0,286715979 |
| Experimental | Control | 0,692638815 | 0,056985507 | -1,25272902 |
| Experimental | Control | 0,772193668 | 0,178077656 | 0,35083638 |
| Experimental | Control | 0,92316567 | 0,540478133 | -0,115698674 |
| Experimental | Control | 0,983491561 | 0,926538898 | 0,046531601 |
| Experimental | Control | 0,772193668 | 0,181351942 | 1,037996113 |
| Experimental | Control | 0,870817871 | 0,359587516 | -0,54104632 |
| Experimental | Control | 0,770427777 | 0,102925518 | -0,628094594 |
| Experimental | Control | 0,789234341 | 0,21091824 | 0,451014671 |
| Experimental | Control | 0,953177433 | 0,656558563 | 0,159567325 |
| Experimental | Control | 0,983491561 | 0,807882046 | -0,174097685 |
| Experimental | Control | 0,790420277 | 0,229753574 | 0,885187047 |
| Experimental | Control | 0,772193668 | 0,17380504 | 0,323313226 |
| Experimental | Control | 0,983491561 | 0,900036991 | -0,054299421 |
| Experimental | Control | 0,942203073 | 0,609483829 | 0,130559565 |
| Experimental | Control | 0,941268639 | 0,574247624 | -0,303747643 |
| Experimental | Control | 0,790420277 | 0,233086805 | -0,378085209 |
| Experimental | Control | 0,959244846 | 0,682213427 | 0,132371824 |
| Experimental | Control | 0,983491561 | 0,828853879 | 0,080770363 |
| Experimental | Control | 0,933825177 | 0,562363235 | 0,214867352 |

| | | | | |
|---|---|---|---|---|
| Experimental | Control | 0,771535076 | 0,10697383 | -0,440520961 |
| Experimental | Control | 0,797872703 | 0,241818211 | 0,235276021 |
| Experimental | Control | 0,858120076 | 0,313998655 | 0,199172339 |
| Experimental | Control | 0,676638733 | 0,040850994 | 0,938668025 |
| Experimental | Control | 0,858120076 | 0,321659594 | 0,280426793 |
| Experimental | Control | 0,692638815 | 0,065826966 | -0,55574665 |
| Experimental | Control | 0,983491561 | 0,871756079 | 0,034953594 |
| Experimental | Control | 0,983491561 | 0,963850061 | -0,016572217 |
| Experimental | Control | 0,98145184 | 0,757811617 | -0,106482726 |
| Experimental | Control | 0,704891642 | 0,071103044 | -1,005456143 |
| Experimental | Control | 0,92316567 | 0,508992315 | 0,119545795 |
| Experimental | Control | 0,772193668 | 0,148074194 | 0,417564684 |
| Experimental | Control | 0,983491561 | 0,832053331 | 0,039972353 |
| Experimental | Control | 0,92316567 | 0,53067002 | 0,169688894 |
| Experimental | Control | 0,772193668 | 0,152474767 | 0,657983216 |
| Experimental | Control | 0,919104202 | 0,471378447 | 0,410484477 |
| Experimental | Control | 0,983491561 | 0,776772169 | -0,091854386 |
| Experimental | Control | 0,885436121 | 0,388606844 | -0,331452676 |
| Experimental | Control | 3,83704E-08 | 7,53838E-11 | 3,337521835 |
| Experimental | Control | 0,98145184 | 0,738326725 | -0,105537658 |
| Experimental | Control | 0,772193668 | 0,116956074 | 0,413883021 |
| Experimental | Control | 0,983491561 | 0,883964756 | 0,078042771 |
| Experimental | Control | 0,98145184 | 0,766756707 | -0,052655483 |
| Experimental | Control | 0,983491561 | 0,956165485 | -0,02281581 |
| Experimental | Control | 0,772193668 | 0,137852727 | -0,515530229 |
| Experimental | Control | 0,783663238 | 0,204686881 | -0,445568025 |
| Experimental | Control | 0,783701121 | 0,207857861 | 0,341324462 |
| Experimental | Control | 0,983491561 | 0,782939097 | -0,077395544 |
| Experimental | Control | 0,885436121 | 0,388356532 | -0,207975915 |
| Experimental | Control | 0,942203073 | 0,617215868 | -0,095869388 |
| Experimental | Control | 0,983491561 | 0,939982767 | 0,019702698 |
| Experimental | Control | 0,772193668 | 0,165719113 | -0,498276076 |
| Experimental | Control | 0,692638815 | 0,059485433 | 0,487264847 |
| Experimental | Control | 0,953177433 | 0,657299173 | 0,190411031 |
| Experimental | Control | 0,983491561 | 0,894685799 | -0,024710224 |
| Experimental | Control | 0,870817871 | 0,348495469 | -0,373847482 |
| Experimental | Control | 0,624192061 | 0,021587708 | 1,294117467 |
| Experimental | Control | 0,983491561 | 0,961107133 | 0,021743332 |
| Experimental | Control | 0,983491561 | 0,773702822 | 0,094522918 |
| Experimental | Control | 0,983491561 | 0,946389385 | -0,01376267 |
| Experimental | Control | 0,959244846 | 0,680149667 | 0,07023106 |
| Experimental | Control | 0,983491561 | 0,952855058 | -0,042678287 |
| Experimental | Control | 0,983491561 | 0,965336556 | 0,009602362 |
| Experimental | Control | 0,68521937 | 0,049744862 | 0,700718444 |
| Experimental | Control | 0,754607597 | 0,096723236 | 1,097032569 |
| Experimental | Control | 0,870817871 | 0,366956354 | -0,444496089 |
| Experimental | Control | 0,98145184 | 0,761758649 | 0,144540405 |
| Experimental | Control | 0,908003536 | 0,426145434 | 0,217688256 |

| | | | | | |
|---|---|---|---|---|---|
| Experimental | Control | | 0,983491561 | 0,798207646 | -0,077282201 |
| Experimental | Control | | 0,942203073 | 0,608010719 | -0,197564385 |
| Experimental | Control | | 0,858120076 | 0,297826816 | 0,296364018 |
| Experimental | Control | | 0,948299071 | 0,636043469 | -0,260484472 |
| Experimental | Control | | 0,983491561 | 0,909284056 | -0,061787953 |
| Experimental | Control | | 0,772193668 | 0,126221277 | -0,590574744 |
| Experimental | Control | | 0,950766093 | 0,648942381 | -0,174305144 |
| Experimental | Control | | 0,754607597 | 0,097846958 | 0,4650607 |
| Experimental | Control | | 0,983491561 | 0,843959769 | 0,084290376 |
| Experimental | Control | | 0,947375571 | 0,628044852 | -0,273777781 |
| Experimental | Control | | 0,915172456 | 0,440925434 | 0,123314474 |
| Experimental | Control | | 0,870817871 | 0,351063413 | 0,470746495 |
| Experimental | Control | | 0,783663238 | 0,196632408 | 0,293619921 |
| Experimental | Control | | 0,964525247 | 0,693548606 | -0,1554113 |
| Experimental | Control | | 0,948299071 | 0,644619801 | 0,247861545 |
| Experimental | Control | | 0,922188811 | 0,505482668 | 0,136751523 |
| Experimental | Control | | 0,870817871 | 0,36400146 | -0,339807428 |
| Experimental | Control | | 0,815638754 | 0,259747165 | -0,390352642 |
| Experimental | Control | | 0,948299071 | 0,640494412 | -0,146732764 |
| Experimental | Control | | 0,983491561 | 0,808668934 | -0,124688778 |
| Experimental | Control | | 0,959244846 | 0,681496982 | -0,110651678 |
| Experimental | Control | | 0,92316567 | 0,512637859 | -0,362727291 |
| Experimental | Control | | 0,840958856 | 0,280870345 | 0,38530731 |
| Experimental | Control | | 0,919104202 | 0,478709662 | 0,118649859 |
| Experimental | Control | | 0,908003536 | 0,430389911 | 0,210189944 |
| Experimental | Control | | 0,858120076 | 0,302958472 | -0,626755501 |
| Experimental | Control | | 0,983491561 | 0,913620992 | -0,031468906 |
| Experimental | Control | | 0,676638733 | 0,042539174 | 1,011532219 |
| Experimental | Control | | 0,917990807 | 0,459598775 | 0,343998134 |
| Experimental | Control | | 0,692638815 | 0,061949257 | -0,942740345 |
| Experimental | Control | | 0,983491561 | 0,906691574 | 0,028549218 |
| Experimental | Control | | 0,844057035 | 0,285221631 | 0,289860998 |
| Experimental | Control | | 0,927175792 | 0,548290596 | -0,21969721 |
| Experimental | Control | | 0,772193668 | 0,167317331 | 0,5171583 |
| Experimental | Control | | 0,870817871 | 0,367830731 | -0,223254085 |
| Experimental | Control | | 0,983491561 | 0,817246419 | 0,168826693 |
| Experimental | Control | | 0,92316567 | 0,529921355 | 0,275357118 |
| Experimental | Control | | 0,98145184 | 0,720619465 | 0,152559952 |
| Experimental | Control | | 0,983491561 | 0,960886317 | 0,016153264 |
| Experimental | Control | | 0,68521937 | 0,047010338 | 0,823548007 |
| Experimental | Control | | 0,964525247 | 0,690201999 | 0,199714431 |
| Experimental | Control | | 0,884905186 | 0,382473754 | -0,276545843 |
| Experimental | Control | | 0,959244846 | 0,667419321 | -0,178499957 |
| Experimental | Control | | 0,92316567 | 0,519890478 | 0,280066241 |
| Experimental | Control | | 0,772193668 | 0,137059911 | 0,477285148 |
| Experimental | Control | | 0,858120076 | 0,325432331 | -0,583704881 |
| Experimental | Control | | 0,98145184 | 0,734414687 | 0,119479137 |
| Experimental | Control | | 0,908003536 | 0,430250775 | 0,36106462 |

| | | | | |
|---|---|---|---|---|
| Experimental | Control | 0,725293596 | 0,076946668 | -0,662984911 |
| Experimental | Control | 0,990728512 | 0,985128955 | -0,009811587 |
| Experimental | Control | 0,915172456 | 0,446787843 | -0,2779679 |
| Experimental | Control | 0,983491561 | 0,890463097 | -0,07297762 |
| Experimental | Control | 0,983491561 | 0,939716835 | 0,027536257 |
| Experimental | Control | 0,789234341 | 0,212922218 | -0,41832082 |
| Experimental | Control | 0,772193668 | 0,161844574 | -0,43918077 |
| Experimental | Control | 0,983491561 | 0,894490958 | -0,06342979 |
| Experimental | Control | 0,983491561 | 0,79098973 | 0,07211279 |
| Experimental | Control | 0,885436121 | 0,409116596 | 0,291515834 |
| Experimental | Control | 0,942203073 | 0,600372267 | -0,325295698 |
| Experimental | Control | 0,772193668 | 0,171205498 | 0,994891629 |
| Experimental | Control | 0,983491561 | 0,806719255 | -0,068290329 |
| Experimental | Control | 0,783663238 | 0,204476438 | -0,347240281 |
| Experimental | Control | 0,942203073 | 0,608413501 | -0,086730715 |
| Experimental | Control | 0,754607597 | 0,097288159 | -0,697375011 |
| Experimental | Control | 0,772193668 | 0,147805739 | 0,368538523 |
| Experimental | Control | 0,983491561 | 0,846739893 | -0,033090572 |
| Experimental | Control | 0,98145184 | 0,757269108 | -0,089994504 |
| Experimental | Control | 0,884905186 | 0,380754524 | 0,282085571 |
| Experimental | Control | 0,858120076 | 0,304515989 | -0,523398841 |
| Experimental | Control | 0,983491561 | 0,955380092 | -0,019002419 |
| Experimental | Control | 0,98145184 | 0,766224152 | -0,136993459 |
| Experimental | Control | 0,983491561 | 0,937089758 | -0,053019512 |
| Experimental | Control | 0,772193668 | 0,171397245 | -0,714508476 |
| Experimental | Control | 0,783701121 | 0,207350818 | -0,539722295 |
| Experimental | Control | 0,98145184 | 0,714839395 | -0,07017043 |
| Experimental | Control | 0,771535076 | 0,108770612 | -0,530178417 |
| Experimental | Control | 0,983491561 | 0,944821038 | 0,033446669 |
| Experimental | Control | 0,942203073 | 0,584433479 | -0,257641026 |
| Experimental | Control | 0,866291532 | 0,3403896 | 0,143499423 |
| Experimental | Control | 0,983491561 | 0,900440434 | -0,037058363 |
| Experimental | Control | 0,983491561 | 0,862326924 | -0,078832489 |
| Experimental | Control | 0,752251491 | 0,084240344 | -0,63264253 |
| Experimental | Control | 0,772193668 | 0,158708389 | 0,965212187 |
| Experimental | Control | 0,813614216 | 0,252556083 | -0,536220566 |
| Experimental | Control | 0,624192061 | 0,016850549 | -0,948461586 |
| Experimental | Control | 0,98145184 | 0,71658101 | -0,136766795 |
| Experimental | Control | 0,815638754 | 0,262799127 | 0,274087825 |
| Experimental | Control | 0,983491561 | 0,932129172 | 0,019802523 |
| Experimental | Control | 0,837851424 | 0,212896673 | 0,349489512 |
| Experimental | Control | 0,96532044 | 0,49659351 | -0,200418096 |
| Experimental | Control | 0,96532044 | 0,40807026 | 0,257715905 |
| Experimental | Control | 0,96532044 | 0,807958935 | -0,108055193 |
| Experimental | Control | 0,96532044 | 0,609152363 | 0,448128825 |
| Experimental | Control | 0,96532044 | 0,886903144 | -0,076878538 |
| Experimental | Control | 0,96532044 | 0,92470743 | 0,038953573 |
| Experimental | Control | 0,96532044 | 0,912402162 | 0,026731792 |

| | | | | |
|---|---|---|---|---|
| Experimental | Control | 0,71152862 | 0,12868639 | -1,078055492 |
| Experimental | Control | 0,96532044 | 0,601567121 | -0,19463906 |
| Experimental | Control | 0,96532044 | 0,685566444 | 0,170949464 |
| Experimental | Control | 0,422845864 | 0,043500191 | -0,834458968 |
| Experimental | Control | 0,96532044 | 0,488525362 | -0,427765615 |
| Experimental | Control | 0,515135012 | 0,076003526 | 0,683526537 |
| Experimental | Control | 0,987064451 | 0,987064451 | -0,005999908 |
| Experimental | Control | 0,96532044 | 0,744193058 | 0,131153536 |
| Experimental | Control | 0,96532044 | 0,816071808 | -0,101495062 |
| Experimental | Control | 0,96532044 | 0,355732549 | 0,371404656 |
| Experimental | Control | 0,96532044 | 0,733596181 | 0,129663073 |
| Experimental | Control | 0,422845864 | 0,045057346 | -0,57431432 |
| Experimental | Control | 0,96532044 | 0,949495515 | -0,018373937 |
| Experimental | Control | 0,71152862 | 0,145805045 | 0,629475845 |
| Experimental | Control | 0,96532044 | 0,74160759 | 0,061488407 |
| Experimental | Control | 0,96532044 | 0,803856608 | -0,075644838 |
| Experimental | Control | 0,646772158 | 0,111329634 | -1,248219001 |
| Experimental | Control | 0,96532044 | 0,838512153 | 0,107014876 |
| Experimental | Control | 0,979963996 | 0,971931504 | 0,008854499 |
| Experimental | Control | 0,96532044 | 0,600201154 | -0,16777924 |
| Experimental | Control | 0,96532044 | 0,903661674 | -0,03585748 |
| Experimental | Control | 0,96532044 | 0,305084534 | 0,387899843 |
| Experimental | Control | 0,452915377 | 0,051973896 | -1,220295086 |
| Experimental | Control | 0,96532044 | 0,616844756 | -0,151465853 |
| Experimental | Control | 0,575644074 | 0,089649487 | 0,689011415 |
| Experimental | Control | 0,71152862 | 0,145004106 | -0,503145235 |
| Experimental | Control | 0,96532044 | 0,330354166 | 0,317598327 |
| Experimental | Control | 0,302511593 | 0,017357223 | -1,043893598 |
| Experimental | Control | 0,96532044 | 0,833295865 | -0,043150531 |
| Experimental | Control | 0,96532044 | 0,310182478 | 0,360972928 |
| Experimental | Control | 0,96532044 | 0,916094333 | -0,060155393 |
| Experimental | Control | 0,96532044 | 0,399948201 | 0,379472381 |
| Experimental | Control | 0,474333398 | 0,062207659 | 0,766840254 |
| Experimental | Control | 0,96532044 | 0,535561468 | 0,205523525 |
| Experimental | Control | 0,96532044 | 0,398425425 | -0,418288995 |
| Experimental | Control | 0,96532044 | 0,445789488 | 0,318208716 |
| Experimental | Control | 0,422845864 | 0,033963617 | 1,63152558 |
| Experimental | Control | 0,96532044 | 0,811330208 | -0,131279629 |
| Experimental | Control | 0,837851424 | 0,207863028 | 0,878514162 |
| Experimental | Control | 0,96532044 | 0,679203618 | -0,192032262 |
| Experimental | Control | 0,96532044 | 0,627699808 | 0,314299733 |
| Experimental | Control | 0,96532044 | 0,290122495 | -0,349279037 |
| Experimental | Control | 0,96532044 | 0,762485353 | -0,230892455 |
| Experimental | Control | 0,96532044 | 0,872541462 | -0,059811074 |
| Experimental | Control | 0,422845864 | 0,044216415 | -0,881109571 |
| Experimental | Control | 0,96532044 | 0,874672434 | 0,037550589 |
| Experimental | Control | 0,96532044 | 0,697589257 | -0,134485931 |
| Experimental | Control | 0,96532044 | 0,594573014 | 0,176515068 |

| | | | | |
|---|---|---|---|---|
| Experimental | Control | 0,96532044 | 0,303209909 | -0,830048729 |
| Experimental | Control | 0,74932928 | 0,165835169 | -0,604950697 |
| Experimental | Control | 0,96532044 | 0,286345517 | -0,690613069 |
| Experimental | Control | 0,474333398 | 0,058331481 | -0,925361424 |
| Experimental | Control | 0,96532044 | 0,810134696 | -0,08061033 |
| Experimental | Control | 0,589019868 | 0,096560634 | 0,901072814 |
| Experimental | Control | 0,422845864 | 0,037479849 | -1,959956326 |
| Experimental | Control | 0,96532044 | 0,841049399 | 0,091360625 |
| Experimental | Control | 0,96532044 | 0,430606127 | 0,338367024 |
| Experimental | Control | 0,493819748 | 0,068810948 | 0,802082025 |
| Experimental | Control | 0,422845864 | 0,040141587 | 0,670652526 |
| Experimental | Control | 0,265523961 | 0,012975283 | -1,340904821 |
| Experimental | Control | 0,96532044 | 0,722869318 | 0,144749392 |
| Experimental | Control | 0,96532044 | 0,370294165 | -0,352685355 |
| Experimental | Control | 0,96532044 | 0,525260099 | 0,398453582 |
| Experimental | Control | 0,96532044 | 0,85879498 | -0,061987937 |
| Experimental | Control | 0,96532044 | 0,667928329 | -0,179425429 |
| Experimental | Control | 0,96532044 | 0,457860953 | -0,413949365 |
| Experimental | Control | 0,96532044 | 0,866198889 | 0,084224682 |
| Experimental | Control | 0,96532044 | 0,729902973 | 0,183471783 |
| Experimental | Control | 0,96532044 | 0,623317073 | 0,102926624 |
| Experimental | Control | 0,96532044 | 0,358489688 | -0,631664569 |
| Experimental | Control | 0,96532044 | 0,94307462 | -0,024579711 |
| Experimental | Control | 0,96532044 | 0,819015459 | -0,048572 |
| Experimental | Control | 0,96532044 | 0,874173231 | -0,110200997 |
| Experimental | Control | 6,06056E-05 | 4,96767E-07 | 2,715309904 |
| Experimental | Control | 0,96532044 | 0,942166052 | 0,023781871 |
| Experimental | Control | 0,96532044 | 0,377617882 | 0,236137569 |
| Experimental | Control | 0,96532044 | 0,472259829 | 0,236314157 |
| Experimental | Control | 0,265523961 | 0,011687004 | -1,13906127 |
| Experimental | Control | 0,96532044 | 0,46068962 | -0,542113819 |
| Experimental | Control | 0,265523961 | 0,013058555 | 1,665368297 |
| Experimental | Control | 0,96532044 | 0,934225521 | 0,050845745 |
| Experimental | Control | 0,71152862 | 0,139090955 | 0,417547947 |
| Experimental | Control | 0,96532044 | 0,718563706 | 0,153659354 |
| Experimental | Control | 0,96532044 | 0,448759852 | 0,314973406 |
| Experimental | Control | 0,96532044 | 0,681945039 | 0,22496009 |
| Experimental | Control | 0,96532044 | 0,885695976 | 0,058163009 |
| Experimental | Control | 0,737236159 | 0,157115903 | 0,310178211 |
| Experimental | Control | 0,96532044 | 0,683027818 | 0,07724492 |
| Experimental | Control | 0,96532044 | 0,569679197 | 0,376870148 |
| Experimental | Control | 0,265523961 | 0,012020837 | 1,072137509 |
| Experimental | Control | 0,96532044 | 0,625744047 | -0,131802037 |
| Experimental | Control | 0,96532044 | 0,807446897 | 0,09102732 |
| Experimental | Control | 0,96532044 | 0,623870246 | 0,340389564 |
| Experimental | Control | 0,96532044 | 0,431350715 | 0,263445494 |
| Experimental | Control | 0,96532044 | 0,923607826 | 0,044725339 |
| Experimental | Control | 0,832111546 | 0,19097642 | -0,742213858 |

| | | | | |
|---|---|---|---|---|
| Experimental | Control | 0,96532044 | 0,505478142 | -0,402345627 |
| Experimental | Control | 0,96532044 | 0,878356363 | 0,08925902 |
| Experimental | Control | 0,96532044 | 0,302010025 | -0,45618472 |
| Experimental | Control | 0,96532044 | 0,925890614 | 0,043427039 |
| Experimental | Control | 0,96532044 | 0,623334075 | 0,216481469 |
| Experimental | Control | 0,96532044 | 0,936363171 | 0,05460961 |
| Experimental | Control | 0,96532044 | 0,723409252 | -0,189238881 |
| Experimental | Control | 0,96532044 | 0,253979837 | -0,803094564 |
| Experimental | Control | 0,265523961 | 0,007434625 | -1,092568019 |
| Experimental | Control | 0,96532044 | 0,775669499 | -0,123650919 |
| Experimental | Control | 0,837851424 | 0,207093794 | -0,625740606 |
| Experimental | Control | 0,96532044 | 0,736681184 | -0,152818202 |
| Experimental | Control | 0,96532044 | 0,448661997 | -0,19024051 |
| Experimental | Control | 0,96532044 | 0,269360243 | -0,533725437 |
| Experimental | Control | 0,96532044 | 0,765981059 | -0,226339627 |
| Experimental | Control | 0,96532044 | 0,392916132 | 0,470878879 |
| Experimental | Control | 0,96532044 | 0,698223762 | -0,272615028 |
| Experimental | Control | 0,96532044 | 0,671257296 | -0,222247103 |
| Experimental | Control | 0,952834963 | 0,4210201 | 0,517978923 |
| Experimental | Control | 0,97102509 | 0,640908398 | -0,135611802 |
| Experimental | Control | 0,97102509 | 0,588886409 | 0,475327076 |
| Experimental | Control | 0,97102509 | 0,925911039 | 0,057493572 |
| Experimental | Control | 0,663258616 | 0,222247585 | 0,455514093 |
| Experimental | Control | 0,97102509 | 0,800138595 | -0,127823363 |
| Experimental | Control | 0,785606885 | 0,293819104 | 0,431031155 |
| Experimental | Control | 0,663258616 | 0,231369285 | -0,490919127 |
| Experimental | Control | 0,97102509 | 0,782997846 | 0,149043118 |
| Experimental | Control | 0,97102509 | 0,925593179 | -0,02851582 |
| Experimental | Control | 0,346490135 | 0,056405371 | -1,577481282 |
| Experimental | Control | 0,97102509 | 0,922484117 | -0,028084453 |
| Experimental | Control | 0,97102509 | 0,7102276 | -0,111598688 |
| Experimental | Control | 0,97102509 | 0,556287471 | 0,111029994 |
| Experimental | Control | 0,663258616 | 0,229819891 | 0,491924108 |
| Experimental | Control | 0,97102509 | 0,630383127 | -0,167263689 |
| Experimental | Control | 0,97102509 | 0,485443536 | 0,354901361 |
| Experimental | Control | 0,236400034 | 0,027488376 | 1,679720798 |
| Experimental | Control | 0,236400034 | 0,023644753 | -0,655064015 |
| Experimental | Control | 0,97102509 | 0,903892314 | -0,049491316 |
| Experimental | Control | 0,97102509 | 0,915954601 | 0,064265506 |
| Experimental | Control | 0,663258616 | 0,197353239 | -0,546964545 |
| Experimental | Control | 0,236400034 | 0,023344044 | -0,537981681 |
| Experimental | Control | 0,006098733 | 0,000141831 | 2,468454515 |
| Experimental | Control | 0,97102509 | 0,848265592 | 0,106674612 |
| Experimental | Control | 0,637127317 | 0,162986058 | 0,391992782 |
| Experimental | Control | 0,356058957 | 0,071741506 | 0,488061027 |
| Experimental | Control | 0,97102509 | 0,889628942 | 0,061516549 |
| Experimental | Control | 0,97102509 | 0,714555663 | -0,094357283 |
| Experimental | Control | 0,356058957 | 0,074523968 | 0,91993359 |

| | | | | |
|---|---|---|---|---|
| Experimental | Control | 0,97102509 | 0,620738091 | -0,18651103 |
| Experimental | Control | 0,423151993 | 0,09840744 | -0,949630331 |
| Experimental | Control | 0,97102509 | 0,707973211 | 0,138248508 |
| Experimental | Control | 0,97102509 | 0,635889376 | 0,321784763 |
| Experimental | Control | 0,97102509 | 0,458940138 | -0,291486224 |
| Experimental | Control | 0,97102509 | 0,948443111 | -0,035529956 |
| Experimental | Control | 0,904032312 | 0,378432131 | -0,61033553 |
| Experimental | Control | 0,173411762 | 0,008065663 | -1,076310493 |
| Experimental | Control | 0,97102509 | 0,507810434 | -0,273465932 |
| Experimental | Control | 0,997364336 | 0,997364336 | -0,001250055 |
| Experimental | Control | 0,97102509 | 0,724213636 | 0,144907495 |
| Experimental | Control | 0,785606885 | 0,310588769 | -0,494373927 |
| Experimental | Control | 0,265397478 | 0,037032206 | 1,15331987 |
| Experimental | Control | 0,778339522 | 0,497824217 | 0,214454185 |
| Experimental | Control | 0,849753109 | 0,849753109 | 0,037329965 |
| Experimental | Control | 0,778339522 | 0,639454394 | 0,182360259 |
| Experimental | Control | 0,511665502 | 0,146190144 | -0,291837586 |
| Experimental | Control | 0,511665502 | 0,07456208 | -0,698071582 |
| Experimental | Control | 0,778339522 | 0,546969156 | 0,260615961 |
| Experimental | Control | 0,778339522 | 0,667148161 | 0,329560518 |
| Experimental | Control | 0,057956521 | 0,00277627 | 1,409240892 |
| Experimental | Control | 0,057956521 | 0,001723478 | 1,360625359 |
| Experimental | Control | 0,174985748 | 0,016787442 | -2,164062143 |
| Experimental | Control | 0,572808567 | 0,196480329 | -0,692041181 |
| Experimental | Control | 0,298114484 | 0,040705471 | -1,43464469 |
| Experimental | Control | 0,121663398 | 0,009170409 | 1,309127913 |
| Experimental | Control | 0,993927207 | 0,993541535 | 0,004157195 |
| Experimental | Control | 0,837515875 | 0,507122219 | 0,393118921 |
| Experimental | Control | 0,63768051 | 0,239861476 | 0,518818034 |
| Experimental | Control | 0,838863437 | 0,514092455 | -0,353792888 |
| Experimental | Control | 0,785579079 | 0,423780274 | -0,462001282 |
| Experimental | Control | 0,993927207 | 0,981400445 | 0,008741022 |
| Experimental | Control | 0,90534009 | 0,70046368 | 0,293183748 |
| Experimental | Control | 0,863880358 | 0,579132729 | -0,297055894 |
| Experimental | Control | 0,451865161 | 0,114417233 | -1,11763514 |
| Experimental | Control | 0,477641458 | 0,122965507 | 1,07303249 |
| Experimental | Control | 0,529467057 | 0,157382868 | -0,924699446 |
| Experimental | Control | 0,554948252 | 0,180010257 | -1,063529539 |
| Experimental | Control | 0,20593209 | 0,021177752 | 1,123220272 |
| Experimental | Control | 0,837079944 | 0,499176113 | 0,359489587 |
| Experimental | Control | 0,041440413 | 0,000716497 | 1,879034643 |
| Experimental | Control | 0,648729218 | 0,252620998 | -0,612573936 |
| Experimental | Control | 0,121663398 | 0,010045602 | 1,189902925 |
| Experimental | Control | 0,522163094 | 0,148017151 | 0,949331418 |
| Experimental | Control | 0,763168505 | 0,393086236 | -0,318167496 |
| Experimental | Control | 0,732971373 | 0,326811089 | -0,711739074 |
| Experimental | Control | 0,958500764 | 0,810447485 | 0,145467416 |
| Experimental | Control | 0,863880358 | 0,597159975 | -0,335391771 |

| | | | | |
|---|---|---|---|---|
| Experimental | Control | 0,897159618 | 0,662499826 | -0,364549265 |
| Experimental | Control | 0,993927207 | 0,978559633 | 0,015088858 |
| Experimental | Control | 0,814252818 | 0,459805857 | -0,546207015 |
| Experimental | Control | 0,733532311 | 0,328909194 | 0,473044016 |
| Experimental | Control | 0,712792386 | 0,305577245 | -0,779006957 |
| Experimental | Control | 0,329988443 | 0,053057768 | -1,286204091 |
| Experimental | Control | 0,572808567 | 0,196541655 | 0,681412228 |
| Experimental | Control | 0,298114484 | 0,042118927 | 0,856972949 |
| Experimental | Control | 0,218578096 | 0,024865765 | -1,154828828 |
| Experimental | Control | 0,988656375 | 0,932364052 | -0,065900788 |
| Experimental | Control | 0,057956521 | 0,003206616 | -2,008626889 |
| Experimental | Control | 0,395419923 | 0,085985857 | -1,363184708 |
| Experimental | Control | 0,742113995 | 0,340460929 | -0,719824576 |
| Experimental | Control | 0,218578096 | 0,02463803 | -1,451013654 |
| Experimental | Control | 0,703604491 | 0,290870921 | -0,602076089 |
| Experimental | Control | 0,930764102 | 0,751979761 | -0,086070276 |
| Experimental | Control | 0,517912006 | 0,1438521 | -0,750878376 |
| Experimental | Control | 0,395419923 | 0,085684664 | -0,604498297 |
| Experimental | Control | 0,851026926 | 0,529522269 | -0,367242853 |
| Experimental | Control | 0,969496774 | 0,852734738 | 0,117835716 |
| Experimental | Control | 0,90221013 | 0,683154272 | -0,117736888 |
| Experimental | Control | 0,686739223 | 0,280996049 | -0,718603923 |
| Experimental | Control | 0,969496774 | 0,857461312 | 0,088651359 |
| Experimental | Control | 0,766140345 | 0,405655059 | 0,282598344 |
| Experimental | Control | 0,513480967 | 0,13783206 | 0,953033805 |
| Experimental | Control | 0,318433538 | 0,046742538 | -1,144257294 |
| Experimental | Control | 0,863880358 | 0,594408589 | 0,25798567 |
| Experimental | Control | 0,993927207 | 0,976662425 | -0,019199163 |
| Experimental | Control | 0,712792386 | 0,304404178 | -0,567854575 |
| Experimental | Control | 0,90221013 | 0,688659475 | 0,288427163 |
| Experimental | Control | 0,81933177 | 0,470552007 | -0,122827841 |
| Experimental | Control | 0,90221013 | 0,68384018 | -0,214443549 |
| Experimental | Control | 0,766015405 | 0,400576863 | -0,678681983 |
| Experimental | Control | 0,657888881 | 0,263155552 | 0,585448134 |
| Experimental | Control | 0,416183377 | 0,101245842 | -0,693158789 |
| Experimental | Control | 0,508976901 | 0,135274311 | -1,191915417 |
| Experimental | Control | 0,72873926 | 0,323586974 | -0,71169923 |
| Experimental | Control | 0,758652695 | 0,365656176 | 0,283076334 |
| Experimental | Control | 0,869288498 | 0,61248951 | 0,288741164 |
| Experimental | Control | 0,135705582 | 0,012201052 | -1,320566313 |
| Experimental | Control | 0,057956521 | 0,00274956 | 1,404289594 |
| Experimental | Control | 0,758652695 | 0,369243997 | -0,480442833 |
| Experimental | Control | 0,536258408 | 0,165305344 | -0,751915256 |
| Experimental | Control | 0,394821831 | 0,084035472 | -0,795104834 |
| Experimental | Control | 0,875638058 | 0,622960301 | -0,230672119 |
| Experimental | Control | 0,778142834 | 0,418242502 | -0,293091487 |
| Experimental | Control | 0,532043431 | 0,161737742 | -0,748542966 |
| Experimental | Control | 0,559060106 | 0,185021148 | -0,475092993 |

| | | | | |
|---|---|---|---|---|
| Experimental | Control | 0,298114484 | 0,041634908 | -0,693460221 |
| Experimental | Control | 0,387976049 | 0,073679369 | 1,209814363 |
| Experimental | Control | 0,942023842 | 0,786319763 | 0,129055632 |
| Experimental | Control | 0,63768051 | 0,238736573 | -0,444718483 |
| Experimental | Control | 0,942023842 | 0,770935791 | 0,18051778 |
| Experimental | Control | 0,442655649 | 0,111273071 | -1,435741082 |
| Experimental | Control | 0,885956934 | 0,643741185 | 0,164788385 |
| Experimental | Control | 0,576705782 | 0,201053392 | -0,741117139 |
| Experimental | Control | 0,919846846 | 0,739253062 | 0,336252977 |
| Experimental | Control | 0,993927207 | 0,958461995 | 0,034094925 |
| Experimental | Control | 0,416183377 | 0,09359265 | 0,477974185 |
| Experimental | Control | 0,789179454 | 0,428618566 | 0,264759867 |
| Experimental | Control | 0,930764102 | 0,753150402 | 0,149865708 |
| Experimental | Control | 0,863880358 | 0,593455776 | -0,232222901 |
| Experimental | Control | 0,960150973 | 0,828678941 | 0,116148763 |
| Experimental | Control | 0,298114484 | 0,039251984 | 1,604383353 |
| Experimental | Control | 0,887212907 | 0,646281696 | 0,194838396 |
| Experimental | Control | 0,416183377 | 0,097412475 | -1,050995479 |
| Experimental | Control | 0,121663398 | 0,009952367 | -2,061357576 |
| Experimental | Control | 0,041440413 | 0,000369879 | 3,405684521 |
| Experimental | Control | 0,645797995 | 0,247654644 | 0,841003041 |
| Experimental | Control | 0,958734741 | 0,819762182 | -0,120805051 |
| Experimental | Control | 0,90221013 | 0,685639185 | -0,295480684 |
| Experimental | Control | 0,057956521 | 0,003138734 | -1,05558398 |
| Experimental | Control | 0,988656375 | 0,932420875 | -0,055559733 |
| Experimental | Control | 0,969496774 | 0,891058195 | -0,058250788 |
| Experimental | Control | 0,837186636 | 0,502836169 | -0,284904043 |
| Experimental | Control | 0,330471664 | 0,058211522 | 1,234481732 |
| Experimental | Control | 0,554948252 | 0,175793699 | -0,684361893 |
| Experimental | Control | 0,90221013 | 0,687110126 | 0,272013561 |
| Experimental | Control | 0,837186636 | 0,503952366 | -0,172250026 |
| Experimental | Control | 0,969496774 | 0,901898833 | -0,054218652 |
| Experimental | Control | 0,899664766 | 0,66690746 | -0,239711233 |
| Experimental | Control | 0,758652695 | 0,365734961 | -0,417775755 |
| Experimental | Control | 0,841882069 | 0,517487143 | 0,301976959 |
| Experimental | Control | 0,703604491 | 0,292117388 | -0,527990644 |
| Experimental | Control | 0,958734741 | 0,813456514 | 0,135466774 |
| Experimental | Control | 0,969496774 | 0,852867887 | -0,049783986 |
| Experimental | Control | 0,330471664 | 0,056581936 | -0,842701625 |
| Experimental | Control | 0,942023842 | 0,769781837 | 0,094527701 |
| Experimental | Control | 0,057956521 | 0,002231549 | -1,518243565 |
| Experimental | Control | 0,969496774 | 0,892302333 | -0,035763466 |
| Experimental | Control | 0,734874952 | 0,331705024 | -0,206529161 |
| Experimental | Control | 0,758652695 | 0,381991762 | 0,267322948 |
| Experimental | Control | 0,552296958 | 0,172556182 | 0,494153094 |
| Experimental | Control | 0,942023842 | 0,782817466 | -0,143344928 |
| Experimental | Control | 0,416183377 | 0,100969006 | -1,705925126 |
| Experimental | Control | 0,879402206 | 0,631941216 | -0,178567779 |

| | | | | |
|---|---|---|---|---|
| Experimental | Control | 0,298114484 | 0,041827054 | -1,318262172 |
| Experimental | Control | 0,90884232 | 0,708116177 | -0,097574813 |
| Experimental | Control | 0,411214661 | 0,091021432 | -1,118201065 |
| Experimental | Control | 0,057956521 | 0,001816162 | -1,505524996 |
| Experimental | Control | 0,993927207 | 0,993927207 | 0,003129775 |
| Experimental | Control | 0,814457542 | 0,46177501 | 0,356684294 |
| Experimental | Control | 0,993927207 | 0,975535197 | -0,018033668 |
| Experimental | Control | 0,973992691 | 0,911442701 | -0,021101069 |
| Experimental | Control | 0,176805734 | 0,017528928 | -0,677257476 |
| Experimental | Control | 0,857987221 | 0,54473702 | -0,454952818 |
| Experimental | Control | 0,532043431 | 0,162499087 | -0,765848978 |
| Experimental | Control | 0,62370955 | 0,226595396 | -0,47203351 |
| Experimental | Control | 0,969496774 | 0,895015082 | -0,0590556 |
| Experimental | Control | 0,121663398 | 0,00983903 | -1,159721717 |
| Experimental | Control | 0,543317864 | 0,168478383 | -0,696422702 |
| Experimental | Control | 0,961029126 | 0,83406748 | 0,138757103 |
| Experimental | Control | 0,969496774 | 0,867627061 | 0,066098255 |
| Experimental | Control | 0,820490176 | 0,475746704 | 0,27202335 |
| Experimental | Control | 0,060227846 | 0,003646824 | -2,933455668 |
| Experimental | Control | 0,069458163 | 0,00433317 | -1,201832392 |
| Experimental | Control | 0,712792386 | 0,308335928 | -0,404429126 |
| Experimental | Control | 0,911914311 | 0,716317382 | 0,199665922 |
| Experimental | Control | 0,960150973 | 0,822783141 | -0,112735358 |
| Experimental | Control | 0,525664202 | 0,15046535 | 0,550783272 |
| Experimental | Control | 0,041440413 | 0,000356456 | 1,126372037 |
| Experimental | Control | 0,837186636 | 0,505384226 | -0,369482146 |
| Experimental | Control | 0,969496774 | 0,88309258 | -0,074283957 |
| Experimental | Control | 0,712792386 | 0,309646185 | 0,838784551 |
| Experimental | Control | 0,969496774 | 0,862612666 | 0,051788868 |
| Experimental | Control | 0,709413773 | 0,299153906 | -0,507219307 |
| Experimental | Control | 0,529545898 | 0,159349591 | 1,121396015 |
| Experimental | Control | 0,837186636 | 0,505366923 | -0,132958238 |
| Experimental | Control | 0,814252818 | 0,460001849 | -0,298376126 |
| Experimental | Control | 0,041440413 | 0,000865905 | -2,315818212 |
| Experimental | Control | 0,969496774 | 0,871594469 | -0,103780063 |
| Experimental | Control | 0,712792386 | 0,305808234 | -0,515441529 |
| Experimental | Control | 0,529467057 | 0,156781726 | 0,662774745 |
| Experimental | Control | 0,742113995 | 0,343978927 | -0,562370373 |
| Experimental | Control | 0,057956521 | 0,001687061 | 3,574125838 |
| Experimental | Control | 0,576326596 | 0,199863719 | -0,381093287 |
| Experimental | Control | 0,792267412 | 0,436110502 | -0,587856775 |
| Experimental | Control | 0,636069717 | 0,236501305 | 0,660184897 |
| Experimental | Control | 0,814252818 | 0,460164896 | -0,171815241 |
| Experimental | Control | 0,825110389 | 0,482954521 | 0,298472143 |
| Experimental | Control | 0,969496774 | 0,873476595 | 0,043048306 |
| Experimental | Control | 0,942023842 | 0,774684595 | 0,100184384 |
| Experimental | Control | 0,969496774 | 0,856414555 | -0,129119311 |
| Experimental | Control | 0,927785783 | 0,747335704 | 0,142388885 |

| | | | | |
|---|---|---|---|---|
| Experimental | Control | 0,742113995 | 0,339273539 | 0,474473193 |
| Experimental | Control | 0,057956521 | 0,003296609 | 1,154401908 |
| Experimental | Control | 0,440573542 | 0,109941288 | -1,082423146 |
| Experimental | Control | 0,899774764 | 0,670290925 | -0,142889054 |
| Experimental | Control | 0,477641458 | 0,124449701 | -0,513532578 |
| Experimental | Control | 0,526624713 | 0,153379965 | -0,956426571 |
| Experimental | Control | 0,559060106 | 0,185669503 | -0,39429847 |
| Experimental | Control | 0,554948252 | 0,178027283 | -0,512100041 |
| Experimental | Control | 0,712792386 | 0,309966597 | -0,601932217 |
| Experimental | Control | 0,471613712 | 0,12028313 | 0,603229971 |
| Experimental | Control | 0,91467373 | 0,729621834 | 0,152637526 |
| Experimental | Control | 0,11915024 | 0,008744972 | -1,497790685 |
| Experimental | Control | 0,581552158 | 0,203810022 | -0,736938121 |
| Experimental | Control | 0,877481963 | 0,626312814 | -0,162365024 |
| Experimental | Control | 0,421631689 | 0,103667241 | -0,428519565 |
| Experimental | Control | 0,416183377 | 0,101151286 | -0,417141069 |
| Experimental | Control | 0,829986385 | 0,489525781 | -0,307896269 |
| Experimental | Control | 0,958734741 | 0,818626248 | -0,065894547 |
| Experimental | Control | 0,829986385 | 0,491900188 | -0,175706826 |
| Experimental | Control | 0,969496774 | 0,884095109 | -0,03657998 |
| Experimental | Control | 0,650951978 | 0,255603162 | -0,52777699 |
| Experimental | Control | 0,877687664 | 0,628070072 | 0,238348609 |
| Experimental | Control | 0,81933177 | 0,466215576 | 0,193478338 |
| Experimental | Control | 0,328429023 | 0,048812387 | -1,07494274 |
| Experimental | Control | 0,969496774 | 0,867783125 | 0,105536586 |
| Experimental | Control | 0,961029126 | 0,833891418 | 0,045239429 |
| Experimental | Control | 0,911914311 | 0,717338129 | -0,090522747 |
| Experimental | Control | 0,969496774 | 0,846170383 | 0,123059708 |
| Experimental | Control | 0,863880358 | 0,560941231 | -0,128945595 |
| Experimental | Control | 0,758652695 | 0,360986924 | 0,176602093 |
| Experimental | Control | 0,757619695 | 0,353092482 | 0,676922084 |
| Experimental | Control | 0,477641458 | 0,123608035 | -0,715297798 |
| Experimental | Control | 0,851026926 | 0,530915881 | -0,469983899 |
| Experimental | Control | 0,941568699 | 0,765348502 | -0,059421083 |
| Experimental | Control | 0,559060106 | 0,184579389 | -0,468394176 |
| Experimental | Control | 0,993927207 | 0,993585443 | -0,002938766 |
| Experimental | Control | 0,703604491 | 0,290653101 | 0,485098345 |
| Experimental | Control | 0,057956521 | 0,002573829 | -1,440167136 |
| Experimental | Control | 0,863880358 | 0,585662634 | 0,248238525 |
| Experimental | Control | 0,993927207 | 0,955047654 | -0,036212213 |
| Experimental | Control | 0,969496774 | 0,898763613 | 0,07521152 |
| Experimental | Control | 0,90221013 | 0,681729536 | -0,159381885 |
| Experimental | Control | 0,857260587 | 0,541096591 | -0,397705867 |
| Experimental | Control | 0,969496774 | 0,87310114 | 0,06956082 |
| Experimental | Control | 0,090375866 | 0,006301437 | 0,862490613 |
| Experimental | Control | 0,71998115 | 0,314633675 | -0,707978792 |
| Experimental | Control | 0,057956521 | 0,002899642 | 1,090829871 |
| Experimental | Control | 0,71998115 | 0,316618535 | -0,402464111 |

| | | | | |
|---|---|---|---|---|
| Experimental | Control | 0,59276098 | 0,208825886 | -0,893150764 |
| Experimental | Control | 0,863880358 | 0,596155137 | -0,360594627 |
| Experimental | Control | 0,81933177 | 0,468930074 | -0,134873137 |
| Experimental | Control | 0,958734741 | 0,819136597 | 0,099527739 |
| Experimental | Control | 0,833767336 | 0,495670857 | -0,147917622 |
| Experimental | Control | 0,90884232 | 0,70873025 | 0,227731002 |
| Experimental | Control | 0,90534009 | 0,702676804 | -0,06476201 |
| Experimental | Control | 0,041440413 | 0,000457632 | -1,404465071 |
| Experimental | Control | 0,758652695 | 0,356420389 | 0,387350991 |
| Experimental | Control | 0,394135539 | 0,082198577 | -0,750164693 |
| Experimental | Control | 0,919665864 | 0,735732691 | 0,089028784 |
| Experimental | Control | 0,942023842 | 0,786043969 | 0,052169756 |
| Experimental | Control | 0,742113995 | 0,344504295 | -0,394503716 |
| Experimental | Control | 0,758652695 | 0,375226667 | -0,271639583 |
| Experimental | Control | 0,703604491 | 0,29435197 | 0,746670703 |
| Experimental | Control | 0,863880358 | 0,596573814 | 0,180247219 |
| Experimental | Control | 0,041440413 | 0,000912449 | 1,140773624 |
| Experimental | Control | 0,763168505 | 0,393157459 | 0,234139191 |
| Experimental | Control | 0,508976901 | 0,135415873 | -0,89186557 |
| Experimental | Control | 0,993927207 | 0,991490511 | 0,002838145 |
| Experimental | Control | 0,209215047 | 0,022265088 | 0,806629622 |
| Experimental | Control | 0,329988443 | 0,051729413 | 0,501796919 |
| Experimental | Control | 0,758652695 | 0,369189661 | -0,389780016 |
| Experimental | Control | 0,672382709 | 0,271420543 | 0,707339265 |
| Experimental | Control | 0,526624713 | 0,153639136 | -0,905101363 |
| Experimental | Control | 0,742113995 | 0,343890613 | -0,164497343 |
| Experimental | Control | 0,90221013 | 0,675969498 | -0,241719996 |
| Experimental | Control | 0,95347105 | 0,803015068 | 0,117227024 |
| Experimental | Control | 0,238560017 | 0,028014387 | -1,054027487 |
| Experimental | Control | 0,041440413 | 0,000433613 | 1,738116156 |
| Experimental | Control | 0,517912006 | 0,14539548 | -0,538425583 |
| Experimental | Control | 0,960150973 | 0,825702998 | 0,164858273 |
| Experimental | Control | 0,394135539 | 0,080004738 | -1,028511823 |
| Experimental | Control | 0,829986385 | 0,490179556 | -0,296529692 |
| Experimental | Control | 0,863880358 | 0,583221251 | -0,315996237 |
| Experimental | Control | 0,627709367 | 0,230352061 | -0,618483194 |
| Experimental | Control | 0,394135539 | 0,08209431 | 0,560516798 |
| Experimental | Control | 0,763168505 | 0,39177822 | -0,326267568 |
| Experimental | Control | 0,919846846 | 0,738727056 | -0,095577097 |
| Experimental | Control | 0,758652695 | 0,378871259 | -0,540568442 |
| Experimental | Control | 0,993927207 | 0,974245585 | -0,016759047 |
| Experimental | Control | 0,646503812 | 0,250297806 | -0,527940717 |
| Experimental | Control | 0,95347105 | 0,800069718 | -0,213627919 |
| Experimental | Control | 0,517912006 | 0,142413701 | -0,595668071 |
| Experimental | Control | 0,942023842 | 0,78595891 | -0,082463227 |
| Experimental | Control | 0,394135539 | 0,083166215 | -0,73534555 |
| Experimental | Control | 0,174985748 | 0,017016963 | -1,545261543 |
| Experimental | Control | 0,969496774 | 0,881436834 | -0,074942855 |

| | | | | |
|---|---|---|---|---|
| Experimental | Control | 0,993927207 | 0,957946006 | 0,013770779 |
| Experimental | Control | 0,554948252 | 0,1802309 | -0,955533523 |
| Experimental | Control | 0,863880358 | 0,589419442 | 0,174743609 |
| Experimental | Control | 0,897159618 | 0,663404268 | 0,159257169 |
| Experimental | Control | 0,394135539 | 0,079889467 | -1,367501389 |
| Experimental | Control | 0,758652695 | 0,368756883 | 0,48307547 |
| Experimental | Control | 0,330471664 | 0,05744981 | 0,358542776 |
| Experimental | Control | 0,330471664 | 0,057612678 | 0,953254382 |
| Experimental | Control | 0,057956521 | 0,003283607 | -1,268231233 |
| Experimental | Control | 0,851026926 | 0,530827213 | 0,20440384 |
| Experimental | Control | 0,71998115 | 0,318376985 | 0,636916149 |
| Experimental | Control | 0,90221013 | 0,68472078 | -0,131867158 |
| Experimental | Control | 0,99096641 | 0,9364178 | -0,018536227 |
| Experimental | Control | 0,562434821 | 0,190873014 | 0,607051416 |
| Experimental | Control | 0,660164871 | 0,26527726 | 0,430515833 |
| Experimental | Control | 0,657777016 | 0,259601516 | -0,487330421 |
| Experimental | Control | 0,709413773 | 0,299385629 | 0,369780816 |
| Experimental | Control | 0,913007919 | 0,725380603 | -0,091903101 |
| Experimental | Control | 0,993927207 | 0,947818715 | 0,015912061 |
| Experimental | Control | 0,712792386 | 0,308656696 | -0,541097636 |
| Experimental | Control | 0,758652695 | 0,380616726 | 0,216315146 |
| Experimental | Control | 0,758652695 | 0,377683283 | -0,487034347 |
| Experimental | Control | 0,673660499 | 0,273172422 | -0,538978872 |
| Experimental | Control | 0,359410474 | 0,065287407 | -0,589905285 |
| Experimental | Control | 0,973937655 | 0,907817117 | 0,030906974 |
| Experimental | Control | 0,387976049 | 0,074035797 | 0,895554987 |
| Experimental | Control | 0,416183377 | 0,100418336 | 1,094192809 |
| Experimental | Control | 0,871121383 | 0,615379325 | 0,070300578 |
| Experimental | Control | 0,394135539 | 0,077914969 | -0,869677546 |
| Experimental | Control | 0,758652695 | 0,378085091 | 0,567443844 |
| Experimental | Control | 0,802180208 | 0,445982758 | 0,129503084 |
| Experimental | Control | 0,766140345 | 0,402690221 | -0,478446491 |
| Experimental | Control | 0,911914311 | 0,717047834 | 0,081884168 |
| Experimental | Control | 0,869288498 | 0,611402106 | -0,323673072 |
| Experimental | Control | 0,763168505 | 0,39142591 | -0,387257636 |
| Experimental | Control | 0,764864297 | 0,397168066 | -0,39301373 |
| Experimental | Control | 0,992179212 | 0,939384355 | -0,027547608 |
| Experimental | Control | 0,882164852 | 0,636129884 | -0,270254578 |
| Experimental | Control | 0,960150973 | 0,82808111 | 0,122243108 |
| Experimental | Control | 0,057956521 | 0,00226993 | -1,097169867 |
| Experimental | Control | 0,858224921 | 0,54800417 | 0,287645291 |
| Experimental | Control | 0,626836806 | 0,228881696 | 0,275908446 |
| Experimental | Control | 0,856198414 | 0,537284143 | 0,36179934 |
| Experimental | Control | 0,911914311 | 0,719492025 | -0,152319373 |
| Experimental | Control | 0,863880358 | 0,575380325 | -0,27154978 |
| Experimental | Control | 0,489434128 | 0,128420331 | 0,422618222 |
| Experimental | Control | 0,863880358 | 0,567930771 | -0,13899169 |
| Experimental | Control | 0,641870357 | 0,244970705 | 0,226792864 |

| | | | | |
|---|---|---|---|---|
| Experimental | Control | 0,758652695 | 0,369128414 | 0,802649015 |
| Experimental | Control | 0,394135539 | 0,080874128 | -0,520013721 |
| Experimental | Control | 0,329988443 | 0,053139078 | 0,972336743 |
| Experimental | Control | 0,992904123 | 0,941892535 | 0,026188336 |
| Experimental | Control | 0,904608978 | 0,692150355 | -0,12228657 |
| Experimental | Control | 0,969496774 | 0,865388006 | -0,061366219 |
| Experimental | Control | 0,529545898 | 0,159344986 | 0,895163567 |
| Experimental | Control | 0,394135539 | 0,082645227 | -0,937741015 |
| Experimental | Control | 0,291305327 | 0,035906109 | 0,604479271 |
| Experimental | Control | 0,648729218 | 0,253540043 | 0,306423034 |
| Experimental | Control | 0,988656375 | 0,929320655 | 0,014384052 |
| Experimental | Control | 0,646335809 | 0,249046825 | 0,509242515 |
| Experimental | Control | 0,371551164 | 0,069538016 | 0,80201795 |
| Experimental | Control | 0,802180208 | 0,44541743 | 0,277207922 |
| Experimental | Control | 0,301277473 | 0,043118611 | -0,796693802 |
| Experimental | Control | 0,073387073 | 0,004982242 | 0,986401335 |
| Experimental | Control | 0,993927207 | 0,967737571 | 0,011909951 |
| Experimental | Control | 0,416183377 | 0,101350899 | 0,403377226 |
| Experimental | Control | 0,993927207 | 0,979844605 | 0,006580072 |
| Experimental | Control | 0,135705582 | 0,012003224 | 0,670151422 |
| Experimental | Control | 0,329988443 | 0,055686583 | 0,90833285 |
| Experimental | Control | 0,758652695 | 0,359797636 | -0,231016268 |
| Experimental | Control | 0,041440413 | 0,000805418 | -1,47880337 |
| Experimental | Control | 0,993927207 | 0,94553179 | 0,01342205 |
| Experimental | Control | 0,875638058 | 0,621459957 | 0,093749006 |
| Experimental | Control | 0,758652695 | 0,372395031 | 0,278032355 |
| Experimental | Control | 0,20593209 | 0,021537852 | -1,10803067 |
| Experimental | Control | 0,857260587 | 0,54096619 | 0,326644515 |
| Experimental | Control | 0,993927207 | 0,988685188 | -0,004083179 |
| Experimental | Control | 0,792267412 | 0,434214889 | -0,170351625 |
| Experimental | Control | 0,073387073 | 0,004972265 | 0,763146138 |
| Experimental | Control | 0,806123861 | 0,450507483 | 0,143755424 |
| Experimental | Control | 0,522163094 | 0,1485051 | -0,475872807 |
| Experimental | Control | 0,291305327 | 0,036346353 | 0,962709906 |
| Experimental | Control | 0,942023842 | 0,783763549 | -0,121550745 |
| Experimental | Control | 0,357558091 | 0,06363878 | -0,730367934 |
| Experimental | Control | 0,758652695 | 0,359140841 | -0,409553291 |
| Experimental | Control | 0,942023842 | 0,78167014 | -0,06215276 |
| Experimental | Control | 0,517912006 | 0,141827093 | -0,558414866 |
| Experimental | Control | 0,057956521 | 0,003059205 | -1,151908667 |
| Experimental | Control | 0,562434821 | 0,190918242 | 0,279570784 |
| Experimental | Control | 0,763168505 | 0,39488719 | -0,320491592 |
| Experimental | Control | 0,395419923 | 0,086339396 | -0,749690035 |
| Experimental | Control | 0,517912006 | 0,143629835 | 0,478459796 |
| Experimental | Control | 0,863880358 | 0,561833856 | 0,224430571 |
| Experimental | Control | 0,883362092 | 0,638614063 | -0,194967173 |
| Experimental | Control | 0,394135539 | 0,076754066 | 0,800577844 |
| Experimental | Control | 0,217858521 | 0,023584684 | 0,742855992 |

| | | | | |
|---|---|---|---|---|
| Experimental | Control | 0,91467373 | 0,730060683 | -0,129910254 |
| Experimental | Control | 0,703604491 | 0,293132733 | 0,195221699 |
| Experimental | Control | 0,96779014 | 0,841711057 | -0,095500728 |
| Experimental | Control | 0,993927207 | 0,956905756 | 0,024948507 |
| Experimental | Control | 0,865761837 | 0,605239009 | -0,284172341 |
| Experimental | Control | 0,942023842 | 0,781039785 | -0,131379147 |
| Experimental | Control | 0,951958206 | 0,796500811 | 0,044567692 |
| Experimental | Control | 0,517912006 | 0,144879453 | -0,394082777 |
| Experimental | Control | 0,616834884 | 0,22070239 | -0,696293858 |
| Experimental | Control | 0,62370955 | 0,225833029 | -0,346181772 |
| Experimental | Control | 0,778142834 | 0,417346932 | -0,301184582 |
| Experimental | Control | 0,857987221 | 0,545646945 | 0,253441892 |
| Experimental | Control | 0,618092173 | 0,22228636 | -0,318588253 |
| Experimental | Control | 0,251610475 | 0,030008589 | 0,983555979 |
| Experimental | Control | 0,969496774 | 0,901812611 | 0,021045389 |
| Experimental | Control | 0,59713352 | 0,211461962 | -0,303237561 |
| Experimental | Control | 0,361430551 | 0,066980708 | -0,780676142 |
| Experimental | Control | 0,863880358 | 0,599168395 | -0,305540329 |
| Experimental | Control | 0,806123861 | 0,451133537 | 0,11606966 |
| Experimental | Control | 0,969496774 | 0,876084473 | -0,094606285 |
| Experimental | Control | 0,640631523 | 0,242146961 | -0,777629962 |
| Experimental | Control | 0,057956521 | 0,002630449 | -1,804349138 |
| Experimental | Control | 0,149288225 | 0,013696167 | -0,844124068 |
| Experimental | Control | 0,969496774 | 0,889582616 | -0,077326963 |
| Experimental | Control | 0,958500764 | 0,810768536 | 0,15982695 |
| Experimental | Control | 0,863880358 | 0,599066565 | -0,21182927 |
| Experimental | Control | 0,993927207 | 0,960004036 | -0,020706423 |
| Experimental | Control | 0,969496774 | 0,886005608 | 0,050853188 |
| Experimental | Control | 0,42651097 | 0,105649506 | -1,07691684 |
| Experimental | Control | 0,969496774 | 0,895327018 | -0,043902795 |
| Experimental | Control | 0,329988443 | 0,0534157 | -0,495035586 |
| Experimental | Control | 0,90534009 | 0,701438061 | 0,161290946 |
| Experimental | Control | 0,329988443 | 0,053200537 | 0,542895408 |
| Experimental | Control | 0,091936922 | 0,006578972 | -2,178288457 |
| Experimental | Control | 0,763168505 | 0,390665849 | -0,515792817 |
| Experimental | Control | 0,605930629 | 0,215689068 | -0,527566798 |
| Experimental | Control | 0,685118193 | 0,279075668 | 0,411839713 |
| Experimental | Control | 0,969496774 | 0,866318028 | 0,072415199 |
| Experimental | Control | 0,765222281 | 0,398758033 | 0,617102461 |
| Experimental | Control | 0,071676165 | 0,004603056 | -2,280856213 |
| Experimental | Control | 0,993927207 | 0,972915566 | -0,005242106 |
| Experimental | Control | 0,416183377 | 0,098021765 | 0,838356828 |
| Experimental | Control | 0,900027447 | 0,672130589 | -0,210508784 |
| Experimental | Control | 0,90534009 | 0,698814311 | 0,152125406 |
| Experimental | Control | 0,985587547 | 0,924101351 | 0,040665106 |
| Experimental | Control | 0,298114484 | 0,040990387 | 0,321487547 |
| Experimental | Control | 0,913007919 | 0,724452271 | -0,149668087 |
| Experimental | Control | 0,416183377 | 0,09821208 | 0,768983715 |

| | | | | |
|---|---|---|---|---|
| Experimental | Control | 0,885956934 | 0,643578419 | 0,218032318 |
| Experimental | Control | 0,562434821 | 0,190266728 | -0,488445988 |
| Experimental | Control | 0,937032606 | 0,75994204 | -0,129136576 |
| Experimental | Control | 0,863880358 | 0,593073071 | 0,146759673 |
| Experimental | Control | 0,837515875 | 0,508656431 | -0,187903289 |
| Experimental | Control | 0,778142834 | 0,417016881 | 0,197766067 |
| Experimental | Control | 0,060227846 | 0,003613154 | -1,079112424 |
| Experimental | Control | 0,857987221 | 0,546278102 | -0,160313898 |
| Experimental | Control | 0,847421357 | 0,52400183 | -0,314040574 |
| Experimental | Control | 0,758652695 | 0,366976037 | 0,184681062 |
| Experimental | Control | 0,820490176 | 0,478520728 | 0,184609266 |
| Experimental | Control | 0,802180208 | 0,443347837 | 0,259998826 |
| Experimental | Control | 0,742113995 | 0,339238433 | -0,254717621 |
| Experimental | Control | 0,829986385 | 0,49085432 | -0,142298483 |
| Experimental | Control | 0,552296958 | 0,173289504 | -0,399591369 |
| Experimental | Control | 0,758652695 | 0,377886045 | 0,201612291 |
| Experimental | Control | 0,942023842 | 0,781237829 | -0,109386463 |
| Experimental | Control | 0,763168505 | 0,393829776 | 0,487761841 |
| Experimental | Control | 0,411214661 | 0,0912972 | -0,581213554 |
| Experimental | Control | 0,969496774 | 0,899942511 | 0,049570985 |
| Experimental | Control | 3,59396E-12 | 6,59443E-15 | 4,227754641 |
| Experimental | Control | 0,856198414 | 0,536614446 | 0,137070415 |
| Experimental | Control | 0,329988443 | 0,05565878 | -0,459743525 |
| Experimental | Control | 0,057956521 | 0,001907853 | -1,707414196 |
| Experimental | Control | 0,27381969 | 0,033159816 | -0,515309243 |
| Experimental | Control | 0,863131153 | 0,552720683 | 0,210807986 |
| Experimental | Control | 0,121663398 | 0,009915013 | -0,833040339 |
| Experimental | Control | 0,298114484 | 0,041079409 | -1,05948377 |
| Experimental | Control | 0,863880358 | 0,56492289 | 0,165507909 |
| Experimental | Control | 0,766140345 | 0,403865934 | -0,194736449 |
| Experimental | Control | 0,960150973 | 0,82978185 | -0,035998382 |
| Experimental | Control | 0,863880358 | 0,564924599 | 0,109670613 |
| Experimental | Control | 0,329988443 | 0,054467466 | -0,662037942 |
| Experimental | Control | 0,993927207 | 0,984517415 | -0,006417194 |
| Experimental | Control | 0,993927207 | 0,97611221 | 0,008719391 |
| Experimental | Control | 0,636069717 | 0,236921381 | -0,492472402 |
| Experimental | Control | 0,71998115 | 0,31798719 | -0,190774666 |
| Experimental | Control | 0,416183377 | 0,101564017 | -0,811468127 |
| Experimental | Control | 0,041440413 | 0,000231231 | 2,555015507 |
| Experimental | Control | 0,628864508 | 0,231929846 | -0,624555616 |
| Experimental | Control | 0,758652695 | 0,378245781 | 0,35409256 |
| Experimental | Control | 0,847421357 | 0,523342166 | 0,09520371 |
| Experimental | Control | 0,863880358 | 0,577031976 | -0,114874004 |
| Experimental | Control | 0,973992691 | 0,910516407 | 0,076627966 |
| Experimental | Control | 0,792267412 | 0,434804893 | 0,297631341 |
| Experimental | Control | 0,298114484 | 0,040070811 | -0,79413301 |
| Experimental | Control | 0,176805734 | 0,01784278 | -1,327440907 |
| Experimental | Control | 0,969496774 | 0,879947261 | 0,061728945 |

| | | | | |
|---|---|---|---|---|
| Experimental | Control | 0,041440413 | 0,000839544 | 1,633410469 |
| Experimental | Control | 0,394135539 | 0,082634205 | 0,358271674 |
| Experimental | Control | 0,899774764 | 0,669152361 | -0,16746037 |
| Experimental | Control | 0,969496774 | 0,861453044 | 0,067051875 |
| Experimental | Control | 0,913007919 | 0,723336388 | -0,080811681 |
| Experimental | Control | 0,742113995 | 0,33672254 | -0,641359327 |
| Experimental | Control | 0,81933177 | 0,469131742 | 0,232748279 |
| Experimental | Control | 0,863880358 | 0,572080644 | 0,218956543 |
| Experimental | Control | 0,865761837 | 0,602986688 | -0,193007295 |
| Experimental | Control | 0,358800439 | 0,064518244 | 0,557129863 |
| Experimental | Control | 0,993927207 | 0,964916751 | 0,019002793 |
| Experimental | Control | 0,837835519 | 0,511849615 | 0,382752935 |
| Experimental | Control | 0,911914311 | 0,718465743 | 0,081940272 |
| Experimental | Control | 0,820490176 | 0,478744727 | -0,347256821 |
| Experimental | Control | 0,969496774 | 0,870730983 | 0,050813755 |
| Experimental | Control | 0,050038837 | 0,001193587 | -1,360435856 |
| Experimental | Control | 0,657777016 | 0,261685064 | 0,567855644 |
| Experimental | Control | 0,879402206 | 0,632524156 | -0,168985813 |
| Experimental | Control | 0,329988443 | 0,055704471 | -1,56073299 |
| Experimental | Control | 0,298114484 | 0,038419037 | 0,575299461 |
| Experimental | Control | 0,863880358 | 0,579284779 | 0,324593239 |
| Experimental | Control | 0,863880358 | 0,597265492 | -0,221715272 |
| Experimental | Control | 0,863880358 | 0,589425872 | 0,141572412 |
| Experimental | Control | 0,416183377 | 0,096521354 | 0,312840349 |
| Experimental | Control | 0,758652695 | 0,382806406 | -0,286955518 |
| Experimental | Control | 0,865761837 | 0,60435316 | -0,343219936 |
| Experimental | Control | 0,90534009 | 0,697932142 | 0,122041237 |
| Experimental | Control | 0,969496774 | 0,898107026 | 0,063504444 |
| Experimental | Control | 0,789179454 | 0,428303824 | 0,388525736 |
| Experimental | Control | 0,863880358 | 0,559572263 | -0,276937326 |
| Experimental | Control | 0,993927207 | 0,993471741 | 0,002267596 |
| Experimental | Control | 0,875638058 | 0,62339003 | 0,206490338 |
| Experimental | Control | 0,863880358 | 0,560674558 | -0,151762122 |
| Experimental | Control | 0,318433538 | 0,046358347 | 0,836708865 |
| Experimental | Control | 0,129384111 | 0,010920494 | -0,934782771 |
| Experimental | Control | 0,057956521 | 0,002109983 | -2,640146094 |
| Experimental | Control | 0,891576681 | 0,652732286 | 0,201269057 |
| Experimental | Control | 0,576326596 | 0,199637299 | -0,287258122 |
| Experimental | Control | 0,733532311 | 0,329753057 | 0,449694536 |
| Experimental | Control | 0,993927207 | 0,950588416 | 0,036082435 |
| Experimental | Control | 0,837835519 | 0,511925189 | 0,298752887 |
| Experimental | Control | 0,554948252 | 0,180066008 | 0,370616037 |
| Experimental | Control | 0,820490176 | 0,478740282 | 0,349249704 |
| Experimental | Control | 0,993927207 | 0,972508165 | 0,015554453 |
| Experimental | Control | 0,360356586 | 0,066120474 | -0,618618216 |
| Experimental | Control | 0,863880358 | 0,59118755 | -0,287474971 |
| Experimental | Control | 0,657777016 | 0,261903876 | 0,377297662 |
| Experimental | Control | 0,562434821 | 0,188866061 | 0,71242243 |

| | | | | |
|---|---|---|---|---|
| Experimental | Control | 0,869288498 | 0,611172158 | -0,174369167 |
| Experimental | Control | 0,218578096 | 0,024636061 | 1,447317213 |
| Experimental | Control | 0,041440413 | 0,000898039 | -1,694568424 |
| Experimental | Control | 0,416183377 | 0,10141254 | 0,963475065 |
| Experimental | Control | 0,863880358 | 0,583079365 | -0,24487346 |
| Experimental | Control | 0,893774272 | 0,655981117 | -0,163901836 |
| Experimental | Control | 0,329988443 | 0,054987807 | -0,686245417 |
| Experimental | Control | 0,057956521 | 0,003155281 | 1,305724749 |
| Experimental | Control | 0,64075595 | 0,243369691 | 0,361333897 |
| Experimental | Control | 0,556427534 | 0,181732295 | 0,609385675 |
| Experimental | Control | 0,942023842 | 0,786460272 | -0,167700522 |
| Experimental | Control | 0,792267412 | 0,432610568 | -0,514897134 |
| Experimental | Control | 0,863880358 | 0,567944065 | 0,155392668 |
| Experimental | Control | 0,958734741 | 0,817730823 | 0,08766029 |
| Experimental | Control | 0,969496774 | 0,870400608 | -0,042096533 |
| Experimental | Control | 0,526624713 | 0,152131141 | 0,417263123 |
| Experimental | Control | 0,90221013 | 0,680990584 | -0,108774493 |
| Experimental | Control | 0,863880358 | 0,598414746 | 0,244165821 |
| Experimental | Control | 0,130507735 | 0,011254796 | -1,281330531 |
| Experimental | Control | 0,820490176 | 0,474609147 | -0,269072286 |
| Experimental | Control | 0,220412032 | 0,025478822 | -0,974716618 |
| Experimental | Control | 0,554948252 | 0,179687231 | 0,884541104 |
| Experimental | Control | 0,993927207 | 0,958011702 | -0,03051296 |
| Experimental | Control | 0,157501496 | 0,014738672 | -1,045996845 |
| Experimental | Control | 0,329988443 | 0,052419388 | -0,673192608 |
| Experimental | Control | 0,993927207 | 0,988905515 | 0,006175341 |
| Experimental | Control | 0,969496774 | 0,861884879 | 0,064274896 |
| Experimental | Control | 0,887720145 | 0,648280033 | 0,076618457 |
| Experimental | Control | 0,513480967 | 0,138498536 | -0,474139748 |
| Experimental | Control | 0,766140345 | 0,406265247 | -0,388049079 |
| Experimental | Control | 0,394135539 | 0,082994376 | -0,790709573 |
| Experimental | Control | 0,778142834 | 0,41834101 | -0,609737043 |
| Experimental | Control | 0,90534009 | 0,696656462 | 0,207523297 |
| Experimental | Control | 0,897159618 | 0,661450823 | -0,171606852 |
| Experimental | Control | 0,529467057 | 0,155582831 | 0,594481596 |
| Experimental | Control | 0,95347105 | 0,801813353 | -0,083812625 |
| Experimental | Control | 0,532043431 | 0,163029822 | -0,258690152 |
| Experimental | Control | 0,9597389 | 0,879088572 | 0,054210444 |
| Experimental | Control | 0,614328094 | 0,314907678 | -0,28387826 |
| Experimental | Control | 0,936720063 | 0,826517703 | -0,059300073 |
| Experimental | Control | 0,935024688 | 0,800779925 | -0,075227273 |
| Experimental | Control | 0,709691399 | 0,43481533 | 0,602445742 |
| Experimental | Control | 0,455706215 | 0,179984807 | 0,76490625 |
| Experimental | Control | 0,058168303 | 0,003799158 | -1,084259567 |
| Experimental | Control | 0,576843266 | 0,269389241 | -0,223416704 |
| Experimental | Control | 0,496231003 | 0,216580025 | -1,120612197 |
| Experimental | Control | 0,255131401 | 0,040735266 | 0,859578412 |
| Experimental | Control | 0,602303154 | 0,298620892 | 0,488378607 |

| | | | | |
|---|---|---|---|---|
| Experimental | Control | 0,496231003 | 0,220491515 | 0,342902385 |
| Experimental | Control | 0,940655603 | 0,85082932 | -0,080851935 |
| Experimental | Control | 0,790012353 | 0,544301518 | 0,450312563 |
| Experimental | Control | 0,162071329 | 0,017705271 | 0,760579139 |
| Experimental | Control | 0,067181439 | 0,006210049 | -1,043842558 |
| Experimental | Control | 0,131981702 | 0,013309079 | 1,039253064 |
| Experimental | Control | 0,935024688 | 0,755411491 | 0,186276327 |
| Experimental | Control | 0,058168303 | 0,004399283 | 1,32136302 |
| Experimental | Control | 0,964945899 | 0,901501205 | -0,055225139 |
| Experimental | Control | 0,421673968 | 0,131108713 | -0,426410347 |
| Experimental | Control | 0,806518916 | 0,586016904 | 0,154417277 |
| Experimental | Control | 0,214321828 | 0,025214333 | 1,154439938 |
| Experimental | Control | 0,935024688 | 0,789244042 | -0,059003431 |
| Experimental | Control | 0,935024688 | 0,76437656 | -0,082950984 |
| Experimental | Control | 0,058168303 | 0,004034873 | -2,885316466 |
| Experimental | Control | 0,062602198 | 0,005260689 | 1,796618084 |
| Experimental | Control | 0,535966654 | 0,243211759 | 0,475946133 |
| Experimental | Control | 0,451974304 | 0,15968099 | -0,439435331 |
| Experimental | Control | 0,455706215 | 0,175451246 | 0,539504536 |
| Experimental | Control | 0,975869894 | 0,958711902 | 0,026891495 |
| Experimental | Control | 0,022618616 | 0,000570217 | -2,587146473 |
| Experimental | Control | 0,685444734 | 0,403202785 | -0,297839326 |
| Experimental | Control | 0,496231003 | 0,221010447 | 0,580140834 |
| Experimental | Control | 0,790012353 | 0,535620207 | 0,28651851 |
| Experimental | Control | 0,576843266 | 0,280632971 | -0,413752837 |
| Experimental | Control | 0,806518916 | 0,58963988 | -0,13298099 |
| Experimental | Control | 0,790012353 | 0,564294538 | -0,250886616 |
| Experimental | Control | 0,996742863 | 0,996742863 | -0,002540864 |
| Experimental | Control | 0,301035244 | 0,060747701 | 0,977463541 |
| Experimental | Control | 0,269622363 | 0,047580417 | 0,542975363 |
| Experimental | Control | 0,255131401 | 0,040042045 | -1,1776085 |
| Experimental | Control | 0,449601171 | 0,147348283 | 0,599589774 |
| Experimental | Control | 0,451974304 | 0,163318446 | -1,005906051 |
| Experimental | Control | 0,646895742 | 0,348661133 | 0,558168784 |
| Experimental | Control | 0,576843266 | 0,274763187 | -0,766702712 |
| Experimental | Control | 0,935024688 | 0,786373217 | 0,121542196 |
| Experimental | Control | 0,935024688 | 0,800616653 | 0,162514938 |
| Experimental | Control | 0,646895742 | 0,36525893 | -0,782249667 |
| Experimental | Control | 0,964945899 | 0,924401954 | -0,041835481 |
| Experimental | Control | 0,255131401 | 0,039600785 | -0,807976653 |
| Experimental | Control | 0,646895742 | 0,344608684 | 0,24050954 |
| Experimental | Control | 0,790012353 | 0,54091339 | 0,201426458 |
| Experimental | Control | 0,463899761 | 0,187119231 | 0,453760038 |
| Experimental | Control | 0,258501644 | 0,043445654 | -1,387210991 |
| Experimental | Control | 0,845432624 | 0,6465073 | -0,263358181 |
| Experimental | Control | 0,613636191 | 0,309396399 | -0,75927612 |
| Experimental | Control | 0,811961081 | 0,600441808 | -0,241832672 |
| Experimental | Control | 0,421673968 | 0,123304619 | -0,550930322 |

| | | | | |
|---|---|---|---|---|
| Experimental | Control | 0,326198004 | 0,076752471 | -0,804098922 |
| Experimental | Control | 0,835257961 | 0,624688727 | -0,530133448 |
| Experimental | Control | 0,841681174 | 0,636565594 | 0,215463407 |
| Experimental | Control | 0,767365703 | 0,487066099 | 0,393769995 |
| Experimental | Control | 0,940655603 | 0,853704244 | 0,083387985 |
| Experimental | Control | 0,964945899 | 0,898074919 | -0,039196552 |
| Experimental | Control | 0,790012353 | 0,552134219 | -0,323836727 |
| Experimental | Control | 0,255131401 | 0,033645867 | 0,792401604 |
| Experimental | Control | 0,391397583 | 0,10524977 | -0,788062669 |
| Experimental | Control | 0,935024688 | 0,809307083 | -0,166617817 |
| Experimental | Control | 0,905600813 | 0,707738451 | 0,144893255 |
| Experimental | Control | 0,614355622 | 0,320084441 | 0,395343405 |
| Experimental | Control | 0,996742863 | 0,995981383 | -0,002181289 |
| Experimental | Control | 0,767365703 | 0,496530749 | 0,355351153 |
| Experimental | Control | 0,326198004 | 0,075664712 | 0,912179085 |
| Experimental | Control | 0,646895742 | 0,36965471 | -0,260755641 |
| Experimental | Control | 0,301035244 | 0,063198904 | -1,249162074 |
| Experimental | Control | 0,774049739 | 0,507360333 | -0,25552278 |
| Experimental | Control | 0,935024688 | 0,805798221 | 0,173971368 |
| Experimental | Control | 1,23994E-08 | 1,04197E-10 | 3,468783118 |
| Experimental | Control | 0,936720063 | 0,825093446 | 0,06054355 |
| Experimental | Control | 0,301120976 | 0,065791138 | 0,392982631 |
| Experimental | Control | 0,32829181 | 0,082762641 | 0,830664751 |
| Experimental | Control | 0,470886195 | 0,193894316 | -0,623259667 |
| Experimental | Control | 0,451974304 | 0,15628875 | -0,930099419 |
| Experimental | Control | 0,935024688 | 0,761628767 | 0,215757979 |
| Experimental | Control | 0,935024688 | 0,783281853 | 0,169438328 |
| Experimental | Control | 0,359854499 | 0,093743609 | -0,444971488 |
| Experimental | Control | 0,451974304 | 0,161521133 | 0,72173189 |
| Experimental | Control | 0,421673968 | 0,130364187 | 1,017707499 |
| Experimental | Control | 0,671568526 | 0,389396876 | 0,493778902 |
| Experimental | Control | 0,455706215 | 0,17679361 | 0,631361489 |
| Experimental | Control | 0,32829181 | 0,082476648 | 0,515512206 |
| Experimental | Control | 0,418818992 | 0,116143082 | 0,463984928 |
| Experimental | Control | 0,646895742 | 0,360680143 | 0,514374271 |
| Experimental | Control | 0,027765826 | 0,000933305 | 1,490986969 |
| Experimental | Control | 0,767365703 | 0,486399713 | -0,25489357 |
| Experimental | Control | 0,936720168 | 0,834389393 | -0,08774261 |
| Experimental | Control | 0,301035244 | 0,063242698 | -0,995171386 |
| Experimental | Control | 0,255131401 | 0,034406868 | -1,014411935 |
| Experimental | Control | 0,964945899 | 0,912513167 | 0,047627269 |
| Experimental | Control | 0,646895742 | 0,367463927 | -0,529857068 |
| Experimental | Control | 0,767365703 | 0,490545847 | -0,460544037 |
| Experimental | Control | 0,058168303 | 0,003069601 | -1,980566092 |
| Experimental | Control | 0,873958874 | 0,675665684 | -0,221568553 |
| Experimental | Control | 0,709691399 | 0,435356908 | 0,343468856 |
| Experimental | Control | 0,576843266 | 0,281150499 | -0,461713139 |
| Experimental | Control | 0,485330744 | 0,203920481 | 0,919252915 |

| | | | | |
|---|---|---|---|---|
| Experimental | Control | 0,301035244 | 0,061104244 | -1,2132611 |
| Experimental | Control | 0,790012353 | 0,552396679 | -0,409749127 |
| Experimental | Control | 0,42500133 | 0,13571471 | -0,801877213 |
| Experimental | Control | 8,72317E-05 | 1,46608E-06 | -2,271885172 |
| Experimental | Control | 0,455706215 | 0,176942637 | -0,889588394 |
| Experimental | Control | 0,421673968 | 0,127730681 | -0,627825975 |
| Experimental | Control | 0,790012353 | 0,561147805 | -0,143835515 |
| Experimental | Control | 0,975869894 | 0,959468719 | 0,025252619 |
| Experimental | Control | 0,964945899 | 0,91970892 | -0,071708476 |
| Experimental | Control | 0,058168303 | 0,00409947 | -1,774535673 |
| Experimental | Control | 0,975869894 | 0,949425271 | -0,04099464 |
| Experimental | Control | 0,703607686 | 0,419799544 | 0,439177869 |
| Experimental | Control | 0,509529902 | 0,248840185 | 0,674975165 |
| Experimental | Control | 0,759620943 | 0,662668517 | -0,116422108 |
| Experimental | Control | 0,544978752 | 0,304174187 | 0,792557364 |
| Experimental | Control | 0,810464525 | 0,79131204 | 0,177019722 |
| Experimental | Control | 0,0123664 | 0,000948574 | 1,288095584 |
| Experimental | Control | 0,56621753 | 0,381867637 | 0,522373364 |
| Experimental | Control | 0,0123664 | 0,001150363 | 1,480319736 |
| Experimental | Control | 0,283914554 | 0,092437297 | -0,757900133 |
| Experimental | Control | 0,023252391 | 0,002703766 | 1,971331687 |
| Experimental | Control | 0,661239618 | 0,552792511 | -0,18317185 |
| Experimental | Control | 0,154900604 | 0,032421057 | -1,991482062 |
| Experimental | Control | 0,241893146 | 0,062291347 | 0,720079448 |
| Experimental | Control | 0,775876751 | 0,71072682 | -0,130169829 |
| Experimental | Control | 0,661239618 | 0,553595959 | 0,122931246 |
| Experimental | Control | 0,509529902 | 0,248159868 | 0,543001013 |
| Experimental | Control | 0,661239618 | 0,493981594 | 0,193647286 |
| Experimental | Control | 0,561186449 | 0,328855049 | -0,4767009 |
| Experimental | Control | 0,509529902 | 0,234967891 | -0,842395473 |
| Experimental | Control | 0,661239618 | 0,531644936 | -0,206965297 |
| Experimental | Control | 0,661239618 | 0,549128881 | 0,235845173 |
| Experimental | Control | 0,561186449 | 0,339322039 | 0,584993139 |
| Experimental | Control | 0,922337234 | 0,922337234 | -0,054112911 |
| Experimental | Control | 0,775876751 | 0,721745815 | -0,080545734 |
| Experimental | Control | 8,86826E-06 | 2,06239E-07 | 3,435820515 |
| Experimental | Control | 0,043275009 | 0,006038373 | -1,768411835 |
| Experimental | Control | 0,648261476 | 0,467351297 | 0,252176438 |
| Experimental | Control | 0,543790166 | 0,290864507 | -0,27311871 |
| Experimental | Control | 0,068057752 | 0,012661907 | 1,31371884 |
| Experimental | Control | 0,582912633 | 0,406683233 | 0,210786067 |
| Experimental | Control | 0,416555 | 0,164684535 | -0,603651678 |
| Experimental | Control | 0,060183506 | 0,009797315 | 1,031861002 |
| Experimental | Control | 0,56621753 | 0,380367591 | -0,443138962 |
| Experimental | Control | 0,810464525 | 0,791616513 | 0,10942684 |
| Experimental | Control | 0,241893146 | 0,067505064 | -0,952495429 |
| Experimental | Control | 0,489307476 | 0,204826385 | -0,603281597 |
| Experimental | Control | 0,321631169 | 0,112196919 | 0,954368128 |

| | | | | |
|---|---|---|---|---|
| Experimental | Control | 0,375099133 | 0,13957177 | -0,977673295 |
| Experimental | Control | 0,529853819 | 0,271088 | -0,575791502 |
| Experimental | Control | 0,000452955 | 2,10677E-05 | -1,811285044 |
| Experimental | Control | 0,56621753 | 0,368305451 | -0,340796255 |
| Experimental | Control | 0,267042863 | 0,080733889 | 0,84844189 |
| Experimental | Control | 0,759620943 | 0,671292926 | 0,213277336 |
| Experimental | Control | 0,165549574 | 0,038499901 | -1,094729479 |
| Experimental | Control | 0,010680887 | 0,001525841 | 0,989648936 |
| Experimental | Control | 0,554084369 | 0,465381647 | -0,15357193 |
| Experimental | Control | 0,73757631 | 0,73757631 | 0,105731962 |
| Experimental | Control | 0,07952014 | 0,02272004 | -0,49981832 |
| Experimental | Control | 0,554084369 | 0,474929459 | -0,340584798 |
| Experimental | Control | 0,158812762 | 0,068062612 | -0,711485921 |
| Experimental | Control | 0,554084369 | 0,43767182 | 0,524525191 |

| Standard error | Sample size | N alternative grou | N reference group | Degrees of freedom |
|---|---|---|---|---|
| 0,467046619 | 142 | 70 | 72 | 139 |
| 0,491735245 | 142 | 70 | 72 | 139 |
| 0,908630377 | 142 | 70 | 72 | 139 |
| 0,321846281 | 142 | 70 | 72 | 139 |
| 0,454057063 | 142 | 70 | 72 | 139 |
| 0,497647978 | 142 | 70 | 72 | 139 |
| 0,434788985 | 142 | 70 | 72 | 139 |
| 0,66612106 | 142 | 70 | 72 | 139 |
| 0,44185375 | 142 | 70 | 72 | 139 |
| 0,556295958 | 142 | 70 | 72 | 139 |
| 0,685991241 | 142 | 70 | 72 | 139 |
| 0,416919232 | 142 | 70 | 72 | 139 |
| 0,747819636 | 142 | 70 | 72 | 139 |
| 0,536823064 | 142 | 70 | 72 | 139 |
| 0,718650886 | 142 | 70 | 72 | 139 |
| 0,569231322 | 142 | 70 | 72 | 139 |
| 0,458654967 | 142 | 70 | 72 | 139 |
| 0,813529669 | 142 | 70 | 72 | 139 |
| 0,540871451 | 142 | 70 | 72 | 139 |
| 0,33764527 | 142 | 70 | 72 | 139 |
| 0,52347921 | 142 | 70 | 72 | 139 |
| 0,538057688 | 142 | 70 | 72 | 139 |
| 0,458883595 | 142 | 70 | 72 | 139 |
| 0,633127759 | 142 | 70 | 72 | 139 |
| 0,312087981 | 142 | 70 | 72 | 139 |
| 0,659842891 | 142 | 70 | 72 | 139 |
| 0,490174794 | 142 | 70 | 72 | 139 |
| 0,678291259 | 142 | 70 | 72 | 139 |
| 0,786906877 | 142 | 70 | 72 | 139 |
| 0,600215352 | 142 | 70 | 72 | 139 |
| 0,570985495 | 142 | 70 | 72 | 139 |
| 0,4294963 | 142 | 70 | 72 | 139 |
| 0,97016173 | 142 | 70 | 72 | 139 |
| 0,677014648 | 142 | 70 | 72 | 139 |
| 0,546611272 | 142 | 70 | 72 | 139 |
| 0,468920033 | 142 | 70 | 72 | 139 |
| 0,479844225 | 142 | 70 | 72 | 139 |
| 0,898536607 | 142 | 70 | 72 | 139 |
| 0,487765302 | 142 | 70 | 72 | 139 |
| 0,680925361 | 142 | 70 | 72 | 139 |
| 0,748061344 | 142 | 70 | 72 | 139 |
| 0,610899907 | 142 | 70 | 72 | 139 |
| 0,525337056 | 142 | 70 | 72 | 139 |
| 0,386175937 | 142 | 70 | 72 | 139 |
| 0,642649202 | 142 | 70 | 72 | 139 |
| 0,320888939 | 142 | 70 | 72 | 139 |
| 0,467475128 | 142 | 70 | 72 | 139 |

| | | | | |
|---|---|---|---|---|
| 0,45664017 | 142 | 70 | 72 | 139 |
| 0,575079882 | 142 | 70 | 72 | 139 |
| 0,445360718 | 142 | 70 | 72 | 139 |
| 0,262018543 | 142 | 70 | 72 | 139 |
| 0,503416803 | 142 | 70 | 72 | 139 |
| 0,715021438 | 142 | 70 | 72 | 139 |
| 0,519367861 | 142 | 70 | 72 | 139 |
| 0,698647608 | 142 | 70 | 72 | 139 |
| 0,446974634 | 142 | 70 | 72 | 139 |
| 0,514963139 | 142 | 70 | 72 | 139 |
| 0,267038348 | 142 | 70 | 72 | 139 |
| 0,59019389 | 142 | 70 | 72 | 139 |
| 0,658066552 | 142 | 70 | 72 | 139 |
| 0,483631128 | 142 | 70 | 72 | 139 |
| 0,458109016 | 142 | 70 | 72 | 139 |
| 0,647133019 | 142 | 70 | 72 | 139 |
| 0,606756145 | 142 | 70 | 72 | 139 |
| 0,648553123 | 142 | 70 | 72 | 139 |
| 0,651727684 | 142 | 70 | 72 | 139 |
| 0,424197315 | 142 | 70 | 72 | 139 |
| 0,533642425 | 142 | 70 | 72 | 139 |
| 0,456983239 | 142 | 70 | 72 | 139 |
| 0,608507108 | 142 | 70 | 72 | 139 |
| 0,359970874 | 142 | 70 | 72 | 139 |
| 0,34471433 | 142 | 70 | 72 | 139 |
| 0,641881637 | 142 | 70 | 72 | 139 |
| 0,20940842 | 142 | 70 | 72 | 139 |
| 0,300990552 | 142 | 70 | 72 | 139 |
| 0,408413328 | 142 | 70 | 72 | 139 |
| 0,779045 | 142 | 70 | 72 | 139 |
| 0,562519574 | 142 | 70 | 72 | 139 |
| 0,508934416 | 142 | 70 | 72 | 139 |
| 0,77658145 | 142 | 70 | 72 | 139 |
| 0,371455883 | 142 | 70 | 72 | 139 |
| 0,626767353 | 142 | 70 | 72 | 139 |
| 0,341904512 | 142 | 70 | 72 | 139 |
| 1,016722114 | 142 | 70 | 72 | 139 |
| 0,663816065 | 142 | 70 | 72 | 139 |
| 0,379221525 | 142 | 70 | 72 | 139 |
| 0,346512911 | 142 | 70 | 72 | 139 |
| 0,516890015 | 142 | 70 | 72 | 139 |
| 0,397292571 | 142 | 70 | 72 | 139 |
| 0,490266063 | 142 | 70 | 72 | 139 |
| 0,687413143 | 142 | 70 | 72 | 139 |
| 0,509588825 | 142 | 70 | 72 | 139 |
| 0,667948563 | 142 | 70 | 72 | 139 |
| 0,725815759 | 142 | 70 | 72 | 139 |
| 1,101479736 | 142 | 70 | 72 | 139 |

| | | | | |
|---|---|---|---|---|
| 0,754823355 | 142 | 70 | 72 | 139 |
| 0,466175104 | 142 | 70 | 72 | 139 |
| 1,014683236 | 142 | 70 | 72 | 139 |
| 0,429372756 | 142 | 70 | 72 | 139 |
| 0,761326023 | 142 | 70 | 72 | 139 |
| 0,554635758 | 142 | 70 | 72 | 139 |
| 0,593604592 | 142 | 70 | 72 | 139 |
| 0,522509549 | 142 | 70 | 72 | 139 |
| 0,427198722 | 142 | 70 | 72 | 139 |
| 0,311977739 | 142 | 70 | 72 | 139 |
| 0,418275712 | 142 | 70 | 72 | 139 |
| 0,345705428 | 142 | 70 | 72 | 139 |
| 0,280766054 | 142 | 70 | 72 | 139 |
| 0,384040973 | 142 | 70 | 72 | 139 |
| 0,435461013 | 142 | 70 | 72 | 139 |
| 0,435828654 | 142 | 70 | 72 | 139 |
| 0,532185895 | 142 | 70 | 72 | 139 |
| 0,373573117 | 142 | 70 | 72 | 139 |
| 0,524314712 | 142 | 70 | 72 | 139 |
| 0,531541794 | 142 | 70 | 72 | 139 |
| 0,416740945 | 142 | 70 | 72 | 139 |
| 0,353542005 | 142 | 70 | 72 | 139 |
| 0,357451127 | 142 | 70 | 72 | 139 |
| 0,382774761 | 142 | 70 | 72 | 139 |
| 0,768976886 | 142 | 70 | 72 | 139 |
| 0,451616514 | 142 | 70 | 72 | 139 |
| 0,48076742 | 142 | 70 | 72 | 139 |
| 0,287088688 | 142 | 70 | 72 | 139 |
| 0,741613143 | 142 | 70 | 72 | 139 |
| 0,509658147 | 142 | 70 | 72 | 139 |
| 0,510331268 | 142 | 70 | 72 | 139 |
| 0,723947237 | 142 | 70 | 72 | 139 |
| 0,662041055 | 142 | 70 | 72 | 139 |
| 0,488323293 | 142 | 70 | 72 | 139 |
| 0,714741818 | 142 | 70 | 72 | 139 |
| 0,583558396 | 142 | 70 | 72 | 139 |
| 0,456916179 | 142 | 70 | 72 | 139 |
| 0,555345962 | 142 | 70 | 72 | 139 |
| 0,468811956 | 142 | 70 | 72 | 139 |
| 0,659292 | 142 | 70 | 72 | 139 |
| 0,436570195 | 142 | 70 | 72 | 139 |
| 0,339083666 | 142 | 70 | 72 | 139 |
| 0,881358852 | 142 | 70 | 72 | 139 |
| 0,479799897 | 142 | 70 | 72 | 139 |
| 0,275309594 | 142 | 70 | 72 | 139 |
| 0,502814549 | 142 | 70 | 72 | 139 |
| 0,503259525 | 142 | 70 | 72 | 139 |
| 0,372364173 | 142 | 70 | 72 | 139 |

| | | | | |
|---|---|---|---|---|
| 0,346590741 | 142 | 70 | 72 | 139 |
| 0,581965529 | 142 | 70 | 72 | 139 |
| 0,416071375 | 142 | 70 | 72 | 139 |
| 0,703581407 | 142 | 70 | 72 | 139 |
| 0,343279054 | 142 | 70 | 72 | 139 |
| 0,503855881 | 142 | 70 | 72 | 139 |
| 0,644256911 | 142 | 70 | 72 | 139 |
| 0,20182308 | 142 | 70 | 72 | 139 |
| 0,495152551 | 142 | 70 | 72 | 139 |
| 0,542796814 | 142 | 70 | 72 | 139 |
| 0,522092244 | 142 | 70 | 72 | 139 |
| 0,67896633 | 142 | 70 | 72 | 139 |
| 0,343258409 | 142 | 70 | 72 | 139 |
| 0,552154958 | 142 | 70 | 72 | 139 |
| 0,610690973 | 142 | 70 | 72 | 139 |
| 0,989322259 | 142 | 70 | 72 | 139 |
| 0,813555514 | 142 | 70 | 72 | 139 |
| 0,67603848 | 142 | 70 | 72 | 139 |
| 0,152512897 | 142 | 70 | 72 | 139 |
| 0,375310733 | 142 | 70 | 72 | 139 |
| 0,249778779 | 142 | 70 | 72 | 139 |
| 0,489610196 | 142 | 70 | 72 | 139 |
| 0,698886123 | 142 | 70 | 72 | 139 |
| 0,551864507 | 142 | 70 | 72 | 139 |
| 0,47403595 | 142 | 70 | 72 | 139 |
| 0,422144495 | 142 | 70 | 72 | 139 |
| 0,626466906 | 142 | 70 | 72 | 139 |
| 0,291209099 | 142 | 70 | 72 | 139 |
| 0,682806308 | 142 | 70 | 72 | 139 |
| 0,347028688 | 142 | 70 | 72 | 139 |
| 0,244891132 | 142 | 70 | 72 | 139 |
| 0,527568087 | 142 | 70 | 72 | 139 |
| 0,437736958 | 142 | 70 | 72 | 139 |
| 0,504562117 | 142 | 70 | 72 | 139 |
| 0,500870383 | 142 | 70 | 72 | 139 |
| 0,442873766 | 142 | 70 | 72 | 139 |
| 0,427963636 | 142 | 70 | 72 | 139 |
| 0,338528254 | 142 | 70 | 72 | 139 |
| 0,304594196 | 142 | 70 | 72 | 139 |
| 0,425795826 | 142 | 70 | 72 | 139 |
| 0,346303637 | 142 | 70 | 72 | 139 |
| 0,290081398 | 142 | 70 | 72 | 139 |
| 0,30813892 | 142 | 70 | 72 | 139 |
| 0,481680164 | 142 | 70 | 72 | 139 |
| 0,524686506 | 142 | 70 | 72 | 139 |
| 0,209240342 | 142 | 70 | 72 | 139 |
| 0,568379591 | 142 | 70 | 72 | 139 |
| 0,320380759 | 142 | 70 | 72 | 139 |

| | | | | |
|---|---|---|---|---|
| 0,231274448 | 142 | 70 | 72 | 139 |
| 0,288047413 | 142 | 70 | 72 | 139 |
| 0,590453153 | 142 | 70 | 72 | 139 |
| 0,173158277 | 142 | 70 | 72 | 139 |
| 0,18189129 | 142 | 70 | 72 | 139 |
| 0,770617661 | 142 | 70 | 72 | 139 |
| 0,402227548 | 142 | 70 | 72 | 139 |
| 0,577064051 | 142 | 70 | 72 | 139 |
| 0,138895314 | 142 | 70 | 72 | 139 |
| 0,489378021 | 142 | 70 | 72 | 139 |
| 0,337362407 | 142 | 70 | 72 | 139 |
| 0,450985475 | 142 | 70 | 72 | 139 |
| 0,48759975 | 142 | 70 | 72 | 139 |
| 0,489778641 | 142 | 70 | 72 | 139 |
| 0,481796107 | 142 | 70 | 72 | 139 |
| 0,783203233 | 142 | 70 | 72 | 139 |
| 0,292206924 | 142 | 70 | 72 | 139 |
| 0,717423941 | 142 | 70 | 72 | 139 |
| 0,376331549 | 142 | 70 | 72 | 139 |
| 0,735965027 | 142 | 70 | 72 | 139 |
| 0,456860924 | 142 | 70 | 72 | 139 |
| 0,432461143 | 142 | 70 | 72 | 139 |
| 0,73386957 | 142 | 70 | 72 | 139 |
| 0,566832471 | 142 | 70 | 72 | 139 |
| 0,281068536 | 142 | 70 | 72 | 139 |
| 0,335922425 | 142 | 70 | 72 | 139 |
| 0,231400901 | 142 | 70 | 72 | 139 |
| 0,460735701 | 142 | 70 | 72 | 139 |
| 0,135660307 | 142 | 70 | 72 | 139 |
| 0,29794428 | 142 | 70 | 72 | 139 |
| 0,520953136 | 142 | 70 | 72 | 139 |
| 0,504482848 | 142 | 70 | 72 | 139 |
| 0,224880544 | 142 | 70 | 72 | 139 |
| 0,454015084 | 142 | 70 | 72 | 139 |
| 0,28796946 | 142 | 70 | 72 | 139 |
| 0,656167603 | 142 | 70 | 72 | 139 |
| 0,252480684 | 142 | 70 | 72 | 139 |
| 0,435951918 | 142 | 70 | 72 | 139 |
| 0,325025791 | 142 | 70 | 72 | 139 |
| 0,543577645 | 142 | 70 | 72 | 139 |
| 0,199175094 | 142 | 70 | 72 | 139 |
| 0,240262601 | 142 | 70 | 72 | 139 |
| 0,359162838 | 142 | 70 | 72 | 139 |
| 0,175634653 | 142 | 70 | 72 | 139 |
| 0,238726974 | 142 | 70 | 72 | 139 |
| 0,608179744 | 142 | 70 | 72 | 139 |
| 0,71116789 | 142 | 70 | 72 | 139 |
| 0,269010566 | 142 | 70 | 72 | 139 |

| | | | | |
|---|---|---|---|---|
| 0,409810828 | 142 | 70 | 72 | 139 |
| 0,397804576 | 142 | 70 | 72 | 139 |
| 0,557727465 | 142 | 70 | 72 | 139 |
| 0,278950331 | 142 | 70 | 72 | 139 |
| 0,448583164 | 142 | 70 | 72 | 139 |
| 0,407823572 | 142 | 70 | 72 | 139 |
| 0,190941457 | 142 | 70 | 72 | 139 |
| 0,812435886 | 142 | 70 | 72 | 139 |
| 0,350537744 | 142 | 70 | 72 | 139 |
| 0,558146712 | 142 | 70 | 72 | 139 |
| 0,387774653 | 142 | 70 | 72 | 139 |
| 0,312883249 | 142 | 70 | 72 | 139 |
| 0,494384708 | 142 | 70 | 72 | 139 |
| 0,366768814 | 142 | 70 | 72 | 139 |
| 0,343696628 | 142 | 70 | 72 | 139 |
| 0,377163811 | 142 | 70 | 72 | 139 |
| 0,500976651 | 142 | 70 | 72 | 139 |
| 0,547733892 | 142 | 70 | 72 | 139 |
| 0,265032479 | 142 | 70 | 72 | 139 |
| 0,907381022 | 142 | 70 | 72 | 139 |
| 0,387964256 | 142 | 70 | 72 | 139 |
| 0,3675115 | 142 | 70 | 72 | 139 |
| 0,191998447 | 142 | 70 | 72 | 139 |
| 0,381896861 | 142 | 70 | 72 | 139 |
| 0,610935056 | 142 | 70 | 72 | 139 |
| 0,489403845 | 142 | 70 | 72 | 139 |
| 0,283634716 | 142 | 70 | 72 | 139 |
| 0,598717776 | 142 | 70 | 72 | 139 |
| 0,275826933 | 142 | 70 | 72 | 139 |
| 0,67155467 | 142 | 70 | 72 | 139 |
| 0,356491545 | 142 | 70 | 72 | 139 |
| 0,273087846 | 142 | 70 | 72 | 139 |
| 0,785809971 | 142 | 70 | 72 | 139 |
| 0,563063851 | 142 | 70 | 72 | 139 |
| 0,552209084 | 142 | 70 | 72 | 139 |
| 0,301269878 | 142 | 70 | 72 | 139 |
| 0,70506869 | 142 | 70 | 72 | 139 |
| 0,260600283 | 142 | 70 | 72 | 139 |
| 0,652168148 | 142 | 70 | 72 | 139 |
| 0,560266356 | 142 | 70 | 72 | 139 |
| 0,348669754 | 142 | 70 | 72 | 139 |
| 0,327061765 | 142 | 70 | 72 | 139 |
| 0,216920591 | 142 | 70 | 72 | 139 |
| 0,639747741 | 142 | 70 | 72 | 139 |
| 0,348127158 | 142 | 70 | 72 | 139 |
| 0,506815943 | 142 | 70 | 72 | 139 |
| 0,433772862 | 142 | 70 | 72 | 139 |
| 0,307620673 | 142 | 70 | 72 | 139 |

| | | | | |
|---|---|---|---|---|
| 0,326497796 | 142 | 70 | 72 | 139 |
| 0,453920922 | 142 | 70 | 72 | 139 |
| 0,621465999 | 142 | 70 | 72 | 139 |
| 0,165399 | 142 | 70 | 72 | 139 |
| 0,360018733 | 142 | 70 | 72 | 139 |
| 0,166034703 | 142 | 70 | 72 | 139 |
| 0,587923436 | 142 | 70 | 72 | 139 |
| 0,681335965 | 142 | 70 | 72 | 139 |
| 0,250217476 | 142 | 70 | 72 | 139 |
| 0,318855294 | 142 | 70 | 72 | 139 |
| 0,623284285 | 142 | 70 | 72 | 139 |
| 0,561833104 | 142 | 70 | 72 | 139 |
| 0,600259351 | 142 | 70 | 72 | 139 |
| 0,28701637 | 142 | 70 | 72 | 139 |
| 0,637187106 | 142 | 70 | 72 | 139 |
| 0,607492282 | 142 | 70 | 72 | 139 |
| 0,498104189 | 142 | 70 | 72 | 139 |
| 0,230903181 | 142 | 70 | 72 | 139 |
| 0,63824795 | 142 | 70 | 72 | 139 |
| 0,380367343 | 142 | 70 | 72 | 139 |
| 0,204709501 | 142 | 70 | 72 | 139 |
| 0,13620204 | 142 | 70 | 72 | 139 |
| 0,522288677 | 142 | 70 | 72 | 139 |
| 0,193917782 | 142 | 70 | 72 | 139 |
| 0,854928317 | 142 | 70 | 72 | 139 |
| 0,321169236 | 142 | 70 | 72 | 139 |
| 0,426415303 | 142 | 70 | 72 | 139 |
| 0,41960933 | 142 | 70 | 72 | 139 |
| 0,432543535 | 142 | 70 | 72 | 139 |
| 0,431439119 | 142 | 70 | 72 | 139 |
| 0,608496865 | 142 | 70 | 72 | 139 |
| 0,568316901 | 142 | 70 | 72 | 139 |
| 0,361479407 | 142 | 70 | 72 | 139 |
| 0,281571643 | 142 | 70 | 72 | 139 |
| 0,549085149 | 142 | 70 | 72 | 139 |
| 0,43601393 | 142 | 70 | 72 | 139 |
| 0,324112364 | 142 | 70 | 72 | 139 |
| 0,420760521 | 142 | 70 | 72 | 139 |
| 0,31335782 | 142 | 70 | 72 | 139 |
| 0,274283502 | 142 | 70 | 72 | 139 |
| 0,202974548 | 142 | 70 | 72 | 139 |
| 0,291699614 | 142 | 70 | 72 | 139 |
| 0,491880778 | 142 | 70 | 72 | 139 |
| 0,155561545 | 142 | 70 | 72 | 139 |
| 0,207409417 | 142 | 70 | 72 | 139 |
| 0,41644819 | 142 | 70 | 72 | 139 |
| 0,226888516 | 142 | 70 | 72 | 139 |
| 0,248967082 | 142 | 70 | 72 | 139 |

| | | | | |
|---|---|---|---|---|
| 0,303069733 | 142 | 70 | 72 | 139 |
| 0,52369144 | 142 | 70 | 72 | 139 |
| 0,534888206 | 142 | 70 | 72 | 139 |
| 0,406382213 | 142 | 70 | 72 | 139 |
| 0,337848689 | 142 | 70 | 72 | 139 |
| 0,242210513 | 142 | 70 | 72 | 139 |
| 0,150042201 | 142 | 70 | 72 | 139 |
| 0,237999073 | 142 | 70 | 72 | 139 |
| 0,439823709 | 142 | 70 | 72 | 139 |
| 0,35596377 | 142 | 70 | 72 | 139 |
| 0,425539567 | 142 | 70 | 72 | 139 |
| 0,393966213 | 142 | 70 | 72 | 139 |
| 0,292081476 | 142 | 70 | 72 | 139 |
| 0,397176702 | 142 | 70 | 72 | 139 |
| 0,397517699 | 142 | 70 | 72 | 139 |
| 0,253033958 | 142 | 70 | 72 | 139 |
| 0,400840454 | 142 | 70 | 72 | 139 |
| 0,503473359 | 142 | 70 | 72 | 139 |
| 0,219105557 | 142 | 70 | 72 | 139 |
| 0,504539535 | 142 | 70 | 72 | 139 |
| 0,357636857 | 142 | 70 | 72 | 139 |
| 0,49515334 | 142 | 70 | 72 | 139 |
| 0,376013318 | 142 | 70 | 72 | 139 |
| 0,442543018 | 142 | 70 | 72 | 139 |
| 0,203136669 | 142 | 70 | 72 | 139 |
| 0,493492989 | 142 | 70 | 72 | 139 |
| 0,351224047 | 142 | 70 | 72 | 139 |
| 0,566224312 | 142 | 70 | 72 | 139 |
| 0,440773411 | 142 | 70 | 72 | 139 |
| 0,243017186 | 142 | 70 | 72 | 139 |
| 0,213163082 | 142 | 70 | 72 | 139 |
| 0,574019956 | 142 | 70 | 72 | 139 |
| 0,278189578 | 142 | 70 | 72 | 139 |
| 0,277535398 | 142 | 70 | 72 | 139 |
| 0,411330338 | 142 | 70 | 72 | 139 |
| 0,172222445 | 142 | 70 | 72 | 139 |
| 0,404533747 | 142 | 70 | 72 | 139 |
| 0,204637197 | 142 | 70 | 72 | 139 |
| 0,263329492 | 142 | 70 | 72 | 139 |
| 0,260514859 | 142 | 70 | 72 | 139 |
| 0,379835808 | 142 | 70 | 72 | 139 |
| 0,474872631 | 142 | 70 | 72 | 139 |
| 0,237193285 | 142 | 70 | 72 | 139 |
| 0,213387615 | 142 | 70 | 72 | 139 |
| 0,189915581 | 142 | 70 | 72 | 139 |
| 0,564303132 | 142 | 70 | 72 | 139 |
| 0,560467097 | 142 | 70 | 72 | 139 |
| 0,487954404 | 142 | 70 | 72 | 139 |

| | | | | |
|---|---|---|---|---|
| 0,397254931 | 142 | 70 | 72 | 139 |
| 0,570341983 | 142 | 70 | 72 | 139 |
| 0,505872058 | 142 | 70 | 72 | 139 |
| 0,49888219 | 142 | 70 | 72 | 139 |
| 0,489657721 | 142 | 70 | 72 | 139 |
| 0,273960537 | 142 | 70 | 72 | 139 |
| 0,707888856 | 142 | 70 | 72 | 139 |
| 0,279401032 | 142 | 70 | 72 | 139 |
| 0,152327123 | 142 | 70 | 72 | 139 |
| 0,418934001 | 142 | 70 | 72 | 139 |
| 0,206285082 | 142 | 70 | 72 | 139 |
| 0,836267255 | 142 | 70 | 72 | 139 |
| 0,638959472 | 142 | 70 | 72 | 139 |
| 0,397862354 | 142 | 70 | 72 | 139 |
| 0,282058211 | 142 | 70 | 72 | 139 |
| 0,436816746 | 142 | 70 | 72 | 139 |
| 0,640715393 | 142 | 70 | 72 | 139 |
| 0,727703947 | 142 | 70 | 72 | 139 |
| 0,369362661 | 142 | 70 | 72 | 139 |
| 0,512764892 | 142 | 70 | 72 | 139 |
| 0,311850524 | 142 | 70 | 72 | 139 |
| 0,653155279 | 142 | 70 | 72 | 139 |
| 0,378346819 | 142 | 70 | 72 | 139 |
| 0,41349225 | 142 | 70 | 72 | 139 |
| 0,153605109 | 142 | 70 | 72 | 139 |
| 0,440153417 | 142 | 70 | 72 | 139 |
| 0,355314238 | 142 | 70 | 72 | 139 |
| 0,260097938 | 142 | 70 | 72 | 139 |
| 0,124854149 | 142 | 70 | 72 | 139 |
| 0,272464742 | 142 | 70 | 72 | 139 |
| 0,532124973 | 142 | 70 | 72 | 139 |
| 0,281943503 | 142 | 70 | 72 | 139 |
| 0,238709123 | 142 | 70 | 72 | 139 |
| 0,253003196 | 142 | 70 | 72 | 139 |
| 0,403895516 | 142 | 70 | 72 | 139 |
| 0,389450187 | 142 | 70 | 72 | 139 |
| 0,58538946 | 142 | 70 | 72 | 139 |
| 0,230821021 | 142 | 70 | 72 | 139 |
| 0,403462554 | 142 | 70 | 72 | 139 |
| 0,249168852 | 142 | 70 | 72 | 139 |
| 0,267798709 | 142 | 70 | 72 | 139 |
| 0,378316445 | 142 | 70 | 72 | 139 |
| 0,242266422 | 142 | 70 | 72 | 139 |
| 0,423653978 | 142 | 70 | 72 | 139 |
| 0,606482793 | 142 | 70 | 72 | 139 |
| 0,343170016 | 142 | 70 | 72 | 139 |
| 0,462630804 | 142 | 70 | 72 | 139 |
| 0,319677351 | 142 | 70 | 72 | 139 |

| | | | | |
|---|---|---|---|---|
| 0,209172179 | 142 | 70 | 72 | 139 |
| 0,567416792 | 142 | 70 | 72 | 139 |
| 0,198402822 | 142 | 70 | 72 | 139 |
| 0,376134411 | 142 | 70 | 72 | 139 |
| 0,289738655 | 142 | 70 | 72 | 139 |
| 0,384499196 | 142 | 70 | 72 | 139 |
| 0,322787641 | 142 | 70 | 72 | 139 |
| 0,401296854 | 142 | 70 | 72 | 139 |
| 0,278611048 | 142 | 70 | 72 | 139 |
| 0,191731389 | 142 | 70 | 72 | 139 |
| 0,246315323 | 142 | 70 | 72 | 139 |
| 0,182915933 | 142 | 70 | 72 | 139 |
| 0,429124341 | 142 | 70 | 72 | 139 |
| 0,19841828 | 142 | 70 | 72 | 139 |
| 0,30344529 | 142 | 70 | 72 | 139 |
| 0,381605073 | 142 | 70 | 72 | 139 |
| 0,300478995 | 142 | 70 | 72 | 139 |
| 0,413367713 | 142 | 70 | 72 | 139 |
| 0,192469171 | 142 | 70 | 72 | 139 |
| 0,510122217 | 142 | 70 | 72 | 139 |
| 0,534331253 | 142 | 70 | 72 | 139 |
| 0,154232021 | 142 | 70 | 72 | 139 |
| 0,566666299 | 142 | 70 | 72 | 139 |
| 0,419552255 | 142 | 70 | 72 | 139 |
| 0,225789416 | 142 | 70 | 72 | 139 |
| 0,273158703 | 142 | 70 | 72 | 139 |
| 0,64616914 | 142 | 70 | 72 | 139 |
| 0,415670056 | 142 | 70 | 72 | 139 |
| 0,309323136 | 142 | 70 | 72 | 139 |
| 0,507269942 | 142 | 70 | 72 | 139 |
| 0,562136252 | 142 | 70 | 72 | 139 |
| 0,531338437 | 142 | 70 | 72 | 139 |
| 0,413641523 | 142 | 70 | 72 | 139 |
| 0,314174231 | 142 | 70 | 72 | 139 |
| 0,349257919 | 142 | 70 | 72 | 139 |
| 0,444468101 | 142 | 70 | 72 | 139 |
| 0,387454549 | 142 | 70 | 72 | 139 |
| 0,665982601 | 142 | 70 | 72 | 139 |
| 0,381333944 | 142 | 70 | 72 | 139 |
| 0,465539115 | 142 | 70 | 72 | 139 |
| 0,34186952 | 142 | 70 | 72 | 139 |
| 0,451303034 | 142 | 70 | 72 | 139 |
| 0,324852307 | 142 | 70 | 72 | 139 |
| 0,222004477 | 142 | 70 | 72 | 139 |
| 0,542199122 | 142 | 70 | 72 | 139 |
| 0,2264636 | 142 | 70 | 72 | 139 |
| 0,445494793 | 142 | 70 | 72 | 139 |
| 0,391024551 | 142 | 70 | 72 | 139 |

| | | | | |
|---|---|---|---|---|
| 0,414248562 | 142 | 70 | 72 | 139 |
| 0,466367161 | 142 | 70 | 72 | 139 |
| 0,382288909 | 142 | 70 | 72 | 139 |
| 0,32686745 | 142 | 70 | 72 | 139 |
| 0,243562331 | 142 | 70 | 72 | 139 |
| 0,47794379 | 142 | 70 | 72 | 139 |
| 0,257083355 | 142 | 70 | 72 | 139 |
| 0,539935413 | 142 | 70 | 72 | 139 |
| 0,335991192 | 142 | 70 | 72 | 139 |
| 0,235643392 | 142 | 70 | 72 | 139 |
| 0,247138879 | 142 | 70 | 72 | 139 |
| 0,372337992 | 142 | 70 | 72 | 139 |
| 0,483018705 | 142 | 70 | 72 | 139 |
| 0,429062194 | 142 | 70 | 72 | 139 |
| 0,598954124 | 142 | 70 | 72 | 139 |
| 0,425911189 | 142 | 70 | 72 | 139 |
| 0,493629845 | 142 | 70 | 72 | 139 |
| 0,32754849 | 142 | 70 | 72 | 139 |
| 0,320723779 | 142 | 70 | 72 | 139 |
| 0,437093944 | 142 | 70 | 72 | 139 |
| 0,401598679 | 142 | 70 | 72 | 139 |
| 0,285458877 | 142 | 70 | 72 | 139 |
| 0,746022981 | 142 | 70 | 72 | 139 |
| 0,479794221 | 142 | 70 | 72 | 139 |
| 0,275142714 | 142 | 70 | 72 | 139 |
| 0,473291094 | 142 | 70 | 72 | 139 |
| 0,40177644 | 142 | 70 | 72 | 139 |
| 0,47510975 | 142 | 70 | 72 | 139 |
| 0,487966259 | 142 | 70 | 72 | 139 |
| 0,433395495 | 142 | 70 | 72 | 139 |
| 0,549108527 | 142 | 70 | 72 | 139 |
| 0,325528874 | 142 | 70 | 72 | 139 |
| 0,592351111 | 142 | 70 | 72 | 139 |
| 0,414936777 | 142 | 70 | 72 | 139 |
| 0,466756062 | 142 | 70 | 72 | 139 |
| 0,331961789 | 142 | 70 | 72 | 139 |
| 0,51144995 | 142 | 70 | 72 | 139 |
| 0,385833206 | 142 | 70 | 72 | 139 |
| 0,569356133 | 142 | 70 | 72 | 139 |
| 0,441869522 | 142 | 70 | 72 | 139 |
| 0,409189138 | 142 | 70 | 72 | 139 |
| 0,388439799 | 142 | 70 | 72 | 139 |
| 0,452668491 | 142 | 70 | 72 | 139 |
| 0,310630991 | 142 | 70 | 72 | 139 |
| 0,413907892 | 142 | 70 | 72 | 139 |
| 0,671889062 | 142 | 70 | 72 | 139 |
| 0,566159596 | 142 | 70 | 72 | 139 |
| 0,333742116 | 142 | 70 | 72 | 139 |

| | | | | |
|---|---|---|---|---|
| 0,252437039 | 142 | 70 | 72 | 139 |
| 0,211016661 | 142 | 70 | 72 | 139 |
| 0,502508496 | 142 | 70 | 72 | 139 |
| 0,262894319 | 142 | 70 | 72 | 139 |
| 0,309351946 | 142 | 70 | 72 | 139 |
| 0,292268067 | 142 | 70 | 72 | 139 |
| 0,386340044 | 142 | 70 | 72 | 139 |
| 0,496602642 | 142 | 70 | 72 | 139 |
| 0,482380256 | 142 | 70 | 72 | 139 |
| 0,365176323 | 142 | 70 | 72 | 139 |
| 0,696563418 | 142 | 70 | 72 | 139 |
| 0,497305295 | 142 | 70 | 72 | 139 |
| 0,425086263 | 142 | 70 | 72 | 139 |
| 0,199829844 | 142 | 70 | 72 | 139 |
| 0,390950899 | 142 | 70 | 72 | 139 |
| 0,519963262 | 142 | 70 | 72 | 139 |
| 0,398523701 | 142 | 70 | 72 | 139 |
| 0,258225081 | 142 | 70 | 72 | 139 |
| 0,318967307 | 142 | 70 | 72 | 139 |
| 0,386570459 | 142 | 70 | 72 | 139 |
| 0,375610036 | 142 | 70 | 72 | 139 |
| 0,551462447 | 142 | 70 | 72 | 139 |
| 0,549290566 | 142 | 70 | 72 | 139 |
| 0,420291887 | 142 | 70 | 72 | 139 |
| 0,490100298 | 142 | 70 | 72 | 139 |
| 0,298813854 | 142 | 70 | 72 | 139 |
| 0,295994275 | 142 | 70 | 72 | 139 |
| 0,289175818 | 142 | 70 | 72 | 139 |
| 0,24425963 | 142 | 70 | 72 | 139 |
| 0,287186744 | 142 | 70 | 72 | 139 |
| 0,366549021 | 142 | 70 | 72 | 139 |
| 0,852576028 | 142 | 70 | 72 | 139 |
| 0,567638707 | 142 | 70 | 72 | 139 |
| 0,500741363 | 142 | 70 | 72 | 139 |
| 0,30021387 | 142 | 70 | 72 | 139 |
| 0,813815873 | 142 | 70 | 72 | 139 |
| 0,434253702 | 142 | 70 | 72 | 139 |
| 0,466436459 | 142 | 70 | 72 | 139 |
| 0,28830061 | 142 | 70 | 72 | 139 |
| 0,406824446 | 142 | 70 | 72 | 139 |
| 0,667464766 | 142 | 70 | 72 | 139 |
| 0,36186885 | 142 | 70 | 72 | 139 |
| 0,311454218 | 142 | 70 | 72 | 139 |
| 0,402444716 | 142 | 70 | 72 | 139 |
| 0,399766524 | 142 | 70 | 72 | 139 |
| 0,5048123 | 142 | 70 | 72 | 139 |
| 0,400988558 | 142 | 70 | 72 | 139 |
| 0,467121971 | 142 | 70 | 72 | 139 |

| | | | | |
|---|---|---|---|---|
| 0,267330921 | 142 | 70 | 72 | 139 |
| 0,383925841 | 142 | 70 | 72 | 139 |
| 0,463243455 | 142 | 70 | 72 | 139 |
| 0,277904346 | 142 | 70 | 72 | 139 |
| 0,292248602 | 142 | 70 | 72 | 139 |
| 0,886212499 | 142 | 70 | 72 | 139 |
| 0,531088532 | 142 | 70 | 72 | 139 |
| 0,21802614 | 142 | 70 | 72 | 139 |
| 0,295200384 | 142 | 70 | 72 | 139 |
| 0,190991907 | 142 | 70 | 72 | 139 |
| 0,291474381 | 142 | 70 | 72 | 139 |
| 0,540874438 | 142 | 70 | 72 | 139 |
| 0,197298037 | 142 | 70 | 72 | 139 |
| 0,666982269 | 142 | 70 | 72 | 139 |
| 0,342023631 | 142 | 70 | 72 | 139 |
| 0,535664808 | 142 | 70 | 72 | 139 |
| 0,377358156 | 142 | 70 | 72 | 139 |
| 0,382988398 | 142 | 70 | 72 | 139 |
| 0,361408669 | 142 | 70 | 72 | 139 |
| 0,178450115 | 142 | 70 | 72 | 139 |
| 0,446780284 | 142 | 70 | 72 | 139 |
| 0,516449587 | 142 | 70 | 72 | 139 |
| 0,552799951 | 142 | 70 | 72 | 139 |
| 0,376906285 | 142 | 70 | 72 | 139 |
| 0,566282179 | 142 | 70 | 72 | 139 |
| 0,50243182 | 142 | 70 | 72 | 139 |
| 0,710547303 | 142 | 70 | 72 | 139 |
| 0,518594112 | 142 | 70 | 72 | 139 |
| 0,566505153 | 142 | 70 | 72 | 139 |
| 0,486582672 | 142 | 70 | 72 | 139 |
| 0,643718805 | 142 | 70 | 72 | 139 |
| 0,326413142 | 142 | 70 | 72 | 139 |
| 0,768156869 | 142 | 70 | 72 | 139 |
| 0,412870594 | 142 | 70 | 72 | 139 |
| 0,409999103 | 142 | 70 | 72 | 139 |
| 0,257259018 | 142 | 70 | 72 | 139 |
| 0,380593531 | 142 | 70 | 72 | 139 |
| 0,350427541 | 142 | 70 | 72 | 139 |
| 0,443546032 | 142 | 70 | 72 | 139 |
| 0,471633696 | 142 | 70 | 72 | 139 |
| 0,956733489 | 142 | 70 | 72 | 139 |
| 0,480884875 | 142 | 70 | 72 | 139 |
| 0,358760723 | 142 | 70 | 72 | 139 |
| 0,539514564 | 142 | 70 | 72 | 139 |
| 1,056715321 | 142 | 70 | 72 | 139 |
| 0,488720605 | 142 | 70 | 72 | 139 |
| 0,43474588 | 142 | 70 | 72 | 139 |
| 0,34748116 | 142 | 70 | 72 | 139 |

| | | | | |
|---|---|---|---|---|
| 0,35961412 | 142 | 70 | 72 | 139 |
| 0,569948639 | 142 | 70 | 72 | 139 |
| 0,429441465 | 142 | 70 | 72 | 139 |
| 0,509577975 | 142 | 70 | 72 | 139 |
| 0,566707503 | 142 | 70 | 72 | 139 |
| 0,428232449 | 142 | 70 | 72 | 139 |
| 0,34261488 | 142 | 70 | 72 | 139 |
| 0,53631249 | 142 | 70 | 72 | 139 |
| 0,505703039 | 142 | 70 | 72 | 139 |
| 0,5699516 | 142 | 70 | 72 | 139 |
| 0,338904793 | 142 | 70 | 72 | 139 |
| 0,761276695 | 142 | 70 | 72 | 139 |
| 0,483517968 | 142 | 70 | 72 | 139 |
| 0,212619812 | 142 | 70 | 72 | 139 |
| 0,882530175 | 142 | 70 | 72 | 139 |
| 0,564201988 | 142 | 70 | 72 | 139 |
| 0,340312791 | 142 | 70 | 72 | 139 |
| 0,307557434 | 142 | 70 | 72 | 139 |
| 0,417611478 | 142 | 70 | 72 | 139 |
| 0,505127462 | 142 | 70 | 72 | 139 |
| 0,684323921 | 142 | 70 | 72 | 139 |
| 0,753967269 | 142 | 70 | 72 | 139 |
| 0,662845144 | 142 | 70 | 72 | 139 |
| 0,232979169 | 142 | 70 | 72 | 139 |
| 0,531865036 | 142 | 70 | 72 | 139 |
| 0,700486805 | 142 | 70 | 72 | 139 |
| 0,604879555 | 142 | 70 | 72 | 139 |
| 0,371448789 | 142 | 70 | 72 | 139 |
| 0,383222536 | 142 | 70 | 72 | 139 |
| 0,307181357 | 142 | 70 | 72 | 139 |
| 0,206591237 | 142 | 70 | 72 | 139 |
| 0,706150156 | 142 | 70 | 72 | 139 |
| 0,423822507 | 142 | 70 | 72 | 139 |
| 0,270487862 | 142 | 70 | 72 | 139 |
| 0,434240699 | 142 | 70 | 72 | 139 |
| 0,63950542 | 142 | 70 | 72 | 139 |
| 0,563779125 | 142 | 70 | 72 | 139 |
| 0,513018407 | 142 | 70 | 72 | 139 |
| 0,599531018 | 142 | 70 | 72 | 139 |
| 0,601688856 | 142 | 70 | 72 | 139 |
| 0,510082063 | 142 | 70 | 72 | 139 |
| 0,489541334 | 142 | 70 | 72 | 139 |
| 0,41180587 | 142 | 70 | 72 | 139 |
| 0,409482257 | 142 | 70 | 72 | 139 |
| 0,777148857 | 142 | 70 | 72 | 139 |
| 0,505482686 | 142 | 70 | 72 | 139 |
| 0,646020709 | 142 | 70 | 72 | 139 |
| 0,424081819 | 142 | 70 | 72 | 139 |

| | | | | |
|---|---|---|---|---|
| 0,388936207 | 142 | 70 | 72 | 139 |
| 0,472001396 | 142 | 70 | 72 | 139 |
| 0,404851967 | 142 | 70 | 72 | 139 |
| 0,248989207 | 142 | 70 | 72 | 139 |
| 0,553290767 | 142 | 70 | 72 | 139 |
| 0,691273837 | 142 | 70 | 72 | 139 |
| 0,501408056 | 142 | 70 | 72 | 139 |
| 0,757966055 | 142 | 70 | 72 | 139 |
| 0,555833788 | 142 | 70 | 72 | 139 |
| 0,712949293 | 142 | 70 | 72 | 139 |
| 0,232045691 | 142 | 70 | 72 | 139 |
| 0,845851841 | 142 | 70 | 72 | 139 |
| 0,589797675 | 142 | 70 | 72 | 139 |
| 0,380060503 | 142 | 70 | 72 | 139 |
| 0,511724046 | 142 | 70 | 72 | 139 |
| 0,393066937 | 142 | 70 | 72 | 139 |
| 0,399442134 | 142 | 70 | 72 | 139 |
| 0,541661286 | 142 | 70 | 72 | 139 |
| 0,286212005 | 142 | 70 | 72 | 139 |
| 0,891271957 | 142 | 70 | 72 | 139 |
| 0,285906827 | 142 | 70 | 72 | 139 |
| 0,33916951 | 142 | 70 | 72 | 139 |
| 0,280593964 | 142 | 70 | 72 | 139 |
| 0,478402411 | 142 | 70 | 72 | 139 |
| 0,31908817 | 142 | 70 | 72 | 139 |
| 0,442731429 | 142 | 70 | 72 | 139 |
| 0,693700892 | 142 | 70 | 72 | 139 |
| 0,301494 | 142 | 70 | 72 | 139 |
| 0,430732418 | 142 | 70 | 72 | 139 |
| 0,657336443 | 142 | 70 | 72 | 139 |
| 0,46087388 | 142 | 70 | 72 | 139 |
| 0,211780483 | 142 | 70 | 72 | 139 |
| 0,841911641 | 142 | 70 | 72 | 139 |
| 0,498449937 | 142 | 70 | 72 | 139 |
| 0,283867464 | 142 | 70 | 72 | 139 |
| 0,227914616 | 142 | 70 | 72 | 139 |
| 0,540702048 | 142 | 70 | 72 | 139 |
| 0,201908277 | 142 | 70 | 72 | 139 |
| 0,529399905 | 142 | 70 | 72 | 139 |
| 0,429218693 | 142 | 70 | 72 | 139 |
| 0,51657773 | 142 | 70 | 72 | 139 |
| 0,426439567 | 142 | 70 | 72 | 139 |
| 0,613423302 | 142 | 70 | 72 | 139 |
| 0,498336856 | 142 | 70 | 72 | 139 |
| 0,580004975 | 142 | 70 | 72 | 139 |
| 0,624524309 | 142 | 70 | 72 | 139 |
| 0,428303387 | 142 | 70 | 72 | 139 |
| 0,359915143 | 142 | 70 | 72 | 139 |

| | | | | |
|---|---|---|---|---|
| 0,361952684 | 142 | 70 | 72 | 139 |
| 0,37506772 | 142 | 70 | 72 | 139 |
| 0,553628205 | 142 | 70 | 72 | 139 |
| 0,488564605 | 142 | 70 | 72 | 139 |
| 0,316747986 | 142 | 70 | 72 | 139 |
| 0,262868271 | 142 | 70 | 72 | 139 |
| 0,38072267 | 142 | 70 | 72 | 139 |
| 0,174017994 | 142 | 70 | 72 | 139 |
| 0,421797202 | 142 | 70 | 72 | 139 |
| 0,390727789 | 142 | 70 | 72 | 139 |
| 0,751007165 | 142 | 70 | 72 | 139 |
| 0,435527909 | 141 | 69 | 72 | 138 |
| 0,482797478 | 141 | 69 | 72 | 138 |
| 0,910274336 | 141 | 69 | 72 | 138 |
| 0,461479156 | 141 | 69 | 72 | 138 |
| 0,81497198 | 141 | 69 | 72 | 138 |
| 0,496685204 | 141 | 69 | 72 | 138 |
| 0,403768342 | 141 | 69 | 72 | 138 |
| 0,604476567 | 141 | 69 | 72 | 138 |
| 0,472883695 | 141 | 69 | 72 | 138 |
| 0,550421714 | 141 | 69 | 72 | 138 |
| 0,659684208 | 141 | 69 | 72 | 138 |
| 0,457284638 | 141 | 69 | 72 | 138 |
| 0,69012561 | 141 | 69 | 72 | 138 |
| 0,554463626 | 141 | 69 | 72 | 138 |
| 0,748791529 | 141 | 69 | 72 | 138 |
| 0,595692609 | 141 | 69 | 72 | 138 |
| 0,48041694 | 141 | 69 | 72 | 138 |
| 0,73516404 | 141 | 69 | 72 | 138 |
| 0,505099197 | 141 | 69 | 72 | 138 |
| 0,404270406 | 141 | 69 | 72 | 138 |
| 0,575203724 | 141 | 69 | 72 | 138 |
| 0,476671724 | 141 | 69 | 72 | 138 |
| 0,484768336 | 141 | 69 | 72 | 138 |
| 0,591183944 | 141 | 69 | 72 | 138 |
| 0,381366376 | 141 | 69 | 72 | 138 |
| 0,677544504 | 141 | 69 | 72 | 138 |
| 0,433214504 | 141 | 69 | 72 | 138 |
| 0,593311198 | 141 | 69 | 72 | 138 |
| 0,819675436 | 141 | 69 | 72 | 138 |
| 0,472342081 | 141 | 69 | 72 | 138 |
| 0,752406686 | 141 | 69 | 72 | 138 |
| 0,348407676 | 141 | 69 | 72 | 138 |
| 0,654935713 | 141 | 69 | 72 | 138 |
| 0,66872998 | 141 | 69 | 72 | 138 |
| 0,440774428 | 141 | 69 | 72 | 138 |
| 0,458725783 | 141 | 69 | 72 | 138 |
| 0,489587832 | 141 | 69 | 72 | 138 |

| | | | | |
|---|---|---|---|---|
| 0,884994198 | 141 | 69 | 72 | 138 |
| 0,714737538 | 141 | 69 | 72 | 138 |
| 0,801626941 | 141 | 69 | 72 | 138 |
| 0,877628967 | 141 | 69 | 72 | 138 |
| 0,608659636 | 141 | 69 | 72 | 138 |
| 0,431825771 | 141 | 69 | 72 | 138 |
| 0,463297929 | 141 | 69 | 72 | 138 |
| 0,289885895 | 141 | 69 | 72 | 138 |
| 0,360828129 | 141 | 69 | 72 | 138 |
| 0,743347746 | 141 | 69 | 72 | 138 |
| 0,274182126 | 141 | 69 | 72 | 138 |
| 0,701677517 | 141 | 69 | 72 | 138 |
| 0,331343488 | 141 | 69 | 72 | 138 |
| 0,730165214 | 141 | 69 | 72 | 138 |
| 0,603472692 | 141 | 69 | 72 | 138 |
| 0,471468264 | 141 | 69 | 72 | 138 |
| 0,675284729 | 141 | 69 | 72 | 138 |
| 0,364698834 | 141 | 69 | 72 | 138 |
| 0,659112929 | 141 | 69 | 72 | 138 |
| 0,180063557 | 141 | 69 | 72 | 138 |
| 0,434605872 | 141 | 69 | 72 | 138 |
| 0,676268608 | 141 | 69 | 72 | 138 |
| 0,462054563 | 141 | 69 | 72 | 138 |
| 0,420785511 | 141 | 69 | 72 | 138 |
| 0,674486804 | 141 | 69 | 72 | 138 |
| 0,58211848 | 141 | 69 | 72 | 138 |
| 0,575542093 | 141 | 69 | 72 | 138 |
| 0,741900161 | 141 | 69 | 72 | 138 |
| 0,407651596 | 141 | 69 | 72 | 138 |
| 0,37128425 | 141 | 69 | 72 | 138 |
| 0,527862962 | 141 | 69 | 72 | 138 |
| 0,519693009 | 141 | 69 | 72 | 138 |
| 0,449336616 | 141 | 69 | 72 | 138 |
| 0,537037034 | 141 | 69 | 72 | 138 |
| 0,298568985 | 141 | 69 | 72 | 138 |
| 0,383589362 | 141 | 69 | 72 | 138 |
| 0,670436117 | 141 | 69 | 72 | 138 |
| 0,625551274 | 141 | 69 | 72 | 138 |
| 0,559577723 | 141 | 69 | 72 | 138 |
| 0,787621222 | 141 | 69 | 72 | 138 |
| 0,375877597 | 141 | 69 | 72 | 138 |
| 0,555226984 | 141 | 69 | 72 | 138 |
| 0,336316422 | 141 | 69 | 72 | 138 |
| 1,03846151 | 141 | 69 | 72 | 138 |
| 0,613135548 | 141 | 69 | 72 | 138 |
| 0,200963723 | 141 | 69 | 72 | 138 |
| 0,230905387 | 141 | 69 | 72 | 138 |
| 0,410139067 | 141 | 69 | 72 | 138 |

| | | | | |
|---|---|---|---|---|
| 0,242367685 | 141 | 69 | 72 | 138 |
| 0,25395184 | 141 | 69 | 72 | 138 |
| 0,6369244 | 141 | 69 | 72 | 138 |
| 0,475087359 | 141 | 69 | 72 | 138 |
| 0,741620532 | 141 | 69 | 72 | 138 |
| 0,780744035 | 141 | 69 | 72 | 138 |
| 0,923036341 | 141 | 69 | 72 | 138 |
| 0,548985304 | 141 | 69 | 72 | 138 |
| 0,319440321 | 141 | 69 | 72 | 138 |
| 0,880787407 | 141 | 69 | 72 | 138 |
| 0,343542997 | 141 | 69 | 72 | 138 |
| 0,721740927 | 141 | 69 | 72 | 138 |
| 0,486629231 | 141 | 69 | 72 | 138 |
| 0,57251013 | 141 | 69 | 72 | 138 |
| 0,500845688 | 141 | 69 | 72 | 138 |
| 0,5942581 | 141 | 69 | 72 | 138 |
| 0,389756299 | 141 | 69 | 72 | 138 |
| 0,371179509 | 141 | 69 | 72 | 138 |
| 0,205965304 | 141 | 69 | 72 | 138 |
| 0,400604498 | 141 | 69 | 72 | 138 |
| 0,494401259 | 141 | 69 | 72 | 138 |
| 0,457785633 | 141 | 69 | 72 | 138 |
| 0,497799492 | 141 | 69 | 72 | 138 |
| 0,252448894 | 141 | 69 | 72 | 138 |
| 0,638559021 | 141 | 69 | 72 | 138 |
| 0,523001577 | 141 | 69 | 72 | 138 |
| 0,406839457 | 141 | 69 | 72 | 138 |
| 0,357687859 | 141 | 69 | 72 | 138 |
| 0,346823532 | 141 | 69 | 72 | 138 |
| 0,350988135 | 141 | 69 | 72 | 138 |
| 0,871439997 | 141 | 69 | 72 | 138 |
| 0,405340958 | 141 | 69 | 72 | 138 |
| 0,511836809 | 141 | 69 | 72 | 138 |
| 0,298665321 | 141 | 69 | 72 | 138 |
| 0,596094083 | 141 | 69 | 72 | 138 |
| 0,559786273 | 141 | 69 | 72 | 138 |
| 0,291473334 | 141 | 69 | 72 | 138 |
| 0,480134476 | 141 | 69 | 72 | 138 |
| 0,558891281 | 141 | 69 | 72 | 138 |
| 0,315716623 | 141 | 69 | 72 | 138 |
| 0,86958355 | 141 | 69 | 72 | 138 |
| 0,45030495 | 141 | 69 | 72 | 138 |
| 0,320565931 | 141 | 69 | 72 | 138 |
| 0,427806745 | 141 | 69 | 72 | 138 |
| 0,615903899 | 141 | 69 | 72 | 138 |
| 0,66538724 | 141 | 69 | 72 | 138 |
| 0,396729232 | 141 | 69 | 72 | 138 |
| 0,226340886 | 141 | 69 | 72 | 138 |

| | | | | |
|---|---|---|---|---|
| 0,777892183 | 141 | 69 | 72 | 138 |
| 0,433736238 | 141 | 69 | 72 | 138 |
| 0,336736105 | 141 | 69 | 72 | 138 |
| 0,510120924 | 141 | 69 | 72 | 138 |
| 0,58727327 | 141 | 69 | 72 | 138 |
| 0,295251356 | 141 | 69 | 72 | 138 |
| 0,306923139 | 141 | 69 | 72 | 138 |
| 0,553699545 | 141 | 69 | 72 | 138 |
| 0,729531188 | 141 | 69 | 72 | 138 |
| 0,357125419 | 141 | 69 | 72 | 138 |
| 0,587881106 | 141 | 69 | 72 | 138 |
| 0,669537198 | 141 | 69 | 72 | 138 |
| 0,396917207 | 141 | 69 | 72 | 138 |
| 0,539622328 | 141 | 69 | 72 | 138 |
| 0,605096157 | 141 | 69 | 72 | 138 |
| 0,589208005 | 141 | 69 | 72 | 138 |
| 0,515934001 | 141 | 69 | 72 | 138 |
| 0,582478147 | 141 | 69 | 72 | 138 |
| 0,966555488 | 141 | 69 | 72 | 138 |
| 0,822850159 | 141 | 69 | 72 | 138 |
| 0,643684229 | 141 | 69 | 72 | 138 |
| 0,17907074 | 141 | 69 | 72 | 138 |
| 0,402614798 | 141 | 69 | 72 | 138 |
| 0,222088427 | 141 | 69 | 72 | 138 |
| 0,556216171 | 141 | 69 | 72 | 138 |
| 0,765322053 | 141 | 69 | 72 | 138 |
| 0,545593186 | 141 | 69 | 72 | 138 |
| 0,489800106 | 141 | 69 | 72 | 138 |
| 0,358428665 | 141 | 69 | 72 | 138 |
| 0,592286819 | 141 | 69 | 72 | 138 |
| 0,266941074 | 141 | 69 | 72 | 138 |
| 0,73253571 | 141 | 69 | 72 | 138 |
| 0,242529798 | 141 | 69 | 72 | 138 |
| 0,436672181 | 141 | 69 | 72 | 138 |
| 0,438148802 | 141 | 69 | 72 | 138 |
| 0,413871707 | 141 | 69 | 72 | 138 |
| 0,362081439 | 141 | 69 | 72 | 138 |
| 0,336922629 | 141 | 69 | 72 | 138 |
| 0,296807363 | 141 | 69 | 72 | 138 |
| 0,404295304 | 141 | 69 | 72 | 138 |
| 0,203438967 | 141 | 69 | 72 | 138 |
| 0,228096944 | 141 | 69 | 72 | 138 |
| 0,438600258 | 141 | 69 | 72 | 138 |
| 0,32348401 | 141 | 69 | 72 | 138 |
| 0,254145735 | 141 | 69 | 72 | 138 |
| 0,477646296 | 141 | 69 | 72 | 138 |
| 0,47558494 | 141 | 69 | 72 | 138 |
| 0,217375212 | 141 | 69 | 72 | 138 |

| | | | | |
|---|---|---|---|---|
| 0,586702664 | 141 | 69 | 72 | 138 |
| 0,450814613 | 141 | 69 | 72 | 138 |
| 0,26141009 | 141 | 69 | 72 | 138 |
| 0,662206609 | 141 | 69 | 72 | 138 |
| 0,61770941 | 141 | 69 | 72 | 138 |
| 0,692088569 | 141 | 69 | 72 | 138 |
| 0,356323466 | 141 | 69 | 72 | 138 |
| 0,644423506 | 141 | 69 | 72 | 138 |
| 0,509386907 | 141 | 69 | 72 | 138 |
| 0,332636985 | 141 | 69 | 72 | 138 |
| 0,38460769 | 141 | 69 | 72 | 138 |
| 0,479828854 | 141 | 69 | 72 | 138 |
| 0,347041856 | 141 | 69 | 72 | 138 |
| 0,681104533 | 141 | 69 | 72 | 138 |
| 0,59225835 | 141 | 69 | 72 | 138 |
| 0,303365859 | 141 | 69 | 72 | 138 |
| 0,543934629 | 141 | 69 | 72 | 138 |
| 0,34860238 | 141 | 69 | 72 | 138 |
| 0,70498439 | 141 | 69 | 72 | 138 |
| 0,391567461 | 141 | 69 | 72 | 138 |
| 0,361433955 | 141 | 69 | 72 | 138 |
| 0,649661364 | 141 | 69 | 72 | 138 |
| 0,636978464 | 141 | 69 | 72 | 138 |
| 0,158818346 | 141 | 69 | 72 | 138 |
| 0,299513503 | 141 | 69 | 72 | 138 |
| 0,405031971 | 141 | 69 | 72 | 138 |
| 0,415173037 | 141 | 69 | 72 | 138 |
| 0,379679261 | 141 | 69 | 72 | 138 |
| 0,523975312 | 141 | 69 | 72 | 138 |
| 0,238917651 | 141 | 69 | 72 | 138 |
| 0,243075079 | 141 | 69 | 72 | 138 |
| 0,694076121 | 141 | 69 | 72 | 138 |
| 0,221073885 | 141 | 69 | 72 | 138 |
| 0,320536979 | 141 | 69 | 72 | 138 |
| 0,213261249 | 141 | 69 | 72 | 138 |
| 0,440202609 | 141 | 69 | 72 | 138 |
| 0,175838765 | 141 | 69 | 72 | 138 |
| 0,240858164 | 141 | 69 | 72 | 138 |
| 0,390408002 | 141 | 69 | 72 | 138 |
| 0,226091127 | 141 | 69 | 72 | 138 |
| 0,225794944 | 141 | 69 | 72 | 138 |
| 0,476256764 | 141 | 69 | 72 | 138 |
| 0,635776287 | 141 | 69 | 72 | 138 |
| 0,178543383 | 141 | 69 | 72 | 138 |
| 0,4543557 | 141 | 69 | 72 | 138 |
| 0,279637334 | 141 | 69 | 72 | 138 |
| 0,309563396 | 141 | 69 | 72 | 138 |
| 0,420475026 | 141 | 69 | 72 | 138 |

| | | | | |
|---|---|---|---|---|
| 0,3899485 | 141 | 69 | 72 | 138 |
| 0,185563131 | 141 | 69 | 72 | 138 |
| 0,789058815 | 141 | 69 | 72 | 138 |
| 0,348785411 | 141 | 69 | 72 | 138 |
| 0,500817267 | 141 | 69 | 72 | 138 |
| 0,375983108 | 141 | 69 | 72 | 138 |
| 0,590903966 | 141 | 69 | 72 | 138 |
| 0,454294667 | 141 | 69 | 72 | 138 |
| 0,336265295 | 141 | 69 | 72 | 138 |
| 0,306722354 | 141 | 69 | 72 | 138 |
| 0,516219265 | 141 | 69 | 72 | 138 |
| 0,381759423 | 141 | 69 | 72 | 138 |
| 0,383764463 | 141 | 69 | 72 | 138 |
| 0,750061087 | 141 | 69 | 72 | 138 |
| 0,423428234 | 141 | 69 | 72 | 138 |
| 0,578505899 | 141 | 69 | 72 | 138 |
| 0,490302864 | 141 | 69 | 72 | 138 |
| 0,282625481 | 141 | 69 | 72 | 138 |
| 0,594674559 | 141 | 69 | 72 | 138 |
| 0,585590467 | 141 | 69 | 72 | 138 |
| 0,314498226 | 141 | 69 | 72 | 138 |
| 0,225031165 | 141 | 69 | 72 | 138 |
| 0,779551769 | 141 | 69 | 72 | 138 |
| 0,57033184 | 141 | 69 | 72 | 138 |
| 0,468081102 | 141 | 69 | 72 | 138 |
| 0,281248196 | 141 | 69 | 72 | 138 |
| 0,150773365 | 141 | 69 | 72 | 138 |
| 0,638176495 | 141 | 69 | 72 | 138 |
| 0,489587578 | 141 | 69 | 72 | 138 |
| 0,456947058 | 141 | 69 | 72 | 138 |
| 0,31914464 | 141 | 69 | 72 | 138 |
| 0,250754993 | 141 | 69 | 72 | 138 |
| 0,266029985 | 141 | 69 | 72 | 138 |
| 0,577728227 | 141 | 69 | 72 | 138 |
| 0,264006217 | 141 | 69 | 72 | 138 |
| 0,51275617 | 141 | 69 | 72 | 138 |
| 0,500658969 | 141 | 69 | 72 | 138 |
| 0,245377348 | 141 | 69 | 72 | 138 |
| 0,328776353 | 141 | 69 | 72 | 138 |
| 0,380890102 | 141 | 69 | 72 | 138 |
| 0,400829427 | 141 | 69 | 72 | 138 |
| 0,142400612 | 141 | 69 | 72 | 138 |
| 0,567931373 | 141 | 69 | 72 | 138 |
| 0,477334955 | 141 | 69 | 72 | 138 |
| 0,527328389 | 141 | 69 | 72 | 138 |
| 0,266763101 | 141 | 69 | 72 | 138 |
| 0,618314643 | 141 | 69 | 72 | 138 |
| 0,467458493 | 141 | 69 | 72 | 138 |

| | | | | |
|---|---|---|---|---|
| 0,471062482 | 141 | 69 | 72 | 138 |
| 0,636819835 | 141 | 69 | 72 | 138 |
| 0,58874537 | 141 | 69 | 72 | 138 |
| 0,406992472 | 141 | 69 | 72 | 138 |
| 0,333433206 | 141 | 69 | 72 | 138 |
| 0,614923092 | 141 | 69 | 72 | 138 |
| 0,347647097 | 141 | 69 | 72 | 138 |
| 0,474004502 | 141 | 69 | 72 | 138 |
| 0,153433404 | 141 | 69 | 72 | 138 |
| 0,854830127 | 141 | 69 | 72 | 138 |
| 0,238202543 | 141 | 69 | 72 | 138 |
| 0,402786122 | 141 | 69 | 72 | 138 |
| 0,31658936 | 141 | 69 | 72 | 138 |
| 0,29674203 | 141 | 69 | 72 | 138 |
| 0,374549454 | 141 | 69 | 72 | 138 |
| 0,616504835 | 141 | 69 | 72 | 138 |
| 0,553786863 | 141 | 69 | 72 | 138 |
| 0,254375613 | 141 | 69 | 72 | 138 |
| 0,307106686 | 141 | 69 | 72 | 138 |
| 0,531138771 | 141 | 69 | 72 | 138 |
| 0,350039715 | 141 | 69 | 72 | 138 |
| 0,349603807 | 141 | 69 | 72 | 138 |
| 0,149190428 | 141 | 69 | 72 | 138 |
| 0,279028856 | 141 | 69 | 72 | 138 |
| 0,274684783 | 141 | 69 | 72 | 138 |
| 0,19244172 | 141 | 69 | 72 | 138 |
| 0,485806811 | 141 | 69 | 72 | 138 |
| 0,215475175 | 141 | 69 | 72 | 138 |
| 0,277023169 | 141 | 69 | 72 | 138 |
| 0,409358954 | 141 | 69 | 72 | 138 |
| 0,201202217 | 141 | 69 | 72 | 138 |
| 0,279989352 | 141 | 69 | 72 | 138 |
| 0,309860301 | 141 | 69 | 72 | 138 |
| 0,397242611 | 141 | 69 | 72 | 138 |
| 0,303015345 | 141 | 69 | 72 | 138 |
| 0,329668316 | 141 | 69 | 72 | 138 |
| 0,227352861 | 141 | 69 | 72 | 138 |
| 0,2882251 | 141 | 69 | 72 | 138 |
| 0,409055169 | 141 | 69 | 72 | 138 |
| 0,443493826 | 141 | 69 | 72 | 138 |
| 0,536799393 | 141 | 69 | 72 | 138 |
| 0,556014205 | 141 | 69 | 72 | 138 |
| 0,317378585 | 141 | 69 | 72 | 138 |
| 0,395893318 | 141 | 69 | 72 | 138 |
| 0,316446715 | 141 | 69 | 72 | 138 |
| 0,37261699 | 141 | 69 | 72 | 138 |
| 0,450174271 | 141 | 69 | 72 | 138 |
| 0,350486487 | 141 | 69 | 72 | 138 |

| | | | | |
|---|---|---|---|---|
| 0,35710067 | 141 | 69 | 72 | 138 |
| 0,417586656 | 141 | 69 | 72 | 138 |
| 0,399326202 | 141 | 69 | 72 | 138 |
| 0,321064691 | 141 | 69 | 72 | 138 |
| 0,159352869 | 141 | 69 | 72 | 138 |
| 0,430624151 | 141 | 69 | 72 | 138 |
| 0,446134774 | 141 | 69 | 72 | 138 |
| 0,498053277 | 141 | 69 | 72 | 138 |
| 0,37114679 | 141 | 69 | 72 | 138 |
| 0,210556673 | 141 | 69 | 72 | 138 |
| 0,232628692 | 141 | 69 | 72 | 138 |
| 0,486080329 | 141 | 69 | 72 | 138 |
| 0,25245468 | 141 | 69 | 72 | 138 |
| 0,268400576 | 141 | 69 | 72 | 138 |
| 0,33944694 | 141 | 69 | 72 | 138 |
| 0,146614631 | 141 | 69 | 72 | 138 |
| 0,362425865 | 141 | 69 | 72 | 138 |
| 0,207207216 | 141 | 69 | 72 | 138 |
| 0,390399288 | 141 | 69 | 72 | 138 |
| 0,428577 | 141 | 69 | 72 | 138 |
| 0,151824033 | 141 | 69 | 72 | 138 |
| 0,539933035 | 141 | 69 | 72 | 138 |
| 0,677412419 | 141 | 69 | 72 | 138 |
| 0,535040611 | 141 | 69 | 72 | 138 |
| 0,303099987 | 141 | 69 | 72 | 138 |
| 0,573515729 | 141 | 69 | 72 | 138 |
| 0,537633396 | 141 | 69 | 72 | 138 |
| 0,518701932 | 141 | 69 | 72 | 138 |
| 0,415337401 | 141 | 69 | 72 | 138 |
| 0,65263441 | 141 | 69 | 72 | 138 |
| 0,259188086 | 141 | 69 | 72 | 138 |
| 0,188551733 | 141 | 69 | 72 | 138 |
| 0,503758265 | 141 | 69 | 72 | 138 |
| 0,772671491 | 141 | 69 | 72 | 138 |
| 0,588596631 | 141 | 69 | 72 | 138 |
| 0,382583786 | 141 | 69 | 72 | 138 |
| 0,358835074 | 141 | 69 | 72 | 138 |
| 0,358066539 | 141 | 69 | 72 | 138 |
| 0,714609541 | 141 | 69 | 72 | 138 |
| 0,733784029 | 141 | 69 | 72 | 138 |
| 0,23648919 | 141 | 69 | 72 | 138 |
| 0,431475168 | 141 | 69 | 72 | 138 |
| 0,255009445 | 141 | 69 | 72 | 138 |
| 0,539373997 | 141 | 69 | 72 | 138 |
| 0,315675959 | 141 | 69 | 72 | 138 |
| 0,322612621 | 141 | 69 | 72 | 138 |
| 0,37293185 | 141 | 69 | 72 | 138 |
| 0,369989232 | 141 | 69 | 72 | 138 |

| | | | | |
|---|---|---|---|---|
| 0,27150373 | 141 | 69 | 72 | 138 |
| 0,200149482 | 141 | 69 | 72 | 138 |
| 0,197092281 | 141 | 69 | 72 | 138 |
| 0,454689525 | 141 | 69 | 72 | 138 |
| 0,281942291 | 141 | 69 | 72 | 138 |
| 0,299702692 | 141 | 69 | 72 | 138 |
| 0,21612492 | 141 | 69 | 72 | 138 |
| 0,364985883 | 141 | 69 | 72 | 138 |
| 0,344639658 | 141 | 69 | 72 | 138 |
| 0,552798914 | 141 | 69 | 72 | 138 |
| 0,180547399 | 141 | 69 | 72 | 138 |
| 0,287081652 | 141 | 69 | 72 | 138 |
| 0,188131108 | 141 | 69 | 72 | 138 |
| 0,2699605 | 141 | 69 | 72 | 138 |
| 0,457317663 | 141 | 69 | 72 | 138 |
| 0,568361216 | 141 | 69 | 72 | 138 |
| 0,323335835 | 141 | 69 | 72 | 138 |
| 0,383236456 | 141 | 69 | 72 | 138 |
| 0,472925679 | 141 | 69 | 72 | 138 |
| 0,315279055 | 141 | 69 | 72 | 138 |
| 0,26235626 | 141 | 69 | 72 | 138 |
| 0,533752227 | 141 | 69 | 72 | 138 |
| 0,177168744 | 141 | 69 | 72 | 138 |
| 0,41433533 | 141 | 69 | 72 | 138 |
| 0,345415625 | 141 | 69 | 72 | 138 |
| 0,34964781 | 141 | 69 | 72 | 138 |
| 0,26973493 | 141 | 69 | 72 | 138 |
| 0,280389668 | 141 | 69 | 72 | 138 |
| 0,240341384 | 141 | 69 | 72 | 138 |
| 0,191381404 | 141 | 69 | 72 | 138 |
| 0,261209151 | 141 | 69 | 72 | 138 |
| 0,357580337 | 141 | 69 | 72 | 138 |
| 0,256420665 | 141 | 69 | 72 | 138 |
| 0,428266684 | 141 | 69 | 72 | 138 |
| 0,186321206 | 141 | 69 | 72 | 138 |
| 0,397408445 | 141 | 69 | 72 | 138 |
| 0,556857407 | 141 | 69 | 72 | 138 |
| 0,445077316 | 141 | 69 | 72 | 138 |
| 0,328091233 | 141 | 69 | 72 | 138 |
| 0,204302863 | 141 | 69 | 72 | 138 |
| 0,169995838 | 141 | 69 | 72 | 138 |
| 0,720558888 | 141 | 69 | 72 | 138 |
| 0,220557342 | 141 | 69 | 72 | 138 |
| 0,353980209 | 141 | 69 | 72 | 138 |
| 0,655986938 | 141 | 69 | 72 | 138 |
| 0,491067182 | 141 | 69 | 72 | 138 |
| 0,475817735 | 141 | 69 | 72 | 138 |
| 0,272735496 | 141 | 69 | 72 | 138 |

| | | | | |
|---|---|---|---|---|
| 0,301693347 | 141 | 69 | 72 | 138 |
| 0,384298582 | 141 | 69 | 72 | 138 |
| 0,283588274 | 141 | 69 | 72 | 138 |
| 0,549210481 | 141 | 69 | 72 | 138 |
| 0,541282935 | 141 | 69 | 72 | 138 |
| 0,383869182 | 141 | 69 | 72 | 138 |
| 0,382056115 | 141 | 69 | 72 | 138 |
| 0,279034879 | 141 | 69 | 72 | 138 |
| 0,427432063 | 141 | 69 | 72 | 138 |
| 0,563832134 | 141 | 69 | 72 | 138 |
| 0,159555496 | 141 | 69 | 72 | 138 |
| 0,503096817 | 141 | 69 | 72 | 138 |
| 0,226300951 | 141 | 69 | 72 | 138 |
| 0,39357604 | 141 | 69 | 72 | 138 |
| 0,536190137 | 141 | 69 | 72 | 138 |
| 0,204830298 | 141 | 69 | 72 | 138 |
| 0,373096539 | 141 | 69 | 72 | 138 |
| 0,344943254 | 141 | 69 | 72 | 138 |
| 0,31350482 | 141 | 69 | 72 | 138 |
| 0,513950665 | 141 | 69 | 72 | 138 |
| 0,269034823 | 141 | 69 | 72 | 138 |
| 0,552572155 | 141 | 69 | 72 | 138 |
| 0,355906473 | 141 | 69 | 72 | 138 |
| 0,167039205 | 141 | 69 | 72 | 138 |
| 0,265778029 | 141 | 69 | 72 | 138 |
| 0,606163757 | 141 | 69 | 72 | 138 |
| 0,289578585 | 141 | 69 | 72 | 138 |
| 0,494113231 | 141 | 69 | 72 | 138 |
| 0,463868556 | 141 | 69 | 72 | 138 |
| 0,50093908 | 141 | 69 | 72 | 138 |
| 0,243120894 | 141 | 69 | 72 | 138 |
| 0,270184524 | 141 | 69 | 72 | 138 |
| 0,365063077 | 141 | 69 | 72 | 138 |
| 0,372542497 | 141 | 69 | 72 | 138 |
| 0,247096928 | 141 | 69 | 72 | 138 |
| 0,729180041 | 141 | 69 | 72 | 138 |
| 0,437271633 | 141 | 69 | 72 | 138 |
| 0,425713745 | 141 | 69 | 72 | 138 |
| 0,328782674 | 141 | 69 | 72 | 138 |
| 0,410919773 | 141 | 69 | 72 | 138 |
| 0,500012393 | 141 | 69 | 72 | 138 |
| 0,315639366 | 141 | 69 | 72 | 138 |
| 0,414522242 | 141 | 69 | 72 | 138 |
| 0,434099182 | 141 | 69 | 72 | 138 |
| 0,319141443 | 141 | 69 | 72 | 138 |
| 0,591469457 | 141 | 69 | 72 | 138 |
| 0,351465914 | 141 | 69 | 72 | 138 |
| 0,456415855 | 141 | 69 | 72 | 138 |

| | | | | |
|---|---|---|---|---|
| 0,372043232 | 141 | 69 | 72 | 138 |
| 0,525443071 | 141 | 69 | 72 | 138 |
| 0,364326939 | 141 | 69 | 72 | 138 |
| 0,528927906 | 141 | 69 | 72 | 138 |
| 0,363449306 | 141 | 69 | 72 | 138 |
| 0,334294356 | 141 | 69 | 72 | 138 |
| 0,312266532 | 141 | 69 | 72 | 138 |
| 0,477387863 | 141 | 69 | 72 | 138 |
| 0,271570293 | 141 | 69 | 72 | 138 |
| 0,352084663 | 141 | 69 | 72 | 138 |
| 0,61952005 | 141 | 69 | 72 | 138 |
| 0,723295209 | 141 | 69 | 72 | 138 |
| 0,278587233 | 141 | 69 | 72 | 138 |
| 0,27236031 | 141 | 69 | 72 | 138 |
| 0,168896731 | 141 | 69 | 72 | 138 |
| 0,417718539 | 141 | 69 | 72 | 138 |
| 0,253206427 | 141 | 69 | 72 | 138 |
| 0,17088412 | 141 | 69 | 72 | 138 |
| 0,290601948 | 141 | 69 | 72 | 138 |
| 0,320798419 | 141 | 69 | 72 | 138 |
| 0,507842226 | 141 | 69 | 72 | 138 |
| 0,339003738 | 141 | 69 | 72 | 138 |
| 0,4598571 | 141 | 69 | 72 | 138 |
| 0,67052045 | 141 | 69 | 72 | 138 |
| 0,519688372 | 141 | 69 | 72 | 138 |
| 0,426042404 | 141 | 69 | 72 | 138 |
| 0,191661892 | 141 | 69 | 72 | 138 |
| 0,328448712 | 141 | 69 | 72 | 138 |
| 0,482372181 | 141 | 69 | 72 | 138 |
| 0,469967813 | 141 | 69 | 72 | 138 |
| 0,149993519 | 141 | 69 | 72 | 138 |
| 0,295673565 | 141 | 69 | 72 | 138 |
| 0,453746691 | 141 | 69 | 72 | 138 |
| 0,363767804 | 141 | 69 | 72 | 138 |
| 0,681119583 | 141 | 69 | 72 | 138 |
| 0,46669694 | 141 | 69 | 72 | 138 |
| 0,392031231 | 141 | 69 | 72 | 138 |
| 0,375962942 | 141 | 69 | 72 | 138 |
| 0,243765557 | 141 | 69 | 72 | 138 |
| 0,232092011 | 141 | 69 | 72 | 138 |
| 0,279273241 | 141 | 69 | 72 | 138 |
| 0,294012798 | 141 | 69 | 72 | 138 |
| 0,310566132 | 141 | 69 | 72 | 138 |
| 0,443709585 | 141 | 69 | 72 | 138 |
| 0,87447578 | 141 | 69 | 72 | 138 |
| 0,539547612 | 141 | 69 | 72 | 138 |
| 0,411429416 | 141 | 69 | 72 | 138 |
| 0,242551056 | 141 | 69 | 72 | 138 |

| | | | | |
|---|---|---|---|---|
| 0,705319718 | 141 | 69 | 72 | 138 |
| 0,371906899 | 141 | 69 | 72 | 138 |
| 0,42133384 | 141 | 69 | 72 | 138 |
| 0,409528752 | 141 | 69 | 72 | 138 |
| 0,615917719 | 141 | 69 | 72 | 138 |
| 0,382329241 | 141 | 69 | 72 | 138 |
| 0,369397378 | 141 | 69 | 72 | 138 |
| 0,401129429 | 141 | 69 | 72 | 138 |
| 0,435517107 | 141 | 69 | 72 | 138 |
| 0,400809033 | 141 | 69 | 72 | 138 |
| 0,380205002 | 141 | 69 | 72 | 138 |
| 0,28396327 | 141 | 69 | 72 | 138 |
| 0,28955568 | 141 | 69 | 72 | 138 |
| 0,430332399 | 141 | 69 | 72 | 138 |
| 0,186109485 | 141 | 69 | 72 | 138 |
| 0,303993193 | 141 | 69 | 72 | 138 |
| 0,778915532 | 141 | 69 | 72 | 138 |
| 0,524115345 | 141 | 69 | 72 | 138 |
| 0,251193011 | 141 | 69 | 72 | 138 |
| 0,319382533 | 141 | 69 | 72 | 138 |
| 0,295707983 | 141 | 69 | 72 | 138 |
| 0,376814996 | 141 | 69 | 72 | 138 |
| 0,622500949 | 141 | 69 | 72 | 138 |
| 0,302048269 | 141 | 69 | 72 | 138 |
| 0,403103664 | 141 | 69 | 72 | 138 |
| 0,343279193 | 141 | 69 | 72 | 138 |
| 0,325125942 | 141 | 69 | 72 | 138 |
| 0,433494697 | 141 | 69 | 72 | 138 |
| 0,204626091 | 141 | 69 | 72 | 138 |
| 0,354386681 | 141 | 69 | 72 | 138 |
| 0,569934565 | 141 | 69 | 72 | 138 |
| 0,449437867 | 141 | 69 | 72 | 138 |
| 0,407880449 | 141 | 69 | 72 | 138 |
| 0,330904981 | 141 | 69 | 72 | 138 |
| 0,493809709 | 141 | 69 | 72 | 138 |
| 0,416153096 | 141 | 69 | 72 | 138 |
| 0,761745877 | 141 | 69 | 72 | 138 |
| 0,548901484 | 141 | 69 | 72 | 138 |
| 0,694262178 | 141 | 69 | 72 | 138 |
| 0,463366171 | 141 | 69 | 72 | 138 |
| 0,646635445 | 141 | 69 | 72 | 138 |
| 0,328909773 | 141 | 69 | 72 | 138 |
| 0,762481963 | 141 | 69 | 72 | 138 |
| 0,372122442 | 141 | 69 | 72 | 138 |
| 0,433915823 | 141 | 69 | 72 | 138 |
| 0,237632645 | 141 | 69 | 72 | 138 |
| 0,345377547 | 141 | 69 | 72 | 138 |
| 0,330886871 | 141 | 69 | 72 | 138 |

| | | | | |
|---|---|---|---|---|
| 0,803197068 | 141 | 69 | 72 | 138 |
| 0,434253666 | 141 | 69 | 72 | 138 |
| 0,645242202 | 141 | 69 | 72 | 138 |
| 0,484723323 | 141 | 69 | 72 | 138 |
| 0,334881298 | 141 | 69 | 72 | 138 |
| 0,53854458 | 141 | 69 | 72 | 138 |
| 0,932969718 | 141 | 69 | 72 | 138 |
| 0,45468954 | 141 | 69 | 72 | 138 |
| 0,428054753 | 141 | 69 | 72 | 138 |
| 0,43734464 | 141 | 69 | 72 | 138 |
| 0,323698028 | 141 | 69 | 72 | 138 |
| 0,532711815 | 141 | 69 | 72 | 138 |
| 0,407342183 | 141 | 69 | 72 | 138 |
| 0,392370635 | 141 | 69 | 72 | 138 |
| 0,62563672 | 141 | 69 | 72 | 138 |
| 0,347775672 | 141 | 69 | 72 | 138 |
| 0,417351326 | 141 | 69 | 72 | 138 |
| 0,5560359 | 141 | 69 | 72 | 138 |
| 0,498952983 | 141 | 69 | 72 | 138 |
| 0,530333391 | 141 | 69 | 72 | 138 |
| 0,209089545 | 141 | 69 | 72 | 138 |
| 0,685606022 | 141 | 69 | 72 | 138 |
| 0,343598431 | 141 | 69 | 72 | 138 |
| 0,211875207 | 141 | 69 | 72 | 138 |
| 0,694598479 | 141 | 69 | 72 | 138 |
| 0,514546322 | 141 | 69 | 72 | 138 |
| 0,327213713 | 141 | 69 | 72 | 138 |
| 0,266778269 | 141 | 69 | 72 | 138 |
| 0,32785575 | 141 | 69 | 72 | 138 |
| 0,445730511 | 141 | 69 | 72 | 138 |
| 0,732803274 | 141 | 69 | 72 | 138 |
| 0,662235051 | 141 | 69 | 72 | 138 |
| 0,614967755 | 141 | 69 | 72 | 138 |
| 0,280652082 | 141 | 69 | 72 | 138 |
| 0,425510456 | 141 | 69 | 72 | 138 |
| 0,414629902 | 141 | 69 | 72 | 138 |
| 0,547776253 | 141 | 69 | 72 | 138 |
| 0,40385795 | 141 | 69 | 72 | 138 |
| 0,218039974 | 141 | 69 | 72 | 138 |
| 0,188770911 | 141 | 69 | 72 | 138 |
| 0,661303837 | 141 | 69 | 72 | 138 |
| 0,421238724 | 141 | 69 | 72 | 138 |
| 0,269632262 | 141 | 69 | 72 | 138 |
| 0,372773597 | 141 | 69 | 72 | 138 |
| 0,692585626 | 141 | 69 | 72 | 138 |
| 0,333814641 | 141 | 69 | 72 | 138 |
| 0,465570741 | 141 | 69 | 72 | 138 |
| 0,564786884 | 141 | 69 | 72 | 138 |

| | | | | |
|---|---|---|---|---|
| 0,602638238 | 141 | 69 | 72 | 138 |
| 0,582109385 | 141 | 69 | 72 | 138 |
| 0,440328998 | 141 | 69 | 72 | 138 |
| 0,466023252 | 141 | 69 | 72 | 138 |
| 0,439791256 | 141 | 69 | 72 | 138 |
| 0,682727823 | 141 | 69 | 72 | 138 |
| 0,533625669 | 141 | 69 | 72 | 138 |
| 0,701079248 | 141 | 69 | 72 | 138 |
| 0,402142404 | 141 | 69 | 72 | 138 |
| 0,433063999 | 141 | 69 | 72 | 138 |
| 0,49366196 | 141 | 69 | 72 | 138 |
| 0,453560061 | 141 | 69 | 72 | 138 |
| 0,250377825 | 141 | 69 | 72 | 138 |
| 0,481268449 | 141 | 69 | 72 | 138 |
| 0,758959478 | 141 | 69 | 72 | 138 |
| 0,549433337 | 141 | 69 | 72 | 138 |
| 0,701660388 | 141 | 69 | 72 | 138 |
| 0,522526349 | 141 | 69 | 72 | 138 |
| 0,641808839 | 141 | 69 | 72 | 138 |
| 0,290103789 | 141 | 69 | 72 | 138 |
| 0,877443895 | 141 | 69 | 72 | 138 |
| 0,617144295 | 141 | 69 | 72 | 138 |
| 0,371513362 | 141 | 69 | 72 | 138 |
| 0,503923414 | 141 | 69 | 72 | 138 |
| 0,409028164 | 141 | 69 | 72 | 138 |
| 0,408372106 | 141 | 69 | 72 | 138 |
| 0,540105489 | 141 | 69 | 72 | 138 |
| 0,304781493 | 141 | 69 | 72 | 138 |
| 0,819870345 | 141 | 69 | 72 | 138 |
| 0,288095314 | 141 | 69 | 72 | 138 |
| 0,299740626 | 141 | 69 | 72 | 138 |
| 0,188249423 | 141 | 69 | 72 | 138 |
| 0,407843417 | 141 | 69 | 72 | 138 |
| 0,34683082 | 141 | 69 | 72 | 138 |
| 0,507395246 | 141 | 69 | 72 | 138 |
| 0,753861448 | 141 | 69 | 72 | 138 |
| 0,286277579 | 141 | 69 | 72 | 138 |
| 0,409127244 | 141 | 69 | 72 | 138 |
| 0,607859199 | 141 | 69 | 72 | 138 |
| 0,422243797 | 141 | 69 | 72 | 138 |
| 0,234592073 | 141 | 69 | 72 | 138 |
| 0,630614827 | 141 | 69 | 72 | 138 |
| 0,556490944 | 141 | 69 | 72 | 138 |
| 0,279478653 | 141 | 69 | 72 | 138 |
| 0,268948275 | 141 | 69 | 72 | 138 |
| 0,442468852 | 141 | 69 | 72 | 138 |
| 0,25745728 | 141 | 69 | 72 | 138 |
| 0,511929815 | 141 | 69 | 72 | 138 |

| | | | | |
|---|---|---|---|---|
| 0,376089182 | 141 | 69 | 72 | 138 |
| 0,570735604 | 141 | 69 | 72 | 138 |
| 0,368314603 | 141 | 69 | 72 | 138 |
| 0,678147388 | 141 | 69 | 72 | 138 |
| 0,392480958 | 141 | 69 | 72 | 138 |
| 0,548473203 | 141 | 69 | 72 | 138 |
| 0,690712959 | 141 | 69 | 72 | 138 |
| 0,400364819 | 141 | 69 | 72 | 138 |
| 0,411858203 | 141 | 69 | 72 | 138 |
| 0,377738903 | 141 | 69 | 72 | 138 |
| 0,409861035 | 141 | 69 | 72 | 138 |
| 0,485762928 | 141 | 69 | 72 | 138 |
| 0,547685022 | 141 | 69 | 72 | 138 |
| 0,315506542 | 141 | 69 | 72 | 138 |
| 0,196691608 | 141 | 69 | 72 | 138 |
| 0,388415066 | 141 | 69 | 72 | 138 |
| 0,19970283 | 141 | 69 | 72 | 138 |
| 0,38851842 | 141 | 69 | 72 | 138 |
| 0,431627689 | 141 | 69 | 72 | 138 |
| 0,764659468 | 141 | 69 | 72 | 138 |
| 0,462418598 | 138 | 69 | 69 | 135 |
| 0,42544243 | 138 | 69 | 69 | 135 |
| 0,893709457 | 138 | 69 | 69 | 135 |
| 0,533135305 | 138 | 69 | 69 | 135 |
| 0,694283908 | 138 | 69 | 69 | 135 |
| 0,495181133 | 138 | 69 | 69 | 135 |
| 0,512627715 | 138 | 69 | 69 | 135 |
| 0,591068843 | 138 | 69 | 69 | 135 |
| 0,439475305 | 138 | 69 | 69 | 135 |
| 0,540797937 | 138 | 69 | 69 | 135 |
| 0,575834517 | 138 | 69 | 69 | 135 |
| 0,374244719 | 138 | 69 | 69 | 135 |
| 0,760501607 | 138 | 69 | 69 | 135 |
| 0,534275348 | 138 | 69 | 69 | 135 |
| 0,703392004 | 138 | 69 | 69 | 135 |
| 0,691316322 | 138 | 69 | 69 | 135 |
| 0,650360371 | 138 | 69 | 69 | 135 |
| 0,789140557 | 138 | 69 | 69 | 135 |
| 0,481645881 | 138 | 69 | 69 | 135 |
| 0,530522659 | 138 | 69 | 69 | 135 |
| 0,542631948 | 138 | 69 | 69 | 135 |
| 0,53317425 | 138 | 69 | 69 | 135 |
| 0,455709081 | 138 | 69 | 69 | 135 |
| 0,652504593 | 138 | 69 | 69 | 135 |
| 0,371353747 | 138 | 69 | 69 | 135 |
| 0,723215045 | 138 | 69 | 69 | 135 |
| 0,605309583 | 138 | 69 | 69 | 135 |
| 0,633121181 | 138 | 69 | 69 | 135 |

| | | | | |
|---|---|---|---|---|
| 0,833392258 | 138 | 69 | 69 | 135 |
| 0,560411615 | 138 | 69 | 69 | 135 |
| 0,73683423 | 138 | 69 | 69 | 135 |
| 0,482768196 | 138 | 69 | 69 | 135 |
| 0,757457151 | 138 | 69 | 69 | 135 |
| 0,659055793 | 138 | 69 | 69 | 135 |
| 0,525019349 | 138 | 69 | 69 | 135 |
| 0,417662211 | 138 | 69 | 69 | 135 |
| 0,508999965 | 138 | 69 | 69 | 135 |
| 0,775036054 | 138 | 69 | 69 | 135 |
| 0,669329478 | 138 | 69 | 69 | 135 |
| 0,788146358 | 138 | 69 | 69 | 135 |
| 0,752453887 | 138 | 69 | 69 | 135 |
| 0,638516504 | 138 | 69 | 69 | 135 |
| 0,567799541 | 138 | 69 | 69 | 135 |
| 0,271794153 | 138 | 69 | 69 | 135 |
| 0,510754855 | 138 | 69 | 69 | 135 |
| 0,34916068 | 138 | 69 | 69 | 135 |
| 0,582588866 | 138 | 69 | 69 | 135 |
| 0,633572294 | 138 | 69 | 69 | 135 |
| 0,28783304 | 138 | 69 | 69 | 135 |
| 0,663887098 | 138 | 69 | 69 | 135 |
| 0,492643208 | 138 | 69 | 69 | 135 |
| 0,338775168 | 138 | 69 | 69 | 135 |
| 0,638435235 | 138 | 69 | 69 | 135 |
| 0,570122637 | 138 | 69 | 69 | 135 |
| 0,483366948 | 138 | 69 | 69 | 135 |
| 0,655088176 | 138 | 69 | 69 | 135 |
| 0,550803031 | 138 | 69 | 69 | 135 |
| 0,718307212 | 138 | 69 | 69 | 135 |
| 0,169741168 | 138 | 69 | 69 | 135 |
| 0,525456531 | 138 | 69 | 69 | 135 |
| 0,804834974 | 138 | 69 | 69 | 135 |
| 0,521023419 | 138 | 69 | 69 | 135 |
| 0,420069032 | 138 | 69 | 69 | 135 |
| 0,793228147 | 138 | 69 | 69 | 135 |
| 0,718354127 | 138 | 69 | 69 | 135 |
| 0,311862406 | 138 | 69 | 69 | 135 |
| 0,568719648 | 138 | 69 | 69 | 135 |
| 0,519804558 | 138 | 69 | 69 | 135 |
| 0,460324609 | 138 | 69 | 69 | 135 |
| 0,533290005 | 138 | 69 | 69 | 135 |
| 0,539005597 | 138 | 69 | 69 | 135 |
| 0,45680415 | 138 | 69 | 69 | 135 |
| 0,46809249 | 138 | 69 | 69 | 135 |
| 0,360966739 | 138 | 69 | 69 | 135 |
| 0,532029195 | 138 | 69 | 69 | 135 |
| 0,35660828 | 138 | 69 | 69 | 135 |

| | | | | |
|---|---|---|---|---|
| 0,337161185 | 138 | 69 | 69 | 135 |
| 0,671143714 | 138 | 69 | 69 | 135 |
| 0,475111632 | 138 | 69 | 69 | 135 |
| 0,375802265 | 138 | 69 | 69 | 135 |
| 0,618769612 | 138 | 69 | 69 | 135 |
| 0,89566198 | 138 | 69 | 69 | 135 |
| 0,355518436 | 138 | 69 | 69 | 135 |
| 0,576823173 | 138 | 69 | 69 | 135 |
| 1,008169477 | 138 | 69 | 69 | 135 |
| 0,653403438 | 138 | 69 | 69 | 135 |
| 0,283048935 | 138 | 69 | 69 | 135 |
| 0,333472187 | 138 | 69 | 69 | 135 |
| 0,475569623 | 138 | 69 | 69 | 135 |
| 0,4339742 | 138 | 69 | 69 | 135 |
| 0,535701636 | 138 | 69 | 69 | 135 |
| 0,770682571 | 138 | 69 | 69 | 135 |
| 0,423594497 | 138 | 69 | 69 | 135 |
| 0,629673133 | 138 | 69 | 69 | 135 |
| 0,788446735 | 138 | 69 | 69 | 135 |
| 0,932303857 | 138 | 69 | 69 | 135 |
| 0,724322642 | 138 | 69 | 69 | 135 |
| 0,529161558 | 138 | 69 | 69 | 135 |
| 0,728406052 | 138 | 69 | 69 | 135 |
| 0,35093668 | 138 | 69 | 69 | 135 |
| 0,653969287 | 138 | 69 | 69 | 135 |
| 0,424494203 | 138 | 69 | 69 | 135 |
| 0,424067806 | 138 | 69 | 69 | 135 |
| 0,64621418 | 138 | 69 | 69 | 135 |
| 0,502852091 | 138 | 69 | 69 | 135 |
| 0,673887811 | 138 | 69 | 69 | 135 |
| 0,257059491 | 138 | 69 | 69 | 135 |
| 0,439035319 | 138 | 69 | 69 | 135 |
| 0,555733095 | 138 | 69 | 69 | 135 |
| 0,460335307 | 138 | 69 | 69 | 135 |
| 0,465347188 | 138 | 69 | 69 | 135 |
| 0,499225123 | 138 | 69 | 69 | 135 |
| 0,572965103 | 138 | 69 | 69 | 135 |
| 0,267920738 | 138 | 69 | 69 | 135 |
| 0,438215587 | 138 | 69 | 69 | 135 |
| 0,322345772 | 138 | 69 | 69 | 135 |
| 0,487055388 | 138 | 69 | 69 | 135 |
| 0,263650817 | 138 | 69 | 69 | 135 |
| 0,212001105 | 138 | 69 | 69 | 135 |
| 0,304781282 | 138 | 69 | 69 | 135 |
| 0,360353432 | 138 | 69 | 69 | 135 |
| 0,518991144 | 138 | 69 | 69 | 135 |
| 1,032980112 | 138 | 69 | 69 | 135 |
| 0,371947393 | 138 | 69 | 69 | 135 |

| | | | | |
|---|---|---|---|---|
| 0,641552818 | 138 | 69 | 69 | 135 |
| 0,260075397 | 138 | 69 | 69 | 135 |
| 0,656919155 | 138 | 69 | 69 | 135 |
| 0,473160365 | 138 | 69 | 69 | 135 |
| 0,410446217 | 138 | 69 | 69 | 135 |
| 0,483290026 | 138 | 69 | 69 | 135 |
| 0,586961106 | 138 | 69 | 69 | 135 |
| 0,189370063 | 138 | 69 | 69 | 135 |
| 0,281605383 | 138 | 69 | 69 | 135 |
| 0,74926149 | 138 | 69 | 69 | 135 |
| 0,545324742 | 138 | 69 | 69 | 135 |
| 0,388596268 | 138 | 69 | 69 | 135 |
| 0,446680462 | 138 | 69 | 69 | 135 |
| 0,442884194 | 138 | 69 | 69 | 135 |
| 0,50300058 | 138 | 69 | 69 | 135 |
| 0,661084387 | 138 | 69 | 69 | 135 |
| 0,395819449 | 138 | 69 | 69 | 135 |
| 0,380368449 | 138 | 69 | 69 | 135 |
| 0,991395057 | 138 | 69 | 69 | 135 |
| 0,414156943 | 138 | 69 | 69 | 135 |
| 0,395499196 | 138 | 69 | 69 | 135 |
| 0,548310164 | 138 | 69 | 69 | 135 |
| 0,502379377 | 138 | 69 | 69 | 135 |
| 0,380873583 | 138 | 69 | 69 | 135 |
| 0,307466328 | 138 | 69 | 69 | 135 |
| 0,553260411 | 138 | 69 | 69 | 135 |
| 0,504206461 | 138 | 69 | 69 | 135 |
| 0,822497002 | 138 | 69 | 69 | 135 |
| 0,298702002 | 138 | 69 | 69 | 135 |
| 0,486653208 | 138 | 69 | 69 | 135 |
| 0,792464983 | 138 | 69 | 69 | 135 |
| 0,199082786 | 138 | 69 | 69 | 135 |
| 0,402686117 | 138 | 69 | 69 | 135 |
| 0,679731668 | 138 | 69 | 69 | 135 |
| 0,64085122 | 138 | 69 | 69 | 135 |
| 0,50142313 | 138 | 69 | 69 | 135 |
| 0,465463332 | 138 | 69 | 69 | 135 |
| 0,592176931 | 138 | 69 | 69 | 135 |
| 1,115252696 | 138 | 69 | 69 | 135 |
| 0,29582323 | 138 | 69 | 69 | 135 |
| 0,75259989 | 138 | 69 | 69 | 135 |
| 0,555213387 | 138 | 69 | 69 | 135 |
| 0,231964977 | 138 | 69 | 69 | 135 |
| 0,424266235 | 138 | 69 | 69 | 135 |
| 0,26981627 | 138 | 69 | 69 | 135 |
| 0,349285144 | 138 | 69 | 69 | 135 |
| 0,712238762 | 138 | 69 | 69 | 135 |
| 0,441086744 | 138 | 69 | 69 | 135 |

| | | | | |
|---|---|---|---|---|
| 0,494758717 | 138 | 69 | 69 | 135 |
| 0,385836555 | 138 | 69 | 69 | 135 |
| 0,672709154 | 138 | 69 | 69 | 135 |
| 0,334884524 | 138 | 69 | 69 | 135 |
| 0,332172212 | 138 | 69 | 69 | 135 |
| 0,66614048 | 138 | 69 | 69 | 135 |
| 0,296403007 | 138 | 69 | 69 | 135 |
| 0,378237868 | 138 | 69 | 69 | 135 |
| 0,590636395 | 138 | 69 | 69 | 135 |
| 0,385827545 | 138 | 69 | 69 | 135 |
| 0,440708067 | 138 | 69 | 69 | 135 |
| 0,562946544 | 138 | 69 | 69 | 135 |
| 0,577105647 | 138 | 69 | 69 | 135 |
| 0,332687546 | 138 | 69 | 69 | 135 |
| 0,261547855 | 138 | 69 | 69 | 135 |
| 0,252725674 | 138 | 69 | 69 | 135 |
| 0,444321153 | 138 | 69 | 69 | 135 |
| 0,286797654 | 138 | 69 | 69 | 135 |
| 0,254954904 | 138 | 69 | 69 | 135 |
| 0,250451878 | 138 | 69 | 69 | 135 |
| 0,462276222 | 138 | 69 | 69 | 135 |
| 0,490879078 | 138 | 69 | 69 | 135 |
| 0,264777132 | 138 | 69 | 69 | 135 |
| 0,540661935 | 138 | 69 | 69 | 135 |
| 0,632742956 | 138 | 69 | 69 | 135 |
| 0,215303629 | 138 | 69 | 69 | 135 |
| 0,249526605 | 138 | 69 | 69 | 135 |
| 0,633087621 | 138 | 69 | 69 | 135 |
| 0,221218559 | 138 | 69 | 69 | 135 |
| 0,192670896 | 138 | 69 | 69 | 135 |
| 0,726450758 | 138 | 69 | 69 | 135 |
| 0,46163797 | 138 | 69 | 69 | 135 |
| 0,748110258 | 138 | 69 | 69 | 135 |
| 0,198687916 | 138 | 69 | 69 | 135 |
| 0,351224324 | 138 | 69 | 69 | 135 |
| 0,364862639 | 138 | 69 | 69 | 135 |
| 0,457274043 | 138 | 69 | 69 | 135 |
| 0,468832494 | 138 | 69 | 69 | 135 |
| 0,454279737 | 138 | 69 | 69 | 135 |
| 0,641218469 | 138 | 69 | 69 | 135 |
| 0,590065448 | 138 | 69 | 69 | 135 |
| 0,387797352 | 138 | 69 | 69 | 135 |
| 0,649089502 | 138 | 69 | 69 | 135 |
| 0,434689175 | 138 | 69 | 69 | 135 |
| 0,310795285 | 138 | 69 | 69 | 135 |
| 0,701451334 | 138 | 69 | 69 | 135 |
| 0,359589539 | 138 | 69 | 69 | 135 |
| 0,40039845 | 138 | 69 | 69 | 135 |

| | | | | |
|---|---|---|---|---|
| 0,707259309 | 138 | 69 | 69 | 135 |
| 0,678834033 | 138 | 69 | 69 | 135 |
| 0,185706388 | 138 | 69 | 69 | 135 |
| 0,434426042 | 138 | 69 | 69 | 135 |
| 0,216516894 | 138 | 69 | 69 | 135 |
| 0,608334852 | 138 | 69 | 69 | 135 |
| 0,169303323 | 138 | 69 | 69 | 135 |
| 0,390940004 | 138 | 69 | 69 | 135 |
| 0,418586631 | 138 | 69 | 69 | 135 |
| 0,428394273 | 138 | 69 | 69 | 135 |
| 0,263237658 | 138 | 69 | 69 | 135 |
| 0,191806459 | 138 | 69 | 69 | 135 |
| 0,415867142 | 138 | 69 | 69 | 135 |
| 0,305326819 | 138 | 69 | 69 | 135 |
| 0,709280583 | 138 | 69 | 69 | 135 |
| 0,339710138 | 138 | 69 | 69 | 135 |
| 0,336372241 | 138 | 69 | 69 | 135 |
| 0,273320156 | 138 | 69 | 69 | 135 |
| 0,593759734 | 138 | 69 | 69 | 135 |
| 0,265618656 | 138 | 69 | 69 | 135 |
| 0,348818987 | 138 | 69 | 69 | 135 |
| 0,255657989 | 138 | 69 | 69 | 135 |
| 0,432605319 | 138 | 69 | 69 | 135 |
| 0,640522581 | 138 | 69 | 69 | 135 |
| 0,630793463 | 138 | 69 | 69 | 135 |
| 0,173184168 | 138 | 69 | 69 | 135 |
| 0,577062576 | 138 | 69 | 69 | 135 |
| 0,469024026 | 138 | 69 | 69 | 135 |
| 0,474564239 | 138 | 69 | 69 | 135 |
| 0,481751757 | 138 | 69 | 69 | 135 |
| 0,367663671 | 138 | 69 | 69 | 135 |
| 0,747168692 | 138 | 69 | 69 | 135 |
| 0,583077698 | 138 | 69 | 69 | 135 |
| 0,428564295 | 138 | 69 | 69 | 135 |
| 0,57452802 | 138 | 69 | 69 | 135 |
| 0,513309092 | 138 | 69 | 69 | 135 |
| 0,319982387 | 138 | 69 | 69 | 135 |
| 0,379754199 | 138 | 69 | 69 | 135 |
| 0,285964768 | 138 | 69 | 69 | 135 |
| 0,612279416 | 138 | 69 | 69 | 135 |
| 0,518151993 | 138 | 69 | 69 | 135 |
| 0,457253596 | 138 | 69 | 69 | 135 |
| 0,841861751 | 138 | 69 | 69 | 135 |
| 0,403713053 | 138 | 69 | 69 | 135 |
| 0,303059411 | 138 | 69 | 69 | 135 |
| 0,421271301 | 138 | 69 | 69 | 135 |
| 0,639521348 | 138 | 69 | 69 | 135 |
| 0,501522646 | 138 | 69 | 69 | 135 |

| | | | | |
|---|---|---|---|---|
| 0,260665537 | 138 | 69 | 69 | 135 |
| 0,709369016 | 138 | 69 | 69 | 135 |
| 0,323018743 | 138 | 69 | 69 | 135 |
| 0,365119621 | 138 | 69 | 69 | 135 |
| 0,774955995 | 138 | 69 | 69 | 135 |
| 0,535665325 | 138 | 69 | 69 | 135 |
| 0,18710862 | 138 | 69 | 69 | 135 |
| 0,497793053 | 138 | 69 | 69 | 135 |
| 0,423699489 | 138 | 69 | 69 | 135 |
| 0,325295325 | 138 | 69 | 69 | 135 |
| 0,635962357 | 138 | 69 | 69 | 135 |
| 0,324072063 | 138 | 69 | 69 | 135 |
| 0,231929537 | 138 | 69 | 69 | 135 |
| 0,461776184 | 138 | 69 | 69 | 135 |
| 0,384854011 | 138 | 69 | 69 | 135 |
| 0,430467941 | 138 | 69 | 69 | 135 |
| 0,354958249 | 138 | 69 | 69 | 135 |
| 0,261081888 | 138 | 69 | 69 | 135 |
| 0,242680052 | 138 | 69 | 69 | 135 |
| 0,529502343 | 138 | 69 | 69 | 135 |
| 0,245912873 | 138 | 69 | 69 | 135 |
| 0,550267017 | 138 | 69 | 69 | 135 |
| 0,489860406 | 138 | 69 | 69 | 135 |
| 0,317423271 | 138 | 69 | 69 | 135 |
| 0,266416786 | 138 | 69 | 69 | 135 |
| 0,497429699 | 138 | 69 | 69 | 135 |
| 0,661479463 | 138 | 69 | 69 | 135 |
| 0,139602417 | 138 | 69 | 69 | 135 |
| 0,489565516 | 138 | 69 | 69 | 135 |
| 0,641658158 | 138 | 69 | 69 | 135 |
| 0,169425346 | 138 | 69 | 69 | 135 |
| 0,569939342 | 138 | 69 | 69 | 135 |
| 0,225472217 | 138 | 69 | 69 | 135 |
| 0,635576567 | 138 | 69 | 69 | 135 |
| 0,45040656 | 138 | 69 | 69 | 135 |
| 0,462701154 | 138 | 69 | 69 | 135 |
| 0,361586254 | 138 | 69 | 69 | 135 |
| 0,569927859 | 138 | 69 | 69 | 135 |
| 0,561818099 | 138 | 69 | 69 | 135 |
| 0,352584617 | 138 | 69 | 69 | 135 |
| 0,477601287 | 138 | 69 | 69 | 135 |
| 0,228264207 | 138 | 69 | 69 | 135 |
| 0,584957063 | 138 | 69 | 69 | 135 |
| 0,423239074 | 138 | 69 | 69 | 135 |
| 0,483605616 | 138 | 69 | 69 | 135 |
| 0,276267566 | 138 | 69 | 69 | 135 |
| 0,242777278 | 138 | 69 | 69 | 135 |
| 0,194215534 | 138 | 69 | 69 | 135 |

| | | | | |
|---|---|---|---|---|
| 0,890722118 | 138 | 69 | 69 | 135 |
| 0,295660832 | 138 | 69 | 69 | 135 |
| 0,498401792 | 138 | 69 | 69 | 135 |
| 0,358610551 | 138 | 69 | 69 | 135 |
| 0,308191207 | 138 | 69 | 69 | 135 |
| 0,361311834 | 138 | 69 | 69 | 135 |
| 0,632584637 | 138 | 69 | 69 | 135 |
| 0,536302222 | 138 | 69 | 69 | 135 |
| 0,285241958 | 138 | 69 | 69 | 135 |
| 0,267225062 | 138 | 69 | 69 | 135 |
| 0,161863541 | 138 | 69 | 69 | 135 |
| 0,439889473 | 138 | 69 | 69 | 135 |
| 0,438391861 | 138 | 69 | 69 | 135 |
| 0,362212197 | 138 | 69 | 69 | 135 |
| 0,390191199 | 138 | 69 | 69 | 135 |
| 0,345487019 | 138 | 69 | 69 | 135 |
| 0,29391991 | 138 | 69 | 69 | 135 |
| 0,244531185 | 138 | 69 | 69 | 135 |
| 0,259973124 | 138 | 69 | 69 | 135 |
| 0,263166049 | 138 | 69 | 69 | 135 |
| 0,470604751 | 138 | 69 | 69 | 135 |
| 0,251410933 | 138 | 69 | 69 | 135 |
| 0,431340221 | 138 | 69 | 69 | 135 |
| 0,196096601 | 138 | 69 | 69 | 135 |
| 0,189421911 | 138 | 69 | 69 | 135 |
| 0,310660438 | 138 | 69 | 69 | 135 |
| 0,476477431 | 138 | 69 | 69 | 135 |
| 0,532939357 | 138 | 69 | 69 | 135 |
| 0,287390328 | 138 | 69 | 69 | 135 |
| 0,217191118 | 138 | 69 | 69 | 135 |
| 0,267228664 | 138 | 69 | 69 | 135 |
| 0,189959669 | 138 | 69 | 69 | 135 |
| 0,3274787 | 138 | 69 | 69 | 135 |
| 0,455377716 | 138 | 69 | 69 | 135 |
| 0,442059083 | 138 | 69 | 69 | 135 |
| 0,390555088 | 138 | 69 | 69 | 135 |
| 0,445098864 | 138 | 69 | 69 | 135 |
| 0,223815396 | 138 | 69 | 69 | 135 |
| 0,377903971 | 138 | 69 | 69 | 135 |
| 0,381903964 | 138 | 69 | 69 | 135 |
| 0,212687709 | 138 | 69 | 69 | 135 |
| 0,375497364 | 138 | 69 | 69 | 135 |
| 0,433938452 | 138 | 69 | 69 | 135 |
| 0,325271116 | 138 | 69 | 69 | 135 |
| 0,385912243 | 138 | 69 | 69 | 135 |
| 0,414206179 | 138 | 69 | 69 | 135 |
| 0,448883737 | 138 | 69 | 69 | 135 |
| 0,324421328 | 138 | 69 | 69 | 135 |

| | | | | |
|---|---|---|---|---|
| 0,375722875 | 138 | 69 | 69 | 135 |
| 0,184976136 | 138 | 69 | 69 | 135 |
| 0,477288141 | 138 | 69 | 69 | 135 |
| 0,460837106 | 138 | 69 | 69 | 135 |
| 0,548493089 | 138 | 69 | 69 | 135 |
| 0,47170544 | 138 | 69 | 69 | 135 |
| 0,172483441 | 138 | 69 | 69 | 135 |
| 0,268751497 | 138 | 69 | 69 | 135 |
| 0,565927538 | 138 | 69 | 69 | 135 |
| 0,284520106 | 138 | 69 | 69 | 135 |
| 0,370219511 | 138 | 69 | 69 | 135 |
| 0,418340454 | 138 | 69 | 69 | 135 |
| 0,259833571 | 138 | 69 | 69 | 135 |
| 0,448466408 | 138 | 69 | 69 | 135 |
| 0,170264535 | 138 | 69 | 69 | 135 |
| 0,241525149 | 138 | 69 | 69 | 135 |
| 0,422741792 | 138 | 69 | 69 | 135 |
| 0,579947532 | 138 | 69 | 69 | 135 |
| 0,153588133 | 138 | 69 | 69 | 135 |
| 0,605554572 | 138 | 69 | 69 | 135 |
| 0,661931349 | 138 | 69 | 69 | 135 |
| 0,588722619 | 138 | 69 | 69 | 135 |
| 0,337940717 | 138 | 69 | 69 | 135 |
| 0,555930414 | 138 | 69 | 69 | 135 |
| 0,666212005 | 138 | 69 | 69 | 135 |
| 0,401961951 | 138 | 69 | 69 | 135 |
| 0,412136139 | 138 | 69 | 69 | 135 |
| 0,354053068 | 138 | 69 | 69 | 135 |
| 0,661100618 | 138 | 69 | 69 | 135 |
| 0,333064197 | 138 | 69 | 69 | 135 |
| 0,25404498 | 138 | 69 | 69 | 135 |
| 0,419814811 | 138 | 69 | 69 | 135 |
| 0,278351013 | 138 | 69 | 69 | 135 |
| 0,789163954 | 138 | 69 | 69 | 135 |
| 0,598937239 | 138 | 69 | 69 | 135 |
| 0,424120515 | 138 | 69 | 69 | 135 |
| 0,378958031 | 138 | 69 | 69 | 135 |
| 0,429360992 | 138 | 69 | 69 | 135 |
| 0,72898317 | 138 | 69 | 69 | 135 |
| 0,791497449 | 138 | 69 | 69 | 135 |
| 0,154112374 | 138 | 69 | 69 | 135 |
| 0,50319967 | 138 | 69 | 69 | 135 |
| 0,496305876 | 138 | 69 | 69 | 135 |
| 0,392332099 | 138 | 69 | 69 | 135 |
| 0,426049257 | 138 | 69 | 69 | 135 |
| 0,155804605 | 138 | 69 | 69 | 135 |
| 0,423689241 | 138 | 69 | 69 | 135 |
| 0,461824587 | 138 | 69 | 69 | 135 |

| | | | | |
|---|---|---|---|---|
| 0,470157192 | 138 | 69 | 69 | 135 |
| 0,371043665 | 138 | 69 | 69 | 135 |
| 0,421771863 | 138 | 69 | 69 | 135 |
| 0,273977767 | 138 | 69 | 69 | 135 |
| 0,2835446 | 138 | 69 | 69 | 135 |
| 0,242923685 | 138 | 69 | 69 | 135 |
| 0,364323458 | 138 | 69 | 69 | 135 |
| 0,265036865 | 138 | 69 | 69 | 135 |
| 0,491570395 | 138 | 69 | 69 | 135 |
| 0,204024072 | 138 | 69 | 69 | 135 |
| 0,259771476 | 138 | 69 | 69 | 135 |
| 0,338182579 | 138 | 69 | 69 | 135 |
| 0,265588392 | 138 | 69 | 69 | 135 |
| 0,205979917 | 138 | 69 | 69 | 135 |
| 0,291897053 | 138 | 69 | 69 | 135 |
| 0,227885166 | 138 | 69 | 69 | 135 |
| 0,393107559 | 138 | 69 | 69 | 135 |
| 0,570196098 | 138 | 69 | 69 | 135 |
| 0,341744739 | 138 | 69 | 69 | 135 |
| 0,393511951 | 138 | 69 | 69 | 135 |
| 0,481770985 | 138 | 69 | 69 | 135 |
| 0,221250662 | 138 | 69 | 69 | 135 |
| 0,238164424 | 138 | 69 | 69 | 135 |
| 0,539216195 | 138 | 69 | 69 | 135 |
| 0,239433067 | 138 | 69 | 69 | 135 |
| 0,354193021 | 138 | 69 | 69 | 135 |
| 0,318464287 | 138 | 69 | 69 | 135 |
| 0,51369418 | 138 | 69 | 69 | 135 |
| 0,286858734 | 138 | 69 | 69 | 135 |
| 0,232557901 | 138 | 69 | 69 | 135 |
| 0,167124702 | 138 | 69 | 69 | 135 |
| 0,190082224 | 138 | 69 | 69 | 135 |
| 0,341260761 | 138 | 69 | 69 | 135 |
| 0,330072988 | 138 | 69 | 69 | 135 |
| 0,290656716 | 138 | 69 | 69 | 135 |
| 0,414540933 | 138 | 69 | 69 | 135 |
| 0,19033504 | 138 | 69 | 69 | 135 |
| 0,492229771 | 138 | 69 | 69 | 135 |
| 0,675230883 | 138 | 69 | 69 | 135 |
| 0,520114453 | 138 | 69 | 69 | 135 |
| 0,400538621 | 138 | 69 | 69 | 135 |
| 0,148786242 | 138 | 69 | 69 | 135 |
| 0,205468939 | 138 | 69 | 69 | 135 |
| 0,68054372 | 138 | 69 | 69 | 135 |
| 0,379956079 | 138 | 69 | 69 | 135 |
| 0,383077872 | 138 | 69 | 69 | 135 |
| 0,553513241 | 138 | 69 | 69 | 135 |
| 0,407929892 | 138 | 69 | 69 | 135 |

| | | | | |
|---|---|---|---|---|
| 0,478147637 | 138 | 69 | 69 | 135 |
| 0,204891243 | 138 | 69 | 69 | 135 |
| 0,391035123 | 138 | 69 | 69 | 135 |
| 0,383464329 | 138 | 69 | 69 | 135 |
| 0,227804052 | 138 | 69 | 69 | 135 |
| 0,665245557 | 138 | 69 | 69 | 135 |
| 0,320616069 | 138 | 69 | 69 | 135 |
| 0,386594296 | 138 | 69 | 69 | 135 |
| 0,370216478 | 138 | 69 | 69 | 135 |
| 0,298917215 | 138 | 69 | 69 | 135 |
| 0,431232889 | 138 | 69 | 69 | 135 |
| 0,581955077 | 138 | 69 | 69 | 135 |
| 0,226814558 | 138 | 69 | 69 | 135 |
| 0,488889093 | 138 | 69 | 69 | 135 |
| 0,311664914 | 138 | 69 | 69 | 135 |
| 0,410911481 | 138 | 69 | 69 | 135 |
| 0,503804722 | 138 | 69 | 69 | 135 |
| 0,352592228 | 138 | 69 | 69 | 135 |
| 0,808671376 | 138 | 69 | 69 | 135 |
| 0,275155136 | 138 | 69 | 69 | 135 |
| 0,584037431 | 138 | 69 | 69 | 135 |
| 0,418653913 | 138 | 69 | 69 | 135 |
| 0,261705327 | 138 | 69 | 69 | 135 |
| 0,186924661 | 138 | 69 | 69 | 135 |
| 0,327726953 | 138 | 69 | 69 | 135 |
| 0,660838878 | 138 | 69 | 69 | 135 |
| 0,313777812 | 138 | 69 | 69 | 135 |
| 0,494990507 | 138 | 69 | 69 | 135 |
| 0,489024422 | 138 | 69 | 69 | 135 |
| 0,473455139 | 138 | 69 | 69 | 135 |
| 0,27662998 | 138 | 69 | 69 | 135 |
| 0,419540511 | 138 | 69 | 69 | 135 |
| 0,260185158 | 138 | 69 | 69 | 135 |
| 0,416146404 | 138 | 69 | 69 | 135 |
| 0,362186174 | 138 | 69 | 69 | 135 |
| 0,84218697 | 138 | 69 | 69 | 135 |
| 0,446299227 | 138 | 69 | 69 | 135 |
| 0,222870851 | 138 | 69 | 69 | 135 |
| 0,45974302 | 138 | 69 | 69 | 135 |
| 0,581195054 | 138 | 69 | 69 | 135 |
| 0,454319035 | 138 | 69 | 69 | 135 |
| 0,275033114 | 138 | 69 | 69 | 135 |
| 0,491689815 | 138 | 69 | 69 | 135 |
| 0,450505317 | 138 | 69 | 69 | 135 |
| 0,333915202 | 138 | 69 | 69 | 135 |
| 0,533943125 | 138 | 69 | 69 | 135 |
| 0,334894905 | 138 | 69 | 69 | 135 |
| 0,53946493 | 138 | 69 | 69 | 135 |

| | | | | |
|---|---|---|---|---|
| 0,342176761 | 138 | 69 | 69 | 135 |
| 0,63688099 | 138 | 69 | 69 | 135 |
| 0,498969268 | 138 | 69 | 69 | 135 |
| 0,584174374 | 138 | 69 | 69 | 135 |
| 0,445048396 | 138 | 69 | 69 | 135 |
| 0,367115169 | 138 | 69 | 69 | 135 |
| 0,354510363 | 138 | 69 | 69 | 135 |
| 0,434344304 | 138 | 69 | 69 | 135 |
| 0,308376698 | 138 | 69 | 69 | 135 |
| 0,453965889 | 138 | 69 | 69 | 135 |
| 0,617797239 | 138 | 69 | 69 | 135 |
| 0,654180735 | 138 | 69 | 69 | 135 |
| 0,271434257 | 138 | 69 | 69 | 135 |
| 0,379622304 | 138 | 69 | 69 | 135 |
| 0,257533933 | 138 | 69 | 69 | 135 |
| 0,289729557 | 138 | 69 | 69 | 135 |
| 0,264013757 | 138 | 69 | 69 | 135 |
| 0,462496763 | 138 | 69 | 69 | 135 |
| 0,498573423 | 138 | 69 | 69 | 135 |
| 0,375272063 | 138 | 69 | 69 | 135 |
| 0,431459014 | 138 | 69 | 69 | 135 |
| 0,65584077 | 138 | 69 | 69 | 135 |
| 0,578480938 | 138 | 69 | 69 | 135 |
| 0,423394745 | 138 | 69 | 69 | 135 |
| 0,344004579 | 138 | 69 | 69 | 135 |
| 0,443277239 | 138 | 69 | 69 | 135 |
| 0,368744154 | 138 | 69 | 69 | 135 |
| 0,167591056 | 138 | 69 | 69 | 135 |
| 0,318167144 | 138 | 69 | 69 | 135 |
| 0,465794713 | 138 | 69 | 69 | 135 |
| 0,452733239 | 138 | 69 | 69 | 135 |
| 0,751101636 | 138 | 69 | 69 | 135 |
| 0,531197488 | 138 | 69 | 69 | 135 |
| 0,391011775 | 138 | 69 | 69 | 135 |
| 0,416285039 | 138 | 69 | 69 | 135 |
| 0,333256753 | 138 | 69 | 69 | 135 |
| 0,184435355 | 138 | 69 | 69 | 135 |
| 0,355680678 | 138 | 69 | 69 | 135 |
| 0,281420841 | 138 | 69 | 69 | 135 |
| 0,270042008 | 138 | 69 | 69 | 135 |
| 0,29753447 | 138 | 69 | 69 | 135 |
| 0,769099564 | 138 | 69 | 69 | 135 |
| 0,567528224 | 138 | 69 | 69 | 135 |
| 0,36811298 | 138 | 69 | 69 | 135 |
| 0,201452459 | 138 | 69 | 69 | 135 |
| 0,902639816 | 138 | 69 | 69 | 135 |
| 0,416048122 | 138 | 69 | 69 | 135 |
| 0,468057791 | 138 | 69 | 69 | 135 |

| | | | | |
|---|---|---|---|---|
| 0,278573143 | 138 | 69 | 69 | 135 |
| 0,429101804 | 138 | 69 | 69 | 135 |
| 0,740816184 | 138 | 69 | 69 | 135 |
| 0,316754041 | 138 | 69 | 69 | 135 |
| 0,375465091 | 138 | 69 | 69 | 135 |
| 0,414296675 | 138 | 69 | 69 | 135 |
| 0,596761066 | 138 | 69 | 69 | 135 |
| 0,456141167 | 138 | 69 | 69 | 135 |
| 0,445370818 | 138 | 69 | 69 | 135 |
| 0,280723474 | 138 | 69 | 69 | 135 |
| 0,282853178 | 138 | 69 | 69 | 135 |
| 0,510074367 | 138 | 69 | 69 | 135 |
| 0,220307247 | 138 | 69 | 69 | 135 |
| 0,276186314 | 138 | 69 | 69 | 135 |
| 0,98624175 | 138 | 69 | 69 | 135 |
| 0,633341318 | 138 | 69 | 69 | 135 |
| 0,406054555 | 138 | 69 | 69 | 135 |
| 0,310787456 | 138 | 69 | 69 | 135 |
| 0,396098159 | 138 | 69 | 69 | 135 |
| 0,518477323 | 138 | 69 | 69 | 135 |
| 0,733044786 | 138 | 69 | 69 | 135 |
| 0,355182685 | 138 | 69 | 69 | 135 |
| 0,471838192 | 138 | 69 | 69 | 135 |
| 0,461352445 | 138 | 69 | 69 | 135 |
| 0,38195871 | 138 | 69 | 69 | 135 |
| 0,245965105 | 138 | 69 | 69 | 135 |
| 0,434135241 | 138 | 69 | 69 | 135 |
| 0,621269382 | 138 | 69 | 69 | 135 |
| 0,516863556 | 138 | 69 | 69 | 135 |
| 0,271579063 | 138 | 69 | 69 | 135 |
| 0,567977331 | 138 | 69 | 69 | 135 |
| 0,411431131 | 138 | 69 | 69 | 135 |
| 0,717664931 | 138 | 69 | 69 | 135 |
| 0,593505803 | 138 | 69 | 69 | 135 |
| 0,699155646 | 138 | 69 | 69 | 135 |
| 0,447566081 | 138 | 69 | 69 | 135 |
| 0,64223169 | 138 | 69 | 69 | 135 |
| 0,861080829 | 138 | 69 | 69 | 135 |
| 0,440059502 | 138 | 69 | 69 | 135 |
| 0,388818785 | 138 | 69 | 69 | 135 |
| 0,253588816 | 138 | 69 | 69 | 135 |
| 0,328595941 | 138 | 69 | 69 | 135 |
| 0,342233666 | 138 | 69 | 69 | 135 |
| 0,680483795 | 138 | 69 | 69 | 135 |
| 0,57295456 | 138 | 69 | 69 | 135 |
| 0,744146702 | 138 | 69 | 69 | 135 |
| 0,460630491 | 138 | 69 | 69 | 135 |
| 0,355268775 | 138 | 69 | 69 | 135 |

| | | | | |
|---|---|---|---|---|
| 0,450855538 | 138 | 69 | 69 | 135 |
| 1,081152945 | 138 | 69 | 69 | 135 |
| 0,454968921 | 138 | 69 | 69 | 135 |
| 0,565033986 | 138 | 69 | 69 | 135 |
| 0,451361617 | 138 | 69 | 69 | 135 |
| 0,305423906 | 138 | 69 | 69 | 135 |
| 0,543296869 | 138 | 69 | 69 | 135 |
| 0,369212672 | 138 | 69 | 69 | 135 |
| 0,483217797 | 138 | 69 | 69 | 135 |
| 0,689091407 | 138 | 69 | 69 | 135 |
| 0,385674316 | 138 | 69 | 69 | 135 |
| 0,396151786 | 138 | 69 | 69 | 135 |
| 0,432285514 | 138 | 69 | 69 | 135 |
| 0,521192306 | 138 | 69 | 69 | 135 |
| 0,509543147 | 138 | 69 | 69 | 135 |
| 0,289687108 | 138 | 69 | 69 | 135 |
| 0,666849379 | 138 | 69 | 69 | 135 |
| 0,384403781 | 138 | 69 | 69 | 135 |
| 0,70624318 | 138 | 69 | 69 | 135 |
| 0,494936952 | 138 | 69 | 69 | 135 |
| 0,273422672 | 138 | 69 | 69 | 135 |
| 0,211861275 | 138 | 69 | 69 | 135 |
| 0,475248003 | 138 | 69 | 69 | 135 |
| 0,477358094 | 138 | 69 | 69 | 135 |
| 0,652421577 | 138 | 69 | 69 | 135 |
| 0,709830977 | 138 | 69 | 69 | 135 |
| 0,61481296 | 138 | 69 | 69 | 135 |
| 0,263627762 | 138 | 69 | 69 | 135 |
| 0,512706308 | 138 | 69 | 69 | 135 |
| 0,668692983 | 138 | 69 | 69 | 135 |
| 0,571837571 | 138 | 69 | 69 | 135 |
| 0,464989955 | 138 | 69 | 69 | 135 |
| 0,294662208 | 138 | 69 | 69 | 135 |
| 0,293414677 | 138 | 69 | 69 | 135 |
| 0,560817062 | 138 | 69 | 69 | 135 |
| 0,440511561 | 138 | 69 | 69 | 135 |
| 0,365195506 | 138 | 69 | 69 | 135 |
| 0,418859285 | 138 | 69 | 69 | 135 |
| 0,53134894 | 138 | 69 | 69 | 135 |
| 0,474705783 | 138 | 69 | 69 | 135 |
| 0,432679205 | 138 | 69 | 69 | 135 |
| 0,585951076 | 138 | 69 | 69 | 135 |
| 0,666171881 | 138 | 69 | 69 | 135 |
| 0,656876514 | 138 | 69 | 69 | 135 |
| 0,528428933 | 138 | 69 | 69 | 135 |
| 0,439000804 | 138 | 69 | 69 | 135 |
| 0,426694367 | 138 | 69 | 69 | 135 |
| 0,720055592 | 138 | 69 | 69 | 135 |

| | | | | |
|---|---|---|---|---|
| 0,642446531 | 138 | 69 | 69 | 135 |
| 0,687885738 | 138 | 69 | 69 | 135 |
| 0,534261555 | 138 | 69 | 69 | 135 |
| 0,45074988 | 138 | 69 | 69 | 135 |
| 0,655397994 | 138 | 69 | 69 | 135 |
| 0,409665124 | 138 | 69 | 69 | 135 |
| 0,246893982 | 138 | 69 | 69 | 135 |
| 0,495978594 | 138 | 69 | 69 | 135 |
| 0,710061318 | 138 | 69 | 69 | 135 |
| 0,6076545 | 138 | 69 | 69 | 135 |
| 0,645111757 | 138 | 69 | 69 | 135 |
| 0,542702313 | 138 | 69 | 69 | 135 |
| 0,58279544 | 138 | 69 | 69 | 135 |
| 0,266293339 | 138 | 69 | 69 | 135 |
| 0,768391476 | 138 | 69 | 69 | 135 |
| 0,667665904 | 138 | 69 | 69 | 135 |
| 0,38111223 | 138 | 69 | 69 | 135 |
| 0,595414635 | 138 | 69 | 69 | 135 |
| 0,445620136 | 138 | 69 | 69 | 135 |
| 0,447217043 | 138 | 69 | 69 | 135 |
| 0,6450602 | 138 | 69 | 69 | 135 |
| 0,307815387 | 138 | 69 | 69 | 135 |
| 0,921345705 | 138 | 69 | 69 | 135 |
| 0,383063024 | 138 | 69 | 69 | 135 |
| 0,350236621 | 138 | 69 | 69 | 135 |
| 0,207001876 | 138 | 69 | 69 | 135 |
| 0,468167651 | 138 | 69 | 69 | 135 |
| 0,282344935 | 138 | 69 | 69 | 135 |
| 0,486445601 | 138 | 69 | 69 | 135 |
| 0,706124375 | 138 | 69 | 69 | 135 |
| 0,330029203 | 138 | 69 | 69 | 135 |
| 0,392697831 | 138 | 69 | 69 | 135 |
| 0,610065276 | 138 | 69 | 69 | 135 |
| 0,554024137 | 138 | 69 | 69 | 135 |
| 0,225700052 | 138 | 69 | 69 | 135 |
| 0,627527369 | 138 | 69 | 69 | 135 |
| 0,633895238 | 138 | 69 | 69 | 135 |
| 0,345988615 | 138 | 69 | 69 | 135 |
| 0,257566462 | 138 | 69 | 69 | 135 |
| 0,519897707 | 138 | 69 | 69 | 135 |
| 0,253243089 | 138 | 69 | 69 | 135 |
| 0,43208392 | 138 | 69 | 69 | 135 |
| 0,393826402 | 138 | 69 | 69 | 135 |
| 0,503508122 | 138 | 69 | 69 | 135 |
| 0,413343059 | 138 | 69 | 69 | 135 |
| 0,516784907 | 138 | 69 | 69 | 135 |
| 0,473506171 | 138 | 69 | 69 | 135 |
| 0,596917704 | 138 | 69 | 69 | 135 |

| | | | | |
|---|---|---|---|---|
| 0,657857537 | 138 | 69 | 69 | 135 |
| 0,521037797 | 138 | 69 | 69 | 135 |
| 0,410868809 | 138 | 69 | 69 | 135 |
| 0,377539787 | 138 | 69 | 69 | 135 |
| 0,482167636 | 138 | 69 | 69 | 135 |
| 0,501471411 | 138 | 69 | 69 | 135 |
| 0,523810879 | 138 | 69 | 69 | 135 |
| 0,305843949 | 138 | 69 | 69 | 135 |
| 0,209771442 | 138 | 69 | 69 | 135 |
| 0,314907386 | 138 | 69 | 69 | 135 |
| 0,216886788 | 138 | 69 | 69 | 135 |
| 0,475354975 | 138 | 69 | 69 | 135 |
| 0,386816545 | 138 | 69 | 69 | 135 |
| 0,673815672 | 138 | 69 | 69 | 135 |

| Test statistic | Model formula fit |
|---|---|
| -0,453690106 | y ~ Intervention + Sex |
| -0,623353512 | y ~ Intervention + Sex |
| -1,092649437 | y ~ Intervention + Sex |
| -0,560703291 | y ~ Intervention + Sex |
| -0,01491497 | y ~ Intervention + Sex |
| -0,950413232 | y ~ Intervention + Sex |
| -1,603331987 | y ~ Intervention + Sex |
| -2,023118555 | y ~ Intervention + Sex |
| -2,187059929 | y ~ Intervention + Sex |
| -0,047834348 | y ~ Intervention + Sex |
| -1,145873945 | y ~ Intervention + Sex |
| -1,796163474 | y ~ Intervention + Sex |
| -0,706431887 | y ~ Intervention + Sex |
| -1,22555141 | y ~ Intervention + Sex |
| -2,541871094 | y ~ Intervention + Sex |
| -0,590910759 | y ~ Intervention + Sex |
| 0,843213562 | y ~ Intervention + Sex |
| -0,680086455 | y ~ Intervention + Sex |
| -0,077668525 | y ~ Intervention + Sex |
| 0,782683846 | y ~ Intervention + Sex |
| 0,782758751 | y ~ Intervention + Sex |
| 1,31868594 | y ~ Intervention + Sex |
| 0,09654112 | y ~ Intervention + Sex |
| -0,348920631 | y ~ Intervention + Sex |
| 0,675674731 | y ~ Intervention + Sex |
| 1,552478966 | y ~ Intervention + Sex |
| -0,975399418 | y ~ Intervention + Sex |
| -1,690542741 | y ~ Intervention + Sex |
| -2,18494258 | y ~ Intervention + Sex |
| -0,389489039 | y ~ Intervention + Sex |
| 0,387214012 | y ~ Intervention + Sex |
| -2,032350022 | y ~ Intervention + Sex |
| -1,632383623 | y ~ Intervention + Sex |
| -1,783250999 | y ~ Intervention + Sex |
| -1,324090846 | y ~ Intervention + Sex |
| -0,383966884 | y ~ Intervention + Sex |
| -4,389643445 | y ~ Intervention + Sex |
| -0,44935143 | y ~ Intervention + Sex |
| 0,108142782 | y ~ Intervention + Sex |
| 0,15273946 | y ~ Intervention + Sex |
| -0,973419279 | y ~ Intervention + Sex |
| -0,632201923 | y ~ Intervention + Sex |
| -0,610084103 | y ~ Intervention + Sex |
| -0,799938459 | y ~ Intervention + Sex |
| -1,314994863 | y ~ Intervention + Sex |
| -1,138555607 | y ~ Intervention + Sex |
| -0,35711209 | y ~ Intervention + Sex |

| | |
|---|---|
| -1,898672535 | y ~ Intervention + Sex |
| -2,051294429 | y ~ Intervention + Sex |
| -1,543556596 | y ~ Intervention + Sex |
| 1,35686082 | y ~ Intervention + Sex |
| 0,078399036 | y ~ Intervention + Sex |
| -1,510346693 | y ~ Intervention + Sex |
| -2,877970318 | y ~ Intervention + Sex |
| -1,995970839 | y ~ Intervention + Sex |
| -1,592782198 | y ~ Intervention + Sex |
| -0,040505932 | y ~ Intervention + Sex |
| 0,461528435 | y ~ Intervention + Sex |
| -0,119128245 | y ~ Intervention + Sex |
| 0,510897675 | y ~ Intervention + Sex |
| -1,776945581 | y ~ Intervention + Sex |
| -1,44963176 | y ~ Intervention + Sex |
| -0,266472198 | y ~ Intervention + Sex |
| -1,276731207 | y ~ Intervention + Sex |
| -1,621675014 | y ~ Intervention + Sex |
| -1,196121566 | y ~ Intervention + Sex |
| 0,488104724 | y ~ Intervention + Sex |
| -0,105573605 | y ~ Intervention + Sex |
| 0,300425525 | y ~ Intervention + Sex |
| -1,60950456 | y ~ Intervention + Sex |
| 0,954151067 | y ~ Intervention + Sex |
| 0,550363898 | y ~ Intervention + Sex |
| -0,595221906 | y ~ Intervention + Sex |
| 1,887693102 | y ~ Intervention + Sex |
| -0,188854159 | y ~ Intervention + Sex |
| -2,46638645 | y ~ Intervention + Sex |
| 0,354366776 | y ~ Intervention + Sex |
| 0,289742709 | y ~ Intervention + Sex |
| -1,172994118 | y ~ Intervention + Sex |
| -0,433933764 | y ~ Intervention + Sex |
| -0,667661554 | y ~ Intervention + Sex |
| -1,394640724 | y ~ Intervention + Sex |
| -1,604847872 | y ~ Intervention + Sex |
| -1,83430437 | y ~ Intervention + Sex |
| -0,669586054 | y ~ Intervention + Sex |
| 0,69697237 | y ~ Intervention + Sex |
| -0,294114705 | y ~ Intervention + Sex |
| -1,071255711 | y ~ Intervention + Sex |
| -1,411337125 | y ~ Intervention + Sex |
| -0,052208915 | y ~ Intervention + Sex |
| -1,46087778 | y ~ Intervention + Sex |
| 0,388882417 | y ~ Intervention + Sex |
| -1,241473528 | y ~ Intervention + Sex |
| -1,474445485 | y ~ Intervention + Sex |
| -0,002290489 | y ~ Intervention + Sex |

| | |
|---|---|
| -0,484049682 | y ~ Intervention + Sex |
| -1,035232361 | y ~ Intervention + Sex |
| -0,985480149 | y ~ Intervention + Sex |
| -2,822588195 | y ~ Intervention + Sex |
| -1,642675189 | y ~ Intervention + Sex |
| 0,216024136 | y ~ Intervention + Sex |
| -0,136689879 | y ~ Intervention + Sex |
| -0,972349509 | y ~ Intervention + Sex |
| 0,810433982 | y ~ Intervention + Sex |
| -0,843572575 | y ~ Intervention + Sex |
| -0,177304993 | y ~ Intervention + Sex |
| -1,205841757 | y ~ Intervention + Sex |
| 0,209513254 | y ~ Intervention + Sex |
| 0,633724289 | y ~ Intervention + Sex |
| 0,77862127 | y ~ Intervention + Sex |
| -0,061228116 | y ~ Intervention + Sex |
| -0,925080963 | y ~ Intervention + Sex |
| -0,821841649 | y ~ Intervention + Sex |
| 0,52827027 | y ~ Intervention + Sex |
| -2,632789969 | y ~ Intervention + Sex |
| 0,47459302 | y ~ Intervention + Sex |
| 1,113333314 | y ~ Intervention + Sex |
| -1,205194587 | y ~ Intervention + Sex |
| -0,418209717 | y ~ Intervention + Sex |
| -0,161510921 | y ~ Intervention + Sex |
| -0,310888133 | y ~ Intervention + Sex |
| -0,820731802 | y ~ Intervention + Sex |
| 1,14302092 | y ~ Intervention + Sex |
| -0,198498084 | y ~ Intervention + Sex |
| -0,020145875 | y ~ Intervention + Sex |
| 0,757831357 | y ~ Intervention + Sex |
| 0,369145577 | y ~ Intervention + Sex |
| 0,215305976 | y ~ Intervention + Sex |
| -0,015039631 | y ~ Intervention + Sex |
| -1,762673685 | y ~ Intervention + Sex |
| 0,452597605 | y ~ Intervention + Sex |
| -0,440328687 | y ~ Intervention + Sex |
| -0,955360915 | y ~ Intervention + Sex |
| -0,592672864 | y ~ Intervention + Sex |
| 2,381979462 | y ~ Intervention + Sex |
| -0,472645467 | y ~ Intervention + Sex |
| -0,614714813 | y ~ Intervention + Sex |
| -0,954242664 | y ~ Intervention + Sex |
| -1,784626407 | y ~ Intervention + Sex |
| -0,68559988 | y ~ Intervention + Sex |
| -0,868391823 | y ~ Intervention + Sex |
| -0,335281803 | y ~ Intervention + Sex |
| -2,032372352 | y ~ Intervention + Sex |

| | |
|---|---|
| 0,446739048 | y ~ Intervention + Sex |
| 0,780776818 | y ~ Intervention + Sex |
| 0,571623468 | y ~ Intervention + Sex |
| -0,258508593 | y ~ Intervention + Sex |
| 0,282962246 | y ~ Intervention + Sex |
| -0,364769577 | y ~ Intervention + Sex |
| -0,227383064 | y ~ Intervention + Sex |
| -0,357496453 | y ~ Intervention + Sex |
| -1,629674737 | y ~ Intervention + Sex |
| -0,689836964 | y ~ Intervention + Sex |
| 1,007582305 | y ~ Intervention + Sex |
| 0,890936529 | y ~ Intervention + Sex |
| 2,046221021 | y ~ Intervention + Sex |
| 0,593852087 | y ~ Intervention + Sex |
| 0,535885685 | y ~ Intervention + Sex |
| 1,459061391 | y ~ Intervention + Sex |
| -0,266255234 | y ~ Intervention + Sex |
| 0,165891451 | y ~ Intervention + Sex |
| -0,910201134 | y ~ Intervention + Sex |
| -1,092416486 | y ~ Intervention + Sex |
| 0,825714456 | y ~ Intervention + Sex |
| -0,16170974 | y ~ Intervention + Sex |
| -1,13359796 | y ~ Intervention + Sex |
| 0,781470582 | y ~ Intervention + Sex |
| -1,791663958 | y ~ Intervention + Sex |
| -0,766419275 | y ~ Intervention + Sex |
| -0,653649893 | y ~ Intervention + Sex |
| -0,949778279 | y ~ Intervention + Sex |
| -2,067720822 | y ~ Intervention + Sex |
| 0,11603649 | y ~ Intervention + Sex |
| -0,316973771 | y ~ Intervention + Sex |
| 1,397680303 | y ~ Intervention + Sex |
| -1,043638544 | y ~ Intervention + Sex |
| 0,910922544 | y ~ Intervention + Sex |
| 0,082512359 | y ~ Intervention + Sex |
| -0,075874349 | y ~ Intervention + Sex |
| -0,408792897 | y ~ Intervention + Sex |
| -0,843699059 | y ~ Intervention + Sex |
| -0,181491708 | y ~ Intervention + Sex |
| -0,905370869 | y ~ Intervention + Sex |
| -0,757877322 | y ~ Intervention + Sex |
| -0,980300133 | y ~ Intervention + Sex |
| -3,051541193 | y ~ Intervention + Sex |
| -0,144186027 | y ~ Intervention + Sex |
| 0,577420987 | y ~ Intervention + Sex |
| -0,9944026 | y ~ Intervention + Sex |
| -0,093845876 | y ~ Intervention + Sex |
| 0,313196155 | y ~ Intervention + Sex |

| | |
|---|---|
| 0,188214694 | y ~ Intervention + Sex |
| -0,421562898 | y ~ Intervention + Sex |
| 1,734572045 | y ~ Intervention + Sex |
| 0,381551932 | y ~ Intervention + Sex |
| 1,179514103 | y ~ Intervention + Sex |
| -0,780919406 | y ~ Intervention + Sex |
| -0,059640086 | y ~ Intervention + Sex |
| -1,852408452 | y ~ Intervention + Sex |
| -0,279830228 | y ~ Intervention + Sex |
| -1,918050183 | y ~ Intervention + Sex |
| 0,687380076 | y ~ Intervention + Sex |
| -1,321316869 | y ~ Intervention + Sex |
| 0,177297635 | y ~ Intervention + Sex |
| -0,644770527 | y ~ Intervention + Sex |
| -0,425149695 | y ~ Intervention + Sex |
| 0,180996133 | y ~ Intervention + Sex |
| -0,881071196 | y ~ Intervention + Sex |
| -1,153182907 | y ~ Intervention + Sex |
| -0,849714427 | y ~ Intervention + Sex |
| -1,014804061 | y ~ Intervention + Sex |
| 0,013675843 | y ~ Intervention + Sex |
| -0,359310829 | y ~ Intervention + Sex |
| 0,332716633 | y ~ Intervention + Sex |
| 0,376765468 | y ~ Intervention + Sex |
| -0,008165121 | y ~ Intervention + Sex |
| -1,049948602 | y ~ Intervention + Sex |
| -0,091599674 | y ~ Intervention + Sex |
| 0,561661284 | y ~ Intervention + Sex |
| 0,856198089 | y ~ Intervention + Sex |
| 0,341986768 | y ~ Intervention + Sex |
| -0,61487969 | y ~ Intervention + Sex |
| -2,053065611 | y ~ Intervention + Sex |
| 0,822923548 | y ~ Intervention + Sex |
| -0,308041865 | y ~ Intervention + Sex |
| -0,110332038 | y ~ Intervention + Sex |
| 0,404161182 | y ~ Intervention + Sex |
| -0,143048115 | y ~ Intervention + Sex |
| 2,162259888 | y ~ Intervention + Sex |
| 0,814103004 | y ~ Intervention + Sex |
| 0,643530704 | y ~ Intervention + Sex |
| 1,195559961 | y ~ Intervention + Sex |
| 0,225167845 | y ~ Intervention + Sex |
| 1,037882137 | y ~ Intervention + Sex |
| 0,235034725 | y ~ Intervention + Sex |
| -0,876493968 | y ~ Intervention + Sex |
| -0,971438058 | y ~ Intervention + Sex |
| -1,393578434 | y ~ Intervention + Sex |
| 0,445758231 | y ~ Intervention + Sex |

| | |
|---|---|
| -0,699102717 | y ~ Intervention + Sex |
| -1,545807322 | y ~ Intervention + Sex |
| -0,013545584 | y ~ Intervention + Sex |
| 0,031135749 | y ~ Intervention + Sex |
| -0,960805412 | y ~ Intervention + Sex |
| -1,198218308 | y ~ Intervention + Sex |
| -0,58865186 | y ~ Intervention + Sex |
| 0,753864193 | y ~ Intervention + Sex |
| 2,125563278 | y ~ Intervention + Sex |
| -0,368393819 | y ~ Intervention + Sex |
| -0,971454994 | y ~ Intervention + Sex |
| -0,609674568 | y ~ Intervention + Sex |
| 0,187733827 | y ~ Intervention + Sex |
| -0,207019197 | y ~ Intervention + Sex |
| -1,142519426 | y ~ Intervention + Sex |
| 0,338335157 | y ~ Intervention + Sex |
| -1,901171588 | y ~ Intervention + Sex |
| 0,087127182 | y ~ Intervention + Sex |
| 0,639959154 | y ~ Intervention + Sex |
| -1,232953124 | y ~ Intervention + Sex |
| 0,304329888 | y ~ Intervention + Sex |
| 0,523917351 | y ~ Intervention + Sex |
| -0,078388592 | y ~ Intervention + Sex |
| 1,067432487 | y ~ Intervention + Sex |
| 0,872796393 | y ~ Intervention + Sex |
| 0,665142942 | y ~ Intervention + Sex |
| 0,726351828 | y ~ Intervention + Sex |
| -0,083579553 | y ~ Intervention + Sex |
| -1,649449405 | y ~ Intervention + Sex |
| -0,813202894 | y ~ Intervention + Sex |
| -0,847717567 | y ~ Intervention + Sex |
| -0,900230198 | y ~ Intervention + Sex |
| 0,77168116 | y ~ Intervention + Sex |
| -0,447293066 | y ~ Intervention + Sex |
| 0,311006863 | y ~ Intervention + Sex |
| -1,393365999 | y ~ Intervention + Sex |
| 0,2673613 | y ~ Intervention + Sex |
| 0,524933098 | y ~ Intervention + Sex |
| -0,626378334 | y ~ Intervention + Sex |
| -0,204630362 | y ~ Intervention + Sex |
| 0,730208337 | y ~ Intervention + Sex |
| 1,020674131 | y ~ Intervention + Sex |
| 1,044430035 | y ~ Intervention + Sex |
| 0,190688921 | y ~ Intervention + Sex |
| -0,938531467 | y ~ Intervention + Sex |
| -0,542898195 | y ~ Intervention + Sex |
| 1,256189216 | y ~ Intervention + Sex |
| 0,421609834 | y ~ Intervention + Sex |

| | |
|---|---|
| -0,961426695 | y ~ Intervention + Sex |
| -0,748291166 | y ~ Intervention + Sex |
| -0,073154226 | y ~ Intervention + Sex |
| -0,210499988 | y ~ Intervention + Sex |
| -1,616843219 | y ~ Intervention + Sex |
| -1,868168513 | y ~ Intervention + Sex |
| 1,137811467 | y ~ Intervention + Sex |
| -0,79078363 | y ~ Intervention + Sex |
| -0,449140255 | y ~ Intervention + Sex |
| 0,590792433 | y ~ Intervention + Sex |
| -0,17980978 | y ~ Intervention + Sex |
| -0,528428557 | y ~ Intervention + Sex |
| -0,797666964 | y ~ Intervention + Sex |
| 0,508503752 | y ~ Intervention + Sex |
| -1,29756439 | y ~ Intervention + Sex |
| 0,0894839 | y ~ Intervention + Sex |
| -0,473925947 | y ~ Intervention + Sex |
| 1,543956136 | y ~ Intervention + Sex |
| 1,60786765 | y ~ Intervention + Sex |
| -0,85494643 | y ~ Intervention + Sex |
| -1,169859768 | y ~ Intervention + Sex |
| 1,582742043 | y ~ Intervention + Sex |
| -0,84005246 | y ~ Intervention + Sex |
| 1,480781498 | y ~ Intervention + Sex |
| 1,37911871 | y ~ Intervention + Sex |
| 0,806254392 | y ~ Intervention + Sex |
| -0,877539341 | y ~ Intervention + Sex |
| -0,415363269 | y ~ Intervention + Sex |
| -1,840790482 | y ~ Intervention + Sex |
| -0,624548365 | y ~ Intervention + Sex |
| -0,653626411 | y ~ Intervention + Sex |
| -0,69083783 | y ~ Intervention + Sex |
| -1,809708523 | y ~ Intervention + Sex |
| 0,33253798 | y ~ Intervention + Sex |
| -0,15127037 | y ~ Intervention + Sex |
| 0,101366107 | y ~ Intervention + Sex |
| -0,070721371 | y ~ Intervention + Sex |
| 1,943099921 | y ~ Intervention + Sex |
| -1,167143665 | y ~ Intervention + Sex |
| -0,970222196 | y ~ Intervention + Sex |
| -1,994172503 | y ~ Intervention + Sex |
| 0,828849679 | y ~ Intervention + Sex |
| 0,345609542 | y ~ Intervention + Sex |
| 0,378806875 | y ~ Intervention + Sex |
| 0,237611877 | y ~ Intervention + Sex |
| 0,000367589 | y ~ Intervention + Sex |
| 1,17377161 | y ~ Intervention + Sex |
| 0,215900728 | y ~ Intervention + Sex |

| | |
|---|---|
| -0,697897381 | y ~ Intervention + Sex |
| -0,183267115 | y ~ Intervention + Sex |
| -0,710630708 | y ~ Intervention + Sex |
| 0,66221551 | y ~ Intervention + Sex |
| 0,465333702 | y ~ Intervention + Sex |
| -0,130368332 | y ~ Intervention + Sex |
| -0,480727759 | y ~ Intervention + Sex |
| -0,594764194 | y ~ Intervention + Sex |
| 0,284185655 | y ~ Intervention + Sex |
| -0,488908383 | y ~ Intervention + Sex |
| -2,001781024 | y ~ Intervention + Sex |
| -2,26977477 | y ~ Intervention + Sex |
| 1,033057113 | y ~ Intervention + Sex |
| -0,962642267 | y ~ Intervention + Sex |
| -0,065503287 | y ~ Intervention + Sex |
| 0,362556163 | y ~ Intervention + Sex |
| -1,071639236 | y ~ Intervention + Sex |
| -0,259853418 | y ~ Intervention + Sex |
| 2,653526146 | y ~ Intervention + Sex |
| 0,761276139 | y ~ Intervention + Sex |
| -2,527978223 | y ~ Intervention + Sex |
| -0,072495721 | y ~ Intervention + Sex |
| -0,530591949 | y ~ Intervention + Sex |
| -0,414494774 | y ~ Intervention + Sex |
| 0,270392976 | y ~ Intervention + Sex |
| -0,282494817 | y ~ Intervention + Sex |
| 0,939004771 | y ~ Intervention + Sex |
| 0,238448115 | y ~ Intervention + Sex |
| -0,406108138 | y ~ Intervention + Sex |
| -0,914147496 | y ~ Intervention + Sex |
| 0,356896531 | y ~ Intervention + Sex |
| -1,065892151 | y ~ Intervention + Sex |
| 0,070037372 | y ~ Intervention + Sex |
| 0,456540432 | y ~ Intervention + Sex |
| -0,193872537 | y ~ Intervention + Sex |
| 0,968413884 | y ~ Intervention + Sex |
| 0,371333789 | y ~ Intervention + Sex |
| 0,394895787 | y ~ Intervention + Sex |
| 0,355508697 | y ~ Intervention + Sex |
| 0,229764588 | y ~ Intervention + Sex |
| -0,741015467 | y ~ Intervention + Sex |
| -1,302686995 | y ~ Intervention + Sex |
| 1,033831109 | y ~ Intervention + Sex |
| 0,19961844 | y ~ Intervention + Sex |
| 0,227489605 | y ~ Intervention + Sex |
| -0,194905322 | y ~ Intervention + Sex |
| -2,076588891 | y ~ Intervention + Sex |
| -0,099456024 | y ~ Intervention + Sex |

| | |
|---|---|
| -0,808367344 | y ~ Intervention + Sex |
| 0,575307268 | y ~ Intervention + Sex |
| 1,683622347 | y ~ Intervention + Sex |
| -0,535658665 | y ~ Intervention + Sex |
| 0,789144362 | y ~ Intervention + Sex |
| -0,53653488 | y ~ Intervention + Sex |
| -0,795430224 | y ~ Intervention + Sex |
| 0,978908141 | y ~ Intervention + Sex |
| 0,303816763 | y ~ Intervention + Sex |
| 1,686068087 | y ~ Intervention + Sex |
| 2,017705839 | y ~ Intervention + Sex |
| -1,05636466 | y ~ Intervention + Sex |
| 0,160086013 | y ~ Intervention + Sex |
| 1,241996367 | y ~ Intervention + Sex |
| -0,811723487 | y ~ Intervention + Sex |
| 0,617315044 | y ~ Intervention + Sex |
| 1,351162557 | y ~ Intervention + Sex |
| -2,461757575 | y ~ Intervention + Sex |
| -0,351353736 | y ~ Intervention + Sex |
| 0,730903243 | y ~ Intervention + Sex |
| -0,222869971 | y ~ Intervention + Sex |
| 0,790064521 | y ~ Intervention + Sex |
| -0,320904336 | y ~ Intervention + Sex |
| -0,274092245 | y ~ Intervention + Sex |
| 0,335684971 | y ~ Intervention + Sex |
| 0,078547132 | y ~ Intervention + Sex |
| 0,724884429 | y ~ Intervention + Sex |
| -0,479060212 | y ~ Intervention + Sex |
| -0,186295672 | y ~ Intervention + Sex |
| -0,569596442 | y ~ Intervention + Sex |
| 0,567056215 | y ~ Intervention + Sex |
| -1,068541414 | y ~ Intervention + Sex |
| 1,239327985 | y ~ Intervention + Sex |
| -1,542942792 | y ~ Intervention + Sex |
| 2,54813429 | y ~ Intervention + Sex |
| 0,557583905 | y ~ Intervention + Sex |
| 2,093159149 | y ~ Intervention + Sex |
| -1,278825291 | y ~ Intervention + Sex |
| -0,142129732 | y ~ Intervention + Sex |
| 0,104718031 | y ~ Intervention + Sex |
| -1,495620447 | y ~ Intervention + Sex |
| -1,452468104 | y ~ Intervention + Sex |
| 1,49838864 | y ~ Intervention + Sex |
| -1,713079361 | y ~ Intervention + Sex |
| 1,715927222 | y ~ Intervention + Sex |
| 0,126580272 | y ~ Intervention + Sex |
| 0,072819461 | y ~ Intervention + Sex |
| 1,016652651 | y ~ Intervention + Sex |

| | |
|---|---|
| -0,136723378 | y ~ Intervention + Sex |
| -0,087453385 | y ~ Intervention + Sex |
| -0,79169556 | y ~ Intervention + Sex |
| -0,580921053 | y ~ Intervention + Sex |
| -0,334581832 | y ~ Intervention + Sex |
| -0,047003787 | y ~ Intervention + Sex |
| -0,036160558 | y ~ Intervention + Sex |
| -0,450495365 | y ~ Intervention + Sex |
| 2,158056491 | y ~ Intervention + Sex |
| 0,707368238 | y ~ Intervention + Sex |
| 0,810090635 | y ~ Intervention + Sex |
| 1,658781519 | y ~ Intervention + Sex |
| -0,49360358 | y ~ Intervention + Sex |
| 0,603890124 | y ~ Intervention + Sex |
| -0,055625839 | y ~ Intervention + Sex |
| 1,295616549 | y ~ Intervention + Sex |
| -1,29028336 | y ~ Intervention + Sex |
| 0,098833193 | y ~ Intervention + Sex |
| 1,071065162 | y ~ Intervention + Sex |
| 1,167957876 | y ~ Intervention + Sex |
| 1,457831741 | y ~ Intervention + Sex |
| 1,003791248 | y ~ Intervention + Sex |
| -0,405695676 | y ~ Intervention + Sex |
| -0,558622764 | y ~ Intervention + Sex |
| 1,024155684 | y ~ Intervention + Sex |
| 1,522872312 | y ~ Intervention + Sex |
| 1,014432758 | y ~ Intervention + Sex |
| 1,641049009 | y ~ Intervention + Sex |
| 0,223566653 | y ~ Intervention + Sex |
| 0,776273523 | y ~ Intervention + Sex |
| -0,572095891 | y ~ Intervention + Sex |
| -0,283474846 | y ~ Intervention + Sex |
| 0,754954159 | y ~ Intervention + Sex |
| -0,559005894 | y ~ Intervention + Sex |
| 0,031917704 | y ~ Intervention + Sex |
| -1,354677457 | y ~ Intervention + Sex |
| 0,7525176 | y ~ Intervention + Sex |
| 0,919312315 | y ~ Intervention + Sex |
| 1,529391825 | y ~ Intervention + Sex |
| -0,533562325 | y ~ Intervention + Sex |
| 0,427064149 | y ~ Intervention + Sex |
| -0,476565122 | y ~ Intervention + Sex |
| -0,017255346 | y ~ Intervention + Sex |
| 0,532990668 | y ~ Intervention + Sex |
| -0,610659285 | y ~ Intervention + Sex |
| -0,096019023 | y ~ Intervention + Sex |
| 1,770432089 | y ~ Intervention + Sex |
| -0,492808611 | y ~ Intervention + Sex |

| | |
|---|---|
| -0,71762324 | y ~ Intervention + Sex |
| 1,028156067 | y ~ Intervention + Sex |
| 1,127024599 | y ~ Intervention + Sex |
| 0,126764363 | y ~ Intervention + Sex |
| 0,173541867 | y ~ Intervention + Sex |
| -0,721836184 | y ~ Intervention + Sex |
| 0,87807467 | y ~ Intervention + Sex |
| 1,482958025 | y ~ Intervention + Sex |
| -1,440906752 | y ~ Intervention + Sex |
| 0,44061994 | y ~ Intervention + Sex |
| 0,135562406 | y ~ Intervention + Sex |
| 0,593861596 | y ~ Intervention + Sex |
| -0,010780111 | y ~ Intervention + Sex |
| -0,513668302 | y ~ Intervention + Sex |
| 0,727801298 | y ~ Intervention + Sex |
| 0,621534567 | y ~ Intervention + Sex |
| -1,452893446 | y ~ Intervention + Sex |
| -0,32615403 | y ~ Intervention + Sex |
| 0,860528999 | y ~ Intervention + Sex |
| 0,66721854 | y ~ Intervention + Sex |
| -0,956113343 | y ~ Intervention + Sex |
| 0,087107725 | y ~ Intervention + Sex |
| -0,286624699 | y ~ Intervention + Sex |
| 1,074480442 | y ~ Intervention + Sex |
| -1,015367271 | y ~ Intervention + Sex |
| 0,847954137 | y ~ Intervention + Sex |
| -0,272484237 | y ~ Intervention + Sex |
| 1,73488479 | y ~ Intervention + Sex |
| 1,296517027 | y ~ Intervention + Sex |
| 1,486307029 | y ~ Intervention + Sex |
| 1,063344679 | y ~ Intervention + Sex |
| -0,362293455 | y ~ Intervention + Sex |
| -0,489980351 | y ~ Intervention + Sex |
| -0,376062901 | y ~ Intervention + Sex |
| 0,543071667 | y ~ Intervention + Sex |
| -0,551354744 | y ~ Intervention + Sex |
| 0,163978865 | y ~ Intervention + Sex |
| 0,312324912 | y ~ Intervention + Sex |
| 0,472054132 | y ~ Intervention + Sex |
| -0,476578456 | y ~ Intervention + Sex |
| 0,365797564 | y ~ Intervention + Sex |
| -0,864884248 | y ~ Intervention + Sex |
| 0,447208828 | y ~ Intervention + Sex |
| -2,006784861 | y ~ Intervention + Sex |
| -0,636518895 | y ~ Intervention + Sex |
| 0,351994284 | y ~ Intervention + Sex |
| 0,426595131 | y ~ Intervention + Sex |
| 2,483868575 | y ~ Intervention + Sex |

| | |
|---|---|
| 0,331614708 | y ~ Intervention + Sex |
| 0,480989493 | y ~ Intervention + Sex |
| 2,388463295 | y ~ Intervention + Sex |
| 0,414040845 | y ~ Intervention + Sex |
| -0,04571396 | y ~ Intervention + Sex |
| 1,038774975 | y ~ Intervention + Sex |
| -0,982725475 | y ~ Intervention + Sex |
| 0,66103187 | y ~ Intervention + Sex |
| 0,902489619 | y ~ Intervention + Sex |
| 0,432890324 | y ~ Intervention + Sex |
| 1,075133395 | y ~ Intervention + Sex |
| 0,430207702 | y ~ Intervention + Sex |
| -0,303610708 | y ~ Intervention + Sex |
| -0,504922704 | y ~ Intervention + Sex |
| -0,571726043 | y ~ Intervention + Sex |
| -1,401245811 | y ~ Intervention + Sex |
| 0,681555981 | y ~ Intervention + Sex |
| 0,656800338 | y ~ Intervention + Sex |
| 0,867151485 | y ~ Intervention + Sex |
| 0,277944387 | y ~ Intervention + Sex |
| -0,846071714 | y ~ Intervention + Sex |
| 0,493477624 | y ~ Intervention + Sex |
| -0,259241557 | y ~ Intervention + Sex |
| -0,186256352 | y ~ Intervention + Sex |
| 0,76173297 | y ~ Intervention + Sex |
| -0,633081876 | y ~ Intervention + Sex |
| 0,082728181 | y ~ Intervention + Sex |
| 0,113820369 | y ~ Intervention + Sex |
| 1,477726365 | y ~ Intervention + Sex |
| 0,983995039 | y ~ Intervention + Sex |
| -0,201539225 | y ~ Intervention + Sex |
| -1,293065659 | y ~ Intervention + Sex |
| -3,075603003 | y ~ Intervention + Sex |
| -0,565611667 | y ~ Intervention + Sex |
| -0,110392594 | y ~ Intervention + Sex |
| -1,851062892 | y ~ Intervention + Sex |
| 0,61723415 | y ~ Intervention + Sex |
| 0,748671963 | y ~ Intervention + Sex |
| 0,17574544 | y ~ Intervention + Sex |
| 0,895692117 | y ~ Intervention + Sex |
| 0,500178975 | y ~ Intervention + Sex |
| -0,841703406 | y ~ Intervention + Sex |
| 0,009557408 | y ~ Intervention + Sex |
| 2,444831196 | y ~ Intervention + Sex |
| -1,167140167 | y ~ Intervention + Sex |
| -0,930802031 | y ~ Intervention + Sex |
| 0,855913436 | y ~ Intervention + Sex |
| -2,476001563 | y ~ Intervention + Sex |

| | |
|---|---|
| 0,38301589 | y ~ Intervention + Sex |
| 0,137809452 | y ~ Intervention + Sex |
| -0,176060907 | y ~ Intervention + Sex |
| 1,328514138 | y ~ Intervention + Sex |
| 0,842529914 | y ~ Intervention + Sex |
| -0,980105941 | y ~ Intervention + Sex |
| -0,044057154 | y ~ Intervention + Sex |
| -0,403921514 | y ~ Intervention + Sex |
| -1,861218271 | y ~ Intervention + Sex |
| 0,158203109 | y ~ Intervention + Sex |
| 0,922128669 | y ~ Intervention + Sex |
| -0,023502833 | y ~ Intervention + Sex |
| -0,915014365 | y ~ Intervention + Sex |
| 0,4118458 | y ~ Intervention + Sex |
| 0,294956183 | y ~ Intervention + Sex |
| -0,418015924 | y ~ Intervention + Sex |
| -2,04373671 | y ~ Intervention + Sex |
| -1,119615016 | y ~ Intervention + Sex |
| -0,429342252 | y ~ Intervention + Sex |
| -0,991746664 | y ~ Intervention + Sex |
| -0,677098856 | y ~ Intervention + Sex |
| 0,700724379 | y ~ Intervention + Sex |
| 0,154738576 | y ~ Intervention + Sex |
| 0,892917563 | y ~ Intervention + Sex |
| -0,887520603 | y ~ Intervention + Sex |
| -1,227900225 | y ~ Intervention + Sex |
| -0,239851034 | y ~ Intervention + Sex |
| 0,713322465 | y ~ Intervention + Sex |
| -2,095283619 | y ~ Intervention + Sex |
| 0,920600075 | y ~ Intervention + Sex |
| 2,369285595 | y ~ Intervention + Sex |
| -0,021287512 | y ~ Intervention + Sex |
| 0,454848744 | y ~ Intervention + Sex |
| -3,100975922 | y ~ Intervention + Sex |
| 1,402782958 | y ~ Intervention + Sex |
| -1,678957727 | y ~ Intervention + Sex |
| -0,83672253 | y ~ Intervention + Sex |
| -1,236982743 | y ~ Intervention + Sex |
| -0,089243355 | y ~ Intervention + Sex |
| -1,432277325 | y ~ Intervention + Sex |
| -2,044927644 | y ~ Intervention + Sex |
| -1,564364497 | y ~ Intervention + Sex |
| -0,006177217 | y ~ Intervention + Sex |
| -2,6592619 | y ~ Intervention + Sex |
| -0,870256781 | y ~ Intervention + Sex |
| 1,321010406 | y ~ Intervention + Sex |
| -1,279758397 | y ~ Intervention + Sex |
| 0,85758222 | y ~ Intervention + Sex |

| | |
|---|---|
| -0,849655259 | y ~ Intervention + Sex |
| 0,68409369 | y ~ Intervention + Sex |
| -1,139161148 | y ~ Intervention + Sex |
| 1,106871827 | y ~ Intervention + Sex |
| 0,287033112 | y ~ Intervention + Sex |
| -0,56189547 | y ~ Intervention + Sex |
| -1,554168196 | y ~ Intervention + Sex |
| -0,935804654 | y ~ Intervention + Sex |
| -1,905934635 | y ~ Intervention + Sex |
| 0,530253698 | y ~ Intervention + Sex |
| 0,264211009 | y ~ Intervention + Sex |
| -0,934575294 | y ~ Intervention + Sex |
| 0,803719884 | y ~ Intervention + Sex |
| -0,759673668 | y ~ Intervention + Sex |
| -0,033607406 | y ~ Intervention + Sex |
| 1,302940144 | y ~ Intervention + Sex |
| -1,054132421 | y ~ Intervention + Sex |
| -0,70595065 | y ~ Intervention + Sex |
| -0,231449906 | y ~ Intervention + Sex |
| -0,288248991 | y ~ Intervention + Sex |
| -2,146870263 | y ~ Intervention + Sex |
| 0,519996714 | y ~ Intervention + Sex |
| 0,488804377 | y ~ Intervention + Sex |
| -0,263550266 | y ~ Intervention + Sex |
| -1,514109586 | y ~ Intervention + Sex |
| -0,196307684 | y ~ Intervention + Sex |
| -2,359604197 | y ~ Intervention + Sex |
| -1,434705301 | y ~ Intervention + Sex |
| 0,561283168 | y ~ Intervention + Sex |
| 1,545365541 | y ~ Intervention + Sex |
| 1,159762525 | y ~ Intervention + Sex |
| 0,354028453 | y ~ Intervention + Sex |
| -0,38004088 | y ~ Intervention + Sex |
| 0,27258543 | y ~ Intervention + Sex |
| -0,011690691 | y ~ Intervention + Sex |
| -2,360633789 | y ~ Intervention + Sex |
| -0,078591338 | y ~ Intervention + Sex |
| -1,479595809 | y ~ Intervention + Sex |
| 0,469893812 | y ~ Intervention + Sex |
| -2,543566873 | y ~ Intervention + Sex |
| -0,293812779 | y ~ Intervention + Sex |
| -0,725548478 | y ~ Intervention + Sex |
| 0,675521315 | y ~ Intervention + Sex |
| -0,007141141 | y ~ Intervention + Sex |
| -0,932828105 | y ~ Intervention + Sex |
| -0,895464093 | y ~ Intervention + Sex |
| -0,590527341 | y ~ Intervention + Sex |
| -0,529366006 | y ~ Intervention + Sex |

| | |
|---|---|
| -0,812026661 | y ~ Intervention + Sex |
| 0,752180001 | y ~ Intervention + Sex |
| -0,094692012 | y ~ Intervention + Sex |
| 0,605097056 | y ~ Intervention + Sex |
| 0,102304962 | y ~ Intervention + Sex |
| -1,194235313 | y ~ Intervention + Sex |
| -0,989003279 | y ~ Intervention + Sex |
| -1,759551766 | y ~ Intervention + Sex |
| -1,909394042 | y ~ Intervention + Sex |
| 0,367229042 | y ~ Intervention + Sex |
| 0,820129773 | y ~ Intervention + Sex |
| -1,481174276 | y ~ Intervention + Sex |
| 0,022364258 | y ~ Intervention + Sex |
| -1,494067956 | y ~ Intervention + Sex |
| -1,19456952 | y ~ Intervention + Sex |
| 0,478301429 | y ~ Intervention + Sex |
| -0,996512833 | y ~ Intervention + Sex |
| -0,37225826 | y ~ Intervention + Sex |
| -2,404256167 | y ~ Intervention + Sex |
| -1,59176563 | y ~ Intervention + Sex |
| 0,256995596 | y ~ Intervention + Sex |
| -0,100707452 | y ~ Intervention + Sex |
| 0,734498096 | y ~ Intervention + Sex |
| -2,545845435 | y ~ Intervention + Sex |
| 0,832774595 | y ~ Intervention + Sex |
| -2,142243157 | y ~ Intervention + Sex |
| -0,462514096 | y ~ Intervention + Sex |
| -0,746864592 | y ~ Intervention + Sex |
| 0,735838766 | y ~ Intervention + Sex |
| 0,096747196 | y ~ Intervention + Sex |
| -1,820839213 | y ~ Intervention + Sex |
| -0,575132158 | y ~ Intervention + Sex |
| 0,226947221 | y ~ Intervention + Sex |
| -0,608368931 | y ~ Intervention + Sex |
| -0,422350599 | y ~ Intervention + Sex |
| -0,950332056 | y ~ Intervention + Sex |
| -1,293130254 | y ~ Intervention + Sex |
| -2,759067227 | y ~ Intervention + Sex |
| -3,031298987 | y ~ Intervention + Sex |
| 0,277701703 | y ~ Intervention + Sex |
| 0,57893705 | y ~ Intervention + Sex |
| -0,361541044 | y ~ Intervention + Sex |
| -2,761365217 | y ~ Intervention + Sex |
| 0,835553507 | y ~ Intervention + Sex |
| -3,301923476 | y ~ Intervention + Sex |
| -2,436868588 | y ~ Intervention + Sex |
| -1,154592413 | y ~ Intervention + Sex |
| -1,441798569 | y ~ Intervention + Sex |

| | |
|---|---|
| 0,244189924 | y ~ Intervention + Sex |
| -1,813149039 | y ~ Intervention + Sex |
| -0,187767456 | y ~ Intervention + Sex |
| -1,440675709 | y ~ Intervention + Sex |
| 2,386260768 | y ~ Intervention + Sex |
| 2,943770769 | y ~ Intervention + Sex |
| -1,626898092 | y ~ Intervention + Sex |
| 1,394670251 | y ~ Intervention + Sex |
| -0,623281099 | y ~ Intervention + Sex |
| -0,925982706 | y ~ Intervention + Sex |
| -0,866822556 | y ~ Intervention + Sex |
| 2,43081002 | y ~ Intervention + Sex |
| 1,812952532 | y ~ Intervention + Sex |
| -1,238317626 | y ~ Intervention + Sex |
| -1,410854328 | y ~ Intervention + Sex |
| -1,013956847 | y ~ Intervention + Sex |
| -0,757737093 | y ~ Intervention + Sex |
| -0,053829793 | y ~ Intervention + Sex |
| 0,831852955 | y ~ Intervention + Sex |
| 0,085037564 | y ~ Intervention + Sex |
| -2,124715987 | y ~ Intervention + Sex |
| -0,914439044 | y ~ Intervention + Sex |
| -1,430849719 | y ~ Intervention + Sex |
| -0,193713306 | y ~ Intervention + Sex |
| -0,138568521 | y ~ Intervention + Sex |
| -1,553430472 | y ~ Intervention + Sex |
| 0,502818947 | y ~ Intervention + Sex |
| -1,328309782 | y ~ Intervention + Sex |
| -1,142313913 | y ~ Intervention + Sex |
| 2,687931159 | y ~ Intervention + Sex |
| -0,840751752 | y ~ Intervention + Sex |
| 1,295757271 | y ~ Intervention + Sex |
| 1,195580976 | y ~ Intervention + Sex |
| 1,030253357 | y ~ Intervention + Sex |
| 0,840982092 | y ~ Intervention + Sex |
| -1,373664081 | y ~ Intervention + Sex |
| -1,848444493 | y ~ Intervention + Sex |
| -0,239191768 | y ~ Intervention + Sex |
| -0,853289952 | y ~ Intervention + Sex |
| -2,310023865 | y ~ Intervention + Sex |
| -1,03099174 | y ~ Intervention + Sex |
| -0,356063255 | y ~ Intervention + Sex |
| 1,286058415 | y ~ Intervention + Sex |
| -0,794918471 | y ~ Intervention + Sex |
| -1,204667977 | y ~ Intervention + Sex |
| 0,216820092 | y ~ Intervention + Sex |
| 1,474036987 | y ~ Intervention + Sex |
| -0,040928532 | y ~ Intervention + Sex |

| | |
|---|---|
| -0,494179548 | y ~ Intervention + Sex |
| -1,450200658 | y ~ Intervention + Sex |
| -1,443668445 | y ~ Intervention + Sex |
| -1,107405497 | y ~ Intervention + Sex |
| -2,764368311 | y ~ Intervention + Sex |
| 0,735234753 | y ~ Intervention + Sex |
| -1,678157839 | y ~ Intervention + Sex |
| -0,351818713 | y ~ Intervention + Sex |
| 1,085435338 | y ~ Intervention + Sex |
| -0,027216374 | y ~ Intervention + Sex |
| -0,627386468 | y ~ Intervention + Sex |
| 1,126751525 | y ~ Intervention + Sex |
| 1,471808785 | y ~ Intervention + Sex |
| 0,628117489 | y ~ Intervention + Sex |
| 0,58619673 | y ~ Intervention + Sex |
| -0,829166784 | y ~ Intervention + Sex |
| -1,207458342 | y ~ Intervention + Sex |
| 0,715773753 | y ~ Intervention + Sex |
| -1,112048399 | y ~ Intervention + Sex |
| -0,32458071 | y ~ Intervention + Sex |
| -0,741091519 | y ~ Intervention + Sex |
| -0,358842549 | y ~ Intervention + Sex |
| -0,067602358 | y ~ Intervention + Sex |
| 1,209041189 | y ~ Intervention + Sex |
| -1,978972053 | y ~ Intervention + Sex |
| -2,35262261 | y ~ Intervention + Sex |
| -0,848037953 | y ~ Intervention + Sex |
| -0,231059888 | y ~ Intervention + Sex |
| 1,140668122 | y ~ Intervention + Sex |
| -0,825458198 | y ~ Intervention + Sex |
| -0,464355052 | y ~ Intervention + Sex |
| -1,938977479 | y ~ Intervention + Sex |
| -0,477987495 | y ~ Intervention + Sex |
| -0,153710171 | y ~ Intervention + Sex |
| 1,911531242 | y ~ Intervention + Sex |
| -0,213917182 | y ~ Intervention + Sex |
| -0,887605052 | y ~ Intervention + Sex |
| -1,196790988 | y ~ Intervention + Sex |
| 0,337465798 | y ~ Intervention + Sex |
| -0,945485682 | y ~ Intervention + Sex |
| -0,062863772 | y ~ Intervention + Sex |
| -0,397401424 | y ~ Intervention + Sex |
| -1,413637733 | y ~ Intervention + Sex |
| -1,39152253 | y ~ Intervention + Sex |
| -0,712513296 | y ~ Intervention + Sex |
| 1,456172116 | y ~ Intervention + Sex |
| -0,83742969 | y ~ Intervention + Sex |
| 0,316223021 | y ~ Intervention + Sex |

| | |
|---|---|
| 0,263228893 | y ~ Intervention + Sex |
| 0,000448708 | y ~ Intervention + Sex |
| -1,30206401 | y ~ Intervention + Sex |
| -0,726177058 | y ~ Intervention + Sex |
| -0,990029083 | y ~ Intervention + Sex |
| -1,463540828 | y ~ Intervention + Sex |
| -0,428272958 | y ~ Intervention + Sex |
| 0,264030665 | y ~ Intervention + Sex |
| 0,040147028 | y ~ Intervention + Sex |
| -2,220326639 | y ~ Intervention + Sex |
| -0,557468533 | y ~ Intervention + Sex |
| -0,498745567 | y ~ Intervention + Sex |
| -0,764893498 | y ~ Intervention + Sex |
| 0,359309131 | y ~ Intervention + Sex |
| 0,100758316 | y ~ Intervention + Sex |
| 1,350778447 | y ~ Intervention + Sex |
| -0,14213888 | y ~ Intervention + Sex |
| 0,296331969 | y ~ Intervention + Sex |
| 2,113626831 | y ~ Intervention + Sex |
| -1,567387436 | y ~ Intervention + Sex |
| 0,088860213 | y ~ Intervention + Sex |
| -0,683178515 | y ~ Intervention + Sex |
| 0,4176874 | y ~ Intervention + Sex |
| -0,992681681 | y ~ Intervention + Sex |
| 0,585031765 | y ~ Intervention + Sex |
| 1,764821672 | y ~ Intervention + Sex |
| -0,132755591 | y ~ Intervention + Sex |
| -0,2018899 | y ~ Intervention + Sex |
| 0,677336098 | y ~ Intervention + Sex |
| 0,889344718 | y ~ Intervention + Sex |
| -0,518912168 | y ~ Intervention + Sex |
| 0,260770304 | y ~ Intervention + Sex |
| 1,183203969 | y ~ Intervention + Sex |
| 0,815649407 | y ~ Intervention + Sex |
| -1,702231437 | y ~ Intervention + Sex |
| -0,229236453 | y ~ Intervention + Sex |
| 0,607061042 | y ~ Intervention + Sex |
| 2,217777186 | y ~ Intervention + Sex |
| -1,865885648 | y ~ Intervention + Sex |
| -0,671892704 | y ~ Intervention + Sex |
| 0,159748119 | y ~ Intervention + Sex |
| -1,35341804 | y ~ Intervention + Sex |
| -0,149651661 | y ~ Intervention + Sex |
| -2,419966734 | y ~ Intervention + Sex |
| 1,409050232 | y ~ Intervention + Sex |
| 0,428198161 | y ~ Intervention + Sex |
| 2,50907762 | y ~ Intervention + Sex |
| 1,398840186 | y ~ Intervention + Sex |

| | |
|---|---|
| -1,630031592 | y ~ Intervention + Sex |
| -1,932117666 | y ~ Intervention + Sex |
| -0,512607289 | y ~ Intervention + Sex |
| 0,317629596 | y ~ Intervention + Sex |
| -0,075182328 | y ~ Intervention + Sex |
| 1,172623265 | y ~ Intervention + Sex |
| 3,024113689 | y ~ Intervention + Sex |
| -2,4236944 | y ~ Intervention + Sex |
| 0,831035723 | y ~ Intervention + Sex |
| 0,054677703 | y ~ Intervention + Sex |
| 0,947527839 | y ~ Intervention + Sex |
| 0,204957007 | y ~ Intervention + Sex |
| 0,992471817 | y ~ Intervention + Sex |
| 1,068997979 | y ~ Intervention + Sex |
| 0,268470384 | y ~ Intervention + Sex |
| -0,6679869 | y ~ Intervention + Sex |
| 0,160043442 | y ~ Intervention + Sex |
| -1,244220256 | y ~ Intervention + Sex |
| 2,225210727 | y ~ Intervention + Sex |
| -1,41356262 | y ~ Intervention + Sex |
| 1,076691826 | y ~ Intervention + Sex |
| -0,314440447 | y ~ Intervention + Sex |
| 0,023605168 | y ~ Intervention + Sex |
| -0,278238742 | y ~ Intervention + Sex |
| 0,159014143 | y ~ Intervention + Sex |
| -0,328288726 | y ~ Intervention + Sex |
| 0,45470965 | y ~ Intervention + Sex |
| -0,519926773 | y ~ Intervention + Sex |
| 0,423817803 | y ~ Intervention + Sex |
| -0,593259607 | y ~ Intervention + Sex |
| -0,226555199 | y ~ Intervention + Sex |
| -0,754588084 | y ~ Intervention + Sex |
| -0,343535246 | y ~ Intervention + Sex |
| 2,136877005 | y ~ Intervention + Sex |
| 0,620930579 | y ~ Intervention + Sex |
| 0,605760049 | y ~ Intervention + Sex |
| 1,757910222 | y ~ Intervention + Sex |
| -1,319457929 | y ~ Intervention + Sex |
| 0,018449594 | y ~ Intervention + Sex |
| 0,542256598 | y ~ Intervention + Sex |
| 0,501761403 | y ~ Intervention + Sex |
| 1,997376691 | y ~ Intervention + Sex |
| -0,302296167 | y ~ Intervention + Sex |
| 2,048589325 | y ~ Intervention + Sex |
| -1,477096472 | y ~ Intervention + Sex |
| -1,318156632 | y ~ Intervention + Sex |
| -1,113204782 | y ~ Intervention + Sex |
| 0,62005455 | y ~ Intervention + Sex |

| | |
|---|---|
| -2,166383572 | y ~ Intervention + Sex |
| 0,234602307 | y ~ Intervention + Sex |
| 2,589116625 | y ~ Intervention + Sex |
| -0,218419062 | y ~ Intervention + Sex |
| 0,77068258 | y ~ Intervention + Sex |
| 0,115589505 | y ~ Intervention + Sex |
| -0,101175107 | y ~ Intervention + Sex |
| 1,287110331 | y ~ Intervention + Sex |
| -0,90813296 | y ~ Intervention + Sex |
| 0,71170569 | y ~ Intervention + Sex |
| -0,846416121 | y ~ Intervention + Sex |
| 1,334365935 | y ~ Intervention + Sex |
| 1,649487585 | y ~ Intervention + Sex |
| 0,015873426 | y ~ Intervention + Sex |
| 0,678738231 | y ~ Intervention + Sex |
| 0,060612141 | y ~ Intervention + Sex |
| -0,961617616 | y ~ Intervention + Sex |
| -0,195676644 | y ~ Intervention + Sex |
| -0,700244456 | y ~ Intervention + Sex |
| 1,677513469 | y ~ Intervention + Sex |
| 0,337030355 | y ~ Intervention + Sex |
| -0,579449696 | y ~ Intervention + Sex |
| 0,361047402 | y ~ Intervention + Sex |
| 0,697663894 | y ~ Intervention + Sex |
| -0,529555882 | y ~ Intervention + Sex |
| -1,212613762 | y ~ Intervention + Sex |
| -1,354457742 | y ~ Intervention + Sex |
| 1,872056048 | y ~ Intervention + Sex |
| 0,692400848 | y ~ Intervention + Sex |
| -0,622145881 | y ~ Intervention + Sex |
| -0,45170914 | y ~ Intervention + Sex |
| 1,194103043 | y ~ Intervention + Sex |
| 1,135678639 | y ~ Intervention + Sex |
| 2,293173688 | y ~ Intervention + Sex |
| -0,175402343 | y ~ Intervention + Sex |
| 0,353137118 | y ~ Intervention + Sex |
| 1,854481721 | y ~ Intervention + Sex |
| 0,183656787 | y ~ Intervention + Sex |
| 1,920302738 | y ~ Intervention + Sex |
| -0,906725103 | y ~ Intervention + Sex |
| 1,506612882 | y ~ Intervention + Sex |
| 0,726335199 | y ~ Intervention + Sex |
| -1,824447628 | y ~ Intervention + Sex |
| -1,015198301 | y ~ Intervention + Sex |
| 0,755740204 | y ~ Intervention + Sex |
| 0,551779754 | y ~ Intervention + Sex |
| 0,85255394 | y ~ Intervention + Sex |
| 0,053515467 | y ~ Intervention + Sex |

| | |
|---|---|
| -0,651461436 | y ~ Intervention + Sex |
| 2,171610881 | y ~ Intervention + Sex |
| -0,519687426 | y ~ Intervention + Sex |
| 1,687433309 | y ~ Intervention + Sex |
| -1,198275477 | y ~ Intervention + Sex |
| 0,57114378 | y ~ Intervention + Sex |
| 1,520979264 | y ~ Intervention + Sex |
| -0,401966807 | y ~ Intervention + Sex |
| 0,357786044 | y ~ Intervention + Sex |
| -0,0704841 | y ~ Intervention + Sex |
| 1,042439641 | y ~ Intervention + Sex |
| 0,9577794 | y ~ Intervention + Sex |
| 0,636397127 | y ~ Intervention + Sex |
| -2,344669412 | y ~ Intervention + Sex |
| -0,938373723 | y ~ Intervention + Sex |
| -0,745574155 | y ~ Intervention + Sex |
| 0,635645167 | y ~ Intervention + Sex |
| 0,080891348 | y ~ Intervention + Sex |
| 0,411225302 | y ~ Intervention + Sex |
| -1,583299573 | y ~ Intervention + Sex |
| 1,502705871 | y ~ Intervention + Sex |
| -0,306780566 | y ~ Intervention + Sex |
| -0,78718 | y ~ Intervention + Sex |
| -1,326451771 | y ~ Intervention + Sex |
| 0,962492743 | y ~ Intervention + Sex |
| 0,268291102 | y ~ Intervention + Sex |
| 1,544502565 | y ~ Intervention + Sex |
| -0,512614687 | y ~ Intervention + Sex |
| 2,352524983 | y ~ Intervention + Sex |
| -0,499713399 | y ~ Intervention + Sex |
| -1,17679597 | y ~ Intervention + Sex |
| 2,791705168 | y ~ Intervention + Sex |
| -0,279573844 | y ~ Intervention + Sex |
| -0,643074333 | y ~ Intervention + Sex |
| -0,066233041 | y ~ Intervention + Sex |
| 0,572249309 | y ~ Intervention + Sex |
| -0,616070869 | y ~ Intervention + Sex |
| 0,546381713 | y ~ Intervention + Sex |
| -0,027908405 | y ~ Intervention + Sex |
| 1,319640591 | y ~ Intervention + Sex |
| 1,004764282 | y ~ Intervention + Sex |
| 0,904266505 | y ~ Intervention + Sex |
| -0,059462602 | y ~ Intervention + Sex |
| -0,157231575 | y ~ Intervention + Sex |
| -0,621332082 | y ~ Intervention + Sex |
| 0,07207329 | y ~ Intervention + Sex |
| 0,959570776 | y ~ Intervention + Sex |
| -1,987038446 | y ~ Intervention + Sex |

| | |
|---|---|
| -0,617321518 | y ~ Intervention + Sex |
| 0,419474106 | y ~ Intervention + Sex |
| -0,204990971 | y ~ Intervention + Sex |
| 0,058903372 | y ~ Intervention + Sex |
| 0,561868575 | y ~ Intervention + Sex |
| -0,014085527 | y ~ Intervention + Sex |
| -0,060460403 | y ~ Intervention + Sex |
| -2,179967934 | y ~ Intervention + Sex |
| 0,466106357 | y ~ Intervention + Sex |
| 1,007027192 | y ~ Intervention + Sex |
| 0,104695199 | y ~ Intervention + Sex |
| 0,785534358 | y ~ Intervention + Sex |
| 2,253476663 | y ~ Intervention + Sex |
| -0,703199158 | y ~ Intervention + Sex |
| 1,692318656 | y ~ Intervention + Sex |
| -0,899766987 | y ~ Intervention + Sex |
| -1,50610316 | y ~ Intervention + Sex |
| 0,711623449 | y ~ Intervention + Sex |
| -0,169027369 | y ~ Intervention + Sex |
| 0,969181746 | y ~ Intervention + Sex |
| 0,693337036 | y ~ Intervention + Sex |
| 1,009634789 | y ~ Intervention + Sex |
| 0,494370164 | y ~ Intervention + Sex |
| -0,830508095 | y ~ Intervention + Sex |
| 0,709551927 | y ~ Intervention + Sex |
| -0,484072055 | y ~ Intervention + Sex |
| -0,210004561 | y ~ Intervention + Sex |
| 0,166407453 | y ~ Intervention + Sex |
| -0,268885827 | y ~ Intervention + Sex |
| -1,518640963 | y ~ Intervention + Sex |
| -0,636139117 | y ~ Intervention + Sex |
| -0,470718387 | y ~ Intervention + Sex |
| -0,428128139 | y ~ Intervention + Sex |
| 0,68205271 | y ~ Intervention + Sex |
| 0,296790219 | y ~ Intervention + Sex |
| 1,150069131 | y ~ Intervention + Sex |
| 1,503123983 | y ~ Intervention + Sex |
| 0,752401244 | y ~ Intervention + Sex |
| -0,336964189 | y ~ Intervention + Sex |
| -0,835782359 | y ~ Intervention + Sex |
| -0,983539013 | y ~ Intervention + Sex |
| 0,767779923 | y ~ Intervention + Sex |
| -0,697533069 | y ~ Intervention + Sex |
| 0,040887462 | y ~ Intervention + Sex |
| 0,553184179 | y ~ Intervention + Sex |
| 0,582356614 | y ~ Intervention + Sex |
| -0,247891819 | y ~ Intervention + Sex |
| 0,296822543 | y ~ Intervention + Sex |

| | |
|---|---|
| 0,153425132 | y ~ Intervention + Sex |
| 1,204838641 | y ~ Intervention + Sex |
| 1,668421492 | y ~ Intervention + Sex |
| 0,921043144 | y ~ Intervention + Sex |
| 1,149421718 | y ~ Intervention + Sex |
| -0,988712585 | y ~ Intervention + Sex |
| 1,802688733 | y ~ Intervention + Sex |
| 0,320671042 | y ~ Intervention + Sex |
| -1,274102521 | y ~ Intervention + Sex |
| 2,68908968 | y ~ Intervention + Sex |
| -1,117691749 | y ~ Intervention + Sex |
| 0,055202977 | y ~ Intervention + Sex |
| -0,509433826 | y ~ Intervention + Sex |
| -0,539343899 | y ~ Intervention + Sex |
| 0,480055814 | y ~ Intervention + Sex |
| -0,16341975 | y ~ Intervention + Sex |
| 2,66773366 | y ~ Intervention + Sex |
| 0,231876926 | y ~ Intervention + Sex |
| 1,429137456 | y ~ Intervention + Sex |
| -1,377894636 | y ~ Intervention + Sex |
| 1,006326085 | y ~ Intervention + Sex |
| 0,99129779 | y ~ Intervention + Sex |
| 1,350202587 | y ~ Intervention + Sex |
| 0,16276216 | y ~ Intervention + Sex |
| 0,929421736 | y ~ Intervention + Sex |
| -0,044667462 | y ~ Intervention + Sex |
| 1,245366957 | y ~ Intervention + Sex |
| -1,612505492 | y ~ Intervention + Sex |
| -0,690320636 | y ~ Intervention + Sex |
| -1,919495818 | y ~ Intervention + Sex |
| 1,353597636 | y ~ Intervention + Sex |
| -0,613617662 | y ~ Intervention + Sex |
| 0,092368908 | y ~ Intervention + Sex |
| 1,343386064 | y ~ Intervention + Sex |
| -0,919214096 | y ~ Intervention + Sex |
| -1,641717752 | y ~ Intervention + Sex |
| 1,256885694 | y ~ Intervention + Sex |
| 0,445635958 | y ~ Intervention + Sex |
| -0,243626309 | y ~ Intervention + Sex |
| 1,206331851 | y ~ Intervention + Sex |
| 1,367137441 | y ~ Intervention + Sex |
| -0,125845993 | y ~ Intervention + Sex |
| 0,51197933 | y ~ Intervention + Sex |
| -0,563148473 | y ~ Intervention + Sex |
| -1,19770036 | y ~ Intervention + Sex |
| 0,410311981 | y ~ Intervention + Sex |
| 0,2165821 | y ~ Intervention + Sex |
| 0,580739476 | y ~ Intervention + Sex |

| | |
|---|---|
| -1,622522685 | y ~ Intervention + Sex |
| 1,175501522 | y ~ Intervention + Sex |
| 1,010553724 | y ~ Intervention + Sex |
| 2,064415327 | y ~ Intervention + Sex |
| 0,994624793 | y ~ Intervention + Sex |
| -1,854326519 | y ~ Intervention + Sex |
| 0,161728663 | y ~ Intervention + Sex |
| -0,045405091 | y ~ Intervention + Sex |
| -0,308968289 | y ~ Intervention + Sex |
| -1,818846089 | y ~ Intervention + Sex |
| 0,662129699 | y ~ Intervention + Sex |
| 1,454515403 | y ~ Intervention + Sex |
| 0,212470727 | y ~ Intervention + Sex |
| 0,628569343 | y ~ Intervention + Sex |
| 1,438788108 | y ~ Intervention + Sex |
| 0,722224644 | y ~ Intervention + Sex |
| -0,284083531 | y ~ Intervention + Sex |
| -0,864877731 | y ~ Intervention + Sex |
| 7,057180401 | y ~ Intervention + Sex |
| -0,334743639 | y ~ Intervention + Sex |
| 1,577561064 | y ~ Intervention + Sex |
| 0,146215354 | y ~ Intervention + Sex |
| -0,297205263 | y ~ Intervention + Sex |
| -0,05506605 | y ~ Intervention + Sex |
| -1,492492499 | y ~ Intervention + Sex |
| -1,274333806 | y ~ Intervention + Sex |
| 1,265406977 | y ~ Intervention + Sex |
| -0,276028516 | y ~ Intervention + Sex |
| -0,865335428 | y ~ Intervention + Sex |
| -0,500933667 | y ~ Intervention + Sex |
| 0,075428818 | y ~ Intervention + Sex |
| -1,393466097 | y ~ Intervention + Sex |
| 1,900255763 | y ~ Intervention + Sex |
| 0,444608554 | y ~ Intervention + Sex |
| -0,13262164 | y ~ Intervention + Sex |
| -0,940713481 | y ~ Intervention + Sex |
| 2,323965615 | y ~ Intervention + Sex |
| 0,048852932 | y ~ Intervention + Sex |
| 0,288099493 | y ~ Intervention + Sex |
| -0,067364056 | y ~ Intervention + Sex |
| 0,413134 | y ~ Intervention + Sex |
| -0,059229423 | y ~ Intervention + Sex |
| 0,043536807 | y ~ Intervention + Sex |
| 1,979541299 | y ~ Intervention + Sex |
| 1,672339044 | y ~ Intervention + Sex |
| -0,9051635 | y ~ Intervention + Sex |
| 0,30377263 | y ~ Intervention + Sex |
| 0,798166205 | y ~ Intervention + Sex |

| | |
|---|---|
| -0,256161437 | y ~ Intervention + Sex |
| -0,514090851 | y ~ Intervention + Sex |
| 1,045050326 | y ~ Intervention + Sex |
| -0,474288967 | y ~ Intervention + Sex |
| -0,114150934 | y ~ Intervention + Sex |
| -1,53847918 | y ~ Intervention + Sex |
| -0,456229168 | y ~ Intervention + Sex |
| 1,666675869 | y ~ Intervention + Sex |
| 0,197201809 | y ~ Intervention + Sex |
| -0,485566119 | y ~ Intervention + Sex |
| 0,772862591 | y ~ Intervention + Sex |
| 0,935697621 | y ~ Intervention + Sex |
| 1,297475418 | y ~ Intervention + Sex |
| -0,394869821 | y ~ Intervention + Sex |
| 0,462264274 | y ~ Intervention + Sex |
| 0,667633277 | y ~ Intervention + Sex |
| -0,910776145 | y ~ Intervention + Sex |
| -1,131643069 | y ~ Intervention + Sex |
| -0,468039898 | y ~ Intervention + Sex |
| -0,242608457 | y ~ Intervention + Sex |
| -0,411291286 | y ~ Intervention + Sex |
| -0,656434254 | y ~ Intervention + Sex |
| 1,082608323 | y ~ Intervention + Sex |
| 0,710311448 | y ~ Intervention + Sex |
| 0,790847706 | y ~ Intervention + Sex |
| -1,033970595 | y ~ Intervention + Sex |
| -0,108671384 | y ~ Intervention + Sex |
| 2,047166837 | y ~ Intervention + Sex |
| 0,741585367 | y ~ Intervention + Sex |
| -1,881946095 | y ~ Intervention + Sex |
| 0,117428074 | y ~ Intervention + Sex |
| 1,072826056 | y ~ Intervention + Sex |
| -0,601806164 | y ~ Intervention + Sex |
| 1,388186056 | y ~ Intervention + Sex |
| -0,903508139 | y ~ Intervention + Sex |
| 0,231529504 | y ~ Intervention + Sex |
| 0,629716398 | y ~ Intervention + Sex |
| 0,358362759 | y ~ Intervention + Sex |
| 0,04913052 | y ~ Intervention + Sex |
| 2,004157649 | y ~ Intervention + Sex |
| 0,399418962 | y ~ Intervention + Sex |
| -0,876144971 | y ~ Intervention + Sex |
| -0,430616114 | y ~ Intervention + Sex |
| 0,645166479 | y ~ Intervention + Sex |
| 1,495528574 | y ~ Intervention + Sex |
| -0,986872398 | y ~ Intervention + Sex |
| 0,339945163 | y ~ Intervention + Sex |
| 0,791086936 | y ~ Intervention + Sex |

| | |
|---|---|
| -1,782010404 | y ~ Intervention + Sex |
| -0,018672979 | y ~ Intervention + Sex |
| -0,762962797 | y ~ Intervention + Sex |
| -0,137972716 | y ~ Intervention + Sex |
| 0,075763681 | y ~ Intervention + Sex |
| -1,251354721 | y ~ Intervention + Sex |
| -1,406429202 | y ~ Intervention + Sex |
| -0,132868461 | y ~ Intervention + Sex |
| 0,265540054 | y ~ Intervention + Sex |
| 0,827970839 | y ~ Intervention + Sex |
| -0,525076949 | y ~ Intervention + Sex |
| 1,375498713 | y ~ Intervention + Sex |
| -0,245130862 | y ~ Intervention + Sex |
| -1,274929822 | y ~ Intervention + Sex |
| -0,513513283 | y ~ Intervention + Sex |
| -1,66948542 | y ~ Intervention + Sex |
| 1,45548645 | y ~ Intervention + Sex |
| -0,193643342 | y ~ Intervention + Sex |
| -0,309683071 | y ~ Intervention + Sex |
| 0,879323444 | y ~ Intervention + Sex |
| -1,030632772 | y ~ Intervention + Sex |
| -0,056053714 | y ~ Intervention + Sex |
| -0,297904413 | y ~ Intervention + Sex |
| -0,079072177 | y ~ Intervention + Sex |
| -1,374878704 | y ~ Intervention + Sex |
| -1,266827643 | y ~ Intervention + Sex |
| -0,366115711 | y ~ Intervention + Sex |
| -1,61418936 | y ~ Intervention + Sex |
| 0,06933789 | y ~ Intervention + Sex |
| -0,54820994 | y ~ Intervention + Sex |
| 0,956704156 | y ~ Intervention + Sex |
| -0,125335396 | y ~ Intervention + Sex |
| -0,17373678 | y ~ Intervention + Sex |
| -1,739138326 | y ~ Intervention + Sex |
| 1,417096515 | y ~ Intervention + Sex |
| -1,148969535 | y ~ Intervention + Sex |
| -2,419352109 | y ~ Intervention + Sex |
| -0,363777329 | y ~ Intervention + Sex |
| 1,124391106 | y ~ Intervention + Sex |
| 0,085321862 | y ~ Intervention + Sex |
| 1,251424985 | y ~ Intervention + Sex |
| -0,68166453 | y ~ Intervention + Sex |
| 0,829826173 | y ~ Intervention + Sex |
| -0,24352684 | y ~ Intervention + Sex |
| 0,512454245 | y ~ Intervention + Sex |
| -0,142487033 | y ~ Intervention + Sex |
| 0,094678628 | y ~ Intervention + Sex |
| 0,11021099 | y ~ Intervention + Sex |

| | |
|---|---|
| -1,528463567 | y ~ Intervention + Sex |
| -0,523354259 | y ~ Intervention + Sex |
| 0,405733999 | y ~ Intervention + Sex |
| -2,03760777 | y ~ Intervention + Sex |
| -0,694517468 | y ~ Intervention + Sex |
| 1,787795607 | y ~ Intervention + Sex |
| -0,016242423 | y ~ Intervention + Sex |
| 0,326960644 | y ~ Intervention + Sex |
| -0,233044948 | y ~ Intervention + Sex |
| 0,926637438 | y ~ Intervention + Sex |
| 0,341034633 | y ~ Intervention + Sex |
| -2,022495093 | y ~ Intervention + Sex |
| -0,063455626 | y ~ Intervention + Sex |
| 1,462766563 | y ~ Intervention + Sex |
| 0,330388358 | y ~ Intervention + Sex |
| -0,248837277 | y ~ Intervention + Sex |
| -1,602508808 | y ~ Intervention + Sex |
| 0,204181917 | y ~ Intervention + Sex |
| 0,035249781 | y ~ Intervention + Sex |
| -0,52532378 | y ~ Intervention + Sex |
| -0,121259765 | y ~ Intervention + Sex |
| 1,029417213 | y ~ Intervention + Sex |
| -1,960310405 | y ~ Intervention + Sex |
| -0,501462409 | y ~ Intervention + Sex |
| 1,709266067 | y ~ Intervention + Sex |
| -1,465702687 | y ~ Intervention + Sex |
| 0,976847081 | y ~ Intervention + Sex |
| -2,408088506 | y ~ Intervention + Sex |
| -0,21087502 | y ~ Intervention + Sex |
| 1,018584916 | y ~ Intervention + Sex |
| -0,105547894 | y ~ Intervention + Sex |
| 0,844326677 | y ~ Intervention + Sex |
| 1,880061315 | y ~ Intervention + Sex |
| 0,621095289 | y ~ Intervention + Sex |
| -0,84706515 | y ~ Intervention + Sex |
| 0,764643394 | y ~ Intervention + Sex |
| 2,141823973 | y ~ Intervention + Sex |
| -0,239167925 | y ~ Intervention + Sex |
| 1,265392513 | y ~ Intervention + Sex |
| -0,414428748 | y ~ Intervention + Sex |
| 0,486053982 | y ~ Intervention + Sex |
| -1,061929641 | y ~ Intervention + Sex |
| -0,30281694 | y ~ Intervention + Sex |
| -0,160729553 | y ~ Intervention + Sex |
| -2,030600229 | y ~ Intervention + Sex |
| 0,158019488 | y ~ Intervention + Sex |
| -0,389388169 | y ~ Intervention + Sex |
| 0,533460476 | y ~ Intervention + Sex |

| | |
|---|---|
| -1,033430975 | y ~ Intervention + Sex |
| -1,393081382 | y ~ Intervention + Sex |
| -1,070316025 | y ~ Intervention + Sex |
| -1,90905075 | y ~ Intervention + Sex |
| -0,240713145 | y ~ Intervention + Sex |
| 1,673162904 | y ~ Intervention + Sex |
| -2,10077164 | y ~ Intervention + Sex |
| 0,200929683 | y ~ Intervention + Sex |
| 0,790476035 | y ~ Intervention + Sex |
| 1,833981607 | y ~ Intervention + Sex |
| 2,071846192 | y ~ Intervention + Sex |
| -2,517129869 | y ~ Intervention + Sex |
| 0,355350854 | y ~ Intervention + Sex |
| -0,898857669 | y ~ Intervention + Sex |
| 0,636876911 | y ~ Intervention + Sex |
| -0,178241154 | y ~ Intervention + Sex |
| -0,429914602 | y ~ Intervention + Sex |
| -0,744465178 | y ~ Intervention + Sex |
| 0,168802843 | y ~ Intervention + Sex |
| 0,345955555 | y ~ Intervention + Sex |
| 0,492260979 | y ~ Intervention + Sex |
| -0,921322958 | y ~ Intervention + Sex |
| -0,071536155 | y ~ Intervention + Sex |
| -0,229248154 | y ~ Intervention + Sex |
| -0,158654244 | y ~ Intervention + Sex |
| 5,277095157 | y ~ Intervention + Sex |
| 0,072679933 | y ~ Intervention + Sex |
| 0,885145442 | y ~ Intervention + Sex |
| 0,720786983 | y ~ Intervention + Sex |
| -2,555493153 | y ~ Intervention + Sex |
| -0,739780836 | y ~ Intervention + Sex |
| 2,514769181 | y ~ Intervention + Sex |
| 0,082680344 | y ~ Intervention + Sex |
| 1,487777834 | y ~ Intervention + Sex |
| 0,361117692 | y ~ Intervention + Sex |
| 0,75964952 | y ~ Intervention + Sex |
| 0,410678792 | y ~ Intervention + Sex |
| 0,144018483 | y ~ Intervention + Sex |
| 1,42257498 | y ~ Intervention + Sex |
| 0,409199275 | y ~ Intervention + Sex |
| 0,569889554 | y ~ Intervention + Sex |
| 2,545201681 | y ~ Intervention + Sex |
| -0,488821465 | y ~ Intervention + Sex |
| 0,24418929 | y ~ Intervention + Sex |
| 0,491476507 | y ~ Intervention + Sex |
| 0,789196943 | y ~ Intervention + Sex |
| 0,096065614 | y ~ Intervention + Sex |
| -1,314148538 | y ~ Intervention + Sex |

| | |
|---|---|
| -0,667640388 | y ~ Intervention + Sex |
| 0,153337194 | y ~ Intervention + Sex |
| -1,036008807 | y ~ Intervention + Sex |
| 0,093186421 | y ~ Intervention + Sex |
| 0,492236864 | y ~ Intervention + Sex |
| 0,079987381 | y ~ Intervention + Sex |
| -0,35462852 | y ~ Intervention + Sex |
| -1,145511818 | y ~ Intervention + Sex |
| -2,716868475 | y ~ Intervention + Sex |
| -0,285525741 | y ~ Intervention + Sex |
| -1,26754876 | y ~ Intervention + Sex |
| -0,336930465 | y ~ Intervention + Sex |
| -0,759813733 | y ~ Intervention + Sex |
| -1,108997355 | y ~ Intervention + Sex |
| -0,298223599 | y ~ Intervention + Sex |
| 0,857026407 | y ~ Intervention + Sex |
| -0,388528457 | y ~ Intervention + Sex |
| -0,425331858 | y ~ Intervention + Sex |
| 0,807061062 | y ~ Intervention + Sex |
| -0,467459603 | y ~ Intervention + Sex |
| 0,541717913 | y ~ Intervention + Sex |
| 0,093160664 | y ~ Intervention + Sex |
| 1,2261042 | y ~ Intervention + Sex |
| -0,253656329 | y ~ Intervention + Sex |
| 1,053793339 | y ~ Intervention + Sex |
| -1,202136776 | y ~ Intervention + Sex |
| 0,275951867 | y ~ Intervention + Sex |
| -0,09356152 | y ~ Intervention + Sex |
| -1,924061884 | y ~ Intervention + Sex |
| -0,097483198 | y ~ Intervention + Sex |
| -0,372317526 | y ~ Intervention + Sex |
| 0,589802576 | y ~ Intervention + Sex |
| 1,206159248 | y ~ Intervention + Sex |
| -0,482263051 | y ~ Intervention + Sex |
| 0,699457403 | y ~ Intervention + Sex |
| 2,228155853 | y ~ Intervention + Sex |
| -2,288212779 | y ~ Intervention + Sex |
| -0,120968028 | y ~ Intervention + Sex |
| 0,105724329 | y ~ Intervention + Sex |
| -1,295376151 | y ~ Intervention + Sex |
| -2,293264529 | y ~ Intervention + Sex |
| 3,914361685 | y ~ Intervention + Sex |
| 0,19169155 | y ~ Intervention + Sex |
| 1,40258577 | y ~ Intervention + Sex |
| 1,814702205 | y ~ Intervention + Sex |
| 0,139030237 | y ~ Intervention + Sex |
| -0,366496855 | y ~ Intervention + Sex |
| 1,796991625 | y ~ Intervention + Sex |

| | |
|---|---|
| -0,495922348 | y ~ Intervention + Sex |
| -1,663870843 | y ~ Intervention + Sex |
| 0,375354402 | y ~ Intervention + Sex |
| 0,474505644 | y ~ Intervention + Sex |
| -0,742676093 | y ~ Intervention + Sex |
| -0,064779749 | y ~ Intervention + Sex |
| -0,883631213 | y ~ Intervention + Sex |
| -2,688324354 | y ~ Intervention + Sex |
| -0,663980784 | y ~ Intervention + Sex |
| -0,003309311 | y ~ Intervention + Sex |
| 0,353552747 | y ~ Intervention + Sex |
| -1,017726752 | y ~ Intervention + Sex |
| 2,105808672 | y ~ Intervention + Sex |
| 0,679713908 | y ~ Intervention + Sex |
| 0,189789311 | y ~ Intervention + Sex |
| 0,469498416 | y ~ Intervention + Sex |
| -1,461359292 | y ~ Intervention + Sex |
| -1,796752859 | y ~ Intervention + Sex |
| 0,603798059 | y ~ Intervention + Sex |
| 0,430989913 | y ~ Intervention + Sex |
| 3,047543711 | y ~ Intervention + Sex |
| 3,198142129 | y ~ Intervention + Sex |
| -2,421438114 | y ~ Intervention + Sex |
| -1,298059186 | y ~ Intervention + Sex |
| -2,066366041 | y ~ Intervention + Sex |
| 2,643735447 | y ~ Intervention + Sex |
| 0,008109579 | y ~ Intervention + Sex |
| 0,665098365 | y ~ Intervention + Sex |
| 1,180539677 | y ~ Intervention + Sex |
| -0,654205322 | y ~ Intervention + Sex |
| -0,802316063 | y ~ Intervention + Sex |
| 0,023356433 | y ~ Intervention + Sex |
| 0,385513647 | y ~ Intervention + Sex |
| -0,555997754 | y ~ Intervention + Sex |
| -1,588922157 | y ~ Intervention + Sex |
| 1,552158478 | y ~ Intervention + Sex |
| -1,421826248 | y ~ Intervention + Sex |
| -1,347706095 | y ~ Intervention + Sex |
| 2,332045839 | y ~ Intervention + Sex |
| 0,677614011 | y ~ Intervention + Sex |
| 3,462816094 | y ~ Intervention + Sex |
| -1,148918831 | y ~ Intervention + Sex |
| 2,611102072 | y ~ Intervention + Sex |
| 1,45490381 | y ~ Intervention + Sex |
| -0,856777394 | y ~ Intervention + Sex |
| -0,984132008 | y ~ Intervention + Sex |
| 0,240319038 | y ~ Intervention + Sex |
| -0,529743406 | y ~ Intervention + Sex |

| | |
|---|---|
| -0,437428188 | y ~ Intervention + Sex |
| 0,026924599 | y ~ Intervention + Sex |
| -0,741288871 | y ~ Intervention + Sex |
| 0,979857455 | y ~ Intervention + Sex |
| -1,028450197 | y ~ Intervention + Sex |
| -1,951586049 | y ~ Intervention + Sex |
| 1,29788022 | y ~ Intervention + Sex |
| 2,051832621 | y ~ Intervention + Sex |
| -2,268819071 | y ~ Intervention + Sex |
| -0,085029319 | y ~ Intervention + Sex |
| -3,000953873 | y ~ Intervention + Sex |
| -1,729608585 | y ~ Intervention + Sex |
| -0,956636132 | y ~ Intervention + Sex |
| -2,272476348 | y ~ Intervention + Sex |
| -1,060367341 | y ~ Intervention + Sex |
| -0,316674496 | y ~ Intervention + Sex |
| -1,470134581 | y ~ Intervention + Sex |
| -1,731289725 | y ~ Intervention + Sex |
| -0,630363666 | y ~ Intervention + Sex |
| 0,18598622 | y ~ Intervention + Sex |
| -0,409045772 | y ~ Intervention + Sex |
| -1,08241887 | y ~ Intervention + Sex |
| 0,179950435 | y ~ Intervention + Sex |
| 0,834176676 | y ~ Intervention + Sex |
| 1,492765049 | y ~ Intervention + Sex |
| -2,007037118 | y ~ Intervention + Sex |
| 0,533726337 | y ~ Intervention + Sex |
| -0,029307753 | y ~ Intervention + Sex |
| -1,030957608 | y ~ Intervention + Sex |
| 0,401537334 | y ~ Intervention + Sex |
| -0,723618447 | y ~ Intervention + Sex |
| -0,408109018 | y ~ Intervention + Sex |
| -0,843256077 | y ~ Intervention + Sex |
| 1,123650325 | y ~ Intervention + Sex |
| -1,650106856 | y ~ Intervention + Sex |
| -1,502613619 | y ~ Intervention + Sex |
| -0,990735911 | y ~ Intervention + Sex |
| 0,90769624 | y ~ Intervention + Sex |
| 0,507703866 | y ~ Intervention + Sex |
| -2,540505449 | y ~ Intervention + Sex |
| 3,05065071 | y ~ Intervention + Sex |
| -0,900903502 | y ~ Intervention + Sex |
| -1,395004544 | y ~ Intervention + Sex |
| -1,74058146 | y ~ Intervention + Sex |
| -0,492791754 | y ~ Intervention + Sex |
| -0,811962585 | y ~ Intervention + Sex |
| -1,40695844 | y ~ Intervention + Sex |
| -1,33225452 | y ~ Intervention + Sex |

| | |
|---|---|
| -2,05676173 | y ~ Intervention + Sex |
| 1,80261595 | y ~ Intervention + Sex |
| 0,27163223 | y ~ Intervention + Sex |
| -1,183384255 | y ~ Intervention + Sex |
| 0,29173666 | y ~ Intervention + Sex |
| -1,602994338 | y ~ Intervention + Sex |
| 0,463515723 | y ~ Intervention + Sex |
| -1,28482553 | y ~ Intervention + Sex |
| 0,333528226 | y ~ Intervention + Sex |
| 0,052180511 | y ~ Intervention + Sex |
| 1,688662725 | y ~ Intervention + Sex |
| 0,793948871 | y ~ Intervention + Sex |
| 0,315128849 | y ~ Intervention + Sex |
| -0,53510762 | y ~ Intervention + Sex |
| 0,216816144 | y ~ Intervention + Sex |
| 2,081769347 | y ~ Intervention + Sex |
| 0,459964417 | y ~ Intervention + Sex |
| -1,669112789 | y ~ Intervention + Sex |
| -2,614453817 | y ~ Intervention + Sex |
| 3,652976972 | y ~ Intervention + Sex |
| 1,161088985 | y ~ Intervention + Sex |
| -0,228295214 | y ~ Intervention + Sex |
| -0,405653801 | y ~ Intervention + Sex |
| -3,007904387 | y ~ Intervention + Sex |
| -0,084957709 | y ~ Intervention + Sex |
| -0,137223989 | y ~ Intervention + Sex |
| -0,671836057 | y ~ Intervention + Sex |
| 1,910329066 | y ~ Intervention + Sex |
| -1,360960618 | y ~ Intervention + Sex |
| 0,40364814 | y ~ Intervention + Sex |
| -0,670078452 | y ~ Intervention + Sex |
| -0,123494967 | y ~ Intervention + Sex |
| -0,431342376 | y ~ Intervention + Sex |
| -0,907546627 | y ~ Intervention + Sex |
| 0,648928298 | y ~ Intervention + Sex |
| -1,057620339 | y ~ Intervention + Sex |
| 0,236431107 | y ~ Intervention + Sex |
| -0,185816097 | y ~ Intervention + Sex |
| -1,923029783 | y ~ Intervention + Sex |
| 0,29324939 | y ~ Intervention + Sex |
| -3,11718873 | y ~ Intervention + Sex |
| -0,135647089 | y ~ Intervention + Sex |
| -0,974189077 | y ~ Intervention + Sex |
| 0,877097656 | y ~ Intervention + Sex |
| 1,371301201 | y ~ Intervention + Sex |
| -0,276199179 | y ~ Intervention + Sex |
| -1,651459796 | y ~ Intervention + Sex |
| -0,480088805 | y ~ Intervention + Sex |

| | |
|---|---|
| -2,05479913 | y ~ Intervention + Sex |
| -0,375178945 | y ~ Intervention + Sex |
| -1,702189769 | y ~ Intervention + Sex |
| -3,181849344 | y ~ Intervention + Sex |
| 0,0076253 | y ~ Intervention + Sex |
| 0,738033633 | y ~ Intervention + Sex |
| -0,030723786 | y ~ Intervention + Sex |
| -0,11142769 | y ~ Intervention + Sex |
| -2,404987673 | y ~ Intervention + Sex |
| -0,607201656 | y ~ Intervention + Sex |
| -1,404390665 | y ~ Intervention + Sex |
| -1,214714469 | y ~ Intervention + Sex |
| -0,132209945 | y ~ Intervention + Sex |
| -2,618566509 | y ~ Intervention + Sex |
| -1,384536579 | y ~ Intervention + Sex |
| 0,209893178 | y ~ Intervention + Sex |
| 0,166990924 | y ~ Intervention + Sex |
| 0,715157503 | y ~ Intervention + Sex |
| -2,95891698 | y ~ Intervention + Sex |
| -2,901876724 | y ~ Intervention + Sex |
| -1,022578881 | y ~ Intervention + Sex |
| 0,364147768 | y ~ Intervention + Sex |
| -0,224402838 | y ~ Intervention + Sex |
| 1,446105207 | y ~ Intervention + Sex |
| 3,663399648 | y ~ Intervention + Sex |
| -0,667826829 | y ~ Intervention + Sex |
| -0,14732845 | y ~ Intervention + Sex |
| 1,019802562 | y ~ Intervention + Sex |
| 0,173379713 | y ~ Intervention + Sex |
| -1,042260276 | y ~ Intervention + Sex |
| 1,415073269 | y ~ Intervention + Sex |
| -0,667854018 | y ~ Intervention + Sex |
| -0,740964522 | y ~ Intervention + Sex |
| -3,406959425 | y ~ Intervention + Sex |
| -0,161940962 | y ~ Intervention + Sex |
| -1,027957224 | y ~ Intervention + Sex |
| 1,423903237 | y ~ Intervention + Sex |
| -0,949666129 | y ~ Intervention + Sex |
| 3,204767719 | y ~ Intervention + Sex |
| -1,288246657 | y ~ Intervention + Sex |
| -0,781101329 | y ~ Intervention + Sex |
| 1,189065165 | y ~ Intervention + Sex |
| -0,740694753 | y ~ Intervention + Sex |
| 0,703501995 | y ~ Intervention + Sex |
| 0,159546739 | y ~ Intervention + Sex |
| 0,286826925 | y ~ Intervention + Sex |
| -0,181286555 | y ~ Intervention + Sex |
| 0,322813793 | y ~ Intervention + Sex |

| | |
|---|---|
| 0,958999158 | y ~ Intervention + Sex |
| 2,991945407 | y ~ Intervention + Sex |
| -1,609050715 | y ~ Intervention + Sex |
| -0,426681569 | y ~ Intervention + Sex |
| -1,545982955 | y ~ Intervention + Sex |
| -1,435773083 | y ~ Intervention + Sex |
| -1,330278241 | y ~ Intervention + Sex |
| -1,353910026 | y ~ Intervention + Sex |
| -1,019124832 | y ~ Intervention + Sex |
| 1,563470466 | y ~ Intervention + Sex |
| 0,346346114 | y ~ Intervention + Sex |
| -2,660626839 | y ~ Intervention + Sex |
| -1,276955312 | y ~ Intervention + Sex |
| -0,488040583 | y ~ Intervention + Sex |
| -1,638398311 | y ~ Intervention + Sex |
| -1,650568631 | y ~ Intervention + Sex |
| -0,692958836 | y ~ Intervention + Sex |
| -0,229759715 | y ~ Intervention + Sex |
| -0,689168252 | y ~ Intervention + Sex |
| -0,14605592 | y ~ Intervention + Sex |
| -1,14169184 | y ~ Intervention + Sex |
| 0,485554629 | y ~ Intervention + Sex |
| 0,730721481 | y ~ Intervention + Sex |
| -1,988197561 | y ~ Intervention + Sex |
| 0,166792195 | y ~ Intervention + Sex |
| 0,210119214 | y ~ Intervention + Sex |
| -0,362777936 | y ~ Intervention + Sex |
| 0,194380215 | y ~ Intervention + Sex |
| -0,582887781 | y ~ Intervention + Sex |
| 0,916599743 | y ~ Intervention + Sex |
| 0,931821017 | y ~ Intervention + Sex |
| -1,549477827 | y ~ Intervention + Sex |
| -0,628228117 | y ~ Intervention + Sex |
| -0,299067422 | y ~ Intervention + Sex |
| -1,333604037 | y ~ Intervention + Sex |
| -0,008054444 | y ~ Intervention + Sex |
| 1,060848199 | y ~ Intervention + Sex |
| -3,071815954 | y ~ Intervention + Sex |
| 0,546444194 | y ~ Intervention + Sex |
| -0,056474065 | y ~ Intervention + Sex |
| 0,127463013 | y ~ Intervention + Sex |
| -0,410992711 | y ~ Intervention + Sex |
| -0,612713449 | y ~ Intervention + Sex |
| 0,160024275 | y ~ Intervention + Sex |
| 2,775108425 | y ~ Intervention + Sex |
| -1,009305645 | y ~ Intervention + Sex |
| 3,033541723 | y ~ Intervention + Sex |
| -1,005159013 | y ~ Intervention + Sex |

| | |
|---|---|
| -1,262833522 | y ~ Intervention + Sex |
| -0,531197036 | y ~ Intervention + Sex |
| -0,726270853 | y ~ Intervention + Sex |
| 0,229101688 | y ~ Intervention + Sex |
| -0,683168962 | y ~ Intervention + Sex |
| 0,374351398 | y ~ Intervention + Sex |
| -0,38252061 | y ~ Intervention + Sex |
| -3,592533529 | y ~ Intervention + Sex |
| 0,925378314 | y ~ Intervention + Sex |
| -1,751108127 | y ~ Intervention + Sex |
| 0,338206867 | y ~ Intervention + Sex |
| 0,271991653 | y ~ Intervention + Sex |
| -0,948629206 | y ~ Intervention + Sex |
| -0,889668272 | y ~ Intervention + Sex |
| 1,052715555 | y ~ Intervention + Sex |
| 0,530591228 | y ~ Intervention + Sex |
| 3,391402398 | y ~ Intervention + Sex |
| 0,856648093 | y ~ Intervention + Sex |
| -1,502064756 | y ~ Intervention + Sex |
| 0,010685036 | y ~ Intervention + Sex |
| 2,312459049 | y ~ Intervention + Sex |
| 1,962766431 | y ~ Intervention + Sex |
| -0,901006064 | y ~ Intervention + Sex |
| 1,104315891 | y ~ Intervention + Sex |
| -1,434861672 | y ~ Intervention + Sex |
| -0,949840534 | y ~ Intervention + Sex |
| -0,418880042 | y ~ Intervention + Sex |
| 0,249938207 | y ~ Intervention + Sex |
| -2,221042802 | y ~ Intervention + Sex |
| 3,607908288 | y ~ Intervention + Sex |
| -1,464451413 | y ~ Intervention + Sex |
| 0,220643979 | y ~ Intervention + Sex |
| -1,76393614 | y ~ Intervention + Sex |
| -0,691914131 | y ~ Intervention + Sex |
| -0,550010141 | y ~ Intervention + Sex |
| -1,204894291 | y ~ Intervention + Sex |
| 1,751711407 | y ~ Intervention + Sex |
| -0,859154602 | y ~ Intervention + Sex |
| -0,334226828 | y ~ Intervention + Sex |
| -0,88287868 | y ~ Intervention + Sex |
| -0,032343882 | y ~ Intervention + Sex |
| -1,154590627 | y ~ Intervention + Sex |
| -0,253756532 | y ~ Intervention + Sex |
| -1,475473894 | y ~ Intervention + Sex |
| -0,272102512 | y ~ Intervention + Sex |
| -1,745539155 | y ~ Intervention + Sex |
| -2,416278279 | y ~ Intervention + Sex |
| -0,14943065 | y ~ Intervention + Sex |

| | |
|---|---|
| 0,052829306 | y ~ Intervention + Sex |
| -1,347018973 | y ~ Intervention + Sex |
| 0,540970495 | y ~ Intervention + Sex |
| 0,436178063 | y ~ Intervention + Sex |
| -1,764618117 | y ~ Intervention + Sex |
| 0,901823299 | y ~ Intervention + Sex |
| 1,916228001 | y ~ Intervention + Sex |
| 1,914961199 | y ~ Intervention + Sex |
| -2,993232859 | y ~ Intervention + Sex |
| 0,628363904 | y ~ Intervention + Sex |
| 1,001499762 | y ~ Intervention + Sex |
| -0,406906899 | y ~ Intervention + Sex |
| -0,079921806 | y ~ Intervention + Sex |
| 1,314600961 | y ~ Intervention + Sex |
| 1,118647126 | y ~ Intervention + Sex |
| -1,132094576 | y ~ Intervention + Sex |
| 1,04175862 | y ~ Intervention + Sex |
| -0,352008719 | y ~ Intervention + Sex |
| 0,065568062 | y ~ Intervention + Sex |
| -1,021898474 | y ~ Intervention + Sex |
| 0,879641409 | y ~ Intervention + Sex |
| -0,885087297 | y ~ Intervention + Sex |
| -1,100270332 | y ~ Intervention + Sex |
| -1,858418518 | y ~ Intervention + Sex |
| 0,116009861 | y ~ Intervention + Sex |
| 1,800364933 | y ~ Intervention + Sex |
| 1,654159908 | y ~ Intervention + Sex |
| 0,503577086 | y ~ Intervention + Sex |
| -1,776427299 | y ~ Intervention + Sex |
| 0,884339795 | y ~ Intervention + Sex |
| 0,764366646 | y ~ Intervention + Sex |
| -0,839469142 | y ~ Intervention + Sex |
| 0,363167439 | y ~ Intervention + Sex |
| -0,509258976 | y ~ Intervention + Sex |
| -0,859795729 | y ~ Intervention + Sex |
| -0,849389993 | y ~ Intervention + Sex |
| -0,076185439 | y ~ Intervention + Sex |
| -0,474190854 | y ~ Intervention + Sex |
| 0,217584852 | y ~ Intervention + Sex |
| -3,111791651 | y ~ Intervention + Sex |
| 0,602270763 | y ~ Intervention + Sex |
| 1,208724093 | y ~ Intervention + Sex |
| 0,618505806 | y ~ Intervention + Sex |
| -0,359889676 | y ~ Intervention + Sex |
| -0,561510808 | y ~ Intervention + Sex |
| 1,529742446 | y ~ Intervention + Sex |
| -0,572506995 | y ~ Intervention + Sex |
| 1,167738024 | y ~ Intervention + Sex |

| | |
|---|---|
| 0,901121684 | y ~ Intervention + Sex |
| -1,758818436 | y ~ Intervention + Sex |
| 1,950909404 | y ~ Intervention + Sex |
| 0,073027231 | y ~ Intervention + Sex |
| -0,396787992 | y ~ Intervention + Sex |
| -0,169842816 | y ~ Intervention + Sex |
| 1,415089008 | y ~ Intervention + Sex |
| -1,748530915 | y ~ Intervention + Sex |
| 2,119180762 | y ~ Intervention + Sex |
| 1,146685242 | y ~ Intervention + Sex |
| 0,088865302 | y ~ Intervention + Sex |
| 1,157660153 | y ~ Intervention + Sex |
| 1,829454471 | y ~ Intervention + Sex |
| 0,765319128 | y ~ Intervention + Sex |
| -2,04180362 | y ~ Intervention + Sex |
| 2,855103903 | y ~ Intervention + Sex |
| 0,040521075 | y ~ Intervention + Sex |
| 1,649594207 | y ~ Intervention + Sex |
| 0,025310586 | y ~ Intervention + Sex |
| 2,546496497 | y ~ Intervention + Sex |
| 1,930139566 | y ~ Intervention + Sex |
| -0,918879166 | y ~ Intervention + Sex |
| -3,428392019 | y ~ Intervention + Sex |
| 0,068446112 | y ~ Intervention + Sex |
| 0,49492166 | y ~ Intervention + Sex |
| 0,894971877 | y ~ Intervention + Sex |
| -2,325463072 | y ~ Intervention + Sex |
| 0,612911227 | y ~ Intervention + Sex |
| -0,014207781 | y ~ Intervention + Sex |
| -0,784339738 | y ~ Intervention + Sex |
| 2,855779494 | y ~ Intervention + Sex |
| 0,756768134 | y ~ Intervention + Sex |
| -1,453141249 | y ~ Intervention + Sex |
| 2,114090945 | y ~ Intervention + Sex |
| -0,274964931 | y ~ Intervention + Sex |
| -1,870076606 | y ~ Intervention + Sex |
| -0,92014005 | y ~ Intervention + Sex |
| -0,277696535 | y ~ Intervention + Sex |
| -1,477663397 | y ~ Intervention + Sex |
| -3,016226005 | y ~ Intervention + Sex |
| 1,314466102 | y ~ Intervention + Sex |
| -0,853512229 | y ~ Intervention + Sex |
| -1,727641398 | y ~ Intervention + Sex |
| 1,470956911 | y ~ Intervention + Sex |
| 0,581558567 | y ~ Intervention + Sex |
| -0,470700783 | y ~ Intervention + Sex |
| 1,783485964 | y ~ Intervention + Sex |
| 2,28978778 | y ~ Intervention + Sex |

| | |
|---|---|
| -0,345760832 | y ~ Intervention + Sex |
| 1,055388567 | y ~ Intervention + Sex |
| -0,200090301 | y ~ Intervention + Sex |
| 0,054137366 | y ~ Intervention + Sex |
| -0,518096485 | y ~ Intervention + Sex |
| -0,278519467 | y ~ Intervention + Sex |
| 0,258388237 | y ~ Intervention + Sex |
| -1,466346348 | y ~ Intervention + Sex |
| -1,230358679 | y ~ Intervention + Sex |
| -1,216721647 | y ~ Intervention + Sex |
| -0,813529738 | y ~ Intervention + Sex |
| 0,60582688 | y ~ Intervention + Sex |
| -1,226124291 | y ~ Intervention + Sex |
| 2,193154183 | y ~ Intervention + Sex |
| 0,123604067 | y ~ Intervention + Sex |
| -1,25551133 | y ~ Intervention + Sex |
| -1,846697338 | y ~ Intervention + Sex |
| -0,526841329 | y ~ Intervention + Sex |
| 0,755720236 | y ~ Intervention + Sex |
| -0,156230816 | y ~ Intervention + Sex |
| -1,174789444 | y ~ Intervention + Sex |
| -3,064854449 | y ~ Intervention + Sex |
| -2,497846588 | y ~ Intervention + Sex |
| -0,139094681 | y ~ Intervention + Sex |
| 0,239904038 | y ~ Intervention + Sex |
| -0,526988362 | y ~ Intervention + Sex |
| -0,050241707 | y ~ Intervention + Sex |
| 0,143631543 | y ~ Intervention + Sex |
| -1,628975695 | y ~ Intervention + Sex |
| -0,131814814 | y ~ Intervention + Sex |
| -1,948613928 | y ~ Intervention + Sex |
| 0,384195464 | y ~ Intervention + Sex |
| 1,950398534 | y ~ Intervention + Sex |
| -2,760248295 | y ~ Intervention + Sex |
| -0,861180076 | y ~ Intervention + Sex |
| -1,243907755 | y ~ Intervention + Sex |
| 1,086768665 | y ~ Intervention + Sex |
| 0,168658077 | y ~ Intervention + Sex |
| 0,846524977 | y ~ Intervention + Sex |
| -2,881697491 | y ~ Intervention + Sex |
| -0,03401483 | y ~ Intervention + Sex |
| 1,666052021 | y ~ Intervention + Sex |
| -0,424151302 | y ~ Intervention + Sex |
| 0,387746521 | y ~ Intervention + Sex |
| 0,09544696 | y ~ Intervention + Sex |
| 2,063402084 | y ~ Intervention + Sex |
| -0,353249676 | y ~ Intervention + Sex |
| 1,665099123 | y ~ Intervention + Sex |

| | |
|---|---|
| 0,463743449 | y ~ Intervention + Sex |
| -1,316411071 | y ~ Intervention + Sex |
| -0,306176365 | y ~ Intervention + Sex |
| 0,535662712 | y ~ Intervention + Sex |
| -0,662693943 | y ~ Intervention + Sex |
| 0,814107799 | y ~ Intervention + Sex |
| -2,961962511 | y ~ Intervention + Sex |
| -0,604873962 | y ~ Intervention + Sex |
| -0,638851682 | y ~ Intervention + Sex |
| 0,905192514 | y ~ Intervention + Sex |
| 0,710660263 | y ~ Intervention + Sex |
| 0,768812004 | y ~ Intervention + Sex |
| -0,959069104 | y ~ Intervention + Sex |
| -0,690836683 | y ~ Intervention + Sex |
| -1,368946225 | y ~ Intervention + Sex |
| 0,884710027 | y ~ Intervention + Sex |
| -0,278260898 | y ~ Intervention + Sex |
| 0,855428234 | y ~ Intervention + Sex |
| -1,700724218 | y ~ Intervention + Sex |
| 0,125970723 | y ~ Intervention + Sex |
| 8,775444702 | y ~ Intervention + Sex |
| 0,619525445 | y ~ Intervention + Sex |
| -1,930361877 | y ~ Intervention + Sex |
| -3,166474246 | y ~ Intervention + Sex |
| -2,152205825 | y ~ Intervention + Sex |
| 0,59517826 | y ~ Intervention + Sex |
| -2,615804579 | y ~ Intervention + Sex |
| -2,062479607 | y ~ Intervention + Sex |
| 0,576966602 | y ~ Intervention + Sex |
| -0,83736759 | y ~ Intervention + Sex |
| -0,215398333 | y ~ Intervention + Sex |
| 0,576964066 | y ~ Intervention + Sex |
| -1,93997675 | y ~ Intervention + Sex |
| -0,019441744 | y ~ Intervention + Sex |
| 0,02999893 | y ~ Intervention + Sex |
| -1,18799463 | y ~ Intervention + Sex |
| -1,00230975 | y ~ Intervention + Sex |
| -1,648555564 | y ~ Intervention + Sex |
| 3,783913873 | y ~ Intervention + Sex |
| -1,200804193 | y ~ Intervention + Sex |
| 0,884040994 | y ~ Intervention + Sex |
| 0,639869043 | y ~ Intervention + Sex |
| -0,559082089 | y ~ Intervention + Sex |
| 0,112598153 | y ~ Intervention + Sex |
| 0,78333091 | y ~ Intervention + Sex |
| -2,073032844 | y ~ Intervention + Sex |
| -2,398209851 | y ~ Intervention + Sex |
| 0,151322437 | y ~ Intervention + Sex |

| | |
|---|---|
| 3,416121597 | y ~ Intervention + Sex |
| 1,748594374 | y ~ Intervention + Sex |
| -0,428248924 | y ~ Intervention + Sex |
| 0,174858181 | y ~ Intervention + Sex |
| -0,354742069 | y ~ Intervention + Sex |
| -0,964094116 | y ~ Intervention + Sex |
| 0,725940779 | y ~ Intervention + Sex |
| 0,566372926 | y ~ Intervention + Sex |
| -0,521336317 | y ~ Intervention + Sex |
| 1,863826621 | y ~ Intervention + Sex |
| 0,044066196 | y ~ Intervention + Sex |
| 0,657701858 | y ~ Intervention + Sex |
| 0,361265488 | y ~ Intervention + Sex |
| -0,710297747 | y ~ Intervention + Sex |
| 0,163039702 | y ~ Intervention + Sex |
| -3,310775967 | y ~ Intervention + Sex |
| 1,127134421 | y ~ Intervention + Sex |
| -0,479266981 | y ~ Intervention + Sex |
| -1,929996581 | y ~ Intervention + Sex |
| 2,090818547 | y ~ Intervention + Sex |
| 0,55577472 | y ~ Intervention + Sex |
| -0,529590827 | y ~ Intervention + Sex |
| 0,54096114 | y ~ Intervention + Sex |
| 1,673617314 | y ~ Intervention + Sex |
| -0,87559328 | y ~ Intervention + Sex |
| -0,519370073 | y ~ Intervention + Sex |
| 0,388941577 | y ~ Intervention + Sex |
| 0,128294266 | y ~ Intervention + Sex |
| 0,794491478 | y ~ Intervention + Sex |
| -0,584928334 | y ~ Intervention + Sex |
| 0,008197218 | y ~ Intervention + Sex |
| 0,49218212 | y ~ Intervention + Sex |
| -0,583285087 | y ~ Intervention + Sex |
| 2,010611785 | y ~ Intervention + Sex |
| -2,580945489 | y ~ Intervention + Sex |
| -3,13486932 | y ~ Intervention + Sex |
| 0,450973349 | y ~ Intervention + Sex |
| -1,28889947 | y ~ Intervention + Sex |
| 0,978143261 | y ~ Intervention + Sex |
| 0,062083177 | y ~ Intervention + Sex |
| 0,657583909 | y ~ Intervention + Sex |
| 1,347532418 | y ~ Intervention + Sex |
| 0,71030494 | y ~ Intervention + Sex |
| 0,03452668 | y ~ Intervention + Sex |
| -1,8526207 | y ~ Intervention + Sex |
| -0,538399985 | y ~ Intervention + Sex |
| 1,126615115 | y ~ Intervention + Sex |
| 1,320609349 | y ~ Intervention + Sex |

| | |
|---|---|
| -0,509587987 | y ~ Intervention + Sex |
| 2,272508108 | y ~ Intervention + Sex |
| -3,396137861 | y ~ Intervention + Sex |
| 1,649293614 | y ~ Intervention + Sex |
| -0,550217599 | y ~ Intervention + Sex |
| -0,446458904 | y ~ Intervention + Sex |
| -1,935755588 | y ~ Intervention + Sex |
| 3,006197474 | y ~ Intervention + Sex |
| 1,171728923 | y ~ Intervention + Sex |
| 1,342360054 | y ~ Intervention + Sex |
| -0,271449128 | y ~ Intervention + Sex |
| -0,787086971 | y ~ Intervention + Sex |
| 0,572487309 | y ~ Intervention + Sex |
| 0,230914489 | y ~ Intervention + Sex |
| -0,163460139 | y ~ Intervention + Sex |
| 1,440181414 | y ~ Intervention + Sex |
| -0,412003124 | y ~ Intervention + Sex |
| 0,527929795 | y ~ Intervention + Sex |
| -2,569993648 | y ~ Intervention + Sex |
| -0,717005908 | y ~ Intervention + Sex |
| -2,259117522 | y ~ Intervention + Sex |
| 1,3487132 | y ~ Intervention + Sex |
| -0,052746699 | y ~ Intervention + Sex |
| -2,470500301 | y ~ Intervention + Sex |
| -1,956929206 | y ~ Intervention + Sex |
| 0,013931102 | y ~ Intervention + Sex |
| 0,174307566 | y ~ Intervention + Sex |
| 0,457175096 | y ~ Intervention + Sex |
| -1,490222223 | y ~ Intervention + Sex |
| -0,833090347 | y ~ Intervention + Sex |
| -1,746524232 | y ~ Intervention + Sex |
| -0,811790327 | y ~ Intervention + Sex |
| 0,390670705 | y ~ Intervention + Sex |
| -0,438878988 | y ~ Intervention + Sex |
| 1,428063804 | y ~ Intervention + Sex |
| -0,251495653 | y ~ Intervention + Sex |
| -1,402606087 | y ~ Intervention + Sex |
| 0,152413237 | y ~ Intervention + Sex |
| -1,008732186 | y ~ Intervention + Sex |
| -0,219595735 | y ~ Intervention + Sex |
| -0,252835487 | y ~ Intervention + Sex |
| 0,783313072 | y ~ Intervention + Sex |
| 1,347785391 | y ~ Intervention + Sex |
| -2,945453232 | y ~ Intervention + Sex |
| -1,109029419 | y ~ Intervention + Sex |
| -1,241483234 | y ~ Intervention + Sex |
| 2,066055261 | y ~ Intervention + Sex |
| 1,043415188 | y ~ Intervention + Sex |

| | |
|---|---|
| 1,230924065 | y ~ Intervention + Sex |
| -0,188421336 | y ~ Intervention + Sex |
| 0,60786005 | y ~ Intervention + Sex |
| 2,401166328 | y ~ Intervention + Sex |
| -2,780132113 | y ~ Intervention + Sex |
| 2,50847551 | y ~ Intervention + Sex |
| 0,312145577 | y ~ Intervention + Sex |
| 2,89682913 | y ~ Intervention + Sex |
| -0,123998109 | y ~ Intervention + Sex |
| -1,518969327 | y ~ Intervention + Sex |
| 0,545927319 | y ~ Intervention + Sex |
| 2,263277695 | y ~ Intervention + Sex |
| -0,267823378 | y ~ Intervention + Sex |
| -0,300344295 | y ~ Intervention + Sex |
| -2,92556715 | y ~ Intervention + Sex |
| 2,836729634 | y ~ Intervention + Sex |
| 1,172123615 | y ~ Intervention + Sex |
| -1,413941662 | y ~ Intervention + Sex |
| 1,362047574 | y ~ Intervention + Sex |
| 0,05186629 | y ~ Intervention + Sex |
| -3,529315699 | y ~ Intervention + Sex |
| -0,838552493 | y ~ Intervention + Sex |
| 1,229533436 | y ~ Intervention + Sex |
| 0,621040407 | y ~ Intervention + Sex |
| -1,083239698 | y ~ Intervention + Sex |
| -0,540649822 | y ~ Intervention + Sex |
| -0,577899678 | y ~ Intervention + Sex |
| -0,004089794 | y ~ Intervention + Sex |
| 1,891144249 | y ~ Intervention + Sex |
| 1,999327036 | y ~ Intervention + Sex |
| -2,07333715 | y ~ Intervention + Sex |
| 1,45732719 | y ~ Intervention + Sex |
| -1,401637459 | y ~ Intervention + Sex |
| 0,940460533 | y ~ Intervention + Sex |
| -1,096612344 | y ~ Intervention + Sex |
| 0,271562571 | y ~ Intervention + Sex |
| 0,253047211 | y ~ Intervention + Sex |
| -0,908450915 | y ~ Intervention + Sex |
| -0,095067783 | y ~ Intervention + Sex |
| -2,078028853 | y ~ Intervention + Sex |
| 0,948423294 | y ~ Intervention + Sex |
| 0,612991316 | y ~ Intervention + Sex |
| 1,325877851 | y ~ Intervention + Sex |
| -2,038565798 | y ~ Intervention + Sex |
| -0,459649333 | y ~ Intervention + Sex |
| -1,020331231 | y ~ Intervention + Sex |
| -0,525003614 | y ~ Intervention + Sex |
| -1,550742315 | y ~ Intervention + Sex |

| | |
|---|---|
| -1,783495718 | y ~ Intervention + Sex |
| -0,490340844 | y ~ Intervention + Sex |
| 0,473578298 | y ~ Intervention + Sex |
| 0,696896124 | y ~ Intervention + Sex |
| 0,184747621 | y ~ Intervention + Sex |
| -0,128334917 | y ~ Intervention + Sex |
| -0,596058518 | y ~ Intervention + Sex |
| 2,14619287 | y ~ Intervention + Sex |
| -1,630864331 | y ~ Intervention + Sex |
| -0,241793491 | y ~ Intervention + Sex |
| 0,37568811 | y ~ Intervention + Sex |
| 0,997959415 | y ~ Intervention + Sex |
| -0,005045946 | y ~ Intervention + Sex |
| 0,681804295 | y ~ Intervention + Sex |
| 1,790190076 | y ~ Intervention + Sex |
| -0,900128564 | y ~ Intervention + Sex |
| -1,873229719 | y ~ Intervention + Sex |
| -0,66472494 | y ~ Intervention + Sex |
| 0,24633352 | y ~ Intervention + Sex |
| 7,008535332 | y ~ Intervention + Sex |
| 0,22142842 | y ~ Intervention + Sex |
| 1,854905437 | y ~ Intervention + Sex |
| 1,747855321 | y ~ Intervention + Sex |
| -1,305643867 | y ~ Intervention + Sex |
| -1,425611065 | y ~ Intervention + Sex |
| 0,303956838 | y ~ Intervention + Sex |
| 0,275593293 | y ~ Intervention + Sex |
| -1,687877955 | y ~ Intervention + Sex |
| 1,407690678 | y ~ Intervention + Sex |
| 1,521935364 | y ~ Intervention + Sex |
| 0,863495032 | y ~ Intervention + Sex |
| 1,357795976 | y ~ Intervention + Sex |
| 1,749502286 | y ~ Intervention + Sex |
| 1,581328285 | y ~ Intervention + Sex |
| 0,917187272 | y ~ Intervention + Sex |
| 3,384671599 | y ~ Intervention + Sex |
| -0,697964696 | y ~ Intervention + Sex |
| -0,209479922 | y ~ Intervention + Sex |
| -1,872914974 | y ~ Intervention + Sex |
| -2,136927695 | y ~ Intervention + Sex |
| 0,110075244 | y ~ Intervention + Sex |
| -0,904268444 | y ~ Intervention + Sex |
| -0,691329146 | y ~ Intervention + Sex |
| -3,015126969 | y ~ Intervention + Sex |
| -0,419296785 | y ~ Intervention + Sex |
| 0,78238776 | y ~ Intervention + Sex |
| -1,082069918 | y ~ Intervention + Sex |
| 1,27664159 | y ~ Intervention + Sex |

| | |
|---|---|
| -1,888501286 | y ~ Intervention + Sex |
| -0,595664519 | y ~ Intervention + Sex |
| -1,50090757 | y ~ Intervention + Sex |
| -5,040234672 | y ~ Intervention + Sex |
| -1,357325475 | y ~ Intervention + Sex |
| -1,532534593 | y ~ Intervention + Sex |
| -0,582580079 | y ~ Intervention + Sex |
| 0,050914736 | y ~ Intervention + Sex |
| -0,100989132 | y ~ Intervention + Sex |
| -2,920303681 | y ~ Intervention + Sex |
| -0,063546571 | y ~ Intervention + Sex |
| 0,80924267 | y ~ Intervention + Sex |
| 1,158168234 | y ~ Intervention + Sex |
| -0,43719497 | y ~ Intervention + Sex |
| 1,031449969 | y ~ Intervention + Sex |
| 0,265132188 | y ~ Intervention + Sex |
| 3,379832717 | y ~ Intervention + Sex |
| 0,877327048 | y ~ Intervention + Sex |
| 3,321931882 | y ~ Intervention + Sex |
| -1,694703155 | y ~ Intervention + Sex |
| 3,056042965 | y ~ Intervention + Sex |
| -0,595070482 | y ~ Intervention + Sex |
| -2,16149275 | y ~ Intervention + Sex |
| 1,879793671 | y ~ Intervention + Sex |
| -0,37166253 | y ~ Intervention + Sex |
| 0,593865369 | y ~ Intervention + Sex |
| 1,159843085 | y ~ Intervention + Sex |
| 0,685853584 | y ~ Intervention + Sex |
| -0,979967541 | y ~ Intervention + Sex |
| -1,192984554 | y ~ Intervention + Sex |
| -0,62711207 | y ~ Intervention + Sex |
| 0,600576715 | y ~ Intervention + Sex |
| 0,958902534 | y ~ Intervention + Sex |
| -0,097672479 | y ~ Intervention + Sex |
| -0,356870694 | y ~ Intervention + Sex |
| 5,475172377 | y ~ Intervention + Sex |
| -2,789754091 | y ~ Intervention + Sex |
| 0,728857617 | y ~ Intervention + Sex |
| -1,060381496 | y ~ Intervention + Sex |
| 2,526879466 | y ~ Intervention + Sex |
| 0,832346769 | y ~ Intervention + Sex |
| -1,397070455 | y ~ Intervention + Sex |
| 2,620090974 | y ~ Intervention + Sex |
| -0,880102907 | y ~ Intervention + Sex |
| 0,264736126 | y ~ Intervention + Sex |
| -1,84311774 | y ~ Intervention + Sex |
| -1,274073357 | y ~ Intervention + Sex |
| 1,598826977 | y ~ Intervention + Sex |

| | |
|---|---|
| -1,486147441 | y ~ Intervention + Sex |
| -1,10508586 | y ~ Intervention + Sex |
| -4,408426743 | y ~ Intervention + Sex |
| -0,902676399 | y ~ Intervention + Sex |
| 1,759640893 | y ~ Intervention + Sex |
| 0,42530308 | y ~ Intervention + Sex |
| -2,089932689 | y ~ Intervention + Sex |
| 3,235797015 | y ~ Intervention + Sex |
| -0,73209169 | y ~ Intervention + Sex |
| 0,335755738 | y ~ Intervention + Sex |
| -2,304512527 | y ~ Intervention + Sex |
| -0,716485187 | y ~ Intervention + Sex |
| -1,839336841 | y ~ Intervention + Sex |
| 0,778440177 | y ~ Intervention + Sex |
