## Supplemental table S10. Comparison of metabolome intensities between EYCF and CM. for "A YOUNG CHILD FORMULA SUPPLEMENTED WITH L. REUTERI AND GALACTO-OLIGOSACCHARIDES MODULATES THE COMPOSITION AND FUNCTION OF THE GUT MICROBIOME SUPPORTING BONE AND MUSCLE DEVELOPMENT IN TODDLERS"

| Visit | Test type | Item | name | ation_level | Feature | Contrast | Alternative | Reference | FDR |
|---|---|---|---|---|---|---|---|---|---|
| baseline | abundance | 2-Methylpr | 2-Methylpr | CFA_panel | Interventio | Experimen | Experimen | Control | 0,97445 |
| baseline | abundance | 3-Methylbu | 3-Methylbu | CFA_panel | Interventio | Experimen | Experimen | Control | 0,957875 |
| baseline | abundance | Acetic acid | Acetic acid | CFA_panel | Interventio | Experimen | Experimen | Control | 0,936399 |
| baseline | abundance | Butanoic a | Butanoic a | CFA_panel | Interventio | Experimen | Experimen | Control | 0,936399 |
| baseline | abundance | Hexanoic a | Hexanoic a | CFA_panel | Interventio | Experimen | Experimen | Control | 0,948031 |
| baseline | abundance | M1 | M1 | IA_module | Interventio | Experimen | Experimen | Control | 0,97445 |
| baseline | abundance | M10 | M10 | IA_module | Interventio | Experimen | Experimen | Control | 0,936399 |
| baseline | abundance | M11 | M11 | IA_module | Interventio | Experimen | Experimen | Control | 0,945105 |
| baseline | abundance | M12 | M12 | IA_module | Interventio | Experimen | Experimen | Control | 0,936399 |
| baseline | abundance | M13 | M13 | IA_module | Interventio | Experimen | Experimen | Control | 0,936399 |
| baseline | abundance | M14 | M14 | IA_module | Interventio | Experimen | Experimen | Control | 0,986763 |
| baseline | abundance | M15 | M15 | IA_module | Interventio | Experimen | Experimen | Control | 0,936399 |
| baseline | abundance | M16 | M16 | IA_module | Interventio | Experimen | Experimen | Control | 0,957875 |
| baseline | abundance | M17 | M17 | IA_module | Interventio | Experimen | Experimen | Control | 0,97445 |
| baseline | abundance | M18 | M18 | IA_module | Interventio | Experimen | Experimen | Control | 0,97445 |
| baseline | abundance | M19 | M19 | IA_module | Interventio | Experimen | Experimen | Control | 0,974375 |
| baseline | abundance | M2 | M2 | IA_module | Interventio | Experimen | Experimen | Control | 0,936399 |
| baseline | abundance | M20 | M20 | IA_module | Interventio | Experimen | Experimen | Control | 0,936399 |
| baseline | abundance | M21 | M21 | IA_module | Interventio | Experimen | Experimen | Control | 0,957875 |
| baseline | abundance | M22 | M22 | IA_module | Interventio | Experimen | Experimen | Control | 0,97445 |
| baseline | abundance | M23 | M23 | IA_module | Interventio | Experimen | Experimen | Control | 0,936399 |
| baseline | abundance | M24 | M24 | IA_module | Interventio | Experimen | Experimen | Control | 0,97445 |
| baseline | abundance | M25 | M25 | IA_module | Interventio | Experimen | Experimen | Control | 0,983461 |
| baseline | abundance | M26 | M26 | IA_module | Interventio | Experimen | Experimen | Control | 0,97445 |
| baseline | abundance | M27 | M27 | IA_module | Interventio | Experimen | Experimen | Control | 0,936399 |
| baseline | abundance | M28 | M28 | IA_module | Interventio | Experimen | Experimen | Control | 0,936399 |
| baseline | abundance | M29 | M29 | IA_module | Interventio | Experimen | Experimen | Control | 0,936399 |
| baseline | abundance | M3 | M3 | IA_module | Interventio | Experimen | Experimen | Control | 0,945105 |
| baseline | abundance | M30 | M30 | IA_module | Interventio | Experimen | Experimen | Control | 0,936399 |
| baseline | abundance | M31 | M31 | IA_module | Interventio | Experimen | Experimen | Control | 0,988948 |
| baseline | abundance | M32 | M32 | IA_module | Interventio | Experimen | Experimen | Control | 0,97445 |
| baseline | abundance | M33 | M33 | IA_module | Interventio | Experimen | Experimen | Control | 0,936399 |
| baseline | abundance | M34 | M34 | IA_module | Interventio | Experimen | Experimen | Control | 0,936399 |
| baseline | abundance | M4 | M4 | IA_module | Interventio | Experimen | Experimen | Control | 0,945105 |
| baseline | abundance | M5 | M5 | IA_module | Interventio | Experimen | Experimen | Control | 0,988948 |
| baseline | abundance | M6 | M6 | IA_module | Interventio | Experimen | Experimen | Control | 0,936399 |
| baseline | abundance | M7 | M7 | IA_module | Interventio | Experimen | Experimen | Control | 0,957875 |
| baseline | abundance | M8 | M8 | IA_module | Interventio | Experimen | Experimen | Control | 0,936399 |
| baseline | abundance | M9 | M9 | IA_module | Interventio | Experimen | Experimen | Control | 0,936399 |
| baseline | abundance | Pentanoic | Pentanoic | CFA_panel | Interventio | Experimen | Experimen | Control | 0,945105 |
| baseline | abundance | Propanoic | Propanoic | CFA_panel | Interventio | Experimen | Experimen | Control | 0,936399 |
| baseline | abundance | SL00001 | Deoxycytid | 2a | Interventio | Experimen | Experimen | Control | 0,948031 |
| baseline | abundance | SL00009 | N-Acetylhi | 2a | Interventio | Experimen | Experimen | Control | 0,97445 |
| baseline | abundance | SL00017 | Aspartic ac | 2a | Interventio | Experimen | Experimen | Control | 0,936399 |
| baseline | abundance | SL00020 | Carnitine | 2a | Interventio | Experimen | Experimen | Control | 0,936399 |
| baseline | abundance | SL00024 | Creatine | 2a | Interventio | Experimen | Experimen | Control | 0,955424 |
| baseline | abundance | SL00035 | Glycylglyci | 2a | Interventio | Experimen | Experimen | Control | 0,936399 |

| | | | | | | | | | |
|---|---|---|---|---|---|---|---|---|---|
| baseline | abundance | SL00040 | Kynurenine | 2a | Interventio | Experimen | Experimen | Control | 0,945105 |
| baseline | abundance | SL00049 | N-Acetylar | 2a | Interventio | Experimen | Experimen | Control | 0,948031 |
| baseline | abundance | SL00052 | N-Isovaler | 2a | Interventio | Experimen | Experimen | Control | 0,936399 |
| baseline | abundance | SL00054 | Ornithine | 1 | Interventio | Experimen | Experimen | Control | 0,945105 |
| baseline | abundance | SL00061 | Proline | 1 | Interventio | Experimen | Experimen | Control | 0,936399 |
| baseline | abundance | SL00063 | Pyridoxal | 1 | Interventio | Experimen | Experimen | Control | 0,97445 |
| baseline | abundance | SL00074 | Valine/ 5-A | 2a | Interventio | Experimen | Experimen | Control | 0,998355 |
| baseline | abundance | SL00081 | 5-Methylcy | 2a | Interventio | Experimen | Experimen | Control | 0,97445 |
| baseline | abundance | SL00082 | Allopurinol | 1 | Interventio | Experimen | Experimen | Control | 0,962445 |
| baseline | abundance | SL00089 | Dissaccari | 2a | Interventio | Experimen | Experimen | Control | 0,936399 |
| baseline | abundance | SL00096 | Hexoses III | 1 | Interventio | Experimen | Experimen | Control | 0,945105 |
| baseline | abundance | SL00097 | Hexoses II | 1 | Interventio | Experimen | Experimen | Control | 0,989213 |
| baseline | abundance | SL00103 | Gulonic ac | 2a | Interventio | Experimen | Experimen | Control | 0,936399 |
| baseline | abundance | SL00104 | Hexoses I | 1 | Interventio | Experimen | Experimen | Control | 0,97445 |
| baseline | abundance | SL00108 | Histidinol | 2a | Interventio | Experimen | Experimen | Control | 0,945105 |
| baseline | abundance | SL00124 | Dissaccari | 2a | Interventio | Experimen | Experimen | Control | 0,97445 |
| baseline | abundance | SL00128 | N-Acetylala | 2a | Interventio | Experimen | Experimen | Control | 0,948031 |
| baseline | abundance | SL00133 | N-Acetylglu | 2a | Interventio | Experimen | Experimen | Control | 0,936399 |
| baseline | abundance | SL00137 | Phenylacet | 2a | Interventio | Experimen | Experimen | Control | 0,945105 |
| baseline | abundance | SL00140 | Pyridoxami | 1 | Interventio | Experimen | Experimen | Control | 0,945105 |
| baseline | abundance | SL00149 | Dissaccari | 2a | Interventio | Experimen | Experimen | Control | 0,945105 |
| baseline | abundance | SL00154 | Trigonelline | 1 | Interventio | Experimen | Experimen | Control | 0,948031 |
| baseline | abundance | SL00159 | Uracil | 1 | Interventio | Experimen | Experimen | Control | 0,936399 |
| baseline | abundance | SL00161 | Uridine | 2a | Interventio | Experimen | Experimen | Control | 0,936399 |
| baseline | abundance | SL00185 | 2-Oxo-3-ph | 1 | Interventio | Experimen | Experimen | Control | 0,945105 |
| baseline | abundance | SL00189 | 3-Hydroxyb | 2a | Interventio | Experimen | Experimen | Control | 0,936399 |
| baseline | abundance | SL00197 | 4-Methyl-2 | 2a | Interventio | Experimen | Experimen | Control | 0,936399 |
| baseline | abundance | SL00199 | 5-Hydroxyi | 2a | Interventio | Experimen | Experimen | Control | 0,936399 |
| baseline | abundance | SL00200 | 8-Aminooc | 2a | Interventio | Experimen | Experimen | Control | 0,936399 |
| baseline | abundance | SL00201 | Pentose II | 2a | Interventio | Experimen | Experimen | Control | 0,937501 |
| baseline | abundance | SL00205 | Citraconic | 2a | Interventio | Experimen | Experimen | Control | 0,945105 |
| baseline | abundance | SL00210 | Galactonic | 2a | Interventio | Experimen | Experimen | Control | 0,948031 |
| baseline | abundance | SL00212 | Galactosar | 2a | Interventio | Experimen | Experimen | Control | 0,936399 |
| baseline | abundance | SL00215 | Glycolic ac | 2a | Interventio | Experimen | Experimen | Control | 0,97445 |
| baseline | abundance | SL00216 | Glyoxylic a | 2a | Interventio | Experimen | Experimen | Control | 0,97445 |
| baseline | abundance | SL00222 | Lactic acid | 1 | Interventio | Experimen | Experimen | Control | 0,97445 |
| baseline | abundance | SL00226 | N-Acetylgly | 2a | Interventio | Experimen | Experimen | Control | 0,945105 |
| baseline | abundance | SL00239 | Pinitol | 2a | Interventio | Experimen | Experimen | Control | 0,97445 |
| baseline | abundance | SL00240 | Pyruvic aci | 1 | Interventio | Experimen | Experimen | Control | 0,983461 |
| baseline | abundance | SL00243 | Deoxysuga | 2a | Interventio | Experimen | Experimen | Control | 0,957875 |
| baseline | abundance | SL00245 | sugar alcol | 2a | Interventio | Experimen | Experimen | Control | 0,936399 |
| baseline | abundance | SL00246 | Pentose III | 2a | Interventio | Experimen | Experimen | Control | 0,937501 |
| baseline | abundance | SL00247 | Shikimic ac | 2a | Interventio | Experimen | Experimen | Control | 0,936399 |
| baseline | abundance | SL00248 | Gluconic a | 2a | Interventio | Experimen | Experimen | Control | 0,936399 |
| baseline | abundance | SL00260 | Pentose I | 2a | Interventio | Experimen | Experimen | Control | 0,945786 |
| baseline | abundance | SL00262 | cis-Aconiti | 2a | Interventio | Experimen | Experimen | Control | 0,99606 |
| baseline | abundance | SL00264 | Methyl ace | 2a | Interventio | Experimen | Experimen | Control | 0,936399 |
| baseline | abundance | SL00265 | Purine | 2a | Interventio | Experimen | Experimen | Control | 0,936399 |

| | | | | | | | | | |
|---|---|---|---|---|---|---|---|---|---|
| baseline | abundance | SL00268 | Thymine | 1 | Interventio | Experimen | Experimen | Control | 0,936399 |
| baseline | abundance | SL00270 | 3-hydroxy- | 2a | Interventio | Experimen | Experimen | Control | 0,936399 |
| baseline | abundance | SL00275 | Dopamine | 2a | Interventio | Experimen | Experimen | Control | 0,936399 |
| baseline | abundance | SL00282 | 3-Hydroxy | 2a | Interventio | Experimen | Experimen | Control | 0,936399 |
| baseline | abundance | SL00285 | Dihydrofer | 2a | Interventio | Experimen | Experimen | Control | 0,936399 |
| baseline | abundance | SL00287 | DOPA | 2a | Interventio | Experimen | Experimen | Control | 0,957875 |
| baseline | abundance | SL00288 | Propionylc | 2a | Interventio | Experimen | Experimen | Control | 0,9737 |
| baseline | abundance | SL00290 | N-Acetylm | 2a | Interventio | Experimen | Experimen | Control | 0,97445 |
| baseline | abundance | SL00293 | Butyrylcarr | 2a | Interventio | Experimen | Experimen | Control | 0,97445 |
| baseline | abundance | SL00295 | 5-Methylur | 2a | Interventio | Experimen | Experimen | Control | 0,936399 |
| baseline | abundance | SL00297 | Acetylmur | 2a | Interventio | Experimen | Experimen | Control | 0,936399 |
| baseline | abundance | SL00299 | β-Murichol | 2a | Interventio | Experimen | Experimen | Control | 0,945105 |
| baseline | abundance | SL00309 | 1-Aminocy | 2a | Interventio | Experimen | Experimen | Control | 0,936399 |
| baseline | abundance | SL00311 | N,N-Dimet | 2a | Interventio | Experimen | Experimen | Control | 0,936399 |
| baseline | abundance | SL00313 | 2-Aminois | 2a | Interventio | Experimen | Experimen | Control | 0,936399 |
| baseline | abundance | SL00315 | Malonic ac | 2a | Interventio | Experimen | Experimen | Control | 0,936399 |
| baseline | abundance | SL00318 | 4-Aminoph | 1 | Interventio | Experimen | Experimen | Control | 0,936399 |
| baseline | abundance | SL00320 | γ-Caprolac | 2a | Interventio | Experimen | Experimen | Control | 0,936399 |
| baseline | abundance | SL00325 | 2-Methylm | 2a | Interventio | Experimen | Experimen | Control | 0,97445 |
| baseline | abundance | SL00328 | 2-(hydroxy | 2a | Interventio | Experimen | Experimen | Control | 0,978464 |
| baseline | abundance | SL00329 | 2-Hydroxy- | 2a | Interventio | Experimen | Experimen | Control | 0,945105 |
| baseline | abundance | SL00334 | Imidazolea | 2a | Interventio | Experimen | Experimen | Control | 0,936399 |
| baseline | abundance | SL00346 | N-Methylni | 2a | Interventio | Experimen | Experimen | Control | 0,97445 |
| baseline | abundance | SL00349 | 1-Aminocy | 2a | Interventio | Experimen | Experimen | Control | 0,948031 |
| baseline | abundance | SL00350 | Stachydrin | 1 | Interventio | Experimen | Experimen | Control | 0,936399 |
| baseline | abundance | SL00353 | Adipic acid | 1 | Interventio | Experimen | Experimen | Control | 0,97445 |
| baseline | abundance | SL00356 | N-(5-Amin | 2a | Interventio | Experimen | Experimen | Control | 0,945105 |
| baseline | abundance | SL00368 | N-Methylty | 1 | Interventio | Experimen | Experimen | Control | 0,936399 |
| baseline | abundance | SL00373 | N1-Methyl- | 2a | Interventio | Experimen | Experimen | Control | 0,97445 |
| baseline | abundance | SL00376 | N-AcetylPr | 2a | Interventio | Experimen | Experimen | Control | 0,936399 |
| baseline | abundance | SL00380 | 2-Aminoad | 2a | Interventio | Experimen | Experimen | Control | 0,936399 |
| baseline | abundance | SL00383 | 3-(2-Hydro | 2a | Interventio | Experimen | Experimen | Control | 0,945105 |
| baseline | abundance | SL00384 | p-Coumari | 1 | Interventio | Experimen | Experimen | Control | 0,936399 |
| baseline | abundance | SL00390 | Gallic acid | 2a | Interventio | Experimen | Experimen | Control | 0,936399 |
| baseline | abundance | SL00391 | N-Acetylle | 1 | Interventio | Experimen | Experimen | Control | 0,936399 |
| baseline | abundance | SL00401 | Theophylli | 1 | Interventio | Experimen | Experimen | Control | 0,936399 |
| baseline | abundance | SL00407 | Homovanil | 2a | Interventio | Experimen | Experimen | Control | 0,936399 |
| baseline | abundance | SL00420 | 4-Hydroxy- | 2a | Interventio | Experimen | Experimen | Control | 0,957875 |
| baseline | abundance | SL00421 | Asymmetri | 2a | Interventio | Experimen | Experimen | Control | 0,978464 |
| baseline | abundance | SL00430 | Homocarn | 2a | Interventio | Experimen | Experimen | Control | 0,936399 |
| baseline | abundance | SL00431 | Tiglylcarnit | 2a | Interventio | Experimen | Experimen | Control | 0,936399 |
| baseline | abundance | SL00433 | Isovaleryl | 2a | Interventio | Experimen | Experimen | Control | 0,948031 |
| baseline | abundance | SL00436 | 1-Carboxy | 2a | Interventio | Experimen | Experimen | Control | 0,936399 |
| baseline | abundance | SL00438 | Daidzein | 2a | Interventio | Experimen | Experimen | Control | 0,957875 |
| baseline | abundance | SL00440 | 5-Methylcy | 2a | Interventio | Experimen | Experimen | Control | 0,936399 |
| baseline | abundance | SL00445 | Glucosami | 2a | Interventio | Experimen | Experimen | Control | 0,945105 |
| baseline | abundance | SL00447 | Apigenin | 2a | Interventio | Experimen | Experimen | Control | 0,945105 |
| baseline | abundance | SL00455 | 12,13-DHC | 2a | Interventio | Experimen | Experimen | Control | 0,936399 |

| | | | | | | | | | |
|---|---|---|---|---|---|---|---|---|---|
| baseline | abundance | SL00467 | Sucralose | 2a | Interventio | Experimen | Experimen | Control | 0,936399 |
| baseline | abundance | SL00502 | 2-Ketobuty | 1 | Interventio | Experimen | Experimen | Control | 0,945105 |
| baseline | abundance | X00011 | Glutamine | 2a | Interventio | Experimen | Experimen | Control | 0,936399 |
| baseline | abundance | X00018 | Succinic ac | 1 | Interventio | Experimen | Experimen | Control | 0,955424 |
| baseline | abundance | X00020 | sugar alcol | 1 | Interventio | Experimen | Experimen | Control | 0,936399 |
| baseline | abundance | X00022 | N-Methylas | 1 | Interventio | Experimen | Experimen | Control | 0,936399 |
| baseline | abundance | X00023 | N-Formylm | 1 | Interventio | Experimen | Experimen | Control | 0,945105 |
| baseline | abundance | X00025 | Acesulfam | 1 | Interventio | Experimen | Experimen | Control | 0,936399 |
| baseline | abundance | X00026 | Inosine | 1 | Interventio | Experimen | Experimen | Control | 0,936399 |
| baseline | abundance | X00027 | N-Acetylglu | 1 | Interventio | Experimen | Experimen | Control | 0,936399 |
| baseline | abundance | X00029 | Urocanic a | 1 | Interventio | Experimen | Experimen | Control | 0,936399 |
| baseline | abundance | X00030 | 3,4-Dihydr | 1 | Interventio | Experimen | Experimen | Control | 0,936399 |
| baseline | abundance | X00032 | Indole-3-pr | 1 | Interventio | Experimen | Experimen | Control | 0,936399 |
| baseline | abundance | X00033 | Deoxycholi | 1 | Interventio | Experimen | Experimen | Control | 0,957875 |
| baseline | abundance | X00034 | Pyroglutam | 1 | Interventio | Experimen | Experimen | Control | 0,945105 |
| baseline | abundance | X00035 | Choline | 1 | Interventio | Experimen | Experimen | Control | 0,97445 |
| baseline | abundance | X00036 | Serine | 1 | Interventio | Experimen | Experimen | Control | 0,936399 |
| baseline | abundance | X00038 | Histamine | 1 | Interventio | Experimen | Experimen | Control | 0,936399 |
| baseline | abundance | X00040 | N-Acetylty | 1 | Interventio | Experimen | Experimen | Control | 0,987751 |
| baseline | abundance | X00042 | Glucuronic | 1 | Interventio | Experimen | Experimen | Control | 0,936399 |
| baseline | abundance | X00045 | Threonine | 1 | Interventio | Experimen | Experimen | Control | 0,97445 |
| baseline | abundance | X00050 | β-Hydroxyi | 1 | Interventio | Experimen | Experimen | Control | 0,955424 |
| baseline | abundance | X00051 | β-D-Gluco | 2b | Interventio | Experimen | Experimen | Control | 0,936399 |
| baseline | abundance | X00056 | 3-(4-hydro | 1 | Interventio | Experimen | Experimen | Control | 0,936399 |
| baseline | abundance | X00057 | N-Acetylor | 1 | Interventio | Experimen | Experimen | Control | 0,948031 |
| baseline | abundance | X00059 | 3,4-Dihydr | 1 | Interventio | Experimen | Experimen | Control | 0,936399 |
| baseline | abundance | X00060 | Ethylmalor | 1 | Interventio | Experimen | Experimen | Control | 0,98802 |
| baseline | abundance | X00061 | Thiamine | 1 | Interventio | Experimen | Experimen | Control | 0,945105 |
| baseline | abundance | X00063 | 2-Hydroxy | 1 | Interventio | Experimen | Experimen | Control | 0,97445 |
| baseline | abundance | X00064 | Traumatic | 1 | Interventio | Experimen | Experimen | Control | 0,957875 |
| baseline | abundance | X00066 | Tryptamine | 1 | Interventio | Experimen | Experimen | Control | 0,945105 |
| baseline | abundance | X00067 | Hypoxanth | 1 | Interventio | Experimen | Experimen | Control | 0,936399 |
| baseline | abundance | X00068 | N-Acetylm | 1 | Interventio | Experimen | Experimen | Control | 0,936399 |
| baseline | abundance | X00070 | α-aminobu | 1 | Interventio | Experimen | Experimen | Control | 0,936399 |
| baseline | abundance | X00071 | Tyrosine | 1 | Interventio | Experimen | Experimen | Control | 0,936399 |
| baseline | abundance | X00072 | 3,5-Dihydr | 1 | Interventio | Experimen | Experimen | Control | 0,936399 |
| baseline | abundance | X00073 | Indole-3-la | 1 | Interventio | Experimen | Experimen | Control | 0,97445 |
| baseline | abundance | X00074 | Methylsuc | 1 | Interventio | Experimen | Experimen | Control | 0,985326 |
| baseline | abundance | X00076 | Isoleucine | 1 | Interventio | Experimen | Experimen | Control | 0,945105 |
| baseline | abundance | X00078 | Acetylagm | 1 | Interventio | Experimen | Experimen | Control | 0,97445 |
| baseline | abundance | X00082 | Cholic acic | 1 | Interventio | Experimen | Experimen | Control | 0,945105 |
| baseline | abundance | X00083 | 1,7-Dimeth | 1 | Interventio | Experimen | Experimen | Control | 0,936399 |
| baseline | abundance | X00084 | Tricarballyl | 1 | Interventio | Experimen | Experimen | Control | 0,936399 |
| baseline | abundance | X00088 | Pantotheni | 1 | Interventio | Experimen | Experimen | Control | 0,948031 |
| baseline | abundance | X00089 | 4-Hydroxyb | 1 | Interventio | Experimen | Experimen | Control | 0,946333 |
| baseline | abundance | X00090 | Malic acid | 1 | Interventio | Experimen | Experimen | Control | 0,992737 |
| baseline | abundance | X00092 | 3-Methylhi | 1 | Interventio | Experimen | Experimen | Control | 0,936399 |
| baseline | abundance | X00093 | Deoxyinosi | 1 | Interventio | Experimen | Experimen | Control | 0,97445 |

| | | | | | | | | | |
|---|---|---|---|---|---|---|---|---|---|
| baseline | abundance | X00094 | Methionine | 1 | Interventio | Experimen | Experimen | Control | 0,945105 |
| baseline | abundance | X00097 | 2,6-Dihydr | 1 | Interventio | Experimen | Experimen | Control | 0,936399 |
| baseline | abundance | X00099 | 7-Methylgu | 1 | Interventio | Experimen | Experimen | Control | 0,936399 |
| baseline | abundance | X00101 | Threonic ac | 1 | Interventio | Experimen | Experimen | Control | 0,986763 |
| baseline | abundance | X00102 | Tryptophan | 1 | Interventio | Experimen | Experimen | Control | 0,936399 |
| baseline | abundance | X00106 | N-Acetyltry | 1 | Interventio | Experimen | Experimen | Control | 0,97445 |
| baseline | abundance | X00107 | N6-Acetylly | 1 | Interventio | Experimen | Experimen | Control | 0,992737 |
| baseline | abundance | X00112 | Xanthosine | 1 | Interventio | Experimen | Experimen | Control | 0,945105 |
| baseline | abundance | X00113 | N-Acetylph | 1 | Interventio | Experimen | Experimen | Control | 0,982558 |
| baseline | abundance | X00114 | Indole-3-m | 1 | Interventio | Experimen | Experimen | Control | 0,936399 |
| baseline | abundance | X00115 | Histidine | 1 | Interventio | Experimen | Experimen | Control | 0,936399 |
| baseline | abundance | X00116 | Cytidine | 1 | Interventio | Experimen | Experimen | Control | 0,945105 |
| baseline | abundance | X00120 | Leucylalan | 1 | Interventio | Experimen | Experimen | Control | 0,936399 |
| baseline | abundance | X00123 | Phenylalar | 1 | Interventio | Experimen | Experimen | Control | 0,945105 |
| baseline | abundance | X00124 | Leucine | 1 | Interventio | Experimen | Experimen | Control | 0,936399 |
| baseline | abundance | X00125 | Taurine | 1 | Interventio | Experimen | Experimen | Control | 0,936399 |
| baseline | abundance | X00127 | Nicotinic a | 1 | Interventio | Experimen | Experimen | Control | 0,936399 |
| baseline | abundance | X00129 | Quinic acid | 1 | Interventio | Experimen | Experimen | Control | 0,936399 |
| baseline | abundance | X00130 | Deoxyuridi | 1 | Interventio | Experimen | Experimen | Control | 0,936399 |
| baseline | abundance | X00132 | Glyceric ac | 1 | Interventio | Experimen | Experimen | Control | 0,936399 |
| baseline | abundance | X00135 | N-Acetylglu | 1 | Interventio | Experimen | Experimen | Control | 0,936399 |
| baseline | abundance | X00136 | leu-gln_a | 3 | Interventio | Experimen | Experimen | Control | 0,936399 |
| baseline | abundance | X00163 | primidone_ | 3 | Interventio | Experimen | Experimen | Control | 0,936399 |
| baseline | abundance | X00180 | 1-(2-Carbo | 3 | Interventio | Experimen | Experimen | Control | 0,945105 |
| baseline | abundance | X00182 | 3-(Butylsul | 3 | Interventio | Experimen | Experimen | Control | 0,945105 |
| baseline | abundance | X00199 | tert-Butyl 3 | 3 | Interventio | Experimen | Experimen | Control | 0,936399 |
| baseline | abundance | X00204 | 4-O-{3-O-[( | 3 | Interventio | Experimen | Experimen | Control | 0,945105 |
| baseline | abundance | X00221 | Astemizole | 3 | Interventio | Experimen | Experimen | Control | 0,948031 |
| baseline | abundance | X00224 | Nifedipine | 3 | Interventio | Experimen | Experimen | Control | 0,945105 |
| baseline | abundance | X00233 | g-Aminobu | 3 | Interventio | Experimen | Experimen | Control | 0,945105 |
| baseline | abundance | X00237 | OI1700000 | 3 | Interventio | Experimen | Experimen | Control | 0,936399 |
| baseline | abundance | X00242 | IN00258 | 3 | Interventio | Experimen | Experimen | Control | 0,989213 |
| baseline | abundance | X00253 | TDP-2_a | 3 | Interventio | Experimen | Experimen | Control | 0,991989 |
| baseline | abundance | X00261 | Mexiletine | 3 | Interventio | Experimen | Experimen | Control | 0,945105 |
| baseline | abundance | X00264 | PEG n12 | 2b | Interventio | Experimen | Experimen | Control | 0,957875 |
| baseline | abundance | X00266 | 2-Oxo-3-(p | 3 | Interventio | Experimen | Experimen | Control | 0,945105 |
| baseline | abundance | X00305 | Formylkynu | 3 | Interventio | Experimen | Experimen | Control | 0,936399 |
| baseline | abundance | X00320 | Panthenol_ | 3 | Interventio | Experimen | Experimen | Control | 0,945105 |
| baseline | abundance | X00327 | 3-Hydroxy- | 3 | Interventio | Experimen | Experimen | Control | 0,936399 |
| baseline | abundance | X00331 | 4-(9H-beta | 3 | Interventio | Experimen | Experimen | Control | 0,936399 |
| baseline | abundance | X00344 | SECONAL_ | 3 | Interventio | Experimen | Experimen | Control | 0,936399 |
| baseline | abundance | X00361 | nitecapone | 3 | Interventio | Experimen | Experimen | Control | 0,97445 |
| baseline | abundance | X00367 | hexobarbit | 3 | Interventio | Experimen | Experimen | Control | 0,959387 |
| baseline | abundance | X00375 | N-[(2S)-2-I | 3 | Interventio | Experimen | Experimen | Control | 0,945105 |
| baseline | abundance | X00383 | pretazettin | 3 | Interventio | Experimen | Experimen | Control | 0,957875 |
| baseline | abundance | X00384 | 3-Formyl-2 | 3 | Interventio | Experimen | Experimen | Control | 0,97445 |
| baseline | abundance | X00403 | 7alpha-Hy | 3 | Interventio | Experimen | Experimen | Control | 0,945105 |
| baseline | abundance | X00404 | piscidic ac | 3 | Interventio | Experimen | Experimen | Control | 0,936399 |

| | | | | | | | | | |
|---|---|---|---|---|---|---|---|---|---|
| baseline | abundance | X00432 | 8-(Methyls | 3 | Interventio | Experimen | Experimen | Control | 0,945105 |
| baseline | abundance | X00450 | Gly-Ser | 3 | Interventio | Experimen | Experimen | Control | 0,957875 |
| baseline | abundance | X00477 | Sparfloxac | 2b | Interventio | Experimen | Experimen | Control | 0,936399 |
| baseline | abundance | X00518 | Nisinic aci | 3 | Interventio | Experimen | Experimen | Control | 0,936399 |
| baseline | abundance | X00528 | R-(+)-Etira | 3 | Interventio | Experimen | Experimen | Control | 0,936399 |
| baseline | abundance | X00545 | 3-hydroxyc | 3 | Interventio | Experimen | Experimen | Control | 0,936399 |
| baseline | abundance | X00549 | 3-[(2Z)-1-C | 3 | Interventio | Experimen | Experimen | Control | 0,945105 |
| baseline | abundance | X00574 | miglustat | 3 | Interventio | Experimen | Experimen | Control | 0,936399 |
| baseline | abundance | X00594 | Nicotinami | 3 | Interventio | Experimen | Experimen | Control | 0,936399 |
| baseline | abundance | X00635 | 4-(5,6-Dihy | 3 | Interventio | Experimen | Experimen | Control | 0,945105 |
| baseline | abundance | X00637 | N~6~,N~6 | 3 | Interventio | Experimen | Experimen | Control | 0,936399 |
| baseline | abundance | X00646 | Deacetyldi | 3 | Interventio | Experimen | Experimen | Control | 0,936399 |
| baseline | abundance | X00670 | 7-Chloro-5 | 3 | Interventio | Experimen | Experimen | Control | 0,97445 |
| baseline | abundance | X00683 | Zinecard_a | 3 | Interventio | Experimen | Experimen | Control | 0,936399 |
| baseline | abundance | X00693 | n-Ribosylh | 3 | Interventio | Experimen | Experimen | Control | 0,936399 |
| baseline | abundance | X00698 | valgancicl | 3 | Interventio | Experimen | Experimen | Control | 0,936399 |
| baseline | abundance | X00702 | N-[(2S)-2-I | 3 | Interventio | Experimen | Experimen | Control | 0,936399 |
| baseline | abundance | X00722 | (3S,5R,6E) | 3 | Interventio | Experimen | Experimen | Control | 0,945105 |
| baseline | abundance | X00723 | Aspartyl-L- | 3 | Interventio | Experimen | Experimen | Control | 0,97445 |
| baseline | abundance | X00742 | MFCD1869 | 3 | Interventio | Experimen | Experimen | Control | 0,989769 |
| baseline | abundance | X00743 | 11beta,13- | 3 | Interventio | Experimen | Experimen | Control | 0,945105 |
| baseline | abundance | X00744 | (1R,2S)-1-( | 3 | Interventio | Experimen | Experimen | Control | 0,936399 |
| baseline | abundance | X00748 | 3-[3-Metho | 3 | Interventio | Experimen | Experimen | Control | 0,945786 |
| baseline | abundance | X00764 | 5-Allyl-5-se | 3 | Interventio | Experimen | Experimen | Control | 0,97445 |
| baseline | abundance | X00807 | Midodrine_ | 3 | Interventio | Experimen | Experimen | Control | 0,945105 |
| baseline | abundance | X00828 | Fenoterol | 3 | Interventio | Experimen | Experimen | Control | 0,957875 |
| baseline | abundance | X00836 | 3-Mercapt | 3 | Interventio | Experimen | Experimen | Control | 0,987642 |
| baseline | abundance | X00853 | 4-O-beta-D | 3 | Interventio | Experimen | Experimen | Control | 0,936399 |
| baseline | abundance | X00889 | LW800000 | 3 | Interventio | Experimen | Experimen | Control | 0,945105 |
| baseline | abundance | X00899 | N-(2,3,4-Tr | 3 | Interventio | Experimen | Experimen | Control | 0,936399 |
| baseline | abundance | X00907 | 1,2,3,4-Tet | 3 | Interventio | Experimen | Experimen | Control | 0,936399 |
| baseline | abundance | X00928 | (7R)-7-(5-c | 3 | Interventio | Experimen | Experimen | Control | 0,945105 |
| baseline | abundance | X00948 | Linamarin | 3 | Interventio | Experimen | Experimen | Control | 0,97445 |
| baseline | abundance | X00949 | primidone_ | 3 | Interventio | Experimen | Experimen | Control | 0,992737 |
| baseline | abundance | X00950 | 2-Phenylet | 3 | Interventio | Experimen | Experimen | Control | 0,936399 |
| baseline | abundance | X00963 | (4S)-4-[(2E | 3 | Interventio | Experimen | Experimen | Control | 0,936399 |
| baseline | abundance | X00985 | 1-(4-Aminc | 3 | Interventio | Experimen | Experimen | Control | 0,97445 |
| baseline | abundance | X00998 | epsilon-(ga | 3 | Interventio | Experimen | Experimen | Control | 0,959387 |
| baseline | abundance | X01007 | pentobarbi | 3 | Interventio | Experimen | Experimen | Control | 0,97445 |
| baseline | abundance | X01011 | DIBEHENIN | 3 | Interventio | Experimen | Experimen | Control | 0,97445 |
| baseline | abundance | X01017 | hexobarbit | 3 | Interventio | Experimen | Experimen | Control | 0,936399 |
| baseline | abundance | X01031 | 3-(14-Ethy | 3 | Interventio | Experimen | Experimen | Control | 0,945105 |
| baseline | abundance | X01045 | Ethyl maltc | 3 | Interventio | Experimen | Experimen | Control | 0,936399 |
| baseline | abundance | X01059 | coronatine | 3 | Interventio | Experimen | Experimen | Control | 0,945105 |
| baseline | abundance | X01067 | g-Aminobu | 3 | Interventio | Experimen | Experimen | Control | 0,936399 |
| baseline | abundance | X01078 | Dihydroura | 3 | Interventio | Experimen | Experimen | Control | 0,936399 |
| baseline | abundance | X01081 | Methyl alpl | 3 | Interventio | Experimen | Experimen | Control | 0,945105 |
| baseline | abundance | X01092 | 17,21-Dihy | 3 | Interventio | Experimen | Experimen | Control | 0,936399 |

| | | | | | | | | | |
|---|---|---|---|---|---|---|---|---|---|
| baseline | abundance | X01098 | (-)-Physost | 3 | Interventio | Experimen | Experimen | Control | 0,945105 |
| baseline | abundance | X01100 | (betaS)-be | 3 | Interventio | Experimen | Experimen | Control | 0,97445 |
| baseline | abundance | X01154 | N-(2,3,4-Tr | 3 | Interventio | Experimen | Experimen | Control | 0,957875 |
| baseline | abundance | X01163 | Prephenic | 3 | Interventio | Experimen | Experimen | Control | 0,948031 |
| baseline | abundance | X01164 | DL-Carboc | 3 | Interventio | Experimen | Experimen | Control | 0,957875 |
| baseline | abundance | X01181 | 2-(3,4-Dim | 3 | Interventio | Experimen | Experimen | Control | 0,936399 |
| baseline | abundance | X01186 | asn-val_b | 3 | Interventio | Experimen | Experimen | Control | 0,936399 |
| baseline | abundance | X01208 | Lys-Pro_c | 3 | Interventio | Experimen | Experimen | Control | 0,945105 |
| baseline | abundance | X01223 | DL-Mevalo | 3 | Interventio | Experimen | Experimen | Control | 0,936399 |
| baseline | abundance | X01235 | 3-[(3-Hydr | 3 | Interventio | Experimen | Experimen | Control | 0,936399 |
| baseline | abundance | X01236 | bis(4-isoth | 3 | Interventio | Experimen | Experimen | Control | 0,936399 |
| baseline | abundance | X01242 | 3-Morpholi | 2b | Interventio | Experimen | Experimen | Control | 0,97445 |
| baseline | abundance | X01246 | (2R)-1-{[(2 | 3 | Interventio | Experimen | Experimen | Control | 0,973066 |
| baseline | abundance | X01252 | (−)-nabilor | 3 | Interventio | Experimen | Experimen | Control | 0,97445 |
| baseline | abundance | X01285 | FB950000( | 3 | Interventio | Experimen | Experimen | Control | 0,97445 |
| baseline | abundance | X01286 | (2,4-Dihyd | 3 | Interventio | Experimen | Experimen | Control | 0,945105 |
| baseline | abundance | X01288 | L-gamma-( | 3 | Interventio | Experimen | Experimen | Control | 0,936399 |
| baseline | abundance | X01316 | 5-Hydanto | 3 | Interventio | Experimen | Experimen | Control | 0,936399 |
| baseline | abundance | X01327 | threonylph | 3 | Interventio | Experimen | Experimen | Control | 0,97445 |
| baseline | abundance | X01340 | Cadralazin | 3 | Interventio | Experimen | Experimen | Control | 0,962445 |
| baseline | abundance | X01341 | metixene | 3 | Interventio | Experimen | Experimen | Control | 0,936399 |
| baseline | abundance | X01346 | IN00260_a | 3 | Interventio | Experimen | Experimen | Control | 0,936399 |
| baseline | abundance | X01363 | Seryltyrosi | 3 | Interventio | Experimen | Experimen | Control | 0,959788 |
| baseline | abundance | X01364 | Oleuropeir | 3 | Interventio | Experimen | Experimen | Control | 0,936399 |
| baseline | abundance | X01367 | MFCD1869 | 3 | Interventio | Experimen | Experimen | Control | 0,945105 |
| baseline | abundance | X01380 | Spermic ac | 3 | Interventio | Experimen | Experimen | Control | 0,936399 |
| baseline | abundance | X01387 | Methyl 4-(4 | 3 | Interventio | Experimen | Experimen | Control | 0,945105 |
| baseline | abundance | X01441 | 1-{3-Carbo | 3 | Interventio | Experimen | Experimen | Control | 0,989213 |
| baseline | abundance | X01463 | 4-Thiapent | 3 | Interventio | Experimen | Experimen | Control | 0,948031 |
| baseline | abundance | X01474 | Glyceroph( | 3 | Interventio | Experimen | Experimen | Control | 0,945105 |
| baseline | abundance | X01518 | L-gamma-( | 3 | Interventio | Experimen | Experimen | Control | 0,97445 |
| baseline | abundance | X01519 | Leucyltrypt | 3 | Interventio | Experimen | Experimen | Control | 0,985326 |
| baseline | abundance | X01528 | butalbital_ | 3 | Interventio | Experimen | Experimen | Control | 0,945105 |
| baseline | abundance | X01530 | INK (Peptic | 2b | Interventio | Experimen | Experimen | Control | 0,957875 |
| baseline | abundance | X01549 | 3-(Sulfooxy | 3 | Interventio | Experimen | Experimen | Control | 0,962445 |
| baseline | abundance | X01553 | uridine 5'-c | 3 | Interventio | Experimen | Experimen | Control | 0,936399 |
| baseline | abundance | X01558 | 2-Furoylgly | 2b | Interventio | Experimen | Experimen | Control | 0,936399 |
| baseline | abundance | X01561 | 2-Ammonio | 3 | Interventio | Experimen | Experimen | Control | 0,97445 |
| baseline | abundance | X01570 | carglumic a | 3 | Interventio | Experimen | Experimen | Control | 0,936399 |
| baseline | abundance | X01574 | N-Acetylpr | 3 | Interventio | Experimen | Experimen | Control | 0,945786 |
| baseline | abundance | X01577 | Prunasin | 3 | Interventio | Experimen | Experimen | Control | 0,945105 |
| baseline | abundance | X01621 | 2-(2,4-Dihy | 3 | Interventio | Experimen | Experimen | Control | 0,945105 |
| baseline | abundance | X01640 | Ro 20-1724 | 3 | Interventio | Experimen | Experimen | Control | 0,989209 |
| baseline | abundance | X01656 | Guanadrel | 3 | Interventio | Experimen | Experimen | Control | 0,945105 |
| baseline | abundance | X01664 | 5-methylth | 3 | Interventio | Experimen | Experimen | Control | 0,936399 |
| baseline | abundance | X01671 | N-(3,5-Dim | 3 | Interventio | Experimen | Experimen | Control | 0,936399 |
| baseline | abundance | X01672 | Triethyl citı | 3 | Interventio | Experimen | Experimen | Control | 0,936399 |
| baseline | abundance | X01679 | meprobam | 3 | Interventio | Experimen | Experimen | Control | 0,945105 |

| | | | | | | | | | |
|---|---|---|---|---|---|---|---|---|---|
| baseline | abundance | X01689 | Ethyl malat | 3 | Interventio | Experimen | Experimen | Control | 0,936399 |
| baseline | abundance | X01732 | 2-Hydroxy- | 3 | Interventio | Experimen | Experimen | Control | 0,936399 |
| baseline | abundance | X01738 | 2-Acetami | 3 | Interventio | Experimen | Experimen | Control | 0,945105 |
| baseline | abundance | X01746 | Astemizole | 3 | Interventio | Experimen | Experimen | Control | 0,936399 |
| baseline | abundance | X01768 | Butabarbit | 3 | Interventio | Experimen | Experimen | Control | 0,936399 |
| baseline | abundance | X01776 | N-(Carboxy | 3 | Interventio | Experimen | Experimen | Control | 0,957875 |
| baseline | abundance | X01793 | Lys-phe_b | 3 | Interventio | Experimen | Experimen | Control | 0,936399 |
| baseline | abundance | X01813 | Erythorbic | 3 | Interventio | Experimen | Experimen | Control | 0,972498 |
| baseline | abundance | X01833 | 2,3,4,9-Tet | 2b | Interventio | Experimen | Experimen | Control | 0,945105 |
| baseline | abundance | X01873 | Methyl 2,3- | 3 | Interventio | Experimen | Experimen | Control | 0,97445 |
| baseline | abundance | X01879 | Tetramethy | 2b | Interventio | Experimen | Experimen | Control | 0,945105 |
| baseline | abundance | X01881 | (2S)-3-Met | 3 | Interventio | Experimen | Experimen | Control | 0,97445 |
| baseline | abundance | X01884 | Roxane | 3 | Interventio | Experimen | Experimen | Control | 0,936399 |
| baseline | abundance | X01887 | lys-tyr_b | 3 | Interventio | Experimen | Experimen | Control | 0,936399 |
| baseline | abundance | X01893 | 4-(METHYL | 3 | Interventio | Experimen | Experimen | Control | 0,955719 |
| baseline | abundance | X01911 | 3-(2,3-Dihy | 3 | Interventio | Experimen | Experimen | Control | 0,97477 |
| baseline | abundance | X01920 | N-(Carboxy | 3 | Interventio | Experimen | Experimen | Control | 0,936399 |
| baseline | abundance | X01932 | asn-val_c | 3 | Interventio | Experimen | Experimen | Control | 0,945105 |
| baseline | abundance | X01943 | Tocainide | 3 | Interventio | Experimen | Experimen | Control | 0,936399 |
| baseline | abundance | X01950 | Histidylgly | 3 | Interventio | Experimen | Experimen | Control | 0,936399 |
| baseline | abundance | X01968 | 7-Chloro-5 | 3 | Interventio | Experimen | Experimen | Control | 0,936399 |
| baseline | abundance | X01996 | 3-Hydroxy- | 3 | Interventio | Experimen | Experimen | Control | 0,936399 |
| baseline | abundance | X02000 | 11-(4-Hydr | 3 | Interventio | Experimen | Experimen | Control | 0,957232 |
| baseline | abundance | X02009 | Arctiopicri | 3 | Interventio | Experimen | Experimen | Control | 0,945105 |
| baseline | abundance | X02013 | Ile-cys | 3 | Interventio | Experimen | Experimen | Control | 0,936399 |
| baseline | abundance | X02020 | Val-Ser_b | 3 | Interventio | Experimen | Experimen | Control | 0,982558 |
| baseline | abundance | X02023 | beta-D-Eth | 3 | Interventio | Experimen | Experimen | Control | 0,957875 |
| baseline | abundance | X02073 | N-acetyl-9- | 3 | Interventio | Experimen | Experimen | Control | 0,936399 |
| baseline | abundance | X02080 | (-)-Aspidos | 3 | Interventio | Experimen | Experimen | Control | 0,986763 |
| baseline | abundance | X02082 | 6-Hydroxyr | 3 | Interventio | Experimen | Experimen | Control | 0,97499 |
| baseline | abundance | X02101 | Succinic ar | 3 | Interventio | Experimen | Experimen | Control | 0,957875 |
| baseline | abundance | X02108 | Homovanil | 3 | Interventio | Experimen | Experimen | Control | 0,936399 |
| baseline | abundance | X02121 | 2-Acetami | 3 | Interventio | Experimen | Experimen | Control | 0,97445 |
| baseline | abundance | X02139 | SECONAL_ | 3 | Interventio | Experimen | Experimen | Control | 0,945105 |
| baseline | abundance | X02145 | 3'-Hydroxy | 3 | Interventio | Experimen | Experimen | Control | 0,936399 |
| baseline | abundance | X02153 | Asarone | 3 | Interventio | Experimen | Experimen | Control | 0,945105 |
| baseline | abundance | X02181 | YWA1 | 3 | Interventio | Experimen | Experimen | Control | 0,941185 |
| baseline | abundance | X02184 | Piperonylo | 2b | Interventio | Experimen | Experimen | Control | 0,936399 |
| baseline | abundance | X02195 | alliin | 3 | Interventio | Experimen | Experimen | Control | 0,97445 |
| baseline | abundance | X02202 | D-2-Amino | 3 | Interventio | Experimen | Experimen | Control | 0,97445 |
| baseline | abundance | X02208 | SECONAL_ | 3 | Interventio | Experimen | Experimen | Control | 0,945105 |
| baseline | abundance | X02214 | (E)-4-Meth | 3 | Interventio | Experimen | Experimen | Control | 0,945105 |
| baseline | abundance | X02219 | Homoanse | 3 | Interventio | Experimen | Experimen | Control | 0,97445 |
| baseline | abundance | X02231 | Ala-Tyr | 3 | Interventio | Experimen | Experimen | Control | 0,945105 |
| baseline | abundance | X02238 | 6-Myoporo | 3 | Interventio | Experimen | Experimen | Control | 0,936399 |
| baseline | abundance | X02251 | Hostmania | 3 | Interventio | Experimen | Experimen | Control | 0,936399 |
| baseline | abundance | X02256 | hexobarbit | 3 | Interventio | Experimen | Experimen | Control | 0,936399 |
| baseline | abundance | X02265 | ophthalmic | 3 | Interventio | Experimen | Experimen | Control | 0,945105 |

| | | | | | | | | | |
|---|---|---|---|---|---|---|---|---|---|
| baseline | abundance | X02268 | 5-Hydroxy- | 3 | Interventio | Experimen | Experimen | Control | 0,97445 |
| baseline | abundance | X02277 | tert-Butyl 3 | 3 | Interventio | Experimen | Experimen | Control | 0,97445 |
| baseline | abundance | X02281 | tert-Butyl 3 | 3 | Interventio | Experimen | Experimen | Control | 0,97445 |
| baseline | abundance | X02288 | 9-Methylur | 2b | Interventio | Experimen | Experimen | Control | 0,97445 |
| baseline | abundance | X02289 | Kynurenic a | 2a | Interventio | Experimen | Experimen | Control | 0,982558 |
| baseline | abundance | X02317 | 2-Aminooc | 2b | Interventio | Experimen | Experimen | Control | 0,955424 |
| baseline | abundance | X02327 | 1H-Pyrazol | 3 | Interventio | Experimen | Experimen | Control | 0,945105 |
| baseline | abundance | X02333 | N-Propiony | 3 | Interventio | Experimen | Experimen | Control | 0,986763 |
| baseline | abundance | X02337 | Methyl 1-hy | 3 | Interventio | Experimen | Experimen | Control | 0,992433 |
| baseline | abundance | X02348 | N-Benzoyla | 3 | Interventio | Experimen | Experimen | Control | 0,936399 |
| baseline | abundance | X02352 | Indole-3-ca | 3 | Interventio | Experimen | Experimen | Control | 0,992433 |
| baseline | abundance | X02380 | Propafenon | 3 | Interventio | Experimen | Experimen | Control | 0,936399 |
| baseline | abundance | X02419 | Glu-Glu | 3 | Interventio | Experimen | Experimen | Control | 0,936399 |
| baseline | abundance | X02426 | Raltitrexed | 3 | Interventio | Experimen | Experimen | Control | 0,936399 |
| baseline | abundance | X02427 | 2-(3,5-dim | 2b | Interventio | Experimen | Experimen | Control | 0,958229 |
| baseline | abundance | X02442 | 3-(Sulfooxy | 3 | Interventio | Experimen | Experimen | Control | 0,936399 |
| baseline | abundance | X02466 | Validamyci | 3 | Interventio | Experimen | Experimen | Control | 0,945105 |
| baseline | abundance | X02468 | N,N-Diethy | 3 | Interventio | Experimen | Experimen | Control | 0,936399 |
| baseline | abundance | X02471 | n-Propyl Ga | 3 | Interventio | Experimen | Experimen | Control | 0,936399 |
| baseline | abundance | X02492 | N-(3,5-Dim | 3 | Interventio | Experimen | Experimen | Control | 0,948031 |
| baseline | abundance | X02494 | Butylphtha | 3 | Interventio | Experimen | Experimen | Control | 0,945105 |
| baseline | abundance | X02504 | DLK (Pepti | 2b | Interventio | Experimen | Experimen | Control | 0,936399 |
| baseline | abundance | X02513 | 7-Methylad | 2b | Interventio | Experimen | Experimen | Control | 0,936399 |
| baseline | abundance | X02514 | 3-Ureidopr | 1 | Interventio | Experimen | Experimen | Control | 0,936399 |
| baseline | abundance | X02521 | 6-(1-Hydro | 3 | Interventio | Experimen | Experimen | Control | 0,945105 |
| baseline | abundance | X02529 | tenivastati | 3 | Interventio | Experimen | Experimen | Control | 0,936399 |
| baseline | abundance | X02537 | Leu-pro_a | 3 | Interventio | Experimen | Experimen | Control | 0,936399 |
| baseline | abundance | X02565 | Dopamine | 3 | Interventio | Experimen | Experimen | Control | 0,97445 |
| baseline | abundance | X02583 | Ethylvanilli | 3 | Interventio | Experimen | Experimen | Control | 0,957875 |
| baseline | abundance | X02599 | Dihydrothy | 3 | Interventio | Experimen | Experimen | Control | 0,936399 |
| baseline | abundance | X02601 | (7E,7'E)-5, | 3 | Interventio | Experimen | Experimen | Control | 0,97445 |
| baseline | abundance | X02606 | 4-Amino-1 | 3 | Interventio | Experimen | Experimen | Control | 0,936399 |
| baseline | abundance | X02615 | 6-APA_a | 3 | Interventio | Experimen | Experimen | Control | 0,936399 |
| baseline | abundance | X02640 | thyronine | 3 | Interventio | Experimen | Experimen | Control | 0,945105 |
| baseline | abundance | X02651 | L-gamma-G | 3 | Interventio | Experimen | Experimen | Control | 0,936399 |
| baseline | abundance | X02652 | Leucyltyros | 3 | Interventio | Experimen | Experimen | Control | 0,936399 |
| baseline | abundance | X02661 | 4-(3-Oxope | 3 | Interventio | Experimen | Experimen | Control | 0,945105 |
| baseline | abundance | X02676 | Val-Trp_a | 3 | Interventio | Experimen | Experimen | Control | 0,936399 |
| baseline | abundance | X02687 | S-Propylcy | 3 | Interventio | Experimen | Experimen | Control | 0,936399 |
| baseline | abundance | X02689 | piscidic ac | 3 | Interventio | Experimen | Experimen | Control | 0,97445 |
| baseline | abundance | X02702 | R-(+)-Etirac | 3 | Interventio | Experimen | Experimen | Control | 0,945105 |
| baseline | abundance | X02720 | ala-ser_b | 3 | Interventio | Experimen | Experimen | Control | 0,936399 |
| baseline | abundance | X02730 | N-Phenylac | 3 | Interventio | Experimen | Experimen | Control | 0,936399 |
| baseline | abundance | X02733 | Calcitriol | 3 | Interventio | Experimen | Experimen | Control | 0,957875 |
| baseline | abundance | X02746 | N,N-Diethy | 3 | Interventio | Experimen | Experimen | Control | 0,970482 |
| baseline | abundance | X02760 | 6-hydroxyp | 3 | Interventio | Experimen | Experimen | Control | 0,957875 |
| baseline | abundance | X02765 | L-Pyrrolysi | 3 | Interventio | Experimen | Experimen | Control | 0,936399 |
| baseline | abundance | X02767 | 6-APA_b | 3 | Interventio | Experimen | Experimen | Control | 0,972498 |

| | | | | | | | | | |
|---|---|---|---|---|---|---|---|---|---|
| baseline | abundance | X02769 | Tetraacety | 3 | Interventio | Experimen | Experimen | Control | 0,945105 |
| baseline | abundance | X02774 | 9-ribosylze | 3 | Interventio | Experimen | Experimen | Control | 0,936399 |
| baseline | abundance | X02793 | 3-Methoxy- | 3 | Interventio | Experimen | Experimen | Control | 0,936399 |
| baseline | abundance | X02820 | FC250500 | 3 | Interventio | Experimen | Experimen | Control | 0,945105 |
| baseline | abundance | X02823 | 6,8-Dimeth | 3 | Interventio | Experimen | Experimen | Control | 0,945105 |
| baseline | abundance | X02824 | Eslicarbaz | 3 | Interventio | Experimen | Experimen | Control | 0,936399 |
| baseline | abundance | X02832 | (+/-)-2-Hyc | 3 | Interventio | Experimen | Experimen | Control | 0,936399 |
| baseline | abundance | X02841 | Kyotorphin | 3 | Interventio | Experimen | Experimen | Control | 0,945105 |
| baseline | abundance | X02843 | 4-(1-Hydro | 3 | Interventio | Experimen | Experimen | Control | 0,936399 |
| baseline | abundance | X02853 | 7alpha-Hy | 3 | Interventio | Experimen | Experimen | Control | 0,945105 |
| baseline | abundance | X02854 | Marimasta | 3 | Interventio | Experimen | Experimen | Control | 0,936399 |
| baseline | abundance | X02868 | 2-(3-Hydro | 3 | Interventio | Experimen | Experimen | Control | 0,957875 |
| baseline | abundance | X02872 | folinic acid | 3 | Interventio | Experimen | Experimen | Control | 0,945105 |
| baseline | abundance | X02878 | N~6~-[5-(1 | 3 | Interventio | Experimen | Experimen | Control | 0,97445 |
| baseline | abundance | X02881 | N-(2,3,4-Tr | 3 | Interventio | Experimen | Experimen | Control | 0,97445 |
| baseline | abundance | X02894 | Toluene_a | 3 | Interventio | Experimen | Experimen | Control | 0,945105 |
| baseline | abundance | X02905 | MFCD0005 | 3 | Interventio | Experimen | Experimen | Control | 0,945105 |
| baseline | abundance | X02920 | L-gamma- | 3 | Interventio | Experimen | Experimen | Control | 0,936399 |
| baseline | abundance | X02922 | (2E)-3-Met | 3 | Interventio | Experimen | Experimen | Control | 0,97445 |
| baseline | abundance | X02928 | N-Acetylpr | 3 | Interventio | Experimen | Experimen | Control | 0,945105 |
| baseline | abundance | X02943 | 3-Methoxy- | 3 | Interventio | Experimen | Experimen | Control | 0,957875 |
| baseline | abundance | X02944 | Valylvaline | 3 | Interventio | Experimen | Experimen | Control | 0,936399 |
| baseline | abundance | X02948 | N-(2-Cyan | 3 | Interventio | Experimen | Experimen | Control | 0,962445 |
| baseline | abundance | X02952 | Vorinostat_ | 3 | Interventio | Experimen | Experimen | Control | 0,948031 |
| baseline | abundance | X02968 | Ro 20-172 | 3 | Interventio | Experimen | Experimen | Control | 0,936399 |
| baseline | abundance | X02972 | 3-(Sulfoox | 3 | Interventio | Experimen | Experimen | Control | 0,936399 |
| baseline | abundance | X02986 | Diacetin_b | 3 | Interventio | Experimen | Experimen | Control | 0,936399 |
| baseline | abundance | X02990 | Pentoxifylli | 3 | Interventio | Experimen | Experimen | Control | 0,945105 |
| baseline | abundance | X03001 | Pseudouric | 2b | Interventio | Experimen | Experimen | Control | 0,945105 |
| baseline | abundance | X03004 | Bicine_b | 3 | Interventio | Experimen | Experimen | Control | 0,936399 |
| baseline | abundance | X03017 | Pro-tyr | 3 | Interventio | Experimen | Experimen | Control | 0,936399 |
| baseline | abundance | X03023 | NPK (Pepti | 2b | Interventio | Experimen | Experimen | Control | 0,936399 |
| baseline | abundance | X03064 | Formimino | 3 | Interventio | Experimen | Experimen | Control | 0,936399 |
| baseline | abundance | X03069 | N-COUMAI | 3 | Interventio | Experimen | Experimen | Control | 0,936399 |
| baseline | abundance | X03070 | 2-Methoxy- | 3 | Interventio | Experimen | Experimen | Control | 0,97445 |
| baseline | abundance | X03077 | nicotianam | 3 | Interventio | Experimen | Experimen | Control | 0,936399 |
| baseline | abundance | X03097 | 6-hydroxyn | 3 | Interventio | Experimen | Experimen | Control | 0,984935 |
| baseline | abundance | X03109 | Guanidino | 3 | Interventio | Experimen | Experimen | Control | 0,936399 |
| baseline | abundance | X03132 | (3S,4S)-7,1 | 3 | Interventio | Experimen | Experimen | Control | 0,936399 |
| baseline | abundance | X03134 | Tetrahydro | 3 | Interventio | Experimen | Experimen | Control | 0,97445 |
| baseline | abundance | X03158 | 5-Hydroxy- | 3 | Interventio | Experimen | Experimen | Control | 0,97445 |
| baseline | abundance | X03223 | 1-(2-Carbo | 3 | Interventio | Experimen | Experimen | Control | 0,958749 |
| baseline | abundance | X03254 | Gly-Trp_b | 3 | Interventio | Experimen | Experimen | Control | 0,945105 |
| baseline | abundance | X03268 | Dihydrouri | 3 | Interventio | Experimen | Experimen | Control | 0,957232 |
| baseline | abundance | X03271 | 1Î²-Hydrox | 3 | Interventio | Experimen | Experimen | Control | 0,957875 |
| baseline | abundance | X03275 | Methdilazi | 3 | Interventio | Experimen | Experimen | Control | 0,957875 |
| baseline | abundance | X03276 | 2'-Deoxyac | 3 | Interventio | Experimen | Experimen | Control | 0,97445 |
| baseline | abundance | X03278 | Hydroxyph | 3 | Interventio | Experimen | Experimen | Control | 0,988867 |

| | | | | | | | | | |
|---|---|---|---|---|---|---|---|---|---|
| baseline | abundance | X03294 | 4,4'-Thiobi | 3 | Interventio | Experimen | Experimen | Control | 0,97445 |
| baseline | abundance | X03340 | His-pro_b | 3 | Interventio | Experimen | Experimen | Control | 0,936399 |
| baseline | abundance | X03343 | L-alpha-As | 3 | Interventio | Experimen | Experimen | Control | 0,97445 |
| baseline | abundance | X03357 | 2-Hydroxyh | 3 | Interventio | Experimen | Experimen | Control | 0,936399 |
| baseline | abundance | X03363 | Choline Alf | 3 | Interventio | Experimen | Experimen | Control | 0,945105 |
| baseline | abundance | X03376 | 4-(9H-beta | 3 | Interventio | Experimen | Experimen | Control | 0,97445 |
| baseline | abundance | X03413 | Val-Trp_b | 3 | Interventio | Experimen | Experimen | Control | 0,936399 |
| baseline | abundance | X03416 | 9-(alpha-D | 3 | Interventio | Experimen | Experimen | Control | 0,97445 |
| baseline | abundance | X03434 | SECONAL_ | 3 | Interventio | Experimen | Experimen | Control | 0,983487 |
| baseline | abundance | X03480 | epsilon-(ga | 3 | Interventio | Experimen | Experimen | Control | 0,945105 |
| baseline | abundance | X03488 | 3,4-Dihydr | 3 | Interventio | Experimen | Experimen | Control | 0,957232 |
| baseline | abundance | X03535 | (1S,3R,4s)- | 3 | Interventio | Experimen | Experimen | Control | 0,97445 |
| baseline | abundance | X03552 | 4-Hydroxya | 3 | Interventio | Experimen | Experimen | Control | 0,957875 |
| baseline | abundance | X03563 | 6-imino-5- | 3 | Interventio | Experimen | Experimen | Control | 0,948031 |
| baseline | abundance | X03568 | 3-[(2Z)-1-C | 3 | Interventio | Experimen | Experimen | Control | 0,936399 |
| baseline | abundance | X03592 | 2-glyceryl 1 | 3 | Interventio | Experimen | Experimen | Control | 0,945105 |
| baseline | abundance | X03595 | Methylol D | 3 | Interventio | Experimen | Experimen | Control | 0,948031 |
| baseline | abundance | X03602 | o-Succinyl | 3 | Interventio | Experimen | Experimen | Control | 0,955424 |
| baseline | abundance | X03604 | Gly-Trp_a | 3 | Interventio | Experimen | Experimen | Control | 0,962445 |
| baseline | abundance | X03629 | Glycylglcy | 3 | Interventio | Experimen | Experimen | Control | 0,936399 |
| baseline | abundance | X03660 | trp-ser | 3 | Interventio | Experimen | Experimen | Control | 0,945786 |
| baseline | abundance | X03675 | asp-gln_a | 3 | Interventio | Experimen | Experimen | Control | 0,945105 |
| baseline | abundance | X03704 | N,N-Dimet | 3 | Interventio | Experimen | Experimen | Control | 0,988867 |
| baseline | abundance | X03707 | 9-[(5R)-5-E | 3 | Interventio | Experimen | Experimen | Control | 0,945105 |
| baseline | abundance | X03709 | Guanfacine | 3 | Interventio | Experimen | Experimen | Control | 0,945105 |
| baseline | abundance | X03714 | Sinapinic a | 3 | Interventio | Experimen | Experimen | Control | 0,97445 |
| baseline | abundance | X03718 | Propanthel | 3 | Interventio | Experimen | Experimen | Control | 0,945105 |
| baseline | abundance | X03719 | Losalen | 3 | Interventio | Experimen | Experimen | Control | 0,936399 |
| baseline | abundance | X03760 | 2-Methoxy- | 3 | Interventio | Experimen | Experimen | Control | 0,97445 |
| baseline | abundance | X03793 | Desonide | 3 | Interventio | Experimen | Experimen | Control | 0,945105 |
| baseline | abundance | X03837 | 2-Methoxy- | 3 | Interventio | Experimen | Experimen | Control | 0,936399 |
| baseline | abundance | X03855 | 2-{(2S,4S)- | 3 | Interventio | Experimen | Experimen | Control | 0,955424 |
| baseline | abundance | X03860 | 2-BUTYL PI | 3 | Interventio | Experimen | Experimen | Control | 0,945105 |
| baseline | abundance | X03892 | shinorine | 3 | Interventio | Experimen | Experimen | Control | 0,945105 |
| baseline | abundance | X03901 | ferrileghen | 3 | Interventio | Experimen | Experimen | Control | 0,936399 |
| baseline | abundance | X03916 | 2-(1-Ethoxy | 3 | Interventio | Experimen | Experimen | Control | 0,936399 |
| baseline | abundance | X03917 | Uramustine | 3 | Interventio | Experimen | Experimen | Control | 0,936399 |
| baseline | abundance | X03934 | 2-Aminom | 3 | Interventio | Experimen | Experimen | Control | 0,936399 |
| baseline | abundance | X03936 | (3R)-2-(3,4 | 3 | Interventio | Experimen | Experimen | Control | 0,936399 |
| baseline | abundance | X03961 | Artesunate | 3 | Interventio | Experimen | Experimen | Control | 0,959387 |
| baseline | abundance | X04021 | Bentazone | 2b | Interventio | Experimen | Experimen | Control | 0,936399 |
| baseline | abundance | X04040 | 3-(3,4-dihy | 3 | Interventio | Experimen | Experimen | Control | 0,987642 |
| baseline | abundance | X04117 | 2,3,4,5-tet | 3 | Interventio | Experimen | Experimen | Control | 0,945105 |
| baseline | abundance | X04119 | Tetraacety | 3 | Interventio | Experimen | Experimen | Control | 0,97445 |
| baseline | abundance | X04146 | L-fucopyra | 3 | Interventio | Experimen | Experimen | Control | 0,957875 |
| baseline | abundance | X04150 | 4-(METHYL | 3 | Interventio | Experimen | Experimen | Control | 0,957875 |
| baseline | abundance | X04157 | 3-Methoxy- | 3 | Interventio | Experimen | Experimen | Control | 0,945105 |
| baseline | abundance | X04181 | 3-Methoxy- | 3 | Interventio | Experimen | Experimen | Control | 0,936399 |

| | | | | | | | | | |
|---|---|---|---|---|---|---|---|---|---|
| baseline | abundance | X04183 | Spaglumic | 3 | Interventio | Experimen | Experimen | Control | 0,992185 |
| baseline | abundance | X04229 | gamma-L-g | 3 | Interventio | Experimen | Experimen | Control | 0,957875 |
| baseline | abundance | X04259 | 3-Hydroxyt | 3 | Interventio | Experimen | Experimen | Control | 0,936399 |
| baseline | abundance | X04274 | 6-[(Z)-2-(3, | 3 | Interventio | Experimen | Experimen | Control | 0,936399 |
| baseline | abundance | X04286 | Zinecard_b | 3 | Interventio | Experimen | Experimen | Control | 0,936399 |
| baseline | abundance | X04310 | Aminohipp | 3 | Interventio | Experimen | Experimen | Control | 0,945105 |
| baseline | abundance | X04315 | Benzamide | 3 | Interventio | Experimen | Experimen | Control | 0,989213 |
| baseline | abundance | X04334 | 3-Benzyl-6 | 3 | Interventio | Experimen | Experimen | Control | 0,936399 |
| baseline | abundance | X04339 | L-Glutamic | 3 | Interventio | Experimen | Experimen | Control | 0,936399 |
| baseline | abundance | X04351 | Pyrimidine | 3 | Interventio | Experimen | Experimen | Control | 0,986763 |
| baseline | abundance | X04377 | Dihydroura | 3 | Interventio | Experimen | Experimen | Control | 0,97445 |
| baseline | abundance | X04378 | 9,11-Dihyc | 3 | Interventio | Experimen | Experimen | Control | 0,936399 |
| baseline | abundance | X04379 | 2-Amino-6- | 3 | Interventio | Experimen | Experimen | Control | 0,945105 |
| baseline | abundance | X04440 | Mono(3-ca | 3 | Interventio | Experimen | Experimen | Control | 0,945105 |
| baseline | abundance | X04450 | 3-Methylac | 3 | Interventio | Experimen | Experimen | Control | 0,936399 |
| baseline | abundance | X04483 | (2E)-5-Hyd | 3 | Interventio | Experimen | Experimen | Control | 0,936399 |
| baseline | abundance | X04493 | Tyrosol | 3 | Interventio | Experimen | Experimen | Control | 0,936399 |
| baseline | abundance | X04526 | N-Nonanoy | 3 | Interventio | Experimen | Experimen | Control | 0,945105 |
| baseline | abundance | X04538 | Valylvaline | 3 | Interventio | Experimen | Experimen | Control | 0,955424 |
| baseline | abundance | X04543 | N-Acetyl-S | 3 | Interventio | Experimen | Experimen | Control | 0,936399 |
| baseline | abundance | X04544 | L-gamma-C | 3 | Interventio | Experimen | Experimen | Control | 0,945105 |
| baseline | abundance | X04553 | N-(4-Amino | 3 | Interventio | Experimen | Experimen | Control | 0,936399 |
| baseline | abundance | X04557 | 1,4-Naphth | 3 | Interventio | Experimen | Experimen | Control | 0,936399 |
| baseline | abundance | X04562 | Lisdexamfe | 3 | Interventio | Experimen | Experimen | Control | 0,945105 |
| baseline | abundance | X04564 | asp-gln_b | 3 | Interventio | Experimen | Experimen | Control | 0,945105 |
| baseline | abundance | X04566 | pterin | 3 | Interventio | Experimen | Experimen | Control | 0,936399 |
| baseline | abundance | X04571 | 13a-Hydro: | 3 | Interventio | Experimen | Experimen | Control | 0,945105 |
| baseline | abundance | X04579 | 6-(alpha-D | 3 | Interventio | Experimen | Experimen | Control | 0,936399 |
| baseline | abundance | X04582 | Dinoseb | 3 | Interventio | Experimen | Experimen | Control | 0,945105 |
| baseline | abundance | X04593 | Methional | 3 | Interventio | Experimen | Experimen | Control | 0,945105 |
| baseline | abundance | X04636 | epsilon-(ga | 3 | Interventio | Experimen | Experimen | Control | 0,936399 |
| baseline | abundance | X04639 | hydroxyhex | 3 | Interventio | Experimen | Experimen | Control | 0,957875 |
| baseline | abundance | X04682 | 1-Methylin | 3 | Interventio | Experimen | Experimen | Control | 0,945105 |
| baseline | abundance | X04684 | AAMU_b | 3 | Interventio | Experimen | Experimen | Control | 0,936399 |
| baseline | abundance | X04688 | Minoxidil | 3 | Interventio | Experimen | Experimen | Control | 0,936399 |
| baseline | abundance | X04695 | N-[(4-Meth | 3 | Interventio | Experimen | Experimen | Control | 0,936399 |
| baseline | abundance | X04697 | Ethosuxim | 3 | Interventio | Experimen | Experimen | Control | 0,945105 |
| baseline | abundance | X04743 | 5-Methoxy- | 3 | Interventio | Experimen | Experimen | Control | 0,936399 |
| baseline | abundance | X04744 | 16alpha-hy | 3 | Interventio | Experimen | Experimen | Control | 0,936399 |
| baseline | abundance | X04759 | 7-Aminome | 3 | Interventio | Experimen | Experimen | Control | 0,945786 |
| baseline | abundance | X04788 | 2-(4-Isopro | 3 | Interventio | Experimen | Experimen | Control | 0,970482 |
| baseline | abundance | X04792 | his-asn | 3 | Interventio | Experimen | Experimen | Control | 0,957875 |
| baseline | abundance | X04814 | meprobam | 3 | Interventio | Experimen | Experimen | Control | 0,987642 |
| baseline | abundance | X04822 | 6-Sulfatoxy | 3 | Interventio | Experimen | Experimen | Control | 0,98802 |
| baseline | abundance | X04852 | Toluene_b | 3 | Interventio | Experimen | Experimen | Control | 0,936399 |
| baseline | abundance | X04877 | Trifluorome | 3 | Interventio | Experimen | Experimen | Control | 0,94488 |
| baseline | abundance | X04897 | Taxifolin | 3 | Interventio | Experimen | Experimen | Control | 0,958229 |
| baseline | abundance | X04898 | 17-Hydroxy | 3 | Interventio | Experimen | Experimen | Control | 0,936399 |

| | | | | | | | | | |
|---|---|---|---|---|---|---|---|---|---|
| baseline | abundance | X04919 | 5-O-alpha- | 3 | Interventio | Experimen | Experimen | Control | 0,957875 |
| baseline | abundance | X04922 | Sinapinic a | 3 | Interventio | Experimen | Experimen | Control | 0,936399 |
| baseline | abundance | X04939 | N-Desalkyl | 3 | Interventio | Experimen | Experimen | Control | 0,97445 |
| baseline | abundance | X04975 | MFCD0995 | 3 | Interventio | Experimen | Experimen | Control | 0,936399 |
| baseline | abundance | X04986 | L-Proline, 4 | 3 | Interventio | Experimen | Experimen | Control | 0,936399 |
| baseline | abundance | X04990 | Cilazapril | 3 | Interventio | Experimen | Experimen | Control | 0,936399 |
| baseline | abundance | X04996 | MFCD0272 | 3 | Interventio | Experimen | Experimen | Control | 0,936399 |
| baseline | abundance | X04999 | 2-Hydroxyp | 1 | Interventio | Experimen | Experimen | Control | 0,936399 |
| baseline | abundance | X05017 | N-(1-{[Metl | 3 | Interventio | Experimen | Experimen | Control | 0,991989 |
| baseline | abundance | X05029 | Hypericin | 3 | Interventio | Experimen | Experimen | Control | 0,936399 |
| baseline | abundance | X05033 | (2E)-N-3,7- | 3 | Interventio | Experimen | Experimen | Control | 0,936399 |
| baseline | abundance | X05046 | Casimiroin | 3 | Interventio | Experimen | Experimen | Control | 0,945105 |
| baseline | abundance | X05068 | pro-gln_c | 3 | Interventio | Experimen | Experimen | Control | 0,936399 |
| baseline | abundance | X05071 | Histidylphe | 3 | Interventio | Experimen | Experimen | Control | 0,97445 |
| baseline | abundance | X05072 | L-gamma-( | 3 | Interventio | Experimen | Experimen | Control | 0,945105 |
| baseline | abundance | X05081 | ELK (Peptic | 2b | Interventio | Experimen | Experimen | Control | 0,936399 |
| baseline | abundance | X05126 | (3aS,5S,6R | 3 | Interventio | Experimen | Experimen | Control | 0,945786 |
| baseline | abundance | X05146 | UQ367500 | 3 | Interventio | Experimen | Experimen | Control | 0,936399 |
| baseline | abundance | X05149 | S-Sulfocys | 3 | Interventio | Experimen | Experimen | Control | 0,936399 |
| baseline | abundance | X05177 | cis-3-Hexe | 3 | Interventio | Experimen | Experimen | Control | 0,988867 |
| baseline | abundance | X05183 | 3,6-Dichlo | 3 | Interventio | Experimen | Experimen | Control | 0,936399 |
| baseline | abundance | X05195 | Lys-phe_a | 3 | Interventio | Experimen | Experimen | Control | 0,936399 |
| baseline | abundance | X05203 | felbamate | 3 | Interventio | Experimen | Experimen | Control | 0,936399 |
| baseline | abundance | X05204 | Urothion | 3 | Interventio | Experimen | Experimen | Control | 0,957875 |
| baseline | abundance | X05214 | [7-Hydroxy | 3 | Interventio | Experimen | Experimen | Control | 0,97445 |
| baseline | abundance | X05225 | 4-(METHYL | 3 | Interventio | Experimen | Experimen | Control | 0,992737 |
| baseline | abundance | X05234 | 3,4-dihydro | 3 | Interventio | Experimen | Experimen | Control | 0,936399 |
| baseline | abundance | X05237 | imazameth | 3 | Interventio | Experimen | Experimen | Control | 0,936399 |
| baseline | abundance | X05240 | Dimeric me | 3 | Interventio | Experimen | Experimen | Control | 0,937501 |
| baseline | abundance | X05243 | N-(4-Hydro | 3 | Interventio | Experimen | Experimen | Control | 0,936399 |
| baseline | abundance | X05275 | H-DL-MET- | 3 | Interventio | Experimen | Experimen | Control | 0,97445 |
| baseline | abundance | X05295 | 3-(4,7-Dim | 3 | Interventio | Experimen | Experimen | Control | 0,936399 |
| baseline | abundance | X05306 | 2-methoxy | 3 | Interventio | Experimen | Experimen | Control | 0,936399 |
| baseline | abundance | X05357 | NPYR | 3 | Interventio | Experimen | Experimen | Control | 0,936399 |
| baseline | abundance | X05391 | 2-Acetamid | 3 | Interventio | Experimen | Experimen | Control | 0,936399 |
| baseline | abundance | X05398 | DIBOA | 3 | Interventio | Experimen | Experimen | Control | 0,936399 |
| baseline | abundance | X05411 | 3-(Sulfooxy | 3 | Interventio | Experimen | Experimen | Control | 0,97445 |
| baseline | abundance | X05415 | Redul | 3 | Interventio | Experimen | Experimen | Control | 0,936399 |
| baseline | abundance | X05417 | (S)-2-hydra | 3 | Interventio | Experimen | Experimen | Control | 0,936399 |
| baseline | abundance | X05421 | S-Allylcyste | 3 | Interventio | Experimen | Experimen | Control | 0,945105 |
| baseline | abundance | X05459 | Scopoletin | 3 | Interventio | Experimen | Experimen | Control | 0,97445 |
| baseline | abundance | X05467 | Azulfidine | 3 | Interventio | Experimen | Experimen | Control | 0,936399 |
| baseline | abundance | X05478 | S(8)-aminc | 3 | Interventio | Experimen | Experimen | Control | 0,957875 |
| baseline | abundance | X05499 | GLY-MET | 3 | Interventio | Experimen | Experimen | Control | 0,945105 |
| baseline | abundance | X05510 | 3-Benzyl-6 | 3 | Interventio | Experimen | Experimen | Control | 0,945105 |
| baseline | abundance | X05514 | (3aS,5S,6R | 3 | Interventio | Experimen | Experimen | Control | 0,97445 |
| baseline | abundance | X05551 | Trolox | 3 | Interventio | Experimen | Experimen | Control | 0,936399 |
| baseline | abundance | X05561 | 1Î²-Hydroxy | 3 | Interventio | Experimen | Experimen | Control | 0,948031 |

| | | | | | | | | | |
|---|---|---|---|---|---|---|---|---|---|
| baseline | abundance | X05580 | Xanthureni | 3 | Interventio | Experimen | Experimen | Control | 0,945105 |
| baseline | abundance | X05581 | N-[(10Z)-7- | 3 | Interventio | Experimen | Experimen | Control | 0,945105 |
| baseline | abundance | X05584 | Serotonin | 1 | Interventio | Experimen | Experimen | Control | 0,987751 |
| baseline | abundance | X05656 | mesifurane | 3 | Interventio | Experimen | Experimen | Control | 0,936399 |
| baseline | abundance | X05709 | 2,3-Dihydr | 3 | Interventio | Experimen | Experimen | Control | 0,945105 |
| baseline | abundance | X05726 | methocarb | 3 | Interventio | Experimen | Experimen | Control | 0,936399 |
| baseline | abundance | X05734 | N-[(2S)-2-H | 3 | Interventio | Experimen | Experimen | Control | 0,97445 |
| baseline | abundance | X05741 | g-Aminobu | 3 | Interventio | Experimen | Experimen | Control | 0,936399 |
| baseline | abundance | X05758 | quinol sulf | 3 | Interventio | Experimen | Experimen | Control | 0,97499 |
| baseline | abundance | X05786 | Nitrendipir | 3 | Interventio | Experimen | Experimen | Control | 0,936399 |
| baseline | abundance | X05812 | (2S)-3-(1H | 3 | Interventio | Experimen | Experimen | Control | 0,957875 |
| baseline | abundance | X05857 | N-D-Gluco | 3 | Interventio | Experimen | Experimen | Control | 0,97445 |
| baseline | abundance | X05859 | 3,7,12,17- | 3 | Interventio | Experimen | Experimen | Control | 0,985326 |
| baseline | abundance | X05878 | 1,2-dihydr | 3 | Interventio | Experimen | Experimen | Control | 0,97445 |
| baseline | abundance | X05892 | (5Z)-2-Ami | 3 | Interventio | Experimen | Experimen | Control | 0,97445 |
| baseline | abundance | X05919 | 2-(2-Amino | 3 | Interventio | Experimen | Experimen | Control | 0,945105 |
| baseline | abundance | X05968 | Tetraacety | 3 | Interventio | Experimen | Experimen | Control | 0,945105 |
| baseline | abundance | X05969 | 4-(METHYL | 3 | Interventio | Experimen | Experimen | Control | 0,936399 |
| baseline | abundance | X05971 | L-gamma-( | 3 | Interventio | Experimen | Experimen | Control | 0,936399 |
| baseline | abundance | X05982 | (19R,25S)- | 3 | Interventio | Experimen | Experimen | Control | 0,97445 |
| baseline | abundance | X05984 | 1-(4-Methy | 2b | Interventio | Experimen | Experimen | Control | 0,936399 |
| baseline | abundance | X05988 | Selsun | 3 | Interventio | Experimen | Experimen | Control | 0,957875 |
| baseline | abundance | X05995 | Diacetin_a | 3 | Interventio | Experimen | Experimen | Control | 0,936399 |
| baseline | abundance | X05997 | FB950000( | 3 | Interventio | Experimen | Experimen | Control | 0,945105 |
| baseline | abundance | X06011 | Coprine_a | 3 | Interventio | Experimen | Experimen | Control | 0,992737 |
| baseline | abundance | X06016 | N,N-Dimet | 3 | Interventio | Experimen | Experimen | Control | 0,955424 |
| baseline | abundance | X06034 | N-[(2S)-2-H | 3 | Interventio | Experimen | Experimen | Control | 0,97445 |
| baseline | abundance | X06039 | gamma-Gl | 3 | Interventio | Experimen | Experimen | Control | 0,948031 |
| baseline | abundance | X06041 | (DL)-3-O-M | 3 | Interventio | Experimen | Experimen | Control | 0,936399 |
| baseline | abundance | X06043 | vinyl sulfid | 3 | Interventio | Experimen | Experimen | Control | 0,945105 |
| baseline | abundance | X06046 | Valylvaline | 3 | Interventio | Experimen | Experimen | Control | 0,987751 |
| baseline | abundance | X06062 | 1-(2,3-Dihy | 3 | Interventio | Experimen | Experimen | Control | 0,948031 |
| baseline | abundance | X06079 | Glycylleuci | 1 | Interventio | Experimen | Experimen | Control | 0,945105 |
| baseline | abundance | X06082 | O-heptano | 3 | Interventio | Experimen | Experimen | Control | 0,936399 |
| baseline | abundance | X06085 | Tetraacety | 3 | Interventio | Experimen | Experimen | Control | 0,945105 |
| baseline | abundance | X06089 | riboprine | 3 | Interventio | Experimen | Experimen | Control | 0,97445 |
| baseline | abundance | X06100 | Midodrine_ | 3 | Interventio | Experimen | Experimen | Control | 0,989769 |
| baseline | abundance | X06107 | Hydroxyca | 3 | Interventio | Experimen | Experimen | Control | 0,97445 |
| baseline | abundance | X06112 | Val-Ser_a | 3 | Interventio | Experimen | Experimen | Control | 0,936399 |
| baseline | abundance | X06124 | MFCD0995 | 3 | Interventio | Experimen | Experimen | Control | 0,936399 |
| baseline | abundance | X06127 | 2-Methoxy- | 3 | Interventio | Experimen | Experimen | Control | 0,987642 |
| baseline | abundance | X06143 | 2-Methylbu | 2a | Interventio | Experimen | Experimen | Control | 0,945105 |
| baseline | abundance | X06146 | S-Methyl-1 | 3 | Interventio | Experimen | Experimen | Control | 0,945105 |
| baseline | abundance | X06149 | Yangonin | 3 | Interventio | Experimen | Experimen | Control | 0,945786 |
| baseline | abundance | X06150 | Zalcitabine | 3 | Interventio | Experimen | Experimen | Control | 0,936399 |
| baseline | abundance | X06152 | alpha-keto | 3 | Interventio | Experimen | Experimen | Control | 0,993031 |
| baseline | abundance | X06167 | N-Acetylas | 2a | Interventio | Experimen | Experimen | Control | 0,936399 |
| baseline | abundance | X06168 | leu-gln_b | 3 | Interventio | Experimen | Experimen | Control | 0,945105 |

| | | | | | | | | | |
|---|---|---|---|---|---|---|---|---|---|
| baseline | abundance | X06183 | Nisinic aci | 3 | Interventio | Experimen | Experimen | Control | 0,936399 |
| baseline | abundance | X06189 | delta-Guar | 3 | Interventio | Experimen | Experimen | Control | 0,994656 |
| baseline | abundance | X06197 | 4-(Nitrosoa | 3 | Interventio | Experimen | Experimen | Control | 0,936399 |
| baseline | abundance | X06220 | Tetrahydro | 3 | Interventio | Experimen | Experimen | Control | 0,936399 |
| baseline | abundance | X06222 | Leucylaspa | 3 | Interventio | Experimen | Experimen | Control | 0,936399 |
| baseline | abundance | X06227 | lys-tyr_a | 3 | Interventio | Experimen | Experimen | Control | 0,936399 |
| baseline | abundance | X06230 | Glu-Gly | 3 | Interventio | Experimen | Experimen | Control | 0,936399 |
| baseline | abundance | X06240 | His-pro_a | 3 | Interventio | Experimen | Experimen | Control | 0,945105 |
| baseline | abundance | X06250 | [3-({3-[(Cy | 2b | Interventio | Experimen | Experimen | Control | 0,945105 |
| baseline | abundance | X06253 | N-(4-Amin | 3 | Interventio | Experimen | Experimen | Control | 0,955719 |
| baseline | abundance | X06254 | D-Alanyl-D | 3 | Interventio | Experimen | Experimen | Control | 0,936399 |
| baseline | abundance | X06259 | (S)-?-glyce | 3 | Interventio | Experimen | Experimen | Control | 0,945105 |
| baseline | abundance | X06268 | mesifurane | 3 | Interventio | Experimen | Experimen | Control | 0,936399 |
| baseline | abundance | X06270 | 9-Methylur | 2b | Interventio | Experimen | Experimen | Control | 0,936399 |
| baseline | abundance | X06276 | asn-pro_b | 3 | Interventio | Experimen | Experimen | Control | 0,936399 |
| baseline | abundance | X06278 | 3-Hydroxy- | 3 | Interventio | Experimen | Experimen | Control | 0,936399 |
| baseline | abundance | X06289 | Nicotinatel | 3 | Interventio | Experimen | Experimen | Control | 0,988948 |
| baseline | abundance | X06291 | Corticoste | 2b | Interventio | Experimen | Experimen | Control | 0,945105 |
| baseline | abundance | X06292 | mesifurane | 3 | Interventio | Experimen | Experimen | Control | 0,936399 |
| baseline | abundance | X06293 | (4R)-4-{[3- | 3 | Interventio | Experimen | Experimen | Control | 0,945105 |
| baseline | abundance | X06313 | Isoquinolir | 2b | Interventio | Experimen | Experimen | Control | 0,95582 |
| baseline | abundance | X06318 | Propamoca | 2b | Interventio | Experimen | Experimen | Control | 0,948031 |
| baseline | abundance | X06320 | 7alpha-Hy | 3 | Interventio | Experimen | Experimen | Control | 0,97445 |
| baseline | abundance | X06329 | Leu-Leu_d | 3 | Interventio | Experimen | Experimen | Control | 0,936399 |
| baseline | abundance | X06334 | Bis-D-fruct | 3 | Interventio | Experimen | Experimen | Control | 0,97445 |
| baseline | abundance | X06337 | Zalcitabine | 3 | Interventio | Experimen | Experimen | Control | 0,945105 |
| baseline | abundance | X06354 | trans-Zeati | 2b | Interventio | Experimen | Experimen | Control | 0,936399 |
| baseline | abundance | X06361 | Oxprenolol | 3 | Interventio | Experimen | Experimen | Control | 0,936399 |
| baseline | abundance | X06368 | TDP-2_b | 3 | Interventio | Experimen | Experimen | Control | 0,936399 |
| baseline | abundance | X06371 | L-gamma-( | 3 | Interventio | Experimen | Experimen | Control | 0,936399 |
| baseline | abundance | X06372 | N,N-dimetl | 2b | Interventio | Experimen | Experimen | Control | 0,945105 |
| baseline | abundance | X06381 | MFCD1869 | 3 | Interventio | Experimen | Experimen | Control | 0,936399 |
| baseline | abundance | X06388 | Caffeic aci | 1 | Interventio | Experimen | Experimen | Control | 0,936399 |
| baseline | abundance | X06400 | Phloionolic | 3 | Interventio | Experimen | Experimen | Control | 0,998245 |
| baseline | abundance | X06401 | Isophthalic | 2b | Interventio | Experimen | Experimen | Control | 0,936399 |
| baseline | abundance | X06404 | Flemichap | 3 | Interventio | Experimen | Experimen | Control | 0,962445 |
| baseline | abundance | X06405 | Asparaginy | 3 | Interventio | Experimen | Experimen | Control | 0,98802 |
| baseline | abundance | X06409 | Glycylproli | 2b | Interventio | Experimen | Experimen | Control | 0,945105 |
| baseline | abundance | X06412 | Butenylcar | 3 | Interventio | Experimen | Experimen | Control | 0,936399 |
| baseline | abundance | X06416 | KYNURAMI | 3 | Interventio | Experimen | Experimen | Control | 0,936399 |
| baseline | abundance | X06419 | Spermic ac | 3 | Interventio | Experimen | Experimen | Control | 0,945105 |
| baseline | abundance | X06433 | 4-(2,5-Diflu | 2b | Interventio | Experimen | Experimen | Control | 0,97445 |
| baseline | abundance | X06434 | butyrin | 3 | Interventio | Experimen | Experimen | Control | 0,97445 |
| baseline | abundance | X06437 | 2-Acetamic | 3 | Interventio | Experimen | Experimen | Control | 0,957875 |
| baseline | abundance | X06448 | 3,8,9-trihy | 2b | Interventio | Experimen | Experimen | Control | 0,936399 |
| baseline | abundance | X06454 | 8-Amino-7- | 3 | Interventio | Experimen | Experimen | Control | 0,936399 |
| baseline | abundance | X06455 | 4-Hydroxyp | 3 | Interventio | Experimen | Experimen | Control | 0,982558 |
| baseline | abundance | X06462 | DNOP_d | 3 | Interventio | Experimen | Experimen | Control | 0,97445 |

| | | | | | | | | | |
|---|---|---|---|---|---|---|---|---|---|
| baseline | abundance | X06463 | 8-Amino-7 | 3 | Interventio | Experimen | Experimen | Control | 0,936399 |
| baseline | abundance | X06472 | N(alpha)-B | 3 | Interventio | Experimen | Experimen | Control | 0,936399 |
| baseline | abundance | X06473 | MFCD0002 | 3 | Interventio | Experimen | Experimen | Control | 0,945105 |
| baseline | abundance | X06477 | APM_b | 3 | Interventio | Experimen | Experimen | Control | 0,936399 |
| baseline | abundance | X06478 | Caffeic aci | 3 | Interventio | Experimen | Experimen | Control | 0,936399 |
| baseline | abundance | X06482 | glu-pro | 3 | Interventio | Experimen | Experimen | Control | 0,957875 |
| baseline | abundance | X06486 | Leu-Val_b | 3 | Interventio | Experimen | Experimen | Control | 0,97445 |
| baseline | abundance | X06495 | 2-Acetami | 3 | Interventio | Experimen | Experimen | Control | 0,945786 |
| baseline | abundance | X06502 | MFCD2836 | 3 | Interventio | Experimen | Experimen | Control | 0,945105 |
| baseline | abundance | X06506 | L-Homocys | 3 | Interventio | Experimen | Experimen | Control | 0,945105 |
| baseline | abundance | X06507 | Gly-Lys | 3 | Interventio | Experimen | Experimen | Control | 0,945105 |
| baseline | abundance | X06508 | Asparaginy | 3 | Interventio | Experimen | Experimen | Control | 0,987751 |
| baseline | abundance | X06510 | N-{4-[(2R,3 | 2b | Interventio | Experimen | Experimen | Control | 0,936399 |
| baseline | abundance | X06529 | Methanesu | 2a | Interventio | Experimen | Experimen | Control | 0,985326 |
| baseline | abundance | X06530 | 3-(1-hydro | 2b | Interventio | Experimen | Experimen | Control | 0,97445 |
| baseline | abundance | X06532 | Leupeptin | 3 | Interventio | Experimen | Experimen | Control | 0,936399 |
| baseline | abundance | X06546 | 4-Acetami | 2b | Interventio | Experimen | Experimen | Control | 0,936399 |
| baseline | abundance | X06549 | Panthenol_ | 3 | Interventio | Experimen | Experimen | Control | 0,936399 |
| baseline | abundance | X06551 | Methyl 1-hy | 3 | Interventio | Experimen | Experimen | Control | 0,97445 |
| baseline | abundance | X06560 | 2,6-Dimeth | 2b | Interventio | Experimen | Experimen | Control | 0,936399 |
| baseline | abundance | X06590 | 3-hydroxy- | 3 | Interventio | Experimen | Experimen | Control | 0,948031 |
| baseline | abundance | X06600 | MFCD0014 | 3 | Interventio | Experimen | Experimen | Control | 0,988948 |
| baseline | abundance | X06606 | Leu-Val_d | 3 | Interventio | Experimen | Experimen | Control | 0,97445 |
| baseline | abundance | X06612 | Spermic ac | 3 | Interventio | Experimen | Experimen | Control | 0,936399 |
| baseline | abundance | X06622 | Lys-Pro_b | 3 | Interventio | Experimen | Experimen | Control | 0,97445 |
| baseline | abundance | X06623 | quinol sulf | 3 | Interventio | Experimen | Experimen | Control | 0,945105 |
| baseline | abundance | X06626 | N-Butyryl-L | 3 | Interventio | Experimen | Experimen | Control | 0,936399 |
| baseline | abundance | X06630 | 4-(METHYL | 3 | Interventio | Experimen | Experimen | Control | 0,936399 |
| baseline | abundance | X06631 | 2-Isopropy | 2b | Interventio | Experimen | Experimen | Control | 0,936399 |
| baseline | abundance | X06634 | Glycitein | 3 | Interventio | Experimen | Experimen | Control | 0,945105 |
| baseline | abundance | X06642 | Homocitru | 2a | Interventio | Experimen | Experimen | Control | 0,936399 |
| baseline | abundance | X06646 | (S)-3-sulfo | 3 | Interventio | Experimen | Experimen | Control | 0,936399 |
| baseline | abundance | X06655 | Hydroxycar | 3 | Interventio | Experimen | Experimen | Control | 0,936399 |
| baseline | abundance | X06656 | Valylvaline | 3 | Interventio | Experimen | Experimen | Control | 0,945105 |
| baseline | abundance | X06663 | Leucyltyros | 3 | Interventio | Experimen | Experimen | Control | 0,945105 |
| baseline | abundance | X06675 | L-gamma-( | 3 | Interventio | Experimen | Experimen | Control | 0,957875 |
| baseline | abundance | X06680 | 7-Hydroxy- | 2b | Interventio | Experimen | Experimen | Control | 0,936399 |
| baseline | abundance | X06681 | Scymnol | 3 | Interventio | Experimen | Experimen | Control | 0,936399 |
| baseline | abundance | X06684 | Bromazine | 3 | Interventio | Experimen | Experimen | Control | 0,945105 |
| baseline | abundance | X06690 | 4-Methylca | 2b | Interventio | Experimen | Experimen | Control | 0,936399 |
| baseline | abundance | X06698 | entecavir | 3 | Interventio | Experimen | Experimen | Control | 0,957875 |
| baseline | abundance | X06703 | N6-METHY | 3 | Interventio | Experimen | Experimen | Control | 0,97445 |
| baseline | abundance | X06704 | feruloylagr | 3 | Interventio | Experimen | Experimen | Control | 0,945105 |
| baseline | abundance | X06709 | Tetraacety | 3 | Interventio | Experimen | Experimen | Control | 0,955424 |
| baseline | abundance | X06722 | LW800000 | 3 | Interventio | Experimen | Experimen | Control | 0,945105 |
| baseline | abundance | X06723 | 3-Hydroxy- | 3 | Interventio | Experimen | Experimen | Control | 0,945105 |
| baseline | abundance | X06727 | MFCD0087 | 3 | Interventio | Experimen | Experimen | Control | 0,948031 |
| baseline | abundance | X06735 | 7-Methylxa | 2b | Interventio | Experimen | Experimen | Control | 0,945105 |

| | | | | | | | | | |
|---|---|---|---|---|---|---|---|---|---|
| baseline | abundance | X06744 | Diethylpyr | 3 | Interventio | Experimen | Experimen | Control | 0,978464 |
| baseline | abundance | X06750 | 1-PYRENYL | 3 | Interventio | Experimen | Experimen | Control | 0,97445 |
| baseline | abundance | X06764 | N-(4-Hydro | 3 | Interventio | Experimen | Experimen | Control | 0,936399 |
| baseline | abundance | X06767 | 2-Hydroxyb | 1 | Interventio | Experimen | Experimen | Control | 0,945105 |
| baseline | abundance | X06768 | Sinapyl alc | 3 | Interventio | Experimen | Experimen | Control | 0,936399 |
| baseline | abundance | X06771 | MFCD0015 | 3 | Interventio | Experimen | Experimen | Control | 0,937501 |
| baseline | abundance | X06783 | N~6~-Octa | 3 | Interventio | Experimen | Experimen | Control | 0,936399 |
| baseline | abundance | X06797 | pro-gln_a | 3 | Interventio | Experimen | Experimen | Control | 0,957232 |
| baseline | abundance | X06805 | (9cis)-O~1 | 3 | Interventio | Experimen | Experimen | Control | 0,957875 |
| baseline | abundance | X06807 | 1,1'-[1,12- | 3 | Interventio | Experimen | Experimen | Control | 0,97445 |
| baseline | abundance | X06812 | 2-Hydroxy- | 3 | Interventio | Experimen | Experimen | Control | 0,936399 |
| baseline | abundance | X06814 | N-Benzoyla | 3 | Interventio | Experimen | Experimen | Control | 0,936399 |
| baseline | abundance | X06818 | Homoanse | 3 | Interventio | Experimen | Experimen | Control | 0,936399 |
| baseline | abundance | X06819 | Primaquine | 3 | Interventio | Experimen | Experimen | Control | 0,945105 |
| baseline | abundance | X06820 | 1-(3,4-dim | 2b | Interventio | Experimen | Experimen | Control | 0,945105 |
| baseline | abundance | X06836 | asn-lys | 3 | Interventio | Experimen | Experimen | Control | 0,982558 |
| baseline | abundance | X06850 | Leu-Val_c | 3 | Interventio | Experimen | Experimen | Control | 0,97445 |
| baseline | abundance | X06856 | Tiglic acid_ | 2b | Interventio | Experimen | Experimen | Control | 0,936399 |
| baseline | abundance | X06858 | tert-Butyl 3 | 3 | Interventio | Experimen | Experimen | Control | 0,945105 |
| baseline | abundance | X06859 | Methylol D | 3 | Interventio | Experimen | Experimen | Control | 0,936399 |
| baseline | abundance | X06861 | 4-(4-Deoxy | 3 | Interventio | Experimen | Experimen | Control | 0,936399 |
| baseline | abundance | X06866 | tert-Butyl 3 | 3 | Interventio | Experimen | Experimen | Control | 0,957875 |
| baseline | abundance | X06870 | Menadiol | 3 | Interventio | Experimen | Experimen | Control | 0,945105 |
| baseline | abundance | X06874 | N-Acetyl-5 | 3 | Interventio | Experimen | Experimen | Control | 0,936399 |
| baseline | abundance | X06875 | Nicotine gl | 3 | Interventio | Experimen | Experimen | Control | 0,936399 |
| baseline | abundance | X06880 | pentobarbi | 3 | Interventio | Experimen | Experimen | Control | 0,945105 |
| baseline | abundance | X06883 | Agomelatir | 3 | Interventio | Experimen | Experimen | Control | 0,962445 |
| baseline | abundance | X06889 | 3,4-Dimeth | 2a | Interventio | Experimen | Experimen | Control | 0,97445 |
| baseline | abundance | X06896 | Pirbuterol | 3 | Interventio | Experimen | Experimen | Control | 0,97445 |
| baseline | abundance | X06902 | Homocyste | 3 | Interventio | Experimen | Experimen | Control | 0,936399 |
| baseline | abundance | X06903 | Esculin | 2b | Interventio | Experimen | Experimen | Control | 0,936399 |
| baseline | abundance | X06905 | Ectoine | 3 | Interventio | Experimen | Experimen | Control | 0,955424 |
| baseline | abundance | X06906 | Arg-pro | 3 | Interventio | Experimen | Experimen | Control | 0,945105 |
| baseline | abundance | X06917 | Alanyltrypt | 3 | Interventio | Experimen | Experimen | Control | 0,936399 |
| baseline | abundance | X06933 | 3-Methoxy- | 3 | Interventio | Experimen | Experimen | Control | 0,968601 |
| baseline | abundance | X06943 | NSC 92778 | 3 | Interventio | Experimen | Experimen | Control | 0,945105 |
| baseline | abundance | X06946 | SECONAL_ | 3 | Interventio | Experimen | Experimen | Control | 0,97445 |
| baseline | abundance | X06948 | (1R,3R,5R) | 3 | Interventio | Experimen | Experimen | Control | 0,945105 |
| baseline | abundance | X06958 | 2-[(carboxy | 2b | Interventio | Experimen | Experimen | Control | 0,936399 |
| baseline | abundance | X06961 | Zalcitabine | 3 | Interventio | Experimen | Experimen | Control | 0,948031 |
| baseline | abundance | X06977 | 4-Hydroxyp | 3 | Interventio | Experimen | Experimen | Control | 0,945786 |
| baseline | abundance | X06985 | N-Ethylpro | 3 | Interventio | Experimen | Experimen | Control | 0,948031 |
| baseline | abundance | X07002 | Paraldehyc | 3 | Interventio | Experimen | Experimen | Control | 0,936399 |
| baseline | abundance | X07013 | pimethixer | 3 | Interventio | Experimen | Experimen | Control | 0,936399 |
| baseline | abundance | X07014 | Octyl benz | 3 | Interventio | Experimen | Experimen | Control | 0,936399 |
| baseline | abundance | X07026 | 7α-Hydroxy | 2b | Interventio | Experimen | Experimen | Control | 0,978198 |
| baseline | abundance | X07027 | Allyl merca | 3 | Interventio | Experimen | Experimen | Control | 0,936399 |
| baseline | abundance | X07028 | N-Pentano | 3 | Interventio | Experimen | Experimen | Control | 0,936399 |

| | | | | | | | | | |
|---|---|---|---|---|---|---|---|---|---|
| baseline | abundance | X07038 | Piperidine_ | 3 | Interventio | Experimen | Experimen | Control | 0,945105 |
| baseline | abundance | X07040 | 6-Hydroxyp | 3 | Interventio | Experimen | Experimen | Control | 0,97445 |
| baseline | abundance | X07050 | 2-Methylth | 3 | Interventio | Experimen | Experimen | Control | 0,945105 |
| baseline | abundance | X07051 | O-succinyl | 3 | Interventio | Experimen | Experimen | Control | 0,97445 |
| baseline | abundance | X07054 | Valylprolin | 2b | Interventio | Experimen | Experimen | Control | 0,987276 |
| baseline | abundance | X07057 | Leu-Leu_c | 3 | Interventio | Experimen | Experimen | Control | 0,968601 |
| baseline | abundance | X07073 | asn-val_a | 3 | Interventio | Experimen | Experimen | Control | 0,936399 |
| baseline | abundance | X07077 | Hyodeoxyc | 1 | Interventio | Experimen | Experimen | Control | 0,945105 |
| baseline | abundance | X07079 | N-Pentano | 3 | Interventio | Experimen | Experimen | Control | 0,936399 |
| baseline | abundance | X07081 | N-LACTOYI | 3 | Interventio | Experimen | Experimen | Control | 0,97445 |
| baseline | abundance | X07084 | heptabarbi | 3 | Interventio | Experimen | Experimen | Control | 0,945105 |
| baseline | abundance | X07089 | Pregabalin | 3 | Interventio | Experimen | Experimen | Control | 0,936399 |
| baseline | abundance | X07092 | Dibutyl ma | 3 | Interventio | Experimen | Experimen | Control | 0,945105 |
| baseline | abundance | X07107 | Lysylvaline | 3 | Interventio | Experimen | Experimen | Control | 0,957875 |
| baseline | abundance | X07112 | MFCD0272 | 3 | Interventio | Experimen | Experimen | Control | 0,954102 |
| baseline | abundance | X07113 | Sulfoaceta | 3 | Interventio | Experimen | Experimen | Control | 0,936399 |
| baseline | abundance | X07125 | Zalcitabine | 3 | Interventio | Experimen | Experimen | Control | 0,936399 |
| baseline | abundance | X07126 | Leu-Val_f | 3 | Interventio | Experimen | Experimen | Control | 0,936399 |
| baseline | abundance | X07127 | N~5~-[P-Ar | 3 | Interventio | Experimen | Experimen | Control | 0,936399 |
| baseline | abundance | X07134 | 4-Phenolsu | 2b | Interventio | Experimen | Experimen | Control | 0,97445 |
| baseline | abundance | X07136 | hexobarbit | 3 | Interventio | Experimen | Experimen | Control | 0,97445 |
| baseline | abundance | X07139 | N,N-Diethy | 3 | Interventio | Experimen | Experimen | Control | 0,936399 |
| baseline | abundance | X07146 | meprobam | 3 | Interventio | Experimen | Experimen | Control | 0,991989 |
| baseline | abundance | X07155 | Naphthale | 3 | Interventio | Experimen | Experimen | Control | 0,945105 |
| baseline | abundance | X07164 | N-Acetyl-5 | 3 | Interventio | Experimen | Experimen | Control | 0,97445 |
| baseline | abundance | X07165 | pro-gln_b | 3 | Interventio | Experimen | Experimen | Control | 0,991989 |
| baseline | abundance | X07168 | L-(+)-Eryth | 3 | Interventio | Experimen | Experimen | Control | 0,948031 |
| baseline | abundance | X07193 | N-(1-Methy | 3 | Interventio | Experimen | Experimen | Control | 0,97445 |
| baseline | abundance | X07201 | 2,4-Quinol | 2b | Interventio | Experimen | Experimen | Control | 0,97445 |
| baseline | abundance | X07216 | pentobarbi | 3 | Interventio | Experimen | Experimen | Control | 0,936399 |
| baseline | abundance | X07219 | 3,3-Dimeth | 2b | Interventio | Experimen | Experimen | Control | 0,97445 |
| baseline | abundance | X07220 | Glycylproly | 3 | Interventio | Experimen | Experimen | Control | 0,936399 |
| baseline | abundance | X07226 | threonylph | 3 | Interventio | Experimen | Experimen | Control | 0,97445 |
| baseline | abundance | X07236 | 2-methylci | 3 | Interventio | Experimen | Experimen | Control | 0,945105 |
| baseline | abundance | X07250 | Leucylaspa | 3 | Interventio | Experimen | Experimen | Control | 0,997375 |
| baseline | abundance | X07260 | (7E,7'E)-5, | 3 | Interventio | Experimen | Experimen | Control | 0,936399 |
| baseline | abundance | X07263 | 3,3-Dimeth | 2b | Interventio | Experimen | Experimen | Control | 0,945105 |
| baseline | abundance | X07269 | (3aR,4R,5R | 3 | Interventio | Experimen | Experimen | Control | 0,962445 |
| baseline | abundance | X07270 | 5-(2-Carbo | 3 | Interventio | Experimen | Experimen | Control | 0,936399 |
| baseline | abundance | X07278 | 5-Phospho | 3 | Interventio | Experimen | Experimen | Control | 0,936399 |
| baseline | abundance | X07283 | 8-Hydroxyh | 3 | Interventio | Experimen | Experimen | Control | 0,957875 |
| baseline | abundance | X07285 | 2-(Carboxy | 3 | Interventio | Experimen | Experimen | Control | 0,936399 |
| baseline | abundance | X07294 | 6-Acetamid | 3 | Interventio | Experimen | Experimen | Control | 0,957875 |
| baseline | abundance | X07303 | Triacetin | 3 | Interventio | Experimen | Experimen | Control | 0,945786 |
| baseline | abundance | X07308 | Nicotinic a | 2a | Interventio | Experimen | Experimen | Control | 0,936399 |
| baseline | abundance | X07309 | 3,4,15-Trih | 3 | Interventio | Experimen | Experimen | Control | 0,988867 |
| baseline | abundance | X07315 | Rivastigmi | 3 | Interventio | Experimen | Experimen | Control | 0,945105 |
| baseline | abundance | X07320 | asn-pro_d | 3 | Interventio | Experimen | Experimen | Control | 0,97445 |

| | | | | | | | | | |
|---|---|---|---|---|---|---|---|---|---|
| baseline | abundance | X07327 | Aspartyl-L- | 3 | Interventio | Experimen | Experimen | Control | 0,936399 |
| baseline | abundance | X07334 | 2,2-Bis(hyd | 3 | Interventio | Experimen | Experimen | Control | 0,972498 |
| baseline | abundance | X07336 | Threonylse | 3 | Interventio | Experimen | Experimen | Control | 0,945105 |
| baseline | abundance | X07344 | (−)-nabilor | 3 | Interventio | Experimen | Experimen | Control | 0,983487 |
| baseline | abundance | X07345 | Phenyl D-g | 3 | Interventio | Experimen | Experimen | Control | 0,936399 |
| baseline | abundance | X07375 | porphobilir | 3 | Interventio | Experimen | Experimen | Control | 0,97445 |
| baseline | abundance | X07381 | Bis-D-fruct | 3 | Interventio | Experimen | Experimen | Control | 0,941185 |
| baseline | abundance | X07394 | Hydroxypro | 3 | Interventio | Experimen | Experimen | Control | 0,936399 |
| baseline | abundance | X07399 | Homoanse | 3 | Interventio | Experimen | Experimen | Control | 0,968601 |
| baseline | abundance | X07400 | Tranexami | 2b | Interventio | Experimen | Experimen | Control | 0,936399 |
| baseline | abundance | X07413 | Chenodeo: | 1 | Interventio | Experimen | Experimen | Control | 0,974375 |
| baseline | abundance | X07414 | 2,2-Bis(hyd | 3 | Interventio | Experimen | Experimen | Control | 0,936399 |
| baseline | abundance | X07417 | δ-Valerolac | 2b | Interventio | Experimen | Experimen | Control | 0,986763 |
| baseline | abundance | X07420 | DNOP_h | 3 | Interventio | Experimen | Experimen | Control | 0,97445 |
| baseline | abundance | X07424 | Rutinose (l | 3 | Interventio | Experimen | Experimen | Control | 0,945105 |
| baseline | abundance | X07425 | N-Acetylas | 2a | Interventio | Experimen | Experimen | Control | 0,936399 |
| baseline | abundance | X07428 | N6-METHY | 3 | Interventio | Experimen | Experimen | Control | 0,936399 |
| baseline | abundance | X07429 | DNOP_c | 3 | Interventio | Experimen | Experimen | Control | 0,97445 |
| baseline | abundance | X07434 | N'-Hydroxy | 2b | Interventio | Experimen | Experimen | Control | 0,97445 |
| baseline | abundance | X07439 | 4-pyridoxic | 1 | Interventio | Experimen | Experimen | Control | 0,936399 |
| baseline | abundance | X07442 | coenzyme | 3 | Interventio | Experimen | Experimen | Control | 0,987642 |
| baseline | abundance | X07443 | Butylparab | 3 | Interventio | Experimen | Experimen | Control | 0,97445 |
| baseline | abundance | X07445 | 3-(Sulfooxy | 3 | Interventio | Experimen | Experimen | Control | 0,948031 |
| baseline | abundance | X07451 | Valylvaline | 3 | Interventio | Experimen | Experimen | Control | 0,936399 |
| baseline | abundance | X07452 | 4-Amino-1- | 3 | Interventio | Experimen | Experimen | Control | 0,936399 |
| baseline | abundance | X07454 | 2,4-Quinol | 2b | Interventio | Experimen | Experimen | Control | 0,945105 |
| baseline | abundance | X07457 | Glucosami | 1 | Interventio | Experimen | Experimen | Control | 0,97445 |
| baseline | abundance | X07460 | 1-pyrroline | 3 | Interventio | Experimen | Experimen | Control | 0,957875 |
| baseline | abundance | X07465 | TO0127900 | 3 | Interventio | Experimen | Experimen | Control | 0,957875 |
| baseline | abundance | X07469 | 1,1'-[1,12-I | 3 | Interventio | Experimen | Experimen | Control | 0,945105 |
| baseline | abundance | X07475 | 2'-Deoxy-5 | 3 | Interventio | Experimen | Experimen | Control | 0,936399 |
| baseline | abundance | X07476 | 4-Hydroxyb | 1 | Interventio | Experimen | Experimen | Control | 0,936399 |
| baseline | abundance | X07480 | Naringenin | 2a | Interventio | Experimen | Experimen | Control | 0,945105 |
| baseline | abundance | X07484 | MFCD0002 | 3 | Interventio | Experimen | Experimen | Control | 0,983461 |
| baseline | abundance | X07487 | Oxypeucec | 3 | Interventio | Experimen | Experimen | Control | 0,989213 |
| baseline | abundance | X07491 | 1-Vinylimic | 2b | Interventio | Experimen | Experimen | Control | 0,936399 |
| baseline | abundance | X07504 | 4-Acetami | 2b | Interventio | Experimen | Experimen | Control | 0,959701 |
| baseline | abundance | X07508 | Procaine_a | 2b | Interventio | Experimen | Experimen | Control | 0,97445 |
| baseline | abundance | X07511 | Piperidine_ | 3 | Interventio | Experimen | Experimen | Control | 0,936399 |
| baseline | abundance | X07512 | 4-{[(3-Hydr | 3 | Interventio | Experimen | Experimen | Control | 0,936399 |
| baseline | abundance | X07513 | (−)-nabilor | 3 | Interventio | Experimen | Experimen | Control | 0,957875 |
| baseline | abundance | X07514 | L-(+)-Eryth | 3 | Interventio | Experimen | Experimen | Control | 0,97445 |
| baseline | abundance | X07518 | (+/-)-2-Hyc | 3 | Interventio | Experimen | Experimen | Control | 0,945105 |
| baseline | abundance | X07519 | Pyrrolidine | 3 | Interventio | Experimen | Experimen | Control | 0,936399 |
| baseline | abundance | X07523 | Ursodeoxy | 1 | Interventio | Experimen | Experimen | Control | 0,945105 |
| baseline | abundance | X07524 | Prolinamid | 2b | Interventio | Experimen | Experimen | Control | 0,936399 |
| baseline | abundance | X07527 | 1,7-Dimeth | 2b | Interventio | Experimen | Experimen | Control | 0,936399 |
| baseline | abundance | X07530 | Alanine | 1 | Interventio | Experimen | Experimen | Control | 0,936399 |

| | | | | | | | | | |
|---|---|---|---|---|---|---|---|---|---|
| baseline | abundance | X07537 | alpha-Cha | 3 | Interventio | Experimen | Experimen | Control | 0,957875 |
| baseline | abundance | X07538 | Homoanse | 3 | Interventio | Experimen | Experimen | Control | 0,936399 |
| baseline | abundance | X07541 | Saccharin | 1 | Interventio | Experimen | Experimen | Control | 0,97445 |
| baseline | abundance | X07544 | Ethyl sulfat | 3 | Interventio | Experimen | Experimen | Control | 0,957875 |
| baseline | abundance | X07558 | Leucylproli | 2b | Interventio | Experimen | Experimen | Control | 0,97445 |
| baseline | abundance | X07563 | Xanthine | 1 | Interventio | Experimen | Experimen | Control | 0,948031 |
| baseline | abundance | X07565 | 5-Hydroxyi | 2b | Interventio | Experimen | Experimen | Control | 0,945105 |
| baseline | abundance | X07566 | 5-Allyl-5-se | 3 | Interventio | Experimen | Experimen | Control | 0,97445 |
| baseline | abundance | X07572 | 8-(3-Furyl)- | 3 | Interventio | Experimen | Experimen | Control | 0,936399 |
| baseline | abundance | X07574 | 7-ketodeox | 3 | Interventio | Experimen | Experimen | Control | 0,957875 |
| baseline | abundance | X07577 | 2-[(Sulfoox | 3 | Interventio | Experimen | Experimen | Control | 0,936399 |
| baseline | abundance | X07578 | 5-Allyl-5-se | 3 | Interventio | Experimen | Experimen | Control | 0,936399 |
| baseline | abundance | X07580 | N-Pentano | 3 | Interventio | Experimen | Experimen | Control | 0,945105 |
| baseline | abundance | X07581 | N-Acetylva | 3 | Interventio | Experimen | Experimen | Control | 0,945105 |
| baseline | abundance | X07582 | 4-methylpy | 2b | Interventio | Experimen | Experimen | Control | 0,97445 |
| baseline | abundance | X07596 | 5-amino-2- | 2b | Interventio | Experimen | Experimen | Control | 0,936399 |
| baseline | abundance | X07597 | p-Cresylsu | 3 | Interventio | Experimen | Experimen | Control | 0,936399 |
| baseline | abundance | X07601 | Asp-lys | 3 | Interventio | Experimen | Experimen | Control | 0,992737 |
| baseline | abundance | X07613 | 1-(4-Aminc | 3 | Interventio | Experimen | Experimen | Control | 0,988948 |
| baseline | abundance | X07618 | N-(3-aceta | 3 | Interventio | Experimen | Experimen | Control | 0,945105 |
| baseline | abundance | X07630 | Methionine | 1 | Interventio | Experimen | Experimen | Control | 0,948059 |
| baseline | abundance | X07634 | 3,8,9-trihy | 2b | Interventio | Experimen | Experimen | Control | 0,945105 |
| baseline | abundance | X07638 | 7-ketodeox | 3 | Interventio | Experimen | Experimen | Control | 0,955424 |
| baseline | abundance | X07645 | O-propeno | 3 | Interventio | Experimen | Experimen | Control | 0,962445 |
| baseline | abundance | X07646 | (2Z)-2-({[(6 | 3 | Interventio | Experimen | Experimen | Control | 0,936399 |
| baseline | abundance | X07647 | Genistein | 1 | Interventio | Experimen | Experimen | Control | 0,936399 |
| baseline | abundance | X07648 | NL8513000 | 3 | Interventio | Experimen | Experimen | Control | 0,936399 |
| baseline | abundance | X07657 | Docosahex | 2b | Interventio | Experimen | Experimen | Control | 0,936399 |
| baseline | abundance | X07658 | δ-Glucono | 1 | Interventio | Experimen | Experimen | Control | 0,945105 |
| baseline | abundance | X07674 | Lysine | 1 | Interventio | Experimen | Experimen | Control | 0,936399 |
| baseline | abundance | X07678 | (2E)-3-(3,4 | 2b | Interventio | Experimen | Experimen | Control | 0,936399 |
| baseline | abundance | X07680 | Indole-3-ac | 1 | Interventio | Experimen | Experimen | Control | 0,978464 |
| baseline | abundance | X07694 | DNOP_a | 3 | Interventio | Experimen | Experimen | Control | 0,97445 |
| baseline | abundance | X07699 | N-(1-Methy | 3 | Interventio | Experimen | Experimen | Control | 0,945105 |
| baseline | abundance | X07701 | 3-Benzyl-6 | 3 | Interventio | Experimen | Experimen | Control | 0,97445 |
| baseline | abundance | X07710 | 3-O-beta-D | 3 | Interventio | Experimen | Experimen | Control | 0,970763 |
| baseline | abundance | X07712 | paracetam | 3 | Interventio | Experimen | Experimen | Control | 0,936399 |
| baseline | abundance | X07731 | 3-[4-methy | 2b | Interventio | Experimen | Experimen | Control | 0,936399 |
| baseline | abundance | X07732 | N-Acetylva | 2b | Interventio | Experimen | Experimen | Control | 0,97499 |
| baseline | abundance | X07743 | D-Alanine r | 2b | Interventio | Experimen | Experimen | Control | 0,936399 |
| baseline | abundance | X07745 | APM_c | 3 | Interventio | Experimen | Experimen | Control | 0,936399 |
| baseline | abundance | X07746 | 3-(2-Oxo-2 | 2b | Interventio | Experimen | Experimen | Control | 0,936399 |
| baseline | abundance | X07747 | Diethylpyrc | 3 | Interventio | Experimen | Experimen | Control | 0,958749 |
| baseline | abundance | X07749 | (DL)-3-O-M | 3 | Interventio | Experimen | Experimen | Control | 0,936399 |
| baseline | abundance | X07753 | Lovastatin_ | 3 | Interventio | Experimen | Experimen | Control | 0,957875 |
| baseline | abundance | X07754 | hexobarbit | 3 | Interventio | Experimen | Experimen | Control | 0,945105 |
| baseline | abundance | X07764 | (S)-2-meth | 3 | Interventio | Experimen | Experimen | Control | 0,957875 |
| baseline | abundance | X07770 | Piperine | 1 | Interventio | Experimen | Experimen | Control | 0,981877 |

| | | | | | | | | | |
|---|---|---|---|---|---|---|---|---|---|
| baseline | abundance | X07771 | Caprolacta | 2b | Interventio | Experimen | Experimen | Control | 0,945105 |
| baseline | abundance | X07774 | 7alpha-Hy | 3 | Interventio | Experimen | Experimen | Control | 0,988948 |
| baseline | abundance | X07777 | Arginine | 1 | Interventio | Experimen | Experimen | Control | 0,97445 |
| baseline | abundance | X07782 | Cytosine | 1 | Interventio | Experimen | Experimen | Control | 0,962445 |
| baseline | abundance | X07783 | 1,3-dimeth | 3 | Interventio | Experimen | Experimen | Control | 0,945105 |
| baseline | abundance | X07792 | N-Acetylhi | 2b | Interventio | Experimen | Experimen | Control | 0,936399 |
| baseline | abundance | X07794 | Prolylleuci | 2b | Interventio | Experimen | Experimen | Control | 0,978464 |
| baseline | abundance | X07798 | N-lauroylgl | 3 | Interventio | Experimen | Experimen | Control | 0,936399 |
| baseline | abundance | X07799 | Capryloylg | 2b | Interventio | Experimen | Experimen | Control | 0,97445 |
| baseline | abundance | X07811 | Metirosine | 3 | Interventio | Experimen | Experimen | Control | 0,936399 |
| baseline | abundance | X07812 | Triethyl cit | 3 | Interventio | Experimen | Experimen | Control | 0,97445 |
| baseline | abundance | X07815 | 3-[2-[(Z)-[3 | 3 | Interventio | Experimen | Experimen | Control | 0,936399 |
| baseline | abundance | X07820 | Creatinine | 1 | Interventio | Experimen | Experimen | Control | 0,945105 |
| baseline | abundance | X07825 | Maltotriose | 1 | Interventio | Experimen | Experimen | Control | 0,936399 |
| baseline | abundance | X07830 | 1,5-Isoquir | 2b | Interventio | Experimen | Experimen | Control | 0,945105 |
| baseline | abundance | X07834 | Choline su | 3 | Interventio | Experimen | Experimen | Control | 0,945105 |
| baseline | abundance | X07835 | 7-ketodeo | 3 | Interventio | Experimen | Experimen | Control | 0,945105 |
| baseline | abundance | X07841 | 2-(5-Benzy | 3 | Interventio | Experimen | Experimen | Control | 0,97445 |
| baseline | abundance | X07843 | Piperidine_ | 3 | Interventio | Experimen | Experimen | Control | 0,936399 |
| baseline | abundance | X07846 | Carbofurar | 3 | Interventio | Experimen | Experimen | Control | 0,945105 |
| baseline | abundance | X07848 | (−)-nabilor | 3 | Interventio | Experimen | Experimen | Control | 0,97445 |
| baseline | abundance | X07849 | Crotamitor | 3 | Interventio | Experimen | Experimen | Control | 0,936399 |
| baseline | abundance | X07852 | N6,N6,N6- | 3 | Interventio | Experimen | Experimen | Control | 0,980969 |
| baseline | abundance | X07854 | TO0127900 | 3 | Interventio | Experimen | Experimen | Control | 0,945105 |
| baseline | abundance | X07856 | (3R,4S,5S, | 3 | Interventio | Experimen | Experimen | Control | 0,97445 |
| baseline | abundance | X07866 | 3,3,5,5-Tet | 2b | Interventio | Experimen | Experimen | Control | 0,936399 |
| baseline | abundance | X07867 | N-Acetylva | 3 | Interventio | Experimen | Experimen | Control | 0,992737 |
| baseline | abundance | X07868 | Biotin | 2b | Interventio | Experimen | Experimen | Control | 0,94488 |
| baseline | abundance | X07869 | Hexamethy | 2a | Interventio | Experimen | Experimen | Control | 0,945105 |
| baseline | abundance | X07873 | (3Z,6Z,9Z,1 | 3 | Interventio | Experimen | Experimen | Control | 0,98802 |
| baseline | abundance | X07874 | δ-Valerolac | 2b | Interventio | Experimen | Experimen | Control | 0,94488 |
| baseline | abundance | X07886 | L-Urobilin | 3 | Interventio | Experimen | Experimen | Control | 0,97445 |
| baseline | abundance | X07889 | UROBILIN, | 3 | Interventio | Experimen | Experimen | Control | 0,936399 |
| baseline | abundance | X07890 | N-(5-aceta | 2b | Interventio | Experimen | Experimen | Control | 0,97445 |
| baseline | abundance | X07891 | Cyclamic a | 2b | Interventio | Experimen | Experimen | Control | 0,936399 |
| baseline | abundance | X07892 | urobilinoge | 3 | Interventio | Experimen | Experimen | Control | 0,936399 |
| baseline | abundance | X07899 | N-Acetylpu | 2b | Interventio | Experimen | Experimen | Control | 0,957875 |
| baseline | abundance | X07900 | presqualer | 3 | Interventio | Experimen | Experimen | Control | 0,945105 |
| baseline | abundance | X07903 | Acetophen | 2b | Interventio | Experimen | Experimen | Control | 0,948031 |
| baseline | abundance | X07905 | Prilocaine | 2b | Interventio | Experimen | Experimen | Control | 0,945105 |
| baseline | abundance | X07907 | 4-Acetami | 1 | Interventio | Experimen | Experimen | Control | 0,988948 |
| baseline | abundance | X07909 | (±)-Albuter | 2b | Interventio | Experimen | Experimen | Control | 0,957875 |
| baseline | abundance | X07914 | (-)-Erythror | 3 | Interventio | Experimen | Experimen | Control | 0,945105 |
| baseline | abundance | X07916 | N6,N6,N6- | 3 | Interventio | Experimen | Experimen | Control | 0,968601 |
| baseline | abundance | X07921 | Pipecolinic | 1 | Interventio | Experimen | Experimen | Control | 0,936399 |
| baseline | abundance | X07925 | DNOP_f | 3 | Interventio | Experimen | Experimen | Control | 0,97445 |
| baseline | abundance | X07930 | N-Methylc | 2b | Interventio | Experimen | Experimen | Control | 0,945105 |
| baseline | abundance | X07935 | Limonin | 3 | Interventio | Experimen | Experimen | Control | 0,936399 |

| | | | | | | | | | | |
|---|---|---|---|---|---|---|---|---|---|---|
| baseline | abundance | X07937 | Atenolol | 1 | Interventio | Experimen | Experimen | Control | | 0,97445 |
| baseline | abundance | X07944 | Styrene | 3 | Interventio | Experimen | Experimen | Control | | 0,97445 |
| baseline | abundance | X07955 | Valylvaline | 3 | Interventio | Experimen | Experimen | Control | | 0,982558 |
| baseline | abundance | X07961 | 2-(Hydroxy | 3 | Interventio | Experimen | Experimen | Control | | 0,936399 |
| baseline | abundance | X07963 | Theobromi | 1 | Interventio | Experimen | Experimen | Control | | 0,936399 |
| baseline | abundance | X07969 | 6-hydroxyp | 3 | Interventio | Experimen | Experimen | Control | | 0,936399 |
| baseline | abundance | X07974 | Isoprene | 3 | Interventio | Experimen | Experimen | Control | | 0,945105 |
| baseline | abundance | X07977 | Tyramine | 2b | Interventio | Experimen | Experimen | Control | | 0,945105 |
| baseline | abundance | X07978 | 1-[(4E)-4-( | 3 | Interventio | Experimen | Experimen | Control | | 0,957875 |
| baseline | abundance | X07981 | Methylimic | 2b | Interventio | Experimen | Experimen | Control | | 0,946624 |
| baseline | abundance | X07983 | butalbital_ | 3 | Interventio | Experimen | Experimen | Control | | 0,936399 |
| baseline | abundance | X07989 | Piperidine_ | 3 | Interventio | Experimen | Experimen | Control | | 0,936399 |
| baseline | abundance | X07993 | UROBILIN, | 3 | Interventio | Experimen | Experimen | Control | | 0,936399 |
| baseline | abundance | X08002 | (2S)-6-Ami | 3 | Interventio | Experimen | Experimen | Control | | 0,97445 |
| baseline | abundance | X08004 | δ-Valerola | 2b | Interventio | Experimen | Experimen | Control | | 0,945105 |
| baseline | abundance | X08007 | MFCD0002 | 3 | Interventio | Experimen | Experimen | Control | | 0,97445 |
| baseline | abundance | X08008 | N-Acetylne | 1 | Interventio | Experimen | Experimen | Control | | 0,936399 |
| baseline | abundance | X08012 | Methylol D | 3 | Interventio | Experimen | Experimen | Control | | 0,936399 |
| baseline | abundance | X08013 | Deoxysuga | 1 | Interventio | Experimen | Experimen | Control | | 0,97477 |
| baseline | abundance | X08014 | DNOP_e | 3 | Interventio | Experimen | Experimen | Control | | 0,970763 |
| baseline | abundance | X08017 | Tropinone | 2b | Interventio | Experimen | Experimen | Control | | 0,936399 |
| baseline | abundance | X08019 | Thymidine | 1 | Interventio | Experimen | Experimen | Control | | 0,936399 |
| baseline | abundance | X08024 | 8-Methyl-8 | 3 | Interventio | Experimen | Experimen | Control | | 0,983461 |
| baseline | abundance | X08028 | Solanidine | 3 | Interventio | Experimen | Experimen | Control | | 0,945105 |
| baseline | abundance | X08035 | Tetrahydro | 3 | Interventio | Experimen | Experimen | Control | | 0,945105 |
| baseline | abundance | X08037 | Crotonic a | 2b | Interventio | Experimen | Experimen | Control | | 0,936399 |
| baseline | abundance | X08038 | N-{6-[(7-C | 2b | Interventio | Experimen | Experimen | Control | | 0,945105 |
| baseline | abundance | X08040 | Prolylleuci | 2b | Interventio | Experimen | Experimen | Control | | 0,97445 |
| baseline | abundance | X08044 | 3-[2-[(Z)-[3 | 3 | Interventio | Experimen | Experimen | Control | | 0,945105 |
| baseline | abundance | X08046 | 3-Hydroxy- | 3 | Interventio | Experimen | Experimen | Control | | 0,945105 |
| baseline | abundance | X08054 | Docosahe | 2b | Interventio | Experimen | Experimen | Control | | 0,97445 |
| baseline | abundance | X08056 | 3-(2-Oxo-2 | 2b | Interventio | Experimen | Experimen | Control | | 0,936399 |
| baseline | abundance | X08057 | N-{3-Carbo | 3 | Interventio | Experimen | Experimen | Control | | 0,936399 |
| baseline | abundance | X08058 | 2-Hydroxy- | 3 | Interventio | Experimen | Experimen | Control | | 0,972498 |
| baseline | abundance | X08059 | Cadaverine | 3 | Interventio | Experimen | Experimen | Control | | 0,936399 |
| baseline | abundance | X08062 | Indole-3-c | 1 | Interventio | Experimen | Experimen | Control | | 0,936399 |
| baseline | abundance | X08072 | 4-Hydroxyp | 3 | Interventio | Experimen | Experimen | Control | | 0,936399 |
| baseline | abundance | X08076 | 3',5,7-Trihy | 2b | Interventio | Experimen | Experimen | Control | | 0,945105 |
| baseline | abundance | X08077 | 6-(alpha-D | 3 | Interventio | Experimen | Experimen | Control | | 0,936399 |
| baseline | abundance | X08078 | Ethyl mala | 3 | Interventio | Experimen | Experimen | Control | | 0,945105 |
| baseline | abundance | X08092 | Safrole | 3 | Interventio | Experimen | Experimen | Control | | 0,945105 |
| baseline | abundance | X08094 | 1,1'-[1,12- | 3 | Interventio | Experimen | Experimen | Control | | 0,945105 |
| baseline | abundance | X08095 | DNOP_g | 3 | Interventio | Experimen | Experimen | Control | | 0,983461 |
| baseline | abundance | X08096 | (2E)-3-Met | 3 | Interventio | Experimen | Experimen | Control | | 0,936399 |
| baseline | abundance | X08098 | Citrulline | 1 | Interventio | Experimen | Experimen | Control | | 0,936399 |
| baseline | abundance | X08099 | g-Butyrobe | 3 | Interventio | Experimen | Experimen | Control | | 0,936399 |
| baseline | abundance | X08100 | Piperidine_ | 3 | Interventio | Experimen | Experimen | Control | | 0,936399 |
| baseline | abundance | X08103 | Isopelletie | 3 | Interventio | Experimen | Experimen | Control | | 0,957875 |

| | | | | | | | | | |
|---|---|---|---|---|---|---|---|---|---|
| baseline | abundance | X08105 | 2-Acetamic | 3 | Interventio | Experimen | Experimen | Control | 0,97445 |
| baseline | abundance | X08106 | 4-Vinylphe | 1 | Interventio | Experimen | Experimen | Control | 0,936399 |
| baseline | abundance | X08111 | 2-Hydroxyc | 2b | Interventio | Experimen | Experimen | Control | 0,945105 |
| baseline | abundance | X08116 | Methyl (2Z, | 3 | Interventio | Experimen | Experimen | Control | 0,936399 |
| baseline | abundance | X08123 | Tiglic acid_ | 2b | Interventio | Experimen | Experimen | Control | 0,936399 |
| baseline | abundance | X08146 | O-Ethyl (4- | 3 | Interventio | Experimen | Experimen | Control | 0,987642 |
| baseline | abundance | X08147 | porphobilir | 3 | Interventio | Experimen | Experimen | Control | 0,945105 |
| baseline | abundance | X08150 | DL-4-Hydro | 2b | Interventio | Experimen | Experimen | Control | 0,945105 |
| baseline | abundance | X08154 | 4-(METHYL | 3 | Interventio | Experimen | Experimen | Control | 0,945105 |
| baseline | abundance | X08157 | Leu-Val_e | 3 | Interventio | Experimen | Experimen | Control | 0,945105 |
| baseline | abundance | X08162 | Valylvaline | 3 | Interventio | Experimen | Experimen | Control | 0,986763 |
| baseline | abundance | X08163 | Phenyl D-g | 3 | Interventio | Experimen | Experimen | Control | 0,945105 |
| baseline | abundance | X08167 | Lanthionin | 3 | Interventio | Experimen | Experimen | Control | 0,97445 |
| baseline | abundance | X08173 | 9-Methylur | 2b | Interventio | Experimen | Experimen | Control | 0,948031 |
| baseline | abundance | X08188 | 2-Hydroxy- | 2b | Interventio | Experimen | Experimen | Control | 0,945105 |
| baseline | abundance | X08193 | Seryltyrosi | 3 | Interventio | Experimen | Experimen | Control | 0,945105 |
| baseline | abundance | X08204 | 2-Amino-4 | 2b | Interventio | Experimen | Experimen | Control | 0,936399 |
| baseline | abundance | X08212 | LU345300 | 3 | Interventio | Experimen | Experimen | Control | 0,993446 |
| baseline | abundance | X08221 | Brilliant blu | 2b | Interventio | Experimen | Experimen | Control | 0,97499 |
| baseline | abundance | X08230 | Coumaron | 3 | Interventio | Experimen | Experimen | Control | 0,936399 |
| baseline | abundance | X08235 | 1-Methylhi | 2b | Interventio | Experimen | Experimen | Control | 0,936399 |
| baseline | abundance | X08244 | Setoclavin | 3 | Interventio | Experimen | Experimen | Control | 0,945105 |
| baseline | abundance | X08270 | Butabarbit | 3 | Interventio | Experimen | Experimen | Control | 0,936399 |
| baseline | abundance | X08274 | Mevalonic | 2b | Interventio | Experimen | Experimen | Control | 0,936399 |
| baseline | abundance | X08277 | adrenaline | 3 | Interventio | Experimen | Experimen | Control | 0,936399 |
| baseline | abundance | X08279 | DNOP_b | 3 | Interventio | Experimen | Experimen | Control | 0,945105 |
| baseline | abundance | X08288 | Capryloylg | 3 | Interventio | Experimen | Experimen | Control | 0,936399 |
| baseline | abundance | X08292 | Lidocaine | 2b | Interventio | Experimen | Experimen | Control | 0,957875 |
| baseline | abundance | X08298 | N6,N6,N6- | 2b | Interventio | Experimen | Experimen | Control | 0,936399 |
| baseline | abundance | X08300 | Indole-3-ac | 3 | Interventio | Experimen | Experimen | Control | 0,987276 |
| baseline | abundance | X08305 | Prolylhydro | 2a | Interventio | Experimen | Experimen | Control | 0,957875 |
| baseline | abundance | X08306 | 1-Methylxa | 2b | Interventio | Experimen | Experimen | Control | 0,945105 |
| baseline | abundance | X08311 | Methyl 3-fo | 3 | Interventio | Experimen | Experimen | Control | 0,985903 |
| baseline | abundance | X08317 | 8-Hydroxyc | 2b | Interventio | Experimen | Experimen | Control | 0,945105 |
| baseline | abundance | X08322 | Glycylproly | 3 | Interventio | Experimen | Experimen | Control | 0,936399 |
| baseline | abundance | X08323 | (15Z)-9,12 | 2b | Interventio | Experimen | Experimen | Control | 0,936399 |
| baseline | abundance | X08326 | 2-(1,3-Ben | 3 | Interventio | Experimen | Experimen | Control | 0,945105 |
| baseline | abundance | X08338 | Methyl [9-( | 3 | Interventio | Experimen | Experimen | Control | 0,936399 |
| baseline | abundance | X08341 | Midodrine_ | 3 | Interventio | Experimen | Experimen | Control | 0,97445 |
| baseline | abundance | X08352 | 7-[4-(tert-b | 2b | Interventio | Experimen | Experimen | Control | 0,936399 |
| baseline | abundance | X08365 | Lysylvaline | 3 | Interventio | Experimen | Experimen | Control | 0,936399 |
| baseline | abundance | X08367 | 11-dehydro | 2b | Interventio | Experimen | Experimen | Control | 0,945105 |
| baseline | abundance | X08373 | 2-Hydroxy- | 3 | Interventio | Experimen | Experimen | Control | 0,936399 |
| baseline | abundance | X08386 | 3b-Hydroxy | 3 | Interventio | Experimen | Experimen | Control | 0,936399 |
| baseline | abundance | X08387 | 4-[(2E,4Z)- | 3 | Interventio | Experimen | Experimen | Control | 0,936399 |
| baseline | abundance | X08412 | 2-Hydroxyp | 2b | Interventio | Experimen | Experimen | Control | 0,957875 |
| baseline | abundance | X08417 | asn-pro_c | 3 | Interventio | Experimen | Experimen | Control | 0,97445 |
| baseline | abundance | X08420 | Dodecane | 1 | Interventio | Experimen | Experimen | Control | 0,945105 |

| | | | | | | | | | |
|---|---|---|---|---|---|---|---|---|---|
| baseline | abundance | X08421 | Cys-tyr | 3 | Interventio | Experimen | Experimen | Control | 0,955424 |
| baseline | abundance | X08422 | N-(3-aceta | 3 | Interventio | Experimen | Experimen | Control | 0,936399 |
| baseline | abundance | X08425 | MFCD1297 | 3 | Interventio | Experimen | Experimen | Control | 0,936399 |
| baseline | abundance | X08436 | N,N-Dimet | 3 | Interventio | Experimen | Experimen | Control | 0,988948 |
| baseline | abundance | X08439 | Leucyltyros | 3 | Interventio | Experimen | Experimen | Control | 0,992737 |
| baseline | abundance | X08451 | L-gamma-( | 3 | Interventio | Experimen | Experimen | Control | 0,936399 |
| baseline | abundance | X08460 | α-Murichol | 1 | Interventio | Experimen | Experimen | Control | 0,936399 |
| baseline | abundance | X08461 | Dihydrouri | 3 | Interventio | Experimen | Experimen | Control | 0,945105 |
| baseline | abundance | X08475 | 1-(4-Amino | 3 | Interventio | Experimen | Experimen | Control | 0,936399 |
| baseline | abundance | X08476 | Coprine_c | 3 | Interventio | Experimen | Experimen | Control | 0,945105 |
| baseline | abundance | X08478 | 1-(beta-D-I | 3 | Interventio | Experimen | Experimen | Control | 0,936399 |
| baseline | abundance | X08485 | Isoprenalir | 3 | Interventio | Experimen | Experimen | Control | 0,945105 |
| baseline | abundance | X08486 | (2E,6E)-9-[ | 3 | Interventio | Experimen | Experimen | Control | 0,936399 |
| baseline | abundance | X08504 | Indole-3-ca | 2a | Interventio | Experimen | Experimen | Control | 0,948031 |
| baseline | abundance | X08509 | Piceid | 3 | Interventio | Experimen | Experimen | Control | 0,936399 |
| baseline | abundance | X08512 | Bile acid I ( | 2b | Interventio | Experimen | Experimen | Control | 0,945105 |
| baseline | abundance | X08514 | N-Ethylpro | 3 | Interventio | Experimen | Experimen | Control | 0,936399 |
| baseline | abundance | X08525 | MFCD1869 | 3 | Interventio | Experimen | Experimen | Control | 0,97445 |
| baseline | abundance | X08529 | 7,8-Diamir | 3 | Interventio | Experimen | Experimen | Control | 0,945105 |
| baseline | abundance | X08533 | tyramine su | 3 | Interventio | Experimen | Experimen | Control | 0,936399 |
| baseline | abundance | X08535 | Nitrosohep | 2b | Interventio | Experimen | Experimen | Control | 0,936399 |
| baseline | abundance | X08546 | 1,3,7-Trime | 2b | Interventio | Experimen | Experimen | Control | 0,957875 |
| baseline | abundance | X08548 | N2-Acetyll | 1 | Interventio | Experimen | Experimen | Control | 0,936399 |
| baseline | abundance | X08549 | 3-Methylsu | 3 | Interventio | Experimen | Experimen | Control | 0,945105 |
| baseline | abundance | X08561 | N-(Carboxy | 3 | Interventio | Experimen | Experimen | Control | 0,97445 |
| baseline | abundance | X08578 | tert-Butyl 3 | 3 | Interventio | Experimen | Experimen | Control | 0,991763 |
| baseline | abundance | X08584 | meticillin | 3 | Interventio | Experimen | Experimen | Control | 0,936399 |
| baseline | abundance | X08588 | 2-Acetamid | 3 | Interventio | Experimen | Experimen | Control | 0,948031 |
| baseline | abundance | X08593 | Uric acid | 1 | Interventio | Experimen | Experimen | Control | 0,936399 |
| baseline | abundance | X08594 | 7-ketodeox | 3 | Interventio | Experimen | Experimen | Control | 0,97445 |
| baseline | abundance | X08600 | N-Propiony | 3 | Interventio | Experimen | Experimen | Control | 0,945105 |
| baseline | abundance | X08606 | Butabarbit | 3 | Interventio | Experimen | Experimen | Control | 0,936399 |
| baseline | abundance | X08610 | Leucyltyros | 3 | Interventio | Experimen | Experimen | Control | 0,936399 |
| baseline | abundance | X08616 | MFCD0002 | 3 | Interventio | Experimen | Experimen | Control | 0,936399 |
| baseline | abundance | X08625 | Hept-2-ulo | 3 | Interventio | Experimen | Experimen | Control | 0,936399 |
| baseline | abundance | X08634 | 1,5-Isoquir | 2b | Interventio | Experimen | Experimen | Control | 0,936399 |
| baseline | abundance | X08639 | hypaphorir | 3 | Interventio | Experimen | Experimen | Control | 0,936399 |
| baseline | abundance | X08641 | meprobam | 3 | Interventio | Experimen | Experimen | Control | 0,936399 |
| baseline | abundance | X08644 | 2,5-Dimeth | 2b | Interventio | Experimen | Experimen | Control | 0,936399 |
| baseline | abundance | X08646 | N-Stearoylt | 3 | Interventio | Experimen | Experimen | Control | 0,945105 |
| baseline | abundance | X08657 | 1-(4-Amino | 3 | Interventio | Experimen | Experimen | Control | 0,97445 |
| baseline | abundance | X08659 | 3,3-Dimeth | 2b | Interventio | Experimen | Experimen | Control | 0,945105 |
| baseline | abundance | X08663 | Pilocarpine | 2b | Interventio | Experimen | Experimen | Control | 0,936399 |
| baseline | abundance | X08670 | Hydroxypro | 3 | Interventio | Experimen | Experimen | Control | 0,945105 |
| baseline | abundance | X08673 | NPC | 3 | Interventio | Experimen | Experimen | Control | 0,97445 |
| baseline | abundance | X08677 | 1-methylhy | 3 | Interventio | Experimen | Experimen | Control | 0,936399 |
| baseline | abundance | X08690 | Ethanoic a | 3 | Interventio | Experimen | Experimen | Control | 0,982558 |
| baseline | abundance | X08693 | 7-Sulfocho | 3 | Interventio | Experimen | Experimen | Control | 0,97445 |

| | | | | | | | | | |
|---|---|---|---|---|---|---|---|---|---|
| baseline | abundance | X08695 | 3b-Hydroxy | 3 | Interventio | Experimen | Experimen | Control | 0,979456 |
| baseline | abundance | X08699 | Aurorix | 3 | Interventio | Experimen | Experimen | Control | 0,936399 |
| baseline | abundance | X08707 | 2-(1-Ethoxy | 3 | Interventio | Experimen | Experimen | Control | 0,948031 |
| baseline | abundance | X08711 | 2-Acetami | 3 | Interventio | Experimen | Experimen | Control | 0,97445 |
| baseline | abundance | X08712 | 4-Iodoanis | 3 | Interventio | Experimen | Experimen | Control | 0,948031 |
| baseline | abundance | X08713 | Phenethyla | 2b | Interventio | Experimen | Experimen | Control | 0,97445 |
| baseline | abundance | X08714 | Arabic acid | 3 | Interventio | Experimen | Experimen | Control | 0,992737 |
| baseline | abundance | X08719 | 6-Methylqu | 2b | Interventio | Experimen | Experimen | Control | 0,945105 |
| baseline | abundance | X08723 | S-Allylcyste | 3 | Interventio | Experimen | Experimen | Control | 0,957875 |
| baseline | abundance | X08726 | YV8195000 | 3 | Interventio | Experimen | Experimen | Control | 0,936399 |
| baseline | abundance | X08733 | N-Methyl-1 | 3 | Interventio | Experimen | Experimen | Control | 0,936399 |
| baseline | abundance | X08739 | 1,1'-[1,12-I | 3 | Interventio | Experimen | Experimen | Control | 0,936399 |
| baseline | abundance | X08758 | Methohexit | 3 | Interventio | Experimen | Experimen | Control | 0,936399 |
| baseline | abundance | X08777 | 3-Aminosa | 2b | Interventio | Experimen | Experimen | Control | 0,936399 |
| baseline | abundance | X08779 | 3-(1-hydro: | 2b | Interventio | Experimen | Experimen | Control | 0,945105 |
| baseline | abundance | X08781 | Procaine_b | 3 | Interventio | Experimen | Experimen | Control | 0,936399 |
| baseline | abundance | X08789 | Acrylic acid | 2b | Interventio | Experimen | Experimen | Control | 0,985903 |
| baseline | abundance | X08792 | 3,4-Methyl | 3 | Interventio | Experimen | Experimen | Control | 0,945105 |
| baseline | abundance | X08795 | N-Acetylva | 3 | Interventio | Experimen | Experimen | Control | 0,936399 |
| baseline | abundance | X08801 | 3,7-Dimeth | 2b | Interventio | Experimen | Experimen | Control | 0,936399 |
| baseline | abundance | X08810 | N~6~,N~6~ | 3 | Interventio | Experimen | Experimen | Control | 0,936399 |
| baseline | abundance | X08822 | CYS-ASP | 3 | Interventio | Experimen | Experimen | Control | 0,945105 |
| baseline | abundance | X08825 | Chenodeox | 3 | Interventio | Experimen | Experimen | Control | 0,936399 |
| baseline | abundance | X08842 | Spermic ac | 3 | Interventio | Experimen | Experimen | Control | 0,936399 |
| baseline | abundance | X08843 | (2R,3S)-3-I | 3 | Interventio | Experimen | Experimen | Control | 0,936399 |
| baseline | abundance | X08844 | 4-Hydroxy- | 3 | Interventio | Experimen | Experimen | Control | 0,97445 |
| baseline | abundance | X08845 | 14-Hydroxy | 3 | Interventio | Experimen | Experimen | Control | 0,945105 |
| baseline | abundance | X08847 | 1-Vinylimic | 2b | Interventio | Experimen | Experimen | Control | 0,936399 |
| baseline | abundance | X08869 | Lanthionin | 3 | Interventio | Experimen | Experimen | Control | 0,945105 |
| baseline | abundance | X08870 | Tetraacety | 3 | Interventio | Experimen | Experimen | Control | 0,945105 |
| baseline | abundance | X08883 | 3-Phenylpr | 3 | Interventio | Experimen | Experimen | Control | 0,957875 |
| baseline | abundance | X08893 | 5beta-Cho | 3 | Interventio | Experimen | Experimen | Control | 0,936399 |
| baseline | abundance | X08895 | pentoxyl | 3 | Interventio | Experimen | Experimen | Control | 0,936399 |
| baseline | abundance | X08908 | Vorinostat_ | 3 | Interventio | Experimen | Experimen | Control | 0,986763 |
| baseline | abundance | X08909 | GLK (Pepti | 2b | Interventio | Experimen | Experimen | Control | 0,936399 |
| baseline | abundance | X08923 | Semilicoise | 3 | Interventio | Experimen | Experimen | Control | 0,945105 |
| baseline | abundance | X08938 | Nonivamid | 3 | Interventio | Experimen | Experimen | Control | 0,936399 |
| baseline | abundance | X08944 | Dipivefrin | 3 | Interventio | Experimen | Experimen | Control | 0,936399 |
| baseline | abundance | X08965 | 1,9-Nonan | 3 | Interventio | Experimen | Experimen | Control | 0,936399 |
| baseline | abundance | X08973 | Aminohipp | 3 | Interventio | Experimen | Experimen | Control | 0,936399 |
| baseline | abundance | X08983 | IN00150 | 3 | Interventio | Experimen | Experimen | Control | 0,957875 |
| baseline | abundance | X08987 | feruloylser | 3 | Interventio | Experimen | Experimen | Control | 0,945105 |
| baseline | abundance | X08990 | Procaine_c | 3 | Interventio | Experimen | Experimen | Control | 0,986763 |
| baseline | abundance | X08992 | N(2)-succi | 3 | Interventio | Experimen | Experimen | Control | 0,936399 |
| baseline | abundance | X09001 | Varanic ac | 3 | Interventio | Experimen | Experimen | Control | 0,957875 |
| baseline | abundance | X09004 | Arenaine | 3 | Interventio | Experimen | Experimen | Control | 0,97445 |
| baseline | abundance | X09008 | 5-guanidin | 3 | Interventio | Experimen | Experimen | Control | 0,97445 |
| baseline | abundance | X09010 | 4-Hydroxyp | 1 | Interventio | Experimen | Experimen | Control | 0,936399 |

| | | | | | | | | | |
|---|---|---|---|---|---|---|---|---|---|
| baseline | abundance | X09055 | N-[(1R,2S,( | 3 | Interventio | Experimen | Experimen | Control | 0,936399 |
| baseline | abundance | X09064 | Tetrahydro | 3 | Interventio | Experimen | Experimen | Control | 0,945105 |
| baseline | abundance | X09068 | N,N-Diethy | 3 | Interventio | Experimen | Experimen | Control | 0,945105 |
| baseline | abundance | X09082 | 1_2-Dihydr | 3 | Interventio | Experimen | Experimen | Control | 0,97445 |
| baseline | abundance | X09091 | N-(3-aceta | 3 | Interventio | Experimen | Experimen | Control | 0,936399 |
| baseline | abundance | X09092 | LysoSM(d1 | 3 | Interventio | Experimen | Experimen | Control | 0,945105 |
| baseline | abundance | X09099 | 1-(2-Hydro | 3 | Interventio | Experimen | Experimen | Control | 0,936399 |
| baseline | abundance | X09100 | Ethyl aceta | 3 | Interventio | Experimen | Experimen | Control | 0,98802 |
| baseline | abundance | X09110 | Valylvaline | 3 | Interventio | Experimen | Experimen | Control | 0,94488 |
| baseline | abundance | X09122 | Valyl-4-hyc | 3 | Interventio | Experimen | Experimen | Control | 0,945105 |
| baseline | abundance | X09129 | Coprine_d | 3 | Interventio | Experimen | Experimen | Control | 0,945105 |
| baseline | abundance | X09152 | Ferulic aci | 1 | Interventio | Experimen | Experimen | Control | 0,936399 |
| baseline | abundance | X09163 | Prenisteine | 3 | Interventio | Experimen | Experimen | Control | 0,97445 |
| baseline | abundance | X09165 | N-[1-Carbo | 3 | Interventio | Experimen | Experimen | Control | 0,936399 |
| baseline | abundance | X09187 | Salicylic ac | 1 | Interventio | Experimen | Experimen | Control | 0,945105 |
| baseline | abundance | X09189 | 4-[(E)-2-(3, | 3 | Interventio | Experimen | Experimen | Control | 0,936399 |
| baseline | abundance | X09207 | AAMU_a | 3 | Interventio | Experimen | Experimen | Control | 0,936399 |
| baseline | abundance | X09208 | IN00260_b | 3 | Interventio | Experimen | Experimen | Control | 0,957875 |
| baseline | abundance | X09222 | L-gamma-( | 3 | Interventio | Experimen | Experimen | Control | 0,945105 |
| baseline | abundance | X09231 | 2-Keto-glut | 3 | Interventio | Experimen | Experimen | Control | 0,936399 |
| baseline | abundance | X09244 | gamma-Gl | 3 | Interventio | Experimen | Experimen | Control | 0,945105 |
| baseline | abundance | X09245 | L-gamma-( | 3 | Interventio | Experimen | Experimen | Control | 0,936399 |
| baseline | abundance | X09256 | Aspartyl-L- | 3 | Interventio | Experimen | Experimen | Control | 0,945105 |
| baseline | abundance | X09260 | (4S)-4-[(2E | 3 | Interventio | Experimen | Experimen | Control | 0,936399 |
| baseline | abundance | X09284 | 4-(Nitrosoa | 3 | Interventio | Experimen | Experimen | Control | 0,957875 |
| baseline | abundance | X09285 | MFCD0272 | 3 | Interventio | Experimen | Experimen | Control | 0,957875 |
| baseline | abundance | X09302 | Sular | 3 | Interventio | Experimen | Experimen | Control | 0,945105 |
| baseline | abundance | X09318 | butalbital_ | 3 | Interventio | Experimen | Experimen | Control | 0,945105 |
| baseline | abundance | X09336 | 1,1'-[1,12-I | 3 | Interventio | Experimen | Experimen | Control | 0,97445 |
| baseline | abundance | X09357 | 5-(5-Methy | 3 | Interventio | Experimen | Experimen | Control | 0,936399 |
| baseline | abundance | X09374 | 3-Deoxy-D- | 3 | Interventio | Experimen | Experimen | Control | 0,936399 |
| baseline | abundance | X09385 | 2-(1-Napht | 3 | Interventio | Experimen | Experimen | Control | 0,936399 |
| baseline | abundance | X09390 | asn-pro_a | 3 | Interventio | Experimen | Experimen | Control | 0,936399 |
| baseline | abundance | X09391 | ophthalmic | 3 | Interventio | Experimen | Experimen | Control | 0,957875 |
| baseline | abundance | X09393 | 2-Isopropy | 3 | Interventio | Experimen | Experimen | Control | 0,957875 |
| baseline | abundance | X09399 | L-gamma-( | 3 | Interventio | Experimen | Experimen | Control | 0,936399 |
| baseline | abundance | X09454 | 2-(2-Hydro | 3 | Interventio | Experimen | Experimen | Control | 0,936399 |
| baseline | abundance | X09476 | N(1),N(8)-I | 3 | Interventio | Experimen | Experimen | Control | 0,936399 |
| baseline | abundance | X09480 | Spermic ac | 3 | Interventio | Experimen | Experimen | Control | 0,982558 |
| baseline | abundance | X09490 | 2-Hydroxy- | 3 | Interventio | Experimen | Experimen | Control | 0,936399 |
| baseline | abundance | X09503 | 4-(METHYL | 3 | Interventio | Experimen | Experimen | Control | 0,936399 |
| baseline | abundance | X09505 | (-)-Physost | 3 | Interventio | Experimen | Experimen | Control | 0,936399 |
| baseline | abundance | X09523 | N-(3,5-Dim | 3 | Interventio | Experimen | Experimen | Control | 0,97445 |
| baseline | abundance | X09525 | Homoanse | 3 | Interventio | Experimen | Experimen | Control | 0,985326 |
| baseline | abundance | X09533 | S(6)-acetyl | 3 | Interventio | Experimen | Experimen | Control | 0,97445 |
| baseline | abundance | X09545 | 4-Methyler | 3 | Interventio | Experimen | Experimen | Control | 0,945105 |
| baseline | abundance | X09546 | asn-phe | 3 | Interventio | Experimen | Experimen | Control | 0,97445 |
| baseline | abundance | X09548 | 2-O-beta-D | 3 | Interventio | Experimen | Experimen | Control | 0,97445 |

| | | | | | | | | | |
|---|---|---|---|---|---|---|---|---|---|
| baseline | abundance | X09554 | N-Propiony | 3 | Interventio | Experimen | Experimen | Control | 0,992737 |
| baseline | abundance | X09555 | N4-(beta-N | 3 | Interventio | Experimen | Experimen | Control | 0,936399 |
| baseline | abundance | X09565 | 3-Methoxy- | 3 | Interventio | Experimen | Experimen | Control | 0,957875 |
| baseline | abundance | X09577 | Spermic ac | 3 | Interventio | Experimen | Experimen | Control | 0,936399 |
| baseline | abundance | X09581 | L-gamma-( | 3 | Interventio | Experimen | Experimen | Control | 0,936399 |
| baseline | abundance | X09607 | N-(1H-Pyrr | 3 | Interventio | Experimen | Experimen | Control | 0,948031 |
| baseline | abundance | X09608 | N-(3,5-Dim | 3 | Interventio | Experimen | Experimen | Control | 0,945105 |
| baseline | abundance | X09609 | N-{3-[(4-Ac | 3 | Interventio | Experimen | Experimen | Control | 0,936399 |
| baseline | abundance | X09612 | 3-hydroxy-: | 3 | Interventio | Experimen | Experimen | Control | 0,936399 |
| baseline | abundance | X09615 | tyramine su | 3 | Interventio | Experimen | Experimen | Control | 0,945105 |
| baseline | abundance | X09622 | N-Hydroxy- | 3 | Interventio | Experimen | Experimen | Control | 0,936399 |
| baseline | abundance | X09623 | Endothal | 2b | Interventio | Experimen | Experimen | Control | 0,936399 |
| baseline | abundance | X09635 | 2,4,6-Triisc | 3 | Interventio | Experimen | Experimen | Control | 0,994656 |
| baseline | abundance | X09643 | N-(3,5-Dim | 3 | Interventio | Experimen | Experimen | Control | 0,97445 |
| baseline | abundance | X09645 | TOLMETIN | 3 | Interventio | Experimen | Experimen | Control | 0,97445 |
| baseline | abundance | X09654 | linatine | 3 | Interventio | Experimen | Experimen | Control | 0,936399 |
| baseline | abundance | X09657 | 2,3,4,5,6-F | 3 | Interventio | Experimen | Experimen | Control | 0,936399 |
| baseline | abundance | X09675 | UK387000 | 3 | Interventio | Experimen | Experimen | Control | 0,97445 |
| baseline | abundance | X09684 | (+)-Etomid | 3 | Interventio | Experimen | Experimen | Control | 0,936399 |
| baseline | abundance | X09703 | 7,8-Didehy | 3 | Interventio | Experimen | Experimen | Control | 0,936399 |
| baseline | abundance | X09710 | LW800000 | 3 | Interventio | Experimen | Experimen | Control | 0,945105 |
| baseline | abundance | X09713 | N(2)-succi | 3 | Interventio | Experimen | Experimen | Control | 0,97445 |
| baseline | abundance | X09721 | 3-(2,3-Dihy | 3 | Interventio | Experimen | Experimen | Control | 0,955424 |
| baseline | abundance | X09724 | Formylkynu | 3 | Interventio | Experimen | Experimen | Control | 0,936399 |
| baseline | abundance | X09726 | indoline-2- | 2b | Interventio | Experimen | Experimen | Control | 0,936399 |
| baseline | abundance | X09728 | {[(15-Hydro | 3 | Interventio | Experimen | Experimen | Control | 0,936399 |
| baseline | abundance | X09738 | (7E,7'E)-5,: | 3 | Interventio | Experimen | Experimen | Control | 0,936399 |
| baseline | abundance | X09740 | MFCD0166 | 3 | Interventio | Experimen | Experimen | Control | 0,945105 |
| baseline | abundance | X09741 | Estrone glu | 3 | Interventio | Experimen | Experimen | Control | 0,982558 |
| baseline | abundance | X09746 | ELK (Peptic | 2b | Interventio | Experimen | Experimen | Control | 0,936399 |
| baseline | abundance | X09765 | Tetrahydro | 3 | Interventio | Experimen | Experimen | Control | 0,945105 |
| baseline | abundance | X09771 | 2-(3-CARB | 3 | Interventio | Experimen | Experimen | Control | 0,936399 |
| baseline | abundance | X09787 | Queuosine | 3 | Interventio | Experimen | Experimen | Control | 0,945105 |
| baseline | abundance | X09805 | Methyl alpl | 3 | Interventio | Experimen | Experimen | Control | 0,945105 |
| baseline | abundance | X09810 | Zalcitabine | 3 | Interventio | Experimen | Experimen | Control | 0,936399 |
| baseline | abundance | X09825 | (2S)-6-Ami | 3 | Interventio | Experimen | Experimen | Control | 0,945105 |
| baseline | abundance | X09849 | 3-Methylac | 3 | Interventio | Experimen | Experimen | Control | 0,988948 |
| baseline | abundance | X09854 | N-Benzyl-3 | 2b | Interventio | Experimen | Experimen | Control | 0,97445 |
| baseline | abundance | X09875 | 4-[(E)-2-(3, | 3 | Interventio | Experimen | Experimen | Control | 0,945786 |
| baseline | abundance | X09882 | Harmane | 3 | Interventio | Experimen | Experimen | Control | 0,936399 |
| baseline | abundance | X09895 | 1-(beta-D-r | 3 | Interventio | Experimen | Experimen | Control | 0,936399 |
| baseline | abundance | X09898 | N(6),N(6)-I | 3 | Interventio | Experimen | Experimen | Control | 0,936399 |
| baseline | abundance | X09912 | MC055530 | 3 | Interventio | Experimen | Experimen | Control | 0,945786 |
| baseline | abundance | X09919 | 4-ethylphe | 3 | Interventio | Experimen | Experimen | Control | 0,936399 |
| baseline | abundance | X09921 | 5-Hydroxy- | 3 | Interventio | Experimen | Experimen | Control | 0,936399 |
| baseline | abundance | X09932 | g-Aminobu | 3 | Interventio | Experimen | Experimen | Control | 0,936399 |
| baseline | abundance | X09940 | pentobarbi | 3 | Interventio | Experimen | Experimen | Control | 0,936399 |
| baseline | abundance | X09942 | Spermic ac | 3 | Interventio | Experimen | Experimen | Control | 0,982558 |

| | | | | | | | | | |
|---|---|---|---|---|---|---|---|---|---|
| baseline | abundance | X09950 | 3-Hydroxyl | 3 | Interventio | Experimen | Experimen | Control | 0,945105 |
| baseline | abundance | X09951 | 5beta-Cypr | 3 | Interventio | Experimen | Experimen | Control | 0,936399 |
| baseline | abundance | X09963 | Iminoglycir | 3 | Interventio | Experimen | Experimen | Control | 0,936399 |
| baseline | abundance | X09965 | Biocytin | 2b | Interventio | Experimen | Experimen | Control | 0,97445 |
| baseline | abundance | X09969 | 3-Methylac | 3 | Interventio | Experimen | Experimen | Control | 0,945105 |
| baseline | abundance | X09977 | TLK (Peptic | 2b | Interventio | Experimen | Experimen | Control | 0,945105 |
| baseline | abundance | X09980 | N-(4-Hepta | 3 | Interventio | Experimen | Experimen | Control | 0,936399 |
| baseline | abundance | X09981 | N-Acetylva | 3 | Interventio | Experimen | Experimen | Control | 0,936399 |
| baseline | abundance | X09998 | N-[(10Z)-7- | 3 | Interventio | Experimen | Experimen | Control | 0,945105 |
| baseline | abundance | X10011 | Midodrine_ | 3 | Interventio | Experimen | Experimen | Control | 0,936399 |
| baseline | abundance | X10025 | 2-[4-(3-Hyc | 3 | Interventio | Experimen | Experimen | Control | 0,945105 |
| baseline | abundance | X10032 | 4,9a-Dimet | 3 | Interventio | Experimen | Experimen | Control | 0,945105 |
| baseline | abundance | X10033 | epsilon-(ga | 3 | Interventio | Experimen | Experimen | Control | 0,962143 |
| baseline | abundance | X10036 | Nimodipin | 3 | Interventio | Experimen | Experimen | Control | 0,978464 |
| baseline | abundance | X10037 | his-gln_a | 3 | Interventio | Experimen | Experimen | Control | 0,936399 |
| baseline | abundance | X10046 | Dehydroac | 2b | Interventio | Experimen | Experimen | Control | 0,936399 |
| baseline | abundance | X10076 | trimethadi | 3 | Interventio | Experimen | Experimen | Control | 0,936399 |
| baseline | abundance | X10091 | (2E,6E)-9-[ | 3 | Interventio | Experimen | Experimen | Control | 0,957875 |
| baseline | abundance | X10097 | Coixol | 3 | Interventio | Experimen | Experimen | Control | 0,97445 |
| baseline | abundance | X10098 | Methyl 2,3- | 3 | Interventio | Experimen | Experimen | Control | 0,945105 |
| baseline | abundance | X10099 | ala-met | 3 | Interventio | Experimen | Experimen | Control | 0,98802 |
| baseline | abundance | X10121 | Leu-pro_b | 3 | Interventio | Experimen | Experimen | Control | 0,945105 |
| baseline | abundance | X10129 | SECONAL_ | 3 | Interventio | Experimen | Experimen | Control | 0,945105 |
| baseline | abundance | X10130 | N-(4-Hydro | 3 | Interventio | Experimen | Experimen | Control | 0,936399 |
| baseline | abundance | X10137 | 1-Methyl-1 | 3 | Interventio | Experimen | Experimen | Control | 0,963031 |
| baseline | abundance | X10144 | imazameth | 3 | Interventio | Experimen | Experimen | Control | 0,986763 |
| baseline | abundance | X10148 | cys-met | 3 | Interventio | Experimen | Experimen | Control | 0,936399 |
| baseline | abundance | X10151 | 2-(3-CARB | 3 | Interventio | Experimen | Experimen | Control | 0,936399 |
| baseline | abundance | X10162 | Quinaldic a | 2a | Interventio | Experimen | Experimen | Control | 0,960702 |
| baseline | abundance | X10177 | Gly-DL-Phe | 3 | Interventio | Experimen | Experimen | Control | 0,957875 |
| baseline | abundance | X10180 | 4-Formyl-2 | 3 | Interventio | Experimen | Experimen | Control | 0,936399 |
| baseline | abundance | X10203 | L-gamma-( | 3 | Interventio | Experimen | Experimen | Control | 0,936399 |
| baseline | abundance | X10224 | Imidazolela | 2b | Interventio | Experimen | Experimen | Control | 0,936399 |
| baseline | abundance | X10246 | Ethynodiol | 3 | Interventio | Experimen | Experimen | Control | 0,957875 |
| baseline | abundance | X10248 | (+)-Etomid | 3 | Interventio | Experimen | Experimen | Control | 0,97445 |
| baseline | abundance | X10265 | 3-methyl-4 | 3 | Interventio | Experimen | Experimen | Control | 0,945786 |
| baseline | abundance | X10276 | Maleamate | 3 | Interventio | Experimen | Experimen | Control | 0,936399 |
| baseline | abundance | X10279 | Lys-Pro_d | 3 | Interventio | Experimen | Experimen | Control | 0,97029 |
| baseline | abundance | X10304 | 2,3,8,9-Tet | 3 | Interventio | Experimen | Experimen | Control | 0,945105 |
| baseline | abundance | X10314 | 4-morpholi | 2b | Interventio | Experimen | Experimen | Control | 0,97445 |
| baseline | abundance | X10319 | 1-Methyl-1 | 3 | Interventio | Experimen | Experimen | Control | 0,970482 |
| baseline | abundance | X10325 | Lovastatin_ | 3 | Interventio | Experimen | Experimen | Control | 0,97499 |
| baseline | abundance | X10330 | Sulfurous a | 3 | Interventio | Experimen | Experimen | Control | 0,936399 |
| baseline | abundance | X10339 | 4H-1-Benz | 2b | Interventio | Experimen | Experimen | Control | 0,936399 |
| baseline | abundance | X10345 | S-Allylcyst | 3 | Interventio | Experimen | Experimen | Control | 0,936399 |
| baseline | abundance | X10353 | L-gamma-( | 3 | Interventio | Experimen | Experimen | Control | 0,945105 |
| baseline | abundance | X10367 | 2-(1-Napht | 3 | Interventio | Experimen | Experimen | Control | 0,941349 |
| baseline | abundance | X10370 | L-Arogenat | 3 | Interventio | Experimen | Experimen | Control | 0,936399 |

| | | | | | | | | | |
|---|---|---|---|---|---|---|---|---|---|
| baseline | abundance | X10375 | 4-Indoleca | 2b | Interventio | Experimen | Experimen | Control | 0,936399 |
| baseline | abundance | X10379 | Phenyl D-g | 3 | Interventio | Experimen | Experimen | Control | 0,945786 |
| baseline | abundance | X10382 | Zalcitabine | 3 | Interventio | Experimen | Experimen | Control | 0,936399 |
| baseline | abundance | X10384 | L-N2-(2-Ca | 3 | Interventio | Experimen | Experimen | Control | 0,957875 |
| baseline | abundance | X10387 | Coenzyme | 3 | Interventio | Experimen | Experimen | Control | 0,936399 |
| baseline | abundance | X10389 | Leucylaspa | 3 | Interventio | Experimen | Experimen | Control | 0,936399 |
| baseline | abundance | X10393 | Ro 20-1724 | 3 | Interventio | Experimen | Experimen | Control | 0,936399 |
| baseline | abundance | X10401 | 3-Methylxa | 1 | Interventio | Experimen | Experimen | Control | 0,936399 |
| baseline | abundance | X10415 | beta-D-Glc | 3 | Interventio | Experimen | Experimen | Control | 0,97445 |
| baseline | abundance | X10417 | L-Sacchar | 2b | Interventio | Experimen | Experimen | Control | 0,936399 |
| baseline | abundance | X10429 | 1,5-Isoquir | 2b | Interventio | Experimen | Experimen | Control | 0,936399 |
| baseline | abundance | X10442 | (6alpha,11 | 3 | Interventio | Experimen | Experimen | Control | 0,97445 |
| baseline | abundance | X10445 | 2-(Carboxy | 3 | Interventio | Experimen | Experimen | Control | 0,97445 |
| baseline | abundance | X10451 | Ro 20-1724 | 3 | Interventio | Experimen | Experimen | Control | 0,945105 |
| baseline | abundance | X10454 | 8-Amino-7 | 3 | Interventio | Experimen | Experimen | Control | 0,936399 |
| baseline | abundance | X10457 | glu-ser | 3 | Interventio | Experimen | Experimen | Control | 0,970772 |
| baseline | abundance | X10476 | GAMMA-HY | 3 | Interventio | Experimen | Experimen | Control | 0,957875 |
| baseline | abundance | X10486 | Coumarin | 2b | Interventio | Experimen | Experimen | Control | 0,94488 |
| baseline | abundance | X10488 | 4-Guanidir | 1 | Interventio | Experimen | Experimen | Control | 0,982558 |
| baseline | abundance | X10494 | 26Q0EO75 | 3 | Interventio | Experimen | Experimen | Control | 0,97445 |
| baseline | abundance | X10497 | 4-Guanidir | 3 | Interventio | Experimen | Experimen | Control | 0,97445 |
| baseline | abundance | X10503 | APM_a | 3 | Interventio | Experimen | Experimen | Control | 0,936399 |
| baseline | abundance | X10511 | dopaquino | 3 | Interventio | Experimen | Experimen | Control | 0,936399 |
| baseline | abundance | X10513 | ophthalmic | 3 | Interventio | Experimen | Experimen | Control | 0,957875 |
| baseline | abundance | X10518 | MFCD0002 | 3 | Interventio | Experimen | Experimen | Control | 0,936399 |
| baseline | abundance | X10534 | L-N2-(2-Ca | 3 | Interventio | Experimen | Experimen | Control | 0,936399 |
| baseline | abundance | X10536 | CMPF | 3 | Interventio | Experimen | Experimen | Control | 0,97445 |
| baseline | abundance | X10549 | Glaucine | 2b | Interventio | Experimen | Experimen | Control | 0,97445 |
| baseline | abundance | X10550 | pentobarbi | 3 | Interventio | Experimen | Experimen | Control | 0,945105 |
| baseline | abundance | X10554 | Glycine | 1 | Interventio | Experimen | Experimen | Control | 0,945105 |
| baseline | abundance | X10564 | 2-(1-Ethoxy | 3 | Interventio | Experimen | Experimen | Control | 0,936399 |
| baseline | abundance | X10580 | Leu-Leu_a | 3 | Interventio | Experimen | Experimen | Control | 0,97445 |
| baseline | abundance | X10588 | (8)-Ginger | 3 | Interventio | Experimen | Experimen | Control | 0,97445 |
| baseline | abundance | X10593 | Vorinostat_ | 3 | Interventio | Experimen | Experimen | Control | 0,936399 |
| baseline | abundance | X10600 | Glycocyam | 3 | Interventio | Experimen | Experimen | Control | 0,936399 |
| baseline | abundance | X10611 | 2-Acrylami | 3 | Interventio | Experimen | Experimen | Control | 0,97445 |
| baseline | abundance | X10614 | Monometh | 2a | Interventio | Experimen | Experimen | Control | 0,957875 |
| baseline | abundance | X10617 | 2-methyl-1 | 2b | Interventio | Experimen | Experimen | Control | 0,936399 |
| baseline | abundance | X10623 | N-Nonanoy | 3 | Interventio | Experimen | Experimen | Control | 0,936399 |
| baseline | abundance | X10636 | 8-hydroxy- | 2b | Interventio | Experimen | Experimen | Control | 0,945105 |
| baseline | abundance | X10647 | 4-Hydroxyp | 3 | Interventio | Experimen | Experimen | Control | 0,955424 |
| baseline | abundance | X10651 | Methyl 1-hy | 3 | Interventio | Experimen | Experimen | Control | 0,936399 |
| baseline | abundance | X10660 | 2-Methylth | 3 | Interventio | Experimen | Experimen | Control | 0,936399 |
| baseline | abundance | X10664 | N1-(5-meth | 2b | Interventio | Experimen | Experimen | Control | 0,985903 |
| baseline | abundance | X10665 | Aprobarbit | 3 | Interventio | Experimen | Experimen | Control | 0,936399 |
| baseline | abundance | X10667 | N-Phenylac | 3 | Interventio | Experimen | Experimen | Control | 0,936399 |
| baseline | abundance | X10676 | 6-(1-Hydro | 3 | Interventio | Experimen | Experimen | Control | 0,945105 |
| baseline | abundance | X10680 | 8-hydroxy-7 | 3 | Interventio | Experimen | Experimen | Control | 0,945105 |

| | | | | | | | | | |
|---|---|---|---|---|---|---|---|---|---|
| baseline | abundance | X10685 | Glycylvalin | 2a | Interventio | Experimen | Experimen | Control | 0,986763 |
| baseline | abundance | X10687 | Gln-Gln | 3 | Interventio | Experimen | Experimen | Control | 0,945105 |
| baseline | abundance | X10691 | (−)-nabilor | 3 | Interventio | Experimen | Experimen | Control | 0,97445 |
| baseline | abundance | X10706 | Furaneol | 3 | Interventio | Experimen | Experimen | Control | 0,936399 |
| baseline | abundance | X10713 | norhaman | 3 | Interventio | Experimen | Experimen | Control | 0,97445 |
| baseline | abundance | X10722 | Acetanilide | 2b | Interventio | Experimen | Experimen | Control | 0,936399 |
| baseline | abundance | X10725 | Ethyl malat | 3 | Interventio | Experimen | Experimen | Control | 0,97445 |
| baseline | abundance | X10736 | Toxopyrimi | 3 | Interventio | Experimen | Experimen | Control | 0,936399 |
| baseline | abundance | X10741 | 5-Nitro-2-p | 3 | Interventio | Experimen | Experimen | Control | 0,988948 |
| baseline | abundance | X10757 | Glycyrin | 3 | Interventio | Experimen | Experimen | Control | 0,945105 |
| baseline | abundance | X10769 | Cystine | 1 | Interventio | Experimen | Experimen | Control | 0,936399 |
| baseline | abundance | X10770 | 2-(4-Isopro | 3 | Interventio | Experimen | Experimen | Control | 0,945105 |
| baseline | abundance | X10772 | Bicine_a | 3 | Interventio | Experimen | Experimen | Control | 0,936399 |
| baseline | abundance | X10776 | Histidylgly | 3 | Interventio | Experimen | Experimen | Control | 0,97445 |
| baseline | abundance | X10793 | Zalcitabine | 3 | Interventio | Experimen | Experimen | Control | 0,945105 |
| baseline | abundance | X10818 | butalbital_ | 3 | Interventio | Experimen | Experimen | Control | 0,97445 |
| baseline | abundance | X10819 | Hyocholic | 2a | Interventio | Experimen | Experimen | Control | 0,936399 |
| baseline | abundance | X10864 | 5-amino-2- | 2b | Interventio | Experimen | Experimen | Control | 0,945105 |
| baseline | abundance | X10865 | Melatonin | 3 | Interventio | Experimen | Experimen | Control | 0,945105 |
| baseline | abundance | X10881 | L-gamma-( | 3 | Interventio | Experimen | Experimen | Control | 0,936399 |
| baseline | abundance | X10886 | L-gamma-( | 3 | Interventio | Experimen | Experimen | Control | 0,936399 |
| baseline | abundance | X10889 | N-acetyl-b | 3 | Interventio | Experimen | Experimen | Control | 0,957875 |
| baseline | abundance | X10896 | 6-Hydroxyr | 2b | Interventio | Experimen | Experimen | Control | 0,945105 |
| baseline | abundance | X10901 | bis-noryan | 3 | Interventio | Experimen | Experimen | Control | 0,945105 |
| baseline | abundance | X10902 | Isoquinolir | 2b | Interventio | Experimen | Experimen | Control | 0,936399 |
| baseline | abundance | X10918 | meprobam | 3 | Interventio | Experimen | Experimen | Control | 0,936399 |
| baseline | abundance | X10929 | Temozolon | 3 | Interventio | Experimen | Experimen | Control | 0,936399 |
| baseline | abundance | X10930 | MFCD0995 | 3 | Interventio | Experimen | Experimen | Control | 0,948031 |
| baseline | abundance | X10936 | clavulanic | 3 | Interventio | Experimen | Experimen | Control | 0,936399 |
| baseline | abundance | X10947 | 1,1'-[1,12- | 3 | Interventio | Experimen | Experimen | Control | 0,945105 |
| baseline | abundance | X10951 | 8-Methyl-8 | 3 | Interventio | Experimen | Experimen | Control | 0,936399 |
| baseline | abundance | X10953 | Vorinostat_ | 3 | Interventio | Experimen | Experimen | Control | 0,936399 |
| baseline | abundance | X10974 | Threonylglu | 3 | Interventio | Experimen | Experimen | Control | 0,941185 |
| baseline | abundance | X10977 | pentobarbi | 3 | Interventio | Experimen | Experimen | Control | 0,97445 |
| baseline | abundance | X10979 | acetyltauri | 3 | Interventio | Experimen | Experimen | Control | 0,936399 |
| baseline | abundance | X10982 | 8-Methyl-8 | 3 | Interventio | Experimen | Experimen | Control | 0,936399 |
| baseline | abundance | X10987 | 2-BUTYL PI | 3 | Interventio | Experimen | Experimen | Control | 0,945105 |
| baseline | abundance | X11001 | 2-[(carboxy | 2b | Interventio | Experimen | Experimen | Control | 0,936399 |
| baseline | abundance | X11012 | Furan | 3 | Interventio | Experimen | Experimen | Control | 0,936399 |
| baseline | abundance | X11028 | butalbital_ | 3 | Interventio | Experimen | Experimen | Control | 0,945105 |
| baseline | abundance | X11032 | 2,4-Bis(3-r | 3 | Interventio | Experimen | Experimen | Control | 0,978464 |
| baseline | abundance | X11033 | N-Acetyl-5 | 3 | Interventio | Experimen | Experimen | Control | 0,983461 |
| baseline | abundance | X11034 | 1-(3-Aminc | 3 | Interventio | Experimen | Experimen | Control | 0,945105 |
| baseline | abundance | X11036 | 3-Hydroxyp | 2b | Interventio | Experimen | Experimen | Control | 0,936399 |
| baseline | abundance | X11038 | (2E)-3-(3,4 | 3 | Interventio | Experimen | Experimen | Control | 0,936399 |
| baseline | abundance | X11042 | Leu-arg | 3 | Interventio | Experimen | Experimen | Control | 0,957875 |
| baseline | abundance | X11059 | Dihydrocor | 3 | Interventio | Experimen | Experimen | Control | 0,945105 |
| baseline | abundance | X11067 | Resveratro | 3 | Interventio | Experimen | Experimen | Control | 0,936399 |

| | | | | | | | | | |
|---|---|---|---|---|---|---|---|---|---|
| baseline | abundance | X11068 | N,N'-Bis[4- | 3 | Interventio | Experimen | Experimen | Control | 0,945786 |
| baseline | abundance | X11110 | Tauropine | 3 | Interventio | Experimen | Experimen | Control | 0,936399 |
| baseline | abundance | X11115 | Hydroxyca | 3 | Interventio | Experimen | Experimen | Control | 0,936399 |
| baseline | abundance | X11117 | Coprine_b | 3 | Interventio | Experimen | Experimen | Control | 0,936399 |
| baseline | abundance | X11125 | 5,8,12-Trih | 3 | Interventio | Experimen | Experimen | Control | 0,936399 |
| baseline | abundance | X11140 | Leu-Leu_e | 3 | Interventio | Experimen | Experimen | Control | 0,936399 |
| baseline | abundance | X11163 | Lys-Pro_a | 3 | Interventio | Experimen | Experimen | Control | 0,936399 |
| baseline | abundance | X11170 | QJ972000 | 3 | Interventio | Experimen | Experimen | Control | 0,936399 |
| baseline | abundance | X11186 | Fluocinolo | 2b | Interventio | Experimen | Experimen | Control | 0,988867 |
| baseline | abundance | X11215 | Leu-Leu_b | 3 | Interventio | Experimen | Experimen | Control | 0,936399 |
| baseline | abundance | X11223 | Ethyl mala | 3 | Interventio | Experimen | Experimen | Control | 0,936399 |
| baseline | abundance | X11245 | MFCD0003 | 3 | Interventio | Experimen | Experimen | Control | 0,936399 |
| baseline | abundance | X11251 | N-Nonano | 3 | Interventio | Experimen | Experimen | Control | 0,958229 |
| baseline | abundance | X11254 | (4R,5S,9S, | 3 | Interventio | Experimen | Experimen | Control | 0,945105 |
| baseline | abundance | X11257 | 1-Methylgu | 2b | Interventio | Experimen | Experimen | Control | 0,955424 |
| baseline | abundance | X11258 | Valyl-4-hyd | 3 | Interventio | Experimen | Experimen | Control | 0,945105 |
| baseline | abundance | X11265 | Sulfurol | 3 | Interventio | Experimen | Experimen | Control | 0,97445 |
| baseline | abundance | X11273 | trp-pro | 3 | Interventio | Experimen | Experimen | Control | 0,936399 |
| baseline | abundance | X11277 | lys-leu | 3 | Interventio | Experimen | Experimen | Control | 0,957875 |
| baseline | abundance | X11281 | Penbutolol | 3 | Interventio | Experimen | Experimen | Control | 0,960702 |
| baseline | abundance | X11301 | (6S)-2-Ami | 3 | Interventio | Experimen | Experimen | Control | 0,97445 |
| baseline | abundance | X11310 | Leu-Val_a | 3 | Interventio | Experimen | Experimen | Control | 0,936399 |
| baseline | abundance | X11319 | Methohexi | 3 | Interventio | Experimen | Experimen | Control | 0,987751 |
| baseline | abundance | X11323 | dihydroxyb | 3 | Interventio | Experimen | Experimen | Control | 0,936399 |
| baseline | abundance | X11331 | N~6~-Octa | 3 | Interventio | Experimen | Experimen | Control | 0,955424 |
| baseline | abundance | X11339 | L-gamma-( | 3 | Interventio | Experimen | Experimen | Control | 0,97445 |
| baseline | abundance | X11347 | Voglibose | 3 | Interventio | Experimen | Experimen | Control | 0,945105 |
| baseline | abundance | X11349 | Butabarbit | 3 | Interventio | Experimen | Experimen | Control | 0,962445 |
| baseline | abundance | X11354 | 3-Oxo-4,6- | 3 | Interventio | Experimen | Experimen | Control | 0,987751 |
| baseline | abundance | X11361 | 1-Methylhi | 3 | Interventio | Experimen | Experimen | Control | 0,936399 |
| baseline | abundance | X11366 | 3-Sulfino-L | 3 | Interventio | Experimen | Experimen | Control | 0,936399 |
| baseline | abundance | X11373 | 3-Succinoy | 2b | Interventio | Experimen | Experimen | Control | 0,945105 |
| baseline | abundance | X11375 | L-gamma-( | 3 | Interventio | Experimen | Experimen | Control | 0,948031 |
| baseline | abundance | X11383 | N-{3-[(4-Ac | 3 | Interventio | Experimen | Experimen | Control | 0,957875 |
| baseline | abundance | X11408 | 5-Amino-6 | 3 | Interventio | Experimen | Experimen | Control | 0,97445 |
| baseline | abundance | X11421 | 8-Amino-7 | 3 | Interventio | Experimen | Experimen | Control | 0,97445 |
| baseline | abundance | X11438 | pro-met | 3 | Interventio | Experimen | Experimen | Control | 0,945105 |
| baseline | abundance | X11441 | beta-D-Eth | 3 | Interventio | Experimen | Experimen | Control | 0,988948 |
| baseline | abundance | X11477 | gamma-Gl | 3 | Interventio | Experimen | Experimen | Control | 0,945105 |
| baseline | abundance | X11481 | his-gln_b | 3 | Interventio | Experimen | Experimen | Control | 0,955424 |
| baseline | abundance | X11494 | 3-Hydroxys | 3 | Interventio | Experimen | Experimen | Control | 0,962445 |
| baseline | abundance | X11498 | Valylvaline | 3 | Interventio | Experimen | Experimen | Control | 0,957875 |
| baseline | abundance | X11505 | FB950000 | 3 | Interventio | Experimen | Experimen | Control | 0,945105 |
| baseline | abundance | X11515 | 13(S)-HOT | 2b | Interventio | Experimen | Experimen | Control | 0,936399 |
| baseline | abundance | X11522 | thr-trp | 3 | Interventio | Experimen | Experimen | Control | 0,988867 |
| baseline | abundance | X11524 | 6-hydroxyp | 3 | Interventio | Experimen | Experimen | Control | 0,936399 |
| baseline | abundance | X11529 | 2-Aminooc | 2b | Interventio | Experimen | Experimen | Control | 0,988948 |
| baseline | abundance | X11549 | 1,1'-[1,12- | 3 | Interventio | Experimen | Experimen | Control | 0,945105 |

| | | | | | | | | | |
|---|---|---|---|---|---|---|---|---|---|
| baseline | abundance | X11556 | N-[(2E)-3-( | 3 | Interventio | Experimen | Experimen | Control | 0,936399 |
| baseline | abundance | X11562 | ALA-PRO | 3 | Interventio | Experimen | Experimen | Control | 0,957875 |
| baseline | abundance | X11563 | threonylph | 3 | Interventio | Experimen | Experimen | Control | 0,936399 |
| baseline | abundance | X11576 | Phenyl D-g | 3 | Interventio | Experimen | Experimen | Control | 0,945105 |
| baseline | abundance | X11605 | 3-Methyl-2 | 3 | Interventio | Experimen | Experimen | Control | 0,957875 |
| baseline | abundance | X11614 | 2-Methylth | 3 | Interventio | Experimen | Experimen | Control | 0,936399 |
| baseline | abundance | X11627 | N-(3-aceta | 3 | Interventio | Experimen | Experimen | Control | 0,936399 |
| baseline | abundance | X11639 | Midodrine_ | 3 | Interventio | Experimen | Experimen | Control | 0,957875 |
| baseline | abundance | X11645 | glu-thr | 3 | Interventio | Experimen | Experimen | Control | 0,945105 |
| baseline | abundance | X11666 | N-(4-Hydro | 3 | Interventio | Experimen | Experimen | Control | 0,987541 |
| baseline | abundance | X11667 | Tetraacety | 3 | Interventio | Experimen | Experimen | Control | 0,962445 |
| baseline | abundance | X11692 | 2_7-Anhyd | 3 | Interventio | Experimen | Experimen | Control | 0,936399 |
| baseline | abundance | X11698 | 1-(beta-D- | 3 | Interventio | Experimen | Experimen | Control | 0,948225 |
| baseline | abundance | X11699 | {2-[2-(Isob | 3 | Interventio | Experimen | Experimen | Control | 0,936399 |
| baseline | abundance | X11716 | ala-ser_a | 3 | Interventio | Experimen | Experimen | Control | 0,945105 |
| baseline | abundance | X11717 | nicotianan | 3 | Interventio | Experimen | Experimen | Control | 0,988948 |
| baseline | abundance | X11733 | 1,3-Dihydr | 3 | Interventio | Experimen | Experimen | Control | 0,97445 |
| baseline | abundance | X11745 | 2-Despipe | 3 | Interventio | Experimen | Experimen | Control | 0,936399 |
| baseline | abundance | X11755 | Furfuranol | 3 | Interventio | Experimen | Experimen | Control | 0,945105 |
| baseline | abundance | p_cresol | p-Cresol | 1 | Interventio | Experimen | Experimen | Control | 0,97445 |
| 3 months | abundance | 2-Methylpr | 2-Methylpr | CFA_panel | Interventio | Experimen | Experimen | Control | 0,106066 |
| 3 months | abundance | 3-Methylbu | 3-Methylbu | CFA_panel | Interventio | Experimen | Experimen | Control | 0,151734 |
| 3 months | abundance | Acetic acid | Acetic acid | CFA_panel | Interventio | Experimen | Experimen | Control | 0,918443 |
| 3 months | abundance | Butanoic a | Butanoic a | CFA_panel | Interventio | Experimen | Experimen | Control | 0,626645 |
| 3 months | abundance | Hexanoic a | Hexanoic a | CFA_panel | Interventio | Experimen | Experimen | Control | 0,108736 |
| 3 months | abundance | M1 | M1 | IA_module | Interventio | Experimen | Experimen | Control | 0,141328 |
| 3 months | abundance | M10 | M10 | IA_module | Interventio | Experimen | Experimen | Control | 0,967096 |
| 3 months | abundance | M11 | M11 | IA_module | Interventio | Experimen | Experimen | Control | 0,787558 |
| 3 months | abundance | M12 | M12 | IA_module | Interventio | Experimen | Experimen | Control | 0,017773 |
| 3 months | abundance | M13 | M13 | IA_module | Interventio | Experimen | Experimen | Control | 0,718544 |
| 3 months | abundance | M14 | M14 | IA_module | Interventio | Experimen | Experimen | Control | 0,581247 |
| 3 months | abundance | M15 | M15 | IA_module | Interventio | Experimen | Experimen | Control | 0,819081 |
| 3 months | abundance | M16 | M16 | IA_module | Interventio | Experimen | Experimen | Control | 0,648353 |
| 3 months | abundance | M17 | M17 | IA_module | Interventio | Experimen | Experimen | Control | 0,953271 |
| 3 months | abundance | M18 | M18 | IA_module | Interventio | Experimen | Experimen | Control | 0,927867 |
| 3 months | abundance | M19 | M19 | IA_module | Interventio | Experimen | Experimen | Control | 0,922757 |
| 3 months | abundance | M2 | M2 | IA_module | Interventio | Experimen | Experimen | Control | 0,136956 |
| 3 months | abundance | M20 | M20 | IA_module | Interventio | Experimen | Experimen | Control | 0,100587 |
| 3 months | abundance | M21 | M21 | IA_module | Interventio | Experimen | Experimen | Control | 0,639019 |
| 3 months | abundance | M22 | M22 | IA_module | Interventio | Experimen | Experimen | Control | 0,55784 |
| 3 months | abundance | M23 | M23 | IA_module | Interventio | Experimen | Experimen | Control | 0,960119 |
| 3 months | abundance | M24 | M24 | IA_module | Interventio | Experimen | Experimen | Control | 0,980029 |
| 3 months | abundance | M25 | M25 | IA_module | Interventio | Experimen | Experimen | Control | 0,174141 |
| 3 months | abundance | M26 | M26 | IA_module | Interventio | Experimen | Experimen | Control | 0,862952 |
| 3 months | abundance | M27 | M27 | IA_module | Interventio | Experimen | Experimen | Control | 0,854716 |
| 3 months | abundance | M28 | M28 | IA_module | Interventio | Experimen | Experimen | Control | 0,633187 |
| 3 months | abundance | M29 | M29 | IA_module | Interventio | Experimen | Experimen | Control | 0,232653 |
| 3 months | abundance | M3 | M3 | IA_module | Interventio | Experimen | Experimen | Control | 0,867514 |

| | | | | | | | | | |
|---|---|---|---|---|---|---|---|---|---|
| 3 months | abundance | M30 | M30 | \A_module | Interventio | Experimen | Experimen | Control | 0,123709 |
| 3 months | abundance | M31 | M31 | \A_module | Interventio | Experimen | Experimen | Control | 0,834977 |
| 3 months | abundance | M32 | M32 | \A_module | Interventio | Experimen | Experimen | Control | 0,59163 |
| 3 months | abundance | M33 | M33 | \A_module | Interventio | Experimen | Experimen | Control | 0,217561 |
| 3 months | abundance | M34 | M34 | \A_module | Interventio | Experimen | Experimen | Control | 0,985356 |
| 3 months | abundance | M4 | M4 | \A_module | Interventio | Experimen | Experimen | Control | 0,541151 |
| 3 months | abundance | M5 | M5 | \A_module | Interventio | Experimen | Experimen | Control | 0,004656 |
| 3 months | abundance | M6 | M6 | \A_module | Interventio | Experimen | Experimen | Control | 0,960119 |
| 3 months | abundance | M7 | M7 | \A_module | Interventio | Experimen | Experimen | Control | 0,651767 |
| 3 months | abundance | M8 | M8 | \A_module | Interventio | Experimen | Experimen | Control | 0,876495 |
| 3 months | abundance | M9 | M9 | \A_module | Interventio | Experimen | Experimen | Control | 0,098744 |
| 3 months | abundance | Pentanoic | Pentanoic | CFA_panel | Interventio | Experimen | Experimen | Control | 0,740178 |
| 3 months | abundance | Propanoic | Propanoic | CFA_panel | Interventio | Experimen | Experimen | Control | 0,400419 |
| 3 months | abundance | SL00001 | Deoxycytid | 2a | Interventio | Experimen | Experimen | Control | 0,997328 |
| 3 months | abundance | SL00009 | N-Acetylhi: | 2a | Interventio | Experimen | Experimen | Control | 0,515877 |
| 3 months | abundance | SL00017 | Aspartic ac | 2a | Interventio | Experimen | Experimen | Control | 0,73207 |
| 3 months | abundance | SL00020 | Carnitine | 2a | Interventio | Experimen | Experimen | Control | 0,916188 |
| 3 months | abundance | SL00024 | Creatine | 2a | Interventio | Experimen | Experimen | Control | 0,808744 |
| 3 months | abundance | SL00035 | Glycylglyci | 2a | Interventio | Experimen | Experimen | Control | 0,808747 |
| 3 months | abundance | SL00040 | Kynurenine | 2a | Interventio | Experimen | Experimen | Control | 0,747665 |
| 3 months | abundance | SL00049 | N-Acetylar | 2a | Interventio | Experimen | Experimen | Control | 0,277347 |
| 3 months | abundance | SL00052 | N-Isovalery | 2a | Interventio | Experimen | Experimen | Control | 0,343109 |
| 3 months | abundance | SL00054 | Ornithine | 1 | Interventio | Experimen | Experimen | Control | 0,513969 |
| 3 months | abundance | SL00061 | Proline | 1 | Interventio | Experimen | Experimen | Control | 0,400419 |
| 3 months | abundance | SL00063 | Pyridoxal | 1 | Interventio | Experimen | Experimen | Control | 0,448346 |
| 3 months | abundance | SL00074 | Valine/ 5-A | 2a | Interventio | Experimen | Experimen | Control | 0,220826 |
| 3 months | abundance | SL00081 | 5-Methylcy | 2a | Interventio | Experimen | Experimen | Control | 0,30026 |
| 3 months | abundance | SL00082 | Allopurinol | 1 | Interventio | Experimen | Experimen | Control | 0,853666 |
| 3 months | abundance | SL00089 | Dissaccari | 2a | Interventio | Experimen | Experimen | Control | 0,235652 |
| 3 months | abundance | SL00096 | Hexoses III | 1 | Interventio | Experimen | Experimen | Control | 0,096259 |
| 3 months | abundance | SL00097 | Hexoses II | 1 | Interventio | Experimen | Experimen | Control | 0,115645 |
| 3 months | abundance | SL00103 | Gulonic ac | 2a | Interventio | Experimen | Experimen | Control | 0,100093 |
| 3 months | abundance | SL00104 | Hexoses I | 1 | Interventio | Experimen | Experimen | Control | 0,099053 |
| 3 months | abundance | SL00108 | Histidinol | 2a | Interventio | Experimen | Experimen | Control | 0,030613 |
| 3 months | abundance | SL00124 | Dissaccari | 2a | Interventio | Experimen | Experimen | Control | 0,503169 |
| 3 months | abundance | SL00128 | N-Acetylala | 2a | Interventio | Experimen | Experimen | Control | 0,778801 |
| 3 months | abundance | SL00133 | N-Acetylglu | 2a | Interventio | Experimen | Experimen | Control | 0,909515 |
| 3 months | abundance | SL00137 | Phenylace | 2a | Interventio | Experimen | Experimen | Control | 0,613297 |
| 3 months | abundance | SL00140 | Pyridoxami | 1 | Interventio | Experimen | Experimen | Control | 0,385109 |
| 3 months | abundance | SL00149 | Dissaccari | 2a | Interventio | Experimen | Experimen | Control | 0,017773 |
| 3 months | abundance | SL00154 | Trigonellin | 1 | Interventio | Experimen | Experimen | Control | 0,449163 |
| 3 months | abundance | SL00159 | Uracil | 1 | Interventio | Experimen | Experimen | Control | 0,98068 |
| 3 months | abundance | SL00161 | Uridine | 2a | Interventio | Experimen | Experimen | Control | 0,993459 |
| 3 months | abundance | SL00185 | 2-Oxo-3-pl | 1 | Interventio | Experimen | Experimen | Control | 0,056989 |
| 3 months | abundance | SL00189 | 3-Hydroxyt | 2a | Interventio | Experimen | Experimen | Control | 0,105225 |
| 3 months | abundance | SL00197 | 4-Methyl-2 | 2a | Interventio | Experimen | Experimen | Control | 0,323676 |
| 3 months | abundance | SL00199 | 5-Hydroxyi | 2a | Interventio | Experimen | Experimen | Control | 0,798192 |
| 3 months | abundance | SL00200 | 8-Aminooc | 2a | Interventio | Experimen | Experimen | Control | 0,143909 |

| | | | | | | | | | |
|---|---|---|---|---|---|---|---|---|---|
| 3 months | abundance | SL00201 | Pentose II | 2a | Interventio | Experimen | Experimen | Control | 0,497383 |
| 3 months | abundance | SL00205 | Citraconic | 2a | Interventio | Experimen | Experimen | Control | 0,985356 |
| 3 months | abundance | SL00210 | Galactonic | 2a | Interventio | Experimen | Experimen | Control | 0,243875 |
| 3 months | abundance | SL00212 | Galactosar | 2a | Interventio | Experimen | Experimen | Control | 0,71076 |
| 3 months | abundance | SL00215 | Glycolic ac | 2a | Interventio | Experimen | Experimen | Control | 0,867667 |
| 3 months | abundance | SL00216 | Glyoxylic a | 2a | Interventio | Experimen | Experimen | Control | 0,584092 |
| 3 months | abundance | SL00222 | Lactic acid | 1 | Interventio | Experimen | Experimen | Control | 0,250053 |
| 3 months | abundance | SL00226 | N-Acetylgly | 2a | Interventio | Experimen | Experimen | Control | 0,198023 |
| 3 months | abundance | SL00239 | Pinitol | 2a | Interventio | Experimen | Experimen | Control | 0,876028 |
| 3 months | abundance | SL00240 | Pyruvic aci | 1 | Interventio | Experimen | Experimen | Control | 0,798192 |
| 3 months | abundance | SL00243 | Deoxysuga | 2a | Interventio | Experimen | Experimen | Control | 0,150298 |
| 3 months | abundance | SL00245 | sugar alcol | 2a | Interventio | Experimen | Experimen | Control | 0,68636 |
| 3 months | abundance | SL00246 | Pentose III | 2a | Interventio | Experimen | Experimen | Control | 0,139689 |
| 3 months | abundance | SL00247 | Shikimic ac | 2a | Interventio | Experimen | Experimen | Control | 0,163182 |
| 3 months | abundance | SL00248 | Gluconic a | 2a | Interventio | Experimen | Experimen | Control | 0,747665 |
| 3 months | abundance | SL00260 | Pentose I | 2a | Interventio | Experimen | Experimen | Control | 0,29731 |
| 3 months | abundance | SL00262 | cis-Aconiti | 2a | Interventio | Experimen | Experimen | Control | 0,365312 |
| 3 months | abundance | SL00264 | Methyl ace | 2a | Interventio | Experimen | Experimen | Control | 0,560103 |
| 3 months | abundance | SL00265 | Purine | 2a | Interventio | Experimen | Experimen | Control | 0,222442 |
| 3 months | abundance | SL00268 | Thymine | 1 | Interventio | Experimen | Experimen | Control | 0,915997 |
| 3 months | abundance | SL00270 | 3-hydroxy- | 2a | Interventio | Experimen | Experimen | Control | 0,98692 |
| 3 months | abundance | SL00275 | Dopamine | 2a | Interventio | Experimen | Experimen | Control | 0,985504 |
| 3 months | abundance | SL00282 | 3-Hydroxyl | 2a | Interventio | Experimen | Experimen | Control | 0,886266 |
| 3 months | abundance | SL00285 | Dihydrofer | 2a | Interventio | Experimen | Experimen | Control | 0,988551 |
| 3 months | abundance | SL00287 | DOPA | 2a | Interventio | Experimen | Experimen | Control | 0,776369 |
| 3 months | abundance | SL00288 | Propionylc | 2a | Interventio | Experimen | Experimen | Control | 0,419167 |
| 3 months | abundance | SL00290 | N-Acetylma | 2a | Interventio | Experimen | Experimen | Control | 0,264089 |
| 3 months | abundance | SL00293 | Butyrylcarr | 2a | Interventio | Experimen | Experimen | Control | 0,960119 |
| 3 months | abundance | SL00295 | 5-Methylur | 2a | Interventio | Experimen | Experimen | Control | 0,254073 |
| 3 months | abundance | SL00297 | Acetylmura | 2a | Interventio | Experimen | Experimen | Control | 0,765006 |
| 3 months | abundance | SL00299 | β-Murichol | 2a | Interventio | Experimen | Experimen | Control | 0,499773 |
| 3 months | abundance | SL00309 | 1-Aminocy | 2a | Interventio | Experimen | Experimen | Control | 0,087526 |
| 3 months | abundance | SL00311 | N,N-Dimet | 2a | Interventio | Experimen | Experimen | Control | 0,836327 |
| 3 months | abundance | SL00313 | 2-Aminoisc | 2a | Interventio | Experimen | Experimen | Control | 0,957479 |
| 3 months | abundance | SL00315 | Malonic ac | 2a | Interventio | Experimen | Experimen | Control | 0,404129 |
| 3 months | abundance | SL00318 | 4-Aminoph | 1 | Interventio | Experimen | Experimen | Control | 0,944871 |
| 3 months | abundance | SL00320 | γ-Caprolac | 2a | Interventio | Experimen | Experimen | Control | 0,437457 |
| 3 months | abundance | SL00325 | 2-Methylm | 2a | Interventio | Experimen | Experimen | Control | 0,375891 |
| 3 months | abundance | SL00328 | 2-(hydroxy | 2a | Interventio | Experimen | Experimen | Control | 0,143745 |
| 3 months | abundance | SL00329 | 2-Hydroxy- | 2a | Interventio | Experimen | Experimen | Control | 0,718544 |
| 3 months | abundance | SL00334 | Imidazolea | 2a | Interventio | Experimen | Experimen | Control | 0,73403 |
| 3 months | abundance | SL00346 | N-Methylni | 2a | Interventio | Experimen | Experimen | Control | 0,9752 |
| 3 months | abundance | SL00349 | 1-Aminocy | 2a | Interventio | Experimen | Experimen | Control | 0,845389 |
| 3 months | abundance | SL00350 | Stachydrin | 1 | Interventio | Experimen | Experimen | Control | 0,561912 |
| 3 months | abundance | SL00353 | Adipic acid | 1 | Interventio | Experimen | Experimen | Control | 0,985504 |
| 3 months | abundance | SL00356 | N-(5-Amin | 2a | Interventio | Experimen | Experimen | Control | 0,066304 |
| 3 months | abundance | SL00368 | N-Methylty | 1 | Interventio | Experimen | Experimen | Control | 0,36547 |
| 3 months | abundance | SL00373 | N1-Methyl- | 2a | Interventio | Experimen | Experimen | Control | 0,004228 |

| | | | | | | | | | |
|---|---|---|---|---|---|---|---|---|---|
| 3 months | abundance | SL00376 | N-AcetylPr | 2a | Interventio | Experimen | Experimen | Control | 0,776369 |
| 3 months | abundance | SL00380 | 2-Aminoad | 2a | Interventio | Experimen | Experimen | Control | 0,325689 |
| 3 months | abundance | SL00383 | 3-(2-Hydro | 2a | Interventio | Experimen | Experimen | Control | 0,655638 |
| 3 months | abundance | SL00384 | p-Coumari | 1 | Interventio | Experimen | Experimen | Control | 0,685901 |
| 3 months | abundance | SL00390 | Gallic acid | 2a | Interventio | Experimen | Experimen | Control | 0,511679 |
| 3 months | abundance | SL00391 | N-Acetylle | 1 | Interventio | Experimen | Experimen | Control | 0,704502 |
| 3 months | abundance | SL00401 | Theophylli | 1 | Interventio | Experimen | Experimen | Control | 0,820887 |
| 3 months | abundance | SL00407 | Homovanil | 2a | Interventio | Experimen | Experimen | Control | 0,691158 |
| 3 months | abundance | SL00420 | 4-Hydroxy- | 2a | Interventio | Experimen | Experimen | Control | 0,84521 |
| 3 months | abundance | SL00421 | Asymmetri | 2a | Interventio | Experimen | Experimen | Control | 0,988551 |
| 3 months | abundance | SL00430 | Homocarn | 2a | Interventio | Experimen | Experimen | Control | 0,419167 |
| 3 months | abundance | SL00431 | Tiglylcarnit | 2a | Interventio | Experimen | Experimen | Control | 0,268012 |
| 3 months | abundance | SL00433 | Isovalerylc | 2a | Interventio | Experimen | Experimen | Control | 0,788303 |
| 3 months | abundance | SL00436 | 1-Carboxy | 2a | Interventio | Experimen | Experimen | Control | 0,666592 |
| 3 months | abundance | SL00438 | Daidzein | 2a | Interventio | Experimen | Experimen | Control | 0,532452 |
| 3 months | abundance | SL00440 | 5-Methylcy | 2a | Interventio | Experimen | Experimen | Control | 0,988198 |
| 3 months | abundance | SL00445 | Glucosami | 2a | Interventio | Experimen | Experimen | Control | 0,860527 |
| 3 months | abundance | SL00447 | Apigenin | 2a | Interventio | Experimen | Experimen | Control | 0,798192 |
| 3 months | abundance | SL00455 | 12,13-DHC | 2a | Interventio | Experimen | Experimen | Control | 0,867514 |
| 3 months | abundance | SL00467 | Sucralose | 2a | Interventio | Experimen | Experimen | Control | 0,997328 |
| 3 months | abundance | SL00502 | 2-Ketobuty | 1 | Interventio | Experimen | Experimen | Control | 0,53654 |
| 3 months | abundance | X00011 | Glutamine | 2a | Interventio | Experimen | Experimen | Control | 0,651289 |
| 3 months | abundance | X00018 | Succinic ac | 1 | Interventio | Experimen | Experimen | Control | 0,511237 |
| 3 months | abundance | X00020 | sugar alcol | 1 | Interventio | Experimen | Experimen | Control | 0,423696 |
| 3 months | abundance | X00022 | N-Methylas | 1 | Interventio | Experimen | Experimen | Control | 0,014777 |
| 3 months | abundance | X00023 | N-Formylm | 1 | Interventio | Experimen | Experimen | Control | 0,142786 |
| 3 months | abundance | X00025 | Acesulfam | 1 | Interventio | Experimen | Experimen | Control | 0,998514 |
| 3 months | abundance | X00026 | Inosine | 1 | Interventio | Experimen | Experimen | Control | 0,364717 |
| 3 months | abundance | X00027 | N-Acetylglu | 1 | Interventio | Experimen | Experimen | Control | 0,104321 |
| 3 months | abundance | X00029 | Urocanic a | 1 | Interventio | Experimen | Experimen | Control | 0,971921 |
| 3 months | abundance | X00030 | 3,4-Dihydr | 1 | Interventio | Experimen | Experimen | Control | 0,587315 |
| 3 months | abundance | X00032 | Indole-3-pr | 1 | Interventio | Experimen | Experimen | Control | 0,002538 |
| 3 months | abundance | X00033 | Deoxycholi | 1 | Interventio | Experimen | Experimen | Control | 0,718544 |
| 3 months | abundance | X00034 | Pyroglutam | 1 | Interventio | Experimen | Experimen | Control | 0,960119 |
| 3 months | abundance | X00035 | Choline | 1 | Interventio | Experimen | Experimen | Control | 0,244993 |
| 3 months | abundance | X00036 | Serine | 1 | Interventio | Experimen | Experimen | Control | 0,261424 |
| 3 months | abundance | X00038 | Histamine | 1 | Interventio | Experimen | Experimen | Control | 0,605819 |
| 3 months | abundance | X00040 | N-Acetylty | 1 | Interventio | Experimen | Experimen | Control | 0,99116 |
| 3 months | abundance | X00042 | Glucuronic | 1 | Interventio | Experimen | Experimen | Control | 0,98336 |
| 3 months | abundance | X00045 | Threonine | 1 | Interventio | Experimen | Experimen | Control | 0,181353 |
| 3 months | abundance | X00050 | β-Hydroxyi | 1 | Interventio | Experimen | Experimen | Control | 0,135093 |
| 3 months | abundance | X00051 | β-D-Gluco | 2b | Interventio | Experimen | Experimen | Control | 0,755448 |
| 3 months | abundance | X00056 | 3-(4-hydro | 1 | Interventio | Experimen | Experimen | Control | 0,823326 |
| 3 months | abundance | X00057 | N-Acetylor | 1 | Interventio | Experimen | Experimen | Control | 0,341701 |
| 3 months | abundance | X00059 | 3,4-Dihydr | 1 | Interventio | Experimen | Experimen | Control | 0,385799 |
| 3 months | abundance | X00060 | Ethylmalor | 1 | Interventio | Experimen | Experimen | Control | 0,468958 |
| 3 months | abundance | X00061 | Thiamine | 1 | Interventio | Experimen | Experimen | Control | 0,94039 |
| 3 months | abundance | X00063 | 2-Hydroxy | 1 | Interventio | Experimen | Experimen | Control | 0,907733 |

| | | | | | | | | | |
|---|---|---|---|---|---|---|---|---|---|
| 3 months | abundance | X00064 | Traumatic | 1 | Interventio | Experimen | Experimen | Control | 0,21397 |
| 3 months | abundance | X00066 | Tryptamine | 1 | Interventio | Experimen | Experimen | Control | 0,98336 |
| 3 months | abundance | X00067 | Hypoxanth | 1 | Interventio | Experimen | Experimen | Control | 0,143745 |
| 3 months | abundance | X00068 | N-Acetylm | 1 | Interventio | Experimen | Experimen | Control | 0,560103 |
| 3 months | abundance | X00070 | α-aminobu | 1 | Interventio | Experimen | Experimen | Control | 0,923057 |
| 3 months | abundance | X00071 | Tyrosine | 1 | Interventio | Experimen | Experimen | Control | 0,833128 |
| 3 months | abundance | X00072 | 3,5-Dihydr | 1 | Interventio | Experimen | Experimen | Control | 0,736809 |
| 3 months | abundance | X00073 | Indole-3-la | 1 | Interventio | Experimen | Experimen | Control | 0,025224 |
| 3 months | abundance | X00074 | Methylsuc | 1 | Interventio | Experimen | Experimen | Control | 0,613297 |
| 3 months | abundance | X00076 | Isoleucine | 1 | Interventio | Experimen | Experimen | Control | 0,560103 |
| 3 months | abundance | X00078 | Acetylagm | 1 | Interventio | Experimen | Experimen | Control | 0,718544 |
| 3 months | abundance | X00082 | Cholic acic | 1 | Interventio | Experimen | Experimen | Control | 0,270355 |
| 3 months | abundance | X00083 | 1,7-Dimeth | 1 | Interventio | Experimen | Experimen | Control | 0,798192 |
| 3 months | abundance | X00084 | Tricarballyl | 1 | Interventio | Experimen | Experimen | Control | 0,761041 |
| 3 months | abundance | X00088 | Pantotheni | 1 | Interventio | Experimen | Experimen | Control | 0,122267 |
| 3 months | abundance | X00089 | 4-Hydroxyb | 1 | Interventio | Experimen | Experimen | Control | 0,840004 |
| 3 months | abundance | X00090 | Malic acid | 1 | Interventio | Experimen | Experimen | Control | 0,387203 |
| 3 months | abundance | X00092 | 3-Methylhi | 1 | Interventio | Experimen | Experimen | Control | 0,339934 |
| 3 months | abundance | X00093 | Deoxyinosi | 1 | Interventio | Experimen | Experimen | Control | 0,000237 |
| 3 months | abundance | X00094 | Methionine | 1 | Interventio | Experimen | Experimen | Control | 0,230723 |
| 3 months | abundance | X00097 | 2,6-Dihydr | 1 | Interventio | Experimen | Experimen | Control | 0,932813 |
| 3 months | abundance | X00099 | 7-Methylgu | 1 | Interventio | Experimen | Experimen | Control | 0,988551 |
| 3 months | abundance | X00101 | Threonic ac | 1 | Interventio | Experimen | Experimen | Control | 0,221894 |
| 3 months | abundance | X00102 | Tryptophar | 1 | Interventio | Experimen | Experimen | Control | 0,141328 |
| 3 months | abundance | X00106 | N-Acetyltry | 1 | Interventio | Experimen | Experimen | Control | 0,161346 |
| 3 months | abundance | X00107 | N6-Acetylly | 1 | Interventio | Experimen | Experimen | Control | 0,017773 |
| 3 months | abundance | X00112 | Xanthosine | 1 | Interventio | Experimen | Experimen | Control | 0,091781 |
| 3 months | abundance | X00113 | N-Acetylph | 1 | Interventio | Experimen | Experimen | Control | 0,361332 |
| 3 months | abundance | X00114 | Indole-3-m | 1 | Interventio | Experimen | Experimen | Control | 0,325689 |
| 3 months | abundance | X00115 | Histidine | 1 | Interventio | Experimen | Experimen | Control | 0,042091 |
| 3 months | abundance | X00116 | Cytidine | 1 | Interventio | Experimen | Experimen | Control | 0,988551 |
| 3 months | abundance | X00120 | Leucylalan | 1 | Interventio | Experimen | Experimen | Control | 0,850869 |
| 3 months | abundance | X00123 | Phenylalar | 1 | Interventio | Experimen | Experimen | Control | 0,836327 |
| 3 months | abundance | X00124 | Leucine | 1 | Interventio | Experimen | Experimen | Control | 0,449163 |
| 3 months | abundance | X00125 | Taurine | 1 | Interventio | Experimen | Experimen | Control | 0,740265 |
| 3 months | abundance | X00127 | Nicotinic a | 1 | Interventio | Experimen | Experimen | Control | 0,04166 |
| 3 months | abundance | X00129 | Quinic acic | 1 | Interventio | Experimen | Experimen | Control | 0,888652 |
| 3 months | abundance | X00130 | Deoxyuridi | 1 | Interventio | Experimen | Experimen | Control | 0,082572 |
| 3 months | abundance | X00132 | Glyceric ac | 1 | Interventio | Experimen | Experimen | Control | 0,957479 |
| 3 months | abundance | X00135 | N-Acetylglu | 1 | Interventio | Experimen | Experimen | Control | 0,235652 |
| 3 months | abundance | X00136 | leu-gln_a | 3 | Interventio | Experimen | Experimen | Control | 0,960119 |
| 3 months | abundance | X00163 | primidone_ | 3 | Interventio | Experimen | Experimen | Control | 0,790121 |
| 3 months | abundance | X00180 | 1-(2-Carbo | 3 | Interventio | Experimen | Experimen | Control | 0,696041 |
| 3 months | abundance | X00182 | 3-(Butylsul | 3 | Interventio | Experimen | Experimen | Control | 0,968857 |
| 3 months | abundance | X00199 | tert-Butyl 3 | 3 | Interventio | Experimen | Experimen | Control | 0,151919 |
| 3 months | abundance | X00204 | 4-O-{3-O-[( | 3 | Interventio | Experimen | Experimen | Control | 0,333178 |
| 3 months | abundance | X00221 | Astemizole | 3 | Interventio | Experimen | Experimen | Control | 0,003817 |
| 3 months | abundance | X00224 | Nifedipine | 3 | Interventio | Experimen | Experimen | Control | 0,765006 |

| | | | | | | | | | |
|---|---|---|---|---|---|---|---|---|---|
| 3 months | abundance | X00233 | g-Aminobu | 3 | Interventio | Experimen | Experimen | Control | 0,212863 |
| 3 months | abundance | X00237 | OI1700000 | 3 | Interventio | Experimen | Experimen | Control | 0,625098 |
| 3 months | abundance | X00242 | IN00258 | 3 | Interventio | Experimen | Experimen | Control | 0,099053 |
| 3 months | abundance | X00253 | TDP-2_a | 3 | Interventio | Experimen | Experimen | Control | 0,684644 |
| 3 months | abundance | X00261 | Mexiletine | 3 | Interventio | Experimen | Experimen | Control | 0,986565 |
| 3 months | abundance | X00264 | PEG n12 | 2b | Interventio | Experimen | Experimen | Control | 0,460268 |
| 3 months | abundance | X00266 | 2-Oxo-3-(p | 3 | Interventio | Experimen | Experimen | Control | 0,423696 |
| 3 months | abundance | X00305 | Formylkynu | 3 | Interventio | Experimen | Experimen | Control | 0,486342 |
| 3 months | abundance | X00320 | Panthenol_ | 3 | Interventio | Experimen | Experimen | Control | 0,277347 |
| 3 months | abundance | X00327 | 3-Hydroxy- | 3 | Interventio | Experimen | Experimen | Control | 0,992999 |
| 3 months | abundance | X00331 | 4-(9H-beta | 3 | Interventio | Experimen | Experimen | Control | 0,520085 |
| 3 months | abundance | X00344 | SECONAL_ | 3 | Interventio | Experimen | Experimen | Control | 0,475034 |
| 3 months | abundance | X00361 | nitecapone | 3 | Interventio | Experimen | Experimen | Control | 0,143745 |
| 3 months | abundance | X00367 | hexobarbit | 3 | Interventio | Experimen | Experimen | Control | 0,174141 |
| 3 months | abundance | X00375 | N-[(2S)-2-H | 3 | Interventio | Experimen | Experimen | Control | 0,021942 |
| 3 months | abundance | X00383 | pretazettin | 3 | Interventio | Experimen | Experimen | Control | 0,405763 |
| 3 months | abundance | X00384 | 3-Formyl-2 | 3 | Interventio | Experimen | Experimen | Control | 0,136644 |
| 3 months | abundance | X00403 | 7alpha-Hyd | 3 | Interventio | Experimen | Experimen | Control | 0,613297 |
| 3 months | abundance | X00404 | piscidic ac | 3 | Interventio | Experimen | Experimen | Control | 0,449163 |
| 3 months | abundance | X00432 | 8-(Methyls | 3 | Interventio | Experimen | Experimen | Control | 0,98336 |
| 3 months | abundance | X00450 | Gly-Ser | 3 | Interventio | Experimen | Experimen | Control | 0,876028 |
| 3 months | abundance | X00477 | Sparfloxac | 2b | Interventio | Experimen | Experimen | Control | 0,63113 |
| 3 months | abundance | X00518 | Nisinic aci | 3 | Interventio | Experimen | Experimen | Control | 0,505325 |
| 3 months | abundance | X00528 | R-(+)-Etira | 3 | Interventio | Experimen | Experimen | Control | 0,787558 |
| 3 months | abundance | X00545 | 3-hydroxyc | 3 | Interventio | Experimen | Experimen | Control | 0,530275 |
| 3 months | abundance | X00549 | 3-[(2Z)-1-C | 3 | Interventio | Experimen | Experimen | Control | 0,798192 |
| 3 months | abundance | X00574 | miglustat | 3 | Interventio | Experimen | Experimen | Control | 0,22934 |
| 3 months | abundance | X00594 | Nicotinami | 3 | Interventio | Experimen | Experimen | Control | 0,988551 |
| 3 months | abundance | X00635 | 4-(5,6-Dihy | 3 | Interventio | Experimen | Experimen | Control | 0,860527 |
| 3 months | abundance | X00637 | N~6~,N~6 | 3 | Interventio | Experimen | Experimen | Control | 0,128284 |
| 3 months | abundance | X00646 | Deacetyldi | 3 | Interventio | Experimen | Experimen | Control | 0,986931 |
| 3 months | abundance | X00670 | 7-Chloro-5 | 3 | Interventio | Experimen | Experimen | Control | 0,894218 |
| 3 months | abundance | X00683 | Zinecard_a | 3 | Interventio | Experimen | Experimen | Control | 0,109824 |
| 3 months | abundance | X00693 | n-Ribosylh | 3 | Interventio | Experimen | Experimen | Control | 0,225072 |
| 3 months | abundance | X00698 | valgancicl | 3 | Interventio | Experimen | Experimen | Control | 0,106496 |
| 3 months | abundance | X00702 | N-[(2S)-2-H | 3 | Interventio | Experimen | Experimen | Control | 0,096259 |
| 3 months | abundance | X00722 | (3S,5R,6E) | 3 | Interventio | Experimen | Experimen | Control | 0,301015 |
| 3 months | abundance | X00723 | Aspartyl-L- | 3 | Interventio | Experimen | Experimen | Control | 0,817686 |
| 3 months | abundance | X00742 | MFCD1869 | 3 | Interventio | Experimen | Experimen | Control | 0,84915 |
| 3 months | abundance | X00743 | 11beta,13- | 3 | Interventio | Experimen | Experimen | Control | 0,835202 |
| 3 months | abundance | X00744 | (1R,2S)-1-( | 3 | Interventio | Experimen | Experimen | Control | 0,713464 |
| 3 months | abundance | X00748 | 3-[3-Metho | 3 | Interventio | Experimen | Experimen | Control | 0,867667 |
| 3 months | abundance | X00764 | 5-Allyl-5-se | 3 | Interventio | Experimen | Experimen | Control | 0,26602 |
| 3 months | abundance | X00807 | Midodrine_ | 3 | Interventio | Experimen | Experimen | Control | 0,667762 |
| 3 months | abundance | X00828 | Fenoterol | 3 | Interventio | Experimen | Experimen | Control | 0,613297 |
| 3 months | abundance | X00836 | 3-Mercapto | 3 | Interventio | Experimen | Experimen | Control | 0,354732 |
| 3 months | abundance | X00853 | 4-O-beta-D | 3 | Interventio | Experimen | Experimen | Control | 0,725009 |
| 3 months | abundance | X00889 | LW800000 | 3 | Interventio | Experimen | Experimen | Control | 0,176658 |

| | | | | | | | | | |
|---|---|---|---|---|---|---|---|---|---|
| 3 months | abundance | X00899 | N-(2,3,4-Tr | 3 | Interventio | Experimen | Experimen | Control | 0,446333 |
| 3 months | abundance | X00907 | 1,2,3,4-Tet | 3 | Interventio | Experimen | Experimen | Control | 0,935319 |
| 3 months | abundance | X00928 | (7R)-7-(5-c | 3 | Interventio | Experimen | Experimen | Control | 0,932813 |
| 3 months | abundance | X00948 | Linamarin | 3 | Interventio | Experimen | Experimen | Control | 0,647023 |
| 3 months | abundance | X00949 | primidone_ | 3 | Interventio | Experimen | Experimen | Control | 0,404129 |
| 3 months | abundance | X00950 | 2-Phenylet | 3 | Interventio | Experimen | Experimen | Control | 0,268483 |
| 3 months | abundance | X00963 | (4S)-4-[(2E | 3 | Interventio | Experimen | Experimen | Control | 0,099053 |
| 3 months | abundance | X00985 | 1-(4-Aminc | 3 | Interventio | Experimen | Experimen | Control | 0,324406 |
| 3 months | abundance | X00998 | epsilon-(ga | 3 | Interventio | Experimen | Experimen | Control | 0,106066 |
| 3 months | abundance | X01007 | pentobarbi | 3 | Interventio | Experimen | Experimen | Control | 0,992999 |
| 3 months | abundance | X01011 | DIBEHENIN | 3 | Interventio | Experimen | Experimen | Control | 0,724969 |
| 3 months | abundance | X01017 | hexobarbit | 3 | Interventio | Experimen | Experimen | Control | 0,424219 |
| 3 months | abundance | X01031 | 3-(14-Ethy | 3 | Interventio | Experimen | Experimen | Control | 0,181353 |
| 3 months | abundance | X01045 | Ethyl maltc | 3 | Interventio | Experimen | Experimen | Control | 0,19078 |
| 3 months | abundance | X01059 | coronatine | 3 | Interventio | Experimen | Experimen | Control | 0,992999 |
| 3 months | abundance | X01067 | g-Aminobu | 3 | Interventio | Experimen | Experimen | Control | 0,918473 |
| 3 months | abundance | X01078 | Dihydroura | 3 | Interventio | Experimen | Experimen | Control | 0,860697 |
| 3 months | abundance | X01081 | Methyl alpl | 3 | Interventio | Experimen | Experimen | Control | 0,927509 |
| 3 months | abundance | X01092 | 17,21-Dihy | 3 | Interventio | Experimen | Experimen | Control | 0,908976 |
| 3 months | abundance | X01098 | (-)-Physost | 3 | Interventio | Experimen | Experimen | Control | 0,758265 |
| 3 months | abundance | X01100 | (betaS)-be | 3 | Interventio | Experimen | Experimen | Control | 0,907733 |
| 3 months | abundance | X01154 | N-(2,3,4-Tr | 3 | Interventio | Experimen | Experimen | Control | 0,262176 |
| 3 months | abundance | X01163 | Prephenic | 3 | Interventio | Experimen | Experimen | Control | 0,944776 |
| 3 months | abundance | X01164 | DL-Carboc | 3 | Interventio | Experimen | Experimen | Control | 0,98692 |
| 3 months | abundance | X01181 | 2-(3,4-Dim | 3 | Interventio | Experimen | Experimen | Control | 0,63113 |
| 3 months | abundance | X01186 | asn-val_b | 3 | Interventio | Experimen | Experimen | Control | 0,698621 |
| 3 months | abundance | X01208 | Lys-Pro_c | 3 | Interventio | Experimen | Experimen | Control | 0,002122 |
| 3 months | abundance | X01223 | DL-Mevalo | 3 | Interventio | Experimen | Experimen | Control | 0,514326 |
| 3 months | abundance | X01235 | 3-[(3-Hydrc | 3 | Interventio | Experimen | Experimen | Control | 0,006023 |
| 3 months | abundance | X01236 | bis(4-isoth | 3 | Interventio | Experimen | Experimen | Control | 0,915997 |
| 3 months | abundance | X01242 | 3-Morpholi | 2b | Interventio | Experimen | Experimen | Control | 0,967096 |
| 3 months | abundance | X01246 | (2R)-1-{[(2 | 3 | Interventio | Experimen | Experimen | Control | 0,002122 |
| 3 months | abundance | X01252 | (−)-nabilor | 3 | Interventio | Experimen | Experimen | Control | 0,808744 |
| 3 months | abundance | X01285 | FB9500000 | 3 | Interventio | Experimen | Experimen | Control | 0,780575 |
| 3 months | abundance | X01286 | (2,4-Dihyd | 3 | Interventio | Experimen | Experimen | Control | 0,76164 |
| 3 months | abundance | X01288 | L-gamma-( | 3 | Interventio | Experimen | Experimen | Control | 0,925953 |
| 3 months | abundance | X01316 | 5-Hydanto | 3 | Interventio | Experimen | Experimen | Control | 0,143745 |
| 3 months | abundance | X01327 | threonylph | 3 | Interventio | Experimen | Experimen | Control | 0,94858 |
| 3 months | abundance | X01340 | Cadralazin | 3 | Interventio | Experimen | Experimen | Control | 0,250053 |
| 3 months | abundance | X01341 | metixene | 3 | Interventio | Experimen | Experimen | Control | 0,36583 |
| 3 months | abundance | X01346 | IN00260_a | 3 | Interventio | Experimen | Experimen | Control | 0,997328 |
| 3 months | abundance | X01363 | Seryltyrosi | 3 | Interventio | Experimen | Experimen | Control | 0,98692 |
| 3 months | abundance | X01364 | Oleuropeir | 3 | Interventio | Experimen | Experimen | Control | 0,862952 |
| 3 months | abundance | X01367 | MFCD1869 | 3 | Interventio | Experimen | Experimen | Control | 0,423696 |
| 3 months | abundance | X01380 | Spermic ac | 3 | Interventio | Experimen | Experimen | Control | 0,96467 |
| 3 months | abundance | X01387 | Methyl 4-(4 | 3 | Interventio | Experimen | Experimen | Control | 0,830617 |
| 3 months | abundance | X01441 | 1-{3-Carbo | 3 | Interventio | Experimen | Experimen | Control | 0,510076 |
| 3 months | abundance | X01463 | 4-Thiapent | 3 | Interventio | Experimen | Experimen | Control | 0,997328 |

| | | | | | | | | | |
|---|---|---|---|---|---|---|---|---|---|
| 3 months | abundance | X01474 | Glyceropho | 3 | Interventio | Experimen | Experimen | Control | 0,004656 |
| 3 months | abundance | X01518 | L-gamma-( | 3 | Interventio | Experimen | Experimen | Control | 0,979909 |
| 3 months | abundance | X01519 | Leucyltrypt | 3 | Interventio | Experimen | Experimen | Control | 0,884928 |
| 3 months | abundance | X01528 | butalbital_ | 3 | Interventio | Experimen | Experimen | Control | 0,588242 |
| 3 months | abundance | X01530 | INK (Peptic | 2b | Interventio | Experimen | Experimen | Control | 0,977246 |
| 3 months | abundance | X01549 | 3-(Sulfooxy | 3 | Interventio | Experimen | Experimen | Control | 0,198011 |
| 3 months | abundance | X01553 | uridine 5'-c | 3 | Interventio | Experimen | Experimen | Control | 0,854716 |
| 3 months | abundance | X01558 | 2-Furoylgly | 2b | Interventio | Experimen | Experimen | Control | 0,778801 |
| 3 months | abundance | X01561 | 2-Ammoni | 3 | Interventio | Experimen | Experimen | Control | 0,680872 |
| 3 months | abundance | X01570 | carglumic a | 3 | Interventio | Experimen | Experimen | Control | 0,949849 |
| 3 months | abundance | X01574 | N-Acetylpr | 3 | Interventio | Experimen | Experimen | Control | 0,960119 |
| 3 months | abundance | X01577 | Prunasin | 3 | Interventio | Experimen | Experimen | Control | 0,883461 |
| 3 months | abundance | X01621 | 2-(2,4-Dihy | 3 | Interventio | Experimen | Experimen | Control | 0,042091 |
| 3 months | abundance | X01640 | Ro 20-1724 | 3 | Interventio | Experimen | Experimen | Control | 0,405692 |
| 3 months | abundance | X01656 | Guanadrel | 3 | Interventio | Experimen | Experimen | Control | 0,015478 |
| 3 months | abundance | X01664 | 5-methylth | 3 | Interventio | Experimen | Experimen | Control | 0,565249 |
| 3 months | abundance | X01671 | N-(3,5-Dim | 3 | Interventio | Experimen | Experimen | Control | 0,9752 |
| 3 months | abundance | X01672 | Triethyl citı | 3 | Interventio | Experimen | Experimen | Control | 0,867667 |
| 3 months | abundance | X01679 | meprobam | 3 | Interventio | Experimen | Experimen | Control | 0,988551 |
| 3 months | abundance | X01689 | Ethyl malat | 3 | Interventio | Experimen | Experimen | Control | 0,003588 |
| 3 months | abundance | X01732 | 2-Hydroxy- | 3 | Interventio | Experimen | Experimen | Control | 0,985356 |
| 3 months | abundance | X01738 | 2-Acetami | 3 | Interventio | Experimen | Experimen | Control | 0,765006 |
| 3 months | abundance | X01746 | Astemizole | 3 | Interventio | Experimen | Experimen | Control | 0,949849 |
| 3 months | abundance | X01768 | Butabarbit | 3 | Interventio | Experimen | Experimen | Control | 2,67E-13 |
| 3 months | abundance | X01776 | N-(Carboxy | 3 | Interventio | Experimen | Experimen | Control | 0,483353 |
| 3 months | abundance | X01793 | Lys-phe_b | 3 | Interventio | Experimen | Experimen | Control | 0,701873 |
| 3 months | abundance | X01813 | Erythorbic | 3 | Interventio | Experimen | Experimen | Control | 0,94878 |
| 3 months | abundance | X01833 | 2,3,4,9-Tet | 2b | Interventio | Experimen | Experimen | Control | 0,808744 |
| 3 months | abundance | X01873 | Methyl 2,3- | 3 | Interventio | Experimen | Experimen | Control | 0,817974 |
| 3 months | abundance | X01879 | Tetramethy | 2b | Interventio | Experimen | Experimen | Control | 0,572678 |
| 3 months | abundance | X01881 | (2S)-3-Met | 3 | Interventio | Experimen | Experimen | Control | 0,867514 |
| 3 months | abundance | X01884 | Roxane | 3 | Interventio | Experimen | Experimen | Control | 5,30E-05 |
| 3 months | abundance | X01887 | lys-tyr_b | 3 | Interventio | Experimen | Experimen | Control | 0,124404 |
| 3 months | abundance | X01893 | 4-(METHYL | 3 | Interventio | Experimen | Experimen | Control | 0,960119 |
| 3 months | abundance | X01911 | 3-(2,3-Dihy | 3 | Interventio | Experimen | Experimen | Control | 0,872597 |
| 3 months | abundance | X01920 | N-(Carboxy | 3 | Interventio | Experimen | Experimen | Control | 0,725856 |
| 3 months | abundance | X01932 | asn-val_c | 3 | Interventio | Experimen | Experimen | Control | 0,26602 |
| 3 months | abundance | X01943 | Tocainide | 3 | Interventio | Experimen | Experimen | Control | 0,859603 |
| 3 months | abundance | X01950 | Histidylgly | 3 | Interventio | Experimen | Experimen | Control | 0,835202 |
| 3 months | abundance | X01968 | 7-Chloro-5 | 3 | Interventio | Experimen | Experimen | Control | 0,835202 |
| 3 months | abundance | X01996 | 3-Hydroxy- | 3 | Interventio | Experimen | Experimen | Control | 0,687176 |
| 3 months | abundance | X02000 | 11-(4-Hydr | 3 | Interventio | Experimen | Experimen | Control | 0,877325 |
| 3 months | abundance | X02009 | Arctiopicri | 3 | Interventio | Experimen | Experimen | Control | 0,416668 |
| 3 months | abundance | X02013 | Ile-cys | 3 | Interventio | Experimen | Experimen | Control | 0,778801 |
| 3 months | abundance | X02020 | Val-Ser_b | 3 | Interventio | Experimen | Experimen | Control | 0,884928 |
| 3 months | abundance | X02023 | beta-D-Eth | 3 | Interventio | Experimen | Experimen | Control | 0,397203 |
| 3 months | abundance | X02073 | N-acetyl-9- | 3 | Interventio | Experimen | Experimen | Control | 0,096437 |
| 3 months | abundance | X02080 | (-)-Aspidos | 3 | Interventio | Experimen | Experimen | Control | 0,210987 |

| | | | | | | | | | |
|---|---|---|---|---|---|---|---|---|---|
| 3 months | abundance | X02082 | 6-Hydroxyr | 3 | Interventio | Experimen | Experimen | Control | 0,647508 |
| 3 months | abundance | X02101 | Succinic ar | 3 | Interventio | Experimen | Experimen | Control | 0,59787 |
| 3 months | abundance | X02108 | Homovanil | 3 | Interventio | Experimen | Experimen | Control | 0,456954 |
| 3 months | abundance | X02121 | 2-Acetamid | 3 | Interventio | Experimen | Experimen | Control | 0,77242 |
| 3 months | abundance | X02139 | SECONAL_ | 3 | Interventio | Experimen | Experimen | Control | 0,150157 |
| 3 months | abundance | X02145 | 3'-Hydroxy | 3 | Interventio | Experimen | Experimen | Control | 0,992999 |
| 3 months | abundance | X02153 | Asarone | 3 | Interventio | Experimen | Experimen | Control | 0,56915 |
| 3 months | abundance | X02181 | YWA1 | 3 | Interventio | Experimen | Experimen | Control | 0,174141 |
| 3 months | abundance | X02184 | Piperonylo | 2b | Interventio | Experimen | Experimen | Control | 0,971921 |
| 3 months | abundance | X02195 | alliin | 3 | Interventio | Experimen | Experimen | Control | 0,988551 |
| 3 months | abundance | X02202 | D-2-Amino | 3 | Interventio | Experimen | Experimen | Control | 0,950648 |
| 3 months | abundance | X02208 | SECONAL_ | 3 | Interventio | Experimen | Experimen | Control | 0,021942 |
| 3 months | abundance | X02214 | (E)-4-Meth | 3 | Interventio | Experimen | Experimen | Control | 0,46054 |
| 3 months | abundance | X02219 | Homoanse | 3 | Interventio | Experimen | Experimen | Control | 0,36547 |
| 3 months | abundance | X02231 | Ala-Tyr | 3 | Interventio | Experimen | Experimen | Control | 0,725339 |
| 3 months | abundance | X02238 | 6-Myoporo | 3 | Interventio | Experimen | Experimen | Control | 0,830535 |
| 3 months | abundance | X02251 | Hostmania | 3 | Interventio | Experimen | Experimen | Control | 0,998514 |
| 3 months | abundance | X02256 | hexobarbit | 3 | Interventio | Experimen | Experimen | Control | 0,052565 |
| 3 months | abundance | X02265 | ophthalmic | 3 | Interventio | Experimen | Experimen | Control | 0,21873 |
| 3 months | abundance | X02268 | 5-Hydroxy- | 3 | Interventio | Experimen | Experimen | Control | 0,640356 |
| 3 months | abundance | X02277 | tert-Butyl 3 | 3 | Interventio | Experimen | Experimen | Control | 0,235652 |
| 3 months | abundance | X02281 | tert-Butyl 3 | 3 | Interventio | Experimen | Experimen | Control | 0,63113 |
| 3 months | abundance | X02288 | 9-Methylur | 2b | Interventio | Experimen | Experimen | Control | 0,392246 |
| 3 months | abundance | X02289 | Kynurenic a | 2a | Interventio | Experimen | Experimen | Control | 0,824813 |
| 3 months | abundance | X02317 | 2-Aminooc | 2b | Interventio | Experimen | Experimen | Control | 0,876083 |
| 3 months | abundance | X02327 | 1H-Pyrazol | 3 | Interventio | Experimen | Experimen | Control | 0,924699 |
| 3 months | abundance | X02333 | N-Propiony | 3 | Interventio | Experimen | Experimen | Control | 0,350171 |
| 3 months | abundance | X02337 | Methyl 1-hy | 3 | Interventio | Experimen | Experimen | Control | 0,698621 |
| 3 months | abundance | X02348 | N-Benzoyla | 3 | Interventio | Experimen | Experimen | Control | 0,63213 |
| 3 months | abundance | X02352 | Indole-3-ca | 3 | Interventio | Experimen | Experimen | Control | 0,251118 |
| 3 months | abundance | X02380 | Propafenor | 3 | Interventio | Experimen | Experimen | Control | 0,142786 |
| 3 months | abundance | X02419 | Glu-Glu | 3 | Interventio | Experimen | Experimen | Control | 0,980029 |
| 3 months | abundance | X02426 | Raltitrexed | 3 | Interventio | Experimen | Experimen | Control | 0,927159 |
| 3 months | abundance | X02427 | 2-(3,5-dim | 2b | Interventio | Experimen | Experimen | Control | 0,690566 |
| 3 months | abundance | X02442 | 3-(Sulfooxy | 3 | Interventio | Experimen | Experimen | Control | 0,655638 |
| 3 months | abundance | X02466 | Validamyci | 3 | Interventio | Experimen | Experimen | Control | 0,59163 |
| 3 months | abundance | X02468 | N,N-Diethy | 3 | Interventio | Experimen | Experimen | Control | 0,819081 |
| 3 months | abundance | X02471 | n-Propyl Ga | 3 | Interventio | Experimen | Experimen | Control | 0,60023 |
| 3 months | abundance | X02492 | N-(3,5-Dim | 3 | Interventio | Experimen | Experimen | Control | 0,88099 |
| 3 months | abundance | X02494 | Butylphtha | 3 | Interventio | Experimen | Experimen | Control | 0,419167 |
| 3 months | abundance | X02504 | DLK (Peptid | 2b | Interventio | Experimen | Experimen | Control | 0,854716 |
| 3 months | abundance | X02513 | 7-Methylac | 2b | Interventio | Experimen | Experimen | Control | 0,012524 |
| 3 months | abundance | X02514 | 3-Ureidopr | 1 | Interventio | Experimen | Experimen | Control | 0,625098 |
| 3 months | abundance | X02521 | 6-(1-Hydro | 3 | Interventio | Experimen | Experimen | Control | 0,837165 |
| 3 months | abundance | X02529 | tenivastati | 3 | Interventio | Experimen | Experimen | Control | 0,920024 |
| 3 months | abundance | X02537 | Leu-pro_a | 3 | Interventio | Experimen | Experimen | Control | 0,968857 |
| 3 months | abundance | X02565 | Dopamine | 3 | Interventio | Experimen | Experimen | Control | 0,680872 |
| 3 months | abundance | X02583 | Ethylvanilli | 3 | Interventio | Experimen | Experimen | Control | 0,276947 |

| | | | | | | | | | |
|---|---|---|---|---|---|---|---|---|---|
| 3 months | abundance | X02599 | Dihydrothy | 3 | Interventio | Experimen | Experimen | Control | 0,141328 |
| 3 months | abundance | X02601 | (7E,7'E)-5, | 3 | Interventio | Experimen | Experimen | Control | 0,15445 |
| 3 months | abundance | X02606 | 4-Amino-1 | 3 | Interventio | Experimen | Experimen | Control | 0,808747 |
| 3 months | abundance | X02615 | 6-APA_a | 3 | Interventio | Experimen | Experimen | Control | 0,860527 |
| 3 months | abundance | X02640 | thyronine | 3 | Interventio | Experimen | Experimen | Control | 0,325689 |
| 3 months | abundance | X02651 | L-gamma-( | 3 | Interventio | Experimen | Experimen | Control | 0,466427 |
| 3 months | abundance | X02652 | Leucyltyros | 3 | Interventio | Experimen | Experimen | Control | 0,244161 |
| 3 months | abundance | X02661 | 4-(3-Oxope | 3 | Interventio | Experimen | Experimen | Control | 0,988551 |
| 3 months | abundance | X02676 | Val-Trp_a | 3 | Interventio | Experimen | Experimen | Control | 0,949849 |
| 3 months | abundance | X02687 | S-Propylcy | 3 | Interventio | Experimen | Experimen | Control | 0,927509 |
| 3 months | abundance | X02689 | piscidic ac | 3 | Interventio | Experimen | Experimen | Control | 0,988551 |
| 3 months | abundance | X02702 | R-(+)-Etira | 3 | Interventio | Experimen | Experimen | Control | 0,639019 |
| 3 months | abundance | X02720 | ala-ser_b | 3 | Interventio | Experimen | Experimen | Control | 0,150157 |
| 3 months | abundance | X02730 | N-Phenyla | 3 | Interventio | Experimen | Experimen | Control | 0,048889 |
| 3 months | abundance | X02733 | Calcitriol | 3 | Interventio | Experimen | Experimen | Control | 0,860527 |
| 3 months | abundance | X02746 | N,N-Diethy | 3 | Interventio | Experimen | Experimen | Control | 0,177953 |
| 3 months | abundance | X02760 | 6-hydroxyp | 3 | Interventio | Experimen | Experimen | Control | 4,83E-14 |
| 3 months | abundance | X02765 | L-Pyrrolysi | 3 | Interventio | Experimen | Experimen | Control | 0,997328 |
| 3 months | abundance | X02767 | 6-APA_b | 3 | Interventio | Experimen | Experimen | Control | 0,181353 |
| 3 months | abundance | X02769 | Tetraacety | 3 | Interventio | Experimen | Experimen | Control | 0,517729 |
| 3 months | abundance | X02774 | 9-ribosylze | 3 | Interventio | Experimen | Experimen | Control | 0,947336 |
| 3 months | abundance | X02793 | 3-Methoxy- | 3 | Interventio | Experimen | Experimen | Control | 0,9752 |
| 3 months | abundance | X02820 | FC250500 | 3 | Interventio | Experimen | Experimen | Control | 0,116825 |
| 3 months | abundance | X02823 | 6,8-Dimeth | 3 | Interventio | Experimen | Experimen | Control | 0,960119 |
| 3 months | abundance | X02824 | Eslicarbaze | 3 | Interventio | Experimen | Experimen | Control | 0,834977 |
| 3 months | abundance | X02832 | (+/-)-2-Hyc | 3 | Interventio | Experimen | Experimen | Control | 0,765006 |
| 3 months | abundance | X02841 | Kyotorphin | 3 | Interventio | Experimen | Experimen | Control | 0,021942 |
| 3 months | abundance | X02843 | 4-(1-Hydro | 3 | Interventio | Experimen | Experimen | Control | 0,631492 |
| 3 months | abundance | X02853 | 7alpha-Hy | 3 | Interventio | Experimen | Experimen | Control | 0,350171 |
| 3 months | abundance | X02854 | Marimasta | 3 | Interventio | Experimen | Experimen | Control | 0,511237 |
| 3 months | abundance | X02868 | 2-(3-Hydro | 3 | Interventio | Experimen | Experimen | Control | 0,729281 |
| 3 months | abundance | X02872 | folinic acid | 3 | Interventio | Experimen | Experimen | Control | 0,059894 |
| 3 months | abundance | X02878 | N~6~-[5-(1 | 3 | Interventio | Experimen | Experimen | Control | 0,924699 |
| 3 months | abundance | X02881 | N-(2,3,4-Tr | 3 | Interventio | Experimen | Experimen | Control | 0,856619 |
| 3 months | abundance | X02894 | Toluene_a | 3 | Interventio | Experimen | Experimen | Control | 0,244993 |
| 3 months | abundance | X02905 | MFCD0005 | 3 | Interventio | Experimen | Experimen | Control | 0,442109 |
| 3 months | abundance | X02920 | L-gamma-( | 3 | Interventio | Experimen | Experimen | Control | 0,552202 |
| 3 months | abundance | X02922 | (2E)-3-Met | 3 | Interventio | Experimen | Experimen | Control | 0,913384 |
| 3 months | abundance | X02928 | N-Acetylpr | 3 | Interventio | Experimen | Experimen | Control | 0,021942 |
| 3 months | abundance | X02943 | 3-Methoxy- | 3 | Interventio | Experimen | Experimen | Control | 0,758687 |
| 3 months | abundance | X02944 | Valylvaline | 3 | Interventio | Experimen | Experimen | Control | 0,255035 |
| 3 months | abundance | X02948 | N-(2-Cyan | 3 | Interventio | Experimen | Experimen | Control | 0,570659 |
| 3 months | abundance | X02952 | Vorinostat_ | 3 | Interventio | Experimen | Experimen | Control | 0,915997 |
| 3 months | abundance | X02968 | Ro 20-172 | 3 | Interventio | Experimen | Experimen | Control | 0,052565 |
| 3 months | abundance | X02972 | 3-(Sulfoox | 3 | Interventio | Experimen | Experimen | Control | 0,445572 |
| 3 months | abundance | X02986 | Diacetin_b | 3 | Interventio | Experimen | Experimen | Control | 0,505325 |
| 3 months | abundance | X02990 | Pentoxifylli | 3 | Interventio | Experimen | Experimen | Control | 0,002026 |
| 3 months | abundance | X03001 | Pseudouric | 2b | Interventio | Experimen | Experimen | Control | 0,988551 |

| | | | | | | | | | |
|---|---|---|---|---|---|---|---|---|---|
| 3 months | abundance | X03004 | Bicine_b | 3 | Interventio | Experimen | Experimen | Control | 0,934681 |
| 3 months | abundance | X03017 | Pro-tyr | 3 | Interventio | Experimen | Experimen | Control | 0,94878 |
| 3 months | abundance | X03023 | NPK (Pepti | 2b | Interventio | Experimen | Experimen | Control | 0,015774 |
| 3 months | abundance | X03064 | Formimino | 3 | Interventio | Experimen | Experimen | Control | 0,465566 |
| 3 months | abundance | X03069 | N-COUMAI | 3 | Interventio | Experimen | Experimen | Control | 0,033766 |
| 3 months | abundance | X03070 | 2-Methoxy- | 3 | Interventio | Experimen | Experimen | Control | 0,801038 |
| 3 months | abundance | X03077 | nicotianam | 3 | Interventio | Experimen | Experimen | Control | 0,593214 |
| 3 months | abundance | X03097 | 6-hydroxyn | 3 | Interventio | Experimen | Experimen | Control | 0,879562 |
| 3 months | abundance | X03109 | Guanidino | 3 | Interventio | Experimen | Experimen | Control | 0,988551 |
| 3 months | abundance | X03132 | (3S,4S)-7,1 | 3 | Interventio | Experimen | Experimen | Control | 0,010133 |
| 3 months | abundance | X03134 | Tetrahydro | 3 | Interventio | Experimen | Experimen | Control | 0,949849 |
| 3 months | abundance | X03158 | 5-Hydroxy- | 3 | Interventio | Experimen | Experimen | Control | 0,004863 |
| 3 months | abundance | X03223 | 1-(2-Carbo | 3 | Interventio | Experimen | Experimen | Control | 0,449163 |
| 3 months | abundance | X03254 | Gly-Trp_b | 3 | Interventio | Experimen | Experimen | Control | 0,971877 |
| 3 months | abundance | X03268 | Dihydrouri | 3 | Interventio | Experimen | Experimen | Control | 0,953271 |
| 3 months | abundance | X03271 | 1Î²-Hydroxy | 3 | Interventio | Experimen | Experimen | Control | 0,71541 |
| 3 months | abundance | X03275 | Methdilazi | 3 | Interventio | Experimen | Experimen | Control | 0,860527 |
| 3 months | abundance | X03276 | 2'-Deoxyac | 3 | Interventio | Experimen | Experimen | Control | 0,577829 |
| 3 months | abundance | X03278 | Hydroxyph | 3 | Interventio | Experimen | Experimen | Control | 0,385347 |
| 3 months | abundance | X03294 | 4,4'-Thiobi | 3 | Interventio | Experimen | Experimen | Control | 0,647023 |
| 3 months | abundance | X03340 | His-pro_b | 3 | Interventio | Experimen | Experimen | Control | 0,232653 |
| 3 months | abundance | X03343 | L-alpha-As | 3 | Interventio | Experimen | Experimen | Control | 0,380548 |
| 3 months | abundance | X03357 | 2-Hydroxyh | 3 | Interventio | Experimen | Experimen | Control | 0,718544 |
| 3 months | abundance | X03363 | Choline Alf | 3 | Interventio | Experimen | Experimen | Control | 0,61536 |
| 3 months | abundance | X03376 | 4-(9H-beta | 3 | Interventio | Experimen | Experimen | Control | 0,817974 |
| 3 months | abundance | X03413 | Val-Trp_b | 3 | Interventio | Experimen | Experimen | Control | 0,657878 |
| 3 months | abundance | X03416 | 9-(alpha-D | 3 | Interventio | Experimen | Experimen | Control | 0,991184 |
| 3 months | abundance | X03434 | SECONAL_ | 3 | Interventio | Experimen | Experimen | Control | 0,907192 |
| 3 months | abundance | X03480 | epsilon-(ga | 3 | Interventio | Experimen | Experimen | Control | 0,836327 |
| 3 months | abundance | X03488 | 3,4-Dihydr | 3 | Interventio | Experimen | Experimen | Control | 0,997328 |
| 3 months | abundance | X03535 | (1S,3R,4s)- | 3 | Interventio | Experimen | Experimen | Control | 0,876028 |
| 3 months | abundance | X03552 | 4-Hydroxya | 3 | Interventio | Experimen | Experimen | Control | 0,88099 |
| 3 months | abundance | X03563 | 6-imino-5- | 3 | Interventio | Experimen | Experimen | Control | 0,244993 |
| 3 months | abundance | X03568 | 3-[(2Z)-1-C | 3 | Interventio | Experimen | Experimen | Control | 0,808744 |
| 3 months | abundance | X03592 | 2-glyceryl 1 | 3 | Interventio | Experimen | Experimen | Control | 0,181353 |
| 3 months | abundance | X03595 | Methylol D | 3 | Interventio | Experimen | Experimen | Control | 0,758265 |
| 3 months | abundance | X03602 | o-Succinyl | 3 | Interventio | Experimen | Experimen | Control | 0,217561 |
| 3 months | abundance | X03604 | Gly-Trp_a | 3 | Interventio | Experimen | Experimen | Control | 1,36E-07 |
| 3 months | abundance | X03629 | Glycylglycy | 3 | Interventio | Experimen | Experimen | Control | 0,931973 |
| 3 months | abundance | X03660 | trp-ser | 3 | Interventio | Experimen | Experimen | Control | 0,949849 |
| 3 months | abundance | X03675 | asp-gln_a | 3 | Interventio | Experimen | Experimen | Control | 0,748275 |
| 3 months | abundance | X03704 | N,N-Dimet | 3 | Interventio | Experimen | Experimen | Control | 0,88099 |
| 3 months | abundance | X03707 | 9-[(5R)-5-E | 3 | Interventio | Experimen | Experimen | Control | 0,019284 |
| 3 months | abundance | X03709 | Guanfacin | 3 | Interventio | Experimen | Experimen | Control | 0,907733 |
| 3 months | abundance | X03714 | Sinapinic a | 3 | Interventio | Experimen | Experimen | Control | 0,718544 |
| 3 months | abundance | X03718 | Propanthel | 3 | Interventio | Experimen | Experimen | Control | 0,808115 |
| 3 months | abundance | X03719 | Losalen | 3 | Interventio | Experimen | Experimen | Control | 0,970457 |
| 3 months | abundance | X03760 | 2-Methoxy- | 3 | Interventio | Experimen | Experimen | Control | 0,834104 |

| | | | | | | | | | |
|---|---|---|---|---|---|---|---|---|---|
| 3 months | abundance | X03793 | Desonide | 3 | Interventio | Experimen | Experimen | Control | 6,33E-05 |
| 3 months | abundance | X03837 | 2-Methoxy- | 3 | Interventio | Experimen | Experimen | Control | 0,448346 |
| 3 months | abundance | X03855 | 2-{(2S,4S)- | 3 | Interventio | Experimen | Experimen | Control | 0,988551 |
| 3 months | abundance | X03860 | 2-BUTYL PI | 3 | Interventio | Experimen | Experimen | Control | 0,968857 |
| 3 months | abundance | X03892 | shinorine | 3 | Interventio | Experimen | Experimen | Control | 0,988551 |
| 3 months | abundance | X03901 | ferrileghen | 3 | Interventio | Experimen | Experimen | Control | 0,899996 |
| 3 months | abundance | X03916 | 2-(1-Ethoxy | 3 | Interventio | Experimen | Experimen | Control | 0,960119 |
| 3 months | abundance | X03917 | Uramustin | 3 | Interventio | Experimen | Experimen | Control | 0,232653 |
| 3 months | abundance | X03934 | 2-Aminom | 3 | Interventio | Experimen | Experimen | Control | 0,845389 |
| 3 months | abundance | X03936 | (3R)-2-(3,4 | 3 | Interventio | Experimen | Experimen | Control | 0,73207 |
| 3 months | abundance | X03961 | Artesunate | 3 | Interventio | Experimen | Experimen | Control | 0,001034 |
| 3 months | abundance | X04021 | Bentazone | 2b | Interventio | Experimen | Experimen | Control | 1,31E-21 |
| 3 months | abundance | X04040 | 3-(3,4-dihy | 3 | Interventio | Experimen | Experimen | Control | 0,98336 |
| 3 months | abundance | X04117 | 2,3,4,5-tet | 3 | Interventio | Experimen | Experimen | Control | 0,604864 |
| 3 months | abundance | X04119 | Tetraacety | 3 | Interventio | Experimen | Experimen | Control | 0,349017 |
| 3 months | abundance | X04146 | L-fucopyra | 3 | Interventio | Experimen | Experimen | Control | 0,571812 |
| 3 months | abundance | X04150 | 4-(METHYL | 3 | Interventio | Experimen | Experimen | Control | 0,61536 |
| 3 months | abundance | X04157 | 3-Methoxy- | 3 | Interventio | Experimen | Experimen | Control | 0,741986 |
| 3 months | abundance | X04181 | 3-Methoxy- | 3 | Interventio | Experimen | Experimen | Control | 0,83895 |
| 3 months | abundance | X04183 | Spaglumic | 3 | Interventio | Experimen | Experimen | Control | 0,99116 |
| 3 months | abundance | X04229 | gamma-L-g | 3 | Interventio | Experimen | Experimen | Control | 0,59787 |
| 3 months | abundance | X04259 | 3-Hydroxyt | 3 | Interventio | Experimen | Experimen | Control | 0,646655 |
| 3 months | abundance | X04274 | 6-[(Z)-2-(3, | 3 | Interventio | Experimen | Experimen | Control | 0,012053 |
| 3 months | abundance | X04286 | Zinecard_b | 3 | Interventio | Experimen | Experimen | Control | 0,761041 |
| 3 months | abundance | X04310 | Aminohipp | 3 | Interventio | Experimen | Experimen | Control | 0,141328 |
| 3 months | abundance | X04315 | Benzamide | 3 | Interventio | Experimen | Experimen | Control | 0,446773 |
| 3 months | abundance | X04334 | 3-Benzyl-6 | 3 | Interventio | Experimen | Experimen | Control | 0,73207 |
| 3 months | abundance | X04339 | L-Glutamic | 3 | Interventio | Experimen | Experimen | Control | 0,385109 |
| 3 months | abundance | X04351 | Pyrimidine | 3 | Interventio | Experimen | Experimen | Control | 0,949849 |
| 3 months | abundance | X04377 | Dihydroura | 3 | Interventio | Experimen | Experimen | Control | 0,582285 |
| 3 months | abundance | X04378 | 9,11-Dihyc | 3 | Interventio | Experimen | Experimen | Control | 0,856135 |
| 3 months | abundance | X04379 | 2-Amino-6 | 3 | Interventio | Experimen | Experimen | Control | 0,820266 |
| 3 months | abundance | X04440 | Mono(3-ca | 3 | Interventio | Experimen | Experimen | Control | 0,867373 |
| 3 months | abundance | X04450 | 3-Methylac | 3 | Interventio | Experimen | Experimen | Control | 0,250532 |
| 3 months | abundance | X04483 | (2E)-5-Hyd | 3 | Interventio | Experimen | Experimen | Control | 0,582285 |
| 3 months | abundance | X04493 | Tyrosol | 3 | Interventio | Experimen | Experimen | Control | 0,722718 |
| 3 months | abundance | X04526 | N-Nonanoy | 3 | Interventio | Experimen | Experimen | Control | 0,198023 |
| 3 months | abundance | X04538 | Valylvaline | 3 | Interventio | Experimen | Experimen | Control | 0,833128 |
| 3 months | abundance | X04543 | N-Acetyl-S | 3 | Interventio | Experimen | Experimen | Control | 0,728208 |
| 3 months | abundance | X04544 | L-gamma-( | 3 | Interventio | Experimen | Experimen | Control | 0,995163 |
| 3 months | abundance | X04553 | N-(4-Amin | 3 | Interventio | Experimen | Experimen | Control | 0,960119 |
| 3 months | abundance | X04557 | 1,4-Naphtl | 3 | Interventio | Experimen | Experimen | Control | 0,431024 |
| 3 months | abundance | X04562 | Lisdexamf | 3 | Interventio | Experimen | Experimen | Control | 0,958777 |
| 3 months | abundance | X04564 | asp-gln_b | 3 | Interventio | Experimen | Experimen | Control | 0,625098 |
| 3 months | abundance | X04566 | pterin | 3 | Interventio | Experimen | Experimen | Control | 0,840004 |
| 3 months | abundance | X04571 | 13a-Hydro | 3 | Interventio | Experimen | Experimen | Control | 0,639019 |
| 3 months | abundance | X04579 | 6-(alpha-D | 3 | Interventio | Experimen | Experimen | Control | 0,861311 |
| 3 months | abundance | X04582 | Dinoseb | 3 | Interventio | Experimen | Experimen | Control | 0,46054 |

| | | | | | | | | | |
|---|---|---|---|---|---|---|---|---|---|
| 3 months | abundance | X04593 | Methional | 3 | Interventio | Experimen | Experimen | Control | 0,619252 |
| 3 months | abundance | X04636 | epsilon-(ga | 3 | Interventio | Experimen | Experimen | Control | 0,725339 |
| 3 months | abundance | X04639 | hydroxyhex | 3 | Interventio | Experimen | Experimen | Control | 0,124404 |
| 3 months | abundance | X04682 | 1-Methylin | 3 | Interventio | Experimen | Experimen | Control | 0,282328 |
| 3 months | abundance | X04684 | AAMU_b | 3 | Interventio | Experimen | Experimen | Control | 0,862952 |
| 3 months | abundance | X04688 | Minoxidil | 3 | Interventio | Experimen | Experimen | Control | 0,393572 |
| 3 months | abundance | X04695 | N-[(4-Meth | 3 | Interventio | Experimen | Experimen | Control | 0,718199 |
| 3 months | abundance | X04697 | Ethosuxim | 3 | Interventio | Experimen | Experimen | Control | 0,820266 |
| 3 months | abundance | X04743 | 5-Methoxy- | 3 | Interventio | Experimen | Experimen | Control | 0,798841 |
| 3 months | abundance | X04744 | 16alpha-hy | 3 | Interventio | Experimen | Experimen | Control | 0,948674 |
| 3 months | abundance | X04759 | 7-Aminom | 3 | Interventio | Experimen | Experimen | Control | 0,960119 |
| 3 months | abundance | X04788 | 2-(4-Isopro | 3 | Interventio | Experimen | Experimen | Control | 0,446309 |
| 3 months | abundance | X04792 | his-asn | 3 | Interventio | Experimen | Experimen | Control | 0,203782 |
| 3 months | abundance | X04814 | meprobam | 3 | Interventio | Experimen | Experimen | Control | 0,927159 |
| 3 months | abundance | X04822 | 6-Sulfatoxy | 3 | Interventio | Experimen | Experimen | Control | 0,141328 |
| 3 months | abundance | X04852 | Toluene_b | 3 | Interventio | Experimen | Experimen | Control | 0,058295 |
| 3 months | abundance | X04877 | Trifluorome | 3 | Interventio | Experimen | Experimen | Control | 0,216079 |
| 3 months | abundance | X04897 | Taxifolin | 3 | Interventio | Experimen | Experimen | Control | 0,082572 |
| 3 months | abundance | X04898 | 17-Hydroxy | 3 | Interventio | Experimen | Experimen | Control | 0,491513 |
| 3 months | abundance | X04919 | 5-O-alpha- | 3 | Interventio | Experimen | Experimen | Control | 0,646655 |
| 3 months | abundance | X04922 | Sinapinic a | 3 | Interventio | Experimen | Experimen | Control | 0,830535 |
| 3 months | abundance | X04939 | N-Desalkyl | 3 | Interventio | Experimen | Experimen | Control | 0,522871 |
| 3 months | abundance | X04975 | MFCD0995 | 3 | Interventio | Experimen | Experimen | Control | 0,639019 |
| 3 months | abundance | X04986 | L-Proline, 4 | 3 | Interventio | Experimen | Experimen | Control | 0,642236 |
| 3 months | abundance | X04990 | Cilazapril | 3 | Interventio | Experimen | Experimen | Control | 0,282368 |
| 3 months | abundance | X04996 | MFCD0272 | 3 | Interventio | Experimen | Experimen | Control | 0,807551 |
| 3 months | abundance | X04999 | 2-Hydroxyp | 1 | Interventio | Experimen | Experimen | Control | 0,819081 |
| 3 months | abundance | X05017 | N-(1-{[Metl | 3 | Interventio | Experimen | Experimen | Control | 0,078386 |
| 3 months | abundance | X05029 | Hypericin | 3 | Interventio | Experimen | Experimen | Control | 0,083526 |
| 3 months | abundance | X05033 | (2E)-N-3,7- | 3 | Interventio | Experimen | Experimen | Control | 1,92E-05 |
| 3 months | abundance | X05046 | Casimiroin | 3 | Interventio | Experimen | Experimen | Control | 0,960119 |
| 3 months | abundance | X05068 | pro-gln_c | 3 | Interventio | Experimen | Experimen | Control | 0,141328 |
| 3 months | abundance | X05071 | Histidylphe | 3 | Interventio | Experimen | Experimen | Control | 0,613297 |
| 3 months | abundance | X05072 | L-gamma-( | 3 | Interventio | Experimen | Experimen | Control | 0,113776 |
| 3 months | abundance | X05081 | ELK (Peptic | 2b | Interventio | Experimen | Experimen | Control | 0,322462 |
| 3 months | abundance | X05126 | (3aS,5S,6F | 3 | Interventio | Experimen | Experimen | Control | 0,008795 |
| 3 months | abundance | X05146 | UQ367500 | 3 | Interventio | Experimen | Experimen | Control | 0,949849 |
| 3 months | abundance | X05149 | S-Sulfocys | 3 | Interventio | Experimen | Experimen | Control | 0,907192 |
| 3 months | abundance | X05177 | cis-3-Hexe | 3 | Interventio | Experimen | Experimen | Control | 0,833128 |
| 3 months | abundance | X05183 | 3,6-Dichlo | 3 | Interventio | Experimen | Experimen | Control | 0,778801 |
| 3 months | abundance | X05195 | Lys-phe_a | 3 | Interventio | Experimen | Experimen | Control | 0,651289 |
| 3 months | abundance | X05203 | felbamate | 3 | Interventio | Experimen | Experimen | Control | 0,820266 |
| 3 months | abundance | X05204 | Urothion | 3 | Interventio | Experimen | Experimen | Control | 0,082572 |
| 3 months | abundance | X05214 | [7-Hydroxy | 3 | Interventio | Experimen | Experimen | Control | 0,423696 |
| 3 months | abundance | X05225 | 4-(METHYL | 3 | Interventio | Experimen | Experimen | Control | 0,503169 |
| 3 months | abundance | X05234 | 3,4-dihydro | 3 | Interventio | Experimen | Experimen | Control | 0,988198 |
| 3 months | abundance | X05237 | imazameth | 3 | Interventio | Experimen | Experimen | Control | 0,985356 |
| 3 months | abundance | X05240 | Dimeric m | 3 | Interventio | Experimen | Experimen | Control | 0,854716 |

| | | | | | | | | | |
|---|---|---|---|---|---|---|---|---|---|
| 3 months | abundance | X05243 | N-(4-Hydro | 3 | Interventio | Experimen | Experimen | Control | 0,971921 |
| 3 months | abundance | X05275 | H-DL-MET- | 3 | Interventio | Experimen | Experimen | Control | 0,042091 |
| 3 months | abundance | X05295 | 3-(4,7-Dim | 3 | Interventio | Experimen | Experimen | Control | 0,78879 |
| 3 months | abundance | X05306 | 2-methoxy | 3 | Interventio | Experimen | Experimen | Control | 0,63213 |
| 3 months | abundance | X05357 | NPYR | 3 | Interventio | Experimen | Experimen | Control | 0,193817 |
| 3 months | abundance | X05391 | 2-Acetami | 3 | Interventio | Experimen | Experimen | Control | 0,206211 |
| 3 months | abundance | X05398 | DIBOA | 3 | Interventio | Experimen | Experimen | Control | 0,424219 |
| 3 months | abundance | X05411 | 3-(Sulfoox | 3 | Interventio | Experimen | Experimen | Control | 0,88266 |
| 3 months | abundance | X05415 | Redul | 3 | Interventio | Experimen | Experimen | Control | 0,923057 |
| 3 months | abundance | X05417 | (S)-2-hydra | 3 | Interventio | Experimen | Experimen | Control | 0,143745 |
| 3 months | abundance | X05421 | S-Allylcyst | 3 | Interventio | Experimen | Experimen | Control | 0,953271 |
| 3 months | abundance | X05459 | Scopoletin | 3 | Interventio | Experimen | Experimen | Control | 0,142786 |
| 3 months | abundance | X05467 | Azulfidine | 3 | Interventio | Experimen | Experimen | Control | 0,333178 |
| 3 months | abundance | X05478 | S(8)-aminc | 3 | Interventio | Experimen | Experimen | Control | 0,385347 |
| 3 months | abundance | X05499 | GLY-MET | 3 | Interventio | Experimen | Experimen | Control | 0,000124 |
| 3 months | abundance | X05510 | 3-Benzyl-6 | 3 | Interventio | Experimen | Experimen | Control | 0,988551 |
| 3 months | abundance | X05514 | (3aS,5S,6F | 3 | Interventio | Experimen | Experimen | Control | 0,936365 |
| 3 months | abundance | X05551 | Trolox | 3 | Interventio | Experimen | Experimen | Control | 0,474449 |
| 3 months | abundance | X05561 | 1Î²-Hydrox | 3 | Interventio | Experimen | Experimen | Control | 0,956306 |
| 3 months | abundance | X05580 | Xanthureni | 3 | Interventio | Experimen | Experimen | Control | 0,948674 |
| 3 months | abundance | X05581 | N-[(10Z)-7- | 3 | Interventio | Experimen | Experimen | Control | 0,656167 |
| 3 months | abundance | X05584 | Serotonin | 1 | Interventio | Experimen | Experimen | Control | 0,815986 |
| 3 months | abundance | X05656 | mesifurane | 3 | Interventio | Experimen | Experimen | Control | 0,947758 |
| 3 months | abundance | X05709 | 2,3-Dihydr | 3 | Interventio | Experimen | Experimen | Control | 0,915552 |
| 3 months | abundance | X05726 | methocarb | 3 | Interventio | Experimen | Experimen | Control | 0,835202 |
| 3 months | abundance | X05734 | N-[(2S)-2-I | 3 | Interventio | Experimen | Experimen | Control | 0,773264 |
| 3 months | abundance | X05741 | g-Aminobu | 3 | Interventio | Experimen | Experimen | Control | 0,001817 |
| 3 months | abundance | X05758 | quinol sulf | 3 | Interventio | Experimen | Experimen | Control | 0,60023 |
| 3 months | abundance | X05786 | Nitrendipir | 3 | Interventio | Experimen | Experimen | Control | 0,988551 |
| 3 months | abundance | X05812 | (2S)-3-(1H | 3 | Interventio | Experimen | Experimen | Control | 0,864576 |
| 3 months | abundance | X05857 | N-D-Gluco | 3 | Interventio | Experimen | Experimen | Control | 0,217561 |
| 3 months | abundance | X05859 | 3,7,12,17- | 3 | Interventio | Experimen | Experimen | Control | 0,960119 |
| 3 months | abundance | X05878 | 1,2-dihydr | 3 | Interventio | Experimen | Experimen | Control | 0,034991 |
| 3 months | abundance | X05892 | (5Z)-2-Ami | 3 | Interventio | Experimen | Experimen | Control | 0,821708 |
| 3 months | abundance | X05919 | 2-(2-Aminc | 3 | Interventio | Experimen | Experimen | Control | 0,651767 |
| 3 months | abundance | X05968 | Tetraacety | 3 | Interventio | Experimen | Experimen | Control | 0,631492 |
| 3 months | abundance | X05969 | 4-(METHYL | 3 | Interventio | Experimen | Experimen | Control | 0,820266 |
| 3 months | abundance | X05971 | L-gamma-( | 3 | Interventio | Experimen | Experimen | Control | 0,681178 |
| 3 months | abundance | X05982 | (19R,25S)- | 3 | Interventio | Experimen | Experimen | Control | 0,10419 |
| 3 months | abundance | X05984 | 1-(4-Methy | 2b | Interventio | Experimen | Experimen | Control | 0,820408 |
| 3 months | abundance | X05988 | Selsun | 3 | Interventio | Experimen | Experimen | Control | 0,179615 |
| 3 months | abundance | X05995 | Diacetin_a | 3 | Interventio | Experimen | Experimen | Control | 0,234997 |
| 3 months | abundance | X05997 | FB950000( | 3 | Interventio | Experimen | Experimen | Control | 0,287585 |
| 3 months | abundance | X06011 | Coprine_a | 3 | Interventio | Experimen | Experimen | Control | 0,392246 |
| 3 months | abundance | X06016 | N,N-Dimet | 3 | Interventio | Experimen | Experimen | Control | 0,161121 |
| 3 months | abundance | X06034 | N-[(2S)-2-I | 3 | Interventio | Experimen | Experimen | Control | 0,59787 |
| 3 months | abundance | X06039 | gamma-Gl | 3 | Interventio | Experimen | Experimen | Control | 1,83E-11 |
| 3 months | abundance | X06041 | (DL)-3-O-M | 3 | Interventio | Experimen | Experimen | Control | 0,798841 |

| | | | | | | | | | |
|---|---|---|---|---|---|---|---|---|---|
| 3 months | abundance | X06043 | vinyl sulfid | 3 | Interventio | Experimen | Experimen | Control | 0,22934 |
| 3 months | abundance | X06046 | Valylvaline | 3 | Interventio | Experimen | Experimen | Control | 0,000496 |
| 3 months | abundance | X06062 | 1-(2,3-Dihy | 3 | Interventio | Experimen | Experimen | Control | 0,957479 |
| 3 months | abundance | X06079 | Glycylleuci | 1 | Interventio | Experimen | Experimen | Control | 0,627129 |
| 3 months | abundance | X06082 | O-heptano | 3 | Interventio | Experimen | Experimen | Control | 0,830535 |
| 3 months | abundance | X06085 | Tetraacety | 3 | Interventio | Experimen | Experimen | Control | 0,992999 |
| 3 months | abundance | X06089 | riboprine | 3 | Interventio | Experimen | Experimen | Control | 0,820408 |
| 3 months | abundance | X06100 | Midodrine_ | 3 | Interventio | Experimen | Experimen | Control | 0,927509 |
| 3 months | abundance | X06107 | Hydroxycar | 3 | Interventio | Experimen | Experimen | Control | 0,787558 |
| 3 months | abundance | X06112 | Val-Ser_a | 3 | Interventio | Experimen | Experimen | Control | 0,724599 |
| 3 months | abundance | X06124 | MFCD0995 | 3 | Interventio | Experimen | Experimen | Control | 0,860527 |
| 3 months | abundance | X06127 | 2-Methoxy- | 3 | Interventio | Experimen | Experimen | Control | 0,625098 |
| 3 months | abundance | X06143 | 2-Methylbu | 2a | Interventio | Experimen | Experimen | Control | 0,938964 |
| 3 months | abundance | X06146 | S-Methyl-1 | 3 | Interventio | Experimen | Experimen | Control | 0,823267 |
| 3 months | abundance | X06149 | Yangonin | 3 | Interventio | Experimen | Experimen | Control | 0,034991 |
| 3 months | abundance | X06150 | Zalcitabine | 3 | Interventio | Experimen | Experimen | Control | 0,707386 |
| 3 months | abundance | X06152 | alpha-keto | 3 | Interventio | Experimen | Experimen | Control | 0,605613 |
| 3 months | abundance | X06167 | N-Acetylas | 2a | Interventio | Experimen | Experimen | Control | 0,198023 |
| 3 months | abundance | X06168 | leu-gln_b | 3 | Interventio | Experimen | Experimen | Control | 0,988551 |
| 3 months | abundance | X06183 | Nisinic aci | 3 | Interventio | Experimen | Experimen | Control | 0,718544 |
| 3 months | abundance | X06189 | delta-Guar | 3 | Interventio | Experimen | Experimen | Control | 0,008037 |
| 3 months | abundance | X06197 | 4-(Nitrosoa | 3 | Interventio | Experimen | Experimen | Control | 0,853633 |
| 3 months | abundance | X06220 | Tetrahydro | 3 | Interventio | Experimen | Experimen | Control | 0,907192 |
| 3 months | abundance | X06222 | Leucylaspa | 3 | Interventio | Experimen | Experimen | Control | 0,739004 |
| 3 months | abundance | X06227 | lys-tyr_a | 3 | Interventio | Experimen | Experimen | Control | 0,698621 |
| 3 months | abundance | X06230 | Glu-Gly | 3 | Interventio | Experimen | Experimen | Control | 0,315213 |
| 3 months | abundance | X06240 | His-pro_a | 3 | Interventio | Experimen | Experimen | Control | 0,105225 |
| 3 months | abundance | X06250 | [3-({3-[(Cy | 2b | Interventio | Experimen | Experimen | Control | 0,10419 |
| 3 months | abundance | X06253 | N-(4-Amino | 3 | Interventio | Experimen | Experimen | Control | 0,718544 |
| 3 months | abundance | X06254 | D-Alanyl-D | 3 | Interventio | Experimen | Experimen | Control | 0,113776 |
| 3 months | abundance | X06259 | (S)-?-glyce | 3 | Interventio | Experimen | Experimen | Control | 0,836327 |
| 3 months | abundance | X06268 | mesifurane | 3 | Interventio | Experimen | Experimen | Control | 0,985356 |
| 3 months | abundance | X06270 | 9-Methylur | 2b | Interventio | Experimen | Experimen | Control | 0,625098 |
| 3 months | abundance | X06276 | asn-pro_b | 3 | Interventio | Experimen | Experimen | Control | 0,431744 |
| 3 months | abundance | X06278 | 3-Hydroxy- | 3 | Interventio | Experimen | Experimen | Control | 0,718544 |
| 3 months | abundance | X06289 | Nicotinatel | 3 | Interventio | Experimen | Experimen | Control | 0,08584 |
| 3 months | abundance | X06291 | Corticoster | 2b | Interventio | Experimen | Experimen | Control | 0,803437 |
| 3 months | abundance | X06292 | mesifurane | 3 | Interventio | Experimen | Experimen | Control | 0,990649 |
| 3 months | abundance | X06293 | (4R)-4-{[3- | 3 | Interventio | Experimen | Experimen | Control | 0,250575 |
| 3 months | abundance | X06313 | Isoquinolir | 2b | Interventio | Experimen | Experimen | Control | 0,881276 |
| 3 months | abundance | X06318 | Propamoca | 2b | Interventio | Experimen | Experimen | Control | 0,819701 |
| 3 months | abundance | X06320 | 7alpha-Hy | 3 | Interventio | Experimen | Experimen | Control | 0,876495 |
| 3 months | abundance | X06329 | Leu-Leu_d | 3 | Interventio | Experimen | Experimen | Control | 0,394274 |
| 3 months | abundance | X06334 | Bis-D-fruct | 3 | Interventio | Experimen | Experimen | Control | 0,980029 |
| 3 months | abundance | X06337 | Zalcitabine | 3 | Interventio | Experimen | Experimen | Control | 0,387209 |
| 3 months | abundance | X06354 | trans-Zeati | 2b | Interventio | Experimen | Experimen | Control | 0,449163 |
| 3 months | abundance | X06361 | Oxprenolol | 3 | Interventio | Experimen | Experimen | Control | 0,124383 |
| 3 months | abundance | X06368 | TDP-2_b | 3 | Interventio | Experimen | Experimen | Control | 0,562487 |

| | | | | | | | | | |
|---|---|---|---|---|---|---|---|---|---|
| 3 months | abundance | X06371 | L-gamma-( | 3 | Interventio | Experimen | Experimen | Control | 0,909315 |
| 3 months | abundance | X06372 | N,N-dimetl | 2b | Interventio | Experimen | Experimen | Control | 0,089222 |
| 3 months | abundance | X06381 | MFCD1869 | 3 | Interventio | Experimen | Experimen | Control | 0,99116 |
| 3 months | abundance | X06388 | Caffeic aci | 1 | Interventio | Experimen | Experimen | Control | 0,71076 |
| 3 months | abundance | X06400 | Phloionolic | 3 | Interventio | Experimen | Experimen | Control | 0,819701 |
| 3 months | abundance | X06401 | Isophthalic | 2b | Interventio | Experimen | Experimen | Control | 0,817715 |
| 3 months | abundance | X06404 | Flemichap | 3 | Interventio | Experimen | Experimen | Control | 0,210209 |
| 3 months | abundance | X06405 | Asparaginy | 3 | Interventio | Experimen | Experimen | Control | 0,322462 |
| 3 months | abundance | X06409 | Glycylproli | 2b | Interventio | Experimen | Experimen | Control | 0,490686 |
| 3 months | abundance | X06412 | Butenylcar | 3 | Interventio | Experimen | Experimen | Control | 0,991184 |
| 3 months | abundance | X06416 | KYNURAMI | 3 | Interventio | Experimen | Experimen | Control | 0,960119 |
| 3 months | abundance | X06419 | Spermic ac | 3 | Interventio | Experimen | Experimen | Control | 0,988551 |
| 3 months | abundance | X06433 | 4-(2,5-Diflu | 2b | Interventio | Experimen | Experimen | Control | 0,935128 |
| 3 months | abundance | X06434 | butyrin | 3 | Interventio | Experimen | Experimen | Control | 0,860304 |
| 3 months | abundance | X06437 | 2-Acetamid | 3 | Interventio | Experimen | Experimen | Control | 0,336689 |
| 3 months | abundance | X06448 | 3,8,9-trihy | 2b | Interventio | Experimen | Experimen | Control | 0,761478 |
| 3 months | abundance | X06454 | 8-Amino-7- | 3 | Interventio | Experimen | Experimen | Control | 0,284646 |
| 3 months | abundance | X06455 | 4-Hydroxyp | 3 | Interventio | Experimen | Experimen | Control | 0,63213 |
| 3 months | abundance | X06462 | DNOP_d | 3 | Interventio | Experimen | Experimen | Control | 0,605613 |
| 3 months | abundance | X06463 | 8-Amino-7- | 3 | Interventio | Experimen | Experimen | Control | 0,696041 |
| 3 months | abundance | X06472 | N(alpha)-B | 3 | Interventio | Experimen | Experimen | Control | 0,927509 |
| 3 months | abundance | X06473 | MFCD0002 | 3 | Interventio | Experimen | Experimen | Control | 0,967096 |
| 3 months | abundance | X06477 | APM_b | 3 | Interventio | Experimen | Experimen | Control | 0,303703 |
| 3 months | abundance | X06478 | Caffeic aci | 3 | Interventio | Experimen | Experimen | Control | 0,860527 |
| 3 months | abundance | X06482 | glu-pro | 3 | Interventio | Experimen | Experimen | Control | 0,967096 |
| 3 months | abundance | X06486 | Leu-Val_b | 3 | Interventio | Experimen | Experimen | Control | 0,096259 |
| 3 months | abundance | X06495 | 2-Acetamid | 3 | Interventio | Experimen | Experimen | Control | 0,633187 |
| 3 months | abundance | X06502 | MFCD2836 | 3 | Interventio | Experimen | Experimen | Control | 0,665073 |
| 3 months | abundance | X06506 | L-Homocys | 3 | Interventio | Experimen | Experimen | Control | 0,758687 |
| 3 months | abundance | X06507 | Gly-Lys | 3 | Interventio | Experimen | Experimen | Control | 0,226642 |
| 3 months | abundance | X06508 | Asparaginy | 3 | Interventio | Experimen | Experimen | Control | 0,475833 |
| 3 months | abundance | X06510 | N-{4-[(2R,3 | 2b | Interventio | Experimen | Experimen | Control | 0,235092 |
| 3 months | abundance | X06529 | Methanesu | 2a | Interventio | Experimen | Experimen | Control | 0,862952 |
| 3 months | abundance | X06530 | 3-(1-hydro: | 2b | Interventio | Experimen | Experimen | Control | 0,998514 |
| 3 months | abundance | X06532 | Leupeptin | 3 | Interventio | Experimen | Experimen | Control | 0,000266 |
| 3 months | abundance | X06546 | 4-Acetamid | 2b | Interventio | Experimen | Experimen | Control | 0,106066 |
| 3 months | abundance | X06549 | Panthenol_ | 3 | Interventio | Experimen | Experimen | Control | 0,853605 |
| 3 months | abundance | X06551 | Methyl 1-hy | 3 | Interventio | Experimen | Experimen | Control | 0,988551 |
| 3 months | abundance | X06560 | 2,6-Dimeth | 2b | Interventio | Experimen | Experimen | Control | 0,150647 |
| 3 months | abundance | X06590 | 3-hydroxy-: | 3 | Interventio | Experimen | Experimen | Control | 0,303703 |
| 3 months | abundance | X06600 | MFCD0014 | 3 | Interventio | Experimen | Experimen | Control | 0,08584 |
| 3 months | abundance | X06606 | Leu-Val_d | 3 | Interventio | Experimen | Experimen | Control | 0,503169 |
| 3 months | abundance | X06612 | Spermic ac | 3 | Interventio | Experimen | Experimen | Control | 0,77242 |
| 3 months | abundance | X06622 | Lys-Pro_b | 3 | Interventio | Experimen | Experimen | Control | 0,152323 |
| 3 months | abundance | X06623 | quinol sulf | 3 | Interventio | Experimen | Experimen | Control | 0,329628 |
| 3 months | abundance | X06626 | N-Butyryl-L | 3 | Interventio | Experimen | Experimen | Control | 0,953271 |
| 3 months | abundance | X06630 | 4-(METHYL | 3 | Interventio | Experimen | Experimen | Control | 0,187996 |
| 3 months | abundance | X06631 | 2-Isopropy | 2b | Interventio | Experimen | Experimen | Control | 0,136002 |

| | | | | | | | | | |
|---|---|---|---|---|---|---|---|---|---|
| 3 months | abundance | X06634 | Glycitein | 3 | Interventio | Experimen | Experimen | Control | 0,143819 |
| 3 months | abundance | X06642 | Homocitru | 2a | Interventio | Experimen | Experimen | Control | 0,798192 |
| 3 months | abundance | X06646 | (S)-3-sulfo | 3 | Interventio | Experimen | Experimen | Control | 0,385799 |
| 3 months | abundance | X06655 | Hydroxyca | 3 | Interventio | Experimen | Experimen | Control | 0,654003 |
| 3 months | abundance | X06656 | Valylvaline | 3 | Interventio | Experimen | Experimen | Control | 0,84521 |
| 3 months | abundance | X06663 | Leucyltyros | 3 | Interventio | Experimen | Experimen | Control | 0,898308 |
| 3 months | abundance | X06675 | L-gamma-( | 3 | Interventio | Experimen | Experimen | Control | 0,430971 |
| 3 months | abundance | X06680 | 7-Hydroxy- | 2b | Interventio | Experimen | Experimen | Control | 0,270117 |
| 3 months | abundance | X06681 | Scymnol | 3 | Interventio | Experimen | Experimen | Control | 0,249675 |
| 3 months | abundance | X06684 | Bromazine | 3 | Interventio | Experimen | Experimen | Control | 0,854716 |
| 3 months | abundance | X06690 | 4-Methylca | 2b | Interventio | Experimen | Experimen | Control | 0,860527 |
| 3 months | abundance | X06698 | entecavir | 3 | Interventio | Experimen | Experimen | Control | 0,997937 |
| 3 months | abundance | X06703 | N6-METHY | 3 | Interventio | Experimen | Experimen | Control | 0,60023 |
| 3 months | abundance | X06704 | feruloylagr | 3 | Interventio | Experimen | Experimen | Control | 0,552202 |
| 3 months | abundance | X06709 | Tetraacety | 3 | Interventio | Experimen | Experimen | Control | 0,798192 |
| 3 months | abundance | X06722 | LW800000 | 3 | Interventio | Experimen | Experimen | Control | 0,146246 |
| 3 months | abundance | X06723 | 3-Hydroxy- | 3 | Interventio | Experimen | Experimen | Control | 0,836327 |
| 3 months | abundance | X06727 | MFCD0087 | 3 | Interventio | Experimen | Experimen | Control | 0,854716 |
| 3 months | abundance | X06735 | 7-Methylxa | 2b | Interventio | Experimen | Experimen | Control | 0,198011 |
| 3 months | abundance | X06744 | Diethylpyr | 3 | Interventio | Experimen | Experimen | Control | 0,150298 |
| 3 months | abundance | X06750 | 1-PYRENYI | 3 | Interventio | Experimen | Experimen | Control | 0,835202 |
| 3 months | abundance | X06764 | N-(4-Hydro | 3 | Interventio | Experimen | Experimen | Control | 0,71541 |
| 3 months | abundance | X06767 | 2-Hydroxyb | 1 | Interventio | Experimen | Experimen | Control | 0,750158 |
| 3 months | abundance | X06768 | Sinapyl alc | 3 | Interventio | Experimen | Experimen | Control | 0,413227 |
| 3 months | abundance | X06771 | MFCD0015 | 3 | Interventio | Experimen | Experimen | Control | 0,198023 |
| 3 months | abundance | X06783 | N~6~-Octa | 3 | Interventio | Experimen | Experimen | Control | 0,385109 |
| 3 months | abundance | X06797 | pro-gln_a | 3 | Interventio | Experimen | Experimen | Control | 0,718544 |
| 3 months | abundance | X06805 | (9cis)-O~1 | 3 | Interventio | Experimen | Experimen | Control | 0,698241 |
| 3 months | abundance | X06807 | 1,1'-[1,12-I | 3 | Interventio | Experimen | Experimen | Control | 0,413564 |
| 3 months | abundance | X06812 | 2-Hydroxy- | 3 | Interventio | Experimen | Experimen | Control | 0,182946 |
| 3 months | abundance | X06814 | N-Benzoyla | 3 | Interventio | Experimen | Experimen | Control | 0,748275 |
| 3 months | abundance | X06818 | Homoanse | 3 | Interventio | Experimen | Experimen | Control | 0,025414 |
| 3 months | abundance | X06819 | Primaquine | 3 | Interventio | Experimen | Experimen | Control | 0,004762 |
| 3 months | abundance | X06820 | 1-(3,4-dim | 2b | Interventio | Experimen | Experimen | Control | 0,993797 |
| 3 months | abundance | X06836 | asn-lys | 3 | Interventio | Experimen | Experimen | Control | 0,194947 |
| 3 months | abundance | X06850 | Leu-Val_c | 3 | Interventio | Experimen | Experimen | Control | 0,01685 |
| 3 months | abundance | X06856 | Tiglic acid_ | 2b | Interventio | Experimen | Experimen | Control | 0,440554 |
| 3 months | abundance | X06858 | tert-Butyl 3 | 3 | Interventio | Experimen | Experimen | Control | 0,056989 |
| 3 months | abundance | X06859 | Methylol D | 3 | Interventio | Experimen | Experimen | Control | 0,697678 |
| 3 months | abundance | X06861 | 4-(4-Deoxy | 3 | Interventio | Experimen | Experimen | Control | 0,894218 |
| 3 months | abundance | X06866 | tert-Butyl 3 | 3 | Interventio | Experimen | Experimen | Control | 0,043633 |
| 3 months | abundance | X06870 | Menadiol | 3 | Interventio | Experimen | Experimen | Control | 0,206211 |
| 3 months | abundance | X06874 | N-Acetyl-5 | 3 | Interventio | Experimen | Experimen | Control | 0,119178 |
| 3 months | abundance | X06875 | Nicotine gl | 3 | Interventio | Experimen | Experimen | Control | 0,909234 |
| 3 months | abundance | X06880 | pentobarbi | 3 | Interventio | Experimen | Experimen | Control | 0,029867 |
| 3 months | abundance | X06883 | Agomelatir | 3 | Interventio | Experimen | Experimen | Control | 0,948674 |
| 3 months | abundance | X06889 | 3,4-Dimetl | 2a | Interventio | Experimen | Experimen | Control | 0,968857 |
| 3 months | abundance | X06896 | Pirbuterol | 3 | Interventio | Experimen | Experimen | Control | 0,642549 |

| | | | | | | | | | |
|---|---|---|---|---|---|---|---|---|---|
| 3 months | abundance | X06902 | Homocyste | 3 | Interventio | Experimen | Experimen | Control | 0,867667 |
| 3 months | abundance | X06903 | Esculin | 2b | Interventio | Experimen | Experimen | Control | 0,515877 |
| 3 months | abundance | X06905 | Ectoine | 3 | Interventio | Experimen | Experimen | Control | 0,997328 |
| 3 months | abundance | X06906 | Arg-pro | 3 | Interventio | Experimen | Experimen | Control | 0,503169 |
| 3 months | abundance | X06917 | Alanyltrypt | 3 | Interventio | Experimen | Experimen | Control | 0,131627 |
| 3 months | abundance | X06933 | 3-Methoxy- | 3 | Interventio | Experimen | Experimen | Control | 0,968857 |
| 3 months | abundance | X06943 | NSC 9277 | 3 | Interventio | Experimen | Experimen | Control | 0,404129 |
| 3 months | abundance | X06946 | SECONAL_ | 3 | Interventio | Experimen | Experimen | Control | 0,58814 |
| 3 months | abundance | X06948 | (1R,3R,5R) | 3 | Interventio | Experimen | Experimen | Control | 0,86519 |
| 3 months | abundance | X06958 | 2-[(carboxy | 2b | Interventio | Experimen | Experimen | Control | 0,000193 |
| 3 months | abundance | X06961 | Zalcitabine | 3 | Interventio | Experimen | Experimen | Control | 0,892548 |
| 3 months | abundance | X06977 | 4-Hydroxyp | 3 | Interventio | Experimen | Experimen | Control | 0,988551 |
| 3 months | abundance | X06985 | N-Ethylpro | 3 | Interventio | Experimen | Experimen | Control | 0,116825 |
| 3 months | abundance | X07002 | Paraldehyc | 3 | Interventio | Experimen | Experimen | Control | 0,980029 |
| 3 months | abundance | X07013 | pimethixer | 3 | Interventio | Experimen | Experimen | Control | 0,21959 |
| 3 months | abundance | X07014 | Octyl benz | 3 | Interventio | Experimen | Experimen | Control | 0,821708 |
| 3 months | abundance | X07026 | 7α-Hydroxy | 2b | Interventio | Experimen | Experimen | Control | 0,820408 |
| 3 months | abundance | X07027 | Allyl merca | 3 | Interventio | Experimen | Experimen | Control | 0,886266 |
| 3 months | abundance | X07028 | N-Pentano | 3 | Interventio | Experimen | Experimen | Control | 0,947336 |
| 3 months | abundance | X07038 | Piperidine_ | 3 | Interventio | Experimen | Experimen | Control | 0,207459 |
| 3 months | abundance | X07040 | 6-Hydroxyp | 3 | Interventio | Experimen | Experimen | Control | 0,988551 |
| 3 months | abundance | X07050 | 2-Methylth | 3 | Interventio | Experimen | Experimen | Control | 0,22934 |
| 3 months | abundance | X07051 | O-succinyl | 3 | Interventio | Experimen | Experimen | Control | 0,655638 |
| 3 months | abundance | X07054 | Valylprolin | 2b | Interventio | Experimen | Experimen | Control | 0,223785 |
| 3 months | abundance | X07057 | Leu-Leu_c | 3 | Interventio | Experimen | Experimen | Control | 0,000943 |
| 3 months | abundance | X07073 | asn-val_a | 3 | Interventio | Experimen | Experimen | Control | 0,468958 |
| 3 months | abundance | X07077 | Hyodeoxyc | 1 | Interventio | Experimen | Experimen | Control | 0,314021 |
| 3 months | abundance | X07079 | N-Pentano | 3 | Interventio | Experimen | Experimen | Control | 0,854716 |
| 3 months | abundance | X07081 | N-LACTOYI | 3 | Interventio | Experimen | Experimen | Control | 0,274678 |
| 3 months | abundance | X07084 | heptabarbi | 3 | Interventio | Experimen | Experimen | Control | 0,867667 |
| 3 months | abundance | X07089 | Pregabalin | 3 | Interventio | Experimen | Experimen | Control | 0,992999 |
| 3 months | abundance | X07092 | Dibutyl ma | 3 | Interventio | Experimen | Experimen | Control | 0,541151 |
| 3 months | abundance | X07107 | Lysylvaline | 3 | Interventio | Experimen | Experimen | Control | 0,004656 |
| 3 months | abundance | X07112 | MFCD0272 | 3 | Interventio | Experimen | Experimen | Control | 0,270117 |
| 3 months | abundance | X07113 | Sulfoaceta | 3 | Interventio | Experimen | Experimen | Control | 0,685901 |
| 3 months | abundance | X07125 | Zalcitabine | 3 | Interventio | Experimen | Experimen | Control | 0,823267 |
| 3 months | abundance | X07126 | Leu-Val_f | 3 | Interventio | Experimen | Experimen | Control | 0,124404 |
| 3 months | abundance | X07127 | N~5~-[P-A | 3 | Interventio | Experimen | Experimen | Control | 0,997328 |
| 3 months | abundance | X07134 | 4-Phenolsu | 2b | Interventio | Experimen | Experimen | Control | 0,44396 |
| 3 months | abundance | X07136 | hexobarbit | 3 | Interventio | Experimen | Experimen | Control | 0,972998 |
| 3 months | abundance | X07139 | N,N-Diethy | 3 | Interventio | Experimen | Experimen | Control | 0,232653 |
| 3 months | abundance | X07146 | meprobam | 3 | Interventio | Experimen | Experimen | Control | 0,988551 |
| 3 months | abundance | X07155 | Naphthale | 3 | Interventio | Experimen | Experimen | Control | 0,952532 |
| 3 months | abundance | X07164 | N-Acetyl-5 | 3 | Interventio | Experimen | Experimen | Control | 0,024021 |
| 3 months | abundance | X07165 | pro-gln_b | 3 | Interventio | Experimen | Experimen | Control | 0,613297 |
| 3 months | abundance | X07168 | L-(+)-Eryth | 3 | Interventio | Experimen | Experimen | Control | 0,172075 |
| 3 months | abundance | X07193 | N-(1-Methy | 3 | Interventio | Experimen | Experimen | Control | 0,651767 |
| 3 months | abundance | X07201 | 2,4-Quinol | 2b | Interventio | Experimen | Experimen | Control | 0,530275 |

| | | | | | | | | | |
|---|---|---|---|---|---|---|---|---|---|
| 3 months | abundance | X07216 | pentobarbi | 3 | Interventio | Experimen | Experimen | Control | 0,988551 |
| 3 months | abundance | X07219 | 3,3-Dimeth | 2b | Interventio | Experimen | Experimen | Control | 0,808747 |
| 3 months | abundance | X07220 | Glycylproly | 3 | Interventio | Experimen | Experimen | Control | 0,836327 |
| 3 months | abundance | X07226 | threonylph | 3 | Interventio | Experimen | Experimen | Control | 0,824533 |
| 3 months | abundance | X07236 | 2-methylci | 3 | Interventio | Experimen | Experimen | Control | 0,988551 |
| 3 months | abundance | X07250 | Leucylaspa | 3 | Interventio | Experimen | Experimen | Control | 0,988551 |
| 3 months | abundance | X07260 | (7E,7'E)-5, | 3 | Interventio | Experimen | Experimen | Control | 0,748275 |
| 3 months | abundance | X07263 | 3,3-Dimeth | 2b | Interventio | Experimen | Experimen | Control | 0,798192 |
| 3 months | abundance | X07269 | (3aR,4R,5R | 3 | Interventio | Experimen | Experimen | Control | 0,433847 |
| 3 months | abundance | X07270 | 5-(2-Carbo | 3 | Interventio | Experimen | Experimen | Control | 0,149754 |
| 3 months | abundance | X07278 | 5-Phospho | 3 | Interventio | Experimen | Experimen | Control | 0,631492 |
| 3 months | abundance | X07283 | 8-Hydroxyh | 3 | Interventio | Experimen | Experimen | Control | 0,102756 |
| 3 months | abundance | X07285 | 2-(Carboxy | 3 | Interventio | Experimen | Experimen | Control | 0,141328 |
| 3 months | abundance | X07294 | 6-Acetamid | 3 | Interventio | Experimen | Experimen | Control | 0,325689 |
| 3 months | abundance | X07303 | Triacetin | 3 | Interventio | Experimen | Experimen | Control | 0,317237 |
| 3 months | abundance | X07308 | Nicotinic a | 2a | Interventio | Experimen | Experimen | Control | 0,409881 |
| 3 months | abundance | X07309 | 3,4,15-Trih | 3 | Interventio | Experimen | Experimen | Control | 0,633187 |
| 3 months | abundance | X07315 | Rivastigmi | 3 | Interventio | Experimen | Experimen | Control | 0,384971 |
| 3 months | abundance | X07320 | asn-pro_d | 3 | Interventio | Experimen | Experimen | Control | 0,456954 |
| 3 months | abundance | X07327 | Aspartyl-L- | 3 | Interventio | Experimen | Experimen | Control | 0,854716 |
| 3 months | abundance | X07334 | 2,2-Bis(hyd | 3 | Interventio | Experimen | Experimen | Control | 0,324406 |
| 3 months | abundance | X07336 | Threonylse | 3 | Interventio | Experimen | Experimen | Control | 0,830535 |
| 3 months | abundance | X07344 | (−)-nabilor | 3 | Interventio | Experimen | Experimen | Control | 0,755448 |
| 3 months | abundance | X07345 | Phenyl D-g | 3 | Interventio | Experimen | Experimen | Control | 0,661083 |
| 3 months | abundance | X07375 | porphobili | 3 | Interventio | Experimen | Experimen | Control | 0,898308 |
| 3 months | abundance | X07381 | Bis-D-fruct | 3 | Interventio | Experimen | Experimen | Control | 0,014081 |
| 3 months | abundance | X07394 | Hydroxypro | 3 | Interventio | Experimen | Experimen | Control | 0,000235 |
| 3 months | abundance | X07399 | Homoanse | 3 | Interventio | Experimen | Experimen | Control | 0,273699 |
| 3 months | abundance | X07400 | Tranexami | 2b | Interventio | Experimen | Experimen | Control | 0,440554 |
| 3 months | abundance | X07413 | Chenodeox | 1 | Interventio | Experimen | Experimen | Control | 0,15162 |
| 3 months | abundance | X07414 | 2,2-Bis(hyd | 3 | Interventio | Experimen | Experimen | Control | 0,115261 |
| 3 months | abundance | X07417 | δ-Valerolac | 2b | Interventio | Experimen | Experimen | Control | 0,836327 |
| 3 months | abundance | X07420 | DNOP_h | 3 | Interventio | Experimen | Experimen | Control | 0,819081 |
| 3 months | abundance | X07424 | Rutinose (b | 3 | Interventio | Experimen | Experimen | Control | 0,860527 |
| 3 months | abundance | X07425 | N-Acetylas | 2a | Interventio | Experimen | Experimen | Control | 0,056989 |
| 3 months | abundance | X07428 | N6-METHY | 3 | Interventio | Experimen | Experimen | Control | 0,216079 |
| 3 months | abundance | X07429 | DNOP_c | 3 | Interventio | Experimen | Experimen | Control | 0,579428 |
| 3 months | abundance | X07434 | N'-Hydroxy | 2b | Interventio | Experimen | Experimen | Control | 0,945538 |
| 3 months | abundance | X07439 | 4-pyridoxic | 1 | Interventio | Experimen | Experimen | Control | 0,052565 |
| 3 months | abundance | X07442 | coenzyme | 3 | Interventio | Experimen | Experimen | Control | 0,817974 |
| 3 months | abundance | X07443 | Butylparab | 3 | Interventio | Experimen | Experimen | Control | 0,514326 |
| 3 months | abundance | X07445 | 3-(Sulfooxy | 3 | Interventio | Experimen | Experimen | Control | 0,988551 |
| 3 months | abundance | X07451 | Valylvaline | 3 | Interventio | Experimen | Experimen | Control | 0,949849 |
| 3 months | abundance | X07452 | 4-Amino-1- | 3 | Interventio | Experimen | Experimen | Control | 0,99116 |
| 3 months | abundance | X07454 | 2,4-Quinol | 2b | Interventio | Experimen | Experimen | Control | 0,988551 |
| 3 months | abundance | X07457 | Glucosami | 1 | Interventio | Experimen | Experimen | Control | 0,14094 |
| 3 months | abundance | X07460 | 1-pyrroline | 3 | Interventio | Experimen | Experimen | Control | 0,404129 |
| 3 months | abundance | X07465 | TO0127900 | 3 | Interventio | Experimen | Experimen | Control | 0,988551 |

| | | | | | | | | | |
|---|---|---|---|---|---|---|---|---|---|
| 3 months | abundance | X07469 | 1,1'-[1,12- | 3 | Interventio | Experimen | Experimen | Control | 0,260224 |
| 3 months | abundance | X07475 | 2'-Deoxy-5 | 3 | Interventio | Experimen | Experimen | Control | 0,935609 |
| 3 months | abundance | X07476 | 4-Hydroxyb | 1 | Interventio | Experimen | Experimen | Control | 0,821708 |
| 3 months | abundance | X07480 | Naringenin | 2a | Interventio | Experimen | Experimen | Control | 0,532452 |
| 3 months | abundance | X07484 | MFCD0002 | 3 | Interventio | Experimen | Experimen | Control | 0,377363 |
| 3 months | abundance | X07487 | Oxypeucec | 3 | Interventio | Experimen | Experimen | Control | 0,762066 |
| 3 months | abundance | X07491 | 1-Vinylimic | 2b | Interventio | Experimen | Experimen | Control | 0,908976 |
| 3 months | abundance | X07504 | 4-Acetamic | 2b | Interventio | Experimen | Experimen | Control | 0,88099 |
| 3 months | abundance | X07508 | Procaine_a | 2b | Interventio | Experimen | Experimen | Control | 0,250053 |
| 3 months | abundance | X07511 | Piperidine_ | 3 | Interventio | Experimen | Experimen | Control | 0,744844 |
| 3 months | abundance | X07512 | 4-{[(3-Hydr | 3 | Interventio | Experimen | Experimen | Control | 0,314021 |
| 3 months | abundance | X07513 | (−)-nabilor | 3 | Interventio | Experimen | Experimen | Control | 0,698315 |
| 3 months | abundance | X07514 | L-(+)-Eryth | 3 | Interventio | Experimen | Experimen | Control | 0,10419 |
| 3 months | abundance | X07518 | (+/-)-2-Hyc | 3 | Interventio | Experimen | Experimen | Control | 0,106066 |
| 3 months | abundance | X07519 | Pyrrolidine | 3 | Interventio | Experimen | Experimen | Control | 0,817974 |
| 3 months | abundance | X07523 | Ursodeoxy | 1 | Interventio | Experimen | Experimen | Control | 0,429858 |
| 3 months | abundance | X07524 | Prolinamid | 2b | Interventio | Experimen | Experimen | Control | 0,593214 |
| 3 months | abundance | X07527 | 1,7-Dimetl | 2b | Interventio | Experimen | Experimen | Control | 0,960119 |
| 3 months | abundance | X07530 | Alanine | 1 | Interventio | Experimen | Experimen | Control | 0,854716 |
| 3 months | abundance | X07537 | alpha-Cha | 3 | Interventio | Experimen | Experimen | Control | 0,135093 |
| 3 months | abundance | X07538 | Homoanse | 3 | Interventio | Experimen | Experimen | Control | 0,052565 |
| 3 months | abundance | X07541 | Saccharin | 1 | Interventio | Experimen | Experimen | Control | 0,503169 |
| 3 months | abundance | X07544 | Ethyl sulfa | 3 | Interventio | Experimen | Experimen | Control | 0,439076 |
| 3 months | abundance | X07558 | Leucylproli | 2b | Interventio | Experimen | Experimen | Control | 0,082572 |
| 3 months | abundance | X07563 | Xanthine | 1 | Interventio | Experimen | Experimen | Control | 0,884928 |
| 3 months | abundance | X07565 | 5-Hydroxyi | 2b | Interventio | Experimen | Experimen | Control | 0,042091 |
| 3 months | abundance | X07566 | 5-Allyl-5-se | 3 | Interventio | Experimen | Experimen | Control | 0,817974 |
| 3 months | abundance | X07572 | 8-(3-Furyl)- | 3 | Interventio | Experimen | Experimen | Control | 0,744844 |
| 3 months | abundance | X07574 | 7-ketodeox | 3 | Interventio | Experimen | Experimen | Control | 0,988551 |
| 3 months | abundance | X07577 | 2-[(Sulfoox | 3 | Interventio | Experimen | Experimen | Control | 0,681513 |
| 3 months | abundance | X07578 | 5-Allyl-5-se | 3 | Interventio | Experimen | Experimen | Control | 0,956306 |
| 3 months | abundance | X07580 | N-Pentano | 3 | Interventio | Experimen | Experimen | Control | 0,333178 |
| 3 months | abundance | X07581 | N-Acetylva | 3 | Interventio | Experimen | Experimen | Control | 0,513969 |
| 3 months | abundance | X07582 | 4-methylpy | 2b | Interventio | Experimen | Experimen | Control | 0,96692 |
| 3 months | abundance | X07596 | 5-amino-2- | 2b | Interventio | Experimen | Experimen | Control | 0,835202 |
| 3 months | abundance | X07597 | p-Cresylsu | 3 | Interventio | Experimen | Experimen | Control | 0,990949 |
| 3 months | abundance | X07601 | Asp-lys | 3 | Interventio | Experimen | Experimen | Control | 0,656167 |
| 3 months | abundance | X07613 | 1-(4-Aminc | 3 | Interventio | Experimen | Experimen | Control | 0,483229 |
| 3 months | abundance | X07618 | N-(3-aceta | 3 | Interventio | Experimen | Experimen | Control | 0,833088 |
| 3 months | abundance | X07630 | Methionine | 1 | Interventio | Experimen | Experimen | Control | 0,617664 |
| 3 months | abundance | X07634 | 3,8,9-trihy | 2b | Interventio | Experimen | Experimen | Control | 0,141328 |
| 3 months | abundance | X07638 | 7-ketodeox | 3 | Interventio | Experimen | Experimen | Control | 0,651767 |
| 3 months | abundance | X07645 | O-propeno | 3 | Interventio | Experimen | Experimen | Control | 0,968857 |
| 3 months | abundance | X07646 | (2Z)-2-({[(6 | 3 | Interventio | Experimen | Experimen | Control | 0,834104 |
| 3 months | abundance | X07647 | Genistein | 1 | Interventio | Experimen | Experimen | Control | 0,023171 |
| 3 months | abundance | X07648 | NL851300 | 3 | Interventio | Experimen | Experimen | Control | 0,886266 |
| 3 months | abundance | X07657 | Docosahex | 2b | Interventio | Experimen | Experimen | Control | 0,323676 |
| 3 months | abundance | X07658 | δ-Glucono | 1 | Interventio | Experimen | Experimen | Control | 0,469877 |

| | | | | | | | | | |
|---|---|---|---|---|---|---|---|---|---|
| 3 months | abundance | X07674 | Lysine | 1 | Interventio | Experimen | Experimen | Control | 0,26602 |
| 3 months | abundance | X07678 | (2E)-3-(3,4 | 2b | Interventio | Experimen | Experimen | Control | 0,322462 |
| 3 months | abundance | X07680 | Indole-3-a | 1 | Interventio | Experimen | Experimen | Control | 0,84521 |
| 3 months | abundance | X07694 | DNOP_a | 3 | Interventio | Experimen | Experimen | Control | 0,985356 |
| 3 months | abundance | X07699 | N-(1-Methy | 3 | Interventio | Experimen | Experimen | Control | 0,915997 |
| 3 months | abundance | X07701 | 3-Benzyl-6 | 3 | Interventio | Experimen | Experimen | Control | 0,505748 |
| 3 months | abundance | X07710 | 3-O-beta-D | 3 | Interventio | Experimen | Experimen | Control | 0,755239 |
| 3 months | abundance | X07712 | paracetam | 3 | Interventio | Experimen | Experimen | Control | 0,635505 |
| 3 months | abundance | X07731 | 3-[4-methy | 2b | Interventio | Experimen | Experimen | Control | 0,124383 |
| 3 months | abundance | X07732 | N-Acetylva | 2b | Interventio | Experimen | Experimen | Control | 0,923057 |
| 3 months | abundance | X07743 | D-Alanine | 2b | Interventio | Experimen | Experimen | Control | 0,694902 |
| 3 months | abundance | X07745 | APM_c | 3 | Interventio | Experimen | Experimen | Control | 0,691158 |
| 3 months | abundance | X07746 | 3-(2-Oxo-2 | 2b | Interventio | Experimen | Experimen | Control | 0,718544 |
| 3 months | abundance | X07747 | Diethylpyr | 3 | Interventio | Experimen | Experimen | Control | 0,108736 |
| 3 months | abundance | X07749 | (DL)-3-O-M | 3 | Interventio | Experimen | Experimen | Control | 0,632225 |
| 3 months | abundance | X07753 | Lovastatin_ | 3 | Interventio | Experimen | Experimen | Control | 0,755448 |
| 3 months | abundance | X07754 | hexobarbit | 3 | Interventio | Experimen | Experimen | Control | 0,787558 |
| 3 months | abundance | X07764 | (S)-2-meth | 3 | Interventio | Experimen | Experimen | Control | 0,833128 |
| 3 months | abundance | X07770 | Piperine | 1 | Interventio | Experimen | Experimen | Control | 0,798841 |
| 3 months | abundance | X07771 | Caprolacta | 2b | Interventio | Experimen | Experimen | Control | 0,60023 |
| 3 months | abundance | X07774 | 7alpha-Hy | 3 | Interventio | Experimen | Experimen | Control | 0,593214 |
| 3 months | abundance | X07777 | Arginine | 1 | Interventio | Experimen | Experimen | Control | 0,980029 |
| 3 months | abundance | X07782 | Cytosine | 1 | Interventio | Experimen | Experimen | Control | 0,172283 |
| 3 months | abundance | X07783 | 1,3-dimeth | 3 | Interventio | Experimen | Experimen | Control | 0,886266 |
| 3 months | abundance | X07792 | N-Acetylhi | 2b | Interventio | Experimen | Experimen | Control | 0,867667 |
| 3 months | abundance | X07794 | Prolylleuci | 2b | Interventio | Experimen | Experimen | Control | 0,867667 |
| 3 months | abundance | X07798 | N-lauroylgl | 3 | Interventio | Experimen | Experimen | Control | 0,713464 |
| 3 months | abundance | X07799 | Capryloylg | 2b | Interventio | Experimen | Experimen | Control | 0,329628 |
| 3 months | abundance | X07811 | Metirosine | 3 | Interventio | Experimen | Experimen | Control | 0,835202 |
| 3 months | abundance | X07812 | Triethyl citr | 3 | Interventio | Experimen | Experimen | Control | 0,718544 |
| 3 months | abundance | X07815 | 3-[2-[(Z)-[3 | 3 | Interventio | Experimen | Experimen | Control | 0,260022 |
| 3 months | abundance | X07820 | Creatinine | 1 | Interventio | Experimen | Experimen | Control | 0,238218 |
| 3 months | abundance | X07825 | Maltotriose | 1 | Interventio | Experimen | Experimen | Control | 0,009306 |
| 3 months | abundance | X07830 | 1,5-Isoquir | 2b | Interventio | Experimen | Experimen | Control | 0,458416 |
| 3 months | abundance | X07834 | Choline su | 3 | Interventio | Experimen | Experimen | Control | 0,907733 |
| 3 months | abundance | X07835 | 7-ketodeox | 3 | Interventio | Experimen | Experimen | Control | 0,659543 |
| 3 months | abundance | X07841 | 2-(5-Benzy | 3 | Interventio | Experimen | Experimen | Control | 0,718544 |
| 3 months | abundance | X07843 | Piperidine_ | 3 | Interventio | Experimen | Experimen | Control | 0,115261 |
| 3 months | abundance | X07846 | Carbofurar | 3 | Interventio | Experimen | Experimen | Control | 0,161121 |
| 3 months | abundance | X07848 | (−)-nabilor | 3 | Interventio | Experimen | Experimen | Control | 0,633187 |
| 3 months | abundance | X07849 | Crotamitor | 3 | Interventio | Experimen | Experimen | Control | 0,142786 |
| 3 months | abundance | X07852 | N6,N6,N6- | 3 | Interventio | Experimen | Experimen | Control | 0,503169 |
| 3 months | abundance | X07854 | TO0127900 | 3 | Interventio | Experimen | Experimen | Control | 4,85E-05 |

| | | | | | | | | | |
|---|---|---|---|---|---|---|---|---|---|
| 3 months | abundance | X07856 | ,6R,7R,9R,11R,12R,13S,14R)-6-{[(2S,3R,4S,6R)-4-(Dimethylamino)-3-hydroxy-6-methyltetrahydro-2H-pyran-2-yl]oxy}-12,13-dihydroxy-14-[(1S)-1-hydroxyethyl]-4-{[(2R,4R,5S,6S)-5-hydroxy-4-methoxy-4,6-dimethyltetrahydro-2H-pyran-2-yl]oxy}-7- | 3 | Interventio | Experimen | Experimen | Control | 0,222442 |
| 3 months | abundance | X07866 | 3,3,5,5-Tet | 2b | Interventio | Experimen | Experimen | Control | 0,001034 |
| 3 months | abundance | X07867 | N-Acetylva | 3 | Interventio | Experimen | Experimen | Control | 0,083448 |
| 3 months | abundance | X07868 | Biotin | 2b | Interventio | Experimen | Experimen | Control | 0,997328 |
| 3 months | abundance | X07869 | Hexamethy | 2a | Interventio | Experimen | Experimen | Control | 0,081509 |
| 3 months | abundance | X07873 | (3Z,6Z,9Z,1 | 3 | Interventio | Experimen | Experimen | Control | 0,392246 |
| 3 months | abundance | X07874 | δ-Valerolac | 2b | Interventio | Experimen | Experimen | Control | 0,985504 |
| 3 months | abundance | X07886 | L-Urobilin | 3 | Interventio | Experimen | Experimen | Control | 0,900417 |
| 3 months | abundance | X07889 | UROBILIN, | 3 | Interventio | Experimen | Experimen | Control | 0,996617 |
| 3 months | abundance | X07890 | N-(5-aceta | 2b | Interventio | Experimen | Experimen | Control | 0,680872 |
| 3 months | abundance | X07891 | Cyclamic a | 2b | Interventio | Experimen | Experimen | Control | 0,867667 |
| 3 months | abundance | X07892 | urobilinoge | 3 | Interventio | Experimen | Experimen | Control | 0,909722 |
| 3 months | abundance | X07899 | N-Acetylpu | 2b | Interventio | Experimen | Experimen | Control | 0,755448 |
| 3 months | abundance | X07900 | presqualer | 3 | Interventio | Experimen | Experimen | Control | 0,021235 |
| 3 months | abundance | X07903 | Acetophen | 2b | Interventio | Experimen | Experimen | Control | 0,985504 |
| 3 months | abundance | X07905 | Prilocaine | 2b | Interventio | Experimen | Experimen | Control | 0,113035 |
| 3 months | abundance | X07907 | 4-Acetami | 1 | Interventio | Experimen | Experimen | Control | 0,821708 |
| 3 months | abundance | X07909 | (±)-Albuter | 2b | Interventio | Experimen | Experimen | Control | 0,620467 |
| 3 months | abundance | X07914 | (-)-Erythror | 3 | Interventio | Experimen | Experimen | Control | 0,231572 |
| 3 months | abundance | X07916 | N6,N6,N6- | 3 | Interventio | Experimen | Experimen | Control | 5,47E-05 |

| | | | | | | | | | |
|---|---|---|---|---|---|---|---|---|---|
| 3 months | abundance | X07921 | Pipecolinic | 1 | Interventio | Experimen | Experimen | Control | 0,000266 |
| 3 months | abundance | X07925 | DNOP_f | 3 | Interventio | Experimen | Experimen | Control | 0,656554 |
| 3 months | abundance | X07930 | N-Methylca | 2b | Interventio | Experimen | Experimen | Control | 0,064289 |
| 3 months | abundance | X07935 | Limonin | 3 | Interventio | Experimen | Experimen | Control | 0,997328 |
| 3 months | abundance | X07937 | Atenolol | 1 | Interventio | Experimen | Experimen | Control | 0,712596 |
| 3 months | abundance | X07944 | Styrene | 3 | Interventio | Experimen | Experimen | Control | 0,725856 |
| 3 months | abundance | X07955 | Valylvaline | 3 | Interventio | Experimen | Experimen | Control | 0,860527 |
| 3 months | abundance | X07961 | 2-(Hydroxy | 3 | Interventio | Experimen | Experimen | Control | 0,560103 |
| 3 months | abundance | X07963 | Theobromi | 1 | Interventio | Experimen | Experimen | Control | 0,817715 |
| 3 months | abundance | X07969 | 6-hydroxyp | 3 | Interventio | Experimen | Experimen | Control | 0,265534 |
| 3 months | abundance | X07974 | Isoprene | 3 | Interventio | Experimen | Experimen | Control | 0,276947 |
| 3 months | abundance | X07977 | Tyramine | 2b | Interventio | Experimen | Experimen | Control | 0,94858 |
| 3 months | abundance | X07978 | 1-[(4E)-4-( | 3 | Interventio | Experimen | Experimen | Control | 0,325689 |
| 3 months | abundance | X07981 | Methylimic | 2b | Interventio | Experimen | Experimen | Control | 0,817974 |
| 3 months | abundance | X07983 | butalbital_ | 3 | Interventio | Experimen | Experimen | Control | 0,297127 |
| 3 months | abundance | X07989 | Piperidine_ | 3 | Interventio | Experimen | Experimen | Control | 0,448277 |
| 3 months | abundance | X07993 | UROBILIN, | 3 | Interventio | Experimen | Experimen | Control | 0,99116 |
| 3 months | abundance | X08002 | (2S)-6-Ami | 3 | Interventio | Experimen | Experimen | Control | 0,506018 |
| 3 months | abundance | X08004 | δ-Valerola | 2b | Interventio | Experimen | Experimen | Control | 0,988551 |
| 3 months | abundance | X08007 | MFCD0002 | 3 | Interventio | Experimen | Experimen | Control | 0,000518 |
| 3 months | abundance | X08008 | N-Acetylne | 1 | Interventio | Experimen | Experimen | Control | 0,086617 |
| 3 months | abundance | X08012 | Methylol D | 3 | Interventio | Experimen | Experimen | Control | 0,242449 |
| 3 months | abundance | X08013 | Deoxysuga | 1 | Interventio | Experimen | Experimen | Control | 0,049638 |
| 3 months | abundance | X08014 | DNOP_e | 3 | Interventio | Experimen | Experimen | Control | 0,703544 |
| 3 months | abundance | X08017 | Tropinone | 2b | Interventio | Experimen | Experimen | Control | 0,750825 |
| 3 months | abundance | X08019 | Thymidine | 1 | Interventio | Experimen | Experimen | Control | 0,082877 |
| 3 months | abundance | X08024 | 8-Methyl-8 | 3 | Interventio | Experimen | Experimen | Control | 0,124404 |
| 3 months | abundance | X08028 | Solanidine | 3 | Interventio | Experimen | Experimen | Control | 0,080062 |
| 3 months | abundance | X08035 | Tetrahydro | 3 | Interventio | Experimen | Experimen | Control | 0,75812 |
| 3 months | abundance | X08037 | Crotonic a | 2b | Interventio | Experimen | Experimen | Control | 0,892974 |
| 3 months | abundance | X08038 | N-{6-[(7-Cl | 2b | Interventio | Experimen | Experimen | Control | 0,85207 |
| 3 months | abundance | X08040 | Prolylleuci | 2b | Interventio | Experimen | Experimen | Control | 0,98336 |
| 3 months | abundance | X08044 | 3-[2-[(Z)-[3 | 3 | Interventio | Experimen | Experimen | Control | 0,633187 |
| 3 months | abundance | X08046 | 3-Hydroxy- | 3 | Interventio | Experimen | Experimen | Control | 0,614079 |
| 3 months | abundance | X08054 | Docosahex | 2b | Interventio | Experimen | Experimen | Control | 0,694902 |
| 3 months | abundance | X08056 | 3-(2-Oxo-2 | 2b | Interventio | Experimen | Experimen | Control | 0,333178 |
| 3 months | abundance | X08057 | N-{3-Carbo | 3 | Interventio | Experimen | Experimen | Control | 0,9752 |
| 3 months | abundance | X08058 | 2-Hydroxy- | 3 | Interventio | Experimen | Experimen | Control | 0,879562 |
| 3 months | abundance | X08059 | Cadaverine | 3 | Interventio | Experimen | Experimen | Control | 0,082877 |
| 3 months | abundance | X08062 | Indole-3-ca | 1 | Interventio | Experimen | Experimen | Control | 0,436028 |
| 3 months | abundance | X08072 | 4-Hydroxyp | 3 | Interventio | Experimen | Experimen | Control | 0,021942 |
| 3 months | abundance | X08076 | 3',5,7-Trihy | 2b | Interventio | Experimen | Experimen | Control | 0,613297 |
| 3 months | abundance | X08077 | 6-(alpha-D | 3 | Interventio | Experimen | Experimen | Control | 0,581247 |
| 3 months | abundance | X08078 | Ethyl mala | 3 | Interventio | Experimen | Experimen | Control | 0,915997 |
| 3 months | abundance | X08092 | Safrole | 3 | Interventio | Experimen | Experimen | Control | 0,94878 |
| 3 months | abundance | X08094 | 1,1'-[1,12- | 3 | Interventio | Experimen | Experimen | Control | 0,503169 |
| 3 months | abundance | X08095 | DNOP_g | 3 | Interventio | Experimen | Experimen | Control | 0,821708 |
| 3 months | abundance | X08096 | (2E)-3-Met | 3 | Interventio | Experimen | Experimen | Control | 0,625098 |

| | | | | | | | | | |
|---|---|---|---|---|---|---|---|---|---|
| 3 months | abundance | X08098 | Citrulline | 1 | Interventio | Experimen | Experimen | Control | 0,30026 |
| 3 months | abundance | X08099 | g-Butyrobe | 3 | Interventio | Experimen | Experimen | Control | 0,702384 |
| 3 months | abundance | X08100 | Piperidine_ | 3 | Interventio | Experimen | Experimen | Control | 0,001936 |
| 3 months | abundance | X08103 | Isopelletier | 3 | Interventio | Experimen | Experimen | Control | 0,399618 |
| 3 months | abundance | X08105 | 2-Acetami | 3 | Interventio | Experimen | Experimen | Control | 0,915366 |
| 3 months | abundance | X08106 | 4-Vinylphe | 1 | Interventio | Experimen | Experimen | Control | 0,581247 |
| 3 months | abundance | X08111 | 2-Hydroxy | 2b | Interventio | Experimen | Experimen | Control | 0,552202 |
| 3 months | abundance | X08116 | Methyl (2Z, | 3 | Interventio | Experimen | Experimen | Control | 0,003352 |
| 3 months | abundance | X08123 | Tiglic acid_ | 2b | Interventio | Experimen | Experimen | Control | 0,765006 |
| 3 months | abundance | X08146 | O-Ethyl (4- | 3 | Interventio | Experimen | Experimen | Control | 0,98719 |
| 3 months | abundance | X08147 | porphobilir | 3 | Interventio | Experimen | Experimen | Control | 0,935319 |
| 3 months | abundance | X08150 | DL-4-Hydro | 2b | Interventio | Experimen | Experimen | Control | 0,404129 |
| 3 months | abundance | X08154 | 4-(METHYL | 3 | Interventio | Experimen | Experimen | Control | 0,752586 |
| 3 months | abundance | X08157 | Leu-Val_e | 3 | Interventio | Experimen | Experimen | Control | 0,787558 |
| 3 months | abundance | X08162 | Valylvaline | 3 | Interventio | Experimen | Experimen | Control | 0,244161 |
| 3 months | abundance | X08163 | Phenyl D-g | 3 | Interventio | Experimen | Experimen | Control | 0,613297 |
| 3 months | abundance | X08167 | Lanthionin | 3 | Interventio | Experimen | Experimen | Control | 0,993803 |
| 3 months | abundance | X08173 | 9-Methylur | 2b | Interventio | Experimen | Experimen | Control | 0,718544 |
| 3 months | abundance | X08188 | 2-Hydroxy- | 2b | Interventio | Experimen | Experimen | Control | 0,681178 |
| 3 months | abundance | X08193 | Seryltyrosi | 3 | Interventio | Experimen | Experimen | Control | 0,761041 |
| 3 months | abundance | X08204 | 2-Amino-4 | 2b | Interventio | Experimen | Experimen | Control | 0,038414 |
| 3 months | abundance | X08212 | LU345300 | 3 | Interventio | Experimen | Experimen | Control | 0,070241 |
| 3 months | abundance | X08221 | Brilliant blu | 2b | Interventio | Experimen | Experimen | Control | 0,238218 |
| 3 months | abundance | X08230 | Coumaron | 3 | Interventio | Experimen | Experimen | Control | 0,46992 |
| 3 months | abundance | X08235 | 1-Methylhi | 2b | Interventio | Experimen | Experimen | Control | 0,821708 |
| 3 months | abundance | X08244 | Setoclavin | 3 | Interventio | Experimen | Experimen | Control | 0,800324 |
| 3 months | abundance | X08270 | Butabarbit | 3 | Interventio | Experimen | Experimen | Control | 0,787558 |
| 3 months | abundance | X08274 | Mevalonic | 2b | Interventio | Experimen | Experimen | Control | 0,244993 |
| 3 months | abundance | X08277 | adrenaline | 3 | Interventio | Experimen | Experimen | Control | 0,703692 |
| 3 months | abundance | X08279 | DNOP_b | 3 | Interventio | Experimen | Experimen | Control | 0,90642 |
| 3 months | abundance | X08288 | Capryloylg | 3 | Interventio | Experimen | Experimen | Control | 0,907733 |
| 3 months | abundance | X08292 | Lidocaine | 2b | Interventio | Experimen | Experimen | Control | 0,778801 |
| 3 months | abundance | X08298 | N6,N6,N6- | 2b | Interventio | Experimen | Experimen | Control | 0,333178 |
| 3 months | abundance | X08300 | Indole-3-a | 3 | Interventio | Experimen | Experimen | Control | 0,808903 |
| 3 months | abundance | X08305 | Prolylhydro | 2a | Interventio | Experimen | Experimen | Control | 0,808744 |
| 3 months | abundance | X08306 | 1-Methylxa | 2b | Interventio | Experimen | Experimen | Control | 0,522871 |
| 3 months | abundance | X08311 | Methyl 3-fo | 3 | Interventio | Experimen | Experimen | Control | 0,755448 |
| 3 months | abundance | X08317 | 8-Hydroxy | 2b | Interventio | Experimen | Experimen | Control | 0,529387 |
| 3 months | abundance | X08322 | Glycylproly | 3 | Interventio | Experimen | Experimen | Control | 0,860527 |
| 3 months | abundance | X08323 | (15Z)-9,12 | 2b | Interventio | Experimen | Experimen | Control | 0,854716 |
| 3 months | abundance | X08326 | 2-(1,3-Ben | 3 | Interventio | Experimen | Experimen | Control | 0,188874 |
| 3 months | abundance | X08338 | Methyl [9-( | 3 | Interventio | Experimen | Experimen | Control | 0,322462 |
| 3 months | abundance | X08341 | Midodrine_ | 3 | Interventio | Experimen | Experimen | Control | 0,892548 |
| 3 months | abundance | X08352 | 7-[4-(tert-b | 2b | Interventio | Experimen | Experimen | Control | 0,953271 |
| 3 months | abundance | X08365 | Lysylvaline | 3 | Interventio | Experimen | Experimen | Control | 0,003817 |
| 3 months | abundance | X08367 | 11-dehydro | 2b | Interventio | Experimen | Experimen | Control | 0,59787 |
| 3 months | abundance | X08373 | 2-Hydroxy- | 3 | Interventio | Experimen | Experimen | Control | 0,854716 |
| 3 months | abundance | X08386 | 3b-Hydroxy | 3 | Interventio | Experimen | Experimen | Control | 0,509355 |

| | | | | | | | | | |
|---|---|---|---|---|---|---|---|---|---|
| 3 months | abundance | X08387 | 4-[(2E,4Z)- | 3 | Interventio | Experimen | Experimen | Control | 0,807551 |
| 3 months | abundance | X08412 | 2-Hydroxyp | 2b | Interventio | Experimen | Experimen | Control | 0,830535 |
| 3 months | abundance | X08417 | asn-pro_c | 3 | Interventio | Experimen | Experimen | Control | 0,798192 |
| 3 months | abundance | X08420 | Dodecane | 1 | Interventio | Experimen | Experimen | Control | 0,047927 |
| 3 months | abundance | X08421 | Cys-tyr | 3 | Interventio | Experimen | Experimen | Control | 0,109563 |
| 3 months | abundance | X08422 | N-(3-aceta | 3 | Interventio | Experimen | Experimen | Control | 0,765006 |
| 3 months | abundance | X08425 | MFCD1297 | 3 | Interventio | Experimen | Experimen | Control | 0,008475 |
| 3 months | abundance | X08436 | N,N-Dimet | 3 | Interventio | Experimen | Experimen | Control | 0,521176 |
| 3 months | abundance | X08439 | Leucyltyros | 3 | Interventio | Experimen | Experimen | Control | 0,070241 |
| 3 months | abundance | X08451 | L-gamma-( | 3 | Interventio | Experimen | Experimen | Control | 0,985356 |
| 3 months | abundance | X08460 | α-Murichol | 1 | Interventio | Experimen | Experimen | Control | 0,985356 |
| 3 months | abundance | X08461 | Dihydrouri | 3 | Interventio | Experimen | Experimen | Control | 0,208353 |
| 3 months | abundance | X08475 | 1-(4-Aminc | 3 | Interventio | Experimen | Experimen | Control | 0,657878 |
| 3 months | abundance | X08476 | Coprine_c | 3 | Interventio | Experimen | Experimen | Control | 0,679693 |
| 3 months | abundance | X08478 | 1-(beta-D-I | 3 | Interventio | Experimen | Experimen | Control | 0,99116 |
| 3 months | abundance | X08485 | Isoprenalir | 3 | Interventio | Experimen | Experimen | Control | 0,980029 |
| 3 months | abundance | X08486 | (2E,6E)-9-[ | 3 | Interventio | Experimen | Experimen | Control | 0,216079 |
| 3 months | abundance | X08504 | Indole-3-ca | 2a | Interventio | Experimen | Experimen | Control | 0,988551 |
| 3 months | abundance | X08509 | Piceid | 3 | Interventio | Experimen | Experimen | Control | 0,412774 |
| 3 months | abundance | X08512 | Bile acid I ( | 2b | Interventio | Experimen | Experimen | Control | 0,645886 |
| 3 months | abundance | X08514 | N-Ethylpro | 3 | Interventio | Experimen | Experimen | Control | 0,242305 |
| 3 months | abundance | X08525 | MFCD1869 | 3 | Interventio | Experimen | Experimen | Control | 0,023171 |
| 3 months | abundance | X08529 | 7,8-Diamir | 3 | Interventio | Experimen | Experimen | Control | 0,014515 |
| 3 months | abundance | X08533 | tyramine su | 3 | Interventio | Experimen | Experimen | Control | 0,598888 |
| 3 months | abundance | X08535 | Nitrosohep | 2b | Interventio | Experimen | Experimen | Control | 0,385109 |
| 3 months | abundance | X08546 | 1,3,7-Trime | 2b | Interventio | Experimen | Experimen | Control | 0,819701 |
| 3 months | abundance | X08548 | N2-Acetylly | 1 | Interventio | Experimen | Experimen | Control | 0,405692 |
| 3 months | abundance | X08549 | 3-Methylsu | 3 | Interventio | Experimen | Experimen | Control | 0,243875 |
| 3 months | abundance | X08561 | N-(Carboxy | 3 | Interventio | Experimen | Experimen | Control | 0,00049 |
| 3 months | abundance | X08578 | tert-Butyl 3 | 3 | Interventio | Experimen | Experimen | Control | 0,625098 |
| 3 months | abundance | X08584 | meticillin | 3 | Interventio | Experimen | Experimen | Control | 0,467786 |
| 3 months | abundance | X08588 | 2-Acetamid | 3 | Interventio | Experimen | Experimen | Control | 0,486342 |
| 3 months | abundance | X08593 | Uric acid | 1 | Interventio | Experimen | Experimen | Control | 0,084515 |
| 3 months | abundance | X08594 | 7-ketodeox | 3 | Interventio | Experimen | Experimen | Control | 0,997328 |
| 3 months | abundance | X08600 | N-Propiony | 3 | Interventio | Experimen | Experimen | Control | 0,60023 |
| 3 months | abundance | X08606 | Butabarbit | 3 | Interventio | Experimen | Experimen | Control | 0,217561 |
| 3 months | abundance | X08610 | Leucyltyros | 3 | Interventio | Experimen | Experimen | Control | 0,833128 |
| 3 months | abundance | X08616 | MFCD0002 | 3 | Interventio | Experimen | Experimen | Control | 0,718544 |
| 3 months | abundance | X08625 | Hept-2-ulo | 3 | Interventio | Experimen | Experimen | Control | 0,322462 |
| 3 months | abundance | X08634 | 1,5-Isoquir | 2b | Interventio | Experimen | Experimen | Control | 0,798571 |
| 3 months | abundance | X08639 | hypaphorir | 3 | Interventio | Experimen | Experimen | Control | 0,99116 |
| 3 months | abundance | X08641 | meprobam | 3 | Interventio | Experimen | Experimen | Control | 0,76643 |
| 3 months | abundance | X08644 | 2,5-Dimeth | 2b | Interventio | Experimen | Experimen | Control | 0,466427 |
| 3 months | abundance | X08646 | N-Stearoyl | 3 | Interventio | Experimen | Experimen | Control | 0,84915 |
| 3 months | abundance | X08657 | 1-(4-Aminc | 3 | Interventio | Experimen | Experimen | Control | 0,99587 |
| 3 months | abundance | X08659 | 3,3-Dimeth | 2b | Interventio | Experimen | Experimen | Control | 0,96467 |
| 3 months | abundance | X08663 | Pilocarpine | 2b | Interventio | Experimen | Experimen | Control | 0,59787 |
| 3 months | abundance | X08670 | Hydroxypro | 3 | Interventio | Experimen | Experimen | Control | 0,004228 |

| | | | | | | | | | |
|---|---|---|---|---|---|---|---|---|---|
| 3 months | abundance | X08673 | NPC | 3 | Interventio | Experimen | Experimen | Control | 0,854716 |
| 3 months | abundance | X08677 | 1-methylhy | 3 | Interventio | Experimen | Experimen | Control | 0,126143 |
| 3 months | abundance | X08690 | Ethanoic a | 3 | Interventio | Experimen | Experimen | Control | 0,146137 |
| 3 months | abundance | X08693 | 7-Sulfocho | 3 | Interventio | Experimen | Experimen | Control | 0,96467 |
| 3 months | abundance | X08695 | 3b-Hydroxy | 3 | Interventio | Experimen | Experimen | Control | 0,231572 |
| 3 months | abundance | X08699 | Aurorix | 3 | Interventio | Experimen | Experimen | Control | 0,124383 |
| 3 months | abundance | X08707 | 2-(1-Ethoxy | 3 | Interventio | Experimen | Experimen | Control | 0,58465 |
| 3 months | abundance | X08711 | 2-Acetamid | 3 | Interventio | Experimen | Experimen | Control | 0,871745 |
| 3 months | abundance | X08712 | 4-Iodoanis | 3 | Interventio | Experimen | Experimen | Control | 0,038414 |
| 3 months | abundance | X08713 | Phenethyla | 2b | Interventio | Experimen | Experimen | Control | 0,838687 |
| 3 months | abundance | X08714 | Arabic acid | 3 | Interventio | Experimen | Experimen | Control | 0,256516 |
| 3 months | abundance | X08719 | 6-Methylqu | 2b | Interventio | Experimen | Experimen | Control | 0,993459 |
| 3 months | abundance | X08723 | S-Allylcyste | 3 | Interventio | Experimen | Experimen | Control | 0,142786 |
| 3 months | abundance | X08726 | YV8195000 | 3 | Interventio | Experimen | Experimen | Control | 0,000211 |
| 3 months | abundance | X08733 | N-Methyl-1 | 3 | Interventio | Experimen | Experimen | Control | 0,977246 |
| 3 months | abundance | X08739 | 1,1'-[1,12-I | 3 | Interventio | Experimen | Experimen | Control | 0,758687 |
| 3 months | abundance | X08758 | Methohexit | 3 | Interventio | Experimen | Experimen | Control | 0,217561 |
| 3 months | abundance | X08777 | 3-Aminosa | 2b | Interventio | Experimen | Experimen | Control | 0,385799 |
| 3 months | abundance | X08779 | 3-(1-hydro | 2b | Interventio | Experimen | Experimen | Control | 0,665073 |
| 3 months | abundance | X08781 | Procaine_b | 3 | Interventio | Experimen | Experimen | Control | 0,174141 |
| 3 months | abundance | X08789 | Acrylic acid | 2b | Interventio | Experimen | Experimen | Control | 0,124404 |
| 3 months | abundance | X08792 | 3,4-Methyl | 3 | Interventio | Experimen | Experimen | Control | 0,967096 |
| 3 months | abundance | X08795 | N-Acetylva | 3 | Interventio | Experimen | Experimen | Control | 0,614079 |
| 3 months | abundance | X08801 | 3,7-Dimeth | 2b | Interventio | Experimen | Experimen | Control | 0,787558 |
| 3 months | abundance | X08810 | N~6~,N~6~ | 3 | Interventio | Experimen | Experimen | Control | 0,011582 |
| 3 months | abundance | X08822 | CYS-ASP | 3 | Interventio | Experimen | Experimen | Control | 0,267725 |
| 3 months | abundance | X08825 | Chenodeox | 3 | Interventio | Experimen | Experimen | Control | 0,584092 |
| 3 months | abundance | X08842 | Spermic ac | 3 | Interventio | Experimen | Experimen | Control | 0,992999 |
| 3 months | abundance | X08843 | (2R,3S)-3-I | 3 | Interventio | Experimen | Experimen | Control | 0,002229 |
| 3 months | abundance | X08844 | 4-Hydroxy- | 3 | Interventio | Experimen | Experimen | Control | 0,321752 |
| 3 months | abundance | X08845 | 14-Hydroxy | 3 | Interventio | Experimen | Experimen | Control | 0,351458 |
| 3 months | abundance | X08847 | 1-Vinylimid | 2b | Interventio | Experimen | Experimen | Control | 0,960119 |
| 3 months | abundance | X08869 | Lanthionin | 3 | Interventio | Experimen | Experimen | Control | 0,9765 |
| 3 months | abundance | X08870 | Tetraacety | 3 | Interventio | Experimen | Experimen | Control | 0,144456 |
| 3 months | abundance | X08883 | 3-Phenylpr | 3 | Interventio | Experimen | Experimen | Control | 0,981401 |
| 3 months | abundance | X08893 | 5beta-Cho | 3 | Interventio | Experimen | Experimen | Control | 0,115645 |
| 3 months | abundance | X08895 | pentoxyl | 3 | Interventio | Experimen | Experimen | Control | 0,565249 |
| 3 months | abundance | X08908 | Vorinostat_ | 3 | Interventio | Experimen | Experimen | Control | 0,191691 |
| 3 months | abundance | X08909 | GLK (Pepti | 2b | Interventio | Experimen | Experimen | Control | 0,442109 |
| 3 months | abundance | X08923 | Semilicois | 3 | Interventio | Experimen | Experimen | Control | 0,28244 |
| 3 months | abundance | X08938 | Nonivamid | 3 | Interventio | Experimen | Experimen | Control | 8,90E-05 |
| 3 months | abundance | X08944 | Dipivefrin | 3 | Interventio | Experimen | Experimen | Control | 0,239969 |
| 3 months | abundance | X08965 | 1,9-Nonan | 3 | Interventio | Experimen | Experimen | Control | 0,820266 |
| 3 months | abundance | X08973 | Aminohipp | 3 | Interventio | Experimen | Experimen | Control | 0,419167 |
| 3 months | abundance | X08983 | IN00150 | 3 | Interventio | Experimen | Experimen | Control | 0,830535 |
| 3 months | abundance | X08987 | feruloylser | 3 | Interventio | Experimen | Experimen | Control | 0,436347 |
| 3 months | abundance | X08990 | Procaine_c | 3 | Interventio | Experimen | Experimen | Control | 0,656554 |
| 3 months | abundance | X08992 | N(2)-succi | 3 | Interventio | Experimen | Experimen | Control | 0,71541 |

| | | | | | | | | | |
|---|---|---|---|---|---|---|---|---|---|
| 3 months | abundance | X09001 | Varanic ac | 3 | Interventio | Experimen | Experimen | Control | 0,957479 |
| 3 months | abundance | X09004 | Arenaine | 3 | Interventio | Experimen | Experimen | Control | 0,070143 |
| 3 months | abundance | X09008 | 5-guanidin | 3 | Interventio | Experimen | Experimen | Control | 0,778801 |
| 3 months | abundance | X09010 | 4-Hydroxyp | 1 | Interventio | Experimen | Experimen | Control | 0,532452 |
| 3 months | abundance | X09055 | N-[(1R,2S,( | 3 | Interventio | Experimen | Experimen | Control | 0,605819 |
| 3 months | abundance | X09064 | Tetrahydro | 3 | Interventio | Experimen | Experimen | Control | 0,518924 |
| 3 months | abundance | X09068 | N,N-Diethy | 3 | Interventio | Experimen | Experimen | Control | 0,000404 |
| 3 months | abundance | X09082 | 1_2-Dihydr | 3 | Interventio | Experimen | Experimen | Control | 0,99116 |
| 3 months | abundance | X09091 | N-(3-aceta | 3 | Interventio | Experimen | Experimen | Control | 0,155891 |
| 3 months | abundance | X09092 | LysoSM(d1 | 3 | Interventio | Experimen | Experimen | Control | 0,787162 |
| 3 months | abundance | X09099 | 1-(2-Hydro | 3 | Interventio | Experimen | Experimen | Control | 0,149795 |
| 3 months | abundance | X09100 | Ethyl aceta | 3 | Interventio | Experimen | Experimen | Control | 0,613297 |
| 3 months | abundance | X09110 | Valylvaline | 3 | Interventio | Experimen | Experimen | Control | 0,100587 |
| 3 months | abundance | X09122 | Valyl-4-hyd | 3 | Interventio | Experimen | Experimen | Control | 0,413564 |
| 3 months | abundance | X09129 | Coprine_d | 3 | Interventio | Experimen | Experimen | Control | 0,655532 |
| 3 months | abundance | X09152 | Ferulic aci | 1 | Interventio | Experimen | Experimen | Control | 0,268018 |
| 3 months | abundance | X09163 | Prenisteine | 3 | Interventio | Experimen | Experimen | Control | 0,938964 |
| 3 months | abundance | X09165 | N-[1-Carbo | 3 | Interventio | Experimen | Experimen | Control | 0,012368 |
| 3 months | abundance | X09187 | Salicylic ac | 1 | Interventio | Experimen | Experimen | Control | 0,76643 |
| 3 months | abundance | X09189 | 4-[(E)-2-(3, | 3 | Interventio | Experimen | Experimen | Control | 0,333863 |
| 3 months | abundance | X09207 | AAMU_a | 3 | Interventio | Experimen | Experimen | Control | 0,333178 |
| 3 months | abundance | X09208 | IN00260_b | 3 | Interventio | Experimen | Experimen | Control | 0,798192 |
| 3 months | abundance | X09222 | L-gamma-( | 3 | Interventio | Experimen | Experimen | Control | 0,619252 |
| 3 months | abundance | X09231 | 2-Keto-glut | 3 | Interventio | Experimen | Experimen | Control | 0,988551 |
| 3 months | abundance | X09244 | gamma-Gl | 3 | Interventio | Experimen | Experimen | Control | 0,409881 |
| 3 months | abundance | X09245 | L-gamma-( | 3 | Interventio | Experimen | Experimen | Control | 0,390151 |
| 3 months | abundance | X09256 | Aspartyl-L- | 3 | Interventio | Experimen | Experimen | Control | 0,451712 |
| 3 months | abundance | X09260 | (4S)-4-[(2E | 3 | Interventio | Experimen | Experimen | Control | 0,23349 |
| 3 months | abundance | X09284 | 4-(Nitrosoa | 3 | Interventio | Experimen | Experimen | Control | 0,913373 |
| 3 months | abundance | X09285 | MFCD0272 | 3 | Interventio | Experimen | Experimen | Control | 0,991184 |
| 3 months | abundance | X09302 | Sular | 3 | Interventio | Experimen | Experimen | Control | 0,836327 |
| 3 months | abundance | X09318 | butalbital_ | 3 | Interventio | Experimen | Experimen | Control | 0,971877 |
| 3 months | abundance | X09336 | 1,1'-[1,12-I | 3 | Interventio | Experimen | Experimen | Control | 0,627129 |
| 3 months | abundance | X09357 | 5-(5-Methy | 3 | Interventio | Experimen | Experimen | Control | 0,469773 |
| 3 months | abundance | X09374 | 3-Deoxy-D- | 3 | Interventio | Experimen | Experimen | Control | 0,46054 |
| 3 months | abundance | X09385 | 2-(1-Napht | 3 | Interventio | Experimen | Experimen | Control | 0,503169 |
| 3 months | abundance | X09390 | asn-pro_a | 3 | Interventio | Experimen | Experimen | Control | 0,142786 |
| 3 months | abundance | X09391 | ophthalmic | 3 | Interventio | Experimen | Experimen | Control | 0,096259 |
| 3 months | abundance | X09393 | 2-Isopropy | 3 | Interventio | Experimen | Experimen | Control | 0,943743 |
| 3 months | abundance | X09399 | L-gamma-( | 3 | Interventio | Experimen | Experimen | Control | 0,718544 |
| 3 months | abundance | X09454 | 2-(2-Hydro | 3 | Interventio | Experimen | Experimen | Control | 0,80026 |
| 3 months | abundance | X09476 | N(1),N(8)-I | 3 | Interventio | Experimen | Experimen | Control | 0,927159 |
| 3 months | abundance | X09480 | Spermic ac | 3 | Interventio | Experimen | Experimen | Control | 0,739091 |
| 3 months | abundance | X09490 | 2-Hydroxy- | 3 | Interventio | Experimen | Experimen | Control | 0,226184 |
| 3 months | abundance | X09503 | 4-(METHYL | 3 | Interventio | Experimen | Experimen | Control | 0,60023 |
| 3 months | abundance | X09505 | (-)-Physost | 3 | Interventio | Experimen | Experimen | Control | 0,991184 |
| 3 months | abundance | X09523 | N-(3,5-Dim | 3 | Interventio | Experimen | Experimen | Control | 0,45097 |
| 3 months | abundance | X09525 | Homoanse | 3 | Interventio | Experimen | Experimen | Control | 0,268483 |

| | | | | | | | | | |
|---|---|---|---|---|---|---|---|---|---|
| 3 months | abundance | X09533 | S(6)-acetyl | 3 | Interventio | Experimen | Experimen | Control | 0,873316 |
| 3 months | abundance | X09545 | 4-Methyler | 3 | Interventio | Experimen | Experimen | Control | 0,691471 |
| 3 months | abundance | X09546 | asn-phe | 3 | Interventio | Experimen | Experimen | Control | 0,625098 |
| 3 months | abundance | X09548 | 2-O-beta-D | 3 | Interventio | Experimen | Experimen | Control | 0,021942 |
| 3 months | abundance | X09554 | N-Propiony | 3 | Interventio | Experimen | Experimen | Control | 0,953482 |
| 3 months | abundance | X09555 | N4-(beta-N | 3 | Interventio | Experimen | Experimen | Control | 0,449163 |
| 3 months | abundance | X09565 | 3-Methoxy- | 3 | Interventio | Experimen | Experimen | Control | 0,30859 |
| 3 months | abundance | X09577 | Spermic ac | 3 | Interventio | Experimen | Experimen | Control | 0,291778 |
| 3 months | abundance | X09581 | L-gamma-( | 3 | Interventio | Experimen | Experimen | Control | 0,76164 |
| 3 months | abundance | X09607 | N-(1H-Pyrr | 3 | Interventio | Experimen | Experimen | Control | 0,244993 |
| 3 months | abundance | X09608 | N-(3,5-Dim | 3 | Interventio | Experimen | Experimen | Control | 0,802409 |
| 3 months | abundance | X09609 | N-{3-[(4-Ac | 3 | Interventio | Experimen | Experimen | Control | 0,886266 |
| 3 months | abundance | X09612 | 3-hydroxy-3 | 3 | Interventio | Experimen | Experimen | Control | 0,60023 |
| 3 months | abundance | X09615 | tyramine su | 3 | Interventio | Experimen | Experimen | Control | 0,691158 |
| 3 months | abundance | X09622 | N-Hydroxy- | 3 | Interventio | Experimen | Experimen | Control | 0,243875 |
| 3 months | abundance | X09623 | Endothal | 2b | Interventio | Experimen | Experimen | Control | 0,161121 |
| 3 months | abundance | X09635 | 2,4,6-Triisc | 3 | Interventio | Experimen | Experimen | Control | 0,712279 |
| 3 months | abundance | X09643 | N-(3,5-Dim | 3 | Interventio | Experimen | Experimen | Control | 0,854716 |
| 3 months | abundance | X09645 | TOLMETIN | 3 | Interventio | Experimen | Experimen | Control | 0,76643 |
| 3 months | abundance | X09654 | linatine | 3 | Interventio | Experimen | Experimen | Control | 0,860527 |
| 3 months | abundance | X09657 | 2,3,4,5,6-F | 3 | Interventio | Experimen | Experimen | Control | 0,771075 |
| 3 months | abundance | X09675 | UK387000 | 3 | Interventio | Experimen | Experimen | Control | 0,821708 |
| 3 months | abundance | X09684 | (+)-Etomid | 3 | Interventio | Experimen | Experimen | Control | 0,598888 |
| 3 months | abundance | X09703 | 7,8-Didehy | 3 | Interventio | Experimen | Experimen | Control | 0,468958 |
| 3 months | abundance | X09710 | LW800000 | 3 | Interventio | Experimen | Experimen | Control | 0,093704 |
| 3 months | abundance | X09713 | N(2)-succi | 3 | Interventio | Experimen | Experimen | Control | 0,581247 |
| 3 months | abundance | X09721 | 3-(2,3-Dihy | 3 | Interventio | Experimen | Experimen | Control | 0,124404 |
| 3 months | abundance | X09724 | Formylkynu | 3 | Interventio | Experimen | Experimen | Control | 0,151919 |
| 3 months | abundance | X09726 | indoline-2- | 2b | Interventio | Experimen | Experimen | Control | 0,143745 |
| 3 months | abundance | X09728 | {[(15-Hydro | 3 | Interventio | Experimen | Experimen | Control | 0,915997 |
| 3 months | abundance | X09738 | (7E,7'E)-5, | 3 | Interventio | Experimen | Experimen | Control | 0,694902 |
| 3 months | abundance | X09740 | MFCD0166 | 3 | Interventio | Experimen | Experimen | Control | 0,365259 |
| 3 months | abundance | X09741 | Estrone glu | 3 | Interventio | Experimen | Experimen | Control | 0,415989 |
| 3 months | abundance | X09746 | ELK (Peptic | 2b | Interventio | Experimen | Experimen | Control | 0,98336 |
| 3 months | abundance | X09765 | Tetrahydro | 3 | Interventio | Experimen | Experimen | Control | 0,657878 |
| 3 months | abundance | X09771 | 2-(3-CARB | 3 | Interventio | Experimen | Experimen | Control | 0,953271 |
| 3 months | abundance | X09787 | Queuosine | 3 | Interventio | Experimen | Experimen | Control | 0,713464 |
| 3 months | abundance | X09805 | Methyl alpl | 3 | Interventio | Experimen | Experimen | Control | 0,787558 |
| 3 months | abundance | X09810 | Zalcitabine | 3 | Interventio | Experimen | Experimen | Control | 0,798192 |
| 3 months | abundance | X09825 | (2S)-6-Ami | 3 | Interventio | Experimen | Experimen | Control | 0,927159 |
| 3 months | abundance | X09849 | 3-Methylac | 3 | Interventio | Experimen | Experimen | Control | 0,002308 |
| 3 months | abundance | X09854 | N-Benzyl-3 | 2b | Interventio | Experimen | Experimen | Control | 0,755448 |
| 3 months | abundance | X09875 | 4-[(E)-2-(3, | 3 | Interventio | Experimen | Experimen | Control | 0,778801 |
| 3 months | abundance | X09882 | Harmane | 3 | Interventio | Experimen | Experimen | Control | 0,986565 |
| 3 months | abundance | X09895 | 1-(beta-D-r | 3 | Interventio | Experimen | Experimen | Control | 0,886266 |
| 3 months | abundance | X09898 | N(6),N(6)-I | 3 | Interventio | Experimen | Experimen | Control | 0,099053 |
| 3 months | abundance | X09912 | MC055530 | 3 | Interventio | Experimen | Experimen | Control | 0,988551 |
| 3 months | abundance | X09919 | 4-ethylphe | 3 | Interventio | Experimen | Experimen | Control | 0,627129 |

| | | | | | | | | | |
|---|---|---|---|---|---|---|---|---|---|
| 3 months | abundance | X09921 | 5-Hydroxy- | 3 | Interventio | Experimen | Experimen | Control | 0,082425 |
| 3 months | abundance | X09932 | g-Aminobu | 3 | Interventio | Experimen | Experimen | Control | 0,001482 |
| 3 months | abundance | X09940 | pentobarbi | 3 | Interventio | Experimen | Experimen | Control | 0,598888 |
| 3 months | abundance | X09942 | Spermic ac | 3 | Interventio | Experimen | Experimen | Control | 0,96467 |
| 3 months | abundance | X09950 | 3-Hydroxyh | 3 | Interventio | Experimen | Experimen | Control | 0,230103 |
| 3 months | abundance | X09951 | 5beta-Cypr | 3 | Interventio | Experimen | Experimen | Control | 0,821759 |
| 3 months | abundance | X09963 | Iminoglycir | 3 | Interventio | Experimen | Experimen | Control | 0,691471 |
| 3 months | abundance | X09965 | Biocytin | 2b | Interventio | Experimen | Experimen | Control | 0,798341 |
| 3 months | abundance | X09969 | 3-Methylac | 3 | Interventio | Experimen | Experimen | Control | 0,005269 |
| 3 months | abundance | X09977 | TLK (Peptic | 2b | Interventio | Experimen | Experimen | Control | 0,898308 |
| 3 months | abundance | X09980 | N-(4-Hepta | 3 | Interventio | Experimen | Experimen | Control | 0,052265 |
| 3 months | abundance | X09981 | N-Acetylva | 3 | Interventio | Experimen | Experimen | Control | 0,503169 |
| 3 months | abundance | X09998 | N-[(10Z)-7- | 3 | Interventio | Experimen | Experimen | Control | 0,766128 |
| 3 months | abundance | X10011 | Midodrine_ | 3 | Interventio | Experimen | Experimen | Control | 0,718544 |
| 3 months | abundance | X10025 | 2-[4-(3-Hyc | 3 | Interventio | Experimen | Experimen | Control | 0,46992 |
| 3 months | abundance | X10032 | 4,9a-Dime | 3 | Interventio | Experimen | Experimen | Control | 0,680872 |
| 3 months | abundance | X10033 | epsilon-(ga | 3 | Interventio | Experimen | Experimen | Control | 0,916051 |
| 3 months | abundance | X10036 | Nimodipin | 3 | Interventio | Experimen | Experimen | Control | 0,9752 |
| 3 months | abundance | X10037 | his-gln_a | 3 | Interventio | Experimen | Experimen | Control | 0,88099 |
| 3 months | abundance | X10046 | Dehydroac | 2b | Interventio | Experimen | Experimen | Control | 0,986931 |
| 3 months | abundance | X10076 | trimethadi | 3 | Interventio | Experimen | Experimen | Control | 0,924699 |
| 3 months | abundance | X10091 | (2E,6E)-9-[ | 3 | Interventio | Experimen | Experimen | Control | 0,977246 |
| 3 months | abundance | X10097 | Coixol | 3 | Interventio | Experimen | Experimen | Control | 0,298368 |
| 3 months | abundance | X10098 | Methyl 2,3- | 3 | Interventio | Experimen | Experimen | Control | 0,583521 |
| 3 months | abundance | X10099 | ala-met | 3 | Interventio | Experimen | Experimen | Control | 0,419167 |
| 3 months | abundance | X10121 | Leu-pro_b | 3 | Interventio | Experimen | Experimen | Control | 0,262176 |
| 3 months | abundance | X10129 | SECONAL_ | 3 | Interventio | Experimen | Experimen | Control | 0,664257 |
| 3 months | abundance | X10130 | N-(4-Hydro | 3 | Interventio | Experimen | Experimen | Control | 0,833128 |
| 3 months | abundance | X10137 | 1-Methyl-1 | 3 | Interventio | Experimen | Experimen | Control | 0,61536 |
| 3 months | abundance | X10144 | imazameth | 3 | Interventio | Experimen | Experimen | Control | 0,907192 |
| 3 months | abundance | X10148 | cys-met | 3 | Interventio | Experimen | Experimen | Control | 0,144997 |
| 3 months | abundance | X10151 | 2-(3-CARB | 3 | Interventio | Experimen | Experimen | Control | 0,741794 |
| 3 months | abundance | X10162 | Quinaldic a | 2a | Interventio | Experimen | Experimen | Control | 0,144456 |
| 3 months | abundance | X10177 | Gly-DL-Phe | 3 | Interventio | Experimen | Experimen | Control | 0,706727 |
| 3 months | abundance | X10180 | 4-Formyl-2 | 3 | Interventio | Experimen | Experimen | Control | 0,328214 |
| 3 months | abundance | X10203 | L-gamma-( | 3 | Interventio | Experimen | Experimen | Control | 0,282689 |
| 3 months | abundance | X10224 | Imidazolela | 2b | Interventio | Experimen | Experimen | Control | 0,433847 |
| 3 months | abundance | X10246 | Ethynodiol | 3 | Interventio | Experimen | Experimen | Control | 0,23701 |
| 3 months | abundance | X10248 | (+)-Etomid | 3 | Interventio | Experimen | Experimen | Control | 0,448962 |
| 3 months | abundance | X10265 | 3-methyl-4 | 3 | Interventio | Experimen | Experimen | Control | 0,099053 |
| 3 months | abundance | X10276 | Maleamate | 3 | Interventio | Experimen | Experimen | Control | 0,718544 |
| 3 months | abundance | X10279 | Lys-Pro_d | 3 | Interventio | Experimen | Experimen | Control | 0,143745 |
| 3 months | abundance | X10304 | 2,3,8,9-Tet | 3 | Interventio | Experimen | Experimen | Control | 0,480445 |
| 3 months | abundance | X10314 | 4-morpholi | 2b | Interventio | Experimen | Experimen | Control | 0,517388 |
| 3 months | abundance | X10319 | 1-Methyl-1 | 3 | Interventio | Experimen | Experimen | Control | 0,232653 |
| 3 months | abundance | X10325 | Lovastatin_ | 3 | Interventio | Experimen | Experimen | Control | 0,633187 |
| 3 months | abundance | X10330 | Sulfurous a | 3 | Interventio | Experimen | Experimen | Control | 0,71076 |
| 3 months | abundance | X10339 | 4H-1-Benz | 2b | Interventio | Experimen | Experimen | Control | 0,965837 |

| | | | | | | | | | |
|---|---|---|---|---|---|---|---|---|---|
| 3 months | abundance | X10345 | S-Allylcyst | 3 | Interventio | Experimen | Experimen | Control | 0,631492 |
| 3 months | abundance | X10353 | L-gamma-( | 3 | Interventio | Experimen | Experimen | Control | 0,468958 |
| 3 months | abundance | X10367 | 2-(1-Napht | 3 | Interventio | Experimen | Experimen | Control | 0,468958 |
| 3 months | abundance | X10370 | L-Arogenat | 3 | Interventio | Experimen | Experimen | Control | 0,613297 |
| 3 months | abundance | X10375 | 4-Indoleca | 2b | Interventio | Experimen | Experimen | Control | 0,143745 |
| 3 months | abundance | X10379 | Phenyl D-g | 3 | Interventio | Experimen | Experimen | Control | 0,625098 |
| 3 months | abundance | X10382 | Zalcitabine | 3 | Interventio | Experimen | Experimen | Control | 0,724259 |
| 3 months | abundance | X10384 | L-N2-(2-Ca | 3 | Interventio | Experimen | Experimen | Control | 0,239969 |
| 3 months | abundance | X10387 | Coenzyme | 3 | Interventio | Experimen | Experimen | Control | 0,886266 |
| 3 months | abundance | X10389 | Leucylaspa | 3 | Interventio | Experimen | Experimen | Control | 0,63213 |
| 3 months | abundance | X10393 | Ro 20-172 | 3 | Interventio | Experimen | Experimen | Control | 0,124404 |
| 3 months | abundance | X10401 | 3-Methylxa | 1 | Interventio | Experimen | Experimen | Control | 0,99116 |
| 3 months | abundance | X10415 | beta-D-Glc | 3 | Interventio | Experimen | Experimen | Control | 0,143745 |
| 3 months | abundance | X10417 | L-Sacchar | 2b | Interventio | Experimen | Experimen | Control | 0,198011 |
| 3 months | abundance | X10429 | 1,5-Isoquir | 2b | Interventio | Experimen | Experimen | Control | 0,246675 |
| 3 months | abundance | X10442 | (6alpha,11 | 3 | Interventio | Experimen | Experimen | Control | 0,055322 |
| 3 months | abundance | X10445 | 2-(Carboxy | 3 | Interventio | Experimen | Experimen | Control | 0,675486 |
| 3 months | abundance | X10451 | Ro 20-172 | 3 | Interventio | Experimen | Experimen | Control | 0,629552 |
| 3 months | abundance | X10454 | 8-Amino-7 | 3 | Interventio | Experimen | Experimen | Control | 0,61536 |
| 3 months | abundance | X10457 | glu-ser | 3 | Interventio | Experimen | Experimen | Control | 0,98719 |
| 3 months | abundance | X10476 | GAMMA-HY | 3 | Interventio | Experimen | Experimen | Control | 0,03075 |
| 3 months | abundance | X10486 | Coumarin | 2b | Interventio | Experimen | Experimen | Control | 0,468958 |
| 3 months | abundance | X10488 | 4-Guanidir | 1 | Interventio | Experimen | Experimen | Control | 0,46992 |
| 3 months | abundance | X10494 | 26Q0EO75 | 3 | Interventio | Experimen | Experimen | Control | 0,835202 |
| 3 months | abundance | X10497 | 4-Guanidir | 3 | Interventio | Experimen | Experimen | Control | 0,490686 |
| 3 months | abundance | X10503 | APM_a | 3 | Interventio | Experimen | Experimen | Control | 0,980029 |
| 3 months | abundance | X10511 | dopaquino | 3 | Interventio | Experimen | Experimen | Control | 0,833128 |
| 3 months | abundance | X10513 | ophthalmi | 3 | Interventio | Experimen | Experimen | Control | 0,297155 |
| 3 months | abundance | X10518 | MFCD0002 | 3 | Interventio | Experimen | Experimen | Control | 0,907733 |
| 3 months | abundance | X10534 | L-N2-(2-Ca | 3 | Interventio | Experimen | Experimen | Control | 0,718544 |
| 3 months | abundance | X10536 | CMPF | 3 | Interventio | Experimen | Experimen | Control | 0,988551 |
| 3 months | abundance | X10549 | Glaucine | 2b | Interventio | Experimen | Experimen | Control | 0,901344 |
| 3 months | abundance | X10550 | pentobarbi | 3 | Interventio | Experimen | Experimen | Control | 0,423696 |
| 3 months | abundance | X10554 | Glycine | 1 | Interventio | Experimen | Experimen | Control | 0,440598 |
| 3 months | abundance | X10564 | 2-(1-Ethox | 3 | Interventio | Experimen | Experimen | Control | 0,206211 |
| 3 months | abundance | X10580 | Leu-Leu_a | 3 | Interventio | Experimen | Experimen | Control | 0,773109 |
| 3 months | abundance | X10588 | (8)-Ginger | 3 | Interventio | Experimen | Experimen | Control | 0,765006 |
| 3 months | abundance | X10593 | Vorinostat_ | 3 | Interventio | Experimen | Experimen | Control | 0,701955 |
| 3 months | abundance | X10600 | Glycocyam | 3 | Interventio | Experimen | Experimen | Control | 0,830535 |
| 3 months | abundance | X10611 | 2-Acrylami | 3 | Interventio | Experimen | Experimen | Control | 0,143745 |
| 3 months | abundance | X10614 | Monometh | 2a | Interventio | Experimen | Experimen | Control | 0,552894 |
| 3 months | abundance | X10617 | 2-methyl-1 | 2b | Interventio | Experimen | Experimen | Control | 0,000106 |
| 3 months | abundance | X10623 | N-Nonano | 3 | Interventio | Experimen | Experimen | Control | 0,960119 |
| 3 months | abundance | X10636 | 8-hydroxy- | 2b | Interventio | Experimen | Experimen | Control | 0,882862 |
| 3 months | abundance | X10647 | 4-Hydroxy | 3 | Interventio | Experimen | Experimen | Control | 0,949849 |
| 3 months | abundance | X10651 | Methyl 1-h | 3 | Interventio | Experimen | Experimen | Control | 0,949849 |
| 3 months | abundance | X10660 | 2-Methylth | 3 | Interventio | Experimen | Experimen | Control | 0,98719 |
| 3 months | abundance | X10664 | N1-(5-metl | 2b | Interventio | Experimen | Experimen | Control | 0,584092 |

| | | | | | | | | | |
|---|---|---|---|---|---|---|---|---|---|
| 3 months | abundance | X10665 | Aprobarbit | 3 | Interventio | Experimen | Experimen | Control | 0,011491 |
| 3 months | abundance | X10667 | N-Phenyla | 3 | Interventio | Experimen | Experimen | Control | 0,997328 |
| 3 months | abundance | X10676 | 6-(1-Hydro | 3 | Interventio | Experimen | Experimen | Control | 0,932175 |
| 3 months | abundance | X10680 | 8-hydroxy- | 3 | Interventio | Experimen | Experimen | Control | 0,714434 |
| 3 months | abundance | X10685 | Glycylvalin | 2a | Interventio | Experimen | Experimen | Control | 0,960119 |
| 3 months | abundance | X10687 | Gln-Gln | 3 | Interventio | Experimen | Experimen | Control | 0,960135 |
| 3 months | abundance | X10691 | (−)-nabilor | 3 | Interventio | Experimen | Experimen | Control | 0,468958 |
| 3 months | abundance | X10706 | Furaneol | 3 | Interventio | Experimen | Experimen | Control | 0,242305 |
| 3 months | abundance | X10713 | norhaman | 3 | Interventio | Experimen | Experimen | Control | 0,748275 |
| 3 months | abundance | X10722 | Acetanilide | 2b | Interventio | Experimen | Experimen | Control | 0,495802 |
| 3 months | abundance | X10725 | Ethyl mala | 3 | Interventio | Experimen | Experimen | Control | 0,354443 |
| 3 months | abundance | X10736 | Toxopyrimi | 3 | Interventio | Experimen | Experimen | Control | 0,217561 |
| 3 months | abundance | X10741 | 5-Nitro-2-p | 3 | Interventio | Experimen | Experimen | Control | 0,915997 |
| 3 months | abundance | X10757 | Glycyrin | 3 | Interventio | Experimen | Experimen | Control | 0,082425 |
| 3 months | abundance | X10769 | Cystine | 1 | Interventio | Experimen | Experimen | Control | 0,63213 |
| 3 months | abundance | X10770 | 2-(4-Isopro | 3 | Interventio | Experimen | Experimen | Control | 0,698621 |
| 3 months | abundance | X10772 | Bicine_a | 3 | Interventio | Experimen | Experimen | Control | 0,042091 |
| 3 months | abundance | X10776 | Histidylgly | 3 | Interventio | Experimen | Experimen | Control | 0,821708 |
| 3 months | abundance | X10793 | Zalcitabine | 3 | Interventio | Experimen | Experimen | Control | 0,122346 |
| 3 months | abundance | X10818 | butalbital_ | 3 | Interventio | Experimen | Experimen | Control | 0,614079 |
| 3 months | abundance | X10819 | Hyocholic | 2a | Interventio | Experimen | Experimen | Control | 0,892772 |
| 3 months | abundance | X10864 | 5-amino-2- | 2b | Interventio | Experimen | Experimen | Control | 0,884928 |
| 3 months | abundance | X10865 | Melatonin | 3 | Interventio | Experimen | Experimen | Control | 0,664511 |
| 3 months | abundance | X10881 | L-gamma- | 3 | Interventio | Experimen | Experimen | Control | 0,94878 |
| 3 months | abundance | X10886 | L-gamma- | 3 | Interventio | Experimen | Experimen | Control | 0,838286 |
| 3 months | abundance | X10889 | N-acetyl-b | 3 | Interventio | Experimen | Experimen | Control | 0,988551 |
| 3 months | abundance | X10896 | 6-Hydroxy | 2b | Interventio | Experimen | Experimen | Control | 0,960119 |
| 3 months | abundance | X10901 | bis-noryan | 3 | Interventio | Experimen | Experimen | Control | 0,143745 |
| 3 months | abundance | X10902 | Isoquinolir | 2b | Interventio | Experimen | Experimen | Control | 0,392246 |
| 3 months | abundance | X10918 | meprobam | 3 | Interventio | Experimen | Experimen | Control | 0,625098 |
| 3 months | abundance | X10929 | Temozolon | 3 | Interventio | Experimen | Experimen | Control | 0,021942 |
| 3 months | abundance | X10930 | MFCD099 | 3 | Interventio | Experimen | Experimen | Control | 0,76643 |
| 3 months | abundance | X10936 | clavulanic | 3 | Interventio | Experimen | Experimen | Control | 0,1745 |
| 3 months | abundance | X10947 | 1,1'-[1,12- | 3 | Interventio | Experimen | Experimen | Control | 0,815986 |
| 3 months | abundance | X10951 | 8-Methyl-8 | 3 | Interventio | Experimen | Experimen | Control | 0,946232 |
| 3 months | abundance | X10953 | Vorinostat_ | 3 | Interventio | Experimen | Experimen | Control | 0,078386 |
| 3 months | abundance | X10974 | Threonylgl | 3 | Interventio | Experimen | Experimen | Control | 0,902223 |
| 3 months | abundance | X10977 | pentobarbi | 3 | Interventio | Experimen | Experimen | Control | 0,952305 |
| 3 months | abundance | X10979 | acetyltauri | 3 | Interventio | Experimen | Experimen | Control | 0,986565 |
| 3 months | abundance | X10982 | 8-Methyl-8 | 3 | Interventio | Experimen | Experimen | Control | 0,330615 |
| 3 months | abundance | X10987 | 2-BUTYL PI | 3 | Interventio | Experimen | Experimen | Control | 0,560103 |
| 3 months | abundance | X11001 | 2-[(carbox | 2b | Interventio | Experimen | Experimen | Control | 0,820266 |
| 3 months | abundance | X11012 | Furan | 3 | Interventio | Experimen | Experimen | Control | 0,765006 |
| 3 months | abundance | X11028 | butalbital_ | 3 | Interventio | Experimen | Experimen | Control | 0,604864 |
| 3 months | abundance | X11032 | 2,4-Bis(3-r | 3 | Interventio | Experimen | Experimen | Control | 0,960119 |
| 3 months | abundance | X11033 | N-Acetyl-5 | 3 | Interventio | Experimen | Experimen | Control | 0,967096 |
| 3 months | abundance | X11034 | 1-(3-Aminc | 3 | Interventio | Experimen | Experimen | Control | 0,565249 |
| 3 months | abundance | X11036 | 3-Hydroxyp | 2b | Interventio | Experimen | Experimen | Control | 0,259613 |

| | | | | | | | | | |
|---|---|---|---|---|---|---|---|---|---|
| 3 months | abundance | X11038 | (2E)-3-(3,4 | 3 | Interventio | Experimen | Experimen | Control | 0,722718 |
| 3 months | abundance | X11042 | Leu-arg | 3 | Interventio | Experimen | Experimen | Control | 0,396367 |
| 3 months | abundance | X11059 | Dihydrocor | 3 | Interventio | Experimen | Experimen | Control | 0,521176 |
| 3 months | abundance | X11067 | Resveratro | 3 | Interventio | Experimen | Experimen | Control | 0,755448 |
| 3 months | abundance | X11068 | N,N'-Bis[4- | 3 | Interventio | Experimen | Experimen | Control | 0,567851 |
| 3 months | abundance | X11110 | Tauropine | 3 | Interventio | Experimen | Experimen | Control | 0,248637 |
| 3 months | abundance | X11115 | Hydroxycar | 3 | Interventio | Experimen | Experimen | Control | 0,80026 |
| 3 months | abundance | X11117 | Coprine_b | 3 | Interventio | Experimen | Experimen | Control | 0,067442 |
| 3 months | abundance | X11125 | 5,8,12-Trih | 3 | Interventio | Experimen | Experimen | Control | 0,029867 |
| 3 months | abundance | X11140 | Leu-Leu_e | 3 | Interventio | Experimen | Experimen | Control | 0,251604 |
| 3 months | abundance | X11163 | Lys-Pro_a | 3 | Interventio | Experimen | Experimen | Control | 0,176658 |
| 3 months | abundance | X11170 | QJ972000( | 3 | Interventio | Experimen | Experimen | Control | 0,510076 |
| 3 months | abundance | X11186 | Fluocinolo | 2b | Interventio | Experimen | Experimen | Control | 0,99116 |
| 3 months | abundance | X11215 | Leu-Leu_b | 3 | Interventio | Experimen | Experimen | Control | 0,099053 |
| 3 months | abundance | X11223 | Ethyl malat | 3 | Interventio | Experimen | Experimen | Control | 0,988551 |
| 3 months | abundance | X11245 | MFCD0003 | 3 | Interventio | Experimen | Experimen | Control | 0,718452 |
| 3 months | abundance | X11251 | N-Nonanoy | 3 | Interventio | Experimen | Experimen | Control | 0,685901 |
| 3 months | abundance | X11254 | (4R,5S,9S, | 3 | Interventio | Experimen | Experimen | Control | 0,614079 |
| 3 months | abundance | X11257 | 1-Methylgu | 2b | Interventio | Experimen | Experimen | Control | 0,191691 |
| 3 months | abundance | X11258 | Valyl-4-hyd | 3 | Interventio | Experimen | Experimen | Control | 0,972998 |
| 3 months | abundance | X11265 | Sulfurol | 3 | Interventio | Experimen | Experimen | Control | 0,342788 |
| 3 months | abundance | X11273 | trp-pro | 3 | Interventio | Experimen | Experimen | Control | 0,191691 |
| 3 months | abundance | X11277 | lys-leu | 3 | Interventio | Experimen | Experimen | Control | 0,000943 |
| 3 months | abundance | X11281 | Penbutolol | 3 | Interventio | Experimen | Experimen | Control | 0,798192 |
| 3 months | abundance | X11301 | (6S)-2-Ami | 3 | Interventio | Experimen | Experimen | Control | 0,552894 |
| 3 months | abundance | X11310 | Leu-Val_a | 3 | Interventio | Experimen | Experimen | Control | 0,633187 |
| 3 months | abundance | X11319 | Methohexit | 3 | Interventio | Experimen | Experimen | Control | 0,248187 |
| 3 months | abundance | X11323 | dihydroxyb | 3 | Interventio | Experimen | Experimen | Control | 0,003846 |
| 3 months | abundance | X11331 | N~6~-Octa | 3 | Interventio | Experimen | Experimen | Control | 0,058295 |
| 3 months | abundance | X11339 | L-gamma-( | 3 | Interventio | Experimen | Experimen | Control | 0,981401 |
| 3 months | abundance | X11347 | Voglibose | 3 | Interventio | Experimen | Experimen | Control | 0,953271 |
| 3 months | abundance | X11349 | Butabarbit | 3 | Interventio | Experimen | Experimen | Control | 0,198469 |
| 3 months | abundance | X11354 | 3-Oxo-4,6- | 3 | Interventio | Experimen | Experimen | Control | 0,78879 |
| 3 months | abundance | X11361 | 1-Methylhi | 3 | Interventio | Experimen | Experimen | Control | 0,150507 |
| 3 months | abundance | X11366 | 3-Sulfino-L | 3 | Interventio | Experimen | Experimen | Control | 0,990745 |
| 3 months | abundance | X11373 | 3-Succinoy | 2b | Interventio | Experimen | Experimen | Control | 0,698621 |
| 3 months | abundance | X11375 | L-gamma-( | 3 | Interventio | Experimen | Experimen | Control | 0,698621 |
| 3 months | abundance | X11383 | N-{3-[(4-Ac | 3 | Interventio | Experimen | Experimen | Control | 0,835202 |
| 3 months | abundance | X11408 | 5-Amino-6- | 3 | Interventio | Experimen | Experimen | Control | 0,767238 |
| 3 months | abundance | X11421 | 8-Amino-7- | 3 | Interventio | Experimen | Experimen | Control | 0,338283 |
| 3 months | abundance | X11438 | pro-met | 3 | Interventio | Experimen | Experimen | Control | 0,748275 |
| 3 months | abundance | X11441 | beta-D-Eth | 3 | Interventio | Experimen | Experimen | Control | 0,958101 |
| 3 months | abundance | X11477 | gamma-Gl | 3 | Interventio | Experimen | Experimen | Control | 0,988198 |
| 3 months | abundance | X11481 | his-gln_b | 3 | Interventio | Experimen | Experimen | Control | 0,000404 |
| 3 months | abundance | X11494 | 3-Hydroxys | 3 | Interventio | Experimen | Experimen | Control | 0,988551 |
| 3 months | abundance | X11498 | Valylvaline | 3 | Interventio | Experimen | Experimen | Control | 0,104321 |
| 3 months | abundance | X11505 | FB950000( | 3 | Interventio | Experimen | Experimen | Control | 0,60023 |
| 3 months | abundance | X11515 | 13(S)-HOT | 2b | Interventio | Experimen | Experimen | Control | 0,992999 |

| | | | | | | | | | |
|---|---|---|---|---|---|---|---|---|---|
| 3 months | abundance | X11522 | thr-trp | 3 | Interventio | Experimen | Experimen | Control | 0,61996 |
| 3 months | abundance | X11524 | 6-hydroxyp | 3 | Interventio | Experimen | Experimen | Control | 0,142786 |
| 3 months | abundance | X11529 | 2-Aminooc | 2b | Interventio | Experimen | Experimen | Control | 0,301015 |
| 3 months | abundance | X11549 | 1,1'-[1,12-I | 3 | Interventio | Experimen | Experimen | Control | 0,797019 |
| 3 months | abundance | X11556 | N-[(2E)-3-( | 3 | Interventio | Experimen | Experimen | Control | 0,960119 |
| 3 months | abundance | X11562 | ALA-PRO | 3 | Interventio | Experimen | Experimen | Control | 0,876028 |
| 3 months | abundance | X11563 | threonylph | 3 | Interventio | Experimen | Experimen | Control | 0,76643 |
| 3 months | abundance | X11576 | Phenyl D-g | 3 | Interventio | Experimen | Experimen | Control | 0,988551 |
| 3 months | abundance | X11605 | 3-Methyl-2 | 3 | Interventio | Experimen | Experimen | Control | 0,985356 |
| 3 months | abundance | X11614 | 2-Methylth | 3 | Interventio | Experimen | Experimen | Control | 0,46054 |
| 3 months | abundance | X11627 | N-(3-aceta | 3 | Interventio | Experimen | Experimen | Control | 0,821708 |
| 3 months | abundance | X11639 | Midodrine_ | 3 | Interventio | Experimen | Experimen | Control | 0,821708 |
| 3 months | abundance | X11645 | glu-thr | 3 | Interventio | Experimen | Experimen | Control | 0,985356 |
| 3 months | abundance | X11666 | N-(4-Hydrc | 3 | Interventio | Experimen | Experimen | Control | 0,801038 |
| 3 months | abundance | X11667 | Tetraacety | 3 | Interventio | Experimen | Experimen | Control | 0,613297 |
| 3 months | abundance | X11692 | 2_7-Anhyd | 3 | Interventio | Experimen | Experimen | Control | 0,05742 |
| 3 months | abundance | X11698 | 1-(beta-D-r | 3 | Interventio | Experimen | Experimen | Control | 1,75E-11 |
| 3 months | abundance | X11699 | {2-[2-(Isob | 3 | Interventio | Experimen | Experimen | Control | 0,511679 |
| 3 months | abundance | X11716 | ala-ser_a | 3 | Interventio | Experimen | Experimen | Control | 0,76643 |
| 3 months | abundance | X11717 | nicotianam | 3 | Interventio | Experimen | Experimen | Control | 0,430971 |
| 3 months | abundance | X11733 | 1,3-Dihydr | 3 | Interventio | Experimen | Experimen | Control | 0,816795 |
| 3 months | abundance | X11745 | 2-Despipei | 3 | Interventio | Experimen | Experimen | Control | 0,834104 |
| 3 months | abundance | X11755 | Furfuranol | 3 | Interventio | Experimen | Experimen | Control | 0,765006 |
| 3 months | abundance | p_cresol | p-Cresol | 1 | Interventio | Experimen | Experimen | Control | 0,575427 |
| 6 months | abundance | X11698 | 1-(beta-D-r | 3 | Interventio | Experimen | Experimen | Control | 1,72E-21 |
| 6 months | abundance | X06039 | gamma-Gl | 3 | Interventio | Experimen | Experimen | Control | 1,72E-21 |
| 6 months | abundance | X08843 | (2R,3S)-3-I | 3 | Interventio | Experimen | Experimen | Control | 6,48E-07 |
| 6 months | abundance | X07921 | Pipecolinic | 1 | Interventio | Experimen | Experimen | Control | 2,83E-06 |
| 6 months | abundance | X03132 | (3S,4S)-7,1 | 3 | Interventio | Experimen | Experimen | Control | 2,83E-06 |
| 6 months | abundance | X10617 | 2-methyl-1 | 2b | Interventio | Experimen | Experimen | Control | 5,94E-06 |
| 6 months | abundance | X08726 | YV819500( | 3 | Interventio | Experimen | Experimen | Control | 3,82E-05 |
| 6 months | abundance | X02928 | N-Acetylpr | 3 | Interventio | Experimen | Experimen | Control | 5,71E-06 |
| 6 months | abundance | X00375 | N-[(2S)-2-I | 3 | Interventio | Experimen | Experimen | Control | 0,000814 |
| 6 months | abundance | X00107 | N6-Acetylly | 1 | Interventio | Experimen | Experimen | Control | 5,65E-05 |
| 6 months | abundance | X08561 | N-(Carboxy | 3 | Interventio | Experimen | Experimen | Control | 8,18E-05 |
| 6 months | abundance | X07782 | Cytosine | 1 | Interventio | Experimen | Experimen | Control | 0,000821 |
| 6 months | abundance | X01768 | Butabarbit | 3 | Interventio | Experimen | Experimen | Control | 0,000235 |
| 6 months | abundance | X00115 | Histidine | 1 | Interventio | Experimen | Experimen | Control | 0,002657 |
| 6 months | abundance | X03023 | NPK (Pepti | 2b | Interventio | Experimen | Experimen | Control | 0,001953 |
| 6 months | abundance | X01689 | Ethyl mala | 3 | Interventio | Experimen | Experimen | Control | 0,002657 |
| 6 months | abundance | X10611 | 2-Acrylami | 3 | Interventio | Experimen | Experimen | Control | 0,003073 |
| 6 months | abundance | X08204 | 2-Amino-4 | 2b | Interventio | Experimen | Experimen | Control | 0,002868 |
| 6 months | abundance | SL00309 | 1-Aminocy | 2a | Interventio | Experimen | Experimen | Control | 0,000833 |
| 6 months | abundance | X07674 | Lysine | 1 | Interventio | Experimen | Experimen | Control | 0,00107 |
| 6 months | abundance | X06046 | Valylvaline | 3 | Interventio | Experimen | Experimen | Control | 0,003235 |
| 6 months | abundance | X00022 | N-Methyla: | 1 | Interventio | Experimen | Experimen | Control | 0,003235 |
| 6 months | abundance | X06477 | APM_b | 3 | Interventio | Experimen | Experimen | Control | 0,002657 |
| 6 months | abundance | X05499 | GLY-MET | 3 | Interventio | Experimen | Experimen | Control | 0,003235 |

| | | | | | | | | | |
|---|---|---|---|---|---|---|---|---|---|
| 6 months | abundance | X07394 | Hydroxypro | 3 | Interventio | Experimen | Experimen | Control | 0,002808 |
| 6 months | abundance | X03961 | Artesunate | 3 | Interventio | Experimen | Experimen | Control | 0,007941 |
| 6 months | abundance | X00032 | Indole-3-pı | 1 | Interventio | Experimen | Experimen | Control | 0,007096 |
| 6 months | abundance | X09256 | Aspartyl-L- | 3 | Interventio | Experimen | Experimen | Control | 0,003479 |
| 6 months | abundance | M12 | M12 | IA_module | Interventio | Experimen | Experimen | Control | 0,002657 |
| 6 months | abundance | X09969 | 3-Methylac | 3 | Interventio | Experimen | Experimen | Control | 0,003399 |
| 6 months | abundance | X00702 | N-[(2S)-2-I | 3 | Interventio | Experimen | Experimen | Control | 0,008517 |
| 6 months | abundance | X05195 | Lys-phe_a | 3 | Interventio | Experimen | Experimen | Control | 0,008517 |
| 6 months | abundance | X06630 | 4-(METHYL | 3 | Interventio | Experimen | Experimen | Control | 0,007523 |
| 6 months | abundance | X09980 | N-(4-Hepta | 3 | Interventio | Experimen | Experimen | Control | 0,002868 |
| 6 months | abundance | M2 | M2 | IA_module | Interventio | Experimen | Experimen | Control | 0,012138 |
| 6 months | abundance | X09165 | N-[1-Carbo | 3 | Interventio | Experimen | Experimen | Control | 0,006049 |
| 6 months | abundance | X00027 | N-Acetylglu | 1 | Interventio | Experimen | Experimen | Control | 0,004511 |
| 6 months | abundance | X06675 | L-gamma-( | 3 | Interventio | Experimen | Experimen | Control | 0,006888 |
| 6 months | abundance | X08944 | Dipivefrin | 3 | Interventio | Experimen | Experimen | Control | 0,004096 |
| 6 months | abundance | X06958 | 2-[(carboxy | 2b | Interventio | Experimen | Experimen | Control | 0,012263 |
| 6 months | abundance | X07825 | Maltotriose | 1 | Interventio | Experimen | Experimen | Control | 0,002657 |
| 6 months | abundance | X05149 | S-Sulfocys | 3 | Interventio | Experimen | Experimen | Control | 0,007941 |
| 6 months | abundance | X07381 | Bis-D-fruct | 3 | Interventio | Experimen | Experimen | Control | 0,002868 |
| 6 months | abundance | X03604 | Gly-Trp_a | 3 | Interventio | Experimen | Experimen | Control | 0,012184 |
| 6 months | abundance | X04526 | N-Nonanoy | 3 | Interventio | Experimen | Experimen | Control | 0,019271 |
| 6 months | abundance | X02843 | 4-(1-Hydro | 3 | Interventio | Experimen | Experimen | Control | 0,009848 |
| 6 months | abundance | X08938 | Nonivamid | 3 | Interventio | Experimen | Experimen | Control | 0,002516 |
| 6 months | abundance | X10901 | bis-noryan | 3 | Interventio | Experimen | Experimen | Control | 0,012138 |
| 6 months | abundance | SL00049 | N-Acetylar | 2a | Interventio | Experimen | Experimen | Control | 0,012184 |
| 6 months | abundance | X01549 | 3-(Sulfooxy | 3 | Interventio | Experimen | Experimen | Control | 0,012184 |
| 6 months | abundance | Hexanoic a | Hexanoic a | CFA_panel | Interventio | Experimen | Experimen | Control | 0,06731 |
| 6 months | abundance | X10929 | Temozolon | 3 | Interventio | Experimen | Experimen | Control | 0,014596 |
| 6 months | abundance | X06866 | tert-Butyl 3 | 3 | Interventio | Experimen | Experimen | Control | 0,013902 |
| 6 months | abundance | X02181 | YWA1 | 3 | Interventio | Experimen | Experimen | Control | 0,012466 |
| 6 months | abundance | X10564 | 2-(1-Ethox | 3 | Interventio | Experimen | Experimen | Control | 0,031241 |
| 6 months | abundance | X00093 | Deoxyinosi | 1 | Interventio | Experimen | Experimen | Control | 0,022391 |
| 6 months | abundance | X08777 | 3-Aminosa | 2b | Interventio | Experimen | Experimen | Control | 0,012184 |
| 6 months | abundance | X06361 | Oxprenolol | 3 | Interventio | Experimen | Experimen | Control | 0,003235 |
| 6 months | abundance | X00045 | Threonine | 1 | Interventio | Experimen | Experimen | Control | 0,025332 |
| 6 months | abundance | X01884 | Roxane | 3 | Interventio | Experimen | Experimen | Control | 0,006888 |
| 6 months | abundance | X11421 | 8-Amino-7 | 3 | Interventio | Experimen | Experimen | Control | 0,005507 |
| 6 months | abundance | X11163 | Lys-Pro_a | 3 | Interventio | Experimen | Experimen | Control | 0,022035 |
| 6 months | abundance | X00114 | Indole-3-m | 1 | Interventio | Experimen | Experimen | Control | 0,017055 |
| 6 months | abundance | X08062 | Indole-3-ca | 1 | Interventio | Experimen | Experimen | Control | 0,013207 |
| 6 months | abundance | X10099 | ala-met | 3 | Interventio | Experimen | Experimen | Control | 0,021862 |
| 6 months | abundance | X00237 | OI170000( | 3 | Interventio | Experimen | Experimen | Control | 0,025324 |
| 6 months | abundance | SL00054 | Ornithine | 1 | Interventio | Experimen | Experimen | Control | 0,009848 |
| 6 months | abundance | X08670 | Hydroxypro | 3 | Interventio | Experimen | Experimen | Control | 0,015345 |
| 6 months | abundance | X10902 | Isoquinolir | 2b | Interventio | Experimen | Experimen | Control | 0,019271 |
| 6 months | abundance | X02513 | 7-Methylac | 2b | Interventio | Experimen | Experimen | Control | 0,024983 |
| 6 months | abundance | X05275 | H-DL-MET- | 3 | Interventio | Experimen | Experimen | Control | 0,012138 |
| 6 months | abundance | X02720 | ala-ser_b | 3 | Interventio | Experimen | Experimen | Control | 0,024328 |

| | | | | | | | | | |
|---|---|---|---|---|---|---|---|---|---|
| 6 months | abundance | X07961 | 2-(Hydroxy | 3 | Interventio | Experimen | Experimen | Control | 0,022718 |
| 6 months | abundance | SL00373 | N1-Methyl- | 2a | Interventio | Experimen | Experimen | Control | 0,027979 |
| 6 months | abundance | X04897 | Taxifolin | 3 | Interventio | Experimen | Experimen | Control | 0,019271 |
| 6 months | abundance | X10736 | Toxopyrimi | 3 | Interventio | Experimen | Experimen | Control | 0,012184 |
| 6 months | abundance | X09622 | N-Hydroxy- | 3 | Interventio | Experimen | Experimen | Control | 0,031688 |
| 6 months | abundance | SL00017 | Aspartic ac | 2a | Interventio | Experimen | Experimen | Control | 0,032825 |
| 6 months | abundance | X11717 | nicotianam | 3 | Interventio | Experimen | Experimen | Control | 0,024334 |
| 6 months | abundance | SL00189 | 3-Hydroxyb | 2a | Interventio | Experimen | Experimen | Control | 0,009848 |
| 6 months | abundance | SL00328 | 2-(hydroxy | 2a | Interventio | Experimen | Experimen | Control | 0,01944 |
| 6 months | abundance | X00320 | Panthenol_ | 3 | Interventio | Experimen | Experimen | Control | 0,034024 |
| 6 months | abundance | X07558 | Leucylproli | 2b | Interventio | Experimen | Experimen | Control | 0,012263 |
| 6 months | abundance | SL00140 | Pyridoxami | 1 | Interventio | Experimen | Experimen | Control | 0,021043 |
| 6 months | abundance | X10554 | Glycine | 1 | Interventio | Experimen | Experimen | Control | 0,030586 |
| 6 months | abundance | X08548 | N2-Acetyll | 1 | Interventio | Experimen | Experimen | Control | 0,021401 |
| 6 months | abundance | X10265 | 3-methyl-4 | 3 | Interventio | Experimen | Experimen | Control | 0,012184 |
| 6 months | abundance | X09963 | Iminoglycir | 3 | Interventio | Experimen | Experimen | Control | 0,040501 |
| 6 months | abundance | X11170 | QJ972000( | 3 | Interventio | Experimen | Experimen | Control | 0,030712 |
| 6 months | abundance | X10687 | Gln-Gln | 3 | Interventio | Experimen | Experimen | Control | 0,030712 |
| 6 months | abundance | X01246 | (2R)-1-{[(2- | 3 | Interventio | Experimen | Experimen | Control | 0,04742 |
| 6 months | abundance | X02380 | Propafenor | 3 | Interventio | Experimen | Experimen | Control | 0,027049 |
| 6 months | abundance | X07630 | Methionine | 1 | Interventio | Experimen | Experimen | Control | 0,020533 |
| 6 months | abundance | X00036 | Serine | 1 | Interventio | Experimen | Experimen | Control | 0,04742 |
| 6 months | abundance | X00127 | Nicotinic a | 1 | Interventio | Experimen | Experimen | Control | 0,035097 |
| 6 months | abundance | X09849 | 3-Methylac | 3 | Interventio | Experimen | Experimen | Control | 0,035097 |
| 6 months | abundance | X05033 | (2E)-N-3,7- | 3 | Interventio | Experimen | Experimen | Control | 0,020208 |
| 6 months | abundance | X01235 | 3-[(3-Hydro | 3 | Interventio | Experimen | Experimen | Control | 0,012184 |
| 6 months | abundance | SL00149 | Dissaccari | 2a | Interventio | Experimen | Experimen | Control | 0,017511 |
| 6 months | abundance | X08712 | 4-Iodoanis | 3 | Interventio | Experimen | Experimen | Control | 0,041133 |
| 6 months | abundance | X10706 | Furaneol | 3 | Interventio | Experimen | Experimen | Control | 0,012552 |
| 6 months | abundance | X11614 | 2-Methylth | 3 | Interventio | Experimen | Experimen | Control | 0,056436 |
| 6 months | abundance | SL00009 | N-Acetylhis | 2a | Interventio | Experimen | Experimen | Control | 0,055668 |
| 6 months | abundance | X08012 | Methylol D | 3 | Interventio | Experimen | Experimen | Control | 0,032515 |
| 6 months | abundance | X10276 | Maleamate | 3 | Interventio | Experimen | Experimen | Control | 0,053894 |
| 6 months | abundance | SL00081 | 5-Methylcy | 2a | Interventio | Experimen | Experimen | Control | 0,031688 |
| 6 months | abundance | X00998 | epsilon-(ga | 3 | Interventio | Experimen | Experimen | Control | 0,046863 |
| 6 months | abundance | X06240 | His-pro_a | 3 | Interventio | Experimen | Experimen | Control | 0,032515 |
| 6 months | abundance | X02760 | 6-hydroxyp | 3 | Interventio | Experimen | Experimen | Control | 0,03552 |
| 6 months | abundance | X08895 | pentoxyl | 3 | Interventio | Experimen | Experimen | Control | 0,04788 |
| 6 months | abundance | X11036 | 3-Hydroxyp | 2b | Interventio | Experimen | Experimen | Control | 0,035388 |
| 6 months | abundance | X09981 | N-Acetylva | 3 | Interventio | Experimen | Experimen | Control | 0,025324 |
| 6 months | abundance | X07125 | Zalcitabine | 3 | Interventio | Experimen | Experimen | Control | 0,035357 |
| 6 months | abundance | X07856 | (3R,4S,5S,( | 3 | Interventio | Experimen | Experimen | Control | 0,032515 |
| 6 months | abundance | X01340 | Cadralazin | 3 | Interventio | Experimen | Experimen | Control | 0,045594 |
| 6 months | abundance | X08116 | Methyl (2Z, | 3 | Interventio | Experimen | Experimen | Control | 0,056436 |
| 6 months | abundance | X10415 | beta-D-Glc | 3 | Interventio | Experimen | Experimen | Control | 0,025324 |
| 6 months | abundance | X06409 | Glycylproli | 2b | Interventio | Experimen | Experimen | Control | 0,035097 |
| 6 months | abundance | X06771 | MFCD0015 | 3 | Interventio | Experimen | Experimen | Control | 0,056436 |
| 6 months | abundance | X08525 | MFCD1869 | 3 | Interventio | Experimen | Experimen | Control | 0,072606 |

| | | | | | | | | | |
|---|---|---|---|---|---|---|---|---|---|
| 6 months | abundance | X08230 | Coumaron | 3 | Interventio | Experimen | Experimen | Control | 0,066068 |
| 6 months | abundance | X00683 | Zinecard_a | 3 | Interventio | Experimen | Experimen | Control | 0,068028 |
| 6 months | abundance | X00057 | N-Acetylor | 1 | Interventio | Experimen | Experimen | Control | 0,068026 |
| 6 months | abundance | X00094 | Methionine | 1 | Interventio | Experimen | Experimen | Control | 0,080527 |
| 6 months | abundance | X06089 | riboprine | 3 | Interventio | Experimen | Experimen | Control | 0,074642 |
| 6 months | abundance | X07460 | 1-pyrroline | 3 | Interventio | Experimen | Experimen | Control | 0,050313 |
| 6 months | abundance | X04339 | L-Glutamic | 3 | Interventio | Experimen | Experimen | Control | 0,055563 |
| 6 months | abundance | X00124 | Leucine | 1 | Interventio | Experimen | Experimen | Control | 0,056436 |
| 6 months | abundance | X10486 | Coumarin | 2b | Interventio | Experimen | Experimen | Control | 0,077439 |
| 6 months | abundance | X10722 | Acetanilide | 2b | Interventio | Experimen | Experimen | Control | 0,06731 |
| 6 months | abundance | X07512 | 4-{[(3-Hydr | 3 | Interventio | Experimen | Experimen | Control | 0,046863 |
| 6 months | abundance | X09724 | Formylkynu | 3 | Interventio | Experimen | Experimen | Control | 0,099094 |
| 6 months | abundance | X04822 | 6-Sulfatoxy | 3 | Interventio | Experimen | Experimen | Control | 0,055312 |
| 6 months | abundance | SL00074 | Valine/ 5-A | 2a | Interventio | Experimen | Experimen | Control | 0,059922 |
| 6 months | abundance | X10713 | norhaman | 3 | Interventio | Experimen | Experimen | Control | 0,065238 |
| 6 months | abundance | X04571 | 13a-Hydro: | 3 | Interventio | Experimen | Experimen | Control | 0,06731 |
| 6 months | abundance | X05029 | Hypericin | 3 | Interventio | Experimen | Experimen | Control | 0,068026 |
| 6 months | abundance | X05459 | Scopoletin | 3 | Interventio | Experimen | Experimen | Control | 0,077865 |
| 6 months | abundance | SL00061 | Proline | 1 | Interventio | Experimen | Experimen | Control | 0,065283 |
| 6 months | abundance | X00076 | Isoleucine | 1 | Interventio | Experimen | Experimen | Control | 0,065793 |
| 6 months | abundance | X06858 | tert-Butyl 3 | 3 | Interventio | Experimen | Experimen | Control | 0,060957 |
| 6 months | abundance | X00073 | Indole-3-la | 1 | Interventio | Experimen | Experimen | Control | 0,047694 |
| 6 months | abundance | X07647 | Genistein | 1 | Interventio | Experimen | Experimen | Control | 0,100431 |
| 6 months | abundance | X07334 | 2,2-Bis(hyc | 3 | Interventio | Experimen | Experimen | Control | 0,080527 |
| 6 months | abundance | X09385 | 2-(1-Napht | 3 | Interventio | Experimen | Experimen | Control | 0,103356 |
| 6 months | abundance | X07989 | Piperidine_ | 3 | Interventio | Experimen | Experimen | Control | 0,071535 |
| 6 months | abundance | X06043 | vinyl sulfid | 3 | Interventio | Experimen | Experimen | Control | 0,103571 |
| 6 months | abundance | X07050 | 2-Methylth | 3 | Interventio | Experimen | Experimen | Control | 0,103571 |
| 6 months | abundance | X09391 | ophthalmic | 3 | Interventio | Experimen | Experimen | Control | 0,110798 |
| 6 months | abundance | X08549 | 3-Methylsu | 3 | Interventio | Experimen | Experimen | Control | 0,106358 |
| 6 months | abundance | X08002 | (2S)-6-Ami | 3 | Interventio | Experimen | Experimen | Control | 0,102658 |
| 6 months | abundance | X06454 | 8-Amino-7- | 3 | Interventio | Experimen | Experimen | Control | 0,103571 |
| 6 months | abundance | X11117 | Coprine_b | 3 | Interventio | Experimen | Experimen | Control | 0,059922 |
| 6 months | abundance | X07974 | Isoprene | 3 | Interventio | Experimen | Experimen | Control | 0,080527 |
| 6 months | abundance | X02442 | 3-(Sulfooxy | 3 | Interventio | Experimen | Experimen | Control | 0,099711 |
| 6 months | abundance | X05995 | Diacetin_a | 3 | Interventio | Experimen | Experimen | Control | 0,094328 |
| 6 months | abundance | X05478 | S(8)-aminc | 3 | Interventio | Experimen | Experimen | Control | 0,074642 |
| 6 months | abundance | X10442 | (6alpha,11 | 3 | Interventio | Experimen | Experimen | Control | 0,113797 |
| 6 months | abundance | X07054 | Valylprolin | 2b | Interventio | Experimen | Experimen | Control | 0,062463 |
| 6 months | abundance | X08699 | Aurorix | 3 | Interventio | Experimen | Experimen | Control | 0,095545 |
| 6 months | abundance | X06812 | 2-Hydroxy- | 3 | Interventio | Experimen | Experimen | Control | 0,127044 |
| 6 months | abundance | X06404 | Flemichap | 3 | Interventio | Experimen | Experimen | Control | 0,12884 |
| 6 months | abundance | X09554 | N-Propiony | 3 | Interventio | Experimen | Experimen | Control | 0,075736 |
| 6 months | abundance | X09726 | indoline-2- | 2b | Interventio | Experimen | Experimen | Control | 0,126909 |
| 6 months | abundance | X10375 | 4-Indoleca | 2b | Interventio | Experimen | Experimen | Control | 0,126909 |
| 6 months | abundance | X00050 | β-Hydroxyi | 1 | Interventio | Experimen | Experimen | Control | 0,143926 |
| 6 months | abundance | X00068 | N-Acetylm | 1 | Interventio | Experimen | Experimen | Control | 0,126909 |
| 6 months | abundance | X08111 | 2-Hydroxyc | 2b | Interventio | Experimen | Experimen | Control | 0,137332 |

| | | | | | | | | | |
|---|---|---|---|---|---|---|---|---|---|
| 6 months | abundance | X07914 | (-)-Erythror | 3 | Interventio | Experimen | Experimen | Control | 0,063697 |
| 6 months | abundance | X05357 | NPYR | 3 | Interventio | Experimen | Experimen | Control | 0,113797 |
| 6 months | abundance | X01621 | 2-(2,4-Dihy | 3 | Interventio | Experimen | Experimen | Control | 0,148418 |
| 6 months | abundance | M9 | M9 | IA_module | Interventio | Experimen | Experimen | Control | 0,118447 |
| 6 months | abundance | X03363 | Choline Alf | 3 | Interventio | Experimen | Experimen | Control | 0,130162 |
| 6 months | abundance | X08421 | Cys-tyr | 3 | Interventio | Experimen | Experimen | Control | 0,133886 |
| 6 months | abundance | X00106 | N-Acetyltry | 1 | Interventio | Experimen | Experimen | Control | 0,097673 |
| 6 months | abundance | X08288 | Capryloylg | 3 | Interventio | Experimen | Experimen | Control | 0,071535 |
| 6 months | abundance | X04877 | Trifluorome | 3 | Interventio | Experimen | Experimen | Control | 0,174109 |
| 6 months | abundance | X06622 | Lys-Pro_b | 3 | Interventio | Experimen | Experimen | Control | 0,105786 |
| 6 months | abundance | SL00247 | Shikimic ac | 2a | Interventio | Experimen | Experimen | Control | 0,068028 |
| 6 months | abundance | X01208 | Lys-Pro_c | 3 | Interventio | Experimen | Experimen | Control | 0,089671 |
| 6 months | abundance | X08677 | 1-methylhy | 3 | Interventio | Experimen | Experimen | Control | 0,126909 |
| 6 months | abundance | X09122 | Valyl-4-hyc | 3 | Interventio | Experimen | Experimen | Control | 0,115704 |
| 6 months | abundance | SL00103 | Gulonic ac | 2a | Interventio | Experimen | Experimen | Control | 0,125436 |
| 6 months | abundance | X08098 | Citrulline | 1 | Interventio | Experimen | Experimen | Control | 0,109405 |
| 6 months | abundance | X05240 | Dimeric me | 3 | Interventio | Experimen | Experimen | Control | 0,157793 |
| 6 months | abundance | X10769 | Cystine | 1 | Interventio | Experimen | Experimen | Control | 0,161409 |
| 6 months | abundance | SL00210 | Galactonic | 2a | Interventio | Experimen | Experimen | Control | 0,146968 |
| 6 months | abundance | SL00334 | Imidazolea | 2a | Interventio | Experimen | Experimen | Control | 0,067529 |
| 6 months | abundance | SL00108 | Histidinol | 2a | Interventio | Experimen | Experimen | Control | 0,110798 |
| 6 months | abundance | X05214 | [7-Hydroxy | 3 | Interventio | Experimen | Experimen | Control | 0,0902 |
| 6 months | abundance | X00130 | Deoxyuridi | 1 | Interventio | Experimen | Experimen | Control | 0,161409 |
| 6 months | abundance | X07476 | 4-Hydroxyb | 1 | Interventio | Experimen | Experimen | Control | 0,145494 |
| 6 months | abundance | SL00215 | Glycolic ac | 2a | Interventio | Experimen | Experimen | Control | 0,142606 |
| 6 months | abundance | X08019 | Thymidine | 1 | Interventio | Experimen | Experimen | Control | 0,149045 |
| 6 months | abundance | X01031 | 3-(14-Ethy | 3 | Interventio | Experimen | Experimen | Control | 0,126909 |
| 6 months | abundance | X07424 | Rutinose (b | 3 | Interventio | Experimen | Experimen | Control | 0,160244 |
| 6 months | abundance | X08822 | CYS-ASP | 3 | Interventio | Experimen | Experimen | Control | 0,170002 |
| 6 months | abundance | X06034 | N-[(2S)-2-H | 3 | Interventio | Experimen | Experimen | Control | 0,170002 |
| 6 months | abundance | X00071 | Tyrosine | 1 | Interventio | Experimen | Experimen | Control | 0,165608 |
| 6 months | abundance | X08150 | DL-4-Hydro | 2b | Interventio | Experimen | Experimen | Control | 0,160244 |
| 6 months | abundance | X08616 | MFCD0002 | 3 | Interventio | Experimen | Experimen | Control | 0,187472 |
| 6 months | abundance | X02080 | (-)-Aspidos | 3 | Interventio | Experimen | Experimen | Control | 0,248374 |
| 6 months | abundance | X10476 | GAMMA-HY | 3 | Interventio | Experimen | Experimen | Control | 0,196868 |
| 6 months | abundance | X00011 | Glutamine | 2a | Interventio | Experimen | Experimen | Control | 0,114299 |
| 6 months | abundance | X09721 | 3-(2,3-Dihy | 3 | Interventio | Experimen | Experimen | Control | 0,22431 |
| 6 months | abundance | X07216 | pentobarbi | 3 | Interventio | Experimen | Experimen | Control | 0,153848 |
| 6 months | abundance | X04788 | 2-(4-Isopro | 3 | Interventio | Experimen | Experimen | Control | 0,172247 |
| 6 months | abundance | X00102 | Tryptophan | 1 | Interventio | Experimen | Experimen | Control | 0,171599 |
| 6 months | abundance | X06870 | Menadiol | 3 | Interventio | Experimen | Experimen | Control | 0,160244 |
| 6 months | abundance | X02101 | Succinic an | 3 | Interventio | Experimen | Experimen | Control | 0,172965 |
| 6 months | abundance | X09741 | Estrone glu | 3 | Interventio | Experimen | Experimen | Control | 0,10858 |
| 6 months | abundance | X02468 | N,N-Diethy | 3 | Interventio | Experimen | Experimen | Control | 0,169648 |
| 6 months | abundance | X07537 | alpha-Cha | 3 | Interventio | Experimen | Experimen | Control | 0,171874 |
| 6 months | abundance | X06507 | Gly-Lys | 3 | Interventio | Experimen | Experimen | Control | 0,201493 |
| 6 months | abundance | X10793 | Zalcitabine | 3 | Interventio | Experimen | Experimen | Control | 0,103211 |
| 6 months | abundance | SL00440 | 5-Methylcy | 2a | Interventio | Experimen | Experimen | Control | 0,162202 |

| | | | | | | | | | |
|---|---|---|---|---|---|---|---|---|---|
| 6 months | abundance | SL00246 | Pentose III | 2a | Interventio | Experimen | Experimen | Control | 0,171874 |
| 6 months | abundance | X00090 | Malic acid | 1 | Interventio | Experimen | Experimen | Control | 0,133886 |
| 6 months | abundance | X06656 | Valylvaline | 3 | Interventio | Experimen | Experimen | Control | 0,20627 |
| 6 months | abundance | X11556 | N-[(2E)-3-( | 3 | Interventio | Experimen | Experimen | Control | 0,148764 |
| 6 months | abundance | X10248 | (+)-Etomid | 3 | Interventio | Experimen | Experimen | Control | 0,167056 |
| 6 months | abundance | X02277 | tert-Butyl 3 | 3 | Interventio | Experimen | Experimen | Control | 0,187192 |
| 6 months | abundance | X01367 | MFCD1869 | 3 | Interventio | Experimen | Experimen | Control | 0,225907 |
| 6 months | abundance | X00060 | Ethylmalor | 1 | Interventio | Experimen | Experimen | Control | 0,186012 |
| 6 months | abundance | SL00436 | 1-Carboxy | 2a | Interventio | Experimen | Experimen | Control | 0,143926 |
| 6 months | abundance | X09654 | linatine | 3 | Interventio | Experimen | Experimen | Control | 0,11721 |
| 6 months | abundance | X08436 | N,N-Dimet | 3 | Interventio | Experimen | Experimen | Control | 0,287584 |
| 6 months | abundance | X10148 | cys-met | 3 | Interventio | Experimen | Experimen | Control | 0,215732 |
| 6 months | abundance | X00088 | Pantotheni | 1 | Interventio | Experimen | Experimen | Control | 0,254807 |
| 6 months | abundance | X00367 | hexobarbit | 3 | Interventio | Experimen | Experimen | Control | 0,179358 |
| 6 months | abundance | X09390 | asn-pro_a | 3 | Interventio | Experimen | Experimen | Control | 0,138188 |
| 6 months | abundance | X06634 | Glycitein | 3 | Interventio | Experimen | Experimen | Control | 0,186692 |
| 6 months | abundance | X11067 | Resveratro | 3 | Interventio | Experimen | Experimen | Control | 0,174109 |
| 6 months | abundance | X02599 | Dihydrothy | 3 | Interventio | Experimen | Experimen | Control | 0,201493 |
| 6 months | abundance | X00204 | 4-O-{3-O-[( | 3 | Interventio | Experimen | Experimen | Control | 0,23628 |
| 6 months | abundance | X11375 | L-gamma-( | 3 | Interventio | Experimen | Experimen | Control | 0,180061 |
| 6 months | abundance | X06631 | 2-Isopropy | 2b | Interventio | Experimen | Experimen | Control | 0,174109 |
| 6 months | abundance | X05878 | 1,2-dihydro | 3 | Interventio | Experimen | Experimen | Control | 0,23628 |
| 6 months | abundance | X09533 | S(6)-acetyl | 3 | Interventio | Experimen | Experimen | Control | 0,234335 |
| 6 months | abundance | X06546 | 4-Acetami | 2b | Interventio | Experimen | Experimen | Control | 0,186094 |
| 6 months | abundance | M25 | M25 | IA_module | Interventio | Experimen | Experimen | Control | 0,246099 |
| 6 months | abundance | X06167 | N-Acetylas | 2a | Interventio | Experimen | Experimen | Control | 0,169034 |
| 6 months | abundance | X07530 | Alanine | 1 | Interventio | Experimen | Experimen | Control | 0,189396 |
| 6 months | abundance | X07873 | (3Z,6Z,9Z,1 | 3 | Interventio | Experimen | Experimen | Control | 0,261581 |
| 6 months | abundance | X02746 | N,N-Diethy | 3 | Interventio | Experimen | Experimen | Control | 0,247541 |
| 6 months | abundance | X02514 | 3-Ureidopr | 1 | Interventio | Experimen | Experimen | Control | 0,220238 |
| 6 months | abundance | X05982 | (19R,25S)- | 3 | Interventio | Experimen | Experimen | Control | 0,25478 |
| 6 months | abundance | X00574 | miglustat | 3 | Interventio | Experimen | Experimen | Control | 0,217996 |
| 6 months | abundance | X00018 | Succinic ac | 1 | Interventio | Experimen | Experimen | Control | 0,196145 |
| 6 months | abundance | X02466 | Validamyci | 3 | Interventio | Experimen | Experimen | Control | 0,186094 |
| 6 months | abundance | X07451 | Valylvaline | 3 | Interventio | Experimen | Experimen | Control | 0,179439 |
| 6 months | abundance | X01045 | Ethyl malto | 3 | Interventio | Experimen | Experimen | Control | 0,134826 |
| 6 months | abundance | X11257 | 1-Methylgu | 2b | Interventio | Experimen | Experimen | Control | 0,287584 |
| 6 months | abundance | X10647 | 4-Hydroxyp | 3 | Interventio | Experimen | Experimen | Control | 0,278551 |
| 6 months | abundance | SL00089 | Dissaccari | 2a | Interventio | Experimen | Experimen | Control | 0,162222 |
| 6 months | abundance | X06254 | D-Alanyl-D | 3 | Interventio | Experimen | Experimen | Control | 0,274433 |
| 6 months | abundance | X05177 | cis-3-Hexe | 3 | Interventio | Experimen | Experimen | Control | 0,284006 |
| 6 months | abundance | X09635 | 2,4,6-Triiso | 3 | Interventio | Experimen | Experimen | Control | 0,301875 |
| 6 months | abundance | X03340 | His-pro_b | 3 | Interventio | Experimen | Experimen | Control | 0,20627 |
| 6 months | abundance | X07270 | 5-(2-Carbo | 3 | Interventio | Experimen | Experimen | Control | 0,250568 |
| 6 months | abundance | X07504 | 4-Acetami | 2b | Interventio | Experimen | Experimen | Control | 0,254963 |
| 6 months | abundance | X10664 | N1-(5-metl | 2b | Interventio | Experimen | Experimen | Control | 0,248374 |
| 6 months | abundance | X08013 | Deoxysuga | 1 | Interventio | Experimen | Experimen | Control | 0,262216 |
| 6 months | abundance | X09609 | N-{3-[(4-Ac | 3 | Interventio | Experimen | Experimen | Control | 0,270217 |

| | | | | | | | | | |
|---|---|---|---|---|---|---|---|---|---|
| 6 months | abundance | X02854 | Marimasta | 3 | Interventio | Experimen | Experimen | Control | 0,301288 |
| 6 months | abundance | X09657 | 2,3,4,5,6-F | 3 | Interventio | Experimen | Experimen | Control | 0,303634 |
| 6 months | abundance | X08028 | Solanidine | 3 | Interventio | Experimen | Experimen | Control | 0,273643 |
| 6 months | abundance | X09555 | N4-(beta-N | 3 | Interventio | Experimen | Experimen | Control | 0,287584 |
| 6 months | abundance | X06977 | 4-Hydroxyp | 3 | Interventio | Experimen | Experimen | Control | 0,287584 |
| 6 months | abundance | X06943 | NSC 92778 | 3 | Interventio | Experimen | Experimen | Control | 0,303982 |
| 6 months | abundance | X04310 | Aminohipp | 3 | Interventio | Experimen | Experimen | Control | 0,22431 |
| 6 months | abundance | X00123 | Phenylalar | 1 | Interventio | Experimen | Experimen | Control | 0,275763 |
| 6 months | abundance | X07524 | Prolinamid | 2b | Interventio | Experimen | Experimen | Control | 0,301288 |
| 6 months | abundance | X09951 | 5beta-Cypr | 3 | Interventio | Experimen | Experimen | Control | 0,310912 |
| 6 months | abundance | X00101 | Threonic ac | 1 | Interventio | Experimen | Experimen | Control | 0,275484 |
| 6 months | abundance | X04682 | 1-Methylin | 3 | Interventio | Experimen | Experimen | Control | 0,334391 |
| 6 months | abundance | X10036 | Nimodipine | 3 | Interventio | Experimen | Experimen | Control | 0,316696 |
| 6 months | abundance | X02606 | 4-Amino-1- | 3 | Interventio | Experimen | Experimen | Control | 0,33088 |
| 6 months | abundance | X07164 | N-Acetyl-5 | 3 | Interventio | Experimen | Experimen | Control | 0,391188 |
| 6 months | abundance | X00344 | SECONAL_ | 3 | Interventio | Experimen | Experimen | Control | 0,324575 |
| 6 months | abundance | X06510 | N-{4-[(2R,3 | 2b | Interventio | Experimen | Experimen | Control | 0,213812 |
| 6 months | abundance | X08870 | Tetraacety | 3 | Interventio | Experimen | Experimen | Control | 0,310448 |
| 6 months | abundance | X07084 | heptabarbi | 3 | Interventio | Experimen | Experimen | Control | 0,33088 |
| 6 months | abundance | X09336 | 1,1'-[1,12-I | 3 | Interventio | Experimen | Experimen | Control | 0,312951 |
| 6 months | abundance | X00182 | 3-(Butylsul | 3 | Interventio | Experimen | Experimen | Control | 0,278551 |
| 6 months | abundance | X00264 | PEG n12 | 2b | Interventio | Experimen | Experimen | Control | 0,373224 |
| 6 months | abundance | X02073 | N-acetyl-9- | 3 | Interventio | Experimen | Experimen | Control | 0,25478 |
| 6 months | abundance | X04553 | N-(4-Amino | 3 | Interventio | Experimen | Experimen | Control | 0,346785 |
| 6 months | abundance | X08714 | Arabic acid | 3 | Interventio | Experimen | Experimen | Control | 0,288871 |
| 6 months | abundance | X01285 | FB9500000 | 3 | Interventio | Experimen | Experimen | Control | 0,365232 |
| 6 months | abundance | X04759 | 7-Aminome | 3 | Interventio | Experimen | Experimen | Control | 0,378791 |
| 6 months | abundance | X09476 | N(1),N(8)-I | 3 | Interventio | Experimen | Experimen | Control | 0,324575 |
| 6 months | abundance | X03097 | 6-hydroxym | 3 | Interventio | Experimen | Experimen | Control | 0,375103 |
| 6 months | abundance | X06906 | Arg-pro | 3 | Interventio | Experimen | Experimen | Control | 0,310912 |
| 6 months | abundance | X06735 | 7-Methylxa | 2b | Interventio | Experimen | Experimen | Control | 0,355054 |
| 6 months | abundance | X01996 | 3-Hydroxy- | 3 | Interventio | Experimen | Experimen | Control | 0,28123 |
| 6 months | abundance | X07420 | DNOP_h | 3 | Interventio | Experimen | Experimen | Control | 0,310912 |
| 6 months | abundance | X07002 | Paraldehyd | 3 | Interventio | Experimen | Experimen | Control | 0,291867 |
| 6 months | abundance | X08412 | 2-Hydroxyp | 2b | Interventio | Experimen | Experimen | Control | 0,320284 |
| 6 months | abundance | X06836 | asn-lys | 3 | Interventio | Experimen | Experimen | Control | 0,358168 |
| 6 months | abundance | X02000 | 11-(4-Hydr | 3 | Interventio | Experimen | Experimen | Control | 0,355054 |
| 6 months | abundance | X09010 | 4-Hydroxyp | 1 | Interventio | Experimen | Experimen | Control | 0,260834 |
| 6 months | abundance | X10896 | 6-Hydroxym | 2b | Interventio | Experimen | Experimen | Control | 0,310448 |
| 6 months | abundance | X09091 | N-(3-aceta | 3 | Interventio | Experimen | Experimen | Control | 0,299663 |
| 6 months | abundance | X00040 | N-Acetyltyr | 1 | Interventio | Experimen | Experimen | Control | 0,346785 |
| 6 months | abundance | X02504 | DLK (Pepti | 2b | Interventio | Experimen | Experimen | Control | 0,30493 |
| 6 months | abundance | X05561 | 1Î²-Hydroxy | 3 | Interventio | Experimen | Experimen | Control | 0,326255 |
| 6 months | abundance | SL00325 | 2-Methylm | 2a | Interventio | Experimen | Experimen | Control | 0,34199 |
| 6 months | abundance | X07907 | 4-Acetamid | 1 | Interventio | Experimen | Experimen | Control | 0,380677 |
| 6 months | abundance | X10224 | Imidazolela | 2b | Interventio | Experimen | Experimen | Control | 0,307119 |
| 6 months | abundance | X11310 | Leu-Val_a | 3 | Interventio | Experimen | Experimen | Control | 0,34021 |
| 6 months | abundance | X00020 | sugar alcol | 1 | Interventio | Experimen | Experimen | Control | 0,381461 |

| | | | | | | | | | |
|---|---|---|---|---|---|---|---|---|---|
| 6 months | abundance | X08096 | (2E)-3-Met | 3 | Interventio | Experimen | Experimen | Control | 0,320854 |
| 6 months | abundance | X06016 | N,N-Dimet | 3 | Interventio | Experimen | Experimen | Control | 0,402458 |
| 6 months | abundance | X07400 | Tranexami | 2b | Interventio | Experimen | Experimen | Control | 0,34199 |
| 6 months | abundance | Acetic acid | Acetic acid | CFA_panel | Interventio | Experimen | Experimen | Control | 0,310448 |
| 6 months | abundance | X00067 | Hypoxanth | 1 | Interventio | Experimen | Experimen | Control | 0,299663 |
| 6 months | abundance | X00403 | 7alpha-Hy | 3 | Interventio | Experimen | Experimen | Control | 0,370881 |
| 6 months | abundance | X06529 | Methanesu | 2a | Interventio | Experimen | Experimen | Control | 0,375103 |
| 6 months | abundance | X06230 | Glu-Gly | 3 | Interventio | Experimen | Experimen | Control | 0,286639 |
| 6 months | abundance | X11258 | Valyl-4-hyc | 3 | Interventio | Experimen | Experimen | Control | 0,378866 |
| 6 months | abundance | X02214 | (E)-4-Meth | 3 | Interventio | Experimen | Experimen | Control | 0,402458 |
| 6 months | abundance | X07073 | asn-val_a | 3 | Interventio | Experimen | Experimen | Control | 0,410744 |
| 6 months | abundance | X11281 | Penbutolol | 3 | Interventio | Experimen | Experimen | Control | 0,294471 |
| 6 months | abundance | SL00265 | Purine | 2a | Interventio | Experimen | Experimen | Control | 0,417706 |
| 6 months | abundance | X10680 | 8-hydroxy- | 3 | Interventio | Experimen | Experimen | Control | 0,452861 |
| 6 months | abundance | X09260 | (4S)-4-[(2E | 3 | Interventio | Experimen | Experimen | Control | 0,303733 |
| 6 months | abundance | SL00315 | Malonic ac | 2a | Interventio | Experimen | Experimen | Control | 0,428209 |
| 6 months | abundance | X04975 | MFCD0995 | 3 | Interventio | Experimen | Experimen | Control | 0,425528 |
| 6 months | abundance | X11319 | Methohexi | 3 | Interventio | Experimen | Experimen | Control | 0,328207 |
| 6 months | abundance | X00242 | IN00258 | 3 | Interventio | Experimen | Experimen | Control | 0,296902 |
| 6 months | abundance | X10367 | 2-(1-Napht | 3 | Interventio | Experimen | Experimen | Control | 0,39359 |
| 6 months | abundance | X08476 | Coprine_c | 3 | Interventio | Experimen | Experimen | Control | 0,401596 |
| 6 months | abundance | X07518 | (+/-)-2-Hyc | 3 | Interventio | Experimen | Experimen | Control | 0,375606 |
| 6 months | abundance | X11186 | Fluocinolo | 2b | Interventio | Experimen | Experimen | Control | 0,433028 |
| 6 months | abundance | X00670 | 7-Chloro-5 | 3 | Interventio | Experimen | Experimen | Control | 0,392683 |
| 6 months | abundance | SL00097 | Hexoses II | 1 | Interventio | Experimen | Experimen | Control | 0,275484 |
| 6 months | abundance | X09895 | 1-(beta-D- | 3 | Interventio | Experimen | Experimen | Control | 0,414019 |
| 6 months | abundance | X09713 | N(2)-succi | 3 | Interventio | Experimen | Experimen | Control | 0,375103 |
| 6 months | abundance | X02317 | 2-Aminooc | 2b | Interventio | Experimen | Experimen | Control | 0,402458 |
| 6 months | abundance | X06372 | N,N-dimet | 2b | Interventio | Experimen | Experimen | Control | 0,42661 |
| 6 months | abundance | M16 | M16 | IA_module | Interventio | Experimen | Experimen | Control | 0,387982 |
| 6 months | abundance | SL00264 | Methyl ace | 2a | Interventio | Experimen | Experimen | Control | 0,411713 |
| 6 months | abundance | X01316 | 5-Hydanto | 3 | Interventio | Experimen | Experimen | Control | 0,371899 |
| 6 months | abundance | X08758 | Methohexi | 3 | Interventio | Experimen | Experimen | Control | 0,375103 |
| 6 months | abundance | X02687 | S-Propylcy | 3 | Interventio | Experimen | Experimen | Control | 0,336351 |
| 6 months | abundance | X10454 | 8-Amino-7 | 3 | Interventio | Experimen | Experimen | Control | 0,428209 |
| 6 months | abundance | X01776 | N-(Carboxy | 3 | Interventio | Experimen | Experimen | Control | 0,425873 |
| 6 months | abundance | X03070 | 2-Methoxy | 3 | Interventio | Experimen | Experimen | Control | 0,429158 |
| 6 months | abundance | X09810 | Zalcitabine | 3 | Interventio | Experimen | Experimen | Control | 0,511339 |
| 6 months | abundance | X02265 | ophthalmi | 3 | Interventio | Experimen | Experimen | Control | 0,452966 |
| 6 months | abundance | X10417 | L-Sacchar | 2b | Interventio | Experimen | Experimen | Control | 0,428209 |
| 6 months | abundance | X01873 | Methyl 2,3 | 3 | Interventio | Experimen | Experimen | Control | 0,472806 |
| 6 months | abundance | X08244 | Setoclavin | 3 | Interventio | Experimen | Experimen | Control | 0,391188 |
| 6 months | abundance | X04593 | Methional | 3 | Interventio | Experimen | Experimen | Control | 0,405943 |
| 6 months | abundance | X00384 | 3-Formyl-2 | 3 | Interventio | Experimen | Experimen | Control | 0,452191 |
| 6 months | abundance | X00807 | Midodrine_ | 3 | Interventio | Experimen | Experimen | Control | 0,279022 |
| 6 months | abundance | X03855 | 2-{(2S,4S)- | 3 | Interventio | Experimen | Experimen | Control | 0,401596 |
| 6 months | abundance | X08323 | (15Z)-9,12 | 2b | Interventio | Experimen | Experimen | Control | 0,441241 |
| 6 months | abundance | X09950 | 3-Hydroxy | 3 | Interventio | Experimen | Experimen | Control | 0,478267 |

| | | | | | | | | | |
|---|---|---|---|---|---|---|---|---|---|
| 6 months | abundance | X06722 | LW800000 | 3 | Interventio | Experimen | Experimen | Control | 0,320854 |
| 6 months | abundance | X10660 | 2-Methylth | 3 | Interventio | Experimen | Experimen | Control | 0,447107 |
| 6 months | abundance | M13 | M13 | IA_module | Interventio | Experimen | Experimen | Control | 0,433028 |
| 6 months | abundance | X02702 | R-(+)-Etira | 3 | Interventio | Experimen | Experimen | Control | 0,459973 |
| 6 months | abundance | X07900 | presqualer | 3 | Interventio | Experimen | Experimen | Control | 0,476616 |
| 6 months | abundance | SL00293 | Butyrylcarr | 2a | Interventio | Experimen | Experimen | Control | 0,416238 |
| 6 months | abundance | SL00240 | Pyruvic aci | 1 | Interventio | Experimen | Experimen | Control | 0,33088 |
| 6 months | abundance | X10772 | Bicine_a | 3 | Interventio | Experimen | Experimen | Control | 0,455427 |
| 6 months | abundance | X00889 | LW800000 | 3 | Interventio | Experimen | Experimen | Control | 0,428209 |
| 6 months | abundance | X08625 | Hept-2-ulo | 3 | Interventio | Experimen | Experimen | Control | 0,463949 |
| 6 months | abundance | X11733 | 1,3-Dihydr | 3 | Interventio | Experimen | Experimen | Control | 0,455427 |
| 6 months | abundance | X02238 | 6-Myoporo | 3 | Interventio | Experimen | Experimen | Control | 0,485832 |
| 6 months | abundance | X08663 | Pilocarpine | 2b | Interventio | Experimen | Experimen | Control | 0,451974 |
| 6 months | abundance | SL00104 | Hexoses l | 1 | Interventio | Experimen | Experimen | Control | 0,403055 |
| 6 months | abundance | X08306 | 1-Methylxa | 2b | Interventio | Experimen | Experimen | Control | 0,474044 |
| 6 months | abundance | X06146 | S-Methyl-1 | 3 | Interventio | Experimen | Experimen | Control | 0,402458 |
| 6 months | abundance | M31 | M31 | IA_module | Interventio | Experimen | Experimen | Control | 0,373224 |
| 6 months | abundance | X07414 | 2,2-Bis(hy | 3 | Interventio | Experimen | Experimen | Control | 0,472806 |
| 6 months | abundance | X08212 | LU345300 | 3 | Interventio | Experimen | Experimen | Control | 0,365141 |
| 6 months | abundance | X04229 | gamma-L-g | 3 | Interventio | Experimen | Experimen | Control | 0,401596 |
| 6 months | abundance | X11273 | trp-pro | 3 | Interventio | Experimen | Experimen | Control | 0,476616 |
| 6 months | abundance | X06642 | Homocitru | 2a | Interventio | Experimen | Experimen | Control | 0,472806 |
| 6 months | abundance | X09222 | L-gamma-( | 3 | Interventio | Experimen | Experimen | Control | 0,462525 |
| 6 months | abundance | X05306 | 2-methoxy | 3 | Interventio | Experimen | Experimen | Control | 0,472806 |
| 6 months | abundance | X01341 | metixene | 3 | Interventio | Experimen | Experimen | Control | 0,474371 |
| 6 months | abundance | X00305 | Formylkyn | 3 | Interventio | Experimen | Experimen | Control | 0,462525 |
| 6 months | abundance | X09008 | 5-guanidin | 3 | Interventio | Experimen | Experimen | Control | 0,452966 |
| 6 months | abundance | X08058 | 2-Hydroxy- | 3 | Interventio | Experimen | Experimen | Control | 0,472806 |
| 6 months | abundance | X10137 | 1-Methyl-1 | 3 | Interventio | Experimen | Experimen | Control | 0,484284 |
| 6 months | abundance | X00163 | primidone_ | 3 | Interventio | Experimen | Experimen | Control | 0,462525 |
| 6 months | abundance | X02601 | (7E,7'E)-5, | 3 | Interventio | Experimen | Experimen | Control | 0,474913 |
| 6 months | abundance | X02184 | Piperonylo | 2b | Interventio | Experimen | Experimen | Control | 0,455918 |
| 6 months | abundance | X06549 | Panthenol_ | 3 | Interventio | Experimen | Experimen | Control | 0,491198 |
| 6 months | abundance | X08154 | 4-(METHYL | 3 | Interventio | Experimen | Experimen | Control | 0,511339 |
| 6 months | abundance | X02767 | 6-APA_b | 3 | Interventio | Experimen | Experimen | Control | 0,487202 |
| 6 months | abundance | X03592 | 2-glyceryl 1 | 3 | Interventio | Experimen | Experimen | Control | 0,547533 |
| 6 months | abundance | X10246 | Ethynodiol | 3 | Interventio | Experimen | Experimen | Control | 0,477197 |
| 6 months | abundance | X08461 | Dihydrouri | 3 | Interventio | Experimen | Experimen | Control | 0,446583 |
| 6 months | abundance | X06107 | Hydroxycar | 3 | Interventio | Experimen | Experimen | Control | 0,495451 |
| 6 months | abundance | SL00243 | Deoxysuga | 2a | Interventio | Experimen | Experimen | Control | 0,444616 |
| 6 months | abundance | X04562 | Lisdexamfe | 3 | Interventio | Experimen | Experimen | Control | 0,539856 |
| 6 months | abundance | SL00063 | Pyridoxal | 1 | Interventio | Experimen | Experimen | Control | 0,452966 |
| 6 months | abundance | X06875 | Nicotine gl | 3 | Interventio | Experimen | Experimen | Control | 0,539856 |
| 6 months | abundance | X02952 | Vorinostat_ | 3 | Interventio | Experimen | Experimen | Control | 0,494948 |
| 6 months | abundance | X10384 | L-N2-(2-Ca | 3 | Interventio | Experimen | Experimen | Control | 0,536933 |
| 6 months | abundance | X06902 | Homocyste | 3 | Interventio | Experimen | Experimen | Control | 0,513677 |
| 6 months | abundance | X07026 | 7α-Hydroxy | 2b | Interventio | Experimen | Experimen | Control | 0,566482 |
| 6 months | abundance | X10345 | S-Allylcyste | 3 | Interventio | Experimen | Experimen | Control | 0,497615 |

| | | | | | | | | | |
|---|---|---|---|---|---|---|---|---|---|
| 6 months | abundance | X05415 | Redul | 3 | Interventio | Experimen | Experimen | Control | 0,462222 |
| 6 months | abundance | X04792 | his-asn | 3 | Interventio | Experimen | Experimen | Control | 0,449538 |
| 6 months | abundance | SL00222 | Lactic acid | 1 | Interventio | Experimen | Experimen | Control | 0,462222 |
| 6 months | abundance | X07457 | Glucosami | 1 | Interventio | Experimen | Experimen | Control | 0,472806 |
| 6 months | abundance | X06727 | MFCD0087 | 3 | Interventio | Experimen | Experimen | Control | 0,516851 |
| 6 months | abundance | X03837 | 2-Methoxy- | 3 | Interventio | Experimen | Experimen | Control | 0,522714 |
| 6 months | abundance | X06478 | Caffeic aci | 3 | Interventio | Experimen | Experimen | Control | 0,536933 |
| 6 months | abundance | X08274 | Mevalonic | 2b | Interventio | Experimen | Experimen | Control | 0,536933 |
| 6 months | abundance | X05581 | N-[(10Z)-7- | 3 | Interventio | Experimen | Experimen | Control | 0,536933 |
| 6 months | abundance | X09244 | gamma-Gl | 3 | Interventio | Experimen | Experimen | Control | 0,452966 |
| 6 months | abundance | X06626 | N-Butyryl-L | 3 | Interventio | Experimen | Experimen | Control | 0,512311 |
| 6 months | abundance | SL00262 | cis-Aconiti | 2a | Interventio | Experimen | Experimen | Control | 0,477197 |
| 6 months | abundance | SL00380 | 2-Aminoad | 2a | Interventio | Experimen | Experimen | Control | 0,535443 |
| 6 months | abundance | SL00020 | Carnitine | 2a | Interventio | Experimen | Experimen | Control | 0,534558 |
| 6 months | abundance | X10304 | 2,3,8,9-Tet | 3 | Interventio | Experimen | Experimen | Control | 0,481144 |
| 6 months | abundance | X01474 | Glyceroph | 3 | Interventio | Experimen | Experimen | Control | 0,547533 |
| 6 months | abundance | SL00128 | N-Acetylal | 2a | Interventio | Experimen | Experimen | Control | 0,507098 |
| 6 months | abundance | M1 | M1 | IA_module | Interventio | Experimen | Experimen | Control | 0,526151 |
| 6 months | abundance | X02121 | 2-Acetami | 3 | Interventio | Experimen | Experimen | Control | 0,550871 |
| 6 months | abundance | X07260 | (7E,7'E)-5, | 3 | Interventio | Experimen | Experimen | Control | 0,530382 |
| 6 months | abundance | X00089 | 4-Hydroxyb | 1 | Interventio | Experimen | Experimen | Control | 0,574199 |
| 6 months | abundance | X07014 | Octyl benz | 3 | Interventio | Experimen | Experimen | Control | 0,574621 |
| 6 months | abundance | X07484 | MFCD0002 | 3 | Interventio | Experimen | Experimen | Control | 0,532547 |
| 6 months | abundance | X08008 | N-Acetylne | 1 | Interventio | Experimen | Experimen | Control | 0,547533 |
| 6 months | abundance | X07815 | 3-[2-[(Z)-[3 | 3 | Interventio | Experimen | Experimen | Control | 0,519242 |
| 6 months | abundance | X07563 | Xanthine | 1 | Interventio | Experimen | Experimen | Control | 0,558374 |
| 6 months | abundance | X04315 | Benzamide | 3 | Interventio | Experimen | Experimen | Control | 0,574621 |
| 6 months | abundance | X07732 | N-Acetylva | 2b | Interventio | Experimen | Experimen | Control | 0,52723 |
| 6 months | abundance | X10097 | Coixol | 3 | Interventio | Experimen | Experimen | Control | 0,584085 |
| 6 months | abundance | X11692 | 2_7-Anhyd | 3 | Interventio | Experimen | Experimen | Control | 0,566785 |
| 6 months | abundance | X00594 | Nicotinami | 3 | Interventio | Experimen | Experimen | Control | 0,596698 |
| 6 months | abundance | X04119 | Tetraacety | 3 | Interventio | Experimen | Experimen | Control | 0,544099 |
| 6 months | abundance | X01968 | 7-Chloro-5 | 3 | Interventio | Experimen | Experimen | Control | 0,539856 |
| 6 months | abundance | SL00124 | Dissaccari | 2a | Interventio | Experimen | Experimen | Control | 0,497615 |
| 6 months | abundance | X11563 | threonylph | 3 | Interventio | Experimen | Experimen | Control | 0,547533 |
| 6 months | abundance | X08659 | 3,3-Dimeth | 2b | Interventio | Experimen | Experimen | Control | 0,622746 |
| 6 months | abundance | X07925 | DNOP_f | 3 | Interventio | Experimen | Experimen | Control | 0,533701 |
| 6 months | abundance | X00635 | 4-(5,6-Dihy | 3 | Interventio | Experimen | Experimen | Control | 0,587711 |
| 6 months | abundance | X07514 | L-(+)-Eryth | 3 | Interventio | Experimen | Experimen | Control | 0,545162 |
| 6 months | abundance | X07155 | Naphthale | 3 | Interventio | Experimen | Experimen | Control | 0,604328 |
| 6 months | abundance | X00035 | Choline | 1 | Interventio | Experimen | Experimen | Control | 0,530382 |
| 6 months | abundance | X08046 | 3-Hydroxy- | 3 | Interventio | Experimen | Experimen | Control | 0,596842 |
| 6 months | abundance | X04990 | Cilazapril | 3 | Interventio | Experimen | Experimen | Control | 0,612948 |
| 6 months | abundance | X08789 | Acrylic acid | 2b | Interventio | Experimen | Experimen | Control | 0,542307 |
| 6 months | abundance | X09152 | Ferulic acid | 1 | Interventio | Experimen | Experimen | Control | 0,573484 |
| 6 months | abundance | X10091 | (2E,6E)-9-[ | 3 | Interventio | Experimen | Experimen | Control | 0,579165 |
| 6 months | abundance | X06473 | MFCD0002 | 3 | Interventio | Experimen | Experimen | Control | 0,588894 |
| 6 months | abundance | X08606 | Butabarbit | 3 | Interventio | Experimen | Experimen | Control | 0,607249 |

| | | | | | | | | | |
|---|---|---|---|---|---|---|---|---|---|
| 6 months | abundance | X02583 | Ethylvanilli | 3 | Interventio | Experimen | Experimen | Control | 0,613206 |
| 6 months | abundance | X09525 | Homoanse | 3 | Interventio | Experimen | Experimen | Control | 0,519176 |
| 6 months | abundance | X02872 | folinic acid | 3 | Interventio | Experimen | Experimen | Control | 0,57055 |
| 6 months | abundance | X06150 | Zalcitabine | 3 | Interventio | Experimen | Experimen | Control | 0,624583 |
| 6 months | abundance | X07596 | 5-amino-2- | 2b | Interventio | Experimen | Experimen | Control | 0,587711 |
| 6 months | abundance | X05786 | Nitrendipir | 3 | Interventio | Experimen | Experimen | Control | 0,615628 |
| 6 months | abundance | X11001 | 2-[(carboxy | 2b | Interventio | Experimen | Experimen | Control | 0,578228 |
| 6 months | abundance | X03294 | 4,4'-Thiobi | 3 | Interventio | Experimen | Experimen | Control | 0,566482 |
| 6 months | abundance | X06183 | Nisinic aci | 3 | Interventio | Experimen | Experimen | Control | 0,614024 |
| 6 months | abundance | X08965 | 1,9-Nonan | 3 | Interventio | Experimen | Experimen | Control | 0,644196 |
| 6 months | abundance | SL00159 | Uracil | 1 | Interventio | Experimen | Experimen | Control | 0,622672 |
| 6 months | abundance | X05017 | N-(1-{[Met | 3 | Interventio | Experimen | Experimen | Control | 0,574621 |
| 6 months | abundance | X08723 | S-Allylcyst | 3 | Interventio | Experimen | Experimen | Control | 0,622672 |
| 6 months | abundance | X06293 | (4R)-4-{[3- | 3 | Interventio | Experimen | Experimen | Control | 0,608921 |
| 6 months | abundance | X06416 | KYNURAMI | 3 | Interventio | Experimen | Experimen | Control | 0,662984 |
| 6 months | abundance | SL00245 | sugar alcol | 2a | Interventio | Experimen | Experimen | Control | 0,559923 |
| 6 months | abundance | X08035 | Tetrahydro | 3 | Interventio | Experimen | Experimen | Control | 0,587711 |
| 6 months | abundance | X02139 | SECONAL_ | 3 | Interventio | Experimen | Experimen | Control | 0,662457 |
| 6 months | abundance | X10518 | MFCD0002 | 3 | Interventio | Experimen | Experimen | Control | 0,538141 |
| 6 months | abundance | SL00288 | Propionylc | 2a | Interventio | Experimen | Experimen | Control | 0,596842 |
| 6 months | abundance | M23 | M23 | IA_module | Interventio | Experimen | Experimen | Control | 0,66921 |
| 6 months | abundance | X07777 | Arginine | 1 | Interventio | Experimen | Experimen | Control | 0,57257 |
| 6 months | abundance | X06337 | Zalcitabine | 3 | Interventio | Experimen | Experimen | Control | 0,622746 |
| 6 months | abundance | X07811 | Metirosine | 3 | Interventio | Experimen | Experimen | Control | 0,66921 |
| 6 months | abundance | X02905 | MFCD0005 | 3 | Interventio | Experimen | Experimen | Control | 0,639371 |
| 6 months | abundance | X01879 | Tetramethy | 2b | Interventio | Experimen | Experimen | Control | 0,67553 |
| 6 months | abundance | X01154 | N-(2,3,4-Tr | 3 | Interventio | Experimen | Experimen | Control | 0,679153 |
| 6 months | abundance | X09882 | Harmane | 3 | Interventio | Experimen | Experimen | Control | 0,67553 |
| 6 months | abundance | X08014 | DNOP_e | 3 | Interventio | Experimen | Experimen | Control | 0,621405 |
| 6 months | abundance | X08099 | g-Butyrobe | 3 | Interventio | Experimen | Experimen | Control | 0,67553 |
| 6 months | abundance | X08279 | DNOP_b | 3 | Interventio | Experimen | Experimen | Control | 0,668366 |
| 6 months | abundance | X07783 | 1,3-dimeth | 3 | Interventio | Experimen | Experimen | Control | 0,660061 |
| 6 months | abundance | X00029 | Urocanic a | 1 | Interventio | Experimen | Experimen | Control | 0,622672 |
| 6 months | abundance | X03278 | Hydroxyph | 3 | Interventio | Experimen | Experimen | Control | 0,660061 |
| 6 months | abundance | X06253 | N-(4-Amin | 3 | Interventio | Experimen | Experimen | Control | 0,660061 |
| 6 months | abundance | X09231 | 2-Keto-glu | 3 | Interventio | Experimen | Experimen | Control | 0,632396 |
| 6 months | abundance | X08425 | MFCD1297 | 3 | Interventio | Experimen | Experimen | Control | 0,621405 |
| 6 months | abundance | X04939 | N-Desalkyl | 3 | Interventio | Experimen | Experimen | Control | 0,613206 |
| 6 months | abundance | X03934 | 2-Aminom | 3 | Interventio | Experimen | Experimen | Control | 0,622746 |
| 6 months | abundance | X10667 | N-Phenyla | 3 | Interventio | Experimen | Experimen | Control | 0,669161 |
| 6 months | abundance | X00072 | 3,5-Dihydr | 1 | Interventio | Experimen | Experimen | Control | 0,62929 |
| 6 months | abundance | X06318 | Propamoca | 2b | Interventio | Experimen | Experimen | Control | 0,660061 |
| 6 months | abundance | M19 | M19 | IA_module | Interventio | Experimen | Experimen | Control | 0,687737 |
| 6 months | abundance | X09612 | 3-hydroxy- | 3 | Interventio | Experimen | Experimen | Control | 0,667479 |
| 6 months | abundance | X08157 | Leu-Val_e | 3 | Interventio | Experimen | Experimen | Control | 0,660061 |
| 6 months | abundance | X06278 | 3-Hydroxy- | 3 | Interventio | Experimen | Experimen | Control | 0,694759 |
| 6 months | abundance | SL00329 | 2-Hydroxy- | 2a | Interventio | Experimen | Experimen | Control | 0,644196 |
| 6 months | abundance | X09805 | Methyl alpl | 3 | Interventio | Experimen | Experimen | Control | 0,641729 |

| | | | | | | | | | |
|---|---|---|---|---|---|---|---|---|---|
| 6 months | abundance | X10691 | (−)-nabilor | 3 | Interventio | Experimen | Experimen | Control | 0,626113 |
| 6 months | abundance | X06463 | 8-Amino-7- | 3 | Interventio | Experimen | Experimen | Control | 0,660061 |
| 6 months | abundance | X11549 | 1,1'-[1,12-l | 3 | Interventio | Experimen | Experimen | Control | 0,672927 |
| 6 months | abundance | X08588 | 2-Acetamid | 3 | Interventio | Experimen | Experimen | Control | 0,660061 |
| 6 months | abundance | X04744 | 16alpha-hy | 3 | Interventio | Experimen | Experimen | Control | 0,682743 |
| 6 months | abundance | X07648 | NL851300( | 3 | Interventio | Experimen | Experimen | Control | 0,679833 |
| 6 months | abundance | X08123 | Tiglic acid_ | 2b | Interventio | Experimen | Experimen | Control | 0,660061 |
| 6 months | abundance | X08146 | O-Ethyl (4- | 3 | Interventio | Experimen | Experimen | Control | 0,696038 |
| 6 months | abundance | X07601 | Asp-lys | 3 | Interventio | Experimen | Experimen | Control | 0,662346 |
| 6 months | abundance | X09921 | 5-Hydroxy- | 3 | Interventio | Experimen | Experimen | Control | 0,663456 |
| 6 months | abundance | X03860 | 2-BUTYL PI | 3 | Interventio | Experimen | Experimen | Control | 0,660061 |
| 6 months | abundance | X07294 | 6-Acetamid | 3 | Interventio | Experimen | Experimen | Control | 0,662984 |
| 6 months | abundance | X08188 | 2-Hydroxy- | 2b | Interventio | Experimen | Experimen | Control | 0,659507 |
| 6 months | abundance | X11605 | 3-Methyl-2 | 3 | Interventio | Experimen | Experimen | Control | 0,662984 |
| 6 months | abundance | X06698 | entecavir | 3 | Interventio | Experimen | Experimen | Control | 0,660061 |
| 6 months | abundance | X08987 | feruloylser | 3 | Interventio | Experimen | Experimen | Control | 0,709598 |
| 6 months | abundance | X04040 | 3-(3,4-dihy | 3 | Interventio | Experimen | Experimen | Control | 0,662984 |
| 6 months | abundance | X08707 | 2-(1-Ethoxy | 3 | Interventio | Experimen | Experimen | Control | 0,696038 |
| 6 months | abundance | X08578 | tert-Butyl 3 | 3 | Interventio | Experimen | Experimen | Control | 0,709124 |
| 6 months | abundance | X03276 | 2'-Deoxyac | 3 | Interventio | Experimen | Experimen | Control | 0,679833 |
| 6 months | abundance | X07434 | N'-Hydroxy | 2b | Interventio | Experimen | Experimen | Control | 0,696038 |
| 6 months | abundance | SL00201 | Pentose II | 2a | Interventio | Experimen | Experimen | Control | 0,660061 |
| 6 months | abundance | X00518 | Nisinic acid | 3 | Interventio | Experimen | Experimen | Control | 0,723284 |
| 6 months | abundance | X01943 | Tocainide | 3 | Interventio | Experimen | Experimen | Control | 0,723552 |
| 6 months | abundance | X01223 | DL-Mevalo | 3 | Interventio | Experimen | Experimen | Control | 0,694971 |
| 6 months | abundance | X11562 | ALA-PRO | 3 | Interventio | Experimen | Experimen | Control | 0,696038 |
| 6 months | abundance | SL00154 | Trigonelline | 1 | Interventio | Experimen | Experimen | Control | 0,682898 |
| 6 months | abundance | X07747 | Diethylpyrc | 3 | Interventio | Experimen | Experimen | Control | 0,65482 |
| 6 months | abundance | X08739 | 1,1'-[1,12-l | 3 | Interventio | Experimen | Experimen | Control | 0,732912 |
| 6 months | abundance | X05510 | 3-Benzyl-6 | 3 | Interventio | Experimen | Experimen | Control | 0,75248 |
| 6 months | abundance | X08733 | N-Methyl-1 | 3 | Interventio | Experimen | Experimen | Control | 0,744735 |
| 6 months | abundance | X11110 | Tauropine | 3 | Interventio | Experimen | Experimen | Control | 0,746475 |
| 6 months | abundance | X01950 | Histidylgly | 3 | Interventio | Experimen | Experimen | Control | 0,741224 |
| 6 months | abundance | SL00216 | Glyoxylic a | 2a | Interventio | Experimen | Experimen | Control | 0,66921 |
| 6 months | abundance | X07442 | coenzyme | 3 | Interventio | Experimen | Experimen | Control | 0,766405 |
| 6 months | abundance | X11524 | 6-hydroxyp | 3 | Interventio | Experimen | Experimen | Control | 0,728006 |
| 6 months | abundance | X08092 | Safrole | 3 | Interventio | Experimen | Experimen | Control | 0,696038 |
| 6 months | abundance | X09854 | N-Benzyl-3 | 2b | Interventio | Experimen | Experimen | Control | 0,71955 |
| 6 months | abundance | X08270 | Butabarbit | 3 | Interventio | Experimen | Experimen | Control | 0,739135 |
| 6 months | abundance | X08095 | DNOP_g | 3 | Interventio | Experimen | Experimen | Control | 0,740789 |
| 6 months | abundance | SL00137 | Phenylacet | 2a | Interventio | Experimen | Experimen | Control | 0,750878 |
| 6 months | abundance | X09675 | UK387000 | 3 | Interventio | Experimen | Experimen | Control | 0,734771 |
| 6 months | abundance | X02730 | N-Phenylac | 3 | Interventio | Experimen | Experimen | Control | 0,67553 |
| 6 months | abundance | X07079 | N-Pentano | 3 | Interventio | Experimen | Experimen | Control | 0,739135 |
| 6 months | abundance | X04538 | Valylvaline | 3 | Interventio | Experimen | Experimen | Control | 0,770082 |
| 6 months | abundance | X10588 | (8)-Ginger( | 3 | Interventio | Experimen | Experimen | Control | 0,766405 |
| 6 months | abundance | X06684 | Bromazine | 3 | Interventio | Experimen | Experimen | Control | 0,731466 |
| 6 months | abundance | X08825 | Chenodeo: | 3 | Interventio | Experimen | Experimen | Control | 0,746475 |

| | | | | | | | | | |
|---|---|---|---|---|---|---|---|---|---|
| 6 months | abundance | X10534 | L-N2-(2-Ca | 3 | Interventio | Experimen | Experimen | Control | 0,751594 |
| 6 months | abundance | X00828 | Fenoterol | 3 | Interventio | Experimen | Experimen | Control | 0,752189 |
| 6 months | abundance | X09998 | N-[(10Z)-7- | 3 | Interventio | Experimen | Experimen | Control | 0,729309 |
| 6 months | abundance | SL00311 | N,N-Dimet | 2a | Interventio | Experimen | Experimen | Control | 0,741993 |
| 6 months | abundance | X09503 | 4-(METHYL | 3 | Interventio | Experimen | Experimen | Control | 0,686987 |
| 6 months | abundance | X05857 | N-D-Gluco | 3 | Interventio | Experimen | Experimen | Control | 0,749793 |
| 6 months | abundance | X09548 | 2-O-beta-D | 3 | Interventio | Experimen | Experimen | Control | 0,756336 |
| 6 months | abundance | X02195 | alliin | 3 | Interventio | Experimen | Experimen | Control | 0,762795 |
| 6 months | abundance | X06334 | Bis-D-fruct | 3 | Interventio | Experimen | Experimen | Control | 0,756336 |
| 6 months | abundance | X03004 | Bicine_b | 3 | Interventio | Experimen | Experimen | Control | 0,723106 |
| 6 months | abundance | SL00430 | Homocarn | 2a | Interventio | Experimen | Experimen | Control | 0,767195 |
| 6 months | abundance | X05146 | UQ367500 | 3 | Interventio | Experimen | Experimen | Control | 0,726086 |
| 6 months | abundance | X03413 | Val-Trp_b | 3 | Interventio | Experimen | Experimen | Control | 0,733178 |
| 6 months | abundance | X07168 | L-(+)-Eryth | 3 | Interventio | Experimen | Experimen | Control | 0,734771 |
| 6 months | abundance | X05859 | 3,7,12,17-T | 3 | Interventio | Experimen | Experimen | Control | 0,755323 |
| 6 months | abundance | SL00383 | 3-(2-Hydro | 2a | Interventio | Experimen | Experimen | Control | 0,682743 |
| 6 months | abundance | X08486 | (2E,6E)-9-[ | 3 | Interventio | Experimen | Experimen | Control | 0,722158 |
| 6 months | abundance | X09684 | (+)-Etomid | 3 | Interventio | Experimen | Experimen | Control | 0,755986 |
| 6 months | abundance | X01463 | 4-Thiapent | 3 | Interventio | Experimen | Experimen | Control | 0,75033 |
| 6 months | abundance | SL00268 | Thymine | 1 | Interventio | Experimen | Experimen | Control | 0,741993 |
| 6 months | abundance | X09393 | 2-Isopropy | 3 | Interventio | Experimen | Experimen | Control | 0,761405 |
| 6 months | abundance | X07812 | Triethyl citı | 3 | Interventio | Experimen | Experimen | Control | 0,709124 |
| 6 months | abundance | SL00096 | Hexoses III | 1 | Interventio | Experimen | Experimen | Control | 0,742624 |
| 6 months | abundance | X10600 | Glycocyam | 3 | Interventio | Experimen | Experimen | Control | 0,770082 |
| 6 months | abundance | X00722 | (3S,5R,6E) | 3 | Interventio | Experimen | Experimen | Control | 0,729456 |
| 6 months | abundance | X11699 | {2-[2-(Isob | 3 | Interventio | Experimen | Experimen | Control | 0,767935 |
| 6 months | abundance | X06329 | Leu-Leu_d | 3 | Interventio | Experimen | Experimen | Control | 0,740789 |
| 6 months | abundance | X02333 | N-Propiony | 3 | Interventio | Experimen | Experimen | Control | 0,769575 |
| 6 months | abundance | M14 | M14 | ۱A_module | Interventio | Experimen | Experimen | Control | 0,761405 |
| 6 months | abundance | X06124 | MFCD0995 | 3 | Interventio | Experimen | Experimen | Control | 0,770082 |
| 6 months | abundance | X03001 | Pseudouric | 2b | Interventio | Experimen | Experimen | Control | 0,756336 |
| 6 months | abundance | X03936 | (3R)-2-(3,4 | 3 | Interventio | Experimen | Experimen | Control | 0,761405 |
| 6 months | abundance | X08594 | 7-ketodeo | 3 | Interventio | Experimen | Experimen | Control | 0,760607 |
| 6 months | abundance | X07544 | Ethyl sulfat | 3 | Interventio | Experimen | Experimen | Control | 0,772129 |
| 6 months | abundance | SL00260 | Pentose I | 2a | Interventio | Experimen | Experimen | Control | 0,767935 |
| 6 months | abundance | SL00433 | Isovalerylc | 2a | Interventio | Experimen | Experimen | Control | 0,662984 |
| 6 months | abundance | X04259 | 3-Hydroxyt | 3 | Interventio | Experimen | Experimen | Control | 0,740789 |
| 6 months | abundance | X02832 | (+/-)-2-Hyd | 3 | Interventio | Experimen | Experimen | Control | 0,766405 |
| 6 months | abundance | X00199 | tert-Butyl 3 | 3 | Interventio | Experimen | Experimen | Control | 0,761405 |
| 6 months | abundance | X07909 | (±)-Albuter | 2b | Interventio | Experimen | Experimen | Control | 0,79043 |
| 6 months | abundance | X00331 | 4-(9H-beta | 3 | Interventio | Experimen | Experimen | Control | 0,772129 |
| 6 months | abundance | X08792 | 3,4-Methyl | 3 | Interventio | Experimen | Experimen | Control | 0,762795 |
| 6 months | abundance | X03760 | 2-Methoxy- | 3 | Interventio | Experimen | Experimen | Control | 0,749793 |
| 6 months | abundance | X07699 | N-(1-Methy | 3 | Interventio | Experimen | Experimen | Control | 0,726086 |
| 6 months | abundance | X08387 | 4-[(2E,4Z)- | 3 | Interventio | Experimen | Experimen | Control | 0,79043 |
| 6 months | abundance | X09374 | 3-Deoxy-D- | 3 | Interventio | Experimen | Experimen | Control | 0,769575 |
| 6 months | abundance | X02023 | beta-D-Eth | 3 | Interventio | Experimen | Experimen | Control | 0,766405 |
| 6 months | abundance | X09001 | Varanic ac | 3 | Interventio | Experimen | Experimen | Control | 0,783629 |

| | | | | | | | | | |
|---|---|---|---|---|---|---|---|---|---|
| 6 months | abundance | X07618 | N-(3-aceta | 3 | Interventio | Experimen | Experimen | Control | 0,79043 |
| 6 months | abundance | X01932 | asn-val_c | 3 | Interventio | Experimen | Experimen | Control | 0,79043 |
| 6 months | abundance | X08634 | 1,5-Isoquir | 2b | Interventio | Experimen | Experimen | Control | 0,780431 |
| 6 months | abundance | X10947 | 1,1'-[1,12-l | 3 | Interventio | Experimen | Experimen | Control | 0,798106 |
| 6 months | abundance | SL00082 | Allopurinol | 1 | Interventio | Experimen | Experimen | Control | 0,77966 |
| 6 months | abundance | X02922 | (2E)-3-Met | 3 | Interventio | Experimen | Experimen | Control | 0,79043 |
| 6 months | abundance | X08893 | 5beta-Cho | 3 | Interventio | Experimen | Experimen | Control | 0,79043 |
| 6 months | abundance | X02009 | Arctiopicri | 3 | Interventio | Experimen | Experimen | Control | 0,790718 |
| 6 months | abundance | X07678 | (2E)-3-(3,4 | 2b | Interventio | Experimen | Experimen | Control | 0,772129 |
| 6 months | abundance | X07774 | 7alpha-Hy | 3 | Interventio | Experimen | Experimen | Control | 0,79043 |
| 6 months | abundance | SL00197 | 4-Methyl-2 | 2a | Interventio | Experimen | Experimen | Control | 0,772129 |
| 6 months | abundance | X01092 | 17,21-Dihy | 3 | Interventio | Experimen | Experimen | Control | 0,79043 |
| 6 months | abundance | X00963 | (4S)-4-[(2E | 3 | Interventio | Experimen | Experimen | Control | 0,782484 |
| 6 months | abundance | X10325 | Lovastatin_ | 3 | Interventio | Experimen | Experimen | Control | 0,792475 |
| 6 months | abundance | X10370 | L-Arogenat | 3 | Interventio | Experimen | Experimen | Control | 0,782484 |
| 6 months | abundance | X08711 | 2-Acetami | 3 | Interventio | Experimen | Experimen | Control | 0,79043 |
| 6 months | abundance | X10623 | N-Nonano | 3 | Interventio | Experimen | Experimen | Control | 0,79043 |
| 6 months | abundance | X00034 | Pyroglutan | 1 | Interventio | Experimen | Experimen | Control | 0,785613 |
| 6 months | abundance | X04564 | asp-gln_b | 3 | Interventio | Experimen | Experimen | Control | 0,79043 |
| 6 months | abundance | X07308 | Nicotinic a | 2a | Interventio | Experimen | Experimen | Control | 0,770082 |
| 6 months | abundance | X01364 | Oleuropeir | 3 | Interventio | Experimen | Experimen | Control | 0,792475 |
| 6 months | abundance | X06905 | Ectoine | 3 | Interventio | Experimen | Experimen | Control | 0,79043 |
| 6 months | abundance | SL00248 | Gluconic a | 2a | Interventio | Experimen | Experimen | Control | 0,783629 |
| 6 months | abundance | X03109 | Guanidino | 3 | Interventio | Experimen | Experimen | Control | 0,828594 |
| 6 months | abundance | X03595 | Methylol D | 3 | Interventio | Experimen | Experimen | Control | 0,821549 |
| 6 months | abundance | X06455 | 4-Hydroxyp | 3 | Interventio | Experimen | Experimen | Control | 0,811341 |
| 6 months | abundance | X10918 | meprobam | 3 | Interventio | Experimen | Experimen | Control | 0,79043 |
| 6 months | abundance | X06368 | TDP-2_b | 3 | Interventio | Experimen | Experimen | Control | 0,825116 |
| 6 months | abundance | X03718 | Propanthel | 3 | Interventio | Experimen | Experimen | Control | 0,823738 |
| 6 months | abundance | X00743 | 11beta,13- | 3 | Interventio | Experimen | Experimen | Control | 0,836775 |
| 6 months | abundance | X06917 | Alanyltrypt | 3 | Interventio | Experimen | Experimen | Control | 0,838691 |
| 6 months | abundance | X06472 | N(alpha)-B | 3 | Interventio | Experimen | Experimen | Control | 0,821549 |
| 6 months | abundance | X10770 | 2-(4-Isoprc | 3 | Interventio | Experimen | Experimen | Control | 0,801 |
| 6 months | abundance | X07165 | pro-gln_b | 3 | Interventio | Experimen | Experimen | Control | 0,838691 |
| 6 months | abundance | SL00455 | 12,13-DHC | 2a | Interventio | Experimen | Experimen | Control | 0,801727 |
| 6 months | abundance | X09581 | L-gamma-( | 3 | Interventio | Experimen | Experimen | Control | 0,825133 |
| 6 months | abundance | X00136 | leu-gln_a | 3 | Interventio | Experimen | Experimen | Control | 0,838691 |
| 6 months | abundance | X01181 | 2-(3,4-Dim | 3 | Interventio | Experimen | Experimen | Control | 0,826734 |
| 6 months | abundance | X10162 | Quinaldic a | 2a | Interventio | Experimen | Experimen | Control | 0,843934 |
| 6 months | abundance | X10379 | Phenyl D-g | 3 | Interventio | Experimen | Experimen | Control | 0,857307 |
| 6 months | abundance | X09919 | 4-ethylphe | 3 | Interventio | Experimen | Experimen | Control | 0,856311 |
| 6 months | abundance | X04150 | 4-(METHYL | 3 | Interventio | Experimen | Experimen | Control | 0,836775 |
| 6 months | abundance | X11494 | 3-Hydroxys | 3 | Interventio | Experimen | Experimen | Control | 0,836775 |
| 6 months | abundance | X07746 | 3-(2-Oxo-2 | 2b | Interventio | Experimen | Experimen | Control | 0,840699 |
| 6 months | abundance | X04440 | Mono(3-ca | 3 | Interventio | Experimen | Experimen | Control | 0,852491 |
| 6 months | abundance | X07899 | N-Acetylpu | 2b | Interventio | Experimen | Experimen | Control | 0,856311 |
| 6 months | abundance | X02268 | 5-Hydroxy- | 3 | Interventio | Experimen | Experimen | Control | 0,856311 |
| 6 months | abundance | X02327 | 1H-Pyrazol | 3 | Interventio | Experimen | Experimen | Control | 0,836775 |

| | | | | | | | | | |
|---|---|---|---|---|---|---|---|---|---|
| 6 months | abundance | X00928 | (7R)-7-(5-c | 3 | Interventio | Experimen | Experimen | Control | 0,847789 |
| 6 months | abundance | X08422 | N-(3-aceta | 3 | Interventio | Experimen | Experimen | Control | 0,856311 |
| 6 months | abundance | X09082 | 1_2-Dihydr | 3 | Interventio | Experimen | Experimen | Control | 0,856186 |
| 6 months | abundance | X07527 | 1,7-Dimetl | 2b | Interventio | Experimen | Experimen | Control | 0,864593 |
| 6 months | abundance | X10076 | trimethadi | 3 | Interventio | Experimen | Experimen | Control | 0,838691 |
| 6 months | abundance | X08844 | 4-Hydroxy- | 3 | Interventio | Experimen | Experimen | Control | 0,852491 |
| 6 months | abundance | X04919 | 5-O-alpha- | 3 | Interventio | Experimen | Experimen | Control | 0,856186 |
| 6 months | abundance | X08690 | Ethanoic a | 3 | Interventio | Experimen | Experimen | Control | 0,848721 |
| 6 months | abundance | M21 | M21 | IA_module | Interventio | Experimen | Experimen | Control | 0,856311 |
| 6 months | abundance | X09898 | N(6),N(6)-I | 3 | Interventio | Experimen | Experimen | Control | 0,856311 |
| 6 months | abundance | X05225 | 4-(METHYL | 3 | Interventio | Experimen | Experimen | Control | 0,864593 |
| 6 months | abundance | SL00239 | Pinitol | 2a | Interventio | Experimen | Experimen | Control | 0,85403 |
| 6 months | abundance | X10725 | Ethyl mala | 3 | Interventio | Experimen | Experimen | Control | 0,863282 |
| 6 months | abundance | X07040 | 6-Hydroxyp | 3 | Interventio | Experimen | Experimen | Control | 0,864593 |
| 6 months | abundance | X00646 | Deacetyldi | 3 | Interventio | Experimen | Experimen | Control | 0,864593 |
| 6 months | abundance | M7 | M7 | IA_module | Interventio | Experimen | Experimen | Control | 0,856311 |
| 6 months | abundance | M27 | M27 | IA_module | Interventio | Experimen | Experimen | Control | 0,864593 |
| 6 months | abundance | X08292 | Lidocaine | 2b | Interventio | Experimen | Experimen | Control | 0,864593 |
| 6 months | abundance | X01346 | IN00260_a | 3 | Interventio | Experimen | Experimen | Control | 0,874528 |
| 6 months | abundance | X09825 | (2S)-6-Ami | 3 | Interventio | Experimen | Experimen | Control | 0,864593 |
| 6 months | abundance | X07375 | porphobilir | 3 | Interventio | Experimen | Experimen | Control | 0,864593 |
| 6 months | abundance | X06388 | Caffeic aci | 1 | Interventio | Experimen | Experimen | Control | 0,864593 |
| 6 months | abundance | X05984 | 1-(4-Methy | 2b | Interventio | Experimen | Experimen | Control | 0,864593 |
| 6 months | abundance | X06268 | mesifurane | 3 | Interventio | Experimen | Experimen | Control | 0,880429 |
| 6 months | abundance | X00135 | N-Acetylgl | 1 | Interventio | Experimen | Experimen | Control | 0,864593 |
| 6 months | abundance | SL00285 | Dihydrofer | 2a | Interventio | Experimen | Experimen | Control | 0,864593 |
| 6 months | abundance | X00061 | Thiamine | 1 | Interventio | Experimen | Experimen | Control | 0,861845 |
| 6 months | abundance | X02820 | FC250500 | 3 | Interventio | Experimen | Experimen | Control | 0,880429 |
| 6 months | abundance | X06681 | Scymnol | 3 | Interventio | Experimen | Experimen | Control | 0,880429 |
| 6 months | abundance | X05126 | (3aS,5S,6F | 3 | Interventio | Experimen | Experimen | Control | 0,864593 |
| 6 months | abundance | X07028 | N-Pentano | 3 | Interventio | Experimen | Experimen | Control | 0,873138 |
| 6 months | abundance | SL00290 | N-Acetylm | 2a | Interventio | Experimen | Experimen | Control | 0,872367 |
| 6 months | abundance | X06502 | MFCD2836 | 3 | Interventio | Experimen | Experimen | Control | 0,872258 |
| 6 months | abundance | X08352 | 7-[4-(tert-b | 2b | Interventio | Experimen | Experimen | Control | 0,885146 |
| 6 months | abundance | X10032 | 4,9a-Dime | 3 | Interventio | Experimen | Experimen | Control | 0,897356 |
| 6 months | abundance | X09710 | LW800000 | 3 | Interventio | Experimen | Experimen | Control | 0,901301 |
| 6 months | abundance | X07770 | Piperine | 1 | Interventio | Experimen | Experimen | Control | 0,885778 |
| 6 months | abundance | SL00384 | p-Coumari | 1 | Interventio | Experimen | Experimen | Control | 0,888946 |
| 6 months | abundance | X01380 | Spermic ac | 3 | Interventio | Experimen | Experimen | Control | 0,880429 |
| 6 months | abundance | X04898 | 17-Hydroxy | 3 | Interventio | Experimen | Experimen | Control | 0,907254 |
| 6 months | abundance | X08460 | α-Murichol | 1 | Interventio | Experimen | Experimen | Control | 0,905579 |
| 6 months | abundance | X07754 | hexobarbit | 3 | Interventio | Experimen | Experimen | Control | 0,907254 |
| 6 months | abundance | X05892 | (5Z)-2-Ami | 3 | Interventio | Experimen | Experimen | Control | 0,905579 |
| 6 months | abundance | X07886 | L-Urobilin | 3 | Interventio | Experimen | Experimen | Control | 0,922666 |
| 6 months | abundance | X03675 | asp-gln_a | 3 | Interventio | Experimen | Experimen | Control | 0,916184 |
| 6 months | abundance | X10445 | 2-(Carboxy | 3 | Interventio | Experimen | Experimen | Control | 0,921647 |
| 6 months | abundance | X06434 | butyrin | 3 | Interventio | Experimen | Experimen | Control | 0,923492 |
| 6 months | abundance | X00907 | 1,2,3,4-Tet | 3 | Interventio | Experimen | Experimen | Control | 0,921679 |

| | | | | | | | | | |
|---|---|---|---|---|---|---|---|---|---|
| 6 months | abundance | X09208 | IN00260_b | 3 | Interventio | Experimen | Experimen | Control | 0,919926 |
| 6 months | abundance | X09546 | asn-phe | 3 | Interventio | Experimen | Experimen | Control | 0,922666 |
| 6 months | abundance | X09055 | N-[(1R,2S,( | 3 | Interventio | Experimen | Experimen | Control | 0,922779 |
| 6 months | abundance | X10451 | Ro 20-1724 | 3 | Interventio | Experimen | Experimen | Control | 0,921679 |
| 6 months | abundance | X07541 | Saccharin | 1 | Interventio | Experimen | Experimen | Control | 0,922779 |
| 6 months | abundance | SL00287 | DOPA | 2a | Interventio | Experimen | Experimen | Control | 0,922674 |
| 6 months | abundance | X11265 | Sulfurol | 3 | Interventio | Experimen | Experimen | Control | 0,916193 |
| 6 months | abundance | X07841 | 2-(5-Benzy | 3 | Interventio | Experimen | Experimen | Control | 0,916184 |
| 6 months | abundance | X07764 | (S)-2-meth | 3 | Interventio | Experimen | Experimen | Control | 0,922666 |
| 6 months | abundance | X00063 | 2-Hydroxy( | 1 | Interventio | Experimen | Experimen | Control | 0,922666 |
| 6 months | abundance | X11438 | pro-met | 3 | Interventio | Experimen | Experimen | Control | 0,922674 |
| 6 months | abundance | X02202 | D-2-Amino | 3 | Interventio | Experimen | Experimen | Control | 0,922779 |
| 6 months | abundance | X08485 | Isoprenalir | 3 | Interventio | Experimen | Experimen | Control | 0,938072 |
| 6 months | abundance | X00078 | Acetylagm | 1 | Interventio | Experimen | Experimen | Control | 0,93201 |
| 6 months | abundance | X00116 | Cytidine | 1 | Interventio | Experimen | Experimen | Control | 0,929246 |
| 6 months | abundance | X06127 | 2-Methoxy- | 3 | Interventio | Experimen | Experimen | Control | 0,938072 |
| 6 months | abundance | X07278 | 5-Phospho | 3 | Interventio | Experimen | Experimen | Control | 0,934752 |
| 6 months | abundance | X11515 | 13(S)-HOT | 2b | Interventio | Experimen | Experimen | Control | 0,939435 |
| 6 months | abundance | X02661 | 4-(3-Oxope | 3 | Interventio | Experimen | Experimen | Control | 0,932328 |
| 6 months | abundance | X07638 | 7-ketodeo | 3 | Interventio | Experimen | Experimen | Control | 0,937075 |
| 6 months | abundance | X11059 | Dihydrocor | 3 | Interventio | Experimen | Experimen | Control | 0,938375 |
| 6 months | abundance | X04286 | Zinecard_b | 3 | Interventio | Experimen | Experimen | Control | 0,939203 |
| 6 months | abundance | X10580 | Leu-Leu_a | 3 | Interventio | Experimen | Experimen | Control | 0,938375 |
| 6 months | abundance | X04183 | Spaglumic | 3 | Interventio | Experimen | Experimen | Control | 0,939525 |
| 6 months | abundance | X03064 | Formimino | 3 | Interventio | Experimen | Experimen | Control | 0,939525 |
| 6 months | abundance | SL00346 | N-Methylni | 2a | Interventio | Experimen | Experimen | Control | 0,940033 |
| 6 months | abundance | X00528 | R-(+)-Etira( | 3 | Interventio | Experimen | Experimen | Control | 0,946138 |
| 6 months | abundance | X07753 | Lovastatin_ | 3 | Interventio | Experimen | Experimen | Control | 0,946963 |
| 6 months | abundance | X10488 | 4-Guanidir | 1 | Interventio | Experimen | Experimen | Control | 0,950361 |
| 6 months | abundance | X01252 | (−)-nabilor | 3 | Interventio | Experimen | Experimen | Control | 0,950361 |
| 6 months | abundance | X08078 | Ethyl mala | 3 | Interventio | Experimen | Experimen | Control | 0,951718 |
| 6 months | abundance | X02537 | Leu-pro_a | 3 | Interventio | Experimen | Experimen | Control | 0,949352 |
| 6 months | abundance | X08317 | 8-Hydroxy( | 2b | Interventio | Experimen | Experimen | Control | 0,962036 |
| 6 months | abundance | X02765 | L-Pyrrolysi | 3 | Interventio | Experimen | Experimen | Control | 0,962302 |
| 6 months | abundance | X09875 | 4-[(E)-2-(3, | 3 | Interventio | Experimen | Experimen | Control | 0,962036 |
| 6 months | abundance | X08024 | 8-Methyl-8 | 3 | Interventio | Experimen | Experimen | Control | 0,954086 |
| 6 months | abundance | X07219 | 3,3-Dimeth | 2b | Interventio | Experimen | Experimen | Control | 0,966723 |
| 6 months | abundance | X05969 | 4-(METHYL | 3 | Interventio | Experimen | Experimen | Control | 0,966723 |
| 6 months | abundance | X10818 | butalbital_ | 3 | Interventio | Experimen | Experimen | Control | 0,968054 |
| 6 months | abundance | X07835 | 7-ketodeo | 3 | Interventio | Experimen | Experimen | Control | 0,969893 |
| 6 months | abundance | X10536 | CMPF | 3 | Interventio | Experimen | Experimen | Control | 0,962036 |
| 6 months | abundance | M4 | M4 | IA_module | Interventio | Experimen | Experimen | Control | 0,9714 |
| 6 months | abundance | SL00133 | N-Acetylgl | 2a | Interventio | Experimen | Experimen | Control | 0,9714 |
| 6 months | abundance | X03552 | 4-Hydroxy | 3 | Interventio | Experimen | Experimen | Control | 0,97202 |
| 6 months | abundance | X08693 | 7-Sulfocho | 3 | Interventio | Experimen | Experimen | Control | 0,9714 |
| 6 months | abundance | X00083 | 1,7-Dimeth | 1 | Interventio | Experimen | Experimen | Control | 0,973253 |
| 6 months | abundance | X06291 | Corticoste | 2b | Interventio | Experimen | Experimen | Control | 0,9714 |
| 6 months | abundance | X11245 | MFCD0003 | 3 | Interventio | Experimen | Experimen | Control | 0,9714 |

| | | | | | | | | | |
|---|---|---|---|---|---|---|---|---|---|
| 6 months | abundance | X05988 | Selsun | 3 | Interventio | Experimen | Experimen | Control | 0,977372 |
| 6 months | abundance | X06462 | DNOP_d | 3 | Interventio | Experimen | Experimen | Control | 0,983736 |
| 6 months | abundance | X02013 | Ile-cys | 3 | Interventio | Experimen | Experimen | Control | 0,98383 |
| 6 months | abundance | X10685 | Glycylvalin | 2a | Interventio | Experimen | Experimen | Control | 0,98383 |
| 6 months | abundance | X04334 | 3-Benzyl-6 | 3 | Interventio | Experimen | Experimen | Control | 0,986192 |
| 6 months | abundance | X04639 | hydroxyhe | 3 | Interventio | Experimen | Experimen | Control | 0,986192 |
| 6 months | abundance | X03343 | L-alpha-As | 3 | Interventio | Experimen | Experimen | Control | 0,989662 |
| 6 months | abundance | X09480 | Spermic ac | 3 | Interventio | Experimen | Experimen | Control | 0,990034 |
| 6 months | abundance | X01679 | meprobam | 3 | Interventio | Experimen | Experimen | Control | 0,990034 |
| 6 months | abundance | X06082 | O-heptano | 3 | Interventio | Experimen | Experimen | Control | 0,991611 |
| 6 months | abundance | X02427 | 2-(3,5-dim | 2b | Interventio | Experimen | Experimen | Control | 0,991713 |
| 6 months | abundance | M22 | M22 | IA_module | Interventio | Experimen | Experimen | Control | 0,991713 |
| 6 months | abundance | X07613 | 1-(4-Aminc | 3 | Interventio | Experimen | Experimen | Control | 0,992985 |
| 6 months | abundance | X02426 | Raltitrexed | 3 | Interventio | Experimen | Experimen | Control | 0,992985 |
| 6 months | abundance | X08167 | Lanthionin | 3 | Interventio | Experimen | Experimen | Control | 0,996256 |
| 6 months | abundance | X08105 | 2-Acetami | 3 | Interventio | Experimen | Experimen | Control | 0,999683 |
| 6 months | abundance | X02769 | Tetraacety | 3 | Interventio | Experimen | Experimen | Control | 0,999683 |
| 6 months | abundance | X09302 | Sular | 3 | Interventio | Experimen | Experimen | Control | 0,999683 |
| 6 months | abundance | X07469 | 1,1'-[1,12-I | 3 | Interventio | Experimen | Experimen | Control | 0,999683 |
| 6 months | abundance | X03535 | (1S,3R,4s)- | 3 | Interventio | Experimen | Experimen | Control | 0,999683 |
| 6 months | abundance | X09245 | L-gamma-( | 3 | Interventio | Experimen | Experimen | Control | 0,996256 |
| 6 months | abundance | X01017 | hexobarbit | 3 | Interventio | Experimen | Experimen | Control | 0,996256 |
| 6 months | abundance | X08646 | N-Stearoyl | 3 | Interventio | Experimen | Experimen | Control | 0,992514 |
| 6 months | abundance | X02853 | 7alpha-Hy | 3 | Interventio | Experimen | Experimen | Control | 0,992514 |
| 6 months | abundance | X02948 | N-(2-Cyan | 3 | Interventio | Experimen | Experimen | Control | 0,989662 |
| 6 months | abundance | X06412 | Butenylcar | 3 | Interventio | Experimen | Experimen | Control | 0,990034 |
| 6 months | abundance | SL00502 | 2-Ketobuty | 1 | Interventio | Experimen | Experimen | Control | 0,989662 |
| 6 months | abundance | X07890 | N-(5-aceta | 2b | Interventio | Experimen | Experimen | Control | 0,989662 |
| 6 months | abundance | X00477 | Sparfloxac | 2b | Interventio | Experimen | Experimen | Control | 0,989662 |
| 6 months | abundance | X07771 | Caprolacta | 2b | Interventio | Experimen | Experimen | Control | 0,989662 |
| 6 months | abundance | X10819 | Hyocholic | 2a | Interventio | Experimen | Experimen | Control | 0,989662 |
| 6 months | abundance | X00253 | TDP-2_a | 3 | Interventio | Experimen | Experimen | Control | 0,989662 |
| 6 months | abundance | X08584 | meticillin | 3 | Interventio | Experimen | Experimen | Control | 0,986192 |
| 6 months | abundance | X09765 | Tetrahydro | 3 | Interventio | Experimen | Experimen | Control | 0,986192 |
| 6 months | abundance | X01664 | 5-methylth | 3 | Interventio | Experimen | Experimen | Control | 0,983955 |
| 6 months | abundance | X08057 | N-{3-Carbc | 3 | Interventio | Experimen | Experimen | Control | 0,986192 |
| 6 months | abundance | X11716 | ala-ser_a | 3 | Interventio | Experimen | Experimen | Control | 0,98383 |
| 6 months | abundance | X08235 | 1-Methylhi | 2b | Interventio | Experimen | Experimen | Control | 0,98383 |
| 6 months | abundance | X11755 | Furfuranol | 3 | Interventio | Experimen | Experimen | Control | 0,977927 |
| 6 months | abundance | X08017 | Tropinone | 2b | Interventio | Experimen | Experimen | Control | 0,973414 |
| 6 months | abundance | X03660 | trp-ser | 3 | Interventio | Experimen | Experimen | Control | 0,975253 |
| 6 months | abundance | X02920 | L-gamma-( | 3 | Interventio | Experimen | Experimen | Control | 0,972448 |
| 6 months | abundance | X01577 | Prunasin | 3 | Interventio | Experimen | Experimen | Control | 0,973253 |
| 6 months | abundance | X10741 | 5-Nitro-2-p | 3 | Interventio | Experimen | Experimen | Control | 0,9714 |
| 6 months | abundance | X09187 | Salicylic ac | 1 | Interventio | Experimen | Experimen | Control | 0,9714 |
| 6 months | abundance | X00099 | 7-Methylgu | 1 | Interventio | Experimen | Experimen | Control | 0,972448 |
| 6 months | abundance | X07345 | Phenyl D-g | 3 | Interventio | Experimen | Experimen | Control | 0,9714 |
| 6 months | abundance | X06011 | Coprine_a | 3 | Interventio | Experimen | Experimen | Control | 0,9714 |

| | | | | | | | | | |
|---|---|---|---|---|---|---|---|---|---|
| 6 months | abundance | X00224 | Nifedipine | 3 | Interventio | Experimen | Experimen | Control | 0,9714 |
| 6 months | abundance | X06805 | (9cis)-O~1 | 3 | Interventio | Experimen | Experimen | Control | 0,96818 |
| 6 months | abundance | X07429 | DNOP_c | 3 | Interventio | Experimen | Experimen | Control | 0,962036 |
| 6 months | abundance | SL00270 | 3-hydroxy- | 2a | Interventio | Experimen | Experimen | Control | 0,95757 |
| 6 months | abundance | X07566 | 5-Allyl-5-se | 3 | Interventio | Experimen | Experimen | Control | 0,952514 |
| 6 months | abundance | SL00313 | 2-Aminoisc | 2a | Interventio | Experimen | Experimen | Control | 0,947377 |
| 6 months | abundance | X08695 | 3b-Hydroxy | 3 | Interventio | Experimen | Experimen | Control | 0,948936 |
| 6 months | abundance | X11529 | 2-Aminooc | 2b | Interventio | Experimen | Experimen | Control | 0,948936 |
| 6 months | abundance | M10 | M10 | IA_module | Interventio | Experimen | Experimen | Control | 0,949352 |
| 6 months | abundance | X03017 | Pro-tyr | 3 | Interventio | Experimen | Experimen | Control | 0,938375 |
| 6 months | abundance | X00836 | 3-Mercapto | 3 | Interventio | Experimen | Experimen | Control | 0,944459 |
| 6 months | abundance | X08094 | 1,1'-[1,12-l | 3 | Interventio | Experimen | Experimen | Control | 0,946138 |
| 6 months | abundance | X02878 | N~6~-[5-(1 | 3 | Interventio | Experimen | Experimen | Control | 0,939435 |
| 6 months | abundance | X04351 | Pyrimidine | 3 | Interventio | Experimen | Experimen | Control | 0,946875 |
| 6 months | abundance | X07634 | 3,8,9-trihy | 2b | Interventio | Experimen | Experimen | Control | 0,939525 |
| 6 months | abundance | X10046 | Dehydroac | 2b | Interventio | Experimen | Experimen | Control | 0,939203 |
| 6 months | abundance | X09545 | 4-Methyler | 3 | Interventio | Experimen | Experimen | Control | 0,937636 |
| 6 months | abundance | X07092 | Dibutyl ma | 3 | Interventio | Experimen | Experimen | Control | 0,938375 |
| 6 months | abundance | X08533 | tyramine su | 3 | Interventio | Experimen | Experimen | Control | 0,93201 |
| 6 months | abundance | X09728 | {[(15-Hydro | 3 | Interventio | Experimen | Experimen | Control | 0,924496 |
| 6 months | abundance | X07226 | threonylph | 3 | Interventio | Experimen | Experimen | Control | 0,922674 |
| 6 months | abundance | X01078 | Dihydroura | 3 | Interventio | Experimen | Experimen | Control | 0,922779 |
| 6 months | abundance | SL00297 | Acetylmura | 2a | Interventio | Experimen | Experimen | Control | 0,922779 |
| 6 months | abundance | X11349 | Butabarbit | 3 | Interventio | Experimen | Experimen | Control | 0,93201 |
| 6 months | abundance | X06606 | Leu-Val_d | 3 | Interventio | Experimen | Experimen | Control | 0,929246 |
| 6 months | abundance | X07743 | D-Alanine r | 2b | Interventio | Experimen | Experimen | Control | 0,923004 |
| 6 months | abundance | X10614 | Monometh | 2a | Interventio | Experimen | Experimen | Control | 0,922779 |
| 6 months | abundance | X07892 | urobilinoge | 3 | Interventio | Experimen | Experimen | Control | 0,929246 |
| 6 months | abundance | X07452 | 4-Amino-1 | 3 | Interventio | Experimen | Experimen | Control | 0,923492 |
| 6 months | abundance | X06313 | Isoquinolir | 2b | Interventio | Experimen | Experimen | Control | 0,922674 |
| 6 months | abundance | X05421 | S-Allylcyste | 3 | Interventio | Experimen | Experimen | Control | 0,919926 |
| 6 months | abundance | X02208 | SECONAL_ | 3 | Interventio | Experimen | Experimen | Control | 0,922666 |
| 6 months | abundance | X08037 | Crotonic ac | 2b | Interventio | Experimen | Experimen | Control | 0,919926 |
| 6 months | abundance | X10339 | 4H-1-Benz | 2b | Interventio | Experimen | Experimen | Control | 0,922666 |
| 6 months | abundance | X09284 | 4-(Nitrosoa | 3 | Interventio | Experimen | Experimen | Control | 0,922779 |
| 6 months | abundance | X07874 | δ-Valerolac | 2b | Interventio | Experimen | Experimen | Control | 0,922666 |
| 6 months | abundance | X11032 | 2,4-Bis(3-r | 3 | Interventio | Experimen | Experimen | Control | 0,922666 |
| 6 months | abundance | X02944 | Valylvaline | 3 | Interventio | Experimen | Experimen | Control | 0,907254 |
| 6 months | abundance | X02256 | hexobarbit | 3 | Interventio | Experimen | Experimen | Control | 0,922779 |
| 6 months | abundance | X10776 | Histidylgly | 3 | Interventio | Experimen | Experimen | Control | 0,905579 |
| 6 months | abundance | X06197 | 4-(Nitrosoa | 3 | Interventio | Experimen | Experimen | Control | 0,922666 |
| 6 months | abundance | X09490 | 2-Hydroxy- | 3 | Interventio | Experimen | Experimen | Control | 0,913548 |
| 6 months | abundance | X03480 | epsilon-(ga | 3 | Interventio | Experimen | Experimen | Control | 0,910919 |
| 6 months | abundance | X07868 | Biotin | 2b | Interventio | Experimen | Experimen | Control | 0,922666 |
| 6 months | abundance | X03416 | 9-(alpha-D | 3 | Interventio | Experimen | Experimen | Control | 0,907254 |
| 6 months | abundance | X05580 | Xanthureni | 3 | Interventio | Experimen | Experimen | Control | 0,906599 |
| 6 months | abundance | X00074 | Methylsucc | 1 | Interventio | Experimen | Experimen | Control | 0,919926 |
| 6 months | abundance | X08923 | Semilicoisc | 3 | Interventio | Experimen | Experimen | Control | 0,911969 |

| | | | | | | | | | |
|---|---|---|---|---|---|---|---|---|---|
| 6 months | abundance | X04814 | meprobam | 3 | Interventio | Experimen | Experimen | Control | 0,913038 |
| 6 months | abundance | X01911 | 3-(2,3-Dihy | 3 | Interventio | Experimen | Experimen | Control | 0,916412 |
| 6 months | abundance | X09100 | Ethyl aceta | 3 | Interventio | Experimen | Experimen | Control | 0,917684 |
| 6 months | abundance | X02492 | N-(3,5-Dim | 3 | Interventio | Experimen | Experimen | Control | 0,8995 |
| 6 months | abundance | X09623 | Endothal | 2b | Interventio | Experimen | Experimen | Control | 0,910969 |
| 6 months | abundance | X07193 | N-(1-Methy | 3 | Interventio | Experimen | Experimen | Control | 0,8995 |
| 6 months | abundance | X10144 | imazameth | 3 | Interventio | Experimen | Experimen | Control | 0,905717 |
| 6 months | abundance | X06767 | 2-Hydroxyb | 1 | Interventio | Experimen | Experimen | Control | 0,902955 |
| 6 months | abundance | M20 | M20 | IA_module | Interventio | Experimen | Experimen | Control | 0,901376 |
| 6 months | abundance | X03077 | nicotianam | 3 | Interventio | Experimen | Experimen | Control | 0,897356 |
| 6 months | abundance | X11115 | Hydroxyca | 3 | Interventio | Experimen | Experimen | Control | 0,902955 |
| 6 months | abundance | Butanoic a | Butanoic a | CFA_panel | Interventio | Experimen | Experimen | Control | 0,878814 |
| 6 months | abundance | X01011 | DIBEHENIN | 3 | Interventio | Experimen | Experimen | Control | 0,897356 |
| 6 months | abundance | SL00438 | Daidzein | 2a | Interventio | Experimen | Experimen | Control | 0,8995 |
| 6 months | abundance | X08847 | 1-Vinylimic | 2b | Interventio | Experimen | Experimen | Control | 0,864906 |
| 6 months | abundance | X11627 | N-(3-aceta | 3 | Interventio | Experimen | Experimen | Control | 0,885146 |
| 6 months | abundance | SL00299 | β-Murichol | 2a | Interventio | Experimen | Experimen | Control | 0,896401 |
| 6 months | abundance | SL00205 | Citraconic | 2a | Interventio | Experimen | Experimen | Control | 0,865759 |
| 6 months | abundance | X07051 | O-succinyl | 3 | Interventio | Experimen | Experimen | Control | 0,885146 |
| 6 months | abundance | X02972 | 3-(Sulfooxy | 3 | Interventio | Experimen | Experimen | Control | 0,885146 |
| 6 months | abundance | X03901 | ferrileghen | 3 | Interventio | Experimen | Experimen | Control | 0,887588 |
| 6 months | abundance | X02881 | N-(2,3,4-Tr | 3 | Interventio | Experimen | Experimen | Control | 0,887588 |
| 6 months | abundance | X04579 | 6-(alpha-D | 3 | Interventio | Experimen | Experimen | Control | 0,889563 |
| 6 months | abundance | X08719 | 6-Methylqu | 2b | Interventio | Experimen | Experimen | Control | 0,880429 |
| 6 months | abundance | X01059 | coronatine | 3 | Interventio | Experimen | Experimen | Control | 0,889563 |
| 6 months | abundance | X05237 | imazameth | 3 | Interventio | Experimen | Experimen | Control | 0,877813 |
| 6 months | abundance | X02529 | tenivastati | 3 | Interventio | Experimen | Experimen | Control | 0,872258 |
| 6 months | abundance | X00033 | Deoxycholi | 1 | Interventio | Experimen | Experimen | Control | 0,877813 |
| 6 months | abundance | X10330 | Sulfurous a | 3 | Interventio | Experimen | Experimen | Control | 0,864593 |
| 6 months | abundance | X09942 | Spermic ac | 3 | Interventio | Experimen | Experimen | Control | 0,864593 |
| 6 months | abundance | X02615 | 6-APA_a | 3 | Interventio | Experimen | Experimen | Control | 0,883824 |
| 6 months | abundance | SL00035 | Glycylglyci | 2a | Interventio | Experimen | Experimen | Control | 0,864593 |
| 6 months | abundance | X06530 | 3-(1-hydro: | 2b | Interventio | Experimen | Experimen | Control | 0,875244 |
| 6 months | abundance | X11223 | Ethyl mala | 3 | Interventio | Experimen | Experimen | Control | 0,864593 |
| 6 months | abundance | X06903 | Esculin | 2b | Interventio | Experimen | Experimen | Control | 0,873138 |
| 6 months | abundance | X06292 | mesifurane | 3 | Interventio | Experimen | Experimen | Control | 0,864593 |
| 6 months | abundance | X08054 | Docosahex | 2b | Interventio | Experimen | Experimen | Control | 0,864906 |
| 6 months | abundance | X07201 | 2,4-Quinol | 2b | Interventio | Experimen | Experimen | Control | 0,867822 |
| 6 months | abundance | X05656 | mesifurane | 3 | Interventio | Experimen | Experimen | Control | 0,864906 |
| 6 months | abundance | X06320 | 7alpha-Hy | 3 | Interventio | Experimen | Experimen | Control | 0,864593 |
| 6 months | abundance | X09746 | ELK (Peptic | 2b | Interventio | Experimen | Experimen | Control | 0,851221 |
| 6 months | abundance | X10497 | 4-Guanidir | 3 | Interventio | Experimen | Experimen | Control | 0,864593 |
| 6 months | abundance | X07694 | DNOP_a | 3 | Interventio | Experimen | Experimen | Control | 0,864593 |
| 6 months | abundance | X00125 | Taurine | 1 | Interventio | Experimen | Experimen | Control | 0,857307 |
| 6 months | abundance | X09740 | MFCD0166 | 3 | Interventio | Experimen | Experimen | Control | 0,864593 |
| 6 months | abundance | X02281 | tert-Butyl 3 | 3 | Interventio | Experimen | Experimen | Control | 0,864593 |
| 6 months | abundance | X00084 | Tricarballyl | 1 | Interventio | Experimen | Experimen | Control | 0,864593 |
| 6 months | abundance | X08600 | N-Propiony | 3 | Interventio | Experimen | Experimen | Control | 0,861845 |

| | | | | | | | | | |
|---|---|---|---|---|---|---|---|---|---|
| 6 months | abundance | X01746 | Astemizole | 3 | Interventio | Experimen | Experimen | Control | 0,860024 |
| 6 months | abundance | X00082 | Cholic acic | 1 | Interventio | Experimen | Experimen | Control | 0,864593 |
| 6 months | abundance | X01833 | 2,3,4,9-Tet | 2b | Interventio | Experimen | Experimen | Control | 0,864593 |
| 6 months | abundance | X04157 | 3-Methoxy- | 3 | Interventio | Experimen | Experimen | Control | 0,864593 |
| 6 months | abundance | X09189 | 4-[(E)-2-(3, | 3 | Interventio | Experimen | Experimen | Control | 0,857746 |
| 6 months | abundance | X06948 | (1R,3R,5R) | 3 | Interventio | Experimen | Experimen | Control | 0,864593 |
| 6 months | abundance | X08004 | δ-Valerolac | 2b | Interventio | Experimen | Experimen | Control | 0,856311 |
| 6 months | abundance | X02145 | 3'-Hydroxy | 3 | Interventio | Experimen | Experimen | Control | 0,850568 |
| 6 months | abundance | X00723 | Aspartyl-L- | 3 | Interventio | Experimen | Experimen | Control | 0,856311 |
| 6 months | abundance | X07523 | Ursodeoxy | 1 | Interventio | Experimen | Experimen | Control | 0,856311 |
| 6 months | abundance | X00764 | 5-Allyl-5-se | 3 | Interventio | Experimen | Experimen | Control | 0,856311 |
| 6 months | abundance | X07799 | Capryloylg | 2b | Interventio | Experimen | Experimen | Control | 0,815129 |
| 6 months | abundance | X06818 | Homoanse | 3 | Interventio | Experimen | Experimen | Control | 0,814559 |
| 6 months | abundance | X07710 | 3-O-beta-D | 3 | Interventio | Experimen | Experimen | Control | 0,860361 |
| 6 months | abundance | X01067 | g-Aminobu | 3 | Interventio | Experimen | Experimen | Control | 0,852621 |
| 6 months | abundance | X07344 | (−)-nabilor | 3 | Interventio | Experimen | Experimen | Control | 0,856311 |
| 6 months | abundance | SL00391 | N-Acetyllei | 1 | Interventio | Experimen | Experimen | Control | 0,82397 |
| 6 months | abundance | X06551 | Methyl 1-hy | 3 | Interventio | Experimen | Experimen | Control | 0,838691 |
| 6 months | abundance | X10974 | Threonylglu | 3 | Interventio | Experimen | Experimen | Control | 0,811341 |
| 6 months | abundance | X00066 | Tryptamine | 1 | Interventio | Experimen | Experimen | Control | 0,842213 |
| 6 months | abundance | X07645 | O-propeno | 3 | Interventio | Experimen | Experimen | Control | 0,838204 |
| 6 months | abundance | X02108 | Homovanil | 3 | Interventio | Experimen | Experimen | Control | 0,825116 |
| 6 months | abundance | SL00052 | N-Isovalery | 2a | Interventio | Experimen | Experimen | Control | 0,81676 |
| 6 months | abundance | X06448 | 3,8,9-trihy | 2b | Interventio | Experimen | Experimen | Control | 0,825116 |
| 6 months | abundance | X00113 | N-Acetylph | 1 | Interventio | Experimen | Experimen | Control | 0,79043 |
| 6 months | abundance | X05204 | Urothion | 3 | Interventio | Experimen | Experimen | Control | 0,836775 |
| 6 months | abundance | SL00199 | 5-Hydroxyi | 2a | Interventio | Experimen | Experimen | Control | 0,851221 |
| 6 months | abundance | SL00226 | N-Acetylgly | 2a | Interventio | Experimen | Experimen | Control | 0,798663 |
| 6 months | abundance | X09645 | TOLMETIN | 3 | Interventio | Experimen | Experimen | Control | 0,836775 |
| 6 months | abundance | X00432 | 8-(Methyls | 3 | Interventio | Experimen | Experimen | Control | 0,838691 |
| 6 months | abundance | X07027 | Allyl merca | 3 | Interventio | Experimen | Experimen | Control | 0,829112 |
| 6 months | abundance | X07978 | 1-[(4E)-4-(4 | 3 | Interventio | Experimen | Experimen | Control | 0,79043 |
| 6 months | abundance | X10353 | L-gamma-C | 3 | Interventio | Experimen | Experimen | Control | 0,79043 |
| 6 months | abundance | M24 | M24 | IA_module | Interventio | Experimen | Experimen | Control | 0,801727 |
| 6 months | abundance | X07578 | 5-Allyl-5-se | 3 | Interventio | Experimen | Experimen | Control | 0,825133 |
| 6 months | abundance | X03709 | Guanfacin | 3 | Interventio | Experimen | Experimen | Control | 0,792475 |
| 6 months | abundance | X11354 | 3-Oxo-4,6- | 3 | Interventio | Experimen | Experimen | Control | 0,814559 |
| 6 months | abundance | X10881 | L-gamma-C | 3 | Interventio | Experimen | Experimen | Control | 0,79043 |
| 6 months | abundance | X08341 | Midodrine_ | 3 | Interventio | Experimen | Experimen | Control | 0,82397 |
| 6 months | abundance | X07658 | δ-Glucono | 1 | Interventio | Experimen | Experimen | Control | 0,822447 |
| 6 months | abundance | X08173 | 9-Methylur | 2b | Interventio | Experimen | Experimen | Control | 0,815129 |
| 6 months | abundance | X08221 | Brilliant blu | 2b | Interventio | Experimen | Experimen | Control | 0,815764 |
| 6 months | abundance | X10098 | Methyl 2,3- | 3 | Interventio | Experimen | Experimen | Control | 0,79043 |
| 6 months | abundance | X00853 | 4-O-beta-D | 3 | Interventio | Experimen | Experimen | Control | 0,767935 |
| 6 months | abundance | X08277 | adrenaline | 3 | Interventio | Experimen | Experimen | Control | 0,81084 |
| 6 months | abundance | X00327 | 3-Hydroxy- | 3 | Interventio | Experimen | Experimen | Control | 0,806712 |
| 6 months | abundance | X06797 | pro-gln_a | 3 | Interventio | Experimen | Experimen | Control | 0,790718 |
| 6 months | abundance | X02733 | Calcitriol | 3 | Interventio | Experimen | Experimen | Control | 0,811341 |

| | | | | | | | | | |
|---|---|---|---|---|---|---|---|---|---|
| 6 months | abundance | X07399 | Homoanse | 3 | Interventio | Experimen | Experimen | Control | 0,79043 |
| 6 months | abundance | X11441 | beta-D-Eth | 3 | Interventio | Experimen | Experimen | Control | 0,772129 |
| 6 months | abundance | X07935 | Limonin | 3 | Interventio | Experimen | Experimen | Control | 0,811341 |
| 6 months | abundance | X03134 | Tetrahydro | 3 | Interventio | Experimen | Experimen | Control | 0,770919 |
| 6 months | abundance | Pentanoic | Pentanoic | CFA_panel | Interventio | Experimen | Experimen | Control | 0,77651 |
| 6 months | abundance | X03916 | 2-(1-Ethox | 3 | Interventio | Experimen | Experimen | Control | 0,79043 |
| 6 months | abundance | X10593 | Vorinostat_ | 3 | Interventio | Experimen | Experimen | Control | 0,79043 |
| 6 months | abundance | X08386 | 3b-Hydrox | 3 | Interventio | Experimen | Experimen | Control | 0,806712 |
| 6 months | abundance | X10930 | MFCD0995 | 3 | Interventio | Experimen | Experimen | Control | 0,79043 |
| 6 months | abundance | X10177 | Gly-DL-Phe | 3 | Interventio | Experimen | Experimen | Control | 0,801 |
| 6 months | abundance | X09940 | pentobarbi | 3 | Interventio | Experimen | Experimen | Control | 0,79043 |
| 6 months | abundance | X08326 | 2-(1,3-Ben | 3 | Interventio | Experimen | Experimen | Control | 0,803444 |
| 6 months | abundance | X07680 | Indole-3-a | 1 | Interventio | Experimen | Experimen | Control | 0,786166 |
| 6 months | abundance | X06623 | quinol sulf | 3 | Interventio | Experimen | Experimen | Control | 0,798346 |
| 6 months | abundance | X07519 | Pyrrolidine | 3 | Interventio | Experimen | Experimen | Control | 0,769775 |
| 6 months | abundance | X06100 | Midodrine_ | 3 | Interventio | Experimen | Experimen | Control | 0,777511 |
| 6 months | abundance | X06723 | 3-Hydroxy- | 3 | Interventio | Experimen | Experimen | Control | 0,790718 |
| 6 months | abundance | SL00349 | 1-Aminocy | 2a | Interventio | Experimen | Experimen | Control | 0,79043 |
| 6 months | abundance | X07320 | asn-pro_d | 3 | Interventio | Experimen | Experimen | Control | 0,790718 |
| 6 months | abundance | X11639 | Midodrine_ | 3 | Interventio | Experimen | Experimen | Control | 0,772129 |
| 6 months | abundance | X11505 | FB950000( | 3 | Interventio | Experimen | Experimen | Control | 0,801 |
| 6 months | abundance | X01738 | 2-Acetami | 3 | Interventio | Experimen | Experimen | Control | 0,796588 |
| 6 months | abundance | X06807 | 1,1'-[1,12-I | 3 | Interventio | Experimen | Experimen | Control | 0,780414 |
| 6 months | abundance | X08795 | N-Acetylva | 3 | Interventio | Experimen | Experimen | Control | 0,780178 |
| 6 months | abundance | X04557 | 1,4-Naphtl | 3 | Interventio | Experimen | Experimen | Control | 0,79043 |
| 6 months | abundance | X06508 | Asparaginy | 3 | Interventio | Experimen | Experimen | Control | 0,714671 |
| 6 months | abundance | X01920 | N-(Carbox | 3 | Interventio | Experimen | Experimen | Control | 0,770082 |
| 6 months | abundance | X01518 | L-gamma-( | 3 | Interventio | Experimen | Experimen | Control | 0,782484 |
| 6 months | abundance | X03271 | 1Î²-Hydrox | 3 | Interventio | Experimen | Experimen | Control | 0,767953 |
| 6 months | abundance | X06861 | 4-(4-Deox | 3 | Interventio | Experimen | Experimen | Control | 0,79043 |
| 6 months | abundance | X11576 | Phenyl D-g | 3 | Interventio | Experimen | Experimen | Control | 0,79043 |
| 6 months | abundance | X10987 | 2-BUTYL PI | 3 | Interventio | Experimen | Experimen | Control | 0,766405 |
| 6 months | abundance | X03434 | SECONAL_ | 3 | Interventio | Experimen | Experimen | Control | 0,79043 |
| 6 months | abundance | X04274 | 6-[(Z)-2-(3, | 3 | Interventio | Experimen | Experimen | Control | 0,79043 |
| 6 months | abundance | X01236 | bis(4-isoth | 3 | Interventio | Experimen | Experimen | Control | 0,767935 |
| 6 months | abundance | X02419 | Glu-Glu | 3 | Interventio | Experimen | Experimen | Control | 0,755323 |
| 6 months | abundance | X08077 | 6-(alpha-D | 3 | Interventio | Experimen | Experimen | Control | 0,769575 |
| 6 months | abundance | X09577 | Spermic ac | 3 | Interventio | Experimen | Experimen | Control | 0,769775 |
| 6 months | abundance | X02288 | 9-Methylur | 2b | Interventio | Experimen | Experimen | Control | 0,770082 |
| 6 months | abundance | X07480 | Naringenin | 2a | Interventio | Experimen | Experimen | Control | 0,772129 |
| 6 months | abundance | X07513 | (−)-nabilor | 3 | Interventio | Experimen | Experimen | Control | 0,769575 |
| 6 months | abundance | X07580 | N-Pentano | 3 | Interventio | Experimen | Experimen | Control | 0,758222 |
| 6 months | abundance | X07220 | Glycylproly | 3 | Interventio | Experimen | Experimen | Control | 0,779289 |
| 6 months | abundance | X11033 | N-Acetyl-5 | 3 | Interventio | Experimen | Experimen | Control | 0,788346 |
| 6 months | abundance | X06985 | N-Ethylpro | 3 | Interventio | Experimen | Experimen | Control | 0,772129 |
| 6 months | abundance | X08992 | N(2)-succi | 3 | Interventio | Experimen | Experimen | Control | 0,758222 |
| 6 months | abundance | X10033 | epsilon-(ga | 3 | Interventio | Experimen | Experimen | Control | 0,767646 |
| 6 months | abundance | X08417 | asn-pro_c | 3 | Interventio | Experimen | Experimen | Control | 0,762795 |

| | | | | | | | | | |
|---|---|---|---|---|---|---|---|---|---|
| 6 months | abundance | X04636 | epsilon-(ga | 3 | Interventio | Experimen | Experimen | Control | 0,756336 |
| 6 months | abundance | X08040 | Prolylleuci | 2b | Interventio | Experimen | Experimen | Control | 0,769775 |
| 6 months | abundance | X10865 | Melatonin | 3 | Interventio | Experimen | Experimen | Control | 0,769775 |
| 6 months | abundance | X05411 | 3-(Sulfooxy | 3 | Interventio | Experimen | Experimen | Control | 0,768316 |
| 6 months | abundance | X09523 | N-(3,5-Dim | 3 | Interventio | Experimen | Experimen | Control | 0,766405 |
| 6 months | abundance | X07413 | Chenodeo: | 1 | Interventio | Experimen | Experimen | Control | 0,767935 |
| 6 months | abundance | SL00421 | Asymmetri | 2a | Interventio | Experimen | Experimen | Control | 0,76776 |
| 6 months | abundance | X00899 | N-(2,3,4-Tr | 3 | Interventio | Experimen | Experimen | Control | 0,767646 |
| 6 months | abundance | X04379 | 2-Amino-6 | 3 | Interventio | Experimen | Experimen | Control | 0,729456 |
| 6 months | abundance | SL00390 | Gallic acid | 2a | Interventio | Experimen | Experimen | Control | 0,728006 |
| 6 months | abundance | X06646 | (S)-3-sulfo | 3 | Interventio | Experimen | Experimen | Control | 0,762795 |
| 6 months | abundance | X01288 | L-gamma-( | 3 | Interventio | Experimen | Experimen | Control | 0,751594 |
| 6 months | abundance | X10387 | Coenzyme | 3 | Interventio | Experimen | Experimen | Control | 0,762795 |
| 6 months | abundance | X09771 | 2-(3-CARB | 3 | Interventio | Experimen | Experimen | Control | 0,770082 |
| 6 months | abundance | X07889 | UROBILIN, | 3 | Interventio | Experimen | Experimen | Control | 0,766405 |
| 6 months | abundance | X07969 | 6-hydroxyp | 3 | Interventio | Experimen | Experimen | Control | 0,769575 |
| 6 months | abundance | X02868 | 2-(3-Hydro | 3 | Interventio | Experimen | Experimen | Control | 0,767935 |
| 6 months | abundance | X07491 | 1-Vinylimic | 2b | Interventio | Experimen | Experimen | Control | 0,735877 |
| 6 months | abundance | X10951 | 8-Methyl-8 | 3 | Interventio | Experimen | Experimen | Control | 0,741993 |
| 6 months | abundance | X09505 | (-)-Physost | 3 | Interventio | Experimen | Experimen | Control | 0,749793 |
| 6 months | abundance | X06437 | 2-Acetamic | 3 | Interventio | Experimen | Experimen | Control | 0,731466 |
| 6 months | abundance | X08305 | Prolylhydrc | 2a | Interventio | Experimen | Experimen | Control | 0,71955 |
| 6 months | abundance | X07597 | p-Cresylsu | 3 | Interventio | Experimen | Experimen | Control | 0,735906 |
| 6 months | abundance | X07136 | hexobarbit | 3 | Interventio | Experimen | Experimen | Control | 0,71955 |
| 6 months | abundance | X07089 | Pregabalin | 3 | Interventio | Experimen | Experimen | Control | 0,766405 |
| 6 months | abundance | X07993 | UROBILIN, | 3 | Interventio | Experimen | Experimen | Control | 0,752328 |
| 6 months | abundance | X10401 | 3-Methylxa | 1 | Interventio | Experimen | Experimen | Control | 0,741993 |
| 6 months | abundance | X00059 | 3,4-Dihydr | 1 | Interventio | Experimen | Experimen | Control | 0,752328 |
| 6 months | abundance | X05971 | L-gamma-( | 3 | Interventio | Experimen | Experimen | Control | 0,733425 |
| 6 months | abundance | X04688 | Minoxidil | 3 | Interventio | Experimen | Experimen | Control | 0,750878 |
| 6 months | abundance | X02352 | Indole-3-ca | 3 | Interventio | Experimen | Experimen | Control | 0,756336 |
| 6 months | abundance | X06371 | L-gamma-( | 3 | Interventio | Experimen | Experimen | Control | 0,697971 |
| 6 months | abundance | X07327 | Aspartyl-L- | 3 | Interventio | Experimen | Experimen | Control | 0,713864 |
| 6 months | abundance | X11645 | glu-thr | 3 | Interventio | Experimen | Experimen | Control | 0,643904 |
| 6 months | abundance | X08883 | 3-Phenylpr | 3 | Interventio | Experimen | Experimen | Control | 0,71955 |
| 6 months | abundance | X06079 | Glycylleuci | 1 | Interventio | Experimen | Experimen | Control | 0,686259 |
| 6 months | abundance | X07127 | N~5~-[P-Ar | 3 | Interventio | Experimen | Experimen | Control | 0,741993 |
| 6 months | abundance | X08162 | Valylvaline | 3 | Interventio | Experimen | Experimen | Control | 0,728006 |
| 6 months | abundance | X07445 | 3-(Sulfooxy | 3 | Interventio | Experimen | Experimen | Control | 0,695875 |
| 6 months | abundance | SL00353 | Adipic acid | 1 | Interventio | Experimen | Experimen | Control | 0,701195 |
| 6 months | abundance | X06085 | Tetraacety | 3 | Interventio | Experimen | Experimen | Control | 0,707314 |
| 6 months | abundance | X07443 | Butylparab | 3 | Interventio | Experimen | Experimen | Control | 0,698931 |
| 6 months | abundance | X05812 | (2S)-3-(1H | 3 | Interventio | Experimen | Experimen | Control | 0,662346 |
| 6 months | abundance | X05758 | quinol sulf | 3 | Interventio | Experimen | Experimen | Control | 0,679833 |
| 6 months | abundance | X06143 | 2-Methylbu | 2a | Interventio | Experimen | Experimen | Control | 0,696038 |
| 6 months | abundance | X06783 | N~6~-Octa | 3 | Interventio | Experimen | Experimen | Control | 0,71955 |
| 6 months | abundance | X00092 | 3-Methylhi | 1 | Interventio | Experimen | Experimen | Control | 0,718213 |
| 6 months | abundance | SL00420 | 4-Hydroxy- | 2a | Interventio | Experimen | Experimen | Control | 0,70518 |

| | | | | | | | | | |
|---|---|---|---|---|---|---|---|---|---|
| 6 months | abundance | X10511 | dopaquino | 3 | Interventio | Experimen | Experimen | Control | 0,694971 |
| 6 months | abundance | X05514 | (3aS,5S,6F | 3 | Interventio | Experimen | Experimen | Control | 0,718952 |
| 6 months | abundance | X11667 | Tetraacety | 3 | Interventio | Experimen | Experimen | Control | 0,660061 |
| 6 months | abundance | X08801 | 3,7-Dimetl | 2b | Interventio | Experimen | Experimen | Control | 0,701195 |
| 6 months | abundance | X11301 | (6S)-2-Ami | 3 | Interventio | Experimen | Experimen | Control | 0,679833 |
| 6 months | abundance | X06690 | 4-Methylca | 2b | Interventio | Experimen | Experimen | Control | 0,714671 |
| 6 months | abundance | X06961 | Zalcitabine | 3 | Interventio | Experimen | Experimen | Control | 0,669742 |
| 6 months | abundance | SL00001 | Deoxycytid | 2a | Interventio | Experimen | Experimen | Control | 0,64451 |
| 6 months | abundance | M32 | M32 | JA_module | Interventio | Experimen | Experimen | Control | 0,679015 |
| 6 months | abundance | X09129 | Coprine_d | 3 | Interventio | Experimen | Experimen | Control | 0,644196 |
| 6 months | abundance | X01671 | N-(3,5-Dim | 3 | Interventio | Experimen | Experimen | Control | 0,707314 |
| 6 months | abundance | X05068 | pro-gln_c | 3 | Interventio | Experimen | Experimen | Control | 0,686987 |
| 6 months | abundance | X08657 | 1-(4-Aminc | 3 | Interventio | Experimen | Experimen | Control | 0,713054 |
| 6 months | abundance | X07963 | Theobromi | 1 | Interventio | Experimen | Experimen | Control | 0,679833 |
| 6 months | abundance | SL00024 | Creatine | 2a | Interventio | Experimen | Experimen | Control | 0,652884 |
| 6 months | abundance | X00132 | Glyceric ac | 1 | Interventio | Experimen | Experimen | Control | 0,679833 |
| 6 months | abundance | X03719 | Losalen | 3 | Interventio | Experimen | Experimen | Control | 0,679494 |
| 6 months | abundance | X07538 | Homoanse | 3 | Interventio | Experimen | Experimen | Control | 0,696038 |
| 6 months | abundance | M26 | M26 | JA_module | Interventio | Experimen | Experimen | Control | 0,696038 |
| 6 months | abundance | X03704 | N,N-Dimet | 3 | Interventio | Experimen | Experimen | Control | 0,716787 |
| 6 months | abundance | X08163 | Phenyl D-g | 3 | Interventio | Experimen | Experimen | Control | 0,709598 |
| 6 months | abundance | X08512 | Bile acid I ( | 2b | Interventio | Experimen | Experimen | Control | 0,659423 |
| 6 months | abundance | X07794 | Prolylleuci | 2b | Interventio | Experimen | Experimen | Control | 0,67553 |
| 6 months | abundance | X02565 | Dopamine | 3 | Interventio | Experimen | Experimen | Control | 0,67778 |
| 6 months | abundance | X09285 | MFCD0272 | 3 | Interventio | Experimen | Experimen | Control | 0,67553 |
| 6 months | abundance | X08983 | IN00150 | 3 | Interventio | Experimen | Experimen | Control | 0,66921 |
| 6 months | abundance | X11012 | Furan | 3 | Interventio | Experimen | Experimen | Control | 0,622746 |
| 6 months | abundance | X06889 | 3,4-Dimetl | 2a | Interventio | Experimen | Experimen | Control | 0,696038 |
| 6 months | abundance | X06112 | Val-Ser_a | 3 | Interventio | Experimen | Experimen | Control | 0,660061 |
| 6 months | abundance | X09092 | LysoSM(d1 | 3 | Interventio | Experimen | Experimen | Control | 0,658755 |
| 6 months | abundance | X06764 | N-(4-Hydrc | 3 | Interventio | Experimen | Experimen | Control | 0,652283 |
| 6 months | abundance | X01793 | Lys-phe_b | 3 | Interventio | Experimen | Experimen | Control | 0,609036 |
| 6 months | abundance | X11347 | Voglibose | 3 | Interventio | Experimen | Experimen | Control | 0,665468 |
| 6 months | abundance | X08367 | 11-dehydr | 2b | Interventio | Experimen | Experimen | Control | 0,66921 |
| 6 months | abundance | X07134 | 4-Phenols | 2b | Interventio | Experimen | Experimen | Control | 0,660061 |
| 6 months | abundance | X03892 | shinorine | 3 | Interventio | Experimen | Experimen | Control | 0,660061 |
| 6 months | abundance | X07792 | N-Acetylhi | 2b | Interventio | Experimen | Experimen | Control | 0,579512 |
| 6 months | abundance | X07263 | 3,3-Dimetl | 2b | Interventio | Experimen | Experimen | Control | 0,662457 |
| 6 months | abundance | SL00401 | Theophylli | 1 | Interventio | Experimen | Experimen | Control | 0,643904 |
| 6 months | abundance | X05072 | L-gamma-( | 3 | Interventio | Experimen | Experimen | Control | 0,66921 |
| 6 months | abundance | X07336 | Threonylse | 3 | Interventio | Experimen | Experimen | Control | 0,574621 |
| 6 months | abundance | X07646 | (2Z)-2-({[(6 | 3 | Interventio | Experimen | Experimen | Control | 0,640371 |
| 6 months | abundance | X06405 | Asparaginy | 3 | Interventio | Experimen | Experimen | Control | 0,535443 |
| 6 months | abundance | X09207 | AAMU_a | 3 | Interventio | Experimen | Experimen | Control | 0,587711 |
| 6 months | abundance | X07712 | paracetam | 3 | Interventio | Experimen | Experimen | Control | 0,644196 |
| 6 months | abundance | X07981 | Methylimic | 2b | Interventio | Experimen | Experimen | Control | 0,66245 |
| 6 months | abundance | X08869 | Lanthionin | 3 | Interventio | Experimen | Experimen | Control | 0,610394 |
| 6 months | abundance | X03917 | Uramustin | 3 | Interventio | Experimen | Experimen | Control | 0,574621 |

| | | | | | | | | | |
|---|---|---|---|---|---|---|---|---|---|
| 6 months | abundance | X07848 | (−)-nabilor | 3 | Interventio | Experimen | Experimen | Control | 0,645111 |
| 6 months | abundance | X06222 | Leucylaspa | 3 | Interventio | Experimen | Experimen | Control | 0,588937 |
| 6 months | abundance | X07798 | N-lauroylgl | 3 | Interventio | Experimen | Experimen | Control | 0,662984 |
| 6 months | abundance | X01164 | DL-Carboc | 3 | Interventio | Experimen | Experimen | Control | 0,607861 |
| 6 months | abundance | X10203 | L-gamma-( | 3 | Interventio | Experimen | Experimen | Control | 0,643904 |
| 6 months | abundance | X10389 | Leucylaspa | 3 | Interventio | Experimen | Experimen | Control | 0,567954 |
| 6 months | abundance | X08300 | Indole-3-a | 3 | Interventio | Experimen | Experimen | Control | 0,646446 |
| 6 months | abundance | X00064 | Traumatic | 1 | Interventio | Experimen | Experimen | Control | 0,608921 |
| 6 months | abundance | X06149 | Yangonin | 3 | Interventio | Experimen | Experimen | Control | 0,644181 |
| 6 months | abundance | X02774 | 9-ribosylze | 3 | Interventio | Experimen | Experimen | Control | 0,622672 |
| 6 months | abundance | X04544 | L-gamma-( | 3 | Interventio | Experimen | Experimen | Control | 0,574621 |
| 6 months | abundance | SL00376 | N-AcetylPr | 2a | Interventio | Experimen | Experimen | Control | 0,547533 |
| 6 months | abundance | X11068 | N,N'-Bis[4- | 3 | Interventio | Experimen | Experimen | Control | 0,622672 |
| 6 months | abundance | X05584 | Serotonin | 1 | Interventio | Experimen | Experimen | Control | 0,628857 |
| 6 months | abundance | X08845 | 14-Hydrox | 3 | Interventio | Experimen | Experimen | Control | 0,621405 |
| 6 months | abundance | X01553 | uridine 5'-( | 3 | Interventio | Experimen | Experimen | Control | 0,628365 |
| 6 months | abundance | X02494 | Butylphtha | 3 | Interventio | Experimen | Experimen | Control | 0,639371 |
| 6 months | abundance | X10429 | 1,5-Isoquir | 2b | Interventio | Experimen | Experimen | Control | 0,613206 |
| 6 months | abundance | X06883 | Agomelatir | 3 | Interventio | Experimen | Experimen | Control | 0,614024 |
| 6 months | abundance | X07657 | Docosahe> | 2b | Interventio | Experimen | Experimen | Control | 0,622672 |
| 6 months | abundance | X05046 | Casimiroin | 3 | Interventio | Experimen | Experimen | Control | 0,622746 |
| 6 months | abundance | X10279 | Lys-Pro_d | 3 | Interventio | Experimen | Experimen | Control | 0,602457 |
| 6 months | abundance | X08673 | NPC | 3 | Interventio | Experimen | Experimen | Control | 0,613827 |
| 6 months | abundance | X05391 | 2-Acetami | 3 | Interventio | Experimen | Experimen | Control | 0,497615 |
| 6 months | abundance | X02153 | Asarone | 3 | Interventio | Experimen | Experimen | Control | 0,607249 |
| 6 months | abundance | M18 | M18 | VA_module | Interventio | Experimen | Experimen | Control | 0,628777 |
| 6 months | abundance | X08909 | GLK (Pepti | 2b | Interventio | Experimen | Experimen | Control | 0,495451 |
| 6 months | abundance | X03254 | Gly-Trp_b | 3 | Interventio | Experimen | Experimen | Control | 0,547533 |
| 6 months | abundance | X06259 | (S)-?-glyce | 3 | Interventio | Experimen | Experimen | Control | 0,612837 |
| 6 months | abundance | X00070 | α-aminobu | 1 | Interventio | Experimen | Experimen | Control | 0,549392 |
| 6 months | abundance | X00744 | (1R,2S)-1-( | 3 | Interventio | Experimen | Experimen | Control | 0,614024 |
| 6 months | abundance | X08044 | 3-[2-[(Z)-[3 | 3 | Interventio | Experimen | Experimen | Control | 0,574621 |
| 6 months | abundance | X08908 | Vorinostat_ | 3 | Interventio | Experimen | Experimen | Control | 0,610394 |
| 6 months | abundance | X03629 | Glycylglyc | 3 | Interventio | Experimen | Experimen | Control | 0,547533 |
| 6 months | abundance | X08593 | Uric acid | 1 | Interventio | Experimen | Experimen | Control | 0,52723 |
| 6 months | abundance | X01186 | asn-val_b | 3 | Interventio | Experimen | Experimen | Control | 0,530545 |
| 6 months | abundance | X06270 | 9-Methylur | 2b | Interventio | Experimen | Experimen | Control | 0,584085 |
| 6 months | abundance | X08973 | Aminohipp | 3 | Interventio | Experimen | Experimen | Control | 0,541353 |
| 6 months | abundance | X06946 | SECONAL_ | 3 | Interventio | Experimen | Experimen | Control | 0,611916 |
| 6 months | abundance | X07574 | 7-ketodeo> | 3 | Interventio | Experimen | Experimen | Control | 0,541353 |
| 6 months | abundance | X11408 | 5-Amino-6 | 3 | Interventio | Experimen | Experimen | Control | 0,587711 |
| 6 months | abundance | X07417 | δ-Valerola | 2b | Interventio | Experimen | Experimen | Control | 0,567461 |
| 6 months | abundance | X11498 | Valylvaline | 3 | Interventio | Experimen | Experimen | Control | 0,547533 |
| 6 months | abundance | X00042 | Glucuronic | 1 | Interventio | Experimen | Experimen | Control | 0,587711 |
| 6 months | abundance | X09965 | Biocytin | 2b | Interventio | Experimen | Experimen | Control | 0,536933 |
| 6 months | abundance | X08546 | 1,3,7-Trim | 2b | Interventio | Experimen | Experimen | Control | 0,60451 |
| 6 months | abundance | X01441 | 1-{3-Carbo | 3 | Interventio | Experimen | Experimen | Control | 0,566785 |
| 6 months | abundance | X10382 | Zalcitabine | 3 | Interventio | Experimen | Experimen | Control | 0,536933 |

| | | | | | | | | | |
|---|---|---|---|---|---|---|---|---|---|
| 6 months | abundance | X09318 | butalbital_ | 3 | Interventio | Experimen | Experimen | Control | 0,536933 |
| 6 months | abundance | X04986 | L-Proline, 4 | 3 | Interventio | Experimen | Experimen | Control | 0,526151 |
| 6 months | abundance | X10977 | pentobarbi | 3 | Interventio | Experimen | Experimen | Control | 0,60451 |
| 6 months | abundance | X00030 | 3,4-Dihydr | 1 | Interventio | Experimen | Experimen | Control | 0,547533 |
| 6 months | abundance | X05071 | Histidylphe | 3 | Interventio | Experimen | Experimen | Control | 0,519242 |
| 6 months | abundance | X06704 | feruloylagr | 3 | Interventio | Experimen | Experimen | Control | 0,621405 |
| 6 months | abundance | X00949 | primidone_ | 3 | Interventio | Experimen | Experimen | Control | 0,547533 |
| 6 months | abundance | X07891 | Cyclamic a | 2b | Interventio | Experimen | Experimen | Control | 0,547533 |
| 6 months | abundance | X10037 | his-gln_a | 3 | Interventio | Experimen | Experimen | Control | 0,536149 |
| 6 months | abundance | SL00350 | Stachydrin | 1 | Interventio | Experimen | Experimen | Control | 0,559923 |
| 6 months | abundance | X00056 | 3-(4-hydro: | 1 | Interventio | Experimen | Experimen | Control | 0,482527 |
| 6 months | abundance | X07511 | Piperidine_ | 3 | Interventio | Experimen | Experimen | Control | 0,496369 |
| 6 months | abundance | X08373 | 2-Hydroxy- | 3 | Interventio | Experimen | Experimen | Control | 0,579512 |
| 6 months | abundance | X02943 | 3-Methoxy- | 3 | Interventio | Experimen | Experimen | Control | 0,552443 |
| 6 months | abundance | X04378 | 9,11-Dihyc | 3 | Interventio | Experimen | Experimen | Control | 0,573484 |
| 6 months | abundance | X08475 | 1-(4-Aminc | 3 | Interventio | Experimen | Experimen | Control | 0,547533 |
| 6 months | abundance | X00742 | MFCD1869 | 3 | Interventio | Experimen | Experimen | Control | 0,581344 |
| 6 months | abundance | X06896 | Pirbuterol | 3 | Interventio | Experimen | Experimen | Control | 0,552645 |
| 6 months | abundance | X11373 | 3-Succinoy | 2b | Interventio | Experimen | Experimen | Control | 0,507626 |
| 6 months | abundance | SL00431 | Tiglylcarnit | 2a | Interventio | Experimen | Experimen | Control | 0,519243 |
| 6 months | abundance | X08779 | 3-(1-hydro: | 2b | Interventio | Experimen | Experimen | Control | 0,448848 |
| 6 months | abundance | M15 | M15 | IA_module | Interventio | Experimen | Experimen | Control | 0,433769 |
| 6 months | abundance | p_cresol | p-Cresol | 1 | Interventio | Experimen | Experimen | Control | 0,475754 |
| 6 months | abundance | X01574 | N-Acetylpr | 3 | Interventio | Experimen | Experimen | Control | 0,458898 |
| 6 months | abundance | X10180 | 4-Formyl-2 | 3 | Interventio | Experimen | Experimen | Control | 0,530382 |
| 6 months | abundance | X02348 | N-Benzoyla | 3 | Interventio | Experimen | Experimen | Control | 0,494948 |
| 6 months | abundance | X07077 | Hyodeoxyc | 1 | Interventio | Experimen | Experimen | Control | 0,536933 |
| 6 months | abundance | X01813 | Erythorbic | 3 | Interventio | Experimen | Experimen | Control | 0,536933 |
| 6 months | abundance | SL00447 | Apigenin | 2a | Interventio | Experimen | Experimen | Control | 0,495451 |
| 6 months | abundance | X07572 | 8-(3-Furyl)- | 3 | Interventio | Experimen | Experimen | Control | 0,514474 |
| 6 months | abundance | X06062 | 1-(2,3-Dihy | 3 | Interventio | Experimen | Experimen | Control | 0,461611 |
| 6 months | abundance | X02841 | Kyotorphin | 3 | Interventio | Experimen | Experimen | Control | 0,402458 |
| 6 months | abundance | X01561 | 2-Ammoni | 3 | Interventio | Experimen | Experimen | Control | 0,547533 |
| 6 months | abundance | X06859 | Methylol D | 3 | Interventio | Experimen | Experimen | Control | 0,522714 |
| 6 months | abundance | X11366 | 3-Sulfino-L | 3 | Interventio | Experimen | Experimen | Control | 0,526151 |
| 6 months | abundance | X01007 | pentobarbi | 3 | Interventio | Experimen | Experimen | Control | 0,487464 |
| 6 months | abundance | X02082 | 6-Hydroxyr | 3 | Interventio | Experimen | Experimen | Control | 0,535443 |
| 6 months | abundance | X02231 | Ala-Tyr | 3 | Interventio | Experimen | Experimen | Control | 0,530382 |
| 6 months | abundance | X07508 | Procaine_a | 2b | Interventio | Experimen | Experimen | Control | 0,481265 |
| 6 months | abundance | X06482 | glu-pro | 3 | Interventio | Experimen | Experimen | Control | 0,458898 |
| 6 months | abundance | X01732 | 2-Hydroxy- | 3 | Interventio | Experimen | Experimen | Control | 0,530382 |
| 6 months | abundance | X03275 | Methdilazi | 3 | Interventio | Experimen | Experimen | Control | 0,474418 |
| 6 months | abundance | X07834 | Choline su | 3 | Interventio | Experimen | Experimen | Control | 0,497529 |
| 6 months | abundance | X06381 | MFCD1869 | 3 | Interventio | Experimen | Experimen | Control | 0,493814 |
| 6 months | abundance | X08509 | Piceid | 3 | Interventio | Experimen | Experimen | Control | 0,495451 |
| 6 months | abundance | X06703 | N6-METHY | 3 | Interventio | Experimen | Experimen | Control | 0,413749 |
| 6 months | abundance | X03568 | 3-[(2Z)-1-C | 3 | Interventio | Experimen | Experimen | Control | 0,494948 |
| 6 months | abundance | X01640 | Ro 20-1724 | 3 | Interventio | Experimen | Experimen | Control | 0,424411 |

| | | | | | | | | | |
|---|---|---|---|---|---|---|---|---|---|
| 6 months | abundance | X08420 | Dodecane | 1 | Interventio | Experimen | Experimen | Control | 0,474418 |
| 6 months | abundance | X10886 | L-gamma-( | 3 | Interventio | Experimen | Experimen | Control | 0,471375 |
| 6 months | abundance | X06856 | Tiglic acid_ | 2b | Interventio | Experimen | Experimen | Control | 0,445492 |
| 6 months | abundance | X09064 | Tetrahydro | 3 | Interventio | Experimen | Experimen | Control | 0,477197 |
| 6 months | abundance | X07081 | N-LACTOYI | 3 | Interventio | Experimen | Experimen | Control | 0,487464 |
| 6 months | abundance | X09787 | Queuosine | 3 | Interventio | Experimen | Experimen | Control | 0,497615 |
| 6 months | abundance | SL00040 | Kynurenine | 2a | Interventio | Experimen | Experimen | Control | 0,474418 |
| 6 months | abundance | X10151 | 2-(3-CARB | 3 | Interventio | Experimen | Experimen | Control | 0,477197 |
| 6 months | abundance | X10549 | Glaucine | 2b | Interventio | Experimen | Experimen | Control | 0,455427 |
| 6 months | abundance | X09912 | MC055530 | 3 | Interventio | Experimen | Experimen | Control | 0,485449 |
| 6 months | abundance | X04543 | N-Acetyl-S | 3 | Interventio | Experimen | Experimen | Control | 0,477197 |
| 6 months | abundance | X10129 | SECONAL_ | 3 | Interventio | Experimen | Experimen | Control | 0,45459 |
| 6 months | abundance | X03069 | N-COUMAI | 3 | Interventio | Experimen | Experimen | Control | 0,475519 |
| 6 months | abundance | X06041 | (DL)-3-O-M | 3 | Interventio | Experimen | Experimen | Control | 0,428209 |
| 6 months | abundance | X07236 | 2-methylci | 3 | Interventio | Experimen | Experimen | Control | 0,495451 |
| 6 months | abundance | X08147 | porphobilir | 3 | Interventio | Experimen | Experimen | Control | 0,472806 |
| 6 months | abundance | X04852 | Toluene_b | 3 | Interventio | Experimen | Experimen | Control | 0,464749 |
| 6 months | abundance | X07112 | MFCD0272 | 3 | Interventio | Experimen | Experimen | Control | 0,462525 |
| 6 months | abundance | X07425 | N-Acetylas | 2a | Interventio | Experimen | Experimen | Control | 0,425825 |
| 6 months | abundance | X02289 | Kynurenic a | 2a | Interventio | Experimen | Experimen | Control | 0,451534 |
| 6 months | abundance | X06709 | Tetraacety | 3 | Interventio | Experimen | Experimen | Control | 0,402821 |
| 6 months | abundance | Propanoic | Propanoic | CFA_panel | Interventio | Experimen | Experimen | Control | 0,446583 |
| 6 months | abundance | X01570 | carglumic a | 3 | Interventio | Experimen | Experimen | Control | 0,472806 |
| 6 months | abundance | X11383 | N-{3-[(4-Ac | 3 | Interventio | Experimen | Experimen | Control | 0,444616 |
| 6 months | abundance | X00450 | Gly-Ser | 3 | Interventio | Experimen | Experimen | Control | 0,421892 |
| 6 months | abundance | X05243 | N-(4-Hydro | 3 | Interventio | Experimen | Experimen | Control | 0,471112 |
| 6 months | abundance | X06506 | L-Homocys | 3 | Interventio | Experimen | Experimen | Control | 0,474951 |
| 6 months | abundance | X07830 | 1,5-Isoquir | 2b | Interventio | Experimen | Experimen | Control | 0,472806 |
| 6 months | abundance | X01519 | Leucyltrypt | 3 | Interventio | Experimen | Experimen | Control | 0,472806 |
| 6 months | abundance | SL00368 | N-Methylty | 1 | Interventio | Experimen | Experimen | Control | 0,472806 |
| 6 months | abundance | X10550 | pentobarbi | 3 | Interventio | Experimen | Experimen | Control | 0,485832 |
| 6 months | abundance | X07701 | 3-Benzyl-6 | 3 | Interventio | Experimen | Experimen | Control | 0,483186 |
| 6 months | abundance | M8 | M8 | IA_module | Interventio | Experimen | Experimen | Control | 0,465501 |
| 6 months | abundance | X07745 | APM_c | 3 | Interventio | Experimen | Experimen | Control | 0,462155 |
| 6 months | abundance | X07565 | 5-Hydroxyi | 2b | Interventio | Experimen | Experimen | Control | 0,444616 |
| 6 months | abundance | X10979 | acetyltauri | 3 | Interventio | Experimen | Experimen | Control | 0,463049 |
| 6 months | abundance | X00051 | β-D-Glucoן | 2b | Interventio | Experimen | Experimen | Control | 0,429158 |
| 6 months | abundance | SL00161 | Uridine | 2a | Interventio | Experimen | Experimen | Control | 0,411713 |
| 6 months | abundance | X07315 | Rivastigmi | 3 | Interventio | Experimen | Experimen | Control | 0,437868 |
| 6 months | abundance | X07846 | Carbofurar | 3 | Interventio | Experimen | Experimen | Control | 0,373897 |
| 6 months | abundance | X09615 | tyramine sı | 3 | Interventio | Experimen | Experimen | Control | 0,476616 |
| 6 months | abundance | X07749 | (DL)-3-O-M | 3 | Interventio | Experimen | Experimen | Control | 0,425825 |
| 6 months | abundance | X02521 | 6-(1-Hydro | 3 | Interventio | Experimen | Experimen | Control | 0,464749 |
| 6 months | abundance | X11034 | 1-(3-Aminc | 3 | Interventio | Experimen | Experimen | Control | 0,416238 |
| 6 months | abundance | X11477 | gamma-Gl | 3 | Interventio | Experimen | Experimen | Control | 0,363159 |
| 6 months | abundance | X06612 | Spermic ac | 3 | Interventio | Experimen | Experimen | Control | 0,315603 |
| 6 months | abundance | X00545 | 3-hydroxyc | 3 | Interventio | Experimen | Experimen | Control | 0,434372 |
| 6 months | abundance | X04684 | AAMU_b | 3 | Interventio | Experimen | Experimen | Control | 0,473588 |

| | | | | | | | | | |
|---|---|---|---|---|---|---|---|---|---|
| 6 months | abundance | X04181 | 3-Methoxy- | 3 | Interventio | Experimen | Experimen | Control | 0,413749 |
| 6 months | abundance | 3-Methylbu | 3-Methylbu | CFA_panel | Interventio | Experimen | Experimen | Control | 0,366113 |
| 6 months | abundance | SL00445 | Glucosami | 2a | Interventio | Experimen | Experimen | Control | 0,462525 |
| 6 months | abundance | SL00185 | 2-Oxo-3-pł | 1 | Interventio | Experimen | Experimen | Control | 0,375103 |
| 6 months | abundance | X04996 | MFCD0272 | 3 | Interventio | Experimen | Experimen | Control | 0,497529 |
| 6 months | abundance | X05551 | Trolox | 3 | Interventio | Experimen | Experimen | Control | 0,435244 |
| 6 months | abundance | X04922 | Sinapinic a | 3 | Interventio | Experimen | Experimen | Control | 0,433769 |
| 6 months | abundance | X08504 | Indole-3-ca | 2a | Interventio | Experimen | Experimen | Control | 0,374505 |
| 6 months | abundance | X09454 | 2-(2-Hydro | 3 | Interventio | Experimen | Experimen | Control | 0,410744 |
| 6 months | abundance | X10457 | glu-ser | 3 | Interventio | Experimen | Experimen | Control | 0,310448 |
| 6 months | abundance | X09004 | Arenaine | 3 | Interventio | Experimen | Experimen | Control | 0,455294 |
| 6 months | abundance | SL00212 | Galactosar | 2a | Interventio | Experimen | Experimen | Control | 0,346393 |
| 6 months | abundance | X08322 | Glycylproly | 3 | Interventio | Experimen | Experimen | Control | 0,402458 |
| 6 months | abundance | X10503 | APM_a | 3 | Interventio | Experimen | Experimen | Control | 0,408947 |
| 6 months | abundance | X06820 | 1-(3,4-dim | 2b | Interventio | Experimen | Experimen | Control | 0,472806 |
| 6 months | abundance | X01528 | butalbital_ | 3 | Interventio | Experimen | Experimen | Control | 0,346785 |
| 6 months | abundance | X04566 | pterin | 3 | Interventio | Experimen | Experimen | Control | 0,464749 |
| 6 months | abundance | X02689 | piscidic ac | 3 | Interventio | Experimen | Experimen | Control | 0,402458 |
| 6 months | abundance | X01881 | (2S)-3-Met | 3 | Interventio | Experimen | Experimen | Control | 0,408947 |
| 6 months | abundance | X07439 | 4-pyridoxic | 1 | Interventio | Experimen | Experimen | Control | 0,328207 |
| 6 months | abundance | X10025 | 2-[4-(3-Hy | 3 | Interventio | Experimen | Experimen | Control | 0,416238 |
| 6 months | abundance | X11038 | (2E)-3-(3,4 | 3 | Interventio | Experimen | Experimen | Control | 0,346785 |
| 6 months | abundance | X00266 | 2-Oxo-3-(p | 3 | Interventio | Experimen | Experimen | Control | 0,411713 |
| 6 months | abundance | 2-Methylpr | 2-Methylpr | CFA_panel | Interventio | Experimen | Experimen | Control | 0,299663 |
| 6 months | abundance | X08076 | 3',5,7-Trihy | 2b | Interventio | Experimen | Experimen | Control | 0,400134 |
| 6 months | abundance | X11028 | butalbital_ | 3 | Interventio | Experimen | Experimen | Control | 0,395346 |
| 6 months | abundance | X08311 | Methyl 3-fc | 3 | Interventio | Experimen | Experimen | Control | 0,421892 |
| 6 months | abundance | X05398 | DIBOA | 3 | Interventio | Experimen | Experimen | Control | 0,373224 |
| 6 months | abundance | X06590 | 3-hydroxy- | 3 | Interventio | Experimen | Experimen | Control | 0,392683 |
| 6 months | abundance | X00383 | pretazettin | 3 | Interventio | Experimen | Experimen | Control | 0,362327 |
| 6 months | abundance | X01100 | (betaS)-be | 3 | Interventio | Experimen | Experimen | Control | 0,408413 |
| 6 months | abundance | X09977 | TLK (Peptic | 2b | Interventio | Experimen | Experimen | Control | 0,297276 |
| 6 months | abundance | X04377 | Dihydroura | 3 | Interventio | Experimen | Experimen | Control | 0,327144 |
| 6 months | abundance | X05417 | (S)-2-hydra | 3 | Interventio | Experimen | Experimen | Control | 0,346785 |
| 6 months | abundance | X08535 | Nitrosohep | 2b | Interventio | Experimen | Experimen | Control | 0,324575 |
| 6 months | abundance | X05734 | N-[(2S)-2-H | 3 | Interventio | Experimen | Experimen | Control | 0,310912 |
| 6 months | abundance | X06220 | Tetrahydro | 3 | Interventio | Experimen | Experimen | Control | 0,40852 |
| 6 months | abundance | X02968 | Ro 20-1724 | 3 | Interventio | Experimen | Experimen | Control | 0,387234 |
| 6 months | abundance | X08990 | Procaine_c | 3 | Interventio | Experimen | Experimen | Control | 0,357085 |
| 6 months | abundance | X05968 | Tetraacety | 3 | Interventio | Experimen | Experimen | Control | 0,303733 |
| 6 months | abundance | X10011 | Midodrine_ | 3 | Interventio | Experimen | Experimen | Control | 0,33088 |
| 6 months | abundance | X02823 | 6,8-Dimetł | 3 | Interventio | Experimen | Experimen | Control | 0,319626 |
| 6 months | abundance | X06486 | Leu-Val_b | 3 | Interventio | Experimen | Experimen | Control | 0,301288 |
| 6 months | abundance | X09357 | 5-(5-Methy | 3 | Interventio | Experimen | Experimen | Control | 0,346785 |
| 6 months | abundance | X06600 | MFCD0014 | 3 | Interventio | Experimen | Experimen | Control | 0,189922 |
| 6 months | abundance | X07849 | Crotamitor | 3 | Interventio | Experimen | Experimen | Control | 0,36404 |
| 6 months | abundance | X06289 | Nicotinatel | 3 | Interventio | Experimen | Experimen | Control | 0,189396 |
| 6 months | abundance | X10936 | clavulanic | 3 | Interventio | Experimen | Experimen | Control | 0,248374 |

| | | | | | | | | | |
|---|---|---|---|---|---|---|---|---|---|
| 6 months | abundance | X04697 | Ethosuxim | 3 | Interventio | Experimen | Experimen | Control | 0,283406 |
| 6 months | abundance | X07250 | Leucylaspa | 3 | Interventio | Experimen | Experimen | Control | 0,250568 |
| 6 months | abundance | X07869 | Hexamethy | 2a | Interventio | Experimen | Experimen | Control | 0,30878 |
| 6 months | abundance | X10864 | 5-amino-2- | 2b | Interventio | Experimen | Experimen | Control | 0,25478 |
| 6 months | abundance | X01530 | INK (Peptic | 2b | Interventio | Experimen | Experimen | Control | 0,20049 |
| 6 months | abundance | X06655 | Hydroxycar | 3 | Interventio | Experimen | Experimen | Control | 0,313913 |
| 6 months | abundance | X06933 | 3-Methoxy- | 3 | Interventio | Experimen | Experimen | Control | 0,316818 |
| 6 months | abundance | X04999 | 2-Hydroxyp | 1 | Interventio | Experimen | Experimen | Control | 0,316818 |
| 6 months | abundance | X07577 | 2-[(Sulfoox | 3 | Interventio | Experimen | Experimen | Control | 0,22808 |
| 6 months | abundance | X06152 | alpha-keto | 3 | Interventio | Experimen | Experimen | Control | 0,330185 |
| 6 months | abundance | M11 | M11 | NA_module | Interventio | Experimen | Experimen | Control | 0,301288 |
| 6 months | abundance | X03714 | Sinapinic a | 3 | Interventio | Experimen | Experimen | Control | 0,375103 |
| 6 months | abundance | SL00407 | Homovanil | 2a | Interventio | Experimen | Experimen | Control | 0,250568 |
| 6 months | abundance | X00120 | Leucylalan | 1 | Interventio | Experimen | Experimen | Control | 0,275681 |
| 6 months | abundance | X04146 | L-fucopyra | 3 | Interventio | Experimen | Experimen | Control | 0,326255 |
| 6 months | abundance | X08439 | Leucyltyros | 3 | Interventio | Experimen | Experimen | Control | 0,292527 |
| 6 months | abundance | X06400 | Phloionolic | 3 | Interventio | Experimen | Experimen | Control | 0,37321 |
| 6 months | abundance | X05726 | methocarb | 3 | Interventio | Experimen | Experimen | Control | 0,27927 |
| 6 months | abundance | X00404 | piscidic ac | 3 | Interventio | Experimen | Experimen | Control | 0,301288 |
| 6 months | abundance | X06495 | 2-Acetamic | 3 | Interventio | Experimen | Experimen | Control | 0,248374 |
| 6 months | abundance | X10757 | Glycyrin | 3 | Interventio | Experimen | Experimen | Control | 0,272268 |
| 6 months | abundance | X06880 | pentobarbi | 3 | Interventio | Experimen | Experimen | Control | 0,327464 |
| 6 months | abundance | X07038 | Piperidine_ | 3 | Interventio | Experimen | Experimen | Control | 0,303488 |
| 6 months | abundance | X00549 | 3-[(2Z)-1-C | 3 | Interventio | Experimen | Experimen | Control | 0,283861 |
| 6 months | abundance | X05081 | ELK (Peptic | 2b | Interventio | Experimen | Experimen | Control | 0,251302 |
| 6 months | abundance | M29 | M29 | NA_module | Interventio | Experimen | Experimen | Control | 0,301288 |
| 6 months | abundance | X00233 | g-Aminobu | 3 | Interventio | Experimen | Experimen | Control | 0,317066 |
| 6 months | abundance | X10636 | 8-hydroxy- | 2b | Interventio | Experimen | Experimen | Control | 0,315603 |
| 6 months | abundance | X07465 | TO0127900 | 3 | Interventio | Experimen | Experimen | Control | 0,246789 |
| 6 months | abundance | X07454 | 2,4-Quinol | 2b | Interventio | Experimen | Experimen | Control | 0,179358 |
| 6 months | abundance | X01672 | Triethyl citı | 3 | Interventio | Experimen | Experimen | Control | 0,254807 |
| 6 months | abundance | X04450 | 3-Methylac | 3 | Interventio | Experimen | Experimen | Control | 0,305574 |
| 6 months | abundance | X07487 | Oxypeucec | 3 | Interventio | Experimen | Experimen | Control | 0,316818 |
| 6 months | abundance | X06768 | Sinapyl alc | 3 | Interventio | Experimen | Experimen | Control | 0,279022 |
| 6 months | abundance | X07820 | Creatinine | 1 | Interventio | Experimen | Experimen | Control | 0,287584 |
| 6 months | abundance | X08072 | 4-Hydroxyp | 3 | Interventio | Experimen | Experimen | Control | 0,303733 |
| 6 months | abundance | X10130 | N-(4-Hydrc | 3 | Interventio | Experimen | Experimen | Control | 0,2532 |
| 6 months | abundance | X07013 | pimethixer | 3 | Interventio | Experimen | Experimen | Control | 0,310448 |
| 6 months | abundance | X07937 | Atenolol | 1 | Interventio | Experimen | Experimen | Control | 0,274405 |
| 6 months | abundance | X10314 | 4-morpholi | 2b | Interventio | Experimen | Experimen | Control | 0,278551 |
| 6 months | abundance | X10665 | Aprobarbit | 3 | Interventio | Experimen | Experimen | Control | 0,309332 |
| 6 months | abundance | X08038 | N-{6-[(7-Cl | 2b | Interventio | Experimen | Experimen | Control | 0,327144 |
| 6 months | abundance | M3 | M3 | NA_module | Interventio | Experimen | Experimen | Control | 0,27927 |
| 6 months | abundance | X05997 | FB9500000 | 3 | Interventio | Experimen | Experimen | Control | 0,118447 |
| 6 months | abundance | M17 | M17 | NA_module | Interventio | Experimen | Experimen | Control | 0,170002 |
| 6 months | abundance | X08451 | L-gamma-( | 3 | Interventio | Experimen | Experimen | Control | 0,162035 |
| 6 months | abundance | X11251 | N-Nonanoy | 3 | Interventio | Experimen | Experimen | Control | 0,246099 |
| 6 months | abundance | X10651 | Methyl 1-hy | 3 | Interventio | Experimen | Experimen | Control | 0,246099 |

| | | | | | | | | | |
|---|---|---|---|---|---|---|---|---|---|
| 6 months | abundance | X08056 | 3-(2-Oxo-2 | 2b | Interventio | Experimen | Experimen | Control | 0,278551 |
| 6 months | abundance | M6 | M6 | NA_module | Interventio | Experimen | Experimen | Control | 0,287584 |
| 6 months | abundance | X10982 | 8-Methyl-8 | 3 | Interventio | Experimen | Experimen | Control | 0,157063 |
| 6 months | abundance | X06354 | trans-Zeati | 2b | Interventio | Experimen | Experimen | Control | 0,245841 |
| 6 months | abundance | X06680 | 7-Hydroxy- | 2b | Interventio | Experimen | Experimen | Control | 0,215732 |
| 6 months | abundance | X07113 | Sulfoaceta | 3 | Interventio | Experimen | Experimen | Control | 0,243384 |
| 6 months | abundance | X06168 | leu-gln_b | 3 | Interventio | Experimen | Experimen | Control | 0,167056 |
| 6 months | abundance | X07428 | N6-METHY | 3 | Interventio | Experimen | Experimen | Control | 0,213235 |
| 6 months | abundance | X02640 | thyronine | 3 | Interventio | Experimen | Experimen | Control | 0,25111 |
| 6 months | abundance | X07303 | Triacetin | 3 | Interventio | Experimen | Experimen | Control | 0,217996 |
| 6 months | abundance | SL00320 | γ-Caprolac | 2a | Interventio | Experimen | Experimen | Control | 0,160244 |
| 6 months | abundance | X02020 | Val-Ser_b | 3 | Interventio | Experimen | Experimen | Control | 0,153848 |
| 6 months | abundance | X07057 | Leu-Leu_c | 3 | Interventio | Experimen | Experimen | Control | 0,276451 |
| 6 months | abundance | X10676 | 6-(1-Hydro | 3 | Interventio | Experimen | Experimen | Control | 0,225907 |
| 6 months | abundance | X06250 | [3-({3-[(Cy | 2b | Interventio | Experimen | Experimen | Control | 0,187192 |
| 6 months | abundance | X00026 | Inosine | 1 | Interventio | Experimen | Experimen | Control | 0,195944 |
| 6 months | abundance | SL00282 | 3-Hydroxyl | 2a | Interventio | Experimen | Experimen | Control | 0,196579 |
| 6 months | abundance | X09163 | Prenisteine | 3 | Interventio | Experimen | Experimen | Control | 0,224766 |
| 6 months | abundance | X07139 | N,N-Diethy | 3 | Interventio | Experimen | Experimen | Control | 0,179358 |
| 6 months | abundance | SL00200 | 8-Aminooc | 2a | Interventio | Experimen | Experimen | Control | 0,246099 |
| 6 months | abundance | X04582 | Dinoseb | 3 | Interventio | Experimen | Experimen | Control | 0,218277 |
| 6 months | abundance | X11666 | N-(4-Hydro | 3 | Interventio | Experimen | Experimen | Control | 0,213812 |
| 6 months | abundance | X06850 | Leu-Val_c | 3 | Interventio | Experimen | Experimen | Control | 0,251476 |
| 6 months | abundance | X01363 | Seryltyrosi | 3 | Interventio | Experimen | Experimen | Control | 0,276451 |
| 6 months | abundance | X00097 | 2,6-Dihydr | 1 | Interventio | Experimen | Experimen | Control | 0,199156 |
| 6 months | abundance | X05183 | 3,6-Dichlo | 3 | Interventio | Experimen | Experimen | Control | 0,248374 |
| 6 months | abundance | X09643 | N-(3,5-Dim | 3 | Interventio | Experimen | Experimen | Control | 0,187115 |
| 6 months | abundance | X05295 | 3-(4,7-Dim | 3 | Interventio | Experimen | Experimen | Control | 0,270217 |
| 6 months | abundance | X10953 | Vorinostat_ | 3 | Interventio | Experimen | Experimen | Control | 0,196579 |
| 6 months | abundance | X01893 | 4-(METHYL | 3 | Interventio | Experimen | Experimen | Control | 0,232202 |
| 6 months | abundance | X10889 | N-acetyl-b | 3 | Interventio | Experimen | Experimen | Control | 0,141367 |
| 6 months | abundance | X11254 | (4R,5S,9S, | 3 | Interventio | Experimen | Experimen | Control | 0,222774 |
| 6 months | abundance | X01163 | Prephenic | 3 | Interventio | Experimen | Experimen | Control | 0,251601 |
| 6 months | abundance | X00748 | 3-[3-Metho | 3 | Interventio | Experimen | Experimen | Control | 0,209548 |
| 6 months | abundance | X09099 | 1-(2-Hydro | 3 | Interventio | Experimen | Experimen | Control | 0,200132 |
| 6 months | abundance | X08106 | 4-Vinylphe | 1 | Interventio | Experimen | Experimen | Control | 0,107899 |
| 6 months | abundance | X11042 | Leu-arg | 3 | Interventio | Experimen | Experimen | Control | 0,08278 |
| 6 months | abundance | X07852 | N6,N6,N6- | 3 | Interventio | Experimen | Experimen | Control | 0,139315 |
| 6 months | abundance | X11125 | 5,8,12-Trih | 3 | Interventio | Experimen | Experimen | Control | 0,143983 |
| 6 months | abundance | X00261 | Mexiletine | 3 | Interventio | Experimen | Experimen | Control | 0,18655 |
| 6 months | abundance | X00948 | Linamarin | 3 | Interventio | Experimen | Experimen | Control | 0,126909 |
| 6 months | abundance | SL00275 | Dopamine | 2a | Interventio | Experimen | Experimen | Control | 0,246789 |
| 6 months | abundance | X04117 | 2,3,4,5-tet | 3 | Interventio | Experimen | Experimen | Control | 0,196471 |
| 6 months | abundance | X00038 | Histamine | 1 | Interventio | Experimen | Experimen | Control | 0,221015 |
| 6 months | abundance | X03357 | 2-Hydroxyl | 3 | Interventio | Experimen | Experimen | Control | 0,12884 |
| 6 months | abundance | X11481 | his-gln_b | 3 | Interventio | Experimen | Experimen | Control | 0,216874 |
| 6 months | abundance | X06189 | delta-Guar | 3 | Interventio | Experimen | Experimen | Control | 0,165482 |
| 6 months | abundance | M28 | M28 | NA_module | Interventio | Experimen | Experimen | Control | 0,136337 |

| | | | | | | | | | |
|---|---|---|---|---|---|---|---|---|---|
| 6 months | abundance | X00221 | Astemizole | 3 | Interventio | Experimen | Experimen | Control | 0,159111 |
| 6 months | abundance | X07475 | 2'-Deoxy-5 | 3 | Interventio | Experimen | Experimen | Control | 0,213235 |
| 6 months | abundance | X06814 | N-Benzoyla | 3 | Interventio | Experimen | Experimen | Control | 0,171599 |
| 6 months | abundance | X06276 | asn-pro_b | 3 | Interventio | Experimen | Experimen | Control | 0,094335 |
| 6 months | abundance | X02824 | Eslicarbaz | 3 | Interventio | Experimen | Experimen | Control | 0,187823 |
| 6 months | abundance | X03268 | Dihydrouri | 3 | Interventio | Experimen | Experimen | Control | 0,171599 |
| 6 months | abundance | X08781 | Procaine_b | 3 | Interventio | Experimen | Experimen | Control | 0,179439 |
| 6 months | abundance | X11215 | Leu-Leu_b | 3 | Interventio | Experimen | Experimen | Control | 0,117512 |
| 6 months | abundance | M34 | M34 | NA_module | Interventio | Experimen | Experimen | Control | 0,196868 |
| 6 months | abundance | X02986 | Diacetin_b | 3 | Interventio | Experimen | Experimen | Control | 0,173912 |
| 6 months | abundance | X08338 | Methyl [9-( | 3 | Interventio | Experimen | Experimen | Control | 0,164527 |
| 6 months | abundance | X05919 | 2-(2-Amino | 3 | Interventio | Experimen | Experimen | Control | 0,175218 |
| 6 months | abundance | X01327 | threonylph | 3 | Interventio | Experimen | Experimen | Control | 0,222739 |
| 6 months | abundance | SL00467 | Sucralose | 2a | Interventio | Experimen | Experimen | Control | 0,20049 |
| 6 months | abundance | X01242 | 3-Morpholi | 2b | Interventio | Experimen | Experimen | Control | 0,080975 |
| 6 months | abundance | X11522 | thr-trp | 3 | Interventio | Experimen | Experimen | Control | 0,098395 |
| 6 months | abundance | X11339 | L-gamma-( | 3 | Interventio | Experimen | Experimen | Control | 0,136337 |
| 6 months | abundance | X01558 | 2-Furoylgly | 2b | Interventio | Experimen | Experimen | Control | 0,145494 |
| 6 months | abundance | X07731 | 3-[4-methy | 2b | Interventio | Experimen | Experimen | Control | 0,113797 |
| 6 months | abundance | X09608 | N-(3,5-Dim | 3 | Interventio | Experimen | Experimen | Control | 0,12884 |
| 6 months | abundance | X06419 | Spermic ac | 3 | Interventio | Experimen | Experimen | Control | 0,068876 |
| 6 months | abundance | X07283 | 8-Hydroxyh | 3 | Interventio | Experimen | Experimen | Control | 0,107899 |
| 6 months | abundance | X01387 | Methyl 4-(4 | 3 | Interventio | Experimen | Experimen | Control | 0,139315 |
| 6 months | abundance | X06227 | lys-tyr_a | 3 | Interventio | Experimen | Experimen | Control | 0,103211 |
| 6 months | abundance | X04695 | N-[(4-Meth | 3 | Interventio | Experimen | Experimen | Control | 0,124409 |
| 6 months | abundance | X00129 | Quinic acic | 1 | Interventio | Experimen | Experimen | Control | 0,115704 |
| 6 months | abundance | X09399 | L-gamma-( | 3 | Interventio | Experimen | Experimen | Control | 0,062463 |
| 6 months | abundance | X05203 | felbamate | 3 | Interventio | Experimen | Experimen | Control | 0,107855 |
| 6 months | abundance | X03158 | 5-Hydroxy- | 3 | Interventio | Experimen | Experimen | Control | 0,103211 |
| 6 months | abundance | M33 | M33 | NA_module | Interventio | Experimen | Experimen | Control | 0,092456 |
| 6 months | abundance | X08713 | Phenethyla | 2b | Interventio | Experimen | Experimen | Control | 0,056436 |
| 6 months | abundance | X07977 | Tyramine | 2b | Interventio | Experimen | Experimen | Control | 0,095967 |
| 6 months | abundance | X08298 | N6,N6,N6- | 2b | Interventio | Experimen | Experimen | Control | 0,095031 |
| 6 months | abundance | X07905 | Prilocaine | 2b | Interventio | Experimen | Experimen | Control | 0,125216 |
| 6 months | abundance | X03563 | 6-imino-5- | 3 | Interventio | Experimen | Experimen | Control | 0,077552 |
| 6 months | abundance | X01887 | lys-tyr_b | 3 | Interventio | Experimen | Experimen | Control | 0,0902 |
| 6 months | abundance | X07146 | meprobam | 3 | Interventio | Experimen | Experimen | Control | 0,055405 |
| 6 months | abundance | X07930 | N-Methylca | 2b | Interventio | Experimen | Experimen | Control | 0,130126 |
| 6 months | abundance | X09607 | N-(1H-Pyrr | 3 | Interventio | Experimen | Experimen | Control | 0,06731 |
| 6 months | abundance | X08639 | hypaphorin | 3 | Interventio | Experimen | Experimen | Control | 0,080527 |
| 6 months | abundance | SL00356 | N-(5-Amin | 2a | Interventio | Experimen | Experimen | Control | 0,126909 |
| 6 months | abundance | X11140 | Leu-Leu_e | 3 | Interventio | Experimen | Experimen | Control | 0,095967 |
| 6 months | abundance | X04483 | (2E)-5-Hyd | 3 | Interventio | Experimen | Experimen | Control | 0,134826 |
| 6 months | abundance | X06401 | Isophthalic | 2b | Interventio | Experimen | Experimen | Control | 0,118447 |
| 6 months | abundance | X00693 | n-Ribosylh | 3 | Interventio | Experimen | Experimen | Control | 0,045594 |
| 6 months | abundance | X00985 | 1-(4-Amino | 3 | Interventio | Experimen | Experimen | Control | 0,068028 |
| 6 months | abundance | M30 | M30 | NA_module | Interventio | Experimen | Experimen | Control | 0,080527 |
| 6 months | abundance | X10513 | ophthalmic | 3 | Interventio | Experimen | Experimen | Control | 0,06242 |

| | | | | | | | | | |
|---|---|---|---|---|---|---|---|---|---|
| 6 months | abundance | X02219 | Homoanse | 3 | Interventio | Experimen | Experimen | Control | 0,110798 |
| 6 months | abundance | X05234 | 3,4-dihydro | 3 | Interventio | Experimen | Experimen | Control | 0,114862 |
| 6 months | abundance | X06433 | 4-(2,5-Diflu | 2b | Interventio | Experimen | Experimen | Control | 0,092456 |
| 6 months | abundance | X07285 | 2-(Carboxy | 3 | Interventio | Experimen | Experimen | Control | 0,113467 |
| 6 months | abundance | X10393 | Ro 20-1724 | 3 | Interventio | Experimen | Experimen | Control | 0,081606 |
| 6 months | abundance | X08478 | 1-(beta-D-I | 3 | Interventio | Experimen | Experimen | Control | 0,074642 |
| 6 months | abundance | X03223 | 1-(2-Carbo | 3 | Interventio | Experimen | Experimen | Control | 0,095967 |
| 6 months | abundance | X00025 | Acesulfam | 1 | Interventio | Experimen | Experimen | Control | 0,077865 |
| 6 months | abundance | X07309 | 3,4,15-Trih | 3 | Interventio | Experimen | Experimen | Control | 0,086596 |
| 6 months | abundance | SL00318 | 4-Aminoph | 1 | Interventio | Experimen | Experimen | Control | 0,097102 |
| 6 months | abundance | X01081 | Methyl alpl | 3 | Interventio | Experimen | Experimen | Control | 0,048201 |
| 6 months | abundance | X08103 | Isopelletier | 3 | Interventio | Experimen | Experimen | Control | 0,053894 |
| 6 months | abundance | X09738 | (7E,7'E)-5, | 3 | Interventio | Experimen | Experimen | Control | 0,060452 |
| 6 months | abundance | X07944 | Styrene | 3 | Interventio | Experimen | Experimen | Control | 0,032515 |
| 6 months | abundance | X03376 | 4-(9H-beta | 3 | Interventio | Experimen | Experimen | Control | 0,080527 |
| 6 months | abundance | X00180 | 1-(2-Carbo | 3 | Interventio | Experimen | Experimen | Control | 0,072309 |
| 6 months | abundance | X07903 | Acetophen | 2b | Interventio | Experimen | Experimen | Control | 0,068026 |
| 6 months | abundance | X02990 | Pentoxifylli | 3 | Interventio | Experimen | Experimen | Control | 0,080527 |
| 6 months | abundance | X03602 | o-Succinyl | 3 | Interventio | Experimen | Experimen | Control | 0,06595 |
| 6 months | abundance | X02337 | Methyl 1-hy | 3 | Interventio | Experimen | Experimen | Control | 0,080527 |
| 6 months | abundance | X07107 | Lysylvaline | 3 | Interventio | Experimen | Experimen | Control | 0,109347 |
| 6 months | abundance | X10121 | Leu-pro_b | 3 | Interventio | Experimen | Experimen | Control | 0,053894 |
| 6 months | abundance | X02793 | 3-Methoxy- | 3 | Interventio | Experimen | Experimen | Control | 0,065283 |
| 6 months | abundance | X07581 | N-Acetylva | 3 | Interventio | Experimen | Experimen | Control | 0,06595 |
| 6 months | abundance | X06663 | Leucyltyros | 3 | Interventio | Experimen | Experimen | Control | 0,095545 |
| 6 months | abundance | X00361 | nitecapone | 3 | Interventio | Experimen | Experimen | Control | 0,085643 |
| 6 months | abundance | X05709 | 2,3-Dihydro | 3 | Interventio | Experimen | Experimen | Control | 0,053894 |
| 6 months | abundance | X03793 | Desonide | 3 | Interventio | Experimen | Experimen | Control | 0,07652 |
| 6 months | abundance | X08193 | Seryltyrosi | 3 | Interventio | Experimen | Experimen | Control | 0,065283 |
| 6 months | abundance | X01656 | Guanadrel | 3 | Interventio | Experimen | Experimen | Control | 0,055563 |
| 6 months | abundance | X04493 | Tyrosol | 3 | Interventio | Experimen | Experimen | Control | 0,080975 |
| 6 months | abundance | X11323 | dihydroxyb | 3 | Interventio | Experimen | Experimen | Control | 0,039418 |
| 6 months | abundance | X06750 | 1-PYRENYL | 3 | Interventio | Experimen | Experimen | Control | 0,080975 |
| 6 months | abundance | X10494 | 26Q0EO75 | 3 | Interventio | Experimen | Experimen | Control | 0,080943 |
| 6 months | abundance | M5 | M5 | IA_module | Interventio | Experimen | Experimen | Control | 0,056436 |
| 6 months | abundance | X06560 | 2,6-Dimeth | 2b | Interventio | Experimen | Experimen | Control | 0,053803 |
| 6 months | abundance | X07582 | 4-methylpy | 2b | Interventio | Experimen | Experimen | Control | 0,074642 |
| 6 months | abundance | X06874 | N-Acetyl-5 | 3 | Interventio | Experimen | Experimen | Control | 0,046544 |
| 6 months | abundance | X11361 | 1-Methylhi | 3 | Interventio | Experimen | Experimen | Control | 0,052526 |
| 6 months | abundance | X02651 | L-gamma-( | 3 | Interventio | Experimen | Experimen | Control | 0,046272 |
| 6 months | abundance | X06744 | Diethylpyro | 3 | Interventio | Experimen | Experimen | Control | 0,053894 |
| 6 months | abundance | X09565 | 3-Methoxy- | 3 | Interventio | Experimen | Experimen | Control | 0,061587 |
| 6 months | abundance | X04743 | 5-Methoxy- | 3 | Interventio | Experimen | Experimen | Control | 0,074608 |
| 6 months | abundance | X01098 | (-)-Physost | 3 | Interventio | Experimen | Experimen | Control | 0,042505 |
| 6 months | abundance | X08610 | Leucyltyros | 3 | Interventio | Experimen | Experimen | Control | 0,060452 |
| 6 months | abundance | X09932 | g-Aminobu | 3 | Interventio | Experimen | Experimen | Control | 0,03552 |
| 6 months | abundance | X00637 | N~6~,N~6 | 3 | Interventio | Experimen | Experimen | Control | 0,023843 |
| 6 months | abundance | X00698 | valganciclo | 3 | Interventio | Experimen | Experimen | Control | 0,046272 |

| | | | | | | | | | |
|---|---|---|---|---|---|---|---|---|---|
| 6 months | abundance | X03488 | 3,4-Dihydr | 3 | Interventio | Experimen | Experimen | Control | 0,046125 |
| 6 months | abundance | X09703 | 7,8-Didehy | 3 | Interventio | Experimen | Experimen | Control | 0,050313 |
| 6 months | abundance | X08644 | 2,5-Dimeth | 2b | Interventio | Experimen | Experimen | Control | 0,040318 |
| 6 months | abundance | X01286 | (2,4-Dihyd | 3 | Interventio | Experimen | Experimen | Control | 0,022499 |
| 6 months | abundance | X07955 | Valylvaline | 3 | Interventio | Experimen | Experimen | Control | 0,012138 |
| 6 months | abundance | X08842 | Spermic ac | 3 | Interventio | Experimen | Experimen | Control | 0,016813 |
| 6 months | abundance | X02652 | Leucyltyros | 3 | Interventio | Experimen | Experimen | Control | 0,023682 |
| 6 months | abundance | X05467 | Azulfidine | 3 | Interventio | Experimen | Experimen | Control | 0,025893 |
| 6 months | abundance | X08641 | meprobam | 3 | Interventio | Experimen | Experimen | Control | 0,017053 |
| 6 months | abundance | X02676 | Val-Trp_a | 3 | Interventio | Experimen | Experimen | Control | 0,060957 |
| 6 months | abundance | X08529 | 7,8-Diamir | 3 | Interventio | Experimen | Experimen | Control | 0,014596 |
| 6 months | abundance | X08514 | N-Ethylpro | 3 | Interventio | Experimen | Experimen | Control | 0,025324 |
| 6 months | abundance | X05741 | g-Aminobu | 3 | Interventio | Experimen | Experimen | Control | 0,023022 |
| 6 months | abundance | X07867 | N-Acetylva | 3 | Interventio | Experimen | Experimen | Control | 0,038212 |
| 6 months | abundance | X11331 | N~6~-Octa | 3 | Interventio | Experimen | Experimen | Control | 0,017053 |
| 6 months | abundance | X00950 | 2-Phenylet | 3 | Interventio | Experimen | Experimen | Control | 0,005134 |
| 6 months | abundance | X11277 | lys-leu | 3 | Interventio | Experimen | Experimen | Control | 0,035097 |
| 6 months | abundance | X10319 | 1-Methyl-1 | 3 | Interventio | Experimen | Experimen | Control | 0,014596 |
| 6 months | abundance | X03707 | 9-[(5R)-5-E | 3 | Interventio | Experimen | Experimen | Control | 0,032515 |
| 6 months | abundance | X02471 | n-Propyl Ga | 3 | Interventio | Experimen | Experimen | Control | 0,013146 |
| 6 months | abundance | X07126 | Leu-Val_f | 3 | Interventio | Experimen | Experimen | Control | 0,012184 |
| 6 months | abundance | X02894 | Toluene_a | 3 | Interventio | Experimen | Experimen | Control | 0,009848 |
| 6 months | abundance | X06532 | Leupeptin | 3 | Interventio | Experimen | Experimen | Control | 0,00969 |
| 6 months | abundance | X07843 | Piperidine_ | 3 | Interventio | Experimen | Experimen | Control | 0,006888 |
| 6 months | abundance | X02251 | Hostmania | 3 | Interventio | Experimen | Experimen | Control | 0,003046 |
| 6 months | abundance | X06819 | Primaquine | 3 | Interventio | Experimen | Experimen | Control | 0,009224 |
| 6 months | abundance | X00023 | N-Formylm | 1 | Interventio | Experimen | Experimen | Control | 0,006049 |
| 6 months | abundance | X08007 | MFCD0002 | 3 | Interventio | Experimen | Experimen | Control | 0,008517 |
| 6 months | abundance | X08100 | Piperidine_ | 3 | Interventio | Experimen | Experimen | Control | 0,007323 |
| 6 months | abundance | X08059 | Cadaverine | 3 | Interventio | Experimen | Experimen | Control | 0,005507 |
| 6 months | abundance | X08810 | N~6~,N~6 | 3 | Interventio | Experimen | Experimen | Control | 0,004096 |
| 6 months | abundance | X07269 | (3aR,4R,5R | 3 | Interventio | Experimen | Experimen | Control | 0,012184 |
| 6 months | abundance | X07866 | 3,3,5,5-Tet | 2b | Interventio | Experimen | Experimen | Control | 0,006888 |
| 6 months | abundance | SL00295 | 5-Methylur | 2a | Interventio | Experimen | Experimen | Control | 0,002657 |
| 6 months | abundance | X09110 | Valylvaline | 3 | Interventio | Experimen | Experimen | Control | 0,002657 |
| 6 months | abundance | X08365 | Lysylvaline | 3 | Interventio | Experimen | Experimen | Control | 0,003479 |
| 6 months | abundance | X11745 | 2-Despiper | 3 | Interventio | Experimen | Experimen | Control | 0,003399 |
| 6 months | abundance | X07854 | TO0127900 | 3 | Interventio | Experimen | Experimen | Control | 0,00194 |
| 6 months | abundance | X09068 | N,N-Diethy | 3 | Interventio | Experimen | Experimen | Control | 0,003525 |
| 6 months | abundance | X07983 | butalbital_ | 3 | Interventio | Experimen | Experimen | Control | 0,002657 |
| 6 months | abundance | X07916 | N6,N6,N6- | 3 | Interventio | Experimen | Experimen | Control | 0,000536 |
| 6 months | abundance | X00112 | Xanthosine | 1 | Interventio | Experimen | Experimen | Control | 5,65E-05 |
| 6 months | abundance | X04021 | Bentazone | 2b | Interventio | Experimen | Experimen | Control | 1,23E-18 |

| P-value | Effect size | Standard e | Sample siz | N alternati | N referenc | Degrees of | Test statist | Model formula fit |
|---|---|---|---|---|---|---|---|---|
| 0,887212 | -0,0202 | 0,142142 | 140 | 69 | 71 | 136 | -0,1421 | y ~ Intervention + Sex + |
| 0,680706 | -0,06074 | 0,147281 | 137 | 68 | 69 | 133 | -0,41241 | y ~ Intervention + Sex + |
| 0,074152 | -0,27675 | 0,153806 | 142 | 70 | 72 | 138 | -1,79933 | y ~ Intervention + Sex + |
| 0,002816 | -0,44677 | 0,146879 | 142 | 70 | 72 | 138 | -3,04176 | y ~ Intervention + Sex + |
| 0,627082 | -0,11769 | 0,241146 | 72 | 31 | 41 | 68 | -0,48805 | y ~ Intervention + Sex + |
| 0,792364 | -0,04171 | 0,158143 | 142 | 70 | 72 | 138 | -0,26375 | y ~ Intervention + Sex + |
| 0,273916 | -0,16803 | 0,152968 | 142 | 70 | 72 | 138 | -1,09846 | y ~ Intervention + Sex + |
| 0,587785 | -0,09077 | 0,167061 | 142 | 70 | 72 | 138 | -0,54332 | y ~ Intervention + Sex + |
| 0,081652 | 0,220251 | 0,125571 | 142 | 70 | 72 | 138 | 1,753992 | y ~ Intervention + Sex + |
| 0,107509 | 0,256055 | 0,158056 | 142 | 70 | 72 | 138 | 1,620029 | y ~ Intervention + Sex + |
| 0,935522 | -0,01382 | 0,170514 | 142 | 70 | 72 | 138 | -0,08105 | y ~ Intervention + Sex + |
| 0,326071 | -0,15097 | 0,153183 | 142 | 70 | 72 | 138 | -0,98557 | y ~ Intervention + Sex + |
| 0,669916 | 0,068003 | 0,159191 | 142 | 70 | 72 | 138 | 0,427177 | y ~ Intervention + Sex + |
| 0,806273 | -0,03563 | 0,145013 | 142 | 70 | 72 | 138 | -0,24571 | y ~ Intervention + Sex + |
| 0,817678 | 0,036569 | 0,158327 | 142 | 70 | 72 | 138 | 0,230973 | y ~ Intervention + Sex + |
| 0,759715 | 0,046793 | 0,152688 | 142 | 70 | 72 | 138 | 0,306461 | y ~ Intervention + Sex + |
| 0,250178 | 0,180989 | 0,156731 | 142 | 70 | 72 | 138 | 1,154776 | y ~ Intervention + Sex + |
| 0,156925 | 0,227993 | 0,160194 | 142 | 70 | 72 | 138 | 1,423234 | y ~ Intervention + Sex + |
| 0,715584 | -0,05696 | 0,156002 | 142 | 70 | 72 | 138 | -0,36512 | y ~ Intervention + Sex + |
| 0,781694 | -0,04056 | 0,14607 | 142 | 70 | 72 | 138 | -0,27765 | y ~ Intervention + Sex + |
| 0,141139 | 0,231593 | 0,156476 | 142 | 70 | 72 | 138 | 1,480053 | y ~ Intervention + Sex + |
| 0,817358 | -0,03787 | 0,163668 | 142 | 70 | 72 | 138 | -0,23139 | y ~ Intervention + Sex + |
| 0,919247 | 0,017117 | 0,168527 | 142 | 70 | 72 | 138 | 0,101569 | y ~ Intervention + Sex + |
| 0,849855 | -0,03079 | 0,162366 | 142 | 70 | 72 | 138 | -0,18966 | y ~ Intervention + Sex + |
| 0,258621 | -0,1727 | 0,152244 | 142 | 70 | 72 | 138 | -1,13433 | y ~ Intervention + Sex + |
| 0,210827 | -0,20755 | 0,1651 | 142 | 70 | 72 | 138 | -1,25714 | y ~ Intervention + Sex + |
| 0,358 | 0,145924 | 0,158223 | 142 | 70 | 72 | 138 | 0,922266 | y ~ Intervention + Sex + |
| 0,539028 | -0,09796 | 0,159066 | 142 | 70 | 72 | 138 | -0,61582 | y ~ Intervention + Sex + |
| 0,038895 | -0,29874 | 0,143269 | 142 | 70 | 72 | 138 | -2,08518 | y ~ Intervention + Sex + |
| 0,964279 | 0,006996 | 0,15592 | 142 | 70 | 72 | 138 | 0,044866 | y ~ Intervention + Sex + |
| 0,876181 | -0,02323 | 0,148797 | 142 | 70 | 72 | 138 | -0,1561 | y ~ Intervention + Sex + |
| 0,109782 | -0,2649 | 0,164581 | 142 | 70 | 72 | 138 | -1,60955 | y ~ Intervention + Sex + |
| 0,304308 | -0,14241 | 0,138117 | 142 | 70 | 72 | 138 | -1,03108 | y ~ Intervention + Sex + |
| 0,485933 | 0,113639 | 0,16265 | 142 | 70 | 72 | 138 | 0,698672 | y ~ Intervention + Sex + |
| 0,966471 | -0,00657 | 0,156001 | 142 | 70 | 72 | 138 | -0,04211 | y ~ Intervention + Sex + |
| 0,304995 | -0,16968 | 0,164804 | 142 | 70 | 72 | 138 | -1,02961 | y ~ Intervention + Sex + |
| 0,705916 | 0,057331 | 0,151619 | 142 | 70 | 72 | 138 | 0,378128 | y ~ Intervention + Sex + |
| 0,09434 | -0,25929 | 0,153922 | 142 | 70 | 72 | 138 | -1,68453 | y ~ Intervention + Sex + |
| 0,321498 | 0,150025 | 0,150785 | 142 | 70 | 72 | 138 | 0,994959 | y ~ Intervention + Sex + |
| 0,567671 | -0,08807 | 0,153726 | 136 | 67 | 69 | 132 | -0,57292 | y ~ Intervention + Sex + |
| 0,128262 | -0,21001 | 0,137247 | 142 | 70 | 72 | 138 | -1,53018 | y ~ Intervention + Sex + |
| 0,637742 | -0,07313 | 0,154965 | 142 | 70 | 72 | 138 | -0,4719 | y ~ Intervention + Sex + |
| 0,783485 | -0,03737 | 0,135727 | 142 | 70 | 72 | 138 | -0,27532 | y ~ Intervention + Sex + |
| 0,169367 | 0,209832 | 0,15189 | 142 | 70 | 72 | 138 | 1,38147 | y ~ Intervention + Sex + |
| 0,371913 | 0,122736 | 0,137011 | 142 | 70 | 72 | 138 | 0,895813 | y ~ Intervention + Sex + |
| 0,654915 | 0,061171 | 0,136567 | 142 | 70 | 72 | 138 | 0,447918 | y ~ Intervention + Sex + |
| 0,373772 | -0,12406 | 0,139031 | 142 | 70 | 72 | 138 | -0,89232 | y ~ Intervention + Sex + |

| | | | | | | | | |
|---|---|---|---|---|---|---|---|---|
| 0,571832 | -0,09384 | 0,165596 | 142 | 70 | 72 | 138 | -0,56671 | y ~ Intervention + Sex + |
| 0,63443 | -0,0781 | 0,163874 | 142 | 70 | 72 | 138 | -0,47656 | y ~ Intervention + Sex + |
| 0,322011 | 0,155308 | 0,156261 | 142 | 70 | 72 | 138 | 0,993901 | y ~ Intervention + Sex + |
| 0,593192 | 0,057069 | 0,10658 | 142 | 70 | 72 | 138 | 0,535463 | y ~ Intervention + Sex + |
| 0,334213 | 0,14214 | 0,146679 | 142 | 70 | 72 | 138 | 0,969055 | y ~ Intervention + Sex + |
| 0,837692 | -0,02798 | 0,136342 | 142 | 70 | 72 | 138 | -0,20523 | y ~ Intervention + Sex + |
| 0,998355 | 0,000345 | 0,166939 | 142 | 70 | 72 | 138 | 0,002066 | y ~ Intervention + Sex + |
| 0,863253 | -0,02496 | 0,144666 | 142 | 70 | 72 | 138 | -0,17256 | y ~ Intervention + Sex + |
| 0,736323 | -0,04822 | 0,142911 | 142 | 70 | 72 | 138 | -0,33741 | y ~ Intervention + Sex + |
| 0,083789 | 0,250961 | 0,14409 | 142 | 70 | 72 | 138 | 1,741698 | y ~ Intervention + Sex + |
| 0,5456 | -0,1004 | 0,165709 | 142 | 70 | 72 | 138 | -0,60587 | y ~ Intervention + Sex + |
| 0,968465 | 0,005249 | 0,13253 | 142 | 70 | 72 | 138 | 0,039605 | y ~ Intervention + Sex + |
| 0,176385 | -0,18705 | 0,137644 | 142 | 70 | 72 | 138 | -1,35893 | y ~ Intervention + Sex + |
| 0,794404 | -0,03541 | 0,135609 | 142 | 70 | 72 | 138 | -0,2611 | y ~ Intervention + Sex + |
| 0,478465 | -0,11008 | 0,154894 | 142 | 70 | 72 | 138 | -0,71071 | y ~ Intervention + Sex + |
| 0,841699 | -0,03168 | 0,158308 | 142 | 70 | 72 | 138 | -0,2001 | y ~ Intervention + Sex + |
| 0,615292 | 0,077871 | 0,154605 | 142 | 70 | 72 | 138 | 0,503676 | y ~ Intervention + Sex + |
| 0,389469 | 0,109753 | 0,127131 | 142 | 70 | 72 | 138 | 0,863303 | y ~ Intervention + Sex + |
| 0,535012 | -0,0867 | 0,139404 | 142 | 70 | 72 | 138 | -0,62193 | y ~ Intervention + Sex + |
| 0,562818 | 0,078146 | 0,13472 | 142 | 70 | 72 | 138 | 0,580063 | y ~ Intervention + Sex + |
| 0,466935 | 0,10384 | 0,142345 | 142 | 70 | 72 | 138 | 0,729495 | y ~ Intervention + Sex + |
| 0,617871 | -0,06748 | 0,134968 | 142 | 70 | 72 | 138 | -0,5 | y ~ Intervention + Sex + |
| 0,292869 | -0,15024 | 0,142285 | 142 | 70 | 72 | 138 | -1,05588 | y ~ Intervention + Sex + |
| 0,075765 | 0,253878 | 0,141889 | 142 | 70 | 72 | 138 | 1,789269 | y ~ Intervention + Sex + |
| 0,580376 | 0,085555 | 0,15439 | 142 | 70 | 72 | 138 | 0,554146 | y ~ Intervention + Sex + |
| 0,185058 | -0,19028 | 0,142855 | 142 | 70 | 72 | 138 | -1,33199 | y ~ Intervention + Sex + |
| 0,141436 | 0,219571 | 0,148465 | 142 | 70 | 72 | 138 | 1,478937 | y ~ Intervention + Sex + |
| 0,164654 | -0,20203 | 0,144616 | 142 | 70 | 72 | 138 | -1,39701 | y ~ Intervention + Sex + |
| 0,366908 | 0,142449 | 0,157358 | 142 | 70 | 72 | 138 | 0,905255 | y ~ Intervention + Sex + |
| 0,398222 | -0,13685 | 0,161489 | 142 | 70 | 72 | 138 | -0,84743 | y ~ Intervention + Sex + |
| 0,509683 | 0,094366 | 0,142752 | 142 | 70 | 72 | 138 | 0,66105 | y ~ Intervention + Sex + |
| 0,636458 | -0,07001 | 0,147794 | 142 | 70 | 72 | 138 | -0,47371 | y ~ Intervention + Sex + |
| 0,354242 | -0,11421 | 0,122869 | 142 | 70 | 72 | 138 | -0,92952 | y ~ Intervention + Sex + |
| 0,788053 | -0,04214 | 0,156439 | 142 | 70 | 72 | 138 | -0,26936 | y ~ Intervention + Sex + |
| 0,76252 | 0,041493 | 0,137045 | 142 | 70 | 72 | 138 | 0,302772 | y ~ Intervention + Sex + |
| 0,882378 | -0,0236 | 0,159234 | 142 | 70 | 72 | 138 | -0,14823 | y ~ Intervention + Sex + |
| 0,43195 | 0,126642 | 0,160678 | 142 | 70 | 72 | 138 | 0,788169 | y ~ Intervention + Sex + |
| 0,881597 | 0,020755 | 0,139089 | 142 | 70 | 72 | 138 | 0,149221 | y ~ Intervention + Sex + |
| 0,920418 | -0,01397 | 0,139579 | 142 | 70 | 72 | 138 | -0,10009 | y ~ Intervention + Sex + |
| 0,716162 | -0,05847 | 0,160489 | 142 | 70 | 72 | 138 | -0,36434 | y ~ Intervention + Sex + |
| 0,196967 | -0,21693 | 0,167317 | 142 | 70 | 72 | 138 | -1,2965 | y ~ Intervention + Sex + |
| 0,398764 | -0,14461 | 0,170847 | 142 | 70 | 72 | 138 | -0,84646 | y ~ Intervention + Sex + |
| 0,169617 | -0,18022 | 0,130534 | 142 | 70 | 72 | 138 | -1,38066 | y ~ Intervention + Sex + |
| 0,30978 | -0,15509 | 0,152133 | 142 | 70 | 72 | 138 | -1,01944 | y ~ Intervention + Sex + |
| 0,607869 | 0,077295 | 0,150293 | 142 | 70 | 72 | 138 | 0,514294 | y ~ Intervention + Sex + |
| 0,994077 | 0,001019 | 0,137044 | 142 | 70 | 72 | 138 | 0,007437 | y ~ Intervention + Sex + |
| 0,188842 | 0,214119 | 0,162146 | 142 | 70 | 72 | 138 | 1,320536 | y ~ Intervention + Sex + |
| 0,32045 | -0,13265 | 0,133031 | 142 | 70 | 72 | 138 | -0,99712 | y ~ Intervention + Sex + |

| | | | | | | | | |
|---|---|---|---|---|---|---|---|---|
| 0,198147 | -0,18109 | 0,140045 | 142 | 70 | 72 | 138 | -1,29307 | y ~ Intervention + Sex + |
| 0,161822 | -0,19428 | 0,138131 | 142 | 70 | 72 | 138 | -1,40651 | y ~ Intervention + Sex + |
| 0,179349 | -0,18457 | 0,136756 | 142 | 70 | 72 | 138 | -1,34962 | y ~ Intervention + Sex + |
| 0,241709 | -0,17646 | 0,150078 | 142 | 70 | 72 | 138 | -1,17578 | y ~ Intervention + Sex + |
| 0,03068 | -0,22814 | 0,104478 | 142 | 70 | 72 | 138 | -2,18362 | y ~ Intervention + Sex + |
| 0,711331 | -0,05965 | 0,160863 | 142 | 70 | 72 | 138 | -0,37083 | y ~ Intervention + Sex + |
| 0,757897 | -0,04098 | 0,132683 | 142 | 70 | 72 | 138 | -0,30886 | y ~ Intervention + Sex + |
| 0,78058 | 0,039614 | 0,141932 | 142 | 70 | 72 | 138 | 0,279107 | y ~ Intervention + Sex + |
| 0,831609 | -0,0299 | 0,140327 | 142 | 70 | 72 | 138 | -0,21304 | y ~ Intervention + Sex + |
| 0,328039 | 0,13358 | 0,136091 | 142 | 70 | 72 | 138 | 0,98155 | y ~ Intervention + Sex + |
| 0,021146 | -0,33744 | 0,144696 | 142 | 70 | 72 | 138 | -2,33204 | y ~ Intervention + Sex + |
| 0,510951 | -0,09828 | 0,149115 | 142 | 70 | 72 | 138 | -0,65907 | y ~ Intervention + Sex + |
| 0,323798 | 0,144666 | 0,146094 | 142 | 70 | 72 | 138 | 0,990222 | y ~ Intervention + Sex + |
| 0,226684 | -0,1696 | 0,139662 | 142 | 70 | 72 | 138 | -1,21436 | y ~ Intervention + Sex + |
| 0,056757 | -0,27494 | 0,143103 | 142 | 70 | 72 | 138 | -1,92129 | y ~ Intervention + Sex + |
| 0,275787 | 0,17477 | 0,159729 | 142 | 70 | 72 | 138 | 1,09417 | y ~ Intervention + Sex + |
| 0,09714 | -0,26129 | 0,156441 | 142 | 70 | 72 | 138 | -1,67023 | y ~ Intervention + Sex + |
| 0,07515 | -0,29319 | 0,163511 | 142 | 70 | 72 | 138 | -1,79308 | y ~ Intervention + Sex + |
| 0,886217 | 0,021843 | 0,152368 | 142 | 70 | 72 | 138 | 0,143358 | y ~ Intervention + Sex + |
| 0,902623 | -0,01922 | 0,15679 | 142 | 70 | 72 | 138 | -0,12257 | y ~ Intervention + Sex + |
| 0,580827 | 0,092019 | 0,166255 | 142 | 70 | 72 | 138 | 0,553485 | y ~ Intervention + Sex + |
| 0,213623 | -0,19309 | 0,154543 | 142 | 70 | 72 | 138 | -1,24943 | y ~ Intervention + Sex + |
| 0,835261 | 0,03152 | 0,151283 | 142 | 70 | 72 | 138 | 0,208352 | y ~ Intervention + Sex + |
| 0,628421 | -0,07629 | 0,157295 | 142 | 70 | 72 | 138 | -0,48503 | y ~ Intervention + Sex + |
| 0,046047 | -0,3054 | 0,151704 | 142 | 70 | 72 | 138 | -2,01312 | y ~ Intervention + Sex + |
| 0,816208 | 0,03776 | 0,16215 | 142 | 70 | 72 | 138 | 0,23287 | y ~ Intervention + Sex + |
| 0,489599 | -0,10131 | 0,146238 | 142 | 70 | 72 | 138 | -0,6928 | y ~ Intervention + Sex + |
| 0,181533 | -0,14709 | 0,109539 | 142 | 70 | 72 | 138 | -1,34283 | y ~ Intervention + Sex + |
| 0,833612 | -0,0313 | 0,148697 | 142 | 70 | 72 | 138 | -0,21047 | y ~ Intervention + Sex + |
| 0,265183 | 0,168567 | 0,150672 | 142 | 70 | 72 | 138 | 1,118768 | y ~ Intervention + Sex + |
| 0,115322 | 0,234092 | 0,14772 | 142 | 70 | 72 | 138 | 1,584707 | y ~ Intervention + Sex + |
| 0,477125 | 0,093609 | 0,131311 | 142 | 70 | 72 | 138 | 0,712877 | y ~ Intervention + Sex + |
| 0,258888 | 0,173055 | 0,152647 | 142 | 70 | 72 | 138 | 1,133696 | y ~ Intervention + Sex + |
| 0,244353 | -0,17583 | 0,150391 | 142 | 70 | 72 | 138 | -1,16916 | y ~ Intervention + Sex + |
| 0,312783 | 0,146902 | 0,145002 | 142 | 70 | 72 | 138 | 1,013105 | y ~ Intervention + Sex + |
| 0,008416 | -0,39018 | 0,145953 | 142 | 70 | 72 | 138 | -2,67335 | y ~ Intervention + Sex + |
| 0,004353 | -0,39653 | 0,136769 | 142 | 70 | 72 | 138 | -2,89926 | y ~ Intervention + Sex + |
| 0,717612 | 0,050522 | 0,139412 | 142 | 70 | 72 | 138 | 0,362395 | y ~ Intervention + Sex + |
| 0,901499 | -0,01984 | 0,160022 | 142 | 70 | 72 | 138 | -0,124 | y ~ Intervention + Sex + |
| 0,373845 | -0,13358 | 0,149722 | 142 | 70 | 72 | 138 | -0,89219 | y ~ Intervention + Sex + |
| 0,393145 | 0,131696 | 0,153741 | 142 | 70 | 72 | 138 | 0,856611 | y ~ Intervention + Sex + |
| 0,637892 | 0,071619 | 0,151833 | 142 | 70 | 72 | 138 | 0,471691 | y ~ Intervention + Sex + |
| 0,34715 | -0,11879 | 0,125923 | 142 | 70 | 72 | 138 | -0,94335 | y ~ Intervention + Sex + |
| 0,702346 | 0,056336 | 0,14711 | 142 | 70 | 72 | 138 | 0,38295 | y ~ Intervention + Sex + |
| 0,311338 | 0,152978 | 0,150547 | 142 | 70 | 72 | 138 | 1,016145 | y ~ Intervention + Sex + |
| 0,421056 | 0,123516 | 0,153056 | 142 | 70 | 72 | 138 | 0,806999 | y ~ Intervention + Sex + |
| 0,550993 | -0,0879 | 0,14706 | 142 | 70 | 72 | 138 | -0,59774 | y ~ Intervention + Sex + |
| 0,329656 | 0,147392 | 0,150667 | 142 | 70 | 72 | 138 | 0,978263 | y ~ Intervention + Sex + |

| | | | | | | | | |
|---|---|---|---|---|---|---|---|---|
| 0,251758 | -0,15097 | 0,131172 | 142 | 70 | 72 | 138 | -1,15091 | y ~ Intervention + Sex + |
| 0,571736 | -0,08931 | 0,157547 | 142 | 70 | 72 | 138 | -0,56685 | y ~ Intervention + Sex + |
| 0,134653 | 0,217591 | 0,144595 | 142 | 70 | 72 | 138 | 1,504829 | y ~ Intervention + Sex + |
| 0,655012 | 0,080706 | 0,180234 | 142 | 70 | 72 | 138 | 0,447783 | y ~ Intervention + Sex + |
| 0,092862 | -0,26934 | 0,159162 | 142 | 70 | 72 | 138 | -1,69221 | y ~ Intervention + Sex + |
| 0,132505 | 0,206607 | 0,136533 | 142 | 70 | 72 | 138 | 1,51324 | y ~ Intervention + Sex + |
| 0,521923 | -0,09471 | 0,14751 | 142 | 70 | 72 | 138 | -0,64202 | y ~ Intervention + Sex + |
| 0,300052 | -0,15382 | 0,147875 | 142 | 70 | 72 | 138 | -1,04023 | y ~ Intervention + Sex + |
| 0,304463 | 0,149174 | 0,144724 | 142 | 70 | 72 | 138 | 1,030747 | y ~ Intervention + Sex + |
| 0,130855 | 0,209335 | 0,13774 | 142 | 70 | 72 | 138 | 1,519777 | y ~ Intervention + Sex + |
| 0,28711 | -0,16469 | 0,154115 | 142 | 70 | 72 | 138 | -1,06861 | y ~ Intervention + Sex + |
| 0,240322 | -0,17247 | 0,146251 | 142 | 70 | 72 | 138 | -1,17927 | y ~ Intervention + Sex + |
| 0,243963 | -0,15593 | 0,133259 | 142 | 70 | 72 | 138 | -1,17014 | y ~ Intervention + Sex + |
| 0,693994 | -0,06306 | 0,159937 | 142 | 70 | 72 | 138 | -0,39426 | y ~ Intervention + Sex + |
| 0,480116 | -0,11776 | 0,166319 | 142 | 70 | 72 | 138 | -0,70804 | y ~ Intervention + Sex + |
| 0,857627 | -0,02993 | 0,166506 | 142 | 70 | 72 | 138 | -0,17973 | y ~ Intervention + Sex + |
| 0,284731 | 0,157875 | 0,147007 | 142 | 70 | 72 | 138 | 1,073924 | y ~ Intervention + Sex + |
| 0,200564 | -0,20558 | 0,159848 | 142 | 70 | 72 | 138 | -1,28609 | y ~ Intervention + Sex + |
| 0,949077 | -0,00876 | 0,13686 | 142 | 70 | 72 | 138 | -0,06398 | y ~ Intervention + Sex + |
| 0,342507 | 0,128585 | 0,134996 | 142 | 70 | 72 | 138 | 0,952504 | y ~ Intervention + Sex + |
| 0,765057 | 0,047638 | 0,159092 | 142 | 70 | 72 | 138 | 0,299437 | y ~ Intervention + Sex + |
| 0,64975 | -0,06892 | 0,151447 | 142 | 70 | 72 | 138 | -0,4551 | y ~ Intervention + Sex + |
| 0,140238 | 0,201693 | 0,135963 | 142 | 70 | 72 | 138 | 1,483438 | y ~ Intervention + Sex + |
| 0,303667 | -0,14853 | 0,143857 | 142 | 70 | 72 | 138 | -1,03245 | y ~ Intervention + Sex + |
| 0,634306 | -0,0655 | 0,137401 | 142 | 70 | 72 | 138 | -0,47673 | y ~ Intervention + Sex + |
| 0,163261 | -0,21752 | 0,155185 | 142 | 70 | 72 | 138 | -1,40166 | y ~ Intervention + Sex + |
| 0,953112 | 0,009134 | 0,155068 | 142 | 70 | 72 | 138 | 0,058906 | y ~ Intervention + Sex + |
| 0,564647 | -0,08588 | 0,148752 | 142 | 70 | 72 | 138 | -0,57735 | y ~ Intervention + Sex + |
| 0,772019 | 0,048054 | 0,165529 | 142 | 70 | 72 | 138 | 0,290304 | y ~ Intervention + Sex + |
| 0,702274 | 0,053739 | 0,140292 | 142 | 70 | 72 | 138 | 0,383047 | y ~ Intervention + Sex + |
| 0,43832 | 0,115543 | 0,148649 | 142 | 70 | 72 | 138 | 0,777286 | y ~ Intervention + Sex + |
| 0,08805 | -0,24043 | 0,139951 | 142 | 70 | 72 | 138 | -1,71794 | y ~ Intervention + Sex + |
| 0,203435 | 0,182995 | 0,143202 | 142 | 70 | 72 | 138 | 1,277885 | y ~ Intervention + Sex + |
| 0,039928 | -0,32227 | 0,155377 | 142 | 70 | 72 | 138 | -2,07411 | y ~ Intervention + Sex + |
| 0,324558 | 0,150811 | 0,15254 | 142 | 70 | 72 | 138 | 0,988663 | y ~ Intervention + Sex + |
| 0,078365 | -0,27871 | 0,157157 | 142 | 70 | 72 | 138 | -1,77342 | y ~ Intervention + Sex + |
| 0,805595 | -0,03865 | 0,156742 | 142 | 70 | 72 | 138 | -0,24659 | y ~ Intervention + Sex + |
| 0,926003 | -0,01525 | 0,163865 | 142 | 70 | 72 | 138 | -0,09304 | y ~ Intervention + Sex + |
| 0,438986 | 0,12511 | 0,161192 | 142 | 70 | 72 | 138 | 0,776154 | y ~ Intervention + Sex + |
| 0,846628 | -0,03157 | 0,162894 | 142 | 70 | 72 | 138 | -0,19379 | y ~ Intervention + Sex + |
| 0,602025 | 0,081432 | 0,155792 | 142 | 70 | 72 | 138 | 0,522695 | y ~ Intervention + Sex + |
| 0,015583 | -0,35813 | 0,146239 | 142 | 70 | 72 | 138 | -2,44891 | y ~ Intervention + Sex + |
| 0,362828 | 0,15801 | 0,173065 | 142 | 70 | 72 | 138 | 0,913012 | y ~ Intervention + Sex + |
| 0,629293 | -0,06141 | 0,126931 | 142 | 70 | 72 | 138 | -0,4838 | y ~ Intervention + Sex + |
| 0,612887 | -0,07318 | 0,144301 | 142 | 70 | 72 | 138 | -0,50711 | y ~ Intervention + Sex + |
| 0,986963 | 0,002677 | 0,163518 | 142 | 70 | 72 | 138 | 0,01637 | y ~ Intervention + Sex + |
| 0,078602 | -0,28066 | 0,158386 | 142 | 70 | 72 | 138 | -1,772 | y ~ Intervention + Sex + |
| 0,830113 | 0,031929 | 0,148532 | 142 | 70 | 72 | 138 | 0,214964 | y ~ Intervention + Sex + |

| | | | | | | | | |
|---|---|---|---|---|---|---|---|---|
| 0,576056 | 0,083855 | 0,149611 | 142 | 70 | 72 | 138 | 0,560487 | y ~ Intervention + Sex + |
| 0,040598 | -0,2974 | 0,143877 | 142 | 70 | 72 | 138 | -2,06706 | y ~ Intervention + Sex + |
| 0,359576 | 0,133048 | 0,144737 | 142 | 70 | 72 | 138 | 0,919237 | y ~ Intervention + Sex + |
| 0,938309 | 0,011695 | 0,150834 | 142 | 70 | 72 | 138 | 0,077537 | y ~ Intervention + Sex + |
| 0,324051 | 0,138855 | 0,1403 | 142 | 70 | 72 | 138 | 0,989703 | y ~ Intervention + Sex + |
| 0,805009 | -0,03886 | 0,157094 | 142 | 70 | 72 | 138 | -0,24734 | y ~ Intervention + Sex + |
| 0,984249 | 0,003055 | 0,154455 | 142 | 70 | 72 | 138 | 0,019778 | y ~ Intervention + Sex + |
| 0,571068 | 0,086795 | 0,152852 | 142 | 70 | 72 | 138 | 0,567838 | y ~ Intervention + Sex + |
| 0,915528 | 0,016589 | 0,156115 | 142 | 70 | 72 | 138 | 0,106263 | y ~ Intervention + Sex + |
| 0,167018 | 0,208431 | 0,15004 | 142 | 70 | 72 | 138 | 1,389174 | y ~ Intervention + Sex + |
| 0,081128 | 0,251709 | 0,143257 | 142 | 70 | 72 | 138 | 1,757047 | y ~ Intervention + Sex + |
| 0,593585 | 0,073822 | 0,138012 | 142 | 70 | 72 | 138 | 0,534892 | y ~ Intervention + Sex + |
| 0,097094 | -0,27211 | 0,162896 | 142 | 70 | 72 | 138 | -1,67046 | y ~ Intervention + Sex + |
| 0,527553 | 0,100961 | 0,159407 | 142 | 70 | 72 | 138 | 0,633351 | y ~ Intervention + Sex + |
| 0,386437 | 0,137869 | 0,158679 | 142 | 70 | 72 | 138 | 0,868851 | y ~ Intervention + Sex + |
| 0,161777 | -0,21271 | 0,15122 | 142 | 70 | 72 | 138 | -1,40666 | y ~ Intervention + Sex + |
| 0,188945 | -0,17892 | 0,135526 | 142 | 70 | 72 | 138 | -1,32023 | y ~ Intervention + Sex + |
| 0,051485 | -0,31541 | 0,160557 | 142 | 70 | 72 | 138 | -1,96447 | y ~ Intervention + Sex + |
| 0,130345 | 0,242223 | 0,159167 | 142 | 70 | 72 | 138 | 1,521809 | y ~ Intervention + Sex + |
| 0,243694 | -0,16798 | 0,143477 | 142 | 70 | 72 | 138 | -1,17081 | y ~ Intervention + Sex + |
| 0,228943 | 0,160414 | 0,132744 | 142 | 70 | 72 | 138 | 1,208443 | y ~ Intervention + Sex + |
| 0,385739 | 0,143897 | 0,165374 | 142 | 70 | 72 | 138 | 0,870133 | y ~ Intervention + Sex + |
| 0,235235 | 0,186572 | 0,156496 | 142 | 70 | 72 | 138 | 1,192184 | y ~ Intervention + Sex + |
| 0,486787 | -0,10935 | 0,156821 | 142 | 70 | 72 | 138 | -0,6973 | y ~ Intervention + Sex + |
| 0,58978 | 0,100217 | 0,185444 | 142 | 70 | 72 | 138 | 0,540418 | y ~ Intervention + Sex + |
| 0,321628 | 0,137428 | 0,138162 | 142 | 70 | 72 | 138 | 0,994689 | y ~ Intervention + Sex + |
| 0,421202 | 0,133621 | 0,16563 | 142 | 70 | 72 | 138 | 0,806745 | y ~ Intervention + Sex + |
| 0,622984 | 0,075713 | 0,153658 | 142 | 70 | 72 | 138 | 0,492733 | y ~ Intervention + Sex + |
| 0,542139 | 0,087769 | 0,143625 | 142 | 70 | 72 | 138 | 0,6111 | y ~ Intervention + Sex + |
| 0,476272 | 0,114222 | 0,159917 | 142 | 70 | 72 | 138 | 0,714261 | y ~ Intervention + Sex + |
| 0,132106 | -0,2316 | 0,152889 | 142 | 70 | 72 | 138 | -1,51482 | y ~ Intervention + Sex + |
| 0,970734 | 0,005704 | 0,155194 | 142 | 70 | 72 | 138 | 0,036754 | y ~ Intervention + Sex + |
| 0,976448 | 0,004281 | 0,144748 | 142 | 70 | 72 | 138 | 0,029576 | y ~ Intervention + Sex + |
| 0,571291 | 0,08943 | 0,157583 | 142 | 70 | 72 | 138 | 0,567508 | y ~ Intervention + Sex + |
| 0,689893 | -0,06736 | 0,168458 | 142 | 70 | 72 | 138 | -0,39984 | y ~ Intervention + Sex + |
| 0,557718 | -0,09938 | 0,169102 | 142 | 70 | 72 | 138 | -0,58766 | y ~ Intervention + Sex + |
| 0,223849 | 0,182961 | 0,149741 | 142 | 70 | 72 | 138 | 1,221847 | y ~ Intervention + Sex + |
| 0,436935 | -0,11632 | 0,149196 | 142 | 70 | 72 | 138 | -0,77964 | y ~ Intervention + Sex + |
| 0,390065 | -0,12711 | 0,147421 | 142 | 70 | 72 | 138 | -0,86221 | y ~ Intervention + Sex + |
| 0,381469 | -0,1422 | 0,161953 | 142 | 70 | 72 | 138 | -0,878 | y ~ Intervention + Sex + |
| 0,305665 | -0,18245 | 0,177445 | 142 | 70 | 72 | 138 | -1,02818 | y ~ Intervention + Sex + |
| 0,765338 | -0,04385 | 0,146636 | 142 | 70 | 72 | 138 | -0,29907 | y ~ Intervention + Sex + |
| 0,723837 | 0,059477 | 0,167988 | 142 | 70 | 72 | 138 | 0,354056 | y ~ Intervention + Sex + |
| 0,55282 | -0,08354 | 0,140397 | 142 | 70 | 72 | 138 | -0,595 | y ~ Intervention + Sex + |
| 0,698698 | 0,053949 | 0,139085 | 142 | 70 | 72 | 138 | 0,387887 | y ~ Intervention + Sex + |
| 0,85075 | -0,02568 | 0,136247 | 142 | 70 | 72 | 138 | -0,18851 | y ~ Intervention + Sex + |
| 0,449255 | 0,128755 | 0,169678 | 142 | 70 | 72 | 138 | 0,758819 | y ~ Intervention + Sex + |
| 0,281372 | -0,18656 | 0,172507 | 142 | 70 | 72 | 138 | -1,08147 | y ~ Intervention + Sex + |

| | | | | | | | | |
|---|---|---|---|---|---|---|---|---|
| 0,600942 | -0,08554 | 0,163168 | 142 | 70 | 72 | 138 | -0,52425 | y ~ Intervention + Sex + |
| 0,698842 | -0,06028 | 0,155491 | 142 | 70 | 72 | 138 | -0,38769 | y ~ Intervention + Sex + |
| 0,125555 | 0,241452 | 0,156664 | 142 | 70 | 72 | 138 | 1,541211 | y ~ Intervention + Sex + |
| 0,043056 | 0,329088 | 0,161159 | 142 | 70 | 72 | 138 | 2,042003 | y ~ Intervention + Sex + |
| 0,136177 | 0,234101 | 0,156179 | 142 | 70 | 72 | 138 | 1,498928 | y ~ Intervention + Sex + |
| 0,287512 | 0,16065 | 0,150461 | 142 | 70 | 72 | 138 | 1,067719 | y ~ Intervention + Sex + |
| 0,411982 | -0,13959 | 0,169632 | 142 | 70 | 72 | 138 | -0,8229 | y ~ Intervention + Sex + |
| 0,172636 | -0,20884 | 0,15234 | 142 | 70 | 72 | 138 | -1,37089 | y ~ Intervention + Sex + |
| 0,068187 | 0,300085 | 0,163252 | 142 | 70 | 72 | 138 | 1,838174 | y ~ Intervention + Sex + |
| 0,558012 | 0,094499 | 0,160925 | 142 | 70 | 72 | 138 | 0,587226 | y ~ Intervention + Sex + |
| 0,047456 | -0,30462 | 0,152303 | 142 | 70 | 72 | 138 | -2,00006 | y ~ Intervention + Sex + |
| 0,257839 | -0,19485 | 0,171491 | 142 | 70 | 72 | 138 | -1,13621 | y ~ Intervention + Sex + |
| 0,814132 | -0,03637 | 0,1544 | 142 | 70 | 72 | 138 | -0,23555 | y ~ Intervention + Sex + |
| 0,237802 | -0,20474 | 0,172684 | 142 | 70 | 72 | 138 | -1,18564 | y ~ Intervention + Sex + |
| 0,050925 | 0,315112 | 0,160014 | 142 | 70 | 72 | 138 | 1,969275 | y ~ Intervention + Sex + |
| 0,166985 | 0,192238 | 0,138372 | 142 | 70 | 72 | 138 | 1,389281 | y ~ Intervention + Sex + |
| 0,280948 | 0,161435 | 0,149141 | 142 | 70 | 72 | 138 | 1,082432 | y ~ Intervention + Sex + |
| 0,523428 | -0,10282 | 0,160731 | 142 | 70 | 72 | 138 | -0,6397 | y ~ Intervention + Sex + |
| 0,864097 | 0,024806 | 0,144659 | 142 | 70 | 72 | 138 | 0,171481 | y ~ Intervention + Sex + |
| 0,972693 | 0,005462 | 0,159264 | 142 | 70 | 72 | 138 | 0,034294 | y ~ Intervention + Sex + |
| 0,527767 | -0,10946 | 0,172915 | 142 | 70 | 72 | 138 | -0,63302 | y ~ Intervention + Sex + |
| 0,33075 | -0,16206 | 0,16604 | 142 | 70 | 72 | 138 | -0,97604 | y ~ Intervention + Sex + |
| 0,606525 | -0,08629 | 0,167162 | 142 | 70 | 72 | 138 | -0,51622 | y ~ Intervention + Sex + |
| 0,859786 | 0,023903 | 0,135062 | 142 | 70 | 72 | 138 | 0,176977 | y ~ Intervention + Sex + |
| 0,536909 | 0,087747 | 0,141747 | 142 | 70 | 72 | 138 | 0,619042 | y ~ Intervention + Sex + |
| 0,690542 | 0,064982 | 0,162879 | 142 | 70 | 72 | 138 | 0,398956 | y ~ Intervention + Sex + |
| 0,944915 | -0,01135 | 0,163958 | 142 | 70 | 72 | 138 | -0,06922 | y ~ Intervention + Sex + |
| 0,211813 | 0,166262 | 0,132542 | 142 | 70 | 72 | 138 | 1,254411 | y ~ Intervention + Sex + |
| 0,534592 | 0,102739 | 0,165024 | 142 | 70 | 72 | 138 | 0,622573 | y ~ Intervention + Sex + |
| 0,375485 | -0,14785 | 0,166292 | 142 | 70 | 72 | 138 | -0,88912 | y ~ Intervention + Sex + |
| 0,250064 | -0,17995 | 0,155792 | 142 | 70 | 72 | 138 | -1,15505 | y ~ Intervention + Sex + |
| 0,485925 | 0,104037 | 0,148904 | 142 | 70 | 72 | 138 | 0,698685 | y ~ Intervention + Sex + |
| 0,79365 | -0,04406 | 0,168101 | 142 | 70 | 72 | 138 | -0,26208 | y ~ Intervention + Sex + |
| 0,987467 | 0,002595 | 0,164929 | 142 | 70 | 72 | 138 | 0,015737 | y ~ Intervention + Sex + |
| 0,221688 | 0,185704 | 0,151274 | 142 | 70 | 72 | 138 | 1,227599 | y ~ Intervention + Sex + |
| 0,023463 | 0,285638 | 0,124664 | 142 | 70 | 72 | 138 | 2,291266 | y ~ Intervention + Sex + |
| 0,794333 | -0,03983 | 0,152477 | 142 | 70 | 72 | 138 | -0,26119 | y ~ Intervention + Sex + |
| 0,722595 | 0,059418 | 0,167036 | 142 | 70 | 72 | 138 | 0,355718 | y ~ Intervention + Sex + |
| 0,822419 | -0,03412 | 0,15174 | 142 | 70 | 72 | 138 | -0,22486 | y ~ Intervention + Sex + |
| 0,864582 | -0,02872 | 0,168105 | 142 | 70 | 72 | 138 | -0,17086 | y ~ Intervention + Sex + |
| 0,195005 | -0,18241 | 0,140077 | 142 | 70 | 72 | 138 | -1,30223 | y ~ Intervention + Sex + |
| 0,452153 | -0,12461 | 0,165278 | 142 | 70 | 72 | 138 | -0,75397 | y ~ Intervention + Sex + |
| 0,326748 | 0,189061 | 0,192099 | 142 | 70 | 72 | 138 | 0,984183 | y ~ Intervention + Sex + |
| 0,446448 | -0,12572 | 0,164661 | 142 | 70 | 72 | 138 | -0,76353 | y ~ Intervention + Sex + |
| 0,315771 | -0,13628 | 0,135356 | 142 | 70 | 72 | 138 | -1,00685 | y ~ Intervention + Sex + |
| 0,317076 | -0,15084 | 0,150224 | 142 | 70 | 72 | 138 | -1,00412 | y ~ Intervention + Sex + |
| 0,558527 | 0,089331 | 0,152324 | 142 | 70 | 72 | 138 | 0,586456 | y ~ Intervention + Sex + |
| 0,125178 | 0,257099 | 0,166648 | 142 | 70 | 72 | 138 | 1,542764 | y ~ Intervention + Sex + |

| | | | | | | | | |
|---|---|---|---|---|---|---|---|---|
| 0,58737 | -0,08269 | 0,152021 | 142 | 70 | 72 | 138 | -0,54393 | y ~ Intervention + Sex + |
| 0,811257 | -0,03558 | 0,148727 | 142 | 70 | 72 | 138 | -0,23926 | y ~ Intervention + Sex + |
| 0,689486 | 0,05652 | 0,14116 | 142 | 70 | 72 | 138 | 0,400393 | y ~ Intervention + Sex + |
| 0,62992 | -0,07113 | 0,14729 | 142 | 70 | 72 | 138 | -0,48292 | y ~ Intervention + Sex + |
| 0,66601 | -0,07074 | 0,163546 | 142 | 70 | 72 | 138 | -0,43256 | y ~ Intervention + Sex + |
| 0,05122 | -0,35036 | 0,178143 | 142 | 70 | 72 | 138 | -1,96673 | y ~ Intervention + Sex + |
| 0,352915 | -0,14257 | 0,152962 | 142 | 70 | 72 | 138 | -0,93209 | y ~ Intervention + Sex + |
| 0,410871 | 0,118562 | 0,143735 | 142 | 70 | 72 | 138 | 0,824867 | y ~ Intervention + Sex + |
| 0,182611 | -0,20634 | 0,154045 | 142 | 70 | 72 | 138 | -1,3395 | y ~ Intervention + Sex + |
| 0,257071 | 0,177465 | 0,155938 | 142 | 70 | 72 | 138 | 1,138051 | y ~ Intervention + Sex + |
| 0,162256 | 0,193495 | 0,137715 | 142 | 70 | 72 | 138 | 1,405041 | y ~ Intervention + Sex + |
| 0,769799 | 0,041872 | 0,142802 | 142 | 70 | 72 | 138 | 0,293214 | y ~ Intervention + Sex + |
| 0,756757 | 0,043721 | 0,140872 | 142 | 70 | 72 | 138 | 0,310358 | y ~ Intervention + Sex + |
| 0,815977 | -0,04 | 0,171547 | 142 | 70 | 72 | 138 | -0,23317 | y ~ Intervention + Sex + |
| 0,870546 | -0,02558 | 0,156646 | 142 | 70 | 72 | 138 | -0,16327 | y ~ Intervention + Sex + |
| 0,431191 | -0,1267 | 0,160486 | 142 | 70 | 72 | 138 | -0,78947 | y ~ Intervention + Sex + |
| 0,228925 | -0,18851 | 0,15599 | 142 | 70 | 72 | 138 | -1,20849 | y ~ Intervention + Sex + |
| 0,384851 | 0,142929 | 0,163953 | 142 | 70 | 72 | 138 | 0,871764 | y ~ Intervention + Sex + |
| 0,793454 | -0,04076 | 0,155368 | 142 | 70 | 72 | 138 | -0,26234 | y ~ Intervention + Sex + |
| 0,733179 | 0,053732 | 0,157299 | 142 | 70 | 72 | 138 | 0,34159 | y ~ Intervention + Sex + |
| 0,324606 | 0,154728 | 0,156518 | 142 | 70 | 72 | 138 | 0,988565 | y ~ Intervention + Sex + |
| 0,180716 | -0,21744 | 0,161623 | 142 | 70 | 72 | 138 | -1,34536 | y ~ Intervention + Sex + |
| 0,725413 | 0,056594 | 0,160803 | 142 | 70 | 72 | 138 | 0,351949 | y ~ Intervention + Sex + |
| 0,045177 | 0,322769 | 0,15968 | 142 | 70 | 72 | 138 | 2,021354 | y ~ Intervention + Sex + |
| 0,589452 | 0,08149 | 0,150657 | 142 | 70 | 72 | 138 | 0,540895 | y ~ Intervention + Sex + |
| 0,071017 | 0,253497 | 0,139329 | 142 | 70 | 72 | 138 | 1,819408 | y ~ Intervention + Sex + |
| 0,559205 | 0,102933 | 0,175821 | 142 | 70 | 72 | 138 | 0,585444 | y ~ Intervention + Sex + |
| 0,970834 | 0,005379 | 0,146858 | 142 | 70 | 72 | 138 | 0,036629 | y ~ Intervention + Sex + |
| 0,630582 | 0,073832 | 0,153183 | 142 | 70 | 72 | 138 | 0,481982 | y ~ Intervention + Sex + |
| 0,518083 | -0,10754 | 0,165971 | 142 | 70 | 72 | 138 | -0,64797 | y ~ Intervention + Sex + |
| 0,794966 | -0,04256 | 0,163468 | 142 | 70 | 72 | 138 | -0,26037 | y ~ Intervention + Sex + |
| 0,926698 | -0,01325 | 0,143773 | 142 | 70 | 72 | 138 | -0,09217 | y ~ Intervention + Sex + |
| 0,574765 | -0,09004 | 0,160101 | 142 | 70 | 72 | 138 | -0,56239 | y ~ Intervention + Sex + |
| 0,710713 | -0,05757 | 0,154887 | 142 | 70 | 72 | 138 | -0,37166 | y ~ Intervention + Sex + |
| 0,7333 | 0,051697 | 0,151413 | 142 | 70 | 72 | 138 | 0,341429 | y ~ Intervention + Sex + |
| 0,029863 | 0,326815 | 0,148916 | 142 | 70 | 72 | 138 | 2,194623 | y ~ Intervention + Sex + |
| 0,349261 | -0,14402 | 0,153344 | 142 | 70 | 72 | 138 | -0,93922 | y ~ Intervention + Sex + |
| 0,796407 | -0,04065 | 0,157262 | 142 | 70 | 72 | 138 | -0,2585 | y ~ Intervention + Sex + |
| 0,117581 | -0,24478 | 0,155431 | 142 | 70 | 72 | 138 | -1,57485 | y ~ Intervention + Sex + |
| 0,610065 | 0,083611 | 0,163576 | 142 | 70 | 72 | 138 | 0,511147 | y ~ Intervention + Sex + |
| 0,45894 | 0,111953 | 0,150742 | 142 | 70 | 72 | 138 | 0,742677 | y ~ Intervention + Sex + |
| 0,449327 | 0,12279 | 0,161843 | 142 | 70 | 72 | 138 | 0,758698 | y ~ Intervention + Sex + |
| 0,967547 | 0,006633 | 0,162731 | 142 | 70 | 72 | 138 | 0,040759 | y ~ Intervention + Sex + |
| 0,543558 | -0,1014 | 0,166516 | 142 | 70 | 72 | 138 | -0,60895 | y ~ Intervention + Sex + |
| 0,232149 | -0,1915 | 0,15957 | 142 | 70 | 72 | 138 | -1,20012 | y ~ Intervention + Sex + |
| 0,361983 | -0,13184 | 0,144143 | 142 | 70 | 72 | 138 | -0,91463 | y ~ Intervention + Sex + |
| 0,162837 | -0,24669 | 0,175822 | 142 | 70 | 72 | 138 | -1,40308 | y ~ Intervention + Sex + |
| 0,558317 | 0,103587 | 0,176538 | 142 | 70 | 72 | 138 | 0,58677 | y ~ Intervention + Sex + |

| | | | | | | | | |
|---|---|---|---|---|---|---|---|---|
| 0,213135 | 0,19047 | 0,152282 | 142 | 70 | 72 | 138 | 1,250768 | y ~ Intervention + Sex + |
| 0,231793 | -0,2136 | 0,177846 | 142 | 70 | 72 | 138 | -1,20104 | y ~ Intervention + Sex + |
| 0,434654 | 0,124782 | 0,159254 | 142 | 70 | 72 | 138 | 0,783538 | y ~ Intervention + Sex + |
| 0,156194 | 0,22944 | 0,160924 | 142 | 70 | 72 | 138 | 1,425767 | y ~ Intervention + Sex + |
| 0,176242 | 0,184895 | 0,136014 | 142 | 70 | 72 | 138 | 1,359385 | y ~ Intervention + Sex + |
| 0,691304 | -0,06667 | 0,167551 | 142 | 70 | 72 | 138 | -0,39792 | y ~ Intervention + Sex + |
| 0,356159 | -0,13928 | 0,150441 | 142 | 70 | 72 | 138 | -0,92581 | y ~ Intervention + Sex + |
| 0,754913 | -0,04086 | 0,130638 | 142 | 70 | 72 | 138 | -0,31279 | y ~ Intervention + Sex + |
| 0,587623 | -0,08248 | 0,151741 | 142 | 70 | 72 | 138 | -0,54356 | y ~ Intervention + Sex + |
| 0,838462 | 0,031806 | 0,155724 | 142 | 70 | 72 | 138 | 0,204246 | y ~ Intervention + Sex + |
| 0,601525 | -0,08463 | 0,161696 | 142 | 70 | 72 | 138 | -0,52342 | y ~ Intervention + Sex + |
| 0,814169 | 0,037338 | 0,158549 | 142 | 70 | 72 | 138 | 0,235501 | y ~ Intervention + Sex + |
| 0,135838 | 0,216841 | 0,144538 | 142 | 70 | 72 | 138 | 1,500235 | y ~ Intervention + Sex + |
| 0,117433 | -0,2384 | 0,151319 | 142 | 70 | 72 | 138 | -1,57549 | y ~ Intervention + Sex + |
| 0,657017 | -0,06684 | 0,150204 | 142 | 70 | 72 | 138 | -0,445 | y ~ Intervention + Sex + |
| 0,891329 | -0,02301 | 0,168104 | 142 | 70 | 72 | 138 | -0,13687 | y ~ Intervention + Sex + |
| 0,241438 | -0,17603 | 0,14963 | 142 | 70 | 72 | 138 | -1,17645 | y ~ Intervention + Sex + |
| 0,514203 | 0,101947 | 0,155884 | 142 | 70 | 72 | 138 | 0,653995 | y ~ Intervention + Sex + |
| 0,119362 | 0,257763 | 0,164475 | 142 | 70 | 72 | 138 | 1,567185 | y ~ Intervention + Sex + |
| 0,388406 | -0,13337 | 0,154136 | 142 | 70 | 72 | 138 | -0,86525 | y ~ Intervention + Sex + |
| 0,322163 | -0,172 | 0,173111 | 142 | 70 | 72 | 138 | -0,99359 | y ~ Intervention + Sex + |
| 0,131649 | 0,251453 | 0,165798 | 142 | 70 | 72 | 138 | 1,516623 | y ~ Intervention + Sex + |
| 0,659475 | -0,07309 | 0,165509 | 142 | 70 | 72 | 138 | -0,44159 | y ~ Intervention + Sex + |
| 0,552981 | 0,101012 | 0,169838 | 142 | 70 | 72 | 138 | 0,594755 | y ~ Intervention + Sex + |
| 0,223374 | -0,18271 | 0,14938 | 142 | 70 | 72 | 138 | -1,22311 | y ~ Intervention + Sex + |
| 0,915529 | -0,01609 | 0,151462 | 142 | 70 | 72 | 138 | -0,10626 | y ~ Intervention + Sex + |
| 0,676618 | -0,06814 | 0,163034 | 142 | 70 | 72 | 138 | -0,41797 | y ~ Intervention + Sex + |
| 0,119797 | -0,24511 | 0,15659 | 142 | 70 | 72 | 138 | -1,56533 | y ~ Intervention + Sex + |
| 0,937786 | 0,012418 | 0,158811 | 142 | 70 | 72 | 138 | 0,078196 | y ~ Intervention + Sex + |
| 0,894234 | 0,022916 | 0,172051 | 142 | 70 | 72 | 138 | 0,133194 | y ~ Intervention + Sex + |
| 0,71745 | 0,067688 | 0,186668 | 142 | 70 | 72 | 138 | 0,362611 | y ~ Intervention + Sex + |
| 0,063124 | -0,32252 | 0,172156 | 142 | 70 | 72 | 138 | -1,87343 | y ~ Intervention + Sex + |
| 0,812157 | -0,03969 | 0,166707 | 142 | 70 | 72 | 138 | -0,2381 | y ~ Intervention + Sex + |
| 0,550997 | 0,081301 | 0,136015 | 142 | 70 | 72 | 138 | 0,597734 | y ~ Intervention + Sex + |
| 0,154606 | 0,220853 | 0,154303 | 142 | 70 | 72 | 138 | 1,431297 | y ~ Intervention + Sex + |
| 0,438134 | 0,126149 | 0,162228 | 142 | 70 | 72 | 138 | 0,777603 | y ~ Intervention + Sex + |
| 0,401662 | 0,128846 | 0,153159 | 142 | 70 | 72 | 138 | 0,841253 | y ~ Intervention + Sex + |
| 0,388289 | 0,123407 | 0,142591 | 142 | 70 | 72 | 138 | 0,865459 | y ~ Intervention + Sex + |
| 0,772503 | -0,04564 | 0,157568 | 142 | 70 | 72 | 138 | -0,28967 | y ~ Intervention + Sex + |
| 0,87958 | 0,024045 | 0,158414 | 142 | 70 | 72 | 138 | 0,151783 | y ~ Intervention + Sex + |
| 0,585922 | -0,08201 | 0,150191 | 142 | 70 | 72 | 138 | -0,54604 | y ~ Intervention + Sex + |
| 0,517586 | 0,105883 | 0,163214 | 142 | 70 | 72 | 138 | 0,648738 | y ~ Intervention + Sex + |
| 0,763996 | 0,049043 | 0,163027 | 142 | 70 | 72 | 138 | 0,300831 | y ~ Intervention + Sex + |
| 0,594493 | -0,07307 | 0,136947 | 142 | 70 | 72 | 138 | -0,53358 | y ~ Intervention + Sex + |
| 0,253443 | -0,18005 | 0,156996 | 142 | 70 | 72 | 138 | -1,14681 | y ~ Intervention + Sex + |
| 0,26986 | -0,17629 | 0,15913 | 142 | 70 | 72 | 138 | -1,10784 | y ~ Intervention + Sex + |
| 0,145566 | 0,21152 | 0,144516 | 142 | 70 | 72 | 138 | 1,46364 | y ~ Intervention + Sex + |
| 0,566205 | 0,097819 | 0,170109 | 142 | 70 | 72 | 138 | 0,575034 | y ~ Intervention + Sex + |

| | | | | | | | | |
|---|---|---|---|---|---|---|---|---|
| 0,77662 | -0,04611 | 0,162212 | 142 | 70 | 72 | 138 | -0,28428 | y ~ Intervention + Sex + |
| 0,805934 | 0,040346 | 0,16391 | 142 | 70 | 72 | 138 | 0,246148 | y ~ Intervention + Sex + |
| 0,816467 | -0,03326 | 0,143039 | 142 | 70 | 72 | 138 | -0,23253 | y ~ Intervention + Sex + |
| 0,846829 | -0,02921 | 0,150921 | 142 | 70 | 72 | 138 | -0,19353 | y ~ Intervention + Sex + |
| 0,91355 | 0,018505 | 0,170144 | 142 | 70 | 72 | 138 | 0,108761 | y ~ Intervention + Sex + |
| 0,647505 | 0,073021 | 0,159352 | 142 | 70 | 72 | 138 | 0,458234 | y ~ Intervention + Sex + |
| 0,547002 | 0,090638 | 0,150125 | 142 | 70 | 72 | 138 | 0,603749 | y ~ Intervention + Sex + |
| 0,932498 | 0,014255 | 0,167989 | 142 | 70 | 72 | 138 | 0,084857 | y ~ Intervention + Sex + |
| 0,980579 | -0,00377 | 0,15447 | 142 | 70 | 72 | 138 | -0,02439 | y ~ Intervention + Sex + |
| 0,147331 | 0,209731 | 0,143926 | 142 | 70 | 72 | 138 | 1,457207 | y ~ Intervention + Sex + |
| 0,980505 | -0,00362 | 0,147782 | 142 | 70 | 72 | 138 | -0,02448 | y ~ Intervention + Sex + |
| 0,324851 | 0,163146 | 0,165117 | 142 | 70 | 72 | 138 | 0,988062 | y ~ Intervention + Sex + |
| 0,20693 | 0,167932 | 0,132438 | 142 | 70 | 72 | 138 | 1,268007 | y ~ Intervention + Sex + |
| 0,294151 | -0,17777 | 0,168816 | 142 | 70 | 72 | 138 | -1,05307 | y ~ Intervention + Sex + |
| 0,719216 | 0,049769 | 0,138153 | 142 | 70 | 72 | 138 | 0,360243 | y ~ Intervention + Sex + |
| 0,222949 | 0,190373 | 0,155503 | 142 | 70 | 72 | 138 | 1,224238 | y ~ Intervention + Sex + |
| 0,515133 | 0,11295 | 0,17309 | 142 | 70 | 72 | 138 | 0,652549 | y ~ Intervention + Sex + |
| 0,169948 | -0,20788 | 0,150687 | 142 | 70 | 72 | 138 | -1,37958 | y ~ Intervention + Sex + |
| 0,165507 | -0,2094 | 0,150197 | 142 | 70 | 72 | 138 | -1,39417 | y ~ Intervention + Sex + |
| 0,618097 | -0,07545 | 0,150988 | 142 | 70 | 72 | 138 | -0,49968 | y ~ Intervention + Sex + |
| 0,496969 | 0,107863 | 0,158373 | 142 | 70 | 72 | 138 | 0,68107 | y ~ Intervention + Sex + |
| 0,061969 | -0,29063 | 0,154443 | 142 | 70 | 72 | 138 | -1,8818 | y ~ Intervention + Sex + |
| 0,368601 | -0,14177 | 0,157164 | 142 | 70 | 72 | 138 | -0,90205 | y ~ Intervention + Sex + |
| 0,182933 | -0,20636 | 0,154169 | 142 | 70 | 72 | 138 | -1,3385 | y ~ Intervention + Sex + |
| 0,433355 | -0,10617 | 0,135119 | 142 | 70 | 72 | 138 | -0,78576 | y ~ Intervention + Sex + |
| 0,263397 | 0,174533 | 0,15542 | 142 | 70 | 72 | 138 | 1,122978 | y ~ Intervention + Sex + |
| 0,358664 | -0,15335 | 0,166503 | 142 | 70 | 72 | 138 | -0,92099 | y ~ Intervention + Sex + |
| 0,802805 | 0,040809 | 0,163107 | 142 | 70 | 72 | 138 | 0,2502 | y ~ Intervention + Sex + |
| 0,694216 | 0,063887 | 0,162164 | 142 | 70 | 72 | 138 | 0,393963 | y ~ Intervention + Sex + |
| 0,321283 | 0,157697 | 0,158426 | 142 | 70 | 72 | 138 | 0,995401 | y ~ Intervention + Sex + |
| 0,819536 | -0,03689 | 0,161411 | 142 | 70 | 72 | 138 | -0,22858 | y ~ Intervention + Sex + |
| 0,211914 | 0,202317 | 0,161321 | 142 | 70 | 72 | 138 | 1,254132 | y ~ Intervention + Sex + |
| 0,143485 | -0,24234 | 0,164712 | 142 | 70 | 72 | 138 | -1,47131 | y ~ Intervention + Sex + |
| 0,503284 | -0,11459 | 0,170758 | 142 | 70 | 72 | 138 | -0,67109 | y ~ Intervention + Sex + |
| 0,369676 | -0,14528 | 0,161414 | 142 | 70 | 72 | 138 | -0,90002 | y ~ Intervention + Sex + |
| 0,157946 | -0,21295 | 0,149997 | 142 | 70 | 72 | 138 | -1,41971 | y ~ Intervention + Sex + |
| 0,409216 | -0,14178 | 0,171279 | 142 | 70 | 72 | 138 | -0,82779 | y ~ Intervention + Sex + |
| 0,11958 | -0,25622 | 0,163586 | 142 | 70 | 72 | 138 | -1,56625 | y ~ Intervention + Sex + |
| 0,329403 | 0,148943 | 0,152173 | 142 | 70 | 72 | 138 | 0,978777 | y ~ Intervention + Sex + |
| 0,871118 | 0,027121 | 0,166855 | 142 | 70 | 72 | 138 | 0,16254 | y ~ Intervention + Sex + |
| 0,506637 | 0,096135 | 0,144386 | 142 | 70 | 72 | 138 | 0,66582 | y ~ Intervention + Sex + |
| 0,216302 | 0,202699 | 0,163189 | 142 | 70 | 72 | 138 | 1,242113 | y ~ Intervention + Sex + |
| 0,220218 | 0,214 | 0,173767 | 142 | 70 | 72 | 138 | 1,231532 | y ~ Intervention + Sex + |
| 0,693037 | -0,05668 | 0,1433 | 142 | 70 | 72 | 138 | -0,39556 | y ~ Intervention + Sex + |
| 0,749574 | -0,05713 | 0,178611 | 142 | 70 | 72 | 138 | -0,31984 | y ~ Intervention + Sex + |
| 0,684912 | 0,060344 | 0,148402 | 142 | 70 | 72 | 138 | 0,406627 | y ~ Intervention + Sex + |
| 0,060196 | 0,293157 | 0,154708 | 142 | 70 | 72 | 138 | 1,894908 | y ~ Intervention + Sex + |
| 0,754441 | -0,04622 | 0,147459 | 142 | 70 | 72 | 138 | -0,31341 | y ~ Intervention + Sex + |

| | | | | | | | | |
|---|---|---|---|---|---|---|---|---|
| 0,476277 | -0,11219 | 0,157073 | 142 | 70 | 72 | 138 | -0,71425 | y ~ Intervention + Sex + |
| 0,240823 | -0,20187 | 0,171368 | 142 | 70 | 72 | 138 | -1,178 | y ~ Intervention + Sex + |
| 0,128786 | -0,23162 | 0,151575 | 142 | 70 | 72 | 138 | -1,52806 | y ~ Intervention + Sex + |
| 0,499616 | -0,1112 | 0,164286 | 142 | 70 | 72 | 138 | -0,67688 | y ~ Intervention + Sex + |
| 0,505104 | -0,10869 | 0,162655 | 142 | 70 | 72 | 138 | -0,66823 | y ~ Intervention + Sex + |
| 0,175238 | -0,2232 | 0,163808 | 142 | 70 | 72 | 138 | -1,36257 | y ~ Intervention + Sex + |
| 0,387759 | 0,127602 | 0,147274 | 142 | 70 | 72 | 138 | 0,866429 | y ~ Intervention + Sex + |
| 0,481497 | 0,107605 | 0,152456 | 142 | 70 | 72 | 138 | 0,705808 | y ~ Intervention + Sex + |
| 0,365971 | 0,144948 | 0,159805 | 142 | 70 | 72 | 138 | 0,907033 | y ~ Intervention + Sex + |
| 0,416364 | 0,127592 | 0,156517 | 142 | 70 | 72 | 138 | 0,815198 | y ~ Intervention + Sex + |
| 0,318807 | -0,1639 | 0,163811 | 142 | 70 | 72 | 138 | -1,00053 | y ~ Intervention + Sex + |
| 0,690138 | -0,06182 | 0,154736 | 142 | 70 | 72 | 138 | -0,39951 | y ~ Intervention + Sex + |
| 0,563706 | 0,084394 | 0,145824 | 142 | 70 | 72 | 138 | 0,578743 | y ~ Intervention + Sex + |
| 0,840179 | 0,02975 | 0,147242 | 142 | 70 | 72 | 138 | 0,202045 | y ~ Intervention + Sex + |
| 0,838666 | -0,0293 | 0,143636 | 142 | 70 | 72 | 138 | -0,20398 | y ~ Intervention + Sex + |
| 0,534496 | -0,09732 | 0,156279 | 142 | 70 | 72 | 138 | -0,62272 | y ~ Intervention + Sex + |
| 0,56315 | -0,09825 | 0,169529 | 142 | 70 | 72 | 138 | -0,57957 | y ~ Intervention + Sex + |
| 0,113114 | 0,262376 | 0,164551 | 142 | 70 | 72 | 138 | 1,594492 | y ~ Intervention + Sex + |
| 0,845073 | -0,02989 | 0,152649 | 142 | 70 | 72 | 138 | -0,19578 | y ~ Intervention + Sex + |
| 0,599551 | 0,086713 | 0,164772 | 142 | 70 | 72 | 138 | 0,526263 | y ~ Intervention + Sex + |
| 0,713732 | 0,058218 | 0,158372 | 142 | 70 | 72 | 138 | 0,367603 | y ~ Intervention + Sex + |
| 0,365186 | 0,151226 | 0,166452 | 142 | 70 | 72 | 138 | 0,908524 | y ~ Intervention + Sex + |
| 0,732282 | 0,047068 | 0,137311 | 142 | 70 | 72 | 138 | 0,342785 | y ~ Intervention + Sex + |
| 0,634862 | -0,07253 | 0,152398 | 142 | 70 | 72 | 138 | -0,47595 | y ~ Intervention + Sex + |
| 0,318334 | -0,15505 | 0,154812 | 142 | 70 | 72 | 138 | -1,00151 | y ~ Intervention + Sex + |
| 0,382521 | -0,1424 | 0,162549 | 142 | 70 | 72 | 138 | -0,87606 | y ~ Intervention + Sex + |
| 0,259091 | -0,17699 | 0,156183 | 142 | 70 | 72 | 138 | -1,13321 | y ~ Intervention + Sex + |
| 0,489148 | 0,111347 | 0,160553 | 142 | 70 | 72 | 138 | 0,693521 | y ~ Intervention + Sex + |
| 0,448423 | -0,10987 | 0,144522 | 142 | 70 | 72 | 138 | -0,76021 | y ~ Intervention + Sex + |
| 0,093502 | 0,2787 | 0,165022 | 142 | 70 | 72 | 138 | 1,688871 | y ~ Intervention + Sex + |
| 0,256975 | 0,152599 | 0,134061 | 142 | 70 | 72 | 138 | 1,138283 | y ~ Intervention + Sex + |
| 0,375443 | 0,140486 | 0,157991 | 142 | 70 | 72 | 138 | 0,8892 | y ~ Intervention + Sex + |
| 0,103914 | 0,243832 | 0,148953 | 142 | 70 | 72 | 138 | 1,636975 | y ~ Intervention + Sex + |
| 0,395769 | 0,14814 | 0,173902 | 142 | 70 | 72 | 138 | 0,851858 | y ~ Intervention + Sex + |
| 0,856243 | -0,02638 | 0,145333 | 142 | 70 | 72 | 138 | -0,1815 | y ~ Intervention + Sex + |
| 0,323065 | -0,1722 | 0,173641 | 142 | 70 | 72 | 138 | -0,99173 | y ~ Intervention + Sex + |
| 0,924152 | -0,01546 | 0,162089 | 142 | 70 | 72 | 138 | -0,09538 | y ~ Intervention + Sex + |
| 0,11886 | -0,25779 | 0,164265 | 142 | 70 | 72 | 138 | -1,56933 | y ~ Intervention + Sex + |
| 0,189817 | 0,204793 | 0,155427 | 142 | 70 | 72 | 138 | 1,317611 | y ~ Intervention + Sex + |
| 0,889648 | -0,02254 | 0,162144 | 142 | 70 | 72 | 138 | -0,13901 | y ~ Intervention + Sex + |
| 0,793735 | -0,03638 | 0,138867 | 142 | 70 | 72 | 138 | -0,26197 | y ~ Intervention + Sex + |
| 0,721268 | -0,05606 | 0,156812 | 142 | 70 | 72 | 138 | -0,35749 | y ~ Intervention + Sex + |
| 0,544032 | -0,09416 | 0,154812 | 142 | 70 | 72 | 138 | -0,60823 | y ~ Intervention + Sex + |
| 0,661162 | 0,067055 | 0,152654 | 142 | 70 | 72 | 138 | 0,439258 | y ~ Intervention + Sex + |
| 0,699837 | -0,05823 | 0,150719 | 142 | 70 | 72 | 138 | -0,38634 | y ~ Intervention + Sex + |
| 0,68202 | -0,06961 | 0,169552 | 142 | 70 | 72 | 138 | -0,41058 | y ~ Intervention + Sex + |
| 0,871516 | 0,027206 | 0,167901 | 142 | 70 | 72 | 138 | 0,162034 | y ~ Intervention + Sex + |
| 0,957692 | -0,00863 | 0,162422 | 142 | 70 | 72 | 138 | -0,05315 | y ~ Intervention + Sex + |

| | | | | | | | | |
|---|---|---|---|---|---|---|---|---|
| 0,822427 | -0,03305 | 0,146979 | 142 | 70 | 72 | 138 | -0,22485 | y ~ Intervention + Sex + |
| 0,39096 | 0,13238 | 0,153825 | 142 | 70 | 72 | 138 | 0,860584 | y ~ Intervention + Sex + |
| 0,813798 | 0,034862 | 0,147733 | 142 | 70 | 72 | 138 | 0,235981 | y ~ Intervention + Sex + |
| 0,298262 | -0,17336 | 0,166039 | 142 | 70 | 72 | 138 | -1,04411 | y ~ Intervention + Sex + |
| 0,587086 | -0,08553 | 0,157123 | 142 | 70 | 72 | 138 | -0,54434 | y ~ Intervention + Sex + |
| 0,855791 | -0,03009 | 0,165287 | 142 | 70 | 72 | 138 | -0,18207 | y ~ Intervention + Sex + |
| 0,278812 | 0,165455 | 0,152174 | 142 | 70 | 72 | 138 | 1,087272 | y ~ Intervention + Sex + |
| 0,787801 | 0,046051 | 0,170753 | 142 | 70 | 72 | 138 | 0,269691 | y ~ Intervention + Sex + |
| 0,922142 | -0,01458 | 0,148933 | 142 | 70 | 72 | 138 | -0,09792 | y ~ Intervention + Sex + |
| 0,516417 | 0,104898 | 0,161245 | 142 | 70 | 72 | 138 | 0,650553 | y ~ Intervention + Sex + |
| 0,65999 | -0,07606 | 0,172524 | 142 | 70 | 72 | 138 | -0,44088 | y ~ Intervention + Sex + |
| 0,885094 | 0,025568 | 0,176596 | 142 | 70 | 72 | 138 | 0,144782 | y ~ Intervention + Sex + |
| 0,668071 | 0,073018 | 0,169921 | 142 | 70 | 72 | 138 | 0,429718 | y ~ Intervention + Sex + |
| 0,624284 | 0,072352 | 0,147389 | 142 | 70 | 72 | 138 | 0,49089 | y ~ Intervention + Sex + |
| 0,2925 | -0,15778 | 0,149317 | 142 | 70 | 72 | 138 | -1,05669 | y ~ Intervention + Sex + |
| 0,505169 | -0,10986 | 0,164436 | 142 | 70 | 72 | 138 | -0,66813 | y ~ Intervention + Sex + |
| 0,621674 | 0,086206 | 0,174297 | 142 | 70 | 72 | 138 | 0,494592 | y ~ Intervention + Sex + |
| 0,64936 | -0,07872 | 0,172763 | 142 | 70 | 72 | 138 | -0,45565 | y ~ Intervention + Sex + |
| 0,730997 | 0,050355 | 0,14617 | 142 | 70 | 72 | 138 | 0,344496 | y ~ Intervention + Sex + |
| 0,281539 | 0,175208 | 0,162065 | 142 | 70 | 72 | 138 | 1,081098 | y ~ Intervention + Sex + |
| 0,607087 | -0,08655 | 0,167918 | 142 | 70 | 72 | 138 | -0,51542 | y ~ Intervention + Sex + |
| 0,489185 | -0,10732 | 0,154763 | 142 | 70 | 72 | 138 | -0,69346 | y ~ Intervention + Sex + |
| 0,956906 | -0,00805 | 0,148671 | 142 | 70 | 72 | 138 | -0,05413 | y ~ Intervention + Sex + |
| 0,572694 | 0,087669 | 0,155046 | 142 | 70 | 72 | 138 | 0,565438 | y ~ Intervention + Sex + |
| 0,569934 | -0,0841 | 0,147672 | 142 | 70 | 72 | 138 | -0,56951 | y ~ Intervention + Sex + |
| 0,828081 | -0,03622 | 0,166473 | 142 | 70 | 72 | 138 | -0,21758 | y ~ Intervention + Sex + |
| 0,544371 | -0,09345 | 0,153765 | 142 | 70 | 72 | 138 | -0,60772 | y ~ Intervention + Sex + |
| 0,071404 | -0,29266 | 0,161078 | 142 | 70 | 72 | 138 | -1,81689 | y ~ Intervention + Sex + |
| 0,869992 | 0,021975 | 0,134015 | 142 | 70 | 72 | 138 | 0,163973 | y ~ Intervention + Sex + |
| 0,471848 | 0,116605 | 0,161623 | 142 | 70 | 72 | 138 | 0,721459 | y ~ Intervention + Sex + |
| 0,286334 | 0,170977 | 0,15974 | 142 | 70 | 72 | 138 | 1,070342 | y ~ Intervention + Sex + |
| 0,653302 | 0,071358 | 0,158517 | 142 | 70 | 72 | 138 | 0,450159 | y ~ Intervention + Sex + |
| 0,554915 | -0,09267 | 0,156574 | 142 | 70 | 72 | 138 | -0,59186 | y ~ Intervention + Sex + |
| 0,584881 | 0,076096 | 0,138974 | 142 | 70 | 72 | 138 | 0,547557 | y ~ Intervention + Sex + |
| 0,014485 | -0,3775 | 0,152444 | 142 | 70 | 72 | 138 | -2,4763 | y ~ Intervention + Sex + |
| 0,338867 | -0,1492 | 0,155456 | 142 | 70 | 72 | 138 | -0,95974 | y ~ Intervention + Sex + |
| 0,361755 | 0,158559 | 0,173276 | 142 | 70 | 72 | 138 | 0,915063 | y ~ Intervention + Sex + |
| 0,024846 | 0,368606 | 0,162483 | 142 | 70 | 72 | 138 | 2,268584 | y ~ Intervention + Sex + |
| 0,346259 | -0,15008 | 0,158799 | 142 | 70 | 72 | 138 | -0,9451 | y ~ Intervention + Sex + |
| 0,72381 | -0,05083 | 0,143557 | 142 | 70 | 72 | 138 | -0,35409 | y ~ Intervention + Sex + |
| 0,187794 | 0,198966 | 0,150311 | 142 | 70 | 72 | 138 | 1,323692 | y ~ Intervention + Sex + |
| 0,944463 | 0,010823 | 0,155082 | 142 | 70 | 72 | 138 | 0,069788 | y ~ Intervention + Sex + |
| 0,54482 | 0,078988 | 0,13012 | 142 | 70 | 72 | 138 | 0,607043 | y ~ Intervention + Sex + |
| 0,799039 | 0,039175 | 0,153576 | 142 | 70 | 72 | 138 | 0,255082 | y ~ Intervention + Sex + |
| 0,680472 | 0,072683 | 0,176119 | 142 | 70 | 72 | 138 | 0,412693 | y ~ Intervention + Sex + |
| 0,704822 | -0,0594 | 0,156475 | 142 | 70 | 72 | 138 | -0,3796 | y ~ Intervention + Sex + |
| 0,553015 | -0,10029 | 0,168637 | 142 | 70 | 72 | 138 | -0,5947 | y ~ Intervention + Sex + |
| 0,069046 | -0,28985 | 0,15818 | 142 | 70 | 72 | 138 | -1,83241 | y ~ Intervention + Sex + |

| | | | | | | | | |
|---|---|---|---|---|---|---|---|---|
| 0,979018 | 0,004221 | 0,16019 | 142 | 70 | 72 | 138 | 0,026348 | y ~ Intervention + Sex + |
| 0,692679 | 0,060974 | 0,153955 | 142 | 70 | 72 | 138 | 0,396051 | y ~ Intervention + Sex + |
| 0,358064 | 0,143188 | 0,155278 | 142 | 70 | 72 | 138 | 0,922141 | y ~ Intervention + Sex + |
| 0,190953 | 0,186355 | 0,141799 | 142 | 70 | 72 | 138 | 1,314218 | y ~ Intervention + Sex + |
| 0,339634 | 0,175556 | 0,183213 | 142 | 70 | 72 | 138 | 0,958207 | y ~ Intervention + Sex + |
| 0,510616 | 0,097594 | 0,147961 | 142 | 70 | 72 | 138 | 0,659591 | y ~ Intervention + Sex + |
| 0,969115 | -0,00556 | 0,143233 | 142 | 70 | 72 | 138 | -0,03879 | y ~ Intervention + Sex + |
| 0,367087 | 0,143976 | 0,159104 | 142 | 70 | 72 | 138 | 0,904917 | y ~ Intervention + Sex + |
| 0,120373 | -0,25916 | 0,165821 | 142 | 70 | 72 | 138 | -1,56287 | y ~ Intervention + Sex + |
| 0,935885 | 0,013062 | 0,162074 | 142 | 70 | 72 | 138 | 0,08059 | y ~ Intervention + Sex + |
| 0,804824 | -0,03676 | 0,148481 | 142 | 70 | 72 | 138 | -0,24758 | y ~ Intervention + Sex + |
| 0,096432 | -0,26414 | 0,157809 | 142 | 70 | 72 | 138 | -1,67381 | y ~ Intervention + Sex + |
| 0,462303 | 0,121213 | 0,164442 | 142 | 70 | 72 | 138 | 0,737117 | y ~ Intervention + Sex + |
| 0,558246 | 0,095753 | 0,163157 | 142 | 70 | 72 | 138 | 0,586875 | y ~ Intervention + Sex + |
| 0,143446 | -0,22853 | 0,155307 | 142 | 70 | 72 | 138 | -1,47145 | y ~ Intervention + Sex + |
| 0,149914 | -0,22633 | 0,156317 | 142 | 70 | 72 | 138 | -1,4479 | y ~ Intervention + Sex + |
| 0,08648 | -0,26313 | 0,152401 | 142 | 70 | 72 | 138 | -1,72659 | y ~ Intervention + Sex + |
| 0,53246 | -0,09516 | 0,15205 | 142 | 70 | 72 | 138 | -0,62583 | y ~ Intervention + Sex + |
| 0,653719 | 0,068596 | 0,152579 | 142 | 70 | 72 | 138 | 0,449579 | y ~ Intervention + Sex + |
| 0,342513 | -0,15656 | 0,164373 | 142 | 70 | 72 | 138 | -0,95249 | y ~ Intervention + Sex + |
| 0,559885 | -0,09837 | 0,168318 | 142 | 70 | 72 | 138 | -0,58443 | y ~ Intervention + Sex + |
| 0,188374 | -0,22056 | 0,166842 | 142 | 70 | 72 | 138 | -1,32194 | y ~ Intervention + Sex + |
| 0,354679 | 0,150468 | 0,162025 | 142 | 70 | 72 | 138 | 0,928675 | y ~ Intervention + Sex + |
| 0,57704 | -0,096 | 0,171722 | 142 | 70 | 72 | 138 | -0,55904 | y ~ Intervention + Sex + |
| 0,593008 | 0,078021 | 0,145635 | 142 | 70 | 72 | 138 | 0,535729 | y ~ Intervention + Sex + |
| 0,353061 | -0,14259 | 0,153024 | 142 | 70 | 72 | 138 | -0,93181 | y ~ Intervention + Sex + |
| 0,430179 | -0,13215 | 0,167019 | 142 | 70 | 72 | 138 | -0,79121 | y ~ Intervention + Sex + |
| 0,275747 | -0,16903 | 0,154468 | 142 | 70 | 72 | 138 | -1,09426 | y ~ Intervention + Sex + |
| 0,591873 | 0,082389 | 0,153317 | 142 | 70 | 72 | 138 | 0,537377 | y ~ Intervention + Sex + |
| 0,50827 | 0,095818 | 0,144466 | 142 | 70 | 72 | 138 | 0,66326 | y ~ Intervention + Sex + |
| 0,293855 | -0,17272 | 0,163919 | 142 | 70 | 72 | 138 | -1,05371 | y ~ Intervention + Sex + |
| 0,699135 | -0,0634 | 0,163711 | 142 | 70 | 72 | 138 | -0,38729 | y ~ Intervention + Sex + |
| 0,574383 | 0,08984 | 0,159588 | 142 | 70 | 72 | 138 | 0,562949 | y ~ Intervention + Sex + |
| 0,368347 | -0,13128 | 0,145453 | 142 | 70 | 72 | 138 | -0,90253 | y ~ Intervention + Sex + |
| 0,263246 | -0,1923 | 0,171191 | 142 | 70 | 72 | 138 | -1,12333 | y ~ Intervention + Sex + |
| 0,24696 | 0,190794 | 0,164096 | 142 | 70 | 72 | 138 | 1,162695 | y ~ Intervention + Sex + |
| 0,543196 | -0,10585 | 0,173668 | 142 | 70 | 72 | 138 | -0,6095 | y ~ Intervention + Sex + |
| 0,335293 | -0,15755 | 0,162948 | 142 | 70 | 72 | 138 | -0,96689 | y ~ Intervention + Sex + |
| 0,00667 | -0,42865 | 0,155614 | 142 | 70 | 72 | 138 | -2,75455 | y ~ Intervention + Sex + |
| 0,611905 | 0,082658 | 0,162548 | 142 | 70 | 72 | 138 | 0,508514 | y ~ Intervention + Sex + |
| 0,748609 | 0,052996 | 0,165037 | 142 | 70 | 72 | 138 | 0,321115 | y ~ Intervention + Sex + |
| 0,698498 | 0,053421 | 0,137626 | 142 | 70 | 72 | 138 | 0,388157 | y ~ Intervention + Sex + |
| 0,944578 | 0,010048 | 0,144277 | 142 | 70 | 72 | 138 | 0,069644 | y ~ Intervention + Sex + |
| 0,951957 | 0,0098 | 0,16237 | 142 | 70 | 72 | 138 | 0,060359 | y ~ Intervention + Sex + |
| 0,304002 | 0,16977 | 0,164549 | 142 | 70 | 72 | 138 | 1,031734 | y ~ Intervention + Sex + |
| 0,406665 | -0,12502 | 0,150212 | 142 | 70 | 72 | 138 | -0,83232 | y ~ Intervention + Sex + |
| 0,719784 | 0,053256 | 0,148147 | 142 | 70 | 72 | 138 | 0,359481 | y ~ Intervention + Sex + |
| 0,287217 | 0,171374 | 0,160406 | 142 | 70 | 72 | 138 | 1,068374 | y ~ Intervention + Sex + |

| | | | | | | | | |
|---|---|---|---|---|---|---|---|---|
| 0,702344 | 0,055377 | 0,144605 | 142 | 70 | 72 | 138 | 0,382953 | y ~ Intervention + Sex + |
| 0,002099 | -0,47833 | 0,15257 | 142 | 70 | 72 | 138 | -3,13515 | y ~ Intervention + Sex + |
| 0,810361 | 0,042335 | 0,176086 | 142 | 70 | 72 | 138 | 0,240421 | y ~ Intervention + Sex + |
| 0,379312 | 0,140155 | 0,158907 | 142 | 70 | 72 | 138 | 0,881998 | y ~ Intervention + Sex + |
| 0,1346 | 0,273509 | 0,181729 | 142 | 70 | 72 | 138 | 1,505037 | y ~ Intervention + Sex + |
| 0,251322 | 0,185889 | 0,161365 | 142 | 70 | 72 | 138 | 1,151978 | y ~ Intervention + Sex + |
| 0,274311 | -0,16864 | 0,153653 | 142 | 70 | 72 | 138 | -1,09755 | y ~ Intervention + Sex + |
| 0,073837 | -0,24968 | 0,138613 | 142 | 70 | 72 | 138 | -1,80131 | y ~ Intervention + Sex + |
| 0,978166 | -0,00445 | 0,162166 | 142 | 70 | 72 | 138 | -0,02742 | y ~ Intervention + Sex + |
| 0,392417 | 0,14073 | 0,164034 | 142 | 70 | 72 | 138 | 0,857932 | y ~ Intervention + Sex + |
| 0,125809 | 0,224388 | 0,14569 | 142 | 70 | 72 | 138 | 1,54017 | y ~ Intervention + Sex + |
| 0,584469 | -0,09573 | 0,174636 | 142 | 70 | 72 | 138 | -0,54816 | y ~ Intervention + Sex + |
| 0,031411 | -0,36511 | 0,167943 | 142 | 70 | 72 | 138 | -2,17399 | y ~ Intervention + Sex + |
| 0,797615 | 0,042585 | 0,165747 | 142 | 70 | 72 | 138 | 0,256931 | y ~ Intervention + Sex + |
| 0,576503 | -0,09014 | 0,161009 | 142 | 70 | 72 | 138 | -0,55983 | y ~ Intervention + Sex + |
| 0,108111 | -0,21467 | 0,132739 | 142 | 70 | 72 | 138 | -1,61723 | y ~ Intervention + Sex + |
| 0,607977 | -0,08153 | 0,158575 | 142 | 70 | 72 | 138 | -0,51414 | y ~ Intervention + Sex + |
| 0,197924 | -0,19891 | 0,153752 | 142 | 70 | 72 | 138 | -1,29372 | y ~ Intervention + Sex + |
| 0,264058 | 0,175176 | 0,15621 | 142 | 70 | 72 | 138 | 1,121416 | y ~ Intervention + Sex + |
| 0,957656 | -0,00801 | 0,150589 | 142 | 70 | 72 | 138 | -0,05319 | y ~ Intervention + Sex + |
| 0,354243 | -0,14178 | 0,152529 | 142 | 70 | 72 | 138 | -0,92952 | y ~ Intervention + Sex + |
| 0,166677 | 0,201474 | 0,144915 | 142 | 70 | 72 | 138 | 1,390296 | y ~ Intervention + Sex + |
| 0,309457 | -0,14084 | 0,138066 | 142 | 70 | 72 | 138 | -1,02012 | y ~ Intervention + Sex + |
| 0,684395 | 0,069395 | 0,170363 | 142 | 70 | 72 | 138 | 0,407333 | y ~ Intervention + Sex + |
| 0,862431 | 0,024998 | 0,143995 | 142 | 70 | 72 | 138 | 0,173605 | y ~ Intervention + Sex + |
| 0,985942 | 0,002848 | 0,161341 | 142 | 70 | 72 | 138 | 0,017653 | y ~ Intervention + Sex + |
| 0,159317 | -0,20133 | 0,142282 | 142 | 70 | 72 | 138 | -1,41502 | y ~ Intervention + Sex + |
| 0,008819 | -0,47268 | 0,177915 | 142 | 70 | 72 | 138 | -2,65677 | y ~ Intervention + Sex + |
| 0,397666 | 0,138441 | 0,163172 | 142 | 70 | 72 | 138 | 0,848434 | y ~ Intervention + Sex + |
| 0,301316 | -0,16561 | 0,159623 | 142 | 70 | 72 | 138 | -1,0375 | y ~ Intervention + Sex + |
| 0,814781 | -0,03155 | 0,134417 | 142 | 70 | 72 | 138 | -0,23471 | y ~ Intervention + Sex + |
| 0,295224 | -0,17636 | 0,167847 | 142 | 70 | 72 | 138 | -1,05072 | y ~ Intervention + Sex + |
| 0,007075 | 0,437121 | 0,159875 | 142 | 70 | 72 | 138 | 2,734149 | y ~ Intervention + Sex + |
| 0,284542 | 0,169908 | 0,15815 | 142 | 70 | 72 | 138 | 1,074348 | y ~ Intervention + Sex + |
| 0,18803 | 0,208733 | 0,157775 | 142 | 70 | 72 | 138 | 1,322979 | y ~ Intervention + Sex + |
| 0,313447 | 0,159696 | 0,157847 | 142 | 70 | 72 | 138 | 1,01171 | y ~ Intervention + Sex + |
| 0,864448 | 0,027403 | 0,160222 | 142 | 70 | 72 | 138 | 0,171033 | y ~ Intervention + Sex + |
| 0,206767 | 0,183108 | 0,144354 | 142 | 70 | 72 | 138 | 1,268467 | y ~ Intervention + Sex + |
| 0,310155 | 0,117593 | 0,115441 | 142 | 70 | 72 | 138 | 1,018643 | y ~ Intervention + Sex + |
| 0,523989 | -0,10414 | 0,163008 | 142 | 70 | 72 | 138 | -0,63884 | y ~ Intervention + Sex + |
| 0,847724 | 0,029717 | 0,154468 | 142 | 70 | 72 | 138 | 0,192384 | y ~ Intervention + Sex + |
| 0,251126 | -0,18154 | 0,157529 | 142 | 70 | 72 | 138 | -1,15246 | y ~ Intervention + Sex + |
| 0,67959 | 0,0578 | 0,139648 | 142 | 70 | 72 | 138 | 0,4139 | y ~ Intervention + Sex + |
| 0,431578 | -0,11824 | 0,149899 | 142 | 70 | 72 | 138 | -0,78881 | y ~ Intervention + Sex + |
| 0,557274 | 0,092129 | 0,156594 | 142 | 70 | 72 | 138 | 0,588328 | y ~ Intervention + Sex + |
| 0,829162 | 0,036301 | 0,167915 | 142 | 70 | 72 | 138 | 0,216186 | y ~ Intervention + Sex + |
| 0,084594 | -0,27463 | 0,158096 | 142 | 70 | 72 | 138 | -1,73714 | y ~ Intervention + Sex + |
| 0,634693 | -0,07368 | 0,154738 | 142 | 70 | 72 | 138 | -0,47619 | y ~ Intervention + Sex + |

| | | | | | | | | |
|---|---|---|---|---|---|---|---|---|
| 0,474345 | 0,121223 | 0,168977 | 142 | 70 | 72 | 138 | 0,717391 | y ~ Intervention + Sex + |
| 0,573396 | 0,090966 | 0,161172 | 142 | 70 | 72 | 138 | 0,564403 | y ~ Intervention + Sex + |
| 0,946732 | 0,010682 | 0,1596 | 142 | 70 | 72 | 138 | 0,066932 | y ~ Intervention + Sex + |
| 0,174268 | 0,222552 | 0,162963 | 142 | 70 | 72 | 138 | 1,365657 | y ~ Intervention + Sex + |
| 0,520571 | -0,10636 | 0,165121 | 142 | 70 | 72 | 138 | -0,64411 | y ~ Intervention + Sex + |
| 0,125087 | -0,24247 | 0,157129 | 142 | 70 | 72 | 138 | -1,54314 | y ~ Intervention + Sex + |
| 0,888881 | -0,02187 | 0,156223 | 142 | 70 | 72 | 138 | -0,13998 | y ~ Intervention + Sex + |
| 0,274567 | 0,183367 | 0,167159 | 142 | 70 | 72 | 138 | 1,096966 | y ~ Intervention + Sex + |
| 0,893063 | 0,023197 | 0,172239 | 142 | 70 | 72 | 138 | 0,134678 | y ~ Intervention + Sex + |
| 0,104881 | 0,258789 | 0,158535 | 142 | 70 | 72 | 138 | 1,632374 | y ~ Intervention + Sex + |
| 0,704684 | -0,06654 | 0,175197 | 142 | 70 | 72 | 138 | -0,37979 | y ~ Intervention + Sex + |
| 0,768121 | -0,04707 | 0,159346 | 142 | 70 | 72 | 138 | -0,29541 | y ~ Intervention + Sex + |
| 0,927343 | -0,01473 | 0,161286 | 142 | 70 | 72 | 138 | -0,09136 | y ~ Intervention + Sex + |
| 0,860516 | 0,027459 | 0,155974 | 142 | 70 | 72 | 138 | 0,176046 | y ~ Intervention + Sex + |
| 0,866353 | 0,027481 | 0,162991 | 142 | 70 | 72 | 138 | 0,168607 | y ~ Intervention + Sex + |
| 0,45116 | -0,12786 | 0,169217 | 142 | 70 | 72 | 138 | -0,75563 | y ~ Intervention + Sex + |
| 0,468945 | -0,11792 | 0,162376 | 142 | 70 | 72 | 138 | -0,7262 | y ~ Intervention + Sex + |
| 0,010091 | -0,43608 | 0,167167 | 142 | 70 | 72 | 138 | -2,60867 | y ~ Intervention + Sex + |
| 0,286644 | -0,15901 | 0,14866 | 142 | 70 | 72 | 138 | -1,06965 | y ~ Intervention + Sex + |
| 0,792816 | -0,04585 | 0,174231 | 142 | 70 | 72 | 138 | -0,26317 | y ~ Intervention + Sex + |
| 0,143802 | 0,220305 | 0,149854 | 142 | 70 | 72 | 138 | 1,470135 | y ~ Intervention + Sex + |
| 0,691091 | 0,059714 | 0,149957 | 142 | 70 | 72 | 138 | 0,39821 | y ~ Intervention + Sex + |
| 0,158912 | -0,23921 | 0,168887 | 142 | 70 | 72 | 138 | -1,4164 | y ~ Intervention + Sex + |
| 0,552088 | -0,0873 | 0,146457 | 142 | 70 | 72 | 138 | -0,5961 | y ~ Intervention + Sex + |
| 0,987232 | -0,00245 | 0,152895 | 142 | 70 | 72 | 138 | -0,01603 | y ~ Intervention + Sex + |
| 0,649655 | 0,068693 | 0,150895 | 142 | 70 | 72 | 138 | 0,455236 | y ~ Intervention + Sex + |
| 0,769455 | 0,040675 | 0,138509 | 142 | 70 | 72 | 138 | 0,293665 | y ~ Intervention + Sex + |
| 0,628774 | 0,070924 | 0,146375 | 142 | 70 | 72 | 138 | 0,484536 | y ~ Intervention + Sex + |
| 0,316498 | -0,14372 | 0,142955 | 142 | 70 | 72 | 138 | -1,00533 | y ~ Intervention + Sex + |
| 0,474664 | 0,105825 | 0,14762 | 142 | 70 | 72 | 138 | 0,716874 | y ~ Intervention + Sex + |
| 0,945959 | 0,010358 | 0,152537 | 142 | 70 | 72 | 138 | 0,067906 | y ~ Intervention + Sex + |
| 0,620363 | 0,087576 | 0,176403 | 142 | 70 | 72 | 138 | 0,496455 | y ~ Intervention + Sex + |
| 0,464356 | -0,10613 | 0,144646 | 142 | 70 | 72 | 138 | -0,73373 | y ~ Intervention + Sex + |
| 0,09982 | -0,25802 | 0,155727 | 142 | 70 | 72 | 138 | -1,65686 | y ~ Intervention + Sex + |
| 0,586928 | -0,08821 | 0,161979 | 142 | 70 | 72 | 138 | -0,54457 | y ~ Intervention + Sex + |
| 0,869768 | 0,025641 | 0,156099 | 142 | 70 | 72 | 138 | 0,164259 | y ~ Intervention + Sex + |
| 0,972508 | 0,00627 | 0,181593 | 142 | 70 | 72 | 138 | 0,034526 | y ~ Intervention + Sex + |
| 0,878994 | 0,025089 | 0,164486 | 142 | 70 | 72 | 138 | 0,152528 | y ~ Intervention + Sex + |
| 0,176945 | 0,230392 | 0,16976 | 142 | 70 | 72 | 138 | 1,357164 | y ~ Intervention + Sex + |
| 0,132068 | 0,194866 | 0,128627 | 142 | 70 | 72 | 138 | 1,514967 | y ~ Intervention + Sex + |
| 0,942808 | 0,012053 | 0,167706 | 142 | 70 | 72 | 138 | 0,071872 | y ~ Intervention + Sex + |
| 0,581757 | -0,08482 | 0,153628 | 142 | 70 | 72 | 138 | -0,55212 | y ~ Intervention + Sex + |
| 0,487222 | 0,109681 | 0,157451 | 142 | 70 | 72 | 138 | 0,696604 | y ~ Intervention + Sex + |
| 0,61134 | -0,0865 | 0,16983 | 142 | 70 | 72 | 138 | -0,50932 | y ~ Intervention + Sex + |
| 0,3933 | -0,13836 | 0,16157 | 142 | 70 | 72 | 138 | -0,85633 | y ~ Intervention + Sex + |
| 0,988418 | 0,00201 | 0,138192 | 142 | 70 | 72 | 138 | 0,014542 | y ~ Intervention + Sex + |
| 0,102739 | -0,26089 | 0,158827 | 142 | 70 | 72 | 138 | -1,64262 | y ~ Intervention + Sex + |
| 0,491712 | -0,11031 | 0,160009 | 142 | 70 | 72 | 138 | -0,68943 | y ~ Intervention + Sex + |

| | | | | | | | | |
|---|---|---|---|---|---|---|---|---|
| 0,16935 | 0,233365 | 0,168918 | 142 | 70 | 72 | 138 | 1,381525 | y ~ Intervention + Sex + |
| 0,992016 | 0,001223 | 0,121952 | 142 | 70 | 72 | 138 | 0,010025 | y ~ Intervention + Sex + |
| 0,122777 | -0,22163 | 0,142738 | 142 | 70 | 72 | 138 | -1,55273 | y ~ Intervention + Sex + |
| 0,348546 | -0,15812 | 0,168101 | 142 | 70 | 72 | 138 | -0,94061 | y ~ Intervention + Sex + |
| 0,241544 | -0,1778 | 0,151164 | 142 | 70 | 72 | 138 | -1,17619 | y ~ Intervention + Sex + |
| 0,184099 | -0,18626 | 0,139525 | 142 | 70 | 72 | 138 | -1,33492 | y ~ Intervention + Sex + |
| 0,394886 | 0,1214 | 0,142245 | 142 | 70 | 72 | 138 | 0,853455 | y ~ Intervention + Sex + |
| 0,410562 | 0,125554 | 0,152111 | 142 | 70 | 72 | 138 | 0,825412 | y ~ Intervention + Sex + |
| 0,467241 | 0,098021 | 0,134461 | 142 | 70 | 72 | 138 | 0,728994 | y ~ Intervention + Sex + |
| 0,656473 | -0,06832 | 0,153276 | 142 | 70 | 72 | 138 | -0,44575 | y ~ Intervention + Sex + |
| 0,030814 | -0,31072 | 0,142413 | 142 | 70 | 72 | 138 | -2,18185 | y ~ Intervention + Sex + |
| 0,565407 | -0,08817 | 0,153023 | 142 | 70 | 72 | 138 | -0,57622 | y ~ Intervention + Sex + |
| 0,296287 | 0,163037 | 0,15551 | 142 | 70 | 72 | 138 | 1,0484 | y ~ Intervention + Sex + |
| 0,349334 | 0,151176 | 0,160984 | 142 | 70 | 72 | 138 | 0,939072 | y ~ Intervention + Sex + |
| 0,291091 | -0,15511 | 0,146362 | 142 | 70 | 72 | 138 | -1,05979 | y ~ Intervention + Sex + |
| 0,250294 | 0,195062 | 0,168959 | 142 | 70 | 72 | 138 | 1,154493 | y ~ Intervention + Sex + |
| 0,963162 | -0,00686 | 0,148225 | 142 | 70 | 72 | 138 | -0,04627 | y ~ Intervention + Sex + |
| 0,418794 | 0,142103 | 0,175231 | 142 | 70 | 72 | 138 | 0,810944 | y ~ Intervention + Sex + |
| 0,146851 | 0,231715 | 0,158823 | 142 | 70 | 72 | 138 | 1,458949 | y ~ Intervention + Sex + |
| 0,49254 | -0,11419 | 0,165953 | 142 | 70 | 72 | 138 | -0,68811 | y ~ Intervention + Sex + |
| 0,657721 | -0,07568 | 0,17045 | 142 | 70 | 72 | 138 | -0,44402 | y ~ Intervention + Sex + |
| 0,618617 | -0,0769 | 0,154129 | 142 | 70 | 72 | 138 | -0,49894 | y ~ Intervention + Sex + |
| 0,805714 | 0,037194 | 0,150929 | 142 | 70 | 72 | 138 | 0,246433 | y ~ Intervention + Sex + |
| 0,324734 | -0,17592 | 0,178006 | 142 | 70 | 72 | 138 | -0,9883 | y ~ Intervention + Sex + |
| 0,792878 | -0,03681 | 0,139899 | 142 | 70 | 72 | 138 | -0,26308 | y ~ Intervention + Sex + |
| 0,463053 | 0,110298 | 0,149886 | 142 | 70 | 72 | 138 | 0,73588 | y ~ Intervention + Sex + |
| 0,201486 | -0,22022 | 0,171585 | 142 | 70 | 72 | 138 | -1,28345 | y ~ Intervention + Sex + |
| 0,019723 | 0,283957 | 0,120369 | 142 | 70 | 72 | 138 | 2,359061 | y ~ Intervention + Sex + |
| 0,182944 | 0,207276 | 0,154861 | 142 | 70 | 72 | 138 | 1,33847 | y ~ Intervention + Sex + |
| 0,070957 | -0,27461 | 0,150901 | 142 | 70 | 72 | 138 | -1,8198 | y ~ Intervention + Sex + |
| 0,466235 | 0,112565 | 0,154063 | 142 | 70 | 72 | 138 | 0,730644 | y ~ Intervention + Sex + |
| 0,36544 | -0,12578 | 0,138514 | 142 | 70 | 72 | 138 | -0,90804 | y ~ Intervention + Sex + |
| 0,309828 | -0,14436 | 0,141621 | 142 | 70 | 72 | 138 | -1,01934 | y ~ Intervention + Sex + |
| 0,997583 | 0,000495 | 0,163141 | 142 | 70 | 72 | 138 | 0,003035 | y ~ Intervention + Sex + |
| 0,066391 | -0,30019 | 0,162226 | 142 | 70 | 72 | 138 | -1,85042 | y ~ Intervention + Sex + |
| 0,735514 | 0,053638 | 0,158466 | 142 | 70 | 72 | 138 | 0,338482 | y ~ Intervention + Sex + |
| 0,950299 | -0,01003 | 0,160566 | 142 | 70 | 72 | 138 | -0,06245 | y ~ Intervention + Sex + |
| 0,443566 | 0,113263 | 0,147402 | 142 | 70 | 72 | 138 | 0,768394 | y ~ Intervention + Sex + |
| 0,295169 | 0,157961 | 0,150319 | 142 | 70 | 72 | 138 | 1,050839 | y ~ Intervention + Sex + |
| 0,183994 | 0,201285 | 0,150748 | 142 | 70 | 72 | 138 | 1,335246 | y ~ Intervention + Sex + |
| 0,584302 | -0,08112 | 0,147928 | 142 | 70 | 72 | 138 | -0,5484 | y ~ Intervention + Sex + |
| 0,857532 | 0,027906 | 0,155158 | 142 | 70 | 72 | 138 | 0,179853 | y ~ Intervention + Sex + |
| 0,807946 | -0,04222 | 0,173357 | 142 | 70 | 72 | 138 | -0,24354 | y ~ Intervention + Sex + |
| 0,666219 | 0,066965 | 0,154914 | 142 | 70 | 72 | 138 | 0,432272 | y ~ Intervention + Sex + |
| 0,240322 | 0,188743 | 0,160051 | 142 | 70 | 72 | 138 | 1,179265 | y ~ Intervention + Sex + |
| 0,102211 | 0,232557 | 0,141357 | 142 | 70 | 72 | 138 | 1,645166 | y ~ Intervention + Sex + |
| 0,915794 | 0,014409 | 0,136026 | 142 | 70 | 72 | 138 | 0,105927 | y ~ Intervention + Sex + |
| 0,855349 | -0,02741 | 0,150081 | 142 | 70 | 72 | 138 | -0,18264 | y ~ Intervention + Sex + |

| | | | | | | | | |
|---|---|---|---|---|---|---|---|---|
| 0,252453 | -0,15867 | 0,138066 | 142 | 70 | 72 | 138 | -1,14922 | y ~ Intervention + Sex + |
| 0,073342 | 0,277726 | 0,153912 | 142 | 70 | 72 | 138 | 1,80445 | y ~ Intervention + Sex + |
| 0,41008 | 0,102836 | 0,124459 | 142 | 70 | 72 | 138 | 0,826266 | y ~ Intervention + Sex + |
| 0,062533 | 0,292216 | 0,155625 | 142 | 70 | 72 | 138 | 1,877697 | y ~ Intervention + Sex + |
| 0,274489 | 0,180976 | 0,164952 | 142 | 70 | 72 | 138 | 1,097145 | y ~ Intervention + Sex + |
| 0,70537 | -0,06241 | 0,164719 | 142 | 70 | 72 | 138 | -0,37887 | y ~ Intervention + Sex + |
| 0,883842 | 0,02457 | 0,167862 | 142 | 70 | 72 | 138 | 0,146371 | y ~ Intervention + Sex + |
| 0,606957 | -0,08407 | 0,163057 | 142 | 70 | 72 | 138 | -0,5156 | y ~ Intervention + Sex + |
| 0,557301 | 0,092993 | 0,158074 | 142 | 70 | 72 | 138 | 0,588287 | y ~ Intervention + Sex + |
| 0,601167 | -0,08665 | 0,165376 | 142 | 70 | 72 | 138 | -0,52393 | y ~ Intervention + Sex + |
| 0,573853 | 0,076448 | 0,135612 | 142 | 70 | 72 | 138 | 0,56373 | y ~ Intervention + Sex + |
| 0,948428 | 0,010655 | 0,164427 | 142 | 70 | 72 | 138 | 0,064798 | y ~ Intervention + Sex + |
| 0,287858 | 0,179482 | 0,16822 | 142 | 70 | 72 | 138 | 1,066949 | y ~ Intervention + Sex + |
| 0,927789 | 0,014288 | 0,157373 | 142 | 70 | 72 | 138 | 0,090793 | y ~ Intervention + Sex + |
| 0,787256 | 0,045391 | 0,167867 | 142 | 70 | 72 | 138 | 0,2704 | y ~ Intervention + Sex + |
| 0,280339 | 0,173443 | 0,160031 | 142 | 70 | 72 | 138 | 1,083811 | y ~ Intervention + Sex + |
| 0,13589 | -0,23986 | 0,1599 | 142 | 70 | 72 | 138 | -1,50003 | y ~ Intervention + Sex + |
| 0,18259 | 0,207864 | 0,155174 | 142 | 70 | 72 | 138 | 1,33956 | y ~ Intervention + Sex + |
| 0,846614 | -0,03505 | 0,180854 | 142 | 70 | 72 | 138 | -0,1938 | y ~ Intervention + Sex + |
| 0,086512 | -0,27392 | 0,158663 | 142 | 70 | 72 | 138 | -1,72641 | y ~ Intervention + Sex + |
| 0,621849 | 0,074865 | 0,151443 | 142 | 70 | 72 | 138 | 0,494344 | y ~ Intervention + Sex + |
| 0,964252 | -0,00666 | 0,148342 | 142 | 70 | 72 | 138 | -0,0449 | y ~ Intervention + Sex + |
| 0,877838 | -0,02185 | 0,141894 | 142 | 70 | 72 | 138 | -0,154 | y ~ Intervention + Sex + |
| 0,154545 | -0,21402 | 0,149508 | 142 | 70 | 72 | 138 | -1,43151 | y ~ Intervention + Sex + |
| 0,829832 | 0,033179 | 0,154087 | 142 | 70 | 72 | 138 | 0,215324 | y ~ Intervention + Sex + |
| 0,56778 | -0,09533 | 0,166455 | 142 | 70 | 72 | 138 | -0,5727 | y ~ Intervention + Sex + |
| 0,395358 | -0,1247 | 0,146257 | 142 | 70 | 72 | 138 | -0,8526 | y ~ Intervention + Sex + |
| 0,044076 | -0,29147 | 0,143442 | 142 | 70 | 72 | 138 | -2,03196 | y ~ Intervention + Sex + |
| 0,347693 | 0,151791 | 0,161088 | 142 | 70 | 72 | 138 | 0,942286 | y ~ Intervention + Sex + |
| 0,456191 | 0,121265 | 0,162284 | 142 | 70 | 72 | 138 | 0,747238 | y ~ Intervention + Sex + |
| 0,366857 | 0,147695 | 0,163135 | 142 | 70 | 72 | 138 | 0,905351 | y ~ Intervention + Sex + |
| 0,251151 | 0,183591 | 0,159313 | 142 | 70 | 72 | 138 | 1,152396 | y ~ Intervention + Sex + |
| 0,333128 | 0,153888 | 0,158445 | 142 | 70 | 72 | 138 | 0,97124 | y ~ Intervention + Sex + |
| 0,566902 | 0,091927 | 0,160151 | 142 | 70 | 72 | 138 | 0,574 | y ~ Intervention + Sex + |
| 0,56509 | -0,08833 | 0,153161 | 142 | 70 | 72 | 138 | -0,57669 | y ~ Intervention + Sex + |
| 0,716474 | -0,06317 | 0,173588 | 142 | 70 | 72 | 138 | -0,36392 | y ~ Intervention + Sex + |
| 0,122262 | 0,215809 | 0,138794 | 142 | 70 | 72 | 138 | 1,554894 | y ~ Intervention + Sex + |
| 0,258277 | -0,1665 | 0,146675 | 142 | 70 | 72 | 138 | -1,13516 | y ~ Intervention + Sex + |
| 0,462336 | 0,107444 | 0,145774 | 142 | 70 | 72 | 138 | 0,737062 | y ~ Intervention + Sex + |
| 0,04903 | -0,28248 | 0,142244 | 142 | 70 | 72 | 138 | -1,98586 | y ~ Intervention + Sex + |
| 0,664951 | -0,06546 | 0,150818 | 142 | 70 | 72 | 138 | -0,43402 | y ~ Intervention + Sex + |
| 0,886838 | 0,021368 | 0,14988 | 142 | 70 | 72 | 138 | 0,14257 | y ~ Intervention + Sex + |
| 0,532023 | -0,10654 | 0,17005 | 142 | 70 | 72 | 138 | -0,6265 | y ~ Intervention + Sex + |
| 0,652089 | -0,07038 | 0,15577 | 142 | 70 | 72 | 138 | -0,45185 | y ~ Intervention + Sex + |
| 0,517198 | 0,11415 | 0,175793 | 142 | 70 | 72 | 138 | 0,649341 | y ~ Intervention + Sex + |
| 0,445056 | 0,135425 | 0,176823 | 142 | 70 | 72 | 138 | 0,765879 | y ~ Intervention + Sex + |
| 0,634819 | 0,067957 | 0,142763 | 142 | 70 | 72 | 138 | 0,476011 | y ~ Intervention + Sex + |
| 0,425244 | -0,12275 | 0,153493 | 142 | 70 | 72 | 138 | -0,79973 | y ~ Intervention + Sex + |

| | | | | | | | | |
|---|---|---|---|---|---|---|---|---|
| 0,900983 | -0,01863 | 0,149461 | 142 | 70 | 72 | 138 | -0,12465 | y ~ Intervention + Sex + |
| 0,80624 | 0,03537 | 0,143926 | 142 | 70 | 72 | 138 | 0,245751 | y ~ Intervention + Sex + |
| 0,296155 | -0,16063 | 0,153172 | 142 | 70 | 72 | 138 | -1,04869 | y ~ Intervention + Sex + |
| 0,459589 | -0,11876 | 0,160144 | 142 | 70 | 72 | 138 | -0,7416 | y ~ Intervention + Sex + |
| 0,385949 | -0,1295 | 0,14889 | 142 | 70 | 72 | 138 | -0,86975 | y ~ Intervention + Sex + |
| 0,397912 | 0,120215 | 0,141765 | 142 | 70 | 72 | 138 | 0,84799 | y ~ Intervention + Sex + |
| 0,373469 | 0,157548 | 0,176447 | 142 | 70 | 72 | 138 | 0,892892 | y ~ Intervention + Sex + |
| 0,661233 | 0,07721 | 0,175814 | 142 | 70 | 72 | 138 | 0,439159 | y ~ Intervention + Sex + |
| 0,698837 | 0,054757 | 0,141236 | 142 | 70 | 72 | 138 | 0,387698 | y ~ Intervention + Sex + |
| 0,794818 | 0,041966 | 0,161058 | 142 | 70 | 72 | 138 | 0,260563 | y ~ Intervention + Sex + |
| 0,371248 | 0,139831 | 0,155877 | 142 | 70 | 72 | 138 | 0,897063 | y ~ Intervention + Sex + |
| 0,140356 | -0,24931 | 0,168113 | 142 | 70 | 72 | 138 | -1,483 | y ~ Intervention + Sex + |
| 0,197408 | 0,177321 | 0,136905 | 142 | 70 | 72 | 138 | 1,295216 | y ~ Intervention + Sex + |
| 0,491003 | 0,114661 | 0,166041 | 142 | 70 | 72 | 138 | 0,690558 | y ~ Intervention + Sex + |
| 0,513837 | -0,09873 | 0,150832 | 142 | 70 | 72 | 138 | -0,65456 | y ~ Intervention + Sex + |
| 0,912865 | -0,01771 | 0,161515 | 142 | 70 | 72 | 138 | -0,10963 | y ~ Intervention + Sex + |
| 0,872644 | -0,02569 | 0,159979 | 142 | 70 | 72 | 138 | -0,1606 | y ~ Intervention + Sex + |
| 0,093056 | 0,267546 | 0,158199 | 142 | 70 | 72 | 138 | 1,691198 | y ~ Intervention + Sex + |
| 0,420032 | 0,124257 | 0,153634 | 142 | 70 | 72 | 138 | 0,808782 | y ~ Intervention + Sex + |
| 0,392357 | 0,150173 | 0,175018 | 142 | 70 | 72 | 138 | 0,858041 | y ~ Intervention + Sex + |
| 0,010648 | 0,374002 | 0,14444 | 142 | 70 | 72 | 138 | 2,589315 | y ~ Intervention + Sex + |
| 0,705059 | 0,054144 | 0,142752 | 142 | 70 | 72 | 138 | 0,379286 | y ~ Intervention + Sex + |
| 0,572615 | -0,08838 | 0,156262 | 142 | 70 | 72 | 138 | -0,56556 | y ~ Intervention + Sex + |
| 0,168115 | 0,203893 | 0,147155 | 142 | 70 | 72 | 138 | 1,385564 | y ~ Intervention + Sex + |
| 0,268728 | 0,16588 | 0,149378 | 142 | 70 | 72 | 138 | 1,110469 | y ~ Intervention + Sex + |
| 0,467277 | 0,108251 | 0,148506 | 142 | 70 | 72 | 138 | 0,728935 | y ~ Intervention + Sex + |
| 0,734398 | -0,05683 | 0,167173 | 142 | 70 | 72 | 138 | -0,33997 | y ~ Intervention + Sex + |
| 0,836381 | -0,02861 | 0,138252 | 142 | 70 | 72 | 138 | -0,20692 | y ~ Intervention + Sex + |
| 0,865405 | -0,02747 | 0,161754 | 142 | 70 | 72 | 138 | -0,16981 | y ~ Intervention + Sex + |
| 0,023598 | -0,30818 | 0,134637 | 142 | 70 | 72 | 138 | -2,289 | y ~ Intervention + Sex + |
| 0,259268 | 0,196695 | 0,173638 | 142 | 70 | 72 | 138 | 1,132786 | y ~ Intervention + Sex + |
| 0,654263 | 0,077747 | 0,173225 | 142 | 70 | 72 | 138 | 0,448824 | y ~ Intervention + Sex + |
| 0,493806 | 0,09975 | 0,145388 | 142 | 70 | 72 | 138 | 0,686092 | y ~ Intervention + Sex + |
| 0,051106 | 0,319545 | 0,162394 | 142 | 70 | 72 | 138 | 1,967711 | y ~ Intervention + Sex + |
| 0,745471 | -0,056 | 0,172167 | 142 | 70 | 72 | 138 | -0,32527 | y ~ Intervention + Sex + |
| 0,426152 | -0,11372 | 0,142481 | 142 | 70 | 72 | 138 | -0,79815 | y ~ Intervention + Sex + |
| 0,839282 | 0,032029 | 0,157629 | 142 | 70 | 72 | 138 | 0,203195 | y ~ Intervention + Sex + |
| 0,563072 | -0,09992 | 0,172373 | 142 | 70 | 72 | 138 | -0,57969 | y ~ Intervention + Sex + |
| 0,027293 | 0,32877 | 0,147362 | 142 | 70 | 72 | 138 | 2,23103 | y ~ Intervention + Sex + |
| 0,625062 | 0,076497 | 0,156185 | 142 | 70 | 72 | 138 | 0,489787 | y ~ Intervention + Sex + |
| 0,611192 | 0,077625 | 0,152346 | 142 | 70 | 72 | 138 | 0,509534 | y ~ Intervention + Sex + |
| 0,625368 | -0,07572 | 0,154725 | 142 | 70 | 72 | 138 | -0,48935 | y ~ Intervention + Sex + |
| 0,241488 | 0,191344 | 0,162662 | 142 | 70 | 72 | 138 | 1,176331 | y ~ Intervention + Sex + |
| 0,282945 | 0,179317 | 0,166353 | 142 | 70 | 72 | 138 | 1,077932 | y ~ Intervention + Sex + |
| 0,210048 | -0,1993 | 0,158266 | 142 | 70 | 72 | 138 | -1,2593 | y ~ Intervention + Sex + |
| 0,898359 | 0,019987 | 0,156186 | 142 | 70 | 72 | 138 | 0,127971 | y ~ Intervention + Sex + |
| 0,290921 | -0,17513 | 0,165195 | 142 | 70 | 72 | 138 | -1,06017 | y ~ Intervention + Sex + |
| 0,223948 | 0,194943 | 0,159582 | 142 | 70 | 72 | 138 | 1,221583 | y ~ Intervention + Sex + |

| | | | | | | | | |
|---|---|---|---|---|---|---|---|---|
| 0,578818 | -0,0861 | 0,154734 | 142 | 70 | 72 | 138 | -0,55643 | y ~ Intervention + Sex + |
| 0,887711 | -0,02429 | 0,171691 | 142 | 70 | 72 | 138 | -0,14146 | y ~ Intervention + Sex + |
| 0,474664 | 0,105825 | 0,14762 | 142 | 70 | 72 | 138 | 0,716874 | y ~ Intervention + Sex + |
| 0,815256 | 0,040982 | 0,175066 | 142 | 70 | 72 | 138 | 0,234097 | y ~ Intervention + Sex + |
| 0,940107 | -0,01169 | 0,155338 | 142 | 70 | 72 | 138 | -0,07527 | y ~ Intervention + Sex + |
| 0,745572 | 0,05312 | 0,163379 | 142 | 70 | 72 | 138 | 0,325134 | y ~ Intervention + Sex + |
| 0,378732 | 0,168996 | 0,191373 | 142 | 70 | 72 | 138 | 0,883074 | y ~ Intervention + Sex + |
| 0,428452 | -0,12658 | 0,159379 | 142 | 70 | 72 | 138 | -0,79418 | y ~ Intervention + Sex + |
| 0,296489 | 0,173341 | 0,165408 | 142 | 70 | 72 | 138 | 1,04796 | y ~ Intervention + Sex + |
| 0,865989 | 0,024154 | 0,142865 | 142 | 70 | 72 | 138 | 0,16907 | y ~ Intervention + Sex + |
| 0,516781 | 0,10386 | 0,159788 | 142 | 70 | 72 | 138 | 0,649988 | y ~ Intervention + Sex + |
| 0,050949 | -0,29228 | 0,148435 | 142 | 70 | 72 | 138 | -1,96906 | y ~ Intervention + Sex + |
| 0,572971 | 0,08929 | 0,158027 | 142 | 70 | 72 | 138 | 0,56503 | y ~ Intervention + Sex + |
| 0,705183 | 0,058091 | 0,153227 | 142 | 70 | 72 | 138 | 0,379118 | y ~ Intervention + Sex + |
| 0,643876 | 0,070709 | 0,152619 | 142 | 70 | 72 | 138 | 0,463304 | y ~ Intervention + Sex + |
| 0,174021 | -0,19304 | 0,141271 | 142 | 70 | 72 | 138 | -1,36645 | y ~ Intervention + Sex + |
| 0,237112 | -0,17803 | 0,14993 | 142 | 70 | 72 | 138 | -1,1874 | y ~ Intervention + Sex + |
| 0,252304 | -0,17446 | 0,15176 | 142 | 70 | 72 | 138 | -1,14958 | y ~ Intervention + Sex + |
| 0,232375 | 0,164283 | 0,136955 | 142 | 70 | 72 | 138 | 1,199537 | y ~ Intervention + Sex + |
| 0,841353 | -0,03327 | 0,165911 | 142 | 70 | 72 | 138 | -0,20054 | y ~ Intervention + Sex + |
| 0,834304 | -0,03621 | 0,172783 | 142 | 70 | 72 | 138 | -0,20958 | y ~ Intervention + Sex + |
| 0,377003 | -0,13822 | 0,15595 | 142 | 70 | 72 | 138 | -0,88629 | y ~ Intervention + Sex + |
| 0,97793 | -0,00389 | 0,140488 | 142 | 70 | 72 | 138 | -0,02771 | y ~ Intervention + Sex + |
| 0,507767 | 0,105069 | 0,158225 | 142 | 70 | 72 | 138 | 0,664049 | y ~ Intervention + Sex + |
| 0,852994 | 0,027419 | 0,147695 | 142 | 70 | 72 | 138 | 0,185647 | y ~ Intervention + Sex + |
| 0,97749 | 0,004574 | 0,161801 | 142 | 70 | 72 | 138 | 0,028267 | y ~ Intervention + Sex + |
| 0,625591 | 0,06356 | 0,129969 | 142 | 70 | 72 | 138 | 0,489039 | y ~ Intervention + Sex + |
| 0,869218 | -0,02451 | 0,148576 | 142 | 70 | 72 | 138 | -0,16496 | y ~ Intervention + Sex + |
| 0,832773 | 0,033753 | 0,159553 | 142 | 70 | 72 | 138 | 0,211546 | y ~ Intervention + Sex + |
| 0,333252 | 0,170181 | 0,175266 | 142 | 70 | 72 | 138 | 0,97099 | y ~ Intervention + Sex + |
| 0,844783 | -0,02983 | 0,152097 | 142 | 70 | 72 | 138 | -0,19615 | y ~ Intervention + Sex + |
| 0,239425 | -0,16896 | 0,143004 | 142 | 70 | 72 | 138 | -1,18153 | y ~ Intervention + Sex + |
| 0,819329 | 0,035265 | 0,154101 | 142 | 70 | 72 | 138 | 0,228844 | y ~ Intervention + Sex + |
| 0,547436 | -0,09335 | 0,154791 | 142 | 70 | 72 | 138 | -0,60309 | y ~ Intervention + Sex + |
| 0,996052 | 0,000696 | 0,140305 | 142 | 70 | 72 | 138 | 0,004957 | y ~ Intervention + Sex + |
| 0,18828 | -0,21101 | 0,159583 | 142 | 70 | 72 | 138 | -1,32223 | y ~ Intervention + Sex + |
| 0,455448 | -0,1095 | 0,146297 | 142 | 70 | 72 | 138 | -0,74847 | y ~ Intervention + Sex + |
| 0,737162 | -0,05052 | 0,150218 | 142 | 70 | 72 | 138 | -0,33629 | y ~ Intervention + Sex + |
| 0,338127 | 0,163102 | 0,169684 | 142 | 70 | 72 | 138 | 0,961211 | y ~ Intervention + Sex + |
| 0,395379 | -0,12818 | 0,150344 | 142 | 70 | 72 | 138 | -0,85256 | y ~ Intervention + Sex + |
| 0,693455 | 0,061656 | 0,156092 | 142 | 70 | 72 | 138 | 0,394996 | y ~ Intervention + Sex + |
| 0,144639 | 0,240335 | 0,163823 | 142 | 70 | 72 | 138 | 1,467044 | y ~ Intervention + Sex + |
| 0,68973 | -0,05591 | 0,139749 | 142 | 70 | 72 | 138 | -0,40006 | y ~ Intervention + Sex + |
| 0,611644 | -0,07819 | 0,153642 | 142 | 70 | 72 | 138 | -0,50889 | y ~ Intervention + Sex + |
| 0,284512 | 0,178139 | 0,165801 | 142 | 70 | 72 | 138 | 1,074414 | y ~ Intervention + Sex + |
| 0,958027 | 0,007312 | 0,13868 | 142 | 70 | 72 | 138 | 0,052726 | y ~ Intervention + Sex + |
| 0,423493 | -0,13213 | 0,164591 | 142 | 70 | 72 | 138 | -0,80276 | y ~ Intervention + Sex + |
| 0,877451 | -0,0233 | 0,150839 | 142 | 70 | 72 | 138 | -0,15449 | y ~ Intervention + Sex + |

| | | | | | | | | |
|---|---|---|---|---|---|---|---|---|
| 0,171394 | -0,18153 | 0,13203 | 142 | 70 | 72 | 138 | -1,37489 | y ~ Intervention + Sex + |
| 0,75567 | -0,05323 | 0,170735 | 142 | 70 | 72 | 138 | -0,31179 | y ~ Intervention + Sex + |
| 0,598502 | -0,08187 | 0,155115 | 142 | 70 | 72 | 138 | -0,52778 | y ~ Intervention + Sex + |
| 0,921664 | -0,01623 | 0,164709 | 142 | 70 | 72 | 138 | -0,09852 | y ~ Intervention + Sex + |
| 0,35306 | 0,149027 | 0,159933 | 142 | 70 | 72 | 138 | 0,931814 | y ~ Intervention + Sex + |
| 0,800037 | -0,04291 | 0,169068 | 142 | 70 | 72 | 138 | -0,25379 | y ~ Intervention + Sex + |
| 0,401823 | -0,13702 | 0,162927 | 142 | 70 | 72 | 138 | -0,84096 | y ~ Intervention + Sex + |
| 0,378608 | -0,12653 | 0,143248 | 142 | 70 | 72 | 138 | -0,88331 | y ~ Intervention + Sex + |
| 0,74401 | -0,05021 | 0,153457 | 142 | 70 | 72 | 138 | -0,3272 | y ~ Intervention + Sex + |
| 0,341937 | 0,151647 | 0,159021 | 142 | 70 | 72 | 138 | 0,953633 | y ~ Intervention + Sex + |
| 0,759573 | 0,049489 | 0,161386 | 142 | 70 | 72 | 138 | 0,306649 | y ~ Intervention + Sex + |
| 0,127047 | -0,2312 | 0,150605 | 142 | 70 | 72 | 138 | -1,53511 | y ~ Intervention + Sex + |
| 0,934966 | 0,013574 | 0,166051 | 142 | 70 | 72 | 138 | 0,081748 | y ~ Intervention + Sex + |
| 0,811721 | 0,039831 | 0,166893 | 142 | 70 | 72 | 138 | 0,238663 | y ~ Intervention + Sex + |
| 0,555649 | -0,09537 | 0,161443 | 142 | 70 | 72 | 138 | -0,59076 | y ~ Intervention + Sex + |
| 0,106801 | 0,236678 | 0,145798 | 142 | 70 | 72 | 138 | 1,623331 | y ~ Intervention + Sex + |
| 0,089962 | -0,2839 | 0,166256 | 142 | 70 | 72 | 138 | -1,70759 | y ~ Intervention + Sex + |
| 0,797003 | 0,044089 | 0,171069 | 142 | 70 | 72 | 138 | 0,257726 | y ~ Intervention + Sex + |
| 0,76214 | 0,049633 | 0,163659 | 142 | 70 | 72 | 138 | 0,303271 | y ~ Intervention + Sex + |
| 0,371707 | 0,151352 | 0,168882 | 142 | 70 | 72 | 138 | 0,896199 | y ~ Intervention + Sex + |
| 0,942861 | 0,012365 | 0,1722 | 142 | 70 | 72 | 138 | 0,071805 | y ~ Intervention + Sex + |
| 0,788602 | 0,041981 | 0,156266 | 142 | 70 | 72 | 138 | 0,268647 | y ~ Intervention + Sex + |
| 0,635344 | 0,068078 | 0,143239 | 142 | 70 | 72 | 138 | 0,475273 | y ~ Intervention + Sex + |
| 0,375449 | -0,12654 | 0,142309 | 142 | 70 | 72 | 138 | -0,88919 | y ~ Intervention + Sex + |
| 0,108415 | -0,27212 | 0,168407 | 142 | 70 | 72 | 138 | -1,61583 | y ~ Intervention + Sex + |
| 0,519596 | 0,106495 | 0,164949 | 142 | 70 | 72 | 138 | 0,645623 | y ~ Intervention + Sex + |
| 0,767618 | -0,04 | 0,135095 | 142 | 70 | 72 | 138 | -0,29607 | y ~ Intervention + Sex + |
| 0,701118 | 0,056966 | 0,148115 | 142 | 70 | 72 | 138 | 0,38461 | y ~ Intervention + Sex + |
| 0,679965 | 0,072178 | 0,174601 | 142 | 70 | 72 | 138 | 0,413386 | y ~ Intervention + Sex + |
| 0,482375 | 0,113512 | 0,161149 | 142 | 70 | 72 | 138 | 0,704393 | y ~ Intervention + Sex + |
| 0,258676 | -0,16152 | 0,142406 | 142 | 70 | 72 | 138 | -1,1342 | y ~ Intervention + Sex + |
| 0,03013 | 0,341657 | 0,155936 | 142 | 70 | 72 | 138 | 2,191005 | y ~ Intervention + Sex + |
| 0,555883 | 0,088379 | 0,149691 | 142 | 70 | 72 | 138 | 0,590407 | y ~ Intervention + Sex + |
| 0,919166 | 0,016659 | 0,163852 | 142 | 70 | 72 | 138 | 0,10167 | y ~ Intervention + Sex + |
| 0,96963 | -0,00529 | 0,138756 | 142 | 70 | 72 | 138 | -0,03814 | y ~ Intervention + Sex + |
| 0,371491 | -0,14776 | 0,164798 | 142 | 70 | 72 | 138 | -0,89661 | y ~ Intervention + Sex + |
| 0,724712 | 0,049466 | 0,140176 | 142 | 70 | 72 | 138 | 0,352887 | y ~ Intervention + Sex + |
| 0,886675 | -0,02308 | 0,161623 | 142 | 70 | 72 | 138 | -0,14278 | y ~ Intervention + Sex + |
| 0,317348 | -0,15942 | 0,158852 | 142 | 70 | 72 | 138 | -1,00356 | y ~ Intervention + Sex + |
| 0,359539 | 0,149656 | 0,162792 | 142 | 70 | 72 | 138 | 0,919307 | y ~ Intervention + Sex + |
| 0,70637 | 0,057866 | 0,153282 | 142 | 70 | 72 | 138 | 0,377516 | y ~ Intervention + Sex + |
| 0,887755 | -0,02023 | 0,143053 | 142 | 70 | 72 | 138 | -0,14141 | y ~ Intervention + Sex + |
| 0,508122 | 0,110046 | 0,165858 | 142 | 70 | 72 | 138 | 0,663493 | y ~ Intervention + Sex + |
| 0,050005 | -0,30766 | 0,155601 | 142 | 70 | 72 | 138 | -1,97726 | y ~ Intervention + Sex + |
| 0,573326 | 0,093441 | 0,165527 | 142 | 70 | 72 | 138 | 0,564506 | y ~ Intervention + Sex + |
| 0,117506 | -0,22982 | 0,145903 | 142 | 70 | 72 | 138 | -1,57518 | y ~ Intervention + Sex + |
| 0,166759 | -0,21981 | 0,158135 | 142 | 70 | 72 | 138 | -1,39003 | y ~ Intervention + Sex + |
| 0,11127 | 0,247167 | 0,154211 | 142 | 70 | 72 | 138 | 1,602779 | y ~ Intervention + Sex + |

| | | | | | | | | |
|---|---|---|---|---|---|---|---|---|
| 0,704945 | 0,064977 | 0,171245 | 142 | 70 | 72 | 138 | 0,379439 | y ~ Intervention + Sex + |
| 0,112679 | 0,221416 | 0,138694 | 142 | 70 | 72 | 138 | 1,596436 | y ~ Intervention + Sex + |
| 0,761755 | 0,047553 | 0,156539 | 142 | 70 | 72 | 138 | 0,303777 | y ~ Intervention + Sex + |
| 0,666146 | -0,05712 | 0,132107 | 142 | 70 | 72 | 138 | -0,43237 | y ~ Intervention + Sex + |
| 0,826082 | 0,034956 | 0,158784 | 142 | 70 | 72 | 138 | 0,220148 | y ~ Intervention + Sex + |
| 0,637652 | -0,0655 | 0,138773 | 142 | 70 | 72 | 138 | -0,47203 | y ~ Intervention + Sex + |
| 0,510429 | -0,10541 | 0,159745 | 142 | 70 | 72 | 138 | -0,65988 | y ~ Intervention + Sex + |
| 0,780549 | 0,03521 | 0,126133 | 142 | 70 | 72 | 138 | 0,279148 | y ~ Intervention + Sex + |
| 0,085565 | -0,25444 | 0,146933 | 142 | 70 | 72 | 138 | -1,73168 | y ~ Intervention + Sex + |
| 0,670223 | -0,06322 | 0,148133 | 142 | 70 | 72 | 138 | -0,42675 | y ~ Intervention + Sex + |
| 0,166141 | -0,21019 | 0,150992 | 142 | 70 | 72 | 138 | -1,39207 | y ~ Intervention + Sex + |
| 0,261523 | -0,15585 | 0,13824 | 142 | 70 | 72 | 138 | -1,12742 | y ~ Intervention + Sex + |
| 0,423241 | 0,121973 | 0,151859 | 142 | 70 | 72 | 138 | 0,803199 | y ~ Intervention + Sex + |
| 0,580436 | 0,066473 | 0,119976 | 142 | 70 | 72 | 138 | 0,554058 | y ~ Intervention + Sex + |
| 0,855506 | -0,02437 | 0,133602 | 142 | 70 | 72 | 138 | -0,18244 | y ~ Intervention + Sex + |
| 0,230437 | 0,186268 | 0,154637 | 142 | 70 | 72 | 138 | 1,204555 | y ~ Intervention + Sex + |
| 0,158673 | 0,207703 | 0,146557 | 142 | 70 | 72 | 138 | 1,417216 | y ~ Intervention + Sex + |
| 0,986173 | 0,002676 | 0,154126 | 142 | 70 | 72 | 138 | 0,017362 | y ~ Intervention + Sex + |
| 0,963328 | 0,007081 | 0,153728 | 142 | 70 | 72 | 138 | 0,046061 | y ~ Intervention + Sex + |
| 0,602057 | 0,079483 | 0,152077 | 142 | 70 | 72 | 138 | 0,522648 | y ~ Intervention + Sex + |
| 0,63854 | 0,068949 | 0,146458 | 142 | 70 | 72 | 138 | 0,470781 | y ~ Intervention + Sex + |
| 0,57721 | -0,09223 | 0,165053 | 142 | 70 | 72 | 138 | -0,55879 | y ~ Intervention + Sex + |
| 0,646452 | 0,06723 | 0,146246 | 142 | 70 | 72 | 138 | 0,459705 | y ~ Intervention + Sex + |
| 0,735402 | -0,05333 | 0,157482 | 142 | 70 | 72 | 138 | -0,33863 | y ~ Intervention + Sex + |
| 0,323361 | -0,16242 | 0,163871 | 142 | 70 | 72 | 138 | -0,99112 | y ~ Intervention + Sex + |
| 0,178888 | 0,218621 | 0,161815 | 142 | 70 | 72 | 138 | 1,351056 | y ~ Intervention + Sex + |
| 0,186556 | 0,206176 | 0,155319 | 142 | 70 | 72 | 138 | 1,327438 | y ~ Intervention + Sex + |
| 0,261368 | -0,17522 | 0,155363 | 142 | 70 | 72 | 138 | -1,12779 | y ~ Intervention + Sex + |
| 0,511595 | 0,089361 | 0,135795 | 142 | 70 | 72 | 138 | 0,658061 | y ~ Intervention + Sex + |
| 0,329992 | 0,161309 | 0,165009 | 142 | 70 | 72 | 138 | 0,977581 | y ~ Intervention + Sex + |
| 0,05723 | -0,32147 | 0,167646 | 142 | 70 | 72 | 138 | -1,91759 | y ~ Intervention + Sex + |
| 0,902449 | -0,02042 | 0,166292 | 142 | 70 | 72 | 138 | -0,12279 | y ~ Intervention + Sex + |
| 0,799368 | 0,042713 | 0,167728 | 142 | 70 | 72 | 138 | 0,254655 | y ~ Intervention + Sex + |
| 0,598867 | -0,08074 | 0,153139 | 142 | 70 | 72 | 138 | -0,52725 | y ~ Intervention + Sex + |
| 0,826262 | 0,036548 | 0,166191 | 142 | 70 | 72 | 138 | 0,219915 | y ~ Intervention + Sex + |
| 0,751101 | -0,04689 | 0,147536 | 142 | 70 | 72 | 138 | -0,31782 | y ~ Intervention + Sex + |
| 0,134644 | -0,24012 | 0,15956 | 142 | 70 | 72 | 138 | -1,50487 | y ~ Intervention + Sex + |
| 0,2282 | 0,159122 | 0,131464 | 142 | 70 | 72 | 138 | 1,210385 | y ~ Intervention + Sex + |
| 0,894765 | 0,02142 | 0,161636 | 142 | 70 | 72 | 138 | 0,132521 | y ~ Intervention + Sex + |
| 0,057021 | -0,30363 | 0,158208 | 142 | 70 | 72 | 138 | -1,91922 | y ~ Intervention + Sex + |
| 0,082684 | -0,24476 | 0,140021 | 142 | 70 | 72 | 138 | -1,74802 | y ~ Intervention + Sex + |
| 0,310565 | -0,15529 | 0,152574 | 142 | 70 | 72 | 138 | -1,01778 | y ~ Intervention + Sex + |
| 0,721447 | -0,04617 | 0,129238 | 142 | 70 | 72 | 138 | -0,35725 | y ~ Intervention + Sex + |
| 0,261022 | -0,1406 | 0,124582 | 142 | 70 | 72 | 138 | -1,12861 | y ~ Intervention + Sex + |
| 0,675152 | 0,059983 | 0,142822 | 142 | 70 | 72 | 138 | 0,419981 | y ~ Intervention + Sex + |
| 0,581883 | -0,08237 | 0,149229 | 142 | 70 | 72 | 138 | -0,55194 | y ~ Intervention + Sex + |
| 0,68146 | 0,064666 | 0,157209 | 142 | 70 | 72 | 138 | 0,411342 | y ~ Intervention + Sex + |
| 0,908253 | 0,018802 | 0,162853 | 142 | 70 | 72 | 138 | 0,115455 | y ~ Intervention + Sex + |

| | | | | | | | | |
|---|---|---|---|---|---|---|---|---|
| 0,524702 | 0,100399 | 0,157431 | 142 | 70 | 72 | 138 | 0,637737 | y ~ Intervention + Sex + |
| 0,965564 | 0,007263 | 0,167928 | 142 | 70 | 72 | 138 | 0,043251 | y ~ Intervention + Sex + |
| 0,763148 | 0,042648 | 0,141244 | 142 | 70 | 72 | 138 | 0,301946 | y ~ Intervention + Sex + |
| 0,73764 | -0,0463 | 0,137947 | 142 | 70 | 72 | 138 | -0,33566 | y ~ Intervention + Sex + |
| 0,480976 | 0,113518 | 0,160643 | 142 | 70 | 72 | 138 | 0,706649 | y ~ Intervention + Sex + |
| 0,07575 | -0,26596 | 0,148631 | 142 | 70 | 72 | 138 | -1,78936 | y ~ Intervention + Sex + |
| 0,903147 | 0,019605 | 0,160812 | 142 | 70 | 72 | 138 | 0,12191 | y ~ Intervention + Sex + |
| 0,062099 | -0,25098 | 0,13344 | 142 | 70 | 72 | 138 | -1,88085 | y ~ Intervention + Sex + |
| 0,863444 | -0,02675 | 0,155245 | 142 | 70 | 72 | 138 | -0,17231 | y ~ Intervention + Sex + |
| 0,231205 | 0,171187 | 0,142351 | 142 | 70 | 72 | 138 | 1,202563 | y ~ Intervention + Sex + |
| 0,793049 | 0,042155 | 0,160369 | 142 | 70 | 72 | 138 | 0,262862 | y ~ Intervention + Sex + |
| 0,341628 | 0,137 | 0,143569 | 142 | 70 | 72 | 138 | 0,954245 | y ~ Intervention + Sex + |
| 0,444819 | -0,11313 | 0,147629 | 142 | 70 | 72 | 138 | -0,76628 | y ~ Intervention + Sex + |
| 0,240005 | -0,17954 | 0,152147 | 142 | 70 | 72 | 138 | -1,18007 | y ~ Intervention + Sex + |
| 0,474322 | 0,108987 | 0,151913 | 142 | 70 | 72 | 138 | 0,71743 | y ~ Intervention + Sex + |
| 0,576962 | -0,08956 | 0,160163 | 142 | 70 | 72 | 138 | -0,55916 | y ~ Intervention + Sex + |
| 0,585049 | 0,077195 | 0,141043 | 142 | 70 | 72 | 138 | 0,547311 | y ~ Intervention + Sex + |
| 0,880632 | -0,0194 | 0,128967 | 142 | 70 | 72 | 138 | -0,15045 | y ~ Intervention + Sex + |
| 0,353328 | -0,14453 | 0,155195 | 142 | 70 | 72 | 138 | -0,93129 | y ~ Intervention + Sex + |
| 0,493063 | 0,104798 | 0,152483 | 142 | 70 | 72 | 138 | 0,687275 | y ~ Intervention + Sex + |
| 0,889743 | 0,022887 | 0,164794 | 142 | 70 | 72 | 138 | 0,138886 | y ~ Intervention + Sex + |
| 0,161458 | 0,233985 | 0,166214 | 142 | 70 | 72 | 138 | 1,407736 | y ~ Intervention + Sex + |
| 0,906762 | 0,020366 | 0,173564 | 142 | 70 | 72 | 138 | 0,117339 | y ~ Intervention + Sex + |
| 0,594533 | -0,08378 | 0,15703 | 142 | 70 | 72 | 138 | -0,53352 | y ~ Intervention + Sex + |
| 0,810718 | 0,038989 | 0,162482 | 142 | 70 | 72 | 138 | 0,239959 | y ~ Intervention + Sex + |
| 0,223179 | -0,17983 | 0,146965 | 142 | 70 | 72 | 138 | -1,22362 | y ~ Intervention + Sex + |
| 0,982215 | -0,00277 | 0,124211 | 142 | 70 | 72 | 138 | -0,02233 | y ~ Intervention + Sex + |
| 0,406662 | 0,132706 | 0,15944 | 142 | 70 | 72 | 138 | 0,832328 | y ~ Intervention + Sex + |
| 0,446078 | -0,11049 | 0,144586 | 142 | 70 | 72 | 138 | -0,76416 | y ~ Intervention + Sex + |
| 0,952697 | 0,010443 | 0,175729 | 142 | 70 | 72 | 138 | 0,059428 | y ~ Intervention + Sex + |
| 0,40752 | -0,13611 | 0,163824 | 142 | 70 | 72 | 138 | -0,8308 | y ~ Intervention + Sex + |
| 0,869445 | -0,02511 | 0,152483 | 142 | 70 | 72 | 138 | -0,16467 | y ~ Intervention + Sex + |
| 0,232674 | 0,198583 | 0,165656 | 142 | 70 | 72 | 138 | 1,198763 | y ~ Intervention + Sex + |
| 0,84853 | -0,02788 | 0,145718 | 142 | 70 | 72 | 138 | -0,19135 | y ~ Intervention + Sex + |
| 0,068022 | -0,29381 | 0,159743 | 142 | 70 | 72 | 138 | -1,83929 | y ~ Intervention + Sex + |
| 0,095084 | 0,245718 | 0,1462 | 142 | 70 | 72 | 138 | 1,680697 | y ~ Intervention + Sex + |
| 0,711834 | 0,059942 | 0,161938 | 142 | 70 | 72 | 138 | 0,370156 | y ~ Intervention + Sex + |
| 0,582771 | -0,0826 | 0,149999 | 142 | 70 | 72 | 138 | -0,55064 | y ~ Intervention + Sex + |
| 0,62624 | 0,077953 | 0,1597 | 142 | 70 | 72 | 138 | 0,488119 | y ~ Intervention + Sex + |
| 0,44603 | 0,117874 | 0,154238 | 142 | 70 | 72 | 138 | 0,764239 | y ~ Intervention + Sex + |
| 0,960362 | -0,00759 | 0,152502 | 142 | 70 | 72 | 138 | -0,04979 | y ~ Intervention + Sex + |
| 0,670626 | 0,065002 | 0,152515 | 142 | 70 | 72 | 138 | 0,426201 | y ~ Intervention + Sex + |
| 0,595445 | 0,076534 | 0,143808 | 142 | 70 | 72 | 138 | 0,532198 | y ~ Intervention + Sex + |
| 0,744907 | -0,04889 | 0,149958 | 142 | 70 | 72 | 138 | -0,32601 | y ~ Intervention + Sex + |
| 0,384974 | 0,123129 | 0,141277 | 142 | 70 | 72 | 138 | 0,871538 | y ~ Intervention + Sex + |
| 0,770365 | 0,046136 | 0,157743 | 142 | 70 | 72 | 138 | 0,292472 | y ~ Intervention + Sex + |
| 0,499879 | -0,09881 | 0,146075 | 142 | 70 | 72 | 138 | -0,67646 | y ~ Intervention + Sex + |
| 0,318646 | -0,15322 | 0,15309 | 142 | 70 | 72 | 138 | -1,00086 | y ~ Intervention + Sex + |

| | | | | | | | | |
|---|---|---|---|---|---|---|---|---|
| 0,851331 | 0,028393 | 0,151211 | 142 | 70 | 72 | 138 | 0,187772 | y ~ Intervention + Sex + |
| 0,849043 | 0,028628 | 0,150122 | 142 | 70 | 72 | 138 | 0,190697 | y ~ Intervention + Sex + |
| 0,913379 | 0,015356 | 0,140912 | 142 | 70 | 72 | 138 | 0,108977 | y ~ Intervention + Sex + |
| 0,353442 | 0,164288 | 0,17645 | 142 | 70 | 72 | 138 | 0,931073 | y ~ Intervention + Sex + |
| 0,036217 | -0,31051 | 0,146804 | 142 | 70 | 72 | 138 | -2,11511 | y ~ Intervention + Sex + |
| 0,329106 | 0,149526 | 0,152674 | 142 | 70 | 72 | 138 | 0,979381 | y ~ Intervention + Sex + |
| 0,460087 | 0,119067 | 0,160732 | 142 | 70 | 72 | 138 | 0,740778 | y ~ Intervention + Sex + |
| 0,570276 | 0,090191 | 0,158506 | 142 | 70 | 72 | 138 | 0,569008 | y ~ Intervention + Sex + |
| 0,696896 | -0,0632 | 0,161904 | 142 | 70 | 72 | 138 | -0,39033 | y ~ Intervention + Sex + |
| 0,613704 | -0,07077 | 0,139884 | 142 | 70 | 72 | 138 | -0,50594 | y ~ Intervention + Sex + |
| 0,173567 | -0,20731 | 0,151551 | 142 | 70 | 72 | 138 | -1,3679 | y ~ Intervention + Sex + |
| 0,336337 | 0,150119 | 0,155597 | 142 | 70 | 72 | 138 | 0,964791 | y ~ Intervention + Sex + |
| 0,337563 | 0,124434 | 0,129304 | 142 | 70 | 72 | 138 | 0,962339 | y ~ Intervention + Sex + |
| 0,82802 | 0,033215 | 0,152606 | 142 | 70 | 72 | 138 | 0,217655 | y ~ Intervention + Sex + |
| 0,467765 | -0,12101 | 0,166193 | 142 | 70 | 72 | 138 | -0,72814 | y ~ Intervention + Sex + |
| 0,866777 | 0,025719 | 0,15303 | 142 | 70 | 72 | 138 | 0,168066 | y ~ Intervention + Sex + |
| 0,186162 | 0,191053 | 0,143797 | 142 | 70 | 72 | 138 | 1,328631 | y ~ Intervention + Sex + |
| 0,206147 | 0,197155 | 0,155215 | 142 | 70 | 72 | 138 | 1,27021 | y ~ Intervention + Sex + |
| 0,890976 | 0,021744 | 0,158344 | 142 | 70 | 72 | 138 | 0,137323 | y ~ Intervention + Sex + |
| 0,750464 | 0,054668 | 0,171554 | 142 | 70 | 72 | 138 | 0,318663 | y ~ Intervention + Sex + |
| 0,252059 | 0,168736 | 0,146704 | 142 | 70 | 72 | 138 | 1,150179 | y ~ Intervention + Sex + |
| 0,319596 | 0,152548 | 0,152717 | 142 | 70 | 72 | 138 | 0,998891 | y ~ Intervention + Sex + |
| 0,919242 | 0,01478 | 0,145508 | 142 | 70 | 72 | 138 | 0,101575 | y ~ Intervention + Sex + |
| 0,444519 | 0,123125 | 0,160573 | 142 | 70 | 72 | 138 | 0,766785 | y ~ Intervention + Sex + |
| 0,546727 | 0,100447 | 0,166258 | 142 | 70 | 72 | 138 | 0,604164 | y ~ Intervention + Sex + |
| 0,030911 | -0,35673 | 0,163595 | 142 | 70 | 72 | 138 | -2,18056 | y ~ Intervention + Sex + |
| 0,454936 | -0,10152 | 0,135479 | 142 | 70 | 72 | 138 | -0,74933 | y ~ Intervention + Sex + |
| 0,844114 | -0,03187 | 0,161789 | 142 | 70 | 72 | 138 | -0,197 | y ~ Intervention + Sex + |
| 0,412083 | 0,140241 | 0,170459 | 142 | 70 | 72 | 138 | 0,822726 | y ~ Intervention + Sex + |
| 0,580091 | 0,082312 | 0,148426 | 142 | 70 | 72 | 138 | 0,554563 | y ~ Intervention + Sex + |
| 0,792129 | -0,04053 | 0,153498 | 142 | 70 | 72 | 138 | -0,26406 | y ~ Intervention + Sex + |
| 0,391086 | 0,127596 | 0,148306 | 142 | 70 | 72 | 138 | 0,860354 | y ~ Intervention + Sex + |
| 0,393657 | -0,13275 | 0,155139 | 142 | 70 | 72 | 138 | -0,85568 | y ~ Intervention + Sex + |
| 0,755472 | -0,04433 | 0,142057 | 142 | 70 | 72 | 138 | -0,31205 | y ~ Intervention + Sex + |
| 0,363072 | -0,14049 | 0,153956 | 142 | 70 | 72 | 138 | -0,91255 | y ~ Intervention + Sex + |
| 0,013172 | 0,371708 | 0,147998 | 142 | 70 | 72 | 138 | 2,511576 | y ~ Intervention + Sex + |
| 0,34822 | 0,140221 | 0,148972 | 142 | 70 | 72 | 138 | 0,941254 | y ~ Intervention + Sex + |
| 0,494726 | -0,1053 | 0,153801 | 142 | 70 | 72 | 138 | -0,68463 | y ~ Intervention + Sex + |
| 0,374436 | -0,14484 | 0,162543 | 142 | 70 | 72 | 138 | -0,89108 | y ~ Intervention + Sex + |
| 0,428034 | 0,129185 | 0,162516 | 142 | 70 | 72 | 138 | 0,794904 | y ~ Intervention + Sex + |
| 0,500308 | -0,09594 | 0,141962 | 142 | 70 | 72 | 138 | -0,67578 | y ~ Intervention + Sex + |
| 0,503212 | 0,113611 | 0,169265 | 142 | 70 | 72 | 138 | 0,671205 | y ~ Intervention + Sex + |
| 0,920811 | -0,01537 | 0,154293 | 142 | 70 | 72 | 138 | -0,09959 | y ~ Intervention + Sex + |
| 0,203692 | -0,17555 | 0,137453 | 142 | 70 | 72 | 138 | -1,27715 | y ~ Intervention + Sex + |
| 0,225191 | 0,191498 | 0,157185 | 142 | 70 | 72 | 138 | 1,218295 | y ~ Intervention + Sex + |
| 0,07671 | 0,292192 | 0,163835 | 142 | 70 | 72 | 138 | 1,783455 | y ~ Intervention + Sex + |
| 0,240607 | -0,17907 | 0,151942 | 142 | 70 | 72 | 138 | -1,17855 | y ~ Intervention + Sex + |
| 0,681598 | -0,06772 | 0,164705 | 142 | 70 | 72 | 138 | -0,41115 | y ~ Intervention + Sex + |

| | | | | | | | | |
|---|---|---|---|---|---|---|---|---|
| 0,871722 | -0,02333 | 0,144231 | 142 | 70 | 72 | 138 | -0,16177 | y ~ Intervention + Sex + |
| 0,163911 | -0,22624 | 0,161659 | 142 | 70 | 72 | 138 | -1,39949 | y ~ Intervention + Sex + |
| 0,489624 | 0,098202 | 0,141754 | 142 | 70 | 72 | 138 | 0,69276 | y ~ Intervention + Sex + |
| 0,012141 | -0,38176 | 0,150206 | 142 | 70 | 72 | 138 | -2,54155 | y ~ Intervention + Sex + |
| 0,351152 | -0,14891 | 0,159172 | 142 | 70 | 72 | 138 | -0,93552 | y ~ Intervention + Sex + |
| 0,945043 | 0,011139 | 0,161306 | 142 | 70 | 72 | 138 | 0,069058 | y ~ Intervention + Sex + |
| 0,562697 | -0,0888 | 0,153031 | 142 | 70 | 72 | 138 | -0,58024 | y ~ Intervention + Sex + |
| 0,486753 | 0,10073 | 0,144446 | 142 | 70 | 72 | 138 | 0,697356 | y ~ Intervention + Sex + |
| 0,475615 | -0,11046 | 0,154421 | 142 | 70 | 72 | 138 | -0,71533 | y ~ Intervention + Sex + |
| 0,560658 | 0,083292 | 0,1428 | 142 | 70 | 72 | 138 | 0,583278 | y ~ Intervention + Sex + |
| 0,937646 | -0,01164 | 0,148529 | 142 | 70 | 72 | 138 | -0,07837 | y ~ Intervention + Sex + |
| 0,545675 | -0,08719 | 0,143934 | 142 | 70 | 72 | 138 | -0,60575 | y ~ Intervention + Sex + |
| 0,886611 | 0,020488 | 0,143416 | 142 | 70 | 72 | 138 | 0,142858 | y ~ Intervention + Sex + |
| 0,635723 | 0,072977 | 0,153719 | 142 | 70 | 72 | 138 | 0,47474 | y ~ Intervention + Sex + |
| 0,523107 | 0,097097 | 0,151667 | 142 | 70 | 72 | 138 | 0,640196 | y ~ Intervention + Sex + |
| 0,58187 | -0,08907 | 0,16138 | 142 | 70 | 72 | 138 | -0,55196 | y ~ Intervention + Sex + |
| 0,07939 | 0,252477 | 0,142861 | 142 | 70 | 72 | 138 | 1,767292 | y ~ Intervention + Sex + |
| 0,989491 | 0,001906 | 0,144412 | 142 | 70 | 72 | 138 | 0,013195 | y ~ Intervention + Sex + |
| 0,894494 | 0,022564 | 0,169829 | 142 | 70 | 72 | 138 | 0,132865 | y ~ Intervention + Sex + |
| 0,307736 | 0,152706 | 0,149161 | 142 | 70 | 72 | 138 | 1,023769 | y ~ Intervention + Sex + |
| 0,29392 | -0,15568 | 0,147763 | 142 | 70 | 72 | 138 | -1,05357 | y ~ Intervention + Sex + |
| 0,424796 | -0,12997 | 0,162364 | 142 | 70 | 72 | 138 | -0,8005 | y ~ Intervention + Sex + |
| 0,333306 | -0,15319 | 0,157785 | 142 | 70 | 72 | 138 | -0,97088 | y ~ Intervention + Sex + |
| 0,099623 | -0,25183 | 0,151906 | 142 | 70 | 72 | 138 | -1,65783 | y ~ Intervention + Sex + |
| 0,090901 | 0,276965 | 0,162675 | 142 | 70 | 72 | 138 | 1,702566 | y ~ Intervention + Sex + |
| 0,413845 | 0,136892 | 0,167018 | 142 | 70 | 72 | 138 | 0,819622 | y ~ Intervention + Sex + |
| 0,12452 | -0,25319 | 0,163823 | 142 | 70 | 72 | 138 | -1,54548 | y ~ Intervention + Sex + |
| 0,713692 | 0,064012 | 0,174107 | 142 | 70 | 72 | 138 | 0,367658 | y ~ Intervention + Sex + |
| 0,216933 | -0,20358 | 0,164122 | 142 | 70 | 72 | 138 | -1,2404 | y ~ Intervention + Sex + |
| 0,940095 | 0,012016 | 0,159598 | 142 | 70 | 72 | 138 | 0,075288 | y ~ Intervention + Sex + |
| 0,698465 | -0,06309 | 0,162518 | 142 | 70 | 72 | 138 | -0,3882 | y ~ Intervention + Sex + |
| 0,529823 | -0,09314 | 0,147877 | 142 | 70 | 72 | 138 | -0,62987 | y ~ Intervention + Sex + |
| 0,929653 | -0,01421 | 0,160688 | 142 | 70 | 72 | 138 | -0,08844 | y ~ Intervention + Sex + |
| 0,54778 | 0,088074 | 0,146163 | 142 | 70 | 72 | 138 | 0,602575 | y ~ Intervention + Sex + |
| 0,389859 | -0,13969 | 0,161942 | 142 | 70 | 72 | 138 | -0,86259 | y ~ Intervention + Sex + |
| 0,144724 | 0,191715 | 0,130709 | 142 | 70 | 72 | 138 | 1,466733 | y ~ Intervention + Sex + |
| 0,590194 | -0,08826 | 0,163502 | 142 | 70 | 72 | 138 | -0,53982 | y ~ Intervention + Sex + |
| 0,08267 | 0,238767 | 0,136586 | 142 | 70 | 72 | 138 | 1,748106 | y ~ Intervention + Sex + |
| 0,789749 | -0,03906 | 0,146199 | 142 | 70 | 72 | 138 | -0,26716 | y ~ Intervention + Sex + |
| 0,283289 | 0,180179 | 0,167272 | 142 | 70 | 72 | 138 | 1,077159 | y ~ Intervention + Sex + |
| 0,332235 | -0,13692 | 0,140714 | 142 | 70 | 72 | 138 | -0,97304 | y ~ Intervention + Sex + |
| 0,480741 | 0,115938 | 0,163979 | 142 | 70 | 72 | 138 | 0,707029 | y ~ Intervention + Sex + |
| 0,354325 | -0,13962 | 0,150236 | 142 | 70 | 72 | 138 | -0,92936 | y ~ Intervention + Sex + |
| 0,325365 | -0,15899 | 0,161084 | 142 | 70 | 72 | 138 | -0,98701 | y ~ Intervention + Sex + |
| 0,38918 | -0,14929 | 0,172821 | 142 | 70 | 72 | 138 | -0,86383 | y ~ Intervention + Sex + |
| 0,711164 | 0,052334 | 0,14104 | 142 | 70 | 72 | 138 | 0,371057 | y ~ Intervention + Sex + |
| 0,820772 | 0,038265 | 0,168579 | 142 | 70 | 72 | 138 | 0,226984 | y ~ Intervention + Sex + |
| 0,564141 | 0,082678 | 0,143018 | 142 | 70 | 72 | 138 | 0,578097 | y ~ Intervention + Sex + |

| | | | | | | | | |
|---|---|---|---|---|---|---|---|---|
| 0,655397 | -0,06435 | 0,143878 | 142 | 70 | 72 | 138 | -0,44725 | y ~ Intervention + Sex + |
| 0,195011 | -0,20135 | 0,154624 | 142 | 70 | 72 | 138 | -1,30222 | y ~ Intervention + Sex + |
| 0,249882 | 0,195337 | 0,16905 | 142 | 70 | 72 | 138 | 1,1555 | y ~ Intervention + Sex + |
| 0,966636 | -0,00658 | 0,157017 | 142 | 70 | 72 | 138 | -0,0419 | y ~ Intervention + Sex + |
| 0,985745 | 0,00262 | 0,146364 | 142 | 70 | 72 | 138 | 0,017899 | y ~ Intervention + Sex + |
| 0,373709 | -0,12199 | 0,136693 | 142 | 70 | 72 | 138 | -0,89244 | y ~ Intervention + Sex + |
| 0,299181 | -0,1737 | 0,166683 | 142 | 70 | 72 | 138 | -1,04211 | y ~ Intervention + Sex + |
| 0,568461 | -0,08256 | 0,144411 | 142 | 70 | 72 | 138 | -0,57169 | y ~ Intervention + Sex + |
| 0,162884 | -0,21587 | 0,153869 | 142 | 70 | 72 | 138 | -1,40293 | y ~ Intervention + Sex + |
| 0,502606 | 0,086269 | 0,128346 | 142 | 70 | 72 | 138 | 0,672159 | y ~ Intervention + Sex + |
| 0,178024 | -0,21921 | 0,161927 | 142 | 70 | 72 | 138 | -1,35377 | y ~ Intervention + Sex + |
| 0,585885 | 0,085485 | 0,15654 | 142 | 70 | 72 | 138 | 0,546092 | y ~ Intervention + Sex + |
| 0,043132 | 0,314072 | 0,153863 | 142 | 70 | 72 | 138 | 2,041247 | y ~ Intervention + Sex + |
| 0,620213 | 0,082859 | 0,16683 | 142 | 70 | 72 | 138 | 0,496669 | y ~ Intervention + Sex + |
| 0,049477 | -0,26979 | 0,136129 | 142 | 70 | 72 | 138 | -1,9819 | y ~ Intervention + Sex + |
| 0,490163 | -0,11227 | 0,162263 | 142 | 70 | 72 | 138 | -0,6919 | y ~ Intervention + Sex + |
| 0,048513 | -0,32712 | 0,164342 | 142 | 70 | 72 | 138 | -1,99048 | y ~ Intervention + Sex + |
| 0,828934 | 0,032653 | 0,150838 | 142 | 70 | 72 | 138 | 0,216479 | y ~ Intervention + Sex + |
| 0,44811 | -0,12909 | 0,169691 | 142 | 70 | 72 | 138 | -0,76074 | y ~ Intervention + Sex + |
| 0,226412 | -0,18846 | 0,155105 | 142 | 70 | 72 | 138 | -1,21508 | y ~ Intervention + Sex + |
| 0,118985 | -0,25978 | 0,165594 | 142 | 70 | 72 | 138 | -1,5688 | y ~ Intervention + Sex + |
| 0,707536 | -0,05133 | 0,136531 | 142 | 70 | 72 | 138 | -0,37594 | y ~ Intervention + Sex + |
| 0,269745 | 0,176657 | 0,159423 | 142 | 70 | 72 | 138 | 1,108102 | y ~ Intervention + Sex + |
| 0,493306 | 0,101798 | 0,148202 | 142 | 70 | 72 | 138 | 0,686888 | y ~ Intervention + Sex + |
| 0,839979 | -0,02936 | 0,145115 | 142 | 70 | 72 | 138 | -0,2023 | y ~ Intervention + Sex + |
| 0,97531 | 0,005685 | 0,183347 | 142 | 70 | 72 | 138 | 0,031005 | y ~ Intervention + Sex + |
| 0,034929 | -0,36092 | 0,169432 | 142 | 70 | 72 | 138 | -2,13021 | y ~ Intervention + Sex + |
| 0,628684 | 0,064885 | 0,133876 | 142 | 70 | 72 | 138 | 0,484662 | y ~ Intervention + Sex + |
| 0,049726 | 0,311584 | 0,157389 | 142 | 70 | 72 | 138 | 1,979711 | y ~ Intervention + Sex + |
| 0,836799 | 0,030838 | 0,149425 | 142 | 70 | 72 | 138 | 0,206379 | y ~ Intervention + Sex + |
| 0,55422 | 0,074672 | 0,125943 | 142 | 70 | 72 | 138 | 0,592898 | y ~ Intervention + Sex + |
| 0,146657 | 0,208684 | 0,142968 | 142 | 70 | 72 | 138 | 1,459656 | y ~ Intervention + Sex + |
| 0,17727 | -0,1796 | 0,132433 | 142 | 70 | 72 | 138 | -1,35614 | y ~ Intervention + Sex + |
| 0,013025 | -0,35898 | 0,142693 | 142 | 70 | 72 | 138 | -2,51573 | y ~ Intervention + Sex + |
| 0,306232 | -0,11842 | 0,115313 | 142 | 70 | 72 | 138 | -1,02697 | y ~ Intervention + Sex + |
| 0,361895 | -0,13973 | 0,152744 | 142 | 70 | 72 | 138 | -0,9148 | y ~ Intervention + Sex + |
| 0,33803 | -0,17182 | 0,178714 | 142 | 70 | 72 | 138 | -0,96141 | y ~ Intervention + Sex + |
| 0,102497 | 0,22842 | 0,13896 | 142 | 70 | 72 | 138 | 1,643786 | y ~ Intervention + Sex + |
| 0,134638 | -0,21983 | 0,146078 | 142 | 70 | 72 | 138 | -1,50489 | y ~ Intervention + Sex + |
| 0,559798 | -0,09559 | 0,163521 | 142 | 70 | 72 | 138 | -0,58456 | y ~ Intervention + Sex + |
| 0,866747 | 0,026874 | 0,159863 | 142 | 70 | 72 | 138 | 0,168104 | y ~ Intervention + Sex + |
| 0,582034 | 0,094768 | 0,171769 | 142 | 70 | 72 | 138 | 0,551718 | y ~ Intervention + Sex + |
| 0,325905 | 0,15357 | 0,155765 | 142 | 70 | 72 | 138 | 0,985905 | y ~ Intervention + Sex + |
| 0,537267 | -0,09979 | 0,161344 | 142 | 70 | 72 | 138 | -0,6185 | y ~ Intervention + Sex + |
| 0,862233 | -0,02409 | 0,13855 | 142 | 70 | 72 | 138 | -0,17386 | y ~ Intervention + Sex + |
| 0,364919 | -0,13948 | 0,153442 | 142 | 70 | 72 | 138 | -0,90903 | y ~ Intervention + Sex + |
| 0,913652 | 0,016342 | 0,150434 | 142 | 70 | 72 | 138 | 0,108632 | y ~ Intervention + Sex + |
| 0,819759 | -0,03571 | 0,156436 | 142 | 70 | 72 | 138 | -0,22829 | y ~ Intervention + Sex + |

| | | | | | | | | |
|---|---|---|---|---|---|---|---|---|
| 0,904713 | 0,020645 | 0,172144 | 142 | 70 | 72 | 138 | 0,11993 | y ~ Intervention + Sex + |
| 0,049185 | 0,331347 | 0,166968 | 142 | 70 | 72 | 138 | 1,98449 | y ~ Intervention + Sex + |
| 0,629984 | -0,07829 | 0,162153 | 142 | 70 | 72 | 138 | -0,48283 | y ~ Intervention + Sex + |
| 0,871001 | -0,02541 | 0,156186 | 142 | 70 | 72 | 138 | -0,16269 | y ~ Intervention + Sex + |
| 0,630753 | 0,071189 | 0,147774 | 142 | 70 | 72 | 138 | 0,481741 | y ~ Intervention + Sex + |
| 0,888158 | 0,021912 | 0,155517 | 142 | 70 | 72 | 138 | 0,140896 | y ~ Intervention + Sex + |
| 0,986536 | 0,002541 | 0,150274 | 142 | 70 | 72 | 138 | 0,016906 | y ~ Intervention + Sex + |
| 0,591462 | 0,077865 | 0,144738 | 142 | 70 | 72 | 138 | 0,537974 | y ~ Intervention + Sex + |
| 0,692098 | 0,061266 | 0,154384 | 142 | 70 | 72 | 138 | 0,39684 | y ~ Intervention + Sex + |
| 0,385295 | 0,12617 | 0,144865 | 142 | 70 | 72 | 138 | 0,870948 | y ~ Intervention + Sex + |
| 0,174087 | 0,224725 | 0,164485 | 142 | 70 | 72 | 138 | 1,366238 | y ~ Intervention + Sex + |
| 0,237847 | 0,190686 | 0,160845 | 142 | 70 | 72 | 138 | 1,185525 | y ~ Intervention + Sex + |
| 0,39581 | 0,133762 | 0,157038 | 142 | 70 | 72 | 138 | 0,851783 | y ~ Intervention + Sex + |
| 0,231543 | 0,182159 | 0,151586 | 142 | 70 | 72 | 138 | 1,201688 | y ~ Intervention + Sex + |
| 0,426152 | -0,13417 | 0,168099 | 142 | 70 | 72 | 138 | -0,79815 | y ~ Intervention + Sex + |
| 0,133583 | 0,240304 | 0,159246 | 142 | 70 | 72 | 138 | 1,509009 | y ~ Intervention + Sex + |
| 0,929312 | -0,01251 | 0,140819 | 142 | 70 | 72 | 138 | -0,08887 | y ~ Intervention + Sex + |
| 0,464758 | 0,102147 | 0,139341 | 142 | 70 | 72 | 138 | 0,733072 | y ~ Intervention + Sex + |
| 0,111733 | -0,23682 | 0,147952 | 142 | 70 | 72 | 138 | -1,60069 | y ~ Intervention + Sex + |
| 0,395446 | -0,13096 | 0,153626 | 142 | 70 | 72 | 138 | -0,85244 | y ~ Intervention + Sex + |
| 0,101866 | -0,25437 | 0,154457 | 142 | 70 | 72 | 138 | -1,64684 | y ~ Intervention + Sex + |
| 0,540435 | -0,08329 | 0,135717 | 142 | 70 | 72 | 138 | -0,61368 | y ~ Intervention + Sex + |
| 0,134426 | 0,238697 | 0,158527 | 142 | 70 | 72 | 138 | 1,505715 | y ~ Intervention + Sex + |
| 0,074487 | 0,238048 | 0,132453 | 142 | 70 | 72 | 138 | 1,797222 | y ~ Intervention + Sex + |
| 0,010529 | 0,378585 | 0,145983 | 142 | 70 | 72 | 138 | 2,593358 | y ~ Intervention + Sex + |
| 0,874613 | 0,021979 | 0,139021 | 142 | 70 | 72 | 138 | 0,158096 | y ~ Intervention + Sex + |
| 0,523329 | -0,10916 | 0,170607 | 142 | 70 | 72 | 138 | -0,63985 | y ~ Intervention + Sex + |
| 0,088022 | -0,26197 | 0,152478 | 142 | 70 | 72 | 138 | -1,7181 | y ~ Intervention + Sex + |
| 0,591769 | -0,0905 | 0,168362 | 142 | 70 | 72 | 138 | -0,53753 | y ~ Intervention + Sex + |
| 0,600304 | 0,078761 | 0,149971 | 142 | 70 | 72 | 138 | 0,525176 | y ~ Intervention + Sex + |
| 0,685217 | 0,064536 | 0,158874 | 142 | 70 | 72 | 138 | 0,406211 | y ~ Intervention + Sex + |
| 0,275358 | -0,17508 | 0,15987 | 142 | 70 | 72 | 138 | -1,09515 | y ~ Intervention + Sex + |
| 0,207608 | -0,21249 | 0,167827 | 142 | 70 | 72 | 138 | -1,26611 | y ~ Intervention + Sex + |
| 0,936306 | -0,01357 | 0,169465 | 142 | 70 | 72 | 138 | -0,08006 | y ~ Intervention + Sex + |
| 0,155973 | -0,24181 | 0,169512 | 142 | 70 | 72 | 138 | -1,42653 | y ~ Intervention + Sex + |
| 0,59421 | 0,08059 | 0,150922 | 142 | 70 | 72 | 138 | 0,533987 | y ~ Intervention + Sex + |
| 0,170298 | 0,195336 | 0,141708 | 142 | 70 | 72 | 138 | 1,378442 | y ~ Intervention + Sex + |
| 0,112345 | 0,201529 | 0,126119 | 142 | 70 | 72 | 138 | 1,597935 | y ~ Intervention + Sex + |
| 0,085724 | 0,277284 | 0,160207 | 142 | 70 | 72 | 138 | 1,730792 | y ~ Intervention + Sex + |
| 0,097837 | -0,22651 | 0,135904 | 142 | 70 | 72 | 138 | -1,66672 | y ~ Intervention + Sex + |
| 0,66708 | -0,07691 | 0,17842 | 142 | 70 | 72 | 138 | -0,43108 | y ~ Intervention + Sex + |
| 0,496029 | -0,12285 | 0,179982 | 142 | 70 | 72 | 138 | -0,68256 | y ~ Intervention + Sex + |
| 0,935128 | 0,012281 | 0,150608 | 142 | 70 | 72 | 138 | 0,081544 | y ~ Intervention + Sex + |
| 0,041344 | -0,31624 | 0,153564 | 142 | 70 | 72 | 138 | -2,05931 | y ~ Intervention + Sex + |
| 0,67764 | -0,06308 | 0,151426 | 142 | 70 | 72 | 138 | -0,41657 | y ~ Intervention + Sex + |
| 0,838849 | 0,034834 | 0,170967 | 142 | 70 | 72 | 138 | 0,20375 | y ~ Intervention + Sex + |
| 0,785859 | -0,03627 | 0,133234 | 142 | 70 | 72 | 138 | -0,27222 | y ~ Intervention + Sex + |
| 0,262661 | -0,17329 | 0,15407 | 142 | 70 | 72 | 138 | -1,12472 | y ~ Intervention + Sex + |

| | | | | | | | | |
|---|---|---|---|---|---|---|---|---|
| 0,355711 | 0,139373 | 0,1504 | 142 | 70 | 72 | 138 | 0,926678 | y ~ Intervention + Sex + |
| 0,419912 | -0,13574 | 0,167786 | 142 | 70 | 72 | 138 | -0,80899 | y ~ Intervention + Sex + |
| 0,426657 | -0,12709 | 0,15941 | 142 | 70 | 72 | 138 | -0,79728 | y ~ Intervention + Sex + |
| 0,874812 | 0,023354 | 0,14796 | 142 | 70 | 72 | 138 | 0,157843 | y ~ Intervention + Sex + |
| 0,057219 | 0,264924 | 0,138149 | 142 | 70 | 72 | 138 | 1,917671 | y ~ Intervention + Sex + |
| 0,490993 | -0,1088 | 0,157547 | 142 | 70 | 72 | 138 | -0,69057 | y ~ Intervention + Sex + |
| 0,177222 | -0,22404 | 0,165185 | 142 | 70 | 72 | 138 | -1,35629 | y ~ Intervention + Sex + |
| 0,953272 | 0,009604 | 0,163606 | 142 | 70 | 72 | 138 | 0,058705 | y ~ Intervention + Sex + |
| 0,405924 | -0,12648 | 0,151722 | 142 | 70 | 72 | 138 | -0,83364 | y ~ Intervention + Sex + |
| 0,506341 | 0,098776 | 0,148249 | 142 | 70 | 72 | 138 | 0,666286 | y ~ Intervention + Sex + |
| 0,460391 | -0,11578 | 0,156396 | 142 | 70 | 72 | 138 | -0,74027 | y ~ Intervention + Sex + |
| 0,086934 | 0,267189 | 0,154976 | 142 | 70 | 72 | 138 | 1,724071 | y ~ Intervention + Sex + |
| 0,839394 | -0,03321 | 0,163536 | 142 | 70 | 72 | 138 | -0,20305 | y ~ Intervention + Sex + |
| 0,24937 | 0,183188 | 0,158363 | 142 | 70 | 72 | 138 | 1,156756 | y ~ Intervention + Sex + |
| 0,461974 | -0,11075 | 0,150142 | 142 | 70 | 72 | 138 | -0,73766 | y ~ Intervention + Sex + |
| 0,244854 | 0,181391 | 0,155311 | 142 | 70 | 72 | 138 | 1,167917 | y ~ Intervention + Sex + |
| 0,314764 | -0,15274 | 0,15139 | 142 | 70 | 72 | 138 | -1,00895 | y ~ Intervention + Sex + |
| 0,680885 | -0,06985 | 0,169475 | 142 | 70 | 72 | 138 | -0,41213 | y ~ Intervention + Sex + |
| 0,52672 | 0,09957 | 0,156895 | 142 | 70 | 72 | 138 | 0,634631 | y ~ Intervention + Sex + |
| 0,003764 | 0,483493 | 0,164042 | 142 | 70 | 72 | 138 | 2,947368 | y ~ Intervention + Sex + |
| 0,589925 | -0,07867 | 0,14563 | 142 | 70 | 72 | 138 | -0,54021 | y ~ Intervention + Sex + |
| 0,014515 | -0,35306 | 0,14262 | 142 | 70 | 72 | 138 | -2,47553 | y ~ Intervention + Sex + |
| 0,419046 | 0,123992 | 0,152982 | 142 | 70 | 72 | 138 | 0,810504 | y ~ Intervention + Sex + |
| 0,080258 | 0,215189 | 0,122117 | 142 | 70 | 72 | 138 | 1,762151 | y ~ Intervention + Sex + |
| 0,688426 | 0,059409 | 0,147844 | 142 | 70 | 72 | 138 | 0,401836 | y ~ Intervention + Sex + |
| 0,694781 | 0,06252 | 0,159003 | 142 | 70 | 72 | 138 | 0,393197 | y ~ Intervention + Sex + |
| 0,499482 | 0,121679 | 0,179708 | 142 | 70 | 72 | 138 | 0,67709 | y ~ Intervention + Sex + |
| 0,576278 | -0,09418 | 0,168123 | 142 | 70 | 72 | 138 | -0,56016 | y ~ Intervention + Sex + |
| 0,855844 | -0,03036 | 0,166796 | 142 | 70 | 72 | 138 | -0,18201 | y ~ Intervention + Sex + |
| 0,197936 | -0,21733 | 0,167993 | 142 | 70 | 72 | 138 | -1,29368 | y ~ Intervention + Sex + |
| 0,362276 | -0,14473 | 0,158332 | 142 | 70 | 72 | 138 | -0,91407 | y ~ Intervention + Sex + |
| 0,150836 | -0,24755 | 0,17136 | 142 | 70 | 72 | 138 | -1,4446 | y ~ Intervention + Sex + |
| 0,338182 | 0,150101 | 0,156176 | 142 | 70 | 72 | 138 | 0,961102 | y ~ Intervention + Sex + |
| 0,680531 | -0,06606 | 0,1601 | 142 | 70 | 72 | 138 | -0,41261 | y ~ Intervention + Sex + |
| 0,680805 | 0,068245 | 0,165548 | 142 | 70 | 72 | 138 | 0,412238 | y ~ Intervention + Sex + |
| 0,208598 | -0,17932 | 0,141939 | 142 | 70 | 72 | 138 | -1,26334 | y ~ Intervention + Sex + |
| 0,243679 | -0,20222 | 0,172711 | 142 | 70 | 72 | 138 | -1,17085 | y ~ Intervention + Sex + |
| 0,204123 | -0,20358 | 0,159555 | 142 | 70 | 72 | 138 | -1,27593 | y ~ Intervention + Sex + |
| 0,916055 | 0,015352 | 0,145378 | 142 | 70 | 72 | 138 | 0,105598 | y ~ Intervention + Sex + |
| 0,321366 | -0,16856 | 0,169368 | 142 | 70 | 72 | 138 | -0,99523 | y ~ Intervention + Sex + |
| 0,219634 | -0,20054 | 0,162633 | 142 | 70 | 72 | 138 | -1,2331 | y ~ Intervention + Sex + |
| 0,266819 | -0,16616 | 0,149034 | 142 | 70 | 72 | 138 | -1,11493 | y ~ Intervention + Sex + |
| 0,867546 | -0,02544 | 0,152238 | 142 | 70 | 72 | 138 | -0,16709 | y ~ Intervention + Sex + |
| 0,9263 | 0,014189 | 0,153107 | 142 | 70 | 72 | 138 | 0,092671 | y ~ Intervention + Sex + |
| 0,824034 | 0,034411 | 0,154461 | 142 | 70 | 72 | 138 | 0,222783 | y ~ Intervention + Sex + |
| 0,586444 | 0,093403 | 0,171295 | 142 | 70 | 72 | 138 | 0,545276 | y ~ Intervention + Sex + |
| 0,824958 | -0,03003 | 0,135506 | 142 | 70 | 72 | 138 | -0,22159 | y ~ Intervention + Sex + |
| 0,830702 | 0,03267 | 0,152515 | 142 | 70 | 72 | 138 | 0,214207 | y ~ Intervention + Sex + |

| | | | | | | | | |
|---|---|---|---|---|---|---|---|---|
| 0,98684 | 0,00254 | 0,153737 | 142 | 70 | 72 | 138 | 0,016524 | y ~ Intervention + Sex + |
| 0,118052 | 0,24102 | 0,153241 | 142 | 70 | 72 | 138 | 1,572813 | y ~ Intervention + Sex + |
| 0,715537 | -0,05513 | 0,150975 | 142 | 70 | 72 | 138 | -0,36518 | y ~ Intervention + Sex + |
| 0,0864 | -0,24659 | 0,142781 | 142 | 70 | 72 | 138 | -1,72703 | y ~ Intervention + Sex + |
| 0,137672 | 0,212797 | 0,142513 | 142 | 70 | 72 | 138 | 1,493182 | y ~ Intervention + Sex + |
| 0,620653 | 0,073104 | 0,147374 | 142 | 70 | 72 | 138 | 0,496043 | y ~ Intervention + Sex + |
| 0,510167 | -0,1024 | 0,155078 | 142 | 70 | 72 | 138 | -0,66029 | y ~ Intervention + Sex + |
| 0,13027 | -0,2267 | 0,148937 | 142 | 70 | 72 | 138 | -1,52211 | y ~ Intervention + Sex + |
| 0,326188 | -0,15397 | 0,156265 | 142 | 70 | 72 | 138 | -0,98533 | y ~ Intervention + Sex + |
| 0,438712 | -0,11676 | 0,150343 | 142 | 70 | 72 | 138 | -0,77662 | y ~ Intervention + Sex + |
| 0,051698 | 0,296196 | 0,150916 | 142 | 70 | 72 | 138 | 1,962651 | y ~ Intervention + Sex + |
| 0,280236 | -0,17462 | 0,161079 | 142 | 70 | 72 | 138 | -1,08404 | y ~ Intervention + Sex + |
| 0,991842 | 0,001727 | 0,168563 | 142 | 70 | 72 | 138 | 0,010243 | y ~ Intervention + Sex + |
| 0,836659 | 0,029762 | 0,144084 | 142 | 70 | 72 | 138 | 0,206558 | y ~ Intervention + Sex + |
| 0,803111 | -0,04266 | 0,170761 | 142 | 70 | 72 | 138 | -0,2498 | y ~ Intervention + Sex + |
| 0,38769 | 0,142579 | 0,164535 | 142 | 70 | 72 | 138 | 0,866555 | y ~ Intervention + Sex + |
| 0,13511 | 0,194252 | 0,129239 | 142 | 70 | 72 | 138 | 1,503053 | y ~ Intervention + Sex + |
| 0,765761 | 0,041852 | 0,140202 | 142 | 70 | 72 | 138 | 0,298513 | y ~ Intervention + Sex + |
| 0,218011 | 0,174668 | 0,141148 | 142 | 70 | 72 | 138 | 1,237478 | y ~ Intervention + Sex + |
| 0,227029 | -0,1997 | 0,164568 | 142 | 70 | 72 | 138 | -1,21346 | y ~ Intervention + Sex + |
| 0,563777 | 0,095093 | 0,164339 | 142 | 70 | 72 | 138 | 0,578638 | y ~ Intervention + Sex + |
| 0,882499 | -0,02391 | 0,161446 | 142 | 70 | 72 | 138 | -0,14808 | y ~ Intervention + Sex + |
| 0,655546 | 0,070509 | 0,157724 | 142 | 70 | 72 | 138 | 0,447041 | y ~ Intervention + Sex + |
| 0,068703 | 0,297717 | 0,162269 | 142 | 70 | 72 | 138 | 1,834706 | y ~ Intervention + Sex + |
| 0,310835 | 0,15181 | 0,149242 | 142 | 70 | 72 | 138 | 1,017207 | y ~ Intervention + Sex + |
| 0,034935 | -0,29732 | 0,13958 | 142 | 70 | 72 | 138 | -2,13013 | y ~ Intervention + Sex + |
| 0,207975 | -0,20593 | 0,162778 | 142 | 70 | 72 | 138 | -1,26508 | y ~ Intervention + Sex + |
| 0,494803 | 0,089487 | 0,130732 | 142 | 70 | 72 | 138 | 0,684507 | y ~ Intervention + Sex + |
| 0,915901 | 0,01533 | 0,144905 | 142 | 70 | 72 | 138 | 0,105792 | y ~ Intervention + Sex + |
| 0,174892 | -0,23137 | 0,169669 | 142 | 70 | 72 | 138 | -1,36367 | y ~ Intervention + Sex + |
| 0,596125 | 0,089244 | 0,168001 | 142 | 70 | 72 | 138 | 0,531214 | y ~ Intervention + Sex + |
| 0,315881 | -0,15661 | 0,155584 | 142 | 70 | 72 | 138 | -1,00662 | y ~ Intervention + Sex + |
| 0,481662 | -0,10891 | 0,154364 | 142 | 70 | 72 | 138 | -0,70554 | y ~ Intervention + Sex + |
| 0,421301 | 0,124748 | 0,154665 | 142 | 70 | 72 | 138 | 0,806572 | y ~ Intervention + Sex + |
| 0,285741 | 0,157249 | 0,146733 | 142 | 70 | 72 | 138 | 1,071666 | y ~ Intervention + Sex + |
| 0,427354 | -0,1299 | 0,163178 | 142 | 70 | 72 | 138 | -0,79608 | y ~ Intervention + Sex + |
| 0,965107 | -0,00632 | 0,14421 | 142 | 70 | 72 | 138 | -0,04382 | y ~ Intervention + Sex + |
| 0,761642 | -0,04961 | 0,163227 | 142 | 70 | 72 | 138 | -0,30393 | y ~ Intervention + Sex + |
| 0,609855 | 0,085461 | 0,167096 | 142 | 70 | 72 | 138 | 0,511448 | y ~ Intervention + Sex + |
| 0,103915 | 0,246988 | 0,150881 | 142 | 70 | 72 | 138 | 1,636971 | y ~ Intervention + Sex + |
| 0,337709 | 0,122114 | 0,126931 | 142 | 70 | 72 | 138 | 0,962047 | y ~ Intervention + Sex + |
| 0,312159 | -0,144 | 0,141954 | 142 | 70 | 72 | 138 | -1,01442 | y ~ Intervention + Sex + |
| 0,607521 | -0,08169 | 0,158684 | 142 | 70 | 72 | 138 | -0,51479 | y ~ Intervention + Sex + |
| 0,142658 | -0,23867 | 0,161877 | 142 | 70 | 72 | 138 | -1,47438 | y ~ Intervention + Sex + |
| 0,023507 | 0,345904 | 0,151015 | 142 | 70 | 72 | 138 | 2,29052 | y ~ Intervention + Sex + |
| 0,281419 | 0,179431 | 0,16593 | 142 | 70 | 72 | 138 | 1,081368 | y ~ Intervention + Sex + |
| 0,0772 | 0,262548 | 0,14746 | 142 | 70 | 72 | 138 | 1,780467 | y ~ Intervention + Sex + |
| 0,9114 | -0,01701 | 0,152609 | 142 | 70 | 72 | 138 | -0,11148 | y ~ Intervention + Sex + |

| | | | | | | | | |
|---|---|---|---|---|---|---|---|---|
| 0,595323 | -0,08801 | 0,165315 | 142 | 70 | 72 | 138 | -0,53237 | y ~ Intervention + Sex + |
| 0,277459 | -0,16842 | 0,154463 | 142 | 70 | 72 | 138 | -1,09035 | y ~ Intervention + Sex + |
| 0,183146 | 0,202268 | 0,151189 | 142 | 70 | 72 | 138 | 1,337851 | y ~ Intervention + Sex + |
| 0,783668 | 0,040721 | 0,148035 | 142 | 70 | 72 | 138 | 0,275077 | y ~ Intervention + Sex + |
| 0,490493 | 0,097444 | 0,140943 | 142 | 70 | 72 | 138 | 0,691373 | y ~ Intervention + Sex + |
| 0,599549 | -0,08294 | 0,157598 | 142 | 70 | 72 | 138 | -0,52626 | y ~ Intervention + Sex + |
| 0,05386 | 0,252357 | 0,129775 | 142 | 70 | 72 | 138 | 1,944573 | y ~ Intervention + Sex + |
| 0,125673 | 0,27505 | 0,178519 | 142 | 70 | 72 | 138 | 1,54073 | y ~ Intervention + Sex + |
| 0,594742 | -0,08382 | 0,157197 | 142 | 70 | 72 | 138 | -0,53322 | y ~ Intervention + Sex + |
| 0,249428 | -0,17734 | 0,153326 | 142 | 70 | 72 | 138 | -1,15661 | y ~ Intervention + Sex + |
| 0,574017 | -0,09505 | 0,168683 | 142 | 70 | 72 | 138 | -0,56349 | y ~ Intervention + Sex + |
| 0,422295 | 0,139651 | 0,173513 | 142 | 70 | 72 | 138 | 0,804842 | y ~ Intervention + Sex + |
| 0,729109 | -0,05571 | 0,160535 | 142 | 70 | 72 | 138 | -0,34701 | y ~ Intervention + Sex + |
| 0,899559 | -0,01972 | 0,155975 | 142 | 70 | 72 | 138 | -0,12645 | y ~ Intervention + Sex + |
| 0,230776 | -0,18981 | 0,157693 | 142 | 70 | 72 | 138 | -1,20368 | y ~ Intervention + Sex + |
| 0,113087 | -0,24335 | 0,15261 | 142 | 70 | 72 | 138 | -1,59461 | y ~ Intervention + Sex + |
| 0,007435 | -0,40137 | 0,147733 | 142 | 70 | 72 | 138 | -2,71686 | y ~ Intervention + Sex + |
| 0,716752 | 0,059776 | 0,164424 | 142 | 70 | 72 | 138 | 0,363547 | y ~ Intervention + Sex + |
| 0,873364 | -0,02312 | 0,144812 | 142 | 70 | 72 | 138 | -0,15968 | y ~ Intervention + Sex + |
| 0,524351 | -0,10705 | 0,167714 | 142 | 70 | 72 | 138 | -0,63828 | y ~ Intervention + Sex + |
| 0,95135 | -0,0093 | 0,152079 | 142 | 70 | 72 | 138 | -0,06112 | y ~ Intervention + Sex + |
| 0,536425 | -0,09926 | 0,160157 | 142 | 70 | 72 | 138 | -0,61978 | y ~ Intervention + Sex + |
| 0,55163 | 0,100551 | 0,168489 | 142 | 70 | 72 | 138 | 0,596784 | y ~ Intervention + Sex + |
| 0,272762 | -0,1811 | 0,164466 | 142 | 70 | 72 | 138 | -1,10112 | y ~ Intervention + Sex + |
| 0,738728 | 0,057375 | 0,171674 | 142 | 70 | 72 | 138 | 0,33421 | y ~ Intervention + Sex + |
| 0,936488 | 0,012804 | 0,16039 | 142 | 70 | 72 | 138 | 0,07983 | y ~ Intervention + Sex + |
| 0,099294 | -0,24405 | 0,147064 | 142 | 70 | 72 | 138 | -1,65946 | y ~ Intervention + Sex + |
| 0,380836 | -0,14431 | 0,164145 | 142 | 70 | 72 | 138 | -0,87917 | y ~ Intervention + Sex + |
| 0,7272 | -0,05616 | 0,160661 | 142 | 70 | 72 | 138 | -0,34956 | y ~ Intervention + Sex + |
| 0,709274 | 0,057917 | 0,155023 | 142 | 70 | 72 | 138 | 0,373601 | y ~ Intervention + Sex + |
| 0,286404 | 0,186782 | 0,174532 | 142 | 70 | 72 | 138 | 1,070186 | y ~ Intervention + Sex + |
| 0,29182 | -0,15357 | 0,14513 | 142 | 70 | 72 | 138 | -1,05818 | y ~ Intervention + Sex + |
| 0,332585 | 0,141731 | 0,145764 | 142 | 70 | 72 | 138 | 0,972335 | y ~ Intervention + Sex + |
| 0,665745 | 0,074205 | 0,171403 | 142 | 70 | 72 | 138 | 0,432926 | y ~ Intervention + Sex + |
| 0,788008 | 0,041854 | 0,155346 | 142 | 70 | 72 | 138 | 0,269421 | y ~ Intervention + Sex + |
| 0,603356 | 0,085288 | 0,16377 | 142 | 70 | 72 | 138 | 0,520779 | y ~ Intervention + Sex + |
| 0,146534 | 0,220161 | 0,150784 | 142 | 70 | 72 | 138 | 1,460106 | y ~ Intervention + Sex + |
| 0,747516 | 0,055842 | 0,173121 | 142 | 70 | 72 | 138 | 0,322561 | y ~ Intervention + Sex + |
| 0,542353 | 0,087517 | 0,143288 | 142 | 70 | 72 | 138 | 0,610776 | y ~ Intervention + Sex + |
| 0,843679 | -0,02931 | 0,148349 | 142 | 70 | 72 | 138 | -0,19756 | y ~ Intervention + Sex + |
| 0,749596 | 0,04453 | 0,139239 | 142 | 70 | 72 | 138 | 0,319811 | y ~ Intervention + Sex + |
| 0,892314 | 0,022013 | 0,162305 | 142 | 70 | 72 | 138 | 0,135627 | y ~ Intervention + Sex + |
| 0,125588 | -0,23841 | 0,154701 | 142 | 70 | 72 | 138 | -1,54108 | y ~ Intervention + Sex + |
| 0,295269 | 0,180784 | 0,172073 | 142 | 70 | 72 | 138 | 1,05062 | y ~ Intervention + Sex + |
| 0,0961 | 0,27699 | 0,165317 | 142 | 70 | 72 | 138 | 1,675503 | y ~ Intervention + Sex + |
| 0,416207 | -0,12865 | 0,157756 | 142 | 70 | 72 | 138 | -0,81547 | y ~ Intervention + Sex + |
| 0,4029 | 0,141219 | 0,168311 | 142 | 70 | 72 | 138 | 0,839036 | y ~ Intervention + Sex + |
| 0,190329 | 0,194714 | 0,14795 | 142 | 70 | 72 | 138 | 1,316081 | y ~ Intervention + Sex + |

| | | | | | | | | |
|---|---|---|---|---|---|---|---|---|
| 0,310835 | 0,15181 | 0,149242 | 142 | 70 | 72 | 138 | 1,017207 | y ~ Intervention + Sex + |
| 0,609215 | 0,08246 | 0,160941 | 142 | 70 | 72 | 138 | 0,512364 | y ~ Intervention + Sex + |
| 0,264103 | -0,18667 | 0,166474 | 142 | 70 | 72 | 138 | -1,12131 | y ~ Intervention + Sex + |
| 0,688152 | 0,06225 | 0,154771 | 142 | 70 | 72 | 138 | 0,402209 | y ~ Intervention + Sex + |
| 0,043493 | 0,346757 | 0,170173 | 142 | 70 | 72 | 138 | 2,037674 | y ~ Intervention + Sex + |
| 0,244618 | -0,16296 | 0,139456 | 142 | 70 | 72 | 138 | -1,1685 | y ~ Intervention + Sex + |
| 0,000735 | -0,50377 | 0,145865 | 142 | 70 | 72 | 138 | -3,45368 | y ~ Intervention + Sex + |
| 0,392739 | -0,1285 | 0,149877 | 142 | 70 | 72 | 138 | -0,85735 | y ~ Intervention + Sex + |
| 0,767909 | 0,036439 | 0,123232 | 142 | 70 | 72 | 138 | 0,295693 | y ~ Intervention + Sex + |
| 0,189273 | 0,20277 | 0,153702 | 142 | 70 | 72 | 138 | 1,319242 | y ~ Intervention + Sex + |
| 0,357955 | 0,15454 | 0,16755 | 142 | 70 | 72 | 138 | 0,922352 | y ~ Intervention + Sex + |
| 0,845557 | 0,030113 | 0,154302 | 142 | 70 | 72 | 138 | 0,195157 | y ~ Intervention + Sex + |
| 0,782473 | 0,042347 | 0,153078 | 142 | 70 | 72 | 138 | 0,276637 | y ~ Intervention + Sex + |
| 0,57196 | 0,089031 | 0,157155 | 142 | 70 | 72 | 138 | 0,566521 | y ~ Intervention + Sex + |
| 0,311504 | -0,16289 | 0,160354 | 142 | 70 | 72 | 138 | -1,0158 | y ~ Intervention + Sex + |
| 0,751752 | -0,04451 | 0,14044 | 142 | 70 | 72 | 138 | -0,31696 | y ~ Intervention + Sex + |
| 0,717084 | -0,05584 | 0,153776 | 142 | 70 | 72 | 138 | -0,3631 | y ~ Intervention + Sex + |
| 0,407546 | 0,121333 | 0,146051 | 142 | 70 | 72 | 138 | 0,830757 | y ~ Intervention + Sex + |
| 0,910581 | -0,01763 | 0,156682 | 142 | 70 | 72 | 138 | -0,11251 | y ~ Intervention + Sex + |
| 0,803675 | 0,035946 | 0,14432 | 142 | 70 | 72 | 138 | 0,249073 | y ~ Intervention + Sex + |
| 0,772229 | 0,036634 | 0,126312 | 142 | 70 | 72 | 138 | 0,29003 | y ~ Intervention + Sex + |
| 0,298812 | -0,14902 | 0,142889 | 142 | 70 | 72 | 138 | -1,04291 | y ~ Intervention + Sex + |
| 0,267035 | -0,19323 | 0,173392 | 142 | 70 | 72 | 138 | -1,11442 | y ~ Intervention + Sex + |
| 0,686458 | -0,05614 | 0,138776 | 142 | 70 | 72 | 138 | -0,40452 | y ~ Intervention + Sex + |
| 0,299998 | -0,14947 | 0,143671 | 142 | 70 | 72 | 138 | -1,04035 | y ~ Intervention + Sex + |
| 0,150959 | 0,232016 | 0,160658 | 142 | 70 | 72 | 138 | 1,444166 | y ~ Intervention + Sex + |
| 0,851289 | 0,029824 | 0,158785 | 142 | 70 | 72 | 138 | 0,187826 | y ~ Intervention + Sex + |
| 0,845699 | -0,03127 | 0,160378 | 142 | 70 | 72 | 138 | -0,19497 | y ~ Intervention + Sex + |
| 0,553471 | -0,09339 | 0,15721 | 142 | 70 | 72 | 138 | -0,59402 | y ~ Intervention + Sex + |
| 0,56886 | 0,092577 | 0,162102 | 142 | 70 | 72 | 138 | 0,571102 | y ~ Intervention + Sex + |
| 0,344433 | -0,15043 | 0,158569 | 142 | 70 | 72 | 138 | -0,9487 | y ~ Intervention + Sex + |
| 0,785431 | 0,046794 | 0,171544 | 142 | 70 | 72 | 138 | 0,272779 | y ~ Intervention + Sex + |
| 0,858372 | 0,029 | 0,162209 | 142 | 70 | 72 | 138 | 0,178781 | y ~ Intervention + Sex + |
| 0,239699 | -0,19078 | 0,16156 | 142 | 70 | 72 | 138 | -1,18084 | y ~ Intervention + Sex + |
| 0,278648 | -0,16415 | 0,150924 | 142 | 70 | 72 | 138 | -1,08765 | y ~ Intervention + Sex + |
| 0,871304 | 0,02323 | 0,143127 | 142 | 70 | 72 | 138 | 0,162304 | y ~ Intervention + Sex + |
| 0,709867 | 0,062955 | 0,16887 | 142 | 70 | 72 | 138 | 0,372803 | y ~ Intervention + Sex + |
| 0,360737 | 0,131817 | 0,143747 | 142 | 70 | 72 | 138 | 0,91701 | y ~ Intervention + Sex + |
| 0,132721 | 0,224997 | 0,148769 | 142 | 70 | 72 | 138 | 1,512392 | y ~ Intervention + Sex + |
| 0,589062 | -0,07546 | 0,139354 | 142 | 70 | 72 | 138 | -0,54146 | y ~ Intervention + Sex + |
| 0,650326 | -0,07125 | 0,156831 | 142 | 70 | 72 | 138 | -0,4543 | y ~ Intervention + Sex + |
| 0,023276 | -0,35608 | 0,155193 | 142 | 70 | 72 | 138 | -2,29442 | y ~ Intervention + Sex + |
| 0,247904 | 0,173562 | 0,149575 | 142 | 70 | 72 | 138 | 1,160364 | y ~ Intervention + Sex + |
| 0,930295 | 0,015553 | 0,177473 | 142 | 70 | 72 | 138 | 0,087634 | y ~ Intervention + Sex + |
| 0,209632 | -0,16161 | 0,128218 | 142 | 70 | 72 | 138 | -1,26046 | y ~ Intervention + Sex + |
| 0,111346 | -0,22644 | 0,141311 | 142 | 70 | 72 | 138 | -1,60244 | y ~ Intervention + Sex + |
| 0,513191 | -0,09857 | 0,150362 | 142 | 70 | 72 | 138 | -0,65557 | y ~ Intervention + Sex + |
| 0,439755 | 0,129305 | 0,166878 | 142 | 70 | 72 | 138 | 0,774847 | y ~ Intervention + Sex + |

| | | | | | | | | |
|---|---|---|---|---|---|---|---|---|
| 0,936147 | 0,012757 | 0,15895 | 142 | 70 | 72 | 138 | 0,080259 | y ~ Intervention + Sex + |
| 0,534347 | 0,11054 | 0,177447 | 142 | 70 | 72 | 138 | 0,622948 | y ~ Intervention + Sex + |
| 0,843941 | 0,034381 | 0,174321 | 142 | 70 | 72 | 138 | 0,197225 | y ~ Intervention + Sex + |
| 0,329753 | 0,133048 | 0,136031 | 142 | 70 | 72 | 138 | 0,978065 | y ~ Intervention + Sex + |
| 0,877793 | 0,022997 | 0,149277 | 142 | 70 | 72 | 138 | 0,154053 | y ~ Intervention + Sex + |
| 0,217621 | 0,185587 | 0,149844 | 142 | 70 | 72 | 138 | 1,238534 | y ~ Intervention + Sex + |
| 0,815168 | -0,04274 | 0,182475 | 142 | 70 | 72 | 138 | -0,23421 | y ~ Intervention + Sex + |
| 0,259488 | -0,14523 | 0,128268 | 142 | 70 | 72 | 138 | -1,13226 | y ~ Intervention + Sex + |
| 0,965455 | 0,006495 | 0,149688 | 142 | 70 | 72 | 138 | 0,043388 | y ~ Intervention + Sex + |
| 0,482873 | -0,09849 | 0,139979 | 142 | 70 | 72 | 138 | -0,70359 | y ~ Intervention + Sex + |
| 0,352282 | 0,149007 | 0,159652 | 142 | 70 | 72 | 138 | 0,933326 | y ~ Intervention + Sex + |
| 0,587614 | -0,08557 | 0,15742 | 142 | 70 | 72 | 138 | -0,54357 | y ~ Intervention + Sex + |
| 0,07375 | 0,283186 | 0,157163 | 142 | 70 | 72 | 138 | 1,80186 | y ~ Intervention + Sex + |
| 0,82599 | -0,03235 | 0,146882 | 142 | 70 | 72 | 138 | -0,22027 | y ~ Intervention + Sex + |
| 0,555558 | 0,093955 | 0,159004 | 142 | 70 | 72 | 138 | 0,590894 | y ~ Intervention + Sex + |
| 0,879943 | -0,02047 | 0,135291 | 142 | 70 | 72 | 138 | -0,15132 | y ~ Intervention + Sex + |
| 0,363162 | 0,15594 | 0,170916 | 142 | 70 | 72 | 138 | 0,912377 | y ~ Intervention + Sex + |
| 0,46597 | -0,11484 | 0,157089 | 142 | 70 | 72 | 138 | -0,73108 | y ~ Intervention + Sex + |
| 0,598266 | 0,085806 | 0,162475 | 142 | 70 | 72 | 138 | 0,528118 | y ~ Intervention + Sex + |
| 0,350694 | -0,14316 | 0,152876 | 142 | 70 | 72 | 138 | -0,93642 | y ~ Intervention + Sex + |
| 0,064138 | -0,25432 | 0,136278 | 142 | 70 | 72 | 138 | -1,86619 | y ~ Intervention + Sex + |
| 0,682195 | 0,059513 | 0,145033 | 142 | 70 | 72 | 138 | 0,410338 | y ~ Intervention + Sex + |
| 0,53374 | -0,10076 | 0,161504 | 142 | 70 | 72 | 138 | -0,62387 | y ~ Intervention + Sex + |
| 0,457036 | 0,118452 | 0,158819 | 142 | 70 | 72 | 138 | 0,745834 | y ~ Intervention + Sex + |
| 0,289246 | 0,162511 | 0,152755 | 142 | 70 | 72 | 138 | 1,063869 | y ~ Intervention + Sex + |
| 0,025528 | 0,354285 | 0,156916 | 142 | 70 | 72 | 138 | 2,2578 | y ~ Intervention + Sex + |
| 0,013769 | -0,34468 | 0,138138 | 142 | 70 | 72 | 138 | -2,49517 | y ~ Intervention + Sex + |
| 0,630886 | -0,07104 | 0,147523 | 142 | 70 | 72 | 138 | -0,48155 | y ~ Intervention + Sex + |
| 0,38956 | -0,1332 | 0,154321 | 142 | 70 | 72 | 138 | -0,86314 | y ~ Intervention + Sex + |
| 0,422436 | 0,126426 | 0,157129 | 142 | 70 | 72 | 138 | 0,804598 | y ~ Intervention + Sex + |
| 0,086527 | 0,305149 | 0,176762 | 142 | 70 | 72 | 138 | 1,726327 | y ~ Intervention + Sex + |
| 0,002415 | -0,45299 | 0,146556 | 142 | 70 | 72 | 138 | -3,09087 | y ~ Intervention + Sex + |
| 0,402205 | -0,13023 | 0,154978 | 142 | 70 | 72 | 138 | -0,84028 | y ~ Intervention + Sex + |
| 0,867802 | 0,026006 | 0,155949 | 142 | 70 | 72 | 138 | 0,166762 | y ~ Intervention + Sex + |
| 0,050565 | -0,30872 | 0,156523 | 142 | 70 | 72 | 138 | -1,97238 | y ~ Intervention + Sex + |
| 0,332938 | 0,164541 | 0,169347 | 142 | 70 | 72 | 138 | 0,971624 | y ~ Intervention + Sex + |
| 0,498055 | -0,09806 | 0,14434 | 142 | 70 | 72 | 138 | -0,67935 | y ~ Intervention + Sex + |
| 0,130165 | 0,209748 | 0,137763 | 142 | 70 | 72 | 138 | 1,522527 | y ~ Intervention + Sex + |
| 0,234208 | -0,16857 | 0,141085 | 142 | 70 | 72 | 138 | -1,19482 | y ~ Intervention + Sex + |
| 0,504933 | -0,09536 | 0,142652 | 142 | 70 | 72 | 138 | -0,6685 | y ~ Intervention + Sex + |
| 0,899942 | 0,021931 | 0,174106 | 142 | 70 | 72 | 138 | 0,125966 | y ~ Intervention + Sex + |
| 0,920409 | 0,01635 | 0,163331 | 142 | 70 | 72 | 138 | 0,100101 | y ~ Intervention + Sex + |
| 0,509453 | -0,1044 | 0,157846 | 142 | 70 | 72 | 138 | -0,66141 | y ~ Intervention + Sex + |
| 0,323935 | 0,151557 | 0,153096 | 142 | 70 | 72 | 138 | 0,989942 | y ~ Intervention + Sex + |
| 0,098698 | -0,24548 | 0,147666 | 142 | 70 | 72 | 138 | -1,66242 | y ~ Intervention + Sex + |
| 0,706962 | -0,05443 | 0,144477 | 142 | 70 | 72 | 138 | -0,37672 | y ~ Intervention + Sex + |
| 0,444404 | 0,135951 | 0,177255 | 142 | 70 | 72 | 138 | 0,766979 | y ~ Intervention + Sex + |
| 0,372005 | 0,141294 | 0,157757 | 142 | 70 | 72 | 138 | 0,895639 | y ~ Intervention + Sex + |

| | | | | | | | | |
|---|---|---|---|---|---|---|---|---|
| 0,603865 | 0,084998 | 0,163443 | 142 | 70 | 72 | 138 | 0,520046 | y ~ Intervention + Sex + |
| 0,004877 | -0,43451 | 0,151861 | 142 | 70 | 72 | 138 | -2,86124 | y ~ Intervention + Sex + |
| 0,216448 | -0,18826 | 0,151616 | 142 | 70 | 72 | 138 | -1,24171 | y ~ Intervention + Sex + |
| 0,133675 | 0,233074 | 0,154492 | 142 | 70 | 72 | 138 | 1,508646 | y ~ Intervention + Sex + |
| 0,030916 | -0,35469 | 0,162667 | 142 | 70 | 72 | 138 | -2,18049 | y ~ Intervention + Sex + |
| 0,343333 | -0,14581 | 0,153341 | 142 | 70 | 72 | 138 | -0,95087 | y ~ Intervention + Sex + |
| 0,063671 | 0,291169 | 0,155746 | 142 | 70 | 72 | 138 | 1,869512 | y ~ Intervention + Sex + |
| 0,015069 | 0,355418 | 0,144391 | 142 | 70 | 72 | 138 | 2,461507 | y ~ Intervention + Sex + |
| 0,956689 | 0,009238 | 0,169787 | 142 | 70 | 72 | 138 | 0,054408 | y ~ Intervention + Sex + |
| 0,247806 | -0,18991 | 0,163633 | 142 | 70 | 72 | 138 | -1,16061 | y ~ Intervention + Sex + |
| 0,197902 | -0,20716 | 0,16012 | 142 | 70 | 72 | 138 | -1,29378 | y ~ Intervention + Sex + |
| 0,044419 | -0,30303 | 0,149376 | 142 | 70 | 72 | 138 | -2,02863 | y ~ Intervention + Sex + |
| 0,718658 | 0,053154 | 0,147244 | 142 | 70 | 72 | 138 | 0,360991 | y ~ Intervention + Sex + |
| 0,579072 | -0,08455 | 0,152058 | 142 | 70 | 72 | 138 | -0,55606 | y ~ Intervention + Sex + |
| 0,650806 | -0,06644 | 0,146454 | 142 | 70 | 72 | 138 | -0,45363 | y ~ Intervention + Sex + |
| 0,453708 | 0,120095 | 0,159834 | 142 | 70 | 72 | 138 | 0,751373 | y ~ Intervention + Sex + |
| 0,843378 | 0,037117 | 0,187512 | 142 | 70 | 72 | 138 | 0,197946 | y ~ Intervention + Sex + |
| 0,227919 | 0,18079 | 0,149275 | 142 | 70 | 72 | 138 | 1,21112 | y ~ Intervention + Sex + |
| 0,684763 | 0,067798 | 0,16665 | 142 | 70 | 72 | 138 | 0,40683 | y ~ Intervention + Sex + |
| 0,727379 | -0,05416 | 0,155044 | 142 | 70 | 72 | 138 | -0,34932 | y ~ Intervention + Sex + |
| 0,797706 | -0,03794 | 0,147721 | 142 | 70 | 72 | 138 | -0,25681 | y ~ Intervention + Sex + |
| 0,081052 | 0,284335 | 0,161785 | 142 | 70 | 72 | 138 | 1,757489 | y ~ Intervention + Sex + |
| 0,94805 | -0,01089 | 0,166776 | 142 | 70 | 72 | 138 | -0,06527 | y ~ Intervention + Sex + |
| 0,268122 | 0,180299 | 0,162156 | 142 | 70 | 72 | 138 | 1,111883 | y ~ Intervention + Sex + |
| 0,647433 | -0,07428 | 0,162062 | 142 | 70 | 72 | 138 | -0,45833 | y ~ Intervention + Sex + |
| 0,82016 | 0,037849 | 0,166169 | 142 | 70 | 72 | 138 | 0,227772 | y ~ Intervention + Sex + |
| 0,558081 | -0,08832 | 0,15043 | 142 | 70 | 72 | 138 | -0,58712 | y ~ Intervention + Sex + |
| 0,736567 | 0,054516 | 0,161728 | 142 | 70 | 72 | 138 | 0,337082 | y ~ Intervention + Sex + |
| 0,94908 | -0,01045 | 0,163286 | 142 | 70 | 72 | 138 | -0,06398 | y ~ Intervention + Sex + |
| 0,085327 | -0,27457 | 0,158433 | 142 | 70 | 72 | 138 | -1,73302 | y ~ Intervention + Sex + |
| 0,067919 | -0,26918 | 0,146294 | 142 | 70 | 72 | 138 | -1,83999 | y ~ Intervention + Sex + |
| 0,444605 | 0,134304 | 0,175186 | 142 | 70 | 72 | 138 | 0,76664 | y ~ Intervention + Sex + |
| 0,635636 | 0,070673 | 0,148828 | 142 | 70 | 72 | 138 | 0,474861 | y ~ Intervention + Sex + |
| 0,71191 | 0,05873 | 0,158708 | 142 | 70 | 72 | 138 | 0,370053 | y ~ Intervention + Sex + |
| 0,875099 | -0,02631 | 0,16708 | 142 | 70 | 72 | 138 | -0,15748 | y ~ Intervention + Sex + |
| 0,793662 | -0,03733 | 0,142429 | 142 | 70 | 72 | 138 | -0,26207 | y ~ Intervention + Sex + |
| 0,577198 | 0,092446 | 0,165434 | 142 | 70 | 72 | 138 | 0,558809 | y ~ Intervention + Sex + |
| 0,963676 | 0,006607 | 0,144808 | 142 | 70 | 72 | 138 | 0,045624 | y ~ Intervention + Sex + |
| 0,426808 | -0,13292 | 0,166766 | 142 | 70 | 72 | 138 | -0,79702 | y ~ Intervention + Sex + |
| 0,647766 | -0,07693 | 0,168019 | 142 | 70 | 72 | 138 | -0,45787 | y ~ Intervention + Sex + |
| 0,732767 | -0,05822 | 0,17017 | 142 | 70 | 72 | 138 | -0,34214 | y ~ Intervention + Sex + |
| 0,704274 | 0,058339 | 0,153383 | 142 | 70 | 72 | 138 | 0,380345 | y ~ Intervention + Sex + |
| 0,551511 | -0,08707 | 0,14585 | 142 | 70 | 72 | 138 | -0,59696 | y ~ Intervention + Sex + |
| 0,163976 | 0,180664 | 0,129113 | 142 | 70 | 72 | 138 | 1,39927 | y ~ Intervention + Sex + |
| 0,954897 | -0,01014 | 0,178988 | 142 | 70 | 72 | 138 | -0,05666 | y ~ Intervention + Sex + |
| 0,291036 | -0,15346 | 0,144786 | 142 | 70 | 72 | 138 | -1,05991 | y ~ Intervention + Sex + |
| 0,959971 | 0,008193 | 0,162936 | 142 | 70 | 72 | 138 | 0,050281 | y ~ Intervention + Sex + |
| 0,491651 | -0,10478 | 0,151962 | 142 | 70 | 72 | 138 | -0,68952 | y ~ Intervention + Sex + |

| | | | | | | | | |
|---|---|---|---|---|---|---|---|---|
| 0,220888 | 0,196693 | 0,159947 | 142 | 70 | 72 | 138 | 1,229738 | y ~ Intervention + Sex + |
| 0,671497 | 0,064959 | 0,152845 | 142 | 70 | 72 | 138 | 0,425002 | y ~ Intervention + Sex + |
| 0,33643 | 0,147163 | 0,152563 | 142 | 70 | 72 | 138 | 0,964606 | y ~ Intervention + Sex + |
| 0,524189 | 0,102536 | 0,160582 | 142 | 70 | 72 | 138 | 0,638527 | y ~ Intervention + Sex + |
| 0,705931 | 0,058598 | 0,154977 | 142 | 70 | 72 | 138 | 0,378108 | y ~ Intervention + Sex + |
| 0,210878 | 0,185869 | 0,147867 | 142 | 70 | 72 | 138 | 1,256996 | y ~ Intervention + Sex + |
| 0,217303 | -0,20808 | 0,167891 | 142 | 70 | 72 | 138 | -1,2394 | y ~ Intervention + Sex + |
| 0,702675 | -0,06165 | 0,161178 | 142 | 70 | 72 | 138 | -0,38251 | y ~ Intervention + Sex + |
| 0,508622 | -0,09512 | 0,14353 | 142 | 70 | 72 | 138 | -0,66271 | y ~ Intervention + Sex + |
| 0,941014 | -0,01162 | 0,156801 | 142 | 70 | 72 | 138 | -0,07413 | y ~ Intervention + Sex + |
| 0,733827 | 0,050569 | 0,148414 | 142 | 70 | 72 | 138 | 0,340727 | y ~ Intervention + Sex + |
| 0,128288 | 0,231009 | 0,150979 | 142 | 70 | 72 | 138 | 1,530072 | y ~ Intervention + Sex + |
| 0,639281 | 0,066554 | 0,141682 | 142 | 70 | 72 | 138 | 0,469742 | y ~ Intervention + Sex + |
| 0,278288 | 0,18778 | 0,172518 | 142 | 70 | 72 | 138 | 1,088464 | y ~ Intervention + Sex + |
| 0,426978 | -0,11497 | 0,144304 | 142 | 70 | 72 | 138 | -0,79673 | y ~ Intervention + Sex + |
| 0,964134 | 0,007482 | 0,166101 | 142 | 70 | 72 | 138 | 0,045048 | y ~ Intervention + Sex + |
| 0,879081 | -0,02611 | 0,171335 | 142 | 70 | 72 | 138 | -0,15242 | y ~ Intervention + Sex + |
| 0,301221 | 0,164853 | 0,158863 | 142 | 70 | 72 | 138 | 1,037708 | y ~ Intervention + Sex + |
| 0,448868 | -0,11255 | 0,148194 | 142 | 70 | 72 | 138 | -0,75947 | y ~ Intervention + Sex + |
| 0,814407 | 0,036123 | 0,15359 | 142 | 70 | 72 | 138 | 0,235193 | y ~ Intervention + Sex + |
| 0,01208 | -0,4175 | 0,1641 | 139 | 68 | 71 | 135 | -2,54417 | y ~ Intervention + Sex + |
| 0,025474 | -0,36409 | 0,161106 | 135 | 65 | 70 | 131 | -2,25997 | y ~ Intervention + Sex + |
| 0,756939 | 0,049375 | 0,159211 | 141 | 69 | 72 | 137 | 0,310123 | y ~ Intervention + Sex + |
| 0,30563 | -0,17339 | 0,168618 | 140 | 69 | 71 | 136 | -1,02831 | y ~ Intervention + Sex + |
| 0,012561 | 0,49606 | 0,194369 | 86 | 42 | 44 | 82 | 2,552152 | y ~ Intervention + Sex + |
| 0,020243 | 0,386427 | 0,164494 | 141 | 69 | 72 | 137 | 2,34919 | y ~ Intervention + Sex + |
| 0,868813 | 0,027618 | 0,166903 | 141 | 69 | 72 | 137 | 0,165476 | y ~ Intervention + Sex + |
| 0,50849 | -0,09815 | 0,148057 | 141 | 69 | 72 | 137 | -0,66293 | y ~ Intervention + Sex + |
| 0,000837 | 0,559172 | 0,163704 | 141 | 69 | 72 | 137 | 3,415753 | y ~ Intervention + Sex + |
| 0,416379 | -0,11699 | 0,143517 | 141 | 69 | 72 | 137 | -0,81519 | y ~ Intervention + Sex + |
| 0,255718 | -0,18456 | 0,161708 | 141 | 69 | 72 | 137 | -1,14134 | y ~ Intervention + Sex + |
| 0,55906 | 0,089399 | 0,152644 | 141 | 69 | 72 | 137 | 0,585672 | y ~ Intervention + Sex + |
| 0,332996 | 0,132341 | 0,136219 | 141 | 69 | 72 | 137 | 0,971531 | y ~ Intervention + Sex + |
| 0,826894 | 0,033363 | 0,152268 | 141 | 69 | 72 | 137 | 0,219106 | y ~ Intervention + Sex + |
| 0,776404 | -0,04159 | 0,14615 | 141 | 69 | 72 | 137 | -0,28457 | y ~ Intervention + Sex + |
| 0,762331 | -0,05151 | 0,169976 | 141 | 69 | 72 | 137 | -0,30302 | y ~ Intervention + Sex + |
| 0,018903 | 0,32949 | 0,138695 | 141 | 69 | 72 | 137 | 2,375646 | y ~ Intervention + Sex + |
| 0,010679 | -0,40953 | 0,158213 | 141 | 69 | 72 | 137 | -2,5885 | y ~ Intervention + Sex + |
| 0,323268 | 0,141605 | 0,142843 | 141 | 69 | 72 | 137 | 0,991339 | y ~ Intervention + Sex + |
| 0,237276 | 0,191743 | 0,161534 | 141 | 69 | 72 | 137 | 1,187014 | y ~ Intervention + Sex + |
| 0,85373 | -0,02835 | 0,153464 | 141 | 69 | 72 | 137 | -0,18471 | y ~ Intervention + Sex + |
| 0,898675 | 0,022818 | 0,178867 | 141 | 69 | 72 | 137 | 0,127572 | y ~ Intervention + Sex + |
| 0,031084 | 0,320875 | 0,147296 | 141 | 69 | 72 | 137 | 2,178438 | y ~ Intervention + Sex + |
| 0,657651 | -0,06306 | 0,141997 | 141 | 69 | 72 | 137 | -0,44413 | y ~ Intervention + Sex + |
| 0,636839 | -0,07797 | 0,164769 | 141 | 69 | 72 | 137 | -0,47318 | y ~ Intervention + Sex + |
| 0,317284 | -0,1627 | 0,162098 | 141 | 69 | 72 | 137 | -1,00372 | y ~ Intervention + Sex + |
| 0,051634 | -0,31325 | 0,159552 | 141 | 69 | 72 | 137 | -1,96332 | y ~ Intervention + Sex + |
| 0,665352 | -0,06138 | 0,14161 | 141 | 69 | 72 | 137 | -0,43347 | y ~ Intervention + Sex + |

| | | | | | | | | |
|---|---|---|---|---|---|---|---|---|
| 0,015597 | -0,37292 | 0,152287 | 141 | 69 | 72 | 137 | -2,44879 | y ~ Intervention + Sex + |
| 0,598391 | 0,079465 | 0,150517 | 141 | 69 | 72 | 137 | 0,527946 | y ~ Intervention + Sex + |
| 0,264878 | 0,177959 | 0,15896 | 141 | 69 | 72 | 137 | 1,119518 | y ~ Intervention + Sex + |
| 0,044837 | -0,31165 | 0,153919 | 141 | 69 | 72 | 137 | -2,02474 | y ~ Intervention + Sex + |
| 0,915518 | 0,019698 | 0,185348 | 141 | 69 | 72 | 137 | 0,106276 | y ~ Intervention + Sex + |
| 0,227749 | 0,195294 | 0,161186 | 141 | 69 | 72 | 137 | 1,211606 | y ~ Intervention + Sex + |
| 0,000148 | -0,61155 | 0,156648 | 141 | 69 | 72 | 137 | -3,90394 | y ~ Intervention + Sex + |
| 0,850683 | -0,02704 | 0,143374 | 141 | 69 | 72 | 137 | -0,1886 | y ~ Intervention + Sex + |
| 0,337334 | -0,16067 | 0,166878 | 141 | 69 | 72 | 137 | -0,96282 | y ~ Intervention + Sex + |
| 0,682619 | 0,062415 | 0,152318 | 141 | 69 | 72 | 137 | 0,409764 | y ~ Intervention + Sex + |
| 0,009894 | 0,400988 | 0,153282 | 141 | 69 | 72 | 137 | 2,616016 | y ~ Intervention + Sex + |
| 0,440079 | -0,1246 | 0,160891 | 135 | 66 | 69 | 131 | -0,77442 | y ~ Intervention + Sex + |
| 0,130993 | -0,22751 | 0,149744 | 141 | 69 | 72 | 137 | -1,51929 | y ~ Intervention + Sex + |
| 0,99468 | 0,000834 | 0,124866 | 141 | 69 | 72 | 137 | 0,006679 | y ~ Intervention + Sex + |
| 0,211554 | 0,183107 | 0,145882 | 141 | 69 | 72 | 137 | 1,25517 | y ~ Intervention + Sex + |
| 0,432799 | 0,126723 | 0,161076 | 141 | 69 | 72 | 137 | 0,78673 | y ~ Intervention + Sex + |
| 0,754472 | 0,050638 | 0,161588 | 141 | 69 | 72 | 137 | 0,313376 | y ~ Intervention + Sex + |
| 0,541904 | 0,087136 | 0,142503 | 141 | 69 | 72 | 137 | 0,611466 | y ~ Intervention + Sex + |
| 0,543637 | 0,087212 | 0,143242 | 141 | 69 | 72 | 137 | 0,608843 | y ~ Intervention + Sex + |
| 0,448004 | -0,10797 | 0,141896 | 141 | 69 | 72 | 137 | -0,76093 | y ~ Intervention + Sex + |
| 0,073432 | 0,260768 | 0,144551 | 141 | 69 | 72 | 137 | 1,80398 | y ~ Intervention + Sex + |
| 0,103138 | -0,25869 | 0,157664 | 141 | 69 | 72 | 137 | -1,64078 | y ~ Intervention + Sex + |
| 0,209245 | 0,175637 | 0,13922 | 141 | 69 | 72 | 137 | 1,261577 | y ~ Intervention + Sex + |
| 0,130914 | 0,197306 | 0,12984 | 141 | 69 | 72 | 137 | 1,519607 | y ~ Intervention + Sex + |
| 0,162142 | 0,258361 | 0,183824 | 141 | 69 | 72 | 137 | 1,405479 | y ~ Intervention + Sex + |
| 0,046451 | 0,269768 | 0,134248 | 141 | 69 | 72 | 137 | 2,009474 | y ~ Intervention + Sex + |
| 0,08191 | 0,276567 | 0,157805 | 141 | 69 | 72 | 137 | 1,752587 | y ~ Intervention + Sex + |
| 0,631611 | -0,06645 | 0,138288 | 141 | 69 | 72 | 137 | -0,48054 | y ~ Intervention + Sex + |
| 0,053479 | 0,291585 | 0,149697 | 141 | 69 | 72 | 137 | 1,947839 | y ~ Intervention + Sex + |
| 0,009517 | 0,459596 | 0,174757 | 141 | 69 | 72 | 137 | 2,629922 | y ~ Intervention + Sex + |
| 0,01412 | 0,33533 | 0,134885 | 141 | 69 | 72 | 137 | 2,486036 | y ~ Intervention + Sex + |
| 0,010494 | 0,404146 | 0,155751 | 141 | 69 | 72 | 137 | 2,594821 | y ~ Intervention + Sex + |
| 0,010303 | 0,396095 | 0,15226 | 141 | 69 | 72 | 137 | 2,601439 | y ~ Intervention + Sex + |
| 0,001788 | 0,587052 | 0,184269 | 141 | 69 | 72 | 137 | 3,185845 | y ~ Intervention + Sex + |
| 0,198676 | 0,190323 | 0,147356 | 141 | 69 | 72 | 137 | 1,291584 | y ~ Intervention + Sex + |
| 0,497204 | 0,095327 | 0,140041 | 141 | 69 | 72 | 137 | 0,680709 | y ~ Intervention + Sex + |
| 0,740527 | -0,05232 | 0,157685 | 141 | 69 | 72 | 137 | -0,33183 | y ~ Intervention + Sex + |
| 0,287773 | 0,157712 | 0,147785 | 141 | 69 | 72 | 137 | 1,067167 | y ~ Intervention + Sex + |
| 0,121035 | 0,255683 | 0,163886 | 141 | 69 | 72 | 137 | 1,560134 | y ~ Intervention + Sex + |
| 0,000827 | 0,498791 | 0,145869 | 141 | 69 | 72 | 137 | 3,41945 | y ~ Intervention + Sex + |
| 0,163444 | 0,220932 | 0,157684 | 141 | 69 | 72 | 137 | 1,401104 | y ~ Intervention + Sex + |
| 0,901939 | -0,02106 | 0,17058 | 141 | 69 | 72 | 137 | -0,12344 | y ~ Intervention + Sex + |
| 0,979615 | 0,003867 | 0,151053 | 141 | 69 | 72 | 137 | 0,025599 | y ~ Intervention + Sex + |
| 0,004271 | -0,39305 | 0,135257 | 141 | 69 | 72 | 137 | -2,90595 | y ~ Intervention + Sex + |
| 0,01173 | 0,474093 | 0,185599 | 141 | 69 | 72 | 137 | 2,554391 | y ~ Intervention + Sex + |
| 0,092141 | -0,24862 | 0,146582 | 141 | 69 | 72 | 137 | -1,69608 | y ~ Intervention + Sex + |
| 0,519661 | -0,10547 | 0,163391 | 141 | 69 | 72 | 137 | -0,64553 | y ~ Intervention + Sex + |
| 0,022728 | -0,37026 | 0,160704 | 141 | 69 | 72 | 137 | -2,30399 | y ~ Intervention + Sex + |

| | | | | | | | | |
|---|---|---|---|---|---|---|---|---|
| 0,195058 | 0,186567 | 0,143279 | 141 | 69 | 72 | 137 | 1,302125 | y ~ Intervention + Sex + |
| 0,912129 | -0,01947 | 0,176099 | 141 | 69 | 72 | 137 | -0,11056 | y ~ Intervention + Sex + |
| 0,057059 | 0,276151 | 0,143901 | 141 | 69 | 72 | 137 | 1,919036 | y ~ Intervention + Sex + |
| 0,399007 | -0,13414 | 0,158546 | 141 | 69 | 72 | 137 | -0,84604 | y ~ Intervention + Sex + |
| 0,669939 | 0,060526 | 0,141696 | 141 | 69 | 72 | 137 | 0,427153 | y ~ Intervention + Sex + |
| 0,259295 | 0,186079 | 0,164271 | 141 | 69 | 72 | 137 | 1,132757 | y ~ Intervention + Sex + |
| 0,060797 | 0,327172 | 0,173057 | 141 | 69 | 72 | 137 | 1,890543 | y ~ Intervention + Sex + |
| 0,038616 | -0,30672 | 0,14687 | 141 | 69 | 72 | 137 | -2,08836 | y ~ Intervention + Sex + |
| 0,680337 | -0,07042 | 0,170562 | 141 | 69 | 72 | 137 | -0,41288 | y ~ Intervention + Sex + |
| 0,522951 | 0,085588 | 0,133638 | 141 | 69 | 72 | 137 | 0,640448 | y ~ Intervention + Sex + |
| 0,024769 | 0,351944 | 0,155041 | 141 | 69 | 72 | 137 | 2,270002 | y ~ Intervention + Sex + |
| 0,370734 | 0,119243 | 0,132779 | 141 | 69 | 72 | 137 | 0,89805 | y ~ Intervention + Sex + |
| 0,019373 | 0,373794 | 0,157973 | 141 | 69 | 72 | 137 | 2,366186 | y ~ Intervention + Sex + |
| 0,028478 | 0,392283 | 0,177179 | 141 | 69 | 72 | 137 | 2,214053 | y ~ Intervention + Sex + |
| 0,447537 | 0,100964 | 0,132548 | 141 | 69 | 72 | 137 | 0,761719 | y ~ Intervention + Sex + |
| 0,08069 | 0,267202 | 0,151845 | 141 | 69 | 72 | 137 | 1,759705 | y ~ Intervention + Sex + |
| 0,112236 | 0,264604 | 0,165533 | 141 | 69 | 72 | 137 | 1,598499 | y ~ Intervention + Sex + |
| 0,23974 | -0,16778 | 0,14209 | 141 | 69 | 72 | 137 | -1,18077 | y ~ Intervention + Sex + |
| 0,047234 | 0,341857 | 0,170738 | 141 | 69 | 72 | 137 | 2,002232 | y ~ Intervention + Sex + |
| 0,752382 | -0,05814 | 0,183921 | 141 | 69 | 72 | 137 | -0,31613 | y ~ Intervention + Sex + |
| 0,925504 | 0,016182 | 0,172744 | 141 | 69 | 72 | 137 | 0,093675 | y ~ Intervention + Sex + |
| 0,919873 | -0,01342 | 0,133128 | 141 | 69 | 72 | 137 | -0,10078 | y ~ Intervention + Sex + |
| 0,702601 | 0,060054 | 0,156957 | 141 | 69 | 72 | 137 | 0,382611 | y ~ Intervention + Sex + |
| 0,951388 | 0,00789 | 0,129193 | 141 | 69 | 72 | 137 | 0,061075 | y ~ Intervention + Sex + |
| 0,492508 | -0,10456 | 0,15194 | 141 | 69 | 72 | 137 | -0,68817 | y ~ Intervention + Sex + |
| 0,142496 | 0,22094 | 0,149786 | 141 | 69 | 72 | 137 | 1,475042 | y ~ Intervention + Sex + |
| 0,066767 | 0,280522 | 0,151802 | 141 | 69 | 72 | 137 | 1,847943 | y ~ Intervention + Sex + |
| 0,844852 | 0,034113 | 0,173989 | 141 | 69 | 72 | 137 | 0,196062 | y ~ Intervention + Sex + |
| 0,062717 | -0,31991 | 0,170484 | 141 | 69 | 72 | 137 | -1,87647 | y ~ Intervention + Sex + |
| 0,474293 | -0,11525 | 0,160633 | 141 | 69 | 72 | 137 | -0,71749 | y ~ Intervention + Sex + |
| 0,196328 | 0,191415 | 0,147423 | 141 | 69 | 72 | 137 | 1,298411 | y ~ Intervention + Sex + |
| 0,008247 | 0,383554 | 0,143076 | 141 | 69 | 72 | 137 | 2,680769 | y ~ Intervention + Sex + |
| 0,607725 | -0,08042 | 0,156308 | 141 | 69 | 72 | 137 | -0,51451 | y ~ Intervention + Sex + |
| 0,836757 | -0,03353 | 0,162412 | 141 | 69 | 72 | 137 | -0,20644 | y ~ Intervention + Sex + |
| 0,132907 | 0,226353 | 0,149731 | 141 | 69 | 72 | 137 | 1,511727 | y ~ Intervention + Sex + |
| 0,800664 | -0,0404 | 0,159699 | 141 | 69 | 72 | 137 | -0,25298 | y ~ Intervention + Sex + |
| 0,15414 | -0,21392 | 0,149281 | 141 | 69 | 72 | 137 | -1,43298 | y ~ Intervention + Sex + |
| 0,116484 | 0,229239 | 0,145117 | 141 | 69 | 72 | 137 | 1,57969 | y ~ Intervention + Sex + |
| 0,021617 | 0,362457 | 0,155987 | 141 | 69 | 72 | 137 | 2,323629 | y ~ Intervention + Sex + |
| 0,415871 | 0,11509 | 0,141027 | 141 | 69 | 72 | 137 | 0,81608 | y ~ Intervention + Sex + |
| 0,434476 | 0,108772 | 0,138764 | 141 | 69 | 72 | 137 | 0,783859 | y ~ Intervention + Sex + |
| 0,888486 | -0,01817 | 0,129308 | 141 | 69 | 72 | 137 | -0,14048 | y ~ Intervention + Sex + |
| 0,621392 | 0,081534 | 0,164714 | 141 | 69 | 72 | 137 | 0,495002 | y ~ Intervention + Sex + |
| 0,241245 | -0,18782 | 0,159576 | 141 | 69 | 72 | 137 | -1,17698 | y ~ Intervention + Sex + |
| 0,919396 | -0,01684 | 0,166152 | 141 | 69 | 72 | 137 | -0,10138 | y ~ Intervention + Sex + |
| 0,005236 | -0,52256 | 0,184152 | 141 | 69 | 72 | 137 | -2,83766 | y ~ Intervention + Sex + |
| 0,112769 | -0,20766 | 0,130103 | 141 | 69 | 72 | 137 | -1,59611 | y ~ Intervention + Sex + |
| 0,000124 | 0,657292 | 0,166379 | 141 | 69 | 72 | 137 | 3,950563 | y ~ Intervention + Sex + |

| | | | | | | | | |
|---|---|---|---|---|---|---|---|---|
| 0,49241 | -0,10677 | 0,155108 | 141 | 69 | 72 | 137 | -0,68833 | y ~ Intervention + Sex + |
| 0,094009 | 0,282217 | 0,167356 | 141 | 69 | 72 | 137 | 1,686328 | y ~ Intervention + Sex + |
| 0,341285 | -0,12737 | 0,133378 | 141 | 69 | 72 | 137 | -0,95495 | y ~ Intervention + Sex + |
| 0,369777 | 0,139787 | 0,155344 | 141 | 69 | 72 | 137 | 0,899854 | y ~ Intervention + Sex + |
| 0,207795 | 0,172746 | 0,136491 | 141 | 69 | 72 | 137 | 1,265626 | y ~ Intervention + Sex + |
| 0,393156 | -0,12889 | 0,150462 | 141 | 69 | 72 | 137 | -0,85661 | y ~ Intervention + Sex + |
| 0,568139 | 0,106855 | 0,186752 | 141 | 69 | 72 | 137 | 0,572179 | y ~ Intervention + Sex + |
| 0,37535 | -0,13118 | 0,147492 | 141 | 69 | 72 | 137 | -0,8894 | y ~ Intervention + Sex + |
| 0,620044 | 0,076363 | 0,153674 | 141 | 69 | 72 | 137 | 0,496918 | y ~ Intervention + Sex + |
| 0,936552 | -0,0137 | 0,171733 | 141 | 69 | 72 | 137 | -0,07975 | y ~ Intervention + Sex + |
| 0,142073 | 0,213534 | 0,14461 | 141 | 69 | 72 | 137 | 1,476618 | y ~ Intervention + Sex + |
| 0,068826 | 0,286125 | 0,156013 | 141 | 69 | 72 | 137 | 1,833987 | y ~ Intervention + Sex + |
| 0,509494 | 0,121283 | 0,183385 | 141 | 69 | 72 | 137 | 0,661358 | y ~ Intervention + Sex + |
| 0,353422 | 0,157192 | 0,168818 | 141 | 69 | 72 | 137 | 0,931134 | y ~ Intervention + Sex + |
| 0,222994 | 0,217918 | 0,178014 | 141 | 69 | 72 | 137 | 1,224158 | y ~ Intervention + Sex + |
| 0,93131 | -0,01334 | 0,154444 | 141 | 69 | 72 | 137 | -0,08636 | y ~ Intervention + Sex + |
| 0,647652 | -0,0805 | 0,175741 | 141 | 69 | 72 | 137 | -0,45804 | y ~ Intervention + Sex + |
| 0,522526 | 0,097789 | 0,152532 | 141 | 69 | 72 | 137 | 0,641104 | y ~ Intervention + Sex + |
| 0,665209 | 0,062406 | 0,143901 | 141 | 69 | 72 | 137 | 0,433671 | y ~ Intervention + Sex + |
| 0,993393 | -0,00155 | 0,186902 | 141 | 69 | 72 | 137 | -0,0083 | y ~ Intervention + Sex + |
| 0,225368 | -0,16817 | 0,138089 | 141 | 69 | 72 | 137 | -1,21787 | y ~ Intervention + Sex + |
| 0,335311 | 0,150307 | 0,155457 | 141 | 69 | 72 | 137 | 0,966873 | y ~ Intervention + Sex + |
| 0,206937 | 0,196139 | 0,15468 | 141 | 69 | 72 | 137 | 1,268032 | y ~ Intervention + Sex + |
| 0,145347 | 0,242675 | 0,165705 | 141 | 69 | 72 | 137 | 1,464504 | y ~ Intervention + Sex + |
| 0,000637 | 0,489705 | 0,140084 | 141 | 69 | 72 | 137 | 3,495797 | y ~ Intervention + Sex + |
| 0,021267 | -0,36879 | 0,15828 | 141 | 69 | 72 | 137 | -2,33 | y ~ Intervention + Sex + |
| 0,998311 | 0,000378 | 0,178304 | 141 | 69 | 72 | 137 | 0,002121 | y ~ Intervention + Sex + |
| 0,111569 | -0,25382 | 0,158487 | 141 | 69 | 72 | 137 | -1,6015 | y ~ Intervention + Sex + |
| 0,011491 | 0,378028 | 0,147556 | 141 | 69 | 72 | 137 | 2,561918 | y ~ Intervention + Sex + |
| 0,880983 | 0,024143 | 0,160949 | 141 | 69 | 72 | 137 | 0,150003 | y ~ Intervention + Sex + |
| 0,261505 | -0,18207 | 0,161485 | 141 | 69 | 72 | 137 | -1,12749 | y ~ Intervention + Sex + |
| 6,40E-05 | 0,68682 | 0,166506 | 141 | 69 | 72 | 137 | 4,124887 | y ~ Intervention + Sex + |
| 0,41033 | -0,13175 | 0,15954 | 141 | 69 | 72 | 137 | -0,82584 | y ~ Intervention + Sex + |
| 0,845161 | 0,025619 | 0,130933 | 141 | 69 | 72 | 137 | 0,195667 | y ~ Intervention + Sex + |
| 0,058267 | 0,326874 | 0,171169 | 141 | 69 | 72 | 137 | 1,909659 | y ~ Intervention + Sex + |
| 0,065573 | 0,272638 | 0,146879 | 141 | 69 | 72 | 137 | 1,856204 | y ~ Intervention + Sex + |
| 0,281308 | -0,17113 | 0,158215 | 141 | 69 | 72 | 137 | -1,08165 | y ~ Intervention + Sex + |
| 0,962358 | -0,00714 | 0,151023 | 141 | 69 | 72 | 137 | -0,04728 | y ~ Intervention + Sex + |
| 0,908549 | -0,01612 | 0,140047 | 141 | 69 | 72 | 137 | -0,11508 | y ~ Intervention + Sex + |
| 0,033292 | 0,300085 | 0,139559 | 141 | 69 | 72 | 137 | 2,150239 | y ~ Intervention + Sex + |
| 0,018377 | 0,390103 | 0,163463 | 141 | 69 | 72 | 137 | 2,386486 | y ~ Intervention + Sex + |
| 0,460421 | -0,11427 | 0,154363 | 141 | 69 | 72 | 137 | -0,74024 | y ~ Intervention + Sex + |
| 0,577476 | -0,07977 | 0,142851 | 141 | 69 | 72 | 137 | -0,55841 | y ~ Intervention + Sex + |
| 0,102261 | 0,279446 | 0,169875 | 141 | 69 | 72 | 137 | 1,645008 | y ~ Intervention + Sex + |
| 0,122435 | 0,22759 | 0,146432 | 141 | 69 | 72 | 137 | 1,554237 | y ~ Intervention + Sex + |
| 0,178188 | 0,235619 | 0,174107 | 141 | 69 | 72 | 137 | 1,353302 | y ~ Intervention + Sex + |
| 0,794995 | 0,036845 | 0,141529 | 141 | 69 | 72 | 137 | 0,260337 | y ~ Intervention + Sex + |
| 0,734421 | 0,043158 | 0,126957 | 141 | 69 | 72 | 137 | 0,339942 | y ~ Intervention + Sex + |

| | | | | | | | | |
|---|---|---|---|---|---|---|---|---|
| 0,043305 | -0,33002 | 0,161801 | 141 | 69 | 72 | 137 | -2,03967 | y ~ Intervention + Sex + |
| 0,908408 | -0,01795 | 0,155757 | 141 | 69 | 72 | 137 | -0,11526 | y ~ Intervention + Sex + |
| 0,022402 | 0,38377 | 0,166158 | 141 | 69 | 72 | 137 | 2,309663 | y ~ Intervention + Sex + |
| 0,240097 | 0,157816 | 0,133757 | 141 | 69 | 72 | 137 | 1,179869 | y ~ Intervention + Sex + |
| 0,763895 | 0,051123 | 0,169863 | 141 | 69 | 72 | 137 | 0,300968 | y ~ Intervention + Sex + |
| 0,593866 | 0,075109 | 0,140523 | 141 | 69 | 72 | 137 | 0,534494 | y ~ Intervention + Sex + |
| 0,43661 | 0,125853 | 0,161306 | 141 | 69 | 72 | 137 | 0,780216 | y ~ Intervention + Sex + |
| 0,001406 | 0,452345 | 0,13876 | 141 | 69 | 72 | 137 | 3,259909 | y ~ Intervention + Sex + |
| 0,289232 | 0,177891 | 0,167202 | 141 | 69 | 72 | 137 | 1,063929 | y ~ Intervention + Sex + |
| 0,238726 | 0,149282 | 0,126154 | 141 | 69 | 72 | 137 | 1,183335 | y ~ Intervention + Sex + |
| 0,41712 | 0,139933 | 0,171931 | 141 | 69 | 72 | 137 | 0,813891 | y ~ Intervention + Sex + |
| 0,070504 | 0,304071 | 0,166809 | 141 | 69 | 72 | 137 | 1,822866 | y ~ Intervention + Sex + |
| 0,519356 | -0,09789 | 0,151532 | 141 | 69 | 72 | 137 | -0,64601 | y ~ Intervention + Sex + |
| 0,468509 | -0,11742 | 0,161533 | 141 | 69 | 72 | 137 | -0,72693 | y ~ Intervention + Sex + |
| 0,015253 | 0,434845 | 0,176969 | 141 | 69 | 72 | 137 | 2,457176 | y ~ Intervention + Sex + |
| 0,614665 | -0,0754 | 0,149432 | 141 | 69 | 72 | 137 | -0,50458 | y ~ Intervention + Sex + |
| 0,123329 | 0,266667 | 0,171988 | 141 | 69 | 72 | 137 | 1,550497 | y ~ Intervention + Sex + |
| 0,101507 | -0,24699 | 0,14981 | 141 | 69 | 72 | 137 | -1,64867 | y ~ Intervention + Sex + |
| 2,83E-06 | 0,715066 | 0,146355 | 141 | 69 | 72 | 137 | 4,885824 | y ~ Intervention + Sex + |
| 0,05037 | 0,272813 | 0,138189 | 141 | 69 | 72 | 137 | 1,9742 | y ~ Intervention + Sex + |
| 0,783018 | -0,0436 | 0,157997 | 141 | 69 | 72 | 137 | -0,27593 | y ~ Intervention + Sex + |
| 0,955914 | 0,010389 | 0,187577 | 141 | 69 | 72 | 137 | 0,055383 | y ~ Intervention + Sex + |
| 0,046823 | 0,30274 | 0,150916 | 141 | 69 | 72 | 137 | 2,006021 | y ~ Intervention + Sex + |
| 0,020444 | 0,341364 | 0,145549 | 141 | 69 | 72 | 137 | 2,345357 | y ~ Intervention + Sex + |
| 0,028051 | 0,330169 | 0,148714 | 141 | 69 | 72 | 137 | 2,220163 | y ~ Intervention + Sex + |
| 0,000823 | 0,487846 | 0,142606 | 141 | 69 | 72 | 137 | 3,420935 | y ~ Intervention + Sex + |
| 0,00877 | -0,44705 | 0,168123 | 141 | 69 | 72 | 137 | -2,65904 | y ~ Intervention + Sex + |
| 0,110294 | -0,2291 | 0,142541 | 141 | 69 | 72 | 137 | -1,60729 | y ~ Intervention + Sex + |
| 0,093524 | 0,257005 | 0,152178 | 141 | 69 | 72 | 137 | 1,688843 | y ~ Intervention + Sex + |
| 0,002728 | 0,495162 | 0,162226 | 141 | 69 | 72 | 137 | 3,052291 | y ~ Intervention + Sex + |
| 0,948917 | -0,00959 | 0,149459 | 141 | 69 | 72 | 137 | -0,06418 | y ~ Intervention + Sex + |
| 0,627283 | 0,077232 | 0,158701 | 141 | 69 | 72 | 137 | 0,486651 | y ~ Intervention + Sex + |
| 0,606813 | 0,066902 | 0,129701 | 141 | 69 | 72 | 137 | 0,515818 | y ~ Intervention + Sex + |
| 0,163651 | 0,183548 | 0,131067 | 141 | 69 | 72 | 137 | 1,400412 | y ~ Intervention + Sex + |
| 0,440622 | 0,131822 | 0,170447 | 141 | 69 | 72 | 137 | 0,773393 | y ~ Intervention + Sex + |
| 0,002626 | 0,509838 | 0,166368 | 141 | 69 | 72 | 137 | 3,064526 | y ~ Intervention + Sex + |
| 0,70644 | -0,0629 | 0,166645 | 141 | 69 | 72 | 137 | -0,37743 | y ~ Intervention + Sex + |
| 0,007191 | 0,422905 | 0,154978 | 141 | 69 | 72 | 137 | 2,728798 | y ~ Intervention + Sex + |
| 0,837397 | -0,03532 | 0,171767 | 141 | 69 | 72 | 137 | -0,20561 | y ~ Intervention + Sex + |
| 0,053277 | 0,291257 | 0,1494 | 141 | 69 | 72 | 137 | 1,949512 | y ~ Intervention + Sex + |
| 0,845192 | -0,03226 | 0,164883 | 141 | 69 | 72 | 137 | -0,19563 | y ~ Intervention + Sex + |
| 0,512242 | 0,097196 | 0,147925 | 141 | 69 | 72 | 137 | 0,657065 | y ~ Intervention + Sex + |
| 0,381362 | -0,13016 | 0,148204 | 141 | 69 | 72 | 137 | -0,87822 | y ~ Intervention + Sex + |
| 0,873825 | -0,02607 | 0,163859 | 141 | 69 | 72 | 137 | -0,1591 | y ~ Intervention + Sex + |
| 0,025622 | 0,342971 | 0,151991 | 141 | 69 | 72 | 137 | 2,256516 | y ~ Intervention + Sex + |
| 0,098037 | 0,278798 | 0,167365 | 141 | 69 | 72 | 137 | 1,665806 | y ~ Intervention + Sex + |
| 0,000106 | -0,61649 | 0,154436 | 141 | 69 | 72 | 137 | -3,99185 | y ~ Intervention + Sex + |
| 0,476112 | -0,12266 | 0,171671 | 141 | 69 | 72 | 137 | -0,71454 | y ~ Intervention + Sex + |

| | | | | | | | | |
|---|---|---|---|---|---|---|---|---|
| 0,04294 | -0,3329 | 0,162924 | 141 | 69 | 72 | 137 | -2,0433 | y ~ Intervention + Sex + |
| 0,304299 | 0,165575 | 0,160577 | 141 | 69 | 72 | 137 | 1,031125 | y ~ Intervention + Sex + |
| 0,010319 | 0,356151 | 0,136935 | 141 | 69 | 72 | 137 | 2,60088 | y ~ Intervention + Sex + |
| 0,367991 | -0,14872 | 0,164649 | 141 | 69 | 72 | 137 | -0,90323 | y ~ Intervention + Sex + |
| 0,923437 | -0,01573 | 0,163425 | 141 | 69 | 72 | 137 | -0,09628 | y ~ Intervention + Sex + |
| 0,170424 | 0,242477 | 0,175952 | 141 | 69 | 72 | 137 | 1,378083 | y ~ Intervention + Sex + |
| 0,144815 | -0,24332 | 0,165925 | 141 | 69 | 72 | 137 | -1,46646 | y ~ Intervention + Sex + |
| 0,188822 | 0,213869 | 0,161943 | 141 | 69 | 72 | 137 | 1,320645 | y ~ Intervention + Sex + |
| 0,073364 | 0,293293 | 0,162542 | 141 | 69 | 72 | 137 | 1,804407 | y ~ Intervention + Sex + |
| 0,974453 | 0,005228 | 0,162963 | 141 | 69 | 72 | 137 | 0,032082 | y ~ Intervention + Sex + |
| 0,21466 | 0,212744 | 0,170655 | 141 | 69 | 72 | 137 | 1,24663 | y ~ Intervention + Sex + |
| 0,182826 | 0,213947 | 0,159795 | 141 | 69 | 72 | 137 | 1,338882 | y ~ Intervention + Sex + |
| 0,022415 | -0,39923 | 0,172868 | 141 | 69 | 72 | 137 | -2,30944 | y ~ Intervention + Sex + |
| 0,030854 | 0,360311 | 0,165168 | 141 | 69 | 72 | 137 | 2,181474 | y ~ Intervention + Sex + |
| 0,001118 | 0,557623 | 0,167474 | 141 | 69 | 72 | 137 | 3,329601 | y ~ Intervention + Sex + |
| 0,134895 | -0,2474 | 0,164499 | 141 | 69 | 72 | 137 | -1,50395 | y ~ Intervention + Sex + |
| 0,018769 | 0,420459 | 0,176784 | 141 | 69 | 72 | 137 | 2,378375 | y ~ Intervention + Sex + |
| 0,288222 | 0,163984 | 0,153807 | 141 | 69 | 72 | 137 | 1,066171 | y ~ Intervention + Sex + |
| 0,164345 | 0,213878 | 0,152979 | 141 | 69 | 72 | 137 | 1,398091 | y ~ Intervention + Sex + |
| 0,908147 | -0,01827 | 0,158037 | 141 | 69 | 72 | 137 | -0,11559 | y ~ Intervention + Sex + |
| 0,68071 | 0,065152 | 0,157992 | 141 | 69 | 72 | 137 | 0,412374 | y ~ Intervention + Sex + |
| 0,310352 | -0,15437 | 0,151607 | 141 | 69 | 72 | 137 | -1,01825 | y ~ Intervention + Sex + |
| 0,201962 | 0,205539 | 0,16031 | 141 | 69 | 72 | 137 | 1,282131 | y ~ Intervention + Sex + |
| 0,506056 | -0,0966 | 0,144878 | 141 | 69 | 72 | 137 | -0,66675 | y ~ Intervention + Sex + |
| 0,221329 | -0,20079 | 0,163433 | 141 | 69 | 72 | 137 | -1,2286 | y ~ Intervention + Sex + |
| 0,522257 | -0,10706 | 0,166886 | 141 | 69 | 72 | 137 | -0,64152 | y ~ Intervention + Sex + |
| 0,049626 | 0,30737 | 0,155181 | 141 | 69 | 72 | 137 | 1,980717 | y ~ Intervention + Sex + |
| 0,952425 | 0,009242 | 0,154615 | 141 | 69 | 72 | 137 | 0,059772 | y ~ Intervention + Sex + |
| 0,653247 | -0,07082 | 0,157295 | 141 | 69 | 72 | 137 | -0,45024 | y ~ Intervention + Sex + |
| 0,017195 | -0,36658 | 0,151989 | 141 | 69 | 72 | 137 | -2,41187 | y ~ Intervention + Sex + |
| 0,927336 | 0,012802 | 0,140116 | 141 | 69 | 72 | 137 | 0,091365 | y ~ Intervention + Sex + |
| 0,714425 | -0,05997 | 0,163546 | 141 | 69 | 72 | 137 | -0,36668 | y ~ Intervention + Sex + |
| 0,012899 | 0,393761 | 0,156282 | 141 | 69 | 72 | 137 | 2,519547 | y ~ Intervention + Sex + |
| 0,048091 | -0,35575 | 0,178371 | 141 | 69 | 72 | 137 | -1,99442 | y ~ Intervention + Sex + |
| 0,012225 | -0,39604 | 0,155965 | 141 | 69 | 72 | 137 | -2,53927 | y ~ Intervention + Sex + |
| 0,00946 | 0,450451 | 0,171138 | 141 | 69 | 72 | 137 | 2,632087 | y ~ Intervention + Sex + |
| 0,082694 | 0,267272 | 0,152897 | 141 | 69 | 72 | 137 | 1,748058 | y ~ Intervention + Sex + |
| 0,552359 | -0,10145 | 0,170308 | 141 | 69 | 72 | 137 | -0,5957 | y ~ Intervention + Sex + |
| 0,625452 | 0,069013 | 0,14106 | 141 | 69 | 72 | 137 | 0,489243 | y ~ Intervention + Sex + |
| 0,601788 | 0,080881 | 0,154634 | 141 | 69 | 72 | 137 | 0,523045 | y ~ Intervention + Sex + |
| 0,402567 | -0,13024 | 0,155107 | 141 | 69 | 72 | 137 | -0,83965 | y ~ Intervention + Sex + |
| 0,669356 | -0,06633 | 0,155 | 141 | 69 | 72 | 137 | -0,42796 | y ~ Intervention + Sex + |
| 0,067727 | -0,35365 | 0,192056 | 141 | 69 | 72 | 137 | -1,84139 | y ~ Intervention + Sex + |
| 0,354486 | 0,137179 | 0,147652 | 141 | 69 | 72 | 137 | 0,929073 | y ~ Intervention + Sex + |
| 0,289352 | -0,16237 | 0,152656 | 141 | 69 | 72 | 137 | -1,06366 | y ~ Intervention + Sex + |
| 0,108044 | -0,25107 | 0,155211 | 141 | 69 | 72 | 137 | -1,61762 | y ~ Intervention + Sex + |
| 0,424325 | 0,121261 | 0,151324 | 141 | 69 | 72 | 137 | 0,801335 | y ~ Intervention + Sex + |
| 0,031885 | -0,32621 | 0,150466 | 141 | 69 | 72 | 137 | -2,16801 | y ~ Intervention + Sex + |

| | | | | | | | | |
|---|---|---|---|---|---|---|---|---|
| 0,16023 | 0,203396 | 0,144052 | 141 | 69 | 72 | 137 | 1,411958 | y ~ Intervention + Sex + |
| 0,787605 | -0,04636 | 0,171733 | 141 | 69 | 72 | 137 | -0,26995 | y ~ Intervention + Sex + |
| 0,78256 | 0,044714 | 0,161696 | 141 | 69 | 72 | 137 | 0,276527 | y ~ Intervention + Sex + |
| 0,331455 | -0,1454 | 0,149178 | 141 | 69 | 72 | 137 | -0,97464 | y ~ Intervention + Sex + |
| 0,132639 | -0,2484 | 0,164201 | 141 | 69 | 72 | 137 | -1,51278 | y ~ Intervention + Sex + |
| 0,069465 | -0,24418 | 0,133449 | 141 | 69 | 72 | 137 | -1,82973 | y ~ Intervention + Sex + |
| 0,010198 | -0,36422 | 0,139809 | 141 | 69 | 72 | 137 | -2,60516 | y ~ Intervention + Sex + |
| 0,09278 | -0,25961 | 0,15337 | 141 | 69 | 72 | 137 | -1,69273 | y ~ Intervention + Sex + |
| 0,012106 | 0,40571 | 0,159548 | 141 | 69 | 72 | 137 | 2,542874 | y ~ Intervention + Sex + |
| 0,97752 | -0,00495 | 0,175354 | 141 | 69 | 72 | 137 | -0,02823 | y ~ Intervention + Sex + |
| 0,423821 | -0,13288 | 0,165638 | 141 | 69 | 72 | 137 | -0,80221 | y ~ Intervention + Sex + |
| 0,146661 | 0,24833 | 0,170124 | 141 | 69 | 72 | 137 | 1,459703 | y ~ Intervention + Sex + |
| 0,033422 | 0,327341 | 0,152349 | 141 | 69 | 72 | 137 | 2,148629 | y ~ Intervention + Sex + |
| 0,0357 | 0,3087 | 0,145526 | 141 | 69 | 72 | 137 | 2,121268 | y ~ Intervention + Sex + |
| 0,977053 | 0,004376 | 0,151844 | 141 | 69 | 72 | 137 | 0,028816 | y ~ Intervention + Sex + |
| 0,757573 | 0,052185 | 0,168725 | 141 | 69 | 72 | 137 | 0,309287 | y ~ Intervention + Sex + |
| 0,653947 | -0,07201 | 0,160292 | 141 | 69 | 72 | 137 | -0,44927 | y ~ Intervention + Sex + |
| 0,775488 | 0,043148 | 0,150989 | 141 | 69 | 72 | 137 | 0,285767 | y ~ Intervention + Sex + |
| 0,737757 | -0,05678 | 0,16923 | 141 | 69 | 72 | 137 | -0,33551 | y ~ Intervention + Sex + |
| 0,463543 | -0,12398 | 0,168661 | 141 | 69 | 72 | 137 | -0,73509 | y ~ Intervention + Sex + |
| 0,735664 | 0,053089 | 0,156934 | 141 | 69 | 72 | 137 | 0,338288 | y ~ Intervention + Sex + |
| 0,066109 | 0,310179 | 0,16744 | 141 | 69 | 72 | 137 | 1,852476 | y ~ Intervention + Sex + |
| 0,799956 | -0,03928 | 0,154717 | 141 | 69 | 72 | 137 | -0,2539 | y ~ Intervention + Sex + |
| 0,926015 | 0,014069 | 0,151232 | 141 | 69 | 72 | 137 | 0,093031 | y ~ Intervention + Sex + |
| 0,310749 | 0,135081 | 0,132769 | 141 | 69 | 72 | 137 | 1,017416 | y ~ Intervention + Sex + |
| 0,38709 | 0,133244 | 0,153565 | 141 | 69 | 72 | 137 | 0,867675 | y ~ Intervention + Sex + |
| 4,93E-05 | 0,661843 | 0,157877 | 141 | 69 | 72 | 137 | 4,192139 | y ~ Intervention + Sex + |
| 0,21023 | 0,222461 | 0,176719 | 141 | 69 | 72 | 137 | 1,258837 | y ~ Intervention + Sex + |
| 0,000208 | 0,519234 | 0,136222 | 141 | 69 | 72 | 137 | 3,811675 | y ~ Intervention + Sex + |
| 0,751713 | -0,04945 | 0,155972 | 141 | 69 | 72 | 137 | -0,31702 | y ~ Intervention + Sex + |
| 0,866341 | 0,024355 | 0,144436 | 141 | 69 | 72 | 137 | 0,168624 | y ~ Intervention + Sex + |
| 4,90E-05 | 0,663808 | 0,158279 | 141 | 69 | 72 | 137 | 4,193897 | y ~ Intervention + Sex + |
| 0,541133 | 0,091606 | 0,149528 | 141 | 69 | 72 | 137 | 0,612635 | y ~ Intervention + Sex + |
| 0,49932 | 0,105509 | 0,155765 | 141 | 69 | 72 | 137 | 0,67736 | y ~ Intervention + Sex + |
| 0,470529 | -0,10336 | 0,14284 | 141 | 69 | 72 | 137 | -0,72363 | y ~ Intervention + Sex + |
| 0,769272 | 0,047975 | 0,163232 | 141 | 69 | 72 | 137 | 0,29391 | y ~ Intervention + Sex + |
| 0,022418 | -0,33413 | 0,144683 | 141 | 69 | 72 | 137 | -2,30938 | y ~ Intervention + Sex + |
| 0,808212 | -0,03428 | 0,140934 | 141 | 69 | 72 | 137 | -0,2432 | y ~ Intervention + Sex + |
| 0,060895 | 0,301796 | 0,159696 | 141 | 69 | 72 | 137 | 1,889815 | y ~ Intervention + Sex + |
| 0,113123 | 0,246824 | 0,154795 | 141 | 69 | 72 | 137 | 1,594523 | y ~ Intervention + Sex + |
| 0,991965 | -0,00163 | 0,161239 | 141 | 69 | 72 | 137 | -0,01009 | y ~ Intervention + Sex + |
| 0,925497 | -0,01426 | 0,152169 | 141 | 69 | 72 | 137 | -0,09368 | y ~ Intervention + Sex + |
| 0,658324 | -0,06588 | 0,148638 | 141 | 69 | 72 | 137 | -0,4432 | y ~ Intervention + Sex + |
| 0,145918 | 0,250458 | 0,171263 | 141 | 69 | 72 | 137 | 1,462415 | y ~ Intervention + Sex + |
| 0,861532 | -0,02813 | 0,160997 | 141 | 69 | 72 | 137 | -0,17475 | y ~ Intervention + Sex + |
| 0,588101 | -0,08362 | 0,15404 | 141 | 69 | 72 | 137 | -0,54287 | y ~ Intervention + Sex + |
| 0,205672 | -0,20379 | 0,160264 | 141 | 69 | 72 | 137 | -1,27159 | y ~ Intervention + Sex + |
| 0,988693 | -0,00202 | 0,142334 | 141 | 69 | 72 | 137 | -0,0142 | y ~ Intervention + Sex + |

| | | | | | | | | |
|---|---|---|---|---|---|---|---|---|
| 0,000146 | 0,607536 | 0,155505 | 141 | 69 | 72 | 137 | 3,906859 | y ~ Intervention + Sex + |
| 0,896028 | -0,02281 | 0,174216 | 141 | 69 | 72 | 137 | -0,13092 | y ~ Intervention + Sex + |
| 0,698782 | 0,063102 | 0,162728 | 141 | 69 | 72 | 137 | 0,387778 | y ~ Intervention + Sex + |
| 0,262699 | 0,165316 | 0,146992 | 141 | 69 | 72 | 137 | 1,124663 | y ~ Intervention + Sex + |
| 0,892521 | -0,02056 | 0,151908 | 141 | 69 | 72 | 137 | -0,13537 | y ~ Intervention + Sex + |
| 0,038104 | 0,324283 | 0,154864 | 141 | 69 | 72 | 137 | 2,093979 | y ~ Intervention + Sex + |
| 0,633012 | -0,07374 | 0,154094 | 141 | 69 | 72 | 137 | -0,47856 | y ~ Intervention + Sex + |
| 0,497668 | -0,11802 | 0,173567 | 141 | 69 | 72 | 137 | -0,67997 | y ~ Intervention + Sex + |
| 0,363247 | -0,16347 | 0,179194 | 141 | 69 | 72 | 137 | -0,91224 | y ~ Intervention + Sex + |
| 0,817203 | -0,03629 | 0,156704 | 141 | 69 | 72 | 137 | -0,23159 | y ~ Intervention + Sex + |
| 0,851716 | -0,02883 | 0,153928 | 141 | 69 | 72 | 137 | -0,18728 | y ~ Intervention + Sex + |
| 0,695279 | 0,062997 | 0,160491 | 141 | 69 | 72 | 137 | 0,392527 | y ~ Intervention + Sex + |
| 0,002793 | 0,518418 | 0,170265 | 141 | 69 | 72 | 137 | 3,044778 | y ~ Intervention + Sex + |
| 0,134603 | -0,22743 | 0,151104 | 141 | 69 | 72 | 137 | -1,50509 | y ~ Intervention + Sex + |
| 0,000678 | -0,5266 | 0,151417 | 141 | 69 | 72 | 137 | -3,47785 | y ~ Intervention + Sex + |
| 0,24406 | 0,167765 | 0,143397 | 141 | 69 | 72 | 137 | 1,16993 | y ~ Intervention + Sex + |
| 0,887962 | 0,0224 | 0,158704 | 141 | 69 | 72 | 137 | 0,141146 | y ~ Intervention + Sex + |
| 0,67 | 0,06611 | 0,154799 | 141 | 69 | 72 | 137 | 0,427068 | y ~ Intervention + Sex + |
| 0,955072 | -0,00857 | 0,151896 | 141 | 69 | 72 | 137 | -0,05644 | y ~ Intervention + Sex + |
| 9,52E-05 | 0,584732 | 0,145416 | 141 | 69 | 72 | 137 | 4,021092 | y ~ Intervention + Sex + |
| 0,916634 | -0,01604 | 0,152918 | 141 | 69 | 72 | 137 | -0,10487 | y ~ Intervention + Sex + |
| 0,477095 | 0,115517 | 0,162029 | 141 | 69 | 72 | 137 | 0,712941 | y ~ Intervention + Sex + |
| 0,819483 | -0,03497 | 0,152929 | 141 | 69 | 72 | 137 | -0,22865 | y ~ Intervention + Sex + |
| 5,31E-16 | 1,408143 | 0,153056 | 141 | 69 | 72 | 137 | 9,200162 | y ~ Intervention + Sex + |
| 0,187311 | 0,221273 | 0,166974 | 141 | 69 | 72 | 137 | 1,325198 | y ~ Intervention + Sex + |
| 0,38936 | -0,13832 | 0,16018 | 141 | 69 | 72 | 137 | -0,86352 | y ~ Intervention + Sex + |
| 0,812716 | 0,036884 | 0,155379 | 141 | 69 | 72 | 137 | 0,237381 | y ~ Intervention + Sex + |
| 0,541687 | 0,090118 | 0,147301 | 141 | 69 | 72 | 137 | 0,611795 | y ~ Intervention + Sex + |
| 0,556172 | -0,10226 | 0,173318 | 141 | 69 | 72 | 137 | -0,58999 | y ~ Intervention + Sex + |
| 0,249288 | 0,182016 | 0,157318 | 141 | 69 | 72 | 137 | 1,156995 | y ~ Intervention + Sex + |
| 0,665459 | 0,060703 | 0,140085 | 141 | 69 | 72 | 137 | 0,433327 | y ~ Intervention + Sex + |
| 3,17E-07 | 0,858172 | 0,159578 | 141 | 69 | 72 | 137 | 5,377746 | y ~ Intervention + Sex + |
| 0,016356 | -0,3719 | 0,152993 | 141 | 69 | 72 | 137 | -2,43085 | y ~ Intervention + Sex + |
| 0,849387 | 0,032832 | 0,172563 | 141 | 69 | 72 | 137 | 0,19026 | y ~ Intervention + Sex + |
| 0,675148 | 0,060271 | 0,143505 | 141 | 69 | 72 | 137 | 0,419993 | y ~ Intervention + Sex + |
| 0,426674 | -0,12873 | 0,161462 | 141 | 69 | 72 | 137 | -0,79727 | y ~ Intervention + Sex + |
| 0,06794 | -0,27259 | 0,148152 | 141 | 69 | 72 | 137 | -1,83995 | y ~ Intervention + Sex + |
| 0,64513 | 0,075047 | 0,162595 | 141 | 69 | 72 | 137 | 0,461558 | y ~ Intervention + Sex + |
| 0,602934 | 0,08262 | 0,158459 | 141 | 69 | 72 | 137 | 0,521394 | y ~ Intervention + Sex + |
| 0,600739 | 0,090356 | 0,172251 | 141 | 69 | 72 | 137 | 0,524557 | y ~ Intervention + Sex + |
| 0,371631 | 0,12967 | 0,144663 | 141 | 69 | 72 | 137 | 0,896363 | y ~ Intervention + Sex + |
| 0,684046 | 0,066918 | 0,164088 | 141 | 69 | 72 | 137 | 0,407816 | y ~ Intervention + Sex + |
| 0,140732 | 0,247009 | 0,166713 | 141 | 69 | 72 | 137 | 1,48164 | y ~ Intervention + Sex + |
| 0,496885 | -0,10472 | 0,153727 | 141 | 69 | 72 | 137 | -0,68122 | y ~ Intervention + Sex + |
| 0,697767 | 0,057936 | 0,148878 | 141 | 69 | 72 | 137 | 0,389153 | y ~ Intervention + Sex + |
| 0,12915 | 0,248421 | 0,162721 | 141 | 69 | 72 | 137 | 1,526662 | y ~ Intervention + Sex + |
| 0,009599 | 0,403701 | 0,153681 | 141 | 69 | 72 | 137 | 2,626867 | y ~ Intervention + Sex + |
| 0,042421 | 0,323137 | 0,157744 | 141 | 69 | 72 | 137 | 2,048491 | y ~ Intervention + Sex + |

| | | | | | | | | |
|---|---|---|---|---|---|---|---|---|
| 0,332133 | 0,159852 | 0,164241 | 141 | 69 | 72 | 137 | 0,973274 | y ~ Intervention + Sex + |
| 0,270923 | 0,164247 | 0,148586 | 141 | 69 | 72 | 137 | 1,105401 | y ~ Intervention + Sex + |
| 0,168411 | 0,220246 | 0,159063 | 141 | 69 | 72 | 137 | 1,384648 | y ~ Intervention + Sex + |
| 0,487952 | 0,112077 | 0,161158 | 141 | 69 | 72 | 137 | 0,695448 | y ~ Intervention + Sex + |
| 0,024611 | 0,380349 | 0,167368 | 141 | 69 | 72 | 137 | 2,272537 | y ~ Intervention + Sex + |
| 0,974694 | 0,004928 | 0,155083 | 141 | 69 | 72 | 137 | 0,031779 | y ~ Intervention + Sex + |
| 0,246619 | 0,188199 | 0,161742 | 141 | 69 | 72 | 137 | 1,163574 | y ~ Intervention + Sex + |
| 0,031051 | 0,324137 | 0,148763 | 141 | 69 | 72 | 137 | 2,178877 | y ~ Intervention + Sex + |
| 0,879796 | -0,02386 | 0,157453 | 141 | 69 | 72 | 137 | -0,15151 | y ~ Intervention + Sex + |
| 0,943467 | 0,010693 | 0,150522 | 141 | 69 | 72 | 137 | 0,071043 | y ~ Intervention + Sex + |
| 0,821329 | 0,035434 | 0,156603 | 141 | 69 | 72 | 137 | 0,226269 | y ~ Intervention + Sex + |
| 0,001135 | 0,54718 | 0,164564 | 141 | 69 | 72 | 137 | 3,325033 | y ~ Intervention + Sex + |
| 0,171748 | 0,211235 | 0,153759 | 141 | 69 | 72 | 137 | 1,3738 | y ~ Intervention + Sex + |
| 0,112543 | -0,26065 | 0,163199 | 141 | 69 | 72 | 137 | -1,59712 | y ~ Intervention + Sex + |
| 0,42514 | -0,13842 | 0,173046 | 141 | 69 | 72 | 137 | -0,79992 | y ~ Intervention + Sex + |
| 0,585827 | 0,093732 | 0,171612 | 141 | 69 | 72 | 137 | 0,546186 | y ~ Intervention + Sex + |
| 0,998514 | 0,000292 | 0,1563 | 141 | 69 | 72 | 137 | 0,001866 | y ~ Intervention + Sex + |
| 0,003761 | -0,49383 | 0,167512 | 141 | 69 | 72 | 137 | -2,94804 | y ~ Intervention + Sex + |
| 0,04572 | 0,314309 | 0,155881 | 141 | 69 | 72 | 137 | 2,016334 | y ~ Intervention + Sex + |
| 0,325065 | -0,1469 | 0,148736 | 141 | 69 | 72 | 137 | -0,98765 | y ~ Intervention + Sex + |
| 0,053419 | 0,294617 | 0,151215 | 141 | 69 | 72 | 137 | 1,948336 | y ~ Intervention + Sex + |
| 0,310512 | -0,16188 | 0,159028 | 141 | 69 | 72 | 137 | -1,01792 | y ~ Intervention + Sex + |
| 0,126332 | -0,25037 | 0,162777 | 141 | 69 | 72 | 137 | -1,53809 | y ~ Intervention + Sex + |
| 0,579613 | 0,086603 | 0,155964 | 141 | 69 | 72 | 137 | 0,555274 | y ~ Intervention + Sex + |
| 0,681333 | 0,069065 | 0,167829 | 141 | 69 | 72 | 137 | 0,411522 | y ~ Intervention + Sex + |
| 0,767617 | 0,047769 | 0,161339 | 141 | 69 | 72 | 137 | 0,296081 | y ~ Intervention + Sex + |
| 0,105794 | 0,253813 | 0,155892 | 141 | 69 | 72 | 137 | 1,628136 | y ~ Intervention + Sex + |
| 0,386572 | -0,13672 | 0,157395 | 141 | 69 | 72 | 137 | -0,86862 | y ~ Intervention + Sex + |
| 0,315016 | -0,15365 | 0,152363 | 141 | 69 | 72 | 137 | -1,00845 | y ~ Intervention + Sex + |
| 0,061655 | -0,31467 | 0,167006 | 141 | 69 | 72 | 137 | -1,88421 | y ~ Intervention + Sex + |
| 0,021313 | 0,34854 | 0,149642 | 141 | 69 | 72 | 137 | 2,329162 | y ~ Intervention + Sex + |
| 0,90043 | 0,020013 | 0,159653 | 141 | 69 | 72 | 137 | 0,125351 | y ~ Intervention + Sex + |
| 0,772324 | 0,043875 | 0,15134 | 141 | 69 | 72 | 137 | 0,28991 | y ~ Intervention + Sex + |
| 0,373923 | -0,13667 | 0,153203 | 141 | 69 | 72 | 137 | -0,89206 | y ~ Intervention + Sex + |
| 0,340844 | 0,144705 | 0,151393 | 141 | 69 | 72 | 137 | 0,955825 | y ~ Intervention + Sex + |
| 0,264997 | -0,16664 | 0,148885 | 141 | 69 | 72 | 137 | -1,11924 | y ~ Intervention + Sex + |
| 0,559823 | 0,10126 | 0,173233 | 141 | 69 | 72 | 137 | 0,584533 | y ~ Intervention + Sex + |
| 0,274812 | -0,18048 | 0,164608 | 141 | 69 | 72 | 137 | -1,09644 | y ~ Intervention + Sex + |
| 0,689864 | -0,06015 | 0,150424 | 141 | 69 | 72 | 137 | -0,39989 | y ~ Intervention + Sex + |
| 0,142967 | 0,217939 | 0,147927 | 141 | 69 | 72 | 137 | 1,473287 | y ~ Intervention + Sex + |
| 0,639761 | -0,0704 | 0,150082 | 141 | 69 | 72 | 137 | -0,46908 | y ~ Intervention + Sex + |
| 0,000515 | 0,530051 | 0,149005 | 141 | 69 | 72 | 137 | 3,557267 | y ~ Intervention + Sex + |
| 0,30124 | 0,183092 | 0,176441 | 141 | 69 | 72 | 137 | 1,037695 | y ~ Intervention + Sex + |
| 0,609958 | -0,09441 | 0,184653 | 141 | 69 | 72 | 137 | -0,51131 | y ~ Intervention + Sex + |
| 0,759463 | 0,04659 | 0,15186 | 141 | 69 | 72 | 137 | 0,306798 | y ~ Intervention + Sex + |
| 0,873424 | 0,025447 | 0,159436 | 141 | 69 | 72 | 137 | 0,159609 | y ~ Intervention + Sex + |
| 0,364156 | 0,145046 | 0,159303 | 141 | 69 | 72 | 137 | 0,910504 | y ~ Intervention + Sex + |
| 0,072958 | -0,30893 | 0,170962 | 141 | 69 | 72 | 137 | -1,80699 | y ~ Intervention + Sex + |

| | | | | | | | | |
|---|---|---|---|---|---|---|---|---|
| 0,020378 | -0,33863 | 0,144306 | 141 | 69 | 72 | 137 | -2,34662 | y ~ Intervention + Sex + |
| 0,02634 | 0,334376 | 0,148911 | 141 | 69 | 72 | 137 | 2,245472 | y ~ Intervention + Sex + |
| 0,543634 | 0,103066 | 0,169281 | 141 | 69 | 72 | 137 | 0,608848 | y ~ Intervention + Sex + |
| 0,653094 | 0,079857 | 0,177281 | 141 | 69 | 72 | 137 | 0,450456 | y ~ Intervention + Sex + |
| 0,094227 | -0,24102 | 0,143022 | 141 | 69 | 72 | 137 | -1,6852 | y ~ Intervention + Sex + |
| 0,174721 | -0,20563 | 0,150723 | 141 | 69 | 72 | 137 | -1,36427 | y ~ Intervention + Sex + |
| 0,057356 | -0,30469 | 0,158964 | 141 | 69 | 72 | 137 | -1,91672 | y ~ Intervention + Sex + |
| 0,935662 | 0,0114 | 0,140958 | 141 | 69 | 72 | 137 | 0,080872 | y ~ Intervention + Sex + |
| 0,817564 | -0,03551 | 0,153656 | 141 | 69 | 72 | 137 | -0,23112 | y ~ Intervention + Sex + |
| 0,773881 | -0,04102 | 0,142484 | 141 | 69 | 72 | 137 | -0,28787 | y ~ Intervention + Sex + |
| 0,951153 | -0,00998 | 0,162617 | 141 | 69 | 72 | 137 | -0,06137 | y ~ Intervention + Sex + |
| 0,322876 | -0,14981 | 0,150994 | 141 | 69 | 72 | 137 | -0,99215 | y ~ Intervention + Sex + |
| 0,024564 | 0,374758 | 0,164852 | 141 | 69 | 72 | 137 | 2,273293 | y ~ Intervention + Sex + |
| 0,003341 | 0,457755 | 0,153262 | 141 | 69 | 72 | 137 | 2,986755 | y ~ Intervention + Sex + |
| 0,653197 | -0,07658 | 0,170067 | 141 | 69 | 72 | 137 | -0,45031 | y ~ Intervention + Sex + |
| 0,032237 | 0,303102 | 0,140098 | 141 | 69 | 72 | 137 | 2,163506 | y ~ Intervention + Sex + |
| 6,41E-17 | 1,28004 | 0,133806 | 141 | 69 | 72 | 137 | 9,566385 | y ~ Intervention + Sex + |
| 0,992655 | -0,00152 | 0,164605 | 141 | 69 | 72 | 137 | -0,00922 | y ~ Intervention + Sex + |
| 0,033455 | 0,34682 | 0,161445 | 141 | 69 | 72 | 137 | 2,148226 | y ~ Intervention + Sex + |
| 0,213001 | -0,18923 | 0,151243 | 141 | 69 | 72 | 137 | -1,25118 | y ~ Intervention + Sex + |
| 0,804939 | -0,03844 | 0,15534 | 141 | 69 | 72 | 137 | -0,24744 | y ~ Intervention + Sex + |
| 0,88634 | -0,02359 | 0,164757 | 141 | 69 | 72 | 137 | -0,1432 | y ~ Intervention + Sex + |
| 0,014419 | 0,397349 | 0,160336 | 141 | 69 | 72 | 137 | 2,478223 | y ~ Intervention + Sex + |
| 0,854359 | -0,02879 | 0,156531 | 141 | 69 | 72 | 137 | -0,18391 | y ~ Intervention + Sex + |
| 0,598057 | -0,08242 | 0,155981 | 141 | 69 | 72 | 137 | -0,52843 | y ~ Intervention + Sex + |
| 0,47549 | 0,1203 | 0,168124 | 141 | 69 | 72 | 137 | 0,715544 | y ~ Intervention + Sex + |
| 0,001148 | -0,5163 | 0,15544 | 141 | 69 | 72 | 137 | -3,32152 | y ~ Intervention + Sex + |
| 0,312095 | 0,157477 | 0,155214 | 141 | 69 | 72 | 137 | 1,014579 | y ~ Intervention + Sex + |
| 0,105957 | -0,24574 | 0,151005 | 141 | 69 | 72 | 137 | -1,62736 | y ~ Intervention + Sex + |
| 0,206721 | 0,192339 | 0,151611 | 141 | 69 | 72 | 137 | 1,268639 | y ~ Intervention + Sex + |
| 0,429729 | 0,12728 | 0,160707 | 141 | 69 | 72 | 137 | 0,792001 | y ~ Intervention + Sex + |
| 0,00465 | 0,457438 | 0,158966 | 141 | 69 | 72 | 137 | 2,877586 | y ~ Intervention + Sex + |
| 0,767257 | 0,051817 | 0,174733 | 141 | 69 | 72 | 137 | 0,296553 | y ~ Intervention + Sex + |
| 0,642322 | 0,071537 | 0,153682 | 141 | 69 | 72 | 137 | 0,465486 | y ~ Intervention + Sex + |
| 0,058374 | -0,29854 | 0,156396 | 141 | 69 | 72 | 137 | -1,90884 | y ~ Intervention + Sex + |
| 0,15754 | 0,225154 | 0,158428 | 141 | 69 | 72 | 137 | 1,421169 | y ~ Intervention + Sex + |
| 0,233779 | -0,1745 | 0,14591 | 141 | 69 | 72 | 137 | -1,19596 | y ~ Intervention + Sex + |
| 0,745496 | 0,048761 | 0,149924 | 141 | 69 | 72 | 137 | 0,325239 | y ~ Intervention + Sex + |
| 0,001143 | 0,52366 | 0,157596 | 141 | 69 | 72 | 137 | 3,322807 | y ~ Intervention + Sex + |
| 0,465202 | -0,1102 | 0,150474 | 141 | 69 | 72 | 137 | -0,73236 | y ~ Intervention + Sex + |
| 0,063124 | -0,25912 | 0,138307 | 141 | 69 | 72 | 137 | -1,87354 | y ~ Intervention + Sex + |
| 0,247652 | 0,204087 | 0,175782 | 141 | 69 | 72 | 137 | 1,161023 | y ~ Intervention + Sex + |
| 0,752768 | 0,052695 | 0,166956 | 141 | 69 | 72 | 137 | 0,315624 | y ~ Intervention + Sex + |
| 0,003802 | -0,46117 | 0,156624 | 141 | 69 | 72 | 137 | -2,94445 | y ~ Intervention + Sex + |
| 0,159365 | 0,242161 | 0,17115 | 141 | 69 | 72 | 137 | 1,414906 | y ~ Intervention + Sex + |
| 0,202197 | -0,21094 | 0,164606 | 141 | 69 | 72 | 137 | -1,28146 | y ~ Intervention + Sex + |
| 4,44E-05 | -0,66796 | 0,158321 | 141 | 69 | 72 | 137 | -4,21902 | y ~ Intervention + Sex + |
| 0,938098 | 0,013093 | 0,16828 | 141 | 69 | 72 | 137 | 0,077803 | y ~ Intervention + Sex + |

| | | | | | | | | |
|---|---|---|---|---|---|---|---|---|
| 0,785207 | 0,041663 | 0,152569 | 141 | 69 | 72 | 137 | 0,273075 | y ~ Intervention + Sex + |
| 0,811673 | -0,038 | 0,159166 | 141 | 69 | 72 | 137 | -0,23873 | y ~ Intervention + Sex + |
| 0,000701 | 0,565246 | 0,162993 | 141 | 69 | 72 | 137 | 3,467912 | y ~ Intervention + Sex + |
| 0,173931 | -0,2034 | 0,148815 | 141 | 69 | 72 | 137 | -1,36679 | y ~ Intervention + Sex + |
| 0,002017 | -0,44492 | 0,141326 | 141 | 69 | 72 | 137 | -3,1482 | y ~ Intervention + Sex + |
| 0,531545 | 0,093764 | 0,149487 | 141 | 69 | 72 | 137 | 0,627242 | y ~ Intervention + Sex + |
| 0,266887 | 0,182822 | 0,163995 | 141 | 69 | 72 | 137 | 1,114802 | y ~ Intervention + Sex + |
| 0,686747 | 0,065502 | 0,162081 | 141 | 69 | 72 | 137 | 0,404131 | y ~ Intervention + Sex + |
| 0,940013 | -0,01236 | 0,163969 | 141 | 69 | 72 | 137 | -0,07539 | y ~ Intervention + Sex + |
| 0,000383 | 0,58317 | 0,160143 | 141 | 69 | 72 | 137 | 3,641566 | y ~ Intervention + Sex + |
| 0,815392 | -0,03342 | 0,14288 | 141 | 69 | 72 | 137 | -0,23393 | y ~ Intervention + Sex + |
| 0,000161 | -0,64103 | 0,165194 | 141 | 69 | 72 | 137 | -3,88045 | y ~ Intervention + Sex + |
| 0,164383 | -0,20976 | 0,15005 | 141 | 69 | 72 | 137 | -1,39796 | y ~ Intervention + Sex + |
| 0,878832 | -0,02508 | 0,164224 | 141 | 69 | 72 | 137 | -0,15274 | y ~ Intervention + Sex + |
| 0,828982 | 0,037342 | 0,172545 | 141 | 69 | 72 | 137 | 0,216421 | y ~ Intervention + Sex + |
| 0,405669 | -0,13868 | 0,166256 | 141 | 69 | 72 | 137 | -0,83411 | y ~ Intervention + Sex + |
| 0,651768 | 0,071263 | 0,157557 | 141 | 69 | 72 | 137 | 0,4523 | y ~ Intervention + Sex + |
| 0,252297 | 0,184032 | 0,160079 | 141 | 69 | 72 | 137 | 1,149636 | y ~ Intervention + Sex + |
| 0,121715 | 0,251756 | 0,161666 | 141 | 69 | 72 | 137 | 1,557262 | y ~ Intervention + Sex + |
| 0,331196 | 0,151965 | 0,155835 | 141 | 69 | 72 | 137 | 0,975168 | y ~ Intervention + Sex + |
| 0,051284 | 0,309078 | 0,157187 | 141 | 69 | 72 | 137 | 1,966312 | y ~ Intervention + Sex + |
| 0,118432 | 0,241643 | 0,15379 | 141 | 69 | 72 | 137 | 1,571247 | y ~ Intervention + Sex + |
| 0,411025 | -0,12972 | 0,157314 | 141 | 69 | 72 | 137 | -0,82461 | y ~ Intervention + Sex + |
| 0,293592 | 0,17207 | 0,163205 | 141 | 69 | 72 | 137 | 1,05432 | y ~ Intervention + Sex + |
| 0,555605 | -0,09827 | 0,166328 | 141 | 69 | 72 | 137 | -0,59083 | y ~ Intervention + Sex + |
| 0,345285 | 0,159016 | 0,167907 | 141 | 69 | 72 | 137 | 0,947043 | y ~ Intervention + Sex + |
| 0,970795 | -0,00567 | 0,154665 | 141 | 69 | 72 | 137 | -0,03668 | y ~ Intervention + Sex + |
| 0,732014 | -0,05255 | 0,153152 | 141 | 69 | 72 | 137 | -0,34315 | y ~ Intervention + Sex + |
| 0,606961 | 0,080854 | 0,156813 | 141 | 69 | 72 | 137 | 0,515606 | y ~ Intervention + Sex + |
| 0,994637 | -0,00102 | 0,152172 | 141 | 69 | 72 | 137 | -0,00673 | y ~ Intervention + Sex + |
| 0,680403 | 0,07263 | 0,175948 | 141 | 69 | 72 | 137 | 0,412794 | y ~ Intervention + Sex + |
| 0,690943 | -0,05851 | 0,146848 | 141 | 69 | 72 | 137 | -0,39842 | y ~ Intervention + Sex + |
| 0,05838 | -0,30335 | 0,158921 | 141 | 69 | 72 | 137 | -1,90879 | y ~ Intervention + Sex + |
| 0,542025 | -0,10016 | 0,163859 | 141 | 69 | 72 | 137 | -0,61128 | y ~ Intervention + Sex + |
| 0,033202 | 0,353832 | 0,164469 | 141 | 69 | 72 | 137 | 2,151355 | y ~ Intervention + Sex + |
| 0,463915 | 0,11585 | 0,157732 | 141 | 69 | 72 | 137 | 0,734474 | y ~ Intervention + Sex + |
| 0,044916 | -0,36429 | 0,179989 | 141 | 69 | 72 | 137 | -2,02399 | y ~ Intervention + Sex + |
| 5,40E-10 | 1,098728 | 0,164388 | 141 | 69 | 72 | 137 | 6,683756 | y ~ Intervention + Sex + |
| 0,780458 | 0,041213 | 0,147575 | 141 | 69 | 72 | 137 | 0,279271 | y ~ Intervention + Sex + |
| 0,819011 | -0,0384 | 0,167508 | 141 | 69 | 72 | 137 | -0,22926 | y ~ Intervention + Sex + |
| 0,450288 | 0,100293 | 0,13247 | 141 | 69 | 72 | 137 | 0,757103 | y ~ Intervention + Sex + |
| 0,690995 | -0,06448 | 0,161868 | 141 | 69 | 72 | 137 | -0,39835 | y ~ Intervention + Sex + |
| 0,000921 | -0,56579 | 0,167027 | 141 | 69 | 72 | 137 | -3,3874 | y ~ Intervention + Sex + |
| 0,735664 | -0,05475 | 0,16184 | 141 | 69 | 72 | 137 | -0,33829 | y ~ Intervention + Sex + |
| 0,415807 | -0,12656 | 0,155062 | 141 | 69 | 72 | 137 | -0,81619 | y ~ Intervention + Sex + |
| 0,538922 | -0,09681 | 0,157164 | 141 | 69 | 72 | 137 | -0,61599 | y ~ Intervention + Sex + |
| 0,876438 | 0,02389 | 0,15336 | 141 | 69 | 72 | 137 | 0,155778 | y ~ Intervention + Sex + |
| 0,596658 | 0,082889 | 0,156262 | 141 | 69 | 72 | 137 | 0,53045 | y ~ Intervention + Sex + |

| | | | | | | | | |
|---|---|---|---|---|---|---|---|---|
| 4,62E-07 | -0,73646 | 0,139091 | 141 | 69 | 72 | 137 | -5,29482 | y ~ Intervention + Sex + |
| 0,161995 | -0,21337 | 0,151757 | 141 | 69 | 72 | 137 | -1,40598 | y ~ Intervention + Sex + |
| 0,934186 | 0,011867 | 0,143441 | 141 | 69 | 72 | 137 | 0,082731 | y ~ Intervention + Sex + |
| 0,874247 | 0,024709 | 0,155831 | 141 | 69 | 72 | 137 | 0,158562 | y ~ Intervention + Sex + |
| 0,945057 | -0,01123 | 0,162608 | 141 | 69 | 72 | 137 | -0,06904 | y ~ Intervention + Sex + |
| 0,72143 | -0,05641 | 0,157885 | 141 | 69 | 72 | 137 | -0,35728 | y ~ Intervention + Sex + |
| 0,846471 | 0,030511 | 0,15728 | 141 | 69 | 72 | 137 | 0,19399 | y ~ Intervention + Sex + |
| 0,051856 | -0,29123 | 0,148476 | 141 | 69 | 72 | 137 | -1,96143 | y ~ Intervention + Sex + |
| 0,62156 | -0,07523 | 0,152059 | 141 | 69 | 72 | 137 | -0,49476 | y ~ Intervention + Sex + |
| 0,43283 | -0,12984 | 0,165047 | 141 | 69 | 72 | 137 | -0,78668 | y ~ Intervention + Sex + |
| 1,99E-05 | 0,655359 | 0,148268 | 141 | 69 | 72 | 137 | 4,420107 | y ~ Intervention + Sex + |
| 8,72E-25 | -1,45305 | 0,114855 | 141 | 69 | 72 | 137 | -12,6512 | y ~ Intervention + Sex + |
| 0,907012 | -0,019 | 0,162321 | 141 | 69 | 72 | 137 | -0,11702 | y ~ Intervention + Sex + |
| 0,279246 | -0,16881 | 0,155398 | 141 | 69 | 72 | 137 | -1,08632 | y ~ Intervention + Sex + |
| 0,105145 | 0,254844 | 0,156231 | 141 | 69 | 72 | 137 | 1,631199 | y ~ Intervention + Sex + |
| 0,248531 | -0,1925 | 0,166112 | 141 | 69 | 72 | 137 | -1,15885 | y ~ Intervention + Sex + |
| 0,292627 | -0,17388 | 0,164593 | 141 | 69 | 72 | 137 | -1,05644 | y ~ Intervention + Sex + |
| 0,442632 | 0,103664 | 0,13463 | 141 | 69 | 72 | 137 | 0,76999 | y ~ Intervention + Sex + |
| 0,612929 | 0,07646 | 0,150792 | 141 | 69 | 72 | 137 | 0,507058 | y ~ Intervention + Sex + |
| 0,966419 | 0,006515 | 0,154476 | 141 | 69 | 72 | 137 | 0,042177 | y ~ Intervention + Sex + |
| 0,270966 | 0,18109 | 0,163837 | 141 | 69 | 72 | 137 | 1,105303 | y ~ Intervention + Sex + |
| 0,330184 | -0,15572 | 0,159353 | 141 | 69 | 72 | 137 | -0,97722 | y ~ Intervention + Sex + |
| 0,00048 | -0,63306 | 0,176949 | 141 | 69 | 72 | 137 | -3,57765 | y ~ Intervention + Sex + |
| 0,468394 | 0,117814 | 0,162028 | 141 | 69 | 72 | 137 | 0,727119 | y ~ Intervention + Sex + |
| 0,020397 | 0,371761 | 0,158448 | 141 | 69 | 72 | 137 | 2,34626 | y ~ Intervention + Sex + |
| 0,160684 | -0,20953 | 0,148562 | 141 | 69 | 72 | 137 | -1,41041 | y ~ Intervention + Sex + |
| 0,432801 | 0,122223 | 0,155356 | 141 | 69 | 72 | 137 | 0,786727 | y ~ Intervention + Sex + |
| 0,120765 | 0,240614 | 0,154113 | 141 | 69 | 72 | 137 | 1,561281 | y ~ Intervention + Sex + |
| 0,820009 | 0,0369 | 0,161861 | 141 | 69 | 72 | 137 | 0,22797 | y ~ Intervention + Sex + |
| 0,256902 | 0,196552 | 0,172642 | 141 | 69 | 72 | 137 | 1,138492 | y ~ Intervention + Sex + |
| 0,641391 | 0,076043 | 0,162907 | 141 | 69 | 72 | 137 | 0,466791 | y ~ Intervention + Sex + |
| 0,565376 | 0,090907 | 0,15775 | 141 | 69 | 72 | 137 | 0,576273 | y ~ Intervention + Sex + |
| 0,663624 | -0,06819 | 0,156457 | 141 | 69 | 72 | 137 | -0,43586 | y ~ Intervention + Sex + |
| 0,061178 | -0,3408 | 0,180536 | 141 | 69 | 72 | 137 | -1,88772 | y ~ Intervention + Sex + |
| 0,256947 | -0,18992 | 0,166837 | 141 | 69 | 72 | 137 | -1,13838 | y ~ Intervention + Sex + |
| 0,421066 | -0,12129 | 0,150295 | 141 | 69 | 72 | 137 | -0,807 | y ~ Intervention + Sex + |
| 0,038544 | 0,366998 | 0,175668 | 141 | 69 | 72 | 137 | 2,089154 | y ~ Intervention + Sex + |
| 0,594095 | -0,07396 | 0,138467 | 141 | 69 | 72 | 137 | -0,53416 | y ~ Intervention + Sex + |
| 0,428614 | -0,12175 | 0,153352 | 141 | 69 | 72 | 137 | -0,79392 | y ~ Intervention + Sex + |
| 0,983276 | 0,003451 | 0,164355 | 141 | 69 | 72 | 137 | 0,021 | y ~ Intervention + Sex + |
| 0,854213 | -0,02835 | 0,154017 | 141 | 69 | 72 | 137 | -0,18409 | y ~ Intervention + Sex + |
| 0,150158 | -0,24559 | 0,169714 | 141 | 69 | 72 | 137 | -1,44708 | y ~ Intervention + Sex + |
| 0,839804 | -0,03397 | 0,167718 | 141 | 69 | 72 | 137 | -0,20253 | y ~ Intervention + Sex + |
| 0,30446 | 0,154317 | 0,149709 | 141 | 69 | 72 | 137 | 1,03078 | y ~ Intervention + Sex + |
| 0,614814 | 0,088375 | 0,17522 | 141 | 69 | 72 | 137 | 0,504367 | y ~ Intervention + Sex + |
| 0,323962 | 0,158336 | 0,15995 | 141 | 69 | 72 | 137 | 0,989913 | y ~ Intervention + Sex + |
| 0,654985 | -0,07336 | 0,163821 | 141 | 69 | 72 | 137 | -0,44783 | y ~ Intervention + Sex + |
| 0,171352 | -0,22086 | 0,160617 | 141 | 69 | 72 | 137 | -1,37508 | y ~ Intervention + Sex + |

| | | | | | | | | |
|---|---|---|---|---|---|---|---|---|
| 0,296682 | 0,150522 | 0,143687 | 141 | 69 | 72 | 137 | 1,047569 | y ~ Intervention + Sex + |
| 0,425481 | 0,134795 | 0,168635 | 141 | 69 | 72 | 137 | 0,799334 | y ~ Intervention + Sex + |
| 0,016272 | -0,375 | 0,154141 | 141 | 69 | 72 | 137 | -2,4328 | y ~ Intervention + Sex + |
| 0,074938 | 0,28953 | 0,161342 | 141 | 69 | 72 | 137 | 1,794505 | y ~ Intervention + Sex + |
| 0,657153 | -0,06947 | 0,156169 | 141 | 69 | 72 | 137 | -0,44482 | y ~ Intervention + Sex + |
| 0,127186 | -0,22803 | 0,148588 | 141 | 69 | 72 | 137 | -1,53461 | y ~ Intervention + Sex + |
| 0,407949 | -0,1371 | 0,165173 | 141 | 69 | 72 | 137 | -0,83006 | y ~ Intervention + Sex + |
| 0,56495 | -0,08785 | 0,152286 | 141 | 69 | 72 | 137 | -0,57691 | y ~ Intervention + Sex + |
| 0,527436 | -0,10071 | 0,15896 | 141 | 69 | 72 | 137 | -0,63354 | y ~ Intervention + Sex + |
| 0,809944 | 0,040173 | 0,166718 | 141 | 69 | 72 | 137 | 0,240963 | y ~ Intervention + Sex + |
| 0,852993 | -0,03005 | 0,161874 | 141 | 69 | 72 | 137 | -0,18565 | y ~ Intervention + Sex + |
| 0,159925 | 0,208459 | 0,14753 | 141 | 69 | 72 | 137 | 1,412996 | y ~ Intervention + Sex + |
| 0,040026 | 0,343052 | 0,165469 | 141 | 69 | 72 | 137 | 2,073211 | y ~ Intervention + Sex + |
| 0,772729 | 0,051055 | 0,17643 | 141 | 69 | 72 | 137 | 0,289379 | y ~ Intervention + Sex + |
| 0,020361 | 0,312251 | 0,133046 | 141 | 69 | 72 | 137 | 2,34694 | y ~ Intervention + Sex + |
| 0,004477 | -0,38397 | 0,132847 | 141 | 69 | 72 | 137 | -2,8903 | y ~ Intervention + Sex + |
| 0,044109 | 0,339979 | 0,16733 | 141 | 69 | 72 | 137 | 2,031782 | y ~ Intervention + Sex + |
| 0,007204 | 0,440925 | 0,16162 | 141 | 69 | 72 | 137 | 2,728166 | y ~ Intervention + Sex + |
| 0,192104 | 0,22148 | 0,16896 | 141 | 69 | 72 | 137 | 1,310841 | y ~ Intervention + Sex + |
| 0,330408 | 0,17454 | 0,178692 | 141 | 69 | 72 | 137 | 0,976764 | y ~ Intervention + Sex + |
| 0,586787 | -0,09201 | 0,168884 | 141 | 69 | 72 | 137 | -0,54478 | y ~ Intervention + Sex + |
| 0,217198 | 0,164374 | 0,132589 | 141 | 69 | 72 | 137 | 1,239721 | y ~ Intervention + Sex + |
| 0,323876 | 0,148255 | 0,149739 | 141 | 69 | 72 | 137 | 0,99009 | y ~ Intervention + Sex + |
| 0,326445 | 0,155228 | 0,157619 | 141 | 69 | 72 | 137 | 0,984827 | y ~ Intervention + Sex + |
| 0,075136 | 0,27661 | 0,154249 | 141 | 69 | 72 | 137 | 1,79327 | y ~ Intervention + Sex + |
| 0,53801 | 0,096639 | 0,156531 | 141 | 69 | 72 | 137 | 0,617378 | y ~ Intervention + Sex + |
| 0,559498 | -0,08622 | 0,147376 | 141 | 69 | 72 | 137 | -0,58502 | y ~ Intervention + Sex + |
| 0,006502 | 0,460055 | 0,166464 | 141 | 69 | 72 | 137 | 2,763685 | y ~ Intervention + Sex + |
| 0,007593 | 0,366144 | 0,135119 | 141 | 69 | 72 | 137 | 2,709788 | y ~ Intervention + Sex + |
| 8,90E-08 | 0,877761 | 0,155336 | 141 | 69 | 72 | 137 | 5,650729 | y ~ Intervention + Sex + |
| 0,852202 | -0,0268 | 0,143557 | 141 | 69 | 72 | 137 | -0,18666 | y ~ Intervention + Sex + |
| 0,020397 | 0,374318 | 0,159538 | 141 | 69 | 72 | 137 | 2,346259 | y ~ Intervention + Sex + |
| 0,287056 | 0,154985 | 0,145013 | 141 | 69 | 72 | 137 | 1,068763 | y ~ Intervention + Sex + |
| 0,013581 | -0,40059 | 0,160206 | 141 | 69 | 72 | 137 | -2,50048 | y ~ Intervention + Sex + |
| 0,091368 | -0,26185 | 0,154014 | 141 | 69 | 72 | 137 | -1,70017 | y ~ Intervention + Sex + |
| 0,000321 | 0,498503 | 0,135043 | 141 | 69 | 72 | 137 | 3,691423 | y ~ Intervention + Sex + |
| 0,817018 | -0,03748 | 0,161664 | 141 | 69 | 72 | 137 | -0,23183 | y ~ Intervention + Sex + |
| 0,731232 | 0,056401 | 0,163867 | 141 | 69 | 72 | 137 | 0,344189 | y ~ Intervention + Sex + |
| 0,592586 | -0,08426 | 0,157091 | 141 | 69 | 72 | 137 | -0,53635 | y ~ Intervention + Sex + |
| 0,496839 | 0,116781 | 0,171412 | 141 | 69 | 72 | 137 | 0,681288 | y ~ Intervention + Sex + |
| 0,335368 | 0,170198 | 0,17605 | 141 | 69 | 72 | 137 | 0,966759 | y ~ Intervention + Sex + |
| 0,565532 | 0,085653 | 0,148693 | 141 | 69 | 72 | 137 | 0,576042 | y ~ Intervention + Sex + |
| 0,007263 | 0,440403 | 0,161596 | 141 | 69 | 72 | 137 | 2,72533 | y ~ Intervention + Sex + |
| 0,145522 | 0,234323 | 0,160072 | 141 | 69 | 72 | 137 | 1,463864 | y ~ Intervention + Sex + |
| 0,200088 | 0,194412 | 0,150998 | 141 | 69 | 72 | 137 | 1,287509 | y ~ Intervention + Sex + |
| 0,932453 | -0,01291 | 0,151979 | 141 | 69 | 72 | 137 | -0,08492 | y ~ Intervention + Sex + |
| 0,916625 | -0,0157 | 0,149694 | 141 | 69 | 72 | 137 | -0,10488 | y ~ Intervention + Sex + |
| 0,638784 | 0,074813 | 0,159025 | 141 | 69 | 72 | 137 | 0,470447 | y ~ Intervention + Sex + |

| | | | | | | | | |
|---|---|---|---|---|---|---|---|---|
| 0,880985 | -0,02037 | 0,1358 | 141 | 69 | 72 | 137 | -0,15 | y ~ Intervention + Sex + |
| 0,002775 | 0,480931 | 0,157847 | 141 | 69 | 72 | 137 | 3,04682 | y ~ Intervention + Sex + |
| 0,510856 | -0,10367 | 0,157266 | 141 | 69 | 72 | 137 | -0,65923 | y ~ Intervention + Sex + |
| 0,313978 | 0,158647 | 0,156979 | 141 | 69 | 72 | 137 | 1,010624 | y ~ Intervention + Sex + |
| 0,036783 | -0,29796 | 0,141297 | 141 | 69 | 72 | 137 | -2,10879 | y ~ Intervention + Sex + |
| 0,040914 | 0,328937 | 0,159376 | 141 | 69 | 72 | 137 | 2,063906 | y ~ Intervention + Sex + |
| 0,146466 | -0,22586 | 0,154656 | 141 | 69 | 72 | 137 | -1,46042 | y ~ Intervention + Sex + |
| 0,693477 | -0,05918 | 0,149833 | 141 | 69 | 72 | 137 | -0,39497 | y ~ Intervention + Sex + |
| 0,764416 | -0,05226 | 0,174033 | 141 | 69 | 72 | 137 | -0,30028 | y ~ Intervention + Sex + |
| 0,022511 | -0,37746 | 0,163563 | 141 | 69 | 72 | 137 | -2,30776 | y ~ Intervention + Sex + |
| 0,828589 | -0,03547 | 0,163511 | 141 | 69 | 72 | 137 | -0,21693 | y ~ Intervention + Sex + |
| 0,020983 | 0,33371 | 0,142902 | 141 | 69 | 72 | 137 | 2,335237 | y ~ Intervention + Sex + |
| 0,098605 | -0,25424 | 0,152881 | 141 | 69 | 72 | 137 | -1,66297 | y ~ Intervention + Sex + |
| 0,121586 | 0,242955 | 0,155959 | 141 | 69 | 72 | 137 | 1,557809 | y ~ Intervention + Sex + |
| 1,15E-06 | 0,799692 | 0,157051 | 141 | 69 | 72 | 137 | 5,091932 | y ~ Intervention + Sex + |
| 0,936269 | -0,0131 | 0,163522 | 141 | 69 | 72 | 137 | -0,08011 | y ~ Intervention + Sex + |
| 0,789728 | 0,035938 | 0,134505 | 141 | 69 | 72 | 137 | 0,267186 | y ~ Intervention + Sex + |
| 0,182287 | -0,20947 | 0,156259 | 141 | 69 | 72 | 137 | -1,34055 | y ~ Intervention + Sex + |
| 0,833833 | -0,03454 | 0,164329 | 141 | 69 | 72 | 137 | -0,21019 | y ~ Intervention + Sex + |
| 0,808967 | -0,03853 | 0,159086 | 141 | 69 | 72 | 137 | -0,24223 | y ~ Intervention + Sex + |
| 0,34267 | 0,173834 | 0,18256 | 141 | 69 | 72 | 137 | 0,952206 | y ~ Intervention + Sex + |
| 0,550127 | -0,08921 | 0,148922 | 141 | 69 | 72 | 137 | -0,59905 | y ~ Intervention + Sex + |
| 0,806254 | -0,03639 | 0,148099 | 141 | 69 | 72 | 137 | -0,24574 | y ~ Intervention + Sex + |
| 0,74848 | 0,051582 | 0,160547 | 141 | 69 | 72 | 137 | 0,32129 | y ~ Intervention + Sex + |
| 0,603367 | -0,08447 | 0,162194 | 141 | 69 | 72 | 137 | -0,52077 | y ~ Intervention + Sex + |
| 0,489511 | -0,10431 | 0,150535 | 141 | 69 | 72 | 137 | -0,69295 | y ~ Intervention + Sex + |
| 3,74E-05 | -0,65221 | 0,153006 | 141 | 69 | 72 | 137 | -4,26264 | y ~ Intervention + Sex + |
| 0,27626 | 0,160512 | 0,146839 | 141 | 69 | 72 | 137 | 1,093118 | y ~ Intervention + Sex + |
| 0,951111 | -0,01025 | 0,166886 | 141 | 69 | 72 | 137 | -0,06142 | y ~ Intervention + Sex + |
| 0,660336 | 0,060821 | 0,138102 | 141 | 69 | 72 | 137 | 0,440407 | y ~ Intervention + Sex + |
| 0,045331 | 0,349097 | 0,172819 | 141 | 69 | 72 | 137 | 2,020019 | y ~ Intervention + Sex + |
| 0,847866 | -0,02922 | 0,152008 | 141 | 69 | 72 | 137 | -0,19221 | y ~ Intervention + Sex + |
| 0,002121 | 0,531586 | 0,169711 | 141 | 69 | 72 | 137 | 3,132304 | y ~ Intervention + Sex + |
| 0,573457 | -0,09594 | 0,170002 | 141 | 69 | 72 | 137 | -0,56432 | y ~ Intervention + Sex + |
| 0,337345 | -0,14599 | 0,151627 | 141 | 69 | 72 | 137 | -0,9628 | y ~ Intervention + Sex + |
| 0,312603 | -0,16808 | 0,165842 | 141 | 69 | 72 | 137 | -1,01351 | y ~ Intervention + Sex + |
| 0,563224 | 0,087335 | 0,150715 | 141 | 69 | 72 | 137 | 0,57947 | y ~ Intervention + Sex + |
| 0,365058 | -0,14589 | 0,160532 | 141 | 69 | 72 | 137 | -0,90879 | y ~ Intervention + Sex + |
| 0,011339 | 0,394792 | 0,153808 | 141 | 69 | 72 | 137 | 2,566775 | y ~ Intervention + Sex + |
| 0,566759 | -0,0892 | 0,155345 | 141 | 69 | 72 | 137 | -0,57422 | y ~ Intervention + Sex + |
| 0,032657 | 0,325662 | 0,150897 | 141 | 69 | 72 | 137 | 2,158176 | y ~ Intervention + Sex + |
| 0,052707 | 0,299012 | 0,153005 | 141 | 69 | 72 | 137 | 1,954264 | y ~ Intervention + Sex + |
| 0,077287 | -0,28011 | 0,157363 | 141 | 69 | 72 | 137 | -1,78003 | y ~ Intervention + Sex + |
| 0,126087 | 0,248702 | 0,161589 | 141 | 69 | 72 | 137 | 1,539098 | y ~ Intervention + Sex + |
| 0,027846 | 0,326625 | 0,146922 | 141 | 69 | 72 | 137 | 2,22312 | y ~ Intervention + Sex + |
| 0,270788 | 0,149248 | 0,134979 | 141 | 69 | 72 | 137 | 1,105716 | y ~ Intervention + Sex + |
| 6,08E-14 | 1,146761 | 0,137066 | 141 | 69 | 72 | 137 | 8,366469 | y ~ Intervention + Sex + |
| 0,527003 | 0,096907 | 0,1528 | 141 | 69 | 72 | 137 | 0,634207 | y ~ Intervention + Sex + |

| | | | | | | | | |
|---|---|---|---|---|---|---|---|---|
| 0,049764 | 0,292762 | 0,147897 | 141 | 69 | 72 | 137 | 1,979502 | y ~ Intervention + Sex + |
| 7,90E-06 | 0,644442 | 0,138751 | 141 | 69 | 72 | 137 | 4,644582 | y ~ Intervention + Sex + |
| 0,836115 | 0,029039 | 0,140112 | 141 | 69 | 72 | 137 | 0,207259 | y ~ Intervention + Sex + |
| 0,306452 | 0,165763 | 0,161479 | 141 | 69 | 72 | 137 | 1,026527 | y ~ Intervention + Sex + |
| 0,58578 | -0,0789 | 0,144432 | 141 | 69 | 72 | 137 | -0,54625 | y ~ Intervention + Sex + |
| 0,977122 | -0,00442 | 0,153953 | 141 | 69 | 72 | 137 | -0,02873 | y ~ Intervention + Sex + |
| 0,567263 | -0,09294 | 0,162059 | 141 | 69 | 72 | 137 | -0,57348 | y ~ Intervention + Sex + |
| 0,774694 | 0,045929 | 0,160141 | 141 | 69 | 72 | 137 | 0,286807 | y ~ Intervention + Sex + |
| 0,507543 | 0,118446 | 0,178272 | 141 | 69 | 72 | 137 | 0,664413 | y ~ Intervention + Sex + |
| 0,423124 | -0,13325 | 0,165848 | 141 | 69 | 72 | 137 | -0,80342 | y ~ Intervention + Sex + |
| 0,649323 | -0,0689 | 0,151189 | 141 | 69 | 72 | 137 | -0,45571 | y ~ Intervention + Sex + |
| 0,303752 | 0,157576 | 0,152646 | 141 | 69 | 72 | 137 | 1,032297 | y ~ Intervention + Sex + |
| 0,793166 | -0,04192 | 0,159561 | 141 | 69 | 72 | 137 | -0,26271 | y ~ Intervention + Sex + |
| 0,576888 | 0,095729 | 0,171166 | 141 | 69 | 72 | 137 | 0,559274 | y ~ Intervention + Sex + |
| 0,002136 | -0,46869 | 0,149739 | 141 | 69 | 72 | 137 | -3,13009 | y ~ Intervention + Sex + |
| 0,395705 | -0,12943 | 0,151918 | 141 | 69 | 72 | 137 | -0,85199 | y ~ Intervention + Sex + |
| 0,280503 | 0,191057 | 0,176338 | 141 | 69 | 72 | 137 | 1,08347 | y ~ Intervention + Sex + |
| 0,038443 | 0,346443 | 0,165742 | 141 | 69 | 72 | 137 | 2,090253 | y ~ Intervention + Sex + |
| 0,940106 | 0,011364 | 0,150967 | 141 | 69 | 72 | 137 | 0,075274 | y ~ Intervention + Sex + |
| 0,409438 | 0,135044 | 0,16321 | 141 | 69 | 72 | 137 | 0,82742 | y ~ Intervention + Sex + |
| 0,000283 | -0,54951 | 0,147446 | 141 | 69 | 72 | 137 | -3,72687 | y ~ Intervention + Sex + |
| 0,63102 | -0,0824 | 0,171186 | 141 | 69 | 72 | 137 | -0,48137 | y ~ Intervention + Sex + |
| 0,731615 | -0,05364 | 0,156074 | 141 | 69 | 72 | 137 | -0,34368 | y ~ Intervention + Sex + |
| 0,438401 | 0,120999 | 0,155693 | 141 | 69 | 72 | 137 | 0,777166 | y ~ Intervention + Sex + |
| 0,385356 | 0,151987 | 0,174526 | 141 | 69 | 72 | 137 | 0,870856 | y ~ Intervention + Sex + |
| 0,08785 | 0,220605 | 0,128324 | 141 | 69 | 72 | 137 | 1,719128 | y ~ Intervention + Sex + |
| 0,011703 | 0,4007 | 0,156814 | 141 | 69 | 72 | 137 | 2,555253 | y ~ Intervention + Sex + |
| 0,011305 | -0,38462 | 0,149783 | 141 | 69 | 72 | 137 | -2,56787 | y ~ Intervention + Sex + |
| 0,41516 | 0,130224 | 0,159329 | 141 | 69 | 72 | 137 | 0,817329 | y ~ Intervention + Sex + |
| 0,01359 | 0,419588 | 0,167818 | 141 | 69 | 72 | 137 | 2,500259 | y ~ Intervention + Sex + |
| 0,608792 | 0,084213 | 0,164164 | 141 | 69 | 72 | 137 | 0,512979 | y ~ Intervention + Sex + |
| 0,914686 | -0,01692 | 0,157629 | 141 | 69 | 72 | 137 | -0,10733 | y ~ Intervention + Sex + |
| 0,303006 | -0,14499 | 0,140233 | 141 | 69 | 72 | 137 | -1,0339 | y ~ Intervention + Sex + |
| 0,150695 | -0,20221 | 0,139921 | 141 | 69 | 72 | 137 | -1,44516 | y ~ Intervention + Sex + |
| 0,417121 | 0,137303 | 0,1687 | 141 | 69 | 72 | 137 | 0,813889 | y ~ Intervention + Sex + |
| 0,007974 | -0,3813 | 0,141608 | 141 | 69 | 72 | 137 | -2,69261 | y ~ Intervention + Sex + |
| 0,534203 | -0,08494 | 0,136299 | 141 | 69 | 72 | 137 | -0,62318 | y ~ Intervention + Sex + |
| 0,959753 | 0,007864 | 0,15555 | 141 | 69 | 72 | 137 | 0,050556 | y ~ Intervention + Sex + |
| 0,061355 | 0,28976 | 0,153603 | 141 | 69 | 72 | 137 | 1,886418 | y ~ Intervention + Sex + |
| 0,691805 | -0,05763 | 0,145084 | 141 | 69 | 72 | 137 | -0,39725 | y ~ Intervention + Sex + |
| 0,561598 | -0,10187 | 0,175061 | 141 | 69 | 72 | 137 | -0,58189 | y ~ Intervention + Sex + |
| 0,682817 | 0,069874 | 0,170635 | 141 | 69 | 72 | 137 | 0,409494 | y ~ Intervention + Sex + |
| 0,127675 | -0,20819 | 0,135836 | 141 | 69 | 72 | 137 | -1,53262 | y ~ Intervention + Sex + |
| 0,897032 | 0,021209 | 0,163583 | 141 | 69 | 72 | 137 | 0,129652 | y ~ Intervention + Sex + |
| 0,123588 | -0,25983 | 0,167693 | 141 | 69 | 72 | 137 | -1,54942 | y ~ Intervention + Sex + |
| 0,164524 | -0,21442 | 0,153434 | 141 | 69 | 72 | 137 | -1,39749 | y ~ Intervention + Sex + |
| 0,01593 | 0,387488 | 0,158752 | 141 | 69 | 72 | 137 | 2,440841 | y ~ Intervention + Sex + |
| 0,241866 | -0,18993 | 0,161583 | 141 | 69 | 72 | 137 | -1,17542 | y ~ Intervention + Sex + |

| | | | | | | | | |
|---|---|---|---|---|---|---|---|---|
| 0,739761 | 0,049654 | 0,149181 | 141 | 69 | 72 | 137 | 0,332844 | y ~ Intervention + Sex + |
| 0,008466 | 0,397691 | 0,148864 | 141 | 69 | 72 | 137 | 2,671517 | y ~ Intervention + Sex + |
| 0,965474 | 0,006295 | 0,145174 | 141 | 69 | 72 | 137 | 0,043364 | y ~ Intervention + Sex + |
| 0,398699 | 0,135887 | 0,16051 | 141 | 69 | 72 | 137 | 0,846591 | y ~ Intervention + Sex + |
| 0,561104 | -0,10077 | 0,172964 | 141 | 69 | 72 | 137 | -0,58262 | y ~ Intervention + Sex + |
| 0,553343 | -0,08746 | 0,147189 | 141 | 69 | 72 | 137 | -0,59422 | y ~ Intervention + Sex + |
| 0,042126 | 0,337281 | 0,164409 | 141 | 69 | 72 | 137 | 2,051479 | y ~ Intervention + Sex + |
| 0,091325 | -0,24205 | 0,142349 | 141 | 69 | 72 | 137 | -1,7004 | y ~ Intervention + Sex + |
| 0,191267 | 0,203009 | 0,154576 | 141 | 69 | 72 | 137 | 1,313329 | y ~ Intervention + Sex + |
| 0,970319 | 0,005722 | 0,153492 | 141 | 69 | 72 | 137 | 0,037276 | y ~ Intervention + Sex + |
| 0,847046 | -0,03111 | 0,16096 | 141 | 69 | 72 | 137 | -0,19325 | y ~ Intervention + Sex + |
| 0,942873 | 0,011409 | 0,158915 | 141 | 69 | 72 | 137 | 0,071791 | y ~ Intervention + Sex + |
| 0,786203 | -0,04358 | 0,160339 | 141 | 69 | 72 | 137 | -0,27178 | y ~ Intervention + Sex + |
| 0,646227 | -0,08159 | 0,177354 | 141 | 69 | 72 | 137 | -0,46003 | y ~ Intervention + Sex + |
| 0,100091 | 0,238041 | 0,143779 | 141 | 69 | 72 | 137 | 1,655604 | y ~ Intervention + Sex + |
| 0,469418 | -0,10401 | 0,14337 | 141 | 69 | 72 | 137 | -0,72544 | y ~ Intervention + Sex + |
| 0,076308 | 0,310706 | 0,173966 | 141 | 69 | 72 | 137 | 1,786017 | y ~ Intervention + Sex + |
| 0,313577 | 0,166993 | 0,1651 | 141 | 69 | 72 | 137 | 1,011465 | y ~ Intervention + Sex + |
| 0,280437 | 0,16324 | 0,150643 | 141 | 69 | 72 | 137 | 1,083619 | y ~ Intervention + Sex + |
| 0,381506 | 0,160218 | 0,182491 | 141 | 69 | 72 | 137 | 0,877954 | y ~ Intervention + Sex + |
| 0,775334 | 0,044998 | 0,157351 | 141 | 69 | 72 | 137 | 0,285968 | y ~ Intervention + Sex + |
| 0,867766 | 0,023426 | 0,140436 | 141 | 69 | 72 | 137 | 0,166809 | y ~ Intervention + Sex + |
| 0,083836 | 0,299095 | 0,171743 | 141 | 69 | 72 | 137 | 1,741527 | y ~ Intervention + Sex + |
| 0,649253 | -0,07496 | 0,164448 | 141 | 69 | 72 | 137 | -0,4558 | y ~ Intervention + Sex + |
| 0,866958 | -0,02444 | 0,145614 | 141 | 69 | 72 | 137 | -0,16784 | y ~ Intervention + Sex + |
| 0,009432 | -0,44101 | 0,167483 | 141 | 69 | 72 | 137 | -2,63313 | y ~ Intervention + Sex + |
| 0,318724 | -0,1582 | 0,158085 | 141 | 69 | 72 | 137 | -1,00073 | y ~ Intervention + Sex + |
| 0,351804 | 0,149621 | 0,160146 | 141 | 69 | 72 | 137 | 0,934278 | y ~ Intervention + Sex + |
| 0,465684 | -0,12587 | 0,172051 | 141 | 69 | 72 | 137 | -0,73157 | y ~ Intervention + Sex + |
| 0,048727 | 0,349005 | 0,175495 | 141 | 69 | 72 | 137 | 1,988694 | y ~ Intervention + Sex + |
| 0,18345 | -0,19209 | 0,143673 | 141 | 69 | 72 | 137 | -1,33697 | y ~ Intervention + Sex + |
| 0,052884 | 0,315836 | 0,161736 | 141 | 69 | 72 | 137 | 1,952782 | y ~ Intervention + Sex + |
| 0,658523 | 0,06236 | 0,140794 | 141 | 69 | 72 | 137 | 0,442919 | y ~ Intervention + Sex + |
| 0,998316 | -0,00031 | 0,148882 | 141 | 69 | 72 | 137 | -0,00211 | y ~ Intervention + Sex + |
| 3,36E-06 | -0,7956 | 0,164175 | 141 | 69 | 72 | 137 | -4,84604 | y ~ Intervention + Sex + |
| 0,011935 | 0,411737 | 0,161588 | 141 | 69 | 72 | 137 | 2,548066 | y ~ Intervention + Sex + |
| 0,630433 | 0,08072 | 0,1674 | 141 | 69 | 72 | 137 | 0,4822 | y ~ Intervention + Sex + |
| 0,952939 | 0,008963 | 0,151595 | 141 | 69 | 72 | 137 | 0,059125 | y ~ Intervention + Sex + |
| 0,025091 | -0,37421 | 0,165226 | 141 | 69 | 72 | 137 | -2,26486 | y ~ Intervention + Sex + |
| 0,083806 | -0,28384 | 0,162966 | 141 | 69 | 72 | 137 | -1,7417 | y ~ Intervention + Sex + |
| 0,00797 | -0,38157 | 0,141701 | 141 | 69 | 72 | 137 | -2,69281 | y ~ Intervention + Sex + |
| 0,200042 | -0,2098 | 0,162937 | 141 | 69 | 72 | 137 | -1,28764 | y ~ Intervention + Sex + |
| 0,48777 | 0,10469 | 0,150472 | 141 | 69 | 72 | 137 | 0,695741 | y ~ Intervention + Sex + |
| 0,025876 | 0,357372 | 0,15865 | 141 | 69 | 72 | 137 | 2,252577 | y ~ Intervention + Sex + |
| 0,096023 | 0,246945 | 0,147344 | 141 | 69 | 72 | 137 | 1,675978 | y ~ Intervention + Sex + |
| 0,828249 | -0,03081 | 0,141754 | 141 | 69 | 72 | 137 | -0,21736 | y ~ Intervention + Sex + |
| 0,03493 | 0,358518 | 0,168291 | 141 | 69 | 72 | 137 | 2,130348 | y ~ Intervention + Sex + |
| 0,018591 | 0,379049 | 0,159128 | 141 | 69 | 72 | 137 | 2,382045 | y ~ Intervention + Sex + |

| | | | | | | | | |
|---|---|---|---|---|---|---|---|---|
| 0,022618 | 0,364262 | 0,15797 | 141 | 69 | 72 | 137 | 2,305894 | y ~ Intervention + Sex + |
| 0,519963 | 0,094012 | 0,14574 | 141 | 69 | 72 | 137 | 0,645066 | y ~ Intervention + Sex + |
| 0,122609 | -0,24226 | 0,155947 | 141 | 69 | 72 | 137 | -1,55351 | y ~ Intervention + Sex + |
| 0,338936 | -0,1502 | 0,156521 | 141 | 69 | 72 | 137 | -0,95962 | y ~ Intervention + Sex + |
| 0,620307 | -0,07538 | 0,151816 | 141 | 69 | 72 | 137 | -0,49654 | y ~ Intervention + Sex + |
| 0,719394 | 0,05484 | 0,152328 | 141 | 69 | 72 | 137 | 0,360011 | y ~ Intervention + Sex + |
| 0,149791 | 0,223666 | 0,154423 | 141 | 69 | 72 | 137 | 1,448396 | y ~ Intervention + Sex + |
| 0,070263 | -0,28985 | 0,158871 | 141 | 69 | 72 | 137 | -1,82445 | y ~ Intervention + Sex + |
| 0,060306 | 0,301955 | 0,15941 | 141 | 69 | 72 | 137 | 1,8942 | y ~ Intervention + Sex + |
| 0,639695 | 0,062259 | 0,132701 | 141 | 69 | 72 | 137 | 0,469168 | y ~ Intervention + Sex + |
| 0,649913 | 0,069106 | 0,151919 | 141 | 69 | 72 | 137 | 0,454883 | y ~ Intervention + Sex + |
| 0,99595 | 0,000851 | 0,167457 | 141 | 69 | 72 | 137 | 0,005085 | y ~ Intervention + Sex + |
| 0,276204 | -0,17867 | 0,163433 | 141 | 69 | 72 | 137 | -1,09325 | y ~ Intervention + Sex + |
| 0,233582 | -0,19681 | 0,164497 | 141 | 69 | 72 | 137 | -1,19646 | y ~ Intervention + Sex + |
| 0,524185 | 0,090892 | 0,142343 | 141 | 69 | 72 | 137 | 0,638545 | y ~ Intervention + Sex + |
| 0,023582 | 0,3536 | 0,154447 | 141 | 69 | 72 | 137 | 2,289453 | y ~ Intervention + Sex + |
| 0,608763 | -0,07711 | 0,150305 | 141 | 69 | 72 | 137 | -0,51302 | y ~ Intervention + Sex + |
| 0,638672 | -0,07323 | 0,155618 | 141 | 69 | 72 | 137 | -0,4706 | y ~ Intervention + Sex + |
| 0,037976 | 0,333273 | 0,15905 | 141 | 69 | 72 | 137 | 2,095397 | y ~ Intervention + Sex + |
| 0,024834 | -0,36709 | 0,161789 | 141 | 69 | 72 | 137 | -2,26896 | y ~ Intervention + Sex + |
| 0,60072 | 0,084507 | 0,161092 | 141 | 69 | 72 | 137 | 0,524584 | y ~ Intervention + Sex + |
| 0,405509 | 0,109766 | 0,131551 | 141 | 69 | 72 | 137 | 0,834398 | y ~ Intervention + Sex + |
| 0,452484 | 0,120314 | 0,159688 | 141 | 69 | 72 | 137 | 0,753431 | y ~ Intervention + Sex + |
| 0,138474 | -0,24841 | 0,166698 | 141 | 69 | 72 | 137 | -1,49019 | y ~ Intervention + Sex + |
| 0,038632 | 0,274906 | 0,131648 | 141 | 69 | 72 | 137 | 2,088186 | y ~ Intervention + Sex + |
| 0,121129 | -0,24718 | 0,158474 | 141 | 69 | 72 | 137 | -1,55974 | y ~ Intervention + Sex + |
| 0,415398 | -0,12031 | 0,14727 | 141 | 69 | 72 | 137 | -0,81691 | y ~ Intervention + Sex + |
| 0,383639 | 0,148364 | 0,169749 | 141 | 69 | 72 | 137 | 0,874017 | y ~ Intervention + Sex + |
| 0,139135 | 0,234861 | 0,157871 | 141 | 69 | 72 | 137 | 1,487673 | y ~ Intervention + Sex + |
| 0,03387 | 0,375048 | 0,175001 | 141 | 69 | 72 | 137 | 2,143124 | y ~ Intervention + Sex + |
| 0,450852 | -0,11508 | 0,152195 | 141 | 69 | 72 | 137 | -0,75616 | y ~ Intervention + Sex + |
| 0,001433 | -0,57732 | 0,177418 | 141 | 69 | 72 | 137 | -3,25399 | y ~ Intervention + Sex + |
| 0,000155 | -0,60518 | 0,15551 | 141 | 69 | 72 | 137 | -3,89157 | y ~ Intervention + Sex + |
| 0,980608 | -0,00377 | 0,154718 | 141 | 69 | 72 | 137 | -0,02435 | y ~ Intervention + Sex + |
| 0,037127 | 0,330069 | 0,15681 | 141 | 69 | 72 | 137 | 2,104893 | y ~ Intervention + Sex + |
| 0,00076 | -0,54669 | 0,158728 | 141 | 69 | 72 | 137 | -3,44421 | y ~ Intervention + Sex + |
| 0,156089 | -0,22879 | 0,160422 | 141 | 69 | 72 | 137 | -1,42619 | y ~ Intervention + Sex + |
| 0,004246 | 0,42785 | 0,147132 | 141 | 69 | 72 | 137 | 2,907938 | y ~ Intervention + Sex + |
| 0,382866 | -0,14024 | 0,160198 | 141 | 69 | 72 | 137 | -0,87544 | y ~ Intervention + Sex + |
| 0,714205 | -0,05724 | 0,15597 | 141 | 69 | 72 | 137 | -0,36697 | y ~ Intervention + Sex + |
| 0,002924 | 0,468586 | 0,154649 | 141 | 69 | 72 | 137 | 3,02999 | y ~ Intervention + Sex + |
| 0,040886 | 0,333473 | 0,161551 | 141 | 69 | 72 | 137 | 2,064194 | y ~ Intervention + Sex + |
| 0,014789 | -0,31936 | 0,12936 | 141 | 69 | 72 | 137 | -2,46877 | y ~ Intervention + Sex + |
| 0,739092 | -0,05286 | 0,158389 | 141 | 69 | 72 | 137 | -0,33373 | y ~ Intervention + Sex + |
| 0,001724 | -0,51939 | 0,162459 | 141 | 69 | 72 | 137 | -3,19706 | y ~ Intervention + Sex + |
| 0,810182 | -0,03492 | 0,145111 | 141 | 69 | 72 | 137 | -0,24066 | y ~ Intervention + Sex + |
| 0,872683 | 0,024038 | 0,14972 | 141 | 69 | 72 | 137 | 0,160552 | y ~ Intervention + Sex + |
| 0,32703 | -0,16796 | 0,170754 | 141 | 69 | 72 | 137 | -0,98363 | y ~ Intervention + Sex + |

| | | | | | | | | |
|---|---|---|---|---|---|---|---|---|
| 0,668411 | 0,0706 | 0,164471 | 141 | 69 | 72 | 137 | 0,429256 | y ~ Intervention + Sex + |
| 0,211503 | 0,188404 | 0,150086 | 141 | 69 | 72 | 137 | 1,25531 | y ~ Intervention + Sex + |
| 0,991309 | 0,001599 | 0,146483 | 141 | 69 | 72 | 137 | 0,010913 | y ~ Intervention + Sex + |
| 0,199526 | 0,193757 | 0,150301 | 141 | 69 | 72 | 137 | 1,289127 | y ~ Intervention + Sex + |
| 0,017731 | 0,350518 | 0,146038 | 141 | 69 | 72 | 137 | 2,400181 | y ~ Intervention + Sex + |
| 0,872084 | -0,02554 | 0,158345 | 141 | 69 | 72 | 137 | -0,16131 | y ~ Intervention + Sex + |
| 0,133092 | 0,279145 | 0,184742 | 141 | 69 | 72 | 137 | 1,510997 | y ~ Intervention + Sex + |
| 0,262263 | -0,18307 | 0,162631 | 141 | 69 | 72 | 137 | -1,1257 | y ~ Intervention + Sex + |
| 0,66138 | -0,06864 | 0,156359 | 141 | 69 | 72 | 137 | -0,43896 | y ~ Intervention + Sex + |
| 1,92E-06 | 0,728475 | 0,146427 | 141 | 69 | 72 | 137 | 4,97501 | y ~ Intervention + Sex + |
| 0,710721 | 0,05217 | 0,140371 | 141 | 69 | 72 | 137 | 0,371659 | y ~ Intervention + Sex + |
| 0,946344 | -0,01028 | 0,152456 | 141 | 69 | 72 | 137 | -0,06742 | y ~ Intervention + Sex + |
| 0,014351 | -0,38734 | 0,156186 | 141 | 69 | 72 | 137 | -2,47998 | y ~ Intervention + Sex + |
| 0,898951 | 0,019333 | 0,151965 | 141 | 69 | 72 | 137 | 0,127223 | y ~ Intervention + Sex + |
| 0,046046 | -0,32947 | 0,163648 | 141 | 69 | 72 | 137 | -2,01327 | y ~ Intervention + Sex + |
| 0,57416 | 0,09824 | 0,174404 | 141 | 69 | 72 | 137 | 0,563288 | y ~ Intervention + Sex + |
| 0,566204 | -0,0947 | 0,164675 | 141 | 69 | 72 | 137 | -0,57505 | y ~ Intervention + Sex + |
| 0,70113 | 0,05914 | 0,153771 | 141 | 69 | 72 | 137 | 0,3846 | y ~ Intervention + Sex + |
| 0,805267 | -0,039 | 0,157896 | 141 | 69 | 72 | 137 | -0,24701 | y ~ Intervention + Sex + |
| 0,041299 | -0,36779 | 0,178546 | 141 | 69 | 72 | 137 | -2,05992 | y ~ Intervention + Sex + |
| 0,951321 | 0,009548 | 0,156122 | 141 | 69 | 72 | 137 | 0,06116 | y ~ Intervention + Sex + |
| 0,049764 | 0,292762 | 0,147897 | 141 | 69 | 72 | 137 | 1,979502 | y ~ Intervention + Sex + |
| 0,341523 | 0,162957 | 0,170729 | 141 | 69 | 72 | 137 | 0,954477 | y ~ Intervention + Sex + |
| 0,047668 | 0,320456 | 0,160367 | 141 | 69 | 72 | 137 | 1,998263 | y ~ Intervention + Sex + |
| 1,63E-05 | -0,64575 | 0,14449 | 141 | 69 | 72 | 137 | -4,46917 | y ~ Intervention + Sex + |
| 0,178229 | 0,206621 | 0,152694 | 141 | 69 | 72 | 137 | 1,353173 | y ~ Intervention + Sex + |
| 0,087309 | -0,26964 | 0,156574 | 141 | 69 | 72 | 137 | -1,7221 | y ~ Intervention + Sex + |
| 0,63923 | 0,073432 | 0,156298 | 141 | 69 | 72 | 137 | 0,46982 | y ~ Intervention + Sex + |
| 0,071996 | -0,28624 | 0,157867 | 141 | 69 | 72 | 137 | -1,81316 | y ~ Intervention + Sex + |
| 0,669739 | 0,073048 | 0,170901 | 141 | 69 | 72 | 137 | 0,427427 | y ~ Intervention + Sex + |
| 0,97533 | -0,00456 | 0,147154 | 141 | 69 | 72 | 137 | -0,03098 | y ~ Intervention + Sex + |
| 0,228023 | -0,18812 | 0,155358 | 141 | 69 | 72 | 137 | -1,21089 | y ~ Intervention + Sex + |
| 0,000148 | -0,6537 | 0,167477 | 141 | 69 | 72 | 137 | -3,90322 | y ~ Intervention + Sex + |
| 0,070096 | -0,28337 | 0,155225 | 141 | 69 | 72 | 137 | -1,82555 | y ~ Intervention + Sex + |
| 0,369125 | -0,12191 | 0,135291 | 141 | 69 | 72 | 137 | -0,90108 | y ~ Intervention + Sex + |
| 0,576373 | 0,075591 | 0,134976 | 141 | 69 | 72 | 137 | 0,560031 | y ~ Intervention + Sex + |
| 0,016399 | -0,37636 | 0,15489 | 141 | 69 | 72 | 137 | -2,42987 | y ~ Intervention + Sex + |
| 0,989553 | -0,00213 | 0,162602 | 141 | 69 | 72 | 137 | -0,01312 | y ~ Intervention + Sex + |
| 0,158494 | -0,23322 | 0,164482 | 141 | 69 | 72 | 137 | -1,41789 | y ~ Intervention + Sex + |
| 0,883252 | 0,021789 | 0,148103 | 141 | 69 | 72 | 137 | 0,147122 | y ~ Intervention + Sex + |
| 0,051872 | -0,32033 | 0,163327 | 141 | 69 | 72 | 137 | -1,9613 | y ~ Intervention + Sex + |
| 0,952586 | -0,00864 | 0,145086 | 141 | 69 | 72 | 137 | -0,05957 | y ~ Intervention + Sex + |
| 0,824222 | 0,034263 | 0,153959 | 141 | 69 | 72 | 137 | 0,222544 | y ~ Intervention + Sex + |
| 0,001323 | 0,53961 | 0,164591 | 141 | 69 | 72 | 137 | 3,278482 | y ~ Intervention + Sex + |
| 0,287594 | -0,18494 | 0,173232 | 141 | 69 | 72 | 137 | -1,06757 | y ~ Intervention + Sex + |
| 0,030145 | 0,33357 | 0,152248 | 141 | 69 | 72 | 137 | 2,190971 | y ~ Intervention + Sex + |
| 0,336881 | -0,16234 | 0,168455 | 141 | 69 | 72 | 137 | -0,96373 | y ~ Intervention + Sex + |
| 0,221113 | 0,209377 | 0,170339 | 141 | 69 | 72 | 137 | 1,229176 | y ~ Intervention + Sex + |

| | | | | | | | | |
|---|---|---|---|---|---|---|---|---|
| 0,95633 | 0,008608 | 0,156911 | 141 | 69 | 72 | 137 | 0,054859 | y ~ Intervention + Sex + |
| 0,543413 | -0,0903 | 0,148226 | 141 | 69 | 72 | 137 | -0,60918 | y ~ Intervention + Sex + |
| 0,606383 | -0,07987 | 0,15466 | 141 | 69 | 72 | 137 | -0,51644 | y ~ Intervention + Sex + |
| 0,57887 | 0,082022 | 0,147425 | 141 | 69 | 72 | 137 | 0,556364 | y ~ Intervention + Sex + |
| 0,957065 | 0,008136 | 0,150854 | 141 | 69 | 72 | 137 | 0,053936 | y ~ Intervention + Sex + |
| 0,956256 | -0,00864 | 0,157134 | 141 | 69 | 72 | 137 | -0,05495 | y ~ Intervention + Sex + |
| 0,450202 | 0,128114 | 0,169183 | 141 | 69 | 72 | 137 | 0,757247 | y ~ Intervention + Sex + |
| 0,520935 | -0,09813 | 0,152479 | 141 | 69 | 72 | 137 | -0,64356 | y ~ Intervention + Sex + |
| 0,151897 | -0,23527 | 0,163278 | 141 | 69 | 72 | 137 | -1,44089 | y ~ Intervention + Sex + |
| 0,024247 | -0,33109 | 0,145314 | 141 | 69 | 72 | 137 | -2,27845 | y ~ Intervention + Sex + |
| 0,312544 | -0,16917 | 0,166894 | 141 | 69 | 72 | 137 | -1,01363 | y ~ Intervention + Sex + |
| 0,010978 | -0,35927 | 0,139331 | 141 | 69 | 72 | 137 | -2,57851 | y ~ Intervention + Sex + |
| 0,02025 | -0,37539 | 0,159805 | 141 | 69 | 72 | 137 | -2,34906 | y ~ Intervention + Sex + |
| 0,094119 | 0,260264 | 0,15439 | 141 | 69 | 72 | 137 | 1,685758 | y ~ Intervention + Sex + |
| 0,088624 | -0,28462 | 0,165972 | 141 | 69 | 72 | 137 | -1,7149 | y ~ Intervention + Sex + |
| 0,136661 | 0,247441 | 0,165277 | 141 | 69 | 72 | 137 | 1,497127 | y ~ Intervention + Sex + |
| 0,318904 | 0,168704 | 0,168645 | 141 | 69 | 72 | 137 | 1,000351 | y ~ Intervention + Sex + |
| 0,120064 | -0,25131 | 0,160656 | 141 | 69 | 72 | 137 | -1,56426 | y ~ Intervention + Sex + |
| 0,168591 | 0,219424 | 0,158537 | 141 | 69 | 72 | 137 | 1,384059 | y ~ Intervention + Sex + |
| 0,636489 | 0,070038 | 0,147862 | 141 | 69 | 72 | 137 | 0,473671 | y ~ Intervention + Sex + |
| 0,092737 | 0,292334 | 0,172677 | 141 | 69 | 72 | 137 | 1,692952 | y ~ Intervention + Sex + |
| 0,586906 | 0,083395 | 0,153127 | 141 | 69 | 72 | 137 | 0,544612 | y ~ Intervention + Sex + |
| 0,459865 | -0,11661 | 0,157338 | 141 | 69 | 72 | 137 | -0,74116 | y ~ Intervention + Sex + |
| 0,348308 | -0,15614 | 0,165913 | 141 | 69 | 72 | 137 | -0,9411 | y ~ Intervention + Sex + |
| 0,719481 | 0,055893 | 0,155303 | 141 | 69 | 72 | 137 | 0,359894 | y ~ Intervention + Sex + |
| 0,000589 | 0,540284 | 0,15354 | 141 | 69 | 72 | 137 | 3,518857 | y ~ Intervention + Sex + |
| 2,65E-06 | 0,734066 | 0,149768 | 141 | 69 | 72 | 137 | 4,901361 | y ~ Intervention + Sex + |
| 0,071558 | 0,307357 | 0,16925 | 141 | 69 | 72 | 137 | 1,815996 | y ~ Intervention + Sex + |
| 0,156109 | 0,233824 | 0,163958 | 141 | 69 | 72 | 137 | 1,426118 | y ~ Intervention + Sex + |
| 0,025354 | 0,367598 | 0,162603 | 141 | 69 | 72 | 137 | 2,260707 | y ~ Intervention + Sex + |
| 0,013884 | 0,4642 | 0,186252 | 141 | 69 | 72 | 137 | 2,492316 | y ~ Intervention + Sex + |
| 0,605138 | -0,08223 | 0,15867 | 141 | 69 | 72 | 137 | -0,51822 | y ~ Intervention + Sex + |
| 0,559707 | -0,10089 | 0,172549 | 141 | 69 | 72 | 137 | -0,58471 | y ~ Intervention + Sex + |
| 0,651169 | 0,076566 | 0,168969 | 141 | 69 | 72 | 137 | 0,453135 | y ~ Intervention + Sex + |
| 0,004273 | -0,43648 | 0,15021 | 141 | 69 | 72 | 137 | -2,90579 | y ~ Intervention + Sex + |
| 0,044092 | -0,32332 | 0,159116 | 141 | 69 | 72 | 137 | -2,03195 | y ~ Intervention + Sex + |
| 0,253379 | 0,187935 | 0,163849 | 141 | 69 | 72 | 137 | 1,147003 | y ~ Intervention + Sex + |
| 0,801857 | -0,04001 | 0,159134 | 141 | 69 | 72 | 137 | -0,25143 | y ~ Intervention + Sex + |
| 0,003708 | -0,41176 | 0,139453 | 141 | 69 | 72 | 137 | -2,95269 | y ~ Intervention + Sex + |
| 0,556201 | 0,098806 | 0,167484 | 141 | 69 | 72 | 137 | 0,589943 | y ~ Intervention + Sex + |
| 0,210236 | -0,22988 | 0,182616 | 141 | 69 | 72 | 137 | -1,25882 | y ~ Intervention + Sex + |
| 0,95093 | -0,01053 | 0,170718 | 141 | 69 | 72 | 137 | -0,06165 | y ~ Intervention + Sex + |
| 0,818431 | -0,037 | 0,160859 | 141 | 69 | 72 | 137 | -0,23001 | y ~ Intervention + Sex + |
| 0,967395 | 0,006181 | 0,150936 | 141 | 69 | 72 | 137 | 0,040951 | y ~ Intervention + Sex + |
| 0,953991 | 0,007475 | 0,129321 | 141 | 69 | 72 | 137 | 0,057802 | y ~ Intervention + Sex + |
| 0,01964 | 0,368849 | 0,156232 | 141 | 69 | 72 | 137 | 2,360901 | y ~ Intervention + Sex + |
| 0,133492 | 0,230259 | 0,152547 | 141 | 69 | 72 | 137 | 1,509431 | y ~ Intervention + Sex + |
| 0,952654 | -0,00961 | 0,161518 | 141 | 69 | 72 | 137 | -0,05948 | y ~ Intervention + Sex + |

| | | | | | | | | |
|---|---|---|---|---|---|---|---|---|
| 0,065099 | 0,302315 | 0,162577 | 141 | 69 | 72 | 137 | 1,859513 | y ~ Intervention + Sex + |
| 0,78847 | 0,048216 | 0,17936 | 141 | 69 | 72 | 137 | 0,268824 | y ~ Intervention + Sex + |
| 0,569453 | -0,07886 | 0,138286 | 141 | 69 | 72 | 137 | -0,57023 | y ~ Intervention + Sex + |
| 0,223298 | 0,200196 | 0,163646 | 141 | 69 | 72 | 137 | 1,223349 | y ~ Intervention + Sex + |
| 0,11719 | -0,20818 | 0,13204 | 141 | 69 | 72 | 137 | -1,57662 | y ~ Intervention + Sex + |
| 0,471298 | 0,116489 | 0,161259 | 141 | 69 | 72 | 137 | 0,722371 | y ~ Intervention + Sex + |
| 0,738279 | -0,05103 | 0,152418 | 141 | 69 | 72 | 137 | -0,33481 | y ~ Intervention + Sex + |
| 0,688997 | 0,073017 | 0,182058 | 141 | 69 | 72 | 137 | 0,401065 | y ~ Intervention + Sex + |
| 0,06058 | -0,29932 | 0,158191 | 141 | 69 | 72 | 137 | -1,89216 | y ~ Intervention + Sex + |
| 0,445306 | -0,11493 | 0,15014 | 141 | 69 | 72 | 137 | -0,76547 | y ~ Intervention + Sex + |
| 0,087265 | 0,232045 | 0,134727 | 141 | 69 | 72 | 137 | 1,722338 | y ~ Intervention + Sex + |
| 0,384143 | 0,14625 | 0,167509 | 141 | 69 | 72 | 137 | 0,873088 | y ~ Intervention + Sex + |
| 0,011237 | 0,396719 | 0,154363 | 141 | 69 | 72 | 137 | 2,570038 | y ~ Intervention + Sex + |
| 0,011973 | 0,418179 | 0,16419 | 141 | 69 | 72 | 137 | 2,546921 | y ~ Intervention + Sex + |
| 0,556896 | 0,088204 | 0,149777 | 141 | 69 | 72 | 137 | 0,588904 | y ~ Intervention + Sex + |
| 0,148896 | 0,2395 | 0,164989 | 141 | 69 | 72 | 137 | 1,451611 | y ~ Intervention + Sex + |
| 0,266589 | 0,175556 | 0,157379 | 141 | 69 | 72 | 137 | 1,1155 | y ~ Intervention + Sex + |
| 0,848626 | -0,03064 | 0,160211 | 141 | 69 | 72 | 137 | -0,19123 | y ~ Intervention + Sex + |
| 0,63599 | 0,062515 | 0,131784 | 141 | 69 | 72 | 137 | 0,474372 | y ~ Intervention + Sex + |
| 0,018353 | 0,377547 | 0,158169 | 141 | 69 | 72 | 137 | 2,386978 | y ~ Intervention + Sex + |
| 0,003776 | -0,50745 | 0,17221 | 141 | 69 | 72 | 137 | -2,94668 | y ~ Intervention + Sex + |
| 0,198836 | 0,21703 | 0,168094 | 141 | 69 | 72 | 137 | 1,291121 | y ~ Intervention + Sex + |
| 0,155002 | 0,232436 | 0,162546 | 141 | 69 | 72 | 137 | 1,42997 | y ~ Intervention + Sex + |
| 0,007287 | 0,426983 | 0,15674 | 141 | 69 | 72 | 137 | 2,724143 | y ~ Intervention + Sex + |
| 0,697423 | 0,058878 | 0,151116 | 141 | 69 | 72 | 137 | 0,389619 | y ~ Intervention + Sex + |
| 0,002744 | -0,48562 | 0,159195 | 141 | 69 | 72 | 137 | -3,05049 | y ~ Intervention + Sex + |
| 0,55648 | -0,09593 | 0,162724 | 141 | 69 | 72 | 137 | -0,58952 | y ~ Intervention + Sex + |
| 0,445325 | -0,11737 | 0,153331 | 141 | 69 | 72 | 137 | -0,76544 | y ~ Intervention + Sex + |
| 0,940406 | -0,01155 | 0,154253 | 141 | 69 | 72 | 137 | -0,0749 | y ~ Intervention + Sex + |
| 0,365855 | -0,15704 | 0,173088 | 141 | 69 | 72 | 137 | -0,90727 | y ~ Intervention + Sex + |
| 0,833819 | 0,031966 | 0,152068 | 141 | 69 | 72 | 137 | 0,210206 | y ~ Intervention + Sex + |
| 0,097867 | -0,26066 | 0,156397 | 141 | 69 | 72 | 137 | -1,66666 | y ~ Intervention + Sex + |
| 0,209407 | -0,21248 | 0,168487 | 141 | 69 | 72 | 137 | -1,26112 | y ~ Intervention + Sex + |
| 0,864903 | 0,026503 | 0,155485 | 141 | 69 | 72 | 137 | 0,170456 | y ~ Intervention + Sex + |
| 0,601261 | 0,0854 | 0,163038 | 141 | 69 | 72 | 137 | 0,523805 | y ~ Intervention + Sex + |
| 0,961359 | 0,007368 | 0,15181 | 141 | 69 | 72 | 137 | 0,048537 | y ~ Intervention + Sex + |
| 0,342628 | 0,15156 | 0,159153 | 141 | 69 | 72 | 137 | 0,952289 | y ~ Intervention + Sex + |
| 0,186943 | 0,194082 | 0,146332 | 141 | 69 | 72 | 137 | 1,326314 | y ~ Intervention + Sex + |
| 0,590403 | 0,095698 | 0,177376 | 141 | 69 | 72 | 137 | 0,539521 | y ~ Intervention + Sex + |
| 0,295101 | 0,155588 | 0,148036 | 141 | 69 | 72 | 137 | 1,051016 | y ~ Intervention + Sex + |
| 0,019956 | -0,40522 | 0,172087 | 141 | 69 | 72 | 137 | -2,35473 | y ~ Intervention + Sex + |
| 0,336176 | 0,146976 | 0,152284 | 141 | 69 | 72 | 137 | 0,965138 | y ~ Intervention + Sex + |
| 0,87435 | 0,024658 | 0,155637 | 141 | 69 | 72 | 137 | 0,158431 | y ~ Intervention + Sex + |
| 0,596074 | 0,081496 | 0,153392 | 141 | 69 | 72 | 137 | 0,531297 | y ~ Intervention + Sex + |
| 0,001261 | 0,540867 | 0,164241 | 141 | 69 | 72 | 137 | 3,293126 | y ~ Intervention + Sex + |
| 0,703016 | -0,05892 | 0,154208 | 141 | 69 | 72 | 137 | -0,38205 | y ~ Intervention + Sex + |
| 0,092113 | -0,26173 | 0,1543 | 141 | 69 | 72 | 137 | -1,69623 | y ~ Intervention + Sex + |
| 0,179283 | 0,242418 | 0,179586 | 141 | 69 | 72 | 137 | 1,349873 | y ~ Intervention + Sex + |

| | | | | | | | | |
|---|---|---|---|---|---|---|---|---|
| 0,067961 | 0,226704 | 0,123222 | 141 | 69 | 72 | 137 | 1,839805 | y ~ Intervention + Sex + |
| 0,090662 | 0,23709 | 0,139144 | 141 | 69 | 72 | 137 | 1,703925 | y ~ Intervention + Sex + |
| 0,620071 | -0,07524 | 0,15143 | 141 | 69 | 72 | 137 | -0,49688 | y ~ Intervention + Sex + |
| 0,916343 | -0,01756 | 0,166834 | 141 | 69 | 72 | 137 | -0,10524 | y ~ Intervention + Sex + |
| 0,751842 | -0,04389 | 0,13852 | 141 | 69 | 72 | 137 | -0,31685 | y ~ Intervention + Sex + |
| 0,202702 | -0,18972 | 0,14822 | 141 | 69 | 72 | 137 | -1,28002 | y ~ Intervention + Sex + |
| 0,457052 | -0,12682 | 0,170041 | 141 | 69 | 72 | 137 | -0,74582 | y ~ Intervention + Sex + |
| 0,320494 | 0,149814 | 0,150256 | 141 | 69 | 72 | 137 | 0,997059 | y ~ Intervention + Sex + |
| 0,015794 | -0,38102 | 0,155896 | 141 | 69 | 72 | 137 | -2,44407 | y ~ Intervention + Sex + |
| 0,764407 | -0,04005 | 0,133364 | 141 | 69 | 72 | 137 | -0,3003 | y ~ Intervention + Sex + |
| 0,379778 | -0,1537 | 0,174427 | 141 | 69 | 72 | 137 | -0,88115 | y ~ Intervention + Sex + |
| 0,375619 | -0,13002 | 0,146274 | 141 | 69 | 72 | 137 | -0,88889 | y ~ Intervention + Sex + |
| 0,412856 | -0,13646 | 0,166136 | 141 | 69 | 72 | 137 | -0,82138 | y ~ Intervention + Sex + |
| 0,012627 | 0,402733 | 0,159347 | 141 | 69 | 72 | 137 | 2,527398 | y ~ Intervention + Sex + |
| 0,315483 | -0,14831 | 0,14721 | 141 | 69 | 72 | 137 | -1,00747 | y ~ Intervention + Sex + |
| 0,459656 | 0,12152 | 0,163883 | 141 | 69 | 72 | 137 | 0,741506 | y ~ Intervention + Sex + |
| 0,507146 | 0,10732 | 0,161375 | 141 | 69 | 72 | 137 | 0,665035 | y ~ Intervention + Sex + |
| 0,594208 | 0,068376 | 0,128045 | 141 | 69 | 72 | 137 | 0,533999 | y ~ Intervention + Sex + |
| 0,52675 | 0,105903 | 0,166882 | 141 | 69 | 72 | 137 | 0,634597 | y ~ Intervention + Sex + |
| 0,275528 | 0,192674 | 0,175991 | 141 | 69 | 72 | 137 | 1,094794 | y ~ Intervention + Sex + |
| 0,266762 | -0,18872 | 0,169241 | 141 | 69 | 72 | 137 | -1,1151 | y ~ Intervention + Sex + |
| 0,899991 | -0,01964 | 0,155956 | 141 | 69 | 72 | 137 | -0,12591 | y ~ Intervention + Sex + |
| 0,030295 | 0,353662 | 0,161568 | 141 | 69 | 72 | 137 | 2,188937 | y ~ Intervention + Sex + |
| 0,70352 | 0,062238 | 0,163197 | 141 | 69 | 72 | 137 | 0,381369 | y ~ Intervention + Sex + |
| 0,670182 | -0,06398 | 0,149904 | 141 | 69 | 72 | 137 | -0,42682 | y ~ Intervention + Sex + |
| 0,667223 | -0,07519 | 0,174495 | 141 | 69 | 72 | 137 | -0,43089 | y ~ Intervention + Sex + |
| 0,402892 | -0,1278 | 0,15231 | 141 | 69 | 72 | 137 | -0,83907 | y ~ Intervention + Sex + |
| 0,095853 | -0,2731 | 0,162868 | 141 | 69 | 72 | 137 | -1,67685 | y ~ Intervention + Sex + |
| 0,602481 | 0,086432 | 0,165563 | 141 | 69 | 72 | 137 | 0,522047 | y ~ Intervention + Sex + |
| 0,410903 | 0,121577 | 0,147396 | 141 | 69 | 72 | 137 | 0,824828 | y ~ Intervention + Sex + |
| 0,064876 | 0,303006 | 0,162812 | 141 | 69 | 72 | 137 | 1,86108 | y ~ Intervention + Sex + |
| 0,054402 | -0,30265 | 0,155982 | 141 | 69 | 72 | 137 | -1,94026 | y ~ Intervention + Sex + |
| 0,000346 | 0,567916 | 0,154722 | 141 | 69 | 72 | 137 | 3,670549 | y ~ Intervention + Sex + |
| 0,169434 | -0,22801 | 0,165068 | 141 | 69 | 72 | 137 | -1,3813 | y ~ Intervention + Sex + |
| 0,733778 | 0,053642 | 0,157402 | 141 | 69 | 72 | 137 | 0,340798 | y ~ Intervention + Sex + |
| 0,347059 | 0,147004 | 0,155799 | 141 | 69 | 72 | 137 | 0,943553 | y ~ Intervention + Sex + |
| 0,417599 | -0,1383 | 0,170106 | 141 | 69 | 72 | 137 | -0,81305 | y ~ Intervention + Sex + |
| 0,01392 | -0,42965 | 0,172458 | 141 | 69 | 72 | 137 | -2,49134 | y ~ Intervention + Sex + |
| 0,027846 | -0,34435 | 0,154894 | 141 | 69 | 72 | 137 | -2,22312 | y ~ Intervention + Sex + |
| 0,318566 | 0,165709 | 0,165534 | 141 | 69 | 72 | 137 | 1,001054 | y ~ Intervention + Sex + |
| 0,02086 | -0,37761 | 0,161541 | 141 | 69 | 72 | 137 | -2,33752 | y ~ Intervention + Sex + |
| 0,200544 | -0,21028 | 0,163492 | 141 | 69 | 72 | 137 | -1,2862 | y ~ Intervention + Sex + |
| 2,57E-07 | -0,75959 | 0,140071 | 141 | 69 | 72 | 137 | -5,42292 | y ~ Intervention + Sex + |

| | | | | | | | | |
|---|---|---|---|---|---|---|---|---|
| 0,0472 | 0,329172 | 0,164377 | 141 | 69 | 72 | 137 | 2,002539 | y ~ Intervention + Sex + |
| 1,94E-05 | -0,75113 | 0,169688 | 141 | 69 | 72 | 137 | -4,42652 | y ~ Intervention + Sex + |
| 0,007531 | -0,40172 | 0,148089 | 141 | 69 | 72 | 137 | -2,71268 | y ~ Intervention + Sex + |
| 0,991267 | 0,001956 | 0,178405 | 141 | 69 | 72 | 137 | 0,010966 | y ~ Intervention + Sex + |
| 0,006869 | -0,44721 | 0,162938 | 141 | 69 | 72 | 137 | -2,74469 | y ~ Intervention + Sex + |
| 0,126497 | 0,245538 | 0,159708 | 141 | 69 | 72 | 137 | 1,537418 | y ~ Intervention + Sex + |
| 0,920763 | 0,017333 | 0,173931 | 141 | 69 | 72 | 137 | 0,099656 | y ~ Intervention + Sex + |
| 0,722365 | -0,05898 | 0,165658 | 141 | 69 | 72 | 137 | -0,35603 | y ~ Intervention + Sex + |
| 0,986036 | -0,00282 | 0,160553 | 141 | 69 | 72 | 137 | -0,01753 | y ~ Intervention + Sex + |
| 0,363779 | -0,15253 | 0,167394 | 141 | 69 | 72 | 137 | -0,91122 | y ~ Intervention + Sex + |
| 0,666588 | 0,078079 | 0,180834 | 141 | 69 | 72 | 137 | 0,431768 | y ~ Intervention + Sex + |
| 0,7413 | -0,0555 | 0,167778 | 141 | 69 | 72 | 137 | -0,3308 | y ~ Intervention + Sex + |
| 0,458143 | 0,135342 | 0,181908 | 141 | 69 | 72 | 137 | 0,744013 | y ~ Intervention + Sex + |
| 0,001029 | 0,459583 | 0,137005 | 141 | 69 | 72 | 137 | 3,354509 | y ~ Intervention + Sex + |
| 0,920142 | -0,01575 | 0,156824 | 141 | 69 | 72 | 137 | -0,10044 | y ~ Intervention + Sex + |
| 0,013351 | -0,39365 | 0,15703 | 141 | 69 | 72 | 137 | -2,50682 | y ~ Intervention + Sex + |
| 0,572353 | 0,097455 | 0,172196 | 141 | 69 | 72 | 137 | 0,565951 | y ~ Intervention + Sex + |
| 0,298088 | 0,174014 | 0,166599 | 141 | 69 | 72 | 137 | 1,044513 | y ~ Intervention + Sex + |
| 0,050863 | 0,318982 | 0,161925 | 141 | 69 | 72 | 137 | 1,969934 | y ~ Intervention + Sex + |
| 3,63E-07 | -0,7986 | 0,14933 | 141 | 69 | 72 | 137 | -5,34787 | y ~ Intervention + Sex + |

| | | | | | | | | |
|---|---|---|---|---|---|---|---|---|
| 3,52E-06 | 0,763247 | 0,157853 | 141 | 69 | 72 | 137 | 4,835165 | y ~ Intervention + Sex + |
| 0,343688 | -0,15906 | 0,167396 | 141 | 69 | 72 | 137 | -0,95019 | y ~ Intervention + Sex + |
| 0,005034 | -0,52466 | 0,18403 | 141 | 69 | 72 | 137 | -2,85093 | y ~ Intervention + Sex + |
| 0,9928 | -0,00149 | 0,164558 | 141 | 69 | 72 | 137 | -0,00904 | y ~ Intervention + Sex + |
| 0,400983 | -0,13887 | 0,164832 | 141 | 69 | 72 | 137 | -0,84249 | y ~ Intervention + Sex + |
| 0,426747 | -0,1222 | 0,153295 | 141 | 69 | 72 | 137 | -0,79714 | y ~ Intervention + Sex + |
| 0,649681 | 0,068499 | 0,150478 | 141 | 69 | 72 | 137 | 0,455207 | y ~ Intervention + Sex + |
| 0,239432 | 0,209044 | 0,176924 | 141 | 69 | 72 | 137 | 1,181548 | y ~ Intervention + Sex + |
| 0,553463 | 0,102046 | 0,171782 | 141 | 69 | 72 | 137 | 0,594042 | y ~ Intervention + Sex + |
| 0,067308 | -0,33663 | 0,182529 | 141 | 69 | 72 | 137 | -1,84424 | y ~ Intervention + Sex + |
| 0,072928 | 0,263113 | 0,145593 | 141 | 69 | 72 | 137 | 1,80718 | y ~ Intervention + Sex + |
| 0,808158 | 0,038837 | 0,159643 | 141 | 69 | 72 | 137 | 0,243273 | y ~ Intervention + Sex + |
| 0,094113 | -0,25416 | 0,150769 | 141 | 69 | 72 | 137 | -1,68579 | y ~ Intervention + Sex + |
| 0,55593 | -0,09027 | 0,152906 | 141 | 69 | 72 | 137 | -0,59035 | y ~ Intervention + Sex + |
| 0,080246 | -0,25335 | 0,143759 | 141 | 69 | 72 | 137 | -1,76232 | y ~ Intervention + Sex + |
| 0,161523 | 0,197329 | 0,140191 | 141 | 69 | 72 | 137 | 1,407572 | y ~ Intervention + Sex + |
| 0,967657 | 0,007021 | 0,172828 | 141 | 69 | 72 | 137 | 0,040622 | y ~ Intervention + Sex + |
| 0,203146 | 0,213555 | 0,167002 | 141 | 69 | 72 | 137 | 1,278753 | y ~ Intervention + Sex + |
| 0,943425 | -0,01163 | 0,163614 | 141 | 69 | 72 | 137 | -0,0711 | y ~ Intervention + Sex + |
| 8,60E-06 | -0,66624 | 0,144073 | 141 | 69 | 72 | 137 | -4,62433 | y ~ Intervention + Sex + |
| 0,008104 | 0,429649 | 0,159903 | 141 | 69 | 72 | 137 | 2,686938 | y ~ Intervention + Sex + |
| 0,056309 | 0,282351 | 0,14668 | 141 | 69 | 72 | 137 | 1,924946 | y ~ Intervention + Sex + |
| 0,003426 | 0,489194 | 0,164234 | 141 | 69 | 72 | 137 | 2,978642 | y ~ Intervention + Sex + |
| 0,391688 | -0,14434 | 0,167979 | 141 | 69 | 72 | 137 | -0,85928 | y ~ Intervention + Sex + |
| 0,453385 | -0,11841 | 0,157471 | 141 | 69 | 72 | 137 | -0,75193 | y ~ Intervention + Sex + |
| 0,007424 | 0,425631 | 0,156617 | 141 | 69 | 72 | 137 | 2,717653 | y ~ Intervention + Sex + |
| 0,01651 | 0,418907 | 0,172581 | 141 | 69 | 72 | 137 | 2,427308 | y ~ Intervention + Sex + |
| 0,006694 | 0,44183 | 0,160454 | 141 | 69 | 72 | 137 | 2,753625 | y ~ Intervention + Sex + |
| 0,462821 | 0,102108 | 0,138682 | 141 | 69 | 72 | 137 | 0,736277 | y ~ Intervention + Sex + |
| 0,712246 | 0,062111 | 0,168046 | 141 | 69 | 72 | 137 | 0,369607 | y ~ Intervention + Sex + |
| 0,628734 | 0,088756 | 0,183153 | 141 | 69 | 72 | 137 | 0,484601 | y ~ Intervention + Sex + |
| 0,909159 | -0,01964 | 0,171855 | 141 | 69 | 72 | 137 | -0,11431 | y ~ Intervention + Sex + |
| 0,31802 | -0,16527 | 0,164905 | 141 | 69 | 72 | 137 | -1,00219 | y ~ Intervention + Sex + |
| 0,291352 | 0,173184 | 0,163497 | 141 | 69 | 72 | 137 | 1,059246 | y ~ Intervention + Sex + |
| 0,379124 | -0,14232 | 0,16129 | 141 | 69 | 72 | 137 | -0,88237 | y ~ Intervention + Sex + |
| 0,098213 | -0,28102 | 0,168787 | 141 | 69 | 72 | 137 | -1,66493 | y ~ Intervention + Sex + |
| 0,888464 | -0,02369 | 0,16859 | 141 | 69 | 72 | 137 | -0,14051 | y ~ Intervention + Sex + |
| 0,686957 | 0,065835 | 0,16302 | 141 | 69 | 72 | 137 | 0,403844 | y ~ Intervention + Sex + |
| 0,007384 | -0,46337 | 0,170383 | 141 | 69 | 72 | 137 | -2,71955 | y ~ Intervention + Sex + |
| 0,153058 | 0,209836 | 0,146045 | 141 | 69 | 72 | 137 | 1,436787 | y ~ Intervention + Sex + |
| 0,001152 | -0,53019 | 0,159676 | 141 | 69 | 72 | 137 | -3,32038 | y ~ Intervention + Sex + |
| 0,286869 | 0,166113 | 0,155365 | 141 | 69 | 72 | 137 | 1,06918 | y ~ Intervention + Sex + |
| 0,255535 | 0,178112 | 0,155994 | 141 | 69 | 72 | 137 | 1,141786 | y ~ Intervention + Sex + |
| 0,753099 | 0,049525 | 0,157129 | 141 | 69 | 72 | 137 | 0,315187 | y ~ Intervention + Sex + |
| 0,812791 | -0,03505 | 0,147723 | 141 | 69 | 72 | 137 | -0,23728 | y ~ Intervention + Sex + |
| 0,198207 | 0,216669 | 0,167578 | 141 | 69 | 72 | 137 | 1,292944 | y ~ Intervention + Sex + |
| 0,571653 | 0,095149 | 0,167815 | 141 | 69 | 72 | 137 | 0,566984 | y ~ Intervention + Sex + |
| 0,303729 | 0,160554 | 0,155524 | 141 | 69 | 72 | 137 | 1,032345 | y ~ Intervention + Sex + |

| | | | | | | | | |
|---|---|---|---|---|---|---|---|---|
| 0,082088 | 0,252854 | 0,14436 | 141 | 69 | 72 | 137 | 1,751555 | y ~ Intervention + Sex + |
| 0,390576 | 0,141566 | 0,164362 | 141 | 69 | 72 | 137 | 0,861304 | y ~ Intervention + Sex + |
| 4,11E-05 | -0,70423 | 0,166154 | 141 | 69 | 72 | 137 | -4,2384 | y ~ Intervention + Sex + |
| 0,130201 | -0,24664 | 0,162001 | 141 | 69 | 72 | 137 | -1,52245 | y ~ Intervention + Sex + |
| 0,747721 | 0,048706 | 0,151123 | 141 | 69 | 72 | 137 | 0,322294 | y ~ Intervention + Sex + |
| 0,255277 | -0,17744 | 0,155324 | 141 | 69 | 72 | 137 | -1,14241 | y ~ Intervention + Sex + |
| 0,233618 | 0,183145 | 0,153084 | 141 | 69 | 72 | 137 | 1,196371 | y ~ Intervention + Sex + |
| 8,67E-05 | 0,594265 | 0,146889 | 141 | 69 | 72 | 137 | 4,045675 | y ~ Intervention + Sex + |
| 0,477324 | 0,116699 | 0,163772 | 141 | 69 | 72 | 137 | 0,71257 | y ~ Intervention + Sex + |
| 0,92919 | 0,014103 | 0,158414 | 141 | 69 | 72 | 137 | 0,089028 | y ~ Intervention + Sex + |
| 0,787102 | 0,040844 | 0,150937 | 141 | 69 | 72 | 137 | 0,270605 | y ~ Intervention + Sex + |
| 0,133548 | 0,211777 | 0,140323 | 141 | 69 | 72 | 137 | 1,509212 | y ~ Intervention + Sex + |
| 0,454947 | 0,12203 | 0,162853 | 141 | 69 | 72 | 137 | 0,749323 | y ~ Intervention + Sex + |
| 0,507169 | -0,10837 | 0,162964 | 141 | 69 | 72 | 137 | -0,665 | y ~ Intervention + Sex + |
| 0,057516 | -0,3217 | 0,167948 | 141 | 69 | 72 | 137 | -1,91547 | y ~ Intervention + Sex + |
| 0,288998 | -0,16058 | 0,150856 | 141 | 69 | 72 | 137 | -1,06445 | y ~ Intervention + Sex + |
| 0,981273 | -0,00365 | 0,155129 | 141 | 69 | 72 | 137 | -0,02352 | y ~ Intervention + Sex + |
| 0,415428 | 0,139836 | 0,171188 | 141 | 69 | 72 | 137 | 0,816857 | y ~ Intervention + Sex + |
| 0,365224 | 0,117843 | 0,129716 | 141 | 69 | 72 | 137 | 0,908473 | y ~ Intervention + Sex + |
| 0,468644 | -0,11174 | 0,153755 | 141 | 69 | 72 | 137 | -0,72671 | y ~ Intervention + Sex + |
| 0,002392 | 0,503059 | 0,162576 | 141 | 69 | 72 | 137 | 3,094289 | y ~ Intervention + Sex + |
| 0,005733 | 0,420177 | 0,149698 | 141 | 69 | 72 | 137 | 2,806836 | y ~ Intervention + Sex + |
| 0,054536 | 0,316307 | 0,163114 | 141 | 69 | 72 | 137 | 1,939174 | y ~ Intervention + Sex + |
| 0,17991 | 0,194016 | 0,143938 | 141 | 69 | 72 | 137 | 1,347915 | y ~ Intervention + Sex + |
| 0,574038 | -0,09508 | 0,168741 | 141 | 69 | 72 | 137 | -0,56347 | y ~ Intervention + Sex + |
| 0,530009 | 0,099019 | 0,157275 | 141 | 69 | 72 | 137 | 0,629594 | y ~ Intervention + Sex + |
| 0,505851 | -0,10596 | 0,158848 | 141 | 69 | 72 | 137 | -0,66707 | y ~ Intervention + Sex + |
| 0,058525 | 0,338143 | 0,177253 | 141 | 69 | 72 | 137 | 1,907681 | y ~ Intervention + Sex + |
| 0,392237 | -0,13287 | 0,154813 | 141 | 69 | 72 | 137 | -0,85828 | y ~ Intervention + Sex + |
| 0,728985 | -0,05527 | 0,159198 | 141 | 69 | 72 | 137 | -0,34718 | y ~ Intervention + Sex + |
| 0,734117 | 0,053321 | 0,156666 | 141 | 69 | 72 | 137 | 0,340347 | y ~ Intervention + Sex + |
| 0,496006 | -0,10975 | 0,160778 | 141 | 69 | 72 | 137 | -0,68261 | y ~ Intervention + Sex + |
| 0,097538 | -0,2592 | 0,155368 | 141 | 69 | 72 | 137 | -1,66831 | y ~ Intervention + Sex + |
| 0,544278 | -0,09026 | 0,148489 | 141 | 69 | 72 | 137 | -0,60787 | y ~ Intervention + Sex + |
| 0,540041 | -0,10281 | 0,167363 | 141 | 69 | 72 | 137 | -0,61429 | y ~ Intervention + Sex + |
| 0,217151 | 0,183277 | 0,147822 | 141 | 69 | 72 | 137 | 1,239847 | y ~ Intervention + Sex + |
| 0,460086 | -0,12291 | 0,165917 | 141 | 69 | 72 | 137 | -0,74079 | y ~ Intervention + Sex + |
| 0,220256 | -0,19672 | 0,15974 | 141 | 69 | 72 | 137 | -1,23147 | y ~ Intervention + Sex + |
| 0,648175 | -0,06977 | 0,152578 | 141 | 69 | 72 | 137 | -0,45731 | y ~ Intervention + Sex + |
| 0,639247 | 0,086349 | 0,1838 | 141 | 69 | 72 | 137 | 0,469797 | y ~ Intervention + Sex + |
| 0,035218 | -0,33332 | 0,156714 | 141 | 69 | 72 | 137 | -2,12693 | y ~ Intervention + Sex + |
| 0,090825 | 0,260797 | 0,153135 | 141 | 69 | 72 | 137 | 1,703056 | y ~ Intervention + Sex + |
| 0,710671 | 0,063551 | 0,170962 | 141 | 69 | 72 | 137 | 0,371726 | y ~ Intervention + Sex + |
| 0,829124 | 0,032012 | 0,148041 | 141 | 69 | 72 | 137 | 0,216237 | y ~ Intervention + Sex + |
| 0,000106 | -0,64514 | 0,161608 | 141 | 69 | 72 | 137 | -3,99203 | y ~ Intervention + Sex + |
| 0,270961 | -0,15451 | 0,139789 | 141 | 69 | 72 | 137 | -1,10531 | y ~ Intervention + Sex + |
| 0,63545 | -0,07369 | 0,155092 | 141 | 69 | 72 | 137 | -0,47513 | y ~ Intervention + Sex + |
| 0,204823 | -0,19803 | 0,155443 | 141 | 69 | 72 | 137 | -1,27399 | y ~ Intervention + Sex + |

| | | | | | | | | |
|---|---|---|---|---|---|---|---|---|
| 0,537958 | -0,09963 | 0,161358 | 141 | 69 | 72 | 137 | -0,61746 | y ~ Intervention + Sex + |
| 0,587341 | 0,099327 | 0,182594 | 141 | 69 | 72 | 137 | 0,543978 | y ~ Intervention + Sex + |
| 0,52436 | -0,09244 | 0,144825 | 141 | 69 | 72 | 137 | -0,63828 | y ~ Intervention + Sex + |
| 0,003244 | -0,44093 | 0,147152 | 141 | 69 | 72 | 137 | -2,9964 | y ~ Intervention + Sex + |
| 0,012796 | 0,407968 | 0,161731 | 141 | 69 | 72 | 137 | 2,522509 | y ~ Intervention + Sex + |
| 0,477685 | 0,11395 | 0,160046 | 141 | 69 | 72 | 137 | 0,711985 | y ~ Intervention + Sex + |
| 0,000304 | 0,506405 | 0,136612 | 141 | 69 | 72 | 137 | 3,706885 | y ~ Intervention + Sex + |
| 0,215775 | 0,236859 | 0,190464 | 141 | 69 | 72 | 137 | 1,243588 | y ~ Intervention + Sex + |
| 0,005724 | -0,43515 | 0,155003 | 141 | 69 | 72 | 137 | -2,80739 | y ~ Intervention + Sex + |
| 0,918009 | -0,01588 | 0,154021 | 141 | 69 | 72 | 137 | -0,10313 | y ~ Intervention + Sex + |
| 0,917483 | 0,016925 | 0,163058 | 141 | 69 | 72 | 137 | 0,103796 | y ~ Intervention + Sex + |
| 0,041615 | 0,356248 | 0,173216 | 141 | 69 | 72 | 137 | 2,056673 | y ~ Intervention + Sex + |
| 0,345373 | -0,15407 | 0,162713 | 141 | 69 | 72 | 137 | -0,94687 | y ~ Intervention + Sex + |
| 0,361721 | 0,118143 | 0,129097 | 141 | 69 | 72 | 137 | 0,91515 | y ~ Intervention + Sex + |
| 0,965505 | -0,00637 | 0,147121 | 141 | 69 | 72 | 137 | -0,04333 | y ~ Intervention + Sex + |
| 0,900173 | 0,019691 | 0,156681 | 141 | 69 | 72 | 137 | 0,125675 | y ~ Intervention + Sex + |
| 0,044162 | -0,28525 | 0,140429 | 141 | 69 | 72 | 137 | -2,03127 | y ~ Intervention + Sex + |
| 0,943978 | 0,010776 | 0,153067 | 141 | 69 | 72 | 137 | 0,070399 | y ~ Intervention + Sex + |
| 0,138048 | -0,23659 | 0,158594 | 141 | 69 | 72 | 137 | -1,49181 | y ~ Intervention + Sex + |
| 0,329158 | -0,15685 | 0,160161 | 141 | 69 | 72 | 137 | -0,9793 | y ~ Intervention + Sex + |
| 0,056086 | -0,29836 | 0,154853 | 141 | 69 | 72 | 137 | -1,92671 | y ~ Intervention + Sex + |
| 0,001248 | 0,493685 | 0,149774 | 141 | 69 | 72 | 137 | 3,296201 | y ~ Intervention + Sex + |
| 0,000616 | -0,48791 | 0,139185 | 141 | 69 | 72 | 137 | -3,5055 | y ~ Intervention + Sex + |
| 0,272619 | -0,16989 | 0,154237 | 141 | 69 | 72 | 137 | -1,10148 | y ~ Intervention + Sex + |
| 0,120428 | -0,24428 | 0,156316 | 141 | 69 | 72 | 137 | -1,56271 | y ~ Intervention + Sex + |
| 0,561878 | -0,09381 | 0,161337 | 141 | 69 | 72 | 137 | -0,58147 | y ~ Intervention + Sex + |
| 0,134365 | 0,207992 | 0,138107 | 141 | 69 | 72 | 137 | 1,506019 | y ~ Intervention + Sex + |
| 0,057125 | 0,28235 | 0,147171 | 141 | 69 | 72 | 137 | 1,918519 | y ~ Intervention + Sex + |
| 7,48E-06 | 0,7952 | 0,170733 | 141 | 69 | 72 | 137 | 4,657568 | y ~ Intervention + Sex + |
| 0,301559 | 0,172937 | 0,166765 | 141 | 69 | 72 | 137 | 1,037009 | y ~ Intervention + Sex + |
| 0,175691 | 0,239049 | 0,175619 | 141 | 69 | 72 | 137 | 1,36118 | y ~ Intervention + Sex + |
| 0,189115 | 0,189554 | 0,143627 | 141 | 69 | 72 | 137 | 1,319763 | y ~ Intervention + Sex + |
| 0,007739 | -0,37893 | 0,140181 | 141 | 69 | 72 | 137 | -2,70312 | y ~ Intervention + Sex + |
| 0,989779 | 0,002195 | 0,171013 | 141 | 69 | 72 | 137 | 0,012834 | y ~ Intervention + Sex + |
| 0,273839 | -0,17667 | 0,160806 | 141 | 69 | 72 | 137 | -1,09867 | y ~ Intervention + Sex + |
| 0,044699 | 0,319362 | 0,157627 | 141 | 69 | 72 | 137 | 2,026066 | y ~ Intervention + Sex + |
| 0,592234 | -0,0811 | 0,151067 | 141 | 69 | 72 | 137 | -0,53686 | y ~ Intervention + Sex + |
| 0,411303 | 0,145284 | 0,176289 | 141 | 69 | 72 | 137 | 0,824122 | y ~ Intervention + Sex + |
| 0,091337 | 0,263518 | 0,15498 | 141 | 69 | 72 | 137 | 1,700336 | y ~ Intervention + Sex + |
| 0,525669 | -0,10688 | 0,167979 | 141 | 69 | 72 | 137 | -0,63626 | y ~ Intervention + Sex + |
| 0,968141 | -0,00703 | 0,175667 | 141 | 69 | 72 | 137 | -0,04001 | y ~ Intervention + Sex + |
| 0,481555 | -0,10833 | 0,153501 | 141 | 69 | 72 | 137 | -0,70573 | y ~ Intervention + Sex + |
| 0,174871 | -0,223 | 0,163515 | 141 | 69 | 72 | 137 | -1,36379 | y ~ Intervention + Sex + |
| 0,625323 | 0,078626 | 0,16065 | 141 | 69 | 72 | 137 | 0,489426 | y ~ Intervention + Sex + |
| 0,984636 | 0,003184 | 0,16502 | 141 | 69 | 72 | 137 | 0,019292 | y ~ Intervention + Sex + |
| 0,860649 | 0,029404 | 0,167185 | 141 | 69 | 72 | 137 | 0,175879 | y ~ Intervention + Sex + |
| 0,269698 | 0,162787 | 0,146887 | 141 | 69 | 72 | 137 | 1,108244 | y ~ Intervention + Sex + |
| 0,000126 | 0,575987 | 0,145956 | 141 | 69 | 72 | 137 | 3,946314 | y ~ Intervention + Sex + |

| | | | | | | | | |
|---|---|---|---|---|---|---|---|---|
| 0,637089 | 0,073262 | 0,154945 | 141 | 69 | 72 | 137 | 0,472828 | y ~ Intervention + Sex + |
| 0,016825 | 0,386124 | 0,159545 | 141 | 69 | 72 | 137 | 2,420153 | y ~ Intervention + Sex + |
| 0,023467 | 0,357405 | 0,155978 | 141 | 69 | 72 | 137 | 2,291375 | y ~ Intervention + Sex + |
| 0,860345 | -0,02872 | 0,162935 | 141 | 69 | 72 | 137 | -0,17627 | y ~ Intervention + Sex + |
| 0,050822 | 0,305112 | 0,154857 | 141 | 69 | 72 | 137 | 1,970287 | y ~ Intervention + Sex + |
| 0,015856 | 0,316303 | 0,129495 | 141 | 69 | 72 | 137 | 2,442583 | y ~ Intervention + Sex + |
| 0,259931 | 0,190177 | 0,168114 | 141 | 69 | 72 | 137 | 1,131239 | y ~ Intervention + Sex + |
| 0,673911 | -0,0731 | 0,173357 | 141 | 69 | 72 | 137 | -0,42169 | y ~ Intervention + Sex + |
| 0,002396 | 0,468512 | 0,151438 | 141 | 69 | 72 | 137 | 3,093758 | y ~ Intervention + Sex + |
| 0,612181 | -0,07947 | 0,1564 | 141 | 69 | 72 | 137 | -0,50813 | y ~ Intervention + Sex + |
| 0,063661 | 0,301375 | 0,16119 | 141 | 69 | 72 | 137 | 1,869692 | y ~ Intervention + Sex + |
| 0,979467 | -0,00401 | 0,155628 | 141 | 69 | 72 | 137 | -0,02578 | y ~ Intervention + Sex + |
| 0,021318 | 0,389878 | 0,167397 | 141 | 69 | 72 | 137 | 2,329061 | y ~ Intervention + Sex + |
| 2,24E-06 | 0,788783 | 0,159681 | 141 | 69 | 72 | 137 | 4,939732 | y ~ Intervention + Sex + |
| 0,892945 | -0,0219 | 0,16243 | 141 | 69 | 72 | 137 | -0,13483 | y ~ Intervention + Sex + |
| 0,465043 | 0,130932 | 0,178718 | 141 | 69 | 72 | 137 | 0,732617 | y ~ Intervention + Sex + |
| 0,045088 | 0,325818 | 0,16111 | 141 | 69 | 72 | 137 | 2,022333 | y ~ Intervention + Sex + |
| 0,122626 | 0,253357 | 0,163095 | 141 | 69 | 72 | 137 | 1,553435 | y ~ Intervention + Sex + |
| 0,352175 | -0,15022 | 0,160913 | 141 | 69 | 72 | 137 | -0,93356 | y ~ Intervention + Sex + |
| 0,030923 | -0,36094 | 0,165526 | 141 | 69 | 72 | 137 | -2,18057 | y ~ Intervention + Sex + |
| 0,016032 | 0,376918 | 0,154575 | 141 | 69 | 72 | 137 | 2,438414 | y ~ Intervention + Sex + |
| 0,86891 | -0,02584 | 0,156245 | 141 | 69 | 72 | 137 | -0,16535 | y ~ Intervention + Sex + |
| 0,291177 | -0,16845 | 0,158966 | 141 | 69 | 72 | 137 | -1,05963 | y ~ Intervention + Sex + |
| 0,505588 | -0,10731 | 0,16077 | 141 | 69 | 72 | 137 | -0,66748 | y ~ Intervention + Sex + |
| 0,000453 | -0,53378 | 0,148527 | 141 | 69 | 72 | 137 | -3,59385 | y ~ Intervention + Sex + |
| 0,068574 | 0,308781 | 0,168211 | 141 | 69 | 72 | 137 | 1,835672 | y ~ Intervention + Sex + |
| 0,259153 | 0,181837 | 0,160478 | 141 | 69 | 72 | 137 | 1,133096 | y ~ Intervention + Sex + |
| 0,977844 | -0,00454 | 0,163163 | 141 | 69 | 72 | 137 | -0,02782 | y ~ Intervention + Sex + |
| 5,33E-05 | 0,699655 | 0,16769 | 141 | 69 | 72 | 137 | 4,172319 | y ~ Intervention + Sex + |
| 0,090099 | 0,264133 | 0,154741 | 141 | 69 | 72 | 137 | 1,706937 | y ~ Intervention + Sex + |
| 0,10658 | -0,25134 | 0,154724 | 141 | 69 | 72 | 137 | -1,62444 | y ~ Intervention + Sex + |
| 0,843998 | 0,030357 | 0,153973 | 141 | 69 | 72 | 137 | 0,197156 | y ~ Intervention + Sex + |
| 0,890319 | 0,019122 | 0,13841 | 141 | 69 | 72 | 137 | 0,138158 | y ~ Intervention + Sex + |
| 0,022997 | 0,36201 | 0,157439 | 141 | 69 | 72 | 137 | 2,299359 | y ~ Intervention + Sex + |
| 0,903905 | -0,01766 | 0,146013 | 141 | 69 | 72 | 137 | -0,12095 | y ~ Intervention + Sex + |
| 0,014055 | 0,399286 | 0,160502 | 141 | 69 | 72 | 137 | 2,48774 | y ~ Intervention + Sex + |
| 0,244179 | 0,20649 | 0,176542 | 141 | 69 | 72 | 137 | 1,169635 | y ~ Intervention + Sex + |
| 0,036017 | -0,34009 | 0,160603 | 141 | 69 | 72 | 137 | -2,11758 | y ~ Intervention + Sex + |
| 0,157524 | 0,215906 | 0,151915 | 141 | 69 | 72 | 137 | 1,421224 | y ~ Intervention + Sex + |
| 0,075342 | 0,274978 | 0,153449 | 141 | 69 | 72 | 137 | 1,791987 | y ~ Intervention + Sex + |
| 7,08E-07 | 0,744818 | 0,143225 | 141 | 69 | 72 | 137 | 5,200321 | y ~ Intervention + Sex + |
| 0,055255 | 0,298196 | 0,154238 | 141 | 69 | 72 | 137 | 1,933355 | y ~ Intervention + Sex + |
| 0,563425 | 0,082075 | 0,141712 | 141 | 69 | 72 | 137 | 0,579171 | y ~ Intervention + Sex + |
| 0,142392 | -0,27966 | 0,189544 | 141 | 69 | 72 | 137 | -1,47543 | y ~ Intervention + Sex + |
| 0,58531 | -0,07839 | 0,143323 | 141 | 69 | 72 | 137 | -0,54694 | y ~ Intervention + Sex + |
| 0,15346 | 0,206245 | 0,143687 | 141 | 69 | 72 | 137 | 1,435374 | y ~ Intervention + Sex + |
| 0,343743 | -0,15641 | 0,164624 | 141 | 69 | 72 | 137 | -0,95008 | y ~ Intervention + Sex + |
| 0,40589 | 0,131257 | 0,157435 | 141 | 69 | 72 | 137 | 0,833721 | y ~ Intervention + Sex + |

| | | | | | | | | |
|---|---|---|---|---|---|---|---|---|
| 0,836954 | 0,035001 | 0,169755 | 141 | 69 | 72 | 137 | 0,206183 | y ~ Intervention + Sex + |
| 0,005632 | -0,45575 | 0,16202 | 141 | 69 | 72 | 137 | -2,8129 | y ~ Intervention + Sex + |
| 0,496225 | -0,11768 | 0,17249 | 141 | 69 | 72 | 137 | -0,68226 | y ~ Intervention + Sex + |
| 0,222882 | 0,184086 | 0,150341 | 141 | 69 | 72 | 137 | 1,224454 | y ~ Intervention + Sex + |
| 0,281402 | 0,157246 | 0,145405 | 141 | 69 | 72 | 137 | 1,081438 | y ~ Intervention + Sex + |
| 0,213837 | -0,18896 | 0,151304 | 141 | 69 | 72 | 137 | -1,24889 | y ~ Intervention + Sex + |
| 5,79E-06 | -0,72664 | 0,153991 | 141 | 69 | 72 | 137 | -4,71874 | y ~ Intervention + Sex + |
| 0,963664 | 0,006819 | 0,149413 | 141 | 69 | 72 | 137 | 0,04564 | y ~ Intervention + Sex + |
| 0,026689 | 0,325061 | 0,145104 | 141 | 69 | 72 | 137 | 2,240194 | y ~ Intervention + Sex + |
| 0,504056 | -0,11557 | 0,172515 | 141 | 69 | 72 | 137 | -0,66989 | y ~ Intervention + Sex + |
| 0,024353 | -0,36299 | 0,159434 | 141 | 69 | 72 | 137 | -2,27672 | y ~ Intervention + Sex + |
| 0,288462 | 0,178856 | 0,16784 | 141 | 69 | 72 | 137 | 1,065639 | y ~ Intervention + Sex + |
| 0,010673 | -0,40533 | 0,156576 | 141 | 69 | 72 | 137 | -2,58871 | y ~ Intervention + Sex + |
| 0,138959 | 0,2354 | 0,158163 | 141 | 69 | 72 | 137 | 1,488343 | y ~ Intervention + Sex + |
| 0,340163 | -0,14213 | 0,148489 | 141 | 69 | 72 | 137 | -0,95718 | y ~ Intervention + Sex + |
| 0,069005 | 0,251536 | 0,137242 | 141 | 69 | 72 | 137 | 1,832787 | y ~ Intervention + Sex + |
| 0,793136 | 0,043449 | 0,165362 | 141 | 69 | 72 | 137 | 0,262753 | y ~ Intervention + Sex + |
| 0,000501 | 0,594436 | 0,166717 | 141 | 69 | 72 | 137 | 3,565533 | y ~ Intervention + Sex + |
| 0,479978 | 0,115302 | 0,162793 | 141 | 69 | 72 | 137 | 0,708275 | y ~ Intervention + Sex + |
| 0,099029 | -0,26243 | 0,158006 | 141 | 69 | 72 | 137 | -1,66086 | y ~ Intervention + Sex + |
| 0,098543 | -0,25937 | 0,155937 | 141 | 69 | 72 | 137 | -1,66327 | y ~ Intervention + Sex + |
| 0,523434 | 0,100621 | 0,157294 | 141 | 69 | 72 | 137 | 0,639703 | y ~ Intervention + Sex + |
| 0,296508 | 0,160571 | 0,153224 | 141 | 69 | 72 | 137 | 1,047948 | y ~ Intervention + Sex + |
| 0,954378 | 0,009366 | 0,16341 | 141 | 69 | 72 | 137 | 0,057315 | y ~ Intervention + Sex + |
| 0,136808 | 0,218222 | 0,145816 | 141 | 69 | 72 | 137 | 1,496559 | y ~ Intervention + Sex + |
| 0,124786 | 0,219936 | 0,142404 | 141 | 69 | 72 | 137 | 1,54445 | y ~ Intervention + Sex + |
| 0,166057 | 0,201382 | 0,144629 | 141 | 69 | 72 | 137 | 1,392399 | y ~ Intervention + Sex + |
| 0,052214 | -0,24985 | 0,127577 | 141 | 69 | 72 | 137 | -1,95841 | y ~ Intervention + Sex + |
| 0,744881 | -0,04828 | 0,14807 | 141 | 69 | 72 | 137 | -0,32605 | y ~ Intervention + Sex + |
| 0,970435 | -0,00634 | 0,170835 | 141 | 69 | 72 | 137 | -0,03713 | y ~ Intervention + Sex + |
| 0,607973 | -0,07145 | 0,138965 | 141 | 69 | 72 | 137 | -0,51415 | y ~ Intervention + Sex + |
| 0,87901 | -0,02339 | 0,153384 | 141 | 69 | 72 | 137 | -0,15251 | y ~ Intervention + Sex + |
| 0,307114 | -0,15544 | 0,151627 | 141 | 69 | 72 | 137 | -1,02512 | y ~ Intervention + Sex + |
| 0,178931 | -0,20326 | 0,150451 | 141 | 69 | 72 | 137 | -1,35097 | y ~ Intervention + Sex + |
| 0,171406 | 0,208391 | 0,151568 | 141 | 69 | 72 | 137 | 1,374901 | y ~ Intervention + Sex + |
| 0,200667 | 0,198344 | 0,154252 | 141 | 69 | 72 | 137 | 1,285844 | y ~ Intervention + Sex + |
| 0,021079 | 0,356215 | 0,152655 | 141 | 69 | 72 | 137 | 2,333465 | y ~ Intervention + Sex + |
| 0,009506 | 0,406396 | 0,154502 | 141 | 69 | 72 | 137 | 2,63036 | y ~ Intervention + Sex + |
| 0,798455 | -0,04045 | 0,158089 | 141 | 69 | 72 | 137 | -0,25584 | y ~ Intervention + Sex + |
| 0,413788 | -0,1246 | 0,152001 | 141 | 69 | 72 | 137 | -0,81974 | y ~ Intervention + Sex + |
| 0,529436 | 0,092363 | 0,146498 | 141 | 69 | 72 | 137 | 0,630473 | y ~ Intervention + Sex + |
| 0,772735 | 0,044632 | 0,154239 | 141 | 69 | 72 | 137 | 0,289371 | y ~ Intervention + Sex + |
| 0,438942 | -0,13194 | 0,169972 | 141 | 69 | 72 | 137 | -0,77625 | y ~ Intervention + Sex + |
| 0,048479 | 0,267877 | 0,134549 | 141 | 69 | 72 | 137 | 1,990922 | y ~ Intervention + Sex + |
| 0,276416 | 0,177913 | 0,16281 | 141 | 69 | 72 | 137 | 1,092762 | y ~ Intervention + Sex + |
| 0,969886 | 0,00619 | 0,163663 | 141 | 69 | 72 | 137 | 0,03782 | y ~ Intervention + Sex + |
| 0,165485 | -0,20503 | 0,14705 | 141 | 69 | 72 | 137 | -1,3943 | y ~ Intervention + Sex + |
| 0,069481 | 0,305966 | 0,16723 | 141 | 69 | 72 | 137 | 1,829616 | y ~ Intervention + Sex + |

| | | | | | | | | |
|---|---|---|---|---|---|---|---|---|
| 0,676284 | 0,07125 | 0,170277 | 141 | 69 | 72 | 137 | 0,418436 | y ~ Intervention + Sex + |
| 0,376609 | -0,13664 | 0,154035 | 141 | 69 | 72 | 137 | -0,88705 | y ~ Intervention + Sex + |
| 0,303064 | 0,162429 | 0,157122 | 141 | 69 | 72 | 137 | 1,033773 | y ~ Intervention + Sex + |
| 0,001088 | 0,502365 | 0,150515 | 141 | 69 | 72 | 137 | 3,337647 | y ~ Intervention + Sex + |
| 0,830105 | 0,030792 | 0,143234 | 141 | 69 | 72 | 137 | 0,214977 | y ~ Intervention + Sex + |
| 0,163896 | 0,206244 | 0,14736 | 141 | 69 | 72 | 137 | 1,399593 | y ~ Intervention + Sex + |
| 0,085389 | -0,26183 | 0,151103 | 141 | 69 | 72 | 137 | -1,73276 | y ~ Intervention + Sex + |
| 0,078608 | -0,28526 | 0,160979 | 141 | 69 | 72 | 137 | -1,77206 | y ~ Intervention + Sex + |
| 0,47017 | 0,10893 | 0,150411 | 141 | 69 | 72 | 137 | 0,724213 | y ~ Intervention + Sex + |
| 0,057982 | -0,30494 | 0,159498 | 141 | 69 | 72 | 137 | -1,91186 | y ~ Intervention + Sex + |
| 0,532987 | -0,09362 | 0,149783 | 141 | 69 | 72 | 137 | -0,62504 | y ~ Intervention + Sex + |
| 0,703955 | -0,06859 | 0,180125 | 141 | 69 | 72 | 137 | -0,38078 | y ~ Intervention + Sex + |
| 0,276209 | -0,1833 | 0,167666 | 141 | 69 | 72 | 137 | -1,09324 | y ~ Intervention + Sex + |
| 0,375056 | -0,14892 | 0,167336 | 141 | 69 | 72 | 137 | -0,88994 | y ~ Intervention + Sex + |
| 0,057052 | 0,296228 | 0,154358 | 141 | 69 | 72 | 137 | 1,919095 | y ~ Intervention + Sex + |
| 0,027905 | 0,377515 | 0,169878 | 141 | 69 | 72 | 137 | 2,222271 | y ~ Intervention + Sex + |
| 0,400332 | 0,120402 | 0,142715 | 141 | 69 | 72 | 137 | 0,843657 | y ~ Intervention + Sex + |
| 0,636356 | -0,06646 | 0,140255 | 141 | 69 | 72 | 137 | -0,47386 | y ~ Intervention + Sex + |
| 0,480334 | 0,107252 | 0,151551 | 141 | 69 | 72 | 137 | 0,707699 | y ~ Intervention + Sex + |
| 0,651856 | -0,07194 | 0,159097 | 141 | 69 | 72 | 137 | -0,45218 | y ~ Intervention + Sex + |
| 0,486079 | -0,11515 | 0,164871 | 141 | 69 | 72 | 137 | -0,69845 | y ~ Intervention + Sex + |
| 0,571056 | -0,08624 | 0,151865 | 141 | 69 | 72 | 137 | -0,56787 | y ~ Intervention + Sex + |
| 0,27261 | 0,171602 | 0,155789 | 141 | 69 | 72 | 137 | 1,101501 | y ~ Intervention + Sex + |
| 0,177034 | -0,2118 | 0,156086 | 141 | 69 | 72 | 137 | -1,35693 | y ~ Intervention + Sex + |
| 0,009016 | 0,424467 | 0,160224 | 141 | 69 | 72 | 137 | 2,649216 | y ~ Intervention + Sex + |
| 0,255287 | 0,160574 | 0,14056 | 141 | 69 | 72 | 137 | 1,142385 | y ~ Intervention + Sex + |
| 0,016432 | 0,396343 | 0,163165 | 141 | 69 | 72 | 137 | 2,4291 | y ~ Intervention + Sex + |
| 0,025706 | 0,303834 | 0,134726 | 141 | 69 | 72 | 137 | 2,255201 | y ~ Intervention + Sex + |
| 0,022116 | 0,360714 | 0,155836 | 141 | 69 | 72 | 137 | 2,314699 | y ~ Intervention + Sex + |
| 0,751113 | -0,05074 | 0,159658 | 141 | 69 | 72 | 137 | -0,31781 | y ~ Intervention + Sex + |
| 0,379959 | -0,1298 | 0,147359 | 141 | 69 | 72 | 137 | -0,88082 | y ~ Intervention + Sex + |
| 0,111977 | 0,294319 | 0,183988 | 141 | 69 | 72 | 137 | 1,599662 | y ~ Intervention + Sex + |
| 0,140227 | 0,237974 | 0,160409 | 141 | 69 | 72 | 137 | 1,483543 | y ~ Intervention + Sex + |
| 0,909625 | 0,01734 | 0,152479 | 141 | 69 | 72 | 137 | 0,113722 | y ~ Intervention + Sex + |
| 0,345746 | -0,16146 | 0,170649 | 141 | 69 | 72 | 137 | -0,94613 | y ~ Intervention + Sex + |
| 0,826884 | -0,03286 | 0,149951 | 141 | 69 | 72 | 137 | -0,21912 | y ~ Intervention + Sex + |
| 0,402424 | -0,12857 | 0,153079 | 141 | 69 | 72 | 137 | -0,83991 | y ~ Intervention + Sex + |
| 0,50837 | 0,104777 | 0,158007 | 141 | 69 | 72 | 137 | 0,663116 | y ~ Intervention + Sex + |
| 0,521602 | 0,112177 | 0,174586 | 141 | 69 | 72 | 137 | 0,642531 | y ~ Intervention + Sex + |
| 0,772154 | 0,044848 | 0,154578 | 141 | 69 | 72 | 137 | 0,290132 | y ~ Intervention + Sex + |
| 5,67E-05 | 0,592487 | 0,14255 | 141 | 69 | 72 | 137 | 4,156334 | y ~ Intervention + Sex + |
| 0,459704 | 0,123056 | 0,165972 | 141 | 69 | 72 | 137 | 0,741426 | y ~ Intervention + Sex + |
| 0,495139 | -0,10906 | 0,15945 | 141 | 69 | 72 | 137 | -0,68399 | y ~ Intervention + Sex + |
| 0,923718 | 0,015663 | 0,163278 | 141 | 69 | 72 | 137 | 0,095928 | y ~ Intervention + Sex + |
| 0,703616 | -0,06265 | 0,164324 | 141 | 69 | 72 | 137 | -0,38124 | y ~ Intervention + Sex + |
| 0,010257 | 0,423721 | 0,162778 | 141 | 69 | 72 | 137 | 2,603057 | y ~ Intervention + Sex + |
| 0,933702 | 0,013147 | 0,157751 | 141 | 69 | 72 | 137 | 0,083342 | y ~ Intervention + Sex + |
| 0,307052 | -0,15841 | 0,154509 | 141 | 69 | 72 | 137 | -1,02525 | y ~ Intervention + Sex + |

| | | | | | | | | |
|---|---|---|---|---|---|---|---|---|
| 0,007043 | 0,397702 | 0,145359 | 141 | 69 | 72 | 137 | 2,735996 | y ~ Intervention + Sex + |
| 2,95E-05 | -0,66178 | 0,153113 | 141 | 69 | 72 | 137 | -4,32215 | y ~ Intervention + Sex + |
| 0,27223 | 0,162913 | 0,147783 | 141 | 69 | 72 | 137 | 1,102379 | y ~ Intervention + Sex + |
| 0,861609 | 0,030561 | 0,174979 | 141 | 69 | 72 | 137 | 0,174654 | y ~ Intervention + Sex + |
| 0,050082 | 0,336537 | 0,170251 | 141 | 69 | 72 | 137 | 1,976712 | y ~ Intervention + Sex + |
| 0,574741 | 0,09175 | 0,16313 | 141 | 69 | 72 | 137 | 0,562432 | y ~ Intervention + Sex + |
| 0,376707 | 0,146407 | 0,165084 | 141 | 69 | 72 | 137 | 0,886862 | y ~ Intervention + Sex + |
| 0,524987 | -0,0984 | 0,154399 | 141 | 69 | 72 | 137 | -0,63731 | y ~ Intervention + Sex + |
| 0,000178 | 0,512178 | 0,132917 | 141 | 69 | 72 | 137 | 3,853371 | y ~ Intervention + Sex + |
| 0,718486 | -0,0536 | 0,148385 | 141 | 69 | 72 | 137 | -0,36123 | y ~ Intervention + Sex + |
| 0,003642 | 0,459622 | 0,15535 | 141 | 69 | 72 | 137 | 2,958627 | y ~ Intervention + Sex + |
| 0,200204 | 0,203648 | 0,158213 | 141 | 69 | 72 | 137 | 1,287175 | y ~ Intervention + Sex + |
| 0,478893 | 0,125334 | 0,176519 | 141 | 69 | 72 | 137 | 0,710028 | y ~ Intervention + Sex + |
| 0,417681 | 0,116649 | 0,143495 | 141 | 69 | 72 | 137 | 0,812909 | y ~ Intervention + Sex + |
| 0,179986 | -0,20707 | 0,153647 | 141 | 69 | 72 | 137 | -1,34768 | y ~ Intervention + Sex + |
| 0,363325 | -0,13705 | 0,150261 | 141 | 69 | 72 | 137 | -0,91209 | y ~ Intervention + Sex + |
| 0,753751 | -0,05302 | 0,168679 | 141 | 69 | 72 | 137 | -0,31433 | y ~ Intervention + Sex + |
| 0,887705 | 0,021141 | 0,149436 | 141 | 69 | 72 | 137 | 0,141472 | y ~ Intervention + Sex + |
| 0,689982 | -0,06424 | 0,1607 | 141 | 69 | 72 | 137 | -0,39972 | y ~ Intervention + Sex + |
| 0,926997 | -0,01303 | 0,141994 | 141 | 69 | 72 | 137 | -0,09179 | y ~ Intervention + Sex + |
| 0,767017 | 0,046675 | 0,157225 | 141 | 69 | 72 | 137 | 0,296868 | y ~ Intervention + Sex + |
| 0,892422 | -0,02087 | 0,154067 | 141 | 69 | 72 | 137 | -0,13549 | y ~ Intervention + Sex + |
| 0,081175 | -0,27017 | 0,153778 | 141 | 69 | 72 | 137 | -1,75686 | y ~ Intervention + Sex + |
| 0,25788 | 0,178529 | 0,157136 | 141 | 69 | 72 | 137 | 1,136144 | y ~ Intervention + Sex + |
| 0,142908 | 0,21641 | 0,146867 | 141 | 69 | 72 | 137 | 1,473509 | y ~ Intervention + Sex + |
| 0,06607 | -0,2642 | 0,142598 | 141 | 69 | 72 | 137 | -1,85275 | y ~ Intervention + Sex + |
| 0,350421 | -0,15013 | 0,16023 | 141 | 69 | 72 | 137 | -0,93697 | y ~ Intervention + Sex + |
| 0,591395 | 0,079945 | 0,148574 | 141 | 69 | 72 | 137 | 0,53808 | y ~ Intervention + Sex + |
| 0,293571 | 0,159375 | 0,151157 | 141 | 69 | 72 | 137 | 1,054367 | y ~ Intervention + Sex + |
| 0,730726 | 0,055531 | 0,161024 | 141 | 69 | 72 | 137 | 0,344864 | y ~ Intervention + Sex + |
| 0,023188 | 0,368448 | 0,160467 | 141 | 69 | 72 | 137 | 2,296097 | y ~ Intervention + Sex + |
| 0,442025 | -0,11852 | 0,153714 | 141 | 69 | 72 | 137 | -0,77102 | y ~ Intervention + Sex + |
| 0,023006 | -0,37038 | 0,161089 | 141 | 69 | 72 | 137 | -2,29921 | y ~ Intervention + Sex + |
| 0,394867 | 0,134428 | 0,1575 | 141 | 69 | 72 | 137 | 0,853509 | y ~ Intervention + Sex + |
| 0,095175 | 0,244485 | 0,1455 | 141 | 69 | 72 | 137 | 1,680312 | y ~ Intervention + Sex + |
| 0,075596 | -0,28648 | 0,160008 | 141 | 69 | 72 | 137 | -1,79041 | y ~ Intervention + Sex + |
| 0,152005 | 0,238936 | 0,165869 | 141 | 69 | 72 | 137 | 1,440509 | y ~ Intervention + Sex + |
| 0,053944 | -0,27683 | 0,142403 | 141 | 69 | 72 | 137 | -1,944 | y ~ Intervention + Sex + |
| 0,162663 | 0,211536 | 0,150696 | 141 | 69 | 72 | 137 | 1,403726 | y ~ Intervention + Sex + |
| 0,010244 | 0,405577 | 0,155781 | 141 | 69 | 72 | 137 | 2,603519 | y ~ Intervention + Sex + |
| 0,412223 | 0,135053 | 0,164199 | 141 | 69 | 72 | 137 | 0,822498 | y ~ Intervention + Sex + |
| 0,022201 | -0,37036 | 0,160108 | 141 | 69 | 72 | 137 | -2,3132 | y ~ Intervention + Sex + |
| 0,185547 | 0,210868 | 0,158482 | 141 | 69 | 72 | 137 | 1,330552 | y ~ Intervention + Sex + |
| 0,212517 | -0,18867 | 0,150635 | 141 | 69 | 72 | 137 | -1,25251 | y ~ Intervention + Sex + |
| 0,051549 | -0,33744 | 0,17181 | 141 | 69 | 72 | 137 | -1,96405 | y ~ Intervention + Sex + |
| 0,318174 | -0,17165 | 0,17133 | 141 | 69 | 72 | 137 | -1,00187 | y ~ Intervention + Sex + |
| 0,39831 | 0,127354 | 0,150307 | 141 | 69 | 72 | 137 | 0,847293 | y ~ Intervention + Sex + |
| 0,863293 | -0,03014 | 0,174707 | 141 | 69 | 72 | 137 | -0,17251 | y ~ Intervention + Sex + |

| | | | | | | | | |
|---|---|---|---|---|---|---|---|---|
| 0,311572 | 0,15608 | 0,15367 | 141 | 69 | 72 | 137 | 1,015681 | y ~ Intervention + Sex + |
| 0,177638 | 0,193191 | 0,142573 | 141 | 69 | 72 | 137 | 1,355031 | y ~ Intervention + Sex + |
| 0,176676 | 0,220364 | 0,162263 | 141 | 69 | 72 | 137 | 1,358064 | y ~ Intervention + Sex + |
| 0,285984 | -0,16324 | 0,152398 | 141 | 69 | 72 | 137 | -1,07115 | y ~ Intervention + Sex + |
| 0,022116 | 0,360714 | 0,155836 | 141 | 69 | 72 | 137 | 2,314699 | y ~ Intervention + Sex + |
| 0,302208 | -0,16248 | 0,156895 | 141 | 69 | 72 | 137 | -1,03561 | y ~ Intervention + Sex + |
| 0,422444 | -0,12998 | 0,16154 | 141 | 69 | 72 | 137 | -0,8046 | y ~ Intervention + Sex + |
| 0,05524 | 0,289675 | 0,149821 | 141 | 69 | 72 | 137 | 1,933478 | y ~ Intervention + Sex + |
| 0,701644 | -0,05992 | 0,156087 | 141 | 69 | 72 | 137 | -0,38391 | y ~ Intervention + Sex + |
| 0,314832 | 0,168951 | 0,167471 | 141 | 69 | 72 | 137 | 1,008835 | y ~ Intervention + Sex + |
| 0,01642 | -0,3887 | 0,159998 | 141 | 69 | 72 | 137 | -2,42939 | y ~ Intervention + Sex + |
| 0,968088 | 0,0069 | 0,172163 | 141 | 69 | 72 | 137 | 0,040079 | y ~ Intervention + Sex + |
| 0,021906 | 0,367507 | 0,158515 | 141 | 69 | 72 | 137 | 2,318444 | y ~ Intervention + Sex + |
| 0,038045 | 0,32044 | 0,152981 | 141 | 69 | 72 | 137 | 2,094639 | y ~ Intervention + Sex + |
| 0,059091 | -0,31348 | 0,164698 | 141 | 69 | 72 | 137 | -1,90336 | y ~ Intervention + Sex + |
| 0,004038 | 0,43629 | 0,149181 | 141 | 69 | 72 | 137 | 2,924559 | y ~ Intervention + Sex + |
| 0,359034 | 0,152544 | 0,165754 | 141 | 69 | 72 | 137 | 0,920299 | y ~ Intervention + Sex + |
| 0,308718 | 0,153658 | 0,150392 | 141 | 69 | 72 | 137 | 1,021711 | y ~ Intervention + Sex + |
| 0,292895 | 0,163783 | 0,15512 | 141 | 69 | 72 | 137 | 1,05585 | y ~ Intervention + Sex + |
| 0,928365 | 0,013503 | 0,149916 | 141 | 69 | 72 | 137 | 0,090068 | y ~ Intervention + Sex + |
| 0,001816 | 0,457605 | 0,143859 | 141 | 69 | 72 | 137 | 3,180934 | y ~ Intervention + Sex + |
| 0,17831 | 0,196532 | 0,145265 | 141 | 69 | 72 | 137 | 1,352919 | y ~ Intervention + Sex + |
| 0,180235 | -0,20465 | 0,151939 | 141 | 69 | 72 | 137 | -1,3469 | y ~ Intervention + Sex + |
| 0,600447 | 0,084781 | 0,161493 | 141 | 69 | 72 | 137 | 0,524979 | y ~ Intervention + Sex + |
| 0,191456 | -0,20256 | 0,154297 | 141 | 69 | 72 | 137 | -1,31277 | y ~ Intervention + Sex + |
| 0,90069 | -0,02097 | 0,167703 | 141 | 69 | 72 | 137 | -0,12502 | y ~ Intervention + Sex + |
| 0,594302 | 0,089439 | 0,167532 | 141 | 69 | 72 | 137 | 0,533863 | y ~ Intervention + Sex + |
| 0,080451 | -0,27886 | 0,158342 | 141 | 69 | 72 | 137 | -1,76111 | y ~ Intervention + Sex + |
| 0,736065 | -0,05681 | 0,168196 | 141 | 69 | 72 | 137 | -0,33776 | y ~ Intervention + Sex + |
| 0,415234 | -0,12027 | 0,14717 | 141 | 69 | 72 | 137 | -0,8172 | y ~ Intervention + Sex + |
| 0,947049 | 0,01063 | 0,159768 | 141 | 69 | 72 | 137 | 0,066535 | y ~ Intervention + Sex + |
| 0,723707 | 0,058806 | 0,166007 | 141 | 69 | 72 | 137 | 0,354236 | y ~ Intervention + Sex + |
| 0,145683 | -0,24382 | 0,166624 | 141 | 69 | 72 | 137 | -1,46327 | y ~ Intervention + Sex + |
| 0,156417 | 0,197525 | 0,138609 | 141 | 69 | 72 | 137 | 1,425051 | y ~ Intervention + Sex + |
| 0,040789 | 0,355381 | 0,172081 | 141 | 69 | 72 | 137 | 2,065201 | y ~ Intervention + Sex + |
| 0,488901 | -0,11106 | 0,16004 | 141 | 69 | 72 | 137 | -0,69393 | y ~ Intervention + Sex + |
| 0,475952 | -0,12038 | 0,168412 | 141 | 69 | 72 | 137 | -0,7148 | y ~ Intervention + Sex + |
| 0,389872 | 0,136101 | 0,157782 | 141 | 69 | 72 | 137 | 0,862588 | y ~ Intervention + Sex + |
| 0,587492 | -0,08635 | 0,158795 | 141 | 69 | 72 | 137 | -0,54376 | y ~ Intervention + Sex + |
| 0,022111 | 0,328326 | 0,141838 | 141 | 69 | 72 | 137 | 2,31479 | y ~ Intervention + Sex + |
| 0,234671 | -0,16037 | 0,134347 | 141 | 69 | 72 | 137 | -1,19367 | y ~ Intervention + Sex + |
| 9,11E-07 | 0,74622 | 0,145063 | 141 | 69 | 72 | 137 | 5,144123 | y ~ Intervention + Sex + |
| 0,844353 | 0,029079 | 0,147832 | 141 | 69 | 72 | 137 | 0,196701 | y ~ Intervention + Sex + |
| 0,694221 | 0,061979 | 0,157323 | 141 | 69 | 72 | 137 | 0,393962 | y ~ Intervention + Sex + |
| 0,816004 | 0,039954 | 0,171376 | 141 | 69 | 72 | 137 | 0,233136 | y ~ Intervention + Sex + |
| 0,819105 | 0,034272 | 0,149572 | 141 | 69 | 72 | 137 | 0,229136 | y ~ Intervention + Sex + |
| 0,929544 | -0,01259 | 0,142133 | 141 | 69 | 72 | 137 | -0,08858 | y ~ Intervention + Sex + |
| 0,258787 | 0,170083 | 0,149989 | 141 | 69 | 72 | 137 | 1,13397 | y ~ Intervention + Sex + |

| | | | | | | | | |
|---|---|---|---|---|---|---|---|---|
| 0,000442 | -0,53605 | 0,148864 | 141 | 69 | 72 | 137 | -3,60096 | y ~ Intervention + Sex + |
| 0,991354 | 0,001722 | 0,158606 | 141 | 69 | 72 | 137 | 0,010856 | y ~ Intervention + Sex + |
| 0,781245 | -0,04527 | 0,162689 | 141 | 69 | 72 | 137 | -0,27824 | y ~ Intervention + Sex + |
| 0,403914 | -0,12819 | 0,153104 | 141 | 69 | 72 | 137 | -0,83724 | y ~ Intervention + Sex + |
| 0,846219 | 0,029819 | 0,153461 | 141 | 69 | 72 | 137 | 0,194312 | y ~ Intervention + Sex + |
| 0,855011 | 0,029937 | 0,163524 | 141 | 69 | 72 | 137 | 0,183074 | y ~ Intervention + Sex + |
| 0,177295 | -0,22072 | 0,162759 | 141 | 69 | 72 | 137 | -1,35611 | y ~ Intervention + Sex + |
| 0,056114 | 0,275396 | 0,142952 | 141 | 69 | 72 | 137 | 1,926486 | y ~ Intervention + Sex + |
| 0,449133 | 0,117927 | 0,155363 | 141 | 69 | 72 | 137 | 0,759039 | y ~ Intervention + Sex + |
| 0,19411 | 0,188914 | 0,144772 | 141 | 69 | 72 | 137 | 1,304914 | y ~ Intervention + Sex + |
| 0,107721 | 0,238921 | 0,147562 | 141 | 69 | 72 | 137 | 1,619123 | y ~ Intervention + Sex + |
| 0,045195 | 0,325938 | 0,161251 | 141 | 69 | 72 | 137 | 2,021311 | y ~ Intervention + Sex + |
| 0,752909 | 0,046471 | 0,147322 | 141 | 69 | 72 | 137 | 0,315438 | y ~ Intervention + Sex + |
| 0,007056 | -0,4244 | 0,15515 | 141 | 69 | 72 | 137 | -2,73539 | y ~ Intervention + Sex + |
| 0,314493 | 0,168655 | 0,16706 | 141 | 69 | 72 | 137 | 1,009546 | y ~ Intervention + Sex + |
| 0,387093 | -0,14483 | 0,166914 | 141 | 69 | 72 | 137 | -0,86767 | y ~ Intervention + Sex + |
| 0,002732 | 0,459647 | 0,150614 | 141 | 69 | 72 | 137 | 3,051814 | y ~ Intervention + Sex + |
| 0,571585 | 0,085839 | 0,151369 | 141 | 69 | 72 | 137 | 0,567084 | y ~ Intervention + Sex + |
| 0,015344 | 0,357617 | 0,145672 | 141 | 69 | 72 | 137 | 2,454943 | y ~ Intervention + Sex + |
| 0,290708 | -0,17471 | 0,16472 | 141 | 69 | 72 | 137 | -1,06067 | y ~ Intervention + Sex + |
| 0,711493 | -0,0593 | 0,160007 | 141 | 69 | 72 | 137 | -0,37062 | y ~ Intervention + Sex + |
| 0,698455 | -0,06287 | 0,161932 | 141 | 69 | 72 | 137 | -0,38822 | y ~ Intervention + Sex + |
| 0,350996 | -0,14567 | 0,155656 | 141 | 69 | 72 | 137 | -0,93585 | y ~ Intervention + Sex + |
| 0,81206 | 0,036568 | 0,1535 | 141 | 69 | 72 | 137 | 0,238228 | y ~ Intervention + Sex + |
| 0,611332 | -0,08275 | 0,162463 | 141 | 69 | 72 | 137 | -0,50934 | y ~ Intervention + Sex + |
| 0,942701 | -0,01158 | 0,160781 | 141 | 69 | 72 | 137 | -0,07201 | y ~ Intervention + Sex + |
| 0,844043 | 0,031086 | 0,157721 | 141 | 69 | 72 | 137 | 0,197098 | y ~ Intervention + Sex + |
| 0,022056 | 0,332754 | 0,143691 | 141 | 69 | 72 | 137 | 2,315768 | y ~ Intervention + Sex + |
| 0,125796 | 0,231143 | 0,150065 | 141 | 69 | 72 | 137 | 1,540289 | y ~ Intervention + Sex + |
| 0,301563 | 0,175099 | 0,168852 | 141 | 69 | 72 | 137 | 1,036999 | y ~ Intervention + Sex + |
| 0,001165 | 0,540283 | 0,162877 | 141 | 69 | 72 | 137 | 3,317114 | y ~ Intervention + Sex + |
| 0,482134 | -0,11636 | 0,165092 | 141 | 69 | 72 | 137 | -0,7048 | y ~ Intervention + Sex + |
| 0,031264 | -0,38847 | 0,17852 | 141 | 69 | 72 | 137 | -2,17608 | y ~ Intervention + Sex + |
| 0,549601 | 0,094446 | 0,157451 | 141 | 69 | 72 | 137 | 0,599844 | y ~ Intervention + Sex + |
| 0,803073 | 0,037034 | 0,148222 | 141 | 69 | 72 | 137 | 0,249856 | y ~ Intervention + Sex + |
| 0,006476 | -0,42079 | 0,152181 | 141 | 69 | 72 | 137 | -2,76504 | y ~ Intervention + Sex + |
| 0,725011 | 0,057268 | 0,162465 | 141 | 69 | 72 | 137 | 0,352492 | y ~ Intervention + Sex + |
| 0,823393 | 0,03364 | 0,15044 | 141 | 69 | 72 | 137 | 0,223611 | y ~ Intervention + Sex + |
| 0,922491 | -0,01568 | 0,160811 | 141 | 69 | 72 | 137 | -0,09748 | y ~ Intervention + Sex + |
| 0,09653 | -0,25408 | 0,151835 | 141 | 69 | 72 | 137 | -1,6734 | y ~ Intervention + Sex + |
| 0,239329 | -0,19073 | 0,161388 | 141 | 69 | 72 | 137 | -1,18181 | y ~ Intervention + Sex + |
| 0,565071 | 0,080291 | 0,139219 | 141 | 69 | 72 | 137 | 0,576726 | y ~ Intervention + Sex + |
| 0,477389 | -0,12355 | 0,173412 | 141 | 69 | 72 | 137 | -0,71246 | y ~ Intervention + Sex + |
| 0,279353 | -0,16875 | 0,155373 | 141 | 69 | 72 | 137 | -1,08607 | y ~ Intervention + Sex + |
| 0,854028 | 0,029691 | 0,161075 | 141 | 69 | 72 | 137 | 0,184328 | y ~ Intervention + Sex + |
| 0,868419 | 0,026054 | 0,156975 | 141 | 69 | 72 | 137 | 0,165978 | y ~ Intervention + Sex + |
| 0,244085 | -0,17376 | 0,148532 | 141 | 69 | 72 | 137 | -1,16987 | y ~ Intervention + Sex + |
| 0,064602 | 0,278315 | 0,14939 | 141 | 69 | 72 | 137 | 1,863013 | y ~ Intervention + Sex + |

| | | | | | | | | |
|---|---|---|---|---|---|---|---|---|
| 0,420765 | -0,148 | 0,183277 | 141 | 69 | 72 | 137 | -0,80752 | y ~ Intervention + Sex + |
| 0,128615 | -0,22971 | 0,150252 | 141 | 69 | 72 | 137 | -1,52882 | y ~ Intervention + Sex + |
| 0,215802 | -0,17613 | 0,141638 | 141 | 69 | 72 | 137 | -1,24351 | y ~ Intervention + Sex + |
| 0,460688 | 0,121968 | 0,164866 | 141 | 69 | 72 | 137 | 0,739799 | y ~ Intervention + Sex + |
| 0,245679 | -0,17734 | 0,152108 | 141 | 69 | 72 | 137 | -1,1659 | y ~ Intervention + Sex + |
| 0,059891 | 0,306877 | 0,161743 | 141 | 69 | 72 | 137 | 1,897317 | y ~ Intervention + Sex + |
| 0,529014 | 0,103347 | 0,163752 | 141 | 69 | 72 | 137 | 0,631119 | y ~ Intervention + Sex + |
| 0,00537 | 0,442877 | 0,156546 | 141 | 69 | 72 | 137 | 2,829061 | y ~ Intervention + Sex + |
| 0,001712 | -0,47974 | 0,149951 | 141 | 69 | 72 | 137 | -3,19931 | y ~ Intervention + Sex + |
| 0,061941 | -0,29951 | 0,159132 | 141 | 69 | 72 | 137 | -1,88212 | y ~ Intervention + Sex + |
| 0,031842 | 0,332981 | 0,153549 | 141 | 69 | 72 | 137 | 2,168568 | y ~ Intervention + Sex + |
| 0,20579 | 0,184397 | 0,145051 | 141 | 69 | 72 | 137 | 1,27126 | y ~ Intervention + Sex + |
| 0,963947 | -0,00726 | 0,160394 | 141 | 69 | 72 | 137 | -0,04528 | y ~ Intervention + Sex + |
| 0,010168 | -0,41751 | 0,160198 | 141 | 69 | 72 | 137 | -2,60621 | y ~ Intervention + Sex + |
| 0,953164 | 0,008473 | 0,143999 | 141 | 69 | 72 | 137 | 0,058842 | y ~ Intervention + Sex + |
| 0,408569 | 0,116641 | 0,140707 | 141 | 69 | 72 | 137 | 0,828961 | y ~ Intervention + Sex + |
| 0,370032 | -0,1512 | 0,168116 | 141 | 69 | 72 | 137 | -0,89937 | y ~ Intervention + Sex + |
| 0,290545 | -0,1636 | 0,154189 | 141 | 69 | 72 | 137 | -1,06103 | y ~ Intervention + Sex + |
| 0,036252 | 0,341858 | 0,161645 | 141 | 69 | 72 | 137 | 2,114866 | y ~ Intervention + Sex + |
| 0,883034 | 0,024877 | 0,168772 | 141 | 69 | 72 | 137 | 0,147399 | y ~ Intervention + Sex + |
| 0,102814 | 0,266705 | 0,162393 | 141 | 69 | 72 | 137 | 1,642337 | y ~ Intervention + Sex + |
| 0,036161 | 0,320413 | 0,15143 | 141 | 69 | 72 | 137 | 2,115911 | y ~ Intervention + Sex + |
| 1,69E-05 | -0,68308 | 0,153137 | 141 | 69 | 72 | 137 | -4,46059 | y ~ Intervention + Sex + |
| 0,523775 | 0,100001 | 0,156452 | 141 | 69 | 72 | 137 | 0,639177 | y ~ Intervention + Sex + |
| 0,234806 | -0,1753 | 0,146902 | 141 | 69 | 72 | 137 | -1,19332 | y ~ Intervention + Sex + |
| 0,317809 | 0,151909 | 0,151511 | 141 | 69 | 72 | 137 | 1,002626 | y ~ Intervention + Sex + |
| 0,059618 | 0,309424 | 0,162908 | 141 | 69 | 72 | 137 | 1,899373 | y ~ Intervention + Sex + |
| 0,00011 | -0,50927 | 0,127842 | 141 | 69 | 72 | 137 | -3,98359 | y ~ Intervention + Sex + |
| 0,004487 | -0,44759 | 0,154903 | 141 | 69 | 72 | 137 | -2,8895 | y ~ Intervention + Sex + |
| 0,903614 | -0,02062 | 0,169938 | 141 | 69 | 72 | 137 | -0,12132 | y ~ Intervention + Sex + |
| 0,829289 | -0,03016 | 0,139625 | 141 | 69 | 72 | 137 | -0,21603 | y ~ Intervention + Sex + |
| 0,038851 | 0,31011 | 0,148676 | 141 | 69 | 72 | 137 | 2,085806 | y ~ Intervention + Sex + |
| 0,510344 | 0,103897 | 0,157413 | 141 | 69 | 72 | 137 | 0,660028 | y ~ Intervention + Sex + |
| 0,024968 | -0,37683 | 0,166237 | 141 | 69 | 72 | 137 | -2,26681 | y ~ Intervention + Sex + |
| 0,960504 | 0,007849 | 0,158213 | 141 | 69 | 72 | 137 | 0,049612 | y ~ Intervention + Sex + |
| 0,387077 | -0,12502 | 0,144084 | 141 | 69 | 72 | 137 | -0,8677 | y ~ Intervention + Sex + |
| 0,385333 | 0,135715 | 0,155833 | 141 | 69 | 72 | 137 | 0,870899 | y ~ Intervention + Sex + |
| 0,60354 | -0,08951 | 0,171965 | 141 | 69 | 72 | 137 | -0,52052 | y ~ Intervention + Sex + |
| 0,483151 | 0,102257 | 0,145425 | 141 | 69 | 72 | 137 | 0,703156 | y ~ Intervention + Sex + |
| 0,100789 | 0,236286 | 0,143015 | 141 | 69 | 72 | 137 | 1,652176 | y ~ Intervention + Sex + |
| 0,450731 | 0,119754 | 0,158329 | 141 | 69 | 72 | 137 | 0,756362 | y ~ Intervention + Sex + |
| 0,838577 | -0,03659 | 0,179268 | 141 | 69 | 72 | 137 | -0,2041 | y ~ Intervention + Sex + |
| 0,93246 | 0,013673 | 0,161031 | 141 | 69 | 72 | 137 | 0,084906 | y ~ Intervention + Sex + |
| 5,89E-06 | -0,67738 | 0,143682 | 141 | 69 | 72 | 137 | -4,71444 | y ~ Intervention + Sex + |
| 0,939073 | 0,011129 | 0,145336 | 141 | 69 | 72 | 137 | 0,076575 | y ~ Intervention + Sex + |
| 0,011491 | -0,41426 | 0,1617 | 141 | 69 | 72 | 137 | -2,56191 | y ~ Intervention + Sex + |
| 0,275496 | -0,17698 | 0,161641 | 141 | 69 | 72 | 137 | -1,09487 | y ~ Intervention + Sex + |
| 0,975032 | 0,004891 | 0,155999 | 141 | 69 | 72 | 137 | 0,031355 | y ~ Intervention + Sex + |

| | | | | | | | | |
|---|---|---|---|---|---|---|---|---|
| 0,297433 | -0,15902 | 0,15204 | 141 | 69 | 72 | 137 | -1,04594 | y ~ Intervention + Sex + |
| 0,021177 | 0,378139 | 0,162176 | 141 | 69 | 72 | 137 | 2,331656 | y ~ Intervention + Sex + |
| 0,082593 | -0,25896 | 0,148091 | 141 | 69 | 72 | 137 | -1,74864 | y ~ Intervention + Sex + |
| 0,517243 | 0,100589 | 0,154923 | 141 | 69 | 72 | 137 | 0,649282 | y ~ Intervention + Sex + |
| 0,848479 | 0,025002 | 0,13061 | 141 | 69 | 72 | 137 | 0,191421 | y ~ Intervention + Sex + |
| 0,680279 | 0,066262 | 0,160455 | 141 | 69 | 72 | 137 | 0,412963 | y ~ Intervention + Sex + |
| 0,480769 | 0,11933 | 0,168784 | 141 | 69 | 72 | 137 | 0,706997 | y ~ Intervention + Sex + |
| 0,956958 | 0,008691 | 0,160732 | 141 | 69 | 72 | 137 | 0,05407 | y ~ Intervention + Sex + |
| 0,917647 | -0,01471 | 0,141992 | 141 | 69 | 72 | 137 | -0,10359 | y ~ Intervention + Sex + |
| 0,171522 | 0,190408 | 0,138526 | 141 | 69 | 72 | 137 | 1,374527 | y ~ Intervention + Sex + |
| 0,572651 | -0,08522 | 0,150694 | 141 | 69 | 72 | 137 | -0,56551 | y ~ Intervention + Sex + |
| 0,570747 | -0,09108 | 0,160257 | 141 | 69 | 72 | 137 | -0,56832 | y ~ Intervention + Sex + |
| 0,917874 | -0,01428 | 0,1382 | 141 | 69 | 72 | 137 | -0,1033 | y ~ Intervention + Sex + |
| 0,531165 | -0,09575 | 0,15251 | 141 | 69 | 72 | 137 | -0,62782 | y ~ Intervention + Sex + |
| 0,286019 | 0,161109 | 0,150418 | 141 | 69 | 72 | 137 | 1,071075 | y ~ Intervention + Sex + |
| 0,004344 | 0,471267 | 0,162486 | 141 | 69 | 72 | 137 | 2,900347 | y ~ Intervention + Sex + |
| 4,65E-14 | 1,181914 | 0,140466 | 141 | 69 | 72 | 137 | 8,414223 | y ~ Intervention + Sex + |
| 0,207667 | -0,20071 | 0,15854 | 141 | 69 | 72 | 137 | -1,26599 | y ~ Intervention + Sex + |
| 0,481675 | 0,107642 | 0,152568 | 141 | 69 | 72 | 137 | 0,705535 | y ~ Intervention + Sex + |
| 0,149853 | 0,232236 | 0,160365 | 141 | 69 | 72 | 137 | 1,448173 | y ~ Intervention + Sex + |
| 0,551214 | -0,09522 | 0,159393 | 141 | 69 | 72 | 137 | -0,59742 | y ~ Intervention + Sex + |
| 0,596259 | -0,08635 | 0,162612 | 141 | 69 | 72 | 137 | -0,53103 | y ~ Intervention + Sex + |
| 0,474527 | -0,12685 | 0,176892 | 141 | 69 | 72 | 137 | -0,71711 | y ~ Intervention + Sex + |
| 0,250867 | -0,20104 | 0,174344 | 141 | 69 | 72 | 137 | -1,15313 | y ~ Intervention + Sex + |
| 1,32E-24 | 1,320778 | 0,104473 | 138 | 69 | 69 | 134 | 12,64228 | y ~ Intervention + Sex + |
| 2,28E-24 | 1,286363 | 0,102514 | 138 | 69 | 69 | 134 | 12,54822 | y ~ Intervention + Sex + |
| 1,72E-09 | 0,998532 | 0,154404 | 138 | 69 | 69 | 134 | 6,466987 | y ~ Intervention + Sex + |
| 1,02E-08 | 0,986572 | 0,161514 | 138 | 69 | 69 | 134 | 6,108277 | y ~ Intervention + Sex + |
| 1,13E-08 | 0,980408 | 0,16103 | 138 | 69 | 69 | 134 | 6,088337 | y ~ Intervention + Sex + |
| 3,15E-08 | 0,850782 | 0,14478 | 138 | 69 | 69 | 134 | 5,876397 | y ~ Intervention + Sex + |
| 2,28E-07 | 0,838897 | 0,153767 | 138 | 69 | 69 | 134 | 5,455629 | y ~ Intervention + Sex + |
| 2,65E-08 | 0,836341 | 0,141463 | 138 | 69 | 69 | 134 | 5,912097 | y ~ Intervention + Sex + |
| 8,11E-06 | 0,816675 | 0,17591 | 138 | 69 | 69 | 134 | 4,642567 | y ~ Intervention + Sex + |
| 4,09E-07 | 0,78665 | 0,14765 | 138 | 69 | 69 | 134 | 5,327819 | y ~ Intervention + Sex + |
| 6,51E-07 | 0,776208 | 0,148561 | 138 | 69 | 69 | 134 | 5,224834 | y ~ Intervention + Sex + |
| 8,71E-06 | 0,712439 | 0,154034 | 138 | 69 | 69 | 134 | 4,625217 | y ~ Intervention + Sex + |
| 2,03E-06 | 0,705348 | 0,141992 | 138 | 69 | 69 | 134 | 4,967532 | y ~ Intervention + Sex + |
| 4,63E-05 | 0,689158 | 0,163647 | 138 | 69 | 69 | 134 | 4,211244 | y ~ Intervention + Sex + |
| 2,59E-05 | 0,686156 | 0,157449 | 138 | 69 | 69 | 134 | 4,357959 | y ~ Intervention + Sex + |
| 4,3E-05 | 0,68525 | 0,161993 | 138 | 69 | 69 | 134 | 4,230125 | y ~ Intervention + Sex + |
| 7,14E-05 | 0,679982 | 0,16587 | 138 | 69 | 69 | 134 | 4,099501 | y ~ Intervention + Sex + |
| 6,18E-05 | 0,678074 | 0,163907 | 138 | 69 | 69 | 134 | 4,136947 | y ~ Intervention + Sex + |
| 9,4E-06 | 0,6667 | 0,144715 | 138 | 69 | 69 | 134 | 4,607004 | y ~ Intervention + Sex + |
| 1,28E-05 | 0,659396 | 0,145478 | 138 | 69 | 69 | 134 | 4,532616 | y ~ Intervention + Sex + |
| 8,07E-05 | 0,652237 | 0,16036 | 138 | 69 | 69 | 134 | 4,067339 | y ~ Intervention + Sex + |
| 8,37E-05 | 0,650966 | 0,160426 | 138 | 69 | 69 | 134 | 4,057733 | y ~ Intervention + Sex + |
| 5,07E-05 | 0,633257 | 0,151206 | 138 | 69 | 69 | 134 | 4,188038 | y ~ Intervention + Sex + |
| 8,21E-05 | 0,621806 | 0,153048 | 138 | 69 | 69 | 134 | 4,062822 | y ~ Intervention + Sex + |

| | | | | | | | | |
|---|---|---|---|---|---|---|---|---|
| 5,59E-05 | 0,618088 | 0,148476 | 138 | 69 | 69 | 134 | 4,162884 | y ~ Intervention + Sex + |
| 0,000321 | 0,616342 | 0,166886 | 138 | 69 | 69 | 134 | 3,69319 | y ~ Intervention + Sex + |
| 0,000268 | 0,613909 | 0,163995 | 138 | 69 | 69 | 134 | 3,743462 | y ~ Intervention + Sex + |
| 9,93E-05 | 0,611897 | 0,152485 | 138 | 69 | 69 | 134 | 4,012843 | y ~ Intervention + Sex + |
| 4,9E-05 | 0,611564 | 0,145717 | 138 | 69 | 69 | 134 | 4,196932 | y ~ Intervention + Sex + |
| 9,25E-05 | 0,609762 | 0,151246 | 138 | 69 | 69 | 134 | 4,031598 | y ~ Intervention + Sex + |
| 0,000358 | 0,602637 | 0,164508 | 138 | 69 | 69 | 134 | 3,663274 | y ~ Intervention + Sex + |
| 0,000362 | 0,601645 | 0,164384 | 138 | 69 | 69 | 134 | 3,659996 | y ~ Intervention + Sex + |
| 0,000295 | 0,59819 | 0,160907 | 138 | 69 | 69 | 134 | 3,717616 | y ~ Intervention + Sex + |
| 6,09E-05 | 0,596912 | 0,144152 | 138 | 69 | 69 | 134 | 4,140846 | y ~ Intervention + Sex + |
| 0,000586 | 0,59136 | 0,167913 | 138 | 69 | 69 | 134 | 3,521833 | y ~ Intervention + Sex + |
| 0,000209 | 0,58924 | 0,154542 | 138 | 69 | 69 | 134 | 3,812807 | y ~ Intervention + Sex + |
| 0,000141 | 0,574872 | 0,146654 | 138 | 69 | 69 | 134 | 3,919917 | y ~ Intervention + Sex + |
| 0,000249 | 0,573138 | 0,152271 | 138 | 69 | 69 | 134 | 3,763921 | y ~ Intervention + Sex + |
| 0,000123 | 0,571918 | 0,144546 | 138 | 69 | 69 | 134 | 3,956652 | y ~ Intervention + Sex + |
| 0,000692 | 0,568272 | 0,163591 | 138 | 69 | 69 | 134 | 3,473744 | y ~ Intervention + Sex + |
| 4,38E-05 | 0,567857 | 0,134398 | 138 | 69 | 69 | 134 | 4,225195 | y ~ Intervention + Sex + |
| 0,000316 | 0,562406 | 0,152097 | 138 | 69 | 69 | 134 | 3,697691 | y ~ Intervention + Sex + |
| 6,28E-05 | 0,561675 | 0,13591 | 138 | 69 | 69 | 134 | 4,132713 | y ~ Intervention + Sex + |
| 0,000668 | 0,5608 | 0,160966 | 138 | 69 | 69 | 134 | 3,483973 | y ~ Intervention + Sex + |
| 0,001292 | 0,555576 | 0,169012 | 138 | 69 | 69 | 134 | 3,287191 | y ~ Intervention + Sex + |
| 0,000457 | 0,555545 | 0,154604 | 138 | 69 | 69 | 134 | 3,593336 | y ~ Intervention + Sex + |
| 3,51E-05 | 0,553618 | 0,12929 | 138 | 69 | 69 | 134 | 4,281984 | y ~ Intervention + Sex + |
| 0,000596 | 0,552842 | 0,157186 | 138 | 69 | 69 | 134 | 3,517127 | y ~ Intervention + Sex + |
| 0,000669 | 0,543593 | 0,156041 | 138 | 69 | 69 | 134 | 3,483657 | y ~ Intervention + Sex + |
| 0,000671 | 0,543199 | 0,155975 | 138 | 69 | 69 | 134 | 3,482613 | y ~ Intervention + Sex + |
| 0,009287 | 0,542055 | 0,20333 | 84 | 45 | 39 | 80 | 2,665892 | y ~ Intervention + Sex + |
| 0,000895 | 0,541746 | 0,159446 | 138 | 69 | 69 | 134 | 3,397689 | y ~ Intervention + Sex + |
| 0,00083 | 0,536595 | 0,156899 | 138 | 69 | 69 | 134 | 3,420007 | y ~ Intervention + Sex + |
| 0,000711 | 0,535588 | 0,154547 | 138 | 69 | 69 | 134 | 3,465529 | y ~ Intervention + Sex + |
| 0,002695 | 0,535073 | 0,175003 | 138 | 69 | 69 | 134 | 3,057515 | y ~ Intervention + Sex + |
| 0,001634 | 0,531991 | 0,165468 | 138 | 69 | 69 | 134 | 3,215078 | y ~ Intervention + Sex + |
| 0,000655 | 0,528321 | 0,151388 | 138 | 69 | 69 | 134 | 3,489856 | y ~ Intervention + Sex + |
| 8,22E-05 | 0,525817 | 0,129431 | 138 | 69 | 69 | 134 | 4,06251 | y ~ Intervention + Sex + |
| 0,002068 | 0,525618 | 0,167303 | 138 | 69 | 69 | 134 | 3,141719 | y ~ Intervention + Sex + |
| 0,000248 | 0,524307 | 0,139244 | 138 | 69 | 69 | 134 | 3,765386 | y ~ Intervention + Sex + |
| 0,000179 | 0,524028 | 0,135956 | 138 | 69 | 69 | 134 | 3,854388 | y ~ Intervention + Sex + |
| 0,001594 | 0,522863 | 0,162235 | 138 | 69 | 69 | 134 | 3,222868 | y ~ Intervention + Sex + |
| 0,001109 | 0,521401 | 0,156412 | 138 | 69 | 69 | 134 | 3,333507 | y ~ Intervention + Sex + |
| 0,00078 | 0,520916 | 0,151497 | 138 | 69 | 69 | 134 | 3,438462 | y ~ Intervention + Sex + |
| 0,001567 | 0,517865 | 0,160422 | 138 | 69 | 69 | 134 | 3,228148 | y ~ Intervention + Sex + |
| 0,002021 | 0,516995 | 0,16418 | 138 | 69 | 69 | 134 | 3,148958 | y ~ Intervention + Sex + |
| 0,000455 | 0,516442 | 0,143662 | 138 | 69 | 69 | 134 | 3,594838 | y ~ Intervention + Sex + |
| 0,000957 | 0,515287 | 0,152555 | 138 | 69 | 69 | 134 | 3,377715 | y ~ Intervention + Sex + |
| 0,001302 | 0,515152 | 0,156825 | 138 | 69 | 69 | 134 | 3,284875 | y ~ Intervention + Sex + |
| 0,001956 | 0,513813 | 0,162645 | 138 | 69 | 69 | 134 | 3,159111 | y ~ Intervention + Sex + |
| 0,000595 | 0,512742 | 0,145759 | 138 | 69 | 69 | 134 | 3,517744 | y ~ Intervention + Sex + |
| 0,001873 | 0,504537 | 0,159021 | 138 | 69 | 69 | 134 | 3,17277 | y ~ Intervention + Sex + |

| | | | | | | | | |
|---|---|---|---|---|---|---|---|---|
| 0,001688 | 0,501848 | 0,156582 | 138 | 69 | 69 | 134 | 3,20501 | y ~ Intervention + Sex + |
| 0,002339 | 0,500192 | 0,161212 | 138 | 69 | 69 | 134 | 3,102691 | y ~ Intervention + Sex + |
| 0,001304 | 0,499581 | 0,15211 | 138 | 69 | 69 | 134 | 3,284349 | y ~ Intervention + Sex + |
| 0,000629 | 0,499339 | 0,142616 | 138 | 69 | 69 | 134 | 3,501291 | y ~ Intervention + Sex + |
| 0,002776 | 0,497149 | 0,163104 | 138 | 69 | 69 | 134 | 3,048046 | y ~ Intervention + Sex + |
| 0,003006 | 0,495809 | 0,164048 | 138 | 69 | 69 | 134 | 3,022333 | y ~ Intervention + Sex + |
| 0,001889 | 0,495579 | 0,156333 | 138 | 69 | 69 | 134 | 3,170016 | y ~ Intervention + Sex + |
| 0,000439 | 0,495172 | 0,137359 | 138 | 69 | 69 | 134 | 3,604961 | y ~ Intervention + Sex + |
| 0,001329 | 0,494718 | 0,150888 | 138 | 69 | 69 | 134 | 3,278713 | y ~ Intervention + Sex + |
| 0,003138 | 0,491877 | 0,163503 | 138 | 69 | 69 | 134 | 3,008363 | y ~ Intervention + Sex + |
| 0,000692 | 0,491788 | 0,141572 | 138 | 69 | 69 | 134 | 3,473762 | y ~ Intervention + Sex + |
| 0,00148 | 0,488881 | 0,150626 | 138 | 69 | 69 | 134 | 3,245662 | y ~ Intervention + Sex + |
| 0,002578 | 0,488021 | 0,158872 | 138 | 69 | 69 | 134 | 3,07178 | y ~ Intervention + Sex + |
| 0,00152 | 0,486826 | 0,150367 | 138 | 69 | 69 | 134 | 3,237575 | y ~ Intervention + Sex + |
| 0,000665 | 0,48032 | 0,13782 | 138 | 69 | 69 | 134 | 3,485114 | y ~ Intervention + Sex + |
| 0,004058 | 0,479015 | 0,16382 | 138 | 69 | 69 | 134 | 2,924036 | y ~ Intervention + Sex + |
| 0,002629 | 0,47761 | 0,155803 | 138 | 69 | 69 | 134 | 3,065468 | y ~ Intervention + Sex + |
| 0,002616 | 0,476502 | 0,155359 | 138 | 69 | 69 | 134 | 3,067106 | y ~ Intervention + Sex + |
| 0,005129 | 0,469055 | 0,164832 | 138 | 69 | 69 | 134 | 2,845654 | y ~ Intervention + Sex + |
| 0,002244 | 0,46773 | 0,150109 | 138 | 69 | 69 | 134 | 3,115934 | y ~ Intervention + Sex + |
| 0,001431 | 0,466817 | 0,143367 | 138 | 69 | 69 | 134 | 3,256095 | y ~ Intervention + Sex + |
| 0,005114 | 0,466328 | 0,163817 | 138 | 69 | 69 | 134 | 2,846646 | y ~ Intervention + Sex + |
| 0,003316 | 0,465357 | 0,155615 | 138 | 69 | 69 | 134 | 2,990434 | y ~ Intervention + Sex + |
| 0,003294 | 0,46469 | 0,15528 | 138 | 69 | 69 | 134 | 2,992591 | y ~ Intervention + Sex + |
| 0,001395 | 0,464134 | 0,142201 | 138 | 69 | 69 | 134 | 3,263926 | y ~ Intervention + Sex + |
| 0,000646 | 0,459641 | 0,131561 | 138 | 69 | 69 | 134 | 3,493734 | y ~ Intervention + Sex + |
| 0,00115 | 0,45694 | 0,13753 | 138 | 69 | 69 | 134 | 3,322479 | y ~ Intervention + Sex + |
| 0,004149 | 0,456556 | 0,156531 | 138 | 69 | 69 | 134 | 2,916713 | y ~ Intervention + Sex + |
| 0,000725 | 0,455517 | 0,131648 | 138 | 69 | 69 | 134 | 3,460117 | y ~ Intervention + Sex + |
| 0,006844 | 0,45396 | 0,165261 | 138 | 69 | 69 | 134 | 2,746924 | y ~ Intervention + Sex + |
| 0,006649 | 0,453083 | 0,164345 | 138 | 69 | 69 | 134 | 2,756907 | y ~ Intervention + Sex + |
| 0,002912 | 0,451897 | 0,149012 | 138 | 69 | 69 | 134 | 3,032613 | y ~ Intervention + Sex + |
| 0,006142 | 0,451758 | 0,162254 | 138 | 69 | 69 | 134 | 2,784257 | y ~ Intervention + Sex + |
| 0,002769 | 0,451672 | 0,148148 | 138 | 69 | 69 | 134 | 3,048795 | y ~ Intervention + Sex + |
| 0,004993 | 0,451311 | 0,158092 | 138 | 69 | 69 | 134 | 2,854735 | y ~ Intervention + Sex + |
| 0,002926 | 0,44951 | 0,1483 | 138 | 69 | 69 | 134 | 3,031082 | y ~ Intervention + Sex + |
| 0,003443 | 0,448708 | 0,150666 | 138 | 69 | 69 | 134 | 2,978172 | y ~ Intervention + Sex + |
| 0,005242 | 0,447422 | 0,157639 | 138 | 69 | 69 | 134 | 2,838265 | y ~ Intervention + Sex + |
| 0,003405 | 0,444463 | 0,149059 | 138 | 69 | 69 | 134 | 2,981801 | y ~ Intervention + Sex + |
| 0,002008 | 0,444308 | 0,14101 | 138 | 69 | 69 | 134 | 3,150891 | y ~ Intervention + Sex + |
| 0,003379 | 0,440465 | 0,147592 | 138 | 69 | 69 | 134 | 2,984341 | y ~ Intervention + Sex + |
| 0,002956 | 0,438022 | 0,144669 | 138 | 69 | 69 | 134 | 3,027753 | y ~ Intervention + Sex + |
| 0,00466 | 0,436104 | 0,151535 | 138 | 69 | 69 | 134 | 2,877913 | y ~ Intervention + Sex + |
| 0,006864 | 0,435907 | 0,158748 | 138 | 69 | 69 | 134 | 2,745901 | y ~ Intervention + Sex + |
| 0,00205 | 0,43473 | 0,138256 | 138 | 69 | 69 | 134 | 3,144387 | y ~ Intervention + Sex + |
| 0,003303 | 0,434299 | 0,145167 | 138 | 69 | 69 | 134 | 2,991728 | y ~ Intervention + Sex + |
| 0,006885 | 0,433023 | 0,15776 | 138 | 69 | 69 | 134 | 2,744818 | y ~ Intervention + Sex + |
| 0,010648 | 0,427039 | 0,164856 | 138 | 69 | 69 | 134 | 2,590384 | y ~ Intervention + Sex + |

| | | | | | | | | |
|---|---|---|---|---|---|---|---|---|
| 0,008987 | 0,426194 | 0,160755 | 138 | 69 | 69 | 134 | 2,651207 | y ~ Intervention + Sex + |
| 0,009727 | 0,425942 | 0,16239 | 138 | 69 | 69 | 134 | 2,622956 | y ~ Intervention + Sex + |
| 0,009615 | 0,421414 | 0,16041 | 138 | 69 | 69 | 134 | 2,627114 | y ~ Intervention + Sex + |
| 0,012763 | 0,420247 | 0,166486 | 138 | 69 | 69 | 134 | 2,524218 | y ~ Intervention + Sex + |
| 0,011154 | 0,418085 | 0,162456 | 138 | 69 | 69 | 134 | 2,573526 | y ~ Intervention + Sex + |
| 0,005607 | 0,414554 | 0,147245 | 138 | 69 | 69 | 134 | 2,815401 | y ~ Intervention + Sex + |
| 0,006574 | 0,413706 | 0,149848 | 138 | 69 | 69 | 134 | 2,760832 | y ~ Intervention + Sex + |
| 0,006891 | 0,413681 | 0,15073 | 138 | 69 | 69 | 134 | 2,744508 | y ~ Intervention + Sex + |
| 0,011767 | 0,413532 | 0,161915 | 138 | 69 | 69 | 134 | 2,554015 | y ~ Intervention + Sex + |
| 0,009335 | 0,413294 | 0,156689 | 138 | 69 | 69 | 134 | 2,63768 | y ~ Intervention + Sex + |
| 0,005007 | 0,413216 | 0,144794 | 138 | 69 | 69 | 134 | 2,853812 | y ~ Intervention + Sex + |
| 0,017425 | 0,412538 | 0,171357 | 138 | 69 | 69 | 134 | 2,407474 | y ~ Intervention + Sex + |
| 0,00646 | 0,412009 | 0,148907 | 138 | 69 | 69 | 134 | 2,766882 | y ~ Intervention + Sex + |
| 0,007454 | 0,411957 | 0,151612 | 138 | 69 | 69 | 134 | 2,717176 | y ~ Intervention + Sex + |
| 0,008571 | 0,409321 | 0,153417 | 138 | 69 | 69 | 134 | 2,668027 | y ~ Intervention + Sex + |
| 0,009307 | 0,408965 | 0,154985 | 138 | 69 | 69 | 134 | 2,638739 | y ~ Intervention + Sex + |
| 0,00958 | 0,405284 | 0,154194 | 138 | 69 | 69 | 134 | 2,628399 | y ~ Intervention + Sex + |
| 0,011987 | 0,404341 | 0,158737 | 138 | 69 | 69 | 134 | 2,547246 | y ~ Intervention + Sex + |
| 0,008707 | 0,403979 | 0,151732 | 138 | 69 | 69 | 134 | 2,66245 | y ~ Intervention + Sex + |
| 0,008819 | 0,403437 | 0,151786 | 138 | 69 | 69 | 134 | 2,657931 | y ~ Intervention + Sex + |
| 0,007736 | 0,395046 | 0,146088 | 138 | 69 | 69 | 134 | 2,704167 | y ~ Intervention + Sex + |
| 0,00519 | 0,394845 | 0,13895 | 138 | 69 | 69 | 134 | 2,84164 | y ~ Intervention + Sex + |
| 0,017794 | 0,393484 | 0,163987 | 138 | 69 | 69 | 134 | 2,399484 | y ~ Intervention + Sex + |
| 0,012518 | 0,393266 | 0,155358 | 138 | 69 | 69 | 134 | 2,531348 | y ~ Intervention + Sex + |
| 0,018655 | 0,393059 | 0,165056 | 138 | 69 | 69 | 134 | 2,381368 | y ~ Intervention + Sex + |
| 0,010374 | 0,392444 | 0,150952 | 138 | 69 | 69 | 134 | 2,599799 | y ~ Intervention + Sex + |
| 0,0189 | 0,390413 | 0,164291 | 138 | 69 | 69 | 134 | 2,376352 | y ~ Intervention + Sex + |
| 0,0189 | 0,390413 | 0,164291 | 138 | 69 | 69 | 134 | 2,376352 | y ~ Intervention + Sex + |
| 0,021027 | 0,387182 | 0,165815 | 138 | 69 | 69 | 134 | 2,335031 | y ~ Intervention + Sex + |
| 0,01955 | 0,386841 | 0,163686 | 138 | 69 | 69 | 134 | 2,363314 | y ~ Intervention + Sex + |
| 0,018256 | 0,385741 | 0,161421 | 138 | 69 | 69 | 134 | 2,389658 | y ~ Intervention + Sex + |
| 0,018785 | 0,384472 | 0,161632 | 138 | 69 | 69 | 134 | 2,378694 | y ~ Intervention + Sex + |
| 0,007475 | 0,382101 | 0,140676 | 138 | 69 | 69 | 134 | 2,716173 | y ~ Intervention + Sex + |
| 0,012661 | 0,381538 | 0,150975 | 138 | 69 | 69 | 134 | 2,527164 | y ~ Intervention + Sex + |
| 0,0176 | 0,381009 | 0,158512 | 138 | 69 | 69 | 134 | 2,403668 | y ~ Intervention + Sex + |
| 0,015899 | 0,377544 | 0,154588 | 138 | 69 | 69 | 134 | 2,442261 | y ~ Intervention + Sex + |
| 0,011074 | 0,377369 | 0,146485 | 138 | 69 | 69 | 134 | 2,576154 | y ~ Intervention + Sex + |
| 0,021898 | 0,375772 | 0,162028 | 138 | 69 | 69 | 134 | 2,319176 | y ~ Intervention + Sex + |
| 0,008092 | 0,374304 | 0,139232 | 138 | 69 | 69 | 134 | 2,688347 | y ~ Intervention + Sex + |
| 0,016357 | 0,373134 | 0,153458 | 138 | 69 | 69 | 134 | 2,431506 | y ~ Intervention + Sex + |
| 0,026134 | 0,371931 | 0,165365 | 138 | 69 | 69 | 134 | 2,249161 | y ~ Intervention + Sex + |
| 0,026683 | 0,370526 | 0,165352 | 138 | 69 | 69 | 134 | 2,240834 | y ~ Intervention + Sex + |
| 0,011408 | 0,368561 | 0,14367 | 138 | 69 | 69 | 134 | 2,565333 | y ~ Intervention + Sex + |
| 0,026022 | 0,367856 | 0,163428 | 138 | 69 | 69 | 134 | 2,25088 | y ~ Intervention + Sex + |
| 0,026022 | 0,367856 | 0,163428 | 138 | 69 | 69 | 134 | 2,25088 | y ~ Intervention + Sex + |
| 0,031421 | 0,367809 | 0,169144 | 138 | 69 | 69 | 134 | 2,17453 | y ~ Intervention + Sex + |
| 0,025556 | 0,364358 | 0,161356 | 138 | 69 | 69 | 134 | 2,258095 | y ~ Intervention + Sex + |
| 0,029344 | 0,363412 | 0,165004 | 138 | 69 | 69 | 134 | 2,202447 | y ~ Intervention + Sex + |

| | | | | | | | | |
|---|---|---|---|---|---|---|---|---|
| 0,008327 | 0,362312 | 0,135278 | 138 | 69 | 69 | 134 | 2,678271 | y ~ Intervention + Sex |
| 0,021866 | 0,360641 | 0,155465 | 138 | 69 | 69 | 134 | 2,319756 | y ~ Intervention + Sex |
| 0,032894 | 0,359546 | 0,166789 | 138 | 69 | 69 | 134 | 2,155691 | y ~ Intervention + Sex |
| 0,0234 | 0,35854 | 0,156357 | 138 | 69 | 69 | 134 | 2,293089 | y ~ Intervention + Sex |
| 0,027207 | 0,357231 | 0,159977 | 138 | 69 | 69 | 134 | 2,23301 | y ~ Intervention + Sex |
| 0,028151 | 0,356018 | 0,160422 | 138 | 69 | 69 | 134 | 2,219262 | y ~ Intervention + Sex |
| 0,017046 | 0,355463 | 0,147137 | 138 | 69 | 69 | 134 | 2,415862 | y ~ Intervention + Sex |
| 0,010396 | 0,353631 | 0,136062 | 138 | 69 | 69 | 134 | 2,599039 | y ~ Intervention + Sex |
| 0,042766 | 0,353269 | 0,172708 | 138 | 69 | 69 | 134 | 2,045467 | y ~ Intervention + Sex |
| 0,019374 | 0,352524 | 0,148946 | 138 | 69 | 69 | 134 | 2,366792 | y ~ Intervention + Sex |
| 0,009695 | 0,35223 | 0,134227 | 138 | 69 | 69 | 134 | 2,624146 | y ~ Intervention + Sex |
| 0,014816 | 0,350511 | 0,141978 | 138 | 69 | 69 | 134 | 2,468774 | y ~ Intervention + Sex |
| 0,026005 | 0,349895 | 0,15543 | 138 | 69 | 69 | 134 | 2,251145 | y ~ Intervention + Sex |
| 0,022511 | 0,34931 | 0,151325 | 138 | 69 | 69 | 134 | 2,308338 | y ~ Intervention + Sex |
| 0,025137 | 0,34621 | 0,152873 | 138 | 69 | 69 | 134 | 2,264689 | y ~ Intervention + Sex |
| 0,020545 | 0,345356 | 0,147333 | 138 | 69 | 69 | 134 | 2,34406 | y ~ Intervention + Sex |
| 0,0356 | 0,343089 | 0,161613 | 138 | 69 | 69 | 134 | 2,122908 | y ~ Intervention + Sex |
| 0,037166 | 0,342738 | 0,162828 | 138 | 69 | 69 | 134 | 2,104914 | y ~ Intervention + Sex |
| 0,032475 | 0,342289 | 0,158396 | 138 | 69 | 69 | 134 | 2,160971 | y ~ Intervention + Sex |
| 0,00941 | 0,341847 | 0,129742 | 138 | 69 | 69 | 134 | 2,634811 | y ~ Intervention + Sex |
| 0,020975 | 0,341548 | 0,146211 | 138 | 69 | 69 | 134 | 2,335997 | y ~ Intervention + Sex |
| 0,015023 | 0,341291 | 0,138534 | 138 | 69 | 69 | 134 | 2,463586 | y ~ Intervention + Sex |
| 0,037102 | 0,341147 | 0,162017 | 138 | 69 | 69 | 134 | 2,105632 | y ~ Intervention + Sex |
| 0,032053 | 0,340779 | 0,157305 | 138 | 69 | 69 | 134 | 2,166357 | y ~ Intervention + Sex |
| 0,030944 | 0,340509 | 0,15614 | 138 | 69 | 69 | 134 | 2,180801 | y ~ Intervention + Sex |
| 0,033231 | 0,340472 | 0,15825 | 138 | 69 | 69 | 134 | 2,151487 | y ~ Intervention + Sex |
| 0,025996 | 0,339023 | 0,150592 | 138 | 69 | 69 | 134 | 2,25127 | y ~ Intervention + Sex |
| 0,036624 | 0,338566 | 0,160377 | 138 | 69 | 69 | 134 | 2,111064 | y ~ Intervention + Sex |
| 0,040551 | 0,337035 | 0,162966 | 138 | 69 | 69 | 134 | 2,068128 | y ~ Intervention + Sex |
| 0,040404 | 0,335566 | 0,162135 | 138 | 69 | 69 | 134 | 2,069668 | y ~ Intervention + Sex |
| 0,038792 | 0,335386 | 0,16071 | 138 | 69 | 69 | 134 | 2,086898 | y ~ Intervention + Sex |
| 0,036685 | 0,332246 | 0,157435 | 138 | 69 | 69 | 134 | 2,110371 | y ~ Intervention + Sex |
| 0,048143 | 0,332213 | 0,166577 | 138 | 69 | 69 | 134 | 1,994349 | y ~ Intervention + Sex |
| 0,07275 | 0,329091 | 0,181956 | 138 | 69 | 69 | 134 | 1,808634 | y ~ Intervention + Sex |
| 0,051988 | 0,327792 | 0,167181 | 138 | 69 | 69 | 134 | 1,960703 | y ~ Intervention + Sex |
| 0,022071 | 0,326241 | 0,140858 | 138 | 69 | 69 | 134 | 2,316093 | y ~ Intervention + Sex |
| 0,062855 | 0,325987 | 0,173783 | 138 | 69 | 69 | 134 | 1,875827 | y ~ Intervention + Sex |
| 0,034506 | 0,325331 | 0,152316 | 138 | 69 | 69 | 134 | 2,135896 | y ~ Intervention + Sex |
| 0,041833 | 0,325163 | 0,158239 | 138 | 69 | 69 | 134 | 2,054885 | y ~ Intervention + Sex |
| 0,041334 | 0,32489 | 0,157714 | 138 | 69 | 69 | 134 | 2,06 | y ~ Intervention + Sex |
| 0,036659 | 0,324669 | 0,153822 | 138 | 69 | 69 | 134 | 2,110672 | y ~ Intervention + Sex |
| 0,042122 | 0,324264 | 0,158027 | 138 | 69 | 69 | 134 | 2,051946 | y ~ Intervention + Sex |
| 0,020246 | 0,323945 | 0,137863 | 138 | 69 | 69 | 134 | 2,349756 | y ~ Intervention + Sex |
| 0,040189 | 0,322636 | 0,155717 | 138 | 69 | 69 | 134 | 2,071939 | y ~ Intervention + Sex |
| 0,041601 | 0,318247 | 0,154694 | 138 | 69 | 69 | 134 | 2,057261 | y ~ Intervention + Sex |
| 0,054017 | 0,317164 | 0,163168 | 138 | 69 | 69 | 134 | 1,943789 | y ~ Intervention + Sex |
| 0,01856 | 0,316465 | 0,132783 | 138 | 69 | 69 | 134 | 2,383323 | y ~ Intervention + Sex |
| 0,037564 | 0,316151 | 0,150516 | 138 | 69 | 69 | 134 | 2,100444 | y ~ Intervention + Sex |

| | | | | | | | | |
|---|---|---|---|---|---|---|---|---|
| 0,041629 | 0,313418 | 0,152368 | 138 | 69 | 69 | 134 | 2,056975 | y ~ Intervention + Sex + |
| 0,028163 | 0,313263 | 0,141168 | 138 | 69 | 69 | 134 | 2,219083 | y ~ Intervention + Sex + |
| 0,055504 | 0,312447 | 0,161745 | 138 | 69 | 69 | 134 | 1,931727 | y ~ Intervention + Sex + |
| 0,03307 | 0,311622 | 0,144705 | 138 | 69 | 69 | 134 | 2,153496 | y ~ Intervention + Sex + |
| 0,039308 | 0,309507 | 0,148707 | 138 | 69 | 69 | 134 | 2,08132 | y ~ Intervention + Sex + |
| 0,047947 | 0,309402 | 0,155001 | 138 | 69 | 69 | 134 | 1,996127 | y ~ Intervention + Sex + |
| 0,06386 | 0,309184 | 0,165461 | 138 | 69 | 69 | 134 | 1,868616 | y ~ Intervention + Sex + |
| 0,046781 | 0,308619 | 0,153785 | 138 | 69 | 69 | 134 | 2,006822 | y ~ Intervention + Sex + |
| 0,031367 | 0,307556 | 0,14139 | 138 | 69 | 69 | 134 | 2,17524 | y ~ Intervention + Sex + |
| 0,022944 | 0,307352 | 0,133582 | 138 | 69 | 69 | 134 | 2,300846 | y ~ Intervention + Sex + |
| 0,094033 | 0,307296 | 0,182213 | 138 | 69 | 69 | 134 | 1,686467 | y ~ Intervention + Sex + |
| 0,059122 | 0,307163 | 0,161369 | 138 | 69 | 69 | 134 | 1,903481 | y ~ Intervention + Sex + |
| 0,07744 | 0,307093 | 0,172581 | 138 | 69 | 69 | 134 | 1,779408 | y ~ Intervention + Sex + |
| 0,044517 | 0,306566 | 0,151149 | 138 | 69 | 69 | 134 | 2,028242 | y ~ Intervention + Sex + |
| 0,029618 | 0,306271 | 0,139299 | 138 | 69 | 69 | 134 | 2,198659 | y ~ Intervention + Sex + |
| 0,047447 | 0,305082 | 0,152489 | 138 | 69 | 69 | 134 | 2,000683 | y ~ Intervention + Sex + |
| 0,04283 | 0,303144 | 0,14825 | 138 | 69 | 69 | 134 | 2,04482 | y ~ Intervention + Sex + |
| 0,053975 | 0,303091 | 0,155901 | 138 | 69 | 69 | 134 | 1,944128 | y ~ Intervention + Sex + |
| 0,067576 | 0,302041 | 0,163908 | 138 | 69 | 69 | 134 | 1,842749 | y ~ Intervention + Sex + |
| 0,045165 | 0,30175 | 0,149232 | 138 | 69 | 69 | 134 | 2,022023 | y ~ Intervention + Sex + |
| 0,042863 | 0,301113 | 0,14728 | 138 | 69 | 69 | 134 | 2,044494 | y ~ Intervention + Sex + |
| 0,067508 | 0,30077 | 0,163177 | 138 | 69 | 69 | 134 | 1,843212 | y ~ Intervention + Sex + |
| 0,066708 | 0,299863 | 0,162204 | 138 | 69 | 69 | 134 | 1,848677 | y ~ Intervention + Sex + |
| 0,046962 | 0,298419 | 0,148826 | 138 | 69 | 69 | 134 | 2,005147 | y ~ Intervention + Sex + |
| 0,071146 | 0,297838 | 0,163738 | 138 | 69 | 69 | 134 | 1,81899 | y ~ Intervention + Sex + |
| 0,039931 | 0,297111 | 0,143209 | 138 | 69 | 69 | 134 | 2,074665 | y ~ Intervention + Sex + |
| 0,048914 | 0,294094 | 0,147977 | 138 | 69 | 69 | 134 | 1,987427 | y ~ Intervention + Sex + |
| 0,080019 | 0,294087 | 0,16672 | 138 | 69 | 69 | 134 | 1,763951 | y ~ Intervention + Sex + |
| 0,072275 | 0,293648 | 0,162086 | 138 | 69 | 69 | 134 | 1,811682 | y ~ Intervention + Sex + |
| 0,061088 | 0,29336 | 0,15532 | 138 | 69 | 69 | 134 | 1,888742 | y ~ Intervention + Sex + |
| 0,077075 | 0,293147 | 0,164539 | 138 | 69 | 69 | 134 | 1,781628 | y ~ Intervention + Sex + |
| 0,060177 | 0,292684 | 0,154408 | 138 | 69 | 69 | 134 | 1,895523 | y ~ Intervention + Sex + |
| 0,051151 | 0,292312 | 0,148545 | 138 | 69 | 69 | 134 | 1,967841 | y ~ Intervention + Sex + |
| 0,047048 | 0,288915 | 0,144144 | 138 | 69 | 69 | 134 | 2,004349 | y ~ Intervention + Sex + |
| 0,04477 | 0,286764 | 0,141556 | 138 | 69 | 69 | 134 | 2,025801 | y ~ Intervention + Sex + |
| 0,028472 | 0,286163 | 0,129212 | 138 | 69 | 69 | 134 | 2,214677 | y ~ Intervention + Sex + |
| 0,093878 | 0,285266 | 0,16907 | 138 | 69 | 69 | 134 | 1,687269 | y ~ Intervention + Sex + |
| 0,08809 | 0,283991 | 0,165295 | 138 | 69 | 69 | 134 | 1,718087 | y ~ Intervention + Sex + |
| 0,037676 | 0,283779 | 0,135185 | 138 | 69 | 69 | 134 | 2,099189 | y ~ Intervention + Sex + |
| 0,085225 | 0,283514 | 0,163507 | 138 | 69 | 69 | 134 | 1,733959 | y ~ Intervention + Sex + |
| 0,09159 | 0,283435 | 0,166799 | 138 | 69 | 69 | 134 | 1,699261 | y ~ Intervention + Sex + |
| 0,10196 | 0,278499 | 0,169124 | 138 | 69 | 69 | 134 | 1,646712 | y ~ Intervention + Sex + |
| 0,055571 | 0,277221 | 0,14355 | 138 | 69 | 69 | 134 | 1,931189 | y ~ Intervention + Sex + |
| 0,074356 | 0,275057 | 0,152941 | 138 | 69 | 69 | 134 | 1,798456 | y ~ Intervention + Sex + |
| 0,077656 | 0,274125 | 0,154168 | 138 | 69 | 69 | 134 | 1,778095 | y ~ Intervention + Sex + |
| 0,073342 | 0,274029 | 0,151828 | 138 | 69 | 69 | 134 | 1,80486 | y ~ Intervention + Sex + |
| 0,080387 | 0,273578 | 0,155285 | 138 | 69 | 69 | 134 | 1,761778 | y ~ Intervention + Sex + |
| 0,083199 | 0,272721 | 0,156247 | 138 | 69 | 69 | 134 | 1,74545 | y ~ Intervention + Sex + |

| | | | | | | | | |
|---|---|---|---|---|---|---|---|---|
| 0,100838 | 0,272099 | 0,16469 | 138 | 69 | 69 | 134 | 1,652188 | y ~ Intervention + Sex + |
| 0,102957 | 0,271681 | 0,165468 | 138 | 69 | 69 | 134 | 1,641891 | y ~ Intervention + Sex + |
| 0,084617 | 0,2708 | 0,155867 | 138 | 69 | 69 | 134 | 1,737384 | y ~ Intervention + Sex + |
| 0,093709 | 0,270126 | 0,160013 | 138 | 69 | 69 | 134 | 1,688149 | y ~ Intervention + Sex + |
| 0,093485 | 0,269033 | 0,159256 | 138 | 69 | 69 | 134 | 1,689311 | y ~ Intervention + Sex + |
| 0,103882 | 0,268112 | 0,163737 | 138 | 69 | 69 | 134 | 1,637452 | y ~ Intervention + Sex + |
| 0,062962 | 0,26785 | 0,142849 | 138 | 69 | 69 | 134 | 1,875054 | y ~ Intervention + Sex + |
| 0,086371 | 0,267397 | 0,154783 | 138 | 69 | 69 | 134 | 1,727563 | y ~ Intervention + Sex + |
| 0,101562 | 0,26586 | 0,161259 | 138 | 69 | 69 | 134 | 1,648649 | y ~ Intervention + Sex + |
| 0,109002 | 0,265678 | 0,164666 | 138 | 69 | 69 | 134 | 1,613437 | y ~ Intervention + Sex + |
| 0,085815 | 0,265306 | 0,153298 | 138 | 69 | 69 | 134 | 1,730656 | y ~ Intervention + Sex + |
| 0,123816 | 0,263902 | 0,170404 | 138 | 69 | 69 | 134 | 1,548687 | y ~ Intervention + Sex + |
| 0,11222 | 0,261491 | 0,163554 | 138 | 69 | 69 | 134 | 1,598801 | y ~ Intervention + Sex + |
| 0,122296 | 0,260887 | 0,167769 | 138 | 69 | 69 | 134 | 1,555037 | y ~ Intervention + Sex + |
| 0,156085 | 0,260819 | 0,182854 | 138 | 69 | 69 | 134 | 1,426378 | y ~ Intervention + Sex + |
| 0,117096 | 0,260331 | 0,165054 | 138 | 69 | 69 | 134 | 1,577249 | y ~ Intervention + Sex + |
| 0,058312 | 0,260326 | 0,13632 | 138 | 69 | 69 | 134 | 1,909675 | y ~ Intervention + Sex + |
| 0,108118 | 0,259927 | 0,160695 | 138 | 69 | 69 | 134 | 1,617518 | y ~ Intervention + Sex + |
| 0,122103 | 0,259224 | 0,166613 | 138 | 69 | 69 | 134 | 1,555846 | y ~ Intervention + Sex + |
| 0,110062 | 0,259199 | 0,161136 | 138 | 69 | 69 | 134 | 1,608578 | y ~ Intervention + Sex + |
| 0,088352 | 0,259017 | 0,150884 | 138 | 69 | 69 | 134 | 1,716657 | y ~ Intervention + Sex + |
| 0,144513 | 0,257306 | 0,175305 | 138 | 69 | 69 | 134 | 1,467759 | y ~ Intervention + Sex + |
| 0,076891 | 0,256611 | 0,143941 | 138 | 69 | 69 | 134 | 1,782751 | y ~ Intervention + Sex + |
| 0,130889 | 0,254389 | 0,167371 | 138 | 69 | 69 | 134 | 1,519912 | y ~ Intervention + Sex + |
| 0,094693 | 0,25423 | 0,151053 | 138 | 69 | 69 | 134 | 1,683056 | y ~ Intervention + Sex + |
| 0,140082 | 0,254157 | 0,171232 | 138 | 69 | 69 | 134 | 1,484283 | y ~ Intervention + Sex + |
| 0,149556 | 0,253902 | 0,175175 | 138 | 69 | 69 | 134 | 1,449423 | y ~ Intervention + Sex + |
| 0,117381 | 0,253094 | 0,160592 | 138 | 69 | 69 | 134 | 1,576011 | y ~ Intervention + Sex + |
| 0,146898 | 0,251845 | 0,172612 | 138 | 69 | 69 | 134 | 1,459026 | y ~ Intervention + Sex + |
| 0,108942 | 0,251643 | 0,15594 | 138 | 69 | 69 | 134 | 1,613713 | y ~ Intervention + Sex + |
| 0,134351 | 0,250187 | 0,166097 | 138 | 69 | 69 | 134 | 1,506271 | y ~ Intervention + Sex + |
| 0,090135 | 0,249914 | 0,146404 | 138 | 69 | 69 | 134 | 1,707015 | y ~ Intervention + Sex + |
| 0,108634 | 0,249108 | 0,154234 | 138 | 69 | 69 | 134 | 1,615132 | y ~ Intervention + Sex + |
| 0,095869 | 0,248592 | 0,148234 | 138 | 69 | 69 | 134 | 1,677025 | y ~ Intervention + Sex + |
| 0,114767 | 0,24851 | 0,156546 | 138 | 69 | 69 | 134 | 1,587455 | y ~ Intervention + Sex + |
| 0,136185 | 0,248452 | 0,165728 | 138 | 69 | 69 | 134 | 1,499159 | y ~ Intervention + Sex + |
| 0,13453 | 0,248334 | 0,164943 | 138 | 69 | 69 | 134 | 1,505577 | y ~ Intervention + Sex + |
| 0,079618 | 0,248096 | 0,140458 | 138 | 69 | 69 | 134 | 1,76633 | y ~ Intervention + Sex + |
| 0,108152 | 0,248026 | 0,153352 | 138 | 69 | 69 | 134 | 1,61736 | y ~ Intervention + Sex + |
| 0,099781 | 0,247874 | 0,149557 | 138 | 69 | 69 | 134 | 1,657383 | y ~ Intervention + Sex + |
| 0,130233 | 0,24513 | 0,161002 | 138 | 69 | 69 | 134 | 1,52253 | y ~ Intervention + Sex + |
| 0,104409 | 0,244285 | 0,149415 | 138 | 69 | 69 | 134 | 1,63494 | y ~ Intervention + Sex + |
| 0,118269 | 0,244258 | 0,155364 | 138 | 69 | 69 | 134 | 1,57217 | y ~ Intervention + Sex + |
| 0,127537 | 0,242623 | 0,158226 | 138 | 69 | 69 | 134 | 1,533391 | y ~ Intervention + Sex + |
| 0,150806 | 0,242288 | 0,167679 | 138 | 69 | 69 | 134 | 1,444952 | y ~ Intervention + Sex + |
| 0,105566 | 0,240394 | 0,14753 | 138 | 69 | 69 | 134 | 1,629453 | y ~ Intervention + Sex + |
| 0,126422 | 0,240197 | 0,156181 | 138 | 69 | 69 | 134 | 1,537939 | y ~ Intervention + Sex + |
| 0,151369 | 0,240059 | 0,166367 | 138 | 69 | 69 | 134 | 1,442946 | y ~ Intervention + Sex + |

| | | | | | | | | |
|---|---|---|---|---|---|---|---|---|
| 0,115397 | 0,238762 | 0,150669 | 138 | 69 | 69 | 134 | 1,584678 | y ~ Intervention + Sex + |
| 0,164775 | 0,238718 | 0,1709 | 138 | 69 | 69 | 134 | 1,396825 | y ~ Intervention + Sex + |
| 0,127505 | 0,237183 | 0,154666 | 138 | 69 | 69 | 134 | 1,533522 | y ~ Intervention + Sex + |
| 0,107419 | 0,237038 | 0,146251 | 138 | 69 | 69 | 134 | 1,620764 | y ~ Intervention + Sex + |
| 0,099821 | 0,236352 | 0,142623 | 138 | 69 | 69 | 134 | 1,657186 | y ~ Intervention + Sex + |
| 0,142741 | 0,23547 | 0,159714 | 138 | 69 | 69 | 134 | 1,47432 | y ~ Intervention + Sex + |
| 0,146865 | 0,234153 | 0,160473 | 138 | 69 | 69 | 134 | 1,459146 | y ~ Intervention + Sex + |
| 0,09263 | 0,232725 | 0,1374 | 138 | 69 | 69 | 134 | 1,693783 | y ~ Intervention + Sex + |
| 0,149837 | 0,23174 | 0,159995 | 138 | 69 | 69 | 134 | 1,448415 | y ~ Intervention + Sex + |
| 0,164196 | 0,231256 | 0,16533 | 138 | 69 | 69 | 134 | 1,398756 | y ~ Intervention + Sex + |
| 0,170419 | 0,23124 | 0,167776 | 138 | 69 | 69 | 134 | 1,378264 | y ~ Intervention + Sex + |
| 0,097115 | 0,231174 | 0,138369 | 138 | 69 | 69 | 134 | 1,670701 | y ~ Intervention + Sex + |
| 0,176285 | 0,231113 | 0,170004 | 138 | 69 | 69 | 134 | 1,359459 | y ~ Intervention + Sex + |
| 0,201939 | 0,23075 | 0,179945 | 138 | 69 | 69 | 134 | 1,282334 | y ~ Intervention + Sex + |
| 0,103596 | 0,230038 | 0,140368 | 138 | 69 | 69 | 134 | 1,638823 | y ~ Intervention + Sex + |
| 0,184411 | 0,228734 | 0,171443 | 138 | 69 | 69 | 134 | 1,334169 | y ~ Intervention + Sex + |
| 0,180715 | 0,228691 | 0,169959 | 138 | 69 | 69 | 134 | 1,345565 | y ~ Intervention + Sex + |
| 0,120219 | 0,227895 | 0,14573 | 138 | 69 | 69 | 134 | 1,563817 | y ~ Intervention + Sex + |
| 0,098114 | 0,227724 | 0,136715 | 138 | 69 | 69 | 134 | 1,665679 | y ~ Intervention + Sex + |
| 0,158011 | 0,227564 | 0,160287 | 138 | 69 | 69 | 134 | 1,419722 | y ~ Intervention + Sex + |
| 0,162557 | 0,227237 | 0,16182 | 138 | 69 | 69 | 134 | 1,404253 | y ~ Intervention + Sex + |
| 0,148049 | 0,226415 | 0,155628 | 138 | 69 | 69 | 134 | 1,454851 | y ~ Intervention + Sex + |
| 0,187571 | 0,225964 | 0,170596 | 138 | 69 | 69 | 134 | 1,324561 | y ~ Intervention + Sex + |
| 0,157386 | 0,225738 | 0,158761 | 138 | 69 | 69 | 134 | 1,421876 | y ~ Intervention + Sex + |
| 0,085917 | 0,225413 | 0,13029 | 138 | 69 | 69 | 134 | 1,730085 | y ~ Intervention + Sex + |
| 0,17363 | 0,224034 | 0,163778 | 138 | 69 | 69 | 134 | 1,36791 | y ~ Intervention + Sex + |
| 0,147602 | 0,221684 | 0,152207 | 138 | 69 | 69 | 134 | 1,456471 | y ~ Intervention + Sex + |
| 0,163237 | 0,221332 | 0,157872 | 138 | 69 | 69 | 134 | 1,401969 | y ~ Intervention + Sex + |
| 0,182307 | 0,221256 | 0,165039 | 138 | 69 | 69 | 134 | 1,340635 | y ~ Intervention + Sex + |
| 0,154472 | 0,221246 | 0,154501 | 138 | 69 | 69 | 134 | 1,432001 | y ~ Intervention + Sex + |
| 0,171573 | 0,221184 | 0,160916 | 138 | 69 | 69 | 134 | 1,374527 | y ~ Intervention + Sex + |
| 0,14338 | 0,220534 | 0,149824 | 138 | 69 | 69 | 134 | 1,471948 | y ~ Intervention + Sex + |
| 0,147376 | 0,220501 | 0,151309 | 138 | 69 | 69 | 134 | 1,457288 | y ~ Intervention + Sex + |
| 0,124764 | 0,2205 | 0,142742 | 138 | 69 | 69 | 134 | 1,544753 | y ~ Intervention + Sex + |
| 0,184048 | 0,220429 | 0,165081 | 138 | 69 | 69 | 134 | 1,335282 | y ~ Intervention + Sex + |
| 0,181709 | 0,220395 | 0,16417 | 138 | 69 | 69 | 134 | 1,342483 | y ~ Intervention + Sex + |
| 0,185389 | 0,220098 | 0,165341 | 138 | 69 | 69 | 134 | 1,331183 | y ~ Intervention + Sex + |
| 0,259251 | 0,219351 | 0,193608 | 138 | 69 | 69 | 134 | 1,132968 | y ~ Intervention + Sex + |
| 0,203189 | 0,218947 | 0,171217 | 138 | 69 | 69 | 134 | 1,278769 | y ~ Intervention + Sex + |
| 0,184231 | 0,217128 | 0,162676 | 138 | 69 | 69 | 134 | 1,33472 | y ~ Intervention + Sex + |
| 0,222286 | 0,216741 | 0,176763 | 138 | 69 | 69 | 134 | 1,226168 | y ~ Intervention + Sex + |
| 0,156267 | 0,216626 | 0,151939 | 138 | 69 | 69 | 134 | 1,425745 | y ~ Intervention + Sex + |
| 0,16701 | 0,216377 | 0,155732 | 138 | 69 | 69 | 134 | 1,389417 | y ~ Intervention + Sex + |
| 0,20134 | 0,215941 | 0,168172 | 138 | 69 | 69 | 134 | 1,284049 | y ~ Intervention + Sex + |
| 0,088872 | 0,215766 | 0,125897 | 138 | 69 | 69 | 134 | 1,71383 | y ~ Intervention + Sex + |
| 0,162361 | 0,215674 | 0,153514 | 138 | 69 | 69 | 134 | 1,404913 | y ~ Intervention + Sex + |
| 0,192952 | 0,214664 | 0,164056 | 138 | 69 | 69 | 134 | 1,308476 | y ~ Intervention + Sex + |
| 0,232628 | 0,212822 | 0,177494 | 138 | 69 | 69 | 134 | 1,199042 | y ~ Intervention + Sex + |

| | | | | | | | | |
|---|---|---|---|---|---|---|---|---|
| 0,115368 | 0,211455 | 0,133427 | 138 | 69 | 69 | 134 | 1,584806 | y ~ Intervention + Sex + |
| 0,197594 | 0,211123 | 0,163047 | 138 | 69 | 69 | 134 | 1,294865 | y ~ Intervention + Sex + |
| 0,187636 | 0,206704 | 0,156078 | 138 | 69 | 69 | 134 | 1,324366 | y ~ Intervention + Sex + |
| 0,209078 | 0,205307 | 0,162661 | 138 | 69 | 69 | 134 | 1,262174 | y ~ Intervention + Sex + |
| 0,229394 | 0,204289 | 0,169193 | 138 | 69 | 69 | 134 | 1,207429 | y ~ Intervention + Sex + |
| 0,174866 | 0,203718 | 0,149357 | 138 | 69 | 69 | 134 | 1,363961 | y ~ Intervention + Sex + |
| 0,122022 | 0,20339 | 0,130698 | 138 | 69 | 69 | 134 | 1,55619 | y ~ Intervention + Sex + |
| 0,205781 | 0,203049 | 0,159702 | 138 | 69 | 69 | 134 | 1,271423 | y ~ Intervention + Sex + |
| 0,183486 | 0,202571 | 0,151511 | 138 | 69 | 69 | 134 | 1,337006 | y ~ Intervention + Sex + |
| 0,214272 | 0,201326 | 0,161342 | 138 | 69 | 69 | 134 | 1,247823 | y ~ Intervention + Sex + |
| 0,205338 | 0,201018 | 0,15795 | 138 | 69 | 69 | 134 | 1,272671 | y ~ Intervention + Sex + |
| 0,238886 | 0,200832 | 0,169759 | 138 | 69 | 69 | 134 | 1,183045 | y ~ Intervention + Sex + |
| 0,200944 | 0,200197 | 0,155773 | 138 | 69 | 69 | 134 | 1,285187 | y ~ Intervention + Sex + |
| 0,165555 | 0,199958 | 0,143418 | 138 | 69 | 69 | 134 | 1,394233 | y ~ Intervention + Sex + |
| 0,225226 | 0,199002 | 0,163336 | 138 | 69 | 69 | 134 | 1,218365 | y ~ Intervention + Sex + |
| 0,164436 | 0,198728 | 0,142156 | 138 | 69 | 69 | 134 | 1,397957 | y ~ Intervention + Sex + |
| 0,144881 | 0,198305 | 0,135232 | 138 | 69 | 69 | 134 | 1,466404 | y ~ Intervention + Sex + |
| 0,223417 | 0,198015 | 0,161889 | 138 | 69 | 69 | 134 | 1,223158 | y ~ Intervention + Sex + |
| 0,139805 | 0,197861 | 0,13321 | 138 | 69 | 69 | 134 | 1,48533 | y ~ Intervention + Sex + |
| 0,16228 | 0,197535 | 0,140575 | 138 | 69 | 69 | 134 | 1,405186 | y ~ Intervention + Sex + |
| 0,229797 | 0,195744 | 0,162257 | 138 | 69 | 69 | 134 | 1,206378 | y ~ Intervention + Sex + |
| 0,22401 | 0,195021 | 0,159646 | 138 | 69 | 69 | 134 | 1,221583 | y ~ Intervention + Sex + |
| 0,212582 | 0,194058 | 0,154941 | 138 | 69 | 69 | 134 | 1,252465 | y ~ Intervention + Sex + |
| 0,221332 | 0,193864 | 0,157778 | 138 | 69 | 69 | 134 | 1,228716 | y ~ Intervention + Sex + |
| 0,225696 | 0,193817 | 0,159242 | 138 | 69 | 69 | 134 | 1,217124 | y ~ Intervention + Sex + |
| 0,213001 | 0,192526 | 0,153859 | 138 | 69 | 69 | 134 | 1,251312 | y ~ Intervention + Sex + |
| 0,202602 | 0,19092 | 0,149105 | 138 | 69 | 69 | 134 | 1,280441 | y ~ Intervention + Sex + |
| 0,222304 | 0,190693 | 0,155526 | 138 | 69 | 69 | 134 | 1,22612 | y ~ Intervention + Sex + |
| 0,237161 | 0,189559 | 0,159639 | 138 | 69 | 69 | 134 | 1,187424 | y ~ Intervention + Sex + |
| 0,212938 | 0,18806 | 0,150269 | 138 | 69 | 69 | 134 | 1,251484 | y ~ Intervention + Sex + |
| 0,227215 | 0,187703 | 0,154726 | 138 | 69 | 69 | 134 | 1,213129 | y ~ Intervention + Sex + |
| 0,206328 | 0,186569 | 0,146918 | 138 | 69 | 69 | 134 | 1,269881 | y ~ Intervention + Sex + |
| 0,242828 | 0,186077 | 0,158617 | 138 | 69 | 69 | 134 | 1,173121 | y ~ Intervention + Sex + |
| 0,259572 | 0,186 | 0,164281 | 138 | 69 | 69 | 134 | 1,132202 | y ~ Intervention + Sex + |
| 0,239883 | 0,185594 | 0,157214 | 138 | 69 | 69 | 134 | 1,180524 | y ~ Intervention + Sex + |
| 0,295457 | 0,184891 | 0,176032 | 138 | 69 | 69 | 134 | 1,05033 | y ~ Intervention + Sex + |
| 0,23179 | 0,184645 | 0,153716 | 138 | 69 | 69 | 134 | 1,201206 | y ~ Intervention + Sex + |
| 0,197066 | 0,184346 | 0,142198 | 138 | 69 | 69 | 134 | 1,296401 | y ~ Intervention + Sex + |
| 0,247136 | 0,18429 | 0,158541 | 138 | 69 | 69 | 134 | 1,162408 | y ~ Intervention + Sex + |
| 0,194805 | 0,182646 | 0,140172 | 138 | 69 | 69 | 134 | 1,303013 | y ~ Intervention + Sex + |
| 0,28805 | 0,182139 | 0,170759 | 138 | 69 | 69 | 134 | 1,066646 | y ~ Intervention + Sex + |
| 0,202961 | 0,181927 | 0,142195 | 138 | 69 | 69 | 134 | 1,279417 | y ~ Intervention + Sex + |
| 0,288735 | 0,181867 | 0,170747 | 138 | 69 | 69 | 134 | 1,065125 | y ~ Intervention + Sex + |
| 0,24569 | 0,18186 | 0,155971 | 138 | 69 | 69 | 134 | 1,165989 | y ~ Intervention + Sex + |
| 0,28549 | 0,181512 | 0,169266 | 138 | 69 | 69 | 134 | 1,07235 | y ~ Intervention + Sex + |
| 0,26144 | 0,18055 | 0,160097 | 138 | 69 | 69 | 134 | 1,127752 | y ~ Intervention + Sex + |
| 0,312845 | 0,179944 | 0,17762 | 138 | 69 | 69 | 134 | 1,013086 | y ~ Intervention + Sex + |
| 0,251002 | 0,179539 | 0,155727 | 138 | 69 | 69 | 134 | 1,152904 | y ~ Intervention + Sex + |

| | | | | | | | | |
|---|---|---|---|---|---|---|---|---|
| 0,211189 | 0,179224 | 0,142659 | 138 | 69 | 69 | 134 | 1,256312 | y ~ Intervention + Sex + |
| 0,199264 | 0,178909 | 0,138687 | 138 | 69 | 69 | 134 | 1,290024 | y ~ Intervention + Sex + |
| 0,211328 | 0,178724 | 0,142305 | 138 | 69 | 69 | 134 | 1,255927 | y ~ Intervention + Sex + |
| 0,221869 | 0,17796 | 0,145004 | 138 | 69 | 69 | 134 | 1,22728 | y ~ Intervention + Sex + |
| 0,263742 | 0,177506 | 0,158163 | 138 | 69 | 69 | 134 | 1,1223 | y ~ Intervention + Sex + |
| 0,268814 | 0,177504 | 0,159856 | 138 | 69 | 69 | 134 | 1,110402 | y ~ Intervention + Sex + |
| 0,283034 | 0,177081 | 0,16429 | 138 | 69 | 69 | 134 | 1,077857 | y ~ Intervention + Sex + |
| 0,283924 | 0,176261 | 0,163833 | 138 | 69 | 69 | 134 | 1,075857 | y ~ Intervention + Sex + |
| 0,284161 | 0,176151 | 0,163812 | 138 | 69 | 69 | 134 | 1,075326 | y ~ Intervention + Sex + |
| 0,203076 | 0,174108 | 0,136119 | 138 | 69 | 69 | 134 | 1,27909 | y ~ Intervention + Sex + |
| 0,260405 | 0,17376 | 0,153741 | 138 | 69 | 69 | 134 | 1,130214 | y ~ Intervention + Sex + |
| 0,231764 | 0,172676 | 0,143744 | 138 | 69 | 69 | 134 | 1,201275 | y ~ Intervention + Sex + |
| 0,281388 | 0,172297 | 0,159304 | 138 | 69 | 69 | 134 | 1,081566 | y ~ Intervention + Sex + |
| 0,279871 | 0,172168 | 0,158681 | 138 | 69 | 69 | 134 | 1,084996 | y ~ Intervention + Sex + |
| 0,234346 | 0,172125 | 0,144084 | 138 | 69 | 69 | 134 | 1,194619 | y ~ Intervention + Sex + |
| 0,298823 | 0,171864 | 0,164777 | 138 | 69 | 69 | 134 | 1,043007 | y ~ Intervention + Sex + |
| 0,256409 | 0,170892 | 0,149933 | 138 | 69 | 69 | 134 | 1,139785 | y ~ Intervention + Sex + |
| 0,271629 | 0,170864 | 0,154786 | 138 | 69 | 69 | 134 | 1,103866 | y ~ Intervention + Sex + |
| 0,301937 | 0,169574 | 0,163637 | 138 | 69 | 69 | 134 | 1,036281 | y ~ Intervention + Sex + |
| 0,275658 | 0,168561 | 0,153994 | 138 | 69 | 69 | 134 | 1,094593 | y ~ Intervention + Sex + |
| 0,32082 | 0,167462 | 0,168056 | 138 | 69 | 69 | 134 | 0,996466 | y ~ Intervention + Sex + |
| 0,323359 | 0,167443 | 0,168924 | 138 | 69 | 69 | 134 | 0,991233 | y ~ Intervention + Sex + |
| 0,278112 | 0,167218 | 0,153553 | 138 | 69 | 69 | 134 | 1,088993 | y ~ Intervention + Sex + |
| 0,297219 | 0,166345 | 0,158955 | 138 | 69 | 69 | 134 | 1,046489 | y ~ Intervention + Sex + |
| 0,265652 | 0,166327 | 0,148799 | 138 | 69 | 69 | 134 | 1,117802 | y ~ Intervention + Sex + |
| 0,307161 | 0,166069 | 0,162002 | 138 | 69 | 69 | 134 | 1,025104 | y ~ Intervention + Sex + |
| 0,322972 | 0,165634 | 0,166964 | 138 | 69 | 69 | 134 | 0,992028 | y ~ Intervention + Sex + |
| 0,272886 | 0,165046 | 0,14991 | 138 | 69 | 69 | 134 | 1,100964 | y ~ Intervention + Sex + |
| 0,331547 | 0,164642 | 0,168945 | 138 | 69 | 69 | 134 | 0,974534 | y ~ Intervention + Sex + |
| 0,314045 | 0,164047 | 0,162332 | 138 | 69 | 69 | 134 | 1,010568 | y ~ Intervention + Sex + |
| 0,342498 | 0,163203 | 0,17132 | 138 | 69 | 69 | 134 | 0,952621 | y ~ Intervention + Sex + |
| 0,292449 | 0,163152 | 0,154365 | 138 | 69 | 69 | 134 | 1,056922 | y ~ Intervention + Sex + |
| 0,288535 | 0,163133 | 0,153095 | 138 | 69 | 69 | 134 | 1,065568 | y ~ Intervention + Sex + |
| 0,251284 | 0,162727 | 0,141229 | 138 | 69 | 69 | 134 | 1,152215 | y ~ Intervention + Sex + |
| 0,29777 | 0,161981 | 0,154962 | 138 | 69 | 69 | 134 | 1,045293 | y ~ Intervention + Sex + |
| 0,373746 | 0,159977 | 0,179254 | 138 | 69 | 69 | 134 | 0,892461 | y ~ Intervention + Sex + |
| 0,279069 | 0,159844 | 0,147075 | 138 | 69 | 69 | 134 | 1,086817 | y ~ Intervention + Sex + |
| 0,335041 | 0,158679 | 0,164011 | 138 | 69 | 69 | 134 | 0,967493 | y ~ Intervention + Sex + |
| 0,293382 | 0,158551 | 0,150303 | 138 | 69 | 69 | 134 | 1,054872 | y ~ Intervention + Sex + |
| 0,348481 | 0,15855 | 0,16852 | 138 | 69 | 69 | 134 | 0,940837 | y ~ Intervention + Sex + |
| 0,276277 | 0,158499 | 0,14499 | 138 | 69 | 69 | 134 | 1,093178 | y ~ Intervention + Sex + |
| 0,343372 | 0,158355 | 0,166534 | 138 | 69 | 69 | 134 | 0,950889 | y ~ Intervention + Sex + |
| 0,358739 | 0,158248 | 0,171835 | 138 | 69 | 69 | 134 | 0,920935 | y ~ Intervention + Sex + |
| 0,291126 | 0,157568 | 0,148672 | 138 | 69 | 69 | 134 | 1,059836 | y ~ Intervention + Sex + |
| 0,32004 | 0,1572 | 0,157503 | 138 | 69 | 69 | 134 | 0,998079 | y ~ Intervention + Sex + |
| 0,327053 | 0,155436 | 0,158018 | 138 | 69 | 69 | 134 | 0,983665 | y ~ Intervention + Sex + |
| 0,337236 | 0,154509 | 0,16043 | 138 | 69 | 69 | 134 | 0,963091 | y ~ Intervention + Sex + |
| 0,351448 | 0,153158 | 0,163798 | 138 | 69 | 69 | 134 | 0,935044 | y ~ Intervention + Sex + |

| | | | | | | | | |
|---|---|---|---|---|---|---|---|---|
| 0,359365 | 0,152628 | 0,165948 | 138 | 69 | 69 | 134 | 0,919734 | y ~ Intervention + Sex |
| 0,265272 | 0,150565 | 0,13459 | 138 | 69 | 69 | 134 | 1,118694 | y ~ Intervention + Sex |
| 0,317267 | 0,150346 | 0,149771 | 138 | 69 | 69 | 134 | 1,003836 | y ~ Intervention + Sex |
| 0,375496 | 0,150217 | 0,168937 | 138 | 69 | 69 | 134 | 0,889188 | y ~ Intervention + Sex |
| 0,335091 | 0,150206 | 0,155269 | 138 | 69 | 69 | 134 | 0,967391 | y ~ Intervention + Sex |
| 0,363576 | 0,149238 | 0,163696 | 138 | 69 | 69 | 134 | 0,911677 | y ~ Intervention + Sex |
| 0,32614 | 0,148783 | 0,150968 | 138 | 69 | 69 | 134 | 0,985529 | y ~ Intervention + Sex |
| 0,313125 | 0,148775 | 0,146939 | 138 | 69 | 69 | 134 | 1,012498 | y ~ Intervention + Sex |
| 0,362216 | 0,145929 | 0,159612 | 138 | 69 | 69 | 134 | 0,914273 | y ~ Intervention + Sex |
| 0,393977 | 0,143471 | 0,167767 | 138 | 69 | 69 | 134 | 0,855183 | y ~ Intervention + Sex |
| 0,371309 | 0,14311 | 0,159537 | 138 | 69 | 69 | 134 | 0,897035 | y ~ Intervention + Sex |
| 0,323725 | 0,142501 | 0,143871 | 138 | 69 | 69 | 134 | 0,99048 | y ~ Intervention + Sex |
| 0,370812 | 0,141463 | 0,157536 | 138 | 69 | 69 | 134 | 0,89797 | y ~ Intervention + Sex |
| 0,353732 | 0,13985 | 0,150278 | 138 | 69 | 69 | 134 | 0,930604 | y ~ Intervention + Sex |
| 0,420176 | 0,138782 | 0,171631 | 138 | 69 | 69 | 134 | 0,808605 | y ~ Intervention + Sex |
| 0,308691 | 0,138677 | 0,135711 | 138 | 69 | 69 | 134 | 1,021854 | y ~ Intervention + Sex |
| 0,335916 | 0,137675 | 0,14256 | 138 | 69 | 69 | 134 | 0,965735 | y ~ Intervention + Sex |
| 0,418774 | 0,137462 | 0,169486 | 138 | 69 | 69 | 134 | 0,811052 | y ~ Intervention + Sex |
| 0,286747 | 0,137271 | 0,128345 | 138 | 69 | 69 | 134 | 1,069546 | y ~ Intervention + Sex |
| 0,343049 | 0,137231 | 0,144222 | 138 | 69 | 69 | 134 | 0,951529 | y ~ Intervention + Sex |
| 0,428555 | 0,137188 | 0,172764 | 138 | 69 | 69 | 134 | 0,794077 | y ~ Intervention + Sex |
| 0,31877 | 0,13665 | 0,136553 | 138 | 69 | 69 | 134 | 1,000712 | y ~ Intervention + Sex |
| 0,373978 | 0,13602 | 0,152484 | 138 | 69 | 69 | 134 | 0,892026 | y ~ Intervention + Sex |
| 0,430746 | 0,135619 | 0,171603 | 138 | 69 | 69 | 134 | 0,790306 | y ~ Intervention + Sex |
| 0,38778 | 0,135508 | 0,156391 | 138 | 69 | 69 | 134 | 0,866472 | y ~ Intervention + Sex |
| 0,437701 | 0,13546 | 0,174021 | 138 | 69 | 69 | 134 | 0,778408 | y ~ Intervention + Sex |
| 0,442103 | 0,134665 | 0,174677 | 138 | 69 | 69 | 134 | 0,770935 | y ~ Intervention + Sex |
| 0,437673 | 0,134262 | 0,172473 | 138 | 69 | 69 | 134 | 0,778455 | y ~ Intervention + Sex |
| 0,368375 | 0,133838 | 0,148286 | 138 | 69 | 69 | 134 | 0,902567 | y ~ Intervention + Sex |
| 0,437634 | 0,131953 | 0,169492 | 138 | 69 | 69 | 134 | 0,778522 | y ~ Intervention + Sex |
| 0,427098 | 0,13161 | 0,165216 | 138 | 69 | 69 | 134 | 0,796591 | y ~ Intervention + Sex |
| 0,410945 | 0,13136 | 0,15926 | 138 | 69 | 69 | 134 | 0,824811 | y ~ Intervention + Sex |
| 0,371103 | 0,131162 | 0,146154 | 138 | 69 | 69 | 134 | 0,897422 | y ~ Intervention + Sex |
| 0,414607 | 0,130953 | 0,160019 | 138 | 69 | 69 | 134 | 0,818356 | y ~ Intervention + Sex |
| 0,412046 | 0,130163 | 0,158182 | 138 | 69 | 69 | 134 | 0,822866 | y ~ Intervention + Sex |
| 0,382711 | 0,130119 | 0,148573 | 138 | 69 | 69 | 134 | 0,875791 | y ~ Intervention + Sex |
| 0,367592 | 0,129547 | 0,143297 | 138 | 69 | 69 | 134 | 0,904048 | y ~ Intervention + Sex |
| 0,360111 | 0,129347 | 0,140855 | 138 | 69 | 69 | 134 | 0,918302 | y ~ Intervention + Sex |
| 0,373187 | 0,129224 | 0,144626 | 138 | 69 | 69 | 134 | 0,893508 | y ~ Intervention + Sex |
| 0,42805 | 0,129222 | 0,162555 | 138 | 69 | 69 | 134 | 0,794947 | y ~ Intervention + Sex |
| 0,380413 | 0,128133 | 0,145599 | 138 | 69 | 69 | 134 | 0,88004 | y ~ Intervention + Sex |
| 0,412765 | 0,127651 | 0,155369 | 138 | 69 | 69 | 134 | 0,821598 | y ~ Intervention + Sex |
| 0,45408 | 0,127384 | 0,169661 | 138 | 69 | 69 | 134 | 0,750816 | y ~ Intervention + Sex |
| 0,426088 | 0,127277 | 0,159427 | 138 | 69 | 69 | 134 | 0,798336 | y ~ Intervention + Sex |
| 0,411999 | 0,126932 | 0,15424 | 138 | 69 | 69 | 134 | 0,822949 | y ~ Intervention + Sex |
| 0,459177 | 0,126639 | 0,170593 | 138 | 69 | 69 | 134 | 0,742346 | y ~ Intervention + Sex |
| 0,394927 | 0,126305 | 0,147992 | 138 | 69 | 69 | 134 | 0,853461 | y ~ Intervention + Sex |
| 0,390062 | 0,125921 | 0,14603 | 138 | 69 | 69 | 134 | 0,862301 | y ~ Intervention + Sex |

| | | | | | | | | |
|---|---|---|---|---|---|---|---|---|
| 0,376831 | 0,125896 | 0,141983 | 138 | 69 | 69 | 134 | 0,886696 | y ~ Intervention + Sex + |
| 0,410284 | 0,125662 | 0,152137 | 138 | 69 | 69 | 134 | 0,825979 | y ~ Intervention + Sex + |
| 0,434031 | 0,125328 | 0,159721 | 138 | 69 | 69 | 134 | 0,784671 | y ~ Intervention + Sex + |
| 0,410334 | 0,125082 | 0,151452 | 138 | 69 | 69 | 134 | 0,82589 | y ~ Intervention + Sex + |
| 0,448289 | 0,125066 | 0,164452 | 138 | 69 | 69 | 134 | 0,760504 | y ~ Intervention + Sex + |
| 0,444816 | 0,124462 | 0,162408 | 138 | 69 | 69 | 134 | 0,76635 | y ~ Intervention + Sex + |
| 0,415221 | 0,124281 | 0,152068 | 138 | 69 | 69 | 134 | 0,817277 | y ~ Intervention + Sex + |
| 0,464128 | 0,124248 | 0,169236 | 138 | 69 | 69 | 134 | 0,734169 | y ~ Intervention + Sex + |
| 0,417261 | 0,123821 | 0,15217 | 138 | 69 | 69 | 134 | 0,8137 | y ~ Intervention + Sex + |
| 0,422639 | 0,123647 | 0,15373 | 138 | 69 | 69 | 134 | 0,804316 | y ~ Intervention + Sex + |
| 0,409945 | 0,12308 | 0,148903 | 138 | 69 | 69 | 134 | 0,82658 | y ~ Intervention + Sex + |
| 0,421539 | 0,121863 | 0,151152 | 138 | 69 | 69 | 134 | 0,80623 | y ~ Intervention + Sex + |
| 0,408308 | 0,12172 | 0,146742 | 138 | 69 | 69 | 134 | 0,829481 | y ~ Intervention + Sex + |
| 0,420255 | 0,121716 | 0,150552 | 138 | 69 | 69 | 134 | 0,808466 | y ~ Intervention + Sex + |
| 0,414151 | 0,121379 | 0,148175 | 138 | 69 | 69 | 134 | 0,819158 | y ~ Intervention + Sex + |
| 0,479723 | 0,119224 | 0,168223 | 138 | 69 | 69 | 134 | 0,70873 | y ~ Intervention + Sex + |
| 0,421899 | 0,119066 | 0,147798 | 138 | 69 | 69 | 134 | 0,805603 | y ~ Intervention + Sex + |
| 0,462629 | 0,118734 | 0,161183 | 138 | 69 | 69 | 134 | 0,736641 | y ~ Intervention + Sex + |
| 0,478381 | 0,117871 | 0,165806 | 138 | 69 | 69 | 134 | 0,710901 | y ~ Intervention + Sex + |
| 0,445153 | 0,117515 | 0,153457 | 138 | 69 | 69 | 134 | 0,765782 | y ~ Intervention + Sex + |
| 0,463561 | 0,117265 | 0,159522 | 138 | 69 | 69 | 134 | 0,735104 | y ~ Intervention + Sex + |
| 0,413692 | 0,116589 | 0,142188 | 138 | 69 | 69 | 134 | 0,819966 | y ~ Intervention + Sex + |
| 0,496268 | 0,114316 | 0,167561 | 138 | 69 | 69 | 134 | 0,682236 | y ~ Intervention + Sex + |
| 0,496932 | 0,114112 | 0,167521 | 138 | 69 | 69 | 134 | 0,681182 | y ~ Intervention + Sex + |
| 0,460118 | 0,113744 | 0,153544 | 138 | 69 | 69 | 134 | 0,740789 | y ~ Intervention + Sex + |
| 0,4628 | 0,113266 | 0,153819 | 138 | 69 | 69 | 134 | 0,736358 | y ~ Intervention + Sex + |
| 0,449073 | 0,112551 | 0,148251 | 138 | 69 | 69 | 134 | 0,75919 | y ~ Intervention + Sex + |
| 0,404103 | 0,112484 | 0,134395 | 138 | 69 | 69 | 134 | 0,836965 | y ~ Intervention + Sex + |
| 0,50871 | 0,111208 | 0,16783 | 138 | 69 | 69 | 134 | 0,662623 | y ~ Intervention + Sex + |
| 0,53877 | 0,108976 | 0,176834 | 138 | 69 | 69 | 134 | 0,616258 | y ~ Intervention + Sex + |
| 0,526306 | 0,108299 | 0,170465 | 138 | 69 | 69 | 134 | 0,635316 | y ~ Intervention + Sex + |
| 0,5285 | 0,107513 | 0,17013 | 138 | 69 | 69 | 134 | 0,631945 | y ~ Intervention + Sex + |
| 0,520382 | 0,107403 | 0,166657 | 138 | 69 | 69 | 134 | 0,644456 | y ~ Intervention + Sex + |
| 0,430305 | 0,107366 | 0,135724 | 138 | 69 | 69 | 134 | 0,791063 | y ~ Intervention + Sex + |
| 0,562591 | 0,106657 | 0,183751 | 138 | 69 | 69 | 134 | 0,580443 | y ~ Intervention + Sex + |
| 0,502123 | 0,106473 | 0,158213 | 138 | 69 | 69 | 134 | 0,672973 | y ~ Intervention + Sex + |
| 0,465484 | 0,106037 | 0,144871 | 138 | 69 | 69 | 134 | 0,731939 | y ~ Intervention + Sex + |
| 0,491524 | 0,10592 | 0,153557 | 138 | 69 | 69 | 134 | 0,689781 | y ~ Intervention + Sex + |
| 0,516499 | 0,105549 | 0,162264 | 138 | 69 | 69 | 134 | 0,650475 | y ~ Intervention + Sex + |
| 0,519261 | 0,105438 | 0,163168 | 138 | 69 | 69 | 134 | 0,646191 | y ~ Intervention + Sex + |
| 0,534633 | 0,105033 | 0,168713 | 138 | 69 | 69 | 134 | 0,622558 | y ~ Intervention + Sex + |
| 0,51195 | 0,10461 | 0,159088 | 138 | 69 | 69 | 134 | 0,657558 | y ~ Intervention + Sex + |
| 0,43821 | 0,104263 | 0,134093 | 138 | 69 | 69 | 134 | 0,777542 | y ~ Intervention + Sex + |
| 0,516953 | 0,103814 | 0,159771 | 138 | 69 | 69 | 134 | 0,64977 | y ~ Intervention + Sex + |
| 0,576953 | 0,103308 | 0,18474 | 138 | 69 | 69 | 134 | 0,559208 | y ~ Intervention + Sex + |
| 0,560938 | 0,102693 | 0,176175 | 138 | 69 | 69 | 134 | 0,582904 | y ~ Intervention + Sex + |
| 0,507172 | 0,101384 | 0,152449 | 138 | 69 | 69 | 134 | 0,665034 | y ~ Intervention + Sex + |
| 0,528526 | 0,101101 | 0,159994 | 138 | 69 | 69 | 134 | 0,631905 | y ~ Intervention + Sex + |

| | | | | | | | | |
|---|---|---|---|---|---|---|---|---|
| 0,53614 | 0,100731 | 0,162401 | 138 | 69 | 69 | 134 | 0,62026 | y ~ Intervention + Sex + |
| 0,537064 | 0,100525 | 0,162437 | 138 | 69 | 69 | 134 | 0,618853 | y ~ Intervention + Sex + |
| 0,503789 | 0,100402 | 0,149776 | 138 | 69 | 69 | 134 | 0,670349 | y ~ Intervention + Sex + |
| 0,5232 | 0,100184 | 0,156512 | 138 | 69 | 69 | 134 | 0,640101 | y ~ Intervention + Sex + |
| 0,452868 | 0,09978 | 0,132539 | 138 | 69 | 69 | 134 | 0,752838 | y ~ Intervention + Sex + |
| 0,531519 | 0,099766 | 0,159036 | 138 | 69 | 69 | 134 | 0,627316 | y ~ Intervention + Sex + |
| 0,544732 | 0,099382 | 0,163667 | 138 | 69 | 69 | 134 | 0,60722 | y ~ Intervention + Sex + |
| 0,555783 | 0,099309 | 0,168149 | 138 | 69 | 69 | 134 | 0,590601 | y ~ Intervention + Sex + |
| 0,545545 | 0,098864 | 0,163143 | 138 | 69 | 69 | 134 | 0,605992 | y ~ Intervention + Sex + |
| 0,495666 | 0,098657 | 0,144406 | 138 | 69 | 69 | 134 | 0,683191 | y ~ Intervention + Sex + |
| 0,564069 | 0,09855 | 0,17043 | 138 | 69 | 69 | 134 | 0,578245 | y ~ Intervention + Sex + |
| 0,499636 | 0,098412 | 0,145386 | 138 | 69 | 69 | 134 | 0,6769 | y ~ Intervention + Sex + |
| 0,509381 | 0,097629 | 0,147571 | 138 | 69 | 69 | 134 | 0,661572 | y ~ Intervention + Sex + |
| 0,511788 | 0,097337 | 0,147972 | 138 | 69 | 69 | 134 | 0,657811 | y ~ Intervention + Sex + |
| 0,541579 | 0,097053 | 0,158586 | 138 | 69 | 69 | 134 | 0,611994 | y ~ Intervention + Sex + |
| 0,448517 | 0,096787 | 0,127331 | 138 | 69 | 69 | 134 | 0,760121 | y ~ Intervention + Sex + |
| 0,494537 | 0,096786 | 0,141296 | 138 | 69 | 69 | 134 | 0,684985 | y ~ Intervention + Sex + |
| 0,542785 | 0,096569 | 0,158266 | 138 | 69 | 69 | 134 | 0,610167 | y ~ Intervention + Sex + |
| 0,533247 | 0,09654 | 0,154544 | 138 | 69 | 69 | 134 | 0,624674 | y ~ Intervention + Sex + |
| 0,523383 | 0,096426 | 0,150709 | 138 | 69 | 69 | 134 | 0,639818 | y ~ Intervention + Sex + |
| 0,552684 | 0,09618 | 0,16158 | 138 | 69 | 69 | 134 | 0,595245 | y ~ Intervention + Sex + |
| 0,478553 | 0,096107 | 0,135243 | 138 | 69 | 69 | 134 | 0,710623 | y ~ Intervention + Sex + |
| 0,524321 | 0,095474 | 0,149559 | 138 | 69 | 69 | 134 | 0,638372 | y ~ Intervention + Sex + |
| 0,578438 | 0,094895 | 0,17036 | 138 | 69 | 69 | 134 | 0,557027 | y ~ Intervention + Sex + |
| 0,504859 | 0,09469 | 0,14161 | 138 | 69 | 69 | 134 | 0,668666 | y ~ Intervention + Sex + |
| 0,569095 | 0,09465 | 0,165821 | 138 | 69 | 69 | 134 | 0,570795 | y ~ Intervention + Sex + |
| 0,519585 | 0,093867 | 0,145374 | 138 | 69 | 69 | 134 | 0,645689 | y ~ Intervention + Sex + |
| 0,573761 | 0,093653 | 0,166079 | 138 | 69 | 69 | 134 | 0,563905 | y ~ Intervention + Sex + |
| 0,5517 | 0,093549 | 0,156771 | 138 | 69 | 69 | 134 | 0,596722 | y ~ Intervention + Sex + |
| 0,578061 | 0,093439 | 0,167579 | 138 | 69 | 69 | 134 | 0,557581 | y ~ Intervention + Sex + |
| 0,54515 | 0,093366 | 0,153919 | 138 | 69 | 69 | 134 | 0,606589 | y ~ Intervention + Sex + |
| 0,551835 | 0,092959 | 0,155836 | 138 | 69 | 69 | 134 | 0,596519 | y ~ Intervention + Sex + |
| 0,550141 | 0,091007 | 0,151916 | 138 | 69 | 69 | 134 | 0,599066 | y ~ Intervention + Sex + |
| 0,584463 | 0,09072 | 0,165485 | 138 | 69 | 69 | 134 | 0,548206 | y ~ Intervention + Sex + |
| 0,567965 | 0,090708 | 0,158452 | 138 | 69 | 69 | 134 | 0,572467 | y ~ Intervention + Sex + |
| 0,421877 | 0,090502 | 0,112336 | 138 | 69 | 69 | 134 | 0,805641 | y ~ Intervention + Sex + |
| 0,519095 | 0,08984 | 0,138974 | 138 | 69 | 69 | 134 | 0,646448 | y ~ Intervention + Sex + |
| 0,559955 | 0,089257 | 0,152742 | 138 | 69 | 69 | 134 | 0,584368 | y ~ Intervention + Sex + |
| 0,552738 | 0,089136 | 0,149767 | 138 | 69 | 69 | 134 | 0,595163 | y ~ Intervention + Sex + |
| 0,614942 | 0,088779 | 0,176075 | 138 | 69 | 69 | 134 | 0,50421 | y ~ Intervention + Sex + |
| 0,586142 | 0,08755 | 0,160421 | 138 | 69 | 69 | 134 | 0,545755 | y ~ Intervention + Sex + |
| 0,554597 | 0,08642 | 0,145888 | 138 | 69 | 69 | 134 | 0,592376 | y ~ Intervention + Sex + |
| 0,532368 | 0,085584 | 0,136712 | 138 | 69 | 69 | 134 | 0,626018 | y ~ Intervention + Sex + |
| 0,499599 | 0,084974 | 0,125523 | 138 | 69 | 69 | 134 | 0,676959 | y ~ Intervention + Sex + |
| 0,622492 | 0,084936 | 0,172123 | 138 | 69 | 69 | 134 | 0,493465 | y ~ Intervention + Sex + |
| 0,57399 | 0,084426 | 0,149806 | 138 | 69 | 69 | 134 | 0,563568 | y ~ Intervention + Sex + |
| 0,56298 | 0,084348 | 0,145462 | 138 | 69 | 69 | 134 | 0,579864 | y ~ Intervention + Sex + |
| 0,601112 | 0,083937 | 0,160171 | 138 | 69 | 69 | 134 | 0,524047 | y ~ Intervention + Sex + |

| | | | | | | | | |
|---|---|---|---|---|---|---|---|---|
| 0,61031 | 0,083719 | 0,163888 | 138 | 69 | 69 | 134 | 0,510832 | y ~ Intervention + Sex + |
| 0,610144 | 0,082732 | 0,16188 | 138 | 69 | 69 | 134 | 0,511069 | y ~ Intervention + Sex + |
| 0,596069 | 0,082642 | 0,155538 | 138 | 69 | 69 | 134 | 0,531332 | y ~ Intervention + Sex + |
| 0,633931 | 0,081569 | 0,170899 | 138 | 69 | 69 | 134 | 0,477294 | y ~ Intervention + Sex + |
| 0,593928 | 0,081547 | 0,152586 | 138 | 69 | 69 | 134 | 0,534432 | y ~ Intervention + Sex + |
| 0,613715 | 0,08101 | 0,160111 | 138 | 69 | 69 | 134 | 0,505961 | y ~ Intervention + Sex + |
| 0,615717 | 0,081002 | 0,161005 | 138 | 69 | 69 | 134 | 0,503104 | y ~ Intervention + Sex + |
| 0,624627 | 0,080326 | 0,163785 | 138 | 69 | 69 | 134 | 0,490437 | y ~ Intervention + Sex + |
| 0,582893 | 0,079831 | 0,145015 | 138 | 69 | 69 | 134 | 0,550501 | y ~ Intervention + Sex + |
| 0,618981 | 0,079763 | 0,160021 | 138 | 69 | 69 | 134 | 0,498455 | y ~ Intervention + Sex + |
| 0,583153 | 0,079593 | 0,144683 | 138 | 69 | 69 | 134 | 0,550119 | y ~ Intervention + Sex + |
| 0,622097 | 0,078271 | 0,158435 | 138 | 69 | 69 | 134 | 0,494025 | y ~ Intervention + Sex + |
| 0,598666 | 0,077554 | 0,147001 | 138 | 69 | 69 | 134 | 0,527576 | y ~ Intervention + Sex + |
| 0,628406 | 0,077072 | 0,158882 | 138 | 69 | 69 | 134 | 0,485088 | y ~ Intervention + Sex + |
| 0,599054 | 0,076534 | 0,145222 | 138 | 69 | 69 | 134 | 0,527016 | y ~ Intervention + Sex + |
| 0,620693 | 0,076529 | 0,154286 | 138 | 69 | 69 | 134 | 0,49602 | y ~ Intervention + Sex + |
| 0,623113 | 0,076367 | 0,155035 | 138 | 69 | 69 | 134 | 0,492583 | y ~ Intervention + Sex + |
| 0,603155 | 0,075566 | 0,145011 | 138 | 69 | 69 | 134 | 0,521103 | y ~ Intervention + Sex + |
| 0,611327 | 0,074964 | 0,147168 | 138 | 69 | 69 | 134 | 0,509375 | y ~ Intervention + Sex + |
| 0,579989 | 0,07432 | 0,13397 | 138 | 69 | 69 | 134 | 0,554752 | y ~ Intervention + Sex + |
| 0,627914 | 0,074012 | 0,152356 | 138 | 69 | 69 | 134 | 0,485783 | y ~ Intervention + Sex + |
| 0,611702 | 0,074006 | 0,145441 | 138 | 69 | 69 | 134 | 0,508839 | y ~ Intervention + Sex + |
| 0,60078 | 0,072987 | 0,139148 | 138 | 69 | 69 | 134 | 0,524524 | y ~ Intervention + Sex + |
| 0,676841 | 0,072604 | 0,173823 | 138 | 69 | 69 | 134 | 0,417692 | y ~ Intervention + Sex + |
| 0,665089 | 0,072182 | 0,166373 | 138 | 69 | 69 | 134 | 0,433858 | y ~ Intervention + Sex + |
| 0,652264 | 0,071206 | 0,157664 | 138 | 69 | 69 | 134 | 0,451633 | y ~ Intervention + Sex + |
| 0,62113 | 0,07073 | 0,142773 | 138 | 69 | 69 | 134 | 0,495399 | y ~ Intervention + Sex + |
| 0,671393 | 0,07033 | 0,165414 | 138 | 69 | 69 | 134 | 0,425172 | y ~ Intervention + Sex + |
| 0,667955 | 0,069271 | 0,16113 | 138 | 69 | 69 | 134 | 0,429906 | y ~ Intervention + Sex + |
| 0,686907 | 0,069148 | 0,171187 | 138 | 69 | 69 | 134 | 0,403931 | y ~ Intervention + Sex + |
| 0,692585 | 0,068759 | 0,173545 | 138 | 69 | 69 | 134 | 0,396204 | y ~ Intervention + Sex + |
| 0,664899 | 0,067661 | 0,155857 | 138 | 69 | 69 | 134 | 0,434121 | y ~ Intervention + Sex + |
| 0,638886 | 0,067423 | 0,143353 | 138 | 69 | 69 | 134 | 0,470327 | y ~ Intervention + Sex + |
| 0,692201 | 0,066449 | 0,167495 | 138 | 69 | 69 | 134 | 0,396725 | y ~ Intervention + Sex + |
| 0,640531 | 0,066349 | 0,141765 | 138 | 69 | 69 | 134 | 0,46802 | y ~ Intervention + Sex + |
| 0,672919 | 0,065417 | 0,154622 | 138 | 69 | 69 | 134 | 0,423075 | y ~ Intervention + Sex + |
| 0,690686 | 0,064241 | 0,161092 | 138 | 69 | 69 | 134 | 0,398785 | y ~ Intervention + Sex + |
| 0,674773 | 0,064113 | 0,152459 | 138 | 69 | 69 | 134 | 0,420528 | y ~ Intervention + Sex + |
| 0,698892 | 0,064026 | 0,165166 | 138 | 69 | 69 | 134 | 0,387648 | y ~ Intervention + Sex + |
| 0,723944 | 0,063451 | 0,179272 | 138 | 69 | 69 | 134 | 0,353935 | y ~ Intervention + Sex + |
| 0,719393 | 0,063022 | 0,175046 | 138 | 69 | 69 | 134 | 0,360028 | y ~ Intervention + Sex + |
| 0,687411 | 0,062867 | 0,155904 | 138 | 69 | 69 | 134 | 0,403245 | y ~ Intervention + Sex + |
| 0,685926 | 0,062432 | 0,154051 | 138 | 69 | 69 | 134 | 0,40527 | y ~ Intervention + Sex + |
| 0,695097 | 0,06159 | 0,156802 | 138 | 69 | 69 | 134 | 0,392792 | y ~ Intervention + Sex + |
| 0,709938 | 0,061109 | 0,163948 | 138 | 69 | 69 | 134 | 0,372731 | y ~ Intervention + Sex + |
| 0,720929 | 0,059121 | 0,165156 | 138 | 69 | 69 | 134 | 0,357971 | y ~ Intervention + Sex + |
| 0,716909 | 0,059085 | 0,162609 | 138 | 69 | 69 | 134 | 0,363359 | y ~ Intervention + Sex + |
| 0,686114 | 0,058794 | 0,145166 | 138 | 69 | 69 | 134 | 0,405013 | y ~ Intervention + Sex + |

| | | | | | | | | |
|---|---|---|---|---|---|---|---|---|
| 0,702647 | 0,05852 | 0,152966 | 138 | 69 | 69 | 134 | 0,382567 | y ~ Intervention + Sex + |
| 0,721 | 0,058505 | 0,163479 | 138 | 69 | 69 | 134 | 0,357875 | y ~ Intervention + Sex + |
| 0,715286 | 0,058276 | 0,159425 | 138 | 69 | 69 | 134 | 0,365538 | y ~ Intervention + Sex + |
| 0,738257 | 0,058126 | 0,173584 | 138 | 69 | 69 | 134 | 0,334856 | y ~ Intervention + Sex + |
| 0,69288 | 0,058075 | 0,146728 | 138 | 69 | 69 | 134 | 0,395803 | y ~ Intervention + Sex + |
| 0,709614 | 0,057499 | 0,154084 | 138 | 69 | 69 | 134 | 0,373166 | y ~ Intervention + Sex + |
| 0,715287 | 0,057322 | 0,156816 | 138 | 69 | 69 | 134 | 0,365537 | y ~ Intervention + Sex + |
| 0,703982 | 0,05698 | 0,149648 | 138 | 69 | 69 | 134 | 0,380763 | y ~ Intervention + Sex + |
| 0,722211 | 0,056486 | 0,158555 | 138 | 69 | 69 | 134 | 0,356254 | y ~ Intervention + Sex + |
| 0,716204 | 0,05631 | 0,154567 | 138 | 69 | 69 | 134 | 0,364306 | y ~ Intervention + Sex + |
| 0,735779 | 0,055994 | 0,165587 | 138 | 69 | 69 | 134 | 0,338151 | y ~ Intervention + Sex + |
| 0,712353 | 0,054963 | 0,148758 | 138 | 69 | 69 | 134 | 0,36948 | y ~ Intervention + Sex + |
| 0,732673 | 0,054419 | 0,158989 | 138 | 69 | 69 | 134 | 0,342285 | y ~ Intervention + Sex + |
| 0,749374 | 0,054419 | 0,169994 | 138 | 69 | 69 | 134 | 0,320122 | y ~ Intervention + Sex + |
| 0,746451 | 0,053951 | 0,166521 | 138 | 69 | 69 | 134 | 0,323989 | y ~ Intervention + Sex + |
| 0,719959 | 0,053697 | 0,149462 | 138 | 69 | 69 | 134 | 0,35927 | y ~ Intervention + Sex + |
| 0,73574 | 0,052788 | 0,156083 | 138 | 69 | 69 | 134 | 0,338202 | y ~ Intervention + Sex + |
| 0,749367 | 0,052445 | 0,163822 | 138 | 69 | 69 | 134 | 0,320132 | y ~ Intervention + Sex + |
| 0,764849 | 0,052375 | 0,174742 | 138 | 69 | 69 | 134 | 0,299728 | y ~ Intervention + Sex + |
| 0,747297 | 0,052281 | 0,161926 | 138 | 69 | 69 | 134 | 0,32287 | y ~ Intervention + Sex + |
| 0,741586 | 0,051881 | 0,157006 | 138 | 69 | 69 | 134 | 0,330436 | y ~ Intervention + Sex + |
| 0,744314 | 0,051803 | 0,158506 | 138 | 69 | 69 | 134 | 0,326821 | y ~ Intervention + Sex + |
| 0,749213 | 0,05129 | 0,160115 | 138 | 69 | 69 | 134 | 0,320336 | y ~ Intervention + Sex + |
| 0,7739 | 0,051106 | 0,177538 | 138 | 69 | 69 | 134 | 0,287859 | y ~ Intervention + Sex + |
| 0,741648 | 0,050939 | 0,154194 | 138 | 69 | 69 | 134 | 0,330354 | y ~ Intervention + Sex + |
| 0,745898 | 0,050608 | 0,15585 | 138 | 69 | 69 | 134 | 0,324722 | y ~ Intervention + Sex + |
| 0,730407 | 0,050445 | 0,146088 | 138 | 69 | 69 | 134 | 0,345305 | y ~ Intervention + Sex + |
| 0,775268 | 0,048576 | 0,169808 | 138 | 69 | 69 | 134 | 0,286067 | y ~ Intervention + Sex + |
| 0,774481 | 0,048484 | 0,168875 | 138 | 69 | 69 | 134 | 0,287098 | y ~ Intervention + Sex + |
| 0,736877 | 0,048112 | 0,142896 | 138 | 69 | 69 | 134 | 0,33669 | y ~ Intervention + Sex + |
| 0,763054 | 0,046879 | 0,155184 | 138 | 69 | 69 | 134 | 0,302087 | y ~ Intervention + Sex + |
| 0,761223 | 0,046798 | 0,153691 | 138 | 69 | 69 | 134 | 0,304496 | y ~ Intervention + Sex + |
| 0,760538 | 0,046441 | 0,152069 | 138 | 69 | 69 | 134 | 0,305397 | y ~ Intervention + Sex + |
| 0,782092 | 0,042998 | 0,155143 | 138 | 69 | 69 | 134 | 0,27715 | y ~ Intervention + Sex + |
| 0,79892 | 0,041535 | 0,16272 | 138 | 69 | 69 | 134 | 0,255252 | y ~ Intervention + Sex + |
| 0,805011 | 0,03979 | 0,16086 | 138 | 69 | 69 | 134 | 0,247356 | y ~ Intervention + Sex + |
| 0,783505 | 0,039584 | 0,14378 | 138 | 69 | 69 | 134 | 0,275306 | y ~ Intervention + Sex + |
| 0,788077 | 0,039527 | 0,14675 | 138 | 69 | 69 | 134 | 0,269347 | y ~ Intervention + Sex + |
| 0,774798 | 0,038854 | 0,13553 | 138 | 69 | 69 | 134 | 0,286682 | y ~ Intervention + Sex + |
| 0,815856 | 0,03869 | 0,165811 | 138 | 69 | 69 | 134 | 0,233337 | y ~ Intervention + Sex + |
| 0,811811 | 0,038385 | 0,160902 | 138 | 69 | 69 | 134 | 0,23856 | y ~ Intervention + Sex + |
| 0,817552 | 0,037604 | 0,162684 | 138 | 69 | 69 | 134 | 0,231148 | y ~ Intervention + Sex + |
| 0,811877 | 0,036281 | 0,152135 | 138 | 69 | 69 | 134 | 0,238475 | y ~ Intervention + Sex + |
| 0,844394 | 0,034332 | 0,17458 | 138 | 69 | 69 | 134 | 0,196656 | y ~ Intervention + Sex + |
| 0,829855 | 0,032898 | 0,152798 | 138 | 69 | 69 | 134 | 0,215307 | y ~ Intervention + Sex + |
| 0,839696 | 0,032533 | 0,16052 | 138 | 69 | 69 | 134 | 0,202676 | y ~ Intervention + Sex + |
| 0,857508 | 0,031781 | 0,176666 | 138 | 69 | 69 | 134 | 0,179894 | y ~ Intervention + Sex + |
| 0,840948 | 0,031728 | 0,157793 | 138 | 69 | 69 | 134 | 0,201071 | y ~ Intervention + Sex + |

| | | | | | | | | |
|---|---|---|---|---|---|---|---|---|
| 0,836852 | 0,031606 | 0,153186 | 138 | 69 | 69 | 134 | 0,206322 | y ~ Intervention + Sex + |
| 0,847664 | 0,030953 | 0,16082 | 138 | 69 | 69 | 134 | 0,192472 | y ~ Intervention + Sex + |
| 0,851215 | 0,030527 | 0,162436 | 138 | 69 | 69 | 134 | 0,187931 | y ~ Intervention + Sex + |
| 0,840582 | 0,029399 | 0,145872 | 138 | 69 | 69 | 134 | 0,201539 | y ~ Intervention + Sex + |
| 0,854994 | 0,029009 | 0,15843 | 138 | 69 | 69 | 134 | 0,183104 | y ~ Intervention + Sex + |
| 0,849701 | 0,029002 | 0,152751 | 138 | 69 | 69 | 134 | 0,189867 | y ~ Intervention + Sex + |
| 0,830471 | 0,028657 | 0,13359 | 138 | 69 | 69 | 134 | 0,214516 | y ~ Intervention + Sex + |
| 0,829374 | 0,028372 | 0,131398 | 138 | 69 | 69 | 134 | 0,215926 | y ~ Intervention + Sex + |
| 0,843989 | 0,02816 | 0,142816 | 138 | 69 | 69 | 134 | 0,197176 | y ~ Intervention + Sex + |
| 0,847971 | 0,026945 | 0,140283 | 138 | 69 | 69 | 134 | 0,192079 | y ~ Intervention + Sex + |
| 0,850428 | 0,02694 | 0,142586 | 138 | 69 | 69 | 134 | 0,188938 | y ~ Intervention + Sex + |
| 0,854343 | 0,026709 | 0,14521 | 138 | 69 | 69 | 134 | 0,183935 | y ~ Intervention + Sex + |
| 0,879062 | 0,026301 | 0,172523 | 138 | 69 | 69 | 134 | 0,152449 | y ~ Intervention + Sex + |
| 0,869871 | 0,026232 | 0,159817 | 138 | 69 | 69 | 134 | 0,164137 | y ~ Intervention + Sex + |
| 0,865734 | 0,026078 | 0,153937 | 138 | 69 | 69 | 134 | 0,169405 | y ~ Intervention + Sex + |
| 0,879559 | 0,025628 | 0,168808 | 138 | 69 | 69 | 134 | 0,151818 | y ~ Intervention + Sex + |
| 0,873966 | 0,024847 | 0,156345 | 138 | 69 | 69 | 134 | 0,158927 | y ~ Intervention + Sex + |
| 0,885523 | 0,024517 | 0,169965 | 138 | 69 | 69 | 134 | 0,144246 | y ~ Intervention + Sex + |
| 0,87108 | 0,024285 | 0,149356 | 138 | 69 | 69 | 134 | 0,162598 | y ~ Intervention + Sex + |
| 0,876759 | 0,02423 | 0,155943 | 138 | 69 | 69 | 134 | 0,155376 | y ~ Intervention + Sex + |
| 0,882152 | 0,024011 | 0,161663 | 138 | 69 | 69 | 134 | 0,148525 | y ~ Intervention + Sex + |
| 0,88403 | 0,023374 | 0,159944 | 138 | 69 | 69 | 134 | 0,146141 | y ~ Intervention + Sex + |
| 0,881387 | 0,023302 | 0,155874 | 138 | 69 | 69 | 134 | 0,149496 | y ~ Intervention + Sex + |
| 0,887006 | 0,023102 | 0,162277 | 138 | 69 | 69 | 134 | 0,142365 | y ~ Intervention + Sex + |
| 0,88778 | 0,02218 | 0,156878 | 138 | 69 | 69 | 134 | 0,141383 | y ~ Intervention + Sex + |
| 0,888883 | 0,02087 | 0,149087 | 138 | 69 | 69 | 134 | 0,139984 | y ~ Intervention + Sex + |
| 0,896539 | 0,019708 | 0,151273 | 138 | 69 | 69 | 134 | 0,130281 | y ~ Intervention + Sex + |
| 0,898578 | 0,019114 | 0,149677 | 138 | 69 | 69 | 134 | 0,1277 | y ~ Intervention + Sex + |
| 0,905732 | 0,018753 | 0,158054 | 138 | 69 | 69 | 134 | 0,118647 | y ~ Intervention + Sex + |
| 0,906217 | 0,018663 | 0,158117 | 138 | 69 | 69 | 134 | 0,118035 | y ~ Intervention + Sex + |
| 0,908142 | 0,018397 | 0,159142 | 138 | 69 | 69 | 134 | 0,1156 | y ~ Intervention + Sex + |
| 0,903994 | 0,018358 | 0,151912 | 138 | 69 | 69 | 134 | 0,120846 | y ~ Intervention + Sex + |
| 0,922457 | 0,016967 | 0,173982 | 138 | 69 | 69 | 134 | 0,097523 | y ~ Intervention + Sex + |
| 0,92335 | 0,016092 | 0,166932 | 138 | 69 | 69 | 134 | 0,096396 | y ~ Intervention + Sex + |
| 0,921697 | 0,016015 | 0,162623 | 138 | 69 | 69 | 134 | 0,098482 | y ~ Intervention + Sex + |
| 0,911668 | 0,01545 | 0,139006 | 138 | 69 | 69 | 134 | 0,111145 | y ~ Intervention + Sex + |
| 0,928835 | 0,014611 | 0,163286 | 138 | 69 | 69 | 134 | 0,089479 | y ~ Intervention + Sex + |
| 0,928875 | 0,013991 | 0,156452 | 138 | 69 | 69 | 134 | 0,089428 | y ~ Intervention + Sex + |
| 0,930797 | 0,01351 | 0,155278 | 138 | 69 | 69 | 134 | 0,087006 | y ~ Intervention + Sex + |
| 0,933852 | 0,013447 | 0,161712 | 138 | 69 | 69 | 134 | 0,083155 | y ~ Intervention + Sex + |
| 0,922139 | 0,013208 | 0,13488 | 138 | 69 | 69 | 134 | 0,097923 | y ~ Intervention + Sex + |
| 0,938933 | 0,012522 | 0,163145 | 138 | 69 | 69 | 134 | 0,076755 | y ~ Intervention + Sex + |
| 0,937225 | 0,012513 | 0,15858 | 138 | 69 | 69 | 134 | 0,078907 | y ~ Intervention + Sex + |
| 0,942995 | 0,012295 | 0,171625 | 138 | 69 | 69 | 134 | 0,07164 | y ~ Intervention + Sex + |
| 0,941297 | 0,011527 | 0,156245 | 138 | 69 | 69 | 134 | 0,073777 | y ~ Intervention + Sex + |
| 0,946775 | 0,011473 | 0,171539 | 138 | 69 | 69 | 134 | 0,066883 | y ~ Intervention + Sex + |
| 0,941749 | 0,01129 | 0,154214 | 138 | 69 | 69 | 134 | 0,073209 | y ~ Intervention + Sex + |
| 0,939334 | 0,009796 | 0,128476 | 138 | 69 | 69 | 134 | 0,07625 | y ~ Intervention + Sex + |

| | | | | | | | | |
|---|---|---|---|---|---|---|---|---|
| 0,952727 | 0,008954 | 0,150753 | 138 | 69 | 69 | 134 | 0,059394 | y ~ Intervention + Sex + |
| 0,960236 | 0,007963 | 0,159411 | 138 | 69 | 69 | 134 | 0,049951 | y ~ Intervention + Sex + |
| 0,96246 | 0,007778 | 0,164952 | 138 | 69 | 69 | 134 | 0,047155 | y ~ Intervention + Sex + |
| 0,962939 | 0,006879 | 0,147777 | 138 | 69 | 69 | 134 | 0,046553 | y ~ Intervention + Sex + |
| 0,96848 | 0,006509 | 0,164413 | 138 | 69 | 69 | 134 | 0,039589 | y ~ Intervention + Sex + |
| 0,969178 | 0,006503 | 0,167989 | 138 | 69 | 69 | 134 | 0,038712 | y ~ Intervention + Sex + |
| 0,976695 | 0,004694 | 0,160381 | 138 | 69 | 69 | 134 | 0,029268 | y ~ Intervention + Sex + |
| 0,979468 | 0,004337 | 0,168219 | 138 | 69 | 69 | 134 | 0,025784 | y ~ Intervention + Sex + |
| 0,98018 | 0,003861 | 0,155139 | 138 | 69 | 69 | 134 | 0,02489 | y ~ Intervention + Sex + |
| 0,982399 | 0,003487 | 0,157787 | 138 | 69 | 69 | 134 | 0,022102 | y ~ Intervention + Sex + |
| 0,983816 | 0,003281 | 0,161425 | 138 | 69 | 69 | 134 | 0,020323 | y ~ Intervention + Sex + |
| 0,983741 | 0,003021 | 0,147945 | 138 | 69 | 69 | 134 | 0,020417 | y ~ Intervention + Sex + |
| 0,987681 | 0,002519 | 0,162841 | 138 | 69 | 69 | 134 | 0,01547 | y ~ Intervention + Sex + |
| 0,987713 | 0,002464 | 0,15969 | 138 | 69 | 69 | 134 | 0,015428 | y ~ Intervention + Sex + |
| 0,991787 | 0,001666 | 0,161561 | 138 | 69 | 69 | 134 | 0,010312 | y ~ Intervention + Sex + |
| 0,998627 | 0,000269 | 0,155849 | 138 | 69 | 69 | 134 | 0,001724 | y ~ Intervention + Sex + |
| 0,998967 | 0,000205 | 0,158257 | 138 | 69 | 69 | 134 | 0,001297 | y ~ Intervention + Sex + |
| 0,999683 | -6,7E-05 | 0,167181 | 138 | 69 | 69 | 134 | -0,0004 | y ~ Intervention + Sex + |
| 0,99929 | -0,00015 | 0,166237 | 138 | 69 | 69 | 134 | -0,00089 | y ~ Intervention + Sex + |
| 0,997527 | -0,00041 | 0,133334 | 138 | 69 | 69 | 134 | -0,00311 | y ~ Intervention + Sex + |
| 0,992951 | -0,00123 | 0,139019 | 138 | 69 | 69 | 134 | -0,00885 | y ~ Intervention + Sex + |
| 0,99259 | -0,00137 | 0,147667 | 138 | 69 | 69 | 134 | -0,00931 | y ~ Intervention + Sex + |
| 0,985928 | -0,00264 | 0,149566 | 138 | 69 | 69 | 134 | -0,01767 | y ~ Intervention + Sex + |
| 0,985741 | -0,00276 | 0,154403 | 138 | 69 | 69 | 134 | -0,0179 | y ~ Intervention + Sex + |
| 0,976508 | -0,00424 | 0,143792 | 138 | 69 | 69 | 134 | -0,0295 | y ~ Intervention + Sex + |
| 0,97953 | -0,00426 | 0,165542 | 138 | 69 | 69 | 134 | -0,02571 | y ~ Intervention + Sex + |
| 0,977841 | -0,00432 | 0,155176 | 138 | 69 | 69 | 134 | -0,02783 | y ~ Intervention + Sex + |
| 0,977324 | -0,00456 | 0,160085 | 138 | 69 | 69 | 134 | -0,02848 | y ~ Intervention + Sex + |
| 0,977017 | -0,00465 | 0,161004 | 138 | 69 | 69 | 134 | -0,02886 | y ~ Intervention + Sex + |
| 0,976619 | -0,00486 | 0,165613 | 138 | 69 | 69 | 134 | -0,02936 | y ~ Intervention + Sex + |
| 0,975193 | -0,00516 | 0,165489 | 138 | 69 | 69 | 134 | -0,03115 | y ~ Intervention + Sex + |
| 0,974408 | -0,00565 | 0,175947 | 138 | 69 | 69 | 134 | -0,03214 | y ~ Intervention + Sex + |
| 0,968917 | -0,00614 | 0,157365 | 138 | 69 | 69 | 134 | -0,03904 | y ~ Intervention + Sex + |
| 0,96905 | -0,00656 | 0,168637 | 138 | 69 | 69 | 134 | -0,03887 | y ~ Intervention + Sex + |
| 0,963714 | -0,00664 | 0,145764 | 138 | 69 | 69 | 134 | -0,04558 | y ~ Intervention + Sex + |
| 0,968752 | -0,00682 | 0,173738 | 138 | 69 | 69 | 134 | -0,03925 | y ~ Intervention + Sex + |
| 0,962645 | -0,00717 | 0,152717 | 138 | 69 | 69 | 134 | -0,04692 | y ~ Intervention + Sex + |
| 0,9626 | -0,00814 | 0,173242 | 138 | 69 | 69 | 134 | -0,04698 | y ~ Intervention + Sex + |
| 0,953917 | -0,00841 | 0,1453 | 138 | 69 | 69 | 134 | -0,0579 | y ~ Intervention + Sex + |
| 0,947577 | -0,0103 | 0,156302 | 138 | 69 | 69 | 134 | -0,06587 | y ~ Intervention + Sex + |
| 0,950014 | -0,01065 | 0,169509 | 138 | 69 | 69 | 134 | -0,06281 | y ~ Intervention + Sex + |
| 0,944701 | -0,01098 | 0,157957 | 138 | 69 | 69 | 134 | -0,06949 | y ~ Intervention + Sex + |
| 0,946603 | -0,01117 | 0,166495 | 138 | 69 | 69 | 134 | -0,0671 | y ~ Intervention + Sex + |
| 0,940566 | -0,01154 | 0,154543 | 138 | 69 | 69 | 134 | -0,0747 | y ~ Intervention + Sex + |
| 0,940462 | -0,01231 | 0,164531 | 138 | 69 | 69 | 134 | -0,07483 | y ~ Intervention + Sex + |
| 0,944092 | -0,01245 | 0,177165 | 138 | 69 | 69 | 134 | -0,07026 | y ~ Intervention + Sex + |
| 0,939477 | -0,01283 | 0,168662 | 138 | 69 | 69 | 134 | -0,07607 | y ~ Intervention + Sex + |
| 0,937362 | -0,0131 | 0,166424 | 138 | 69 | 69 | 134 | -0,07873 | y ~ Intervention + Sex + |

| | | | | | | | | |
|---|---|---|---|---|---|---|---|---|
| 0,938943 | -0,01312 | 0,17099 | 138 | 69 | 69 | 134 | -0,07674 | y ~ Intervention + Sex + |
| 0,93156 | -0,01382 | 0,160637 | 138 | 69 | 69 | 134 | -0,08604 | y ~ Intervention + Sex + |
| 0,920663 | -0,01612 | 0,16154 | 138 | 69 | 69 | 134 | -0,09979 | y ~ Intervention + Sex + |
| 0,915633 | -0,01668 | 0,15714 | 138 | 69 | 69 | 134 | -0,10614 | y ~ Intervention + Sex + |
| 0,909534 | -0,01697 | 0,149029 | 138 | 69 | 69 | 134 | -0,11384 | y ~ Intervention + Sex + |
| 0,899599 | -0,01825 | 0,144375 | 138 | 69 | 69 | 134 | -0,12641 | y ~ Intervention + Sex + |
| 0,90234 | -0,01904 | 0,154872 | 138 | 69 | 69 | 134 | -0,12294 | y ~ Intervention + Sex + |
| 0,902202 | -0,01913 | 0,155353 | 138 | 69 | 69 | 134 | -0,12311 | y ~ Intervention + Sex + |
| 0,903448 | -0,0193 | 0,158803 | 138 | 69 | 69 | 134 | -0,12154 | y ~ Intervention + Sex + |
| 0,882334 | -0,02022 | 0,136327 | 138 | 69 | 69 | 134 | -0,14829 | y ~ Intervention + Sex + |
| 0,893695 | -0,02114 | 0,157911 | 138 | 69 | 69 | 134 | -0,13388 | y ~ Intervention + Sex + |
| 0,895985 | -0,02136 | 0,163111 | 138 | 69 | 69 | 134 | -0,13098 | y ~ Intervention + Sex + |
| 0,885824 | -0,022 | 0,152919 | 138 | 69 | 69 | 134 | -0,14386 | y ~ Intervention + Sex + |
| 0,897866 | -0,02215 | 0,172208 | 138 | 69 | 69 | 134 | -0,1286 | y ~ Intervention + Sex + |
| 0,887586 | -0,02339 | 0,16513 | 138 | 69 | 69 | 134 | -0,14163 | y ~ Intervention + Sex + |
| 0,884359 | -0,02354 | 0,161531 | 138 | 69 | 69 | 134 | -0,14572 | y ~ Intervention + Sex + |
| 0,877906 | -0,0249 | 0,161749 | 138 | 69 | 69 | 134 | -0,15392 | y ~ Intervention + Sex + |
| 0,880963 | -0,02549 | 0,169899 | 138 | 69 | 69 | 134 | -0,15003 | y ~ Intervention + Sex + |
| 0,870164 | -0,02558 | 0,156226 | 138 | 69 | 69 | 134 | -0,16376 | y ~ Intervention + Sex + |
| 0,859469 | -0,02588 | 0,145906 | 138 | 69 | 69 | 134 | -0,17739 | y ~ Intervention + Sex + |
| 0,850206 | -0,02597 | 0,137234 | 138 | 69 | 69 | 134 | -0,18922 | y ~ Intervention + Sex + |
| 0,854624 | -0,02652 | 0,144444 | 138 | 69 | 69 | 134 | -0,18357 | y ~ Intervention + Sex + |
| 0,855423 | -0,02685 | 0,147093 | 138 | 69 | 69 | 134 | -0,18256 | y ~ Intervention + Sex + |
| 0,868978 | -0,027 | 0,163374 | 138 | 69 | 69 | 134 | -0,16527 | y ~ Intervention + Sex + |
| 0,865441 | -0,02719 | 0,160158 | 138 | 69 | 69 | 134 | -0,16978 | y ~ Intervention + Sex + |
| 0,856244 | -0,02762 | 0,152174 | 138 | 69 | 69 | 134 | -0,18151 | y ~ Intervention + Sex + |
| 0,853908 | -0,02821 | 0,152927 | 138 | 69 | 69 | 134 | -0,18449 | y ~ Intervention + Sex + |
| 0,864815 | -0,02828 | 0,16582 | 138 | 69 | 69 | 134 | -0,17058 | y ~ Intervention + Sex + |
| 0,857922 | -0,02943 | 0,164052 | 138 | 69 | 69 | 134 | -0,17936 | y ~ Intervention + Sex + |
| 0,84943 | -0,02959 | 0,155573 | 138 | 69 | 69 | 134 | -0,19021 | y ~ Intervention + Sex + |
| 0,83724 | -0,0304 | 0,147714 | 138 | 69 | 69 | 134 | -0,20583 | y ~ Intervention + Sex + |
| 0,847786 | -0,03044 | 0,158294 | 138 | 69 | 69 | 134 | -0,19232 | y ~ Intervention + Sex + |
| 0,837517 | -0,03083 | 0,150062 | 138 | 69 | 69 | 134 | -0,20547 | y ~ Intervention + Sex + |
| 0,846604 | -0,03091 | 0,159482 | 138 | 69 | 69 | 134 | -0,19383 | y ~ Intervention + Sex + |
| 0,852373 | -0,03098 | 0,166147 | 138 | 69 | 69 | 134 | -0,18645 | y ~ Intervention + Sex + |
| 0,846661 | -0,03099 | 0,15996 | 138 | 69 | 69 | 134 | -0,19376 | y ~ Intervention + Sex + |
| 0,843143 | -0,03137 | 0,158232 | 138 | 69 | 69 | 134 | -0,19826 | y ~ Intervention + Sex + |
| 0,817134 | -0,03191 | 0,13773 | 138 | 69 | 69 | 134 | -0,23169 | y ~ Intervention + Sex + |
| 0,852946 | -0,03203 | 0,172489 | 138 | 69 | 69 | 134 | -0,18572 | y ~ Intervention + Sex + |
| 0,812437 | -0,03214 | 0,135185 | 138 | 69 | 69 | 134 | -0,23775 | y ~ Intervention + Sex + |
| 0,84637 | -0,03217 | 0,165704 | 138 | 69 | 69 | 134 | -0,19413 | y ~ Intervention + Sex + |
| 0,826255 | -0,03217 | 0,146259 | 138 | 69 | 69 | 134 | -0,21994 | y ~ Intervention + Sex + |
| 0,821459 | -0,03317 | 0,146682 | 138 | 69 | 69 | 134 | -0,22611 | y ~ Intervention + Sex + |
| 0,844311 | -0,03423 | 0,173971 | 138 | 69 | 69 | 134 | -0,19676 | y ~ Intervention + Sex + |
| 0,816474 | -0,03441 | 0,147981 | 138 | 69 | 69 | 134 | -0,23254 | y ~ Intervention + Sex + |
| 0,814555 | -0,0364 | 0,154863 | 138 | 69 | 69 | 134 | -0,23502 | y ~ Intervention + Sex + |
| 0,835721 | -0,03734 | 0,179691 | 138 | 69 | 69 | 134 | -0,20777 | y ~ Intervention + Sex + |
| 0,823617 | -0,03741 | 0,167523 | 138 | 69 | 69 | 134 | -0,22333 | y ~ Intervention + Sex + |

| | | | | | | | | |
|---|---|---|---|---|---|---|---|---|
| 0,825187 | -0,03755 | 0,169662 | 138 | 69 | 69 | 134 | -0,22131 | y ~ Intervention + Sex + |
| 0,831277 | -0,03777 | 0,176942 | 138 | 69 | 69 | 134 | -0,21348 | y ~ Intervention + Sex + |
| 0,83304 | -0,03819 | 0,180822 | 138 | 69 | 69 | 134 | -0,21122 | y ~ Intervention + Sex + |
| 0,802806 | -0,03877 | 0,154947 | 138 | 69 | 69 | 134 | -0,25021 | y ~ Intervention + Sex + |
| 0,822109 | -0,03881 | 0,172272 | 138 | 69 | 69 | 134 | -0,22527 | y ~ Intervention + Sex + |
| 0,802431 | -0,03891 | 0,155219 | 138 | 69 | 69 | 134 | -0,2507 | y ~ Intervention + Sex + |
| 0,813162 | -0,03892 | 0,164361 | 138 | 69 | 69 | 134 | -0,23681 | y ~ Intervention + Sex + |
| 0,80795 | -0,03917 | 0,160845 | 138 | 69 | 69 | 134 | -0,24355 | y ~ Intervention + Sex + |
| 0,805676 | -0,03975 | 0,161249 | 138 | 69 | 69 | 134 | -0,2465 | y ~ Intervention + Sex + |
| 0,799105 | -0,03976 | 0,155896 | 138 | 69 | 69 | 134 | -0,25501 | y ~ Intervention + Sex + |
| 0,808286 | -0,03989 | 0,164064 | 138 | 69 | 69 | 134 | -0,24312 | y ~ Intervention + Sex + |
| 0,77093 | -0,04039 | 0,138432 | 138 | 69 | 69 | 134 | -0,29175 | y ~ Intervention + Sex + |
| 0,798191 | -0,04049 | 0,158056 | 138 | 69 | 69 | 134 | -0,2562 | y ~ Intervention + Sex + |
| 0,801719 | -0,04091 | 0,16258 | 138 | 69 | 69 | 134 | -0,25162 | y ~ Intervention + Sex + |
| 0,751842 | -0,04155 | 0,131129 | 138 | 69 | 69 | 134 | -0,31686 | y ~ Intervention + Sex + |
| 0,781513 | -0,04155 | 0,14952 | 138 | 69 | 69 | 134 | -0,27791 | y ~ Intervention + Sex + |
| 0,79647 | -0,04159 | 0,160951 | 138 | 69 | 69 | 134 | -0,25843 | y ~ Intervention + Sex + |
| 0,753159 | -0,04163 | 0,132102 | 138 | 69 | 69 | 134 | -0,31512 | y ~ Intervention + Sex + |
| 0,781969 | -0,04206 | 0,151674 | 138 | 69 | 69 | 134 | -0,27731 | y ~ Intervention + Sex + |
| 0,782358 | -0,04366 | 0,157716 | 138 | 69 | 69 | 134 | -0,2768 | y ~ Intervention + Sex + |
| 0,786256 | -0,04375 | 0,161026 | 138 | 69 | 69 | 134 | -0,27172 | y ~ Intervention + Sex + |
| 0,786284 | -0,04387 | 0,161482 | 138 | 69 | 69 | 134 | -0,27168 | y ~ Intervention + Sex + |
| 0,789805 | -0,0451 | 0,168852 | 138 | 69 | 69 | 134 | -0,2671 | y ~ Intervention + Sex + |
| 0,773528 | -0,04555 | 0,157977 | 138 | 69 | 69 | 134 | -0,28835 | y ~ Intervention + Sex + |
| 0,789415 | -0,04558 | 0,17034 | 138 | 69 | 69 | 134 | -0,26761 | y ~ Intervention + Sex + |
| 0,76947 | -0,04584 | 0,156114 | 138 | 69 | 69 | 134 | -0,29366 | y ~ Intervention + Sex + |
| 0,760549 | -0,04648 | 0,152206 | 138 | 69 | 69 | 134 | -0,30538 | y ~ Intervention + Sex + |
| 0,768986 | -0,0468 | 0,159025 | 138 | 69 | 69 | 134 | -0,2943 | y ~ Intervention + Sex + |
| 0,744498 | -0,04698 | 0,143853 | 138 | 69 | 69 | 134 | -0,32658 | y ~ Intervention + Sex + |
| 0,743828 | -0,04702 | 0,143591 | 138 | 69 | 69 | 134 | -0,32746 | y ~ Intervention + Sex + |
| 0,778844 | -0,04722 | 0,167793 | 138 | 69 | 69 | 134 | -0,28139 | y ~ Intervention + Sex + |
| 0,742442 | -0,04756 | 0,144441 | 138 | 69 | 69 | 134 | -0,3293 | y ~ Intervention + Sex + |
| 0,766057 | -0,04808 | 0,161266 | 138 | 69 | 69 | 134 | -0,29814 | y ~ Intervention + Sex + |
| 0,746333 | -0,04829 | 0,148965 | 138 | 69 | 69 | 134 | -0,32415 | y ~ Intervention + Sex + |
| 0,762791 | -0,04843 | 0,160121 | 138 | 69 | 69 | 134 | -0,30243 | y ~ Intervention + Sex + |
| 0,748015 | -0,04868 | 0,151222 | 138 | 69 | 69 | 134 | -0,32192 | y ~ Intervention + Sex + |
| 0,75143 | -0,04991 | 0,157236 | 138 | 69 | 69 | 134 | -0,3174 | y ~ Intervention + Sex + |
| 0,755529 | -0,05117 | 0,164001 | 138 | 69 | 69 | 134 | -0,312 | y ~ Intervention + Sex + |
| 0,751199 | -0,05128 | 0,161413 | 138 | 69 | 69 | 134 | -0,31771 | y ~ Intervention + Sex + |
| 0,743507 | -0,05143 | 0,156855 | 138 | 69 | 69 | 134 | -0,32789 | y ~ Intervention + Sex + |
| 0,70775 | -0,05155 | 0,13721 | 138 | 69 | 69 | 134 | -0,37568 | y ~ Intervention + Sex + |
| 0,735299 | -0,05159 | 0,15227 | 138 | 69 | 69 | 134 | -0,33879 | y ~ Intervention + Sex + |
| 0,74847 | -0,05174 | 0,161016 | 138 | 69 | 69 | 134 | -0,32132 | y ~ Intervention + Sex + |
| 0,724189 | -0,05212 | 0,147393 | 138 | 69 | 69 | 134 | -0,35361 | y ~ Intervention + Sex + |
| 0,738266 | -0,0527 | 0,157382 | 138 | 69 | 69 | 134 | -0,33484 | y ~ Intervention + Sex + |
| 0,737097 | -0,05302 | 0,157602 | 138 | 69 | 69 | 134 | -0,3364 | y ~ Intervention + Sex + |
| 0,749849 | -0,05348 | 0,167387 | 138 | 69 | 69 | 134 | -0,31949 | y ~ Intervention + Sex + |
| 0,730881 | -0,05348 | 0,155172 | 138 | 69 | 69 | 134 | -0,34467 | y ~ Intervention + Sex + |

| | | | | | | | | |
|---|---|---|---|---|---|---|---|---|
| 0,727625 | -0,0537 | 0,153872 | 138 | 69 | 69 | 134 | -0,34902 | y ~ Intervention + Sex + |
| 0,745146 | -0,05437 | 0,166916 | 138 | 69 | 69 | 134 | -0,32572 | y ~ Intervention + Sex + |
| 0,744477 | -0,05502 | 0,168468 | 138 | 69 | 69 | 134 | -0,3266 | y ~ Intervention + Sex + |
| 0,74107 | -0,05549 | 0,167591 | 138 | 69 | 69 | 134 | -0,33112 | y ~ Intervention + Sex + |
| 0,725128 | -0,05553 | 0,15759 | 138 | 69 | 69 | 134 | -0,35235 | y ~ Intervention + Sex + |
| 0,745936 | -0,05573 | 0,171638 | 138 | 69 | 69 | 134 | -0,32467 | y ~ Intervention + Sex + |
| 0,721297 | -0,05642 | 0,157828 | 138 | 69 | 69 | 134 | -0,35748 | y ~ Intervention + Sex + |
| 0,706079 | -0,05715 | 0,151222 | 138 | 69 | 69 | 134 | -0,37793 | y ~ Intervention + Sex + |
| 0,721852 | -0,05743 | 0,160987 | 138 | 69 | 69 | 134 | -0,35673 | y ~ Intervention + Sex + |
| 0,720158 | -0,05851 | 0,162989 | 138 | 69 | 69 | 134 | -0,359 | y ~ Intervention + Sex + |
| 0,717841 | -0,05874 | 0,162214 | 138 | 69 | 69 | 134 | -0,36211 | y ~ Intervention + Sex + |
| 0,657729 | -0,05914 | 0,133176 | 138 | 69 | 69 | 134 | -0,44404 | y ~ Intervention + Sex + |
| 0,656188 | -0,06039 | 0,135352 | 138 | 69 | 69 | 134 | -0,44618 | y ~ Intervention + Sex + |
| 0,728481 | -0,06039 | 0,173611 | 138 | 69 | 69 | 134 | -0,34787 | y ~ Intervention + Sex + |
| 0,710611 | -0,06041 | 0,162457 | 138 | 69 | 69 | 134 | -0,37182 | y ~ Intervention + Sex + |
| 0,719517 | -0,06044 | 0,167961 | 138 | 69 | 69 | 134 | -0,35986 | y ~ Intervention + Sex + |
| 0,669237 | -0,06082 | 0,142052 | 138 | 69 | 69 | 134 | -0,42814 | y ~ Intervention + Sex + |
| 0,692853 | -0,06143 | 0,155187 | 138 | 69 | 69 | 134 | -0,39584 | y ~ Intervention + Sex + |
| 0,651253 | -0,06237 | 0,137675 | 138 | 69 | 69 | 134 | -0,45304 | y ~ Intervention + Sex + |
| 0,696908 | -0,06285 | 0,161022 | 138 | 69 | 69 | 134 | -0,39034 | y ~ Intervention + Sex + |
| 0,68914 | -0,06288 | 0,156861 | 138 | 69 | 69 | 134 | -0,40089 | y ~ Intervention + Sex + |
| 0,67181 | -0,06395 | 0,150619 | 138 | 69 | 69 | 134 | -0,4246 | y ~ Intervention + Sex + |
| 0,660128 | -0,06446 | 0,146256 | 138 | 69 | 69 | 134 | -0,44072 | y ~ Intervention + Sex + |
| 0,671732 | -0,06453 | 0,151931 | 138 | 69 | 69 | 134 | -0,42471 | y ~ Intervention + Sex + |
| 0,610384 | -0,06586 | 0,128961 | 138 | 69 | 69 | 134 | -0,51072 | y ~ Intervention + Sex + |
| 0,684686 | -0,06667 | 0,163835 | 138 | 69 | 69 | 134 | -0,40696 | y ~ Intervention + Sex + |
| 0,707246 | -0,06737 | 0,179004 | 138 | 69 | 69 | 134 | -0,37636 | y ~ Intervention + Sex + |
| 0,635432 | -0,06778 | 0,142643 | 138 | 69 | 69 | 134 | -0,47518 | y ~ Intervention + Sex + |
| 0,687006 | -0,06787 | 0,16809 | 138 | 69 | 69 | 134 | -0,4038 | y ~ Intervention + Sex + |
| 0,691995 | -0,06788 | 0,170968 | 138 | 69 | 69 | 134 | -0,39701 | y ~ Intervention + Sex + |
| 0,677814 | -0,06906 | 0,165876 | 138 | 69 | 69 | 134 | -0,41636 | y ~ Intervention + Sex + |
| 0,612339 | -0,06927 | 0,13638 | 138 | 69 | 69 | 134 | -0,50793 | y ~ Intervention + Sex + |
| 0,613857 | -0,07006 | 0,138523 | 138 | 69 | 69 | 134 | -0,50576 | y ~ Intervention + Sex + |
| 0,640311 | -0,07016 | 0,14982 | 138 | 69 | 69 | 134 | -0,46833 | y ~ Intervention + Sex + |
| 0,672692 | -0,07035 | 0,166153 | 138 | 69 | 69 | 134 | -0,42339 | y ~ Intervention + Sex + |
| 0,627694 | -0,07079 | 0,145623 | 138 | 69 | 69 | 134 | -0,48609 | y ~ Intervention + Sex + |
| 0,656072 | -0,07106 | 0,159206 | 138 | 69 | 69 | 134 | -0,44634 | y ~ Intervention + Sex + |
| 0,620944 | -0,07113 | 0,143501 | 138 | 69 | 69 | 134 | -0,49566 | y ~ Intervention + Sex + |
| 0,668708 | -0,07114 | 0,165886 | 138 | 69 | 69 | 134 | -0,42887 | y ~ Intervention + Sex + |
| 0,666362 | -0,07149 | 0,165456 | 138 | 69 | 69 | 134 | -0,4321 | y ~ Intervention + Sex + |
| 0,657454 | -0,07184 | 0,161655 | 138 | 69 | 69 | 134 | -0,44442 | y ~ Intervention + Sex + |
| 0,658782 | -0,07187 | 0,162398 | 138 | 69 | 69 | 134 | -0,44258 | y ~ Intervention + Sex + |
| 0,616443 | -0,07211 | 0,143618 | 138 | 69 | 69 | 134 | -0,50207 | y ~ Intervention + Sex + |
| 0,568896 | -0,07321 | 0,128198 | 138 | 69 | 69 | 134 | -0,57109 | y ~ Intervention + Sex + |
| 0,649963 | -0,07409 | 0,162884 | 138 | 69 | 69 | 134 | -0,45484 | y ~ Intervention + Sex + |
| 0,646119 | -0,07449 | 0,161864 | 138 | 69 | 69 | 134 | -0,4602 | y ~ Intervention + Sex + |
| 0,624142 | -0,07473 | 0,152167 | 138 | 69 | 69 | 134 | -0,49112 | y ~ Intervention + Sex + |
| 0,652519 | -0,07476 | 0,165658 | 138 | 69 | 69 | 134 | -0,45128 | y ~ Intervention + Sex + |

| | | | | | | | | |
|---|---|---|---|---|---|---|---|---|
| 0,61865 | -0,07549 | 0,151298 | 138 | 69 | 69 | 134 | -0,49893 | y ~ Intervention + Sex + |
| 0,585151 | -0,07653 | 0,13985 | 138 | 69 | 69 | 134 | -0,5472 | y ~ Intervention + Sex + |
| 0,652203 | -0,07699 | 0,170429 | 138 | 69 | 69 | 134 | -0,45172 | y ~ Intervention + Sex + |
| 0,581131 | -0,07701 | 0,139236 | 138 | 69 | 69 | 134 | -0,55308 | y ~ Intervention + Sex + |
| 0,589982 | -0,07707 | 0,142676 | 134 | 67 | 67 | 130 | -0,5402 | y ~ Intervention + Sex + |
| 0,61145 | -0,07716 | 0,15154 | 138 | 69 | 69 | 134 | -0,5092 | y ~ Intervention + Sex + |
| 0,617896 | -0,0773 | 0,15459 | 138 | 69 | 69 | 134 | -0,5 | y ~ Intervention + Sex + |
| 0,645814 | -0,07758 | 0,168432 | 138 | 69 | 69 | 134 | -0,46063 | y ~ Intervention + Sex + |
| 0,610577 | -0,07762 | 0,152066 | 138 | 69 | 69 | 134 | -0,51045 | y ~ Intervention + Sex + |
| 0,638613 | -0,07854 | 0,166857 | 138 | 69 | 69 | 134 | -0,47071 | y ~ Intervention + Sex + |
| 0,615767 | -0,07889 | 0,156824 | 138 | 69 | 69 | 134 | -0,50303 | y ~ Intervention + Sex + |
| 0,642435 | -0,07909 | 0,169967 | 138 | 69 | 69 | 134 | -0,46535 | y ~ Intervention + Sex + |
| 0,604101 | -0,07914 | 0,152276 | 138 | 69 | 69 | 134 | -0,51974 | y ~ Intervention + Sex + |
| 0,634651 | -0,07921 | 0,166315 | 138 | 69 | 69 | 134 | -0,47628 | y ~ Intervention + Sex + |
| 0,575791 | -0,07929 | 0,141352 | 138 | 69 | 69 | 134 | -0,56092 | y ~ Intervention + Sex + |
| 0,591259 | -0,07999 | 0,148601 | 138 | 69 | 69 | 134 | -0,53831 | y ~ Intervention + Sex + |
| 0,625278 | -0,08044 | 0,164318 | 138 | 69 | 69 | 134 | -0,48951 | y ~ Intervention + Sex + |
| 0,623112 | -0,08063 | 0,163679 | 138 | 69 | 69 | 134 | -0,49258 | y ~ Intervention + Sex + |
| 0,625438 | -0,08091 | 0,165363 | 138 | 69 | 69 | 134 | -0,48929 | y ~ Intervention + Sex + |
| 0,585938 | -0,08099 | 0,148311 | 138 | 69 | 69 | 134 | -0,54605 | y ~ Intervention + Sex + |
| 0,63866 | -0,08109 | 0,172288 | 138 | 69 | 69 | 134 | -0,47064 | y ~ Intervention + Sex + |
| 0,632196 | -0,08138 | 0,169644 | 138 | 69 | 69 | 134 | -0,47974 | y ~ Intervention + Sex + |
| 0,595538 | -0,08162 | 0,153389 | 138 | 69 | 69 | 134 | -0,5321 | y ~ Intervention + Sex + |
| 0,59484 | -0,08308 | 0,155836 | 138 | 69 | 69 | 134 | -0,53311 | y ~ Intervention + Sex + |
| 0,621609 | -0,08309 | 0,167963 | 138 | 69 | 69 | 134 | -0,49472 | y ~ Intervention + Sex + |
| 0,485142 | -0,0832 | 0,118856 | 138 | 69 | 69 | 134 | -0,7 | y ~ Intervention + Sex + |
| 0,579707 | -0,08338 | 0,150184 | 138 | 69 | 69 | 134 | -0,55517 | y ~ Intervention + Sex + |
| 0,599195 | -0,08371 | 0,158898 | 138 | 69 | 69 | 134 | -0,52681 | y ~ Intervention + Sex + |
| 0,569722 | -0,08399 | 0,147378 | 138 | 69 | 69 | 134 | -0,56987 | y ~ Intervention + Sex + |
| 0,620494 | -0,08477 | 0,170794 | 138 | 69 | 69 | 134 | -0,4963 | y ~ Intervention + Sex + |
| 0,617193 | -0,08522 | 0,170092 | 138 | 69 | 69 | 134 | -0,501 | y ~ Intervention + Sex + |
| 0,559663 | -0,0856 | 0,146367 | 138 | 69 | 69 | 134 | -0,5848 | y ~ Intervention + Sex + |
| 0,618645 | -0,08609 | 0,172555 | 138 | 69 | 69 | 134 | -0,49893 | y ~ Intervention + Sex + |
| 0,616936 | -0,08657 | 0,172658 | 138 | 69 | 69 | 134 | -0,50137 | y ~ Intervention + Sex + |
| 0,569199 | -0,08661 | 0,15178 | 138 | 69 | 69 | 134 | -0,57064 | y ~ Intervention + Sex + |
| 0,541808 | -0,08682 | 0,141946 | 138 | 69 | 69 | 134 | -0,61165 | y ~ Intervention + Sex + |
| 0,572208 | -0,08687 | 0,153423 | 138 | 69 | 69 | 134 | -0,5662 | y ~ Intervention + Sex + |
| 0,574688 | -0,08698 | 0,154622 | 138 | 69 | 69 | 134 | -0,56254 | y ~ Intervention + Sex + |
| 0,57931 | -0,08727 | 0,157038 | 138 | 69 | 69 | 134 | -0,55575 | y ~ Intervention + Sex + |
| 0,585851 | -0,08741 | 0,160034 | 138 | 69 | 69 | 134 | -0,54618 | y ~ Intervention + Sex + |
| 0,572872 | -0,08741 | 0,154655 | 138 | 69 | 69 | 134 | -0,56522 | y ~ Intervention + Sex + |
| 0,547757 | -0,08782 | 0,145718 | 138 | 69 | 69 | 134 | -0,60265 | y ~ Intervention + Sex + |
| 0,593128 | -0,08789 | 0,164102 | 138 | 69 | 69 | 134 | -0,53559 | y ~ Intervention + Sex + |
| 0,606299 | -0,08793 | 0,170216 | 138 | 69 | 69 | 134 | -0,51658 | y ~ Intervention + Sex + |
| 0,583649 | -0,08858 | 0,161231 | 138 | 69 | 69 | 134 | -0,54939 | y ~ Intervention + Sex + |
| 0,547912 | -0,08897 | 0,147686 | 138 | 69 | 69 | 134 | -0,60242 | y ~ Intervention + Sex + |
| 0,565293 | -0,08915 | 0,154668 | 138 | 69 | 69 | 134 | -0,57643 | y ~ Intervention + Sex + |
| 0,555786 | -0,09028 | 0,152866 | 138 | 69 | 69 | 134 | -0,5906 | y ~ Intervention + Sex + |

| | | | | | | | | |
|---|---|---|---|---|---|---|---|---|
| 0,545224 | -0,09067 | 0,149496 | 138 | 69 | 69 | 134 | -0,60648 | y ~ Intervention + Sex + |
| 0,576182 | -0,0912 | 0,162763 | 138 | 69 | 69 | 134 | -0,56034 | y ~ Intervention + Sex + |
| 0,575221 | -0,09239 | 0,164468 | 138 | 69 | 69 | 134 | -0,56176 | y ~ Intervention + Sex + |
| 0,570501 | -0,09242 | 0,162506 | 138 | 69 | 69 | 134 | -0,56872 | y ~ Intervention + Sex + |
| 0,562292 | -0,09271 | 0,159604 | 138 | 69 | 69 | 134 | -0,58089 | y ~ Intervention + Sex + |
| 0,568052 | -0,09302 | 0,162532 | 138 | 69 | 69 | 134 | -0,57234 | y ~ Intervention + Sex + |
| 0,566013 | -0,09345 | 0,162427 | 138 | 69 | 69 | 134 | -0,57536 | y ~ Intervention + Sex + |
| 0,565419 | -0,09388 | 0,162915 | 138 | 69 | 69 | 134 | -0,57624 | y ~ Intervention + Sex + |
| 0,504697 | -0,09539 | 0,142606 | 138 | 69 | 69 | 134 | -0,66892 | y ~ Intervention + Sex + |
| 0,501644 | -0,09568 | 0,142013 | 138 | 69 | 69 | 134 | -0,67373 | y ~ Intervention + Sex + |
| 0,556279 | -0,09602 | 0,162793 | 138 | 69 | 69 | 134 | -0,58986 | y ~ Intervention + Sex + |
| 0,536038 | -0,09607 | 0,154841 | 138 | 69 | 69 | 134 | -0,62042 | y ~ Intervention + Sex + |
| 0,555462 | -0,09632 | 0,162961 | 138 | 69 | 69 | 134 | -0,59108 | y ~ Intervention + Sex + |
| 0,579698 | -0,09651 | 0,173831 | 138 | 69 | 69 | 134 | -0,55518 | y ~ Intervention + Sex + |
| 0,562333 | -0,0966 | 0,166319 | 138 | 69 | 69 | 134 | -0,58083 | y ~ Intervention + Sex + |
| 0,573556 | -0,09674 | 0,17147 | 138 | 69 | 69 | 134 | -0,56421 | y ~ Intervention + Sex + |
| 0,567261 | -0,09698 | 0,169092 | 138 | 69 | 69 | 134 | -0,57351 | y ~ Intervention + Sex + |
| 0,51321 | -0,09707 | 0,148069 | 138 | 69 | 69 | 134 | -0,65559 | y ~ Intervention + Sex + |
| 0,52303 | -0,09715 | 0,151711 | 138 | 69 | 69 | 134 | -0,64036 | y ~ Intervention + Sex + |
| 0,532219 | -0,09725 | 0,155283 | 138 | 69 | 69 | 134 | -0,62625 | y ~ Intervention + Sex + |
| 0,507221 | -0,09917 | 0,149142 | 138 | 69 | 69 | 134 | -0,66496 | y ~ Intervention + Sex + |
| 0,491886 | -0,10025 | 0,14546 | 138 | 69 | 69 | 134 | -0,68921 | y ~ Intervention + Sex + |
| 0,513718 | -0,10061 | 0,153655 | 138 | 69 | 69 | 134 | -0,6548 | y ~ Intervention + Sex + |
| 0,49164 | -0,10109 | 0,14659 | 138 | 69 | 69 | 134 | -0,6896 | y ~ Intervention + Sex + |
| 0,561279 | -0,10165 | 0,174542 | 138 | 69 | 69 | 134 | -0,5824 | y ~ Intervention + Sex + |
| 0,538162 | -0,1019 | 0,165103 | 138 | 69 | 69 | 134 | -0,61718 | y ~ Intervention + Sex + |
| 0,523367 | -0,10256 | 0,16029 | 138 | 69 | 69 | 134 | -0,63984 | y ~ Intervention + Sex + |
| 0,537948 | -0,1035 | 0,167614 | 138 | 69 | 69 | 134 | -0,61751 | y ~ Intervention + Sex + |
| 0,51004 | -0,10388 | 0,15726 | 138 | 69 | 69 | 134 | -0,66054 | y ~ Intervention + Sex + |
| 0,534435 | -0,10505 | 0,168655 | 138 | 69 | 69 | 134 | -0,62286 | y ~ Intervention + Sex + |
| 0,544887 | -0,10554 | 0,173879 | 138 | 69 | 69 | 134 | -0,60699 | y ~ Intervention + Sex + |
| 0,467321 | -0,10627 | 0,145797 | 138 | 69 | 69 | 134 | -0,72892 | y ~ Intervention + Sex + |
| 0,483646 | -0,10672 | 0,151931 | 138 | 69 | 69 | 134 | -0,7024 | y ~ Intervention + Sex + |
| 0,39264 | -0,10778 | 0,12568 | 138 | 69 | 69 | 134 | -0,85761 | y ~ Intervention + Sex + |
| 0,491334 | -0,10795 | 0,156431 | 138 | 69 | 69 | 134 | -0,69008 | y ~ Intervention + Sex + |
| 0,451738 | -0,10798 | 0,143069 | 138 | 69 | 69 | 134 | -0,75473 | y ~ Intervention + Sex + |
| 0,523298 | -0,10802 | 0,168791 | 138 | 69 | 69 | 134 | -0,63995 | y ~ Intervention + Sex + |
| 0,502406 | -0,10803 | 0,160636 | 138 | 69 | 69 | 134 | -0,67253 | y ~ Intervention + Sex + |
| 0,4613 | -0,11007 | 0,148984 | 138 | 69 | 69 | 134 | -0,73883 | y ~ Intervention + Sex + |
| 0,470876 | -0,11093 | 0,153408 | 138 | 69 | 69 | 134 | -0,7231 | y ~ Intervention + Sex + |
| 0,476136 | -0,11102 | 0,15537 | 138 | 69 | 69 | 134 | -0,71454 | y ~ Intervention + Sex + |
| 0,468427 | -0,11159 | 0,153477 | 138 | 69 | 69 | 134 | -0,72711 | y ~ Intervention + Sex + |
| 0,417537 | -0,11187 | 0,137567 | 138 | 69 | 69 | 134 | -0,81321 | y ~ Intervention + Sex + |
| 0,445649 | -0,11205 | 0,146483 | 138 | 69 | 69 | 134 | -0,76495 | y ~ Intervention + Sex + |
| 0,465565 | -0,11232 | 0,153482 | 138 | 69 | 69 | 134 | -0,73181 | y ~ Intervention + Sex + |
| 0,492273 | -0,11286 | 0,163898 | 138 | 69 | 69 | 134 | -0,68859 | y ~ Intervention + Sex + |
| 0,488499 | -0,11294 | 0,162594 | 138 | 69 | 69 | 134 | -0,69461 | y ~ Intervention + Sex + |
| 0,474019 | -0,11295 | 0,157311 | 138 | 69 | 69 | 134 | -0,71798 | y ~ Intervention + Sex + |

| | | | | | | | | |
|---|---|---|---|---|---|---|---|---|
| 0,460239 | -0,113 | 0,152578 | 138 | 69 | 69 | 134 | -0,74059 | y ~ Intervention + Sex + |
| 0,489479 | -0,11366 | 0,164 | 138 | 69 | 69 | 134 | -0,69305 | y ~ Intervention + Sex + |
| 0,413999 | -0,11443 | 0,139649 | 138 | 69 | 69 | 134 | -0,81942 | y ~ Intervention + Sex + |
| 0,470773 | -0,11542 | 0,159587 | 138 | 69 | 69 | 134 | -0,72327 | y ~ Intervention + Sex + |
| 0,444021 | -0,11704 | 0,152458 | 138 | 69 | 69 | 134 | -0,76769 | y ~ Intervention + Sex + |
| 0,485085 | -0,11768 | 0,168093 | 138 | 69 | 69 | 134 | -0,70009 | y ~ Intervention + Sex + |
| 0,431532 | -0,11811 | 0,149699 | 138 | 69 | 69 | 134 | -0,78895 | y ~ Intervention + Sex + |
| 0,395602 | -0,11819 | 0,13868 | 138 | 69 | 69 | 134 | -0,85224 | y ~ Intervention + Sex + |
| 0,441563 | -0,11846 | 0,153479 | 138 | 69 | 69 | 134 | -0,77185 | y ~ Intervention + Sex + |
| 0,394878 | -0,11859 | 0,138934 | 138 | 69 | 69 | 134 | -0,85355 | y ~ Intervention + Sex + |
| 0,476393 | -0,11861 | 0,166088 | 138 | 69 | 69 | 134 | -0,71412 | y ~ Intervention + Sex + |
| 0,453129 | -0,11877 | 0,157848 | 138 | 69 | 69 | 134 | -0,7524 | y ~ Intervention + Sex + |
| 0,482624 | -0,11982 | 0,170193 | 138 | 69 | 69 | 134 | -0,70405 | y ~ Intervention + Sex + |
| 0,444244 | -0,12024 | 0,156697 | 138 | 69 | 69 | 134 | -0,76732 | y ~ Intervention + Sex + |
| 0,402475 | -0,12047 | 0,143442 | 138 | 69 | 69 | 134 | -0,83987 | y ~ Intervention + Sex + |
| 0,445704 | -0,12115 | 0,158395 | 138 | 69 | 69 | 134 | -0,76485 | y ~ Intervention + Sex + |
| 0,442776 | -0,12145 | 0,157769 | 138 | 69 | 69 | 134 | -0,7698 | y ~ Intervention + Sex + |
| 0,465558 | -0,12206 | 0,166793 | 138 | 69 | 69 | 134 | -0,73182 | y ~ Intervention + Sex + |
| 0,465469 | -0,12348 | 0,168702 | 138 | 69 | 69 | 134 | -0,73196 | y ~ Intervention + Sex + |
| 0,487053 | -0,12392 | 0,17781 | 138 | 69 | 69 | 134 | -0,69693 | y ~ Intervention + Sex + |
| 0,479815 | -0,12525 | 0,176767 | 138 | 69 | 69 | 134 | -0,70858 | y ~ Intervention + Sex + |
| 0,407818 | -0,12544 | 0,151069 | 138 | 69 | 69 | 134 | -0,83035 | y ~ Intervention + Sex + |
| 0,437172 | -0,12651 | 0,162342 | 138 | 69 | 69 | 134 | -0,77931 | y ~ Intervention + Sex + |
| 0,44031 | -0,12656 | 0,163517 | 138 | 69 | 69 | 134 | -0,77397 | y ~ Intervention + Sex + |
| 0,438399 | -0,12667 | 0,162972 | 138 | 69 | 69 | 134 | -0,77722 | y ~ Intervention + Sex + |
| 0,429946 | -0,12733 | 0,160838 | 138 | 69 | 69 | 134 | -0,79168 | y ~ Intervention + Sex + |
| 0,372694 | -0,12756 | 0,142611 | 138 | 69 | 69 | 134 | -0,89443 | y ~ Intervention + Sex + |
| 0,463551 | -0,12846 | 0,17475 | 138 | 69 | 69 | 134 | -0,73512 | y ~ Intervention + Sex + |
| 0,413777 | -0,12942 | 0,157863 | 138 | 69 | 69 | 134 | -0,81982 | y ~ Intervention + Sex + |
| 0,406968 | -0,12996 | 0,156229 | 138 | 69 | 69 | 134 | -0,83186 | y ~ Intervention + Sex + |
| 0,401671 | -0,13032 | 0,154895 | 138 | 69 | 69 | 134 | -0,84131 | y ~ Intervention + Sex + |
| 0,354429 | -0,13109 | 0,141068 | 138 | 69 | 69 | 134 | -0,92925 | y ~ Intervention + Sex + |
| 0,424363 | -0,13128 | 0,163824 | 138 | 69 | 69 | 134 | -0,80132 | y ~ Intervention + Sex + |
| 0,429443 | -0,13144 | 0,165839 | 138 | 69 | 69 | 134 | -0,79255 | y ~ Intervention + Sex + |
| 0,415071 | -0,13195 | 0,161398 | 138 | 69 | 69 | 134 | -0,81754 | y ~ Intervention + Sex + |
| 0,414369 | -0,13291 | 0,162331 | 138 | 69 | 69 | 134 | -0,81877 | y ~ Intervention + Sex + |
| 0,328018 | -0,13295 | 0,135425 | 138 | 69 | 69 | 134 | -0,9817 | y ~ Intervention + Sex + |
| 0,418926 | -0,13297 | 0,164001 | 138 | 69 | 69 | 134 | -0,81079 | y ~ Intervention + Sex + |
| 0,391841 | -0,13308 | 0,154913 | 138 | 69 | 69 | 134 | -0,85906 | y ~ Intervention + Sex + |
| 0,429432 | -0,13347 | 0,168405 | 138 | 69 | 69 | 134 | -0,79257 | y ~ Intervention + Sex + |
| 0,322524 | -0,13358 | 0,134531 | 138 | 69 | 69 | 134 | -0,99295 | y ~ Intervention + Sex + |
| 0,388812 | -0,13363 | 0,154558 | 138 | 69 | 69 | 134 | -0,86458 | y ~ Intervention + Sex + |
| 0,281401 | -0,13372 | 0,12364 | 138 | 69 | 69 | 134 | -1,08154 | y ~ Intervention + Sex + |
| 0,335891 | -0,13531 | 0,1401 | 138 | 69 | 69 | 134 | -0,96579 | y ~ Intervention + Sex + |
| 0,394982 | -0,13561 | 0,158917 | 138 | 69 | 69 | 134 | -0,85336 | y ~ Intervention + Sex + |
| 0,418043 | -0,13574 | 0,167103 | 138 | 69 | 69 | 134 | -0,81233 | y ~ Intervention + Sex + |
| 0,355978 | -0,13669 | 0,147573 | 138 | 69 | 69 | 134 | -0,92626 | y ~ Intervention + Sex + |
| 0,322147 | -0,13717 | 0,138036 | 138 | 69 | 69 | 134 | -0,99373 | y ~ Intervention + Sex + |

| | | | | | | | | |
|---|---|---|---|---|---|---|---|---|
| 0,396399 | -0,13898 | 0,163348 | 138 | 69 | 69 | 134 | -0,8508 | y ~ Intervention + Sex + |
| 0,337652 | -0,139 | 0,144454 | 138 | 69 | 69 | 134 | -0,96226 | y ~ Intervention + Sex + |
| 0,421038 | -0,13952 | 0,172866 | 138 | 69 | 69 | 134 | -0,8071 | y ~ Intervention + Sex + |
| 0,352535 | -0,14067 | 0,150781 | 138 | 69 | 69 | 134 | -0,93293 | y ~ Intervention + Sex + |
| 0,392666 | -0,14103 | 0,164456 | 138 | 69 | 69 | 134 | -0,85756 | y ~ Intervention + Sex + |
| 0,315446 | -0,14115 | 0,140085 | 138 | 69 | 69 | 134 | -1,00763 | y ~ Intervention + Sex + |
| 0,397648 | -0,14143 | 0,166676 | 138 | 69 | 69 | 134 | -0,84854 | y ~ Intervention + Sex + |
| 0,353958 | -0,1422 | 0,152875 | 138 | 69 | 69 | 134 | -0,93017 | y ~ Intervention + Sex + |
| 0,393262 | -0,14311 | 0,167096 | 138 | 69 | 69 | 134 | -0,85648 | y ~ Intervention + Sex + |
| 0,371868 | -0,14329 | 0,159929 | 138 | 69 | 69 | 134 | -0,89598 | y ~ Intervention + Sex + |
| 0,323622 | -0,14439 | 0,145751 | 138 | 69 | 69 | 134 | -0,99069 | y ~ Intervention + Sex + |
| 0,29837 | -0,14571 | 0,139571 | 138 | 69 | 69 | 134 | -1,04399 | y ~ Intervention + Sex + |
| 0,371831 | -0,14619 | 0,163149 | 138 | 69 | 69 | 134 | -0,89605 | y ~ Intervention + Sex + |
| 0,379735 | -0,1466 | 0,166348 | 138 | 69 | 69 | 134 | -0,8813 | y ~ Intervention + Sex + |
| 0,368637 | -0,14716 | 0,163134 | 138 | 69 | 69 | 134 | -0,90207 | y ~ Intervention + Sex + |
| 0,378603 | -0,14742 | 0,166882 | 138 | 69 | 69 | 134 | -0,8834 | y ~ Intervention + Sex + |
| 0,387438 | -0,14891 | 0,171736 | 138 | 69 | 69 | 134 | -0,8671 | y ~ Intervention + Sex + |
| 0,359915 | -0,14897 | 0,16216 | 138 | 69 | 69 | 134 | -0,91868 | y ~ Intervention + Sex + |
| 0,362221 | -0,14912 | 0,163099 | 138 | 69 | 69 | 134 | -0,91426 | y ~ Intervention + Sex + |
| 0,370434 | -0,15202 | 0,169164 | 138 | 69 | 69 | 134 | -0,89868 | y ~ Intervention + Sex + |
| 0,373566 | -0,15228 | 0,17056 | 138 | 69 | 69 | 134 | -0,8928 | y ~ Intervention + Sex + |
| 0,347002 | -0,15254 | 0,161631 | 138 | 69 | 69 | 134 | -0,94374 | y ~ Intervention + Sex + |
| 0,360883 | -0,15287 | 0,166734 | 138 | 69 | 69 | 134 | -0,91682 | y ~ Intervention + Sex + |
| 0,251083 | -0,15335 | 0,133033 | 138 | 69 | 69 | 134 | -1,15271 | y ~ Intervention + Sex + |
| 0,351778 | -0,15411 | 0,164933 | 138 | 69 | 69 | 134 | -0,9344 | y ~ Intervention + Sex + |
| 0,379269 | -0,15412 | 0,174711 | 138 | 69 | 69 | 134 | -0,88216 | y ~ Intervention + Sex + |
| 0,2473 | -0,15462 | 0,133065 | 138 | 69 | 69 | 134 | -1,162 | y ~ Intervention + Sex + |
| 0,298327 | -0,15465 | 0,148119 | 138 | 69 | 69 | 134 | -1,04408 | y ~ Intervention + Sex + |
| 0,358268 | -0,15519 | 0,168352 | 138 | 69 | 69 | 134 | -0,92184 | y ~ Intervention + Sex + |
| 0,300762 | -0,15523 | 0,149428 | 138 | 69 | 69 | 134 | -1,03881 | y ~ Intervention + Sex + |
| 0,361654 | -0,15542 | 0,169798 | 138 | 69 | 69 | 134 | -0,91535 | y ~ Intervention + Sex + |
| 0,323567 | -0,15587 | 0,157314 | 138 | 69 | 69 | 134 | -0,9908 | y ~ Intervention + Sex + |
| 0,356029 | -0,15607 | 0,168511 | 138 | 69 | 69 | 134 | -0,92616 | y ~ Intervention + Sex + |
| 0,297789 | -0,15635 | 0,149585 | 138 | 69 | 69 | 134 | -1,04525 | y ~ Intervention + Sex + |
| 0,272766 | -0,15678 | 0,142368 | 138 | 69 | 69 | 134 | -1,10124 | y ~ Intervention + Sex + |
| 0,276714 | -0,15688 | 0,143636 | 138 | 69 | 69 | 134 | -1,09218 | y ~ Intervention + Sex + |
| 0,33177 | -0,1578 | 0,162002 | 138 | 69 | 69 | 134 | -0,97408 | y ~ Intervention + Sex + |
| 0,289956 | -0,15838 | 0,149075 | 138 | 69 | 69 | 134 | -1,06242 | y ~ Intervention + Sex + |
| 0,357323 | -0,15876 | 0,171884 | 138 | 69 | 69 | 134 | -0,92366 | y ~ Intervention + Sex + |
| 0,290254 | -0,15938 | 0,150109 | 138 | 69 | 69 | 134 | -1,06176 | y ~ Intervention + Sex + |
| 0,336169 | -0,15939 | 0,165133 | 138 | 69 | 69 | 134 | -0,96523 | y ~ Intervention + Sex + |
| 0,314796 | -0,15951 | 0,158083 | 138 | 69 | 69 | 134 | -1,00899 | y ~ Intervention + Sex + |
| 0,298258 | -0,1596 | 0,152837 | 138 | 69 | 69 | 134 | -1,04423 | y ~ Intervention + Sex + |
| 0,335225 | -0,16019 | 0,165631 | 138 | 69 | 69 | 134 | -0,96712 | y ~ Intervention + Sex + |
| 0,28369 | -0,16223 | 0,150716 | 138 | 69 | 69 | 134 | -1,07638 | y ~ Intervention + Sex + |
| 0,349351 | -0,16295 | 0,173515 | 138 | 69 | 69 | 134 | -0,93914 | y ~ Intervention + Sex + |
| 0,313712 | -0,16309 | 0,161276 | 138 | 69 | 69 | 134 | -1,01127 | y ~ Intervention + Sex + |
| 0,285656 | -0,16342 | 0,152446 | 138 | 69 | 69 | 134 | -1,07198 | y ~ Intervention + Sex + |

| | | | | | | | | |
|---|---|---|---|---|---|---|---|---|
| 0,285747 | -0,16351 | 0,152559 | 138 | 69 | 69 | 134 | -1,07178 | y ~ Intervention + Sex + |
| 0,271592 | -0,16352 | 0,148122 | 138 | 69 | 69 | 134 | -1,10395 | y ~ Intervention + Sex + |
| 0,349388 | -0,16368 | 0,174299 | 138 | 69 | 69 | 134 | -0,93906 | y ~ Intervention + Sex + |
| 0,296011 | -0,16376 | 0,15609 | 138 | 69 | 69 | 134 | -1,04912 | y ~ Intervention + Sex + |
| 0,265995 | -0,16379 | 0,146632 | 138 | 69 | 69 | 134 | -1,117 | y ~ Intervention + Sex + |
| 0,368375 | -0,16418 | 0,181901 | 138 | 69 | 69 | 134 | -0,90257 | y ~ Intervention + Sex + |
| 0,299381 | -0,16477 | 0,158163 | 138 | 69 | 69 | 134 | -1,0418 | y ~ Intervention + Sex + |
| 0,299028 | -0,16551 | 0,158755 | 138 | 69 | 69 | 134 | -1,04256 | y ~ Intervention + Sex + |
| 0,282127 | -0,16569 | 0,153432 | 138 | 69 | 69 | 134 | -1,0799 | y ~ Intervention + Sex + |
| 0,308757 | -0,16665 | 0,163107 | 138 | 69 | 69 | 134 | -1,02172 | y ~ Intervention + Sex + |
| 0,23566 | -0,1677 | 0,140777 | 138 | 69 | 69 | 134 | -1,19125 | y ~ Intervention + Sex + |
| 0,248679 | -0,16807 | 0,145063 | 138 | 69 | 69 | 134 | -1,1586 | y ~ Intervention + Sex + |
| 0,327912 | -0,1699 | 0,17303 | 138 | 69 | 69 | 134 | -0,98191 | y ~ Intervention + Sex + |
| 0,303165 | -0,17052 | 0,164966 | 138 | 69 | 69 | 134 | -1,03364 | y ~ Intervention + Sex + |
| 0,319665 | -0,17104 | 0,171236 | 138 | 69 | 69 | 134 | -0,99886 | y ~ Intervention + Sex + |
| 0,297531 | -0,17228 | 0,164731 | 138 | 69 | 69 | 134 | -1,04581 | y ~ Intervention + Sex + |
| 0,329441 | -0,1727 | 0,176444 | 138 | 69 | 69 | 134 | -0,9788 | y ~ Intervention + Sex + |
| 0,303643 | -0,1729 | 0,167441 | 138 | 69 | 69 | 134 | -1,03262 | y ~ Intervention + Sex + |
| 0,257013 | -0,174 | 0,152856 | 138 | 69 | 69 | 134 | -1,13833 | y ~ Intervention + Sex + |
| 0,26634 | -0,17527 | 0,157022 | 138 | 69 | 69 | 134 | -1,11619 | y ~ Intervention + Sex + |
| 0,198661 | -0,17713 | 0,137122 | 138 | 69 | 69 | 134 | -1,29177 | y ~ Intervention + Sex + |
| 0,188533 | -0,17724 | 0,134108 | 138 | 69 | 69 | 134 | -1,32166 | y ~ Intervention + Sex + |
| 0,228564 | -0,17795 | 0,147111 | 138 | 69 | 69 | 134 | -1,20959 | y ~ Intervention + Sex + |
| 0,208286 | -0,17932 | 0,141822 | 138 | 69 | 69 | 134 | -1,26439 | y ~ Intervention + Sex + |
| 0,275925 | -0,17969 | 0,164256 | 138 | 69 | 69 | 134 | -1,09398 | y ~ Intervention + Sex + |
| 0,245996 | -0,18029 | 0,154727 | 138 | 69 | 69 | 134 | -1,16523 | y ~ Intervention + Sex + |
| 0,284708 | -0,18071 | 0,168247 | 138 | 69 | 69 | 134 | -1,0741 | y ~ Intervention + Sex + |
| 0,285572 | -0,18142 | 0,169204 | 138 | 69 | 69 | 134 | -1,07217 | y ~ Intervention + Sex + |
| 0,24789 | -0,18158 | 0,156464 | 138 | 69 | 69 | 134 | -1,16055 | y ~ Intervention + Sex + |
| 0,262187 | -0,18243 | 0,162023 | 138 | 69 | 69 | 134 | -1,12598 | y ~ Intervention + Sex + |
| 0,210129 | -0,18271 | 0,145092 | 138 | 69 | 69 | 134 | -1,25925 | y ~ Intervention + Sex + |
| 0,164532 | -0,18336 | 0,13119 | 138 | 69 | 69 | 134 | -1,39764 | y ~ Intervention + Sex + |
| 0,29525 | -0,18471 | 0,175787 | 138 | 69 | 69 | 134 | -1,05078 | y ~ Intervention + Sex + |
| 0,268578 | -0,18602 | 0,167442 | 138 | 69 | 69 | 134 | -1,11095 | y ~ Intervention + Sex + |
| 0,271369 | -0,18675 | 0,169087 | 138 | 69 | 69 | 134 | -1,10447 | y ~ Intervention + Sex + |
| 0,240556 | -0,18678 | 0,158446 | 138 | 69 | 69 | 134 | -1,17883 | y ~ Intervention + Sex + |
| 0,280981 | -0,18682 | 0,172589 | 138 | 69 | 69 | 134 | -1,08249 | y ~ Intervention + Sex + |
| 0,275275 | -0,18684 | 0,170557 | 138 | 69 | 69 | 134 | -1,09547 | y ~ Intervention + Sex + |
| 0,234724 | -0,187 | 0,156658 | 138 | 69 | 69 | 134 | -1,19365 | y ~ Intervention + Sex + |
| 0,20807 | -0,18743 | 0,148166 | 138 | 69 | 69 | 134 | -1,26499 | y ~ Intervention + Sex + |
| 0,275693 | -0,1896 | 0,173224 | 138 | 69 | 69 | 134 | -1,09451 | y ~ Intervention + Sex + |
| 0,226663 | -0,18976 | 0,156234 | 138 | 69 | 69 | 134 | -1,21458 | y ~ Intervention + Sex + |
| 0,24992 | -0,18983 | 0,164272 | 138 | 69 | 69 | 134 | -1,15555 | y ~ Intervention + Sex + |
| 0,244449 | -0,18995 | 0,162481 | 138 | 69 | 69 | 134 | -1,16907 | y ~ Intervention + Sex + |
| 0,247864 | -0,19031 | 0,163974 | 138 | 69 | 69 | 134 | -1,16061 | y ~ Intervention + Sex + |
| 0,173012 | -0,19087 | 0,139334 | 138 | 69 | 69 | 134 | -1,36989 | y ~ Intervention + Sex + |
| 0,245629 | -0,19213 | 0,164761 | 138 | 69 | 69 | 134 | -1,16614 | y ~ Intervention + Sex + |
| 0,179959 | -0,19236 | 0,142709 | 138 | 69 | 69 | 134 | -1,34792 | y ~ Intervention + Sex + |

| | | | | | | | | |
|---|---|---|---|---|---|---|---|---|
| 0,226473 | -0,19288 | 0,158735 | 138 | 69 | 69 | 134 | -1,21508 | y ~ Intervention + Sex + |
| 0,219579 | -0,19315 | 0,156599 | 138 | 69 | 69 | 134 | -1,23342 | y ~ Intervention + Sex + |
| 0,195993 | -0,19344 | 0,148852 | 138 | 69 | 69 | 134 | -1,29953 | y ~ Intervention + Sex + |
| 0,231588 | -0,19425 | 0,161645 | 138 | 69 | 69 | 134 | -1,20173 | y ~ Intervention + Sex + |
| 0,240659 | -0,19429 | 0,164854 | 138 | 69 | 69 | 134 | -1,17857 | y ~ Intervention + Sex + |
| 0,250713 | -0,19438 | 0,168494 | 138 | 69 | 69 | 134 | -1,15361 | y ~ Intervention + Sex + |
| 0,226307 | -0,19441 | 0,15994 | 138 | 69 | 69 | 134 | -1,21551 | y ~ Intervention + Sex + |
| 0,231464 | -0,19658 | 0,163539 | 138 | 69 | 69 | 134 | -1,20205 | y ~ Intervention + Sex + |
| 0,205803 | -0,19665 | 0,154679 | 138 | 69 | 69 | 134 | -1,27136 | y ~ Intervention + Sex + |
| 0,238054 | -0,19676 | 0,166017 | 138 | 69 | 69 | 134 | -1,18516 | y ~ Intervention + Sex + |
| 0,230827 | -0,19731 | 0,16392 | 138 | 69 | 69 | 134 | -1,2037 | y ~ Intervention + Sex + |
| 0,204219 | -0,19802 | 0,155205 | 138 | 69 | 69 | 134 | -1,27584 | y ~ Intervention + Sex + |
| 0,228135 | -0,19837 | 0,163842 | 138 | 69 | 69 | 134 | -1,21072 | y ~ Intervention + Sex + |
| 0,183978 | -0,19861 | 0,148716 | 138 | 69 | 69 | 134 | -1,3355 | y ~ Intervention + Sex + |
| 0,246586 | -0,19887 | 0,170883 | 138 | 69 | 69 | 134 | -1,16377 | y ~ Intervention + Sex + |
| 0,223111 | -0,19925 | 0,162787 | 138 | 69 | 69 | 134 | -1,22397 | y ~ Intervention + Sex + |
| 0,215321 | -0,20019 | 0,1608 | 138 | 69 | 69 | 134 | -1,24496 | y ~ Intervention + Sex + |
| 0,212953 | -0,20048 | 0,160202 | 138 | 69 | 69 | 134 | -1,25144 | y ~ Intervention + Sex + |
| 0,181406 | -0,20066 | 0,149362 | 138 | 69 | 69 | 134 | -1,34342 | y ~ Intervention + Sex + |
| 0,200449 | -0,20085 | 0,15611 | 138 | 69 | 69 | 134 | -1,28661 | y ~ Intervention + Sex + |
| 0,165191 | -0,20094 | 0,144 | 138 | 69 | 69 | 134 | -1,39544 | y ~ Intervention + Sex + |
| 0,196942 | -0,201 | 0,155003 | 138 | 69 | 69 | 134 | -1,29676 | y ~ Intervention + Sex + |
| 0,223872 | -0,20104 | 0,164526 | 138 | 69 | 69 | 134 | -1,22195 | y ~ Intervention + Sex + |
| 0,195232 | -0,20123 | 0,154586 | 138 | 69 | 69 | 134 | -1,30176 | y ~ Intervention + Sex + |
| 0,178611 | -0,20133 | 0,1489 | 138 | 69 | 69 | 134 | -1,35213 | y ~ Intervention + Sex + |
| 0,219144 | -0,20136 | 0,163102 | 138 | 69 | 69 | 134 | -1,23459 | y ~ Intervention + Sex + |
| 0,227548 | -0,20185 | 0,166512 | 138 | 69 | 69 | 134 | -1,21226 | y ~ Intervention + Sex + |
| 0,222911 | -0,20216 | 0,165096 | 138 | 69 | 69 | 134 | -1,2245 | y ~ Intervention + Sex + |
| 0,222132 | -0,2031 | 0,165579 | 138 | 69 | 69 | 134 | -1,22658 | y ~ Intervention + Sex + |
| 0,223891 | -0,20333 | 0,166407 | 138 | 69 | 69 | 134 | -1,2219 | y ~ Intervention + Sex + |
| 0,238813 | -0,20347 | 0,171963 | 138 | 69 | 69 | 134 | -1,18323 | y ~ Intervention + Sex + |
| 0,236303 | -0,20356 | 0,171118 | 138 | 69 | 69 | 134 | -1,18961 | y ~ Intervention + Sex + |
| 0,216225 | -0,20405 | 0,164228 | 138 | 69 | 69 | 134 | -1,24249 | y ~ Intervention + Sex + |
| 0,210684 | -0,20457 | 0,162651 | 138 | 69 | 69 | 134 | -1,25771 | y ~ Intervention + Sex + |
| 0,195313 | -0,20576 | 0,158091 | 138 | 69 | 69 | 134 | -1,30152 | y ~ Intervention + Sex + |
| 0,213549 | -0,20676 | 0,165436 | 138 | 69 | 69 | 134 | -1,2498 | y ~ Intervention + Sex + |
| 0,185154 | -0,20709 | 0,155486 | 138 | 69 | 69 | 134 | -1,3319 | y ~ Intervention + Sex + |
| 0,171795 | -0,20762 | 0,151126 | 138 | 69 | 69 | 134 | -1,37381 | y ~ Intervention + Sex + |
| 0,191186 | -0,20867 | 0,15884 | 138 | 69 | 69 | 134 | -1,31372 | y ~ Intervention + Sex + |
| 0,14539 | -0,20874 | 0,142532 | 138 | 69 | 69 | 134 | -1,46453 | y ~ Intervention + Sex + |
| 0,229927 | -0,20879 | 0,173124 | 138 | 69 | 69 | 134 | -1,20604 | y ~ Intervention + Sex + |
| 0,181356 | -0,20894 | 0,155509 | 138 | 69 | 69 | 134 | -1,34358 | y ~ Intervention + Sex + |
| 0,215567 | -0,20936 | 0,168256 | 138 | 69 | 69 | 134 | -1,24428 | y ~ Intervention + Sex + |
| 0,175389 | -0,2098 | 0,154004 | 138 | 69 | 69 | 134 | -1,3623 | y ~ Intervention + Sex + |
| 0,138564 | -0,21018 | 0,141056 | 138 | 69 | 69 | 134 | -1,49004 | y ~ Intervention + Sex + |
| 0,111487 | -0,21018 | 0,131191 | 138 | 69 | 69 | 134 | -1,6021 | y ~ Intervention + Sex + |
| 0,189083 | -0,21113 | 0,159949 | 138 | 69 | 69 | 134 | -1,32001 | y ~ Intervention + Sex + |
| 0,224695 | -0,21179 | 0,173631 | 138 | 69 | 69 | 134 | -1,21977 | y ~ Intervention + Sex + |

| | | | | | | | | |
|---|---|---|---|---|---|---|---|---|
| 0,173242 | -0,21274 | 0,15538 | 138 | 69 | 69 | 134 | -1,36915 | y ~ Intervention + Sex + |
| 0,140663 | -0,2137 | 0,144183 | 137 | 68 | 69 | 133 | -1,48216 | y ~ Intervention + Sex + |
| 0,211834 | -0,21374 | 0,170371 | 138 | 69 | 69 | 134 | -1,25453 | y ~ Intervention + Sex + |
| 0,146785 | -0,21455 | 0,147006 | 138 | 69 | 69 | 134 | -1,45944 | y ~ Intervention + Sex + |
| 0,249906 | -0,21548 | 0,186472 | 138 | 69 | 69 | 134 | -1,15559 | y ~ Intervention + Sex + |
| 0,189751 | -0,21616 | 0,164004 | 138 | 69 | 69 | 134 | -1,318 | y ~ Intervention + Sex + |
| 0,188493 | -0,21672 | 0,163961 | 138 | 69 | 69 | 134 | -1,32178 | y ~ Intervention + Sex + |
| 0,145876 | -0,21828 | 0,149222 | 138 | 69 | 69 | 134 | -1,46276 | y ~ Intervention + Sex + |
| 0,170621 | -0,21864 | 0,158713 | 138 | 69 | 69 | 134 | -1,37761 | y ~ Intervention + Sex + |
| 0,108086 | -0,21905 | 0,135409 | 138 | 69 | 69 | 134 | -1,61767 | y ~ Intervention + Sex + |
| 0,204837 | -0,2191 | 0,171966 | 138 | 69 | 69 | 134 | -1,27409 | y ~ Intervention + Sex + |
| 0,129409 | -0,21944 | 0,143816 | 138 | 69 | 69 | 134 | -1,52583 | y ~ Intervention + Sex + |
| 0,164771 | -0,22008 | 0,157559 | 138 | 69 | 69 | 134 | -1,39684 | y ~ Intervention + Sex + |
| 0,169177 | -0,22105 | 0,159914 | 138 | 69 | 69 | 134 | -1,38231 | y ~ Intervention + Sex + |
| 0,223234 | -0,22125 | 0,180814 | 138 | 69 | 69 | 134 | -1,22364 | y ~ Intervention + Sex + |
| 0,129786 | -0,22139 | 0,145239 | 138 | 69 | 69 | 134 | -1,52432 | y ~ Intervention + Sex + |
| 0,215047 | -0,22233 | 0,178475 | 138 | 69 | 69 | 134 | -1,2457 | y ~ Intervention + Sex + |
| 0,164541 | -0,22354 | 0,159942 | 138 | 69 | 69 | 134 | -1,39761 | y ~ Intervention + Sex + |
| 0,169332 | -0,22475 | 0,162648 | 138 | 69 | 69 | 134 | -1,3818 | y ~ Intervention + Sex + |
| 0,120209 | -0,22748 | 0,145458 | 138 | 69 | 69 | 134 | -1,56386 | y ~ Intervention + Sex + |
| 0,175336 | -0,2275 | 0,166975 | 138 | 69 | 69 | 134 | -1,36247 | y ~ Intervention + Sex + |
| 0,130325 | -0,22885 | 0,150343 | 138 | 69 | 69 | 134 | -1,52216 | y ~ Intervention + Sex + |
| 0,171843 | -0,23111 | 0,168243 | 138 | 69 | 69 | 134 | -1,37365 | y ~ Intervention + Sex + |
| 0,09968 | -0,2316 | 0,139696 | 138 | 69 | 69 | 134 | -1,65788 | y ~ Intervention + Sex + |
| 0,161169 | -0,23211 | 0,16474 | 138 | 69 | 69 | 134 | -1,40894 | y ~ Intervention + Sex + |
| 0,158978 | -0,23276 | 0,164328 | 138 | 69 | 69 | 134 | -1,4164 | y ~ Intervention + Sex + |
| 0,178564 | -0,23368 | 0,172802 | 138 | 69 | 69 | 134 | -1,35228 | y ~ Intervention + Sex + |
| 0,144753 | -0,23651 | 0,161231 | 138 | 69 | 69 | 134 | -1,46687 | y ~ Intervention + Sex + |
| 0,157338 | -0,23655 | 0,166344 | 138 | 69 | 69 | 134 | -1,42204 | y ~ Intervention + Sex + |
| 0,138006 | -0,23832 | 0,159716 | 138 | 69 | 69 | 134 | -1,49217 | y ~ Intervention + Sex + |
| 0,168298 | -0,23911 | 0,172622 | 138 | 69 | 69 | 134 | -1,38518 | y ~ Intervention + Sex + |
| 0,098434 | -0,24061 | 0,144589 | 138 | 69 | 69 | 134 | -1,66407 | y ~ Intervention + Sex + |
| 0,11909 | -0,24062 | 0,153395 | 138 | 69 | 69 | 134 | -1,56864 | y ~ Intervention + Sex + |
| 0,130886 | -0,24077 | 0,158412 | 138 | 69 | 69 | 134 | -1,51992 | y ~ Intervention + Sex + |
| 0,117301 | -0,24134 | 0,153099 | 138 | 69 | 69 | 134 | -1,57636 | y ~ Intervention + Sex + |
| 0,109139 | -0,2414 | 0,149677 | 138 | 69 | 69 | 134 | -1,61281 | y ~ Intervention + Sex + |
| 0,168613 | -0,2448 | 0,17686 | 138 | 69 | 69 | 134 | -1,38415 | y ~ Intervention + Sex + |
| 0,153917 | -0,24496 | 0,170826 | 138 | 69 | 69 | 134 | -1,43395 | y ~ Intervention + Sex + |
| 0,135536 | -0,24519 | 0,16328 | 138 | 69 | 69 | 134 | -1,50167 | y ~ Intervention + Sex + |
| 0,103365 | -0,24554 | 0,149723 | 138 | 69 | 69 | 134 | -1,63993 | y ~ Intervention + Sex + |
| 0,121706 | -0,24649 | 0,15826 | 138 | 69 | 69 | 134 | -1,55752 | y ~ Intervention + Sex + |
| 0,114319 | -0,24662 | 0,155161 | 138 | 69 | 69 | 134 | -1,58943 | y ~ Intervention + Sex + |
| 0,101275 | -0,2467 | 0,149511 | 138 | 69 | 69 | 134 | -1,65005 | y ~ Intervention + Sex + |
| 0,130936 | -0,24694 | 0,162491 | 138 | 69 | 69 | 134 | -1,51973 | y ~ Intervention + Sex + |
| 0,049276 | -0,24742 | 0,124695 | 138 | 69 | 69 | 134 | -1,9842 | y ~ Intervention + Sex + |
| 0,139142 | -0,24766 | 0,166457 | 138 | 69 | 69 | 134 | -1,48784 | y ~ Intervention + Sex + |
| 0,049014 | -0,24786 | 0,124771 | 138 | 69 | 69 | 134 | -1,98653 | y ~ Intervention + Sex + |
| 0,073187 | -0,24848 | 0,137596 | 138 | 69 | 69 | 134 | -1,80585 | y ~ Intervention + Sex + |

| | | | | | | | | |
|---|---|---|---|---|---|---|---|---|
| 0,091021 | -0,25201 | 0,14804 | 138 | 69 | 69 | 134 | -1,70228 | y ~ Intervention + Sex + |
| 0,074245 | -0,25217 | 0,140159 | 138 | 69 | 69 | 134 | -1,79915 | y ~ Intervention + Sex + |
| 0,106342 | -0,25221 | 0,15513 | 138 | 69 | 69 | 134 | -1,6258 | y ~ Intervention + Sex + |
| 0,077093 | -0,25367 | 0,14239 | 138 | 69 | 69 | 134 | -1,78152 | y ~ Intervention + Sex + |
| 0,053452 | -0,25371 | 0,130214 | 138 | 69 | 69 | 134 | -1,94844 | y ~ Intervention + Sex + |
| 0,110609 | -0,25464 | 0,158547 | 138 | 69 | 69 | 134 | -1,60609 | y ~ Intervention + Sex + |
| 0,112511 | -0,25532 | 0,159827 | 138 | 69 | 69 | 134 | -1,5975 | y ~ Intervention + Sex + |
| 0,112894 | -0,25546 | 0,160082 | 138 | 69 | 69 | 134 | -1,59578 | y ~ Intervention + Sex + |
| 0,064625 | -0,25649 | 0,13766 | 138 | 69 | 69 | 134 | -1,86319 | y ~ Intervention + Sex + |
| 0,121163 | -0,25649 | 0,164438 | 138 | 69 | 69 | 134 | -1,55981 | y ~ Intervention + Sex + |
| 0,101398 | -0,25672 | 0,155641 | 138 | 69 | 69 | 134 | -1,64945 | y ~ Intervention + Sex + |
| 0,147541 | -0,25784 | 0,177001 | 138 | 69 | 69 | 134 | -1,45669 | y ~ Intervention + Sex + |
| 0,074489 | -0,25885 | 0,143993 | 138 | 69 | 69 | 134 | -1,79762 | y ~ Intervention + Sex + |
| 0,086162 | -0,26207 | 0,151596 | 138 | 69 | 69 | 134 | -1,72872 | y ~ Intervention + Sex + |
| 0,118422 | -0,26216 | 0,166817 | 138 | 69 | 69 | 134 | -1,57151 | y ~ Intervention + Sex + |
| 0,09628 | -0,26397 | 0,157598 | 138 | 69 | 69 | 134 | -1,67493 | y ~ Intervention + Sex + |
| 0,144133 | -0,26403 | 0,179713 | 138 | 69 | 69 | 134 | -1,46916 | y ~ Intervention + Sex + |
| 0,08927 | -0,26472 | 0,154653 | 138 | 69 | 69 | 134 | -1,71168 | y ~ Intervention + Sex + |
| 0,101414 | -0,26582 | 0,161162 | 138 | 69 | 69 | 134 | -1,64937 | y ~ Intervention + Sex + |
| 0,073226 | -0,26607 | 0,147359 | 138 | 69 | 69 | 134 | -1,8056 | y ~ Intervention + Sex + |
| 0,084011 | -0,26699 | 0,153368 | 138 | 69 | 69 | 134 | -1,74082 | y ~ Intervention + Sex + |
| 0,119512 | -0,2688 | 0,171558 | 138 | 69 | 69 | 134 | -1,56683 | y ~ Intervention + Sex + |
| 0,102707 | -0,26886 | 0,16363 | 138 | 69 | 69 | 134 | -1,6431 | y ~ Intervention + Sex + |
| 0,091355 | -0,26988 | 0,158705 | 138 | 69 | 69 | 134 | -1,70051 | y ~ Intervention + Sex + |
| 0,075041 | -0,27014 | 0,150565 | 138 | 69 | 69 | 134 | -1,79417 | y ~ Intervention + Sex + |
| 0,100799 | -0,27136 | 0,164226 | 138 | 69 | 69 | 134 | -1,65238 | y ~ Intervention + Sex + |
| 0,113193 | -0,27146 | 0,170254 | 138 | 69 | 69 | 134 | -1,59444 | y ~ Intervention + Sex + |
| 0,111623 | -0,2721 | 0,169907 | 138 | 69 | 69 | 134 | -1,60149 | y ~ Intervention + Sex + |
| 0,071891 | -0,27237 | 0,150136 | 138 | 69 | 69 | 134 | -1,81415 | y ~ Intervention + Sex + |
| 0,0446 | -0,27311 | 0,134708 | 138 | 69 | 69 | 134 | -2,02745 | y ~ Intervention + Sex + |
| 0,077293 | -0,27368 | 0,153729 | 138 | 69 | 69 | 134 | -1,7803 | y ~ Intervention + Sex + |
| 0,104832 | -0,27371 | 0,167622 | 138 | 69 | 69 | 134 | -1,63293 | y ~ Intervention + Sex + |
| 0,112829 | -0,2741 | 0,171734 | 138 | 69 | 69 | 134 | -1,59607 | y ~ Intervention + Sex + |
| 0,088744 | -0,27411 | 0,159876 | 138 | 69 | 69 | 134 | -1,71453 | y ~ Intervention + Sex + |
| 0,09408 | -0,27612 | 0,163748 | 138 | 69 | 69 | 134 | -1,68622 | y ~ Intervention + Sex + |
| 0,10353 | -0,27716 | 0,169086 | 138 | 69 | 69 | 134 | -1,63914 | y ~ Intervention + Sex + |
| 0,076111 | -0,27779 | 0,155403 | 138 | 69 | 69 | 134 | -1,78754 | y ~ Intervention + Sex + |
| 0,107825 | -0,27836 | 0,171945 | 138 | 69 | 69 | 134 | -1,61888 | y ~ Intervention + Sex + |
| 0,085035 | -0,28108 | 0,162002 | 138 | 69 | 69 | 134 | -1,73503 | y ~ Intervention + Sex + |
| 0,088164 | -0,282 | 0,164177 | 138 | 69 | 69 | 134 | -1,71769 | y ~ Intervention + Sex + |
| 0,106737 | -0,28324 | 0,174413 | 138 | 69 | 69 | 134 | -1,62395 | y ~ Intervention + Sex + |
| 0,119179 | -0,28404 | 0,18112 | 138 | 69 | 69 | 134 | -1,56826 | y ~ Intervention + Sex + |
| 0,089322 | -0,28531 | 0,166711 | 138 | 69 | 69 | 134 | -1,7114 | y ~ Intervention + Sex + |
| 0,023501 | -0,28706 | 0,125279 | 138 | 69 | 69 | 134 | -2,29139 | y ~ Intervention + Sex + |
| 0,040611 | -0,28748 | 0,139045 | 138 | 69 | 69 | 134 | -2,0675 | y ~ Intervention + Sex + |
| 0,037418 | -0,28891 | 0,13744 | 138 | 69 | 69 | 134 | -2,10208 | y ~ Intervention + Sex + |
| 0,071054 | -0,29069 | 0,159755 | 138 | 69 | 69 | 134 | -1,81959 | y ~ Intervention + Sex + |
| 0,071352 | -0,29156 | 0,160408 | 138 | 69 | 69 | 134 | -1,81764 | y ~ Intervention + Sex + |

| | | | | | | | | |
|---|---|---|---|---|---|---|---|---|
| 0,088196 | -0,29327 | 0,170753 | 138 | 69 | 69 | 134 | -1,71751 | y ~ Intervention + Sex + |
| 0,093148 | -0,29383 | 0,173755 | 138 | 69 | 69 | 134 | -1,69107 | y ~ Intervention + Sex + |
| 0,035331 | -0,29437 | 0,138456 | 138 | 69 | 69 | 134 | -2,12607 | y ~ Intervention + Sex + |
| 0,070637 | -0,29465 | 0,161689 | 138 | 69 | 69 | 134 | -1,82232 | y ~ Intervention + Sex + |
| 0,059086 | -0,29488 | 0,154891 | 138 | 69 | 69 | 134 | -1,90376 | y ~ Intervention + Sex + |
| 0,069769 | -0,29653 | 0,162211 | 138 | 69 | 69 | 134 | -1,82803 | y ~ Intervention + Sex + |
| 0,039353 | -0,29748 | 0,14296 | 138 | 69 | 69 | 134 | -2,08084 | y ~ Intervention + Sex + |
| 0,057872 | -0,2979 | 0,155719 | 138 | 69 | 69 | 134 | -1,91307 | y ~ Intervention + Sex + |
| 0,074816 | -0,29848 | 0,166229 | 138 | 69 | 69 | 134 | -1,79557 | y ~ Intervention + Sex + |
| 0,060096 | -0,29916 | 0,157776 | 138 | 69 | 69 | 134 | -1,89613 | y ~ Intervention + Sex + |
| 0,036398 | -0,29943 | 0,141663 | 138 | 69 | 69 | 134 | -2,11365 | y ~ Intervention + Sex + |
| 0,034404 | -0,30038 | 0,140555 | 138 | 69 | 69 | 134 | -2,13712 | y ~ Intervention + Sex + |
| 0,086926 | -0,30064 | 0,174338 | 138 | 69 | 69 | 134 | -1,72448 | y ~ Intervention + Sex + |
| 0,063732 | -0,30189 | 0,16148 | 138 | 69 | 69 | 134 | -1,86953 | y ~ Intervention + Sex + |
| 0,047937 | -0,30257 | 0,151573 | 138 | 69 | 69 | 134 | -1,99621 | y ~ Intervention + Sex + |
| 0,050969 | -0,30356 | 0,15414 | 138 | 69 | 69 | 134 | -1,96941 | y ~ Intervention + Sex + |
| 0,051642 | -0,30365 | 0,154636 | 138 | 69 | 69 | 134 | -1,96364 | y ~ Intervention + Sex + |
| 0,063239 | -0,30399 | 0,162295 | 138 | 69 | 69 | 134 | -1,87306 | y ~ Intervention + Sex + |
| 0,044631 | -0,30399 | 0,149959 | 138 | 69 | 69 | 134 | -2,02714 | y ~ Intervention + Sex + |
| 0,071364 | -0,30444 | 0,167498 | 138 | 69 | 69 | 134 | -1,81757 | y ~ Intervention + Sex + |
| 0,060399 | -0,30449 | 0,160776 | 138 | 69 | 69 | 134 | -1,89386 | y ~ Intervention + Sex + |
| 0,058306 | -0,30675 | 0,160625 | 138 | 69 | 69 | 134 | -1,90972 | y ~ Intervention + Sex + |
| 0,075259 | -0,30702 | 0,171249 | 138 | 69 | 69 | 134 | -1,79281 | y ~ Intervention + Sex + |
| 0,086953 | -0,30747 | 0,178312 | 138 | 69 | 69 | 134 | -1,72434 | y ~ Intervention + Sex + |
| 0,052729 | -0,30908 | 0,158142 | 138 | 69 | 69 | 134 | -1,95446 | y ~ Intervention + Sex + |
| 0,073224 | -0,30911 | 0,171192 | 138 | 69 | 69 | 134 | -1,80561 | y ~ Intervention + Sex + |
| 0,047679 | -0,3092 | 0,154713 | 138 | 69 | 69 | 134 | -1,99857 | y ~ Intervention + Sex + |
| 0,083144 | -0,30954 | 0,177311 | 138 | 69 | 69 | 134 | -1,74576 | y ~ Intervention + Sex + |
| 0,051656 | -0,31053 | 0,158149 | 138 | 69 | 69 | 134 | -1,96352 | y ~ Intervention + Sex + |
| 0,065947 | -0,31095 | 0,167724 | 138 | 69 | 69 | 134 | -1,85394 | y ~ Intervention + Sex + |
| 0,030581 | -0,31107 | 0,142328 | 138 | 69 | 69 | 134 | -2,18562 | y ~ Intervention + Sex + |
| 0,062235 | -0,31966 | 0,170001 | 138 | 69 | 69 | 134 | -1,88032 | y ~ Intervention + Sex + |
| 0,075464 | -0,32074 | 0,179029 | 138 | 69 | 69 | 134 | -1,79154 | y ~ Intervention + Sex + |
| 0,056593 | -0,32249 | 0,167698 | 138 | 69 | 69 | 134 | -1,92306 | y ~ Intervention + Sex + |
| 0,053121 | -0,32333 | 0,165711 | 138 | 69 | 69 | 134 | -1,95119 | y ~ Intervention + Sex + |
| 0,02003 | -0,32387 | 0,137587 | 138 | 69 | 69 | 134 | -2,35392 | y ~ Intervention + Sex + |
| 0,013513 | -0,32488 | 0,129792 | 138 | 69 | 69 | 134 | -2,50309 | y ~ Intervention + Sex + |
| 0,030045 | -0,32532 | 0,148356 | 138 | 69 | 69 | 134 | -2,19284 | y ~ Intervention + Sex + |
| 0,031529 | -0,32588 | 0,149961 | 138 | 69 | 69 | 134 | -2,17312 | y ~ Intervention + Sex + |
| 0,047287 | -0,32631 | 0,16298 | 138 | 69 | 69 | 134 | -2,00215 | y ~ Intervention + Sex + |
| 0,025875 | -0,32668 | 0,14499 | 138 | 69 | 69 | 134 | -2,25314 | y ~ Intervention + Sex + |
| 0,071856 | -0,32716 | 0,180317 | 138 | 69 | 69 | 134 | -1,81438 | y ~ Intervention + Sex + |
| 0,051367 | -0,32782 | 0,166746 | 138 | 69 | 69 | 134 | -1,96599 | y ~ Intervention + Sex + |
| 0,06145 | -0,32799 | 0,173904 | 138 | 69 | 69 | 134 | -1,88607 | y ~ Intervention + Sex + |
| 0,026606 | -0,32818 | 0,14638 | 138 | 69 | 69 | 134 | -2,24198 | y ~ Intervention + Sex + |
| 0,059579 | -0,32921 | 0,173268 | 138 | 69 | 69 | 134 | -1,90002 | y ~ Intervention + Sex + |
| 0,038653 | -0,33035 | 0,158183 | 138 | 69 | 69 | 134 | -2,08842 | y ~ Intervention + Sex + |
| 0,029041 | -0,33544 | 0,152011 | 138 | 69 | 69 | 134 | -2,20666 | y ~ Intervention + Sex + |

| | | | | | | | | |
|---|---|---|---|---|---|---|---|---|
| 0,036003 | -0,33545 | 0,158364 | 138 | 69 | 69 | 134 | -2,11821 | y ~ Intervention + Sex + |
| 0,057775 | -0,33663 | 0,175894 | 138 | 69 | 69 | 134 | -1,91382 | y ~ Intervention + Sex + |
| 0,041281 | -0,33843 | 0,164241 | 138 | 69 | 69 | 134 | -2,06055 | y ~ Intervention + Sex + |
| 0,015963 | -0,34171 | 0,140004 | 138 | 69 | 69 | 134 | -2,44075 | y ~ Intervention + Sex + |
| 0,048358 | -0,3422 | 0,171752 | 138 | 69 | 69 | 134 | -1,99241 | y ~ Intervention + Sex + |
| 0,041332 | -0,34278 | 0,166395 | 138 | 69 | 69 | 134 | -2,06001 | y ~ Intervention + Sex + |
| 0,04489 | -0,34409 | 0,169951 | 138 | 69 | 69 | 134 | -2,02466 | y ~ Intervention + Sex + |
| 0,023081 | -0,34436 | 0,149822 | 138 | 69 | 69 | 134 | -2,2985 | y ~ Intervention + Sex + |
| 0,051993 | -0,34665 | 0,176804 | 138 | 69 | 69 | 134 | -1,96066 | y ~ Intervention + Sex + |
| 0,042468 | -0,34692 | 0,169358 | 138 | 69 | 69 | 134 | -2,04845 | y ~ Intervention + Sex + |
| 0,038321 | -0,34847 | 0,166569 | 138 | 69 | 69 | 134 | -2,09205 | y ~ Intervention + Sex + |
| 0,043252 | -0,34871 | 0,170886 | 138 | 69 | 69 | 134 | -2,04062 | y ~ Intervention + Sex + |
| 0,062077 | -0,34945 | 0,185732 | 138 | 69 | 69 | 134 | -1,88147 | y ~ Intervention + Sex + |
| 0,053482 | -0,35032 | 0,179817 | 138 | 69 | 69 | 134 | -1,9482 | y ~ Intervention + Sex + |
| 0,013111 | -0,35322 | 0,140485 | 138 | 69 | 69 | 134 | -2,51428 | y ~ Intervention + Sex + |
| 0,017237 | -0,35521 | 0,147291 | 138 | 69 | 69 | 134 | -2,41161 | y ~ Intervention + Sex + |
| 0,028959 | -0,35597 | 0,161235 | 138 | 69 | 69 | 134 | -2,2078 | y ~ Intervention + Sex + |
| 0,032016 | -0,35908 | 0,165715 | 138 | 69 | 69 | 134 | -2,16684 | y ~ Intervention + Sex + |
| 0,021899 | -0,35947 | 0,154998 | 138 | 69 | 69 | 134 | -2,31917 | y ~ Intervention + Sex + |
| 0,02676 | -0,35969 | 0,160601 | 138 | 69 | 69 | 134 | -2,23967 | y ~ Intervention + Sex + |
| 0,009918 | -0,3628 | 0,138686 | 138 | 69 | 69 | 134 | -2,61598 | y ~ Intervention + Sex + |
| 0,020048 | -0,36462 | 0,154922 | 138 | 69 | 69 | 134 | -2,35358 | y ~ Intervention + Sex + |
| 0,030029 | -0,36487 | 0,166378 | 138 | 69 | 69 | 134 | -2,19305 | y ~ Intervention + Sex + |
| 0,018495 | -0,36512 | 0,153111 | 138 | 69 | 69 | 134 | -2,38467 | y ~ Intervention + Sex + |
| 0,024766 | -0,3664 | 0,161367 | 138 | 69 | 69 | 134 | -2,27061 | y ~ Intervention + Sex + |
| 0,022573 | -0,36943 | 0,160114 | 138 | 69 | 69 | 134 | -2,30727 | y ~ Intervention + Sex + |
| 0,008124 | -0,37003 | 0,137711 | 138 | 69 | 69 | 134 | -2,68697 | y ~ Intervention + Sex + |
| 0,019896 | -0,37115 | 0,1575 | 138 | 69 | 69 | 134 | -2,35651 | y ~ Intervention + Sex + |
| 0,018531 | -0,37254 | 0,15627 | 138 | 69 | 69 | 134 | -2,38393 | y ~ Intervention + Sex + |
| 0,015522 | -0,37505 | 0,153 | 138 | 69 | 69 | 134 | -2,4513 | y ~ Intervention + Sex + |
| 0,006966 | -0,37549 | 0,137003 | 138 | 69 | 69 | 134 | -2,74079 | y ~ Intervention + Sex + |
| 0,016577 | -0,37735 | 0,155517 | 138 | 69 | 69 | 134 | -2,42645 | y ~ Intervention + Sex + |
| 0,016143 | -0,37812 | 0,155189 | 138 | 69 | 69 | 134 | -2,43649 | y ~ Intervention + Sex + |
| 0,02501 | -0,37819 | 0,166845 | 138 | 69 | 69 | 134 | -2,26671 | y ~ Intervention + Sex + |
| 0,011836 | -0,37831 | 0,148249 | 138 | 69 | 69 | 134 | -2,55189 | y ~ Intervention + Sex + |
| 0,015023 | -0,37872 | 0,153729 | 138 | 69 | 69 | 134 | -2,46357 | y ~ Intervention + Sex + |
| 0,006507 | -0,37962 | 0,137327 | 138 | 69 | 69 | 134 | -2,76435 | y ~ Intervention + Sex + |
| 0,027113 | -0,38003 | 0,170082 | 138 | 69 | 69 | 134 | -2,2344 | y ~ Intervention + Sex + |
| 0,009318 | -0,38362 | 0,145402 | 138 | 69 | 69 | 134 | -2,63833 | y ~ Intervention + Sex + |
| 0,012824 | -0,38439 | 0,152387 | 138 | 69 | 69 | 134 | -2,52243 | y ~ Intervention + Sex + |
| 0,025824 | -0,38469 | 0,170675 | 138 | 69 | 69 | 134 | -2,25393 | y ~ Intervention + Sex + |
| 0,016508 | -0,38525 | 0,158667 | 138 | 69 | 69 | 134 | -2,42803 | y ~ Intervention + Sex + |
| 0,02854 | -0,38569 | 0,174226 | 138 | 69 | 69 | 134 | -2,21371 | y ~ Intervention + Sex + |
| 0,023461 | -0,38693 | 0,168813 | 138 | 69 | 69 | 134 | -2,29206 | y ~ Intervention + Sex + |
| 0,004689 | -0,38803 | 0,134928 | 138 | 69 | 69 | 134 | -2,87583 | y ~ Intervention + Sex + |
| 0,009751 | -0,38819 | 0,148046 | 138 | 69 | 69 | 134 | -2,62209 | y ~ Intervention + Sex + |
| 0,012807 | -0,38828 | 0,153901 | 138 | 69 | 69 | 134 | -2,52294 | y ~ Intervention + Sex + |
| 0,008036 | -0,39208 | 0,14571 | 138 | 69 | 69 | 134 | -2,69082 | y ~ Intervention + Sex + |

| | | | | | | | | |
|---|---|---|---|---|---|---|---|---|
| 0,020987 | -0,39277 | 0,168155 | 138 | 69 | 69 | 134 | -2,33579 | y ~ Intervention + Sex + |
| 0,022256 | -0,39371 | 0,170228 | 138 | 69 | 69 | 134 | -2,31282 | y ~ Intervention + Sex + |
| 0,015477 | -0,39394 | 0,160635 | 138 | 69 | 69 | 134 | -2,4524 | y ~ Intervention + Sex + |
| 0,021609 | -0,39502 | 0,169948 | 138 | 69 | 69 | 134 | -2,32437 | y ~ Intervention + Sex + |
| 0,013267 | -0,39541 | 0,15754 | 138 | 69 | 69 | 134 | -2,5099 | y ~ Intervention + Sex + |
| 0,011194 | -0,39832 | 0,154855 | 138 | 69 | 69 | 134 | -2,57224 | y ~ Intervention + Sex + |
| 0,016621 | -0,39834 | 0,164231 | 138 | 69 | 69 | 134 | -2,42545 | y ~ Intervention + Sex + |
| 0,01195 | -0,39964 | 0,15682 | 138 | 69 | 69 | 134 | -2,54838 | y ~ Intervention + Sex + |
| 0,014251 | -0,4005 | 0,161278 | 138 | 69 | 69 | 134 | -2,48332 | y ~ Intervention + Sex + |
| 0,016882 | -0,40179 | 0,166059 | 138 | 69 | 69 | 134 | -2,41954 | y ~ Intervention + Sex + |
| 0,005309 | -0,40353 | 0,14239 | 138 | 69 | 69 | 134 | -2,83395 | y ~ Intervention + Sex + |
| 0,006166 | -0,40384 | 0,145117 | 138 | 69 | 69 | 134 | -2,78288 | y ~ Intervention + Sex + |
| 0,0076 | -0,40751 | 0,150349 | 138 | 69 | 69 | 134 | -2,71041 | y ~ Intervention + Sex + |
| 0,002951 | -0,40809 | 0,134758 | 138 | 69 | 69 | 134 | -3,02833 | y ~ Intervention + Sex + |
| 0,012469 | -0,40814 | 0,161142 | 138 | 69 | 69 | 134 | -2,53279 | y ~ Intervention + Sex + |
| 0,010556 | -0,40833 | 0,157444 | 138 | 69 | 69 | 134 | -2,5935 | y ~ Intervention + Sex + |
| 0,009576 | -0,4103 | 0,156094 | 138 | 69 | 69 | 134 | -2,62856 | y ~ Intervention + Sex + |
| 0,01275 | -0,41119 | 0,162874 | 138 | 69 | 69 | 134 | -2,52458 | y ~ Intervention + Sex + |
| 0,008886 | -0,41281 | 0,155467 | 138 | 69 | 69 | 134 | -2,65526 | y ~ Intervention + Sex + |
| 0,012671 | -0,4146 | 0,164075 | 138 | 69 | 69 | 134 | -2,52687 | y ~ Intervention + Sex + |
| 0,020462 | -0,41524 | 0,177025 | 138 | 69 | 69 | 134 | -2,34564 | y ~ Intervention + Sex + |
| 0,006258 | -0,41771 | 0,150375 | 138 | 69 | 69 | 134 | -2,77779 | y ~ Intervention + Sex + |
| 0,008641 | -0,418 | 0,156837 | 138 | 69 | 69 | 134 | -2,66517 | y ~ Intervention + Sex + |
| 0,008927 | -0,41892 | 0,157868 | 138 | 69 | 69 | 134 | -2,65358 | y ~ Intervention + Sex + |
| 0,016314 | -0,42141 | 0,173242 | 138 | 69 | 69 | 134 | -2,4325 | y ~ Intervention + Sex + |
| 0,014037 | -0,42282 | 0,169879 | 138 | 69 | 69 | 134 | -2,48895 | y ~ Intervention + Sex + |
| 0,006254 | -0,43152 | 0,155331 | 138 | 69 | 69 | 134 | -2,77805 | y ~ Intervention + Sex + |
| 0,011577 | -0,4319 | 0,168711 | 138 | 69 | 69 | 134 | -2,55998 | y ~ Intervention + Sex + |
| 0,008674 | -0,43442 | 0,163084 | 138 | 69 | 69 | 134 | -2,66379 | y ~ Intervention + Sex + |
| 0,0066 | -0,43494 | 0,157616 | 138 | 69 | 69 | 134 | -2,75949 | y ~ Intervention + Sex + |
| 0,013075 | -0,43802 | 0,174144 | 138 | 69 | 69 | 134 | -2,51528 | y ~ Intervention + Sex + |
| 0,003897 | -0,43884 | 0,149396 | 138 | 69 | 69 | 134 | -2,93742 | y ~ Intervention + Sex + |
| 0,01305 | -0,44195 | 0,175655 | 138 | 69 | 69 | 134 | -2,51599 | y ~ Intervention + Sex + |
| 0,012945 | -0,44222 | 0,175555 | 138 | 69 | 69 | 134 | -2,51899 | y ~ Intervention + Sex + |
| 0,006957 | -0,44364 | 0,16184 | 138 | 69 | 69 | 134 | -2,74123 | y ~ Intervention + Sex + |
| 0,006069 | -0,44417 | 0,159297 | 138 | 69 | 69 | 134 | -2,78833 | y ~ Intervention + Sex + |
| 0,011178 | -0,44437 | 0,172721 | 138 | 69 | 69 | 134 | -2,57276 | y ~ Intervention + Sex + |
| 0,004911 | -0,4448 | 0,155506 | 138 | 69 | 69 | 134 | -2,86033 | y ~ Intervention + Sex + |
| 0,00589 | -0,4469 | 0,159687 | 138 | 69 | 69 | 134 | -2,79858 | y ~ Intervention + Sex + |
| 0,004851 | -0,4471 | 0,156087 | 138 | 69 | 69 | 134 | -2,86442 | y ~ Intervention + Sex + |
| 0,006218 | -0,45037 | 0,162002 | 138 | 69 | 69 | 134 | -2,78001 | y ~ Intervention + Sex + |
| 0,007887 | -0,45417 | 0,168375 | 138 | 69 | 69 | 134 | -2,69737 | y ~ Intervention + Sex + |
| 0,010991 | -0,45461 | 0,176283 | 138 | 69 | 69 | 134 | -2,57889 | y ~ Intervention + Sex + |
| 0,004315 | -0,45644 | 0,157196 | 138 | 69 | 69 | 134 | -2,90362 | y ~ Intervention + Sex + |
| 0,007622 | -0,45803 | 0,169053 | 138 | 69 | 69 | 134 | -2,70939 | y ~ Intervention + Sex + |
| 0,003465 | -0,46058 | 0,154762 | 138 | 69 | 69 | 134 | -2,9761 | y ~ Intervention + Sex + |
| 0,001819 | -0,46225 | 0,145283 | 138 | 69 | 69 | 134 | -3,18176 | y ~ Intervention + Sex + |
| 0,004832 | -0,46565 | 0,16249 | 138 | 69 | 69 | 134 | -2,86574 | y ~ Intervention + Sex + |

| | | | | | | | | |
|---|---|---|---|---|---|---|---|---|
| 0,004775 | -0,47096 | 0,164109 | 138 | 69 | 69 | 134 | -2,86978 | y ~ Intervention + Sex + |
| 0,005609 | -0,47237 | 0,167787 | 138 | 69 | 69 | 134 | -2,81531 | y ~ Intervention + Sex + |
| 0,004013 | -0,47409 | 0,161932 | 138 | 69 | 69 | 134 | -2,92774 | y ~ Intervention + Sex + |
| 0,001657 | -0,47854 | 0,14904 | 138 | 69 | 69 | 134 | -3,21079 | y ~ Intervention + Sex + |
| 0,000584 | -0,48361 | 0,137267 | 138 | 69 | 69 | 134 | -3,52315 | y ~ Intervention + Sex + |
| 0,00106 | -0,48374 | 0,144523 | 138 | 69 | 69 | 134 | -3,34716 | y ~ Intervention + Sex + |
| 0,001792 | -0,4856 | 0,15239 | 138 | 69 | 69 | 134 | -3,18659 | y ~ Intervention + Sex + |
| 0,002131 | -0,48603 | 0,155168 | 138 | 69 | 69 | 134 | -3,13227 | y ~ Intervention + Sex + |
| 0,001088 | -0,48844 | 0,146269 | 138 | 69 | 69 | 134 | -3,33931 | y ~ Intervention + Sex + |
| 0,007766 | -0,48848 | 0,180731 | 138 | 69 | 69 | 134 | -2,70281 | y ~ Intervention + Sex + |
| 0,000901 | -0,49244 | 0,145013 | 138 | 69 | 69 | 134 | -3,39582 | y ~ Intervention + Sex + |
| 0,002049 | -0,49384 | 0,157044 | 138 | 69 | 69 | 134 | -3,14463 | y ~ Intervention + Sex + |
| 0,001726 | -0,50071 | 0,156565 | 138 | 69 | 69 | 134 | -3,19813 | y ~ Intervention + Sex + |
| 0,003753 | -0,50204 | 0,170189 | 138 | 69 | 69 | 134 | -2,94989 | y ~ Intervention + Sex + |
| 0,001098 | -0,50241 | 0,150574 | 138 | 69 | 69 | 134 | -3,33663 | y ~ Intervention + Sex + |
| 0,000164 | -0,51024 | 0,131529 | 138 | 69 | 69 | 134 | -3,87932 | y ~ Intervention + Sex + |
| 0,00333 | -0,51259 | 0,17149 | 138 | 69 | 69 | 134 | -2,98903 | y ~ Intervention + Sex + |
| 0,000884 | -0,51649 | 0,151847 | 138 | 69 | 69 | 134 | -3,40136 | y ~ Intervention + Sex + |
| 0,002892 | -0,5172 | 0,170422 | 138 | 69 | 69 | 134 | -3,03484 | y ~ Intervention + Sex + |
| 0,000768 | -0,52119 | 0,151371 | 138 | 69 | 69 | 134 | -3,44314 | y ~ Intervention + Sex + |
| 0,000637 | -0,5271 | 0,150699 | 138 | 69 | 69 | 134 | -3,49771 | y ~ Intervention + Sex + |
| 0,000457 | -0,53866 | 0,149897 | 138 | 69 | 69 | 134 | -3,59354 | y ~ Intervention + Sex + |
| 0,000424 | -0,56575 | 0,156513 | 138 | 69 | 69 | 134 | -3,61472 | y ~ Intervention + Sex + |
| 0,000255 | -0,56882 | 0,151355 | 138 | 69 | 69 | 134 | -3,7582 | y ~ Intervention + Sex + |
| 6,87E-05 | -0,56971 | 0,138637 | 138 | 69 | 69 | 134 | -4,10934 | y ~ Intervention + Sex + |
| 0,000398 | -0,57032 | 0,156981 | 138 | 69 | 69 | 134 | -3,63304 | y ~ Intervention + Sex + |
| 0,000208 | -0,5787 | 0,151739 | 138 | 69 | 69 | 134 | -3,81376 | y ~ Intervention + Sex + |
| 0,000355 | -0,58054 | 0,158406 | 138 | 69 | 69 | 134 | -3,66488 | y ~ Intervention + Sex + |
| 0,000282 | -0,58118 | 0,155818 | 138 | 69 | 69 | 134 | -3,72989 | y ~ Intervention + Sex + |
| 0,000183 | -0,58908 | 0,153039 | 138 | 69 | 69 | 134 | -3,84919 | y ~ Intervention + Sex + |
| 0,000125 | -0,59101 | 0,149565 | 138 | 69 | 69 | 134 | -3,9515 | y ~ Intervention + Sex + |
| 0,000636 | -0,59103 | 0,168939 | 138 | 69 | 69 | 134 | -3,49849 | y ~ Intervention + Sex + |
| 0,000256 | -0,61888 | 0,164744 | 138 | 69 | 69 | 134 | -3,75665 | y ~ Intervention + Sex + |
| 4,54E-05 | -0,63768 | 0,151249 | 138 | 69 | 69 | 134 | -4,21609 | y ~ Intervention + Sex + |
| 4,98E-05 | -0,64057 | 0,152784 | 138 | 69 | 69 | 134 | -4,19265 | y ~ Intervention + Sex + |
| 9,89E-05 | -0,6433 | 0,160266 | 138 | 69 | 69 | 134 | -4,01397 | y ~ Intervention + Sex + |
| 9,05E-05 | -0,64481 | 0,159713 | 138 | 69 | 69 | 134 | -4,03732 | y ~ Intervention + Sex + |
| 2,45E-05 | -0,64498 | 0,147509 | 138 | 69 | 69 | 134 | -4,37248 | y ~ Intervention + Sex + |
| 0,000103 | -0,6739 | 0,168336 | 138 | 69 | 69 | 134 | -4,00329 | y ~ Intervention + Sex + |
| 5,11E-05 | -0,69057 | 0,164978 | 138 | 69 | 69 | 134 | -4,18581 | y ~ Intervention + Sex + |
| 4,98E-06 | -0,72042 | 0,151392 | 138 | 69 | 69 | 134 | -4,75862 | y ~ Intervention + Sex + |
| 4,12E-07 | -0,76718 | 0,144041 | 138 | 69 | 69 | 134 | -5,32613 | y ~ Intervention + Sex + |
| 2,45E-21 | -1,47592 | 0,130046 | 138 | 69 | 69 | 134 | -11,3492 | y ~ Intervention + Sex + |

⊦ storage_time
⊦ storage_time
⊦ storage_time
⊦ storage_time
⊦ storage_time
⊦ storage_time
⊦ storage_time
⊦ storage_time
⊦ storage_time
⊦ storage_time
⊦ storage_time
⊦ storage_time
⊦ storage_time
⊦ storage_time
⊦ storage_time
⊦ storage_time
⊦ storage_time
⊦ storage_time
⊦ storage_time
⊦ storage_time
⊦ storage_time
⊦ storage_time
⊦ storage_time
⊦ storage_time
⊦ storage_time
⊦ storage_time
⊦ storage_time
⊦ storage_time
⊦ storage_time
⊦ storage_time
⊦ storage_time
⊦ storage_time
⊦ storage_time
⊦ storage_time
⊦ storage_time
⊦ storage_time
⊦ storage_time
⊦ storage_time
⊦ storage_time
⊦ storage_time
⊦ storage_time
⊦ storage_time
⊦ storage_time
⊦ storage_time
⊦ storage_time
⊦ storage_time
⊦ storage_time

├ storage_time
├ storage_time
├ storage_time
├ storage_time
├ storage_time
├ storage_time
├ storage_time
├ storage_time
├ storage_time
├ storage_time
├ storage_time
├ storage_time
├ storage_time
├ storage_time
├ storage_time
├ storage_time
├ storage_time
├ storage_time
├ storage_time
├ storage_time
├ storage_time
├ storage_time
├ storage_time
├ storage_time
├ storage_time
├ storage_time
├ storage_time
├ storage_time
├ storage_time
├ storage_time
├ storage_time
├ storage_time
├ storage_time
├ storage_time
├ storage_time
├ storage_time
├ storage_time
├ storage_time
├ storage_time
├ storage_time
├ storage_time
├ storage_time
├ storage_time
├ storage_time
├ storage_time
├ storage_time
├ storage_time
├ storage_time

⊦ storage_time
⊦ storage_time
⊦ storage_time
⊦ storage_time
⊦ storage_time
⊦ storage_time
⊦ storage_time
⊦ storage_time
⊦ storage_time
⊦ storage_time
⊦ storage_time
⊦ storage_time
⊦ storage_time
⊦ storage_time
⊦ storage_time
⊦ storage_time
⊦ storage_time
⊦ storage_time
⊦ storage_time
⊦ storage_time
⊦ storage_time
⊦ storage_time
⊦ storage_time
⊦ storage_time
⊦ storage_time
⊦ storage_time
⊦ storage_time
⊦ storage_time
⊦ storage_time
⊦ storage_time
⊦ storage_time
⊦ storage_time
⊦ storage_time
⊦ storage_time
⊦ storage_time
⊦ storage_time
⊦ storage_time
⊦ storage_time
⊦ storage_time
⊦ storage_time
⊦ storage_time
⊦ storage_time
⊦ storage_time
⊦ storage_time
⊦ storage_time
⊦ storage_time
⊦ storage_time
⊦ storage_time

⊦ storage_time
⊦ storage_time
⊦ storage_time
⊦ storage_time
⊦ storage_time
⊦ storage_time
⊦ storage_time
⊦ storage_time
⊦ storage_time
⊦ storage_time
⊦ storage_time
⊦ storage_time
⊦ storage_time
⊦ storage_time
⊦ storage_time
⊦ storage_time
⊦ storage_time
⊦ storage_time
⊦ storage_time
⊦ storage_time
⊦ storage_time
⊦ storage_time
⊦ storage_time
⊦ storage_time
⊦ storage_time
⊦ storage_time
⊦ storage_time
⊦ storage_time
⊦ storage_time
⊦ storage_time
⊦ storage_time
⊦ storage_time
⊦ storage_time
⊦ storage_time
⊦ storage_time
⊦ storage_time
⊦ storage_time
⊦ storage_time
⊦ storage_time
⊦ storage_time
⊦ storage_time
⊦ storage_time
⊦ storage_time
⊦ storage_time
⊦ storage_time
⊦ storage_time
⊦ storage_time
⊦ storage_time

⊦ storage_time
⊦ storage_time
⊦ storage_time
⊦ storage_time
⊦ storage_time
⊦ storage_time
⊦ storage_time
⊦ storage_time
⊦ storage_time
⊦ storage_time
⊦ storage_time
⊦ storage_time
⊦ storage_time
⊦ storage_time
⊦ storage_time
⊦ storage_time
⊦ storage_time
⊦ storage_time
⊦ storage_time
⊦ storage_time
⊦ storage_time
⊦ storage_time
⊦ storage_time
⊦ storage_time
⊦ storage_time
⊦ storage_time
⊦ storage_time
⊦ storage_time
⊦ storage_time
⊦ storage_time
⊦ storage_time
⊦ storage_time
⊦ storage_time
⊦ storage_time
⊦ storage_time
⊦ storage_time
⊦ storage_time
⊦ storage_time
⊦ storage_time
⊦ storage_time
⊦ storage_time
⊦ storage_time
⊦ storage_time
⊦ storage_time
⊦ storage_time
⊦ storage_time
⊦ storage_time
⊦ storage_time

⊦ storage_time
⊦ storage_time
⊦ storage_time
⊦ storage_time
⊦ storage_time
⊦ storage_time
⊦ storage_time
⊦ storage_time
⊦ storage_time
⊦ storage_time
⊦ storage_time
⊦ storage_time
⊦ storage_time
⊦ storage_time
⊦ storage_time
⊦ storage_time
⊦ storage_time
⊦ storage_time
⊦ storage_time
⊦ storage_time
⊦ storage_time
⊦ storage_time
⊦ storage_time
⊦ storage_time
⊦ storage_time
⊦ storage_time
⊦ storage_time
⊦ storage_time
⊦ storage_time
⊦ storage_time
⊦ storage_time
⊦ storage_time
⊦ storage_time
⊦ storage_time
⊦ storage_time
⊦ storage_time
⊦ storage_time
⊦ storage_time
⊦ storage_time
⊦ storage_time
⊦ storage_time
⊦ storage_time
⊦ storage_time
⊦ storage_time
⊦ storage_time
⊦ storage_time
⊦ storage_time
⊦ storage_time

├ storage_time
├ storage_time
├ storage_time
├ storage_time
├ storage_time
├ storage_time
├ storage_time
├ storage_time
├ storage_time
├ storage_time
├ storage_time
├ storage_time
├ storage_time
├ storage_time
├ storage_time
├ storage_time
├ storage_time
├ storage_time
├ storage_time
├ storage_time
├ storage_time
├ storage_time
├ storage_time
├ storage_time
├ storage_time
├ storage_time
├ storage_time
├ storage_time
├ storage_time
├ storage_time
├ storage_time
├ storage_time
├ storage_time
├ storage_time
├ storage_time
├ storage_time
├ storage_time
├ storage_time
├ storage_time
├ storage_time
├ storage_time
├ storage_time
├ storage_time
├ storage_time
├ storage_time
├ storage_time
├ storage_time
├ storage_time

├ storage_time
├ storage_time
├ storage_time
├ storage_time
├ storage_time
├ storage_time
├ storage_time
├ storage_time
├ storage_time
├ storage_time
├ storage_time
├ storage_time
├ storage_time
├ storage_time
├ storage_time
├ storage_time
├ storage_time
├ storage_time
├ storage_time
├ storage_time
├ storage_time
├ storage_time
├ storage_time
├ storage_time
├ storage_time
├ storage_time
├ storage_time
├ storage_time
├ storage_time
├ storage_time
├ storage_time
├ storage_time
├ storage_time
├ storage_time
├ storage_time
├ storage_time
├ storage_time
├ storage_time
├ storage_time
├ storage_time
├ storage_time
├ storage_time
├ storage_time
├ storage_time
├ storage_time
├ storage_time
├ storage_time
├ storage_time

├ storage_time
├ storage_time
├ storage_time
├ storage_time
├ storage_time
├ storage_time
├ storage_time
├ storage_time
├ storage_time
├ storage_time
├ storage_time
├ storage_time
├ storage_time
├ storage_time
├ storage_time
├ storage_time
├ storage_time
├ storage_time
├ storage_time
├ storage_time
├ storage_time
├ storage_time
├ storage_time
├ storage_time
├ storage_time
├ storage_time
├ storage_time
├ storage_time
├ storage_time
├ storage_time
├ storage_time
├ storage_time
├ storage_time
├ storage_time
├ storage_time
├ storage_time
├ storage_time
├ storage_time
├ storage_time
├ storage_time
├ storage_time
├ storage_time
├ storage_time
├ storage_time
├ storage_time
├ storage_time
├ storage_time
├ storage_time

├ storage_time
├ storage_time
├ storage_time
├ storage_time
├ storage_time
├ storage_time
├ storage_time
├ storage_time
├ storage_time
├ storage_time
├ storage_time
├ storage_time
├ storage_time
├ storage_time
├ storage_time
├ storage_time
├ storage_time
├ storage_time
├ storage_time
├ storage_time
├ storage_time
├ storage_time
├ storage_time
├ storage_time
├ storage_time
├ storage_time
├ storage_time
├ storage_time
├ storage_time
├ storage_time
├ storage_time
├ storage_time
├ storage_time
├ storage_time
├ storage_time
├ storage_time
├ storage_time
├ storage_time
├ storage_time
├ storage_time
├ storage_time
├ storage_time
├ storage_time
├ storage_time
├ storage_time
├ storage_time
├ storage_time
├ storage_time

├ storage_time
├ storage_time
├ storage_time
├ storage_time
├ storage_time
├ storage_time
├ storage_time
├ storage_time
├ storage_time
├ storage_time
├ storage_time
├ storage_time
├ storage_time
├ storage_time
├ storage_time
├ storage_time
├ storage_time
├ storage_time
├ storage_time
├ storage_time
├ storage_time
├ storage_time
├ storage_time
├ storage_time
├ storage_time
├ storage_time
├ storage_time
├ storage_time
├ storage_time
├ storage_time
├ storage_time
├ storage_time
├ storage_time
├ storage_time
├ storage_time
├ storage_time
├ storage_time
├ storage_time
├ storage_time
├ storage_time
├ storage_time
├ storage_time
├ storage_time
├ storage_time
├ storage_time
├ storage_time
├ storage_time
├ storage_time

⊦ storage_time
⊦ storage_time
⊦ storage_time
⊦ storage_time
⊦ storage_time
⊦ storage_time
⊦ storage_time
⊦ storage_time
⊦ storage_time
⊦ storage_time
⊦ storage_time
⊦ storage_time
⊦ storage_time
⊦ storage_time
⊦ storage_time
⊦ storage_time
⊦ storage_time
⊦ storage_time
⊦ storage_time
⊦ storage_time
⊦ storage_time
⊦ storage_time
⊦ storage_time
⊦ storage_time
⊦ storage_time
⊦ storage_time
⊦ storage_time
⊦ storage_time
⊦ storage_time
⊦ storage_time
⊦ storage_time
⊦ storage_time
⊦ storage_time
⊦ storage_time
⊦ storage_time
⊦ storage_time
⊦ storage_time
⊦ storage_time
⊦ storage_time
⊦ storage_time
⊦ storage_time
⊦ storage_time
⊦ storage_time
⊦ storage_time
⊦ storage_time
⊦ storage_time
⊦ storage_time
⊦ storage_time

├ storage_time
├ storage_time
├ storage_time
├ storage_time
├ storage_time
├ storage_time
├ storage_time
├ storage_time
├ storage_time
├ storage_time
├ storage_time
├ storage_time
├ storage_time
├ storage_time
├ storage_time
├ storage_time
├ storage_time
├ storage_time
├ storage_time
├ storage_time
├ storage_time
├ storage_time
├ storage_time
├ storage_time
├ storage_time
├ storage_time
├ storage_time
├ storage_time
├ storage_time
├ storage_time
├ storage_time
├ storage_time
├ storage_time
├ storage_time
├ storage_time
├ storage_time
├ storage_time
├ storage_time
├ storage_time
├ storage_time
├ storage_time
├ storage_time
├ storage_time
├ storage_time
├ storage_time
├ storage_time
├ storage_time
├ storage_time

├ storage_time
├ storage_time
├ storage_time
├ storage_time
├ storage_time
├ storage_time
├ storage_time
├ storage_time
├ storage_time
├ storage_time
├ storage_time
├ storage_time
├ storage_time
├ storage_time
├ storage_time
├ storage_time
├ storage_time
├ storage_time
├ storage_time
├ storage_time
├ storage_time
├ storage_time
├ storage_time
├ storage_time
├ storage_time
├ storage_time
├ storage_time
├ storage_time
├ storage_time
├ storage_time
├ storage_time
├ storage_time
├ storage_time
├ storage_time
├ storage_time
├ storage_time
├ storage_time
├ storage_time
├ storage_time
├ storage_time
├ storage_time
├ storage_time
├ storage_time
├ storage_time
├ storage_time
├ storage_time
├ storage_time
├ storage_time

⊦ storage_time
⊦ storage_time
⊦ storage_time
⊦ storage_time
⊦ storage_time
⊦ storage_time
⊦ storage_time
⊦ storage_time
⊦ storage_time
⊦ storage_time
⊦ storage_time
⊦ storage_time
⊦ storage_time
⊦ storage_time
⊦ storage_time
⊦ storage_time
⊦ storage_time
⊦ storage_time
⊦ storage_time
⊦ storage_time
⊦ storage_time
⊦ storage_time
⊦ storage_time
⊦ storage_time
⊦ storage_time
⊦ storage_time
⊦ storage_time
⊦ storage_time
⊦ storage_time
⊦ storage_time
⊦ storage_time
⊦ storage_time
⊦ storage_time
⊦ storage_time
⊦ storage_time
⊦ storage_time
⊦ storage_time
⊦ storage_time
⊦ storage_time
⊦ storage_time
⊦ storage_time
⊦ storage_time
⊦ storage_time
⊦ storage_time
⊦ storage_time
⊦ storage_time
⊦ storage_time
⊦ storage_time

⊦ storage_time
⊦ storage_time
⊦ storage_time
⊦ storage_time
⊦ storage_time
⊦ storage_time
⊦ storage_time
⊦ storage_time
⊦ storage_time
⊦ storage_time
⊦ storage_time
⊦ storage_time
⊦ storage_time
⊦ storage_time
⊦ storage_time
⊦ storage_time
⊦ storage_time
⊦ storage_time
⊦ storage_time
⊦ storage_time
⊦ storage_time
⊦ storage_time
⊦ storage_time
⊦ storage_time
⊦ storage_time
⊦ storage_time
⊦ storage_time
⊦ storage_time
⊦ storage_time
⊦ storage_time
⊦ storage_time
⊦ storage_time
⊦ storage_time
⊦ storage_time
⊦ storage_time
⊦ storage_time
⊦ storage_time
⊦ storage_time
⊦ storage_time
⊦ storage_time
⊦ storage_time
⊦ storage_time
⊦ storage_time
⊦ storage_time
⊦ storage_time
⊦ storage_time
⊦ storage_time
⊦ storage_time

⊦ storage_time
⊦ storage_time
⊦ storage_time
⊦ storage_time
⊦ storage_time
⊦ storage_time
⊦ storage_time
⊦ storage_time
⊦ storage_time
⊦ storage_time
⊦ storage_time
⊦ storage_time
⊦ storage_time
⊦ storage_time
⊦ storage_time
⊦ storage_time
⊦ storage_time
⊦ storage_time
⊦ storage_time
⊦ storage_time
⊦ storage_time
⊦ storage_time
⊦ storage_time
⊦ storage_time
⊦ storage_time
⊦ storage_time
⊦ storage_time
⊦ storage_time
⊦ storage_time
⊦ storage_time
⊦ storage_time
⊦ storage_time
⊦ storage_time
⊦ storage_time
⊦ storage_time
⊦ storage_time
⊦ storage_time
⊦ storage_time
⊦ storage_time
⊦ storage_time
⊦ storage_time
⊦ storage_time
⊦ storage_time
⊦ storage_time
⊦ storage_time
⊦ storage_time
⊦ storage_time
⊦ storage_time

├ storage_time
├ storage_time
├ storage_time
├ storage_time
├ storage_time
├ storage_time
├ storage_time
├ storage_time
├ storage_time
├ storage_time
├ storage_time
├ storage_time
├ storage_time
├ storage_time
├ storage_time
├ storage_time
├ storage_time
├ storage_time
├ storage_time
├ storage_time
├ storage_time
├ storage_time
├ storage_time
├ storage_time
├ storage_time
├ storage_time
├ storage_time
├ storage_time
├ storage_time
├ storage_time
├ storage_time
├ storage_time
├ storage_time
├ storage_time
├ storage_time
├ storage_time
├ storage_time
├ storage_time
├ storage_time
├ storage_time
├ storage_time
├ storage_time
├ storage_time
├ storage_time
├ storage_time
├ storage_time
├ storage_time
├ storage_time

⊦ storage_time
⊦ storage_time
⊦ storage_time
⊦ storage_time
⊦ storage_time
⊦ storage_time
⊦ storage_time
⊦ storage_time
⊦ storage_time
⊦ storage_time
⊦ storage_time
⊦ storage_time
⊦ storage_time
⊦ storage_time
⊦ storage_time
⊦ storage_time
⊦ storage_time
⊦ storage_time
⊦ storage_time
⊦ storage_time
⊦ storage_time
⊦ storage_time
⊦ storage_time
⊦ storage_time
⊦ storage_time
⊦ storage_time
⊦ storage_time
⊦ storage_time
⊦ storage_time
⊦ storage_time
⊦ storage_time
⊦ storage_time
⊦ storage_time
⊦ storage_time
⊦ storage_time
⊦ storage_time
⊦ storage_time
⊦ storage_time
⊦ storage_time
⊦ storage_time
⊦ storage_time
⊦ storage_time
⊦ storage_time
⊦ storage_time
⊦ storage_time
⊦ storage_time
⊦ storage_time
⊦ storage_time

├ storage_time
├ storage_time
├ storage_time
├ storage_time
├ storage_time
├ storage_time
├ storage_time
├ storage_time
├ storage_time
├ storage_time
├ storage_time
├ storage_time
├ storage_time
├ storage_time
├ storage_time
├ storage_time
├ storage_time
├ storage_time
├ storage_time
├ storage_time
├ storage_time
├ storage_time
├ storage_time
├ storage_time
├ storage_time
├ storage_time
├ storage_time
├ storage_time
├ storage_time
├ storage_time
├ storage_time
├ storage_time
├ storage_time
├ storage_time
├ storage_time
├ storage_time
├ storage_time
├ storage_time
├ storage_time
├ storage_time
├ storage_time
├ storage_time
├ storage_time
├ storage_time
├ storage_time
├ storage_time
├ storage_time
├ storage_time

├ storage_time
├ storage_time
├ storage_time
├ storage_time
├ storage_time
├ storage_time
├ storage_time
├ storage_time
├ storage_time
├ storage_time
├ storage_time
├ storage_time
├ storage_time
├ storage_time
├ storage_time
├ storage_time
├ storage_time
├ storage_time
├ storage_time
├ storage_time
├ storage_time
├ storage_time
├ storage_time
├ storage_time
├ storage_time
├ storage_time
├ storage_time
├ storage_time
├ storage_time
├ storage_time
├ storage_time
├ storage_time
├ storage_time
├ storage_time
├ storage_time
├ storage_time
├ storage_time
├ storage_time
├ storage_time
├ storage_time
├ storage_time
├ storage_time
├ storage_time
├ storage_time
├ storage_time
├ storage_time
├ storage_time
├ storage_time

⊦ storage_time
⊦ storage_time
⊦ storage_time
⊦ storage_time
⊦ storage_time
⊦ storage_time
⊦ storage_time
⊦ storage_time
⊦ storage_time
⊦ storage_time
⊦ storage_time
⊦ storage_time
⊦ storage_time
⊦ storage_time
⊦ storage_time
⊦ storage_time
⊦ storage_time
⊦ storage_time
⊦ storage_time
⊦ storage_time
⊦ storage_time
⊦ storage_time
⊦ storage_time
⊦ storage_time
⊦ storage_time
⊦ storage_time
⊦ storage_time
⊦ storage_time
⊦ storage_time
⊦ storage_time
⊦ storage_time
⊦ storage_time
⊦ storage_time
⊦ storage_time
⊦ storage_time
⊦ storage_time
⊦ storage_time
⊦ storage_time
⊦ storage_time
⊦ storage_time
⊦ storage_time
⊦ storage_time
⊦ storage_time
⊦ storage_time
⊦ storage_time
⊦ storage_time
⊦ storage_time
⊦ storage_time

⊦ storage_time
⊦ storage_time
⊦ storage_time
⊦ storage_time
⊦ storage_time
⊦ storage_time
⊦ storage_time
⊦ storage_time
⊦ storage_time
⊦ storage_time
⊦ storage_time
⊦ storage_time
⊦ storage_time
⊦ storage_time
⊦ storage_time
⊦ storage_time
⊦ storage_time
⊦ storage_time
⊦ storage_time
⊦ storage_time
⊦ storage_time
⊦ storage_time
⊦ storage_time
⊦ storage_time
⊦ storage_time
⊦ storage_time
⊦ storage_time
⊦ storage_time
⊦ storage_time
⊦ storage_time
⊦ storage_time
⊦ storage_time
⊦ storage_time
⊦ storage_time
⊦ storage_time
⊦ storage_time
⊦ storage_time
⊦ storage_time
⊦ storage_time
⊦ storage_time
⊦ storage_time
⊦ storage_time
⊦ storage_time
⊦ storage_time
⊦ storage_time
⊦ storage_time
⊦ storage_time
⊦ storage_time

⊦ storage_time
⊦ storage_time
⊦ storage_time
⊦ storage_time
⊦ storage_time
⊦ storage_time
⊦ storage_time
⊦ storage_time
⊦ storage_time
⊦ storage_time
⊦ storage_time
⊦ storage_time
⊦ storage_time
⊦ storage_time
⊦ storage_time
⊦ storage_time
⊦ storage_time
⊦ storage_time
⊦ storage_time
⊦ storage_time
⊦ storage_time
⊦ storage_time
⊦ storage_time
⊦ storage_time
⊦ storage_time
⊦ storage_time
⊦ storage_time
⊦ storage_time
⊦ storage_time
⊦ storage_time
⊦ storage_time
⊦ storage_time
⊦ storage_time
⊦ storage_time
⊦ storage_time
⊦ storage_time
⊦ storage_time
⊦ storage_time
⊦ storage_time
⊦ storage_time
⊦ storage_time
⊦ storage_time
⊦ storage_time
⊦ storage_time
⊦ storage_time
⊦ storage_time
⊦ storage_time
⊦ storage_time

⊦ storage_time
⊦ storage_time
⊦ storage_time
⊦ storage_time
⊦ storage_time
⊦ storage_time
⊦ storage_time
⊦ storage_time
⊦ storage_time
⊦ storage_time
⊦ storage_time
⊦ storage_time
⊦ storage_time
⊦ storage_time
⊦ storage_time
⊦ storage_time
⊦ storage_time
⊦ storage_time
⊦ storage_time
⊦ storage_time
⊦ storage_time
⊦ storage_time
⊦ storage_time
⊦ storage_time
⊦ storage_time
⊦ storage_time
⊦ storage_time
⊦ storage_time
⊦ storage_time
⊦ storage_time
⊦ storage_time
⊦ storage_time
⊦ storage_time
⊦ storage_time
⊦ storage_time
⊦ storage_time
⊦ storage_time
⊦ storage_time
⊦ storage_time
⊦ storage_time
⊦ storage_time
⊦ storage_time
⊦ storage_time
⊦ storage_time
⊦ storage_time
⊦ storage_time
⊦ storage_time
⊦ storage_time

├ storage_time
├ storage_time
├ storage_time
├ storage_time
├ storage_time
├ storage_time
├ storage_time
├ storage_time
├ storage_time
├ storage_time
├ storage_time
├ storage_time
├ storage_time
├ storage_time
├ storage_time
├ storage_time
├ storage_time
├ storage_time
├ storage_time
├ storage_time
├ storage_time
├ storage_time
├ storage_time
├ storage_time
├ storage_time
├ storage_time
├ storage_time
├ storage_time
├ storage_time
├ storage_time
├ storage_time
├ storage_time
├ storage_time
├ storage_time
├ storage_time
├ storage_time
├ storage_time
├ storage_time
├ storage_time
├ storage_time
├ storage_time
├ storage_time
├ storage_time
├ storage_time
├ storage_time
├ storage_time
├ storage_time
├ storage_time

⊦ storage_time
⊦ storage_time
⊦ storage_time
⊦ storage_time
⊦ storage_time
⊦ storage_time
⊦ storage_time
⊦ storage_time
⊦ storage_time
⊦ storage_time
⊦ storage_time
⊦ storage_time
⊦ storage_time
⊦ storage_time
⊦ storage_time
⊦ storage_time
⊦ storage_time
⊦ storage_time
⊦ storage_time
⊦ storage_time
⊦ storage_time
⊦ storage_time
⊦ storage_time
⊦ storage_time
⊦ storage_time
⊦ storage_time
⊦ storage_time
⊦ storage_time
⊦ storage_time
⊦ storage_time
⊦ storage_time
⊦ storage_time
⊦ storage_time
⊦ storage_time
⊦ storage_time
⊦ storage_time
⊦ storage_time
⊦ storage_time
⊦ storage_time
⊦ storage_time
⊦ storage_time
⊦ storage_time
⊦ storage_time
⊦ storage_time
⊦ storage_time
⊦ storage_time
⊦ storage_time
⊦ storage_time

⊦ storage_time
⊦ storage_time
⊦ storage_time
⊦ storage_time
⊦ storage_time
⊦ storage_time
⊦ storage_time
⊦ storage_time
⊦ storage_time
⊦ storage_time
⊦ storage_time
⊦ storage_time
⊦ storage_time
⊦ storage_time
⊦ storage_time
⊦ storage_time
⊦ storage_time
⊦ storage_time
⊦ storage_time
⊦ storage_time
⊦ storage_time
⊦ storage_time
⊦ storage_time
⊦ storage_time
⊦ storage_time
⊦ storage_time
⊦ storage_time
⊦ storage_time
⊦ storage_time
⊦ storage_time
⊦ storage_time
⊦ storage_time
⊦ storage_time
⊦ storage_time
⊦ storage_time
⊦ storage_time
⊦ storage_time
⊦ storage_time
⊦ storage_time
⊦ storage_time
⊦ storage_time
⊦ storage_time
⊦ storage_time
⊦ storage_time
⊦ storage_time
⊦ storage_time
⊦ storage_time
⊦ storage_time

⊦ storage_time
⊦ storage_time
⊦ storage_time
⊦ storage_time
⊦ storage_time
⊦ storage_time
⊦ storage_time
⊦ storage_time
⊦ storage_time
⊦ storage_time
⊦ storage_time
⊦ storage_time
⊦ storage_time
⊦ storage_time
⊦ storage_time
⊦ storage_time
⊦ storage_time
⊦ storage_time
⊦ storage_time
⊦ storage_time
⊦ storage_time
⊦ storage_time
⊦ storage_time
⊦ storage_time
⊦ storage_time
⊦ storage_time
⊦ storage_time
⊦ storage_time
⊦ storage_time
⊦ storage_time
⊦ storage_time
⊦ storage_time
⊦ storage_time
⊦ storage_time
⊦ storage_time
⊦ storage_time
⊦ storage_time
⊦ storage_time
⊦ storage_time
⊦ storage_time
⊦ storage_time
⊦ storage_time
⊦ storage_time
⊦ storage_time
⊦ storage_time
⊦ storage_time
⊦ storage_time
⊦ storage_time

⊦ storage_time
⊦ storage_time
⊦ storage_time
⊦ storage_time
⊦ storage_time
⊦ storage_time
⊦ storage_time
⊦ storage_time
⊦ storage_time
⊦ storage_time
⊦ storage_time
⊦ storage_time
⊦ storage_time
⊦ storage_time
⊦ storage_time
⊦ storage_time
⊦ storage_time
⊦ storage_time
⊦ storage_time
⊦ storage_time
⊦ storage_time
⊦ storage_time
⊦ storage_time
⊦ storage_time
⊦ storage_time
⊦ storage_time
⊦ storage_time
⊦ storage_time
⊦ storage_time
⊦ storage_time
⊦ storage_time
⊦ storage_time
⊦ storage_time
⊦ storage_time
⊦ storage_time
⊦ storage_time
⊦ storage_time
⊦ storage_time
⊦ storage_time
⊦ storage_time
⊦ storage_time
⊦ storage_time
⊦ storage_time
⊦ storage_time
⊦ storage_time
⊦ storage_time
⊦ storage_time
⊦ storage_time

├ storage_time
├ storage_time
├ storage_time
├ storage_time
├ storage_time
├ storage_time
├ storage_time
├ storage_time
├ storage_time
├ storage_time
├ storage_time
├ storage_time
├ storage_time
├ storage_time
├ storage_time
├ storage_time
├ storage_time
├ storage_time
├ storage_time
├ storage_time
├ storage_time
├ storage_time
├ storage_time
├ storage_time
├ storage_time
├ storage_time
├ storage_time
├ storage_time
├ storage_time
├ storage_time
├ storage_time
├ storage_time
├ storage_time
├ storage_time
├ storage_time
├ storage_time
├ storage_time
├ storage_time
├ storage_time
├ storage_time
├ storage_time
├ storage_time
├ storage_time
├ storage_time
├ storage_time
├ storage_time
├ storage_time
├ storage_time

├ storage_time
├ storage_time
├ storage_time
├ storage_time
├ storage_time
├ storage_time
├ storage_time
├ storage_time
├ storage_time
├ storage_time
├ storage_time
├ storage_time
├ storage_time
├ storage_time
├ storage_time
├ storage_time
├ storage_time
├ storage_time
├ storage_time
├ storage_time
├ storage_time
├ storage_time
├ storage_time
├ storage_time
├ storage_time
├ storage_time
├ storage_time
├ storage_time
├ storage_time
├ storage_time
├ storage_time
├ storage_time
├ storage_time
├ storage_time
├ storage_time
├ storage_time
├ storage_time
├ storage_time
├ storage_time
├ storage_time
├ storage_time
├ storage_time
├ storage_time
├ storage_time
├ storage_time
├ storage_time
├ storage_time
├ storage_time

├ storage_time
├ storage_time
├ storage_time
├ storage_time
├ storage_time
├ storage_time
├ storage_time
├ storage_time
├ storage_time
├ storage_time
├ storage_time
├ storage_time
├ storage_time
├ storage_time
├ storage_time
├ storage_time
├ storage_time
├ storage_time
├ storage_time
├ storage_time
├ storage_time
├ storage_time
├ storage_time
├ storage_time
├ storage_time
├ storage_time
├ storage_time
├ storage_time
├ storage_time
├ storage_time
├ storage_time
├ storage_time
├ storage_time
├ storage_time
├ storage_time
├ storage_time
├ storage_time
├ storage_time
├ storage_time
├ storage_time
├ storage_time
├ storage_time
├ storage_time
├ storage_time
├ storage_time
├ storage_time
├ storage_time
├ storage_time

├ storage_time
├ storage_time
├ storage_time
├ storage_time
├ storage_time
├ storage_time
├ storage_time
├ storage_time
├ storage_time
├ storage_time
├ storage_time
├ storage_time
├ storage_time
├ storage_time
├ storage_time
├ storage_time
├ storage_time
├ storage_time
├ storage_time
├ storage_time
├ storage_time
├ storage_time
├ storage_time
├ storage_time
├ storage_time
├ storage_time
├ storage_time
├ storage_time
├ storage_time
├ storage_time
├ storage_time
├ storage_time
├ storage_time
├ storage_time
├ storage_time
├ storage_time
├ storage_time
├ storage_time
├ storage_time
├ storage_time
├ storage_time
├ storage_time
├ storage_time
├ storage_time
├ storage_time
├ storage_time
├ storage_time
├ storage_time

├ storage_time
├ storage_time
├ storage_time
├ storage_time
├ storage_time
├ storage_time
├ storage_time
├ storage_time
├ storage_time
├ storage_time
├ storage_time
├ storage_time
├ storage_time
├ storage_time
├ storage_time
├ storage_time
├ storage_time
├ storage_time
├ storage_time
├ storage_time
├ storage_time
├ storage_time
├ storage_time
├ storage_time
├ storage_time
├ storage_time
├ storage_time
├ storage_time
├ storage_time
├ storage_time
├ storage_time
├ storage_time
├ storage_time
├ storage_time
├ storage_time
├ storage_time
├ storage_time
├ storage_time
├ storage_time
├ storage_time
├ storage_time
├ storage_time
├ storage_time
├ storage_time
├ storage_time
├ storage_time
├ storage_time
├ storage_time

⊦ storage_time
⊦ storage_time
⊦ storage_time
⊦ storage_time
⊦ storage_time
⊦ storage_time
⊦ storage_time
⊦ storage_time
⊦ storage_time
⊦ storage_time
⊦ storage_time
⊦ storage_time
⊦ storage_time
⊦ storage_time
⊦ storage_time
⊦ storage_time
⊦ storage_time
⊦ storage_time
⊦ storage_time
⊦ storage_time
⊦ storage_time
⊦ storage_time
⊦ storage_time
⊦ storage_time
⊦ storage_time
⊦ storage_time
⊦ storage_time
⊦ storage_time
⊦ storage_time
⊦ storage_time
⊦ storage_time
⊦ storage_time
⊦ storage_time
⊦ storage_time
⊦ storage_time
⊦ storage_time
⊦ storage_time
⊦ storage_time
⊦ storage_time
⊦ storage_time
⊦ storage_time
⊦ storage_time
⊦ storage_time
⊦ storage_time
⊦ storage_time
⊦ storage_time
⊦ storage_time
⊦ storage_time

├ storage_time
├ storage_time
├ storage_time
├ storage_time
├ storage_time
├ storage_time
├ storage_time
├ storage_time
├ storage_time
├ storage_time
├ storage_time
├ storage_time
├ storage_time
├ storage_time
├ storage_time
├ storage_time
├ storage_time
├ storage_time
├ storage_time
├ storage_time
├ storage_time
├ storage_time
├ storage_time
├ storage_time
├ storage_time
├ storage_time
├ storage_time
├ storage_time
├ storage_time
├ storage_time
├ storage_time
├ storage_time
├ storage_time
├ storage_time
├ storage_time
├ storage_time
├ storage_time
├ storage_time
├ storage_time
├ storage_time
├ storage_time
├ storage_time
├ storage_time
├ storage_time
├ storage_time
├ storage_time
├ storage_time
├ storage_time

├ storage_time
├ storage_time
├ storage_time
├ storage_time
├ storage_time
├ storage_time
├ storage_time
├ storage_time
├ storage_time
├ storage_time
├ storage_time
├ storage_time
├ storage_time
├ storage_time
├ storage_time
├ storage_time
├ storage_time
├ storage_time
├ storage_time
├ storage_time
├ storage_time
├ storage_time
├ storage_time
├ storage_time
├ storage_time
├ storage_time
├ storage_time
├ storage_time
├ storage_time
├ storage_time
├ storage_time
├ storage_time
├ storage_time
├ storage_time
├ storage_time
├ storage_time
├ storage_time
├ storage_time
├ storage_time
├ storage_time
├ storage_time
├ storage_time
├ storage_time
├ storage_time
├ storage_time
├ storage_time
├ storage_time
├ storage_time

⊦ storage_time
⊦ storage_time
⊦ storage_time
⊦ storage_time
⊦ storage_time
⊦ storage_time
⊦ storage_time
⊦ storage_time
⊦ storage_time
⊦ storage_time
⊦ storage_time
⊦ storage_time
⊦ storage_time
⊦ storage_time
⊦ storage_time
⊦ storage_time
⊦ storage_time
⊦ storage_time
⊦ storage_time
⊦ storage_time
⊦ storage_time
⊦ storage_time
⊦ storage_time
⊦ storage_time
⊦ storage_time
⊦ storage_time
⊦ storage_time
⊦ storage_time
⊦ storage_time
⊦ storage_time
⊦ storage_time
⊦ storage_time
⊦ storage_time
⊦ storage_time
⊦ storage_time
⊦ storage_time
⊦ storage_time
⊦ storage_time
⊦ storage_time
⊦ storage_time
⊦ storage_time
⊦ storage_time
⊦ storage_time
⊦ storage_time
⊦ storage_time
⊦ storage_time
⊦ storage_time
⊦ storage_time

⊦ storage_time
⊦ storage_time
⊦ storage_time
⊦ storage_time
⊦ storage_time
⊦ storage_time
⊦ storage_time
⊦ storage_time
⊦ storage_time
⊦ storage_time
⊦ storage_time
⊦ storage_time
⊦ storage_time
⊦ storage_time
⊦ storage_time
⊦ storage_time
⊦ storage_time
⊦ storage_time
⊦ storage_time
⊦ storage_time
⊦ storage_time
⊦ storage_time
⊦ storage_time
⊦ storage_time
⊦ storage_time
⊦ storage_time
⊦ storage_time
⊦ storage_time
⊦ storage_time
⊦ storage_time
⊦ storage_time
⊦ storage_time
⊦ storage_time
⊦ storage_time
⊦ storage_time
⊦ storage_time
⊦ storage_time
⊦ storage_time
⊦ storage_time
⊦ storage_time
⊦ storage_time
⊦ storage_time
⊦ storage_time
⊦ storage_time
⊦ storage_time
⊦ storage_time
⊦ storage_time
⊦ storage_time

├ storage_time
├ storage_time
├ storage_time
├ storage_time
├ storage_time
├ storage_time
├ storage_time
├ storage_time
├ storage_time
├ storage_time
├ storage_time
├ storage_time
├ storage_time
├ storage_time
├ storage_time
├ storage_time
├ storage_time
├ storage_time
├ storage_time
├ storage_time
├ storage_time
├ storage_time
├ storage_time
├ storage_time
├ storage_time
├ storage_time
├ storage_time
├ storage_time
├ storage_time
├ storage_time
├ storage_time
├ storage_time
├ storage_time
├ storage_time
├ storage_time
├ storage_time
├ storage_time
├ storage_time
├ storage_time
├ storage_time
├ storage_time
├ storage_time
├ storage_time
├ storage_time
├ storage_time
├ storage_time
├ storage_time
├ storage_time

├ storage_time
├ storage_time
├ storage_time
├ storage_time
├ storage_time
├ storage_time
├ storage_time
├ storage_time
├ storage_time
├ storage_time
├ storage_time
├ storage_time
├ storage_time
├ storage_time
├ storage_time
├ storage_time
├ storage_time
├ storage_time
├ storage_time
├ storage_time
├ storage_time
├ storage_time
├ storage_time
├ storage_time
├ storage_time
├ storage_time
├ storage_time
├ storage_time
├ storage_time
├ storage_time
├ storage_time
├ storage_time
├ storage_time
├ storage_time
├ storage_time
├ storage_time
├ storage_time
├ storage_time
├ storage_time
├ storage_time
├ storage_time
├ storage_time
├ storage_time
├ storage_time
├ storage_time
├ storage_time
├ storage_time
├ storage_time

├ storage_time
├ storage_time
├ storage_time
├ storage_time
├ storage_time
├ storage_time
├ storage_time
├ storage_time
├ storage_time
├ storage_time
├ storage_time
├ storage_time
├ storage_time
├ storage_time
├ storage_time
├ storage_time
├ storage_time
├ storage_time
├ storage_time
├ storage_time
├ storage_time
├ storage_time
├ storage_time
├ storage_time
├ storage_time
├ storage_time
├ storage_time
├ storage_time
├ storage_time
├ storage_time
├ storage_time
├ storage_time
├ storage_time
├ storage_time
├ storage_time
├ storage_time
├ storage_time
├ storage_time
├ storage_time
├ storage_time
├ storage_time
├ storage_time
├ storage_time
├ storage_time
├ storage_time
├ storage_time
├ storage_time
├ storage_time

⊦ storage_time
⊦ storage_time
⊦ storage_time
⊦ storage_time
⊦ storage_time
⊦ storage_time
⊦ storage_time
⊦ storage_time
⊦ storage_time
⊦ storage_time
⊦ storage_time
⊦ storage_time
⊦ storage_time
⊦ storage_time
⊦ storage_time
⊦ storage_time
⊦ storage_time
⊦ storage_time
⊦ storage_time
⊦ storage_time
⊦ storage_time
⊦ storage_time
⊦ storage_time
⊦ storage_time
⊦ storage_time
⊦ storage_time
⊦ storage_time
⊦ storage_time
⊦ storage_time
⊦ storage_time
⊦ storage_time
⊦ storage_time
⊦ storage_time
⊦ storage_time
⊦ storage_time
⊦ storage_time
⊦ storage_time
⊦ storage_time
⊦ storage_time
⊦ storage_time
⊦ storage_time
⊦ storage_time
⊦ storage_time
⊦ storage_time
⊦ storage_time
⊦ storage_time
⊦ storage_time
⊦ storage_time

⊦ storage_time
⊦ storage_time
⊦ storage_time
⊦ storage_time
⊦ storage_time
⊦ storage_time
⊦ storage_time
⊦ storage_time
⊦ storage_time
⊦ storage_time
⊦ storage_time
⊦ storage_time
⊦ storage_time
⊦ storage_time
⊦ storage_time
⊦ storage_time
⊦ storage_time
⊦ storage_time
⊦ storage_time
⊦ storage_time
⊦ storage_time
⊦ storage_time
⊦ storage_time
⊦ storage_time
⊦ storage_time
⊦ storage_time
⊦ storage_time
⊦ storage_time
⊦ storage_time
⊦ storage_time
⊦ storage_time
⊦ storage_time
⊦ storage_time
⊦ storage_time
⊦ storage_time
⊦ storage_time
⊦ storage_time
⊦ storage_time
⊦ storage_time
⊦ storage_time
⊦ storage_time
⊦ storage_time
⊦ storage_time
⊦ storage_time
⊦ storage_time
⊦ storage_time
⊦ storage_time
⊦ storage_time

⊦ storage_time
⊦ storage_time
⊦ storage_time
⊦ storage_time
⊦ storage_time
⊦ storage_time
⊦ storage_time
⊦ storage_time
⊦ storage_time
⊦ storage_time
⊦ storage_time
⊦ storage_time
⊦ storage_time
⊦ storage_time
⊦ storage_time
⊦ storage_time
⊦ storage_time
⊦ storage_time
⊦ storage_time
⊦ storage_time
⊦ storage_time
⊦ storage_time
⊦ storage_time
⊦ storage_time
⊦ storage_time
⊦ storage_time
⊦ storage_time
⊦ storage_time
⊦ storage_time
⊦ storage_time
⊦ storage_time
⊦ storage_time
⊦ storage_time
⊦ storage_time
⊦ storage_time
⊦ storage_time
⊦ storage_time
⊦ storage_time
⊦ storage_time
⊦ storage_time
⊦ storage_time
⊦ storage_time
⊦ storage_time
⊦ storage_time
⊦ storage_time
⊦ storage_time
⊦ storage_time
⊦ storage_time

⊦ storage_time
⊦ storage_time
⊦ storage_time
⊦ storage_time
⊦ storage_time
⊦ storage_time
⊦ storage_time
⊦ storage_time
⊦ storage_time
⊦ storage_time
⊦ storage_time
⊦ storage_time
⊦ storage_time
⊦ storage_time
⊦ storage_time
⊦ storage_time
⊦ storage_time
⊦ storage_time
⊦ storage_time
⊦ storage_time
⊦ storage_time
⊦ storage_time
⊦ storage_time
⊦ storage_time
⊦ storage_time
⊦ storage_time
⊦ storage_time
⊦ storage_time
⊦ storage_time
⊦ storage_time
⊦ storage_time
⊦ storage_time
⊦ storage_time
⊦ storage_time
⊦ storage_time
⊦ storage_time
⊦ storage_time
⊦ storage_time
⊦ storage_time
⊦ storage_time
⊦ storage_time
⊦ storage_time
⊦ storage_time
⊦ storage_time
⊦ storage_time
⊦ storage_time
⊦ storage_time
⊦ storage_time

├ storage_time
├ storage_time
├ storage_time
├ storage_time
├ storage_time
├ storage_time
├ storage_time
├ storage_time
├ storage_time
├ storage_time
├ storage_time
├ storage_time
├ storage_time
├ storage_time
├ storage_time
├ storage_time
├ storage_time
├ storage_time
├ storage_time
├ storage_time
├ storage_time
├ storage_time
├ storage_time
├ storage_time
├ storage_time
├ storage_time
├ storage_time
├ storage_time
├ storage_time
├ storage_time
├ storage_time
├ storage_time
├ storage_time
├ storage_time
├ storage_time
├ storage_time
├ storage_time
├ storage_time
├ storage_time
├ storage_time
├ storage_time
├ storage_time
├ storage_time
├ storage_time
├ storage_time
├ storage_time
├ storage_time
├ storage_time

├ storage_time
├ storage_time
├ storage_time
├ storage_time
├ storage_time
├ storage_time
├ storage_time
├ storage_time
├ storage_time
├ storage_time
├ storage_time
├ storage_time
├ storage_time
├ storage_time
├ storage_time
├ storage_time
├ storage_time
├ storage_time
├ storage_time
├ storage_time
├ storage_time
├ storage_time
├ storage_time
├ storage_time
├ storage_time
├ storage_time
├ storage_time
├ storage_time
├ storage_time
├ storage_time
├ storage_time
├ storage_time
├ storage_time
├ storage_time
├ storage_time
├ storage_time
├ storage_time
├ storage_time
├ storage_time
├ storage_time
├ storage_time
├ storage_time
├ storage_time
├ storage_time
├ storage_time
├ storage_time
├ storage_time
├ storage_time

├ storage_time
├ storage_time
├ storage_time
├ storage_time
├ storage_time
├ storage_time
├ storage_time
├ storage_time
├ storage_time
├ storage_time
├ storage_time
├ storage_time
├ storage_time
├ storage_time
├ storage_time
├ storage_time
├ storage_time
├ storage_time
├ storage_time
├ storage_time
├ storage_time
├ storage_time
├ storage_time
├ storage_time
├ storage_time
├ storage_time
├ storage_time
├ storage_time
├ storage_time
├ storage_time
├ storage_time
├ storage_time
├ storage_time
├ storage_time
├ storage_time
├ storage_time
├ storage_time
├ storage_time
├ storage_time
├ storage_time
├ storage_time
├ storage_time
├ storage_time
├ storage_time
├ storage_time
├ storage_time
├ storage_time
├ storage_time

⊦ storage_time
⊦ storage_time
⊦ storage_time
⊦ storage_time
⊦ storage_time
⊦ storage_time
⊦ storage_time
⊦ storage_time
⊦ storage_time
⊦ storage_time
⊦ storage_time
⊦ storage_time
⊦ storage_time
⊦ storage_time
⊦ storage_time
⊦ storage_time
⊦ storage_time
⊦ storage_time
⊦ storage_time
⊦ storage_time
⊦ storage_time
⊦ storage_time
⊦ storage_time
⊦ storage_time
⊦ storage_time
⊦ storage_time
⊦ storage_time
⊦ storage_time
⊦ storage_time
⊦ storage_time
⊦ storage_time
⊦ storage_time
⊦ storage_time
⊦ storage_time
⊦ storage_time
⊦ storage_time
⊦ storage_time
⊦ storage_time
⊦ storage_time
⊦ storage_time
⊦ storage_time
⊦ storage_time
⊦ storage_time
⊦ storage_time
⊦ storage_time
⊦ storage_time
⊦ storage_time
⊦ storage_time

├ storage_time
├ storage_time
├ storage_time
├ storage_time
├ storage_time
├ storage_time
├ storage_time
├ storage_time
├ storage_time
├ storage_time
├ storage_time
├ storage_time
├ storage_time
├ storage_time
├ storage_time
├ storage_time
├ storage_time
├ storage_time
├ storage_time
├ storage_time
├ storage_time
├ storage_time
├ storage_time
├ storage_time
├ storage_time
├ storage_time
├ storage_time
├ storage_time
├ storage_time
├ storage_time
├ storage_time
├ storage_time
├ storage_time
├ storage_time
├ storage_time
├ storage_time
├ storage_time
├ storage_time
├ storage_time
├ storage_time
├ storage_time
├ storage_time
├ storage_time

⊦ storage_time
⊦ storage_time
⊦ storage_time
⊦ storage_time
⊦ storage_time
⊦ storage_time
⊦ storage_time
⊦ storage_time
⊦ storage_time
⊦ storage_time
⊦ storage_time
⊦ storage_time
⊦ storage_time
⊦ storage_time
⊦ storage_time
⊦ storage_time
⊦ storage_time
⊦ storage_time
⊦ storage_time
⊦ storage_time

├ storage_time
├ storage_time
├ storage_time
├ storage_time
├ storage_time
├ storage_time
├ storage_time
├ storage_time
├ storage_time
├ storage_time
├ storage_time
├ storage_time
├ storage_time
├ storage_time
├ storage_time
├ storage_time
├ storage_time
├ storage_time
├ storage_time
├ storage_time
├ storage_time
├ storage_time
├ storage_time
├ storage_time
├ storage_time
├ storage_time
├ storage_time
├ storage_time
├ storage_time
├ storage_time
├ storage_time
├ storage_time
├ storage_time
├ storage_time
├ storage_time
├ storage_time
├ storage_time
├ storage_time
├ storage_time
├ storage_time
├ storage_time
├ storage_time
├ storage_time
├ storage_time
├ storage_time
├ storage_time
├ storage_time
├ storage_time

⊦ storage_time
⊦ storage_time
⊦ storage_time
⊦ storage_time
⊦ storage_time
⊦ storage_time
⊦ storage_time
⊦ storage_time
⊦ storage_time
⊦ storage_time
⊦ storage_time
⊦ storage_time
⊦ storage_time
⊦ storage_time
⊦ storage_time
⊦ storage_time
⊦ storage_time
⊦ storage_time
⊦ storage_time
⊦ storage_time
⊦ storage_time
⊦ storage_time
⊦ storage_time
⊦ storage_time
⊦ storage_time
⊦ storage_time
⊦ storage_time
⊦ storage_time
⊦ storage_time
⊦ storage_time
⊦ storage_time
⊦ storage_time
⊦ storage_time
⊦ storage_time
⊦ storage_time
⊦ storage_time
⊦ storage_time
⊦ storage_time
⊦ storage_time
⊦ storage_time
⊦ storage_time
⊦ storage_time
⊦ storage_time
⊦ storage_time
⊦ storage_time
⊦ storage_time
⊦ storage_time
⊦ storage_time

├ storage_time
├ storage_time
├ storage_time
├ storage_time
├ storage_time
├ storage_time
├ storage_time
├ storage_time
├ storage_time
├ storage_time
├ storage_time
├ storage_time
├ storage_time
├ storage_time
├ storage_time
├ storage_time
├ storage_time
├ storage_time
├ storage_time
├ storage_time
├ storage_time
├ storage_time
├ storage_time
├ storage_time
├ storage_time
├ storage_time
├ storage_time
├ storage_time
├ storage_time
├ storage_time
├ storage_time
├ storage_time
├ storage_time
├ storage_time
├ storage_time
├ storage_time
├ storage_time
├ storage_time
├ storage_time
├ storage_time
├ storage_time
├ storage_time
├ storage_time
├ storage_time
├ storage_time
├ storage_time
├ storage_time
├ storage_time

⊦ storage_time
⊦ storage_time
⊦ storage_time
⊦ storage_time
⊦ storage_time
⊦ storage_time
⊦ storage_time
⊦ storage_time
⊦ storage_time
⊦ storage_time
⊦ storage_time
⊦ storage_time
⊦ storage_time
⊦ storage_time
⊦ storage_time
⊦ storage_time
⊦ storage_time
⊦ storage_time
⊦ storage_time
⊦ storage_time
⊦ storage_time
⊦ storage_time
⊦ storage_time
⊦ storage_time
⊦ storage_time
⊦ storage_time
⊦ storage_time
⊦ storage_time
⊦ storage_time
⊦ storage_time
⊦ storage_time
⊦ storage_time
⊦ storage_time
⊦ storage_time
⊦ storage_time
⊦ storage_time
⊦ storage_time
⊦ storage_time
⊦ storage_time
⊦ storage_time
⊦ storage_time
⊦ storage_time
⊦ storage_time
⊦ storage_time
⊦ storage_time
⊦ storage_time
⊦ storage_time
⊦ storage_time

⊦ storage_time
⊦ storage_time
⊦ storage_time
⊦ storage_time
⊦ storage_time
⊦ storage_time
⊦ storage_time
⊦ storage_time
⊦ storage_time
⊦ storage_time
⊦ storage_time
⊦ storage_time
⊦ storage_time
⊦ storage_time
⊦ storage_time
⊦ storage_time
⊦ storage_time
⊦ storage_time
⊦ storage_time
⊦ storage_time
⊦ storage_time
⊦ storage_time
⊦ storage_time
⊦ storage_time
⊦ storage_time
⊦ storage_time
⊦ storage_time
⊦ storage_time
⊦ storage_time
⊦ storage_time
⊦ storage_time
⊦ storage_time
⊦ storage_time
⊦ storage_time
⊦ storage_time
⊦ storage_time
⊦ storage_time
⊦ storage_time
⊦ storage_time
⊦ storage_time
⊦ storage_time
⊦ storage_time
⊦ storage_time
⊦ storage_time
⊦ storage_time
⊦ storage_time
⊦ storage_time
⊦ storage_time

⊢ storage_time
⊢ storage_time
⊢ storage_time
⊢ storage_time
⊢ storage_time
⊢ storage_time
⊢ storage_time
⊢ storage_time
⊢ storage_time
⊢ storage_time
⊢ storage_time
⊢ storage_time
⊢ storage_time
⊢ storage_time
⊢ storage_time
⊢ storage_time
⊢ storage_time
⊢ storage_time
⊢ storage_time
⊢ storage_time
⊢ storage_time
⊢ storage_time
⊢ storage_time
⊢ storage_time
⊢ storage_time
⊢ storage_time
⊢ storage_time
⊢ storage_time
⊢ storage_time
⊢ storage_time
⊢ storage_time
⊢ storage_time
⊢ storage_time
⊢ storage_time
⊢ storage_time
⊢ storage_time
⊢ storage_time
⊢ storage_time
⊢ storage_time
⊢ storage_time
⊢ storage_time
⊢ storage_time
⊢ storage_time
⊢ storage_time
⊢ storage_time
⊢ storage_time
⊢ storage_time
⊢ storage_time

⊦ storage_time
⊦ storage_time
⊦ storage_time
⊦ storage_time
⊦ storage_time
⊦ storage_time
⊦ storage_time
⊦ storage_time
⊦ storage_time
⊦ storage_time
⊦ storage_time
⊦ storage_time
⊦ storage_time
⊦ storage_time
⊦ storage_time
⊦ storage_time
⊦ storage_time
⊦ storage_time
⊦ storage_time
⊦ storage_time
⊦ storage_time
⊦ storage_time
⊦ storage_time
⊦ storage_time
⊦ storage_time
⊦ storage_time
⊦ storage_time
⊦ storage_time
⊦ storage_time
⊦ storage_time
⊦ storage_time
⊦ storage_time
⊦ storage_time
⊦ storage_time
⊦ storage_time
⊦ storage_time
⊦ storage_time
⊦ storage_time
⊦ storage_time
⊦ storage_time
⊦ storage_time
⊦ storage_time
⊦ storage_time
⊦ storage_time
⊦ storage_time
⊦ storage_time
⊦ storage_time
⊦ storage_time

⊦ storage_time
⊦ storage_time
⊦ storage_time
⊦ storage_time
⊦ storage_time
⊦ storage_time
⊦ storage_time
⊦ storage_time
⊦ storage_time
⊦ storage_time
⊦ storage_time
⊦ storage_time
⊦ storage_time
⊦ storage_time
⊦ storage_time
⊦ storage_time
⊦ storage_time
⊦ storage_time
⊦ storage_time
⊦ storage_time
⊦ storage_time
⊦ storage_time
⊦ storage_time
⊦ storage_time
⊦ storage_time
⊦ storage_time
⊦ storage_time
⊦ storage_time
⊦ storage_time
⊦ storage_time
⊦ storage_time
⊦ storage_time
⊦ storage_time
⊦ storage_time
⊦ storage_time
⊦ storage_time
⊦ storage_time
⊦ storage_time
⊦ storage_time
⊦ storage_time
⊦ storage_time
⊦ storage_time
⊦ storage_time
⊦ storage_time
⊦ storage_time
⊦ storage_time
⊦ storage_time
⊦ storage_time

⊦ storage_time
⊦ storage_time
⊦ storage_time
⊦ storage_time
⊦ storage_time
⊦ storage_time
⊦ storage_time
⊦ storage_time
⊦ storage_time
⊦ storage_time
⊦ storage_time
⊦ storage_time
⊦ storage_time
⊦ storage_time
⊦ storage_time
⊦ storage_time
⊦ storage_time
⊦ storage_time
⊦ storage_time
⊦ storage_time
⊦ storage_time
⊦ storage_time
⊦ storage_time
⊦ storage_time
⊦ storage_time
⊦ storage_time
⊦ storage_time
⊦ storage_time
⊦ storage_time
⊦ storage_time
⊦ storage_time
⊦ storage_time
⊦ storage_time
⊦ storage_time
⊦ storage_time
⊦ storage_time
⊦ storage_time
⊦ storage_time
⊦ storage_time
⊦ storage_time
⊦ storage_time
⊦ storage_time
⊦ storage_time
⊦ storage_time
⊦ storage_time
⊦ storage_time
⊦ storage_time
⊦ storage_time

├ storage_time
├ storage_time
├ storage_time
├ storage_time
├ storage_time
├ storage_time
├ storage_time
├ storage_time
├ storage_time
├ storage_time
├ storage_time
├ storage_time
├ storage_time
├ storage_time
├ storage_time
├ storage_time
├ storage_time
├ storage_time
├ storage_time
├ storage_time
├ storage_time
├ storage_time
├ storage_time
├ storage_time
├ storage_time
├ storage_time
├ storage_time
├ storage_time
├ storage_time
├ storage_time
├ storage_time
├ storage_time
├ storage_time
├ storage_time
├ storage_time
├ storage_time
├ storage_time
├ storage_time
├ storage_time
├ storage_time
├ storage_time
├ storage_time
├ storage_time
├ storage_time
├ storage_time
├ storage_time
├ storage_time
├ storage_time

⊦ storage_time
⊦ storage_time
⊦ storage_time
⊦ storage_time
⊦ storage_time
⊦ storage_time
⊦ storage_time
⊦ storage_time
⊦ storage_time
⊦ storage_time
⊦ storage_time
⊦ storage_time
⊦ storage_time
⊦ storage_time
⊦ storage_time
⊦ storage_time
⊦ storage_time
⊦ storage_time
⊦ storage_time
⊦ storage_time
⊦ storage_time
⊦ storage_time
⊦ storage_time
⊦ storage_time
⊦ storage_time
⊦ storage_time
⊦ storage_time
⊦ storage_time
⊦ storage_time
⊦ storage_time
⊦ storage_time
⊦ storage_time
⊦ storage_time
⊦ storage_time
⊦ storage_time
⊦ storage_time
⊦ storage_time
⊦ storage_time
⊦ storage_time
⊦ storage_time
⊦ storage_time
⊦ storage_time
⊦ storage_time
⊦ storage_time
⊦ storage_time
⊦ storage_time
⊦ storage_time
⊦ storage_time

⊦ storage_time
⊦ storage_time
⊦ storage_time
⊦ storage_time
⊦ storage_time
⊦ storage_time
⊦ storage_time
⊦ storage_time
⊦ storage_time
⊦ storage_time
⊦ storage_time
⊦ storage_time
⊦ storage_time
⊦ storage_time
⊦ storage_time
⊦ storage_time
⊦ storage_time
⊦ storage_time
⊦ storage_time
⊦ storage_time
⊦ storage_time
⊦ storage_time
⊦ storage_time
⊦ storage_time
⊦ storage_time
⊦ storage_time
⊦ storage_time
⊦ storage_time
⊦ storage_time
⊦ storage_time
⊦ storage_time
⊦ storage_time
⊦ storage_time
⊦ storage_time
⊦ storage_time
⊦ storage_time
⊦ storage_time
⊦ storage_time
⊦ storage_time
⊦ storage_time
⊦ storage_time
⊦ storage_time
⊦ storage_time
⊦ storage_time
⊦ storage_time
⊦ storage_time
⊦ storage_time
⊦ storage_time

├ storage_time
├ storage_time
├ storage_time
├ storage_time
├ storage_time
├ storage_time
├ storage_time
├ storage_time
├ storage_time
├ storage_time
├ storage_time
├ storage_time
├ storage_time
├ storage_time
├ storage_time
├ storage_time
├ storage_time
├ storage_time
├ storage_time
├ storage_time
├ storage_time
├ storage_time
├ storage_time
├ storage_time
├ storage_time
├ storage_time
├ storage_time
├ storage_time
├ storage_time
├ storage_time
├ storage_time
├ storage_time
├ storage_time
├ storage_time
├ storage_time
├ storage_time
├ storage_time
├ storage_time
├ storage_time
├ storage_time
├ storage_time
├ storage_time
├ storage_time
├ storage_time
├ storage_time
├ storage_time
├ storage_time
├ storage_time

⊦ storage_time
⊦ storage_time
⊦ storage_time
⊦ storage_time
⊦ storage_time
⊦ storage_time
⊦ storage_time
⊦ storage_time
⊦ storage_time
⊦ storage_time
⊦ storage_time
⊦ storage_time
⊦ storage_time
⊦ storage_time
⊦ storage_time
⊦ storage_time
⊦ storage_time
⊦ storage_time
⊦ storage_time
⊦ storage_time
⊦ storage_time
⊦ storage_time
⊦ storage_time
⊦ storage_time
⊦ storage_time
⊦ storage_time
⊦ storage_time
⊦ storage_time
⊦ storage_time
⊦ storage_time
⊦ storage_time
⊦ storage_time
⊦ storage_time
⊦ storage_time
⊦ storage_time
⊦ storage_time
⊦ storage_time
⊦ storage_time
⊦ storage_time
⊦ storage_time
⊦ storage_time
⊦ storage_time
⊦ storage_time
⊦ storage_time
⊦ storage_time
⊦ storage_time
⊦ storage_time
⊦ storage_time

⊦ storage_time
⊦ storage_time
⊦ storage_time
⊦ storage_time
⊦ storage_time
⊦ storage_time
⊦ storage_time
⊦ storage_time
⊦ storage_time
⊦ storage_time
⊦ storage_time
⊦ storage_time
⊦ storage_time
⊦ storage_time
⊦ storage_time
⊦ storage_time
⊦ storage_time
⊦ storage_time
⊦ storage_time
⊦ storage_time
⊦ storage_time
⊦ storage_time
⊦ storage_time
⊦ storage_time
⊦ storage_time
⊦ storage_time
⊦ storage_time
⊦ storage_time
⊦ storage_time
⊦ storage_time
⊦ storage_time
⊦ storage_time
⊦ storage_time
⊦ storage_time
⊦ storage_time
⊦ storage_time
⊦ storage_time
⊦ storage_time
⊦ storage_time
⊦ storage_time
⊦ storage_time
⊦ storage_time
⊦ storage_time
⊦ storage_time
⊦ storage_time
⊦ storage_time
⊦ storage_time
⊦ storage_time

├ storage_time
├ storage_time
├ storage_time
├ storage_time
├ storage_time
├ storage_time
├ storage_time
├ storage_time
├ storage_time
├ storage_time
├ storage_time
├ storage_time
├ storage_time
├ storage_time
├ storage_time
├ storage_time
├ storage_time
├ storage_time
├ storage_time
├ storage_time
├ storage_time
├ storage_time
├ storage_time
├ storage_time
├ storage_time
├ storage_time
├ storage_time
├ storage_time
├ storage_time
├ storage_time
├ storage_time
├ storage_time
├ storage_time
├ storage_time
├ storage_time
├ storage_time
├ storage_time
├ storage_time
├ storage_time
├ storage_time
├ storage_time
├ storage_time
├ storage_time
├ storage_time
├ storage_time
├ storage_time
├ storage_time
├ storage_time

⊦ storage_time
⊦ storage_time
⊦ storage_time
⊦ storage_time
⊦ storage_time
⊦ storage_time
⊦ storage_time
⊦ storage_time
⊦ storage_time
⊦ storage_time
⊦ storage_time
⊦ storage_time
⊦ storage_time
⊦ storage_time
⊦ storage_time
⊦ storage_time
⊦ storage_time
⊦ storage_time
⊦ storage_time
⊦ storage_time
⊦ storage_time
⊦ storage_time
⊦ storage_time
⊦ storage_time
⊦ storage_time
⊦ storage_time
⊦ storage_time
⊦ storage_time
⊦ storage_time
⊦ storage_time
⊦ storage_time
⊦ storage_time
⊦ storage_time
⊦ storage_time
⊦ storage_time
⊦ storage_time
⊦ storage_time
⊦ storage_time
⊦ storage_time
⊦ storage_time
⊦ storage_time
⊦ storage_time
⊦ storage_time
⊦ storage_time
⊦ storage_time
⊦ storage_time
⊦ storage_time
⊦ storage_time

├ storage_time
├ storage_time
├ storage_time
├ storage_time
├ storage_time
├ storage_time
├ storage_time
├ storage_time
├ storage_time
├ storage_time
├ storage_time
├ storage_time
├ storage_time
├ storage_time
├ storage_time
├ storage_time
├ storage_time
├ storage_time
├ storage_time
├ storage_time
├ storage_time
├ storage_time
├ storage_time
├ storage_time
├ storage_time
├ storage_time
├ storage_time
├ storage_time
├ storage_time
├ storage_time
├ storage_time
├ storage_time
├ storage_time
├ storage_time
├ storage_time
├ storage_time
├ storage_time
├ storage_time
├ storage_time
├ storage_time
├ storage_time
├ storage_time
├ storage_time
├ storage_time
├ storage_time
├ storage_time
├ storage_time
├ storage_time

├ storage_time
├ storage_time
├ storage_time
├ storage_time
├ storage_time
├ storage_time
├ storage_time
├ storage_time
├ storage_time
├ storage_time
├ storage_time
├ storage_time
├ storage_time
├ storage_time
├ storage_time
├ storage_time
├ storage_time
├ storage_time
├ storage_time
├ storage_time
├ storage_time
├ storage_time
├ storage_time
├ storage_time
├ storage_time
├ storage_time
├ storage_time
├ storage_time
├ storage_time
├ storage_time
├ storage_time
├ storage_time
├ storage_time
├ storage_time
├ storage_time
├ storage_time
├ storage_time
├ storage_time
├ storage_time
├ storage_time
├ storage_time
├ storage_time
├ storage_time
├ storage_time
├ storage_time
├ storage_time
├ storage_time
├ storage_time

├ storage_time
├ storage_time
├ storage_time
├ storage_time
├ storage_time
├ storage_time
├ storage_time
├ storage_time
├ storage_time
├ storage_time
├ storage_time
├ storage_time
├ storage_time
├ storage_time
├ storage_time
├ storage_time
├ storage_time
├ storage_time
├ storage_time
├ storage_time
├ storage_time
├ storage_time
├ storage_time
├ storage_time
├ storage_time
├ storage_time
├ storage_time
├ storage_time
├ storage_time
├ storage_time
├ storage_time
├ storage_time
├ storage_time
├ storage_time
├ storage_time
├ storage_time
├ storage_time
├ storage_time
├ storage_time
├ storage_time
├ storage_time
├ storage_time
├ storage_time
├ storage_time
├ storage_time
├ storage_time
├ storage_time
├ storage_time

├ storage_time
├ storage_time
├ storage_time
├ storage_time
├ storage_time
├ storage_time
├ storage_time
├ storage_time
├ storage_time
├ storage_time
├ storage_time
├ storage_time
├ storage_time
├ storage_time
├ storage_time
├ storage_time
├ storage_time
├ storage_time
├ storage_time
├ storage_time
├ storage_time
├ storage_time
├ storage_time
├ storage_time
├ storage_time
├ storage_time
├ storage_time
├ storage_time
├ storage_time
├ storage_time
├ storage_time
├ storage_time
├ storage_time
├ storage_time
├ storage_time
├ storage_time
├ storage_time
├ storage_time
├ storage_time
├ storage_time
├ storage_time
├ storage_time
├ storage_time
├ storage_time
├ storage_time
├ storage_time
├ storage_time
├ storage_time

⊦ storage_time
⊦ storage_time
⊦ storage_time
⊦ storage_time
⊦ storage_time
⊦ storage_time
⊦ storage_time
⊦ storage_time
⊦ storage_time
⊦ storage_time
⊦ storage_time
⊦ storage_time
⊦ storage_time
⊦ storage_time
⊦ storage_time
⊦ storage_time
⊦ storage_time
⊦ storage_time
⊦ storage_time
⊦ storage_time
⊦ storage_time
⊦ storage_time
⊦ storage_time
⊦ storage_time
⊦ storage_time
⊦ storage_time
⊦ storage_time
⊦ storage_time
⊦ storage_time
⊦ storage_time
⊦ storage_time
⊦ storage_time
⊦ storage_time
⊦ storage_time
⊦ storage_time
⊦ storage_time
⊦ storage_time
⊦ storage_time
⊦ storage_time
⊦ storage_time
⊦ storage_time
⊦ storage_time
⊦ storage_time
⊦ storage_time
⊦ storage_time
⊦ storage_time
⊦ storage_time
⊦ storage_time

⊦ storage_time
⊦ storage_time
⊦ storage_time
⊦ storage_time
⊦ storage_time
⊦ storage_time
⊦ storage_time
⊦ storage_time
⊦ storage_time
⊦ storage_time
⊦ storage_time
⊦ storage_time
⊦ storage_time
⊦ storage_time
⊦ storage_time
⊦ storage_time
⊦ storage_time
⊦ storage_time
⊦ storage_time
⊦ storage_time
⊦ storage_time
⊦ storage_time
⊦ storage_time
⊦ storage_time
⊦ storage_time
⊦ storage_time
⊦ storage_time
⊦ storage_time
⊦ storage_time
⊦ storage_time
⊦ storage_time
⊦ storage_time
⊦ storage_time
⊦ storage_time
⊦ storage_time
⊦ storage_time
⊦ storage_time
⊦ storage_time
⊦ storage_time
⊦ storage_time
⊦ storage_time
⊦ storage_time
⊦ storage_time
⊦ storage_time
⊦ storage_time
⊦ storage_time
⊦ storage_time
⊦ storage_time

⊦ storage_time
⊦ storage_time
⊦ storage_time
⊦ storage_time
⊦ storage_time
⊦ storage_time
⊦ storage_time
⊦ storage_time
⊦ storage_time
⊦ storage_time
⊦ storage_time
⊦ storage_time
⊦ storage_time
⊦ storage_time
⊦ storage_time
⊦ storage_time
⊦ storage_time
⊦ storage_time
⊦ storage_time
⊦ storage_time
⊦ storage_time
⊦ storage_time
⊦ storage_time
⊦ storage_time
⊦ storage_time
⊦ storage_time
⊦ storage_time
⊦ storage_time
⊦ storage_time
⊦ storage_time
⊦ storage_time
⊦ storage_time
⊦ storage_time
⊦ storage_time
⊦ storage_time
⊦ storage_time
⊦ storage_time
⊦ storage_time
⊦ storage_time
⊦ storage_time
⊦ storage_time
⊦ storage_time
⊦ storage_time
⊦ storage_time
⊦ storage_time
⊦ storage_time
⊦ storage_time
⊦ storage_time

⊦ storage_time
⊦ storage_time
⊦ storage_time
⊦ storage_time
⊦ storage_time
⊦ storage_time
⊦ storage_time
⊦ storage_time
⊦ storage_time
⊦ storage_time
⊦ storage_time
⊦ storage_time
⊦ storage_time
⊦ storage_time
⊦ storage_time
⊦ storage_time
⊦ storage_time
⊦ storage_time
⊦ storage_time
⊦ storage_time
⊦ storage_time
⊦ storage_time
⊦ storage_time
⊦ storage_time
⊦ storage_time
⊦ storage_time
⊦ storage_time
⊦ storage_time
⊦ storage_time
⊦ storage_time
⊦ storage_time
⊦ storage_time
⊦ storage_time
⊦ storage_time
⊦ storage_time
⊦ storage_time
⊦ storage_time
⊦ storage_time
⊦ storage_time
⊦ storage_time
⊦ storage_time
⊦ storage_time
⊦ storage_time
⊦ storage_time
⊦ storage_time
⊦ storage_time
⊦ storage_time
⊦ storage_time

⊦ storage_time
⊦ storage_time
⊦ storage_time
⊦ storage_time
⊦ storage_time
⊦ storage_time
⊦ storage_time
⊦ storage_time
⊦ storage_time
⊦ storage_time
⊦ storage_time
⊦ storage_time
⊦ storage_time
⊦ storage_time
⊦ storage_time
⊦ storage_time
⊦ storage_time
⊦ storage_time
⊦ storage_time
⊦ storage_time
⊦ storage_time
⊦ storage_time
⊦ storage_time
⊦ storage_time
⊦ storage_time
⊦ storage_time
⊦ storage_time
⊦ storage_time
⊦ storage_time
⊦ storage_time
⊦ storage_time
⊦ storage_time
⊦ storage_time
⊦ storage_time
⊦ storage_time
⊦ storage_time
⊦ storage_time
⊦ storage_time
⊦ storage_time
⊦ storage_time
⊦ storage_time
⊦ storage_time
⊦ storage_time
⊦ storage_time
⊦ storage_time
⊦ storage_time
⊦ storage_time
⊦ storage_time

⊦ storage_time
⊦ storage_time
⊦ storage_time
⊦ storage_time
⊦ storage_time
⊦ storage_time
⊦ storage_time
⊦ storage_time
⊦ storage_time
⊦ storage_time
⊦ storage_time
⊦ storage_time
⊦ storage_time
⊦ storage_time
⊦ storage_time
⊦ storage_time
⊦ storage_time
⊦ storage_time
⊦ storage_time
⊦ storage_time
⊦ storage_time
⊦ storage_time
⊦ storage_time
⊦ storage_time
⊦ storage_time
⊦ storage_time
⊦ storage_time
⊦ storage_time
⊦ storage_time
⊦ storage_time
⊦ storage_time
⊦ storage_time
⊦ storage_time
⊦ storage_time
⊦ storage_time
⊦ storage_time
⊦ storage_time
⊦ storage_time
⊦ storage_time
⊦ storage_time
⊦ storage_time
⊦ storage_time
⊦ storage_time
⊦ storage_time
⊦ storage_time
⊦ storage_time
⊦ storage_time
⊦ storage_time

⊦ storage_time
⊦ storage_time
⊦ storage_time
⊦ storage_time
⊦ storage_time
⊦ storage_time
⊦ storage_time
⊦ storage_time
⊦ storage_time
⊦ storage_time
⊦ storage_time
⊦ storage_time
⊦ storage_time
⊦ storage_time
⊦ storage_time
⊦ storage_time
⊦ storage_time
⊦ storage_time
⊦ storage_time
⊦ storage_time
⊦ storage_time
⊦ storage_time
⊦ storage_time
⊦ storage_time
⊦ storage_time
⊦ storage_time
⊦ storage_time
⊦ storage_time
⊦ storage_time
⊦ storage_time
⊦ storage_time
⊦ storage_time
⊦ storage_time
⊦ storage_time
⊦ storage_time
⊦ storage_time
⊦ storage_time
⊦ storage_time
⊦ storage_time
⊦ storage_time
⊦ storage_time
⊦ storage_time
⊦ storage_time
⊦ storage_time
⊦ storage_time
⊦ storage_time
⊦ storage_time
⊦ storage_time

⊦ storage_time
⊦ storage_time
⊦ storage_time
⊦ storage_time
⊦ storage_time
⊦ storage_time
⊦ storage_time
⊦ storage_time
⊦ storage_time
⊦ storage_time
⊦ storage_time
⊦ storage_time
⊦ storage_time
⊦ storage_time
⊦ storage_time
⊦ storage_time
⊦ storage_time
⊦ storage_time
⊦ storage_time
⊦ storage_time
⊦ storage_time
⊦ storage_time
⊦ storage_time
⊦ storage_time
⊦ storage_time
⊦ storage_time
⊦ storage_time
⊦ storage_time
⊦ storage_time
⊦ storage_time
⊦ storage_time
⊦ storage_time
⊦ storage_time
⊦ storage_time
⊦ storage_time
⊦ storage_time
⊦ storage_time
⊦ storage_time
⊦ storage_time
⊦ storage_time
⊦ storage_time
⊦ storage_time
⊦ storage_time
⊦ storage_time
⊦ storage_time
⊦ storage_time
⊦ storage_time
⊦ storage_time

⊦ storage_time
⊦ storage_time
⊦ storage_time
⊦ storage_time
⊦ storage_time
⊦ storage_time
⊦ storage_time
⊦ storage_time
⊦ storage_time
⊦ storage_time
⊦ storage_time
⊦ storage_time
⊦ storage_time
⊦ storage_time
⊦ storage_time
⊦ storage_time
⊦ storage_time
⊦ storage_time
⊦ storage_time
⊦ storage_time
⊦ storage_time
⊦ storage_time
⊦ storage_time
⊦ storage_time
⊦ storage_time
⊦ storage_time
⊦ storage_time
⊦ storage_time
⊦ storage_time
⊦ storage_time
⊦ storage_time
⊦ storage_time
⊦ storage_time
⊦ storage_time
⊦ storage_time
⊦ storage_time
⊦ storage_time
⊦ storage_time
⊦ storage_time
⊦ storage_time
⊦ storage_time
⊦ storage_time
⊦ storage_time
⊦ storage_time
⊦ storage_time
⊦ storage_time
⊦ storage_time
⊦ storage_time

⊦ storage_time
⊦ storage_time
⊦ storage_time
⊦ storage_time
⊦ storage_time
⊦ storage_time
⊦ storage_time
⊦ storage_time
⊦ storage_time
⊦ storage_time
⊦ storage_time
⊦ storage_time
⊦ storage_time
⊦ storage_time
⊦ storage_time
⊦ storage_time
⊦ storage_time
⊦ storage_time
⊦ storage_time
⊦ storage_time
⊦ storage_time
⊦ storage_time
⊦ storage_time
⊦ storage_time
⊦ storage_time
⊦ storage_time
⊦ storage_time
⊦ storage_time
⊦ storage_time
⊦ storage_time
⊦ storage_time
⊦ storage_time
⊦ storage_time
⊦ storage_time
⊦ storage_time
⊦ storage_time
⊦ storage_time
⊦ storage_time
⊦ storage_time
⊦ storage_time
⊦ storage_time
⊦ storage_time
⊦ storage_time
⊦ storage_time
⊦ storage_time
⊦ storage_time
⊦ storage_time
⊦ storage_time

├ storage_time
├ storage_time
├ storage_time
├ storage_time
├ storage_time
├ storage_time
├ storage_time
├ storage_time
├ storage_time
├ storage_time
├ storage_time
├ storage_time
├ storage_time
├ storage_time
├ storage_time
├ storage_time
├ storage_time
├ storage_time
├ storage_time
├ storage_time
├ storage_time
├ storage_time
├ storage_time
├ storage_time
├ storage_time
├ storage_time
├ storage_time
├ storage_time
├ storage_time
├ storage_time
├ storage_time
├ storage_time
├ storage_time
├ storage_time
├ storage_time
├ storage_time
├ storage_time
├ storage_time
├ storage_time
├ storage_time
├ storage_time
├ storage_time
├ storage_time
├ storage_time
├ storage_time
├ storage_time
├ storage_time
├ storage_time

⊦ storage_time
⊦ storage_time
⊦ storage_time
⊦ storage_time
⊦ storage_time
⊦ storage_time
⊦ storage_time
⊦ storage_time
⊦ storage_time
⊦ storage_time
⊦ storage_time
⊦ storage_time
⊦ storage_time
⊦ storage_time
⊦ storage_time
⊦ storage_time
⊦ storage_time
⊦ storage_time
⊦ storage_time
⊦ storage_time
⊦ storage_time
⊦ storage_time
⊦ storage_time
⊦ storage_time
⊦ storage_time
⊦ storage_time
⊦ storage_time
⊦ storage_time
⊦ storage_time
⊦ storage_time
⊦ storage_time
⊦ storage_time
⊦ storage_time
⊦ storage_time
⊦ storage_time
⊦ storage_time
⊦ storage_time
⊦ storage_time
⊦ storage_time
⊦ storage_time
⊦ storage_time
⊦ storage_time
⊦ storage_time
⊦ storage_time
⊦ storage_time
⊦ storage_time
⊦ storage_time
⊦ storage_time

⊦ storage_time
⊦ storage_time
⊦ storage_time
⊦ storage_time
⊦ storage_time
⊦ storage_time
⊦ storage_time
⊦ storage_time
⊦ storage_time
⊦ storage_time
⊦ storage_time
⊦ storage_time
⊦ storage_time
⊦ storage_time
⊦ storage_time
⊦ storage_time
⊦ storage_time
⊦ storage_time
⊦ storage_time
⊦ storage_time
⊦ storage_time
⊦ storage_time
⊦ storage_time
⊦ storage_time
⊦ storage_time
⊦ storage_time
⊦ storage_time
⊦ storage_time
⊦ storage_time
⊦ storage_time
⊦ storage_time
⊦ storage_time
⊦ storage_time
⊦ storage_time
⊦ storage_time
⊦ storage_time
⊦ storage_time
⊦ storage_time
⊦ storage_time
⊦ storage_time
⊦ storage_time
⊦ storage_time
⊦ storage_time
⊦ storage_time
⊦ storage_time
⊦ storage_time
⊦ storage_time
⊦ storage_time

├ storage_time
├ storage_time
├ storage_time
├ storage_time
├ storage_time
├ storage_time
├ storage_time
├ storage_time
├ storage_time
├ storage_time
├ storage_time
├ storage_time
├ storage_time
├ storage_time
├ storage_time
├ storage_time
├ storage_time
├ storage_time
├ storage_time
├ storage_time
├ storage_time
├ storage_time
├ storage_time
├ storage_time
├ storage_time
├ storage_time
├ storage_time
├ storage_time
├ storage_time
├ storage_time
├ storage_time
├ storage_time
├ storage_time
├ storage_time
├ storage_time
├ storage_time
├ storage_time
├ storage_time
├ storage_time
├ storage_time
├ storage_time
├ storage_time
├ storage_time
├ storage_time
├ storage_time
├ storage_time
├ storage_time
├ storage_time

⊦ storage_time
⊦ storage_time
⊦ storage_time
⊦ storage_time
⊦ storage_time
⊦ storage_time
⊦ storage_time
⊦ storage_time
⊦ storage_time
⊦ storage_time
⊦ storage_time
⊦ storage_time
⊦ storage_time
⊦ storage_time
⊦ storage_time
⊦ storage_time
⊦ storage_time
⊦ storage_time
⊦ storage_time
⊦ storage_time
⊦ storage_time
⊦ storage_time
⊦ storage_time
⊦ storage_time
⊦ storage_time
⊦ storage_time
⊦ storage_time
⊦ storage_time
⊦ storage_time
⊦ storage_time
⊦ storage_time
⊦ storage_time
⊦ storage_time
⊦ storage_time
⊦ storage_time
⊦ storage_time
⊦ storage_time
⊦ storage_time
⊦ storage_time
⊦ storage_time
⊦ storage_time
⊦ storage_time
⊦ storage_time
⊦ storage_time
⊦ storage_time
⊦ storage_time
⊦ storage_time
⊦ storage_time

├ storage_time
├ storage_time
├ storage_time
├ storage_time
├ storage_time
├ storage_time
├ storage_time
├ storage_time
├ storage_time
├ storage_time
├ storage_time
├ storage_time
├ storage_time
├ storage_time
├ storage_time
├ storage_time
├ storage_time
├ storage_time
├ storage_time
├ storage_time
├ storage_time
├ storage_time
├ storage_time
├ storage_time
├ storage_time
├ storage_time
├ storage_time
├ storage_time
├ storage_time
├ storage_time
├ storage_time
├ storage_time
├ storage_time
├ storage_time
├ storage_time
├ storage_time
├ storage_time
├ storage_time
├ storage_time
├ storage_time
├ storage_time
├ storage_time
├ storage_time
├ storage_time
├ storage_time
├ storage_time
├ storage_time
├ storage_time

├ storage_time
├ storage_time
├ storage_time
├ storage_time
├ storage_time
├ storage_time
├ storage_time
├ storage_time
├ storage_time
├ storage_time
├ storage_time
├ storage_time
├ storage_time
├ storage_time
├ storage_time
├ storage_time
├ storage_time
├ storage_time
├ storage_time
├ storage_time
├ storage_time
├ storage_time
├ storage_time
├ storage_time
├ storage_time
├ storage_time
├ storage_time
├ storage_time
├ storage_time
├ storage_time
├ storage_time
├ storage_time
├ storage_time
├ storage_time
├ storage_time
├ storage_time
├ storage_time
├ storage_time
├ storage_time
├ storage_time
├ storage_time
├ storage_time
├ storage_time
├ storage_time
├ storage_time
├ storage_time
├ storage_time
├ storage_time

⊦ storage_time
⊦ storage_time
⊦ storage_time
⊦ storage_time
⊦ storage_time
⊦ storage_time
⊦ storage_time
⊦ storage_time
⊦ storage_time
⊦ storage_time
⊦ storage_time
⊦ storage_time
⊦ storage_time
⊦ storage_time
⊦ storage_time
⊦ storage_time
⊦ storage_time
⊦ storage_time
⊦ storage_time
⊦ storage_time
⊦ storage_time
⊦ storage_time
⊦ storage_time
⊦ storage_time
⊦ storage_time
⊦ storage_time
⊦ storage_time
⊦ storage_time
⊦ storage_time
⊦ storage_time
⊦ storage_time
⊦ storage_time
⊦ storage_time
⊦ storage_time
⊦ storage_time
⊦ storage_time
⊦ storage_time
⊦ storage_time
⊦ storage_time
⊦ storage_time
⊦ storage_time
⊦ storage_time
⊦ storage_time
⊦ storage_time
⊦ storage_time
⊦ storage_time
⊦ storage_time
⊦ storage_time

⊦ storage_time
⊦ storage_time
⊦ storage_time
⊦ storage_time
⊦ storage_time
⊦ storage_time
⊦ storage_time
⊦ storage_time
⊦ storage_time
⊦ storage_time
⊦ storage_time
⊦ storage_time
⊦ storage_time
⊦ storage_time
⊦ storage_time
⊦ storage_time
⊦ storage_time
⊦ storage_time
⊦ storage_time
⊦ storage_time
⊦ storage_time
⊦ storage_time
⊦ storage_time
⊦ storage_time
⊦ storage_time
⊦ storage_time
⊦ storage_time
⊦ storage_time
⊦ storage_time
⊦ storage_time
⊦ storage_time
⊦ storage_time
⊦ storage_time
⊦ storage_time
⊦ storage_time
⊦ storage_time
⊦ storage_time
⊦ storage_time
⊦ storage_time
⊦ storage_time
⊦ storage_time
⊦ storage_time
⊦ storage_time
⊦ storage_time
⊦ storage_time
⊦ storage_time
⊦ storage_time
⊦ storage_time

⊦ storage_time
⊦ storage_time
⊦ storage_time
⊦ storage_time
⊦ storage_time
⊦ storage_time
⊦ storage_time
⊦ storage_time
⊦ storage_time
⊦ storage_time
⊦ storage_time
⊦ storage_time
⊦ storage_time
⊦ storage_time
⊦ storage_time
⊦ storage_time
⊦ storage_time
⊦ storage_time
⊦ storage_time
⊦ storage_time
⊦ storage_time
⊦ storage_time
⊦ storage_time
⊦ storage_time
⊦ storage_time
⊦ storage_time
⊦ storage_time
⊦ storage_time
⊦ storage_time
⊦ storage_time
⊦ storage_time
⊦ storage_time
⊦ storage_time
⊦ storage_time
⊦ storage_time
⊦ storage_time
⊦ storage_time
⊦ storage_time
⊦ storage_time
⊦ storage_time
⊦ storage_time
⊦ storage_time
⊦ storage_time
⊦ storage_time
⊦ storage_time
⊦ storage_time
⊦ storage_time
⊦ storage_time

⊦ storage_time
⊦ storage_time
⊦ storage_time
⊦ storage_time
⊦ storage_time
⊦ storage_time
⊦ storage_time
⊦ storage_time
⊦ storage_time
⊦ storage_time
⊦ storage_time
⊦ storage_time
⊦ storage_time
⊦ storage_time
⊦ storage_time
⊦ storage_time
⊦ storage_time
⊦ storage_time
⊦ storage_time
⊦ storage_time
⊦ storage_time
⊦ storage_time
⊦ storage_time
⊦ storage_time
⊦ storage_time
⊦ storage_time
⊦ storage_time
⊦ storage_time
⊦ storage_time
⊦ storage_time
⊦ storage_time
⊦ storage_time
⊦ storage_time
⊦ storage_time
⊦ storage_time
⊦ storage_time
⊦ storage_time
⊦ storage_time
⊦ storage_time
⊦ storage_time
⊦ storage_time
⊦ storage_time
⊦ storage_time
