## Supplemental table S11. Procrustes analysis of microbiome-metabolome associations. for "A YOUNG CHILD FORMULA SUPPLEMENTED WITH L. REUTERI AND GALACTO-OLIGOSACCHARIDES MODULATES THE COMPOSITION AND FUNCTION OF THE GUT MICROBIOME SUPPORTING BONE AND MUSCLE DEVELOPMENT IN TODDLERS"

| Intervention | Visit | P-value | Correlation coefficient |
| --- | --- | --- | --- |
| Pooled | All | 0,0001 | 0,342 |
| Pooled | V1 | 0,0001 | 0,405 |
| Pooled | V2 | 0,0001 | 0,296 |
| Pooled | V3 | 0,0001 | 0,322 |
| Experimental | All | 0,0001 | 0,42 |
| Experimental | V1 | 0,0001 | 0,462 |
| Experimental | V2 | 0,0004 | 0,339 |
| Experimental | V3 | 0,0001 | 0,469 |
| Control | All | 0,0001 | 0,323 |
| Control | V1 | 0,0001 | 0,425 |
| Control | V2 | 0,012 | 0,265 |
| Control | V3 | 0,0007 | 0,335 |
| Reference | All | 0,0001 | 0,27 |
| Reference | V1 | 0,0001 | 0,363 |
| Reference | V2 | 0,0001 | 0,276 |
| Reference | V3 | 0,0947 | 0,191 |
