## Supplemental table S12. Associations between microbiome taxonomic composition and clinical outcomes. for "A YOUNG CHILD FORMULA SUPPLEMENTED WITH L. REUTERI AND GALACTO-OLIGOSACCHARIDES MODULATES THE COMPOSITION AND FUNCTION OF THE GUT MICROBIOME SUPPORTING BONE AND MUSCLE DEVELOPMENT IN TODDLERS"

| Category | Test type | Item | Feature |
|---|---|---|---|
| MGS | abundance | Hgn3C.0001 - Phocaeicola vulgatus | Tibia_length |
| MGS | abundance | Hgn3C.0001 - Phocaeicola vulgatus | Radius_length |
| MGS | abundance | Hgn3C.0001 - Phocaeicola vulgatus | Radius_SOS |
| MGS | abundance | Hgn3C.0001 - Phocaeicola vulgatus | Tibia_SOS |
| MGS | abundance | Hgn3C.0001 - Phocaeicola vulgatus | Handgrip |
| MGS | abundance | Hgn3C.0002 - Bacteroides uniformis | Tibia_length |
| MGS | abundance | Hgn3C.0002 - Bacteroides uniformis | Radius_length |
| MGS | abundance | Hgn3C.0002 - Bacteroides uniformis | Radius_SOS |
| MGS | abundance | Hgn3C.0002 - Bacteroides uniformis | Tibia_SOS |
| MGS | abundance | Hgn3C.0002 - Bacteroides uniformis | Handgrip |
| MGS | abundance | Hgn3C.0003 - Lachnospiraceae sp. | Tibia_length |
| MGS | abundance | Hgn3C.0003 - Lachnospiraceae sp. | Radius_length |
| MGS | abundance | Hgn3C.0003 - Lachnospiraceae sp. | Radius_SOS |
| MGS | abundance | Hgn3C.0003 - Lachnospiraceae sp. | Tibia_SOS |
| MGS | abundance | Hgn3C.0003 - Lachnospiraceae sp. | Handgrip |
| MGS | abundance | Hgn3C.0004 - Blautia wexlerae | Tibia_length |
| MGS | abundance | Hgn3C.0004 - Blautia wexlerae | Radius_length |
| MGS | abundance | Hgn3C.0004 - Blautia wexlerae | Radius_SOS |
| MGS | abundance | Hgn3C.0004 - Blautia wexlerae | Tibia_SOS |
| MGS | abundance | Hgn3C.0004 - Blautia wexlerae | Handgrip |
| MGS | abundance | Hgn3C.0005 - Fusicatenibacter saccharivorans | Tibia_length |
| MGS | abundance | Hgn3C.0005 - Fusicatenibacter saccharivorans | Radius_length |
| MGS | abundance | Hgn3C.0005 - Fusicatenibacter saccharivorans | Radius_SOS |
| MGS | abundance | Hgn3C.0005 - Fusicatenibacter saccharivorans | Tibia_SOS |
| MGS | abundance | Hgn3C.0005 - Fusicatenibacter saccharivorans | Handgrip |
| MGS | abundance | Hgn3C.0006 - Bacteroides ovatus | Tibia_length |
| MGS | abundance | Hgn3C.0006 - Bacteroides ovatus | Radius_length |
| MGS | abundance | Hgn3C.0006 - Bacteroides ovatus | Radius_SOS |
| MGS | abundance | Hgn3C.0006 - Bacteroides ovatus | Tibia_SOS |
| MGS | abundance | Hgn3C.0006 - Bacteroides ovatus | Handgrip |
| MGS | abundance | Hgn3C.0007 - Parabacteroides distasonis | Tibia_length |
| MGS | abundance | Hgn3C.0007 - Parabacteroides distasonis | Radius_length |
| MGS | abundance | Hgn3C.0007 - Parabacteroides distasonis | Radius_SOS |
| MGS | abundance | Hgn3C.0007 - Parabacteroides distasonis | Tibia_SOS |
| MGS | abundance | Hgn3C.0007 - Parabacteroides distasonis | Handgrip |
| MGS | abundance | Hgn3C.0008 - Alistipes putredinis | Tibia_length |
| MGS | abundance | Hgn3C.0008 - Alistipes putredinis | Radius_length |
| MGS | abundance | Hgn3C.0008 - Alistipes putredinis | Radius_SOS |
| MGS | abundance | Hgn3C.0008 - Alistipes putredinis | Tibia_SOS |
| MGS | abundance | Hgn3C.0008 - Alistipes putredinis | Handgrip |
| MGS | abundance | Hgn3C.0009 - Faecalibacterium longum CLA-AA-H243 | Tibia_length |
| MGS | abundance | Hgn3C.0009 - Faecalibacterium longum CLA-AA-H243 | Radius_length |
| MGS | abundance | Hgn3C.0009 - Faecalibacterium longum CLA-AA-H243 | Radius_SOS |
| MGS | abundance | Hgn3C.0009 - Faecalibacterium longum CLA-AA-H243 | Tibia_SOS |
| MGS | abundance | Hgn3C.0009 - Faecalibacterium longum CLA-AA-H243 | Handgrip |
| MGS | abundance | Hgn3C.0010 - Anaerostipes hadrus | Tibia_length |
| MGS | abundance | Hgn3C.0010 - Anaerostipes hadrus | Radius_length |

| | | | |
|---|---|---|---|
| MGS | abundance | Hgn3C.0010 - Anaerostipes hadrus | Radius_SOS |
| MGS | abundance | Hgn3C.0010 - Anaerostipes hadrus | Tibia_SOS |
| MGS | abundance | Hgn3C.0010 - Anaerostipes hadrus | Handgrip |
| MGS | abundance | Hgn3C.0011 - Roseburia faecis | Tibia_length |
| MGS | abundance | Hgn3C.0011 - Roseburia faecis | Radius_length |
| MGS | abundance | Hgn3C.0011 - Roseburia faecis | Radius_SOS |
| MGS | abundance | Hgn3C.0011 - Roseburia faecis | Tibia_SOS |
| MGS | abundance | Hgn3C.0011 - Roseburia faecis | Handgrip |
| MGS | abundance | Hgn3C.0012 - Lachnospira sp. | Tibia_length |
| MGS | abundance | Hgn3C.0012 - Lachnospira sp. | Radius_length |
| MGS | abundance | Hgn3C.0012 - Lachnospira sp. | Radius_SOS |
| MGS | abundance | Hgn3C.0012 - Lachnospira sp. | Tibia_SOS |
| MGS | abundance | Hgn3C.0012 - Lachnospira sp. | Handgrip |
| MGS | abundance | Hgn3C.0013 - Ruminococcus bromii | Tibia_length |
| MGS | abundance | Hgn3C.0013 - Ruminococcus bromii | Radius_length |
| MGS | abundance | Hgn3C.0013 - Ruminococcus bromii | Radius_SOS |
| MGS | abundance | Hgn3C.0013 - Ruminococcus bromii | Tibia_SOS |
| MGS | abundance | Hgn3C.0013 - Ruminococcus bromii | Handgrip |
| MGS | abundance | Hgn3C.0014 - Lacrimispora saccharolytica | Tibia_length |
| MGS | abundance | Hgn3C.0014 - Lacrimispora saccharolytica | Radius_length |
| MGS | abundance | Hgn3C.0014 - Lacrimispora saccharolytica | Radius_SOS |
| MGS | abundance | Hgn3C.0014 - Lacrimispora saccharolytica | Tibia_SOS |
| MGS | abundance | Hgn3C.0014 - Lacrimispora saccharolytica | Handgrip |
| MGS | abundance | Hgn3C.0015 - Faecalibacterium sp. Marseille-Q4896 | Tibia_length |
| MGS | abundance | Hgn3C.0015 - Faecalibacterium sp. Marseille-Q4896 | Radius_length |
| MGS | abundance | Hgn3C.0015 - Faecalibacterium sp. Marseille-Q4896 | Radius_SOS |
| MGS | abundance | Hgn3C.0015 - Faecalibacterium sp. Marseille-Q4896 | Tibia_SOS |
| MGS | abundance | Hgn3C.0015 - Faecalibacterium sp. Marseille-Q4896 | Handgrip |
| MGS | abundance | Hgn3C.0016 - Parabacteroides merdae | Tibia_length |
| MGS | abundance | Hgn3C.0016 - Parabacteroides merdae | Radius_length |
| MGS | abundance | Hgn3C.0016 - Parabacteroides merdae | Radius_SOS |
| MGS | abundance | Hgn3C.0016 - Parabacteroides merdae | Tibia_SOS |
| MGS | abundance | Hgn3C.0016 - Parabacteroides merdae | Handgrip |
| MGS | abundance | Hgn3C.0017 - Subdoligranulum sp. APC924/74 | Tibia_length |
| MGS | abundance | Hgn3C.0017 - Subdoligranulum sp. APC924/74 | Radius_length |
| MGS | abundance | Hgn3C.0017 - Subdoligranulum sp. APC924/74 | Radius_SOS |
| MGS | abundance | Hgn3C.0017 - Subdoligranulum sp. APC924/74 | Tibia_SOS |
| MGS | abundance | Hgn3C.0017 - Subdoligranulum sp. APC924/74 | Handgrip |
| MGS | abundance | Hgn3C.0018 - Faecalibacterium prausnitzii | Tibia_length |
| MGS | abundance | Hgn3C.0018 - Faecalibacterium prausnitzii | Radius_length |
| MGS | abundance | Hgn3C.0018 - Faecalibacterium prausnitzii | Radius_SOS |
| MGS | abundance | Hgn3C.0018 - Faecalibacterium prausnitzii | Tibia_SOS |
| MGS | abundance | Hgn3C.0018 - Faecalibacterium prausnitzii | Handgrip |
| MGS | abundance | Hgn3C.0019 - Phocaeicola dorei | Tibia_length |
| MGS | abundance | Hgn3C.0019 - Phocaeicola dorei | Radius_length |
| MGS | abundance | Hgn3C.0019 - Phocaeicola dorei | Radius_SOS |
| MGS | abundance | Hgn3C.0019 - Phocaeicola dorei | Tibia_SOS |
| MGS | abundance | Hgn3C.0019 - Phocaeicola dorei | Handgrip |

| | | | |
|---|---|---|---|
| MGS | abundance | Hgn3C.0020 - Collinsella aerofaciens | Tibia_length |
| MGS | abundance | Hgn3C.0020 - Collinsella aerofaciens | Radius_length |
| MGS | abundance | Hgn3C.0020 - Collinsella aerofaciens | Radius_SOS |
| MGS | abundance | Hgn3C.0020 - Collinsella aerofaciens | Tibia_SOS |
| MGS | abundance | Hgn3C.0020 - Collinsella aerofaciens | Handgrip |
| MGS | abundance | Hgn3C.0021 - Bifidobacterium longum subsp. longum | Tibia_length |
| MGS | abundance | Hgn3C.0021 - Bifidobacterium longum subsp. longum | Radius_length |
| MGS | abundance | Hgn3C.0021 - Bifidobacterium longum subsp. longum | Radius_SOS |
| MGS | abundance | Hgn3C.0021 - Bifidobacterium longum subsp. longum | Tibia_SOS |
| MGS | abundance | Hgn3C.0021 - Bifidobacterium longum subsp. longum | Handgrip |
| MGS | abundance | Hgn3C.0022 - Alistipes shahii | Tibia_length |
| MGS | abundance | Hgn3C.0022 - Alistipes shahii | Radius_length |
| MGS | abundance | Hgn3C.0022 - Alistipes shahii | Radius_SOS |
| MGS | abundance | Hgn3C.0022 - Alistipes shahii | Tibia_SOS |
| MGS | abundance | Hgn3C.0022 - Alistipes shahii | Handgrip |
| MGS | abundance | Hgn3C.0023 - Bacteroides xylanisolvens | Tibia_length |
| MGS | abundance | Hgn3C.0023 - Bacteroides xylanisolvens | Radius_length |
| MGS | abundance | Hgn3C.0023 - Bacteroides xylanisolvens | Radius_SOS |
| MGS | abundance | Hgn3C.0023 - Bacteroides xylanisolvens | Tibia_SOS |
| MGS | abundance | Hgn3C.0023 - Bacteroides xylanisolvens | Handgrip |
| MGS | abundance | Hgn3C.0024 - Bacteroides caccae | Tibia_length |
| MGS | abundance | Hgn3C.0024 - Bacteroides caccae | Radius_length |
| MGS | abundance | Hgn3C.0024 - Bacteroides caccae | Radius_SOS |
| MGS | abundance | Hgn3C.0024 - Bacteroides caccae | Tibia_SOS |
| MGS | abundance | Hgn3C.0024 - Bacteroides caccae | Handgrip |
| MGS | abundance | Hgn3C.0025 - Ruminococcus bicirculans | Tibia_length |
| MGS | abundance | Hgn3C.0025 - Ruminococcus bicirculans | Radius_length |
| MGS | abundance | Hgn3C.0025 - Ruminococcus bicirculans | Radius_SOS |
| MGS | abundance | Hgn3C.0025 - Ruminococcus bicirculans | Tibia_SOS |
| MGS | abundance | Hgn3C.0025 - Ruminococcus bicirculans | Handgrip |
| MGS | abundance | Hgn3C.0026 - Dorea longicatena | Tibia_length |
| MGS | abundance | Hgn3C.0026 - Dorea longicatena | Radius_length |
| MGS | abundance | Hgn3C.0026 - Dorea longicatena | Radius_SOS |
| MGS | abundance | Hgn3C.0026 - Dorea longicatena | Tibia_SOS |
| MGS | abundance | Hgn3C.0026 - Dorea longicatena | Handgrip |
| MGS | abundance | Hgn3C.0027 - Barnesiella intestinihominis | Tibia_length |
| MGS | abundance | Hgn3C.0027 - Barnesiella intestinihominis | Radius_length |
| MGS | abundance | Hgn3C.0027 - Barnesiella intestinihominis | Radius_SOS |
| MGS | abundance | Hgn3C.0027 - Barnesiella intestinihominis | Tibia_SOS |
| MGS | abundance | Hgn3C.0027 - Barnesiella intestinihominis | Handgrip |
| MGS | abundance | Hgn3C.0028 - Oscillibacter sp. ER4 | Tibia_length |
| MGS | abundance | Hgn3C.0028 - Oscillibacter sp. ER4 | Radius_length |
| MGS | abundance | Hgn3C.0028 - Oscillibacter sp. ER4 | Radius_SOS |
| MGS | abundance | Hgn3C.0028 - Oscillibacter sp. ER4 | Tibia_SOS |
| MGS | abundance | Hgn3C.0028 - Oscillibacter sp. ER4 | Handgrip |
| MGS | abundance | Hgn3C.0029 - Anaerobutyricum hallii | Tibia_length |
| MGS | abundance | Hgn3C.0029 - Anaerobutyricum hallii | Radius_length |
| MGS | abundance | Hgn3C.0029 - Anaerobutyricum hallii | Radius_SOS |

| | | | |
|---|---|---|---|
| MGS | abundance | Hgn3C.0029 - Anaerobutyricum hallii | Tibia_SOS |
| MGS | abundance | Hgn3C.0029 - Anaerobutyricum hallii | Handgrip |
| MGS | abundance | Hgn3C.0030 - Faecalibacterium prausnitzii | Tibia_length |
| MGS | abundance | Hgn3C.0030 - Faecalibacterium prausnitzii | Radius_length |
| MGS | abundance | Hgn3C.0030 - Faecalibacterium prausnitzii | Radius_SOS |
| MGS | abundance | Hgn3C.0030 - Faecalibacterium prausnitzii | Tibia_SOS |
| MGS | abundance | Hgn3C.0030 - Faecalibacterium prausnitzii | Handgrip |
| MGS | abundance | Hgn3C.0031 - Blautia massiliensis | Tibia_length |
| MGS | abundance | Hgn3C.0031 - Blautia massiliensis | Radius_length |
| MGS | abundance | Hgn3C.0031 - Blautia massiliensis | Radius_SOS |
| MGS | abundance | Hgn3C.0031 - Blautia massiliensis | Tibia_SOS |
| MGS | abundance | Hgn3C.0031 - Blautia massiliensis | Handgrip |
| MGS | abundance | Hgn3C.0032 - Alistipes onderdonkii subsp. vulgaris | Tibia_length |
| MGS | abundance | Hgn3C.0032 - Alistipes onderdonkii subsp. vulgaris | Radius_length |
| MGS | abundance | Hgn3C.0032 - Alistipes onderdonkii subsp. vulgaris | Radius_SOS |
| MGS | abundance | Hgn3C.0032 - Alistipes onderdonkii subsp. vulgaris | Tibia_SOS |
| MGS | abundance | Hgn3C.0032 - Alistipes onderdonkii subsp. vulgaris | Handgrip |
| MGS | abundance | Hgn3C.0033 - Gemmiger formicilis | Tibia_length |
| MGS | abundance | Hgn3C.0033 - Gemmiger formicilis | Radius_length |
| MGS | abundance | Hgn3C.0033 - Gemmiger formicilis | Radius_SOS |
| MGS | abundance | Hgn3C.0033 - Gemmiger formicilis | Tibia_SOS |
| MGS | abundance | Hgn3C.0033 - Gemmiger formicilis | Handgrip |
| MGS | abundance | Hgn3C.0034 - Roseburia inulinivorans | Tibia_length |
| MGS | abundance | Hgn3C.0034 - Roseburia inulinivorans | Radius_length |
| MGS | abundance | Hgn3C.0034 - Roseburia inulinivorans | Radius_SOS |
| MGS | abundance | Hgn3C.0034 - Roseburia inulinivorans | Tibia_SOS |
| MGS | abundance | Hgn3C.0034 - Roseburia inulinivorans | Handgrip |
| MGS | abundance | Hgn3C.0035 - Odoribacter splanchnicus | Tibia_length |
| MGS | abundance | Hgn3C.0035 - Odoribacter splanchnicus | Radius_length |
| MGS | abundance | Hgn3C.0035 - Odoribacter splanchnicus | Radius_SOS |
| MGS | abundance | Hgn3C.0035 - Odoribacter splanchnicus | Tibia_SOS |
| MGS | abundance | Hgn3C.0035 - Odoribacter splanchnicus | Handgrip |
| MGS | abundance | Hgn3C.0036 - Bacteroides thetaiotaomicron | Tibia_length |
| MGS | abundance | Hgn3C.0036 - Bacteroides thetaiotaomicron | Radius_length |
| MGS | abundance | Hgn3C.0036 - Bacteroides thetaiotaomicron | Radius_SOS |
| MGS | abundance | Hgn3C.0036 - Bacteroides thetaiotaomicron | Tibia_SOS |
| MGS | abundance | Hgn3C.0036 - Bacteroides thetaiotaomicron | Handgrip |
| MGS | abundance | Hgn3C.0037 - Roseburia intestinalis | Tibia_length |
| MGS | abundance | Hgn3C.0037 - Roseburia intestinalis | Radius_length |
| MGS | abundance | Hgn3C.0037 - Roseburia intestinalis | Radius_SOS |
| MGS | abundance | Hgn3C.0037 - Roseburia intestinalis | Tibia_SOS |
| MGS | abundance | Hgn3C.0037 - Roseburia intestinalis | Handgrip |
| MGS | abundance | Hgn3C.0038 - Bifidobacterium adolescentis | Tibia_length |
| MGS | abundance | Hgn3C.0038 - Bifidobacterium adolescentis | Radius_length |
| MGS | abundance | Hgn3C.0038 - Bifidobacterium adolescentis | Radius_SOS |
| MGS | abundance | Hgn3C.0038 - Bifidobacterium adolescentis | Tibia_SOS |
| MGS | abundance | Hgn3C.0038 - Bifidobacterium adolescentis | Handgrip |
| MGS | abundance | Hgn3C.0039 - Coprococcus comes | Tibia_length |

| | | | |
|---|---|---|---|
| MGS | abundance | Hgn3C.0039 - Coprococcus comes | Radius_length |
| MGS | abundance | Hgn3C.0039 - Coprococcus comes | Radius_SOS |
| MGS | abundance | Hgn3C.0039 - Coprococcus comes | Tibia_SOS |
| MGS | abundance | Hgn3C.0039 - Coprococcus comes | Handgrip |
| MGS | abundance | Hgn3C.0040 - Eubacteriales sp. | Tibia_length |
| MGS | abundance | Hgn3C.0040 - Eubacteriales sp. | Radius_length |
| MGS | abundance | Hgn3C.0040 - Eubacteriales sp. | Radius_SOS |
| MGS | abundance | Hgn3C.0040 - Eubacteriales sp. | Tibia_SOS |
| MGS | abundance | Hgn3C.0040 - Eubacteriales sp. | Handgrip |
| MGS | abundance | Hgn3C.0041 - Ruminococcus sp. | Tibia_length |
| MGS | abundance | Hgn3C.0041 - Ruminococcus sp. | Radius_length |
| MGS | abundance | Hgn3C.0041 - Ruminococcus sp. | Radius_SOS |
| MGS | abundance | Hgn3C.0041 - Ruminococcus sp. | Tibia_SOS |
| MGS | abundance | Hgn3C.0041 - Ruminococcus sp. | Handgrip |
| MGS | abundance | Hgn3C.0042 - Blautia faecis | Tibia_length |
| MGS | abundance | Hgn3C.0042 - Blautia faecis | Radius_length |
| MGS | abundance | Hgn3C.0042 - Blautia faecis | Radius_SOS |
| MGS | abundance | Hgn3C.0042 - Blautia faecis | Tibia_SOS |
| MGS | abundance | Hgn3C.0042 - Blautia faecis | Handgrip |
| MGS | abundance | Hgn3C.0043 - Lachnoclostridium sp. 210928-DFI.6.3 | Tibia_length |
| MGS | abundance | Hgn3C.0043 - Lachnoclostridium sp. 210928-DFI.6.3 | Radius_length |
| MGS | abundance | Hgn3C.0043 - Lachnoclostridium sp. 210928-DFI.6.3 | Radius_SOS |
| MGS | abundance | Hgn3C.0043 - Lachnoclostridium sp. 210928-DFI.6.3 | Tibia_SOS |
| MGS | abundance | Hgn3C.0043 - Lachnoclostridium sp. 210928-DFI.6.3 | Handgrip |
| MGS | abundance | Hgn3C.0044 - Lachnospira pectinoschiza | Tibia_length |
| MGS | abundance | Hgn3C.0044 - Lachnospira pectinoschiza | Radius_length |
| MGS | abundance | Hgn3C.0044 - Lachnospira pectinoschiza | Radius_SOS |
| MGS | abundance | Hgn3C.0044 - Lachnospira pectinoschiza | Tibia_SOS |
| MGS | abundance | Hgn3C.0044 - Lachnospira pectinoschiza | Handgrip |
| MGS | abundance | Hgn3C.0045 - Faecalibacterium sp. | Tibia_length |
| MGS | abundance | Hgn3C.0045 - Faecalibacterium sp. | Radius_length |
| MGS | abundance | Hgn3C.0045 - Faecalibacterium sp. | Radius_SOS |
| MGS | abundance | Hgn3C.0045 - Faecalibacterium sp. | Tibia_SOS |
| MGS | abundance | Hgn3C.0045 - Faecalibacterium sp. | Handgrip |
| MGS | abundance | Hgn3C.0046 - Clostridium sp. AF37-5 | Tibia_length |
| MGS | abundance | Hgn3C.0046 - Clostridium sp. AF37-5 | Radius_length |
| MGS | abundance | Hgn3C.0046 - Clostridium sp. AF37-5 | Radius_SOS |
| MGS | abundance | Hgn3C.0046 - Clostridium sp. AF37-5 | Tibia_SOS |
| MGS | abundance | Hgn3C.0046 - Clostridium sp. AF37-5 | Handgrip |
| MGS | abundance | Hgn3C.0047 - Dysosmobacter sp. BX15 | Tibia_length |
| MGS | abundance | Hgn3C.0047 - Dysosmobacter sp. BX15 | Radius_length |
| MGS | abundance | Hgn3C.0047 - Dysosmobacter sp. BX15 | Radius_SOS |
| MGS | abundance | Hgn3C.0047 - Dysosmobacter sp. BX15 | Tibia_SOS |
| MGS | abundance | Hgn3C.0047 - Dysosmobacter sp. BX15 | Handgrip |
| MGS | abundance | Hgn3C.0048 - Akkermansia muciniphila | Tibia_length |
| MGS | abundance | Hgn3C.0048 - Akkermansia muciniphila | Radius_length |
| MGS | abundance | Hgn3C.0048 - Akkermansia muciniphila | Radius_SOS |
| MGS | abundance | Hgn3C.0048 - Akkermansia muciniphila | Tibia_SOS |

| | | | |
|---|---|---|---|
| MGS | abundance | Hgn3C.0048 - Akkermansia muciniphila | Handgrip |
| MGS | abundance | Hgn3C.0049 - Eubacteriales sp. | Tibia_length |
| MGS | abundance | Hgn3C.0049 - Eubacteriales sp. | Radius_length |
| MGS | abundance | Hgn3C.0049 - Eubacteriales sp. | Radius_SOS |
| MGS | abundance | Hgn3C.0049 - Eubacteriales sp. | Tibia_SOS |
| MGS | abundance | Hgn3C.0049 - Eubacteriales sp. | Handgrip |
| MGS | abundance | Hgn3C.0050 - Escherichia coli | Tibia_length |
| MGS | abundance | Hgn3C.0050 - Escherichia coli | Radius_length |
| MGS | abundance | Hgn3C.0050 - Escherichia coli | Radius_SOS |
| MGS | abundance | Hgn3C.0050 - Escherichia coli | Tibia_SOS |
| MGS | abundance | Hgn3C.0050 - Escherichia coli | Handgrip |
| MGS | abundance | Hgn3C.0051 - Alistipes communis | Tibia_length |
| MGS | abundance | Hgn3C.0051 - Alistipes communis | Radius_length |
| MGS | abundance | Hgn3C.0051 - Alistipes communis | Radius_SOS |
| MGS | abundance | Hgn3C.0051 - Alistipes communis | Tibia_SOS |
| MGS | abundance | Hgn3C.0051 - Alistipes communis | Handgrip |
| MGS | abundance | Hgn3C.0052 - Phocaeicola massiliensis | Tibia_length |
| MGS | abundance | Hgn3C.0052 - Phocaeicola massiliensis | Radius_length |
| MGS | abundance | Hgn3C.0052 - Phocaeicola massiliensis | Radius_SOS |
| MGS | abundance | Hgn3C.0052 - Phocaeicola massiliensis | Tibia_SOS |
| MGS | abundance | Hgn3C.0052 - Phocaeicola massiliensis | Handgrip |
| MGS | abundance | Hgn3C.0053 - Eubacteriales sp. | Tibia_length |
| MGS | abundance | Hgn3C.0053 - Eubacteriales sp. | Radius_length |
| MGS | abundance | Hgn3C.0053 - Eubacteriales sp. | Radius_SOS |
| MGS | abundance | Hgn3C.0053 - Eubacteriales sp. | Tibia_SOS |
| MGS | abundance | Hgn3C.0053 - Eubacteriales sp. | Handgrip |
| MGS | abundance | Hgn3C.0054 - Prevotella copri | Tibia_length |
| MGS | abundance | Hgn3C.0054 - Prevotella copri | Radius_length |
| MGS | abundance | Hgn3C.0054 - Prevotella copri | Radius_SOS |
| MGS | abundance | Hgn3C.0054 - Prevotella copri | Tibia_SOS |
| MGS | abundance | Hgn3C.0054 - Prevotella copri | Handgrip |
| MGS | abundance | Hgn3C.0055 - Alistipes finegoldii | Tibia_length |
| MGS | abundance | Hgn3C.0055 - Alistipes finegoldii | Radius_length |
| MGS | abundance | Hgn3C.0055 - Alistipes finegoldii | Radius_SOS |
| MGS | abundance | Hgn3C.0055 - Alistipes finegoldii | Tibia_SOS |
| MGS | abundance | Hgn3C.0055 - Alistipes finegoldii | Handgrip |
| MGS | abundance | Hgn3C.0056 - Bacteroides fragilis | Tibia_length |
| MGS | abundance | Hgn3C.0056 - Bacteroides fragilis | Radius_length |
| MGS | abundance | Hgn3C.0056 - Bacteroides fragilis | Radius_SOS |
| MGS | abundance | Hgn3C.0056 - Bacteroides fragilis | Tibia_SOS |
| MGS | abundance | Hgn3C.0056 - Bacteroides fragilis | Handgrip |
| MGS | abundance | Hgn3C.0057 - Oscillibacter sp. MSJ-31 | Tibia_length |
| MGS | abundance | Hgn3C.0057 - Oscillibacter sp. MSJ-31 | Radius_length |
| MGS | abundance | Hgn3C.0057 - Oscillibacter sp. MSJ-31 | Radius_SOS |
| MGS | abundance | Hgn3C.0057 - Oscillibacter sp. MSJ-31 | Tibia_SOS |
| MGS | abundance | Hgn3C.0057 - Oscillibacter sp. MSJ-31 | Handgrip |
| MGS | abundance | Hgn3C.0058 - Lachnospiraceae sp. | Tibia_length |
| MGS | abundance | Hgn3C.0058 - Lachnospiraceae sp. | Radius_length |

| | | | |
|---|---|---|---|
| MGS | abundance | Hgn3C.0058 - Lachnospiraceae sp. | Radius_SOS |
| MGS | abundance | Hgn3C.0058 - Lachnospiraceae sp. | Tibia_SOS |
| MGS | abundance | Hgn3C.0058 - Lachnospiraceae sp. | Handgrip |
| MGS | abundance | Hgn3C.0059 - Bacteroides cellulosilyticus | Tibia_length |
| MGS | abundance | Hgn3C.0059 - Bacteroides cellulosilyticus | Radius_length |
| MGS | abundance | Hgn3C.0059 - Bacteroides cellulosilyticus | Radius_SOS |
| MGS | abundance | Hgn3C.0059 - Bacteroides cellulosilyticus | Tibia_SOS |
| MGS | abundance | Hgn3C.0059 - Bacteroides cellulosilyticus | Handgrip |
| MGS | abundance | Hgn3C.0060 - Eubacteriales sp. | Tibia_length |
| MGS | abundance | Hgn3C.0060 - Eubacteriales sp. | Radius_length |
| MGS | abundance | Hgn3C.0060 - Eubacteriales sp. | Radius_SOS |
| MGS | abundance | Hgn3C.0060 - Eubacteriales sp. | Tibia_SOS |
| MGS | abundance | Hgn3C.0060 - Eubacteriales sp. | Handgrip |
| MGS | abundance | Hgn3C.0061 - Faecalibacillus sp. TM498 | Tibia_length |
| MGS | abundance | Hgn3C.0061 - Faecalibacillus sp. TM498 | Radius_length |
| MGS | abundance | Hgn3C.0061 - Faecalibacillus sp. TM498 | Radius_SOS |
| MGS | abundance | Hgn3C.0061 - Faecalibacillus sp. TM498 | Tibia_SOS |
| MGS | abundance | Hgn3C.0061 - Faecalibacillus sp. TM498 | Handgrip |
| MGS | abundance | Hgn3C.0062 - Waltera intestinalis | Tibia_length |
| MGS | abundance | Hgn3C.0062 - Waltera intestinalis | Radius_length |
| MGS | abundance | Hgn3C.0062 - Waltera intestinalis | Radius_SOS |
| MGS | abundance | Hgn3C.0062 - Waltera intestinalis | Tibia_SOS |
| MGS | abundance | Hgn3C.0062 - Waltera intestinalis | Handgrip |
| MGS | abundance | Hgn3C.0063 - Eubacteriales sp. | Tibia_length |
| MGS | abundance | Hgn3C.0063 - Eubacteriales sp. | Radius_length |
| MGS | abundance | Hgn3C.0063 - Eubacteriales sp. | Radius_SOS |
| MGS | abundance | Hgn3C.0063 - Eubacteriales sp. | Tibia_SOS |
| MGS | abundance | Hgn3C.0063 - Eubacteriales sp. | Handgrip |
| MGS | abundance | Hgn3C.0064 - Blautia sp. DFI.9.9 | Tibia_length |
| MGS | abundance | Hgn3C.0064 - Blautia sp. DFI.9.9 | Radius_length |
| MGS | abundance | Hgn3C.0064 - Blautia sp. DFI.9.9 | Radius_SOS |
| MGS | abundance | Hgn3C.0064 - Blautia sp. DFI.9.9 | Tibia_SOS |
| MGS | abundance | Hgn3C.0064 - Blautia sp. DFI.9.9 | Handgrip |
| MGS | abundance | Hgn3C.0065 - Blautia obeum | Tibia_length |
| MGS | abundance | Hgn3C.0065 - Blautia obeum | Radius_length |
| MGS | abundance | Hgn3C.0065 - Blautia obeum | Radius_SOS |
| MGS | abundance | Hgn3C.0065 - Blautia obeum | Tibia_SOS |
| MGS | abundance | Hgn3C.0065 - Blautia obeum | Handgrip |
| MGS | abundance | Hgn3C.0066 - Oscillospiraceae sp. | Tibia_length |
| MGS | abundance | Hgn3C.0066 - Oscillospiraceae sp. | Radius_length |
| MGS | abundance | Hgn3C.0066 - Oscillospiraceae sp. | Radius_SOS |
| MGS | abundance | Hgn3C.0066 - Oscillospiraceae sp. | Tibia_SOS |
| MGS | abundance | Hgn3C.0066 - Oscillospiraceae sp. | Handgrip |
| MGS | abundance | Hgn3C.0067 - Roseburia hominis | Tibia_length |
| MGS | abundance | Hgn3C.0067 - Roseburia hominis | Radius_length |
| MGS | abundance | Hgn3C.0067 - Roseburia hominis | Radius_SOS |
| MGS | abundance | Hgn3C.0067 - Roseburia hominis | Tibia_SOS |
| MGS | abundance | Hgn3C.0067 - Roseburia hominis | Handgrip |

| | | | |
|---|---|---|---|
| MGS | abundance | Hgn3C.0068 - [Ruminococcus] lactaris | Tibia_length |
| MGS | abundance | Hgn3C.0068 - [Ruminococcus] lactaris | Radius_length |
| MGS | abundance | Hgn3C.0068 - [Ruminococcus] lactaris | Radius_SOS |
| MGS | abundance | Hgn3C.0068 - [Ruminococcus] lactaris | Tibia_SOS |
| MGS | abundance | Hgn3C.0068 - [Ruminococcus] lactaris | Handgrip |
| MGS | abundance | Hgn3C.0069 - Oscillibacter sp. KLE 1728 | Tibia_length |
| MGS | abundance | Hgn3C.0069 - Oscillibacter sp. KLE 1728 | Radius_length |
| MGS | abundance | Hgn3C.0069 - Oscillibacter sp. KLE 1728 | Radius_SOS |
| MGS | abundance | Hgn3C.0069 - Oscillibacter sp. KLE 1728 | Tibia_SOS |
| MGS | abundance | Hgn3C.0069 - Oscillibacter sp. KLE 1728 | Handgrip |
| MGS | abundance | Hgn3C.0070 - Faecalibacterium sp. OF04-11AC | Tibia_length |
| MGS | abundance | Hgn3C.0070 - Faecalibacterium sp. OF04-11AC | Radius_length |
| MGS | abundance | Hgn3C.0070 - Faecalibacterium sp. OF04-11AC | Radius_SOS |
| MGS | abundance | Hgn3C.0070 - Faecalibacterium sp. OF04-11AC | Tibia_SOS |
| MGS | abundance | Hgn3C.0070 - Faecalibacterium sp. OF04-11AC | Handgrip |
| MGS | abundance | Hgn3C.0071 - Faecalicatena fissicatena | Tibia_length |
| MGS | abundance | Hgn3C.0071 - Faecalicatena fissicatena | Radius_length |
| MGS | abundance | Hgn3C.0071 - Faecalicatena fissicatena | Radius_SOS |
| MGS | abundance | Hgn3C.0071 - Faecalicatena fissicatena | Tibia_SOS |
| MGS | abundance | Hgn3C.0071 - Faecalicatena fissicatena | Handgrip |
| MGS | abundance | Hgn3C.0072 - Clostridium fessum | Tibia_length |
| MGS | abundance | Hgn3C.0072 - Clostridium fessum | Radius_length |
| MGS | abundance | Hgn3C.0072 - Clostridium fessum | Radius_SOS |
| MGS | abundance | Hgn3C.0072 - Clostridium fessum | Tibia_SOS |
| MGS | abundance | Hgn3C.0072 - Clostridium fessum | Handgrip |
| MGS | abundance | Hgn3C.0073 - Blautia sp. | Tibia_length |
| MGS | abundance | Hgn3C.0073 - Blautia sp. | Radius_length |
| MGS | abundance | Hgn3C.0073 - Blautia sp. | Radius_SOS |
| MGS | abundance | Hgn3C.0073 - Blautia sp. | Tibia_SOS |
| MGS | abundance | Hgn3C.0073 - Blautia sp. | Handgrip |
| MGS | abundance | Hgn3C.0074 - Clostridium sp. D43t1_170807_D5 | Tibia_length |
| MGS | abundance | Hgn3C.0074 - Clostridium sp. D43t1_170807_D5 | Radius_length |
| MGS | abundance | Hgn3C.0074 - Clostridium sp. D43t1_170807_D5 | Radius_SOS |
| MGS | abundance | Hgn3C.0074 - Clostridium sp. D43t1_170807_D5 | Tibia_SOS |
| MGS | abundance | Hgn3C.0074 - Clostridium sp. D43t1_170807_D5 | Handgrip |
| MGS | abundance | Hgn3C.0075 - Agathobaculum butyriciproducens | Tibia_length |
| MGS | abundance | Hgn3C.0075 - Agathobaculum butyriciproducens | Radius_length |
| MGS | abundance | Hgn3C.0075 - Agathobaculum butyriciproducens | Radius_SOS |
| MGS | abundance | Hgn3C.0075 - Agathobaculum butyriciproducens | Tibia_SOS |
| MGS | abundance | Hgn3C.0075 - Agathobaculum butyriciproducens | Handgrip |
| MGS | abundance | Hgn3C.0076 - Lachnospira pectinoschiza | Tibia_length |
| MGS | abundance | Hgn3C.0076 - Lachnospira pectinoschiza | Radius_length |
| MGS | abundance | Hgn3C.0076 - Lachnospira pectinoschiza | Radius_SOS |
| MGS | abundance | Hgn3C.0076 - Lachnospira pectinoschiza | Tibia_SOS |
| MGS | abundance | Hgn3C.0076 - Lachnospira pectinoschiza | Handgrip |
| MGS | abundance | Hgn3C.0077 - Sutterella wadsworthensis | Tibia_length |
| MGS | abundance | Hgn3C.0077 - Sutterella wadsworthensis | Radius_length |
| MGS | abundance | Hgn3C.0077 - Sutterella wadsworthensis | Radius_SOS |

| | | | |
|---|---|---|---|
| MGS | abundance | Hgn3C.0077 - Sutterella wadsworthensis | Tibia_SOS |
| MGS | abundance | Hgn3C.0077 - Sutterella wadsworthensis | Handgrip |
| MGS | abundance | Hgn3C.0078 - Clostridium sp. MCC328 | Tibia_length |
| MGS | abundance | Hgn3C.0078 - Clostridium sp. MCC328 | Radius_length |
| MGS | abundance | Hgn3C.0078 - Clostridium sp. MCC328 | Radius_SOS |
| MGS | abundance | Hgn3C.0078 - Clostridium sp. MCC328 | Tibia_SOS |
| MGS | abundance | Hgn3C.0078 - Clostridium sp. MCC328 | Handgrip |
| MGS | abundance | Hgn3C.0079 - [Ruminococcus] torques | Tibia_length |
| MGS | abundance | Hgn3C.0079 - [Ruminococcus] torques | Radius_length |
| MGS | abundance | Hgn3C.0079 - [Ruminococcus] torques | Radius_SOS |
| MGS | abundance | Hgn3C.0079 - [Ruminococcus] torques | Tibia_SOS |
| MGS | abundance | Hgn3C.0079 - [Ruminococcus] torques | Handgrip |
| MGS | abundance | Hgn3C.0080 - Phascolarctobacterium faecium | Tibia_length |
| MGS | abundance | Hgn3C.0080 - Phascolarctobacterium faecium | Radius_length |
| MGS | abundance | Hgn3C.0080 - Phascolarctobacterium faecium | Radius_SOS |
| MGS | abundance | Hgn3C.0080 - Phascolarctobacterium faecium | Tibia_SOS |
| MGS | abundance | Hgn3C.0080 - Phascolarctobacterium faecium | Handgrip |
| MGS | abundance | Hgn3C.0081 - Coprococcus eutactus | Tibia_length |
| MGS | abundance | Hgn3C.0081 - Coprococcus eutactus | Radius_length |
| MGS | abundance | Hgn3C.0081 - Coprococcus eutactus | Radius_SOS |
| MGS | abundance | Hgn3C.0081 - Coprococcus eutactus | Tibia_SOS |
| MGS | abundance | Hgn3C.0081 - Coprococcus eutactus | Handgrip |
| MGS | abundance | Hgn3C.0082 - Roseburia sp. CLA-AA-H204 | Tibia_length |
| MGS | abundance | Hgn3C.0082 - Roseburia sp. CLA-AA-H204 | Radius_length |
| MGS | abundance | Hgn3C.0082 - Roseburia sp. CLA-AA-H204 | Radius_SOS |
| MGS | abundance | Hgn3C.0082 - Roseburia sp. CLA-AA-H204 | Tibia_SOS |
| MGS | abundance | Hgn3C.0082 - Roseburia sp. CLA-AA-H204 | Handgrip |
| MGS | abundance | Hgn3C.0083 - Eubacteriales sp. | Tibia_length |
| MGS | abundance | Hgn3C.0083 - Eubacteriales sp. | Radius_length |
| MGS | abundance | Hgn3C.0083 - Eubacteriales sp. | Radius_SOS |
| MGS | abundance | Hgn3C.0083 - Eubacteriales sp. | Tibia_SOS |
| MGS | abundance | Hgn3C.0083 - Eubacteriales sp. | Handgrip |
| MGS | abundance | Hgn3C.0084 - Flavonifractor plautii | Tibia_length |
| MGS | abundance | Hgn3C.0084 - Flavonifractor plautii | Radius_length |
| MGS | abundance | Hgn3C.0084 - Flavonifractor plautii | Radius_SOS |
| MGS | abundance | Hgn3C.0084 - Flavonifractor plautii | Tibia_SOS |
| MGS | abundance | Hgn3C.0084 - Flavonifractor plautii | Handgrip |
| MGS | abundance | Hgn3C.0085 - Faecalibacterium sp. | Tibia_length |
| MGS | abundance | Hgn3C.0085 - Faecalibacterium sp. | Radius_length |
| MGS | abundance | Hgn3C.0085 - Faecalibacterium sp. | Radius_SOS |
| MGS | abundance | Hgn3C.0085 - Faecalibacterium sp. | Tibia_SOS |
| MGS | abundance | Hgn3C.0085 - Faecalibacterium sp. | Handgrip |
| MGS | abundance | Hgn3C.0086 - Dialister invisus | Tibia_length |
| MGS | abundance | Hgn3C.0086 - Dialister invisus | Radius_length |
| MGS | abundance | Hgn3C.0086 - Dialister invisus | Radius_SOS |
| MGS | abundance | Hgn3C.0086 - Dialister invisus | Tibia_SOS |
| MGS | abundance | Hgn3C.0086 - Dialister invisus | Handgrip |
| MGS | abundance | Hgn3C.0087 - Eubacteriales sp. | Tibia_length |

| | | | |
|---|---|---|---|
| MGS | abundanc | Hgn3C.0087 - Eubacteriales sp. | Radius_length |
| MGS | abundanc | Hgn3C.0087 - Eubacteriales sp. | Radius_SOS |
| MGS | abundanc | Hgn3C.0087 - Eubacteriales sp. | Tibia_SOS |
| MGS | abundanc | Hgn3C.0087 - Eubacteriales sp. | Handgrip |
| MGS | abundanc | Hgn3C.0088 - [Ruminococcus] gnavus | Tibia_length |
| MGS | abundanc | Hgn3C.0088 - [Ruminococcus] gnavus | Radius_length |
| MGS | abundanc | Hgn3C.0088 - [Ruminococcus] gnavus | Radius_SOS |
| MGS | abundanc | Hgn3C.0088 - [Ruminococcus] gnavus | Tibia_SOS |
| MGS | abundanc | Hgn3C.0088 - [Ruminococcus] gnavus | Handgrip |
| MGS | abundanc | Hgn3C.0089 - Paraprevotella clara | Tibia_length |
| MGS | abundanc | Hgn3C.0089 - Paraprevotella clara | Radius_length |
| MGS | abundanc | Hgn3C.0089 - Paraprevotella clara | Radius_SOS |
| MGS | abundanc | Hgn3C.0089 - Paraprevotella clara | Tibia_SOS |
| MGS | abundanc | Hgn3C.0089 - Paraprevotella clara | Handgrip |
| MGS | abundanc | Hgn3C.0091 - Lachnospira sp. NSJ-43 | Tibia_length |
| MGS | abundanc | Hgn3C.0091 - Lachnospira sp. NSJ-43 | Radius_length |
| MGS | abundanc | Hgn3C.0091 - Lachnospira sp. NSJ-43 | Radius_SOS |
| MGS | abundanc | Hgn3C.0091 - Lachnospira sp. NSJ-43 | Tibia_SOS |
| MGS | abundanc | Hgn3C.0091 - Lachnospira sp. NSJ-43 | Handgrip |
| MGS | abundanc | Hgn3C.0092 - Lachnospiraceae sp. | Tibia_length |
| MGS | abundanc | Hgn3C.0092 - Lachnospiraceae sp. | Radius_length |
| MGS | abundanc | Hgn3C.0092 - Lachnospiraceae sp. | Radius_SOS |
| MGS | abundanc | Hgn3C.0092 - Lachnospiraceae sp. | Tibia_SOS |
| MGS | abundanc | Hgn3C.0092 - Lachnospiraceae sp. | Handgrip |
| MGS | abundanc | Hgn3C.0093 - Bilophila wadsworthia | Tibia_length |
| MGS | abundanc | Hgn3C.0093 - Bilophila wadsworthia | Radius_length |
| MGS | abundanc | Hgn3C.0093 - Bilophila wadsworthia | Radius_SOS |
| MGS | abundanc | Hgn3C.0093 - Bilophila wadsworthia | Tibia_SOS |
| MGS | abundanc | Hgn3C.0093 - Bilophila wadsworthia | Handgrip |
| MGS | abundanc | Hgn3C.0094 - Eubacteriales sp. | Tibia_length |
| MGS | abundanc | Hgn3C.0094 - Eubacteriales sp. | Radius_length |
| MGS | abundanc | Hgn3C.0094 - Eubacteriales sp. | Radius_SOS |
| MGS | abundanc | Hgn3C.0094 - Eubacteriales sp. | Tibia_SOS |
| MGS | abundanc | Hgn3C.0094 - Eubacteriales sp. | Handgrip |
| MGS | abundanc | Hgn3C.0095 - Eubacteriales sp. | Tibia_length |
| MGS | abundanc | Hgn3C.0095 - Eubacteriales sp. | Radius_length |
| MGS | abundanc | Hgn3C.0095 - Eubacteriales sp. | Radius_SOS |
| MGS | abundanc | Hgn3C.0095 - Eubacteriales sp. | Tibia_SOS |
| MGS | abundanc | Hgn3C.0095 - Eubacteriales sp. | Handgrip |
| MGS | abundanc | Hgn3C.0096 - Eubacteriales sp. | Tibia_length |
| MGS | abundanc | Hgn3C.0096 - Eubacteriales sp. | Radius_length |
| MGS | abundanc | Hgn3C.0096 - Eubacteriales sp. | Radius_SOS |
| MGS | abundanc | Hgn3C.0096 - Eubacteriales sp. | Tibia_SOS |
| MGS | abundanc | Hgn3C.0096 - Eubacteriales sp. | Handgrip |
| MGS | abundanc | Hgn3C.0097 - Eubacterium ventriosum | Tibia_length |
| MGS | abundanc | Hgn3C.0097 - Eubacterium ventriosum | Radius_length |
| MGS | abundanc | Hgn3C.0097 - Eubacterium ventriosum | Radius_SOS |
| MGS | abundanc | Hgn3C.0097 - Eubacterium ventriosum | Tibia_SOS |

| | | | |
|---|---|---|---|
| MGS | abundance | Hgn3C.0097 - Eubacterium ventriosum | Handgrip |
| MGS | abundance | Hgn3C.0098 - Coprococcus catus | Tibia_length |
| MGS | abundance | Hgn3C.0098 - Coprococcus catus | Radius_length |
| MGS | abundance | Hgn3C.0098 - Coprococcus catus | Radius_SOS |
| MGS | abundance | Hgn3C.0098 - Coprococcus catus | Tibia_SOS |
| MGS | abundance | Hgn3C.0098 - Coprococcus catus | Handgrip |
| MGS | abundance | Hgn3C.0099 - Dorea longicatena | Tibia_length |
| MGS | abundance | Hgn3C.0099 - Dorea longicatena | Radius_length |
| MGS | abundance | Hgn3C.0099 - Dorea longicatena | Radius_SOS |
| MGS | abundance | Hgn3C.0099 - Dorea longicatena | Tibia_SOS |
| MGS | abundance | Hgn3C.0099 - Dorea longicatena | Handgrip |
| MGS | abundance | Hgn3C.0100 - Bifidobacterium bifidum | Tibia_length |
| MGS | abundance | Hgn3C.0100 - Bifidobacterium bifidum | Radius_length |
| MGS | abundance | Hgn3C.0100 - Bifidobacterium bifidum | Radius_SOS |
| MGS | abundance | Hgn3C.0100 - Bifidobacterium bifidum | Tibia_SOS |
| MGS | abundance | Hgn3C.0100 - Bifidobacterium bifidum | Handgrip |
| MGS | abundance | Hgn3C.0101 - Bifidobacterium pseudocatenulatum | Tibia_length |
| MGS | abundance | Hgn3C.0101 - Bifidobacterium pseudocatenulatum | Radius_length |
| MGS | abundance | Hgn3C.0101 - Bifidobacterium pseudocatenulatum | Radius_SOS |
| MGS | abundance | Hgn3C.0101 - Bifidobacterium pseudocatenulatum | Tibia_SOS |
| MGS | abundance | Hgn3C.0101 - Bifidobacterium pseudocatenulatum | Handgrip |
| MGS | abundance | Hgn3C.0102 - Eubacteriales sp. | Tibia_length |
| MGS | abundance | Hgn3C.0102 - Eubacteriales sp. | Radius_length |
| MGS | abundance | Hgn3C.0102 - Eubacteriales sp. | Radius_SOS |
| MGS | abundance | Hgn3C.0102 - Eubacteriales sp. | Tibia_SOS |
| MGS | abundance | Hgn3C.0102 - Eubacteriales sp. | Handgrip |
| MGS | abundance | Hgn3C.0103 - Holdemanella porci | Tibia_length |
| MGS | abundance | Hgn3C.0103 - Holdemanella porci | Radius_length |
| MGS | abundance | Hgn3C.0103 - Holdemanella porci | Radius_SOS |
| MGS | abundance | Hgn3C.0103 - Holdemanella porci | Tibia_SOS |
| MGS | abundance | Hgn3C.0103 - Holdemanella porci | Handgrip |
| MGS | abundance | Hgn3C.0104 - Clostridium sp. AM32-2 | Tibia_length |
| MGS | abundance | Hgn3C.0104 - Clostridium sp. AM32-2 | Radius_length |
| MGS | abundance | Hgn3C.0104 - Clostridium sp. AM32-2 | Radius_SOS |
| MGS | abundance | Hgn3C.0104 - Clostridium sp. AM32-2 | Tibia_SOS |
| MGS | abundance | Hgn3C.0104 - Clostridium sp. AM32-2 | Handgrip |
| MGS | abundance | Hgn3C.0105 - Vescimonas coprocola | Tibia_length |
| MGS | abundance | Hgn3C.0105 - Vescimonas coprocola | Radius_length |
| MGS | abundance | Hgn3C.0105 - Vescimonas coprocola | Radius_SOS |
| MGS | abundance | Hgn3C.0105 - Vescimonas coprocola | Tibia_SOS |
| MGS | abundance | Hgn3C.0105 - Vescimonas coprocola | Handgrip |
| MGS | abundance | Hgn3C.0106 - Faecalibacterium sp. CLA-AA-H233 | Tibia_length |
| MGS | abundance | Hgn3C.0106 - Faecalibacterium sp. CLA-AA-H233 | Radius_length |
| MGS | abundance | Hgn3C.0106 - Faecalibacterium sp. CLA-AA-H233 | Radius_SOS |
| MGS | abundance | Hgn3C.0106 - Faecalibacterium sp. CLA-AA-H233 | Tibia_SOS |
| MGS | abundance | Hgn3C.0106 - Faecalibacterium sp. CLA-AA-H233 | Handgrip |
| MGS | abundance | Hgn3C.0107 - Parasutterella excrementihominis | Tibia_length |
| MGS | abundance | Hgn3C.0107 - Parasutterella excrementihominis | Radius_length |

| | | | |
|---|---|---|---|
| MGS | abundance | Hgn3C.0107 - Parasutterella excrementihominis | Radius_SOS |
| MGS | abundance | Hgn3C.0107 - Parasutterella excrementihominis | Tibia_SOS |
| MGS | abundance | Hgn3C.0107 - Parasutterella excrementihominis | Handgrip |
| MGS | abundance | Hgn3C.0108 - Eubacteriales sp. | Tibia_length |
| MGS | abundance | Hgn3C.0108 - Eubacteriales sp. | Radius_length |
| MGS | abundance | Hgn3C.0108 - Eubacteriales sp. | Radius_SOS |
| MGS | abundance | Hgn3C.0108 - Eubacteriales sp. | Tibia_SOS |
| MGS | abundance | Hgn3C.0108 - Eubacteriales sp. | Handgrip |
| MGS | abundance | Hgn3C.0109 - Bacteroides eggerthii | Tibia_length |
| MGS | abundance | Hgn3C.0109 - Bacteroides eggerthii | Radius_length |
| MGS | abundance | Hgn3C.0109 - Bacteroides eggerthii | Radius_SOS |
| MGS | abundance | Hgn3C.0109 - Bacteroides eggerthii | Tibia_SOS |
| MGS | abundance | Hgn3C.0109 - Bacteroides eggerthii | Handgrip |
| MGS | abundance | Hgn3C.0110 - Eubacteriales sp. | Tibia_length |
| MGS | abundance | Hgn3C.0110 - Eubacteriales sp. | Radius_length |
| MGS | abundance | Hgn3C.0110 - Eubacteriales sp. | Radius_SOS |
| MGS | abundance | Hgn3C.0110 - Eubacteriales sp. | Tibia_SOS |
| MGS | abundance | Hgn3C.0110 - Eubacteriales sp. | Handgrip |
| MGS | abundance | Hgn3C.0111 - Eubacteriales sp. | Tibia_length |
| MGS | abundance | Hgn3C.0111 - Eubacteriales sp. | Radius_length |
| MGS | abundance | Hgn3C.0111 - Eubacteriales sp. | Radius_SOS |
| MGS | abundance | Hgn3C.0111 - Eubacteriales sp. | Tibia_SOS |
| MGS | abundance | Hgn3C.0111 - Eubacteriales sp. | Handgrip |
| MGS | abundance | Hgn3C.0113 - Clostridium sp. MCC345 | Tibia_length |
| MGS | abundance | Hgn3C.0113 - Clostridium sp. MCC345 | Radius_length |
| MGS | abundance | Hgn3C.0113 - Clostridium sp. MCC345 | Radius_SOS |
| MGS | abundance | Hgn3C.0113 - Clostridium sp. MCC345 | Tibia_SOS |
| MGS | abundance | Hgn3C.0113 - Clostridium sp. MCC345 | Handgrip |
| MGS | abundance | Hgn3C.0114 - Eubacteriales sp. | Tibia_length |
| MGS | abundance | Hgn3C.0114 - Eubacteriales sp. | Radius_length |
| MGS | abundance | Hgn3C.0114 - Eubacteriales sp. | Radius_SOS |
| MGS | abundance | Hgn3C.0114 - Eubacteriales sp. | Tibia_SOS |
| MGS | abundance | Hgn3C.0114 - Eubacteriales sp. | Handgrip |
| MGS | abundance | Hgn3C.0115 - Eubacteriales sp. | Tibia_length |
| MGS | abundance | Hgn3C.0115 - Eubacteriales sp. | Radius_length |
| MGS | abundance | Hgn3C.0115 - Eubacteriales sp. | Radius_SOS |
| MGS | abundance | Hgn3C.0115 - Eubacteriales sp. | Tibia_SOS |
| MGS | abundance | Hgn3C.0115 - Eubacteriales sp. | Handgrip |
| MGS | abundance | Hgn3C.0116 - Anaerostipes hadrus | Tibia_length |
| MGS | abundance | Hgn3C.0116 - Anaerostipes hadrus | Radius_length |
| MGS | abundance | Hgn3C.0116 - Anaerostipes hadrus | Radius_SOS |
| MGS | abundance | Hgn3C.0116 - Anaerostipes hadrus | Tibia_SOS |
| MGS | abundance | Hgn3C.0116 - Anaerostipes hadrus | Handgrip |
| MGS | abundance | Hgn3C.0117 - Methanobrevibacter smithii | Tibia_length |
| MGS | abundance | Hgn3C.0117 - Methanobrevibacter smithii | Radius_length |
| MGS | abundance | Hgn3C.0117 - Methanobrevibacter smithii | Radius_SOS |
| MGS | abundance | Hgn3C.0117 - Methanobrevibacter smithii | Tibia_SOS |
| MGS | abundance | Hgn3C.0117 - Methanobrevibacter smithii | Handgrip |

| | | | |
|---|---|---|---|
| MGS | abundance | Hgn3C.0118 - Eubacteriales sp. | Tibia_length |
| MGS | abundance | Hgn3C.0118 - Eubacteriales sp. | Radius_length |
| MGS | abundance | Hgn3C.0118 - Eubacteriales sp. | Radius_SOS |
| MGS | abundance | Hgn3C.0118 - Eubacteriales sp. | Tibia_SOS |
| MGS | abundance | Hgn3C.0118 - Eubacteriales sp. | Handgrip |
| MGS | abundance | Hgn3C.0119 - Bacteroides faecis | Tibia_length |
| MGS | abundance | Hgn3C.0119 - Bacteroides faecis | Radius_length |
| MGS | abundance | Hgn3C.0119 - Bacteroides faecis | Radius_SOS |
| MGS | abundance | Hgn3C.0119 - Bacteroides faecis | Tibia_SOS |
| MGS | abundance | Hgn3C.0119 - Bacteroides faecis | Handgrip |
| MGS | abundance | Hgn3C.0120 - Flavonifractor plautii | Tibia_length |
| MGS | abundance | Hgn3C.0120 - Flavonifractor plautii | Radius_length |
| MGS | abundance | Hgn3C.0120 - Flavonifractor plautii | Radius_SOS |
| MGS | abundance | Hgn3C.0120 - Flavonifractor plautii | Tibia_SOS |
| MGS | abundance | Hgn3C.0120 - Flavonifractor plautii | Handgrip |
| MGS | abundance | Hgn3C.0121 - Oscillospiraceae sp. | Tibia_length |
| MGS | abundance | Hgn3C.0121 - Oscillospiraceae sp. | Radius_length |
| MGS | abundance | Hgn3C.0121 - Oscillospiraceae sp. | Radius_SOS |
| MGS | abundance | Hgn3C.0121 - Oscillospiraceae sp. | Tibia_SOS |
| MGS | abundance | Hgn3C.0121 - Oscillospiraceae sp. | Handgrip |
| MGS | abundance | Hgn3C.0122 - Bacteroides finegoldii | Tibia_length |
| MGS | abundance | Hgn3C.0122 - Bacteroides finegoldii | Radius_length |
| MGS | abundance | Hgn3C.0122 - Bacteroides finegoldii | Radius_SOS |
| MGS | abundance | Hgn3C.0122 - Bacteroides finegoldii | Tibia_SOS |
| MGS | abundance | Hgn3C.0122 - Bacteroides finegoldii | Handgrip |
| MGS | abundance | Hgn3C.0123 - Eubacteriales sp. | Tibia_length |
| MGS | abundance | Hgn3C.0123 - Eubacteriales sp. | Radius_length |
| MGS | abundance | Hgn3C.0123 - Eubacteriales sp. | Radius_SOS |
| MGS | abundance | Hgn3C.0123 - Eubacteriales sp. | Tibia_SOS |
| MGS | abundance | Hgn3C.0123 - Eubacteriales sp. | Handgrip |
| MGS | abundance | Hgn3C.0124 - Ruthenibacterium lactatiformans | Tibia_length |
| MGS | abundance | Hgn3C.0124 - Ruthenibacterium lactatiformans | Radius_length |
| MGS | abundance | Hgn3C.0124 - Ruthenibacterium lactatiformans | Radius_SOS |
| MGS | abundance | Hgn3C.0124 - Ruthenibacterium lactatiformans | Tibia_SOS |
| MGS | abundance | Hgn3C.0124 - Ruthenibacterium lactatiformans | Handgrip |
| MGS | abundance | Hgn3C.0125 - Sutterella sp. KLE1602 | Tibia_length |
| MGS | abundance | Hgn3C.0125 - Sutterella sp. KLE1602 | Radius_length |
| MGS | abundance | Hgn3C.0125 - Sutterella sp. KLE1602 | Radius_SOS |
| MGS | abundance | Hgn3C.0125 - Sutterella sp. KLE1602 | Tibia_SOS |
| MGS | abundance | Hgn3C.0125 - Sutterella sp. KLE1602 | Handgrip |
| MGS | abundance | Hgn3C.0126 - Ruminococcus sp. BSD2780120874_150323_B10 | Tibia_length |
| MGS | abundance | Hgn3C.0126 - Ruminococcus sp. BSD2780120874_150323_B10 | Radius_length |
| MGS | abundance | Hgn3C.0126 - Ruminococcus sp. BSD2780120874_150323_B10 | Radius_SOS |
| MGS | abundance | Hgn3C.0126 - Ruminococcus sp. BSD2780120874_150323_B10 | Tibia_SOS |
| MGS | abundance | Hgn3C.0126 - Ruminococcus sp. BSD2780120874_150323_B10 | Handgrip |
| MGS | abundance | Hgn3C.0127 - Faecalibacterium sp. | Tibia_length |
| MGS | abundance | Hgn3C.0127 - Faecalibacterium sp. | Radius_length |
| MGS | abundance | Hgn3C.0127 - Faecalibacterium sp. | Radius_SOS |

| | | | |
|---|---|---|---|
| MGS | abundance | Hgn3C.0127 - Faecalibacterium sp. | Tibia_SOS |
| MGS | abundance | Hgn3C.0127 - Faecalibacterium sp. | Handgrip |
| MGS | abundance | Hgn3C.0128 - Eubacteriales sp. | Tibia_length |
| MGS | abundance | Hgn3C.0128 - Eubacteriales sp. | Radius_length |
| MGS | abundance | Hgn3C.0128 - Eubacteriales sp. | Radius_SOS |
| MGS | abundance | Hgn3C.0128 - Eubacteriales sp. | Tibia_SOS |
| MGS | abundance | Hgn3C.0128 - Eubacteriales sp. | Handgrip |
| MGS | abundance | Hgn3C.0129 - Haemophilus parainfluenzae | Tibia_length |
| MGS | abundance | Hgn3C.0129 - Haemophilus parainfluenzae | Radius_length |
| MGS | abundance | Hgn3C.0129 - Haemophilus parainfluenzae | Radius_SOS |
| MGS | abundance | Hgn3C.0129 - Haemophilus parainfluenzae | Tibia_SOS |
| MGS | abundance | Hgn3C.0129 - Haemophilus parainfluenzae | Handgrip |
| MGS | abundance | Hgn3C.0130 - Phocaeicola plebeius | Tibia_length |
| MGS | abundance | Hgn3C.0130 - Phocaeicola plebeius | Radius_length |
| MGS | abundance | Hgn3C.0130 - Phocaeicola plebeius | Radius_SOS |
| MGS | abundance | Hgn3C.0130 - Phocaeicola plebeius | Tibia_SOS |
| MGS | abundance | Hgn3C.0130 - Phocaeicola plebeius | Handgrip |
| MGS | abundance | Hgn3C.0131 - Streptococcus thermophilus | Tibia_length |
| MGS | abundance | Hgn3C.0131 - Streptococcus thermophilus | Radius_length |
| MGS | abundance | Hgn3C.0131 - Streptococcus thermophilus | Radius_SOS |
| MGS | abundance | Hgn3C.0131 - Streptococcus thermophilus | Tibia_SOS |
| MGS | abundance | Hgn3C.0131 - Streptococcus thermophilus | Handgrip |
| MGS | abundance | Hgn3C.0132 - Eubacteriales sp. | Tibia_length |
| MGS | abundance | Hgn3C.0132 - Eubacteriales sp. | Radius_length |
| MGS | abundance | Hgn3C.0132 - Eubacteriales sp. | Radius_SOS |
| MGS | abundance | Hgn3C.0132 - Eubacteriales sp. | Tibia_SOS |
| MGS | abundance | Hgn3C.0132 - Eubacteriales sp. | Handgrip |
| MGS | abundance | Hgn3C.0134 - Eubacteriales sp. | Tibia_length |
| MGS | abundance | Hgn3C.0134 - Eubacteriales sp. | Radius_length |
| MGS | abundance | Hgn3C.0134 - Eubacteriales sp. | Radius_SOS |
| MGS | abundance | Hgn3C.0134 - Eubacteriales sp. | Tibia_SOS |
| MGS | abundance | Hgn3C.0134 - Eubacteriales sp. | Handgrip |
| MGS | abundance | Hgn3C.0135 - Firmicutes sp. | Tibia_length |
| MGS | abundance | Hgn3C.0135 - Firmicutes sp. | Radius_length |
| MGS | abundance | Hgn3C.0135 - Firmicutes sp. | Radius_SOS |
| MGS | abundance | Hgn3C.0135 - Firmicutes sp. | Tibia_SOS |
| MGS | abundance | Hgn3C.0135 - Firmicutes sp. | Handgrip |
| MGS | abundance | Hgn3C.0136 - Alistipes ihumii | Tibia_length |
| MGS | abundance | Hgn3C.0136 - Alistipes ihumii | Radius_length |
| MGS | abundance | Hgn3C.0136 - Alistipes ihumii | Radius_SOS |
| MGS | abundance | Hgn3C.0136 - Alistipes ihumii | Tibia_SOS |
| MGS | abundance | Hgn3C.0136 - Alistipes ihumii | Handgrip |
| MGS | abundance | Hgn3C.0137 - Clostridium sp. OF03-18AA | Tibia_length |
| MGS | abundance | Hgn3C.0137 - Clostridium sp. OF03-18AA | Radius_length |
| MGS | abundance | Hgn3C.0137 - Clostridium sp. OF03-18AA | Radius_SOS |
| MGS | abundance | Hgn3C.0137 - Clostridium sp. OF03-18AA | Tibia_SOS |
| MGS | abundance | Hgn3C.0137 - Clostridium sp. OF03-18AA | Handgrip |
| MGS | abundance | Hgn3C.0138 - Anaerobutyricum soehngenii | Tibia_length |

| | | | |
|---|---|---|---|
| MGS | abundance | Hgn3C.0138 - Anaerobutyricum soehngenii | Radius_length |
| MGS | abundance | Hgn3C.0138 - Anaerobutyricum soehngenii | Radius_SOS |
| MGS | abundance | Hgn3C.0138 - Anaerobutyricum soehngenii | Tibia_SOS |
| MGS | abundance | Hgn3C.0138 - Anaerobutyricum soehngenii | Handgrip |
| MGS | abundance | Hgn3C.0139 - Ruminococcus callidus | Tibia_length |
| MGS | abundance | Hgn3C.0139 - Ruminococcus callidus | Radius_length |
| MGS | abundance | Hgn3C.0139 - Ruminococcus callidus | Radius_SOS |
| MGS | abundance | Hgn3C.0139 - Ruminococcus callidus | Tibia_SOS |
| MGS | abundance | Hgn3C.0139 - Ruminococcus callidus | Handgrip |
| MGS | abundance | Hgn3C.0140 - Veillonella parvula | Tibia_length |
| MGS | abundance | Hgn3C.0140 - Veillonella parvula | Radius_length |
| MGS | abundance | Hgn3C.0140 - Veillonella parvula | Radius_SOS |
| MGS | abundance | Hgn3C.0140 - Veillonella parvula | Tibia_SOS |
| MGS | abundance | Hgn3C.0140 - Veillonella parvula | Handgrip |
| MGS | abundance | Hgn3C.0141 - Adlercreutzia equolifaciens subsp. celatus | Tibia_length |
| MGS | abundance | Hgn3C.0141 - Adlercreutzia equolifaciens subsp. celatus | Radius_length |
| MGS | abundance | Hgn3C.0141 - Adlercreutzia equolifaciens subsp. celatus | Radius_SOS |
| MGS | abundance | Hgn3C.0141 - Adlercreutzia equolifaciens subsp. celatus | Tibia_SOS |
| MGS | abundance | Hgn3C.0141 - Adlercreutzia equolifaciens subsp. celatus | Handgrip |
| MGS | abundance | Hgn3C.0142 - Eubacterium ramulus | Tibia_length |
| MGS | abundance | Hgn3C.0142 - Eubacterium ramulus | Radius_length |
| MGS | abundance | Hgn3C.0142 - Eubacterium ramulus | Radius_SOS |
| MGS | abundance | Hgn3C.0142 - Eubacterium ramulus | Tibia_SOS |
| MGS | abundance | Hgn3C.0142 - Eubacterium ramulus | Handgrip |
| MGS | abundance | Hgn3C.0143 - Streptococcus salivarius | Tibia_length |
| MGS | abundance | Hgn3C.0143 - Streptococcus salivarius | Radius_length |
| MGS | abundance | Hgn3C.0143 - Streptococcus salivarius | Radius_SOS |
| MGS | abundance | Hgn3C.0143 - Streptococcus salivarius | Tibia_SOS |
| MGS | abundance | Hgn3C.0143 - Streptococcus salivarius | Handgrip |
| MGS | abundance | Hgn3C.0144 - Eubacteriales sp. | Tibia_length |
| MGS | abundance | Hgn3C.0144 - Eubacteriales sp. | Radius_length |
| MGS | abundance | Hgn3C.0144 - Eubacteriales sp. | Radius_SOS |
| MGS | abundance | Hgn3C.0144 - Eubacteriales sp. | Tibia_SOS |
| MGS | abundance | Hgn3C.0144 - Eubacteriales sp. | Handgrip |
| MGS | abundance | Hgn3C.0146 - Phascolarctobacterium succinatutens | Tibia_length |
| MGS | abundance | Hgn3C.0146 - Phascolarctobacterium succinatutens | Radius_length |
| MGS | abundance | Hgn3C.0146 - Phascolarctobacterium succinatutens | Radius_SOS |
| MGS | abundance | Hgn3C.0146 - Phascolarctobacterium succinatutens | Tibia_SOS |
| MGS | abundance | Hgn3C.0146 - Phascolarctobacterium succinatutens | Handgrip |
| MGS | abundance | Hgn3C.0147 - Oscillospiraceae sp. | Tibia_length |
| MGS | abundance | Hgn3C.0147 - Oscillospiraceae sp. | Radius_length |
| MGS | abundance | Hgn3C.0147 - Oscillospiraceae sp. | Radius_SOS |
| MGS | abundance | Hgn3C.0147 - Oscillospiraceae sp. | Tibia_SOS |
| MGS | abundance | Hgn3C.0147 - Oscillospiraceae sp. | Handgrip |
| MGS | abundance | Hgn3C.0148 - Agathobaculum sp. M2 | Tibia_length |
| MGS | abundance | Hgn3C.0148 - Agathobaculum sp. M2 | Radius_length |
| MGS | abundance | Hgn3C.0148 - Agathobaculum sp. M2 | Radius_SOS |
| MGS | abundance | Hgn3C.0148 - Agathobaculum sp. M2 | Tibia_SOS |

| | | | |
|---|---|---|---|
| MGS | abundance | Hgn3C.0148 - Agathobaculum sp. M2 | Handgrip |
| MGS | abundance | Hgn3C.0149 - Eggerthellales sp. | Tibia_length |
| MGS | abundance | Hgn3C.0149 - Eggerthellales sp. | Radius_length |
| MGS | abundance | Hgn3C.0149 - Eggerthellales sp. | Radius_SOS |
| MGS | abundance | Hgn3C.0149 - Eggerthellales sp. | Tibia_SOS |
| MGS | abundance | Hgn3C.0149 - Eggerthellales sp. | Handgrip |
| MGS | abundance | Hgn3C.0151 - Clostridium sp. AF34-13 | Tibia_length |
| MGS | abundance | Hgn3C.0151 - Clostridium sp. AF34-13 | Radius_length |
| MGS | abundance | Hgn3C.0151 - Clostridium sp. AF34-13 | Radius_SOS |
| MGS | abundance | Hgn3C.0151 - Clostridium sp. AF34-13 | Tibia_SOS |
| MGS | abundance | Hgn3C.0151 - Clostridium sp. AF34-13 | Handgrip |
| MGS | abundance | Hgn3C.0152 - Bacteroides salyersiae | Tibia_length |
| MGS | abundance | Hgn3C.0152 - Bacteroides salyersiae | Radius_length |
| MGS | abundance | Hgn3C.0152 - Bacteroides salyersiae | Radius_SOS |
| MGS | abundance | Hgn3C.0152 - Bacteroides salyersiae | Tibia_SOS |
| MGS | abundance | Hgn3C.0152 - Bacteroides salyersiae | Handgrip |
| MGS | abundance | Hgn3C.0153 - Lachnospiraceae sp. | Tibia_length |
| MGS | abundance | Hgn3C.0153 - Lachnospiraceae sp. | Radius_length |
| MGS | abundance | Hgn3C.0153 - Lachnospiraceae sp. | Radius_SOS |
| MGS | abundance | Hgn3C.0153 - Lachnospiraceae sp. | Tibia_SOS |
| MGS | abundance | Hgn3C.0153 - Lachnospiraceae sp. | Handgrip |
| MGS | abundance | Hgn3C.0154 - Prevotella copri | Tibia_length |
| MGS | abundance | Hgn3C.0154 - Prevotella copri | Radius_length |
| MGS | abundance | Hgn3C.0154 - Prevotella copri | Radius_SOS |
| MGS | abundance | Hgn3C.0154 - Prevotella copri | Tibia_SOS |
| MGS | abundance | Hgn3C.0154 - Prevotella copri | Handgrip |
| MGS | abundance | Hgn3C.0155 - Blautia sp. | Tibia_length |
| MGS | abundance | Hgn3C.0155 - Blautia sp. | Radius_length |
| MGS | abundance | Hgn3C.0155 - Blautia sp. | Radius_SOS |
| MGS | abundance | Hgn3C.0155 - Blautia sp. | Tibia_SOS |
| MGS | abundance | Hgn3C.0155 - Blautia sp. | Handgrip |
| MGS | abundance | Hgn3C.0157 - Eubacteriales sp. | Tibia_length |
| MGS | abundance | Hgn3C.0157 - Eubacteriales sp. | Radius_length |
| MGS | abundance | Hgn3C.0157 - Eubacteriales sp. | Radius_SOS |
| MGS | abundance | Hgn3C.0157 - Eubacteriales sp. | Tibia_SOS |
| MGS | abundance | Hgn3C.0157 - Eubacteriales sp. | Handgrip |
| MGS | abundance | Hgn3C.0159 - Firmicutes sp. | Tibia_length |
| MGS | abundance | Hgn3C.0159 - Firmicutes sp. | Radius_length |
| MGS | abundance | Hgn3C.0159 - Firmicutes sp. | Radius_SOS |
| MGS | abundance | Hgn3C.0159 - Firmicutes sp. | Tibia_SOS |
| MGS | abundance | Hgn3C.0159 - Firmicutes sp. | Handgrip |
| MGS | abundance | Hgn3C.0161 - Blautia sp. | Tibia_length |
| MGS | abundance | Hgn3C.0161 - Blautia sp. | Radius_length |
| MGS | abundance | Hgn3C.0161 - Blautia sp. | Radius_SOS |
| MGS | abundance | Hgn3C.0161 - Blautia sp. | Tibia_SOS |
| MGS | abundance | Hgn3C.0161 - Blautia sp. | Handgrip |
| MGS | abundance | Hgn3C.0162 - Prevotella sp. | Tibia_length |
| MGS | abundance | Hgn3C.0162 - Prevotella sp. | Radius_length |

| | | | |
|---|---|---|---|
| MGS | abundance | Hgn3C.0162 - Prevotella sp. | Radius_SOS |
| MGS | abundance | Hgn3C.0162 - Prevotella sp. | Tibia_SOS |
| MGS | abundance | Hgn3C.0162 - Prevotella sp. | Handgrip |
| MGS | abundance | Hgn3C.0164 - Kineothrix sp. | Tibia_length |
| MGS | abundance | Hgn3C.0164 - Kineothrix sp. | Radius_length |
| MGS | abundance | Hgn3C.0164 - Kineothrix sp. | Radius_SOS |
| MGS | abundance | Hgn3C.0164 - Kineothrix sp. | Tibia_SOS |
| MGS | abundance | Hgn3C.0164 - Kineothrix sp. | Handgrip |
| MGS | abundance | Hgn3C.0166 - Eubacteriales sp. | Tibia_length |
| MGS | abundance | Hgn3C.0166 - Eubacteriales sp. | Radius_length |
| MGS | abundance | Hgn3C.0166 - Eubacteriales sp. | Radius_SOS |
| MGS | abundance | Hgn3C.0166 - Eubacteriales sp. | Tibia_SOS |
| MGS | abundance | Hgn3C.0166 - Eubacteriales sp. | Handgrip |
| MGS | abundance | Hgn3C.0167 - Lachnospiraceae sp. | Tibia_length |
| MGS | abundance | Hgn3C.0167 - Lachnospiraceae sp. | Radius_length |
| MGS | abundance | Hgn3C.0167 - Lachnospiraceae sp. | Radius_SOS |
| MGS | abundance | Hgn3C.0167 - Lachnospiraceae sp. | Tibia_SOS |
| MGS | abundance | Hgn3C.0167 - Lachnospiraceae sp. | Handgrip |
| MGS | abundance | Hgn3C.0168 - Evtepia gabavorous | Tibia_length |
| MGS | abundance | Hgn3C.0168 - Evtepia gabavorous | Radius_length |
| MGS | abundance | Hgn3C.0168 - Evtepia gabavorous | Radius_SOS |
| MGS | abundance | Hgn3C.0168 - Evtepia gabavorous | Tibia_SOS |
| MGS | abundance | Hgn3C.0168 - Evtepia gabavorous | Handgrip |
| MGS | abundance | Hgn3C.0169 - Bacteroides intestinalis | Tibia_length |
| MGS | abundance | Hgn3C.0169 - Bacteroides intestinalis | Radius_length |
| MGS | abundance | Hgn3C.0169 - Bacteroides intestinalis | Radius_SOS |
| MGS | abundance | Hgn3C.0169 - Bacteroides intestinalis | Tibia_SOS |
| MGS | abundance | Hgn3C.0169 - Bacteroides intestinalis | Handgrip |
| MGS | abundance | Hgn3C.0170 - Oscillospiraceae sp. | Tibia_length |
| MGS | abundance | Hgn3C.0170 - Oscillospiraceae sp. | Radius_length |
| MGS | abundance | Hgn3C.0170 - Oscillospiraceae sp. | Radius_SOS |
| MGS | abundance | Hgn3C.0170 - Oscillospiraceae sp. | Tibia_SOS |
| MGS | abundance | Hgn3C.0170 - Oscillospiraceae sp. | Handgrip |
| MGS | abundance | Hgn3C.0171 - Catenibacterium mitsuokai | Tibia_length |
| MGS | abundance | Hgn3C.0171 - Catenibacterium mitsuokai | Radius_length |
| MGS | abundance | Hgn3C.0171 - Catenibacterium mitsuokai | Radius_SOS |
| MGS | abundance | Hgn3C.0171 - Catenibacterium mitsuokai | Tibia_SOS |
| MGS | abundance | Hgn3C.0171 - Catenibacterium mitsuokai | Handgrip |
| MGS | abundance | Hgn3C.0174 - Lachnospiraceae sp. | Tibia_length |
| MGS | abundance | Hgn3C.0174 - Lachnospiraceae sp. | Radius_length |
| MGS | abundance | Hgn3C.0174 - Lachnospiraceae sp. | Radius_SOS |
| MGS | abundance | Hgn3C.0174 - Lachnospiraceae sp. | Tibia_SOS |
| MGS | abundance | Hgn3C.0174 - Lachnospiraceae sp. | Handgrip |
| MGS | abundance | Hgn3C.0175 - Oscillospiraceae sp. | Tibia_length |
| MGS | abundance | Hgn3C.0175 - Oscillospiraceae sp. | Radius_length |
| MGS | abundance | Hgn3C.0175 - Oscillospiraceae sp. | Radius_SOS |
| MGS | abundance | Hgn3C.0175 - Oscillospiraceae sp. | Tibia_SOS |
| MGS | abundance | Hgn3C.0175 - Oscillospiraceae sp. | Handgrip |

| | | | |
|---|---|---|---|
| MGS | abundance | Hgn3C.0176 - Alistipes indistinctus | Tibia_length |
| MGS | abundance | Hgn3C.0176 - Alistipes indistinctus | Radius_length |
| MGS | abundance | Hgn3C.0176 - Alistipes indistinctus | Radius_SOS |
| MGS | abundance | Hgn3C.0176 - Alistipes indistinctus | Tibia_SOS |
| MGS | abundance | Hgn3C.0176 - Alistipes indistinctus | Handgrip |
| MGS | abundance | Hgn3C.0178 - Clostridia sp. | Tibia_length |
| MGS | abundance | Hgn3C.0178 - Clostridia sp. | Radius_length |
| MGS | abundance | Hgn3C.0178 - Clostridia sp. | Radius_SOS |
| MGS | abundance | Hgn3C.0178 - Clostridia sp. | Tibia_SOS |
| MGS | abundance | Hgn3C.0178 - Clostridia sp. | Handgrip |
| MGS | abundance | Hgn3C.0179 - Bacteroidales sp. | Tibia_length |
| MGS | abundance | Hgn3C.0179 - Bacteroidales sp. | Radius_length |
| MGS | abundance | Hgn3C.0179 - Bacteroidales sp. | Radius_SOS |
| MGS | abundance | Hgn3C.0179 - Bacteroidales sp. | Tibia_SOS |
| MGS | abundance | Hgn3C.0179 - Bacteroidales sp. | Handgrip |
| MGS | abundance | Hgn3C.0180 - Bacteria sp. | Tibia_length |
| MGS | abundance | Hgn3C.0180 - Bacteria sp. | Radius_length |
| MGS | abundance | Hgn3C.0180 - Bacteria sp. | Radius_SOS |
| MGS | abundance | Hgn3C.0180 - Bacteria sp. | Tibia_SOS |
| MGS | abundance | Hgn3C.0180 - Bacteria sp. | Handgrip |
| MGS | abundance | Hgn3C.0181 - Alistipes onderdonkii | Tibia_length |
| MGS | abundance | Hgn3C.0181 - Alistipes onderdonkii | Radius_length |
| MGS | abundance | Hgn3C.0181 - Alistipes onderdonkii | Radius_SOS |
| MGS | abundance | Hgn3C.0181 - Alistipes onderdonkii | Tibia_SOS |
| MGS | abundance | Hgn3C.0181 - Alistipes onderdonkii | Handgrip |
| MGS | abundance | Hgn3C.0182 - Butyricimonas faecihominis | Tibia_length |
| MGS | abundance | Hgn3C.0182 - Butyricimonas faecihominis | Radius_length |
| MGS | abundance | Hgn3C.0182 - Butyricimonas faecihominis | Radius_SOS |
| MGS | abundance | Hgn3C.0182 - Butyricimonas faecihominis | Tibia_SOS |
| MGS | abundance | Hgn3C.0182 - Butyricimonas faecihominis | Handgrip |
| MGS | abundance | Hgn3C.0183 - Intestinibacter bartlettii | Tibia_length |
| MGS | abundance | Hgn3C.0183 - Intestinibacter bartlettii | Radius_length |
| MGS | abundance | Hgn3C.0183 - Intestinibacter bartlettii | Radius_SOS |
| MGS | abundance | Hgn3C.0183 - Intestinibacter bartlettii | Tibia_SOS |
| MGS | abundance | Hgn3C.0183 - Intestinibacter bartlettii | Handgrip |
| MGS | abundance | Hgn3C.0184 - Eubacteriales sp. | Tibia_length |
| MGS | abundance | Hgn3C.0184 - Eubacteriales sp. | Radius_length |
| MGS | abundance | Hgn3C.0184 - Eubacteriales sp. | Radius_SOS |
| MGS | abundance | Hgn3C.0184 - Eubacteriales sp. | Tibia_SOS |
| MGS | abundance | Hgn3C.0184 - Eubacteriales sp. | Handgrip |
| MGS | abundance | Hgn3C.0185 - Bifidobacterium catenulatum subsp. kashiwanohense | Tibia_length |
| MGS | abundance | Hgn3C.0185 - Bifidobacterium catenulatum subsp. kashiwanohense | Radius_length |
| MGS | abundance | Hgn3C.0185 - Bifidobacterium catenulatum subsp. kashiwanohense | Radius_SOS |
| MGS | abundance | Hgn3C.0185 - Bifidobacterium catenulatum subsp. kashiwanohense | Tibia_SOS |
| MGS | abundance | Hgn3C.0185 - Bifidobacterium catenulatum subsp. kashiwanohense | Handgrip |
| MGS | abundance | Hgn3C.0186 - Butyricimonas virosa | Tibia_length |
| MGS | abundance | Hgn3C.0186 - Butyricimonas virosa | Radius_length |
| MGS | abundance | Hgn3C.0186 - Butyricimonas virosa | Radius_SOS |

| | | | |
|---|---|---|---|
| MGS | abundance | Hgn3C.0186 - Butyricimonas virosa | Tibia_SOS |
| MGS | abundance | Hgn3C.0186 - Butyricimonas virosa | Handgrip |
| MGS | abundance | Hgn3C.0191 - Prevotella sp. | Tibia_length |
| MGS | abundance | Hgn3C.0191 - Prevotella sp. | Radius_length |
| MGS | abundance | Hgn3C.0191 - Prevotella sp. | Radius_SOS |
| MGS | abundance | Hgn3C.0191 - Prevotella sp. | Tibia_SOS |
| MGS | abundance | Hgn3C.0191 - Prevotella sp. | Handgrip |
| MGS | abundance | Hgn3C.0192 - Erysipelatoclostridium ramosum | Tibia_length |
| MGS | abundance | Hgn3C.0192 - Erysipelatoclostridium ramosum | Radius_length |
| MGS | abundance | Hgn3C.0192 - Erysipelatoclostridium ramosum | Radius_SOS |
| MGS | abundance | Hgn3C.0192 - Erysipelatoclostridium ramosum | Tibia_SOS |
| MGS | abundance | Hgn3C.0192 - Erysipelatoclostridium ramosum | Handgrip |
| MGS | abundance | Hgn3C.0193 - Eubacteriales sp. | Tibia_length |
| MGS | abundance | Hgn3C.0193 - Eubacteriales sp. | Radius_length |
| MGS | abundance | Hgn3C.0193 - Eubacteriales sp. | Radius_SOS |
| MGS | abundance | Hgn3C.0193 - Eubacteriales sp. | Tibia_SOS |
| MGS | abundance | Hgn3C.0193 - Eubacteriales sp. | Handgrip |
| MGS | abundance | Hgn3C.0194 - Faecalibacillus faecis | Tibia_length |
| MGS | abundance | Hgn3C.0194 - Faecalibacillus faecis | Radius_length |
| MGS | abundance | Hgn3C.0194 - Faecalibacillus faecis | Radius_SOS |
| MGS | abundance | Hgn3C.0194 - Faecalibacillus faecis | Tibia_SOS |
| MGS | abundance | Hgn3C.0194 - Faecalibacillus faecis | Handgrip |
| MGS | abundance | Hgn3C.0195 - Eubacteriales sp. | Tibia_length |
| MGS | abundance | Hgn3C.0195 - Eubacteriales sp. | Radius_length |
| MGS | abundance | Hgn3C.0195 - Eubacteriales sp. | Radius_SOS |
| MGS | abundance | Hgn3C.0195 - Eubacteriales sp. | Tibia_SOS |
| MGS | abundance | Hgn3C.0195 - Eubacteriales sp. | Handgrip |
| MGS | abundance | Hgn3C.0196 - Phocaeicola coprocola | Tibia_length |
| MGS | abundance | Hgn3C.0196 - Phocaeicola coprocola | Radius_length |
| MGS | abundance | Hgn3C.0196 - Phocaeicola coprocola | Radius_SOS |
| MGS | abundance | Hgn3C.0196 - Phocaeicola coprocola | Tibia_SOS |
| MGS | abundance | Hgn3C.0196 - Phocaeicola coprocola | Handgrip |
| MGS | abundance | Hgn3C.0197 - Bacteroides clarus | Tibia_length |
| MGS | abundance | Hgn3C.0197 - Bacteroides clarus | Radius_length |
| MGS | abundance | Hgn3C.0197 - Bacteroides clarus | Radius_SOS |
| MGS | abundance | Hgn3C.0197 - Bacteroides clarus | Tibia_SOS |
| MGS | abundance | Hgn3C.0197 - Bacteroides clarus | Handgrip |
| MGS | abundance | Hgn3C.0199 - Bacteroidales sp. | Tibia_length |
| MGS | abundance | Hgn3C.0199 - Bacteroidales sp. | Radius_length |
| MGS | abundance | Hgn3C.0199 - Bacteroidales sp. | Radius_SOS |
| MGS | abundance | Hgn3C.0199 - Bacteroidales sp. | Tibia_SOS |
| MGS | abundance | Hgn3C.0199 - Bacteroidales sp. | Handgrip |
| MGS | abundance | Hgn3C.0201 - Veillonella dispar | Tibia_length |
| MGS | abundance | Hgn3C.0201 - Veillonella dispar | Radius_length |
| MGS | abundance | Hgn3C.0201 - Veillonella dispar | Radius_SOS |
| MGS | abundance | Hgn3C.0201 - Veillonella dispar | Tibia_SOS |
| MGS | abundance | Hgn3C.0201 - Veillonella dispar | Handgrip |
| MGS | abundance | Hgn3C.0202 - Eggerthella lenta | Tibia_length |

| | | | |
|---|---|---|---|
| MGS | abundance | Hgn3C.0202 - Eggerthella lenta | Radius_length |
| MGS | abundance | Hgn3C.0202 - Eggerthella lenta | Radius_SOS |
| MGS | abundance | Hgn3C.0202 - Eggerthella lenta | Tibia_SOS |
| MGS | abundance | Hgn3C.0202 - Eggerthella lenta | Handgrip |
| MGS | abundance | Hgn3C.0203 - Faecalibacterium sp. Marseille-Q3530 | Tibia_length |
| MGS | abundance | Hgn3C.0203 - Faecalibacterium sp. Marseille-Q3530 | Radius_length |
| MGS | abundance | Hgn3C.0203 - Faecalibacterium sp. Marseille-Q3530 | Radius_SOS |
| MGS | abundance | Hgn3C.0203 - Faecalibacterium sp. Marseille-Q3530 | Tibia_SOS |
| MGS | abundance | Hgn3C.0203 - Faecalibacterium sp. Marseille-Q3530 | Handgrip |
| MGS | abundance | Hgn3C.0204 - Coprobacter fastidiosus | Tibia_length |
| MGS | abundance | Hgn3C.0204 - Coprobacter fastidiosus | Radius_length |
| MGS | abundance | Hgn3C.0204 - Coprobacter fastidiosus | Radius_SOS |
| MGS | abundance | Hgn3C.0204 - Coprobacter fastidiosus | Tibia_SOS |
| MGS | abundance | Hgn3C.0204 - Coprobacter fastidiosus | Handgrip |
| MGS | abundance | Hgn3C.0206 - Gemmiger formicilis | Tibia_length |
| MGS | abundance | Hgn3C.0206 - Gemmiger formicilis | Radius_length |
| MGS | abundance | Hgn3C.0206 - Gemmiger formicilis | Radius_SOS |
| MGS | abundance | Hgn3C.0206 - Gemmiger formicilis | Tibia_SOS |
| MGS | abundance | Hgn3C.0206 - Gemmiger formicilis | Handgrip |
| MGS | abundance | Hgn3C.0207 - Eubacteriales sp. | Tibia_length |
| MGS | abundance | Hgn3C.0207 - Eubacteriales sp. | Radius_length |
| MGS | abundance | Hgn3C.0207 - Eubacteriales sp. | Radius_SOS |
| MGS | abundance | Hgn3C.0207 - Eubacteriales sp. | Tibia_SOS |
| MGS | abundance | Hgn3C.0207 - Eubacteriales sp. | Handgrip |
| MGS | abundance | Hgn3C.0208 - Romboutsia timonensis | Tibia_length |
| MGS | abundance | Hgn3C.0208 - Romboutsia timonensis | Radius_length |
| MGS | abundance | Hgn3C.0208 - Romboutsia timonensis | Radius_SOS |
| MGS | abundance | Hgn3C.0208 - Romboutsia timonensis | Tibia_SOS |
| MGS | abundance | Hgn3C.0208 - Romboutsia timonensis | Handgrip |
| MGS | abundance | Hgn3C.0209 - Bifidobacterium breve | Tibia_length |
| MGS | abundance | Hgn3C.0209 - Bifidobacterium breve | Radius_length |
| MGS | abundance | Hgn3C.0209 - Bifidobacterium breve | Radius_SOS |
| MGS | abundance | Hgn3C.0209 - Bifidobacterium breve | Tibia_SOS |
| MGS | abundance | Hgn3C.0209 - Bifidobacterium breve | Handgrip |
| MGS | abundance | Hgn3C.0211 - Eubacteriales sp. | Tibia_length |
| MGS | abundance | Hgn3C.0211 - Eubacteriales sp. | Radius_length |
| MGS | abundance | Hgn3C.0211 - Eubacteriales sp. | Radius_SOS |
| MGS | abundance | Hgn3C.0211 - Eubacteriales sp. | Tibia_SOS |
| MGS | abundance | Hgn3C.0211 - Eubacteriales sp. | Handgrip |
| MGS | abundance | Hgn3C.0212 - Holdemanella biformis | Tibia_length |
| MGS | abundance | Hgn3C.0212 - Holdemanella biformis | Radius_length |
| MGS | abundance | Hgn3C.0212 - Holdemanella biformis | Radius_SOS |
| MGS | abundance | Hgn3C.0212 - Holdemanella biformis | Tibia_SOS |
| MGS | abundance | Hgn3C.0212 - Holdemanella biformis | Handgrip |
| MGS | abundance | Hgn3C.0213 - Akkermansia sp. GGCC_0220 | Tibia_length |
| MGS | abundance | Hgn3C.0213 - Akkermansia sp. GGCC_0220 | Radius_length |
| MGS | abundance | Hgn3C.0213 - Akkermansia sp. GGCC_0220 | Radius_SOS |
| MGS | abundance | Hgn3C.0213 - Akkermansia sp. GGCC_0220 | Tibia_SOS |

| | | | |
|---|---|---|---|
| MGS | abundance | Hgn3C.0213 - Akkermansia sp. GGCC_0220 | Handgrip |
| MGS | abundance | Hgn3C.0214 - Duodenibacillus massiliensis | Tibia_length |
| MGS | abundance | Hgn3C.0214 - Duodenibacillus massiliensis | Radius_length |
| MGS | abundance | Hgn3C.0214 - Duodenibacillus massiliensis | Radius_SOS |
| MGS | abundance | Hgn3C.0214 - Duodenibacillus massiliensis | Tibia_SOS |
| MGS | abundance | Hgn3C.0214 - Duodenibacillus massiliensis | Handgrip |
| MGS | abundance | Hgn3C.0215 - Alistipes sp. cv1 | Tibia_length |
| MGS | abundance | Hgn3C.0215 - Alistipes sp. cv1 | Radius_length |
| MGS | abundance | Hgn3C.0215 - Alistipes sp. cv1 | Radius_SOS |
| MGS | abundance | Hgn3C.0215 - Alistipes sp. cv1 | Tibia_SOS |
| MGS | abundance | Hgn3C.0215 - Alistipes sp. cv1 | Handgrip |
| MGS | abundance | Hgn3C.0217 - Eubacteriales sp. | Tibia_length |
| MGS | abundance | Hgn3C.0217 - Eubacteriales sp. | Radius_length |
| MGS | abundance | Hgn3C.0217 - Eubacteriales sp. | Radius_SOS |
| MGS | abundance | Hgn3C.0217 - Eubacteriales sp. | Tibia_SOS |
| MGS | abundance | Hgn3C.0217 - Eubacteriales sp. | Handgrip |
| MGS | abundance | Hgn3C.0218 - Eubacteriales sp. | Tibia_length |
| MGS | abundance | Hgn3C.0218 - Eubacteriales sp. | Radius_length |
| MGS | abundance | Hgn3C.0218 - Eubacteriales sp. | Radius_SOS |
| MGS | abundance | Hgn3C.0218 - Eubacteriales sp. | Tibia_SOS |
| MGS | abundance | Hgn3C.0218 - Eubacteriales sp. | Handgrip |
| MGS | abundance | Hgn3C.0219 - Tyzzerella nexilis | Tibia_length |
| MGS | abundance | Hgn3C.0219 - Tyzzerella nexilis | Radius_length |
| MGS | abundance | Hgn3C.0219 - Tyzzerella nexilis | Radius_SOS |
| MGS | abundance | Hgn3C.0219 - Tyzzerella nexilis | Tibia_SOS |
| MGS | abundance | Hgn3C.0219 - Tyzzerella nexilis | Handgrip |
| MGS | abundance | Hgn3C.0220 - Clostridium sp. MCC334 | Tibia_length |
| MGS | abundance | Hgn3C.0220 - Clostridium sp. MCC334 | Radius_length |
| MGS | abundance | Hgn3C.0220 - Clostridium sp. MCC334 | Radius_SOS |
| MGS | abundance | Hgn3C.0220 - Clostridium sp. MCC334 | Tibia_SOS |
| MGS | abundance | Hgn3C.0220 - Clostridium sp. MCC334 | Handgrip |
| MGS | abundance | Hgn3C.0221 - Clostridium sp. | Tibia_length |
| MGS | abundance | Hgn3C.0221 - Clostridium sp. | Radius_length |
| MGS | abundance | Hgn3C.0221 - Clostridium sp. | Radius_SOS |
| MGS | abundance | Hgn3C.0221 - Clostridium sp. | Tibia_SOS |
| MGS | abundance | Hgn3C.0221 - Clostridium sp. | Handgrip |
| MGS | abundance | Hgn3C.0222 - Parabacteroides merdae | Tibia_length |
| MGS | abundance | Hgn3C.0222 - Parabacteroides merdae | Radius_length |
| MGS | abundance | Hgn3C.0222 - Parabacteroides merdae | Radius_SOS |
| MGS | abundance | Hgn3C.0222 - Parabacteroides merdae | Tibia_SOS |
| MGS | abundance | Hgn3C.0222 - Parabacteroides merdae | Handgrip |
| MGS | abundance | Hgn3C.0224 - Veillonellales sp. | Tibia_length |
| MGS | abundance | Hgn3C.0224 - Veillonellales sp. | Radius_length |
| MGS | abundance | Hgn3C.0224 - Veillonellales sp. | Radius_SOS |
| MGS | abundance | Hgn3C.0224 - Veillonellales sp. | Tibia_SOS |
| MGS | abundance | Hgn3C.0224 - Veillonellales sp. | Handgrip |
| MGS | abundance | Hgn3C.0226 - Eubacteriales sp. | Tibia_length |
| MGS | abundance | Hgn3C.0226 - Eubacteriales sp. | Radius_length |

| | | | |
|---|---|---|---|
| MGS | abundance | Hgn3C.0226 - Eubacteriales sp. | Radius_SOS |
| MGS | abundance | Hgn3C.0226 - Eubacteriales sp. | Tibia_SOS |
| MGS | abundance | Hgn3C.0226 - Eubacteriales sp. | Handgrip |
| MGS | abundance | Hgn3C.0228 - Coprococcus eutactus | Tibia_length |
| MGS | abundance | Hgn3C.0228 - Coprococcus eutactus | Radius_length |
| MGS | abundance | Hgn3C.0228 - Coprococcus eutactus | Radius_SOS |
| MGS | abundance | Hgn3C.0228 - Coprococcus eutactus | Tibia_SOS |
| MGS | abundance | Hgn3C.0228 - Coprococcus eutactus | Handgrip |
| MGS | abundance | Hgn3C.0230 - Gemmiger sp. | Tibia_length |
| MGS | abundance | Hgn3C.0230 - Gemmiger sp. | Radius_length |
| MGS | abundance | Hgn3C.0230 - Gemmiger sp. | Radius_SOS |
| MGS | abundance | Hgn3C.0230 - Gemmiger sp. | Tibia_SOS |
| MGS | abundance | Hgn3C.0230 - Gemmiger sp. | Handgrip |
| MGS | abundance | Hgn3C.0232 - Megamonas funiformis | Tibia_length |
| MGS | abundance | Hgn3C.0232 - Megamonas funiformis | Radius_length |
| MGS | abundance | Hgn3C.0232 - Megamonas funiformis | Radius_SOS |
| MGS | abundance | Hgn3C.0232 - Megamonas funiformis | Tibia_SOS |
| MGS | abundance | Hgn3C.0232 - Megamonas funiformis | Handgrip |
| MGS | abundance | Hgn3C.0236 - Oscillospiraceae sp. | Tibia_length |
| MGS | abundance | Hgn3C.0236 - Oscillospiraceae sp. | Radius_length |
| MGS | abundance | Hgn3C.0236 - Oscillospiraceae sp. | Radius_SOS |
| MGS | abundance | Hgn3C.0236 - Oscillospiraceae sp. | Tibia_SOS |
| MGS | abundance | Hgn3C.0236 - Oscillospiraceae sp. | Handgrip |
| MGS | abundance | Hgn3C.0238 - Oscillospiraceae sp. | Tibia_length |
| MGS | abundance | Hgn3C.0238 - Oscillospiraceae sp. | Radius_length |
| MGS | abundance | Hgn3C.0238 - Oscillospiraceae sp. | Radius_SOS |
| MGS | abundance | Hgn3C.0238 - Oscillospiraceae sp. | Tibia_SOS |
| MGS | abundance | Hgn3C.0238 - Oscillospiraceae sp. | Handgrip |
| MGS | abundance | Hgn3C.0239 - Blautia sp. BIOML-A1 | Tibia_length |
| MGS | abundance | Hgn3C.0239 - Blautia sp. BIOML-A1 | Radius_length |
| MGS | abundance | Hgn3C.0239 - Blautia sp. BIOML-A1 | Radius_SOS |
| MGS | abundance | Hgn3C.0239 - Blautia sp. BIOML-A1 | Tibia_SOS |
| MGS | abundance | Hgn3C.0239 - Blautia sp. BIOML-A1 | Handgrip |
| MGS | abundance | Hgn3C.0242 - Enterocloster bolteae | Tibia_length |
| MGS | abundance | Hgn3C.0242 - Enterocloster bolteae | Radius_length |
| MGS | abundance | Hgn3C.0242 - Enterocloster bolteae | Radius_SOS |
| MGS | abundance | Hgn3C.0242 - Enterocloster bolteae | Tibia_SOS |
| MGS | abundance | Hgn3C.0242 - Enterocloster bolteae | Handgrip |
| MGS | abundance | Hgn3C.0244 - Desulfovibrio piger | Tibia_length |
| MGS | abundance | Hgn3C.0244 - Desulfovibrio piger | Radius_length |
| MGS | abundance | Hgn3C.0244 - Desulfovibrio piger | Radius_SOS |
| MGS | abundance | Hgn3C.0244 - Desulfovibrio piger | Tibia_SOS |
| MGS | abundance | Hgn3C.0244 - Desulfovibrio piger | Handgrip |
| MGS | abundance | Hgn3C.0245 - Blautia stercoris | Tibia_length |
| MGS | abundance | Hgn3C.0245 - Blautia stercoris | Radius_length |
| MGS | abundance | Hgn3C.0245 - Blautia stercoris | Radius_SOS |
| MGS | abundance | Hgn3C.0245 - Blautia stercoris | Tibia_SOS |
| MGS | abundance | Hgn3C.0245 - Blautia stercoris | Handgrip |

| | | | |
|---|---|---|---|
| MGS | abundanc | Hgn3C.0248 - Coprococcus catus | Tibia_length |
| MGS | abundanc | Hgn3C.0248 - Coprococcus catus | Radius_length |
| MGS | abundanc | Hgn3C.0248 - Coprococcus catus | Radius_SOS |
| MGS | abundanc | Hgn3C.0248 - Coprococcus catus | Tibia_SOS |
| MGS | abundanc | Hgn3C.0248 - Coprococcus catus | Handgrip |
| MGS | abundanc | Hgn3C.0249 - Clostridium sp. AM33-3 | Tibia_length |
| MGS | abundanc | Hgn3C.0249 - Clostridium sp. AM33-3 | Radius_length |
| MGS | abundanc | Hgn3C.0249 - Clostridium sp. AM33-3 | Radius_SOS |
| MGS | abundanc | Hgn3C.0249 - Clostridium sp. AM33-3 | Tibia_SOS |
| MGS | abundanc | Hgn3C.0249 - Clostridium sp. AM33-3 | Handgrip |
| MGS | abundanc | Hgn3C.0250 - Eubacteriales sp. | Tibia_length |
| MGS | abundanc | Hgn3C.0250 - Eubacteriales sp. | Radius_length |
| MGS | abundanc | Hgn3C.0250 - Eubacteriales sp. | Radius_SOS |
| MGS | abundanc | Hgn3C.0250 - Eubacteriales sp. | Tibia_SOS |
| MGS | abundanc | Hgn3C.0250 - Eubacteriales sp. | Handgrip |
| MGS | abundanc | Hgn3C.0251 - Alistipes senegalensis | Tibia_length |
| MGS | abundanc | Hgn3C.0251 - Alistipes senegalensis | Radius_length |
| MGS | abundanc | Hgn3C.0251 - Alistipes senegalensis | Radius_SOS |
| MGS | abundanc | Hgn3C.0251 - Alistipes senegalensis | Tibia_SOS |
| MGS | abundanc | Hgn3C.0251 - Alistipes senegalensis | Handgrip |
| MGS | abundanc | Hgn3C.0253 - Waltera sp. | Tibia_length |
| MGS | abundanc | Hgn3C.0253 - Waltera sp. | Radius_length |
| MGS | abundanc | Hgn3C.0253 - Waltera sp. | Radius_SOS |
| MGS | abundanc | Hgn3C.0253 - Waltera sp. | Tibia_SOS |
| MGS | abundanc | Hgn3C.0253 - Waltera sp. | Handgrip |
| MGS | abundanc | Hgn3C.0260 - Hydrogenoanaerobacterium saccharovorans | Tibia_length |
| MGS | abundanc | Hgn3C.0260 - Hydrogenoanaerobacterium saccharovorans | Radius_length |
| MGS | abundanc | Hgn3C.0260 - Hydrogenoanaerobacterium saccharovorans | Radius_SOS |
| MGS | abundanc | Hgn3C.0260 - Hydrogenoanaerobacterium saccharovorans | Tibia_SOS |
| MGS | abundanc | Hgn3C.0260 - Hydrogenoanaerobacterium saccharovorans | Handgrip |
| MGS | abundanc | Hgn3C.0262 - Oscillospiraceae sp. | Tibia_length |
| MGS | abundanc | Hgn3C.0262 - Oscillospiraceae sp. | Radius_length |
| MGS | abundanc | Hgn3C.0262 - Oscillospiraceae sp. | Radius_SOS |
| MGS | abundanc | Hgn3C.0262 - Oscillospiraceae sp. | Tibia_SOS |
| MGS | abundanc | Hgn3C.0262 - Oscillospiraceae sp. | Handgrip |
| MGS | abundanc | Hgn3C.0264 - Clostridium sp. AF27-2AA | Tibia_length |
| MGS | abundanc | Hgn3C.0264 - Clostridium sp. AF27-2AA | Radius_length |
| MGS | abundanc | Hgn3C.0264 - Clostridium sp. AF27-2AA | Radius_SOS |
| MGS | abundanc | Hgn3C.0264 - Clostridium sp. AF27-2AA | Tibia_SOS |
| MGS | abundanc | Hgn3C.0264 - Clostridium sp. AF27-2AA | Handgrip |
| MGS | abundanc | Hgn3C.0266 - Dorea sp. AF36-15AT | Tibia_length |
| MGS | abundanc | Hgn3C.0266 - Dorea sp. AF36-15AT | Radius_length |
| MGS | abundanc | Hgn3C.0266 - Dorea sp. AF36-15AT | Radius_SOS |
| MGS | abundanc | Hgn3C.0266 - Dorea sp. AF36-15AT | Tibia_SOS |
| MGS | abundanc | Hgn3C.0266 - Dorea sp. AF36-15AT | Handgrip |
| MGS | abundanc | Hgn3C.0267 - Oscillospiraceae sp. | Tibia_length |
| MGS | abundanc | Hgn3C.0267 - Oscillospiraceae sp. | Radius_length |
| MGS | abundanc | Hgn3C.0267 - Oscillospiraceae sp. | Radius_SOS |

| | | | |
|---|---|---|---|
| MGS | abundance | Hgn3C.0267 - Oscillospiraceae sp. | Tibia_SOS |
| MGS | abundance | Hgn3C.0267 - Oscillospiraceae sp. | Handgrip |
| MGS | abundance | Hgn3C.0269 - Flintibacter sp. NSJ-23 | Tibia_length |
| MGS | abundance | Hgn3C.0269 - Flintibacter sp. NSJ-23 | Radius_length |
| MGS | abundance | Hgn3C.0269 - Flintibacter sp. NSJ-23 | Radius_SOS |
| MGS | abundance | Hgn3C.0269 - Flintibacter sp. NSJ-23 | Tibia_SOS |
| MGS | abundance | Hgn3C.0269 - Flintibacter sp. NSJ-23 | Handgrip |
| MGS | abundance | Hgn3C.0271 - Oscillibacter sp. | Tibia_length |
| MGS | abundance | Hgn3C.0271 - Oscillibacter sp. | Radius_length |
| MGS | abundance | Hgn3C.0271 - Oscillibacter sp. | Radius_SOS |
| MGS | abundance | Hgn3C.0271 - Oscillibacter sp. | Tibia_SOS |
| MGS | abundance | Hgn3C.0271 - Oscillibacter sp. | Handgrip |
| MGS | abundance | Hgn3C.0272 - Blautia faecicola | Tibia_length |
| MGS | abundance | Hgn3C.0272 - Blautia faecicola | Radius_length |
| MGS | abundance | Hgn3C.0272 - Blautia faecicola | Radius_SOS |
| MGS | abundance | Hgn3C.0272 - Blautia faecicola | Tibia_SOS |
| MGS | abundance | Hgn3C.0272 - Blautia faecicola | Handgrip |
| MGS | abundance | Hgn3C.0273 - Slackia isoflavoniconvertens | Tibia_length |
| MGS | abundance | Hgn3C.0273 - Slackia isoflavoniconvertens | Radius_length |
| MGS | abundance | Hgn3C.0273 - Slackia isoflavoniconvertens | Radius_SOS |
| MGS | abundance | Hgn3C.0273 - Slackia isoflavoniconvertens | Tibia_SOS |
| MGS | abundance | Hgn3C.0273 - Slackia isoflavoniconvertens | Handgrip |
| MGS | abundance | Hgn3C.0275 - Lachnospiraceae sp. | Tibia_length |
| MGS | abundance | Hgn3C.0275 - Lachnospiraceae sp. | Radius_length |
| MGS | abundance | Hgn3C.0275 - Lachnospiraceae sp. | Radius_SOS |
| MGS | abundance | Hgn3C.0275 - Lachnospiraceae sp. | Tibia_SOS |
| MGS | abundance | Hgn3C.0275 - Lachnospiraceae sp. | Handgrip |
| MGS | abundance | Hgn3C.0279 - Oscillospiraceae sp. | Tibia_length |
| MGS | abundance | Hgn3C.0279 - Oscillospiraceae sp. | Radius_length |
| MGS | abundance | Hgn3C.0279 - Oscillospiraceae sp. | Radius_SOS |
| MGS | abundance | Hgn3C.0279 - Oscillospiraceae sp. | Tibia_SOS |
| MGS | abundance | Hgn3C.0279 - Oscillospiraceae sp. | Handgrip |
| MGS | abundance | Hgn3C.0281 - Eubacteriales sp. | Tibia_length |
| MGS | abundance | Hgn3C.0281 - Eubacteriales sp. | Radius_length |
| MGS | abundance | Hgn3C.0281 - Eubacteriales sp. | Radius_SOS |
| MGS | abundance | Hgn3C.0281 - Eubacteriales sp. | Tibia_SOS |
| MGS | abundance | Hgn3C.0281 - Eubacteriales sp. | Handgrip |
| MGS | abundance | Hgn3C.0284 - Erysipelotrichaceae sp. | Tibia_length |
| MGS | abundance | Hgn3C.0284 - Erysipelotrichaceae sp. | Radius_length |
| MGS | abundance | Hgn3C.0284 - Erysipelotrichaceae sp. | Radius_SOS |
| MGS | abundance | Hgn3C.0284 - Erysipelotrichaceae sp. | Tibia_SOS |
| MGS | abundance | Hgn3C.0284 - Erysipelotrichaceae sp. | Handgrip |
| MGS | abundance | Hgn3C.0285 - Colidextribacter sp. | Tibia_length |
| MGS | abundance | Hgn3C.0285 - Colidextribacter sp. | Radius_length |
| MGS | abundance | Hgn3C.0285 - Colidextribacter sp. | Radius_SOS |
| MGS | abundance | Hgn3C.0285 - Colidextribacter sp. | Tibia_SOS |
| MGS | abundance | Hgn3C.0285 - Colidextribacter sp. | Handgrip |
| MGS | abundance | Hgn3C.0286 - Clostridia sp. | Tibia_length |

| | | | |
|---|---|---|---|
| MGS | abundance | Hgn3C.0286 - Clostridia sp. | Radius_length |
| MGS | abundance | Hgn3C.0286 - Clostridia sp. | Radius_SOS |
| MGS | abundance | Hgn3C.0286 - Clostridia sp. | Tibia_SOS |
| MGS | abundance | Hgn3C.0286 - Clostridia sp. | Handgrip |
| MGS | abundance | Hgn3C.0288 - Butyricimonas paravirosa | Tibia_length |
| MGS | abundance | Hgn3C.0288 - Butyricimonas paravirosa | Radius_length |
| MGS | abundance | Hgn3C.0288 - Butyricimonas paravirosa | Radius_SOS |
| MGS | abundance | Hgn3C.0288 - Butyricimonas paravirosa | Tibia_SOS |
| MGS | abundance | Hgn3C.0288 - Butyricimonas paravirosa | Handgrip |
| MGS | abundance | Hgn3C.0289 - Bacteroides cellulosilyticus | Tibia_length |
| MGS | abundance | Hgn3C.0289 - Bacteroides cellulosilyticus | Radius_length |
| MGS | abundance | Hgn3C.0289 - Bacteroides cellulosilyticus | Radius_SOS |
| MGS | abundance | Hgn3C.0289 - Bacteroides cellulosilyticus | Tibia_SOS |
| MGS | abundance | Hgn3C.0289 - Bacteroides cellulosilyticus | Handgrip |
| MGS | abundance | Hgn3C.0290 - Sellimonas intestinalis | Tibia_length |
| MGS | abundance | Hgn3C.0290 - Sellimonas intestinalis | Radius_length |
| MGS | abundance | Hgn3C.0290 - Sellimonas intestinalis | Radius_SOS |
| MGS | abundance | Hgn3C.0290 - Sellimonas intestinalis | Tibia_SOS |
| MGS | abundance | Hgn3C.0290 - Sellimonas intestinalis | Handgrip |
| MGS | abundance | Hgn3C.0291 - Parabacteroides johnsonii | Tibia_length |
| MGS | abundance | Hgn3C.0291 - Parabacteroides johnsonii | Radius_length |
| MGS | abundance | Hgn3C.0291 - Parabacteroides johnsonii | Radius_SOS |
| MGS | abundance | Hgn3C.0291 - Parabacteroides johnsonii | Tibia_SOS |
| MGS | abundance | Hgn3C.0291 - Parabacteroides johnsonii | Handgrip |
| MGS | abundance | Hgn3C.0295 - Eubacteriales sp. | Tibia_length |
| MGS | abundance | Hgn3C.0295 - Eubacteriales sp. | Radius_length |
| MGS | abundance | Hgn3C.0295 - Eubacteriales sp. | Radius_SOS |
| MGS | abundance | Hgn3C.0295 - Eubacteriales sp. | Tibia_SOS |
| MGS | abundance | Hgn3C.0295 - Eubacteriales sp. | Handgrip |
| MGS | abundance | Hgn3C.0298 - Monoglobus pectinilyticus | Tibia_length |
| MGS | abundance | Hgn3C.0298 - Monoglobus pectinilyticus | Radius_length |
| MGS | abundance | Hgn3C.0298 - Monoglobus pectinilyticus | Radius_SOS |
| MGS | abundance | Hgn3C.0298 - Monoglobus pectinilyticus | Tibia_SOS |
| MGS | abundance | Hgn3C.0298 - Monoglobus pectinilyticus | Handgrip |
| MGS | abundance | Hgn3C.0299 - Escherichia coli | Tibia_length |
| MGS | abundance | Hgn3C.0299 - Escherichia coli | Radius_length |
| MGS | abundance | Hgn3C.0299 - Escherichia coli | Radius_SOS |
| MGS | abundance | Hgn3C.0299 - Escherichia coli | Tibia_SOS |
| MGS | abundance | Hgn3C.0299 - Escherichia coli | Handgrip |
| MGS | abundance | Hgn3C.0300 - Clostridia sp. | Tibia_length |
| MGS | abundance | Hgn3C.0300 - Clostridia sp. | Radius_length |
| MGS | abundance | Hgn3C.0300 - Clostridia sp. | Radius_SOS |
| MGS | abundance | Hgn3C.0300 - Clostridia sp. | Tibia_SOS |
| MGS | abundance | Hgn3C.0300 - Clostridia sp. | Handgrip |
| MGS | abundance | Hgn3C.0301 - Prevotella hominis | Tibia_length |
| MGS | abundance | Hgn3C.0301 - Prevotella hominis | Radius_length |
| MGS | abundance | Hgn3C.0301 - Prevotella hominis | Radius_SOS |
| MGS | abundance | Hgn3C.0301 - Prevotella hominis | Tibia_SOS |

| | | | |
|---|---|---|---|
| MGS | abundance | Hgn3C.0301 - Prevotella hominis | Handgrip |
| MGS | abundance | Hgn3C.0302 - Lachnospiraceae sp. | Tibia_length |
| MGS | abundance | Hgn3C.0302 - Lachnospiraceae sp. | Radius_length |
| MGS | abundance | Hgn3C.0302 - Lachnospiraceae sp. | Radius_SOS |
| MGS | abundance | Hgn3C.0302 - Lachnospiraceae sp. | Tibia_SOS |
| MGS | abundance | Hgn3C.0302 - Lachnospiraceae sp. | Handgrip |
| MGS | abundance | Hgn3C.0304 - Eubacteriales sp. | Tibia_length |
| MGS | abundance | Hgn3C.0304 - Eubacteriales sp. | Radius_length |
| MGS | abundance | Hgn3C.0304 - Eubacteriales sp. | Radius_SOS |
| MGS | abundance | Hgn3C.0304 - Eubacteriales sp. | Tibia_SOS |
| MGS | abundance | Hgn3C.0304 - Eubacteriales sp. | Handgrip |
| MGS | abundance | Hgn3C.0305 - Candidatus Borkfalkia ceftriaxoniphila | Tibia_length |
| MGS | abundance | Hgn3C.0305 - Candidatus Borkfalkia ceftriaxoniphila | Radius_length |
| MGS | abundance | Hgn3C.0305 - Candidatus Borkfalkia ceftriaxoniphila | Radius_SOS |
| MGS | abundance | Hgn3C.0305 - Candidatus Borkfalkia ceftriaxoniphila | Tibia_SOS |
| MGS | abundance | Hgn3C.0305 - Candidatus Borkfalkia ceftriaxoniphila | Handgrip |
| MGS | abundance | Hgn3C.0306 - Bacteroides nordii | Tibia_length |
| MGS | abundance | Hgn3C.0306 - Bacteroides nordii | Radius_length |
| MGS | abundance | Hgn3C.0306 - Bacteroides nordii | Radius_SOS |
| MGS | abundance | Hgn3C.0306 - Bacteroides nordii | Tibia_SOS |
| MGS | abundance | Hgn3C.0306 - Bacteroides nordii | Handgrip |
| MGS | abundance | Hgn3C.0307 - Veillonella atypica | Tibia_length |
| MGS | abundance | Hgn3C.0307 - Veillonella atypica | Radius_length |
| MGS | abundance | Hgn3C.0307 - Veillonella atypica | Radius_SOS |
| MGS | abundance | Hgn3C.0307 - Veillonella atypica | Tibia_SOS |
| MGS | abundance | Hgn3C.0307 - Veillonella atypica | Handgrip |
| MGS | abundance | Hgn3C.0308 - [Clostridium] symbiosum | Tibia_length |
| MGS | abundance | Hgn3C.0308 - [Clostridium] symbiosum | Radius_length |
| MGS | abundance | Hgn3C.0308 - [Clostridium] symbiosum | Radius_SOS |
| MGS | abundance | Hgn3C.0308 - [Clostridium] symbiosum | Tibia_SOS |
| MGS | abundance | Hgn3C.0308 - [Clostridium] symbiosum | Handgrip |
| MGS | abundance | Hgn3C.0309 - Eubacteriales sp. | Tibia_length |
| MGS | abundance | Hgn3C.0309 - Eubacteriales sp. | Radius_length |
| MGS | abundance | Hgn3C.0309 - Eubacteriales sp. | Radius_SOS |
| MGS | abundance | Hgn3C.0309 - Eubacteriales sp. | Tibia_SOS |
| MGS | abundance | Hgn3C.0309 - Eubacteriales sp. | Handgrip |
| MGS | abundance | Hgn3C.0310 - Ligilactobacillus ruminis | Tibia_length |
| MGS | abundance | Hgn3C.0310 - Ligilactobacillus ruminis | Radius_length |
| MGS | abundance | Hgn3C.0310 - Ligilactobacillus ruminis | Radius_SOS |
| MGS | abundance | Hgn3C.0310 - Ligilactobacillus ruminis | Tibia_SOS |
| MGS | abundance | Hgn3C.0310 - Ligilactobacillus ruminis | Handgrip |
| MGS | abundance | Hgn3C.0313 - Ruminococcus bromii | Tibia_length |
| MGS | abundance | Hgn3C.0313 - Ruminococcus bromii | Radius_length |
| MGS | abundance | Hgn3C.0313 - Ruminococcus bromii | Radius_SOS |
| MGS | abundance | Hgn3C.0313 - Ruminococcus bromii | Tibia_SOS |
| MGS | abundance | Hgn3C.0313 - Ruminococcus bromii | Handgrip |
| MGS | abundance | Hgn3C.0314 - Ruminococcus sp. | Tibia_length |
| MGS | abundance | Hgn3C.0314 - Ruminococcus sp. | Radius_length |

| | | | |
|---|---|---|---|
| MGS | abundance | Hgn3C.0314 - Ruminococcus sp. | Radius_SOS |
| MGS | abundance | Hgn3C.0314 - Ruminococcus sp. | Tibia_SOS |
| MGS | abundance | Hgn3C.0314 - Ruminococcus sp. | Handgrip |
| MGS | abundance | Hgn3C.0315 - Bacteroidia sp. | Tibia_length |
| MGS | abundance | Hgn3C.0315 - Bacteroidia sp. | Radius_length |
| MGS | abundance | Hgn3C.0315 - Bacteroidia sp. | Radius_SOS |
| MGS | abundance | Hgn3C.0315 - Bacteroidia sp. | Tibia_SOS |
| MGS | abundance | Hgn3C.0315 - Bacteroidia sp. | Handgrip |
| MGS | abundance | Hgn3C.0317 - Intestinimonas butyriciproducens | Tibia_length |
| MGS | abundance | Hgn3C.0317 - Intestinimonas butyriciproducens | Radius_length |
| MGS | abundance | Hgn3C.0317 - Intestinimonas butyriciproducens | Radius_SOS |
| MGS | abundance | Hgn3C.0317 - Intestinimonas butyriciproducens | Tibia_SOS |
| MGS | abundance | Hgn3C.0317 - Intestinimonas butyriciproducens | Handgrip |
| MGS | abundance | Hgn3C.0319 - Clostridiaceae sp. | Tibia_length |
| MGS | abundance | Hgn3C.0319 - Clostridiaceae sp. | Radius_length |
| MGS | abundance | Hgn3C.0319 - Clostridiaceae sp. | Radius_SOS |
| MGS | abundance | Hgn3C.0319 - Clostridiaceae sp. | Tibia_SOS |
| MGS | abundance | Hgn3C.0319 - Clostridiaceae sp. | Handgrip |
| MGS | abundance | Hgn3C.0323 - Bacteroides sp. | Tibia_length |
| MGS | abundance | Hgn3C.0323 - Bacteroides sp. | Radius_length |
| MGS | abundance | Hgn3C.0323 - Bacteroides sp. | Radius_SOS |
| MGS | abundance | Hgn3C.0323 - Bacteroides sp. | Tibia_SOS |
| MGS | abundance | Hgn3C.0323 - Bacteroides sp. | Handgrip |
| MGS | abundance | Hgn3C.0324 - Prevotella stercorea | Tibia_length |
| MGS | abundance | Hgn3C.0324 - Prevotella stercorea | Radius_length |
| MGS | abundance | Hgn3C.0324 - Prevotella stercorea | Radius_SOS |
| MGS | abundance | Hgn3C.0324 - Prevotella stercorea | Tibia_SOS |
| MGS | abundance | Hgn3C.0324 - Prevotella stercorea | Handgrip |
| MGS | abundance | Hgn3C.0325 - Roseburia sp. BX0805 | Tibia_length |
| MGS | abundance | Hgn3C.0325 - Roseburia sp. BX0805 | Radius_length |
| MGS | abundance | Hgn3C.0325 - Roseburia sp. BX0805 | Radius_SOS |
| MGS | abundance | Hgn3C.0325 - Roseburia sp. BX0805 | Tibia_SOS |
| MGS | abundance | Hgn3C.0325 - Roseburia sp. BX0805 | Handgrip |
| MGS | abundance | Hgn3C.0328 - Eubacteriales sp. | Tibia_length |
| MGS | abundance | Hgn3C.0328 - Eubacteriales sp. | Radius_length |
| MGS | abundance | Hgn3C.0328 - Eubacteriales sp. | Radius_SOS |
| MGS | abundance | Hgn3C.0328 - Eubacteriales sp. | Tibia_SOS |
| MGS | abundance | Hgn3C.0328 - Eubacteriales sp. | Handgrip |
| MGS | abundance | Hgn3C.0329 - Eubacteriales sp. | Tibia_length |
| MGS | abundance | Hgn3C.0329 - Eubacteriales sp. | Radius_length |
| MGS | abundance | Hgn3C.0329 - Eubacteriales sp. | Radius_SOS |
| MGS | abundance | Hgn3C.0329 - Eubacteriales sp. | Tibia_SOS |
| MGS | abundance | Hgn3C.0329 - Eubacteriales sp. | Handgrip |
| MGS | abundance | Hgn3C.0330 - Faecalibacillus intestinalis | Tibia_length |
| MGS | abundance | Hgn3C.0330 - Faecalibacillus intestinalis | Radius_length |
| MGS | abundance | Hgn3C.0330 - Faecalibacillus intestinalis | Radius_SOS |
| MGS | abundance | Hgn3C.0330 - Faecalibacillus intestinalis | Tibia_SOS |
| MGS | abundance | Hgn3C.0330 - Faecalibacillus intestinalis | Handgrip |

| | | | |
|---|---|---|---|
| MGS | abundance | Hgn3C.0332 - Klebsiella pneumoniae subsp. pneumoniae | Tibia_length |
| MGS | abundance | Hgn3C.0332 - Klebsiella pneumoniae subsp. pneumoniae | Radius_length |
| MGS | abundance | Hgn3C.0332 - Klebsiella pneumoniae subsp. pneumoniae | Radius_SOS |
| MGS | abundance | Hgn3C.0332 - Klebsiella pneumoniae subsp. pneumoniae | Tibia_SOS |
| MGS | abundance | Hgn3C.0332 - Klebsiella pneumoniae subsp. pneumoniae | Handgrip |
| MGS | abundance | Hgn3C.0334 - Bacteroides cellulosilyticus | Tibia_length |
| MGS | abundance | Hgn3C.0334 - Bacteroides cellulosilyticus | Radius_length |
| MGS | abundance | Hgn3C.0334 - Bacteroides cellulosilyticus | Radius_SOS |
| MGS | abundance | Hgn3C.0334 - Bacteroides cellulosilyticus | Tibia_SOS |
| MGS | abundance | Hgn3C.0334 - Bacteroides cellulosilyticus | Handgrip |
| MGS | abundance | Hgn3C.0335 - Alistipes sp. Marseille-P2263 | Tibia_length |
| MGS | abundance | Hgn3C.0335 - Alistipes sp. Marseille-P2263 | Radius_length |
| MGS | abundance | Hgn3C.0335 - Alistipes sp. Marseille-P2263 | Radius_SOS |
| MGS | abundance | Hgn3C.0335 - Alistipes sp. Marseille-P2263 | Tibia_SOS |
| MGS | abundance | Hgn3C.0335 - Alistipes sp. Marseille-P2263 | Handgrip |
| MGS | abundance | Hgn3C.0336 - Parabacteroides goldsteinii | Tibia_length |
| MGS | abundance | Hgn3C.0336 - Parabacteroides goldsteinii | Radius_length |
| MGS | abundance | Hgn3C.0336 - Parabacteroides goldsteinii | Radius_SOS |
| MGS | abundance | Hgn3C.0336 - Parabacteroides goldsteinii | Tibia_SOS |
| MGS | abundance | Hgn3C.0336 - Parabacteroides goldsteinii | Handgrip |
| MGS | abundance | Hgn3C.0337 - Eubacteriales sp. | Tibia_length |
| MGS | abundance | Hgn3C.0337 - Eubacteriales sp. | Radius_length |
| MGS | abundance | Hgn3C.0337 - Eubacteriales sp. | Radius_SOS |
| MGS | abundance | Hgn3C.0337 - Eubacteriales sp. | Tibia_SOS |
| MGS | abundance | Hgn3C.0337 - Eubacteriales sp. | Handgrip |
| MGS | abundance | Hgn3C.0338 - Eubacteriales sp. | Tibia_length |
| MGS | abundance | Hgn3C.0338 - Eubacteriales sp. | Radius_length |
| MGS | abundance | Hgn3C.0338 - Eubacteriales sp. | Radius_SOS |
| MGS | abundance | Hgn3C.0338 - Eubacteriales sp. | Tibia_SOS |
| MGS | abundance | Hgn3C.0338 - Eubacteriales sp. | Handgrip |
| MGS | abundance | Hgn3C.0345 - Oscillospiraceae sp. | Tibia_length |
| MGS | abundance | Hgn3C.0345 - Oscillospiraceae sp. | Radius_length |
| MGS | abundance | Hgn3C.0345 - Oscillospiraceae sp. | Radius_SOS |
| MGS | abundance | Hgn3C.0345 - Oscillospiraceae sp. | Tibia_SOS |
| MGS | abundance | Hgn3C.0345 - Oscillospiraceae sp. | Handgrip |
| MGS | abundance | Hgn3C.0348 - Streptococcus sp. | Tibia_length |
| MGS | abundance | Hgn3C.0348 - Streptococcus sp. | Radius_length |
| MGS | abundance | Hgn3C.0348 - Streptococcus sp. | Radius_SOS |
| MGS | abundance | Hgn3C.0348 - Streptococcus sp. | Tibia_SOS |
| MGS | abundance | Hgn3C.0348 - Streptococcus sp. | Handgrip |
| MGS | abundance | Hgn3C.0349 - Eubacteriales sp. | Tibia_length |
| MGS | abundance | Hgn3C.0349 - Eubacteriales sp. | Radius_length |
| MGS | abundance | Hgn3C.0349 - Eubacteriales sp. | Radius_SOS |
| MGS | abundance | Hgn3C.0349 - Eubacteriales sp. | Tibia_SOS |
| MGS | abundance | Hgn3C.0349 - Eubacteriales sp. | Handgrip |
| MGS | abundance | Hgn3C.0350 - [Clostridium] symbiosum | Tibia_length |
| MGS | abundance | Hgn3C.0350 - [Clostridium] symbiosum | Radius_length |
| MGS | abundance | Hgn3C.0350 - [Clostridium] symbiosum | Radius_SOS |

| | | | |
|---|---|---|---|
| MGS | abundance | Hgn3C.0350 - [Clostridium] symbiosum | Tibia_SOS |
| MGS | abundance | Hgn3C.0350 - [Clostridium] symbiosum | Handgrip |
| MGS | abundance | Hgn3C.0351 - Prevotella sp. | Tibia_length |
| MGS | abundance | Hgn3C.0351 - Prevotella sp. | Radius_length |
| MGS | abundance | Hgn3C.0351 - Prevotella sp. | Radius_SOS |
| MGS | abundance | Hgn3C.0351 - Prevotella sp. | Tibia_SOS |
| MGS | abundance | Hgn3C.0351 - Prevotella sp. | Handgrip |
| MGS | abundance | Hgn3C.0352 - Eubacteriales sp. | Tibia_length |
| MGS | abundance | Hgn3C.0352 - Eubacteriales sp. | Radius_length |
| MGS | abundance | Hgn3C.0352 - Eubacteriales sp. | Radius_SOS |
| MGS | abundance | Hgn3C.0352 - Eubacteriales sp. | Tibia_SOS |
| MGS | abundance | Hgn3C.0352 - Eubacteriales sp. | Handgrip |
| MGS | abundance | Hgn3C.0353 - Prevotella sp. P4-67 | Tibia_length |
| MGS | abundance | Hgn3C.0353 - Prevotella sp. P4-67 | Radius_length |
| MGS | abundance | Hgn3C.0353 - Prevotella sp. P4-67 | Radius_SOS |
| MGS | abundance | Hgn3C.0353 - Prevotella sp. P4-67 | Tibia_SOS |
| MGS | abundance | Hgn3C.0353 - Prevotella sp. P4-67 | Handgrip |
| MGS | abundance | Hgn3C.0354 - Sutterella seckii | Tibia_length |
| MGS | abundance | Hgn3C.0354 - Sutterella seckii | Radius_length |
| MGS | abundance | Hgn3C.0354 - Sutterella seckii | Radius_SOS |
| MGS | abundance | Hgn3C.0354 - Sutterella seckii | Tibia_SOS |
| MGS | abundance | Hgn3C.0354 - Sutterella seckii | Handgrip |
| MGS | abundance | Hgn3C.0356 - Blautia sp. M29 | Tibia_length |
| MGS | abundance | Hgn3C.0356 - Blautia sp. M29 | Radius_length |
| MGS | abundance | Hgn3C.0356 - Blautia sp. M29 | Radius_SOS |
| MGS | abundance | Hgn3C.0356 - Blautia sp. M29 | Tibia_SOS |
| MGS | abundance | Hgn3C.0356 - Blautia sp. M29 | Handgrip |
| MGS | abundance | Hgn3C.0357 - Eubacteriales sp. | Tibia_length |
| MGS | abundance | Hgn3C.0357 - Eubacteriales sp. | Radius_length |
| MGS | abundance | Hgn3C.0357 - Eubacteriales sp. | Radius_SOS |
| MGS | abundance | Hgn3C.0357 - Eubacteriales sp. | Tibia_SOS |
| MGS | abundance | Hgn3C.0357 - Eubacteriales sp. | Handgrip |
| MGS | abundance | Hgn3C.0359 - Eubacterium sp. BX4 | Tibia_length |
| MGS | abundance | Hgn3C.0359 - Eubacterium sp. BX4 | Radius_length |
| MGS | abundance | Hgn3C.0359 - Eubacterium sp. BX4 | Radius_SOS |
| MGS | abundance | Hgn3C.0359 - Eubacterium sp. BX4 | Tibia_SOS |
| MGS | abundance | Hgn3C.0359 - Eubacterium sp. BX4 | Handgrip |
| MGS | abundance | Hgn3C.0361 - Dorea sp. AF24-7LB | Tibia_length |
| MGS | abundance | Hgn3C.0361 - Dorea sp. AF24-7LB | Radius_length |
| MGS | abundance | Hgn3C.0361 - Dorea sp. AF24-7LB | Radius_SOS |
| MGS | abundance | Hgn3C.0361 - Dorea sp. AF24-7LB | Tibia_SOS |
| MGS | abundance | Hgn3C.0361 - Dorea sp. AF24-7LB | Handgrip |
| MGS | abundance | Hgn3C.0362 - Dysosmobacter sp. NSJ-60 | Tibia_length |
| MGS | abundance | Hgn3C.0362 - Dysosmobacter sp. NSJ-60 | Radius_length |
| MGS | abundance | Hgn3C.0362 - Dysosmobacter sp. NSJ-60 | Radius_SOS |
| MGS | abundance | Hgn3C.0362 - Dysosmobacter sp. NSJ-60 | Tibia_SOS |
| MGS | abundance | Hgn3C.0362 - Dysosmobacter sp. NSJ-60 | Handgrip |
| MGS | abundance | Hgn3C.0364 - Eubacteriales sp. | Tibia_length |

| | | | |
|---|---|---|---|
| MGS | abundanc | Hgn3C.0364 - Eubacteriales sp. | Radius_length |
| MGS | abundanc | Hgn3C.0364 - Eubacteriales sp. | Radius_SOS |
| MGS | abundanc | Hgn3C.0364 - Eubacteriales sp. | Tibia_SOS |
| MGS | abundanc | Hgn3C.0364 - Eubacteriales sp. | Handgrip |
| MGS | abundanc | Hgn3C.0365 - Eubacteriales sp. | Tibia_length |
| MGS | abundanc | Hgn3C.0365 - Eubacteriales sp. | Radius_length |
| MGS | abundanc | Hgn3C.0365 - Eubacteriales sp. | Radius_SOS |
| MGS | abundanc | Hgn3C.0365 - Eubacteriales sp. | Tibia_SOS |
| MGS | abundanc | Hgn3C.0365 - Eubacteriales sp. | Handgrip |
| MGS | abundanc | Hgn3C.0367 - Lachnospiraceae sp. | Tibia_length |
| MGS | abundanc | Hgn3C.0367 - Lachnospiraceae sp. | Radius_length |
| MGS | abundanc | Hgn3C.0367 - Lachnospiraceae sp. | Radius_SOS |
| MGS | abundanc | Hgn3C.0367 - Lachnospiraceae sp. | Tibia_SOS |
| MGS | abundanc | Hgn3C.0367 - Lachnospiraceae sp. | Handgrip |
| MGS | abundanc | Hgn3C.0368 - Eubacteriales sp. | Tibia_length |
| MGS | abundanc | Hgn3C.0368 - Eubacteriales sp. | Radius_length |
| MGS | abundanc | Hgn3C.0368 - Eubacteriales sp. | Radius_SOS |
| MGS | abundanc | Hgn3C.0368 - Eubacteriales sp. | Tibia_SOS |
| MGS | abundanc | Hgn3C.0368 - Eubacteriales sp. | Handgrip |
| MGS | abundanc | Hgn3C.0369 - Prevotella sp. | Tibia_length |
| MGS | abundanc | Hgn3C.0369 - Prevotella sp. | Radius_length |
| MGS | abundanc | Hgn3C.0369 - Prevotella sp. | Radius_SOS |
| MGS | abundanc | Hgn3C.0369 - Prevotella sp. | Tibia_SOS |
| MGS | abundanc | Hgn3C.0369 - Prevotella sp. | Handgrip |
| MGS | abundanc | Hgn3C.0371 - Lachnospiraceae sp. | Tibia_length |
| MGS | abundanc | Hgn3C.0371 - Lachnospiraceae sp. | Radius_length |
| MGS | abundanc | Hgn3C.0371 - Lachnospiraceae sp. | Radius_SOS |
| MGS | abundanc | Hgn3C.0371 - Lachnospiraceae sp. | Tibia_SOS |
| MGS | abundanc | Hgn3C.0371 - Lachnospiraceae sp. | Handgrip |
| MGS | abundanc | Hgn3C.0373 - Enterocloster clostridioformis | Tibia_length |
| MGS | abundanc | Hgn3C.0373 - Enterocloster clostridioformis | Radius_length |
| MGS | abundanc | Hgn3C.0373 - Enterocloster clostridioformis | Radius_SOS |
| MGS | abundanc | Hgn3C.0373 - Enterocloster clostridioformis | Tibia_SOS |
| MGS | abundanc | Hgn3C.0373 - Enterocloster clostridioformis | Handgrip |
| MGS | abundanc | Hgn3C.0375 - Ellagibacter isourolithinifaciens | Tibia_length |
| MGS | abundanc | Hgn3C.0375 - Ellagibacter isourolithinifaciens | Radius_length |
| MGS | abundanc | Hgn3C.0375 - Ellagibacter isourolithinifaciens | Radius_SOS |
| MGS | abundanc | Hgn3C.0375 - Ellagibacter isourolithinifaciens | Tibia_SOS |
| MGS | abundanc | Hgn3C.0375 - Ellagibacter isourolithinifaciens | Handgrip |
| MGS | abundanc | Hgn3C.0378 - Firmicutes sp. | Tibia_length |
| MGS | abundanc | Hgn3C.0378 - Firmicutes sp. | Radius_length |
| MGS | abundanc | Hgn3C.0378 - Firmicutes sp. | Radius_SOS |
| MGS | abundanc | Hgn3C.0378 - Firmicutes sp. | Tibia_SOS |
| MGS | abundanc | Hgn3C.0378 - Firmicutes sp. | Handgrip |
| MGS | abundanc | Hgn3C.0379 - Eubacteriales sp. | Tibia_length |
| MGS | abundanc | Hgn3C.0379 - Eubacteriales sp. | Radius_length |
| MGS | abundanc | Hgn3C.0379 - Eubacteriales sp. | Radius_SOS |
| MGS | abundanc | Hgn3C.0379 - Eubacteriales sp. | Tibia_SOS |

| | | | |
|---|---|---|---|
| MGS | abundance | Hgn3C.0379 - Eubacteriales sp. | Handgrip |
| MGS | abundance | Hgn3C.0381 - Clostridium sp. AM30-24 | Tibia_length |
| MGS | abundance | Hgn3C.0381 - Clostridium sp. AM30-24 | Radius_length |
| MGS | abundance | Hgn3C.0381 - Clostridium sp. AM30-24 | Radius_SOS |
| MGS | abundance | Hgn3C.0381 - Clostridium sp. AM30-24 | Tibia_SOS |
| MGS | abundance | Hgn3C.0381 - Clostridium sp. AM30-24 | Handgrip |
| MGS | abundance | Hgn3C.0385 - Veillonella rogosae | Tibia_length |
| MGS | abundance | Hgn3C.0385 - Veillonella rogosae | Radius_length |
| MGS | abundance | Hgn3C.0385 - Veillonella rogosae | Radius_SOS |
| MGS | abundance | Hgn3C.0385 - Veillonella rogosae | Tibia_SOS |
| MGS | abundance | Hgn3C.0385 - Veillonella rogosae | Handgrip |
| MGS | abundance | Hgn3C.0386 - Clostridium sp. AT4 | Tibia_length |
| MGS | abundance | Hgn3C.0386 - Clostridium sp. AT4 | Radius_length |
| MGS | abundance | Hgn3C.0386 - Clostridium sp. AT4 | Radius_SOS |
| MGS | abundance | Hgn3C.0386 - Clostridium sp. AT4 | Tibia_SOS |
| MGS | abundance | Hgn3C.0386 - Clostridium sp. AT4 | Handgrip |
| MGS | abundance | Hgn3C.0388 - Clostridium sp. | Tibia_length |
| MGS | abundance | Hgn3C.0388 - Clostridium sp. | Radius_length |
| MGS | abundance | Hgn3C.0388 - Clostridium sp. | Radius_SOS |
| MGS | abundance | Hgn3C.0388 - Clostridium sp. | Tibia_SOS |
| MGS | abundance | Hgn3C.0388 - Clostridium sp. | Handgrip |
| MGS | abundance | Hgn3C.0389 - Vescimonas fastidiosa | Tibia_length |
| MGS | abundance | Hgn3C.0389 - Vescimonas fastidiosa | Radius_length |
| MGS | abundance | Hgn3C.0389 - Vescimonas fastidiosa | Radius_SOS |
| MGS | abundance | Hgn3C.0389 - Vescimonas fastidiosa | Tibia_SOS |
| MGS | abundance | Hgn3C.0389 - Vescimonas fastidiosa | Handgrip |
| MGS | abundance | Hgn3C.0392 - Clostridia sp. | Tibia_length |
| MGS | abundance | Hgn3C.0392 - Clostridia sp. | Radius_length |
| MGS | abundance | Hgn3C.0392 - Clostridia sp. | Radius_SOS |
| MGS | abundance | Hgn3C.0392 - Clostridia sp. | Tibia_SOS |
| MGS | abundance | Hgn3C.0392 - Clostridia sp. | Handgrip |
| MGS | abundance | Hgn3C.0394 - Hungatella hathewayi | Tibia_length |
| MGS | abundance | Hgn3C.0394 - Hungatella hathewayi | Radius_length |
| MGS | abundance | Hgn3C.0394 - Hungatella hathewayi | Radius_SOS |
| MGS | abundance | Hgn3C.0394 - Hungatella hathewayi | Tibia_SOS |
| MGS | abundance | Hgn3C.0394 - Hungatella hathewayi | Handgrip |
| MGS | abundance | Hgn3C.0395 - Bacteria sp. | Tibia_length |
| MGS | abundance | Hgn3C.0395 - Bacteria sp. | Radius_length |
| MGS | abundance | Hgn3C.0395 - Bacteria sp. | Radius_SOS |
| MGS | abundance | Hgn3C.0395 - Bacteria sp. | Tibia_SOS |
| MGS | abundance | Hgn3C.0395 - Bacteria sp. | Handgrip |
| MGS | abundance | Hgn3C.0396 - Bacteroidales sp. | Tibia_length |
| MGS | abundance | Hgn3C.0396 - Bacteroidales sp. | Radius_length |
| MGS | abundance | Hgn3C.0396 - Bacteroidales sp. | Radius_SOS |
| MGS | abundance | Hgn3C.0396 - Bacteroidales sp. | Tibia_SOS |
| MGS | abundance | Hgn3C.0396 - Bacteroidales sp. | Handgrip |
| MGS | abundance | Hgn3C.0397 - Parabacteroides sp. | Tibia_length |
| MGS | abundance | Hgn3C.0397 - Parabacteroides sp. | Radius_length |

| | | | |
|---|---|---|---|
| MGS | abundance | Hgn3C.0397 - Parabacteroides sp. | Radius_SOS |
| MGS | abundance | Hgn3C.0397 - Parabacteroides sp. | Tibia_SOS |
| MGS | abundance | Hgn3C.0397 - Parabacteroides sp. | Handgrip |
| MGS | abundance | Hgn3C.0400 - Prevotellamassilia timonensis | Tibia_length |
| MGS | abundance | Hgn3C.0400 - Prevotellamassilia timonensis | Radius_length |
| MGS | abundance | Hgn3C.0400 - Prevotellamassilia timonensis | Radius_SOS |
| MGS | abundance | Hgn3C.0400 - Prevotellamassilia timonensis | Tibia_SOS |
| MGS | abundance | Hgn3C.0400 - Prevotellamassilia timonensis | Handgrip |
| MGS | abundance | Hgn3C.0401 - Eubacteriales sp. | Tibia_length |
| MGS | abundance | Hgn3C.0401 - Eubacteriales sp. | Radius_length |
| MGS | abundance | Hgn3C.0401 - Eubacteriales sp. | Radius_SOS |
| MGS | abundance | Hgn3C.0401 - Eubacteriales sp. | Tibia_SOS |
| MGS | abundance | Hgn3C.0401 - Eubacteriales sp. | Handgrip |
| MGS | abundance | Hgn3C.0404 - Eubacteriales sp. | Tibia_length |
| MGS | abundance | Hgn3C.0404 - Eubacteriales sp. | Radius_length |
| MGS | abundance | Hgn3C.0404 - Eubacteriales sp. | Radius_SOS |
| MGS | abundance | Hgn3C.0404 - Eubacteriales sp. | Tibia_SOS |
| MGS | abundance | Hgn3C.0404 - Eubacteriales sp. | Handgrip |
| MGS | abundance | Hgn3C.0405 - Bacteroides ovatus | Tibia_length |
| MGS | abundance | Hgn3C.0405 - Bacteroides ovatus | Radius_length |
| MGS | abundance | Hgn3C.0405 - Bacteroides ovatus | Radius_SOS |
| MGS | abundance | Hgn3C.0405 - Bacteroides ovatus | Tibia_SOS |
| MGS | abundance | Hgn3C.0405 - Bacteroides ovatus | Handgrip |
| MGS | abundance | Hgn3C.0407 - Eubacteriales sp. | Tibia_length |
| MGS | abundance | Hgn3C.0407 - Eubacteriales sp. | Radius_length |
| MGS | abundance | Hgn3C.0407 - Eubacteriales sp. | Radius_SOS |
| MGS | abundance | Hgn3C.0407 - Eubacteriales sp. | Tibia_SOS |
| MGS | abundance | Hgn3C.0407 - Eubacteriales sp. | Handgrip |
| MGS | abundance | Hgn3C.0408 - Eubacteriales sp. | Tibia_length |
| MGS | abundance | Hgn3C.0408 - Eubacteriales sp. | Radius_length |
| MGS | abundance | Hgn3C.0408 - Eubacteriales sp. | Radius_SOS |
| MGS | abundance | Hgn3C.0408 - Eubacteriales sp. | Tibia_SOS |
| MGS | abundance | Hgn3C.0408 - Eubacteriales sp. | Handgrip |
| MGS | abundance | Hgn3C.0412 - Eubacteriales sp. | Tibia_length |
| MGS | abundance | Hgn3C.0412 - Eubacteriales sp. | Radius_length |
| MGS | abundance | Hgn3C.0412 - Eubacteriales sp. | Radius_SOS |
| MGS | abundance | Hgn3C.0412 - Eubacteriales sp. | Tibia_SOS |
| MGS | abundance | Hgn3C.0412 - Eubacteriales sp. | Handgrip |
| MGS | abundance | Hgn3C.0413 - Blautia caecimuris | Tibia_length |
| MGS | abundance | Hgn3C.0413 - Blautia caecimuris | Radius_length |
| MGS | abundance | Hgn3C.0413 - Blautia caecimuris | Radius_SOS |
| MGS | abundance | Hgn3C.0413 - Blautia caecimuris | Tibia_SOS |
| MGS | abundance | Hgn3C.0413 - Blautia caecimuris | Handgrip |
| MGS | abundance | Hgn3C.0415 - Clostridium sp. C5-48 | Tibia_length |
| MGS | abundance | Hgn3C.0415 - Clostridium sp. C5-48 | Radius_length |
| MGS | abundance | Hgn3C.0415 - Clostridium sp. C5-48 | Radius_SOS |
| MGS | abundance | Hgn3C.0415 - Clostridium sp. C5-48 | Tibia_SOS |
| MGS | abundance | Hgn3C.0415 - Clostridium sp. C5-48 | Handgrip |

| | | | |
|---|---|---|---|
| MGS | abundance | Hgn3C.0417 - Eubacteriales sp. | Tibia_length |
| MGS | abundance | Hgn3C.0417 - Eubacteriales sp. | Radius_length |
| MGS | abundance | Hgn3C.0417 - Eubacteriales sp. | Radius_SOS |
| MGS | abundance | Hgn3C.0417 - Eubacteriales sp. | Tibia_SOS |
| MGS | abundance | Hgn3C.0417 - Eubacteriales sp. | Handgrip |
| MGS | abundance | Hgn3C.0418 - Blautia sp. | Tibia_length |
| MGS | abundance | Hgn3C.0418 - Blautia sp. | Radius_length |
| MGS | abundance | Hgn3C.0418 - Blautia sp. | Radius_SOS |
| MGS | abundance | Hgn3C.0418 - Blautia sp. | Tibia_SOS |
| MGS | abundance | Hgn3C.0418 - Blautia sp. | Handgrip |
| MGS | abundance | Hgn3C.0419 - Prevotella sp. 885 | Tibia_length |
| MGS | abundance | Hgn3C.0419 - Prevotella sp. 885 | Radius_length |
| MGS | abundance | Hgn3C.0419 - Prevotella sp. 885 | Radius_SOS |
| MGS | abundance | Hgn3C.0419 - Prevotella sp. 885 | Tibia_SOS |
| MGS | abundance | Hgn3C.0419 - Prevotella sp. 885 | Handgrip |
| MGS | abundance | Hgn3C.0420 - Clostridium sp. 1001270J_160509_D11 | Tibia_length |
| MGS | abundance | Hgn3C.0420 - Clostridium sp. 1001270J_160509_D11 | Radius_length |
| MGS | abundance | Hgn3C.0420 - Clostridium sp. 1001270J_160509_D11 | Radius_SOS |
| MGS | abundance | Hgn3C.0420 - Clostridium sp. 1001270J_160509_D11 | Tibia_SOS |
| MGS | abundance | Hgn3C.0420 - Clostridium sp. 1001270J_160509_D11 | Handgrip |
| MGS | abundance | Hgn3C.0428 - Eubacteriales sp. | Tibia_length |
| MGS | abundance | Hgn3C.0428 - Eubacteriales sp. | Radius_length |
| MGS | abundance | Hgn3C.0428 - Eubacteriales sp. | Radius_SOS |
| MGS | abundance | Hgn3C.0428 - Eubacteriales sp. | Tibia_SOS |
| MGS | abundance | Hgn3C.0428 - Eubacteriales sp. | Handgrip |
| MGS | abundance | Hgn3C.0430 - Enterocloster aldenensis | Tibia_length |
| MGS | abundance | Hgn3C.0430 - Enterocloster aldenensis | Radius_length |
| MGS | abundance | Hgn3C.0430 - Enterocloster aldenensis | Radius_SOS |
| MGS | abundance | Hgn3C.0430 - Enterocloster aldenensis | Tibia_SOS |
| MGS | abundance | Hgn3C.0430 - Enterocloster aldenensis | Handgrip |
| MGS | abundance | Hgn3C.0434 - Firmicutes sp. | Tibia_length |
| MGS | abundance | Hgn3C.0434 - Firmicutes sp. | Radius_length |
| MGS | abundance | Hgn3C.0434 - Firmicutes sp. | Radius_SOS |
| MGS | abundance | Hgn3C.0434 - Firmicutes sp. | Tibia_SOS |
| MGS | abundance | Hgn3C.0434 - Firmicutes sp. | Handgrip |
| MGS | abundance | Hgn3C.0435 - Eubacteriales sp. | Tibia_length |
| MGS | abundance | Hgn3C.0435 - Eubacteriales sp. | Radius_length |
| MGS | abundance | Hgn3C.0435 - Eubacteriales sp. | Radius_SOS |
| MGS | abundance | Hgn3C.0435 - Eubacteriales sp. | Tibia_SOS |
| MGS | abundance | Hgn3C.0435 - Eubacteriales sp. | Handgrip |
| MGS | abundance | Hgn3C.0436 - Eubacteriales sp. | Tibia_length |
| MGS | abundance | Hgn3C.0436 - Eubacteriales sp. | Radius_length |
| MGS | abundance | Hgn3C.0436 - Eubacteriales sp. | Radius_SOS |
| MGS | abundance | Hgn3C.0436 - Eubacteriales sp. | Tibia_SOS |
| MGS | abundance | Hgn3C.0436 - Eubacteriales sp. | Handgrip |
| MGS | abundance | Hgn3C.0437 - Prevotella sp. | Tibia_length |
| MGS | abundance | Hgn3C.0437 - Prevotella sp. | Radius_length |
| MGS | abundance | Hgn3C.0437 - Prevotella sp. | Radius_SOS |

| | | | |
|---|---|---|---|
| MGS | abundance | Hgn3C.0437 - Prevotella sp. | Tibia_SOS |
| MGS | abundance | Hgn3C.0437 - Prevotella sp. | Handgrip |
| MGS | abundance | Hgn3C.0438 - Coprobacillus cateniformis | Tibia_length |
| MGS | abundance | Hgn3C.0438 - Coprobacillus cateniformis | Radius_length |
| MGS | abundance | Hgn3C.0438 - Coprobacillus cateniformis | Radius_SOS |
| MGS | abundance | Hgn3C.0438 - Coprobacillus cateniformis | Tibia_SOS |
| MGS | abundance | Hgn3C.0438 - Coprobacillus cateniformis | Handgrip |
| MGS | abundance | Hgn3C.0439 - Enterococcus faecalis | Tibia_length |
| MGS | abundance | Hgn3C.0439 - Enterococcus faecalis | Radius_length |
| MGS | abundance | Hgn3C.0439 - Enterococcus faecalis | Radius_SOS |
| MGS | abundance | Hgn3C.0439 - Enterococcus faecalis | Tibia_SOS |
| MGS | abundance | Hgn3C.0439 - Enterococcus faecalis | Handgrip |
| MGS | abundance | Hgn3C.0442 - Holdemanella sp. | Tibia_length |
| MGS | abundance | Hgn3C.0442 - Holdemanella sp. | Radius_length |
| MGS | abundance | Hgn3C.0442 - Holdemanella sp. | Radius_SOS |
| MGS | abundance | Hgn3C.0442 - Holdemanella sp. | Tibia_SOS |
| MGS | abundance | Hgn3C.0442 - Holdemanella sp. | Handgrip |
| MGS | abundance | Hgn3C.0443 - Prevotella stercorea | Tibia_length |
| MGS | abundance | Hgn3C.0443 - Prevotella stercorea | Radius_length |
| MGS | abundance | Hgn3C.0443 - Prevotella stercorea | Radius_SOS |
| MGS | abundance | Hgn3C.0443 - Prevotella stercorea | Tibia_SOS |
| MGS | abundance | Hgn3C.0443 - Prevotella stercorea | Handgrip |
| MGS | abundance | Hgn3C.0444 - Eubacteriales sp. | Tibia_length |
| MGS | abundance | Hgn3C.0444 - Eubacteriales sp. | Radius_length |
| MGS | abundance | Hgn3C.0444 - Eubacteriales sp. | Radius_SOS |
| MGS | abundance | Hgn3C.0444 - Eubacteriales sp. | Tibia_SOS |
| MGS | abundance | Hgn3C.0444 - Eubacteriales sp. | Handgrip |
| MGS | abundance | Hgn3C.0445 - Aeromonadales sp. | Tibia_length |
| MGS | abundance | Hgn3C.0445 - Aeromonadales sp. | Radius_length |
| MGS | abundance | Hgn3C.0445 - Aeromonadales sp. | Radius_SOS |
| MGS | abundance | Hgn3C.0445 - Aeromonadales sp. | Tibia_SOS |
| MGS | abundance | Hgn3C.0445 - Aeromonadales sp. | Handgrip |
| MGS | abundance | Hgn3C.0448 - Firmicutes sp. | Tibia_length |
| MGS | abundance | Hgn3C.0448 - Firmicutes sp. | Radius_length |
| MGS | abundance | Hgn3C.0448 - Firmicutes sp. | Radius_SOS |
| MGS | abundance | Hgn3C.0448 - Firmicutes sp. | Tibia_SOS |
| MGS | abundance | Hgn3C.0448 - Firmicutes sp. | Handgrip |
| MGS | abundance | Hgn3C.0451 - Roseburia sp. AM16-25 | Tibia_length |
| MGS | abundance | Hgn3C.0451 - Roseburia sp. AM16-25 | Radius_length |
| MGS | abundance | Hgn3C.0451 - Roseburia sp. AM16-25 | Radius_SOS |
| MGS | abundance | Hgn3C.0451 - Roseburia sp. AM16-25 | Tibia_SOS |
| MGS | abundance | Hgn3C.0451 - Roseburia sp. AM16-25 | Handgrip |
| MGS | abundance | Hgn3C.0452 - Oscillospiraceae sp. | Tibia_length |
| MGS | abundance | Hgn3C.0452 - Oscillospiraceae sp. | Radius_length |
| MGS | abundance | Hgn3C.0452 - Oscillospiraceae sp. | Radius_SOS |
| MGS | abundance | Hgn3C.0452 - Oscillospiraceae sp. | Tibia_SOS |
| MGS | abundance | Hgn3C.0452 - Oscillospiraceae sp. | Handgrip |
| MGS | abundance | Hgn3C.0453 - Blautia sp. Marseille-P3087 | Tibia_length |

| | | | |
|---|---|---|---|
| MGS | abundance | Hgn3C.0453 - Blautia sp. Marseille-P3087 | Radius_length |
| MGS | abundance | Hgn3C.0453 - Blautia sp. Marseille-P3087 | Radius_SOS |
| MGS | abundance | Hgn3C.0453 - Blautia sp. Marseille-P3087 | Tibia_SOS |
| MGS | abundance | Hgn3C.0453 - Blautia sp. Marseille-P3087 | Handgrip |
| MGS | abundance | Hgn3C.0455 - Peptostreptococcaceae sp. | Tibia_length |
| MGS | abundance | Hgn3C.0455 - Peptostreptococcaceae sp. | Radius_length |
| MGS | abundance | Hgn3C.0455 - Peptostreptococcaceae sp. | Radius_SOS |
| MGS | abundance | Hgn3C.0455 - Peptostreptococcaceae sp. | Tibia_SOS |
| MGS | abundance | Hgn3C.0455 - Peptostreptococcaceae sp. | Handgrip |
| MGS | abundance | Hgn3C.0456 - Alistipes sp. | Tibia_length |
| MGS | abundance | Hgn3C.0456 - Alistipes sp. | Radius_length |
| MGS | abundance | Hgn3C.0456 - Alistipes sp. | Radius_SOS |
| MGS | abundance | Hgn3C.0456 - Alistipes sp. | Tibia_SOS |
| MGS | abundance | Hgn3C.0456 - Alistipes sp. | Handgrip |
| MGS | abundance | Hgn3C.0457 - Coprobacter secundus subsp. similis | Tibia_length |
| MGS | abundance | Hgn3C.0457 - Coprobacter secundus subsp. similis | Radius_length |
| MGS | abundance | Hgn3C.0457 - Coprobacter secundus subsp. similis | Radius_SOS |
| MGS | abundance | Hgn3C.0457 - Coprobacter secundus subsp. similis | Tibia_SOS |
| MGS | abundance | Hgn3C.0457 - Coprobacter secundus subsp. similis | Handgrip |
| MGS | abundance | Hgn3C.0458 - Eubacteriales sp. | Tibia_length |
| MGS | abundance | Hgn3C.0458 - Eubacteriales sp. | Radius_length |
| MGS | abundance | Hgn3C.0458 - Eubacteriales sp. | Radius_SOS |
| MGS | abundance | Hgn3C.0458 - Eubacteriales sp. | Tibia_SOS |
| MGS | abundance | Hgn3C.0458 - Eubacteriales sp. | Handgrip |
| MGS | abundance | Hgn3C.0459 - Eubacteriales sp. | Tibia_length |
| MGS | abundance | Hgn3C.0459 - Eubacteriales sp. | Radius_length |
| MGS | abundance | Hgn3C.0459 - Eubacteriales sp. | Radius_SOS |
| MGS | abundance | Hgn3C.0459 - Eubacteriales sp. | Tibia_SOS |
| MGS | abundance | Hgn3C.0459 - Eubacteriales sp. | Handgrip |
| MGS | abundance | Hgn3C.0464 - Bifidobacterium longum subsp. infantis | Tibia_length |
| MGS | abundance | Hgn3C.0464 - Bifidobacterium longum subsp. infantis | Radius_length |
| MGS | abundance | Hgn3C.0464 - Bifidobacterium longum subsp. infantis | Radius_SOS |
| MGS | abundance | Hgn3C.0464 - Bifidobacterium longum subsp. infantis | Tibia_SOS |
| MGS | abundance | Hgn3C.0464 - Bifidobacterium longum subsp. infantis | Handgrip |
| MGS | abundance | Hgn3C.0465 - Eisenbergiella massiliensis | Tibia_length |
| MGS | abundance | Hgn3C.0465 - Eisenbergiella massiliensis | Radius_length |
| MGS | abundance | Hgn3C.0465 - Eisenbergiella massiliensis | Radius_SOS |
| MGS | abundance | Hgn3C.0465 - Eisenbergiella massiliensis | Tibia_SOS |
| MGS | abundance | Hgn3C.0465 - Eisenbergiella massiliensis | Handgrip |
| MGS | abundance | Hgn3C.0468 - Clostridium sp. NSJ-42 | Tibia_length |
| MGS | abundance | Hgn3C.0468 - Clostridium sp. NSJ-42 | Radius_length |
| MGS | abundance | Hgn3C.0468 - Clostridium sp. NSJ-42 | Radius_SOS |
| MGS | abundance | Hgn3C.0468 - Clostridium sp. NSJ-42 | Tibia_SOS |
| MGS | abundance | Hgn3C.0468 - Clostridium sp. NSJ-42 | Handgrip |
| MGS | abundance | Hgn3C.0470 - Eubacteriales sp. | Tibia_length |
| MGS | abundance | Hgn3C.0470 - Eubacteriales sp. | Radius_length |
| MGS | abundance | Hgn3C.0470 - Eubacteriales sp. | Radius_SOS |
| MGS | abundance | Hgn3C.0470 - Eubacteriales sp. | Tibia_SOS |

| | | | |
|---|---|---|---|
| MGS | abundance | Hgn3C.0470 - Eubacteriales sp. | Handgrip |
| MGS | abundance | Hgn3C.0472 - Parabacteroides goldsteinii | Tibia_length |
| MGS | abundance | Hgn3C.0472 - Parabacteroides goldsteinii | Radius_length |
| MGS | abundance | Hgn3C.0472 - Parabacteroides goldsteinii | Radius_SOS |
| MGS | abundance | Hgn3C.0472 - Parabacteroides goldsteinii | Tibia_SOS |
| MGS | abundance | Hgn3C.0472 - Parabacteroides goldsteinii | Handgrip |
| MGS | abundance | Hgn3C.0475 - Coriobacteriia sp. | Tibia_length |
| MGS | abundance | Hgn3C.0475 - Coriobacteriia sp. | Radius_length |
| MGS | abundance | Hgn3C.0475 - Coriobacteriia sp. | Radius_SOS |
| MGS | abundance | Hgn3C.0475 - Coriobacteriia sp. | Tibia_SOS |
| MGS | abundance | Hgn3C.0475 - Coriobacteriia sp. | Handgrip |
| MGS | abundance | Hgn3C.0476 - Streptococcus sp. | Tibia_length |
| MGS | abundance | Hgn3C.0476 - Streptococcus sp. | Radius_length |
| MGS | abundance | Hgn3C.0476 - Streptococcus sp. | Radius_SOS |
| MGS | abundance | Hgn3C.0476 - Streptococcus sp. | Tibia_SOS |
| MGS | abundance | Hgn3C.0476 - Streptococcus sp. | Handgrip |
| MGS | abundance | Hgn3C.0477 - Eubacteriales sp. | Tibia_length |
| MGS | abundance | Hgn3C.0477 - Eubacteriales sp. | Radius_length |
| MGS | abundance | Hgn3C.0477 - Eubacteriales sp. | Radius_SOS |
| MGS | abundance | Hgn3C.0477 - Eubacteriales sp. | Tibia_SOS |
| MGS | abundance | Hgn3C.0477 - Eubacteriales sp. | Handgrip |
| MGS | abundance | Hgn3C.0482 - Lachnospiraceae sp. | Tibia_length |
| MGS | abundance | Hgn3C.0482 - Lachnospiraceae sp. | Radius_length |
| MGS | abundance | Hgn3C.0482 - Lachnospiraceae sp. | Radius_SOS |
| MGS | abundance | Hgn3C.0482 - Lachnospiraceae sp. | Tibia_SOS |
| MGS | abundance | Hgn3C.0482 - Lachnospiraceae sp. | Handgrip |
| MGS | abundance | Hgn3C.0489 - Eubacteriales sp. | Tibia_length |
| MGS | abundance | Hgn3C.0489 - Eubacteriales sp. | Radius_length |
| MGS | abundance | Hgn3C.0489 - Eubacteriales sp. | Radius_SOS |
| MGS | abundance | Hgn3C.0489 - Eubacteriales sp. | Tibia_SOS |
| MGS | abundance | Hgn3C.0489 - Eubacteriales sp. | Handgrip |
| MGS | abundance | Hgn3C.0492 - Prevotellaceae sp. | Tibia_length |
| MGS | abundance | Hgn3C.0492 - Prevotellaceae sp. | Radius_length |
| MGS | abundance | Hgn3C.0492 - Prevotellaceae sp. | Radius_SOS |
| MGS | abundance | Hgn3C.0492 - Prevotellaceae sp. | Tibia_SOS |
| MGS | abundance | Hgn3C.0492 - Prevotellaceae sp. | Handgrip |
| MGS | abundance | Hgn3C.0493 - Eisenbergiella tayi | Tibia_length |
| MGS | abundance | Hgn3C.0493 - Eisenbergiella tayi | Radius_length |
| MGS | abundance | Hgn3C.0493 - Eisenbergiella tayi | Radius_SOS |
| MGS | abundance | Hgn3C.0493 - Eisenbergiella tayi | Tibia_SOS |
| MGS | abundance | Hgn3C.0493 - Eisenbergiella tayi | Handgrip |
| MGS | abundance | Hgn3C.0494 - Prevotellaceae sp. | Tibia_length |
| MGS | abundance | Hgn3C.0494 - Prevotellaceae sp. | Radius_length |
| MGS | abundance | Hgn3C.0494 - Prevotellaceae sp. | Radius_SOS |
| MGS | abundance | Hgn3C.0494 - Prevotellaceae sp. | Tibia_SOS |
| MGS | abundance | Hgn3C.0494 - Prevotellaceae sp. | Handgrip |
| MGS | abundance | Hgn3C.0495 - Ruminococcus sp. | Tibia_length |
| MGS | abundance | Hgn3C.0495 - Ruminococcus sp. | Radius_length |

| | | | |
|---|---|---|---|
| MGS | abundance | Hgn3C.0495 - Ruminococcus sp. | Radius_SOS |
| MGS | abundance | Hgn3C.0495 - Ruminococcus sp. | Tibia_SOS |
| MGS | abundance | Hgn3C.0495 - Ruminococcus sp. | Handgrip |
| MGS | abundance | Hgn3C.0498 - Prevotella sp. | Tibia_length |
| MGS | abundance | Hgn3C.0498 - Prevotella sp. | Radius_length |
| MGS | abundance | Hgn3C.0498 - Prevotella sp. | Radius_SOS |
| MGS | abundance | Hgn3C.0498 - Prevotella sp. | Tibia_SOS |
| MGS | abundance | Hgn3C.0498 - Prevotella sp. | Handgrip |
| MGS | abundance | Hgn3C.0499 - Bacteroides sp. | Tibia_length |
| MGS | abundance | Hgn3C.0499 - Bacteroides sp. | Radius_length |
| MGS | abundance | Hgn3C.0499 - Bacteroides sp. | Radius_SOS |
| MGS | abundance | Hgn3C.0499 - Bacteroides sp. | Tibia_SOS |
| MGS | abundance | Hgn3C.0499 - Bacteroides sp. | Handgrip |
| MGS | abundance | Hgn3C.0500 - Anaerotignum sp. | Tibia_length |
| MGS | abundance | Hgn3C.0500 - Anaerotignum sp. | Radius_length |
| MGS | abundance | Hgn3C.0500 - Anaerotignum sp. | Radius_SOS |
| MGS | abundance | Hgn3C.0500 - Anaerotignum sp. | Tibia_SOS |
| MGS | abundance | Hgn3C.0500 - Anaerotignum sp. | Handgrip |
| MGS | abundance | Hgn3C.0501 - Oscillospiraceae sp. | Tibia_length |
| MGS | abundance | Hgn3C.0501 - Oscillospiraceae sp. | Radius_length |
| MGS | abundance | Hgn3C.0501 - Oscillospiraceae sp. | Radius_SOS |
| MGS | abundance | Hgn3C.0501 - Oscillospiraceae sp. | Tibia_SOS |
| MGS | abundance | Hgn3C.0501 - Oscillospiraceae sp. | Handgrip |
| MGS | abundance | Hgn3C.0503 - Eubacteriales sp. | Tibia_length |
| MGS | abundance | Hgn3C.0503 - Eubacteriales sp. | Radius_length |
| MGS | abundance | Hgn3C.0503 - Eubacteriales sp. | Radius_SOS |
| MGS | abundance | Hgn3C.0503 - Eubacteriales sp. | Tibia_SOS |
| MGS | abundance | Hgn3C.0503 - Eubacteriales sp. | Handgrip |
| MGS | abundance | Hgn3C.0504 - Megasphaera sp. BL7 | Tibia_length |
| MGS | abundance | Hgn3C.0504 - Megasphaera sp. BL7 | Radius_length |
| MGS | abundance | Hgn3C.0504 - Megasphaera sp. BL7 | Radius_SOS |
| MGS | abundance | Hgn3C.0504 - Megasphaera sp. BL7 | Tibia_SOS |
| MGS | abundance | Hgn3C.0504 - Megasphaera sp. BL7 | Handgrip |
| MGS | abundance | Hgn3C.0507 - [Clostridium] spiroforme | Tibia_length |
| MGS | abundance | Hgn3C.0507 - [Clostridium] spiroforme | Radius_length |
| MGS | abundance | Hgn3C.0507 - [Clostridium] spiroforme | Radius_SOS |
| MGS | abundance | Hgn3C.0507 - [Clostridium] spiroforme | Tibia_SOS |
| MGS | abundance | Hgn3C.0507 - [Clostridium] spiroforme | Handgrip |
| MGS | abundance | Hgn3C.0514 - Blautia glucerasea | Tibia_length |
| MGS | abundance | Hgn3C.0514 - Blautia glucerasea | Radius_length |
| MGS | abundance | Hgn3C.0514 - Blautia glucerasea | Radius_SOS |
| MGS | abundance | Hgn3C.0514 - Blautia glucerasea | Tibia_SOS |
| MGS | abundance | Hgn3C.0514 - Blautia glucerasea | Handgrip |
| MGS | abundance | Hgn3C.0518 - Eubacteriales sp. | Tibia_length |
| MGS | abundance | Hgn3C.0518 - Eubacteriales sp. | Radius_length |
| MGS | abundance | Hgn3C.0518 - Eubacteriales sp. | Radius_SOS |
| MGS | abundance | Hgn3C.0518 - Eubacteriales sp. | Tibia_SOS |
| MGS | abundance | Hgn3C.0518 - Eubacteriales sp. | Handgrip |

| | | | |
|---|---|---|---|
| MGS | abundance | Hgn3C.0522 - Roseburia sp. BX1005 | Tibia_length |
| MGS | abundance | Hgn3C.0522 - Roseburia sp. BX1005 | Radius_length |
| MGS | abundance | Hgn3C.0522 - Roseburia sp. BX1005 | Radius_SOS |
| MGS | abundance | Hgn3C.0522 - Roseburia sp. BX1005 | Tibia_SOS |
| MGS | abundance | Hgn3C.0522 - Roseburia sp. BX1005 | Handgrip |
| MGS | abundance | Hgn3C.0523 - Phascolarctobacterium succinatutens | Tibia_length |
| MGS | abundance | Hgn3C.0523 - Phascolarctobacterium succinatutens | Radius_length |
| MGS | abundance | Hgn3C.0523 - Phascolarctobacterium succinatutens | Radius_SOS |
| MGS | abundance | Hgn3C.0523 - Phascolarctobacterium succinatutens | Tibia_SOS |
| MGS | abundance | Hgn3C.0523 - Phascolarctobacterium succinatutens | Handgrip |
| MGS | abundance | Hgn3C.0524 - Butyricicoccus sp. | Tibia_length |
| MGS | abundance | Hgn3C.0524 - Butyricicoccus sp. | Radius_length |
| MGS | abundance | Hgn3C.0524 - Butyricicoccus sp. | Radius_SOS |
| MGS | abundance | Hgn3C.0524 - Butyricicoccus sp. | Tibia_SOS |
| MGS | abundance | Hgn3C.0524 - Butyricicoccus sp. | Handgrip |
| MGS | abundance | Hgn3C.0526 - Anaerostipes sp. | Tibia_length |
| MGS | abundance | Hgn3C.0526 - Anaerostipes sp. | Radius_length |
| MGS | abundance | Hgn3C.0526 - Anaerostipes sp. | Radius_SOS |
| MGS | abundance | Hgn3C.0526 - Anaerostipes sp. | Tibia_SOS |
| MGS | abundance | Hgn3C.0526 - Anaerostipes sp. | Handgrip |
| MGS | abundance | Hgn3C.0527 - Sutterella sp. | Tibia_length |
| MGS | abundance | Hgn3C.0527 - Sutterella sp. | Radius_length |
| MGS | abundance | Hgn3C.0527 - Sutterella sp. | Radius_SOS |
| MGS | abundance | Hgn3C.0527 - Sutterella sp. | Tibia_SOS |
| MGS | abundance | Hgn3C.0527 - Sutterella sp. | Handgrip |
| MGS | abundance | Hgn3C.0528 - Coprococcus sp. AM27-12LB | Tibia_length |
| MGS | abundance | Hgn3C.0528 - Coprococcus sp. AM27-12LB | Radius_length |
| MGS | abundance | Hgn3C.0528 - Coprococcus sp. AM27-12LB | Radius_SOS |
| MGS | abundance | Hgn3C.0528 - Coprococcus sp. AM27-12LB | Tibia_SOS |
| MGS | abundance | Hgn3C.0528 - Coprococcus sp. AM27-12LB | Handgrip |
| MGS | abundance | Hgn3C.0532 - Paraprevotella xylaniphila | Tibia_length |
| MGS | abundance | Hgn3C.0532 - Paraprevotella xylaniphila | Radius_length |
| MGS | abundance | Hgn3C.0532 - Paraprevotella xylaniphila | Radius_SOS |
| MGS | abundance | Hgn3C.0532 - Paraprevotella xylaniphila | Tibia_SOS |
| MGS | abundance | Hgn3C.0532 - Paraprevotella xylaniphila | Handgrip |
| MGS | abundance | Hgn3C.0533 - Flintibacter sp. KGMB00164 | Tibia_length |
| MGS | abundance | Hgn3C.0533 - Flintibacter sp. KGMB00164 | Radius_length |
| MGS | abundance | Hgn3C.0533 - Flintibacter sp. KGMB00164 | Radius_SOS |
| MGS | abundance | Hgn3C.0533 - Flintibacter sp. KGMB00164 | Tibia_SOS |
| MGS | abundance | Hgn3C.0533 - Flintibacter sp. KGMB00164 | Handgrip |
| MGS | abundance | Hgn3C.0535 - Victivallis vadensis | Tibia_length |
| MGS | abundance | Hgn3C.0535 - Victivallis vadensis | Radius_length |
| MGS | abundance | Hgn3C.0535 - Victivallis vadensis | Radius_SOS |
| MGS | abundance | Hgn3C.0535 - Victivallis vadensis | Tibia_SOS |
| MGS | abundance | Hgn3C.0535 - Victivallis vadensis | Handgrip |
| MGS | abundance | Hgn3C.0537 - Bifidobacterium dentium | Tibia_length |
| MGS | abundance | Hgn3C.0537 - Bifidobacterium dentium | Radius_length |
| MGS | abundance | Hgn3C.0537 - Bifidobacterium dentium | Radius_SOS |

| | | | |
|---|---|---|---|
| MGS | abundance | Hgn3C.0537 - Bifidobacterium dentium | Tibia_SOS |
| MGS | abundance | Hgn3C.0537 - Bifidobacterium dentium | Handgrip |
| MGS | abundance | Hgn3C.0540 - Firmicutes sp. | Tibia_length |
| MGS | abundance | Hgn3C.0540 - Firmicutes sp. | Radius_length |
| MGS | abundance | Hgn3C.0540 - Firmicutes sp. | Radius_SOS |
| MGS | abundance | Hgn3C.0540 - Firmicutes sp. | Tibia_SOS |
| MGS | abundance | Hgn3C.0540 - Firmicutes sp. | Handgrip |
| MGS | abundance | Hgn3C.0541 - Phocaeicola plebeius | Tibia_length |
| MGS | abundance | Hgn3C.0541 - Phocaeicola plebeius | Radius_length |
| MGS | abundance | Hgn3C.0541 - Phocaeicola plebeius | Radius_SOS |
| MGS | abundance | Hgn3C.0541 - Phocaeicola plebeius | Tibia_SOS |
| MGS | abundance | Hgn3C.0541 - Phocaeicola plebeius | Handgrip |
| MGS | abundance | Hgn3C.0542 - [Clostridium] innocuum | Tibia_length |
| MGS | abundance | Hgn3C.0542 - [Clostridium] innocuum | Radius_length |
| MGS | abundance | Hgn3C.0542 - [Clostridium] innocuum | Radius_SOS |
| MGS | abundance | Hgn3C.0542 - [Clostridium] innocuum | Tibia_SOS |
| MGS | abundance | Hgn3C.0542 - [Clostridium] innocuum | Handgrip |
| MGS | abundance | Hgn3C.0543 - Oscillospiraceae sp. | Tibia_length |
| MGS | abundance | Hgn3C.0543 - Oscillospiraceae sp. | Radius_length |
| MGS | abundance | Hgn3C.0543 - Oscillospiraceae sp. | Radius_SOS |
| MGS | abundance | Hgn3C.0543 - Oscillospiraceae sp. | Tibia_SOS |
| MGS | abundance | Hgn3C.0543 - Oscillospiraceae sp. | Handgrip |
| MGS | abundance | Hgn3C.0544 - Eubacteriales sp. | Tibia_length |
| MGS | abundance | Hgn3C.0544 - Eubacteriales sp. | Radius_length |
| MGS | abundance | Hgn3C.0544 - Eubacteriales sp. | Radius_SOS |
| MGS | abundance | Hgn3C.0544 - Eubacteriales sp. | Tibia_SOS |
| MGS | abundance | Hgn3C.0544 - Eubacteriales sp. | Handgrip |
| MGS | abundance | Hgn3C.0546 - Bacteroidia sp. | Tibia_length |
| MGS | abundance | Hgn3C.0546 - Bacteroidia sp. | Radius_length |
| MGS | abundance | Hgn3C.0546 - Bacteroidia sp. | Radius_SOS |
| MGS | abundance | Hgn3C.0546 - Bacteroidia sp. | Tibia_SOS |
| MGS | abundance | Hgn3C.0546 - Bacteroidia sp. | Handgrip |
| MGS | abundance | Hgn3C.0549 - Longicatena caecimuris | Tibia_length |
| MGS | abundance | Hgn3C.0549 - Longicatena caecimuris | Radius_length |
| MGS | abundance | Hgn3C.0549 - Longicatena caecimuris | Radius_SOS |
| MGS | abundance | Hgn3C.0549 - Longicatena caecimuris | Tibia_SOS |
| MGS | abundance | Hgn3C.0549 - Longicatena caecimuris | Handgrip |
| MGS | abundance | Hgn3C.0557 - Enterocloster bolteae | Tibia_length |
| MGS | abundance | Hgn3C.0557 - Enterocloster bolteae | Radius_length |
| MGS | abundance | Hgn3C.0557 - Enterocloster bolteae | Radius_SOS |
| MGS | abundance | Hgn3C.0557 - Enterocloster bolteae | Tibia_SOS |
| MGS | abundance | Hgn3C.0557 - Enterocloster bolteae | Handgrip |
| MGS | abundance | Hgn3C.0558 - Olsenella sp. | Tibia_length |
| MGS | abundance | Hgn3C.0558 - Olsenella sp. | Radius_length |
| MGS | abundance | Hgn3C.0558 - Olsenella sp. | Radius_SOS |
| MGS | abundance | Hgn3C.0558 - Olsenella sp. | Tibia_SOS |
| MGS | abundance | Hgn3C.0558 - Olsenella sp. | Handgrip |
| MGS | abundance | Hgn3C.0560 - Oscillospiraceae sp. | Tibia_length |

| | | | |
|---|---|---|---|
| MGS | abundance | Hgn3C.0560 - Oscillospiraceae sp. | Radius_length |
| MGS | abundance | Hgn3C.0560 - Oscillospiraceae sp. | Radius_SOS |
| MGS | abundance | Hgn3C.0560 - Oscillospiraceae sp. | Tibia_SOS |
| MGS | abundance | Hgn3C.0560 - Oscillospiraceae sp. | Handgrip |
| MGS | abundance | Hgn3C.0561 - Enterocloster clostridioformis | Tibia_length |
| MGS | abundance | Hgn3C.0561 - Enterocloster clostridioformis | Radius_length |
| MGS | abundance | Hgn3C.0561 - Enterocloster clostridioformis | Radius_SOS |
| MGS | abundance | Hgn3C.0561 - Enterocloster clostridioformis | Tibia_SOS |
| MGS | abundance | Hgn3C.0561 - Enterocloster clostridioformis | Handgrip |
| MGS | abundance | Hgn3C.0570 - Eubacteriales sp. | Tibia_length |
| MGS | abundance | Hgn3C.0570 - Eubacteriales sp. | Radius_length |
| MGS | abundance | Hgn3C.0570 - Eubacteriales sp. | Radius_SOS |
| MGS | abundance | Hgn3C.0570 - Eubacteriales sp. | Tibia_SOS |
| MGS | abundance | Hgn3C.0570 - Eubacteriales sp. | Handgrip |
| MGS | abundance | Hgn3C.0573 - Prevotella sp. Marseille-P4119 | Tibia_length |
| MGS | abundance | Hgn3C.0573 - Prevotella sp. Marseille-P4119 | Radius_length |
| MGS | abundance | Hgn3C.0573 - Prevotella sp. Marseille-P4119 | Radius_SOS |
| MGS | abundance | Hgn3C.0573 - Prevotella sp. Marseille-P4119 | Tibia_SOS |
| MGS | abundance | Hgn3C.0573 - Prevotella sp. Marseille-P4119 | Handgrip |
| MGS | abundance | Hgn3C.0575 - Bacteria sp. | Tibia_length |
| MGS | abundance | Hgn3C.0575 - Bacteria sp. | Radius_length |
| MGS | abundance | Hgn3C.0575 - Bacteria sp. | Radius_SOS |
| MGS | abundance | Hgn3C.0575 - Bacteria sp. | Tibia_SOS |
| MGS | abundance | Hgn3C.0575 - Bacteria sp. | Handgrip |
| MGS | abundance | Hgn3C.0579 - Eubacteriales sp. | Tibia_length |
| MGS | abundance | Hgn3C.0579 - Eubacteriales sp. | Radius_length |
| MGS | abundance | Hgn3C.0579 - Eubacteriales sp. | Radius_SOS |
| MGS | abundance | Hgn3C.0579 - Eubacteriales sp. | Tibia_SOS |
| MGS | abundance | Hgn3C.0579 - Eubacteriales sp. | Handgrip |
| MGS | abundance | Hgn3C.0581 - Negativibacillus massiliensis | Tibia_length |
| MGS | abundance | Hgn3C.0581 - Negativibacillus massiliensis | Radius_length |
| MGS | abundance | Hgn3C.0581 - Negativibacillus massiliensis | Radius_SOS |
| MGS | abundance | Hgn3C.0581 - Negativibacillus massiliensis | Tibia_SOS |
| MGS | abundance | Hgn3C.0581 - Negativibacillus massiliensis | Handgrip |
| MGS | abundance | Hgn3C.0582 - Bacteroides sp. | Tibia_length |
| MGS | abundance | Hgn3C.0582 - Bacteroides sp. | Radius_length |
| MGS | abundance | Hgn3C.0582 - Bacteroides sp. | Radius_SOS |
| MGS | abundance | Hgn3C.0582 - Bacteroides sp. | Tibia_SOS |
| MGS | abundance | Hgn3C.0582 - Bacteroides sp. | Handgrip |
| MGS | abundance | Hgn3C.0584 - Bacteroides fragilis | Tibia_length |
| MGS | abundance | Hgn3C.0584 - Bacteroides fragilis | Radius_length |
| MGS | abundance | Hgn3C.0584 - Bacteroides fragilis | Radius_SOS |
| MGS | abundance | Hgn3C.0584 - Bacteroides fragilis | Tibia_SOS |
| MGS | abundance | Hgn3C.0584 - Bacteroides fragilis | Handgrip |
| MGS | abundance | Hgn3C.0590 - Clostridia sp. | Tibia_length |
| MGS | abundance | Hgn3C.0590 - Clostridia sp. | Radius_length |
| MGS | abundance | Hgn3C.0590 - Clostridia sp. | Radius_SOS |
| MGS | abundance | Hgn3C.0590 - Clostridia sp. | Tibia_SOS |

| | | | |
|---|---|---|---|
| MGS | abundance | Hgn3C.0590 - Clostridia sp. | Handgrip |
| MGS | abundance | Hgn3C.0595 - Eubacteriales sp. | Tibia_length |
| MGS | abundance | Hgn3C.0595 - Eubacteriales sp. | Radius_length |
| MGS | abundance | Hgn3C.0595 - Eubacteriales sp. | Radius_SOS |
| MGS | abundance | Hgn3C.0595 - Eubacteriales sp. | Tibia_SOS |
| MGS | abundance | Hgn3C.0595 - Eubacteriales sp. | Handgrip |
| MGS | abundance | Hgn3C.0596 - Butyricimonas sp. | Tibia_length |
| MGS | abundance | Hgn3C.0596 - Butyricimonas sp. | Radius_length |
| MGS | abundance | Hgn3C.0596 - Butyricimonas sp. | Radius_SOS |
| MGS | abundance | Hgn3C.0596 - Butyricimonas sp. | Tibia_SOS |
| MGS | abundance | Hgn3C.0596 - Butyricimonas sp. | Handgrip |
| MGS | abundance | Hgn3C.0598 - Eubacteriales sp. | Tibia_length |
| MGS | abundance | Hgn3C.0598 - Eubacteriales sp. | Radius_length |
| MGS | abundance | Hgn3C.0598 - Eubacteriales sp. | Radius_SOS |
| MGS | abundance | Hgn3C.0598 - Eubacteriales sp. | Tibia_SOS |
| MGS | abundance | Hgn3C.0598 - Eubacteriales sp. | Handgrip |
| MGS | abundance | Hgn3C.0601 - Bacteroidales sp. | Tibia_length |
| MGS | abundance | Hgn3C.0601 - Bacteroidales sp. | Radius_length |
| MGS | abundance | Hgn3C.0601 - Bacteroidales sp. | Radius_SOS |
| MGS | abundance | Hgn3C.0601 - Bacteroidales sp. | Tibia_SOS |
| MGS | abundance | Hgn3C.0601 - Bacteroidales sp. | Handgrip |
| MGS | abundance | Hgn3C.0604 - Megasphaera elsdenii | Tibia_length |
| MGS | abundance | Hgn3C.0604 - Megasphaera elsdenii | Radius_length |
| MGS | abundance | Hgn3C.0604 - Megasphaera elsdenii | Radius_SOS |
| MGS | abundance | Hgn3C.0604 - Megasphaera elsdenii | Tibia_SOS |
| MGS | abundance | Hgn3C.0604 - Megasphaera elsdenii | Handgrip |
| MGS | abundance | Hgn3C.0607 - Eubacteriales sp. | Tibia_length |
| MGS | abundance | Hgn3C.0607 - Eubacteriales sp. | Radius_length |
| MGS | abundance | Hgn3C.0607 - Eubacteriales sp. | Radius_SOS |
| MGS | abundance | Hgn3C.0607 - Eubacteriales sp. | Tibia_SOS |
| MGS | abundance | Hgn3C.0607 - Eubacteriales sp. | Handgrip |
| MGS | abundance | Hgn3C.0608 - Lachnospiraceae sp. | Tibia_length |
| MGS | abundance | Hgn3C.0608 - Lachnospiraceae sp. | Radius_length |
| MGS | abundance | Hgn3C.0608 - Lachnospiraceae sp. | Radius_SOS |
| MGS | abundance | Hgn3C.0608 - Lachnospiraceae sp. | Tibia_SOS |
| MGS | abundance | Hgn3C.0608 - Lachnospiraceae sp. | Handgrip |
| MGS | abundance | Hgn3C.0610 - Eubacteriales sp. | Tibia_length |
| MGS | abundance | Hgn3C.0610 - Eubacteriales sp. | Radius_length |
| MGS | abundance | Hgn3C.0610 - Eubacteriales sp. | Radius_SOS |
| MGS | abundance | Hgn3C.0610 - Eubacteriales sp. | Tibia_SOS |
| MGS | abundance | Hgn3C.0610 - Eubacteriales sp. | Handgrip |
| MGS | abundance | Hgn3C.0612 - Bacteroidales sp. | Tibia_length |
| MGS | abundance | Hgn3C.0612 - Bacteroidales sp. | Radius_length |
| MGS | abundance | Hgn3C.0612 - Bacteroidales sp. | Radius_SOS |
| MGS | abundance | Hgn3C.0612 - Bacteroidales sp. | Tibia_SOS |
| MGS | abundance | Hgn3C.0612 - Bacteroidales sp. | Handgrip |
| MGS | abundance | Hgn3C.0613 - Clostridiaceae sp. | Tibia_length |
| MGS | abundance | Hgn3C.0613 - Clostridiaceae sp. | Radius_length |

| | | | |
|---|---|---|---|
| MGS | abundanc | Hgn3C.0613 - Clostridiaceae sp. | Radius_SOS |
| MGS | abundanc | Hgn3C.0613 - Clostridiaceae sp. | Tibia_SOS |
| MGS | abundanc | Hgn3C.0613 - Clostridiaceae sp. | Handgrip |
| MGS | abundanc | Hgn3C.0616 - Lachnospiraceae sp. | Tibia_length |
| MGS | abundanc | Hgn3C.0616 - Lachnospiraceae sp. | Radius_length |
| MGS | abundanc | Hgn3C.0616 - Lachnospiraceae sp. | Radius_SOS |
| MGS | abundanc | Hgn3C.0616 - Lachnospiraceae sp. | Tibia_SOS |
| MGS | abundanc | Hgn3C.0616 - Lachnospiraceae sp. | Handgrip |
| MGS | abundanc | Hgn3C.0617 - Prevotella sp. | Tibia_length |
| MGS | abundanc | Hgn3C.0617 - Prevotella sp. | Radius_length |
| MGS | abundanc | Hgn3C.0617 - Prevotella sp. | Radius_SOS |
| MGS | abundanc | Hgn3C.0617 - Prevotella sp. | Tibia_SOS |
| MGS | abundanc | Hgn3C.0617 - Prevotella sp. | Handgrip |
| MGS | abundanc | Hgn3C.0629 - Collinsella intestinalis | Tibia_length |
| MGS | abundanc | Hgn3C.0629 - Collinsella intestinalis | Radius_length |
| MGS | abundanc | Hgn3C.0629 - Collinsella intestinalis | Radius_SOS |
| MGS | abundanc | Hgn3C.0629 - Collinsella intestinalis | Tibia_SOS |
| MGS | abundanc | Hgn3C.0629 - Collinsella intestinalis | Handgrip |
| MGS | abundanc | Hgn3C.0630 - Allisonella histaminiformans | Tibia_length |
| MGS | abundanc | Hgn3C.0630 - Allisonella histaminiformans | Radius_length |
| MGS | abundanc | Hgn3C.0630 - Allisonella histaminiformans | Radius_SOS |
| MGS | abundanc | Hgn3C.0630 - Allisonella histaminiformans | Tibia_SOS |
| MGS | abundanc | Hgn3C.0630 - Allisonella histaminiformans | Handgrip |
| MGS | abundanc | Hgn3C.0634 - Blautia schinkii | Tibia_length |
| MGS | abundanc | Hgn3C.0634 - Blautia schinkii | Radius_length |
| MGS | abundanc | Hgn3C.0634 - Blautia schinkii | Radius_SOS |
| MGS | abundanc | Hgn3C.0634 - Blautia schinkii | Tibia_SOS |
| MGS | abundanc | Hgn3C.0634 - Blautia schinkii | Handgrip |
| MGS | abundanc | Hgn3C.0635 - Eubacteriales sp. | Tibia_length |
| MGS | abundanc | Hgn3C.0635 - Eubacteriales sp. | Radius_length |
| MGS | abundanc | Hgn3C.0635 - Eubacteriales sp. | Radius_SOS |
| MGS | abundanc | Hgn3C.0635 - Eubacteriales sp. | Tibia_SOS |
| MGS | abundanc | Hgn3C.0635 - Eubacteriales sp. | Handgrip |
| MGS | abundanc | Hgn3C.0636 - Anaerostipes caccae | Tibia_length |
| MGS | abundanc | Hgn3C.0636 - Anaerostipes caccae | Radius_length |
| MGS | abundanc | Hgn3C.0636 - Anaerostipes caccae | Radius_SOS |
| MGS | abundanc | Hgn3C.0636 - Anaerostipes caccae | Tibia_SOS |
| MGS | abundanc | Hgn3C.0636 - Anaerostipes caccae | Handgrip |
| MGS | abundanc | Hgn3C.0640 - Eubacterium sp. AF22-8LB | Tibia_length |
| MGS | abundanc | Hgn3C.0640 - Eubacterium sp. AF22-8LB | Radius_length |
| MGS | abundanc | Hgn3C.0640 - Eubacterium sp. AF22-8LB | Radius_SOS |
| MGS | abundanc | Hgn3C.0640 - Eubacterium sp. AF22-8LB | Tibia_SOS |
| MGS | abundanc | Hgn3C.0640 - Eubacterium sp. AF22-8LB | Handgrip |
| MGS | abundanc | Hgn3C.0643 - Eubacteriales sp. | Tibia_length |
| MGS | abundanc | Hgn3C.0643 - Eubacteriales sp. | Radius_length |
| MGS | abundanc | Hgn3C.0643 - Eubacteriales sp. | Radius_SOS |
| MGS | abundanc | Hgn3C.0643 - Eubacteriales sp. | Tibia_SOS |
| MGS | abundanc | Hgn3C.0643 - Eubacteriales sp. | Handgrip |

| | | | |
|---|---|---|---|
| MGS | abundance | Hgn3C.0644 - Bacteroides cellulosilyticus | Tibia_length |
| MGS | abundance | Hgn3C.0644 - Bacteroides cellulosilyticus | Radius_length |
| MGS | abundance | Hgn3C.0644 - Bacteroides cellulosilyticus | Radius_SOS |
| MGS | abundance | Hgn3C.0644 - Bacteroides cellulosilyticus | Tibia_SOS |
| MGS | abundance | Hgn3C.0644 - Bacteroides cellulosilyticus | Handgrip |
| MGS | abundance | Hgn3C.0645 - Eubacteriales sp. | Tibia_length |
| MGS | abundance | Hgn3C.0645 - Eubacteriales sp. | Radius_length |
| MGS | abundance | Hgn3C.0645 - Eubacteriales sp. | Radius_SOS |
| MGS | abundance | Hgn3C.0645 - Eubacteriales sp. | Tibia_SOS |
| MGS | abundance | Hgn3C.0645 - Eubacteriales sp. | Handgrip |
| MGS | abundance | Hgn3C.0649 - Lachnospiraceae sp. | Tibia_length |
| MGS | abundance | Hgn3C.0649 - Lachnospiraceae sp. | Radius_length |
| MGS | abundance | Hgn3C.0649 - Lachnospiraceae sp. | Radius_SOS |
| MGS | abundance | Hgn3C.0649 - Lachnospiraceae sp. | Tibia_SOS |
| MGS | abundance | Hgn3C.0649 - Lachnospiraceae sp. | Handgrip |
| MGS | abundance | Hgn3C.0651 - Eubacteriales sp. | Tibia_length |
| MGS | abundance | Hgn3C.0651 - Eubacteriales sp. | Radius_length |
| MGS | abundance | Hgn3C.0651 - Eubacteriales sp. | Radius_SOS |
| MGS | abundance | Hgn3C.0651 - Eubacteriales sp. | Tibia_SOS |
| MGS | abundance | Hgn3C.0651 - Eubacteriales sp. | Handgrip |
| MGS | abundance | Hgn3C.0657 - Parolsenella catena | Tibia_length |
| MGS | abundance | Hgn3C.0657 - Parolsenella catena | Radius_length |
| MGS | abundance | Hgn3C.0657 - Parolsenella catena | Radius_SOS |
| MGS | abundance | Hgn3C.0657 - Parolsenella catena | Tibia_SOS |
| MGS | abundance | Hgn3C.0657 - Parolsenella catena | Handgrip |
| MGS | abundance | Hgn3C.0659 - Oscillospiraceae sp. | Tibia_length |
| MGS | abundance | Hgn3C.0659 - Oscillospiraceae sp. | Radius_length |
| MGS | abundance | Hgn3C.0659 - Oscillospiraceae sp. | Radius_SOS |
| MGS | abundance | Hgn3C.0659 - Oscillospiraceae sp. | Tibia_SOS |
| MGS | abundance | Hgn3C.0659 - Oscillospiraceae sp. | Handgrip |
| MGS | abundance | Hgn3C.0660 - Eubacteriales sp. | Tibia_length |
| MGS | abundance | Hgn3C.0660 - Eubacteriales sp. | Radius_length |
| MGS | abundance | Hgn3C.0660 - Eubacteriales sp. | Radius_SOS |
| MGS | abundance | Hgn3C.0660 - Eubacteriales sp. | Tibia_SOS |
| MGS | abundance | Hgn3C.0660 - Eubacteriales sp. | Handgrip |
| MGS | abundance | Hgn3C.0661 - Alistipes sp. | Tibia_length |
| MGS | abundance | Hgn3C.0661 - Alistipes sp. | Radius_length |
| MGS | abundance | Hgn3C.0661 - Alistipes sp. | Radius_SOS |
| MGS | abundance | Hgn3C.0661 - Alistipes sp. | Tibia_SOS |
| MGS | abundance | Hgn3C.0661 - Alistipes sp. | Handgrip |
| MGS | abundance | Hgn3C.0664 - Blautia hansenii | Tibia_length |
| MGS | abundance | Hgn3C.0664 - Blautia hansenii | Radius_length |
| MGS | abundance | Hgn3C.0664 - Blautia hansenii | Radius_SOS |
| MGS | abundance | Hgn3C.0664 - Blautia hansenii | Tibia_SOS |
| MGS | abundance | Hgn3C.0664 - Blautia hansenii | Handgrip |
| MGS | abundance | Hgn3C.0665 - Desulfovibrionales sp. | Tibia_length |
| MGS | abundance | Hgn3C.0665 - Desulfovibrionales sp. | Radius_length |
| MGS | abundance | Hgn3C.0665 - Desulfovibrionales sp. | Radius_SOS |

| | | | |
|---|---|---|---|
| MGS | abundance | Hgn3C.0665 - Desulfovibrionales sp. | Tibia_SOS |
| MGS | abundance | Hgn3C.0665 - Desulfovibrionales sp. | Handgrip |
| MGS | abundance | Hgn3C.0669 - Burkholderiales sp. | Tibia_length |
| MGS | abundance | Hgn3C.0669 - Burkholderiales sp. | Radius_length |
| MGS | abundance | Hgn3C.0669 - Burkholderiales sp. | Radius_SOS |
| MGS | abundance | Hgn3C.0669 - Burkholderiales sp. | Tibia_SOS |
| MGS | abundance | Hgn3C.0669 - Burkholderiales sp. | Handgrip |
| MGS | abundance | Hgn3C.0675 - Eubacteriales sp. | Tibia_length |
| MGS | abundance | Hgn3C.0675 - Eubacteriales sp. | Radius_length |
| MGS | abundance | Hgn3C.0675 - Eubacteriales sp. | Radius_SOS |
| MGS | abundance | Hgn3C.0675 - Eubacteriales sp. | Tibia_SOS |
| MGS | abundance | Hgn3C.0675 - Eubacteriales sp. | Handgrip |
| MGS | abundance | Hgn3C.0676 - Bacteroides sp. | Tibia_length |
| MGS | abundance | Hgn3C.0676 - Bacteroides sp. | Radius_length |
| MGS | abundance | Hgn3C.0676 - Bacteroides sp. | Radius_SOS |
| MGS | abundance | Hgn3C.0676 - Bacteroides sp. | Tibia_SOS |
| MGS | abundance | Hgn3C.0676 - Bacteroides sp. | Handgrip |
| MGS | abundance | Hgn3C.0677 - Bacteroides sp. | Tibia_length |
| MGS | abundance | Hgn3C.0677 - Bacteroides sp. | Radius_length |
| MGS | abundance | Hgn3C.0677 - Bacteroides sp. | Radius_SOS |
| MGS | abundance | Hgn3C.0677 - Bacteroides sp. | Tibia_SOS |
| MGS | abundance | Hgn3C.0677 - Bacteroides sp. | Handgrip |
| MGS | abundance | Hgn3C.0679 - Holdemania filiformis | Tibia_length |
| MGS | abundance | Hgn3C.0679 - Holdemania filiformis | Radius_length |
| MGS | abundance | Hgn3C.0679 - Holdemania filiformis | Radius_SOS |
| MGS | abundance | Hgn3C.0679 - Holdemania filiformis | Tibia_SOS |
| MGS | abundance | Hgn3C.0679 - Holdemania filiformis | Handgrip |
| MGS | abundance | Hgn3C.0680 - Eubacteriales sp. | Tibia_length |
| MGS | abundance | Hgn3C.0680 - Eubacteriales sp. | Radius_length |
| MGS | abundance | Hgn3C.0680 - Eubacteriales sp. | Radius_SOS |
| MGS | abundance | Hgn3C.0680 - Eubacteriales sp. | Tibia_SOS |
| MGS | abundance | Hgn3C.0680 - Eubacteriales sp. | Handgrip |
| MGS | abundance | Hgn3C.0681 - Pseudoflavonifractor sp. | Tibia_length |
| MGS | abundance | Hgn3C.0681 - Pseudoflavonifractor sp. | Radius_length |
| MGS | abundance | Hgn3C.0681 - Pseudoflavonifractor sp. | Radius_SOS |
| MGS | abundance | Hgn3C.0681 - Pseudoflavonifractor sp. | Tibia_SOS |
| MGS | abundance | Hgn3C.0681 - Pseudoflavonifractor sp. | Handgrip |
| MGS | abundance | Hgn3C.0682 - [Clostridium] scindens | Tibia_length |
| MGS | abundance | Hgn3C.0682 - [Clostridium] scindens | Radius_length |
| MGS | abundance | Hgn3C.0682 - [Clostridium] scindens | Radius_SOS |
| MGS | abundance | Hgn3C.0682 - [Clostridium] scindens | Tibia_SOS |
| MGS | abundance | Hgn3C.0682 - [Clostridium] scindens | Handgrip |
| MGS | abundance | Hgn3C.0683 - Clostridium disporicum | Tibia_length |
| MGS | abundance | Hgn3C.0683 - Clostridium disporicum | Radius_length |
| MGS | abundance | Hgn3C.0683 - Clostridium disporicum | Radius_SOS |
| MGS | abundance | Hgn3C.0683 - Clostridium disporicum | Tibia_SOS |
| MGS | abundance | Hgn3C.0683 - Clostridium disporicum | Handgrip |
| MGS | abundance | Hgn3C.0685 - Lachnospiraceae sp. | Tibia_length |

| | | | |
|---|---|---|---|
| MGS | abundance | Hgn3C.0685 - Lachnospiraceae sp. | Radius_length |
| MGS | abundance | Hgn3C.0685 - Lachnospiraceae sp. | Radius_SOS |
| MGS | abundance | Hgn3C.0685 - Lachnospiraceae sp. | Tibia_SOS |
| MGS | abundance | Hgn3C.0685 - Lachnospiraceae sp. | Handgrip |
| MGS | abundance | Hgn3C.0690 - Eubacteriales sp. | Tibia_length |
| MGS | abundance | Hgn3C.0690 - Eubacteriales sp. | Radius_length |
| MGS | abundance | Hgn3C.0690 - Eubacteriales sp. | Radius_SOS |
| MGS | abundance | Hgn3C.0690 - Eubacteriales sp. | Tibia_SOS |
| MGS | abundance | Hgn3C.0690 - Eubacteriales sp. | Handgrip |
| MGS | abundance | Hgn3C.0693 - Hungatella hathewayi | Tibia_length |
| MGS | abundance | Hgn3C.0693 - Hungatella hathewayi | Radius_length |
| MGS | abundance | Hgn3C.0693 - Hungatella hathewayi | Radius_SOS |
| MGS | abundance | Hgn3C.0693 - Hungatella hathewayi | Tibia_SOS |
| MGS | abundance | Hgn3C.0693 - Hungatella hathewayi | Handgrip |
| MGS | abundance | Hgn3C.0694 - Anaeromassilibacillus sp. An250 | Tibia_length |
| MGS | abundance | Hgn3C.0694 - Anaeromassilibacillus sp. An250 | Radius_length |
| MGS | abundance | Hgn3C.0694 - Anaeromassilibacillus sp. An250 | Radius_SOS |
| MGS | abundance | Hgn3C.0694 - Anaeromassilibacillus sp. An250 | Tibia_SOS |
| MGS | abundance | Hgn3C.0694 - Anaeromassilibacillus sp. An250 | Handgrip |
| MGS | abundance | Hgn3C.0700 - Eubacteriales sp. | Tibia_length |
| MGS | abundance | Hgn3C.0700 - Eubacteriales sp. | Radius_length |
| MGS | abundance | Hgn3C.0700 - Eubacteriales sp. | Radius_SOS |
| MGS | abundance | Hgn3C.0700 - Eubacteriales sp. | Tibia_SOS |
| MGS | abundance | Hgn3C.0700 - Eubacteriales sp. | Handgrip |
| MGS | abundance | Hgn3C.0701 - Bacteroides sp. | Tibia_length |
| MGS | abundance | Hgn3C.0701 - Bacteroides sp. | Radius_length |
| MGS | abundance | Hgn3C.0701 - Bacteroides sp. | Radius_SOS |
| MGS | abundance | Hgn3C.0701 - Bacteroides sp. | Tibia_SOS |
| MGS | abundance | Hgn3C.0701 - Bacteroides sp. | Handgrip |
| MGS | abundance | Hgn3C.0704 - [Clostridium] scindens | Tibia_length |
| MGS | abundance | Hgn3C.0704 - [Clostridium] scindens | Radius_length |
| MGS | abundance | Hgn3C.0704 - [Clostridium] scindens | Radius_SOS |
| MGS | abundance | Hgn3C.0704 - [Clostridium] scindens | Tibia_SOS |
| MGS | abundance | Hgn3C.0704 - [Clostridium] scindens | Handgrip |
| MGS | abundance | Hgn3C.0706 - Eubacteriales sp. | Tibia_length |
| MGS | abundance | Hgn3C.0706 - Eubacteriales sp. | Radius_length |
| MGS | abundance | Hgn3C.0706 - Eubacteriales sp. | Radius_SOS |
| MGS | abundance | Hgn3C.0706 - Eubacteriales sp. | Tibia_SOS |
| MGS | abundance | Hgn3C.0706 - Eubacteriales sp. | Handgrip |
| MGS | abundance | Hgn3C.0711 - Enterocloster asparagiformis | Tibia_length |
| MGS | abundance | Hgn3C.0711 - Enterocloster asparagiformis | Radius_length |
| MGS | abundance | Hgn3C.0711 - Enterocloster asparagiformis | Radius_SOS |
| MGS | abundance | Hgn3C.0711 - Enterocloster asparagiformis | Tibia_SOS |
| MGS | abundance | Hgn3C.0711 - Enterocloster asparagiformis | Handgrip |
| MGS | abundance | Hgn3C.0712 - Megasphaera micronuciformis | Tibia_length |
| MGS | abundance | Hgn3C.0712 - Megasphaera micronuciformis | Radius_length |
| MGS | abundance | Hgn3C.0712 - Megasphaera micronuciformis | Radius_SOS |
| MGS | abundance | Hgn3C.0712 - Megasphaera micronuciformis | Tibia_SOS |

| | | | |
|---|---|---|---|
| MGS | abundance | Hgn3C.0712 - Megasphaera micronuciformis | Handgrip |
| MGS | abundance | Hgn3C.0713 - Veillonellales sp. | Tibia_length |
| MGS | abundance | Hgn3C.0713 - Veillonellales sp. | Radius_length |
| MGS | abundance | Hgn3C.0713 - Veillonellales sp. | Radius_SOS |
| MGS | abundance | Hgn3C.0713 - Veillonellales sp. | Tibia_SOS |
| MGS | abundance | Hgn3C.0713 - Veillonellales sp. | Handgrip |
| MGS | abundance | Hgn3C.0714 - Intestinimonas sp. MSJ-38 | Tibia_length |
| MGS | abundance | Hgn3C.0714 - Intestinimonas sp. MSJ-38 | Radius_length |
| MGS | abundance | Hgn3C.0714 - Intestinimonas sp. MSJ-38 | Radius_SOS |
| MGS | abundance | Hgn3C.0714 - Intestinimonas sp. MSJ-38 | Tibia_SOS |
| MGS | abundance | Hgn3C.0714 - Intestinimonas sp. MSJ-38 | Handgrip |
| MGS | abundance | Hgn3C.0715 - Desulfovibrio fairfieldensis | Tibia_length |
| MGS | abundance | Hgn3C.0715 - Desulfovibrio fairfieldensis | Radius_length |
| MGS | abundance | Hgn3C.0715 - Desulfovibrio fairfieldensis | Radius_SOS |
| MGS | abundance | Hgn3C.0715 - Desulfovibrio fairfieldensis | Tibia_SOS |
| MGS | abundance | Hgn3C.0715 - Desulfovibrio fairfieldensis | Handgrip |
| MGS | abundance | Hgn3C.0716 - Eubacteriales sp. | Tibia_length |
| MGS | abundance | Hgn3C.0716 - Eubacteriales sp. | Radius_length |
| MGS | abundance | Hgn3C.0716 - Eubacteriales sp. | Radius_SOS |
| MGS | abundance | Hgn3C.0716 - Eubacteriales sp. | Tibia_SOS |
| MGS | abundance | Hgn3C.0716 - Eubacteriales sp. | Handgrip |
| MGS | abundance | Hgn3C.0717 - Eubacteriales sp. | Tibia_length |
| MGS | abundance | Hgn3C.0717 - Eubacteriales sp. | Radius_length |
| MGS | abundance | Hgn3C.0717 - Eubacteriales sp. | Radius_SOS |
| MGS | abundance | Hgn3C.0717 - Eubacteriales sp. | Tibia_SOS |
| MGS | abundance | Hgn3C.0717 - Eubacteriales sp. | Handgrip |
| MGS | abundance | Hgn3C.0718 - Eubacteriales sp. | Tibia_length |
| MGS | abundance | Hgn3C.0718 - Eubacteriales sp. | Radius_length |
| MGS | abundance | Hgn3C.0718 - Eubacteriales sp. | Radius_SOS |
| MGS | abundance | Hgn3C.0718 - Eubacteriales sp. | Tibia_SOS |
| MGS | abundance | Hgn3C.0718 - Eubacteriales sp. | Handgrip |
| MGS | abundance | Hgn3C.0729 - Eubacteriales sp. | Tibia_length |
| MGS | abundance | Hgn3C.0729 - Eubacteriales sp. | Radius_length |
| MGS | abundance | Hgn3C.0729 - Eubacteriales sp. | Radius_SOS |
| MGS | abundance | Hgn3C.0729 - Eubacteriales sp. | Tibia_SOS |
| MGS | abundance | Hgn3C.0729 - Eubacteriales sp. | Handgrip |
| MGS | abundance | Hgn3C.0731 - Eubacteriales sp. | Tibia_length |
| MGS | abundance | Hgn3C.0731 - Eubacteriales sp. | Radius_length |
| MGS | abundance | Hgn3C.0731 - Eubacteriales sp. | Radius_SOS |
| MGS | abundance | Hgn3C.0731 - Eubacteriales sp. | Tibia_SOS |
| MGS | abundance | Hgn3C.0731 - Eubacteriales sp. | Handgrip |
| MGS | abundance | Hgn3C.0732 - Eubacterium ramulus | Tibia_length |
| MGS | abundance | Hgn3C.0732 - Eubacterium ramulus | Radius_length |
| MGS | abundance | Hgn3C.0732 - Eubacterium ramulus | Radius_SOS |
| MGS | abundance | Hgn3C.0732 - Eubacterium ramulus | Tibia_SOS |
| MGS | abundance | Hgn3C.0732 - Eubacterium ramulus | Handgrip |
| MGS | abundance | Hgn3C.0736 - Coprococcus sp. | Tibia_length |
| MGS | abundance | Hgn3C.0736 - Coprococcus sp. | Radius_length |

| | | | |
|---|---|---|---|
| MGS | abundance | Hgn3C.0736 - Coprococcus sp. | Radius_SOS |
| MGS | abundance | Hgn3C.0736 - Coprococcus sp. | Tibia_SOS |
| MGS | abundance | Hgn3C.0736 - Coprococcus sp. | Handgrip |
| MGS | abundance | Hgn3C.0737 - Alistipes timonensis | Tibia_length |
| MGS | abundance | Hgn3C.0737 - Alistipes timonensis | Radius_length |
| MGS | abundance | Hgn3C.0737 - Alistipes timonensis | Radius_SOS |
| MGS | abundance | Hgn3C.0737 - Alistipes timonensis | Tibia_SOS |
| MGS | abundance | Hgn3C.0737 - Alistipes timonensis | Handgrip |
| MGS | abundance | Hgn3C.0740 - Anaerostipes sp. NSJ-7 | Tibia_length |
| MGS | abundance | Hgn3C.0740 - Anaerostipes sp. NSJ-7 | Radius_length |
| MGS | abundance | Hgn3C.0740 - Anaerostipes sp. NSJ-7 | Radius_SOS |
| MGS | abundance | Hgn3C.0740 - Anaerostipes sp. NSJ-7 | Tibia_SOS |
| MGS | abundance | Hgn3C.0740 - Anaerostipes sp. NSJ-7 | Handgrip |
| MGS | abundance | Hgn3C.0741 - Blautia sp. OF01-4LB | Tibia_length |
| MGS | abundance | Hgn3C.0741 - Blautia sp. OF01-4LB | Radius_length |
| MGS | abundance | Hgn3C.0741 - Blautia sp. OF01-4LB | Radius_SOS |
| MGS | abundance | Hgn3C.0741 - Blautia sp. OF01-4LB | Tibia_SOS |
| MGS | abundance | Hgn3C.0741 - Blautia sp. OF01-4LB | Handgrip |
| MGS | abundance | Hgn3C.0742 - Eubacteriales sp. | Tibia_length |
| MGS | abundance | Hgn3C.0742 - Eubacteriales sp. | Radius_length |
| MGS | abundance | Hgn3C.0742 - Eubacteriales sp. | Radius_SOS |
| MGS | abundance | Hgn3C.0742 - Eubacteriales sp. | Tibia_SOS |
| MGS | abundance | Hgn3C.0742 - Eubacteriales sp. | Handgrip |
| MGS | abundance | Hgn3C.0743 - Enterobacter sp. | Tibia_length |
| MGS | abundance | Hgn3C.0743 - Enterobacter sp. | Radius_length |
| MGS | abundance | Hgn3C.0743 - Enterobacter sp. | Radius_SOS |
| MGS | abundance | Hgn3C.0743 - Enterobacter sp. | Tibia_SOS |
| MGS | abundance | Hgn3C.0743 - Enterobacter sp. | Handgrip |
| MGS | abundance | Hgn3C.0749 - Eubacteriales sp. | Tibia_length |
| MGS | abundance | Hgn3C.0749 - Eubacteriales sp. | Radius_length |
| MGS | abundance | Hgn3C.0749 - Eubacteriales sp. | Radius_SOS |
| MGS | abundance | Hgn3C.0749 - Eubacteriales sp. | Tibia_SOS |
| MGS | abundance | Hgn3C.0749 - Eubacteriales sp. | Handgrip |
| MGS | abundance | Hgn3C.0750 - Merdimonas faecis | Tibia_length |
| MGS | abundance | Hgn3C.0750 - Merdimonas faecis | Radius_length |
| MGS | abundance | Hgn3C.0750 - Merdimonas faecis | Radius_SOS |
| MGS | abundance | Hgn3C.0750 - Merdimonas faecis | Tibia_SOS |
| MGS | abundance | Hgn3C.0750 - Merdimonas faecis | Handgrip |
| MGS | abundance | Hgn3C.0754 - Collinsella bouchesdurhonensis | Tibia_length |
| MGS | abundance | Hgn3C.0754 - Collinsella bouchesdurhonensis | Radius_length |
| MGS | abundance | Hgn3C.0754 - Collinsella bouchesdurhonensis | Radius_SOS |
| MGS | abundance | Hgn3C.0754 - Collinsella bouchesdurhonensis | Tibia_SOS |
| MGS | abundance | Hgn3C.0754 - Collinsella bouchesdurhonensis | Handgrip |
| MGS | abundance | Hgn3C.0755 - Oscillibacter sp. | Tibia_length |
| MGS | abundance | Hgn3C.0755 - Oscillibacter sp. | Radius_length |
| MGS | abundance | Hgn3C.0755 - Oscillibacter sp. | Radius_SOS |
| MGS | abundance | Hgn3C.0755 - Oscillibacter sp. | Tibia_SOS |
| MGS | abundance | Hgn3C.0755 - Oscillibacter sp. | Handgrip |

| | | | |
|---|---|---|---|
| MGS | abundance | Hgn3C.0756 - Oscillibacter valericigenes | Tibia_length |
| MGS | abundance | Hgn3C.0756 - Oscillibacter valericigenes | Radius_length |
| MGS | abundance | Hgn3C.0756 - Oscillibacter valericigenes | Radius_SOS |
| MGS | abundance | Hgn3C.0756 - Oscillibacter valericigenes | Tibia_SOS |
| MGS | abundance | Hgn3C.0756 - Oscillibacter valericigenes | Handgrip |
| MGS | abundance | Hgn3C.0758 - Bacteroides sp. | Tibia_length |
| MGS | abundance | Hgn3C.0758 - Bacteroides sp. | Radius_length |
| MGS | abundance | Hgn3C.0758 - Bacteroides sp. | Radius_SOS |
| MGS | abundance | Hgn3C.0758 - Bacteroides sp. | Tibia_SOS |
| MGS | abundance | Hgn3C.0758 - Bacteroides sp. | Handgrip |
| MGS | abundance | Hgn3C.0760 - Klebsiella oxytoca | Tibia_length |
| MGS | abundance | Hgn3C.0760 - Klebsiella oxytoca | Radius_length |
| MGS | abundance | Hgn3C.0760 - Klebsiella oxytoca | Radius_SOS |
| MGS | abundance | Hgn3C.0760 - Klebsiella oxytoca | Tibia_SOS |
| MGS | abundance | Hgn3C.0760 - Klebsiella oxytoca | Handgrip |
| MGS | abundance | Hgn3C.0761 - Bacteroides sp. | Tibia_length |
| MGS | abundance | Hgn3C.0761 - Bacteroides sp. | Radius_length |
| MGS | abundance | Hgn3C.0761 - Bacteroides sp. | Radius_SOS |
| MGS | abundance | Hgn3C.0761 - Bacteroides sp. | Tibia_SOS |
| MGS | abundance | Hgn3C.0761 - Bacteroides sp. | Handgrip |
| MGS | abundance | Hgn3C.0762 - Enterocloster lavalensis | Tibia_length |
| MGS | abundance | Hgn3C.0762 - Enterocloster lavalensis | Radius_length |
| MGS | abundance | Hgn3C.0762 - Enterocloster lavalensis | Radius_SOS |
| MGS | abundance | Hgn3C.0762 - Enterocloster lavalensis | Tibia_SOS |
| MGS | abundance | Hgn3C.0762 - Enterocloster lavalensis | Handgrip |
| MGS | abundance | Hgn3C.0767 - Citrobacter freundii | Tibia_length |
| MGS | abundance | Hgn3C.0767 - Citrobacter freundii | Radius_length |
| MGS | abundance | Hgn3C.0767 - Citrobacter freundii | Radius_SOS |
| MGS | abundance | Hgn3C.0767 - Citrobacter freundii | Tibia_SOS |
| MGS | abundance | Hgn3C.0767 - Citrobacter freundii | Handgrip |
| MGS | abundance | Hgn3C.0777 - Holdemanella sp. | Tibia_length |
| MGS | abundance | Hgn3C.0777 - Holdemanella sp. | Radius_length |
| MGS | abundance | Hgn3C.0777 - Holdemanella sp. | Radius_SOS |
| MGS | abundance | Hgn3C.0777 - Holdemanella sp. | Tibia_SOS |
| MGS | abundance | Hgn3C.0777 - Holdemanella sp. | Handgrip |
| MGS | abundance | Hgn3C.0781 - Collinsella sp. | Tibia_length |
| MGS | abundance | Hgn3C.0781 - Collinsella sp. | Radius_length |
| MGS | abundance | Hgn3C.0781 - Collinsella sp. | Radius_SOS |
| MGS | abundance | Hgn3C.0781 - Collinsella sp. | Tibia_SOS |
| MGS | abundance | Hgn3C.0781 - Collinsella sp. | Handgrip |
| MGS | abundance | Hgn3C.0782 - Streptococcus sp. | Tibia_length |
| MGS | abundance | Hgn3C.0782 - Streptococcus sp. | Radius_length |
| MGS | abundance | Hgn3C.0782 - Streptococcus sp. | Radius_SOS |
| MGS | abundance | Hgn3C.0782 - Streptococcus sp. | Tibia_SOS |
| MGS | abundance | Hgn3C.0782 - Streptococcus sp. | Handgrip |
| MGS | abundance | Hgn3C.0790 - Oscillospiraceae sp. | Tibia_length |
| MGS | abundance | Hgn3C.0790 - Oscillospiraceae sp. | Radius_length |
| MGS | abundance | Hgn3C.0790 - Oscillospiraceae sp. | Radius_SOS |

| | | | |
|---|---|---|---|
| MGS | abundance | Hgn3C.0790 - Oscillospiraceae sp. | Tibia_SOS |
| MGS | abundance | Hgn3C.0790 - Oscillospiraceae sp. | Handgrip |
| MGS | abundance | Hgn3C.0793 - Prevotellaceae sp. | Tibia_length |
| MGS | abundance | Hgn3C.0793 - Prevotellaceae sp. | Radius_length |
| MGS | abundance | Hgn3C.0793 - Prevotellaceae sp. | Radius_SOS |
| MGS | abundance | Hgn3C.0793 - Prevotellaceae sp. | Tibia_SOS |
| MGS | abundance | Hgn3C.0793 - Prevotellaceae sp. | Handgrip |
| MGS | abundance | Hgn3C.0794 - Bacteroides sp. | Tibia_length |
| MGS | abundance | Hgn3C.0794 - Bacteroides sp. | Radius_length |
| MGS | abundance | Hgn3C.0794 - Bacteroides sp. | Radius_SOS |
| MGS | abundance | Hgn3C.0794 - Bacteroides sp. | Tibia_SOS |
| MGS | abundance | Hgn3C.0794 - Bacteroides sp. | Handgrip |
| MGS | abundance | Hgn3C.0797 - Senegalimassilia sp. | Tibia_length |
| MGS | abundance | Hgn3C.0797 - Senegalimassilia sp. | Radius_length |
| MGS | abundance | Hgn3C.0797 - Senegalimassilia sp. | Radius_SOS |
| MGS | abundance | Hgn3C.0797 - Senegalimassilia sp. | Tibia_SOS |
| MGS | abundance | Hgn3C.0797 - Senegalimassilia sp. | Handgrip |
| MGS | abundance | Hgn3C.0798 - Streptococcus vestibularis | Tibia_length |
| MGS | abundance | Hgn3C.0798 - Streptococcus vestibularis | Radius_length |
| MGS | abundance | Hgn3C.0798 - Streptococcus vestibularis | Radius_SOS |
| MGS | abundance | Hgn3C.0798 - Streptococcus vestibularis | Tibia_SOS |
| MGS | abundance | Hgn3C.0798 - Streptococcus vestibularis | Handgrip |
| MGS | abundance | Hgn3C.0807 - Enorma massiliensis | Tibia_length |
| MGS | abundance | Hgn3C.0807 - Enorma massiliensis | Radius_length |
| MGS | abundance | Hgn3C.0807 - Enorma massiliensis | Radius_SOS |
| MGS | abundance | Hgn3C.0807 - Enorma massiliensis | Tibia_SOS |
| MGS | abundance | Hgn3C.0807 - Enorma massiliensis | Handgrip |
| MGS | abundance | Hgn3C.0814 - Massilimicrobiota timonensis | Tibia_length |
| MGS | abundance | Hgn3C.0814 - Massilimicrobiota timonensis | Radius_length |
| MGS | abundance | Hgn3C.0814 - Massilimicrobiota timonensis | Radius_SOS |
| MGS | abundance | Hgn3C.0814 - Massilimicrobiota timonensis | Tibia_SOS |
| MGS | abundance | Hgn3C.0814 - Massilimicrobiota timonensis | Handgrip |
| MGS | abundance | Hgn3C.0815 - Eubacteriales sp. | Tibia_length |
| MGS | abundance | Hgn3C.0815 - Eubacteriales sp. | Radius_length |
| MGS | abundance | Hgn3C.0815 - Eubacteriales sp. | Radius_SOS |
| MGS | abundance | Hgn3C.0815 - Eubacteriales sp. | Tibia_SOS |
| MGS | abundance | Hgn3C.0815 - Eubacteriales sp. | Handgrip |
| MGS | abundance | Hgn3C.0817 - Prevotella sp. | Tibia_length |
| MGS | abundance | Hgn3C.0817 - Prevotella sp. | Radius_length |
| MGS | abundance | Hgn3C.0817 - Prevotella sp. | Radius_SOS |
| MGS | abundance | Hgn3C.0817 - Prevotella sp. | Tibia_SOS |
| MGS | abundance | Hgn3C.0817 - Prevotella sp. | Handgrip |
| MGS | abundance | Hgn3C.0818 - Rothia sp. | Tibia_length |
| MGS | abundance | Hgn3C.0818 - Rothia sp. | Radius_length |
| MGS | abundance | Hgn3C.0818 - Rothia sp. | Radius_SOS |
| MGS | abundance | Hgn3C.0818 - Rothia sp. | Tibia_SOS |
| MGS | abundance | Hgn3C.0818 - Rothia sp. | Handgrip |
| MGS | abundance | Hgn3C.0823 - Anaerotignum lactatifermentans | Tibia_length |

| | | | |
|---|---|---|---|
| MGS | abundance | Hgn3C.0823 - Anaerotignum lactatifermentans | Radius_length |
| MGS | abundance | Hgn3C.0823 - Anaerotignum lactatifermentans | Radius_SOS |
| MGS | abundance | Hgn3C.0823 - Anaerotignum lactatifermentans | Tibia_SOS |
| MGS | abundance | Hgn3C.0823 - Anaerotignum lactatifermentans | Handgrip |
| MGS | abundance | Hgn3C.0828 - Bacteroides congonensis | Tibia_length |
| MGS | abundance | Hgn3C.0828 - Bacteroides congonensis | Radius_length |
| MGS | abundance | Hgn3C.0828 - Bacteroides congonensis | Radius_SOS |
| MGS | abundance | Hgn3C.0828 - Bacteroides congonensis | Tibia_SOS |
| MGS | abundance | Hgn3C.0828 - Bacteroides congonensis | Handgrip |
| MGS | abundance | Hgn3C.0829 - Dorea phocaeensis | Tibia_length |
| MGS | abundance | Hgn3C.0829 - Dorea phocaeensis | Radius_length |
| MGS | abundance | Hgn3C.0829 - Dorea phocaeensis | Radius_SOS |
| MGS | abundance | Hgn3C.0829 - Dorea phocaeensis | Tibia_SOS |
| MGS | abundance | Hgn3C.0829 - Dorea phocaeensis | Handgrip |
| MGS | abundance | Hgn3C.0832 - Veillonellales sp. | Tibia_length |
| MGS | abundance | Hgn3C.0832 - Veillonellales sp. | Radius_length |
| MGS | abundance | Hgn3C.0832 - Veillonellales sp. | Radius_SOS |
| MGS | abundance | Hgn3C.0832 - Veillonellales sp. | Tibia_SOS |
| MGS | abundance | Hgn3C.0832 - Veillonellales sp. | Handgrip |
| MGS | abundance | Hgn3C.0833 - Eisenbergiella tayi | Tibia_length |
| MGS | abundance | Hgn3C.0833 - Eisenbergiella tayi | Radius_length |
| MGS | abundance | Hgn3C.0833 - Eisenbergiella tayi | Radius_SOS |
| MGS | abundance | Hgn3C.0833 - Eisenbergiella tayi | Tibia_SOS |
| MGS | abundance | Hgn3C.0833 - Eisenbergiella tayi | Handgrip |
| MGS | abundance | Hgn3C.0836 - Lachnospiraceae sp. | Tibia_length |
| MGS | abundance | Hgn3C.0836 - Lachnospiraceae sp. | Radius_length |
| MGS | abundance | Hgn3C.0836 - Lachnospiraceae sp. | Radius_SOS |
| MGS | abundance | Hgn3C.0836 - Lachnospiraceae sp. | Tibia_SOS |
| MGS | abundance | Hgn3C.0836 - Lachnospiraceae sp. | Handgrip |
| MGS | abundance | Hgn3C.0837 - Anaerotruncus colihominis | Tibia_length |
| MGS | abundance | Hgn3C.0837 - Anaerotruncus colihominis | Radius_length |
| MGS | abundance | Hgn3C.0837 - Anaerotruncus colihominis | Radius_SOS |
| MGS | abundance | Hgn3C.0837 - Anaerotruncus colihominis | Tibia_SOS |
| MGS | abundance | Hgn3C.0837 - Anaerotruncus colihominis | Handgrip |
| MGS | abundance | Hgn3C.0840 - Eubacteriales sp. | Tibia_length |
| MGS | abundance | Hgn3C.0840 - Eubacteriales sp. | Radius_length |
| MGS | abundance | Hgn3C.0840 - Eubacteriales sp. | Radius_SOS |
| MGS | abundance | Hgn3C.0840 - Eubacteriales sp. | Tibia_SOS |
| MGS | abundance | Hgn3C.0840 - Eubacteriales sp. | Handgrip |
| MGS | abundance | Hgn3C.0851 - Oscillospiraceae sp. | Tibia_length |
| MGS | abundance | Hgn3C.0851 - Oscillospiraceae sp. | Radius_length |
| MGS | abundance | Hgn3C.0851 - Oscillospiraceae sp. | Radius_SOS |
| MGS | abundance | Hgn3C.0851 - Oscillospiraceae sp. | Tibia_SOS |
| MGS | abundance | Hgn3C.0851 - Oscillospiraceae sp. | Handgrip |
| MGS | abundance | Hgn3C.0855 - Eubacteriales sp. | Tibia_length |
| MGS | abundance | Hgn3C.0855 - Eubacteriales sp. | Radius_length |
| MGS | abundance | Hgn3C.0855 - Eubacteriales sp. | Radius_SOS |
| MGS | abundance | Hgn3C.0855 - Eubacteriales sp. | Tibia_SOS |

| | | | |
|---|---|---|---|
| MGS | abundance | Hgn3C.0855 - Eubacteriales sp. | Handgrip |
| MGS | abundance | Hgn3C.0858 - Turicibacter sanguinis | Tibia_length |
| MGS | abundance | Hgn3C.0858 - Turicibacter sanguinis | Radius_length |
| MGS | abundance | Hgn3C.0858 - Turicibacter sanguinis | Radius_SOS |
| MGS | abundance | Hgn3C.0858 - Turicibacter sanguinis | Tibia_SOS |
| MGS | abundance | Hgn3C.0858 - Turicibacter sanguinis | Handgrip |
| MGS | abundance | Hgn3C.0859 - Hydrogeniiclostidium mannosilyticum | Tibia_length |
| MGS | abundance | Hgn3C.0859 - Hydrogeniiclostidium mannosilyticum | Radius_length |
| MGS | abundance | Hgn3C.0859 - Hydrogeniiclostidium mannosilyticum | Radius_SOS |
| MGS | abundance | Hgn3C.0859 - Hydrogeniiclostidium mannosilyticum | Tibia_SOS |
| MGS | abundance | Hgn3C.0859 - Hydrogeniiclostidium mannosilyticum | Handgrip |
| MGS | abundance | Hgn3C.0873 - Clostridium perfringens | Tibia_length |
| MGS | abundance | Hgn3C.0873 - Clostridium perfringens | Radius_length |
| MGS | abundance | Hgn3C.0873 - Clostridium perfringens | Radius_SOS |
| MGS | abundance | Hgn3C.0873 - Clostridium perfringens | Tibia_SOS |
| MGS | abundance | Hgn3C.0873 - Clostridium perfringens | Handgrip |
| MGS | abundance | Hgn3C.0875 - Lachnospiraceae sp. | Tibia_length |
| MGS | abundance | Hgn3C.0875 - Lachnospiraceae sp. | Radius_length |
| MGS | abundance | Hgn3C.0875 - Lachnospiraceae sp. | Radius_SOS |
| MGS | abundance | Hgn3C.0875 - Lachnospiraceae sp. | Tibia_SOS |
| MGS | abundance | Hgn3C.0875 - Lachnospiraceae sp. | Handgrip |
| MGS | abundance | Hgn3C.0882 - Eubacteriales sp. | Tibia_length |
| MGS | abundance | Hgn3C.0882 - Eubacteriales sp. | Radius_length |
| MGS | abundance | Hgn3C.0882 - Eubacteriales sp. | Radius_SOS |
| MGS | abundance | Hgn3C.0882 - Eubacteriales sp. | Tibia_SOS |
| MGS | abundance | Hgn3C.0882 - Eubacteriales sp. | Handgrip |
| MGS | abundance | Hgn3C.0885 - Eubacteriales sp. | Tibia_length |
| MGS | abundance | Hgn3C.0885 - Eubacteriales sp. | Radius_length |
| MGS | abundance | Hgn3C.0885 - Eubacteriales sp. | Radius_SOS |
| MGS | abundance | Hgn3C.0885 - Eubacteriales sp. | Tibia_SOS |
| MGS | abundance | Hgn3C.0885 - Eubacteriales sp. | Handgrip |
| MGS | abundance | Hgn3C.0886 - Bacteroides sp. | Tibia_length |
| MGS | abundance | Hgn3C.0886 - Bacteroides sp. | Radius_length |
| MGS | abundance | Hgn3C.0886 - Bacteroides sp. | Radius_SOS |
| MGS | abundance | Hgn3C.0886 - Bacteroides sp. | Tibia_SOS |
| MGS | abundance | Hgn3C.0886 - Bacteroides sp. | Handgrip |
| MGS | abundance | Hgn3C.0889 - Clostridium paraputrificum | Tibia_length |
| MGS | abundance | Hgn3C.0889 - Clostridium paraputrificum | Radius_length |
| MGS | abundance | Hgn3C.0889 - Clostridium paraputrificum | Radius_SOS |
| MGS | abundance | Hgn3C.0889 - Clostridium paraputrificum | Tibia_SOS |
| MGS | abundance | Hgn3C.0889 - Clostridium paraputrificum | Handgrip |
| MGS | abundance | Hgn3C.0890 - Enterobacter roggenkampii | Tibia_length |
| MGS | abundance | Hgn3C.0890 - Enterobacter roggenkampii | Radius_length |
| MGS | abundance | Hgn3C.0890 - Enterobacter roggenkampii | Radius_SOS |
| MGS | abundance | Hgn3C.0890 - Enterobacter roggenkampii | Tibia_SOS |
| MGS | abundance | Hgn3C.0890 - Enterobacter roggenkampii | Handgrip |
| MGS | abundance | Hgn3C.0891 - Eubacteriales sp. | Tibia_length |
| MGS | abundance | Hgn3C.0891 - Eubacteriales sp. | Radius_length |

| | | | |
|---|---|---|---|
| MGS | abundance | Hgn3C.0891 - Eubacteriales sp. | Radius_SOS |
| MGS | abundance | Hgn3C.0891 - Eubacteriales sp. | Tibia_SOS |
| MGS | abundance | Hgn3C.0891 - Eubacteriales sp. | Handgrip |
| MGS | abundance | Hgn3C.0893 - Phocea massiliensis | Tibia_length |
| MGS | abundance | Hgn3C.0893 - Phocea massiliensis | Radius_length |
| MGS | abundance | Hgn3C.0893 - Phocea massiliensis | Radius_SOS |
| MGS | abundance | Hgn3C.0893 - Phocea massiliensis | Tibia_SOS |
| MGS | abundance | Hgn3C.0893 - Phocea massiliensis | Handgrip |
| MGS | abundance | Hgn3C.0904 - Bacteroidia sp. | Tibia_length |
| MGS | abundance | Hgn3C.0904 - Bacteroidia sp. | Radius_length |
| MGS | abundance | Hgn3C.0904 - Bacteroidia sp. | Radius_SOS |
| MGS | abundance | Hgn3C.0904 - Bacteroidia sp. | Tibia_SOS |
| MGS | abundance | Hgn3C.0904 - Bacteroidia sp. | Handgrip |
| MGS | abundance | Hgn3C.0908 - Eubacteriales sp. | Tibia_length |
| MGS | abundance | Hgn3C.0908 - Eubacteriales sp. | Radius_length |
| MGS | abundance | Hgn3C.0908 - Eubacteriales sp. | Radius_SOS |
| MGS | abundance | Hgn3C.0908 - Eubacteriales sp. | Tibia_SOS |
| MGS | abundance | Hgn3C.0908 - Eubacteriales sp. | Handgrip |
| MGS | abundance | Hgn3C.0913 - Erysipelotrichaceae sp. | Tibia_length |
| MGS | abundance | Hgn3C.0913 - Erysipelotrichaceae sp. | Radius_length |
| MGS | abundance | Hgn3C.0913 - Erysipelotrichaceae sp. | Radius_SOS |
| MGS | abundance | Hgn3C.0913 - Erysipelotrichaceae sp. | Tibia_SOS |
| MGS | abundance | Hgn3C.0913 - Erysipelotrichaceae sp. | Handgrip |
| MGS | abundance | Hgn3C.0921 - Bacteroides sp. | Tibia_length |
| MGS | abundance | Hgn3C.0921 - Bacteroides sp. | Radius_length |
| MGS | abundance | Hgn3C.0921 - Bacteroides sp. | Radius_SOS |
| MGS | abundance | Hgn3C.0921 - Bacteroides sp. | Tibia_SOS |
| MGS | abundance | Hgn3C.0921 - Bacteroides sp. | Handgrip |
| MGS | abundance | Hgn3C.0922 - Eubacteriales sp. | Tibia_length |
| MGS | abundance | Hgn3C.0922 - Eubacteriales sp. | Radius_length |
| MGS | abundance | Hgn3C.0922 - Eubacteriales sp. | Radius_SOS |
| MGS | abundance | Hgn3C.0922 - Eubacteriales sp. | Tibia_SOS |
| MGS | abundance | Hgn3C.0922 - Eubacteriales sp. | Handgrip |
| MGS | abundance | Hgn3C.0924 - Clostridioides difficile | Tibia_length |
| MGS | abundance | Hgn3C.0924 - Clostridioides difficile | Radius_length |
| MGS | abundance | Hgn3C.0924 - Clostridioides difficile | Radius_SOS |
| MGS | abundance | Hgn3C.0924 - Clostridioides difficile | Tibia_SOS |
| MGS | abundance | Hgn3C.0924 - Clostridioides difficile | Handgrip |
| MGS | abundance | Hgn3C.0940 - Eubacteriales sp. | Tibia_length |
| MGS | abundance | Hgn3C.0940 - Eubacteriales sp. | Radius_length |
| MGS | abundance | Hgn3C.0940 - Eubacteriales sp. | Radius_SOS |
| MGS | abundance | Hgn3C.0940 - Eubacteriales sp. | Tibia_SOS |
| MGS | abundance | Hgn3C.0940 - Eubacteriales sp. | Handgrip |
| MGS | abundance | Hgn3C.0943 - Lactiplantibacillus plantarum subsp. plantarum | Tibia_length |
| MGS | abundance | Hgn3C.0943 - Lactiplantibacillus plantarum subsp. plantarum | Radius_length |
| MGS | abundance | Hgn3C.0943 - Lactiplantibacillus plantarum subsp. plantarum | Radius_SOS |
| MGS | abundance | Hgn3C.0943 - Lactiplantibacillus plantarum subsp. plantarum | Tibia_SOS |
| MGS | abundance | Hgn3C.0943 - Lactiplantibacillus plantarum subsp. plantarum | Handgrip |

| | | | |
|---|---|---|---|
| MGS | abundance | Hgn3C.0946 - Eubacteriales sp. | Tibia_length |
| MGS | abundance | Hgn3C.0946 - Eubacteriales sp. | Radius_length |
| MGS | abundance | Hgn3C.0946 - Eubacteriales sp. | Radius_SOS |
| MGS | abundance | Hgn3C.0946 - Eubacteriales sp. | Tibia_SOS |
| MGS | abundance | Hgn3C.0946 - Eubacteriales sp. | Handgrip |
| MGS | abundance | Hgn3C.0948 - Enterobacter sp. | Tibia_length |
| MGS | abundance | Hgn3C.0948 - Enterobacter sp. | Radius_length |
| MGS | abundance | Hgn3C.0948 - Enterobacter sp. | Radius_SOS |
| MGS | abundance | Hgn3C.0948 - Enterobacter sp. | Tibia_SOS |
| MGS | abundance | Hgn3C.0948 - Enterobacter sp. | Handgrip |
| MGS | abundance | Hgn3C.0951 - Eubacteriales sp. | Tibia_length |
| MGS | abundance | Hgn3C.0951 - Eubacteriales sp. | Radius_length |
| MGS | abundance | Hgn3C.0951 - Eubacteriales sp. | Radius_SOS |
| MGS | abundance | Hgn3C.0951 - Eubacteriales sp. | Tibia_SOS |
| MGS | abundance | Hgn3C.0951 - Eubacteriales sp. | Handgrip |
| MGS | abundance | Hgn3C.0973 - Desulfovibrio piger | Tibia_length |
| MGS | abundance | Hgn3C.0973 - Desulfovibrio piger | Radius_length |
| MGS | abundance | Hgn3C.0973 - Desulfovibrio piger | Radius_SOS |
| MGS | abundance | Hgn3C.0973 - Desulfovibrio piger | Tibia_SOS |
| MGS | abundance | Hgn3C.0973 - Desulfovibrio piger | Handgrip |
| MGS | abundance | Hgn3C.0978 - Veillonella sp. | Tibia_length |
| MGS | abundance | Hgn3C.0978 - Veillonella sp. | Radius_length |
| MGS | abundance | Hgn3C.0978 - Veillonella sp. | Radius_SOS |
| MGS | abundance | Hgn3C.0978 - Veillonella sp. | Tibia_SOS |
| MGS | abundance | Hgn3C.0978 - Veillonella sp. | Handgrip |
| MGS | abundance | Hgn3C.0986 - Desulfovibrio sp. | Tibia_length |
| MGS | abundance | Hgn3C.0986 - Desulfovibrio sp. | Radius_length |
| MGS | abundance | Hgn3C.0986 - Desulfovibrio sp. | Radius_SOS |
| MGS | abundance | Hgn3C.0986 - Desulfovibrio sp. | Tibia_SOS |
| MGS | abundance | Hgn3C.0986 - Desulfovibrio sp. | Handgrip |
| MGS | abundance | Hgn3C.0987 - Fusobacterium mortiferum | Tibia_length |
| MGS | abundance | Hgn3C.0987 - Fusobacterium mortiferum | Radius_length |
| MGS | abundance | Hgn3C.0987 - Fusobacterium mortiferum | Radius_SOS |
| MGS | abundance | Hgn3C.0987 - Fusobacterium mortiferum | Tibia_SOS |
| MGS | abundance | Hgn3C.0987 - Fusobacterium mortiferum | Handgrip |
| MGS | abundance | Hgn3C.0988 - Eubacteriales sp. | Tibia_length |
| MGS | abundance | Hgn3C.0988 - Eubacteriales sp. | Radius_length |
| MGS | abundance | Hgn3C.0988 - Eubacteriales sp. | Radius_SOS |
| MGS | abundance | Hgn3C.0988 - Eubacteriales sp. | Tibia_SOS |
| MGS | abundance | Hgn3C.0988 - Eubacteriales sp. | Handgrip |
| MGS | abundance | Hgn3C.0993 - Faecalitalea cylindroides | Tibia_length |
| MGS | abundance | Hgn3C.0993 - Faecalitalea cylindroides | Radius_length |
| MGS | abundance | Hgn3C.0993 - Faecalitalea cylindroides | Radius_SOS |
| MGS | abundance | Hgn3C.0993 - Faecalitalea cylindroides | Tibia_SOS |
| MGS | abundance | Hgn3C.0993 - Faecalitalea cylindroides | Handgrip |
| MGS | abundance | Hgn3C.0996 - Oscillospiraceae sp. | Tibia_length |
| MGS | abundance | Hgn3C.0996 - Oscillospiraceae sp. | Radius_length |
| MGS | abundance | Hgn3C.0996 - Oscillospiraceae sp. | Radius_SOS |

| | | | |
|---|---|---|---|
| MGS | abundance | Hgn3C.0996 - Oscillospiraceae sp. | Tibia_SOS |
| MGS | abundance | Hgn3C.0996 - Oscillospiraceae sp. | Handgrip |
| MGS | abundance | Hgn3C.0998 - Eubacteriales sp. | Tibia_length |
| MGS | abundance | Hgn3C.0998 - Eubacteriales sp. | Radius_length |
| MGS | abundance | Hgn3C.0998 - Eubacteriales sp. | Radius_SOS |
| MGS | abundance | Hgn3C.0998 - Eubacteriales sp. | Tibia_SOS |
| MGS | abundance | Hgn3C.0998 - Eubacteriales sp. | Handgrip |
| MGS | abundance | Hgn3C.1012 - Erysipelotrichaceae sp. | Tibia_length |
| MGS | abundance | Hgn3C.1012 - Erysipelotrichaceae sp. | Radius_length |
| MGS | abundance | Hgn3C.1012 - Erysipelotrichaceae sp. | Radius_SOS |
| MGS | abundance | Hgn3C.1012 - Erysipelotrichaceae sp. | Tibia_SOS |
| MGS | abundance | Hgn3C.1012 - Erysipelotrichaceae sp. | Handgrip |
| MGS | abundance | Hgn3C.1017 - Emergencia timonensis | Tibia_length |
| MGS | abundance | Hgn3C.1017 - Emergencia timonensis | Radius_length |
| MGS | abundance | Hgn3C.1017 - Emergencia timonensis | Radius_SOS |
| MGS | abundance | Hgn3C.1017 - Emergencia timonensis | Tibia_SOS |
| MGS | abundance | Hgn3C.1017 - Emergencia timonensis | Handgrip |
| MGS | abundance | Hgn3C.1035 - Klebsiella sp. | Tibia_length |
| MGS | abundance | Hgn3C.1035 - Klebsiella sp. | Radius_length |
| MGS | abundance | Hgn3C.1035 - Klebsiella sp. | Radius_SOS |
| MGS | abundance | Hgn3C.1035 - Klebsiella sp. | Tibia_SOS |
| MGS | abundance | Hgn3C.1035 - Klebsiella sp. | Handgrip |
| MGS | abundance | Hgn3C.1036 - Escherichia coli | Tibia_length |
| MGS | abundance | Hgn3C.1036 - Escherichia coli | Radius_length |
| MGS | abundance | Hgn3C.1036 - Escherichia coli | Radius_SOS |
| MGS | abundance | Hgn3C.1036 - Escherichia coli | Tibia_SOS |
| MGS | abundance | Hgn3C.1036 - Escherichia coli | Handgrip |
| MGS | abundance | Hgn3C.1037 - Rothia sp. | Tibia_length |
| MGS | abundance | Hgn3C.1037 - Rothia sp. | Radius_length |
| MGS | abundance | Hgn3C.1037 - Rothia sp. | Radius_SOS |
| MGS | abundance | Hgn3C.1037 - Rothia sp. | Tibia_SOS |
| MGS | abundance | Hgn3C.1037 - Rothia sp. | Handgrip |
| MGS | abundance | Hgn3C.1038 - Veillonella sp. | Tibia_length |
| MGS | abundance | Hgn3C.1038 - Veillonella sp. | Radius_length |
| MGS | abundance | Hgn3C.1038 - Veillonella sp. | Radius_SOS |
| MGS | abundance | Hgn3C.1038 - Veillonella sp. | Tibia_SOS |
| MGS | abundance | Hgn3C.1038 - Veillonella sp. | Handgrip |
| MGS | abundance | Hgn3C.1040 - Lachnospiraceae sp. | Tibia_length |
| MGS | abundance | Hgn3C.1040 - Lachnospiraceae sp. | Radius_length |
| MGS | abundance | Hgn3C.1040 - Lachnospiraceae sp. | Radius_SOS |
| MGS | abundance | Hgn3C.1040 - Lachnospiraceae sp. | Tibia_SOS |
| MGS | abundance | Hgn3C.1040 - Lachnospiraceae sp. | Handgrip |
| MGS | abundance | Hgn3C.1041 - Bacteroides stercoris | Tibia_length |
| MGS | abundance | Hgn3C.1041 - Bacteroides stercoris | Radius_length |
| MGS | abundance | Hgn3C.1041 - Bacteroides stercoris | Radius_SOS |
| MGS | abundance | Hgn3C.1041 - Bacteroides stercoris | Tibia_SOS |
| MGS | abundance | Hgn3C.1041 - Bacteroides stercoris | Handgrip |
| MGS | abundance | Hgn3C.1042 - Dorea formicigenerans | Tibia_length |

| | | | |
|---|---|---|---|
| MGS | abundance | Hgn3C.1042 - Dorea formicigenerans | Radius_length |
| MGS | abundance | Hgn3C.1042 - Dorea formicigenerans | Radius_SOS |
| MGS | abundance | Hgn3C.1042 - Dorea formicigenerans | Tibia_SOS |
| MGS | abundance | Hgn3C.1042 - Dorea formicigenerans | Handgrip |
| MGS | abundance | Hgn3C.1043 - Clostridium phoceensis | Tibia_length |
| MGS | abundance | Hgn3C.1043 - Clostridium phoceensis | Radius_length |
| MGS | abundance | Hgn3C.1043 - Clostridium phoceensis | Radius_SOS |
| MGS | abundance | Hgn3C.1043 - Clostridium phoceensis | Tibia_SOS |
| MGS | abundance | Hgn3C.1043 - Clostridium phoceensis | Handgrip |
| MGS | abundance | Hgn3C.1044 - Eubacteriales sp. | Tibia_length |
| MGS | abundance | Hgn3C.1044 - Eubacteriales sp. | Radius_length |
| MGS | abundance | Hgn3C.1044 - Eubacteriales sp. | Radius_SOS |
| MGS | abundance | Hgn3C.1044 - Eubacteriales sp. | Tibia_SOS |
| MGS | abundance | Hgn3C.1044 - Eubacteriales sp. | Handgrip |
| MGS | abundance | Hgn3C.1045 - Lactococcus lactis subsp. lactis | Tibia_length |
| MGS | abundance | Hgn3C.1045 - Lactococcus lactis subsp. lactis | Radius_length |
| MGS | abundance | Hgn3C.1045 - Lactococcus lactis subsp. lactis | Radius_SOS |
| MGS | abundance | Hgn3C.1045 - Lactococcus lactis subsp. lactis | Tibia_SOS |
| MGS | abundance | Hgn3C.1045 - Lactococcus lactis subsp. lactis | Handgrip |
| MGS | abundance | Hgn3C.1046 - Bacteroidales sp. | Tibia_length |
| MGS | abundance | Hgn3C.1046 - Bacteroidales sp. | Radius_length |
| MGS | abundance | Hgn3C.1046 - Bacteroidales sp. | Radius_SOS |
| MGS | abundance | Hgn3C.1046 - Bacteroidales sp. | Tibia_SOS |
| MGS | abundance | Hgn3C.1046 - Bacteroidales sp. | Handgrip |
| MGS | abundance | Hgn3C.1047 - Eubacteriales sp. | Tibia_length |
| MGS | abundance | Hgn3C.1047 - Eubacteriales sp. | Radius_length |
| MGS | abundance | Hgn3C.1047 - Eubacteriales sp. | Radius_SOS |
| MGS | abundance | Hgn3C.1047 - Eubacteriales sp. | Tibia_SOS |
| MGS | abundance | Hgn3C.1047 - Eubacteriales sp. | Handgrip |
| MGS | abundance | Hgn3C.1049 - Enterococcus faecium | Tibia_length |
| MGS | abundance | Hgn3C.1049 - Enterococcus faecium | Radius_length |
| MGS | abundance | Hgn3C.1049 - Enterococcus faecium | Radius_SOS |
| MGS | abundance | Hgn3C.1049 - Enterococcus faecium | Tibia_SOS |
| MGS | abundance | Hgn3C.1049 - Enterococcus faecium | Handgrip |
| MGS | abundance | Hgn3C.1050 - Staphylococcus epidermidis | Tibia_length |
| MGS | abundance | Hgn3C.1050 - Staphylococcus epidermidis | Radius_length |
| MGS | abundance | Hgn3C.1050 - Staphylococcus epidermidis | Radius_SOS |
| MGS | abundance | Hgn3C.1050 - Staphylococcus epidermidis | Tibia_SOS |
| MGS | abundance | Hgn3C.1050 - Staphylococcus epidermidis | Handgrip |
| MGS | abundance | Hgn3C.1051 - Holdemania massiliensis | Tibia_length |
| MGS | abundance | Hgn3C.1051 - Holdemania massiliensis | Radius_length |
| MGS | abundance | Hgn3C.1051 - Holdemania massiliensis | Radius_SOS |
| MGS | abundance | Hgn3C.1051 - Holdemania massiliensis | Tibia_SOS |
| MGS | abundance | Hgn3C.1051 - Holdemania massiliensis | Handgrip |
| MGS | abundance | Hgn3C.1052 - Enterococcus gallinarum | Tibia_length |
| MGS | abundance | Hgn3C.1052 - Enterococcus gallinarum | Radius_length |
| MGS | abundance | Hgn3C.1052 - Enterococcus gallinarum | Radius_SOS |
| MGS | abundance | Hgn3C.1052 - Enterococcus gallinarum | Tibia_SOS |

| | | | |
|---|---|---|---|
| MGS | abundance | Hgn3C.1052 - Enterococcus gallinarum | Handgrip |
| MGS | abundance | Hgn3C.1054 - Streptococcus lutetiensis | Tibia_length |
| MGS | abundance | Hgn3C.1054 - Streptococcus lutetiensis | Radius_length |
| MGS | abundance | Hgn3C.1054 - Streptococcus lutetiensis | Radius_SOS |
| MGS | abundance | Hgn3C.1054 - Streptococcus lutetiensis | Tibia_SOS |
| MGS | abundance | Hgn3C.1054 - Streptococcus lutetiensis | Handgrip |
| MGS | abundance | Hgn3C.1058 - Mediterraneibacter glycyrrhizinilyticus | Tibia_length |
| MGS | abundance | Hgn3C.1058 - Mediterraneibacter glycyrrhizinilyticus | Radius_length |
| MGS | abundance | Hgn3C.1058 - Mediterraneibacter glycyrrhizinilyticus | Radius_SOS |
| MGS | abundance | Hgn3C.1058 - Mediterraneibacter glycyrrhizinilyticus | Tibia_SOS |
| MGS | abundance | Hgn3C.1058 - Mediterraneibacter glycyrrhizinilyticus | Handgrip |
| MGS | abundance | Hgn3C.1059 - Faecalicatena sp. | Tibia_length |
| MGS | abundance | Hgn3C.1059 - Faecalicatena sp. | Radius_length |
| MGS | abundance | Hgn3C.1059 - Faecalicatena sp. | Radius_SOS |
| MGS | abundance | Hgn3C.1059 - Faecalicatena sp. | Tibia_SOS |
| MGS | abundance | Hgn3C.1059 - Faecalicatena sp. | Handgrip |
| MGS | abundance | Hgn3C.1063 - Enterococcus avium | Tibia_length |
| MGS | abundance | Hgn3C.1063 - Enterococcus avium | Radius_length |
| MGS | abundance | Hgn3C.1063 - Enterococcus avium | Radius_SOS |
| MGS | abundance | Hgn3C.1063 - Enterococcus avium | Tibia_SOS |
| MGS | abundance | Hgn3C.1063 - Enterococcus avium | Handgrip |
| MGS | abundance | Hgn3C.1065 - Klebsiella variicola subsp. variicola | Tibia_length |
| MGS | abundance | Hgn3C.1065 - Klebsiella variicola subsp. variicola | Radius_length |
| MGS | abundance | Hgn3C.1065 - Klebsiella variicola subsp. variicola | Radius_SOS |
| MGS | abundance | Hgn3C.1065 - Klebsiella variicola subsp. variicola | Tibia_SOS |
| MGS | abundance | Hgn3C.1065 - Klebsiella variicola subsp. variicola | Handgrip |
| MGS | abundance | Hgn3C.1067 - Lachnospiraceae sp. | Tibia_length |
| MGS | abundance | Hgn3C.1067 - Lachnospiraceae sp. | Radius_length |
| MGS | abundance | Hgn3C.1067 - Lachnospiraceae sp. | Radius_SOS |
| MGS | abundance | Hgn3C.1067 - Lachnospiraceae sp. | Tibia_SOS |
| MGS | abundance | Hgn3C.1067 - Lachnospiraceae sp. | Handgrip |
| MGS | abundance | Hgn3C.1071 - Lactobacillus gasseri | Tibia_length |
| MGS | abundance | Hgn3C.1071 - Lactobacillus gasseri | Radius_length |
| MGS | abundance | Hgn3C.1071 - Lactobacillus gasseri | Radius_SOS |
| MGS | abundance | Hgn3C.1071 - Lactobacillus gasseri | Tibia_SOS |
| MGS | abundance | Hgn3C.1071 - Lactobacillus gasseri | Handgrip |
| MGS | abundance | Hgn3C.1073 - Streptococcus gallolyticus subsp. gallolyticus | Tibia_length |
| MGS | abundance | Hgn3C.1073 - Streptococcus gallolyticus subsp. gallolyticus | Radius_length |
| MGS | abundance | Hgn3C.1073 - Streptococcus gallolyticus subsp. gallolyticus | Radius_SOS |
| MGS | abundance | Hgn3C.1073 - Streptococcus gallolyticus subsp. gallolyticus | Tibia_SOS |
| MGS | abundance | Hgn3C.1073 - Streptococcus gallolyticus subsp. gallolyticus | Handgrip |
| MGS | abundance | Hgn3C.1081 - Lacticaseibacillus rhamnosus | Tibia_length |
| MGS | abundance | Hgn3C.1081 - Lacticaseibacillus rhamnosus | Radius_length |
| MGS | abundance | Hgn3C.1081 - Lacticaseibacillus rhamnosus | Radius_SOS |
| MGS | abundance | Hgn3C.1081 - Lacticaseibacillus rhamnosus | Tibia_SOS |
| MGS | abundance | Hgn3C.1081 - Lacticaseibacillus rhamnosus | Handgrip |
| MGS | abundance | Hgn3C.1082 - Lacticaseibacillus paracasei subsp. paracasei | Tibia_length |
| MGS | abundance | Hgn3C.1082 - Lacticaseibacillus paracasei subsp. paracasei | Radius_length |

| | | | |
|---|---|---|---|
| MGS | abundanc | Hgn3C.1082 - Lacticaseibacillus paracasei subsp. paracasei | Radius_SOS |
| MGS | abundanc | Hgn3C.1082 - Lacticaseibacillus paracasei subsp. paracasei | Tibia_SOS |
| MGS | abundanc | Hgn3C.1082 - Lacticaseibacillus paracasei subsp. paracasei | Handgrip |
| MGS | abundanc | Hgn3C.1084 - Massilimicrobiota timonensis | Tibia_length |
| MGS | abundanc | Hgn3C.1084 - Massilimicrobiota timonensis | Radius_length |
| MGS | abundanc | Hgn3C.1084 - Massilimicrobiota timonensis | Radius_SOS |
| MGS | abundanc | Hgn3C.1084 - Massilimicrobiota timonensis | Tibia_SOS |
| MGS | abundanc | Hgn3C.1084 - Massilimicrobiota timonensis | Handgrip |
| MGS | abundanc | Hgn3C.1087 - Ligilactobacillus salivarius | Tibia_length |
| MGS | abundanc | Hgn3C.1087 - Ligilactobacillus salivarius | Radius_length |
| MGS | abundanc | Hgn3C.1087 - Ligilactobacillus salivarius | Radius_SOS |
| MGS | abundanc | Hgn3C.1087 - Ligilactobacillus salivarius | Tibia_SOS |
| MGS | abundanc | Hgn3C.1087 - Ligilactobacillus salivarius | Handgrip |
| MGS | abundanc | Hgn3C.1088 - Enterococcus casseliflavus | Tibia_length |
| MGS | abundanc | Hgn3C.1088 - Enterococcus casseliflavus | Radius_length |
| MGS | abundanc | Hgn3C.1088 - Enterococcus casseliflavus | Radius_SOS |
| MGS | abundanc | Hgn3C.1088 - Enterococcus casseliflavus | Tibia_SOS |
| MGS | abundanc | Hgn3C.1088 - Enterococcus casseliflavus | Handgrip |
| MGS | abundanc | Hgn3C.1090 - Turicibacter sanguinis | Tibia_length |
| MGS | abundanc | Hgn3C.1090 - Turicibacter sanguinis | Radius_length |
| MGS | abundanc | Hgn3C.1090 - Turicibacter sanguinis | Radius_SOS |
| MGS | abundanc | Hgn3C.1090 - Turicibacter sanguinis | Tibia_SOS |
| MGS | abundanc | Hgn3C.1090 - Turicibacter sanguinis | Handgrip |
| MGS | abundanc | Hgn3C.1093 - Enterobacter kobei | Tibia_length |
| MGS | abundanc | Hgn3C.1093 - Enterobacter kobei | Radius_length |
| MGS | abundanc | Hgn3C.1093 - Enterobacter kobei | Radius_SOS |
| MGS | abundanc | Hgn3C.1093 - Enterobacter kobei | Tibia_SOS |
| MGS | abundanc | Hgn3C.1093 - Enterobacter kobei | Handgrip |
| MGS | abundanc | Hgn3C.1097 - Alistipes provencensis | Tibia_length |
| MGS | abundanc | Hgn3C.1097 - Alistipes provencensis | Radius_length |
| MGS | abundanc | Hgn3C.1097 - Alistipes provencensis | Radius_SOS |
| MGS | abundanc | Hgn3C.1097 - Alistipes provencensis | Tibia_SOS |
| MGS | abundanc | Hgn3C.1097 - Alistipes provencensis | Handgrip |
| MGS | abundanc | Hgn3C.1102 - Eubacteriales sp. | Tibia_length |
| MGS | abundanc | Hgn3C.1102 - Eubacteriales sp. | Radius_length |
| MGS | abundanc | Hgn3C.1102 - Eubacteriales sp. | Radius_SOS |
| MGS | abundanc | Hgn3C.1102 - Eubacteriales sp. | Tibia_SOS |
| MGS | abundanc | Hgn3C.1102 - Eubacteriales sp. | Handgrip |
| MGS | abundanc | Hgn3C.1106 - Clostridium butyricum | Tibia_length |
| MGS | abundanc | Hgn3C.1106 - Clostridium butyricum | Radius_length |
| MGS | abundanc | Hgn3C.1106 - Clostridium butyricum | Radius_SOS |
| MGS | abundanc | Hgn3C.1106 - Clostridium butyricum | Tibia_SOS |
| MGS | abundanc | Hgn3C.1106 - Clostridium butyricum | Handgrip |
| MGS | abundanc | Hgn3C.1107 - Eubacteriales sp. | Tibia_length |
| MGS | abundanc | Hgn3C.1107 - Eubacteriales sp. | Radius_length |
| MGS | abundanc | Hgn3C.1107 - Eubacteriales sp. | Radius_SOS |
| MGS | abundanc | Hgn3C.1107 - Eubacteriales sp. | Tibia_SOS |
| MGS | abundanc | Hgn3C.1107 - Eubacteriales sp. | Handgrip |

| | | | |
|---|---|---|---|
| MGS | abundance | Hgn3C.1110 - Porphyromonas sp. | Tibia_length |
| MGS | abundance | Hgn3C.1110 - Porphyromonas sp. | Radius_length |
| MGS | abundance | Hgn3C.1110 - Porphyromonas sp. | Radius_SOS |
| MGS | abundance | Hgn3C.1110 - Porphyromonas sp. | Tibia_SOS |
| MGS | abundance | Hgn3C.1110 - Porphyromonas sp. | Handgrip |
| MGS | abundance | Hgn3C.1117 - Atopobiaceae sp. | Tibia_length |
| MGS | abundance | Hgn3C.1117 - Atopobiaceae sp. | Radius_length |
| MGS | abundance | Hgn3C.1117 - Atopobiaceae sp. | Radius_SOS |
| MGS | abundance | Hgn3C.1117 - Atopobiaceae sp. | Tibia_SOS |
| MGS | abundance | Hgn3C.1117 - Atopobiaceae sp. | Handgrip |
| MGS | abundance | Hgn3C.1120 - Actinomycetaceae sp. | Tibia_length |
| MGS | abundance | Hgn3C.1120 - Actinomycetaceae sp. | Radius_length |
| MGS | abundance | Hgn3C.1120 - Actinomycetaceae sp. | Radius_SOS |
| MGS | abundance | Hgn3C.1120 - Actinomycetaceae sp. | Tibia_SOS |
| MGS | abundance | Hgn3C.1120 - Actinomycetaceae sp. | Handgrip |
| MGS | abundance | Hgn3C.1122 - Ruminococcus sp. | Tibia_length |
| MGS | abundance | Hgn3C.1122 - Ruminococcus sp. | Radius_length |
| MGS | abundance | Hgn3C.1122 - Ruminococcus sp. | Radius_SOS |
| MGS | abundance | Hgn3C.1122 - Ruminococcus sp. | Tibia_SOS |
| MGS | abundance | Hgn3C.1122 - Ruminococcus sp. | Handgrip |
| MGS | abundance | Hgn3C.1124 - Clostridium porci | Tibia_length |
| MGS | abundance | Hgn3C.1124 - Clostridium porci | Radius_length |
| MGS | abundance | Hgn3C.1124 - Clostridium porci | Radius_SOS |
| MGS | abundance | Hgn3C.1124 - Clostridium porci | Tibia_SOS |
| MGS | abundance | Hgn3C.1124 - Clostridium porci | Handgrip |
| MGS | abundance | Hgn3C.1126 - Phocaeicola sartorii | Tibia_length |
| MGS | abundance | Hgn3C.1126 - Phocaeicola sartorii | Radius_length |
| MGS | abundance | Hgn3C.1126 - Phocaeicola sartorii | Radius_SOS |
| MGS | abundance | Hgn3C.1126 - Phocaeicola sartorii | Tibia_SOS |
| MGS | abundance | Hgn3C.1126 - Phocaeicola sartorii | Handgrip |
| MGS | abundance | Hgn3C.1146 - Bacteria sp. | Tibia_length |
| MGS | abundance | Hgn3C.1146 - Bacteria sp. | Radius_length |
| MGS | abundance | Hgn3C.1146 - Bacteria sp. | Radius_SOS |
| MGS | abundance | Hgn3C.1146 - Bacteria sp. | Tibia_SOS |
| MGS | abundance | Hgn3C.1146 - Bacteria sp. | Handgrip |
| MGS | abundance | Hgn3C.1148 - Eggerthellales sp. | Tibia_length |
| MGS | abundance | Hgn3C.1148 - Eggerthellales sp. | Radius_length |
| MGS | abundance | Hgn3C.1148 - Eggerthellales sp. | Radius_SOS |
| MGS | abundance | Hgn3C.1148 - Eggerthellales sp. | Tibia_SOS |
| MGS | abundance | Hgn3C.1148 - Eggerthellales sp. | Handgrip |
| MGS | abundance | Hgn3C.1153 - Eubacterium sp. OM08-24 | Tibia_length |
| MGS | abundance | Hgn3C.1153 - Eubacterium sp. OM08-24 | Radius_length |
| MGS | abundance | Hgn3C.1153 - Eubacterium sp. OM08-24 | Radius_SOS |
| MGS | abundance | Hgn3C.1153 - Eubacterium sp. OM08-24 | Tibia_SOS |
| MGS | abundance | Hgn3C.1153 - Eubacterium sp. OM08-24 | Handgrip |
| MGS | abundance | Hgn3C.1154 - Lactococcus garvieae | Tibia_length |
| MGS | abundance | Hgn3C.1154 - Lactococcus garvieae | Radius_length |
| MGS | abundance | Hgn3C.1154 - Lactococcus garvieae | Radius_SOS |

| | | | |
|---|---|---|---|
| MGS | abundance | Hgn3C.1154 - Lactococcus garvieae | Tibia_SOS |
| MGS | abundance | Hgn3C.1154 - Lactococcus garvieae | Handgrip |
| MGS | abundance | Hgn3C.1169 - Oscillospiraceae sp. | Tibia_length |
| MGS | abundance | Hgn3C.1169 - Oscillospiraceae sp. | Radius_length |
| MGS | abundance | Hgn3C.1169 - Oscillospiraceae sp. | Radius_SOS |
| MGS | abundance | Hgn3C.1169 - Oscillospiraceae sp. | Tibia_SOS |
| MGS | abundance | Hgn3C.1169 - Oscillospiraceae sp. | Handgrip |
| MGS | abundance | Hgn3C.1180 - Enterocloster citroniae | Tibia_length |
| MGS | abundance | Hgn3C.1180 - Enterocloster citroniae | Radius_length |
| MGS | abundance | Hgn3C.1180 - Enterocloster citroniae | Radius_SOS |
| MGS | abundance | Hgn3C.1180 - Enterocloster citroniae | Tibia_SOS |
| MGS | abundance | Hgn3C.1180 - Enterocloster citroniae | Handgrip |
| MGS | abundance | Hgn3C.1182 - Comamonas kerstersii | Tibia_length |
| MGS | abundance | Hgn3C.1182 - Comamonas kerstersii | Radius_length |
| MGS | abundance | Hgn3C.1182 - Comamonas kerstersii | Radius_SOS |
| MGS | abundance | Hgn3C.1182 - Comamonas kerstersii | Tibia_SOS |
| MGS | abundance | Hgn3C.1182 - Comamonas kerstersii | Handgrip |
| MGS | abundance | Hgn3C.1185 - Acinetobacter ursingii | Tibia_length |
| MGS | abundance | Hgn3C.1185 - Acinetobacter ursingii | Radius_length |
| MGS | abundance | Hgn3C.1185 - Acinetobacter ursingii | Radius_SOS |
| MGS | abundance | Hgn3C.1185 - Acinetobacter ursingii | Tibia_SOS |
| MGS | abundance | Hgn3C.1185 - Acinetobacter ursingii | Handgrip |
| MGS | abundance | Hgn3C.1188 - Actinomyces oris | Tibia_length |
| MGS | abundance | Hgn3C.1188 - Actinomyces oris | Radius_length |
| MGS | abundance | Hgn3C.1188 - Actinomyces oris | Radius_SOS |
| MGS | abundance | Hgn3C.1188 - Actinomyces oris | Tibia_SOS |
| MGS | abundance | Hgn3C.1188 - Actinomyces oris | Handgrip |
| MGS | abundance | Hgn3C.1190 - Fenollaria massiliensis | Tibia_length |
| MGS | abundance | Hgn3C.1190 - Fenollaria massiliensis | Radius_length |
| MGS | abundance | Hgn3C.1190 - Fenollaria massiliensis | Radius_SOS |
| MGS | abundance | Hgn3C.1190 - Fenollaria massiliensis | Tibia_SOS |
| MGS | abundance | Hgn3C.1190 - Fenollaria massiliensis | Handgrip |
| MGS | abundance | Hgn3C.1193 - Atopobium sp. | Tibia_length |
| MGS | abundance | Hgn3C.1193 - Atopobium sp. | Radius_length |
| MGS | abundance | Hgn3C.1193 - Atopobium sp. | Radius_SOS |
| MGS | abundance | Hgn3C.1193 - Atopobium sp. | Tibia_SOS |
| MGS | abundance | Hgn3C.1193 - Atopobium sp. | Handgrip |
| MGS | abundance | Hgn3C.1196 - Blautia marasmi | Tibia_length |
| MGS | abundance | Hgn3C.1196 - Blautia marasmi | Radius_length |
| MGS | abundance | Hgn3C.1196 - Blautia marasmi | Radius_SOS |
| MGS | abundance | Hgn3C.1196 - Blautia marasmi | Tibia_SOS |
| MGS | abundance | Hgn3C.1196 - Blautia marasmi | Handgrip |
| MGS | abundance | Hgn3C.1197 - Blautia hansenii | Tibia_length |
| MGS | abundance | Hgn3C.1197 - Blautia hansenii | Radius_length |
| MGS | abundance | Hgn3C.1197 - Blautia hansenii | Radius_SOS |
| MGS | abundance | Hgn3C.1197 - Blautia hansenii | Tibia_SOS |
| MGS | abundance | Hgn3C.1197 - Blautia hansenii | Handgrip |
| MGS | abundance | Hgn3C.1203 - Citrobacter sp. CRE-46 | Tibia_length |

| | | | |
|---|---|---|---|
| MGS | abundance | Hgn3C.1203 - Citrobacter sp. CRE-46 | Radius_length |
| MGS | abundance | Hgn3C.1203 - Citrobacter sp. CRE-46 | Radius_SOS |
| MGS | abundance | Hgn3C.1203 - Citrobacter sp. CRE-46 | Tibia_SOS |
| MGS | abundance | Hgn3C.1203 - Citrobacter sp. CRE-46 | Handgrip |
| MGS | abundance | Hgn3C.1206 - Citrobacter pasteurii | Tibia_length |
| MGS | abundance | Hgn3C.1206 - Citrobacter pasteurii | Radius_length |
| MGS | abundance | Hgn3C.1206 - Citrobacter pasteurii | Radius_SOS |
| MGS | abundance | Hgn3C.1206 - Citrobacter pasteurii | Tibia_SOS |
| MGS | abundance | Hgn3C.1206 - Citrobacter pasteurii | Handgrip |
| MGS | abundance | Hgn3C.1207 - Citrobacter sp. | Tibia_length |
| MGS | abundance | Hgn3C.1207 - Citrobacter sp. | Radius_length |
| MGS | abundance | Hgn3C.1207 - Citrobacter sp. | Radius_SOS |
| MGS | abundance | Hgn3C.1207 - Citrobacter sp. | Tibia_SOS |
| MGS | abundance | Hgn3C.1207 - Citrobacter sp. | Handgrip |
| MGS | abundance | Hgn3C.1210 - Dielma sp. | Tibia_length |
| MGS | abundance | Hgn3C.1210 - Dielma sp. | Radius_length |
| MGS | abundance | Hgn3C.1210 - Dielma sp. | Radius_SOS |
| MGS | abundance | Hgn3C.1210 - Dielma sp. | Tibia_SOS |
| MGS | abundance | Hgn3C.1210 - Dielma sp. | Handgrip |
| MGS | abundance | Hgn3C.1211 - Lachnoclostridium pacaense | Tibia_length |
| MGS | abundance | Hgn3C.1211 - Lachnoclostridium pacaense | Radius_length |
| MGS | abundance | Hgn3C.1211 - Lachnoclostridium pacaense | Radius_SOS |
| MGS | abundance | Hgn3C.1211 - Lachnoclostridium pacaense | Tibia_SOS |
| MGS | abundance | Hgn3C.1211 - Lachnoclostridium pacaense | Handgrip |
| MGS | abundance | Hgn3C.1214 - Corynebacterium tuberculostearicum | Tibia_length |
| MGS | abundance | Hgn3C.1214 - Corynebacterium tuberculostearicum | Radius_length |
| MGS | abundance | Hgn3C.1214 - Corynebacterium tuberculostearicum | Radius_SOS |
| MGS | abundance | Hgn3C.1214 - Corynebacterium tuberculostearicum | Tibia_SOS |
| MGS | abundance | Hgn3C.1214 - Corynebacterium tuberculostearicum | Handgrip |
| MGS | abundance | Hgn3C.1215 - Cutibacterium acnes subsp. acnes | Tibia_length |
| MGS | abundance | Hgn3C.1215 - Cutibacterium acnes subsp. acnes | Radius_length |
| MGS | abundance | Hgn3C.1215 - Cutibacterium acnes subsp. acnes | Radius_SOS |
| MGS | abundance | Hgn3C.1215 - Cutibacterium acnes subsp. acnes | Tibia_SOS |
| MGS | abundance | Hgn3C.1215 - Cutibacterium acnes subsp. acnes | Handgrip |
| MGS | abundance | Hgn3C.1218 - Enterobacter bugandensis | Tibia_length |
| MGS | abundance | Hgn3C.1218 - Enterobacter bugandensis | Radius_length |
| MGS | abundance | Hgn3C.1218 - Enterobacter bugandensis | Radius_SOS |
| MGS | abundance | Hgn3C.1218 - Enterobacter bugandensis | Tibia_SOS |
| MGS | abundance | Hgn3C.1218 - Enterobacter bugandensis | Handgrip |
| MGS | abundance | Hgn3C.1219 - Enterobacter sp. NFIX58 | Tibia_length |
| MGS | abundance | Hgn3C.1219 - Enterobacter sp. NFIX58 | Radius_length |
| MGS | abundance | Hgn3C.1219 - Enterobacter sp. NFIX58 | Radius_SOS |
| MGS | abundance | Hgn3C.1219 - Enterobacter sp. NFIX58 | Tibia_SOS |
| MGS | abundance | Hgn3C.1219 - Enterobacter sp. NFIX58 | Handgrip |
| MGS | abundance | Hgn3C.1221 - Phytobacter diazotrophicus | Tibia_length |
| MGS | abundance | Hgn3C.1221 - Phytobacter diazotrophicus | Radius_length |
| MGS | abundance | Hgn3C.1221 - Phytobacter diazotrophicus | Radius_SOS |
| MGS | abundance | Hgn3C.1221 - Phytobacter diazotrophicus | Tibia_SOS |

| | | | |
|---|---|---|---|
| MGS | abundance | Hgn3C.1221 - Phytobacter diazotrophicus | Handgrip |
| MGS | abundance | Hgn3C.1224 - Enterococcus raffinosus | Tibia_length |
| MGS | abundance | Hgn3C.1224 - Enterococcus raffinosus | Radius_length |
| MGS | abundance | Hgn3C.1224 - Enterococcus raffinosus | Radius_SOS |
| MGS | abundance | Hgn3C.1224 - Enterococcus raffinosus | Tibia_SOS |
| MGS | abundance | Hgn3C.1224 - Enterococcus raffinosus | Handgrip |
| MGS | abundance | Hgn3C.1225 - Enterococcus faecium | Tibia_length |
| MGS | abundance | Hgn3C.1225 - Enterococcus faecium | Radius_length |
| MGS | abundance | Hgn3C.1225 - Enterococcus faecium | Radius_SOS |
| MGS | abundance | Hgn3C.1225 - Enterococcus faecium | Tibia_SOS |
| MGS | abundance | Hgn3C.1225 - Enterococcus faecium | Handgrip |
| MGS | abundance | Hgn3C.1228 - Escherichia marmotae | Tibia_length |
| MGS | abundance | Hgn3C.1228 - Escherichia marmotae | Radius_length |
| MGS | abundance | Hgn3C.1228 - Escherichia marmotae | Radius_SOS |
| MGS | abundance | Hgn3C.1228 - Escherichia marmotae | Tibia_SOS |
| MGS | abundance | Hgn3C.1228 - Escherichia marmotae | Handgrip |
| MGS | abundance | Hgn3C.1229 - Escherichia sp. 93.0750 | Tibia_length |
| MGS | abundance | Hgn3C.1229 - Escherichia sp. 93.0750 | Radius_length |
| MGS | abundance | Hgn3C.1229 - Escherichia sp. 93.0750 | Radius_SOS |
| MGS | abundance | Hgn3C.1229 - Escherichia sp. 93.0750 | Tibia_SOS |
| MGS | abundance | Hgn3C.1229 - Escherichia sp. 93.0750 | Handgrip |
| MGS | abundance | Hgn3C.1230 - Eubacterium limosum | Tibia_length |
| MGS | abundance | Hgn3C.1230 - Eubacterium limosum | Radius_length |
| MGS | abundance | Hgn3C.1230 - Eubacterium limosum | Radius_SOS |
| MGS | abundance | Hgn3C.1230 - Eubacterium limosum | Tibia_SOS |
| MGS | abundance | Hgn3C.1230 - Eubacterium limosum | Handgrip |
| MGS | abundance | Hgn3C.1234 - Granulicatella adiacens | Tibia_length |
| MGS | abundance | Hgn3C.1234 - Granulicatella adiacens | Radius_length |
| MGS | abundance | Hgn3C.1234 - Granulicatella adiacens | Radius_SOS |
| MGS | abundance | Hgn3C.1234 - Granulicatella adiacens | Tibia_SOS |
| MGS | abundance | Hgn3C.1234 - Granulicatella adiacens | Handgrip |
| MGS | abundance | Hgn3C.1238 - Klebsiella quasivariicola | Tibia_length |
| MGS | abundance | Hgn3C.1238 - Klebsiella quasivariicola | Radius_length |
| MGS | abundance | Hgn3C.1238 - Klebsiella quasivariicola | Radius_SOS |
| MGS | abundance | Hgn3C.1238 - Klebsiella quasivariicola | Tibia_SOS |
| MGS | abundance | Hgn3C.1238 - Klebsiella quasivariicola | Handgrip |
| MGS | abundance | Hgn3C.1241 - Limosilactobacillus reuteri subsp. reuteri | Tibia_length |
| MGS | abundance | Hgn3C.1241 - Limosilactobacillus reuteri subsp. reuteri | Radius_length |
| MGS | abundance | Hgn3C.1241 - Limosilactobacillus reuteri subsp. reuteri | Radius_SOS |
| MGS | abundance | Hgn3C.1241 - Limosilactobacillus reuteri subsp. reuteri | Tibia_SOS |
| MGS | abundance | Hgn3C.1241 - Limosilactobacillus reuteri subsp. reuteri | Handgrip |
| MGS | abundance | Hgn3C.1242 - Limosilactobacillus vaginalis | Tibia_length |
| MGS | abundance | Hgn3C.1242 - Limosilactobacillus vaginalis | Radius_length |
| MGS | abundance | Hgn3C.1242 - Limosilactobacillus vaginalis | Radius_SOS |
| MGS | abundance | Hgn3C.1242 - Limosilactobacillus vaginalis | Tibia_SOS |
| MGS | abundance | Hgn3C.1242 - Limosilactobacillus vaginalis | Handgrip |
| MGS | abundance | Hgn3C.1248 - Neisseria sp. HMSC061E12 | Tibia_length |
| MGS | abundance | Hgn3C.1248 - Neisseria sp. HMSC061E12 | Radius_length |

| | | | |
|---|---|---|---|
| MGS | abundance | Hgn3C.1248 - Neisseria sp. HMSC061E12 | Radius_SOS |
| MGS | abundance | Hgn3C.1248 - Neisseria sp. HMSC061E12 | Tibia_SOS |
| MGS | abundance | Hgn3C.1248 - Neisseria sp. HMSC061E12 | Handgrip |
| MGS | abundance | Hgn3C.1250 - Parabacteroides gordonii | Tibia_length |
| MGS | abundance | Hgn3C.1250 - Parabacteroides gordonii | Radius_length |
| MGS | abundance | Hgn3C.1250 - Parabacteroides gordonii | Radius_SOS |
| MGS | abundance | Hgn3C.1250 - Parabacteroides gordonii | Tibia_SOS |
| MGS | abundance | Hgn3C.1250 - Parabacteroides gordonii | Handgrip |
| MGS | abundance | Hgn3C.1258 - Rothia sp. HMSC072E10 | Tibia_length |
| MGS | abundance | Hgn3C.1258 - Rothia sp. HMSC072E10 | Radius_length |
| MGS | abundance | Hgn3C.1258 - Rothia sp. HMSC072E10 | Radius_SOS |
| MGS | abundance | Hgn3C.1258 - Rothia sp. HMSC072E10 | Tibia_SOS |
| MGS | abundance | Hgn3C.1258 - Rothia sp. HMSC072E10 | Handgrip |
| MGS | abundance | Hgn3C.1264 - Staphylococcus hominis subsp. hominis | Tibia_length |
| MGS | abundance | Hgn3C.1264 - Staphylococcus hominis subsp. hominis | Radius_length |
| MGS | abundance | Hgn3C.1264 - Staphylococcus hominis subsp. hominis | Radius_SOS |
| MGS | abundance | Hgn3C.1264 - Staphylococcus hominis subsp. hominis | Tibia_SOS |
| MGS | abundance | Hgn3C.1264 - Staphylococcus hominis subsp. hominis | Handgrip |
| MGS | abundance | Hgn3C.1268 - Streptococcus sp. | Tibia_length |
| MGS | abundance | Hgn3C.1268 - Streptococcus sp. | Radius_length |
| MGS | abundance | Hgn3C.1268 - Streptococcus sp. | Radius_SOS |
| MGS | abundance | Hgn3C.1268 - Streptococcus sp. | Tibia_SOS |
| MGS | abundance | Hgn3C.1268 - Streptococcus sp. | Handgrip |
| MGS | abundance | Hgn3C.1269 - Streptococcus constellatus subsp. constellatus | Tibia_length |
| MGS | abundance | Hgn3C.1269 - Streptococcus constellatus subsp. constellatus | Radius_length |
| MGS | abundance | Hgn3C.1269 - Streptococcus constellatus subsp. constellatus | Radius_SOS |
| MGS | abundance | Hgn3C.1269 - Streptococcus constellatus subsp. constellatus | Tibia_SOS |
| MGS | abundance | Hgn3C.1269 - Streptococcus constellatus subsp. constellatus | Handgrip |
| MGS | abundance | Hgn3C.1270 - Streptococcus sp. | Tibia_length |
| MGS | abundance | Hgn3C.1270 - Streptococcus sp. | Radius_length |
| MGS | abundance | Hgn3C.1270 - Streptococcus sp. | Radius_SOS |
| MGS | abundance | Hgn3C.1270 - Streptococcus sp. | Tibia_SOS |
| MGS | abundance | Hgn3C.1270 - Streptococcus sp. | Handgrip |
| MGS | abundance | Hgn3C.1271 - Streptococcus agalactiae | Tibia_length |
| MGS | abundance | Hgn3C.1271 - Streptococcus agalactiae | Radius_length |
| MGS | abundance | Hgn3C.1271 - Streptococcus agalactiae | Radius_SOS |
| MGS | abundance | Hgn3C.1271 - Streptococcus agalactiae | Tibia_SOS |
| MGS | abundance | Hgn3C.1271 - Streptococcus agalactiae | Handgrip |
| MGS | abundance | Hgn3C.1275 - Lachnospiraceae sp. | Tibia_length |
| MGS | abundance | Hgn3C.1275 - Lachnospiraceae sp. | Radius_length |
| MGS | abundance | Hgn3C.1275 - Lachnospiraceae sp. | Radius_SOS |
| MGS | abundance | Hgn3C.1275 - Lachnospiraceae sp. | Tibia_SOS |
| MGS | abundance | Hgn3C.1275 - Lachnospiraceae sp. | Handgrip |
| MGS | abundance | Hgn3C.1277 - Lachnospiraceae sp. | Tibia_length |
| MGS | abundance | Hgn3C.1277 - Lachnospiraceae sp. | Radius_length |
| MGS | abundance | Hgn3C.1277 - Lachnospiraceae sp. | Radius_SOS |
| MGS | abundance | Hgn3C.1277 - Lachnospiraceae sp. | Tibia_SOS |
| MGS | abundance | Hgn3C.1277 - Lachnospiraceae sp. | Handgrip |

| | | | |
|---|---|---|---|
| MGS | abundance | Hgn3C.1278 - Clostridium sp. NSJ-6 | Tibia_length |
| MGS | abundance | Hgn3C.1278 - Clostridium sp. NSJ-6 | Radius_length |
| MGS | abundance | Hgn3C.1278 - Clostridium sp. NSJ-6 | Radius_SOS |
| MGS | abundance | Hgn3C.1278 - Clostridium sp. NSJ-6 | Tibia_SOS |
| MGS | abundance | Hgn3C.1278 - Clostridium sp. NSJ-6 | Handgrip |
| MGS | abundance | Hgn3C.1282 - Terrisporobacter othiniensis | Tibia_length |
| MGS | abundance | Hgn3C.1282 - Terrisporobacter othiniensis | Radius_length |
| MGS | abundance | Hgn3C.1282 - Terrisporobacter othiniensis | Radius_SOS |
| MGS | abundance | Hgn3C.1282 - Terrisporobacter othiniensis | Tibia_SOS |
| MGS | abundance | Hgn3C.1282 - Terrisporobacter othiniensis | Handgrip |
| MGS | abundance | Hgn3C.1283 - Clostridium baratii | Tibia_length |
| MGS | abundance | Hgn3C.1283 - Clostridium baratii | Radius_length |
| MGS | abundance | Hgn3C.1283 - Clostridium baratii | Radius_SOS |
| MGS | abundance | Hgn3C.1283 - Clostridium baratii | Tibia_SOS |
| MGS | abundance | Hgn3C.1283 - Clostridium baratii | Handgrip |
| MGS | abundance | Hgn3C.1285 - Enterobacteriaceae sp. | Tibia_length |
| MGS | abundance | Hgn3C.1285 - Enterobacteriaceae sp. | Radius_length |
| MGS | abundance | Hgn3C.1285 - Enterobacteriaceae sp. | Radius_SOS |
| MGS | abundance | Hgn3C.1285 - Enterobacteriaceae sp. | Tibia_SOS |
| MGS | abundance | Hgn3C.1285 - Enterobacteriaceae sp. | Handgrip |
| MGS | abundance | Hgn3C.1286 - Corynebacterium variabile | Tibia_length |
| MGS | abundance | Hgn3C.1286 - Corynebacterium variabile | Radius_length |
| MGS | abundance | Hgn3C.1286 - Corynebacterium variabile | Radius_SOS |
| MGS | abundance | Hgn3C.1286 - Corynebacterium variabile | Tibia_SOS |
| MGS | abundance | Hgn3C.1286 - Corynebacterium variabile | Handgrip |
| MGS | abundance | Hgn3C.1288 - Enterobacter hormaechei | Tibia_length |
| MGS | abundance | Hgn3C.1288 - Enterobacter hormaechei | Radius_length |
| MGS | abundance | Hgn3C.1288 - Enterobacter hormaechei | Radius_SOS |
| MGS | abundance | Hgn3C.1288 - Enterobacter hormaechei | Tibia_SOS |
| MGS | abundance | Hgn3C.1288 - Enterobacter hormaechei | Handgrip |
| MGS | abundance | Hgn3C.1292 - Limosilactobacillus portuensis | Tibia_length |
| MGS | abundance | Hgn3C.1292 - Limosilactobacillus portuensis | Radius_length |
| MGS | abundance | Hgn3C.1292 - Limosilactobacillus portuensis | Radius_SOS |
| MGS | abundance | Hgn3C.1292 - Limosilactobacillus portuensis | Tibia_SOS |
| MGS | abundance | Hgn3C.1292 - Limosilactobacillus portuensis | Handgrip |
| MGS | abundance | Hgn3C.1295 - Butyricicoccus pullicaecorum | Tibia_length |
| MGS | abundance | Hgn3C.1295 - Butyricicoccus pullicaecorum | Radius_length |
| MGS | abundance | Hgn3C.1295 - Butyricicoccus pullicaecorum | Radius_SOS |
| MGS | abundance | Hgn3C.1295 - Butyricicoccus pullicaecorum | Tibia_SOS |
| MGS | abundance | Hgn3C.1295 - Butyricicoccus pullicaecorum | Handgrip |
| MGS | abundance | Hgn3C.1297 - Clostridium sp. | Tibia_length |
| MGS | abundance | Hgn3C.1297 - Clostridium sp. | Radius_length |
| MGS | abundance | Hgn3C.1297 - Clostridium sp. | Radius_SOS |
| MGS | abundance | Hgn3C.1297 - Clostridium sp. | Tibia_SOS |
| MGS | abundance | Hgn3C.1297 - Clostridium sp. | Handgrip |
| MGS | abundance | Hgn3C.1298 - Firmicutes sp. | Tibia_length |
| MGS | abundance | Hgn3C.1298 - Firmicutes sp. | Radius_length |
| MGS | abundance | Hgn3C.1298 - Firmicutes sp. | Radius_SOS |

| | | | |
|---|---|---|---|
| MGS | abundance | Hgn3C.1298 - Firmicutes sp. | Tibia_SOS |
| MGS | abundance | Hgn3C.1298 - Firmicutes sp. | Handgrip |
| MGS | abundance | Hgn3C.1299 - Erysipelotrichaceae sp. | Tibia_length |
| MGS | abundance | Hgn3C.1299 - Erysipelotrichaceae sp. | Radius_length |
| MGS | abundance | Hgn3C.1299 - Erysipelotrichaceae sp. | Radius_SOS |
| MGS | abundance | Hgn3C.1299 - Erysipelotrichaceae sp. | Tibia_SOS |
| MGS | abundance | Hgn3C.1299 - Erysipelotrichaceae sp. | Handgrip |
| MGS | abundance | Hgn3C.1300 - Rothia sp. | Tibia_length |
| MGS | abundance | Hgn3C.1300 - Rothia sp. | Radius_length |
| MGS | abundance | Hgn3C.1300 - Rothia sp. | Radius_SOS |
| MGS | abundance | Hgn3C.1300 - Rothia sp. | Tibia_SOS |
| MGS | abundance | Hgn3C.1300 - Rothia sp. | Handgrip |
| MGS | abundance | Hgn3C.1302 - Schaalia sp. | Tibia_length |
| MGS | abundance | Hgn3C.1302 - Schaalia sp. | Radius_length |
| MGS | abundance | Hgn3C.1302 - Schaalia sp. | Radius_SOS |
| MGS | abundance | Hgn3C.1302 - Schaalia sp. | Tibia_SOS |
| MGS | abundance | Hgn3C.1302 - Schaalia sp. | Handgrip |
| MGS | abundance | Hgn3C.1303 - Clostridium paraputrificum | Tibia_length |
| MGS | abundance | Hgn3C.1303 - Clostridium paraputrificum | Radius_length |
| MGS | abundance | Hgn3C.1303 - Clostridium paraputrificum | Radius_SOS |
| MGS | abundance | Hgn3C.1303 - Clostridium paraputrificum | Tibia_SOS |
| MGS | abundance | Hgn3C.1303 - Clostridium paraputrificum | Handgrip |
| MGS | abundance | Hgn3C.1305 - Isoptericola variabilis | Tibia_length |
| MGS | abundance | Hgn3C.1305 - Isoptericola variabilis | Radius_length |
| MGS | abundance | Hgn3C.1305 - Isoptericola variabilis | Radius_SOS |
| MGS | abundance | Hgn3C.1305 - Isoptericola variabilis | Tibia_SOS |
| MGS | abundance | Hgn3C.1305 - Isoptericola variabilis | Handgrip |
| MGS | abundance | Hgn3C.1308 - Eisenbergiella sp. | Tibia_length |
| MGS | abundance | Hgn3C.1308 - Eisenbergiella sp. | Radius_length |
| MGS | abundance | Hgn3C.1308 - Eisenbergiella sp. | Radius_SOS |
| MGS | abundance | Hgn3C.1308 - Eisenbergiella sp. | Tibia_SOS |
| MGS | abundance | Hgn3C.1308 - Eisenbergiella sp. | Handgrip |
| MGS | abundance | Hgn3C.1309 - Enterobacter cloacae subsp. cloacae | Tibia_length |
| MGS | abundance | Hgn3C.1309 - Enterobacter cloacae subsp. cloacae | Radius_length |
| MGS | abundance | Hgn3C.1309 - Enterobacter cloacae subsp. cloacae | Radius_SOS |
| MGS | abundance | Hgn3C.1309 - Enterobacter cloacae subsp. cloacae | Tibia_SOS |
| MGS | abundance | Hgn3C.1309 - Enterobacter cloacae subsp. cloacae | Handgrip |
| MGS | abundance | Hgn3C.1310 - Erysipelotrichaceae sp. | Tibia_length |
| MGS | abundance | Hgn3C.1310 - Erysipelotrichaceae sp. | Radius_length |
| MGS | abundance | Hgn3C.1310 - Erysipelotrichaceae sp. | Radius_SOS |
| MGS | abundance | Hgn3C.1310 - Erysipelotrichaceae sp. | Tibia_SOS |
| MGS | abundance | Hgn3C.1310 - Erysipelotrichaceae sp. | Handgrip |
| MGS | abundance | Hgn3C.1311 - Lachnospiraceae sp. | Tibia_length |
| MGS | abundance | Hgn3C.1311 - Lachnospiraceae sp. | Radius_length |
| MGS | abundance | Hgn3C.1311 - Lachnospiraceae sp. | Radius_SOS |
| MGS | abundance | Hgn3C.1311 - Lachnospiraceae sp. | Tibia_SOS |
| MGS | abundance | Hgn3C.1311 - Lachnospiraceae sp. | Handgrip |
| MGS | abundance | Hgn3C.1312 - Candida tropicalis | Tibia_length |

| | | | |
|---|---|---|---|
| MGS | abundance | Hgn3C.1312 - Candida tropicalis | Radius_length |
| MGS | abundance | Hgn3C.1312 - Candida tropicalis | Radius_SOS |
| MGS | abundance | Hgn3C.1312 - Candida tropicalis | Tibia_SOS |
| MGS | abundance | Hgn3C.1312 - Candida tropicalis | Handgrip |
| MGS | abundance | Hgn3C.1313 - Klebsiella aerogenes | Tibia_length |
| MGS | abundance | Hgn3C.1313 - Klebsiella aerogenes | Radius_length |
| MGS | abundance | Hgn3C.1313 - Klebsiella aerogenes | Radius_SOS |
| MGS | abundance | Hgn3C.1313 - Klebsiella aerogenes | Tibia_SOS |
| MGS | abundance | Hgn3C.1313 - Klebsiella aerogenes | Handgrip |
| MGS | abundance | Hgn3C.1314 - Eubacteriales sp. | Tibia_length |
| MGS | abundance | Hgn3C.1314 - Eubacteriales sp. | Radius_length |
| MGS | abundance | Hgn3C.1314 - Eubacteriales sp. | Radius_SOS |
| MGS | abundance | Hgn3C.1314 - Eubacteriales sp. | Tibia_SOS |
| MGS | abundance | Hgn3C.1314 - Eubacteriales sp. | Handgrip |
| MGS | abundance | Hgn3C.1315 - Eubacteriales sp. | Tibia_length |
| MGS | abundance | Hgn3C.1315 - Eubacteriales sp. | Radius_length |
| MGS | abundance | Hgn3C.1315 - Eubacteriales sp. | Radius_SOS |
| MGS | abundance | Hgn3C.1315 - Eubacteriales sp. | Tibia_SOS |
| MGS | abundance | Hgn3C.1315 - Eubacteriales sp. | Handgrip |
| MGS | abundance | Hgn3C.1316 - Eubacteriales sp. | Tibia_length |
| MGS | abundance | Hgn3C.1316 - Eubacteriales sp. | Radius_length |
| MGS | abundance | Hgn3C.1316 - Eubacteriales sp. | Radius_SOS |
| MGS | abundance | Hgn3C.1316 - Eubacteriales sp. | Tibia_SOS |
| MGS | abundance | Hgn3C.1316 - Eubacteriales sp. | Handgrip |
| MGS | abundance | Hgn3C.1319 - Atlantibacter hermannii | Tibia_length |
| MGS | abundance | Hgn3C.1319 - Atlantibacter hermannii | Radius_length |
| MGS | abundance | Hgn3C.1319 - Atlantibacter hermannii | Radius_SOS |
| MGS | abundance | Hgn3C.1319 - Atlantibacter hermannii | Tibia_SOS |
| MGS | abundance | Hgn3C.1319 - Atlantibacter hermannii | Handgrip |
| MGS | abundance | Hgn3C.1321 - Veillonellaceae sp. | Tibia_length |
| MGS | abundance | Hgn3C.1321 - Veillonellaceae sp. | Radius_length |
| MGS | abundance | Hgn3C.1321 - Veillonellaceae sp. | Radius_SOS |
| MGS | abundance | Hgn3C.1321 - Veillonellaceae sp. | Tibia_SOS |
| MGS | abundance | Hgn3C.1321 - Veillonellaceae sp. | Handgrip |
| MGS | abundance | Hgn3C.1322 - Enterococcus hirae | Tibia_length |
| MGS | abundance | Hgn3C.1322 - Enterococcus hirae | Radius_length |
| MGS | abundance | Hgn3C.1322 - Enterococcus hirae | Radius_SOS |
| MGS | abundance | Hgn3C.1322 - Enterococcus hirae | Tibia_SOS |
| MGS | abundance | Hgn3C.1322 - Enterococcus hirae | Handgrip |
| MGS | abundance | Hgn3C.1323 - Lachnospiraceae sp. | Tibia_length |
| MGS | abundance | Hgn3C.1323 - Lachnospiraceae sp. | Radius_length |
| MGS | abundance | Hgn3C.1323 - Lachnospiraceae sp. | Radius_SOS |
| MGS | abundance | Hgn3C.1323 - Lachnospiraceae sp. | Tibia_SOS |
| MGS | abundance | Hgn3C.1323 - Lachnospiraceae sp. | Handgrip |
| MGS | abundance | Hgn3C.1324 - Eubacterium sp. c-25 | Tibia_length |
| MGS | abundance | Hgn3C.1324 - Eubacterium sp. c-25 | Radius_length |
| MGS | abundance | Hgn3C.1324 - Eubacterium sp. c-25 | Radius_SOS |
| MGS | abundance | Hgn3C.1324 - Eubacterium sp. c-25 | Tibia_SOS |

| | | | |
|---|---|---|---|
| MGS | abundance | Hgn3C.1324 - Eubacterium sp. c-25 | Handgrip |
| MGS | abundance | Hgn3C.1325 - Luxibacter massiliensis | Tibia_length |
| MGS | abundance | Hgn3C.1325 - Luxibacter massiliensis | Radius_length |
| MGS | abundance | Hgn3C.1325 - Luxibacter massiliensis | Radius_SOS |
| MGS | abundance | Hgn3C.1325 - Luxibacter massiliensis | Tibia_SOS |
| MGS | abundance | Hgn3C.1325 - Luxibacter massiliensis | Handgrip |
| MGS | abundance | Hgn3C.1326 - Eggerthella guodeyinii | Tibia_length |
| MGS | abundance | Hgn3C.1326 - Eggerthella guodeyinii | Radius_length |
| MGS | abundance | Hgn3C.1326 - Eggerthella guodeyinii | Radius_SOS |
| MGS | abundance | Hgn3C.1326 - Eggerthella guodeyinii | Tibia_SOS |
| MGS | abundance | Hgn3C.1326 - Eggerthella guodeyinii | Handgrip |
| MGS | abundance | Hgn3C.1328 - Lacrimispora saccharolytica | Tibia_length |
| MGS | abundance | Hgn3C.1328 - Lacrimispora saccharolytica | Radius_length |
| MGS | abundance | Hgn3C.1328 - Lacrimispora saccharolytica | Radius_SOS |
| MGS | abundance | Hgn3C.1328 - Lacrimispora saccharolytica | Tibia_SOS |
| MGS | abundance | Hgn3C.1328 - Lacrimispora saccharolytica | Handgrip |
| MGS | abundance | Hgn3C.1329 - Eubacteriales sp. | Tibia_length |
| MGS | abundance | Hgn3C.1329 - Eubacteriales sp. | Radius_length |
| MGS | abundance | Hgn3C.1329 - Eubacteriales sp. | Radius_SOS |
| MGS | abundance | Hgn3C.1329 - Eubacteriales sp. | Tibia_SOS |
| MGS | abundance | Hgn3C.1329 - Eubacteriales sp. | Handgrip |
| MGS | abundance | Hgn3C.1330 - Lachnospiraceae sp. | Tibia_length |
| MGS | abundance | Hgn3C.1330 - Lachnospiraceae sp. | Radius_length |
| MGS | abundance | Hgn3C.1330 - Lachnospiraceae sp. | Radius_SOS |
| MGS | abundance | Hgn3C.1330 - Lachnospiraceae sp. | Tibia_SOS |
| MGS | abundance | Hgn3C.1330 - Lachnospiraceae sp. | Handgrip |
| MGS | abundance | Hgn3C.1331 - Lachnospiraceae sp. | Tibia_length |
| MGS | abundance | Hgn3C.1331 - Lachnospiraceae sp. | Radius_length |
| MGS | abundance | Hgn3C.1331 - Lachnospiraceae sp. | Radius_SOS |
| MGS | abundance | Hgn3C.1331 - Lachnospiraceae sp. | Tibia_SOS |
| MGS | abundance | Hgn3C.1331 - Lachnospiraceae sp. | Handgrip |
| MGS | abundance | Hgn3C.1334 - Eubacteriales sp. | Tibia_length |
| MGS | abundance | Hgn3C.1334 - Eubacteriales sp. | Radius_length |
| MGS | abundance | Hgn3C.1334 - Eubacteriales sp. | Radius_SOS |
| MGS | abundance | Hgn3C.1334 - Eubacteriales sp. | Tibia_SOS |
| MGS | abundance | Hgn3C.1334 - Eubacteriales sp. | Handgrip |
| MGS | abundance | Hgn3C.1335 - Eubacteriales sp. | Tibia_length |
| MGS | abundance | Hgn3C.1335 - Eubacteriales sp. | Radius_length |
| MGS | abundance | Hgn3C.1335 - Eubacteriales sp. | Radius_SOS |
| MGS | abundance | Hgn3C.1335 - Eubacteriales sp. | Tibia_SOS |
| MGS | abundance | Hgn3C.1335 - Eubacteriales sp. | Handgrip |
| MGS | abundance | Hgn3C.1336 - Eubacteriales sp. | Tibia_length |
| MGS | abundance | Hgn3C.1336 - Eubacteriales sp. | Radius_length |
| MGS | abundance | Hgn3C.1336 - Eubacteriales sp. | Radius_SOS |
| MGS | abundance | Hgn3C.1336 - Eubacteriales sp. | Tibia_SOS |
| MGS | abundance | Hgn3C.1336 - Eubacteriales sp. | Handgrip |
| MGS | abundance | Hgn3C.1338 - Erysipelatoclostridium sp. | Tibia_length |
| MGS | abundance | Hgn3C.1338 - Erysipelatoclostridium sp. | Radius_length |

| | | | |
|---|---|---|---|
| MGS | abundance | Hgn3C.1338 - Erysipelatoclostridium sp. | Radius_SOS |
| MGS | abundance | Hgn3C.1338 - Erysipelatoclostridium sp. | Tibia_SOS |
| MGS | abundance | Hgn3C.1338 - Erysipelatoclostridium sp. | Handgrip |
| MGS | abundance | Hgn3C.1340 - Clostridiaceae sp. | Tibia_length |
| MGS | abundance | Hgn3C.1340 - Clostridiaceae sp. | Radius_length |
| MGS | abundance | Hgn3C.1340 - Clostridiaceae sp. | Radius_SOS |
| MGS | abundance | Hgn3C.1340 - Clostridiaceae sp. | Tibia_SOS |
| MGS | abundance | Hgn3C.1340 - Clostridiaceae sp. | Handgrip |
| MGS | abundance | Hgn3C.1341 - Lachnospiraceae sp. | Tibia_length |
| MGS | abundance | Hgn3C.1341 - Lachnospiraceae sp. | Radius_length |
| MGS | abundance | Hgn3C.1341 - Lachnospiraceae sp. | Radius_SOS |
| MGS | abundance | Hgn3C.1341 - Lachnospiraceae sp. | Tibia_SOS |
| MGS | abundance | Hgn3C.1341 - Lachnospiraceae sp. | Handgrip |
| MGS | abundance | Hgn3C.1342 - Eubacteriales sp. | Tibia_length |
| MGS | abundance | Hgn3C.1342 - Eubacteriales sp. | Radius_length |
| MGS | abundance | Hgn3C.1342 - Eubacteriales sp. | Radius_SOS |
| MGS | abundance | Hgn3C.1342 - Eubacteriales sp. | Tibia_SOS |
| MGS | abundance | Hgn3C.1342 - Eubacteriales sp. | Handgrip |
| MGS | abundance | Hgn3C.1343 - Enterococcus italicus | Tibia_length |
| MGS | abundance | Hgn3C.1343 - Enterococcus italicus | Radius_length |
| MGS | abundance | Hgn3C.1343 - Enterococcus italicus | Radius_SOS |
| MGS | abundance | Hgn3C.1343 - Enterococcus italicus | Tibia_SOS |
| MGS | abundance | Hgn3C.1343 - Enterococcus italicus | Handgrip |
| MGS | abundance | Hgn3C.1345 - Limosilactobacillus mucosae | Tibia_length |
| MGS | abundance | Hgn3C.1345 - Limosilactobacillus mucosae | Radius_length |
| MGS | abundance | Hgn3C.1345 - Limosilactobacillus mucosae | Radius_SOS |
| MGS | abundance | Hgn3C.1345 - Limosilactobacillus mucosae | Tibia_SOS |
| MGS | abundance | Hgn3C.1345 - Limosilactobacillus mucosae | Handgrip |
| MGS | abundance | Hgn3C.1346 - Erysipelotrichales sp. | Tibia_length |
| MGS | abundance | Hgn3C.1346 - Erysipelotrichales sp. | Radius_length |
| MGS | abundance | Hgn3C.1346 - Erysipelotrichales sp. | Radius_SOS |
| MGS | abundance | Hgn3C.1346 - Erysipelotrichales sp. | Tibia_SOS |
| MGS | abundance | Hgn3C.1346 - Erysipelotrichales sp. | Handgrip |
| MGS | abundance | Hgn3C.1347 - [Clostridium] hylemonae | Tibia_length |
| MGS | abundance | Hgn3C.1347 - [Clostridium] hylemonae | Radius_length |
| MGS | abundance | Hgn3C.1347 - [Clostridium] hylemonae | Radius_SOS |
| MGS | abundance | Hgn3C.1347 - [Clostridium] hylemonae | Tibia_SOS |
| MGS | abundance | Hgn3C.1347 - [Clostridium] hylemonae | Handgrip |
| MGS | abundance | Hgn3C.1348 - Eubacteriales sp. | Tibia_length |
| MGS | abundance | Hgn3C.1348 - Eubacteriales sp. | Radius_length |
| MGS | abundance | Hgn3C.1348 - Eubacteriales sp. | Radius_SOS |
| MGS | abundance | Hgn3C.1348 - Eubacteriales sp. | Tibia_SOS |
| MGS | abundance | Hgn3C.1348 - Eubacteriales sp. | Handgrip |
| MGS | abundance | Hgn3C.1349 - Corynebacterium falsenii | Tibia_length |
| MGS | abundance | Hgn3C.1349 - Corynebacterium falsenii | Radius_length |
| MGS | abundance | Hgn3C.1349 - Corynebacterium falsenii | Radius_SOS |
| MGS | abundance | Hgn3C.1349 - Corynebacterium falsenii | Tibia_SOS |
| MGS | abundance | Hgn3C.1349 - Corynebacterium falsenii | Handgrip |

| | | | |
|---|---|---|---|
| MGS | abundance | Hgn3C.1351 - Actinomyces sp. ICM47 | Tibia_length |
| MGS | abundance | Hgn3C.1351 - Actinomyces sp. ICM47 | Radius_length |
| MGS | abundance | Hgn3C.1351 - Actinomyces sp. ICM47 | Radius_SOS |
| MGS | abundance | Hgn3C.1351 - Actinomyces sp. ICM47 | Tibia_SOS |
| MGS | abundance | Hgn3C.1351 - Actinomyces sp. ICM47 | Handgrip |
| MGS | abundance | Hgn3C.1352 - Akkermansia sp. | Tibia_length |
| MGS | abundance | Hgn3C.1352 - Akkermansia sp. | Radius_length |
| MGS | abundance | Hgn3C.1352 - Akkermansia sp. | Radius_SOS |
| MGS | abundance | Hgn3C.1352 - Akkermansia sp. | Tibia_SOS |
| MGS | abundance | Hgn3C.1352 - Akkermansia sp. | Handgrip |
| MGS | abundance | Hgn3C.1353 - Beduini sp. | Tibia_length |
| MGS | abundance | Hgn3C.1353 - Beduini sp. | Radius_length |
| MGS | abundance | Hgn3C.1353 - Beduini sp. | Radius_SOS |
| MGS | abundance | Hgn3C.1353 - Beduini sp. | Tibia_SOS |
| MGS | abundance | Hgn3C.1353 - Beduini sp. | Handgrip |
| MGS | abundance | Hgn3C.1354 - Actinomyces sp. | Tibia_length |
| MGS | abundance | Hgn3C.1354 - Actinomyces sp. | Radius_length |
| MGS | abundance | Hgn3C.1354 - Actinomyces sp. | Radius_SOS |
| MGS | abundance | Hgn3C.1354 - Actinomyces sp. | Tibia_SOS |
| MGS | abundance | Hgn3C.1354 - Actinomyces sp. | Handgrip |
| MGS | abundance | Hgn3C.1355 - Collinsella stercoris | Tibia_length |
| MGS | abundance | Hgn3C.1355 - Collinsella stercoris | Radius_length |
| MGS | abundance | Hgn3C.1355 - Collinsella stercoris | Radius_SOS |
| MGS | abundance | Hgn3C.1355 - Collinsella stercoris | Tibia_SOS |
| MGS | abundance | Hgn3C.1355 - Collinsella stercoris | Handgrip |
| MGS | abundance | Hgn3C.1356 - Eubacteriales sp. | Tibia_length |
| MGS | abundance | Hgn3C.1356 - Eubacteriales sp. | Radius_length |
| MGS | abundance | Hgn3C.1356 - Eubacteriales sp. | Radius_SOS |
| MGS | abundance | Hgn3C.1356 - Eubacteriales sp. | Tibia_SOS |
| MGS | abundance | Hgn3C.1356 - Eubacteriales sp. | Handgrip |
| MGS | abundance | Hgn3C.1357 - Bacteroidales sp. | Tibia_length |
| MGS | abundance | Hgn3C.1357 - Bacteroidales sp. | Radius_length |
| MGS | abundance | Hgn3C.1357 - Bacteroidales sp. | Radius_SOS |
| MGS | abundance | Hgn3C.1357 - Bacteroidales sp. | Tibia_SOS |
| MGS | abundance | Hgn3C.1357 - Bacteroidales sp. | Handgrip |
| MGS | abundance | Hgn3C.1358 - Lachnospiraceae sp. | Tibia_length |
| MGS | abundance | Hgn3C.1358 - Lachnospiraceae sp. | Radius_length |
| MGS | abundance | Hgn3C.1358 - Lachnospiraceae sp. | Radius_SOS |
| MGS | abundance | Hgn3C.1358 - Lachnospiraceae sp. | Tibia_SOS |
| MGS | abundance | Hgn3C.1358 - Lachnospiraceae sp. | Handgrip |
| MGS | abundance | Hgn3C.1359 - Extibacter muris | Tibia_length |
| MGS | abundance | Hgn3C.1359 - Extibacter muris | Radius_length |
| MGS | abundance | Hgn3C.1359 - Extibacter muris | Radius_SOS |
| MGS | abundance | Hgn3C.1359 - Extibacter muris | Tibia_SOS |
| MGS | abundance | Hgn3C.1359 - Extibacter muris | Handgrip |
| MGS | abundance | Hgn3C.1360 - Bifidobacterium crudilactis | Tibia_length |
| MGS | abundance | Hgn3C.1360 - Bifidobacterium crudilactis | Radius_length |
| MGS | abundance | Hgn3C.1360 - Bifidobacterium crudilactis | Radius_SOS |

| | | | |
|---|---|---|---|
| MGS | abundance | Hgn3C.1360 - Bifidobacterium crudilactis | Tibia_SOS |
| MGS | abundance | Hgn3C.1360 - Bifidobacterium crudilactis | Handgrip |
| MGS | abundance | Hgn3C.1361 - Senegalimassilia anaerobia | Tibia_length |
| MGS | abundance | Hgn3C.1361 - Senegalimassilia anaerobia | Radius_length |
| MGS | abundance | Hgn3C.1361 - Senegalimassilia anaerobia | Radius_SOS |
| MGS | abundance | Hgn3C.1361 - Senegalimassilia anaerobia | Tibia_SOS |
| MGS | abundance | Hgn3C.1361 - Senegalimassilia anaerobia | Handgrip |
| MGS | abundance | Hgn3C.1362 - Christensenella intestinihominis | Tibia_length |
| MGS | abundance | Hgn3C.1362 - Christensenella intestinihominis | Radius_length |
| MGS | abundance | Hgn3C.1362 - Christensenella intestinihominis | Radius_SOS |
| MGS | abundance | Hgn3C.1362 - Christensenella intestinihominis | Tibia_SOS |
| MGS | abundance | Hgn3C.1362 - Christensenella intestinihominis | Handgrip |
| MGS | abundance | Hgn3C.1366 - Lachnospiraceae sp. | Tibia_length |
| MGS | abundance | Hgn3C.1366 - Lachnospiraceae sp. | Radius_length |
| MGS | abundance | Hgn3C.1366 - Lachnospiraceae sp. | Radius_SOS |
| MGS | abundance | Hgn3C.1366 - Lachnospiraceae sp. | Tibia_SOS |
| MGS | abundance | Hgn3C.1366 - Lachnospiraceae sp. | Handgrip |
| MGS | abundance | Hgn3C.1369 - Lachnospiraceae sp. | Tibia_length |
| MGS | abundance | Hgn3C.1369 - Lachnospiraceae sp. | Radius_length |
| MGS | abundance | Hgn3C.1369 - Lachnospiraceae sp. | Radius_SOS |
| MGS | abundance | Hgn3C.1369 - Lachnospiraceae sp. | Tibia_SOS |
| MGS | abundance | Hgn3C.1369 - Lachnospiraceae sp. | Handgrip |
| MGS | abundance | Hgn3C.1370 - Kluyvera genomosp. 3 | Tibia_length |
| MGS | abundance | Hgn3C.1370 - Kluyvera genomosp. 3 | Radius_length |
| MGS | abundance | Hgn3C.1370 - Kluyvera genomosp. 3 | Radius_SOS |
| MGS | abundance | Hgn3C.1370 - Kluyvera genomosp. 3 | Tibia_SOS |
| MGS | abundance | Hgn3C.1370 - Kluyvera genomosp. 3 | Handgrip |
| MGS | abundance | Hgn3C.1371 - Lachnospiraceae sp. | Tibia_length |
| MGS | abundance | Hgn3C.1371 - Lachnospiraceae sp. | Radius_length |
| MGS | abundance | Hgn3C.1371 - Lachnospiraceae sp. | Radius_SOS |
| MGS | abundance | Hgn3C.1371 - Lachnospiraceae sp. | Tibia_SOS |
| MGS | abundance | Hgn3C.1371 - Lachnospiraceae sp. | Handgrip |
| MGS | abundance | Hgn3C.1372 - Eubacteriales sp. | Tibia_length |
| MGS | abundance | Hgn3C.1372 - Eubacteriales sp. | Radius_length |
| MGS | abundance | Hgn3C.1372 - Eubacteriales sp. | Radius_SOS |
| MGS | abundance | Hgn3C.1372 - Eubacteriales sp. | Tibia_SOS |
| MGS | abundance | Hgn3C.1372 - Eubacteriales sp. | Handgrip |
| MGS | abundance | Hgn3C.1373 - Eubacteriales sp. | Tibia_length |
| MGS | abundance | Hgn3C.1373 - Eubacteriales sp. | Radius_length |
| MGS | abundance | Hgn3C.1373 - Eubacteriales sp. | Radius_SOS |
| MGS | abundance | Hgn3C.1373 - Eubacteriales sp. | Tibia_SOS |
| MGS | abundance | Hgn3C.1373 - Eubacteriales sp. | Handgrip |
| MGS | abundance | Hgn3C.1375 - Atopobiaceae sp. | Tibia_length |
| MGS | abundance | Hgn3C.1375 - Atopobiaceae sp. | Radius_length |
| MGS | abundance | Hgn3C.1375 - Atopobiaceae sp. | Radius_SOS |
| MGS | abundance | Hgn3C.1375 - Atopobiaceae sp. | Tibia_SOS |
| MGS | abundance | Hgn3C.1375 - Atopobiaceae sp. | Handgrip |
| MGS | abundance | Hgn3C.1376 - Kluyvera sp. CRP | Tibia_length |

| | | | |
|---|---|---|---|
| MGS | abundance | Hgn3C.1376 - Kluyvera sp. CRP | Radius_length |
| MGS | abundance | Hgn3C.1376 - Kluyvera sp. CRP | Radius_SOS |
| MGS | abundance | Hgn3C.1376 - Kluyvera sp. CRP | Tibia_SOS |
| MGS | abundance | Hgn3C.1376 - Kluyvera sp. CRP | Handgrip |
| MGS | abundance | Hgn3C.1377 - Collinsella tanakaei | Tibia_length |
| MGS | abundance | Hgn3C.1377 - Collinsella tanakaei | Radius_length |
| MGS | abundance | Hgn3C.1377 - Collinsella tanakaei | Radius_SOS |
| MGS | abundance | Hgn3C.1377 - Collinsella tanakaei | Tibia_SOS |
| MGS | abundance | Hgn3C.1377 - Collinsella tanakaei | Handgrip |
| MGS | abundance | Hgn3C.1378 - Collinsella sp. AF08-23 | Tibia_length |
| MGS | abundance | Hgn3C.1378 - Collinsella sp. AF08-23 | Radius_length |
| MGS | abundance | Hgn3C.1378 - Collinsella sp. AF08-23 | Radius_SOS |
| MGS | abundance | Hgn3C.1378 - Collinsella sp. AF08-23 | Tibia_SOS |
| MGS | abundance | Hgn3C.1378 - Collinsella sp. AF08-23 | Handgrip |
| MGS | abundance | Hgn3C.1379 - Gordonibacter urolithinfaciens | Tibia_length |
| MGS | abundance | Hgn3C.1379 - Gordonibacter urolithinfaciens | Radius_length |
| MGS | abundance | Hgn3C.1379 - Gordonibacter urolithinfaciens | Radius_SOS |
| MGS | abundance | Hgn3C.1379 - Gordonibacter urolithinfaciens | Tibia_SOS |
| MGS | abundance | Hgn3C.1379 - Gordonibacter urolithinfaciens | Handgrip |
| MGS | abundance | Hgn3C.1380 - Kluyvera georgiana | Tibia_length |
| MGS | abundance | Hgn3C.1380 - Kluyvera georgiana | Radius_length |
| MGS | abundance | Hgn3C.1380 - Kluyvera georgiana | Radius_SOS |
| MGS | abundance | Hgn3C.1380 - Kluyvera georgiana | Tibia_SOS |
| MGS | abundance | Hgn3C.1380 - Kluyvera georgiana | Handgrip |
| MGS | abundance | Hgn3C.1381 - Eubacteriales sp. | Tibia_length |
| MGS | abundance | Hgn3C.1381 - Eubacteriales sp. | Radius_length |
| MGS | abundance | Hgn3C.1381 - Eubacteriales sp. | Radius_SOS |
| MGS | abundance | Hgn3C.1381 - Eubacteriales sp. | Tibia_SOS |
| MGS | abundance | Hgn3C.1381 - Eubacteriales sp. | Handgrip |
| MGS | abundance | Hgn3C.1382 - Oscillospiraceae sp. | Tibia_length |
| MGS | abundance | Hgn3C.1382 - Oscillospiraceae sp. | Radius_length |
| MGS | abundance | Hgn3C.1382 - Oscillospiraceae sp. | Radius_SOS |
| MGS | abundance | Hgn3C.1382 - Oscillospiraceae sp. | Tibia_SOS |
| MGS | abundance | Hgn3C.1382 - Oscillospiraceae sp. | Handgrip |
| MGS | abundance | Hgn3C.1383 - Sellimonas monacensis | Tibia_length |
| MGS | abundance | Hgn3C.1383 - Sellimonas monacensis | Radius_length |
| MGS | abundance | Hgn3C.1383 - Sellimonas monacensis | Radius_SOS |
| MGS | abundance | Hgn3C.1383 - Sellimonas monacensis | Tibia_SOS |
| MGS | abundance | Hgn3C.1383 - Sellimonas monacensis | Handgrip |
| MGS | abundance | Hgn3C.1384 - Anaerococcus sp. | Tibia_length |
| MGS | abundance | Hgn3C.1384 - Anaerococcus sp. | Radius_length |
| MGS | abundance | Hgn3C.1384 - Anaerococcus sp. | Radius_SOS |
| MGS | abundance | Hgn3C.1384 - Anaerococcus sp. | Tibia_SOS |
| MGS | abundance | Hgn3C.1384 - Anaerococcus sp. | Handgrip |
| MGS | abundance | Hgn3C.1385 - Eubacteriales sp. | Tibia_length |
| MGS | abundance | Hgn3C.1385 - Eubacteriales sp. | Radius_length |
| MGS | abundance | Hgn3C.1385 - Eubacteriales sp. | Radius_SOS |
| MGS | abundance | Hgn3C.1385 - Eubacteriales sp. | Tibia_SOS |

| | | | |
|---|---|---|---|
| MGS | abundance | Hgn3C.1385 - Eubacteriales sp. | Handgrip |
| MGS | abundance | Hgn3C.1386 - Veillonella sp. | Tibia_length |
| MGS | abundance | Hgn3C.1386 - Veillonella sp. | Radius_length |
| MGS | abundance | Hgn3C.1386 - Veillonella sp. | Radius_SOS |
| MGS | abundance | Hgn3C.1386 - Veillonella sp. | Tibia_SOS |
| MGS | abundance | Hgn3C.1386 - Veillonella sp. | Handgrip |
| MGS | abundance | Hgn3C.1387 - Alistipes sp. | Tibia_length |
| MGS | abundance | Hgn3C.1387 - Alistipes sp. | Radius_length |
| MGS | abundance | Hgn3C.1387 - Alistipes sp. | Radius_SOS |
| MGS | abundance | Hgn3C.1387 - Alistipes sp. | Tibia_SOS |
| MGS | abundance | Hgn3C.1387 - Alistipes sp. | Handgrip |
| MGS | abundance | Hgn3C.1388 - Streptococcus infantarius subsp. infantarius | Tibia_length |
| MGS | abundance | Hgn3C.1388 - Streptococcus infantarius subsp. infantarius | Radius_length |
| MGS | abundance | Hgn3C.1388 - Streptococcus infantarius subsp. infantarius | Radius_SOS |
| MGS | abundance | Hgn3C.1388 - Streptococcus infantarius subsp. infantarius | Tibia_SOS |
| MGS | abundance | Hgn3C.1388 - Streptococcus infantarius subsp. infantarius | Handgrip |
| MGS | abundance | Hgn3C.1390 - Erysipelatoclostridium sp. | Tibia_length |
| MGS | abundance | Hgn3C.1390 - Erysipelatoclostridium sp. | Radius_length |
| MGS | abundance | Hgn3C.1390 - Erysipelatoclostridium sp. | Radius_SOS |
| MGS | abundance | Hgn3C.1390 - Erysipelatoclostridium sp. | Tibia_SOS |
| MGS | abundance | Hgn3C.1390 - Erysipelatoclostridium sp. | Handgrip |
| MGS | abundance | Hgn3C.1392 - Enterobacter sp. JBIWA005 | Tibia_length |
| MGS | abundance | Hgn3C.1392 - Enterobacter sp. JBIWA005 | Radius_length |
| MGS | abundance | Hgn3C.1392 - Enterobacter sp. JBIWA005 | Radius_SOS |
| MGS | abundance | Hgn3C.1392 - Enterobacter sp. JBIWA005 | Tibia_SOS |
| MGS | abundance | Hgn3C.1392 - Enterobacter sp. JBIWA005 | Handgrip |
| MGS | abundance | Hgn3C.1393 - Anaerofustis stercorihominis | Tibia_length |
| MGS | abundance | Hgn3C.1393 - Anaerofustis stercorihominis | Radius_length |
| MGS | abundance | Hgn3C.1393 - Anaerofustis stercorihominis | Radius_SOS |
| MGS | abundance | Hgn3C.1393 - Anaerofustis stercorihominis | Tibia_SOS |
| MGS | abundance | Hgn3C.1393 - Anaerofustis stercorihominis | Handgrip |
| MGS | abundance | Hgn3C.1394 - Eubacteriales sp. | Tibia_length |
| MGS | abundance | Hgn3C.1394 - Eubacteriales sp. | Radius_length |
| MGS | abundance | Hgn3C.1394 - Eubacteriales sp. | Radius_SOS |
| MGS | abundance | Hgn3C.1394 - Eubacteriales sp. | Tibia_SOS |
| MGS | abundance | Hgn3C.1394 - Eubacteriales sp. | Handgrip |
| MGS | abundance | Hgn3C.1397 - Erysipelotrichales sp. | Tibia_length |
| MGS | abundance | Hgn3C.1397 - Erysipelotrichales sp. | Radius_length |
| MGS | abundance | Hgn3C.1397 - Erysipelotrichales sp. | Radius_SOS |
| MGS | abundance | Hgn3C.1397 - Erysipelotrichales sp. | Tibia_SOS |
| MGS | abundance | Hgn3C.1397 - Erysipelotrichales sp. | Handgrip |
| MGS | abundance | Hgn3C.1398 - Actinomyces sp. | Tibia_length |
| MGS | abundance | Hgn3C.1398 - Actinomyces sp. | Radius_length |
| MGS | abundance | Hgn3C.1398 - Actinomyces sp. | Radius_SOS |
| MGS | abundance | Hgn3C.1398 - Actinomyces sp. | Tibia_SOS |
| MGS | abundance | Hgn3C.1398 - Actinomyces sp. | Handgrip |
| MGS | abundance | Hgn3C.1400 - Eubacterium sp. | Tibia_length |
| MGS | abundance | Hgn3C.1400 - Eubacterium sp. | Radius_length |

| | | | |
|---|---|---|---|
| MGS | abundance | Hgn3C.1400 - Eubacterium sp. | Radius_SOS |
| MGS | abundance | Hgn3C.1400 - Eubacterium sp. | Tibia_SOS |
| MGS | abundance | Hgn3C.1400 - Eubacterium sp. | Handgrip |
| MGS | abundance | Hgn3C.1401 - Eubacteriales sp. | Tibia_length |
| MGS | abundance | Hgn3C.1401 - Eubacteriales sp. | Radius_length |
| MGS | abundance | Hgn3C.1401 - Eubacteriales sp. | Radius_SOS |
| MGS | abundance | Hgn3C.1401 - Eubacteriales sp. | Tibia_SOS |
| MGS | abundance | Hgn3C.1401 - Eubacteriales sp. | Handgrip |
| MGS | abundance | Hgn3C.1402 - Collinsella phocaeensis | Tibia_length |
| MGS | abundance | Hgn3C.1402 - Collinsella phocaeensis | Radius_length |
| MGS | abundance | Hgn3C.1402 - Collinsella phocaeensis | Radius_SOS |
| MGS | abundance | Hgn3C.1402 - Collinsella phocaeensis | Tibia_SOS |
| MGS | abundance | Hgn3C.1402 - Collinsella phocaeensis | Handgrip |
| MGS | abundance | Hgn3C.1403 - Eubacteriaceae sp. | Tibia_length |
| MGS | abundance | Hgn3C.1403 - Eubacteriaceae sp. | Radius_length |
| MGS | abundance | Hgn3C.1403 - Eubacteriaceae sp. | Radius_SOS |
| MGS | abundance | Hgn3C.1403 - Eubacteriaceae sp. | Tibia_SOS |
| MGS | abundance | Hgn3C.1403 - Eubacteriaceae sp. | Handgrip |
| MGS | abundance | Hgn3C.1404 - Clostridium sp. | Tibia_length |
| MGS | abundance | Hgn3C.1404 - Clostridium sp. | Radius_length |
| MGS | abundance | Hgn3C.1404 - Clostridium sp. | Radius_SOS |
| MGS | abundance | Hgn3C.1404 - Clostridium sp. | Tibia_SOS |
| MGS | abundance | Hgn3C.1404 - Clostridium sp. | Handgrip |
| MGS | abundance | Hgn3C.1405 - Eubacteriales sp. | Tibia_length |
| MGS | abundance | Hgn3C.1405 - Eubacteriales sp. | Radius_length |
| MGS | abundance | Hgn3C.1405 - Eubacteriales sp. | Radius_SOS |
| MGS | abundance | Hgn3C.1405 - Eubacteriales sp. | Tibia_SOS |
| MGS | abundance | Hgn3C.1405 - Eubacteriales sp. | Handgrip |
| MGS | abundance | Hgn3C.1406 - Slackia piriformis | Tibia_length |
| MGS | abundance | Hgn3C.1406 - Slackia piriformis | Radius_length |
| MGS | abundance | Hgn3C.1406 - Slackia piriformis | Radius_SOS |
| MGS | abundance | Hgn3C.1406 - Slackia piriformis | Tibia_SOS |
| MGS | abundance | Hgn3C.1406 - Slackia piriformis | Handgrip |
| MGS | abundance | Hgn3C.1407 - Christensenella minuta | Tibia_length |
| MGS | abundance | Hgn3C.1407 - Christensenella minuta | Radius_length |
| MGS | abundance | Hgn3C.1407 - Christensenella minuta | Radius_SOS |
| MGS | abundance | Hgn3C.1407 - Christensenella minuta | Tibia_SOS |
| MGS | abundance | Hgn3C.1407 - Christensenella minuta | Handgrip |
| MGS | abundance | Hgn3C.1408 - Eubacterium callanderi | Tibia_length |
| MGS | abundance | Hgn3C.1408 - Eubacterium callanderi | Radius_length |
| MGS | abundance | Hgn3C.1408 - Eubacterium callanderi | Radius_SOS |
| MGS | abundance | Hgn3C.1408 - Eubacterium callanderi | Tibia_SOS |
| MGS | abundance | Hgn3C.1408 - Eubacterium callanderi | Handgrip |
| MGS | abundance | Hgn3C.1409 - Eggerthella timonensis | Tibia_length |
| MGS | abundance | Hgn3C.1409 - Eggerthella timonensis | Radius_length |
| MGS | abundance | Hgn3C.1409 - Eggerthella timonensis | Radius_SOS |
| MGS | abundance | Hgn3C.1409 - Eggerthella timonensis | Tibia_SOS |
| MGS | abundance | Hgn3C.1409 - Eggerthella timonensis | Handgrip |

| | | | |
|---|---|---|---|
| MGS | abundance | Hgn3C.1411 - Eubacteriales sp. | Tibia_length |
| MGS | abundance | Hgn3C.1411 - Eubacteriales sp. | Radius_length |
| MGS | abundance | Hgn3C.1411 - Eubacteriales sp. | Radius_SOS |
| MGS | abundance | Hgn3C.1411 - Eubacteriales sp. | Tibia_SOS |
| MGS | abundance | Hgn3C.1411 - Eubacteriales sp. | Handgrip |
| MGS | abundance | Hgn3C.1412 - Christensenella hongkongensis | Tibia_length |
| MGS | abundance | Hgn3C.1412 - Christensenella hongkongensis | Radius_length |
| MGS | abundance | Hgn3C.1412 - Christensenella hongkongensis | Radius_SOS |
| MGS | abundance | Hgn3C.1412 - Christensenella hongkongensis | Tibia_SOS |
| MGS | abundance | Hgn3C.1412 - Christensenella hongkongensis | Handgrip |
| MGS | abundance | Hgn3C.1413 - Streptococcus sp. | Tibia_length |
| MGS | abundance | Hgn3C.1413 - Streptococcus sp. | Radius_length |
| MGS | abundance | Hgn3C.1413 - Streptococcus sp. | Radius_SOS |
| MGS | abundance | Hgn3C.1413 - Streptococcus sp. | Tibia_SOS |
| MGS | abundance | Hgn3C.1413 - Streptococcus sp. | Handgrip |
| MGS | abundance | Hgn3C.1414 - Collinsella sp. | Tibia_length |
| MGS | abundance | Hgn3C.1414 - Collinsella sp. | Radius_length |
| MGS | abundance | Hgn3C.1414 - Collinsella sp. | Radius_SOS |
| MGS | abundance | Hgn3C.1414 - Collinsella sp. | Tibia_SOS |
| MGS | abundance | Hgn3C.1414 - Collinsella sp. | Handgrip |
| MGS | abundance | Hgn3C.1415 - Klebsiella quasipneumoniae subsp. quasipneumoniae | Tibia_length |
| MGS | abundance | Hgn3C.1415 - Klebsiella quasipneumoniae subsp. quasipneumoniae | Radius_length |
| MGS | abundance | Hgn3C.1415 - Klebsiella quasipneumoniae subsp. quasipneumoniae | Radius_SOS |
| MGS | abundance | Hgn3C.1415 - Klebsiella quasipneumoniae subsp. quasipneumoniae | Tibia_SOS |
| MGS | abundance | Hgn3C.1415 - Klebsiella quasipneumoniae subsp. quasipneumoniae | Handgrip |
| MGS | abundance | Hgn3C.1416 - Limosilactobacillus fermentum | Tibia_length |
| MGS | abundance | Hgn3C.1416 - Limosilactobacillus fermentum | Radius_length |
| MGS | abundance | Hgn3C.1416 - Limosilactobacillus fermentum | Radius_SOS |
| MGS | abundance | Hgn3C.1416 - Limosilactobacillus fermentum | Tibia_SOS |
| MGS | abundance | Hgn3C.1416 - Limosilactobacillus fermentum | Handgrip |
| MGS | abundance | Hgn3C.1418 - Carnobacteriaceae sp. | Tibia_length |
| MGS | abundance | Hgn3C.1418 - Carnobacteriaceae sp. | Radius_length |
| MGS | abundance | Hgn3C.1418 - Carnobacteriaceae sp. | Radius_SOS |
| MGS | abundance | Hgn3C.1418 - Carnobacteriaceae sp. | Tibia_SOS |
| MGS | abundance | Hgn3C.1418 - Carnobacteriaceae sp. | Handgrip |
| MGS | abundance | Hgn3C.1419 - Agathobaculum sp. NSJ-28 | Tibia_length |
| MGS | abundance | Hgn3C.1419 - Agathobaculum sp. NSJ-28 | Radius_length |
| MGS | abundance | Hgn3C.1419 - Agathobaculum sp. NSJ-28 | Radius_SOS |
| MGS | abundance | Hgn3C.1419 - Agathobaculum sp. NSJ-28 | Tibia_SOS |
| MGS | abundance | Hgn3C.1419 - Agathobaculum sp. NSJ-28 | Handgrip |
| MGS | abundance | Hgn3C.1420 - Eubacteriales sp. | Tibia_length |
| MGS | abundance | Hgn3C.1420 - Eubacteriales sp. | Radius_length |
| MGS | abundance | Hgn3C.1420 - Eubacteriales sp. | Radius_SOS |
| MGS | abundance | Hgn3C.1420 - Eubacteriales sp. | Tibia_SOS |
| MGS | abundance | Hgn3C.1420 - Eubacteriales sp. | Handgrip |
| MGS | abundance | Hgn3C.1423 - Lachnospiraceae sp. | Tibia_length |
| MGS | abundance | Hgn3C.1423 - Lachnospiraceae sp. | Radius_length |
| MGS | abundance | Hgn3C.1423 - Lachnospiraceae sp. | Radius_SOS |

| | | | |
|---|---|---|---|
| MGS | abundance | Hgn3C.1423 - Lachnospiraceae sp. | Tibia_SOS |
| MGS | abundance | Hgn3C.1423 - Lachnospiraceae sp. | Handgrip |
| MGS | abundance | Hgn3C.1424 - Mogibacterium sp. NSJ-24 | Tibia_length |
| MGS | abundance | Hgn3C.1424 - Mogibacterium sp. NSJ-24 | Radius_length |
| MGS | abundance | Hgn3C.1424 - Mogibacterium sp. NSJ-24 | Radius_SOS |
| MGS | abundance | Hgn3C.1424 - Mogibacterium sp. NSJ-24 | Tibia_SOS |
| MGS | abundance | Hgn3C.1424 - Mogibacterium sp. NSJ-24 | Handgrip |
| MGS | abundance | Hgn3C.1425 - Eubacteriales sp. | Tibia_length |
| MGS | abundance | Hgn3C.1425 - Eubacteriales sp. | Radius_length |
| MGS | abundance | Hgn3C.1425 - Eubacteriales sp. | Radius_SOS |
| MGS | abundance | Hgn3C.1425 - Eubacteriales sp. | Tibia_SOS |
| MGS | abundance | Hgn3C.1425 - Eubacteriales sp. | Handgrip |
| MGS | abundance | Hgn3C.1426 - Limosilactobacillus oris | Tibia_length |
| MGS | abundance | Hgn3C.1426 - Limosilactobacillus oris | Radius_length |
| MGS | abundance | Hgn3C.1426 - Limosilactobacillus oris | Radius_SOS |
| MGS | abundance | Hgn3C.1426 - Limosilactobacillus oris | Tibia_SOS |
| MGS | abundance | Hgn3C.1426 - Limosilactobacillus oris | Handgrip |
| MGS | abundance | Hgn3C.1427 - Erysipelotrichales sp. | Tibia_length |
| MGS | abundance | Hgn3C.1427 - Erysipelotrichales sp. | Radius_length |
| MGS | abundance | Hgn3C.1427 - Erysipelotrichales sp. | Radius_SOS |
| MGS | abundance | Hgn3C.1427 - Erysipelotrichales sp. | Tibia_SOS |
| MGS | abundance | Hgn3C.1427 - Erysipelotrichales sp. | Handgrip |
| MGS | abundance | Hgn3C.1429 - Lachnospiraceae sp. | Tibia_length |
| MGS | abundance | Hgn3C.1429 - Lachnospiraceae sp. | Radius_length |
| MGS | abundance | Hgn3C.1429 - Lachnospiraceae sp. | Radius_SOS |
| MGS | abundance | Hgn3C.1429 - Lachnospiraceae sp. | Tibia_SOS |
| MGS | abundance | Hgn3C.1429 - Lachnospiraceae sp. | Handgrip |
| MGS | abundance | Hgn3C.1430 - Corynebacterium provencense | Tibia_length |
| MGS | abundance | Hgn3C.1430 - Corynebacterium provencense | Radius_length |
| MGS | abundance | Hgn3C.1430 - Corynebacterium provencense | Radius_SOS |
| MGS | abundance | Hgn3C.1430 - Corynebacterium provencense | Tibia_SOS |
| MGS | abundance | Hgn3C.1430 - Corynebacterium provencense | Handgrip |
| MGS | abundance | Hgn3C.1431 - Bacteroidales sp. | Tibia_length |
| MGS | abundance | Hgn3C.1431 - Bacteroidales sp. | Radius_length |
| MGS | abundance | Hgn3C.1431 - Bacteroidales sp. | Radius_SOS |
| MGS | abundance | Hgn3C.1431 - Bacteroidales sp. | Tibia_SOS |
| MGS | abundance | Hgn3C.1431 - Bacteroidales sp. | Handgrip |
| MGS | abundance | Hgn3C.1432 - Eggerthella timonensis | Tibia_length |
| MGS | abundance | Hgn3C.1432 - Eggerthella timonensis | Radius_length |
| MGS | abundance | Hgn3C.1432 - Eggerthella timonensis | Radius_SOS |
| MGS | abundance | Hgn3C.1432 - Eggerthella timonensis | Tibia_SOS |
| MGS | abundance | Hgn3C.1432 - Eggerthella timonensis | Handgrip |
| MGS | abundance | Hgn3C.1434 - Blautia sp. | Tibia_length |
| MGS | abundance | Hgn3C.1434 - Blautia sp. | Radius_length |
| MGS | abundance | Hgn3C.1434 - Blautia sp. | Radius_SOS |
| MGS | abundance | Hgn3C.1434 - Blautia sp. | Tibia_SOS |
| MGS | abundance | Hgn3C.1434 - Blautia sp. | Handgrip |
| MGS | abundance | Hgn3C.1435 - Acinetobacter baumannii | Tibia_length |

| | | | |
|---|---|---|---|
| MGS | abundance | Hgn3C.1435 - Acinetobacter baumannii | Radius_length |
| MGS | abundance | Hgn3C.1435 - Acinetobacter baumannii | Radius_SOS |
| MGS | abundance | Hgn3C.1435 - Acinetobacter baumannii | Tibia_SOS |
| MGS | abundance | Hgn3C.1435 - Acinetobacter baumannii | Handgrip |
| MGS | abundance | Hgn3C.1436 - Bacteria sp. | Tibia_length |
| MGS | abundance | Hgn3C.1436 - Bacteria sp. | Radius_length |
| MGS | abundance | Hgn3C.1436 - Bacteria sp. | Radius_SOS |
| MGS | abundance | Hgn3C.1436 - Bacteria sp. | Tibia_SOS |
| MGS | abundance | Hgn3C.1436 - Bacteria sp. | Handgrip |
| MGS | abundance | Hgn3C.1437 - Candidatus Stoquefichus sp. SB1 | Tibia_length |
| MGS | abundance | Hgn3C.1437 - Candidatus Stoquefichus sp. SB1 | Radius_length |
| MGS | abundance | Hgn3C.1437 - Candidatus Stoquefichus sp. SB1 | Radius_SOS |
| MGS | abundance | Hgn3C.1437 - Candidatus Stoquefichus sp. SB1 | Tibia_SOS |
| MGS | abundance | Hgn3C.1437 - Candidatus Stoquefichus sp. SB1 | Handgrip |
| MGS | abundance | Hgn3C.1438 - Dielma fastidiosa | Tibia_length |
| MGS | abundance | Hgn3C.1438 - Dielma fastidiosa | Radius_length |
| MGS | abundance | Hgn3C.1438 - Dielma fastidiosa | Radius_SOS |
| MGS | abundance | Hgn3C.1438 - Dielma fastidiosa | Tibia_SOS |
| MGS | abundance | Hgn3C.1438 - Dielma fastidiosa | Handgrip |
| MGS | abundance | Hgn3C.1439 - Streptococcus sp. IMAU 99161 | Tibia_length |
| MGS | abundance | Hgn3C.1439 - Streptococcus sp. IMAU 99161 | Radius_length |
| MGS | abundance | Hgn3C.1439 - Streptococcus sp. IMAU 99161 | Radius_SOS |
| MGS | abundance | Hgn3C.1439 - Streptococcus sp. IMAU 99161 | Tibia_SOS |
| MGS | abundance | Hgn3C.1439 - Streptococcus sp. IMAU 99161 | Handgrip |
| MGS | abundance | Hgn3C.1440 - Eubacteriales sp. | Tibia_length |
| MGS | abundance | Hgn3C.1440 - Eubacteriales sp. | Radius_length |
| MGS | abundance | Hgn3C.1440 - Eubacteriales sp. | Radius_SOS |
| MGS | abundance | Hgn3C.1440 - Eubacteriales sp. | Tibia_SOS |
| MGS | abundance | Hgn3C.1440 - Eubacteriales sp. | Handgrip |
| MGS | abundance | Hgn3C.1441 - Lancefieldella sp. | Tibia_length |
| MGS | abundance | Hgn3C.1441 - Lancefieldella sp. | Radius_length |
| MGS | abundance | Hgn3C.1441 - Lancefieldella sp. | Radius_SOS |
| MGS | abundance | Hgn3C.1441 - Lancefieldella sp. | Tibia_SOS |
| MGS | abundance | Hgn3C.1441 - Lancefieldella sp. | Handgrip |
| MGS | abundance | Hgn3C.1443 - Streptococcus sp. | Tibia_length |
| MGS | abundance | Hgn3C.1443 - Streptococcus sp. | Radius_length |
| MGS | abundance | Hgn3C.1443 - Streptococcus sp. | Radius_SOS |
| MGS | abundance | Hgn3C.1443 - Streptococcus sp. | Tibia_SOS |
| MGS | abundance | Hgn3C.1443 - Streptococcus sp. | Handgrip |
| MGS | abundance | Hgn3C.1444 - Ruminococcus sp. 1001175B_160314_E1 | Tibia_length |
| MGS | abundance | Hgn3C.1444 - Ruminococcus sp. 1001175B_160314_E1 | Radius_length |
| MGS | abundance | Hgn3C.1444 - Ruminococcus sp. 1001175B_160314_E1 | Radius_SOS |
| MGS | abundance | Hgn3C.1444 - Ruminococcus sp. 1001175B_160314_E1 | Tibia_SOS |
| MGS | abundance | Hgn3C.1444 - Ruminococcus sp. 1001175B_160314_E1 | Handgrip |
| MGS | abundance | Hgn3C.1445 - Oscillospiraceae sp. | Tibia_length |
| MGS | abundance | Hgn3C.1445 - Oscillospiraceae sp. | Radius_length |
| MGS | abundance | Hgn3C.1445 - Oscillospiraceae sp. | Radius_SOS |
| MGS | abundance | Hgn3C.1445 - Oscillospiraceae sp. | Tibia_SOS |

| | | | |
|---|---|---|---|
| MGS | abundance | Hgn3C.1445 - Oscillospiraceae sp. | Handgrip |
| MGS | abundance | Hgn3C.1446 - Eubacteriales sp. | Tibia_length |
| MGS | abundance | Hgn3C.1446 - Eubacteriales sp. | Radius_length |
| MGS | abundance | Hgn3C.1446 - Eubacteriales sp. | Radius_SOS |
| MGS | abundance | Hgn3C.1446 - Eubacteriales sp. | Tibia_SOS |
| MGS | abundance | Hgn3C.1446 - Eubacteriales sp. | Handgrip |
| MGS | abundance | Hgn3C.1447 - Bacteria sp. | Tibia_length |
| MGS | abundance | Hgn3C.1447 - Bacteria sp. | Radius_length |
| MGS | abundance | Hgn3C.1447 - Bacteria sp. | Radius_SOS |
| MGS | abundance | Hgn3C.1447 - Bacteria sp. | Tibia_SOS |
| MGS | abundance | Hgn3C.1447 - Bacteria sp. | Handgrip |
| MGS | abundance | Hgn3C.1448 - Leuconostoc falkenbergense | Tibia_length |
| MGS | abundance | Hgn3C.1448 - Leuconostoc falkenbergense | Radius_length |
| MGS | abundance | Hgn3C.1448 - Leuconostoc falkenbergense | Radius_SOS |
| MGS | abundance | Hgn3C.1448 - Leuconostoc falkenbergense | Tibia_SOS |
| MGS | abundance | Hgn3C.1448 - Leuconostoc falkenbergense | Handgrip |
| MGS | abundance | Hgn3C.1450 - Lactococcus sp. NH2-7C | Tibia_length |
| MGS | abundance | Hgn3C.1450 - Lactococcus sp. NH2-7C | Radius_length |
| MGS | abundance | Hgn3C.1450 - Lactococcus sp. NH2-7C | Radius_SOS |
| MGS | abundance | Hgn3C.1450 - Lactococcus sp. NH2-7C | Tibia_SOS |
| MGS | abundance | Hgn3C.1450 - Lactococcus sp. NH2-7C | Handgrip |
| MGS | abundance | Hgn3C.1451 - Faecalicatena orotica | Tibia_length |
| MGS | abundance | Hgn3C.1451 - Faecalicatena orotica | Radius_length |
| MGS | abundance | Hgn3C.1451 - Faecalicatena orotica | Radius_SOS |
| MGS | abundance | Hgn3C.1451 - Faecalicatena orotica | Tibia_SOS |
| MGS | abundance | Hgn3C.1451 - Faecalicatena orotica | Handgrip |
| MGS | abundance | Hgn3C.1452 - Raoultibacter timonensis | Tibia_length |
| MGS | abundance | Hgn3C.1452 - Raoultibacter timonensis | Radius_length |
| MGS | abundance | Hgn3C.1452 - Raoultibacter timonensis | Radius_SOS |
| MGS | abundance | Hgn3C.1452 - Raoultibacter timonensis | Tibia_SOS |
| MGS | abundance | Hgn3C.1452 - Raoultibacter timonensis | Handgrip |
| MGS | abundance | Hgn3C.1453 - Collinsella sp. | Tibia_length |
| MGS | abundance | Hgn3C.1453 - Collinsella sp. | Radius_length |
| MGS | abundance | Hgn3C.1453 - Collinsella sp. | Radius_SOS |
| MGS | abundance | Hgn3C.1453 - Collinsella sp. | Tibia_SOS |
| MGS | abundance | Hgn3C.1453 - Collinsella sp. | Handgrip |
| MGS | abundance | Hgn3C.1454 - Escherichia fergusonii | Tibia_length |
| MGS | abundance | Hgn3C.1454 - Escherichia fergusonii | Radius_length |
| MGS | abundance | Hgn3C.1454 - Escherichia fergusonii | Radius_SOS |
| MGS | abundance | Hgn3C.1454 - Escherichia fergusonii | Tibia_SOS |
| MGS | abundance | Hgn3C.1454 - Escherichia fergusonii | Handgrip |
| MGS | abundance | Hgn3C.1455 - Enterocloster sp. | Tibia_length |
| MGS | abundance | Hgn3C.1455 - Enterocloster sp. | Radius_length |
| MGS | abundance | Hgn3C.1455 - Enterocloster sp. | Radius_SOS |
| MGS | abundance | Hgn3C.1455 - Enterocloster sp. | Tibia_SOS |
| MGS | abundance | Hgn3C.1455 - Enterocloster sp. | Handgrip |
| MGS | abundance | Hgn3C.1456 - Eubacteriales sp. | Tibia_length |
| MGS | abundance | Hgn3C.1456 - Eubacteriales sp. | Radius_length |

| | | | |
|---|---|---|---|
| MGS | abundance | Hgn3C.1456 - Eubacteriales sp. | Radius_SOS |
| MGS | abundance | Hgn3C.1456 - Eubacteriales sp. | Tibia_SOS |
| MGS | abundance | Hgn3C.1456 - Eubacteriales sp. | Handgrip |
| MGS | abundance | Hgn3C.1458 - Intestinimonas timonensis | Tibia_length |
| MGS | abundance | Hgn3C.1458 - Intestinimonas timonensis | Radius_length |
| MGS | abundance | Hgn3C.1458 - Intestinimonas timonensis | Radius_SOS |
| MGS | abundance | Hgn3C.1458 - Intestinimonas timonensis | Tibia_SOS |
| MGS | abundance | Hgn3C.1458 - Intestinimonas timonensis | Handgrip |
| MGS | abundance | Hgn3C.1460 - Blautia sp. | Tibia_length |
| MGS | abundance | Hgn3C.1460 - Blautia sp. | Radius_length |
| MGS | abundance | Hgn3C.1460 - Blautia sp. | Radius_SOS |
| MGS | abundance | Hgn3C.1460 - Blautia sp. | Tibia_SOS |
| MGS | abundance | Hgn3C.1460 - Blautia sp. | Handgrip |
| MGS | abundance | Hgn3C.1461 - Eubacteriales sp. | Tibia_length |
| MGS | abundance | Hgn3C.1461 - Eubacteriales sp. | Radius_length |
| MGS | abundance | Hgn3C.1461 - Eubacteriales sp. | Radius_SOS |
| MGS | abundance | Hgn3C.1461 - Eubacteriales sp. | Tibia_SOS |
| MGS | abundance | Hgn3C.1461 - Eubacteriales sp. | Handgrip |
| MGS | abundance | Hgn3C.1463 - Enterobacter sp. | Tibia_length |
| MGS | abundance | Hgn3C.1463 - Enterobacter sp. | Radius_length |
| MGS | abundance | Hgn3C.1463 - Enterobacter sp. | Radius_SOS |
| MGS | abundance | Hgn3C.1463 - Enterobacter sp. | Tibia_SOS |
| MGS | abundance | Hgn3C.1463 - Enterobacter sp. | Handgrip |
| MGS | abundance | Hgn3C.1465 - Clostridia sp. | Tibia_length |
| MGS | abundance | Hgn3C.1465 - Clostridia sp. | Radius_length |
| MGS | abundance | Hgn3C.1465 - Clostridia sp. | Radius_SOS |
| MGS | abundance | Hgn3C.1465 - Clostridia sp. | Tibia_SOS |
| MGS | abundance | Hgn3C.1465 - Clostridia sp. | Handgrip |
| MGS | abundance | Hgn3C.1467 - Eubacteriales sp. | Tibia_length |
| MGS | abundance | Hgn3C.1467 - Eubacteriales sp. | Radius_length |
| MGS | abundance | Hgn3C.1467 - Eubacteriales sp. | Radius_SOS |
| MGS | abundance | Hgn3C.1467 - Eubacteriales sp. | Tibia_SOS |
| MGS | abundance | Hgn3C.1467 - Eubacteriales sp. | Handgrip |
| MGS | abundance | Hgn3C.1468 - Staphylococcus aureus | Tibia_length |
| MGS | abundance | Hgn3C.1468 - Staphylococcus aureus | Radius_length |
| MGS | abundance | Hgn3C.1468 - Staphylococcus aureus | Radius_SOS |
| MGS | abundance | Hgn3C.1468 - Staphylococcus aureus | Tibia_SOS |
| MGS | abundance | Hgn3C.1468 - Staphylococcus aureus | Handgrip |
| MGS | abundance | Hgn3C.1469 - Lachnospiraceae sp. | Tibia_length |
| MGS | abundance | Hgn3C.1469 - Lachnospiraceae sp. | Radius_length |
| MGS | abundance | Hgn3C.1469 - Lachnospiraceae sp. | Radius_SOS |
| MGS | abundance | Hgn3C.1469 - Lachnospiraceae sp. | Tibia_SOS |
| MGS | abundance | Hgn3C.1469 - Lachnospiraceae sp. | Handgrip |
| MGS | abundance | Hgn3C.1471 - Eggerthella sp. | Tibia_length |
| MGS | abundance | Hgn3C.1471 - Eggerthella sp. | Radius_length |
| MGS | abundance | Hgn3C.1471 - Eggerthella sp. | Radius_SOS |
| MGS | abundance | Hgn3C.1471 - Eggerthella sp. | Tibia_SOS |
| MGS | abundance | Hgn3C.1471 - Eggerthella sp. | Handgrip |

| | | | |
|---|---|---|---|
| MGS | abundance | Hgn3C.1472 - Amedibacillus dolichus | Tibia_length |
| MGS | abundance | Hgn3C.1472 - Amedibacillus dolichus | Radius_length |
| MGS | abundance | Hgn3C.1472 - Amedibacillus dolichus | Radius_SOS |
| MGS | abundance | Hgn3C.1472 - Amedibacillus dolichus | Tibia_SOS |
| MGS | abundance | Hgn3C.1472 - Amedibacillus dolichus | Handgrip |

| Effect size | Standard e | Bias mode | Test statist | Sample siz | Degrees of | P-value | FDR | Model formula fit |
|---|---|---|---|---|---|---|---|---|
| 0,00423 | 0,049202 | -0,07874 | 0,085979 | 421 | 413,0724 | 0,931525 | 0,991839 | y ~ current_feature + S |
| 0,024423 | 0,061765 | 0,034007 | 0,395421 | 421 | 261,9694 | 0,692854 | 0,916569 | y ~ current_feature + S |
| 0,101956 | 0,049514 | -0,04751 | 2,059146 | 420 | 403,6533 | 0,040121 | 0,612508 | y ~ current_feature + S |
| -0,01113 | 0,054585 | 0,061498 | -0,20396 | 367 | 335,5832 | 0,838509 | 0,996179 | y ~ current_feature + S |
| 0,009443 | 0,060435 | -0,02429 | 0,156243 | 280 | 273,7698 | 0,875956 | 0,984468 | y ~ current_feature + S |
| 0,01462 | 0,050486 | -0,07874 | 0,289595 | 415 | 392,254 | 0,772279 | 0,965962 | y ~ current_feature + S |
| 0,027272 | 0,065364 | 0,034007 | 0,417237 | 415 | 233,8815 | 0,676887 | 0,912427 | y ~ current_feature + S |
| 0,043118 | 0,051355 | -0,04751 | 0,839599 | 414 | 378,4587 | 0,401663 | 0,803665 | y ~ current_feature + S |
| 0,045009 | 0,054776 | 0,061498 | 0,821707 | 361 | 332,6186 | 0,411832 | 0,934833 | y ~ current_feature + S |
| 0,032157 | 0,06081 | -0,02429 | 0,528806 | 276 | 270,1439 | 0,597374 | 0,924682 | y ~ current_feature + S |
| -0,04648 | 0,05257 | -0,07874 | -0,8841 | 388 | 361,0588 | 0,377229 | 0,789252 | y ~ current_feature + S |
| -0,14828 | 0,06754 | 0,034007 | -2,19542 | 388 | 214,3972 | 0,029206 | 0,347616 | y ~ current_feature + S |
| -0,04404 | 0,053134 | -0,04751 | -0,82886 | 387 | 353,5153 | 0,407742 | 0,803665 | y ~ current_feature + S |
| 0,016179 | 0,058231 | 0,061498 | 0,27784 | 335 | 294,8309 | 0,78133 | 0,978149 | y ~ current_feature + S |
| -0,10497 | 0,062276 | -0,02429 | -1,68562 | 258 | 255 | 0,093092 | 0,605474 | y ~ current_feature + S |
| 0,056146 | 0,051914 | -0,07874 | 1,081504 | 421 | 369,8727 | 0,280178 | 0,71741 | y ~ current_feature + S |
| 0,071604 | 0,068417 | 0,034007 | 1,046583 | 421 | 212,5409 | 0,296481 | 0,718742 | y ~ current_feature + S |
| 0,090739 | 0,053434 | -0,04751 | 1,698145 | 420 | 347,3555 | 0,090376 | 0,640171 | y ~ current_feature + S |
| 0,015664 | 0,056697 | 0,061498 | 0,276277 | 367 | 311,0055 | 0,782519 | 0,978149 | y ~ current_feature + S |
| 0,155065 | 0,059357 | -0,02429 | 2,612392 | 280 | 277 | 0,009482 | 0,295269 | y ~ current_feature + S |
| 0,045947 | 0,05055 | -0,07874 | 0,908944 | 403 | 390,5224 | 0,36394 | 0,773896 | y ~ current_feature + S |
| -0,04476 | 0,063625 | 0,034007 | -0,70353 | 403 | 246,5305 | 0,482391 | 0,831402 | y ~ current_feature + S |
| -0,03818 | 0,051093 | -0,04751 | -0,74724 | 402 | 382,5131 | 0,455375 | 0,832044 | y ~ current_feature + S |
| -0,01836 | 0,054228 | 0,061498 | -0,33853 | 349 | 339,9453 | 0,735175 | 0,972573 | y ~ current_feature + S |
| 0,013528 | 0,062863 | -0,02429 | 0,215196 | 268 | 253,0027 | 0,829788 | 0,976969 | y ~ current_feature + S |
| -0,02123 | 0,051389 | -0,07874 | -0,41306 | 395 | 378,4938 | 0,679797 | 0,934653 | y ~ current_feature + S |
| 0,005939 | 0,065397 | 0,034007 | 0,090814 | 395 | 233,8104 | 0,927718 | 0,9732 | y ~ current_feature + S |
| -0,02164 | 0,052277 | -0,04751 | -0,414 | 394 | 365,7453 | 0,679114 | 0,92829 | y ~ current_feature + S |
| -0,01992 | 0,056462 | 0,061498 | -0,35273 | 343 | 313,5582 | 0,724529 | 0,972573 | y ~ current_feature + S |
| -0,01512 | 0,063085 | -0,02429 | -0,23964 | 262 | 251,2185 | 0,810808 | 0,974251 | y ~ current_feature + S |
| 0,029819 | 0,05051 | -0,07874 | 0,590367 | 418 | 391,6176 | 0,555285 | 0,891348 | y ~ current_feature + S |
| -0,04913 | 0,065719 | 0,034007 | -0,74762 | 418 | 230,9758 | 0,455451 | 0,821488 | y ~ current_feature + S |
| -0,0321 | 0,05145 | -0,04751 | -0,62393 | 417 | 377,3792 | 0,533051 | 0,883994 | y ~ current_feature + S |
| -0,0579 | 0,055981 | 0,061498 | -1,03436 | 365 | 318,0232 | 0,301756 | 0,893818 | y ~ current_feature + S |
| 0,057939 | 0,060218 | -0,02429 | 0,96216 | 278 | 274,849 | 0,336815 | 0,785829 | y ~ current_feature + S |
| -0,01021 | 0,056058 | -0,07874 | -0,18215 | 338 | 318,1874 | 0,855584 | 0,983548 | y ~ current_feature + S |
| 0,011115 | 0,073101 | 0,034007 | 0,152045 | 338 | 187,1093 | 0,879315 | 0,96591 | y ~ current_feature + S |
| 0,096416 | 0,057229 | -0,04751 | 1,684759 | 337 | 302,4954 | 0,093066 | 0,640171 | y ~ current_feature + S |
| 0,009498 | 0,059531 | 0,061498 | 0,15954 | 293 | 282,1464 | 0,873358 | 0,996179 | y ~ current_feature + S |
| 0,066181 | 0,068457 | -0,02429 | 0,966753 | 225 | 212,4478 | 0,334767 | 0,785829 | y ~ current_feature + S |
| 0,008659 | 0,049947 | -0,07874 | 0,173369 | 421 | 400,8257 | 0,862449 | 0,985453 | y ~ current_feature + S |
| 0,023878 | 0,064311 | 0,034007 | 0,371283 | 421 | 241,6481 | 0,710752 | 0,920814 | y ~ current_feature + S |
| 0,047071 | 0,050744 | -0,04751 | 0,927614 | 420 | 387,4983 | 0,354185 | 0,795089 | y ~ current_feature + S |
| -0,00941 | 0,055588 | 0,061498 | -0,16936 | 367 | 323,5944 | 0,86562 | 0,996179 | y ~ current_feature + S |
| 0,04392 | 0,060214 | -0,02429 | 0,729394 | 280 | 275,2754 | 0,466381 | 0,876224 | y ~ current_feature + S |
| 0,064893 | 0,051208 | -0,07874 | 1,267243 | 415 | 379,7486 | 0,205845 | 0,622938 | y ~ current_feature + S |
| 0,086323 | 0,067115 | 0,034007 | 1,286197 | 415 | 220,352 | 0,199724 | 0,647595 | y ~ current_feature + S |

| | | | | | | | | |
|---|---|---|---|---|---|---|---|---|
| 0,088952 | 0,052428 | -0,04751 | 1,69666 | 414 | 360,9328 | 0,090623 | 0,640171 | y ~ current_feature + S |
| 0,06274 | 0,055771 | 0,061498 | 1,124967 | 361 | 320,2409 | 0,261445 | 0,893818 | y ~ current_feature + S |
| 0,149647 | 0,061257 | -0,02429 | 2,442932 | 276 | 260,5251 | 0,015233 | 0,417502 | y ~ current_feature + S |
| -0,05607 | 0,054469 | -0,07874 | -1,02932 | 339 | 336 | 0,304069 | 0,718687 | y ~ current_feature + S |
| -0,10861 | 0,054232 | 0,034007 | -2,00276 | 339 | 336 | 0,046006 | 0,400524 | y ~ current_feature + S |
| -0,05682 | 0,054548 | -0,04751 | -1,04164 | 338 | 335 | 0,29833 | 0,753461 | y ~ current_feature + S |
| -0,13668 | 0,058373 | 0,061498 | -2,34144 | 291 | 288 | 0,019891 | 0,609646 | y ~ current_feature + S |
| -0,0973 | 0,066647 | -0,02429 | -1,45989 | 226 | 223 | 0,145728 | 0,619762 | y ~ current_feature + S |
| -0,081 | 0,05904 | -0,07874 | -1,37195 | 288 | 285 | 0,171158 | 0,550681 | y ~ current_feature + S |
| -0,11002 | 0,058875 | 0,034007 | -1,8687 | 288 | 285 | 0,062691 | 0,437643 | y ~ current_feature + S |
| 0,001515 | 0,059339 | -0,04751 | 0,025537 | 287 | 284 | 0,979644 | 0,992311 | y ~ current_feature + S |
| -0,03373 | 0,063721 | 0,061498 | -0,52926 | 249 | 246 | 0,597102 | 0,945257 | y ~ current_feature + S |
| -0,01922 | 0,072726 | -0,02429 | -0,2643 | 192 | 189 | 0,791835 | 0,967756 | y ~ current_feature + S |
| -0,02246 | 0,063872 | -0,07874 | -0,35165 | 248 | 245 | 0,725402 | 0,955155 | y ~ current_feature + S |
| 0,133254 | 0,063318 | 0,034007 | 2,104525 | 248 | 245 | 0,036352 | 0,384289 | y ~ current_feature + S |
| 0,041277 | 0,063833 | -0,04751 | 0,646644 | 248 | 245 | 0,518467 | 0,873954 | y ~ current_feature + S |
| -0,02168 | 0,067558 | 0,061498 | -0,32097 | 222 | 219 | 0,748542 | 0,972573 | y ~ current_feature + S |
| 0,110687 | 0,078085 | -0,02429 | 1,41753 | 165 | 162 | 0,158249 | 0,623497 | y ~ current_feature + S |
| 0,031857 | 0,052006 | -0,07874 | 0,612564 | 389 | 369,3679 | 0,540542 | 0,882344 | y ~ current_feature + S |
| -0,02603 | 0,066591 | 0,034007 | -0,39095 | 389 | 225,3586 | 0,696204 | 0,918206 | y ~ current_feature + S |
| 0,046829 | 0,052746 | -0,04751 | 0,887822 | 389 | 358,6482 | 0,375232 | 0,803665 | y ~ current_feature + S |
| 0,118537 | 0,057733 | 0,061498 | 2,053191 | 336 | 295,8037 | 0,040933 | 0,658491 | y ~ current_feature + S |
| 0,095521 | 0,062882 | -0,02429 | 1,519038 | 259 | 250,5888 | 0,130013 | 0,617591 | y ~ current_feature + S |
| 0,003856 | 0,055888 | -0,07874 | 0,068988 | 347 | 320,1479 | 0,945042 | 0,994607 | y ~ current_feature + S |
| -0,05808 | 0,072054 | 0,034007 | -0,80605 | 347 | 191,9622 | 0,421212 | 0,791109 | y ~ current_feature + S |
| 0,046247 | 0,056075 | -0,04751 | 0,824745 | 346 | 317,3493 | 0,410136 | 0,803665 | y ~ current_feature + S |
| -0,06022 | 0,06074 | 0,061498 | -0,99137 | 298 | 270,0692 | 0,322392 | 0,893818 | y ~ current_feature + S |
| -0,04808 | 0,067013 | -0,02429 | -0,7175 | 231 | 222,1673 | 0,473817 | 0,876561 | y ~ current_feature + S |
| -0,04313 | 0,05343 | -0,07874 | -0,80723 | 377 | 349,6365 | 0,420081 | 0,811887 | y ~ current_feature + S |
| -0,03943 | 0,069184 | 0,034007 | -0,56994 | 377 | 208,5995 | 0,569333 | 0,863333 | y ~ current_feature + S |
| 0,124461 | 0,055114 | -0,04751 | 2,258246 | 376 | 324,1139 | 0,024595 | 0,568759 | y ~ current_feature + S |
| 0,006877 | 0,057329 | 0,061498 | 0,119959 | 328 | 304,2476 | 0,904595 | 0,996179 | y ~ current_feature + S |
| 0,079123 | 0,063766 | -0,02429 | 1,240844 | 251 | 244,3973 | 0,215853 | 0,682612 | y ~ current_feature + S |
| 0,008179 | 0,071426 | -0,07874 | 0,114508 | 199 | 196 | 0,908953 | 0,987929 | y ~ current_feature + S |
| -0,12397 | 0,070878 | 0,034007 | -1,74905 | 199 | 196 | 0,081847 | 0,473527 | y ~ current_feature + S |
| -0,17694 | 0,070482 | -0,04751 | -2,51046 | 198 | 195 | 0,01287 | 0,526293 | y ~ current_feature + S |
| -0,06962 | 0,082972 | 0,061498 | -0,83903 | 164 | 144,5521 | 0,402836 | 0,93433 | y ~ current_feature + S |
| -0,17429 | 0,086697 | -0,02429 | -2,01035 | 132 | 129 | 0,046479 | 0,525585 | y ~ current_feature + S |
| -0,01745 | 0,056069 | -0,07874 | -0,31123 | 321 | 318 | 0,75583 | 0,964335 | y ~ current_feature + S |
| -0,03924 | 0,056034 | 0,034007 | -0,70033 | 321 | 318 | 0,484235 | 0,831402 | y ~ current_feature + S |
| 0,095482 | 0,055821 | -0,04751 | 1,710509 | 321 | 318 | 0,088147 | 0,640171 | y ~ current_feature + S |
| -0,01578 | 0,060738 | 0,061498 | -0,2598 | 274 | 271 | 0,795212 | 0,984075 | y ~ current_feature + S |
| -0,10518 | 0,068461 | -0,02429 | -1,53639 | 214 | 211 | 0,125942 | 0,617591 | y ~ current_feature + S |
| -0,00379 | 0,05194 | -0,07874 | -0,07289 | 374 | 370,6763 | 0,941933 | 0,994607 | y ~ current_feature + S |
| -0,06038 | 0,062688 | 0,034007 | -0,96317 | 374 | 253,5397 | 0,33638 | 0,73474 | y ~ current_feature + S |
| 0,043731 | 0,052184 | -0,04751 | 0,838023 | 374 | 366,5236 | 0,402564 | 0,803665 | y ~ current_feature + S |
| 0,022577 | 0,05659 | 0,061498 | 0,398954 | 322 | 312,1068 | 0,6902 | 0,971004 | y ~ current_feature + S |
| -0,0787 | 0,06547 | -0,02429 | -1,20208 | 249 | 231,8541 | 0,23056 | 0,695919 | y ~ current_feature + S |

| | | | | | | | | |
|---|---|---|---|---|---|---|---|---|
| -0,00645 | 0,049744 | -0,07874 | -0,12964 | 418 | 404,116 | 0,896916 | 0,987929 | y ~ current_feature + S |
| 0,049664 | 0,063271 | 0,034007 | 0,784935 | 418 | 249,1829 | 0,433237 | 0,796709 | y ~ current_feature + S |
| 0,028905 | 0,050399 | -0,04751 | 0,573525 | 417 | 393,3659 | 0,566617 | 0,903656 | y ~ current_feature + S |
| 0,089076 | 0,055059 | 0,061498 | 1,617827 | 364 | 327,2508 | 0,106663 | 0,8197 | y ~ current_feature + S |
| 0,024455 | 0,062381 | -0,02429 | 0,392033 | 278 | 256,8262 | 0,695359 | 0,928819 | y ~ current_feature + S |
| 0,146297 | 0,050073 | -0,07874 | 2,921669 | 421 | 390,2971 | 0,003684 | 0,069632 | y ~ current_feature + S |
| 0,229413 | 0,06443 | 0,034007 | 3,560683 | 421 | 228,2173 | 4,50E-04 | 0,067708 | y ~ current_feature + S |
| 0,094593 | 0,051466 | -0,04751 | 1,837992 | 420 | 374,1632 | 0,066856 | 0,640171 | y ~ current_feature + S |
| -0,00812 | 0,056506 | 0,061498 | -0,14377 | 367 | 313,1697 | 0,885774 | 0,996179 | y ~ current_feature + S |
| 0,144817 | 0,060025 | -0,02429 | 2,412625 | 280 | 271,7285 | 0,016501 | 0,428958 | y ~ current_feature + S |
| 0,133796 | 0,059053 | -0,07874 | 2,265715 | 321 | 281,6286 | 0,024228 | 0,197019 | y ~ current_feature + S |
| 0,054279 | 0,078542 | 0,034007 | 0,691086 | 321 | 161,6276 | 0,490503 | 0,837983 | y ~ current_feature + S |
| 0,157421 | 0,059105 | -0,04751 | 2,663414 | 320 | 279,1598 | 0,008185 | 0,432614 | y ~ current_feature + S |
| 0,037046 | 0,063 | 0,061498 | 0,588036 | 277 | 251,6101 | 0,557035 | 0,937665 | y ~ current_feature + S |
| 0,090102 | 0,068563 | -0,02429 | 1,31415 | 214 | 211 | 0,190223 | 0,65347 | y ~ current_feature + S |
| 0,046425 | 0,050061 | -0,07874 | 0,927366 | 418 | 398,1673 | 0,354298 | 0,765762 | y ~ current_feature + S |
| -0,0301 | 0,064391 | 0,034007 | -0,46747 | 418 | 240,9649 | 0,640584 | 0,899624 | y ~ current_feature + S |
| 0,004264 | 0,051018 | -0,04751 | 0,083569 | 417 | 384,1917 | 0,933443 | 0,988194 | y ~ current_feature + S |
| 0,04176 | 0,055191 | 0,061498 | 0,756645 | 365 | 327,7222 | 0,449806 | 0,937665 | y ~ current_feature + S |
| -0,0233 | 0,061203 | -0,02429 | -0,38069 | 278 | 266,8181 | 0,703736 | 0,93118 | y ~ current_feature + S |
| 0,076012 | 0,053767 | -0,07874 | 1,413726 | 367 | 343,9168 | 0,158347 | 0,527823 | y ~ current_feature + S |
| 0,024445 | 0,068828 | 0,034007 | 0,355167 | 367 | 210,965 | 0,722819 | 0,920814 | y ~ current_feature + S |
| 0,019455 | 0,054754 | -0,04751 | 0,355312 | 367 | 333,4332 | 0,722581 | 0,93314 | y ~ current_feature + S |
| -0,03516 | 0,057169 | 0,061498 | -0,61505 | 321 | 305,5936 | 0,53898 | 0,937665 | y ~ current_feature + S |
| 0,074341 | 0,064752 | -0,02429 | 1,148089 | 245 | 237,1848 | 0,252089 | 0,717483 | y ~ current_feature + S |
| 0,040621 | 0,084146 | -0,07874 | 0,482743 | 144 | 141 | 0,630027 | 0,92504 | y ~ current_feature + S |
| -0,04034 | 0,084147 | 0,034007 | -0,47944 | 144 | 141 | 0,632369 | 0,899624 | y ~ current_feature + S |
| -0,05826 | 0,084072 | -0,04751 | -0,69302 | 144 | 141 | 0,489436 | 0,860292 | y ~ current_feature + S |
| -0,07301 | 0,090294 | 0,061498 | -0,80853 | 125 | 122 | 0,420358 | 0,937665 | y ~ current_feature + S |
| -0,12516 | 0,10288 | -0,02429 | -1,21661 | 96 | 93 | 0,226833 | 0,695919 | y ~ current_feature + S |
| 0,004643 | 0,055265 | -0,07874 | 0,084016 | 346 | 327,4043 | 0,933095 | 0,992084 | y ~ current_feature + S |
| -0,00704 | 0,070757 | 0,034007 | -0,09949 | 346 | 199,7282 | 0,920852 | 0,9732 | y ~ current_feature + S |
| -0,07437 | 0,056063 | -0,04751 | -1,3266 | 346 | 316,3962 | 0,185597 | 0,704811 | y ~ current_feature + S |
| -0,06865 | 0,059217 | 0,061498 | -1,15938 | 300 | 283,8319 | 0,247274 | 0,893818 | y ~ current_feature + S |
| -0,03681 | 0,067033 | -0,02429 | -0,54914 | 230 | 222,2451 | 0,583459 | 0,924682 | y ~ current_feature + S |
| 0,139526 | 0,063654 | -0,07874 | 2,191951 | 245 | 242 | 0,029336 | 0,219281 | y ~ current_feature + S |
| 0,065354 | 0,064145 | 0,034007 | 1,018845 | 245 | 242 | 0,309294 | 0,718742 | y ~ current_feature + S |
| 0,120015 | 0,06395 | -0,04751 | 1,876691 | 244 | 241 | 0,061769 | 0,640171 | y ~ current_feature + S |
| 0,120692 | 0,069164 | 0,061498 | 1,745008 | 209 | 206 | 0,082475 | 0,753473 | y ~ current_feature + S |
| 0,126236 | 0,078425 | -0,02429 | 1,609648 | 163 | 160 | 0,109446 | 0,605474 | y ~ current_feature + S |
| 0,051181 | 0,065133 | -0,07874 | 0,785799 | 272 | 235,1069 | 0,432777 | 0,814177 | y ~ current_feature + S |
| -0,07469 | 0,087642 | 0,034007 | -0,85224 | 272 | 129,4642 | 0,395656 | 0,775482 | y ~ current_feature + S |
| 0,040947 | 0,06593 | -0,04751 | 0,621062 | 271 | 229,6687 | 0,535175 | 0,883994 | y ~ current_feature + S |
| 0,066306 | 0,06873 | 0,061498 | 0,96473 | 232 | 210,7647 | 0,335786 | 0,893818 | y ~ current_feature + S |
| -0,0826 | 0,075427 | -0,02429 | -1,09504 | 181 | 174,5719 | 0,275006 | 0,734108 | y ~ current_feature + S |
| 0,033861 | 0,055541 | -0,07874 | 0,609648 | 376 | 323,7965 | 0,542522 | 0,882344 | y ~ current_feature + S |
| 0,010155 | 0,072483 | 0,034007 | 0,140108 | 376 | 190,3204 | 0,888723 | 0,96591 | y ~ current_feature + S |
| 0,021319 | 0,057398 | -0,04751 | 0,371427 | 375 | 303,3978 | 0,710579 | 0,93314 | y ~ current_feature + S |

| | | | | | | | | |
|---|---|---|---|---|---|---|---|---|
| 0,036049 | 0,061519 | 0,061498 | 0,585976 | 326 | 263,883 | 0,558393 | 0,937665 | y ~ current_feature + S |
| 0,062229 | 0,063505 | -0,02429 | 0,979902 | 250 | 247 | 0,328093 | 0,775683 | y ~ current_feature + S |
| 0,067826 | 0,069012 | -0,07874 | 0,982807 | 212 | 209 | 0,326839 | 0,726391 | y ~ current_feature + S |
| -0,03828 | 0,069121 | 0,034007 | -0,5538 | 212 | 209 | 0,58031 | 0,869291 | y ~ current_feature + S |
| -0,06848 | 0,069009 | -0,04751 | -0,99228 | 212 | 209 | 0,322208 | 0,774585 | y ~ current_feature + S |
| -0,07339 | 0,074751 | 0,061498 | -0,98177 | 181 | 178 | 0,327546 | 0,893818 | y ~ current_feature + S |
| -0,03692 | 0,085068 | -0,02429 | -0,43398 | 141 | 138 | 0,664982 | 0,926342 | y ~ current_feature + S |
| 0,101502 | 0,053945 | -0,07874 | 1,881563 | 377 | 340,0885 | 0,060749 | 0,325854 | y ~ current_feature + S |
| -0,04308 | 0,069857 | 0,034007 | -0,61671 | 377 | 204,5375 | 0,538113 | 0,850862 | y ~ current_feature + S |
| 0,10383 | 0,053697 | -0,04751 | 1,93362 | 376 | 343,0754 | 0,053982 | 0,640171 | y ~ current_feature + S |
| 0,057469 | 0,057363 | 0,061498 | 1,001842 | 326 | 302,8957 | 0,317219 | 0,893818 | y ~ current_feature + S |
| 0,051372 | 0,064895 | -0,02429 | 0,791621 | 251 | 236,8271 | 0,429374 | 0,849564 | y ~ current_feature + S |
| 0,171304 | 0,071206 | -0,07874 | 2,405741 | 211 | 191,4376 | 0,017091 | 0,176938 | y ~ current_feature + S |
| 0,137604 | 0,094045 | 0,034007 | 1,463168 | 211 | 110,9233 | 0,146249 | 0,594724 | y ~ current_feature + S |
| 0,077269 | 0,077633 | -0,04751 | 0,995304 | 210 | 164,9309 | 0,321047 | 0,774585 | y ~ current_feature + S |
| -0,00979 | 0,077544 | 0,061498 | -0,12627 | 180 | 166,2876 | 0,899671 | 0,996179 | y ~ current_feature + S |
| 0,052733 | 0,085317 | -0,02429 | 0,618086 | 140 | 137 | 0,537545 | 0,906112 | y ~ current_feature + S |
| 0,032766 | 0,053611 | -0,07874 | 0,611184 | 380 | 347,5507 | 0,541477 | 0,882344 | y ~ current_feature + S |
| -3,07E-04 | 0,069086 | 0,034007 | -0,00445 | 380 | 209,5202 | 0,996453 | 0,997989 | y ~ current_feature + S |
| 0,096914 | 0,054331 | -0,04751 | 1,783775 | 380 | 335,593 | 0,075364 | 0,640171 | y ~ current_feature + S |
| 0,028918 | 0,057752 | 0,061498 | 0,500729 | 328 | 299,5712 | 0,61693 | 0,948092 | y ~ current_feature + S |
| 9,79E-04 | 0,06355 | -0,02429 | 0,015406 | 253 | 247,6082 | 0,98772 | 0,99444 | y ~ current_feature + S |
| -0,01147 | 0,050273 | -0,07874 | -0,22815 | 404 | 395,6195 | 0,819649 | 0,979871 | y ~ current_feature + S |
| -0,16999 | 0,062758 | 0,034007 | -2,70873 | 404 | 246,5636 | 0,007227 | 0,205704 | y ~ current_feature + S |
| -0,05735 | 0,050667 | -0,04751 | -1,13194 | 403 | 388,261 | 0,258358 | 0,742928 | y ~ current_feature + S |
| -0,02922 | 0,055772 | 0,061498 | -0,52385 | 350 | 321,2142 | 0,600742 | 0,945257 | y ~ current_feature + S |
| -0,11705 | 0,063684 | -0,02429 | -1,83801 | 269 | 243,195 | 0,06728 | 0,592706 | y ~ current_feature + S |
| -0,00132 | 0,05517 | -0,07874 | -0,02395 | 368 | 328,5474 | 0,980904 | 0,996318 | y ~ current_feature + S |
| -0,01533 | 0,069757 | 0,034007 | -0,21982 | 368 | 205,4557 | 0,826233 | 0,953842 | y ~ current_feature + S |
| 0,090133 | 0,056288 | -0,04751 | 1,601303 | 367 | 313,0642 | 0,110318 | 0,640171 | y ~ current_feature + S |
| -0,0173 | 0,057864 | 0,061498 | -0,299 | 319 | 298,5787 | 0,765151 | 0,972573 | y ~ current_feature + S |
| 0,120285 | 0,064424 | -0,02429 | 1,867069 | 245 | 237,449 | 0,063124 | 0,569659 | y ~ current_feature + S |
| 0,029436 | 0,049508 | -0,07874 | 0,59458 | 415 | 407,6361 | 0,552454 | 0,889995 | y ~ current_feature + S |
| -0,03534 | 0,061958 | 0,034007 | -0,57035 | 415 | 260,1755 | 0,568932 | 0,863333 | y ~ current_feature + S |
| 0,009662 | 0,049982 | -0,04751 | 0,193316 | 414 | 400,2445 | 0,84681 | 0,980218 | y ~ current_feature + S |
| 0,024787 | 0,053472 | 0,061498 | 0,463553 | 362 | 349,5264 | 0,643256 | 0,956496 | y ~ current_feature + S |
| 0,042812 | 0,061574 | -0,02429 | 0,695301 | 276 | 263,2748 | 0,48748 | 0,888665 | y ~ current_feature + S |
| -0,01679 | 0,055208 | -0,07874 | -0,30418 | 331 | 328 | 0,761186 | 0,965008 | y ~ current_feature + S |
| -0,05867 | 0,055121 | 0,034007 | -1,06448 | 331 | 328 | 0,287894 | 0,718742 | y ~ current_feature + S |
| 0,087046 | 0,05509 | -0,04751 | 1,580069 | 330 | 327 | 0,115058 | 0,640171 | y ~ current_feature + S |
| -0,05849 | 0,059237 | 0,061498 | -0,98736 | 287 | 284 | 0,324304 | 0,893818 | y ~ current_feature + S |
| -0,01709 | 0,067875 | -0,02429 | -0,25185 | 220 | 217 | 0,801392 | 0,968297 | y ~ current_feature + S |
| -0,11221 | 0,056898 | -0,07874 | -1,97221 | 308 | 305 | 0,049489 | 0,29534 | y ~ current_feature + S |
| -0,07935 | 0,057079 | 0,034007 | -1,39013 | 308 | 305 | 0,165504 | 0,612363 | y ~ current_feature + S |
| -0,00315 | 0,05726 | -0,04751 | -0,05507 | 308 | 305 | 0,956122 | 0,990156 | y ~ current_feature + S |
| -0,09867 | 0,060787 | 0,061498 | -1,62315 | 271 | 268 | 0,105733 | 0,8197 | y ~ current_feature + S |
| -0,15758 | 0,069481 | -0,02429 | -2,268 | 205 | 202 | 0,024387 | 0,485321 | y ~ current_feature + S |
| 0,059304 | 0,055554 | -0,07874 | 1,067495 | 359 | 322,8767 | 0,286546 | 0,718687 | y ~ current_feature + S |

| | | | | | | | | |
|---|---|---|---|---|---|---|---|---|
| -0,07735 | 0,073023 | 0,034007 | -1,05918 | 359 | 186,4109 | 0,290887 | 0,718742 | y ~ current_feature + S |
| -0,02665 | 0,056983 | -0,04751 | -0,46771 | 358 | 307,7521 | 0,640324 | 0,925756 | y ~ current_feature + S |
| -0,035 | 0,059862 | 0,061498 | -0,58474 | 310 | 278,7221 | 0,559198 | 0,937665 | y ~ current_feature + S |
| -0,04748 | 0,065525 | -0,02429 | -0,7246 | 239 | 232,3822 | 0,469424 | 0,876224 | y ~ current_feature + S |
| -0,02505 | 0,050326 | -0,07874 | -0,49778 | 410 | 394,5809 | 0,618918 | 0,919937 | y ~ current_feature + S |
| -0,00943 | 0,065108 | 0,034007 | -0,14477 | 410 | 235,883 | 0,88502 | 0,96591 | y ~ current_feature + S |
| 0,082642 | 0,051073 | -0,04751 | 1,618134 | 409 | 380,7567 | 0,106462 | 0,640171 | y ~ current_feature + S |
| -0,02437 | 0,054733 | 0,061498 | -0,44527 | 356 | 333,612 | 0,656415 | 0,957207 | y ~ current_feature + S |
| -0,04011 | 0,062516 | -0,02429 | -0,64162 | 273 | 255,4593 | 0,521692 | 0,899808 | y ~ current_feature + S |
| 0,090898 | 0,052198 | -0,07874 | 1,741416 | 401 | 363,9958 | 0,082456 | 0,388948 | y ~ current_feature + S |
| -0,01541 | 0,067466 | 0,034007 | -0,22848 | 401 | 219,6476 | 0,819488 | 0,952501 | y ~ current_feature + S |
| 0,116319 | 0,052528 | -0,04751 | 2,214391 | 400 | 357,515 | 0,027432 | 0,588545 | y ~ current_feature + S |
| 0,016219 | 0,05697 | 0,061498 | 0,284691 | 348 | 308,0302 | 0,776072 | 0,975031 | y ~ current_feature + S |
| -0,00317 | 0,062637 | -0,02429 | -0,05065 | 266 | 254,8826 | 0,959642 | 0,99444 | y ~ current_feature + S |
| 0,067043 | 0,05429 | -0,07874 | 1,234898 | 374 | 337,7541 | 0,217727 | 0,639357 | y ~ current_feature + S |
| -0,03816 | 0,07161 | 0,034007 | -0,53284 | 374 | 194,7217 | 0,594753 | 0,878477 | y ~ current_feature + S |
| -0,00768 | 0,055041 | -0,04751 | -0,13945 | 373 | 330,0627 | 0,889179 | 0,985217 | y ~ current_feature + S |
| 0,002409 | 0,060359 | 0,061498 | 0,039918 | 324 | 274,4847 | 0,968188 | 0,996179 | y ~ current_feature + S |
| -0,03917 | 0,063709 | -0,02429 | -0,6149 | 249 | 246 | 0,539192 | 0,906823 | y ~ current_feature + S |
| -0,0085 | 0,054634 | -0,07874 | -0,15552 | 338 | 335 | 0,876505 | 0,987929 | y ~ current_feature + S |
| -0,05888 | 0,054541 | 0,034007 | -1,07953 | 338 | 335 | 0,281127 | 0,715049 | y ~ current_feature + S |
| 0,067268 | 0,054594 | -0,04751 | 1,232162 | 337 | 334 | 0,218755 | 0,742928 | y ~ current_feature + S |
| -0,04374 | 0,058565 | 0,061498 | -0,7469 | 294 | 291 | 0,455729 | 0,937665 | y ~ current_feature + S |
| 0,030025 | 0,067085 | -0,02429 | 0,447559 | 225 | 222 | 0,654908 | 0,924682 | y ~ current_feature + S |
| -0,01867 | 0,078554 | -0,07874 | -0,23763 | 165 | 162 | 0,81247 | 0,979871 | y ~ current_feature + S |
| -0,2389 | 0,076293 | 0,034007 | -3,13131 | 165 | 162 | 0,002065 | 0,133248 | y ~ current_feature + S |
| -0,01791 | 0,078555 | -0,04751 | -0,228 | 165 | 162 | 0,819932 | 0,972405 | y ~ current_feature + S |
| -0,01994 | 0,084198 | 0,061498 | -0,23683 | 144 | 141 | 0,813136 | 0,995348 | y ~ current_feature + S |
| -0,14576 | 0,095641 | -0,02429 | -1,52408 | 110 | 107 | 0,130438 | 0,617591 | y ~ current_feature + S |
| -0,14206 | 0,063111 | -0,07874 | -2,251 | 249 | 246 | 0,025269 | 0,201065 | y ~ current_feature + S |
| -0,22018 | 0,062193 | 0,034007 | -3,54023 | 249 | 246 | 4,78E-04 | 0,067708 | y ~ current_feature + S |
| -0,01765 | 0,063748 | -0,04751 | -0,27684 | 249 | 246 | 0,782136 | 0,94882 | y ~ current_feature + S |
| -0,04988 | 0,068757 | 0,061498 | -0,72551 | 214 | 211 | 0,468943 | 0,937665 | y ~ current_feature + S |
| -0,20313 | 0,076693 | -0,02429 | -2,64857 | 166 | 163 | 0,008878 | 0,295269 | y ~ current_feature + S |
| -0,03726 | 0,083275 | -0,07874 | -0,44743 | 147 | 144 | 0,655235 | 0,932255 | y ~ current_feature + S |
| -0,04993 | 0,083229 | 0,034007 | -0,5999 | 147 | 144 | 0,549519 | 0,854294 | y ~ current_feature + S |
| -0,05952 | 0,083186 | -0,04751 | -0,71548 | 147 | 144 | 0,47547 | 0,842057 | y ~ current_feature + S |
| -0,13212 | 0,089015 | 0,061498 | -1,48429 | 127 | 124 | 0,140269 | 0,8197 | y ~ current_feature + S |
| 0,04913 | 0,102474 | -0,02429 | 0,479443 | 98 | 95 | 0,632726 | 0,924682 | y ~ current_feature + S |
| 0,060997 | 0,061845 | -0,07874 | 0,98629 | 298 | 260,4785 | 0,324906 | 0,726376 | y ~ current_feature + S |
| 0,008941 | 0,080804 | 0,034007 | 0,110647 | 298 | 153,1437 | 0,912041 | 0,9732 | y ~ current_feature + S |
| 0,029545 | 0,064445 | -0,04751 | 0,458452 | 298 | 240,571 | 0,647041 | 0,925756 | y ~ current_feature + S |
| 0,034691 | 0,067542 | 0,061498 | 0,513627 | 258 | 218,9438 | 0,60803 | 0,945257 | y ~ current_feature + S |
| 0,02427 | 0,071408 | -0,02429 | 0,339876 | 199 | 196 | 0,734314 | 0,944216 | y ~ current_feature + S |
| 0,071884 | 0,071795 | -0,07874 | 1,001235 | 196 | 193 | 0,317967 | 0,723052 | y ~ current_feature + S |
| 0,065876 | 0,071825 | 0,034007 | 0,917177 | 196 | 193 | 0,360195 | 0,7498 | y ~ current_feature + S |
| 0,008696 | 0,072166 | -0,04751 | 0,120502 | 195 | 192 | 0,904212 | 0,985217 | y ~ current_feature + S |
| -0,04881 | 0,07729 | 0,061498 | -0,63148 | 170 | 167 | 0,52859 | 0,937665 | y ~ current_feature + S |

| | | | | | | | | |
|---|---|---|---|---|---|---|---|---|
| 0,136693 | 0,087903 | -0,02429 | 1,555044 | 130 | 127 | 0,122424 | 0,617591 | y ~ current_feature + S |
| -0,03933 | 0,097982 | -0,07874 | -0,4014 | 107 | 104 | 0,688949 | 0,93638 | y ~ current_feature + S |
| 0,109092 | 0,097473 | 0,034007 | 1,1192 | 107 | 104 | 0,265633 | 0,71095 | y ~ current_feature + S |
| 0,073796 | 0,097791 | -0,04751 | 0,754631 | 107 | 104 | 0,452176 | 0,832044 | y ~ current_feature + S |
| -0,1143 | 0,107754 | 0,061498 | -1,06078 | 88 | 85 | 0,291794 | 0,893818 | y ~ current_feature + S |
| 0,018881 | 0,121246 | -0,02429 | 0,155728 | 71 | 68 | 0,876709 | 0,984468 | y ~ current_feature + S |
| -0,02793 | 0,049934 | -0,07874 | -0,55933 | 421 | 400,7485 | 0,57625 | 0,899631 | y ~ current_feature + S |
| -0,07971 | 0,063929 | 0,034007 | -1,24682 | 421 | 243,1296 | 0,213663 | 0,664036 | y ~ current_feature + S |
| -0,0219 | 0,050827 | -0,04751 | -0,43086 | 420 | 386,8972 | 0,66681 | 0,92822 | y ~ current_feature + S |
| -0,04421 | 0,055146 | 0,061498 | -0,80177 | 367 | 328,1921 | 0,423268 | 0,937665 | y ~ current_feature + S |
| 0,008246 | 0,061593 | -0,02429 | 0,133872 | 280 | 263,5805 | 0,893606 | 0,99444 | y ~ current_feature + S |
| -0,0915 | 0,068392 | -0,07874 | -1,33788 | 215 | 212 | 0,18237 | 0,581699 | y ~ current_feature + S |
| -0,0666 | 0,068528 | 0,034007 | -0,97184 | 215 | 212 | 0,33224 | 0,734392 | y ~ current_feature + S |
| 0,132781 | 0,068072 | -0,04751 | 1,950598 | 215 | 212 | 0,052423 | 0,640171 | y ~ current_feature + S |
| 0,078983 | 0,07493 | 0,061498 | 1,054089 | 180 | 177 | 0,293279 | 0,893818 | y ~ current_feature + S |
| 0,016858 | 0,084503 | -0,02429 | 0,199491 | 143 | 140 | 0,842168 | 0,977734 | y ~ current_feature + S |
| -0,00157 | 0,095346 | -0,07874 | -0,01647 | 113 | 110 | 0,986893 | 0,996318 | y ~ current_feature + S |
| -0,09946 | 0,094873 | 0,034007 | -1,04834 | 113 | 110 | 0,296781 | 0,718742 | y ~ current_feature + S |
| 0,125936 | 0,09502 | -0,04751 | 1,325363 | 112 | 109 | 0,187822 | 0,709124 | y ~ current_feature + S |
| 0,163396 | 0,104221 | 0,061498 | 1,567792 | 95 | 89,60662 | 0,120454 | 0,8197 | y ~ current_feature + S |
| -0,0414 | 0,11775 | -0,02429 | -0,35157 | 75 | 72 | 0,726191 | 0,942774 | y ~ current_feature + S |
| 0,017915 | 0,056784 | -0,07874 | 0,315498 | 337 | 310,0368 | 0,752596 | 0,963531 | y ~ current_feature + S |
| 0,104123 | 0,075444 | 0,034007 | 1,380132 | 337 | 173,7862 | 0,169319 | 0,61875 | y ~ current_feature + S |
| 0,0639 | 0,059309 | -0,04751 | 1,077414 | 337 | 283,1313 | 0,282213 | 0,747915 | y ~ current_feature + S |
| 0,153869 | 0,061209 | 0,061498 | 2,51381 | 292 | 260,5907 | 0,012547 | 0,55988 | y ~ current_feature + S |
| 0,121318 | 0,066674 | -0,02429 | 1,819569 | 225 | 221,6377 | 0,070174 | 0,605474 | y ~ current_feature + S |
| 0,017291 | 0,05117 | -0,07874 | 0,337909 | 394 | 381,8067 | 0,735618 | 0,959225 | y ~ current_feature + S |
| -0,04136 | 0,065687 | 0,034007 | -0,62968 | 394 | 231,3622 | 0,529523 | 0,848387 | y ~ current_feature + S |
| 0,090154 | 0,052192 | -0,04751 | 1,727344 | 393 | 364,1217 | 0,084954 | 0,640171 | y ~ current_feature + S |
| 0,029325 | 0,055869 | 0,061498 | 0,524881 | 341 | 320,1009 | 0,600029 | 0,945257 | y ~ current_feature + S |
| 0,086709 | 0,062425 | -0,02429 | 1,38901 | 262 | 254,6835 | 0,166043 | 0,624945 | y ~ current_feature + S |
| 0,031012 | 0,059733 | -0,07874 | 0,519173 | 283 | 280 | 0,604051 | 0,908531 | y ~ current_feature + S |
| -0,04349 | 0,059705 | 0,034007 | -0,72842 | 283 | 280 | 0,466968 | 0,831402 | y ~ current_feature + S |
| 9,34E-04 | 0,059868 | -0,04751 | 0,015606 | 282 | 279 | 0,98756 | 0,994138 | y ~ current_feature + S |
| 0,024168 | 0,069591 | 0,061498 | 0,347286 | 243 | 206,3672 | 0,72873 | 0,972573 | y ~ current_feature + S |
| -0,07809 | 0,073366 | -0,02429 | -1,0644 | 188 | 184,6503 | 0,288536 | 0,734108 | y ~ current_feature + S |
| -0,03758 | 0,052133 | -0,07874 | -0,72084 | 379 | 367,4114 | 0,471468 | 0,840689 | y ~ current_feature + S |
| -0,12924 | 0,067201 | 0,034007 | -1,92323 | 379 | 217,7347 | 0,055756 | 0,420946 | y ~ current_feature + S |
| -0,12438 | 0,05277 | -0,04751 | -2,35698 | 378 | 353,549 | 0,018969 | 0,54566 | y ~ current_feature + S |
| -0,16462 | 0,055592 | 0,061498 | -2,96116 | 328 | 314,8076 | 0,003298 | 0,305054 | y ~ current_feature + S |
| -0,00713 | 0,065427 | -0,02429 | -0,10896 | 252 | 233,5987 | 0,913329 | 0,99444 | y ~ current_feature + S |
| 0,139968 | 0,064454 | -0,07874 | 2,171612 | 239 | 236 | 0,030882 | 0,224043 | y ~ current_feature + S |
| 0,039684 | 0,065043 | 0,034007 | 0,610112 | 239 | 236 | 0,542375 | 0,852506 | y ~ current_feature + S |
| 0,11231 | 0,064683 | -0,04751 | 1,736321 | 239 | 236 | 0,083812 | 0,640171 | y ~ current_feature + S |
| 0,002896 | 0,075514 | 0,061498 | 0,038352 | 201 | 175,3643 | 0,969451 | 0,996179 | y ~ current_feature + S |
| 0,046186 | 0,080846 | -0,02429 | 0,571285 | 159 | 152,6719 | 0,568647 | 0,924682 | y ~ current_feature + S |
| -0,09255 | 0,054979 | -0,07874 | -1,6834 | 331 | 328 | 0,093248 | 0,42595 | y ~ current_feature + S |
| -0,10533 | 0,054909 | 0,034007 | -1,91836 | 331 | 328 | 0,055934 | 0,420946 | y ~ current_feature + S |

| | | | | | | | | |
|---|---|---|---|---|---|---|---|---|
| -0,01728 | 0,055208 | -0,04751 | -0,31296 | 331 | 328 | 0,754508 | 0,9394 | y ~ current_feature + S |
| -0,00441 | 0,063015 | 0,061498 | -0,07005 | 287 | 251,8253 | 0,944211 | 0,996179 | y ~ current_feature + S |
| -0,10637 | 0,067344 | -0,02429 | -1,57948 | 221 | 218 | 0,115675 | 0,614731 | y ~ current_feature + S |
| -0,03877 | 0,081862 | -0,07874 | -0,47359 | 152 | 149 | 0,636489 | 0,927169 | y ~ current_feature + S |
| -0,18839 | 0,080456 | 0,034007 | -2,34154 | 152 | 149 | 0,020529 | 0,31947 | y ~ current_feature + S |
| -0,01191 | 0,081917 | -0,04751 | -0,14537 | 152 | 149 | 0,884616 | 0,985217 | y ~ current_feature + S |
| -0,16225 | 0,088258 | 0,061498 | -1,83833 | 128 | 125 | 0,068387 | 0,715249 | y ~ current_feature + S |
| -0,00629 | 0,101013 | -0,02429 | -0,06225 | 101 | 98 | 0,950492 | 0,99444 | y ~ current_feature + S |
| 0,007869 | 0,072355 | -0,07874 | 0,108756 | 194 | 191 | 0,91351 | 0,987929 | y ~ current_feature + S |
| -0,12442 | 0,071795 | 0,034007 | -1,73294 | 194 | 191 | 0,08472 | 0,478574 | y ~ current_feature + S |
| -0,02802 | 0,072329 | -0,04751 | -0,38735 | 194 | 191 | 0,698926 | 0,931902 | y ~ current_feature + S |
| -0,06637 | 0,077915 | 0,061498 | -0,85185 | 167 | 164 | 0,395537 | 0,93433 | y ~ current_feature + S |
| -0,08744 | 0,088746 | -0,02429 | -0,98532 | 129 | 126 | 0,326353 | 0,774043 | y ~ current_feature + S |
| -0,05919 | 0,057992 | -0,07874 | -1,02062 | 337 | 296,3059 | 0,308269 | 0,718687 | y ~ current_feature + S |
| -0,08226 | 0,07583 | 0,034007 | -1,08475 | 337 | 172,7329 | 0,279545 | 0,715049 | y ~ current_feature + S |
| -0,0475 | 0,059983 | -0,04751 | -0,79197 | 337 | 277,3074 | 0,429057 | 0,819126 | y ~ current_feature + S |
| -0,04093 | 0,064435 | 0,061498 | -0,63519 | 294 | 240,4521 | 0,525911 | 0,937665 | y ~ current_feature + S |
| -0,01107 | 0,067263 | -0,02429 | -0,16458 | 224 | 221 | 0,869421 | 0,984468 | y ~ current_feature + S |
| -0,04207 | 0,061169 | -0,07874 | -0,68769 | 319 | 266,7861 | 0,492244 | 0,853069 | y ~ current_feature + S |
| -0,0364 | 0,078986 | 0,034007 | -0,46081 | 319 | 160,0757 | 0,645563 | 0,900251 | y ~ current_feature + S |
| 0,10738 | 0,061647 | -0,04751 | 1,741855 | 318 | 260,0981 | 0,082716 | 0,640171 | y ~ current_feature + S |
| 0,099486 | 0,063268 | 0,061498 | 1,572459 | 275 | 247,3516 | 0,117122 | 0,8197 | y ~ current_feature + S |
| 0,030517 | 0,069286 | -0,02429 | 0,440454 | 212 | 208,1159 | 0,660065 | 0,924682 | y ~ current_feature + S |
| -0,03538 | 0,066923 | -0,07874 | -0,52865 | 226 | 223 | 0,597575 | 0,90386 | y ~ current_feature + S |
| -0,04286 | 0,066903 | 0,034007 | -0,64068 | 226 | 223 | 0,52239 | 0,845883 | y ~ current_feature + S |
| -0,01286 | 0,066959 | -0,04751 | -0,19202 | 226 | 223 | 0,847898 | 0,980218 | y ~ current_feature + S |
| 0,029665 | 0,072137 | 0,061498 | 0,411229 | 195 | 192 | 0,681363 | 0,962245 | y ~ current_feature + S |
| -0,0837 | 0,082189 | -0,02429 | -1,01834 | 150 | 147 | 0,310189 | 0,762591 | y ~ current_feature + S |
| -0,00829 | 0,056886 | -0,07874 | -0,14574 | 312 | 309 | 0,884219 | 0,987929 | y ~ current_feature + S |
| -0,02009 | 0,056877 | 0,034007 | -0,35316 | 312 | 309 | 0,724208 | 0,920814 | y ~ current_feature + S |
| -0,05634 | 0,05689 | -0,04751 | -0,99029 | 311 | 308 | 0,322807 | 0,774585 | y ~ current_feature + S |
| 0,068751 | 0,061054 | 0,061498 | 1,126058 | 270 | 267 | 0,261152 | 0,893818 | y ~ current_feature + S |
| -0,00645 | 0,070013 | -0,02429 | -0,09213 | 207 | 204 | 0,926685 | 0,99444 | y ~ current_feature + S |
| 0,051311 | 0,05697 | -0,07874 | 0,90066 | 346 | 307,2964 | 0,368474 | 0,77906 | y ~ current_feature + S |
| -0,04151 | 0,075681 | 0,034007 | -0,54851 | 346 | 174,2911 | 0,584045 | 0,871357 | y ~ current_feature + S |
| -3,43E-04 | 0,057981 | -0,04751 | -0,00592 | 345 | 297,4593 | 0,995282 | 0,995481 | y ~ current_feature + S |
| -0,01589 | 0,060174 | 0,061498 | -0,26404 | 299 | 276,1036 | 0,791943 | 0,983887 | y ~ current_feature + S |
| 0,023253 | 0,067488 | -0,02429 | 0,344554 | 230 | 219,4367 | 0,73076 | 0,944216 | y ~ current_feature + S |
| -0,01515 | 0,103683 | -0,07874 | -0,14612 | 96 | 93 | 0,88414 | 0,987929 | y ~ current_feature + S |
| 0,296826 | 0,099022 | 0,034007 | 2,997581 | 96 | 93 | 0,00349 | 0,151911 | y ~ current_feature + S |
| 0,007356 | 0,103692 | -0,04751 | 0,070937 | 96 | 93 | 0,9436 | 0,990156 | y ~ current_feature + S |
| -0,03136 | 0,111748 | 0,061498 | -0,28067 | 83 | 80 | 0,779684 | 0,977909 | y ~ current_feature + S |
| 0,197741 | 0,125509 | -0,02429 | 1,575517 | 64 | 61 | 0,12031 | 0,615361 | y ~ current_feature + S |
| -0,06331 | 0,051062 | -0,07874 | -1,23996 | 385 | 382 | 0,215752 | 0,639357 | y ~ current_feature + S |
| -0,07288 | 0,051028 | 0,034007 | -1,42826 | 385 | 382 | 0,154035 | 0,60427 | y ~ current_feature + S |
| 0,043637 | 0,051183 | -0,04751 | 0,852582 | 384 | 381 | 0,394427 | 0,803665 | y ~ current_feature + S |
| 0,063467 | 0,060122 | 0,061498 | 1,05564 | 334 | 275,5365 | 0,292058 | 0,893818 | y ~ current_feature + S |
| 0,049434 | 0,063001 | -0,02429 | 0,784655 | 256 | 251,3319 | 0,433395 | 0,852958 | y ~ current_feature + S |

| | | | | | | | | |
|---|---|---|---|---|---|---|---|---|
| -0,0888 | 0,062498 | -0,07874 | -1,42084 | 257 | 254 | 0,15659 | 0,524327 | y ~ current_feature + S |
| -0,19031 | 0,061599 | 0,034007 | -3,08951 | 257 | 254 | 0,002228 | 0,133248 | y ~ current_feature + S |
| -0,11584 | 0,062323 | -0,04751 | -1,85877 | 257 | 254 | 0,064216 | 0,640171 | y ~ current_feature + S |
| -0,07368 | 0,066783 | 0,061498 | -1,10321 | 226 | 223 | 0,271125 | 0,893818 | y ~ current_feature + S |
| -0,09595 | 0,076796 | -0,02429 | -1,24948 | 171 | 168 | 0,213229 | 0,679441 | y ~ current_feature + S |
| 0,083019 | 0,053055 | -0,07874 | 1,56476 | 390 | 352,8107 | 0,118536 | 0,474142 | y ~ current_feature + S |
| 0,235206 | 0,068611 | 0,034007 | 3,428092 | 390 | 200,6748 | 7,37E-04 | 0,068218 | y ~ current_feature + S |
| 0,092753 | 0,055455 | -0,04751 | 1,672583 | 389 | 322,3821 | 0,09538 | 0,640171 | y ~ current_feature + S |
| 0,034432 | 0,059176 | 0,061498 | 0,581856 | 339 | 285,2321 | 0,561123 | 0,937665 | y ~ current_feature + S |
| 0,183571 | 0,061633 | -0,02429 | 2,978434 | 260 | 254,3791 | 0,003177 | 0,18344 | y ~ current_feature + S |
| 0,016105 | 0,069162 | -0,07874 | 0,232863 | 212 | 209 | 0,816096 | 0,979871 | y ~ current_feature + S |
| -0,06376 | 0,069031 | 0,034007 | -0,92371 | 212 | 209 | 0,356705 | 0,7498 | y ~ current_feature + S |
| -0,06351 | 0,069032 | -0,04751 | -0,91994 | 212 | 209 | 0,358664 | 0,798092 | y ~ current_feature + S |
| 0,022232 | 0,074517 | 0,061498 | 0,298343 | 183 | 180 | 0,765785 | 0,972573 | y ~ current_feature + S |
| -0,06588 | 0,084941 | -0,02429 | -0,77561 | 141 | 138 | 0,439307 | 0,858965 | y ~ current_feature + S |
| -0,01299 | 0,069499 | -0,07874 | -0,18696 | 210 | 207 | 0,851873 | 0,983548 | y ~ current_feature + S |
| -0,14959 | 0,068723 | 0,034007 | -2,17672 | 210 | 207 | 0,030633 | 0,347616 | y ~ current_feature + S |
| -0,02513 | 0,069483 | -0,04751 | -0,36165 | 210 | 207 | 0,71798 | 0,93314 | y ~ current_feature + S |
| -0,04879 | 0,07572 | 0,061498 | -0,6444 | 177 | 174 | 0,520165 | 0,937665 | y ~ current_feature + S |
| -0,10136 | 0,084996 | -0,02429 | -1,19255 | 140 | 137 | 0,235108 | 0,695919 | y ~ current_feature + S |
| -0,01599 | 0,052681 | -0,07874 | -0,3036 | 381 | 360,2311 | 0,761604 | 0,965008 | y ~ current_feature + S |
| -0,21262 | 0,069351 | 0,034007 | -3,06584 | 381 | 198,5221 | 0,002473 | 0,133248 | y ~ current_feature + S |
| -0,0241 | 0,05435 | -0,04751 | -0,44339 | 381 | 338,3301 | 0,657767 | 0,925756 | y ~ current_feature + S |
| -0,09213 | 0,0571 | 0,061498 | -1,61344 | 330 | 304,1096 | 0,107687 | 0,8197 | y ~ current_feature + S |
| -0,04882 | 0,063492 | -0,02429 | -0,76891 | 254 | 247,4684 | 0,442679 | 0,858965 | y ~ current_feature + S |
| 0,115448 | 0,056903 | -0,07874 | 2,02886 | 332 | 304,7228 | 0,043342 | 0,277028 | y ~ current_feature + S |
| -0,05323 | 0,0753 | 0,034007 | -0,70693 | 332 | 175,8621 | 0,480543 | 0,831402 | y ~ current_feature + S |
| -0,00729 | 0,058428 | -0,04751 | -0,12476 | 331 | 292,9125 | 0,900797 | 0,985217 | y ~ current_feature + S |
| 0,035172 | 0,062727 | 0,061498 | 0,560719 | 284 | 253,8332 | 0,575484 | 0,937665 | y ~ current_feature + S |
| 0,036818 | 0,068685 | -0,02429 | 0,536042 | 221 | 211,6828 | 0,592492 | 0,924682 | y ~ current_feature + S |
| 0,082775 | 0,084834 | -0,07874 | 0,975734 | 141 | 138 | 0,330903 | 0,730951 | y ~ current_feature + S |
| 0,020668 | 0,085107 | 0,034007 | 0,242845 | 141 | 138 | 0,808486 | 0,952501 | y ~ current_feature + S |
| -0,23493 | 0,082743 | -0,04751 | -2,83922 | 141 | 138 | 0,005206 | 0,422981 | y ~ current_feature + S |
| -0,05582 | 0,090767 | 0,061498 | -0,615 | 124 | 121 | 0,539713 | 0,937665 | y ~ current_feature + S |
| -0,00409 | 0,104828 | -0,02429 | -0,03904 | 94 | 91 | 0,968947 | 0,99444 | y ~ current_feature + S |
| -0,02792 | 0,050947 | -0,07874 | -0,54807 | 401 | 384,9627 | 0,583963 | 0,902886 | y ~ current_feature + S |
| -0,04719 | 0,065591 | 0,034007 | -0,71946 | 401 | 231,9232 | 0,472579 | 0,831402 | y ~ current_feature + S |
| 0,006715 | 0,052037 | -0,04751 | 0,129047 | 400 | 369,286 | 0,897391 | 0,985217 | y ~ current_feature + S |
| 0,038952 | 0,055845 | 0,061498 | 0,697497 | 347 | 320,1641 | 0,485998 | 0,937665 | y ~ current_feature + S |
| -0,01765 | 0,062905 | -0,02429 | -0,28066 | 267 | 252,6377 | 0,779203 | 0,961068 | y ~ current_feature + S |
| -0,00999 | 0,092843 | -0,07874 | -0,10764 | 119 | 116 | 0,914466 | 0,987929 | y ~ current_feature + S |
| -0,10016 | 0,092381 | 0,034007 | -1,08426 | 119 | 116 | 0,280499 | 0,715049 | y ~ current_feature + S |
| -0,03387 | 0,092794 | -0,04751 | -0,36502 | 119 | 116 | 0,71576 | 0,93314 | y ~ current_feature + S |
| -0,21191 | 0,096766 | 0,061498 | -2,18987 | 105 | 102 | 0,030812 | 0,658491 | y ~ current_feature + S |
| 0,058333 | 0,114513 | -0,02429 | 0,509405 | 79 | 76 | 0,611944 | 0,924682 | y ~ current_feature + S |
| 0,067998 | 0,061993 | -0,07874 | 1,096856 | 262 | 259 | 0,273723 | 0,705864 | y ~ current_feature + S |
| 0,050939 | 0,062056 | 0,034007 | 0,820855 | 262 | 259 | 0,412484 | 0,785427 | y ~ current_feature + S |
| 0,009004 | 0,062134 | -0,04751 | 0,144913 | 262 | 259 | 0,884892 | 0,985217 | y ~ current_feature + S |

| | | | | | | | | |
|---|---|---|---|---|---|---|---|---|
| -0,0507 | 0,067029 | 0,061498 | -0,75633 | 225 | 222 | 0,45025 | 0,937665 | y ~ current_feature + S |
| 0,017741 | 0,076237 | -0,02429 | 0,232709 | 175 | 172 | 0,816264 | 0,974251 | y ~ current_feature + S |
| -0,00932 | 0,052292 | -0,07874 | -0,17827 | 385 | 365,6684 | 0,858611 | 0,983548 | y ~ current_feature + S |
| -0,09005 | 0,069025 | 0,034007 | -1,30455 | 385 | 208,1883 | 0,193485 | 0,6438 | y ~ current_feature + S |
| -0,03037 | 0,053501 | -0,04751 | -0,56771 | 384 | 349,0444 | 0,570599 | 0,903741 | y ~ current_feature + S |
| -0,08719 | 0,05782 | 0,061498 | -1,50787 | 333 | 296,84 | 0,132652 | 0,8197 | y ~ current_feature + S |
| -0,01327 | 0,063065 | -0,02429 | -0,21043 | 256 | 251,3861 | 0,833503 | 0,977484 | y ~ current_feature + S |
| 0,082032 | 0,053783 | -0,07874 | 1,525228 | 402 | 343,3762 | 0,128123 | 0,481525 | y ~ current_feature + S |
| -0,00428 | 0,069743 | 0,034007 | -0,06141 | 402 | 205,5871 | 0,951095 | 0,988497 | y ~ current_feature + S |
| 0,032036 | 0,054545 | -0,04751 | 0,587327 | 401 | 335,7694 | 0,557379 | 0,897386 | y ~ current_feature + S |
| -0,07062 | 0,056647 | 0,061498 | -1,24664 | 348 | 310,0843 | 0,213469 | 0,893818 | y ~ current_feature + S |
| 0,016106 | 0,063 | -0,02429 | 0,255644 | 267 | 251,8847 | 0,798434 | 0,968297 | y ~ current_feature + S |
| -0,08 | 0,064655 | -0,07874 | -1,23737 | 286 | 237,6887 | 0,21717 | 0,639357 | y ~ current_feature + S |
| -0,18558 | 0,084445 | 0,034007 | -2,19766 | 286 | 135,4033 | 0,029673 | 0,347616 | y ~ current_feature + S |
| -0,02859 | 0,062676 | -0,04751 | -0,45618 | 285 | 254,3601 | 0,648652 | 0,925756 | y ~ current_feature + S |
| -0,14426 | 0,066674 | 0,061498 | -2,16364 | 242 | 220,2706 | 0,031568 | 0,658491 | y ~ current_feature + S |
| -0,21321 | 0,071446 | -0,02429 | -2,98423 | 190 | 186,9975 | 0,003223 | 0,18344 | y ~ current_feature + S |
| -0,02919 | 0,115421 | -0,07874 | -0,25292 | 78 | 75 | 0,801023 | 0,979102 | y ~ current_feature + S |
| -0,03959 | 0,11538 | 0,034007 | -0,3431 | 78 | 75 | 0,732483 | 0,922572 | y ~ current_feature + S |
| 0,257753 | 0,111568 | -0,04751 | 2,310273 | 78 | 75 | 0,023625 | 0,563956 | y ~ current_feature + S |
| -0,12319 | 0,12309 | 0,061498 | -1,00079 | 68 | 65 | 0,320639 | 0,893818 | y ~ current_feature + S |
| -0,09876 | 0,142159 | -0,02429 | -0,69472 | 52 | 49 | 0,490512 | 0,889654 | y ~ current_feature + S |
| 0,107707 | 0,065707 | -0,07874 | 1,639191 | 281 | 228,9327 | 0,102547 | 0,449022 | y ~ current_feature + S |
| 0,020591 | 0,059963 | 0,034007 | 0,343386 | 281 | 278 | 0,731567 | 0,922572 | y ~ current_feature + S |
| -0,01142 | 0,06008 | -0,04751 | -0,19016 | 280 | 277 | 0,849322 | 0,980218 | y ~ current_feature + S |
| 0,076092 | 0,071138 | 0,061498 | 1,06964 | 243 | 196,463 | 0,286094 | 0,893818 | y ~ current_feature + S |
| -0,06063 | 0,073786 | -0,02429 | -0,82173 | 186 | 183 | 0,4123 | 0,837702 | y ~ current_feature + S |
| -0,03554 | 0,056578 | -0,07874 | -0,62813 | 315 | 312 | 0,530378 | 0,880735 | y ~ current_feature + S |
| -0,00152 | 0,056614 | 0,034007 | -0,02687 | 315 | 312 | 0,978579 | 0,989859 | y ~ current_feature + S |
| -0,00754 | 0,056612 | -0,04751 | -0,13317 | 315 | 312 | 0,894142 | 0,985217 | y ~ current_feature + S |
| 0,022423 | 0,06073 | 0,061498 | 0,369223 | 274 | 271 | 0,71225 | 0,972444 | y ~ current_feature + S |
| -0,00976 | 0,06967 | -0,02429 | -0,14007 | 209 | 206 | 0,888738 | 0,992711 | y ~ current_feature + S |
| 0,115658 | 0,048583 | -0,07874 | 2,380604 | 421 | 418 | 0,017732 | 0,177316 | y ~ current_feature + S |
| 0,049021 | 0,048853 | 0,034007 | 1,003446 | 421 | 418 | 0,316226 | 0,718742 | y ~ current_feature + S |
| 0,022926 | 0,048957 | -0,04751 | 0,46828 | 420 | 417 | 0,639829 | 0,925756 | y ~ current_feature + S |
| -0,02771 | 0,052394 | 0,061498 | -0,52887 | 367 | 364 | 0,597216 | 0,945257 | y ~ current_feature + S |
| 0,083094 | 0,059876 | -0,02429 | 1,387751 | 280 | 277 | 0,166328 | 0,624945 | y ~ current_feature + S |
| -0,01076 | 0,059192 | -0,07874 | -0,18185 | 309 | 285,3801 | 0,855828 | 0,983548 | y ~ current_feature + S |
| -0,10433 | 0,078369 | 0,034007 | -1,33121 | 309 | 161,0496 | 0,185002 | 0,638805 | y ~ current_feature + S |
| -0,02447 | 0,0607 | -0,04751 | -0,4032 | 309 | 271,2456 | 0,687117 | 0,929555 | y ~ current_feature + S |
| 0,030297 | 0,064048 | 0,061498 | 0,473044 | 267 | 243,5517 | 0,636605 | 0,955689 | y ~ current_feature + S |
| -0,07068 | 0,070011 | -0,02429 | -1,00953 | 206 | 203 | 0,313922 | 0,765236 | y ~ current_feature + S |
| -0,18151 | 0,089399 | -0,07874 | -2,03039 | 124 | 121 | 0,044509 | 0,279123 | y ~ current_feature + S |
| -0,10369 | 0,090419 | 0,034007 | -1,14677 | 124 | 121 | 0,25374 | 0,704827 | y ~ current_feature + S |
| -0,07618 | 0,091022 | -0,04751 | -0,83692 | 123 | 120 | 0,404299 | 0,803665 | y ~ current_feature + S |
| -0,10572 | 0,09846 | 0,061498 | -1,07374 | 105 | 102 | 0,285473 | 0,893818 | y ~ current_feature + S |
| -0,20971 | 0,110007 | -0,02429 | -1,90632 | 82 | 79 | 0,060246 | 0,558147 | y ~ current_feature + S |
| -0,00908 | 0,056794 | -0,07874 | -0,15988 | 313 | 310 | 0,873084 | 0,987929 | y ~ current_feature + S |

| | | | | | | | | |
|---|---|---|---|---|---|---|---|---|
| -0,05763 | 0,056702 | 0,034007 | -1,01641 | 313 | 310 | 0,310226 | 0,718742 | y ~ current_feature + S |
| -0,01742 | 0,056879 | -0,04751 | -0,30631 | 312 | 309 | 0,759579 | 0,9394 | y ~ current_feature + S |
| 0,003558 | 0,060745 | 0,061498 | 0,058568 | 274 | 271 | 0,95334 | 0,996179 | y ~ current_feature + S |
| -0,06291 | 0,069705 | -0,02429 | -0,90246 | 208 | 205 | 0,36787 | 0,807787 | y ~ current_feature + S |
| 0,051411 | 0,050357 | -0,07874 | 1,020942 | 421 | 393,3087 | 0,307909 | 0,718687 | y ~ current_feature + S |
| 0,052183 | 0,065351 | 0,034007 | 0,798502 | 421 | 233,5108 | 0,425391 | 0,791701 | y ~ current_feature + S |
| 0,046661 | 0,05134 | -0,04751 | 0,908858 | 420 | 378,5697 | 0,364003 | 0,801674 | y ~ current_feature + S |
| -0,06747 | 0,055153 | 0,061498 | -1,22335 | 367 | 327,2501 | 0,222078 | 0,893818 | y ~ current_feature + S |
| -0,02062 | 0,060472 | -0,02429 | -0,34095 | 280 | 273,3434 | 0,7334 | 0,944216 | y ~ current_feature + S |
| -0,03394 | 0,093605 | -0,07874 | -0,36258 | 117 | 114 | 0,717593 | 0,948276 | y ~ current_feature + S |
| -0,12556 | 0,092917 | 0,034007 | -1,35134 | 117 | 114 | 0,179262 | 0,630217 | y ~ current_feature + S |
| -0,01547 | 0,093647 | -0,04751 | -0,16524 | 117 | 114 | 0,86905 | 0,985217 | y ~ current_feature + S |
| 0,03106 | 0,103092 | 0,061498 | 0,301282 | 97 | 94 | 0,763865 | 0,972573 | y ~ current_feature + S |
| 0,057235 | 0,115281 | -0,02429 | 0,496483 | 78 | 75 | 0,621006 | 0,924682 | y ~ current_feature + S |
| -0,15625 | 0,14724 | -0,07874 | -1,06118 | 48 | 45 | 0,294273 | 0,718687 | y ~ current_feature + S |
| -0,13345 | 0,147738 | 0,034007 | -0,9033 | 48 | 45 | 0,371175 | 0,750317 | y ~ current_feature + S |
| 0,03499 | 0,14898 | -0,04751 | 0,234861 | 48 | 45 | 0,815382 | 0,972405 | y ~ current_feature + S |
| 0,091001 | 0,161548 | 0,061498 | 0,563304 | 41 | 38 | 0,576537 | 0,937665 | y ~ current_feature + S |
| 0,117869 | 0,184401 | -0,02429 | 0,639201 | 32 | 29 | 0,527709 | 0,899808 | y ~ current_feature + S |
| 0,013799 | 0,074118 | -0,07874 | 0,186171 | 185 | 182 | 0,852518 | 0,983548 | y ~ current_feature + S |
| 0,050882 | 0,074029 | 0,034007 | 0,687323 | 185 | 182 | 0,492754 | 0,838248 | y ~ current_feature + S |
| 0,084952 | 0,073857 | -0,04751 | 1,150219 | 185 | 182 | 0,251563 | 0,742928 | y ~ current_feature + S |
| -0,00972 | 0,080578 | 0,061498 | -0,1206 | 157 | 154 | 0,904162 | 0,996179 | y ~ current_feature + S |
| -0,00361 | 0,091286 | -0,02429 | -0,03953 | 123 | 120 | 0,968533 | 0,99444 | y ~ current_feature + S |
| 0,028698 | 0,05017 | -0,07874 | 0,572009 | 410 | 396,9642 | 0,56764 | 0,898449 | y ~ current_feature + S |
| 0,049649 | 0,063637 | 0,034007 | 0,780195 | 410 | 246,3282 | 0,436025 | 0,79866 | y ~ current_feature + S |
| 0,017572 | 0,050825 | -0,04751 | 0,345736 | 409 | 386,9961 | 0,729729 | 0,93314 | y ~ current_feature + S |
| -0,03447 | 0,054498 | 0,061498 | -0,63244 | 359 | 336,3011 | 0,527526 | 0,937665 | y ~ current_feature + S |
| 0,070604 | 0,062041 | -0,02429 | 1,138022 | 272 | 258,505 | 0,256165 | 0,718089 | y ~ current_feature + S |
| 0,126944 | 0,081811 | -0,07874 | 1,55167 | 150 | 147 | 0,122891 | 0,477837 | y ~ current_feature + S |
| 0,050055 | 0,082375 | 0,034007 | 0,607648 | 150 | 147 | 0,544358 | 0,852506 | y ~ current_feature + S |
| 0,025225 | 0,082452 | -0,04751 | 0,30594 | 150 | 147 | 0,760083 | 0,9394 | y ~ current_feature + S |
| -0,00834 | 0,090533 | 0,061498 | -0,09209 | 125 | 122 | 0,926778 | 0,996179 | y ~ current_feature + S |
| 0,044564 | 0,101434 | -0,02429 | 0,439342 | 100 | 97 | 0,661391 | 0,924682 | y ~ current_feature + S |
| -0,01311 | 0,084208 | -0,07874 | -0,15565 | 144 | 141 | 0,876532 | 0,987929 | y ~ current_feature + S |
| 0,044408 | 0,084132 | 0,034007 | 0,527833 | 144 | 141 | 0,598445 | 0,878669 | y ~ current_feature + S |
| -0,05255 | 0,084099 | -0,04751 | -0,62487 | 144 | 141 | 0,533068 | 0,883994 | y ~ current_feature + S |
| 0,030292 | 0,091628 | 0,061498 | 0,330595 | 122 | 119 | 0,741531 | 0,972573 | y ~ current_feature + S |
| 0,070923 | 0,103434 | -0,02429 | 0,685687 | 96 | 93 | 0,494616 | 0,892722 | y ~ current_feature + S |
| 0,132422 | 0,063068 | -0,07874 | 2,099668 | 250 | 247 | 0,036773 | 0,251962 | y ~ current_feature + S |
| 0,023378 | 0,063611 | 0,034007 | 0,367516 | 250 | 247 | 0,713549 | 0,920814 | y ~ current_feature + S |
| -0,00359 | 0,063757 | -0,04751 | -0,05625 | 249 | 246 | 0,955192 | 0,990156 | y ~ current_feature + S |
| 0,026151 | 0,068657 | 0,061498 | 0,380887 | 215 | 212 | 0,703668 | 0,972444 | y ~ current_feature + S |
| 0,24718 | 0,075896 | -0,02429 | 3,256848 | 166 | 163 | 0,001371 | 0,144895 | y ~ current_feature + S |
| -0,0546 | 0,06315 | -0,07874 | -0,86468 | 277 | 250,0103 | 0,388044 | 0,789252 | y ~ current_feature + S |
| -0,08319 | 0,08151 | 0,034007 | -1,02063 | 277 | 149,4721 | 0,309081 | 0,718742 | y ~ current_feature + S |
| 0,019379 | 0,067021 | -0,04751 | 0,289147 | 276 | 222,5468 | 0,772738 | 0,945414 | y ~ current_feature + S |
| 0,070647 | 0,072432 | 0,061498 | 0,975349 | 239 | 189,6544 | 0,330629 | 0,893818 | y ~ current_feature + S |

| | | | | | | | | |
|---|---|---|---|---|---|---|---|---|
| -0,08102 | 0,075009 | -0,02429 | -1,08021 | 184 | 176,5706 | 0,281523 | 0,734108 | y ~ current_feature + S |
| 0,047485 | 0,061593 | -0,07874 | 0,770946 | 266 | 263 | 0,441431 | 0,818694 | y ~ current_feature + S |
| 3,47E-04 | 0,061663 | 0,034007 | 0,00562 | 266 | 263 | 0,99552 | 0,997989 | y ~ current_feature + S |
| -0,05768 | 0,06156 | -0,04751 | -0,93696 | 266 | 263 | 0,349641 | 0,795089 | y ~ current_feature + S |
| -0,0455 | 0,066746 | 0,061498 | -0,68165 | 227 | 224 | 0,496162 | 0,937665 | y ~ current_feature + S |
| -0,08026 | 0,075565 | -0,02429 | -1,06207 | 177 | 174 | 0,289675 | 0,734108 | y ~ current_feature + S |
| 0,009713 | 0,052431 | -0,07874 | 0,18525 | 386 | 363,7374 | 0,853136 | 0,983548 | y ~ current_feature + S |
| -0,0374 | 0,068102 | 0,034007 | -0,54915 | 386 | 215,3119 | 0,583469 | 0,871357 | y ~ current_feature + S |
| 0,06301 | 0,053652 | -0,04751 | 1,174411 | 386 | 346,0166 | 0,241038 | 0,742928 | y ~ current_feature + S |
| 0,021198 | 0,058117 | 0,061498 | 0,364743 | 334 | 295,9392 | 0,715564 | 0,972573 | y ~ current_feature + S |
| -0,06079 | 0,063223 | -0,02429 | -0,96146 | 257 | 249,2541 | 0,337253 | 0,785829 | y ~ current_feature + S |
| -0,05518 | 0,055586 | -0,07874 | -0,99262 | 370 | 322,656 | 0,321639 | 0,725761 | y ~ current_feature + S |
| 0,101761 | 0,071603 | 0,034007 | 1,421184 | 370 | 193,0279 | 0,156877 | 0,60427 | y ~ current_feature + S |
| 0,050098 | 0,056609 | -0,04751 | 0,884985 | 369 | 311,2752 | 0,376848 | 0,803665 | y ~ current_feature + S |
| 0,135231 | 0,062633 | 0,061498 | 2,159093 | 320 | 250,2506 | 0,031794 | 0,658491 | y ~ current_feature + S |
| 0,036176 | 0,064108 | -0,02429 | 0,564305 | 246 | 243 | 0,573067 | 0,924682 | y ~ current_feature + S |
| 0,12623 | 0,051791 | -0,07874 | 2,43732 | 415 | 366,8788 | 0,015271 | 0,174676 | y ~ current_feature + S |
| 0,191026 | 0,067212 | 0,034007 | 2,842162 | 415 | 213,289 | 0,004916 | 0,167881 | y ~ current_feature + S |
| 0,056813 | 0,053431 | -0,04751 | 1,063303 | 414 | 349,1476 | 0,28838 | 0,747915 | y ~ current_feature + S |
| 0,060721 | 0,057824 | 0,061498 | 1,050087 | 361 | 297,9722 | 0,294529 | 0,893818 | y ~ current_feature + S |
| 0,100496 | 0,060482 | -0,02429 | 1,661589 | 276 | 270,6107 | 0,097753 | 0,605474 | y ~ current_feature + S |
| 0,175653 | 0,076639 | -0,07874 | 2,291934 | 168 | 165 | 0,023173 | 0,194866 | y ~ current_feature + S |
| 0,070901 | 0,077654 | 0,034007 | 0,913033 | 168 | 165 | 0,362557 | 0,7498 | y ~ current_feature + S |
| 0,253689 | 0,075303 | -0,04751 | 3,368903 | 168 | 165 | 9,39E-04 | 0,132163 | y ~ current_feature + S |
| -0,00494 | 0,084214 | 0,061498 | -0,0586 | 144 | 141 | 0,953352 | 0,996179 | y ~ current_feature + S |
| 0,131466 | 0,094951 | -0,02429 | 1,384559 | 112 | 109 | 0,169015 | 0,625356 | y ~ current_feature + S |
| -0,02955 | 0,054209 | -0,07874 | -0,54505 | 343 | 340 | 0,586074 | 0,902886 | y ~ current_feature + S |
| -0,0863 | 0,05403 | 0,034007 | -1,59732 | 343 | 340 | 0,111124 | 0,540997 | y ~ current_feature + S |
| 0,011222 | 0,054309 | -0,04751 | 0,20663 | 342 | 339 | 0,836423 | 0,979827 | y ~ current_feature + S |
| 0,024693 | 0,058106 | 0,061498 | 0,424972 | 299 | 296 | 0,671166 | 0,958606 | y ~ current_feature + S |
| 0,076723 | 0,06647 | -0,02429 | 1,154247 | 228 | 225 | 0,249623 | 0,717451 | y ~ current_feature + S |
| -0,1943 | 0,06204 | -0,07874 | -3,13189 | 253 | 250 | 0,001944 | 0,055318 | y ~ current_feature + S |
| -0,17902 | 0,062224 | 0,034007 | -2,877 | 253 | 250 | 0,004361 | 0,167881 | y ~ current_feature + S |
| -0,10033 | 0,063053 | -0,04751 | -1,59121 | 252 | 249 | 0,112831 | 0,640171 | y ~ current_feature + S |
| -0,03844 | 0,067991 | 0,061498 | -0,56542 | 219 | 216 | 0,572375 | 0,937665 | y ~ current_feature + S |
| -0,13205 | 0,076935 | -0,02429 | -1,7164 | 169 | 166 | 0,087954 | 0,605474 | y ~ current_feature + S |
| -0,02399 | 0,053743 | -0,07874 | -0,44646 | 379 | 346,0236 | 0,655542 | 0,932255 | y ~ current_feature + S |
| -0,05608 | 0,069506 | 0,034007 | -0,80677 | 379 | 206,3397 | 0,420729 | 0,791109 | y ~ current_feature + S |
| 0,069484 | 0,054137 | -0,04751 | 1,283491 | 378 | 339,5592 | 0,200195 | 0,723562 | y ~ current_feature + S |
| 0,077126 | 0,057274 | 0,061498 | 1,346615 | 328 | 303,0392 | 0,179111 | 0,855109 | y ~ current_feature + S |
| -0,01308 | 0,064859 | -0,02429 | -0,20168 | 252 | 237,6755 | 0,840338 | 0,977734 | y ~ current_feature + S |
| -0,0705 | 0,067252 | -0,07874 | -1,04831 | 223 | 220 | 0,295645 | 0,718687 | y ~ current_feature + S |
| -0,11431 | 0,066978 | 0,034007 | -1,70661 | 223 | 220 | 0,089305 | 0,487845 | y ~ current_feature + S |
| -0,13509 | 0,066802 | -0,04751 | -2,02231 | 223 | 220 | 0,044355 | 0,63563 | y ~ current_feature + S |
| 0,026042 | 0,071587 | 0,061498 | 0,363785 | 198 | 195 | 0,716413 | 0,972573 | y ~ current_feature + S |
| -0,05972 | 0,082897 | -0,02429 | -0,72038 | 148 | 145 | 0,472451 | 0,876224 | y ~ current_feature + S |
| -0,36916 | 0,131432 | -0,07874 | -2,80875 | 53 | 50 | 0,007077 | 0,109106 | y ~ current_feature + S |
| -0,0374 | 0,141322 | 0,034007 | -0,26465 | 53 | 50 | 0,792369 | 0,945731 | y ~ current_feature + S |

| | | | | | | | | |
|---|---|---|---|---|---|---|---|---|
| 0,008527 | 0,141416 | -0,04751 | 0,060296 | 53 | 50 | 0,95216 | 0,990156 | y ~ current_feature + S |
| 0,12745 | 0,149526 | 0,061498 | 0,852357 | 47 | 44 | 0,398632 | 0,93433 | y ~ current_feature + S |
| -0,37203 | 0,161583 | -0,02429 | -2,30238 | 36 | 33 | 0,02776 | 0,485321 | y ~ current_feature + S |
| 4,44E-04 | 0,105409 | -0,07874 | 0,004215 | 93 | 90 | 0,996647 | 0,996647 | y ~ current_feature + S |
| 0,118842 | 0,104662 | 0,034007 | 1,135483 | 93 | 90 | 0,259188 | 0,71095 | y ~ current_feature + S |
| 0,102735 | 0,104852 | -0,04751 | 0,979818 | 93 | 90 | 0,329803 | 0,779476 | y ~ current_feature + S |
| -0,0367 | 0,111728 | 0,061498 | -0,32849 | 83 | 80 | 0,743401 | 0,972573 | y ~ current_feature + S |
| -0,04019 | 0,130084 | -0,02429 | -0,30894 | 62 | 59 | 0,758457 | 0,951727 | y ~ current_feature + S |
| -0,2172 | 0,111903 | -0,07874 | -1,94094 | 81 | 76,09054 | 0,055969 | 0,312071 | y ~ current_feature + S |
| 0,019392 | 0,159132 | 0,034007 | 0,121859 | 81 | 39,4748 | 0,903629 | 0,970516 | y ~ current_feature + S |
| -0,13108 | 0,117893 | -0,04751 | -1,11188 | 81 | 70,71222 | 0,269957 | 0,747915 | y ~ current_feature + S |
| 0,217818 | 0,121853 | 0,061498 | 1,787552 | 69 | 64,15347 | 0,078572 | 0,738262 | y ~ current_feature + S |
| -0,18191 | 0,137692 | -0,02429 | -1,32115 | 54 | 51 | 0,192349 | 0,65347 | y ~ current_feature + S |
| -0,0515 | 0,148873 | -0,07874 | -0,34591 | 48 | 45 | 0,731025 | 0,957449 | y ~ current_feature + S |
| 0,130298 | 0,1478 | 0,034007 | 0,88158 | 48 | 45 | 0,382689 | 0,759222 | y ~ current_feature + S |
| 0,333845 | 0,140519 | -0,04751 | 2,375806 | 48 | 45 | 0,021829 | 0,557019 | y ~ current_feature + S |
| 0,013991 | 0,162206 | 0,061498 | 0,086255 | 41 | 38 | 0,931717 | 0,996179 | y ~ current_feature + S |
| 0,013795 | 0,185678 | -0,02429 | 0,074294 | 32 | 29 | 0,941286 | 0,99444 | y ~ current_feature + S |
| 0,092975 | 0,053918 | -0,07874 | 1,724395 | 367 | 341,0103 | 0,085543 | 0,400643 | y ~ current_feature + S |
| 0,016593 | 0,071188 | 0,034007 | 0,233083 | 367 | 197,274 | 0,815939 | 0,952501 | y ~ current_feature + S |
| 0,00621 | 0,055476 | -0,04751 | 0,111949 | 366 | 324,9168 | 0,910933 | 0,985217 | y ~ current_feature + S |
| -0,03716 | 0,058491 | 0,061498 | -0,6353 | 318 | 291,8962 | 0,525728 | 0,937665 | y ~ current_feature + S |
| 0,100228 | 0,064335 | -0,02429 | 1,557907 | 244 | 239,177 | 0,120578 | 0,615361 | y ~ current_feature + S |
| 0,012201 | 0,058617 | -0,07874 | 0,208142 | 294 | 291 | 0,835264 | 0,982764 | y ~ current_feature + S |
| -0,0248 | 0,058603 | 0,034007 | -0,42316 | 294 | 291 | 0,672494 | 0,910826 | y ~ current_feature + S |
| 0,032136 | 0,058591 | -0,04751 | 0,548479 | 294 | 291 | 0,583784 | 0,907563 | y ~ current_feature + S |
| 5,20E-04 | 0,065703 | 0,061498 | 0,007918 | 254 | 231,6502 | 0,99369 | 0,999005 | y ~ current_feature + S |
| -0,11382 | 0,071514 | -0,02429 | -1,59156 | 196 | 193 | 0,113121 | 0,614731 | y ~ current_feature + S |
| 0,189294 | 0,078617 | -0,07874 | 2,407811 | 159 | 156 | 0,017216 | 0,176938 | y ~ current_feature + S |
| -0,0425 | 0,079992 | 0,034007 | -0,53126 | 159 | 156 | 0,595994 | 0,878557 | y ~ current_feature + S |
| 0,108737 | 0,079589 | -0,04751 | 1,366223 | 159 | 156 | 0,173835 | 0,700235 | y ~ current_feature + S |
| 0,089811 | 0,086361 | 0,061498 | 1,039955 | 136 | 133 | 0,300247 | 0,893818 | y ~ current_feature + S |
| 0,184857 | 0,096835 | -0,02429 | 1,908996 | 106 | 103 | 0,059046 | 0,558147 | y ~ current_feature + S |
| 0,069031 | 0,089952 | -0,07874 | 0,767418 | 126 | 123 | 0,444303 | 0,819912 | y ~ current_feature + S |
| 0,002776 | 0,090167 | 0,034007 | 0,030793 | 126 | 123 | 0,975485 | 0,989859 | y ~ current_feature + S |
| 0,048438 | 0,090061 | -0,04751 | 0,537835 | 126 | 123 | 0,591663 | 0,91049 | y ~ current_feature + S |
| -0,1441 | 0,097505 | 0,061498 | -1,47788 | 106 | 103 | 0,14249 | 0,8197 | y ~ current_feature + S |
| -0,04808 | 0,110983 | -0,02429 | -0,43321 | 84 | 81 | 0,666013 | 0,926342 | y ~ current_feature + S |
| -0,06292 | 0,153998 | -0,07874 | -0,40856 | 45 | 42 | 0,684941 | 0,93638 | y ~ current_feature + S |
| -0,17359 | 0,151961 | 0,034007 | -1,14237 | 45 | 42 | 0,259773 | 0,71095 | y ~ current_feature + S |
| 0,055046 | 0,154069 | -0,04751 | 0,357282 | 45 | 42 | 0,72267 | 0,93314 | y ~ current_feature + S |
| 0,225229 | 0,160175 | 0,061498 | 1,406145 | 40 | 37 | 0,168024 | 0,834482 | y ~ current_feature + S |
| -0,07838 | 0,191858 | -0,02429 | -0,40854 | 30 | 27 | 0,686098 | 0,926342 | y ~ current_feature + S |
| 0,134596 | 0,143024 | -0,07874 | 0,941073 | 51 | 48 | 0,35138 | 0,765193 | y ~ current_feature + S |
| 0,305217 | 0,13745 | 0,034007 | 2,220565 | 51 | 48 | 0,031135 | 0,347616 | y ~ current_feature + S |
| -0,18548 | 0,141833 | -0,04751 | -1,30771 | 51 | 48 | 0,1972 | 0,723562 | y ~ current_feature + S |
| -0,07104 | 0,157714 | 0,061498 | -0,45045 | 43 | 40 | 0,654816 | 0,957207 | y ~ current_feature + S |
| -0,10149 | 0,178678 | -0,02429 | -0,56799 | 34 | 31 | 0,574131 | 0,924682 | y ~ current_feature + S |

| | | | | | | | | |
|---|---|---|---|---|---|---|---|---|
| 0,162828 | 0,127377 | -0,07874 | 1,278324 | 63 | 60 | 0,206058 | 0,622938 | y ~ current_feature + S |
| 0,24295 | 0,125231 | 0,034007 | 1,940005 | 63 | 60 | 0,057083 | 0,420946 | y ~ current_feature + S |
| -0,03368 | 0,129026 | -0,04751 | -0,26103 | 63 | 60 | 0,794962 | 0,959661 | y ~ current_feature + S |
| 0,104642 | 0,142073 | 0,061498 | 0,736535 | 52 | 49 | 0,464917 | 0,937665 | y ~ current_feature + S |
| 0,21167 | 0,1565 | -0,02429 | 1,352525 | 42 | 39 | 0,183998 | 0,649807 | y ~ current_feature + S |
| -0,11277 | 0,107145 | -0,07874 | -1,05245 | 89 | 86 | 0,29554 | 0,718687 | y ~ current_feature + S |
| -0,14767 | 0,10665 | 0,034007 | -1,38466 | 89 | 86 | 0,169738 | 0,61875 | y ~ current_feature + S |
| -0,11855 | 0,107072 | -0,04751 | -1,10723 | 89 | 86 | 0,271281 | 0,747915 | y ~ current_feature + S |
| -0,08928 | 0,113505 | 0,061498 | -0,7866 | 80 | 77 | 0,433933 | 0,937665 | y ~ current_feature + S |
| -0,136 | 0,132389 | -0,02429 | -1,02731 | 59 | 56 | 0,308691 | 0,761472 | y ~ current_feature + S |
| 0,090937 | 0,04997 | -0,07874 | 1,819837 | 421 | 397,1683 | 0,069537 | 0,352445 | y ~ current_feature + S |
| 0,042221 | 0,064683 | 0,034007 | 0,652743 | 421 | 238,5886 | 0,51455 | 0,841996 | y ~ current_feature + S |
| -0,01789 | 0,051026 | -0,04751 | -0,35053 | 420 | 383,9504 | 0,726135 | 0,93314 | y ~ current_feature + S |
| 0,019735 | 0,056546 | 0,061498 | 0,34901 | 367 | 312,6242 | 0,727317 | 0,972573 | y ~ current_feature + S |
| 0,080155 | 0,059891 | -0,02429 | 1,338355 | 280 | 277 | 0,181878 | 0,647067 | y ~ current_feature + S |
| 0,191158 | 0,078841 | -0,07874 | 2,424611 | 158 | 155 | 0,016475 | 0,174676 | y ~ current_feature + S |
| 0,089046 | 0,080003 | 0,034007 | 1,113033 | 158 | 155 | 0,267417 | 0,71095 | y ~ current_feature + S |
| 0,018462 | 0,080308 | -0,04751 | 0,22989 | 158 | 155 | 0,81848 | 0,972405 | y ~ current_feature + S |
| 0,22963 | 0,084078 | 0,061498 | 2,731139 | 137 | 134 | 0,007162 | 0,417322 | y ~ current_feature + S |
| -0,03595 | 0,098469 | -0,02429 | -0,36507 | 106 | 103 | 0,71581 | 0,936058 | y ~ current_feature + S |
| -0,12957 | 0,116705 | -0,07874 | -1,11023 | 83 | 72,18831 | 0,270586 | 0,702573 | y ~ current_feature + S |
| -0,10042 | 0,126714 | 0,034007 | -0,79252 | 83 | 61,65206 | 0,431094 | 0,795535 | y ~ current_feature + S |
| -0,00981 | 0,113556 | -0,04751 | -0,08641 | 82 | 77,54157 | 0,931361 | 0,987403 | y ~ current_feature + S |
| -0,2176 | 0,116257 | 0,061498 | -1,87174 | 74 | 70,48528 | 0,065392 | 0,715249 | y ~ current_feature + S |
| -0,19129 | 0,150529 | -0,02429 | -1,27081 | 55 | 42,5177 | 0,210704 | 0,675518 | y ~ current_feature + S |
| 0,004167 | 0,084214 | -0,07874 | 0,049483 | 144 | 141 | 0,960605 | 0,994607 | y ~ current_feature + S |
| -0,08831 | 0,083886 | 0,034007 | -1,05279 | 144 | 141 | 0,29424 | 0,718742 | y ~ current_feature + S |
| -0,05528 | 0,084086 | -0,04751 | -0,65747 | 144 | 141 | 0,511952 | 0,868909 | y ~ current_feature + S |
| -0,06585 | 0,090712 | 0,061498 | -0,72597 | 124 | 121 | 0,469258 | 0,937665 | y ~ current_feature + S |
| -0,15519 | 0,102439 | -0,02429 | -1,51497 | 96 | 93 | 0,133171 | 0,617591 | y ~ current_feature + S |
| 0,114045 | 0,049461 | -0,07874 | 2,305741 | 421 | 403,4468 | 0,021632 | 0,19062 | y ~ current_feature + S |
| 0,15349 | 0,064961 | 0,034007 | 2,362819 | 421 | 231,3906 | 0,018965 | 0,311868 | y ~ current_feature + S |
| 0,033361 | 0,050582 | -0,04751 | 0,659533 | 420 | 390,4124 | 0,509942 | 0,868909 | y ~ current_feature + S |
| 0,064569 | 0,05559 | 0,061498 | 1,161533 | 367 | 322,252 | 0,246285 | 0,893818 | y ~ current_feature + S |
| 0,078567 | 0,060092 | -0,02429 | 1,30745 | 280 | 275,2184 | 0,192151 | 0,65347 | y ~ current_feature + S |
| 0,119861 | 0,052882 | -0,07874 | 2,266567 | 374 | 352,4479 | 0,024022 | 0,197019 | y ~ current_feature + S |
| 0,040341 | 0,069744 | 0,034007 | 0,578416 | 374 | 205,2487 | 0,563617 | 0,861729 | y ~ current_feature + S |
| 0,094032 | 0,055777 | -0,04751 | 1,685862 | 373 | 318,5915 | 0,092801 | 0,640171 | y ~ current_feature + S |
| 0,011687 | 0,059344 | 0,061498 | 0,196936 | 326 | 283,9133 | 0,844019 | 0,996179 | y ~ current_feature + S |
| 0,115044 | 0,064604 | -0,02429 | 1,780754 | 248 | 236,425 | 0,076237 | 0,605474 | y ~ current_feature + S |
| 0,16884 | 0,072271 | -0,07874 | 2,336211 | 189 | 186 | 0,020546 | 0,189455 | y ~ current_feature + S |
| 0,016349 | 0,073314 | 0,034007 | 0,222995 | 189 | 186 | 0,823784 | 0,952501 | y ~ current_feature + S |
| -0,00169 | 0,073323 | -0,04751 | -0,02311 | 189 | 186 | 0,981584 | 0,992311 | y ~ current_feature + S |
| -0,00233 | 0,078811 | 0,061498 | -0,02961 | 164 | 161 | 0,976417 | 0,996179 | y ~ current_feature + S |
| 0,057628 | 0,090017 | -0,02429 | 0,64019 | 126 | 123 | 0,52324 | 0,899808 | y ~ current_feature + S |
| 0,093405 | 0,075696 | -0,07874 | 1,233945 | 176 | 173 | 0,218897 | 0,639583 | y ~ current_feature + S |
| -0,02143 | 0,076011 | 0,034007 | -0,28187 | 176 | 173 | 0,77838 | 0,942718 | y ~ current_feature + S |
| -0,12981 | 0,075385 | -0,04751 | -1,7219 | 176 | 173 | 0,086875 | 0,640171 | y ~ current_feature + S |

| | | | | | | | | |
|---|---|---|---|---|---|---|---|---|
| -0,06619 | 0,082298 | 0,061498 | -0,80424 | 150 | 147 | 0,42256 | 0,937665 | y ~ current_feature + S |
| -0,08406 | 0,093327 | -0,02429 | -0,90066 | 117 | 114 | 0,36967 | 0,809337 | y ~ current_feature + S |
| 0,019203 | 0,082463 | -0,07874 | 0,232867 | 150 | 147 | 0,816188 | 0,979871 | y ~ current_feature + S |
| -0,04988 | 0,082376 | 0,034007 | -0,60551 | 150 | 147 | 0,545773 | 0,852506 | y ~ current_feature + S |
| 0,007666 | 0,082476 | -0,04751 | 0,092948 | 150 | 147 | 0,926071 | 0,985217 | y ~ current_feature + S |
| 0,031507 | 0,090122 | 0,061498 | 0,349604 | 126 | 123 | 0,727234 | 0,972573 | y ~ current_feature + S |
| -0,11549 | 0,100855 | -0,02429 | -1,14511 | 100 | 97 | 0,254981 | 0,718089 | y ~ current_feature + S |
| -0,10709 | 0,050475 | -0,07874 | -2,12153 | 391 | 388 | 0,034511 | 0,238671 | y ~ current_feature + S |
| -0,12694 | 0,050357 | 0,034007 | -2,52091 | 391 | 388 | 0,012105 | 0,235731 | y ~ current_feature + S |
| 0,061805 | 0,050736 | -0,04751 | 1,218176 | 390 | 387 | 0,223899 | 0,742928 | y ~ current_feature + S |
| -0,00877 | 0,054471 | 0,061498 | -0,16107 | 340 | 337 | 0,872139 | 0,996179 | y ~ current_feature + S |
| -0,10912 | 0,062006 | -0,02429 | -1,75987 | 260 | 257 | 0,079619 | 0,605474 | y ~ current_feature + S |
| 0,252212 | 0,122894 | -0,07874 | 2,052263 | 65 | 62 | 0,044372 | 0,279123 | y ~ current_feature + S |
| 0,10806 | 0,162355 | 0,034007 | 0,665581 | 65 | 37,49448 | 0,509753 | 0,841996 | y ~ current_feature + S |
| 0,104911 | 0,126299 | -0,04751 | 0,83065 | 65 | 62 | 0,409359 | 0,803665 | y ~ current_feature + S |
| 0,204638 | 0,169877 | 0,061498 | 1,204625 | 56 | 33,20111 | 0,236865 | 0,893818 | y ~ current_feature + S |
| 0,332484 | 0,149119 | -0,02429 | 2,22966 | 43 | 40 | 0,03145 | 0,485321 | y ~ current_feature + S |
| -0,16136 | 0,050478 | -0,07874 | -3,19664 | 418 | 382,2453 | 0,001506 | 0,048726 | y ~ current_feature + S |
| -0,08783 | 0,066389 | 0,034007 | -1,3229 | 418 | 225,1389 | 0,18721 | 0,638805 | y ~ current_feature + S |
| -0,02916 | 0,052544 | -0,04751 | -0,55496 | 417 | 361,8939 | 0,579266 | 0,904733 | y ~ current_feature + S |
| -0,0819 | 0,057138 | 0,061498 | -1,43331 | 364 | 304,2436 | 0,152796 | 0,8197 | y ~ current_feature + S |
| -0,14182 | 0,060008 | -0,02429 | -2,36338 | 278 | 272,1195 | 0,018812 | 0,428958 | y ~ current_feature + S |
| -0,07043 | 0,101809 | -0,07874 | -0,69174 | 99 | 96 | 0,490768 | 0,852508 | y ~ current_feature + S |
| -0,05659 | 0,101899 | 0,034007 | -0,55532 | 99 | 96 | 0,579966 | 0,869291 | y ~ current_feature + S |
| 0,00318 | 0,102062 | -0,04751 | 0,031161 | 99 | 96 | 0,975206 | 0,992311 | y ~ current_feature + S |
| -0,03937 | 0,111025 | 0,061498 | -0,35461 | 84 | 81 | 0,723802 | 0,972573 | y ~ current_feature + S |
| -0,07664 | 0,125618 | -0,02429 | -0,6101 | 66 | 63 | 0,543988 | 0,911914 | y ~ current_feature + S |
| 0,287093 | 0,104516 | -0,07874 | 2,746885 | 87 | 84 | 0,00736 | 0,110197 | y ~ current_feature + S |
| 0,173329 | 0,107457 | 0,034007 | 1,613005 | 87 | 84 | 0,110494 | 0,540997 | y ~ current_feature + S |
| -0,04558 | 0,108996 | -0,04751 | -0,4182 | 87 | 84 | 0,676866 | 0,928258 | y ~ current_feature + S |
| -0,23275 | 0,114615 | 0,061498 | -2,0307 | 75 | 72 | 0,045979 | 0,700056 | y ~ current_feature + S |
| 0,078904 | 0,13442 | -0,02429 | 0,587 | 58 | 55 | 0,559606 | 0,92024 | y ~ current_feature + S |
| 0,075613 | 0,099219 | -0,07874 | 0,762083 | 104 | 101 | 0,447786 | 0,822238 | y ~ current_feature + S |
| 0,064418 | 0,099297 | 0,034007 | 0,648739 | 104 | 101 | 0,517979 | 0,841996 | y ~ current_feature + S |
| -0,18345 | 0,098303 | -0,04751 | -1,86614 | 103 | 100 | 0,064952 | 0,640171 | y ~ current_feature + S |
| -0,04969 | 0,1077 | 0,061498 | -0,4614 | 89 | 86 | 0,645679 | 0,956496 | y ~ current_feature + S |
| 0,269332 | 0,118543 | -0,02429 | 2,272022 | 69 | 66 | 0,02635 | 0,485321 | y ~ current_feature + S |
| 0,009053 | 0,077379 | -0,07874 | 0,117 | 170 | 167 | 0,907001 | 0,987929 | y ~ current_feature + S |
| 0,089855 | 0,077069 | 0,034007 | 1,165894 | 170 | 167 | 0,24532 | 0,702074 | y ~ current_feature + S |
| 0,073856 | 0,077403 | -0,04751 | 0,95418 | 169 | 166 | 0,34138 | 0,789051 | y ~ current_feature + S |
| 0,130583 | 0,08764 | 0,061498 | 1,489983 | 144 | 127,9742 | 0,138689 | 0,8197 | y ~ current_feature + S |
| 0,165805 | 0,095433 | -0,02429 | 1,737405 | 113 | 106,7821 | 0,085199 | 0,605474 | y ~ current_feature + S |
| -0,03783 | 0,057125 | -0,07874 | -0,66214 | 309 | 306 | 0,508378 | 0,863164 | y ~ current_feature + S |
| -0,01847 | 0,057156 | 0,034007 | -0,32314 | 309 | 306 | 0,746807 | 0,928802 | y ~ current_feature + S |
| 0,040502 | 0,057119 | -0,04751 | 0,709077 | 309 | 306 | 0,478817 | 0,845643 | y ~ current_feature + S |
| -0,02603 | 0,061877 | 0,061498 | -0,42073 | 264 | 261 | 0,674302 | 0,958606 | y ~ current_feature + S |
| 0,058763 | 0,070238 | -0,02429 | 0,836619 | 205 | 202 | 0,403795 | 0,830024 | y ~ current_feature + S |
| 0,011515 | 0,055358 | -0,07874 | 0,208012 | 387 | 326,2689 | 0,835349 | 0,982764 | y ~ current_feature + S |

| | | | | | | | | |
|---|---|---|---|---|---|---|---|---|
| 0,073273 | 0,072975 | 0,034007 | 1,004088 | 387 | 186,7743 | 0,316635 | 0,718742 | y ~ current_feature + S |
| 0,038196 | 0,056778 | -0,04751 | 0,672727 | 386 | 309,7411 | 0,501623 | 0,868909 | y ~ current_feature + S |
| 0,00391 | 0,058865 | 0,061498 | 0,066421 | 336 | 288,5871 | 0,947088 | 0,996179 | y ~ current_feature + S |
| 0,034126 | 0,062668 | -0,02429 | 0,544547 | 258 | 254,3337 | 0,586542 | 0,924682 | y ~ current_feature + S |
| 0,009534 | 0,080579 | -0,07874 | 0,11832 | 157 | 154 | 0,905969 | 0,987929 | y ~ current_feature + S |
| -0,04106 | 0,080514 | 0,034007 | -0,51002 | 157 | 154 | 0,61077 | 0,889068 | y ~ current_feature + S |
| 0,067982 | 0,080658 | -0,04751 | 0,842838 | 156 | 153 | 0,400635 | 0,803665 | y ~ current_feature + S |
| -0,10671 | 0,085894 | 0,061498 | -1,24241 | 137 | 134 | 0,216257 | 0,893818 | y ~ current_feature + S |
| 0,015855 | 0,099491 | -0,02429 | 0,159356 | 104 | 101 | 0,873706 | 0,984468 | y ~ current_feature + S |
| -0,15828 | 0,05231 | -0,07874 | -3,02582 | 415 | 356,2919 | 0,00266 | 0,065908 | y ~ current_feature + S |
| -0,16757 | 0,068464 | 0,034007 | -2,44755 | 415 | 207,3512 | 0,015215 | 0,268069 | y ~ current_feature + S |
| -0,057 | 0,054653 | -0,04751 | -1,04293 | 414 | 333,7028 | 0,297734 | 0,753461 | y ~ current_feature + S |
| -0,06859 | 0,059046 | 0,061498 | -1,16166 | 361 | 285,4785 | 0,246343 | 0,893818 | y ~ current_feature + S |
| -0,02547 | 0,060533 | -0,02429 | -0,42074 | 276 | 272,734 | 0,674275 | 0,926342 | y ~ current_feature + S |
| -0,02114 | 0,088025 | -0,07874 | -0,24014 | 132 | 129 | 0,810604 | 0,979871 | y ~ current_feature + S |
| -0,06687 | 0,087848 | 0,034007 | -0,76116 | 132 | 129 | 0,44795 | 0,814299 | y ~ current_feature + S |
| 0,117611 | 0,087434 | -0,04751 | 1,345145 | 132 | 129 | 0,180938 | 0,704707 | y ~ current_feature + S |
| 0,051075 | 0,099016 | 0,061498 | 0,515821 | 111 | 101,7313 | 0,607099 | 0,945257 | y ~ current_feature + S |
| 0,042188 | 0,110831 | -0,02429 | 0,380656 | 88 | 81,26514 | 0,704452 | 0,93118 | y ~ current_feature + S |
| 0,033818 | 0,05879 | -0,07874 | 0,57523 | 292 | 289 | 0,565583 | 0,898449 | y ~ current_feature + S |
| 0,031601 | 0,058794 | 0,034007 | 0,537488 | 292 | 289 | 0,591344 | 0,876338 | y ~ current_feature + S |
| 0,007155 | 0,058822 | -0,04751 | 0,121637 | 292 | 289 | 0,903271 | 0,985217 | y ~ current_feature + S |
| 0,019947 | 0,063233 | 0,061498 | 0,315455 | 253 | 250 | 0,752679 | 0,972573 | y ~ current_feature + S |
| -0,0346 | 0,072126 | -0,02429 | -0,47973 | 195 | 192 | 0,631967 | 0,924682 | y ~ current_feature + S |
| 0,02466 | 0,049437 | -0,07874 | 0,498823 | 421 | 408,9081 | 0,618172 | 0,919937 | y ~ current_feature + S |
| -0,01606 | 0,0628 | 0,034007 | -0,25571 | 421 | 253,4937 | 0,798383 | 0,94832 | y ~ current_feature + S |
| 0,024673 | 0,050117 | -0,04751 | 0,492304 | 420 | 397,8867 | 0,622776 | 0,925756 | y ~ current_feature + S |
| -0,02692 | 0,054771 | 0,061498 | -0,49143 | 367 | 333,1073 | 0,623446 | 0,950526 | y ~ current_feature + S |
| 0,007388 | 0,062081 | -0,02429 | 0,119007 | 280 | 259,4523 | 0,905362 | 0,99444 | y ~ current_feature + S |
| 0,039015 | 0,077093 | -0,07874 | 0,506082 | 171 | 168 | 0,613462 | 0,918951 | y ~ current_feature + S |
| 0,011574 | 0,077147 | 0,034007 | 0,150026 | 171 | 168 | 0,880924 | 0,96591 | y ~ current_feature + S |
| -0,05979 | 0,077014 | -0,04751 | -0,77635 | 171 | 168 | 0,438637 | 0,823749 | y ~ current_feature + S |
| 0,199861 | 0,083108 | 0,061498 | 2,404848 | 142 | 139 | 0,017496 | 0,562908 | y ~ current_feature + S |
| -0,10715 | 0,094369 | -0,02429 | -1,13541 | 114 | 111 | 0,25865 | 0,719554 | y ~ current_feature + S |
| -0,04367 | 0,064895 | -0,07874 | -0,67287 | 240 | 237 | 0,501688 | 0,863164 | y ~ current_feature + S |
| -0,02608 | 0,064935 | 0,034007 | -0,40162 | 240 | 237 | 0,688322 | 0,916569 | y ~ current_feature + S |
| 0,113204 | 0,064539 | -0,04751 | 1,754022 | 240 | 237 | 0,080719 | 0,640171 | y ~ current_feature + S |
| 0,107978 | 0,071937 | 0,061498 | 1,50101 | 209 | 190,9857 | 0,135004 | 0,8197 | y ~ current_feature + S |
| -0,00413 | 0,080525 | -0,02429 | -0,05129 | 160 | 154,2164 | 0,959159 | 0,99444 | y ~ current_feature + S |
| -0,00805 | 0,068197 | -0,07874 | -0,11806 | 218 | 215 | 0,906133 | 0,987929 | y ~ current_feature + S |
| -0,08108 | 0,067975 | 0,034007 | -1,19284 | 218 | 215 | 0,234246 | 0,685147 | y ~ current_feature + S |
| -0,10416 | 0,067828 | -0,04751 | -1,53567 | 218 | 215 | 0,126091 | 0,654671 | y ~ current_feature + S |
| -0,13307 | 0,080589 | 0,061498 | -1,65116 | 187 | 151,2481 | 0,100781 | 0,8197 | y ~ current_feature + S |
| -0,04432 | 0,084229 | -0,02429 | -0,52617 | 145 | 140,6773 | 0,5996 | 0,924682 | y ~ current_feature + S |
| 0,197028 | 0,146149 | -0,07874 | 1,348127 | 48 | 45 | 0,184365 | 0,585536 | y ~ current_feature + S |
| 0,135418 | 0,147698 | 0,034007 | 0,916855 | 48 | 45 | 0,364106 | 0,7498 | y ~ current_feature + S |
| 0,290058 | 0,142662 | -0,04751 | 2,033177 | 48 | 45 | 0,047958 | 0,640171 | y ~ current_feature + S |
| 0,058161 | 0,159857 | 0,061498 | 0,363831 | 42 | 39 | 0,71795 | 0,972573 | y ~ current_feature + S |

| | | | | | | | | |
|---|---|---|---|---|---|---|---|---|
| 0,241747 | 0,180187 | -0,02429 | 1,341642 | 32 | 29 | 0,190124 | 0,65347 | y ~ current_feature + S |
| 0,219543 | 0,111899 | -0,07874 | 1,961984 | 81 | 76,01447 | 0,053425 | 0,30647 | y ~ current_feature + S |
| 0,176781 | 0,157753 | 0,034007 | 1,120619 | 81 | 38,92753 | 0,269316 | 0,71095 | y ~ current_feature + S |
| -0,21869 | 0,131083 | -0,04751 | -1,66834 | 81 | 55,41461 | 0,10089 | 0,640171 | y ~ current_feature + S |
| -0,13274 | 0,130623 | 0,061498 | -1,01617 | 72 | 57,57574 | 0,3138 | 0,893818 | y ~ current_feature + S |
| 0,12483 | 0,144592 | -0,02429 | 0,863327 | 54 | 47,08584 | 0,392334 | 0,821036 | y ~ current_feature + S |
| 0,272486 | 0,148464 | -0,07874 | 1,835358 | 45 | 42 | 0,07354 | 0,362799 | y ~ current_feature + S |
| -0,04257 | 0,154163 | 0,034007 | -0,27611 | 45 | 42 | 0,783819 | 0,945731 | y ~ current_feature + S |
| 0,321282 | 0,146123 | -0,04751 | 2,198714 | 45 | 42 | 0,033459 | 0,603897 | y ~ current_feature + S |
| 0,174825 | 0,157662 | 0,061498 | 1,108862 | 42 | 39 | 0,274281 | 0,893818 | y ~ current_feature + S |
| 0,09882 | 0,194004 | -0,02429 | 0,50937 | 30 | 26,30968 | 0,614739 | 0,924682 | y ~ current_feature + S |
| -0,44557 | 0,121827 | -0,07874 | -3,65742 | 57 | 54 | 5,79E-04 | 0,031617 | y ~ current_feature + S |
| -0,25756 | 0,131492 | 0,034007 | -1,95877 | 57 | 54 | 0,055312 | 0,420946 | y ~ current_feature + S |
| -0,21628 | 0,132862 | -0,04751 | -1,62786 | 57 | 54 | 0,109376 | 0,640171 | y ~ current_feature + S |
| -0,03069 | 0,145796 | 0,061498 | -0,21048 | 50 | 47 | 0,8342 | 0,996179 | y ~ current_feature + S |
| -0,21082 | 0,165232 | -0,02429 | -1,27588 | 38 | 35 | 0,2104 | 0,675518 | y ~ current_feature + S |
| -0,06756 | 0,067574 | -0,07874 | -0,99976 | 221 | 218 | 0,318534 | 0,723052 | y ~ current_feature + S |
| -0,10405 | 0,067361 | 0,034007 | -1,54467 | 221 | 218 | 0,123876 | 0,572145 | y ~ current_feature + S |
| 0,0358 | 0,067685 | -0,04751 | 0,528914 | 221 | 218 | 0,597403 | 0,911502 | y ~ current_feature + S |
| 0,082185 | 0,072686 | 0,061498 | 1,130685 | 191 | 188 | 0,259628 | 0,893818 | y ~ current_feature + S |
| -0,06333 | 0,083166 | -0,02429 | -0,76148 | 147 | 144 | 0,447616 | 0,858965 | y ~ current_feature + S |
| -0,03374 | 0,056835 | -0,07874 | -0,59356 | 343 | 309,2236 | 0,55324 | 0,889995 | y ~ current_feature + S |
| 0,048127 | 0,075463 | 0,034007 | 0,637756 | 343 | 175,1952 | 0,524465 | 0,84739 | y ~ current_feature + S |
| -0,00108 | 0,057637 | -0,04751 | -0,01875 | 342 | 301,0169 | 0,985055 | 0,993835 | y ~ current_feature + S |
| -0,05668 | 0,059922 | 0,061498 | -0,94593 | 295 | 277,6028 | 0,345008 | 0,907332 | y ~ current_feature + S |
| 0,068342 | 0,066781 | -0,02429 | 1,023368 | 228 | 223,1819 | 0,307242 | 0,761472 | y ~ current_feature + S |
| 0,013646 | 0,062864 | -0,07874 | 0,21708 | 256 | 253 | 0,828321 | 0,981192 | y ~ current_feature + S |
| -0,09313 | 0,062596 | 0,034007 | -1,48787 | 256 | 253 | 0,138031 | 0,58036 | y ~ current_feature + S |
| -0,07493 | 0,062693 | -0,04751 | -1,19526 | 256 | 253 | 0,233104 | 0,742928 | y ~ current_feature + S |
| -0,09084 | 0,067917 | 0,061498 | -1,33756 | 218 | 215 | 0,182453 | 0,857147 | y ~ current_feature + S |
| -0,12929 | 0,076733 | -0,02429 | -1,68488 | 170 | 167 | 0,09388 | 0,605474 | y ~ current_feature + S |
| 0,177053 | 0,058437 | -0,07874 | 3,029797 | 325 | 283,6538 | 0,002673 | 0,065908 | y ~ current_feature + S |
| 0,183088 | 0,078358 | 0,034007 | 2,336559 | 325 | 157,4071 | 0,020722 | 0,31947 | y ~ current_feature + S |
| 0,167799 | 0,060227 | -0,04751 | 2,786089 | 325 | 267,9223 | 0,005716 | 0,422981 | y ~ current_feature + S |
| -0,00723 | 0,06451 | 0,061498 | -0,11201 | 277 | 240,2799 | 0,91091 | 0,996179 | y ~ current_feature + S |
| 0,265591 | 0,066118 | -0,02429 | 4,016928 | 216 | 212,6149 | 8,18E-05 | 0,020175 | y ~ current_feature + S |
| 0,36762 | 0,093942 | -0,07874 | 3,91328 | 101 | 98 | 1,68E-04 | 0,015552 | y ~ current_feature + S |
| 0,152483 | 0,099834 | 0,034007 | 1,527361 | 101 | 98 | 0,129892 | 0,572145 | y ~ current_feature + S |
| 0,123538 | 0,100757 | -0,04751 | 1,226097 | 100 | 97 | 0,22313 | 0,742928 | y ~ current_feature + S |
| -0,01074 | 0,112502 | 0,061498 | -0,09542 | 82 | 79 | 0,924221 | 0,996179 | y ~ current_feature + S |
| 0,263132 | 0,120595 | -0,02429 | 2,181946 | 67 | 64 | 0,032792 | 0,485321 | y ~ current_feature + S |
| -0,07605 | 0,067225 | -0,07874 | -1,13128 | 223 | 220 | 0,259168 | 0,689873 | y ~ current_feature + S |
| -0,10114 | 0,067074 | 0,034007 | -1,50789 | 223 | 220 | 0,133017 | 0,572281 | y ~ current_feature + S |
| -0,03591 | 0,067377 | -0,04751 | -0,53293 | 223 | 220 | 0,594617 | 0,91049 | y ~ current_feature + S |
| -0,00215 | 0,072739 | 0,061498 | -0,02955 | 192 | 189 | 0,976458 | 0,996179 | y ~ current_feature + S |
| -0,13815 | 0,082249 | -0,02429 | -1,67961 | 148 | 145 | 0,095187 | 0,605474 | y ~ current_feature + S |
| -0,11777 | 0,079002 | -0,07874 | -1,49066 | 161 | 158 | 0,138045 | 0,498495 | y ~ current_feature + S |
| -0,04267 | 0,079483 | 0,034007 | -0,53686 | 161 | 158 | 0,59212 | 0,876338 | y ~ current_feature + S |

| | | | | | | | | |
|---|---|---|---|---|---|---|---|---|
| -0,07367 | 0,07934 | -0,04751 | -0,9285 | 161 | 158 | 0,354567 | 0,795089 | y ~ current_feature + S |
| 0,133543 | 0,085613 | 0,061498 | 1,559847 | 137 | 134 | 0,121155 | 0,8197 | y ~ current_feature + S |
| -0,17249 | 0,096588 | -0,02429 | -1,78579 | 107 | 104 | 0,077047 | 0,605474 | y ~ current_feature + S |
| 0,084763 | 0,119953 | -0,07874 | 0,706639 | 72 | 69 | 0,48217 | 0,845512 | y ~ current_feature + S |
| 0,003392 | 0,120385 | 0,034007 | 0,028179 | 72 | 69 | 0,977601 | 0,989859 | y ~ current_feature + S |
| -0,02843 | 0,120337 | -0,04751 | -0,23627 | 72 | 69 | 0,813924 | 0,972405 | y ~ current_feature + S |
| -0,03563 | 0,134754 | 0,061498 | -0,26444 | 58 | 55 | 0,792428 | 0,983887 | y ~ current_feature + S |
| -0,01052 | 0,149063 | -0,02429 | -0,0706 | 48 | 45 | 0,944032 | 0,99444 | y ~ current_feature + S |
| 0,311853 | 0,116953 | -0,07874 | 2,666485 | 69 | 66 | 0,00963 | 0,129567 | y ~ current_feature + S |
| 0,043585 | 0,122975 | 0,034007 | 0,35442 | 69 | 66 | 0,724154 | 0,920814 | y ~ current_feature + S |
| 0,059376 | 0,122874 | -0,04751 | 0,483224 | 69 | 66 | 0,630536 | 0,925756 | y ~ current_feature + S |
| 0,138987 | 0,134762 | 0,061498 | 1,031351 | 57 | 54 | 0,306974 | 0,893818 | y ~ current_feature + S |
| 0,051358 | 0,152297 | -0,02429 | 0,337223 | 46 | 43 | 0,737589 | 0,944216 | y ~ current_feature + S |
| 0,032277 | 0,139955 | -0,07874 | 0,230621 | 54 | 51 | 0,818532 | 0,979871 | y ~ current_feature + S |
| 0,047314 | 0,139871 | 0,034007 | 0,338267 | 54 | 51 | 0,73655 | 0,923809 | y ~ current_feature + S |
| 0,008649 | 0,140023 | -0,04751 | 0,06177 | 54 | 51 | 0,950987 | 0,990156 | y ~ current_feature + S |
| -0,04699 | 0,15233 | 0,061498 | -0,3085 | 46 | 43 | 0,759195 | 0,972573 | y ~ current_feature + S |
| 0,03367 | 0,173979 | -0,02429 | 0,193532 | 36 | 33 | 0,847729 | 0,977734 | y ~ current_feature + S |
| 0,081727 | 0,054814 | -0,07874 | 1,490969 | 385 | 330,5978 | 0,136923 | 0,498495 | y ~ current_feature + S |
| 0,043574 | 0,072484 | 0,034007 | 0,60115 | 385 | 189,9723 | 0,548456 | 0,854294 | y ~ current_feature + S |
| 0,00766 | 0,056547 | -0,04751 | 0,135456 | 384 | 312,7196 | 0,892338 | 0,985217 | y ~ current_feature + S |
| -0,00228 | 0,059418 | 0,061498 | -0,03836 | 333 | 283,2437 | 0,969429 | 0,996179 | y ~ current_feature + S |
| 0,107321 | 0,062796 | -0,02429 | 1,709027 | 256 | 250,668 | 0,088684 | 0,605474 | y ~ current_feature + S |
| -0,09194 | 0,073015 | -0,07874 | -1,25917 | 222 | 185,9907 | 0,209547 | 0,630344 | y ~ current_feature + S |
| -0,03785 | 0,095317 | 0,034007 | -0,39713 | 222 | 109,9107 | 0,692044 | 0,916569 | y ~ current_feature + S |
| -0,00232 | 0,07646 | -0,04751 | -0,0304 | 222 | 171,0533 | 0,97578 | 0,992311 | y ~ current_feature + S |
| 0,069906 | 0,081661 | 0,061498 | 0,856051 | 192 | 149,2266 | 0,393342 | 0,93433 | y ~ current_feature + S |
| 0,010025 | 0,083045 | -0,02429 | 0,120719 | 148 | 144,9878 | 0,904081 | 0,99444 | y ~ current_feature + S |
| 0,083334 | 0,082192 | -0,07874 | 1,013898 | 150 | 147 | 0,312298 | 0,718687 | y ~ current_feature + S |
| 0,106753 | 0,082007 | 0,034007 | 1,301745 | 150 | 147 | 0,19504 | 0,6438 | y ~ current_feature + S |
| -0,08121 | 0,082206 | -0,04751 | -0,98789 | 150 | 147 | 0,32483 | 0,775402 | y ~ current_feature + S |
| -2,20E-04 | 0,088045 | 0,061498 | -0,0025 | 132 | 129 | 0,998006 | 0,999005 | y ~ current_feature + S |
| -0,06662 | 0,101309 | -0,02429 | -0,65755 | 100 | 97 | 0,512383 | 0,899808 | y ~ current_feature + S |
| 0,052293 | 0,056446 | -0,07874 | 0,926421 | 316 | 313 | 0,354941 | 0,765762 | y ~ current_feature + S |
| -0,09937 | 0,056244 | 0,034007 | -1,76673 | 316 | 313 | 0,078249 | 0,470764 | y ~ current_feature + S |
| 0,038136 | 0,056573 | -0,04751 | 0,674099 | 315 | 312 | 0,500748 | 0,868909 | y ~ current_feature + S |
| 0,006272 | 0,061084 | 0,061498 | 0,102683 | 271 | 268 | 0,918291 | 0,996179 | y ~ current_feature + S |
| -0,04123 | 0,069446 | -0,02429 | -0,5937 | 210 | 207 | 0,553364 | 0,918138 | y ~ current_feature + S |
| -0,1316 | 0,061478 | -0,07874 | -2,14063 | 263 | 260 | 0,033234 | 0,236472 | y ~ current_feature + S |
| -0,19211 | 0,060862 | 0,034007 | -3,15652 | 263 | 260 | 0,001785 | 0,132069 | y ~ current_feature + S |
| 0,149978 | 0,061316 | -0,04751 | 2,445991 | 263 | 260 | 0,015109 | 0,526293 | y ~ current_feature + S |
| -0,00495 | 0,066518 | 0,061498 | -0,07439 | 229 | 226 | 0,940765 | 0,996179 | y ~ current_feature + S |
| -0,17574 | 0,075063 | -0,02429 | -2,34124 | 175 | 172 | 0,020366 | 0,443263 | y ~ current_feature + S |
| 0,280645 | 0,084836 | -0,07874 | 3,308083 | 131 | 128 | 0,00122 | 0,047511 | y ~ current_feature + S |
| 0,142703 | 0,087484 | 0,034007 | 1,631199 | 131 | 128 | 0,105306 | 0,526532 | y ~ current_feature + S |
| 0,046045 | 0,088295 | -0,04751 | 0,521497 | 131 | 128 | 0,602922 | 0,91263 | y ~ current_feature + S |
| -0,08718 | 0,094983 | 0,061498 | -0,91786 | 113 | 110 | 0,3607 | 0,920619 | y ~ current_feature + S |
| 0,21893 | 0,106462 | -0,02429 | 2,056419 | 87 | 84 | 0,042846 | 0,525585 | y ~ current_feature + S |

| | | | | | | | | |
|---|---|---|---|---|---|---|---|---|
| -0,06038 | 0,06147 | -0,07874 | -0,98225 | 295 | 263,6904 | 0,326876 | 0,726391 | y ~ current_feature + S |
| -0,09855 | 0,07973 | 0,034007 | -1,23608 | 295 | 155,7829 | 0,218288 | 0,664036 | y ~ current_feature + S |
| -0,0015 | 0,063512 | -0,04751 | -0,02367 | 294 | 247,906 | 0,981133 | 0,992311 | y ~ current_feature + S |
| 0,028544 | 0,065398 | 0,061498 | 0,43646 | 249 | 233,6215 | 0,662906 | 0,957207 | y ~ current_feature + S |
| 0,044493 | 0,07191 | -0,02429 | 0,618728 | 196 | 193 | 0,536825 | 0,906112 | y ~ current_feature + S |
| 0,395577 | 0,115712 | -0,07874 | 3,418648 | 66 | 63 | 0,001108 | 0,045561 | y ~ current_feature + S |
| 0,189 | 0,123717 | 0,034007 | 1,527675 | 66 | 63 | 0,131599 | 0,572281 | y ~ current_feature + S |
| 0,117528 | 0,125115 | -0,04751 | 0,939359 | 66 | 63 | 0,351134 | 0,795089 | y ~ current_feature + S |
| -0,0082 | 0,13867 | 0,061498 | -0,0591 | 55 | 52 | 0,953098 | 0,996179 | y ~ current_feature + S |
| 0,101071 | 0,155374 | -0,02429 | 0,650501 | 44 | 41 | 0,518998 | 0,899808 | y ~ current_feature + S |
| 0,019597 | 0,077946 | -0,07874 | 0,251417 | 203 | 164,5307 | 0,801805 | 0,979102 | y ~ current_feature + S |
| 0,030972 | 0,070677 | 0,034007 | 0,438215 | 203 | 200 | 0,661704 | 0,900251 | y ~ current_feature + S |
| 0,072344 | 0,079039 | -0,04751 | 0,915296 | 203 | 159,2341 | 0,36142 | 0,798361 | y ~ current_feature + S |
| -0,05461 | 0,081111 | 0,061498 | -0,6733 | 176 | 151,5449 | 0,501781 | 0,937665 | y ~ current_feature + S |
| -0,04985 | 0,087842 | -0,02429 | -0,56745 | 135 | 129,2762 | 0,571394 | 0,924682 | y ~ current_feature + S |
| 0,076726 | 0,133237 | -0,07874 | 0,575863 | 59 | 56 | 0,567015 | 0,898449 | y ~ current_feature + S |
| -0,00575 | 0,133628 | 0,034007 | -0,04302 | 59 | 56 | 0,965839 | 0,989859 | y ~ current_feature + S |
| 0,102832 | 0,132922 | -0,04751 | 0,773623 | 59 | 56 | 0,442408 | 0,824639 | y ~ current_feature + S |
| -0,06802 | 0,145527 | 0,061498 | -0,46737 | 50 | 47 | 0,642393 | 0,956496 | y ~ current_feature + S |
| 0,009684 | 0,166659 | -0,02429 | 0,058105 | 39 | 36 | 0,953986 | 0,99444 | y ~ current_feature + S |
| 0,155135 | 0,0754 | -0,07874 | 2,057502 | 181 | 171,6655 | 0,04115 | 0,274334 | y ~ current_feature + S |
| 0,055772 | 0,096122 | 0,034007 | 0,580215 | 181 | 107,8946 | 0,562979 | 0,861729 | y ~ current_feature + S |
| 0,07073 | 0,078678 | -0,04751 | 0,898984 | 180 | 160,7366 | 0,370006 | 0,802018 | y ~ current_feature + S |
| -0,07074 | 0,081056 | 0,061498 | -0,8727 | 155 | 151,4424 | 0,384209 | 0,93433 | y ~ current_feature + S |
| 0,121529 | 0,094178 | -0,02429 | 1,290415 | 120 | 111,0808 | 0,199586 | 0,660578 | y ~ current_feature + S |
| 0,155631 | 0,07554 | -0,07874 | 2,060244 | 174 | 171 | 0,04089 | 0,274334 | y ~ current_feature + S |
| 0,124411 | 0,075878 | 0,034007 | 1,639619 | 174 | 171 | 0,102923 | 0,525261 | y ~ current_feature + S |
| 0,075692 | 0,076253 | -0,04751 | 0,992654 | 174 | 171 | 0,322281 | 0,774585 | y ~ current_feature + S |
| -0,04427 | 0,089839 | 0,061498 | -0,4928 | 149 | 123,6574 | 0,623026 | 0,950526 | y ~ current_feature + S |
| 0,225315 | 0,093501 | -0,02429 | 2,409761 | 116 | 108,5781 | 0,017644 | 0,428958 | y ~ current_feature + S |
| 0,157043 | 0,050551 | -0,07874 | 3,106641 | 418 | 381,683 | 0,002034 | 0,055741 | y ~ current_feature + S |
| -0,03777 | 0,06744 | 0,034007 | -0,55999 | 418 | 219,5579 | 0,576058 | 0,869291 | y ~ current_feature + S |
| 0,04131 | 0,05242 | -0,04751 | 0,788062 | 417 | 363,2968 | 0,431175 | 0,820106 | y ~ current_feature + S |
| -0,04804 | 0,055718 | 0,061498 | -0,86219 | 364 | 321,3747 | 0,389226 | 0,93433 | y ~ current_feature + S |
| -0,03963 | 0,061235 | -0,02429 | -0,64715 | 278 | 266,2707 | 0,518094 | 0,899808 | y ~ current_feature + S |
| -0,03079 | 0,08244 | -0,07874 | -0,37343 | 150 | 147 | 0,709368 | 0,9454 | y ~ current_feature + S |
| -0,03287 | 0,082434 | 0,034007 | -0,39872 | 150 | 147 | 0,690675 | 0,916569 | y ~ current_feature + S |
| -0,04395 | 0,082399 | -0,04751 | -0,53339 | 150 | 147 | 0,594569 | 0,91049 | y ~ current_feature + S |
| 0,0309 | 0,090493 | 0,061498 | 0,341465 | 125 | 122 | 0,733341 | 0,972573 | y ~ current_feature + S |
| -0,06595 | 0,101314 | -0,02429 | -0,65093 | 100 | 97 | 0,516632 | 0,899808 | y ~ current_feature + S |
| 0,06849 | 0,049151 | -0,07874 | 1,393464 | 415 | 412 | 0,164231 | 0,535378 | y ~ current_feature + S |
| 0,077241 | 0,049119 | 0,034007 | 1,572512 | 415 | 412 | 0,1166 | 0,552558 | y ~ current_feature + S |
| 0,045135 | 0,049276 | -0,04751 | 0,91597 | 414 | 411 | 0,36022 | 0,798092 | y ~ current_feature + S |
| 0,087904 | 0,058238 | 0,061498 | 1,509396 | 361 | 292,5666 | 0,132277 | 0,8197 | y ~ current_feature + S |
| 0,115977 | 0,060567 | -0,02429 | 1,914858 | 276 | 268,9353 | 0,056571 | 0,558147 | y ~ current_feature + S |
| 0,148459 | 0,077754 | -0,07874 | 1,909358 | 187 | 161,7635 | 0,057986 | 0,318142 | y ~ current_feature + S |
| -0,06472 | 0,099942 | 0,034007 | -0,64757 | 187 | 99,69689 | 0,518753 | 0,841996 | y ~ current_feature + S |
| 0,089204 | 0,083924 | -0,04751 | 1,06291 | 187 | 140,8494 | 0,289642 | 0,747915 | y ~ current_feature + S |

| | | | | | | | | |
|---|---|---|---|---|---|---|---|---|
| 0,175935 | 0,084547 | 0,061498 | 2,080925 | 162 | 135,566 | 0,039323 | 0,658491 | y ~ current_feature + S |
| 0,035443 | 0,090852 | -0,02429 | 0,390116 | 124 | 121 | 0,697137 | 0,929516 | y ~ current_feature + S |
| -0,10318 | 0,06706 | -0,07874 | -1,53864 | 223 | 220 | 0,12533 | 0,478063 | y ~ current_feature + S |
| -0,14328 | 0,09654 | 0,034007 | -1,48412 | 223 | 105,0938 | 0,140771 | 0,588533 | y ~ current_feature + S |
| -0,11242 | 0,074023 | -0,04751 | -1,51871 | 222 | 180,1928 | 0,130588 | 0,654671 | y ~ current_feature + S |
| 0,019002 | 0,073151 | 0,061498 | 0,259767 | 190 | 186,8113 | 0,79533 | 0,984075 | y ~ current_feature + S |
| -0,20629 | 0,081456 | -0,02429 | -2,53255 | 148 | 144,2993 | 0,012393 | 0,366843 | y ~ current_feature + S |
| 0,101476 | 0,051011 | -0,07874 | 1,989314 | 421 | 380,3491 | 0,047382 | 0,289576 | y ~ current_feature + S |
| 0,158052 | 0,066468 | 0,034007 | 2,377848 | 421 | 220,69 | 0,018267 | 0,307222 | y ~ current_feature + S |
| 0,067772 | 0,051901 | -0,04751 | 1,305786 | 420 | 369,5291 | 0,192438 | 0,717449 | y ~ current_feature + S |
| -0,00608 | 0,056377 | 0,061498 | -0,10789 | 367 | 314,6106 | 0,914154 | 0,996179 | y ~ current_feature + S |
| 0,036417 | 0,060044 | -0,02429 | 0,606494 | 280 | 277 | 0,544684 | 0,911914 | y ~ current_feature + S |
| 0,200336 | 0,097487 | -0,07874 | 2,055012 | 104 | 101 | 0,042459 | 0,275611 | y ~ current_feature + S |
| -0,0977 | 0,099028 | 0,034007 | -0,98657 | 104 | 101 | 0,326212 | 0,728793 | y ~ current_feature + S |
| 0,132288 | 0,098629 | -0,04751 | 1,341261 | 104 | 101 | 0,182844 | 0,704811 | y ~ current_feature + S |
| -0,01319 | 0,107823 | 0,061498 | -0,12231 | 89 | 86 | 0,902942 | 0,996179 | y ~ current_feature + S |
| -0,03394 | 0,123021 | -0,02429 | -0,27592 | 69 | 66 | 0,783468 | 0,964545 | y ~ current_feature + S |
| -0,33314 | 0,128309 | -0,07874 | -2,59638 | 57 | 54 | 0,012111 | 0,144555 | y ~ current_feature + S |
| -0,18446 | 0,133748 | 0,034007 | -1,37914 | 57 | 54 | 0,173538 | 0,619418 | y ~ current_feature + S |
| -0,27806 | 0,130716 | -0,04751 | -2,12719 | 57 | 54 | 0,037988 | 0,611115 | y ~ current_feature + S |
| 0,007713 | 0,154299 | 0,061498 | 0,049987 | 45 | 42 | 0,96037 | 0,996179 | y ~ current_feature + S |
| 0,036159 | 0,16892 | -0,02429 | 0,214058 | 38 | 35 | 0,831744 | 0,976969 | y ~ current_feature + S |
| -0,08811 | 0,056214 | -0,07874 | -1,56742 | 317 | 314 | 0,118024 | 0,474142 | y ~ current_feature + S |
| -0,07458 | 0,056276 | 0,034007 | -1,32532 | 317 | 314 | 0,186027 | 0,638805 | y ~ current_feature + S |
| -0,07103 | 0,056291 | -0,04751 | -1,2619 | 317 | 314 | 0,207923 | 0,725768 | y ~ current_feature + S |
| -0,05725 | 0,066339 | 0,061498 | -0,86292 | 272 | 226,4809 | 0,389093 | 0,93433 | y ~ current_feature + S |
| -0,07725 | 0,069257 | -0,02429 | -1,11537 | 211 | 207,2387 | 0,265984 | 0,723659 | y ~ current_feature + S |
| 0,005866 | 0,15108 | -0,07874 | 0,038825 | 47 | 43,80964 | 0,969206 | 0,994607 | y ~ current_feature + S |
| 0,249239 | 0,197012 | 0,034007 | 1,265097 | 47 | 24,16372 | 0,217901 | 0,664036 | y ~ current_feature + S |
| -0,03514 | 0,162965 | -0,04751 | -0,21563 | 46 | 37,60743 | 0,830441 | 0,975438 | y ~ current_feature + S |
| 0,273003 | 0,177892 | 0,061498 | 1,534657 | 44 | 29,24494 | 0,135617 | 0,8197 | y ~ current_feature + S |
| 0,20889 | 0,184813 | -0,02429 | 1,130276 | 31 | 28 | 0,267949 | 0,723659 | y ~ current_feature + S |
| -0,11198 | 0,128287 | -0,07874 | -0,87288 | 63 | 60 | 0,386207 | 0,789252 | y ~ current_feature + S |
| -0,20779 | 0,126282 | 0,034007 | -1,64546 | 63 | 60 | 0,105104 | 0,526532 | y ~ current_feature + S |
| -0,1856 | 0,126857 | -0,04751 | -1,46303 | 63 | 60 | 0,148678 | 0,661823 | y ~ current_feature + S |
| -0,11478 | 0,136453 | 0,061498 | -0,84117 | 56 | 53 | 0,404035 | 0,93433 | y ~ current_feature + S |
| 0,064267 | 0,159797 | -0,02429 | 0,402177 | 42 | 39 | 0,689749 | 0,926342 | y ~ current_feature + S |
| -0,01405 | 0,137347 | -0,07874 | -0,10229 | 56 | 53 | 0,918912 | 0,989804 | y ~ current_feature + S |
| 0,367897 | 0,127727 | 0,034007 | 2,880343 | 56 | 53 | 0,005721 | 0,170187 | y ~ current_feature + S |
| 0,488785 | 0,120981 | -0,04751 | 4,040184 | 55 | 52 | 1,77E-04 | 0,067562 | y ~ current_feature + S |
| -0,1175 | 0,146421 | 0,061498 | -0,80252 | 49 | 46 | 0,426382 | 0,937665 | y ~ current_feature + S |
| 0,033408 | 0,172467 | -0,02429 | 0,193708 | 37 | 33,58171 | 0,847572 | 0,977734 | y ~ current_feature + S |
| -0,07129 | 0,049503 | -0,07874 | -1,44009 | 409 | 406 | 0,150612 | 0,513608 | y ~ current_feature + S |
| -0,10439 | 0,049358 | 0,034007 | -2,11492 | 409 | 406 | 0,035045 | 0,375847 | y ~ current_feature + S |
| 0,002571 | 0,04969 | -0,04751 | 0,051735 | 408 | 405 | 0,958766 | 0,990156 | y ~ current_feature + S |
| -0,0013 | 0,0606 | 0,061498 | -0,02138 | 356 | 272,3059 | 0,982959 | 0,997791 | y ~ current_feature + S |
| -0,02609 | 0,06095 | -0,02429 | -0,4281 | 272 | 269 | 0,668917 | 0,926342 | y ~ current_feature + S |
| 0,143452 | 0,049683 | -0,07874 | 2,887364 | 421 | 396,7904 | 0,004097 | 0,073642 | y ~ current_feature + S |

| | | | | | | | | |
|---|---|---|---|---|---|---|---|---|
| 0,168382 | 0,063963 | 0,034007 | 2,632507 | 421 | 237,4967 | 0,009032 | 0,215592 | y ~ current_feature + S |
| 0,060533 | 0,050679 | -0,04751 | 1,194454 | 420 | 387,9304 | 0,23303 | 0,742928 | y ~ current_feature + S |
| -0,05354 | 0,054864 | 0,061498 | -0,97578 | 367 | 331,2657 | 0,329886 | 0,893818 | y ~ current_feature + S |
| 0,078447 | 0,060049 | -0,02429 | 1,306391 | 280 | 275,618 | 0,192509 | 0,65347 | y ~ current_feature + S |
| -0,10387 | 0,079377 | -0,07874 | -1,30856 | 160 | 157 | 0,192595 | 0,598826 | y ~ current_feature + S |
| -0,07292 | 0,079596 | 0,034007 | -0,91612 | 160 | 157 | 0,361007 | 0,7498 | y ~ current_feature + S |
| -0,15183 | 0,078883 | -0,04751 | -1,9247 | 160 | 157 | 0,056075 | 0,640171 | y ~ current_feature + S |
| -0,07143 | 0,086166 | 0,061498 | -0,82895 | 137 | 134 | 0,408604 | 0,934833 | y ~ current_feature + S |
| -0,09913 | 0,098048 | -0,02429 | -1,01104 | 106 | 103 | 0,314367 | 0,765236 | y ~ current_feature + S |
| -0,00744 | 0,140024 | -0,07874 | -0,0531 | 54 | 51 | 0,957858 | 0,994607 | y ~ current_feature + S |
| 0,269583 | 0,134844 | 0,034007 | 1,999223 | 54 | 51 | 0,050926 | 0,420946 | y ~ current_feature + S |
| 0,050601 | 0,139849 | -0,04751 | 0,361829 | 54 | 51 | 0,718975 | 0,93314 | y ~ current_feature + S |
| 0,112513 | 0,148125 | 0,061498 | 0,759585 | 48 | 45 | 0,451464 | 0,937665 | y ~ current_feature + S |
| 0,493781 | 0,151376 | -0,02429 | 3,261956 | 36 | 33 | 0,002574 | 0,18344 | y ~ current_feature + S |
| -0,20124 | 0,092974 | -0,07874 | -2,16451 | 114 | 111 | 0,032568 | 0,233981 | y ~ current_feature + S |
| -0,17362 | 0,093474 | 0,034007 | -1,85744 | 114 | 111 | 0,065899 | 0,439525 | y ~ current_feature + S |
| -0,11002 | 0,09434 | -0,04751 | -1,16626 | 114 | 111 | 0,246009 | 0,742928 | y ~ current_feature + S |
| -0,13174 | 0,103348 | 0,061498 | -1,27475 | 95 | 92 | 0,205609 | 0,893818 | y ~ current_feature + S |
| -0,1875 | 0,114965 | -0,02429 | -1,63095 | 76 | 73 | 0,107208 | 0,605474 | y ~ current_feature + S |
| 0,003574 | 0,055222 | -0,07874 | 0,064727 | 351 | 327,9198 | 0,948431 | 0,994607 | y ~ current_feature + S |
| 0,052104 | 0,072355 | 0,034007 | 0,720114 | 351 | 190,4954 | 0,472337 | 0,831402 | y ~ current_feature + S |
| -0,00148 | 0,056188 | -0,04751 | -0,0263 | 350 | 316,7468 | 0,979036 | 0,992311 | y ~ current_feature + S |
| 0,009462 | 0,059467 | 0,061498 | 0,159107 | 310 | 282,7584 | 0,873699 | 0,996179 | y ~ current_feature + S |
| -0,0046 | 0,06616 | -0,02429 | -0,06949 | 233 | 228,4541 | 0,944664 | 0,99444 | y ~ current_feature + S |
| 0,029225 | 0,051764 | -0,07874 | 0,564583 | 416 | 372,8847 | 0,572697 | 0,899533 | y ~ current_feature + S |
| -0,00712 | 0,066982 | 0,034007 | -0,1063 | 416 | 222,8756 | 0,915444 | 0,9732 | y ~ current_feature + S |
| 0,065474 | 0,052287 | -0,04751 | 1,252189 | 415 | 364,2024 | 0,211305 | 0,733915 | y ~ current_feature + S |
| -0,02168 | 0,05605 | 0,061498 | -0,38674 | 362 | 318,1562 | 0,699204 | 0,972444 | y ~ current_feature + S |
| 0,011865 | 0,061228 | -0,02429 | 0,193787 | 276 | 266,706 | 0,84649 | 0,977734 | y ~ current_feature + S |
| 0,120085 | 0,051954 | -0,07874 | 2,311357 | 397 | 365,1299 | 0,021369 | 0,19062 | y ~ current_feature + S |
| 0,146844 | 0,06631 | 0,034007 | 2,214514 | 397 | 222,5257 | 0,027808 | 0,347616 | y ~ current_feature + S |
| 0,046161 | 0,053003 | -0,04751 | 0,870914 | 396 | 355,195 | 0,384389 | 0,803665 | y ~ current_feature + S |
| -0,03259 | 0,056895 | 0,061498 | -0,57287 | 345 | 308,598 | 0,567151 | 0,937665 | y ~ current_feature + S |
| 0,088377 | 0,062381 | -0,02429 | 1,416733 | 264 | 254,9731 | 0,157782 | 0,623497 | y ~ current_feature + S |
| 0,390503 | 0,104238 | -0,07874 | 3,746278 | 81 | 78 | 3,42E-04 | 0,023018 | y ~ current_feature + S |
| 0,240091 | 0,109916 | 0,034007 | 2,184314 | 81 | 78 | 0,031943 | 0,347616 | y ~ current_feature + S |
| 0,046298 | 0,113106 | -0,04751 | 0,409334 | 81 | 78 | 0,683417 | 0,928814 | y ~ current_feature + S |
| 0,001741 | 0,120386 | 0,061498 | 0,01446 | 72 | 69 | 0,988505 | 0,997945 | y ~ current_feature + S |
| -0,07448 | 0,139639 | -0,02429 | -0,53339 | 54 | 51 | 0,596078 | 0,924682 | y ~ current_feature + S |
| -0,01256 | 0,05717 | -0,07874 | -0,21972 | 323 | 305,9105 | 0,82624 | 0,981192 | y ~ current_feature + S |
| 0,050766 | 0,071041 | 0,034007 | 0,714593 | 323 | 197,6321 | 0,475704 | 0,831402 | y ~ current_feature + S |
| 0,016917 | 0,056741 | -0,04751 | 0,298146 | 322 | 310,5128 | 0,765791 | 0,941338 | y ~ current_feature + S |
| -0,09207 | 0,060592 | 0,061498 | -1,51942 | 283 | 270,0634 | 0,129826 | 0,8197 | y ~ current_feature + S |
| -0,04658 | 0,072026 | -0,02429 | -0,64677 | 215 | 192,3416 | 0,518549 | 0,899808 | y ~ current_feature + S |
| 0,024251 | 0,065214 | -0,07874 | 0,371866 | 238 | 235 | 0,710327 | 0,9454 | y ~ current_feature + S |
| -0,00351 | 0,065232 | 0,034007 | -0,05376 | 238 | 235 | 0,957175 | 0,989859 | y ~ current_feature + S |
| -0,02574 | 0,065211 | -0,04751 | -0,39474 | 238 | 235 | 0,693395 | 0,930326 | y ~ current_feature + S |
| -0,06203 | 0,069709 | 0,061498 | -0,88986 | 208 | 205 | 0,374586 | 0,932536 | y ~ current_feature + S |

| | | | | | | | | |
|---|---|---|---|---|---|---|---|---|
| -0,02683 | 0,080035 | -0,02429 | -0,33517 | 159 | 156 | 0,737949 | 0,944216 | y ~ current_feature + S |
| 0,468594 | 0,07493 | -0,07874 | 6,253737 | 142 | 139 | 4,63E-09 | 3,42E-06 | y ~ current_feature + S |
| 0,081515 | 0,084537 | 0,034007 | 0,964259 | 142 | 139 | 0,33659 | 0,73474 | y ~ current_feature + S |
| 0,03018 | 0,08478 | -0,04751 | 0,355978 | 142 | 139 | 0,722397 | 0,93314 | y ~ current_feature + S |
| -0,12687 | 0,089077 | 0,061498 | -1,42428 | 127 | 124 | 0,156877 | 0,8197 | y ~ current_feature + S |
| 0,211048 | 0,101909 | -0,02429 | 2,070946 | 95 | 92 | 0,041165 | 0,525585 | y ~ current_feature + S |
| 0,001741 | 0,09759 | -0,07874 | 0,017839 | 108 | 105 | 0,985801 | 0,996318 | y ~ current_feature + S |
| 0,04326 | 0,145022 | 0,034007 | 0,298297 | 108 | 47,45907 | 0,766778 | 0,93943 | y ~ current_feature + S |
| 0,042298 | 0,113624 | -0,04751 | 0,372266 | 108 | 77,3185 | 0,710713 | 0,93314 | y ~ current_feature + S |
| -0,10928 | 0,117467 | 0,061498 | -0,93026 | 93 | 71,60574 | 0,35536 | 0,920619 | y ~ current_feature + S |
| -0,04341 | 0,120272 | -0,02429 | -0,36095 | 72 | 69 | 0,719243 | 0,937042 | y ~ current_feature + S |
| 0,309994 | 0,072283 | -0,07874 | 4,288594 | 176 | 173 | 2,98E-05 | 0,005519 | y ~ current_feature + S |
| 0,131337 | 0,07537 | 0,034007 | 1,742562 | 176 | 173 | 0,083187 | 0,473527 | y ~ current_feature + S |
| 0,149925 | 0,075169 | -0,04751 | 1,994498 | 176 | 173 | 0,047668 | 0,640171 | y ~ current_feature + S |
| 0,01441 | 0,083616 | 0,061498 | 0,172336 | 146 | 143 | 0,863417 | 0,996179 | y ~ current_feature + S |
| 0,082416 | 0,093341 | -0,02429 | 0,882951 | 117 | 113,9963 | 0,379121 | 0,815551 | y ~ current_feature + S |
| 0,173603 | 0,05809 | -0,07874 | 2,988518 | 331 | 287,4152 | 0,003045 | 0,066821 | y ~ current_feature + S |
| 0,126773 | 0,076706 | 0,034007 | 1,652712 | 331 | 167,2271 | 0,100266 | 0,518857 | y ~ current_feature + S |
| 0,026613 | 0,060212 | -0,04751 | 0,441987 | 331 | 275,631 | 0,658845 | 0,925756 | y ~ current_feature + S |
| 0,082772 | 0,06216 | 0,061498 | 1,331602 | 282 | 257,0365 | 0,184171 | 0,857147 | y ~ current_feature + S |
| 0,097767 | 0,067559 | -0,02429 | 1,447133 | 220 | 217 | 0,149303 | 0,622449 | y ~ current_feature + S |
| 0,058938 | 0,055822 | -0,07874 | 1,055817 | 390 | 319,7989 | 0,291849 | 0,718687 | y ~ current_feature + S |
| 0,036575 | 0,072425 | 0,034007 | 0,505006 | 390 | 190,3897 | 0,614138 | 0,889359 | y ~ current_feature + S |
| -0,01712 | 0,056398 | -0,04751 | -0,30354 | 389 | 314,3013 | 0,761675 | 0,9394 | y ~ current_feature + S |
| -0,014 | 0,060508 | 0,061498 | -0,23145 | 338 | 273,0816 | 0,817138 | 0,996179 | y ~ current_feature + S |
| -0,05544 | 0,062718 | -0,02429 | -0,88395 | 259 | 253,4416 | 0,37756 | 0,815551 | y ~ current_feature + S |
| 0,083133 | 0,051393 | -0,07874 | 1,617599 | 379 | 376 | 0,106588 | 0,449749 | y ~ current_feature + S |
| 0,095999 | 0,051333 | 0,034007 | 1,870124 | 379 | 376 | 0,062243 | 0,437643 | y ~ current_feature + S |
| 0,043974 | 0,05159 | -0,04751 | 0,852368 | 378 | 375 | 0,394554 | 0,803665 | y ~ current_feature + S |
| -0,03731 | 0,062375 | 0,061498 | -0,59821 | 328 | 256,6674 | 0,550227 | 0,937665 | y ~ current_feature + S |
| 0,107655 | 0,063049 | -0,02429 | 1,707473 | 252 | 248,6449 | 0,088982 | 0,605474 | y ~ current_feature + S |
| -0,07765 | 0,050702 | -0,07874 | -1,53159 | 409 | 386,6566 | 0,12644 | 0,479825 | y ~ current_feature + S |
| -0,05758 | 0,065713 | 0,034007 | -0,87624 | 409 | 230,8137 | 0,381811 | 0,759222 | y ~ current_feature + S |
| -0,09418 | 0,051696 | -0,04751 | -1,82174 | 408 | 370,868 | 0,069299 | 0,640171 | y ~ current_feature + S |
| -0,06317 | 0,055957 | 0,061498 | -1,1289 | 356 | 318,0943 | 0,259791 | 0,893818 | y ~ current_feature + S |
| -0,01871 | 0,061272 | -0,02429 | -0,30536 | 272 | 266,2748 | 0,760332 | 0,951727 | y ~ current_feature + S |
| 0,01743 | 0,055969 | -0,07874 | 0,311416 | 341 | 319,13 | 0,755687 | 0,964335 | y ~ current_feature + S |
| 0,032122 | 0,073318 | 0,034007 | 0,438123 | 341 | 185,837 | 0,661806 | 0,900251 | y ~ current_feature + S |
| 0,096228 | 0,057902 | -0,04751 | 1,661917 | 340 | 295,5132 | 0,09759 | 0,640171 | y ~ current_feature + S |
| 0,034778 | 0,060469 | 0,061498 | 0,575134 | 295 | 273,1586 | 0,565674 | 0,937665 | y ~ current_feature + S |
| 0,119735 | 0,067413 | -0,02429 | 1,776137 | 227 | 216,8905 | 0,077112 | 0,605474 | y ~ current_feature + S |
| -0,05295 | 0,060401 | -0,07874 | -0,87657 | 326 | 273,3363 | 0,38149 | 0,789252 | y ~ current_feature + S |
| -0,03494 | 0,079312 | 0,034007 | -0,44056 | 326 | 158,778 | 0,660133 | 0,900251 | y ~ current_feature + S |
| -0,05142 | 0,061471 | -0,04751 | -0,83642 | 326 | 263,9444 | 0,403673 | 0,803665 | y ~ current_feature + S |
| -0,08731 | 0,062007 | 0,061498 | -1,40813 | 279 | 258,1057 | 0,160295 | 0,8197 | y ~ current_feature + S |
| 0,005887 | 0,068834 | -0,02429 | 0,085521 | 217 | 211,0439 | 0,931928 | 0,99444 | y ~ current_feature + S |
| -5,82E-04 | 0,077152 | -0,07874 | -0,00754 | 171 | 168 | 0,993995 | 0,996647 | y ~ current_feature + S |
| -0,0094 | 0,077148 | 0,034007 | -0,1219 | 171 | 168 | 0,903124 | 0,970516 | y ~ current_feature + S |

| | | | | | | | | |
|---|---|---|---|---|---|---|---|---|
| 0,021939 | 0,077133 | -0,04751 | 0,284429 | 171 | 168 | 0,776432 | 0,948119 | y ~ current_feature + S |
| -0,07122 | 0,083705 | 0,061498 | -0,85082 | 145 | 142 | 0,396303 | 0,93433 | y ~ current_feature + S |
| -0,03523 | 0,094857 | -0,02429 | -0,3714 | 114 | 111 | 0,71105 | 0,934596 | y ~ current_feature + S |
| 0,291249 | 0,123503 | -0,07874 | 2,358243 | 63 | 60 | 0,021638 | 0,19062 | y ~ current_feature + S |
| 0,103642 | 0,128404 | 0,034007 | 0,807154 | 63 | 60 | 0,422766 | 0,791701 | y ~ current_feature + S |
| -0,04325 | 0,128979 | -0,04751 | -0,3353 | 63 | 60 | 0,738568 | 0,933594 | y ~ current_feature + S |
| 0,127835 | 0,140261 | 0,061498 | 0,911406 | 53 | 50 | 0,366455 | 0,925517 | y ~ current_feature + S |
| 0,071923 | 0,159713 | -0,02429 | 0,450327 | 42 | 39 | 0,654968 | 0,924682 | y ~ current_feature + S |
| 0,17984 | 0,0974 | -0,07874 | 1,846403 | 105 | 102 | 0,067733 | 0,34663 | y ~ current_feature + S |
| 0,103737 | 0,098481 | 0,034007 | 1,053375 | 105 | 102 | 0,294658 | 0,718742 | y ~ current_feature + S |
| 0,019259 | 0,098996 | -0,04751 | 0,194538 | 105 | 102 | 0,846142 | 0,980218 | y ~ current_feature + S |
| 0,050461 | 0,106465 | 0,061498 | 0,47397 | 91 | 88 | 0,636695 | 0,955689 | y ~ current_feature + S |
| -0,00255 | 0,122169 | -0,02429 | -0,02088 | 70 | 67 | 0,983404 | 0,99444 | y ~ current_feature + S |
| -0,04156 | 0,052805 | -0,07874 | -0,78709 | 388 | 358,0103 | 0,431752 | 0,814177 | y ~ current_feature + S |
| -0,05815 | 0,069632 | 0,034007 | -0,83516 | 388 | 205,5493 | 0,4046 | 0,779245 | y ~ current_feature + S |
| 0,02045 | 0,054981 | -0,04751 | 0,37195 | 387 | 330,6664 | 0,710168 | 0,93314 | y ~ current_feature + S |
| -0,07265 | 0,058363 | 0,061498 | -1,24474 | 335 | 292,0254 | 0,214224 | 0,893818 | y ~ current_feature + S |
| -0,03965 | 0,062791 | -0,02429 | -0,63144 | 258 | 253,2322 | 0,528324 | 0,899808 | y ~ current_feature + S |
| 0,007546 | 0,064474 | -0,07874 | 0,117045 | 269 | 240,5504 | 0,906922 | 0,987929 | y ~ current_feature + S |
| -0,03968 | 0,084822 | 0,034007 | -0,46778 | 269 | 138,7724 | 0,640678 | 0,899624 | y ~ current_feature + S |
| 0,081323 | 0,066209 | -0,04751 | 1,228268 | 269 | 226,6106 | 0,220621 | 0,742928 | y ~ current_feature + S |
| 0,007594 | 0,069065 | 0,061498 | 0,109951 | 229 | 209,6347 | 0,912554 | 0,996179 | y ~ current_feature + S |
| -0,1034 | 0,075657 | -0,02429 | -1,36665 | 179 | 172,8359 | 0,173511 | 0,626332 | y ~ current_feature + S |
| 0,119582 | 0,076598 | -0,07874 | 1,561157 | 171 | 168 | 0,120368 | 0,477837 | y ~ current_feature + S |
| -0,02005 | 0,077136 | 0,034007 | -0,25987 | 171 | 168 | 0,795279 | 0,946875 | y ~ current_feature + S |
| -0,03074 | 0,077115 | -0,04751 | -0,39867 | 171 | 168 | 0,690644 | 0,930326 | y ~ current_feature + S |
| -0,02535 | 0,085098 | 0,061498 | -0,2979 | 141 | 138 | 0,766229 | 0,972573 | y ~ current_feature + S |
| -0,19184 | 0,093153 | -0,02429 | -2,05944 | 114 | 110,9997 | 0,041792 | 0,525585 | y ~ current_feature + S |
| 0,007824 | 0,074951 | -0,07874 | 0,104387 | 181 | 178 | 0,91698 | 0,989162 | y ~ current_feature + S |
| 0,023861 | 0,074932 | 0,034007 | 0,318434 | 181 | 178 | 0,750529 | 0,931865 | y ~ current_feature + S |
| -0,03122 | 0,074917 | -0,04751 | -0,41673 | 181 | 178 | 0,677378 | 0,928258 | y ~ current_feature + S |
| 0,062458 | 0,080165 | 0,061498 | 0,779111 | 158 | 155 | 0,437102 | 0,937665 | y ~ current_feature + S |
| 0,078187 | 0,091776 | -0,02429 | 0,851936 | 121 | 118 | 0,395975 | 0,823699 | y ~ current_feature + S |
| -3,48E-04 | 0,054404 | -0,07874 | -0,0064 | 385 | 337,8655 | 0,994898 | 0,996647 | y ~ current_feature + S |
| -0,04711 | 0,072882 | 0,034007 | -0,64634 | 385 | 187,8441 | 0,518846 | 0,841996 | y ~ current_feature + S |
| -0,06699 | 0,056837 | -0,04751 | -1,17862 | 384 | 308,1699 | 0,239457 | 0,742928 | y ~ current_feature + S |
| -0,11766 | 0,061155 | 0,061498 | -1,92403 | 335 | 263,6863 | 0,055427 | 0,715249 | y ~ current_feature + S |
| -0,08093 | 0,062991 | -0,02429 | -1,2848 | 256 | 250,3734 | 0,200049 | 0,660578 | y ~ current_feature + S |
| 0,077584 | 0,070006 | -0,07874 | 1,10824 | 222 | 202,8177 | 0,269071 | 0,702573 | y ~ current_feature + S |
| 0,059067 | 0,097529 | 0,034007 | 0,605639 | 222 | 104,7643 | 0,546065 | 0,852506 | y ~ current_feature + S |
| -0,0789 | 0,07164 | -0,04751 | -1,10127 | 222 | 193,6297 | 0,272146 | 0,747915 | y ~ current_feature + S |
| -0,0994 | 0,074466 | 0,061498 | -1,3348 | 191 | 178,5526 | 0,183641 | 0,857147 | y ~ current_feature + S |
| -0,06911 | 0,083064 | -0,02429 | -0,832 | 148 | 144,2416 | 0,406786 | 0,831551 | y ~ current_feature + S |
| 0,193169 | 0,079323 | -0,07874 | 2,435231 | 156 | 153 | 0,016032 | 0,174676 | y ~ current_feature + S |
| -0,07552 | 0,080614 | 0,034007 | -0,93686 | 156 | 153 | 0,350308 | 0,749214 | y ~ current_feature + S |
| 0,114345 | 0,080315 | -0,04751 | 1,423708 | 156 | 153 | 0,156567 | 0,673603 | y ~ current_feature + S |
| 0,075059 | 0,088485 | 0,061498 | 0,84826 | 130 | 127 | 0,397889 | 0,93433 | y ~ current_feature + S |
| 0,062292 | 0,09931 | -0,02429 | 0,627247 | 104 | 101 | 0,531912 | 0,901895 | y ~ current_feature + S |

| | | | | | | | | |
|---|---|---|---|---|---|---|---|---|
| 0,053898 | 0,071507 | -0,07874 | 0,753743 | 198 | 195 | 0,451913 | 0,82458 | y ~ current_feature + S |
| 0,029638 | 0,07158 | 0,034007 | 0,414047 | 198 | 195 | 0,679294 | 0,912427 | y ~ current_feature + S |
| 0,067408 | 0,071449 | -0,04751 | 0,943443 | 198 | 195 | 0,346622 | 0,795089 | y ~ current_feature + S |
| 0,382207 | 0,07692 | 0,061498 | 4,968861 | 171 | 144,3222 | 1,88E-06 | 0,001391 | y ~ current_feature + S |
| 0,042735 | 0,087965 | -0,02429 | 0,485823 | 132 | 129 | 0,627917 | 0,924682 | y ~ current_feature + S |
| 0,043725 | 0,056024 | -0,07874 | 0,780468 | 321 | 318 | 0,435696 | 0,81418 | y ~ current_feature + S |
| -0,05944 | 0,055978 | 0,034007 | -1,06187 | 321 | 318 | 0,289099 | 0,718742 | y ~ current_feature + S |
| -0,03647 | 0,05604 | -0,04751 | -0,65073 | 321 | 318 | 0,515688 | 0,871254 | y ~ current_feature + S |
| -0,13923 | 0,06513 | 0,061498 | -2,13775 | 277 | 231,1735 | 0,033587 | 0,658491 | y ~ current_feature + S |
| -0,08254 | 0,068608 | -0,02429 | -1,20314 | 214 | 211 | 0,23027 | 0,695919 | y ~ current_feature + S |
| -0,01757 | 0,089163 | -0,07874 | -0,19701 | 168 | 125,7468 | 0,844139 | 0,983548 | y ~ current_feature + S |
| 0,038266 | 0,100262 | 0,034007 | 0,381664 | 168 | 99,33279 | 0,703526 | 0,920814 | y ~ current_feature + S |
| 0,22618 | 0,085559 | -0,04751 | 2,643556 | 168 | 129,6171 | 0,009216 | 0,454673 | y ~ current_feature + S |
| -0,05664 | 0,090274 | 0,061498 | -0,62742 | 142 | 122,3158 | 0,531551 | 0,937665 | y ~ current_feature + S |
| 0,131857 | 0,096091 | -0,02429 | 1,372213 | 112 | 106,4192 | 0,172883 | 0,626332 | y ~ current_feature + S |
| 0,013531 | 0,086703 | -0,07874 | 0,156059 | 136 | 133 | 0,876224 | 0,987929 | y ~ current_feature + S |
| -0,0893 | 0,086365 | 0,034007 | -1,03403 | 136 | 133 | 0,302999 | 0,718742 | y ~ current_feature + S |
| 0,06869 | 0,086506 | -0,04751 | 0,794046 | 136 | 133 | 0,428583 | 0,819126 | y ~ current_feature + S |
| -0,0542 | 0,095206 | 0,061498 | -0,56926 | 113 | 110 | 0,57034 | 0,937665 | y ~ current_feature + S |
| -0,14199 | 0,106125 | -0,02429 | -1,33793 | 90 | 87 | 0,184405 | 0,649807 | y ~ current_feature + S |
| 0,075575 | 0,063004 | -0,07874 | 1,199528 | 275 | 250,4814 | 0,231456 | 0,657483 | y ~ current_feature + S |
| 0,057648 | 0,08524 | 0,034007 | 0,676305 | 275 | 137,1719 | 0,499986 | 0,841996 | y ~ current_feature + S |
| 0,067403 | 0,068105 | -0,04751 | 0,989687 | 275 | 214,6174 | 0,323442 | 0,774585 | y ~ current_feature + S |
| 0,041666 | 0,067352 | 0,061498 | 0,618621 | 239 | 220,0598 | 0,536806 | 0,937665 | y ~ current_feature + S |
| 0,02973 | 0,074687 | -0,02429 | 0,398063 | 183 | 179,1106 | 0,691058 | 0,926418 | y ~ current_feature + S |
| -0,00293 | 0,078811 | -0,07874 | -0,03715 | 164 | 161 | 0,970414 | 0,994607 | y ~ current_feature + S |
| -0,05264 | 0,078702 | 0,034007 | -0,66886 | 164 | 161 | 0,504545 | 0,841996 | y ~ current_feature + S |
| 0,065529 | 0,078887 | -0,04751 | 0,83067 | 163 | 160 | 0,407398 | 0,803665 | y ~ current_feature + S |
| -0,04831 | 0,085336 | 0,061498 | -0,56607 | 140 | 137 | 0,572269 | 0,937665 | y ~ current_feature + S |
| -0,06998 | 0,09689 | -0,02429 | -0,72223 | 109 | 106 | 0,471745 | 0,876224 | y ~ current_feature + S |
| 0,110151 | 0,106746 | -0,07874 | 1,031905 | 93 | 86,69588 | 0,304987 | 0,718687 | y ~ current_feature + S |
| 0,020691 | 0,152218 | 0,034007 | 0,135929 | 93 | 43,14031 | 0,89251 | 0,96591 | y ~ current_feature + S |
| -0,07409 | 0,115072 | -0,04751 | -0,64387 | 93 | 75,10476 | 0,521619 | 0,875279 | y ~ current_feature + S |
| 0,010661 | 0,128176 | 0,061498 | 0,083173 | 82 | 60,86082 | 0,933987 | 0,996179 | y ~ current_feature + S |
| 0,024585 | 0,131098 | -0,02429 | 0,18753 | 62 | 58,1495 | 0,851898 | 0,980411 | y ~ current_feature + S |
| -0,01083 | 0,076692 | -0,07874 | -0,14115 | 173 | 170 | 0,887915 | 0,987929 | y ~ current_feature + S |
| -0,08028 | 0,076449 | 0,034007 | -1,0501 | 173 | 170 | 0,295162 | 0,718742 | y ~ current_feature + S |
| -0,08295 | 0,076432 | -0,04751 | -1,08531 | 173 | 170 | 0,279322 | 0,747915 | y ~ current_feature + S |
| -0,04096 | 0,081854 | 0,061498 | -0,50039 | 152 | 149 | 0,617541 | 0,948092 | y ~ current_feature + S |
| -0,11341 | 0,093882 | -0,02429 | -1,208 | 115 | 112 | 0,229593 | 0,695919 | y ~ current_feature + S |
| 0,072048 | 0,062955 | -0,07874 | 1,144422 | 254 | 251 | 0,253539 | 0,679778 | y ~ current_feature + S |
| 0,02102 | 0,063105 | 0,034007 | 0,333098 | 254 | 251 | 0,739339 | 0,925737 | y ~ current_feature + S |
| 0,046005 | 0,063053 | -0,04751 | 0,729636 | 254 | 251 | 0,466293 | 0,842057 | y ~ current_feature + S |
| 0,031524 | 0,068166 | 0,061498 | 0,462456 | 218 | 215 | 0,644222 | 0,956496 | y ~ current_feature + S |
| 0,039445 | 0,077555 | -0,02429 | 0,508611 | 169 | 166 | 0,6117 | 0,924682 | y ~ current_feature + S |
| 0,076641 | 0,050359 | -0,07874 | 1,5219 | 395 | 392 | 0,12884 | 0,481525 | y ~ current_feature + S |
| 0,07696 | 0,050358 | 0,034007 | 1,528268 | 395 | 392 | 0,127253 | 0,572145 | y ~ current_feature + S |
| 0,060703 | 0,050479 | -0,04751 | 1,202552 | 394 | 391 | 0,229878 | 0,742928 | y ~ current_feature + S |

| | | | | | | | | |
|---|---|---|---|---|---|---|---|---|
| -0,08399 | 0,054041 | 0,061498 | -1,55419 | 343 | 340 | 0,121069 | 0,8197 | y ~ current_feature + S |
| 0,085421 | 0,061791 | -0,02429 | 1,382419 | 263 | 260 | 0,168029 | 0,624945 | y ~ current_feature + S |
| 0,066079 | 0,076529 | -0,07874 | 0,863447 | 173 | 170 | 0,389108 | 0,789252 | y ~ current_feature + S |
| 0,08021 | 0,076449 | 0,034007 | 1,049196 | 173 | 170 | 0,295577 | 0,718742 | y ~ current_feature + S |
| 0,085634 | 0,076641 | -0,04751 | 1,117347 | 172 | 169 | 0,265432 | 0,747915 | y ~ current_feature + S |
| -0,01027 | 0,083041 | 0,061498 | -0,12366 | 148 | 145 | 0,901759 | 0,996179 | y ~ current_feature + S |
| 0,157079 | 0,093318 | -0,02429 | 1,683262 | 115 | 112 | 0,09511 | 0,605474 | y ~ current_feature + S |
| 0,255912 | 0,144107 | -0,07874 | 1,775849 | 48 | 45 | 0,08252 | 0,388948 | y ~ current_feature + S |
| 0,040821 | 0,148947 | 0,034007 | 0,274065 | 48 | 45 | 0,785289 | 0,945731 | y ~ current_feature + S |
| -0,07966 | 0,148598 | -0,04751 | -0,53606 | 48 | 45 | 0,59456 | 0,91049 | y ~ current_feature + S |
| 0,048268 | 0,164207 | 0,061498 | 0,293948 | 40 | 37 | 0,770441 | 0,973246 | y ~ current_feature + S |
| -0,05247 | 0,18544 | -0,02429 | -0,28292 | 32 | 29 | 0,779245 | 0,961068 | y ~ current_feature + S |
| 0,028468 | 0,063534 | -0,07874 | 0,448077 | 259 | 247,5361 | 0,65449 | 0,932255 | y ~ current_feature + S |
| -0,11177 | 0,081457 | 0,034007 | -1,3721 | 259 | 148,8271 | 0,172097 | 0,618964 | y ~ current_feature + S |
| -0,02742 | 0,065105 | -0,04751 | -0,42118 | 258 | 235,7475 | 0,674007 | 0,928258 | y ~ current_feature + S |
| -0,00532 | 0,06825 | 0,061498 | -0,07789 | 226 | 214,677 | 0,937986 | 0,996179 | y ~ current_feature + S |
| -0,12924 | 0,077412 | -0,02429 | -1,66949 | 172 | 164,0867 | 0,096927 | 0,605474 | y ~ current_feature + S |
| -0,13715 | 0,062774 | -0,07874 | -2,18483 | 252 | 249 | 0,029834 | 0,220772 | y ~ current_feature + S |
| -0,11514 | 0,062951 | 0,034007 | -1,82909 | 252 | 249 | 0,068582 | 0,441313 | y ~ current_feature + S |
| 0,05914 | 0,063389 | -0,04751 | 0,932966 | 251 | 248 | 0,351745 | 0,795089 | y ~ current_feature + S |
| -0,07095 | 0,06787 | 0,061498 | -1,04537 | 219 | 216 | 0,297019 | 0,893818 | y ~ current_feature + S |
| -0,07815 | 0,077612 | -0,02429 | -1,00697 | 168 | 165 | 0,315425 | 0,765294 | y ~ current_feature + S |
| 0,050505 | 0,079454 | -0,07874 | 0,635648 | 161 | 158 | 0,525925 | 0,876542 | y ~ current_feature + S |
| 0,087927 | 0,079248 | 0,034007 | 1,109521 | 161 | 158 | 0,268892 | 0,71095 | y ~ current_feature + S |
| 0,168283 | 0,078421 | -0,04751 | 2,145891 | 161 | 158 | 0,033409 | 0,603897 | y ~ current_feature + S |
| 0,007719 | 0,092185 | 0,061498 | 0,083737 | 133 | 117,6675 | 0,933408 | 0,996179 | y ~ current_feature + S |
| 0,023004 | 0,098032 | -0,02429 | 0,234654 | 107 | 104 | 0,814939 | 0,974251 | y ~ current_feature + S |
| 0,264963 | 0,109887 | -0,07874 | 2,411222 | 80 | 77 | 0,018282 | 0,180385 | y ~ current_feature + S |
| 0,152498 | 0,112628 | 0,034007 | 1,354 | 80 | 77 | 0,179697 | 0,630217 | y ~ current_feature + S |
| -0,0429 | 0,113856 | -0,04751 | -0,3768 | 80 | 77 | 0,707357 | 0,93314 | y ~ current_feature + S |
| -0,06117 | 0,168162 | 0,061498 | -0,36376 | 67 | 35,23012 | 0,718213 | 0,972573 | y ~ current_feature + S |
| 0,110724 | 0,145781 | -0,02429 | 0,75952 | 53 | 46,4774 | 0,451377 | 0,861794 | y ~ current_feature + S |
| 0,0321 | 0,085082 | -0,07874 | 0,37728 | 141 | 138 | 0,706545 | 0,94376 | y ~ current_feature + S |
| 0,038742 | 0,085062 | 0,034007 | 0,455454 | 141 | 138 | 0,649499 | 0,900251 | y ~ current_feature + S |
| 0,066904 | 0,084935 | -0,04751 | 0,787709 | 141 | 138 | 0,432218 | 0,820106 | y ~ current_feature + S |
| 0,125064 | 0,09095 | 0,061498 | 1,375089 | 122 | 119 | 0,171687 | 0,840578 | y ~ current_feature + S |
| -0,17577 | 0,103196 | -0,02429 | -1,70326 | 94 | 91 | 0,091932 | 0,605474 | y ~ current_feature + S |
| 0,068127 | 0,051997 | -0,07874 | 1,310223 | 418 | 368,1549 | 0,190937 | 0,596175 | y ~ current_feature + S |
| 0,115343 | 0,068216 | 0,034007 | 1,690866 | 418 | 212,0394 | 0,092332 | 0,495112 | y ~ current_feature + S |
| 0,034646 | 0,053201 | -0,04751 | 0,651234 | 417 | 352,8872 | 0,515319 | 0,871254 | y ~ current_feature + S |
| -0,03735 | 0,057985 | 0,061498 | -0,64416 | 364 | 297,0007 | 0,519967 | 0,937665 | y ~ current_feature + S |
| -0,07036 | 0,060153 | -0,02429 | -1,16975 | 278 | 275 | 0,243115 | 0,711086 | y ~ current_feature + S |
| 0,107556 | 0,091914 | -0,07874 | 1,170186 | 120 | 117 | 0,244304 | 0,67447 | y ~ current_feature + S |
| 0,00501 | 0,092449 | 0,034007 | 0,054188 | 120 | 117 | 0,956878 | 0,989859 | y ~ current_feature + S |
| -0,0453 | 0,092355 | -0,04751 | -0,49051 | 120 | 117 | 0,62469 | 0,925756 | y ~ current_feature + S |
| 0,108763 | 0,098913 | 0,061498 | 1,099579 | 104 | 101 | 0,274129 | 0,893818 | y ~ current_feature + S |
| -0,14387 | 0,112775 | -0,02429 | -1,27571 | 80 | 77 | 0,205893 | 0,674164 | y ~ current_feature + S |
| -0,10973 | 0,100921 | -0,07874 | -1,08733 | 100 | 97 | 0,279586 | 0,71741 | y ~ current_feature + S |

| | | | | | | | | |
|---|---|---|---|---|---|---|---|---|
| 0,063869 | 0,101327 | 0,034007 | 0,630322 | 100 | 97 | 0,529966 | 0,848387 | y ~ current_feature + S |
| -0,01105 | 0,101528 | -0,04751 | -0,10882 | 100 | 97 | 0,913574 | 0,985217 | y ~ current_feature + S |
| -0,05253 | 0,108315 | 0,061498 | -0,48496 | 88 | 85 | 0,628952 | 0,952306 | y ~ current_feature + S |
| -0,05955 | 0,125765 | -0,02429 | -0,47352 | 66 | 63 | 0,637479 | 0,924682 | y ~ current_feature + S |
| -0,06095 | 0,113749 | -0,07874 | -0,53581 | 80 | 77 | 0,593637 | 0,903286 | y ~ current_feature + S |
| 0,108981 | 0,113282 | 0,034007 | 0,962032 | 80 | 77 | 0,339045 | 0,737921 | y ~ current_feature + S |
| 0,119148 | 0,113149 | -0,04751 | 1,053019 | 80 | 77 | 0,295625 | 0,752909 | y ~ current_feature + S |
| 0,017806 | 0,12215 | 0,061498 | 0,145773 | 70 | 67 | 0,884538 | 0,996179 | y ~ current_feature + S |
| 0,326983 | 0,133647 | -0,02429 | 2,446609 | 53 | 50 | 0,01798 | 0,428958 | y ~ current_feature + S |
| -0,04971 | 0,154113 | -0,07874 | -0,32255 | 45 | 42 | 0,748637 | 0,962676 | y ~ current_feature + S |
| -0,25271 | 0,149295 | 0,034007 | -1,69272 | 45 | 42 | 0,097916 | 0,517555 | y ~ current_feature + S |
| 0,099687 | 0,153535 | -0,04751 | 0,649277 | 45 | 42 | 0,519695 | 0,874032 | y ~ current_feature + S |
| -0,13582 | 0,158644 | 0,061498 | -0,85616 | 42 | 39 | 0,397141 | 0,93433 | y ~ current_feature + S |
| -0,22887 | 0,18736 | -0,02429 | -1,22152 | 30 | 26,99481 | 0,232448 | 0,695919 | y ~ current_feature + S |
| 0,03911 | 0,061499 | -0,07874 | 0,635943 | 267 | 264 | 0,525365 | 0,876542 | y ~ current_feature + S |
| 0,06015 | 0,061434 | 0,034007 | 0,979092 | 267 | 264 | 0,328431 | 0,729847 | y ~ current_feature + S |
| 0,050985 | 0,061582 | -0,04751 | 0,827911 | 266 | 263 | 0,408471 | 0,803665 | y ~ current_feature + S |
| 0,042138 | 0,074284 | 0,061498 | 0,567257 | 230 | 180,9015 | 0,571243 | 0,937665 | y ~ current_feature + S |
| 0,082945 | 0,075332 | -0,02429 | 1,101047 | 178 | 175 | 0,272388 | 0,732972 | y ~ current_feature + S |
| 0,119235 | 0,12926 | -0,07874 | 0,922445 | 62 | 59 | 0,360053 | 0,770057 | y ~ current_feature + S |
| -0,31152 | 0,12371 | 0,034007 | -2,51818 | 62 | 59 | 0,014529 | 0,262237 | y ~ current_feature + S |
| -0,03771 | 0,130096 | -0,04751 | -0,28986 | 62 | 59 | 0,77294 | 0,945414 | y ~ current_feature + S |
| 0,014722 | 0,141406 | 0,061498 | 0,104109 | 53 | 50 | 0,917499 | 0,996179 | y ~ current_feature + S |
| -0,13906 | 0,160645 | -0,02429 | -0,86563 | 41 | 38 | 0,392124 | 0,821036 | y ~ current_feature + S |
| 0,129906 | 0,069937 | -0,07874 | 1,857481 | 204 | 201 | 0,064706 | 0,339589 | y ~ current_feature + S |
| 0,064233 | 0,070389 | 0,034007 | 0,912548 | 204 | 201 | 0,362574 | 0,7498 | y ~ current_feature + S |
| 0,154407 | 0,069689 | -0,04751 | 2,215667 | 204 | 201 | 0,027837 | 0,588545 | y ~ current_feature + S |
| 0,080627 | 0,075781 | 0,061498 | 1,063953 | 176 | 173 | 0,288833 | 0,893818 | y ~ current_feature + S |
| -0,01208 | 0,086705 | -0,02429 | -0,13931 | 136 | 133 | 0,889416 | 0,992711 | y ~ current_feature + S |
| -0,11489 | 0,135182 | -0,07874 | -0,8499 | 57 | 54 | 0,399132 | 0,791912 | y ~ current_feature + S |
| -0,19462 | 0,133481 | 0,034007 | -1,45805 | 57 | 54 | 0,150619 | 0,604094 | y ~ current_feature + S |
| -0,19294 | 0,133526 | -0,04751 | -1,445 | 57 | 54 | 0,154236 | 0,672434 | y ~ current_feature + S |
| -0,03417 | 0,144253 | 0,061498 | -0,23687 | 51 | 48 | 0,813764 | 0,995348 | y ~ current_feature + S |
| -0,07832 | 0,168512 | -0,02429 | -0,46475 | 38 | 35 | 0,644992 | 0,924682 | y ~ current_feature + S |
| -8,67E-04 | 0,051109 | -0,07874 | -0,01697 | 412 | 382,8314 | 0,986473 | 0,996318 | y ~ current_feature + S |
| -0,07414 | 0,06611 | 0,034007 | -1,12141 | 412 | 227,5464 | 0,263295 | 0,71095 | y ~ current_feature + S |
| -0,02782 | 0,052352 | -0,04751 | -0,53134 | 411 | 364,5853 | 0,59551 | 0,91049 | y ~ current_feature + S |
| -0,06434 | 0,056721 | 0,061498 | -1,13427 | 359 | 309,5388 | 0,257558 | 0,893818 | y ~ current_feature + S |
| 0,019596 | 0,06132 | -0,02429 | 0,319566 | 274 | 265,8431 | 0,749548 | 0,94977 | y ~ current_feature + S |
| 0,040697 | 0,091594 | -0,07874 | 0,44432 | 122 | 119 | 0,657618 | 0,932255 | y ~ current_feature + S |
| 0,025864 | 0,091639 | 0,034007 | 0,282234 | 122 | 119 | 0,778255 | 0,942718 | y ~ current_feature + S |
| 0,174215 | 0,090268 | -0,04751 | 1,92998 | 122 | 119 | 0,055989 | 0,640171 | y ~ current_feature + S |
| -0,19537 | 0,094371 | 0,061498 | -2,07027 | 111 | 108 | 0,040812 | 0,658491 | y ~ current_feature + S |
| 0,022312 | 0,112481 | -0,02429 | 0,198359 | 82 | 79 | 0,843274 | 0,977734 | y ~ current_feature + S |
| -0,2084 | 0,06594 | -0,07874 | -3,16049 | 223 | 220 | 0,001797 | 0,053181 | y ~ current_feature + S |
| -0,13699 | 0,066784 | 0,034007 | -2,05119 | 223 | 220 | 0,041434 | 0,398193 | y ~ current_feature + S |
| -0,04572 | 0,067349 | -0,04751 | -0,67882 | 223 | 220 | 0,497965 | 0,868909 | y ~ current_feature + S |
| 0,008601 | 0,07789 | 0,061498 | 0,110419 | 192 | 164,8168 | 0,912212 | 0,996179 | y ~ current_feature + S |

| | | | | | | | | |
|---|---|---|---|---|---|---|---|---|
| -0,13765 | 0,081983 | -0,02429 | -1,67897 | 149 | 145,9644 | 0,095297 | 0,605474 | y ~ current_feature + S |
| -0,00141 | 0,11547 | -0,07874 | -0,01223 | 78 | 75 | 0,990278 | 0,996647 | y ~ current_feature + S |
| -0,23219 | 0,112314 | 0,034007 | -2,06729 | 78 | 75 | 0,042158 | 0,399959 | y ~ current_feature + S |
| -0,09543 | 0,114943 | -0,04751 | -0,83027 | 78 | 75 | 0,409023 | 0,803665 | y ~ current_feature + S |
| -0,0469 | 0,122956 | 0,061498 | -0,38141 | 69 | 66 | 0,704126 | 0,972444 | y ~ current_feature + S |
| 0,136241 | 0,141525 | -0,02429 | 0,962662 | 52 | 49 | 0,340444 | 0,785829 | y ~ current_feature + S |
| 0,120188 | 0,07526 | -0,07874 | 1,596962 | 177 | 174 | 0,112089 | 0,465986 | y ~ current_feature + S |
| 0,117595 | 0,075284 | 0,034007 | 1,562023 | 177 | 174 | 0,1201 | 0,558954 | y ~ current_feature + S |
| 0,138944 | 0,075074 | -0,04751 | 1,850749 | 177 | 174 | 0,065901 | 0,640171 | y ~ current_feature + S |
| 0,127698 | 0,080447 | 0,061498 | 1,587366 | 155 | 152 | 0,114508 | 0,8197 | y ~ current_feature + S |
| 0,163192 | 0,092 | -0,02429 | 1,773819 | 118 | 115 | 0,07874 | 0,605474 | y ~ current_feature + S |
| 0,129742 | 0,147811 | -0,07874 | 0,877757 | 48 | 45 | 0,384738 | 0,789252 | y ~ current_feature + S |
| 0,130748 | 0,147792 | 0,034007 | 0,88468 | 48 | 45 | 0,381032 | 0,759222 | y ~ current_feature + S |
| 0,015205 | 0,149054 | -0,04751 | 0,102009 | 48 | 45 | 0,919203 | 0,985217 | y ~ current_feature + S |
| 0,29635 | 0,157014 | 0,061498 | 1,887413 | 40 | 37 | 0,066968 | 0,715249 | y ~ current_feature + S |
| 0,346119 | 0,174218 | -0,02429 | 1,986707 | 32 | 29 | 0,056476 | 0,558147 | y ~ current_feature + S |
| -0,00852 | 0,060488 | -0,07874 | -0,14078 | 318 | 273,2951 | 0,88815 | 0,987929 | y ~ current_feature + S |
| -0,10617 | 0,077451 | 0,034007 | -1,37078 | 318 | 164,8232 | 0,172306 | 0,618964 | y ~ current_feature + S |
| -0,01218 | 0,061057 | -0,04751 | -0,19942 | 317 | 268,2046 | 0,842084 | 0,980218 | y ~ current_feature + S |
| -0,01323 | 0,06641 | 0,061498 | -0,19919 | 272 | 226,7035 | 0,84229 | 0,996179 | y ~ current_feature + S |
| -0,00346 | 0,069337 | -0,02429 | -0,0499 | 211 | 208 | 0,96025 | 0,99444 | y ~ current_feature + S |
| -0,09595 | 0,049957 | -0,07874 | -1,92067 | 400 | 397 | 0,05549 | 0,312071 | y ~ current_feature + S |
| -0,09651 | 0,049954 | 0,034007 | -1,93198 | 400 | 397 | 0,054073 | 0,420946 | y ~ current_feature + S |
| -0,04195 | 0,050208 | -0,04751 | -0,83552 | 399 | 396 | 0,403928 | 0,803665 | y ~ current_feature + S |
| -0,02418 | 0,053901 | 0,061498 | -0,44862 | 347 | 344 | 0,653988 | 0,957207 | y ~ current_feature + S |
| -0,06059 | 0,061549 | -0,02429 | -0,98445 | 266 | 263 | 0,325801 | 0,774043 | y ~ current_feature + S |
| 0,023324 | 0,054287 | -0,07874 | 0,429637 | 386 | 339,131 | 0,667732 | 0,934653 | y ~ current_feature + S |
| -0,0321 | 0,071234 | 0,034007 | -0,45065 | 386 | 196,8688 | 0,652741 | 0,900251 | y ~ current_feature + S |
| -0,06439 | 0,056155 | -0,04751 | -1,14671 | 385 | 315,8066 | 0,252368 | 0,742928 | y ~ current_feature + S |
| -0,02904 | 0,063563 | 0,061498 | -0,45693 | 337 | 247,2979 | 0,648126 | 0,957207 | y ~ current_feature + S |
| 0,014668 | 0,062739 | -0,02429 | 0,233795 | 257 | 254 | 0,815332 | 0,974251 | y ~ current_feature + S |
| 0,169527 | 0,066444 | -0,07874 | 2,551427 | 223 | 220 | 0,011407 | 0,140685 | y ~ current_feature + S |
| 0,119393 | 0,066938 | 0,034007 | 1,783646 | 223 | 220 | 0,07586 | 0,464477 | y ~ current_feature + S |
| 0,140167 | 0,066907 | -0,04751 | 2,09497 | 222 | 219 | 0,037323 | 0,611115 | y ~ current_feature + S |
| 0,083969 | 0,072102 | 0,061498 | 1,164592 | 194 | 191 | 0,245637 | 0,893818 | y ~ current_feature + S |
| 0,203443 | 0,081309 | -0,02429 | 2,502103 | 148 | 145 | 0,013456 | 0,382986 | y ~ current_feature + S |
| -0,05063 | 0,079203 | -0,07874 | -0,63923 | 162 | 159 | 0,523595 | 0,876542 | y ~ current_feature + S |
| -0,10731 | 0,078847 | 0,034007 | -1,361 | 162 | 159 | 0,17544 | 0,621173 | y ~ current_feature + S |
| -0,09575 | 0,07919 | -0,04751 | -1,20911 | 161 | 158 | 0,228429 | 0,742928 | y ~ current_feature + S |
| -0,11703 | 0,083637 | 0,061498 | -1,39924 | 144 | 141 | 0,163937 | 0,8197 | y ~ current_feature + S |
| -0,1959 | 0,095699 | -0,02429 | -2,04707 | 108 | 105 | 0,043148 | 0,525585 | y ~ current_feature + S |
| -0,07884 | 0,074511 | -0,07874 | -1,05813 | 182 | 179 | 0,291421 | 0,718687 | y ~ current_feature + S |
| 0,127883 | 0,07413 | 0,034007 | 1,725128 | 182 | 179 | 0,08623 | 0,479777 | y ~ current_feature + S |
| 0,079038 | 0,07451 | -0,04751 | 1,060769 | 182 | 179 | 0,290224 | 0,747915 | y ~ current_feature + S |
| 0,041889 | 0,078742 | 0,061498 | 0,531983 | 164 | 161 | 0,595471 | 0,945257 | y ~ current_feature + S |
| 0,087695 | 0,091703 | -0,02429 | 0,956292 | 121 | 118 | 0,34088 | 0,785829 | y ~ current_feature + S |
| 0,043069 | 0,05287 | -0,07874 | 0,814627 | 400 | 357,0904 | 0,415829 | 0,811887 | y ~ current_feature + S |
| -0,08606 | 0,068778 | 0,034007 | -1,25123 | 400 | 209,8294 | 0,212245 | 0,664036 | y ~ current_feature + S |

| | | | | | | | | |
|---|---|---|---|---|---|---|---|---|
| 0,130506 | 0,053154 | -0,04751 | 2,455242 | 400 | 347,9118 | 0,014568 | 0,526293 | y ~ current_feature + S |
| -0,00587 | 0,057817 | 0,061498 | -0,10151 | 348 | 299,1399 | 0,919211 | 0,996179 | y ~ current_feature + S |
| -0,02621 | 0,062613 | -0,02429 | -0,41868 | 266 | 254,9002 | 0,675806 | 0,926342 | y ~ current_feature + S |
| 0,004356 | 0,109763 | -0,07874 | 0,039682 | 86 | 83 | 0,968442 | 0,994607 | y ~ current_feature + S |
| 0,116433 | 0,109018 | 0,034007 | 1,068015 | 86 | 83 | 0,28861 | 0,718742 | y ~ current_feature + S |
| 0,043034 | 0,109663 | -0,04751 | 0,392424 | 86 | 83 | 0,695751 | 0,931023 | y ~ current_feature + S |
| 0,081162 | 0,119129 | 0,061498 | 0,681294 | 73 | 70 | 0,497933 | 0,937665 | y ~ current_feature + S |
| 0,187228 | 0,133676 | -0,02429 | 1,400606 | 57 | 54 | 0,167052 | 0,624945 | y ~ current_feature + S |
| 0,010751 | 0,060189 | -0,07874 | 0,178612 | 279 | 276 | 0,858374 | 0,983548 | y ~ current_feature + S |
| -0,07304 | 0,060032 | 0,034007 | -1,21669 | 279 | 276 | 0,224762 | 0,678874 | y ~ current_feature + S |
| -0,0255 | 0,060283 | -0,04751 | -0,42303 | 278 | 275 | 0,672604 | 0,928258 | y ~ current_feature + S |
| 0,063916 | 0,064961 | 0,061498 | 0,983906 | 239 | 236 | 0,326169 | 0,893818 | y ~ current_feature + S |
| 0,147469 | 0,073114 | -0,02429 | 2,01697 | 186 | 183 | 0,04516 | 0,525585 | y ~ current_feature + S |
| -0,01794 | 0,051907 | -0,07874 | -0,34554 | 401 | 371,0319 | 0,729883 | 0,957449 | y ~ current_feature + S |
| -0,07842 | 0,06767 | 0,034007 | -1,15889 | 401 | 217,0338 | 0,247774 | 0,702074 | y ~ current_feature + S |
| 0,074778 | 0,052008 | -0,04751 | 1,437832 | 400 | 367,6464 | 0,151332 | 0,666583 | y ~ current_feature + S |
| 0,112476 | 0,0581 | 0,061498 | 1,935897 | 350 | 292,4904 | 0,053844 | 0,715249 | y ~ current_feature + S |
| 0,003511 | 0,062009 | -0,02429 | 0,056628 | 267 | 260,0631 | 0,954885 | 0,99444 | y ~ current_feature + S |
| 0,041109 | 0,058361 | -0,07874 | 0,704396 | 327 | 293,1028 | 0,481746 | 0,845512 | y ~ current_feature + S |
| 0,048024 | 0,078774 | 0,034007 | 0,609645 | 327 | 160,7812 | 0,542958 | 0,852506 | y ~ current_feature + S |
| -0,06923 | 0,061207 | -0,04751 | -1,13112 | 326 | 265,649 | 0,259023 | 0,742928 | y ~ current_feature + S |
| -7,89E-05 | 0,063151 | 0,061498 | -0,00125 | 282 | 250,7455 | 0,999005 | 0,999005 | y ~ current_feature + S |
| -8,73E-04 | 0,068199 | -0,02429 | -0,0128 | 218 | 215 | 0,989803 | 0,995182 | y ~ current_feature + S |
| -0,08763 | 0,086377 | -0,07874 | -1,01445 | 136 | 133 | 0,31221 | 0,718687 | y ~ current_feature + S |
| -0,15979 | 0,085597 | 0,034007 | -1,86678 | 136 | 133 | 0,064134 | 0,439434 | y ~ current_feature + S |
| -0,06217 | 0,086543 | -0,04751 | -0,71831 | 136 | 133 | 0,473825 | 0,842057 | y ~ current_feature + S |
| 0,110847 | 0,097667 | 0,061498 | 1,134948 | 118 | 103,5472 | 0,259017 | 0,893818 | y ~ current_feature + S |
| -0,17524 | 0,104951 | -0,02429 | -1,66972 | 91 | 88 | 0,098529 | 0,605474 | y ~ current_feature + S |
| 0,519514 | 0,124636 | -0,07874 | 4,168252 | 50 | 47 | 1,31E-04 | 0,013812 | y ~ current_feature + S |
| 0,279166 | 0,140066 | 0,034007 | 1,993104 | 50 | 47 | 0,052073 | 0,420946 | y ~ current_feature + S |
| 0,260339 | 0,140835 | -0,04751 | 1,848537 | 50 | 47 | 0,070821 | 0,640171 | y ~ current_feature + S |
| 0,064448 | 0,155849 | 0,061498 | 0,41353 | 44 | 41 | 0,681374 | 0,962245 | y ~ current_feature + S |
| 0,367222 | 0,169818 | -0,02429 | 2,162437 | 33 | 30 | 0,038684 | 0,520476 | y ~ current_feature + S |
| 0,050486 | 0,128935 | -0,07874 | 0,391562 | 63 | 60 | 0,696769 | 0,939646 | y ~ current_feature + S |
| 0,179064 | 0,127013 | 0,034007 | 1,40981 | 63 | 60 | 0,163759 | 0,61196 | y ~ current_feature + S |
| 0,057653 | 0,128885 | -0,04751 | 0,44732 | 63 | 60 | 0,656255 | 0,925756 | y ~ current_feature + S |
| 0,216782 | 0,13946 | 0,061498 | 1,554439 | 52 | 49 | 0,126516 | 0,8197 | y ~ current_feature + S |
| 0,277424 | 0,153843 | -0,02429 | 1,8033 | 42 | 39 | 0,079071 | 0,605474 | y ~ current_feature + S |
| 0,104383 | 0,114839 | -0,07874 | 0,908945 | 78 | 75 | 0,366291 | 0,776663 | y ~ current_feature + S |
| 0,218034 | 0,112692 | 0,034007 | 1,934777 | 78 | 75 | 0,056789 | 0,420946 | y ~ current_feature + S |
| -0,1711 | 0,113767 | -0,04751 | -1,50394 | 78 | 75 | 0,136798 | 0,657012 | y ~ current_feature + S |
| -0,12772 | 0,123976 | 0,061498 | -1,03016 | 67 | 64 | 0,306813 | 0,893818 | y ~ current_feature + S |
| 0,048049 | 0,142692 | -0,02429 | 0,336731 | 52 | 49 | 0,737758 | 0,944216 | y ~ current_feature + S |
| -0,08931 | 0,055248 | -0,07874 | -1,61645 | 328 | 325 | 0,106967 | 0,449749 | y ~ current_feature + S |
| -0,06842 | 0,05534 | 0,034007 | -1,2363 | 328 | 325 | 0,21724 | 0,664036 | y ~ current_feature + S |
| -0,02899 | 0,055447 | -0,04751 | -0,52282 | 328 | 325 | 0,601454 | 0,91263 | y ~ current_feature + S |
| -0,05483 | 0,066257 | 0,061498 | -0,82759 | 285 | 227,1069 | 0,408771 | 0,934833 | y ~ current_feature + S |
| -0,00425 | 0,068199 | -0,02429 | -0,06235 | 218 | 215 | 0,950344 | 0,99444 | y ~ current_feature + S |

| | | | | | | | | |
|---|---|---|---|---|---|---|---|---|
| -0,03414 | 0,051202 | -0,07874 | -0,66681 | 421 | 381,0016 | 0,505296 | 0,863164 | y ~ current_feature + S |
| -0,06037 | 0,067236 | 0,034007 | -0,89785 | 421 | 220,4002 | 0,370245 | 0,750317 | y ~ current_feature + S |
| -0,02639 | 0,052602 | -0,04751 | -0,50164 | 420 | 361,1513 | 0,616223 | 0,924009 | y ~ current_feature + S |
| -0,01667 | 0,05716 | 0,061498 | -0,29171 | 367 | 305,9762 | 0,770706 | 0,973246 | y ~ current_feature + S |
| -0,05762 | 0,06029 | -0,02429 | -0,95579 | 280 | 274,1998 | 0,340023 | 0,785829 | y ~ current_feature + S |
| -0,10724 | 0,083142 | -0,07874 | -1,28987 | 146 | 143 | 0,199179 | 0,611596 | y ~ current_feature + S |
| -0,21035 | 0,081753 | 0,034007 | -2,57295 | 146 | 143 | 0,011102 | 0,233774 | y ~ current_feature + S |
| -0,0263 | 0,083595 | -0,04751 | -0,31457 | 146 | 143 | 0,753549 | 0,9394 | y ~ current_feature + S |
| 0,01391 | 0,090527 | 0,061498 | 0,153652 | 125 | 122 | 0,878138 | 0,996179 | y ~ current_feature + S |
| -0,04749 | 0,103026 | -0,02429 | -0,46094 | 97 | 94 | 0,645904 | 0,924682 | y ~ current_feature + S |
| 0,075677 | 0,133247 | -0,07874 | 0,567947 | 59 | 56 | 0,572341 | 0,899533 | y ~ current_feature + S |
| 0,207718 | 0,130716 | 0,034007 | 1,589076 | 59 | 56 | 0,117674 | 0,552558 | y ~ current_feature + S |
| -0,11681 | 0,133917 | -0,04751 | -0,87225 | 58 | 55 | 0,386865 | 0,803665 | y ~ current_feature + S |
| 0,113204 | 0,157097 | 0,061498 | 0,7206 | 43 | 40 | 0,475346 | 0,937665 | y ~ current_feature + S |
| 0,095127 | 0,165911 | -0,02429 | 0,57336 | 39 | 36 | 0,569965 | 0,924682 | y ~ current_feature + S |
| -0,12887 | 0,064827 | -0,07874 | -1,98792 | 237 | 234 | 0,047986 | 0,289576 | y ~ current_feature + S |
| -0,1517 | 0,064615 | 0,034007 | -2,34771 | 237 | 234 | 0,019723 | 0,317291 | y ~ current_feature + S |
| -0,08756 | 0,065121 | -0,04751 | -1,34461 | 237 | 234 | 0,180053 | 0,704707 | y ~ current_feature + S |
| 0,046475 | 0,069598 | 0,061498 | 0,667758 | 209 | 206 | 0,505036 | 0,937665 | y ~ current_feature + S |
| -0,12179 | 0,079982 | -0,02429 | -1,5227 | 157 | 154 | 0,129884 | 0,617591 | y ~ current_feature + S |
| 0,089004 | 0,088038 | -0,07874 | 1,010982 | 131 | 128 | 0,313932 | 0,718687 | y ~ current_feature + S |
| 0,021339 | 0,088368 | 0,034007 | 0,24148 | 131 | 128 | 0,809569 | 0,952501 | y ~ current_feature + S |
| 0,080992 | 0,088098 | -0,04751 | 0,919345 | 131 | 128 | 0,359645 | 0,798092 | y ~ current_feature + S |
| -0,04294 | 0,095694 | 0,061498 | -0,44873 | 112 | 109 | 0,65452 | 0,957207 | y ~ current_feature + S |
| -0,00883 | 0,109105 | -0,02429 | -0,08093 | 87 | 84 | 0,935688 | 0,99444 | y ~ current_feature + S |
| 0,125282 | 0,067979 | -0,07874 | 1,842956 | 216 | 213 | 0,066725 | 0,346137 | y ~ current_feature + S |
| 0,089465 | 0,068244 | 0,034007 | 1,31096 | 216 | 213 | 0,191283 | 0,643406 | y ~ current_feature + S |
| 0,111824 | 0,068089 | -0,04751 | 1,642325 | 216 | 213 | 0,101998 | 0,640171 | y ~ current_feature + S |
| 0,097626 | 0,073771 | 0,061498 | 1,323365 | 185 | 182 | 0,187374 | 0,866606 | y ~ current_feature + S |
| 0,13494 | 0,083445 | -0,02429 | 1,617112 | 144 | 141 | 0,108089 | 0,605474 | y ~ current_feature + S |
| -0,02227 | 0,111084 | -0,07874 | -0,20046 | 84 | 81 | 0,841623 | 0,983548 | y ~ current_feature + S |
| 0,084691 | 0,110712 | 0,034007 | 0,764966 | 84 | 81 | 0,446515 | 0,813845 | y ~ current_feature + S |
| 0,225317 | 0,108254 | -0,04751 | 2,081375 | 84 | 81 | 0,040558 | 0,612508 | y ~ current_feature + S |
| 0,031617 | 0,123973 | 0,061498 | 0,255032 | 68 | 65 | 0,799503 | 0,984413 | y ~ current_feature + S |
| -0,02945 | 0,137301 | -0,02429 | -0,21451 | 56 | 53 | 0,83097 | 0,976969 | y ~ current_feature + S |
| -0,01636 | 0,049602 | -0,07874 | -0,32978 | 418 | 406,3361 | 0,741736 | 0,959588 | y ~ current_feature + S |
| -0,028 | 0,063231 | 0,034007 | -0,44283 | 418 | 249,9155 | 0,658269 | 0,900251 | y ~ current_feature + S |
| -0,00897 | 0,050414 | -0,04751 | -0,17796 | 417 | 393,4194 | 0,858849 | 0,983821 | y ~ current_feature + S |
| -0,08201 | 0,055131 | 0,061498 | -1,4876 | 364 | 326,8001 | 0,137821 | 0,8197 | y ~ current_feature + S |
| -0,05789 | 0,061141 | -0,02429 | -0,94687 | 278 | 266,6141 | 0,344564 | 0,789404 | y ~ current_feature + S |
| 0,09416 | 0,135478 | -0,07874 | 0,695022 | 57 | 54 | 0,490022 | 0,852508 | y ~ current_feature + S |
| 0,618827 | 0,106897 | 0,034007 | 5,789018 | 57 | 54 | 3,71E-07 | 2,74E-04 | y ~ current_feature + S |
| 0,146674 | 0,134611 | -0,04751 | 1,089617 | 57 | 54 | 0,280721 | 0,747915 | y ~ current_feature + S |
| 0,393827 | 0,134077 | 0,061498 | 2,937322 | 50 | 47 | 0,005114 | 0,378455 | y ~ current_feature + S |
| 0,3367 | 0,17174 | -0,02429 | 1,960515 | 38 | 30,06068 | 0,05926 | 0,558147 | y ~ current_feature + S |
| -0,02464 | 0,051419 | -0,07874 | -0,47922 | 381 | 378 | 0,632062 | 0,92619 | y ~ current_feature + S |
| -0,06155 | 0,051337 | 0,034007 | -1,1989 | 381 | 378 | 0,23132 | 0,681978 | y ~ current_feature + S |
| -0,07969 | 0,051339 | -0,04751 | -1,55222 | 380 | 377 | 0,121448 | 0,651244 | y ~ current_feature + S |

| | | | | | | | | |
|---|---|---|---|---|---|---|---|---|
| -0,00979 | 0,055046 | 0,061498 | -0,17782 | 333 | 330 | 0,858974 | 0,996179 | y ~ current_feature + S |
| 0,028258 | 0,06322 | -0,02429 | 0,44697 | 253 | 250 | 0,655284 | 0,924682 | y ~ current_feature + S |
| -0,05431 | 0,052595 | -0,07874 | -1,03254 | 379 | 360,4322 | 0,302512 | 0,718687 | y ~ current_feature + S |
| -0,11854 | 0,06755 | 0,034007 | -1,75484 | 379 | 216,0769 | 0,080704 | 0,473527 | y ~ current_feature + S |
| 0,024029 | 0,053503 | -0,04751 | 0,449114 | 378 | 349,1395 | 0,653628 | 0,925756 | y ~ current_feature + S |
| 0,025974 | 0,057282 | 0,061498 | 0,453452 | 327 | 304,5624 | 0,650546 | 0,957207 | y ~ current_feature + S |
| -0,01979 | 0,065403 | -0,02429 | -0,30264 | 252 | 233,6869 | 0,76243 | 0,951727 | y ~ current_feature + S |
| 0,00662 | 0,113225 | -0,07874 | 0,058464 | 81 | 78 | 0,953529 | 0,994607 | y ~ current_feature + S |
| -0,07318 | 0,112924 | 0,034007 | -0,64806 | 81 | 78 | 0,518851 | 0,841996 | y ~ current_feature + S |
| 0,063144 | 0,113002 | -0,04751 | 0,558792 | 81 | 78 | 0,577904 | 0,904733 | y ~ current_feature + S |
| -0,26089 | 0,116217 | 0,061498 | -2,24489 | 72 | 69 | 0,027983 | 0,658491 | y ~ current_feature + S |
| 0,064163 | 0,144216 | -0,02429 | 0,444906 | 54 | 47,88314 | 0,658392 | 0,924682 | y ~ current_feature + S |
| 0,072525 | 0,108822 | -0,07874 | 0,666458 | 87 | 84 | 0,506945 | 0,863164 | y ~ current_feature + S |
| -0,04836 | 0,108981 | 0,034007 | -0,44375 | 87 | 84 | 0,658363 | 0,900251 | y ~ current_feature + S |
| -0,01375 | 0,109099 | -0,04751 | -0,12601 | 87 | 84 | 0,900028 | 0,985217 | y ~ current_feature + S |
| -0,08781 | 0,117396 | 0,061498 | -0,74802 | 75 | 72 | 0,456887 | 0,937665 | y ~ current_feature + S |
| 0,054817 | 0,134637 | -0,02429 | 0,407146 | 58 | 55 | 0,685481 | 0,926342 | y ~ current_feature + S |
| -0,14765 | 0,08242 | -0,07874 | -1,79141 | 147 | 144 | 0,075329 | 0,366731 | y ~ current_feature + S |
| -5,31E-04 | 0,083333 | 0,034007 | -0,00638 | 147 | 144 | 0,994921 | 0,997989 | y ~ current_feature + S |
| 0,070627 | 0,083125 | -0,04751 | 0,849647 | 147 | 144 | 0,396932 | 0,803665 | y ~ current_feature + S |
| -0,10977 | 0,091245 | 0,061498 | -1,20298 | 124 | 118,6646 | 0,231379 | 0,893818 | y ~ current_feature + S |
| 0,061051 | 0,102435 | -0,02429 | 0,595996 | 98 | 94,94784 | 0,552596 | 0,918138 | y ~ current_feature + S |
| 0,053723 | 0,074222 | -0,07874 | 0,723818 | 184 | 181 | 0,470112 | 0,840689 | y ~ current_feature + S |
| -0,05165 | 0,07423 | 0,034007 | -0,69587 | 184 | 181 | 0,487401 | 0,834899 | y ~ current_feature + S |
| -0,05908 | 0,0742 | -0,04751 | -0,79619 | 184 | 181 | 0,426964 | 0,819126 | y ~ current_feature + S |
| -0,09617 | 0,080209 | 0,061498 | -1,199 | 157 | 154 | 0,232368 | 0,893818 | y ~ current_feature + S |
| -0,11721 | 0,090658 | -0,02429 | -1,29285 | 123 | 120 | 0,198547 | 0,660578 | y ~ current_feature + S |
| 0,091557 | 0,114985 | -0,07874 | 0,796254 | 78 | 75 | 0,428398 | 0,814177 | y ~ current_feature + S |
| 0,016119 | 0,115455 | 0,034007 | 0,13961 | 78 | 75 | 0,889342 | 0,96591 | y ~ current_feature + S |
| -0,11242 | 0,114738 | -0,04751 | -0,97981 | 78 | 75 | 0,330328 | 0,779476 | y ~ current_feature + S |
| -0,45487 | 0,11132 | 0,061498 | -4,08615 | 67 | 64 | 1,25E-04 | 0,023038 | y ~ current_feature + S |
| -0,196 | 0,140086 | -0,02429 | -1,39917 | 52 | 49 | 0,168059 | 0,624945 | y ~ current_feature + S |
| 0,138223 | 0,093169 | -0,07874 | 1,483576 | 116 | 113 | 0,140705 | 0,498495 | y ~ current_feature + S |
| -0,02196 | 0,094049 | 0,034007 | -0,23346 | 116 | 113 | 0,815824 | 0,952501 | y ~ current_feature + S |
| 0,136408 | 0,093608 | -0,04751 | 1,45723 | 115 | 112 | 0,147851 | 0,661823 | y ~ current_feature + S |
| -0,01837 | 0,101517 | 0,061498 | -0,18094 | 100 | 97 | 0,856794 | 0,996179 | y ~ current_feature + S |
| 0,189914 | 0,114132 | -0,02429 | 1,663983 | 77 | 74 | 0,100345 | 0,605474 | y ~ current_feature + S |
| -0,09388 | 0,064399 | -0,07874 | -1,45772 | 242 | 239 | 0,146229 | 0,506121 | y ~ current_feature + S |
| -0,04549 | 0,064618 | 0,034007 | -0,70402 | 242 | 239 | 0,482103 | 0,831402 | y ~ current_feature + S |
| -0,03925 | 0,064635 | -0,04751 | -0,6073 | 242 | 239 | 0,544227 | 0,892966 | y ~ current_feature + S |
| 0,127212 | 0,069616 | 0,061498 | 1,827345 | 206 | 203 | 0,069116 | 0,715249 | y ~ current_feature + S |
| -0,09002 | 0,079233 | -0,02429 | -1,13621 | 161 | 158 | 0,257591 | 0,71931 | y ~ current_feature + S |
| 0,214205 | 0,058796 | -0,07874 | 3,643196 | 279 | 276 | 3,22E-04 | 0,023018 | y ~ current_feature + S |
| 0,12057 | 0,059754 | 0,034007 | 2,017774 | 279 | 276 | 0,044582 | 0,400524 | y ~ current_feature + S |
| 0,136288 | 0,05974 | -0,04751 | 2,281364 | 278 | 275 | 0,023291 | 0,563956 | y ~ current_feature + S |
| -0,05708 | 0,064988 | 0,061498 | -0,87833 | 239 | 236 | 0,380657 | 0,93433 | y ~ current_feature + S |
| 0,203664 | 0,072373 | -0,02429 | 2,814096 | 186 | 183 | 0,005427 | 0,236215 | y ~ current_feature + S |
| 0,206472 | 0,137011 | -0,07874 | 1,506974 | 54 | 51 | 0,137987 | 0,498495 | y ~ current_feature + S |

| | | | | | | | | |
|---|---|---|---|---|---|---|---|---|
| -0,09834 | 0,139349 | 0,034007 | -0,70571 | 54 | 51 | 0,483582 | 0,831402 | y ~ current_feature + S |
| -0,05551 | 0,139812 | -0,04751 | -0,39705 | 54 | 51 | 0,692991 | 0,930326 | y ~ current_feature + S |
| 0,136034 | 0,151081 | 0,061498 | 0,900406 | 46 | 43 | 0,372919 | 0,932536 | y ~ current_feature + S |
| 0,085878 | 0,173435 | -0,02429 | 0,49516 | 36 | 33 | 0,623768 | 0,924682 | y ~ current_feature + S |
| 0,078342 | 0,062925 | -0,07874 | 1,244992 | 254 | 251 | 0,214296 | 0,639357 | y ~ current_feature + S |
| 0,088418 | 0,062872 | 0,034007 | 1,406318 | 254 | 251 | 0,160867 | 0,60427 | y ~ current_feature + S |
| 0,064695 | 0,06781 | -0,04751 | 0,95407 | 253 | 216,5665 | 0,341112 | 0,789051 | y ~ current_feature + S |
| -0,00676 | 0,07023 | 0,061498 | -0,09628 | 222 | 202,7387 | 0,923393 | 0,996179 | y ~ current_feature + S |
| 0,052374 | 0,077509 | -0,02429 | 0,675722 | 169 | 166 | 0,500157 | 0,89399 | y ~ current_feature + S |
| -0,04019 | 0,093996 | -0,07874 | -0,42757 | 116 | 113 | 0,669774 | 0,934653 | y ~ current_feature + S |
| -0,14846 | 0,09303 | 0,034007 | -1,5958 | 116 | 113 | 0,113327 | 0,548117 | y ~ current_feature + S |
| 0,063784 | 0,094299 | -0,04751 | 0,676402 | 115 | 112 | 0,50018 | 0,868909 | y ~ current_feature + S |
| 0,118148 | 0,101347 | 0,061498 | 1,16577 | 99 | 96 | 0,246596 | 0,893818 | y ~ current_feature + S |
| 0,09244 | 0,11575 | -0,02429 | 0,79862 | 77 | 74 | 0,427067 | 0,847264 | y ~ current_feature + S |
| -0,02338 | 0,121235 | -0,07874 | -0,19287 | 71 | 68 | 0,847638 | 0,983548 | y ~ current_feature + S |
| 0,04115 | 0,121165 | 0,034007 | 0,339617 | 71 | 68 | 0,735192 | 0,923671 | y ~ current_feature + S |
| -0,05056 | 0,122013 | -0,04751 | -0,41439 | 70 | 67 | 0,67991 | 0,92829 | y ~ current_feature + S |
| -0,08034 | 0,129768 | 0,061498 | -0,6191 | 62 | 59 | 0,538236 | 0,937665 | y ~ current_feature + S |
| -0,01229 | 0,150744 | -0,02429 | -0,08151 | 47 | 44 | 0,935408 | 0,99444 | y ~ current_feature + S |
| -0,04043 | 0,075316 | -0,07874 | -0,53683 | 179 | 176 | 0,59206 | 0,903286 | y ~ current_feature + S |
| -0,07801 | 0,075148 | 0,034007 | -1,03804 | 179 | 176 | 0,300675 | 0,718742 | y ~ current_feature + S |
| -0,09828 | 0,075013 | -0,04751 | -1,31024 | 179 | 176 | 0,191822 | 0,717449 | y ~ current_feature + S |
| -0,03933 | 0,081316 | 0,061498 | -0,48371 | 154 | 151 | 0,629294 | 0,952306 | y ~ current_feature + S |
| 0,02057 | 0,092828 | -0,02429 | 0,221589 | 119 | 116 | 0,825024 | 0,976828 | y ~ current_feature + S |
| -0,01532 | 0,104919 | -0,07874 | -0,14597 | 123 | 90,82179 | 0,884266 | 0,987929 | y ~ current_feature + S |
| 0,061794 | 0,135483 | 0,034007 | 0,456102 | 123 | 54,271 | 0,650136 | 0,900251 | y ~ current_feature + S |
| 0,097676 | 0,09563 | -0,04751 | 1,021398 | 123 | 108,3051 | 0,309342 | 0,76529 | y ~ current_feature + S |
| 0,066483 | 0,100912 | 0,061498 | 0,658826 | 104 | 97,76654 | 0,511556 | 0,937665 | y ~ current_feature + S |
| 0,053705 | 0,113644 | -0,02429 | 0,472574 | 82 | 77,20606 | 0,63785 | 0,924682 | y ~ current_feature + S |
| 0,20698 | 0,086138 | -0,07874 | 2,40287 | 132 | 129 | 0,017691 | 0,177316 | y ~ current_feature + S |
| 0,041264 | 0,08797 | 0,034007 | 0,469064 | 132 | 129 | 0,639816 | 0,899624 | y ~ current_feature + S |
| 0,074291 | 0,087802 | -0,04751 | 0,84612 | 132 | 129 | 0,399053 | 0,803665 | y ~ current_feature + S |
| -0,0496 | 0,095229 | 0,061498 | -0,52083 | 113 | 110 | 0,603529 | 0,945257 | y ~ current_feature + S |
| 0,064924 | 0,108236 | -0,02429 | 0,599835 | 88 | 85 | 0,550212 | 0,91702 | y ~ current_feature + S |
| -0,13072 | 0,088675 | -0,07874 | -1,47419 | 128 | 125 | 0,142944 | 0,501016 | y ~ current_feature + S |
| -0,11925 | 0,088804 | 0,034007 | -1,34289 | 128 | 125 | 0,181741 | 0,634381 | y ~ current_feature + S |
| 0,049628 | 0,089333 | -0,04751 | 0,55554 | 128 | 125 | 0,579518 | 0,904733 | y ~ current_feature + S |
| -0,04845 | 0,096112 | 0,061498 | -0,50407 | 111 | 108 | 0,615236 | 0,948092 | y ~ current_feature + S |
| 0,108634 | 0,109778 | -0,02429 | 0,98958 | 85 | 82 | 0,32529 | 0,774043 | y ~ current_feature + S |
| -0,02785 | 0,109067 | -0,07874 | -0,25537 | 87 | 84 | 0,79906 | 0,979102 | y ~ current_feature + S |
| -0,08321 | 0,108731 | 0,034007 | -0,76528 | 87 | 84 | 0,446249 | 0,813845 | y ~ current_feature + S |
| 0,06431 | 0,108883 | -0,04751 | 0,590631 | 87 | 84 | 0,556353 | 0,897386 | y ~ current_feature + S |
| 0,119451 | 0,117007 | 0,061498 | 1,020883 | 75 | 72 | 0,310728 | 0,893818 | y ~ current_feature + S |
| -0,0812 | 0,134395 | -0,02429 | -0,60421 | 58 | 55 | 0,54819 | 0,915712 | y ~ current_feature + S |
| 0,109287 | 0,153379 | -0,07874 | 0,712529 | 45 | 42 | 0,480077 | 0,845512 | y ~ current_feature + S |
| 0,15766 | 0,152374 | 0,034007 | 1,034697 | 45 | 42 | 0,306732 | 0,718742 | y ~ current_feature + S |
| -0,1592 | 0,152336 | -0,04751 | -1,04503 | 45 | 42 | 0,301987 | 0,754967 | y ~ current_feature + S |
| -0,1031 | 0,175835 | 0,061498 | -0,58636 | 35 | 32 | 0,561744 | 0,937665 | y ~ current_feature + S |

| | | | | | | | | |
|---|---|---|---|---|---|---|---|---|
| 0,016513 | 0,192424 | -0,02429 | 0,085815 | 30 | 27 | 0,932247 | 0,99444 | y ~ current_feature + S |
| 0,244613 | 0,090027 | -0,07874 | 2,71711 | 119 | 116 | 0,007595 | 0,110197 | y ~ current_feature + S |
| 0,054882 | 0,092708 | 0,034007 | 0,591988 | 119 | 116 | 0,55501 | 0,857986 | y ~ current_feature + S |
| 0,039212 | 0,092776 | -0,04751 | 0,422653 | 119 | 116 | 0,673331 | 0,928258 | y ~ current_feature + S |
| 0,060987 | 0,099814 | 0,061498 | 0,611007 | 103 | 100 | 0,542581 | 0,937665 | y ~ current_feature + S |
| -0,00362 | 0,114707 | -0,02429 | -0,0316 | 79 | 76 | 0,974875 | 0,99444 | y ~ current_feature + S |
| 0,05842 | 0,067925 | -0,07874 | 0,860061 | 219 | 216 | 0,390709 | 0,789252 | y ~ current_feature + S |
| -0,00389 | 0,068041 | 0,034007 | -0,05718 | 219 | 216 | 0,954457 | 0,989859 | y ~ current_feature + S |
| -0,019 | 0,068029 | -0,04751 | -0,27933 | 219 | 216 | 0,780262 | 0,94882 | y ~ current_feature + S |
| -0,03103 | 0,073288 | 0,061498 | -0,42346 | 189 | 186 | 0,672446 | 0,958606 | y ~ current_feature + S |
| -0,01375 | 0,083616 | -0,02429 | -0,1645 | 146 | 143 | 0,869573 | 0,984468 | y ~ current_feature + S |
| 0,095614 | 0,049585 | -0,07874 | 1,928273 | 406 | 403 | 0,054523 | 0,310364 | y ~ current_feature + S |
| 0,110379 | 0,049509 | 0,034007 | 2,229459 | 406 | 403 | 0,026334 | 0,347616 | y ~ current_feature + S |
| 0,066927 | 0,049764 | -0,04751 | 1,344892 | 405 | 402 | 0,179418 | 0,704707 | y ~ current_feature + S |
| -0,05756 | 0,060216 | 0,061498 | -0,95598 | 352 | 274,8787 | 0,339924 | 0,901591 | y ~ current_feature + S |
| 0,11684 | 0,060796 | -0,02429 | 1,921839 | 270 | 266,8605 | 0,055691 | 0,558147 | y ~ current_feature + S |
| -0,05922 | 0,053656 | -0,07874 | -1,10368 | 402 | 346,1297 | 0,2705 | 0,702573 | y ~ current_feature + S |
| -0,11908 | 0,069412 | 0,034007 | -1,71551 | 402 | 204,6139 | 0,087766 | 0,484679 | y ~ current_feature + S |
| 0,096863 | 0,053442 | -0,04751 | 1,812476 | 401 | 346,8447 | 0,070777 | 0,640171 | y ~ current_feature + S |
| 0,034889 | 0,058038 | 0,061498 | 0,601136 | 350 | 296,5128 | 0,548209 | 0,937665 | y ~ current_feature + S |
| -0,03347 | 0,062024 | -0,02429 | -0,53956 | 267 | 259,6492 | 0,589963 | 0,924682 | y ~ current_feature + S |
| 0,054871 | 0,122906 | -0,07874 | 0,446443 | 69 | 66 | 0,656738 | 0,932255 | y ~ current_feature + S |
| 0,03606 | 0,123011 | 0,034007 | 0,29314 | 69 | 66 | 0,770335 | 0,940673 | y ~ current_feature + S |
| -0,0426 | 0,12298 | -0,04751 | -0,34643 | 69 | 66 | 0,730119 | 0,93314 | y ~ current_feature + S |
| -0,09894 | 0,135415 | 0,061498 | -0,73064 | 57 | 54 | 0,468157 | 0,937665 | y ~ current_feature + S |
| -0,2923 | 0,145838 | -0,02429 | -2,00431 | 46 | 43 | 0,051361 | 0,558147 | y ~ current_feature + S |
| -0,07901 | 0,092161 | -0,07874 | -0,85731 | 120 | 117 | 0,393026 | 0,789252 | y ~ current_feature + S |
| -0,20305 | 0,090524 | 0,034007 | -2,24304 | 120 | 117 | 0,026777 | 0,347616 | y ~ current_feature + S |
| -0,1366 | 0,091583 | -0,04751 | -1,49156 | 120 | 117 | 0,138505 | 0,657012 | y ~ current_feature + S |
| -0,09082 | 0,099092 | 0,061498 | -0,91654 | 104 | 101 | 0,361568 | 0,920619 | y ~ current_feature + S |
| -0,32253 | 0,10787 | -0,02429 | -2,99002 | 80 | 77 | 0,003744 | 0,184706 | y ~ current_feature + S |
| -0,04627 | 0,052721 | -0,07874 | -0,87758 | 362 | 359 | 0,380761 | 0,789252 | y ~ current_feature + S |
| -0,17502 | 0,072679 | 0,034007 | -2,40808 | 362 | 183,5137 | 0,017027 | 0,293019 | y ~ current_feature + S |
| -0,09138 | 0,059154 | -0,04751 | -1,54484 | 361 | 283,393 | 0,1235 | 0,652786 | y ~ current_feature + S |
| -0,08255 | 0,064873 | 0,061498 | -1,27249 | 315 | 235,9981 | 0,20445 | 0,893818 | y ~ current_feature + S |
| -0,09534 | 0,064974 | -0,02429 | -1,46739 | 240 | 234,726 | 0,143607 | 0,618941 | y ~ current_feature + S |
| 0,064757 | 0,11299 | -0,07874 | 0,573125 | 81 | 78 | 0,568208 | 0,898449 | y ~ current_feature + S |
| 0,079586 | 0,112869 | 0,034007 | 0,705125 | 81 | 78 | 0,482833 | 0,831402 | y ~ current_feature + S |
| -0,04467 | 0,113115 | -0,04751 | -0,39493 | 81 | 78 | 0,693973 | 0,930326 | y ~ current_feature + S |
| -0,21361 | 0,115939 | 0,061498 | -1,8424 | 74 | 71 | 0,069592 | 0,715249 | y ~ current_feature + S |
| -0,11795 | 0,139051 | -0,02429 | -0,84827 | 54 | 51 | 0,40025 | 0,825029 | y ~ current_feature + S |
| -0,26039 | 0,135198 | -0,07874 | -1,92597 | 54 | 51 | 0,059691 | 0,324792 | y ~ current_feature + S |
| 0,006435 | 0,140025 | 0,034007 | 0,045956 | 54 | 51 | 0,963525 | 0,989859 | y ~ current_feature + S |
| -0,17692 | 0,137819 | -0,04751 | -1,28369 | 54 | 51 | 0,205051 | 0,723562 | y ~ current_feature + S |
| 0,152985 | 0,156253 | 0,061498 | 0,979087 | 43 | 40 | 0,333422 | 0,893818 | y ~ current_feature + S |
| 0,038841 | 0,173946 | -0,02429 | 0,223292 | 36 | 33 | 0,824685 | 0,976828 | y ~ current_feature + S |
| 0,030565 | 0,056407 | -0,07874 | 0,541866 | 317 | 314 | 0,588295 | 0,902886 | y ~ current_feature + S |
| -0,05675 | 0,056342 | 0,034007 | -1,00727 | 317 | 314 | 0,314583 | 0,718742 | y ~ current_feature + S |

| | | | | | | | | |
|---|---|---|---|---|---|---|---|---|
| 0,032394 | 0,056404 | -0,04751 | 0,574318 | 317 | 314 | 0,566164 | 0,903656 | y ~ current_feature + S |
| -0,02601 | 0,060502 | 0,061498 | -0,4299 | 276 | 273 | 0,667604 | 0,958606 | y ~ current_feature + S |
| 0,032278 | 0,069301 | -0,02429 | 0,46576 | 211 | 208 | 0,641875 | 0,924682 | y ~ current_feature + S |
| -0,04485 | 0,078732 | -0,07874 | -0,56965 | 164 | 161 | 0,56971 | 0,898903 | y ~ current_feature + S |
| -0,01761 | 0,078799 | 0,034007 | -0,22345 | 164 | 161 | 0,823467 | 0,952501 | y ~ current_feature + S |
| -0,00714 | 0,078809 | -0,04751 | -0,09054 | 164 | 161 | 0,927968 | 0,985217 | y ~ current_feature + S |
| 0,064944 | 0,093031 | 0,061498 | 0,69809 | 141 | 115,0553 | 0,486529 | 0,937665 | y ~ current_feature + S |
| -0,03672 | 0,097063 | -0,02429 | -0,37827 | 109 | 106 | 0,705987 | 0,93118 | y ~ current_feature + S |
| -0,05889 | 0,109463 | -0,07874 | -0,538 | 87 | 83,16845 | 0,592011 | 0,903286 | y ~ current_feature + S |
| 0,021913 | 0,165309 | 0,034007 | 0,132559 | 87 | 36,57635 | 0,895269 | 0,96591 | y ~ current_feature + S |
| -0,17593 | 0,113686 | -0,04751 | -1,54748 | 87 | 74,97713 | 0,125958 | 0,654671 | y ~ current_feature + S |
| 0,078066 | 0,141085 | 0,061498 | 0,55333 | 70 | 49,93257 | 0,582507 | 0,945257 | y ~ current_feature + S |
| -0,14433 | 0,133428 | -0,02429 | -1,08168 | 58 | 55 | 0,284115 | 0,734108 | y ~ current_feature + S |
| 0,258526 | 0,140977 | -0,07874 | 1,833821 | 50 | 46,95283 | 0,073023 | 0,362665 | y ~ current_feature + S |
| -0,09174 | 0,209613 | 0,034007 | -0,43766 | 50 | 22,56807 | 0,665792 | 0,904011 | y ~ current_feature + S |
| -0,06131 | 0,145894 | -0,04751 | -0,42021 | 50 | 46,80485 | 0,676258 | 0,928258 | y ~ current_feature + S |
| -0,17537 | 0,194549 | 0,061498 | -0,9014 | 42 | 25,60797 | 0,375776 | 0,932536 | y ~ current_feature + S |
| 0,008241 | 0,184317 | -0,02429 | 0,04471 | 33 | 29,43348 | 0,96464 | 0,99444 | y ~ current_feature + S |
| -0,03917 | 0,05328 | -0,07874 | -0,73513 | 383 | 351,7216 | 0,462748 | 0,835203 | y ~ current_feature + S |
| -0,08764 | 0,067909 | 0,034007 | -1,29062 | 383 | 215,1796 | 0,198222 | 0,646185 | y ~ current_feature + S |
| -0,04155 | 0,053803 | -0,04751 | -0,77233 | 382 | 344,8515 | 0,440449 | 0,823749 | y ~ current_feature + S |
| 0,030076 | 0,058412 | 0,061498 | 0,514895 | 333 | 292,8222 | 0,607015 | 0,945257 | y ~ current_feature + S |
| -0,01493 | 0,063349 | -0,02429 | -0,23575 | 254 | 249,1272 | 0,813821 | 0,974251 | y ~ current_feature + S |
| 0,108459 | 0,057882 | -0,07874 | 1,873794 | 332 | 294,9678 | 0,061947 | 0,329792 | y ~ current_feature + S |
| 0,036457 | 0,0727 | 0,034007 | 0,501471 | 332 | 188,9524 | 0,616623 | 0,889725 | y ~ current_feature + S |
| 0,069154 | 0,05986 | -0,04751 | 1,155264 | 331 | 277,7472 | 0,248975 | 0,742928 | y ~ current_feature + S |
| -0,00671 | 0,063341 | 0,061498 | -0,10588 | 287 | 249,235 | 0,915762 | 0,996179 | y ~ current_feature + S |
| 0,128848 | 0,06753 | -0,02429 | 1,908015 | 221 | 215,6435 | 0,057718 | 0,558147 | y ~ current_feature + S |
| -0,04945 | 0,093957 | -0,07874 | -0,52628 | 116 | 113 | 0,599723 | 0,90386 | y ~ current_feature + S |
| -0,08012 | 0,09377 | 0,034007 | -0,85446 | 116 | 113 | 0,394659 | 0,775482 | y ~ current_feature + S |
| 0,014848 | 0,094062 | -0,04751 | 0,157854 | 116 | 113 | 0,874854 | 0,985217 | y ~ current_feature + S |
| 0,06463 | 0,099791 | 0,061498 | 0,647654 | 103 | 100 | 0,518693 | 0,937665 | y ~ current_feature + S |
| -0,07759 | 0,115897 | -0,02429 | -0,6695 | 77 | 74 | 0,505263 | 0,894767 | y ~ current_feature + S |
| 0,173382 | 0,130447 | -0,07874 | 1,329136 | 60 | 57 | 0,189099 | 0,592937 | y ~ current_feature + S |
| 0,235348 | 0,128733 | 0,034007 | 1,828189 | 60 | 57 | 0,072756 | 0,460165 | y ~ current_feature + S |
| 0,00916 | 0,132448 | -0,04751 | 0,069159 | 60 | 57 | 0,945105 | 0,990156 | y ~ current_feature + S |
| -0,12268 | 0,144763 | 0,061498 | -0,84748 | 50 | 47 | 0,401025 | 0,93433 | y ~ current_feature + S |
| -0,02699 | 0,164339 | -0,02429 | -0,16426 | 40 | 37 | 0,870421 | 0,984468 | y ~ current_feature + S |
| 0,153748 | 0,051209 | -0,07874 | 3,002377 | 412 | 372,3271 | 0,00286 | 0,066128 | y ~ current_feature + S |
| 0,197335 | 0,066801 | 0,034007 | 2,954063 | 412 | 215,3687 | 0,003485 | 0,151911 | y ~ current_feature + S |
| 0,125197 | 0,052705 | -0,04751 | 2,375456 | 411 | 354,3583 | 0,018059 | 0,54566 | y ~ current_feature + S |
| 0,029884 | 0,055784 | 0,061498 | 0,535709 | 359 | 321,0656 | 0,592531 | 0,945257 | y ~ current_feature + S |
| 0,091 | 0,061245 | -0,02429 | 1,485823 | 274 | 264,3886 | 0,138517 | 0,618941 | y ~ current_feature + S |
| -0,03819 | 0,091603 | -0,07874 | -0,41693 | 122 | 119 | 0,677481 | 0,934653 | y ~ current_feature + S |
| 0,017796 | 0,091655 | 0,034007 | 0,194168 | 122 | 119 | 0,846376 | 0,959445 | y ~ current_feature + S |
| 0,152458 | 0,090598 | -0,04751 | 1,682796 | 122 | 119 | 0,095037 | 0,640171 | y ~ current_feature + S |
| 0,036754 | 0,098466 | 0,061498 | 0,373261 | 106 | 103 | 0,709721 | 0,972444 | y ~ current_feature + S |
| -0,00473 | 0,113226 | -0,02429 | -0,04173 | 81 | 78 | 0,96682 | 0,99444 | y ~ current_feature + S |

| | | | | | | | | |
|---|---|---|---|---|---|---|---|---|
| 0,033041 | 0,057042 | -0,07874 | 0,579246 | 310 | 307 | 0,562848 | 0,897646 | y ~ current_feature + S |
| 0,033577 | 0,057041 | 0,034007 | 0,588646 | 310 | 307 | 0,556531 | 0,857986 | y ~ current_feature + S |
| 0,092037 | 0,056831 | -0,04751 | 1,619498 | 310 | 307 | 0,106367 | 0,640171 | y ~ current_feature + S |
| 0,040373 | 0,068104 | 0,061498 | 0,592809 | 267 | 215,252 | 0,553931 | 0,937665 | y ~ current_feature + S |
| -0,06051 | 0,069886 | -0,02429 | -0,86585 | 207 | 204 | 0,387591 | 0,819479 | y ~ current_feature + S |
| -0,03533 | 0,052291 | -0,07874 | -0,67572 | 421 | 365,2638 | 0,499649 | 0,861866 | y ~ current_feature + S |
| -0,0164 | 0,068501 | 0,034007 | -0,23946 | 421 | 213,0547 | 0,810976 | 0,952501 | y ~ current_feature + S |
| 0,091643 | 0,05355 | -0,04751 | 1,711359 | 420 | 345,7982 | 0,087912 | 0,640171 | y ~ current_feature + S |
| 0,019087 | 0,058311 | 0,061498 | 0,327327 | 367 | 293,9961 | 0,743654 | 0,972573 | y ~ current_feature + S |
| 0,12715 | 0,059597 | -0,02429 | 2,133513 | 280 | 277 | 0,033761 | 0,489869 | y ~ current_feature + S |
| 0,054078 | 0,070084 | -0,07874 | 0,771626 | 206 | 203 | 0,441233 | 0,818694 | y ~ current_feature + S |
| -0,00794 | 0,070184 | 0,034007 | -0,11309 | 206 | 203 | 0,910069 | 0,9732 | y ~ current_feature + S |
| 0,006373 | 0,070185 | -0,04751 | 0,090805 | 206 | 203 | 0,927737 | 0,985217 | y ~ current_feature + S |
| 0,045408 | 0,079784 | 0,061498 | 0,569146 | 173 | 156,7752 | 0,570071 | 0,937665 | y ~ current_feature + S |
| -0,01058 | 0,087664 | -0,02429 | -0,12069 | 138 | 130,1093 | 0,904126 | 0,99444 | y ~ current_feature + S |
| 0,19113 | 0,055199 | -0,07874 | 3,462577 | 337 | 316,2139 | 6,09E-04 | 0,031617 | y ~ current_feature + S |
| -0,0025 | 0,077616 | 0,034007 | -0,0322 | 337 | 165,9947 | 0,974348 | 0,989859 | y ~ current_feature + S |
| -0,00739 | 0,062104 | -0,04751 | -0,11893 | 336 | 259,2602 | 0,905426 | 0,985217 | y ~ current_feature + S |
| 0,011281 | 0,065028 | 0,061498 | 0,173472 | 296 | 236,4516 | 0,862429 | 0,996179 | y ~ current_feature + S |
| 0,063873 | 0,067818 | -0,02429 | 0,94183 | 224 | 216,5366 | 0,347329 | 0,790842 | y ~ current_feature + S |
| -0,01373 | 0,149057 | -0,07874 | -0,09214 | 48 | 45 | 0,926997 | 0,990519 | y ~ current_feature + S |
| 0,169198 | 0,146922 | 0,034007 | 1,15162 | 48 | 45 | 0,255559 | 0,705649 | y ~ current_feature + S |
| 0,138527 | 0,147634 | -0,04751 | 0,938313 | 48 | 45 | 0,353093 | 0,795089 | y ~ current_feature + S |
| 0,174415 | 0,159735 | 0,061498 | 1,091905 | 41 | 38 | 0,281749 | 0,893818 | y ~ current_feature + S |
| -0,109 | 0,184589 | -0,02429 | -0,59049 | 32 | 29 | 0,559437 | 0,92024 | y ~ current_feature + S |
| -0,00653 | 0,056432 | -0,07874 | -0,11577 | 317 | 314 | 0,907909 | 0,987929 | y ~ current_feature + S |
| -9,48E-04 | 0,056433 | 0,034007 | -0,01681 | 317 | 314 | 0,986602 | 0,994667 | y ~ current_feature + S |
| -0,02031 | 0,056512 | -0,04751 | -0,35938 | 316 | 313 | 0,719556 | 0,93314 | y ~ current_feature + S |
| -0,06326 | 0,060291 | 0,061498 | -1,04929 | 277 | 274 | 0,294968 | 0,893818 | y ~ current_feature + S |
| -0,03106 | 0,069471 | -0,02429 | -0,44705 | 210 | 207 | 0,655308 | 0,924682 | y ~ current_feature + S |
| 0,113859 | 0,108399 | -0,07874 | 1,050366 | 87 | 84 | 0,296562 | 0,718687 | y ~ current_feature + S |
| 0,234632 | 0,106063 | 0,034007 | 2,212198 | 87 | 84 | 0,029667 | 0,347616 | y ~ current_feature + S |
| 0,112826 | 0,108412 | -0,04751 | 1,040709 | 87 | 84 | 0,300997 | 0,754967 | y ~ current_feature + S |
| 0,130454 | 0,115254 | 0,061498 | 1,131877 | 77 | 74 | 0,261341 | 0,893818 | y ~ current_feature + S |
| 0,220061 | 0,131535 | -0,02429 | 1,673031 | 58 | 55 | 0,100001 | 0,605474 | y ~ current_feature + S |
| -0,04078 | 0,093994 | -0,07874 | -0,43388 | 116 | 113 | 0,6652 | 0,934653 | y ~ current_feature + S |
| 0,054722 | 0,093931 | 0,034007 | 0,582577 | 116 | 113 | 0,561339 | 0,861729 | y ~ current_feature + S |
| 0,022457 | 0,094048 | -0,04751 | 0,238779 | 116 | 113 | 0,811709 | 0,972405 | y ~ current_feature + S |
| 0,070536 | 0,100764 | 0,061498 | 0,700014 | 101 | 98 | 0,485577 | 0,937665 | y ~ current_feature + S |
| 0,005649 | 0,116246 | -0,02429 | 0,048591 | 77 | 74 | 0,961376 | 0,99444 | y ~ current_feature + S |
| 0,138945 | 0,068997 | -0,07874 | 2,013772 | 209 | 206 | 0,045334 | 0,281911 | y ~ current_feature + S |
| -0,00132 | 0,069673 | 0,034007 | -0,01891 | 209 | 206 | 0,984932 | 0,994338 | y ~ current_feature + S |
| 0,019385 | 0,06966 | -0,04751 | 0,278281 | 209 | 206 | 0,781076 | 0,94882 | y ~ current_feature + S |
| -0,03101 | 0,079069 | 0,061498 | -0,39217 | 181 | 159,7991 | 0,695454 | 0,972444 | y ~ current_feature + S |
| 0,013778 | 0,085741 | -0,02429 | 0,160697 | 139 | 136 | 0,872571 | 0,984468 | y ~ current_feature + S |
| -0,02636 | 0,066204 | -0,07874 | -0,39814 | 231 | 228 | 0,690897 | 0,93638 | y ~ current_feature + S |
| -0,05389 | 0,06613 | 0,034007 | -0,81487 | 231 | 228 | 0,415997 | 0,787309 | y ~ current_feature + S |
| -0,02249 | 0,066356 | -0,04751 | -0,33886 | 230 | 227 | 0,735027 | 0,933491 | y ~ current_feature + S |

| | | | | | | | | |
|---|---|---|---|---|---|---|---|---|
| -0,04509 | 0,080109 | 0,061498 | -0,56288 | 201 | 155,5065 | 0,574324 | 0,937665 | y ~ current_feature + S |
| -0,08641 | 0,081147 | -0,02429 | -1,06486 | 154 | 150,7284 | 0,28864 | 0,734108 | y ~ current_feature + S |
| -0,10835 | 0,059839 | -0,07874 | -1,81062 | 279 | 276 | 0,071287 | 0,356433 | y ~ current_feature + S |
| -0,05274 | 0,060109 | 0,034007 | -0,87742 | 279 | 276 | 0,381019 | 0,759222 | y ~ current_feature + S |
| -0,06648 | 0,060169 | -0,04751 | -1,10495 | 278 | 275 | 0,270146 | 0,747915 | y ~ current_feature + S |
| 4,94E-04 | 0,06455 | 0,061498 | 0,007655 | 243 | 240 | 0,993899 | 0,999005 | y ~ current_feature + S |
| -0,01613 | 0,074115 | -0,02429 | -0,21768 | 185 | 182 | 0,827924 | 0,976969 | y ~ current_feature + S |
| 0,179352 | 0,069219 | -0,07874 | 2,591082 | 205 | 202 | 0,010265 | 0,135649 | y ~ current_feature + S |
| -0,01008 | 0,070356 | 0,034007 | -0,14329 | 205 | 202 | 0,886204 | 0,96591 | y ~ current_feature + S |
| 0,076876 | 0,070326 | -0,04751 | 1,093145 | 204 | 201 | 0,275639 | 0,747915 | y ~ current_feature + S |
| -0,1817 | 0,073913 | 0,061498 | -2,4583 | 180 | 177 | 0,014922 | 0,55988 | y ~ current_feature + S |
| 0,126819 | 0,086011 | -0,02429 | 1,474455 | 136 | 133 | 0,142722 | 0,618941 | y ~ current_feature + S |
| -0,01824 | 0,054224 | -0,07874 | -0,33634 | 343 | 340 | 0,736821 | 0,959225 | y ~ current_feature + S |
| -0,0809 | 0,054055 | 0,034007 | -1,49669 | 343 | 340 | 0,135401 | 0,576909 | y ~ current_feature + S |
| 0,036081 | 0,054277 | -0,04751 | 0,664748 | 342 | 339 | 0,506664 | 0,868909 | y ~ current_feature + S |
| 0,056404 | 0,058031 | 0,061498 | 0,971961 | 299 | 296 | 0,331864 | 0,893818 | y ~ current_feature + S |
| 0,095833 | 0,06636 | -0,02429 | 1,444147 | 228 | 225 | 0,150089 | 0,622449 | y ~ current_feature + S |
| 0,184005 | 0,0993 | -0,07874 | 1,853022 | 101 | 97,98083 | 0,066889 | 0,346137 | y ~ current_feature + S |
| 0,018132 | 0,135434 | 0,034007 | 0,133883 | 101 | 54,50041 | 0,893988 | 0,96591 | y ~ current_feature + S |
| 0,296172 | 0,113503 | -0,04751 | 2,609384 | 101 | 70,81371 | 0,01106 | 0,481414 | y ~ current_feature + S |
| 0,043558 | 0,109687 | 0,061498 | 0,397111 | 86 | 82,95917 | 0,692306 | 0,972118 | y ~ current_feature + S |
| 0,06997 | 0,125398 | -0,02429 | 0,557986 | 67 | 63,28333 | 0,578823 | 0,924682 | y ~ current_feature + S |
| 0,037942 | 0,125897 | -0,07874 | 0,301369 | 66 | 63 | 0,764126 | 0,965008 | y ~ current_feature + S |
| -0,08545 | 0,125527 | 0,034007 | -0,68076 | 66 | 63 | 0,498519 | 0,841996 | y ~ current_feature + S |
| -0,04819 | 0,125842 | -0,04751 | -0,38298 | 66 | 63 | 0,703027 | 0,93314 | y ~ current_feature + S |
| 0,103937 | 0,137924 | 0,061498 | 0,753583 | 55 | 52 | 0,454499 | 0,937665 | y ~ current_feature + S |
| 0,01273 | 0,156161 | -0,02429 | 0,081519 | 44 | 41 | 0,935426 | 0,99444 | y ~ current_feature + S |
| 0,039299 | 0,095273 | -0,07874 | 0,412493 | 113 | 110 | 0,680781 | 0,934653 | y ~ current_feature + S |
| 0,139486 | 0,094414 | 0,034007 | 1,477387 | 113 | 110 | 0,142429 | 0,588814 | y ~ current_feature + S |
| 0,101379 | 0,094855 | -0,04751 | 1,068773 | 113 | 110 | 0,287512 | 0,747915 | y ~ current_feature + S |
| -0,04267 | 0,100412 | 0,061498 | -0,42492 | 102 | 99 | 0,671814 | 0,958606 | y ~ current_feature + S |
| -0,0019 | 0,117851 | -0,02429 | -0,01611 | 75 | 72 | 0,987193 | 0,99444 | y ~ current_feature + S |
| 0,054122 | 0,154077 | -0,07874 | 0,351264 | 45 | 42 | 0,727146 | 0,955752 | y ~ current_feature + S |
| -0,03868 | 0,154188 | 0,034007 | -0,25087 | 45 | 42 | 0,803138 | 0,952439 | y ~ current_feature + S |
| 0,137493 | 0,152838 | -0,04751 | 0,899597 | 45 | 42 | 0,373464 | 0,803381 | y ~ current_feature + S |
| -0,10148 | 0,16355 | 0,061498 | -0,62046 | 40 | 37 | 0,538756 | 0,937665 | y ~ current_feature + S |
| -0,26649 | 0,185491 | -0,02429 | -1,43665 | 30 | 27 | 0,162304 | 0,623497 | y ~ current_feature + S |
| -0,00849 | 0,129095 | -0,07874 | -0,06573 | 63 | 60 | 0,947808 | 0,994607 | y ~ current_feature + S |
| -0,10256 | 0,128419 | 0,034007 | -0,79861 | 63 | 60 | 0,427664 | 0,791701 | y ~ current_feature + S |
| -0,10871 | 0,128334 | -0,04751 | -0,84712 | 63 | 60 | 0,400297 | 0,803665 | y ~ current_feature + S |
| 0,048143 | 0,15157 | 0,061498 | 0,317629 | 50 | 43,42769 | 0,752288 | 0,972573 | y ~ current_feature + S |
| 0,177738 | 0,181554 | -0,02429 | 0,978987 | 42 | 29,37985 | 0,33558 | 0,785829 | y ~ current_feature + S |
| 0,121595 | 0,102926 | -0,07874 | 1,181382 | 96 | 93 | 0,240463 | 0,668958 | y ~ current_feature + S |
| 0,039834 | 0,103613 | 0,034007 | 0,384454 | 96 | 93 | 0,70152 | 0,920814 | y ~ current_feature + S |
| -0,09988 | 0,103177 | -0,04751 | -0,96803 | 96 | 93 | 0,335541 | 0,785761 | y ~ current_feature + S |
| -0,12652 | 0,121993 | 0,061498 | -1,03712 | 79 | 66,11877 | 0,303458 | 0,893818 | y ~ current_feature + S |
| 0,110743 | 0,127249 | -0,02429 | 0,870283 | 64 | 61 | 0,387558 | 0,819479 | y ~ current_feature + S |
| 0,026024 | 0,062848 | -0,07874 | 0,414073 | 256 | 253 | 0,679171 | 0,934653 | y ~ current_feature + S |

| | | | | | | | | |
|---|---|---|---|---|---|---|---|---|
| -0,11496 | 0,062453 | 0,034007 | -1,84079 | 256 | 253 | 0,066823 | 0,439525 | y ~ current_feature + S |
| -0,03036 | 0,06284 | -0,04751 | -0,48309 | 256 | 253 | 0,62945 | 0,925756 | y ~ current_feature + S |
| 0,020502 | 0,068027 | 0,061498 | 0,301386 | 219 | 216 | 0,76341 | 0,972573 | y ~ current_feature + S |
| -0,05046 | 0,077284 | -0,02429 | -0,65288 | 170 | 167 | 0,514731 | 0,899808 | y ~ current_feature + S |
| 0,083328 | 0,053943 | -0,07874 | 1,54475 | 390 | 341,2762 | 0,123334 | 0,477837 | y ~ current_feature + S |
| -0,08071 | 0,070287 | 0,034007 | -1,14833 | 390 | 201,0973 | 0,252198 | 0,704252 | y ~ current_feature + S |
| 0,032372 | 0,05518 | -0,04751 | 0,586662 | 389 | 328,0834 | 0,557834 | 0,897386 | y ~ current_feature + S |
| 0,048185 | 0,060749 | 0,061498 | 0,793178 | 340 | 270,3439 | 0,42837 | 0,937665 | y ~ current_feature + S |
| 0,04269 | 0,062712 | -0,02429 | 0,680729 | 259 | 253,807 | 0,496664 | 0,89399 | y ~ current_feature + S |
| 0,23883 | 0,150962 | -0,07874 | 1,582049 | 45 | 41,37678 | 0,121253 | 0,477837 | y ~ current_feature + S |
| 0,007452 | 0,190293 | 0,034007 | 0,039159 | 45 | 27,61393 | 0,969046 | 0,989859 | y ~ current_feature + S |
| 0,206761 | 0,159002 | -0,04751 | 1,300372 | 45 | 37,86367 | 0,201333 | 0,723562 | y ~ current_feature + S |
| -0,13147 | 0,161258 | 0,061498 | -0,81529 | 41 | 37,79088 | 0,420014 | 0,937665 | y ~ current_feature + S |
| -0,08487 | 0,191756 | -0,02429 | -0,44257 | 30 | 27 | 0,661601 | 0,924682 | y ~ current_feature + S |
| -0,36171 | 0,121374 | -0,07874 | -2,98013 | 62 | 59 | 0,00418 | 0,073642 | y ~ current_feature + S |
| -0,04622 | 0,13005 | 0,034007 | -0,35537 | 62 | 59 | 0,723579 | 0,920814 | y ~ current_feature + S |
| -0,01422 | 0,130176 | -0,04751 | -0,1092 | 62 | 59 | 0,913414 | 0,985217 | y ~ current_feature + S |
| 0,089854 | 0,147036 | 0,061498 | 0,6111 | 49 | 45,88062 | 0,544149 | 0,937665 | y ~ current_feature + S |
| 0,140042 | 0,16378 | -0,02429 | 0,85506 | 41 | 36,54916 | 0,398088 | 0,823699 | y ~ current_feature + S |
| -0,03266 | 0,070149 | -0,07874 | -0,46554 | 206 | 203 | 0,642044 | 0,928694 | y ~ current_feature + S |
| -0,00643 | 0,070185 | 0,034007 | -0,09167 | 206 | 203 | 0,927054 | 0,9732 | y ~ current_feature + S |
| -0,06955 | 0,070189 | -0,04751 | -0,99092 | 205 | 202 | 0,322909 | 0,774585 | y ~ current_feature + S |
| -0,14391 | 0,078666 | 0,061498 | -1,82938 | 173 | 158,2489 | 0,069225 | 0,715249 | y ~ current_feature + S |
| -0,06022 | 0,08623 | -0,02429 | -0,69839 | 137 | 134 | 0,486141 | 0,888665 | y ~ current_feature + S |
| 0,122987 | 0,072767 | -0,07874 | 1,690155 | 189 | 186 | 0,092674 | 0,42595 | y ~ current_feature + S |
| 0,067677 | 0,073155 | 0,034007 | 0,925116 | 189 | 186 | 0,356104 | 0,7498 | y ~ current_feature + S |
| 0,007099 | 0,073322 | -0,04751 | 0,096823 | 189 | 186 | 0,922971 | 0,985217 | y ~ current_feature + S |
| -0,08969 | 0,078986 | 0,061498 | -1,13555 | 162 | 159 | 0,257853 | 0,893818 | y ~ current_feature + S |
| -0,01944 | 0,090247 | -0,02429 | -0,21539 | 126 | 122,7343 | 0,829824 | 0,976969 | y ~ current_feature + S |
| 0,036447 | 0,085692 | -0,07874 | 0,425328 | 139 | 136 | 0,67127 | 0,934653 | y ~ current_feature + S |
| 0,03069 | 0,085709 | 0,034007 | 0,358075 | 139 | 136 | 0,720842 | 0,920814 | y ~ current_feature + S |
| 0,026744 | 0,085719 | -0,04751 | 0,312003 | 139 | 136 | 0,755516 | 0,9394 | y ~ current_feature + S |
| -0,04893 | 0,092736 | 0,061498 | -0,52765 | 119 | 116 | 0,59875 | 0,945257 | y ~ current_feature + S |
| -0,07849 | 0,105084 | -0,02429 | -0,74695 | 93 | 90 | 0,457038 | 0,866856 | y ~ current_feature + S |
| -0,07083 | 0,100762 | -0,07874 | -0,70294 | 101 | 98 | 0,483758 | 0,846291 | y ~ current_feature + S |
| 0,062896 | 0,100815 | 0,034007 | 0,623877 | 101 | 98 | 0,534158 | 0,849319 | y ~ current_feature + S |
| 0,096448 | 0,101061 | -0,04751 | 0,954352 | 100 | 97 | 0,342278 | 0,789051 | y ~ current_feature + S |
| 0,008121 | 0,111107 | 0,061498 | 0,073094 | 84 | 81 | 0,941911 | 0,996179 | y ~ current_feature + S |
| -0,02071 | 0,124973 | -0,02429 | -0,16575 | 67 | 64 | 0,868875 | 0,984468 | y ~ current_feature + S |
| -0,04281 | 0,054243 | -0,07874 | -0,78922 | 395 | 339,2445 | 0,430537 | 0,814177 | y ~ current_feature + S |
| -0,12903 | 0,050085 | 0,034007 | -2,57613 | 395 | 392 | 0,010357 | 0,232238 | y ~ current_feature + S |
| 0,031198 | 0,055095 | -0,04751 | 0,56627 | 394 | 329,1236 | 0,571596 | 0,903741 | y ~ current_feature + S |
| -3,13E-04 | 0,059316 | 0,061498 | -0,00529 | 343 | 284,2256 | 0,995787 | 0,999005 | y ~ current_feature + S |
| -0,00988 | 0,062014 | -0,02429 | -0,15929 | 263 | 260 | 0,87356 | 0,984468 | y ~ current_feature + S |
| 0,065751 | 0,053557 | -0,07874 | 1,227686 | 408 | 347,1218 | 0,220397 | 0,639583 | y ~ current_feature + S |
| -0,00907 | 0,069769 | 0,034007 | -0,12995 | 408 | 205,4197 | 0,89673 | 0,96591 | y ~ current_feature + S |
| 0,008906 | 0,05547 | -0,04751 | 0,160553 | 407 | 324,9797 | 0,872546 | 0,985217 | y ~ current_feature + S |
| -0,04101 | 0,059045 | 0,061498 | -0,69453 | 354 | 286,3581 | 0,487915 | 0,937665 | y ~ current_feature + S |

| | | | | | | | | |
|---|---|---|---|---|---|---|---|---|
| 0,085336 | 0,060862 | -0,02429 | 1,402128 | 271 | 268 | 0,162034 | 0,623497 | y ~ current_feature + S |
| -0,13958 | 0,070194 | -0,07874 | -1,98845 | 202 | 199 | 0,048132 | 0,289576 | y ~ current_feature + S |
| -0,15215 | 0,070063 | 0,034007 | -2,17157 | 202 | 199 | 0,031071 | 0,347616 | y ~ current_feature + S |
| -0,06149 | 0,070754 | -0,04751 | -0,8691 | 202 | 199 | 0,385837 | 0,803665 | y ~ current_feature + S |
| 0,102689 | 0,075193 | 0,061498 | 1,36567 | 178 | 175 | 0,173795 | 0,840578 | y ~ current_feature + S |
| -0,15293 | 0,086343 | -0,02429 | -1,77124 | 134 | 131 | 0,078847 | 0,605474 | y ~ current_feature + S |
| 0,072757 | 0,120946 | -0,07874 | 0,601565 | 71 | 68 | 0,549464 | 0,888464 | y ~ current_feature + S |
| 0,122588 | 0,120353 | 0,034007 | 1,018571 | 71 | 68 | 0,312017 | 0,718742 | y ~ current_feature + S |
| 0,171956 | 0,119461 | -0,04751 | 1,439423 | 71 | 68 | 0,154618 | 0,672434 | y ~ current_feature + S |
| -0,18767 | 0,128973 | 0,061498 | -1,45513 | 61 | 58 | 0,151023 | 0,8197 | y ~ current_feature + S |
| 0,125634 | 0,149561 | -0,02429 | 0,840014 | 47 | 44 | 0,405442 | 0,8311 | y ~ current_feature + S |
| -0,0879 | 0,052424 | -0,07874 | -1,67663 | 385 | 361,0536 | 0,09448 | 0,42893 | y ~ current_feature + S |
| -0,06883 | 0,06995 | 0,034007 | -0,98403 | 385 | 203,4078 | 0,326272 | 0,728793 | y ~ current_feature + S |
| -0,07667 | 0,054735 | -0,04751 | -1,40069 | 384 | 331,8299 | 0,162242 | 0,693983 | y ~ current_feature + S |
| -0,10151 | 0,057559 | 0,061498 | -1,76364 | 335 | 298,7285 | 0,078814 | 0,738262 | y ~ current_feature + S |
| -0,00144 | 0,063209 | -0,02429 | -0,02274 | 256 | 250,2879 | 0,981875 | 0,99444 | y ~ current_feature + S |
| 0,096039 | 0,11571 | -0,07874 | 0,829991 | 77 | 74 | 0,409213 | 0,806796 | y ~ current_feature + S |
| 0,084567 | 0,115831 | 0,034007 | 0,730092 | 77 | 74 | 0,467639 | 0,831402 | y ~ current_feature + S |
| -0,0499 | 0,116103 | -0,04751 | -0,42983 | 77 | 74 | 0,668569 | 0,92822 | y ~ current_feature + S |
| -0,02598 | 0,126957 | 0,061498 | -0,20462 | 65 | 62 | 0,838539 | 0,996179 | y ~ current_feature + S |
| -0,03938 | 0,144226 | -0,02429 | -0,27307 | 51 | 48 | 0,785974 | 0,964545 | y ~ current_feature + S |
| 0,050082 | 0,052785 | -0,07874 | 0,948784 | 361 | 358 | 0,343371 | 0,751758 | y ~ current_feature + S |
| 5,97E-04 | 0,052852 | 0,034007 | 0,011296 | 361 | 358 | 0,990993 | 0,997735 | y ~ current_feature + S |
| 0,003218 | 0,052851 | -0,04751 | 0,060885 | 361 | 358 | 0,951485 | 0,990156 | y ~ current_feature + S |
| -0,06504 | 0,056585 | 0,061498 | -1,14949 | 314 | 311 | 0,251238 | 0,893818 | y ~ current_feature + S |
| -0,04389 | 0,064894 | -0,02429 | -0,67628 | 240 | 237 | 0,499525 | 0,89399 | y ~ current_feature + S |
| -0,0878 | 0,106797 | -0,07874 | -0,82207 | 90 | 87 | 0,413283 | 0,811218 | y ~ current_feature + S |
| 0,118599 | 0,106455 | 0,034007 | 1,114083 | 90 | 87 | 0,268312 | 0,71095 | y ~ current_feature + S |
| -0,08479 | 0,106825 | -0,04751 | -0,79377 | 90 | 87 | 0,429488 | 0,819126 | y ~ current_feature + S |
| 0,028108 | 0,113916 | 0,061498 | 0,246746 | 80 | 77 | 0,805761 | 0,988828 | y ~ current_feature + S |
| 0,085569 | 0,131967 | -0,02429 | 0,648407 | 60 | 57 | 0,519325 | 0,899808 | y ~ current_feature + S |
| 0,236903 | 0,125424 | -0,07874 | 1,888811 | 63 | 60 | 0,063753 | 0,336982 | y ~ current_feature + S |
| -0,07038 | 0,128779 | 0,034007 | -0,54651 | 63 | 60 | 0,586745 | 0,87187 | y ~ current_feature + S |
| 0,164969 | 0,127331 | -0,04751 | 1,295593 | 63 | 60 | 0,200078 | 0,723562 | y ~ current_feature + S |
| -0,29734 | 0,132403 | 0,061498 | -2,24569 | 55 | 52 | 0,029 | 0,658491 | y ~ current_feature + S |
| 0,017789 | 0,160103 | -0,02429 | 0,111107 | 42 | 39 | 0,912101 | 0,99444 | y ~ current_feature + S |
| 0,024282 | 0,053911 | -0,07874 | 0,450412 | 392 | 343,8666 | 0,652697 | 0,932255 | y ~ current_feature + S |
| 0,084742 | 0,0714 | 0,034007 | 1,186853 | 392 | 194,7466 | 0,236731 | 0,68969 | y ~ current_feature + S |
| 0,04902 | 0,054643 | -0,04751 | 0,897101 | 391 | 334,1121 | 0,370311 | 0,802018 | y ~ current_feature + S |
| -0,01066 | 0,058758 | 0,061498 | -0,1814 | 342 | 289,6107 | 0,85618 | 0,996179 | y ~ current_feature + S |
| 0,088451 | 0,062134 | -0,02429 | 1,423552 | 260 | 257 | 0,155789 | 0,623156 | y ~ current_feature + S |
| 0,015225 | 0,102586 | -0,07874 | 0,148411 | 98 | 95 | 0,882333 | 0,987929 | y ~ current_feature + S |
| -0,01983 | 0,102578 | 0,034007 | -0,19327 | 98 | 95 | 0,847159 | 0,959445 | y ~ current_feature + S |
| -0,07189 | 0,102332 | -0,04751 | -0,70253 | 98 | 95 | 0,484065 | 0,852876 | y ~ current_feature + S |
| 0,017129 | 0,109093 | 0,061498 | 0,157015 | 87 | 84 | 0,87561 | 0,996179 | y ~ current_feature + S |
| -0,10474 | 0,126302 | -0,02429 | -0,8293 | 65 | 62 | 0,410115 | 0,836046 | y ~ current_feature + S |
| 0,078661 | 0,099195 | -0,07874 | 0,792987 | 104 | 101 | 0,429644 | 0,814177 | y ~ current_feature + S |
| -0,05175 | 0,09937 | 0,034007 | -0,52073 | 104 | 101 | 0,603691 | 0,883662 | y ~ current_feature + S |

| | | | | | | | | |
|---|---|---|---|---|---|---|---|---|
| -0,173 | 0,098003 | -0,04751 | -1,7652 | 104 | 101 | 0,080553 | 0,640171 | y ~ current_feature + S |
| 0,135138 | 0,105027 | 0,061498 | 1,28669 | 92 | 89 | 0,201539 | 0,893818 | y ~ current_feature + S |
| -0,12306 | 0,122156 | -0,02429 | -1,00743 | 69 | 66 | 0,317405 | 0,766329 | y ~ current_feature + S |
| 0,09461 | 0,153611 | -0,07874 | 0,615906 | 45 | 42 | 0,54128 | 0,882344 | y ~ current_feature + S |
| -0,03496 | 0,154209 | 0,034007 | -0,22673 | 45 | 42 | 0,821734 | 0,952501 | y ~ current_feature + S |
| -0,10372 | 0,153471 | -0,04751 | -0,67583 | 45 | 42 | 0,502854 | 0,868909 | y ~ current_feature + S |
| 0,186742 | 0,166057 | 0,061498 | 1,124564 | 38 | 35 | 0,268427 | 0,893818 | y ~ current_feature + S |
| 0,086969 | 0,191721 | -0,02429 | 0,453622 | 30 | 27 | 0,653726 | 0,924682 | y ~ current_feature + S |
| 0,079551 | 0,105664 | -0,07874 | 0,752864 | 92 | 89 | 0,453518 | 0,82458 | y ~ current_feature + S |
| 0,051104 | 0,105861 | 0,034007 | 0,482748 | 92 | 89 | 0,63046 | 0,899624 | y ~ current_feature + S |
| 0,166702 | 0,104517 | -0,04751 | 1,594978 | 92 | 89 | 0,114262 | 0,640171 | y ~ current_feature + S |
| -0,15591 | 0,113305 | 0,061498 | -1,37601 | 79 | 76 | 0,172859 | 0,840578 | y ~ current_feature + S |
| 0,031323 | 0,131242 | -0,02429 | 0,238666 | 61 | 58 | 0,812206 | 0,974251 | y ~ current_feature + S |
| 0,038985 | 0,050729 | -0,07874 | 0,768508 | 391 | 388 | 0,442653 | 0,818908 | y ~ current_feature + S |
| 0,096854 | 0,050529 | 0,034007 | 1,916809 | 391 | 388 | 0,055996 | 0,420946 | y ~ current_feature + S |
| 0,011149 | 0,05083 | -0,04751 | 0,219333 | 390 | 387 | 0,826507 | 0,975438 | y ~ current_feature + S |
| 0,010204 | 0,063445 | 0,061498 | 0,160832 | 339 | 248,4026 | 0,872356 | 0,996179 | y ~ current_feature + S |
| 0,078084 | 0,062188 | -0,02429 | 1,255615 | 260 | 257 | 0,210396 | 0,675518 | y ~ current_feature + S |
| 0,13848 | 0,094001 | -0,07874 | 1,473174 | 114 | 111 | 0,143534 | 0,501016 | y ~ current_feature + S |
| -0,05781 | 0,094757 | 0,034007 | -0,61011 | 114 | 111 | 0,543034 | 0,852506 | y ~ current_feature + S |
| 5,39E-04 | 0,094916 | -0,04751 | 0,005677 | 114 | 111 | 0,995481 | 0,995481 | y ~ current_feature + S |
| -0,13701 | 0,107375 | 0,061498 | -1,276 | 99 | 85,10716 | 0,205426 | 0,893818 | y ~ current_feature + S |
| -0,13906 | 0,115904 | -0,02429 | -1,19974 | 76 | 73 | 0,234118 | 0,695919 | y ~ current_feature + S |
| 0,197909 | 0,100043 | -0,07874 | 1,978234 | 99 | 96 | 0,050769 | 0,30055 | y ~ current_feature + S |
| 0,086286 | 0,101681 | 0,034007 | 0,84859 | 99 | 96 | 0,398221 | 0,775482 | y ~ current_feature + S |
| 0,06303 | 0,101859 | -0,04751 | 0,618795 | 99 | 96 | 0,537517 | 0,885885 | y ~ current_feature + S |
| -0,02566 | 0,112472 | 0,061498 | -0,2281 | 82 | 79 | 0,820154 | 0,996179 | y ~ current_feature + S |
| -0,02857 | 0,125937 | -0,02429 | -0,2269 | 66 | 63 | 0,821237 | 0,976828 | y ~ current_feature + S |
| 0,003667 | 0,071795 | -0,07874 | 0,051079 | 197 | 194 | 0,959316 | 0,994607 | y ~ current_feature + S |
| -0,04099 | 0,071735 | 0,034007 | -0,57143 | 197 | 194 | 0,568372 | 0,863333 | y ~ current_feature + S |
| -7,51E-04 | 0,071796 | -0,04751 | -0,01046 | 197 | 194 | 0,991663 | 0,99446 | y ~ current_feature + S |
| -0,08641 | 0,076863 | 0,061498 | -1,12418 | 171 | 168 | 0,26254 | 0,893818 | y ~ current_feature + S |
| -0,00712 | 0,088386 | -0,02429 | -0,08056 | 131 | 128 | 0,935915 | 0,99444 | y ~ current_feature + S |
| 0,058917 | 0,069384 | -0,07874 | 0,849147 | 210 | 207 | 0,39678 | 0,791422 | y ~ current_feature + S |
| -0,0308 | 0,069472 | 0,034007 | -0,44337 | 210 | 207 | 0,657962 | 0,900251 | y ~ current_feature + S |
| 0,134491 | 0,068873 | -0,04751 | 1,952722 | 210 | 207 | 0,052201 | 0,640171 | y ~ current_feature + S |
| -0,04847 | 0,074448 | 0,061498 | -0,65109 | 183 | 180 | 0,515819 | 0,937665 | y ~ current_feature + S |
| -0,13308 | 0,084676 | -0,02429 | -1,57158 | 140 | 137 | 0,118354 | 0,615361 | y ~ current_feature + S |
| 0,062904 | 0,070047 | -0,07874 | 0,898016 | 206 | 203 | 0,370241 | 0,780565 | y ~ current_feature + S |
| -0,11534 | 0,069718 | 0,034007 | -1,6544 | 206 | 203 | 0,099592 | 0,518857 | y ~ current_feature + S |
| -0,02625 | 0,070162 | -0,04751 | -0,37416 | 206 | 203 | 0,708677 | 0,93314 | y ~ current_feature + S |
| 0,024559 | 0,076006 | 0,061498 | 0,323116 | 176 | 173 | 0,746997 | 0,972573 | y ~ current_feature + S |
| -0,10375 | 0,085921 | -0,02429 | -1,20748 | 137 | 134 | 0,229376 | 0,695919 | y ~ current_feature + S |
| -0,1509 | 0,144195 | -0,07874 | -1,04653 | 50 | 47 | 0,300667 | 0,718687 | y ~ current_feature + S |
| -0,17552 | 0,143601 | 0,034007 | -1,22226 | 50 | 47 | 0,227703 | 0,681978 | y ~ current_feature + S |
| -0,30134 | 0,140589 | -0,04751 | -2,1434 | 49 | 46 | 0,037402 | 0,611115 | y ~ current_feature + S |
| 0,178848 | 0,155565 | 0,061498 | 1,149673 | 43 | 40 | 0,257106 | 0,893818 | y ~ current_feature + S |
| -0,12215 | 0,181207 | -0,02429 | -0,67408 | 33 | 30 | 0,505423 | 0,894767 | y ~ current_feature + S |

| | | | | | | | | |
|---|---|---|---|---|---|---|---|---|
| -0,03805 | 0,135984 | -0,07874 | -0,27978 | 57 | 54 | 0,780717 | 0,97261 | y ~ current_feature + S |
| -0,09637 | 0,135449 | 0,034007 | -0,71149 | 57 | 54 | 0,479841 | 0,831402 | y ~ current_feature + S |
| 0,004147 | 0,136082 | -0,04751 | 0,030472 | 57 | 54 | 0,975803 | 0,992311 | y ~ current_feature + S |
| 0,045626 | 0,147288 | 0,061498 | 0,309775 | 49 | 46 | 0,758131 | 0,972573 | y ~ current_feature + S |
| -0,04386 | 0,168868 | -0,02429 | -0,25971 | 38 | 35 | 0,796608 | 0,968297 | y ~ current_feature + S |
| 0,028555 | 0,073892 | -0,07874 | 0,386449 | 186 | 183 | 0,699613 | 0,939646 | y ~ current_feature + S |
| -0,02766 | 0,073894 | 0,034007 | -0,37429 | 186 | 183 | 0,708623 | 0,920814 | y ~ current_feature + S |
| 0,111003 | 0,073465 | -0,04751 | 1,510958 | 186 | 183 | 0,132524 | 0,657012 | y ~ current_feature + S |
| 0,045379 | 0,084405 | 0,061498 | 0,53763 | 161 | 140,0773 | 0,591685 | 0,945257 | y ~ current_feature + S |
| -0,00609 | 0,09092 | -0,02429 | -0,06703 | 124 | 120,9674 | 0,946665 | 0,99444 | y ~ current_feature + S |
| 0,319973 | 0,132666 | -0,07874 | 2,411861 | 54 | 51 | 0,019509 | 0,187487 | y ~ current_feature + S |
| -0,1632 | 0,138151 | 0,034007 | -1,18134 | 54 | 51 | 0,242948 | 0,702074 | y ~ current_feature + S |
| 0,018245 | 0,140005 | -0,04751 | 0,130314 | 54 | 51 | 0,89683 | 0,985217 | y ~ current_feature + S |
| -0,07138 | 0,15211 | 0,061498 | -0,46928 | 46 | 43 | 0,641244 | 0,956496 | y ~ current_feature + S |
| -0,64636 | 0,132828 | -0,02429 | -4,86612 | 36 | 33 | 2,74E-05 | 0,010121 | y ~ current_feature + S |
| 0,049261 | 0,100893 | -0,07874 | 0,488256 | 101 | 98 | 0,626459 | 0,92163 | y ~ current_feature + S |
| 0,277217 | 0,097056 | 0,034007 | 2,856256 | 101 | 98 | 0,005234 | 0,168399 | y ~ current_feature + S |
| 0,13952 | 0,100542 | -0,04751 | 1,387683 | 100 | 97 | 0,168413 | 0,700235 | y ~ current_feature + S |
| -0,00958 | 0,110426 | 0,061498 | -0,08678 | 85 | 82 | 0,931056 | 0,996179 | y ~ current_feature + S |
| 0,249193 | 0,121057 | -0,02429 | 2,058481 | 67 | 64 | 0,04362 | 0,525585 | y ~ current_feature + S |
| -0,01016 | 0,078322 | -0,07874 | -0,12977 | 166 | 163 | 0,896912 | 0,987929 | y ~ current_feature + S |
| -0,08299 | 0,078056 | 0,034007 | -1,06322 | 166 | 163 | 0,289254 | 0,718742 | y ~ current_feature + S |
| 0,06562 | 0,078157 | -0,04751 | 0,839589 | 166 | 163 | 0,402369 | 0,803665 | y ~ current_feature + S |
| -0,12664 | 0,084136 | 0,061498 | -1,50517 | 142 | 139 | 0,134549 | 0,8197 | y ~ current_feature + S |
| -0,04693 | 0,096567 | -0,02429 | -0,48597 | 110 | 107 | 0,62798 | 0,924682 | y ~ current_feature + S |
| 0,014584 | 0,07254 | -0,07874 | 0,201047 | 193 | 190 | 0,840877 | 0,983548 | y ~ current_feature + S |
| -0,14831 | 0,071745 | 0,034007 | -2,06712 | 193 | 190 | 0,040078 | 0,396441 | y ~ current_feature + S |
| 0,001867 | 0,072739 | -0,04751 | 0,025664 | 192 | 189 | 0,979552 | 0,992311 | y ~ current_feature + S |
| -0,01664 | 0,078557 | 0,061498 | -0,21187 | 165 | 162 | 0,832474 | 0,996179 | y ~ current_feature + S |
| -0,1041 | 0,089219 | -0,02429 | -1,16673 | 128 | 124,2658 | 0,245552 | 0,715387 | y ~ current_feature + S |
| -0,05056 | 0,139849 | -0,07874 | -0,36155 | 54 | 51 | 0,71918 | 0,948651 | y ~ current_feature + S |
| -0,26581 | 0,134991 | 0,034007 | -1,96909 | 54 | 51 | 0,054387 | 0,420946 | y ~ current_feature + S |
| 0,026891 | 0,139977 | -0,04751 | 0,192109 | 54 | 51 | 0,84842 | 0,980218 | y ~ current_feature + S |
| 0,035065 | 0,156078 | 0,061498 | 0,224663 | 44 | 41 | 0,823357 | 0,996179 | y ~ current_feature + S |
| 0,089164 | 0,173384 | -0,02429 | 0,514256 | 36 | 33 | 0,6105 | 0,924682 | y ~ current_feature + S |
| -0,00424 | 0,090908 | -0,07874 | -0,04664 | 124 | 121 | 0,962876 | 0,994607 | y ~ current_feature + S |
| 0,077061 | 0,090639 | 0,034007 | 0,850203 | 124 | 121 | 0,396891 | 0,775482 | y ~ current_feature + S |
| 0,044085 | 0,091198 | -0,04751 | 0,483401 | 123 | 120 | 0,629692 | 0,925756 | y ~ current_feature + S |
| 0,062134 | 0,097401 | 0,061498 | 0,637913 | 108 | 105 | 0,524919 | 0,937665 | y ~ current_feature + S |
| 0,008074 | 0,1118 | -0,02429 | 0,072215 | 83 | 80 | 0,942611 | 0,99444 | y ~ current_feature + S |
| 0,444551 | 0,121897 | -0,07874 | 3,646952 | 57 | 54 | 5,98E-04 | 0,031617 | y ~ current_feature + S |
| 0,269277 | 0,131056 | 0,034007 | 2,054667 | 57 | 54 | 0,044763 | 0,400524 | y ~ current_feature + S |
| 0,062489 | 0,135817 | -0,04751 | 0,460096 | 57 | 54 | 0,647295 | 0,925756 | y ~ current_feature + S |
| -0,09611 | 0,14519 | 0,061498 | -0,66195 | 50 | 47 | 0,511235 | 0,937665 | y ~ current_feature + S |
| 0,239171 | 0,164125 | -0,02429 | 1,45725 | 38 | 35 | 0,153962 | 0,623156 | y ~ current_feature + S |
| 0,139478 | 0,060971 | -0,07874 | 2,287613 | 304 | 263,7675 | 0,022952 | 0,194866 | y ~ current_feature + S |
| 0,097962 | 0,081044 | 0,034007 | 1,20875 | 304 | 150,7879 | 0,228651 | 0,681978 | y ~ current_feature + S |
| 0,073159 | 0,062661 | -0,04751 | 1,167536 | 304 | 253,324 | 0,244092 | 0,742928 | y ~ current_feature + S |

| | | | | | | | | |
|---|---|---|---|---|---|---|---|---|
| -0,13171 | 0,066869 | 0,061498 | -1,96961 | 259 | 219,7618 | 0,050139 | 0,700056 | y ~ current_feature + S |
| 0,107165 | 0,070692 | -0,02429 | 1,515934 | 203 | 197,8049 | 0,131132 | 0,617591 | y ~ current_feature + S |
| 0,165013 | 0,121404 | -0,07874 | 1,359202 | 69 | 66 | 0,178709 | 0,572488 | y ~ current_feature + S |
| 0,152106 | 0,121659 | 0,034007 | 1,25026 | 69 | 66 | 0,215619 | 0,664036 | y ~ current_feature + S |
| 0,020367 | 0,123066 | -0,04751 | 0,165493 | 69 | 66 | 0,869062 | 0,985217 | y ~ current_feature + S |
| -0,16056 | 0,129603 | 0,061498 | -1,23889 | 61 | 58 | 0,220377 | 0,893818 | y ~ current_feature + S |
| -0,22073 | 0,148737 | -0,02429 | -1,48405 | 46 | 43 | 0,145089 | 0,619762 | y ~ current_feature + S |
| 0,165064 | 0,14227 | -0,07874 | 1,160215 | 59 | 48,05913 | 0,251693 | 0,679754 | y ~ current_feature + S |
| 0,160062 | 0,15743 | 0,034007 | 1,016714 | 59 | 39,31432 | 0,315506 | 0,718742 | y ~ current_feature + S |
| 0,208548 | 0,140683 | -0,04751 | 1,482398 | 59 | 48,32859 | 0,144726 | 0,661823 | y ~ current_feature + S |
| -0,18802 | 0,184285 | 0,061498 | -1,02025 | 51 | 28,40453 | 0,316223 | 0,893818 | y ~ current_feature + S |
| 0,58531 | 0,135162 | -0,02429 | 4,330449 | 39 | 35,98578 | 1,14E-04 | 0,021049 | y ~ current_feature + S |
| 0,058516 | 0,051919 | -0,07874 | 1,127058 | 421 | 369,7019 | 0,260449 | 0,690796 | y ~ current_feature + S |
| 0,043417 | 0,068324 | 0,034007 | 0,63546 | 421 | 213,8108 | 0,525808 | 0,847708 | y ~ current_feature + S |
| 0,01224 | 0,053045 | -0,04751 | 0,230743 | 420 | 355,3474 | 0,817647 | 0,972405 | y ~ current_feature + S |
| -0,09259 | 0,057747 | 0,061498 | -1,60331 | 367 | 297,3071 | 0,109927 | 0,8197 | y ~ current_feature + S |
| -0,10265 | 0,059767 | -0,02429 | -1,71756 | 280 | 277 | 0,086994 | 0,605474 | y ~ current_feature + S |
| -0,0905 | 0,073821 | -0,07874 | -1,22596 | 185 | 182 | 0,221799 | 0,641138 | y ~ current_feature + S |
| -0,06937 | 0,073946 | 0,034007 | -0,93807 | 185 | 182 | 0,34945 | 0,749214 | y ~ current_feature + S |
| -0,03453 | 0,074081 | -0,04751 | -0,46605 | 185 | 182 | 0,641734 | 0,925756 | y ~ current_feature + S |
| 0,168354 | 0,080217 | 0,061498 | 2,098721 | 154 | 151 | 0,037506 | 0,658491 | y ~ current_feature + S |
| -0,07422 | 0,091035 | -0,02429 | -0,81524 | 123 | 120 | 0,416553 | 0,839968 | y ~ current_feature + S |
| -0,01497 | 0,129085 | -0,07874 | -0,11598 | 63 | 60 | 0,908056 | 0,987929 | y ~ current_feature + S |
| 0,164779 | 0,127335 | 0,034007 | 1,294062 | 63 | 60 | 0,200603 | 0,647595 | y ~ current_feature + S |
| -0,17076 | 0,127203 | -0,04751 | -1,34243 | 63 | 60 | 0,184515 | 0,704811 | y ~ current_feature + S |
| 0,107409 | 0,143503 | 0,061498 | 0,748482 | 51 | 48 | 0,457822 | 0,937665 | y ~ current_feature + S |
| -0,0846 | 0,159554 | -0,02429 | -0,53023 | 42 | 39 | 0,598961 | 0,924682 | y ~ current_feature + S |
| -0,02711 | 0,123046 | -0,07874 | -0,22029 | 69 | 66 | 0,826328 | 0,981192 | y ~ current_feature + S |
| 0,03042 | 0,123035 | 0,034007 | 0,24725 | 69 | 66 | 0,805482 | 0,952501 | y ~ current_feature + S |
| 0,009313 | 0,123086 | -0,04751 | 0,075666 | 69 | 66 | 0,939913 | 0,990156 | y ~ current_feature + S |
| 0,085394 | 0,133143 | 0,061498 | 0,641371 | 59 | 56 | 0,523898 | 0,937665 | y ~ current_feature + S |
| 0,132109 | 0,151162 | -0,02429 | 0,873958 | 46 | 43 | 0,386996 | 0,819479 | y ~ current_feature + S |
| -0,01005 | 0,082474 | -0,07874 | -0,12188 | 150 | 147 | 0,903161 | 0,987929 | y ~ current_feature + S |
| 0,015548 | 0,082469 | 0,034007 | 0,188538 | 150 | 147 | 0,850715 | 0,959445 | y ~ current_feature + S |
| 0,073682 | 0,082254 | -0,04751 | 0,895779 | 150 | 147 | 0,371835 | 0,80221 | y ~ current_feature + S |
| 0,022947 | 0,089064 | 0,061498 | 0,257651 | 129 | 126 | 0,797097 | 0,984075 | y ~ current_feature + S |
| 0,153038 | 0,100339 | -0,02429 | 1,525212 | 100 | 97 | 0,13046 | 0,617591 | y ~ current_feature + S |
| -0,07597 | 0,052191 | -0,07874 | -1,45559 | 368 | 365 | 0,146365 | 0,506121 | y ~ current_feature + S |
| -0,00492 | 0,052342 | 0,034007 | -0,09404 | 368 | 365 | 0,925128 | 0,9732 | y ~ current_feature + S |
| -0,05084 | 0,052346 | -0,04751 | -0,97113 | 367 | 364 | 0,332126 | 0,780234 | y ~ current_feature + S |
| -0,00811 | 0,056164 | 0,061498 | -0,14442 | 320 | 317 | 0,885261 | 0,996179 | y ~ current_feature + S |
| -0,05938 | 0,064169 | -0,02429 | -0,92539 | 245 | 242 | 0,355687 | 0,800026 | y ~ current_feature + S |
| 0,025774 | 0,067093 | -0,07874 | 0,384153 | 225 | 222 | 0,701233 | 0,940058 | y ~ current_feature + S |
| 0,012098 | 0,067111 | 0,034007 | 0,180266 | 225 | 222 | 0,857108 | 0,961 | y ~ current_feature + S |
| -0,00826 | 0,067113 | -0,04751 | -0,12312 | 225 | 222 | 0,902126 | 0,985217 | y ~ current_feature + S |
| -0,04788 | 0,072656 | 0,061498 | -0,65905 | 192 | 189 | 0,510665 | 0,937665 | y ~ current_feature + S |
| -0,00289 | 0,082478 | -0,02429 | -0,03501 | 150 | 147 | 0,972118 | 0,99444 | y ~ current_feature + S |
| 0,11119 | 0,086173 | -0,07874 | 1,290309 | 136 | 133 | 0,199182 | 0,611596 | y ~ current_feature + S |

| | | | | | | | | |
|---|---|---|---|---|---|---|---|---|
| 0,02696 | 0,086679 | 0,034007 | 0,311026 | 136 | 133 | 0,756267 | 0,93416 | y ~ current_feature + S |
| 0,077949 | 0,086774 | -0,04751 | 0,898298 | 135 | 132 | 0,370662 | 0,802018 | y ~ current_feature + S |
| 0,056657 | 0,093101 | 0,061498 | 0,608557 | 118 | 115 | 0,544018 | 0,937665 | y ~ current_feature + S |
| 0,159964 | 0,105228 | -0,02429 | 1,520173 | 91 | 88 | 0,132053 | 0,617591 | y ~ current_feature + S |
| 0,113397 | 0,115498 | -0,07874 | 0,981814 | 77 | 74 | 0,329391 | 0,729788 | y ~ current_feature + S |
| -0,07585 | 0,115913 | 0,034007 | -0,6544 | 77 | 74 | 0,514886 | 0,841996 | y ~ current_feature + S |
| -0,02503 | 0,116211 | -0,04751 | -0,21538 | 77 | 74 | 0,830067 | 0,975438 | y ~ current_feature + S |
| -0,19051 | 0,124674 | 0,061498 | -1,52807 | 65 | 62 | 0,131581 | 0,8197 | y ~ current_feature + S |
| -0,02934 | 0,144275 | -0,02429 | -0,20339 | 51 | 48 | 0,839694 | 0,977734 | y ~ current_feature + S |
| 0,052255 | 0,113073 | -0,07874 | 0,462131 | 81 | 78 | 0,645274 | 0,930804 | y ~ current_feature + S |
| -0,02135 | 0,113202 | 0,034007 | -0,18862 | 81 | 78 | 0,850878 | 0,959445 | y ~ current_feature + S |
| -0,04064 | 0,113134 | -0,04751 | -0,35923 | 81 | 78 | 0,720391 | 0,93314 | y ~ current_feature + S |
| 0,063972 | 0,12674 | 0,061498 | 0,504749 | 65 | 62 | 0,615524 | 0,948092 | y ~ current_feature + S |
| -0,06661 | 0,139717 | -0,02429 | -0,47676 | 54 | 51 | 0,635569 | 0,924682 | y ~ current_feature + S |
| 0,021176 | 0,086367 | -0,07874 | 0,24518 | 137 | 134 | 0,806692 | 0,979871 | y ~ current_feature + S |
| -0,06126 | 0,086225 | 0,034007 | -0,71051 | 137 | 134 | 0,478626 | 0,831402 | y ~ current_feature + S |
| -0,03751 | 0,086326 | -0,04751 | -0,43451 | 137 | 134 | 0,664615 | 0,927953 | y ~ current_feature + S |
| -0,11609 | 0,099361 | 0,061498 | -1,16835 | 116 | 99,9245 | 0,245446 | 0,893818 | y ~ current_feature + S |
| -0,12832 | 0,110236 | -0,02429 | -1,16406 | 91 | 80,93679 | 0,24782 | 0,717451 | y ~ current_feature + S |
| 0,01146 | 0,138666 | -0,07874 | 0,082645 | 55 | 52 | 0,934451 | 0,9921 | y ~ current_feature + S |
| -0,12571 | 0,137575 | 0,034007 | -0,91375 | 55 | 52 | 0,365064 | 0,7498 | y ~ current_feature + S |
| -0,18967 | 0,137486 | -0,04751 | -1,37954 | 54 | 51 | 0,173747 | 0,700235 | y ~ current_feature + S |
| 0,067548 | 0,153951 | 0,061498 | 0,438764 | 45 | 42 | 0,663081 | 0,957207 | y ~ current_feature + S |
| -0,05762 | 0,173788 | -0,02429 | -0,33154 | 36 | 33 | 0,74233 | 0,945481 | y ~ current_feature + S |
| 0,015786 | 0,136066 | -0,07874 | 0,116019 | 57 | 54 | 0,908067 | 0,987929 | y ~ current_feature + S |
| -0,06118 | 0,135828 | 0,034007 | -0,4504 | 57 | 54 | 0,654224 | 0,900251 | y ~ current_feature + S |
| 0,109079 | 0,135271 | -0,04751 | 0,806375 | 57 | 54 | 0,423564 | 0,819126 | y ~ current_feature + S |
| 0,030195 | 0,150687 | 0,061498 | 0,200383 | 47 | 44 | 0,842104 | 0,996179 | y ~ current_feature + S |
| -0,25657 | 0,163373 | -0,02429 | -1,57046 | 38 | 35 | 0,125305 | 0,617591 | y ~ current_feature + S |
| 0,019257 | 0,140002 | -0,07874 | 0,137547 | 54 | 51 | 0,89114 | 0,987929 | y ~ current_feature + S |
| 0,144539 | 0,138558 | 0,034007 | 1,043168 | 54 | 51 | 0,30179 | 0,718742 | y ~ current_feature + S |
| 0,229773 | 0,136281 | -0,04751 | 1,686021 | 54 | 51 | 0,097901 | 0,640171 | y ~ current_feature + S |
| -0,00654 | 0,147439 | 0,061498 | -0,04437 | 49 | 46 | 0,964802 | 0,996179 | y ~ current_feature + S |
| -0,0912 | 0,173352 | -0,02429 | -0,52609 | 36 | 33 | 0,602347 | 0,924682 | y ~ current_feature + S |
| -0,11603 | 0,109023 | -0,07874 | -1,06431 | 86 | 83 | 0,290274 | 0,718687 | y ~ current_feature + S |
| -0,11633 | 0,109019 | 0,034007 | -1,06702 | 86 | 83 | 0,289057 | 0,718742 | y ~ current_feature + S |
| -0,15244 | 0,108481 | -0,04751 | -1,40521 | 86 | 83 | 0,163691 | 0,694976 | y ~ current_feature + S |
| -0,23956 | 0,126327 | 0,061498 | -1,89632 | 77 | 59,06604 | 0,062812 | 0,715249 | y ~ current_feature + S |
| -0,01474 | 0,136068 | -0,02429 | -0,10835 | 57 | 54 | 0,914122 | 0,99444 | y ~ current_feature + S |
| -0,01667 | 0,063491 | -0,07874 | -0,26255 | 251 | 248 | 0,793117 | 0,979102 | y ~ current_feature + S |
| -0,11808 | 0,063056 | 0,034007 | -1,87264 | 251 | 248 | 0,062295 | 0,437643 | y ~ current_feature + S |
| 0,001709 | 0,0635 | -0,04751 | 0,026913 | 251 | 248 | 0,97855 | 0,992311 | y ~ current_feature + S |
| 0,051121 | 0,067952 | 0,061498 | 0,75231 | 219 | 216 | 0,452684 | 0,937665 | y ~ current_feature + S |
| -0,02639 | 0,07806 | -0,02429 | -0,33804 | 167 | 164 | 0,735769 | 0,944216 | y ~ current_feature + S |
| 0,117288 | 0,143341 | -0,07874 | 0,818242 | 51 | 48 | 0,417262 | 0,811887 | y ~ current_feature + S |
| 0,047887 | 0,144172 | 0,034007 | 0,332148 | 51 | 48 | 0,741223 | 0,926529 | y ~ current_feature + S |
| 0,226466 | 0,140588 | -0,04751 | 1,610854 | 51 | 48 | 0,113769 | 0,640171 | y ~ current_feature + S |
| 0,045278 | 0,156014 | 0,061498 | 0,290219 | 44 | 41 | 0,773112 | 0,974621 | y ~ current_feature + S |

| | | | | | | | | |
|---|---|---|---|---|---|---|---|---|
| -0,02351 | 0,179556 | -0,02429 | -0,13094 | 34 | 31 | 0,896665 | 0,99444 | y ~ current_feature + S |
| -0,04876 | 0,116109 | -0,07874 | -0,41997 | 77 | 74 | 0,675723 | 0,934653 | y ~ current_feature + S |
| 0,107018 | 0,11558 | 0,034007 | 0,925918 | 77 | 74 | 0,357498 | 0,7498 | y ~ current_feature + S |
| 0,139271 | 0,1159 | -0,04751 | 1,201645 | 76 | 73 | 0,233385 | 0,742928 | y ~ current_feature + S |
| 0,020037 | 0,129074 | 0,061498 | 0,155239 | 63 | 60 | 0,877154 | 0,996179 | y ~ current_feature + S |
| 0,161588 | 0,142441 | -0,02429 | 1,134421 | 51 | 48 | 0,262251 | 0,723659 | y ~ current_feature + S |
| -0,00405 | 0,11785 | -0,07874 | -0,03435 | 75 | 72 | 0,972694 | 0,995565 | y ~ current_feature + S |
| 0,131923 | 0,116821 | 0,034007 | 1,129272 | 75 | 72 | 0,262532 | 0,71095 | y ~ current_feature + S |
| 0,190536 | 0,115692 | -0,04751 | 1,64692 | 75 | 72 | 0,103933 | 0,640171 | y ~ current_feature + S |
| 0,051407 | 0,135258 | 0,061498 | 0,380068 | 63 | 54,51612 | 0,705371 | 0,972444 | y ~ current_feature + S |
| 0,310279 | 0,138666 | -0,02429 | 2,237602 | 50 | 47 | 0,030026 | 0,485321 | y ~ current_feature + S |
| 0,037862 | 0,052888 | -0,07874 | 0,715891 | 360 | 357 | 0,474526 | 0,84411 | y ~ current_feature + S |
| 0,043249 | 0,052876 | 0,034007 | 0,817933 | 360 | 357 | 0,413941 | 0,785427 | y ~ current_feature + S |
| 0,091335 | 0,052778 | -0,04751 | 1,730545 | 359 | 356 | 0,084399 | 0,640171 | y ~ current_feature + S |
| 0,035893 | 0,061989 | 0,061498 | 0,579011 | 314 | 259,8996 | 0,563083 | 0,937665 | y ~ current_feature + S |
| 0,194216 | 0,063813 | -0,02429 | 3,043526 | 240 | 236,3125 | 0,002603 | 0,18344 | y ~ current_feature + S |
| 0,044221 | 0,121149 | -0,07874 | 0,365015 | 71 | 68 | 0,716233 | 0,948276 | y ~ current_feature + S |
| 0,087292 | 0,120805 | 0,034007 | 0,722586 | 71 | 68 | 0,472412 | 0,831402 | y ~ current_feature + S |
| -0,21311 | 0,118482 | -0,04751 | -1,79864 | 71 | 68 | 0,076514 | 0,640171 | y ~ current_feature + S |
| 0,073286 | 0,134477 | 0,061498 | 0,544967 | 58 | 55 | 0,587979 | 0,945257 | y ~ current_feature + S |
| -0,13715 | 0,149331 | -0,02429 | -0,91841 | 47 | 44 | 0,363412 | 0,805981 | y ~ current_feature + S |
| -0,00255 | 0,102062 | -0,07874 | -0,02501 | 99 | 96 | 0,980101 | 0,996318 | y ~ current_feature + S |
| 0,023061 | 0,102035 | 0,034007 | 0,226009 | 99 | 96 | 0,821675 | 0,952501 | y ~ current_feature + S |
| 0,088602 | 0,102194 | -0,04751 | 0,866995 | 98 | 95 | 0,388129 | 0,803665 | y ~ current_feature + S |
| -0,13819 | 0,112681 | 0,061498 | -1,22639 | 88 | 77,2552 | 0,223776 | 0,893818 | y ~ current_feature + S |
| -0,00819 | 0,126996 | -0,02429 | -0,06451 | 65 | 62 | 0,948769 | 0,99444 | y ~ current_feature + S |
| -0,1553 | 0,097813 | -0,07874 | -1,58776 | 105 | 102 | 0,115436 | 0,469356 | y ~ current_feature + S |
| -0,01317 | 0,099006 | 0,034007 | -0,13307 | 105 | 102 | 0,894403 | 0,96591 | y ~ current_feature + S |
| 0,106009 | 0,098457 | -0,04751 | 1,076703 | 105 | 102 | 0,284153 | 0,747915 | y ~ current_feature + S |
| -0,09794 | 0,107314 | 0,061498 | -0,9126 | 89 | 86 | 0,364002 | 0,92247 | y ~ current_feature + S |
| 0,012787 | 0,122159 | -0,02429 | 0,104674 | 70 | 67 | 0,916947 | 0,99444 | y ~ current_feature + S |
| -0,00741 | 0,052264 | -0,07874 | -0,14169 | 403 | 366,0763 | 0,887405 | 0,987929 | y ~ current_feature + S |
| -0,03897 | 0,068341 | 0,034007 | -0,57023 | 403 | 213,7846 | 0,569118 | 0,863333 | y ~ current_feature + S |
| -0,05965 | 0,05273 | -0,04751 | -1,13132 | 402 | 358,3706 | 0,258678 | 0,742928 | y ~ current_feature + S |
| -0,11004 | 0,05575 | 0,061498 | -1,97387 | 353 | 317,8444 | 0,049263 | 0,700056 | y ~ current_feature + S |
| -0,03836 | 0,061384 | -0,02429 | -0,62485 | 268 | 265 | 0,532606 | 0,901895 | y ~ current_feature + S |
| -0,03625 | 0,059152 | -0,07874 | -0,61275 | 296 | 285,4264 | 0,540528 | 0,882344 | y ~ current_feature + S |
| 0,094568 | 0,078152 | 0,034007 | 1,210064 | 296 | 162,2644 | 0,228014 | 0,681978 | y ~ current_feature + S |
| -0,03223 | 0,060103 | -0,04751 | -0,5363 | 295 | 276,5356 | 0,592181 | 0,91049 | y ~ current_feature + S |
| -0,12898 | 0,065171 | 0,061498 | -1,97906 | 255 | 231,5265 | 0,048995 | 0,700056 | y ~ current_feature + S |
| 0,020961 | 0,072261 | -0,02429 | 0,290073 | 197 | 191,4259 | 0,772074 | 0,957298 | y ~ current_feature + S |
| -0,00682 | 0,140025 | -0,07874 | -0,04868 | 54 | 51 | 0,961366 | 0,994607 | y ~ current_feature + S |
| -0,10076 | 0,139315 | 0,034007 | -0,72326 | 54 | 51 | 0,472825 | 0,831402 | y ~ current_feature + S |
| -0,10377 | 0,139272 | -0,04751 | -0,74508 | 54 | 51 | 0,459642 | 0,837772 | y ~ current_feature + S |
| -0,14249 | 0,15458 | 0,061498 | -0,9218 | 44 | 41 | 0,362027 | 0,920619 | y ~ current_feature + S |
| -0,28119 | 0,167054 | -0,02429 | -1,68325 | 36 | 33 | 0,101766 | 0,605474 | y ~ current_feature + S |
| 0,071515 | 0,056052 | -0,07874 | 1,275863 | 370 | 316,6576 | 0,202939 | 0,620557 | y ~ current_feature + S |
| -0,00961 | 0,07175 | 0,034007 | -0,13399 | 370 | 194,2287 | 0,893552 | 0,96591 | y ~ current_feature + S |

| | | | | | | | | |
|---|---|---|---|---|---|---|---|---|
| 0,035778 | 0,05694 | -0,04751 | 0,628338 | 369 | 308,0358 | 0,530248 | 0,883747 | y ~ current_feature + S |
| -0,11825 | 0,059769 | 0,061498 | -1,9784 | 322 | 276,0135 | 0,048877 | 0,700056 | y ~ current_feature + S |
| 0,026727 | 0,064403 | -0,02429 | 0,414991 | 246 | 240,9248 | 0,678518 | 0,926342 | y ~ current_feature + S |
| -0,07911 | 0,105079 | -0,07874 | -0,75283 | 93 | 90 | 0,453519 | 0,82458 | y ~ current_feature + S |
| 0,036752 | 0,105338 | 0,034007 | 0,3489 | 93 | 90 | 0,727979 | 0,922439 | y ~ current_feature + S |
| -0,11425 | 0,104719 | -0,04751 | -1,09102 | 93 | 90 | 0,278177 | 0,747915 | y ~ current_feature + S |
| -0,04598 | 0,123811 | 0,061498 | -0,3714 | 78 | 65,09683 | 0,711542 | 0,972444 | y ~ current_feature + S |
| 0,195117 | 0,127687 | -0,02429 | 1,528092 | 62 | 59 | 0,131834 | 0,617591 | y ~ current_feature + S |
| -0,08191 | 0,056362 | -0,07874 | -1,45329 | 352 | 312,6795 | 0,147145 | 0,506453 | y ~ current_feature + S |
| -0,03686 | 0,075361 | 0,034007 | -0,48906 | 352 | 175,8396 | 0,625409 | 0,896904 | y ~ current_feature + S |
| -0,02111 | 0,057267 | -0,04751 | -0,36857 | 351 | 304,788 | 0,712701 | 0,93314 | y ~ current_feature + S |
| -0,02943 | 0,06002 | 0,061498 | -0,49036 | 305 | 277,3538 | 0,624264 | 0,950526 | y ~ current_feature + S |
| -0,02934 | 0,066934 | -0,02429 | -0,43842 | 234 | 223,0132 | 0,661509 | 0,924682 | y ~ current_feature + S |
| -0,03117 | 0,072897 | -0,07874 | -0,42756 | 191 | 188 | 0,66946 | 0,934653 | y ~ current_feature + S |
| 0,028893 | 0,072902 | 0,034007 | 0,396333 | 191 | 188 | 0,692309 | 0,916569 | y ~ current_feature + S |
| 0,032795 | 0,072893 | -0,04751 | 0,449901 | 191 | 188 | 0,6533 | 0,925756 | y ~ current_feature + S |
| 0,139275 | 0,077091 | 0,061498 | 1,806629 | 168 | 165 | 0,072642 | 0,721272 | y ~ current_feature + S |
| 0,078171 | 0,089528 | -0,02429 | 0,87315 | 127 | 124 | 0,384269 | 0,819479 | y ~ current_feature + S |
| -0,06258 | 0,059432 | -0,07874 | -1,053 | 285 | 282 | 0,293241 | 0,718687 | y ~ current_feature + S |
| -0,10835 | 0,059199 | 0,034007 | -1,83031 | 285 | 282 | 0,068259 | 0,441313 | y ~ current_feature + S |
| 0,001064 | 0,059655 | -0,04751 | 0,017843 | 284 | 281 | 0,985776 | 0,993835 | y ~ current_feature + S |
| -0,18466 | 0,07286 | 0,061498 | -2,53446 | 246 | 181,9515 | 0,012105 | 0,55988 | y ~ current_feature + S |
| -0,19584 | 0,073545 | -0,02429 | -2,66281 | 189 | 177,7904 | 0,00846 | 0,295269 | y ~ current_feature + S |
| 0,121798 | 0,092156 | -0,07874 | 1,321641 | 119 | 116 | 0,188888 | 0,592937 | y ~ current_feature + S |
| 0,047145 | 0,092744 | 0,034007 | 0,508336 | 119 | 116 | 0,612183 | 0,889068 | y ~ current_feature + S |
| -0,0404 | 0,092772 | -0,04751 | -0,43548 | 119 | 116 | 0,664026 | 0,927953 | y ~ current_feature + S |
| 0,113763 | 0,098858 | 0,061498 | 1,150772 | 104 | 101 | 0,252543 | 0,893818 | y ~ current_feature + S |
| -0,10096 | 0,114122 | -0,02429 | -0,88468 | 79 | 76 | 0,379118 | 0,815551 | y ~ current_feature + S |
| 0,116276 | 0,093434 | -0,07874 | 1,244472 | 116 | 113 | 0,215901 | 0,639357 | y ~ current_feature + S |
| 0,012334 | 0,094065 | 0,034007 | 0,131119 | 116 | 113 | 0,895915 | 0,96591 | y ~ current_feature + S |
| -0,10011 | 0,094016 | -0,04751 | -1,06479 | 115 | 112 | 0,289261 | 0,747915 | y ~ current_feature + S |
| -0,12841 | 0,101217 | 0,061498 | -1,26865 | 99 | 96 | 0,207633 | 0,893818 | y ~ current_feature + S |
| 0,123737 | 0,115354 | -0,02429 | 1,072669 | 77 | 74 | 0,286905 | 0,734108 | y ~ current_feature + S |
| 0,069949 | 0,060097 | -0,07874 | 1,163934 | 308 | 275,525 | 0,245458 | 0,67447 | y ~ current_feature + S |
| 0,052392 | 0,075991 | 0,034007 | 0,68945 | 308 | 172,697 | 0,491466 | 0,837983 | y ~ current_feature + S |
| 0,016795 | 0,060476 | -0,04751 | 0,277713 | 307 | 273,3446 | 0,781443 | 0,94882 | y ~ current_feature + S |
| -0,06113 | 0,06276 | 0,061498 | -0,974 | 273 | 252,9319 | 0,330987 | 0,893818 | y ~ current_feature + S |
| 0,001231 | 0,070892 | -0,02429 | 0,017369 | 205 | 198,9781 | 0,98616 | 0,99444 | y ~ current_feature + S |
| 0,142303 | 0,062762 | -0,07874 | 2,267368 | 280 | 248,7301 | 0,024226 | 0,197019 | y ~ current_feature + S |
| 0,143961 | 0,079359 | 0,034007 | 1,814034 | 280 | 155,4919 | 0,0716 | 0,456761 | y ~ current_feature + S |
| 0,046846 | 0,065432 | -0,04751 | 0,71595 | 279 | 233,0565 | 0,474739 | 0,842057 | y ~ current_feature + S |
| 0,051249 | 0,06921 | 0,061498 | 0,74049 | 241 | 208,2196 | 0,459837 | 0,937665 | y ~ current_feature + S |
| 0,086329 | 0,074822 | -0,02429 | 1,153793 | 186 | 177,2937 | 0,250138 | 0,717451 | y ~ current_feature + S |
| 0,197088 | 0,10006 | -0,07874 | 1,969689 | 99 | 96 | 0,051756 | 0,303964 | y ~ current_feature + S |
| 0,217402 | 0,099621 | 0,034007 | 2,18229 | 99 | 96 | 0,031528 | 0,347616 | y ~ current_feature + S |
| 0,156947 | 0,100797 | -0,04751 | 1,557053 | 99 | 96 | 0,122749 | 0,652786 | y ~ current_feature + S |
| 0,202158 | 0,106856 | 0,061498 | 1,891867 | 87 | 84 | 0,061954 | 0,715249 | y ~ current_feature + S |
| -0,06598 | 0,125714 | -0,02429 | -0,52481 | 66 | 63 | 0,601556 | 0,924682 | y ~ current_feature + S |

| | | | | | | | | |
|---|---|---|---|---|---|---|---|---|
| -0,00108 | 0,120386 | -0,07874 | -0,00895 | 72 | 69 | 0,992884 | 0,996647 | y ~ current_feature + S |
| -0,27076 | 0,115889 | 0,034007 | -2,33638 | 72 | 69 | 0,022382 | 0,331255 | y ~ current_feature + S |
| -0,03672 | 0,120305 | -0,04751 | -0,30521 | 72 | 69 | 0,761126 | 0,9394 | y ~ current_feature + S |
| 0,023182 | 0,130154 | 0,061498 | 0,178111 | 62 | 59 | 0,859246 | 0,996179 | y ~ current_feature + S |
| -0,12096 | 0,148031 | -0,02429 | -0,81714 | 48 | 44,967 | 0,418154 | 0,840853 | y ~ current_feature + S |
| 0,160451 | 0,148802 | -0,07874 | 1,078281 | 47 | 44 | 0,286784 | 0,718687 | y ~ current_feature + S |
| -0,01143 | 0,150746 | 0,034007 | -0,07582 | 47 | 44 | 0,939906 | 0,981002 | y ~ current_feature + S |
| -0,02348 | 0,152457 | -0,04751 | -0,15398 | 46 | 43 | 0,878346 | 0,985217 | y ~ current_feature + S |
| -0,18064 | 0,16625 | 0,061498 | -1,08656 | 38 | 35 | 0,284659 | 0,893818 | y ~ current_feature + S |
| -0,27973 | 0,181438 | -0,02429 | -1,54172 | 31 | 28 | 0,134368 | 0,617591 | y ~ current_feature + S |
| 0,077282 | 0,066467 | -0,07874 | 1,162704 | 228 | 225 | 0,246181 | 0,67447 | y ~ current_feature + S |
| -0,07716 | 0,066468 | 0,034007 | -1,16086 | 228 | 225 | 0,24693 | 0,702074 | y ~ current_feature + S |
| -0,04391 | 0,066602 | -0,04751 | -0,65925 | 228 | 225 | 0,51041 | 0,868909 | y ~ current_feature + S |
| 0,003654 | 0,077671 | 0,061498 | 0,047043 | 194 | 165,7597 | 0,962536 | 0,996179 | y ~ current_feature + S |
| -0,06309 | 0,08176 | -0,02429 | -0,7717 | 152 | 149 | 0,441515 | 0,858965 | y ~ current_feature + S |
| 0,263176 | 0,145419 | -0,07874 | 1,809785 | 48 | 44,01372 | 0,077161 | 0,370448 | y ~ current_feature + S |
| 0,432439 | 0,161435 | 0,034007 | 2,678719 | 48 | 31,19554 | 0,011689 | 0,233774 | y ~ current_feature + S |
| -0,07975 | 0,171167 | -0,04751 | -0,4659 | 48 | 33,91496 | 0,644263 | 0,925756 | y ~ current_feature + S |
| 0,090709 | 0,166733 | 0,061498 | 0,544033 | 41 | 35,67523 | 0,589803 | 0,945257 | y ~ current_feature + S |
| 0,644876 | 0,177531 | -0,02429 | 3,632464 | 32 | 18,53378 | 0,001832 | 0,169424 | y ~ current_feature + S |
| 0,012348 | 0,09016 | -0,07874 | 0,136957 | 126 | 123 | 0,891288 | 0,987929 | y ~ current_feature + S |
| -0,02721 | 0,090134 | 0,034007 | -0,30186 | 126 | 123 | 0,763268 | 0,93668 | y ~ current_feature + S |
| 0,090687 | 0,089795 | -0,04751 | 1,009928 | 126 | 123 | 0,314512 | 0,77066 | y ~ current_feature + S |
| -0,44101 | 0,091018 | 0,061498 | -4,84528 | 111 | 97,23336 | 4,78E-06 | 0,001769 | y ~ current_feature + S |
| -0,05208 | 0,11096 | -0,02429 | -0,46934 | 84 | 81 | 0,640089 | 0,924682 | y ~ current_feature + S |
| -0,12336 | 0,153125 | -0,07874 | -0,80562 | 45 | 42 | 0,425002 | 0,814177 | y ~ current_feature + S |
| -0,08604 | 0,153731 | 0,034007 | -0,55966 | 45 | 42 | 0,578682 | 0,869291 | y ~ current_feature + S |
| 0,080044 | 0,153808 | -0,04751 | 0,520413 | 45 | 42 | 0,605509 | 0,914441 | y ~ current_feature + S |
| -0,18695 | 0,159361 | 0,061498 | -1,17315 | 41 | 38 | 0,248037 | 0,893818 | y ~ current_feature + S |
| -0,10161 | 0,191454 | -0,02429 | -0,53074 | 30 | 27 | 0,599935 | 0,924682 | y ~ current_feature + S |
| -0,07713 | 0,105095 | -0,07874 | -0,73394 | 93 | 90 | 0,464893 | 0,837033 | y ~ current_feature + S |
| -0,15267 | 0,104174 | 0,034007 | -1,4655 | 93 | 90 | 0,14627 | 0,594724 | y ~ current_feature + S |
| -0,08065 | 0,105066 | -0,04751 | -0,76758 | 93 | 90 | 0,444749 | 0,824847 | y ~ current_feature + S |
| 0,004811 | 0,113226 | 0,061498 | 0,042486 | 81 | 78 | 0,96622 | 0,996179 | y ~ current_feature + S |
| -0,24828 | 0,126112 | -0,02429 | -1,96872 | 62 | 59 | 0,053688 | 0,558147 | y ~ current_feature + S |
| 0,16642 | 0,110244 | -0,07874 | 1,509556 | 83 | 80 | 0,135096 | 0,49737 | y ~ current_feature + S |
| 0,018679 | 0,111784 | 0,034007 | 0,167098 | 83 | 80 | 0,867714 | 0,96591 | y ~ current_feature + S |
| -0,14103 | 0,110686 | -0,04751 | -1,27411 | 83 | 80 | 0,206313 | 0,723562 | y ~ current_feature + S |
| -0,09407 | 0,119852 | 0,061498 | -0,78492 | 72 | 69 | 0,43519 | 0,937665 | y ~ current_feature + S |
| 0,207742 | 0,139086 | -0,02429 | 1,493626 | 55 | 49,46225 | 0,141626 | 0,618941 | y ~ current_feature + S |
| 0,101056 | 0,052146 | -0,07874 | 1,937953 | 367 | 364 | 0,053402 | 0,30647 | y ~ current_feature + S |
| 0,093099 | 0,052187 | 0,034007 | 1,78397 | 367 | 364 | 0,075261 | 0,464477 | y ~ current_feature + S |
| 0,054839 | 0,052407 | -0,04751 | 1,046392 | 366 | 363 | 0,296076 | 0,752909 | y ~ current_feature + S |
| -0,04498 | 0,062726 | 0,061498 | -0,71711 | 319 | 253,6479 | 0,473967 | 0,937665 | y ~ current_feature + S |
| -0,00954 | 0,064413 | -0,02429 | -0,14818 | 244 | 241 | 0,882325 | 0,987777 | y ~ current_feature + S |
| 0,031243 | 0,117794 | -0,07874 | 0,265231 | 75 | 72 | 0,791589 | 0,979102 | y ~ current_feature + S |
| 0,187955 | 0,115751 | 0,034007 | 1,623794 | 75 | 72 | 0,10879 | 0,536698 | y ~ current_feature + S |
| -0,05425 | 0,117678 | -0,04751 | -0,46096 | 75 | 72 | 0,646213 | 0,925756 | y ~ current_feature + S |

| | | | | | | | | |
|---|---|---|---|---|---|---|---|---|
| 0,069838 | 0,150148 | 0,061498 | 0,465132 | 64 | 44,14075 | 0,644124 | 0,956496 | y ~ current_feature + S |
| 0,234878 | 0,141784 | -0,02429 | 1,656584 | 50 | 47 | 0,104265 | 0,605474 | y ~ current_feature + S |
| -0,0432 | 0,128979 | -0,07874 | -0,33491 | 63 | 60 | 0,738863 | 0,959225 | y ~ current_feature + S |
| -0,19295 | 0,126673 | 0,034007 | -1,52322 | 63 | 60 | 0,132957 | 0,572281 | y ~ current_feature + S |
| 0,139182 | 0,127843 | -0,04751 | 1,088696 | 63 | 60 | 0,280642 | 0,747915 | y ~ current_feature + S |
| 0,065826 | 0,142547 | 0,061498 | 0,461783 | 52 | 49 | 0,646281 | 0,956496 | y ~ current_feature + S |
| 0,025613 | 0,160076 | -0,02429 | 0,160008 | 42 | 39 | 0,873701 | 0,984468 | y ~ current_feature + S |
| -0,15147 | 0,080175 | -0,07874 | -1,88922 | 155 | 152 | 0,060767 | 0,325854 | y ~ current_feature + S |
| -0,12264 | 0,080498 | 0,034007 | -1,52348 | 155 | 152 | 0,129716 | 0,572145 | y ~ current_feature + S |
| -0,10247 | 0,080684 | -0,04751 | -1,26998 | 155 | 152 | 0,206031 | 0,723562 | y ~ current_feature + S |
| 0,006001 | 0,087369 | 0,061498 | 0,068687 | 134 | 131 | 0,945344 | 0,996179 | y ~ current_feature + S |
| -0,23056 | 0,096823 | -0,02429 | -2,38123 | 104 | 101 | 0,019129 | 0,428958 | y ~ current_feature + S |
| -0,02826 | 0,050371 | -0,07874 | -0,56107 | 406 | 393,8134 | 0,575072 | 0,899631 | y ~ current_feature + S |
| -0,08983 | 0,063289 | 0,034007 | -1,41937 | 406 | 247,6449 | 0,157048 | 0,60427 | y ~ current_feature + S |
| -0,01504 | 0,050963 | -0,04751 | -0,29516 | 405 | 384,9326 | 0,76803 | 0,942525 | y ~ current_feature + S |
| -0,05808 | 0,054322 | 0,061498 | -1,06917 | 353 | 337,7405 | 0,285757 | 0,893818 | y ~ current_feature + S |
| -0,01912 | 0,062116 | -0,02429 | -0,30789 | 270 | 259,0823 | 0,758414 | 0,951727 | y ~ current_feature + S |
| 0,064893 | 0,062368 | -0,07874 | 1,040482 | 259 | 256 | 0,299098 | 0,718687 | y ~ current_feature + S |
| -0,0313 | 0,062469 | 0,034007 | -0,50101 | 259 | 256 | 0,616796 | 0,889725 | y ~ current_feature + S |
| 0,0721 | 0,062337 | -0,04751 | 1,156616 | 259 | 256 | 0,248507 | 0,742928 | y ~ current_feature + S |
| -0,20714 | 0,072329 | 0,061498 | -2,8638 | 227 | 182,9491 | 0,004675 | 0,378455 | y ~ current_feature + S |
| 0,113378 | 0,076241 | -0,02429 | 1,487093 | 173 | 169,8256 | 0,138845 | 0,618941 | y ~ current_feature + S |
| -0,02204 | 0,06665 | -0,07874 | -0,33063 | 228 | 225 | 0,741231 | 0,959588 | y ~ current_feature + S |
| -0,01769 | 0,066656 | 0,034007 | -0,26541 | 228 | 225 | 0,790935 | 0,945731 | y ~ current_feature + S |
| 0,043955 | 0,066602 | -0,04751 | 0,659964 | 228 | 225 | 0,509952 | 0,868909 | y ~ current_feature + S |
| 0,022622 | 0,072339 | 0,061498 | 0,312725 | 194 | 191 | 0,754831 | 0,972573 | y ~ current_feature + S |
| 0,059035 | 0,08178 | -0,02429 | 0,721873 | 152 | 149 | 0,471504 | 0,876224 | y ~ current_feature + S |
| -0,06964 | 0,062606 | -0,07874 | -1,11243 | 278 | 253,8973 | 0,267007 | 0,702573 | y ~ current_feature + S |
| 0,038917 | 0,083126 | 0,034007 | 0,468172 | 278 | 144,5008 | 0,640367 | 0,899624 | y ~ current_feature + S |
| 0,048474 | 0,067542 | -0,04751 | 0,717678 | 277 | 218,6889 | 0,473722 | 0,842057 | y ~ current_feature + S |
| -0,06376 | 0,065966 | 0,061498 | -0,96653 | 243 | 228,87 | 0,334801 | 0,893818 | y ~ current_feature + S |
| 0,131095 | 0,073485 | -0,02429 | 1,78396 | 185 | 182 | 0,076096 | 0,605474 | y ~ current_feature + S |
| -0,07059 | 0,15038 | -0,07874 | -0,46941 | 47 | 44 | 0,641097 | 0,928694 | y ~ current_feature + S |
| 0,003961 | 0,150754 | 0,034007 | 0,026274 | 47 | 44 | 0,979158 | 0,989859 | y ~ current_feature + S |
| -0,25058 | 0,147633 | -0,04751 | -1,69728 | 46 | 43 | 0,096871 | 0,640171 | y ~ current_feature + S |
| -0,2328 | 0,15377 | 0,061498 | -1,51397 | 43 | 40 | 0,137897 | 0,8197 | y ~ current_feature + S |
| 0,205111 | 0,184964 | -0,02429 | 1,108924 | 31 | 28 | 0,276897 | 0,734108 | y ~ current_feature + S |
| 0,083069 | 0,051325 | -0,07874 | 1,61851 | 380 | 377 | 0,106389 | 0,449749 | y ~ current_feature + S |
| 0,097029 | 0,05126 | 0,034007 | 1,892898 | 380 | 377 | 0,059137 | 0,424869 | y ~ current_feature + S |
| 0,056918 | 0,051487 | -0,04751 | 1,105465 | 379 | 376 | 0,269665 | 0,747915 | y ~ current_feature + S |
| -0,009 | 0,060721 | 0,061498 | -0,14827 | 330 | 271,2016 | 0,882239 | 0,996179 | y ~ current_feature + S |
| 0,139146 | 0,062786 | -0,02429 | 2,216191 | 253 | 248,7608 | 0,027584 | 0,485321 | y ~ current_feature + S |
| 0,047194 | 0,055901 | -0,07874 | 0,844242 | 381 | 319,2924 | 0,399166 | 0,791912 | y ~ current_feature + S |
| -0,05146 | 0,072378 | 0,034007 | -0,71103 | 381 | 190,385 | 0,477936 | 0,831402 | y ~ current_feature + S |
| 0,038637 | 0,056755 | -0,04751 | 0,680773 | 380 | 309,9889 | 0,496523 | 0,868909 | y ~ current_feature + S |
| -0,01068 | 0,060986 | 0,061498 | -0,17507 | 332 | 268,8377 | 0,861153 | 0,996179 | y ~ current_feature + S |
| 0,057754 | 0,063475 | -0,02429 | 0,909863 | 253 | 247,3655 | 0,363781 | 0,805981 | y ~ current_feature + S |
| -0,12738 | 0,153046 | -0,07874 | -0,83231 | 45 | 42 | 0,40994 | 0,806796 | y ~ current_feature + S |

| | | | | | | | | |
|---|---|---|---|---|---|---|---|---|
| -0,18988 | 0,151496 | 0,034007 | -1,25339 | 45 | 42 | 0,216998 | 0,664036 | y ~ current_feature + S |
| -0,13878 | 0,15281 | -0,04751 | -0,90816 | 45 | 42 | 0,368977 | 0,802018 | y ~ current_feature + S |
| 0,129778 | 0,172622 | 0,061498 | 0,751806 | 36 | 32,99385 | 0,457497 | 0,937665 | y ~ current_feature + S |
| 0,001943 | 0,19245 | -0,02429 | 0,010096 | 30 | 27 | 0,992019 | 0,996057 | y ~ current_feature + S |
| -0,01479 | 0,136068 | -0,07874 | -0,10867 | 57 | 54 | 0,913868 | 0,987929 | y ~ current_feature + S |
| 0,056459 | 0,135866 | 0,034007 | 0,415547 | 57 | 54 | 0,679388 | 0,912427 | y ~ current_feature + S |
| -0,16125 | 0,134302 | -0,04751 | -1,20067 | 57 | 54 | 0,235118 | 0,742928 | y ~ current_feature + S |
| -0,06305 | 0,14405 | 0,061498 | -0,43769 | 51 | 48 | 0,663577 | 0,957207 | y ~ current_feature + S |
| 0,124896 | 0,167707 | -0,02429 | 0,744727 | 38 | 35 | 0,461408 | 0,871026 | y ~ current_feature + S |
| 0,028773 | 0,056773 | -0,07874 | 0,506819 | 313 | 310 | 0,612642 | 0,918951 | y ~ current_feature + S |
| -0,00158 | 0,056796 | 0,034007 | -0,02785 | 313 | 310 | 0,977798 | 0,989859 | y ~ current_feature + S |
| -0,01348 | 0,056883 | -0,04751 | -0,23705 | 312 | 309 | 0,812774 | 0,972405 | y ~ current_feature + S |
| -0,07551 | 0,060572 | 0,061498 | -1,24669 | 274 | 271 | 0,213589 | 0,893818 | y ~ current_feature + S |
| 0,053884 | 0,069742 | -0,02429 | 0,772624 | 208 | 205 | 0,440635 | 0,858965 | y ~ current_feature + S |
| -0,0567 | 0,092698 | -0,07874 | -0,61164 | 119 | 116 | 0,541973 | 0,882344 | y ~ current_feature + S |
| -0,18414 | 0,09126 | 0,034007 | -2,01772 | 119 | 116 | 0,045928 | 0,400524 | y ~ current_feature + S |
| 0,0093 | 0,093246 | -0,04751 | 0,099736 | 118 | 115 | 0,920727 | 0,985217 | y ~ current_feature + S |
| -0,26402 | 0,096452 | 0,061498 | -2,73738 | 103 | 100 | 0,007331 | 0,417322 | y ~ current_feature + S |
| 0,055766 | 0,114529 | -0,02429 | 0,486918 | 79 | 76 | 0,627718 | 0,924682 | y ~ current_feature + S |
| 0,16901 | 0,102757 | -0,07874 | 1,644751 | 95 | 92 | 0,103434 | 0,449749 | y ~ current_feature + S |
| 0,098526 | 0,10375 | 0,034007 | 0,949648 | 95 | 92 | 0,344779 | 0,746014 | y ~ current_feature + S |
| 0,277679 | 0,100706 | -0,04751 | 2,757321 | 94 | 91 | 0,007042 | 0,432614 | y ~ current_feature + S |
| -0,05538 | 0,108942 | 0,061498 | -0,50833 | 87 | 84 | 0,612555 | 0,948092 | y ~ current_feature + S |
| 0,274799 | 0,124129 | -0,02429 | 2,213809 | 63 | 60 | 0,030654 | 0,485321 | y ~ current_feature + S |
| -0,0153 | 0,058715 | -0,07874 | -0,26055 | 293 | 290 | 0,794627 | 0,979102 | y ~ current_feature + S |
| -0,08262 | 0,058521 | 0,034007 | -1,41183 | 293 | 290 | 0,159072 | 0,60427 | y ~ current_feature + S |
| -0,02869 | 0,058698 | -0,04751 | -0,48876 | 293 | 290 | 0,625378 | 0,925756 | y ~ current_feature + S |
| -0,04412 | 0,062933 | 0,061498 | -0,70114 | 255 | 252 | 0,483863 | 0,937665 | y ~ current_feature + S |
| -0,03155 | 0,072133 | -0,02429 | -0,43746 | 195 | 192 | 0,662272 | 0,924682 | y ~ current_feature + S |
| 0,138368 | 0,062066 | -0,07874 | 2,229356 | 315 | 254,6211 | 0,026663 | 0,204832 | y ~ current_feature + S |
| 0,077378 | 0,056444 | 0,034007 | 1,370878 | 315 | 312 | 0,171398 | 0,618964 | y ~ current_feature + S |
| 0,041209 | 0,056657 | -0,04751 | 0,727354 | 314 | 311 | 0,467556 | 0,842057 | y ~ current_feature + S |
| -0,06716 | 0,066227 | 0,061498 | -1,01416 | 274 | 226,9713 | 0,311585 | 0,893818 | y ~ current_feature + S |
| 0,07075 | 0,069332 | -0,02429 | 1,020453 | 210 | 206,9904 | 0,308705 | 0,761472 | y ~ current_feature + S |
| -0,02658 | 0,132406 | -0,07874 | -0,20073 | 60 | 57 | 0,841622 | 0,983548 | y ~ current_feature + S |
| -0,12806 | 0,131363 | 0,034007 | -0,97487 | 60 | 57 | 0,333746 | 0,734392 | y ~ current_feature + S |
| -0,00818 | 0,132449 | -0,04751 | -0,06176 | 60 | 57 | 0,950967 | 0,990156 | y ~ current_feature + S |
| -0,08141 | 0,140952 | 0,061498 | -0,57758 | 53 | 50 | 0,566141 | 0,937665 | y ~ current_feature + S |
| -0,12535 | 0,163102 | -0,02429 | -0,76854 | 40 | 37 | 0,447046 | 0,858965 | y ~ current_feature + S |
| 0,007134 | 0,051707 | -0,07874 | 0,137963 | 377 | 374 | 0,890344 | 0,987929 | y ~ current_feature + S |
| -1,31E-04 | 0,051709 | 0,034007 | -0,00253 | 377 | 374 | 0,997984 | 0,997989 | y ~ current_feature + S |
| -0,01121 | 0,051775 | -0,04751 | -0,2166 | 376 | 373 | 0,828635 | 0,975438 | y ~ current_feature + S |
| -0,08442 | 0,055357 | 0,061498 | -1,52504 | 327 | 324 | 0,128224 | 0,8197 | y ~ current_feature + S |
| -0,03655 | 0,063458 | -0,02429 | -0,57597 | 251 | 248 | 0,56516 | 0,924682 | y ~ current_feature + S |
| 0,036249 | 0,056577 | -0,07874 | 0,640712 | 315 | 312 | 0,52218 | 0,876542 | y ~ current_feature + S |
| -0,02599 | 0,056595 | 0,034007 | -0,45927 | 315 | 312 | 0,646357 | 0,900251 | y ~ current_feature + S |
| -0,09571 | 0,056354 | -0,04751 | -1,6983 | 315 | 312 | 0,090448 | 0,640171 | y ~ current_feature + S |
| -0,15464 | 0,059905 | 0,061498 | -2,58139 | 275 | 272 | 0,010364 | 0,547829 | y ~ current_feature + S |

| | | | | | | | | |
|---|---|---|---|---|---|---|---|---|
| -0,06919 | 0,069338 | -0,02429 | -0,99791 | 210 | 207 | 0,31949 | 0,767607 | y ~ current_feature + S |
| 0,011045 | 0,064546 | -0,07874 | 0,171126 | 243 | 240 | 0,864269 | 0,985453 | y ~ current_feature + S |
| 0,022679 | 0,064533 | 0,034007 | 0,351439 | 243 | 240 | 0,725567 | 0,92096 | y ~ current_feature + S |
| 0,065228 | 0,064412 | -0,04751 | 1,012666 | 243 | 240 | 0,312239 | 0,767632 | y ~ current_feature + S |
| 0,020174 | 0,068993 | 0,061498 | 0,292405 | 213 | 210 | 0,770265 | 0,973246 | y ~ current_feature + S |
| 0,05639 | 0,079179 | -0,02429 | 0,712187 | 162 | 159 | 0,477392 | 0,878782 | y ~ current_feature + S |
| 0,075837 | 0,075161 | -0,07874 | 1,008991 | 179 | 176 | 0,314364 | 0,718687 | y ~ current_feature + S |
| -0,07444 | 0,075169 | 0,034007 | -0,9903 | 179 | 176 | 0,323387 | 0,728793 | y ~ current_feature + S |
| 0,063595 | 0,07544 | -0,04751 | 0,842985 | 178 | 175 | 0,400387 | 0,803665 | y ~ current_feature + S |
| -0,01726 | 0,081911 | 0,061498 | -0,21069 | 152 | 149 | 0,83342 | 0,996179 | y ~ current_feature + S |
| -0,07056 | 0,092616 | -0,02429 | -0,76191 | 119 | 116 | 0,447663 | 0,858965 | y ~ current_feature + S |
| -0,00855 | 0,089084 | -0,07874 | -0,09597 | 129 | 126 | 0,923695 | 0,990519 | y ~ current_feature + S |
| -0,01348 | 0,089079 | 0,034007 | -0,15131 | 129 | 126 | 0,87997 | 0,96591 | y ~ current_feature + S |
| -0,01271 | 0,08908 | -0,04751 | -0,14272 | 129 | 126 | 0,886736 | 0,985217 | y ~ current_feature + S |
| 0,209506 | 0,094528 | 0,061498 | 2,216333 | 110 | 107 | 0,028785 | 0,658491 | y ~ current_feature + S |
| 0,035773 | 0,109694 | -0,02429 | 0,326119 | 86 | 83 | 0,745155 | 0,947448 | y ~ current_feature + S |
| 0,249916 | 0,107585 | -0,07874 | 2,322958 | 84 | 81 | 0,022691 | 0,194866 | y ~ current_feature + S |
| 0,114964 | 0,110374 | 0,034007 | 1,041586 | 84 | 81 | 0,300703 | 0,718742 | y ~ current_feature + S |
| 0,25148 | 0,10754 | -0,04751 | 2,33847 | 84 | 81 | 0,021829 | 0,557019 | y ~ current_feature + S |
| 0,167031 | 0,118695 | 0,061498 | 1,407233 | 72 | 69 | 0,163847 | 0,8197 | y ~ current_feature + S |
| 0,177194 | 0,135187 | -0,02429 | 1,310732 | 56 | 53 | 0,1956 | 0,657926 | y ~ current_feature + S |
| 0,064361 | 0,117607 | -0,07874 | 0,547255 | 75 | 72 | 0,585896 | 0,902886 | y ~ current_feature + S |
| 0,122796 | 0,116959 | 0,034007 | 1,049903 | 75 | 72 | 0,297274 | 0,718742 | y ~ current_feature + S |
| -0,07856 | 0,117487 | -0,04751 | -0,66865 | 75 | 72 | 0,505855 | 0,868909 | y ~ current_feature + S |
| -0,12905 | 0,12802 | 0,061498 | -1,00803 | 63 | 60 | 0,31749 | 0,893818 | y ~ current_feature + S |
| -0,00972 | 0,145858 | -0,02429 | -0,06663 | 50 | 47 | 0,94716 | 0,99444 | y ~ current_feature + S |
| 0,13278 | 0,08347 | -0,07874 | 1,590764 | 144 | 141 | 0,113902 | 0,467692 | y ~ current_feature + S |
| 0,11972 | 0,083609 | 0,034007 | 1,431901 | 144 | 141 | 0,154385 | 0,60427 | y ~ current_feature + S |
| 0,002623 | 0,084215 | -0,04751 | 0,031148 | 144 | 141 | 0,975195 | 0,992311 | y ~ current_feature + S |
| 0,027981 | 0,092414 | 0,061498 | 0,302777 | 120 | 117 | 0,762597 | 0,972573 | y ~ current_feature + S |
| 0,198716 | 0,101627 | -0,02429 | 1,955342 | 96 | 93 | 0,053545 | 0,558147 | y ~ current_feature + S |
| 0,079029 | 0,054828 | -0,07874 | 1,441382 | 370 | 330,5736 | 0,150423 | 0,513608 | y ~ current_feature + S |
| 0,138545 | 0,07032 | 0,034007 | 1,970209 | 370 | 198,346 | 0,050206 | 0,420946 | y ~ current_feature + S |
| 0,081481 | 0,055151 | -0,04751 | 1,477405 | 369 | 326,5831 | 0,140531 | 0,660782 | y ~ current_feature + S |
| -0,00579 | 0,059111 | 0,061498 | -0,09796 | 318 | 286,1873 | 0,922036 | 0,996179 | y ~ current_feature + S |
| 0,087737 | 0,06461 | -0,02429 | 1,357951 | 246 | 237,7075 | 0,175767 | 0,631395 | y ~ current_feature + S |
| 0,003596 | 0,090535 | -0,07874 | 0,039719 | 125 | 122 | 0,968382 | 0,994607 | y ~ current_feature + S |
| 0,02966 | 0,090496 | 0,034007 | 0,327751 | 125 | 122 | 0,743661 | 0,927328 | y ~ current_feature + S |
| -0,01287 | 0,090528 | -0,04751 | -0,14217 | 125 | 122 | 0,887183 | 0,985217 | y ~ current_feature + S |
| -0,1411 | 0,097077 | 0,061498 | -1,45351 | 107 | 104 | 0,149092 | 0,8197 | y ~ current_feature + S |
| -0,1341 | 0,110794 | -0,02429 | -1,21037 | 83 | 80 | 0,2297 | 0,695919 | y ~ current_feature + S |
| -0,13861 | 0,098543 | -0,07874 | -1,40663 | 104 | 101 | 0,162605 | 0,532425 | y ~ current_feature + S |
| -0,20228 | 0,097447 | 0,034007 | -2,0758 | 104 | 101 | 0,040452 | 0,396441 | y ~ current_feature + S |
| -0,15634 | 0,09828 | -0,04751 | -1,59079 | 104 | 101 | 0,114783 | 0,640171 | y ~ current_feature + S |
| 0,078643 | 0,113606 | 0,061498 | 0,692248 | 84 | 77,00274 | 0,490864 | 0,937665 | y ~ current_feature + S |
| 0,062077 | 0,132073 | -0,02429 | 0,470017 | 69 | 57,1079 | 0,640133 | 0,924682 | y ~ current_feature + S |
| 0,058169 | 0,057498 | -0,07874 | 1,01167 | 344 | 301,4535 | 0,312507 | 0,718687 | y ~ current_feature + S |
| -0,0993 | 0,07645 | 0,034007 | -1,29888 | 344 | 169,4119 | 0,19575 | 0,6438 | y ~ current_feature + S |

| | | | | | | | | |
|---|---|---|---|---|---|---|---|---|
| -0,08204 | 0,059939 | -0,04751 | -1,36873 | 343 | 276,4727 | 0,172193 | 0,700235 | y ~ current_feature + S |
| -0,06964 | 0,061008 | 0,061498 | -1,14155 | 298 | 267,3674 | 0,254661 | 0,893818 | y ~ current_feature + S |
| -0,07992 | 0,067063 | -0,02429 | -1,19176 | 229 | 220,925 | 0,234634 | 0,695919 | y ~ current_feature + S |
| 0,021505 | 0,099481 | -0,07874 | 0,216169 | 104 | 101 | 0,829292 | 0,981192 | y ~ current_feature + S |
| 0,087428 | 0,099123 | 0,034007 | 0,882021 | 104 | 101 | 0,37986 | 0,759222 | y ~ current_feature + S |
| -0,15506 | 0,09879 | -0,04751 | -1,56963 | 103 | 100 | 0,119661 | 0,651097 | y ~ current_feature + S |
| -0,19629 | 0,105126 | 0,061498 | -1,86722 | 90 | 87 | 0,065239 | 0,715249 | y ~ current_feature + S |
| -0,05912 | 0,122876 | -0,02429 | -0,48114 | 69 | 66 | 0,632008 | 0,924682 | y ~ current_feature + S |
| 0,044916 | 0,098915 | -0,07874 | 0,454091 | 105 | 102 | 0,650728 | 0,932255 | y ~ current_feature + S |
| 0,00486 | 0,099014 | 0,034007 | 0,049085 | 105 | 102 | 0,960948 | 0,989859 | y ~ current_feature + S |
| 0,112774 | 0,098383 | -0,04751 | 1,146274 | 105 | 102 | 0,254364 | 0,742928 | y ~ current_feature + S |
| 0,156645 | 0,124387 | 0,061498 | 1,259333 | 89 | 63,04657 | 0,212552 | 0,893818 | y ~ current_feature + S |
| -0,05139 | 0,124962 | -0,02429 | -0,41122 | 70 | 63,86965 | 0,682284 | 0,926342 | y ~ current_feature + S |
| 0,015121 | 0,095335 | -0,07874 | 0,158605 | 113 | 110 | 0,874271 | 0,987929 | y ~ current_feature + S |
| -0,00387 | 0,095346 | 0,034007 | -0,04056 | 113 | 110 | 0,96772 | 0,989859 | y ~ current_feature + S |
| 0,292855 | 0,091583 | -0,04751 | 3,197696 | 112 | 109 | 0,001814 | 0,191719 | y ~ current_feature + S |
| -0,11804 | 0,100825 | 0,061498 | -1,17078 | 100 | 97 | 0,244555 | 0,893818 | y ~ current_feature + S |
| 0,074509 | 0,117524 | -0,02429 | 0,633991 | 75 | 72 | 0,528096 | 0,899808 | y ~ current_feature + S |
| -0,00737 | 0,141418 | -0,07874 | -0,05211 | 53 | 50 | 0,958649 | 0,994607 | y ~ current_feature + S |
| -0,10387 | 0,140656 | 0,034007 | -0,73846 | 53 | 50 | 0,463688 | 0,830821 | y ~ current_feature + S |
| 0,008216 | 0,142852 | -0,04751 | 0,057514 | 52 | 49 | 0,95437 | 0,990156 | y ~ current_feature + S |
| -0,05709 | 0,15225 | 0,061498 | -0,37495 | 46 | 43 | 0,70954 | 0,972444 | y ~ current_feature + S |
| 0,215543 | 0,172621 | -0,02429 | 1,248647 | 35 | 32 | 0,220852 | 0,69545 | y ~ current_feature + S |
| -0,01703 | 0,052844 | -0,07874 | -0,32221 | 361 | 358 | 0,74748 | 0,962676 | y ~ current_feature + S |
| -0,04834 | 0,05279 | 0,034007 | -0,91569 | 361 | 358 | 0,360446 | 0,7498 | y ~ current_feature + S |
| -0,01685 | 0,052918 | -0,04751 | -0,31847 | 360 | 357 | 0,750315 | 0,9394 | y ~ current_feature + S |
| -0,00218 | 0,05698 | 0,061498 | -0,03819 | 311 | 308 | 0,969562 | 0,996179 | y ~ current_feature + S |
| -0,12869 | 0,064417 | -0,02429 | -1,99784 | 240 | 237 | 0,046876 | 0,525585 | y ~ current_feature + S |
| 0,080835 | 0,105653 | -0,07874 | 0,765104 | 92 | 89 | 0,446234 | 0,821425 | y ~ current_feature + S |
| 0,010523 | 0,105994 | 0,034007 | 0,099276 | 92 | 89 | 0,921142 | 0,9732 | y ~ current_feature + S |
| -0,05989 | 0,105809 | -0,04751 | -0,56606 | 92 | 89 | 0,572776 | 0,903741 | y ~ current_feature + S |
| 0,083463 | 0,112833 | 0,061498 | 0,739702 | 81 | 78 | 0,461701 | 0,937665 | y ~ current_feature + S |
| 0,068987 | 0,130994 | -0,02429 | 0,526644 | 61 | 58 | 0,600449 | 0,924682 | y ~ current_feature + S |
| -0,01133 | 0,113953 | -0,07874 | -0,0994 | 80 | 77 | 0,921082 | 0,990519 | y ~ current_feature + S |
| 2,88E-04 | 0,113961 | 0,034007 | 0,002528 | 80 | 77 | 0,997989 | 0,997989 | y ~ current_feature + S |
| -0,02015 | 0,114685 | -0,04751 | -0,17569 | 79 | 76 | 0,861005 | 0,984767 | y ~ current_feature + S |
| -0,13043 | 0,12204 | 0,061498 | -1,06876 | 69 | 66 | 0,289072 | 0,893818 | y ~ current_feature + S |
| 0,15167 | 0,139785 | -0,02429 | 1,08502 | 53 | 50 | 0,283117 | 0,734108 | y ~ current_feature + S |
| 0,019825 | 0,076681 | -0,07874 | 0,258539 | 173 | 170 | 0,796304 | 0,979102 | y ~ current_feature + S |
| 0,021628 | 0,076679 | 0,034007 | 0,282058 | 173 | 170 | 0,778242 | 0,942718 | y ~ current_feature + S |
| -0,01906 | 0,076683 | -0,04751 | -0,24861 | 173 | 170 | 0,80396 | 0,968942 | y ~ current_feature + S |
| -0,10689 | 0,084516 | 0,061498 | -1,26469 | 148 | 138,3972 | 0,208108 | 0,893818 | y ~ current_feature + S |
| -0,06819 | 0,094293 | -0,02429 | -0,72319 | 115 | 111,9488 | 0,471074 | 0,876224 | y ~ current_feature + S |
| -0,13825 | 0,108524 | -0,07874 | -1,2739 | 90 | 83,28586 | 0,206243 | 0,622938 | y ~ current_feature + S |
| -0,16926 | 0,151737 | 0,034007 | -1,1155 | 90 | 42,18825 | 0,27095 | 0,71095 | y ~ current_feature + S |
| 0,109013 | 0,106805 | -0,04751 | 1,02067 | 90 | 86,6212 | 0,310253 | 0,76529 | y ~ current_feature + S |
| 0,125346 | 0,11618 | 0,061498 | 1,078901 | 76 | 72,92249 | 0,284188 | 0,893818 | y ~ current_feature + S |
| -0,06949 | 0,133499 | -0,02429 | -0,52053 | 60 | 55,83949 | 0,604749 | 0,924682 | y ~ current_feature + S |

| | | | | | | | | |
|---|---|---|---|---|---|---|---|---|
| 0,00749 | 0,150751 | -0,07874 | 0,049683 | 47 | 44 | 0,960599 | 0,994607 | y ~ current_feature + S |
| -0,1264 | 0,149546 | 0,034007 | -0,84524 | 47 | 44 | 0,402552 | 0,778997 | y ~ current_feature + S |
| 0,148357 | 0,150811 | -0,04751 | 0,983731 | 46 | 43 | 0,330751 | 0,779476 | y ~ current_feature + S |
| -0,263 | 0,152548 | 0,061498 | -1,72404 | 43 | 40 | 0,092423 | 0,795265 | y ~ current_feature + S |
| 0,14908 | 0,18687 | -0,02429 | 0,797772 | 31 | 28 | 0,431717 | 0,851922 | y ~ current_feature + S |
| 0,030371 | 0,12593 | -0,07874 | 0,241175 | 66 | 63 | 0,810202 | 0,979871 | y ~ current_feature + S |
| 0,180503 | 0,123919 | 0,034007 | 1,456626 | 66 | 63 | 0,150186 | 0,604094 | y ~ current_feature + S |
| 0,07454 | 0,125638 | -0,04751 | 0,593297 | 66 | 63 | 0,555106 | 0,897386 | y ~ current_feature + S |
| -0,07908 | 0,133212 | 0,061498 | -0,59366 | 59 | 56 | 0,555128 | 0,937665 | y ~ current_feature + S |
| 0,316638 | 0,148138 | -0,02429 | 2,137449 | 44 | 41 | 0,038573 | 0,520476 | y ~ current_feature + S |
| 0,427882 | 0,139465 | -0,07874 | 3,068031 | 45 | 42 | 0,003764 | 0,069632 | y ~ current_feature + S |
| 0,026127 | 0,154251 | 0,034007 | 0,169379 | 45 | 42 | 0,866311 | 0,96591 | y ~ current_feature + S |
| 0,058483 | 0,154039 | -0,04751 | 0,379666 | 45 | 42 | 0,706107 | 0,93314 | y ~ current_feature + S |
| 0,07356 | 0,166215 | 0,061498 | 0,44256 | 39 | 36 | 0,660732 | 0,957207 | y ~ current_feature + S |
| -0,08077 | 0,191821 | -0,02429 | -0,42109 | 30 | 27 | 0,677025 | 0,926342 | y ~ current_feature + S |
| -0,03496 | 0,062063 | -0,07874 | -0,56324 | 297 | 259,2979 | 0,573756 | 0,899533 | y ~ current_feature + S |
| -0,0731 | 0,082583 | 0,034007 | -0,88515 | 297 | 145,8459 | 0,377534 | 0,759171 | y ~ current_feature + S |
| -0,13233 | 0,064233 | -0,04751 | -2,0601 | 297 | 238,1319 | 0,040476 | 0,612508 | y ~ current_feature + S |
| -0,16813 | 0,069191 | 0,061498 | -2,43002 | 261 | 202,9781 | 0,015968 | 0,55988 | y ~ current_feature + S |
| -0,1028 | 0,071232 | -0,02429 | -1,44321 | 198 | 195 | 0,150565 | 0,622449 | y ~ current_feature + S |
| -0,01241 | 0,065648 | -0,07874 | -0,18906 | 235 | 232 | 0,850209 | 0,983548 | y ~ current_feature + S |
| -0,05552 | 0,065552 | 0,034007 | -0,84694 | 235 | 232 | 0,397901 | 0,775482 | y ~ current_feature + S |
| -0,01242 | 0,06579 | -0,04751 | -0,18881 | 234 | 231 | 0,850405 | 0,980218 | y ~ current_feature + S |
| -0,21177 | 0,069107 | 0,061498 | -3,06431 | 203 | 200 | 0,002483 | 0,305054 | y ~ current_feature + S |
| -0,1337 | 0,080119 | -0,02429 | -1,66874 | 156 | 153 | 0,097214 | 0,605474 | y ~ current_feature + S |
| 0,011791 | 0,09534 | -0,07874 | 0,123677 | 113 | 110 | 0,901797 | 0,987929 | y ~ current_feature + S |
| -0,07587 | 0,095071 | 0,034007 | -0,79806 | 113 | 110 | 0,426554 | 0,791701 | y ~ current_feature + S |
| -0,02296 | 0,095757 | -0,04751 | -0,23974 | 112 | 109 | 0,810984 | 0,972405 | y ~ current_feature + S |
| 0,075124 | 0,103402 | 0,061498 | 0,72652 | 96 | 93 | 0,469345 | 0,937665 | y ~ current_feature + S |
| 0,088794 | 0,117386 | -0,02429 | 0,756429 | 75 | 72 | 0,45186 | 0,861794 | y ~ current_feature + S |
| -0,02864 | 0,071399 | -0,07874 | -0,40117 | 199 | 196 | 0,68873 | 0,93638 | y ~ current_feature + S |
| -0,14255 | 0,070699 | 0,034007 | -2,01624 | 199 | 196 | 0,04514 | 0,400524 | y ~ current_feature + S |
| 0,007073 | 0,071427 | -0,04751 | 0,099021 | 199 | 196 | 0,921223 | 0,985217 | y ~ current_feature + S |
| -0,09265 | 0,076592 | 0,061498 | -1,2097 | 172 | 169 | 0,228084 | 0,893818 | y ~ current_feature + S |
| -0,18642 | 0,086172 | -0,02429 | -2,16338 | 133 | 129,9891 | 0,032341 | 0,485321 | y ~ current_feature + S |
| -0,01913 | 0,05024 | -0,07874 | -0,38086 | 418 | 396,036 | 0,703512 | 0,941409 | y ~ current_feature + S |
| 0,058014 | 0,064627 | 0,034007 | 0,897685 | 418 | 238,6234 | 0,370258 | 0,750317 | y ~ current_feature + S |
| 0,037431 | 0,051094 | -0,04751 | 0,732592 | 417 | 382,5106 | 0,464256 | 0,842057 | y ~ current_feature + S |
| 0,052518 | 0,056117 | 0,061498 | 0,935867 | 364 | 316,6787 | 0,350055 | 0,915337 | y ~ current_feature + S |
| 0,032605 | 0,061064 | -0,02429 | 0,53395 | 278 | 267,9008 | 0,593819 | 0,924682 | y ~ current_feature + S |
| 0,037848 | 0,05429 | -0,07874 | 0,697144 | 372 | 338,7953 | 0,486191 | 0,84854 | y ~ current_feature + S |
| -0,02412 | 0,070634 | 0,034007 | -0,34152 | 372 | 200,3192 | 0,733071 | 0,922572 | y ~ current_feature + S |
| -0,01137 | 0,05585 | -0,04751 | -0,20364 | 372 | 320,5535 | 0,838765 | 0,980218 | y ~ current_feature + S |
| -0,03937 | 0,059972 | 0,061498 | -0,65653 | 325 | 277,6094 | 0,512029 | 0,937665 | y ~ current_feature + S |
| -0,12872 | 0,063801 | -0,02429 | -2,0175 | 248 | 241,5994 | 0,044749 | 0,525585 | y ~ current_feature + S |
| 0,029091 | 0,053603 | -0,07874 | 0,542703 | 377 | 347,7346 | 0,587682 | 0,902886 | y ~ current_feature + S |
| 0,003106 | 0,069048 | 0,034007 | 0,044983 | 377 | 209,7488 | 0,964164 | 0,989859 | y ~ current_feature + S |
| 0,087361 | 0,053745 | -0,04751 | 1,625494 | 377 | 343,5614 | 0,104974 | 0,640171 | y ~ current_feature + S |

| | | | | | | | | |
|---|---|---|---|---|---|---|---|---|
| 0,035545 | 0,058396 | 0,061498 | 0,608687 | 325 | 292,8723 | 0,543204 | 0,937665 | y ~ current_feature + S |
| -0,04023 | 0,063811 | -0,02429 | -0,63052 | 251 | 245,1905 | 0,528941 | 0,899808 | y ~ current_feature + S |
| -0,06566 | 0,05539 | -0,07874 | -1,18537 | 358 | 324,5374 | 0,236739 | 0,663586 | y ~ current_feature + S |
| -0,10567 | 0,07204 | 0,034007 | -1,46688 | 358 | 190,5351 | 0,144058 | 0,592239 | y ~ current_feature + S |
| 0,009247 | 0,056288 | -0,04751 | 0,164272 | 357 | 315,5954 | 0,869622 | 0,985217 | y ~ current_feature + S |
| 0,005694 | 0,061827 | 0,061498 | 0,092104 | 309 | 261,5993 | 0,926686 | 0,996179 | y ~ current_feature + S |
| -0,02747 | 0,065435 | -0,02429 | -0,4198 | 238 | 233,3743 | 0,675017 | 0,926342 | y ~ current_feature + S |
| 0,032309 | 0,083581 | -0,07874 | 0,386562 | 146 | 143 | 0,699655 | 0,939646 | y ~ current_feature + S |
| -0,22829 | 0,081416 | 0,034007 | -2,80398 | 146 | 143 | 0,00575 | 0,170187 | y ~ current_feature + S |
| 0,090114 | 0,083284 | -0,04751 | 1,082011 | 146 | 143 | 0,281069 | 0,747915 | y ~ current_feature + S |
| 0,011633 | 0,090161 | 0,061498 | 0,129021 | 126 | 123 | 0,897552 | 0,996179 | y ~ current_feature + S |
| -0,0231 | 0,103115 | -0,02429 | -0,22398 | 97 | 94 | 0,823256 | 0,976828 | y ~ current_feature + S |
| 0,150897 | 0,061952 | -0,07874 | 2,435718 | 281 | 254,6187 | 0,015549 | 0,174676 | y ~ current_feature + S |
| -0,03568 | 0,088731 | 0,034007 | -0,4021 | 281 | 126,8511 | 0,68829 | 0,916569 | y ~ current_feature + S |
| 0,075817 | 0,06714 | -0,04751 | 1,129236 | 281 | 220,5655 | 0,260025 | 0,742928 | y ~ current_feature + S |
| -0,04523 | 0,067905 | 0,061498 | -0,66602 | 244 | 216,4244 | 0,506108 | 0,937665 | y ~ current_feature + S |
| 0,029495 | 0,073689 | -0,02429 | 0,400264 | 187 | 184 | 0,689426 | 0,926342 | y ~ current_feature + S |
| -0,04 | 0,051139 | -0,07874 | -0,78218 | 407 | 381,7646 | 0,434595 | 0,814177 | y ~ current_feature + S |
| -0,07159 | 0,065912 | 0,034007 | -1,08617 | 407 | 229,0004 | 0,278545 | 0,715049 | y ~ current_feature + S |
| -0,03034 | 0,052129 | -0,04751 | -0,58205 | 406 | 367,6531 | 0,56089 | 0,900344 | y ~ current_feature + S |
| -0,0846 | 0,057097 | 0,061498 | -1,48168 | 353 | 304,5423 | 0,13946 | 0,8197 | y ~ current_feature + S |
| -0,00162 | 0,061541 | -0,02429 | -0,02626 | 271 | 264,0364 | 0,979067 | 0,99444 | y ~ current_feature + S |
| 0,268666 | 0,103868 | -0,07874 | 2,586611 | 89 | 86 | 0,011374 | 0,140685 | y ~ current_feature + S |
| 0,020675 | 0,10781 | 0,034007 | 0,191776 | 89 | 86 | 0,848369 | 0,959445 | y ~ current_feature + S |
| 0,156733 | 0,107125 | -0,04751 | 1,463089 | 88 | 85 | 0,147131 | 0,661823 | y ~ current_feature + S |
| -0,04022 | 0,116946 | 0,061498 | -0,34393 | 76 | 73 | 0,731887 | 0,972573 | y ~ current_feature + S |
| 0,005816 | 0,133628 | -0,02429 | 0,043521 | 59 | 56 | 0,965441 | 0,99444 | y ~ current_feature + S |
| -0,12082 | 0,101846 | -0,07874 | -1,1863 | 98 | 95 | 0,238464 | 0,6659 | y ~ current_feature + S |
| -0,03897 | 0,10252 | 0,034007 | -0,38011 | 98 | 95 | 0,70471 | 0,920814 | y ~ current_feature + S |
| -0,0213 | 0,103119 | -0,04751 | -0,20653 | 97 | 94 | 0,836825 | 0,979827 | y ~ current_feature + S |
| 0,040588 | 0,107744 | 0,061498 | 0,37671 | 89 | 86 | 0,707317 | 0,972444 | y ~ current_feature + S |
| 0,116551 | 0,126135 | -0,02429 | 0,924023 | 65 | 62 | 0,359057 | 0,800662 | y ~ current_feature + S |
| 0,089604 | 0,148472 | -0,07874 | 0,60351 | 48 | 45 | 0,5492 | 0,888464 | y ~ current_feature + S |
| -0,26018 | 0,143937 | 0,034007 | -1,80757 | 48 | 45 | 0,077361 | 0,469239 | y ~ current_feature + S |
| 0,34053 | 0,140162 | -0,04751 | 2,429551 | 48 | 45 | 0,019172 | 0,54566 | y ~ current_feature + S |
| 0,184996 | 0,155385 | 0,061498 | 1,190565 | 43 | 40 | 0,240842 | 0,893818 | y ~ current_feature + S |
| 0,087055 | 0,18499 | -0,02429 | 0,470593 | 32 | 29 | 0,641452 | 0,924682 | y ~ current_feature + S |
| -0,03964 | 0,062267 | -0,07874 | -0,63668 | 276 | 257,5156 | 0,5249 | 0,876542 | y ~ current_feature + S |
| -0,00791 | 0,083251 | 0,034007 | -0,09499 | 276 | 144,2749 | 0,924458 | 0,9732 | y ~ current_feature + S |
| -0,02169 | 0,062623 | -0,04751 | -0,34634 | 276 | 254,876 | 0,729375 | 0,93314 | y ~ current_feature + S |
| -0,03908 | 0,066091 | 0,061498 | -0,59133 | 241 | 228,5892 | 0,554885 | 0,937665 | y ~ current_feature + S |
| -0,01906 | 0,076436 | -0,02429 | -0,2493 | 183 | 171,0993 | 0,803425 | 0,968297 | y ~ current_feature + S |
| -0,18852 | 0,058698 | -0,07874 | -3,21175 | 316 | 279,9268 | 0,001473 | 0,048726 | y ~ current_feature + S |
| -0,28696 | 0,077044 | 0,034007 | -3,72459 | 316 | 154,597 | 2,74E-04 | 0,067708 | y ~ current_feature + S |
| -0,07245 | 0,062508 | -0,04751 | -1,15908 | 315 | 254,5912 | 0,247509 | 0,742928 | y ~ current_feature + S |
| -0,0596 | 0,064854 | 0,061498 | -0,919 | 272 | 236,9121 | 0,359031 | 0,920619 | y ~ current_feature + S |
| -0,15158 | 0,068812 | -0,02429 | -2,20276 | 210 | 206,3342 | 0,028718 | 0,485321 | y ~ current_feature + S |
| -0,08147 | 0,080576 | -0,07874 | -1,0111 | 156 | 153 | 0,313564 | 0,718687 | y ~ current_feature + S |

| | | | | | | | | |
|---|---|---|---|---|---|---|---|---|
| -0,10456 | 0,080402 | 0,034007 | -1,30045 | 156 | 153 | 0,195401 | 0,6438 | y ~ current_feature + S |
| -0,07137 | 0,080639 | -0,04751 | -0,88507 | 156 | 153 | 0,377508 | 0,803665 | y ~ current_feature + S |
| -0,25323 | 0,082954 | 0,061498 | -3,05259 | 139 | 136 | 0,002729 | 0,305054 | y ~ current_feature + S |
| 0,008365 | 0,0995 | -0,02429 | 0,084067 | 104 | 101 | 0,93317 | 0,99444 | y ~ current_feature + S |
| 0,192615 | 0,121712 | -0,07874 | 1,582543 | 68 | 65 | 0,11838 | 0,474142 | y ~ current_feature + S |
| -0,0484 | 0,123889 | 0,034007 | -0,39064 | 68 | 65 | 0,69734 | 0,918206 | y ~ current_feature + S |
| -0,05641 | 0,124801 | -0,04751 | -0,45198 | 67 | 64 | 0,652811 | 0,925756 | y ~ current_feature + S |
| -0,05863 | 0,132225 | 0,061498 | -0,44342 | 60 | 57 | 0,659142 | 0,957207 | y ~ current_feature + S |
| 0,045776 | 0,154142 | -0,02429 | 0,296977 | 45 | 42 | 0,767948 | 0,955095 | y ~ current_feature + S |
| 0,079765 | 0,076452 | -0,07874 | 1,043339 | 173 | 170 | 0,298273 | 0,718687 | y ~ current_feature + S |
| 0,069667 | 0,07651 | 0,034007 | 0,910561 | 173 | 170 | 0,363816 | 0,7498 | y ~ current_feature + S |
| 0,066285 | 0,076754 | -0,04751 | 0,863608 | 172 | 169 | 0,389027 | 0,803665 | y ~ current_feature + S |
| -0,12644 | 0,081817 | 0,061498 | -1,54542 | 150 | 147 | 0,124395 | 0,8197 | y ~ current_feature + S |
| 0,13425 | 0,093636 | -0,02429 | 1,433749 | 115 | 112 | 0,15443 | 0,623156 | y ~ current_feature + S |
| -0,03094 | 0,066566 | -0,07874 | -0,46475 | 251 | 225,4672 | 0,642556 | 0,928694 | y ~ current_feature + S |
| -0,0126 | 0,085384 | 0,034007 | -0,14762 | 251 | 137,1436 | 0,882861 | 0,96591 | y ~ current_feature + S |
| 0,102745 | 0,070202 | -0,04751 | 1,463549 | 250 | 200,7653 | 0,144881 | 0,661823 | y ~ current_feature + S |
| 0,05465 | 0,073326 | 0,061498 | 0,745296 | 217 | 185,4313 | 0,457036 | 0,937665 | y ~ current_feature + S |
| -0,11665 | 0,077791 | -0,02429 | -1,49957 | 166 | 163 | 0,135661 | 0,618941 | y ~ current_feature + S |
| 0,016157 | 0,054819 | -0,07874 | 0,29474 | 383 | 332,6814 | 0,768377 | 0,965616 | y ~ current_feature + S |
| 0,070477 | 0,072747 | 0,034007 | 0,968806 | 383 | 188,023 | 0,333887 | 0,734392 | y ~ current_feature + S |
| 0,033388 | 0,055532 | -0,04751 | 0,601244 | 382 | 323,9126 | 0,548098 | 0,895709 | y ~ current_feature + S |
| -0,00612 | 0,060051 | 0,061498 | -0,10196 | 333 | 277,2929 | 0,91886 | 0,996179 | y ~ current_feature + S |
| 0,078404 | 0,062925 | -0,02429 | 1,245983 | 254 | 251 | 0,213932 | 0,679441 | y ~ current_feature + S |
| -0,08112 | 0,103353 | -0,07874 | -0,7849 | 96 | 93 | 0,434508 | 0,814177 | y ~ current_feature + S |
| -0,20221 | 0,101553 | 0,034007 | -1,99114 | 96 | 93 | 0,0494 | 0,420946 | y ~ current_feature + S |
| 0,02742 | 0,103656 | -0,04751 | 0,264529 | 96 | 93 | 0,791958 | 0,957596 | y ~ current_feature + S |
| -0,22809 | 0,109543 | 0,061498 | -2,08223 | 82 | 79 | 0,040559 | 0,658491 | y ~ current_feature + S |
| 0,060206 | 0,127805 | -0,02429 | 0,47108 | 64 | 61 | 0,639264 | 0,924682 | y ~ current_feature + S |
| -0,01233 | 0,053952 | -0,07874 | -0,22851 | 381 | 343,4932 | 0,819385 | 0,979871 | y ~ current_feature + S |
| -0,02533 | 0,067545 | 0,034007 | -0,37497 | 381 | 219,0486 | 0,708046 | 0,920814 | y ~ current_feature + S |
| -5,51E-04 | 0,053403 | -0,04751 | -0,01032 | 380 | 350,6493 | 0,991772 | 0,99446 | y ~ current_feature + S |
| -0,06918 | 0,05868 | 0,061498 | -1,1789 | 330 | 289,029 | 0,239408 | 0,893818 | y ~ current_feature + S |
| -0,01166 | 0,063241 | -0,02429 | -0,18439 | 253 | 250 | 0,853859 | 0,981143 | y ~ current_feature + S |
| -0,03324 | 0,113165 | -0,07874 | -0,29377 | 81 | 78 | 0,769717 | 0,965616 | y ~ current_feature + S |
| -0,0582 | 0,113036 | 0,034007 | -0,51492 | 81 | 78 | 0,608062 | 0,887506 | y ~ current_feature + S |
| -0,09769 | 0,112686 | -0,04751 | -0,86693 | 81 | 78 | 0,388639 | 0,803665 | y ~ current_feature + S |
| 0,105202 | 0,123346 | 0,061498 | 0,852901 | 68 | 65 | 0,396846 | 0,93433 | y ~ current_feature + S |
| 0,042545 | 0,139901 | -0,02429 | 0,304108 | 54 | 51 | 0,762283 | 0,951727 | y ~ current_feature + S |
| 0,084297 | 0,053262 | -0,07874 | 1,582691 | 353 | 350 | 0,114395 | 0,467692 | y ~ current_feature + S |
| 0,063772 | 0,053343 | 0,034007 | 1,195502 | 353 | 350 | 0,2327 | 0,683327 | y ~ current_feature + S |
| 0,098021 | 0,053271 | -0,04751 | 1,840052 | 352 | 349 | 0,06661 | 0,640171 | y ~ current_feature + S |
| 0,006174 | 0,057638 | 0,061498 | 0,107117 | 304 | 301 | 0,914767 | 0,996179 | y ~ current_feature + S |
| 0,169048 | 0,064708 | -0,02429 | 2,612464 | 235 | 232 | 0,009576 | 0,295269 | y ~ current_feature + S |
| 0,00546 | 0,094914 | -0,07874 | 0,057524 | 114 | 111 | 0,954231 | 0,994607 | y ~ current_feature + S |
| 0,076533 | 0,094637 | 0,034007 | 0,808697 | 114 | 111 | 0,420419 | 0,791109 | y ~ current_feature + S |
| 0,332915 | 0,089502 | -0,04751 | 3,719657 | 114 | 111 | 3,15E-04 | 0,077615 | y ~ current_feature + S |
| 0,033862 | 0,100957 | 0,061498 | 0,335414 | 101 | 98 | 0,73803 | 0,972573 | y ~ current_feature + S |

| | | | | | | | | |
|---|---|---|---|---|---|---|---|---|
| 0,063302 | 0,116806 | -0,02429 | 0,541938 | 76 | 73 | 0,589511 | 0,924682 | y ~ current_feature + S |
| 0,05064 | 0,054404 | -0,07874 | 0,930827 | 340 | 337 | 0,352609 | 0,765193 | y ~ current_feature + S |
| -0,08507 | 0,054276 | 0,034007 | -1,56732 | 340 | 337 | 0,117979 | 0,552558 | y ~ current_feature + S |
| 0,082663 | 0,054368 | -0,04751 | 1,520444 | 339 | 336 | 0,12934 | 0,654671 | y ~ current_feature + S |
| -0,08065 | 0,066019 | 0,061498 | -1,22164 | 300 | 227,9454 | 0,223107 | 0,893818 | y ~ current_feature + S |
| 0,006485 | 0,066964 | -0,02429 | 0,096847 | 226 | 223 | 0,922935 | 0,99444 | y ~ current_feature + S |
| 0,007569 | 0,058221 | -0,07874 | 0,130006 | 298 | 295 | 0,89665 | 0,987929 | y ~ current_feature + S |
| 0,05996 | 0,058117 | 0,034007 | 1,031696 | 298 | 295 | 0,30306 | 0,718742 | y ~ current_feature + S |
| 0,116795 | 0,057922 | -0,04751 | 2,016413 | 297 | 294 | 0,044666 | 0,63563 | y ~ current_feature + S |
| -0,04582 | 0,062313 | 0,061498 | -0,73532 | 260 | 257 | 0,462816 | 0,937665 | y ~ current_feature + S |
| 0,091937 | 0,071126 | -0,02429 | 1,292593 | 199 | 196 | 0,197674 | 0,660578 | y ~ current_feature + S |
| 0,044453 | 0,050652 | -0,07874 | 0,877627 | 392 | 389 | 0,380688 | 0,789252 | y ~ current_feature + S |
| -0,08595 | 0,050514 | 0,034007 | -1,70145 | 392 | 389 | 0,089658 | 0,487845 | y ~ current_feature + S |
| 0,070554 | 0,050641 | -0,04751 | 1,393215 | 391 | 388 | 0,164352 | 0,694976 | y ~ current_feature + S |
| 0,004693 | 0,060532 | 0,061498 | 0,077531 | 340 | 272,9105 | 0,938258 | 0,996179 | y ~ current_feature + S |
| -0,0098 | 0,062375 | -0,02429 | -0,15712 | 260 | 257 | 0,875271 | 0,984468 | y ~ current_feature + S |
| 0,043167 | 0,050232 | -0,07874 | 0,859356 | 418 | 395,5787 | 0,390665 | 0,789252 | y ~ current_feature + S |
| 0,029163 | 0,06494 | 0,034007 | 0,449072 | 418 | 236,9226 | 0,65379 | 0,900251 | y ~ current_feature + S |
| 0,083265 | 0,05104 | -0,04751 | 1,631389 | 417 | 381,2093 | 0,103634 | 0,640171 | y ~ current_feature + S |
| -0,02664 | 0,055424 | 0,061498 | -0,48065 | 365 | 325,3098 | 0,631092 | 0,953078 | y ~ current_feature + S |
| 0,049071 | 0,060313 | -0,02429 | 0,813603 | 278 | 274,2364 | 0,416579 | 0,839968 | y ~ current_feature + S |
| -0,05052 | 0,120232 | -0,07874 | -0,42022 | 72 | 69 | 0,67563 | 0,934653 | y ~ current_feature + S |
| -0,10074 | 0,119773 | 0,034007 | -0,84112 | 72 | 69 | 0,403184 | 0,778997 | y ~ current_feature + S |
| 0,021866 | 0,120357 | -0,04751 | 0,181673 | 72 | 69 | 0,856372 | 0,983239 | y ~ current_feature + S |
| -0,18171 | 0,125905 | 0,061498 | -1,4432 | 64 | 61 | 0,15408 | 0,8197 | y ~ current_feature + S |
| -0,16898 | 0,146927 | -0,02429 | -1,15009 | 48 | 45 | 0,256183 | 0,718089 | y ~ current_feature + S |
| 0,355841 | 0,123784 | -0,07874 | 2,874697 | 60 | 57 | 0,005676 | 0,092669 | y ~ current_feature + S |
| 0,290214 | 0,126753 | 0,034007 | 2,289608 | 60 | 57 | 0,025767 | 0,347616 | y ~ current_feature + S |
| -0,19393 | 0,129939 | -0,04751 | -1,49249 | 60 | 57 | 0,141086 | 0,660782 | y ~ current_feature + S |
| 0,015543 | 0,14284 | 0,061498 | 0,108812 | 52 | 49 | 0,913796 | 0,996179 | y ~ current_feature + S |
| 0,176519 | 0,161817 | -0,02429 | 1,090855 | 40 | 37 | 0,28239 | 0,734108 | y ~ current_feature + S |
| -0,10112 | 0,082056 | -0,07874 | -1,23233 | 150 | 147 | 0,219793 | 0,639583 | y ~ current_feature + S |
| -0,04039 | 0,082411 | 0,034007 | -0,49016 | 150 | 147 | 0,624752 | 0,896904 | y ~ current_feature + S |
| -0,14691 | 0,081584 | -0,04751 | -1,80074 | 150 | 147 | 0,073794 | 0,640171 | y ~ current_feature + S |
| -0,06166 | 0,098899 | 0,061498 | -0,6235 | 134 | 101,8502 | 0,534352 | 0,937665 | y ~ current_feature + S |
| -0,01329 | 0,101526 | -0,02429 | -0,1309 | 100 | 97 | 0,896124 | 0,99444 | y ~ current_feature + S |
| 0,068192 | 0,07375 | -0,07874 | 0,924635 | 186 | 183 | 0,356373 | 0,766617 | y ~ current_feature + S |
| 0,019673 | 0,073908 | 0,034007 | 0,266179 | 186 | 183 | 0,790401 | 0,945731 | y ~ current_feature + S |
| -0,01337 | 0,074118 | -0,04751 | -0,18043 | 185 | 182 | 0,857012 | 0,983239 | y ~ current_feature + S |
| -0,04304 | 0,078984 | 0,061498 | -0,5449 | 163 | 160 | 0,586581 | 0,945257 | y ~ current_feature + S |
| -0,11661 | 0,090664 | -0,02429 | -1,28619 | 123 | 120 | 0,200852 | 0,660578 | y ~ current_feature + S |
| 0,020266 | 0,051767 | -0,07874 | 0,391486 | 376 | 373 | 0,695661 | 0,939646 | y ~ current_feature + S |
| -0,01584 | 0,051772 | 0,034007 | -0,30595 | 376 | 373 | 0,759814 | 0,935544 | y ~ current_feature + S |
| -0,06858 | 0,051726 | -0,04751 | -1,32578 | 375 | 372 | 0,185727 | 0,704811 | y ~ current_feature + S |
| 0,018753 | 0,055546 | 0,061498 | 0,337605 | 327 | 324 | 0,73588 | 0,972573 | y ~ current_feature + S |
| -0,03044 | 0,063599 | -0,02429 | -0,47858 | 250 | 247 | 0,632659 | 0,924682 | y ~ current_feature + S |
| -0,02293 | 0,079284 | -0,07874 | -0,28925 | 162 | 159 | 0,77277 | 0,965962 | y ~ current_feature + S |
| 0,175496 | 0,078074 | 0,034007 | 2,247804 | 162 | 159 | 0,025964 | 0,347616 | y ~ current_feature + S |

| | | | | | | | | |
|---|---|---|---|---|---|---|---|---|
| 0,114148 | 0,078787 | -0,04751 | 1,448819 | 162 | 159 | 0,149357 | 0,661823 | y ~ current_feature + S |
| -0,00727 | 0,087037 | 0,061498 | -0,08349 | 135 | 132 | 0,933586 | 0,996179 | y ~ current_feature + S |
| 0,108423 | 0,097015 | -0,02429 | 1,117596 | 108 | 105 | 0,26629 | 0,723659 | y ~ current_feature + S |
| 0,018258 | 0,056156 | -0,07874 | 0,325122 | 320 | 317 | 0,745303 | 0,96252 | y ~ current_feature + S |
| -0,07387 | 0,056012 | 0,034007 | -1,3188 | 320 | 317 | 0,188189 | 0,638805 | y ~ current_feature + S |
| -0,01788 | 0,056245 | -0,04751 | -0,3178 | 319 | 316 | 0,750843 | 0,9394 | y ~ current_feature + S |
| -0,04296 | 0,060578 | 0,061498 | -0,70914 | 275 | 272 | 0,478845 | 0,937665 | y ~ current_feature + S |
| -0,02627 | 0,068983 | -0,02429 | -0,3808 | 213 | 210 | 0,703738 | 0,93118 | y ~ current_feature + S |
| 0,138823 | 0,127849 | -0,07874 | 1,085829 | 63 | 60 | 0,281898 | 0,718628 | y ~ current_feature + S |
| 0,023996 | 0,129062 | 0,034007 | 0,185928 | 63 | 60 | 0,853128 | 0,959445 | y ~ current_feature + S |
| 0,073173 | 0,128753 | -0,04751 | 0,568317 | 63 | 60 | 0,57194 | 0,903741 | y ~ current_feature + S |
| 0,03567 | 0,138587 | 0,061498 | 0,257383 | 55 | 52 | 0,797899 | 0,984075 | y ~ current_feature + S |
| 0,074135 | 0,159688 | -0,02429 | 0,464252 | 42 | 39 | 0,645048 | 0,924682 | y ~ current_feature + S |
| 0,101839 | 0,066767 | -0,07874 | 1,525298 | 225 | 222 | 0,128608 | 0,481525 | y ~ current_feature + S |
| 0,085707 | 0,066869 | 0,034007 | 1,281715 | 225 | 222 | 0,20128 | 0,647595 | y ~ current_feature + S |
| 0,064362 | 0,066976 | -0,04751 | 0,960971 | 225 | 222 | 0,337612 | 0,788117 | y ~ current_feature + S |
| 0,10642 | 0,081028 | 0,061498 | 1,313376 | 196 | 150,5857 | 0,191054 | 0,878136 | y ~ current_feature + S |
| 0,120581 | 0,081877 | -0,02429 | 1,472709 | 150 | 147 | 0,142967 | 0,618941 | y ~ current_feature + S |
| 0,024502 | 0,083599 | -0,07874 | 0,293084 | 146 | 143 | 0,769883 | 0,965616 | y ~ current_feature + S |
| 0,03682 | 0,083567 | 0,034007 | 0,440597 | 146 | 143 | 0,66017 | 0,900251 | y ~ current_feature + S |
| 0,130381 | 0,08291 | -0,04751 | 1,572559 | 146 | 143 | 0,118031 | 0,646984 | y ~ current_feature + S |
| -0,04756 | 0,091184 | 0,061498 | -0,52162 | 123 | 120 | 0,602898 | 0,945257 | y ~ current_feature + S |
| 0,121923 | 0,102373 | -0,02429 | 1,190969 | 97 | 94 | 0,236665 | 0,697311 | y ~ current_feature + S |
| -0,03275 | 0,0522 | -0,07874 | -0,62735 | 383 | 366,6025 | 0,530821 | 0,880735 | y ~ current_feature + S |
| -0,06065 | 0,066483 | 0,034007 | -0,9122 | 383 | 225,4133 | 0,362639 | 0,7498 | y ~ current_feature + S |
| 0,087244 | 0,053389 | -0,04751 | 1,634133 | 382 | 348,1625 | 0,103135 | 0,640171 | y ~ current_feature + S |
| 0,058715 | 0,057066 | 0,061498 | 1,028895 | 334 | 306,0117 | 0,304341 | 0,893818 | y ~ current_feature + S |
| -0,04393 | 0,064016 | -0,02429 | -0,68622 | 255 | 243,5489 | 0,493225 | 0,892387 | y ~ current_feature + S |
| -0,07686 | 0,097301 | -0,07874 | -0,78989 | 108 | 105 | 0,431372 | 0,814177 | y ~ current_feature + S |
| -0,20089 | 0,0956 | 0,034007 | -2,10139 | 108 | 105 | 0,038 | 0,390551 | y ~ current_feature + S |
| -0,02632 | 0,097556 | -0,04751 | -0,26984 | 108 | 105 | 0,787811 | 0,954141 | y ~ current_feature + S |
| -0,02891 | 0,105955 | 0,061498 | -0,27285 | 92 | 89 | 0,785598 | 0,979725 | y ~ current_feature + S |
| -0,06053 | 0,120165 | -0,02429 | -0,50375 | 72 | 69 | 0,616039 | 0,924682 | y ~ current_feature + S |
| 0,314776 | 0,111091 | -0,07874 | 2,833487 | 76 | 73 | 0,00595 | 0,093673 | y ~ current_feature + S |
| 0,18982 | 0,114913 | 0,034007 | 1,651854 | 76 | 73 | 0,10286 | 0,525261 | y ~ current_feature + S |
| 0,186142 | 0,114996 | -0,04751 | 1,618684 | 76 | 73 | 0,109828 | 0,640171 | y ~ current_feature + S |
| -0,26718 | 0,123382 | 0,061498 | -2,1655 | 64 | 61 | 0,034269 | 0,658491 | y ~ current_feature + S |
| 0,098356 | 0,145158 | -0,02429 | 0,677581 | 50 | 47 | 0,501359 | 0,89399 | y ~ current_feature + S |
| -0,18842 | 0,109801 | -0,07874 | -1,71601 | 83 | 80 | 0,09003 | 0,419008 | y ~ current_feature + S |
| -0,16619 | 0,110249 | 0,034007 | -1,50738 | 83 | 80 | 0,135652 | 0,576909 | y ~ current_feature + S |
| -0,10085 | 0,111935 | -0,04751 | -0,901 | 82 | 79 | 0,370328 | 0,802018 | y ~ current_feature + S |
| -0,18282 | 0,119224 | 0,061498 | -1,5334 | 71 | 68 | 0,129816 | 0,8197 | y ~ current_feature + S |
| -0,1295 | 0,137507 | -0,02429 | -0,9418 | 55 | 52 | 0,35065 | 0,795954 | y ~ current_feature + S |
| 0,118309 | 0,126108 | -0,07874 | 0,938153 | 65 | 62 | 0,351806 | 0,765193 | y ~ current_feature + S |
| -0,02865 | 0,126948 | 0,034007 | -0,22572 | 65 | 62 | 0,822161 | 0,952501 | y ~ current_feature + S |
| 0,187361 | 0,124751 | -0,04751 | 1,501877 | 65 | 62 | 0,138205 | 0,657012 | y ~ current_feature + S |
| -0,04102 | 0,135968 | 0,061498 | -0,30167 | 57 | 54 | 0,764059 | 0,972573 | y ~ current_feature + S |
| 0,186414 | 0,153436 | -0,02429 | 1,21493 | 44 | 41 | 0,231343 | 0,695919 | y ~ current_feature + S |

| | | | | | | | | |
|---|---|---|---|---|---|---|---|---|
| 0,091993 | 0,139434 | -0,07874 | 0,659757 | 54 | 51 | 0,512378 | 0,867643 | y ~ current_feature + S |
| -0,07114 | 0,139673 | 0,034007 | -0,5093 | 54 | 51 | 0,612736 | 0,889068 | y ~ current_feature + S |
| 0,057625 | 0,139795 | -0,04751 | 0,412209 | 54 | 51 | 0,681914 | 0,928814 | y ~ current_feature + S |
| 0,215222 | 0,150687 | 0,061498 | 1,428267 | 45 | 42 | 0,16061 | 0,8197 | y ~ current_feature + S |
| 0,017605 | 0,174051 | -0,02429 | 0,101151 | 36 | 33 | 0,920042 | 0,99444 | y ~ current_feature + S |
| -0,07198 | 0,072743 | -0,07874 | -0,98944 | 191 | 188 | 0,323718 | 0,725913 | y ~ current_feature + S |
| -0,14499 | 0,072162 | 0,034007 | -2,00921 | 191 | 188 | 0,045945 | 0,400524 | y ~ current_feature + S |
| -0,13312 | 0,072283 | -0,04751 | -1,8417 | 191 | 188 | 0,067095 | 0,640171 | y ~ current_feature + S |
| -0,13373 | 0,078593 | 0,061498 | -1,70154 | 162 | 159 | 0,090797 | 0,795265 | y ~ current_feature + S |
| -0,03197 | 0,089397 | -0,02429 | -0,35762 | 128 | 125 | 0,72123 | 0,937979 | y ~ current_feature + S |
| 0,100089 | 0,113388 | -0,07874 | 0,882714 | 80 | 77 | 0,380138 | 0,789252 | y ~ current_feature + S |
| -0,01692 | 0,113944 | 0,034007 | -0,14849 | 80 | 77 | 0,882348 | 0,96591 | y ~ current_feature + S |
| 0,040397 | 0,114614 | -0,04751 | 0,352457 | 79 | 76 | 0,725471 | 0,93314 | y ~ current_feature + S |
| -0,29236 | 0,116832 | 0,061498 | -2,50239 | 70 | 67 | 0,014783 | 0,55988 | y ~ current_feature + S |
| -0,01312 | 0,141409 | -0,02429 | -0,09281 | 53 | 50 | 0,926424 | 0,99444 | y ~ current_feature + S |
| 0,101122 | 0,135385 | -0,07874 | 0,746923 | 57 | 54 | 0,45835 | 0,829288 | y ~ current_feature + S |
| 0,042969 | 0,135957 | 0,034007 | 0,316047 | 57 | 54 | 0,753185 | 0,933597 | y ~ current_feature + S |
| 0,230918 | 0,132405 | -0,04751 | 1,744033 | 57 | 54 | 0,086844 | 0,640171 | y ~ current_feature + S |
| -0,02368 | 0,145824 | 0,061498 | -0,1624 | 50 | 47 | 0,87169 | 0,996179 | y ~ current_feature + S |
| 0,135023 | 0,167483 | -0,02429 | 0,806187 | 38 | 35 | 0,425579 | 0,847264 | y ~ current_feature + S |
| -0,08556 | 0,052806 | -0,07874 | -1,62037 | 359 | 356 | 0,106039 | 0,449749 | y ~ current_feature + S |
| -0,1163 | 0,05264 | 0,034007 | -2,20927 | 359 | 356 | 0,027792 | 0,347616 | y ~ current_feature + S |
| -0,03522 | 0,052967 | -0,04751 | -0,66486 | 359 | 356 | 0,506569 | 0,868909 | y ~ current_feature + S |
| -0,02509 | 0,05687 | 0,061498 | -0,44112 | 312 | 309 | 0,659438 | 0,957207 | y ~ current_feature + S |
| -0,10576 | 0,064729 | -0,02429 | -1,63381 | 239 | 236 | 0,103631 | 0,605474 | y ~ current_feature + S |
| 0,281396 | 0,113089 | -0,07874 | 2,488267 | 75 | 72 | 0,015151 | 0,174676 | y ~ current_feature + S |
| 0,017211 | 0,117834 | 0,034007 | 0,146058 | 75 | 72 | 0,884283 | 0,96591 | y ~ current_feature + S |
| 0,05388 | 0,11768 | -0,04751 | 0,457856 | 75 | 72 | 0,648435 | 0,925756 | y ~ current_feature + S |
| -0,36929 | 0,119974 | 0,061498 | -3,07811 | 63 | 60 | 0,003139 | 0,305054 | y ~ current_feature + S |
| 0,216987 | 0,14239 | -0,02429 | 1,523892 | 50 | 47 | 0,134237 | 0,617591 | y ~ current_feature + S |
| 0,198903 | 0,05403 | -0,07874 | 3,681334 | 332 | 329 | 2,71E-04 | 0,022288 | y ~ current_feature + S |
| -0,02037 | 0,05512 | 0,034007 | -0,36949 | 332 | 329 | 0,712002 | 0,920814 | y ~ current_feature + S |
| 0,024844 | 0,055115 | -0,04751 | 0,45076 | 332 | 329 | 0,65246 | 0,925756 | y ~ current_feature + S |
| -0,01458 | 0,059229 | 0,061498 | -0,24618 | 288 | 285 | 0,805723 | 0,988828 | y ~ current_feature + S |
| 0,007092 | 0,067572 | -0,02429 | 0,10495 | 222 | 219 | 0,916512 | 0,99444 | y ~ current_feature + S |
| 0,241758 | 0,144649 | -0,07874 | 1,671343 | 48 | 45 | 0,101595 | 0,447502 | y ~ current_feature + S |
| 0,030197 | 0,149003 | 0,034007 | 0,202663 | 48 | 45 | 0,840312 | 0,958985 | y ~ current_feature + S |
| -0,07395 | 0,148663 | -0,04751 | -0,49742 | 48 | 45 | 0,621318 | 0,925756 | y ~ current_feature + S |
| -0,10331 | 0,168126 | 0,061498 | -0,61448 | 38 | 35 | 0,542872 | 0,937665 | y ~ current_feature + S |
| 0,06656 | 0,204326 | -0,02429 | 0,325755 | 32 | 23,8464 | 0,747451 | 0,948737 | y ~ current_feature + S |
| 0,007294 | 0,141418 | -0,07874 | 0,051581 | 53 | 50 | 0,959068 | 0,994607 | y ~ current_feature + S |
| -0,0056 | 0,141419 | 0,034007 | -0,03961 | 53 | 50 | 0,968566 | 0,989859 | y ~ current_feature + S |
| 0,078894 | 0,142412 | -0,04751 | 0,553988 | 52 | 49 | 0,582107 | 0,906861 | y ~ current_feature + S |
| -0,22232 | 0,150442 | 0,061498 | -1,47775 | 45 | 42 | 0,146936 | 0,8197 | y ~ current_feature + S |
| -0,27196 | 0,170114 | -0,02429 | -1,5987 | 35 | 32 | 0,119717 | 0,615361 | y ~ current_feature + S |
| 0,023656 | 0,096647 | -0,07874 | 0,244764 | 110 | 107 | 0,807108 | 0,979871 | y ~ current_feature + S |
| -0,05372 | 0,096534 | 0,034007 | -0,55649 | 110 | 107 | 0,579035 | 0,869291 | y ~ current_feature + S |
| -0,22172 | 0,094711 | -0,04751 | -2,34107 | 109 | 106 | 0,021099 | 0,557019 | y ~ current_feature + S |

| | | | | | | | | |
|---|---|---|---|---|---|---|---|---|
| -0,2158 | 0,100712 | 0,061498 | -2,14278 | 97 | 94 | 0,034714 | 0,658491 | y ~ current_feature + S |
| -0,09557 | 0,118976 | -0,02429 | -0,80329 | 73 | 70 | 0,424524 | 0,847264 | y ~ current_feature + S |
| 0,030358 | 0,090494 | -0,07874 | 0,335471 | 125 | 122 | 0,737846 | 0,959225 | y ~ current_feature + S |
| -0,07823 | 0,090258 | 0,034007 | -0,86674 | 125 | 122 | 0,387784 | 0,767272 | y ~ current_feature + S |
| -0,11072 | 0,089979 | -0,04751 | -1,23046 | 125 | 122 | 0,220892 | 0,742928 | y ~ current_feature + S |
| 0,00251 | 0,1 | 0,061498 | 0,025101 | 103 | 100 | 0,980025 | 0,996179 | y ~ current_feature + S |
| -0,11803 | 0,111022 | -0,02429 | -1,0631 | 83 | 80 | 0,290933 | 0,734779 | y ~ current_feature + S |
| 0,068139 | 0,115977 | -0,07874 | 0,587519 | 77 | 74 | 0,558644 | 0,894753 | y ~ current_feature + S |
| -0,16106 | 0,11473 | 0,034007 | -1,40378 | 77 | 74 | 0,164568 | 0,61196 | y ~ current_feature + S |
| -0,21402 | 0,114329 | -0,04751 | -1,87193 | 76 | 73 | 0,065222 | 0,640171 | y ~ current_feature + S |
| -0,11923 | 0,121298 | 0,061498 | -0,98296 | 70 | 67 | 0,329161 | 0,893818 | y ~ current_feature + S |
| 0,036215 | 0,144243 | -0,02429 | 0,251067 | 51 | 48 | 0,802834 | 0,968297 | y ~ current_feature + S |
| 6,84E-04 | 0,127 | -0,07874 | 0,005388 | 65 | 62 | 0,995718 | 0,996647 | y ~ current_feature + S |
| -0,0502 | 0,12684 | 0,034007 | -0,39579 | 65 | 62 | 0,69362 | 0,916569 | y ~ current_feature + S |
| -0,25651 | 0,123753 | -0,04751 | -2,07272 | 64 | 61 | 0,042431 | 0,627977 | y ~ current_feature + S |
| 0,003472 | 0,13736 | 0,061498 | 0,025277 | 56 | 53 | 0,979929 | 0,996179 | y ~ current_feature + S |
| 0,136354 | 0,156637 | -0,02429 | 0,87051 | 43 | 40 | 0,389216 | 0,82057 | y ~ current_feature + S |
| -0,04436 | 0,050572 | -0,07874 | -0,87709 | 418 | 390,2291 | 0,38098 | 0,789252 | y ~ current_feature + S |
| -0,13179 | 0,065127 | 0,034007 | -2,02363 | 418 | 231,6656 | 0,044157 | 0,400524 | y ~ current_feature + S |
| -0,10033 | 0,051505 | -0,04751 | -1,94797 | 417 | 373,1719 | 0,052168 | 0,640171 | y ~ current_feature + S |
| -0,02962 | 0,056034 | 0,061498 | -0,52866 | 365 | 318,2085 | 0,597413 | 0,945257 | y ~ current_feature + S |
| -0,08875 | 0,061903 | -0,02429 | -1,43376 | 278 | 258,9056 | 0,152848 | 0,623156 | y ~ current_feature + S |
| -0,04039 | 0,050058 | -0,07874 | -0,8069 | 421 | 398,423 | 0,420206 | 0,811887 | y ~ current_feature + S |
| -0,10459 | 0,064163 | 0,034007 | -1,63013 | 421 | 240,2432 | 0,104384 | 0,526532 | y ~ current_feature + S |
| -0,02747 | 0,051024 | -0,04751 | -0,53829 | 420 | 383,8236 | 0,590691 | 0,91049 | y ~ current_feature + S |
| -0,05376 | 0,055398 | 0,061498 | -0,97048 | 367 | 324,9004 | 0,33253 | 0,893818 | y ~ current_feature + S |
| -0,00439 | 0,061726 | -0,02429 | -0,07107 | 280 | 262,459 | 0,943395 | 0,99444 | y ~ current_feature + S |
| -0,04968 | 0,050173 | -0,07874 | -0,99022 | 412 | 396,2713 | 0,322669 | 0,725761 | y ~ current_feature + S |
| -0,09014 | 0,063605 | 0,034007 | -1,4172 | 412 | 245,1778 | 0,157693 | 0,60427 | y ~ current_feature + S |
| -0,05803 | 0,05083 | -0,04751 | -1,14171 | 411 | 385,7327 | 0,254282 | 0,742928 | y ~ current_feature + S |
| -0,01303 | 0,055801 | 0,061498 | -0,23344 | 358 | 321,0981 | 0,815568 | 0,995908 | y ~ current_feature + S |
| 0,029476 | 0,060753 | -0,02429 | 0,485183 | 274 | 270,6987 | 0,627939 | 0,924682 | y ~ current_feature + S |
| -0,11505 | 0,056238 | -0,07874 | -2,04572 | 315 | 312 | 0,041622 | 0,275003 | y ~ current_feature + S |
| -0,08669 | 0,056401 | 0,034007 | -1,53703 | 315 | 312 | 0,125299 | 0,572145 | y ~ current_feature + S |
| -0,01999 | 0,056603 | -0,04751 | -0,35324 | 315 | 312 | 0,724146 | 0,93314 | y ~ current_feature + S |
| -1,16E-04 | 0,060746 | 0,061498 | -0,00192 | 274 | 271 | 0,998473 | 0,999005 | y ~ current_feature + S |
| -0,1242 | 0,068967 | -0,02429 | -1,80087 | 210 | 207 | 0,073179 | 0,605474 | y ~ current_feature + S |
| 0,062437 | 0,053842 | -0,07874 | 1,159637 | 395 | 343,6081 | 0,247002 | 0,67447 | y ~ current_feature + S |
| 0,007009 | 0,070176 | 0,034007 | 0,099881 | 395 | 203,0477 | 0,920538 | 0,9732 | y ~ current_feature + S |
| 0,005018 | 0,0551 | -0,04751 | 0,091075 | 394 | 329,3676 | 0,927489 | 0,985217 | y ~ current_feature + S |
| -0,10779 | 0,05719 | 0,061498 | -1,8847 | 343 | 302,1894 | 0,06043 | 0,715249 | y ~ current_feature + S |
| -0,05378 | 0,062826 | -0,02429 | -0,85608 | 263 | 252,6157 | 0,392766 | 0,821036 | y ~ current_feature + S |
| 0,079708 | 0,056763 | -0,07874 | 1,404232 | 345 | 308,391 | 0,161256 | 0,532425 | y ~ current_feature + S |
| 0,038018 | 0,073224 | 0,034007 | 0,519206 | 345 | 186,2386 | 0,604234 | 0,883662 | y ~ current_feature + S |
| 0,044556 | 0,058823 | -0,04751 | 0,757449 | 344 | 288,4283 | 0,449399 | 0,831388 | y ~ current_feature + S |
| -0,03317 | 0,062266 | 0,061498 | -0,53274 | 299 | 257,6476 | 0,594672 | 0,945257 | y ~ current_feature + S |
| 0,02018 | 0,066746 | -0,02429 | 0,302347 | 230 | 224,377 | 0,762667 | 0,951727 | y ~ current_feature + S |
| 0,08506 | 0,050958 | -0,07874 | 1,669238 | 415 | 382,3224 | 0,095889 | 0,431938 | y ~ current_feature + S |

| | | | | | | | | |
|---|---|---|---|---|---|---|---|---|
| 0,070469 | 0,066977 | 0,034007 | 1,05214 | 415 | 221,8114 | 0,29388 | 0,718742 | y ~ current_feature + S |
| 0,031254 | 0,052435 | -0,04751 | 0,596061 | 414 | 363,3627 | 0,551506 | 0,897386 | y ~ current_feature + S |
| 0,034953 | 0,055933 | 0,061498 | 0,624901 | 361 | 319,2487 | 0,532482 | 0,937665 | y ~ current_feature + S |
| 0,090135 | 0,061486 | -0,02429 | 1,465941 | 276 | 262,3633 | 0,143862 | 0,618941 | y ~ current_feature + S |
| -0,03723 | 0,055603 | -0,07874 | -0,66959 | 326 | 323 | 0,503594 | 0,863164 | y ~ current_feature + S |
| -0,00396 | 0,055641 | 0,034007 | -0,07114 | 326 | 323 | 0,943334 | 0,98181 | y ~ current_feature + S |
| 0,02529 | 0,05571 | -0,04751 | 0,453955 | 325 | 322 | 0,650167 | 0,925756 | y ~ current_feature + S |
| 0,001741 | 0,064886 | 0,061498 | 0,026839 | 280 | 237,5155 | 0,978611 | 0,996179 | y ~ current_feature + S |
| 0,068369 | 0,068996 | -0,02429 | 0,990911 | 217 | 209,082 | 0,322874 | 0,773227 | y ~ current_feature + S |
| 0,068981 | 0,087162 | -0,07874 | 0,791413 | 134 | 131 | 0,430134 | 0,814177 | y ~ current_feature + S |
| 0,031571 | 0,087327 | 0,034007 | 0,361524 | 134 | 131 | 0,71829 | 0,920814 | y ~ current_feature + S |
| 0,01026 | 0,087366 | -0,04751 | 0,117434 | 134 | 131 | 0,906696 | 0,985217 | y ~ current_feature + S |
| 0,066956 | 0,095132 | 0,061498 | 0,703816 | 113 | 110 | 0,483034 | 0,937665 | y ~ current_feature + S |
| 0,018342 | 0,107815 | -0,02429 | 0,170123 | 89 | 86 | 0,865313 | 0,984468 | y ~ current_feature + S |
| -0,00671 | 0,054078 | -0,07874 | -0,12416 | 389 | 341,9334 | 0,901259 | 0,987929 | y ~ current_feature + S |
| -0,04646 | 0,071316 | 0,034007 | -0,65153 | 389 | 196,195 | 0,515464 | 0,841996 | y ~ current_feature + S |
| -0,06358 | 0,055999 | -0,04751 | -1,1354 | 388 | 317,6031 | 0,257062 | 0,742928 | y ~ current_feature + S |
| 0,020866 | 0,059697 | 0,061498 | 0,349539 | 337 | 280,4812 | 0,726947 | 0,972573 | y ~ current_feature + S |
| -0,0046 | 0,062499 | -0,02429 | -0,07363 | 259 | 256 | 0,941359 | 0,99444 | y ~ current_feature + S |
| 0,112207 | 0,140528 | -0,07874 | 0,798466 | 53 | 50 | 0,428376 | 0,814177 | y ~ current_feature + S |
| -0,01226 | 0,141411 | 0,034007 | -0,08671 | 53 | 50 | 0,93125 | 0,974557 | y ~ current_feature + S |
| 0,042626 | 0,141293 | -0,04751 | 0,301683 | 53 | 50 | 0,764145 | 0,940878 | y ~ current_feature + S |
| -0,09716 | 0,150042 | 0,061498 | -0,64753 | 47 | 44 | 0,520652 | 0,937665 | y ~ current_feature + S |
| 0,201062 | 0,173167 | -0,02429 | 1,161087 | 35 | 32 | 0,254195 | 0,718089 | y ~ current_feature + S |
| 0,308431 | 0,129448 | -0,07874 | 2,382658 | 57 | 54 | 0,020738 | 0,189455 | y ~ current_feature + S |
| 0,108524 | 0,135279 | 0,034007 | 0,802224 | 57 | 54 | 0,42594 | 0,791701 | y ~ current_feature + S |
| 0,073213 | 0,135718 | -0,04751 | 0,539451 | 57 | 54 | 0,591793 | 0,91049 | y ~ current_feature + S |
| 0,110412 | 0,149834 | 0,061498 | 0,736898 | 47 | 44 | 0,465095 | 0,937665 | y ~ current_feature + S |
| -0,00976 | 0,169023 | -0,02429 | -0,05777 | 38 | 35 | 0,954262 | 0,99444 | y ~ current_feature + S |
| 0,051913 | 0,073622 | -0,07874 | 0,705132 | 187 | 184 | 0,48162 | 0,845512 | y ~ current_feature + S |
| 0,013534 | 0,073714 | 0,034007 | 0,183599 | 187 | 184 | 0,85453 | 0,959563 | y ~ current_feature + S |
| 0,085724 | 0,07345 | -0,04751 | 1,167108 | 187 | 184 | 0,244677 | 0,742928 | y ~ current_feature + S |
| 0,025755 | 0,077589 | 0,061498 | 0,331938 | 169 | 166 | 0,740355 | 0,972573 | y ~ current_feature + S |
| -0,03694 | 0,090847 | -0,02429 | -0,40663 | 124 | 121 | 0,684999 | 0,926342 | y ~ current_feature + S |
| 0,121077 | 0,081872 | -0,07874 | 1,478859 | 150 | 147 | 0,141318 | 0,498495 | y ~ current_feature + S |
| 0,094939 | 0,082106 | 0,034007 | 1,156303 | 150 | 147 | 0,249433 | 0,702074 | y ~ current_feature + S |
| 0,074048 | 0,082252 | -0,04751 | 0,900258 | 150 | 147 | 0,369455 | 0,802018 | y ~ current_feature + S |
| -0,08259 | 0,089496 | 0,061498 | -0,92278 | 127 | 124 | 0,357913 | 0,920619 | y ~ current_feature + S |
| -0,02676 | 0,101498 | -0,02429 | -0,26367 | 100 | 97 | 0,79259 | 0,967756 | y ~ current_feature + S |
| 0,148652 | 0,050719 | -0,07874 | 2,930921 | 407 | 380,1567 | 0,003584 | 0,069632 | y ~ current_feature + S |
| 0,144226 | 0,065615 | 0,034007 | 2,198067 | 407 | 227,4383 | 0,028953 | 0,347616 | y ~ current_feature + S |
| 0,050538 | 0,051724 | -0,04751 | 0,977074 | 406 | 372,8218 | 0,329166 | 0,779476 | y ~ current_feature + S |
| 0,06752 | 0,05532 | 0,061498 | 1,220536 | 353 | 325,2761 | 0,223146 | 0,893818 | y ~ current_feature + S |
| 0,084382 | 0,061537 | -0,02429 | 1,371244 | 271 | 262,1944 | 0,171471 | 0,626332 | y ~ current_feature + S |
| 0,143473 | 0,092286 | -0,07874 | 1,554659 | 118 | 115 | 0,122775 | 0,477837 | y ~ current_feature + S |
| 0,126504 | 0,092501 | 0,034007 | 1,367592 | 118 | 115 | 0,174107 | 0,619418 | y ~ current_feature + S |
| 0,201976 | 0,091728 | -0,04751 | 2,201889 | 117 | 114 | 0,029687 | 0,603897 | y ~ current_feature + S |
| -0,05011 | 0,100888 | 0,061498 | -0,49673 | 101 | 98 | 0,620493 | 0,950526 | y ~ current_feature + S |

| | | | | | | | | |
|---|---|---|---|---|---|---|---|---|
| 0,148794 | 0,113431 | -0,02429 | 1,311755 | 79 | 76 | 0,193552 | 0,65401 | y ~ current_feature + S |
| -0,17838 | 0,052226 | -0,07874 | -3,41561 | 386 | 354,959 | 7,10E-04 | 0,031617 | y ~ current_feature + S |
| -0,17305 | 0,069401 | 0,034007 | -2,49343 | 386 | 201,4 | 0,013457 | 0,255339 | y ~ current_feature + S |
| -0,0347 | 0,054448 | -0,04751 | -0,63736 | 385 | 336,9115 | 0,524323 | 0,876197 | y ~ current_feature + S |
| 0,101585 | 0,058034 | 0,061498 | 1,750419 | 339 | 293,8486 | 0,08109 | 0,750079 | y ~ current_feature + S |
| -0,12799 | 0,062905 | -0,02429 | -2,03465 | 256 | 248,577 | 0,042948 | 0,525585 | y ~ current_feature + S |
| 0,168316 | 0,052074 | -0,07874 | 3,232285 | 415 | 358,3312 | 0,001342 | 0,048726 | y ~ current_feature + S |
| 0,179864 | 0,069009 | 0,034007 | 2,606391 | 415 | 203,1926 | 0,009828 | 0,227273 | y ~ current_feature + S |
| 0,129207 | 0,053193 | -0,04751 | 2,429011 | 414 | 347,5166 | 0,015647 | 0,526293 | y ~ current_feature + S |
| -0,06553 | 0,057818 | 0,061498 | -1,13345 | 361 | 297,8542 | 0,257935 | 0,893818 | y ~ current_feature + S |
| 0,116471 | 0,060111 | -0,02429 | 1,937603 | 276 | 273 | 0,053703 | 0,558147 | y ~ current_feature + S |
| 0,102415 | 0,050262 | -0,07874 | 2,037623 | 421 | 391,6883 | 0,042259 | 0,275611 | y ~ current_feature + S |
| 0,097914 | 0,066433 | 0,034007 | 1,473874 | 421 | 224,4139 | 0,141917 | 0,588814 | y ~ current_feature + S |
| 0,064531 | 0,051643 | -0,04751 | 1,249567 | 420 | 373,398 | 0,21224 | 0,733915 | y ~ current_feature + S |
| 0,001462 | 0,056583 | 0,061498 | 0,025834 | 367 | 312,3406 | 0,979406 | 0,996179 | y ~ current_feature + S |
| 0,034623 | 0,060459 | -0,02429 | 0,572668 | 280 | 273,2523 | 0,567341 | 0,924682 | y ~ current_feature + S |
| 3,81E-04 | 0,08737 | -0,07874 | 0,004361 | 134 | 131 | 0,996527 | 0,996647 | y ~ current_feature + S |
| -0,01886 | 0,087355 | 0,034007 | -0,2159 | 134 | 131 | 0,829402 | 0,956008 | y ~ current_feature + S |
| 0,057449 | 0,087226 | -0,04751 | 0,658627 | 134 | 131 | 0,511291 | 0,868909 | y ~ current_feature + S |
| -0,07999 | 0,092154 | 0,061498 | -0,868 | 120 | 117 | 0,38717 | 0,93433 | y ~ current_feature + S |
| 0,009388 | 0,107828 | -0,02429 | 0,087066 | 89 | 86 | 0,930821 | 0,99444 | y ~ current_feature + S |
| -0,05596 | 0,054068 | -0,07874 | -1,035 | 344 | 341 | 0,301404 | 0,718687 | y ~ current_feature + S |
| -0,0599 | 0,054056 | 0,034007 | -1,10806 | 344 | 341 | 0,268619 | 0,71095 | y ~ current_feature + S |
| -0,03206 | 0,054125 | -0,04751 | -0,59236 | 344 | 341 | 0,554001 | 0,897386 | y ~ current_feature + S |
| -0,0386 | 0,058179 | 0,061498 | -0,66349 | 298 | 295 | 0,507533 | 0,937665 | y ~ current_feature + S |
| -0,02694 | 0,066495 | -0,02429 | -0,40514 | 229 | 226 | 0,685759 | 0,926342 | y ~ current_feature + S |
| 0,038744 | 0,071573 | -0,07874 | 0,541324 | 254 | 194,9147 | 0,588902 | 0,902886 | y ~ current_feature + S |
| 0,114213 | 0,086565 | 0,034007 | 1,319394 | 254 | 131,7088 | 0,189327 | 0,639734 | y ~ current_feature + S |
| 0,026735 | 0,067706 | -0,04751 | 0,394863 | 253 | 217,9896 | 0,69333 | 0,930326 | y ~ current_feature + S |
| -0,10793 | 0,076384 | 0,061498 | -1,413 | 218 | 169,3979 | 0,159491 | 0,8197 | y ~ current_feature + S |
| 0,148684 | 0,076989 | -0,02429 | 1,931235 | 169 | 164,9808 | 0,055168 | 0,558147 | y ~ current_feature + S |
| 0,052896 | 0,122919 | -0,07874 | 0,430329 | 69 | 66 | 0,668358 | 0,934653 | y ~ current_feature + S |
| 0,011104 | 0,123084 | 0,034007 | 0,090211 | 69 | 66 | 0,928393 | 0,9732 | y ~ current_feature + S |
| 0,163028 | 0,121445 | -0,04751 | 1,342405 | 69 | 66 | 0,184064 | 0,704811 | y ~ current_feature + S |
| -0,14393 | 0,128833 | 0,061498 | -1,1172 | 62 | 59 | 0,268437 | 0,893818 | y ~ current_feature + S |
| 0,156231 | 0,150626 | -0,02429 | 1,037214 | 46 | 43 | 0,305434 | 0,761472 | y ~ current_feature + S |
| -0,12614 | 0,057562 | -0,07874 | -2,1914 | 300 | 297 | 0,029199 | 0,219281 | y ~ current_feature + S |
| -0,15303 | 0,057342 | 0,034007 | -2,66867 | 300 | 297 | 0,008034 | 0,212321 | y ~ current_feature + S |
| -0,02032 | 0,058112 | -0,04751 | -0,3497 | 299 | 296 | 0,72681 | 0,93314 | y ~ current_feature + S |
| 0,048963 | 0,068475 | 0,061498 | 0,715049 | 264 | 212,7608 | 0,475363 | 0,937665 | y ~ current_feature + S |
| -0,12365 | 0,07088 | -0,02429 | -1,74456 | 199 | 196 | 0,082629 | 0,605474 | y ~ current_feature + S |
| 0,038433 | 0,086323 | -0,07874 | 0,445224 | 137 | 134 | 0,656876 | 0,932255 | y ~ current_feature + S |
| 0,029735 | 0,086349 | 0,034007 | 0,344364 | 137 | 134 | 0,731113 | 0,922572 | y ~ current_feature + S |
| -0,03882 | 0,086322 | -0,04751 | -0,44975 | 137 | 134 | 0,653619 | 0,925756 | y ~ current_feature + S |
| -0,14621 | 0,092248 | 0,061498 | -1,585 | 118 | 115 | 0,115713 | 0,8197 | y ~ current_feature + S |
| -0,14732 | 0,104843 | -0,02429 | -1,40515 | 92 | 89 | 0,163457 | 0,623497 | y ~ current_feature + S |
| 0,09159 | 0,06938 | -0,07874 | 1,32011 | 209 | 206 | 0,188263 | 0,592937 | y ~ current_feature + S |
| 0,183991 | 0,068484 | 0,034007 | 2,68663 | 209 | 206 | 0,007807 | 0,212321 | y ~ current_feature + S |

| | | | | | | | | |
|---|---|---|---|---|---|---|---|---|
| 0,12709 | 0,069108 | -0,04751 | 1,839001 | 209 | 206 | 0,067355 | 0,640171 | y ~ current_feature + S |
| 0,004972 | 0,075164 | 0,061498 | 0,066152 | 180 | 177 | 0,947332 | 0,996179 | y ~ current_feature + S |
| 0,059583 | 0,085597 | -0,02429 | 0,696083 | 139 | 136 | 0,487565 | 0,888665 | y ~ current_feature + S |
| 0,165158 | 0,101189 | -0,07874 | 1,632173 | 98 | 95 | 0,105954 | 0,449749 | y ~ current_feature + S |
| 0,02945 | 0,102553 | 0,034007 | 0,287165 | 98 | 95 | 0,774612 | 0,942718 | y ~ current_feature + S |
| 0,152936 | 0,101929 | -0,04751 | 1,500422 | 97 | 94 | 0,136857 | 0,657012 | y ~ current_feature + S |
| 0,202857 | 0,105591 | 0,061498 | 1,921163 | 89 | 86 | 0,058024 | 0,715249 | y ~ current_feature + S |
| 0,1011 | 0,126349 | -0,02429 | 0,800161 | 65 | 62 | 0,426673 | 0,847264 | y ~ current_feature + S |
| -0,00865 | 0,062135 | -0,07874 | -0,13926 | 262 | 259 | 0,889354 | 0,987929 | y ~ current_feature + S |
| 0,011714 | 0,062133 | 0,034007 | 0,188526 | 262 | 259 | 0,850612 | 0,959445 | y ~ current_feature + S |
| 0,049264 | 0,062182 | -0,04751 | 0,792263 | 261 | 258 | 0,428935 | 0,819126 | y ~ current_feature + S |
| -0,04463 | 0,0672 | 0,061498 | -0,66412 | 224 | 221 | 0,507308 | 0,937665 | y ~ current_feature + S |
| 0,015137 | 0,076463 | -0,02429 | 0,197965 | 174 | 171 | 0,843308 | 0,977734 | y ~ current_feature + S |
| -0,03945 | 0,139919 | -0,07874 | -0,28196 | 54 | 51 | 0,779118 | 0,972255 | y ~ current_feature + S |
| 0,093534 | 0,139414 | 0,034007 | 0,670905 | 54 | 51 | 0,505308 | 0,841996 | y ~ current_feature + S |
| 0,26073 | 0,135185 | -0,04751 | 1,928698 | 54 | 51 | 0,059343 | 0,640171 | y ~ current_feature + S |
| -0,09647 | 0,159381 | 0,061498 | -0,60529 | 42 | 39 | 0,548492 | 0,937665 | y ~ current_feature + S |
| 0,163978 | 0,171721 | -0,02429 | 0,954906 | 36 | 33 | 0,346568 | 0,790842 | y ~ current_feature + S |
| 0,104163 | 0,05881 | -0,07874 | 1,771191 | 289 | 286 | 0,077594 | 0,370448 | y ~ current_feature + S |
| -0,01561 | 0,059124 | 0,034007 | -0,264 | 289 | 286 | 0,791968 | 0,945731 | y ~ current_feature + S |
| 0,085619 | 0,059017 | -0,04751 | 1,450735 | 288 | 285 | 0,147953 | 0,661823 | y ~ current_feature + S |
| 9,26E-04 | 0,0635 | 0,061498 | 0,014583 | 251 | 248 | 0,988376 | 0,997945 | y ~ current_feature + S |
| 0,08172 | 0,072496 | -0,02429 | 1,127237 | 192 | 189 | 0,261071 | 0,723568 | y ~ current_feature + S |
| 0,036778 | 0,057792 | -0,07874 | 0,636378 | 302 | 299 | 0,525017 | 0,876542 | y ~ current_feature + S |
| -0,02402 | 0,057815 | 0,034007 | -0,41554 | 302 | 299 | 0,678044 | 0,912427 | y ~ current_feature + S |
| -0,08762 | 0,057706 | -0,04751 | -1,51838 | 301 | 298 | 0,129979 | 0,654671 | y ~ current_feature + S |
| -0,05088 | 0,061818 | 0,061498 | -0,82312 | 264 | 261 | 0,41119 | 0,934833 | y ~ current_feature + S |
| 0,018595 | 0,071055 | -0,02429 | 0,261705 | 201 | 198 | 0,793821 | 0,967756 | y ~ current_feature + S |
| 0,177942 | 0,127039 | -0,07874 | 1,400689 | 63 | 60 | 0,166458 | 0,540258 | y ~ current_feature + S |
| 0,141891 | 0,127793 | 0,034007 | 1,110314 | 63 | 60 | 0,271294 | 0,71095 | y ~ current_feature + S |
| -0,06479 | 0,128828 | -0,04751 | -0,50295 | 63 | 60 | 0,616838 | 0,924009 | y ~ current_feature + S |
| 0,132089 | 0,138801 | 0,061498 | 0,951642 | 54 | 51 | 0,345767 | 0,907332 | y ~ current_feature + S |
| 0,253588 | 0,154894 | -0,02429 | 1,637171 | 42 | 39 | 0,10964 | 0,605474 | y ~ current_feature + S |
| 0,075853 | 0,139625 | -0,07874 | 0,543263 | 54 | 51 | 0,589316 | 0,902886 | y ~ current_feature + S |
| 0,334977 | 0,131938 | 0,034007 | 2,538896 | 54 | 51 | 0,014214 | 0,262237 | y ~ current_feature + S |
| 0,17696 | 0,137818 | -0,04751 | 1,284008 | 54 | 51 | 0,204942 | 0,723562 | y ~ current_feature + S |
| -0,16808 | 0,145344 | 0,061498 | -1,15646 | 49 | 46 | 0,253464 | 0,893818 | y ~ current_feature + S |
| 0,52122 | 0,148562 | -0,02429 | 3,508444 | 36 | 33 | 0,001325 | 0,144895 | y ~ current_feature + S |
| 0,111263 | 0,121411 | -0,07874 | 0,916415 | 70 | 67 | 0,362736 | 0,773558 | y ~ current_feature + S |
| -0,02464 | 0,122132 | 0,034007 | -0,20174 | 70 | 67 | 0,840733 | 0,958985 | y ~ current_feature + S |
| 0,091361 | 0,121659 | -0,04751 | 0,750962 | 70 | 67 | 0,455305 | 0,832044 | y ~ current_feature + S |
| -0,01623 | 0,130172 | 0,061498 | -0,12472 | 62 | 59 | 0,90117 | 0,996179 | y ~ current_feature + S |
| -0,15573 | 0,150638 | -0,02429 | -1,03381 | 46 | 43 | 0,307005 | 0,761472 | y ~ current_feature + S |
| 0,113232 | 0,100366 | -0,07874 | 1,128193 | 101 | 98 | 0,261993 | 0,692409 | y ~ current_feature + S |
| 0,046468 | 0,100906 | 0,034007 | 0,46051 | 101 | 98 | 0,646171 | 0,900251 | y ~ current_feature + S |
| 0,117205 | 0,100835 | -0,04751 | 1,162343 | 100 | 97 | 0,247948 | 0,742928 | y ~ current_feature + S |
| -0,14557 | 0,103712 | 0,061498 | -1,40356 | 94 | 91 | 0,163854 | 0,8197 | y ~ current_feature + S |
| 0,280522 | 0,119981 | -0,02429 | 2,33805 | 67 | 64 | 0,022522 | 0,467656 | y ~ current_feature + S |

| | | | | | | | | |
|---|---|---|---|---|---|---|---|---|
| -0,0258 | 0,058676 | -0,07874 | -0,43966 | 338 | 290,2662 | 0,660511 | 0,934566 | y ~ current_feature + S |
| -0,01611 | 0,077711 | 0,034007 | -0,20731 | 338 | 165,5476 | 0,836022 | 0,958985 | y ~ current_feature + S |
| 0,115823 | 0,060636 | -0,04751 | 1,910131 | 338 | 268,3332 | 0,057182 | 0,640171 | y ~ current_feature + S |
| 0,063626 | 0,064099 | 0,061498 | 0,99261 | 292 | 242,3988 | 0,32189 | 0,893818 | y ~ current_feature + S |
| 0,039927 | 0,067783 | -0,02429 | 0,589034 | 225 | 217,3023 | 0,55645 | 0,92024 | y ~ current_feature + S |
| 0,043596 | 0,088304 | -0,07874 | 0,4937 | 131 | 128 | 0,622364 | 0,919937 | y ~ current_feature + S |
| 0,041808 | 0,088311 | 0,034007 | 0,473418 | 131 | 128 | 0,636722 | 0,899624 | y ~ current_feature + S |
| 0,019177 | 0,088719 | -0,04751 | 0,216153 | 130 | 127 | 0,829215 | 0,975438 | y ~ current_feature + S |
| -0,1331 | 0,09365 | 0,061498 | -1,42119 | 115 | 112 | 0,158039 | 0,8197 | y ~ current_feature + S |
| 0,061473 | 0,108903 | -0,02429 | 0,564474 | 87 | 84 | 0,573936 | 0,924682 | y ~ current_feature + S |
| 0,169539 | 0,062963 | -0,07874 | 2,692679 | 248 | 245 | 0,007577 | 0,110197 | y ~ current_feature + S |
| 0,166112 | 0,063 | 0,034007 | 2,636697 | 248 | 245 | 0,008907 | 0,215592 | y ~ current_feature + S |
| 0,054522 | 0,063793 | -0,04751 | 0,85467 | 248 | 245 | 0,393569 | 0,803665 | y ~ current_feature + S |
| 0,045859 | 0,068608 | 0,061498 | 0,668418 | 215 | 212 | 0,504594 | 0,937665 | y ~ current_feature + S |
| 0,123269 | 0,077729 | -0,02429 | 1,585884 | 166 | 163 | 0,114704 | 0,614731 | y ~ current_feature + S |
| 0,24162 | 0,149732 | -0,07874 | 1,613686 | 45 | 42 | 0,114085 | 0,467692 | y ~ current_feature + S |
| -0,01316 | 0,15429 | 0,034007 | -0,08532 | 45 | 42 | 0,932414 | 0,974557 | y ~ current_feature + S |
| 0,171602 | 0,152014 | -0,04751 | 1,128852 | 45 | 42 | 0,265367 | 0,747915 | y ~ current_feature + S |
| -0,32114 | 0,151647 | 0,061498 | -2,11767 | 42 | 39 | 0,040632 | 0,658491 | y ~ current_feature + S |
| 0,230524 | 0,187267 | -0,02429 | 1,230991 | 30 | 27 | 0,228942 | 0,695919 | y ~ current_feature + S |
| 0,429982 | 0,119584 | -0,07874 | 3,59566 | 60 | 57 | 6,77E-04 | 0,031617 | y ~ current_feature + S |
| 0,267713 | 0,127619 | 0,034007 | 2,097757 | 60 | 57 | 0,040372 | 0,396441 | y ~ current_feature + S |
| 0,003496 | 0,132452 | -0,04751 | 0,026396 | 60 | 57 | 0,979034 | 0,992311 | y ~ current_feature + S |
| 0,115028 | 0,140483 | 0,061498 | 0,818805 | 53 | 50 | 0,416783 | 0,937665 | y ~ current_feature + S |
| 0,064929 | 0,164052 | -0,02429 | 0,395782 | 40 | 37 | 0,694539 | 0,928819 | y ~ current_feature + S |
| -0,14507 | 0,139925 | -0,07874 | -1,03676 | 53 | 50 | 0,304835 | 0,718687 | y ~ current_feature + S |
| -0,14213 | 0,139986 | 0,034007 | -1,01531 | 53 | 50 | 0,314847 | 0,718742 | y ~ current_feature + S |
| 0,020743 | 0,141391 | -0,04751 | 0,146708 | 53 | 50 | 0,883952 | 0,985217 | y ~ current_feature + S |
| -0,10324 | 0,148275 | 0,061498 | -0,69627 | 48 | 45 | 0,489843 | 0,937665 | y ~ current_feature + S |
| 0,085141 | 0,173446 | -0,02429 | 0,49088 | 36 | 33 | 0,62676 | 0,924682 | y ~ current_feature + S |
| -0,00735 | 0,120383 | -0,07874 | -0,06107 | 72 | 69 | 0,95148 | 0,994607 | y ~ current_feature + S |
| -0,06306 | 0,166472 | 0,034007 | -0,37877 | 72 | 35,94068 | 0,707085 | 0,920814 | y ~ current_feature + S |
| 0,104851 | 0,13437 | -0,04751 | 0,780314 | 72 | 54,7762 | 0,438565 | 0,823749 | y ~ current_feature + S |
| -0,23367 | 0,16073 | 0,061498 | -1,45382 | 60 | 36,59491 | 0,154519 | 0,8197 | y ~ current_feature + S |
| -0,0727 | 0,148677 | -0,02429 | -0,48899 | 48 | 45 | 0,627225 | 0,924682 | y ~ current_feature + S |
| -0,01028 | 0,090531 | -0,07874 | -0,11355 | 125 | 122 | 0,909778 | 0,987929 | y ~ current_feature + S |
| -0,12972 | 0,089771 | 0,034007 | -1,44499 | 125 | 122 | 0,151023 | 0,604094 | y ~ current_feature + S |
| -0,04039 | 0,090462 | -0,04751 | -0,44647 | 125 | 122 | 0,656051 | 0,925756 | y ~ current_feature + S |
| 0,029225 | 0,097548 | 0,061498 | 0,2996 | 108 | 105 | 0,765075 | 0,972573 | y ~ current_feature + S |
| -0,12875 | 0,110873 | -0,02429 | -1,16127 | 83 | 80 | 0,248987 | 0,717451 | y ~ current_feature + S |
| 0,103356 | 0,09571 | -0,07874 | 1,079893 | 111 | 108 | 0,282596 | 0,718628 | y ~ current_feature + S |
| 0,025647 | 0,096193 | 0,034007 | 0,266619 | 111 | 108 | 0,790271 | 0,945731 | y ~ current_feature + S |
| 0,09944 | 0,095748 | -0,04751 | 1,038563 | 111 | 108 | 0,301328 | 0,754967 | y ~ current_feature + S |
| -0,00858 | 0,104253 | 0,061498 | -0,08226 | 95 | 92 | 0,934619 | 0,996179 | y ~ current_feature + S |
| 0,034051 | 0,118609 | -0,02429 | 0,287087 | 74 | 71 | 0,774882 | 0,958884 | y ~ current_feature + S |
| 0,01193 | 0,061658 | -0,07874 | 0,193479 | 266 | 263 | 0,846734 | 0,983548 | y ~ current_feature + S |
| 0,013033 | 0,061657 | 0,034007 | 0,211372 | 266 | 263 | 0,832761 | 0,958387 | y ~ current_feature + S |
| 0,098105 | 0,061365 | -0,04751 | 1,598714 | 266 | 263 | 0,111085 | 0,640171 | y ~ current_feature + S |

| | | | | | | | | |
|---|---|---|---|---|---|---|---|---|
| -0,08049 | 0,066012 | 0,061498 | -1,21937 | 231 | 228 | 0,223964 | 0,893818 | y ~ current_feature + S |
| 0,020145 | 0,075578 | -0,02429 | 0,266541 | 178 | 175 | 0,790136 | 0,967756 | y ~ current_feature + S |
| 0,17527 | 0,124038 | -0,07874 | 1,413038 | 66 | 63 | 0,162566 | 0,532425 | y ~ current_feature + S |
| 0,223084 | 0,122813 | 0,034007 | 1,816453 | 66 | 63 | 0,074059 | 0,464437 | y ~ current_feature + S |
| 0,1358 | 0,124821 | -0,04751 | 1,087959 | 66 | 63 | 0,280758 | 0,747915 | y ~ current_feature + S |
| 0,226338 | 0,133796 | 0,061498 | 1,691665 | 56 | 53 | 0,096582 | 0,8197 | y ~ current_feature + S |
| 0,073976 | 0,156674 | -0,02429 | 0,472169 | 44 | 40,51573 | 0,63934 | 0,924682 | y ~ current_feature + S |
| -0,01063 | 0,05634 | -0,07874 | -0,18868 | 318 | 315 | 0,850466 | 0,983548 | y ~ current_feature + S |
| -0,15849 | 0,055631 | 0,034007 | -2,849 | 318 | 315 | 0,004674 | 0,167881 | y ~ current_feature + S |
| -0,0103 | 0,05643 | -0,04751 | -0,18256 | 317 | 314 | 0,85526 | 0,983239 | y ~ current_feature + S |
| -0,10794 | 0,060059 | 0,061498 | -1,79723 | 277 | 274 | 0,073401 | 0,721272 | y ~ current_feature + S |
| -0,02525 | 0,069315 | -0,02429 | -0,36429 | 211 | 208 | 0,716014 | 0,936058 | y ~ current_feature + S |
| 0,115492 | 0,118723 | -0,07874 | 0,972789 | 73 | 70 | 0,334008 | 0,735613 | y ~ current_feature + S |
| -0,21864 | 0,116631 | 0,034007 | -1,87461 | 73 | 70 | 0,065018 | 0,439525 | y ~ current_feature + S |
| -0,16336 | 0,117917 | -0,04751 | -1,38536 | 73 | 70 | 0,170341 | 0,700235 | y ~ current_feature + S |
| -0,19786 | 0,130989 | 0,061498 | -1,51049 | 59 | 56 | 0,13654 | 0,8197 | y ~ current_feature + S |
| -0,13148 | 0,147777 | -0,02429 | -0,88968 | 48 | 45 | 0,378367 | 0,815551 | y ~ current_feature + S |
| -0,06404 | 0,150446 | -0,07874 | -0,42566 | 47 | 44 | 0,672434 | 0,934653 | y ~ current_feature + S |
| -0,10188 | 0,149971 | 0,034007 | -0,67931 | 47 | 44 | 0,500497 | 0,841996 | y ~ current_feature + S |
| -0,20127 | 0,14767 | -0,04751 | -1,363 | 47 | 44 | 0,179818 | 0,704707 | y ~ current_feature + S |
| -0,14073 | 0,160607 | 0,061498 | -0,87621 | 41 | 38 | 0,386419 | 0,93433 | y ~ current_feature + S |
| -0,12593 | 0,184217 | -0,02429 | -0,68359 | 32 | 29 | 0,499659 | 0,89399 | y ~ current_feature + S |
| 0,091063 | 0,051204 | -0,07874 | 1,778426 | 421 | 378,2431 | 0,076137 | 0,368245 | y ~ current_feature + S |
| 0,056587 | 0,067106 | 0,034007 | 0,843243 | 421 | 221,3521 | 0,400003 | 0,776908 | y ~ current_feature + S |
| 0,085494 | 0,052587 | -0,04751 | 1,625764 | 420 | 358,9723 | 0,104878 | 0,640171 | y ~ current_feature + S |
| -0,02195 | 0,057207 | 0,061498 | -0,38373 | 367 | 305,4143 | 0,701442 | 0,972444 | y ~ current_feature + S |
| 0,168631 | 0,059244 | -0,02429 | 2,846383 | 280 | 276,8107 | 0,004753 | 0,219812 | y ~ current_feature + S |
| -0,01908 | 0,068994 | -0,07874 | -0,27655 | 213 | 210 | 0,7824 | 0,973069 | y ~ current_feature + S |
| 0,069492 | 0,06884 | 0,034007 | 1,009482 | 213 | 210 | 0,313905 | 0,718742 | y ~ current_feature + S |
| 0,078842 | 0,068792 | -0,04751 | 1,146094 | 213 | 210 | 0,253061 | 0,742928 | y ~ current_feature + S |
| -0,00478 | 0,073323 | 0,061498 | -0,06517 | 189 | 186 | 0,94811 | 0,996179 | y ~ current_feature + S |
| 0,053825 | 0,084696 | -0,02429 | 0,635511 | 142 | 139 | 0,52614 | 0,899808 | y ~ current_feature + S |
| 0,090666 | 0,050667 | -0,07874 | 1,789447 | 412 | 386,337 | 0,074326 | 0,364247 | y ~ current_feature + S |
| 0,027435 | 0,064984 | 0,034007 | 0,422186 | 412 | 236,6263 | 0,673273 | 0,910826 | y ~ current_feature + S |
| 0,062805 | 0,052169 | -0,04751 | 1,203868 | 411 | 365,9744 | 0,229419 | 0,742928 | y ~ current_feature + S |
| -0,01292 | 0,056354 | 0,061498 | -0,22922 | 359 | 314,8288 | 0,818843 | 0,996179 | y ~ current_feature + S |
| 0,056968 | 0,061781 | -0,02429 | 0,922097 | 274 | 261,1396 | 0,357329 | 0,800662 | y ~ current_feature + S |
| -0,06556 | 0,076307 | -0,07874 | -0,85913 | 174 | 171 | 0,391472 | 0,789252 | y ~ current_feature + S |
| -0,219 | 0,074616 | 0,034007 | -2,93506 | 174 | 171 | 0,003793 | 0,15595 | y ~ current_feature + S |
| -0,0585 | 0,076341 | -0,04751 | -0,76629 | 174 | 171 | 0,444557 | 0,824847 | y ~ current_feature + S |
| -0,22392 | 0,080384 | 0,061498 | -2,78565 | 150 | 147 | 0,006046 | 0,406759 | y ~ current_feature + S |
| -0,1862 | 0,092427 | -0,02429 | -2,01454 | 116 | 113 | 0,046328 | 0,525585 | y ~ current_feature + S |
| 0,097674 | 0,063453 | -0,07874 | 1,539319 | 249 | 246 | 0,125012 | 0,478063 | y ~ current_feature + S |
| 0,068838 | 0,063606 | 0,034007 | 1,082245 | 249 | 246 | 0,280203 | 0,715049 | y ~ current_feature + S |
| 0,087362 | 0,063643 | -0,04751 | 1,372685 | 248 | 245 | 0,171105 | 0,700235 | y ~ current_feature + S |
| -0,11704 | 0,080235 | 0,061498 | -1,45869 | 212 | 153,2085 | 0,146698 | 0,8197 | y ~ current_feature + S |
| 0,016229 | 0,078316 | -0,02429 | 0,207222 | 166 | 163 | 0,836095 | 0,977734 | y ~ current_feature + S |
| 0,173713 | 0,11448 | -0,07874 | 1,517407 | 77 | 74 | 0,133425 | 0,493672 | y ~ current_feature + S |

| | | | | | | | | |
|---|---|---|---|---|---|---|---|---|
| 0,200416 | 0,113889 | 0,034007 | 1,759751 | 77 | 74 | 0,082583 | 0,473527 | y ~ current_feature + S |
| 0,070411 | 0,116751 | -0,04751 | 0,603089 | 76 | 73 | 0,548319 | 0,895709 | y ~ current_feature + S |
| 0,096449 | 0,126408 | 0,061498 | 0,762997 | 65 | 62 | 0,448358 | 0,937665 | y ~ current_feature + S |
| 0,192291 | 0,141644 | -0,02429 | 1,357564 | 51 | 48 | 0,180951 | 0,646876 | y ~ current_feature + S |
| -0,00549 | 0,130187 | -0,07874 | -0,0422 | 62 | 59 | 0,966485 | 0,994607 | y ~ current_feature + S |
| 0,047216 | 0,130044 | 0,034007 | 0,363076 | 62 | 59 | 0,717846 | 0,920814 | y ~ current_feature + S |
| -0,18873 | 0,128947 | -0,04751 | -1,46366 | 61 | 58 | 0,148687 | 0,661823 | y ~ current_feature + S |
| 0,115272 | 0,136445 | 0,061498 | 0,844821 | 56 | 53 | 0,402008 | 0,93433 | y ~ current_feature + S |
| 0,244562 | 0,157295 | -0,02429 | 1,554793 | 41 | 38 | 0,128286 | 0,617591 | y ~ current_feature + S |
| 0,193318 | 0,147912 | -0,07874 | 1,30698 | 47 | 44 | 0,198009 | 0,611596 | y ~ current_feature + S |
| 0,15919 | 0,148833 | 0,034007 | 1,069584 | 47 | 44 | 0,290639 | 0,718742 | y ~ current_feature + S |
| -0,19772 | 0,149488 | -0,04751 | -1,32267 | 46 | 43 | 0,192936 | 0,717449 | y ~ current_feature + S |
| -0,20271 | 0,165522 | 0,061498 | -1,22466 | 38 | 35 | 0,228883 | 0,893818 | y ~ current_feature + S |
| 0,292848 | 0,180697 | -0,02429 | 1,620657 | 31 | 28 | 0,1163 | 0,614731 | y ~ current_feature + S |
| 0,172295 | 0,065524 | -0,07874 | 2,629488 | 229 | 226 | 0,009139 | 0,125233 | y ~ current_feature + S |
| 0,091362 | 0,066241 | 0,034007 | 1,379246 | 229 | 226 | 0,169182 | 0,61875 | y ~ current_feature + S |
| 0,055617 | 0,066563 | -0,04751 | 0,835551 | 228 | 225 | 0,404294 | 0,803665 | y ~ current_feature + S |
| -0,09002 | 0,070958 | 0,061498 | -1,26864 | 200 | 197 | 0,206066 | 0,893818 | y ~ current_feature + S |
| 0,169452 | 0,080738 | -0,02429 | 2,098774 | 152 | 149 | 0,037524 | 0,520476 | y ~ current_feature + S |
| -0,09123 | 0,054006 | -0,07874 | -1,68919 | 343 | 340 | 0,092101 | 0,42595 | y ~ current_feature + S |
| -0,16124 | 0,053523 | 0,034007 | -3,01259 | 343 | 340 | 0,002785 | 0,137369 | y ~ current_feature + S |
| -0,09042 | 0,05409 | -0,04751 | -1,67163 | 342 | 339 | 0,09552 | 0,640171 | y ~ current_feature + S |
| -0,12323 | 0,057487 | 0,061498 | -2,14367 | 301 | 298 | 0,032868 | 0,658491 | y ~ current_feature + S |
| -0,17438 | 0,065645 | -0,02429 | -2,6564 | 228 | 225 | 0,008464 | 0,295269 | y ~ current_feature + S |
| 0,017647 | 0,076237 | -0,07874 | 0,231479 | 175 | 172 | 0,817218 | 0,979871 | y ~ current_feature + S |
| -0,06807 | 0,076072 | 0,034007 | -0,89485 | 175 | 172 | 0,372117 | 0,750317 | y ~ current_feature + S |
| 0,012244 | 0,076466 | -0,04751 | 0,160128 | 174 | 171 | 0,872969 | 0,985217 | y ~ current_feature + S |
| -0,15933 | 0,080607 | 0,061498 | -1,97658 | 153 | 150 | 0,049923 | 0,700056 | y ~ current_feature + S |
| 0,127937 | 0,093299 | -0,02429 | 1,371262 | 116 | 113 | 0,17301 | 0,626332 | y ~ current_feature + S |
| 0,215579 | 0,100185 | -0,07874 | 2,151805 | 98 | 95 | 0,033948 | 0,236995 | y ~ current_feature + S |
| 0,14358 | 0,101535 | 0,034007 | 1,414095 | 98 | 95 | 0,160603 | 0,60427 | y ~ current_feature + S |
| 0,22003 | 0,100083 | -0,04751 | 2,198464 | 98 | 95 | 0,030345 | 0,603897 | y ~ current_feature + S |
| -0,2302 | 0,108801 | 0,061498 | -2,11579 | 83 | 80 | 0,037473 | 0,658491 | y ~ current_feature + S |
| 0,206279 | 0,124269 | -0,02429 | 1,659945 | 65 | 62 | 0,101976 | 0,605474 | y ~ current_feature + S |
| -0,00224 | 0,084819 | -0,07874 | -0,02637 | 142 | 139 | 0,978998 | 0,996318 | y ~ current_feature + S |
| -0,07061 | 0,084607 | 0,034007 | -0,83452 | 142 | 139 | 0,405418 | 0,779245 | y ~ current_feature + S |
| -0,01927 | 0,08511 | -0,04751 | -0,22644 | 141 | 138 | 0,821193 | 0,972405 | y ~ current_feature + S |
| 0,002722 | 0,090909 | 0,061498 | 0,029946 | 124 | 121 | 0,97616 | 0,996179 | y ~ current_feature + S |
| -0,0933 | 0,104371 | -0,02429 | -0,89396 | 94 | 91 | 0,373699 | 0,815551 | y ~ current_feature + S |
| -0,02675 | 0,066495 | -0,07874 | -0,4023 | 229 | 226 | 0,687843 | 0,93638 | y ~ current_feature + S |
| -0,17331 | 0,065512 | 0,034007 | -2,64549 | 229 | 226 | 0,00873 | 0,215592 | y ~ current_feature + S |
| -0,07573 | 0,066328 | -0,04751 | -1,14182 | 229 | 226 | 0,254739 | 0,742928 | y ~ current_feature + S |
| 0,036906 | 0,071199 | 0,061498 | 0,518354 | 200 | 197 | 0,604793 | 0,945257 | y ~ current_feature + S |
| -0,03633 | 0,081596 | -0,02429 | -0,44518 | 153 | 150 | 0,656829 | 0,924682 | y ~ current_feature + S |
| 0,092889 | 0,150104 | -0,07874 | 0,618831 | 47 | 44 | 0,539218 | 0,882344 | y ~ current_feature + S |
| -0,04708 | 0,150588 | 0,034007 | -0,31267 | 47 | 44 | 0,756007 | 0,93416 | y ~ current_feature + S |
| 0,05134 | 0,150557 | -0,04751 | 0,341002 | 47 | 44 | 0,734725 | 0,933491 | y ~ current_feature + S |
| 0,17641 | 0,159677 | 0,061498 | 1,104792 | 41 | 38 | 0,276196 | 0,893818 | y ~ current_feature + S |

| | | | | | | | | |
|---|---|---|---|---|---|---|---|---|
| -0,08017 | 0,188374 | -0,02429 | -0,42557 | 31 | 28 | 0,673671 | 0,926342 | y ~ current_feature + S |
| 0,074697 | 0,150334 | -0,07874 | 0,496874 | 47 | 44 | 0,621752 | 0,919937 | y ~ current_feature + S |
| 0,136478 | 0,149345 | 0,034007 | 0,913841 | 47 | 44 | 0,365781 | 0,7498 | y ~ current_feature + S |
| -0,16873 | 0,148594 | -0,04751 | -1,1355 | 47 | 44 | 0,262312 | 0,74658 | y ~ current_feature + S |
| 0,027729 | 0,160067 | 0,061498 | 0,173233 | 42 | 39 | 0,863364 | 0,996179 | y ~ current_feature + S |
| -0,49158 | 0,164572 | -0,02429 | -2,98702 | 31 | 28 | 0,005801 | 0,238467 | y ~ current_feature + S |
| 0,03964 | 0,087637 | -0,07874 | 0,45232 | 133 | 130 | 0,651793 | 0,932255 | y ~ current_feature + S |
| 0,041786 | 0,087629 | 0,034007 | 0,476855 | 133 | 130 | 0,634266 | 0,899624 | y ~ current_feature + S |
| 0,021576 | 0,087685 | -0,04751 | 0,24606 | 133 | 130 | 0,806024 | 0,96985 | y ~ current_feature + S |
| -0,01485 | 0,094062 | 0,061498 | -0,15785 | 116 | 113 | 0,874856 | 0,996179 | y ~ current_feature + S |
| 0,024952 | 0,108431 | -0,02429 | 0,23012 | 88 | 85 | 0,818552 | 0,975408 | y ~ current_feature + S |
| -0,12948 | 0,113001 | -0,07874 | -1,14586 | 80 | 77 | 0,255404 | 0,682306 | y ~ current_feature + S |
| -0,06368 | 0,113729 | 0,034007 | -0,55995 | 80 | 77 | 0,577139 | 0,869291 | y ~ current_feature + S |
| -0,01798 | 0,113942 | -0,04751 | -0,15778 | 80 | 77 | 0,875046 | 0,985217 | y ~ current_feature + S |
| -0,04096 | 0,122988 | 0,061498 | -0,33302 | 69 | 66 | 0,740173 | 0,972573 | y ~ current_feature + S |
| -0,07072 | 0,141067 | -0,02429 | -0,50132 | 53 | 50 | 0,618346 | 0,924682 | y ~ current_feature + S |
| -0,00175 | 0,055132 | -0,07874 | -0,03181 | 332 | 329 | 0,974641 | 0,99618 | y ~ current_feature + S |
| -0,04372 | 0,055079 | 0,034007 | -0,79369 | 332 | 329 | 0,427947 | 0,791701 | y ~ current_feature + S |
| -0,0265 | 0,055112 | -0,04751 | -0,48079 | 332 | 329 | 0,630986 | 0,925756 | y ~ current_feature + S |
| -0,05551 | 0,06976 | 0,061498 | -0,79579 | 289 | 204,8543 | 0,427074 | 0,937665 | y ~ current_feature + S |
| -0,00271 | 0,067838 | -0,02429 | -0,03999 | 221 | 217,2952 | 0,968138 | 0,99444 | y ~ current_feature + S |
| 0,115356 | 0,051295 | -0,07874 | 2,248871 | 378 | 375 | 0,025101 | 0,201065 | y ~ current_feature + S |
| 0,032379 | 0,051613 | 0,034007 | 0,627345 | 378 | 375 | 0,530815 | 0,848387 | y ~ current_feature + S |
| -0,01644 | 0,051702 | -0,04751 | -0,31803 | 377 | 374 | 0,750643 | 0,9394 | y ~ current_feature + S |
| -0,08816 | 0,055169 | 0,061498 | -1,59806 | 329 | 326 | 0,110998 | 0,8197 | y ~ current_feature + S |
| -0,02415 | 0,063354 | -0,02429 | -0,38121 | 252 | 249 | 0,703369 | 0,93118 | y ~ current_feature + S |
| -0,03744 | 0,051535 | -0,07874 | -0,7265 | 379 | 376 | 0,467984 | 0,840554 | y ~ current_feature + S |
| -0,02725 | 0,051552 | 0,034007 | -0,5285 | 379 | 376 | 0,597462 | 0,878669 | y ~ current_feature + S |
| -0,01746 | 0,051632 | -0,04751 | -0,3381 | 378 | 375 | 0,735473 | 0,933491 | y ~ current_feature + S |
| 0,012483 | 0,061937 | 0,061498 | 0,201552 | 329 | 260,6368 | 0,840424 | 0,996179 | y ~ current_feature + S |
| -0,07264 | 0,063272 | -0,02429 | -1,14799 | 252 | 248,4754 | 0,252078 | 0,717483 | y ~ current_feature + S |
| -0,01874 | 0,061535 | -0,07874 | -0,30453 | 267 | 264 | 0,760964 | 0,965008 | y ~ current_feature + S |
| -0,08153 | 0,061341 | 0,034007 | -1,32919 | 267 | 264 | 0,184932 | 0,638805 | y ~ current_feature + S |
| 0,006996 | 0,061544 | -0,04751 | 0,113673 | 267 | 264 | 0,909584 | 0,985217 | y ~ current_feature + S |
| 0,001041 | 0,066372 | 0,061498 | 0,015684 | 230 | 227 | 0,9875 | 0,997945 | y ~ current_feature + S |
| -0,0804 | 0,075564 | -0,02429 | -1,06399 | 177 | 174 | 0,288807 | 0,734108 | y ~ current_feature + S |
| 0,270663 | 0,063477 | -0,07874 | 4,263961 | 233 | 230 | 2,93E-05 | 0,005519 | y ~ current_feature + S |
| 0,220686 | 0,064312 | 0,034007 | 3,431474 | 233 | 230 | 7,12E-04 | 0,068218 | y ~ current_feature + S |
| 0,243284 | 0,063957 | -0,04751 | 3,803866 | 233 | 230 | 1,83E-04 | 0,067562 | y ~ current_feature + S |
| 0,056542 | 0,070598 | 0,061498 | 0,800911 | 203 | 200 | 0,424133 | 0,937665 | y ~ current_feature + S |
| 0,33435 | 0,076192 | -0,02429 | 4,388235 | 156 | 153 | 2,12E-05 | 0,010121 | y ~ current_feature + S |
| -0,02391 | 0,074722 | -0,07874 | -0,32002 | 182 | 179 | 0,749326 | 0,962676 | y ~ current_feature + S |
| -0,05011 | 0,07465 | 0,034007 | -0,67126 | 182 | 179 | 0,502923 | 0,841996 | y ~ current_feature + S |
| 0,06154 | 0,074602 | -0,04751 | 0,82491 | 182 | 179 | 0,410521 | 0,803665 | y ~ current_feature + S |
| -0,12159 | 0,079726 | 0,061498 | -1,52505 | 158 | 155 | 0,129284 | 0,8197 | y ~ current_feature + S |
| 0,002162 | 0,09167 | -0,02429 | 0,023588 | 122 | 119 | 0,981221 | 0,99444 | y ~ current_feature + S |
| 0,00178 | 0,088388 | -0,07874 | 0,020135 | 131 | 128 | 0,983967 | 0,996318 | y ~ current_feature + S |
| 0,042772 | 0,088307 | 0,034007 | 0,484359 | 131 | 128 | 0,628959 | 0,899624 | y ~ current_feature + S |

| | | | | | | | | |
|---|---|---|---|---|---|---|---|---|
| -0,0094 | 0,088384 | -0,04751 | -0,10637 | 131 | 128 | 0,915452 | 0,985217 | y ~ current_feature + S |
| 0,030484 | 0,094028 | 0,061498 | 0,324203 | 116 | 113 | 0,746384 | 0,972573 | y ~ current_feature + S |
| 0,032604 | 0,109051 | -0,02429 | 0,298977 | 87 | 84 | 0,765696 | 0,953898 | y ~ current_feature + S |
| -0,04647 | 0,116122 | -0,07874 | -0,40015 | 77 | 74 | 0,690195 | 0,93638 | y ~ current_feature + S |
| -0,03539 | 0,116175 | 0,034007 | -0,30466 | 77 | 74 | 0,761483 | 0,936042 | y ~ current_feature + S |
| -0,04895 | 0,116108 | -0,04751 | -0,42159 | 77 | 74 | 0,674547 | 0,928258 | y ~ current_feature + S |
| -0,12086 | 0,143502 | 0,061498 | -0,84224 | 69 | 47,85124 | 0,403841 | 0,93433 | y ~ current_feature + S |
| 0,009284 | 0,144331 | -0,02429 | 0,064322 | 51 | 48 | 0,948981 | 0,99444 | y ~ current_feature + S |
| -0,03483 | 0,052636 | -0,07874 | -0,66173 | 394 | 360,5014 | 0,508567 | 0,863164 | y ~ current_feature + S |
| -0,13251 | 0,069514 | 0,034007 | -1,9063 | 394 | 203,3119 | 0,058022 | 0,420946 | y ~ current_feature + S |
| -0,06616 | 0,054599 | -0,04751 | -1,21174 | 393 | 333,9873 | 0,226469 | 0,742928 | y ~ current_feature + S |
| -0,00451 | 0,05995 | 0,061498 | -0,07521 | 341 | 278,2356 | 0,940104 | 0,996179 | y ~ current_feature + S |
| -0,03311 | 0,062103 | -0,02429 | -0,53307 | 262 | 259 | 0,594441 | 0,924682 | y ~ current_feature + S |
| 0,02812 | 0,083591 | -0,07874 | 0,336398 | 146 | 143 | 0,737064 | 0,959225 | y ~ current_feature + S |
| 0,084317 | 0,083326 | 0,034007 | 1,011894 | 146 | 143 | 0,313298 | 0,718742 | y ~ current_feature + S |
| 0,137952 | 0,083116 | -0,04751 | 1,659757 | 145 | 142 | 0,09917 | 0,640171 | y ~ current_feature + S |
| -0,08755 | 0,09056 | 0,061498 | -0,96675 | 124 | 121 | 0,335598 | 0,893818 | y ~ current_feature + S |
| 0,228609 | 0,100411 | -0,02429 | 2,276735 | 97 | 94 | 0,025071 | 0,485321 | y ~ current_feature + S |
| 0,029285 | 0,055722 | -0,07874 | 0,525562 | 365 | 321,7946 | 0,599554 | 0,90386 | y ~ current_feature + S |
| 0,048212 | 0,073871 | 0,034007 | 0,652643 | 365 | 182,8255 | 0,514806 | 0,841996 | y ~ current_feature + S |
| 0,022866 | 0,059045 | -0,04751 | 0,387259 | 364 | 286,6822 | 0,698852 | 0,931902 | y ~ current_feature + S |
| -0,06308 | 0,063229 | 0,061498 | -0,99771 | 314 | 249,1355 | 0,319387 | 0,893818 | y ~ current_feature + S |
| -0,00397 | 0,064549 | -0,02429 | -0,06157 | 243 | 240 | 0,950954 | 0,99444 | y ~ current_feature + S |
| -0,01333 | 0,052082 | -0,07874 | -0,25597 | 412 | 368,5973 | 0,798121 | 0,979102 | y ~ current_feature + S |
| -0,07751 | 0,067815 | 0,034007 | -1,14298 | 412 | 216,1402 | 0,254309 | 0,704827 | y ~ current_feature + S |
| 0,01794 | 0,053314 | -0,04751 | 0,336491 | 411 | 351,7055 | 0,736701 | 0,933491 | y ~ current_feature + S |
| -0,11498 | 0,057164 | 0,061498 | -2,01147 | 358 | 301,98 | 0,045164 | 0,700056 | y ~ current_feature + S |
| -0,01551 | 0,061164 | -0,02429 | -0,25358 | 274 | 267,24 | 0,800017 | 0,968297 | y ~ current_feature + S |
| 0,03599 | 0,050932 | -0,07874 | 0,706624 | 388 | 385 | 0,480227 | 0,845512 | y ~ current_feature + S |
| -0,00503 | 0,050964 | 0,034007 | -0,09873 | 388 | 385 | 0,921401 | 0,9732 | y ~ current_feature + S |
| -0,0055 | 0,05103 | -0,04751 | -0,1077 | 387 | 384 | 0,914293 | 0,985217 | y ~ current_feature + S |
| -0,07346 | 0,062085 | 0,061498 | -1,18319 | 338 | 258,0305 | 0,237824 | 0,893818 | y ~ current_feature + S |
| -0,06818 | 0,062482 | -0,02429 | -1,09125 | 258 | 254,9591 | 0,276193 | 0,734108 | y ~ current_feature + S |
| 0,094168 | 0,093242 | -0,07874 | 1,009929 | 117 | 114 | 0,314668 | 0,718687 | y ~ current_feature + S |
| -0,05882 | 0,093496 | 0,034007 | -0,62914 | 117 | 114 | 0,530516 | 0,848387 | y ~ current_feature + S |
| -0,10121 | 0,093589 | -0,04751 | -1,08141 | 116 | 113 | 0,281819 | 0,747915 | y ~ current_feature + S |
| -0,16178 | 0,101246 | 0,061498 | -1,59793 | 98 | 95 | 0,113379 | 0,8197 | y ~ current_feature + S |
| 0,008382 | 0,115466 | -0,02429 | 0,072593 | 78 | 75 | 0,942323 | 0,99444 | y ~ current_feature + S |
| -0,06444 | 0,056227 | -0,07874 | -1,14603 | 318 | 315 | 0,252651 | 0,679778 | y ~ current_feature + S |
| -0,12876 | 0,055875 | 0,034007 | -2,30447 | 318 | 315 | 0,021847 | 0,329927 | y ~ current_feature + S |
| -0,06149 | 0,056237 | -0,04751 | -1,0934 | 318 | 315 | 0,275052 | 0,747915 | y ~ current_feature + S |
| -0,12797 | 0,059483 | 0,061498 | -2,15137 | 281 | 278 | 0,032309 | 0,658491 | y ~ current_feature + S |
| -0,14857 | 0,068404 | -0,02429 | -2,17189 | 212 | 209 | 0,03099 | 0,485321 | y ~ current_feature + S |
| 0,035243 | 0,090853 | -0,07874 | 0,387913 | 124 | 121 | 0,698762 | 0,939646 | y ~ current_feature + S |
| -0,07447 | 0,090657 | 0,034007 | -0,82142 | 124 | 121 | 0,413023 | 0,785427 | y ~ current_feature + S |
| 0,045811 | 0,090814 | -0,04751 | 0,504453 | 124 | 121 | 0,61486 | 0,924009 | y ~ current_feature + S |
| -0,09426 | 0,096696 | 0,061498 | -0,97476 | 109 | 106 | 0,331896 | 0,893818 | y ~ current_feature + S |
| -0,08617 | 0,111388 | -0,02429 | -0,77358 | 83 | 80 | 0,441457 | 0,858965 | y ~ current_feature + S |

| | | | | | | | | |
|---|---|---|---|---|---|---|---|---|
| 0,004693 | 0,103694 | -0,07874 | 0,045254 | 96 | 93 | 0,964002 | 0,994607 | y ~ current_feature + S |
| -0,2858 | 0,09937 | 0,034007 | -2,87616 | 96 | 93 | 0,004991 | 0,167881 | y ~ current_feature + S |
| 0,006626 | 0,104255 | -0,04751 | 0,063554 | 95 | 92 | 0,949463 | 0,990156 | y ~ current_feature + S |
| 0,007722 | 0,1118 | 0,061498 | 0,069066 | 83 | 80 | 0,945109 | 0,996179 | y ~ current_feature + S |
| -0,16191 | 0,127396 | -0,02429 | -1,27094 | 63 | 60 | 0,208657 | 0,675518 | y ~ current_feature + S |
| 0,123635 | 0,07417 | -0,07874 | 1,666919 | 182 | 179 | 0,097279 | 0,431938 | y ~ current_feature + S |
| 0,010313 | 0,07474 | 0,034007 | 0,137987 | 182 | 179 | 0,890405 | 0,96591 | y ~ current_feature + S |
| 0,053502 | 0,074846 | -0,04751 | 0,714833 | 181 | 178 | 0,475649 | 0,842057 | y ~ current_feature + S |
| 0,030513 | 0,080027 | 0,061498 | 0,38129 | 159 | 156 | 0,703507 | 0,972444 | y ~ current_feature + S |
| 0,001665 | 0,092057 | -0,02429 | 0,018082 | 121 | 118 | 0,985604 | 0,99444 | y ~ current_feature + S |
| 0,396241 | 0,0942 | -0,07874 | 4,206392 | 98 | 95 | 5,88E-05 | 0,008233 | y ~ current_feature + S |
| 0,122781 | 0,101822 | 0,034007 | 1,20584 | 98 | 95 | 0,230874 | 0,681978 | y ~ current_feature + S |
| 0,154493 | 0,101366 | -0,04751 | 1,524113 | 98 | 95 | 0,130803 | 0,654671 | y ~ current_feature + S |
| 0,012748 | 0,1091 | 0,061498 | 0,116847 | 87 | 84 | 0,907261 | 0,996179 | y ~ current_feature + S |
| 0,024396 | 0,126962 | -0,02429 | 0,192153 | 65 | 62 | 0,84825 | 0,977734 | y ~ current_feature + S |
| 0,329037 | 0,12619 | -0,07874 | 2,607477 | 59 | 56 | 0,01167 | 0,141568 | y ~ current_feature + S |
| 0,099753 | 0,132964 | 0,034007 | 0,750227 | 59 | 56 | 0,456259 | 0,821488 | y ~ current_feature + S |
| -0,02594 | 0,134795 | -0,04751 | -0,19247 | 58 | 55 | 0,848084 | 0,980218 | y ~ current_feature + S |
| -0,26433 | 0,142198 | 0,061498 | -1,85892 | 49 | 46 | 0,069442 | 0,715249 | y ~ current_feature + S |
| 0,27035 | 0,16046 | -0,02429 | 1,68484 | 39 | 36 | 0,100671 | 0,605474 | y ~ current_feature + S |
| 0,175416 | 0,086016 | -0,07874 | 2,039351 | 134 | 131 | 0,043426 | 0,277028 | y ~ current_feature + S |
| 0,096765 | 0,08696 | 0,034007 | 1,112747 | 134 | 131 | 0,267855 | 0,71095 | y ~ current_feature + S |
| 0,008867 | 0,087367 | -0,04751 | 0,101488 | 134 | 131 | 0,919318 | 0,985217 | y ~ current_feature + S |
| -0,05738 | 0,095189 | 0,061498 | -0,60279 | 113 | 110 | 0,547891 | 0,937665 | y ~ current_feature + S |
| 0,02935 | 0,107165 | -0,02429 | 0,273875 | 90 | 87 | 0,78483 | 0,964545 | y ~ current_feature + S |
| -0,00497 | 0,064149 | -0,07874 | -0,0775 | 246 | 243 | 0,938287 | 0,994607 | y ~ current_feature + S |
| -0,01189 | 0,064145 | 0,034007 | -0,18536 | 246 | 243 | 0,8531 | 0,959445 | y ~ current_feature + S |
| -0,00398 | 0,06415 | -0,04751 | -0,06202 | 246 | 243 | 0,950595 | 0,990156 | y ~ current_feature + S |
| -0,0156 | 0,069329 | 0,061498 | -0,22501 | 211 | 208 | 0,822192 | 0,996179 | y ~ current_feature + S |
| -0,07119 | 0,078611 | -0,02429 | -0,90561 | 164 | 161 | 0,366494 | 0,807159 | y ~ current_feature + S |
| -0,32263 | 0,104526 | -0,07874 | -3,08664 | 85 | 82 | 0,002761 | 0,065908 | y ~ current_feature + S |
| -0,1377 | 0,10938 | 0,034007 | -1,25894 | 85 | 82 | 0,211623 | 0,664036 | y ~ current_feature + S |
| -0,02133 | 0,111086 | -0,04751 | -0,192 | 84 | 81 | 0,848221 | 0,980218 | y ~ current_feature + S |
| 0,008331 | 0,119519 | 0,061498 | 0,069708 | 73 | 70 | 0,944625 | 0,996179 | y ~ current_feature + S |
| 0,012403 | 0,136072 | -0,02429 | 0,091147 | 57 | 54 | 0,927713 | 0,99444 | y ~ current_feature + S |
| -0,00643 | 0,097588 | -0,07874 | -0,06589 | 108 | 105 | 0,94759 | 0,994607 | y ~ current_feature + S |
| -0,0028 | 0,09759 | 0,034007 | -0,02869 | 108 | 105 | 0,977165 | 0,989859 | y ~ current_feature + S |
| 0,085073 | 0,097236 | -0,04751 | 0,874914 | 108 | 105 | 0,383617 | 0,803665 | y ~ current_feature + S |
| 0,112009 | 0,104169 | 0,061498 | 1,075264 | 94 | 91 | 0,285099 | 0,893818 | y ~ current_feature + S |
| 0,15031 | 0,119018 | -0,02429 | 1,262914 | 72 | 69 | 0,210871 | 0,675518 | y ~ current_feature + S |
| -0,08853 | 0,103848 | -0,07874 | -0,85252 | 95 | 92 | 0,396138 | 0,791422 | y ~ current_feature + S |
| -0,05691 | 0,104088 | 0,034007 | -0,5467 | 95 | 92 | 0,585906 | 0,87187 | y ~ current_feature + S |
| 0,141523 | 0,103773 | -0,04751 | 1,363767 | 94 | 91 | 0,176005 | 0,700235 | y ~ current_feature + S |
| -0,1557 | 0,109756 | 0,061498 | -1,41856 | 84 | 81 | 0,159863 | 0,8197 | y ~ current_feature + S |
| -0,15225 | 0,127594 | -0,02429 | -1,19326 | 63 | 60 | 0,237463 | 0,697311 | y ~ current_feature + S |
| 0,102816 | 0,128415 | -0,07874 | 0,800654 | 63 | 60 | 0,426491 | 0,814177 | y ~ current_feature + S |
| 0,058689 | 0,128877 | 0,034007 | 0,455391 | 63 | 60 | 0,650472 | 0,900251 | y ~ current_feature + S |
| 0,149302 | 0,127652 | -0,04751 | 1,169598 | 63 | 60 | 0,246788 | 0,742928 | y ~ current_feature + S |

| | | | | | | | | |
|---|---|---|---|---|---|---|---|---|
| -0,30876 | 0,129434 | 0,061498 | -2,38545 | 57 | 54 | 0,020596 | 0,609646 | y ~ current_feature + S |
| -0,11651 | 0,159038 | -0,02429 | -0,7326 | 42 | 39 | 0,468185 | 0,876224 | y ~ current_feature + S |
| 0,087949 | 0,06238 | -0,07874 | 1,409893 | 258 | 255 | 0,15979 | 0,530244 | y ~ current_feature + S |
| 0,088062 | 0,062379 | 0,034007 | 1,411722 | 258 | 255 | 0,159251 | 0,60427 | y ~ current_feature + S |
| 0,032107 | 0,06259 | -0,04751 | 0,512966 | 258 | 255 | 0,608419 | 0,916966 | y ~ current_feature + S |
| -0,08501 | 0,066278 | 0,061498 | -1,2826 | 229 | 226 | 0,200945 | 0,893818 | y ~ current_feature + S |
| 0,042001 | 0,076855 | -0,02429 | 0,546494 | 172 | 169 | 0,585447 | 0,924682 | y ~ current_feature + S |
| 0,081544 | 0,068291 | -0,07874 | 1,194079 | 216 | 213 | 0,233775 | 0,657771 | y ~ current_feature + S |
| -0,03388 | 0,06848 | 0,034007 | -0,49475 | 216 | 213 | 0,621284 | 0,894455 | y ~ current_feature + S |
| 0,152429 | 0,067718 | -0,04751 | 2,250938 | 216 | 213 | 0,025411 | 0,569815 | y ~ current_feature + S |
| -0,00523 | 0,074328 | 0,061498 | -0,07036 | 184 | 181 | 0,943985 | 0,996179 | y ~ current_feature + S |
| 0,071277 | 0,084001 | -0,02429 | 0,848524 | 144 | 141 | 0,397585 | 0,823699 | y ~ current_feature + S |
| 0,154309 | 0,052841 | -0,07874 | 2,920256 | 396 | 349,6166 | 0,003724 | 0,069632 | y ~ current_feature + S |
| 0,098426 | 0,069095 | 0,034007 | 1,424502 | 396 | 207,4353 | 0,155804 | 0,60427 | y ~ current_feature + S |
| 0,061236 | 0,05404 | -0,04751 | 1,133149 | 395 | 341,1423 | 0,257947 | 0,742928 | y ~ current_feature + S |
| -0,02193 | 0,059145 | 0,061498 | -0,3708 | 343 | 285,7334 | 0,711063 | 0,972444 | y ~ current_feature + S |
| 0,087753 | 0,062517 | -0,02429 | 1,403669 | 263 | 253,8906 | 0,161639 | 0,623497 | y ~ current_feature + S |
| 0,089043 | 0,087023 | -0,07874 | 1,023211 | 134 | 131 | 0,308094 | 0,718687 | y ~ current_feature + S |
| -0,09076 | 0,08701 | 0,034007 | -1,04316 | 134 | 131 | 0,298796 | 0,718742 | y ~ current_feature + S |
| -0,11948 | 0,086745 | -0,04751 | -1,37733 | 134 | 131 | 0,170758 | 0,700235 | y ~ current_feature + S |
| -0,04402 | 0,091968 | 0,061498 | -0,4786 | 121 | 118 | 0,633112 | 0,95418 | y ~ current_feature + S |
| -0,20105 | 0,105631 | -0,02429 | -1,90335 | 89 | 86 | 0,06034 | 0,558147 | y ~ current_feature + S |
| 0,144332 | 0,051963 | -0,07874 | 2,777617 | 404 | 362,6407 | 0,005761 | 0,092669 | y ~ current_feature + S |
| 0,056516 | 0,068302 | 0,034007 | 0,827441 | 404 | 213,6684 | 0,40891 | 0,783921 | y ~ current_feature + S |
| 0,021849 | 0,053759 | -0,04751 | 0,406424 | 403 | 345,8498 | 0,684682 | 0,928814 | y ~ current_feature + S |
| 0,010828 | 0,058118 | 0,061498 | 0,186318 | 350 | 296,0216 | 0,852323 | 0,996179 | y ~ current_feature + S |
| 0,102514 | 0,060991 | -0,02429 | 1,680804 | 269 | 266 | 0,093975 | 0,605474 | y ~ current_feature + S |
| 0,157508 | 0,105873 | -0,07874 | 1,487706 | 90 | 87 | 0,140444 | 0,498495 | y ~ current_feature + S |
| -0,02781 | 0,10717 | 0,034007 | -0,25947 | 90 | 87 | 0,795887 | 0,946875 | y ~ current_feature + S |
| -0,04612 | 0,107097 | -0,04751 | -0,4306 | 90 | 87 | 0,667821 | 0,92822 | y ~ current_feature + S |
| -0,05135 | 0,113078 | 0,061498 | -0,45408 | 81 | 78 | 0,651029 | 0,957207 | y ~ current_feature + S |
| -6,69E-04 | 0,132453 | -0,02429 | -0,00505 | 60 | 57 | 0,995989 | 0,998688 | y ~ current_feature + S |
| -0,03037 | 0,050744 | -0,07874 | -0,59845 | 391 | 388 | 0,549887 | 0,888464 | y ~ current_feature + S |
| -0,06269 | 0,050667 | 0,034007 | -1,23725 | 391 | 388 | 0,216741 | 0,664036 | y ~ current_feature + S |
| -0,06174 | 0,050736 | -0,04751 | -1,21685 | 390 | 387 | 0,224402 | 0,742928 | y ~ current_feature + S |
| -0,03993 | 0,06066 | 0,061498 | -0,65821 | 342 | 271,3292 | 0,510964 | 0,937665 | y ~ current_feature + S |
| -0,04441 | 0,062317 | -0,02429 | -0,71273 | 260 | 257 | 0,47666 | 0,878782 | y ~ current_feature + S |
| 0,026761 | 0,073837 | -0,07874 | 0,362439 | 200 | 183,2901 | 0,717442 | 0,948276 | y ~ current_feature + S |
| 0,032601 | 0,090078 | 0,034007 | 0,361917 | 200 | 123,1108 | 0,718035 | 0,920814 | y ~ current_feature + S |
| 0,083691 | 0,078081 | -0,04751 | 1,071843 | 199 | 162,8748 | 0,285376 | 0,747915 | y ~ current_feature + S |
| 0,072157 | 0,078068 | 0,061498 | 0,924283 | 175 | 163,223 | 0,356703 | 0,920619 | y ~ current_feature + S |
| 0,04808 | 0,089476 | -0,02429 | 0,537358 | 133 | 124,6195 | 0,591979 | 0,924682 | y ~ current_feature + S |
| 0,008448 | 0,078565 | -0,07874 | 0,107529 | 165 | 162 | 0,914502 | 0,987929 | y ~ current_feature + S |
| -0,14333 | 0,077756 | 0,034007 | -1,84327 | 165 | 162 | 0,067117 | 0,439525 | y ~ current_feature + S |
| -0,10618 | 0,078123 | -0,04751 | -1,35916 | 165 | 162 | 0,175984 | 0,700235 | y ~ current_feature + S |
| -0,15642 | 0,082595 | 0,061498 | -1,89376 | 146 | 143 | 0,060276 | 0,715249 | y ~ current_feature + S |
| -0,13822 | 0,095746 | -0,02429 | -1,44362 | 110 | 107 | 0,151767 | 0,623156 | y ~ current_feature + S |
| -0,06423 | 0,076538 | -0,07874 | -0,8392 | 173 | 170 | 0,402533 | 0,796456 | y ~ current_feature + S |

| | | | | | | | | |
|---|---|---|---|---|---|---|---|---|
| -0,01253 | 0,07669 | 0,034007 | -0,16334 | 173 | 170 | 0,870444 | 0,96591 | y ~ current_feature + S |
| 0,059199 | 0,076562 | -0,04751 | 0,773221 | 173 | 170 | 0,440466 | 0,823749 | y ~ current_feature + S |
| 0,002754 | 0,081923 | 0,061498 | 0,033621 | 152 | 149 | 0,973224 | 0,996179 | y ~ current_feature + S |
| 0,065111 | 0,093872 | -0,02429 | 0,693608 | 116 | 113 | 0,489351 | 0,889654 | y ~ current_feature + S |
| 0,109131 | 0,105367 | -0,07874 | 1,035725 | 92 | 89 | 0,303136 | 0,718687 | y ~ current_feature + S |
| -0,03801 | 0,105923 | 0,034007 | -0,35881 | 92 | 89 | 0,720588 | 0,920814 | y ~ current_feature + S |
| 0,017345 | 0,105984 | -0,04751 | 0,163661 | 92 | 89 | 0,870369 | 0,985217 | y ~ current_feature + S |
| -0,16253 | 0,114702 | 0,061498 | -1,41699 | 77 | 74 | 0,160684 | 0,8197 | y ~ current_feature + S |
| 0,145621 | 0,129907 | -0,02429 | 1,120963 | 61 | 58 | 0,266923 | 0,723659 | y ~ current_feature + S |
| 0,013919 | 0,102588 | -0,07874 | 0,135682 | 98 | 95 | 0,89236 | 0,987929 | y ~ current_feature + S |
| -0,00927 | 0,102593 | 0,034007 | -0,0904 | 98 | 95 | 0,928159 | 0,9732 | y ~ current_feature + S |
| 0,096424 | 0,102662 | -0,04751 | 0,939238 | 97 | 94 | 0,350016 | 0,795089 | y ~ current_feature + S |
| 0,06606 | 0,111559 | 0,061498 | 0,59215 | 83 | 80 | 0,55542 | 0,937665 | y ~ current_feature + S |
| 0,17344 | 0,125075 | -0,02429 | 1,386687 | 65 | 62 | 0,170503 | 0,626332 | y ~ current_feature + S |
| 0,113094 | 0,058958 | -0,07874 | 1,918198 | 287 | 284 | 0,056088 | 0,312071 | y ~ current_feature + S |
| 0,083639 | 0,059131 | 0,034007 | 1,414457 | 287 | 284 | 0,158323 | 0,60427 | y ~ current_feature + S |
| 0,054312 | 0,059251 | -0,04751 | 0,916643 | 287 | 284 | 0,360108 | 0,798092 | y ~ current_feature + S |
| -0,03101 | 0,063598 | 0,061498 | -0,48753 | 250 | 247 | 0,626317 | 0,951693 | y ~ current_feature + S |
| -0,00953 | 0,072736 | -0,02429 | -0,13103 | 192 | 189 | 0,895887 | 0,99444 | y ~ current_feature + S |
| 0,037463 | 0,090472 | -0,07874 | 0,414087 | 125 | 122 | 0,679537 | 0,934653 | y ~ current_feature + S |
| 0,008142 | 0,090533 | 0,034007 | 0,089936 | 125 | 122 | 0,928486 | 0,9732 | y ~ current_feature + S |
| 0,001283 | 0,090909 | -0,04751 | 0,014111 | 124 | 121 | 0,988765 | 0,994138 | y ~ current_feature + S |
| -0,0117 | 0,099008 | 0,061498 | -0,11818 | 105 | 102 | 0,906156 | 0,996179 | y ~ current_feature + S |
| -0,10712 | 0,11116 | -0,02429 | -0,96369 | 83 | 80 | 0,338105 | 0,785829 | y ~ current_feature + S |
| -0,01581 | 0,087695 | -0,07874 | -0,18034 | 133 | 130 | 0,857167 | 0,983548 | y ~ current_feature + S |
| -0,05854 | 0,087555 | 0,034007 | -0,66858 | 133 | 130 | 0,504949 | 0,841996 | y ~ current_feature + S |
| -0,20569 | 0,086162 | -0,04751 | -2,38725 | 132 | 129 | 0,018426 | 0,54566 | y ~ current_feature + S |
| -0,03196 | 0,094867 | 0,061498 | -0,33685 | 114 | 111 | 0,736863 | 0,972573 | y ~ current_feature + S |
| -0,05859 | 0,107648 | -0,02429 | -0,54423 | 89 | 86 | 0,58769 | 0,924682 | y ~ current_feature + S |
| -0,08762 | 0,054024 | -0,07874 | -1,62195 | 343 | 340 | 0,105742 | 0,449749 | y ~ current_feature + S |
| -0,09609 | 0,053982 | 0,034007 | -1,78013 | 343 | 340 | 0,075948 | 0,464477 | y ~ current_feature + S |
| -0,0694 | 0,054182 | -0,04751 | -1,28096 | 342 | 339 | 0,201082 | 0,723562 | y ~ current_feature + S |
| -0,03178 | 0,058391 | 0,061498 | -0,5443 | 296 | 293 | 0,586652 | 0,945257 | y ~ current_feature + S |
| -0,09469 | 0,066367 | -0,02429 | -1,42669 | 228 | 225 | 0,155055 | 0,623156 | y ~ current_feature + S |
| -0,09086 | 0,07873 | -0,07874 | -1,1541 | 163 | 160 | 0,25018 | 0,679754 | y ~ current_feature + S |
| -0,07445 | 0,078838 | 0,034007 | -0,94431 | 163 | 160 | 0,346436 | 0,747412 | y ~ current_feature + S |
| -0,05347 | 0,078944 | -0,04751 | -0,67737 | 163 | 160 | 0,499148 | 0,868909 | y ~ current_feature + S |
| -0,06709 | 0,095376 | 0,061498 | -0,70345 | 144 | 109,437 | 0,483268 | 0,937665 | y ~ current_feature + S |
| 2,28E-05 | 0,09759 | -0,02429 | 2,33E-04 | 108 | 105 | 0,999814 | 0,999814 | y ~ current_feature + S |
| 0,025455 | 0,063479 | -0,07874 | 0,400999 | 251 | 248 | 0,688766 | 0,93638 | y ~ current_feature + S |
| 0,012747 | 0,063495 | 0,034007 | 0,200753 | 251 | 248 | 0,841056 | 0,958985 | y ~ current_feature + S |
| 0,070007 | 0,063472 | -0,04751 | 1,102956 | 250 | 247 | 0,27112 | 0,747915 | y ~ current_feature + S |
| 0,07258 | 0,068499 | 0,061498 | 1,059572 | 215 | 212 | 0,290545 | 0,893818 | y ~ current_feature + S |
| 0,084046 | 0,077811 | -0,02429 | 1,080141 | 167 | 164 | 0,281665 | 0,734108 | y ~ current_feature + S |
| 0,12232 | 0,085376 | -0,07874 | 1,432719 | 153 | 135,1385 | 0,154248 | 0,518833 | y ~ current_feature + S |
| 0,025521 | 0,113536 | 0,034007 | 0,224786 | 153 | 77,52606 | 0,822737 | 0,952501 | y ~ current_feature + S |
| 0,149015 | 0,083367 | -0,04751 | 1,787455 | 152 | 140,6878 | 0,076017 | 0,640171 | y ~ current_feature + S |
| -0,06812 | 0,093844 | 0,061498 | -0,72592 | 137 | 113,0233 | 0,469388 | 0,937665 | y ~ current_feature + S |

| | | | | | | | | |
|---|---|---|---|---|---|---|---|---|
| 0,006673 | 0,100502 | -0,02429 | 0,066396 | 102 | 99 | 0,947196 | 0,99444 | y ~ current_feature + S |
| -0,01744 | 0,071785 | -0,07874 | -0,24301 | 197 | 194 | 0,808251 | 0,979871 | y ~ current_feature + S |
| -0,05055 | 0,071704 | 0,034007 | -0,70495 | 197 | 194 | 0,481686 | 0,831402 | y ~ current_feature + S |
| -0,05194 | 0,071699 | -0,04751 | -0,72436 | 197 | 194 | 0,469719 | 0,842057 | y ~ current_feature + S |
| -0,01281 | 0,077376 | 0,061498 | -0,16551 | 170 | 167 | 0,868745 | 0,996179 | y ~ current_feature + S |
| 0,027719 | 0,088354 | -0,02429 | 0,313721 | 131 | 128 | 0,754243 | 0,951432 | y ~ current_feature + S |
| 0,127392 | 0,079668 | -0,07874 | 1,599049 | 158 | 155 | 0,111846 | 0,465986 | y ~ current_feature + S |
| -0,00904 | 0,080319 | 0,034007 | -0,11257 | 158 | 155 | 0,910516 | 0,9732 | y ~ current_feature + S |
| 0,166944 | 0,079451 | -0,04751 | 2,101202 | 157 | 154 | 0,037252 | 0,611115 | y ~ current_feature + S |
| -0,10046 | 0,091831 | 0,061498 | -1,09398 | 135 | 117,3868 | 0,276202 | 0,893818 | y ~ current_feature + S |
| 0,103956 | 0,0991 | -0,02429 | 1,049001 | 105 | 100,7233 | 0,296688 | 0,744235 | y ~ current_feature + S |
| 0,370654 | 0,135475 | -0,07874 | 2,735955 | 50 | 47 | 0,008749 | 0,122156 | y ~ current_feature + S |
| 0,276209 | 0,14019 | 0,034007 | 1,970243 | 50 | 47 | 0,054718 | 0,420946 | y ~ current_feature + S |
| 0,111168 | 0,146528 | -0,04751 | 0,758681 | 49 | 46 | 0,451914 | 0,832044 | y ~ current_feature + S |
| 0,17853 | 0,153665 | 0,061498 | 1,161814 | 44 | 41 | 0,25203 | 0,893818 | y ~ current_feature + S |
| 0,135985 | 0,180878 | -0,02429 | 0,751802 | 33 | 30 | 0,458028 | 0,866856 | y ~ current_feature + S |
| -0,10702 | 0,072131 | -0,07874 | -1,4837 | 193 | 190 | 0,139546 | 0,498495 | y ~ current_feature + S |
| -0,0922 | 0,072239 | 0,034007 | -1,2763 | 193 | 190 | 0,203406 | 0,65103 | y ~ current_feature + S |
| -0,02421 | 0,072718 | -0,04751 | -0,33298 | 192 | 189 | 0,739518 | 0,933594 | y ~ current_feature + S |
| 0,059696 | 0,078186 | 0,061498 | 0,763508 | 166 | 163 | 0,446264 | 0,937665 | y ~ current_feature + S |
| -0,14212 | 0,088183 | -0,02429 | -1,61163 | 129 | 126 | 0,109545 | 0,605474 | y ~ current_feature + S |
| 0,023829 | 0,094045 | -0,07874 | 0,25338 | 116 | 113 | 0,800435 | 0,979102 | y ~ current_feature + S |
| 0,037518 | 0,094006 | 0,034007 | 0,399103 | 116 | 113 | 0,690571 | 0,916569 | y ~ current_feature + S |
| 0,010056 | 0,094486 | -0,04751 | 0,106424 | 115 | 112 | 0,915436 | 0,985217 | y ~ current_feature + S |
| -0,01942 | 0,102043 | 0,061498 | -0,19034 | 99 | 96 | 0,849448 | 0,996179 | y ~ current_feature + S |
| 0,047445 | 0,116117 | -0,02429 | 0,408601 | 77 | 74 | 0,684013 | 0,926342 | y ~ current_feature + S |
| 0,16757 | 0,0585 | -0,07874 | 2,864441 | 287 | 284 | 0,004489 | 0,077255 | y ~ current_feature + S |
| 0,134077 | 0,058803 | 0,034007 | 2,280087 | 287 | 284 | 0,023344 | 0,338715 | y ~ current_feature + S |
| 0,067655 | 0,059308 | -0,04751 | 1,140739 | 286 | 283 | 0,254943 | 0,742928 | y ~ current_feature + S |
| 0,013532 | 0,073447 | 0,061498 | 0,184236 | 248 | 185,3432 | 0,85403 | 0,996179 | y ~ current_feature + S |
| 0,12343 | 0,072377 | -0,02429 | 1,705386 | 191 | 187,9896 | 0,089774 | 0,605474 | y ~ current_feature + S |
| 0,18306 | 0,078211 | -0,07874 | 2,34058 | 161 | 158 | 0,020503 | 0,189455 | y ~ current_feature + S |
| 0,092572 | 0,079214 | 0,034007 | 1,168628 | 161 | 158 | 0,244314 | 0,702074 | y ~ current_feature + S |
| 0,125161 | 0,079181 | -0,04751 | 1,580697 | 160 | 157 | 0,11596 | 0,640375 | y ~ current_feature + S |
| 0,008702 | 0,097039 | 0,061498 | 0,089673 | 139 | 106,1882 | 0,928716 | 0,996179 | y ~ current_feature + S |
| -0,1525 | 0,097166 | -0,02429 | -1,56944 | 107 | 103,4559 | 0,119598 | 0,615361 | y ~ current_feature + S |
| 0,285021 | 0,124789 | -0,07874 | 2,284025 | 62 | 59 | 0,025985 | 0,204564 | y ~ current_feature + S |
| 0,127632 | 0,129124 | 0,034007 | 0,988443 | 62 | 59 | 0,326972 | 0,728793 | y ~ current_feature + S |
| 0,177392 | 0,129224 | -0,04751 | 1,37275 | 61 | 58 | 0,175115 | 0,700235 | y ~ current_feature + S |
| 0,204454 | 0,139839 | 0,061498 | 1,462065 | 52 | 49 | 0,150108 | 0,8197 | y ~ current_feature + S |
| 0,079148 | 0,161713 | -0,02429 | 0,489438 | 41 | 38 | 0,627345 | 0,924682 | y ~ current_feature + S |
| 0,215477 | 0,088409 | -0,07874 | 2,437275 | 125 | 122 | 0,016241 | 0,174676 | y ~ current_feature + S |
| 0,08089 | 0,090239 | 0,034007 | 0,896391 | 125 | 122 | 0,371809 | 0,750317 | y ~ current_feature + S |
| 0,136947 | 0,090053 | -0,04751 | 1,52074 | 124 | 121 | 0,130934 | 0,654671 | y ~ current_feature + S |
| -0,05684 | 0,097432 | 0,061498 | -0,58335 | 108 | 105 | 0,56091 | 0,937665 | y ~ current_feature + S |
| 0,084933 | 0,111399 | -0,02429 | 0,762414 | 83 | 80 | 0,448055 | 0,858965 | y ~ current_feature + S |
| 0,034805 | 0,073279 | -0,07874 | 0,474962 | 189 | 186 | 0,635371 | 0,927169 | y ~ current_feature + S |
| -0,02394 | 0,073303 | 0,034007 | -0,32656 | 189 | 186 | 0,744369 | 0,927328 | y ~ current_feature + S |

| | | | | | | | | |
|---|---|---|---|---|---|---|---|---|
| -0,00534 | 0,073323 | -0,04751 | -0,0728 | 189 | 186 | 0,942046 | 0,990156 | y ~ current_feature + S |
| -0,15872 | 0,076401 | 0,061498 | -2,07745 | 170 | 167 | 0,03929 | 0,658491 | y ~ current_feature + S |
| 0,073832 | 0,089921 | -0,02429 | 0,821077 | 126 | 123 | 0,413191 | 0,837702 | y ~ current_feature + S |
| 0,179255 | 0,056541 | -0,07874 | 3,170346 | 364 | 302,7515 | 0,001679 | 0,051754 | y ~ current_feature + S |
| 0,027475 | 0,073457 | 0,034007 | 0,374031 | 364 | 185,1835 | 0,708809 | 0,920814 | y ~ current_feature + S |
| 0,070151 | 0,058647 | -0,04751 | 1,196154 | 363 | 289,3098 | 0,232616 | 0,742928 | y ~ current_feature + S |
| 0,127151 | 0,065123 | 0,061498 | 1,952477 | 315 | 231,9832 | 0,052084 | 0,71375 | y ~ current_feature + S |
| 0,114671 | 0,064124 | -0,02429 | 1,788269 | 243 | 240 | 0,074994 | 0,605474 | y ~ current_feature + S |
| 0,242295 | 0,101151 | -0,07874 | 2,395387 | 95 | 92 | 0,018629 | 0,181384 | y ~ current_feature + S |
| -0,09471 | 0,103789 | 0,034007 | -0,91252 | 95 | 92 | 0,363882 | 0,7498 | y ~ current_feature + S |
| 0,178066 | 0,103153 | -0,04751 | 1,726225 | 94 | 91 | 0,087701 | 0,640171 | y ~ current_feature + S |
| -0,03433 | 0,112442 | 0,061498 | -0,30531 | 82 | 79 | 0,760935 | 0,972573 | y ~ current_feature + S |
| 0,021578 | 0,129069 | -0,02429 | 0,167185 | 63 | 60 | 0,867787 | 0,984468 | y ~ current_feature + S |
| -0,1172 | 0,149717 | -0,07874 | -0,78284 | 47 | 44 | 0,437917 | 0,816268 | y ~ current_feature + S |
| -0,11442 | 0,149766 | 0,034007 | -0,76397 | 47 | 44 | 0,448965 | 0,814299 | y ~ current_feature + S |
| 0,220778 | 0,148736 | -0,04751 | 1,484367 | 46 | 43 | 0,145006 | 0,661823 | y ~ current_feature + S |
| -0,18264 | 0,157435 | 0,061498 | -1,16011 | 42 | 39 | 0,253057 | 0,893818 | y ~ current_feature + S |
| 0,30016 | 0,180268 | -0,02429 | 1,665073 | 31 | 28 | 0,10705 | 0,605474 | y ~ current_feature + S |
| -0,02734 | 0,058802 | -0,07874 | -0,46502 | 292 | 289 | 0,642266 | 0,928694 | y ~ current_feature + S |
| -0,07415 | 0,058662 | 0,034007 | -1,26403 | 292 | 289 | 0,207237 | 0,655365 | y ~ current_feature + S |
| -0,06739 | 0,058792 | -0,04751 | -1,14625 | 291 | 288 | 0,252644 | 0,742928 | y ~ current_feature + S |
| 0,064985 | 0,068715 | 0,061498 | 0,945715 | 251 | 210,8914 | 0,345376 | 0,907332 | y ~ current_feature + S |
| -0,08643 | 0,072114 | -0,02429 | -1,19858 | 195 | 190,8569 | 0,232177 | 0,695919 | y ~ current_feature + S |
| -0,00969 | 0,102057 | -0,07874 | -0,09491 | 99 | 96 | 0,924582 | 0,990519 | y ~ current_feature + S |
| 0,003855 | 0,102061 | 0,034007 | 0,037771 | 99 | 96 | 0,969949 | 0,989859 | y ~ current_feature + S |
| -0,08121 | 0,101725 | -0,04751 | -0,79835 | 99 | 96 | 0,426639 | 0,819126 | y ~ current_feature + S |
| -0,09453 | 0,110614 | 0,061498 | -0,85464 | 84 | 81 | 0,395272 | 0,93433 | y ~ current_feature + S |
| -0,20203 | 0,124381 | -0,02429 | -1,62424 | 65 | 62 | 0,109398 | 0,605474 | y ~ current_feature + S |
| -0,02052 | 0,091268 | -0,07874 | -0,22483 | 123 | 120 | 0,822496 | 0,981192 | y ~ current_feature + S |
| -0,15341 | 0,090207 | 0,034007 | -1,70065 | 123 | 120 | 0,0916 | 0,494772 | y ~ current_feature + S |
| -0,14468 | 0,090327 | -0,04751 | -1,60176 | 123 | 120 | 0,111839 | 0,640171 | y ~ current_feature + S |
| 0,117448 | 0,097851 | 0,061498 | 1,200272 | 106 | 103 | 0,232787 | 0,893818 | y ~ current_feature + S |
| -0,10181 | 0,111924 | -0,02429 | -0,90961 | 82 | 79 | 0,365793 | 0,807159 | y ~ current_feature + S |
| 0,064222 | 0,053572 | -0,07874 | 1,1988 | 350 | 347 | 0,231424 | 0,657483 | y ~ current_feature + S |
| 0,035664 | 0,053649 | 0,034007 | 0,664777 | 350 | 347 | 0,506635 | 0,841996 | y ~ current_feature + S |
| 0,06928 | 0,053631 | -0,04751 | 1,291785 | 349 | 346 | 0,197294 | 0,723562 | y ~ current_feature + S |
| -0,07035 | 0,057496 | 0,061498 | -1,22363 | 304 | 301 | 0,222048 | 0,893818 | y ~ current_feature + S |
| 0,05569 | 0,065836 | -0,02429 | 0,845892 | 233 | 230 | 0,398492 | 0,823699 | y ~ current_feature + S |
| -0,08587 | 0,100133 | -0,07874 | -0,85756 | 102 | 99 | 0,393208 | 0,789252 | y ~ current_feature + S |
| -0,1125 | 0,099866 | 0,034007 | -1,12656 | 102 | 99 | 0,262653 | 0,71095 | y ~ current_feature + S |
| -0,14893 | 0,099383 | -0,04751 | -1,49859 | 102 | 99 | 0,137161 | 0,657012 | y ~ current_feature + S |
| -0,09672 | 0,107957 | 0,061498 | -0,89595 | 88 | 85 | 0,372809 | 0,932536 | y ~ current_feature + S |
| -0,02488 | 0,123996 | -0,02429 | -0,20069 | 68 | 65 | 0,841569 | 0,977734 | y ~ current_feature + S |
| 0,199876 | 0,093001 | -0,07874 | 2,149189 | 114 | 111 | 0,033791 | 0,236995 | y ~ current_feature + S |
| 0,088167 | 0,094546 | 0,034007 | 0,932533 | 114 | 111 | 0,353085 | 0,7498 | y ~ current_feature + S |
| -0,02149 | 0,094894 | -0,04751 | -0,22642 | 114 | 111 | 0,821288 | 0,972405 | y ~ current_feature + S |
| 0,004981 | 0,099999 | 0,061498 | 0,04981 | 103 | 100 | 0,960374 | 0,996179 | y ~ current_feature + S |
| 0,006164 | 0,117039 | -0,02429 | 0,052668 | 76 | 73 | 0,95814 | 0,99444 | y ~ current_feature + S |

| | | | | | | | | |
|---|---|---|---|---|---|---|---|---|
| 0,222465 | 0,052464 | -0,07874 | 4,240336 | 393 | 345,3285 | 2,87E-05 | 0,005519 | y ~ current_feature + S |
| 0,142971 | 0,068215 | 0,034007 | 2,095907 | 393 | 210,5117 | 0,037285 | 0,388609 | y ~ current_feature + S |
| 0,139465 | 0,054207 | -0,04751 | 2,572827 | 392 | 333,7012 | 0,010519 | 0,481414 | y ~ current_feature + S |
| 0,010312 | 0,058899 | 0,061498 | 0,175084 | 341 | 288,2289 | 0,861137 | 0,996179 | y ~ current_feature + S |
| 0,111595 | 0,062733 | -0,02429 | 1,778901 | 261 | 250,9399 | 0,076466 | 0,605474 | y ~ current_feature + S |
| 0,101924 | 0,08233 | -0,07874 | 1,238001 | 149 | 146 | 0,217703 | 0,639357 | y ~ current_feature + S |
| -0,04878 | 0,082662 | 0,034007 | -0,59008 | 149 | 146 | 0,556048 | 0,857986 | y ~ current_feature + S |
| -0,01041 | 0,082756 | -0,04751 | -0,12575 | 149 | 146 | 0,900105 | 0,985217 | y ~ current_feature + S |
| -0,15025 | 0,088427 | 0,061498 | -1,69908 | 128 | 125 | 0,091791 | 0,795265 | y ~ current_feature + S |
| -0,06567 | 0,101315 | -0,02429 | -0,64816 | 100 | 97 | 0,518411 | 0,899808 | y ~ current_feature + S |
| -0,37481 | 0,12982 | -0,07874 | -2,88714 | 54 | 51 | 0,005689 | 0,092669 | y ~ current_feature + S |
| -0,47529 | 0,123201 | 0,034007 | -3,85782 | 54 | 51 | 3,23E-04 | 0,067708 | y ~ current_feature + S |
| 0,079911 | 0,13958 | -0,04751 | 0,572507 | 54 | 51 | 0,569494 | 0,903741 | y ~ current_feature + S |
| -0,19877 | 0,146097 | 0,061498 | -1,36053 | 48 | 45 | 0,180437 | 0,85592 | y ~ current_feature + S |
| -0,27225 | 0,167502 | -0,02429 | -1,62538 | 36 | 33 | 0,113596 | 0,614731 | y ~ current_feature + S |
| -0,01126 | 0,052844 | -0,07874 | -0,21311 | 403 | 358,0589 | 0,831361 | 0,981192 | y ~ current_feature + S |
| -0,05115 | 0,069495 | 0,034007 | -0,736 | 403 | 206,519 | 0,462565 | 0,830821 | y ~ current_feature + S |
| 0,028474 | 0,054633 | -0,04751 | 0,521191 | 402 | 334,7623 | 0,602578 | 0,91263 | y ~ current_feature + S |
| -0,06037 | 0,058545 | 0,061498 | -1,03117 | 351 | 290,6903 | 0,303317 | 0,893818 | y ~ current_feature + S |
| 0,001104 | 0,06145 | -0,02429 | 0,01796 | 268 | 264,821 | 0,985684 | 0,99444 | y ~ current_feature + S |
| 0,044044 | 0,062439 | -0,07874 | 0,70539 | 259 | 256 | 0,481209 | 0,845512 | y ~ current_feature + S |
| 0,01721 | 0,062491 | 0,034007 | 0,275399 | 259 | 256 | 0,783232 | 0,945731 | y ~ current_feature + S |
| 0,05466 | 0,062529 | -0,04751 | 0,87415 | 258 | 255 | 0,382859 | 0,803665 | y ~ current_feature + S |
| -0,00581 | 0,066666 | 0,061498 | -0,08718 | 228 | 225 | 0,930603 | 0,996179 | y ~ current_feature + S |
| 0,037457 | 0,076869 | -0,02429 | 0,48728 | 172 | 169 | 0,626691 | 0,924682 | y ~ current_feature + S |
| 0,214868 | 0,115906 | -0,07874 | 1,853808 | 74 | 71 | 0,067921 | 0,34663 | y ~ current_feature + S |
| 0,080227 | 0,118296 | 0,034007 | 0,678192 | 74 | 71 | 0,499855 | 0,841996 | y ~ current_feature + S |
| 0,13876 | 0,118367 | -0,04751 | 1,172291 | 73 | 70 | 0,245055 | 0,742928 | y ~ current_feature + S |
| -0,29163 | 0,118643 | 0,061498 | -2,45802 | 68 | 65 | 0,016645 | 0,55988 | y ~ current_feature + S |
| 3,09E-04 | 0,147442 | -0,02429 | 0,002093 | 49 | 46 | 0,998339 | 0,99969 | y ~ current_feature + S |
| -0,40964 | 0,133065 | -0,07874 | -3,07853 | 50 | 47 | 0,003467 | 0,069632 | y ~ current_feature + S |
| -0,2419 | 0,141533 | 0,034007 | -1,70916 | 50 | 47 | 0,094017 | 0,500522 | y ~ current_feature + S |
| -0,23183 | 0,141891 | -0,04751 | -1,63384 | 50 | 47 | 0,108975 | 0,640171 | y ~ current_feature + S |
| 0,0081 | 0,162216 | 0,061498 | 0,049934 | 41 | 38 | 0,960437 | 0,996179 | y ~ current_feature + S |
| 0,314337 | 0,17332 | -0,02429 | 1,813627 | 33 | 30 | 0,07975 | 0,605474 | y ~ current_feature + S |
| 0,106477 | 0,077407 | -0,07874 | 1,375537 | 168 | 165 | 0,170829 | 0,550681 | y ~ current_feature + S |
| 0,048361 | 0,077759 | 0,034007 | 0,621938 | 168 | 165 | 0,534841 | 0,849319 | y ~ current_feature + S |
| 0,074485 | 0,077634 | -0,04751 | 0,959442 | 168 | 165 | 0,33874 | 0,788263 | y ~ current_feature + S |
| -0,13845 | 0,084002 | 0,061498 | -1,6482 | 142 | 139 | 0,101571 | 0,8197 | y ~ current_feature + S |
| 0,203681 | 0,093775 | -0,02429 | 2,172025 | 112 | 109 | 0,032021 | 0,485321 | y ~ current_feature + S |
| 0,022234 | 0,081359 | -0,07874 | 0,273278 | 154 | 151 | 0,785013 | 0,974681 | y ~ current_feature + S |
| -0,01067 | 0,081374 | 0,034007 | -0,13107 | 154 | 151 | 0,895897 | 0,96591 | y ~ current_feature + S |
| -0,1582 | 0,080621 | -0,04751 | -1,9623 | 153 | 150 | 0,051578 | 0,640171 | y ~ current_feature + S |
| -0,07298 | 0,08781 | 0,061498 | -0,8311 | 132 | 129 | 0,407451 | 0,934833 | y ~ current_feature + S |
| -0,1399 | 0,099515 | -0,02429 | -1,4058 | 102 | 99 | 0,162913 | 0,623497 | y ~ current_feature + S |
| 0,008349 | 0,095343 | -0,07874 | 0,087572 | 113 | 110 | 0,930376 | 0,991839 | y ~ current_feature + S |
| -0,01481 | 0,095336 | 0,034007 | -0,15535 | 113 | 110 | 0,87683 | 0,96591 | y ~ current_feature + S |
| -0,07731 | 0,095496 | -0,04751 | -0,80956 | 112 | 109 | 0,419959 | 0,819126 | y ~ current_feature + S |

| | | | | | | | | |
|---|---|---|---|---|---|---|---|---|
| -0,07154 | 0,10399 | 0,061498 | -0,68794 | 95 | 92 | 0,493222 | 0,937665 | y ~ current_feature + S |
| -0,17406 | 0,116052 | -0,02429 | -1,49982 | 75 | 72 | 0,138032 | 0,618941 | y ~ current_feature + S |
| 0,080121 | 0,055985 | -0,07874 | 1,431123 | 320 | 317 | 0,15338 | 0,518271 | y ~ current_feature + S |
| 0,022196 | 0,056152 | 0,034007 | 0,39529 | 320 | 317 | 0,692895 | 0,916569 | y ~ current_feature + S |
| 0,024779 | 0,056148 | -0,04751 | 0,441312 | 320 | 317 | 0,659288 | 0,925756 | y ~ current_feature + S |
| 0,017405 | 0,064164 | 0,061498 | 0,271252 | 274 | 242,8208 | 0,786428 | 0,979725 | y ~ current_feature + S |
| 0,022919 | 0,068988 | -0,02429 | 0,332211 | 213 | 210 | 0,740061 | 0,944216 | y ~ current_feature + S |
| 0,126275 | 0,085377 | -0,07874 | 1,479019 | 138 | 135 | 0,141465 | 0,498495 | y ~ current_feature + S |
| 0,014318 | 0,086057 | 0,034007 | 0,16638 | 138 | 135 | 0,868107 | 0,96591 | y ~ current_feature + S |
| 0,029305 | 0,08635 | -0,04751 | 0,339371 | 137 | 134 | 0,734862 | 0,933491 | y ~ current_feature + S |
| 0,071073 | 0,094856 | 0,061498 | 0,749272 | 115 | 110,5795 | 0,455284 | 0,937665 | y ~ current_feature + S |
| 0,218732 | 0,103608 | -0,02429 | 2,111151 | 92 | 88,70006 | 0,037573 | 0,520476 | y ~ current_feature + S |
| 0,134635 | 0,065152 | -0,07874 | 2,066467 | 272 | 231,3103 | 0,039898 | 0,270866 | y ~ current_feature + S |
| 0,137277 | 0,060394 | 0,034007 | 2,273023 | 272 | 269 | 0,023814 | 0,338887 | y ~ current_feature + S |
| -0,02888 | 0,065751 | -0,04751 | -0,43916 | 271 | 231,1166 | 0,660957 | 0,926341 | y ~ current_feature + S |
| -0,11286 | 0,072412 | 0,061498 | -1,55864 | 234 | 188,2813 | 0,120761 | 0,8197 | y ~ current_feature + S |
| 0,035182 | 0,074907 | -0,02429 | 0,469678 | 181 | 177,9998 | 0,63916 | 0,924682 | y ~ current_feature + S |
| 0,022155 | 0,052759 | -0,07874 | 0,419925 | 421 | 359,08 | 0,674792 | 0,934653 | y ~ current_feature + S |
| -0,00496 | 0,069097 | 0,034007 | -0,07171 | 421 | 209,4453 | 0,942897 | 0,98181 | y ~ current_feature + S |
| 0,050789 | 0,054303 | -0,04751 | 0,935283 | 420 | 338,2462 | 0,35031 | 0,795089 | y ~ current_feature + S |
| 0,018413 | 0,059214 | 0,061498 | 0,310962 | 367 | 285,1058 | 0,756057 | 0,972573 | y ~ current_feature + S |
| 0,090282 | 0,059839 | -0,02429 | 1,508756 | 280 | 277 | 0,132501 | 0,617591 | y ~ current_feature + S |
| 0,056002 | 0,084383 | -0,07874 | 0,663672 | 143 | 140 | 0,507992 | 0,863164 | y ~ current_feature + S |
| -0,02615 | 0,084487 | 0,034007 | -0,30947 | 143 | 140 | 0,757427 | 0,93416 | y ~ current_feature + S |
| -0,03995 | 0,084751 | -0,04751 | -0,4714 | 142 | 139 | 0,638095 | 0,925756 | y ~ current_feature + S |
| 1,61E-04 | 0,090536 | 0,061498 | 0,001773 | 125 | 122 | 0,998588 | 0,999005 | y ~ current_feature + S |
| 0,006512 | 0,104255 | -0,02429 | 0,062461 | 95 | 92 | 0,950331 | 0,99444 | y ~ current_feature + S |
| 0,090942 | 0,139448 | -0,07874 | 0,652157 | 54 | 51 | 0,517228 | 0,873856 | y ~ current_feature + S |
| 0,024613 | 0,139986 | 0,034007 | 0,175824 | 54 | 51 | 0,861129 | 0,964047 | y ~ current_feature + S |
| 0,147744 | 0,138491 | -0,04751 | 1,066813 | 54 | 51 | 0,291081 | 0,747915 | y ~ current_feature + S |
| -0,15039 | 0,152548 | 0,061498 | -0,98584 | 45 | 42 | 0,329856 | 0,893818 | y ~ current_feature + S |
| 0,139978 | 0,172364 | -0,02429 | 0,812107 | 36 | 33 | 0,42255 | 0,847264 | y ~ current_feature + S |
| 0,089603 | 0,085092 | -0,07874 | 1,05301 | 140 | 137 | 0,29419 | 0,718687 | y ~ current_feature + S |
| 0,110177 | 0,084916 | 0,034007 | 1,297489 | 140 | 137 | 0,196644 | 0,643878 | y ~ current_feature + S |
| -0,06428 | 0,085572 | -0,04751 | -0,75119 | 139 | 136 | 0,453836 | 0,832044 | y ~ current_feature + S |
| 0,094816 | 0,09283 | 0,061498 | 1,021391 | 118 | 115 | 0,309213 | 0,893818 | y ~ current_feature + S |
| 0,195994 | 0,103365 | -0,02429 | 1,896141 | 93 | 90 | 0,06115 | 0,558651 | y ~ current_feature + S |
| 0,056699 | 0,070073 | -0,07874 | 0,809136 | 206 | 203 | 0,419384 | 0,811887 | y ~ current_feature + S |
| 0,003055 | 0,070186 | 0,034007 | 0,043526 | 206 | 203 | 0,965325 | 0,989859 | y ~ current_feature + S |
| 0,051216 | 0,070267 | -0,04751 | 0,728866 | 205 | 202 | 0,466928 | 0,842057 | y ~ current_feature + S |
| 0,061552 | 0,074602 | 0,061498 | 0,825071 | 182 | 179 | 0,410429 | 0,934833 | y ~ current_feature + S |
| 0,02504 | 0,08636 | -0,02429 | 0,289945 | 137 | 134 | 0,772307 | 0,957298 | y ~ current_feature + S |
| 0,05037 | 0,074316 | -0,07874 | 0,677774 | 185 | 180,6036 | 0,498783 | 0,861866 | y ~ current_feature + S |
| -0,03695 | 0,101348 | 0,034007 | -0,36458 | 185 | 97,22553 | 0,716215 | 0,920814 | y ~ current_feature + S |
| 0,004707 | 0,077969 | -0,04751 | 0,060369 | 184 | 164,4924 | 0,951935 | 0,990156 | y ~ current_feature + S |
| -0,31853 | 0,076521 | 0,061498 | -4,16261 | 159 | 153,4517 | 5,23E-05 | 0,012893 | y ~ current_feature + S |
| -0,05193 | 0,092223 | -0,02429 | -0,5631 | 123 | 117,259 | 0,574441 | 0,924682 | y ~ current_feature + S |
| -0,03874 | 0,063452 | -0,07874 | -0,61048 | 251 | 248 | 0,542106 | 0,882344 | y ~ current_feature + S |

| | | | | | | | | |
|---|---|---|---|---|---|---|---|---|
| -0,04203 | 0,063444 | 0,034007 | -0,66253 | 251 | 248 | 0,508247 | 0,841996 | y ~ current_feature + S |
| -0,06802 | 0,063481 | -0,04751 | -1,07149 | 250 | 247 | 0,284997 | 0,747915 | y ~ current_feature + S |
| 0,058869 | 0,067767 | 0,061498 | 0,8687 | 220 | 217 | 0,385971 | 0,93433 | y ~ current_feature + S |
| -0,07799 | 0,077849 | -0,02429 | -1,00179 | 167 | 164 | 0,317923 | 0,766329 | y ~ current_feature + S |
| 0,191287 | 0,081232 | -0,07874 | 2,354811 | 149 | 146 | 0,019863 | 0,188446 | y ~ current_feature + S |
| -0,08232 | 0,08248 | 0,034007 | -0,998 | 149 | 146 | 0,319929 | 0,723998 | y ~ current_feature + S |
| 0,275786 | 0,079825 | -0,04751 | 3,454889 | 148 | 145 | 7,22E-04 | 0,132163 | y ~ current_feature + S |
| -0,11289 | 0,088518 | 0,061498 | -1,27537 | 129 | 126 | 0,204526 | 0,893818 | y ~ current_feature + S |
| 0,002603 | 0,102062 | -0,02429 | 0,025508 | 99 | 96 | 0,979703 | 0,99444 | y ~ current_feature + S |
| 0,065093 | 0,080152 | -0,07874 | 0,812118 | 158 | 155 | 0,417969 | 0,811887 | y ~ current_feature + S |
| 0,050147 | 0,080221 | 0,034007 | 0,62511 | 158 | 155 | 0,532818 | 0,849319 | y ~ current_feature + S |
| 0,152835 | 0,079636 | -0,04751 | 1,919182 | 157 | 154 | 0,056811 | 0,640171 | y ~ current_feature + S |
| 0,075897 | 0,085818 | 0,061498 | 0,884396 | 138 | 135 | 0,378055 | 0,932536 | y ~ current_feature + S |
| 0,103785 | 0,09848 | -0,02429 | 1,053867 | 105 | 102 | 0,294434 | 0,741092 | y ~ current_feature + S |
| 0,089637 | 0,053789 | -0,07874 | 1,666463 | 390 | 342,8544 | 0,096534 | 0,431938 | y ~ current_feature + S |
| 0,21149 | 0,069125 | 0,034007 | 3,059519 | 390 | 199,9194 | 0,002521 | 0,133248 | y ~ current_feature + S |
| 0,114133 | 0,05471 | -0,04751 | 2,086132 | 389 | 329,7367 | 0,037735 | 0,611115 | y ~ current_feature + S |
| 0,025 | 0,058893 | 0,061498 | 0,424503 | 338 | 288,1407 | 0,671516 | 0,958606 | y ~ current_feature + S |
| 0,107939 | 0,062519 | -0,02429 | 1,726507 | 260 | 252,8664 | 0,085478 | 0,605474 | y ~ current_feature + S |
| -0,06245 | 0,071472 | -0,07874 | -0,87379 | 198 | 195 | 0,383306 | 0,789252 | y ~ current_feature + S |
| -0,06913 | 0,07144 | 0,034007 | -0,96759 | 198 | 195 | 0,334446 | 0,734392 | y ~ current_feature + S |
| -0,04561 | 0,071537 | -0,04751 | -0,63752 | 198 | 195 | 0,524534 | 0,876197 | y ~ current_feature + S |
| 0,058123 | 0,07612 | 0,061498 | 0,763565 | 175 | 172 | 0,446173 | 0,937665 | y ~ current_feature + S |
| -0,02927 | 0,088007 | -0,02429 | -0,3326 | 132 | 129 | 0,739978 | 0,944216 | y ~ current_feature + S |
| 0,051546 | 0,092327 | -0,07874 | 0,558302 | 120 | 117 | 0,577705 | 0,900004 | y ~ current_feature + S |
| 0,072408 | 0,092207 | 0,034007 | 0,785269 | 120 | 117 | 0,433883 | 0,796709 | y ~ current_feature + S |
| 0,142809 | 0,091502 | -0,04751 | 1,560711 | 120 | 117 | 0,121293 | 0,651244 | y ~ current_feature + S |
| 0,056133 | 0,099347 | 0,061498 | 0,565025 | 104 | 101 | 0,57331 | 0,937665 | y ~ current_feature + S |
| 0,047542 | 0,113832 | -0,02429 | 0,417648 | 80 | 77 | 0,677367 | 0,926342 | y ~ current_feature + S |
| 0,012396 | 0,072352 | -0,07874 | 0,171333 | 194 | 191 | 0,864144 | 0,985453 | y ~ current_feature + S |
| -0,01486 | 0,072349 | 0,034007 | -0,20533 | 194 | 191 | 0,837532 | 0,958985 | y ~ current_feature + S |
| 0,072062 | 0,072169 | -0,04751 | 0,998518 | 194 | 191 | 0,319291 | 0,774585 | y ~ current_feature + S |
| -0,11061 | 0,076908 | 0,061498 | -1,4382 | 170 | 167 | 0,152248 | 0,8197 | y ~ current_feature + S |
| 0,098272 | 0,088306 | -0,02429 | 1,112855 | 130 | 127 | 0,267873 | 0,723659 | y ~ current_feature + S |
| 0,151168 | 0,125541 | -0,07874 | 1,204136 | 65 | 62 | 0,233114 | 0,657771 | y ~ current_feature + S |
| 0,060255 | 0,126769 | 0,034007 | 0,475313 | 65 | 62 | 0,636234 | 0,899624 | y ~ current_feature + S |
| -0,21084 | 0,125159 | -0,04751 | -1,68456 | 64 | 61 | 0,097184 | 0,640171 | y ~ current_feature + S |
| -0,19646 | 0,132212 | 0,061498 | -1,48595 | 58 | 55 | 0,143002 | 0,8197 | y ~ current_feature + S |
| 0,118092 | 0,157008 | -0,02429 | 0,75214 | 43 | 40 | 0,45637 | 0,866856 | y ~ current_feature + S |
| 0,07918 | 0,051409 | -0,07874 | 1,5402 | 379 | 376 | 0,124353 | 0,478063 | y ~ current_feature + S |
| 0,090311 | 0,05136 | 0,034007 | 1,758373 | 379 | 376 | 0,079498 | 0,473527 | y ~ current_feature + S |
| 0,039762 | 0,05153 | -0,04751 | 0,771632 | 379 | 376 | 0,440817 | 0,823749 | y ~ current_feature + S |
| -0,00274 | 0,060821 | 0,061498 | -0,04505 | 328 | 270,3281 | 0,964097 | 0,996179 | y ~ current_feature + S |
| 0,067371 | 0,063389 | -0,02429 | 1,062806 | 252 | 247,7376 | 0,288905 | 0,734108 | y ~ current_feature + S |
| -0,04918 | 0,154117 | -0,07874 | -0,31912 | 45 | 42 | 0,751218 | 0,963434 | y ~ current_feature + S |
| -0,20247 | 0,151108 | 0,034007 | -1,33989 | 45 | 42 | 0,187485 | 0,638805 | y ~ current_feature + S |
| -0,3945 | 0,141788 | -0,04751 | -2,78235 | 45 | 42 | 0,00805 | 0,432614 | y ~ current_feature + S |
| -0,24189 | 0,155373 | 0,061498 | -1,55683 | 42 | 39 | 0,127589 | 0,8197 | y ~ current_feature + S |

| | | | | | | | | |
|---|---|---|---|---|---|---|---|---|
| -0,26689 | 0,185469 | -0,02429 | -1,43901 | 30 | 27 | 0,16164 | 0,623497 | y ~ current_feature + S |
| -0,14612 | 0,054875 | -0,07874 | -2,66278 | 328 | 325 | 0,008136 | 0,115775 | y ~ current_feature + S |
| -0,14088 | 0,054917 | 0,034007 | -2,56534 | 328 | 325 | 0,010756 | 0,233774 | y ~ current_feature + S |
| -0,06406 | 0,055441 | -0,04751 | -1,15554 | 327 | 324 | 0,248721 | 0,742928 | y ~ current_feature + S |
| 0,022144 | 0,05922 | 0,061498 | 0,373926 | 288 | 285 | 0,708737 | 0,972444 | y ~ current_feature + S |
| -0,10396 | 0,06783 | -0,02429 | -1,53269 | 218 | 215 | 0,126821 | 0,617591 | y ~ current_feature + S |
| 0,018132 | 0,096209 | -0,07874 | 0,188469 | 111 | 108 | 0,850863 | 0,983548 | y ~ current_feature + S |
| -0,06308 | 0,096033 | 0,034007 | -0,65681 | 111 | 108 | 0,512703 | 0,841996 | y ~ current_feature + S |
| -0,0031 | 0,096225 | -0,04751 | -0,03223 | 111 | 108 | 0,974348 | 0,992311 | y ~ current_feature + S |
| -0,00278 | 0,102597 | 0,061498 | -0,02713 | 98 | 95 | 0,978417 | 0,996179 | y ~ current_feature + S |
| 0,126514 | 0,117725 | -0,02429 | 1,074663 | 74 | 71 | 0,286164 | 0,734108 | y ~ current_feature + S |
| -0,13371 | 0,052273 | -0,07874 | -2,55797 | 388 | 359,4333 | 0,010938 | 0,140685 | y ~ current_feature + S |
| -0,08005 | 0,069321 | 0,034007 | -1,15476 | 388 | 206,7665 | 0,249521 | 0,702074 | y ~ current_feature + S |
| -0,02404 | 0,054445 | -0,04751 | -0,44147 | 387 | 337,1572 | 0,659158 | 0,925756 | y ~ current_feature + S |
| 0,051432 | 0,058163 | 0,061498 | 0,884279 | 341 | 294,823 | 0,377266 | 0,932536 | y ~ current_feature + S |
| -0,1168 | 0,062886 | -0,02429 | -1,85724 | 258 | 249,4142 | 0,064456 | 0,574667 | y ~ current_feature + S |
| 0,132606 | 0,069566 | -0,07874 | 1,90618 | 206 | 203 | 0,058039 | 0,318142 | y ~ current_feature + S |
| 0,020683 | 0,070171 | 0,034007 | 0,294752 | 206 | 203 | 0,768484 | 0,939964 | y ~ current_feature + S |
| 0,138683 | 0,06968 | -0,04751 | 1,990295 | 205 | 202 | 0,047907 | 0,640171 | y ~ current_feature + S |
| 0,08524 | 0,075534 | 0,061498 | 1,128504 | 177 | 174 | 0,260661 | 0,893818 | y ~ current_feature + S |
| 0,231063 | 0,084049 | -0,02429 | 2,74914 | 137 | 134 | 0,0068 | 0,26484 | y ~ current_feature + S |
| 0,073326 | 0,084896 | -0,07874 | 0,863707 | 141 | 138 | 0,389247 | 0,789252 | y ~ current_feature + S |
| -0,06355 | 0,084954 | 0,034007 | -0,74804 | 141 | 138 | 0,455707 | 0,821488 | y ~ current_feature + S |
| -0,15942 | 0,084037 | -0,04751 | -1,89705 | 141 | 138 | 0,059911 | 0,640171 | y ~ current_feature + S |
| -0,17169 | 0,091469 | 0,061498 | -1,87701 | 119 | 116 | 0,063029 | 0,715249 | y ~ current_feature + S |
| -0,01561 | 0,104816 | -0,02429 | -0,14891 | 94 | 91 | 0,881955 | 0,987777 | y ~ current_feature + S |
| 0,003047 | 0,068519 | -0,07874 | 0,044468 | 216 | 213 | 0,964573 | 0,994607 | y ~ current_feature + S |
| 0,006476 | 0,068517 | 0,034007 | 0,094509 | 216 | 213 | 0,924793 | 0,9732 | y ~ current_feature + S |
| 0,184876 | 0,067338 | -0,04751 | 2,745507 | 216 | 213 | 0,006558 | 0,432614 | y ~ current_feature + S |
| 0,008976 | 0,075691 | 0,061498 | 0,118587 | 188 | 174,5307 | 0,905739 | 0,996179 | y ~ current_feature + S |
| 0,093783 | 0,083844 | -0,02429 | 1,118544 | 144 | 141 | 0,265237 | 0,723659 | y ~ current_feature + S |
| -0,05993 | 0,102413 | -0,07874 | -0,58516 | 98 | 95 | 0,559825 | 0,894753 | y ~ current_feature + S |
| -0,08454 | 0,102231 | 0,034007 | -0,82695 | 98 | 95 | 0,41034 | 0,784629 | y ~ current_feature + S |
| -0,08225 | 0,102793 | -0,04751 | -0,80015 | 97 | 94 | 0,425643 | 0,819126 | y ~ current_feature + S |
| -0,06875 | 0,106958 | 0,061498 | -0,64274 | 90 | 87 | 0,522081 | 0,937665 | y ~ current_feature + S |
| 0,010615 | 0,126993 | -0,02429 | 0,083586 | 65 | 62 | 0,933655 | 0,99444 | y ~ current_feature + S |
| 0,288602 | 0,123606 | -0,07874 | 2,334849 | 63 | 60 | 0,022915 | 0,194866 | y ~ current_feature + S |
| 0,222473 | 0,125864 | 0,034007 | 1,767565 | 63 | 60 | 0,082218 | 0,473527 | y ~ current_feature + S |
| 0,07204 | 0,128764 | -0,04751 | 0,559471 | 63 | 60 | 0,577923 | 0,904733 | y ~ current_feature + S |
| 0,092847 | 0,135495 | 0,061498 | 0,68524 | 57 | 54 | 0,496124 | 0,937665 | y ~ current_feature + S |
| 0,138725 | 0,15858 | -0,02429 | 0,874799 | 42 | 39 | 0,387039 | 0,819479 | y ~ current_feature + S |
| 0,188257 | 0,055161 | -0,07874 | 3,412838 | 320 | 317 | 7,26E-04 | 0,031617 | y ~ current_feature + S |
| 0,179095 | 0,055257 | 0,034007 | 3,241102 | 320 | 317 | 0,001317 | 0,108307 | y ~ current_feature + S |
| 0,065591 | 0,056045 | -0,04751 | 1,170339 | 320 | 317 | 0,242744 | 0,742928 | y ~ current_feature + S |
| 0,066012 | 0,06028 | 0,061498 | 1,095076 | 277 | 274 | 0,274445 | 0,893818 | y ~ current_feature + S |
| 0,225656 | 0,067227 | -0,02429 | 3,356646 | 213 | 210 | 9,36E-04 | 0,138591 | y ~ current_feature + S |
| 0,060358 | 0,059759 | -0,07874 | 1,01002 | 282 | 279 | 0,313361 | 0,718687 | y ~ current_feature + S |
| -0,06491 | 0,059742 | 0,034007 | -1,08654 | 282 | 279 | 0,278179 | 0,715049 | y ~ current_feature + S |

| | | | | | | | | |
|---|---|---|---|---|---|---|---|---|
| -0,06762 | 0,059839 | -0,04751 | -1,13008 | 281 | 278 | 0,259418 | 0,742928 | y ~ current_feature + S |
| -0,06513 | 0,063883 | 0,061498 | -1,01956 | 247 | 244 | 0,308945 | 0,893818 | y ~ current_feature + S |
| -0,11923 | 0,072997 | -0,02429 | -1,63336 | 188 | 185 | 0,104093 | 0,605474 | y ~ current_feature + S |
| -0,11588 | 0,07618 | -0,07874 | -1,52108 | 173 | 170 | 0,130099 | 0,483784 | y ~ current_feature + S |
| -0,14193 | 0,07592 | 0,034007 | -1,86944 | 173 | 170 | 0,063281 | 0,437643 | y ~ current_feature + S |
| -0,11471 | 0,076415 | -0,04751 | -1,50116 | 172 | 169 | 0,135179 | 0,657012 | y ~ current_feature + S |
| -0,02374 | 0,083022 | 0,061498 | -0,28593 | 148 | 145 | 0,775341 | 0,975031 | y ~ current_feature + S |
| -0,00245 | 0,094491 | -0,02429 | -0,02596 | 115 | 112 | 0,979336 | 0,99444 | y ~ current_feature + S |
| 0,212052 | 0,096763 | -0,07874 | 2,191455 | 105 | 102 | 0,030694 | 0,224043 | y ~ current_feature + S |
| 0,016997 | 0,099 | 0,034007 | 0,171688 | 105 | 102 | 0,864023 | 0,965827 | y ~ current_feature + S |
| 0,003863 | 0,099014 | -0,04751 | 0,039012 | 105 | 102 | 0,968957 | 0,992311 | y ~ current_feature + S |
| 0,14724 | 0,103686 | 0,061498 | 1,420058 | 94 | 91 | 0,159008 | 0,8197 | y ~ current_feature + S |
| 0,175339 | 0,120302 | -0,02429 | 1,457482 | 70 | 66,97149 | 0,149657 | 0,622449 | y ~ current_feature + S |
| 0,183264 | 0,057236 | -0,07874 | 3,201889 | 298 | 295 | 0,001514 | 0,048726 | y ~ current_feature + S |
| 0,089055 | 0,057991 | 0,034007 | 1,535671 | 298 | 295 | 0,125691 | 0,572145 | y ~ current_feature + S |
| 0,067152 | 0,058091 | -0,04751 | 1,15598 | 298 | 295 | 0,248625 | 0,742928 | y ~ current_feature + S |
| -0,11229 | 0,062226 | 0,061498 | -1,80461 | 258 | 255 | 0,072316 | 0,721272 | y ~ current_feature + S |
| 0,099989 | 0,071071 | -0,02429 | 1,406898 | 199 | 196 | 0,161041 | 0,623497 | y ~ current_feature + S |
| 0,042623 | 0,056292 | -0,07874 | 0,757165 | 318 | 315 | 0,449517 | 0,823372 | y ~ current_feature + S |
| -0,0332 | 0,056313 | 0,034007 | -0,58949 | 318 | 315 | 0,555955 | 0,857986 | y ~ current_feature + S |
| 0,045075 | 0,056376 | -0,04751 | 0,799543 | 317 | 314 | 0,42458 | 0,819126 | y ~ current_feature + S |
| -0,00578 | 0,060633 | 0,061498 | -0,09525 | 275 | 272 | 0,92419 | 0,996179 | y ~ current_feature + S |
| 0,16245 | 0,068253 | -0,02429 | 2,380134 | 212 | 209 | 0,018205 | 0,428958 | y ~ current_feature + S |
| 0,054136 | 0,113062 | -0,07874 | 0,47882 | 81 | 78 | 0,633406 | 0,926325 | y ~ current_feature + S |
| 0,113457 | 0,112497 | 0,034007 | 1,008536 | 81 | 78 | 0,316316 | 0,718742 | y ~ current_feature + S |
| 0,070439 | 0,112946 | -0,04751 | 0,623648 | 81 | 78 | 0,534678 | 0,883994 | y ~ current_feature + S |
| 0,013806 | 0,11784 | 0,061498 | 0,11716 | 75 | 72 | 0,907059 | 0,996179 | y ~ current_feature + S |
| 0,122877 | 0,138967 | -0,02429 | 0,884215 | 54 | 51 | 0,38073 | 0,816637 | y ~ current_feature + S |
| 0,065282 | 0,067584 | -0,07874 | 0,965933 | 221 | 218 | 0,335148 | 0,735933 | y ~ current_feature + S |
| -0,10589 | 0,067348 | 0,034007 | -1,57225 | 221 | 218 | 0,117343 | 0,552558 | y ~ current_feature + S |
| 0,052829 | 0,06779 | -0,04751 | 0,779309 | 220 | 217 | 0,436647 | 0,823749 | y ~ current_feature + S |
| 0,013102 | 0,073121 | 0,061498 | 0,179187 | 190 | 187 | 0,857985 | 0,996179 | y ~ current_feature + S |
| 0,002579 | 0,083333 | -0,02429 | 0,030951 | 147 | 144 | 0,975351 | 0,99444 | y ~ current_feature + S |
| -0,02042 | 0,083628 | -0,07874 | -0,24419 | 158 | 142,9257 | 0,807437 | 0,979871 | y ~ current_feature + S |
| 0,099612 | 0,098702 | 0,034007 | 1,009223 | 158 | 101,6297 | 0,315265 | 0,718742 | y ~ current_feature + S |
| 0,219995 | 0,078354 | -0,04751 | 2,807703 | 158 | 155 | 0,005631 | 0,422981 | y ~ current_feature + S |
| 0,098623 | 0,091199 | 0,061498 | 1,081404 | 140 | 119,062 | 0,281703 | 0,893818 | y ~ current_feature + S |
| 0,017132 | 0,100612 | -0,02429 | 0,17028 | 105 | 98,75906 | 0,865138 | 0,984468 | y ~ current_feature + S |
| 0,034669 | 0,063462 | -0,07874 | 0,546294 | 251 | 248 | 0,585355 | 0,902886 | y ~ current_feature + S |
| -0,01458 | 0,063493 | 0,034007 | -0,22956 | 251 | 248 | 0,818623 | 0,952501 | y ~ current_feature + S |
| 0,003237 | 0,0635 | -0,04751 | 0,050982 | 251 | 248 | 0,959381 | 0,990156 | y ~ current_feature + S |
| -0,00285 | 0,068358 | 0,061498 | -0,04162 | 217 | 214 | 0,966837 | 0,996179 | y ~ current_feature + S |
| 0,003809 | 0,077849 | -0,02429 | 0,048924 | 168 | 165 | 0,961039 | 0,99444 | y ~ current_feature + S |
| -0,00112 | 0,055902 | -0,07874 | -0,01997 | 323 | 320 | 0,98408 | 0,996318 | y ~ current_feature + S |
| -0,00512 | 0,055901 | 0,034007 | -0,09154 | 323 | 320 | 0,927119 | 0,9732 | y ~ current_feature + S |
| 0,137694 | 0,055456 | -0,04751 | 2,482942 | 322 | 319 | 0,013544 | 0,526293 | y ~ current_feature + S |
| 0,001049 | 0,066318 | 0,061498 | 0,01581 | 279 | 227,375 | 0,987399 | 0,997945 | y ~ current_feature + S |
| 0,109309 | 0,068856 | -0,02429 | 1,5875 | 215 | 208,3979 | 0,113915 | 0,614731 | y ~ current_feature + S |

| | | | | | | | | |
|---|---|---|---|---|---|---|---|---|
| -0,04029 | 0,111021 | -0,07874 | -0,36291 | 84 | 81 | 0,717614 | 0,948276 | y ~ current_feature + S |
| 0,023275 | 0,111081 | 0,034007 | 0,209531 | 84 | 81 | 0,83456 | 0,958966 | y ~ current_feature + S |
| -0,16868 | 0,109519 | -0,04751 | -1,54021 | 84 | 81 | 0,127407 | 0,654671 | y ~ current_feature + S |
| -0,06793 | 0,115203 | 0,061498 | -0,58965 | 78 | 75 | 0,557195 | 0,937665 | y ~ current_feature + S |
| -0,02349 | 0,137323 | -0,02429 | -0,17106 | 56 | 53 | 0,864831 | 0,984468 | y ~ current_feature + S |
| 0,015373 | 0,052193 | -0,07874 | 0,294545 | 382 | 366,999 | 0,768508 | 0,965616 | y ~ current_feature + S |
| -0,01903 | 0,066921 | 0,034007 | -0,2843 | 382 | 223,2114 | 0,776446 | 0,942718 | y ~ current_feature + S |
| 0,038189 | 0,053084 | -0,04751 | 0,719404 | 381 | 354,3579 | 0,472366 | 0,842057 | y ~ current_feature + S |
| 0,028183 | 0,055988 | 0,061498 | 0,503383 | 332 | 318,764 | 0,615043 | 0,948092 | y ~ current_feature + S |
| 0,020592 | 0,065082 | -0,02429 | 0,316394 | 254 | 235,9871 | 0,751983 | 0,951227 | y ~ current_feature + S |
| -0,06655 | 0,05469 | -0,07874 | -1,21687 | 370 | 332,8561 | 0,224517 | 0,646468 | y ~ current_feature + S |
| -0,11561 | 0,072818 | 0,034007 | -1,58758 | 370 | 186,0687 | 0,114079 | 0,548173 | y ~ current_feature + S |
| -0,03261 | 0,058264 | -0,04751 | -0,55965 | 369 | 294,2637 | 0,576144 | 0,904733 | y ~ current_feature + S |
| -0,1122 | 0,063085 | 0,061498 | -1,77858 | 320 | 248,1103 | 0,076533 | 0,735511 | y ~ current_feature + S |
| -0,07763 | 0,063956 | -0,02429 | -1,21383 | 246 | 243 | 0,225993 | 0,695919 | y ~ current_feature + S |
| 0,026537 | 0,049971 | -0,07874 | 0,53105 | 412 | 400,1772 | 0,595679 | 0,903286 | y ~ current_feature + S |
| 0,028837 | 0,063552 | 0,034007 | 0,453752 | 412 | 247,3917 | 0,650405 | 0,900251 | y ~ current_feature + S |
| -0,07539 | 0,050714 | -0,04751 | -1,48666 | 411 | 386,6077 | 0,137919 | 0,657012 | y ~ current_feature + S |
| -0,0117 | 0,054192 | 0,061498 | -0,21592 | 359 | 340,4669 | 0,829177 | 0,996179 | y ~ current_feature + S |
| 0,041203 | 0,063112 | -0,02429 | 0,652855 | 274 | 250,6363 | 0,514448 | 0,899808 | y ~ current_feature + S |
| -0,0032 | 0,055048 | -0,07874 | -0,05821 | 333 | 330 | 0,953617 | 0,994607 | y ~ current_feature + S |
| -0,10461 | 0,054746 | 0,034007 | -1,91076 | 333 | 330 | 0,056902 | 0,420946 | y ~ current_feature + S |
| -0,01704 | 0,05504 | -0,04751 | -0,30962 | 333 | 330 | 0,757045 | 0,9394 | y ~ current_feature + S |
| 0,008116 | 0,059233 | 0,061498 | 0,137011 | 288 | 285 | 0,891119 | 0,996179 | y ~ current_feature + S |
| -0,09812 | 0,067402 | -0,02429 | -1,45567 | 221 | 218 | 0,146921 | 0,621266 | y ~ current_feature + S |
| -0,05758 | 0,062519 | -0,07874 | -0,92094 | 258 | 255 | 0,357952 | 0,767781 | y ~ current_feature + S |
| -0,04395 | 0,062562 | 0,034007 | -0,70246 | 258 | 255 | 0,483033 | 0,831402 | y ~ current_feature + S |
| -0,06384 | 0,062495 | -0,04751 | -1,02152 | 258 | 255 | 0,307974 | 0,76529 | y ~ current_feature + S |
| -0,04059 | 0,067673 | 0,061498 | -0,59983 | 221 | 218 | 0,549242 | 0,937665 | y ~ current_feature + S |
| -0,03088 | 0,076886 | -0,02429 | -0,40158 | 172 | 169 | 0,688504 | 0,926342 | y ~ current_feature + S |
| -0,0785 | 0,054795 | -0,07874 | -1,43264 | 334 | 331 | 0,152905 | 0,518271 | y ~ current_feature + S |
| -0,05143 | 0,054892 | 0,034007 | -0,93692 | 334 | 331 | 0,349485 | 0,749214 | y ~ current_feature + S |
| -0,10566 | 0,05474 | -0,04751 | -1,93015 | 333 | 330 | 0,054446 | 0,640171 | y ~ current_feature + S |
| -0,10104 | 0,06908 | 0,061498 | -1,46263 | 289 | 207,4116 | 0,145083 | 0,8197 | y ~ current_feature + S |
| -0,02118 | 0,067706 | -0,02429 | -0,31282 | 222 | 218,0471 | 0,754717 | 0,951432 | y ~ current_feature + S |
| 0,111295 | 0,050087 | -0,07874 | 2,222022 | 410 | 393,6732 | 0,02685 | 0,204832 | y ~ current_feature + S |
| 0,118201 | 0,064176 | 0,034007 | 1,841834 | 410 | 239,4121 | 0,066736 | 0,439525 | y ~ current_feature + S |
| 0,045083 | 0,051119 | -0,04751 | 0,881911 | 409 | 381,8981 | 0,378379 | 0,803665 | y ~ current_feature + S |
| 0,087865 | 0,054556 | 0,061498 | 1,610565 | 357 | 333,3928 | 0,108221 | 0,8197 | y ~ current_feature + S |
| 0,101922 | 0,062227 | -0,02429 | 1,637899 | 273 | 255,5664 | 0,102673 | 0,605474 | y ~ current_feature + S |
| 0,079838 | 0,066602 | -0,07874 | 1,198739 | 227 | 224 | 0,231896 | 0,657483 | y ~ current_feature + S |
| 0,126474 | 0,066279 | 0,034007 | 1,908219 | 227 | 224 | 0,057641 | 0,420946 | y ~ current_feature + S |
| 0,070083 | 0,0668 | -0,04751 | 1,049145 | 226 | 223 | 0,295247 | 0,752909 | y ~ current_feature + S |
| 0,044426 | 0,072286 | 0,061498 | 0,61459 | 194 | 191 | 0,539557 | 0,937665 | y ~ current_feature + S |
| 0,241856 | 0,080267 | -0,02429 | 3,013144 | 151 | 146,1333 | 0,003048 | 0,18344 | y ~ current_feature + S |
| 0,070392 | 0,060044 | -0,07874 | 1,172346 | 279 | 276 | 0,242069 | 0,670903 | y ~ current_feature + S |
| 0,105035 | 0,05986 | 0,034007 | 1,754683 | 279 | 276 | 0,080423 | 0,473527 | y ~ current_feature + S |
| 0,0269 | 0,06028 | -0,04751 | 0,446243 | 278 | 275 | 0,655773 | 0,925756 | y ~ current_feature + S |

| | | | | | | | | |
|---|---|---|---|---|---|---|---|---|
| 0,027153 | 0,064933 | 0,061498 | 0,418164 | 240 | 237 | 0,676206 | 0,958606 | y ~ current_feature + S |
| 0,097627 | 0,073771 | -0,02429 | 1,323375 | 185 | 182 | 0,187371 | 0,65347 | y ~ current_feature + S |
| 0,091943 | 0,05532 | -0,07874 | 1,66201 | 327 | 324 | 0,097478 | 0,431938 | y ~ current_feature + S |
| 0,11341 | 0,055197 | 0,034007 | 2,054639 | 327 | 324 | 0,040716 | 0,396441 | y ~ current_feature + S |
| 0,032637 | 0,055526 | -0,04751 | 0,58777 | 327 | 324 | 0,557096 | 0,897386 | y ~ current_feature + S |
| 0,012898 | 0,06571 | 0,061498 | 0,196293 | 281 | 231,5606 | 0,844553 | 0,996179 | y ~ current_feature + S |
| 0,089807 | 0,068082 | -0,02429 | 1,319092 | 217 | 214 | 0,188548 | 0,65347 | y ~ current_feature + S |
| 0,152467 | 0,063141 | -0,07874 | 2,414714 | 248 | 245 | 0,016481 | 0,174676 | y ~ current_feature + S |
| 0,076451 | 0,063701 | 0,034007 | 1,200155 | 248 | 245 | 0,231238 | 0,681978 | y ~ current_feature + S |
| 0,001928 | 0,064018 | -0,04751 | 0,03011 | 247 | 244 | 0,976004 | 0,992311 | y ~ current_feature + S |
| 0,043493 | 0,068454 | 0,061498 | 0,635364 | 216 | 213 | 0,525873 | 0,937665 | y ~ current_feature + S |
| 0,034507 | 0,078521 | -0,02429 | 0,439463 | 165 | 162 | 0,660911 | 0,924682 | y ~ current_feature + S |
| -0,00183 | 0,081111 | -0,07874 | -0,02255 | 155 | 152 | 0,982037 | 0,996318 | y ~ current_feature + S |
| 0,08841 | 0,080793 | 0,034007 | 1,094273 | 155 | 152 | 0,275566 | 0,715049 | y ~ current_feature + S |
| -0,01018 | 0,081107 | -0,04751 | -0,12549 | 155 | 152 | 0,9003 | 0,985217 | y ~ current_feature + S |
| -0,1572 | 0,087289 | 0,061498 | -1,80088 | 131 | 128 | 0,074077 | 0,721272 | y ~ current_feature + S |
| 0,108092 | 0,098921 | -0,02429 | 1,092713 | 104 | 101 | 0,277119 | 0,734108 | y ~ current_feature + S |
| 0,071863 | 0,145488 | -0,07874 | 0,493944 | 50 | 47 | 0,623647 | 0,919937 | y ~ current_feature + S |
| 0,178814 | 0,143514 | 0,034007 | 1,245965 | 50 | 47 | 0,218952 | 0,664036 | y ~ current_feature + S |
| 0,202113 | 0,144399 | -0,04751 | 1,39968 | 49 | 46 | 0,168317 | 0,700235 | y ~ current_feature + S |
| -0,20934 | 0,150885 | 0,061498 | -1,38739 | 45 | 42 | 0,172641 | 0,840578 | y ~ current_feature + S |
| 0,337314 | 0,171874 | -0,02429 | 1,962565 | 33 | 30 | 0,05903 | 0,558147 | y ~ current_feature + S |
| -0,06208 | 0,0859 | -0,07874 | -0,72269 | 138 | 135 | 0,471122 | 0,840689 | y ~ current_feature + S |
| -0,05562 | 0,085933 | 0,034007 | -0,64726 | 138 | 135 | 0,518563 | 0,841996 | y ~ current_feature + S |
| -0,03821 | 0,086003 | -0,04751 | -0,44432 | 138 | 135 | 0,657521 | 0,925756 | y ~ current_feature + S |
| -0,00339 | 0,09245 | 0,061498 | -0,03663 | 120 | 117 | 0,970839 | 0,996179 | y ~ current_feature + S |
| 0,10023 | 0,105466 | -0,02429 | 0,950353 | 92 | 89 | 0,344507 | 0,789404 | y ~ current_feature + S |
| 0,08032 | 0,150269 | -0,07874 | 0,534509 | 47 | 44 | 0,59568 | 0,903286 | y ~ current_feature + S |
| -0,03713 | 0,150652 | 0,034007 | -0,2465 | 47 | 44 | 0,806445 | 0,952501 | y ~ current_feature + S |
| -0,24019 | 0,148034 | -0,04751 | -1,62252 | 46 | 43 | 0,112003 | 0,640171 | y ~ current_feature + S |
| 0,065757 | 0,159782 | 0,061498 | 0,411541 | 42 | 39 | 0,682929 | 0,962605 | y ~ current_feature + S |
| 0,414536 | 0,17198 | -0,02429 | 2,410372 | 31 | 28 | 0,022751 | 0,467656 | y ~ current_feature + S |
| 0,097387 | 0,093625 | -0,07874 | 1,040181 | 116 | 113 | 0,300476 | 0,718687 | y ~ current_feature + S |
| -0,1193 | 0,0934 | 0,034007 | -1,27732 | 116 | 113 | 0,204107 | 0,65103 | y ~ current_feature + S |
| 0,003668 | 0,09449 | -0,04751 | 0,038821 | 115 | 112 | 0,969103 | 0,992311 | y ~ current_feature + S |
| 0,071208 | 0,100759 | 0,061498 | 0,706713 | 101 | 98 | 0,481422 | 0,937665 | y ~ current_feature + S |
| -0,0295 | 0,116197 | -0,02429 | -0,25385 | 77 | 74 | 0,800315 | 0,968297 | y ~ current_feature + S |
| 0,527493 | 0,12106 | -0,07874 | 4,357278 | 57 | 49,24744 | 6,68E-05 | 0,008233 | y ~ current_feature + S |
| 0,279304 | 0,182817 | 0,034007 | 1,52778 | 57 | 27,58621 | 0,137953 | 0,58036 | y ~ current_feature + S |
| 0,074223 | 0,141827 | -0,04751 | 0,523339 | 57 | 49,44074 | 0,603076 | 0,91263 | y ~ current_feature + S |
| -0,0812 | 0,142451 | 0,061498 | -0,57 | 52 | 48,9549 | 0,571286 | 0,937665 | y ~ current_feature + S |
| 0,467127 | 0,149455 | -0,02429 | 3,125526 | 38 | 35 | 0,003559 | 0,184706 | y ~ current_feature + S |
| 0,080661 | 0,073481 | -0,07874 | 1,097721 | 187 | 184 | 0,273761 | 0,705864 | y ~ current_feature + S |
| 0,063155 | 0,073574 | 0,034007 | 0,858387 | 187 | 184 | 0,391795 | 0,773143 | y ~ current_feature + S |
| 0,00395 | 0,073922 | -0,04751 | 0,053433 | 186 | 183 | 0,957445 | 0,990156 | y ~ current_feature + S |
| -0,0736 | 0,07934 | 0,061498 | -0,9277 | 161 | 158 | 0,354981 | 0,920619 | y ~ current_feature + S |
| 0,032848 | 0,090487 | -0,02429 | 0,363014 | 125 | 122 | 0,717223 | 0,936058 | y ~ current_feature + S |
| -0,01464 | 0,068351 | -0,07874 | -0,2142 | 217 | 214 | 0,830593 | 0,981192 | y ~ current_feature + S |

| | | | | | | | | |
|---|---|---|---|---|---|---|---|---|
| -0,00189 | 0,068358 | 0,034007 | -0,02758 | 217 | 214 | 0,978022 | 0,989859 | y ~ current_feature + S |
| 0,11563 | 0,0679 | -0,04751 | 1,70295 | 217 | 214 | 0,090029 | 0,640171 | y ~ current_feature + S |
| -0,0028 | 0,073127 | 0,061498 | -0,03831 | 190 | 187 | 0,96948 | 0,996179 | y ~ current_feature + S |
| 0,030672 | 0,083879 | -0,02429 | 0,365665 | 145 | 142 | 0,715159 | 0,936058 | y ~ current_feature + S |
| 0,073464 | 0,066193 | -0,07874 | 1,109839 | 230 | 227 | 0,268243 | 0,702573 | y ~ current_feature + S |
| -0,03118 | 0,06634 | 0,034007 | -0,46995 | 230 | 227 | 0,638839 | 0,899624 | y ~ current_feature + S |
| -0,03817 | 0,066471 | -0,04751 | -0,5742 | 229 | 226 | 0,566404 | 0,903656 | y ~ current_feature + S |
| -0,13285 | 0,071344 | 0,061498 | -1,86213 | 196 | 193 | 0,064105 | 0,715249 | y ~ current_feature + S |
| -0,02797 | 0,081618 | -0,02429 | -0,34269 | 153 | 150 | 0,732311 | 0,944216 | y ~ current_feature + S |
| 0,017338 | 0,079293 | -0,07874 | 0,218655 | 162 | 159 | 0,827199 | 0,981192 | y ~ current_feature + S |
| -0,0909 | 0,078977 | 0,034007 | -1,15091 | 162 | 159 | 0,251496 | 0,704252 | y ~ current_feature + S |
| -0,01239 | 0,079299 | -0,04751 | -0,15627 | 162 | 159 | 0,876017 | 0,985217 | y ~ current_feature + S |
| -0,11541 | 0,084865 | 0,061498 | -1,35987 | 140 | 137 | 0,176104 | 0,846213 | y ~ current_feature + S |
| -0,1177 | 0,096912 | -0,02429 | -1,21451 | 108 | 105 | 0,227277 | 0,695919 | y ~ current_feature + S |
| 0,032504 | 0,052243 | -0,07874 | 0,62216 | 369 | 366 | 0,534224 | 0,882344 | y ~ current_feature + S |
| 0,060381 | 0,052175 | 0,034007 | 1,157277 | 369 | 366 | 0,247914 | 0,702074 | y ~ current_feature + S |
| 0,021222 | 0,052331 | -0,04751 | 0,405546 | 368 | 365 | 0,685314 | 0,928814 | y ~ current_feature + S |
| -0,0389 | 0,056123 | 0,061498 | -0,69308 | 320 | 317 | 0,488769 | 0,937665 | y ~ current_feature + S |
| 0,136939 | 0,063546 | -0,02429 | 2,154966 | 246 | 243 | 0,032147 | 0,485321 | y ~ current_feature + S |
| -0,11814 | 0,049083 | -0,07874 | -2,40706 | 418 | 409,2989 | 0,016523 | 0,174676 | y ~ current_feature + S |
| -0,10728 | 0,062239 | 0,034007 | -1,72369 | 418 | 255,1828 | 0,085975 | 0,479777 | y ~ current_feature + S |
| -0,05806 | 0,050245 | -0,04751 | -1,15564 | 417 | 394,7808 | 0,248528 | 0,742928 | y ~ current_feature + S |
| -0,12909 | 0,053226 | 0,061498 | -2,42538 | 364 | 347,1053 | 0,015802 | 0,55988 | y ~ current_feature + S |
| -0,06311 | 0,062336 | -0,02429 | -1,01235 | 278 | 256,3273 | 0,312324 | 0,765236 | y ~ current_feature + S |
| 0,06732 | 0,153953 | -0,07874 | 0,437277 | 45 | 42 | 0,664151 | 0,934653 | y ~ current_feature + S |
| 0,166113 | 0,15216 | 0,034007 | 1,091699 | 45 | 42 | 0,281188 | 0,715049 | y ~ current_feature + S |
| 0,0949 | 0,153607 | -0,04751 | 0,617813 | 45 | 42 | 0,540034 | 0,888057 | y ~ current_feature + S |
| 0,048619 | 0,157927 | 0,061498 | 0,307856 | 43 | 40 | 0,75979 | 0,972573 | y ~ current_feature + S |
| 0,014621 | 0,19243 | -0,02429 | 0,075982 | 30 | 27 | 0,939994 | 0,99444 | y ~ current_feature + S |
| -0,11398 | 0,058746 | -0,07874 | -1,94016 | 289 | 286 | 0,053343 | 0,30647 | y ~ current_feature + S |
| -0,12867 | 0,05864 | 0,034007 | -2,19431 | 289 | 286 | 0,029018 | 0,347616 | y ~ current_feature + S |
| -0,06025 | 0,059127 | -0,04751 | -1,019 | 288 | 285 | 0,309066 | 0,76529 | y ~ current_feature + S |
| -0,10806 | 0,063384 | 0,061498 | -1,70488 | 249 | 246 | 0,089479 | 0,795265 | y ~ current_feature + S |
| -0,12619 | 0,072158 | -0,02429 | -1,74877 | 192 | 189 | 0,081955 | 0,605474 | y ~ current_feature + S |
| 0,015205 | 0,049795 | -0,07874 | 0,305351 | 418 | 403,2004 | 0,760256 | 0,965008 | y ~ current_feature + S |
| -0,00865 | 0,06338 | 0,034007 | -0,13652 | 418 | 248,9187 | 0,891524 | 0,96591 | y ~ current_feature + S |
| 0,015921 | 0,050493 | -0,04751 | 0,315306 | 417 | 392,1214 | 0,752697 | 0,9394 | y ~ current_feature + S |
| -0,02347 | 0,054299 | 0,061498 | -0,43215 | 364 | 338,9865 | 0,665908 | 0,958606 | y ~ current_feature + S |
| -0,09236 | 0,062067 | -0,02429 | -1,48806 | 278 | 257,3697 | 0,137958 | 0,618941 | y ~ current_feature + S |
| 0,156191 | 0,052394 | -0,07874 | 2,981093 | 396 | 355,394 | 0,00307 | 0,066821 | y ~ current_feature + S |
| 0,238673 | 0,067974 | 0,034007 | 3,511212 | 396 | 204,0968 | 5,49E-04 | 0,067708 | y ~ current_feature + S |
| 0,068988 | 0,054235 | -0,04751 | 1,272017 | 395 | 338,35 | 0,20424 | 0,723562 | y ~ current_feature + S |
| -0,00955 | 0,057842 | 0,061498 | -0,16511 | 343 | 298,8631 | 0,868968 | 0,996179 | y ~ current_feature + S |
| 0,03276 | 0,062371 | -0,02429 | 0,525239 | 264 | 256,7865 | 0,599871 | 0,924682 | y ~ current_feature + S |
| -0,04711 | 0,156 | -0,07874 | -0,302 | 44 | 41 | 0,764182 | 0,965008 | y ~ current_feature + S |
| -0,03034 | 0,156102 | 0,034007 | -0,19439 | 44 | 41 | 0,846832 | 0,959445 | y ~ current_feature + S |
| 0,294329 | 0,15111 | -0,04751 | 1,947778 | 43 | 40 | 0,058484 | 0,640171 | y ~ current_feature + S |
| -0,09955 | 0,173213 | 0,061498 | -0,57473 | 36 | 33 | 0,56937 | 0,937665 | y ~ current_feature + S |

| | | | | | | | | |
|---|---|---|---|---|---|---|---|---|
| 0,180057 | 0,192911 | -0,02429 | 0,933372 | 29 | 26 | 0,359216 | 0,800662 | y ~ current_feature + S |
| 0,082443 | 0,055799 | -0,07874 | 1,477506 | 322 | 319 | 0,140527 | 0,498495 | y ~ current_feature + S |
| 0,140721 | 0,055432 | 0,034007 | 2,538619 | 322 | 319 | 0,011604 | 0,233774 | y ~ current_feature + S |
| 0,006373 | 0,056076 | -0,04751 | 0,113645 | 321 | 318 | 0,909591 | 0,985217 | y ~ current_feature + S |
| -0,05352 | 0,060107 | 0,061498 | -0,89046 | 279 | 276 | 0,373994 | 0,932536 | y ~ current_feature + S |
| 0,074615 | 0,068651 | -0,02429 | 1,086869 | 214 | 211 | 0,278335 | 0,734108 | y ~ current_feature + S |
| -0,02426 | 0,049162 | -0,07874 | -0,49349 | 418 | 413,513 | 0,621925 | 0,919937 | y ~ current_feature + S |
| -0,09291 | 0,060559 | 0,034007 | -1,53411 | 418 | 270,3157 | 0,126171 | 0,572145 | y ~ current_feature + S |
| -0,00476 | 0,049508 | -0,04751 | -0,09606 | 417 | 407,9831 | 0,923522 | 0,985217 | y ~ current_feature + S |
| -0,13103 | 0,05393 | 0,061498 | -2,42967 | 364 | 337,925 | 0,015633 | 0,55988 | y ~ current_feature + S |
| -0,03604 | 0,061351 | -0,02429 | -0,58751 | 278 | 265,3297 | 0,557363 | 0,92024 | y ~ current_feature + S |
| -0,07168 | 0,095536 | -0,07874 | -0,75028 | 112 | 109 | 0,454703 | 0,824707 | y ~ current_feature + S |
| 0,025431 | 0,095752 | 0,034007 | 0,265598 | 112 | 109 | 0,791051 | 0,945731 | y ~ current_feature + S |
| -0,00739 | 0,096222 | -0,04751 | -0,07675 | 111 | 108 | 0,938963 | 0,990156 | y ~ current_feature + S |
| 0,042654 | 0,101442 | 0,061498 | 0,420478 | 100 | 97 | 0,675066 | 0,958606 | y ~ current_feature + S |
| -0,05983 | 0,118466 | -0,02429 | -0,50504 | 74 | 71 | 0,615097 | 0,924682 | y ~ current_feature + S |
| 0,035069 | 0,070845 | -0,07874 | 0,495018 | 202 | 199 | 0,621133 | 0,919937 | y ~ current_feature + S |
| -0,08172 | 0,070651 | 0,034007 | -1,15667 | 202 | 199 | 0,248795 | 0,702074 | y ~ current_feature + S |
| -0,11641 | 0,070584 | -0,04751 | -1,64918 | 201 | 198 | 0,100697 | 0,640171 | y ~ current_feature + S |
| -0,04761 | 0,075942 | 0,061498 | -0,62693 | 176 | 173 | 0,53153 | 0,937665 | y ~ current_feature + S |
| -0,03275 | 0,086992 | -0,02429 | -0,37644 | 135 | 132 | 0,707194 | 0,93118 | y ~ current_feature + S |
| -0,10513 | 0,122409 | -0,07874 | -0,8588 | 69 | 66 | 0,393559 | 0,789252 | y ~ current_feature + S |
| -0,18581 | 0,120948 | 0,034007 | -1,53626 | 69 | 66 | 0,129254 | 0,572145 | y ~ current_feature + S |
| -0,021 | 0,123064 | -0,04751 | -0,17063 | 69 | 66 | 0,86504 | 0,985217 | y ~ current_feature + S |
| -0,01811 | 0,129078 | 0,061498 | -0,14028 | 63 | 60 | 0,888907 | 0,996179 | y ~ current_feature + S |
| -0,07567 | 0,155476 | -0,02429 | -0,48669 | 46 | 41,13176 | 0,62906 | 0,924682 | y ~ current_feature + S |
| 0,170014 | 0,057192 | -0,07874 | 2,972668 | 315 | 296,8832 | 0,003194 | 0,067531 | y ~ current_feature + S |
| 0,126891 | 0,076696 | 0,034007 | 1,654452 | 315 | 167,2629 | 0,099911 | 0,518857 | y ~ current_feature + S |
| 0,197213 | 0,059639 | -0,04751 | 3,30678 | 314 | 270,2169 | 0,001072 | 0,132163 | y ~ current_feature + S |
| 0,091514 | 0,060987 | 0,061498 | 1,500554 | 271 | 266,6084 | 0,134654 | 0,8197 | y ~ current_feature + S |
| -0,03196 | 0,07198 | -0,02429 | -0,44397 | 209 | 192,8136 | 0,657561 | 0,924682 | y ~ current_feature + S |
| 0,049044 | 0,049526 | -0,07874 | 0,990281 | 418 | 406,7206 | 0,322626 | 0,725761 | y ~ current_feature + S |
| 0,059222 | 0,062492 | 0,034007 | 0,947668 | 418 | 255,1641 | 0,344195 | 0,746014 | y ~ current_feature + S |
| -0,00499 | 0,050223 | -0,04751 | -0,09941 | 417 | 396,4479 | 0,920866 | 0,985217 | y ~ current_feature + S |
| 0,036555 | 0,05424 | 0,061498 | 0,67396 | 364 | 339,4556 | 0,500795 | 0,937665 | y ~ current_feature + S |
| 0,043587 | 0,062317 | -0,02429 | 0,699439 | 278 | 257,0169 | 0,48491 | 0,888665 | y ~ current_feature + S |
| 0,134228 | 0,126878 | -0,07874 | 1,057926 | 64 | 61 | 0,29426 | 0,718687 | y ~ current_feature + S |
| -0,12608 | 0,127015 | 0,034007 | -0,99262 | 64 | 61 | 0,324818 | 0,728793 | y ~ current_feature + S |
| -0,04528 | 0,127906 | -0,04751 | -0,35397 | 64 | 61 | 0,72458 | 0,93314 | y ~ current_feature + S |
| 0,049077 | 0,135919 | 0,061498 | 0,361074 | 57 | 54 | 0,719453 | 0,972573 | y ~ current_feature + S |
| -0,20777 | 0,156634 | -0,02429 | -1,32646 | 42 | 39 | 0,192402 | 0,65347 | y ~ current_feature + S |
| 0,230983 | 0,075745 | -0,07874 | 3,049498 | 168 | 165 | 0,002671 | 0,065908 | y ~ current_feature + S |
| 0,048122 | 0,07776 | 0,034007 | 0,618857 | 168 | 165 | 0,536864 | 0,850706 | y ~ current_feature + S |
| 0,083885 | 0,077576 | -0,04751 | 1,081335 | 168 | 165 | 0,281126 | 0,747915 | y ~ current_feature + S |
| -0,15181 | 0,082367 | 0,061498 | -1,84312 | 147 | 144 | 0,067367 | 0,715249 | y ~ current_feature + S |
| 0,039425 | 0,095708 | -0,02429 | 0,411931 | 112 | 109 | 0,681199 | 0,926342 | y ~ current_feature + S |
| 0,129251 | 0,064548 | -0,07874 | 2,002393 | 239 | 236 | 0,046387 | 0,286052 | y ~ current_feature + S |
| 0,084241 | 0,064863 | 0,034007 | 1,298747 | 239 | 236 | 0,195298 | 0,6438 | y ~ current_feature + S |

| | | | | | | | | |
|---|---|---|---|---|---|---|---|---|
| 0,019857 | 0,065082 | -0,04751 | 0,305103 | 239 | 236 | 0,760556 | 0,9394 | y ~ current_feature + S |
| -0,09758 | 0,069851 | 0,061498 | -1,397 | 206 | 203 | 0,16394 | 0,8197 | y ~ current_feature + S |
| 0,060733 | 0,079661 | -0,02429 | 0,762388 | 160 | 157 | 0,446972 | 0,858965 | y ~ current_feature + S |
| 0,055989 | 0,113782 | -0,07874 | 0,492077 | 80 | 77 | 0,624065 | 0,919937 | y ~ current_feature + S |
| -0,10189 | 0,113367 | 0,034007 | -0,89877 | 80 | 77 | 0,371576 | 0,750317 | y ~ current_feature + S |
| 0,241056 | 0,1106 | -0,04751 | 2,179527 | 80 | 77 | 0,032349 | 0,603897 | y ~ current_feature + S |
| -0,20528 | 0,116977 | 0,061498 | -1,75487 | 73 | 70 | 0,083658 | 0,754962 | y ~ current_feature + S |
| 0,12944 | 0,13885 | -0,02429 | 0,932227 | 54 | 51 | 0,355612 | 0,800026 | y ~ current_feature + S |
| 0,130755 | 0,051229 | -0,07874 | 2,55238 | 421 | 374,5281 | 0,011095 | 0,140685 | y ~ current_feature + S |
| 0,085689 | 0,067737 | 0,034007 | 1,265022 | 421 | 216,3454 | 0,207224 | 0,655365 | y ~ current_feature + S |
| 0,088359 | 0,052703 | -0,04751 | 1,676549 | 420 | 357,2125 | 0,094506 | 0,640171 | y ~ current_feature + S |
| -0,04459 | 0,056178 | 0,061498 | -0,79376 | 367 | 316,2322 | 0,427932 | 0,937665 | y ~ current_feature + S |
| 0,08813 | 0,05985 | -0,02429 | 1,472505 | 280 | 277 | 0,14202 | 0,618941 | y ~ current_feature + S |
| 0,01025 | 0,111798 | -0,07874 | 0,09168 | 83 | 80 | 0,927182 | 0,990519 | y ~ current_feature + S |
| 0,026512 | 0,111764 | 0,034007 | 0,237217 | 83 | 80 | 0,813095 | 0,952501 | y ~ current_feature + S |
| -0,08069 | 0,112142 | -0,04751 | -0,71953 | 82 | 79 | 0,473938 | 0,842057 | y ~ current_feature + S |
| 0,198098 | 0,128032 | 0,061498 | 1,54725 | 71 | 58,61036 | 0,127186 | 0,8197 | y ~ current_feature + S |
| 0,123003 | 0,137622 | -0,02429 | 0,893777 | 55 | 52 | 0,375559 | 0,815551 | y ~ current_feature + S |
| -0,01301 | 0,050375 | -0,07874 | -0,25821 | 397 | 394 | 0,796382 | 0,979102 | y ~ current_feature + S |
| -0,07628 | 0,050232 | 0,034007 | -1,51863 | 397 | 394 | 0,129659 | 0,572145 | y ~ current_feature + S |
| -0,08402 | 0,050265 | -0,04751 | -1,67153 | 396 | 393 | 0,095412 | 0,640171 | y ~ current_feature + S |
| -0,03958 | 0,059416 | 0,061498 | -0,66608 | 346 | 282,8199 | 0,505903 | 0,937665 | y ~ current_feature + S |
| -0,04144 | 0,061941 | -0,02429 | -0,66896 | 264 | 260,1902 | 0,504116 | 0,894767 | y ~ current_feature + S |
| 0,141362 | 0,063246 | -0,07874 | 2,235104 | 248 | 245 | 0,026312 | 0,204832 | y ~ current_feature + S |
| 0,096003 | 0,063593 | 0,034007 | 1,509657 | 248 | 245 | 0,13242 | 0,572281 | y ~ current_feature + S |
| 0,029594 | 0,06399 | -0,04751 | 0,462475 | 247 | 244 | 0,644153 | 0,925756 | y ~ current_feature + S |
| 0,069093 | 0,068516 | 0,061498 | 1,008412 | 215 | 212 | 0,314406 | 0,893818 | y ~ current_feature + S |
| 0,032967 | 0,078525 | -0,02429 | 0,419831 | 165 | 162 | 0,675165 | 0,926342 | y ~ current_feature + S |
| 0,062868 | 0,051815 | -0,07874 | 1,213321 | 374 | 371 | 0,225779 | 0,647583 | y ~ current_feature + S |
| -0,03033 | 0,051894 | 0,034007 | -0,58447 | 374 | 371 | 0,559257 | 0,860396 | y ~ current_feature + S |
| -0,01722 | 0,05198 | -0,04751 | -0,33135 | 373 | 370 | 0,740567 | 0,933594 | y ~ current_feature + S |
| -0,06956 | 0,064077 | 0,061498 | -1,08561 | 322 | 242,3721 | 0,27873 | 0,893818 | y ~ current_feature + S |
| -0,03341 | 0,063759 | -0,02429 | -0,52402 | 249 | 245,7138 | 0,600736 | 0,924682 | y ~ current_feature + S |
| 0,102589 | 0,056588 | -0,07874 | 1,812914 | 312 | 309 | 0,070815 | 0,356433 | y ~ current_feature + S |
| 0,080388 | 0,056704 | 0,034007 | 1,417676 | 312 | 309 | 0,157293 | 0,60427 | y ~ current_feature + S |
| 0,072539 | 0,05683 | -0,04751 | 1,276422 | 311 | 308 | 0,202768 | 0,723562 | y ~ current_feature + S |
| -0,12766 | 0,060812 | 0,061498 | -2,09922 | 269 | 266 | 0,03674 | 0,658491 | y ~ current_feature + S |
| -0,02641 | 0,06999 | -0,02429 | -0,37731 | 207 | 204 | 0,706334 | 0,93118 | y ~ current_feature + S |
| 0,125084 | 0,080474 | -0,07874 | 1,554352 | 155 | 152 | 0,12218 | 0,477837 | y ~ current_feature + S |
| 0,130295 | 0,080419 | 0,034007 | 1,620191 | 155 | 152 | 0,107264 | 0,532721 | y ~ current_feature + S |
| 0,115424 | 0,080835 | -0,04751 | 1,427896 | 154 | 151 | 0,155387 | 0,672434 | y ~ current_feature + S |
| 0,033089 | 0,08834 | 0,061498 | 0,374559 | 131 | 128 | 0,708608 | 0,972444 | y ~ current_feature + S |
| 0,162478 | 0,098671 | -0,02429 | 1,646663 | 103 | 100 | 0,102767 | 0,605474 | y ~ current_feature + S |
| 0,006689 | 0,07352 | -0,07874 | 0,090979 | 188 | 185 | 0,927608 | 0,990519 | y ~ current_feature + S |
| -0,08076 | 0,073281 | 0,034007 | -1,10201 | 188 | 185 | 0,27189 | 0,71095 | y ~ current_feature + S |
| 0,157582 | 0,0728 | -0,04751 | 2,164588 | 187 | 184 | 0,031707 | 0,603897 | y ~ current_feature + S |
| -0,17059 | 0,07839 | 0,061498 | -2,17618 | 161 | 158 | 0,031027 | 0,658491 | y ~ current_feature + S |
| 0,083752 | 0,090218 | -0,02429 | 0,92833 | 125 | 122 | 0,35507 | 0,800026 | y ~ current_feature + S |

| | | | | | | | | |
|---|---|---|---|---|---|---|---|---|
| -0,10027 | 0,08693 | -0,07874 | -1,15342 | 134 | 131 | 0,250838 | 0,679754 | y ~ current_feature + S |
| -0,16642 | 0,086152 | 0,034007 | -1,93168 | 134 | 131 | 0,055559 | 0,420946 | y ~ current_feature + S |
| -0,01292 | 0,087698 | -0,04751 | -0,14732 | 133 | 130 | 0,883105 | 0,985217 | y ~ current_feature + S |
| 0,053309 | 0,094357 | 0,061498 | 0,564969 | 115 | 112 | 0,573225 | 0,937665 | y ~ current_feature + S |
| -0,06866 | 0,107578 | -0,02429 | -0,6382 | 89 | 86 | 0,525037 | 0,899808 | y ~ current_feature + S |

ex + (1 | Subject)
ex + (1 | Subject)
ex + (1 | Subject)
ex + (1 | Subject)
ex + (1 | Subject)
ex + (1 | Subject)
ex + (1 | Subject)
ex + (1 | Subject)
ex + (1 | Subject)
ex + (1 | Subject)
ex + (1 | Subject)
ex + (1 | Subject)
ex + (1 | Subject)
ex + (1 | Subject)
ex + (1 | Subject)
ex + (1 | Subject)
ex + (1 | Subject)
ex + (1 | Subject)
ex + (1 | Subject)
ex + (1 | Subject)
ex + (1 | Subject)
ex + (1 | Subject)
ex + (1 | Subject)
ex + (1 | Subject)
ex + (1 | Subject)
ex + (1 | Subject)
ex + (1 | Subject)
ex + (1 | Subject)
ex + (1 | Subject)
ex + (1 | Subject)
ex + (1 | Subject)
ex + (1 | Subject)
ex + (1 | Subject)
ex + (1 | Subject)
ex + (1 | Subject)
ex + (1 | Subject)
ex + (1 | Subject)
ex + (1 | Subject)
ex + (1 | Subject)
ex + (1 | Subject)
ex + (1 | Subject)
ex + (1 | Subject)
ex + (1 | Subject)
ex + (1 | Subject)
ex + (1 | Subject)
ex + (1 | Subject)
ex + (1 | Subject)

ex + (1 | Subject)
ex + (1 | Subject)
ex + (1 | Subject)
ex + (1 | Subject)
ex + (1 | Subject)
ex + (1 | Subject)
ex + (1 | Subject)
ex + (1 | Subject)
ex + (1 | Subject)
ex + (1 | Subject)
ex + (1 | Subject)
ex + (1 | Subject)
ex + (1 | Subject)
ex + (1 | Subject)
ex + (1 | Subject)
ex + (1 | Subject)
ex + (1 | Subject)
ex + (1 | Subject)
ex + (1 | Subject)
ex + (1 | Subject)
ex + (1 | Subject)
ex + (1 | Subject)
ex + (1 | Subject)
ex + (1 | Subject)
ex + (1 | Subject)
ex + (1 | Subject)
ex + (1 | Subject)
ex + (1 | Subject)
ex + (1 | Subject)
ex + (1 | Subject)
ex + (1 | Subject)
ex + (1 | Subject)
ex + (1 | Subject)
ex + (1 | Subject)
ex + (1 | Subject)
ex + (1 | Subject)
ex + (1 | Subject)
ex + (1 | Subject)
ex + (1 | Subject)
ex + (1 | Subject)
ex + (1 | Subject)
ex + (1 | Subject)
ex + (1 | Subject)
ex + (1 | Subject)
ex + (1 | Subject)
ex + (1 | Subject)
ex + (1 | Subject)
ex + (1 | Subject)

ex + (1 | Subject)
ex + (1 | Subject)
ex + (1 | Subject)
ex + (1 | Subject)
ex + (1 | Subject)
ex + (1 | Subject)
ex + (1 | Subject)
ex + (1 | Subject)
ex + (1 | Subject)
ex + (1 | Subject)
ex + (1 | Subject)
ex + (1 | Subject)
ex + (1 | Subject)
ex + (1 | Subject)
ex + (1 | Subject)
ex + (1 | Subject)
ex + (1 | Subject)
ex + (1 | Subject)
ex + (1 | Subject)
ex + (1 | Subject)
ex + (1 | Subject)
ex + (1 | Subject)
ex + (1 | Subject)
ex + (1 | Subject)
ex + (1 | Subject)
ex + (1 | Subject)
ex + (1 | Subject)
ex + (1 | Subject)
ex + (1 | Subject)
ex + (1 | Subject)
ex + (1 | Subject)
ex + (1 | Subject)
ex + (1 | Subject)
ex + (1 | Subject)
ex + (1 | Subject)
ex + (1 | Subject)
ex + (1 | Subject)
ex + (1 | Subject)
ex + (1 | Subject)
ex + (1 | Subject)
ex + (1 | Subject)
ex + (1 | Subject)
ex + (1 | Subject)
ex + (1 | Subject)
ex + (1 | Subject)
ex + (1 | Subject)
ex + (1 | Subject)
ex + (1 | Subject)

ex + (1 | Subject)
ex + (1 | Subject)
ex + (1 | Subject)
ex + (1 | Subject)
ex + (1 | Subject)
ex + (1 | Subject)
ex + (1 | Subject)
ex + (1 | Subject)
ex + (1 | Subject)
ex + (1 | Subject)
ex + (1 | Subject)
ex + (1 | Subject)
ex + (1 | Subject)
ex + (1 | Subject)
ex + (1 | Subject)
ex + (1 | Subject)
ex + (1 | Subject)
ex + (1 | Subject)
ex + (1 | Subject)
ex + (1 | Subject)
ex + (1 | Subject)
ex + (1 | Subject)
ex + (1 | Subject)
ex + (1 | Subject)
ex + (1 | Subject)
ex + (1 | Subject)
ex + (1 | Subject)
ex + (1 | Subject)
ex + (1 | Subject)
ex + (1 | Subject)
ex + (1 | Subject)
ex + (1 | Subject)
ex + (1 | Subject)
ex + (1 | Subject)
ex + (1 | Subject)
ex + (1 | Subject)
ex + (1 | Subject)
ex + (1 | Subject)
ex + (1 | Subject)
ex + (1 | Subject)
ex + (1 | Subject)
ex + (1 | Subject)
ex + (1 | Subject)
ex + (1 | Subject)
ex + (1 | Subject)
ex + (1 | Subject)
ex + (1 | Subject)
ex + (1 | Subject)

ex + (1 | Subject)
ex + (1 | Subject)
ex + (1 | Subject)
ex + (1 | Subject)
ex + (1 | Subject)
ex + (1 | Subject)
ex + (1 | Subject)
ex + (1 | Subject)
ex + (1 | Subject)
ex + (1 | Subject)
ex + (1 | Subject)
ex + (1 | Subject)
ex + (1 | Subject)
ex + (1 | Subject)
ex + (1 | Subject)
ex + (1 | Subject)
ex + (1 | Subject)
ex + (1 | Subject)
ex + (1 | Subject)
ex + (1 | Subject)
ex + (1 | Subject)
ex + (1 | Subject)
ex + (1 | Subject)
ex + (1 | Subject)
ex + (1 | Subject)
ex + (1 | Subject)
ex + (1 | Subject)
ex + (1 | Subject)
ex + (1 | Subject)
ex + (1 | Subject)
ex + (1 | Subject)
ex + (1 | Subject)
ex + (1 | Subject)
ex + (1 | Subject)
ex + (1 | Subject)
ex + (1 | Subject)
ex + (1 | Subject)
ex + (1 | Subject)
ex + (1 | Subject)
ex + (1 | Subject)
ex + (1 | Subject)
ex + (1 | Subject)
ex + (1 | Subject)
ex + (1 | Subject)
ex + (1 | Subject)
ex + (1 | Subject)
ex + (1 | Subject)
ex + (1 | Subject)

ex + (1 | Subject)
ex + (1 | Subject)
ex + (1 | Subject)
ex + (1 | Subject)
ex + (1 | Subject)
ex + (1 | Subject)
ex + (1 | Subject)
ex + (1 | Subject)
ex + (1 | Subject)
ex + (1 | Subject)
ex + (1 | Subject)
ex + (1 | Subject)
ex + (1 | Subject)
ex + (1 | Subject)
ex + (1 | Subject)
ex + (1 | Subject)
ex + (1 | Subject)
ex + (1 | Subject)
ex + (1 | Subject)
ex + (1 | Subject)
ex + (1 | Subject)
ex + (1 | Subject)
ex + (1 | Subject)
ex + (1 | Subject)
ex + (1 | Subject)
ex + (1 | Subject)
ex + (1 | Subject)
ex + (1 | Subject)
ex + (1 | Subject)
ex + (1 | Subject)
ex + (1 | Subject)
ex + (1 | Subject)
ex + (1 | Subject)
ex + (1 | Subject)
ex + (1 | Subject)
ex + (1 | Subject)
ex + (1 | Subject)
ex + (1 | Subject)
ex + (1 | Subject)
ex + (1 | Subject)
ex + (1 | Subject)
ex + (1 | Subject)
ex + (1 | Subject)
ex + (1 | Subject)
ex + (1 | Subject)
ex + (1 | Subject)
ex + (1 | Subject)
ex + (1 | Subject)

ex + (1 | Subject)
ex + (1 | Subject)
ex + (1 | Subject)
ex + (1 | Subject)
ex + (1 | Subject)
ex + (1 | Subject)
ex + (1 | Subject)
ex + (1 | Subject)
ex + (1 | Subject)
ex + (1 | Subject)
ex + (1 | Subject)
ex + (1 | Subject)
ex + (1 | Subject)
ex + (1 | Subject)
ex + (1 | Subject)
ex + (1 | Subject)
ex + (1 | Subject)
ex + (1 | Subject)
ex + (1 | Subject)
ex + (1 | Subject)
ex + (1 | Subject)
ex + (1 | Subject)
ex + (1 | Subject)
ex + (1 | Subject)
ex + (1 | Subject)
ex + (1 | Subject)
ex + (1 | Subject)
ex + (1 | Subject)
ex + (1 | Subject)
ex + (1 | Subject)
ex + (1 | Subject)
ex + (1 | Subject)
ex + (1 | Subject)
ex + (1 | Subject)
ex + (1 | Subject)
ex + (1 | Subject)
ex + (1 | Subject)
ex + (1 | Subject)
ex + (1 | Subject)
ex + (1 | Subject)
ex + (1 | Subject)
ex + (1 | Subject)
ex + (1 | Subject)
ex + (1 | Subject)
ex + (1 | Subject)
ex + (1 | Subject)
ex + (1 | Subject)
ex + (1 | Subject)

ex + (1 | Subject)
ex + (1 | Subject)
ex + (1 | Subject)
ex + (1 | Subject)
ex + (1 | Subject)
ex + (1 | Subject)
ex + (1 | Subject)
ex + (1 | Subject)
ex + (1 | Subject)
ex + (1 | Subject)
ex + (1 | Subject)
ex + (1 | Subject)
ex + (1 | Subject)
ex + (1 | Subject)
ex + (1 | Subject)
ex + (1 | Subject)
ex + (1 | Subject)
ex + (1 | Subject)
ex + (1 | Subject)
ex + (1 | Subject)
ex + (1 | Subject)
ex + (1 | Subject)
ex + (1 | Subject)
ex + (1 | Subject)
ex + (1 | Subject)
ex + (1 | Subject)
ex + (1 | Subject)
ex + (1 | Subject)
ex + (1 | Subject)
ex + (1 | Subject)
ex + (1 | Subject)
ex + (1 | Subject)
ex + (1 | Subject)
ex + (1 | Subject)
ex + (1 | Subject)
ex + (1 | Subject)
ex + (1 | Subject)
ex + (1 | Subject)
ex + (1 | Subject)
ex + (1 | Subject)
ex + (1 | Subject)
ex + (1 | Subject)
ex + (1 | Subject)
ex + (1 | Subject)
ex + (1 | Subject)
ex + (1 | Subject)
ex + (1 | Subject)
ex + (1 | Subject)

ex + (1 | Subject)
ex + (1 | Subject)
ex + (1 | Subject)
ex + (1 | Subject)
ex + (1 | Subject)
ex + (1 | Subject)
ex + (1 | Subject)
ex + (1 | Subject)
ex + (1 | Subject)
ex + (1 | Subject)
ex + (1 | Subject)
ex + (1 | Subject)
ex + (1 | Subject)
ex + (1 | Subject)
ex + (1 | Subject)
ex + (1 | Subject)
ex + (1 | Subject)
ex + (1 | Subject)
ex + (1 | Subject)
ex + (1 | Subject)
ex + (1 | Subject)
ex + (1 | Subject)
ex + (1 | Subject)
ex + (1 | Subject)
ex + (1 | Subject)
ex + (1 | Subject)
ex + (1 | Subject)
ex + (1 | Subject)
ex + (1 | Subject)
ex + (1 | Subject)
ex + (1 | Subject)
ex + (1 | Subject)
ex + (1 | Subject)
ex + (1 | Subject)
ex + (1 | Subject)
ex + (1 | Subject)
ex + (1 | Subject)
ex + (1 | Subject)
ex + (1 | Subject)
ex + (1 | Subject)
ex + (1 | Subject)
ex + (1 | Subject)
ex + (1 | Subject)
ex + (1 | Subject)
ex + (1 | Subject)
ex + (1 | Subject)
ex + (1 | Subject)
ex + (1 | Subject)

ex + (1 | Subject)
ex + (1 | Subject)
ex + (1 | Subject)
ex + (1 | Subject)
ex + (1 | Subject)
ex + (1 | Subject)
ex + (1 | Subject)
ex + (1 | Subject)
ex + (1 | Subject)
ex + (1 | Subject)
ex + (1 | Subject)
ex + (1 | Subject)
ex + (1 | Subject)
ex + (1 | Subject)
ex + (1 | Subject)
ex + (1 | Subject)
ex + (1 | Subject)
ex + (1 | Subject)
ex + (1 | Subject)
ex + (1 | Subject)
ex + (1 | Subject)
ex + (1 | Subject)
ex + (1 | Subject)
ex + (1 | Subject)
ex + (1 | Subject)
ex + (1 | Subject)
ex + (1 | Subject)
ex + (1 | Subject)
ex + (1 | Subject)
ex + (1 | Subject)
ex + (1 | Subject)
ex + (1 | Subject)
ex + (1 | Subject)
ex + (1 | Subject)
ex + (1 | Subject)
ex + (1 | Subject)
ex + (1 | Subject)
ex + (1 | Subject)
ex + (1 | Subject)
ex + (1 | Subject)
ex + (1 | Subject)
ex + (1 | Subject)
ex + (1 | Subject)
ex + (1 | Subject)
ex + (1 | Subject)
ex + (1 | Subject)
ex + (1 | Subject)
ex + (1 | Subject)

ex + (1 | Subject)
ex + (1 | Subject)
ex + (1 | Subject)
ex + (1 | Subject)
ex + (1 | Subject)
ex + (1 | Subject)
ex + (1 | Subject)
ex + (1 | Subject)
ex + (1 | Subject)
ex + (1 | Subject)
ex + (1 | Subject)
ex + (1 | Subject)
ex + (1 | Subject)
ex + (1 | Subject)
ex + (1 | Subject)
ex + (1 | Subject)
ex + (1 | Subject)
ex + (1 | Subject)
ex + (1 | Subject)
ex + (1 | Subject)
ex + (1 | Subject)
ex + (1 | Subject)
ex + (1 | Subject)
ex + (1 | Subject)
ex + (1 | Subject)
ex + (1 | Subject)
ex + (1 | Subject)
ex + (1 | Subject)
ex + (1 | Subject)
ex + (1 | Subject)
ex + (1 | Subject)
ex + (1 | Subject)
ex + (1 | Subject)
ex + (1 | Subject)
ex + (1 | Subject)
ex + (1 | Subject)
ex + (1 | Subject)
ex + (1 | Subject)
ex + (1 | Subject)
ex + (1 | Subject)
ex + (1 | Subject)
ex + (1 | Subject)
ex + (1 | Subject)
ex + (1 | Subject)
ex + (1 | Subject)
ex + (1 | Subject)
ex + (1 | Subject)
ex + (1 | Subject)

ex + (1 | Subject)
ex + (1 | Subject)
ex + (1 | Subject)
ex + (1 | Subject)
ex + (1 | Subject)
ex + (1 | Subject)
ex + (1 | Subject)
ex + (1 | Subject)
ex + (1 | Subject)
ex + (1 | Subject)
ex + (1 | Subject)
ex + (1 | Subject)
ex + (1 | Subject)
ex + (1 | Subject)
ex + (1 | Subject)
ex + (1 | Subject)
ex + (1 | Subject)
ex + (1 | Subject)
ex + (1 | Subject)
ex + (1 | Subject)
ex + (1 | Subject)
ex + (1 | Subject)
ex + (1 | Subject)
ex + (1 | Subject)
ex + (1 | Subject)
ex + (1 | Subject)
ex + (1 | Subject)
ex + (1 | Subject)
ex + (1 | Subject)
ex + (1 | Subject)
ex + (1 | Subject)
ex + (1 | Subject)
ex + (1 | Subject)
ex + (1 | Subject)
ex + (1 | Subject)
ex + (1 | Subject)
ex + (1 | Subject)
ex + (1 | Subject)
ex + (1 | Subject)
ex + (1 | Subject)
ex + (1 | Subject)
ex + (1 | Subject)
ex + (1 | Subject)
ex + (1 | Subject)
ex + (1 | Subject)
ex + (1 | Subject)
ex + (1 | Subject)
ex + (1 | Subject)

ex + (1 | Subject)
ex + (1 | Subject)
ex + (1 | Subject)
ex + (1 | Subject)
ex + (1 | Subject)
ex + (1 | Subject)
ex + (1 | Subject)
ex + (1 | Subject)
ex + (1 | Subject)
ex + (1 | Subject)
ex + (1 | Subject)
ex + (1 | Subject)
ex + (1 | Subject)
ex + (1 | Subject)
ex + (1 | Subject)
ex + (1 | Subject)
ex + (1 | Subject)
ex + (1 | Subject)
ex + (1 | Subject)
ex + (1 | Subject)
ex + (1 | Subject)
ex + (1 | Subject)
ex + (1 | Subject)
ex + (1 | Subject)
ex + (1 | Subject)
ex + (1 | Subject)
ex + (1 | Subject)
ex + (1 | Subject)
ex + (1 | Subject)
ex + (1 | Subject)
ex + (1 | Subject)
ex + (1 | Subject)
ex + (1 | Subject)
ex + (1 | Subject)
ex + (1 | Subject)
ex + (1 | Subject)
ex + (1 | Subject)
ex + (1 | Subject)
ex + (1 | Subject)
ex + (1 | Subject)
ex + (1 | Subject)
ex + (1 | Subject)
ex + (1 | Subject)
ex + (1 | Subject)
ex + (1 | Subject)
ex + (1 | Subject)
ex + (1 | Subject)
ex + (1 | Subject)

ex + (1 | Subject)
ex + (1 | Subject)
ex + (1 | Subject)
ex + (1 | Subject)
ex + (1 | Subject)
ex + (1 | Subject)
ex + (1 | Subject)
ex + (1 | Subject)
ex + (1 | Subject)
ex + (1 | Subject)
ex + (1 | Subject)
ex + (1 | Subject)
ex + (1 | Subject)
ex + (1 | Subject)
ex + (1 | Subject)
ex + (1 | Subject)
ex + (1 | Subject)
ex + (1 | Subject)
ex + (1 | Subject)
ex + (1 | Subject)
ex + (1 | Subject)
ex + (1 | Subject)
ex + (1 | Subject)
ex + (1 | Subject)
ex + (1 | Subject)
ex + (1 | Subject)
ex + (1 | Subject)
ex + (1 | Subject)
ex + (1 | Subject)
ex + (1 | Subject)
ex + (1 | Subject)
ex + (1 | Subject)
ex + (1 | Subject)
ex + (1 | Subject)
ex + (1 | Subject)
ex + (1 | Subject)
ex + (1 | Subject)
ex + (1 | Subject)
ex + (1 | Subject)
ex + (1 | Subject)
ex + (1 | Subject)
ex + (1 | Subject)
ex + (1 | Subject)
ex + (1 | Subject)
ex + (1 | Subject)
ex + (1 | Subject)
ex + (1 | Subject)
ex + (1 | Subject)

ex + (1 | Subject)
ex + (1 | Subject)
ex + (1 | Subject)
ex + (1 | Subject)
ex + (1 | Subject)
ex + (1 | Subject)
ex + (1 | Subject)
ex + (1 | Subject)
ex + (1 | Subject)
ex + (1 | Subject)
ex + (1 | Subject)
ex + (1 | Subject)
ex + (1 | Subject)
ex + (1 | Subject)
ex + (1 | Subject)
ex + (1 | Subject)
ex + (1 | Subject)
ex + (1 | Subject)
ex + (1 | Subject)
ex + (1 | Subject)
ex + (1 | Subject)
ex + (1 | Subject)
ex + (1 | Subject)
ex + (1 | Subject)
ex + (1 | Subject)
ex + (1 | Subject)
ex + (1 | Subject)
ex + (1 | Subject)
ex + (1 | Subject)
ex + (1 | Subject)
ex + (1 | Subject)
ex + (1 | Subject)
ex + (1 | Subject)
ex + (1 | Subject)
ex + (1 | Subject)
ex + (1 | Subject)
ex + (1 | Subject)
ex + (1 | Subject)
ex + (1 | Subject)
ex + (1 | Subject)
ex + (1 | Subject)
ex + (1 | Subject)
ex + (1 | Subject)
ex + (1 | Subject)
ex + (1 | Subject)
ex + (1 | Subject)
ex + (1 | Subject)
ex + (1 | Subject)

ex + (1 | Subject)
ex + (1 | Subject)
ex + (1 | Subject)
ex + (1 | Subject)
ex + (1 | Subject)
ex + (1 | Subject)
ex + (1 | Subject)
ex + (1 | Subject)
ex + (1 | Subject)
ex + (1 | Subject)
ex + (1 | Subject)
ex + (1 | Subject)
ex + (1 | Subject)
ex + (1 | Subject)
ex + (1 | Subject)
ex + (1 | Subject)
ex + (1 | Subject)
ex + (1 | Subject)
ex + (1 | Subject)
ex + (1 | Subject)
ex + (1 | Subject)
ex + (1 | Subject)
ex + (1 | Subject)
ex + (1 | Subject)
ex + (1 | Subject)
ex + (1 | Subject)
ex + (1 | Subject)
ex + (1 | Subject)
ex + (1 | Subject)
ex + (1 | Subject)
ex + (1 | Subject)
ex + (1 | Subject)
ex + (1 | Subject)
ex + (1 | Subject)
ex + (1 | Subject)
ex + (1 | Subject)
ex + (1 | Subject)
ex + (1 | Subject)
ex + (1 | Subject)
ex + (1 | Subject)
ex + (1 | Subject)
ex + (1 | Subject)
ex + (1 | Subject)
ex + (1 | Subject)
ex + (1 | Subject)
ex + (1 | Subject)
ex + (1 | Subject)
ex + (1 | Subject)

ex + (1 | Subject)
ex + (1 | Subject)
ex + (1 | Subject)
ex + (1 | Subject)
ex + (1 | Subject)
ex + (1 | Subject)
ex + (1 | Subject)
ex + (1 | Subject)
ex + (1 | Subject)
ex + (1 | Subject)
ex + (1 | Subject)
ex + (1 | Subject)
ex + (1 | Subject)
ex + (1 | Subject)
ex + (1 | Subject)
ex + (1 | Subject)
ex + (1 | Subject)
ex + (1 | Subject)
ex + (1 | Subject)
ex + (1 | Subject)
ex + (1 | Subject)
ex + (1 | Subject)
ex + (1 | Subject)
ex + (1 | Subject)
ex + (1 | Subject)
ex + (1 | Subject)
ex + (1 | Subject)
ex + (1 | Subject)
ex + (1 | Subject)
ex + (1 | Subject)
ex + (1 | Subject)
ex + (1 | Subject)
ex + (1 | Subject)
ex + (1 | Subject)
ex + (1 | Subject)
ex + (1 | Subject)
ex + (1 | Subject)
ex + (1 | Subject)
ex + (1 | Subject)
ex + (1 | Subject)
ex + (1 | Subject)
ex + (1 | Subject)
ex + (1 | Subject)
ex + (1 | Subject)
ex + (1 | Subject)
ex + (1 | Subject)
ex + (1 | Subject)
ex + (1 | Subject)

ex + (1 | Subject)
ex + (1 | Subject)
ex + (1 | Subject)
ex + (1 | Subject)
ex + (1 | Subject)
ex + (1 | Subject)
ex + (1 | Subject)
ex + (1 | Subject)
ex + (1 | Subject)
ex + (1 | Subject)
ex + (1 | Subject)
ex + (1 | Subject)
ex + (1 | Subject)
ex + (1 | Subject)
ex + (1 | Subject)
ex + (1 | Subject)
ex + (1 | Subject)
ex + (1 | Subject)
ex + (1 | Subject)
ex + (1 | Subject)
ex + (1 | Subject)
ex + (1 | Subject)
ex + (1 | Subject)
ex + (1 | Subject)
ex + (1 | Subject)
ex + (1 | Subject)
ex + (1 | Subject)
ex + (1 | Subject)
ex + (1 | Subject)
ex + (1 | Subject)
ex + (1 | Subject)
ex + (1 | Subject)
ex + (1 | Subject)
ex + (1 | Subject)
ex + (1 | Subject)
ex + (1 | Subject)
ex + (1 | Subject)
ex + (1 | Subject)
ex + (1 | Subject)
ex + (1 | Subject)
ex + (1 | Subject)
ex + (1 | Subject)
ex + (1 | Subject)
ex + (1 | Subject)
ex + (1 | Subject)
ex + (1 | Subject)
ex + (1 | Subject)
ex + (1 | Subject)

ex + (1 | Subject)
ex + (1 | Subject)
ex + (1 | Subject)
ex + (1 | Subject)
ex + (1 | Subject)
ex + (1 | Subject)
ex + (1 | Subject)
ex + (1 | Subject)
ex + (1 | Subject)
ex + (1 | Subject)
ex + (1 | Subject)
ex + (1 | Subject)
ex + (1 | Subject)
ex + (1 | Subject)
ex + (1 | Subject)
ex + (1 | Subject)
ex + (1 | Subject)
ex + (1 | Subject)
ex + (1 | Subject)
ex + (1 | Subject)
ex + (1 | Subject)
ex + (1 | Subject)
ex + (1 | Subject)
ex + (1 | Subject)
ex + (1 | Subject)
ex + (1 | Subject)
ex + (1 | Subject)
ex + (1 | Subject)
ex + (1 | Subject)
ex + (1 | Subject)
ex + (1 | Subject)
ex + (1 | Subject)
ex + (1 | Subject)
ex + (1 | Subject)
ex + (1 | Subject)
ex + (1 | Subject)
ex + (1 | Subject)
ex + (1 | Subject)
ex + (1 | Subject)
ex + (1 | Subject)
ex + (1 | Subject)
ex + (1 | Subject)
ex + (1 | Subject)
ex + (1 | Subject)
ex + (1 | Subject)
ex + (1 | Subject)
ex + (1 | Subject)
ex + (1 | Subject)

ex + (1 | Subject)
ex + (1 | Subject)
ex + (1 | Subject)
ex + (1 | Subject)
ex + (1 | Subject)
ex + (1 | Subject)
ex + (1 | Subject)
ex + (1 | Subject)
ex + (1 | Subject)
ex + (1 | Subject)
ex + (1 | Subject)
ex + (1 | Subject)
ex + (1 | Subject)
ex + (1 | Subject)
ex + (1 | Subject)
ex + (1 | Subject)
ex + (1 | Subject)
ex + (1 | Subject)
ex + (1 | Subject)
ex + (1 | Subject)
ex + (1 | Subject)
ex + (1 | Subject)
ex + (1 | Subject)
ex + (1 | Subject)
ex + (1 | Subject)
ex + (1 | Subject)
ex + (1 | Subject)
ex + (1 | Subject)
ex + (1 | Subject)
ex + (1 | Subject)
ex + (1 | Subject)
ex + (1 | Subject)
ex + (1 | Subject)
ex + (1 | Subject)
ex + (1 | Subject)
ex + (1 | Subject)
ex + (1 | Subject)
ex + (1 | Subject)
ex + (1 | Subject)
ex + (1 | Subject)
ex + (1 | Subject)
ex + (1 | Subject)
ex + (1 | Subject)
ex + (1 | Subject)
ex + (1 | Subject)
ex + (1 | Subject)
ex + (1 | Subject)
ex + (1 | Subject)

ex + (1 | Subject)
ex + (1 | Subject)
ex + (1 | Subject)
ex + (1 | Subject)
ex + (1 | Subject)
ex + (1 | Subject)
ex + (1 | Subject)
ex + (1 | Subject)
ex + (1 | Subject)
ex + (1 | Subject)
ex + (1 | Subject)
ex + (1 | Subject)
ex + (1 | Subject)
ex + (1 | Subject)
ex + (1 | Subject)
ex + (1 | Subject)
ex + (1 | Subject)
ex + (1 | Subject)
ex + (1 | Subject)
ex + (1 | Subject)
ex + (1 | Subject)
ex + (1 | Subject)
ex + (1 | Subject)
ex + (1 | Subject)
ex + (1 | Subject)
ex + (1 | Subject)
ex + (1 | Subject)
ex + (1 | Subject)
ex + (1 | Subject)
ex + (1 | Subject)
ex + (1 | Subject)
ex + (1 | Subject)
ex + (1 | Subject)
ex + (1 | Subject)
ex + (1 | Subject)
ex + (1 | Subject)
ex + (1 | Subject)
ex + (1 | Subject)
ex + (1 | Subject)
ex + (1 | Subject)
ex + (1 | Subject)
ex + (1 | Subject)
ex + (1 | Subject)
ex + (1 | Subject)
ex + (1 | Subject)
ex + (1 | Subject)
ex + (1 | Subject)
ex + (1 | Subject)

ex + (1 | Subject)
ex + (1 | Subject)
ex + (1 | Subject)
ex + (1 | Subject)
ex + (1 | Subject)
ex + (1 | Subject)
ex + (1 | Subject)
ex + (1 | Subject)
ex + (1 | Subject)
ex + (1 | Subject)
ex + (1 | Subject)
ex + (1 | Subject)
ex + (1 | Subject)
ex + (1 | Subject)
ex + (1 | Subject)
ex + (1 | Subject)
ex + (1 | Subject)
ex + (1 | Subject)
ex + (1 | Subject)
ex + (1 | Subject)
ex + (1 | Subject)
ex + (1 | Subject)
ex + (1 | Subject)
ex + (1 | Subject)
ex + (1 | Subject)
ex + (1 | Subject)
ex + (1 | Subject)
ex + (1 | Subject)
ex + (1 | Subject)
ex + (1 | Subject)
ex + (1 | Subject)
ex + (1 | Subject)
ex + (1 | Subject)
ex + (1 | Subject)
ex + (1 | Subject)
ex + (1 | Subject)
ex + (1 | Subject)
ex + (1 | Subject)
ex + (1 | Subject)
ex + (1 | Subject)
ex + (1 | Subject)
ex + (1 | Subject)
ex + (1 | Subject)
ex + (1 | Subject)
ex + (1 | Subject)
ex + (1 | Subject)
ex + (1 | Subject)
ex + (1 | Subject)

ex + (1 | Subject)
ex + (1 | Subject)
ex + (1 | Subject)
ex + (1 | Subject)
ex + (1 | Subject)
ex + (1 | Subject)
ex + (1 | Subject)
ex + (1 | Subject)
ex + (1 | Subject)
ex + (1 | Subject)
ex + (1 | Subject)
ex + (1 | Subject)
ex + (1 | Subject)
ex + (1 | Subject)
ex + (1 | Subject)
ex + (1 | Subject)
ex + (1 | Subject)
ex + (1 | Subject)
ex + (1 | Subject)
ex + (1 | Subject)
ex + (1 | Subject)
ex + (1 | Subject)
ex + (1 | Subject)
ex + (1 | Subject)
ex + (1 | Subject)
ex + (1 | Subject)
ex + (1 | Subject)
ex + (1 | Subject)
ex + (1 | Subject)
ex + (1 | Subject)
ex + (1 | Subject)
ex + (1 | Subject)
ex + (1 | Subject)
ex + (1 | Subject)
ex + (1 | Subject)
ex + (1 | Subject)
ex + (1 | Subject)
ex + (1 | Subject)
ex + (1 | Subject)
ex + (1 | Subject)
ex + (1 | Subject)
ex + (1 | Subject)
ex + (1 | Subject)
ex + (1 | Subject)
ex + (1 | Subject)
ex + (1 | Subject)
ex + (1 | Subject)
ex + (1 | Subject)

ex + (1 | Subject)
ex + (1 | Subject)
ex + (1 | Subject)
ex + (1 | Subject)
ex + (1 | Subject)
ex + (1 | Subject)
ex + (1 | Subject)
ex + (1 | Subject)
ex + (1 | Subject)
ex + (1 | Subject)
ex + (1 | Subject)
ex + (1 | Subject)
ex + (1 | Subject)
ex + (1 | Subject)
ex + (1 | Subject)
ex + (1 | Subject)
ex + (1 | Subject)
ex + (1 | Subject)
ex + (1 | Subject)
ex + (1 | Subject)
ex + (1 | Subject)
ex + (1 | Subject)
ex + (1 | Subject)
ex + (1 | Subject)
ex + (1 | Subject)
ex + (1 | Subject)
ex + (1 | Subject)
ex + (1 | Subject)
ex + (1 | Subject)
ex + (1 | Subject)
ex + (1 | Subject)
ex + (1 | Subject)
ex + (1 | Subject)
ex + (1 | Subject)
ex + (1 | Subject)
ex + (1 | Subject)
ex + (1 | Subject)
ex + (1 | Subject)
ex + (1 | Subject)
ex + (1 | Subject)
ex + (1 | Subject)
ex + (1 | Subject)
ex + (1 | Subject)
ex + (1 | Subject)
ex + (1 | Subject)
ex + (1 | Subject)
ex + (1 | Subject)
ex + (1 | Subject)

ex + (1 | Subject)
ex + (1 | Subject)
ex + (1 | Subject)
ex + (1 | Subject)
ex + (1 | Subject)
ex + (1 | Subject)
ex + (1 | Subject)
ex + (1 | Subject)
ex + (1 | Subject)
ex + (1 | Subject)
ex + (1 | Subject)
ex + (1 | Subject)
ex + (1 | Subject)
ex + (1 | Subject)
ex + (1 | Subject)
ex + (1 | Subject)
ex + (1 | Subject)
ex + (1 | Subject)
ex + (1 | Subject)
ex + (1 | Subject)
ex + (1 | Subject)
ex + (1 | Subject)
ex + (1 | Subject)
ex + (1 | Subject)
ex + (1 | Subject)
ex + (1 | Subject)
ex + (1 | Subject)
ex + (1 | Subject)
ex + (1 | Subject)
ex + (1 | Subject)
ex + (1 | Subject)
ex + (1 | Subject)
ex + (1 | Subject)
ex + (1 | Subject)
ex + (1 | Subject)
ex + (1 | Subject)
ex + (1 | Subject)
ex + (1 | Subject)
ex + (1 | Subject)
ex + (1 | Subject)
ex + (1 | Subject)
ex + (1 | Subject)
ex + (1 | Subject)
ex + (1 | Subject)
ex + (1 | Subject)
ex + (1 | Subject)
ex + (1 | Subject)
ex + (1 | Subject)

ex + (1 | Subject)
ex + (1 | Subject)
ex + (1 | Subject)
ex + (1 | Subject)
ex + (1 | Subject)
ex + (1 | Subject)
ex + (1 | Subject)
ex + (1 | Subject)
ex + (1 | Subject)
ex + (1 | Subject)
ex + (1 | Subject)
ex + (1 | Subject)
ex + (1 | Subject)
ex + (1 | Subject)
ex + (1 | Subject)
ex + (1 | Subject)
ex + (1 | Subject)
ex + (1 | Subject)
ex + (1 | Subject)
ex + (1 | Subject)
ex + (1 | Subject)
ex + (1 | Subject)
ex + (1 | Subject)
ex + (1 | Subject)
ex + (1 | Subject)
ex + (1 | Subject)
ex + (1 | Subject)
ex + (1 | Subject)
ex + (1 | Subject)
ex + (1 | Subject)
ex + (1 | Subject)
ex + (1 | Subject)
ex + (1 | Subject)
ex + (1 | Subject)
ex + (1 | Subject)
ex + (1 | Subject)
ex + (1 | Subject)
ex + (1 | Subject)
ex + (1 | Subject)
ex + (1 | Subject)
ex + (1 | Subject)
ex + (1 | Subject)
ex + (1 | Subject)
ex + (1 | Subject)
ex + (1 | Subject)
ex + (1 | Subject)
ex + (1 | Subject)
ex + (1 | Subject)

ex + (1 | Subject)
ex + (1 | Subject)
ex + (1 | Subject)
ex + (1 | Subject)
ex + (1 | Subject)
ex + (1 | Subject)
ex + (1 | Subject)
ex + (1 | Subject)
ex + (1 | Subject)
ex + (1 | Subject)
ex + (1 | Subject)
ex + (1 | Subject)
ex + (1 | Subject)
ex + (1 | Subject)
ex + (1 | Subject)
ex + (1 | Subject)
ex + (1 | Subject)
ex + (1 | Subject)
ex + (1 | Subject)
ex + (1 | Subject)
ex + (1 | Subject)
ex + (1 | Subject)
ex + (1 | Subject)
ex + (1 | Subject)
ex + (1 | Subject)
ex + (1 | Subject)
ex + (1 | Subject)
ex + (1 | Subject)
ex + (1 | Subject)
ex + (1 | Subject)
ex + (1 | Subject)
ex + (1 | Subject)
ex + (1 | Subject)
ex + (1 | Subject)
ex + (1 | Subject)
ex + (1 | Subject)
ex + (1 | Subject)
ex + (1 | Subject)
ex + (1 | Subject)
ex + (1 | Subject)
ex + (1 | Subject)
ex + (1 | Subject)
ex + (1 | Subject)
ex + (1 | Subject)
ex + (1 | Subject)
ex + (1 | Subject)
ex + (1 | Subject)
ex + (1 | Subject)

ex + (1 | Subject)
ex + (1 | Subject)
ex + (1 | Subject)
ex + (1 | Subject)
ex + (1 | Subject)
ex + (1 | Subject)
ex + (1 | Subject)
ex + (1 | Subject)
ex + (1 | Subject)
ex + (1 | Subject)
ex + (1 | Subject)
ex + (1 | Subject)
ex + (1 | Subject)
ex + (1 | Subject)
ex + (1 | Subject)
ex + (1 | Subject)
ex + (1 | Subject)
ex + (1 | Subject)
ex + (1 | Subject)
ex + (1 | Subject)
ex + (1 | Subject)
ex + (1 | Subject)
ex + (1 | Subject)
ex + (1 | Subject)
ex + (1 | Subject)
ex + (1 | Subject)
ex + (1 | Subject)
ex + (1 | Subject)
ex + (1 | Subject)
ex + (1 | Subject)
ex + (1 | Subject)
ex + (1 | Subject)
ex + (1 | Subject)
ex + (1 | Subject)
ex + (1 | Subject)
ex + (1 | Subject)
ex + (1 | Subject)
ex + (1 | Subject)
ex + (1 | Subject)
ex + (1 | Subject)
ex + (1 | Subject)
ex + (1 | Subject)
ex + (1 | Subject)
ex + (1 | Subject)
ex + (1 | Subject)
ex + (1 | Subject)
ex + (1 | Subject)
ex + (1 | Subject)

ex + (1 | Subject)
ex + (1 | Subject)
ex + (1 | Subject)
ex + (1 | Subject)
ex + (1 | Subject)
ex + (1 | Subject)
ex + (1 | Subject)
ex + (1 | Subject)
ex + (1 | Subject)
ex + (1 | Subject)
ex + (1 | Subject)
ex + (1 | Subject)
ex + (1 | Subject)
ex + (1 | Subject)
ex + (1 | Subject)
ex + (1 | Subject)
ex + (1 | Subject)
ex + (1 | Subject)
ex + (1 | Subject)
ex + (1 | Subject)
ex + (1 | Subject)
ex + (1 | Subject)
ex + (1 | Subject)
ex + (1 | Subject)
ex + (1 | Subject)
ex + (1 | Subject)
ex + (1 | Subject)
ex + (1 | Subject)
ex + (1 | Subject)
ex + (1 | Subject)
ex + (1 | Subject)
ex + (1 | Subject)
ex + (1 | Subject)
ex + (1 | Subject)
ex + (1 | Subject)
ex + (1 | Subject)
ex + (1 | Subject)
ex + (1 | Subject)
ex + (1 | Subject)
ex + (1 | Subject)
ex + (1 | Subject)
ex + (1 | Subject)
ex + (1 | Subject)
ex + (1 | Subject)
ex + (1 | Subject)
ex + (1 | Subject)
ex + (1 | Subject)
ex + (1 | Subject)

ex + (1 | Subject)
ex + (1 | Subject)
ex + (1 | Subject)
ex + (1 | Subject)
ex + (1 | Subject)
ex + (1 | Subject)
ex + (1 | Subject)
ex + (1 | Subject)
ex + (1 | Subject)
ex + (1 | Subject)
ex + (1 | Subject)
ex + (1 | Subject)
ex + (1 | Subject)
ex + (1 | Subject)
ex + (1 | Subject)
ex + (1 | Subject)
ex + (1 | Subject)
ex + (1 | Subject)
ex + (1 | Subject)
ex + (1 | Subject)
ex + (1 | Subject)
ex + (1 | Subject)
ex + (1 | Subject)
ex + (1 | Subject)
ex + (1 | Subject)
ex + (1 | Subject)
ex + (1 | Subject)
ex + (1 | Subject)
ex + (1 | Subject)
ex + (1 | Subject)
ex + (1 | Subject)
ex + (1 | Subject)
ex + (1 | Subject)
ex + (1 | Subject)
ex + (1 | Subject)
ex + (1 | Subject)
ex + (1 | Subject)
ex + (1 | Subject)
ex + (1 | Subject)
ex + (1 | Subject)
ex + (1 | Subject)
ex + (1 | Subject)
ex + (1 | Subject)
ex + (1 | Subject)
ex + (1 | Subject)
ex + (1 | Subject)
ex + (1 | Subject)
ex + (1 | Subject)

ex + (1 | Subject)
ex + (1 | Subject)
ex + (1 | Subject)
ex + (1 | Subject)
ex + (1 | Subject)
ex + (1 | Subject)
ex + (1 | Subject)
ex + (1 | Subject)
ex + (1 | Subject)
ex + (1 | Subject)
ex + (1 | Subject)
ex + (1 | Subject)
ex + (1 | Subject)
ex + (1 | Subject)
ex + (1 | Subject)
ex + (1 | Subject)
ex + (1 | Subject)
ex + (1 | Subject)
ex + (1 | Subject)
ex + (1 | Subject)
ex + (1 | Subject)
ex + (1 | Subject)
ex + (1 | Subject)
ex + (1 | Subject)
ex + (1 | Subject)
ex + (1 | Subject)
ex + (1 | Subject)
ex + (1 | Subject)
ex + (1 | Subject)
ex + (1 | Subject)
ex + (1 | Subject)
ex + (1 | Subject)
ex + (1 | Subject)
ex + (1 | Subject)
ex + (1 | Subject)
ex + (1 | Subject)
ex + (1 | Subject)
ex + (1 | Subject)
ex + (1 | Subject)
ex + (1 | Subject)
ex + (1 | Subject)
ex + (1 | Subject)
ex + (1 | Subject)
ex + (1 | Subject)
ex + (1 | Subject)
ex + (1 | Subject)
ex + (1 | Subject)
ex + (1 | Subject)

ex + (1 | Subject)
ex + (1 | Subject)
ex + (1 | Subject)
ex + (1 | Subject)
ex + (1 | Subject)
ex + (1 | Subject)
ex + (1 | Subject)
ex + (1 | Subject)
ex + (1 | Subject)
ex + (1 | Subject)
ex + (1 | Subject)
ex + (1 | Subject)
ex + (1 | Subject)
ex + (1 | Subject)
ex + (1 | Subject)
ex + (1 | Subject)
ex + (1 | Subject)
ex + (1 | Subject)
ex + (1 | Subject)
ex + (1 | Subject)
ex + (1 | Subject)
ex + (1 | Subject)
ex + (1 | Subject)
ex + (1 | Subject)
ex + (1 | Subject)
ex + (1 | Subject)
ex + (1 | Subject)
ex + (1 | Subject)
ex + (1 | Subject)
ex + (1 | Subject)
ex + (1 | Subject)
ex + (1 | Subject)
ex + (1 | Subject)
ex + (1 | Subject)
ex + (1 | Subject)
ex + (1 | Subject)
ex + (1 | Subject)
ex + (1 | Subject)
ex + (1 | Subject)
ex + (1 | Subject)
ex + (1 | Subject)
ex + (1 | Subject)
ex + (1 | Subject)
ex + (1 | Subject)
ex + (1 | Subject)
ex + (1 | Subject)
ex + (1 | Subject)
ex + (1 | Subject)

ex + (1 | Subject)
ex + (1 | Subject)
ex + (1 | Subject)
ex + (1 | Subject)
ex + (1 | Subject)
ex + (1 | Subject)
ex + (1 | Subject)
ex + (1 | Subject)
ex + (1 | Subject)
ex + (1 | Subject)
ex + (1 | Subject)
ex + (1 | Subject)
ex + (1 | Subject)
ex + (1 | Subject)
ex + (1 | Subject)
ex + (1 | Subject)
ex + (1 | Subject)
ex + (1 | Subject)
ex + (1 | Subject)
ex + (1 | Subject)
ex + (1 | Subject)
ex + (1 | Subject)
ex + (1 | Subject)
ex + (1 | Subject)
ex + (1 | Subject)
ex + (1 | Subject)
ex + (1 | Subject)
ex + (1 | Subject)
ex + (1 | Subject)
ex + (1 | Subject)
ex + (1 | Subject)
ex + (1 | Subject)
ex + (1 | Subject)
ex + (1 | Subject)
ex + (1 | Subject)
ex + (1 | Subject)
ex + (1 | Subject)
ex + (1 | Subject)
ex + (1 | Subject)
ex + (1 | Subject)
ex + (1 | Subject)
ex + (1 | Subject)
ex + (1 | Subject)
ex + (1 | Subject)
ex + (1 | Subject)
ex + (1 | Subject)
ex + (1 | Subject)
ex + (1 | Subject)

ex + (1 | Subject)
ex + (1 | Subject)
ex + (1 | Subject)
ex + (1 | Subject)
ex + (1 | Subject)
ex + (1 | Subject)
ex + (1 | Subject)
ex + (1 | Subject)
ex + (1 | Subject)
ex + (1 | Subject)
ex + (1 | Subject)
ex + (1 | Subject)
ex + (1 | Subject)
ex + (1 | Subject)
ex + (1 | Subject)
ex + (1 | Subject)
ex + (1 | Subject)
ex + (1 | Subject)
ex + (1 | Subject)
ex + (1 | Subject)
ex + (1 | Subject)
ex + (1 | Subject)
ex + (1 | Subject)
ex + (1 | Subject)
ex + (1 | Subject)
ex + (1 | Subject)
ex + (1 | Subject)
ex + (1 | Subject)
ex + (1 | Subject)
ex + (1 | Subject)
ex + (1 | Subject)
ex + (1 | Subject)
ex + (1 | Subject)
ex + (1 | Subject)
ex + (1 | Subject)
ex + (1 | Subject)
ex + (1 | Subject)
ex + (1 | Subject)
ex + (1 | Subject)
ex + (1 | Subject)
ex + (1 | Subject)
ex + (1 | Subject)
ex + (1 | Subject)
ex + (1 | Subject)
ex + (1 | Subject)
ex + (1 | Subject)
ex + (1 | Subject)
ex + (1 | Subject)

ex + (1 | Subject)
ex + (1 | Subject)
ex + (1 | Subject)
ex + (1 | Subject)
ex + (1 | Subject)
ex + (1 | Subject)
ex + (1 | Subject)
ex + (1 | Subject)
ex + (1 | Subject)
ex + (1 | Subject)
ex + (1 | Subject)
ex + (1 | Subject)
ex + (1 | Subject)
ex + (1 | Subject)
ex + (1 | Subject)
ex + (1 | Subject)
ex + (1 | Subject)
ex + (1 | Subject)
ex + (1 | Subject)
ex + (1 | Subject)
ex + (1 | Subject)
ex + (1 | Subject)
ex + (1 | Subject)
ex + (1 | Subject)
ex + (1 | Subject)
ex + (1 | Subject)
ex + (1 | Subject)
ex + (1 | Subject)
ex + (1 | Subject)
ex + (1 | Subject)
ex + (1 | Subject)
ex + (1 | Subject)
ex + (1 | Subject)
ex + (1 | Subject)
ex + (1 | Subject)
ex + (1 | Subject)
ex + (1 | Subject)
ex + (1 | Subject)
ex + (1 | Subject)
ex + (1 | Subject)
ex + (1 | Subject)
ex + (1 | Subject)
ex + (1 | Subject)
ex + (1 | Subject)
ex + (1 | Subject)
ex + (1 | Subject)
ex + (1 | Subject)
ex + (1 | Subject)

ex + (1 | Subject)
ex + (1 | Subject)
ex + (1 | Subject)
ex + (1 | Subject)
ex + (1 | Subject)
ex + (1 | Subject)
ex + (1 | Subject)
ex + (1 | Subject)
ex + (1 | Subject)
ex + (1 | Subject)
ex + (1 | Subject)
ex + (1 | Subject)
ex + (1 | Subject)
ex + (1 | Subject)
ex + (1 | Subject)
ex + (1 | Subject)
ex + (1 | Subject)
ex + (1 | Subject)
ex + (1 | Subject)
ex + (1 | Subject)
ex + (1 | Subject)
ex + (1 | Subject)
ex + (1 | Subject)
ex + (1 | Subject)
ex + (1 | Subject)
ex + (1 | Subject)
ex + (1 | Subject)
ex + (1 | Subject)
ex + (1 | Subject)
ex + (1 | Subject)
ex + (1 | Subject)
ex + (1 | Subject)
ex + (1 | Subject)
ex + (1 | Subject)
ex + (1 | Subject)
ex + (1 | Subject)
ex + (1 | Subject)
ex + (1 | Subject)
ex + (1 | Subject)
ex + (1 | Subject)
ex + (1 | Subject)
ex + (1 | Subject)
ex + (1 | Subject)
ex + (1 | Subject)
ex + (1 | Subject)
ex + (1 | Subject)
ex + (1 | Subject)
ex + (1 | Subject)

ex + (1 | Subject)
ex + (1 | Subject)
ex + (1 | Subject)
ex + (1 | Subject)
ex + (1 | Subject)
ex + (1 | Subject)
ex + (1 | Subject)
ex + (1 | Subject)
ex + (1 | Subject)
ex + (1 | Subject)
ex + (1 | Subject)
ex + (1 | Subject)
ex + (1 | Subject)
ex + (1 | Subject)
ex + (1 | Subject)
ex + (1 | Subject)
ex + (1 | Subject)
ex + (1 | Subject)
ex + (1 | Subject)
ex + (1 | Subject)
ex + (1 | Subject)
ex + (1 | Subject)
ex + (1 | Subject)
ex + (1 | Subject)
ex + (1 | Subject)
ex + (1 | Subject)
ex + (1 | Subject)
ex + (1 | Subject)
ex + (1 | Subject)
ex + (1 | Subject)
ex + (1 | Subject)
ex + (1 | Subject)
ex + (1 | Subject)
ex + (1 | Subject)
ex + (1 | Subject)
ex + (1 | Subject)
ex + (1 | Subject)
ex + (1 | Subject)
ex + (1 | Subject)
ex + (1 | Subject)
ex + (1 | Subject)
ex + (1 | Subject)
ex + (1 | Subject)
ex + (1 | Subject)
ex + (1 | Subject)
ex + (1 | Subject)
ex + (1 | Subject)
ex + (1 | Subject)

ex + (1 | Subject)
ex + (1 | Subject)
ex + (1 | Subject)
ex + (1 | Subject)
ex + (1 | Subject)
ex + (1 | Subject)
ex + (1 | Subject)
ex + (1 | Subject)
ex + (1 | Subject)
ex + (1 | Subject)
ex + (1 | Subject)
ex + (1 | Subject)
ex + (1 | Subject)
ex + (1 | Subject)
ex + (1 | Subject)
ex + (1 | Subject)
ex + (1 | Subject)
ex + (1 | Subject)
ex + (1 | Subject)
ex + (1 | Subject)
ex + (1 | Subject)
ex + (1 | Subject)
ex + (1 | Subject)
ex + (1 | Subject)
ex + (1 | Subject)
ex + (1 | Subject)
ex + (1 | Subject)
ex + (1 | Subject)
ex + (1 | Subject)
ex + (1 | Subject)
ex + (1 | Subject)
ex + (1 | Subject)
ex + (1 | Subject)
ex + (1 | Subject)
ex + (1 | Subject)
ex + (1 | Subject)
ex + (1 | Subject)
ex + (1 | Subject)
ex + (1 | Subject)
ex + (1 | Subject)
ex + (1 | Subject)
ex + (1 | Subject)
ex + (1 | Subject)
ex + (1 | Subject)
ex + (1 | Subject)
ex + (1 | Subject)
ex + (1 | Subject)
ex + (1 | Subject)

ex + (1 | Subject)
ex + (1 | Subject)
ex + (1 | Subject)
ex + (1 | Subject)
ex + (1 | Subject)
ex + (1 | Subject)
ex + (1 | Subject)
ex + (1 | Subject)
ex + (1 | Subject)
ex + (1 | Subject)
ex + (1 | Subject)
ex + (1 | Subject)
ex + (1 | Subject)
ex + (1 | Subject)
ex + (1 | Subject)
ex + (1 | Subject)
ex + (1 | Subject)
ex + (1 | Subject)
ex + (1 | Subject)
ex + (1 | Subject)
ex + (1 | Subject)
ex + (1 | Subject)
ex + (1 | Subject)
ex + (1 | Subject)
ex + (1 | Subject)
ex + (1 | Subject)
ex + (1 | Subject)
ex + (1 | Subject)
ex + (1 | Subject)
ex + (1 | Subject)
ex + (1 | Subject)
ex + (1 | Subject)
ex + (1 | Subject)
ex + (1 | Subject)
ex + (1 | Subject)
ex + (1 | Subject)
ex + (1 | Subject)
ex + (1 | Subject)
ex + (1 | Subject)
ex + (1 | Subject)
ex + (1 | Subject)
ex + (1 | Subject)
ex + (1 | Subject)
ex + (1 | Subject)
ex + (1 | Subject)
ex + (1 | Subject)
ex + (1 | Subject)
ex + (1 | Subject)

ex + (1 | Subject)
ex + (1 | Subject)
ex + (1 | Subject)
ex + (1 | Subject)
ex + (1 | Subject)
ex + (1 | Subject)
ex + (1 | Subject)
ex + (1 | Subject)
ex + (1 | Subject)
ex + (1 | Subject)
ex + (1 | Subject)
ex + (1 | Subject)
ex + (1 | Subject)
ex + (1 | Subject)
ex + (1 | Subject)
ex + (1 | Subject)
ex + (1 | Subject)
ex + (1 | Subject)
ex + (1 | Subject)
ex + (1 | Subject)
ex + (1 | Subject)
ex + (1 | Subject)
ex + (1 | Subject)
ex + (1 | Subject)
ex + (1 | Subject)
ex + (1 | Subject)
ex + (1 | Subject)
ex + (1 | Subject)
ex + (1 | Subject)
ex + (1 | Subject)
ex + (1 | Subject)
ex + (1 | Subject)
ex + (1 | Subject)
ex + (1 | Subject)
ex + (1 | Subject)
ex + (1 | Subject)
ex + (1 | Subject)
ex + (1 | Subject)
ex + (1 | Subject)
ex + (1 | Subject)
ex + (1 | Subject)
ex + (1 | Subject)
ex + (1 | Subject)
ex + (1 | Subject)
ex + (1 | Subject)
ex + (1 | Subject)
ex + (1 | Subject)
ex + (1 | Subject)

ex + (1 | Subject)
ex + (1 | Subject)
ex + (1 | Subject)
ex + (1 | Subject)
ex + (1 | Subject)
ex + (1 | Subject)
ex + (1 | Subject)
ex + (1 | Subject)
ex + (1 | Subject)
ex + (1 | Subject)
ex + (1 | Subject)
ex + (1 | Subject)
ex + (1 | Subject)
ex + (1 | Subject)
ex + (1 | Subject)
ex + (1 | Subject)
ex + (1 | Subject)
ex + (1 | Subject)
ex + (1 | Subject)
ex + (1 | Subject)
ex + (1 | Subject)
ex + (1 | Subject)
ex + (1 | Subject)
ex + (1 | Subject)
ex + (1 | Subject)
ex + (1 | Subject)
ex + (1 | Subject)
ex + (1 | Subject)
ex + (1 | Subject)
ex + (1 | Subject)
ex + (1 | Subject)
ex + (1 | Subject)
ex + (1 | Subject)
ex + (1 | Subject)
ex + (1 | Subject)
ex + (1 | Subject)
ex + (1 | Subject)
ex + (1 | Subject)
ex + (1 | Subject)
ex + (1 | Subject)
ex + (1 | Subject)
ex + (1 | Subject)
ex + (1 | Subject)
ex + (1 | Subject)
ex + (1 | Subject)
ex + (1 | Subject)
ex + (1 | Subject)
ex + (1 | Subject)

ex + (1 | Subject)
ex + (1 | Subject)
ex + (1 | Subject)
ex + (1 | Subject)
ex + (1 | Subject)
ex + (1 | Subject)
ex + (1 | Subject)
ex + (1 | Subject)
ex + (1 | Subject)
ex + (1 | Subject)
ex + (1 | Subject)
ex + (1 | Subject)
ex + (1 | Subject)
ex + (1 | Subject)
ex + (1 | Subject)
ex + (1 | Subject)
ex + (1 | Subject)
ex + (1 | Subject)
ex + (1 | Subject)
ex + (1 | Subject)
ex + (1 | Subject)
ex + (1 | Subject)
ex + (1 | Subject)
ex + (1 | Subject)
ex + (1 | Subject)
ex + (1 | Subject)
ex + (1 | Subject)
ex + (1 | Subject)
ex + (1 | Subject)
ex + (1 | Subject)
ex + (1 | Subject)
ex + (1 | Subject)
ex + (1 | Subject)
ex + (1 | Subject)
ex + (1 | Subject)
ex + (1 | Subject)
ex + (1 | Subject)
ex + (1 | Subject)
ex + (1 | Subject)
ex + (1 | Subject)
ex + (1 | Subject)
ex + (1 | Subject)
ex + (1 | Subject)
ex + (1 | Subject)
ex + (1 | Subject)
ex + (1 | Subject)
ex + (1 | Subject)
ex + (1 | Subject)

ex + (1 | Subject)
ex + (1 | Subject)
ex + (1 | Subject)
ex + (1 | Subject)
ex + (1 | Subject)
ex + (1 | Subject)
ex + (1 | Subject)
ex + (1 | Subject)
ex + (1 | Subject)
ex + (1 | Subject)
ex + (1 | Subject)
ex + (1 | Subject)
ex + (1 | Subject)
ex + (1 | Subject)
ex + (1 | Subject)
ex + (1 | Subject)
ex + (1 | Subject)
ex + (1 | Subject)
ex + (1 | Subject)
ex + (1 | Subject)
ex + (1 | Subject)
ex + (1 | Subject)
ex + (1 | Subject)
ex + (1 | Subject)
ex + (1 | Subject)
ex + (1 | Subject)
ex + (1 | Subject)
ex + (1 | Subject)
ex + (1 | Subject)
ex + (1 | Subject)
ex + (1 | Subject)
ex + (1 | Subject)
ex + (1 | Subject)
ex + (1 | Subject)
ex + (1 | Subject)
ex + (1 | Subject)
ex + (1 | Subject)
ex + (1 | Subject)
ex + (1 | Subject)
ex + (1 | Subject)
ex + (1 | Subject)
ex + (1 | Subject)
ex + (1 | Subject)
ex + (1 | Subject)
ex + (1 | Subject)
ex + (1 | Subject)
ex + (1 | Subject)
ex + (1 | Subject)

ex + (1 | Subject)
ex + (1 | Subject)
ex + (1 | Subject)
ex + (1 | Subject)
ex + (1 | Subject)
ex + (1 | Subject)
ex + (1 | Subject)
ex + (1 | Subject)
ex + (1 | Subject)
ex + (1 | Subject)
ex + (1 | Subject)
ex + (1 | Subject)
ex + (1 | Subject)
ex + (1 | Subject)
ex + (1 | Subject)
ex + (1 | Subject)
ex + (1 | Subject)
ex + (1 | Subject)
ex + (1 | Subject)
ex + (1 | Subject)
ex + (1 | Subject)
ex + (1 | Subject)
ex + (1 | Subject)
ex + (1 | Subject)
ex + (1 | Subject)
ex + (1 | Subject)
ex + (1 | Subject)
ex + (1 | Subject)
ex + (1 | Subject)
ex + (1 | Subject)
ex + (1 | Subject)
ex + (1 | Subject)
ex + (1 | Subject)
ex + (1 | Subject)
ex + (1 | Subject)
ex + (1 | Subject)
ex + (1 | Subject)
ex + (1 | Subject)
ex + (1 | Subject)
ex + (1 | Subject)
ex + (1 | Subject)
ex + (1 | Subject)
ex + (1 | Subject)
ex + (1 | Subject)
ex + (1 | Subject)
ex + (1 | Subject)
ex + (1 | Subject)
ex + (1 | Subject)

ex + (1 | Subject)
ex + (1 | Subject)
ex + (1 | Subject)
ex + (1 | Subject)
ex + (1 | Subject)
ex + (1 | Subject)
ex + (1 | Subject)
ex + (1 | Subject)
ex + (1 | Subject)
ex + (1 | Subject)
ex + (1 | Subject)
ex + (1 | Subject)
ex + (1 | Subject)
ex + (1 | Subject)
ex + (1 | Subject)
ex + (1 | Subject)
ex + (1 | Subject)
ex + (1 | Subject)
ex + (1 | Subject)
ex + (1 | Subject)
ex + (1 | Subject)
ex + (1 | Subject)
ex + (1 | Subject)
ex + (1 | Subject)
ex + (1 | Subject)
ex + (1 | Subject)
ex + (1 | Subject)
ex + (1 | Subject)
ex + (1 | Subject)
ex + (1 | Subject)
ex + (1 | Subject)
ex + (1 | Subject)
ex + (1 | Subject)
ex + (1 | Subject)
ex + (1 | Subject)
ex + (1 | Subject)
ex + (1 | Subject)
ex + (1 | Subject)
ex + (1 | Subject)
ex + (1 | Subject)
ex + (1 | Subject)
ex + (1 | Subject)
ex + (1 | Subject)
ex + (1 | Subject)
ex + (1 | Subject)
ex + (1 | Subject)
ex + (1 | Subject)
ex + (1 | Subject)

ex + (1 | Subject)
ex + (1 | Subject)
ex + (1 | Subject)
ex + (1 | Subject)
ex + (1 | Subject)
ex + (1 | Subject)
ex + (1 | Subject)
ex + (1 | Subject)
ex + (1 | Subject)
ex + (1 | Subject)
ex + (1 | Subject)
ex + (1 | Subject)
ex + (1 | Subject)
ex + (1 | Subject)
ex + (1 | Subject)
ex + (1 | Subject)
ex + (1 | Subject)
ex + (1 | Subject)
ex + (1 | Subject)
ex + (1 | Subject)
ex + (1 | Subject)
ex + (1 | Subject)
ex + (1 | Subject)
ex + (1 | Subject)
ex + (1 | Subject)
ex + (1 | Subject)
ex + (1 | Subject)
ex + (1 | Subject)
ex + (1 | Subject)
ex + (1 | Subject)
ex + (1 | Subject)
ex + (1 | Subject)
ex + (1 | Subject)
ex + (1 | Subject)
ex + (1 | Subject)
ex + (1 | Subject)
ex + (1 | Subject)
ex + (1 | Subject)
ex + (1 | Subject)
ex + (1 | Subject)
ex + (1 | Subject)
ex + (1 | Subject)
ex + (1 | Subject)
ex + (1 | Subject)
ex + (1 | Subject)
ex + (1 | Subject)
ex + (1 | Subject)
ex + (1 | Subject)

ex + (1 | Subject)
ex + (1 | Subject)
ex + (1 | Subject)
ex + (1 | Subject)
ex + (1 | Subject)
ex + (1 | Subject)
ex + (1 | Subject)
ex + (1 | Subject)
ex + (1 | Subject)
ex + (1 | Subject)
ex + (1 | Subject)
ex + (1 | Subject)
ex + (1 | Subject)
ex + (1 | Subject)
ex + (1 | Subject)
ex + (1 | Subject)
ex + (1 | Subject)
ex + (1 | Subject)
ex + (1 | Subject)
ex + (1 | Subject)
ex + (1 | Subject)
ex + (1 | Subject)
ex + (1 | Subject)
ex + (1 | Subject)
ex + (1 | Subject)
ex + (1 | Subject)
ex + (1 | Subject)
ex + (1 | Subject)
ex + (1 | Subject)
ex + (1 | Subject)
ex + (1 | Subject)
ex + (1 | Subject)
ex + (1 | Subject)
ex + (1 | Subject)
ex + (1 | Subject)
ex + (1 | Subject)
ex + (1 | Subject)
ex + (1 | Subject)
ex + (1 | Subject)
ex + (1 | Subject)
ex + (1 | Subject)
ex + (1 | Subject)
ex + (1 | Subject)
ex + (1 | Subject)
ex + (1 | Subject)
ex + (1 | Subject)
ex + (1 | Subject)
ex + (1 | Subject)

ex + (1 | Subject)
ex + (1 | Subject)
ex + (1 | Subject)
ex + (1 | Subject)
ex + (1 | Subject)
ex + (1 | Subject)
ex + (1 | Subject)
ex + (1 | Subject)
ex + (1 | Subject)
ex + (1 | Subject)
ex + (1 | Subject)
ex + (1 | Subject)
ex + (1 | Subject)
ex + (1 | Subject)
ex + (1 | Subject)
ex + (1 | Subject)
ex + (1 | Subject)
ex + (1 | Subject)
ex + (1 | Subject)
ex + (1 | Subject)
ex + (1 | Subject)
ex + (1 | Subject)
ex + (1 | Subject)
ex + (1 | Subject)
ex + (1 | Subject)
ex + (1 | Subject)
ex + (1 | Subject)
ex + (1 | Subject)
ex + (1 | Subject)
ex + (1 | Subject)
ex + (1 | Subject)
ex + (1 | Subject)
ex + (1 | Subject)
ex + (1 | Subject)
ex + (1 | Subject)
ex + (1 | Subject)
ex + (1 | Subject)
ex + (1 | Subject)
ex + (1 | Subject)
ex + (1 | Subject)
ex + (1 | Subject)
ex + (1 | Subject)
ex + (1 | Subject)
ex + (1 | Subject)
ex + (1 | Subject)
ex + (1 | Subject)
ex + (1 | Subject)
ex + (1 | Subject)

ex + (1 | Subject)
ex + (1 | Subject)
ex + (1 | Subject)
ex + (1 | Subject)
ex + (1 | Subject)
ex + (1 | Subject)
ex + (1 | Subject)
ex + (1 | Subject)
ex + (1 | Subject)
ex + (1 | Subject)
ex + (1 | Subject)
ex + (1 | Subject)
ex + (1 | Subject)
ex + (1 | Subject)
ex + (1 | Subject)
ex + (1 | Subject)
ex + (1 | Subject)
ex + (1 | Subject)
ex + (1 | Subject)
ex + (1 | Subject)
ex + (1 | Subject)
ex + (1 | Subject)
ex + (1 | Subject)
ex + (1 | Subject)
ex + (1 | Subject)
ex + (1 | Subject)
ex + (1 | Subject)
ex + (1 | Subject)
ex + (1 | Subject)
ex + (1 | Subject)
ex + (1 | Subject)
ex + (1 | Subject)
ex + (1 | Subject)
ex + (1 | Subject)
ex + (1 | Subject)
ex + (1 | Subject)
ex + (1 | Subject)
ex + (1 | Subject)
ex + (1 | Subject)
ex + (1 | Subject)
ex + (1 | Subject)
ex + (1 | Subject)
ex + (1 | Subject)
ex + (1 | Subject)
ex + (1 | Subject)
ex + (1 | Subject)
ex + (1 | Subject)
ex + (1 | Subject)

ex + (1 | Subject)
ex + (1 | Subject)
ex + (1 | Subject)
ex + (1 | Subject)
ex + (1 | Subject)
ex + (1 | Subject)
ex + (1 | Subject)
ex + (1 | Subject)
ex + (1 | Subject)
ex + (1 | Subject)
ex + (1 | Subject)
ex + (1 | Subject)
ex + (1 | Subject)
ex + (1 | Subject)
ex + (1 | Subject)
ex + (1 | Subject)
ex + (1 | Subject)
ex + (1 | Subject)
ex + (1 | Subject)
ex + (1 | Subject)
ex + (1 | Subject)
ex + (1 | Subject)
ex + (1 | Subject)
ex + (1 | Subject)
ex + (1 | Subject)
ex + (1 | Subject)
ex + (1 | Subject)
ex + (1 | Subject)
ex + (1 | Subject)
ex + (1 | Subject)
ex + (1 | Subject)
ex + (1 | Subject)
ex + (1 | Subject)
ex + (1 | Subject)
ex + (1 | Subject)
ex + (1 | Subject)
ex + (1 | Subject)
ex + (1 | Subject)
ex + (1 | Subject)
ex + (1 | Subject)
ex + (1 | Subject)
ex + (1 | Subject)
ex + (1 | Subject)
ex + (1 | Subject)
ex + (1 | Subject)
ex + (1 | Subject)
ex + (1 | Subject)
ex + (1 | Subject)

ex + (1 | Subject)
ex + (1 | Subject)
ex + (1 | Subject)
ex + (1 | Subject)
ex + (1 | Subject)
ex + (1 | Subject)
ex + (1 | Subject)
ex + (1 | Subject)
ex + (1 | Subject)
ex + (1 | Subject)
ex + (1 | Subject)
ex + (1 | Subject)
ex + (1 | Subject)
ex + (1 | Subject)
ex + (1 | Subject)
ex + (1 | Subject)
ex + (1 | Subject)
ex + (1 | Subject)
ex + (1 | Subject)
ex + (1 | Subject)
ex + (1 | Subject)
ex + (1 | Subject)
ex + (1 | Subject)
ex + (1 | Subject)
ex + (1 | Subject)
ex + (1 | Subject)
ex + (1 | Subject)
ex + (1 | Subject)
ex + (1 | Subject)
ex + (1 | Subject)
ex + (1 | Subject)
ex + (1 | Subject)
ex + (1 | Subject)
ex + (1 | Subject)
ex + (1 | Subject)
ex + (1 | Subject)
ex + (1 | Subject)
ex + (1 | Subject)
ex + (1 | Subject)
ex + (1 | Subject)
ex + (1 | Subject)
ex + (1 | Subject)
ex + (1 | Subject)
ex + (1 | Subject)
ex + (1 | Subject)
ex + (1 | Subject)
ex + (1 | Subject)
ex + (1 | Subject)

ex + (1 | Subject)
ex + (1 | Subject)
ex + (1 | Subject)
ex + (1 | Subject)
ex + (1 | Subject)
ex + (1 | Subject)
ex + (1 | Subject)
ex + (1 | Subject)
ex + (1 | Subject)
ex + (1 | Subject)
ex + (1 | Subject)
ex + (1 | Subject)
ex + (1 | Subject)
ex + (1 | Subject)
ex + (1 | Subject)
ex + (1 | Subject)
ex + (1 | Subject)
ex + (1 | Subject)
ex + (1 | Subject)
ex + (1 | Subject)
ex + (1 | Subject)
ex + (1 | Subject)
ex + (1 | Subject)
ex + (1 | Subject)
ex + (1 | Subject)
ex + (1 | Subject)
ex + (1 | Subject)
ex + (1 | Subject)
ex + (1 | Subject)
ex + (1 | Subject)
ex + (1 | Subject)
ex + (1 | Subject)
ex + (1 | Subject)
ex + (1 | Subject)
ex + (1 | Subject)
ex + (1 | Subject)
ex + (1 | Subject)
ex + (1 | Subject)
ex + (1 | Subject)
ex + (1 | Subject)
ex + (1 | Subject)
ex + (1 | Subject)
ex + (1 | Subject)
ex + (1 | Subject)
ex + (1 | Subject)
ex + (1 | Subject)
ex + (1 | Subject)
ex + (1 | Subject)

ex + (1 | Subject)
ex + (1 | Subject)
ex + (1 | Subject)
ex + (1 | Subject)
ex + (1 | Subject)
ex + (1 | Subject)
ex + (1 | Subject)
ex + (1 | Subject)
ex + (1 | Subject)
ex + (1 | Subject)
ex + (1 | Subject)
ex + (1 | Subject)
ex + (1 | Subject)
ex + (1 | Subject)
ex + (1 | Subject)
ex + (1 | Subject)
ex + (1 | Subject)
ex + (1 | Subject)
ex + (1 | Subject)
ex + (1 | Subject)
ex + (1 | Subject)
ex + (1 | Subject)
ex + (1 | Subject)
ex + (1 | Subject)
ex + (1 | Subject)
ex + (1 | Subject)
ex + (1 | Subject)
ex + (1 | Subject)
ex + (1 | Subject)
ex + (1 | Subject)
ex + (1 | Subject)
ex + (1 | Subject)
ex + (1 | Subject)
ex + (1 | Subject)
ex + (1 | Subject)
ex + (1 | Subject)
ex + (1 | Subject)
ex + (1 | Subject)
ex + (1 | Subject)
ex + (1 | Subject)
ex + (1 | Subject)
ex + (1 | Subject)
ex + (1 | Subject)
ex + (1 | Subject)
ex + (1 | Subject)
ex + (1 | Subject)
ex + (1 | Subject)
ex + (1 | Subject)

ex + (1 | Subject)
ex + (1 | Subject)
ex + (1 | Subject)
ex + (1 | Subject)
ex + (1 | Subject)
ex + (1 | Subject)
ex + (1 | Subject)
ex + (1 | Subject)
ex + (1 | Subject)
ex + (1 | Subject)
ex + (1 | Subject)
ex + (1 | Subject)
ex + (1 | Subject)
ex + (1 | Subject)
ex + (1 | Subject)
ex + (1 | Subject)
ex + (1 | Subject)
ex + (1 | Subject)
ex + (1 | Subject)
ex + (1 | Subject)
ex + (1 | Subject)
ex + (1 | Subject)
ex + (1 | Subject)
ex + (1 | Subject)
ex + (1 | Subject)
ex + (1 | Subject)
ex + (1 | Subject)
ex + (1 | Subject)
ex + (1 | Subject)
ex + (1 | Subject)
ex + (1 | Subject)
ex + (1 | Subject)
ex + (1 | Subject)
ex + (1 | Subject)
ex + (1 | Subject)
ex + (1 | Subject)
ex + (1 | Subject)
ex + (1 | Subject)
ex + (1 | Subject)
ex + (1 | Subject)
ex + (1 | Subject)
ex + (1 | Subject)
ex + (1 | Subject)
ex + (1 | Subject)
ex + (1 | Subject)
ex + (1 | Subject)
ex + (1 | Subject)
ex + (1 | Subject)

ex + (1 | Subject)
ex + (1 | Subject)
ex + (1 | Subject)
ex + (1 | Subject)
ex + (1 | Subject)
ex + (1 | Subject)
ex + (1 | Subject)
ex + (1 | Subject)
ex + (1 | Subject)
ex + (1 | Subject)
ex + (1 | Subject)
ex + (1 | Subject)
ex + (1 | Subject)
ex + (1 | Subject)
ex + (1 | Subject)
ex + (1 | Subject)
ex + (1 | Subject)
ex + (1 | Subject)
ex + (1 | Subject)
ex + (1 | Subject)
ex + (1 | Subject)
ex + (1 | Subject)
ex + (1 | Subject)
ex + (1 | Subject)
ex + (1 | Subject)
ex + (1 | Subject)
ex + (1 | Subject)
ex + (1 | Subject)
ex + (1 | Subject)
ex + (1 | Subject)
ex + (1 | Subject)
ex + (1 | Subject)
ex + (1 | Subject)
ex + (1 | Subject)
ex + (1 | Subject)
ex + (1 | Subject)
ex + (1 | Subject)
ex + (1 | Subject)
ex + (1 | Subject)
ex + (1 | Subject)
ex + (1 | Subject)
ex + (1 | Subject)
ex + (1 | Subject)
ex + (1 | Subject)
ex + (1 | Subject)
ex + (1 | Subject)
ex + (1 | Subject)
ex + (1 | Subject)

ex + (1 | Subject)
ex + (1 | Subject)
ex + (1 | Subject)
ex + (1 | Subject)
ex + (1 | Subject)
ex + (1 | Subject)
ex + (1 | Subject)
ex + (1 | Subject)
ex + (1 | Subject)
ex + (1 | Subject)
ex + (1 | Subject)
ex + (1 | Subject)
ex + (1 | Subject)
ex + (1 | Subject)
ex + (1 | Subject)
ex + (1 | Subject)
ex + (1 | Subject)
ex + (1 | Subject)
ex + (1 | Subject)
ex + (1 | Subject)
ex + (1 | Subject)
ex + (1 | Subject)
ex + (1 | Subject)
ex + (1 | Subject)
ex + (1 | Subject)
ex + (1 | Subject)
ex + (1 | Subject)
ex + (1 | Subject)
ex + (1 | Subject)
ex + (1 | Subject)
ex + (1 | Subject)
ex + (1 | Subject)
ex + (1 | Subject)
ex + (1 | Subject)
ex + (1 | Subject)
ex + (1 | Subject)
ex + (1 | Subject)
ex + (1 | Subject)
ex + (1 | Subject)
ex + (1 | Subject)
ex + (1 | Subject)
ex + (1 | Subject)
ex + (1 | Subject)
ex + (1 | Subject)
ex + (1 | Subject)
ex + (1 | Subject)
ex + (1 | Subject)
ex + (1 | Subject)

ex + (1 | Subject)
ex + (1 | Subject)
ex + (1 | Subject)
ex + (1 | Subject)
ex + (1 | Subject)
ex + (1 | Subject)
ex + (1 | Subject)
ex + (1 | Subject)
ex + (1 | Subject)
ex + (1 | Subject)
ex + (1 | Subject)
ex + (1 | Subject)
ex + (1 | Subject)
ex + (1 | Subject)
ex + (1 | Subject)
ex + (1 | Subject)
ex + (1 | Subject)
ex + (1 | Subject)
ex + (1 | Subject)
ex + (1 | Subject)
ex + (1 | Subject)
ex + (1 | Subject)
ex + (1 | Subject)
ex + (1 | Subject)
ex + (1 | Subject)
ex + (1 | Subject)
ex + (1 | Subject)
ex + (1 | Subject)
ex + (1 | Subject)
ex + (1 | Subject)
ex + (1 | Subject)
ex + (1 | Subject)
ex + (1 | Subject)
ex + (1 | Subject)
ex + (1 | Subject)
ex + (1 | Subject)
ex + (1 | Subject)
ex + (1 | Subject)
ex + (1 | Subject)
ex + (1 | Subject)
ex + (1 | Subject)
ex + (1 | Subject)
ex + (1 | Subject)
ex + (1 | Subject)
ex + (1 | Subject)
ex + (1 | Subject)
ex + (1 | Subject)
ex + (1 | Subject)

ex + (1 | Subject)
ex + (1 | Subject)
ex + (1 | Subject)
ex + (1 | Subject)
ex + (1 | Subject)
ex + (1 | Subject)
ex + (1 | Subject)
ex + (1 | Subject)
ex + (1 | Subject)
ex + (1 | Subject)
ex + (1 | Subject)
ex + (1 | Subject)
ex + (1 | Subject)
ex + (1 | Subject)
ex + (1 | Subject)
ex + (1 | Subject)
ex + (1 | Subject)
ex + (1 | Subject)
ex + (1 | Subject)
ex + (1 | Subject)
ex + (1 | Subject)
ex + (1 | Subject)
ex + (1 | Subject)
ex + (1 | Subject)
ex + (1 | Subject)
ex + (1 | Subject)
ex + (1 | Subject)
ex + (1 | Subject)
ex + (1 | Subject)
ex + (1 | Subject)
ex + (1 | Subject)
ex + (1 | Subject)
ex + (1 | Subject)
ex + (1 | Subject)
ex + (1 | Subject)
ex + (1 | Subject)
ex + (1 | Subject)
ex + (1 | Subject)
ex + (1 | Subject)
ex + (1 | Subject)
ex + (1 | Subject)
ex + (1 | Subject)
ex + (1 | Subject)
ex + (1 | Subject)
ex + (1 | Subject)
ex + (1 | Subject)
ex + (1 | Subject)
ex + (1 | Subject)

ex + (1 | Subject)
ex + (1 | Subject)
ex + (1 | Subject)
ex + (1 | Subject)
ex + (1 | Subject)
ex + (1 | Subject)
ex + (1 | Subject)
ex + (1 | Subject)
ex + (1 | Subject)
ex + (1 | Subject)
ex + (1 | Subject)
ex + (1 | Subject)
ex + (1 | Subject)
ex + (1 | Subject)
ex + (1 | Subject)
ex + (1 | Subject)
ex + (1 | Subject)
ex + (1 | Subject)
ex + (1 | Subject)
ex + (1 | Subject)
ex + (1 | Subject)
ex + (1 | Subject)
ex + (1 | Subject)
ex + (1 | Subject)
ex + (1 | Subject)
ex + (1 | Subject)
ex + (1 | Subject)
ex + (1 | Subject)
ex + (1 | Subject)
ex + (1 | Subject)
ex + (1 | Subject)
ex + (1 | Subject)
ex + (1 | Subject)
ex + (1 | Subject)
ex + (1 | Subject)
ex + (1 | Subject)
ex + (1 | Subject)
ex + (1 | Subject)
ex + (1 | Subject)
ex + (1 | Subject)
ex + (1 | Subject)
ex + (1 | Subject)
ex + (1 | Subject)
ex + (1 | Subject)
ex + (1 | Subject)
ex + (1 | Subject)
ex + (1 | Subject)
ex + (1 | Subject)

ex + (1 | Subject)
ex + (1 | Subject)
ex + (1 | Subject)
ex + (1 | Subject)
ex + (1 | Subject)
ex + (1 | Subject)
ex + (1 | Subject)
ex + (1 | Subject)
ex + (1 | Subject)
ex + (1 | Subject)
ex + (1 | Subject)
ex + (1 | Subject)
ex + (1 | Subject)
ex + (1 | Subject)
ex + (1 | Subject)
ex + (1 | Subject)
ex + (1 | Subject)
ex + (1 | Subject)
ex + (1 | Subject)
ex + (1 | Subject)
ex + (1 | Subject)
ex + (1 | Subject)
ex + (1 | Subject)
ex + (1 | Subject)
ex + (1 | Subject)
ex + (1 | Subject)
ex + (1 | Subject)
ex + (1 | Subject)
ex + (1 | Subject)
ex + (1 | Subject)
ex + (1 | Subject)
ex + (1 | Subject)
ex + (1 | Subject)
ex + (1 | Subject)
ex + (1 | Subject)
ex + (1 | Subject)
ex + (1 | Subject)
ex + (1 | Subject)
ex + (1 | Subject)
ex + (1 | Subject)
ex + (1 | Subject)
ex + (1 | Subject)
ex + (1 | Subject)
ex + (1 | Subject)
ex + (1 | Subject)
ex + (1 | Subject)
ex + (1 | Subject)
ex + (1 | Subject)

ex + (1 | Subject)
ex + (1 | Subject)
ex + (1 | Subject)
ex + (1 | Subject)
ex + (1 | Subject)
ex + (1 | Subject)
ex + (1 | Subject)
ex + (1 | Subject)
ex + (1 | Subject)
ex + (1 | Subject)
ex + (1 | Subject)
ex + (1 | Subject)
ex + (1 | Subject)
ex + (1 | Subject)
ex + (1 | Subject)
ex + (1 | Subject)
ex + (1 | Subject)
ex + (1 | Subject)
ex + (1 | Subject)
ex + (1 | Subject)
ex + (1 | Subject)
ex + (1 | Subject)
ex + (1 | Subject)
ex + (1 | Subject)
ex + (1 | Subject)
ex + (1 | Subject)
ex + (1 | Subject)
ex + (1 | Subject)
ex + (1 | Subject)
ex + (1 | Subject)
ex + (1 | Subject)
ex + (1 | Subject)
ex + (1 | Subject)
ex + (1 | Subject)
ex + (1 | Subject)
ex + (1 | Subject)
ex + (1 | Subject)
ex + (1 | Subject)
ex + (1 | Subject)
ex + (1 | Subject)
ex + (1 | Subject)
ex + (1 | Subject)
ex + (1 | Subject)
ex + (1 | Subject)
ex + (1 | Subject)
ex + (1 | Subject)
ex + (1 | Subject)
ex + (1 | Subject)

ex + (1 | Subject)
ex + (1 | Subject)
ex + (1 | Subject)
ex + (1 | Subject)
ex + (1 | Subject)
ex + (1 | Subject)
ex + (1 | Subject)
ex + (1 | Subject)
ex + (1 | Subject)
ex + (1 | Subject)
ex + (1 | Subject)
ex + (1 | Subject)
ex + (1 | Subject)
ex + (1 | Subject)
ex + (1 | Subject)
ex + (1 | Subject)
ex + (1 | Subject)
ex + (1 | Subject)
ex + (1 | Subject)
ex + (1 | Subject)
ex + (1 | Subject)
ex + (1 | Subject)
ex + (1 | Subject)
ex + (1 | Subject)
ex + (1 | Subject)
ex + (1 | Subject)
ex + (1 | Subject)
ex + (1 | Subject)
ex + (1 | Subject)
ex + (1 | Subject)
ex + (1 | Subject)
ex + (1 | Subject)
ex + (1 | Subject)
ex + (1 | Subject)
ex + (1 | Subject)
ex + (1 | Subject)
ex + (1 | Subject)
ex + (1 | Subject)
ex + (1 | Subject)
ex + (1 | Subject)
ex + (1 | Subject)
ex + (1 | Subject)
ex + (1 | Subject)
ex + (1 | Subject)
ex + (1 | Subject)
ex + (1 | Subject)
ex + (1 | Subject)
ex + (1 | Subject)

ex + (1 | Subject)
ex + (1 | Subject)
ex + (1 | Subject)
ex + (1 | Subject)
ex + (1 | Subject)
ex + (1 | Subject)
ex + (1 | Subject)
ex + (1 | Subject)
ex + (1 | Subject)
ex + (1 | Subject)
ex + (1 | Subject)
ex + (1 | Subject)
ex + (1 | Subject)
ex + (1 | Subject)
ex + (1 | Subject)
ex + (1 | Subject)
ex + (1 | Subject)
ex + (1 | Subject)
ex + (1 | Subject)
ex + (1 | Subject)
ex + (1 | Subject)
ex + (1 | Subject)
ex + (1 | Subject)
ex + (1 | Subject)
ex + (1 | Subject)
ex + (1 | Subject)
ex + (1 | Subject)
ex + (1 | Subject)
ex + (1 | Subject)
ex + (1 | Subject)
ex + (1 | Subject)
ex + (1 | Subject)
ex + (1 | Subject)
ex + (1 | Subject)
ex + (1 | Subject)
ex + (1 | Subject)
ex + (1 | Subject)
ex + (1 | Subject)
ex + (1 | Subject)
ex + (1 | Subject)
ex + (1 | Subject)
ex + (1 | Subject)
ex + (1 | Subject)
ex + (1 | Subject)
ex + (1 | Subject)
ex + (1 | Subject)
ex + (1 | Subject)
ex + (1 | Subject)

ex + (1 | Subject)
ex + (1 | Subject)
ex + (1 | Subject)
ex + (1 | Subject)
ex + (1 | Subject)
ex + (1 | Subject)
ex + (1 | Subject)
ex + (1 | Subject)
ex + (1 | Subject)
ex + (1 | Subject)
ex + (1 | Subject)
ex + (1 | Subject)
ex + (1 | Subject)
ex + (1 | Subject)
ex + (1 | Subject)
ex + (1 | Subject)
ex + (1 | Subject)
ex + (1 | Subject)
ex + (1 | Subject)
ex + (1 | Subject)
ex + (1 | Subject)
ex + (1 | Subject)
ex + (1 | Subject)
ex + (1 | Subject)
ex + (1 | Subject)
ex + (1 | Subject)
ex + (1 | Subject)
ex + (1 | Subject)
ex + (1 | Subject)
ex + (1 | Subject)
ex + (1 | Subject)
ex + (1 | Subject)
ex + (1 | Subject)
ex + (1 | Subject)
ex + (1 | Subject)
ex + (1 | Subject)
ex + (1 | Subject)
ex + (1 | Subject)
ex + (1 | Subject)
ex + (1 | Subject)
ex + (1 | Subject)
ex + (1 | Subject)
ex + (1 | Subject)
ex + (1 | Subject)
ex + (1 | Subject)
ex + (1 | Subject)
ex + (1 | Subject)
ex + (1 | Subject)

ex + (1 | Subject)
ex + (1 | Subject)
ex + (1 | Subject)
ex + (1 | Subject)
ex + (1 | Subject)
ex + (1 | Subject)
ex + (1 | Subject)
ex + (1 | Subject)
ex + (1 | Subject)
ex + (1 | Subject)
ex + (1 | Subject)
ex + (1 | Subject)
ex + (1 | Subject)
ex + (1 | Subject)
ex + (1 | Subject)
ex + (1 | Subject)
ex + (1 | Subject)
ex + (1 | Subject)
ex + (1 | Subject)
ex + (1 | Subject)
ex + (1 | Subject)
ex + (1 | Subject)
ex + (1 | Subject)
ex + (1 | Subject)
ex + (1 | Subject)
ex + (1 | Subject)
ex + (1 | Subject)
ex + (1 | Subject)
ex + (1 | Subject)
ex + (1 | Subject)
ex + (1 | Subject)
ex + (1 | Subject)
ex + (1 | Subject)
ex + (1 | Subject)
ex + (1 | Subject)
ex + (1 | Subject)
ex + (1 | Subject)
ex + (1 | Subject)
ex + (1 | Subject)
ex + (1 | Subject)
ex + (1 | Subject)
ex + (1 | Subject)
ex + (1 | Subject)
ex + (1 | Subject)
ex + (1 | Subject)
ex + (1 | Subject)
ex + (1 | Subject)
ex + (1 | Subject)

ex + (1 | Subject)
ex + (1 | Subject)
ex + (1 | Subject)
ex + (1 | Subject)
ex + (1 | Subject)
ex + (1 | Subject)
ex + (1 | Subject)
ex + (1 | Subject)
ex + (1 | Subject)
ex + (1 | Subject)
ex + (1 | Subject)
ex + (1 | Subject)
ex + (1 | Subject)
ex + (1 | Subject)
ex + (1 | Subject)
ex + (1 | Subject)
ex + (1 | Subject)
ex + (1 | Subject)
ex + (1 | Subject)
ex + (1 | Subject)
ex + (1 | Subject)
ex + (1 | Subject)
ex + (1 | Subject)
ex + (1 | Subject)
ex + (1 | Subject)
ex + (1 | Subject)
ex + (1 | Subject)
ex + (1 | Subject)
ex + (1 | Subject)
ex + (1 | Subject)
ex + (1 | Subject)
ex + (1 | Subject)
ex + (1 | Subject)
ex + (1 | Subject)
ex + (1 | Subject)
ex + (1 | Subject)
ex + (1 | Subject)
ex + (1 | Subject)
ex + (1 | Subject)
ex + (1 | Subject)
ex + (1 | Subject)
ex + (1 | Subject)
ex + (1 | Subject)
ex + (1 | Subject)
ex + (1 | Subject)
ex + (1 | Subject)
ex + (1 | Subject)
ex + (1 | Subject)

ex + (1 | Subject)
ex + (1 | Subject)
ex + (1 | Subject)
ex + (1 | Subject)
ex + (1 | Subject)
ex + (1 | Subject)
ex + (1 | Subject)
ex + (1 | Subject)
ex + (1 | Subject)
ex + (1 | Subject)
ex + (1 | Subject)
ex + (1 | Subject)
ex + (1 | Subject)
ex + (1 | Subject)
ex + (1 | Subject)
ex + (1 | Subject)
ex + (1 | Subject)
ex + (1 | Subject)
ex + (1 | Subject)
ex + (1 | Subject)
ex + (1 | Subject)
ex + (1 | Subject)
ex + (1 | Subject)
ex + (1 | Subject)
ex + (1 | Subject)
ex + (1 | Subject)
ex + (1 | Subject)
ex + (1 | Subject)
ex + (1 | Subject)
ex + (1 | Subject)
ex + (1 | Subject)
ex + (1 | Subject)
ex + (1 | Subject)
ex + (1 | Subject)
ex + (1 | Subject)
ex + (1 | Subject)
ex + (1 | Subject)
ex + (1 | Subject)
ex + (1 | Subject)
ex + (1 | Subject)
ex + (1 | Subject)
ex + (1 | Subject)
ex + (1 | Subject)
ex + (1 | Subject)
ex + (1 | Subject)
ex + (1 | Subject)
ex + (1 | Subject)
ex + (1 | Subject)

ex + (1 | Subject)
ex + (1 | Subject)
ex + (1 | Subject)
ex + (1 | Subject)
ex + (1 | Subject)
ex + (1 | Subject)
ex + (1 | Subject)
ex + (1 | Subject)
ex + (1 | Subject)
ex + (1 | Subject)
ex + (1 | Subject)
ex + (1 | Subject)
ex + (1 | Subject)
ex + (1 | Subject)
ex + (1 | Subject)
ex + (1 | Subject)
ex + (1 | Subject)
ex + (1 | Subject)
ex + (1 | Subject)
ex + (1 | Subject)
ex + (1 | Subject)
ex + (1 | Subject)
ex + (1 | Subject)
ex + (1 | Subject)
ex + (1 | Subject)
ex + (1 | Subject)
ex + (1 | Subject)
ex + (1 | Subject)
ex + (1 | Subject)
ex + (1 | Subject)
ex + (1 | Subject)
ex + (1 | Subject)
ex + (1 | Subject)
ex + (1 | Subject)
ex + (1 | Subject)
ex + (1 | Subject)
ex + (1 | Subject)
ex + (1 | Subject)
ex + (1 | Subject)
ex + (1 | Subject)
ex + (1 | Subject)
ex + (1 | Subject)
ex + (1 | Subject)
ex + (1 | Subject)
ex + (1 | Subject)
ex + (1 | Subject)
ex + (1 | Subject)
ex + (1 | Subject)

ex + (1 | Subject)
ex + (1 | Subject)
ex + (1 | Subject)
ex + (1 | Subject)
ex + (1 | Subject)
ex + (1 | Subject)
ex + (1 | Subject)
ex + (1 | Subject)
ex + (1 | Subject)
ex + (1 | Subject)
ex + (1 | Subject)
ex + (1 | Subject)
ex + (1 | Subject)
ex + (1 | Subject)
ex + (1 | Subject)
ex + (1 | Subject)
ex + (1 | Subject)
ex + (1 | Subject)
ex + (1 | Subject)
ex + (1 | Subject)
ex + (1 | Subject)
ex + (1 | Subject)
ex + (1 | Subject)
ex + (1 | Subject)
ex + (1 | Subject)
ex + (1 | Subject)
ex + (1 | Subject)
ex + (1 | Subject)
ex + (1 | Subject)
ex + (1 | Subject)
ex + (1 | Subject)
ex + (1 | Subject)
ex + (1 | Subject)
ex + (1 | Subject)
ex + (1 | Subject)
ex + (1 | Subject)
ex + (1 | Subject)
ex + (1 | Subject)
ex + (1 | Subject)
ex + (1 | Subject)
ex + (1 | Subject)
ex + (1 | Subject)
ex + (1 | Subject)
ex + (1 | Subject)
ex + (1 | Subject)
ex + (1 | Subject)
ex + (1 | Subject)
ex + (1 | Subject)

ex + (1 | Subject)
ex + (1 | Subject)
ex + (1 | Subject)
ex + (1 | Subject)
ex + (1 | Subject)
ex + (1 | Subject)
ex + (1 | Subject)
ex + (1 | Subject)
ex + (1 | Subject)
ex + (1 | Subject)
ex + (1 | Subject)
ex + (1 | Subject)
ex + (1 | Subject)
ex + (1 | Subject)
ex + (1 | Subject)
ex + (1 | Subject)
ex + (1 | Subject)
ex + (1 | Subject)
ex + (1 | Subject)
ex + (1 | Subject)
ex + (1 | Subject)
ex + (1 | Subject)
ex + (1 | Subject)
ex + (1 | Subject)
ex + (1 | Subject)
ex + (1 | Subject)
ex + (1 | Subject)
ex + (1 | Subject)
ex + (1 | Subject)
ex + (1 | Subject)
ex + (1 | Subject)
ex + (1 | Subject)
ex + (1 | Subject)
ex + (1 | Subject)
ex + (1 | Subject)
ex + (1 | Subject)
ex + (1 | Subject)
ex + (1 | Subject)
ex + (1 | Subject)
ex + (1 | Subject)
ex + (1 | Subject)
ex + (1 | Subject)
ex + (1 | Subject)
ex + (1 | Subject)
ex + (1 | Subject)
ex + (1 | Subject)
ex + (1 | Subject)
ex + (1 | Subject)

ex + (1 | Subject)
ex + (1 | Subject)
ex + (1 | Subject)
ex + (1 | Subject)
ex + (1 | Subject)
ex + (1 | Subject)
ex + (1 | Subject)
ex + (1 | Subject)
ex + (1 | Subject)
ex + (1 | Subject)
ex + (1 | Subject)
ex + (1 | Subject)
ex + (1 | Subject)
ex + (1 | Subject)
ex + (1 | Subject)
ex + (1 | Subject)
ex + (1 | Subject)
ex + (1 | Subject)
ex + (1 | Subject)
ex + (1 | Subject)
ex + (1 | Subject)
ex + (1 | Subject)
ex + (1 | Subject)
ex + (1 | Subject)
ex + (1 | Subject)
ex + (1 | Subject)
ex + (1 | Subject)
ex + (1 | Subject)
ex + (1 | Subject)
ex + (1 | Subject)
ex + (1 | Subject)
ex + (1 | Subject)
ex + (1 | Subject)
ex + (1 | Subject)
ex + (1 | Subject)
ex + (1 | Subject)
ex + (1 | Subject)
ex + (1 | Subject)
ex + (1 | Subject)
ex + (1 | Subject)
ex + (1 | Subject)
ex + (1 | Subject)
ex + (1 | Subject)
ex + (1 | Subject)
ex + (1 | Subject)
ex + (1 | Subject)
ex + (1 | Subject)
ex + (1 | Subject)

ex + (1 | Subject)
ex + (1 | Subject)
ex + (1 | Subject)
ex + (1 | Subject)
ex + (1 | Subject)
ex + (1 | Subject)
ex + (1 | Subject)
ex + (1 | Subject)
ex + (1 | Subject)
ex + (1 | Subject)
ex + (1 | Subject)
ex + (1 | Subject)
ex + (1 | Subject)
ex + (1 | Subject)
ex + (1 | Subject)
ex + (1 | Subject)
ex + (1 | Subject)
ex + (1 | Subject)
ex + (1 | Subject)
ex + (1 | Subject)
ex + (1 | Subject)
ex + (1 | Subject)
ex + (1 | Subject)
ex + (1 | Subject)
ex + (1 | Subject)
ex + (1 | Subject)
ex + (1 | Subject)
ex + (1 | Subject)
ex + (1 | Subject)
ex + (1 | Subject)
ex + (1 | Subject)
ex + (1 | Subject)
ex + (1 | Subject)
ex + (1 | Subject)
ex + (1 | Subject)
ex + (1 | Subject)
ex + (1 | Subject)
ex + (1 | Subject)
ex + (1 | Subject)
ex + (1 | Subject)
ex + (1 | Subject)
ex + (1 | Subject)
ex + (1 | Subject)
ex + (1 | Subject)
ex + (1 | Subject)
ex + (1 | Subject)
ex + (1 | Subject)
ex + (1 | Subject)

ex + (1 | Subject)
ex + (1 | Subject)
ex + (1 | Subject)
ex + (1 | Subject)
ex + (1 | Subject)
ex + (1 | Subject)
ex + (1 | Subject)
ex + (1 | Subject)
ex + (1 | Subject)
ex + (1 | Subject)
ex + (1 | Subject)
ex + (1 | Subject)
ex + (1 | Subject)
ex + (1 | Subject)
ex + (1 | Subject)
ex + (1 | Subject)
ex + (1 | Subject)
ex + (1 | Subject)
ex + (1 | Subject)
ex + (1 | Subject)
ex + (1 | Subject)
ex + (1 | Subject)
ex + (1 | Subject)
ex + (1 | Subject)
ex + (1 | Subject)
ex + (1 | Subject)
ex + (1 | Subject)
ex + (1 | Subject)
ex + (1 | Subject)
ex + (1 | Subject)
ex + (1 | Subject)
ex + (1 | Subject)
ex + (1 | Subject)
ex + (1 | Subject)
ex + (1 | Subject)
ex + (1 | Subject)
ex + (1 | Subject)
ex + (1 | Subject)
ex + (1 | Subject)
ex + (1 | Subject)
ex + (1 | Subject)
ex + (1 | Subject)
ex + (1 | Subject)
ex + (1 | Subject)
ex + (1 | Subject)
ex + (1 | Subject)
ex + (1 | Subject)
ex + (1 | Subject)

ex + (1 | Subject)
ex + (1 | Subject)
ex + (1 | Subject)
ex + (1 | Subject)
ex + (1 | Subject)
ex + (1 | Subject)
ex + (1 | Subject)
ex + (1 | Subject)
ex + (1 | Subject)
ex + (1 | Subject)
ex + (1 | Subject)
ex + (1 | Subject)
ex + (1 | Subject)
ex + (1 | Subject)
ex + (1 | Subject)
ex + (1 | Subject)
ex + (1 | Subject)
ex + (1 | Subject)
ex + (1 | Subject)
ex + (1 | Subject)
ex + (1 | Subject)
ex + (1 | Subject)
ex + (1 | Subject)
ex + (1 | Subject)
ex + (1 | Subject)
ex + (1 | Subject)
ex + (1 | Subject)
ex + (1 | Subject)
ex + (1 | Subject)
ex + (1 | Subject)
ex + (1 | Subject)
ex + (1 | Subject)
ex + (1 | Subject)
ex + (1 | Subject)
ex + (1 | Subject)
ex + (1 | Subject)
ex + (1 | Subject)
ex + (1 | Subject)
ex + (1 | Subject)
ex + (1 | Subject)
ex + (1 | Subject)
ex + (1 | Subject)
ex + (1 | Subject)
ex + (1 | Subject)
ex + (1 | Subject)
ex + (1 | Subject)
ex + (1 | Subject)
ex + (1 | Subject)

ex + (1 | Subject)
ex + (1 | Subject)
ex + (1 | Subject)
ex + (1 | Subject)
ex + (1 | Subject)
ex + (1 | Subject)
ex + (1 | Subject)
ex + (1 | Subject)
ex + (1 | Subject)
ex + (1 | Subject)
ex + (1 | Subject)
ex + (1 | Subject)
ex + (1 | Subject)
ex + (1 | Subject)
ex + (1 | Subject)
ex + (1 | Subject)
ex + (1 | Subject)
ex + (1 | Subject)
ex + (1 | Subject)
ex + (1 | Subject)
ex + (1 | Subject)
ex + (1 | Subject)
ex + (1 | Subject)
ex + (1 | Subject)
ex + (1 | Subject)
ex + (1 | Subject)
ex + (1 | Subject)
ex + (1 | Subject)
ex + (1 | Subject)
ex + (1 | Subject)
ex + (1 | Subject)
ex + (1 | Subject)
ex + (1 | Subject)
ex + (1 | Subject)
ex + (1 | Subject)
ex + (1 | Subject)
ex + (1 | Subject)
ex + (1 | Subject)
ex + (1 | Subject)
ex + (1 | Subject)
ex + (1 | Subject)
ex + (1 | Subject)
ex + (1 | Subject)
ex + (1 | Subject)
ex + (1 | Subject)
ex + (1 | Subject)
ex + (1 | Subject)
ex + (1 | Subject)

ex + (1 | Subject)
ex + (1 | Subject)
ex + (1 | Subject)
ex + (1 | Subject)
ex + (1 | Subject)
ex + (1 | Subject)
ex + (1 | Subject)
ex + (1 | Subject)
ex + (1 | Subject)
ex + (1 | Subject)
ex + (1 | Subject)
ex + (1 | Subject)
ex + (1 | Subject)
ex + (1 | Subject)
ex + (1 | Subject)
ex + (1 | Subject)
ex + (1 | Subject)
ex + (1 | Subject)
ex + (1 | Subject)
ex + (1 | Subject)
ex + (1 | Subject)
ex + (1 | Subject)
ex + (1 | Subject)
ex + (1 | Subject)
ex + (1 | Subject)
ex + (1 | Subject)
ex + (1 | Subject)
ex + (1 | Subject)
ex + (1 | Subject)
ex + (1 | Subject)
ex + (1 | Subject)
ex + (1 | Subject)
ex + (1 | Subject)
ex + (1 | Subject)
ex + (1 | Subject)
ex + (1 | Subject)
ex + (1 | Subject)
ex + (1 | Subject)
ex + (1 | Subject)
ex + (1 | Subject)
ex + (1 | Subject)
ex + (1 | Subject)
ex + (1 | Subject)
ex + (1 | Subject)
ex + (1 | Subject)
ex + (1 | Subject)
ex + (1 | Subject)
ex + (1 | Subject)

ex + (1 | Subject)
ex + (1 | Subject)
ex + (1 | Subject)
ex + (1 | Subject)
ex + (1 | Subject)
ex + (1 | Subject)
ex + (1 | Subject)
ex + (1 | Subject)
ex + (1 | Subject)
ex + (1 | Subject)
ex + (1 | Subject)
ex + (1 | Subject)
ex + (1 | Subject)
ex + (1 | Subject)
ex + (1 | Subject)
ex + (1 | Subject)
ex + (1 | Subject)
ex + (1 | Subject)
ex + (1 | Subject)
ex + (1 | Subject)
ex + (1 | Subject)
ex + (1 | Subject)
ex + (1 | Subject)
ex + (1 | Subject)
ex + (1 | Subject)
ex + (1 | Subject)
ex + (1 | Subject)
ex + (1 | Subject)
ex + (1 | Subject)
ex + (1 | Subject)
ex + (1 | Subject)
ex + (1 | Subject)
ex + (1 | Subject)
ex + (1 | Subject)
ex + (1 | Subject)
ex + (1 | Subject)
ex + (1 | Subject)
ex + (1 | Subject)
ex + (1 | Subject)
ex + (1 | Subject)
ex + (1 | Subject)
ex + (1 | Subject)
ex + (1 | Subject)
ex + (1 | Subject)
ex + (1 | Subject)
ex + (1 | Subject)
ex + (1 | Subject)
ex + (1 | Subject)

[illegible]ex + (1 | Subject)
[illegible]ex + (1 | Subject)
[illegible]ex + (1 | Subject)
[illegible]ex + (1 | Subject)
[illegible]ex + (1 | Subject)
